# Rapid and Accurate Identification of SARS-CoV-2 Omicron Variants Using Droplet Digital PCR (RT-ddPCR)

**DOI:** 10.1101/2022.01.11.22268981

**Authors:** Margaret G. Mills, Pooneh Hajian, Shah Mohamed Bakhash, Hong Xie, Derrek Mantzke, Haiying Zhu, Garrett A. Perchetti, Meei-Li Huang, Gregory Pepper, Keith R. Jerome, Pavitra Roychoudhury, Alexander L. Greninger

## Abstract

**Background:** Mutations in the receptor binding domain of the SARS-CoV-2 Spike protein are associated with increased transmission or substantial reductions in vaccine efficacy, including in the recently described Omicron variant. The changing frequencies of these mutations combined with their differing susceptibility to available therapies have posed significant problems for clinicians and public health professionals.

**Objective:** To develop an assay capable of rapidly and accurately identifying variants including Omicron in clinical specimens to enable case tracking and/or selection of appropriate clinical treatment.

**Study Design:** Using three duplex RT-ddPCR reactions targeting four amino acids, we tested 419 positive clinical specimens from February to December 2021 during a period of rapidly shifting variant prevalences and compared genotyping results to genome sequences for each sample, determining the sensitivity and specificity of the assay for each variant.

**Results:** Mutation determinations for 99.7% of detected samples agree with NGS data for those samples, and are accurate despite wide variation in RNA concentration and potential confounding factors like transport medium, presence of additional respiratory viruses, and additional mutations in primer and probe sequences. The assay accurately identified the first 15 Omicron variants in our laboratory including the first Omicron in Washington State and discriminated against an S-gene dropout Delta specimen.

**Conclusion:** We describe an accurate, precise, and specific RT-ddPCR assay for variant detection that remains robust despite being designed prior to the emergence of Delta and Omicron variants. The assay can quickly identify mutations in current and past SARS-CoV-2 variants, and can be adapted to future mutations.

## 1. Background

The evolution of SARS-CoV-2 enabled by two years and nearly 300 million cases of human-to-human transmission has resulted in numerous mutations in the receptor binding domain (RBD) of the spike protein. This is the region that binds to the human cell receptor Angiotensin-converting enzyme 2 (ACE2) to enable viral invasion of the host cell[1-3] and it is the region targeted by most antibodies, both illness- and vaccine-derived.[4,5] Consequently, a number of the amino acid changes observed in the RBD of SARS-CoV-2 variants have been predicted or demonstrated to correlate with increased transmissibility and/or reduced plasma neutralization and vaccine efficacy,[5-11] including N501Y, K417N/T, L452R, and E484K/Q/A. These RBD mutations were detected in a parade of lineages identified as Variants of Concern (VOC) or Interest (VOI) through the first half of 2021: Alpha (B.1.1.7, N501Y);[12-14] Beta (B.1.351, K417N/E484K/N501Y); [15,16] Gamma (P.1, K417T/E484K/N501Y);[17-19] Delta (B.1.617.2 and AY.x, L452R);[20,21] Kappa (B.1.617.1, L452R/E484Q);[22] and more. Before the approval of vaccines against SARS-CoV-2, monoclonal antibodies (mAbs) were the primary tool available to protect patients at risk of severe COVID-19 from the worst effects of the disease,[23,24] and even with the availability of vaccines, mAbs remain important treatment options for vulnerable patients. [25,26] These drugs need to be administered within a limited time after infection in order to provide protection.[26-28] But because of the kaleidoscope of RBD amino acid combinations presented by the circulating variants, because of the varying effectiveness with which different mAbs neutralize different variants,[21,23,29,30] and because of the constantly changing frequencies of the variants themselves in different areas of the world, selection of mAb or mAb cocktail was challenging for large parts of 2021, prompting calls for clinical tests capable of rapidly identifying SARS-CoV-2 variant in clinical specimens. With the recent appearance and rise of the Omicron variant (B.1.1.529, K417N/E484A/G496S/Q498R/N501Y)[31] this need is again growing.[32-34]

## 2. Objective

Single nucleotide mutations, such as those encoding these amino acid changes, are challenging to identify with routine RT-PCR. Efforts to identify variants using other larger changes elsewhere in the viral genome, such as the S-gene target failure (SGTF) used to identify both Alpha and Omicron, have been extremely useful for surveillance purposes by us and others, [35-37] but are not accurate enough for making clinical decisions. Sequencing identifies mutations definitively, but not quickly enough to allow for treatment decisions.

Droplet digital (dd)PCR enables rapid and accurate genotyping of small-but-critical mutations.[38-40] Building on our earlier assay,[36] we sought to develop an assay that could identify these key functional mutations in Spike, regardless of genetic background, quickly enough to be of use to clinicians.

## 3. Study Design

### 3.1. Sample extraction

Total nucleic acids were extracted from nasal/pharyngeal and nasal swabs using either Roche MagNA Pure 96 instrument and DNA & Viral NA Small Volume kit or ThermoFisher KingFisher according to manufacturer instructions. All MagNA Pure extractions used 200 µl of input volume and 100 µl elution; all KingFisher extractions used 400 µl input volume and 50 µl elution.

### 3.2. Viral whole genome sequencing

Sequencing and genomic analyses were performed as previously described. [41,42] Sequencing libraries were prepared using multiplexed amplicon panels from Swift Biosciences or Illumina COVIDSeq. Consensus sequences were assembled using a custom bioinformatics pipeline (https://github.com/greninger-lab/covid_swift_pipeline, [42]). Consensus sequences with >10% Ns were excluded, and phylogenetic lineage was assigned using the PANGOLIN (Phylogenetic Assignment of Named Global Outbreak LINeages, https://pangolin.cog-uk.io/) and NextClade (https://clades.nextstrain.org/) tools.

### 3.3. RT-ddPCR

RT-ddPCR was carried out using the One-Step RT-ddPCR Advanced Kit for Probes (Bio-Rad) according to manufacturer instructions. Three reactions were set up for each specimen, using the primers and probes in Table 1. Droplet generation, PCR, and droplet reading were performed as previously described.[36] RNA from Reference Specimens (Supplement 1) were included as positive controls in each RT-ddPCR run. Data analysis was conducted with QuantaSoft Pro 1.0.596 version software, using two methods. Mutation identification: designating all assays as Amplitude Multiplex (Table 2), using 2D amplitude of positive controls as guides for droplet cluster selection, as in Figure 2. (Note that in all droplet amplitude figures, droplets are colored for ease of visualization.) All concentration calculations were exported to Excel and collated for each specimen: for each amino acid, the allele with the highest concentration of droplets (at least 5-10x the concentration of the next allele) was identified as the allele for that specimen. Droplet amplitudes: designating all assays as Simplex/Duplex and selecting all droplets other than empty (water) droplets as expressing all probes, then exporting all Cluster Data to Excel. For each specimen, the mean amplitude and standard deviation of amplitude for each probe was provided, along with the number of droplets used to generate those calculations. In both analysis methods, samples were only included if they had a minimum of 10,000 measured droplets and a minimum of 3 droplets in a cluster.

**Table 1.**
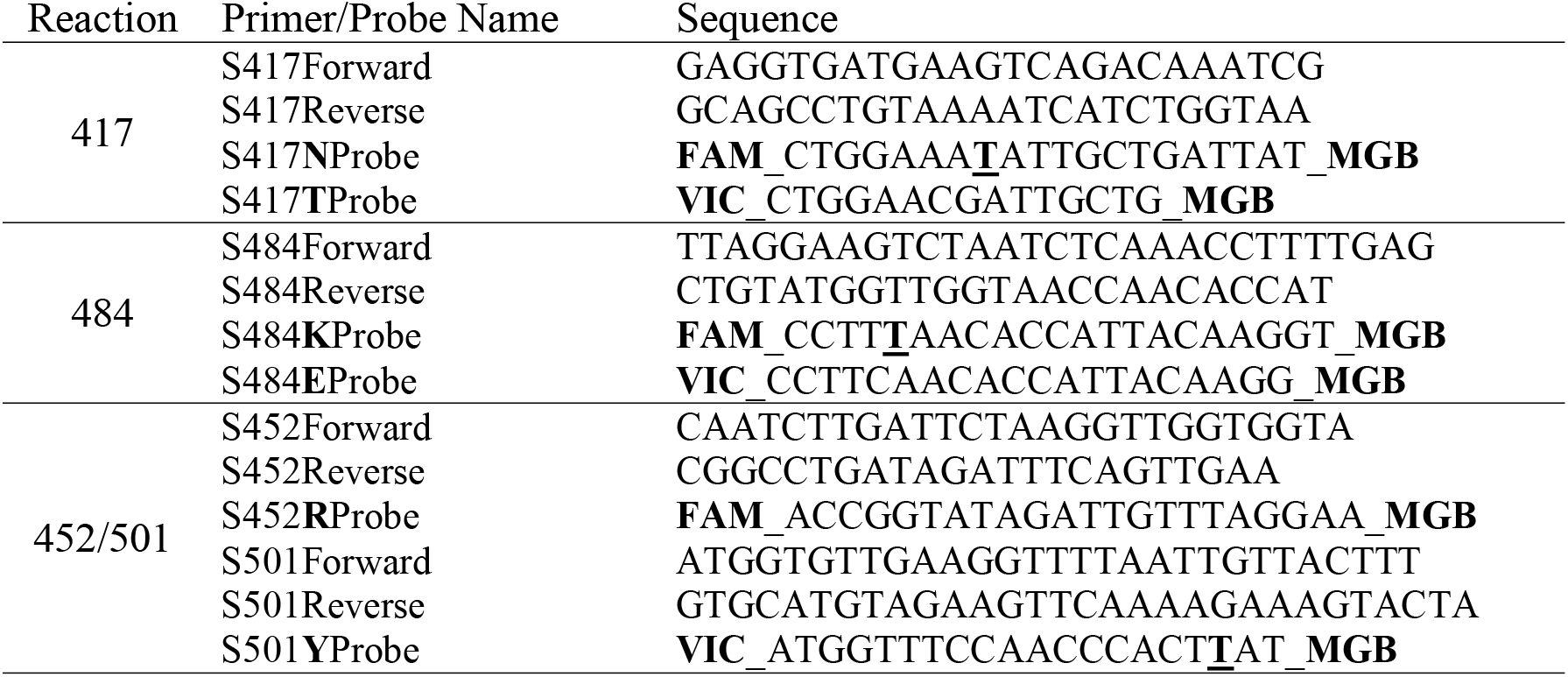
Primer and Probe Sequences for RT-ddPCR assays. Each assay is identified by the amino acid(s) in Spike RBD it targets. For each probe name, bold letters indicate the amino acid detected by that probe. For each probe sequence, bold/underlined letters indicate the mutation that results in the amino acid change. Primer and probe sequences for 501Y are the same as the S1B set listed in [36]. Abbreviations: FAM, 6-carboxyfluorescein; MGB, Minor Groove Binder; VIC, 2’-chloro-7’-phenyl-1,4-dichloro-6-carboxy-fluoroscein.

**Table 2.**
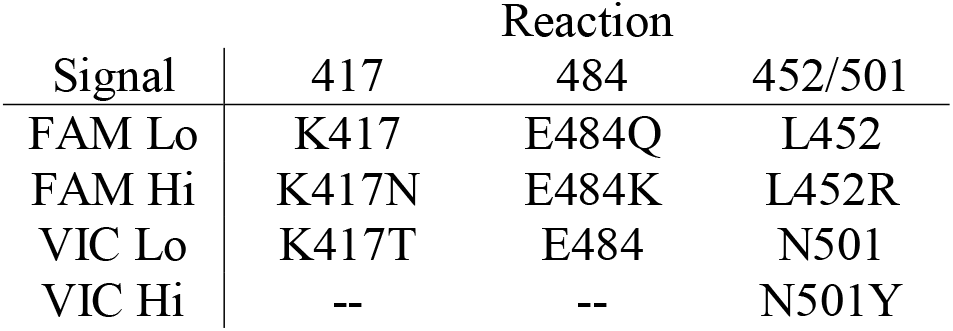
QuantaSoft analysis settings for RT-ddPCR assays. To identify alleles in RT-ddPCR results, the assay type Amplitude Multiplex was selected with allele identifiers for each reaction.

### 3.4. Clinical specimens

Reference Specimens: Four lineages (D614G, Beta, Gamma, and Kappa) were selected to represent the amino acids at the four RBD sites present at the time (Table 3). One high-concentration clinical specimen from each lineage was identified in the UWVL SARS-CoV-2 repository based on NGS results, and diluted in PBS into high-concentration (∼5000 copies/µl) mutation controls.

**Table 3.** Amino Acid Target

| Lineage | Amino Acid Target |  |  |  |
| --- | --- | --- | --- | --- |
|  | 417 | 452 | 484 | 501 |
| D614G | K | L | E | N |
| Beta | N | L | K | Y |
| Gamma | T | L | K | Y |
| Kappa | K | R | Q | N |

Validation Specimens gathered from UWVL: 419 SARS-CoV-2 positive clinical specimens collected between 1/29/2021 and 6/17/2021; 16 SARS-CoV-2 negative clinical specimens (8 collected in PBS and 8 in UTM); and 24 samples positive for other respiratory viruses.

Omicron Specimens: From 11/29/21 to 12/8/21, 2657 positive clinical specimens were screened by TaqPath assay as previously described.[36] Sixteen of these were identified as S-gene dropouts and were tested in the RT-ddPCR assay.

This study was approved under a waiver of consent by the University of Washington institutional review board. GISAD IDs for all specimens are listed in Supplement 1.

## 4. Results

### 4.1. How the Assay Works

Droplet digital (dd)PCR reactions take place inside oil-separated droplets, using TaqMan probe detection: when a probe is bound to the template, amplification separates the dye on the 5’ end of the probe from the quencher on the 3’ end of the probe, causing a release of fluorescent dye. However, when the probe binds poorly to the template because of differences in probe and template sequence, the dye is cleaved less frequently and less fluorescence is released inside that droplet. At the end of the PCR reaction, the fluorescence within each droplet is measured. The amplitude (brightness) of fluorescence within each droplet indicates how well the probe bound to the template and therefore can be used to determine the mutation state of that template.

Using four high-concentration specimens with each allele (Supplement 2), we diluted RNA to the point where each droplet contained at most one copy of template, ∼Ct 28-30. We measured the amplitude of all non-empty droplets (identifiable by comparison to a water-only control) from each specimen and determined that all alleles were clearly identifiable with three combinations of primers and probes (Fig. 1).

**Fig. 1.**
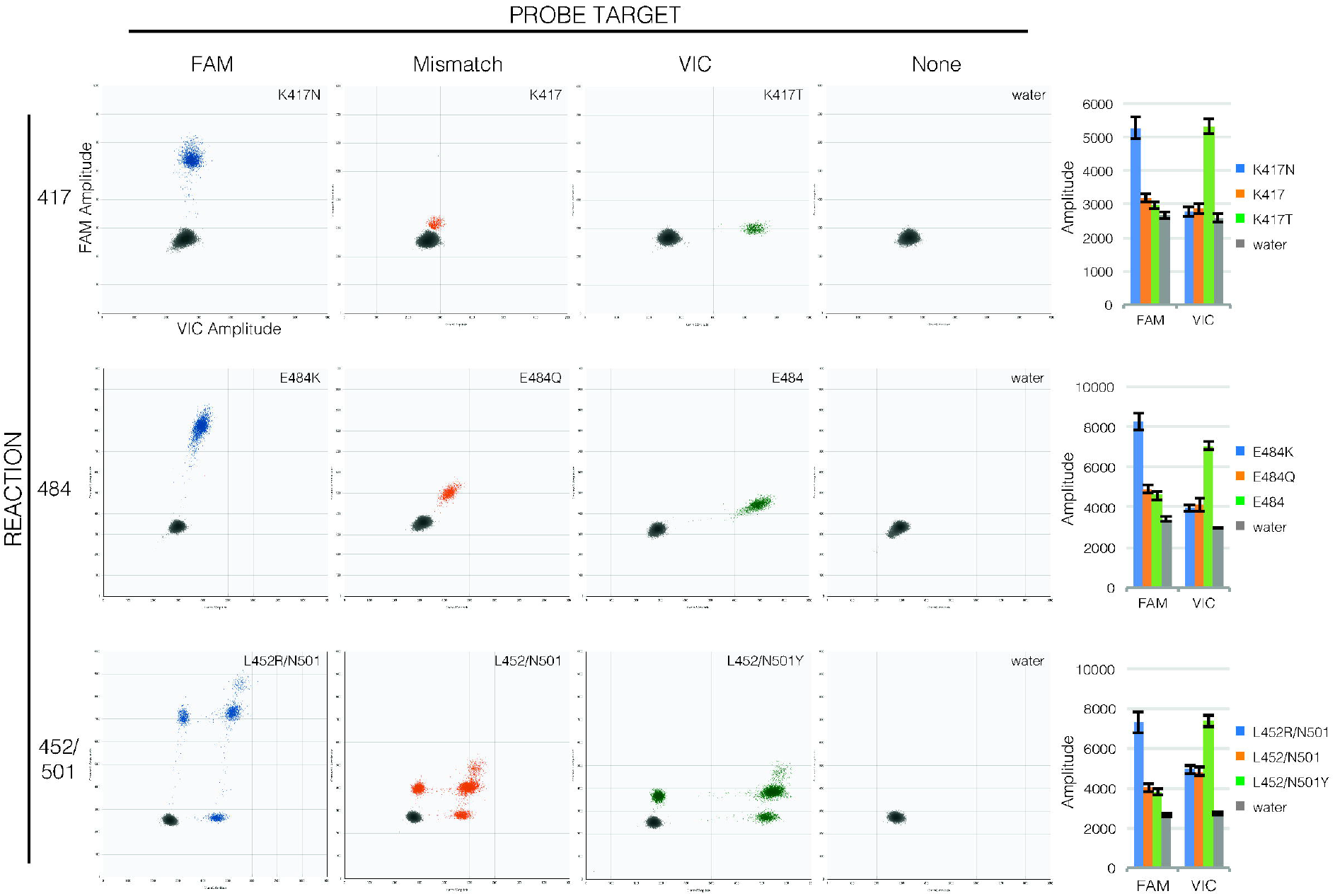
Probe binding clearly distinguishes mutations in RT-ddPCR. Droplet amplitude plots (n=1 per allele) illustrate how FAM fluorescence (X axis) and VIC fluorescence (Y axis) are diagnostic of templates matching the FAM probe (first column, blue droplets) or VIC probe (third column, green droplets) compared to templates with mutations in probe sequences (second column, orange droplets) and to droplets that lack template (fourth column, grey droplets) for each reaction (rows). Bar graphs (final column) show how consistent amplitudes are between specimens (average mean amplitude ± average standard deviation of 190-1900 positive droplets each, n=4 per allele). Note that in the 452/501 reaction, unlike the 417 and 484 reactions, the two probes have separate targets so droplets may show fluorescence from only one or the other target (droplets lower along the axes) or from both targets (droplets higher along the axes).

### 4.2. Template Concentration Range

To confirm that extra copies of template do not increase fluorescence amplitude within a droplet, we tested the four high-concentration specimens along with serial 10-fold dilutions of each, and compared amplitudes (Fig. 2, Supplement 2). RT-ddPCR assays contain 10k+ droplets each, so template concentration ranged from 1 to >50 copies per droplet. Increased concentration did in some cases change amplitude (e.g., 417N and 452R), but these are still readily distinguishable from high-and low-concentration amplitudes from other mutations. All concentrations above LoD are accurately identified with each assay.

**Fig. 2.**
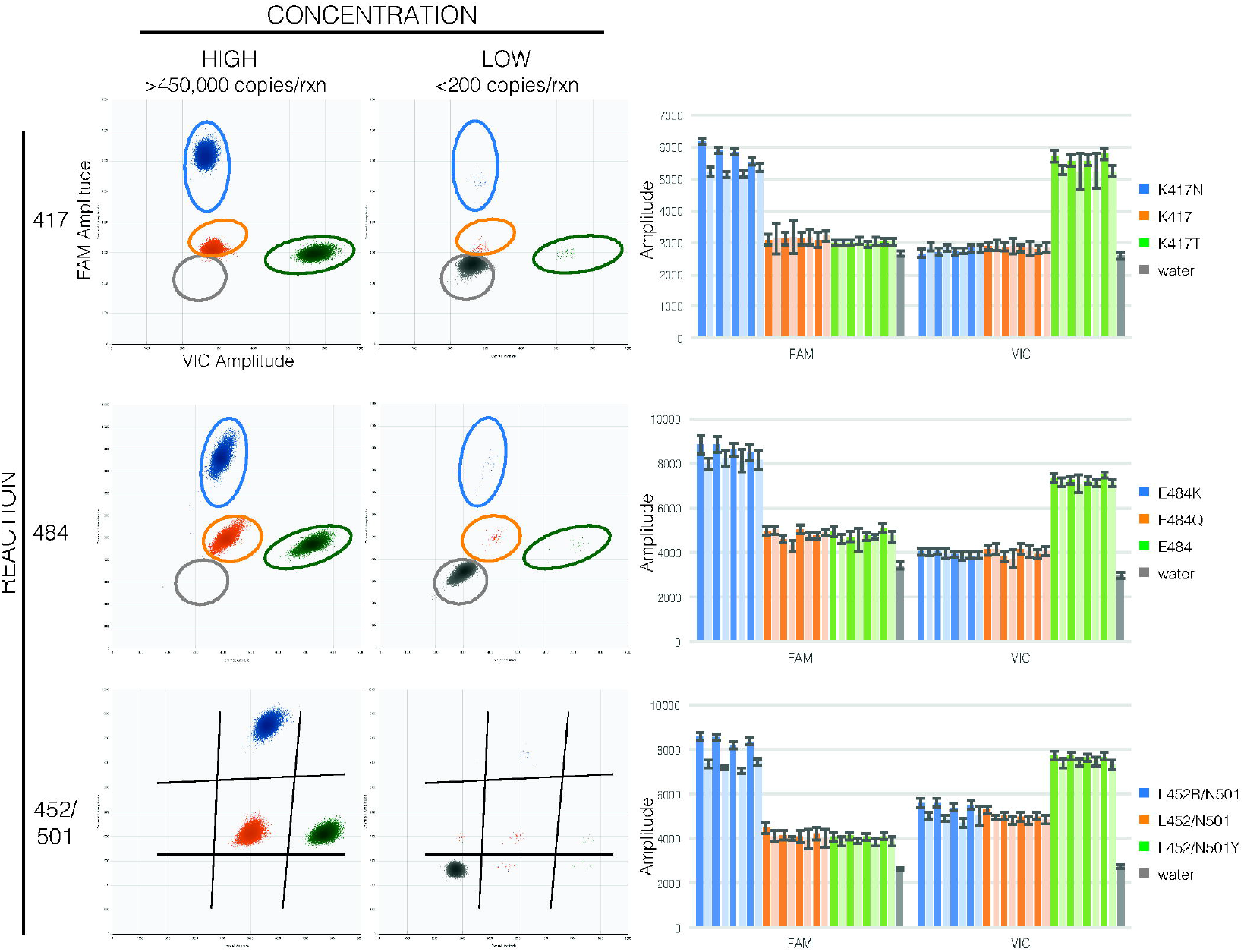
Template concentration does not affect assay accuracy. For each reaction (rows), composite droplet amplitude plots showing the highest-concentration sample of each allele (first column) and lowest-concentration dilution with >10 positive droplets of the same samples (second column) illustrate that amplitudes are diagnostic of template sequences despite wide differences in copy number per droplet. Circles or lines separating droplets from different alleles are for reference on the low-concentration plots. Bar graphs (final column) show this is consistent between specimens (mean amplitude ± standard deviation of highest [dark] and lowest [light] concentration of each specimen, n=4 per allele).

We determined a rough lower limit of detection (LoD) using serial 10-fold dilutions of one specimen per mutation, with four replicates per concentration (Table 4). Each dilution was also measured in RT-PCR with E gene primers and probe[43] in duplicate. Higher template concentrations were necessary to obtain at least 3 positive droplets for mismatch alleles (i.e., 417K and 484Q), but for the targets definitively identified by a probe, lower LoD ranged from 0.7 copies / µL RNA to 1.4 copies / µL RNA.

**Table 4.** Rough Limit of Detection (LoD) for each RT-ddPCR reaction. Dilutions were measured in quadruplicate in RT-ddPCR and in duplicate in RT-PCR (using primer/probe set from [43] as described in [44]). Targets with mismatches to both FAM and VIC probes are indicated with blue type. Mean concentrations are those measured in RT-ddPCR replicates at the LoD; for the dilution beyond LoD, concentrations are calculated from the LoD.

| Reaction | Amino Acid(s) | E Ct | Mean Copies/μL | Mean Copies/Rxn | Positive Replicates | |
| --- | --- | --- | --- | --- | --- | --- |
| 417 | K | 33.4 | 2.22 | 22.2 | 4/4 | At LoD |
|  | N | 35.7 | 0.77 | 7.7 | 4/4 |  |
|  | T | 35.1 | 1.39 | 13.9 | 4/4 |  |
|  | K | 37.1 | 0.22 | 2.2 | 1/4 | Beyond LoD |
|  | N | 37.3 | 0.07 | 0.8 | 0/4 |  |
|  | T | NDET | 0.14 | 1.4 | 0/4 |  |
| 484 | E | 33.4 | 0.65 | 6.5 | 4/4 | At LoD |
|  | K | 35.1 | 0.88 | 8.8 | 4/4 |  |
|  | Q | 30.8 | 13.10 | 131.0 | 4/4 |  |
|  | E | 37.1 | 0.07 | 0.7 | 0/4 | Beyond LoD |
|  | K | NDET | 0.09 | 0.9 | 0/4 |  |
|  | Q | 34.1 | 1.31 | 13.1 | 3/4 |  |
| 452/501 | L/N | 33.4 | 1.17 | 11.7 | 4/4 | At LoD |
|  | L/Y | 35.1 | 0.86 | 8.6 | 4/4 |  |
|  | R/N | 34.1 | 0.91 | 9.1 | 4/4 |  |
|  | L/N | 37.1 | 0.11 | 1.2 | 1/4 | Beyond LoD |
|  | L/Y | NDET | 0.09 | 0.9 | 0/4 |  |
|  | R/N | NDET | 0.09 | 0.9 | 2/4 |  |

### 4.3 Accuracy

We tested 390 additional clinical specimens with the assay, and compared ddPCR results to Whole Genome sequencing (WGS) results for each. Mutation determination for 99.0% of detected samples agree with WGS for those samples (Table 5, Supplement 1).

**Table 5.**
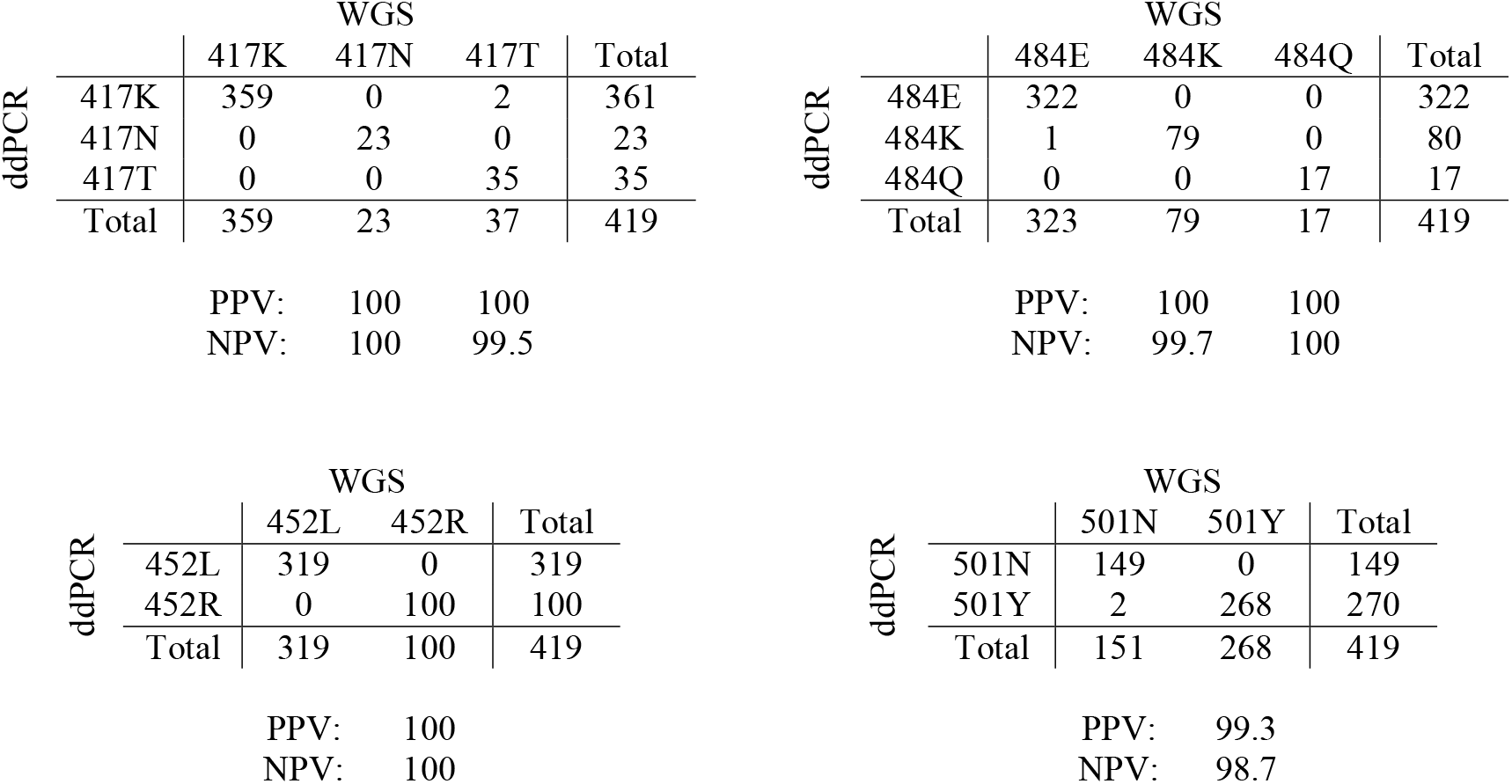
Comparison of RT-ddPCR and WGS results for clinical specimens at each of four amino acids. The number of specimens with a given genotype based on RT-ddPCR (rows) and WGS (columns) is listed for each assay. For each mutation, the Positive Predictive Value (PPV) and Negative Predictive Value (NPV) are calculated based on this comparison.

During the course of testing these specimens, we found two additional mutations in our probe regions that affected droplet amplitude. A search of all sequences in GISAID deposited from UWVL conducted in late July 2021 showed that these were the only two mutations in probe regions that occurred at greater than 1% frequency. Both happened to be synonymous mutations within the codon for the mutation of interest for that probe (Fig. 3). In the 484E probe, 1.1% of Alpha sequences included GAG instead of GAA (Fig. 3A), resulting in a reduction in amplitude that was still easily distinguishable from the reductions caused by mutations to CAA (Q) or AAA (K). In the 417T probe, 4.0% of Gamma sequences included ACA instead of ACG (Fig. 3B), resulting in amplitude almost indistinguishable from AAG (K). A search of all Spike_K417T sequences in GISAID completed on 12/14/2021 (Supplement 3) revealed that even though UWVL deposited only a tiny percentage of Spike_K417T sequences (Fig. 3C), this mutation was greatly enriched in samples sequenced by UWVL, and its presence lasted only a little over a month (Fig. 3D). The last sequence deposited by UWVL with this mutation was collected before 5/17/2021.

**Fig. 3.**
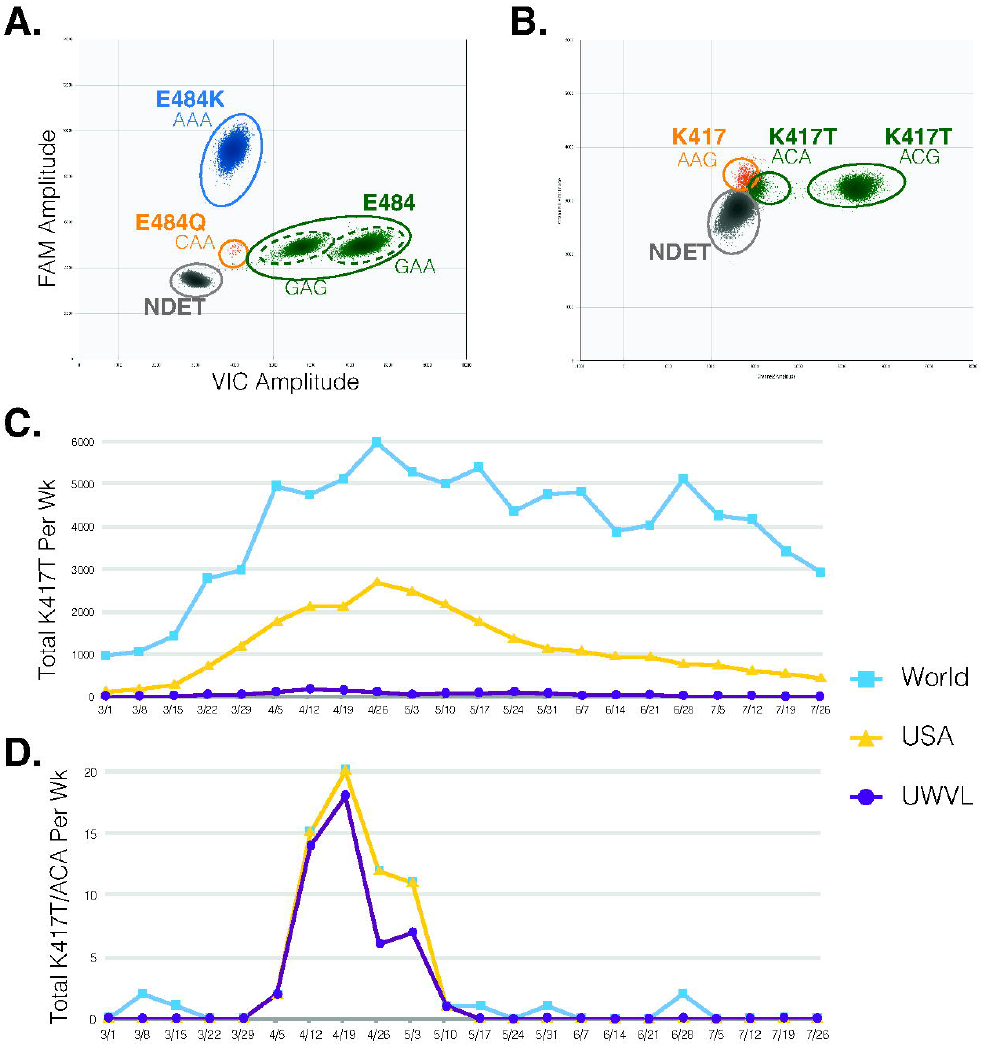
Additional UWVL-identified mutations in probe sequence have variable effects on assay accuracy. (A) Mutation A->G in Spike_E484 results in decreased droplet amplitude that is still distinguishable from other alleles. (B) Mutation G->A in Spike_K417T results in decreased amplitude that is barely distinguishable from K417. (C-D) K417T sequences from samples collected within the week beginning each listed date in the world as a whole, the USA as a whole, or by UWVL: (C) total K417T sequences; (D) sequences with K417T encoded by ACA instead of ACG codon.

### 4.4. Collection media equivalency and Specificity

Negative clinical specimens collected in PBS and in VTM were analyzed alone and with 1/100 dilution of mutation controls spiked in. All negative samples were undetected with the assay, all spiked samples were detected accurately, and the medium into which the samples were spiked did not affect the concentration at which they were detected (Table 6).

To measure cross-reactivity, RNA from 24 individual specimens with high copy number of 10 different respiratory viruses (AdV, BoV, two other human CoV, FluA, MPV, PIV1, PIV4, RhV, and RSV) were analyzed using the assay. To measure microbial interference, these RNA samples were spiked with 1/100 dilution of each mutation control. No amplification of other viruses was detected, and the presence of those viruses did not affect the accuracy of mutation determination for spiked-in controls (Table 6).

**Table 6.** RT-ddPCR reactions are specific for SARS-CoV-2. Sixteen SARS-CoV-2-negative clinical specimens and 24 specimens positive for additional respiratory viruses were tested both alone and with spiked-in RNA from each allele in all assay reactions. Allele determinations were made based on droplet amplitudes.

### 4.5. Accurate Identification of Omicron from SGTF Specimens

Between November 29 and December 8, 2021, we tested 2657 SARS-CoV-2 positive clinical specimens by TaqPath assay, with 16 clear SGTF results. These 16 specimens were tested by the RT-ddPCR assay along with five non-SGTF specimens (Fig. 4). All non-SGTF specimens were clearly Delta. The first SGTF was identified as Delta, and the subsequent 15 were identified as not-Delta, with a combination of droplet amplitudes that matched published sequences for Omicron (Supplement 3): K417N; L452; and mutations in the regions of both 484 and 501 that we had not seen in any previous variants. Whole genome sequencing confirmed the identification of all 16 SGTF specimens: one Delta with the 69-70 deletion associated with SGTF; and 15 Omicron with E484A and G496S/Q498R/N501Y.

**Fig. 4.**
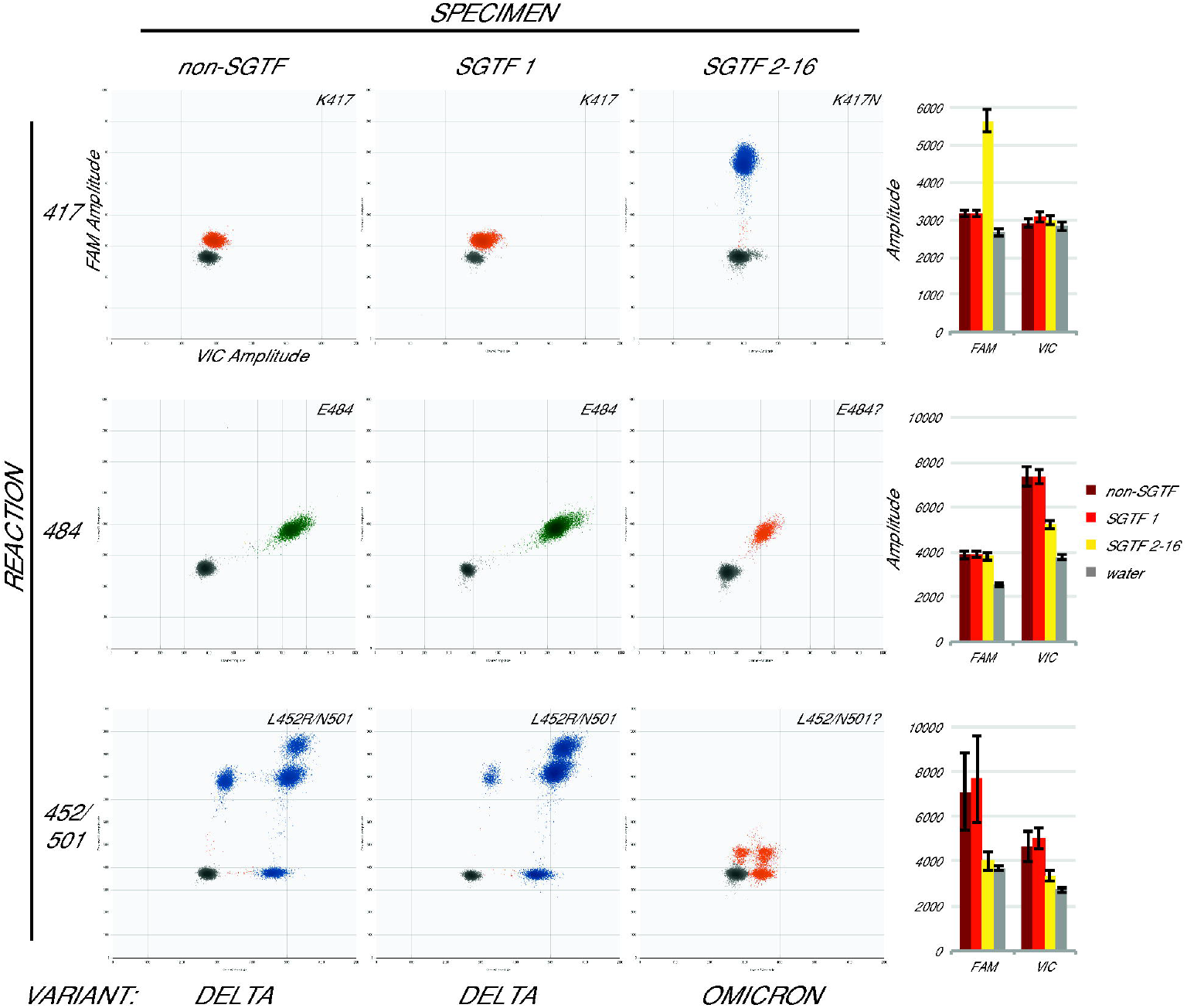
Omicron specimens are accurately identified with the assay. Droplet amplitude plots (n=1 per specimen type) illustrate the different assay results for three categories of newly-collected specimen: non-SGTF (first column), the first SGTF identified at UWVL (second column), and all subsequent SGTF (third column) for each reaction (rows). Bar graphs (final column) show average mean amplitude (± average standard deviation) for samples from each category (n=5, n=1, and n=7 respectively). Variant determination (bottom) based on the assay was confirmed in all cases by whole-genome sequencing.

## 4. Discussion

Through the spring and summer of 2021, numerous SARS-CoV-2 VOC appeared and spread and changed in frequency across the globe in a way that complicated efforts by public health professionals. Some of the mutations carried by these variants were associated with increased transmissibility, which was of concern to those working to contain the epidemic. Others were associated with significant immune evasion, which was of great concern to doctors looking to treat vulnerable COVID-19 patients with monoclonal antibodies, some of the only drugs available for combatting SARS-CoV-2 at the time. [23,24]

When the Delta variant overtook all other variants, accounting for over 99% of all cases sequenced by UWVL by 8/22/21, it was tempting to think that the need to rapidly identify variants had ended. Certainly, the complexity of mAb selection decisions appeared to be reduced. But as the emergence and rapid spread of the Omicron variant demonstrates, viral evolution continues and so does our need to track it.

Omicron was first identified in late November 2021, with a substantial number of mutations resulting in amino acid changes in its Spike and Nucleocapsid proteins in particular.[31,45] Many of the previously concerning mutations are back: K417N, N501Y, a change at E484 (E484A), along with an impressive number of novel mutations. Early evidence from modeling and reports of breakthrough infections show that these mutations result in a substantial reduction in the efficacy of endogenous as well as many mAbs,[46-49] while sotrovimab and related mAbs may retain the ability to neutralize Omicron.[32,49] Once again, clinicians may need to identify variant before prescribing mAbs for their patients, and once again this is a task that must be accomplished rapidly in order for mAb treatment to be effective in preventing severe disease. Because this RT-ddPCR assay targets the sequences directly related to the antigen escape of SARS-CoV-2 variants, and does so rapidly, it is a useful tool for clinical decision-making.

There are other advantages to a method that allows rapid and conclusive identification of virus variant for a particular clinical specimen. Limited resources for case tracking and tracing can be focused on mutations/variants of greater concern, such as current efforts being put into finding and following patients carrying the Omicron variant. Rapid variant identification can also allow selection of specimens for scientific analysis without the delay and added expense of whole genome sequencing. For example, use of this assay in March and August of 2021 allowed us to rapidly select Alpha, Epsilon, and Delta variant samples,[11] enabling a comparison of variant growth in culture that would have been much more tenuous if the specimens needed to be held for sequencing (either subjected to lengthy storage at 4°C or to additional freeze-thaw cycles) before selection.

As is the case for all PCR-based genotyping methods, there are limitations to the utility of this RT-ddPCR assay. The example of ACG->ACA mutation in K417T illustrates that assay accuracy is subject to change with additional mutations. There is also no guarantee that the particular set of mutations of concern detected by the assay will continue to be mutations of concern in the future. However, direct targeting of those mutations of concern, rather than relying on larger-but-less-relevant genomic changes, increases the likelihood that the assay will continue to be useful to researchers and clinicians alike. Reliance on other changes like the 69-70 deletion increases the chances of false positives (as in the case of the SGTF-Delta specimen we identified) or false negatives (as in the case of the non-SGTF Omicron specimens that have been identified in several countries[50]).

We have previously used RT-ddPCR to identify SARS-CoV-2 Alpha variant in clinical specimens, but here we expand both the number of mutational lineages that can be tracked with the assay and our understanding of how robust the assay is. It can accommodate a wide range of sample concentrations, coinfections, and other confounds, and still yields an accurate determination of SARS-CoV-2 mutations quickly enough to enable clinical decision-making.

## Data Availability

All data produced in the present work are contained in the manuscript

## 5. Acknowledgements

We gratefully acknowledge all the laboratories around the world involved in the generation and deposition of SARS-CoV-2 sequence data that we obtained from GISAID (Supplement 3).

## Notes

### Competing Interest Statement

ALG and KRJ report contract testing from Abbot and ALG research support from Merck and
Gilead. The other authors declare no conflicts of interest.

### Funding Statement

This study did not receive any funding

### Author Declarations

This study was approved under a waiver of consent by the University of Washington institutional review board

