## Supplement 3 for "Rapid and Accurate Identification of SARS-CoV-2 Omicron Variants Using Droplet Digital PCR (RT-ddPCR)": All_0301-0307.pdf

We gratefully acknowledge the following Authors from the Originating laboratories responsible for obtaining the specimens, as well as the Submitting laboratories where the genome data were generated and shared via GISAID, on which this research is based.

All Submitters of data may be contacted directly via www.gisaid.org

Authors are sorted alphabetically.

| Accession ID | Originating Laboratory | Submitting Laboratory | Authors |
|---|---|---|---|
| EPI_ISL_1447987 | 20210300728 | TIGEM | Antonio Grimaldi Patrizia Annunziata Francesco Panariello Biancamaria Pierri Claudia Tiberio Valentina Bouche Chiara Colantuono Maria Concetta Cuomo Denise Di Concilio Lucio Di Filippo Anna Manfredi Marcello Salvi Antonio Limone Luigi Atripaldi Pellegrino Cerino Andrea Ballabio Davide Cacchiarelli |
| EPI_ISL_5915124, EPI_ISL_5915219, EPI_ISL_5915328, EPI_ISL_6573815, EPI_ISL_6573817, EPI_ISL_6573852, EPI_ISL_6573962 | | | |
| see above | AMALAB/FACISA/UFRN | WallauLab on behalf of Fiocruz COVID-19 Genomic Surveillance Network | Alexandre Freitas da Silva; Allan Robert Dias Nunes; Antonio Marinho da Silva Neto; Cassia Docena; Constância Flávia Junqueira Ayres; Filipe Zimmer Dezordi; Gabriel Luz Wallau; Gustavo Barbosa de Lima; Joana Cristina Medeiros Tavares Marques; Katya Anaya Jacinto; Lais Ceschini Machado; Lilian Caroliny Amorim Silva; Marcelo Henrique dos Santos Paiva; Mariane dos Santos Duarte; Matheus Filgueira Bezerra; Sinval Pinto Brandão Filho |
| EPI_ISL_1169173, EPI_ISL_1219105, EPI_ISL_1219130, EPI_ISL_1229261, EPI_ISL_1229280, EPI_ISL_1229340, EPI_ISL_1229343, EPI_ISL_1229605, EPI_ISL_1229701, EPI_ISL_1229889, EPI_ISL_1298510, EPI_ISL_1298522, EPI_ISL_1298527, EPI_ISL_1298636, EPI_ISL_1298651, EPI_ISL_1298658, EPI_ISL_1298664, EPI_ISL_1298670, EPI_ISL_1298676, EPI_ISL_1298690, EPI_ISL_1298704, EPI_ISL_1298739, EPI_ISL_1298749, EPI_ISL_1298763, EPI_ISL_1298946, EPI_ISL_1298978, EPI_ISL_1299060, EPI_ISL_1299111, EPI_ISL_1299124 | | | |
| see above | ASL Napoli 1 Centro | AMES Centro Polidiagnostico Strumentale S.r.l. | "Giovanni Savarese; Antonella Di Carlo; Antonio Fico"; Eloisa Evangelista; Luigi D'Amore; Luisa Circelli; Maurizio D'Amora; Monica Ianniello; Nadia Petrillo; Raffaella Ruggiero; Roberto Sirica |
| EPI_ISL_1675317 | AYUDAS DIAGNOSTICAS SURA | Universidad Nacional de Colombia - Laboratorio Genómico One Health | Andres F. Cardona-Rios; Carlos Franco-Muñoz; Daniel O. Maldonado-Perez; Diego A. Álvarez-Díaz; Hector Alejandro Ruiz-Moreno; Idabely Betancur Ortiz; Jorge E. Osorio; Juan P. Hernandez-Ortiz; Karl A Ciuoderis; Katherine Laiton-Donato; Laura Silvana Perez; Lina M. Hurtado; Marcela Mercado-Reyes; Maria Angélica Maya; Maria Stella López; Rita Almanza Payares; Sandra Ines Cano; Simón Villegas Velásquez |
| EPI_ISL_1222737 | AZDelta | AZDelta | Dieter De Smet; Geert Martens |
| EPI_ISL_1493499, EPI_ISL_1493500, EPI_ISL_1493501, EPI_ISL_1493552, EPI_ISL_1493553, EPI_ISL_1550407, EPI_ISL_2186024, EPI_ISL_2186039, EPI_ISL_2241697, EPI_ISL_2241753, EPI_ISL_2241758, EPI_ISL_2241761, EPI_ISL_2241780, EPI_ISL_2242903, EPI_ISL_4960920 | | | |
| see above | Aegis Sciences Corporation | Centers for Disease Control and Prevention Division of Viral Diseases, Pathogen Discovery | Adrian Paskey; Alec Vest; Benjamin Rambo-Martin; Christopher Gulvick; Clinton Paden; Clinton R. Paden; Cyndi Clark; Dakota Howard; Darlene Wagner; Dhwani Batra; Dillon Nall; Duncan MacCannell; Erisa Sula; Ethan Sanders; Holly Houdeshell; Jason Caravas; Kara Moser; Kristine Lacek; Matthew Hardison; Matthew Schmerer; Ola Kvalvaag; Patrick Campbell; Peter Cook; Peter W. Cook; Rob Case; Scott Sammons; Shatavia Morrison; Shaun Westlund; Tymeckia Kendall; Victoria Caban Figueroa; Vikramsinha Ghorpade; Yvette Unoarumhi |
| EPI_ISL_1652201 | Alaska State Virology Laboratory | Alaska State Virology Laboratory | Elva House; Jack Chen; Lisa Smith; Ph.D.; Stephanie DeRonde |
| EPI_ISL_2166896 | Alberta Precision Labs (APL) | Public Health Agency of Canada (PHAC) National Microbiology Laboratory | Buss; Croxen M; Deo A; Dieu P; E; Ferrato C; Gill K; Khan F; Koleva P; Li V; Lloyd C; Lynch T; Ma R; Murphy S; Pabbaraju K; Shokoples S; Thayer J; Tipples G; Whitehouse M; Wong A; Yu C; Zelyas N |
| EPI_ISL_1121976 | Area of Virology, Serology and Virology Division (SAViD), New South Wales Health Pathology Randwick | Virology Research Laboratory; Area of Virology, Serology and Virology Division (SAViD), New South Wales Health Pathology Randwick | Au, J.; Bull, R.; Deveson, I.; Foster, C.; Rawlinson, W.; Ruiz Silva, M.; Van Hal, S. |
| EPI_ISL_1321759, EPI_ISL_1321761, EPI_ISL_1321765, EPI_ISL_1321778, EPI_ISL_1321781, EPI_ISL_1321790, EPI_ISL_1336178 | | | |
| see above | Azienda Ospedaliera Terni | Istituto Zooprofilattico Sperimentale dell'Abruzzo e Molise "G. Caporale" | Ancora M; Calistri P; Cammà C; Curini V; Di Domenico M; Di Pasquale A; Lorusso A; Mangone I; Marcacci M; Palumbo M; Puglia I; Rinaldi A; Savini G; Scaccetti A; Scialabba S |
| EPI_ISL_2522555, EPI_ISL_2522648, EPI_ISL_2524856, EPI_ISL_2524879, EPI_ISL_2525008, EPI_ISL_2525010, EPI_ISL_2525012, EPI_ISL_2525013, EPI_ISL_2525015, EPI_ISL_2525018, EPI_ISL_2525020, EPI_ISL_2525168, EPI_ISL_2525197, EPI_ISL_2525205, EPI_ISL_2525379, EPI_ISL_2525388, EPI_ISL_2525400, EPI_ISL_2525419, EPI_ISL_2525504, EPI_ISL_2525516, EPI_ISL_2525595 | | | |
| see above | BCCDC Public Health Laboratory | BCCDC Public Health Laboratory | Ana Pacagnella; Corinne Ng; Dan Fornika; John Tyson; Kim Macdonald; Kimia Kamelian; Linda Hoang; Loretta Janz; Mel Krajden; Prystajecky Natalie; Robert Azana; Shannon Russell |
| EPI_ISL_7671942, EPI_ISL_7672031 | Bambino Gesù Pediatric Hospital | Microbiology and Immunology Diagnosis Bambino Gesù Pediatric Hospital | Carlo Federico Perno; Claudia Alteri; Luna Colagrossi; Rossana Scutari; Valentino Costabile |
| EPI_ISL_4053500, EPI_ISL_4053520, EPI_ISL_4053522 | Biolab Diagnostic Laboratories | Biolab Diagnostic Laboratories | Ahmad Tibi; Amid Abdelnour; Badia Saddedin; Eiad Atwa; Issa Abu-Dayyeh; Lama Hussein; Shaima Ali |
| EPI_ISL_7743987, EPI_ISL_7744019, EPI_ISL_7744020, EPI_ISL_7744021, EPI_ISL_7744050, EPI_ISL_7744105 | Biome/SENAI/Chapecó | Laboratório de Bioinformática - Universidade Federal de Santa Catarina | "Aline Daina Schlindwein"; "Ana Paula Christoff"; "Antuani Baptistela"; "Carolina Leite Martins"; "Darcita Buerger Rovaris"; "Dayane Azevedo Padilha"; "Doris Sobral Marques SouzaSobral"; "Edmundo Carlos Grisard"; "Eric Kazuo Kawagoe"; "Fernanda Luiza Ferrari"; "Fernanda Roesene Melo"; "Fernando Hartmann Barazzetti"; "Gislaine Fongaro"; "Glauber Wagner"; "Guilherme Augusto Maia"; "Guilherme Razzera"; "Guilherme Toledo e Silva"; "Julia Kinetz Wachter"; "Luiz Felipe de Oliveira"; "Marcos André Schörne"; "Marcus Vinícius Duarte Rodrigues"; "Maria Luiza Bazzo"; "Marlei Pickler Debiasi dos Anjos"; "Milene Moehr de Moraes"; "Nestor Wendt"; "Patrícia Hermes Stoco"; "Paula Sacchet"; "Renato Simões Moreira"; "Rodrigo de Paula Baptista"; "Tamela Zamboni Madaloz"; "Tatiany Aparecida Teixeira Soratto"; "Vilmar Benetti Filho" |
| EPI_ISL_3494243 | Boston University CTL | Boston University/National Emerging Infectious Disease Laboratories | Catherine Klapperich; Jacquelyn Turcinovic; John H. Connor; Lena Landeverde; Lynn Doucette-Stamm |
| EPI_ISL_1701128 | CA DPH Viral and Rickettsial Disease Laboratory | Chan-Zuckerberg Biohub | CZB Cliahub Consortium |
| EPI_ISL_2891189 | CDPH VBL | California Department of Public Health | CDPH-COVIDNet |
| EPI_ISL_5530148 | CENTRO DE ATENCAO ESPECIALIZADA | Analytical Competence Molecular Epidemiology Lab/ACME, Oswaldo Cruz Foundation, Ceara (FIOCRUZ CE) | Carlos Leonardo de Aragao Araujo; Cecilia Leite Costa & Eduardo Ruback dos Santos on behalf of COVID-19 FIOCRUZ Genomic Network; Cleber Furtado Aksenen; Fabio Miyajima; Fernando Braga Stehling; Francisco Eder de Moura Lopes; Igor Oliveira Duarte; Jamille Maria Mendes Bezerra; Joaquim Cesar do Nascimento Sousa Junior; Pedro Miguel Carneiro Jeronimo; Suzana Porto Almeida; Thais Ferreira de Oliveira; Thais de Oliveira Costa; Ticiane Cavalcante de Souza; Veridiana Pessoa Miyajima |
| EPI_ISL_5530067 | CENTRO DE SAUDE DE MOMBACA | Analytical Competence Molecular Epidemiology Lab/ACME, Oswaldo Cruz Foundation, Ceara (FIOCRUZ CE) | Carlos Leonardo de Aragao Araujo; Cecilia Leite Costa & Eduardo Ruback dos Santos on behalf of COVID-19 FIOCRUZ Genomic Network; Cleber Furtado Aksenen; Fabio Miyajima; Fernando Braga Stehling; Francisco Eder de Moura Lopes; Igor Oliveira Duarte; Jamille Maria Mendes Bezerra; Joaquim Cesar do Nascimento Sousa Junior; Pedro Miguel Carneiro Jeronimo; Suzana Porto Almeida; Thais Ferreira de Oliveira; Thais de Oliveira Costa; Ticiane Cavalcante de Souza; Veridiana Pessoa Miyajima |
| EPI_ISL_5530179 | CENTRO DE SAUDE DE PENTECOSTE | Analytical Competence Molecular Epidemiology Lab/ACME, Oswaldo Cruz Foundation, Ceara (FIOCRUZ CE) | Carlos Leonardo de Aragao Araujo; Cecilia Leite Costa & Eduardo Ruback dos Santos on behalf of COVID-19 FIOCRUZ Genomic Network; Cleber Furtado Aksenen; Fabio Miyajima; Fernando Braga Stehling; Francisco Eder de Moura Lopes; Igor Oliveira Duarte; Jamille Maria Mendes Bezerra; Joaquim Cesar do Nascimento Sousa Junior; Pedro Miguel Carneiro Jeronimo; Suzana Porto Almeida; Thais Ferreira de Oliveira; Thais de Oliveira Costa; Ticiane Cavalcante de Souza; Veridiana Pessoa Miyajima |
| EPI_ISL_5529949, EPI_ISL_5529950 | CENTRO DE SAUDE DR MIRANDA TAVARES | Analytical Competence Molecular Epidemiology Lab/ACME, Oswaldo Cruz Foundation, Ceara (FIOCRUZ CE) | Carlos Leonardo de Aragao Araujo; Cecilia Leite Costa & Eduardo Ruback dos Santos on behalf of COVID-19 FIOCRUZ Genomic Network; Cleber Furtado Aksenen; Fabio Miyajima; Fernando Braga Stehling; Francisco Eder de Moura Lopes; Igor Oliveira Duarte; Jamille Maria Mendes Bezerra; Joaquim Cesar do Nascimento Sousa Junior; Pedro Miguel Carneiro Jeronimo; Suzana Porto Almeida; Thais Ferreira de Oliveira; Thais de Oliveira Costa; Ticiane Cavalcante de Souza; Veridiana Pessoa Miyajima |
| EPI_ISL_1399630 | CH Porto - H Sto Antonio | Instituto Nacional de Saude (INSA) and BioSystems & Integrative Sciences Institute (BioISI) Genomics Unit, FCUL | Borges et al |
| EPI_ISL_1696250 | CH. CHARLES NICOLLE | Department of Virology, Henri Mondor University Hospital, Assistance Publique Hôpitaux de Paris, Université Paris-Est Créteil, INSERM U955 | Alexandre Soulier; Christophe Rodriguez; Elisabeth Trawinski; Guillaume Gricourt; Jean-Michel Pawlotsky; Melissa N'Debi; Slim Fourati; Vanessa Demontant |
| EPI_ISL_1672730, EPI_ISL_1672741, EPI_ISL_1696254, EPI_ISL_1706628 | CH.INTERCOMMUNAL DE CRETEIL | Department of Virology, Henri Mondor University Hospital, Assistance Publique Hôpitaux de Paris, Université Paris-Est Créteil, INSERM U955 | Alexandre Soulier; Christophe Rodriguez; Elisabeth Trawinski; Guillaume Gricourt; Jean-Michel Pawlotsky; Melissa N'Debi; Slim Fourati; Vanessa Demontant |
| EPI_ISL_1593980 | CHC Andrée Rosemon | Institut Pasteur de la Guyane | Anne Lavergne; Dominique Rousset |
| EPI_ISL_1399631 | CHU Sao Joao, Porto | Instituto Nacional de Saude (INSA) and Instituto Gulbenkian de Ciencia (IGC) | Borges et al |
| EPI_ISL_5530096 | CSF BARACHO ANTUNINO HERCULANO DE MESQUITA | Analytical Competence Molecular Epidemiology Lab/ACME, Oswaldo Cruz Foundation, Ceara (FIOCRUZ CE) | Carlos Leonardo de Aragao Araujo; Cecilia Leite Costa & Eduardo Ruback dos Santos on behalf of COVID-19 FIOCRUZ Genomic Network; Cleber Furtado Aksenen; Fabio Miyajima; Fernando Braga Stehling; Francisco Eder de Moura Lopes; Igor Oliveira Duarte; Jamille Maria Mendes Bezerra; Joaquim Cesar do Nascimento Sousa Junior; Pedro Miguel Carneiro Jeronimo; Suzana Porto Almeida; Thais Ferreira de Oliveira; Thais de Oliveira Costa; Ticiane Cavalcante de Souza; Veridiana Pessoa Miyajima |
| EPI_ISL_5530162 | CSF TAPERUABA | Analytical Competence Molecular Epidemiology Lab/ACME, Oswaldo Cruz Foundation, Ceara (FIOCRUZ CE) | Carlos Leonardo de Aragao Araujo; Cecilia Leite Costa & Eduardo Ruback dos Santos on behalf of COVID-19 FIOCRUZ Genomic Network; Cleber Furtado Aksenen; Fabio Miyajima; Fernando Braga Stehling; Francisco Eder de Moura Lopes; Igor Oliveira Duarte; Jamille Maria Mendes Bezerra; Joaquim Cesar do Nascimento Sousa Junior; Pedro Miguel Carneiro Jeronimo; Suzana Porto Almeida; Thais Ferreira de Oliveira; Thais de Oliveira Costa; Ticiane Cavalcante de Souza; Veridiana Pessoa Miyajima |
| EPI_ISL_2697909, EPI_ISL_2697920, EPI_ISL_2697972, EPI_ISL_2697974, EPI_ISL_2698010, EPI_ISL_2698012, EPI_ISL_2698014 | | | |
| see above | CTvacinas | CTvacinas | A.P.; B.L.; Coelho; D.B.; Dorlass; Durigon; E.G.; E.L.; F.G.; Fernandes; Fiorini, A.; Fonseca; G.P.; H.P.; K.L.; L.M.; Lourenco; Magalhaes; Oliveira; Ometto, T.; Peixoto, R.; R.D.; Sato, H.; Scagion; Teixeira, S.; Telezynski; Thomazelli |
| EPI_ISL_1583672, EPI_ISL_1583673, EPI_ISL_1583676, EPI_ISL_1583679, EPI_ISL_1583681, EPI_ISL_1583716, EPI_ISL_1583725, EPI_ISL_1583730, EPI_ISL_3255134, EPI_ISL_3266087, EPI_ISL_3266110, EPI_ISL_3266111 | | | |

| | | | |
|---|---|---|---|
| see above | Central Public Health Laboratory - LACEN - Bahia, Salvador, Brazil | Central Public Health Laboratory - LACEN - Bahia, Salvador, Brazil | Arabela Leal; Breno Dominguez; Felicidade Pereira; Jaqueline Gomes; Luciana Oliveira; Luiz Alcantara; Marcela Gómez; Marta Giovanetti; Patrícia Cajado; Stephane Tosta; Vagner Fonseca; Vanessa Nardy |
| EPI_ISL_4169886 | Central Virology Laboratory, Israel Ministry of Health | Israel Institute for Biological Research | Adi Beth-Din; Anat Zvi; Inbar Cohen-Gihon; Nir Paran; Ofir Israeli; Yfat Yahalom Ronen |
| EPI_ISL_2612316 | Centro de Infectologia Charles Mérieux/ Laboratório Rodolphe Mérieux, FUNDHACRE | Bioinformatics Laboratory / LNCC | Alessandra P Lamarca; Alexandra L Gerber; Ana Paula de C Guimarães; Ana Tereza R Vasconcelos; Andreas Stocker; Cirley Maria de Oliveira Lobato; Douglas Terra Machado; Luiz Fellype Alves de Souza; Luiz G P de Almeida; Ronaldo da Silva F Jr |
| EPI_ISL_2031707, EPI_ISL_2031708, EPI_ISL_2031709, EPI_ISL_2031710, EPI_ISL_2031711, EPI_ISL_2031712, EPI_ISL_2031713, EPI_ISL_2031714, EPI_ISL_2031724 | | | |
| see above | Centro de Innovación en Vigilancia Epidemiológica (CiVE), Institut Pasteur Montevideo, Uruguay | Centro de Innovación en Vigilancia Epidemiológica (CiVE), Institut Pasteur Montevideo, Uruguay | Alicia Costábile; Alvaro Fajardo; Andrés Lizosaín; Belén González; Bernardina Rivera; Cecilia Alonso; Cecilia Salazar; Gonzalo Moratorio; Gregorio Iraola; Henry Albornoz; Ignacio Ferrés; Inés Bellini; Juan Zanetti; Julio Medina; Lucia Bilbao; Luciana Griffero; Lucía Spangenberg; Ma Noel Bentancor; Ma Pía Techera; Mailen Arleo; Martina Alonso; María José Benítez; Matías Maidana; Mauricio Méndez; Melissa Duquía; Mercedes Paz; Natalia Rego; Natalia Reyes; Odhille Chappos; Paula Perbolianachis; Pilar Moreno; Rodney Colina; Rodrigo Arce; Tamara Fernández; Tania Possi |
| EPI_ISL_1381068 | Conjunto Hospitalar do Mandaqui de Sao Paulo | Instituto Adolfo Lutz, Interdiciplinary Procedures Center, Strategic Laboratory | Caio Vinicius Dias Lopes; Claudia Regina Gonçalves; Claudio Tavares Sacchi; Erica Valessa Ramos Gomes; Karoline Rodrigues Campos |
| EPI_ISL_1583675 | DNA Laboratory | Central Public Health Laboratory - LACEN - Bahia, Salvador, Brazil | Arabela Leal; Breno Dominguez; Felicidade Pereira; Jaqueline Gomes; Luciana Oliveira; Luiz Alcantara; Marcela Gómez; Marta Giovanetti; Patrícia Cajado; Stephane Tosta; Vagner Fonseca; Vanessa Nardy |
| EPI_ISL_1860143 | Department of Clinical Microbiology and Center for Genomic Medicine, Rigshospitalet, Copenhagen, Denmark | Aalborg University | Danish Covid-19 Genome Consortium |
| EPI_ISL_1259194, EPI_ISL_1259195 | Department of Virology, Istituto Zooprofilattico Sperimentale del Lazio e della Toscana (IZSLT) | Department of General Diagnostics; Department of Virology; Istituto Zooprofilattico Sperimentale del Lazio e della Toscana (IZSLT) | Alessia Franco; Antonella Cersini; Antonio Battisti.; Elena L. Diaconu; Fabiola Feltrin; Giuseppe Manna; Patricia Alba; Raffaela Conti; Teresa Scicluna; Virginia Carfora |
| EPI_ISL_1821212 | Diagnóstico da America S/A | Instituto Adolfo Lutz, Interdiciplinary Procedures Center, Strategic Laboratory | Caio Vinicius Dias Lopes; Claudia Regina Gonçalves; Claudio Tavares Sacchi; Erica Valessa Ramos Gomes; Karoline Rodrigues Campos; Leonardo Jose Tadeu de Araujo |
| EPI_ISL_1311010, EPI_ISL_2145481 | Dutch COVID-19 response team | Erasmus Medical Center | Anne van der Linden; Annemiek van der Eijk; Bas Oude Munnink; Corine GeurtsvanKessel; David Nieuwenhuijse; Emmanuelle Munger; Irina Chestakova; Marion Koopmans; Marjan Boter; Reina Sikkema; Richard Molenkamp; on behalf of the Dutch national COVID-19 respo; on behalf of the Dutch national COVID-19 response team. |
| EPI_ISL_1289218, EPI_ISL_1289220, EPI_ISL_1289221, EPI_ISL_1289222, EPI_ISL_1289224, EPI_ISL_1370615, EPI_ISL_1371081, EPI_ISL_1371282, EPI_ISL_1371610, EPI_ISL_1456642, EPI_ISL_1521300, EPI_ISL_1521301, EPI_ISL_1521302, EPI_ISL_1521324, EPI_ISL_1521325, EPI_ISL_1596027, EPI_ISL_1597037, EPI_ISL_1597126, EPI_ISL_1597139, EPI_ISL_1597162, EPI_ISL_1597220, EPI_ISL_1597302, EPI_ISL_1597424, EPI_ISL_1597436, EPI_ISL_1597446, EPI_ISL_1792430 | | | |
| see above | Dutch COVID-19 response team | National Institute for Public Health and the Environment (RIVM) | Adam Meijer; AnneMarie van den Brandt; Annelies Kroneman; Bas van der Veer; Chantal Reusken; Dennis Schmitz; Dirk Eggink; Euníce Then; Florian Zwagemaker; Harry Vennema; James Groot; Jeroen Cremer; Jolienke Hardeman; Karim Hajji; Kim Freriks; Linda van de Nes; Lisa Wijsman; Lynn Aarts; Melissa van Tuil; Robert Kohl; Ryanne Jaarsma; Sanne Bos; Sharon van den Brink; Sjoerd Kuiling; on behalf of the national COVID-19 response team |
| EPI_ISL_1795098, EPI_ISL_2344544 | ESALQ | Instituto Butantan / ESALQ-Piracicaba | Antonio Jorge Martins; Bianca Cechetto Carlos. Mendelics: Bibiana Santos; Claudia Renata dos Santos Barros; David Schlesinger; David Schlesinger. Hemocentro Ribeirão Preto: Simone Kashima; Debora Botequio Moretti; Debora Botequio Moretti. Centro de Genômica Funcional da ESALQ: Luiz Lehmann Coutinho; Dimas Tadeu Covas; Elaine Cristina Marqueze; Elaine Vieira Santos; Elaine Vieira dos Santos; Elisangela Chicaroni Mattos; Erika Freitas; Evandra Strazza Rodrigues; Felipe Allan da Silva da Costa; Flavia Aburjaile; Guilherme Targino Valente; Heidge Fukumasu; Heidge Fukumasu. USP-Botucatu: Rejane Maria Tommasini Grotto; Instituto Butantan: Alexander Roberto Precioso; Jayme A. Souza-Neto; Jayme Augusto de Souza-Neto; Jessika Cristina Chagas Lesbon; José Salvatore Leister Patané; João Paulo Kitajima; Luiz Alcantara; Luiz Carlos Junior de Alcantara; Luiz Lehmann Coutinho; Maria Carolina Elias; Marta Giovanetti; Maurício Lacerda Nogueira; Patricia Akemi Assato; Rafael dos Santos Bezerra; Raquel de Lello Rocha Campos Cassano. NGS Soluções Genômicas: Pilar Drummond Sampaio Corrêa Mariani. FZEA-USP Pirassununga: Mirele Daiana Poleti; Raul Machado Neto; Rejane Maria Tommasini Grotto; Ricardo Augusto Brassaloti; Ricardo Haddad; Rodrigo Tocantins Calado.; Sandra Coccuzzo Sampaio; Sandra Coccuzzo Sampaio Vessoni; Simone Kashima; Svetoslav Nanev Slavov; Vagner Fonseca; Vincent Louis Viala |
| EPI_ISL_1813476, EPI_ISL_1814101, EPI_ISL_1815018, EPI_ISL_1815577, EPI_ISL_1815751, EPI_ISL_1815988, EPI_ISL_1816366 | | | |
| see above | EXCITE Lab | Andersen lab at Scripps Research | Alexandre Bolze; Alice Summerfield; Celena Andrade; Charlotte Rivera-Garcia; David Becker; Efren Sandoval; Elizabeth Cirulli; Francisco Tanudjaja; Geraint Levan; James Lu + SEARCH; Jason Nguyen; Jimmy Ramirez; Kelly Schiabor Barrett; Magnus Isaksson; Marc Laurent; Nicole L Washington; Ryan Cho; Sherry Wang; Simon White; Tyler Cassens; William Lee |
| EPI_ISL_2112527 | Epidemiology of Microbial Diseases, Yale School of Public Health | Epidemiology of Microbial Diseases, Yale School of Public Health | A.E.; A.F.; Alpert, T.; Breban; Brito; C.B.; C.C.; Fauver; Grubaugh; I.M.; Iwasaki, A.; J.E.; J.R.; Kalinich; Landry; Lucas, C.; M.E.; M.I.; M.L.; N.D.; Ott; Petrone; Rothman; Vogels; Watkins |
| EPI_ISL_6565798 | Florida Bureau of Public Health Laboratories | Florida Bureau of Public Health Laboratories | Jason Blanton; Namratha Tarigopula; Sarah Schmedes; Tiffany Splatt |
| EPI_ISL_1555280, EPI_ISL_1555282, EPI_ISL_1555283, EPI_ISL_1555326, EPI_ISL_1555330, EPI_ISL_1555351, EPI_ISL_1555406, EPI_ISL_1555411, EPI_ISL_1555418, EPI_ISL_1555473, EPI_ISL_1555884, EPI_ISL_1556238 | | | |
| see above | Fulgent Genetics | Centers for Disease Control and Prevention Division of Viral Diseases, Pathogen Discovery | Adrian Paskey; Becky Tsai; Benafsh Sapra; Benjamin Rambo-Martin; Christopher Gulvick; Clinton R. Paden; Dakota Howard; Darlene Wagner; Dhwani Batra; Doreen Ng; Duncan MacCannell; Harry Gao; James Xie; Jason Caravas; John Gao; Joseph Fierro; Kara Moser; Matthew Schmerer; Mickey Li; Peter W. Cook; Scott Sammons; Shatavia Morrison; Yan Meng; Yvette Unoarumhi |
| EPI_ISL_1231495 | Fulgent Genetics | Fulgent Genetics | Becky Tsai; Benafsh Sapra; Doreen Ng; Harry Gao; James Xie; John Gao; Joseph Fierro; Mickey Li; Yan Meng |
| EPI_ISL_1239137, EPI_ISL_1239138 | Fundação Ezequiel Dias | Coordenação Geral de Laboratórios de Saúde Pública (CGLAB) | ; Vagner Fonseca et al |
| EPI_ISL_1399624, EPI_ISL_1399625, EPI_ISL_1399626 | Germano de Sousa | Instituto Nacional de Saude (INSA) | Borges et al |
| EPI_ISL_1696296 | Groupe LCD | Department of Virology, Henri Mondor University Hospital, Assistance Publique Hôpitaux de Paris, Université Paris-Est Créteil, INSERM U955 | Alexandre Soulier; Christophe Rodriguez; Elisabeth Trawinski; Guillaume Gricourt; Jean-Michel Pawlotsky; Melissa N'Debi; Slim Fourati; Vanessa Demontant |
| EPI_ISL_3102238 | H J M A HOSPITAL JOSE MARTINIANO DE ALENCAR | Analytical Competence Molecular Epidemiology Lab/ACME, Oswaldo Cruz Foundation, Ceara (FIOCRUZ CE) | Cleber Furtado Aksenen; Fabio Miyajima; Fernando Braga Stehling; Francisco Eder de Moura Lopes; Jamille Maria Mendes Bezerra; Joaquim César do Nascimento Sousa Junior; Pedro Miguel Carneiro Jeronimo; Suzana Porto Almeida e Lucas Delerino; Thais Ferreira de Oliveira; Thais de Oliveira Costa; Ticiane Cavalcante de Souza; Veridiana Pessoa Miyajima |
| EPI_ISL_3102219 | HEMOCE CENTRO DE HEMATOLOGIA E HEMOTERAPIA | Analytical Competence Molecular Epidemiology Lab/ACME, Oswaldo Cruz Foundation, Ceara (FIOCRUZ CE) | Cleber Furtado Aksenen; Fabio Miyajima; Fernando Braga Stehling; Francisco Eder de Moura Lopes; Jamille Maria Mendes Bezerra; Joaquim César do Nascimento Sousa Junior; Pedro Miguel Carneiro Jeronimo; Suzana Porto Almeida e Lucas Delerino; Thais Ferreira de Oliveira; Thais de Oliveira Costa; Ticiane Cavalcante de Souza; Veridiana Pessoa Miyajima |
| EPI_ISL_2017286, EPI_ISL_2017294, EPI_ISL_2017471, EPI_ISL_2017480, EPI_ISL_2102518, EPI_ISL_2187826, EPI_ISL_2187829, EPI_ISL_2187830, EPI_ISL_2187832, EPI_ISL_2187833, EPI_ISL_2187834, EPI_ISL_2187835, EPI_ISL_2187836, EPI_ISL_2187837, EPI_ISL_2187838, EPI_ISL_2187839, EPI_ISL_2187840, EPI_ISL_2187841, EPI_ISL_2187842, EPI_ISL_2187844, EPI_ISL_2187845, EPI_ISL_2187846, EPI_ISL_2187847, EPI_ISL_2187848, EPI_ISL_2187849, EPI_ISL_2187850, EPI_ISL_2187851, EPI_ISL_2187852, EPI_ISL_2187853 | | | |
| see above | HLAGYN - Laboratorio de Imunologia de Transplantes de Goias | HLAGYN - Laboratorio de Imunologia de Transplantes de Goias | Alessandro Leonardo Alvares Magalhaes; Daniel Ferreira de Sousa; Danielle de Paiva Rezende; Erika Lopes Rocha Batista; Fernando Antonio Vinhal dos Santos; Frederico Rodrigues Vinhal; Lucas Carlos Gomes Pereira; Paola Cristina Resende Silva; Raphael Bessa Parmigiane; Sabrina Sara Moreira Duarte |
| EPI_ISL_3102228, EPI_ISL_3102229, EPI_ISL_3102230, EPI_ISL_3102231, EPI_ISL_3102413, EPI_ISL_3102447 | HM HOSPITAL DE MESSEJANA DR CARLOS ALBERTO STUDART GOMES | Analytical Competence Molecular Epidemiology Lab/ACME, Oswaldo Cruz Foundation, Ceara (FIOCRUZ CE) | Cleber Furtado Aksenen; Fabio Miyajima; Fernando Braga Stehling; Francisco Eder de Moura Lopes; Jamille Maria Mendes Bezerra; Joaquim César do Nascimento Sousa Junior; Pedro Miguel Carneiro Jeronimo; Suzana Porto Almeida e Lucas Delerino; Thais Ferreira de Oliveira; Thais de Oliveira Costa; Ticiane Cavalcante de Souza; Veridiana Pessoa Miyajima |
| EPI_ISL_3102259 | HOSP MATERN SANTA IZABEL ARACOIABA | Analytical Competence Molecular Epidemiology Lab/ACME, Oswaldo Cruz Foundation, Ceara (FIOCRUZ CE) | Cleber Furtado Aksenen; Fabio Miyajima; Fernando Braga Stehling; Francisco Eder de Moura Lopes; Jamille Maria Mendes Bezerra; Joaquim César do Nascimento Sousa Junior; Pedro Miguel Carneiro Jeronimo; Suzana Porto Almeida e Lucas Delerino; Thais Ferreira de Oliveira; Thais de Oliveira Costa; Ticiane Cavalcante de Souza; Veridiana Pessoa Miyajima |
| EPI_ISL_5529959, EPI_ISL_5530117, EPI_ISL_5530174 | HOSP MUN ABELARDO GADELHA DA ROCHA | Analytical Competence Molecular Epidemiology Lab/ACME, Oswaldo Cruz Foundation, Ceara (FIOCRUZ CE) | Carlos Leonardo de Aragao Araujo; Cecilia Leite Costa & Eduardo Ruback dos Santos on behalf of COVID-19 FIOCRUZ Genomic Network; Cleber Furtado Aksenen; Fabio Miyajima; Fernando Braga Stehling; Francisco Eder de Moura Lopes; Igor Oliveira Duarte; Jamille Maria Mendes Bezerra; Joaquim Cesar do Nascimento Sousa Junior; Pedro Miguel Carneiro Jeronimo; Suzana Porto Almeida; Thais Ferreira de Oliveira; Thais de Oliveira Costa; Ticiane Cavalcante de Souza; Veridiana Pessoa Miyajima |
| EPI_ISL_5529951 | HOSPITAL DE PEQUENO PORTE DE CARIDADE | Analytical Competence Molecular Epidemiology Lab/ACME, Oswaldo Cruz Foundation, Ceara (FIOCRUZ CE) | Carlos Leonardo de Aragao Araujo; Cecilia Leite Costa & Eduardo Ruback dos Santos on behalf of COVID-19 FIOCRUZ Genomic Network; Cleber Furtado Aksenen; Fabio Miyajima; Fernando Braga Stehling; Francisco Eder de Moura Lopes; Igor Oliveira Duarte; Jamille Maria Mendes Bezerra; Joaquim Cesar do Nascimento Sousa Junior; Pedro Miguel Carneiro Jeronimo; Suzana Porto Almeida; Thais Ferreira de Oliveira; Thais de Oliveira Costa; Ticiane Cavalcante de Souza; Veridiana Pessoa Miyajima |
| EPI_ISL_5529985, EPI_ISL_5529986, EPI_ISL_5529996, EPI_ISL_5529997, EPI_ISL_5529998 | HOSPITAL DISTRITAL GONZAGA MOTA BARRA DO CEARA | Analytical Competence Molecular Epidemiology Lab/ACME, Oswaldo Cruz Foundation, Ceara (FIOCRUZ CE) | Carlos Leonardo de Aragao Araujo; Cecilia Leite Costa & Eduardo Ruback dos Santos on behalf of COVID-19 FIOCRUZ Genomic Network; Cleber Furtado Aksenen; Fabio Miyajima; Fernando Braga Stehling; Francisco Eder de Moura Lopes; Igor Oliveira Duarte; Jamille Maria Mendes Bezerra; Joaquim Cesar do Nascimento Sousa Junior; Pedro Miguel Carneiro Jeronimo; Suzana Porto Almeida; Thais Ferreira de Oliveira; Thais de Oliveira Costa; Ticiane Cavalcante de Souza; Veridiana Pessoa Miyajima |

| | | | |
|---|---|---|---|
| EPI_ISL_5530172 | HOSPITAL DISTRITAL NOSSA SENHORA DA CONCEICAO | Analytical Competence Molecular Epidemiology Lab/ACME, Oswaldo Cruz Foundation, Ceara (FIOCRUZ CE) | Carlos Leonardo de Aragao Araujo; Cecilia Leite Costa & Eduardo Ruback dos Santos on behalf of COVID-19 FIOCRUZ Genomic Network; Cleber Furtado Aksenen; Fabio Miyajima; Fernando Braga Stehling; Francisco Eder de Moura Lopes; Igor Oliveira Duarte; Jamille Maria Mendes Bezerra; Joaquim Cesar do Nascimento Sousa Junior; Pedro Miguel Carneiro Jeronimo; Suzana Porto Almeida; Thais Ferreira de Oliveira; Thais de Oliveira Costa; Ticiane Cavalcante de Souza; Veridiana Pessoa Miyajima |
| EPI_ISL_5530118 | HOSPITAL DR ESTEVAM | Analytical Competence Molecular Epidemiology Lab/ACME, Oswaldo Cruz Foundation, Ceara (FIOCRUZ CE) | Carlos Leonardo de Aragao Araujo; Cecilia Leite Costa & Eduardo Ruback dos Santos on behalf of COVID-19 FIOCRUZ Genomic Network; Cleber Furtado Aksenen; Fabio Miyajima; Fernando Braga Stehling; Francisco Eder de Moura Lopes; Igor Oliveira Duarte; Jamille Maria Mendes Bezerra; Joaquim Cesar do Nascimento Sousa Junior; Pedro Miguel Carneiro Jeronimo; Suzana Porto Almeida; Thais Ferreira de Oliveira; Thais de Oliveira Costa; Ticiane Cavalcante de Souza; Veridiana Pessoa Miyajima |
| EPI_ISL_1712404 | HOSPITAL DR. ENRIQUE BALTODANO BRICEÑO | Inciensa, Instituto Costarricense de Investigación y Enseñanza en Nutrición y Salud | Adriana Godínez; Claudio Soto-Garita; Estela Cordero; Francisco Duarte; Hebleen Porras; Joselyn Prado & Adriana Bermúdez-Espinoza; José Luis Vargas; Mariela Gutiérrez; Melany Calderón |
| EPI_ISL_3102479 | HOSPITAL E MATERNIDADE ADOLFO BEZERRA DE MENEZES | Analytical Competence Molecular Epidemiology Lab/ACME, Oswaldo Cruz Foundation, Ceara (FIOCRUZ CE) | Cleber Furtado Aksenen; Fabio Miyajima; Fernando Braga Stehling; Francisco Eder de Moura Lopes; Jamille Maria Mendes Bezerra; Joaquim César do Nascimento Sousa Junior; Pedro Miguel Carneiro Jeronimo; Suzana Porto Almeida e Lucas Delerino; Thais Ferreira de Oliveira; Thais de Oliveira Costa; Ticiane Cavalcante de Souza; Veridiana Pessoa Miyajima |
| EPI_ISL_3102221, EPI_ISL_3102222, EPI_ISL_3102390 | HOSPITAL E MATERNIDADE DRA ZILDA ARNS NEUMANN | Analytical Competence Molecular Epidemiology Lab/ACME, Oswaldo Cruz Foundation, Ceara (FIOCRUZ CE) | Cleber Furtado Aksenen; Fabio Miyajima; Fernando Braga Stehling; Francisco Eder de Moura Lopes; Jamille Maria Mendes Bezerra; Joaquim César do Nascimento Sousa Junior; Pedro Miguel Carneiro Jeronimo; Suzana Porto Almeida e Lucas Delerino; Thais Ferreira de Oliveira; Thais de Oliveira Costa; Ticiane Cavalcante de Souza; Veridiana Pessoa Miyajima |
| EPI_ISL_3102478 | HOSPITAL E MATERNIDADE GASTROCLINICA | Analytical Competence Molecular Epidemiology Lab/ACME, Oswaldo Cruz Foundation, Ceara (FIOCRUZ CE) | Cleber Furtado Aksenen; Fabio Miyajima; Fernando Braga Stehling; Francisco Eder de Moura Lopes; Jamille Maria Mendes Bezerra; Joaquim César do Nascimento Sousa Junior; Pedro Miguel Carneiro Jeronimo; Suzana Porto Almeida e Lucas Delerino; Thais Ferreira de Oliveira; Thais de Oliveira Costa; Ticiane Cavalcante de Souza; Veridiana Pessoa Miyajima |
| EPI_ISL_5530095 | HOSPITAL MUNICIPAL ANTONIO RIBEIRO DA SILVA | Analytical Competence Molecular Epidemiology Lab/ACME, Oswaldo Cruz Foundation, Ceara (FIOCRUZ CE) | Carlos Leonardo de Aragao Araujo; Cecilia Leite Costa & Eduardo Ruback dos Santos on behalf of COVID-19 FIOCRUZ Genomic Network; Cleber Furtado Aksenen; Fabio Miyajima; Fernando Braga Stehling; Francisco Eder de Moura Lopes; Igor Oliveira Duarte; Jamille Maria Mendes Bezerra; Joaquim Cesar do Nascimento Sousa Junior; Pedro Miguel Carneiro Jeronimo; Suzana Porto Almeida; Thais Ferreira de Oliveira; Thais de Oliveira Costa; Ticiane Cavalcante de Souza; Veridiana Pessoa Miyajima |
| EPI_ISL_5530168 | HOSPITAL MUNICIPAL JOAO LEOPOLDO PINHEIRO LANDIM | Analytical Competence Molecular Epidemiology Lab/ACME, Oswaldo Cruz Foundation, Ceara (FIOCRUZ CE) | Carlos Leonardo de Aragao Araujo; Cecilia Leite Costa & Eduardo Ruback dos Santos on behalf of COVID-19 FIOCRUZ Genomic Network; Cleber Furtado Aksenen; Fabio Miyajima; Fernando Braga Stehling; Francisco Eder de Moura Lopes; Igor Oliveira Duarte; Jamille Maria Mendes Bezerra; Joaquim Cesar do Nascimento Sousa Junior; Pedro Miguel Carneiro Jeronimo; Suzana Porto Almeida; Thais Ferreira de Oliveira; Thais de Oliveira Costa; Ticiane Cavalcante de Souza; Veridiana Pessoa Miyajima |
| EPI_ISL_5529933 | HOSPITAL MUNICIPAL MARIA IDALINA RODRIGUES DE MEDEIROS | Analytical Competence Molecular Epidemiology Lab/ACME, Oswaldo Cruz Foundation, Ceara (FIOCRUZ CE) | Carlos Leonardo de Aragao Araujo; Cecilia Leite Costa & Eduardo Ruback dos Santos on behalf of COVID-19 FIOCRUZ Genomic Network; Cleber Furtado Aksenen; Fabio Miyajima; Fernando Braga Stehling; Francisco Eder de Moura Lopes; Igor Oliveira Duarte; Jamille Maria Mendes Bezerra; Joaquim Cesar do Nascimento Sousa Junior; Pedro Miguel Carneiro Jeronimo; Suzana Porto Almeida; Thais Ferreira de Oliveira; Thais de Oliveira Costa; Ticiane Cavalcante de Souza; Veridiana Pessoa Miyajima |
| EPI_ISL_3102242, EPI_ISL_3102243, EPI_ISL_3102244, EPI_ISL_3102245, EPI_ISL_3102246, EPI_ISL_3102414, EPI_ISL_3102421 | | | |
| see above | HOSPITAL SAO JOSE DE DOENCAS INFECCIOSAS | Analytical Competence Molecular Epidemiology Lab/ACME, Oswaldo Cruz Foundation, Ceara (FIOCRUZ CE) | Cleber Furtado Aksenen; Fabio Miyajima; Fernando Braga Stehling; Francisco Eder de Moura Lopes; Jamille Maria Mendes Bezerra; Joaquim César do Nascimento Sousa Junior; Pedro Miguel Carneiro Jeronimo; Suzana Porto Almeida e Lucas Delerino; Thais Ferreira de Oliveira; Thais de Oliveira Costa; Ticiane Cavalcante de Souza; Veridiana Pessoa Miyajima |
| EPI_ISL_2801313 | HOSPITAL SÃO JOSE DE DOENÇAS INFECCIOSAS | Analytical Competence Molecular Epidemiology Lab/ACME, Oswaldo Cruz Foundation, Ceara (FIOCRUZ CE) | Cleber Furtado Aksenen e Suzana Porto Almeida; Fabio Miyajima; Fernando Braga Stehling; Francisco Eder de Moura Lopes; Jamille Maria Mendes Bezerra; Joaquim César do Nascimento Sousa Junior; Pedro Miguel Carneiro Jeronimo; Thais Ferreira de Oliveira; Thais de Oliveira Costa; Ticiane Cavalcante de Souza; Veridiana Pessoa Miyajima |
| EPI_ISL_1421440, EPI_ISL_1421740, EPI_ISL_1480020, EPI_ISL_1480106, EPI_ISL_1480124, EPI_ISL_1480193, EPI_ISL_1480227, EPI_ISL_1480288, EPI_ISL_1480289, EPI_ISL_1480453, EPI_ISL_1480464 | | | |
| see above | Helix/Illumina | Centers for Disease Control and Prevention Division of Viral Diseases, Pathogen Discovery | Adrian Paskey; Alexandre Bolze; Ary Ascencio; Ben L. Rambo-Martin; Benjamin Rambo-Martin; Brad Sickler; Charlotte Rivera-Garcia; Christine Tran; Christopher Gulvick; Clinton R. Paden; Dakota Howard; Darlene Wagner; David Becker; Dhwani Batra; Duncan MacCannell; Efren Sandoval; Eileen de Feo; Elizabeth Cirulli; Eric Allen; Geraint Levan; James Lu; Jan Antico; Jason Caravas; Jason Nguyen; Jimmy Ramirez; Jingtao Liu; Kara Moser; Kelly Schiabor Barrett; Kim Gietzen; Magnus Isaksson; Marc Laurent; Matthew Schmerer; Matthew Tolentino; Nicole L. Washington; Peter W. Cook; Phil Febbo; Ryan Cho; Scott Sammons; Shannon Wickline; Shatavia Morrison; Sherry Wang; Simon White; Summer Galloway; Suxiang Tong; Tyler Cassens; William Lee; Yvette Unoarumhi |
| EPI_ISL_3149978, EPI_ISL_3149979, EPI_ISL_3149982, EPI_ISL_3149984, EPI_ISL_3149994, EPI_ISL_3149997, EPI_ISL_3149998, EPI_ISL_3149999, EPI_ISL_3150013 | | | |
| see above | Hermes Pardini | Universidade Federal de Ciencias da Saude de Porto Alegre | Andrea Roberto de Souza; Andy Goren; Carlos Gustavo Wambier; Claudia Elizabeth Thompson; Daniel do Nascimento Fonseca; Emilyn Oliveira Guerreiro; Flávio Adsuara Cadegiani; Liane Nanci Rotta; Patrícia Aline Gröhs Ferrareze; Ricardo Ariel Zimerman; Zhihua Ren |
| EPI_ISL_1262988 | Hopital | National Reference Center for Viruses of Respiratory Infections, Institut Pasteur, Paris | Angela Brisebarre; Camille Capel; Etienne Simon-Lorière; Marion Barbet; Maud Vanpeene; Méline Bizard; Sylvie Behillil; Sylvie van der Werf; Vincent Enouf; Woercel Isabelle |
| EPI_ISL_1381216, EPI_ISL_1381217 | Hospital | National Reference Center for Viruses of Respiratory Infections, Institut Pasteur, Paris | Angela Brisebarre; Camille Capel; Etienne Simon-Lorière; Louise Lefrançois; Marion Barbet; Maud Vanpeene; Méline Bizard; Rousset Dominique; Sylvie Behillil; Sylvie van der Werf; Vincent Enouf |
| EPI_ISL_1533726 | Hospital Estadual de Campanha Covid 19 Barradas | Instituto Adolfo Lutz, Interdiciplinary Procedures Center, Strategic Laboratory | Caio Vinicius Dias Lopes; Claudia Regina Gonçalves; Claudio Tavares Sacchi; Erica Valessa Ramos Gomes; Karoline Rodrigues Campos; Leonardo Jose Tadeu de Araujo |
| EPI_ISL_1533709, EPI_ISL_1533710 | Hospital Estadual de Vila Alpina Org Social Seconci Sao Paulo | Instituto Adolfo Lutz, Interdiciplinary Procedures Center, Strategic Laboratory | Caio Vinicius Dias Lopes; Claudia Regina Gonçalves; Claudio Tavares Sacchi; Erica Valessa Ramos Gomes; Karoline Rodrigues Campos; Leonardo Jose Tadeu de Araujo |
| EPI_ISL_1307715 | Hospital General Universitario Gregorio Marañón | Hospital General Universitario Gregorio Marañón | Cristina Rodriguez-Grande; Darío García de Viedma; Laura Pérez-Lago; Patricia Muñoz; Pedro Sola Campoy; Pilar Catalán; Sergio Buenestado Serrano |
| EPI_ISL_1533725 | Hospital Geral de Guarulhos | Instituto Adolfo Lutz, Interdiciplinary Procedures Center, Strategic Laboratory | Caio Vinicius Dias Lopes; Claudia Regina Gonçalves; Claudio Tavares Sacchi; Erica Valessa Ramos Gomes; Karoline Rodrigues Campos; Leonardo Jose Tadeu de Araujo |
| EPI_ISL_1381069 | Hospital Heliopolis | Instituto Adolfo Lutz, Interdiciplinary Procedures Center, Strategic Laboratory | Caio Vinicius Dias Lopes; Claudia Regina Gonçalves; Claudio Tavares Sacchi; Erica Valessa Ramos Gomes; Karoline Rodrigues Campos |
| EPI_ISL_1675296 | Hospital Pablo Tobón Uribe | Universidad Nacional de Colombia - Laboratorio Genómico One Health | Andres F. Cardona-Rios; Carlos Franco-Muñoz; Daniel O. Maldonado-Perez; Diego A. Álvarez-Díaz; Hector Alejandro Ruiz-Moreno; Idabely Betancur Ortiz; Jorge E. Osorio; Juan P. Hernandez-Ortiz; Karl A Ciuoderis; Katherine Laiton-Donato; Laura Silvana Perez; Lina M. Hurtado; Marcela Mercado-Reyes; Maria Angélica Maya; Maria Stella López; Rita Almanza Payares; Sandra Ines Cano; Simón Villegas Velásquez |
| EPI_ISL_1264408 | Hospital Universitari Arnau de Vilanova | Hospital Universitari Vall d'Hebron - Vall d'Hebron Institut de Recerca | Andrés Antón; Ariadna Rando; Carla Castillo; Cristina Andrés; Damir Garcia-Cehic; Josep F Abril; Josep Quer; Juliana Esperalba; Maria Carmen Martin; Maria Gema Codina; Maria Piñana; Tomàs Pumarola |
| EPI_ISL_1250798 | Hospital Universitari Vall d'Hebron - Vall d'Hebron Institut de Recerca | Hospital Universitari Vall d'Hebron - Vall d'Hebron Institut de Recerca | Andrés Antón; Ariadna Rando; Carla Castillo; Cristina Andrés; Damir Garcia-Cehic; Josep F Abril; Josep Quer; Juliana Esperalba; Maria Carmen Martin; Maria Gema Codina; Maria Piñana; Tomàs Pumarola |
| EPI_ISL_1358288, EPI_ISL_1358289, EPI_ISL_1358290 | IAL Regional de Aracatuba | Instituto Adolfo Lutz, Interdiciplinary Procedures Center, Strategic Laboratory | Caio Vinicius Dias Lopes; Claudia Regina Gonçalves; Claudio Tavares Sacchi; Erica Valessa Ramos Gomes; Karoline Rodrigues Campos |
| EPI_ISL_3368626, EPI_ISL_3373373 | IEC- Instituto Evandro Chagas | ITV-Vale Institute of Technology | Amanda Vidal; Guilherme Oliveira; Mirleide Cordeiro dos Santos; Tatianne Costa Negri |
| EPI_ISL_2444776, EPI_ISL_2444778, EPI_ISL_2444779, EPI_ISL_2444798, EPI_ISL_2444799 | IICS-UNA | IICS-UNA | Adriana Valenzuela; Alejandra Rojas; Chyntia Diaz; Eva Nara; Fatima Cardozo; Florencia del Puerto; Joel Ortiz; Jonas Fernandez; Laura Franco; Laura Mendoza; Leticia Rojas; Magaly Martinez; Maria Eugenia Galeano. |
| EPI_ISL_1509925 | IL Department of Public Health Chicago Laboratory | Genomics and Discovery, Respiratory Viruses Branch, Division of Viral Diseases, Centers for Disease Control and Prevention | Adam Retchless; Anna Kelleher; Anna Montmayeur; Anna Uehara; Brian Lynch; Clinton R. Paden; Haibin Wang; Han Jia Justin Ng; Jing Zhang; Justin Lee; Krista Queen; Mark Burroughs; Peter Cook; Rachel Marine; Suxiang Tong; Yan Li; Ying Tao |
| EPI_ISL_1399627 | IMM | Instituto Nacional de Saude (INSA) | Borges et al |
| EPI_ISL_1547381 | INSA | Instituto Nacional de Saude (INSA) | Borges et al |
| EPI_ISL_1670941, EPI_ISL_2576947, EPI_ISL_2576976, EPI_ISL_2576980 | IRCCS San Gallicano Dermatological Institute | IRCCS Regina Elena National Cancer Institute | Aldo Morrone; Alice Massacci; Antonio Federico; Eleonora Sperandio; Elisabetta Trento; Fabrizio Ensoli; Francesca De Nicola; Frauke Goeman; Fulvia Pimpinelli; Gennaro Ciliberto; Giovanni Blandino; Giulia Orlandi; Grazia Prignano; Matteo Pallocca; Maurizio Fanciulli; Sabrina Strano; Sara Donzelli; di Domenico Enea Gino |
| EPI_ISL_1447712, EPI_ISL_1447713, EPI_ISL_1447714, EPI_ISL_1447715, EPI_ISL_1447716, EPI_ISL_1447717, EPI_ISL_1447718, EPI_ISL_1447719, EPI_ISL_1447720, EPI_ISL_1447721, EPI_ISL_1447722, EPI_ISL_1447723, EPI_ISL_1447724, EPI_ISL_1447725, EPI_ISL_1447726, EPI_ISL_1447727, EPI_ISL_1447728, EPI_ISL_1447729, EPI_ISL_1447730, EPI_ISL_1447731 | | | |
| see above | IZSM | TIGEM | Antonio Grimaldi Patrizia Annunziata Francesco Panariello Biancamaria Pierri Claudia Tiberio Valentina Bouche Chiara Colantuono Maria Concetta Cuomo Denise Di Concilio Lucio Di Filippo Anna Manfredi Marcello Salvi Antonio Limone Luigi Atripaldi Pellegrino Cerino Andrea Ballabio Davide Cacchiarelli |
| EPI_ISL_2691096 | Instituto Adolfo Lutz - Regional de Marilia | Instituto Adolfo Lutz, Interdiciplinary Procedures Center, Strategic Laboratory | Caio Vinicius Dias Lopes; Claudia Regina Gonçalves; Claudio Tavares Sacchi; Erica Valessa Ramos Gomes; Karoline Rodrigues Campos; Leonardo Jose Tadeu de Araujo |
| EPI_ISL_2756443, EPI_ISL_2756475, | Instituto Adolfo Lutz Central | Instituto Adolfo Lutz, Interdiciplinary Procedures Center, Strategic Laboratory | Caio Vinicius Dias Lopes; Claudia Regina Gonçalves; Claudio Tavares Sacchi; Erica Valessa Ramos Gomes; Karoline Rodrigues Campos; Leonardo Jose Tadeu de Araujo; Leonardo Tadeu de Araujo; Marlon Benedito Nascimento Santos |

| | | | |
|---|---|---|---|
| EPI_ISL_2756479, EPI_ISL_4888108 | | | |
| EPI_ISL_2614603, EPI_ISL_2614604, EPI_ISL_2614605, EPI_ISL_2614606, EPI_ISL_2614608, EPI_ISL_2614609 | Instituto Biologico | Instituto Adolfo Lutz, Interdiciplinary Procedures Center, Strategic Laboratory | Caio Vinicius Dias Lopes; Claudia Regina Gonçalves; Claudio Tavares Sacchi; Erica Valessa Ramos Gomes; Karoline Rodrigues Campos; Leonardo Jose Tadeu de Araujo |
| EPI_ISL_3418674, EPI_ISL_3418675, EPI_ISL_3418676, EPI_ISL_3418678, EPI_ISL_3418679, EPI_ISL_3418680, EPI_ISL_3418681, EPI_ISL_3418682, EPI_ISL_3418683, EPI_ISL_3418684 | | | |
| see above | Instituto Carlos Chagas - ICC, FIOCRUZ Parana | Instituto Carlos Chagas - ICC, FIOCRUZ Parana | A.A.; A.M.; A.R.; Aguiar; Albrecht, L.; Alves; Avila; Balsanelli, E.; Becker, G.; Blanes, L.; Dallagiovanna, B.; Debur; E.M.; F.K.; F.O.; Faoro, H.; Graef, T.; H.G.; I.N.; L.G.; L.R.; M.M.; M.O.; Marchini; Md.C.; Morello; Nardeli; Oliveira; P.C.; Passetti, F.; Pedrosa; Resende; Riediger; S.C.; Schemberger; Suzukawa; V.A.; Zanette, D.; de Baura; de Souza; dos Santos |
| EPI_ISL_2894869, EPI_ISL_2894872, EPI_ISL_2894873 | Instituto de Medicina Tropical de Sao Paulo | Instituto de Medicina Tropical de Sao Paulo | Brazil-UK Centre for Arbovirus Discovery Diagnosis Genomics and Epidemiology (CADDE) Genomic Network - Instituto de Medicina Tropical |
| EPI_ISL_2037442 | Instituto de Virologia "Dr. J. M. Vanella", Facultad de Ciencias Médicas, Universidad Nacional de Córdoba. | Centro de Investigaciones Agropecuarias (CIAP), Instituto Nacional de Tecnologia Agropecuaria (INTA) Cordoba, Argentina, on behalf of 'Proyecto Argentino Interinstitucional de Genomica de SARS-CoV-2 (PAIS Consortium) | Adrian Diaz; Brenda Konigheim; Franco Fernandez; Gabriela Barbas; Gonzalo Castro; Humberto Debat; Javier Aguilar; Lorena Spinsanti; Maria Belen Pisano; Maria Elisa Rivarola; Mauricio Beranek; Nathalie Marquez; Sandra Gallego.; Sebastian Blanco; Viviana Re |
| EPI_ISL_1299536, EPI_ISL_1299540, EPI_ISL_1299544 | Istituto Zooprofilattico Sperimentale Umbria e Marche "Togo Rosati" | Istituto Zooprofilattico Sperimentale dell'Abruzzo e Molise "G. Caporale" | Ancora M; Biagetti M; Calistri P; Cammà C; Curini V; Di Domenico M; Di Pasquale A; Giammarioli M; Lorusso A; Mangone I; Marcacci M; Puglia I; Rinaldi A; Savini G; Scialabba S |
| EPI_ISL_1380752, EPI_ISL_1380774, EPI_ISL_1380839, EPI_ISL_1380845, EPI_ISL_1380896, EPI_ISL_1382589, EPI_ISL_1382630, EPI_ISL_1382681, EPI_ISL_1382682, EPI_ISL_1382683, EPI_ISL_1382684, EPI_ISL_1382685, EPI_ISL_1382688, EPI_ISL_1382689, EPI_ISL_1382702, EPI_ISL_1382709, EPI_ISL_1382714, EPI_ISL_1382721, EPI_ISL_2404086 | | | |
| see above | KU Leuven, Rega Institute, Clinical and Epidemiological Virology | KU Leuven, Rega Institute, Clinical and Epidemiological Virology | Bert Vanmechelen; Joan Marti-Carerras; Piet Maes; Tony Wawina-Bokalanga |
| EPI_ISL_3102477 | LABORATORIO MUN ANALISES CLINICAS | Analytical Competence Molecular Epidemiology Lab/ACME, Oswaldo Cruz Foundation, Ceara (FIOCRUZ CE) | Cleber Furtado Aksenen; Fabio Miyajima; Fernando Braga Stehling; Francisco Eder de Moura Lopes; Jamille Maria Mendes Bezerra; Joaquim César do Nascimento Sousa Junior; Pedro Miguel Carneiro Jeronimo; Suzana Porto Almeida e Lucas Delerino; Thais Ferreira de Oliveira; Thais de Oliveira Costa; Ticiane Cavalcante de Souza; Veridiana Pessoa Miyajima |
| EPI_ISL_1716458 | LACEN (Laboratorio de Saude Publica Dr. Giovanni Cysneiros) | LGBio (Laboratorio de Genetica & Biodiversidade) | Amanda Alves de Melo; Cintia Pelegrineti Targueta de Azevedo Brito; Daniela de Melo e Silva; Elisangela de Paula Silveira Lacerda; Francyelli Mello Andrade; Mariana Pires de Campos Telles; Ramilla dos Santos Braga; Renata de Oliveira Dias; Rhewter Nunes; Thais Guimarães Castro; Thays Millena Alves Pedroso |
| EPI_ISL_2488804 | LACEN - Laboratório Central de Saúde Pública do Amapá | Evandro Chagas Institute | A.M.; Barbagelata; E.C.; E.M.A.; Ferreira; J.A.; Junior; K.C.; L.C.; L.S.; M.C.; P.S.; Pinheiro; Santos; Silva; Sousa; Sousa Junior; W.D.C.; da Silva |
| EPI_ISL_2488799 | LACEN - Laboratório Central de Saúde Pública do Ceará | Evandro Chagas Institute | A.M.; Barbagelata; E.C.; E.M.A.; Ferreira; J.A.; Junior; K.C.; L.C.; L.S.; M.C.; P.S.; Pinheiro; Santos; Silva; Sousa; Sousa Junior; W.D.C.; da Silva |
| EPI_ISL_2919229 | LACEN do Estado de Goias | Instituto Adolfo Lutz, Interdiciplinary Procedures Center, Strategic Laboratory | Caio Vinicius Dias Lopes; Claudia Regina Gonçalves; Claudio Tavares Sacchi; Erica Valessa Ramos Gomes; Karoline Rodrigues Campos |
| EPI_ISL_1520108, EPI_ISL_1520109 | LACEN do Estado de Rondonia | Instituto Adolfo Lutz, Interdiciplinary Procedures Center, Strategic Laboratory | Caio Vinicius Dias Lopes; Claudia Regina Gonçalves; Claudio Tavares Sacchi; Erica Valessa Ramos Gomes; Karoline Rodrigues Campos |
| EPI_ISL_3671908, EPI_ISL_3671909, EPI_ISL_3671911, EPI_ISL_3671912 | LACEN do Estado do Mato Grosso do Sul | Instituto Adolfo Lutz, Interdiciplinary Procedures Center, Strategic Laboratory | Caio Vinicius Dias Lopes; Claudia Regina Gonçalves; Claudio Tavares Sacchi; Karoline Rodrigues Campos; Leonardo Tadeu de Araujo; Marlon Benedito Nascomento Santos |
| EPI_ISL_2761936 | LACEN do Mato Grosso do Sul | Instituto Adolfo Lutz, Interdiciplinary Procedures Center, Strategic Laboratory | Caio Vinicius Dias Lopes; Claudia Regina Gonçalves; Claudio Tavares Sacchi; Erica Valessa Ramos Gomes; Karoline Rodrigues Campos; Leonardo Jose Tadeu de Araujo |
| EPI_ISL_2221885, EPI_ISL_6573811, EPI_ISL_6573812, EPI_ISL_6573813, EPI_ISL_6573814, EPI_ISL_6573816, EPI_ISL_6573844, EPI_ISL_6573845, EPI_ISL_6573846, EPI_ISL_6573847, EPI_ISL_6573848, EPI_ISL_6573849 | | | |
| see above | LACEN/PE | WallauLab on behalf of Fiocruz COVID-19 Genomic Surveillance Network | Alexandre Freitas da Silva; Antonio Marinho da Silva Neto; Antonio Mauro Rezende; Cassia Docena; Constância Flávia Junqueira Ayres; Cássia Docena; Duschinka Ribeiro Duarte Guedes; Elisama Helvecio; Filipe Zimmer Dezordi; Gabriel Luz Wallau; Gustavo Barbosa de Lima; Lais Ceschini Machado; Larissa Krokovsky; Laís Ceschini Machado; Lilian Caroliny Amorim Silva; Marcelo Henrique dos Santos Paiva; Matheus Filgueira Bezerra; Sinval Pinto Brandão Filho |
| EPI_ISL_1322021, EPI_ISL_1369117, EPI_ISL_1369125, EPI_ISL_1369135, EPI_ISL_1369137, EPI_ISL_1404193, EPI_ISL_1404234 | | | |
| see above | Lab voor klinische biologie | Lab voor klinische biologie | Bruno Verhasselt; Hannelore Hamerlinck; Marija Janevska |
| EPI_ISL_1250700 | LabPLUS | Institute of Environmental Science and Research (ESR) | Anja Werno; Antje van der Linden; Arlo Upton; Chris Mansell; David Hammer; Dragana Drinkovic; Erasmus Smit; Gary McAuliffe; Hana Sofia Andersson; Hermes Perez; James Ussher; Jill Sherwood; Jing Wang; Joep de Ligt; Josh Freeman; Julia Howard; Juliet Elvy; Lauren Jelly; Mary DeAlmeida; Matt Blakiston; Matt Storey; Matthew Rogers; Max Bloomfield; Michael Addidle; Michelle Balm; Muhammad Faisal; Nikki Freed; Olin Silander; Olivia Stroeven; Rachel Boyle; Sally Roberts; SallyAnn Harbison; Sarah Jefferies; Sharmini Muttaiyah; Susan Morpeth; Susan Taylor; Timothy Blackmore; Vani Sathyendran; Veronica Playle; Virginia Hope; Xiaoyun Ren |
| EPI_ISL_1381222 | Labo Analyses Med | National Reference Center for Viruses of Respiratory Infections, Institut Pasteur, Paris | Angela Brisebarre; Camille Capel; Etienne Simon-Lorière; Louise Lefrançois; Marion Barbet; Maud Vanpeene; Méline Bizard; Rousset Dominique; Sylvie Behillil; Sylvie van der Werf; Vincent Enouf |
| EPI_ISL_1593984 | Labo Carage | Institut Pasteur de la Guyane | Anne Lavergne; Dominique Rousset |
| EPI_ISL_1568558 | Labor Becker & Kollegen (Standort MÃ¼nchen) | Robert Koch Institute | |
| EPI_ISL_1570442 | Labor Dr. Spranger | Robert Koch Institute | |
| EPI_ISL_1384478 | Laboratoires d'analyses medicales - Ketterthill | Laboratoire national de sante, Microbiology, Microbial Genomics Platform | Anke Wienecke-Baldacchino; Caroline Scheiber; Catherine Ragimbeau; Fatu Djabi; Jessica Tapp; Lise Pignon; Raoul Salmon; Serge Vedy; Tamir Abdelrahman |
| EPI_ISL_2196270, EPI_ISL_2196272 | Laboratorio Central de Saude Publica do Estado de Minas Gerais (LACEN/MG) | Laboratory of Respiratory Viruses and Measles, Oswaldo Cruz Institute, FIOCRUZ | Alice Sampaio Rocha; Ana Carolina Mendonca; Andre Felipe Leal Bernardes; Anna Carolina Paixao; Elisa Cavalcante Pereira; Fernando Motta; Luciana Appolinario; Marilda Siqueira on behalf of the Fiocruz COVID-19 Genomic Surveillance Network; Paola Resende; Renata Serrano Lopes; Taina Venas |
| EPI_ISL_2645888, EPI_ISL_2645889, EPI_ISL_2645890 | Laboratorio Central de Saude Publica do Estado do Para (LACEN/PA) | Laboratory of Respiratory Viruses and Measles, Oswaldo Cruz Institute, FIOCRUZ | Alice Sampaio Rocha; Ana Carolina Mendonca; Anna Carolina Paixao; Elisa Cavalcante Pereira; Fernando Motta; Luciana Appolinario; Marilda Siqueira on behalf of the Fiocruz COVID-19 Genomic Surveillance Network; Paola Resende; Renata Serrano Lopes; Taina Venas; Valnete Andrade |
| EPI_ISL_3982719, EPI_ISL_3982722, EPI_ISL_3982728, EPI_ISL_3982731 | Laboratorio Antonello, Pelotas, Rio Grande do Sul | Hemocentro de Ribeirao Preto FMRP USP | Antonio Jorge Martins; Claudia Renata dos Santos Barros; David Schlesinger; Debora Botequio Moretti; Dimas Tadeu Covas; Elaine Cristina Marqueze; Elaine Vieira Santos; Evandra Strazza Rodrigues; Heidge Fukumasu; Jayme Augusto de Souza-Neto; José Salvatore Leister Patané; Luiz Alcantara; Luiz Lehmann Coutinho; Maria Carolina Elias; Maurício Lacerda Nogueira; Rafael dos Santos Bezerra; Raul Machado Neto; Rejane Maria Tommasini Grotto; Ricardo Haddad; Rodrigo Proto de Siqueira; Sandra Coccuzzo Sampaio Vessoni; Simone Kashima; Svetoslav Nanev Slavov; VV Cantarelli; Vincent Louis Viala |
| EPI_ISL_3553588, EPI_ISL_3553614, EPI_ISL_3553615, EPI_ISL_3553634, EPI_ISL_3553654, EPI_ISL_3553655, EPI_ISL_3553665, EPI_ISL_3553666, EPI_ISL_3553678, EPI_ISL_3553684 | | | |
| see above | Laboratorio Central de Saude Publica de Mato Grosso (LACEN-MT) | Fundação Ezequiel Dias | Andre Leal; Cynthia Vazquez; Elaine Cristina; Felipe Iani; Flavia Aburjaile; Gislene Garcia de Castro Lichs; Glauco Carvalho; Hegger Fritsch; Joilson Xavier; Luiz Alcantara.; Luiz Henrique Ferraz Demarchi; Luiz Takao Watanabe; Marina Castilhos Souza Umaki Zardin; Marta Giovanetti; Natalia Guimaraes; Raquel da Silva Ferreira; Talita Adelino; Vagner Fonseca; de Oliveira |
| EPI_ISL_4030368, EPI_ISL_4030369, EPI_ISL_4030370, EPI_ISL_4030371, EPI_ISL_4030372, EPI_ISL_4030373, EPI_ISL_4030374, EPI_ISL_4030375, EPI_ISL_4030376, EPI_ISL_4030378, EPI_ISL_4030380, EPI_ISL_4030381, EPI_ISL_4030382 | | | |
| see above | Laboratorio Central de Saude Publica do Amazonas - LACEN-AM | Laboratorio de Ecologia de Doencas Transmissiveis na Amazonia, Instituto Leonidas e Maria Deane - Fiocruz Amazonia | André Corado; Debora Duarte; Felipe Naveca; Fernanda Nascimento; George Silva; Karina Pessoa; Luciana Gonçalves; Maria Júlia Brandão; Matilde Mejía; Michele Jesus; Valdinete Nascimento; Victor Souza; Ágatha Costa |
| EPI_ISL_2157412, EPI_ISL_2157414, EPI_ISL_2157415 | Laboratorio Central de Saude Publica do Esatado de Alagoas (LACEN/AL) | Laboratory of Respiratory Viruses and Measles, Oswaldo Cruz Institute, FIOCRUZ | Alice Sampaio Rocha; Ana Carolina Mendonca; Anderson Brandao Leite; Anna Carolina Paixao; Elisa Cavalcante Pereira; Fernando Motta; Luciana Appolinario; Marilda Siqueira on behalf of the Fiocruz COVID-19 Genomic Surveillance Network; Paola Resende; Renata Serrano Lopes; Taina Venas |

| | | | |
|---|---|---|---|
| EPI_ISL_2274080, EPI_ISL_2274082, EPI_ISL_2274087, EPI_ISL_2443556 | Laboratorio Central de Saude Publica do Estado Maranhao (LACEN-MA) | Laboratory of Respiratory Viruses and Measles, Oswaldo Cruz Institute, FIOCRUZ | Alice Sampaio Rocha; Ana Carolina Mendonca; Anna Carolina Paixao; Elisa Cavalcante Pereira; Fernando Motta; Lidio Gonçalves Lima Neto; Luciana Appolinario; Marilda Siqueira on behalf of the Fiocruz COVID-19 Genomic Surveillance Network; Paola Resende; Renata Serrano Lopes; Taina Venas |
| EPI_ISL_2157394, EPI_ISL_2157395, EPI_ISL_2157401, EPI_ISL_2274093, EPI_ISL_2536310 | Laboratorio Central de Saude Publica do Estado da Paraiba (LACEN-PB) | Laboratory of Respiratory Viruses and Measles, Oswaldo Cruz Institute, FIOCRUZ | Alice Sampaio Rocha; Ana Carolina Mendonca; Anna Carolina Paixao; Dalane Loudal Florentino Teixeira; Elisa Cavalcante Pereira; Fernando Motta; Irina Riediger; Joao Felipe Bezerra; Luciana Appolinario; Marilda Siqueira on behalf of the Fiocruz COVID-19 Genomic Surveillance Network; Paola Resende; Renata Serrano Lopes; Taina Venas |
| EPI_ISL_2274062 | Laboratorio Central de Saude Publica do Estado de Alagoas (LACEN/AL) | Laboratory of Respiratory Viruses and Measles, Oswaldo Cruz Institute, FIOCRUZ | Alice Sampaio Rocha; Ana Carolina Mendonca; Anderson Brandao Leite; Anna Carolina Paixao; Elisa Cavalcante Pereira; Fernando Motta; Luciana Appolinario; Marilda Siqueira on behalf of the Fiocruz COVID-19 Genomic Surveillance Network; Paola Resende; Renata Serrano Lopes; Taina Venas |
| EPI_ISL_2660471, EPI_ISL_2660472 | Laboratorio Central de Saude Publica do Estado de Minas Gerais (LACEN/MG) | Laboratory of Respiratory Viruses and Measles, Oswaldo Cruz Institute, FIOCRUZ | Alice Sampaio Rocha; Ana Carolina Mendonca; Andre Felipe Leal Bernardes; Anna Carolina Paixao; Elisa Cavalcante Pereira; Fernando Motta; Luciana Appolinario; Marilda Siqueira on behalf of the Fiocruz COVID-19 Genomic Surveillance Network; Paola Resende; Renata Serrano Lopes; Taina Venas |
| EPI_ISL_3190166 | Laboratorio Central de Saude Publica do Estado de Santa Catarina (LACEN/SC) | Laboratory of Respiratory Viruses and Measles, Oswaldo Cruz Institute, FIOCRUZ | Alice Sampaio Rocha; Ana Carolina Mendonca; Anna Carolina Paixao; Darcita Buerger Rovaris; Elisa Cavalcante Pereira; Fernando Motta; Luciana Appolinario; Marilda Siqueira on behalf of the Fiocruz COVID-19 Genomic Surveillance Network; Paola Resende; Renata Serrano Lopes; Sandra Bianchini Fernandes; Taina Venas |
| EPI_ISL_2983164, EPI_ISL_2983165, EPI_ISL_2983167, EPI_ISL_2983168, EPI_ISL_2983169, EPI_ISL_2983170, EPI_ISL_2983171, EPI_ISL_2983172 | | | |
| see above | Laboratorio Central de Saude Publica do Estado do Amapa (LACEN/AP) | Laboratory of Respiratory Viruses and Measles, Oswaldo Cruz Institute, FIOCRUZ | Agatha Cristinne Prudencio; Alice Sampaio Rocha; Ana Carolina Mendonca; Andreia Santos Costa; Anna Carolina Paixao; Anne Caroline da Silva Soledade; Elisa Cavalcante Pereira; Fernando Motta; Ighor Leonardo Arantes Gomes; Lindomar dos Anjos Silva; Luciana Appolinario; Marcia Socorro Pereira Cavalcante; Marilda Siqueira on behalf of the Fiocruz COVID-19 Genomic Surveillance Network; Paola Resende; Renata Serrano Lopes; Taina Venas |
| EPI_ISL_2038968 | Laboratorio Central de Saude Publica do Estado do Espitito Santo (LACEN-ES) | Laboratory of Respiratory Viruses and Measles, Oswaldo Cruz Institute, FIOCRUZ | Alice Sampaio Rocha; Ana Carolina Mendonca; Anna Carolina Paixao; Fernando Motta; Luciana Appolinario; Marilda Siqueira on behalf of the Fiocruz COVID-19 Genomic Surveillance Network; Paola Resende; Renata Serrano Lopes; Rodrigo Ribeiro Rodrigues |
| EPI_ISL_2758943, EPI_ISL_2758944, EPI_ISL_2758945, EPI_ISL_2758947, EPI_ISL_2758948, EPI_ISL_2758949, EPI_ISL_2758950, EPI_ISL_2758951, EPI_ISL_2758952, EPI_ISL_2758953, EPI_ISL_2758954, EPI_ISL_2758957, EPI_ISL_2758958, EPI_ISL_2758959, EPI_ISL_2758961, EPI_ISL_2758962, EPI_ISL_2758964, EPI_ISL_2758968, EPI_ISL_2758970, EPI_ISL_2758971, EPI_ISL_2758972, EPI_ISL_2758975, EPI_ISL_2758979, EPI_ISL_2758981, EPI_ISL_2775390, EPI_ISL_2775391, EPI_ISL_2775393, EPI_ISL_2775394, EPI_ISL_2775395, EPI_ISL_2775399, EPI_ISL_2775400, EPI_ISL_2775403, EPI_ISL_2775404, EPI_ISL_2775410, EPI_ISL_2775412, EPI_ISL_2775413, EPI_ISL_2775415 | | | |
| see above | Laboratorio Central de Saude Publica do Estado do Parana (Instituto de Biologia Molecular do Paraná (LAC)EN-PR) | Instituto Carlos Chagas - Fiocruz | Alessandra De Melo Aguiar; Andreia Akemi Suzukawa; Andréa Rodrigues Ávila; Bruno Dallagiovanna; Dalila Zanette; Eduardo Balsanelli; Emanuel Maltempi de Souza; Fabio Passetti; Fabricio Klerynton Marchini; Fábio de Oliveira Pedrosa; Guilherme Becker; Helisson Faoro; Hellen Geremias dos Santos; Irina Nastassja Riediger; Letusa Albrecht; Lucas Blanes; Luis Gustavo Morello; Lysangela Ronalte Alves; Maria do Carmo Debur; Mauro de Medeiros Oliveira; Michelle Orane Schemberger; Paola Cristina Resende; Sheila Cristina Nardeli; Tiago Gräf; Valter Antônio de Baura |
| EPI_ISL_3061892 | Laboratorio Central de Saude Publica do Estado do Parana (LACEN/PR) | Laboratory of Respiratory Viruses and Measles, Oswaldo Cruz Institute, FIOCRUZ | Alice Sampaio Rocha; Ana Carolina Mendonca; Anna Carolina Paixao; Elisa Cavalcante Pereira; Fernando Motta; Irina Riediger; Luciana Appolinario; Marilda Siqueira on behalf of the Fiocruz COVID-19 Genomic Surveillance Network; Paola Resende; Renata Serrano Lopes; Taina Venas |
| EPI_ISL_2038959, EPI_ISL_2274122, EPI_ISL_2466136, EPI_ISL_2661752, EPI_ISL_2661754, EPI_ISL_2661755, EPI_ISL_2661756, EPI_ISL_2661779, EPI_ISL_2661781 | | | |
| see above | Laboratorio Central de Saude Publica do Estado do Rio Grande do Sul (LACEN-RS) | Laboratory of Respiratory Viruses and Measles, Oswaldo Cruz Institute, FIOCRUZ | Alice Sampaio Rocha; Ana Carolina Mendonca; Anna Carolina Paixao; Elisa Cavalcante Pereira; Fernando Motta; Luciana Appolinario; Marilda Siqueira on behalf of the Fiocruz COVID-19 Genomic Surveillance Network; Paola Resende; Renata Serrano Lopes; Richard Salvato; Taina Venas; Tatiana Schaffer Gregianini |
| EPI_ISL_3048779 | Laboratorio Central de Saude Publica do Estado do Rio Grande do Sul (LACEN-RS) | Laboratório de Biologia Molecular da Universidade Federal de Ciências da Saúde de Porto Alegre | Adriana Seixas; Ana B. G. Veiga; Ana Paula Muterle Varela; Fabiana Quoos Mayer; Fernando Hayashi Sant'Anna; Janira Prichula; Letícia Garay Martins; Richard Steiner Salvato; Tatiana Schäffer Gregianini |
| EPI_ISL_1716397, EPI_ISL_1716398, EPI_ISL_1716400, EPI_ISL_1716401, EPI_ISL_1716402, EPI_ISL_1716403, EPI_ISL_1716404, EPI_ISL_1716405, EPI_ISL_1716406, EPI_ISL_1716407, EPI_ISL_1716408, EPI_ISL_1716409, EPI_ISL_1716410, EPI_ISL_1716411, EPI_ISL_1716412, EPI_ISL_1716413, EPI_ISL_1716414, EPI_ISL_1716416, EPI_ISL_1716417, EPI_ISL_1716418, EPI_ISL_1716419, EPI_ISL_1716420, EPI_ISL_1716421, EPI_ISL_1716422, EPI_ISL_1716423, EPI_ISL_1716424, EPI_ISL_1716425, EPI_ISL_1716427, EPI_ISL_1716428, EPI_ISL_1716429, EPI_ISL_1716430, EPI_ISL_1716431, EPI_ISL_1716432, EPI_ISL_1716433, EPI_ISL_1716434, EPI_ISL_1716435, EPI_ISL_1716436, EPI_ISL_1716437, EPI_ISL_1716438, EPI_ISL_1716439, EPI_ISL_1716440, EPI_ISL_1716441, EPI_ISL_1716442, EPI_ISL_1716443, EPI_ISL_1716444, EPI_ISL_1716445, EPI_ISL_1716446, EPI_ISL_1716447, EPI_ISL_1716448, EPI_ISL_1716449, EPI_ISL_1716450, EPI_ISL_1716451, EPI_ISL_1716452, EPI_ISL_1716453, EPI_ISL_1716454, EPI_ISL_1716455, EPI_ISL_1716456 | | | |
| see above | Laboratorio Saude | LGBio (Laboratorio de Genetica & Biodiversidade) | Amanda Alves de Melo; Cintia Pelegrineti Targueta de Azevedo Brito; Daniela de Melo e Silva; Elisangela de Paula Silveira Lacerda; Francyelli Mello Andrade; Mariana Pires de Campos Telles; Ramilla dos Santos Braga; Renata de Oliveira Dias; Rhewter Nunes; Thais Guimarães Castro; Thays Millena Alves Pedroso |
| EPI_ISL_2427556, EPI_ISL_2427562, EPI_ISL_2427588, EPI_ISL_2427595, EPI_ISL_2427604 | Laboratorio de Biología Molecular Médica Uruguaya | Departments of Pathology and Medicine, New York University School of Medicine | Adriana Heguy; Cecilia Sorhouet; Christian Marier; Dacia Dimartino; Gonzalo Manrique; Maria Cristina Mogdasy; Maria Noel Zubillaga; Maria Victoria Elizondo; Paul Zappile |
| EPI_ISL_2777487 | Laboratorio de Ecologia de Doencas Transmissiveis na Amazonia, Instituto Leonidas e Maria Deane - Fiocruz Amazonia | Laboratorio de Ecologia de Doencas Transmissiveis na Amazonia, Instituto Leonidas e Maria Deane - Fiocruz Amazonia | André Corado; Debora Duarte; Felipe Naveca; Fernanda Nascimento; George Silva; Karina Pessoa; Luciana Gonçalves; Maria Júlia Brandão; Matilde Mejía; Michele Jesus; Valdinete Nascimento; Victor Souza; Ágatha Costa |
| EPI_ISL_2728599, EPI_ISL_2728600 | Laboratorio de Infectologia y Virologia Molecular | Laboratory of Molecular Virology, School of Medicine, Pontificia Universidad Catolica de Chile | Ana Maria Contreras; Andres E. Munoz-Marcos; Carlos Palma; Catalina Pardo-Roa; Constanza Maldonado; Constanza Martinez-Valdevenito; Eileen Serrano; Erick Salinas; Estefany Poblete; Francisco Melo; Jennifer Angulo; Jorge Levican; Leonardo I. Almonacid; M. Belen Leyton; Marcela Ferres; Maria Jose Avendano; Rafael A. Medina; Tamara Garcia-Salum |
| EPI_ISL_3761750, EPI_ISL_3761751 | Laboratorio de Pesquisa em Virologia, FAMERP, SJRP | Laboratorio de Pesquisa em Virologia, FAMERP, SJRP | Beatriz de Carvalho Marques; Cecília Artico Banho; Cíntia Bittar; Fábio Sossai Possebon; Guilherme Campos; Helena Lage Ferreira; Jorge A. Petroli Marchesi; João Pessoa Araújo Jr.; Leila Sabrina Ullmann; Lívia Sacchetto; Maisa C. Pereira Parra; Marília Moraes; Maurício L. Nogueira.; Paula Rahal; Paulo Inacio da Costa |
| EPI_ISL_3401597 | Laboratorio de Referencia Nacional de Virus Respiratorios. Centro Nacional de Salud Publica. Instituto Nacional de Salud Peru. | Laboratorio de Referencia Nacional de Virus Respiratorios. Centro Nacional de Salud Publica. Instituto Nacional de Salud Peru. | Carlos Padilla Rojas; Henri Bailon Calderon; Iris Silva Molina; Joseph Huayra Niquen; Lely Solari Zerpa; Luis Barcena Flores; Marco Galarza Perez; Nancy Rojas Serrano; Nieves Sevilla Castañeda; Omar Caceres Rey; Orson Mestanza Millones; Princesa Medrano Alhuay; Priscila Lope Pari; Sandra Morales Ruiz; Sara Gordillo Vilchez; Steve Acedo Lazo; Veronica Hurtado Vela; Victor Jimenez Vasquez; Wendy Lizarraga Olivares |
| EPI_ISL_3023410, EPI_ISL_3375970, EPI_ISL_3375978, EPI_ISL_3375979, EPI_ISL_3375988, EPI_ISL_3375990, EPI_ISL_3375994, EPI_ISL_3376047, EPI_ISL_3376394 | | | |
| see above | Laboratorio de Referencial Nacional de Virus Respiratorios | Laboratorio de Referencial Nacional de Virus Respiratorios | Carlos Padilla Rojas; Henri Bailon Calderon; Iris Silva Molina; Joseph Huayra Niquen; Lely Solari Zerpa; Luis Barcena Flores; Marco Galarza Perez; Nancy Rojas Serrano; Omar Caceres Rey; Orson Mestanza Millones; Priscila Lope Pari; Sandra Morales Ruiz; Steve Acedo Lazo; Veronica Hurtado Vela |
| EPI_ISL_1786560, EPI_ISL_1786561, EPI_ISL_1786562, EPI_ISL_1786564, EPI_ISL_1786565, EPI_ISL_1786567, EPI_ISL_1786568 | | | |
| see above | Laboratorio de Virologia Clinica do HCFMRP-USP | Hocentro de Ribeirao Preto - Laboratório de Biologia Molecular | Aparecida Yulie Yamamoto; Diego Villa Clé; Dimas Tadeu Covas; Elaine Vieira Santos; Evandra Strazza Rodrigues; Glauco de Carvalho Pereira; Joilson Xavier; Josiane Serrano Borges; Luiz Carlos Junior Alcantara; Mariane Evaristo; Marta Giovanetti; Rafael dos Santos Bezerra; Rodrigo Tocantins Calado; Simone Kashima; Svetoslav Nanev Slavov; Talita Adelino; Vagner Fonseca |
| EPI_ISL_2007528 | Laboratorio de Virología del Hospital de Niños Dr. Ricardo Gutierrez | Área de Secuenciación del Laboratorio de Virología del Hospital de Niños Dr. Ricardo Gutierrez on behalf of 'Proyecto Argentino Interinstitucional de genomica de SARS-CoV-2' (PAIS Consortium) | A; Acevedo; Acuña; Alexay; Alvarez Lopez; Barreda Frank; C; D; E; G; Goya; Grandis; Jacques; LE; Labarta; Lusso; M; ME; MI; Medina; Mistchenko; N; Nabaes Jodar; Natale; O; S; Streitenberger; Thomas; Valinotto; Viegas, M.; Villegas |
| EPI_ISL_1319247, EPI_ISL_1319463, EPI_ISL_1421775, EPI_ISL_1421776, EPI_ISL_1421968, EPI_ISL_1422127 | Laboratory Corporation of America | Centers for Disease Control and Prevention Division of Viral Diseases, Pathogen Discovery | Amanda Douglas; Amanda Suchanek; Andrea Throop; Ayla Burns; Ben L. Rambo-Martin; Bobbi Croy; Brian Krueger; Brian Norvell; Christos Petropoulos; Clinton R. Paden; Craig Lukasik; Dakota Howard; Debbie Boles; Dhwani Batra; Duncan MacCannell; Eyad Almasri; Goran Stevovic; Howard Engler; Hrushikesh Deshmukh; Jake Humphrey; Jana Schroth; Joe Voshell; John Pruitt; Jonathan Meltzer; Jonathan Williams; Kimberly Wagner; Lax Iyer; Lyndon Tilson; Manoj Jain; Marcia Eisenberg; Mary Ann Cristobal; Mary Williamson; Michael Levandoski; Mike Sapeta; Mindy Nye; Minoo Agarwal; Mohan Kolli; Nuthawin Charoensri; Oren Cohen; Peter W. Cook; Prashant Gupta; Qian Zeng; Rama Ghatti; Scott Parker; Scott Ryan; Stanley Letovsky; Steven Ragan; Suresh Babu Selvaraju; Susan Countryman; Susan Hicks; Suxiang Tong; Suzanne Dale; Thomas Urban; Tim Kuphal; Tricia Zwiefelhofer; Vincent Drouillon |
| EPI_ISL_2614081, | Laboratory of Molecular | Laboratory of Respiratory Viruses and | Alice Sampaio Rocha; Amilcar Tanuri; Ana Carolina Mendonca; Anna Carolina Paixao; Elisa Cavalcante Pereira; Fernando Motta; Luciana Appolinario; Marilda Siqueira on behalf of the Fiocruz COVID-19 Genomic Surveillance Network; Paola Resende; Renata Serrano Lopes; Taina Venas |

| | | | |
|---|---|---|---|
| EPI_ISL_2614082, EPI_ISL_2614088, EPI_ISL_2614089 | Virology, Federal University of Rio de Janeito, UFRJ | Measles, Oswaldo Cruz Institute, FIOCRUZ | |
| EPI_ISL_2196185, EPI_ISL_2196186, EPI_ISL_2196187, EPI_ISL_2196190, EPI_ISL_2196191, EPI_ISL_2196194, EPI_ISL_2196195, EPI_ISL_2196196, EPI_ISL_2614300, EPI_ISL_2614301, EPI_ISL_2614302, EPI_ISL_2614303, EPI_ISL_2614304, EPI_ISL_2614305, EPI_ISL_2614306, EPI_ISL_2614307, EPI_ISL_2614308, EPI_ISL_2614309 | | | |
| see above | Laboratory of Respiratory Viruses and Measles, Oswaldo Cruz Institute, FIOCRUZ | Laboratory of Respiratory Viruses and Measles, Oswaldo Cruz Institute, FIOCRUZ | Alice Sampaio Rocha; Ana Carolina Mendonca; Anna Carolina Paixao; Elisa Cavalcante Pereira; Fernando Motta; Luciana Appolinario; Marilda Siqueira on behalf of the Fiocruz COVID-19 Genomic Surveillance Network; Paola Resende; Renata Serrano Lopes; Taina Venas |
| EPI_ISL_2157391, EPI_ISL_2157392, EPI_ISL_2157393, EPI_ISL_2157396, EPI_ISL_2157398, EPI_ISL_2157399, EPI_ISL_2157400, EPI_ISL_2157405, EPI_ISL_2157406, EPI_ISL_2157407, EPI_ISL_2157409, EPI_ISL_2157410, EPI_ISL_2157411, EPI_ISL_2157547 | | | |
| see above | Laboratório Central de Saude Publica do Estado de Santa Catarina (LACEN/SC) | Laboratory of Respiratory Viruses and Measles, Oswaldo Cruz Institute, FIOCRUZ | Alice Sampaio Rocha; Ana Carolina Mendonca; Anna Carolina Paixao; Darcita Buerger Rovaris; Elisa Cavalcante Pereira; Fernando Motta; Luciana Appolinario; Marilda Siqueira on behalf of the Fiocruz COVID-19 Genomic Surveillance Network; Paola Resende; Renata Serrano Lopes; Sandra Bianchini Fernandes; Taina Venas |
| EPI_ISL_2241559, EPI_ISL_2241576, EPI_ISL_2241586 | Laboratório Central de Saúde Pública da Paraiba | Coordenação Geral de Laboratórios de Saúde Pública (CGLAB/DAEVS/SVS/MS) | Vagner Fonseca; et al. |
| EPI_ISL_2298847, EPI_ISL_2298849 | Laboratório Central de Saúde Pública de Roraima | Coordenação Geral de Laboratórios de Saúde Pública (CGLAB/DAEVS/SVS/MS) | Vagner Fonseca; et al. |
| EPI_ISL_2249395 | Laboratório Central de Saúde Pública de Santa Catarina | Coordenação Geral de Laboratórios de Saúde Pública (CGLAB/DAEVS/SVS/MS) | Vagner Fonseca; et al. |
| EPI_ISL_2298828, EPI_ISL_2298830, EPI_ISL_2298832 | Laboratório Central de Saúde Pública do Amapá | Coordenação Geral de Laboratórios de Saúde Pública (CGLAB/DAEVS/SVS/MS) | Vagner Fonseca; et al. |
| EPI_ISL_2777237, EPI_ISL_2777265, EPI_ISL_2777266, EPI_ISL_2777267, EPI_ISL_2777306, EPI_ISL_2777309, EPI_ISL_2777328, EPI_ISL_2777340, EPI_ISL_2777505, EPI_ISL_2777534, EPI_ISL_2777538, EPI_ISL_2777544, EPI_ISL_2777549, EPI_ISL_2777710, EPI_ISL_2777738, EPI_ISL_2777743, EPI_ISL_2777744, EPI_ISL_2777745, EPI_ISL_2777746, EPI_ISL_2777747, EPI_ISL_2777748, EPI_ISL_2777762, EPI_ISL_2777763, EPI_ISL_2777767, EPI_ISL_2777768, EPI_ISL_2777769, EPI_ISL_2777771 | | | |
| see above | Laboratório Central de Saúde Pública do Amazonas - LACEN-AM | Laboratorio de Ecologia de Doencas Transmissiveis na Amazonia, Instituto Leonidas e Maria Deane - Fiocruz Amazonia | André Corado; Debora Duarte; Felipe Naveca; Fernanda Nascimento; George Silva; Karina Pessoa; Luciana Gonçalves; Maria Júlia Brandão; Matilde Mejía; Michele Jesus; Valdinete Nascimento; Victor Souza; Ágatha Costa |
| EPI_ISL_4945103, EPI_ISL_4945104, EPI_ISL_4945108 | Laboratório Central de Saúde Pública do Distrito Federal – LACEN-DF | Laboratory of Baculovirus, University of Brasília | Agenor de Castro Moreira dos Santos Junior; Alessandra Pinheiro Medeiros; Aline Belmok; Anamélia Lorenzetti Bocca; Bergmann Morais Ribeiro; Brenno Vinicius Henrique; Fabiano José Queiroz Costa; Fernando Melo; Jordan Barros Silva; Lucas Luiz Vieira; Renato de Oliveira Resende |
| EPI_ISL_2298842, EPI_ISL_2298844, EPI_ISL_2298845 | Laboratório Central de Saúde Pública do Maranhão | Coordenação Geral de Laboratórios de Saúde Pública (CGLAB/DAEVS/SVS/MS) | Vagner Fonseca; et al. |
| EPI_ISL_4600498, EPI_ISL_4600501 | Laboratório Central de Saúde Pública do Paraná | Coordenação Geral de Laboratórios de Saúde Pública (CGLAB/DAEVS/SVS/MS) | Vagner Fonseca; et al. |
| EPI_ISL_2298810, EPI_ISL_2298812, EPI_ISL_2298814 | Laboratório Central de Saúde Pública do Pará | Coordenação Geral de Laboratórios de Saúde Pública (CGLAB/DAEVS/SVS/MS) | Vagner Fonseca; et al. |
| EPI_ISL_2249443 | Laboratório Central de Saúde Pública do Rio de Janeiro | Coordenação Geral de Laboratórios de Saúde Pública (CGLAB/DAEVS/SVS/MS) | Vagner Fonseca; et al. |
| EPI_ISL_1592227, EPI_ISL_1592228, EPI_ISL_1592229 | Laboratório Hermes Pardini | Laboratório de Biologia Integrativa, Instituto de Ciências Biológicas, Universidade Federal de Minas Gerais | Alessandro Clayton de Souza Ferreira; Aline Brito de Lima; Carolina Moreira Voloch; Daniel Costa Queiroz; Danielle Alves Gomes Zauli; Diego Menezes Bonfim; Filipe Romero Rebello Moreira; Frederico Scott Varella Malta; Joice do Prado Silva; Rafael Marques de Souza; Renan Pedra de Souza; Renato Santana Aguiar.; Rennan Garcias Moreira; Victor Cavalcanti Pardini; Victor Emmanuel Viana Geddes; Wagner Carlos Santos Magalhaes; Walyson Coelho Costa |
| EPI_ISL_3031298, EPI_ISL_3031309 | Laboratório Municipal de Biologia Molecular | Instituto René Rachou / Fiocruz Minas | André Menezes; Anna Salim; Eneida Oliveira; Gabriel Fernandes; Pedro Alves; Rubens do Monte Neto; Thaís Silva |
| EPI_ISL_1495010, EPI_ISL_1495015, EPI_ISL_1495016, EPI_ISL_1495019, EPI_ISL_1495023, EPI_ISL_1495031 | Laboratório de Biologia Integrativa | Laboratório de Biologia Integrativa | Alessandro Clayton de Souza Ferreira; Aline Brito de Lima; Carolina Moreira Voloch; Daniel Costa Queiroz; Danielle Alves Gomes Zauli; Diego Menezes Bonfim; Filipe Romero Rebello Moreira; Frederico Scott Varella Malta; Joice do Prado Silva; Lucyene Miguita Luiz; Nuno Rodrigues Faria; Paula Luize Camargos Fonseca; Rafael Marques de Souza; Renan Pedra de Souza; Renato Santana Aguiar; Rennan Garcias Moreira; Victor Cavalcanti Pardini; Victor Emmanuel Viana Geddes |
| EPI_ISL_6508504, EPI_ISL_6508569, EPI_ISL_6514132, EPI_ISL_6514157, EPI_ISL_6514265, EPI_ISL_6514276 | Laboratório de Biologia Integrativa/ UFMG | Laboratório de Biologia Integrativa/ UFMG | Adriana Aparecida Ribeiro; Alana Vitor Barbosa Costa; Alessandro Luís Gonçalves; Aline de Brito Lima; Ana Paula De Battisti Ribeiro; Ana Paula Salles Moura Fernandes; Andre Luiz Menezes; Bruna Walker Ferreira; Carolina Senra Alves de Souza; Cristiane P. T. Brito Mendonça; Daniel Costa Queiroz; Danielle Alves Gomes Zauli; Diego Menezes; Eneida Santos de Oliveira; Eva Lídia Arcoverde Medeiros; Felipe Campos de Melo Iani; Fernanda Gil de Souza; Fernanda Santos Mendes; Filipe Romero Rebello Moreira; Flávio Guimarães da Fonseca; Frederico Scott Varella Malta; Hugo Itaru Sato; Hugo José Alves; Igor Pereira Godinho; Jaqueline Silva de Oliveira; Joice do Prado Silva; José Nélio Januario; Juliana Wilke Saliba; Karine Lima Lourenço; Lucyene Miguita; Luige Biciati Alvim; Nara Oliveira Carvalho; Natiely Pereira Silva; Natália Rocha Guimarães; Paula Luize Camargos Fonseca; Pedro Henrique Barbosa de Paula Mendes; Rafael Marques de Souza; Renan Pedra de Souza; Renata Barbosa Peixoto Peixoto; Renato Santana de Aguiar; Rennan Garcias Moreira; Rillery Calixto Dias; Rubens Daniel Miserani Magalhães; Santuza Maria Ribeiro Teixeira; Talita Emile Ribeiro Adelino; Victor Emmanuel Viana Geddes; Walyson Coelho Costa |
| EPI_ISL_2466208, EPI_ISL_2466209, EPI_ISL_2466210, EPI_ISL_2466211, EPI_ISL_2466212, EPI_ISL_2466213, EPI_ISL_2466214, EPI_ISL_2466215, EPI_ISL_2466216 | | | |
| see above | Laboratório de Biologia Molecular de Doenças Infecciosas e do Câncer (LADIC - UFRN) | Laboratory of Respiratory Viruses and Measles, Oswaldo Cruz Institute, FIOCRUZ | Alice Sampaio Rocha; Ana Carolina Mendonca; Anna Carolina Paixao; Elisa Cavalcante Pereira; Fernando Motta; Josélio Araújo; Luciana Appolinario; Marilda Siqueira on behalf of the Fiocruz COVID-19 Genomic Surveillance Network; Paola Resende; Renata Serrano Lopes; Taina Venas |
| EPI_ISL_2431439, EPI_ISL_2431853, EPI_ISL_4051951 | Laboratório de Microbiologia Molecular - Universidade FEEVALE | Molecular Microbiology Laboratory | Alana Witt Hansen; Fernando Rosado Spilki; Flávio Silveira; Fágner Henrique Heldt; Juliana Schons Gularte; Juliane Deise Fleck; Mariana Soares da Silva; Matheus Nunes Weber; Meriane Demoliner; Micheli Filippi; Micheli Filippi.; Paula Rodrigues de Almeida; Vyctoria Malayhka de Abreu Góes Pereira. |
| EPI_ISL_1464630, EPI_ISL_1464631, EPI_ISL_1464654, EPI_ISL_1464655, EPI_ISL_1464656, EPI_ISL_1464657, EPI_ISL_1464658, EPI_ISL_1464659, EPI_ISL_1464660, EPI_ISL_1464661, EPI_ISL_1464662, EPI_ISL_1464663, EPI_ISL_1464664, EPI_ISL_1464665, EPI_ISL_1464666, EPI_ISL_1464667, EPI_ISL_1464668, EPI_ISL_1464669, EPI_ISL_1464670, EPI_ISL_1464671 | | | |
| see above | Laboratório de Virologia - UNIFESP | Laboratory of Respiratory Viruses and Measles, Oswaldo Cruz Institute, FIOCRUZ | Alice Sampaio Rocha; Ana Carolina Mendonca; Anna Carolina Paixao; Fernando Motta; Luciana Appolinario; Marilda Siqueira on behalf of the Fiocruz COVID-19 Genomic Surveillance Network; Nancy Belei; Paola Resende; Renata Serrano Lopes |
| EPI_ISL_2629734, EPI_ISL_2629736, EPI_ISL_2629738, EPI_ISL_2629739, EPI_ISL_2629740, EPI_ISL_2629741, EPI_ISL_2629742 | | | |
| see above | Laboratório de Virologia Molecular - Universidade Federal do Rio de Janeiro | Laboratório de Virologia Molecular - Universidade Federal do Rio de Janeiro | ; Alice Laschuk Herlinger; Amilcar Tanuri; André Felipe Andrade dos Santos; Carolina Moreira Voloch; Cássia Cristina Alves Gonçalves; Diana Mariani; Débora Souza Faffe; Filipe Romero Rebello Moreira; Francine Bittencourt Schiffler; Isabela de Carvalho Leitão; Marcelo Calado de Paula Tôrres; Matheus Augusto Calvano Cosentino; Mirela D'arc; Orlando da Costa Ferreira Junior; Rafael Mello Galliez; Raíssa Mirella dos Santos Cunha da Costa; Renato Santana de Aguiar; Terezinha Marta Pereira Pinto Castineiras; Thamiris dos Santos Miranda; Átila Duque Rossi |
| EPI_ISL_2196254, EPI_ISL_2196255, EPI_ISL_2196256, EPI_ISL_2536265, EPI_ISL_2536266, EPI_ISL_2677139, EPI_ISL_2677141, EPI_ISL_2677148 | | | |
| see above | Labortorio Central de Saude Publica do Estado de Santa Catarina (LACEN/SC) | Laboratory of Respiratory Viruses and Measles, Oswaldo Cruz Institute, FIOCRUZ | Alice Sampaio Rocha; Ana Carolina Mendonca; Anna Carolina Paixao; Darcita Buerger Rovaris; Elisa Cavalcante Pereira; Fernando Motta; Luciana Appolinario; Marilda Siqueira on behalf of the Fiocruz COVID-19 Genomic Surveillance Network; Paola Resende; Renata Serrano Lopes; Sandra Bianchini Fernandes; Taina Venas |
| EPI_ISL_2614359, EPI_ISL_2614360 | Labortorio Central de Saude Publica do Estado do Rio de Janeiro (LACEN/RJ) | Laboratory of Respiratory Viruses and Measles, Oswaldo Cruz Institute, FIOCRUZ | Alice Sampaio Rocha; Ana Carolina Mendonca; Andrea Cony Cavalcanti; Anna Carolina Paixao; Elisa Cavalcante Pereira; Fernando Motta; Luciana Appolinario; Marilda Siqueira on behalf of the Fiocruz COVID-19 Genomic Surveillance Network; Paola Resende; Renata Serrano Lopes; Taina Venas |
| EPI_ISL_2157397, EPI_ISL_2157402, EPI_ISL_2157403, EPI_ISL_2157404, EPI_ISL_2157408, EPI_ISL_2157413, EPI_ISL_2157416, EPI_ISL_2157417, EPI_ISL_2274094 | | | |
| see above | Lboratorio Central de Saude Publica do Estado do Parana (LACEN/PR) | Laboratory of Respiratory Viruses and Measles, Oswaldo Cruz Institute, FIOCRUZ | Alice Sampaio Rocha; Ana Carolina Mendonca; Anna Carolina Paixao; Elisa Cavalcante Pereira; Fernando Motta; Irina Riediger; Luciana Appolinario; Marilda Siqueira on behalf of the Fiocruz COVID-19 Genomic Surveillance Network; Paola Resende; Renata Serrano Lopes; Taina Venas |
| EPI_ISL_1329280, EPI_ISL_1333304 | Lighthouse Lab in Cambridge | Wellcome Sanger Institute for the COVID-19 Genomics UK (COG-UK) Consortium | Cordelia Langford; David K. Jackson; Dominic Kwiatkowski; Ewan Harrison; Ian Johnston; Jeffrey Barrett; John Sillitoe on behalf of the Wellcome Sanger Institute COVID-19 Surveillance Team; Rob Howes; Roberto Amato; Sonia Goncalves; The Lighthouse Lab in Cambridge and Alex Alderton |

| | | | |
|---|---|---|---|
| EPI_ISL_1294650 | Lighthouse Lab in Glasgow | Wellcome Sanger Institute for the COVID-19 Genomics UK (COG-UK) Consortium | Anna Dominiczak and Alex Alderton; Carol Clugston; Cordelia Langford; David Gray; David K. Jackson; Dominic Kwiatkowski; Ewan Harrison; Harper VanSteenhouse; Ian Johnston; Jeffrey Barrett; John Sillitoe on behalf of the Wellcome Sanger Institute COVID-19 Surveillance Team; Roberto Amato; Sonia Goncalves; Yumi Kasai |
| EPI_ISL_1350466, EPI_ISL_1350502 | Limbach - MVZ Humangenetik Ulm | Robert Koch Institute | |
| EPI_ISL_1502027 | Lurie Children's Hospital of Chicago | Northwestern University - Ozer Lab | Egon A. Ozer; Judd F. Hultquist; Lacy M. Simons; Larry K. Kociolek; Michael G. Ison; Ramon Lorenzo-Redondo; Taylor J. Dean; William J. Muller; Xiaotian; Zheng |
| EPI_ISL_5825553 | MARCOS AURELIO SABOIA LEITAO | Analytical Competence Molecular Epidemiology Lab/ACME, Oswaldo Cruz Foundation, Ceara (FIOCRUZ CE) | Carlos Leonardo de Aragao Araujo; Cecilia Leite Costa & Eduardo Ruback dos Santos on behalf of COVID-19 FIOCRUZ Genomic Network; Cleber Furtado Aksenen; Fabio Miyajima; Fernando Braga Stehling; Francisco Eder de Moura Lopes; Igor Oliveira Duarte; Jamille Maria Mendes Bezerra; Joaquim Cesar do Nascimento Sousa Junior; Pedro Miguel Carneiro Jeronimo; Suzana Porto Almeida; Thais Ferreira de Oliveira; Thais de Oliveira Costa; Ticiane Cavalcante de Souza; Veridiana Pessoa Miyajima |
| EPI_ISL_1300819 | MSHS Clinical Microbiology Laboratories | MSHS Pathogen Surveillance Program | Adolfo García-Sastre; Adriana van de Guchte; Ajay Obla; Alberto Paniz-Mondolfi; Ana S. Gonzalez-Reiche; Angela Amoako; Ashley Salimbangon; Betsaida Salom Melo; Bremy Alburquerque; Brianne Ciferri; Charles Gleason; Daniel Floda; Deena R. Altman; Denise Jurczyszak; Emilia Mia Sordillo; Gintaras Deikus; Giulio Kleiner; Gopi Patel; Hala Alshammary; Harm van Bakel; Irina Oussenko; Jayeeta Dutta; Juan Soto; Julia Matthews; Katherine Beach; Kathryn Twyman; Kayla Russo; Komal Srivastava; Levy Sominsky; Mahmoud Awawda; Marta Luksza; Matthew M. Hernandez; Melissa Gitman; Michael D. Nowak; Mitchell J. Sullivan; Nancy Francoeur; Robert Sebra; Sarah Schaefer; Shelcie Fabre; Shwetha Hara Sridhar; Viviana Simon; Ying-Chih Wang; Zenab Khan |
| EPI_ISL_1372646 | Maine Health and Environmental Testing Laboratory | Tewhey Lab, The Jackson Laboratory | Barter, M.; Dewey, H.; H. and Tewhey, R.; Iosue, F.; Lynch, R.; Matluk, N.; Munger |
| EPI_ISL_1255064 | Marche en Famenne | Plateforme de testing Namuroise | ; Degosserie Jonathan; Denis Olivier; Mullier François; Otto Gaetan |
| EPI_ISL_1503468 | Massachusetts State Public Health Laboratory | Massachusetts State Public Health Laboratory | Andrew Lang; Glen Gallagher; Sandra Smole; Timelia Fink |
| EPI_ISL_1752110 | Max von Pettenkofer Institute, Virology, National Reference Center for Retroviruses, LMU Munich | Laboratory for Functional Genome Analysis; Dept. Genomics; Gene Center of the LMU Munich | Alexander Graf; Helmut Blum; Max Muenchhoff; Oliver Keppler; Stefan Krebs |
| EPI_ISL_1340191, EPI_ISL_1340222 | NJDOH, Public Health and Environmental Laboratories | New Jersey Public Health and Environmental Laboratories (PHEL) | Byeong Jeong; Dana Woell; Lindsey Bodnar; Shiv K. Verma |
| EPI_ISL_3046157 | NUPIT/UFPE | WallauLab on behalf of Fiocruz COVID-19 Genomic Surveillance Network | Alexandre Freitas da Silva; Cassia Docena; Constância Flávia Junqueira Ayres; Filipe Zimmer Dezordi; Gabriel Luz Wallau; Gustavo Barbosa de Lima; Lais Ceschini Machado; Lilian Caroliny Amorim Silva; Maira Galdino da Rocha Pitta; Marcelo Henrique dos Santos Paiva; Matheus Filgueira Bezerra; Michelly Cristiny Pereira; Rômulo Pessoa e Silva; Sinval Pinto Brandão Filho |
| EPI_ISL_1184826 | National Institute of Infectious Diseases-Prof. Dr. Matei Bals Molecular Diagnostics Laboratory | National Institute of Infectious Diseases-Prof. Dr. Matei Bals Molecular Diagnostics Laboratory | Andreea Tudor; Corina Casangiu; Dan Otelea; Leontina Banica; Marius Surleac; Petre Milu; Simona Paraschiv |
| EPI_ISL_1301964, EPI_ISL_1357775, EPI_ISL_1357776, EPI_ISL_1358253 | National Virus Reference Laboratory | National Virus Reference Laboratory | Calum Walsh; Charlene Bennet; Charlene Bennett; Cillian F De Gascun; Fiona Crispie; Gabriel Gonzalez; Jonathan Dean; Michael Carr; Paul Cotter; Zoe Yandle |
| EPI_ISL_1502062 | Northwestern Memorial Hospital | Northwestern University - Ozer Lab | Chad J. Achenbach; Chao Qi; Egon A. Ozer; Judd F. Hultquist; Lacy M. Simons; Lawrence J. Jennings; Michael G. Ison; Ramon Lorenzo-Redondo; Taylor J. Dean |
| EPI_ISL_2246699 | Northwestern Memorial Hospital | RIPHL at Rush University Medical Center | Amber Kimble; Chao Qi; Felix Araujo Perez; Kevin Kunstman; Laura Furtado; Marieta Hyde; Max Kolton; Stefan Green |
| EPI_ISL_1229141 | Ospedale "F. Spaziani" Frosinone | INMI Lazzaro Spallanzani IRCCS | A Di Caro; B Bartolini; C Gargiulo; C Sias; CEM Gruber; E Giombini; F Messina; M Rueca; MR Capobianchi; O Butera; R Pulselli |
| EPI_ISL_1929417 | Pathogen Genomics Center, National Institute of Infectious Diseases | Pathogen Genomics Center, National Institute of Infectious Diseases | Hidemasa Izumiya; Ken Shimuta; Kentaro Itokawa; Makoto Kuroda; Masanori Hashino; Nobuo Koizumi; Rina Tanaka; Sunao Iyoda; Tsuyoshi Sekizuka |
| EPI_ISL_2663257, EPI_ISL_2663262, EPI_ISL_2663273, EPI_ISL_2663274, EPI_ISL_2663275, EPI_ISL_2663276, EPI_ISL_2663277, EPI_ISL_2663278, EPI_ISL_2663282, EPI_ISL_2663283, EPI_ISL_2663285, EPI_ISL_2663287, EPI_ISL_2663288, EPI_ISL_2663289, EPI_ISL_2663290, EPI_ISL_2663291, EPI_ISL_2663293, EPI_ISL_2663294 | | | |
| see above | Plataforma de Vigilancia Molecular (PVM) - FIOCRUZ/BA | Plataforma de Vigilancia Molecular (PVM) - FIOCRUZ/BA | Bruno Bezerril Andrade; Camila I. de Oliveira on behalf of the Fiocruz COVID-19 Genomic Surveillance Network.; Clarissa Araújo Gurgel; Leonardo Paiva Farias; Marina Cucco; Ricardo Khouri; Tiago Graf |
| EPI_ISL_1220089, EPI_ISL_1220091, EPI_ISL_1220092 | Plateforme de testing Namuroise | Plateforme de testing Namuroise | ; Degosserie Jonathan; Denis Olivier; Mullier François; Otto Gaetan |
| EPI_ISL_1192385, EPI_ISL_1192387 | Plateforme de testing namuroise | Plateforme de testing Namuroise | Céline Maschietto; Degosserie Jonathan; Denis Olivier; Mullier François; Otto Gaetan |
| EPI_ISL_7406395 | Programa Laboratorio de Salud Pública "Dr Dalmiro Pérez Laborda" | Área de Secuenciación del Laboratorio de Virología del Hospital de Niños Dr. Ricardo Gutierrez on behalf of 'Proyecto Argentino Interinstitucional de genomica de SARS-CoV-2' (PAIS Consortium) | A; AM; Acuña; Aguilera; BH; Bohn; Cabral Bombardieri; Campos; Carrizo; D; E; Esteves; F; G; Goya; J; JM; JP; Jofré; L; LE; Lacaze; Lusso; M; MA; MI; MS; Mastrodonato; Molina; Nabaes Jodar; Natale; Olivera; Perez Diaz; Peñalva; Quijano; Rivero; Rosales; S; Talia; Valinotto; Viegas, M. |
| EPI_ISL_1608118, EPI_ISL_1608120, EPI_ISL_1608121, EPI_ISL_1608122, EPI_ISL_2375878, EPI_ISL_2375879 | Programa de Oncovirologia, Instituto Nacional de Câncer | Programa de Oncovirologia, Instituto Nacional de Câncer | Ana Cristina P. M. Pereira; Brunna M. Alves; Claudia Cicala; James Arthos; João P.B. Viola; Juliana D. Siqueira; Livia R. Goes; Marcelo A. Soares; Marianne M. Garrido |
| EPI_ISL_1533714 | Pronto Socorro Dr Osmar Mesquita | Instituto Adolfo Lutz, Interdiciplinary Procedures Center, Strategic Laboratory | Caio Vinicius Dias Lopes; Claudia Regina Gonçalves; Claudio Tavares Sacchi; Erica Valessa Ramos Gomes; Karoline Rodrigues Campos; Leonardo Jose Tadeu de Araujo |
| EPI_ISL_1374280 | Randox Laboratories | Wellcome Sanger Institute for the COVID-19 Genomics UK (COG-UK) Consortium | Cordelia Langford; David K. Jackson; Dominic Kwiatkowski; Ewan Harrison; Ian Johnston; Jeffrey Barrett; John Sillitoe on behalf of the Wellcome Sanger Institute COVID-19 Surveillance Team; Randox Laboratories and Alex Alderton; Roberto Amato; Sonia Goncalves |
| EPI_ISL_1580755, EPI_ISL_1580756, EPI_ISL_1580758, EPI_ISL_1580761, EPI_ISL_1580767, EPI_ISL_1580768, EPI_ISL_1580773, EPI_ISL_1580775 | | | |
| see above | Reditus Laboratories | Reditus Laboratories | Alexa Eichelberger; Cassy Philips; Joshua J. Geltz; M.S.; Ph.D.; Robert M. Sgambelluri |
| EPI_ISL_2739913 | Respiratory Virus Unit, Microbiology Services Colindale, Public Health England | COVID-19 Genomics UK (COG-UK) Consortium | PHE Covid Sequencing Team |
| EPI_ISL_2458108, EPI_ISL_2458109 | Retrovirus Laboratory Adolfo Lutz Institute_Sao Paulo | Dr Arnaldo 355 Sao Paulo City, State of Sao Paulo CEP 01246-902, Brazil | Audrey Cilli; Cintia Ahagon; Gabriela Bastos Cabral; Giselle I S Lopez-Lopes; Igor Mohamed Hussein; Luis Brigido; Paula Morena Guimaraes |
| EPI_ISL_1407415 | Rhode Island Department of Health | Infectious Disease Program, Broad Institute of Harvard and MIT | Adams, G.; Azevedo, K.; B.L.; B.W.; Bauer, M.; Birren; Carter, A.; Chaluvadi, S.; D.J.; DeRuff, K.; Gladden-Young, A.; Huard, R.; J.E.; K.J.; King, E.; Lagerborg, K.; Lemieux; Loreth, C.; Miller, A.; Normandin, E.; P.C.; Park; Pearlman, L.; Reilly, S.; Rudy, M.; Sabeti; Siddle; Tomkins-Tinch, C.; and MacInnis |
| EPI_ISL_2534064, EPI_ISL_2534065, EPI_ISL_2534066 | S.C. Patologia Clinica, Ospedale Sant'Andrea, ASL 5 | U.O. Igiene, Ospedale Policlinico San Martino | Battolla Enrico; Bruzzone Bianca; Caligiuri Patrizia; De Pace Vanessa; Domnich Alexander; Icardi Giancarlo; Orsi Andrea; Ricucci Valentina |
| EPI_ISL_3102253 | SAO CARLOS DIAGNOSTICO POR IMAGEM | Analytical Competence Molecular Epidemiology Lab/ACME, Oswaldo Cruz Foundation, Ceara (FIOCRUZ CE) | Cleber Furtado Aksenen; Fabio Miyajima; Fernando Braga Stehling; Francisco Eder de Moura Lopes; Jamille Maria Mendes Bezerra; Joaquim César do Nascimento Sousa Junior; Pedro Miguel Carneiro Jeronimo; Suzana Porto Almeida e Lucas Delerino; Thais Ferreira de Oliveira; Thais de Oliveira Costa; Ticiane Cavalcante de Souza; Veridiana Pessoa Miyajima |
| EPI_ISL_2346069, EPI_ISL_2346072 | SERRANA | Instituto Butantan / Mendelics | Antonio Jorge Martins; Claudia Renata dos Santos Barros; David Schlesinger; Debora Botequio Moretti; Dimas Tadeu Covas; Elaine Cristina Marqueze; Elaine Vieira Santos; Evandra Strazza Rodrigues; Heidge Fukumasu; Jayme Augusto de Souza-Neto; José Salvatore Leister Patané; Luiz Alcantara; Luiz Lehmann Coutinho; Maria Carolina Elias; Maurício Lacerda Nogueira; Rafael dos Santos Bezerra; Raul Machado Neto; Rejane Maria Tommasini Grotto; Ricardo Haddad; Sandra Coccuzzo Sampaio Vessoni; Simone Kashima; Svetoslav Nanev Slavov; Vincent Louis Viala |
| EPI_ISL_1251115 | SIESP L'AQUILA | Istituto Zooprofilattico Sperimentale dell'Abruzzo e Molise "G. Caporale" | Ancora M; Calistri P; Cammà C; Curini V; Di Domenico M; Di Pasquale A; Lorusso A; Mangone I; Marcacci M; Puglia I; Rinaldi A; Savini G; Scialabba S |
| EPI_ISL_1241716, | SYNLAB | GIGA Medical Genomics | Bouchra Boujemla; Cécile Meex; Keith Durkin; Maria Artesi; Marie-Pierre Hayette; Nathalie Renotte; Pierrette Melin; Raphaël Boreux; Sébastien Bontems; Vincent Bours |

| | | | |
|---|---|---|---|
| EPI_ISL_1241789, EPI_ISL_1241802 | | | |
| EPI_ISL_1260858 | SYNLAB | Instituto Nacional de Saude (INSA) | Borges et al |
| EPI_ISL_1348076 | SYNLAB MVZ Leinfelden-Echterdingen | Robert Koch Institute | |
| EPI_ISL_1215159, EPI_ISL_1281024, EPI_ISL_1281074 | SYNLAB MVZ Weiden | Robert Koch Institute | |
| EPI_ISL_1430698 | Saitama Prefectural Institute of Public Health | Pathogen Genomics Center, National Institute of Infectious Diseases | Chang-Kweng Lim; Eri Nakayama; Kentaro Itokawa; Makoto Kuroda; Masanori Hashino; Motohiko Ogawa; Rina Tanaka; Shigeru Tanjima; Takahiro Maeki; Tsuyoshi Sekizuka |
| EPI_ISL_1969267, EPI_ISL_1969269 | San Diego County Public Health Laboratory | Andersen lab at Scripps Research | Brett Austin; Jovan Shephard; SEARCH Alliance San Diego with Tracy Basler |
| EPI_ISL_1628374 | Santa Casa De Cravinhos | Instituto Adolfo Lutz, Interdiciplinary Procedures Center, Strategic Laboratory | Caio Vinicius Dias Lopes; Claudia Regina Gonçalves; Claudio Tavares Sacchi; Erica Valessa Ramos Gomes; Karoline Rodrigues Campos; Katia Correa de Oliveira Santos; Leonardo Jose Tadeu de Araujo |
| EPI_ISL_1533720 | Santa Casa de Penapolis | Instituto Adolfo Lutz, Interdiciplinary Procedures Center, Strategic Laboratory | Caio Vinicius Dias Lopes; Claudia Regina Gonçalves; Claudio Tavares Sacchi; Erica Valessa Ramos Gomes; Karoline Rodrigues Campos; Leonardo Jose Tadeu de Araujo |
| EPI_ISL_2011571 | Seattle Flu Study | Seattle Flu Study | Amanda Adler; Barry R. Lutz; Benjamin Pelle; Caitlin R. Wolf; Chris D. Frazar; Deborah A. Nickerson; Elisabeth Brandstetter; Erica Ryke; Helen Y. Chu; Janet A. Englund; Jay Shendure; Jover Lee; Kairsten Fay; Kirsten Lacombe; Lea M. Starita; Mark J. Rieder; Matthew Richardson; Matthew Thompson; Melissa Truong; Michael Boeckh; Michael Famulare; Misja Ilcisin; Peter D. Han; Thomas R. Sibley; Trevor Bedford |
| EPI_ISL_5316566 | Shared Hospital Laboratory | Shared Hospital Laboratory | Christie Vermeiren; Finlay Maguire; Kevin Katz; Patryk Aftanas; Robert Kozak; Samira Mubareka |
| EPI_ISL_1254643, EPI_ISL_1254644 | Sonora Quest Laboratories | TGen North | "Jolene Bowers; Ashlyn Pfeiffer; Chris French; Darrin Lemmer; Dave Engelthaler; Hayley Yaglom; Heather Centner; The Arizona COVID Genomics Union (ACGU)" |
| EPI_ISL_1604430, EPI_ISL_1808214, EPI_ISL_1808549, EPI_ISL_1808983, EPI_ISL_1809023 | Swedish national genomic surveillance program of SARS-CoV-2 | The Public Health Agency of Sweden | Alma Brolund; Maria Lind Karlberg; Maximilian Riess; Swedish national genomic surveillance program of SARS-CoV-2 |
| EPI_ISL_1215088, EPI_ISL_1215122, EPI_ISL_1215136 | Synlab MVZ Augsburg | Robert Koch Institute | |
| EPI_ISL_5529993 | UAPS AIDA SANTOS | Analytical Competence Molecular Epidemiology Lab/ACME, Oswaldo Cruz Foundation, Ceara (FIOCRUZ CE) | Carlos Leonardo de Aragao Araujo; Cecilia Leite Costa & Eduardo Ruback dos Santos on behalf of COVID-19 FIOCRUZ Genomic Network; Cleber Furtado Aksenen; Fabio Miyajima; Fernando Braga Stehling; Francisco Eder de Moura Lopes; Igor Oliveira Duarte; Jamille Maria Mendes Bezerra; Joaquim Cesar do Nascimento Sousa Junior; Pedro Miguel Carneiro Jeronimo; Suzana Porto Almeida; Thais Ferreira de Oliveira; Thais de Oliveira Costa; Ticiane Cavalcante de Souza; Veridiana Pessoa Miyajima |
| EPI_ISL_5603293 | UAPS ALARICO LEITE | Analytical Competence Molecular Epidemiology Lab/ACME, Oswaldo Cruz Foundation, Ceara (FIOCRUZ CE) | Carlos Leonardo de Aragao Araujo; Cecilia Leite Costa & Eduardo Ruback dos Santos on behalf of COVID-19 FIOCRUZ Genomic Network; Cleber Furtado Aksenen; Fabio Miyajima; Fernando Braga Stehling; Francisco Eder de Moura Lopes; Igor Oliveira Duarte; Jamille Maria Mendes Bezerra; Joaquim Cesar do Nascimento Sousa Junior; Pedro Miguel Carneiro Jeronimo; Suzana Porto Almeida; Thais Ferreira de Oliveira; Thais de Oliveira Costa; Ticiane Cavalcante de Souza; Veridiana Pessoa Miyajima |
| EPI_ISL_5530132 | UAPS ARGEU HERBSTER | Analytical Competence Molecular Epidemiology Lab/ACME, Oswaldo Cruz Foundation, Ceara (FIOCRUZ CE) | Carlos Leonardo de Aragao Araujo; Cecilia Leite Costa & Eduardo Ruback dos Santos on behalf of COVID-19 FIOCRUZ Genomic Network; Cleber Furtado Aksenen; Fabio Miyajima; Fernando Braga Stehling; Francisco Eder de Moura Lopes; Igor Oliveira Duarte; Jamille Maria Mendes Bezerra; Joaquim Cesar do Nascimento Sousa Junior; Pedro Miguel Carneiro Jeronimo; Suzana Porto Almeida; Thais Ferreira de Oliveira; Thais de Oliveira Costa; Ticiane Cavalcante de Souza; Veridiana Pessoa Miyajima |
| EPI_ISL_5529991 | UAPS EDMAR FUJITA | Analytical Competence Molecular Epidemiology Lab/ACME, Oswaldo Cruz Foundation, Ceara (FIOCRUZ CE) | Carlos Leonardo de Aragao Araujo; Cecilia Leite Costa & Eduardo Ruback dos Santos on behalf of COVID-19 FIOCRUZ Genomic Network; Cleber Furtado Aksenen; Fabio Miyajima; Fernando Braga Stehling; Francisco Eder de Moura Lopes; Igor Oliveira Duarte; Jamille Maria Mendes Bezerra; Joaquim Cesar do Nascimento Sousa Junior; Pedro Miguel Carneiro Jeronimo; Suzana Porto Almeida; Thais Ferreira de Oliveira; Thais de Oliveira Costa; Ticiane Cavalcante de Souza; Veridiana Pessoa Miyajima |
| EPI_ISL_5529995 | UAPS FLORESTA | Analytical Competence Molecular Epidemiology Lab/ACME, Oswaldo Cruz Foundation, Ceara (FIOCRUZ CE) | Carlos Leonardo de Aragao Araujo; Cecilia Leite Costa & Eduardo Ruback dos Santos on behalf of COVID-19 FIOCRUZ Genomic Network; Cleber Furtado Aksenen; Fabio Miyajima; Fernando Braga Stehling; Francisco Eder de Moura Lopes; Igor Oliveira Duarte; Jamille Maria Mendes Bezerra; Joaquim Cesar do Nascimento Sousa Junior; Pedro Miguel Carneiro Jeronimo; Suzana Porto Almeida; Thais Ferreira de Oliveira; Thais de Oliveira Costa; Ticiane Cavalcante de Souza; Veridiana Pessoa Miyajima |
| EPI_ISL_5529994 | UAPS LUIS FRANKLIN | Analytical Competence Molecular Epidemiology Lab/ACME, Oswaldo Cruz Foundation, Ceara (FIOCRUZ CE) | Carlos Leonardo de Aragao Araujo; Cecilia Leite Costa & Eduardo Ruback dos Santos on behalf of COVID-19 FIOCRUZ Genomic Network; Cleber Furtado Aksenen; Fabio Miyajima; Fernando Braga Stehling; Francisco Eder de Moura Lopes; Igor Oliveira Duarte; Jamille Maria Mendes Bezerra; Joaquim Cesar do Nascimento Sousa Junior; Pedro Miguel Carneiro Jeronimo; Suzana Porto Almeida; Thais Ferreira de Oliveira; Thais de Oliveira Costa; Ticiane Cavalcante de Souza; Veridiana Pessoa Miyajima |
| EPI_ISL_5529999 | UAPS MACIEL DE BRITO | Analytical Competence Molecular Epidemiology Lab/ACME, Oswaldo Cruz Foundation, Ceara (FIOCRUZ CE) | Carlos Leonardo de Aragao Araujo; Cecilia Leite Costa & Eduardo Ruback dos Santos on behalf of COVID-19 FIOCRUZ Genomic Network; Cleber Furtado Aksenen; Fabio Miyajima; Fernando Braga Stehling; Francisco Eder de Moura Lopes; Igor Oliveira Duarte; Jamille Maria Mendes Bezerra; Joaquim Cesar do Nascimento Sousa Junior; Pedro Miguel Carneiro Jeronimo; Suzana Porto Almeida; Thais Ferreira de Oliveira; Thais de Oliveira Costa; Ticiane Cavalcante de Souza; Veridiana Pessoa Miyajima |
| EPI_ISL_5603291 | UAPS MELO JABORANDI | Analytical Competence Molecular Epidemiology Lab/ACME, Oswaldo Cruz Foundation, Ceara (FIOCRUZ CE) | Carlos Leonardo de Aragao Araujo; Cecilia Leite Costa & Eduardo Ruback dos Santos on behalf of COVID-19 FIOCRUZ Genomic Network; Cleber Furtado Aksenen; Fabio Miyajima; Fernando Braga Stehling; Francisco Eder de Moura Lopes; Igor Oliveira Duarte; Jamille Maria Mendes Bezerra; Joaquim Cesar do Nascimento Sousa Junior; Pedro Miguel Carneiro Jeronimo; Suzana Porto Almeida; Thais Ferreira de Oliveira; Thais de Oliveira Costa; Ticiane Cavalcante de Souza; Veridiana Pessoa Miyajima |
| EPI_ISL_5529984, EPI_ISL_5530195 | UAPS PAULO MARCELO | Analytical Competence Molecular Epidemiology Lab/ACME, Oswaldo Cruz Foundation, Ceara (FIOCRUZ CE) | Carlos Leonardo de Aragao Araujo; Cecilia Leite Costa & Eduardo Ruback dos Santos on behalf of COVID-19 FIOCRUZ Genomic Network; Cleber Furtado Aksenen; Fabio Miyajima; Fernando Braga Stehling; Francisco Eder de Moura Lopes; Igor Oliveira Duarte; Jamille Maria Mendes Bezerra; Joaquim Cesar do Nascimento Sousa Junior; Pedro Miguel Carneiro Jeronimo; Suzana Porto Almeida; Thais Ferreira de Oliveira; Thais de Oliveira Costa; Ticiane Cavalcante de Souza; Veridiana Pessoa Miyajima |
| EPI_ISL_5530000 | UAPS PONTES NETO | Analytical Competence Molecular Epidemiology Lab/ACME, Oswaldo Cruz Foundation, Ceara (FIOCRUZ CE) | Carlos Leonardo de Aragao Araujo; Cecilia Leite Costa & Eduardo Ruback dos Santos on behalf of COVID-19 FIOCRUZ Genomic Network; Cleber Furtado Aksenen; Fabio Miyajima; Fernando Braga Stehling; Francisco Eder de Moura Lopes; Igor Oliveira Duarte; Jamille Maria Mendes Bezerra; Joaquim Cesar do Nascimento Sousa Junior; Pedro Miguel Carneiro Jeronimo; Suzana Porto Almeida; Thais Ferreira de Oliveira; Thais de Oliveira Costa; Ticiane Cavalcante de Souza; Veridiana Pessoa Miyajima |
| EPI_ISL_5529992 | UAPS RONALDO DE ALBUQUERQUE RIBEIRO | Analytical Competence Molecular Epidemiology Lab/ACME, Oswaldo Cruz Foundation, Ceara (FIOCRUZ CE) | Carlos Leonardo de Aragao Araujo; Cecilia Leite Costa & Eduardo Ruback dos Santos on behalf of COVID-19 FIOCRUZ Genomic Network; Cleber Furtado Aksenen; Fabio Miyajima; Fernando Braga Stehling; Francisco Eder de Moura Lopes; Igor Oliveira Duarte; Jamille Maria Mendes Bezerra; Joaquim Cesar do Nascimento Sousa Junior; Pedro Miguel Carneiro Jeronimo; Suzana Porto Almeida; Thais Ferreira de Oliveira; Thais de Oliveira Costa; Ticiane Cavalcante de Souza; Veridiana Pessoa Miyajima |
| EPI_ISL_5529940 | UBS ANTONIO ROCHA FREIRE | Analytical Competence Molecular Epidemiology Lab/ACME, Oswaldo Cruz Foundation, Ceara (FIOCRUZ CE) | Carlos Leonardo de Aragao Araujo; Cecilia Leite Costa & Eduardo Ruback dos Santos on behalf of COVID-19 FIOCRUZ Genomic Network; Cleber Furtado Aksenen; Fabio Miyajima; Fernando Braga Stehling; Francisco Eder de Moura Lopes; Igor Oliveira Duarte; Jamille Maria Mendes Bezerra; Joaquim Cesar do Nascimento Sousa Junior; Pedro Miguel Carneiro Jeronimo; Suzana Porto Almeida; Thais Ferreira de Oliveira; Thais de Oliveira Costa; Ticiane Cavalcante de Souza; Veridiana Pessoa Miyajima |
| EPI_ISL_5530058, EPI_ISL_5530061 | UBS CENTRO | Analytical Competence Molecular Epidemiology Lab/ACME, Oswaldo Cruz Foundation, Ceara (FIOCRUZ CE) | Carlos Leonardo de Aragao Araujo; Cecilia Leite Costa & Eduardo Ruback dos Santos on behalf of COVID-19 FIOCRUZ Genomic Network; Cleber Furtado Aksenen; Fabio Miyajima; Fernando Braga Stehling; Francisco Eder de Moura Lopes; Igor Oliveira Duarte; Jamille Maria Mendes Bezerra; Joaquim Cesar do Nascimento Sousa Junior; Pedro Miguel Carneiro Jeronimo; Suzana Porto Almeida; Thais Ferreira de Oliveira; Thais de Oliveira Costa; Ticiane Cavalcante de Souza; Veridiana Pessoa Miyajima |
| EPI_ISL_5530116 | UBS DE LAGES | Analytical Competence Molecular Epidemiology Lab/ACME, Oswaldo Cruz Foundation, Ceara (FIOCRUZ CE) | Carlos Leonardo de Aragao Araujo; Cecilia Leite Costa & Eduardo Ruback dos Santos on behalf of COVID-19 FIOCRUZ Genomic Network; Cleber Furtado Aksenen; Fabio Miyajima; Fernando Braga Stehling; Francisco Eder de Moura Lopes; Igor Oliveira Duarte; Jamille Maria Mendes Bezerra; Joaquim Cesar do Nascimento Sousa Junior; Pedro Miguel Carneiro Jeronimo; Suzana Porto Almeida; Thais Ferreira de Oliveira; Thais de Oliveira Costa; Ticiane Cavalcante de Souza; Veridiana Pessoa Miyajima |
| EPI_ISL_5530130 | UBS MARIA LIDIA DE MOURA | Analytical Competence Molecular Epidemiology Lab/ACME, Oswaldo Cruz Foundation, Ceara (FIOCRUZ CE) | Carlos Leonardo de Aragao Araujo; Cecilia Leite Costa & Eduardo Ruback dos Santos on behalf of COVID-19 FIOCRUZ Genomic Network; Cleber Furtado Aksenen; Fabio Miyajima; Fernando Braga Stehling; Francisco Eder de Moura Lopes; Igor Oliveira Duarte; Jamille Maria Mendes Bezerra; Joaquim Cesar do Nascimento Sousa Junior; Pedro Miguel Carneiro Jeronimo; Suzana Porto Almeida; Thais Ferreira de Oliveira; Thais de Oliveira Costa; Ticiane Cavalcante de Souza; Veridiana Pessoa Miyajima |
| EPI_ISL_5530171 | UBS MARIA LUCINEIDE COSTA | Analytical Competence Molecular Epidemiology Lab/ACME, Oswaldo Cruz Foundation, Ceara (FIOCRUZ CE) | Carlos Leonardo de Aragao Araujo; Cecilia Leite Costa & Eduardo Ruback dos Santos on behalf of COVID-19 FIOCRUZ Genomic Network; Cleber Furtado Aksenen; Fabio Miyajima; Fernando Braga Stehling; Francisco Eder de Moura Lopes; Igor Oliveira Duarte; Jamille Maria Mendes Bezerra; Joaquim Cesar do Nascimento Sousa Junior; Pedro Miguel Carneiro Jeronimo; Suzana Porto Almeida; Thais Ferreira de Oliveira; Thais de Oliveira Costa; Ticiane Cavalcante de Souza; Veridiana Pessoa Miyajima |
| EPI_ISL_1675303 | UDEA | Universidad Nacional de Colombia - Laboratorio Genómico One Health | Andres F. Cardona-Rios; Carlos Franco-Muñoz; Daniel O. Maldonado-Perez; Diego A. Álvarez-Díaz; Hector Alejandro Ruiz-Moreno; Idabely Betancur Ortiz; Jorge E. Osorio; Juan P. Hernandez-Ortiz; Karl A Ciuoderis; Katherine Laiton-Donato; Laura Silvana Perez; Lina M. Hurtado; Marcela Mercado-Reyes; Maria Angélica Maya; Maria Stella López; Rita Almanza Payares; Sandra Ines Cano; Simón Villegas Velásquez |
| EPI_ISL_5530072 | UNID MISTA DE MORAUJO | Analytical Competence Molecular Epidemiology Lab/ACME, Oswaldo Cruz Foundation, Ceara (FIOCRUZ CE) | Carlos Leonardo de Aragao Araujo; Cecilia Leite Costa & Eduardo Ruback dos Santos on behalf of COVID-19 FIOCRUZ Genomic Network; Cleber Furtado Aksenen; Fabio Miyajima; Fernando Braga Stehling; Francisco Eder de Moura Lopes; Igor Oliveira Duarte; Jamille Maria Mendes Bezerra; Joaquim Cesar do Nascimento Sousa Junior; Pedro Miguel Carneiro Jeronimo; Suzana Porto Almeida; Thais Ferreira de Oliveira; Thais de Oliveira Costa; Ticiane Cavalcante de Souza; Veridiana Pessoa Miyajima |
| EPI_ISL_5529941 | UNIDADE BASICA DE SAUDE CENTRO DE SAUDE DE AMONTADA | Analytical Competence Molecular Epidemiology Lab/ACME, Oswaldo Cruz Foundation, Ceara (FIOCRUZ CE) | Carlos Leonardo de Aragao Araujo; Cecilia Leite Costa & Eduardo Ruback dos Santos on behalf of COVID-19 FIOCRUZ Genomic Network; Cleber Furtado Aksenen; Fabio Miyajima; Fernando Braga Stehling; Francisco Eder de Moura Lopes; Igor Oliveira Duarte; Jamille Maria Mendes Bezerra; Joaquim Cesar do Nascimento Sousa Junior; Pedro Miguel Carneiro Jeronimo; Suzana Porto Almeida; Thais Ferreira de Oliveira; Thais de Oliveira Costa; Ticiane Cavalcante de Souza; Veridiana Pessoa Miyajima |
| EPI_ISL_5529944 | UNIDADE DE PRONTO ATENDIMENTO ARACATI UPA ARACATI | Analytical Competence Molecular Epidemiology Lab/ACME, Oswaldo Cruz Foundation, Ceara (FIOCRUZ CE) | Carlos Leonardo de Aragao Araujo; Cecilia Leite Costa & Eduardo Ruback dos Santos on behalf of COVID-19 FIOCRUZ Genomic Network; Cleber Furtado Aksenen; Fabio Miyajima; Fernando Braga Stehling; Francisco Eder de Moura Lopes; Igor Oliveira Duarte; Jamille Maria Mendes Bezerra; Joaquim Cesar do Nascimento Sousa Junior; Pedro Miguel Carneiro Jeronimo; Suzana Porto Almeida; Thais Ferreira de Oliveira; Thais de Oliveira Costa; Ticiane Cavalcante de Souza; Veridiana Pessoa Miyajima |
| EPI_ISL_5529960 | UNIDADE DE PRONTO ATENDIMENTO DA JUREMA | Analytical Competence Molecular Epidemiology Lab/ACME, Oswaldo Cruz Foundation, Ceara (FIOCRUZ CE) | Carlos Leonardo de Aragao Araujo; Cecilia Leite Costa & Eduardo Ruback dos Santos on behalf of COVID-19 FIOCRUZ Genomic Network; Cleber Furtado Aksenen; Fabio Miyajima; Fernando Braga Stehling; Francisco Eder de Moura Lopes; Igor Oliveira Duarte; Jamille Maria Mendes Bezerra; Joaquim Cesar do Nascimento Sousa Junior; Pedro Miguel Carneiro Jeronimo; Suzana Porto Almeida; Thais Ferreira de Oliveira; Thais de Oliveira Costa; Ticiane Cavalcante de Souza; Veridiana Pessoa Miyajima |
| EPI_ISL_5530092 | UNIDADE DE PRONTO | Analytical Competence Molecular | Carlos Leonardo de Aragao Araujo; Cecilia Leite Costa & Eduardo Ruback dos Santos on behalf of COVID-19 FIOCRUZ Genomic Network; Cleber Furtado Aksenen; Fabio Miyajima; Fernando Braga Stehling; Francisco Eder de Moura Lopes; Igor Oliveira Duarte; Jamille Maria Mendes Bezerra; Joaquim Cesar do |

| | | | |
|---|---|---|---|
| | ATENDIMENTO DE RUSSAS | Epidemiology Lab/ACME, Oswaldo Cruz Foundation, Ceara (FIOCRUZ CE) | Nascimento Sousa Junior; Pedro Miguel Carneiro Jeronimo; Suzana Porto Almeida; Thais Ferreira de Oliveira; Thais de Oliveira Costa; Ticiane Cavalcante de Souza; Veridiana Pessoa Miyajima |
| EPI_ISL_5529982, EPI_ISL_5529983 | UNIDADE PRONTO ATENDIMENTO CANINDEZINHO | Analytical Competence Molecular Epidemiology Lab/ACME, Oswaldo Cruz Foundation, Ceara (FIOCRUZ CE) | Carlos Leonardo de Aragao Araujo; Cecilia Leite Costa & Eduardo Ruback dos Santos on behalf of COVID-19 FIOCRUZ Genomic Network; Cleber Furtado Aksenen; Fabio Miyajima; Fernando Braga Stehling; Francisco Eder de Moura Lopes; Igor Oliveira Duarte; Jamille Maria Mendes Bezerra; Joaquim Cesar do Nascimento Sousa Junior; Pedro Miguel Carneiro Jeronimo; Suzana Porto Almeida; Thais Ferreira de Oliveira; Thais de Oliveira Costa; Ticiane Cavalcante de Souza; Veridiana Pessoa Miyajima |
| EPI_ISL_3102480 | UNIDADE PRONTO ATENDIMENTO JANGURUSSU | Analytical Competence Molecular Epidemiology Lab/ACME, Oswaldo Cruz Foundation, Ceara (FIOCRUZ CE) | Cleber Furtado Aksenen; Fabio Miyajima; Fernando Braga Stehling; Francisco Eder de Moura Lopes; Jamille Maria Mendes Bezerra; Joaquim César do Nascimento Sousa Junior; Pedro Miguel Carneiro Jeronimo; Suzana Porto Almeida e Lucas Delerino; Thais Ferreira de Oliveira; Thais de Oliveira Costa; Ticiane Cavalcante de Souza; Veridiana Pessoa Miyajima |
| EPI_ISL_3102476 | UNIDADE PRONTO ATENDIMENTO PRAIA DO FUTURO | Analytical Competence Molecular Epidemiology Lab/ACME, Oswaldo Cruz Foundation, Ceara (FIOCRUZ CE) | Cleber Furtado Aksenen; Fabio Miyajima; Fernando Braga Stehling; Francisco Eder de Moura Lopes; Jamille Maria Mendes Bezerra; Joaquim César do Nascimento Sousa Junior; Pedro Miguel Carneiro Jeronimo; Suzana Porto Almeida e Lucas Delerino; Thais Ferreira de Oliveira; Thais de Oliveira Costa; Ticiane Cavalcante de Souza; Veridiana Pessoa Miyajima |
| EPI_ISL_2801318 | UNIDADE REFERENCIA COVID 19 CRATO | Analytical Competence Molecular Epidemiology Lab/ACME, Oswaldo Cruz Foundation, Ceara (FIOCRUZ CE) | Cleber Furtado Aksenen e Suzana Porto Almeida; Fabio Miyajima; Fernando Braga Stehling; Francisco Eder de Moura Lopes; Jamille Maria Mendes Bezerra; Joaquim César do Nascimento Sousa Junior; Pedro Miguel Carneiro Jeronimo; Thais Ferreira de Oliveira; Thais de Oliveira Costa; Ticiane Cavalcante de Souza; Veridiana Pessoa Miyajima |
| EPI_ISL_5530043, EPI_ISL_5530045 | UPA ANTONIA COELHO DE OLIVEIRA | Analytical Competence Molecular Epidemiology Lab/ACME, Oswaldo Cruz Foundation, Ceara (FIOCRUZ CE) | Carlos Leonardo de Aragao Araujo; Cecilia Leite Costa & Eduardo Ruback dos Santos on behalf of COVID-19 FIOCRUZ Genomic Network; Cleber Furtado Aksenen; Fabio Miyajima; Fernando Braga Stehling; Francisco Eder de Moura Lopes; Igor Oliveira Duarte; Jamille Maria Mendes Bezerra; Joaquim Cesar do Nascimento Sousa Junior; Pedro Miguel Carneiro Jeronimo; Suzana Porto Almeida; Thais Ferreira de Oliveira; Thais de Oliveira Costa; Ticiane Cavalcante de Souza; Veridiana Pessoa Miyajima |
| EPI_ISL_5530079 | UPA I UNIDADE DE PRONTO ATENDIMENTO | Analytical Competence Molecular Epidemiology Lab/ACME, Oswaldo Cruz Foundation, Ceara (FIOCRUZ CE) | Carlos Leonardo de Aragao Araujo; Cecilia Leite Costa & Eduardo Ruback dos Santos on behalf of COVID-19 FIOCRUZ Genomic Network; Cleber Furtado Aksenen; Fabio Miyajima; Fernando Braga Stehling; Francisco Eder de Moura Lopes; Igor Oliveira Duarte; Jamille Maria Mendes Bezerra; Joaquim Cesar do Nascimento Sousa Junior; Pedro Miguel Carneiro Jeronimo; Suzana Porto Almeida; Thais Ferreira de Oliveira; Thais de Oliveira Costa; Ticiane Cavalcante de Souza; Veridiana Pessoa Miyajima |
| EPI_ISL_5530131 | USF GERARDO GUALBERTO DE ARAUJO | Analytical Competence Molecular Epidemiology Lab/ACME, Oswaldo Cruz Foundation, Ceara (FIOCRUZ CE) | Carlos Leonardo de Aragao Araujo; Cecilia Leite Costa & Eduardo Ruback dos Santos on behalf of COVID-19 FIOCRUZ Genomic Network; Cleber Furtado Aksenen; Fabio Miyajima; Fernando Braga Stehling; Francisco Eder de Moura Lopes; Igor Oliveira Duarte; Jamille Maria Mendes Bezerra; Joaquim Cesar do Nascimento Sousa Junior; Pedro Miguel Carneiro Jeronimo; Suzana Porto Almeida; Thais Ferreira de Oliveira; Thais de Oliveira Costa; Ticiane Cavalcante de Souza; Veridiana Pessoa Miyajima |
| EPI_ISL_1263461, EPI_ISL_1400378, EPI_ISL_1405128, EPI_ISL_1405162, EPI_ISL_1405283 | UW Virology Lab | UW Virology Lab | Alexander Greninger; Hong Xie; Keith R Jerome; Lasata Shrestha; Meei-Li Huang; Michelle Lin; Noah Baker; Noah R. Baker; Pavitra Roychoudhury; Saraswathi Sathees; Sean Ellis; Shah Mohamed Bakhash |
| EPI_ISL_2660555, EPI_ISL_2660556, EPI_ISL_2660558, EPI_ISL_2660559, EPI_ISL_2660560, EPI_ISL_2660561 | Universidade Federal de Viçosa (UFV) | Laboratory of Respiratory Viruses and Measles, Oswaldo Cruz Institute, FIOCRUZ | Alice Sampaio Rocha; Ana Carolina Mendonca; Anna Carolina Paixao; Elisa Cavalcante Pereira; Fernando Motta; Luciana Appolinario; Marilda Siqueira on behalf of the Fiocruz COVID-19 Genomic Surveillance Network; Paola Resende; Renata Serrano Lopes; Rubens Pasa; Taina Venas |
| EPI_ISL_4413337 | Universidade Federal do Rio de Janeiro | Abbott | Amilcar Atanuri; Ana Olivo; Ana Vallari; Barbara Harris; Gavin Cloherty; Mary Rodgers; Todd Meyer |
| EPI_ISL_2365970 | University of Bari Biomedical Sciences and Human Oncology, Policlinico | University of Bari Biomedical Sciences and Human Oncology | Anna Sallustio; Daniela Loconsole; Maria Chironna; Marisa Accogli |
| EPI_ISL_1577565, EPI_ISL_1577569, EPI_ISL_1577575 | University of Rome Tor Vergata: Departm Experim Medicine Chair of Virology | University of Rome Tor Vergata: Departm Experim Medicine Chair of Virology | Francesca Ceccherini-Silberstein; Loredana Sarmati; Lorenzo Piermatteo; Luca Carioti; Marco Iannetta; Maria Botticelli; Maria Concetta Bellocchi; Massimo Andreoni |
| EPI_ISL_1633102 | Università Federico II - Dipartimento di scienze mediche traslazionali - Napoli | Telethon Institute of Genetics and Medicine (TIGEM) | Antonio Grimaldi Patrizia Annunziata Francesco Panariello Teresa Giuliano Michele Cennamo Valentina Bouche Chiara Colantuono Lucio Di Filippo Mariano Fiorenza Anna Manfredi Marcello Salvi Giuseppe Portella Andrea Ballabio Davide Cacchiarelli |
| EPI_ISL_1299562, EPI_ISL_1299564, EPI_ISL_1299565, EPI_ISL_1299566, EPI_ISL_1321734, EPI_ISL_1321735, EPI_ISL_1321736, EPI_ISL_1321737, EPI_ISL_1321738, EPI_ISL_1321739, EPI_ISL_1321740, EPI_ISL_1321741, EPI_ISL_1321742, EPI_ISL_1321805, EPI_ISL_1321806 | | | |
| see above | Università degli Studi di Perugia | Istituto Zooprofilattico Sperimentale dell’Abruzzo e Molise “G. Caporale” | Ancora M; Calistri P; Camilloni B; Cammà C; Curini V; Di Domenico M; Di Pasquale A; Lorusso A; Mangone I; Marcacci M; Mencacci A; Puglia I; Rinaldi A; Savini G; Scialabba S |
| EPI_ISL_7045642 | Urbino | Microbiology University Politecnica delle Marche | Anna Valenza; Carla Acciarri; Katia Marinelli; Monica Lucia Ferreri; Patrizia Bagnarelli; Roberta Longo; Sara Caucci; Stefano Menzo |
| EPI_ISL_1366655 | Usansolo-Galdakao University Hospital | Cruces University Hospital | Ana Belén de la Hoz; Ana Gual-de-Torrella; Izaskun Alejo-Cancho; mikel Gallego |
| EPI_ISL_1966090 | VIGILANCIA EPIDEMIOLOGICA | Instituto Butantan / Mendelics | Antonio Jorge Martins; Bianca Cechetto Carlos. Mendelics: Bibiana Santos; Claudia Renata dos Santos Barros; Cíntia Bittar; David Schlesinger. Hemocentro Ribeirão Preto: Simone Kashima; Debora Botequio Moretti; Elaine Cristina Marqueze; Elaine Vieira dos Santos; Elisangela Chicaroni Mattos; Erika Freitas; Evandra Strazza Rodrigues; Felipe Allan da Silva da Costa; Flavia Aburjaile; Fábio Sossai Possebon; Guilherme Campos; Guilherme Targino Valente; Heidge Fukumasu. USP-Botucatu: Rejane Maria Tommasini Grotto; Helena Lage Ferreira; Instituto Butantan: Dimas Tadeu Covas; Jardelina de Souza Todao Bernardino; Jayme A. Souza-Neto; Jessika Cristina Chagas Lesbon; Jorge A. Petroli Marchesi; José Salvatore Leister Patané; João Paulo Kitajima; João Pessoa Araújo Jr.; Leila Sabrina Ullmann; Loyze Paola Oliveira de Lima; Luiz Aurelio de Campos Crispin. Centro de Genômica Funcional da ESALQ: Luiz Lehmann Coutinho; Luiz Carlos Junior de Alcantara; Lívia Sacchetto; Maisa C. Pereira Parra; Maria Carolina Elias; Marta Giovanetti; Marília Moraes; Maurício Lacerda Nogueira. Prefeitura de Sao Paulo: Melissa Palmieri.; Patricia Akemi Assato; Paula Rahal; Paulo Inacio da Costa; Rafael dos Santos Bezerra; Raquel de Lello Rocha Campos Cassano. NGS Soluções Genômicas: Pilar Drummond Sampaio Corrêa Mariani. FZEA-USP Pirassununga: Mirele Daiana Poleti; Raul Machado Neto; Ricardo Augusto Brassaloti; Ricardo Haddad; Rodrigo Tocantins Calado. FAMERP-SJRP: Cecília Artico Banho; Sandra Coccuzzo Sampaio; Svetoslav Nanev Slavov; Vagner Fonseca; Vincent Louis Viala |
| EPI_ISL_2344589 | VIGILANCIA EPIDEMIOLOGICA E CONTROLE DE VETORES PIRASSUNUN | Instituto Butantan / FZEA-USP-Pirassununga | Antonio Jorge Martins; Claudia Renata dos Santos Barros; David Schlesinger; Debora Botequio Moretti; Dimas Tadeu Covas; Elaine Cristina Marqueze; Elaine Vieira Santos; Evandra Strazza Rodrigues; Heidge Fukumasu; Jayme Augusto de Souza-Neto; José Salvatore Leister Patané; Luiz Alcantara; Luiz Lehmann Coutinho; Maria Carolina Elias; Maurício Lacerda Nogueira; Rafael dos Santos Bezerra; Raul Machado Neto; Rejane Maria Tommasini Grotto; Ricardo Haddad; Sandra Coccuzzo Sampaio Vessoni; Simone Kashima; Svetoslav Nanev Slavov; Vincent Louis Viala |
| EPI_ISL_2036257 | VIROLOGY LABORATORY-CHU NICE | VIROLOGY LABORATORY-CHU NICE | Aicha El Yakine; Geraldine Gonfrier; Jean Machowiak; Sebastien Vitale; Valerie Giordanengo; Virginie Flahou |
| EPI_ISL_5801754 | Vigilancia Epidemiologica | Instituto Butantan | Antonio Jorge Martins; Claudia Renata dos Santos Barros; David Schlesinger; Debora Botequio Moretti; Dimas Tadeu Covas; Elaine Cristina Marqueze; Elaine Vieira Santos; Evandra Strazza Rodrigues; Heidge Fukumasu; Jayme Augusto de Souza-Neto; José Salvatore Leister Patané; Luiz Alcantara; Luiz Lehmann Coutinho; Maria Carolina Elias; Maurício Lacerda Nogueira; Rafael dos Santos Bezerra; Raul Machado Neto; Rejane Maria Tommasini Grotto; Ricardo Haddad; Sandra Coccuzzo Sampaio Vessoni; Simone Kashima; Svetoslav Nanev Slavov; Vincent Louis Viala |
| EPI_ISL_5782547 | Vigilancia Epidemiologica E Controle De Vetores Pirassununga | Instituto Butantan | Antonio Jorge Martins; Claudia Renata dos Santos Barros; David Schlesinger; Debora Botequio Moretti; Dimas Tadeu Covas; Elaine Cristina Marqueze; Elaine Vieira Santos; Evandra Strazza Rodrigues; Heidge Fukumasu; Jayme Augusto de Souza-Neto; José Salvatore Leister Patané; Luiz Alcantara; Luiz Lehmann Coutinho; Maria Carolina Elias; Maurício Lacerda Nogueira; Rafael dos Santos Bezerra; Raul Machado Neto; Rejane Maria Tommasini Grotto; Ricardo Haddad; Sandra Coccuzzo Sampaio Vessoni; Simone Kashima; Svetoslav Nanev Slavov; Vincent Louis Viala |
| EPI_ISL_2036877, EPI_ISL_2036883 | Virology Laboratory, Scientific Department, Army Medical Center | Virology Laboratory, Scientific Department, Army Medical Center | Anella Monte; Anna Anselmo; Antonella Fortunato; Filippo Molinari; Florigio Lista; Francesco Giordani; Giancarlo Petralito; Giandomenico Cerreto; Giulia Campoli; Lucia Nicosia; Marzia Cavalli; Riccardo De Sanctis; Rossella Brandi; Silvia Fillo; Vanessa Vera Fain |
| EPI_ISL_1420978 | Wisconsin State Laboratory of Hygiene Communicable Disease Division | Wisconsin State Laboratory of Hygiene Communicable Disease Division | Abigail C. Shockey; Kelsey R. Florek |
| EPI_ISL_1293215 | Yale Clinical Virology Lab | Grubaugh Lab - Yale School of Public Health | Anderson Brito; Annie Watkins; Chaney Kalinich; Chantal Vogels; Isabell Ott; Jessica Rothman; Joseph Fauver; Mallery Breban; Marie L. Landry; Mary Petrone; Nathan Grubaugh; Tara Alpert |
| EPI_ISL_1262813 | hopital | National Reference Center for Viruses of Respiratory Infections, Institut Pasteur, Paris | Angela Brisebarre; Camille Capel; Etienne Simon-Lorière; Leruez-Ville Marianne; Marion Barbet; Maud Vanpeene; Méline Bizard; Sylvie Behillil; Sylvie van der Werf; Vincent Enouf |
| EPI_ISL_1799013 | unknown | Instituto Nacional de Saude (INSA) | Borges et al |
