## Supplement 3 for "Rapid and Accurate Identification of SARS-CoV-2 Omicron Variants Using Droplet Digital PCR (RT-ddPCR)": All_0322-0328.pdf

We gratefully acknowledge the following Authors from the Originating laboratories responsible for obtaining the specimens, as well as the Submitting laboratories where the genome data were generated and shared via GISAID, on which this research is based.

All Submitters of data may be contacted directly via www.gisaid.org

Authors are sorted alphabetically.

| Accession ID | Originating Laboratory | Submitting Laboratory | Authors |
|---|---|---|---|
| EPI_ISL_3354677 | AL Dept. of Public Health Bureau of Clinical Laboratories | Centers for Disease Control and Prevention Division of Viral Diseases, Pathogen Discovery | Alex Burgin; Ben Rambo-Martin; Clinton Paden; Dakota Howard; Dave Wentworth; Dhwani Batra; Jasmine Padilla; Justin Lee; Krista Queen; Kristen Knipe; Kristine Lacek; Mark Burroughs; Matthew Schmerer; Meghan Bentz; Mili Sheth; Peter Cook; Sam Shepard; Sarah Nobles; Suxiang Tong; Vivien Dugan; Yvette Unoarumhi |
| EPI_ISL_5915190, EPI_ISL_5915204, EPI_ISL_5915246, EPI_ISL_5915291, EPI_ISL_6573797, EPI_ISL_6573799, EPI_ISL_6573833, EPI_ISL_6573834, EPI_ISL_6573876, EPI_ISL_6573915 | | | |
| see above | AMALAB/FACISA/UFRN | WallauLab on behalf of Fiocruz COVID-19 Genomic Surveillance Network | Alexandre Freitas da Silva; Allan Roberto Dias Nunes; Antonio Marinho da Silva Neto; Cassia Docena; Constância Flávia Junqueira Ayres; Filipe Zimmer Dezordi; Gabriel Luz Wallau; Gustavo Barbosa de Lima; Joana Cristina Medeiros Tavares Marques; Katya Anaya Jacinto; Lais Ceschini Machado; Lilian Caroliny Amorim Silva; Marcelo Henrique dos Santos Paiva; Mariane dos Santos Duarte; Matheus Filgueira Bezerra; Sinval Pinto Brandão Filho |
| EPI_ISL_1795113, EPI_ISL_2345312 | AMBULATORIO DE ATENDIMENTO DE DST DE GUARIBA | Instituto Butantan / ESALQ-Piracicaba | Antonio Jorge Martins; Bianca Cechetto Carlos. Mendelics: Bibiana Santos; Claudia Renata dos Santos Barros; David Schlesinger; David Schlesinger. Hemocentro Ribeirão Preto: Simone Kashima; Debora Botequio Moretti; Debora Botequio Moretti. Centro de Genômica Funcional da ESALQ: Luiz Lehmann Coutinho; Dimas Tadeu Covas; Elaine Cristina Marqueze; Elaine Vieira Santos; Elaine Vieira dos Santos; Elisangela Chicaroni Mattos; Erika Freitas; Evandra Strazza Rodrigues; Felipe Allan da Silva da Costa; Flavia Aburjaile; Guilherme Targino Valente; Heidge Fukumasu; Heidge Fukumasu. USP-Botucatu: Rejane Maria Tommasini Grotto; Instituto Butantan: Alexander Roberto Precioso; Jayme A. Souza-Neto; Jayme Augusto de Souza-Neto; Jessika Cristina Chagas Lesbon; José Salvatore Leister Patané; João Paulo Kitajima; Luiz Alcantara; Luiz Carlos Junior de Alcantara; Luiz Lehmann Coutinho; Maria Carolina Elias; Marta Giovanetti; Maurício Lacerda Nogueira; Patricia Akemi Assato; Rafael dos Santos Bezerra; Raquel de Lello Rocha Campos Cassano. NGS Soluções Genômicas: Pilar Drummond Sampaio Corrêa Mariani. FZEA-USP Pirassununga: Mirele Daiana Poleti; Raul Machado Neto; Rejane Maria Tommasini Grotto; Ricardo Augusto Brassaloti; Ricardo Haddad; Rodrigo Tocantins Calado.; Sandra Coccuzzo Sampaio; Sandra Coccuzzo Sampaio Vessoni; Simone Kashima; Svetoslav Nanev Slavov; Vagner Fonseca; Vincent Louis Viala |
| EPI_ISL_1795367, EPI_ISL_1795369, EPI_ISL_2345940, EPI_ISL_2345942 | AMBULATORIO MEDICO MUNICIPAL DE AGUDOS | Instituto Butantan / ESALQ-Piracicaba | Antonio Jorge Martins; Bianca Cechetto Carlos. Mendelics: Bibiana Santos; Claudia Renata dos Santos Barros; David Schlesinger; David Schlesinger. Hemocentro Ribeirão Preto: Simone Kashima; Debora Botequio Moretti; Debora Botequio Moretti. Centro de Genômica Funcional da ESALQ: Luiz Lehmann Coutinho; Dimas Tadeu Covas; Elaine Cristina Marqueze; Elaine Vieira Santos; Elaine Vieira dos Santos; Elisangela Chicaroni Mattos; Erika Freitas; Evandra Strazza Rodrigues; Felipe Allan da Silva da Costa; Flavia Aburjaile; Guilherme Targino Valente; Heidge Fukumasu; Heidge Fukumasu. USP-Botucatu: Rejane Maria Tommasini Grotto; Instituto Butantan: Alexander Roberto Precioso; Jayme A. Souza-Neto; Jayme Augusto de Souza-Neto; Jessika Cristina Chagas Lesbon; José Salvatore Leister Patané; João Paulo Kitajima; Luiz Alcantara; Luiz Carlos Junior de Alcantara; Luiz Lehmann Coutinho; Maria Carolina Elias; Marta Giovanetti; Maurício Lacerda Nogueira; Patricia Akemi Assato; Rafael dos Santos Bezerra; Raquel de Lello Rocha Campos Cassano. NGS Soluções Genômicas: Pilar Drummond Sampaio Corrêa Mariani. FZEA-USP Pirassununga: Mirele Daiana Poleti; Raul Machado Neto; Rejane Maria Tommasini Grotto; Ricardo Augusto Brassaloti; Ricardo Haddad; Rodrigo Tocantins Calado.; Sandra Coccuzzo Sampaio; Sandra Coccuzzo Sampaio Vessoni; Simone Kashima; Svetoslav Nanev Slavov; Vagner Fonseca; Vincent Louis Viala |
| EPI_ISL_1557818 | ASL Napoli 1 Centro | AMES Centro Polidiagnostico Strumentale S.r.l. | "Giovanni Savarese; Antonella Di Carlo; Antonio Fico"; Eloisa Evangelista; Luigi D'Amore; Luisa Circelli; Maurizio D'Amora; Monica Ianniello; Nadia Petrillo; Raffaella Ruggiero; Roberto Sirica |
| EPI_ISL_1403623, EPI_ISL_1456450, EPI_ISL_1457196, EPI_ISL_1457409, EPI_ISL_1457775 | AZ Klina | AZ Klina | Carl Vael - Lynsey Berckmans |
| EPI_ISL_1404433, EPI_ISL_1469127 | AZDelta | AZDelta | Dieter De Smet; Geert Martens |
| EPI_ISL_1551112, EPI_ISL_1551186, EPI_ISL_1551279, EPI_ISL_1551325, EPI_ISL_1551327, EPI_ISL_1551331, EPI_ISL_1551340, EPI_ISL_1551366, EPI_ISL_1551415, EPI_ISL_1551573, EPI_ISL_1551581, EPI_ISL_1551585, EPI_ISL_1551610, EPI_ISL_1559994, EPI_ISL_1560472, EPI_ISL_1560507, EPI_ISL_1560534, EPI_ISL_1560587, EPI_ISL_1560646, EPI_ISL_1560649, EPI_ISL_1560669, EPI_ISL_1560762, EPI_ISL_1560783, EPI_ISL_1560788, EPI_ISL_1560789, EPI_ISL_1560846, EPI_ISL_1560857, EPI_ISL_1560865, EPI_ISL_1560964, EPI_ISL_1561024, EPI_ISL_1649929, EPI_ISL_1649947, EPI_ISL_1649978, EPI_ISL_1649984, EPI_ISL_1650057, EPI_ISL_1650105, EPI_ISL_1650172, EPI_ISL_1650219, EPI_ISL_1650262, EPI_ISL_1650314, EPI_ISL_1650348, EPI_ISL_1650376, EPI_ISL_1650389, EPI_ISL_1650398, EPI_ISL_1650399, EPI_ISL_1650441, EPI_ISL_1650469, EPI_ISL_1650489, EPI_ISL_1650587, EPI_ISL_1650596, EPI_ISL_1650624, EPI_ISL_1650644, EPI_ISL_1650652, EPI_ISL_1650668, EPI_ISL_1650741, EPI_ISL_1650742, EPI_ISL_1650750, EPI_ISL_1650978, EPI_ISL_1651013, EPI_ISL_1651035, EPI_ISL_1651095, EPI_ISL_1651186, EPI_ISL_1667827, EPI_ISL_1685809, EPI_ISL_1685870, EPI_ISL_1686319, EPI_ISL_1686339, EPI_ISL_1686355, EPI_ISL_1686356, EPI_ISL_1686362, EPI_ISL_1686391, EPI_ISL_1686412, EPI_ISL_1686554, EPI_ISL_1686605, EPI_ISL_1686609, EPI_ISL_1686626, EPI_ISL_1686628, EPI_ISL_1686670, EPI_ISL_1686678, EPI_ISL_1686683, EPI_ISL_1686692, EPI_ISL_1686835, EPI_ISL_1687097, EPI_ISL_1687984, EPI_ISL_1695330, EPI_ISL_1695385, EPI_ISL_1925151, EPI_ISL_1925476, EPI_ISL_1925517, EPI_ISL_2242070, EPI_ISL_2242079, EPI_ISL_2242099, EPI_ISL_2242148, EPI_ISL_2242150, EPI_ISL_2242265, EPI_ISL_2242313, EPI_ISL_2242344, EPI_ISL_2242361, EPI_ISL_2242362, EPI_ISL_2482383, EPI_ISL_2482422, EPI_ISL_4360553, EPI_ISL_4360603, EPI_ISL_4360650, EPI_ISL_4361521, EPI_ISL_4362023, EPI_ISL_4362327, EPI_ISL_4362642, EPI_ISL_4363067, EPI_ISL_4363272, EPI_ISL_4363783, EPI_ISL_4363901, EPI_ISL_4363992, EPI_ISL_4364694, EPI_ISL_4365918 | | | |
| see above | Aegis Sciences Corporation | Centers for Disease Control and Prevention Division of Viral Diseases, Pathogen Discovery | Adrian Paskey; Alec Vest; Benjamin Rambo-Martin; Christopher Gulvick; Clinton Paden; Clinton R. Paden; Cyndi Clark; Dakota Howard; Darlene Wagner; Dhwani Batra; Dillon Nall; Duncan MacCannell; Erisa Sula; Ethan Sanders; Holly Houdeshell; Jason Caravas; Kara Moser; Kristine Lacek; Matthew Hardison; Matthew Schmerer; Ola Kvalvaag; Patrick Campbell; Peter Cook; Peter W. Cook; Rob Case; Scott Sammons; Shatavia Morrison; Shaun Westlund; Tymeckia Kendall; Victoria Caban Figueroa; Vikramsinha Ghorpade; Yvette Unoarumhi |
| EPI_ISL_2168197, EPI_ISL_2168198, EPI_ISL_2168199, EPI_ISL_2168200, EPI_ISL_2168201, EPI_ISL_2168202, EPI_ISL_2168203, EPI_ISL_2168204, EPI_ISL_2168205, EPI_ISL_2168206, EPI_ISL_2168207, EPI_ISL_2168208, EPI_ISL_2168209, EPI_ISL_2168210, EPI_ISL_2168211, EPI_ISL_2168212, EPI_ISL_2168213, EPI_ISL_2168417, EPI_ISL_2168434, EPI_ISL_2168435, EPI_ISL_2168438, EPI_ISL_2168465, EPI_ISL_2168466, EPI_ISL_2168468, EPI_ISL_2168477, EPI_ISL_2168487, EPI_ISL_2168501, EPI_ISL_2168549, EPI_ISL_2168583, EPI_ISL_2168586, EPI_ISL_2168587, EPI_ISL_2168633, EPI_ISL_2168663, EPI_ISL_2168690, EPI_ISL_2168710, EPI_ISL_2168726, EPI_ISL_2168735, EPI_ISL_2168743, EPI_ISL_2168755, EPI_ISL_2168805, EPI_ISL_2168815, EPI_ISL_2168828, EPI_ISL_2168833 | | | |
| see above | Alberta Precision Labs (APL) | Public Health Agency of Canada (PHAC) National Microbiology Laboratory | Buss; Croxen M; Deo A; Dieu P; E; Ferrato C; Gill K; Khan F; Koleva P; Li V; Lloyd C; Lynch T; Ma R; Murphy S; Pabbaraju K; Shokoples S; Thayer J; Tipples G; Whitehouse M; Wong A; Yu C; Zelyas N |
| EPI_ISL_1636499, EPI_ISL_1636502 | Althaia. Xarxa Assistencial Universitària de Manresa | IrsiCaixa | Bonaventura Clotet; Bonaventura Clotet Gloria Trujillo; Carolina Gonzalez Fernandez; Eulalia Grau; Francesc Catala-Moll; Jaume Trape Pujol; Marc Noguera-Julian; Maria Casadellà; Mariona Parera; Pilar Armengol; Rafel Perez Vidal; Roger Paredes |
| EPI_ISL_1587218, EPI_ISL_1587221, EPI_ISL_1587237, EPI_ISL_1587250, EPI_ISL_1587251, EPI_ISL_1822830, EPI_ISL_1822831, EPI_ISL_1822865, EPI_ISL_1822877, EPI_ISL_1822944, EPI_ISL_1823052, EPI_ISL_1823090 | | | |
| see above | Altius Institute | Seattle Flu Study | Alex Nguyen; Amanda Adler; Andrew Meuser; Barry R. Lutz; Benjamin Pelle; Caitlin R. Wolf; Chris D. Frazar; Clem Green; Daniel Bates; Deborah A. Nickerson; Elisabeth Brandstetter; Erica Ryke; Hannah Petersen; Helen Y. Chu; Jacob Rodriguez; Janet A. Englund; Jay Shendure; Jessica Halow; John Stamatoyannopoulos; Joshua Richards; Jover Lee; Julia Wald; Kairsten Fay; Kirsten Lacombe; Kneshay Harper; Lea M. Starita; Mark J. Rieder; Matt Hartman; Matthew Richardson; Matthew Thompson; Melissa Truong; Michael Boeckh; Michael Famulare; Misja Ilcisin; Muhammad Halimun; Olivia Waltner; Peter D. Han; Rebecca Bruders; Ryan Alexander; Sadie Patraw; Sofia Olsson; Stephanie DeBaun; Thomas R. Sibley; Tobias Ragoczy; Trevor Bedford; Truong Nguyen |
| EPI_ISL_5801956, EPI_ISL_5801957, EPI_ISL_5801962, EPI_ISL_5801993, EPI_ISL_5801996, EPI_ISL_5802041 | Ambulatorio Ib | Instituto Butantan | Antonio Jorge Martins; Claudia Renata dos Santos Barros; David Schlesinger; Debora Botequio Moretti; Dimas Tadeu Covas; Elaine Cristina Marqueze; Elaine Vieira Santos; Evandra Strazza Rodrigues; Heidge Fukumasu; Jayme Augusto de Souza-Neto; José Salvatore Leister Patané; Luiz Alcantara; Luiz Lehmann Coutinho; Maria Carolina Elias; Maurício Lacerda Nogueira; Rafael dos Santos Bezerra; Raul Machado Neto; Rejane Maria Tommasini Grotto; Ricardo Haddad; Sandra Coccuzzo Sampaio Vessoni; Simone Kashima; Svetoslav Nanev Slavov; Vincent Louis Viala |
| EPI_ISL_5799777, EPI_ISL_5799782 | Ambulatorio Medico Municipal De Agudos | Instituto Butantan | Antonio Jorge Martins; Claudia Renata dos Santos Barros; David Schlesinger; Debora Botequio Moretti; Dimas Tadeu Covas; Elaine Cristina Marqueze; Elaine Vieira Santos; Evandra Strazza Rodrigues; Heidge Fukumasu; Jayme Augusto de Souza-Neto; José Salvatore Leister Patané; Luiz Alcantara; Luiz Lehmann Coutinho; Maria Carolina Elias; Maurício Lacerda Nogueira; Rafael dos Santos Bezerra; Raul Machado Neto; Rejane Maria Tommasini Grotto; Ricardo Haddad; Sandra Coccuzzo Sampaio Vessoni; Simone Kashima; Svetoslav Nanev Slavov; Vincent Louis Viala |
| EPI_ISL_1966324, EPI_ISL_1966325, EPI_ISL_1966328, EPI_ISL_1966331, EPI_ISL_1966340, EPI_ISL_1966341 | Ambulatório IB | Instituto Butantan / Mendelics | Antonio Jorge Martins; Bianca Cechetto Carlos. Mendelics: Bibiana Santos; Claudia Renata dos Santos Barros; Cíntia Bittar; David Schlesinger. Hemocentro Ribeirão Preto: Simone Kashima; Debora Botequio Moretti; Elaine Cristina Marqueze; Elaine Vieira dos Santos; Elisangela Chicaroni Mattos; Erika Freitas; Evandra Strazza Rodrigues; Felipe Allan da Silva da Costa; Flavia Aburjaile; Fábio Sossai Possebon; Guilherme Campos; Guilherme Targino Valente; Heidge Fukumasu. USP-Botucatu: Rejane Maria Tommasini Grotto; Helena Lage Ferreira; Instituto Butantan: Dimas Tadeu Covas; Jardelina de Souza Todao Bernardino; Jayme A. Souza-Neto; Jessika Cristina Chagas Lesbon; Jorge A. Petroli Marchesi; José Salvatore Leister Patané; João Paulo Kitajima; João Pessoa Araújo Jr.; Leila Sabrina Ullmann; Loyze Paola Oliveira de Lima; Luiz Aurelio de Campos Crispin. Centro de Genômica Funcional da ESALQ: Luiz Lehmann Coutinho; Luiz Carlos Junior de Alcantara; Lívia Sacchetto; Maisa C. Pereira Parra; Maria Carolina Elias; Marta Giovanetti; Marília Moraes; Maurício Lacerda Nogueira. Prefeitura de Sao Paulo: Melissa Palmieri.; Patricia Akemi Assato; Paula Rahal; Paulo Inacio da Costa; Rafael dos Santos Bezerra; Raquel de Lello Rocha Campos Cassano. NGS Soluções Genômicas: Pilar Drummond Sampaio Corrêa Mariani. FZEA-USP Pirassununga: Mirele Daiana Poleti; Raul Machado Neto; Ricardo Augusto Brassaloti; Ricardo Haddad; Rodrigo Tocantins Calado. FAMERP-SJRP: Cecília Artico Banho; Sandra Coccuzzo Sampaio; Svetoslav Nanev Slavov; Vagner Fonseca; Vincent Louis Viala |
| EPI_ISL_1574966 | American Indian Health Service of Chicago | Illinois Department of Public Health - Chicago Lab | Ira Heimler; Vineet K. Dhiman |
| EPI_ISL_1406435 | Area of Virology, Serology and Virology Division (SAViD), New South Wales Health Pathology Randwick | Virology Research Laboratory; Area of Virology, Serology and Virology Division (SAViD), New South Wales Health Pathology Randwick | Au, J.; Bull, R.; Deveson, I.; Foster, C.; Rawlinson, W.; Ruiz Silva, M.; Van Hal, S.; Wong, M. |
| EPI_ISL_2716683 | Arizona State University | Arizona State University | Efrem S. Lim; Joshua LaBaer; Joy M. Blain; LaRinda A. Holland; Matthew F. Smith; Nicholas J. Mellor; Peter T. Skidmore; Rabia Maqsood; Valerie Harris; Vel Murugan |
| EPI_ISL_2105735 | Arión Genética | Instituto Nacional de Medicina Genomica | Cedro-Tanda A; Escobar-Arrazola M; Gonzalez-Barrera D; Herrera-Montalvo LA.; Hidalgo-Miranda A; Mendoza-Vargas A; Munguia-Garza P; Ramirez-Vega O; Rangel-DeLeon D; Reyes-Grajeda JP; Roldan-Castillo Magaly; Uribe-Figueroa Laura; Verea Jazmin |
| EPI_ISL_1492573 | Azienda Ospedaliera San Giovanni Addolorata | INMI Lazzaro Spallanzani IRCCS | A Di Caro; B Bartolini; CEM Gruber; E Giombini; F Messina; F Santini; G Bonfiglio; M Gaudio; M Rueca; MR Capobianchi; O Butera; PM Placanica |
| EPI_ISL_1669916, EPI_ISL_1669917, EPI_ISL_1669948, EPI_ISL_1669953 | Azienda Ospedaliera Terni | Istituto Zooprofilattico Sperimentale dell'Abruzzo e Molise "G. Caporale" | Ancora M; Calistri P; Cammà C; Curini V; Di Domenico M; Di Pasquale A; Lorusso A; Mangone I; Marcacci M; Palumbo M; Puglia I; Rinaldi A; Savini G; Scaccetti A; Scialabba S |
| EPI_ISL_1545715 | Azienda Sanitaria dell'Alto Adige Laboratorio Aziendale di Microbiologia e Virologia | Istituto di Genomica Applicata | Davide Scaglione; Eleonora Paparelli; Elisa Masi; Elisabetta Giacobazzi; Elisabetta Pagani; Gabriele Magris; Irena Jurman; Irene Bianconi; Michele Morgante; Stefanie Wieser; Vera Vendramin |
| EPI_ISL_2528424, EPI_ISL_2528431, EPI_ISL_2528759, EPI_ISL_2528763, EPI_ISL_2528765, EPI_ISL_2528767, EPI_ISL_2528769, EPI_ISL_2528778, EPI_ISL_2528792, EPI_ISL_2528799, EPI_ISL_2528840, EPI_ISL_2529669, EPI_ISL_2529670, EPI_ISL_2529673, EPI_ISL_2529676, EPI_ISL_2529677, EPI_ISL_2529678, EPI_ISL_2529679, EPI_ISL_2529681, EPI_ISL_2529682, EPI_ISL_2529683, EPI_ISL_2529686, EPI_ISL_2529688, EPI_ISL_2529689, EPI_ISL_2529691, EPI_ISL_2529693, EPI_ISL_2529695, EPI_ISL_2529696, EPI_ISL_2529701, EPI_ISL_2529702, EPI_ISL_2529707, EPI_ISL_2529713, EPI_ISL_2529716, EPI_ISL_2529723, EPI_ISL_2529724, EPI_ISL_2529729, EPI_ISL_2529733, EPI_ISL_2529736, EPI_ISL_2529738, EPI_ISL_2529740, EPI_ISL_2529746, EPI_ISL_2529751, EPI_ISL_2529752, EPI_ISL_2529755, EPI_ISL_2529760, EPI_ISL_2529762, EPI_ISL_2529764, EPI_ISL_2529765, EPI_ISL_2529766, EPI_ISL_2529767, EPI_ISL_2529770, EPI_ISL_2529771, EPI_ISL_2529773, EPI_ISL_2529775, EPI_ISL_2529776, EPI_ISL_2529777, EPI_ISL_2529778, EPI_ISL_2529779, EPI_ISL_2529780, EPI_ISL_2529781, EPI_ISL_2529782, EPI_ISL_2529783, EPI_ISL_2529785, EPI_ISL_2529787, EPI_ISL_2529788, EPI_ISL_2529791, EPI_ISL_2529792, EPI_ISL_2529793, EPI_ISL_2529794, EPI_ISL_2529796, EPI_ISL_2529799, EPI_ISL_2529800, EPI_ISL_2529801, EPI_ISL_2529803, EPI_ISL_2529805, EPI_ISL_2529806, EPI_ISL_2529810, EPI_ISL_2529822, EPI_ISL_2529824, EPI_ISL_2529825, EPI_ISL_2529826, EPI_ISL_2529828, EPI_ISL_2529835, EPI_ISL_2529836, EPI_ISL_2529841, EPI_ISL_2529842, EPI_ISL_2529843, EPI_ISL_2529845, EPI_ISL_2529846, EPI_ISL_2529847, EPI_ISL_2529848, EPI_ISL_2529855, EPI_ISL_2529857, EPI_ISL_2529862, EPI_ISL_2529863, EPI_ISL_2529868, EPI_ISL_2529875, EPI_ISL_2529882, EPI_ISL_2529889, EPI_ISL_2529891, EPI_ISL_2529892, EPI_ISL_2529901, EPI_ISL_2529902, EPI_ISL_2529903, EPI_ISL_2529907, EPI_ISL_2529908, EPI_ISL_2529909, EPI_ISL_2529912, EPI_ISL_2529915, EPI_ISL_2529916, EPI_ISL_2529919, EPI_ISL_2529924, EPI_ISL_2529925, EPI_ISL_2529926, EPI_ISL_2529927, EPI_ISL_2529931, EPI_ISL_2529938, EPI_ISL_2529939, EPI_ISL_2529940, EPI_ISL_2529942, EPI_ISL_2529947, EPI_ISL_2529949, EPI_ISL_2529956, EPI_ISL_2529958, EPI_ISL_2529963, EPI_ISL_2529965, EPI_ISL_2529967, EPI_ISL_2529972, EPI_ISL_2529996, EPI_ISL_2530009, EPI_ISL_2530016, EPI_ISL_2530018, | | | |

| | | | |
|---|---|---|---|
| EPI_ISL_2530021, EPI_ISL_2530023, EPI_ISL_2530024, EPI_ISL_2530026, EPI_ISL_2530028, EPI_ISL_2530029, EPI_ISL_2530030, EPI_ISL_2530031, EPI_ISL_2530033, EPI_ISL_2530034, EPI_ISL_2530035, EPI_ISL_2530038, EPI_ISL_2530040, EPI_ISL_2530041, EPI_ISL_2530044, EPI_ISL_2530046, EPI_ISL_2530047, EPI_ISL_2530051, EPI_ISL_2530054, EPI_ISL_2530055, EPI_ISL_2530058, EPI_ISL_2530060, EPI_ISL_2530062, EPI_ISL_2530064, EPI_ISL_2530065, EPI_ISL_2530066, EPI_ISL_2530067, EPI_ISL_2530068, EPI_ISL_2530072, EPI_ISL_2530073, EPI_ISL_2530074, EPI_ISL_2530079, EPI_ISL_2530088, EPI_ISL_2530093, EPI_ISL_2530094, EPI_ISL_2530098, EPI_ISL_2530099, EPI_ISL_2530102, EPI_ISL_2530108, EPI_ISL_2530112, EPI_ISL_2530123, EPI_ISL_2530130, EPI_ISL_2530138, EPI_ISL_2530142, EPI_ISL_2530150, EPI_ISL_2530164, EPI_ISL_2530171, EPI_ISL_2530184, EPI_ISL_2530197, EPI_ISL_2530200, EPI_ISL_2530203, EPI_ISL_2530211, EPI_ISL_2530215, EPI_ISL_2530225, EPI_ISL_2530234, EPI_ISL_2530244, EPI_ISL_2530254, EPI_ISL_2530266, EPI_ISL_2530271, EPI_ISL_2530274, EPI_ISL_2530282, EPI_ISL_2530284, EPI_ISL_2530287, EPI_ISL_2530291, EPI_ISL_2530293, EPI_ISL_2530294, EPI_ISL_2530300, EPI_ISL_2530303, EPI_ISL_2530306, EPI_ISL_2530310, EPI_ISL_2530311, EPI_ISL_2530312, EPI_ISL_2530320, EPI_ISL_2530327, EPI_ISL_2530331, EPI_ISL_2530342, EPI_ISL_2530347, EPI_ISL_2530350, EPI_ISL_2530351, EPI_ISL_2530358, EPI_ISL_2530361, EPI_ISL_2530372, EPI_ISL_2530375, EPI_ISL_2530377, EPI_ISL_2530380, EPI_ISL_2530381, EPI_ISL_2530397, EPI_ISL_2530403, EPI_ISL_2530406, EPI_ISL_2530408, EPI_ISL_2530411, EPI_ISL_2530419, EPI_ISL_2530420, EPI_ISL_2530425, EPI_ISL_2530427, EPI_ISL_2530428, EPI_ISL_2530431, EPI_ISL_2530433, EPI_ISL_2530436, EPI_ISL_2530437, EPI_ISL_2530440, EPI_ISL_2530441, EPI_ISL_2530442, EPI_ISL_2530444, EPI_ISL_2530446, EPI_ISL_2530447, EPI_ISL_2530448, EPI_ISL_2530449, EPI_ISL_2530450, EPI_ISL_2530451, EPI_ISL_2530454, EPI_ISL_2530455, EPI_ISL_2530456, EPI_ISL_2530458, EPI_ISL_2530460, EPI_ISL_2530463, EPI_ISL_2530466, EPI_ISL_2530467, EPI_ISL_2530468, EPI_ISL_2530469, EPI_ISL_2530470, EPI_ISL_2530473, EPI_ISL_2530474, EPI_ISL_2530475, EPI_ISL_2530477, EPI_ISL_2530479, EPI_ISL_2530480, EPI_ISL_2530481, EPI_ISL_2530482, EPI_ISL_2530485, EPI_ISL_2530486, EPI_ISL_2530487, EPI_ISL_2530489, EPI_ISL_2530492, EPI_ISL_2530498, EPI_ISL_2530500, EPI_ISL_2530503, EPI_ISL_2530504, EPI_ISL_2530507, EPI_ISL_2530509, EPI_ISL_2530510, EPI_ISL_2530512, EPI_ISL_2530517, EPI_ISL_2530521, EPI_ISL_2530522, EPI_ISL_2530523, EPI_ISL_2530526, EPI_ISL_2530530, EPI_ISL_2530534, EPI_ISL_2530538, EPI_ISL_2530539, EPI_ISL_2530540, EPI_ISL_2530541, EPI_ISL_2530549, EPI_ISL_2530554, EPI_ISL_2530555, EPI_ISL_2530556, EPI_ISL_2530565, EPI_ISL_2530566, EPI_ISL_2530571, EPI_ISL_2530573, EPI_ISL_2530574, EPI_ISL_2530581, EPI_ISL_2530582, EPI_ISL_2530588, EPI_ISL_2530590, EPI_ISL_2530593, EPI_ISL_2530600, EPI_ISL_2530602, EPI_ISL_2530603, EPI_ISL_2530606, EPI_ISL_2530610, EPI_ISL_2530611, EPI_ISL_2530614, EPI_ISL_2530615, EPI_ISL_2530616, EPI_ISL_2530617, EPI_ISL_2530620, EPI_ISL_2530623, EPI_ISL_2530625, EPI_ISL_2530627, EPI_ISL_2530628, EPI_ISL_2530632, EPI_ISL_2530633, EPI_ISL_2530636, EPI_ISL_2530644, EPI_ISL_2530653, EPI_ISL_2530654, EPI_ISL_2530658, EPI_ISL_2530661, EPI_ISL_2530667, EPI_ISL_2530674, EPI_ISL_2530675, EPI_ISL_2530679, EPI_ISL_2530683, EPI_ISL_2530684, EPI_ISL_2530687, EPI_ISL_2530688, EPI_ISL_2530689, EPI_ISL_2530691, EPI_ISL_2530696, EPI_ISL_2530699, EPI_ISL_2530700, EPI_ISL_2530701, EPI_ISL_2530703, EPI_ISL_2530704, EPI_ISL_2530706, EPI_ISL_2530708, EPI_ISL_2530718, EPI_ISL_2530719, EPI_ISL_2530720, EPI_ISL_2530722, EPI_ISL_2530725, EPI_ISL_2530726, EPI_ISL_2530728, EPI_ISL_2530733, EPI_ISL_2530735, EPI_ISL_2530736, EPI_ISL_2530740, EPI_ISL_2530741, EPI_ISL_2530744, EPI_ISL_2530748, EPI_ISL_2530751, EPI_ISL_2530753, EPI_ISL_2530754, EPI_ISL_2530758, EPI_ISL_2530765, EPI_ISL_2530767, EPI_ISL_2530770, EPI_ISL_2530771, EPI_ISL_2530772, EPI_ISL_2530773, EPI_ISL_2530774, EPI_ISL_2530777, EPI_ISL_2530778, EPI_ISL_2530781, EPI_ISL_2530783, EPI_ISL_2530785, EPI_ISL_2530787, EPI_ISL_2530790, EPI_ISL_2530792, EPI_ISL_2530796, EPI_ISL_2530798, EPI_ISL_2530799, EPI_ISL_2530800, EPI_ISL_2530801, EPI_ISL_2530804, EPI_ISL_2530806, EPI_ISL_2530810, EPI_ISL_2530812, EPI_ISL_2530813, EPI_ISL_2530814, EPI_ISL_2530815, EPI_ISL_2530818, EPI_ISL_2530820, EPI_ISL_2530821, EPI_ISL_2530822, EPI_ISL_2530823, EPI_ISL_2531117, EPI_ISL_2531119, EPI_ISL_2531120, EPI_ISL_2531121, EPI_ISL_2531124, EPI_ISL_2531125, EPI_ISL_2531126, EPI_ISL_2531127, EPI_ISL_2531128, EPI_ISL_2531129, EPI_ISL_2531130, EPI_ISL_2531131, EPI_ISL_2531132, EPI_ISL_2531133, EPI_ISL_2531134, EPI_ISL_2531135, EPI_ISL_2531136, EPI_ISL_2531137, EPI_ISL_2531138, EPI_ISL_2531139, EPI_ISL_2531140, EPI_ISL_2531141, EPI_ISL_2531142, EPI_ISL_2531143, EPI_ISL_2531144, EPI_ISL_2531145, EPI_ISL_2531147, EPI_ISL_2531161, EPI_ISL_2531162, EPI_ISL_2531163, EPI_ISL_2531164, EPI_ISL_2531165, EPI_ISL_2531166, EPI_ISL_2531167, EPI_ISL_2531168, EPI_ISL_2531169, EPI_ISL_2531170, EPI_ISL_2531171, EPI_ISL_2531172, EPI_ISL_2531173, EPI_ISL_2531174, EPI_ISL_2531175, EPI_ISL_2531176, EPI_ISL_2531177, EPI_ISL_2531178, EPI_ISL_2531179, EPI_ISL_2531180, EPI_ISL_2531192, EPI_ISL_2531194, EPI_ISL_2531195, EPI_ISL_2531197, EPI_ISL_2531198, EPI_ISL_2531199, EPI_ISL_2531200, EPI_ISL_2531201, EPI_ISL_2531202, EPI_ISL_2531203, EPI_ISL_2531204, EPI_ISL_2531205, EPI_ISL_2531206, EPI_ISL_2531207, EPI_ISL_2531224, EPI_ISL_2531228, EPI_ISL_2531233, EPI_ISL_2531238, EPI_ISL_2531245, EPI_ISL_2531247, EPI_ISL_2531248, EPI_ISL_2531249, EPI_ISL_2531250, EPI_ISL_2531251, EPI_ISL_2531252, EPI_ISL_2531253, EPI_ISL_2531254, EPI_ISL_2531255, EPI_ISL_2531256, EPI_ISL_2531260, EPI_ISL_2531261, EPI_ISL_2531262, EPI_ISL_2531263, EPI_ISL_2531264, EPI_ISL_2531265, EPI_ISL_2531266, EPI_ISL_2531271, EPI_ISL_2531282, EPI_ISL_2531297, EPI_ISL_2531298, EPI_ISL_2531300, EPI_ISL_2531307, EPI_ISL_2531315, EPI_ISL_2531317, EPI_ISL_2531318, EPI_ISL_2531320, EPI_ISL_2531321, EPI_ISL_2531322, EPI_ISL_2531323, EPI_ISL_2531324, EPI_ISL_2531327, EPI_ISL_2531329, EPI_ISL_2531330, EPI_ISL_2531331, EPI_ISL_2531332, EPI_ISL_2531333, EPI_ISL_2531334, EPI_ISL_2531336, EPI_ISL_2531340, EPI_ISL_2531346, EPI_ISL_2531361, EPI_ISL_2531362, EPI_ISL_2531363, EPI_ISL_2531364, EPI_ISL_2531365, EPI_ISL_2531366, EPI_ISL_2531367, EPI_ISL_2531368, EPI_ISL_2531369, EPI_ISL_2531370, EPI_ISL_2531373, EPI_ISL_2531374, EPI_ISL_2531375, EPI_ISL_2531376, EPI_ISL_2531377, EPI_ISL_2531380, EPI_ISL_2531383, EPI_ISL_2531393, EPI_ISL_2531399, EPI_ISL_2531414, EPI_ISL_2531415, EPI_ISL_2531416, EPI_ISL_2531417, EPI_ISL_2531418, EPI_ISL_2531419, EPI_ISL_2531420, EPI_ISL_2531421, EPI_ISL_2531422, EPI_ISL_2531423, EPI_ISL_2531425, EPI_ISL_2531426, EPI_ISL_2531427, EPI_ISL_2531428, EPI_ISL_2531435, EPI_ISL_2531436, EPI_ISL_2531438, EPI_ISL_2531440, EPI_ISL_2531442, EPI_ISL_2531443, EPI_ISL_2531444, EPI_ISL_2531445, EPI_ISL_2531446, EPI_ISL_2531456, EPI_ISL_2531460, EPI_ISL_2531463, EPI_ISL_2531466, EPI_ISL_2531467, EPI_ISL_2531468, EPI_ISL_2531469, EPI_ISL_2531474, EPI_ISL_2531475, EPI_ISL_2531476, EPI_ISL_2531484, EPI_ISL_2531489, EPI_ISL_2531493, EPI_ISL_2531495, EPI_ISL_2531506, EPI_ISL_2531507, EPI_ISL_2531510, EPI_ISL_2531520, EPI_ISL_2531522, EPI_ISL_2531545, EPI_ISL_2531546, EPI_ISL_2531558, EPI_ISL_2531561, EPI_ISL_2531566, EPI_ISL_2531567, EPI_ISL_2531568, EPI_ISL_2531569, EPI_ISL_2531571, EPI_ISL_2531573, EPI_ISL_2531575, EPI_ISL_2531580, EPI_ISL_2531583, EPI_ISL_2531585, EPI_ISL_2531586, EPI_ISL_2531589, EPI_ISL_2531592, EPI_ISL_2531594, EPI_ISL_2531596, EPI_ISL_2531598, EPI_ISL_2531602, EPI_ISL_2531606, EPI_ISL_2531607, EPI_ISL_2531610, EPI_ISL_2531619, EPI_ISL_2531624, EPI_ISL_2531627, EPI_ISL_2531632, EPI_ISL_2531634, EPI_ISL_2531636, EPI_ISL_2531637, EPI_ISL_2531639, EPI_ISL_2531641, EPI_ISL_2531647, EPI_ISL_2531666, EPI_ISL_2531667, EPI_ISL_2531668, EPI_ISL_2531669, EPI_ISL_2531670, EPI_ISL_2531671, EPI_ISL_2531692, EPI_ISL_2531736, EPI_ISL_2531739, EPI_ISL_2531741, EPI_ISL_2531742, EPI_ISL_2531743, EPI_ISL_2531745, EPI_ISL_2531752, EPI_ISL_2531756, EPI_ISL_2531763, EPI_ISL_2531779, EPI_ISL_2531780, EPI_ISL_2531781, EPI_ISL_2531871, EPI_ISL_2531875, EPI_ISL_2531876, EPI_ISL_2531880, EPI_ISL_2531882, EPI_ISL_2531883, EPI_ISL_2531886, EPI_ISL_2531887, EPI_ISL_2531891, EPI_ISL_2531892, EPI_ISL_2531895, EPI_ISL_2531899, EPI_ISL_2531903, EPI_ISL_2531904, EPI_ISL_2531910, EPI_ISL_2531916, EPI_ISL_2531918, EPI_ISL_2531921, EPI_ISL_2531923, EPI_ISL_2531924, EPI_ISL_2531925, EPI_ISL_2531927, EPI_ISL_2531928, EPI_ISL_2531929, EPI_ISL_2531932, EPI_ISL_2531933, EPI_ISL_2531935, EPI_ISL_2531936, EPI_ISL_2531937, EPI_ISL_2531938, EPI_ISL_2531941, EPI_ISL_2531946, EPI_ISL_2531956, EPI_ISL_2531960, EPI_ISL_2531961, EPI_ISL_2531968, EPI_ISL_2531969, EPI_ISL_2531973, EPI_ISL_2531977, EPI_ISL_2531979, EPI_ISL_2531981, EPI_ISL_2531983, EPI_ISL_2531990, EPI_ISL_2531994, EPI_ISL_2532007, EPI_ISL_2532009, EPI_ISL_2532012, EPI_ISL_2532018, EPI_ISL_2532037, EPI_ISL_2532038, EPI_ISL_2532118, EPI_ISL_2532123, EPI_ISL_2532124, EPI_ISL_2532127, EPI_ISL_2532128, EPI_ISL_2532129, EPI_ISL_2532131, EPI_ISL_2532132, EPI_ISL_2532133, EPI_ISL_2532135, EPI_ISL_2532137, EPI_ISL_2532139, EPI_ISL_2532141, EPI_ISL_2532142, EPI_ISL_2532175, EPI_ISL_2532210, EPI_ISL_2532212, EPI_ISL_2532213, EPI_ISL_2532214, EPI_ISL_2532224, EPI_ISL_2532226, EPI_ISL_2532228, EPI_ISL_2532229, EPI_ISL_2532231, EPI_ISL_2532233, EPI_ISL_2532235, EPI_ISL_2532236, EPI_ISL_2532239, EPI_ISL_2532242, EPI_ISL_2532244, EPI_ISL_2532245, EPI_ISL_2532246, EPI_ISL_2532247, EPI_ISL_2532253, EPI_ISL_2532257, EPI_ISL_2532258, EPI_ISL_2532259, EPI_ISL_2532260, EPI_ISL_2532261, EPI_ISL_2532262, EPI_ISL_2532263, EPI_ISL_2653076, EPI_ISL_2653078, EPI_ISL_2653079, EPI_ISL_2653080, EPI_ISL_2653082, EPI_ISL_2654820, EPI_ISL_2654826, EPI_ISL_2654828, EPI_ISL_2654831, EPI_ISL_2654834, EPI_ISL_2654835, EPI_ISL_2654839, EPI_ISL_2654840, EPI_ISL_2654841 | | | |
| see above | BCCDC Public Health Laboratory | BCCDC Public Health Laboratory | Ana Pacagnella; Corrinne Ng; Dan Fornika; John Tyson; Kim Macdonald; Kimia Kamelian; Linda Hoang; Loretta Janz; Mel Krajden; Prystajecky Natalie; Robert Azana; Shannon Russell |
| EPI_ISL_2321578 | BIOREFERENCIA | Universidad Nacional de Colombia - Laboratorio Genómico One Health | Andres F. Cardona-Rios; Carlos Franco-Muñoz; Carolina Muñoz-Arango; Celeny Ortiz; Daniel O. Maldonado-Perez; Diego A. Álvarez-Díaz; Hector Alejandro Ruiz-Moreno; Idabely Betancur Ortiz; Jorge E. Osorio; Juan P. Hernandez-Ortiz; Karl A Ciuoderis; Katherine Laiton-Donato; Laura Silvana Perez; Lina M. Hurtado; Marcela Mercado-Reyes; Maria Angélica Maya; Maria Stella López; Rita Almanza Payares; Sandra Ines Cano; Simón Villegas Velásquez |
| EPI_ISL_7672399, EPI_ISL_7672402 | Bambino Gesù Pediatric Hospital | Microbiology and Immunology Diagnosis Bambino Gesù Pediatric Hospital | Carlo Federico Perno; Claudia Alteri; Luna Colagrossi; Rossana Scutari; Valentino Costabile |
| EPI_ISL_1749270 | Biology Lab, HIA PERCY | IRBA, 2MI | CHAPUS C.; DEPEILLE A.; FOISSAUD V.; GORGE O.; GRANDPERRET V.; JARJAVAL F.; NOLENT F.; SARILAR V.; VERGUET N. |
| EPI_ISL_3479230, EPI_ISL_3479287, EPI_ISL_4422919, EPI_ISL_4423182 | British Columbia Centre For Disease Control | BCCDC Public Health Laboratory | Ana Pacagnella; Corrinne Ng; Dan Fornika; John Tyson; Kim Macdonald; Kimia Kamelian; Linda Hoang; Loretta Janz; Mel Krajden; Prystajecky Natalie; Robert Azana; Shannon Russell |
| EPI_ISL_1516684, EPI_ISL_1516700, EPI_ISL_1516701, EPI_ISL_1516720, EPI_ISL_1516732, EPI_ISL_1578230, EPI_ISL_1578340, EPI_ISL_1578343, EPI_ISL_1578344, EPI_ISL_1578345, EPI_ISL_1578346, EPI_ISL_1578347, EPI_ISL_1578348, EPI_ISL_1710255, EPI_ISL_1710257, EPI_ISL_1710310, EPI_ISL_1710318, EPI_ISL_1710324, EPI_ISL_1710339, EPI_ISL_1710345, EPI_ISL_1710500, EPI_ISL_1710501 | | | |
| see above | Broad Institute Clinical Research Sequencing Platform | Infectious Disease Program, Broad Institute of Harvard and MIT | Adams, G.; B.L.; B.W.; Bauer, M.; Birren; Blumenstiel, B.; Brown, C.; Carter, A.; Chaluvadi, S.; D.J.; DeFelice, M.; DeRuff, K.; Dodge, S.; Gabriel, S.; Gallagher, G.; Gladden-Young, A.; Granger, B.; J.E.; K.J.; Lagerborg, K.; Larkin, K.; Lee, M.; Lemieux; Lennon, N.; Loreth, C.; Madoff, L.; McGovern, S.; Meldrim, J.; Normandin, E.; P.C.; Park; Pearlman, L.; Reilly, S.; Rudy, M.; Sabeti; Siddle; Smole, S.; Tomkins-Tinch, C.; Vicente, G.; and MacInnis |
| EPI_ISL_2103128 | CA DPH Viral and Rickettsial Disease Laboratory | Chan-Zuckerberg Biohub | CZB Cliahub Consortium |
| EPI_ISL_5530101, EPI_ISL_5530102 | CASA DE SAUDE MATERNIDADE CELESTINA COLARES | Analytical Competence Molecular Epidemiology Lab/ACME, Oswaldo Cruz Foundation, Ceara (FIOCRUZ CE) | Carlos Leonardo de Aragao Araujo; Cecilia Leite Costa & Eduardo Ruback dos Santos on behalf of COVID-19 FIOCRUZ Genomic Network; Cleber Furtado Aksenen; Fabio Miyajima; Fernando Braga Stehling; Francisco Eder de Moura Lopes; Igor Oliveira Duarte; Jamille Maria Mendes Bezerra; Joaquim Cesar do Nascimento Sousa Junior; Pedro Miguel Carneiro Jeronimo; Suzana Porto Almeida; Thais Ferreira de Oliveira; Thais de Oliveira Costa; Ticiane Cavalcante de Souza; Veridiana Pessoa Miyajima |
| EPI_ISL_2825736, EPI_ISL_2889938, EPI_ISL_2890065, EPI_ISL_2921695, EPI_ISL_2923353, EPI_ISL_5696451, EPI_ISL_5696453, EPI_ISL_5696467, EPI_ISL_7700146, EPI_ISL_7700347 | | | |
| see above | CDPH VBL | California Department of Public Health | CDPH-COVIDNet; Emily Smith on behalf of CDPH-COVIDNet; Emily Smith on behalf of CDPH-COVIDNet and Priva Path Labs Inc.; UCLA Technology Center for Genomics & Bioinformatics |
| EPI_ISL_5530170 | CENTRO DE ATENDIMENTO COVID 19 TEREZINHA NUNES DA ROCHA | Analytical Competence Molecular Epidemiology Lab/ACME, Oswaldo Cruz Foundation, Ceara (FIOCRUZ CE) | Carlos Leonardo de Aragao Araujo; Cecilia Leite Costa & Eduardo Ruback dos Santos on behalf of COVID-19 FIOCRUZ Genomic Network; Cleber Furtado Aksenen; Fabio Miyajima; Fernando Braga Stehling; Francisco Eder de Moura Lopes; Igor Oliveira Duarte; Jamille Maria Mendes Bezerra; Joaquim Cesar do Nascimento Sousa Junior; Pedro Miguel Carneiro Jeronimo; Suzana Porto Almeida; Thais Ferreira de Oliveira; Thais de Oliveira Costa; Ticiane Cavalcante de Souza; Veridiana Pessoa Miyajima |
| EPI_ISL_1795325, EPI_ISL_1795329, EPI_ISL_1795333, EPI_ISL_2345586, EPI_ISL_2345590, EPI_ISL_2345594 | CENTRO DE ESPECIALIDADES DE PRIMAVERA | Instituto Butantan / ESALQ-Piracicaba | Antonio Jorge Martins; Bianca Cechetto Carlos. Mendelics: Bibiana Santos; Claudia Renata dos Santos Barros; David Schlesinger; David Schlesinger. Hemocentro Ribeirão Preto: Simone Kashima; Debora Botequio Moretti; Debora Botequio Moretti. Centro de Genômica Funcional da ESALQ: Luiz Lehmann Coutinho; Dimas Tadeu Covas; Elaine Cristina Marqueze; Elaine Vieira Santos; Elaine Vieira dos Santos; Elisangela Chicaroni Mattos; Erika Freitas; Evandra Strazza Rodrigues; Felipe Allan da Silva da Costa; Flavia Aburjaile; Guilherme Targino Valente; Heidge Fukumasu; Heidge Fukumasu. USP-Botucatu: Rejane Maria Tommasini Grotto; Instituto Butantan: Alexander Roberto Precioso; Jayme A. Souza-Neto; Jayme Augusto de Souza-Neto; Jessika Cristina Chagas Lesbon; José Salvatore Leister Patané; João Paulo Kitajima; Luiz Alcantara; Luiz Carlos Junior de Alcantara; Luiz Lehmann Coutinho; Maria Carolina Elias; Marta Giovanetti; Maurício Lacerda Nogueira; Patricia Akemi Assato; Rafael dos Santos Bezerra; Raquel de Lello Rocha Campos Cassano. NGS Soluções Genômicas: Pilar Drummond Sampaio Corrêa Mariani. FZEA-USP Pirassununga: Mirele Daiana Poleti; Raul Machado Neto; Rejane Maria Tommasini Grotto; Ricardo Augusto Brassaloti; Ricardo Haddad; Rodrigo Tocantins Calado.; Sandra Coccuzzo Sampaio; Sandra Coccuzzo Sampaio Vessoni; Simone Kashima; Svetoslav Nanev Slavov; Vagner Fonseca; Vincent Louis Viala |
| EPI_ISL_1795109, EPI_ISL_2344683 | CENTRO DE ESPECIALIDDS MEDICAS IRMA LEOPOLDINA PIRASSUNUNGA | Instituto Butantan / ESALQ-Piracicaba | Antonio Jorge Martins; Bianca Cechetto Carlos. Mendelics: Bibiana Santos; Claudia Renata dos Santos Barros; David Schlesinger; David Schlesinger. Hemocentro Ribeirão Preto: Simone Kashima; Debora Botequio Moretti; Debora Botequio Moretti. Centro de Genômica Funcional da ESALQ: Luiz Lehmann Coutinho; Dimas Tadeu Covas; Elaine Cristina Marqueze; Elaine Vieira Santos; Elaine Vieira dos Santos; Elisangela Chicaroni Mattos; Erika Freitas; Evandra Strazza Rodrigues; Felipe Allan da Silva da Costa; Flavia Aburjaile; Guilherme Targino Valente; Heidge Fukumasu; Heidge Fukumasu. USP-Botucatu: Rejane Maria Tommasini Grotto; Instituto Butantan: Alexander Roberto Precioso; Jayme A. Souza-Neto; Jayme Augusto de Souza-Neto; Jessika Cristina Chagas Lesbon; José Salvatore Leister Patané; João Paulo Kitajima; Luiz Alcantara; Luiz Carlos Junior de Alcantara; Luiz Lehmann Coutinho; Maria Carolina Elias; Marta Giovanetti; Maurício Lacerda Nogueira; Patricia Akemi Assato; Rafael dos Santos Bezerra; Raquel de Lello Rocha Campos Cassano. NGS Soluções Genômicas: Pilar Drummond Sampaio Corrêa Mariani. FZEA-USP Pirassununga: Mirele Daiana Poleti; Raul Machado Neto; Rejane Maria Tommasini Grotto; Ricardo Augusto Brassaloti; Ricardo Haddad; Rodrigo Tocantins Calado.; Sandra Coccuzzo Sampaio; Sandra Coccuzzo Sampaio Vessoni; Simone Kashima; Svetoslav Nanev Slavov; Vagner Fonseca; Vincent Louis Viala |
| EPI_ISL_2344665, EPI_ISL_2344666, EPI_ISL_2344678, EPI_ISL_2344679, EPI_ISL_2344680 | CENTRO DE ESPECIALIDDS MEDICAS IRMA LEOPOLDINA PIRASSUNUNGA | Instituto Butantan / FZEA-USP-Pirassununga | Antonio Jorge Martins; Claudia Renata dos Santos Barros; David Schlesinger; Debora Botequio Moretti; Dimas Tadeu Covas; Elaine Cristina Marqueze; Elaine Vieira Santos; Evandra Strazza Rodrigues; Heidge Fukumasu; Jayme Augusto de Souza-Neto; José Salvatore Leister Patané; Luiz Alcantara; Luiz Lehmann Coutinho; Maria Carolina Elias; Maurício Lacerda Nogueira; Rafael dos Santos Bezerra; Raul Machado Neto; Rejane Maria Tommasini Grotto; Ricardo Haddad; Sandra Coccuzzo Sampaio Vessoni; Simone Kashima; Svetoslav Nanev Slavov; Vincent Louis Viala |
| EPI_ISL_1795228, EPI_ISL_1795229, EPI_ISL_2345459, EPI_ISL_2345460 | CENTRO DE REFERENCIA DO IDOSO DR HUMBERTO MENDES DE CARVALHO | Instituto Butantan / ESALQ-Piracicaba | Antonio Jorge Martins; Bianca Cechetto Carlos. Mendelics: Bibiana Santos; Claudia Renata dos Santos Barros; David Schlesinger; David Schlesinger. Hemocentro Ribeirão Preto: Simone Kashima; Debora Botequio Moretti; Debora Botequio Moretti. Centro de Genômica Funcional da ESALQ: Luiz Lehmann Coutinho; Dimas Tadeu Covas; Elaine Cristina Marqueze; Elaine Vieira Santos; Elaine Vieira dos Santos; Elisangela Chicaroni Mattos; Erika Freitas; Evandra Strazza Rodrigues; Felipe Allan da Silva da Costa; Flavia Aburjaile; Guilherme Targino Valente; Heidge Fukumasu; Heidge Fukumasu. USP-Botucatu: Rejane Maria Tommasini Grotto; Instituto Butantan: Alexander Roberto Precioso; Jayme A. Souza-Neto; Jayme Augusto de Souza-Neto; Jessika Cristina Chagas Lesbon; José Salvatore Leister Patané; João Paulo Kitajima; Luiz Alcantara; Luiz Carlos Junior de Alcantara; Luiz Lehmann Coutinho; Maria Carolina Elias; Marta Giovanetti; Maurício Lacerda Nogueira; Patricia Akemi Assato; Rafael dos Santos Bezerra; Raquel de Lello Rocha Campos Cassano. NGS Soluções Genômicas: Pilar Drummond Sampaio Corrêa Mariani. FZEA-USP Pirassununga: Mirele Daiana Poleti; Raul Machado Neto; Rejane Maria Tommasini Grotto; Ricardo Augusto Brassaloti; Ricardo Haddad; Rodrigo Tocantins Calado.; Sandra Coccuzzo Sampaio; Sandra Coccuzzo Sampaio Vessoni; Simone Kashima; Svetoslav Nanev Slavov; Vagner Fonseca; Vincent Louis Viala |
| EPI_ISL_1795120, EPI_ISL_1795121, EPI_ISL_1795151, EPI_ISL_2345320, EPI_ISL_2345321, EPI_ISL_2345360 | CENTRO DE SAUDE DE CAJOBI | Instituto Butantan / ESALQ-Piracicaba | Antonio Jorge Martins; Bianca Cechetto Carlos. Mendelics: Bibiana Santos; Claudia Renata dos Santos Barros; David Schlesinger; David Schlesinger. Hemocentro Ribeirão Preto: Simone Kashima; Debora Botequio Moretti; Debora Botequio Moretti. Centro de Genômica Funcional da ESALQ: Luiz Lehmann Coutinho; Dimas Tadeu Covas; Elaine Cristina Marqueze; Elaine Vieira Santos; Elaine Vieira dos Santos; Elisangela Chicaroni Mattos; Erika Freitas; Evandra Strazza Rodrigues; Felipe Allan da Silva da Costa; Flavia Aburjaile; Guilherme Targino Valente; Heidge Fukumasu; Heidge Fukumasu. USP-Botucatu: Rejane Maria Tommasini Grotto; Instituto Butantan: Alexander Roberto Precioso; Jayme A. Souza-Neto; Jayme Augusto de Souza-Neto; Jessika Cristina Chagas Lesbon; José Salvatore Leister Patané; João Paulo Kitajima; Luiz Alcantara; Luiz Carlos Junior de Alcantara; Luiz Lehmann Coutinho; Maria Carolina Elias; Marta Giovanetti; Maurício Lacerda Nogueira; Patricia Akemi Assato; Rafael dos Santos Bezerra; Raquel de Lello Rocha Campos Cassano. NGS Soluções Genômicas: Pilar Drummond Sampaio Corrêa Mariani. FZEA-USP Pirassununga: Mirele Daiana Poleti; Raul Machado Neto; Rejane Maria Tommasini Grotto; Ricardo Augusto Brassaloti; Ricardo Haddad; Rodrigo Tocantins Calado.; Sandra Coccuzzo Sampaio; Sandra Coccuzzo Sampaio Vessoni; Simone Kashima; Svetoslav Nanev Slavov; Vagner Fonseca; Vincent Louis Viala |
| EPI_ISL_1795167, EPI_ISL_1795168, EPI_ISL_1795169, EPI_ISL_2345378, EPI_ISL_2345379, EPI_ISL_2345380 | CENTRO DE SAUDE DE FLORINEA | Instituto Butantan / ESALQ-Piracicaba | Antonio Jorge Martins; Bianca Cechetto Carlos. Mendelics: Bibiana Santos; Claudia Renata dos Santos Barros; David Schlesinger; David Schlesinger. Hemocentro Ribeirão Preto: Simone Kashima; Debora Botequio Moretti; Debora Botequio Moretti. Centro de Genômica Funcional da ESALQ: Luiz Lehmann Coutinho; Dimas Tadeu Covas; Elaine Cristina Marqueze; Elaine Vieira Santos; Elaine Vieira dos Santos; Elisangela Chicaroni Mattos; Erika Freitas; Evandra Strazza Rodrigues; Felipe Allan da Silva da Costa; Flavia Aburjaile; Guilherme Targino Valente; Heidge Fukumasu; Heidge Fukumasu. USP-Botucatu: Rejane Maria Tommasini Grotto; Instituto Butantan: Alexander Roberto Precioso; Jayme A. Souza-Neto; Jayme Augusto de Souza-Neto; Jessika Cristina Chagas Lesbon; José Salvatore Leister Patané; João Paulo Kitajima; Luiz Alcantara; Luiz Carlos Junior de Alcantara; Luiz Lehmann Coutinho; Maria Carolina Elias; Marta Giovanetti; Maurício Lacerda Nogueira; Patricia Akemi Assato; Rafael dos Santos Bezerra; Raquel de Lello Rocha Campos Cassano. NGS Soluções Genômicas: Pilar Drummond Sampaio Corrêa Mariani. FZEA-USP Pirassununga: Mirele Daiana Poleti; Raul Machado Neto; Rejane Maria Tommasini Grotto; Ricardo Augusto Brassaloti; Ricardo Haddad; Rodrigo Tocantins Calado.; Sandra Coccuzzo Sampaio; Sandra Coccuzzo Sampaio Vessoni; Simone Kashima; Svetoslav Nanev Slavov; Vagner Fonseca; Vincent Louis Viala |
| EPI_ISL_1795170, EPI_ISL_1795172, EPI_ISL_2345381, EPI_ISL_2345387 | CENTRO DE SAUDE DE MARACAI | Instituto Butantan / ESALQ-Piracicaba | Antonio Jorge Martins; Bianca Cechetto Carlos. Mendelics: Bibiana Santos; Claudia Renata dos Santos Barros; David Schlesinger; David Schlesinger. Hemocentro Ribeirão Preto: Simone Kashima; Debora Botequio Moretti; Debora Botequio Moretti. Centro de Genômica Funcional da ESALQ: Luiz Lehmann Coutinho; Dimas Tadeu Covas; Elaine Cristina Marqueze; Elaine Vieira Santos; Elaine Vieira dos Santos; Elisangela Chicaroni Mattos; Erika Freitas; Evandra Strazza Rodrigues; Felipe Allan da Silva da Costa; Flavia Aburjaile; Guilherme Targino Valente; Heidge Fukumasu; Heidge Fukumasu. USP-Botucatu: Rejane Maria Tommasini Grotto; Instituto Butantan: Alexander Roberto Precioso; Jayme A. Souza-Neto; Jayme Augusto de Souza-Neto; Jessika Cristina Chagas Lesbon; José Salvatore Leister Patané; João Paulo Kitajima; Luiz Alcantara; Luiz Carlos Junior de Alcantara; Luiz Lehmann Coutinho; Maria Carolina Elias; Marta Giovanetti; Maurício Lacerda Nogueira; Patricia Akemi Assato; Rafael dos Santos Bezerra; Raquel de Lello Rocha Campos Cassano. NGS Soluções Genômicas: Pilar Drummond Sampaio Corrêa Mariani. FZEA-USP Pirassununga: Mirele Daiana Poleti; Raul Machado Neto; Rejane Maria Tommasini Grotto; Ricardo Augusto Brassaloti; Ricardo Haddad; Rodrigo Tocantins Calado.; Sandra Coccuzzo Sampaio; Sandra Coccuzzo Sampaio Vessoni; Simone Kashima; Svetoslav Nanev Slavov; Vagner Fonseca; Vincent Louis Viala |
| EPI_ISL_5530051, EPI_ISL_5530053, EPI_ISL_5530054 | CENTRO DE SAUDE DONA TEREZA HOLANDA DE OLIVEIRA | Analytical Competence Molecular Epidemiology Lab/ACME, Oswaldo Cruz Foundation, Ceara (FIOCRUZ CE) | Carlos Leonardo de Aragao Araujo; Cecilia Leite Costa & Eduardo Ruback dos Santos on behalf of COVID-19 FIOCRUZ Genomic Network; Cleber Furtado Aksenen; Fabio Miyajima; Fernando Braga Stehling; Francisco Eder de Moura Lopes; Igor Oliveira Duarte; Jamille Maria Mendes Bezerra; Joaquim Cesar do Nascimento Sousa Junior; Pedro Miguel Carneiro Jeronimo; Suzana Porto Almeida; Thais Ferreira de Oliveira; Thais de Oliveira Costa; Ticiane Cavalcante de Souza; Veridiana Pessoa Miyajima |
| EPI_ISL_1795162, EPI_ISL_1795163, EPI_ISL_1795164, EPI_ISL_1795166, EPI_ISL_2345373, EPI_ISL_2345374, EPI_ISL_2345375, EPI_ISL_2345377 | | | |
| see above | CENTRO DE SAUDE DR PLINIO ALBERS DE GALIA | Instituto Butantan / ESALQ-Piracicaba | Antonio Jorge Martins; Bianca Cechetto Carlos. Mendelics: Bibiana Santos; Claudia Renata dos Santos Barros; David Schlesinger; David Schlesinger. Hemocentro Ribeirão Preto: Simone Kashima; Debora Botequio Moretti; Debora Botequio Moretti. Centro de Genômica Funcional da ESALQ: Luiz Lehmann Coutinho; Dimas Tadeu Covas; Elaine Cristina Marqueze; Elaine Vieira Santos; Elaine Vieira dos Santos; Elisangela Chicaroni Mattos; Erika Freitas; Evandra Strazza Rodrigues; Felipe Allan da Silva da Costa; Flavia Aburjaile; Guilherme Targino Valente; Heidge Fukumasu; Heidge Fukumasu. USP-Botucatu: Rejane Maria Tommasini Grotto; Instituto Butantan: Alexander Roberto Precioso; Jayme A. Souza-Neto; Jayme Augusto de Souza-Neto; Jessika Cristina Chagas Lesbon; José Salvatore Leister Patané; João Paulo Kitajima; Luiz Alcantara; Luiz Carlos Junior de Alcantara; Luiz Lehmann Coutinho; Maria Carolina Elias; Marta Giovanetti; Maurício Lacerda Nogueira; Patricia Akemi Assato; Rafael dos Santos Bezerra; Raquel de Lello Rocha Campos Cassano. NGS Soluções Genômicas: Pilar Drummond Sampaio Corrêa Mariani. FZEA-USP Pirassununga: Mirele Daiana Poleti; Raul Machado Neto; Rejane Maria Tommasini Grotto; Ricardo Augusto |

| | | | |
|---|---|---|---|
| | | | Brassaloti; Ricardo Haddad; Rodrigo Tocantins Calado.; Sandra Coccuzzo Sampaio; Sandra Coccuzzo Sampaio Vessoni; Simone Kashima; Svetoslav Nanev Slavov; Vagner Fonseca; Vincent Louis Viala |
| EPI_ISL_1795327, EPI_ISL_1795331, EPI_ISL_2345588, EPI_ISL_2345592 | CENTRO DE SAUDE II JUNQUEIROPOLIS | Instituto Butantan / ESALQ-Piracicaba | Antonio Jorge Martins; Bianca Cechetto Carlos. Mendelics: Bibiana Santos; Claudia Renata dos Santos Barros; David Schlesinger; David Schlesinger. Hemocentro Ribeirão Preto: Simone Kashima; Debora Botequio Moretti; Debora Botequio Moretti. Centro de Genômica Funcional da ESALQ: Luiz Lehmann Coutinho; Dimas Tadeu Covas; Elaine Cristina Marqueze; Elaine Vieira Santos; Elaine Vieira dos Santos; Elisangela Chicaroni Mattos; Erika Freitas; Evandra Strazza Rodrigues; Felipe Allan da Silva da Costa; Flavia Aburjaile; Guilherme Targino Valente; Heidge Fukumasu; Heidge Fukumasu. USP-Botucatu: Rejane Maria Tommasini Grotto; Instituto Butantan: Alexander Roberto Precioso; Jayme A. Souza-Neto; Jayme Augusto de Souza-Neto; Jessika Cristina Chagas Lesbon; José Salvatore Leister Patané; João Paulo Kitajima; Luiz Alcantara; Luiz Carlos Junior de Alcantara; Luiz Lehmann Coutinho; Maria Carolina Elias; Marta Giovanetti; Maurício Lacerda Nogueira; Patricia Akemi Assato; Rafael dos Santos Bezerra; Raquel de Lello Rocha Campos Cassano. NGS Soluções Genômicas: Pilar Drummond Sampaio Corrêa Mariani. FZEA-USP Pirassununga: Mirele Daiana Poleti; Raul Machado Neto; Rejane Maria Tommasini Grotto; Ricardo Augusto Brassaloti; Ricardo Haddad; Rodrigo Tocantins Calado.; Sandra Coccuzzo Sampaio; Sandra Coccuzzo Sampaio Vessoni; Simone Kashima; Svetoslav Nanev Slavov; Vagner Fonseca; Vincent Louis Viala |
| EPI_ISL_1795081, EPI_ISL_1795296, EPI_ISL_1795297, EPI_ISL_1795299, EPI_ISL_2345538, EPI_ISL_2345539, EPI_ISL_2345542, EPI_ISL_2345546 | | | |
| see above | CENTRO DE SAUDE III AFFONSO LUZZI SANTA CRUZ DAS PALMEIRAS | Instituto Butantan / ESALQ-Piracicaba | Antonio Jorge Martins; Bianca Cechetto Carlos. Mendelics: Bibiana Santos; Claudia Renata dos Santos Barros; David Schlesinger; David Schlesinger. Hemocentro Ribeirão Preto: Simone Kashima; Debora Botequio Moretti; Debora Botequio Moretti. Centro de Genômica Funcional da ESALQ: Luiz Lehmann Coutinho; Dimas Tadeu Covas; Elaine Cristina Marqueze; Elaine Vieira Santos; Elaine Vieira dos Santos; Elisangela Chicaroni Mattos; Erika Freitas; Evandra Strazza Rodrigues; Felipe Allan da Silva da Costa; Flavia Aburjaile; Guilherme Targino Valente; Heidge Fukumasu; Heidge Fukumasu. USP-Botucatu: Rejane Maria Tommasini Grotto; Instituto Butantan: Alexander Roberto Precioso; Jayme A. Souza-Neto; Jayme Augusto de Souza-Neto; Jessika Cristina Chagas Lesbon; José Salvatore Leister Patané; João Paulo Kitajima; Luiz Alcantara; Luiz Carlos Junior de Alcantara; Luiz Lehmann Coutinho; Maria Carolina Elias; Marta Giovanetti; Maurício Lacerda Nogueira; Patricia Akemi Assato; Rafael dos Santos Bezerra; Raquel de Lello Rocha Campos Cassano. NGS Soluções Genômicas: Pilar Drummond Sampaio Corrêa Mariani. FZEA-USP Pirassununga: Mirele Daiana Poleti; Raul Machado Neto; Rejane Maria Tommasini Grotto; Ricardo Augusto Brassaloti; Ricardo Haddad; Rodrigo Tocantins Calado.; Sandra Coccuzzo Sampaio; Sandra Coccuzzo Sampaio Vessoni; Simone Kashima; Svetoslav Nanev Slavov; Vagner Fonseca; Vincent Louis Viala |
| EPI_ISL_1795114, EPI_ISL_1795115, EPI_ISL_2345313, EPI_ISL_2345314 | CENTRO DE SAUDE III BORBOREMA | Instituto Butantan / ESALQ-Piracicaba | Antonio Jorge Martins; Bianca Cechetto Carlos. Mendelics: Bibiana Santos; Claudia Renata dos Santos Barros; David Schlesinger; David Schlesinger. Hemocentro Ribeirão Preto: Simone Kashima; Debora Botequio Moretti; Debora Botequio Moretti. Centro de Genômica Funcional da ESALQ: Luiz Lehmann Coutinho; Dimas Tadeu Covas; Elaine Cristina Marqueze; Elaine Vieira Santos; Elaine Vieira dos Santos; Elisangela Chicaroni Mattos; Erika Freitas; Evandra Strazza Rodrigues; Felipe Allan da Silva da Costa; Flavia Aburjaile; Guilherme Targino Valente; Heidge Fukumasu; Heidge Fukumasu. USP-Botucatu: Rejane Maria Tommasini Grotto; Instituto Butantan: Alexander Roberto Precioso; Jayme A. Souza-Neto; Jayme Augusto de Souza-Neto; Jessika Cristina Chagas Lesbon; José Salvatore Leister Patané; João Paulo Kitajima; Luiz Alcantara; Luiz Carlos Junior de Alcantara; Luiz Lehmann Coutinho; Maria Carolina Elias; Marta Giovanetti; Maurício Lacerda Nogueira; Patricia Akemi Assato; Rafael dos Santos Bezerra; Raquel de Lello Rocha Campos Cassano. NGS Soluções Genômicas: Pilar Drummond Sampaio Corrêa Mariani. FZEA-USP Pirassununga: Mirele Daiana Poleti; Raul Machado Neto; Rejane Maria Tommasini Grotto; Ricardo Augusto Brassaloti; Ricardo Haddad; Rodrigo Tocantins Calado.; Sandra Coccuzzo Sampaio; Sandra Coccuzzo Sampaio Vessoni; Simone Kashima; Svetoslav Nanev Slavov; Vagner Fonseca; Vincent Louis Viala |
| EPI_ISL_1795126, EPI_ISL_2345328 | CENTRO DE SAUDE III SALES OLIVEIRA | Instituto Butantan / ESALQ-Piracicaba | Antonio Jorge Martins; Bianca Cechetto Carlos. Mendelics: Bibiana Santos; Claudia Renata dos Santos Barros; David Schlesinger; David Schlesinger. Hemocentro Ribeirão Preto: Simone Kashima; Debora Botequio Moretti; Debora Botequio Moretti. Centro de Genômica Funcional da ESALQ: Luiz Lehmann Coutinho; Dimas Tadeu Covas; Elaine Cristina Marqueze; Elaine Vieira Santos; Elaine Vieira dos Santos; Elisangela Chicaroni Mattos; Erika Freitas; Evandra Strazza Rodrigues; Felipe Allan da Silva da Costa; Flavia Aburjaile; Guilherme Targino Valente; Heidge Fukumasu; Heidge Fukumasu. USP-Botucatu: Rejane Maria Tommasini Grotto; Instituto Butantan: Alexander Roberto Precioso; Jayme A. Souza-Neto; Jayme Augusto de Souza-Neto; Jessika Cristina Chagas Lesbon; José Salvatore Leister Patané; João Paulo Kitajima; Luiz Alcantara; Luiz Carlos Junior de Alcantara; Luiz Lehmann Coutinho; Maria Carolina Elias; Marta Giovanetti; Maurício Lacerda Nogueira; Patricia Akemi Assato; Rafael dos Santos Bezerra; Raquel de Lello Rocha Campos Cassano. NGS Soluções Genômicas: Pilar Drummond Sampaio Corrêa Mariani. FZEA-USP Pirassununga: Mirele Daiana Poleti; Raul Machado Neto; Rejane Maria Tommasini Grotto; Ricardo Augusto Brassaloti; Ricardo Haddad; Rodrigo Tocantins Calado.; Sandra Coccuzzo Sampaio; Sandra Coccuzzo Sampaio Vessoni; Simone Kashima; Svetoslav Nanev Slavov; Vagner Fonseca; Vincent Louis Viala |
| EPI_ISL_1966345 | CENTRO DE SAUDE III TABATINGA | Instituto Butantan / Mendelics | Antonio Jorge Martins; Bianca Cechetto Carlos. Mendelics: Bibiana Santos; Claudia Renata dos Santos Barros; Cíntia Bittar; David Schlesinger. Hemocentro Ribeirão Preto: Simone Kashima; Debora Botequio Moretti; Elaine Cristina Marqueze; Elaine Vieira dos Santos; Elisangela Chicaroni Mattos; Erika Freitas; Evandra Strazza Rodrigues; Felipe Allan da Silva da Costa; Flavia Aburjaile; Fábio Sossai Possebon; Guilherme Campos; Guilherme Targino Valente; Heidge Fukumasu. USP-Botucatu: Rejane Maria Tommasini Grotto; Helena Lage Ferreira; Instituto Butantan: Dimas Tadeu Covas; Jardelina de Souza Todao Bernardino; Jayme A. Souza-Neto; Jessika Cristina Chagas Lesbon; Jorge A. Petroli Marchesi; José Salvatore Leister Patané; João Paulo Kitajima; João Pessoa Araújo Jr.; Leila Sabrina Ullmann; Loyze Paola Oliveira de Lima; Luiz Aurelio de Campos Crispin. Centro de Genômica Funcional da ESALQ: Luiz Lehmann Coutinho; Luiz Carlos Junior de Alcantara; Lívia Sacchetto; Maisa C. Pereira Parra; Maria Carolina Elias; Marta Giovanetti; Marília Moraes; Maurício Lacerda Nogueira. Prefeitura de Sao Paulo: Melissa Palmieri.; Patricia Akemi Assato; Paula Rahal; Paulo Inacio da Costa; Rafael dos Santos Bezerra; Raquel de Lello Rocha Campos Cassano. NGS Soluções Genômicas: Pilar Drummond Sampaio Corrêa Mariani. FZEA-USP Pirassununga: Mirele Daiana Poleti; Raul Machado Neto; Ricardo Augusto Brassaloti; Ricardo Haddad; Rodrigo Tocantins Calado. FAMERP-SJRP: Cecília Artico Banho; Sandra Coccuzzo Sampaio; Svetoslav Nanev Slavov; Vagner Fonseca; Vincent Louis Viala |
| EPI_ISL_1795093, EPI_ISL_1795368, EPI_ISL_1795370, EPI_ISL_2345939, EPI_ISL_2345941, EPI_ISL_2345944 | CENTRO INTEGRADO DE SAUDE | Instituto Butantan / ESALQ-Piracicaba | Antonio Jorge Martins; Bianca Cechetto Carlos. Mendelics: Bibiana Santos; Claudia Renata dos Santos Barros; David Schlesinger; David Schlesinger. Hemocentro Ribeirão Preto: Simone Kashima; Debora Botequio Moretti; Debora Botequio Moretti. Centro de Genômica Funcional da ESALQ: Luiz Lehmann Coutinho; Dimas Tadeu Covas; Elaine Cristina Marqueze; Elaine Vieira Santos; Elaine Vieira dos Santos; Elisangela Chicaroni Mattos; Erika Freitas; Evandra Strazza Rodrigues; Felipe Allan da Silva da Costa; Flavia Aburjaile; Guilherme Targino Valente; Heidge Fukumasu; Heidge Fukumasu. USP-Botucatu: Rejane Maria Tommasini Grotto; Instituto Butantan: Alexander Roberto Precioso; Jayme A. Souza-Neto; Jayme Augusto de Souza-Neto; Jessika Cristina Chagas Lesbon; José Salvatore Leister Patané; João Paulo Kitajima; Luiz Alcantara; Luiz Carlos Junior de Alcantara; Luiz Lehmann Coutinho; Maria Carolina Elias; Marta Giovanetti; Maurício Lacerda Nogueira; Patricia Akemi Assato; Rafael dos Santos Bezerra; Raquel de Lello Rocha Campos Cassano. NGS Soluções Genômicas: Pilar Drummond Sampaio Corrêa Mariani. FZEA-USP Pirassununga: Mirele Daiana Poleti; Raul Machado Neto; Rejane Maria Tommasini Grotto; Ricardo Augusto Brassaloti; Ricardo Haddad; Rodrigo Tocantins Calado.; Sandra Coccuzzo Sampaio; Sandra Coccuzzo Sampaio Vessoni; Simone Kashima; Svetoslav Nanev Slavov; Vagner Fonseca; Vincent Louis Viala |
| EPI_ISL_1795274, EPI_ISL_1795275, EPI_ISL_2345510, EPI_ISL_2345513 | CENTRO MEDICO PMESP | Instituto Butantan / ESALQ-Piracicaba | Antonio Jorge Martins; Bianca Cechetto Carlos. Mendelics: Bibiana Santos; Claudia Renata dos Santos Barros; David Schlesinger; David Schlesinger. Hemocentro Ribeirão Preto: Simone Kashima; Debora Botequio Moretti; Debora Botequio Moretti. Centro de Genômica Funcional da ESALQ: Luiz Lehmann Coutinho; Dimas Tadeu Covas; Elaine Cristina Marqueze; Elaine Vieira Santos; Elaine Vieira dos Santos; Elisangela Chicaroni Mattos; Erika Freitas; Evandra Strazza Rodrigues; Felipe Allan da Silva da Costa; Flavia Aburjaile; Guilherme Targino Valente; Heidge Fukumasu; Heidge Fukumasu. USP-Botucatu: Rejane Maria Tommasini Grotto; Instituto Butantan: Alexander Roberto Precioso; Jayme A. Souza-Neto; Jayme Augusto de Souza-Neto; Jessika Cristina Chagas Lesbon; José Salvatore Leister Patané; João Paulo Kitajima; Luiz Alcantara; Luiz Carlos Junior de Alcantara; Luiz Lehmann Coutinho; Maria Carolina Elias; Marta Giovanetti; Maurício Lacerda Nogueira; Patricia Akemi Assato; Rafael dos Santos Bezerra; Raquel de Lello Rocha Campos Cassano. NGS Soluções Genômicas: Pilar Drummond Sampaio Corrêa Mariani. FZEA-USP Pirassununga: Mirele Daiana Poleti; Raul Machado Neto; Rejane Maria Tommasini Grotto; Ricardo Augusto Brassaloti; Ricardo Haddad; Rodrigo Tocantins Calado.; Sandra Coccuzzo Sampaio; Sandra Coccuzzo Sampaio Vessoni; Simone Kashima; Svetoslav Nanev Slavov; Vagner Fonseca; Vincent Louis Viala |
| EPI_ISL_2004295 | CH Setubal | Instituto Nacional de Saude (INSA) | Borges et al |
| EPI_ISL_1910892, EPI_ISL_1910893, EPI_ISL_1910907 | CH Tourcoing | CHU Lille | AIT YAHYA Emilie; ALIDJINOU Enagnon Kazali; BOCKET Laurence; CREPIN Michel; DEMAY Christophe; ENGELMANN Ilka; GEFFROY Sandrine; GUIGON Aurélie; LAMBERT Valérie; LAZREK Mouna; NOBILLIAUX Florian; PREVOST Brigitte; THUILLIER Caroline; TINEZ Claire |
| EPI_ISL_3133968 | CH de l'Ouest Guyanais Frank Joly | Institut Pasteur de la Guyane | Anne Lavergne; Dominique Rousset |
| EPI_ISL_3133961, EPI_ISL_3133962, EPI_ISL_3133970 | CHC Andrée Rosemon | Institut Pasteur de la Guyane | Anne Lavergne; Dominique Rousset |
| EPI_ISL_1451467, EPI_ISL_1451470, EPI_ISL_1451473, EPI_ISL_1451476, EPI_ISL_1545445, EPI_ISL_1545446, EPI_ISL_1545447, EPI_ISL_1545448, EPI_ISL_1545449, EPI_ISL_1545450, EPI_ISL_1545646, EPI_ISL_1545647 | | | |
| see above | CHWAPI - SITE NOTRE DAME | Institut de Pathologie et Genetique (IPG) | Jérémie Gras; Pascale Hilbert |
| EPI_ISL_3133960, EPI_ISL_3133963, EPI_ISL_3133964, EPI_ISL_3133966, EPI_ISL_3133967 | CNR Institut Pasteur de la Guyane | Institut Pasteur de la Guyane | Anne Lavergne; Dominique Rousset |
| EPI_ISL_1795107, EPI_ISL_2344660 | COORDENADORIA MUNICIPAL DE SAUDE DE IRACEMAPOLIS | Instituto Butantan / ESALQ-Piracicaba | Antonio Jorge Martins; Bianca Cechetto Carlos. Mendelics: Bibiana Santos; Claudia Renata dos Santos Barros; David Schlesinger; David Schlesinger. Hemocentro Ribeirão Preto: Simone Kashima; Debora Botequio Moretti; Debora Botequio Moretti. Centro de Genômica Funcional da ESALQ: Luiz Lehmann Coutinho; Dimas Tadeu Covas; Elaine Cristina Marqueze; Elaine Vieira Santos; Elaine Vieira dos Santos; Elisangela Chicaroni Mattos; Erika Freitas; Evandra Strazza Rodrigues; Felipe Allan da Silva da Costa; Flavia Aburjaile; Guilherme Targino Valente; Heidge Fukumasu; Heidge Fukumasu. USP-Botucatu: Rejane Maria Tommasini Grotto; Instituto Butantan: Alexander Roberto Precioso; Jayme A. Souza-Neto; Jayme Augusto de Souza-Neto; Jessika Cristina Chagas Lesbon; José Salvatore Leister Patané; João Paulo Kitajima; Luiz Alcantara; Luiz Carlos Junior de Alcantara; Luiz Lehmann Coutinho; Maria Carolina Elias; Marta Giovanetti; Maurício Lacerda Nogueira; Patricia Akemi Assato; Rafael dos Santos Bezerra; Raquel de Lello Rocha Campos Cassano. NGS Soluções Genômicas: Pilar Drummond Sampaio Corrêa Mariani. FZEA-USP Pirassununga: Mirele Daiana Poleti; Raul Machado Neto; Rejane Maria Tommasini Grotto; Ricardo Augusto Brassaloti; Ricardo Haddad; Rodrigo Tocantins Calado.; Sandra Coccuzzo Sampaio; Sandra Coccuzzo Sampaio Vessoni; Simone Kashima; Svetoslav Nanev Slavov; Vagner Fonseca; Vincent Louis Viala |
| EPI_ISL_1795165, EPI_ISL_2345376 | CS DE ALVARO CARVALHO | Instituto Butantan / ESALQ-Piracicaba | Antonio Jorge Martins; Bianca Cechetto Carlos. Mendelics: Bibiana Santos; Claudia Renata dos Santos Barros; David Schlesinger; David Schlesinger. Hemocentro Ribeirão Preto: Simone Kashima; Debora Botequio Moretti; Debora Botequio Moretti. Centro de Genômica Funcional da ESALQ: Luiz Lehmann Coutinho; Dimas Tadeu Covas; Elaine Cristina Marqueze; Elaine Vieira Santos; Elaine Vieira dos Santos; Elisangela Chicaroni Mattos; Erika Freitas; Evandra Strazza Rodrigues; Felipe Allan da Silva da Costa; Flavia Aburjaile; Guilherme Targino Valente; Heidge Fukumasu; Heidge Fukumasu. USP-Botucatu: Rejane Maria Tommasini Grotto; Instituto Butantan: Alexander Roberto Precioso; Jayme A. Souza-Neto; Jayme Augusto de Souza-Neto; Jessika Cristina Chagas Lesbon; José Salvatore Leister Patané; João Paulo Kitajima; Luiz Alcantara; Luiz Carlos Junior de Alcantara; Luiz Lehmann Coutinho; Maria Carolina Elias; Marta Giovanetti; Maurício Lacerda Nogueira; Patricia Akemi Assato; Rafael dos Santos Bezerra; Raquel de Lello Rocha Campos Cassano. NGS Soluções Genômicas: Pilar Drummond Sampaio Corrêa Mariani. FZEA-USP Pirassununga: Mirele Daiana Poleti; Raul Machado Neto; Rejane Maria Tommasini Grotto; Ricardo Augusto Brassaloti; Ricardo Haddad; Rodrigo Tocantins Calado.; Sandra Coccuzzo Sampaio; Sandra Coccuzzo Sampaio Vessoni; Simone Kashima; Svetoslav Nanev Slavov; Vagner Fonseca; Vincent Louis Viala |
| EPI_ISL_1795234, EPI_ISL_1795235, EPI_ISL_1795236, EPI_ISL_1795237, EPI_ISL_1795238, EPI_ISL_1795241, EPI_ISL_1795389, EPI_ISL_2345466, EPI_ISL_2345467, EPI_ISL_2345468, EPI_ISL_2345469, EPI_ISL_2345470, EPI_ISL_2345473, EPI_ISL_2345475 | | | |
| see above | CS DE BALSAMO | Instituto Butantan / ESALQ-Piracicaba | Antonio Jorge Martins; Bianca Cechetto Carlos. Mendelics: Bibiana Santos; Claudia Renata dos Santos Barros; David Schlesinger; David Schlesinger. Hemocentro Ribeirão Preto: Simone Kashima; Debora Botequio Moretti; Debora Botequio Moretti. Centro de Genômica Funcional da ESALQ: Luiz Lehmann Coutinho; Dimas Tadeu Covas; Elaine Cristina Marqueze; Elaine Vieira Santos; Elaine Vieira dos Santos; Elisangela Chicaroni Mattos; Erika Freitas; Evandra Strazza Rodrigues; Felipe Allan da Silva da Costa; Flavia Aburjaile; Guilherme Targino Valente; Heidge Fukumasu; Heidge Fukumasu. USP-Botucatu: Rejane Maria Tommasini Grotto; Instituto Butantan: Alexander Roberto Precioso; Jayme A. Souza-Neto; Jayme Augusto de Souza-Neto; Jessika Cristina Chagas Lesbon; José Salvatore Leister Patané; João Paulo Kitajima; Luiz Alcantara; Luiz Carlos Junior de Alcantara; Luiz Lehmann Coutinho; Maria Carolina Elias; Marta Giovanetti; Maurício Lacerda Nogueira; Patricia Akemi Assato; Rafael dos Santos Bezerra; Raquel de Lello Rocha Campos Cassano. NGS Soluções Genômicas: Pilar Drummond Sampaio Corrêa Mariani. FZEA-USP Pirassununga: Mirele Daiana Poleti; Raul Machado Neto; Rejane Maria Tommasini Grotto; Ricardo Augusto Brassaloti; Ricardo Haddad; Rodrigo Tocantins Calado.; Sandra Coccuzzo Sampaio; Sandra Coccuzzo Sampaio Vessoni; Simone Kashima; Svetoslav Nanev Slavov; Vagner Fonseca; Vincent Louis Viala |
| EPI_ISL_1795259, EPI_ISL_1795260, EPI_ISL_1795261, EPI_ISL_1795262, EPI_ISL_1795263, EPI_ISL_1795265, EPI_ISL_2345493, EPI_ISL_2345494, EPI_ISL_2345495, EPI_ISL_2345496, EPI_ISL_2345497, EPI_ISL_2345499 | | | |
| see above | CS DE ICEM | Instituto Butantan / ESALQ-Piracicaba | Antonio Jorge Martins; Bianca Cechetto Carlos. Mendelics: Bibiana Santos; Claudia Renata dos Santos Barros; David Schlesinger; David Schlesinger. Hemocentro Ribeirão Preto: Simone Kashima; Debora Botequio Moretti; Debora Botequio Moretti. Centro de Genômica Funcional da ESALQ: Luiz Lehmann Coutinho; Dimas Tadeu Covas; Elaine Cristina Marqueze; Elaine Vieira Santos; Elaine Vieira dos Santos; Elisangela Chicaroni Mattos; Erika Freitas; Evandra Strazza Rodrigues; Felipe Allan da Silva da Costa; Flavia Aburjaile; Guilherme Targino Valente; Heidge Fukumasu; Heidge Fukumasu. USP-Botucatu: Rejane Maria Tommasini Grotto; Instituto Butantan: Alexander Roberto Precioso; Jayme A. Souza-Neto; Jayme Augusto de Souza-Neto; Jessika Cristina Chagas Lesbon; José Salvatore Leister Patané; João Paulo Kitajima; Luiz Alcantara; Luiz Carlos Junior de Alcantara; Luiz Lehmann Coutinho; Maria Carolina Elias; Marta Giovanetti; Maurício Lacerda Nogueira; Patricia Akemi Assato; Rafael dos Santos Bezerra; Raquel de Lello Rocha Campos Cassano. NGS Soluções Genômicas: Pilar Drummond Sampaio Corrêa Mariani. FZEA-USP Pirassununga: Mirele Daiana Poleti; Raul Machado Neto; Rejane Maria Tommasini Grotto; Ricardo Augusto Brassaloti; Ricardo Haddad; Rodrigo Tocantins Calado.; Sandra Coccuzzo Sampaio; Sandra Coccuzzo Sampaio Vessoni; Simone Kashima; Svetoslav Nanev Slavov; Vagner Fonseca; Vincent Louis Viala |
| EPI_ISL_1795288, EPI_ISL_2345526 | CS DE NIPOA | Instituto Butantan / ESALQ-Piracicaba | Antonio Jorge Martins; Bianca Cechetto Carlos. Mendelics: Bibiana Santos; Claudia Renata dos Santos Barros; David Schlesinger; David Schlesinger. Hemocentro Ribeirão Preto: Simone Kashima; Debora Botequio Moretti; Debora Botequio Moretti. Centro de Genômica Funcional da ESALQ: Luiz Lehmann Coutinho; Dimas Tadeu Covas; Elaine Cristina Marqueze; Elaine Vieira Santos; Elaine Vieira dos Santos; Elisangela Chicaroni Mattos; Erika Freitas; Evandra Strazza Rodrigues; Felipe Allan da Silva da Costa; Flavia Aburjaile; Guilherme Targino Valente; Heidge Fukumasu; Heidge Fukumasu. USP-Botucatu: Rejane Maria Tommasini Grotto; Instituto Butantan: Alexander Roberto Precioso; Jayme A. Souza-Neto; Jayme Augusto de Souza-Neto; Jessika Cristina Chagas Lesbon; José Salvatore Leister Patané; João Paulo Kitajima; Luiz Alcantara; Luiz Carlos Junior de Alcantara; Luiz Lehmann Coutinho; Maria Carolina Elias; Marta Giovanetti; Maurício Lacerda Nogueira; Patricia Akemi Assato; Rafael dos Santos Bezerra; Raquel de Lello Rocha Campos Cassano. NGS Soluções Genômicas: Pilar Drummond Sampaio Corrêa Mariani. FZEA-USP Pirassununga: Mirele Daiana Poleti; Raul Machado Neto; Rejane Maria Tommasini Grotto; Ricardo Augusto Brassaloti; Ricardo Haddad; Rodrigo Tocantins Calado.; Sandra Coccuzzo Sampaio; Sandra Coccuzzo Sampaio Vessoni; Simone Kashima; Svetoslav Nanev Slavov; Vagner Fonseca; Vincent Louis Viala |
| EPI_ISL_1795171, EPI_ISL_1795173, EPI_ISL_1795390, EPI_ISL_2345383, EPI_ISL_2345388, EPI_ISL_2345389 | CS DE OSCAR BRESSANE PSF | Instituto Butantan / ESALQ-Piracicaba | Antonio Jorge Martins; Bianca Cechetto Carlos. Mendelics: Bibiana Santos; Claudia Renata dos Santos Barros; David Schlesinger; David Schlesinger. Hemocentro Ribeirão Preto: Simone Kashima; Debora Botequio Moretti; Debora Botequio Moretti. Centro de Genômica Funcional da ESALQ: Luiz Lehmann Coutinho; Dimas Tadeu Covas; Elaine Cristina Marqueze; Elaine Vieira Santos; Elaine Vieira dos Santos; Elisangela Chicaroni Mattos; Erika Freitas; Evandra Strazza Rodrigues; Felipe Allan da Silva da Costa; Flavia Aburjaile; Guilherme Targino Valente; Heidge Fukumasu; Heidge Fukumasu. USP-Botucatu: Rejane Maria Tommasini Grotto; Instituto Butantan: Alexander Roberto Precioso; Jayme A. Souza-Neto; Jayme Augusto de Souza-Neto; Jessika Cristina Chagas Lesbon; José Salvatore Leister Patané; João Paulo Kitajima; Luiz Alcantara; Luiz Carlos Junior de Alcantara; Luiz Lehmann Coutinho; Maria Carolina Elias; Marta Giovanetti; Maurício Lacerda Nogueira; Patricia Akemi Assato; Rafael dos Santos Bezerra; Raquel de Lello Rocha Campos Cassano. NGS Soluções Genômicas: Pilar Drummond Sampaio Corrêa Mariani. FZEA-USP Pirassununga: Mirele Daiana Poleti; Raul Machado Neto; Rejane Maria Tommasini Grotto; Ricardo Augusto Brassaloti; Ricardo Haddad; Rodrigo Tocantins Calado.; Sandra Coccuzzo Sampaio; Sandra Coccuzzo Sampaio Vessoni; Simone Kashima; Svetoslav Nanev Slavov; Vagner Fonseca; Vincent Louis Viala |
| EPI_ISL_1795277, EPI_ISL_1795278, EPI_ISL_1795279, EPI_ISL_1795280, EPI_ISL_1795281, EPI_ISL_1795282, EPI_ISL_1795283, EPI_ISL_1795284, EPI_ISL_1795285, EPI_ISL_1795286, EPI_ISL_2345515, EPI_ISL_2345516, EPI_ISL_2345517, EPI_ISL_2345518, EPI_ISL_2345519, EPI_ISL_2345520, EPI_ISL_2345521, EPI_ISL_2345522, EPI_ISL_2345523, EPI_ISL_2345524 | | | |
| see above | CS DE PALESTINA | Instituto Butantan / ESALQ-Piracicaba | Antonio Jorge Martins; Bianca Cechetto Carlos. Mendelics: Bibiana Santos; Claudia Renata dos Santos Barros; David Schlesinger; David Schlesinger. Hemocentro Ribeirão Preto: Simone Kashima; Debora Botequio Moretti; Debora Botequio Moretti. Centro de Genômica Funcional da ESALQ: Luiz Lehmann Coutinho; Dimas Tadeu Covas; Elaine Cristina Marqueze; Elaine Vieira Santos; Elaine Vieira dos Santos; Elisangela Chicaroni Mattos; Erika Freitas; Evandra Strazza Rodrigues; Felipe Allan da Silva da Costa; Flavia Aburjaile; Guilherme Targino Valente; Heidge Fukumasu; Heidge Fukumasu. USP-Botucatu: Rejane Maria Tommasini Grotto; Instituto Butantan: Alexander Roberto Precioso; Jayme A. Souza-Neto; Jayme Augusto de Souza-Neto; Jessika Cristina Chagas Lesbon; José Salvatore Leister Patané; João Paulo Kitajima; Luiz Alcantara; Luiz Carlos Junior de Alcantara; Luiz Lehmann Coutinho; Maria Carolina Elias; Marta Giovanetti; Maurício Lacerda Nogueira; Patricia Akemi Assato; Rafael dos Santos Bezerra; Raquel de Lello Rocha Campos Cassano. NGS Soluções Genômicas: Pilar Drummond Sampaio Corrêa Mariani. FZEA-USP Pirassununga: Mirele Daiana Poleti; Raul Machado Neto; Rejane Maria Tommasini Grotto; Ricardo Augusto Brassaloti; Ricardo Haddad; Rodrigo Tocantins Calado.; Sandra Coccuzzo Sampaio; Sandra Coccuzzo Sampaio Vessoni; Simone Kashima; Svetoslav Nanev Slavov; Vagner Fonseca; Vincent Louis Viala |
| EPI_ISL_1795264, EPI_ISL_1795271, EPI_ISL_2345498, EPI_ISL_2345506 | CS DE PARANAPUA | Instituto Butantan / ESALQ-Piracicaba | Antonio Jorge Martins; Bianca Cechetto Carlos. Mendelics: Bibiana Santos; Claudia Renata dos Santos Barros; David Schlesinger; David Schlesinger. Hemocentro Ribeirão Preto: Simone Kashima; Debora Botequio Moretti; Debora Botequio Moretti. Centro de Genômica Funcional da ESALQ: Luiz Lehmann Coutinho; Dimas Tadeu Covas; Elaine Cristina Marqueze; Elaine Vieira Santos; Elaine Vieira dos Santos; Elisangela Chicaroni Mattos; Erika Freitas; Evandra Strazza Rodrigues; Felipe Allan da Silva da Costa; Flavia Aburjaile; Guilherme Targino Valente; Heidge Fukumasu; Heidge Fukumasu. USP-Botucatu: Rejane Maria Tommasini Grotto; Instituto Butantan: Alexander Roberto Precioso; Jayme A. Souza-Neto; Jayme Augusto de Souza-Neto; Jessika Cristina Chagas Lesbon; José Salvatore Leister Patané; João Paulo Kitajima; Luiz Alcantara; Luiz Carlos Junior de Alcantara; Luiz Lehmann Coutinho; Maria Carolina Elias; Marta Giovanetti; Maurício Lacerda Nogueira; Patricia Akemi Assato; Rafael dos Santos Bezerra; Raquel de Lello Rocha Campos Cassano. NGS Soluções Genômicas: Pilar Drummond Sampaio Corrêa Mariani. FZEA-USP Pirassununga: Mirele Daiana Poleti; Raul Machado Neto; Rejane Maria Tommasini Grotto; Ricardo Augusto |

| | | | |
|---|---|---|---|
| | | | Brassaloti; Ricardo Haddad; Rodrigo Tocantins Calado.; Sandra Coccuzzo Sampaio; Sandra Coccuzzo Sampaio Vessoni; Simone Kashima; Svetoslav Nanev Slavov; Vagner Fonseca; Vincent Louis Viala |
| EPI_ISL_1795258, EPI_ISL_2345492 | CS DE PLANALTO | Instituto Butantan / ESALQ-Piracicaba | Antonio Jorge Martins; Bianca Cechetto Carlos. Mendelics: Bibiana Santos; Claudia Renata dos Santos Barros; David Schlesinger; David Schlesinger. Hemocentro Ribeirão Preto: Simone Kashima; Debora Botequio Moretti; Debora Botequio Moretti. Centro de Genômica Funcional da ESALQ: Luiz Lehmann Coutinho; Dimas Tadeu Covas; Elaine Cristina Marqueze; Elaine Vieira Santos; Elaine Vieira dos Santos; Elisangela Chicaroni Mattos; Erika Freitas; Evandra Strazza Rodrigues; Felipe Allan da Silva da Costa; Flavia Aburjaile; Guilherme Targino Valente; Heidge Fukumasu; Heidge Fukumasu. USP-Botucatu: Rejane Maria Tommasini Grotto; Instituto Butantan: Alexander Roberto Precioso; Jayme A. Souza-Neto; Jayme Augusto de Souza-Neto; Jessika Cristina Chagas Lesbon; José Salvatore Leister Patané; João Paulo Kitajima; Luiz Alcantara; Luiz Carlos Junior de Alcantara; Luiz Lehmann Coutinho; Maria Carolina Elias; Marta Giovanetti; Maurício Lacerda Nogueira; Patricia Akemi Assato; Rafael dos Santos Bezerra; Raquel de Lello Rocha Campos Cassano. NGS Soluções Genômicas: Pilar Drummond Sampaio Corrêa Mariani. FZEA-USP Pirassununga: Mirele Daiana Poleti; Raul Machado Neto; Rejane Maria Tommasini Grotto; Ricardo Augusto Brassaloti; Ricardo Haddad; Rodrigo Tocantins Calado.; Sandra Coccuzzo Sampaio; Sandra Coccuzzo Sampaio Vessoni; Simone Kashima; Svetoslav Nanev Slavov; Vagner Fonseca; Vincent Louis Viala |
| EPI_ISL_1795240, EPI_ISL_2345472 | CS DE SEBASTIANOPOLIS DO SUL | Instituto Butantan / ESALQ-Piracicaba | Antonio Jorge Martins; Bianca Cechetto Carlos. Mendelics: Bibiana Santos; Claudia Renata dos Santos Barros; David Schlesinger; David Schlesinger. Hemocentro Ribeirão Preto: Simone Kashima; Debora Botequio Moretti; Debora Botequio Moretti. Centro de Genômica Funcional da ESALQ: Luiz Lehmann Coutinho; Dimas Tadeu Covas; Elaine Cristina Marqueze; Elaine Vieira Santos; Elaine Vieira dos Santos; Elisangela Chicaroni Mattos; Erika Freitas; Evandra Strazza Rodrigues; Felipe Allan da Silva da Costa; Flavia Aburjaile; Guilherme Targino Valente; Heidge Fukumasu; Heidge Fukumasu. USP-Botucatu: Rejane Maria Tommasini Grotto; Instituto Butantan: Alexander Roberto Precioso; Jayme A. Souza-Neto; Jayme Augusto de Souza-Neto; Jessika Cristina Chagas Lesbon; José Salvatore Leister Patané; João Paulo Kitajima; Luiz Alcantara; Luiz Carlos Junior de Alcantara; Luiz Lehmann Coutinho; Maria Carolina Elias; Marta Giovanetti; Maurício Lacerda Nogueira; Patricia Akemi Assato; Rafael dos Santos Bezerra; Raquel de Lello Rocha Campos Cassano. NGS Soluções Genômicas: Pilar Drummond Sampaio Corrêa Mariani. FZEA-USP Pirassununga: Mirele Daiana Poleti; Raul Machado Neto; Rejane Maria Tommasini Grotto; Ricardo Augusto Brassaloti; Ricardo Haddad; Rodrigo Tocantins Calado.; Sandra Coccuzzo Sampaio; Sandra Coccuzzo Sampaio Vessoni; Simone Kashima; Svetoslav Nanev Slavov; Vagner Fonseca; Vincent Louis Viala |
| EPI_ISL_1795232, EPI_ISL_1795239, EPI_ISL_1795242, EPI_ISL_1795243, EPI_ISL_1795244, EPI_ISL_2345464, EPI_ISL_2345471, EPI_ISL_2345474, EPI_ISL_2345476, EPI_ISL_2345477 | | | |
| see above | CS DE URUPES | Instituto Butantan / ESALQ-Piracicaba | Antonio Jorge Martins; Bianca Cechetto Carlos. Mendelics: Bibiana Santos; Claudia Renata dos Santos Barros; David Schlesinger; David Schlesinger. Hemocentro Ribeirão Preto: Simone Kashima; Debora Botequio Moretti; Debora Botequio Moretti. Centro de Genômica Funcional da ESALQ: Luiz Lehmann Coutinho; Dimas Tadeu Covas; Elaine Cristina Marqueze; Elaine Vieira Santos; Elaine Vieira dos Santos; Elisangela Chicaroni Mattos; Erika Freitas; Evandra Strazza Rodrigues; Felipe Allan da Silva da Costa; Flavia Aburjaile; Guilherme Targino Valente; Heidge Fukumasu; Heidge Fukumasu. USP-Botucatu: Rejane Maria Tommasini Grotto; Instituto Butantan: Alexander Roberto Precioso; Jayme A. Souza-Neto; Jayme Augusto de Souza-Neto; Jessika Cristina Chagas Lesbon; José Salvatore Leister Patané; João Paulo Kitajima; Luiz Alcantara; Luiz Carlos Junior de Alcantara; Luiz Lehmann Coutinho; Maria Carolina Elias; Marta Giovanetti; Maurício Lacerda Nogueira; Patricia Akemi Assato; Rafael dos Santos Bezerra; Raquel de Lello Rocha Campos Cassano. NGS Soluções Genômicas: Pilar Drummond Sampaio Corrêa Mariani. FZEA-USP Pirassununga: Mirele Daiana Poleti; Raul Machado Neto; Rejane Maria Tommasini Grotto; Ricardo Augusto Brassaloti; Ricardo Haddad; Rodrigo Tocantins Calado.; Sandra Coccuzzo Sampaio; Sandra Coccuzzo Sampaio Vessoni; Simone Kashima; Svetoslav Nanev Slavov; Vagner Fonseca; Vincent Louis Viala |
| EPI_ISL_1795161, EPI_ISL_2345372 | CS III VILA ODILON | Instituto Butantan / ESALQ-Piracicaba | Antonio Jorge Martins; Bianca Cechetto Carlos. Mendelics: Bibiana Santos; Claudia Renata dos Santos Barros; David Schlesinger; David Schlesinger. Hemocentro Ribeirão Preto: Simone Kashima; Debora Botequio Moretti; Debora Botequio Moretti. Centro de Genômica Funcional da ESALQ: Luiz Lehmann Coutinho; Dimas Tadeu Covas; Elaine Cristina Marqueze; Elaine Vieira Santos; Elaine Vieira dos Santos; Elisangela Chicaroni Mattos; Erika Freitas; Evandra Strazza Rodrigues; Felipe Allan da Silva da Costa; Flavia Aburjaile; Guilherme Targino Valente; Heidge Fukumasu; Heidge Fukumasu. USP-Botucatu: Rejane Maria Tommasini Grotto; Instituto Butantan: Alexander Roberto Precioso; Jayme A. Souza-Neto; Jayme Augusto de Souza-Neto; Jessika Cristina Chagas Lesbon; José Salvatore Leister Patané; João Paulo Kitajima; Luiz Alcantara; Luiz Carlos Junior de Alcantara; Luiz Lehmann Coutinho; Maria Carolina Elias; Marta Giovanetti; Maurício Lacerda Nogueira; Patricia Akemi Assato; Rafael dos Santos Bezerra; Raquel de Lello Rocha Campos Cassano. NGS Soluções Genômicas: Pilar Drummond Sampaio Corrêa Mariani. FZEA-USP Pirassununga: Mirele Daiana Poleti; Raul Machado Neto; Rejane Maria Tommasini Grotto; Ricardo Augusto Brassaloti; Ricardo Haddad; Rodrigo Tocantins Calado.; Sandra Coccuzzo Sampaio; Sandra Coccuzzo Sampaio Vessoni; Simone Kashima; Svetoslav Nanev Slavov; Vagner Fonseca; Vincent Louis Viala |
| EPI_ISL_1715142 | CS III de Patrocinio Paulista | Instituto Adolfo Lutz, Interdiciplinary Procedures Center, Strategic Laboratory | Caio Vinicius Dias Lopes; Claudia Regina Gonçalves; Claudio Tavares Sacchi; Erica Valessa Ramos Gomes; Karoline Rodrigues Campos; Katia Correa de Oliveira Santos; Leonardo Jose Tadeu de Araujo |
| EPI_ISL_5530099 | CSF CARACARA | Analytical Competence Molecular Epidemiology Lab/ACME, Oswaldo Cruz Foundation, Ceara (FIOCRUZ CE) | Carlos Leonardo de Aragao Araujo; Cecilia Leite Costa & Eduardo Ruback dos Santos on behalf of COVID-19 FIOCRUZ Genomic Network; Cleber Furtado Aksenen; Fabio Miyajima; Fernando Braga Stehling; Francisco Eder de Moura Lopes; Igor Oliveira Duarte; Jamille Maria Mendes Bezerra; Joaquim Cesar do Nascimento Sousa Junior; Pedro Miguel Carneiro Jeronimo; Suzana Porto Almeida; Thais Ferreira de Oliveira; Thais de Oliveira Costa; Ticiane Cavalcante de Souza; Veridiana Pessoa Miyajima |
| EPI_ISL_5530100 | CSF TAPERUABA | Analytical Competence Molecular Epidemiology Lab/ACME, Oswaldo Cruz Foundation, Ceara (FIOCRUZ CE) | Carlos Leonardo de Aragao Araujo; Cecilia Leite Costa & Eduardo Ruback dos Santos on behalf of COVID-19 FIOCRUZ Genomic Network; Cleber Furtado Aksenen; Fabio Miyajima; Fernando Braga Stehling; Francisco Eder de Moura Lopes; Igor Oliveira Duarte; Jamille Maria Mendes Bezerra; Joaquim Cesar do Nascimento Sousa Junior; Pedro Miguel Carneiro Jeronimo; Suzana Porto Almeida; Thais Ferreira de Oliveira; Thais de Oliveira Costa; Ticiane Cavalcante de Souza; Veridiana Pessoa Miyajima |
| EPI_ISL_2036752 | CT Department of Public Health | CT Department of Public Health | Claire_Pearson; Tu_N_Nguyen |
| EPI_ISL_2697914, EPI_ISL_2698024, EPI_ISL_2698025, EPI_ISL_2698054, EPI_ISL_2698060 | CTvacinas | CTvacinas | A.P.; B.L.; Coelho; D.B.; Dorlass; Durigon; E.G.; E.L.; F.G.; Fernandes; Fiorini, A.; Fonseca; G.P.; H.P.; K.L.; L.M.; Lourenco; Magalhaes; Oliveira; Ometto, T.; Peixoto, R.; R.D.; Sato, H.; Scagion; Teixeira, S.; Telezynski; Thomazelli |
| EPI_ISL_2778688 | California Department of Public Health Valencia Branch Laboratory (CDPH VBL) | California Department of Public Health | CDPH-COVIDNet; UCLA Technology Center for Genomics & Bioinformatics |
| EPI_ISL_1760267 | Centers for Disease Control and Prevention, Dengue Branch | Centers for Disease Control and Prevention, Dengue Branch | Betzabel Flores; Gabriela Paz-Bailey; Gilberto A. Santiago; Glenda Gonzalez; Jorge L. Munoz-Jordan; Keyla Charriez |
| EPI_ISL_2502410 | Central Laboratory, Bureau of Public Health (BOG) and Academic Hospital Paramaribo | Erasmus Medical Center | Bas B Oude Munnink; Cherise Beek; Consuella Partowidjojo; Dion Gajadin; Ed PF IJzerman; Emmanuelle Munger; Gary Gummels; Ingrid SK Krishnadath; Lycke Woittiez; Marion PG Koopmans; Mireille Van de Veer; Phyllis Pinas; Princes Wongsowidjojo; Radjesh Ori; Ranisha Doerbalie; Rohma Banwari; Soeradj Harkisoen; Stephen Vreden; Tilotmadebie Ramlal; Verne Nanhoe |
| EPI_ISL_3255187, EPI_ISL_3255193, EPI_ISL_3266095, EPI_ISL_3266099 | Central Public Health Laboratory - LACEN - Bahia, Salvador, Brazil | Central Public Health Laboratory - LACEN -Bahia, Salvador, Brazil | Arabela Leal; Breno Dominguez; Felicidade Pereira; Jaqueline Gomes; Luciana Oliveira; Luiz Alcantara; Marcela Gómez; Marta Giovanetti; Patrícia Cajado; Stephane Tosta; Vagner Fonseca; Vanessa Nardy |
| EPI_ISL_1706506 | Centre Hospitalier Universitaire de Rouen Laboratoire de Virologie | Centre Hospitalier Universitaire de Rouen Laboratoire de Virologie | Alice Moisan; Fabienne De Oliveira; Marie Leoz |
| EPI_ISL_5782665, EPI_ISL_5782666, EPI_ISL_5782676, EPI_ISL_5782677, EPI_ISL_5782678, EPI_ISL_5799801 | Centro De Especialidds Medicas Irma Leopoldina Pirassununga | Instituto Butantan | Antonio Jorge Martins; Claudia Renata dos Santos Barros; David Schlesinger; Debora Botequio Moretti; Dimas Tadeu Covas; Elaine Cristina Marqueze; Elaine Vieira Santos; Evandra Strazza Rodrigues; Heidge Fukumasu; Jayme Augusto de Souza-Neto; José Salvatore Leister Patané; Luiz Alcantara; Luiz Lehmann Coutinho; Maria Carolina Elias; Maurício Lacerda Nogueira; Rafael dos Santos Bezerra; Raul Machado Neto; Rejane Maria Tommasini Grotto; Ricardo Haddad; Sandra Coccuzzo Sampaio Vessoni; Simone Kashima; Svetoslav Nanev Slavov; Vincent Louis Viala |
| EPI_ISL_5802003 | Centro De Saude Iii Tabatinga | Instituto Butantan | Antonio Jorge Martins; Claudia Renata dos Santos Barros; David Schlesinger; Debora Botequio Moretti; Dimas Tadeu Covas; Elaine Cristina Marqueze; Elaine Vieira Santos; Evandra Strazza Rodrigues; Heidge Fukumasu; Jayme Augusto de Souza-Neto; José Salvatore Leister Patané; Luiz Alcantara; Luiz Lehmann Coutinho; Maria Carolina Elias; Maurício Lacerda Nogueira; Rafael dos Santos Bezerra; Raul Machado Neto; Rejane Maria Tommasini Grotto; Ricardo Haddad; Sandra Coccuzzo Sampaio Vessoni; Simone Kashima; Svetoslav Nanev Slavov; Vincent Louis Viala |
| EPI_ISL_5799787, EPI_ISL_5799789, EPI_ISL_5799791 | Centro Integrado De Saude | Instituto Butantan | Antonio Jorge Martins; Claudia Renata dos Santos Barros; David Schlesinger; Debora Botequio Moretti; Dimas Tadeu Covas; Elaine Cristina Marqueze; Elaine Vieira Santos; Evandra Strazza Rodrigues; Heidge Fukumasu; Jayme Augusto de Souza-Neto; José Salvatore Leister Patané; Luiz Alcantara; Luiz Lehmann Coutinho; Maria Carolina Elias; Maurício Lacerda Nogueira; Rafael dos Santos Bezerra; Raul Machado Neto; Rejane Maria Tommasini Grotto; Ricardo Haddad; Sandra Coccuzzo Sampaio Vessoni; Simone Kashima; Svetoslav Nanev Slavov; Vincent Louis Viala |
| EPI_ISL_2274030, EPI_ISL_2274031, EPI_ISL_2274032, EPI_ISL_2274034, EPI_ISL_2274035, EPI_ISL_2274037, EPI_ISL_2274039 | | | |
| see above | Centro Nacional de Enfermidades Tropicales (CENETROP) | Laboratory of Respiratory Viruses and Measles, Oswaldo Cruz Institute, FIOCRUZ | Alice Sampaio Rocha; Ana Carolina Mendonca; Anna Carolina Paixao; Cinthia Avila; Elisa Cavalcante Pereira; Fernando Motta; Luciana Appolinario; Marilda Siqueira on behalf of the Fiocruz COVID-19 Genomic Surveillance Network; Paola Resende; Renata Serrano Lopes; Roxana Loayza; Taina Venas |
| EPI_ISL_2612335, EPI_ISL_2612379, EPI_ISL_2612380, EPI_ISL_2612381, EPI_ISL_2612383, EPI_ISL_2612384, EPI_ISL_2612385, EPI_ISL_2612386 | | | |
| see above | Centro de Infectologia Charles Mérieux/ Laboratório Rodolphe Mérieux, FUNDHACRE | Bioinformatics Laboratory / LNCC | Alessandra P Lamarca; Alexandra L Gerber; Ana Paula de C Guimarães; Ana Tereza R Vasconcelos; Andreas Stocker; Cirley Maria de Oliveira Lobato; Douglas Terra Machado; Luiz Fellype Alves de Souza; Luiz G P de Almeida; Ronaldo da Silva F Jr |
| EPI_ISL_2031732, EPI_ISL_2031733, EPI_ISL_2031759, EPI_ISL_2031760, EPI_ISL_2031761, EPI_ISL_2031762 | Centro de Innovación en Vigilancia Epidemiológica (CiVE), Institut Pasteur Montevideo, Uruguay | Centro de Innovación en Vigilancia Epidemiológica (CiVE), Institut Pasteur Montevideo, Uruguay | Alicia Costábile; Alvaro Fajardo; Andrés Lizosaín; Belén González; Bernardina Rivera; Cecilia Alonso; Cecilia Salazar; Gonzalo Moratorio; Gregorio Iraola; Henry Albornoz; Ignacio Ferrés; Inés Bellini; Juan Zanetti; Julio Medina; Lucia Bilbao; Luciana Griffero; Lucía Spangenberg; Ma Noel Bentancor; Ma Pía Techera; Mailen Arleo; Martina Alonso; María José Benítez; Matías Maidana; Mauricio Méndez; Melissa Duquía; Mercedes Paz; Natalia Rego; Natalia Reyes; Odhille Chappos; Paula Perbolianachis; Pilar Moreno; Rodney Colina; Rodrigo Arce; Tamara Fernández; Tania Possi |
| EPI_ISL_1752545, EPI_ISL_1752551 | Centro de Investigación Biomédica de La Rioja - Hospital San Pedro Logroño | SeqCOVID-SPAIN consortium/IBV(CSIC) | José Manuel Azcona Gutiérrez; María Pilar Bea Escudero; María de Toro; Miriam Blasco Alberdi and SeqCOVID-SPAIN consortium |
| EPI_ISL_1931618 | Chiba Prefectural Institute of Public Health | Pathogen Genomics Center, National Institute of Infectious Diseases | Hazuka Y Furihata; Kentaro Itokawa; Makoto Kuroda; Masanori Hashino; Masumichi Saito; Naomi Nojiri; Nozomu Hanaoka; Rina Tanaka; Sana Uchikoba; Tsuguto Fujimoto; Tsuyoshi Sekizuka |
| EPI_ISL_4498198 | Children's Hospital of Philadelphia | Planet | Ahmed M. Moustafa; Alex Arvanitis; Azad Ahmed; Brandy Neide; Colleen Bianco; Josh Chang Mell; Lidiya Denu; Paul J. Planet; Rebecca M. Harris; Susan Coffin; Swetha Rajagopal |
| EPI_ISL_1738812, EPI_ISL_1738813 | Cintramedica | Instituto Nacional de Saude (INSA) | Borges et al |
| EPI_ISL_2341524 | Contra Costa County Public Health Lab | Chan-Zuckerberg Biohub | CZB Cliahub Consortium |
| EPI_ISL_5799795 | Coordenadoria Municipal De Saude De Iracemapolis | Instituto Butantan | Antonio Jorge Martins; Claudia Renata dos Santos Barros; David Schlesinger; Debora Botequio Moretti; Dimas Tadeu Covas; Elaine Cristina Marqueze; Elaine Vieira Santos; Evandra Strazza Rodrigues; Heidge Fukumasu; Jayme Augusto de Souza-Neto; José Salvatore Leister Patané; Luiz Alcantara; Luiz Lehmann Coutinho; Maria Carolina Elias; Maurício Lacerda Nogueira; Rafael dos Santos Bezerra; Raul Machado Neto; Rejane Maria Tommasini Grotto; Ricardo Haddad; Sandra Coccuzzo Sampaio Vessoni; Simone Kashima; Svetoslav Nanev Slavov; Vincent Louis Viala |

| | | | |
|---|---|---|---|
| EPI_ISL_1805734, EPI_ISL_1805735, EPI_ISL_1805737 | County of San Diego Health and Human Services Agency | Pathogen Discovery, Respiratory Viruses Branch, Division of Viral Diseases, Centers for Disease Control and Prevention | Adam Retchless; Anna Kelleher; Anna Montmayeur; Anna Uehara; Brian Lynch; Clinton R. Paden; Haibin Wang; Han Jia Justin Ng; Jing Zhang; Justin Lee; Krista Queen; Mark Burroughs; Peter Cook; Rachel Marine; Suxiang Tong; Yan Li; Ying Tao |
| EPI_ISL_1482644, EPI_ISL_2854090, EPI_ISL_3482011 | DC Public Health Lab/ Dept. of Forensic Sciences | DC Public Health Lab/ Dept. of Forensic Sciences | Brittany Hamilton; Connie Maza; David Payne; Elizabeth Zelaya; Janis Doss; Jocelyn Hauser; Monica Mann; Sarah Scott; Scott Nguyen |
| EPI_ISL_1502478, EPI_ISL_1580506 | DIP. PREV. AVEZZANO SERVIZIO DI IGIENE EPIDEMIOLOGIA E SANITA' PUBBLICA AVEZZANO(L'AQUILA) | Istituto Zooprofilattico Sperimentale dell'Abruzzo e Molise "G. Caporale" | Ancora M; Calistri P; Cammà C; Caporale M; Curini V; Delli Compagni E; Di Domenico M; Di Lollo Valeria; Di Pasquale A; Lorusso A; Mangone I; Marcacci M; Puglia I; Rinaldi A; Savini G; Scialabba S |
| EPI_ISL_1595624 | Department of Clinical Microbiology | GIGA Medical Genomics | Bouchra Boujemla; Cécile Meex; Keith Durkin; Maria Artesi; Marie-Pierre Hayette; Nathalie Renotte; Pierrette Melin; Raphaël Boreux; Sébastien Bontems; Vincent Bours |
| EPI_ISL_1749429 | Dolomiti srl - Casa di Cura Nuova ITOR | Eurofins Genoma Group | Ettore Cotroneo (Eurofins Genoma Group); Fabrizio Carletti (I.R.C.C.S. I.N.M.I. Lazzaro Spallanzani); Francesca Spinella; Luca Musella (Dolomiti srl - Casa di Cura Nuova ITOR); Riccardo Giannico; Valentina Andrioletti |
| EPI_ISL_2145333, EPI_ISL_2145334, EPI_ISL_2145428, EPI_ISL_2145479, EPI_ISL_2145630, EPI_ISL_2145632 | Dutch COVID-19 response team | Erasmus Medical Center | Anne van der Linden; Annemiek van der Eijk; Bas Oude Munnink; Corine GeurtsvanKessel; David Nieuwenhuijse; Emmanuelle Munger; Irina Chestakova; Marion Koopmans; Marjan Boter; Reina Sikkema; Richard Molenkamp; on behalf of the Dutch national COVID-19 respo |
| EPI_ISL_1521304, EPI_ISL_1521311, EPI_ISL_1521317, EPI_ISL_1521318, EPI_ISL_1521319, EPI_ISL_1521320, EPI_ISL_1521322, EPI_ISL_1521327, EPI_ISL_1521328, EPI_ISL_1521331, EPI_ISL_1521332, EPI_ISL_1521335, EPI_ISL_1521337, EPI_ISL_1521338, EPI_ISL_1521339, EPI_ISL_1596280, EPI_ISL_1596319, EPI_ISL_1596609, EPI_ISL_1597233, EPI_ISL_1597244, EPI_ISL_1597256, EPI_ISL_1597291, EPI_ISL_1597313, EPI_ISL_1597352, EPI_ISL_1597384, EPI_ISL_1597522, EPI_ISL_1597525, EPI_ISL_1597590, EPI_ISL_1703183, EPI_ISL_1703215, EPI_ISL_1703251, EPI_ISL_1703252, EPI_ISL_1703260, EPI_ISL_1703270, EPI_ISL_1703313, EPI_ISL_1703336, EPI_ISL_1705130, EPI_ISL_1705414, EPI_ISL_1705510, EPI_ISL_1705522, EPI_ISL_1962274, EPI_ISL_2475704 | | | |
| see above | Dutch COVID-19 response team | National Institute for Public Health and the Environment (RIVM) | Adam Meijer; AnneMarie van den Brandt; Annelies Kroneman; Bas van der Veer; Chantal Reusken; Dennis Schmitz; Dirk Eggink; Euníce Then; Florian Zwagemaker; Harry Vennema; James Groot; Jeroen Cremer; Jolienke Hardeman; Karim Hajji; Kim Freriks; Linda van de Nes; Lisa Wijsman; Lynn Aarts; Melissa van Tuil; Robert Kohl; Ryanne Jaarsma; Sanne Bos; Sharon van den Brink; Sjoerd Kuiling; on behalf of the national COVID-19 response team |
| EPI_ISL_1795245, EPI_ISL_1795246, EPI_ISL_1795247, EPI_ISL_2345478, EPI_ISL_2345480, EPI_ISL_2345481 | EMERGENCIA RESPIRATORIA DE NOVA GRANADA | Instituto Butantan / ESALQ-Piracicaba | Antonio Jorge Martins; Bianca Cechetto Carlos. Mendelics: Bibiana Santos; Claudia Renata dos Santos Barros; David Schlesinger; David Schlesinger. Hemocentro Ribeirão Preto: Simone Kashima; Debora Botequio Moretti; Debora Botequio Moretti. Centro de Genômica Funcional da ESALQ: Luiz Lehmann Coutinho; Dimas Tadeu Covas; Elaine Cristina Marqueze; Elaine Vieira Santos; Elaine Vieira dos Santos; Elisangela Chicaroni Mattos; Erika Freitas; Evandra Strazza Rodrigues; Felipe Allan da Silva da Costa; Flavia Aburjaile; Guilherme Targino Valente; Heidge Fukumasu; Heidge Fukumasu. USP-Botucatu: Rejane Maria Tommasini Grotto; Instituto Butantan: Alexander Roberto Precioso; Jayme A. Souza-Neto; Jayme Augusto de Souza-Neto; Jessika Cristina Chagas Lesbon; José Salvatore Leister Patané; João Paulo Kitajima; Luiz Alcantara; Luiz Carlos Junior de Alcantara; Luiz Lehmann Coutinho; Maria Carolina Elias; Marta Giovanetti; Maurício Lacerda Nogueira; Patricia Akemi Assato; Rafael dos Santos Bezerra; Raquel de Lello Rocha Campos Cassano. NGS Soluções Genômicas: Pilar Drummond Sampaio Corrêa Mariani. FZEA-USP Pirassununga: Mirele Daiana Poleti; Raul Machado Neto; Rejane Maria Tommasini Grotto; Ricardo Augusto Brassaloti; Ricardo Haddad; Rodrigo Tocantins Calado.; Sandra Coccuzzo Sampaio; Sandra Coccuzzo Sampaio Vessoni; Simone Kashima; Svetoslav Nanev Slavov; Vagner Fonseca; Vincent Louis Viala |
| EPI_ISL_1795376, EPI_ISL_1795377, EPI_ISL_1795378, EPI_ISL_2345952, EPI_ISL_2345953, EPI_ISL_2345954 | ESF MINEIROS DO TIETE | Instituto Butantan / ESALQ-Piracicaba | Antonio Jorge Martins; Bianca Cechetto Carlos. Mendelics: Bibiana Santos; Claudia Renata dos Santos Barros; David Schlesinger; David Schlesinger. Hemocentro Ribeirão Preto: Simone Kashima; Debora Botequio Moretti; Debora Botequio Moretti. Centro de Genômica Funcional da ESALQ: Luiz Lehmann Coutinho; Dimas Tadeu Covas; Elaine Cristina Marqueze; Elaine Vieira Santos; Elaine Vieira dos Santos; Elisangela Chicaroni Mattos; Erika Freitas; Evandra Strazza Rodrigues; Felipe Allan da Silva da Costa; Flavia Aburjaile; Guilherme Targino Valente; Heidge Fukumasu; Heidge Fukumasu. USP-Botucatu: Rejane Maria Tommasini Grotto; Instituto Butantan: Alexander Roberto Precioso; Jayme A. Souza-Neto; Jayme Augusto de Souza-Neto; Jessika Cristina Chagas Lesbon; José Salvatore Leister Patané; João Paulo Kitajima; Luiz Alcantara; Luiz Carlos Junior de Alcantara; Luiz Lehmann Coutinho; Maria Carolina Elias; Marta Giovanetti; Maurício Lacerda Nogueira; Patricia Akemi Assato; Rafael dos Santos Bezerra; Raquel de Lello Rocha Campos Cassano. NGS Soluções Genômicas: Pilar Drummond Sampaio Corrêa Mariani. FZEA-USP Pirassununga: Mirele Daiana Poleti; Raul Machado Neto; Rejane Maria Tommasini Grotto; Ricardo Augusto Brassaloti; Ricardo Haddad; Rodrigo Tocantins Calado.; Sandra Coccuzzo Sampaio; Sandra Coccuzzo Sampaio Vessoni; Simone Kashima; Svetoslav Nanev Slavov; Vagner Fonseca; Vincent Louis Viala |
| EPI_ISL_1795248, EPI_ISL_1795249, EPI_ISL_1795250, EPI_ISL_2345482, EPI_ISL_2345483, EPI_ISL_2345484 | ESF NOVA TANABI II | Instituto Butantan / ESALQ-Piracicaba | Antonio Jorge Martins; Bianca Cechetto Carlos. Mendelics: Bibiana Santos; Claudia Renata dos Santos Barros; David Schlesinger; David Schlesinger. Hemocentro Ribeirão Preto: Simone Kashima; Debora Botequio Moretti; Debora Botequio Moretti. Centro de Genômica Funcional da ESALQ: Luiz Lehmann Coutinho; Dimas Tadeu Covas; Elaine Cristina Marqueze; Elaine Vieira Santos; Elaine Vieira dos Santos; Elisangela Chicaroni Mattos; Erika Freitas; Evandra Strazza Rodrigues; Felipe Allan da Silva da Costa; Flavia Aburjaile; Guilherme Targino Valente; Heidge Fukumasu; Heidge Fukumasu. USP-Botucatu: Rejane Maria Tommasini Grotto; Instituto Butantan: Alexander Roberto Precioso; Jayme A. Souza-Neto; Jayme Augusto de Souza-Neto; Jessika Cristina Chagas Lesbon; José Salvatore Leister Patané; João Paulo Kitajima; Luiz Alcantara; Luiz Carlos Junior de Alcantara; Luiz Lehmann Coutinho; Maria Carolina Elias; Marta Giovanetti; Maurício Lacerda Nogueira; Patricia Akemi Assato; Rafael dos Santos Bezerra; Raquel de Lello Rocha Campos Cassano. NGS Soluções Genômicas: Pilar Drummond Sampaio Corrêa Mariani. FZEA-USP Pirassununga: Mirele Daiana Poleti; Raul Machado Neto; Rejane Maria Tommasini Grotto; Ricardo Augusto Brassaloti; Ricardo Haddad; Rodrigo Tocantins Calado.; Sandra Coccuzzo Sampaio; Sandra Coccuzzo Sampaio Vessoni; Simone Kashima; Svetoslav Nanev Slavov; Vagner Fonseca; Vincent Louis Viala |
| EPI_ISL_1812763, EPI_ISL_3215359, EPI_ISL_3215730 | EXCITE Lab | Andersen lab at Scripps Research | Abigail Schnapper; Angela Scioscia; Cheryl Anderson; Chip Schooley; David Pride; Greg Humphrey; Helena Tubb; Natasha Martin; Natasha Martin Cheryl Anderson; Sawyer Farmer; Sharon Reed; Smruthi Karthikeyan; Tommy Valles + SEARCH |
| EPI_ISL_5802040 | Emergencia Respiratoria De Nova Granada | Instituto Butantan | Antonio Jorge Martins; Claudia Renata dos Santos Barros; David Schlesinger; Debora Botequio Moretti; Dimas Tadeu Covas; Elaine Cristina Marqueze; Elaine Vieira Santos; Evandra Strazza Rodrigues; Heidge Fukumasu; Jayme Augusto de Souza-Neto; José Salvatore Leister Patané; Luiz Alcantara; Luiz Lehmann Coutinho; Maria Carolina Elias; Maurício Lacerda Nogueira; Rafael dos Santos Bezerra; Raul Machado Neto; Rejane Maria Tommasini Grotto; Ricardo Haddad; Sandra Coccuzzo Sampaio Vessoni; Simone Kashima; Svetoslav Nanev Slavov; Vincent Louis Viala |
| EPI_ISL_5799776, EPI_ISL_5799778, EPI_ISL_5799779 | Esf Mineiros Do Tiete | Instituto Butantan | Antonio Jorge Martins; Claudia Renata dos Santos Barros; David Schlesinger; Debora Botequio Moretti; Dimas Tadeu Covas; Elaine Cristina Marqueze; Elaine Vieira Santos; Evandra Strazza Rodrigues; Heidge Fukumasu; Jayme Augusto de Souza-Neto; José Salvatore Leister Patané; Luiz Alcantara; Luiz Lehmann Coutinho; Maria Carolina Elias; Maurício Lacerda Nogueira; Rafael dos Santos Bezerra; Raul Machado Neto; Rejane Maria Tommasini Grotto; Ricardo Haddad; Sandra Coccuzzo Sampaio Vessoni; Simone Kashima; Svetoslav Nanev Slavov; Vincent Louis Viala |
| EPI_ISL_3354635 | FL Bur. of Public Health Laboratories-Jacksonville | Centers for Disease Control and Prevention Division of Viral Diseases, Pathogen Discovery | Alex Burgin; Ben Rambo-Martin; Clinton Paden; Dakota Howard; Dave Wentworth; Dhwani Batra; Jasmine Padilla; Justin Lee; Krista Queen; Kristen Knipe; Kristine Lacek; Mark Burroughs; Matthew Schmerer; Meghan Bentz; Mili Sheth; Peter Cook; Sam Shepard; Sarah Nobles; Suxiang Tong; Vivien Dugan; Yvette Unoarumhi |
| EPI_ISL_3354607 | FL Bureau of Public Health Laboratories-Miami | Centers for Disease Control and Prevention Division of Viral Diseases, Pathogen Discovery | Alex Burgin; Ben Rambo-Martin; Clinton Paden; Dakota Howard; Dave Wentworth; Dhwani Batra; Jasmine Padilla; Justin Lee; Krista Queen; Kristen Knipe; Kristine Lacek; Mark Burroughs; Matthew Schmerer; Meghan Bentz; Mili Sheth; Peter Cook; Sam Shepard; Sarah Nobles; Suxiang Tong; Vivien Dugan; Yvette Unoarumhi |
| EPI_ISL_1620639 | Fleury | Instituto Butantan / Mendelics | Alexander Roberto Precioso; Antonio Jorge Martins; Bibiana Santos; Claudia Renata dos Santos Barros; David Schlesinger; Debora Botequio Moretti; Dimas Tadeu Covas; Elaine Cristina Marqueze; Elaine Vieira dos Santos; Erika Freitas; Evandra Strazza Rodrigues; Flavia Aburjaile; José Salvatore Leister Patané; João Paulo Kitajima; Luiz Carlos Junior de Alcantara; Maria Carolina Elias; Marta Giovanetti; Rafael dos Santos Bezerra; Raul Machado Neto; Ricardo Haddad; Rodrigo Tocantins Calado.; Sandra Coccuzzo Sampaio; Simone Kashima; Svetoslav Nanev Slavov; Vagner Fonseca; Vincent Louis Viala |
| EPI_ISL_1531212, EPI_ISL_1531237, EPI_ISL_1531355, EPI_ISL_1651341, EPI_ISL_6366957, EPI_ISL_6367039 | Florida Bureau of Public Health Laboratories | Florida Bureau of Public Health Laboratories | Jason Blanton; Namratha Tarigopula; Sarah Schmedes; Tiffany Splatt |
| EPI_ISL_1578327 | Flow Health | Infectious Disease Program, Broad Institute of Harvard and MIT | Adams, G.; B.L.; B.W.; Bauer, M.; Birren; Carter, A.; Chaluvadi, S.; D.J.; DeRuff, K.; Gallagher, G.; Gladden-Young, A.; J.E.; K.J.; Lagerborg, K.; Lemieux; Loreth, C.; MacInnis; Normandin, E.; P.C.; Park; Reilly, S.; Rudy, M.; Siddle; Smole, S.; Tomkins-Tinch, C.; and Sabeti |
| EPI_ISL_2621519 | Fondazione IRCCS Ca' Granda Ospedale Maggiore Policlinico | Fondazione IRCCS Ca' Granda Ospedale Maggiore Policlinico | Ferruccio Ceriotti; Sara Uceda Renteria |
| EPI_ISL_4926619 | Fort Belvoir Community Hospital | Naval Medical Research Center Biological Defense Research Directorate | Andrea E. Luquette; Andrew J. Bennett; Britta Babel; Catherine E. Arnold; Emily Hackett; Francisco J. Malagon; Gregory K. Rice; Haven L. Miner; Kimberly A. Bishop-Lilly; Kyle A. Long; Lindsay A. Glang; Logan J. Voegtly; Raven Stone; Regina Z. Cer; Robin H. Miller; Tasheka Pearcey |
| EPI_ISL_1525813, EPI_ISL_1555587, EPI_ISL_1555591, EPI_ISL_1555604, EPI_ISL_1555606, EPI_ISL_1555955, EPI_ISL_1556080, EPI_ISL_1556087, EPI_ISL_1556095, EPI_ISL_1556101, EPI_ISL_1556115, EPI_ISL_1556441, EPI_ISL_1556448, EPI_ISL_1556452, EPI_ISL_1556457, EPI_ISL_1556486, EPI_ISL_1556646, EPI_ISL_1556647, EPI_ISL_1556671, EPI_ISL_1556672, EPI_ISL_1556682, EPI_ISL_1556720, EPI_ISL_1556729, EPI_ISL_1556737, EPI_ISL_1556744, EPI_ISL_1556758, EPI_ISL_1556762, EPI_ISL_1556787, EPI_ISL_1556802, EPI_ISL_1556805, EPI_ISL_1556836, EPI_ISL_1556859, EPI_ISL_1556892, EPI_ISL_1556907, EPI_ISL_1556918, EPI_ISL_1556919, EPI_ISL_1556921, EPI_ISL_1556924, EPI_ISL_1556932, EPI_ISL_1556935, EPI_ISL_1556937, EPI_ISL_1556944, EPI_ISL_1556947, EPI_ISL_1556969, EPI_ISL_1556993, EPI_ISL_1556998, EPI_ISL_1556999, EPI_ISL_1557000, EPI_ISL_1557003, EPI_ISL_1557030, EPI_ISL_1557033, EPI_ISL_1557050, EPI_ISL_1557060, EPI_ISL_1557066, EPI_ISL_1557076, EPI_ISL_1557095, EPI_ISL_1557096, EPI_ISL_1557146, EPI_ISL_1557203, EPI_ISL_1612874, EPI_ISL_1612881, EPI_ISL_1612906, EPI_ISL_1612914, EPI_ISL_1612915, EPI_ISL_1612916, EPI_ISL_1612924, EPI_ISL_1612932, EPI_ISL_1612943, EPI_ISL_1613112, EPI_ISL_1614238, EPI_ISL_1614255, EPI_ISL_1614354, EPI_ISL_1840157 | | | |
| see above | Fulgent Genetics | Centers for Disease Control and Prevention Division of Viral Diseases, Pathogen Discovery | Adrian Paskey; Becky Tsai; Benafsh Sapra; Benjamin Rambo-Martin; Christopher Gulvick; Clinton R. Paden; Dakota Howard; Darlene Wagner; Dhwani Batra; Doreen Ng; Duncan MacCannell; Harry Gao; James Xie; Jason Caravas; John Gao; Joseph Fierro; Kara Moser; Matthew Schmerer; Mickey Li; Peter W. Cook; Scott Sammons; Shatavia Morrison; Yan Meng; Yvette Unoarumhi |
| EPI_ISL_1678367 | GA Department of Public Health Laboratory | Centers for Disease Control and Prevention Division of Viral Diseases, Pathogen Discovery | Alison Laufer Halpin; Ben L. Rambo-Martin; Clinton R. Paden; Dakota Howard; Darlene Wagner; Dave Wentworth; Dhwani Batra; Jasmine Padilla; Justin Lee; Katie Dillon; Krista Queen; Kristen Knipe; Kristine Lacek; Mark Burroughs; Matthew Schmerer; Mili Sheth; Peter Cook; Sam Shepard; Sarah Nobles; Shoshona Le; Suxiang Tong; Vivien Dugan; Yvette Unoarumhi |
| EPI_ISL_1754726 | GH A.CHENEVIER-H.MONDOR | Department of Virology, Henri Mondor University Hospital, Assistance Publique Hôpitaux de Paris, Université Paris-Est Créteil, INSERM U955 | Alexandre Soulier; Christophe Rodriguez; Elisabeth Trawinski; Guillaume Gricourt; Jean-Michel Pawlotsky; Melissa N'Debi; Slim Fourati; Vanessa Demontant |
| EPI_ISL_1755627 | GH de l'Est Francilien | Department of Virology, Henri Mondor University Hospital, Assistance Publique Hôpitaux de Paris, Université Paris-Est Créteil, INSERM U955 | Alexandre Soulier; Christophe Rodriguez; Elisabeth Trawinski; Guillaume Gricourt; Jean-Michel Pawlotsky; Melissa N'Debi; Slim Fourati; Vanessa Demontant |
| EPI_ISL_1755025 | GROUPE HOSPITALIER SUD ILE DE FRANCE | Department of Virology, Henri Mondor University Hospital, Assistance Publique Hôpitaux de Paris, Université Paris-Est Créteil, INSERM U955 | Alexandre Soulier; Christophe Rodriguez; Elisabeth Trawinski; Guillaume Gricourt; Jean-Michel Pawlotsky; Melissa N'Debi; Slim Fourati; Vanessa Demontant |
| EPI_ISL_1626613, EPI_ISL_1626615 | Gencore - Universidad de los Andes | Gencore - Universidad de los Andes | Ana Maria Palacio; Cristian Barrera; David Gonzalez; Erica Salguero; Gabriela Ariza; Luisa Sacristan; Marcela Guevara; Silvia Restrepo |

| | | | |
|---|---|---|---|
| EPI_ISL_1470451, EPI_ISL_1470454, EPI_ISL_1470463, EPI_ISL_1470465, EPI_ISL_1470477, EPI_ISL_1470522, EPI_ISL_1470523, EPI_ISL_1470542, EPI_ISL_1470550, EPI_ISL_1534575, EPI_ISL_2009227 | | | |
| see above | Genetica Molecular and Subdepartamento de Virologia ISP Chile | Instituto de Salud Publica de Chile | Andres Castillo; Barbara Parra; Gisselle Barra; Jaime Lagos; Javier Tognarelli; Jorge Fernandez; Karen Orostica; Loredana Arata; Patricia Bustos; Rodrigo Fasce; Soledad Ulloa |
| EPI_ISL_4744597, EPI_ISL_4744608 | Gorgas Memorial Laboratory of Health Studies | Gorgas Memorial Laboratory of Health Studies | Castillo Jorge; Chen Maria; Franco Danilo; Gonzalez Claudia; Jessica Gondola; Leyda Abrego; Lopez-Verges Sandra; Marlenne Castillo; Martinez Alexander; Menacho Abdiel; Moreno Ambar; Moreno Brechla; Oris Chavarria; Ortiz Alma; Salazar Jacqueline |
| EPI_ISL_3102368, EPI_ISL_3102402 | HEMOCE CENTRO DE HEMATOLOGIA E HEMOTERAPIA DO CEARA | Analytical Competence Molecular Epidemiology Lab/ACME, Oswaldo Cruz Foundation, Ceara (FIOCRUZ CE) | Cleber Furtado Aksenen; Fabio Miyajima; Fernando Braga Stehling; Francisco Eder de Moura Lopes; Jamille Maria Mendes Bezerra; Joaquim César do Nascimento Sousa Junior; Pedro Miguel Carneiro Jeronimo; Suzana Porto Almeida e Lucas Delerino; Thais Ferreira de Oliveira; Thais de Oliveira Costa; Ticiane Cavalcante de Souza; Veridiana Pessoa Miyajima |
| EPI_ISL_3102380, EPI_ISL_3102388 | HGCC HOSPITAL GERAL DR CESAR CALS | Analytical Competence Molecular Epidemiology Lab/ACME, Oswaldo Cruz Foundation, Ceara (FIOCRUZ CE) | Cleber Furtado Aksenen; Fabio Miyajima; Fernando Braga Stehling; Francisco Eder de Moura Lopes; Jamille Maria Mendes Bezerra; Joaquim César do Nascimento Sousa Junior; Pedro Miguel Carneiro Jeronimo; Suzana Porto Almeida e Lucas Delerino; Thais Ferreira de Oliveira; Thais de Oliveira Costa; Ticiane Cavalcante de Souza; Veridiana Pessoa Miyajima |
| EPI_ISL_5530021, EPI_ISL_5530023, EPI_ISL_5530024, EPI_ISL_5530025 | HGF HOSPITAL GERAL DE FORTALEZA | Analytical Competence Molecular Epidemiology Lab/ACME, Oswaldo Cruz Foundation, Ceara (FIOCRUZ CE) | Carlos Leonardo de Aragao Araujo; Cecilia Leite Costa & Eduardo Ruback dos Santos on behalf of COVID-19 FIOCRUZ Genomic Network; Cleber Furtado Aksenen; Fabio Miyajima; Fernando Braga Stehling; Francisco Eder de Moura Lopes; Igor Oliveira Duarte; Jamille Maria Mendes Bezerra; Joaquim Cesar do Nascimento Sousa Junior; Pedro Miguel Carneiro Jeronimo; Suzana Porto Almeida; Thais Ferreira de Oliveira; Thais de Oliveira Costa; Ticiane Cavalcante de Souza; Veridiana Pessoa Miyajima |
| EPI_ISL_2017412, EPI_ISL_2017413, EPI_ISL_2017414, EPI_ISL_2017415, EPI_ISL_2017416, EPI_ISL_2017417, EPI_ISL_2017418, EPI_ISL_2017419, EPI_ISL_2017420, EPI_ISL_2017421, EPI_ISL_2017423, EPI_ISL_2017424, EPI_ISL_2017425, EPI_ISL_2017426, EPI_ISL_2017427, EPI_ISL_2017428, EPI_ISL_2017429, EPI_ISL_2017452, EPI_ISL_2017472, EPI_ISL_2017476, EPI_ISL_2017483, EPI_ISL_2102519, EPI_ISL_2222898, EPI_ISL_2348603, EPI_ISL_2466445 | | | |
| see above | HLAGYN - Laboratorio de Imunologia de Transplantes de Goias | HLAGYN - Laboratorio de Imunologia de Transplantes de Goias | Alessandro Leonardo Alvares Magalhaes; Daniel Ferreira de Sousa; Danielle de Paiva Rezende; Erika Lopes Rocha Batista; Fernando Antonio Vinhal dos Santos; Frederico Rodrigues Vinhal; Lucas Carlos Gomes Pereira; Paola Cristina Resende Silva; Sabrina Sara Moreira Duarte |
| EPI_ISL_3102415, EPI_ISL_3102418, EPI_ISL_3102452 | HM HOSPITAL DE MESSEJANA DR CARLOS ALBERTO STUDART GOMES | Analytical Competence Molecular Epidemiology Lab/ACME, Oswaldo Cruz Foundation, Ceara (FIOCRUZ CE) | Cleber Furtado Aksenen; Fabio Miyajima; Fernando Braga Stehling; Francisco Eder de Moura Lopes; Jamille Maria Mendes Bezerra; Joaquim César do Nascimento Sousa Junior; Pedro Miguel Carneiro Jeronimo; Suzana Porto Almeida e Lucas Delerino; Thais Ferreira de Oliveira; Thais de Oliveira Costa; Ticiane Cavalcante de Souza; Veridiana Pessoa Miyajima |
| EPI_ISL_5529888, EPI_ISL_5530138 | HOSP GERAL LUIZA ALCANTARA SILVA | Analytical Competence Molecular Epidemiology Lab/ACME, Oswaldo Cruz Foundation, Ceara (FIOCRUZ CE) | Carlos Leonardo de Aragao Araujo; Cecilia Leite Costa & Eduardo Ruback dos Santos on behalf of COVID-19 FIOCRUZ Genomic Network; Cleber Furtado Aksenen; Fabio Miyajima; Fernando Braga Stehling; Francisco Eder de Moura Lopes; Igor Oliveira Duarte; Jamille Maria Mendes Bezerra; Joaquim Cesar do Nascimento Sousa Junior; Pedro Miguel Carneiro Jeronimo; Suzana Porto Almeida; Thais Ferreira de Oliveira; Thais de Oliveira Costa; Ticiane Cavalcante de Souza; Veridiana Pessoa Miyajima |
| EPI_ISL_1795345, EPI_ISL_2345612 | HOSPITAL DE CAMPANHA COVID 19 MUNICIPIO DE TAUBATE | Instituto Butantan / ESALQ-Piracicaba | Antonio Jorge Martins; Bianca Cechetto Carlos. Mendelics: Bibiana Santos; Claudia Renata dos Santos Barros; David Schlesinger; David Schlesinger. Hemocentro Ribeirão Preto: Simone Kashima; Debora Botequio Moretti; Debora Botequio Moretti. Centro de Genômica Funcional da ESALQ: Luiz Lehmann Coutinho; Dimas Tadeu Covas; Elaine Cristina Marqueze; Elaine Vieira Santos; Elaine Vieira dos Santos; Elisangela Chicaroni Mattos; Erika Freitas; Evandra Strazza Rodrigues; Felipe Allan da Silva da Costa; Flavia Aburjaile; Guilherme Targino Valente; Heidge Fukumasu; Heidge Fukumasu. USP-Botucatu: Rejane Maria Tommasini Grotto; Instituto Butantan: Alexander Roberto Precioso; Jayme A. Souza-Neto; Jayme Augusto de Souza-Neto; Jessika Cristina Chagas Lesbon; José Salvatore Leister Patané; João Paulo Kitajima; Luiz Alcantara; Luiz Carlos Junior de Alcantara; Luiz Lehmann Coutinho; Maria Carolina Elias; Marta Giovanetti; Maurício Lacerda Nogueira; Patricia Akemi Assato; Rafael dos Santos Bezerra; Raquel de Lello Rocha Campos Cassano. NGS Soluções Genômicas: Pilar Drummond Sampaio Corrêa Mariani. FZEA-USP Pirassununga: Mirele Daiana Poleti; Raul Machado Neto; Rejane Maria Tommasini Grotto; Ricardo Augusto Brassaloti; Ricardo Haddad; Rodrigo Tocantins Calado.; Sandra Coccuzzo Sampaio; Sandra Coccuzzo Sampaio Vessoni; Simone Kashima; Svetoslav Nanev Slavov; Vagner Fonseca; Vincent Louis Viala |
| EPI_ISL_3102379 | HOSPITAL E MATERNIDADE DRA ZILDA ARNS NEUMANN | Analytical Competence Molecular Epidemiology Lab/ACME, Oswaldo Cruz Foundation, Ceara (FIOCRUZ CE) | Cleber Furtado Aksenen; Fabio Miyajima; Fernando Braga Stehling; Francisco Eder de Moura Lopes; Jamille Maria Mendes Bezerra; Joaquim César do Nascimento Sousa Junior; Pedro Miguel Carneiro Jeronimo; Suzana Porto Almeida e Lucas Delerino; Thais Ferreira de Oliveira; Thais de Oliveira Costa; Ticiane Cavalcante de Souza; Veridiana Pessoa Miyajima |
| EPI_ISL_1795344, EPI_ISL_2345610 | HOSPITAL E MATERNIDADE NOSSA SENHORA DA AJUDA | Instituto Butantan / ESALQ-Piracicaba | Antonio Jorge Martins; Bianca Cechetto Carlos. Mendelics: Bibiana Santos; Claudia Renata dos Santos Barros; David Schlesinger; David Schlesinger. Hemocentro Ribeirão Preto: Simone Kashima; Debora Botequio Moretti; Debora Botequio Moretti. Centro de Genômica Funcional da ESALQ: Luiz Lehmann Coutinho; Dimas Tadeu Covas; Elaine Cristina Marqueze; Elaine Vieira Santos; Elaine Vieira dos Santos; Elisangela Chicaroni Mattos; Erika Freitas; Evandra Strazza Rodrigues; Felipe Allan da Silva da Costa; Flavia Aburjaile; Guilherme Targino Valente; Heidge Fukumasu; Heidge Fukumasu. USP-Botucatu: Rejane Maria Tommasini Grotto; Instituto Butantan: Alexander Roberto Precioso; Jayme A. Souza-Neto; Jayme Augusto de Souza-Neto; Jessika Cristina Chagas Lesbon; José Salvatore Leister Patané; João Paulo Kitajima; Luiz Alcantara; Luiz Carlos Junior de Alcantara; Luiz Lehmann Coutinho; Maria Carolina Elias; Marta Giovanetti; Maurício Lacerda Nogueira; Patricia Akemi Assato; Rafael dos Santos Bezerra; Raquel de Lello Rocha Campos Cassano. NGS Soluções Genômicas: Pilar Drummond Sampaio Corrêa Mariani. FZEA-USP Pirassununga: Mirele Daiana Poleti; Raul Machado Neto; Rejane Maria Tommasini Grotto; Ricardo Augusto Brassaloti; Ricardo Haddad; Rodrigo Tocantins Calado.; Sandra Coccuzzo Sampaio; Sandra Coccuzzo Sampaio Vessoni; Simone Kashima; Svetoslav Nanev Slavov; Vagner Fonseca; Vincent Louis Viala |
| EPI_ISL_3102529 | HOSPITAL ESTADUAL LEONARDO DA VINCI | Analytical Competence Molecular Epidemiology Lab/ACME, Oswaldo Cruz Foundation, Ceara (FIOCRUZ CE) | Cleber Furtado Aksenen; Fabio Miyajima; Fernando Braga Stehling; Francisco Eder de Moura Lopes; Jamille Maria Mendes Bezerra; Joaquim César do Nascimento Sousa Junior; Pedro Miguel Carneiro Jeronimo; Suzana Porto Almeida e Lucas Delerino; Thais Ferreira de Oliveira; Thais de Oliveira Costa; Ticiane Cavalcante de Souza; Veridiana Pessoa Miyajima |
| EPI_ISL_5530075 | HOSPITAL JOSE MARIA PHILOMENO GOMES | Analytical Competence Molecular Epidemiology Lab/ACME, Oswaldo Cruz Foundation, Ceara (FIOCRUZ CE) | Carlos Leonardo de Aragao Araujo; Cecilia Leite Costa & Eduardo Ruback dos Santos on behalf of COVID-19 FIOCRUZ Genomic Network; Cleber Furtado Aksenen; Fabio Miyajima; Fernando Braga Stehling; Francisco Eder de Moura Lopes; Igor Oliveira Duarte; Jamille Maria Mendes Bezerra; Joaquim Cesar do Nascimento Sousa Junior; Pedro Miguel Carneiro Jeronimo; Suzana Porto Almeida; Thais Ferreira de Oliveira; Thais de Oliveira Costa; Ticiane Cavalcante de Souza; Veridiana Pessoa Miyajima |
| EPI_ISL_1795380, EPI_ISL_2345957 | HOSPITAL MATERNIDADE SAO JOSE ITAPUI | Instituto Butantan / ESALQ-Piracicaba | Antonio Jorge Martins; Bianca Cechetto Carlos. Mendelics: Bibiana Santos; Claudia Renata dos Santos Barros; David Schlesinger; David Schlesinger. Hemocentro Ribeirão Preto: Simone Kashima; Debora Botequio Moretti; Debora Botequio Moretti. Centro de Genômica Funcional da ESALQ: Luiz Lehmann Coutinho; Dimas Tadeu Covas; Elaine Cristina Marqueze; Elaine Vieira Santos; Elaine Vieira dos Santos; Elisangela Chicaroni Mattos; Erika Freitas; Evandra Strazza Rodrigues; Felipe Allan da Silva da Costa; Flavia Aburjaile; Guilherme Targino Valente; Heidge Fukumasu; Heidge Fukumasu. USP-Botucatu: Rejane Maria Tommasini Grotto; Instituto Butantan: Alexander Roberto Precioso; Jayme A. Souza-Neto; Jayme Augusto de Souza-Neto; Jessika Cristina Chagas Lesbon; José Salvatore Leister Patané; João Paulo Kitajima; Luiz Alcantara; Luiz Carlos Junior de Alcantara; Luiz Lehmann Coutinho; Maria Carolina Elias; Marta Giovanetti; Maurício Lacerda Nogueira; Patricia Akemi Assato; Rafael dos Santos Bezerra; Raquel de Lello Rocha Campos Cassano. NGS Soluções Genômicas: Pilar Drummond Sampaio Corrêa Mariani. FZEA-USP Pirassununga: Mirele Daiana Poleti; Raul Machado Neto; Rejane Maria Tommasini Grotto; Ricardo Augusto Brassaloti; Ricardo Haddad; Rodrigo Tocantins Calado.; Sandra Coccuzzo Sampaio; Sandra Coccuzzo Sampaio Vessoni; Simone Kashima; Svetoslav Nanev Slavov; Vagner Fonseca; Vincent Louis Viala |
| EPI_ISL_5530086 | HOSPITAL MUNICIPAL JOAO MUNIZ | Analytical Competence Molecular Epidemiology Lab/ACME, Oswaldo Cruz Foundation, Ceara (FIOCRUZ CE) | Carlos Leonardo de Aragao Araujo; Cecilia Leite Costa & Eduardo Ruback dos Santos on behalf of COVID-19 FIOCRUZ Genomic Network; Cleber Furtado Aksenen; Fabio Miyajima; Fernando Braga Stehling; Francisco Eder de Moura Lopes; Igor Oliveira Duarte; Jamille Maria Mendes Bezerra; Joaquim Cesar do Nascimento Sousa Junior; Pedro Miguel Carneiro Jeronimo; Suzana Porto Almeida; Thais Ferreira de Oliveira; Thais de Oliveira Costa; Ticiane Cavalcante de Souza; Veridiana Pessoa Miyajima |
| EPI_ISL_1795337, EPI_ISL_1795338, EPI_ISL_1795341, EPI_ISL_2345603, EPI_ISL_2345604, EPI_ISL_2345607 | HOSPITAL MUNICIPAL REYNALDO GUERRA CAJATI | Instituto Butantan / ESALQ-Piracicaba | Antonio Jorge Martins; Bianca Cechetto Carlos. Mendelics: Bibiana Santos; Claudia Renata dos Santos Barros; David Schlesinger; David Schlesinger. Hemocentro Ribeirão Preto: Simone Kashima; Debora Botequio Moretti; Debora Botequio Moretti. Centro de Genômica Funcional da ESALQ: Luiz Lehmann Coutinho; Dimas Tadeu Covas; Elaine Cristina Marqueze; Elaine Vieira Santos; Elaine Vieira dos Santos; Elisangela Chicaroni Mattos; Erika Freitas; Evandra Strazza Rodrigues; Felipe Allan da Silva da Costa; Flavia Aburjaile; Guilherme Targino Valente; Heidge Fukumasu; Heidge Fukumasu. USP-Botucatu: Rejane Maria Tommasini Grotto; Instituto Butantan: Alexander Roberto Precioso; Jayme A. Souza-Neto; Jayme Augusto de Souza-Neto; Jessika Cristina Chagas Lesbon; José Salvatore Leister Patané; João Paulo Kitajima; Luiz Alcantara; Luiz Carlos Junior de Alcantara; Luiz Lehmann Coutinho; Maria Carolina Elias; Marta Giovanetti; Maurício Lacerda Nogueira; Patricia Akemi Assato; Rafael dos Santos Bezerra; Raquel de Lello Rocha Campos Cassano. NGS Soluções Genômicas: Pilar Drummond Sampaio Corrêa Mariani. FZEA-USP Pirassununga: Mirele Daiana Poleti; Raul Machado Neto; Rejane Maria Tommasini Grotto; Ricardo Augusto Brassaloti; Ricardo Haddad; Rodrigo Tocantins Calado.; Sandra Coccuzzo Sampaio; Sandra Coccuzzo Sampaio Vessoni; Simone Kashima; Svetoslav Nanev Slavov; Vagner Fonseca; Vincent Louis Viala |
| EPI_ISL_3102323 | HOSPITAL OTOCLINICA | Analytical Competence Molecular Epidemiology Lab/ACME, Oswaldo Cruz Foundation, Ceara (FIOCRUZ CE) | Cleber Furtado Aksenen; Fabio Miyajima; Fernando Braga Stehling; Francisco Eder de Moura Lopes; Jamille Maria Mendes Bezerra; Joaquim César do Nascimento Sousa Junior; Pedro Miguel Carneiro Jeronimo; Suzana Porto Almeida e Lucas Delerino; Thais Ferreira de Oliveira; Thais de Oliveira Costa; Ticiane Cavalcante de Souza; Veridiana Pessoa Miyajima |
| EPI_ISL_1795371, EPI_ISL_1795372, EPI_ISL_2345945, EPI_ISL_2345946 | HOSPITAL SANTA THEREZINHA BROTAS | Instituto Butantan / ESALQ-Piracicaba | Antonio Jorge Martins; Bianca Cechetto Carlos. Mendelics: Bibiana Santos; Claudia Renata dos Santos Barros; David Schlesinger; David Schlesinger. Hemocentro Ribeirão Preto: Simone Kashima; Debora Botequio Moretti; Debora Botequio Moretti. Centro de Genômica Funcional da ESALQ: Luiz Lehmann Coutinho; Dimas Tadeu Covas; Elaine Cristina Marqueze; Elaine Vieira Santos; Elaine Vieira dos Santos; Elisangela Chicaroni Mattos; Erika Freitas; Evandra Strazza Rodrigues; Felipe Allan da Silva da Costa; Flavia Aburjaile; Guilherme Targino Valente; Heidge Fukumasu; Heidge Fukumasu. USP-Botucatu: Rejane Maria Tommasini Grotto; Instituto Butantan: Alexander Roberto Precioso; Jayme A. Souza-Neto; Jayme Augusto de Souza-Neto; Jessika Cristina Chagas Lesbon; José Salvatore Leister Patané; João Paulo Kitajima; Luiz Alcantara; Luiz Carlos Junior de Alcantara; Luiz Lehmann Coutinho; Maria Carolina Elias; Marta Giovanetti; Maurício Lacerda Nogueira; Patricia Akemi Assato; Rafael dos Santos Bezerra; Raquel de Lello Rocha Campos Cassano. NGS Soluções Genômicas: Pilar Drummond Sampaio Corrêa Mariani. FZEA-USP Pirassununga: Mirele Daiana Poleti; Raul Machado Neto; Rejane Maria Tommasini Grotto; Ricardo Augusto Brassaloti; Ricardo Haddad; Rodrigo Tocantins Calado.; Sandra Coccuzzo Sampaio; Sandra Coccuzzo Sampaio Vessoni; Simone Kashima; Svetoslav Nanev Slavov; Vagner Fonseca; Vincent Louis Viala |
| EPI_ISL_5529942, EPI_ISL_5529943 | HOSPITAL SAO SEBASTIAO | Analytical Competence Molecular Epidemiology Lab/ACME, Oswaldo Cruz Foundation, Ceara (FIOCRUZ CE) | Carlos Leonardo de Aragao Araujo; Cecilia Leite Costa & Eduardo Ruback dos Santos on behalf of COVID-19 FIOCRUZ Genomic Network; Cleber Furtado Aksenen; Fabio Miyajima; Fernando Braga Stehling; Francisco Eder de Moura Lopes; Igor Oliveira Duarte; Jamille Maria Mendes Bezerra; Joaquim Cesar do Nascimento Sousa Junior; Pedro Miguel Carneiro Jeronimo; Suzana Porto Almeida; Thais Ferreira de Oliveira; Thais de Oliveira Costa; Ticiane Cavalcante de Souza; Veridiana Pessoa Miyajima |
| EPI_ISL_2150635 | HOSPITAL UNIVERSITARIO VIRGEN DE LA ARRIXACA | Instituto de Salud Carlos III | A. Monzón; F. Casas; I. Jiménez; I. MORENO PARRADO; LAURA; M. Camarero; P. Zaballos; S. Cuesta; S. Iglesias-Caballero; S. Pozo; S. Varona; Sandonís; V. Vázquez-Morón |
| EPI_ISL_2483427, EPI_ISL_2483438 | Hackensack Medical Center | New York Genome Center | Andre Corvelo; Barry Kreiswirth; David Perlin; Dayna M. Oschwald; Jose Mediavilla; Kaelea Composto; Kar Chow; Liang Chen; Marcus Cunningham; Michael Zody; Samantha Fennessey; Tom Maniatis |
| EPI_ISL_2179166 | HealthQuest Esoterics | New York City Public Health Laboratory | Jade Wang; et al. |
| EPI_ISL_1554659, EPI_ISL_1554791, EPI_ISL_1554801, EPI_ISL_1554811, EPI_ISL_1554823, EPI_ISL_1554839, EPI_ISL_1554845, EPI_ISL_1554859, EPI_ISL_1575524, EPI_ISL_1575552, EPI_ISL_1575623, EPI_ISL_1575663, EPI_ISL_1575765, EPI_ISL_1575797, EPI_ISL_1575872, EPI_ISL_1575934, EPI_ISL_1576009, EPI_ISL_1576051, EPI_ISL_1576082, EPI_ISL_1576086, EPI_ISL_1576104, EPI_ISL_1576112, EPI_ISL_1576115, EPI_ISL_1576122, EPI_ISL_1576139, EPI_ISL_1576191, EPI_ISL_1576345, EPI_ISL_1576376, EPI_ISL_1576393, EPI_ISL_1576395, EPI_ISL_1576405, EPI_ISL_1576414, EPI_ISL_1576440, EPI_ISL_1576530, EPI_ISL_1576599, EPI_ISL_1576618, EPI_ISL_1576637, EPI_ISL_1576647, EPI_ISL_1576683, EPI_ISL_1576714, EPI_ISL_1576731, EPI_ISL_1576735, EPI_ISL_1580837, EPI_ISL_1580982, EPI_ISL_1580989, EPI_ISL_1580990, EPI_ISL_1580995, EPI_ISL_1581042, EPI_ISL_1581249, EPI_ISL_1581263, EPI_ISL_1581305, EPI_ISL_1581378, EPI_ISL_1581435, EPI_ISL_1581448, EPI_ISL_1581490, EPI_ISL_1581537, EPI_ISL_1581546, EPI_ISL_1581567, EPI_ISL_1581626, EPI_ISL_1581731, EPI_ISL_1581760, EPI_ISL_1581778, EPI_ISL_1581793, EPI_ISL_1581803, EPI_ISL_1581809, EPI_ISL_1581848, EPI_ISL_1592498, EPI_ISL_1592505, EPI_ISL_1592525, EPI_ISL_1592526, EPI_ISL_1592539, EPI_ISL_1592556, EPI_ISL_1592727, EPI_ISL_1592761, EPI_ISL_1615106, EPI_ISL_1615156, EPI_ISL_1615324 | | | |
| see above | Helix/Illumina | Centers for Disease Control and Prevention Division of Viral Diseases, Pathogen Discovery | Adrian Paskey; Alexandre Bolze; Ary Ascencio; Benjamin Rambo-Martin; Brad Sickler; Charlotte Rivera-Garcia; Christine Tran; Christopher Gulvick; Clinton R. Paden; Dakota Howard; Darlene Wagner; David Becker; Dhwani Batra; Duncan MacCannell; Efren Sandoval; Eileen de Feo; Elizabeth Cirulli; Eric Allen; Geraint Levan; James Lu; Jan Antico; Jason Caravas; Jason Nguyen; Jimmy Ramirez; Jingtao Liu; Kara Moser; Kelly Schiabor Barrett; Kim Gietzen; Magnus Isaksson; Marc Laurent; Matthew Schmerer; Matthew Tolentino; Nicole L. Washington; Peter W. Cook; Phil Febbo; Ryan Cho; Scott Sammons; Shannon Wickline; Shatavia Morrison; Sherry Wang; Simon White; Tyler Cassens; William Lee; Yvette Unoarumhi |
| EPI_ISL_5771863 | Hospital Central Mendoza | Nodo de Secuenciación Tierra del Fuego - Hospital Regional Ushuaia - Centro Austral De Investigaciones Científicas - Universidad Nacional De Tierra Del Fuego | AE; CF; Ceballos; F; Gallego; Gramundi; ID; Nardi; Rojas; SG |
| EPI_ISL_1401466, EPI_ISL_1401467, EPI_ISL_1476981, EPI_ISL_1477052, EPI_ISL_1477053, EPI_ISL_1477055, EPI_ISL_1647951 | | | |
| see above | Hospital General Universitario Gregorio Marañón | Hospital General Universitario Gregorio Marañón | Cristina Rodriguez-Grande; Darío García de Viedma; Laura Pérez-Lago; Patricia Muñoz; Pedro Sola Campoy; Pilar Catalán; Sergio Buenestado Serrano |
| EPI_ISL_2363535 | Hospital Jaime Ferre - | Grupo de Genómica y Bioinformática | AF; Amadio; C; Eberhardt; Irazoqui; Isaia; JF; JM; MF; Pandolfi; Quaranta; V |

| | | | |
|---|---|---|---|
| | SAMCO Rafaela | del Instituto de Investigación de la Cadena Láctea CONICET-INTA on behalf of 'Proyecto Argentino Interinstitucional de genomica de SARS-CoV-2' (PAIS Consortium) | |
| EPI_ISL_5799794 | Hospital Maternidade Sao Jose Itapui | Instituto Butantan | Antonio Jorge Martins; Claudia Renata dos Santos Barros; David Schlesinger; Debora Botequio Moretti; Dimas Tadeu Covas; Elaine Cristina Marqueze; Elaine Vieira Santos; Evandra Strazza Rodrigues; Heidge Fukumasu; Jayme Augusto de Souza-Neto; José Salvatore Leister Patané; Luiz Alcantara; Luiz Lehmann Coutinho; Maria Carolina Elias; Maurício Lacerda Nogueira; Rafael dos Santos Bezerra; Raul Machado Neto; Rejane Maria Tommasini Grotto; Ricardo Haddad; Sandra Coccuzzo Sampaio Vessoni; Simone Kashima; Svetoslav Nanev Slavov; Vincent Louis Viala |
| EPI_ISL_1620638 | Hospital Municipal Dr. Ignacio Proença de Gouvea | Instituto Butantan / Mendelics | Alexander Roberto Precioso; Antonio Jorge Martins; Bibiana Santos; Claudia Renata dos Santos Barros; David Schlesinger; Debora Botequio Moretti; Dimas Tadeu Covas; Elaine Cristina Marqueze; Elaine Vieira dos Santos; Erika Freitas; Evandra Strazza Rodrigues; Flavia Aburjaile; José Salvatore Leister Patané; João Paulo Kitajima; Luiz Carlos Junior de Alcantara; Maria Carolina Elias; Marta Giovanetti; Rafael dos Santos Bezerra; Raul Machado Neto; Ricardo Haddad; Rodrigo Tocantins Calado.; Sandra Coccuzzo Sampaio; Simone Kashima; Svetoslav Nanev Slavov; Vagner Fonseca; Vincent Louis Viala |
| EPI_ISL_5799792, EPI_ISL_5799793 | Hospital Santa Therezinha Brotas | Instituto Butantan | Antonio Jorge Martins; Claudia Renata dos Santos Barros; David Schlesinger; Debora Botequio Moretti; Dimas Tadeu Covas; Elaine Cristina Marqueze; Elaine Vieira dos Santos; Evandra Strazza Rodrigues; Heidge Fukumasu; Jayme Augusto de Souza-Neto; José Salvatore Leister Patané; Luiz Alcantara; Luiz Lehmann Coutinho; Maria Carolina Elias; Maurício Lacerda Nogueira; Rafael dos Santos Bezerra; Raul Machado Neto; Rejane Maria Tommasini Grotto; Ricardo Haddad; Sandra Coccuzzo Sampaio Vessoni; Simone Kashima; Svetoslav Nanev Slavov; Vincent Louis Viala |
| EPI_ISL_1854462 | Hospital Universitari Vall d'Hebron - Vall d'Hebron Institut de Recerca | Hospital Universitari Vall d'Hebron - Vall d'Hebron Institut de Recerca | Andrés Antón; Ariadna Rando; Carla Castillo; Cristina Andrés; Damir Garcia-Cehic; Josep Quer; Juliana Esperalba; Maria Carmen Martin; Maria Gema Codina; Maria Piñana; Tomàs Pumarola |
| EPI_ISL_7307288 | Hospital Universitario San Ignacio | Centro de Investigaciones en Microbiología y Biotecnología-UR (CIMBIUR), Facultad de Ciencias Naturales, Universidad del Rosario, Bogotá, Colombia | Alberto Paniz-Mondolfi; Angie Ramírez; Beatriz Ariza; Camilo A. Correa-Cárdenas; Carlos Gómez-Restrepo; Claudia Cardozo-Romero; Claudia Méndez; David-Santiago Quevedo; Guido España; Hernando Díaz; Juan David Ramírez; Juliana Cuervo-Rojas; Julie Pérez; Luz H. Patiño; Manuel-Antonio Franco; Maria-Clara Duque; Marina Muñoz; Nathalia Ballesteros; Nicolas Luna; Sergio Castañeda; Zulma M. Cucunubá |
| EPI_ISL_2003938, EPI_ISL_2003966, EPI_ISL_2597518, EPI_ISL_2653024, EPI_ISL_3022639 | Hospital of the University of Pennsylvania Molecular Pathology Lab | Bushman Lab - University of Pennsylvania | Abigail Glascock; Aoife M. Roche; Arupa Ganguly; Ayannah S. Fitzgerald; Brendan Kelly; Jevon Graham-Wooten; John Everett; John K. Everett; Kyle Rodino; Layla A. Khatib; Mike Feldman; Pascha Hokama; Ronald G. Collman and Frederic Bushan; Ronald G. Collman and Frederic Bushman; Samantha A. Whiteside; Scott Sherrill-Mix; Shantan Reddy; Young Hwang |
| EPI_ISL_1479113 | Houston Health Dept. | Houston Health Dept. | Adolpho Lara; Pamela Brown; Ryker Penn; Yanlai Lai |
| EPI_ISL_2201494, EPI_ISL_2201524, EPI_ISL_2203377 | Houston Methodist Hospital | Houston Methodist Hospital | Ilya J. Finkelstein; James J. Davis; Jessica Cambric; Jimmy Gollihar; Kristina Reppond; Layne Pruitt; Madison N. Shyer; Marcus Nguyen; Matthew Ojeda Saavedra; Paul A. Christensen; Prasanti Yerramilli; Randall J. Olsen; Robert Olson; Ryan Gadd; S. Wesley Long; Sishir Subedi; and James M. Musser |
| EPI_ISL_1754996, EPI_ISL_1755133 | Hôpital Avicenne | Department of Virology, Henri Mondor University Hospital, Assistance Publique Hôpitaux de Paris, Université Paris-Est Créteil, INSERM U955 | Alexandre Soulier; Christophe Rodriguez; Elisabeth Trawinski; Guillaume Gricourt; Jean-Michel Pawlotsky; Melissa N'Debi; Slim Fourati; Vanessa Demontant |
| EPI_ISL_2614545, EPI_ISL_2614546, EPI_ISL_2614549, EPI_ISL_2614550, EPI_ISL_2614551, EPI_ISL_2614553 | IAL Presidente Prudente | Instituto Adolfo Lutz, Interdiciplinary Procedures Center, Strategic Laboratory | Caio Vinicius Dias Lopes; Claudia Regina Gonçalves; Claudio Tavares Sacchi; Erica Valessa Ramos Gomes; Karoline Rodrigues Campos; Leonardo Jose Tadeu de Araujo |
| EPI_ISL_2444804, EPI_ISL_2444805, EPI_ISL_2444808 | IICS-UNA | IICS-UNA | Adriana Valenzuela; Alejandra Rojas; Chyntia Diaz; Eva Nara; Fatima Cardozo; Florencia del Puerto; Joel Ortiz; Jonas Fernandez; Laura Franco; Laura Mendoza; Leticia Rojas; Magaly Martinez; Maria Eugenia Galeano. |
| EPI_ISL_3354527, EPI_ISL_3354530, EPI_ISL_3354532, EPI_ISL_3354536, EPI_ISL_3354545 | IL Dept. of Public Health Springfield Laboratory | Centers for Disease Control and Prevention Division of Viral Diseases, Pathogen Discovery | Alex Burgin; Ben Rambo-Martin; Clinton Paden; Dakota Howard; Dave Wentworth; Dhwani Batra; Jasmine Padilla; Justin Lee; Krista Queen; Kristen Knipe; Kristine Lacek; Mark Burroughs; Matthew Schmerer; Meghan Bentz; Mili Sheth; Peter Cook; Sam Shepard; Sarah Nobles; Suxiang Tong; Vivien Dugan; Yvette Unoarumhi |
| EPI_ISL_1639313, EPI_ISL_1639325 | IMD - Medizinisches Labor Rostock | Robert Koch Institute | |
| EPI_ISL_1563643 | IN State Department of Health Laboratory Services | IN State Department of Health Laboratory Services | Ankita Kashikar; Brian Pope; Cassandra Campion; Jamie Yeadon; Kyle Brownlee; Lixia Liu; Mark Glazier; Melissa Hindenlang |
| EPI_ISL_1524727, EPI_ISL_1524728, EPI_ISL_1524738, EPI_ISL_1524746, EPI_ISL_1524747 | INMI Lazzaro Spallanzani IRCCS | INMI Lazzaro Spallanzani IRCCS | A Di Caro; B Bartolini; CEM Gruber; E Giombini; F Messina; F Santini; G Bonfiglio; M Rueca; MR Capobianchi; O Butera |
| EPI_ISL_1589741, EPI_ISL_1589853 | INSACOG-WB | National Institute of Biomedical Genomics – INSACOG | Ajay Chakraborti; Arindam Maitra; Bhaswati Bandyopadhyay; Nidhan Kumar Biswas; Saumitra Das; Sreedhar Chinnaswamy; Tamal Ghosh |
| EPI_ISL_1670935, EPI_ISL_1670948, EPI_ISL_1670949, EPI_ISL_1670952, EPI_ISL_2576975 | IRCCS San Gallicano Dermatological Institute | IRCCS Regina Elena National Cancer Institute | Aldo Morrone; Fabrizio Ensoli; Francesca De Nicola; Fulvia Pimpinelli; Gennaro Ciliberto; Giovanni Blandino; Grazia Prignano; Ludovica Ciuffreda; Matteo Pallocca; Maurizio Fanciulli; Sabrina Strano; Sara Donzelli |
| EPI_ISL_1509455 | IZSM | TIGEM | Antonio Grimaldi Patrizia Annunziata Francesco Panariello Biancamaria Pierri Claudia Tiberio Valentina Bouche Chiara Colantuono Maria Concetta Cuomo Denise Di Concilio Lucio Di Filippo Anna Manfredi Marcello Salvi Antonio Limone Luigi Atripaldi Pellegrino Cerino Andrea Ballabio Davide Cacchiarelli |
| EPI_ISL_1663067, EPI_ISL_1663082 | Illinois Department of Public Health | Illinois Department of Public Health - Chicago Lab | Ira Heimler; Vineet K. Dhiman |
| EPI_ISL_1478850, EPI_ISL_1478861, EPI_ISL_1478879, EPI_ISL_1478882, EPI_ISL_1478902, EPI_ISL_1478906, EPI_ISL_1478920, EPI_ISL_1500153, EPI_ISL_1500177, EPI_ISL_1500184, EPI_ISL_1553196, EPI_ISL_1553205, EPI_ISL_1553207, EPI_ISL_1553209, EPI_ISL_1553215 | | | |
| see above | Illinois Department of Public Health - Springfield Lab | Illinois Department of Public Health - Springfield Lab | Bryan Sim; Gordon McCall |
| EPI_ISL_1464607 | Imeda Hospital | Imelda Hospital | Dagmar Obbels; Hanne Valgaeren; Johan Frans |
| EPI_ISL_1373360 | Imelda Hospital | Imelda Hospital | Dagmar Obbels; Hanne Valgaeren; Johan Frans |
| EPI_ISL_1692730, EPI_ISL_1693117, EPI_ISL_1693319, EPI_ISL_1693420, EPI_ISL_1693672, EPI_ISL_1702287, EPI_ISL_1702296, EPI_ISL_3305285, EPI_ISL_4384179 | | | |
| see above | Infinity Biologix | Centers for Disease Control and Prevention Division of Viral Diseases, Pathogen Discovery | Adrian Paskey; Benjamin Rambo-Martin; Chirayu Goswami; Christian Bixby; Christopher Gulvick; Clinton Paden; Clinton R. Paden; Dakota Howard; Darlene Wagner; Dhwani Batra; Duncan MacCannell; Erisa Sula; Jason Caravas; Jonathan Schultz; Kara Moser; Kristine Lacek; Matthew Schmerer; Peter Cook; Peter W. Cook; Robin Grimwood; Russ Hager; Scott Sammons; Shatavia Morrison; Tymeckia Kendall; Victoria Caban Figueroa; Yihe Wang; Yvette Unoarumhi |
| EPI_ISL_3556914, EPI_ISL_3563989 | InnovoLab Chile | InnovoLab Chile | Alejandro Zuñiga; Harry Bohle |
| EPI_ISL_5736452 | Institut Louis Malardé | Institut Louis Malardé | Dr Van-Mai Cao-Lormeau; Paoaafaite Tuterarii; Teissier Anita |
| EPI_ISL_1663016 | Institut de Virologie du CHU de Strasbourg | Institut de Virologie du CHU de Strasbourg | Fafi-Kremer Samira; Gallais Floriane; Gantner Pierre; Laugel Elodie; Solis Morgane; Velay Aurélie; Wendling Marie-Josée |
| EPI_ISL_2919235, EPI_ISL_2919236 | Instituto Adolfo Lutz - Regional de Bauru | Instituto Adolfo Lutz, Interdiciplinary Procedures Center, Strategic Laboratory | Caio Vinicius Dias Lopes; Claudia Regina Gonçalves; Claudio Tavares Sacchi; Erica Valessa Ramos Gomes; Karoline Rodrigues Campos |
| EPI_ISL_2003166, EPI_ISL_2614521, EPI_ISL_2614522, EPI_ISL_2614523, EPI_ISL_2614524, EPI_ISL_2614525, EPI_ISL_2614526, EPI_ISL_2614529, EPI_ISL_2614530, EPI_ISL_2614531, EPI_ISL_2614532, EPI_ISL_2614533, EPI_ISL_2614534, EPI_ISL_2614535, EPI_ISL_2614537 | | | |
| see above | Instituto Adolfo Lutz - Regional de Campinas | Instituto Adolfo Lutz, Interdiciplinary Procedures Center, Strategic Laboratory | Caio Vinicius Dias Lopes; Claudia Regina Gonçalves; Claudio Tavares Sacchi; Erica Valessa Ramos Gomes; Karoline Rodrigues Campos; Leonardo Jose Tadeu de Araujo |
| EPI_ISL_1821242, EPI_ISL_2691103, EPI_ISL_2691104, EPI_ISL_2691105 | Instituto Adolfo Lutz - Regional de Marilia | Instituto Adolfo Lutz, Interdiciplinary Procedures Center, Strategic Laboratory | Caio Vinicius Dias Lopes; Claudia Regina Gonçalves; Claudio Tavares Sacchi; Erica Valessa Ramos Gomes; Karoline Rodrigues Campos; Leonardo Jose Tadeu de Araujo |
| EPI_ISL_1731581, EPI_ISL_1821250, EPI_ISL_1821253, EPI_ISL_1821256, EPI_ISL_1821257, EPI_ISL_1821260, EPI_ISL_1821263, EPI_ISL_1821265 | | | |
| see above | Instituto Adolfo Lutz - | Instituto Adolfo Lutz, Interdiciplinary | Caio Vinicius Dias Lopes; Claudia Regina Gonçalves; Claudio Tavares Sacchi; Erica Valessa Ramos Gomes; Karoline Rodrigues Campos; Katia Correa de Oliveira Santos; Leonardo Jose Tadeu de Araujo |

| | | | |
|---|---|---|---|
| | Regional de Ribeirao Preto | Procedures Center, Strategic Laboratory | |
| EPI_ISL_2003141, EPI_ISL_2003142, EPI_ISL_2003143, EPI_ISL_2003146, EPI_ISL_2003148, EPI_ISL_2003149 | Instituto Adolfo Lutz - Regional de Santos | Instituto Adolfo Lutz, Interdiciplinary Procedures Center, Strategic Laboratory | Caio Vinicius Dias Lopes; Claudia Regina Gonçalves; Claudio Tavares Sacchi; Erica Valessa Ramos Gomes; Karoline Rodrigues Campos; Leonardo Jose Tadeu de Araujo |
| EPI_ISL_1731598, EPI_ISL_1731599, EPI_ISL_1731600, EPI_ISL_1731601, EPI_ISL_1731602, EPI_ISL_1731603, EPI_ISL_1752668, EPI_ISL_2003114, EPI_ISL_2003115, EPI_ISL_2003117, EPI_ISL_2003131, EPI_ISL_2756445, EPI_ISL_2756446, EPI_ISL_2756447, EPI_ISL_2756462, EPI_ISL_2756463, EPI_ISL_2756464, EPI_ISL_2756465, EPI_ISL_2756476, EPI_ISL_2756488, EPI_ISL_2756489, EPI_ISL_2756490, EPI_ISL_2756491, EPI_ISL_2756492 | | | |
| see above | Instituto Adolfo Lutz Central | Instituto Adolfo Lutz, Interdiciplinary Procedures Center, Strategic Laboratory | Caio Vinicius Dias Lopes; Claudia Regina Gonçalves; Claudio Tavares Sacchi; Erica Valessa Ramos Gomes; Karoline Rodrigues Campos; Katia Correa de Oliveira Santos; Leonardo Jose Tadeu de Araujo |
| EPI_ISL_4104690 | Instituto Butantan | Instituto Butantan | Antonio Jorge Martins; Claudia Renata dos Santos Barros; David Schlesinger; Debora Botequio Moretti; Dimas Tadeu Covas; Elaine Cristina Marqueze; Elaine Vieira Santos; Evandra Strazza Rodrigues; Heidge Fukumasu; Jayme Augusto de Souza-Neto; José Salvatore Leister Patané; Luiz Alcantara; Luiz Lehmann Coutinho; Maria Carolina Elias; Maurício Lacerda Nogueira; Rafael dos Santos Bezerra; Raul Machado Neto; Rejane Maria Tommasini Grotto; Ricardo Haddad; Sandra Coccuzzo Sampaio Vessoni; Simone Kashima; Svetoslav Nanev Slavov; Vincent Louis Viala |
| EPI_ISL_2493026 | Instituto Nacional de Investigação em Saúde | CERI, Centre for Epidemic Response and Innvoation, Stellenbosch University and KRISP, KZN Research Innovation and Sequencing Platform, UKZN. | Afonso P; David K; Emmanuel SJ; Freitas RH; Giandhari J; Inglês L; Lutucuta S; Miranda J; Morais J; Mufinda M; Naidoo Y; Neto Z; Paulo A Carralero RR Paixão JP; Pereira A; Pillay S; Tegally H; Wilkinson E; de Oliveira T |
| EPI_ISL_3118800, EPI_ISL_3118801, EPI_ISL_3118804, EPI_ISL_3118805, EPI_ISL_3118808 | Instituto de Biotecnologia - UNESP-Botucatu-SP | Instituto de Biotecnologia - UNESP-Botucatu-SP | Cecília Artico Banho; Cíntia Bittar; Fábio Sossai Possebon; Guilherme Campos; Helena Lage Ferreira; Jorge A. Petroli Marchesi; João Pessoa Araújo Jr.; Leila Sabrina Ullmann; Lívia Sacchetto; Maisa C. Pereira Parra; Marília Moraes; Maurício L. Nogueira; Paula Rahal; Paulo Inacio da Costa |
| EPI_ISL_2894886 | Instituto de Medicina Tropical de Sao Paulo | Instituto de Medicina Tropical de Sao Paulo | Brazil-UK Centre for Arbovirus Discovery Diagnosis Genomics and Epidemiology (CADDE) Genomic Network - Instituto de Medicina Tropical |
| EPI_ISL_2404060, EPI_ISL_2404082, EPI_ISL_2404190, EPI_ISL_2404256, EPI_ISL_2404338, EPI_ISL_2404362, EPI_ISL_2404372, EPI_ISL_2404380, EPI_ISL_2404382, EPI_ISL_2404392, EPI_ISL_2404597, EPI_ISL_2425144, EPI_ISL_2425147, EPI_ISL_2425148, EPI_ISL_2833096, EPI_ISL_2840493, EPI_ISL_2840496, EPI_ISL_2840506, EPI_ISL_2840508, EPI_ISL_2840514, EPI_ISL_2840521, EPI_ISL_2840523 | | | |
| see above | KU Leuven, Rega Institute, Clinical and Epidemiological Virology | KU Leuven, Rega Institute, Clinical and Epidemiological Virology | Bert Vanmechelen; Joan Marti-Carerras; Piet Maes; Tony Wawina-Bokalanga |
| EPI_ISL_1599531 | Klinisch Laboratorium GZA | Klinisch Laboratorium ZNA | Verstrepen et al. |
| EPI_ISL_3102494, EPI_ISL_3102526 | LABORATORIO CLEMENTINO FRAGA | Analytical Competence Molecular Epidemiology Lab/ACME, Oswaldo Cruz Foundation, Ceara (FIOCRUZ CE) | Cleber Furtado Aksenen; Fabio Miyajima; Fernando Braga Stehling; Francisco Eder de Moura Lopes; Jamille Maria Mendes Bezerra; Joaquim César do Nascimento Sousa Junior; Pedro Miguel Carneiro Jeronimo; Suzana Porto Almeida e Lucas Delerino; Thais Ferreira de Oliveira; Thais de Oliveira Costa; Ticiane Cavalcante de Souza; Veridiana Pessoa Miyajima |
| EPI_ISL_1795303, EPI_ISL_1795306, EPI_ISL_1795307, EPI_ISL_1795308, EPI_ISL_1795309, EPI_ISL_1795310, EPI_ISL_1795311, EPI_ISL_1795313, EPI_ISL_1795314, EPI_ISL_1795315, EPI_ISL_1795316, EPI_ISL_1795317, EPI_ISL_1795319, EPI_ISL_1795320, EPI_ISL_1795321, EPI_ISL_1795322, EPI_ISL_1795323, EPI_ISL_2345549, EPI_ISL_2345554, EPI_ISL_2345555, EPI_ISL_2345557, EPI_ISL_2345558, EPI_ISL_2345559, EPI_ISL_2345560, EPI_ISL_2345563, EPI_ISL_2345565, EPI_ISL_2345566, EPI_ISL_2345568, EPI_ISL_2345569, EPI_ISL_2345570, EPI_ISL_2345576, EPI_ISL_2345578, EPI_ISL_2345579, EPI_ISL_2345582, EPI_ISL_2345584 | | | |
| see above | LABORATORIO DE FRANCA | Instituto Butantan / ESALQ-Piracicaba | Antonio Jorge Martins; Bianca Cechetto Carlos. Mendelics: Bibiana Santos; Claudia Renata dos Santos Barros; David Schlesinger; David Schlesinger. Hemocentro Ribeirão Preto: Simone Kashima; Debora Botequio Moretti; Debora Botequio Moretti. Centro de Genômica Funcional da ESALQ: Luiz Lehmann Coutinho; Dimas Tadeu Covas; Elaine Cristina Marqueze; Elaine Vieira Santos; Elaine Vieira dos Santos; Elisangela Chicaroni Mattos; Erika Freitas; Evandra Strazza Rodrigues; Felipe Allan da Silva da Costa; Flavia Aburjaile; Guilherme Targino Valente; Heidge Fukumasu; Heidge Fukumasu. USP-Botucatu: Rejane Maria Tommasini Grotto; Instituto Butantan: Alexander Roberto Precioso; Jayme A. Souza-Neto; Jayme Augusto de Souza-Neto; Jessika Cristina Chagas Lesbon; José Salvatore Leister Patané; João Paulo Kitajima; Luiz Alcantara; Luiz Carlos Junior de Alcantara; Luiz Lehmann Coutinho; Maria Carolina Elias; Marta Giovanetti; Maurício Lacerda Nogueira; Patricia Akemi Assato; Rafael dos Santos Bezerra; Raquel de Lello Rocha Campos Cassano. NGS Soluções Genômicas: Pilar Drummond Sampaio Corrêa Mariani. FZEA-USP Pirassununga: Mirele Daiana Poleti; Raul Machado Neto; Rejane Maria Tommasini Grotto; Ricardo Augusto Brassaloti; Ricardo Haddad; Rodrigo Tocantins Calado.; Sandra Coccuzzo Sampaio; Sandra Coccuzzo Sampaio Vessoni; Simone Kashima; Svetoslav Nanev Slavov; Vagner Fonseca; Vincent Louis Viala |
| EPI_ISL_1795347, EPI_ISL_1795355, EPI_ISL_1795365, EPI_ISL_1795366, EPI_ISL_2345615, EPI_ISL_2345628, EPI_ISL_2345640, EPI_ISL_2345641 | | | |
| see above | LABORATORIO DR PAULO EMILIO DALESSANDRO PINDAMONHANGABA | Instituto Butantan / ESALQ-Piracicaba | Antonio Jorge Martins; Bianca Cechetto Carlos. Mendelics: Bibiana Santos; Claudia Renata dos Santos Barros; David Schlesinger; David Schlesinger. Hemocentro Ribeirão Preto: Simone Kashima; Debora Botequio Moretti; Debora Botequio Moretti. Centro de Genômica Funcional da ESALQ: Luiz Lehmann Coutinho; Dimas Tadeu Covas; Elaine Cristina Marqueze; Elaine Vieira Santos; Elaine Vieira dos Santos; Elisangela Chicaroni Mattos; Erika Freitas; Evandra Strazza Rodrigues; Felipe Allan da Silva da Costa; Flavia Aburjaile; Guilherme Targino Valente; Heidge Fukumasu; Heidge Fukumasu. USP-Botucatu: Rejane Maria Tommasini Grotto; Instituto Butantan: Alexander Roberto Precioso; Jayme A. Souza-Neto; Jayme Augusto de Souza-Neto; Jessika Cristina Chagas Lesbon; José Salvatore Leister Patané; João Paulo Kitajima; Luiz Alcantara; Luiz Carlos Junior de Alcantara; Luiz Lehmann Coutinho; Maria Carolina Elias; Marta Giovanetti; Maurício Lacerda Nogueira; Patricia Akemi Assato; Rafael dos Santos Bezerra; Raquel de Lello Rocha Campos Cassano. NGS Soluções Genômicas: Pilar Drummond Sampaio Corrêa Mariani. FZEA-USP Pirassununga: Mirele Daiana Poleti; Raul Machado Neto; Rejane Maria Tommasini Grotto; Ricardo Augusto Brassaloti; Ricardo Haddad; Rodrigo Tocantins Calado.; Sandra Coccuzzo Sampaio; Sandra Coccuzzo Sampaio Vessoni; Simone Kashima; Svetoslav Nanev Slavov; Vagner Fonseca; Vincent Louis Viala |
| EPI_ISL_1675322 | LABORATORIO ECHAVARRIA | Universidad Nacional de Colombia - Laboratorio Genómico One Health | Andres F. Cardona-Rios; Carlos Franco-Muñoz; Daniel O. Maldonado-Perez; Diego A. Álvarez-Díaz; Hector Alejandro Ruiz-Moreno; Idabely Betancur Ortiz; Jorge E. Osorio; Juan P. Hernandez-Ortiz; Karl A Ciuoderis; Katherine Laiton-Donato; Laura Silvana Perez; Lina M. Hurtado; Marcela Mercado-Reyes; Maria Angélica Maya; Maria Stella López; Rita Almanza Payares; Sandra Ines Cano; Simón Villegas Velásquez |
| EPI_ISL_7744031, EPI_ISL_7744036, EPI_ISL_7744043 | LACEN | Laboratório de Bioinformática - Universidade Federal de Santa Catarina | "Aline Daina Schlindwein"; "Ana Paula Christoff"; "Antuani Baptistela"; "Carolina Leite Martins"; "Darcita Buerger Rovaris"; "Dayane Azevedo Padilha"; "Doris Sobral Marques SouzaSobral"; "Edmundo Carlos Grisard"; "Eric Kazuo Kawagoe"; "Fernanda Luiza Ferrari"; "Fernanda Roesene Melo"; "Fernando Hartmann Barazzetti"; "Gislaine Fongaro"; "Glauber Wagner"; "Guilherme Augusto Maia"; "Guilherme Razzera"; "Guilherme Toledo e Silva"; "Julia Kinetz Wachter"; "Luiz Felipe de Oliveira"; "Marcos André Schörne"; "Marcus Vinícius Duarte Rodrigues"; "Maria Luiza Bazzo"; "Marlei Pickler Debiasi dos Anjos"; "Milene Moehr de Moraes"; "Nestor Wendt"; "Patrícia Hermes Stoco"; "Paula Sacchet"; "Renato Simões Moreira"; "Rodrigo de Paula Baptista"; "Tamela Zamboni Madaloz"; "Tatiany Aparecida Teixeira Soratto"; "Vilmar Benetti Filho" |
| EPI_ISL_2488802 | LACEN - Laboratório Central de Saúde Pública do Ceará | Evandro Chagas Institute | A.M.; Barbagelata; E.C.; E.M.A.; Ferreira; J.A.; Junior; K.C.; L.C.; L.S.; M.C.; P.S.; Pinheiro; Santos; Silva; Sousa; Sousa Junior; W.D.C.; da Silva |
| EPI_ISL_2958885, EPI_ISL_2958886, EPI_ISL_2958888, EPI_ISL_3316191 | LACEN do Estado de Mato Grosso | Instituto Adolfo Lutz, Interdiciplinary Procedures Center, Strategic Laboratory | Caio Vinicius Dias Lopes; Claudia Regina Gonçalves; Claudio Tavares Sacchi; Erica Valessa Ramos Gomes; Karoline Rodrigues Campos; Leonardo Tadeu de Araujo; Marlon Benedito Nascomento Santos |
| EPI_ISL_3691387, EPI_ISL_3691388, EPI_ISL_3691389 | LACEN do Estado do Mato Grosso do Sul | Instituto Adolfo Lutz, Interdiciplinary Procedures Center, Strategic Laboratory | Caio Vinicius Dias Lopes; Claudia Regina Gonçalves; Claudio Tavares Sacchi; Karoline Rodrigues Campos; Leonardo Tadeu de Araujo; Marlon Benedito Nascomento Santos |
| EPI_ISL_2958855 | LACEN do Mato Grosso do Sul | Instituto Adolfo Lutz, Interdiciplinary Procedures Center, Strategic Laboratory | Caio Vinicius Dias Lopes; Claudia Regina Gonçalves; Claudio Tavares Sacchi; Erica Valessa Ramos Gomes; Karoline Rodrigues Campos |
| EPI_ISL_3703554, EPI_ISL_6573784, EPI_ISL_6573794, EPI_ISL_6573795, EPI_ISL_6573796, EPI_ISL_6573798, EPI_ISL_6573831, EPI_ISL_6573832, EPI_ISL_6573835 | | | |
| see above | LACEN/PE | WallauLab on behalf of Fiocruz COVID-19 Genomic Surveillance Network | Alexandre Freitas da Silva; Antonio Marinho da Silva Neto; Cassia Docena; Constância Flávia Junqueira Ayres; Filipe Zimmer Dezordi; Gabriel Luz Wallau; Gustavo Barbosa de Lima; Lais Ceschini Machado; Lilian Caroliny Amorim Silva; Marcelo Henrique dos Santos Paiva; Matheus Filgueira Bezerra; Sinval Pinto Brandão Filho |
| EPI_ISL_4488044, EPI_ISL_4488047 | LACEN/PI | WallauLab on behalf of Fiocruz COVID-19 Genomic Surveillance Network | Adelino Soares Lima Neto; Alexandre Freitas da Silva; Antônio Marinho; Cassia Docena; Constância Flávia Junqueira Ayres; Filipe Zimmer Dezordi; Gabriel Luz Wallau; Gustavo Barbosa de Lima; Hellen de Oliveira Amaral; Jacenir Reis dos Santos Mallet; Joana Carolina Viana Lima; Lais Ceschini Machado; Leandro de Mattos; Lilian Caroliny Amorim Silva; Marcela de Lacerda Valença Queiroz; Marcelo Adriano da Cunha e Silva Vieira; Marcelo Henrique dos Santos Paiva; Matheus Filgueira Bezerra; Sinval Pinto Brandão Filho; Túlio de Lima Campos; Vladimir Costa Silva; Walterlene de Carvalho Goçalves |
| EPI_ISL_6229761 | LACLIM | ACME Lab, Oswaldo Cruz Foundation, FIOCRUZ/CE | Carlos Leonardo de Aragao Araujo; Cecilia Leite Costa & Eduardo Ruback dos Santos on behalf of COVID-19 FIOCRUZ Genomic Network; Cleber Furtado Aksenen; Fabio Miyajima; Fernando Braga Stehling; Francisco Eder de Moura Lopes; Igor Oliveira Duarte; Jamille Maria Mendes Bezerra; Joaquim Cesar do Nascimento Sousa Junior; Pedro Miguel Carneiro Jeronimo; Suzana Porto Almeida; Thais Ferreira de Oliveira; Thais de Oliveira Costa; Ticiane Cavalcante de Souza; Veridiana Pessoa Miyajima |
| EPI_ISL_1626821 | LESP Quintana Roo | Instituto de Diagnostico y Referencia Epidemiologicos (INDRE) | Abril Rodriguez-Maldonado; Ariadna Medina-Benitez; Claudia Wong-Arambula; Ernesto Ramirez-Gonzalez.; Gisela Barrera-Badillo; Irma Lopez-Martinez; Joaquin Quiroz-Mercado; Lucia Hernandez-Rivas; Natividad Cruz-Ortiz; Sergio Rangel-Guerrero; Tatiana Nunez-Garcia; Vanessa Rivero-Arredondo |
| EPI_ISL_1524938, EPI_ISL_1524947, EPI_ISL_1591183, EPI_ISL_1591189, EPI_ISL_1591191, EPI_ISL_1608054 | LHUB-ULB | Labo Klinische Biologie, UZA | Basil Britto Xavier; Christine Lammens; Herman Goossens; Jasmine Coppens; Marie Le Mercier; Veerle Matheeussen |
| EPI_ISL_1689303, EPI_ISL_1689402, EPI_ISL_1689642, EPI_ISL_1689660, EPI_ISL_1689673, EPI_ISL_1689697 | Lab voor klinische biologie | Lab voor klinische biologie | Bruno Verhasselt; Hannelore Hamerlinck; Marija Janevska |
| EPI_ISL_1643837 | Labor Dr. Spranger | Robert Koch Institute | |
| EPI_ISL_3133971 | Laboratoire Carage | Institut Pasteur de la Guyane | Anne Lavergne; Dominique Rousset |
| EPI_ISL_2975372, EPI_ISL_3143930, EPI_ISL_3458270, EPI_ISL_3458272, EPI_ISL_3458273, EPI_ISL_3458274, EPI_ISL_5873171, EPI_ISL_5891521 | | | |
| see above | Laboratoire de santé publique du Québec | Laboratoire de santé publique du Québec | Guillaume Bourque; Ioannis Ragoussis; Jesse Shapiro; Mark Lathrop and Michel Roger on behalf of the CoVSeQ research group; Sandrine Moreira |
| EPI_ISL_2196257, EPI_ISL_2196276, EPI_ISL_2196278, | Laboratorio Central de Saude Publica do Estado de Minas Gerais | Laboratory of Respiratory Viruses and Measles, Oswaldo Cruz Institute, FIOCRUZ | Alice Sampaio Rocha; Ana Carolina Mendonca; Andre Felipe Leal Bernardes; Anna Carolina Paixao; Elisa Cavalcante Pereira; Fernando Motta; Luciana Appolinario; Marilda Siqueira on behalf of the Fiocruz COVID-19 Genomic Surveillance Network; Paola Resende; Renata Serrano Lopes; Taina Venas |

| | | | |
|---|---|---|---|
| EPI_ISL_2196279, EPI_ISL_2557348 | (LACEN/MG) | | |
| EPI_ISL_2196281, EPI_ISL_2443563, EPI_ISL_2443564, EPI_ISL_2645823, EPI_ISL_2645828, EPI_ISL_2645848, EPI_ISL_2645872, EPI_ISL_2645873, EPI_ISL_2645894, EPI_ISL_2645898, EPI_ISL_2645918, EPI_ISL_2645919, EPI_ISL_2645920, EPI_ISL_2645931, EPI_ISL_4080768 | | | |
| see above | Laboratorio Central de Saude Publica do Estado do Para (LACEN/PA) | Laboratory of Respiratory Viruses and Measles, Oswaldo Cruz Institute, FIOCRUZ | Agatha Soares; Alice Sampaio Rocha; Ana Carolina Mendonca; Anna Carolina Paixao; Elisa Cavalcante Pereira; Fernando Motta; Ighor Arantes; Luciana Appolinario; Marilda Siqueira on behalf of the Fiocruz COVID-19 Genomic Surveillance Network; Paola Resende; Renata Serrano Lopes; Taina Venas; Valnete Andrade |
| EPI_ISL_1670644 | Laboratorio Analisi Osp. Città di Castello - Azienda USL Umbria1 | Istituto Zooprofilattico Sperimentale dell'Abruzzo e Molise "G. Caporale" | Ancora M; Calistri P; Cammà C; Caporale M; Curini V; Delli Compagni E; Di Domenico M; Di Lollo Valeria; Di Pasquale A; Lorusso A; Malagigi V; Mangone I; Marcacci M; Puglia I; Rinaldi A; Savini G; Scialabba S; Tacconi P |
| EPI_ISL_1664113, EPI_ISL_1664114, EPI_ISL_1664115, EPI_ISL_1664116, EPI_ISL_1664117, EPI_ISL_1664118, EPI_ISL_1664119, EPI_ISL_1664120, EPI_ISL_1664121, EPI_ISL_1664122, EPI_ISL_1664123, EPI_ISL_1664124, EPI_ISL_1664125, EPI_ISL_1664127, EPI_ISL_1664128, EPI_ISL_1664129, EPI_ISL_1664130, EPI_ISL_1664131, EPI_ISL_1664132, EPI_ISL_1664133, EPI_ISL_1664134, EPI_ISL_1664135, EPI_ISL_1664136, EPI_ISL_1664137, EPI_ISL_1664139, EPI_ISL_1664140, EPI_ISL_1664141, EPI_ISL_1664142, EPI_ISL_1664143, EPI_ISL_1664144, EPI_ISL_1664145, EPI_ISL_1664146, EPI_ISL_1664147, EPI_ISL_1664148, EPI_ISL_1664149, EPI_ISL_1664150, EPI_ISL_1664151, EPI_ISL_1664152, EPI_ISL_1664153, EPI_ISL_1664154, EPI_ISL_1664155, EPI_ISL_1664156, EPI_ISL_1664157, EPI_ISL_1664158, EPI_ISL_1664159, EPI_ISL_1664160, EPI_ISL_1664161, EPI_ISL_1664162, EPI_ISL_1664163, EPI_ISL_1664164, EPI_ISL_1664165, EPI_ISL_1664166, EPI_ISL_1664167, EPI_ISL_1664168, EPI_ISL_1664169, EPI_ISL_1664170, EPI_ISL_1664171, EPI_ISL_1664172, EPI_ISL_1664173, EPI_ISL_1664174, EPI_ISL_1664176, EPI_ISL_1664177, EPI_ISL_1664178, EPI_ISL_1664179, EPI_ISL_1664180, EPI_ISL_1664181, EPI_ISL_1664182, EPI_ISL_1664183, EPI_ISL_1664187, EPI_ISL_1664188, EPI_ISL_1664189, EPI_ISL_1664190, EPI_ISL_1664191, EPI_ISL_1664192, EPI_ISL_1664193, EPI_ISL_1664194, EPI_ISL_1664195, EPI_ISL_1664196, EPI_ISL_1664197, EPI_ISL_1664198, EPI_ISL_1664199, EPI_ISL_1664200, EPI_ISL_1664201, EPI_ISL_1664202, EPI_ISL_1858267, EPI_ISL_2101485, EPI_ISL_2101541, EPI_ISL_2101621 | | | |
| see above | Laboratorio Central Noel Nutels | Bioinformatics Laboratory / LNCC | Alessandra P Lamarca; Alexandra L Gerber; Amilcar Tanuri; Ana Paula de C Guimaraes; Ana Paula de C Guimarães; Ana Tereza R Vasconcelos; Andrea Cony Cavalcanti; Andréa Cony Cavalcanti; Caio Luiz Pereira Ribeiro; Cassia Alves; Cintia Policarpo; Claudia Maria Braga de Mello; Cristiane Gomes da Silva; Diana Mariani; Douglas Terra Machado; Flavio Dias da Silva; Flávio Dias da Silva; Gleidson da Silva de Oliveira; Leandro Magalhaes de Souza; Leandro Magalhães de Souza; Liliane Cavalcante; Luiz G P de Almeida; Marcio Henrique de Oliveira Garcia; Mario Sergio Ribeiro; Ronaldo da Silva F Jr; Silvia Carvalho; Thais Felix Cruz |
| EPI_ISL_2157509, EPI_ISL_2157512, EPI_ISL_2157514 | Laboratorio Central de Saude Publica do Esatado de Alagoas (LACEN/AL) | Laboratory of Respiratory Viruses and Measles, Oswaldo Cruz Institute, FIOCRUZ | Alice Sampaio Rocha; Ana Carolina Mendonca; Anderson Brandao Leite; Anna Carolina Paixao; Elisa Cavalcante Pereira; Fernando Motta; Luciana Appolinario; Marilda Siqueira on behalf of the Fiocruz COVID-19 Genomic Surveillance Network; Paola Resende; Renata Serrano Lopes; Taina Venas |
| EPI_ISL_2196355, EPI_ISL_2274076, EPI_ISL_2536355 | Laboratorio Central de Saude Publica do Estado Maranhao (LACEN-MA) | Laboratory of Respiratory Viruses and Measles, Oswaldo Cruz Institute, FIOCRUZ | Alice Sampaio Rocha; Ana Carolina Mendonca; Anna Carolina Paixao; Elisa Cavalcante Pereira; Fernando Motta; Lidio Gonçalves Lima Neto; Luciana Appolinario; Marilda Siqueira on behalf of the Fiocruz COVID-19 Genomic Surveillance Network; Paola Resende; Renata Serrano Lopes; Taina Venas |
| EPI_ISL_2491706, EPI_ISL_2491707, EPI_ISL_2491708, EPI_ISL_2491709, EPI_ISL_2491710, EPI_ISL_2491711, EPI_ISL_2491712 | | | |
| see above | Laboratorio Central de Saude Publica do Estado da Bahia (LACEN/BA) | Laboratory of Respiratory Viruses and Measles, Oswaldo Cruz Institute, FIOCRUZ | Alice Sampaio Rocha; Ana Carolina Mendonca; Anna Carolina Paixao; Elisa Cavalcante Pereira; Felicidade Pereira; Fernando Motta; Luciana Appolinario; Marilda Siqueira on behalf of the Fiocruz COVID-19 Genomic Surveillance Network; Paola Resende; Renata Serrano Lopes; Taina Venas |
| EPI_ISL_2536333, EPI_ISL_2536351 | Laboratorio Central de Saude Publica do Estado da Paraiba (LACEN-PB) | Laboratory of Respiratory Viruses and Measles, Oswaldo Cruz Institute, FIOCRUZ | Alice Sampaio Rocha; Ana Carolina Mendonca; Anna Carolina Paixao; Dalane Loudal Florentino Teixeira; Elisa Cavalcante Pereira; Fernando Motta; Joao Felipe Bezerra; Luciana Appolinario; Marilda Siqueira on behalf of the Fiocruz COVID-19 Genomic Surveillance Network; Paola Resende; Renata Serrano Lopes; Taina Venas |
| EPI_ISL_2274058 | Laboratorio Central de Saude Publica do Estado de Alagoas (LACEN/AL) | Laboratory of Respiratory Viruses and Measles, Oswaldo Cruz Institute, FIOCRUZ | Alice Sampaio Rocha; Ana Carolina Mendonca; Anderson Brandao Leite; Anna Carolina Paixao; Elisa Cavalcante Pereira; Fernando Motta; Luciana Appolinario; Marilda Siqueira on behalf of the Fiocruz COVID-19 Genomic Surveillance Network; Paola Resende; Renata Serrano Lopes; Taina Venas |
| EPI_ISL_2274111, EPI_ISL_2274112, EPI_ISL_2274126, EPI_ISL_2274130 | Laboratorio Central de Saude Publica do Estado do Rio Grande do Sul (LACEN-RS) | Laboratory of Respiratory Viruses and Measles, Oswaldo Cruz Institute, FIOCRUZ | Alice Sampaio Rocha; Ana Carolina Mendonca; Anna Carolina Paixao; Elisa Cavalcante Pereira; Fernando Motta; Luciana Appolinario; Marilda Siqueira on behalf of the Fiocruz COVID-19 Genomic Surveillance Network; Paola Resende; Renata Serrano Lopes; Richard Salvato; Taina Venas; Tatiana Schaffer Gregianini |
| EPI_ISL_3048784, EPI_ISL_3048786, EPI_ISL_3048787, EPI_ISL_3048788, EPI_ISL_3048789 | Laboratorio Central de Saude Publica do Estado do Rio Grande do Sul (LACEN-RS) | Laboratório de Biologia Molecular da Universidade Federal de Ciências da Saúde de Porto Alegre | Adriana Seixas; Ana B. G. Veiga; Ana Paula Muterle Varela; Fabiana Quoos Mayer; Fernando Hayashi Sant'Anna; Janira Prichula; Letícia Garay Martins; Richard Steiner Salvato; Tatiana Schäffer Gregianini |
| EPI_ISL_2196201, EPI_ISL_2196203 | Laboratorio Central de Saude Publica do Estado do Rio de Janeiro (LACEN-RJ) | Laboratory of Respiratory Viruses and Measles, Oswaldo Cruz Institute, FIOCRUZ | Alice Sampaio Rocha; Ana Carolina Mendonca; Andrea Cony Cavalcanti; Anna Carolina Paixao; Elisa Cavalcante Pereira; Fernando Motta; Luciana Appolinario; Marilda Siqueira on behalf of the Fiocruz COVID-19 Genomic Surveillance Network; Paola Resende; Renata Serrano Lopes; Taina Venas |
| EPI_ISL_2363545 | Laboratorio Central de la Ciudad de Santa Fe | Grupo de Genómica y Bioinformática del Instituto de Investigación de la Cadena Láctea CONICET-INTA on behalf of 'Proyecto Argentino Interinstitucional de genomica de SARS-CoV-2' (PAIS Consortium) | AF; Amadio; C; Eberhardt; G; Irazoqui; JM; MF; Mugna; Ojeda; Pastor; Rompato; V |
| EPI_ISL_2427510, EPI_ISL_2427518, EPI_ISL_2427526, EPI_ISL_2427533, EPI_ISL_2427534, EPI_ISL_2427541, EPI_ISL_2427570, EPI_ISL_2427586, EPI_ISL_2427594 | | | |
| see above | Laboratorio de Biología Molecular Médica Uruguaya | Departments of Pathology and Medicine, New York University School of Medicine | Adriana Heguy; Cecilia Sorhouet; Christian Marier; Dacia Dimartino; Gonzalo Manrique; Maria Cristina Mogdasy; Maria Noel Zubillaga; Maria Victoria Elizondo; Paul Zappile |
| EPI_ISL_2777428, EPI_ISL_2777429, EPI_ISL_2777430 | Laboratorio de Ecologia de Doencas Transmissiveis na Amazonia, Instituto Leonidas e Maria Deane - Fiocruz Amazonia | Laboratorio de Ecologia de Doencas Transmissiveis na Amazonia, Instituto Leonidas e Maria Deane - Fiocruz Amazonia | André Corado; Debora Duarte; Felipe Naveca; Fernanda Nascimento; George Silva; Karina Pessoa; Luciana Gonçalves; Maria Júlia Brandão; Matilde Mejía; Michele Jesus; Valdinete Nascimento; Victor Souza; Ágatha Costa |
| EPI_ISL_2008941 | Laboratorio de Pesquisa em Virologia, FAMERP, SJRP | Laboratorio de Pesquisa em Virologia, FAMERP, SJRP | Cecília Artico Banho; Cíntia Bittar; Fábio Sossai Possebon; Guilherme Campos; Helena Lage Ferreira; Jorge A. Petroli Marchesi; João Pessoa Araújo Jr.; Leila Sabrina Ullmann; Lívia Sacchetto; Maisa C. Pereira Parra; Marília Moraes; Maurício L. Nogueira.; Paula Rahal; Paulo Inacio da Costa |
| EPI_ISL_3401587, EPI_ISL_3401588 | Laboratorio de Referencia Nacional de Virus Respiratorios. Centro Nacional de Salud Publica. Instituto Nacional de Salud Peru. | Laboratorio de Referencia Nacional de Virus Respiratorios. Centro Nacional de Salud Publica. Instituto Nacional de Salud Peru. | Carlos Padilla Rojas; Henri Bailon Calderon; Iris Silva Molina; Joseph Huayra Niquen; Lely Solari Zerpa; Luis Barcena Flores; Marco Galarza Perez; Nancy Rojas Serrano; Nieves Sevilla Castañeda; Omar Caceres Rey; Orson Mestanza Millones; Princesa Medrano Alhuay; Priscila Lope Pari; Sandra Morales Ruiz; Sara Gordillo Vilchez; Steve Acedo Lazo; Veronica Hurtado Vela; Victor Jimenez Vasquez; Wendy Lizarraga Olivares |
| EPI_ISL_3375998, EPI_ISL_3376036, EPI_ISL_3376038, EPI_ISL_3376044, EPI_ISL_3376176, EPI_ISL_3376177, EPI_ISL_3376376, EPI_ISL_3376404, EPI_ISL_3376406 | | | |
| see above | Laboratorio de Referencial Nacional de Virus Respiratorios | Laboratorio de Referencial Nacional de Virus Respiratorios | Carlos Padilla Rojas; Henri Bailon Calderon; Iris Silva Molina; Joseph Huayra Niquen; Lely Solari Zerpa; Luis Barcena Flores; Marco Galarza Perez; Nancy Rojas Serrano; Omar Caceres Rey; Orson Mestanza Millones; Priscila Lope Pari; Sandra Morales Ruiz; Steve Acedo Lazo; Veronica Hurtado Vela |
| EPI_ISL_1626618 | Laboratorio de Salud Pública Bogota | Gencore - Universidad de los Andes | Alejandro Gomez; Ana Maria Palacio; David Gonzalez; Gabriela Delgado; Johana Hernandez; Luisa Sacristan; Marcela Guevara; Silvia Restrepo |
| EPI_ISL_1673278, EPI_ISL_1673279 | Laboratorio de Virología HUCA | Laboratorio de Virología HUCA | Abreu F; Alvarez-Arguelles ME; Boga JA; Castelló C; Costales I; Coto E; Gómez de Oña J; Martín-Rodríguez G; Melón S; Perez-Martínez Z; Rojo S; Sandoval M |
| EPI_ISL_2007472, EPI_ISL_2007475, EPI_ISL_2007477, EPI_ISL_2007488, EPI_ISL_2007490, EPI_ISL_2007520, EPI_ISL_2007531, EPI_ISL_2007533, EPI_ISL_2007534 | | | |
| see above | Laboratorio de Virología del Hospital de Niños Dr. Ricardo Gutierrez | Área de Secuenciación del Laboratorio de Virología del Hospital de Niños Dr. Ricardo Gutierrez on behalf of 'Proyecto Argentino Interinstitucional de genomica de SARS-CoV-2' (PAIS Consortium) | A; Acevedo; Acuña; Alexay; Alvarez Lopez; Barreda Frank; C; D; E; G; Goya; Grandis; Jacques; LE; Labarta; Lusso; M; ME; MI; Medina; Mistchenko; N; Nabaes Jodar; Natale; O; S; Streitenberger; Thomas; Valinotto; Viegas, M.; Villegas |
| EPI_ISL_5771862 | Laboratorio del Hospital Regional Ushuaia Gdor. Ernesto Campos | Nodo de Secuenciación Tierra del Fuego - Hospital Regional Ushuaia - Centro Austral De Investigaciones Científicas - Universidad Nacional De Tierra Del Fuego | AE; Boutureira, MF.; CA; CB; CF; Castro; Ceballos; Cáceres; De Roccis; F; G; Gallego; Gramundi; ID; Nardi; Rojas; SB; SG; Yulan |
| EPI_ISL_1463438, EPI_ISL_1514006, EPI_ISL_1549125, EPI_ISL_1549272, EPI_ISL_1549343, EPI_ISL_1549382, EPI_ISL_1549387, EPI_ISL_1549546, EPI_ISL_1549554, EPI_ISL_1549568, EPI_ISL_1549580, EPI_ISL_1550088, EPI_ISL_1550102, EPI_ISL_1550146, EPI_ISL_1550147, EPI_ISL_1550151, EPI_ISL_1550152, EPI_ISL_1550153, EPI_ISL_1550192, EPI_ISL_1550204, EPI_ISL_1550261, EPI_ISL_1550317, EPI_ISL_1550322, EPI_ISL_1608842, EPI_ISL_1608844, EPI_ISL_1608858, EPI_ISL_1608860, EPI_ISL_1608903, EPI_ISL_1608904, EPI_ISL_1608940, EPI_ISL_1608950, EPI_ISL_1608987, EPI_ISL_1609004, EPI_ISL_1609017, EPI_ISL_1609020, EPI_ISL_1609051, EPI_ISL_1609112, EPI_ISL_1609136, EPI_ISL_1609142, EPI_ISL_1609208, EPI_ISL_1609219, EPI_ISL_1609340, EPI_ISL_1609395, EPI_ISL_1609449, EPI_ISL_1609458, EPI_ISL_1609534, EPI_ISL_1609566, EPI_ISL_1609567, EPI_ISL_1609575, EPI_ISL_1609578, EPI_ISL_1609799, EPI_ISL_1609848, EPI_ISL_1609873, EPI_ISL_1609906, EPI_ISL_1609990, EPI_ISL_1609994, EPI_ISL_1610024, EPI_ISL_1610310, EPI_ISL_1610604, EPI_ISL_1610610, EPI_ISL_1610619, EPI_ISL_1610644, EPI_ISL_1611785, EPI_ISL_1612269, EPI_ISL_4373341, EPI_ISL_4376050, EPI_ISL_4377413 | | | |
| see above | Laboratory Corporation of America | Centers for Disease Control and Prevention Division of Viral Diseases, | Adrian Paskey; Amanda Douglas; Amanda Suchanek; Andrea Throop; Ayla Burns; Benjamin Rambo-Martin; Bobbi Croy; Brian Krueger; Brian Norvell; Christopher Gulvick; Christos Petropoulos; Clinton Paden; Clinton R. Paden; Craig Lukasik; Dakota Howard; Darlene Wagner; Debbie Boles; Dhwani Batra; Duncan MacCannell; Eyad Almasri; Goran Stevovic; Howard Engler; Hrushikesh Deshmukh; Jake Humphrey; Jana Schroth; Jason Caravas; Joe Voshell; John Pruitt; Jonathan Meltzer; Jonathan Williams; Kara Moser; Kimberly Wagner; Kristine Lacek; Lax Iyer; Lisa Pfefferle; Lyndon Tilson; Manoj Jain; Marcia Eisenberg; Mary Ann |

| | | | |
|---|---|---|---|
| | | Pathogen Discovery | Cristobal; Mary Cristobal; Mary Williamson; Matthew Robinson; Matthew Schmerer; Michael Levandoski; Mike Sapeta; Mindy Nye; Minoo Agarwal; Mohan Kolli; Nuthawin Charoensri; Oren Cohen; Peter Cook; Peter W. Cook; Prashant Gupta; Qian Zeng; Rama Ghatti; Scott Parker; Scott Ryan; Scott Sammons; Shatavia Morrison; Stanley Letovsky; Steven Ragan; Suresh Babu Selvaraju; Suresh Selvaraju; Susan Countryman; Susan Hicks; Suzanne Dale; Thomas Urban; Tim Kuphal; Tricia Zwiefelhofer; Tymeckia Kendall; Victoria Caban Figueroa; Vincent Drouillon; Yvette Unoarumhi |
| EPI_ISL_1909230 | Laboratory of Clinical Microbiology, Virology and Bioemergencies, ASST Fatebenefratelli Sacco - Sacco Hospital | Laboratory of Clinical Microbiology, Virology and Bioemergencies, ASST Fatebenefratelli Sacco - Sacco Hospital | Alberto Rizzo; Alessandro Mancon; Fiorenza Bracchitta; Luca Rizzuto; Maria Rita Gismondo; Valeria Micheli |
| EPI_ISL_1534015, EPI_ISL_1534016, EPI_ISL_2443587, EPI_ISL_2443588, EPI_ISL_2443589, EPI_ISL_2443590, EPI_ISL_2443591, EPI_ISL_2536271, EPI_ISL_2557392, EPI_ISL_2614334, EPI_ISL_2614335, EPI_ISL_2614336, EPI_ISL_2614337, EPI_ISL_2614338, EPI_ISL_2614339, EPI_ISL_2614340, EPI_ISL_2614341, EPI_ISL_2614342, EPI_ISL_2614343, EPI_ISL_2614344, EPI_ISL_2614345, EPI_ISL_2614346, EPI_ISL_2614347, EPI_ISL_2614348, EPI_ISL_2614349, EPI_ISL_2614350, EPI_ISL_6898986, EPI_ISL_6899005, EPI_ISL_7111314, EPI_ISL_7111330 | | | |
| see above | Laboratory of Respiratory Viruses and Measles, Oswaldo Cruz Institute, FIOCRUZ | Laboratory of Respiratory Viruses and Measles, Oswaldo Cruz Institute, FIOCRUZ | Alice Sampaio Rocha; Ana Carolina Mendonca; Anna Carolina Paixao; Bruna Mendonça da Silva; Elisa Cavalcante Pereira; Fernando Motta; Ighor Arantes; Jéssica Graça Macedo de Carvalho; Larissa Macedo Pinto; Luciana Appolinario; Marilda Siqueira on behalf of the Fiocruz COVID-19 Genomic Surveillance Network; Paola Resende; Patricia Brasil; Renata Serrano Lopes; Taina Venas; Victor Guimaraes |
| EPI_ISL_2157510, EPI_ISL_2157511, EPI_ISL_2157513, EPI_ISL_2157515, EPI_ISL_2157518 | Laboratório Central de Saude Publica do Estado de Santa Catarina (LACEN/SC) | Laboratory of Respiratory Viruses and Measles, Oswaldo Cruz Institute, FIOCRUZ | Alice Sampaio Rocha; Ana Carolina Mendonca; Anna Carolina Paixao; Darcita Buerger Rovaris; Elisa Cavalcante Pereira; Fernando Motta; Luciana Appolinario; Marilda Siqueira on behalf of the Fiocruz COVID-19 Genomic Surveillance Network; Paola Resende; Renata Serrano Lopes; Sandra Bianchini Fernandes; Taina Venas |
| EPI_ISL_2157516, EPI_ISL_2157517 | Laboratório Central de Saude Publica do Estado do Paraná (LACEN/PR) | Laboratory of Respiratory Viruses and Measles, Oswaldo Cruz Institute, FIOCRUZ | Alice Sampaio Rocha; Ana Carolina Mendonca; Anna Carolina Paixao; Elisa Cavalcante Pereira; Fernando Motta; Irina Nastassja Riediger; Luciana Appolinario; Maria do Carmo Debur; Marilda Siqueira on behalf of the Fiocruz COVID-19 Genomic Surveillance Network; Paola Resende; Renata Serrano Lopes; Taina Venas |
| EPI_ISL_2293001, EPI_ISL_4600577 | Laboratório Central de Saúde Pública de Santa Catarina | Coordenação Geral de Laboratórios de Saúde Pública (CGLAB/DAEVS/SVS/MS) | Vagner Fonseca; et al. |
| EPI_ISL_2777272, EPI_ISL_2777273, EPI_ISL_2777274, EPI_ISL_2777275, EPI_ISL_2777501, EPI_ISL_2777502, EPI_ISL_2777506, EPI_ISL_2777568, EPI_ISL_2777569, EPI_ISL_2777596, EPI_ISL_2777638, EPI_ISL_2777730, EPI_ISL_2777731, EPI_ISL_2777735, EPI_ISL_2777736, EPI_ISL_2777737, EPI_ISL_2777781, EPI_ISL_2777782, EPI_ISL_2777784, EPI_ISL_2777786, EPI_ISL_2777790, EPI_ISL_2777791, EPI_ISL_2777792, EPI_ISL_2777793, EPI_ISL_2777794, EPI_ISL_2777795, EPI_ISL_2777796, EPI_ISL_2777797, EPI_ISL_2777838, EPI_ISL_2777840, EPI_ISL_2777841, EPI_ISL_2777842, EPI_ISL_2777843, EPI_ISL_2777847, EPI_ISL_2777894 | | | |
| see above | Laboratório Central de Saúde Pública do Amazonas - LACEN-AM | Laboratorio de Ecologia de Doencas Transmissiveis na Amazonia, Instituto Leonidas e Maria Deane - Fiocruz Amazonia | André Corado; Debora Duarte; Felipe Naveca; Fernanda Nascimento; George Silva; Karina Pessoa; Luciana Gonçalves; Maria Júlia Brandão; Matilde Mejía; Michele Jesus; Valdinete Nascimento; Victor Souza; Ágatha Costa |
| EPI_ISL_4945109 | Laboratório Central de Saúde Pública do Distrito Federal – LACEN-DF | Laboratory of Baculovirus, University of Brasília | Agenor de Castro Moreira dos Santos Junior; Alessandra Pinheiro Medeiros; Aline Belmok; Anamélia Lorenzetti Bocca; Bergmann Morais Ribeiro; Brenno Vinicius Henrique; Fabiano José Queiroz Costa; Fernando Melo; Jordan Barros Silva; Lucas Luiz Vieira; Renato de Oliveira Resende |
| EPI_ISL_2102018, EPI_ISL_2102063 | Laboratório Central de Saúde Pública do Estado do Amazonas (LACEN-AM) | Laboratorio de Ecologia de Doencas Transmissiveis na Amazonia, Instituto Leonidas e Maria Deane - Fiocruz Amazonia | André Corado; Debora Duarte; Felipe Naveca; Fernanda Nascimento; George Silva; Karina Pessoa; Luciana Gonçalves; Maria Júlia Brandão; Matilde Mejía; Valdinete Nascimento; Victor Souza; Ágatha Costa |
| EPI_ISL_3031297, EPI_ISL_3031303, EPI_ISL_3031304, EPI_ISL_3031305 | Laboratório Municipal de Biologia Molecular | Instituto René Rachou / Fiocruz Minas | André Menezes; Anna Salim; Eneida Oliveira; Gabriel Fernandes; Pedro Alves; Rubens do Monte Neto; Thaís Silva |
| EPI_ISL_4422311 | Laboratório de Baculovirus, Universidade de Brasília (UnB), Instituto de Ciências Biológicas (IB) | Laboratório de Virologia, Faculdade de Medicina, Universidade Federal de Mato Grosso (UFMT) | Bergman Morais Ribeiro; Fernando Lucas Melo; Francisco Scoffoni Kennedy de Azevedo; Gessica Fernanda Colnago de Lima; Renata Dezengrini Slhessarenko; Thais Campos Cruz |
| EPI_ISL_6508500, EPI_ISL_6508507, EPI_ISL_6508520, EPI_ISL_6508521, EPI_ISL_6508531, EPI_ISL_6508532, EPI_ISL_6508552, EPI_ISL_6508557, EPI_ISL_6508562, EPI_ISL_6508563, EPI_ISL_6508572, EPI_ISL_6508574, EPI_ISL_6513957, EPI_ISL_6513981, EPI_ISL_6513987, EPI_ISL_6514038, EPI_ISL_6514068, EPI_ISL_6514074, EPI_ISL_6514083, EPI_ISL_6514093, EPI_ISL_6514122, EPI_ISL_6514143, EPI_ISL_6514167, EPI_ISL_6514178, EPI_ISL_6514181, EPI_ISL_6514190, EPI_ISL_6514207, EPI_ISL_6514217, EPI_ISL_6514233, EPI_ISL_6514242, EPI_ISL_6514249, EPI_ISL_6514259, EPI_ISL_6514279, EPI_ISL_6514282, EPI_ISL_6514291 | | | |
| see above | Laboratório de Biologia Integrativa/ UFMG | Laboratório de Biologia Integrativa/ UFMG | Adriana Aparecida Ribeiro; Alana Vitor Barbosa Costa; Alessandro Luís Gonçalves; Aline de Brito Lima; Ana Paula De Battisti Ribeiro; Ana Paula Salles Moura Fernandes; Andre Luiz Menezes; Bruna Walker Ferreira; Carolina Senra Alves de Souza; Cristiane P. T. Brito Mendonça; Daniel Costa Queiroz; Danielle Alves Gomes Zauli; Diego Menezes; Eneida Santos de Oliveira; Eva Lídia Arcoverde Medeiros; Felipe Campos de Melo Iani; Fernanda Gil de Souza; Fernanda Santos Mendes; Filipe Romero Rebello Moreira; Flávio Guimarães da Fonseca; Frederico Scott Varella Malta; Hugo Itaru Sato; Hugo José Alves; Igor Pereira Godinho; Jaqueline Silva de Oliveira; Joice do Prado Silva; José Nélio Januario; Juliana Wilke Saliba; Karine Lima Lourenço; Lucyene Miguita; Luige Biciati Alvim; Nara Oliveira Carvalho; Natiely Pereira Silva; Natália Rocha Guimarães; Paula Luize Camargos Fonseca; Pedro Henrique Barbosa de Paula Mendes; Rafael Marques de Souza; Renan Pedra de Souza; Renata Barbosa Peixoto Peixoto; Renato Santana de Aguiar; Rennan Garcias Moreira; Rillery Calixto Dias; Rubens Daniel Miserani Magalhães; Santuza Maria Ribeiro Teixeira; Talita Emile Ribeiro Adelino; Victor Emmanuel Viana Geddes; Walyson Coelho Costa |
| EPI_ISL_2375505, EPI_ISL_4051954 | Laboratório de Microbiologia Molecular - Universidade FEEVALE | Molecular Microbiology Laboratory | Alana Witt Hansen; Fernando Rosado Spilki; Flávio Silveira; Fágner Henrique Heldt; Juliana Schons Gularte; Juliane Deise Fleck; Mariana Soares da Silva; Matheus Nunes Weber; Meriane Demoliner; Micheli Filippi; Micheli Filippi.; Paula Rodrigues de Almeida; Vyctoria Malayhka de Abreu Góes Pereira. |
| EPI_ISL_2629770, EPI_ISL_2629771, EPI_ISL_2629772, EPI_ISL_2629773, EPI_ISL_2629774, EPI_ISL_2629775, EPI_ISL_2629776, EPI_ISL_2629777, EPI_ISL_2629778, EPI_ISL_2629779, EPI_ISL_2629780, EPI_ISL_2629781, EPI_ISL_2629782, EPI_ISL_2629783, EPI_ISL_2629784, EPI_ISL_2629785, EPI_ISL_2629786, EPI_ISL_2629787, EPI_ISL_2629788, EPI_ISL_2629789 | | | |
| see above | Laboratório de Virologia Molecular - Universidade Federal do Rio de Janeiro | Laboratório de Virologia Molecular - Universidade Federal do Rio de Janeiro | ; Alice Laschuk Herlinger; Amilcar Tanuri; André Felipe Andrade dos Santos; Carolina Moreira Voloch; Cássia Cristina Alves Gonçalves; Diana Mariani; Débora Souza Faffe; Filipe Romero Rebello Moreira; Francine Bittencourt Schiffler; Isabela de Carvalho Leitão; Marcelo Calado de Paula Tôrres; Matheus Augusto Calvano Cosentino; Mirela D'arc; Orlando da Costa Ferreira Junior; Rafael Mello Galliez; Raíssa Mirella dos Santos Cunha da Costa; Renato Santana de Aguiar; Terezinha Marta Pereira Pinto Castineiras; Thamiris dos Santos Miranda; Átila Duque Rossi |
| EPI_ISL_2443711, EPI_ISL_2677225, EPI_ISL_2677226, EPI_ISL_2677227, EPI_ISL_3061900 | Laboratorio Central de Saude Publica do Estado de Santa Catarina (LACEN/SC) | Laboratory of Respiratory Viruses and Measles, Oswaldo Cruz Institute, FIOCRUZ | Alice Sampaio Rocha; Ana Carolina Mendonca; Anna Carolina Paixao; Darcita Buerger Rovaris; Elisa Cavalcante Pereira; Fernando Motta; Luciana Appolinario; Marilda Siqueira on behalf of the Fiocruz COVID-19 Genomic Surveillance Network; Paola Resende; Renata Serrano Lopes; Sandra Bianchini Fernandes; Taina Venas |
| EPI_ISL_2614367, EPI_ISL_2614368, EPI_ISL_2614369, EPI_ISL_2614370, EPI_ISL_2614371, EPI_ISL_2614372, EPI_ISL_2614373, EPI_ISL_2614374, EPI_ISL_2614375 | | | |
| see above | Labortorio Central de Saude Publica do Estado do Rio de Janeiro (LACEN/RJ) | Laboratory of Respiratory Viruses and Measles, Oswaldo Cruz Institute, FIOCRUZ | Alice Sampaio Rocha; Ana Carolina Mendonca; Andrea Cony Cavalcanti; Anna Carolina Paixao; Elisa Cavalcante Pereira; Fernando Motta; Luciana Appolinario; Marilda Siqueira on behalf of the Fiocruz COVID-19 Genomic Surveillance Network; Paola Resende; Renata Serrano Lopes; Taina Venas |
| EPI_ISL_2196259, EPI_ISL_2443530 | Lboratorio Central de Saude Publica do Estado do Parana (LACEN/PR) | Laboratory of Respiratory Viruses and Measles, Oswaldo Cruz Institute, FIOCRUZ | Alice Sampaio Rocha; Ana Carolina Mendonca; Anna Carolina Paixao; Elisa Cavalcante Pereira; Fernando Motta; Irina Riediger; Luciana Appolinario; Marilda Siqueira on behalf of the Fiocruz COVID-19 Genomic Surveillance Network; Paola Resende; Renata Serrano Lopes; Taina Venas |
| EPI_ISL_1449193 | Lighthouse Lab in Alderley Park | Wellcome Sanger Institute for the COVID-19 Genomics UK (COG-UK) Consortium | Cordelia Langford; David K. Jackson; Dominic Kwiatkowski; Ewan Harrison; Ian Johnston; Jacquelyn Wynn; Jeffrey Barrett; John Sillitoe on behalf of the Wellcome Sanger Institute COVID-19 Surveillance Team; Mairead Hyland; Roberto Amato; Sonia Goncalves; The Lighthouse Lab in Alderley Park and Alex Alderton |
| EPI_ISL_1409655, EPI_ISL_1454248 | Lighthouse Lab in Cambridge | Wellcome Sanger Institute for the COVID-19 Genomics UK (COG-UK) Consortium | Cordelia Langford; David K. Jackson; Dominic Kwiatkowski; Ewan Harrison; Ian Johnston; Jeffrey Barrett; John Sillitoe on behalf of the Wellcome Sanger Institute COVID-19 Surveillance Team; Rob Howes; Roberto Amato; Sonia Goncalves; The Lighthouse Lab in Cambridge and Alex Alderton |
| EPI_ISL_1507943 | Lighthouse Lab in Glasgow | Wellcome Sanger Institute for the COVID-19 Genomics UK (COG-UK) Consortium | Anna Dominiczak and Alex Alderton; Carol Clugston; Cordelia Langford; David Gray; David K. Jackson; Dominic Kwiatkowski; Ewan Harrison; Harper VanSteenhouse; Ian Johnston; Jeffrey Barrett; John Sillitoe on behalf of the Wellcome Sanger Institute COVID-19 Surveillance Team; Roberto Amato; Sonia Goncalves; Yumi Kasai |
| EPI_ISL_1574983 | Loretto | Illinois Department of Public Health - Chicago Lab | Ira Heimler; Vineet K. Dhiman |
| EPI_ISL_1704772 | Lurie Children's Hospital of Chicago | Northwestern University - Ozer Lab | Egon A. Ozer; Judd F. Hultquist; Lacy M. Simons; Larry K. Kociolek; Michael G. Ison; Ramon Lorenzo-Redondo; Taylor J. Dean; William J. Muller; Xiaotian; Zheng |
| EPI_ISL_1531712 | M Health Fairview | Minnesota Department of Health, Public Health Laboratory | Alexandra Lorentz; Jacob Garfin; Matt Plumb; and Xiong Wang |
| EPI_ISL_1709625, EPI_ISL_1709626 | MSHS Clinical Microbiology Laboratories | MSHS Pathogen Surveillance Program | Adolfo García-Sastre; Adriana van de Guchte; Ajay Obla; Alberto Paniz-Mondolfi; Ana S. Gonzalez-Reiche; Angela Amoako; Ashley Salimbangon; Betsaida Salom Melo; Bremy Alburquerque; Brianne Ciferri; Charles Gleason; Daniel Floda; Deena R. Altman; Denise Jurczyszak; Emilia Mia Sordillo; Gintaras Deikus; Giulio Kleiner; Gopi Patel; Hala Alshammary; Harm van Bakel; Irina Oussenko; Jayeeta Dutta; Juan Soto; Julia Matthews; Katherine Beach; Kathryn Twyman; Kayla Russo; Komal Srivastava; Levy Sominsky; Mahmoud Awawda; Marta Luksza; Matthew M. Hernandez; Melissa Gitman; Michael D. Nowak; Mitchell J. Sullivan; Nancy Francoeur; Robert Sebra; Sarah Schaefer; Shelcie Fabre; Shwetha Hara Sridhar; Viviana Simon; Ying-Chih Wang; Zenab Khan |
| EPI_ISL_1442902 | MVZ Labor Krone GbR | Robert Koch Institute | |
| EPI_ISL_1706155, EPI_ISL_1706156 | Maryland Genomics, Institute for Genome Sciences, University of Maryland School of Medicine | Maryland Genomics, Institute for Genome Sciences, University of Maryland School of Medicine | Aditya; Claire M; Fraser; Holly; Humphrys; Jacques; Kranthi; Lisa D; Luke J; Mehta; Mike; Ott; Ravel; Roussey; Sadzewicz; Sandra; Tallon; Vavikolanu |

| | | | |
|---|---|---|---|
| EPI_ISL_1404623, EPI_ISL_1527218, EPI_ISL_1528147 | Massachusetts State Public Health Laboratory | Massachusetts State Public Health Laboratory | Andrew Lang; Glen Gallagher; Sandra Smole; Timelia Fink |
| EPI_ISL_1906135, EPI_ISL_1906185 | Michigan Department of Health and Human Services, Bureau of Laboratories | Michigan Department of Health and Human Services, Bureau of Laboratories | Blankenship HM; Riner D; Soehnlen MK |
| EPI_ISL_1503906, EPI_ISL_1547769, EPI_ISL_1547780, EPI_ISL_1583556, EPI_ISL_1583601, EPI_ISL_1583612, EPI_ISL_1636419, EPI_ISL_1636431, EPI_ISL_1636443, EPI_ISL_1636455 | | | |
| see above | Microbiology Department, Laboratori Clínic Metropolitana Nord. Hospital Universitari Germans Trias i Pujol. | Can Ruti SARS-CoV-2 Sequencing Hub (HUGTiP/IrsiCaixa/IGTP) | Alba Sánchez; Anna Not; Antoni E Bordoy; Bonaventura Clotet; Cristina Casañ; Cristina Esteban; Francesc Catala-Moll; Gemma Clara; Ignacio Blanco; Marc Noguera-Julian; Maria Casadellà; Mariona Parera; Mercedes Guerrero; Montserrat Giménez; Pere-Joan Cardona; Pilar Armengol; Roger Paredes; Verónica Saludes; and Elisa Martró on behalf of the Can Ruti SARS-CoV-2 Sequencing Hub. |
| EPI_ISL_1478843, EPI_ISL_2086122 | Microbiology Department. Complexo Hospitalario Universitario de Vigo | Microbiology Department. Complexo Hospitalario Universitario de Vigo | Alfaia N; Alfaya N; Alonso I; Alvarez M; Cabrera JJ; Carballo R; Cores O; Cortizo S; Martinez L; Mediero G; Perez S; Potel C; Regueiro B; Rey S; Vasallo FJ; del-Campo V |
| EPI_ISL_2090619 | Microbiology, University of Pennsylvania | Microbiology, University of Pennsylvania | A.M.; A.S.; Collman; Everett, J.; Feldman, M.; Fitzgerald; Ganguly, A.; Glascock, A.; Graham-Wooten, J.; Hokama, P.; Hwang, Y.; Kelly, B.; Khatib; L.A.; R.G. and Bushman, F.; Reddy, S.; Roche; Rodino, K.; S.A.; Sherrill-Mix, S.; Whiteside |
| EPI_ISL_1661393, EPI_ISL_1661416 | Microvida | Microvida | Jaco Verweij; Joep Stöhr; Suzan D. Pas |
| EPI_ISL_1760136, EPI_ISL_1760137, EPI_ISL_2139609, EPI_ISL_2139610, EPI_ISL_2139611 | Ministry of Health Turkey | Ministry of Health Turkey | Fatma Bayrakdar; Gulay Korukluoglu; Suleyman Yalcin; Yasemin Cosgun |
| EPI_ISL_1798542 | NE Public Health Laboratory | Centers for Disease Control and Prevention Division of Viral Diseases, Pathogen Discovery | Alison Laufer Halpin; Ben L. Rambo-Martin; Clinton R. Paden; Dakota Howard; Darlene Wagner; Dave Wentworth; Dhwani Batra; Jasmine Padilla; Justin Lee; Katie Dillon; Krista Queen; Kristen Knipe; Kristine Lacek; Mark Burroughs; Matthew Schmerer; Mili Sheth; Peter Cook; Sam Shepard; Sarah Nobles; Shoshona Le; Suxiang Tong; Vivien Dugan; Yvette Unoarumhi |
| EPI_ISL_3102314 | NIDADE MISTA DE SAUDE DE MISSAO VELHA | Analytical Competence Molecular Epidemiology Lab/ACME, Oswaldo Cruz Foundation, Ceara (FIOCRUZ CE) | Cleber Furtado Aksenen; Fabio Miyajima; Fernando Braga Stehling; Francisco Eder de Moura Lopes; Jamille Maria Mendes Bezerra; Joaquim César do Nascimento Sousa Junior; Pedro Miguel Carneiro Jeronimo; Suzana Porto Almeida e Lucas Delerino; Thais Ferreira de Oliveira; Thais de Oliveira Costa; Ticiane Cavalcante de Souza; Veridiana Pessoa Miyajima |
| EPI_ISL_1530943 | NJDOH, Public Health and Environmental Laboratories | New Jersey Public Health and Environmental Laboratories (NJPHEL) | Byeong Jeong; Dana Woell; Jacquelyn Deverell; Lindsey Bodnar; Matthew Scarnati; Mohammad M Ali; Shiv K. Verma |
| EPI_ISL_1795379, EPI_ISL_2345956 | NUCLEO DE SAUDE VILA FALCAO DE BAURU | Instituto Butantan / ESALQ-Piracicaba | Antonio Jorge Martins; Bianca Cechetto Carlos. Mendelics: Bibiana Santos; Claudia Renata dos Santos Barros; David Schlesinger; David Schlesinger. Hemocentro Ribeirão Preto: Simone Kashima; Debora Botequio Moretti; Debora Botequio Moretti. Centro de Genômica Funcional da ESALQ: Luiz Lehmann Coutinho; Dimas Tadeu Covas; Elaine Cristina Marqueze; Elaine Vieira Santos; Elaine Vieira dos Santos; Elisangela Chicaroni Mattos; Erika Freitas; Evandra Strazza Rodrigues; Felipe Allan da Silva da Costa; Flavia Aburjaile; Guilherme Targino Valente; Heidge Fukumasu; Heidge Fukumasu. USP-Botucatu: Rejane Maria Tommasini Grotto; Instituto Butantan: Alexander Roberto Precioso; Jayme A. Souza-Neto; Jayme Augusto de Souza-Neto; Jessika Cristina Chagas Lesbon; José Salvatore Leister Patané; João Paulo Kitajima; Luiz Alcantara; Luiz Carlos Junior de Alcantara; Luiz Lehmann Coutinho; Maria Carolina Elias; Marta Giovanetti; Maurício Lacerda Nogueira; Patricia Akemi Assato; Rafael dos Santos Bezerra; Raquel de Lello Rocha Campos Cassano. NGS Soluções Genômicas: Pilar Drummond Sampaio Corrêa Mariani. FZEA-USP Pirassununga: Mirele Daiana Poleti; Raul Machado Neto; Rejane Maria Tommasini Grotto; Ricardo Augusto Brassaloti; Ricardo Haddad; Rodrigo Tocantins Calado.; Sandra Coccuzzo Sampaio; Sandra Coccuzzo Sampaio Vessoni; Simone Kashima; Svetoslav Nanev Slavov; Vagner Fonseca; Vincent Louis Viala |
| EPI_ISL_1711773 | NV State Public Health Laboratory | Centers for Disease Control and Prevention Division of Viral Diseases, Pathogen Discovery | Alison Laufer Halpin; Ben L. Rambo-Martin; Clinton R. Paden; Dakota Howard; Darlene Wagner; Dave Wentworth; Dhwani Batra; Jasmine Padilla; Justin Lee; Katie Dillon; Krista Queen; Kristen Knipe; Kristine Lacek; Mark Burroughs; Matthew Schmerer; Mili Sheth; Peter Cook; Sam Shepard; Sarah Nobles; Shoshona Le; Suxiang Tong; Vivien Dugan; Yvette Unoarumhi |
| EPI_ISL_1654584 | NYU Langone Health | Departments of Pathology and Medicine, New York University School of Medicine | Adriana Heguy; Christian Marier; Dacia Dimartino; Emily Guzman; Gael Westby; Guiqing Wang; Paolo Cotzia; Paul Zappile; Peter Meyn; Sitharam Ramaswami; Yutong Zhang |
| EPI_ISL_1623665, EPI_ISL_1623675, EPI_ISL_1665214 | National Platform bis UMONS/Jolimont | National Platform bis UMONS/Jolimont | François Dufrasne; Gautier Detry; Guillaume Bayon-Vicente; Ruddy Wattiez |
| EPI_ISL_1577415, EPI_ISL_1651764, EPI_ISL_1651781, EPI_ISL_1651814 | National Virus Reference Laboratory | National Virus Reference Laboratory | Calum Walsh; Charlene Bennet; Charlene Bennett; Cillian F De Gascun; Fiona Crispie; Gabriel Gonzalez; Jonathan Dean; Matthew McCabe; Michael Carr; Paul Cotter; Zoe Yandle |
| EPI_ISL_1574981 | Near North Health Service | Illinois Department of Public Health - Chicago Lab | Ira Heimler; Vineet K. Dhiman |
| EPI_ISL_2692030 | Northwestern Memorial Hospital | RIPHL at Rush University Medical Center | Amber Kimble; Chao Qi; Felix Araujo Perez; Kevin Kunstman; Laura Furtado; Marieta Hyde; Max Kolton; Stefan Green |
| EPI_ISL_5799781 | Nucleo De Saude Vila Falcao De Bauru | Instituto Butantan | Antonio Jorge Martins; Claudia Renata dos Santos Barros; David Schlesinger; Debora Botequio Moretti; Dimas Tadeu Covas; Elaine Cristina Marqueze; Elaine Vieira Santos; Evandra Strazza Rodrigues; Heidge Fukumasu; Jayme Augusto de Souza-Neto; José Salvatore Leister Patané; Luiz Alcantara; Luiz Lehmann Coutinho; Maria Carolina Elias; Maurício Lacerda Nogueira; Rafael dos Santos Bezerra; Raul Machado Neto; Rejane Maria Tommasini Grotto; Ricardo Haddad; Sandra Coccuzzo Sampaio Vessoni; Simone Kashima; Svetoslav Nanev Slavov; Vincent Louis Viala |
| EPI_ISL_1542030 | OHSU Lab Services Molecular Microbiology Lab | Oregon SARS-CoV-2 Genome Sequencing Center | Alec J. Hirsch; Andrew C. Adey; Benjamin N. Bimber; Brendan L. O'Connell; Brian J. O'Roak; Daniel N. Streblow; Donna Hansel; Guang Fan; Kayla Carter; Ruth V. Nichols; Sally Grindstaff; Sonia Acharya; William B. Messer; Xuan Qin |
| EPI_ISL_7274019, EPI_ISL_7274163, EPI_ISL_7274176, EPI_ISL_7274504, EPI_ISL_7274525, EPI_ISL_7274546, EPI_ISL_7274632, EPI_ISL_7274644, EPI_ISL_7274687, EPI_ISL_7274738, EPI_ISL_7274875 | | | |
| see above | Ontario's COVID-19 Genomics Rapid Response Coalition | McMaster University | Ahmed Draia; Allison McGeer; Andrew G. McArthur; Angel Li; Emily Panousis; Hooman Derakhshani; Jalees Nasir; Kuganya Nirmalarajah; Michael Surette; Patryk Aftanas; Samira Mubareka; Sheridan Baker |
| EPI_ISL_1931621 | Osaka Institute of Public Health, Morinomiya Center | Pathogen Genomics Center, National Institute of Infectious Diseases | Hazuka Y Furihata; Kentaro Itokawa; Makoto Kuroda; Masanori Hashino; Masumichi Saito; Naomi Nojiri; Nozomu Hanaoka; Rina Tanaka; Sana Uchikoba; Tsuguto Fujimoto; Tsuyoshi Sekizuka |
| EPI_ISL_1795336, EPI_ISL_2345602 | PAS JOAO ANTONIO DO NASCIMENTO | Instituto Butantan / ESALQ-Piracicaba | Antonio Jorge Martins; Bianca Cechetto Carlos. Mendelics: Bibiana Santos; Claudia Renata dos Santos Barros; David Schlesinger; David Schlesinger. Hemocentro Ribeirão Preto: Simone Kashima; Debora Botequio Moretti; Debora Botequio Moretti. Centro de Genômica Funcional da ESALQ: Luiz Lehmann Coutinho; Dimas Tadeu Covas; Elaine Cristina Marqueze; Elaine Vieira Santos; Elaine Vieira dos Santos; Elisangela Chicaroni Mattos; Erika Freitas; Evandra Strazza Rodrigues; Felipe Allan da Silva da Costa; Flavia Aburjaile; Guilherme Targino Valente; Heidge Fukumasu; Heidge Fukumasu. USP-Botucatu: Rejane Maria Tommasini Grotto; Instituto Butantan: Alexander Roberto Precioso; Jayme A. Souza-Neto; Jayme Augusto de Souza-Neto; Jessika Cristina Chagas Lesbon; José Salvatore Leister Patané; João Paulo Kitajima; Luiz Alcantara; Luiz Carlos Junior de Alcantara; Luiz Lehmann Coutinho; Maria Carolina Elias; Marta Giovanetti; Maurício Lacerda Nogueira; Patricia Akemi Assato; Rafael dos Santos Bezerra; Raquel de Lello Rocha Campos Cassano. NGS Soluções Genômicas: Pilar Drummond Sampaio Corrêa Mariani. FZEA-USP Pirassununga: Mirele Daiana Poleti; Raul Machado Neto; Rejane Maria Tommasini Grotto; Ricardo Augusto Brassaloti; Ricardo Haddad; Rodrigo Tocantins Calado.; Sandra Coccuzzo Sampaio; Sandra Coccuzzo Sampaio Vessoni; Simone Kashima; Svetoslav Nanev Slavov; Vagner Fonseca; Vincent Louis Viala |
| EPI_ISL_5530052 | POSTO DE SAUDE FRANCISCA ROMANA DE OLIVEIRA | Analytical Competence Molecular Epidemiology Lab/ACME, Oswaldo Cruz Foundation, Ceara (FIOCRUZ CE) | Carlos Leonardo de Aragao Araujo; Cecilia Leite Costa & Eduardo Ruback dos Santos on behalf of COVID-19 FIOCRUZ Genomic Network; Cleber Furtado Aksenen; Fabio Miyajima; Fernando Braga Stehling; Francisco Eder de Moura Lopes; Igor Oliveira Duarte; Jamille Maria Mendes Bezerra; Joaquim Cesar do Nascimento Sousa Junior; Pedro Miguel Carneiro Jeronimo; Suzana Porto Almeida; Thais Ferreira de Oliveira; Thais de Oliveira Costa; Ticiane Cavalcante de Souza; Veridiana Pessoa Miyajima |
| EPI_ISL_1966315, EPI_ISL_1966320 | PRONTO SOCORRO DE AGENOR DE CAMPOS MONGAGUA | Instituto Butantan / Mendelics | Antonio Jorge Martins; Bianca Cechetto Carlos. Mendelics: Bibiana Santos; Claudia Renata dos Santos Barros; Cíntia Bittar; David Schlesinger. Hemocentro Ribeirão Preto: Simone Kashima; Debora Botequio Moretti; Elaine Cristina Marqueze; Elaine Vieira dos Santos; Elisangela Chicaroni Mattos; Erika Freitas; Evandra Strazza Rodrigues; Felipe Allan da Silva da Costa; Flavia Aburjaile; Fábio Sossai Possebon; Guilherme Campos; Guilherme Targino Valente; Heidge Fukumasu. USP-Botucatu: Rejane Maria Tommasini Grotto; Helena Lage Ferreira; Instituto Butantan: Dimas Tadeu Covas; Jardelina de Souza Todao Bernardino; Jayme A. Souza-Neto; Jessika Cristina Chagas Lesbon; Jorge A. Petroli Marchesi; José Salvatore Leister Patané; João Paulo Kitajima; João Pessoa Araújo Jr.; Leila Sabrina Ullmann; Loyze Paola Oliveira de Lima; Luiz Aurelio de Campos Crispin. Centro de Genômica Funcional da ESALQ: Luiz Lehmann Coutinho; Luiz Carlos Junior de Alcantara; Lívia Sacchetto; Maisa C. Pereira Parra; Maria Carolina Elias; Marta Giovanetti; Marília Moraes; Maurício Lacerda Nogueira. Prefeitura de Sao Paulo: Melissa Palmieri.; Patricia Akemi Assato; Paula Rahal; Paulo Inacio da Costa; Rafael dos Santos Bezerra; Raquel de Lello Rocha Campos Cassano. NGS Soluções Genômicas: Pilar Drummond Sampaio Corrêa Mariani. FZEA-USP Pirassununga: Mirele Daiana Poleti; Raul Machado Neto; Ricardo Augusto Brassaloti; Ricardo Haddad; Rodrigo Tocantins Calado. FAMERP-SJRP: Cecília Artico Banho; Sandra Coccuzzo Sampaio; Svetoslav Nanev Slavov; Vagner Fonseca; Vincent Louis Viala |
| EPI_ISL_1795213, EPI_ISL_1795214, EPI_ISL_1795215, EPI_ISL_1795216, EPI_ISL_2345441, EPI_ISL_2345442, EPI_ISL_2345443, EPI_ISL_2345444 | | | |
| see above | PRONTO SOCORRO MUNICIPAL DE SEVERINIA | Instituto Butantan / ESALQ-Piracicaba | Antonio Jorge Martins; Bianca Cechetto Carlos. Mendelics: Bibiana Santos; Claudia Renata dos Santos Barros; David Schlesinger; David Schlesinger. Hemocentro Ribeirão Preto: Simone Kashima; Debora Botequio Moretti; Debora Botequio Moretti. Centro de Genômica Funcional da ESALQ: Luiz Lehmann Coutinho; Dimas Tadeu Covas; Elaine Cristina Marqueze; Elaine Vieira Santos; Elaine Vieira dos Santos; Elisangela Chicaroni Mattos; Erika Freitas; Evandra Strazza Rodrigues; Felipe Allan da Silva da Costa; Flavia Aburjaile; Guilherme Targino Valente; Heidge Fukumasu; Heidge Fukumasu. USP-Botucatu: Rejane Maria Tommasini Grotto; Instituto Butantan: Alexander Roberto Precioso; Jayme A. Souza-Neto; Jayme Augusto de Souza-Neto; Jessika Cristina Chagas Lesbon; José Salvatore Leister Patané; João Paulo Kitajima; Luiz Alcantara; Luiz Carlos Junior de Alcantara; Luiz Lehmann Coutinho; Maria Carolina Elias; Marta Giovanetti; Maurício Lacerda Nogueira; Patricia Akemi Assato; Rafael dos Santos Bezerra; Raquel de Lello Rocha Campos Cassano. NGS Soluções Genômicas: Pilar Drummond Sampaio Corrêa Mariani. FZEA-USP Pirassununga: Mirele Daiana Poleti; Raul Machado Neto; Rejane Maria Tommasini Grotto; Ricardo Augusto Brassaloti; Ricardo Haddad; Rodrigo Tocantins Calado.; Sandra Coccuzzo Sampaio; Sandra Coccuzzo Sampaio Vessoni; Simone Kashima; Svetoslav Nanev Slavov; Vagner Fonseca; Vincent Louis Viala |
| EPI_ISL_1795300, EPI_ISL_1795301, EPI_ISL_1795302, EPI_ISL_1795304, EPI_ISL_2345543, EPI_ISL_2345545, EPI_ISL_2345548, EPI_ISL_2345550 | | | |
| see above | PRONTO SOCORRO MUNICIPAL TAMBAU | Instituto Butantan / ESALQ-Piracicaba | Antonio Jorge Martins; Bianca Cechetto Carlos. Mendelics: Bibiana Santos; Claudia Renata dos Santos Barros; David Schlesinger; David Schlesinger. Hemocentro Ribeirão Preto: Simone Kashima; Debora Botequio Moretti; Debora Botequio Moretti. Centro de Genômica Funcional da ESALQ: Luiz Lehmann Coutinho; Dimas Tadeu Covas; Elaine Cristina Marqueze; Elaine Vieira Santos; Elaine Vieira dos Santos; Elisangela Chicaroni Mattos; Erika Freitas; Evandra Strazza Rodrigues; Felipe Allan da Silva da Costa; Flavia Aburjaile; Guilherme Targino Valente; Heidge Fukumasu; Heidge Fukumasu. USP-Botucatu: Rejane Maria Tommasini Grotto; Instituto Butantan: Alexander Roberto Precioso; Jayme A. Souza-Neto; Jayme Augusto de Souza-Neto; Jessika Cristina Chagas Lesbon; José Salvatore Leister Patané; João Paulo Kitajima; Luiz Alcantara; Luiz Carlos Junior de Alcantara; Luiz Lehmann Coutinho; Maria Carolina Elias; Marta Giovanetti; Maurício Lacerda Nogueira; Patricia Akemi Assato; Rafael dos Santos Bezerra; Raquel de Lello Rocha Campos Cassano. NGS Soluções Genômicas: Pilar Drummond Sampaio Corrêa Mariani. FZEA-USP Pirassununga: Mirele Daiana Poleti; Raul Machado Neto; Rejane Maria Tommasini Grotto; Ricardo Augusto Brassaloti; Ricardo Haddad; Rodrigo Tocantins Calado.; Sandra Coccuzzo Sampaio; Sandra Coccuzzo Sampaio Vessoni; Simone Kashima; Svetoslav Nanev Slavov; Vagner Fonseca; Vincent Louis Viala |
| EPI_ISL_1385316, EPI_ISL_1385397, EPI_ISL_1385576, EPI_ISL_1385656, EPI_ISL_1385740, EPI_ISL_1385758, EPI_ISL_1385771, EPI_ISL_1470859, EPI_ISL_1470879, EPI_ISL_1470885, EPI_ISL_1470898, EPI_ISL_1471084, EPI_ISL_1471160, EPI_ISL_1471230, EPI_ISL_1471246, EPI_ISL_1471407, EPI_ISL_1471500, EPI_ISL_1471583 | | | |

| | | | |
|---|---|---|---|
| see above | Pandemic Response Lab - NYC | Pandemic Response Lab, R&D | Cybill del Castillo; Dylan Law; Haiping Hao; Henry Lee; Jon Laurent; Melissa Hopkins; Michael Hammerling; Pradeep Bugga; Shinyoung Clair Kang; Sol Rey; William Ward |
| EPI_ISL_1416322 | PathWest Laboratory Medicine WA | PathWest Laboratory Medicine WA Microbial Surveillance Unit | PathWest Laboratory Medicine WA Microbial Surveillance Unit |
| EPI_ISL_1929419 | Pathogen Genomics Center, National Institute of Infectious Diseases | Pathogen Genomics Center, National Institute of Infectious Diseases | Hidemasa Izumiya; Ken-ichi Lee; Kentaro Itokawa; Makoto Kuroda; Masanori Hashino; Masatomo Morita; Nobuo Koizumi; Rina Tanaka; Shouji Yamamoto; Sunao Iyoda; Tsuyoshi Sekizuka |
| EPI_ISL_2663303, EPI_ISL_2663304, EPI_ISL_2663305, EPI_ISL_2663306, EPI_ISL_2663307 | Plataforma de Vigilancia Molecular (PVM) - FIOCRUZ/BA | Plataforma de Vigilancia Molecular (PVM) - FIOCRUZ/BA | Bruno Bezerril Andrade; Camila I. de Oliveira on behalf of the Fiocruz COVID-19 Genomic Surveillance Network.; Clarissa Araújo Gurgel; Leonardo Paiva Farias; Marina Cucco; Ricardo Khouri; Tiago Graf |
| EPI_ISL_1788109 | Plateforme de testing Namuroise | Plateforme de testing Namuroise | Céline Maschietto; Degosserie Jonathan; Denis Olivier; Mullier François; Otto Gaetan |
| EPI_ISL_1443701, EPI_ISL_1443720, EPI_ISL_1498358, EPI_ISL_1498359, EPI_ISL_1498360, EPI_ISL_1498363, EPI_ISL_1498364, EPI_ISL_1498365, EPI_ISL_1498367 | | | |
| see above | Platform BIS UZA/UAntwerpen | Labo Klinische Biologie, UZA | Basil Britto Xavier; Christine Lammens; Herman Goossens; Jasmine Coppens; Marie Le Mercier; Veerle Matheeussen |
| EPI_ISL_1416805 | Platform BIS UZA/UAntwerpen | UAntwerp, Laboratory of Medical Microbiology | Basil Britto Xavier; Christine Lammens; Herman Goossens; Jasmine Coppens; Marie Le Mercier; Veerle Matheeussen |
| EPI_ISL_2759571 | Population Medicine and Diagnostic Sciences, Cornell University | Population Medicine and Diagnostic Sciences, Cornell University | Anderson; B.D.; Caserta; Cronk; D.G.; Diel; L.C.; Laverack, M.; Mitchell; P.K.; Plocharczyk, E.; R.R.; Venugopalan, R. |
| EPI_ISL_5802024, EPI_ISL_5802032 | Pronto Socorro De Agenor De Campos Mongagua | Instituto Butantan | Antonio Jorge Martins; Claudia Renata dos Santos Barros; David Schlesinger; Debora Botequio Moretti; Dimas Tadeu Covas; Elaine Cristina Marqueze; Elaine Vieira Santos; Evandra Strazza Rodrigues; Heidge Fukumasu; Jayme Augusto de Souza-Neto; José Salvatore Leister Patané; Luiz Alcantara; Luiz Lehmann Coutinho; Maria Carolina Elias; Maurício Lacerda Nogueira; Rafael dos Santos Bezerra; Raul Machado Neto; Rejane Maria Tommasini Grotto; Ricardo Haddad; Sandra Coccuzzo Sampaio Vessoni; Simone Kashima; Svetoslav Nanev Slavov; Vincent Louis Viala |
| EPI_ISL_2551514 | Providence Oregon Regional Laboratories | Providence St. Joseph Health Molecular Genomics Laboratory | Alexa K Dowdell; Brian D Piening; Carlo B Bifulco; Fred L Robinson; Mary Campbell; Rogan Rattray |
| EPI_ISL_2551272, EPI_ISL_2551274 | Providence Regional Medical Center Everett | Providence St. Joseph Health Molecular Genomics Laboratory | Alexa K Dowdell; Brian D Piening; Carlo B Bifulco; Fred L Robinson; Mary Campbell; Rogan Rattray |
| EPI_ISL_1559414, EPI_ISL_1559467, EPI_ISL_1559538, EPI_ISL_1559585, EPI_ISL_1581990, EPI_ISL_1582080, EPI_ISL_1694690, EPI_ISL_1694691, EPI_ISL_1694692, EPI_ISL_1694693, EPI_ISL_1694694, EPI_ISL_1694695, EPI_ISL_1694946, EPI_ISL_1694947, EPI_ISL_1694948, EPI_ISL_1694949, EPI_ISL_1694950, EPI_ISL_1694951, EPI_ISL_1694952, EPI_ISL_1694953, EPI_ISL_1694954, EPI_ISL_1694955, EPI_ISL_1694956, EPI_ISL_4369570 | | | |
| see above | Quest Diagnostics Incorporated | Centers for Disease Control and Prevention Division of Viral Diseases, Pathogen Discovery | A. Gerasimova; A. Perez; Adrian Paskey; B. Anderson; Benjamin Rambo-Martin; Christopher Gulvick; Clinton Paden; Clinton R. Paden; Dakota Howard; Darlene Wagner; Dhwani Batra; Duncan MacCannell; Erisa Sula; F. Lacbawan; I. A. Shlyakhter; I. Shlyakhter; Jason Caravas; K. Livingston; K.E. Livingston; Kara Moser; Kristine Lacek; L. Bernstein; L.E. Bernstein; M. Hua; Matthew Schmerer; P. Tanpaiboon; Peter Cook; Peter W. Cook; R. Kagan; R. M. Kagan; R. Owen; R. Rolando; R. V. Rolando; S. H. Rosenthal; S. Rosenthal; Scott Sammons; Shatavia Morrison; Tymeckia Kendall; Victoria Caban Figueroa; Y. Liu; Yvette Unoarumhi |
| EPI_ISL_1976763 | Regional Virus Laboratory, Belfast Health and Social Care Trust | COVID-19 Genomics UK (COG-UK) Consortium | Alison Watt; Ciara Cox; Conall McCaughey; David Simpson; Derek Fairley; James McKenna; Mairead Connor; Susan Feeney; Tanya Curran; Zoltan Molnar |
| EPI_ISL_7156280, EPI_ISL_7156284, EPI_ISL_7156288, EPI_ISL_7156289, EPI_ISL_7156290, EPI_ISL_7156291, EPI_ISL_7156292, EPI_ISL_7156293, EPI_ISL_7156294, EPI_ISL_7156295, EPI_ISL_7156296, EPI_ISL_7156297 | | | |
| see above | Research Education in Disease Diagnosis and Intervention (REDDI) Lab, Clemson University | Research Education in Disease Diagnosis and Intervention (REDDI) Lab, Clemson University | Adib Shafi; Brian Krueger; Chloe Emerson; Christopher Parkinson; Christopher Saski; Congyue Peng; Delphine Dean; John Pruitt; Justin Napolitano; Kaitlyn Williams; Keegan Sell; Kylie King; Lax Iyer; Rachel Dango; Rachel Ham; Scott Parker; Stevin Wilson; Sujata Srikanth |
| EPI_ISL_1578134, EPI_ISL_1578138, EPI_ISL_1578145, EPI_ISL_1578151, EPI_ISL_1578349 | Rhode Island Department of Health | Infectious Disease Program, Broad Institute of Harvard and MIT | Adams, G.; Azevedo, K.; B.L.; B.W.; Bauer, M.; Birren; Carter, A.; Chaluvadi, S.; D.J.; DeRuff, K.; Gladden-Young, A.; Huard, R.; J.E.; K.J.; King, E.; Lagerborg, K.; Lemieux; Loreth, C.; Miller, A.; Normandin, E.; P.C.; Park; Pearlman, L.; Reilly, S.; Rudy, M.; Sabeti; Siddle; Tomkins-Tinch, C.; and MacInnis |
| EPI_ISL_2603858 | Rhode Island State Health Laboratory | Rhode Island State Health Laboratory | Ewa King; Kristin Carpenter-Azevedo; Richard C. Huard |
| EPI_ISL_1795361, EPI_ISL_2345636 | SANTA CASA DE MISERICORDIA DE UBATUBA | Instituto Butantan / ESALQ-Piracicaba | Antonio Jorge Martins; Bianca Cechetto Carlos. Mendelics: Bibiana Santos; Claudia Renata dos Santos Barros; David Schlesinger. Hemocentro Ribeirão Preto: Simone Kashima; Debora Botequio Moretti; Debora Botequio Moretti. Centro de Genômica Funcional da ESALQ: Luiz Lehmann Coutinho; Dimas Tadeu Covas; Elaine Cristina Marqueze; Elaine Vieira Santos; Elaine Vieira dos Santos; Elisangela Chicaroni Mattos; Erika Freitas; Evandra Strazza Rodrigues; Felipe Allan da Silva da Costa; Flavia Aburjaile; Guilherme Targino Valente; Heidge Fukumasu; Heidge Fukumasu. USP-Botucatu: Rejane Maria Tommasini Grotto; Instituto Butantan: Alexander Roberto Precioso; Jayme A. Souza-Neto; Jayme Augusto de Souza-Neto; Jessika Cristina Chagas Lesbon; José Salvatore Leister Patané; João Paulo Kitajima; Luiz Alcantara; Luiz Carlos Junior de Alcantara; Luiz Lehmann Coutinho; Maria Carolina Elias; Marta Giovanetti; Maurício Lacerda Nogueira; Patricia Akemi Assato; Rafael dos Santos Bezerra; Raquel de Lello Rocha Campos Cassano. NGS Soluções Genômicas: Pilar Drummond Sampaio Corrêa Mariani. FZEA-USP Pirassununga: Mirele Daiana Poleti; Raul Machado Neto; Rejane Maria Tommasini Grotto; Ricardo Augusto Brassaloti; Ricardo Haddad; Rodrigo Tocantins Calado.; Sandra Coccuzzo Sampaio; Sandra Coccuzzo Sampaio Vessoni; Simone Kashima; Svetoslav Nanev Slavov; Vagner Fonseca; Vincent Louis Viala |
| EPI_ISL_2327224 | SARS-CoV-2 testing team, National Institute of Infectious Diseases | Pathogen Genomics Center, National Institute of Infectious Diseases | Hazuka Y Furihata; Kentaro Itokawa; Makoto Kuroda; Masanori Hashino; Masumichi Saito; Naomi Nojiri; Nozomu Hanaoka; Rina Tanaka; Sana Uchikoba; Tsuguto Fujimoto; Tsuyoshi Sekizuka |
| EPI_ISL_1795364, EPI_ISL_2345639 | SECRETARIA DE SAUDE | Instituto Butantan / ESALQ-Piracicaba | Antonio Jorge Martins; Bianca Cechetto Carlos. Mendelics: Bibiana Santos; Claudia Renata dos Santos Barros; David Schlesinger; David Schlesinger. Hemocentro Ribeirão Preto: Simone Kashima; Debora Botequio Moretti; Debora Botequio Moretti. Centro de Genômica Funcional da ESALQ: Luiz Lehmann Coutinho; Dimas Tadeu Covas; Elaine Cristina Marqueze; Elaine Vieira Santos; Elaine Vieira dos Santos; Elisangela Chicaroni Mattos; Erika Freitas; Evandra Strazza Rodrigues; Felipe Allan da Silva da Costa; Flavia Aburjaile; Guilherme Targino Valente; Heidge Fukumasu; Heidge Fukumasu. USP-Botucatu: Rejane Maria Tommasini Grotto; Instituto Butantan: Alexander Roberto Precioso; Jayme A. Souza-Neto; Jayme Augusto de Souza-Neto; Jessika Cristina Chagas Lesbon; José Salvatore Leister Patané; João Paulo Kitajima; Luiz Alcantara; Luiz Carlos Junior de Alcantara; Luiz Lehmann Coutinho; Maria Carolina Elias; Marta Giovanetti; Maurício Lacerda Nogueira; Patricia Akemi Assato; Rafael dos Santos Bezerra; Raquel de Lello Rocha Campos Cassano. NGS Soluções Genômicas: Pilar Drummond Sampaio Corrêa Mariani. FZEA-USP Pirassununga: Mirele Daiana Poleti; Raul Machado Neto; Rejane Maria Tommasini Grotto; Ricardo Augusto Brassaloti; Ricardo Haddad; Rodrigo Tocantins Calado.; Sandra Coccuzzo Sampaio; Sandra Coccuzzo Sampaio Vessoni; Simone Kashima; Svetoslav Nanev Slavov; Vagner Fonseca; Vincent Louis Viala |
| EPI_ISL_1795108, EPI_ISL_2344662 | SECRETARIA DE SAUDE DE SAO PEDRO | Instituto Butantan / ESALQ-Piracicaba | Antonio Jorge Martins; Bianca Cechetto Carlos. Mendelics: Bibiana Santos; Claudia Renata dos Santos Barros; David Schlesinger; David Schlesinger. Hemocentro Ribeirão Preto: Simone Kashima; Debora Botequio Moretti; Debora Botequio Moretti. Centro de Genômica Funcional da ESALQ: Luiz Lehmann Coutinho; Dimas Tadeu Covas; Elaine Cristina Marqueze; Elaine Vieira Santos; Elaine Vieira dos Santos; Elisangela Chicaroni Mattos; Erika Freitas; Evandra Strazza Rodrigues; Felipe Allan da Silva da Costa; Flavia Aburjaile; Guilherme Targino Valente; Heidge Fukumasu; Heidge Fukumasu. USP-Botucatu: Rejane Maria Tommasini Grotto; Instituto Butantan: Alexander Roberto Precioso; Jayme A. Souza-Neto; Jayme Augusto de Souza-Neto; Jessika Cristina Chagas Lesbon; José Salvatore Leister Patané; João Paulo Kitajima; Luiz Alcantara; Luiz Carlos Junior de Alcantara; Luiz Lehmann Coutinho; Maria Carolina Elias; Marta Giovanetti; Maurício Lacerda Nogueira; Patricia Akemi Assato; Rafael dos Santos Bezerra; Raquel de Lello Rocha Campos Cassano. NGS Soluções Genômicas: Pilar Drummond Sampaio Corrêa Mariani. FZEA-USP Pirassununga: Mirele Daiana Poleti; Raul Machado Neto; Rejane Maria Tommasini Grotto; Ricardo Augusto Brassaloti; Ricardo Haddad; Rodrigo Tocantins Calado.; Sandra Coccuzzo Sampaio; Sandra Coccuzzo Sampaio Vessoni; Simone Kashima; Svetoslav Nanev Slavov; Vagner Fonseca; Vincent Louis Viala |
| EPI_ISL_1795111, EPI_ISL_1795112, EPI_ISL_2345310, EPI_ISL_2345311 | SECRETARIA MUNICIPAL DA SAUDE DE GUARIBA | Instituto Butantan / ESALQ-Piracicaba | Antonio Jorge Martins; Bianca Cechetto Carlos. Mendelics: Bibiana Santos; Claudia Renata dos Santos Barros; David Schlesinger; David Schlesinger. Hemocentro Ribeirão Preto: Simone Kashima; Debora Botequio Moretti; Debora Botequio Moretti. Centro de Genômica Funcional da ESALQ: Luiz Lehmann Coutinho; Dimas Tadeu Covas; Elaine Cristina Marqueze; Elaine Vieira Santos; Elaine Vieira dos Santos; Elisangela Chicaroni Mattos; Erika Freitas; Evandra Strazza Rodrigues; Felipe Allan da Silva da Costa; Flavia Aburjaile; Guilherme Targino Valente; Heidge Fukumasu; Heidge Fukumasu. USP-Botucatu: Rejane Maria Tommasini Grotto; Instituto Butantan: Alexander Roberto Precioso; Jayme A. Souza-Neto; Jayme Augusto de Souza-Neto; Jessika Cristina Chagas Lesbon; José Salvatore Leister Patané; João Paulo Kitajima; Luiz Alcantara; Luiz Carlos Junior de Alcantara; Luiz Lehmann Coutinho; Maria Carolina Elias; Marta Giovanetti; Maurício Lacerda Nogueira; Patricia Akemi Assato; Rafael dos Santos Bezerra; Raquel de Lello Rocha Campos Cassano. NGS Soluções Genômicas: Pilar Drummond Sampaio Corrêa Mariani. FZEA-USP Pirassununga: Mirele Daiana Poleti; Raul Machado Neto; Rejane Maria Tommasini Grotto; Ricardo Augusto Brassaloti; Ricardo Haddad; Rodrigo Tocantins Calado.; Sandra Coccuzzo Sampaio; Sandra Coccuzzo Sampaio Vessoni; Simone Kashima; Svetoslav Nanev Slavov; Vagner Fonseca; Vincent Louis Viala |
| EPI_ISL_2344672, EPI_ISL_2344675 | SECRETARIA MUNICIPAL DE SAUDE DE CORDEIROPOLIS | Instituto Butantan / FZEA-USP-Pirassununga | Antonio Jorge Martins; Claudia Renata dos Santos Barros; David Schlesinger; Debora Botequio Moretti; Dimas Tadeu Covas; Elaine Cristina Marqueze; Elaine Vieira Santos; Evandra Strazza Rodrigues; Heidge Fukumasu; Jayme Augusto de Souza-Neto; José Salvatore Leister Patané; Luiz Alcantara; Luiz Lehmann Coutinho; Maria Carolina Elias; Maurício Lacerda Nogueira; Rafael dos Santos Bezerra; Raul Machado Neto; Rejane Maria Tommasini Grotto; Ricardo Haddad; Sandra Coccuzzo Sampaio Vessoni; Simone Kashima; Svetoslav Nanev Slavov; Vincent Louis Viala |
| EPI_ISL_3102301 | SECRETARIA MUNICIPAL DE SAUDE DE JAGUARIBE | Analytical Competence Molecular Epidemiology Lab/ACME, Oswaldo Cruz Foundation, Ceara (FIOCRUZ CE) | Cleber Furtado Aksenen; Fabio Miyajima; Fernando Braga Stehling; Francisco Eder de Moura Lopes; Jamille Maria Mendes Bezerra; Joaquim César do Nascimento Sousa Junior; Pedro Miguel Carneiro Jeronimo; Suzana Porto Almeida e Lucas Delerino; Thais Ferreira de Oliveira; Thais de Oliveira Costa; Ticiane Cavalcante de Souza; Veridiana Pessoa Miyajima |
| EPI_ISL_1795381, EPI_ISL_2345958 | SECRETARIA MUNICIPAL DE SAUDE DE MACATUBA | Instituto Butantan / ESALQ-Piracicaba | Antonio Jorge Martins; Bianca Cechetto Carlos. Mendelics: Bibiana Santos; Claudia Renata dos Santos Barros; David Schlesinger; David Schlesinger. Hemocentro Ribeirão Preto: Simone Kashima; Debora Botequio Moretti; Debora Botequio Moretti. Centro de Genômica Funcional da ESALQ: Luiz Lehmann Coutinho; Dimas Tadeu Covas; Elaine Cristina Marqueze; Elaine Vieira Santos; Elaine Vieira dos Santos; Elisangela Chicaroni Mattos; Erika Freitas; Evandra Strazza Rodrigues; Felipe Allan da Silva da Costa; Flavia Aburjaile; Guilherme Targino Valente; Heidge Fukumasu; Heidge Fukumasu. USP-Botucatu: Rejane Maria Tommasini Grotto; Instituto Butantan: Alexander Roberto Precioso; Jayme A. Souza-Neto; Jayme Augusto de Souza-Neto; Jessika Cristina Chagas Lesbon; José Salvatore Leister Patané; João Paulo Kitajima; Luiz Alcantara; Luiz Carlos Junior de Alcantara; Luiz Lehmann Coutinho; Maria Carolina Elias; Marta Giovanetti; Maurício Lacerda Nogueira; Patricia Akemi Assato; Rafael dos Santos Bezerra; Raquel de Lello Rocha Campos Cassano. NGS Soluções Genômicas: Pilar Drummond Sampaio Corrêa Mariani. FZEA-USP Pirassununga: Mirele Daiana Poleti; Raul Machado Neto; Rejane Maria Tommasini Grotto; Ricardo Augusto Brassaloti; Ricardo Haddad; Rodrigo Tocantins Calado.; Sandra Coccuzzo Sampaio; Sandra Coccuzzo Sampaio Vessoni; Simone Kashima; Svetoslav Nanev Slavov; Vagner Fonseca; Vincent Louis Viala |
| EPI_ISL_1795373, EPI_ISL_2345947 | SECRETARIA MUNICIPAL DE SAUDE DE PEDERNEIRAS | Instituto Butantan / ESALQ-Piracicaba | Antonio Jorge Martins; Bianca Cechetto Carlos. Mendelics: Bibiana Santos; Claudia Renata dos Santos Barros; David Schlesinger; David Schlesinger. Hemocentro Ribeirão Preto: Simone Kashima; Debora Botequio Moretti; Debora Botequio Moretti. Centro de Genômica Funcional da ESALQ: Luiz Lehmann Coutinho; Dimas Tadeu Covas; Elaine Cristina Marqueze; Elaine Vieira Santos; Elaine Vieira dos Santos; Elisangela Chicaroni Mattos; Erika Freitas; Evandra Strazza Rodrigues; Felipe Allan da Silva da Costa; Flavia Aburjaile; Guilherme Targino Valente; Heidge Fukumasu; Heidge Fukumasu. USP-Botucatu: Rejane Maria Tommasini Grotto; Instituto Butantan: Alexander Roberto Precioso; Jayme A. Souza-Neto; Jayme Augusto de Souza-Neto; Jessika Cristina Chagas Lesbon; José Salvatore Leister Patané; João Paulo Kitajima; Luiz Alcantara; Luiz Carlos Junior de Alcantara; Luiz Lehmann Coutinho; Maria Carolina Elias; Marta Giovanetti; Maurício Lacerda Nogueira; Patricia Akemi Assato; Rafael dos Santos Bezerra; Raquel de Lello Rocha Campos Cassano. NGS Soluções Genômicas: Pilar Drummond Sampaio Corrêa Mariani. FZEA-USP Pirassununga: Mirele Daiana Poleti; Raul Machado Neto; Rejane Maria Tommasini Grotto; Ricardo Augusto Brassaloti; Ricardo Haddad; Rodrigo Tocantins Calado.; Sandra Coccuzzo Sampaio; Sandra Coccuzzo Sampaio Vessoni; Simone Kashima; Svetoslav Nanev Slavov; Vagner Fonseca; Vincent Louis Viala |
| EPI_ISL_3102282, EPI_ISL_3102527 | SECRETARIA MUNICIPAL DE SAUDE DE TIANGUA | Analytical Competence Molecular Epidemiology Lab/ACME, Oswaldo Cruz Foundation, Ceara (FIOCRUZ CE) | Cleber Furtado Aksenen; Fabio Miyajima; Fernando Braga Stehling; Francisco Eder de Moura Lopes; Jamille Maria Mendes Bezerra; Joaquim César do Nascimento Sousa Junior; Pedro Miguel Carneiro Jeronimo; Suzana Porto Almeida e Lucas Delerino; Thais Ferreira de Oliveira; Thais de Oliveira Costa; Ticiane Cavalcante de Souza; Veridiana Pessoa Miyajima |
| EPI_ISL_1580529 | SIESP CHIETI - DRIVE IN LANCIANO LANCIANO(CHIETI) | Istituto Zooprofilattico Sperimentale dell'Abruzzo e Molise "G. Caporale" | Ancora M; Calistri P; Cammà C; Caporale M; Curini V; Delli Compagni E; Di Domenico M; Di Lollo Valeria; Di Pasquale A; Lorusso A; Mangone I; Marcacci M; Puglia I; Rinaldi A; Savini G; Scialabba S |

| | | | |
|---|---|---|---|
| EPI_ISL_5155897 | SUNY Upstate Medical University | 505 Irving Avenue, Syracuse, NY 13210 - Frank Middleton Lab | Brian Pavlovitz; Frank Middleton |
| EPI_ISL_6512556 | SURA | Laboratorio Departamental de Salud Publica de Antioquia | Ana Victoria Valencia Duarte; Cristian Arbey Velarde Hoyos; Gloria Isabel Escobar; Idabely Betancur Ortiz; Juan P. Hernandez-Ortiz; Juan Pablo Isaza Agudelo; Maria Stella López |
| EPI_ISL_1492926, EPI_ISL_1492937, EPI_ISL_1533853, EPI_ISL_1533854, EPI_ISL_1533855, EPI_ISL_1533865, EPI_ISL_1533875, EPI_ISL_1533902, EPI_ISL_1533924, EPI_ISL_1533930, EPI_ISL_1595697, EPI_ISL_1595702, EPI_ISL_1595704, EPI_ISL_1595708 | | | |
| see above | SYNLAB | GIGA Medical Genomics | Bouchra Boujemla; Cécile Meex; Keith Durkin; Maria Artesi; Marie-Pierre Hayette; Nathalie Renotte; Pierrette Melin; Raphaël Boreux; Sébastien Bontems; Vincent Bours |
| EPI_ISL_1738830, EPI_ISL_1738831 | SYNLAB | Instituto Nacional de Saude (INSA) | Borges et al |
| EPI_ISL_1639917 | SYNLAB MVZ Heidelberg | Robert Koch Institute | |
| EPI_ISL_1566557, EPI_ISL_1566580, EPI_ISL_1566632, EPI_ISL_1566637 | SYNLAB MVZ Trier | Robert Koch Institute | |
| EPI_ISL_1791420, EPI_ISL_1791421, EPI_ISL_1791422, EPI_ISL_1791423, EPI_ISL_1791424 | San Diego County Public Health Laboratory | Andersen lab at Scripps Research | Brett Austin; Jovan Shephard; SEARCH Alliance San Diego with Ashleigh Murphy |
| EPI_ISL_1587116, EPI_ISL_2736355, EPI_ISL_2736357 | Seattle Flu Study | Seattle Flu Study | Amanda Adler; Barry R. Lutz; Benjamin Pelle; Caitlin R. Wolf; Chris D. Frazar; Deborah A. Nickerson; Elisabeth Brandstetter; Erica Ryke; Helen Y. Chu; Janet A. Englund; Jay Shendure; Jeff Duchin; Jover Lee; Kairsten Fay; Karen Cowgill; Kirsten Lacombe; Lea M. Starita; Mark J. Rieder; Matthew Richardson; Matthew Thompson; Melissa Truong; Michael Boeckh; Michael Famulare; Misja Ilcisin; Peter D. Han; Stephanie Schrag; Thomas R. Sibley; Trevor Bedford |
| EPI_ISL_5799796 | Secretaria De Saude De Sao Pedro | Instituto Butantan | Antonio Jorge Martins; Claudia Renata dos Santos Barros; David Schlesinger; Debora Botequio Moretti; Dimas Tadeu Covas; Elaine Cristina Marqueze; Elaine Vieira Santos; Evandra Strazza Rodrigues; Heidge Fukumasu; Jayme Augusto de Souza-Neto; José Salvatore Leister Patané; Luiz Alcantara; Luiz Lehmann Coutinho; Maria Carolina Elias; Maurício Lacerda Nogueira; Rafael dos Santos Bezerra; Raul Machado Neto; Rejane Maria Tommasini Grotto; Ricardo Haddad; Sandra Coccuzzo Sampaio Vessoni; Simone Kashima; Svetoslav Nanev Slavov; Vincent Louis Viala |
| EPI_ISL_5782671, EPI_ISL_5782674 | Secretaria Municipal De Saude De Cordeiropolis | Instituto Butantan | Antonio Jorge Martins; Claudia Renata dos Santos Barros; David Schlesinger; Debora Botequio Moretti; Dimas Tadeu Covas; Elaine Cristina Marqueze; Elaine Vieira Santos; Evandra Strazza Rodrigues; Heidge Fukumasu; Jayme Augusto de Souza-Neto; José Salvatore Leister Patané; Luiz Alcantara; Luiz Lehmann Coutinho; Maria Carolina Elias; Maurício Lacerda Nogueira; Rafael dos Santos Bezerra; Raul Machado Neto; Rejane Maria Tommasini Grotto; Ricardo Haddad; Sandra Coccuzzo Sampaio Vessoni; Simone Kashima; Svetoslav Nanev Slavov; Vincent Louis Viala |
| EPI_ISL_5799783 | Secretaria Municipal De Saude De Macatuba | Instituto Butantan | Antonio Jorge Martins; Claudia Renata dos Santos Barros; David Schlesinger; Debora Botequio Moretti; Dimas Tadeu Covas; Elaine Cristina Marqueze; Elaine Vieira Santos; Evandra Strazza Rodrigues; Heidge Fukumasu; Jayme Augusto de Souza-Neto; José Salvatore Leister Patané; Luiz Alcantara; Luiz Lehmann Coutinho; Maria Carolina Elias; Maurício Lacerda Nogueira; Rafael dos Santos Bezerra; Raul Machado Neto; Rejane Maria Tommasini Grotto; Ricardo Haddad; Sandra Coccuzzo Sampaio Vessoni; Simone Kashima; Svetoslav Nanev Slavov; Vincent Louis Viala |
| EPI_ISL_5799785 | Secretaria Municipal De Saude De Pederneiras | Instituto Butantan | Antonio Jorge Martins; Claudia Renata dos Santos Barros; David Schlesinger; Debora Botequio Moretti; Dimas Tadeu Covas; Elaine Cristina Marqueze; Elaine Vieira Santos; Evandra Strazza Rodrigues; Heidge Fukumasu; Jayme Augusto de Souza-Neto; José Salvatore Leister Patané; Luiz Alcantara; Luiz Lehmann Coutinho; Maria Carolina Elias; Maurício Lacerda Nogueira; Rafael dos Santos Bezerra; Raul Machado Neto; Rejane Maria Tommasini Grotto; Ricardo Haddad; Sandra Coccuzzo Sampaio Vessoni; Simone Kashima; Svetoslav Nanev Slavov; Vincent Louis Viala |
| EPI_ISL_2136065, EPI_ISL_2136066, EPI_ISL_2136067, EPI_ISL_2136077 | Servicio Virosis Respiratorias-Departamento Virología-INEI | Instituto Nacional Enfermedades Infecciosas C.G.Malbran | Avaro M.; Baumeister E.; Benedetti E.; Campos J.; Cisterna D.; Dattero ME; Lorenzo F.; Molina V.; Perandones C.; Poklepovich T.; Pontoriero A.; Russo M.; Tuduri E. |
| EPI_ISL_2004015 | Servicio de Microbiología. Hospital Clínico Universitario de Valencia | SeqCOVID-SPAIN consortium/IBV(CSIC) | David Navarro Ortega; Eliseo Albert Vicent; Ignacio Torres and SeqCOVID-SPAIN consortium |
| EPI_ISL_5316560, EPI_ISL_5316561, EPI_ISL_5316563, EPI_ISL_5316565 | Shared Hospital Laboratory | Shared Hospital Laboratory | Christie Vermeiren; Finlay Maguire; Kevin Katz; Patryk Aftanas; Robert Kozak; Samira Mubareka |
| EPI_ISL_1794689, EPI_ISL_1794713 | Sharp HealthCare Laboratory | Andersen lab at Scripps Research | Art Mendoza; Cathy Woerle; Jacquelyn Berumen; Liam McGinnis; Omid Bakhtar; SEARCH Alliance San Diego with Aaron Harding |
| EPI_ISL_1652509, EPI_ISL_1652510, EPI_ISL_1652511, EPI_ISL_1652514 | Simple Laboratories | Gagnon Lab, Southern Illinois University | Keith Gagnon |
| EPI_ISL_1761663, EPI_ISL_1761710, EPI_ISL_1761773, EPI_ISL_1761803 | Sonora Quest Laboratories | TGen North | "Jolene Bowers; Ashlyn Pfeiffer; Chris French; Darrin Lemmer; Dave Engelthaler; Hayley Yaglom; Heather Centner; Jolene Bowers; The Arizona COVID Genomics Union (ACGU); The Arizona COVID Genomics Union (ACGU)" |
| EPI_ISL_1620463 | Stanford Clinical Virology Laboratory | Santa Clara County Public Health Laboratory | Santa Clara County Public Health Department |
| EPI_ISL_3050285, EPI_ISL_3387311 | Stanford Health Care | Stanford University School of Medicine, Clinical Virology Laboratory | Becky Jiang; Bernadette Troung; Daniel Solis; Fiona Yamamoto; James Zehnder; Malaya K. Sahoo; Mamdouh Sibai; Nathan Hammond; Selamawit Bihon; and Benjamin A. Pinsky |
| EPI_ISL_1770730 | State Laboratories Division, Hawaii State Department of Health | State Laboratories Division, Hawaii State Department of Health | Ayana Garnet; Drew Kuwazaki; Edward Desmond; Pamela O'Brien; Razvan Sultana |
| EPI_ISL_1605767, EPI_ISL_1606420, EPI_ISL_1808650, EPI_ISL_2220583, EPI_ISL_2221389, EPI_ISL_2221847, EPI_ISL_2222188, EPI_ISL_2255538, EPI_ISL_2255651, EPI_ISL_2255694 | | | |
| see above | Swedish national genomic surveillance program of SARS-CoV-2 | The Public Health Agency of Sweden | Alma Brolund; Maria Lind Karlberg; Maximilian Riess; Swedish national genomic surveillance program of SARS-CoV-2 |
| EPI_ISL_1582240, EPI_ISL_1582241, EPI_ISL_1582358 | TEMPUS LABS INC | Wadsworth Center, New York State Department of Health | Alexis Russell; Catharine Prussing; Daryl M. Lamson; Erasmus Schneider; Erica Lasek-Nesselquist; John Kelly; Jonathan Plitnick; Kirsten St. George; Matthew Shudt; Melissa A Leisner; Navjot Singh |
| EPI_ISL_1591935 | TGen North | TGen North | Ashlyn Pfeiffer; Chris French; Darrin Lemmer; Dave Engelthaler; Hayley Yaglom; Heather Centner; Jolene Bowers; The Arizona COVID Genomics Union (ACGU) |
| EPI_ISL_2200101, EPI_ISL_2200109, EPI_ISL_2200110, EPI_ISL_2200131, EPI_ISL_2200135, EPI_ISL_2200145, EPI_ISL_2200147, EPI_ISL_2382781, EPI_ISL_2382783, EPI_ISL_2382786, EPI_ISL_2382789 | | | |
| see above | The Ohio State University Applied Microbiology Services Laboratory | The Ohio State University Applied Microbiology Services Laboratory | Seth A. Faith PhD |
| EPI_ISL_1770444 | Tirol/Ralf Herwig | Elling and Cochella laboratories, IMBA/IMP | Alexander Stark; Ezgi Oezkan; Luisa Cochella and Ulrich Elling; Marcus Strobl; Maria Novatchkova; Ramesh Yelagandula; Tanino Albanese |
| EPI_ISL_3177036, EPI_ISL_3177063, EPI_ISL_3184335, EPI_ISL_3184420 | U.O. Microbiologia Laboratorio Unico Centro Servizi - AUSL della Romagna | U.O. Microbiologia, Laboratorio Unico Centro Servizi - AUSL della Romagna | Giorgio Dirani |
| EPI_ISL_5530014 | UAPS ABEL PINTO | Analytical Competence Molecular Epidemiology Lab/ACME, Oswaldo Cruz Foundation, Ceara (FIOCRUZ CE) | Carlos Leonardo de Aragao Araujo; Cecilia Leite Costa & Eduardo Ruback dos Santos on behalf of COVID-19 FIOCRUZ Genomic Network; Cleber Furtado Aksenen; Fabio Miyajima; Fernando Braga Stehling; Francisco Eder de Moura Lopes; Igor Oliveira Duarte; Jamille Maria Mendes Bezerra; Joaquim Cesar do Nascimento Sousa Junior; Pedro Miguel Carneiro Jeronimo; Suzana Porto Almeida; Thais Ferreira de Oliveira; Thais de Oliveira Costa; Ticiane Cavalcante de Souza; Veridiana Pessoa Miyajima |
| EPI_ISL_5530153 | UAPS AIDA SANTOS | Analytical Competence Molecular Epidemiology Lab/ACME, Oswaldo Cruz Foundation, Ceara (FIOCRUZ CE) | Carlos Leonardo de Aragao Araujo; Cecilia Leite Costa & Eduardo Ruback dos Santos on behalf of COVID-19 FIOCRUZ Genomic Network; Cleber Furtado Aksenen; Fabio Miyajima; Fernando Braga Stehling; Francisco Eder de Moura Lopes; Igor Oliveira Duarte; Jamille Maria Mendes Bezerra; Joaquim Cesar do Nascimento Sousa Junior; Pedro Miguel Carneiro Jeronimo; Suzana Porto Almeida; Thais Ferreira de Oliveira; Thais de Oliveira Costa; Ticiane Cavalcante de Souza; Veridiana Pessoa Miyajima |
| EPI_ISL_5530016 | UAPS ANTONIO CIRIACO DE HOLANDA NETO | Analytical Competence Molecular Epidemiology Lab/ACME, Oswaldo Cruz Foundation, Ceara (FIOCRUZ CE) | Carlos Leonardo de Aragao Araujo; Cecilia Leite Costa & Eduardo Ruback dos Santos on behalf of COVID-19 FIOCRUZ Genomic Network; Cleber Furtado Aksenen; Fabio Miyajima; Fernando Braga Stehling; Francisco Eder de Moura Lopes; Igor Oliveira Duarte; Jamille Maria Mendes Bezerra; Joaquim Cesar do Nascimento Sousa Junior; Pedro Miguel Carneiro Jeronimo; Suzana Porto Almeida; Thais Ferreira de Oliveira; Thais de Oliveira Costa; Ticiane Cavalcante de Souza; Veridiana Pessoa Miyajima |
| EPI_ISL_5530015, EPI_ISL_5530019, EPI_ISL_5530020, EPI_ISL_5530022 | UAPS DOM ALOISIO LORSCHEIDER | Analytical Competence Molecular Epidemiology Lab/ACME, Oswaldo Cruz Foundation, Ceara (FIOCRUZ CE) | Carlos Leonardo de Aragao Araujo; Cecilia Leite Costa & Eduardo Ruback dos Santos on behalf of COVID-19 FIOCRUZ Genomic Network; Cleber Furtado Aksenen; Fabio Miyajima; Fernando Braga Stehling; Francisco Eder de Moura Lopes; Igor Oliveira Duarte; Jamille Maria Mendes Bezerra; Joaquim Cesar do Nascimento Sousa Junior; Pedro Miguel Carneiro Jeronimo; Suzana Porto Almeida; Thais Ferreira de Oliveira; Thais de Oliveira Costa; Ticiane Cavalcante de Souza; Veridiana Pessoa Miyajima |
| EPI_ISL_5530137 | UAPS FRANCISCO PEREIRA DE ALMEIDA | Analytical Competence Molecular Epidemiology Lab/ACME, Oswaldo Cruz Foundation, Ceara (FIOCRUZ CE) | Carlos Leonardo de Aragao Araujo; Cecilia Leite Costa & Eduardo Ruback dos Santos on behalf of COVID-19 FIOCRUZ Genomic Network; Cleber Furtado Aksenen; Fabio Miyajima; Fernando Braga Stehling; Francisco Eder de Moura Lopes; Igor Oliveira Duarte; Jamille Maria Mendes Bezerra; Joaquim Cesar do Nascimento Sousa Junior; Pedro Miguel Carneiro Jeronimo; Suzana Porto Almeida; Thais Ferreira de Oliveira; Thais de Oliveira Costa; Ticiane Cavalcante de Souza; Veridiana Pessoa Miyajima |
| EPI_ISL_5530187 | UAPS JOAO XXIII | Analytical Competence Molecular | Carlos Leonardo de Aragao Araujo; Cecilia Leite Costa & Eduardo Ruback dos Santos on behalf of COVID-19 FIOCRUZ Genomic Network; Cleber Furtado Aksenen; Fabio Miyajima; Fernando Braga Stehling; Francisco Eder de Moura Lopes; Igor Oliveira Duarte; Jamille Maria Mendes Bezerra; Joaquim Cesar do |

| | | | |
|---|---|---|---|
| | | Epidemiology Lab/ACME, Oswaldo Cruz Foundation, Ceara (FIOCRUZ CE) | Nascimento Sousa Junior; Pedro Miguel Carneiro Jeronimo; Suzana Porto Almeida; Thais Ferreira de Oliveira; Thais de Oliveira Costa; Ticiane Cavalcante de Souza; Veridiana Pessoa Miyajima |
| EPI_ISL_5530180 | UAPS RECAMONDE CAPELO | Analytical Competence Molecular Epidemiology Lab/ACME, Oswaldo Cruz Foundation, Ceara (FIOCRUZ CE) | Carlos Leonardo de Aragao Araujo; Cecilia Leite Costa & Eduardo Ruback dos Santos on behalf of COVID-19 FIOCRUZ Genomic Network; Cleber Furtado Aksenen; Fabio Miyajima; Fernando Braga Stehling; Francisco Eder de Moura Lopes; Igor Oliveira Duarte; Jamille Maria Mendes Bezerra; Joaquim Cesar do Nascimento Sousa Junior; Pedro Miguel Carneiro Jeronimo; Suzana Porto Almeida; Thais Ferreira de Oliveira; Thais de Oliveira Costa; Ticiane Cavalcante de Souza; Veridiana Pessoa Miyajima |
| EPI_ISL_3102312 | UBASF AGUAS BELAS | Analytical Competence Molecular Epidemiology Lab/ACME, Oswaldo Cruz Foundation, Ceara (FIOCRUZ CE) | Cleber Furtado Aksenen; Fabio Miyajima; Fernando Braga Stehling; Francisco Eder de Moura Lopes; Jamille Maria Mendes Bezerra; Joaquim César do Nascimento Sousa Junior; Pedro Miguel Carneiro Jeronimo; Suzana Porto Almeida e Lucas Delerino; Thais Ferreira de Oliveira; Thais de Oliveira Costa; Ticiane Cavalcante de Souza; Veridiana Pessoa Miyajima |
| EPI_ISL_1752637 | UBDS Dr Marco Antonio Sahao Vila Virginia | Instituto Adolfo Lutz, Interdiciplinary Procedures Center, Strategic Laboratory | Caio Vinicius Dias Lopes; Claudia Regina Gonçalves; Claudio Tavares Sacchi; Erica Valessa Ramos Gomes; Karoline Rodrigues Campos; Katia Correa de Oliveira Santos; Leonardo Jose Tadeu de Araujo |
| EPI_ISL_1795287, EPI_ISL_1795289, EPI_ISL_2345525, EPI_ISL_2345527 | UBS ALCIMINIO DE ASSIS LOURENCO BADY BASSITT | Instituto Butantan / ESALQ-Piracicaba | Antonio Jorge Martins; Bianca Cechetto Carlos. Mendelics: Bibiana Santos; Claudia Renata dos Santos Barros; David Schlesinger; David Schlesinger. Hemocentro Ribeirão Preto: Simone Kashima; Debora Botequio Moretti; Debora Botequio Moretti. Centro de Genômica Funcional da ESALQ: Luiz Lehmann Coutinho; Dimas Tadeu Covas; Elaine Cristina Marqueze; Elaine Vieira Santos; Elaine Vieira dos Santos; Elisangela Chicaroni Mattos; Erika Freitas; Evandra Strazza Rodrigues; Felipe Allan da Silva da Costa; Flavia Aburjaile; Guilherme Targino Valente; Heidge Fukumasu; Heidge Fukumasu. USP-Botucatu: Rejane Maria Tommasini Grotto; Instituto Butantan: Alexander Roberto Precioso; Jayme A. Souza-Neto; Jayme Augusto de Souza-Neto; Jessika Cristina Chagas Lesbon; José Salvatore Leister Patané; João Paulo Kitajima; Luiz Alcantara; Luiz Carlos Junior de Alcantara; Luiz Lehmann Coutinho; Maria Carolina Elias; Marta Giovanetti; Maurício Lacerda Nogueira; Patricia Akemi Assato; Rafael dos Santos Bezerra; Raquel de Lello Rocha Campos Cassano. NGS Soluções Genômicas: Pilar Drummond Sampaio Corrêa Mariani. FZEA-USP Pirassununga: Mirele Daiana Poleti; Raul Machado Neto; Rejane Maria Tommasini Grotto; Ricardo Augusto Brassaloti; Ricardo Haddad; Rodrigo Tocantins Calado.; Sandra Coccuzzo Sampaio; Sandra Coccuzzo Sampaio Vessoni; Simone Kashima; Svetoslav Nanev Slavov; Vagner Fonseca; Vincent Louis Viala |
| EPI_ISL_1795324, EPI_ISL_2345585 | UBS II DE NARANDIBA | Instituto Butantan / ESALQ-Piracicaba | Antonio Jorge Martins; Bianca Cechetto Carlos. Mendelics: Bibiana Santos; Claudia Renata dos Santos Barros; David Schlesinger; David Schlesinger. Hemocentro Ribeirão Preto: Simone Kashima; Debora Botequio Moretti; Debora Botequio Moretti. Centro de Genômica Funcional da ESALQ: Luiz Lehmann Coutinho; Dimas Tadeu Covas; Elaine Cristina Marqueze; Elaine Vieira Santos; Elaine Vieira dos Santos; Elisangela Chicaroni Mattos; Erika Freitas; Evandra Strazza Rodrigues; Felipe Allan da Silva da Costa; Flavia Aburjaile; Guilherme Targino Valente; Heidge Fukumasu; Heidge Fukumasu. USP-Botucatu: Rejane Maria Tommasini Grotto; Instituto Butantan: Alexander Roberto Precioso; Jayme A. Souza-Neto; Jayme Augusto de Souza-Neto; Jessika Cristina Chagas Lesbon; José Salvatore Leister Patané; João Paulo Kitajima; Luiz Alcantara; Luiz Carlos Junior de Alcantara; Luiz Lehmann Coutinho; Maria Carolina Elias; Marta Giovanetti; Maurício Lacerda Nogueira; Patricia Akemi Assato; Rafael dos Santos Bezerra; Raquel de Lello Rocha Campos Cassano. NGS Soluções Genômicas: Pilar Drummond Sampaio Corrêa Mariani. FZEA-USP Pirassununga: Mirele Daiana Poleti; Raul Machado Neto; Rejane Maria Tommasini Grotto; Ricardo Augusto Brassaloti; Ricardo Haddad; Rodrigo Tocantins Calado.; Sandra Coccuzzo Sampaio; Sandra Coccuzzo Sampaio Vessoni; Simone Kashima; Svetoslav Nanev Slavov; Vagner Fonseca; Vincent Louis Viala |
| EPI_ISL_1795326, EPI_ISL_2345587 | UBS II DE REGENTE FEIJO | Instituto Butantan / ESALQ-Piracicaba | Antonio Jorge Martins; Bianca Cechetto Carlos. Mendelics: Bibiana Santos; Claudia Renata dos Santos Barros; David Schlesinger; David Schlesinger. Hemocentro Ribeirão Preto: Simone Kashima; Debora Botequio Moretti; Debora Botequio Moretti. Centro de Genômica Funcional da ESALQ: Luiz Lehmann Coutinho; Dimas Tadeu Covas; Elaine Cristina Marqueze; Elaine Vieira Santos; Elaine Vieira dos Santos; Elisangela Chicaroni Mattos; Erika Freitas; Evandra Strazza Rodrigues; Felipe Allan da Silva da Costa; Flavia Aburjaile; Guilherme Targino Valente; Heidge Fukumasu; Heidge Fukumasu. USP-Botucatu: Rejane Maria Tommasini Grotto; Instituto Butantan: Alexander Roberto Precioso; Jayme A. Souza-Neto; Jayme Augusto de Souza-Neto; Jessika Cristina Chagas Lesbon; José Salvatore Leister Patané; João Paulo Kitajima; Luiz Alcantara; Luiz Carlos Junior de Alcantara; Luiz Lehmann Coutinho; Maria Carolina Elias; Marta Giovanetti; Maurício Lacerda Nogueira; Patricia Akemi Assato; Rafael dos Santos Bezerra; Raquel de Lello Rocha Campos Cassano. NGS Soluções Genômicas: Pilar Drummond Sampaio Corrêa Mariani. FZEA-USP Pirassununga: Mirele Daiana Poleti; Raul Machado Neto; Rejane Maria Tommasini Grotto; Ricardo Augusto Brassaloti; Ricardo Haddad; Rodrigo Tocantins Calado.; Sandra Coccuzzo Sampaio; Sandra Coccuzzo Sampaio Vessoni; Simone Kashima; Svetoslav Nanev Slavov; Vagner Fonseca; Vincent Louis Viala |
| EPI_ISL_1795251, EPI_ISL_1795252, EPI_ISL_1795253, EPI_ISL_1795254, EPI_ISL_1795255, EPI_ISL_1795256, EPI_ISL_1795257, EPI_ISL_2345485, EPI_ISL_2345486, EPI_ISL_2345487, EPI_ISL_2345488, EPI_ISL_2345489, EPI_ISL_2345490, EPI_ISL_2345491 | | | |
| see above | UBS II DE TANABI MILTON MARTINS PERCHES | Instituto Butantan / ESALQ-Piracicaba | Antonio Jorge Martins; Bianca Cechetto Carlos. Mendelics: Bibiana Santos; Claudia Renata dos Santos Barros; David Schlesinger; David Schlesinger. Hemocentro Ribeirão Preto: Simone Kashima; Debora Botequio Moretti; Debora Botequio Moretti. Centro de Genômica Funcional da ESALQ: Luiz Lehmann Coutinho; Dimas Tadeu Covas; Elaine Cristina Marqueze; Elaine Vieira Santos; Elaine Vieira dos Santos; Elisangela Chicaroni Mattos; Erika Freitas; Evandra Strazza Rodrigues; Felipe Allan da Silva da Costa; Flavia Aburjaile; Guilherme Targino Valente; Heidge Fukumasu; Heidge Fukumasu. USP-Botucatu: Rejane Maria Tommasini Grotto; Instituto Butantan: Alexander Roberto Precioso; Jayme A. Souza-Neto; Jayme Augusto de Souza-Neto; Jessika Cristina Chagas Lesbon; José Salvatore Leister Patané; João Paulo Kitajima; Luiz Alcantara; Luiz Carlos Junior de Alcantara; Luiz Lehmann Coutinho; Maria Carolina Elias; Marta Giovanetti; Maurício Lacerda Nogueira; Patricia Akemi Assato; Rafael dos Santos Bezerra; Raquel de Lello Rocha Campos Cassano. NGS Soluções Genômicas: Pilar Drummond Sampaio Corrêa Mariani. FZEA-USP Pirassununga: Mirele Daiana Poleti; Raul Machado Neto; Rejane Maria Tommasini Grotto; Ricardo Augusto Brassaloti; Ricardo Haddad; Rodrigo Tocantins Calado.; Sandra Coccuzzo Sampaio; Sandra Coccuzzo Sampaio Vessoni; Simone Kashima; Svetoslav Nanev Slavov; Vagner Fonseca; Vincent Louis Viala |
| EPI_ISL_1795328, EPI_ISL_1795330, EPI_ISL_2345589, EPI_ISL_2345591 | UBS II DR EXPEDITO SHIZUO KUROCE | Instituto Butantan / ESALQ-Piracicaba | Antonio Jorge Martins; Bianca Cechetto Carlos. Mendelics: Bibiana Santos; Claudia Renata dos Santos Barros; David Schlesinger; David Schlesinger. Hemocentro Ribeirão Preto: Simone Kashima; Debora Botequio Moretti; Debora Botequio Moretti. Centro de Genômica Funcional da ESALQ: Luiz Lehmann Coutinho; Dimas Tadeu Covas; Elaine Cristina Marqueze; Elaine Vieira Santos; Elaine Vieira dos Santos; Elisangela Chicaroni Mattos; Erika Freitas; Evandra Strazza Rodrigues; Felipe Allan da Silva da Costa; Flavia Aburjaile; Guilherme Targino Valente; Heidge Fukumasu; Heidge Fukumasu. USP-Botucatu: Rejane Maria Tommasini Grotto; Instituto Butantan: Alexander Roberto Precioso; Jayme A. Souza-Neto; Jayme Augusto de Souza-Neto; Jessika Cristina Chagas Lesbon; José Salvatore Leister Patané; João Paulo Kitajima; Luiz Alcantara; Luiz Carlos Junior de Alcantara; Luiz Lehmann Coutinho; Maria Carolina Elias; Marta Giovanetti; Maurício Lacerda Nogueira; Patricia Akemi Assato; Rafael dos Santos Bezerra; Raquel de Lello Rocha Campos Cassano. NGS Soluções Genômicas: Pilar Drummond Sampaio Corrêa Mariani. FZEA-USP Pirassununga: Mirele Daiana Poleti; Raul Machado Neto; Rejane Maria Tommasini Grotto; Ricardo Augusto Brassaloti; Ricardo Haddad; Rodrigo Tocantins Calado.; Sandra Coccuzzo Sampaio; Sandra Coccuzzo Sampaio Vessoni; Simone Kashima; Svetoslav Nanev Slavov; Vagner Fonseca; Vincent Louis Viala |
| EPI_ISL_1795332, EPI_ISL_1795335, EPI_ISL_2345593, EPI_ISL_2345599 | UBS III DE RANCHARIA | Instituto Butantan / ESALQ-Piracicaba | Antonio Jorge Martins; Bianca Cechetto Carlos. Mendelics: Bibiana Santos; Claudia Renata dos Santos Barros; David Schlesinger; David Schlesinger. Hemocentro Ribeirão Preto: Simone Kashima; Debora Botequio Moretti; Debora Botequio Moretti. Centro de Genômica Funcional da ESALQ: Luiz Lehmann Coutinho; Dimas Tadeu Covas; Elaine Cristina Marqueze; Elaine Vieira Santos; Elaine Vieira dos Santos; Elisangela Chicaroni Mattos; Erika Freitas; Evandra Strazza Rodrigues; Felipe Allan da Silva da Costa; Flavia Aburjaile; Guilherme Targino Valente; Heidge Fukumasu; Heidge Fukumasu. USP-Botucatu: Rejane Maria Tommasini Grotto; Instituto Butantan: Alexander Roberto Precioso; Jayme A. Souza-Neto; Jayme Augusto de Souza-Neto; Jessika Cristina Chagas Lesbon; José Salvatore Leister Patané; João Paulo Kitajima; Luiz Alcantara; Luiz Carlos Junior de Alcantara; Luiz Lehmann Coutinho; Maria Carolina Elias; Marta Giovanetti; Maurício Lacerda Nogueira; Patricia Akemi Assato; Rafael dos Santos Bezerra; Raquel de Lello Rocha Campos Cassano. NGS Soluções Genômicas: Pilar Drummond Sampaio Corrêa Mariani. FZEA-USP Pirassununga: Mirele Daiana Poleti; Raul Machado Neto; Rejane Maria Tommasini Grotto; Ricardo Augusto Brassaloti; Ricardo Haddad; Rodrigo Tocantins Calado.; Sandra Coccuzzo Sampaio; Sandra Coccuzzo Sampaio Vessoni; Simone Kashima; Svetoslav Nanev Slavov; Vagner Fonseca; Vincent Louis Viala |
| EPI_ISL_1795160, EPI_ISL_2345371 | UBS JARDIM ITAMARATY | Instituto Butantan / ESALQ-Piracicaba | Antonio Jorge Martins; Bianca Cechetto Carlos. Mendelics: Bibiana Santos; Claudia Renata dos Santos Barros; David Schlesinger; David Schlesinger. Hemocentro Ribeirão Preto: Simone Kashima; Debora Botequio Moretti; Debora Botequio Moretti. Centro de Genômica Funcional da ESALQ: Luiz Lehmann Coutinho; Dimas Tadeu Covas; Elaine Cristina Marqueze; Elaine Vieira Santos; Elaine Vieira dos Santos; Elisangela Chicaroni Mattos; Erika Freitas; Evandra Strazza Rodrigues; Felipe Allan da Silva da Costa; Flavia Aburjaile; Guilherme Targino Valente; Heidge Fukumasu; Heidge Fukumasu. USP-Botucatu: Rejane Maria Tommasini Grotto; Instituto Butantan: Alexander Roberto Precioso; Jayme A. Souza-Neto; Jayme Augusto de Souza-Neto; Jessika Cristina Chagas Lesbon; José Salvatore Leister Patané; João Paulo Kitajima; Luiz Alcantara; Luiz Carlos Junior de Alcantara; Luiz Lehmann Coutinho; Maria Carolina Elias; Marta Giovanetti; Maurício Lacerda Nogueira; Patricia Akemi Assato; Rafael dos Santos Bezerra; Raquel de Lello Rocha Campos Cassano. NGS Soluções Genômicas: Pilar Drummond Sampaio Corrêa Mariani. FZEA-USP Pirassununga: Mirele Daiana Poleti; Raul Machado Neto; Rejane Maria Tommasini Grotto; Ricardo Augusto Brassaloti; Ricardo Haddad; Rodrigo Tocantins Calado.; Sandra Coccuzzo Sampaio; Sandra Coccuzzo Sampaio Vessoni; Simone Kashima; Svetoslav Nanev Slavov; Vagner Fonseca; Vincent Louis Viala |
| EPI_ISL_1795266, EPI_ISL_1795267, EPI_ISL_1795268, EPI_ISL_2345500, EPI_ISL_2345501, EPI_ISL_2345502 | UBS LUIS FACHIN IPIGUA | Instituto Butantan / ESALQ-Piracicaba | Antonio Jorge Martins; Bianca Cechetto Carlos. Mendelics: Bibiana Santos; Claudia Renata dos Santos Barros; David Schlesinger; David Schlesinger. Hemocentro Ribeirão Preto: Simone Kashima; Debora Botequio Moretti; Debora Botequio Moretti. Centro de Genômica Funcional da ESALQ: Luiz Lehmann Coutinho; Dimas Tadeu Covas; Elaine Cristina Marqueze; Elaine Vieira Santos; Elaine Vieira dos Santos; Elisangela Chicaroni Mattos; Erika Freitas; Evandra Strazza Rodrigues; Felipe Allan da Silva da Costa; Flavia Aburjaile; Guilherme Targino Valente; Heidge Fukumasu; Heidge Fukumasu. USP-Botucatu: Rejane Maria Tommasini Grotto; Instituto Butantan: Alexander Roberto Precioso; Jayme A. Souza-Neto; Jayme Augusto de Souza-Neto; Jessika Cristina Chagas Lesbon; José Salvatore Leister Patané; João Paulo Kitajima; Luiz Alcantara; Luiz Carlos Junior de Alcantara; Luiz Lehmann Coutinho; Maria Carolina Elias; Marta Giovanetti; Maurício Lacerda Nogueira; Patricia Akemi Assato; Rafael dos Santos Bezerra; Raquel de Lello Rocha Campos Cassano. NGS Soluções Genômicas: Pilar Drummond Sampaio Corrêa Mariani. FZEA-USP Pirassununga: Mirele Daiana Poleti; Raul Machado Neto; Rejane Maria Tommasini Grotto; Ricardo Augusto Brassaloti; Ricardo Haddad; Rodrigo Tocantins Calado.; Sandra Coccuzzo Sampaio; Sandra Coccuzzo Sampaio Vessoni; Simone Kashima; Svetoslav Nanev Slavov; Vagner Fonseca; Vincent Louis Viala |
| EPI_ISL_1795334, EPI_ISL_2345596 | UBS MARCIA CRISTIANE DA SILVA DE OURO VERDE | Instituto Butantan / ESALQ-Piracicaba | Antonio Jorge Martins; Bianca Cechetto Carlos. Mendelics: Bibiana Santos; Claudia Renata dos Santos Barros; David Schlesinger; David Schlesinger. Hemocentro Ribeirão Preto: Simone Kashima; Debora Botequio Moretti; Debora Botequio Moretti. Centro de Genômica Funcional da ESALQ: Luiz Lehmann Coutinho; Dimas Tadeu Covas; Elaine Cristina Marqueze; Elaine Vieira Santos; Elaine Vieira dos Santos; Elisangela Chicaroni Mattos; Erika Freitas; Evandra Strazza Rodrigues; Felipe Allan da Silva da Costa; Flavia Aburjaile; Guilherme Targino Valente; Heidge Fukumasu; Heidge Fukumasu. USP-Botucatu: Rejane Maria Tommasini Grotto; Instituto Butantan: Alexander Roberto Precioso; Jayme A. Souza-Neto; Jayme Augusto de Souza-Neto; Jessika Cristina Chagas Lesbon; José Salvatore Leister Patané; João Paulo Kitajima; Luiz Alcantara; Luiz Carlos Junior de Alcantara; Luiz Lehmann Coutinho; Maria Carolina Elias; Marta Giovanetti; Maurício Lacerda Nogueira; Patricia Akemi Assato; Rafael dos Santos Bezerra; Raquel de Lello Rocha Campos Cassano. NGS Soluções Genômicas: Pilar Drummond Sampaio Corrêa Mariani. FZEA-USP Pirassununga: Mirele Daiana Poleti; Raul Machado Neto; Rejane Maria Tommasini Grotto; Ricardo Augusto Brassaloti; Ricardo Haddad; Rodrigo Tocantins Calado.; Sandra Coccuzzo Sampaio; Sandra Coccuzzo Sampaio Vessoni; Simone Kashima; Svetoslav Nanev Slavov; Vagner Fonseca; Vincent Louis Viala |
| EPI_ISL_1795269, EPI_ISL_1795270, EPI_ISL_1795272, EPI_ISL_1795273, EPI_ISL_2345503, EPI_ISL_2345504, EPI_ISL_2345507, EPI_ISL_2345509 | | | |
| see above | UBS TEREZA GALLO UCHOA | Instituto Butantan / ESALQ-Piracicaba | Antonio Jorge Martins; Bianca Cechetto Carlos. Mendelics: Bibiana Santos; Claudia Renata dos Santos Barros; David Schlesinger; David Schlesinger. Hemocentro Ribeirão Preto: Simone Kashima; Debora Botequio Moretti; Debora Botequio Moretti. Centro de Genômica Funcional da ESALQ: Luiz Lehmann Coutinho; Dimas Tadeu Covas; Elaine Cristina Marqueze; Elaine Vieira Santos; Elaine Vieira dos Santos; Elisangela Chicaroni Mattos; Erika Freitas; Evandra Strazza Rodrigues; Felipe Allan da Silva da Costa; Flavia Aburjaile; Guilherme Targino Valente; Heidge Fukumasu; Heidge Fukumasu. USP-Botucatu: Rejane Maria Tommasini Grotto; Instituto Butantan: Alexander Roberto Precioso; Jayme A. Souza-Neto; Jayme Augusto de Souza-Neto; Jessika Cristina Chagas Lesbon; José Salvatore Leister Patané; João Paulo Kitajima; Luiz Alcantara; Luiz Carlos Junior de Alcantara; Luiz Lehmann Coutinho; Maria Carolina Elias; Marta Giovanetti; Maurício Lacerda Nogueira; Patricia Akemi Assato; Rafael dos Santos Bezerra; Raquel de Lello Rocha Campos Cassano. NGS Soluções Genômicas: Pilar Drummond Sampaio Corrêa Mariani. FZEA-USP Pirassununga: Mirele Daiana Poleti; Raul Machado Neto; Rejane Maria Tommasini Grotto; Ricardo Augusto Brassaloti; Ricardo Haddad; Rodrigo Tocantins Calado.; Sandra Coccuzzo Sampaio; Sandra Coccuzzo Sampaio Vessoni; Simone Kashima; Svetoslav Nanev Slavov; Vagner Fonseca; Vincent Louis Viala |
| EPI_ISL_2325016, EPI_ISL_2365879 | ULSS 3 Venezia | Istituto Zooprofilattico Sperimentale delle Venezie | Adelaide Milani; Alessia Schivo; Alice Fusaro; Ambra Pastori; Annalisa Salviato; Antonia Ricci; Calogero Terregino; Edoardo Giussani; Elisa Palumbo; Erika Giorgia Quaranta; Isabella Monne; Luca Tassoni |
| EPI_ISL_1511626 | UMC Groningen, Clinical Virology, Department of Medical Microbiology and Infection Prevention | UMC Groningen, Clinical Virology, Department of Medical Microbiology and Infection Prevention | Alexander Friedrich; Coretta Van Leer-Buter; Erley Lizarazo-Forero; Hubert Niesters; Lilli Gard; Marjolein Knoester; Monika Fliss; Sigrid Rosema; Xuewei Zhou |
| EPI_ISL_3409302, EPI_ISL_3410074, EPI_ISL_3410075 | UMass Memorial Medical Center | Center for Microbiome Research | Doyle V. Ward; Karl J. Simon; Richard T. Ellison III |
| EPI_ISL_1795362, EPI_ISL_1795363, EPI_ISL_2345637, EPI_ISL_2345638 | UNIDADE BASICA DE SAUDE DE PIQUETE | Instituto Butantan / ESALQ-Piracicaba | Antonio Jorge Martins; Bianca Cechetto Carlos. Mendelics: Bibiana Santos; Claudia Renata dos Santos Barros; David Schlesinger; David Schlesinger. Hemocentro Ribeirão Preto: Simone Kashima; Debora Botequio Moretti; Debora Botequio Moretti. Centro de Genômica Funcional da ESALQ: Luiz Lehmann Coutinho; Dimas Tadeu Covas; Elaine Cristina Marqueze; Elaine Vieira Santos; Elaine Vieira dos Santos; Elisangela Chicaroni Mattos; Erika Freitas; Evandra Strazza Rodrigues; Felipe Allan da Silva da Costa; Flavia Aburjaile; Guilherme Targino Valente; Heidge Fukumasu; Heidge Fukumasu. USP-Botucatu: Rejane Maria Tommasini Grotto; Instituto Butantan: Alexander Roberto Precioso; Jayme A. Souza-Neto; Jayme Augusto de Souza-Neto; Jessika Cristina Chagas Lesbon; José Salvatore Leister Patané; João Paulo Kitajima; Luiz Alcantara; Luiz Carlos Junior de Alcantara; Luiz Lehmann Coutinho; Maria Carolina Elias; Marta Giovanetti; Maurício Lacerda Nogueira; Patricia Akemi Assato; Rafael dos Santos Bezerra; Raquel de Lello Rocha Campos Cassano. NGS Soluções Genômicas: Pilar Drummond Sampaio Corrêa Mariani. FZEA-USP Pirassununga: Mirele Daiana Poleti; Raul Machado Neto; Rejane Maria Tommasini Grotto; Ricardo Augusto Brassaloti; Ricardo Haddad; Rodrigo Tocantins Calado.; Sandra Coccuzzo Sampaio; Sandra Coccuzzo Sampaio Vessoni; Simone Kashima; Svetoslav Nanev Slavov; Vagner Fonseca; Vincent Louis Viala |
| EPI_ISL_1795224, EPI_ISL_2345455 | UNIDADE BASICA DE SAUDE DR APOLONIO MORAES E SOUZA | Instituto Butantan / ESALQ-Piracicaba | Antonio Jorge Martins; Bianca Cechetto Carlos. Mendelics: Bibiana Santos; Claudia Renata dos Santos Barros; David Schlesinger; David Schlesinger. Hemocentro Ribeirão Preto: Simone Kashima; Debora Botequio Moretti; Debora Botequio Moretti. Centro de Genômica Funcional da ESALQ: Luiz Lehmann Coutinho; Dimas Tadeu Covas; Elaine Cristina Marqueze; Elaine Vieira Santos; Elaine Vieira dos Santos; Elisangela Chicaroni Mattos; Erika Freitas; Evandra Strazza Rodrigues; Felipe Allan da Silva da Costa; Flavia Aburjaile; Guilherme Targino Valente; Heidge Fukumasu; Heidge Fukumasu. USP-Botucatu: Rejane Maria Tommasini Grotto; Instituto Butantan: Alexander Roberto Precioso; Jayme A. Souza-Neto; Jayme Augusto de Souza-Neto; Jessika Cristina Chagas Lesbon; José Salvatore Leister Patané; João Paulo Kitajima; Luiz Alcantara; Luiz Carlos Junior de Alcantara; Luiz Lehmann Coutinho; Maria Carolina Elias; Marta Giovanetti; Maurício Lacerda Nogueira; Patricia Akemi Assato; Rafael dos Santos Bezerra; Raquel de Lello Rocha Campos Cassano. NGS Soluções Genômicas: Pilar Drummond Sampaio Corrêa Mariani. FZEA-USP Pirassununga: Mirele Daiana Poleti; Raul Machado Neto; Rejane Maria Tommasini Grotto; Ricardo Augusto Brassaloti; Ricardo Haddad; Rodrigo Tocantins Calado.; Sandra Coccuzzo Sampaio; Sandra Coccuzzo Sampaio Vessoni; Simone Kashima; Svetoslav Nanev Slavov; Vagner Fonseca; Vincent Louis Viala |
| EPI_ISL_1795227, EPI_ISL_2345458 | UNIDADE BASICA DE SAUDE DR LOTFALLAH MIZIARA | Instituto Butantan / ESALQ-Piracicaba | Antonio Jorge Martins; Bianca Cechetto Carlos. Mendelics: Bibiana Santos; Claudia Renata dos Santos Barros; David Schlesinger; David Schlesinger. Hemocentro Ribeirão Preto: Simone Kashima; Debora Botequio Moretti; Debora Botequio Moretti. Centro de Genômica Funcional da ESALQ: Luiz Lehmann Coutinho; Dimas Tadeu Covas; Elaine Cristina Marqueze; Elaine Vieira Santos; Elaine Vieira dos Santos; Elisangela Chicaroni Mattos; Erika Freitas; Evandra Strazza Rodrigues; Felipe Allan da Silva da Costa; Flavia Aburjaile; Guilherme Targino Valente; Heidge Fukumasu; Heidge Fukumasu. USP-Botucatu: Rejane Maria Tommasini Grotto; Instituto Butantan: Alexander Roberto Precioso; Jayme A. Souza-Neto; Jayme Augusto de Souza-Neto; Jessika Cristina Chagas Lesbon; José Salvatore Leister Patané; João Paulo Kitajima; Luiz Alcantara; Luiz Carlos Junior de Alcantara; Luiz Lehmann Coutinho; Maria Carolina Elias; Marta Giovanetti; Maurício Lacerda Nogueira; Patricia Akemi Assato; Rafael dos Santos Bezerra; Raquel de Lello Rocha Campos Cassano. NGS Soluções Genômicas: Pilar Drummond Sampaio Corrêa Mariani. FZEA-USP Pirassununga: Mirele Daiana Poleti; Raul Machado Neto; Rejane Maria Tommasini Grotto; Ricardo Augusto Brassaloti; Ricardo Haddad; Rodrigo Tocantins Calado.; Sandra Coccuzzo Sampaio; Sandra Coccuzzo Sampaio Vessoni; Simone Kashima; Svetoslav Nanev Slavov; Vagner Fonseca; Vincent Louis Viala |
| EPI_ISL_1795225, EPI_ISL_2345456 | UNIDADE BASICA DE SAUDE DR WILSON HAYEK SAIHG | Instituto Butantan / ESALQ-Piracicaba | Antonio Jorge Martins; Bianca Cechetto Carlos. Mendelics: Bibiana Santos; Claudia Renata dos Santos Barros; David Schlesinger; David Schlesinger. Hemocentro Ribeirão Preto: Simone Kashima; Debora Botequio Moretti; Debora Botequio Moretti. Centro de Genômica Funcional da ESALQ: Luiz Lehmann Coutinho; Dimas Tadeu Covas; Elaine Cristina Marqueze; Elaine Vieira Santos; Elaine Vieira dos Santos; Elisangela Chicaroni Mattos; Erika Freitas; Evandra Strazza Rodrigues; Felipe Allan da Silva da Costa; Flavia Aburjaile; Guilherme Targino Valente; Heidge Fukumasu; Heidge Fukumasu. USP-Botucatu: Rejane Maria Tommasini Grotto; Instituto Butantan: Alexander Roberto Precioso; Jayme A. Souza-Neto; Jayme Augusto de Souza-Neto; Jessika Cristina Chagas Lesbon; José Salvatore Leister Patané; João Paulo Kitajima; Luiz Alcantara; Luiz Carlos Junior de Alcantara; Luiz Lehmann Coutinho; Maria Carolina Elias; Marta Giovanetti; Maurício Lacerda Nogueira; Patricia Akemi Assato; Rafael dos Santos Bezerra; Raquel de Lello Rocha Campos Cassano. NGS Soluções Genômicas: Pilar Drummond Sampaio Corrêa Mariani. FZEA-USP Pirassununga: Mirele Daiana Poleti; Raul Machado Neto; Rejane Maria Tommasini Grotto; Ricardo Augusto Brassaloti; Ricardo Haddad; Rodrigo Tocantins Calado.; Sandra Coccuzzo Sampaio; Sandra Coccuzzo Sampaio Vessoni; Simone Kashima; Svetoslav Nanev Slavov; Vagner Fonseca; Vincent Louis Viala |
| EPI_ISL_5530047 | UNIDADE BASICA DE SAUDE HUGO PORFIRIO DA MOTA | Analytical Competence Molecular Epidemiology Lab/ACME, Oswaldo Cruz Foundation, Ceara (FIOCRUZ CE) | Carlos Leonardo de Aragao Araujo; Cecilia Leite Costa & Eduardo Ruback dos Santos on behalf of COVID-19 FIOCRUZ Genomic Network; Cleber Furtado Aksenen; Fabio Miyajima; Fernando Braga Stehling; Francisco Eder de Moura Lopes; Igor Oliveira Duarte; Jamille Maria Mendes Bezerra; Joaquim Cesar do Nascimento Sousa Junior; Pedro Miguel Carneiro Jeronimo; Suzana Porto Almeida; Thais Ferreira de Oliveira; Thais de Oliveira Costa; Ticiane Cavalcante de Souza; Veridiana Pessoa Miyajima |
| EPI_ISL_1795226, EPI_ISL_2345457 | UNIDADE BASICA DR JOSE PARASSU CARVALHO | Instituto Butantan / ESALQ-Piracicaba | Antonio Jorge Martins; Bianca Cechetto Carlos. Mendelics: Bibiana Santos; Claudia Renata dos Santos Barros; David Schlesinger; David Schlesinger. Hemocentro Ribeirão Preto: Simone Kashima; Debora Botequio Moretti; Debora Botequio Moretti. Centro de Genômica Funcional da ESALQ: Luiz Lehmann Coutinho; Dimas Tadeu Covas; Elaine Cristina Marqueze; Elaine Vieira Santos; Elaine Vieira dos Santos; Elisangela Chicaroni Mattos; Erika Freitas; Evandra Strazza Rodrigues; Felipe Allan da Silva da Costa; Flavia Aburjaile; Guilherme Targino Valente; Heidge Fukumasu; Heidge Fukumasu. USP-Botucatu: Rejane Maria Tommasini Grotto; Instituto Butantan: Alexander Roberto Precioso; Jayme A. Souza-Neto; Jayme Augusto de Souza-Neto; Jessika Cristina Chagas Lesbon; José Salvatore Leister Patané; João Paulo Kitajima; Luiz Alcantara; Luiz Carlos Junior de Alcantara; Luiz Lehmann Coutinho; Maria Carolina Elias; Marta Giovanetti; Maurício Lacerda Nogueira; Patricia Akemi Assato; Rafael dos Santos Bezerra; Raquel de Lello Rocha Campos Cassano. NGS Soluções Genômicas: Pilar Drummond Sampaio Corrêa Mariani. FZEA-USP Pirassununga: Mirele Daiana Poleti; Raul Machado Neto; Rejane Maria Tommasini Grotto; Ricardo Augusto |

| | | | |
|---|---|---|---|
| | | | Brassaloti; Ricardo Haddad; Rodrigo Tocantins Calado.; Sandra Coccuzzo Sampaio; Sandra Coccuzzo Sampaio Vessoni; Simone Kashima; Svetoslav Nanev Slavov; Vagner Fonseca; Vincent Louis Viala |
| EPI_ISL_1795305, EPI_ISL_2345553 | UNIDADE DA SAUDE DO ADULTO CASA BRANCA PREFEITURA | Instituto Butantan / ESALQ-Piracicaba | Antonio Jorge Martins; Bianca Cechetto Carlos. Mendelics: Bibiana Santos; Claudia Renata dos Santos Barros; David Schlesinger; David Schlesinger. Hemocentro Ribeirão Preto: Simone Kashima; Debora Botequio Moretti; Debora Botequio Moretti. Centro de Genômica Funcional da ESALQ: Luiz Lehmann Coutinho; Dimas Tadeu Covas; Elaine Cristina Marqueze; Elaine Vieira Santos; Elaine Vieira dos Santos; Elisangela Chicaroni Mattos; Erika Freitas; Evandra Strazza Rodrigues; Felipe Allan da Silva da Costa; Flavia Aburjaile; Guilherme Targino Valente; Heidge Fukumasu; Heidge Fukumasu. USP-Botucatu: Rejane Maria Tommasini Grotto; Instituto Butantan: Alexander Roberto Precioso; Jayme A. Souza-Neto; Jayme Augusto de Souza-Neto; Jessika Cristina Chagas Lesbon; José Salvatore Leister Patané; João Paulo Kitajima; Luiz Alcantara; Luiz Carlos Junior de Alcantara; Luiz Lehmann Coutinho; Maria Carolina Elias; Marta Giovanetti; Maurício Lacerda Nogueira; Patricia Akemi Assato; Rafael dos Santos Bezerra; Raquel de Lello Rocha Campos Cassano. NGS Soluções Genômicas: Pilar Drummond Sampaio Corrêa Mariani. FZEA-USP Pirassununga: Mirele Daiana Poleti; Raul Machado Neto; Rejane Maria Tommasini Grotto; Ricardo Augusto Brassaloti; Ricardo Haddad; Rodrigo Tocantins Calado.; Sandra Coccuzzo Sampaio; Sandra Coccuzzo Sampaio Vessoni; Simone Kashima; Svetoslav Nanev Slavov; Vagner Fonseca; Vincent Louis Viala |
| EPI_ISL_5529963 | UNIDADE DE PRONTO ATENDIMENTO DA JUREMA | Analytical Competence Molecular Epidemiology Lab/ACME, Oswaldo Cruz Foundation, Ceara (FIOCRUZ CE) | Carlos Leonardo de Aragao Araujo; Cecilia Leite Costa & Eduardo Ruback dos Santos on behalf of COVID-19 FIOCRUZ Genomic Network; Cleber Furtado Aksenen; Fabio Miyajima; Fernando Braga Stehling; Francisco Eder de Moura Lopes; Igor Oliveira Duarte; Jamille Maria Mendes Bezerra; Joaquim Cesar do Nascimento Sousa Junior; Pedro Miguel Carneiro Jeronimo; Suzana Porto Almeida; Thais Ferreira de Oliveira; Thais de Oliveira Costa; Ticiane Cavalcante de Souza; Veridiana Pessoa Miyajima |
| EPI_ISL_1795230, EPI_ISL_1795231, EPI_ISL_2345461, EPI_ISL_2345463 | UNIDADE DE SAUDE DA FAMILIA JOSE ADALBERTO LELLIS GARCIA | Instituto Butantan / ESALQ-Piracicaba | Antonio Jorge Martins; Bianca Cechetto Carlos. Mendelics: Bibiana Santos; Claudia Renata dos Santos Barros; David Schlesinger; David Schlesinger. Hemocentro Ribeirão Preto: Simone Kashima; Debora Botequio Moretti; Debora Botequio Moretti. Centro de Genômica Funcional da ESALQ: Luiz Lehmann Coutinho; Dimas Tadeu Covas; Elaine Cristina Marqueze; Elaine Vieira Santos; Elaine Vieira dos Santos; Elisangela Chicaroni Mattos; Erika Freitas; Evandra Strazza Rodrigues; Felipe Allan da Silva da Costa; Flavia Aburjaile; Guilherme Targino Valente; Heidge Fukumasu; Heidge Fukumasu. USP-Botucatu: Rejane Maria Tommasini Grotto; Instituto Butantan: Alexander Roberto Precioso; Jayme A. Souza-Neto; Jayme Augusto de Souza-Neto; Jessika Cristina Chagas Lesbon; José Salvatore Leister Patané; João Paulo Kitajima; Luiz Alcantara; Luiz Carlos Junior de Alcantara; Luiz Lehmann Coutinho; Maria Carolina Elias; Marta Giovanetti; Maurício Lacerda Nogueira; Patricia Akemi Assato; Rafael dos Santos Bezerra; Raquel de Lello Rocha Campos Cassano. NGS Soluções Genômicas: Pilar Drummond Sampaio Corrêa Mariani. FZEA-USP Pirassununga: Mirele Daiana Poleti; Raul Machado Neto; Rejane Maria Tommasini Grotto; Ricardo Augusto Brassaloti; Ricardo Haddad; Rodrigo Tocantins Calado.; Sandra Coccuzzo Sampaio; Sandra Coccuzzo Sampaio Vessoni; Simone Kashima; Svetoslav Nanev Slavov; Vagner Fonseca; Vincent Louis Viala |
| EPI_ISL_1795293, EPI_ISL_1795294, EPI_ISL_1795298, EPI_ISL_2345534, EPI_ISL_2345535, EPI_ISL_2345540 | UNIDADE DE SAUDE DE ITOBI ALCIBIADES PIRES | Instituto Butantan / ESALQ-Piracicaba | Antonio Jorge Martins; Bianca Cechetto Carlos. Mendelics: Bibiana Santos; Claudia Renata dos Santos Barros; David Schlesinger; David Schlesinger. Hemocentro Ribeirão Preto: Simone Kashima; Debora Botequio Moretti; Debora Botequio Moretti. Centro de Genômica Funcional da ESALQ: Luiz Lehmann Coutinho; Dimas Tadeu Covas; Elaine Cristina Marqueze; Elaine Vieira Santos; Elaine Vieira dos Santos; Elisangela Chicaroni Mattos; Erika Freitas; Evandra Strazza Rodrigues; Felipe Allan da Silva da Costa; Flavia Aburjaile; Guilherme Targino Valente; Heidge Fukumasu; Heidge Fukumasu. USP-Botucatu: Rejane Maria Tommasini Grotto; Instituto Butantan: Alexander Roberto Precioso; Jayme A. Souza-Neto; Jayme Augusto de Souza-Neto; Jessika Cristina Chagas Lesbon; José Salvatore Leister Patané; João Paulo Kitajima; Luiz Alcantara; Luiz Carlos Junior de Alcantara; Luiz Lehmann Coutinho; Maria Carolina Elias; Marta Giovanetti; Maurício Lacerda Nogueira; Patricia Akemi Assato; Rafael dos Santos Bezerra; Raquel de Lello Rocha Campos Cassano. NGS Soluções Genômicas: Pilar Drummond Sampaio Corrêa Mariani. FZEA-USP Pirassununga: Mirele Daiana Poleti; Raul Machado Neto; Rejane Maria Tommasini Grotto; Ricardo Augusto Brassaloti; Ricardo Haddad; Rodrigo Tocantins Calado.; Sandra Coccuzzo Sampaio; Sandra Coccuzzo Sampaio Vessoni; Simone Kashima; Svetoslav Nanev Slavov; Vagner Fonseca; Vincent Louis Viala |
| EPI_ISL_3102324, EPI_ISL_3102528 | UNIDADE MISTA DE SAUDE DE MISSAO VELHA | Analytical Competence Molecular Epidemiology Lab/ACME, Oswaldo Cruz Foundation, Ceara (FIOCRUZ CE) | Cleber Furtado Aksenen; Fabio Miyajima; Fernando Braga Stehling; Francisco Eder de Moura Lopes; Jamille Maria Mendes Bezerra; Joaquim César do Nascimento Sousa Junior; Pedro Miguel Carneiro Jeronimo; Suzana Porto Almeida e Lucas Delerino; Thais Ferreira de Oliveira; Thais de Oliveira Costa; Ticiane Cavalcante de Souza; Veridiana Pessoa Miyajima |
| EPI_ISL_1795136, EPI_ISL_1795137, EPI_ISL_1795138, EPI_ISL_1795139, EPI_ISL_1795140, EPI_ISL_1795149, EPI_ISL_1795150, EPI_ISL_2345339, EPI_ISL_2345340, EPI_ISL_2345341, EPI_ISL_2345342, EPI_ISL_2345343, EPI_ISL_2345358, EPI_ISL_2345359 | | | |
| see above | UNIDADE MISTA DE VISTA ALEGRE DO ALTO VISTA ALEGRE DO ALTO | Instituto Butantan / ESALQ-Piracicaba | Antonio Jorge Martins; Bianca Cechetto Carlos. Mendelics: Bibiana Santos; Claudia Renata dos Santos Barros; David Schlesinger; David Schlesinger. Hemocentro Ribeirão Preto: Simone Kashima; Debora Botequio Moretti; Debora Botequio Moretti. Centro de Genômica Funcional da ESALQ: Luiz Lehmann Coutinho; Dimas Tadeu Covas; Elaine Cristina Marqueze; Elaine Vieira Santos; Elaine Vieira dos Santos; Elisangela Chicaroni Mattos; Erika Freitas; Evandra Strazza Rodrigues; Felipe Allan da Silva da Costa; Flavia Aburjaile; Guilherme Targino Valente; Heidge Fukumasu; Heidge Fukumasu. USP-Botucatu: Rejane Maria Tommasini Grotto; Instituto Butantan: Alexander Roberto Precioso; Jayme A. Souza-Neto; Jayme Augusto de Souza-Neto; Jessika Cristina Chagas Lesbon; José Salvatore Leister Patané; João Paulo Kitajima; Luiz Alcantara; Luiz Carlos Junior de Alcantara; Luiz Lehmann Coutinho; Maria Carolina Elias; Marta Giovanetti; Maurício Lacerda Nogueira; Patricia Akemi Assato; Rafael dos Santos Bezerra; Raquel de Lello Rocha Campos Cassano. NGS Soluções Genômicas: Pilar Drummond Sampaio Corrêa Mariani. FZEA-USP Pirassununga: Mirele Daiana Poleti; Raul Machado Neto; Rejane Maria Tommasini Grotto; Ricardo Augusto Brassaloti; Ricardo Haddad; Rodrigo Tocantins Calado.; Sandra Coccuzzo Sampaio; Sandra Coccuzzo Sampaio Vessoni; Simone Kashima; Svetoslav Nanev Slavov; Vagner Fonseca; Vincent Louis Viala |
| EPI_ISL_3102530 | UNIDADE PRONTO ATENDIMENTO CONJUNTO CEARA | Analytical Competence Molecular Epidemiology Lab/ACME, Oswaldo Cruz Foundation, Ceara (FIOCRUZ CE) | Cleber Furtado Aksenen; Fabio Miyajima; Fernando Braga Stehling; Francisco Eder de Moura Lopes; Jamille Maria Mendes Bezerra; Joaquim César do Nascimento Sousa Junior; Pedro Miguel Carneiro Jeronimo; Suzana Porto Almeida e Lucas Delerino; Thais Ferreira de Oliveira; Thais de Oliveira Costa; Ticiane Cavalcante de Souza; Veridiana Pessoa Miyajima |
| EPI_ISL_5529969, EPI_ISL_5530146 | UNIDADE REFERENCIA COVID 19 | Analytical Competence Molecular Epidemiology Lab/ACME, Oswaldo Cruz Foundation, Ceara (FIOCRUZ CE) | Carlos Leonardo de Aragao Araujo; Cecilia Leite Costa & Eduardo Ruback dos Santos on behalf of COVID-19 FIOCRUZ Genomic Network; Cleber Furtado Aksenen; Fabio Miyajima; Fernando Braga Stehling; Francisco Eder de Moura Lopes; Igor Oliveira Duarte; Jamille Maria Mendes Bezerra; Joaquim Cesar do Nascimento Sousa Junior; Pedro Miguel Carneiro Jeronimo; Suzana Porto Almeida; Thais Ferreira de Oliveira; Thais de Oliveira Costa; Ticiane Cavalcante de Souza; Veridiana Pessoa Miyajima |
| EPI_ISL_5530056 | UPA 24 HORA MARACANAU | Analytical Competence Molecular Epidemiology Lab/ACME, Oswaldo Cruz Foundation, Ceara (FIOCRUZ CE) | Carlos Leonardo de Aragao Araujo; Cecilia Leite Costa & Eduardo Ruback dos Santos on behalf of COVID-19 FIOCRUZ Genomic Network; Cleber Furtado Aksenen; Fabio Miyajima; Fernando Braga Stehling; Francisco Eder de Moura Lopes; Igor Oliveira Duarte; Jamille Maria Mendes Bezerra; Joaquim Cesar do Nascimento Sousa Junior; Pedro Miguel Carneiro Jeronimo; Suzana Porto Almeida; Thais Ferreira de Oliveira; Thais de Oliveira Costa; Ticiane Cavalcante de Souza; Veridiana Pessoa Miyajima |
| EPI_ISL_1628373 | UPA De Bebedouro | Instituto Adolfo Lutz, Interdiciplinary Procedures Center, Strategic Laboratory | Caio Vinicius Dias Lopes; Claudia Regina Gonçalves; Claudio Tavares Sacchi; Erica Valessa Ramos Gomes; Karoline Rodrigues Campos; Katia Correa de Oliveira Santos; Leonardo Jose Tadeu de Araujo |
| EPI_ISL_1508923 | USC Clinical Lab | Los Angeles County Public Health Laboratories | P. Hemarajata et al. |
| EPI_ISL_1577924, EPI_ISL_1577953 | USCA Avezzano AVEZZANO(L'AQUILA) | Istituto Zooprofilattico Sperimentale dell'Abruzzo e Molise "G. Caporale" | Ancora M; Calistri P; Cammà C; Curini V; Di Domenico M; Di Pasquale A; Lorusso A; Mangone I; Marcacci M; Puglia I; Rinaldi A; Savini G; Scialabba S |
| EPI_ISL_1577927, EPI_ISL_1580507 | USCA Tagliacozzo TAGLIACOZZO(L'AQUILA) | Istituto Zooprofilattico Sperimentale dell'Abruzzo e Molise "G. Caporale" | Ancora M; Calistri P; Cammà C; Caporale M; Curini V; Delli Compagni E; Di Domenico M; Di Lollo Valeria; Di Pasquale A; Lorusso A; Mangone I; Marcacci M; Puglia I; Rinaldi A; Savini G; Scialabba S |
| EPI_ISL_1795290, EPI_ISL_2345529 | USF EUCALIPTOS | Instituto Butantan / ESALQ-Piracicaba | Antonio Jorge Martins; Bianca Cechetto Carlos. Mendelics: Bibiana Santos; Claudia Renata dos Santos Barros; David Schlesinger; David Schlesinger. Hemocentro Ribeirão Preto: Simone Kashima; Debora Botequio Moretti; Debora Botequio Moretti. Centro de Genômica Funcional da ESALQ: Luiz Lehmann Coutinho; Dimas Tadeu Covas; Elaine Cristina Marqueze; Elaine Vieira Santos; Elaine Vieira dos Santos; Elisangela Chicaroni Mattos; Erika Freitas; Evandra Strazza Rodrigues; Felipe Allan da Silva da Costa; Flavia Aburjaile; Guilherme Targino Valente; Heidge Fukumasu; Heidge Fukumasu. USP-Botucatu: Rejane Maria Tommasini Grotto; Instituto Butantan: Alexander Roberto Precioso; Jayme A. Souza-Neto; Jayme Augusto de Souza-Neto; Jessika Cristina Chagas Lesbon; José Salvatore Leister Patané; João Paulo Kitajima; Luiz Alcantara; Luiz Carlos Junior de Alcantara; Luiz Lehmann Coutinho; Maria Carolina Elias; Marta Giovanetti; Maurício Lacerda Nogueira; Patricia Akemi Assato; Rafael dos Santos Bezerra; Raquel de Lello Rocha Campos Cassano. NGS Soluções Genômicas: Pilar Drummond Sampaio Corrêa Mariani. FZEA-USP Pirassununga: Mirele Daiana Poleti; Raul Machado Neto; Rejane Maria Tommasini Grotto; Ricardo Augusto Brassaloti; Ricardo Haddad; Rodrigo Tocantins Calado.; Sandra Coccuzzo Sampaio; Sandra Coccuzzo Sampaio Vessoni; Simone Kashima; Svetoslav Nanev Slavov; Vagner Fonseca; Vincent Louis Viala |
| EPI_ISL_1795295, EPI_ISL_2345536 | USF GUACUANO | Instituto Butantan / ESALQ-Piracicaba | Antonio Jorge Martins; Bianca Cechetto Carlos. Mendelics: Bibiana Santos; Claudia Renata dos Santos Barros; David Schlesinger; David Schlesinger. Hemocentro Ribeirão Preto: Simone Kashima; Debora Botequio Moretti; Debora Botequio Moretti. Centro de Genômica Funcional da ESALQ: Luiz Lehmann Coutinho; Dimas Tadeu Covas; Elaine Cristina Marqueze; Elaine Vieira Santos; Elaine Vieira dos Santos; Elisangela Chicaroni Mattos; Erika Freitas; Evandra Strazza Rodrigues; Felipe Allan da Silva da Costa; Flavia Aburjaile; Guilherme Targino Valente; Heidge Fukumasu; Heidge Fukumasu. USP-Botucatu: Rejane Maria Tommasini Grotto; Instituto Butantan: Alexander Roberto Precioso; Jayme A. Souza-Neto; Jayme Augusto de Souza-Neto; Jessika Cristina Chagas Lesbon; José Salvatore Leister Patané; João Paulo Kitajima; Luiz Alcantara; Luiz Carlos Junior de Alcantara; Luiz Lehmann Coutinho; Maria Carolina Elias; Marta Giovanetti; Maurício Lacerda Nogueira; Patricia Akemi Assato; Rafael dos Santos Bezerra; Raquel de Lello Rocha Campos Cassano. NGS Soluções Genômicas: Pilar Drummond Sampaio Corrêa Mariani. FZEA-USP Pirassununga: Mirele Daiana Poleti; Raul Machado Neto; Rejane Maria Tommasini Grotto; Ricardo Augusto Brassaloti; Ricardo Haddad; Rodrigo Tocantins Calado.; Sandra Coccuzzo Sampaio; Sandra Coccuzzo Sampaio Vessoni; Simone Kashima; Svetoslav Nanev Slavov; Vagner Fonseca; Vincent Louis Viala |
| EPI_ISL_1795391, EPI_ISL_2345528 | USF ROSA CRUZ | Instituto Butantan / ESALQ-Piracicaba | Antonio Jorge Martins; Bianca Cechetto Carlos. Mendelics: Bibiana Santos; Claudia Renata dos Santos Barros; David Schlesinger; David Schlesinger. Hemocentro Ribeirão Preto: Simone Kashima; Debora Botequio Moretti; Debora Botequio Moretti. Centro de Genômica Funcional da ESALQ: Luiz Lehmann Coutinho; Dimas Tadeu Covas; Elaine Cristina Marqueze; Elaine Vieira Santos; Elaine Vieira dos Santos; Elisangela Chicaroni Mattos; Erika Freitas; Evandra Strazza Rodrigues; Felipe Allan da Silva da Costa; Flavia Aburjaile; Guilherme Targino Valente; Heidge Fukumasu; Heidge Fukumasu. USP-Botucatu: Rejane Maria Tommasini Grotto; Instituto Butantan: Alexander Roberto Precioso; Jayme A. Souza-Neto; Jayme Augusto de Souza-Neto; Jessika Cristina Chagas Lesbon; José Salvatore Leister Patané; João Paulo Kitajima; Luiz Alcantara; Luiz Carlos Junior de Alcantara; Luiz Lehmann Coutinho; Maria Carolina Elias; Marta Giovanetti; Maurício Lacerda Nogueira; Patricia Akemi Assato; Rafael dos Santos Bezerra; Raquel de Lello Rocha Campos Cassano. NGS Soluções Genômicas: Pilar Drummond Sampaio Corrêa Mariani. FZEA-USP Pirassununga: Mirele Daiana Poleti; Raul Machado Neto; Rejane Maria Tommasini Grotto; Ricardo Augusto Brassaloti; Ricardo Haddad; Rodrigo Tocantins Calado.; Sandra Coccuzzo Sampaio; Sandra Coccuzzo Sampaio Vessoni; Simone Kashima; Svetoslav Nanev Slavov; Vagner Fonseca; Vincent Louis Viala |
| EPI_ISL_1497850, EPI_ISL_1497853, EPI_ISL_1497872, EPI_ISL_1497879, EPI_ISL_1497884, EPI_ISL_1497886, EPI_ISL_1498016, EPI_ISL_1498029, EPI_ISL_1498030, EPI_ISL_1498033, EPI_ISL_1523801, EPI_ISL_1601315, EPI_ISL_1601340, EPI_ISL_1601342, EPI_ISL_1601344, EPI_ISL_1601353, EPI_ISL_1601358, EPI_ISL_1601409, EPI_ISL_1601421, EPI_ISL_1601426, EPI_ISL_1601432, EPI_ISL_1616677 | | | |
| see above | UW Virology Lab | UW Virology Lab | Alexander Greninger; Hong Xie; Keith R Jerome; Lasata Shrestha; Meei-Li Huang; Michelle Lin; Noah R. Baker; Pavitra Roychoudhury; Saraswathi Sathees; Sean Ellis; Shah Mohamed Bakhash |
| EPI_ISL_1662185 | Unidad de Investigacion Medica de Yucatan (UIMY) | Unidad de Genomica Avanzada | Alejandro Sanchez-Flores; Alfredo Herrera-Estrella; Alicia Ocana-Mondragon; Angel Gustavo Salas-Lais; Bernardo Martinez-Miguel; Blanca Taboada; Brenda Irasema Maldonado-Meza; Carla Ivon Herrera-Najera; Carlos F. Arias; Celia Boukadida; Celida Duque Molina; Clara Esperanza Santacruz-Tinoco; Concepcion Grajales-Muniz; Consorcio Mexicano de Vigilancia Genomica (CoViGen-Mex). Authors (in alphabetical order): Julio Elias Alvarado-Yaah; Fernando Fontove-Herrera; Francisco Pulido; Gloria Elena Espinosa-Ayala; Gloria Maria Molina-Salinas; Gloria Vazquez; Hector Esteban Paz-Juarez; Hector Montoya-Fuentes; Helen Haydee Fernanda Ramirez-Plascencia; Jose Antonio Enciso-Moreno; Jose Esteban Munoz-Medina; Jose de Jesus Nunez-Contreras; Juan Bautista Chale-Dzul; Luis Alberto Ochoa-Carrera; Margarita Matias-Florentino; Maria Guadalupe Santiago-Mauricio; Maria Guadalupe de Jesus Mireles-Rivera; Nelly Selem-Mojica; Pavel Isa; Ricardo Grande; Santiago Ávila-Rios; Victor Eduardo Garcia-Arias; Victor Hugo Borja-Aburto |
| EPI_ISL_1858758, EPI_ISL_1858761, EPI_ISL_1858763, EPI_ISL_1858765, EPI_ISL_1858767, EPI_ISL_1858769, EPI_ISL_1858771, EPI_ISL_1858773, EPI_ISL_1858775, EPI_ISL_1858777, EPI_ISL_1858779, EPI_ISL_1858781, EPI_ISL_1858783, EPI_ISL_1858786, EPI_ISL_1858788, EPI_ISL_1858790, EPI_ISL_1858792, EPI_ISL_1858794, EPI_ISL_1858796, EPI_ISL_1858798, EPI_ISL_1858800, EPI_ISL_1858802, EPI_ISL_1858803, EPI_ISL_1858805, EPI_ISL_1858807, EPI_ISL_1858809, EPI_ISL_1858811, EPI_ISL_1858814, EPI_ISL_1858816, EPI_ISL_1858818, EPI_ISL_1858819, EPI_ISL_1858822, EPI_ISL_1858824, EPI_ISL_1858826, EPI_ISL_1858828, EPI_ISL_1858830, EPI_ISL_1858832, EPI_ISL_1858834, EPI_ISL_1858836, EPI_ISL_1858838, EPI_ISL_1858840, EPI_ISL_1858842, EPI_ISL_1858843, EPI_ISL_1858846, EPI_ISL_1858847, EPI_ISL_1858850, EPI_ISL_1858852, EPI_ISL_1858853, EPI_ISL_1858856, EPI_ISL_1858858, EPI_ISL_1858860, EPI_ISL_1858862, EPI_ISL_1858864, EPI_ISL_1858866, EPI_ISL_1858868, EPI_ISL_1858870, EPI_ISL_1858872, EPI_ISL_1858874, EPI_ISL_1858880, EPI_ISL_1858882, EPI_ISL_1858884, EPI_ISL_1858886, EPI_ISL_1858888, EPI_ISL_1858890, EPI_ISL_1858892, EPI_ISL_1858894, EPI_ISL_1858896, EPI_ISL_1858898, EPI_ISL_1858900, EPI_ISL_1858902, EPI_ISL_1858904, EPI_ISL_1858906, EPI_ISL_1858908, EPI_ISL_1858910, EPI_ISL_1858912, EPI_ISL_1858914, EPI_ISL_1858916, EPI_ISL_1858918, EPI_ISL_1858920, EPI_ISL_1858922, EPI_ISL_1858924, EPI_ISL_1858926, EPI_ISL_1858928, EPI_ISL_1858930, EPI_ISL_1858933, EPI_ISL_1858934, EPI_ISL_1858936, EPI_ISL_1858938, EPI_ISL_1858940, EPI_ISL_1858942, EPI_ISL_1858944, EPI_ISL_1858946, EPI_ISL_1858948, EPI_ISL_1858950, EPI_ISL_1858952, EPI_ISL_1858954, EPI_ISL_1858957, EPI_ISL_1858958, EPI_ISL_1858960, EPI_ISL_1858963, EPI_ISL_1858964, EPI_ISL_1858966, EPI_ISL_1858968, EPI_ISL_1858970, EPI_ISL_1858973, EPI_ISL_1858975, EPI_ISL_1858977, EPI_ISL_1858979, EPI_ISL_1858981, EPI_ISL_1858984, EPI_ISL_1858986, EPI_ISL_1858988, EPI_ISL_1858990, EPI_ISL_1858992, EPI_ISL_1858994, EPI_ISL_1858996, EPI_ISL_1858998, EPI_ISL_1859000, EPI_ISL_1859002, EPI_ISL_1859005, EPI_ISL_1859007, EPI_ISL_1859008 | | | |
| see above | Unidade de apoio ao diagnóstico da COVID – UNADIG | Bioinformatics Laboratory / LNCC | Alessandra P Lamarca; Alexandra L Gerber; Amilcar Tanuri; Ana Paula de C Guimarães; Ana Tereza R Vasconcelos; Andréa Cony Cavalcanti; Caio Luiz Pereira Ribeiro; Cassia Alves; Claudia Maria Braga de Mello; Cristiane Gomes da Silva; Diana Mariani; Douglas Terra Machado; Flávio Dias da Silva; Leandro Magalhães de Souza; Liliane Cavalcante; Luiz G P de Almeida; Marcio Henrique de Oliveira Garcia; Mario Sergio Ribeiro; Ronaldo da Silva F Jr; Silvia Carvalho; Thais Felix Cruz |
| EPI_ISL_1675333 | Universidad Nacional de Colombia - Laboratorio Genómico One Health | Universidad Nacional de Colombia - Laboratorio Genómico One Health | Andres F. Cardona-Rios; Carlos Franco-Muñoz; Daniel O. Maldonado-Perez; Diego A. Álvarez-Díaz; Hector Alejandro Ruiz-Moreno; Idabely Betancur Ortiz; Jorge E. Osorio; Juan P. Hernandez-Ortiz; Karl A Ciuoderis; Katherine Laiton-Donato; Laura Silvana Perez; Lina M. Hurtado; Marcela Mercado-Reyes; Maria Angélica Maya; Maria Stella López; Rita Almanza Payares; Sandra Ines Cano; Simón Villegas Velásquez |
| EPI_ISL_1472387, EPI_ISL_1472400 | University Hospitals Translational Laboratory (UHTL), University Hospitals | University Hospitals Translational Laboratory (UHTL), University Hospitals | Alouani, D.; Sadri, N.; Song, X. |
| EPI_ISL_1533405, EPI_ISL_1533406 | University Hospitals of Geneva, Laboratory of Virology | HUG, Laboratory of Virology and the Health2030 Genome Center | Ana Rita Goncalves; Deborah Penet; Emmanouil Dermitzakis; Henri Pegeot; Ioannis Xenarios; Keith Harshman; Laurent Kaiser; Lorenzo Cerutti; Melyssa Elies; Samuel Cordey |
| EPI_ISL_2246635 | University of Chicago Medicine | RIPHL at Rush University Medical Center | Cindy Bethel; Felix Araujo Perez; Kevin Kunstman; Laura Furtado; Lauren Megger; Marieta Hyde; Max Kolton; Stefan Green |
| EPI_ISL_1492779, EPI_ISL_1492989 | University of Liège COVID-19 testing center | GIGA Medical Genomics | Bouchra Boujemla; Cécile Meex; Keith Durkin; Maria Artesi; Marie-Pierre Hayette; Nathalie Renotte; Pierrette Melin; Raphaël Boreux; Sébastien Bontems; Vincent Bours |
| EPI_ISL_1491426, EPI_ISL_1491429, EPI_ISL_1491508, EPI_ISL_1580479, EPI_ISL_1580498, EPI_ISL_1855929, EPI_ISL_1855930 | | | |
| see above | University of Michigan Clinical Microbiology Laboratory | Lauring Lab, University of Michigan, Department of Microbiology and Immunology | Valesano |
| EPI_ISL_1527417 | University of Mississippi | University of Mississippi Medical | Ashley C. Johnson; D. Ashley Robinson; Ithiel J. Frame; Krishna K. Ayyalasomayajula; Michael R. Garrett |

| | | | |
|---|---|---|---|
| | Medical Center, Department of Pathology | Center, Molecular and Genomics Core Facility | |
| EPI_ISL_1582628, EPI_ISL_1594028 | University of Wisconsin-Madison AIDS Vaccine Research Laboratories | University of Wisconsin-Madison AIDS Vaccine Research Laboratories | Gage Moreno; Katarina Braun; et al. AIDS Vaccine Research Laboratories |
| EPI_ISL_2465224, EPI_ISL_2465230, EPI_ISL_2465234, EPI_ISL_2465241 | Università Federico II - Dipartimento di scienze mediche traslazionali - Napoli | TIGEM | Antonio Grimaldi Patrizia Annunziata Francesco Panariello Teresa Giuliano Michele Cennamo Valentina Bouche Chiara Colantuono Lucio Di Filippo Mariano Fiorenza Anna Manfredi Marcello Salvi Giuseppe Portella Andrea Ballabio Davide Cacchiarelli |
| EPI_ISL_1499628, EPI_ISL_1499629, EPI_ISL_1499632, EPI_ISL_1499635, EPI_ISL_1499639, EPI_ISL_1499640, EPI_ISL_1520098, EPI_ISL_1669931, EPI_ISL_1669932, EPI_ISL_1669933, EPI_ISL_1669934, EPI_ISL_1669935, EPI_ISL_1669936, EPI_ISL_1669937, EPI_ISL_1669938 | | | |
| see above | Università degli Studi di Perugia | Istituto Zooprofilattico Sperimentale dell'Abruzzo e Molise "G. Caporale" | Ancora M; Calistri P; Camilloni B; Cammà C; Caporale M; Curini V; Delli Compagni E; Di Domenico M; Di Lollo Valeria; Di Pasquale A; Lorusso A; Mangone I; Marcacci M; Mencacci A; Puglia I; Rinaldi A; Savini G; Scialabba S |
| EPI_ISL_1743716 | Usansolo-Galdakao University Hospital | Cruces University Hospital | Ana Belén de la Hoz; Ana Gual-de-Torrella; Izaskun Alejo-Cancho; Mikel Gallego |
| EPI_ISL_2290229, EPI_ISL_2290232, EPI_ISL_2290289, EPI_ISL_2290348, EPI_ISL_2290361, EPI_ISL_2290459, EPI_ISL_2292463, EPI_ISL_2292472, EPI_ISL_2292497, EPI_ISL_2292599, EPI_ISL_2458488 | | | |
| see above | Utah Public Health Laboratory | Utah Public Health Laboratory | Erin L. Young; Kelly F. Oakeson; Tara Gallagher |
| EPI_ISL_2283916 | VA Connecticut Healthcare System | Yale Center for Genomic Analysis | Brooke Sullivan; Curt Scharfe; Irina Tikhonova; Kaya Bilguvar; Shrikant Mane |
| EPI_ISL_1795339, EPI_ISL_1795340, EPI_ISL_1795342, EPI_ISL_1795343, EPI_ISL_2345605, EPI_ISL_2345606, EPI_ISL_2345608, EPI_ISL_2345609 | | | |
| see above | VIGILANCIA EM SAUDE | Instituto Butantan / ESALQ-Piracicaba | Antonio Jorge Martins; Bianca Cechetto Carlos. Mendelics: Bibiana Santos; Claudia Renata dos Santos Barros; David Schlesinger; David Schlesinger. Hemocentro Ribeirão Preto: Simone Kashima; Debora Botequio Moretti; Debora Botequio Moretti. Centro de Genômica Funcional da ESALQ: Luiz Lehmann Coutinho; Dimas Tadeu Covas; Elaine Cristina Marqueze; Elaine Vieira Santos; Elaine Vieira dos Santos; Elisangela Chicaroni Mattos; Erika Freitas; Evandra Strazza Rodrigues; Felipe Allan da Silva da Costa; Flavia Aburjaile; Guilherme Targino Valente; Heidge Fukumasu; Heidge Fukumasu. USP-Botucatu: Rejane Maria Tommasini Grotto; Instituto Butantan: Alexander Roberto Precioso; Jayme A. Souza-Neto; Jayme Augusto de Souza-Neto; Jessika Cristina Chagas Lesbon; José Salvatore Leister Patané; João Paulo Kitajima; Luiz Alcantara; Luiz Carlos Junior de Alcantara; Luiz Lehmann Coutinho; Maria Carolina Elias; Marta Giovanetti; Maurício Lacerda Nogueira; Patricia Akemi Assato; Rafael dos Santos Bezerra; Raquel de Lello Rocha Campos Cassano. NGS Soluções Genômicas: Pilar Drummond Sampaio Corrêa Mariani. FZEA-USP Pirassununga: Mirele Daiana Poleti; Raul Machado Neto; Rejane Maria Tommasini Grotto; Ricardo Augusto Brassaloti; Ricardo Haddad; Rodrigo Tocantins Calado.; Sandra Coccuzzo Sampaio; Sandra Coccuzzo Sampaio Vessoni; Simone Kashima; Svetoslav Nanev Slavov; Vagner Fonseca; Vincent Louis Viala |
| EPI_ISL_1966296, EPI_ISL_1966300 | VIGILANCIA EPIDEMIOLOGICA DE IBATE | Instituto Butantan / Mendelics | Antonio Jorge Martins; Bianca Cechetto Carlos. Mendelics: Bibiana Santos; Claudia Renata dos Santos Barros; Cíntia Bittar; David Schlesinger. Hemocentro Ribeirão Preto: Simone Kashima; Debora Botequio Moretti; Elaine Cristina Marqueze; Elaine Vieira dos Santos; Elisangela Chicaroni Mattos; Erika Freitas; Evandra Strazza Rodrigues; Felipe Allan da Silva da Costa; Flavia Aburjaile; Fábio Sossai Possebon; Guilherme Campos; Guilherme Targino Valente; Heidge Fukumasu. USP-Botucatu: Rejane Maria Tommasini Grotto; Helena Lage Ferreira; Instituto Butantan: Dimas Tadeu Covas; Jardelina de Souza Todao Bernardino; Jayme A. Souza-Neto; Jessika Cristina Chagas Lesbon; Jorge A. Petroli Marchesi; José Salvatore Leister Patané; João Paulo Kitajima; João Pessoa Araújo Jr.; Leila Sabrina Ullmann; Loyze Paola Oliveira de Lima; Luiz Aurelio de Campos Crispin. Centro de Genômica Funcional da ESALQ: Luiz Lehmann Coutinho; Luiz Carlos Junior de Alcantara; Lívia Sacchetto; Maisa C. Pereira Parra; Maria Carolina Elias; Marta Giovanetti; Marília Moraes; Maurício Lacerda Nogueira. Prefeitura de Sao Paulo: Melissa Palmieri.; Patricia Akemi Assato; Paula Rahal; Paulo Inacio da Costa; Rafael dos Santos Bezerra; Raquel de Lello Rocha Campos Cassano. NGS Soluções Genômicas: Pilar Drummond Sampaio Corrêa Mariani. FZEA-USP Pirassununga: Mirele Daiana Poleti; Raul Machado Neto; Ricardo Augusto Brassaloti; Ricardo Haddad; Rodrigo Tocantins Calado. FAMERP-SJRP: Cecília Artico Banho; Sandra Coccuzzo Sampaio; Svetoslav Nanev Slavov; Vagner Fonseca; Vincent Louis Viala |
| EPI_ISL_1795094, EPI_ISL_2344682 | VIGILANCIA EPIDEMIOLOGICA DE LEME | Instituto Butantan / ESALQ-Piracicaba | Antonio Jorge Martins; Bianca Cechetto Carlos. Mendelics: Bibiana Santos; Claudia Renata dos Santos Barros; David Schlesinger; David Schlesinger. Hemocentro Ribeirão Preto: Simone Kashima; Debora Botequio Moretti; Debora Botequio Moretti. Centro de Genômica Funcional da ESALQ: Luiz Lehmann Coutinho; Dimas Tadeu Covas; Elaine Cristina Marqueze; Elaine Vieira Santos; Elaine Vieira dos Santos; Elisangela Chicaroni Mattos; Erika Freitas; Evandra Strazza Rodrigues; Felipe Allan da Silva da Costa; Flavia Aburjaile; Guilherme Targino Valente; Heidge Fukumasu; Heidge Fukumasu. USP-Botucatu: Rejane Maria Tommasini Grotto; Instituto Butantan: Alexander Roberto Precioso; Jayme A. Souza-Neto; Jayme Augusto de Souza-Neto; Jessika Cristina Chagas Lesbon; José Salvatore Leister Patané; João Paulo Kitajima; Luiz Alcantara; Luiz Carlos Junior de Alcantara; Luiz Lehmann Coutinho; Maria Carolina Elias; Marta Giovanetti; Maurício Lacerda Nogueira; Patricia Akemi Assato; Rafael dos Santos Bezerra; Raquel de Lello Rocha Campos Cassano. NGS Soluções Genômicas: Pilar Drummond Sampaio Corrêa Mariani. FZEA-USP Pirassununga: Mirele Daiana Poleti; Raul Machado Neto; Rejane Maria Tommasini Grotto; Ricardo Augusto Brassaloti; Ricardo Haddad; Rodrigo Tocantins Calado.; Sandra Coccuzzo Sampaio; Sandra Coccuzzo Sampaio Vessoni; Simone Kashima; Svetoslav Nanev Slavov; Vagner Fonseca; Vincent Louis Viala |
| EPI_ISL_2210189 | VIGILANCIA EPIDEMIOLOGICA E CONTROLE DE VETORES PIRASSUNUN | Instituto Butantan | Antonio Jorge Martins; Claudia Renata dos Santos Barros; David Schlesinger; Debora Botequio Moretti; Dimas Tadeu Covas; Elaine Cristina Marqueze; Elaine Vieira Santos; Evandra Strazza Rodrigues; Heidge Fukumasu; Jayme Augusto de Souza-Neto; José Salvatore Leister Patané; Luiz Alcantara; Luiz Lehmann Coutinho; Maria Carolina Elias; Maurício Lacerda Nogueira; Rafael dos Santos Bezerra; Raul Machado Neto; Rejane Maria Tommasini Grotto; Ricardo Haddad; Sandra Coccuzzo Sampaio Vessoni; Simone Kashima; Svetoslav Nanev Slavov; Vincent Louis Viala. |
| EPI_ISL_2344663, EPI_ISL_2344664, EPI_ISL_2344667, EPI_ISL_2344668, EPI_ISL_2344669, EPI_ISL_2344670 | VIGILANCIA EPIDEMIOLOGICA E CONTROLE DE VETORES PIRASSUNUN | Instituto Butantan / FZEA-USP-Pirassununga | Antonio Jorge Martins; Claudia Renata dos Santos Barros; David Schlesinger; Debora Botequio Moretti; Dimas Tadeu Covas; Elaine Cristina Marqueze; Elaine Vieira Santos; Evandra Strazza Rodrigues; Heidge Fukumasu; Jayme Augusto de Souza-Neto; José Salvatore Leister Patané; Luiz Alcantara; Luiz Lehmann Coutinho; Maria Carolina Elias; Maurício Lacerda Nogueira; Rafael dos Santos Bezerra; Raul Machado Neto; Rejane Maria Tommasini Grotto; Ricardo Haddad; Sandra Coccuzzo Sampaio Vessoni; Simone Kashima; Svetoslav Nanev Slavov; Vincent Louis Viala |
| EPI_ISL_1795217, EPI_ISL_2345445 | VIGILANCIA EPIDEMIOLOGICA JARDINOPOLIS SP | Instituto Butantan / ESALQ-Piracicaba | Antonio Jorge Martins; Bianca Cechetto Carlos. Mendelics: Bibiana Santos; Claudia Renata dos Santos Barros; David Schlesinger; David Schlesinger. Hemocentro Ribeirão Preto: Simone Kashima; Debora Botequio Moretti; Debora Botequio Moretti. Centro de Genômica Funcional da ESALQ: Luiz Lehmann Coutinho; Dimas Tadeu Covas; Elaine Cristina Marqueze; Elaine Vieira Santos; Elaine Vieira dos Santos; Elisangela Chicaroni Mattos; Erika Freitas; Evandra Strazza Rodrigues; Felipe Allan da Silva da Costa; Flavia Aburjaile; Guilherme Targino Valente; Heidge Fukumasu; Heidge Fukumasu. USP-Botucatu: Rejane Maria Tommasini Grotto; Instituto Butantan: Alexander Roberto Precioso; Jayme A. Souza-Neto; Jayme Augusto de Souza-Neto; Jessika Cristina Chagas Lesbon; José Salvatore Leister Patané; João Paulo Kitajima; Luiz Alcantara; Luiz Carlos Junior de Alcantara; Luiz Lehmann Coutinho; Maria Carolina Elias; Marta Giovanetti; Maurício Lacerda Nogueira; Patricia Akemi Assato; Rafael dos Santos Bezerra; Raquel de Lello Rocha Campos Cassano. NGS Soluções Genômicas: Pilar Drummond Sampaio Corrêa Mariani. FZEA-USP Pirassununga: Mirele Daiana Poleti; Raul Machado Neto; Rejane Maria Tommasini Grotto; Ricardo Augusto Brassaloti; Ricardo Haddad; Rodrigo Tocantins Calado.; Sandra Coccuzzo Sampaio; Sandra Coccuzzo Sampaio Vessoni; Simone Kashima; Svetoslav Nanev Slavov; Vagner Fonseca; Vincent Louis Viala |
| EPI_ISL_2674281 | VIROLOGIA INS DRSP | Instituto Nacional de Salud- Dirección de Investigación en Salud Pública | Carlos Franco-Muñoz; Carmen Osorio; Diana Malo; Diego A. Álvarez-Díaz; Diego Andrés Prada; Gerardo Santamaría; Hector Alejandro Ruiz-Moreno; Jhonnatan Reales-González; Jorge Rivera; Juan Camilo Martinez; Julian Naizaque; Katherine Laiton-Donato; Lisseth Pardo; Magdalena Wiesner; Marcela Mercado-Reyes; Maria T. Herrera-Sepúlveda; Marta Lopez Blanco; Martha Lucia Ospina Martinez; Paola Rojas; Sergio Gomez; Sheryll Corchuelo; Ángela Alarcon Cruz |
| EPI_ISL_2036236, EPI_ISL_2293028 | VIROLOGY LABORATORY-CHU NICE | VIROLOGY LABORATORY-CHU NICE | Aicha El Yakine; Geraldine Gonfrier; Jean Machowiak; Sebastien Vitale; Valerie Giordanengo; Virginie Flahou |
| EPI_ISL_5802000, EPI_ISL_5802005 | Vigilancia Epidemiologica De Ibate | Instituto Butantan | Antonio Jorge Martins; Claudia Renata dos Santos Barros; David Schlesinger; Debora Botequio Moretti; Dimas Tadeu Covas; Elaine Cristina Marqueze; Elaine Vieira Santos; Evandra Strazza Rodrigues; Heidge Fukumasu; Jayme Augusto de Souza-Neto; José Salvatore Leister Patané; Luiz Alcantara; Luiz Lehmann Coutinho; Maria Carolina Elias; Maurício Lacerda Nogueira; Rafael dos Santos Bezerra; Raul Machado Neto; Rejane Maria Tommasini Grotto; Ricardo Haddad; Sandra Coccuzzo Sampaio Vessoni; Simone Kashima; Svetoslav Nanev Slavov; Vincent Louis Viala |
| EPI_ISL_5799799 | Vigilancia Epidemiologica De Leme | Instituto Butantan | Antonio Jorge Martins; Claudia Renata dos Santos Barros; David Schlesinger; Debora Botequio Moretti; Dimas Tadeu Covas; Elaine Cristina Marqueze; Elaine Vieira Santos; Evandra Strazza Rodrigues; Heidge Fukumasu; Jayme Augusto de Souza-Neto; José Salvatore Leister Patané; Luiz Alcantara; Luiz Lehmann Coutinho; Maria Carolina Elias; Maurício Lacerda Nogueira; Rafael dos Santos Bezerra; Raul Machado Neto; Rejane Maria Tommasini Grotto; Ricardo Haddad; Sandra Coccuzzo Sampaio Vessoni; Simone Kashima; Svetoslav Nanev Slavov; Vincent Louis Viala |
| EPI_ISL_5782662, EPI_ISL_5782663, EPI_ISL_5782664, EPI_ISL_5782667, EPI_ISL_5782668, EPI_ISL_5782669, EPI_ISL_5782670 | | | |
| see above | Vigilancia Epidemiologica E Controle De Vetores Pirassununga | Instituto Butantan | Antonio Jorge Martins; Claudia Renata dos Santos Barros; David Schlesinger; Debora Botequio Moretti; Dimas Tadeu Covas; Elaine Cristina Marqueze; Elaine Vieira Santos; Evandra Strazza Rodrigues; Heidge Fukumasu; Jayme Augusto de Souza-Neto; José Salvatore Leister Patané; Luiz Alcantara; Luiz Lehmann Coutinho; Maria Carolina Elias; Maurício Lacerda Nogueira; Rafael dos Santos Bezerra; Raul Machado Neto; Rejane Maria Tommasini Grotto; Ricardo Haddad; Sandra Coccuzzo Sampaio Vessoni; Simone Kashima; Svetoslav Nanev Slavov; Vincent Louis Viala |
| EPI_ISL_1598772 | Viollier AG | Department of Biosystems Science and Engineering, ETH Zürich | Chaoran Chen; Christian Beisel; Christiane Beckmann; Christoph Noppen; David Dreifuss; Elodie Burcklen; Ina Nissen; Ivan Topolsky; Katharina Jahn; Lara Fuhrmann; Maurice Redondo; Mirjam Feldkamp; Natascha Santacroce; Niko Beerenwinkel; Noemie Santamaria de Souza; Olivier Kobel; Philipp Jablonski; Rebecca Denes; Sarah Nadeau; Sophie Seidel; Tanja Stadler |
| EPI_ISL_1805663 | Virginia Division of Consolidated Laboratory Services | Virginia Division of Consolidated Laboratory Services | Virginia DCLS |
| EPI_ISL_2884187 | Virology Laboratory, Scientific Department, Army Medical Center | Virology Laboratory, Scientific Department, Army Medical Center | Anella Monte; Anna Anselmo; Antonella Fortunato; Filippo Molinari; Florigio Lista; Francesco Giordani; Giancarlo Petralito; Giandomenico Cerreto; Giulia Campoli; Lucia Nicosia; Marzia Cavalli; Riccardo De Sanctis; Rossella Brandi; Silvia Fillo; Vanessa Vera Fain |
| EPI_ISL_1626455, EPI_ISL_1626464, EPI_ISL_1793915, EPI_ISL_1793994, EPI_ISL_1794199, EPI_ISL_1794205, EPI_ISL_1794519 | | | |
| see above | Wisconsin State Laboratory of Hygiene Communicable Disease Division | Wisconsin State Laboratory of Hygiene Communicable Disease Division | Abigail C. Shockey; Alicia J. Mooney; Kelsey R. Florek; Sara Wagner |
| EPI_ISL_1587277, EPI_ISL_1587301, EPI_ISL_1674553, EPI_ISL_2023066 | Yale Clinical Virology Lab | Grubaugh Lab - Yale School of Public Health | Anderson Brito; Annie Watkins; Chaney Kalinich; Chantal Vogels; Isabel Ott; Jessica Rothman; Joseph Fauver; Mallery Breban; Marie L. Landry; Mary Petrone; Nathan Grubaugh; Tara Alpert |
| EPI_ISL_2488796 | Evandro Chagas Institute | Evandro Chagas Institute | A.M.; Barbagelata; E.C.; E.M.A.; Ferreira; J.A.; Junior; K.C.; L.C.; L.S.; M.C.; P.S.; Pinheiro; Santos; Silva; Sousa; Sousa Junior; W.D.C.; da Silva |
