## Supplement 3 for "Rapid and Accurate Identification of SARS-CoV-2 Omicron Variants Using Droplet Digital PCR (RT-ddPCR)": All_0329-0404.pdf

We gratefully acknowledge the following Authors from the Originating laboratories responsible for obtaining the specimens, as well as the Submitting laboratories where the genome data were generated and shared via GISAID, on which this research is based.

All Submitters of data may be contacted directly via www.gisaid.org

Authors are sorted alphabetically.

| Accession ID | Originating Laboratory | Submitting Laboratory | Authors |
|---|---|---|---|
| EPI_ISL_2645372 | "PA Department of Health, Bureau of Laboratories" | Centers for Disease Control and Prevention Division of Viral Diseases, Pathogen Discovery | Alison Laufer Halpin; Ben L. Rambo-Martin; Clinton R. Paden; Dakota Howard; Darlene Wagner; Dave Wentworth; Dhwani Batra; Jasmine Padilla; Justin Lee; Katie Dillon; Krista Queen; Kristen Knipe; Kristine Lacek; Mark Burroughs; Matthew Schmerer; Mili Sheth; Peter Cook; Sam Shepard; Sarah Nobles; Shoshona Le; Suxiang Tong; Vivien Dugan; Yvette Unoarumhi |
| EPI_ISL_2720456 | A.S.L. VERCELLI | Fondazione del Piemonte per l'Oncologia IRCCS | Antonino Sottile; Giorgio Giardina; Paola Marino; Silvia Brossa |
| EPI_ISL_2614566 | AMA Dr. Cesar Antunes da Rocha | Instituto Adolfo Lutz, Interdiciplinary Procedures Center, Strategic Laboratory | Caio Vinicius Dias Lopes; Claudia Regina Gonçalves; Claudio Tavares Sacchi; Erica Valessa Ramos Gomes; Karoline Rodrigues Campos; Leonardo Jose Tadeu de Araujo |
| EPI_ISL_5915247, EPI_ISL_5915248, EPI_ISL_5915285, EPI_ISL_6573869, EPI_ISL_6573878, EPI_ISL_6573946 | AMALAB/FACISA/UFRN | WallauLab on behalf of Fiocruz COVID-19 Genomic Surveillance Network | Alexandre Freitas da Silva; Allan Roberto Dias Nunes; Antonio Marinho da Silva Neto; Cassia Docena; Constância Flávia Junqueira Ayres; Filipe Zimmer Dezordi; Gabriel Luz Wallau; Gustavo Barbosa de Lima; Joana Cristina Medeiros Tavares Marques; Katya Anaya Jacinto; Lais Ceschini Machado; Lilian Caroliny Amorim Silva; Marcelo Henrique dos Santos Paiva; Mariane dos Santos Duarte; Matheus Filgueira Bezerra; Sinval Pinto Brandão Filho |
| EPI_ISL_1557527, EPI_ISL_1557546, EPI_ISL_1557955, EPI_ISL_1558010, EPI_ISL_1940206, EPI_ISL_1940230 | ASL Napoli 1 Centro | AMES Centro Polidiagnostico Strumentale S.r.l. | "Giovanni Savarese; Antonella Di Carlo; Antonio Fico; Antonio Fico"; Eloisa Evangelista; Giovanni Savarese; Luigi D'Amore; Luisa Circelli; Maurizio D'Amora; Monica Ianniello; Nadia Petrillo; Raffaella Ruggiero; Roberto Sirica |
| EPI_ISL_1599525, EPI_ISL_1599527 | AZ Klina | AZ Klina | Carl Vael - Lynsey Berckmans |
| EPI_ISL_1667450, EPI_ISL_1667793, EPI_ISL_1667799, EPI_ISL_1667800, EPI_ISL_1667935, EPI_ISL_1667953, EPI_ISL_1668005, EPI_ISL_1668007, EPI_ISL_1668010, EPI_ISL_1668012, EPI_ISL_1668078, EPI_ISL_1668099, EPI_ISL_1668107, EPI_ISL_1668208, EPI_ISL_1668233, EPI_ISL_1668276, EPI_ISL_1668327, EPI_ISL_1668426, EPI_ISL_1668442, EPI_ISL_1685515, EPI_ISL_1685525, EPI_ISL_1685548, EPI_ISL_1685551, EPI_ISL_1685615, EPI_ISL_1685625, EPI_ISL_1685650, EPI_ISL_1685679, EPI_ISL_1685686, EPI_ISL_1685696, EPI_ISL_1685697, EPI_ISL_1685896, EPI_ISL_1685935, EPI_ISL_1685956, EPI_ISL_1685971, EPI_ISL_1685973, EPI_ISL_1686002, EPI_ISL_1686005, EPI_ISL_1686069, EPI_ISL_1686166, EPI_ISL_1686186, EPI_ISL_1686243, EPI_ISL_1686249, EPI_ISL_1686252, EPI_ISL_1686715, EPI_ISL_1686858, EPI_ISL_1686866, EPI_ISL_1686870, EPI_ISL_1686946, EPI_ISL_1686996, EPI_ISL_1687001, EPI_ISL_1687015, EPI_ISL_1687059, EPI_ISL_1687245, EPI_ISL_1687247, EPI_ISL_1687296, EPI_ISL_1687298, EPI_ISL_1687312, EPI_ISL_1687322, EPI_ISL_1687354, EPI_ISL_1687425, EPI_ISL_1687428, EPI_ISL_1687429, EPI_ISL_1687436, EPI_ISL_1687449, EPI_ISL_1687464, EPI_ISL_1687541, EPI_ISL_1687693, EPI_ISL_1687700, EPI_ISL_1687733, EPI_ISL_1687791, EPI_ISL_1687792, EPI_ISL_1687828, EPI_ISL_1687846, EPI_ISL_1687873, EPI_ISL_1687884, EPI_ISL_1687937, EPI_ISL_1687939, EPI_ISL_1687941, EPI_ISL_1687947, EPI_ISL_1688102, EPI_ISL_1688118, EPI_ISL_1688171, EPI_ISL_1688174, EPI_ISL_1688263, EPI_ISL_1688305, EPI_ISL_1688353, EPI_ISL_1688362, EPI_ISL_1688422, EPI_ISL_1688450, EPI_ISL_1688455, EPI_ISL_1688570, EPI_ISL_1688611, EPI_ISL_1688647, EPI_ISL_1688668, EPI_ISL_1688712, EPI_ISL_1688770, EPI_ISL_1688775, EPI_ISL_1688803, EPI_ISL_1688843, EPI_ISL_1688846, EPI_ISL_1688852, EPI_ISL_1688887, EPI_ISL_1688970, EPI_ISL_1689035, EPI_ISL_1689068, EPI_ISL_1689070, EPI_ISL_1689089, EPI_ISL_1689214, EPI_ISL_1689333, EPI_ISL_1689544, EPI_ISL_1689795, EPI_ISL_1689810, EPI_ISL_1689933, EPI_ISL_1690137, EPI_ISL_1690158, EPI_ISL_1690251, EPI_ISL_1701897, EPI_ISL_1701962, EPI_ISL_1701964, EPI_ISL_1702103, EPI_ISL_1702109, EPI_ISL_1702112, EPI_ISL_1702120, EPI_ISL_1702722, EPI_ISL_1702738, EPI_ISL_1702809, EPI_ISL_1702841, EPI_ISL_1702872, EPI_ISL_1702924, EPI_ISL_1702935, EPI_ISL_1702936, EPI_ISL_1702937, EPI_ISL_1702943, EPI_ISL_1702959, EPI_ISL_1702962, EPI_ISL_1711975, EPI_ISL_1735697, EPI_ISL_1735706, EPI_ISL_1735708, EPI_ISL_1735737, EPI_ISL_1735739, EPI_ISL_1735779, EPI_ISL_1735796, EPI_ISL_1735838, EPI_ISL_1735874, EPI_ISL_1735883, EPI_ISL_1735889, EPI_ISL_1735953, EPI_ISL_1735979, EPI_ISL_1735987, EPI_ISL_1736085, EPI_ISL_1736123, EPI_ISL_1736133, EPI_ISL_1736134, EPI_ISL_1736135, EPI_ISL_1736148, EPI_ISL_1736181, EPI_ISL_1736214, EPI_ISL_1736221, EPI_ISL_1736224, EPI_ISL_1736247, EPI_ISL_1736312, EPI_ISL_1737030, EPI_ISL_1737142, EPI_ISL_1737163, EPI_ISL_1737249, EPI_ISL_1737358, EPI_ISL_1737385, EPI_ISL_1737387, EPI_ISL_1737414, EPI_ISL_1737459, EPI_ISL_1737476, EPI_ISL_1737492, EPI_ISL_1737531, EPI_ISL_1737532, EPI_ISL_1737575, EPI_ISL_1737587, EPI_ISL_1834789, EPI_ISL_1834822, EPI_ISL_1834839, EPI_ISL_1834853, EPI_ISL_1834865, EPI_ISL_1834866, EPI_ISL_1834874, EPI_ISL_1834876, EPI_ISL_1834887, EPI_ISL_1834899, EPI_ISL_1834944, EPI_ISL_1834993, EPI_ISL_1835043, EPI_ISL_1835044, EPI_ISL_1835045, EPI_ISL_1835046, EPI_ISL_1835092, EPI_ISL_1835153, EPI_ISL_1835185, EPI_ISL_1835193, EPI_ISL_1835258, EPI_ISL_1835267, EPI_ISL_1835310, EPI_ISL_1900233, EPI_ISL_1900275, EPI_ISL_1900320, EPI_ISL_1900372, EPI_ISL_1925109, EPI_ISL_1925233, EPI_ISL_1925312, EPI_ISL_1925313, EPI_ISL_1925339, EPI_ISL_2421304, EPI_ISL_2421336, EPI_ISL_2421341, EPI_ISL_2421490, EPI_ISL_2421516, EPI_ISL_2421540, EPI_ISL_2421551, EPI_ISL_2421556, EPI_ISL_2421586, EPI_ISL_2421598, EPI_ISL_2421614, EPI_ISL_2421616, EPI_ISL_2421641, EPI_ISL_2421733, EPI_ISL_2421740, EPI_ISL_2421767, EPI_ISL_2421769, EPI_ISL_2442316, EPI_ISL_2442319, EPI_ISL_2442321, EPI_ISL_2599964, EPI_ISL_2599996, EPI_ISL_2600021, EPI_ISL_2600032, EPI_ISL_2649903, EPI_ISL_2649904, EPI_ISL_4360321, EPI_ISL_4361083, EPI_ISL_4364221, EPI_ISL_4364368, EPI_ISL_4365132 | | | |
| see above | Aegis Sciences Corporation | Centers for Disease Control and Prevention Division of Viral Diseases, Pathogen Discovery | Adrian Paskey; Alec Vest; Benjamin Rambo-Martin; Christopher Gulvick; Clinton Paden; Clinton R. Paden; Cyndi Clark; Dakota Howard; Darlene Wagner; Dhwani Batra; Dillon Nall; Duncan MacCannell; Erisa Sula; Ethan Sanders; Holly Houdeshell; Jason Caravas; Kara Moser; Kristine Lacek; Matthew Hardison; Matthew Schmerer; Ola Kvalvaag; Patrick Campbell; Peter Cook; Peter W. Cook; Rob Case; Scott Sammons; Shatavia Morrison; Shaun Westlund; Tymeckia Kendall; Victoria Caban Figueroa; Vikramsinha Ghorpade; Yvette Unoarumhi |
| EPI_ISL_1653895, EPI_ISL_1675103, EPI_ISL_1789630, EPI_ISL_1893796 | Alaska State Virology Laboratory | Alaska State Virology Laboratory | Elva House; Jack Chen; Jacob Zidek; Lisa Smith; Ph.D.; Stephanie DeRonde |
| EPI_ISL_2168277, EPI_ISL_2168278, EPI_ISL_2168279, EPI_ISL_2168280, EPI_ISL_2168281, EPI_ISL_2168282, EPI_ISL_2168283, EPI_ISL_2168284, EPI_ISL_2168285, EPI_ISL_2168286, EPI_ISL_2168287, EPI_ISL_2168288, EPI_ISL_2168289, EPI_ISL_2168290, EPI_ISL_2168291, EPI_ISL_2168292, EPI_ISL_2168293, EPI_ISL_2168294, EPI_ISL_2168295, EPI_ISL_2168296, EPI_ISL_2168297, EPI_ISL_2168298, EPI_ISL_2168299, EPI_ISL_2168300, EPI_ISL_2168301, EPI_ISL_2168302, EPI_ISL_2168303, EPI_ISL_2168304, EPI_ISL_2168305, EPI_ISL_2168306, EPI_ISL_2168307, EPI_ISL_2168308, EPI_ISL_2168309, EPI_ISL_2168310, EPI_ISL_2168311, EPI_ISL_2168312, EPI_ISL_2168313, EPI_ISL_2168314, EPI_ISL_2168315, EPI_ISL_2168316, EPI_ISL_2168317, EPI_ISL_2168318, EPI_ISL_2168319, EPI_ISL_2168320, EPI_ISL_2168321, EPI_ISL_2168322, EPI_ISL_2168323, EPI_ISL_2168324, EPI_ISL_2168325, EPI_ISL_2168686, EPI_ISL_2168704, EPI_ISL_2168706, EPI_ISL_2168722, EPI_ISL_2168753, EPI_ISL_2168762, EPI_ISL_2168764, EPI_ISL_2168786, EPI_ISL_2169243, EPI_ISL_2169249, EPI_ISL_2169265, EPI_ISL_2169268, EPI_ISL_2169271, EPI_ISL_2169272, EPI_ISL_2169273, EPI_ISL_2169300, EPI_ISL_2169301, EPI_ISL_2169302, EPI_ISL_2169307, EPI_ISL_2169310, EPI_ISL_2169312, EPI_ISL_2169313, EPI_ISL_2169315, EPI_ISL_2169403, EPI_ISL_2169404, EPI_ISL_2169405, EPI_ISL_2169406, EPI_ISL_2169407, EPI_ISL_2169408, EPI_ISL_2169409, EPI_ISL_2169498, EPI_ISL_2169500, EPI_ISL_2169506 | | | |
| see above | Alberta Precision Labs (APL) | Public Health Agency of Canada (PHAC) National Microbiology Laboratory | Buss; Croxen M; Deo A; Dieu P; E; Ferrato C; Gill K; Khan F; Koleva P; Li V; Lloyd C; Lynch T; Ma R; Murphy S; Pabbaraju K; Shokoples S; Thayer J; Tipples G; Whitehouse M; Wong A; Yu C; Zelyas N |
| EPI_ISL_1591222 | Algemeen Medisch Labo | Labo Klinische Biologie, UZA | Basil Britto Xavier; Christine Lammens; Herman Goossens; Jasmine Coppens; Marie Le Mercier; Veerle Matheeussen |
| EPI_ISL_1636515 | Althaia. Xarxa Assistencial Universitària de Manresa | IrsiCaixa | Bonaventura Clotet; Bonaventura Clotet Gloria Trujillo; Carolina Gonzalez Fernandez; Eulalia Grau; Francesc Catala-Moll; Jaume Trape Pujol; Marc Noguera-Julian; Maria Casadellà; Mariona Parera; Pilar Armengol; Rafel Perez Vidal; Roger Paredes |
| EPI_ISL_5802052, EPI_ISL_5802054, EPI_ISL_5802110, EPI_ISL_5802148 | Ambulatorio Ib | Instituto Butantan | Antonio Jorge Martins; Claudia Renata dos Santos Barros; David Schlesinger; Debora Botequio Moretti; Dimas Tadeu Covas; Elaine Cristina Marqueze; Elaine Vieira Santos; Evandra Strazza Rodrigues; Heidge Fukumasu; Jayme Augusto de Souza-Neto; José Salvatore Leister Patané; Luiz Alcantara; Luiz Lehmann Coutinho; Maria Carolina Elias; Maurício Lacerda Nogueira; Rafael dos Santos Bezerra; Raul Machado Neto; Rejane Maria Tommasini Grotto; Ricardo Haddad; Sandra Coccuzzo Sampaio Vessoni; Simone Kashima; Svetoslav Nanev Slavov; Vincent Louis Viala |
| EPI_ISL_1966499, EPI_ISL_1966500, EPI_ISL_1966501, EPI_ISL_1966506 | Ambulatório IB | Instituto Butantan / Mendelics | Antonio Jorge Martins; Bianca Cechetto Carlos. Mendelics: Bibiana Santos; Claudia Renata dos Santos Barros; Cíntia Bittar; David Schlesinger. Hemocentro Ribeirão Preto: Simone Kashima; Debora Botequio Moretti; Elaine Cristina Marqueze; Elaine Vieira dos Santos; Elisangela Chicaroni Mattos; Erika Freitas; Evandra Strazza Rodrigues; Felipe Allan da Silva da Costa; Flavia Aburjaile; Fábio Sossai Possebon; Guilherme Campos; Guilherme Targino Valente; Heidge Fukumasu. USP-Botucatu: Rejane Maria Tommasini Grotto; Helena Lage Ferreira; Instituto Butantan: Dimas Tadeu Covas; Jardelina de Souza Todao Bernardino; Jayme A. Souza-Neto; Jessika Cristina Chagas Lesbon; Jorge A. Petroli Marchesi; José Salvatore Leister Patané; João Paulo Kitajima; João Pessoa Araújo Jr.; Leila Sabrina Ullmann; Loyze Paola Oliveira de Lima; Luiz Aurelio de Campos Crispin. Centro de Genômica Funcional da ESALQ: Luiz Lehmann Coutinho; Luiz Carlos Junior de Alcantara; Lívia Sacchetto; Maisa C. Pereira Parra; Maria Carolina Elias; Marta Giovanetti; Marília Moraes; Maurício Lacerda Nogueira. Prefeitura de Sao Paulo: Melissa Palmieri.; Patricia Akemi Assato; Paula Rahal; Paulo Inacio da Costa; Rafael dos Santos Bezerra; Raquel de Lello Rocha Campos Cassano. NGS Soluções Genômicas: Pilar Drummond Sampaio Corrêa Mariani. FZEA-USP Pirassununga: Mirele Daiana Poleti; Raul Machado Neto; Ricardo Augusto Brassaloti; Ricardo Haddad; Rodrigo Tocantins Calado. FAMERP-SJRP: Cecília Artico Banho; Sandra Coccuzzo Sampaio; Svetoslav Nanev Slavov; Vagner Fonseca; Vincent Louis Viala |
| EPI_ISL_3243027 | Applied DNA Sciences Inc | Applied DNA Sciences Inc | Huan Dong; Lucas Liu; Spencer William; Yuhua Sun |
| EPI_ISL_1789682 | Arizona State University | Arizona State University | Efrem S. Lim; Joshua LaBaer; Joy M. Blain; LaRinda A. Holland; Nicholas J. Mellor; Peter T. Skidmore; Rabia Maqsood; Valerie Harris; Vel Murugan |
| EPI_ISL_1711459, EPI_ISL_1711624, EPI_ISL_1823028 | Atlas Genomics | Seattle Flu Study | Amanda Adler; Barry R. Lutz; Benjamin Pelle; Brian Hiatt; Caitlin R. Wolf; Chris D. Frazar; Deborah A. Nickerson; Elisabeth Brandstetter; Erica Ryke; Geoff Melly; Helen Y. Chu; Janet A. Englund; Jay Shendure; Jover Lee; Kairsten Fay; Kirsten Lacombe; Lea M. Starita; Mark J. Rieder; Matthew Richardson; Matthew Thompson; Melissa Truong; Michael Boeckh; Michael Famulare; Misja Ilcisin; Peter D. Han; Philip Dykema; Romesh Gautom; Scott Lindquist; Thomas R. Sibley; Trevor Bedford |
| EPI_ISL_3267033, EPI_ISL_3276561, EPI_ISL_3276562, EPI_ISL_3276602, EPI_ISL_3276603 | Ayudas Diagnosticas Sura | Universidad Nacional de Colombia - Laboratorio Genómico One Health | Andres F. Cardona-Rios; Carlos Franco-Muñoz; Carolina Muñoz-Arango; Celeny Ortiz; Daniel O. Maldonado-Perez; Diego A. Álvarez-Díaz; Hector Alejandro Ruiz-Moreno; Idabely Betancur Ortiz; Jorge E. Osorio; Juan P. Hernandez-Ortiz; Karl A Ciuoderis; Katherine Laiton-Donato; Laura Silvana Perez; Lina M. Hurtado; Marcela Mercado-Reyes; Maria Angélica Maya; Maria Stella López; Rita Almanza Payares; Sandra Ines Cano; Simón Villegas Velásquez |
| EPI_ISL_1670638 | Azienda Ospedaliera Terni | Istituto Zooprofilattico Sperimentale dell'Abruzzo e Molise "G. Caporale" | Ancora M; Calistri P; Cammà C; Caporale M; Curini V; Delli Compagni E; Di Domenico M; Di Lollo Valeria; Di Pasquale A; Lorusso A; Mangone I; Marcacci M; Palumbo M; Puglia I; Rinaldi A; Savini G; Scaccetti A; Scialabba S |
| EPI_ISL_1670664, EPI_ISL_1670665, EPI_ISL_2047868 | Azienda USL Umbria 2 | Istituto Zooprofilattico Sperimentale dell'Abruzzo e Molise "G. Caporale" | Ancora M; Calistri P; Cammà C; Caporale M; Curini V; Delli Compagni E; Di Domenico M; Di Lollo Valeria; Di Pasquale A; Lorusso A; Mangone I; Marcacci M; Pistoni E; Proietti A; Puglia I; Rinaldi A; Savini G; Scialabba S |
| EPI_ISL_2531914, EPI_ISL_2532003, EPI_ISL_2532004, EPI_ISL_2532031, EPI_ISL_2532043, EPI_ISL_2532061, EPI_ISL_2532062, EPI_ISL_2532067, EPI_ISL_2532068, EPI_ISL_2532072, EPI_ISL_2532112, EPI_ISL_2532113, EPI_ISL_2532116, EPI_ISL_2532117, EPI_ISL_2532120, EPI_ISL_2532143, EPI_ISL_2532145, EPI_ISL_2532149, EPI_ISL_2532154, EPI_ISL_2532161, EPI_ISL_2532168, EPI_ISL_2532169, EPI_ISL_2532174, EPI_ISL_2653093, EPI_ISL_2653094, EPI_ISL_2653096, EPI_ISL_2653097, EPI_ISL_2653098, EPI_ISL_2653099, EPI_ISL_2653100, EPI_ISL_2653101, EPI_ISL_2653102, EPI_ISL_2653104, EPI_ISL_2653106, EPI_ISL_2653107, EPI_ISL_2653108, EPI_ISL_2653109, EPI_ISL_2653111, EPI_ISL_2653112, EPI_ISL_2653113, EPI_ISL_2653114, EPI_ISL_2653115, EPI_ISL_2653117, EPI_ISL_2653118, EPI_ISL_2653120, EPI_ISL_2653121, EPI_ISL_2653122, EPI_ISL_2653123, EPI_ISL_2653124, EPI_ISL_2653125, EPI_ISL_2653126, EPI_ISL_2653127, EPI_ISL_2653129, EPI_ISL_2653131, EPI_ISL_2653133, EPI_ISL_2653144, EPI_ISL_2653145, EPI_ISL_2653146, EPI_ISL_2653147, EPI_ISL_2653151, EPI_ISL_2653152, EPI_ISL_2653153, EPI_ISL_2653155, EPI_ISL_2653156, EPI_ISL_2653157, EPI_ISL_2653158, EPI_ISL_2653162, EPI_ISL_2653163, EPI_ISL_2653165, EPI_ISL_2653167, EPI_ISL_2653170, EPI_ISL_2653172, EPI_ISL_2653173, EPI_ISL_2653175, EPI_ISL_2653176, EPI_ISL_2653177, EPI_ISL_2653180, EPI_ISL_2653181, EPI_ISL_2653183, EPI_ISL_2653184, EPI_ISL_2653186, EPI_ISL_2653193, EPI_ISL_2653194, EPI_ISL_2653196, EPI_ISL_2653226, EPI_ISL_2653227, EPI_ISL_2653228, EPI_ISL_2653229, EPI_ISL_2653230, EPI_ISL_2653232, EPI_ISL_2653233, EPI_ISL_2653234, EPI_ISL_2653236, EPI_ISL_2653237, EPI_ISL_2653238, EPI_ISL_2653239, EPI_ISL_2653240, EPI_ISL_2653241, EPI_ISL_2653242, EPI_ISL_2653244, EPI_ISL_2653245, EPI_ISL_2653246, EPI_ISL_2653247, EPI_ISL_2653248, EPI_ISL_2653249, EPI_ISL_2653250, EPI_ISL_2653252, EPI_ISL_2653253, EPI_ISL_2653254, EPI_ISL_2653255, EPI_ISL_2653256, EPI_ISL_2653257, EPI_ISL_2653258, EPI_ISL_2653259, EPI_ISL_2653260, EPI_ISL_2653262, EPI_ISL_2653264, EPI_ISL_2653265, EPI_ISL_2653268, EPI_ISL_2653269, EPI_ISL_2653271, EPI_ISL_2653272, EPI_ISL_2653273, EPI_ISL_2653274, EPI_ISL_2653282, EPI_ISL_2653285, EPI_ISL_2653289, EPI_ISL_2653290, EPI_ISL_2653291, EPI_ISL_2653292, EPI_ISL_2653295, EPI_ISL_2653296, EPI_ISL_2653297, EPI_ISL_2653299, EPI_ISL_2653301, EPI_ISL_2653302, EPI_ISL_2653303, EPI_ISL_2653304, EPI_ISL_2653305, EPI_ISL_2653306, EPI_ISL_2653308, EPI_ISL_2653309, EPI_ISL_2653311, EPI_ISL_2653313, EPI_ISL_2653316, EPI_ISL_2653317, EPI_ISL_2653318, EPI_ISL_2653320, EPI_ISL_2653322, EPI_ISL_2653323, EPI_ISL_2653325, EPI_ISL_2653326, EPI_ISL_2653331, EPI_ISL_2653334, EPI_ISL_2653337, EPI_ISL_2653339, EPI_ISL_2653341, EPI_ISL_2653352, EPI_ISL_2653353, EPI_ISL_2653354, EPI_ISL_2653356, EPI_ISL_2653357, EPI_ISL_2653358, EPI_ISL_2653359, EPI_ISL_2653362, EPI_ISL_2653363, EPI_ISL_2653364, EPI_ISL_2653365, EPI_ISL_2653367, EPI_ISL_2653368, EPI_ISL_2653370, EPI_ISL_2653371, EPI_ISL_2653373, EPI_ISL_2653375, EPI_ISL_2653376, EPI_ISL_2653377, EPI_ISL_2653378, EPI_ISL_2653379, EPI_ISL_2653380, EPI_ISL_2653381, EPI_ISL_2653384, EPI_ISL_2653385, EPI_ISL_2653387, EPI_ISL_2653389, EPI_ISL_2653390, EPI_ISL_2653391, EPI_ISL_2653393, EPI_ISL_2653394, EPI_ISL_2653395, EPI_ISL_2653396, EPI_ISL_2653397, EPI_ISL_2653399, EPI_ISL_2653400, EPI_ISL_2653402, EPI_ISL_2653403, EPI_ISL_2653404, EPI_ISL_2653406, EPI_ISL_2653411, EPI_ISL_2653412, EPI_ISL_2653413, EPI_ISL_2653414, EPI_ISL_2653415, EPI_ISL_2653416, EPI_ISL_2653417, EPI_ISL_2653418, EPI_ISL_2653420, EPI_ISL_2653427, EPI_ISL_2653428, EPI_ISL_2653429, EPI_ISL_2653430, EPI_ISL_2653431, EPI_ISL_2653432, EPI_ISL_2653434, EPI_ISL_2653437, EPI_ISL_2653447, EPI_ISL_2653454, EPI_ISL_2653455, EPI_ISL_2653456, EPI_ISL_2653457, EPI_ISL_2653460, EPI_ISL_2653465, EPI_ISL_2653466, EPI_ISL_2653467, EPI_ISL_2653468, EPI_ISL_2653471, EPI_ISL_2653472, EPI_ISL_2653474, EPI_ISL_2653475, EPI_ISL_2653476, EPI_ISL_2653477, EPI_ISL_2653478, EPI_ISL_2653479, EPI_ISL_2653480, EPI_ISL_2653481, EPI_ISL_2653487, EPI_ISL_2653488, EPI_ISL_2653502, EPI_ISL_2653505, EPI_ISL_2653507, EPI_ISL_2653508, EPI_ISL_2653509, EPI_ISL_2653510, EPI_ISL_2653511, EPI_ISL_2653512, EPI_ISL_2653514, EPI_ISL_2653515, EPI_ISL_2653527, EPI_ISL_2653528, EPI_ISL_2653530, EPI_ISL_2653531, EPI_ISL_2653532, EPI_ISL_2653533, EPI_ISL_2653534, EPI_ISL_2653536, EPI_ISL_2653537, EPI_ISL_2653539, EPI_ISL_2653540, EPI_ISL_2653541, EPI_ISL_2653542, EPI_ISL_2653544, EPI_ISL_2653545, EPI_ISL_2653550, EPI_ISL_2653552, EPI_ISL_2653553, EPI_ISL_2653554, EPI_ISL_2653555, EPI_ISL_2653556, EPI_ISL_2653557, EPI_ISL_2653560, EPI_ISL_2653576, EPI_ISL_2653580, EPI_ISL_2653581, EPI_ISL_2653583, EPI_ISL_2653584, EPI_ISL_2653585, EPI_ISL_2653586, EPI_ISL_2653587, EPI_ISL_2653588, EPI_ISL_2653589, EPI_ISL_2653590, EPI_ISL_2653591, EPI_ISL_2653592, EPI_ISL_2653593, EPI_ISL_2653594, EPI_ISL_2653595, EPI_ISL_2653596, EPI_ISL_2653599, EPI_ISL_2653600, EPI_ISL_2653601, EPI_ISL_2653603, EPI_ISL_2653604, EPI_ISL_2653611, EPI_ISL_2653612, EPI_ISL_2653613, EPI_ISL_2653614, EPI_ISL_2653617, EPI_ISL_2653618, EPI_ISL_2653622, EPI_ISL_2653624, EPI_ISL_2653625, EPI_ISL_2653626, EPI_ISL_2653627, EPI_ISL_2653630, EPI_ISL_2653633, EPI_ISL_2653638, EPI_ISL_2653639, EPI_ISL_2653641, EPI_ISL_2653643, | | | |

| | | | |
|---|---|---|---|
| EPI_ISL_2653691, EPI_ISL_2653693, EPI_ISL_2653694, EPI_ISL_2653697, EPI_ISL_2653698, EPI_ISL_2653719, EPI_ISL_2653720, EPI_ISL_2653722, EPI_ISL_2653734, EPI_ISL_2653735, EPI_ISL_2653736, EPI_ISL_2653740, EPI_ISL_2653743, EPI_ISL_2653744, EPI_ISL_2653747, EPI_ISL_2653748, EPI_ISL_2653749, EPI_ISL_2653750, EPI_ISL_2653752, EPI_ISL_2653753, EPI_ISL_2653755, EPI_ISL_2653805, EPI_ISL_2653811, EPI_ISL_2653826, EPI_ISL_2653827, EPI_ISL_2653828, EPI_ISL_2653830, EPI_ISL_2653833, EPI_ISL_2653834, EPI_ISL_2653835, EPI_ISL_2653837, EPI_ISL_2653838, EPI_ISL_2653839, EPI_ISL_2653840, EPI_ISL_2653845, EPI_ISL_2653846, EPI_ISL_2653847, EPI_ISL_2653848, EPI_ISL_2653850, EPI_ISL_2653855, EPI_ISL_2653856, EPI_ISL_2653858, EPI_ISL_2653859, EPI_ISL_2653861, EPI_ISL_2653862, EPI_ISL_2653878, EPI_ISL_2653879, EPI_ISL_2653885, EPI_ISL_2653888, EPI_ISL_2653889, EPI_ISL_2653906, EPI_ISL_2653907, EPI_ISL_2653910, EPI_ISL_2653911, EPI_ISL_2653940, EPI_ISL_2653942, EPI_ISL_2653944, EPI_ISL_2653945, EPI_ISL_2653947, EPI_ISL_2653950, EPI_ISL_2653956, EPI_ISL_2653962, EPI_ISL_2653963, EPI_ISL_2653967, EPI_ISL_2653968, EPI_ISL_2653970, EPI_ISL_2653973, EPI_ISL_2653977, EPI_ISL_2654016, EPI_ISL_2654017, EPI_ISL_2654022, EPI_ISL_2654028, EPI_ISL_2654029, EPI_ISL_2654033, EPI_ISL_2654041, EPI_ISL_2654042, EPI_ISL_2654043, EPI_ISL_2654044, EPI_ISL_2654046, EPI_ISL_2654049, EPI_ISL_2654052, EPI_ISL_2654056, EPI_ISL_2654062, EPI_ISL_2654065, EPI_ISL_2654118, EPI_ISL_2654119, EPI_ISL_2654120, EPI_ISL_2654135, EPI_ISL_2654138, EPI_ISL_2654141, EPI_ISL_2654147, EPI_ISL_2654151, EPI_ISL_2654152, EPI_ISL_2654153, EPI_ISL_2654154, EPI_ISL_2654164, EPI_ISL_2655675, EPI_ISL_2655677, EPI_ISL_2655679, EPI_ISL_2655683, EPI_ISL_2655693, EPI_ISL_2655694, EPI_ISL_2655696, EPI_ISL_2656111, EPI_ISL_2656129, EPI_ISL_2656136, EPI_ISL_2718639, EPI_ISL_2718640, EPI_ISL_2718641, EPI_ISL_2718642, EPI_ISL_2718644, EPI_ISL_2718645, EPI_ISL_2718646, EPI_ISL_2718647, EPI_ISL_2718648, EPI_ISL_2718649, EPI_ISL_2718652, EPI_ISL_2718653, EPI_ISL_2718654, EPI_ISL_2718657, EPI_ISL_2718658, EPI_ISL_2718660, EPI_ISL_2718662, EPI_ISL_2718663, EPI_ISL_2718664, EPI_ISL_2718665, EPI_ISL_2718666, EPI_ISL_2718667, EPI_ISL_2718675, EPI_ISL_2718687, EPI_ISL_2718691, EPI_ISL_2718839, EPI_ISL_2718841, EPI_ISL_2718843, EPI_ISL_2718844, EPI_ISL_2718846, EPI_ISL_2718847, EPI_ISL_2719281 | | | |
| see above | BCCDC Public Health Laboratory | BCCDC Public Health Laboratory | Ana Pacagnella; Corrinne Ng; Dan Fornika; John Tyson; Kim Macdonald; Kimia Kamelian; Linda Hoang; Loretta Janz; Mel Krajden; Prystajecky Natalie; Robert Azana; Shannon Russell |
| EPI_ISL_7672051, EPI_ISL_7672052 | Bambino Gesù Pediatric Hospital | Microbiology and Immunology Diagnosis Bambino Gesù Pediatric Hospital | Carlo Federico Perno; Claudia Alteri; Luna Colagrossi; Rossana Scutari; Valentino Costabile |
| EPI_ISL_1755515, EPI_ISL_1755613 | Biogroup Bio Lam-LCD Saint-Denis | Department of Virology, Henri Mondor University Hospital, Assistance Publique Hôpitaux de Paris, Université Paris-Est Créteil, INSERM U955 | Alexandre Soulier; Christophe Rodriguez; Elisabeth Trawinski; Guillaume Gricourt; Jean-Michel Pawlotsky; Melissa N'Debi; Slim Fourati; Vanessa Demontant |
| EPI_ISL_2698101 | Biology, UFLA, Universidade Federal de Lavras | Biology, UFLA | Barcante, J.; Cherem, J.; Fernandes, G.; Luciano, P.; Melo, D.; Pylro, V. |
| EPI_ISL_3334239, EPI_ISL_3334248, EPI_ISL_3334260 | BioneXt Lab | Laboratoire national de sante, Microbiology, Microbial Genomics Platform | Anke Wienecke-Baldacchino; Catherine Ragimbeau; Elodie Solarino; Fatu Djabi; Jessica Tapp; Lise Pignon; Raoul Salmon; Tamir Abdelrahman; Thibault Ferrandon; Virginie Jover |
| EPI_ISL_3494215, EPI_ISL_3494226, EPI_ISL_3494230, EPI_ISL_3494231, EPI_ISL_3494238, EPI_ISL_3494239, EPI_ISL_3494241 | | | |
| see above | Boston University CTL | Boston University/National Emerging Infectious Disease Laboratories | Catherine Klapperich; Jacquelyn Turcinovic; John H. Connor; Lena Landeverde; Lynn Doucette-Stamm |
| EPI_ISL_3478912, EPI_ISL_3478915, EPI_ISL_3478917, EPI_ISL_3479246, EPI_ISL_3479249, EPI_ISL_3479265, EPI_ISL_3479279, EPI_ISL_3479293, EPI_ISL_3479298, EPI_ISL_3479302, EPI_ISL_3479303, EPI_ISL_3479305, EPI_ISL_3479309, EPI_ISL_3479310, EPI_ISL_3479312, EPI_ISL_3479313, EPI_ISL_3479319, EPI_ISL_3479320, EPI_ISL_3479321, EPI_ISL_3479322, EPI_ISL_3479325, EPI_ISL_3479326, EPI_ISL_3479327, EPI_ISL_3479333, EPI_ISL_3479334, EPI_ISL_3479335, EPI_ISL_3479336, EPI_ISL_3479340, EPI_ISL_3479341, EPI_ISL_3479343, EPI_ISL_3479345, EPI_ISL_3479346, EPI_ISL_3479350, EPI_ISL_3479351, EPI_ISL_3479355, EPI_ISL_3479357, EPI_ISL_3479358, EPI_ISL_3479359, EPI_ISL_3479360, EPI_ISL_3479361, EPI_ISL_3479364, EPI_ISL_3479369, EPI_ISL_3479371, EPI_ISL_3479372, EPI_ISL_3479374, EPI_ISL_3479376, EPI_ISL_3479378, EPI_ISL_3479381, EPI_ISL_3479382, EPI_ISL_3479385, EPI_ISL_3479388, EPI_ISL_3479391, EPI_ISL_3479393, EPI_ISL_3479394, EPI_ISL_3479395, EPI_ISL_3479396, EPI_ISL_3479397, EPI_ISL_3479398, EPI_ISL_3479399, EPI_ISL_3479400, EPI_ISL_3479401, EPI_ISL_3479406, EPI_ISL_3479407, EPI_ISL_3479408, EPI_ISL_3479411, EPI_ISL_3479412, EPI_ISL_3479413, EPI_ISL_3479414, EPI_ISL_3479423, EPI_ISL_3479425, EPI_ISL_3479660, EPI_ISL_3479710, EPI_ISL_3479720, EPI_ISL_3479721, EPI_ISL_3479722, EPI_ISL_3479723, EPI_ISL_3479727, EPI_ISL_3479729, EPI_ISL_3479732, EPI_ISL_3479733, EPI_ISL_3479734, EPI_ISL_3479738, EPI_ISL_3479739, EPI_ISL_3479740, EPI_ISL_3479741, EPI_ISL_3479742, EPI_ISL_3479743, EPI_ISL_3479764, EPI_ISL_3479766, EPI_ISL_3479838, EPI_ISL_3479870, EPI_ISL_3479883, EPI_ISL_3597593, EPI_ISL_3597596, EPI_ISL_3597601, EPI_ISL_3597605, EPI_ISL_3597608, EPI_ISL_3597610, EPI_ISL_3597620, EPI_ISL_3597639, EPI_ISL_3597669, EPI_ISL_3597676, EPI_ISL_3597684, EPI_ISL_3597686, EPI_ISL_3597689, EPI_ISL_3597694, EPI_ISL_3597721, EPI_ISL_3597734, EPI_ISL_3597761, EPI_ISL_3597764, EPI_ISL_3597769, EPI_ISL_3597788, EPI_ISL_3597790, EPI_ISL_3597794, EPI_ISL_3597797, EPI_ISL_3597804, EPI_ISL_3597822, EPI_ISL_3597840, EPI_ISL_3597845, EPI_ISL_3597847, EPI_ISL_3597850, EPI_ISL_3597868, EPI_ISL_3597878, EPI_ISL_3597889, EPI_ISL_3597899, EPI_ISL_3597912, EPI_ISL_3597921, EPI_ISL_3597932, EPI_ISL_3597937, EPI_ISL_3597941, EPI_ISL_3597944, EPI_ISL_3597945, EPI_ISL_3597955, EPI_ISL_3597957, EPI_ISL_3597958, EPI_ISL_3597959, EPI_ISL_3597960, EPI_ISL_3597964, EPI_ISL_3597965, EPI_ISL_3597968, EPI_ISL_3597969, EPI_ISL_3597971, EPI_ISL_3597974, EPI_ISL_3597978, EPI_ISL_3597985, EPI_ISL_3597986, EPI_ISL_3598011, EPI_ISL_3598014, EPI_ISL_3598015, EPI_ISL_3598016, EPI_ISL_3598022, EPI_ISL_3598030, EPI_ISL_3598032, EPI_ISL_3598033, EPI_ISL_3598034, EPI_ISL_3598035, EPI_ISL_3598036, EPI_ISL_3598037, EPI_ISL_3598046 | | | |
| see above | British Columbia Centre For Disease Control | BCCDC Public Health Laboratory | Ana Pacagnella; Corrinne Ng; Dan Fornika; John Tyson; Kim Macdonald; Kimia Kamelian; Linda Hoang; Loretta Janz; Mel Krajden; Prystajecky Natalie; Robert Azana; Shannon Russell |
| EPI_ISL_1710053, EPI_ISL_1710066, EPI_ISL_1710081, EPI_ISL_1710136, EPI_ISL_1710138, EPI_ISL_1710163, EPI_ISL_1710270, EPI_ISL_1710355, EPI_ISL_1710356, EPI_ISL_1710358, EPI_ISL_1710362, EPI_ISL_1710365, EPI_ISL_1710375, EPI_ISL_1710390, EPI_ISL_1710394, EPI_ISL_1710411, EPI_ISL_1710412, EPI_ISL_1710413, EPI_ISL_1710414, EPI_ISL_1710425, EPI_ISL_1710434, EPI_ISL_1710450, EPI_ISL_1710460, EPI_ISL_1710466, EPI_ISL_1710490, EPI_ISL_1710492, EPI_ISL_1757029, EPI_ISL_1757039 | | | |
| see above | Broad Institute Clinical Research Sequencing Platform | Infectious Disease Program, Broad Institute of Harvard and MIT | Adams, G.; B.L.; B.W.; Bauer, M.; Birren; Blumenstiel, B.; Brown, C.; Carter, A.; Chaluvadi, S.; D.J.; DeFelice, M.; DeRuff, K.; Dodge, S.; Gabriel, S.; Gallagher, G.; Gladden-Young, A.; Granger, B.; J.E.; K.J.; Lagerborg, K.; Larkin, K.; Lee, M.; Lemieux; Lennon, N.; Loreth, C.; Madoff, L.; McGovern, S.; Meldrim, J.; Normandin, E.; P.C.; Park; Pearlman, L.; Reilly, S.; Rudy, M.; Sabeti; Siddle; Smole, S.; Tomkins-Tinch, C.; Vicente, G.; and MacInnis |
| EPI_ISL_1661103, EPI_ISL_1661120, EPI_ISL_1661146 | Brussels Academic Hospital Lab (LHUB-ULB) | Labo Klinische Biologie, UZA | Basil Britto Xavier; Christine Lammens; Herman Goossens; Jasmine Coppens; Marie Le Mercier; Veerle Matheeussen |
| EPI_ISL_1937829 | CA-Los Angeles County Public Health Laboratory | Centers for Disease Control and Prevention Division of Viral Diseases, Pathogen Discovery | Alison Laufer Halpin; Ben L. Rambo-Martin; Clinton R. Paden; Dakota Howard; Darlene Wagner; Dave Wentworth; Dhwani Batra; Jasmine Padilla; Justin Lee; Katie Dillon; Krista Queen; Kristen Knipe; Kristine Lacek; Mark Burroughs; Matthew Schmerer; Mili Sheth; Peter Cook; Sam Shepard; Sarah Nobles; Shoshona Le; Suxiang Tong; Vivien Dugan; Yvette Unoarumhi |
| EPI_ISL_5530105, EPI_ISL_5530173 | CASA DE SAUDE MATERNIDADE CELESTINA COLARES | Analytical Competence Molecular Epidemiology Lab/ACME, Oswaldo Cruz Foundation, Ceara (FIOCRUZ CE) | Carlos Leonardo de Aragao Araujo; Cecilia Leite Costa & Eduardo Ruback dos Santos on behalf of COVID-19 FIOCRUZ Genomic Network; Cleber Furtado Aksenen; Fabio Miyajima; Fernando Braga Stehling; Francisco Eder de Moura Lopes; Igor Oliveira Duarte; Jamille Maria Mendes Bezerra; Joaquim Cesar do Nascimento Sousa Junior; Pedro Miguel Carneiro Jeronimo; Suzana Porto Almeida; Thais Ferreira de Oliveira; Thais de Oliveira Costa; Ticiane Cavalcante de Souza; Veridiana Pessoa Miyajima |
| EPI_ISL_2825770, EPI_ISL_2858421, EPI_ISL_2858457, EPI_ISL_2858492, EPI_ISL_2858570, EPI_ISL_2882784, EPI_ISL_2882785, EPI_ISL_2882851, EPI_ISL_2882858, EPI_ISL_2882859, EPI_ISL_2882880, EPI_ISL_2882927, EPI_ISL_2883013, EPI_ISL_2883024, EPI_ISL_2883050, EPI_ISL_2922007, EPI_ISL_5443780, EPI_ISL_5443781, EPI_ISL_5443782, EPI_ISL_7700356, EPI_ISL_7700484, EPI_ISL_7700643 | | | |
| see above | CDPH VBL | California Department of Public Health | CDPH-COVIDNet; Emily Smith on behalf of CDPH-COVIDNet; Emily Smith on behalf of CDPH-COVIDNet and UCSF Center for Advanced Technology; UCLA Technology Center for Genomics & Bioinformatics |
| EPI_ISL_1798770 | CDPH, Viral and Rickettsial Disease Laboratory | Centers for Disease Control and Prevention Division of Viral Diseases, Pathogen Discovery | Alison Laufer Halpin; Ben L. Rambo-Martin; Clinton R. Paden; Dakota Howard; Darlene Wagner; Dave Wentworth; Dhwani Batra; Jasmine Padilla; Justin Lee; Katie Dillon; Krista Queen; Kristen Knipe; Kristine Lacek; Mark Burroughs; Matthew Schmerer; Mili Sheth; Peter Cook; Sam Shepard; Sarah Nobles; Shoshona Le; Suxiang Tong; Vivien Dugan; Yvette Unoarumhi |
| EPI_ISL_3102322 | CENTRO DE ATENDIMENTO PARA ENFRENTAMENTO AO COVID 19 | Analytical Competence Molecular Epidemiology Lab/ACME, Oswaldo Cruz Foundation, Ceara (FIOCRUZ CE) | Cleber Furtado Aksenen; Fabio Miyajima; Fernando Braga Stehling; Francisco Eder de Moura Lopes; Jamille Maria Mendes Bezerra; Joaquim César do Nascimento Sousa Junior; Pedro Miguel Carneiro Jeronimo; Suzana Porto Almeida e Lucas Delerino; Thais Ferreira de Oliveira; Thais de Oliveira Costa; Ticiane Cavalcante de Souza; Veridiana Pessoa Miyajima |
| EPI_ISL_1966522 | CENTRO DE SAUDE DE ADAMANTINA | Instituto Butantan / FZEA-USP (Pirassununga) | Antonio Jorge Martins; Bianca Cechetto Carlos. Mendelics: Bibiana Santos; Claudia Renata dos Santos Barros; Cíntia Bittar; David Schlesinger. Hemocentro Ribeirão Preto: Simone Kashima; Debora Botequio Moretti; Elaine Cristina Marqueze; Elaine Vieira dos Santos; Elisangela Chicaroni Mattos; Erika Freitas; Evandra Strazza Rodrigues; Felipe Allan da Silva da Costa; Flavia Aburjaile; Fábio Sossai Possebon; Guilherme Campos; Guilherme Targino Valente; Heidge Fukumasu. USP-Botucatu: Rejane Maria Tommasini Grotto; Helena Lage Ferreira; Instituto Butantan: Dimas Tadeu Covas; Jardelina de Souza Todao Bernardino; Jayme A. Souza-Neto; Jessika Cristina Chagas Lesbon; Jorge A. Petroli Marchesi; José Salvatore Leister Patané; João Paulo Kitajima; João Pessoa Araújo Jr.; Leila Sabrina Ullmann; Loyze Paola Oliveira de Lima; Luiz Aurelio de Campos Crispin. Centro de Genômica Funcional da ESALQ: Luiz Lehmann Coutinho; Luiz Carlos Junior de Alcantara; Lívia Sacchetto; Maisa C. Pereira Parra; Maria Carolina Elias; Marta Giovanetti; Marília Moraes; Maurício Lacerda Nogueira. Prefeitura de Sao Paulo: Melissa Palmieri.; Patricia Akemi Assato; Paula Rahal; Paulo Inacio da Costa; Rafael dos Santos Bezerra; Raquel de Lello Rocha Campos Cassano. NGS Soluções Genômicas: Pilar Drummond Sampaio Corrêa Mariani. FZEA-USP Pirassununga: Mirele Daiana Poleti; Raul Machado Neto; Ricardo Augusto Brassaloti; Ricardo Haddad; Rodrigo Tocantins Calado. FAMERP-SJRP: Cecília Artico Banho; Sandra Coccuzzo Sampaio; Svetoslav Nanev Slavov; Vagner Fonseca; Vincent Louis Viala |
| EPI_ISL_5530055 | CENTRO DE SAUDE DONA TEREZA HOLANDA DE OLIVEIRA | Analytical Competence Molecular Epidemiology Lab/ACME, Oswaldo Cruz Foundation, Ceara (FIOCRUZ CE) | Carlos Leonardo de Aragao Araujo; Cecilia Leite Costa & Eduardo Ruback dos Santos on behalf of COVID-19 FIOCRUZ Genomic Network; Cleber Furtado Aksenen; Fabio Miyajima; Fernando Braga Stehling; Francisco Eder de Moura Lopes; Igor Oliveira Duarte; Jamille Maria Mendes Bezerra; Joaquim Cesar do Nascimento Sousa Junior; Pedro Miguel Carneiro Jeronimo; Suzana Porto Almeida; Thais Ferreira de Oliveira; Thais de Oliveira Costa; Ticiane Cavalcante de Souza; Veridiana Pessoa Miyajima |
| EPI_ISL_2208950 | CENTRO DE SAUDE DR RENATO DE CARVALHO RIBEIRO ANGATUBA | Instituto Butantan | Antonio Jorge Martins; Claudia Renata dos Santos Barros; David Schlesinger; Debora Botequio Moretti; Dimas Tadeu Covas; Elaine Cristina Marqueze; Elaine Vieira Santos; Evandra Strazza Rodrigues; Heidge Fukumasu; Jayme Augusto de Souza-Neto; José Salvatore Leister Patané; Luiz Alcantara; Luiz Lehmann Coutinho; Maria Carolina Elias; Maurício Lacerda Nogueira; Rafael dos Santos Bezerra; Raul Machado Neto; Rejane Maria Tommasini Grotto; Ricardo Haddad; Sandra Coccuzzo Sampaio Vessoni; Simone Kashima; Svetoslav Nanev Slavov; Vincent Louis Viala. |
| EPI_ISL_2344587 | CENTRO DE SAUDE DR RENATO DE CARVALHO RIBEIRO ANGATUBA | Instituto Butantan / UNESP-Botucatu | Antonio Jorge Martins; Claudia Renata dos Santos Barros; David Schlesinger; Debora Botequio Moretti; Dimas Tadeu Covas; Elaine Cristina Marqueze; Elaine Vieira Santos; Evandra Strazza Rodrigues; Heidge Fukumasu; Jayme Augusto de Souza-Neto; José Salvatore Leister Patané; Luiz Alcantara; Luiz Lehmann Coutinho; Maria Carolina Elias; Maurício Lacerda Nogueira; Rafael dos Santos Bezerra; Raul Machado Neto; Rejane Maria Tommasini Grotto; Ricardo Haddad; Sandra Coccuzzo Sampaio Vessoni; Simone Kashima; Svetoslav Nanev Slavov; Vincent Louis Viala |
| EPI_ISL_1966536, EPI_ISL_1966537, EPI_ISL_1966538, EPI_ISL_1966539, EPI_ISL_1966540 | CENTRO DE SAUDE I TACITO LEITE DE CARVALHO E SILVA | Instituto Butantan / FZEA-USP (Pirassununga) | Antonio Jorge Martins; Bianca Cechetto Carlos. Mendelics: Bibiana Santos; Claudia Renata dos Santos Barros; Cíntia Bittar; David Schlesinger. Hemocentro Ribeirão Preto: Simone Kashima; Debora Botequio Moretti; Elaine Cristina Marqueze; Elaine Vieira dos Santos; Elisangela Chicaroni Mattos; Erika Freitas; Evandra Strazza Rodrigues; Felipe Allan da Silva da Costa; Flavia Aburjaile; Fábio Sossai Possebon; Guilherme Campos; Guilherme Targino Valente; Heidge Fukumasu. USP-Botucatu: Rejane Maria Tommasini Grotto; Helena Lage Ferreira; Instituto Butantan: Dimas Tadeu Covas; Jardelina de Souza Todao Bernardino; Jayme A. Souza-Neto; Jessika Cristina Chagas Lesbon; Jorge A. Petroli Marchesi; José Salvatore Leister Patané; João Paulo Kitajima; João Pessoa Araújo Jr.; Leila Sabrina Ullmann; Loyze Paola Oliveira de Lima; Luiz Aurelio de Campos Crispin. Centro de Genômica Funcional da ESALQ: Luiz Lehmann Coutinho; Luiz Carlos Junior de Alcantara; Lívia Sacchetto; Maisa C. Pereira Parra; Maria Carolina Elias; Marta Giovanetti; Marília Moraes; Maurício Lacerda Nogueira. Prefeitura de Sao Paulo: Melissa Palmieri.; Patricia Akemi Assato; Paula Rahal; Paulo Inacio da Costa; Rafael dos Santos Bezerra; Raquel de Lello Rocha Campos Cassano. NGS Soluções Genômicas: Pilar Drummond Sampaio Corrêa Mariani. FZEA-USP Pirassununga: Mirele Daiana Poleti; Raul Machado Neto; Ricardo Augusto Brassaloti; Ricardo Haddad; Rodrigo Tocantins Calado. FAMERP-SJRP: Cecília Artico Banho; Sandra Coccuzzo Sampaio; Svetoslav Nanev Slavov; Vagner Fonseca; Vincent Louis Viala |
| EPI_ISL_1966472 | CENTRO DE SAUDE II DR ALCIDES FACUNDO ARROYO | Instituto Butantan / Mendelics | Antonio Jorge Martins; Bianca Cechetto Carlos. Mendelics: Bibiana Santos; Claudia Renata dos Santos Barros; Cíntia Bittar; David Schlesinger. Hemocentro Ribeirão Preto: Simone Kashima; Debora Botequio Moretti; Elaine Cristina Marqueze; Elaine Vieira dos Santos; Elisangela Chicaroni Mattos; Erika Freitas; Evandra Strazza Rodrigues; Felipe Allan da Silva da Costa; Flavia Aburjaile; Fábio Sossai Possebon; Guilherme Campos; Guilherme Targino Valente; Heidge Fukumasu. USP-Botucatu: Rejane Maria Tommasini Grotto; Helena Lage Ferreira; Instituto Butantan: Dimas Tadeu Covas; Jardelina de Souza Todao Bernardino; Jayme A. Souza-Neto; Jessika Cristina Chagas Lesbon; Jorge A. Petroli Marchesi; José Salvatore Leister Patané; João Paulo Kitajima; João Pessoa Araújo Jr.; Leila Sabrina Ullmann; Loyze Paola Oliveira de Lima; Luiz Aurelio de Campos Crispin. Centro de Genômica Funcional da ESALQ: Luiz Lehmann Coutinho; Luiz Carlos Junior de Alcantara; Lívia Sacchetto; Maisa C. Pereira Parra; Maria Carolina Elias; Marta Giovanetti; Marília Moraes; Maurício Lacerda Nogueira. Prefeitura de Sao Paulo: Melissa Palmieri.; Patricia Akemi Assato; Paula Rahal; Paulo Inacio da Costa; Rafael dos Santos Bezerra; Raquel de Lello Rocha Campos Cassano. NGS Soluções Genômicas: Pilar Drummond Sampaio Corrêa Mariani. FZEA-USP Pirassununga: Mirele Daiana Poleti; Raul Machado Neto; Ricardo Augusto Brassaloti; Ricardo Haddad; Rodrigo Tocantins Calado. FAMERP-SJRP: Cecília Artico Banho; Sandra Coccuzzo Sampaio; Svetoslav Nanev Slavov; Vagner Fonseca; Vincent Louis Viala |
| EPI_ISL_1966471 | CENTRO DE SAUDE II MAIRINQUE MAIRINQUE | Instituto Butantan / Mendelics | Antonio Jorge Martins; Bianca Cechetto Carlos. Mendelics: Bibiana Santos; Claudia Renata dos Santos Barros; Cíntia Bittar; David Schlesinger. Hemocentro Ribeirão Preto: Simone Kashima; Debora Botequio Moretti; Elaine Cristina Marqueze; Elaine Vieira dos Santos; Elisangela Chicaroni Mattos; Erika Freitas; Evandra Strazza Rodrigues; Felipe Allan da Silva da Costa; Flavia Aburjaile; Fábio Sossai Possebon; Guilherme Campos; Guilherme Targino Valente; Heidge Fukumasu. USP-Botucatu: Rejane Maria Tommasini Grotto; Helena Lage Ferreira; Instituto Butantan: Dimas Tadeu Covas; Jardelina de Souza Todao Bernardino; Jayme A. Souza-Neto; Jessika Cristina Chagas Lesbon; Jorge A. Petroli Marchesi; José Salvatore Leister Patané; João Paulo Kitajima; João Pessoa Araújo Jr.; Leila Sabrina Ullmann; Loyze Paola Oliveira de Lima; Luiz Aurelio de Campos Crispin. Centro de Genômica Funcional da ESALQ: Luiz Lehmann Coutinho; Luiz Carlos Junior de Alcantara; Lívia Sacchetto; Maisa C. Pereira Parra; Maria Carolina Elias; Marta Giovanetti; Marília Moraes; Maurício Lacerda Nogueira. Prefeitura de Sao Paulo: Melissa Palmieri.; Patricia Akemi Assato; Paula Rahal; Paulo Inacio da Costa; Rafael dos Santos Bezerra; Raquel de Lello Rocha Campos Cassano. NGS Soluções Genômicas: Pilar Drummond Sampaio Corrêa Mariani. FZEA-USP Pirassununga: Mirele Daiana Poleti; Raul Machado Neto; Ricardo Augusto Brassaloti; Ricardo Haddad; Rodrigo Tocantins Calado. FAMERP-SJRP: Cecília Artico Banho; Sandra Coccuzzo Sampaio; Svetoslav Nanev Slavov; Vagner Fonseca; Vincent Louis Viala |
| EPI_ISL_2344689 | CENTRO DE SAUDE III DE AGUAS DE SAO PEDRO | Instituto Butantan / FZEA-USP-Pirassununga | Antonio Jorge Martins; Claudia Renata dos Santos Barros; David Schlesinger; Debora Botequio Moretti; Dimas Tadeu Covas; Elaine Cristina Marqueze; Elaine Vieira Santos; Evandra Strazza Rodrigues; Heidge Fukumasu; Jayme Augusto de Souza-Neto; José Salvatore Leister Patané; Luiz Alcantara; Luiz Lehmann Coutinho; Maria Carolina Elias; Maurício Lacerda Nogueira; Rafael dos Santos Bezerra; Raul Machado Neto; Rejane Maria Tommasini Grotto; Ricardo Haddad; Sandra Coccuzzo Sampaio Vessoni; Simone Kashima; Svetoslav Nanev Slavov; Vincent Louis Viala |
| EPI_ISL_2208998 | CENTRO DE SAUDE SAO ROQUE DR JOSE CARVALHO BRITO | Instituto Butantan | Antonio Jorge Martins; Claudia Renata dos Santos Barros; David Schlesinger; Debora Botequio Moretti; Dimas Tadeu Covas; Elaine Cristina Marqueze; Elaine Vieira Santos; Evandra Strazza Rodrigues; Heidge Fukumasu; Jayme Augusto de Souza-Neto; José Salvatore Leister Patané; Luiz Alcantara; Luiz Lehmann Coutinho; Maria Carolina Elias; Maurício Lacerda Nogueira; Rafael dos Santos Bezerra; Raul Machado Neto; Rejane Maria Tommasini Grotto; Ricardo Haddad; Sandra Coccuzzo Sampaio Vessoni; Simone Kashima; Svetoslav Nanev Slavov; Vincent Louis Viala. |
| EPI_ISL_2344542 | CENTRO DE SAUDE SAO ROQUE DR JOSE CARVALHO BRITO | Instituto Butantan / UNESP-Botucatu | Antonio Jorge Martins; Claudia Renata dos Santos Barros; David Schlesinger; Debora Botequio Moretti; Dimas Tadeu Covas; Elaine Cristina Marqueze; Elaine Vieira Santos; Evandra Strazza Rodrigues; Heidge Fukumasu; Jayme Augusto de Souza-Neto; José Salvatore Leister Patané; Luiz Alcantara; Luiz Lehmann Coutinho; Maria Carolina Elias; Maurício Lacerda Nogueira; Rafael dos Santos Bezerra; Raul Machado Neto; Rejane Maria Tommasini Grotto; Ricardo Haddad; Sandra Coccuzzo Sampaio Vessoni; Simone Kashima; Svetoslav Nanev Slavov; Vincent Louis Viala |
| EPI_ISL_2344690 | CENTRO MEDICO DR NELSON SALOME DE CONCHAL | Instituto Butantan / FZEA-USP-Pirassununga | Antonio Jorge Martins; Claudia Renata dos Santos Barros; David Schlesinger; Debora Botequio Moretti; Dimas Tadeu Covas; Elaine Cristina Marqueze; Elaine Vieira Santos; Evandra Strazza Rodrigues; Heidge Fukumasu; Jayme Augusto de Souza-Neto; José Salvatore Leister Patané; Luiz Alcantara; Luiz Lehmann Coutinho; Maria Carolina Elias; Maurício Lacerda Nogueira; Rafael dos Santos Bezerra; Raul Machado Neto; Rejane Maria Tommasini Grotto; Ricardo Haddad; Sandra Coccuzzo Sampaio Vessoni; Simone Kashima; Svetoslav Nanev Slavov; Vincent Louis Viala |
| EPI_ISL_1545645, | CHWAPI - SITE NOTRE DAME | Institut de Pathologie et Genetique | Jérémie Gras; Pascale Hilbert |

| | | | |
|---|---|---|---|
| EPI_ISL_1573892 | | (IPG) | |
| EPI_ISL_2209080 | COORDENADORIA MUNICIPAL DE SAUDE DE IRACEMAPOLIS | Instituto Butantan | Antonio Jorge Martins; Claudia Renata dos Santos Barros; David Schlesinger; Debora Botequio Moretti; Dimas Tadeu Covas; Elaine Cristina Marqueze; Elaine Vieira Santos; Evandra Strazza Rodrigues; Heidge Fukumasu; Jayme Augusto de Souza-Neto; José Salvatore Leister Patané; Luiz Alcantara; Luiz Lehmann Coutinho; Maria Carolina Elias; Maurício Lacerda Nogueira; Rafael dos Santos Bezerra; Raul Machado Neto; Rejane Maria Tommasini Grotto; Ricardo Haddad; Sandra Coccuzzo Sampaio Vessoni; Simone Kashima; Svetoslav Nanev Slavov; Vincent Louis Viala. |
| EPI_ISL_2344685 | COORDENADORIA MUNICIPAL DE SAUDE DE IRACEMAPOLIS | Instituto Butantan / FZEA-USP-Pirassununga | Antonio Jorge Martins; Claudia Renata dos Santos Barros; David Schlesinger; Debora Botequio Moretti; Dimas Tadeu Covas; Elaine Cristina Marqueze; Elaine Vieira Santos; Evandra Strazza Rodrigues; Heidge Fukumasu; Jayme Augusto de Souza-Neto; José Salvatore Leister Patané; Luiz Alcantara; Luiz Lehmann Coutinho; Maria Carolina Elias; Maurício Lacerda Nogueira; Rafael dos Santos Bezerra; Raul Machado Neto; Rejane Maria Tommasini Grotto; Ricardo Haddad; Sandra Coccuzzo Sampaio Vessoni; Simone Kashima; Svetoslav Nanev Slavov; Vincent Louis Viala |
| EPI_ISL_1804963 | CQRC_QUALITY CONTROL CHEMICAL BIOLOGICAL RISK_AOOR Villa Sofia Cervello Palermo | CQRC_QUALITY CONTROL CHEMICAL BIOLOGICAL RISK_AOOR Villa Sofia Cervello Palermo | ; Brunacci, G.; Contino, F.; Di Gaudio, F. |
| EPI_ISL_2209116 | CS DE BALSAMO | Instituto Butantan | Antonio Jorge Martins; Claudia Renata dos Santos Barros; David Schlesinger; Debora Botequio Moretti; Dimas Tadeu Covas; Elaine Cristina Marqueze; Elaine Vieira Santos; Evandra Strazza Rodrigues; Heidge Fukumasu; Jayme Augusto de Souza-Neto; José Salvatore Leister Patané; Luiz Alcantara; Luiz Lehmann Coutinho; Maria Carolina Elias; Maurício Lacerda Nogueira; Rafael dos Santos Bezerra; Raul Machado Neto; Rejane Maria Tommasini Grotto; Ricardo Haddad; Sandra Coccuzzo Sampaio Vessoni; Simone Kashima; Svetoslav Nanev Slavov; Vincent Louis Viala. |
| EPI_ISL_1966523, EPI_ISL_1966524 | CS DE BALSAMO | Instituto Butantan / FZEA-USP (Pirassununga) | Antonio Jorge Martins; Bianca Cechetto Carlos. Mendelics: Bibiana Santos; Claudia Renata dos Santos Barros; Cíntia Bittar; David Schlesinger. Hemocentro Ribeirão Preto: Simone Kashima; Debora Botequio Moretti; Elaine Cristina Marqueze; Elaine Vieira dos Santos; Elisangela Chicaroni Mattos; Erika Freitas; Evandra Strazza Rodrigues; Felipe Allan da Silva da Costa; Flavia Aburjaile; Fábio Sossai Possebon; Guilherme Campos; Guilherme Targino Valente; Heidge Fukumasu. USP-Botucatu: Rejane Maria Tommasini Grotto; Helena Lage Ferreira; Instituto Butantan: Dimas Tadeu Covas; Jardelina de Souza Todao Bernardino; Jayme A. Souza-Neto; Jessika Cristina Chagas Lesbon; Jorge A. Petroli Marchesi; José Salvatore Leister Patané; João Paulo Kitajima; João Pessoa Araújo Jr.; Leila Sabrina Ullmann; Loyze Paola Oliveira de Lima; Luiz Aurelio de Campos Crispin. Centro de Genômica Funcional da ESALQ: Luiz Lehmann Coutinho; Luiz Carlos Junior de Alcantara; Lívia Sacchetto; Maisa C. Pereira Parra; Maria Carolina Elias; Marta Giovanetti; Marília Moraes; Maurício Lacerda Nogueira. Prefeitura de Sao Paulo: Melissa Palmieri.; Patricia Akemi Assato; Paula Rahal; Paulo Inacio da Costa; Rafael dos Santos Bezerra; Raquel de Lello Rocha Campos Cassano. NGS Soluções Genômicas: Pilar Drummond Sampaio Corrêa Mariani. FZEA-USP Pirassununga: Mirele Daiana Poleti; Raul Machado Neto; Ricardo Augusto Brassaloti; Ricardo Haddad; Rodrigo Tocantins Calado. FAMERP-SJRP: Cecília Artico Banho; Sandra Coccuzzo Sampaio; Svetoslav Nanev Slavov; Vagner Fonseca; Vincent Louis Viala |
| EPI_ISL_2345303 | CS DE BALSAMO | Instituto Butantan / UNESP-Botucatu | Antonio Jorge Martins; Claudia Renata dos Santos Barros; David Schlesinger; Debora Botequio Moretti; Dimas Tadeu Covas; Elaine Cristina Marqueze; Elaine Vieira Santos; Evandra Strazza Rodrigues; Heidge Fukumasu; Jayme Augusto de Souza-Neto; José Salvatore Leister Patané; Luiz Alcantara; Luiz Lehmann Coutinho; Maria Carolina Elias; Maurício Lacerda Nogueira; Rafael dos Santos Bezerra; Raul Machado Neto; Rejane Maria Tommasini Grotto; Ricardo Haddad; Sandra Coccuzzo Sampaio Vessoni; Simone Kashima; Svetoslav Nanev Slavov; Vincent Louis Viala |
| EPI_ISL_1966547 | CS DE SANTA RITA DOESTE | Instituto Butantan / FZEA-USP (Pirassununga) | Antonio Jorge Martins; Bianca Cechetto Carlos. Mendelics: Bibiana Santos; Claudia Renata dos Santos Barros; Cíntia Bittar; David Schlesinger. Hemocentro Ribeirão Preto: Simone Kashima; Debora Botequio Moretti; Elaine Cristina Marqueze; Elaine Vieira dos Santos; Elisangela Chicaroni Mattos; Erika Freitas; Evandra Strazza Rodrigues; Felipe Allan da Silva da Costa; Flavia Aburjaile; Fábio Sossai Possebon; Guilherme Campos; Guilherme Targino Valente; Heidge Fukumasu. USP-Botucatu: Rejane Maria Tommasini Grotto; Helena Lage Ferreira; Instituto Butantan: Dimas Tadeu Covas; Jardelina de Souza Todao Bernardino; Jayme A. Souza-Neto; Jessika Cristina Chagas Lesbon; Jorge A. Petroli Marchesi; José Salvatore Leister Patané; João Paulo Kitajima; João Pessoa Araújo Jr.; Leila Sabrina Ullmann; Loyze Paola Oliveira de Lima; Luiz Aurelio de Campos Crispin. Centro de Genômica Funcional da ESALQ: Luiz Lehmann Coutinho; Luiz Carlos Junior de Alcantara; Lívia Sacchetto; Maisa C. Pereira Parra; Maria Carolina Elias; Marta Giovanetti; Marília Moraes; Maurício Lacerda Nogueira. Prefeitura de Sao Paulo: Melissa Palmieri.; Patricia Akemi Assato; Paula Rahal; Paulo Inacio da Costa; Rafael dos Santos Bezerra; Raquel de Lello Rocha Campos Cassano. NGS Soluções Genômicas: Pilar Drummond Sampaio Corrêa Mariani. FZEA-USP Pirassununga: Mirele Daiana Poleti; Raul Machado Neto; Ricardo Augusto Brassaloti; Ricardo Haddad; Rodrigo Tocantins Calado. FAMERP-SJRP: Cecília Artico Banho; Sandra Coccuzzo Sampaio; Svetoslav Nanev Slavov; Vagner Fonseca; Vincent Louis Viala |
| EPI_ISL_1966480 | CS III SANTOPOLIS DO AGUAPEI | Instituto Butantan / Mendelics | Antonio Jorge Martins; Bianca Cechetto Carlos. Mendelics: Bibiana Santos; Claudia Renata dos Santos Barros; Cíntia Bittar; David Schlesinger. Hemocentro Ribeirão Preto: Simone Kashima; Debora Botequio Moretti; Elaine Cristina Marqueze; Elaine Vieira dos Santos; Elisangela Chicaroni Mattos; Erika Freitas; Evandra Strazza Rodrigues; Felipe Allan da Silva da Costa; Flavia Aburjaile; Fábio Sossai Possebon; Guilherme Campos; Guilherme Targino Valente; Heidge Fukumasu. USP-Botucatu: Rejane Maria Tommasini Grotto; Helena Lage Ferreira; Instituto Butantan: Dimas Tadeu Covas; Jardelina de Souza Todao Bernardino; Jayme A. Souza-Neto; Jessika Cristina Chagas Lesbon; Jorge A. Petroli Marchesi; José Salvatore Leister Patané; João Paulo Kitajima; João Pessoa Araújo Jr.; Leila Sabrina Ullmann; Loyze Paola Oliveira de Lima; Luiz Aurelio de Campos Crispin. Centro de Genômica Funcional da ESALQ: Luiz Lehmann Coutinho; Luiz Carlos Junior de Alcantara; Lívia Sacchetto; Maisa C. Pereira Parra; Maria Carolina Elias; Marta Giovanetti; Marília Moraes; Maurício Lacerda Nogueira. Prefeitura de Sao Paulo: Melissa Palmieri.; Patricia Akemi Assato; Paula Rahal; Paulo Inacio da Costa; Rafael dos Santos Bezerra; Raquel de Lello Rocha Campos Cassano. NGS Soluções Genômicas: Pilar Drummond Sampaio Corrêa Mariani. FZEA-USP Pirassununga: Mirele Daiana Poleti; Raul Machado Neto; Ricardo Augusto Brassaloti; Ricardo Haddad; Rodrigo Tocantins Calado. FAMERP-SJRP: Cecília Artico Banho; Sandra Coccuzzo Sampaio; Svetoslav Nanev Slavov; Vagner Fonseca; Vincent Louis Viala |
| EPI_ISL_2697888, EPI_ISL_2697907, EPI_ISL_2697915, EPI_ISL_2697938, EPI_ISL_2697980, EPI_ISL_2698057, EPI_ISL_2698061, EPI_ISL_2698075, EPI_ISL_2698081, EPI_ISL_2698090 | | | |
| see above | CTvacinas | CTvacinas | A.P.; B.L.; Coelho; D.B.; Dorlass; Durigon; E.G.; E.L.; F.G.; Fernandes; Fiorini, A.; Fonseca; G.P.; H.P.; K.L.; L.M.; Lourenco; Magalhaes; Oliveira; Ometto, T.; Peixoto, R.; R.D.; Sato, H.; Scagion; Teixeira, S.; Telezynski; Thomazelli |
| EPI_ISL_2234338, EPI_ISL_2537465 | California Department of Public Health | California Department of Public Health | CDPH IDLB COVIDNet et al |
| EPI_ISL_2778735, EPI_ISL_2838026 | California Department of Public Health Valencia Branch Laboratory (CDPH VBL) | California Department of Public Health | CDPH-COVIDNet; UCLA Technology Center for Genomics & Bioinformatics |
| EPI_ISL_2497148 | Cayuga Medical Center | Cornell Covid-19 Testing Laboratory | Brittany Cronk; Diego Diel; Elizabeth Plocharczyk; Leonardo Caserta; Melissa Laverack; Patrick Mitchell; Renee Anderson; Roopa Venugopalan |
| EPI_ISL_1760280, EPI_ISL_1760281, EPI_ISL_1760284 | Centers for Disease Control and Prevention, Dengue Branch | Centers for Disease Control and Prevention, Dengue Branch | Betzabel Flores; Gabriela Paz-Bailey; Gilberto A. Santiago; Glenda Gonzalez; Jorge L. Munoz-Jordan; Keyla Charriez |
| EPI_ISL_2502375, EPI_ISL_2502384, EPI_ISL_2502389, EPI_ISL_2502390, EPI_ISL_2502411, EPI_ISL_2502412, EPI_ISL_2502413, EPI_ISL_2502447, EPI_ISL_2502448, EPI_ISL_2502449, EPI_ISL_2502450, EPI_ISL_2502451, EPI_ISL_2502452, EPI_ISL_2502453, EPI_ISL_2502454, EPI_ISL_2502455, EPI_ISL_2502456 | | | |
| see above | Central Laboratory, Bureau of Public Health (BOG) and Academic Hospital Paramaribo | Erasmus Medical Center | Bas B Oude Munnink; Cherise Beek; Consuella Partowidjojo; Dion Gajadin; Ed PF IJzerman; Emmanuelle Munger; Gary Gummels; Ingrid SK Krishnadath; Lycke Woittiez; Marion PG Koopmans; Mireille Van de Veer; Phyllis Pinas; Princes Wongsowidjojo; Radjesh Ori; Ranisha Doerbalie; Rohma Banwari; Soeradj Harkisoen; Stephen Vreden; Tilotmadebie Ramlal; Verne Nanhoe |
| EPI_ISL_3255058, EPI_ISL_3255077, EPI_ISL_3255079, EPI_ISL_3255195, EPI_ISL_3266103 | Central Public Health Laboratory - LACEN -Bahia, Salvador, Brazil | Central Public Health Laboratory - LACEN -Bahia, Salvador, Brazil | Arabela Leal; Breno Dominguez; Felicidade Pereira; Jaqueline Gomes; Luciana Oliveira; Luiz Alcantara; Marcela Gómez; Marta Giovanetti; Patrícia Cajado; Stephane Tosta; Vagner Fonseca; Vanessa Nardy |
| EPI_ISL_1547454 | Centre De Prelevement COVID RIOM | CHU Clermont-Ferrand, service de virologie | Bisseux Maxime; Combes Patricia; Henquell Cécile; Mirand Audrey |
| EPI_ISL_1706429 | Centre Hospitalier Eure Seine | Centre Hospitalier Universitaire de Rouen Laboratoire de Virologie | Alice Moisan; Fabienne De Oliveira; Marie Leoz |
| EPI_ISL_1917208, EPI_ISL_1917221 | Centre Hospitalier du Nord | Laboratoire national de sante, Microbiology, Microbial Genomics Platform | Anke Wienecke-Baldacchino; Catherine Ragimbeau; Fatiha Boulmerka; Fatu Djabi; Jacqueline Parmentier; Jessica Tapp; Lise Pignon; Raoul Salmon; Tamir Abdelrahman |
| EPI_ISL_5802056 | Centro De Saude De Adamantina | Instituto Butantan | Antonio Jorge Martins; Claudia Renata dos Santos Barros; David Schlesinger; Debora Botequio Moretti; Dimas Tadeu Covas; Elaine Cristina Marqueze; Elaine Vieira Santos; Evandra Strazza Rodrigues; Heidge Fukumasu; Jayme Augusto de Souza-Neto; José Salvatore Leister Patané; Luiz Alcantara; Luiz Lehmann Coutinho; Maria Carolina Elias; Maurício Lacerda Nogueira; Rafael dos Santos Bezerra; Raul Machado Neto; Rejane Maria Tommasini Grotto; Ricardo Haddad; Sandra Coccuzzo Sampaio Vessoni; Simone Kashima; Svetoslav Nanev Slavov; Vincent Louis Viala |
| EPI_ISL_5802078, EPI_ISL_5802079, EPI_ISL_5802080, EPI_ISL_5802081, EPI_ISL_5802082 | Centro De Saude I Tacito Leite De Carvalho E Silva | Instituto Butantan | Antonio Jorge Martins; Claudia Renata dos Santos Barros; David Schlesinger; Debora Botequio Moretti; Dimas Tadeu Covas; Elaine Cristina Marqueze; Elaine Vieira Santos; Evandra Strazza Rodrigues; Heidge Fukumasu; Jayme Augusto de Souza-Neto; José Salvatore Leister Patané; Luiz Alcantara; Luiz Lehmann Coutinho; Maria Carolina Elias; Maurício Lacerda Nogueira; Rafael dos Santos Bezerra; Raul Machado Neto; Rejane Maria Tommasini Grotto; Ricardo Haddad; Sandra Coccuzzo Sampaio Vessoni; Simone Kashima; Svetoslav Nanev Slavov; Vincent Louis Viala |
| EPI_ISL_5802157 | Centro De Saude Ii Dr Alcides Facundo Arroyo | Instituto Butantan | Antonio Jorge Martins; Claudia Renata dos Santos Barros; David Schlesinger; Debora Botequio Moretti; Dimas Tadeu Covas; Elaine Cristina Marqueze; Elaine Vieira Santos; Evandra Strazza Rodrigues; Heidge Fukumasu; Jayme Augusto de Souza-Neto; José Salvatore Leister Patané; Luiz Alcantara; Luiz Lehmann Coutinho; Maria Carolina Elias; Maurício Lacerda Nogueira; Rafael dos Santos Bezerra; Raul Machado Neto; Rejane Maria Tommasini Grotto; Ricardo Haddad; Sandra Coccuzzo Sampaio Vessoni; Simone Kashima; Svetoslav Nanev Slavov; Vincent Louis Viala |
| EPI_ISL_5802201 | Centro De Saude Ii Mairinque Mairinque | Instituto Butantan | Antonio Jorge Martins; Claudia Renata dos Santos Barros; David Schlesinger; Debora Botequio Moretti; Dimas Tadeu Covas; Elaine Cristina Marqueze; Elaine Vieira Santos; Evandra Strazza Rodrigues; Heidge Fukumasu; Jayme Augusto de Souza-Neto; José Salvatore Leister Patané; Luiz Alcantara; Luiz Lehmann Coutinho; Maria Carolina Elias; Maurício Lacerda Nogueira; Rafael dos Santos Bezerra; Raul Machado Neto; Rejane Maria Tommasini Grotto; Ricardo Haddad; Sandra Coccuzzo Sampaio Vessoni; Simone Kashima; Svetoslav Nanev Slavov; Vincent Louis Viala |
| EPI_ISL_5782682 | Centro De Saude Iii De Aguas De Sao Pedro | Instituto Butantan | Antonio Jorge Martins; Claudia Renata dos Santos Barros; David Schlesinger; Debora Botequio Moretti; Dimas Tadeu Covas; Elaine Cristina Marqueze; Elaine Vieira Santos; Evandra Strazza Rodrigues; Heidge Fukumasu; Jayme Augusto de Souza-Neto; José Salvatore Leister Patané; Luiz Alcantara; Luiz Lehmann Coutinho; Maria Carolina Elias; Maurício Lacerda Nogueira; Rafael dos Santos Bezerra; Raul Machado Neto; Rejane Maria Tommasini Grotto; Ricardo Haddad; Sandra Coccuzzo Sampaio Vessoni; Simone Kashima; Svetoslav Nanev Slavov; Vincent Louis Viala |
| EPI_ISL_5782683 | Centro Medico Dr Nelson Salome De Conchal | Instituto Butantan | Antonio Jorge Martins; Claudia Renata dos Santos Barros; David Schlesinger; Debora Botequio Moretti; Dimas Tadeu Covas; Elaine Cristina Marqueze; Elaine Vieira Santos; Evandra Strazza Rodrigues; Heidge Fukumasu; Jayme Augusto de Souza-Neto; José Salvatore Leister Patané; Luiz Alcantara; Luiz Lehmann Coutinho; Maria Carolina Elias; Maurício Lacerda Nogueira; Rafael dos Santos Bezerra; Raul Machado Neto; Rejane Maria Tommasini Grotto; Ricardo Haddad; Sandra Coccuzzo Sampaio Vessoni; Simone Kashima; Svetoslav Nanev Slavov; Vincent Louis Viala |
| EPI_ISL_4405258 | Centro Provincial de Referencia VIH/SIDA y Hepatitis Virales – Instituto Biológico Dr Tomás Perón | Área de Secuenciación del Laboratorio de Virología del Hospital de Niños Dr. Ricardo Gutierrez on behalf of 'Proyecto Argentino Interinstitucional de genomica de SARS-CoV-2' (PAIS Consortium) | A; Acuña; C; Carlos; Corazza; D; Dehaut; Dolcini; F; Fresina; Gatti; Gimenez; González; Goya; Gutierrez; K; L; LE; Lusso; M; MI; MS; Nabaes Jodar; Nardone; Natale; Nogueira Laspuir; Pagliari; R; Real; Romanko; S; Sanchez; Suarez; Szymanowski; Talavera; Ten Huver; V; Valinotto; Valle; Viegas, M. |
| EPI_ISL_2612338, EPI_ISL_2612339, EPI_ISL_2612340, EPI_ISL_2612387, EPI_ISL_2612388, EPI_ISL_2612389 | Centro de Infectologia Charles Mérieux/ Laboratório Rodolphe Mérieux, FUNDHACRE | Bioinformatics Laboratory / LNCC | Alessandra P Lamarca; Alexandra L Gerber; Ana Paula de C Guimarães; Ana Tereza R Vasconcelos; Andreas Stocker; Cirley Maria de Oliveira Lobato; Douglas Terra Machado; Luiz Fellype Alves de Souza; Luiz G P de Almeida; Ronaldo da Silva F Jr |
| EPI_ISL_1662101 | Centro de Investigacion Biomedica de Occidente (CIBO) | Unidad de Genomica Avanzada | Alejandro Sanchez-Flores; Alfredo Herrera-Estrella; Alicia Ocana-Mondragon; Angel Gustavo Salas-Lais; Bernardo Martinez-Miguel; Blanca Taboada; Brenda Irasema Maldonado-Meza; Carla Ivon Herrera-Najera; Carlos F. Arias; Celia Boukadida; Celida Duque Molina; Clara Esperanza Santacruz-Tinoco; Concepcion Grajales-Muniz; Consorcio Mexicano de Vigilancia Genomica (CoViGen-Mex). Authors (in alphabetical order): Julio Elias Alvarado-Yaah; Fernando Fontove-Herrera; Francisco Pulido; Gloria Elena Espinosa-Ayala; Gloria Maria Molina-Salinas; Gloria Vazquez; Hector Esteban Paz-Juarez; Hector Montoya-Fuentes; Helen Haydee Fernanda Ramirez-Plascencia; Jose Antonio Enciso-Moreno; Jose Esteban Munoz-Medina; Jose de Jesus Nunez-Contreras; Juan Bautista Chale-Dzul; Luis Alberto Ochoa-Carrera; Margarita Matias-Florentino; Maria Guadalupe Santiago-Mauricio; Maria Guadalupe de Jesus Mireles-Rivera; Nelly Selem-Mojica; Pavel Isa; Ricardo Grande; Santiago Ávila-Rios; Victor Eduardo Garcia-Arias; Victor Hugo Borja-Aburto |
| EPI_ISL_1662047 | Centro de Investigacion Biomedica del Noreste (CIBIN) | Unidad de Genomica Avanzada | Alejandro Sanchez-Flores; Alfredo Herrera-Estrella; Alicia Ocana-Mondragon; Angel Gustavo Salas-Lais; Bernardo Martinez-Miguel; Blanca Taboada; Brenda Irasema Maldonado-Meza; Carla Ivon Herrera-Najera; Carlos F. Arias; Celia Boukadida; Celida Duque Molina; Clara Esperanza Santacruz-Tinoco; Concepcion Grajales-Muniz; Consorcio Mexicano de Vigilancia Genomica (CoViGen-Mex). Authors (in alphabetical order): Julio Elias Alvarado-Yaah; Fernando Fontove-Herrera; Francisco Pulido; Gloria Elena Espinosa-Ayala; Gloria Maria Molina-Salinas; Gloria Vazquez; Hector Esteban Paz-Juarez; Hector Montoya-Fuentes; Helen Haydee Fernanda Ramirez-Plascencia; Jose Antonio Enciso-Moreno; Jose Esteban Munoz-Medina; Jose de Jesus Nunez-Contreras; Juan Bautista Chale-Dzul; Luis Alberto Ochoa-Carrera; Margarita Matias-Florentino; Maria Guadalupe Santiago-Mauricio; Maria Guadalupe de Jesus Mireles-Rivera; Nelly Selem-Mojica; Pavel Isa; Ricardo Grande; Santiago Ávila-Rios; Victor Eduardo Garcia-Arias; Victor Hugo Borja-Aburto |
| EPI_ISL_1971059 | Centro de Investigación Biomédica de La Rioja - Hospital San Pedro Logroño | SeqCOVID-SPAIN consortium/IBV(CSIC) | José Manuel Azcona Gutiérrez; María Pilar Bea Escudero; María de Toro; Miriam Blasco Alberdi and SeqCOVID-SPAIN consortium |

| | | | |
|---|---|---|---|
| EPI_ISL_1933725, EPI_ISL_1933726, EPI_ISL_1933727, EPI_ISL_1933728, EPI_ISL_1933729, EPI_ISL_1933730, EPI_ISL_1933731, EPI_ISL_1933732, EPI_ISL_1933733, EPI_ISL_1933734, EPI_ISL_1933735 | | | |
| see above | Chiba Prefectural Institute of Public Health | Pathogen Genomics Center, National Institute of Infectious Diseases | Hazuka Y Furihata; Kentaro Itokawa; Makoto Kuroda; Masanori Hashino; Masumichi Saito; Naomi Nojiri; Nozomu Hanaoka; Rina Tanaka; Sana Uchikoba; Tsuguto Fujimoto; Tsuyoshi Sekizuka |
| EPI_ISL_4498200 | Children's Hospital of Philadelphia | Planet | Ahmed M. Moustafa; Alex Arvanitis; Azad Ahmed; Brandy Neide; Colleen Bianco; Josh Chang Mell; Lidiya Denu; Paul J. Planet; Rebecca M. Harris; Susan Coffin; Swetha Rajagopal |
| EPI_ISL_2234845 | City of Milwaukee Health Department Laboratory | City of Milwaukee Health Department Laboratory | Amy Bauer; Jennifer Lentz; Manjeet Khubbar; Samantha Scott; Sanjib Bhattacharyya |
| EPI_ISL_1599178 | Cliniques universitaires Saint-Luc | UCLouvain/IREC/MBLG | Benoit Kabamba Mukadi; Jean Ruelle; Lysa Pinsmaye |
| EPI_ISL_1617331, EPI_ISL_1617332, EPI_ISL_2195081 | Colorado Department of Public Health and Environment | Colorado Department of Public Health and Environment | Alexandria Rossheim; Diana Ir; Emily A. Travanty; Laura Bankers; Molly C. Hetherington-Rauth; Sarah Elizabeth Totten; Shannon Ely; Shannon R. Matzinger |
| EPI_ISL_1595844 | Compass Laboratory Services, LLC | Compass Laboratory Services | PhD; William Budd |
| EPI_ISL_2034730 | Connecticut DPH | Yale Center for Genomic Analysis | Anderson Brito; Annie Watkins; Anthony Muyombwe; Brooke Sullivan; Chaney Kalinich; Chantal Vogels; Curt Scharfe; Irina Tikhonova; Isabel Ott; Jafar Razeq; Jessica Rothman; Joseph Fauver; Kaya Bilguvar; Mallery Breban; Mary Petrone; Nathan Grubaugh; Randy Downing; Shrikant Mane; Stephen M. Bart; Tara Alpert |
| EPI_ISL_5782679 | Coordenadoria Municipal De Saude De Iracemapolis | Instituto Butantan | Antonio Jorge Martins; Claudia Renata dos Santos Barros; David Schlesinger; Debora Botequio Moretti; Dimas Tadeu Covas; Elaine Cristina Marqueze; Elaine Vieira Santos; Evandra Strazza Rodrigues; Heidge Fukumasu; Jayme Augusto de Souza-Neto; José Salvatore Leister Patané; Luiz Alcantara; Luiz Lehmann Coutinho; Maria Carolina Elias; Maurício Lacerda Nogueira; Rafael dos Santos Bezerra; Raul Machado Neto; Rejane Maria Tommasini Grotto; Ricardo Haddad; Sandra Coccuzzo Sampaio Vessoni; Simone Kashima; Svetoslav Nanev Slavov; Vincent Louis Viala |
| EPI_ISL_7727195, EPI_ISL_7727328, EPI_ISL_7727332 | Covid Laboratory Biogem | Covid Laboratory Biogem | Alessandra Fucci and Michele Caraglia; Alessia Maria Cossu; Cinzia Miarelli; Clara Iannarone; Egildo Luca D'andrea; Federica Melisi; Giovambattista Capasso; Marco Bocchetti; Marianna Scrima; Michele Ceccarelli; Piera Grisolia; Teresa Maria Rosaria Noviello; Ylenia Abruzzese |
| EPI_ISL_1575080 | Cruces University Hospital | Biocruces Bizkaia | Ana Belen de la Hoz Rastrollo; Mikel Gallego Rodrigo |
| EPI_ISL_5802167, EPI_ISL_5802172, EPI_ISL_5802173 | Cs De Balsamo | Instituto Butantan | Antonio Jorge Martins; Claudia Renata dos Santos Barros; David Schlesinger; Debora Botequio Moretti; Dimas Tadeu Covas; Elaine Cristina Marqueze; Elaine Vieira Santos; Evandra Strazza Rodrigues; Heidge Fukumasu; Jayme Augusto de Souza-Neto; José Salvatore Leister Patané; Luiz Alcantara; Luiz Lehmann Coutinho; Maria Carolina Elias; Maurício Lacerda Nogueira; Rafael dos Santos Bezerra; Raul Machado Neto; Rejane Maria Tommasini Grotto; Ricardo Haddad; Sandra Coccuzzo Sampaio Vessoni; Simone Kashima; Svetoslav Nanev Slavov; Vincent Louis Viala |
| EPI_ISL_5802166 | Cs De Santa Rita Doeste | Instituto Butantan | Antonio Jorge Martins; Claudia Renata dos Santos Barros; David Schlesinger; Debora Botequio Moretti; Dimas Tadeu Covas; Elaine Cristina Marqueze; Elaine Vieira Santos; Evandra Strazza Rodrigues; Heidge Fukumasu; Jayme Augusto de Souza-Neto; José Salvatore Leister Patané; Luiz Alcantara; Luiz Lehmann Coutinho; Maria Carolina Elias; Maurício Lacerda Nogueira; Rafael dos Santos Bezerra; Raul Machado Neto; Rejane Maria Tommasini Grotto; Ricardo Haddad; Sandra Coccuzzo Sampaio Vessoni; Simone Kashima; Svetoslav Nanev Slavov; Vincent Louis Viala |
| EPI_ISL_5802126 | Cs Iii Santopolis Do Aguapei | Instituto Butantan | Antonio Jorge Martins; Claudia Renata dos Santos Barros; David Schlesinger; Debora Botequio Moretti; Dimas Tadeu Covas; Elaine Cristina Marqueze; Elaine Vieira Santos; Evandra Strazza Rodrigues; Heidge Fukumasu; Jayme Augusto de Souza-Neto; José Salvatore Leister Patané; Luiz Alcantara; Luiz Lehmann Coutinho; Maria Carolina Elias; Maurício Lacerda Nogueira; Rafael dos Santos Bezerra; Raul Machado Neto; Rejane Maria Tommasini Grotto; Ricardo Haddad; Sandra Coccuzzo Sampaio Vessoni; Simone Kashima; Svetoslav Nanev Slavov; Vincent Louis Viala |
| EPI_ISL_2754989, EPI_ISL_2755817 | Curative Labs | Curative Labs | Elias L. Salfati; Eugenia Khorosheva; George Way; J.Cesar Ignacio-Espinoza; Janet Chen; Mikhail Hanewich-Hollatz; Nabjot Sandhu; Sophia Quasem; Vladimir Slepnev; Zhiyi Xie |
| EPI_ISL_3243122, EPI_ISL_3243124, EPI_ISL_3243158, EPI_ISL_3243184, EPI_ISL_3452096, EPI_ISL_3452313 | Curative Labs | New Mexico Department of Health Scientific Laboratory | Anastacia Griego-Fisher; D'eldra Malone; Ellie Johnson; Jennifer Benoit; Justin Griego; Linda Salazar; Linda Salzar; Michael Sam; Monica Manginell; Ratheesh Rajan; T. Mark Willmon |
| EPI_ISL_2209152 | DEPARTAMENTO DE SAUDE COLETIVA | Instituto Butantan | Antonio Jorge Martins; Claudia Renata dos Santos Barros; David Schlesinger; Debora Botequio Moretti; Dimas Tadeu Covas; Elaine Cristina Marqueze; Elaine Vieira Santos; Evandra Strazza Rodrigues; Heidge Fukumasu; Jayme Augusto de Souza-Neto; José Salvatore Leister Patané; Luiz Alcantara; Luiz Lehmann Coutinho; Maria Carolina Elias; Maurício Lacerda Nogueira; Rafael dos Santos Bezerra; Raul Machado Neto; Rejane Maria Tommasini Grotto; Ricardo Haddad; Sandra Coccuzzo Sampaio Vessoni; Simone Kashima; Svetoslav Nanev Slavov; Vincent Louis Viala. |
| EPI_ISL_2344545 | DEPARTAMENTO DE SAUDE COLETIVA | Instituto Butantan / UNESP-Botucatu | Antonio Jorge Martins; Claudia Renata dos Santos Barros; David Schlesinger; Debora Botequio Moretti; Dimas Tadeu Covas; Elaine Cristina Marqueze; Elaine Vieira Santos; Evandra Strazza Rodrigues; Heidge Fukumasu; Jayme Augusto de Souza-Neto; José Salvatore Leister Patané; Luiz Alcantara; Luiz Lehmann Coutinho; Maria Carolina Elias; Maurício Lacerda Nogueira; Rafael dos Santos Bezerra; Raul Machado Neto; Rejane Maria Tommasini Grotto; Ricardo Haddad; Sandra Coccuzzo Sampaio Vessoni; Simone Kashima; Svetoslav Nanev Slavov; Vincent Louis Viala |
| EPI_ISL_1577989, EPI_ISL_1580554, EPI_ISL_1580560, EPI_ISL_1580591, EPI_ISL_1580625, EPI_ISL_1580626, EPI_ISL_1580651, EPI_ISL_1580656 | | | |
| see above | DIP. PREV. AVEZZANO SERVIZIO DI IGIENE EPIDEMIOLOGIA E SANITA' PUBBLICA AVEZZANO(L'AQUILA) | Istituto Zooprofilattico Sperimentale dell'Abruzzo e Molise "G. Caporale" | Ancora M; Calistri P; Cammà C; Caporale M; Curini V; Delli Compagni E; Di Domenico M; Di Lollo Valeria; Di Pasquale A; Lorusso A; Mangone I; Marcacci M; Puglia I; Rinaldi A; Savini G; Scialabba S |
| EPI_ISL_1908995 | DPHL | Delaware Public Health Lab | Rebecca Savage |
| EPI_ISL_5802187 | Departamento De Saude Coletiva | Instituto Butantan | Antonio Jorge Martins; Claudia Renata dos Santos Barros; David Schlesinger; Debora Botequio Moretti; Dimas Tadeu Covas; Elaine Cristina Marqueze; Elaine Vieira Santos; Evandra Strazza Rodrigues; Heidge Fukumasu; Jayme Augusto de Souza-Neto; José Salvatore Leister Patané; Luiz Alcantara; Luiz Lehmann Coutinho; Maria Carolina Elias; Maurício Lacerda Nogueira; Rafael dos Santos Bezerra; Raul Machado Neto; Rejane Maria Tommasini Grotto; Ricardo Haddad; Sandra Coccuzzo Sampaio Vessoni; Simone Kashima; Svetoslav Nanev Slavov; Vincent Louis Viala |
| EPI_ISL_1595645 | Department of Clinical Microbiology | GIGA Medical Genomics | Bouchra Boujemla; Cécile Meex; Keith Durkin; Maria Artesi; Marie-Pierre Hayette; Nathalie Renotte; Pierrette Melin; Raphaël Boreux; Sébastien Bontems; Vincent Bours |
| EPI_ISL_2145424, EPI_ISL_2145438, EPI_ISL_2145478 | Dutch COVID-19 response team | Erasmus Medical Center | Anne van der Linden; Annemiek van der Eijk; Bas Oude Munnink; Corine GeurtsvanKessel; David Nieuwenhuijse; Emmanuelle Munger; Irina Chestakova; Marion Koopmans; Marjan Boter; Reina Sikkema; Richard Molenkamp; on behalf of the Dutch national COVID-19 respo |
| EPI_ISL_1596573, EPI_ISL_1596777, EPI_ISL_1596936, EPI_ISL_1597044, EPI_ISL_1597245, EPI_ISL_1597268, EPI_ISL_1703330, EPI_ISL_1703620, EPI_ISL_1705099, EPI_ISL_1705115, EPI_ISL_1705398, EPI_ISL_1705409, EPI_ISL_1705413, EPI_ISL_1705443, EPI_ISL_1705502, EPI_ISL_1705513, EPI_ISL_1705525, EPI_ISL_1961155, EPI_ISL_1962270 | | | |
| see above | Dutch COVID-19 response team | National Institute for Public Health and the Environment (RIVM) | Adam Meijer; AnneMarie van den Brandt; Annelies Kroneman; Bas van der Veer; Chantal Reusken; Dennis Schmitz; Dirk Eggink; Euníce Then; Florian Zwagemaker; Harry Vennema; James Groot; Jeroen Cremer; Jolienke Hardeman; Karim Hajji; Kim Freriks; Linda van de Nes; Lisa Wijsman; Lynn Aarts; Melissa van Tuil; Robert Kohl; Ryanne Jaarsma; Sanne Bos; Sharon van den Brink; Sjoerd Kuiling; on behalf of the national COVID-19 response team |
| EPI_ISL_1966532, EPI_ISL_1966533 | EMERGENCIA RESPIRATORIA DE NOVA GRANADA | Instituto Butantan / FZEA-USP (Pirassununga) | Antonio Jorge Martins; Bianca Cechetto Carlos. Mendelics: Bibiana Santos; Claudia Renata dos Santos Barros; Cíntia Bittar; David Schlesinger. Hemocentro Ribeirão Preto: Simone Kashima; Debora Botequio Moretti; Elaine Cristina Marqueze; Elaine Vieira dos Santos; Elisangela Chicaroni Mattos; Erika Freitas; Evandra Strazza Rodrigues; Felipe Allan da Silva da Costa; Flavia Aburjaile; Fábio Sossai Possebon; Guilherme Campos; Guilherme Targino Valente; Heidge Fukumasu. USP-Botucatu: Rejane Maria Tommasini Grotto; Helena Lage Ferreira; Instituto Butantan: Dimas Tadeu Covas; Jardelina de Souza Todao Bernardino; Jayme A. Souza-Neto; Jessika Cristina Chagas Lesbon; Jorge A. Petroli Marchesi; José Salvatore Leister Patané; João Paulo Kitajima; João Pessoa Araújo Jr.; Leila Sabrina Ullmann; Loyze Paola Oliveira de Lima; Luiz Aurelio de Campos Crispin. Centro de Genômica Funcional da ESALQ: Luiz Lehmann Coutinho; Luiz Carlos Junior de Alcantara; Lívia Sacchetto; Maisa C. Pereira Parra; Maria Carolina Elias; Marta Giovanetti; Marília Moraes; Maurício Lacerda Nogueira. Prefeitura de Sao Paulo: Melissa Palmieri.; Patricia Akemi Assato; Paula Rahal; Paulo Inacio da Costa; Rafael dos Santos Bezerra; Raquel de Lello Rocha Campos Cassano. NGS Soluções Genômicas: Pilar Drummond Sampaio Corrêa Mariani. FZEA-USP Pirassununga: Mirele Daiana Poleti; Raul Machado Neto; Ricardo Augusto Brassaloti; Ricardo Haddad; Rodrigo Tocantins Calado. FAMERP-SJRP: Cecília Artico Banho; Sandra Coccuzzo Sampaio; Svetoslav Nanev Slavov; Vagner Fonseca; Vincent Louis Viala |
| EPI_ISL_1795099, EPI_ISL_2344547 | ESALQ | Instituto Butantan / ESALQ-Piracicaba | Antonio Jorge Martins; Bianca Cechetto Carlos. Mendelics: Bibiana Santos; Claudia Renata dos Santos Barros; David Schlesinger; David Schlesinger. Hemocentro Ribeirão Preto: Simone Kashima; Debora Botequio Moretti; Debora Botequio Moretti. Centro de Genômica Funcional da ESALQ: Luiz Lehmann Coutinho; Dimas Tadeu Covas; Elaine Cristina Marqueze; Elaine Vieira Santos; Elaine Vieira dos Santos; Elisangela Chicaroni Mattos; Erika Freitas; Evandra Strazza Rodrigues; Felipe Allan da Silva da Costa; Flavia Aburjaile; Guilherme Targino Valente; Heidge Fukumasu; Heidge Fukumasu. USP-Botucatu: Rejane Maria Tommasini Grotto; Instituto Butantan: Alexander Roberto Precioso; Jayme A. Souza-Neto; Jayme Augusto de Souza-Neto; Jessika Cristina Chagas Lesbon; José Salvatore Leister Patané; João Paulo Kitajima; Luiz Alcantara; Luiz Carlos Junior de Alcantara; Luiz Lehmann Coutinho; Maria Carolina Elias; Marta Giovanetti; Maurício Lacerda Nogueira; Patricia Akemi Assato; Rafael dos Santos Bezerra; Raquel de Lello Rocha Campos Cassano. NGS Soluções Genômicas: Pilar Drummond Sampaio Corrêa Mariani. FZEA-USP Pirassununga: Mirele Daiana Poleti; Raul Machado Neto; Rejane Maria Tommasini Grotto; Ricardo Augusto Brassaloti; Ricardo Haddad; Rodrigo Tocantins Calado.; Sandra Coccuzzo Sampaio; Sandra Coccuzzo Sampaio Vessoni; Simone Kashima; Svetoslav Nanev Slavov; Vagner Fonseca; Vincent Louis Viala |
| EPI_ISL_3215498 | EXCITE Lab | Andersen lab at Scripps Research | Christine Mitchell + SEARCH; Fred Wu |
| EPI_ISL_2477644, EPI_ISL_2477647, EPI_ISL_2477650, EPI_ISL_2477651, EPI_ISL_2477657, EPI_ISL_2477658, EPI_ISL_2477659, EPI_ISL_2477666, EPI_ISL_2477672, EPI_ISL_2477673, EPI_ISL_2477677, EPI_ISL_2477680, EPI_ISL_2477681, EPI_ISL_2477687, EPI_ISL_2478307, EPI_ISL_2478326, EPI_ISL_2478328 | | | |
| see above | Edmonton Provincial Lab | Public Health Agency of Canada (PHAC) National Microbiology Laboratory | Buss; Croxen M; Deo A; Dieu P; E; Ferrato C; Gill K; Khan F; Koleva P; Li V; Lloyd C; Lynch T; Ma R; Murphy S; Pabbaraju K; Shokoples S; Thayer J; Tipples G; Whitehouse M; Wong A; Yu C; Zelyas N |
| EPI_ISL_5802169, EPI_ISL_5802170 | Emergencia Respiratoria De Nova Granada | Instituto Butantan | Antonio Jorge Martins; Claudia Renata dos Santos Barros; David Schlesinger; Debora Botequio Moretti; Dimas Tadeu Covas; Elaine Cristina Marqueze; Elaine Vieira Santos; Evandra Strazza Rodrigues; Heidge Fukumasu; Jayme Augusto de Souza-Neto; José Salvatore Leister Patané; Luiz Alcantara; Luiz Lehmann Coutinho; Maria Carolina Elias; Maurício Lacerda Nogueira; Rafael dos Santos Bezerra; Raul Machado Neto; Rejane Maria Tommasini Grotto; Ricardo Haddad; Sandra Coccuzzo Sampaio Vessoni; Simone Kashima; Svetoslav Nanev Slavov; Vincent Louis Viala |
| EPI_ISL_2107410 | Emory Molecular Diagnostics Laboratory, Emory Healthcare | Piantadosi Lab, Emory Department of Pathology | Ahmed Babiker; Anne Piantadosi |
| EPI_ISL_1571326, EPI_ISL_1643893, EPI_ISL_1722713, EPI_ISL_1722729 | Eurofins LifeCodexx GmbH | Robert Koch Institute | |
| EPI_ISL_1823684 | FL Bureau of Public Health Laboratories-Tampa | Centers for Disease Control and Prevention Division of Viral Diseases, Pathogen Discovery | Alison Laufer Halpin; Ben L. Rambo-Martin; Clinton R. Paden; Dakota Howard; Darlene Wagner; Dave Wentworth; Dhwani Batra; Jasmine Padilla; Justin Lee; Katie Dillon; Krista Queen; Kristen Knipe; Kristine Lacek; Mark Burroughs; Matthew Schmerer; Mili Sheth; Peter Cook; Sam Shepard; Sarah Nobles; Shoshona Le; Suxiang Tong; Vivien Dugan; Yvette Unoarumhi |
| EPI_ISL_1651446, EPI_ISL_1651454, EPI_ISL_1651496, EPI_ISL_1651523, EPI_ISL_1651531, EPI_ISL_2008410, EPI_ISL_2008573, EPI_ISL_2229962, EPI_ISL_2442426, EPI_ISL_6050659, EPI_ISL_6366955, EPI_ISL_6366958, EPI_ISL_6366959, EPI_ISL_6366960, EPI_ISL_6366995, EPI_ISL_6367503 | | | |
| see above | Florida Bureau of Public Health Laboratories | Florida Bureau of Public Health Laboratories | Jason Blanton; Namratha Tarigopula; Sarah Schmedes; Tiffany Splatt |
| EPI_ISL_1578330, EPI_ISL_1578333 | Flow Health | Infectious Disease Program, Broad Institute of Harvard and MIT | Adams, G.; B.L.; B.W.; Bauer, M.; Birren; Carter, A.; Chaluvadi, S.; D.J.; DeRuff, K.; Gallagher, G.; Gladden-Young, A.; J.E.; K.J.; Lagerborg, K.; Lemieux; Loreth, C.; MacInnis; Normandin, E.; P.C.; Park; Reilly, S.; Rudy, M.; Siddle; Smole, S.; Tomkins-Tinch, C.; and Sabeti |
| EPI_ISL_7500768 | Fort Belvoir Community Hospital | Naval Medical Research Center Biological Defense Research | Andrea E. Luquette; Andrew J. Bennett; Britta Babel; Catherine E. Arnold; Emily Hackett; Francisco J. Malagon; Gregory K. Rice; Haven L. Miner; Kimberly A. Bishop-Lilly; Kyle A. Long; Lindsay A. Glang; Logan J. Voegtly; Raven Stone; Regina Z. Cer; Robin H. Miller; Tasheka Pearcey |

| | | | |
|---|---|---|---|
| | | Directorate | |
| EPI_ISL_1556395, EPI_ISL_1556396, EPI_ISL_1556399, EPI_ISL_1556410, EPI_ISL_1557006, EPI_ISL_1612961, EPI_ISL_1612964, EPI_ISL_1612965, EPI_ISL_1613001, EPI_ISL_1613005, EPI_ISL_1613006, EPI_ISL_1613055, EPI_ISL_1613068, EPI_ISL_1613185, EPI_ISL_1613209, EPI_ISL_1613339, EPI_ISL_1613410, EPI_ISL_1613525, EPI_ISL_1613534, EPI_ISL_1613543, EPI_ISL_1613731, EPI_ISL_1613749, EPI_ISL_1613836, EPI_ISL_1613849, EPI_ISL_1613883, EPI_ISL_1613957, EPI_ISL_1613983, EPI_ISL_1614004, EPI_ISL_1614012, EPI_ISL_1614024, EPI_ISL_1614028, EPI_ISL_1614034, EPI_ISL_1614045, EPI_ISL_1614056, EPI_ISL_1614057, EPI_ISL_1614069, EPI_ISL_1614083, EPI_ISL_1614101, EPI_ISL_1614117, EPI_ISL_1614119, EPI_ISL_1614154, EPI_ISL_1614179, EPI_ISL_1614186, EPI_ISL_1614197, EPI_ISL_1614213, EPI_ISL_1614247, EPI_ISL_1614257, EPI_ISL_1614393, EPI_ISL_1614396, EPI_ISL_1614415, EPI_ISL_1614436, EPI_ISL_1614445, EPI_ISL_1614497, EPI_ISL_1614525, EPI_ISL_1614561, EPI_ISL_1614569, EPI_ISL_1665906, EPI_ISL_1665913, EPI_ISL_1666575, EPI_ISL_1666581, EPI_ISL_1666584, EPI_ISL_1666585, EPI_ISL_1666637, EPI_ISL_1666649, EPI_ISL_1666657, EPI_ISL_1666746, EPI_ISL_1666793, EPI_ISL_1666802, EPI_ISL_1666808, EPI_ISL_1666820, EPI_ISL_1666873, EPI_ISL_1666934, EPI_ISL_1667016, EPI_ISL_1667025, EPI_ISL_1667066, EPI_ISL_1667068, EPI_ISL_1733768, EPI_ISL_1733769, EPI_ISL_1733818, EPI_ISL_1733829, EPI_ISL_1733832, EPI_ISL_1734048, EPI_ISL_1734526, EPI_ISL_1734530, EPI_ISL_4368746, EPI_ISL_4370310 | | | |
| see above | Fulgent Genetics | Centers for Disease Control and Prevention Division of Viral Diseases, Pathogen Discovery | Adrian Paskey; Becky Tsai; Benafsh Sapra; Benjamin Rambo-Martin; Christopher Gulvick; Clinton Paden; Clinton R. Paden; Dakota Howard; Darlene Wagner; Dhwani Batra; Doreen Ng; Duncan MacCannell; Erisa Sula; Harry Gao; James Xie; Jason Caravas; John Gao; Joseph Fierro; Kara Moser; Kristine Lacek; Matthew Schmerer; Mickey Li; Peter Cook; Peter W. Cook; Scott Sammons; Shatavia Morrison; Tymeckia Kendall; Victoria Caban Figueroa; Yan Meng; Yvette Unoarumhi |
| EPI_ISL_3553536, EPI_ISL_3553537, EPI_ISL_3553547, EPI_ISL_3553555, EPI_ISL_3553559, EPI_ISL_3553567, EPI_ISL_3553568, EPI_ISL_3553569, EPI_ISL_3553570, EPI_ISL_3553575, EPI_ISL_3553576, EPI_ISL_3553577, EPI_ISL_3553581, EPI_ISL_3553582, EPI_ISL_3553650, EPI_ISL_3553688, EPI_ISL_3553694 | | | |
| see above | Fundação Ezequiel Dias (FUNED) | Fundação Ezequiel Dias | Andre Leal; Cynthia Vazquez; Elaine Cristina; Felipe Iani; Flavia Aburjaile; Gislene Garcia de Castro Lichs; Glauco Carvalho; Hegger Fritsch; Joilson Xavier; Luiz Alcantara.; Luiz Henrique Ferraz Demarchi; Luiz Takao Watanabe; Marina Castilhos Souza Umaki Zardin; Marta Giovanetti; Natalia Guimaraes; Raquel da Silva Ferreira; Talita Adelino; Vagner Fonseca; de Oliveira |
| EPI_ISL_1755321 | GH A.CHENEVIER-H.MONDOR | Department of Virology, Henri Mondor University Hospital, Assistance Publique Hôpitaux de Paris, Université Paris-Est Créteil, INSERM U955 | Alexandre Soulier; Christophe Rodriguez; Elisabeth Trawinski; Guillaume Gricourt; Jean-Michel Pawlotsky; Melissa N'Debi; Slim Fourati; Vanessa Demontant |
| EPI_ISL_1756134 | GH de l'Est Francilien | Department of Virology, Henri Mondor University Hospital, Assistance Publique Hôpitaux de Paris, Université Paris-Est Créteil, INSERM U955 | Alexandre Soulier; Christophe Rodriguez; Elisabeth Trawinski; Guillaume Gricourt; Jean-Michel Pawlotsky; Melissa N'Debi; Slim Fourati; Vanessa Demontant |
| EPI_ISL_1626616 | Gencore - Universidad de los Andes | Gencore - Universidad de los Andes | Ana Maria Palacio; Cristian Barrera; David Gonzalez; Erica Salguero; Gabriela Ariza; Luisa Sacristan; Marcela Guevara; Silvia Restrepo |
| EPI_ISL_1470548, EPI_ISL_1470554, EPI_ISL_1534596, EPI_ISL_1534603, EPI_ISL_1541015, EPI_ISL_1541016, EPI_ISL_1541042, EPI_ISL_1633502, EPI_ISL_2009237, EPI_ISL_2009243 | | | |
| see above | Genetica Molecular and Subdepartamento de Virologia ISP Chile | Instituto de Salud Publica de Chile | Andres Castillo; Barbara Parra; Gisselle Barra; Jaime Lagos; Javier Tognarelli; Jorge Fernandez; Karen Orostica; Loredana Arata; Patricia Bustos; Rodrigo Fasce; Soledad Ulloa |
| EPI_ISL_5196295, EPI_ISL_5196345 | Gorgas Memorial Institute of Health Studies | Gorgas Memorial Institute of Health Studies | Castillo Jorge; Chen Maria; Franco Danilo; Gonzalez Claudia; Jessica Gondola; Leyda Abrego; Lopez-Verges Sandra; Marlenne Castillo; Martinez Alexander; Menacho Abdiel; Moreno Ambar; Moreno Brechla; Oris Chavarria; Ortiz Alma; Salazar Jacqueline |
| EPI_ISL_4744603, EPI_ISL_4744607, EPI_ISL_4744626, EPI_ISL_4744627 | Gorgas Memorial Laboratory of Health Studies | Gorgas Memorial Laboratory of Health Studies | Castillo Jorge; Chen Maria; Franco Danilo; Gonzalez Claudia; Jessica Gondola; Leyda Abrego; Lopez-Verges Sandra; Marlenne Castillo; Martinez Alexander; Menacho Abdiel; Moreno Ambar; Moreno Brechla; Oris Chavarria; Ortiz Alma; Salazar Jacqueline |
| EPI_ISL_7307284, EPI_ISL_7307290 | Grupo de Investigación en Enfermedades Tropicales del Ejército (GINETEJ), Laboratorio de Referencia e Investigación, Dirección de Sanidad Ejército, Bogotá, Colombia | Centro de Investigaciones en Microbiología y Biotecnología-UR (CIMBIUR), Facultad de Ciencias Naturales, Universidad del Rosario, Bogotá, Colombia | Alberto Paniz-Mondolfi; Angie Ramírez; Beatriz Ariza; Camilo A. Correa-Cárdenas; Carlos Gómez-Restrepo; Claudia Cardozo-Romero; Claudia Méndez; David-Santiago Quevedo; Guido España; Hernando Díaz; Juan David Ramírez; Juliana Cuervo-Rojas; Julie Pérez; Luz H. Patiño; Manuel-Antonio Franco; Maria-Clara Duque; Marina Muñoz; Nathalia Ballesteros; Nicolas Luna; Sergio Castañeda; Zulma M. Cucunubá |
| EPI_ISL_3912160 | H J M A HOSPITAL JOSE MARTINIANO DE ALENCAR | Analytical Competence Molecular Epidemiology Lab/ACME, Oswaldo Cruz Foundation, Ceara (FIOCRUZ CE) | Cleber Furtado Aksenen; Fabio Miyajima; Fernando Braga Stehling; Francisco Eder de Moura Lopes; Jamille Maria Mendes Bezerra; Joaquim Cesar do Nascimento Sousa Junior; Pedro Miguel Carneiro Jeronimo; Suzana Porto Almeida & Lucas Delerino on behalf of COVID-19 FIOCRUZ Genomic Network; Thais Ferreira de Oliveira; Thais de Oliveira Costa; Ticiane Cavalcante de Souza; Veridiana Pessoa Miyajima |
| EPI_ISL_5529884, EPI_ISL_5530028, EPI_ISL_5530034, EPI_ISL_5530035 | HGF HOSPITAL GERAL DE FORTALEZA | Analytical Competence Molecular Epidemiology Lab/ACME, Oswaldo Cruz Foundation, Ceara (FIOCRUZ CE) | Carlos Leonardo de Aragao Araujo; Cecilia Leite Costa & Eduardo Ruback dos Santos on behalf of COVID-19 FIOCRUZ Genomic Network; Cleber Furtado Aksenen; Fabio Miyajima; Fernando Braga Stehling; Francisco Eder de Moura Lopes; Igor Oliveira Duarte; Jamille Maria Mendes Bezerra; Joaquim Cesar do Nascimento Sousa Junior; Pedro Miguel Carneiro Jeronimo; Suzana Porto Almeida; Thais Ferreira de Oliveira; Thais de Oliveira Costa; Ticiane Cavalcante de Souza; Veridiana Pessoa Miyajima |
| EPI_ISL_3102366 | HIAS HOSPITAL INFANTIL ALBERT SABIN | Analytical Competence Molecular Epidemiology Lab/ACME, Oswaldo Cruz Foundation, Ceara (FIOCRUZ CE) | Cleber Furtado Aksenen; Fabio Miyajima; Fernando Braga Stehling; Francisco Eder de Moura Lopes; Jamille Maria Mendes Bezerra; Joaquim César do Nascimento Sousa Junior; Pedro Miguel Carneiro Jeronimo; Suzana Porto Almeida e Lucas Delerino; Thais Ferreira de Oliveira; Thais de Oliveira Costa; Ticiane Cavalcante de Souza; Veridiana Pessoa Miyajima |
| EPI_ISL_2017282, EPI_ISL_2017391, EPI_ISL_2017392, EPI_ISL_2017393, EPI_ISL_2017394, EPI_ISL_2017395, EPI_ISL_2017396, EPI_ISL_2017397, EPI_ISL_2017398, EPI_ISL_2017399, EPI_ISL_2017400, EPI_ISL_2017401, EPI_ISL_2017402, EPI_ISL_2017403, EPI_ISL_2017404, EPI_ISL_2017405, EPI_ISL_2017406, EPI_ISL_2017407, EPI_ISL_2017408, EPI_ISL_2017409, EPI_ISL_2017411, EPI_ISL_2017453, EPI_ISL_2017454, EPI_ISL_2017456, EPI_ISL_2017477, EPI_ISL_2187911, EPI_ISL_2187912, EPI_ISL_2187913, EPI_ISL_2187914, EPI_ISL_2187915, EPI_ISL_2187919, EPI_ISL_2187920, EPI_ISL_2187921, EPI_ISL_2187925, EPI_ISL_2187926, EPI_ISL_2187927, EPI_ISL_2187928, EPI_ISL_2187929, EPI_ISL_2187931, EPI_ISL_2187932, EPI_ISL_2187933, EPI_ISL_2187934, EPI_ISL_2187935, EPI_ISL_2187936, EPI_ISL_2187937, EPI_ISL_2187938, EPI_ISL_2187939, EPI_ISL_2187940, EPI_ISL_2187941, EPI_ISL_2187942, EPI_ISL_2187944, EPI_ISL_2187945, EPI_ISL_2187946 | | | |
| see above | HLAGYN - Laboratorio de Imunologia de Transplantes de Goias | HLAGYN - Laboratorio de Imunologia de Transplantes de Goias | Alessandro Leonardo Alvares Magalhaes; Daniel Ferreira de Sousa; Danielle de Paiva Rezende; Erika Lopes Rocha Batista; Fernando Antonio Vinhal dos Santos; Frederico Rodrigues Vinhal; Lucas Carlos Gomes Pereira; Paola Cristina Resende Silva; Raphael Bessa Parmigiane; Sabrina Sara Moreira Duarte |
| EPI_ISL_5529875, EPI_ISL_5529953 | HOSP DR THADEU DE PAULO BRITO | Analytical Competence Molecular Epidemiology Lab/ACME, Oswaldo Cruz Foundation, Ceara (FIOCRUZ CE) | Carlos Leonardo de Aragao Araujo; Cecilia Leite Costa & Eduardo Ruback dos Santos on behalf of COVID-19 FIOCRUZ Genomic Network; Cleber Furtado Aksenen; Fabio Miyajima; Fernando Braga Stehling; Francisco Eder de Moura Lopes; Igor Oliveira Duarte; Jamille Maria Mendes Bezerra; Joaquim Cesar do Nascimento Sousa Junior; Pedro Miguel Carneiro Jeronimo; Suzana Porto Almeida; Thais Ferreira de Oliveira; Thais de Oliveira Costa; Ticiane Cavalcante de Souza; Veridiana Pessoa Miyajima |
| EPI_ISL_2209204 | HOSP E MATERNIDADE MUNICIPAL N SRA MONTE SERRAT | Instituto Butantan | Antonio Jorge Martins; Claudia Renata dos Santos Barros; David Schlesinger; Debora Botequio Moretti; Dimas Tadeu Covas; Elaine Cristina Marqueze; Elaine Vieira Santos; Evandra Strazza Rodrigues; Heidge Fukumasu; Jayme Augusto de Souza-Neto; José Salvatore Leister Patané; Luiz Alcantara; Luiz Lehmann Coutinho; Maria Carolina Elias; Maurício Lacerda Nogueira; Rafael dos Santos Bezerra; Raul Machado Neto; Rejane Maria Tommasini Grotto; Ricardo Haddad; Sandra Coccuzzo Sampaio Vessoni; Simone Kashima; Svetoslav Nanev Slavov; Vincent Louis Viala. |
| EPI_ISL_2170937 | HOSP E MATERNIDADE MUNICIPAL N SRA MONTE SERRAT | Instituto Butantan / Mendelics | Antonio Jorge Martins; Bianca Cechetto Carlos. Mendelics: Bibiana Santos; Claudia Renata dos Santos Barros; Cíntia Bittar; David Schlesinger. Hemocentro Ribeirão Preto: Simone Kashima; Debora Botequio Moretti; Elaine Cristina Marqueze; Elaine Vieira dos Santos; Elisangela Chicaroni Mattos; Erika Freitas; Evandra Strazza Rodrigues; Felipe Allan da Silva da Costa; Flavia Aburjaile; Fábio Sossai Possebon; Guilherme Campos; Guilherme Targino Valente; Heidge Fukumasu. USP-Botucatu: Rejane Maria Tommasini Grotto; Helena Lage Ferreira; Instituto Butantan: Dimas Tadeu Covas; Jardelina de Souza Todao Bernardino; Jayme A. Souza-Neto; Jessika Cristina Chagas Lesbon; Jorge A. Petroli Marchesi; José Salvatore Leister Patané; João Paulo Kitajima; João Pessoa Araújo Jr.; Leila Sabrina Ullmann; Loyze Paola Oliveira de Lima; Luiz Aurelio de Campos Crispin. Centro de Genômica Funcional da ESALQ: Luiz Lehmann Coutinho; Luiz Carlos Junior de Alcantara; Lívia Sacchetto; Maisa C. Pereira Parra; Maria Carolina Elias; Marta Giovanetti; Marília Moraes; Maurício Lacerda Nogueira. Prefeitura de Sao Paulo: Melissa Palmieri.; Patricia Akemi Assato; Paula Rahal; Paulo Inacio da Costa; Rafael dos Santos Bezerra; Raquel de Lello Rocha Campos Cassano. NGS Soluções Genômicas: Pilar Drummond Sampaio Corrêa Mariani. FZEA-USP Pirassununga: Mirele Daiana Poleti; Raul Machado Neto; Ricardo Augusto Brassaloti; Ricardo Haddad; Rodrigo Tocantins Calado. FAMERP-SJRP: Cecília Artico Banho; Sandra Coccuzzo Sampaio; Svetoslav Nanev Slavov; Vagner Fonseca; Vincent Louis Viala |
| EPI_ISL_2344546 | HOSP E MATERNIDADE MUNICIPAL N SRA MONTE SERRAT | Instituto Butantan / UNESP-Botucatu | Antonio Jorge Martins; Claudia Renata dos Santos Barros; David Schlesinger; Debora Botequio Moretti; Dimas Tadeu Covas; Elaine Cristina Marqueze; Elaine Vieira Santos; Evandra Strazza Rodrigues; Heidge Fukumasu; Jayme Augusto de Souza-Neto; José Salvatore Leister Patané; Luiz Alcantara; Luiz Lehmann Coutinho; Maria Carolina Elias; Maurício Lacerda Nogueira; Rafael dos Santos Bezerra; Raul Machado Neto; Rejane Maria Tommasini Grotto; Ricardo Haddad; Sandra Coccuzzo Sampaio Vessoni; Simone Kashima; Svetoslav Nanev Slavov; Vincent Louis Viala |
| EPI_ISL_5530135 | HOSP MATERN MAE TOTONHA | Analytical Competence Molecular Epidemiology Lab/ACME, Oswaldo Cruz Foundation, Ceara (FIOCRUZ CE) | Carlos Leonardo de Aragao Araujo; Cecilia Leite Costa & Eduardo Ruback dos Santos on behalf of COVID-19 FIOCRUZ Genomic Network; Cleber Furtado Aksenen; Fabio Miyajima; Fernando Braga Stehling; Francisco Eder de Moura Lopes; Igor Oliveira Duarte; Jamille Maria Mendes Bezerra; Joaquim Cesar do Nascimento Sousa Junior; Pedro Miguel Carneiro Jeronimo; Suzana Porto Almeida; Thais Ferreira de Oliveira; Thais de Oliveira Costa; Ticiane Cavalcante de Souza; Veridiana Pessoa Miyajima |
| EPI_ISL_5530093, EPI_ISL_5530094 | HOSP MATERN SENHORA SANTANA | Analytical Competence Molecular Epidemiology Lab/ACME, Oswaldo Cruz Foundation, Ceara (FIOCRUZ CE) | Carlos Leonardo de Aragao Araujo; Cecilia Leite Costa & Eduardo Ruback dos Santos on behalf of COVID-19 FIOCRUZ Genomic Network; Cleber Furtado Aksenen; Fabio Miyajima; Fernando Braga Stehling; Francisco Eder de Moura Lopes; Igor Oliveira Duarte; Jamille Maria Mendes Bezerra; Joaquim Cesar do Nascimento Sousa Junior; Pedro Miguel Carneiro Jeronimo; Suzana Porto Almeida; Thais Ferreira de Oliveira; Thais de Oliveira Costa; Ticiane Cavalcante de Souza; Veridiana Pessoa Miyajima |
| EPI_ISL_5529964, EPI_ISL_5530194 | HOSP MUN ABELARDO GADELHA DA ROCHA | Analytical Competence Molecular Epidemiology Lab/ACME, Oswaldo Cruz Foundation, Ceara (FIOCRUZ CE) | Carlos Leonardo de Aragao Araujo; Cecilia Leite Costa & Eduardo Ruback dos Santos on behalf of COVID-19 FIOCRUZ Genomic Network; Cleber Furtado Aksenen; Fabio Miyajima; Fernando Braga Stehling; Francisco Eder de Moura Lopes; Igor Oliveira Duarte; Jamille Maria Mendes Bezerra; Joaquim Cesar do Nascimento Sousa Junior; Pedro Miguel Carneiro Jeronimo; Suzana Porto Almeida; Thais Ferreira de Oliveira; Thais de Oliveira Costa; Ticiane Cavalcante de Souza; Veridiana Pessoa Miyajima |
| EPI_ISL_5530110, EPI_ISL_5530186 | HOSP MUNIC ANTONIO NERY FILHO | Analytical Competence Molecular Epidemiology Lab/ACME, Oswaldo Cruz Foundation, Ceara (FIOCRUZ CE) | Carlos Leonardo de Aragao Araujo; Cecilia Leite Costa & Eduardo Ruback dos Santos on behalf of COVID-19 FIOCRUZ Genomic Network; Cleber Furtado Aksenen; Fabio Miyajima; Fernando Braga Stehling; Francisco Eder de Moura Lopes; Igor Oliveira Duarte; Jamille Maria Mendes Bezerra; Joaquim Cesar do Nascimento Sousa Junior; Pedro Miguel Carneiro Jeronimo; Suzana Porto Almeida; Thais Ferreira de Oliveira; Thais de Oliveira Costa; Ticiane Cavalcante de Souza; Veridiana Pessoa Miyajima |
| EPI_ISL_2801321 | HOSPITAL ANTONIO ROSENO DE MATOS | Analytical Competence Molecular Epidemiology Lab/ACME, Oswaldo Cruz Foundation, Ceara (FIOCRUZ CE) | Cleber Furtado Aksenen e Suzana Porto Almeida; Fabio Miyajima; Fernando Braga Stehling; Francisco Eder de Moura Lopes; Jamille Maria Mendes Bezerra; Joaquim César do Nascimento Sousa Junior; Pedro Miguel Carneiro Jeronimo; Thais Ferreira de Oliveira; Thais de Oliveira Costa; Ticiane Cavalcante de Souza; Veridiana Pessoa Miyajima |
| EPI_ISL_1795101, EPI_ISL_1795103, EPI_ISL_2344550, EPI_ISL_2344552 | HOSPITAL DOS FORNCEDORES | Instituto Butantan / ESALQ-Piracicaba | Antonio Jorge Martins; Bianca Cechetto Carlos. Mendelics: Bibiana Santos; Claudia Renata dos Santos Barros; David Schlesinger; David Schlesinger. Hemocentro Ribeirão Preto: Simone Kashima; Debora Botequio Moretti; Debora Botequio Moretti. Centro de Genômica Funcional da ESALQ: Luiz Lehmann Coutinho; Dimas Tadeu Covas; Elaine Cristina Marqueze; Elaine Vieira Santos; Elaine Vieira dos Santos; Elisangela Chicaroni Mattos; Erika Freitas; Evandra Strazza Rodrigues; Felipe Allan da Silva da Costa; Flavia Aburjaile; Guilherme Targino Valente; Heidge Fukumasu; Heidge Fukumasu. USP-Botucatu: Rejane Maria Tommasini Grotto; Instituto Butantan: Alexander Roberto Precioso; Jayme A. Souza-Neto; Jayme Augusto de Souza-Neto; Jessika Cristina Chagas Lesbon; José Salvatore Leister Patané; João Paulo Kitajima; Luiz Alcantara; Luiz Carlos Junior de Alcantara; Luiz Lehmann Coutinho; Maria Carolina Elias; Marta Giovanetti; Maurício Lacerda Nogueira; Patricia Akemi Assato; Rafael dos Santos Bezerra; Raquel de Lello Rocha Campos Cassano. NGS Soluções Genômicas: Pilar Drummond Sampaio Corrêa Mariani. FZEA-USP Pirassununga: Mirele Daiana Poleti; Raul Machado Neto; Rejane Maria Tommasini Grotto; Ricardo Augusto Brassaloti; Ricardo Haddad; Rodrigo Tocantins Calado.; Sandra Coccuzzo Sampaio; Sandra Coccuzzo Sampaio Vessoni; Simone Kashima; Svetoslav Nanev Slavov; Vagner Fonseca; Vincent Louis Viala |
| EPI_ISL_3102389, EPI_ISL_3102532, EPI_ISL_3912209, EPI_ISL_3912210 | HOSPITAL ESTADUAL LEONARDO DA VINCI | Analytical Competence Molecular Epidemiology Lab/ACME, Oswaldo Cruz Foundation, Ceara (FIOCRUZ CE) | Cleber Furtado Aksenen; Fabio Miyajima; Fernando Braga Stehling; Francisco Eder de Moura Lopes; Jamille Maria Mendes Bezerra; Joaquim Cesar do Nascimento Sousa Junior; Joaquim César do Nascimento Sousa Junior; Pedro Miguel Carneiro Jeronimo; Suzana Porto Almeida & Lucas Delerino on behalf of COVID-19 FIOCRUZ Genomic Network; Suzana Porto Almeida e Lucas Delerino; Thais Ferreira de Oliveira; Thais de Oliveira Costa; Ticiane Cavalcante de Souza; Veridiana Pessoa Miyajima |
| EPI_ISL_2150636 | HOSPITAL GENERAL UNIVERSITARIO DE GUADALAJARA | Instituto de Salud Carlos III | A. Monzón; ALEJANDRO; F. Casas; I. Jiménez; I.GONZALEZ PRAETORIUS; M. Sandonís; P. Zaballos; S. Cuesta; S. Iglesias-Caballero; S. Pozo; S. Varona; V. Camarero; Vázquez-Morón |
| EPI_ISL_5530076, EPI_ISL_5530077 | HOSPITAL JOSE MARIA PHILOMENO GOMES | Analytical Competence Molecular Epidemiology Lab/ACME, Oswaldo | Carlos Leonardo de Aragao Araujo; Cecilia Leite Costa & Eduardo Ruback dos Santos on behalf of COVID-19 FIOCRUZ Genomic Network; Cleber Furtado Aksenen; Fabio Miyajima; Fernando Braga Stehling; Francisco Eder de Moura Lopes; Igor Oliveira Duarte; Jamille Maria Mendes Bezerra; Joaquim Cesar do Nascimento Sousa Junior; Pedro Miguel Carneiro Jeronimo; Suzana Porto Almeida; Thais Ferreira de Oliveira; Thais de Oliveira Costa; Ticiane Cavalcante de Souza; Veridiana Pessoa Miyajima |

| | | | |
|---|---|---|---|
| | | Cruz Foundation, Ceara (FIOCRUZ CE) | |
| EPI_ISL_1966530 | HOSPITAL MUNICIPAL DR TABAJARA RAMOS | Instituto Butantan / FZEA-USP (Pirassununga) | Antonio Jorge Martins; Bianca Cechetto Carlos. Mendelics: Bibiana Santos; Claudia Renata dos Santos Barros; Cíntia Bittar; David Schlesinger. Hemocentro Ribeirão Preto: Simone Kashima; Debora Botequio Moretti; Elaine Cristina Marqueze; Elaine Vieira dos Santos; Elisangela Chicaroni Mattos; Erika Freitas; Evandra Strazza Rodrigues; Felipe Allan da Silva da Costa; Flavia Aburjaile; Fábio Sossai Possebon; Guilherme Campos; Guilherme Targino Valente; Heidge Fukumasu. USP-Botucatu: Rejane Maria Tommasini Grotto; Helena Lage Ferreira; Instituto Butantan: Dimas Tadeu Covas; Jardelina de Souza Todao Bernardino; Jayme A. Souza-Neto; Jessika Cristina Chagas Lesbon; Jorge A. Petroli Marchesi; José Salvatore Leister Patané; João Paulo Kitajima; João Pessoa Araújo Jr.; Leila Sabrina Ullmann; Loyze Paola Oliveira de Lima; Luiz Aurelio de Campos Crispin. Centro de Genômica Funcional da ESALQ: Luiz Lehmann Coutinho; Luiz Carlos Junior de Alcantara; Lívia Sacchetto; Maisa C. Pereira Parra; Maria Carolina Elias; Marta Giovanetti; Marília Moraes; Maurício Lacerda Nogueira. Prefeitura de Sao Paulo: Melissa Palmieri.; Patricia Akemi Assato; Paula Rahal; Paulo Inacio da Costa; Rafael dos Santos Bezerra; Raquel de Lello Rocha Campos Cassano. NGS Soluções Genômicas: Pilar Drummond Sampaio Corrêa Mariani. FZEA-USP Pirassununga: Mirele Daiana Poleti; Raul Machado Neto; Ricardo Augusto Brassaloti; Ricardo Haddad; Rodrigo Tocantins Calado. FAMERP-SJRP: Cecília Artico Banho; Sandra Coccuzzo Sampaio; Svetoslav Nanev Slavov; Vagner Fonseca; Vincent Louis Viala |
| EPI_ISL_5530080, EPI_ISL_5530081, EPI_ISL_5530119, EPI_ISL_5530181 | HOSPITAL RAIMUNDO CELIO RODRIGUES | Analytical Competence Molecular Epidemiology Lab/ACME, Oswaldo Cruz Foundation, Ceara (FIOCRUZ CE) | Carlos Leonardo de Aragao Araujo; Cecilia Leite Costa & Eduardo Ruback dos Santos on behalf of COVID-19 FIOCRUZ Genomic Network; Cleber Furtado Aksenen; Fabio Miyajima; Fernando Braga Stehling; Francisco Eder de Moura Lopes; Igor Oliveira Duarte; Jamille Maria Mendes Bezerra; Joaquim Cesar do Nascimento Sousa Junior; Pedro Miguel Carneiro Jeronimo; Suzana Porto Almeida; Thais Ferreira de Oliveira; Thais de Oliveira Costa; Ticiane Cavalcante de Souza; Veridiana Pessoa Miyajima |
| EPI_ISL_3102423 | HOSPITAL SAO JOSE DE DOENCAS INFECCIOSAS | Analytical Competence Molecular Epidemiology Lab/ACME, Oswaldo Cruz Foundation, Ceara (FIOCRUZ CE) | Cleber Furtado Aksenen; Fabio Miyajima; Fernando Braga Stehling; Francisco Eder de Moura Lopes; Jamille Maria Mendes Bezerra; Joaquim César do Nascimento Sousa Junior; Pedro Miguel Carneiro Jeronimo; Suzana Porto Almeida e Lucas Delerino; Thais Ferreira de Oliveira; Thais de Oliveira Costa; Ticiane Cavalcante de Souza; Veridiana Pessoa Miyajima |
| EPI_ISL_3102283 | HOSPITAL SAO LUCAS | Analytical Competence Molecular Epidemiology Lab/ACME, Oswaldo Cruz Foundation, Ceara (FIOCRUZ CE) | Cleber Furtado Aksenen; Fabio Miyajima; Fernando Braga Stehling; Francisco Eder de Moura Lopes; Jamille Maria Mendes Bezerra; Joaquim César do Nascimento Sousa Junior; Pedro Miguel Carneiro Jeronimo; Suzana Porto Almeida e Lucas Delerino; Thais Ferreira de Oliveira; Thais de Oliveira Costa; Ticiane Cavalcante de Souza; Veridiana Pessoa Miyajima |
| EPI_ISL_3532494 | HOSPITAL UNIVERSITARIO 12 DE OCTUBRE | HOSPITAL UNIVERSITARIO 12 DE OCTUBRE | Carmen Martín-Higuera; Esther Viedma; Irene Múñoz-Gallego; M.ª Dolores Folgueira; Mar Aguilera; Noelia Moral; Rafael Delgado; Sagrario Zurita |
| EPI_ISL_2020179, EPI_ISL_2100319 | HOSPITAL UNIVERSITARIO SON ESPASES | HOSPITAL UNIVERSITARIO SON ESPASES | Antonio Oliver; Carla López-Causapé; Pablo Fraile-Ribot; SeqCovid |
| EPI_ISL_2081109, EPI_ISL_2483430 | Hackensack Medical Center | New York Genome Center | Andre Corvelo; Barry Kreiswirth; David Perlin; Dayna M. Oschwald; Jose Mediavilla; Kaelea Composto; Kar Chow; Liang Chen; Marcus Cunningham; Michael Zody; Samantha Fennessey; Tom Maniatis |
| EPI_ISL_1580904, EPI_ISL_1581105, EPI_ISL_1581109, EPI_ISL_1592620, EPI_ISL_1592626, EPI_ISL_1592630, EPI_ISL_1592644, EPI_ISL_1592650, EPI_ISL_1592678, EPI_ISL_1592791, EPI_ISL_1592803, EPI_ISL_1592815, EPI_ISL_1592825, EPI_ISL_1592849, EPI_ISL_1592942, EPI_ISL_1593008, EPI_ISL_1593020, EPI_ISL_1593063, EPI_ISL_1593159, EPI_ISL_1593163, EPI_ISL_1593172, EPI_ISL_1615255, EPI_ISL_1615308, EPI_ISL_1615322, EPI_ISL_1615326, EPI_ISL_1615327, EPI_ISL_1615338, EPI_ISL_1615350, EPI_ISL_1615371, EPI_ISL_1615495, EPI_ISL_1615502, EPI_ISL_1615524, EPI_ISL_1678891, EPI_ISL_1678906, EPI_ISL_1678910, EPI_ISL_1678958, EPI_ISL_1678959, EPI_ISL_1678970, EPI_ISL_1678978, EPI_ISL_1679006, EPI_ISL_1679025, EPI_ISL_1679026, EPI_ISL_1679027, EPI_ISL_1679031, EPI_ISL_1679040, EPI_ISL_1679074, EPI_ISL_1679111, EPI_ISL_1679117, EPI_ISL_1679121, EPI_ISL_1679139, EPI_ISL_1679177, EPI_ISL_1679340, EPI_ISL_1679365, EPI_ISL_1679498, EPI_ISL_1679505, EPI_ISL_1679506, EPI_ISL_1679551, EPI_ISL_1679573, EPI_ISL_1679579, EPI_ISL_1679595, EPI_ISL_1679599, EPI_ISL_1679623, EPI_ISL_1679674, EPI_ISL_1679764, EPI_ISL_1679841, EPI_ISL_1679862, EPI_ISL_1679868, EPI_ISL_1679881, EPI_ISL_1679942, EPI_ISL_1679959, EPI_ISL_1679973, EPI_ISL_1680144, EPI_ISL_1680153, EPI_ISL_1680155, EPI_ISL_1680182, EPI_ISL_1680266, EPI_ISL_1680325, EPI_ISL_1690332, EPI_ISL_1690338, EPI_ISL_1690405, EPI_ISL_1690441, EPI_ISL_1690458, EPI_ISL_1690498, EPI_ISL_1690564, EPI_ISL_1690580, EPI_ISL_1690619, EPI_ISL_1690638, EPI_ISL_1690718, EPI_ISL_1690748, EPI_ISL_1690815, EPI_ISL_1690877, EPI_ISL_1690934, EPI_ISL_1690982, EPI_ISL_1690996, EPI_ISL_1691026, EPI_ISL_1691032, EPI_ISL_1691069, EPI_ISL_1691074, EPI_ISL_1691081, EPI_ISL_1691095, EPI_ISL_1691114, EPI_ISL_1691123, EPI_ISL_1691133, EPI_ISL_1691196, EPI_ISL_1691236, EPI_ISL_1691254, EPI_ISL_1691348, EPI_ISL_1691353, EPI_ISL_1701835, EPI_ISL_1701843, EPI_ISL_1701864, EPI_ISL_1734824, EPI_ISL_1734912, EPI_ISL_1734916, EPI_ISL_1734946, EPI_ISL_1734985, EPI_ISL_1734993, EPI_ISL_1753502, EPI_ISL_4346653 | | | |
| see above | Helix/Illumina | Centers for Disease Control and Prevention Division of Viral Diseases, Pathogen Discovery | Adrian Paskey; Alexandre Bolze; Ary Ascencio; Benjamin Rambo-Martin; Brad Sickler; Charlotte Rivera-Garcia; Christine Tran; Christopher Gulvick; Chrstine Tran; Clinton Paden; Clinton R. Paden; Dakota Howard; Darlene Wagner; David Becker; Dhwani Batra; Duncan MacCannell; Efren Sandoval; Eileen De Feo; Eileen de Feo; Elizabeth Cirulli; Eric Allen; Geraint Levan; James Lu; Jan Antico; Jason Caravas; Jason Nguyen; Jimmy Ramirez; Jingtao Liu; Kara Moser; Kelly Barrett; Kelly Schiabor Barrett; Kim Gietzen; Kristine Lacek; Magnus Isaksson; Marc Laurent; Matthew Schmerer; Matthew Tolentino; Nicole L. Washington; Nicole Washington; Peter Cook; Peter W. Cook; Phil Febbo; Ryan Cho; Scott Sammons; Shannon Wickline; Shatavia Morrison; Sherry Wang; Simon White; Tyler Cassens; William Lee; Yvette Unoarumhi |
| EPI_ISL_5802193 | Hosp E Maternidade Municipal N Sra Monte Serrat | Instituto Butantan | Antonio Jorge Martins; Claudia Renata dos Santos Barros; David Schlesinger; Debora Botequio Moretti; Dimas Tadeu Covas; Elaine Cristina Marqueze; Elaine Vieira Santos; Evandra Strazza Rodrigues; Heidge Fukumasu; Jayme Augusto de Souza-Neto; José Salvatore Leister Patané; Luiz Alcantara; Luiz Lehmann Coutinho; Maria Carolina Elias; Maurício Lacerda Nogueira; Rafael dos Santos Bezerra; Raul Machado Neto; Rejane Maria Tommasini Grotto; Ricardo Haddad; Sandra Coccuzzo Sampaio Vessoni; Simone Kashima; Svetoslav Nanev Slavov; Vincent Louis Viala |
| EPI_ISL_1583049, EPI_ISL_1621822, EPI_ISL_1675020 | Hospital | National Reference Center for Viruses of Respiratory Infections, Institut Pasteur, Paris | Adrien Pain; Angela Brisebarre; Camille Capel; Christophe Malabat; Corinne Maufrais; Eleha Guillotel; Etienne Simon-Lorière; Frédéric Lemoine; JéRome Guinard; Laurence Louvet; Louise Lefrançois; Marion Barbet; Maud Vanpeene; Méline Bizard; Pierre Lechat; Sylvie Behillil; Sylvie Van der Werf; Sylvie van der Werf; Vincent Enouf |
| EPI_ISL_1673295 | Hospital Alvarez Buylla | Laboratorio de Virología HUCA | Abreu F; Alvarez-Arguelles ME; Boga JA; Castelló C; Costales I; Coto E; Gómez de Oña J; Martín-Rodríguez G; Melón S; Perez-Martínez Z; Rojo S; Sandoval M |
| EPI_ISL_1511207, EPI_ISL_1524837, EPI_ISL_1524838, EPI_ISL_1524839, EPI_ISL_1524840, EPI_ISL_1524841, EPI_ISL_1524842 | | | |
| see above | Hospital General Universitario Gregorio Marañón | Hospital General Universitario Gregorio Marañón | Cristina Rodriguez-Grande; Darío García de Viedma; Laura Pérez-Lago; Patricia Muñoz; Pedro Sola Campoy; Pilar Catalán; Sergio Buenestado Serrano |
| EPI_ISL_2363562 | Hospital Jaime Ferre - SAMCO Rafaela | Grupo de Genómica y Bioinformática del Instituto de Investigación de la Cadena Láctea CONICET-INTA on behalf of 'Proyecto Argentino Interinstitucional de genomica de SARS-CoV-2' (PAIS Consortium) | AF; Amadio; C; Eberhardt; Irazoqui; Isaia; JF; JM; MF; Pandolfi; Quaranta; V |
| EPI_ISL_3031345, EPI_ISL_3031346 | Hospital Metropolitano Dr. Célio de Castro | Instituto René Rachou / Fiocruz Minas | Anna Salim; Cristina Fonseca; Gabriel Fernandes; Mariana Melo; Núbia Fernandes; Pedro Alves; Rosiane Pereira; Rubens do Monte Neto; Sandra Gava; Thaís Santos; Thaís Silva; Wilma Patrícia Bernardes |
| EPI_ISL_1731577 | Hospital Municipal Dr Mario Gatti Campinas | Instituto Adolfo Lutz, Interdiciplinary Procedures Center, Strategic Laboratory | Caio Vinicius Dias Lopes; Claudia Regina Gonçalves; Claudio Tavares Sacchi; Erica Valessa Ramos Gomes; Karoline Rodrigues Campos; Katia Correa de Oliveira Santos; Leonardo Jose Tadeu de Araujo |
| EPI_ISL_5802117 | Hospital Municipal Dr Tabajara Ramos | Instituto Butantan | Antonio Jorge Martins; Claudia Renata dos Santos Barros; David Schlesinger; Debora Botequio Moretti; Dimas Tadeu Covas; Elaine Cristina Marqueze; Elaine Vieira Santos; Evandra Strazza Rodrigues; Heidge Fukumasu; Jayme Augusto de Souza-Neto; José Salvatore Leister Patané; Luiz Alcantara; Luiz Lehmann Coutinho; Maria Carolina Elias; Maurício Lacerda Nogueira; Rafael dos Santos Bezerra; Raul Machado Neto; Rejane Maria Tommasini Grotto; Ricardo Haddad; Sandra Coccuzzo Sampaio Vessoni; Simone Kashima; Svetoslav Nanev Slavov; Vincent Louis Viala |
| EPI_ISL_1675330 | Hospital Pablo Tobón Uribe | Universidad Nacional de Colombia - Laboratorio Genómico One Health | Andres F. Cardona-Rios; Carlos Franco-Muñoz; Daniel O. Maldonado-Perez; Diego A. Álvarez-Díaz; Hector Alejandro Ruiz-Moreno; Idabely Betancur Ortiz; Jorge E. Osorio; Juan P. Hernandez-Ortiz; Karl A Ciuoderis; Katherine Laiton-Donato; Laura Silvana Perez; Lina M. Hurtado; Marcela Mercado-Reyes; Maria Angélica Maya; Maria Stella López; Rita Almanza Payares; Sandra Ines Cano; Simón Villegas Velásquez |
| EPI_ISL_3022632, EPI_ISL_3022646, EPI_ISL_3022647, EPI_ISL_3022655, EPI_ISL_3022745, EPI_ISL_3022749, EPI_ISL_3022750 | | | |
| see above | Hospital of the University of Pennsylvania Molecular Pathology Lab | Bushman Lab - University of Pennsylvania | Aoife M. Roche; Arupa Ganguly; Ayannah S. Fitzgerald; Brendan Kelly; Jevon Graham-Wooten; John K. Everett; Kyle Rodino; Layla A. Khatib; Mike Feldman; Ronald G. Collman and Frederic Bushan; Samantha A. Whiteside; Scott Sherrill-Mix; Shantan Reddy; Young Hwang |
| EPI_ISL_2205669, EPI_ISL_2212042 | Houston Methodist Hospital | Houston Methodist Hospital | Ilya J. Finkelstein; James J. Davis; Jessica Cambric; Jimmy Gollihar; Kristina Reppond; Layne Pruitt; Madison N. Shyer; Marcus Nguyen; Matthew Ojeda Saavedra; Paul A. Christensen; Prasanti Yerramilli; Randall J. Olsen; Robert Olson; Ryan Gadd; S. Wesley Long; Sishir Subedi; and James M. Musser |
| EPI_ISL_1755809 | Hôpital Avicenne | Department of Virology, Henri Mondor University Hospital, Assistance Publique Hôpitaux de Paris, Université Paris-Est Créteil, INSERM U955 | Alexandre Soulier; Christophe Rodriguez; Elisabeth Trawinski; Guillaume Gricourt; Jean-Michel Pawlotsky; Melissa N'Debi; Slim Fourati; Vanessa Demontant |
| EPI_ISL_1755783 | Hôpital Paul Brousse | Department of Virology, Henri Mondor University Hospital, Assistance Publique Hôpitaux de Paris, Université Paris-Est Créteil, INSERM U955 | Alexandre Soulier; Christophe Rodriguez; Elisabeth Trawinski; Guillaume Gricourt; Jean-Michel Pawlotsky; Melissa N'Debi; Slim Fourati; Vanessa Demontant |
| EPI_ISL_2614555 | IAL Presidente Prudente | Instituto Adolfo Lutz, Interdiciplinary Procedures Center, Strategic Laboratory | Caio Vinicius Dias Lopes; Claudia Regina Gonçalves; Claudio Tavares Sacchi; Erica Valessa Ramos Gomes; Karoline Rodrigues Campos; Leonardo Jose Tadeu de Araujo |
| EPI_ISL_3368625, EPI_ISL_3464691 | IEC- Instituto Evandro Chagas | ITV-Vale Institute of Technology | Amanda Vidal; Guilherme Oliveira; Mirleide Cordeiro dos Santos; Tatianne Costa Negri |
| EPI_ISL_2234873, EPI_ISL_2234874, EPI_ISL_2234875, EPI_ISL_2234904, EPI_ISL_2444801 | IICS-UNA | IICS-UNA | Adriana Valenzuela; Alejandra Rojas; Chyntia Diaz; Eva Nara; Fatima Cardozo; Florencia del Puerto; Joel Ortiz; Jonas Fernandez; Laura Franco; Laura Mendoza; Leticia Rojas; Magaly Martinez; Maria Eugenia Galeano. |
| EPI_ISL_1798496, EPI_ISL_1798508 | IL Dept. of Public Health Springfield Laboratory | Centers for Disease Control and Prevention Division of Viral Diseases, Pathogen Discovery | Alison Laufer Halpin; Ben L. Rambo-Martin; Clinton R. Paden; Dakota Howard; Darlene Wagner; Dave Wentworth; Dhwani Batra; Jasmine Padilla; Justin Lee; Katie Dillon; Krista Queen; Kristen Knipe; Kristine Lacek; Mark Burroughs; Matthew Schmerer; Mili Sheth; Peter Cook; Sam Shepard; Sarah Nobles; Shoshona Le; Suxiang Tong; Vivien Dugan; Yvette Unoarumhi |
| EPI_ISL_1626585 | IN State Department of Health Laboratory Services | IN State Department of Health Laboratory Services | Ankita Kashikar; Brian Pope; Cassandra Campion; Jamie Yeadon; Kyle Brownlee; Lixia Liu; Mark Glazier; Melissa Hindenlang |
| EPI_ISL_2716751 | IRCCS San Gallicano Dermatological Institute | IRCCS Regina Elena National Cancer Institute | Aldo Morrone; Alice Massacci; Fabrizio Ensoli; Francesca Sivori; Fulvia Pimpinelli; Gennaro Ciliberto; Giovanni Blandino; Maurizio Fanciulli; Sabrina Strano; Sara Donzelli |
| EPI_ISL_2335108 | IVY3 Central Lab, Vanderbilt University Medical Center | Lauring Lab, University of Michigan, Department of Microbiology and Immunology | Gilbert |
| EPI_ISL_1663079, EPI_ISL_1663095, EPI_ISL_1663107, EPI_ISL_1663111, EPI_ISL_1663114 | Illinois Department of Public Health | Illinois Department of Public Health - Chicago Lab | Ira Heimler; Vineet K. Dhiman |

| | | | |
|---|---|---|---|
| EPI_ISL_1478912, EPI_ISL_1500174, EPI_ISL_1500175, EPI_ISL_1500178, EPI_ISL_1500190, EPI_ISL_1500192, EPI_ISL_1500193, EPI_ISL_1500200, EPI_ISL_1500209, EPI_ISL_1500210, EPI_ISL_1500220, EPI_ISL_1500221, EPI_ISL_1500223, EPI_ISL_1500224, EPI_ISL_1500225, EPI_ISL_1500228, EPI_ISL_1500233, EPI_ISL_1500234, EPI_ISL_1500238, EPI_ISL_1500243, EPI_ISL_1500244, EPI_ISL_1553174, EPI_ISL_1553175, EPI_ISL_1553176, EPI_ISL_1553179, EPI_ISL_1553180, EPI_ISL_1553184, EPI_ISL_1553185, EPI_ISL_1553227, EPI_ISL_1553228, EPI_ISL_1553230, EPI_ISL_1553234, EPI_ISL_1553236, EPI_ISL_1553237, EPI_ISL_1553238, EPI_ISL_1553239, EPI_ISL_1553240, EPI_ISL_1553241, EPI_ISL_1553242, EPI_ISL_1553243, EPI_ISL_1595558, EPI_ISL_1595559, EPI_ISL_1595560, EPI_ISL_1624098, EPI_ISL_1624104, EPI_ISL_1624111, EPI_ISL_1624112, EPI_ISL_1624115, EPI_ISL_1624117, EPI_ISL_1624121, EPI_ISL_1624122, EPI_ISL_1624123, EPI_ISL_1624124, EPI_ISL_1624125, EPI_ISL_1624126, EPI_ISL_1624142, EPI_ISL_1708270, EPI_ISL_1708274, EPI_ISL_1708280, EPI_ISL_1708281, EPI_ISL_1708282, EPI_ISL_1708284, EPI_ISL_1708285, EPI_ISL_1708286, EPI_ISL_1708287, EPI_ISL_1732601, EPI_ISL_1732606, EPI_ISL_1732609, EPI_ISL_1732616, EPI_ISL_1789200, EPI_ISL_1938521 | | | |
| see above | Illinois Department of Public Health - Springfield Lab | Illinois Department of Public Health - Springfield Lab | Bryan Sim; Gordon McCall |
| EPI_ISL_1628603 | InDRE | Instituto Nacional de Medicina Genomica | Alcaraz N; Canseco Mendez JC; Cedro-Tanda A; Garcia-Cardenas FJ; Gisela Barrera-Badillo; Gonzalez-Barrera D; Gonzalez-Woge MA; Herrera-Montalvo LA.; Hidalgo-Miranda A; Irma Lopez-Martinez; Jose Ernesto Ramirez González; Mendoza-Vargas A; Miranda-Ortiz H; Munguia-Garza P; Ramirez-Vega O; Rangel-DeLeon D; Reyes-Grajeda JP; Rosas-Escobar P |
| EPI_ISL_1517432, EPI_ISL_1517433 | Inciensa, Instituto Costarricense de Investigación y Enseñanza en Nutrición y Salud | Inciensa, Instituto Costarricense de Investigación y Enseñanza en Nutrición y Salud | Barboza-Arguedas E & Centeno-Miranda M; Cristian Peréz-Corrales; Cristian Peréz-Corrales & Barboza-Arguedas E |
| EPI_ISL_1587699, EPI_ISL_1587982, EPI_ISL_1588021, EPI_ISL_1588171, EPI_ISL_1588286, EPI_ISL_1693958, EPI_ISL_1693959, EPI_ISL_1694006, EPI_ISL_1694045, EPI_ISL_1694068, EPI_ISL_1694128, EPI_ISL_1694246, EPI_ISL_1694277, EPI_ISL_1694289, EPI_ISL_1694297, EPI_ISL_1694376, EPI_ISL_1694404, EPI_ISL_1694474, EPI_ISL_1694530, EPI_ISL_1732753, EPI_ISL_1732776, EPI_ISL_1732822, EPI_ISL_1732836, EPI_ISL_1732892, EPI_ISL_1733024, EPI_ISL_1753468, EPI_ISL_1802805, EPI_ISL_4383480, EPI_ISL_4383881, EPI_ISL_4384121, EPI_ISL_4384499, EPI_ISL_4384688, EPI_ISL_4385336, EPI_ISL_4385433, EPI_ISL_4385451, EPI_ISL_4386847, EPI_ISL_4387368, EPI_ISL_4387388, EPI_ISL_4387428, EPI_ISL_4387434, EPI_ISL_4388259, EPI_ISL_4388312, EPI_ISL_4389529, EPI_ISL_4390341 | | | |
| see above | Infinity Biologix | Centers for Disease Control and Prevention Division of Viral Diseases, Pathogen Discovery | Adrian Paskey; Benjamin Rambo-Martin; Chirayu Goswami; Christian Bixby; Christopher Gulvick; Clinton Paden; Clinton R. Paden; Dakota Howard; Darlene Wagner; Dhwani Batra; Duncan MacCannell; Erisa Sula; Jason Caravas; Jonathan Schultz; Kara Moser; Kristine Lacek; Matthew Schmerer; Peter Cook; Peter W. Cook; Robin Grimwood; Russ Hager; Scott Sammons; Shatavia Morrison; Tymeckia Kendall; Victoria Caban Figueroa; Yihe Wang; Yvette Unoarumhi |
| EPI_ISL_1656147, EPI_ISL_1805707, EPI_ISL_2361456, EPI_ISL_2361457 | Institute of Microbiology, Universidad San Francisco de Quito | Institute of Microbiology, Universidad San Francisco de Quito | Belén Prado-Vivar; Bernardo Gutiérrez; Fernada Zurita; Freddy Saldarriaga Mera; Freddy Saldarriaga-Mera; Gabriel Trueba; Guzmán Bernabéu Lorenzo; Juan José Guadalupe; Mayra Perero Intriago; Michelle Grunauer; Miguel Sacoto Mazini; Monica Becerra-Wong; Patricio Rojas-Silva; Paúl Cárdenas; Sully Márquez; Verónica Barragán |
| EPI_ISL_1715146, EPI_ISL_1715147, EPI_ISL_1731578, EPI_ISL_1731579, EPI_ISL_1731580, EPI_ISL_1821214, EPI_ISL_1821215, EPI_ISL_1821216, EPI_ISL_1821217, EPI_ISL_1821218, EPI_ISL_1821219, EPI_ISL_1821220, EPI_ISL_1821221, EPI_ISL_1821222, EPI_ISL_1821223, EPI_ISL_1821224 | | | |
| see above | Instituto Adolfo Lutz - Regional de Aracatuba | Instituto Adolfo Lutz, Interdiciplinary Procedures Center, Strategic Laboratory | Caio Vinicius Dias Lopes; Claudia Regina Gonçalves; Claudio Tavares Sacchi; Erica Valessa Ramos Gomes; Karoline Rodrigues Campos; Katia Correa de Oliveira Santos; Leonardo Jose Tadeu de Araujo |
| EPI_ISL_2691121 | Instituto Adolfo Lutz - Regional de Bauru | Instituto Adolfo Lutz, Interdiciplinary Procedures Center, Strategic Laboratory | Caio Vinicius Dias Lopes; Claudia Regina Gonçalves; Claudio Tavares Sacchi; Erica Valessa Ramos Gomes; Karoline Rodrigues Campos; Leonardo Jose Tadeu de Araujo |
| EPI_ISL_2003162, EPI_ISL_2003163, EPI_ISL_2003164, EPI_ISL_2003167, EPI_ISL_2003168 | Instituto Adolfo Lutz - Regional de Campinas | Instituto Adolfo Lutz, Interdiciplinary Procedures Center, Strategic Laboratory | Caio Vinicius Dias Lopes; Claudia Regina Gonçalves; Claudio Tavares Sacchi; Erica Valessa Ramos Gomes; Karoline Rodrigues Campos; Leonardo Jose Tadeu de Araujo |
| EPI_ISL_2691106 | Instituto Adolfo Lutz - Regional de Marilia | Instituto Adolfo Lutz, Interdiciplinary Procedures Center, Strategic Laboratory | Caio Vinicius Dias Lopes; Claudia Regina Gonçalves; Claudio Tavares Sacchi; Erica Valessa Ramos Gomes; Karoline Rodrigues Campos; Leonardo Jose Tadeu de Araujo |
| EPI_ISL_1715148, EPI_ISL_1731582, EPI_ISL_1821246, EPI_ISL_1821247, EPI_ISL_1821248, EPI_ISL_1821249, EPI_ISL_1821251, EPI_ISL_1821252, EPI_ISL_1821254, EPI_ISL_1821255, EPI_ISL_1821258, EPI_ISL_1821259, EPI_ISL_1821261, EPI_ISL_1821262, EPI_ISL_1821264 | | | |
| see above | Instituto Adolfo Lutz - Regional de Ribeirao Preto | Instituto Adolfo Lutz, Interdiciplinary Procedures Center, Strategic Laboratory | Caio Vinicius Dias Lopes; Claudia Regina Gonçalves; Claudio Tavares Sacchi; Erica Valessa Ramos Gomes; Karoline Rodrigues Campos; Katia Correa de Oliveira Santos; Leonardo Jose Tadeu de Araujo |
| EPI_ISL_2003144, EPI_ISL_2003145 | Instituto Adolfo Lutz - Regional de Santos | Instituto Adolfo Lutz, Interdiciplinary Procedures Center, Strategic Laboratory | Caio Vinicius Dias Lopes; Claudia Regina Gonçalves; Claudio Tavares Sacchi; Erica Valessa Ramos Gomes; Karoline Rodrigues Campos; Leonardo Jose Tadeu de Araujo |
| EPI_ISL_2958840 | Instituto Adolfo Lutz - Regional de Sorocaba | Instituto Adolfo Lutz, Interdiciplinary Procedures Center, Strategic Laboratory | Caio Vinicius Dias Lopes; Claudia Regina Gonçalves; Claudio Tavares Sacchi; Erica Valessa Ramos Gomes; Karoline Rodrigues Campos |
| EPI_ISL_1715149, EPI_ISL_1715150, EPI_ISL_1715151, EPI_ISL_1715152, EPI_ISL_1715153, EPI_ISL_1731583, EPI_ISL_1731584, EPI_ISL_1731585, EPI_ISL_1731586, EPI_ISL_1731587, EPI_ISL_1731588, EPI_ISL_1731604, EPI_ISL_1731605, EPI_ISL_1731607, EPI_ISL_1731608, EPI_ISL_1731609, EPI_ISL_1731610, EPI_ISL_1731611, EPI_ISL_1731612, EPI_ISL_1821266, EPI_ISL_1821267, EPI_ISL_1821268, EPI_ISL_1821270, EPI_ISL_1821271, EPI_ISL_1821272, EPI_ISL_1821273, EPI_ISL_2003119, EPI_ISL_2003120, EPI_ISL_2003121, EPI_ISL_2003122, EPI_ISL_2003123, EPI_ISL_2003124, EPI_ISL_2003125, EPI_ISL_2003126, EPI_ISL_2003127, EPI_ISL_2003128, EPI_ISL_2003129, EPI_ISL_2003130, EPI_ISL_2756459, EPI_ISL_2756461, EPI_ISL_2756466, EPI_ISL_2756472 | | | |
| see above | Instituto Adolfo Lutz Central | Instituto Adolfo Lutz, Interdiciplinary Procedures Center, Strategic Laboratory | Caio Vinicius Dias Lopes; Claudia Regina Gonçalves; Claudio Tavares Sacchi; Erica Valessa Ramos Gomes; Karoline Rodrigues Campos; Katia Correa de Oliveira Santos; Leonardo Jose Tadeu de Araujo |
| EPI_ISL_1585916, EPI_ISL_1585918, EPI_ISL_1585919 | Instituto Nacional de Medicina Genomica | Instituto Nacional de Medicina Genomica | Alcaraz N; Canseco Mendez JC; Cedro-Tanda A; Garcia-Cardenas FJ; Gonzalez-Barrera D; Gonzalez-Woge MA; Herrera-Montalvo LA; Hidalgo-Miranda A; Mendoza-Vargas A; Miranda-Ortiz H; Munguia-Garza P; Ramirez-Vega O; Rangel-DeLeon D; Reyes-Grajeda JP; Rosas-Escobar P |
| EPI_ISL_1734843, EPI_ISL_1734844, EPI_ISL_1911958, EPI_ISL_2295388, EPI_ISL_2295389, EPI_ISL_2308609, EPI_ISL_3118809, EPI_ISL_3118811, EPI_ISL_3118813, EPI_ISL_3118814 | | | |
| see above | Instituto de Biotecnologia - UNESP-Botucatu-SP | Instituto de Biotecnologia - UNESP-Botucatu-SP | Cecília Artico Banho; Cíntia Bittar; Fábio Sossai Possebon; Guilherme Campos; Helena Lage Ferreira; Jorge A. Petroli Marchesi; João Pessoa Araújo Jr.; Leila Sabrina Ullmann; Lívia Sacchetto; Maisa C. Pereira Parra; Marília Moraes; Maurício L. Nogueira; Paula Rahal; Paulo Inacio da Costa |
| EPI_ISL_2776166 | Instituto de Medicina Tropical & Salud Global (IMTSAG) | Grubaugh Lab - Yale School of Public Health | Alejandaro Vallejo Degaudenzi; Anderson Brito; Annie Watkins; Chaney Kalinich; Chantal Vogels; Elisa Contreras; Esperanza Mendoza; Isabel Ott; Jessica Rothman; Joseph Fauver; Kendall Billig; Mallery Breban; Mary Petrone; Nathan Grubaugh; Robert Paulino-Ramirez; Tara Alpert; Tobias Koch; Victor Virgilio Calderon |
| EPI_ISL_2691149 | Intituto Adolfo Lutz - Regional de Sorocaba | Instituto Adolfo Lutz, Interdiciplinary Procedures Center, Strategic Laboratory | Caio Vinicius Dias Lopes; Claudia Regina Gonçalves; Claudio Tavares Sacchi; Erica Valessa Ramos Gomes; Karoline Rodrigues Campos; Leonardo Jose Tadeu de Araujo |
| EPI_ISL_1669921, EPI_ISL_1670662 | Istituto Zooprofilattico Sperimentale Umbria e Marche "Togo Rosati" | Istituto Zooprofilattico Sperimentale dell'Abruzzo e Molise "G. Caporale" | Ancora M; Biagetti M; Calistri P; Cammà C; Caporale M; Curini V; Delli Compagni E; Di Domenico M; Di Lollo Valeria; Di Pasquale A; Giammarioli M; Lorusso A; Mangone I; Marcacci M; Puglia I; Rinaldi A; Savini G; Scialabba S |
| EPI_ISL_1630112, EPI_ISL_1630135, EPI_ISL_1630146, EPI_ISL_1656404, EPI_ISL_1656405, EPI_ISL_1673539, EPI_ISL_1673540, EPI_ISL_1673541, EPI_ISL_1673542, EPI_ISL_1673543, EPI_ISL_1673544, EPI_ISL_1673595 | | | |
| see above | Istituto Zooprofilattico Sperimentale del Mezzogiorno | TIGEM | Antonio Grimaldi Patrizia Annunziata Francesco Panariello Biancamaria Pierri Claudia Tiberio Teresa Giuliano Valentina Bouche Chiara Colantuono Maria Concetta Cuomo Denise Di Concilio Lucio Di Filippo Anna Manfredi Marcello Salvi Antonio Limone Luigi Atripaldi Pellegrino Cerino Andrea Ballabio Davide Cacchiarelli |
| EPI_ISL_1632915 | Istituto Zooprofilattico Sperimentale del Mezzogiorno | Telethon Institute of Genetics and Medicine (TIGEM) | Antonio Grimaldi Patrizia Annunziata Francesco Panariello Biancamaria Pierri Claudia Tiberio Teresa Giuliano Valentina Bouche Chiara Colantuono Maria Concetta Cuomo Denise Di Concilio Lucio Di Filippo Anna Manfredi Marcello Salvi Antonio Limone Luigi Atripaldi Pellegrino Cerino Andrea Ballabio Davide Cacchiarelli |
| EPI_ISL_1759563 | Istituto Zooprofilattico Sperimentale del Mezzogiorno - Azienda Ospedaliera "Pugliese Ciaccio" di Catanzaro | TIGEM | Antonio Grimaldi Patrizia Annunziata Francesco Panariello Biancamaria Pierri Claudia Tiberio Teresa Giuliano Valentina Bouche Chiara Colantuono Maria Concetta Cuomo Denise Di Concilio Lucio Di Filippo Anna Manfredi Pasquale Minchella Marcello Salvi Antonio Limone Luigi Atripaldi Pellegrino Cerino Andrea Ballabio Davide Cacchiarelli |
| EPI_ISL_1577672, EPI_ISL_1577741 | Jessa | Jessa | Cruys et al. on behalf of the Jessa_cmdLab |
| EPI_ISL_1468644 | Johns Hopkins Hospital Department of Pathology | Johns Hopkins Hospital Department of Pathology | Adannaya Amadi; C. Paul Morris; Chun Huai Luo; Heba H. Mostafa; Matthew Schwartz |
| EPI_ISL_2404398, EPI_ISL_2404519, EPI_ISL_2404520, EPI_ISL_2404596, EPI_ISL_2404615, EPI_ISL_2404633, EPI_ISL_2404644, EPI_ISL_2404649, EPI_ISL_2404654, EPI_ISL_2424327, EPI_ISL_2424360, EPI_ISL_2424405, EPI_ISL_2424410, EPI_ISL_2424461, EPI_ISL_2425111, EPI_ISL_2425112, EPI_ISL_2425117, EPI_ISL_2425124, EPI_ISL_2425143, EPI_ISL_2425146, EPI_ISL_2425151, EPI_ISL_2425161, EPI_ISL_2425166, EPI_ISL_2425167, EPI_ISL_2425170, EPI_ISL_2425171, EPI_ISL_2425175, EPI_ISL_2425185, EPI_ISL_2425188, EPI_ISL_2425190, EPI_ISL_2425194, EPI_ISL_2425199, EPI_ISL_2425204, EPI_ISL_2425205, EPI_ISL_2425212, EPI_ISL_2425223, EPI_ISL_2425228, EPI_ISL_2425305, EPI_ISL_2425307, EPI_ISL_2840559 | | | |
| see above | KU Leuven, Rega Institute, Clinical and Epidemiological Virology | KU Leuven, Rega Institute, Clinical and Epidemiological Virology | Bert Vanmechelen; Joan Marti-Carerras; Piet Maes; Tony Wawina-Bokalanga |
| EPI_ISL_1732156, EPI_ISL_1732157 | Klinisch Laboratorium GZA | Klinisch Laboratorium ZNA | Verstrepen et al. |
| EPI_ISL_1843740 | Klinisch Laboratorium ZNA | Klinisch Laboratorium ZNA | Verstrepen et al. |
| EPI_ISL_1795100, EPI_ISL_1795105, EPI_ISL_2344549, EPI_ISL_2344554 | LABORATORIO MUNICIPAL DE PIRACICABA | Instituto Butantan / ESALQ-Piracicaba | Antonio Jorge Martins; Bianca Cechetto Carlos. Mendelics: Bibiana Santos; Claudia Renata dos Santos Barros; David Schlesinger; David Schlesinger. Hemocentro Ribeirão Preto: Simone Kashima; Debora Botequio Moretti; Debora Botequio Moretti. Centro de Genômica Funcional da ESALQ: Luiz Lehmann Coutinho; Dimas Tadeu Covas; Elaine Cristina Marqueze; Elaine Vieira Santos; Elaine Vieira dos Santos; Elisangela Chicaroni Mattos; Erika Freitas; Evandra Strazza Rodrigues; Felipe Allan da Silva da Costa; Flavia Aburjaile; Guilherme Targino Valente; Heidge Fukumasu; Heidge Fukumasu. USP-Botucatu: Rejane Maria Tommasini Grotto; Instituto Butantan: Alexander Roberto Precioso; Jayme A. Souza-Neto; Jayme Augusto de Souza-Neto; Jessika Cristina Chagas Lesbon; José Salvatore Leister Patané; João Paulo Kitajima; Luiz Alcantara; Luiz Carlos Junior de Alcantara; Luiz Lehmann Coutinho; Maria Carolina Elias; Marta Giovanetti; Maurício Lacerda Nogueira; Patricia Akemi Assato; Rafael dos Santos Bezerra; Raquel de Lello Rocha Campos Cassano. NGS Soluções Genômicas: Pilar Drummond Sampaio Corrêa Mariani. FZEA-USP Pirassununga: Mirele Daiana Poleti; Raul Machado Neto; Rejane Maria Tommasini Grotto; Ricardo Augusto Brassaloti; Ricardo Haddad; Rodrigo Tocantins Calado.; Sandra Coccuzzo Sampaio; Sandra Coccuzzo Sampaio Vessoni; Simone Kashima; Svetoslav Nanev Slavov; Vagner Fonseca; Vincent Louis Viala |
| EPI_ISL_1966996, EPI_ISL_1966997, EPI_ISL_1966998 | LABORATORIO MUNICIPAL DE PIRACICABA | Instituto Butantan / ESALQ-USP (Piracicaba) | Antonio Jorge Martins; Bianca Cechetto Carlos. Mendelics: Bibiana Santos; Claudia Renata dos Santos Barros; Cíntia Bittar; David Schlesinger. Hemocentro Ribeirão Preto: Simone Kashima; Debora Botequio Moretti; Elaine Cristina Marqueze; Elaine Vieira dos Santos; Elisangela Chicaroni Mattos; Erika Freitas; Evandra Strazza Rodrigues; Felipe Allan da Silva da Costa; Flavia Aburjaile; Fábio Sossai Possebon; Guilherme Campos; Guilherme Targino Valente; Heidge Fukumasu. USP-Botucatu: Rejane Maria Tommasini Grotto; Helena Lage Ferreira; Instituto Butantan: Dimas Tadeu Covas; Jardelina de Souza Todao Bernardino; Jayme A. Souza-Neto; Jessika Cristina Chagas Lesbon; Jorge A. Petroli Marchesi; José Salvatore Leister Patané; João Paulo Kitajima; João Pessoa Araújo Jr.; Leila Sabrina Ullmann; Loyze Paola Oliveira de Lima; Luiz Aurelio de Campos Crispin. Centro de Genômica Funcional da ESALQ: Luiz Lehmann Coutinho; Luiz Carlos Junior de Alcantara; Lívia Sacchetto; Maisa C. Pereira Parra; Maria Carolina Elias; Marta Giovanetti; Marília Moraes; Maurício Lacerda Nogueira. Prefeitura de Sao Paulo: Melissa Palmieri.; Patricia Akemi Assato; Paula Rahal; Paulo Inacio da Costa; Rafael dos Santos Bezerra; Raquel de Lello Rocha Campos Cassano. NGS Soluções Genômicas: Pilar Drummond Sampaio Corrêa Mariani. FZEA-USP Pirassununga: Mirele Daiana Poleti; Raul Machado Neto; Ricardo Augusto Brassaloti; Ricardo Haddad; Rodrigo Tocantins Calado. FAMERP-SJRP: Cecília Artico Banho; Sandra Coccuzzo Sampaio; Svetoslav |

| | | | |
|---|---|---|---|
| | | | Nanev Slavov; Vagner Fonseca; Vincent Louis Viala |
| EPI_ISL_7744028, EPI_ISL_7744032, EPI_ISL_7744042, EPI_ISL_7744044, EPI_ISL_7744116 | LACEN | Laboratório de Bioinformática - Universidade Federal de Santa Catarina | "Aline Daina Schlindwein"; "Ana Paula Christoff"; "Antuani Baptistela"; "Carolina Leite Martins"; "Darcita Buerger Rovaris"; "Dayane Azevedo Padilha"; "Doris Sobral Marques SouzaSobral"; "Edmundo Carlos Grisard"; "Eric Kazuo Kawagoe"; "Fernanda Luiza Ferrari"; "Fernanda Roesene Melo"; "Fernando Hartmann Barazzetti"; "Gislaine Fongaro"; "Glauber Wagner"; "Guilherme Augusto Maia"; "Guilherme Razzera"; "Guilherme Toledo e Silva"; "Julia Kinetz Wachter"; "Luiz Felipe de Oliveira"; "Marcos André Schörne"; "Marcus Vinícius Duarte Rodrigues"; "Maria Luiza Bazzo"; "Marlei Pickler Debiasi dos Anjos"; "Milene Moehr de Moraes"; "Nestor Wendt"; "Patrícia Hermes Stoco"; "Paula Sacchet"; "Renato Simões Moreira"; "Rodrigo de Paula Baptista"; "Tamela Zamboni Madaloz"; "Tatiany Aparecida Teixeira Soratto"; "Vilmar Benetti Filho" |
| EPI_ISL_2488800, EPI_ISL_2488801, EPI_ISL_2488803 | LACEN - Laboratório Central de Saúde Pública do Ceará | Evandro Chagas Institute | A.M.; Barbagelata; E.C.; E.M.A.; Ferreira; J.A.; Junior; K.C.; L.C.; L.S.; M.C.; P.S.; Pinheiro; Santos; Silva; Sousa; Sousa Junior; W.D.C.; da Silva |
| EPI_ISL_2488812 | LACEN - Laboratório Central de Saúde Pública do Rio Grande do Norte | Evandro Chagas Institute | A.M.; Barbagelata; E.C.; E.M.A.; Ferreira; J.A.; Junior; K.C.; L.C.; L.S.; M.C.; P.S.; Pinheiro; Santos; Silva; Sousa; Sousa Junior; W.D.C.; da Silva |
| EPI_ISL_2919228, EPI_ISL_2919230 | LACEN do Estado de Goias | Instituto Adolfo Lutz, Interdiciplinary Procedures Center, Strategic Laboratory | Caio Vinicius Dias Lopes; Claudia Regina Gonçalves; Claudio Tavares Sacchi; Erica Valessa Ramos Gomes; Karoline Rodrigues Campos |
| EPI_ISL_2958889, EPI_ISL_2958890, EPI_ISL_3316173, EPI_ISL_3316187, EPI_ISL_3316209, EPI_ISL_3316210, EPI_ISL_3316218, EPI_ISL_3316224 | | | |
| see above | LACEN do Estado de Mato Grosso | Instituto Adolfo Lutz, Interdiciplinary Procedures Center, Strategic Laboratory | Caio Vinicius Dias Lopes; Claudia Regina Gonçalves; Claudio Tavares Sacchi; Erica Valessa Ramos Gomes; Karoline Rodrigues Campos; Leonardo Tadeu de Araujo; Marlon Benedito Nascomento Santos |
| EPI_ISL_3671917, EPI_ISL_3671918, EPI_ISL_3691386 | LACEN do Estado do Mato Grosso do Sul | Instituto Adolfo Lutz, Interdiciplinary Procedures Center, Strategic Laboratory | Caio Vinicius Dias Lopes; Claudia Regina Gonçalves; Claudio Tavares Sacchi; Karoline Rodrigues Campos; Leonardo Tadeu de Araujo; Marlon Benedito Nascomento Santos |
| EPI_ISL_1960058 | LAS AMERICAS | Universidad Nacional de Colombia - Laboratorio Genómico One Health | Andres F. Cardona-Rios; Carlos Franco-Muñoz; Carolina Muñoz-Arango; Celeny Ortiz; Daniel O. Maldonado-Perez; Diego A. Álvarez-Díaz; Hector Alejandro Ruiz-Moreno; Idabely Betancur Ortiz; Jorge E. Osorio; Juan P. Hernandez-Ortiz; Karl A Ciuoderis; Katherine Laiton-Donato; Laura Silvana Perez; Lina M. Hurtado; Marcela Mercado-Reyes; Maria Angélica Maya; Maria Stella López; Rita Almanza Payares; Sandra Ines Cano; Simón Villegas Velásquez |
| EPI_ISL_3268021 | LATE - Laboratório de Técnicas Especiais - Hospital Israelita Albert Einstein | LATE - Laboratório de Técnicas Especiais - Hospital Israelita Albert Einstein | Alexandre Hideaki Takara; Ana Paula Moreira Salles; Anelisie da Silva Santos; Deyvid Amgarten; Erick Gustavo Dorlass; Fernanda de Mello Malta; João Renato Rebello Pinho; Marcio Anunciacao Menezes; Pedro Henrique Sebe Rodrigues; Raquel Riyuzo |
| EPI_ISL_1960051 | LDSP | Universidad Nacional de Colombia - Laboratorio Genómico One Health | Andres F. Cardona-Rios; Carlos Franco-Muñoz; Carolina Muñoz-Arango; Celeny Ortiz; Daniel O. Maldonado-Perez; Diego A. Álvarez-Díaz; Hector Alejandro Ruiz-Moreno; Idabely Betancur Ortiz; Jorge E. Osorio; Juan P. Hernandez-Ortiz; Karl A Ciuoderis; Katherine Laiton-Donato; Laura Silvana Perez; Lina M. Hurtado; Marcela Mercado-Reyes; Maria Angélica Maya; Maria Stella López; Rita Almanza Payares; Sandra Ines Cano; Simón Villegas Velásquez |
| EPI_ISL_1805503, EPI_ISL_1805504 | LESP Nuevo Leon | Instituto de Diagnostico y Referencia Epidemiologicos (INDRE) | Abril Rodriguez-Maldonado; Ariadna Medina-Benitez; Claudia Wong-Arambula; Ernesto Ramirez-Gonzalez.; Gisela Barrera-Badillo; Irma Lopez-Martinez; Joaquin Quiroz-Mercado; Lucia Hernandez-Rivas; Natividad Cruz-Ortiz; Sergio Rangel-Guerrero; Tatiana Nunez-Garcia; Vanessa Rivero-Arredondo |
| EPI_ISL_2105693 | LESP Quintana Roo | Instituto de Diagnostico y Referencia Epidemiologicos (INDRE) | Abril Rodriguez-Maldonado; Ariadna Medina-Benitez; Claudia Wong-Arambula; Ernesto Ramirez-Gonzalez.; Gisela Barrera-Badillo; Irma Lopez-Martinez; Joaquin Quiroz-Mercado; Lucia Hernandez-Rivas; Natividad Cruz-Ortiz; Sergio Rangel-Guerrero; Tatiana Nunez-Garcia; Vanessa Rivero-Arredondo |
| EPI_ISL_1600188 | LHUB-ULB | Labo Klinische Biologie, UZA | Basil Britto Xavier; Christine Lammens; Herman Goossens; Jasmine Coppens; Marie Le Mercier; Veerle Matheeussen |
| EPI_ISL_1689767, EPI_ISL_1689785, EPI_ISL_1827029, EPI_ISL_1827032, EPI_ISL_1827033, EPI_ISL_1827053, EPI_ISL_1827055, EPI_ISL_1827091, EPI_ISL_1827102, EPI_ISL_1827117, EPI_ISL_1827153 | | | |
| see above | Lab voor klinische biologie | Lab voor klinische biologie | Bruno Verhasselt; Hannelore Hamerlinck; Marija Janevska |
| EPI_ISL_1594450, EPI_ISL_1821633 | Labo Analyses Med | National Reference Center for Viruses of Respiratory Infections, Institut Pasteur, Paris | Amaury Vaysse; Angela Brisebarre; Camille Capel; Christophe Malabat; Corinne Maufrais; Dominique Rousset; Emmanuelle Permal; Etienne Simon-Lorière; Frédéric Lemoine; Louise Lefrançois; Marion Barbet; Maud Vanpeene; Méline Bizard; Pierre-Yves Leonard; Sylvie Behillil; Sylvie Van der Werf; Sylvie van der Werf; Vincent Enouf |
| EPI_ISL_1846786 | Labor Becker & Kollegen (Standort MÃ¼nchen) | Robert Koch Institute | |
| EPI_ISL_1643459 | Labor LÃ¼beck bzw. LaborÃ¤rztliche Gemeinschaftspraxis LÃ¼beck | Robert Koch Institute | |
| EPI_ISL_1627106 | Laboratoire Carage | Institut Pasteur de la Guyane | Anne Lavergne; Dominique Rousset |
| EPI_ISL_2374084, EPI_ISL_2374085 | Laboratoire Virologie Saint Louis APHP | Laboratoire Virologie Saint Louis APHP | Constance Delaugerre; Jérome Le Goff; Linda Feghoul; Marie Laure Chaix; Marie Laure Néré; Maud Salmona; Severine Mercier Delarue; Sophia Achaibou |
| EPI_ISL_3143995, EPI_ISL_3144000, EPI_ISL_3144001, EPI_ISL_3144423, EPI_ISL_3458280, EPI_ISL_3458287, EPI_ISL_3458288, EPI_ISL_3458290, EPI_ISL_3458291, EPI_ISL_3458298, EPI_ISL_3458374, EPI_ISL_3458379, EPI_ISL_3458381, EPI_ISL_3508438, EPI_ISL_3508439, EPI_ISL_3508440, EPI_ISL_5872716, EPI_ISL_5872721, EPI_ISL_5873188, EPI_ISL_5891523, EPI_ISL_5891524, EPI_ISL_5891526, EPI_ISL_5891528, EPI_ISL_5891529, EPI_ISL_5891531, EPI_ISL_5897808, EPI_ISL_7713419, EPI_ISL_7713423, EPI_ISL_7713434, EPI_ISL_7713435, EPI_ISL_7715291, EPI_ISL_7715320 | | | |
| see above | Laboratoire de santé publique du Québec | Laboratoire de santé publique du Québec | Guillaume Bourque; Ioannis Ragoussis; Jesse Shapiro; Mark Lathrop and Judith Fafard on behalf of the CoVSeQ research group; Mark Lathrop and Michel Roger on behalf of the CoVSeQ research group; Sandrine Moreira |
| EPI_ISL_1918027, EPI_ISL_1918248, EPI_ISL_1918271, EPI_ISL_1918272 | Laboratoire national de sante, Microbiology, Virology | Laboratoire national de sante, Microbiology, Microbial Genomics Platform | Anke Wienecke-Baldacchino; Catherine Ragimbeau; Fatu Djabi; Jessica Tapp; Lise Pignon; Raoul Salmon; Tamir Abdelrahman; Trung Nguyen Nguyen |
| EPI_ISL_1917601 | Laboratoires d'analyses medicales - Ketterthill | Laboratoire national de sante, Microbiology, Microbial Genomics Platform | Anke Wienecke-Baldacchino; Caroline Scheiber; Catherine Ragimbeau; Fatu Djabi; Jessica Tapp; Lise Pignon; Raoul Salmon; Serge Vedy; Tamir Abdelrahman |
| EPI_ISL_2289143 | Laboratori de Referencia de Catalunya | Laboratori de Referencia de Catalunya | Bellosillo B.; Canal M.; Hernandez JJ.; Padilla E.; Ramirez A.; Vilas A. |
| EPI_ISL_2557354 | Laboratorio Central de Saude Publica do Estado de Minas Gerais (LACEN/MG) | Laboratory of Respiratory Viruses and Measles, Oswaldo Cruz Institute, FIOCRUZ | Alice Sampaio Rocha; Ana Carolina Mendonca; Andre Felipe Leal Bernardes; Anna Carolina Paixao; Elisa Cavalcante Pereira; Fernando Motta; Luciana Appolinario; Marilda Siqueira on behalf of the Fiocruz COVID-19 Genomic Surveillance Network; Paola Resende; Renata Serrano Lopes; Taina Venas |
| EPI_ISL_2196282, EPI_ISL_2196283, EPI_ISL_2196284, EPI_ISL_2196285, EPI_ISL_2196286 | Laboratorio Central de Saude Publica do Estado do Para (LACEN/PA) | Laboratory of Respiratory Viruses and Measles, Oswaldo Cruz Institute, FIOCRUZ | Alice Sampaio Rocha; Ana Carolina Mendonca; Anna Carolina Paixao; Elisa Cavalcante Pereira; Fernando Motta; Luciana Appolinario; Marilda Siqueira on behalf of the Fiocruz COVID-19 Genomic Surveillance Network; Paola Resende; Renata Serrano Lopes; Taina Venas; Valnete Andrade |
| EPI_ISL_1670639, EPI_ISL_1670640, EPI_ISL_1670641, EPI_ISL_1670642, EPI_ISL_1670643 | Laboratorio Analisi Osp. Città di Castello - Azienda USL Umbria1 | Istituto Zooprofilattico Sperimentale dell'Abruzzo e Molise "G. Caporale" | Ancora M; Calistri P; Cammà C; Caporale M; Curini V; Delli Compagni E; Di Domenico M; Di Lollo Valeria; Di Pasquale A; Lorusso A; Malagigi V; Mangone I; Marcacci M; Puglia I; Rinaldi A; Savini G; Scialabba S; Tacconi P |
| EPI_ISL_2884631 | Laboratorio Biologia molecolare dell'Istituto di Medicina Aerospaziale di Roma | Virology Laboratory, Scientific Department, Army Medical Center | Anella Monte; Anna Anselmo; Antonella Fortunato; Carmelo Campanella; Carmen Nigro; Filippo Molinari; Florigio Lista; Francesco Giordani; Giancarlo Petralito; Giandomenico Cerreto; Giulia Campoli; Lucia Nicosia; Maria Gravina; Marzia Cavalli; Mattia Rencricca; Mirko Tavernese; Raffaele Cresta; Riccardo De Sanctis; Rossella Brandi; Sara Felici.; Silvia Fillo; Tania Pistoni; Vanessa Vera Fain |
| EPI_ISL_1662023 | Laboratorio Central de Epidemiologia (LCE) | Unidad de Genomica Avanzada | Alejandro Sanchez-Flores; Alfredo Herrera-Estrella; Alicia Ocana-Mondragon; Angel Gustavo Salas-Lais; Bernardo Martinez-Miguel; Blanca Taboada; Brenda Irasema Maldonado-Meza; Carla Ivon Herrera-Najera; Carlos F. Arias; Celia Boukadida; Celida Duque Molina; Clara Esperanza Santacruz-Tinoco; Concepcion Grajales-Muniz; Consorcio Mexicano de Vigilancia Genomica (CoViGen-Mex). Authors (in alphabetical order): Julio Elias Alvarado-Yaah; Fernando Fontove-Herrera; Francisco Pulido; Gloria Elena Espinosa-Ayala; Gloria Maria Molina-Salinas; Gloria Vazquez; Hector Esteban Paz-Juarez; Hector Montoya-Fuentes; Helen Haydee Fernanda Ramirez-Plascencia; Jose Antonio Enciso-Moreno; Jose Esteban Munoz-Medina; Jose de Jesus Nunez-Contreras; Juan Bautista Chale-Dzul; Luis Alberto Ochoa-Carrera; Margarita Matias-Florentino; Maria Guadalupe Santiago-Mauricio; Maria Guadalupe de Jesus Mireles-Rivera; Nelly Selem-Mojica; Pavel Isa; Ricardo Grande; Santiago Ávila-Rios; Victor Eduardo Garcia-Arias; Victor Hugo Borja-Aburto |
| EPI_ISL_4518738 | Laboratorio Central de Salud Publica de Paraguay | Fundação Ezequiel Dias | Andre Leal; Andrea Gómez de la Fuente; Cynthia Vazquez; Elaine Cristina; Felipe Iani; Flavia Aburjaile; Gislene Garcia de Castro Lichs; Glauco Carvalho; Hegger Fritsch; Joilson Xavier; Juan Torales; Luiz Alcantara.; Luiz Henrique Ferraz Demarchi; Luiz Takao Watanabe; Marina Castilhos Souza Umaki Zardin; Marta Giovanetti; María José Ortega; María Liz Gamarra; Natalia Guimaraes; Raquel da Silva Ferreira; Shirley Villalba; Talita Adelino; Vagner Fonseca; de Oliveira |
| EPI_ISL_2157519, EPI_ISL_2157520, EPI_ISL_2157521, EPI_ISL_2157522, EPI_ISL_2157523 | Laboratorio Central de Saude Publica do Esatado de Alagoas (LACEN/AL) | Laboratory of Respiratory Viruses and Measles, Oswaldo Cruz Institute, FIOCRUZ | Alice Sampaio Rocha; Ana Carolina Mendonca; Anderson Brandao Leite; Anna Carolina Paixao; Elisa Cavalcante Pereira; Fernando Motta; Luciana Appolinario; Marilda Siqueira on behalf of the Fiocruz COVID-19 Genomic Surveillance Network; Paola Resende; Renata Serrano Lopes; Taina Venas |
| EPI_ISL_2536326, EPI_ISL_2536328, EPI_ISL_2536329, EPI_ISL_2536331, EPI_ISL_2536346, EPI_ISL_2536347, EPI_ISL_2536348, EPI_ISL_3061884 | | | |
| see above | Laboratorio Central de Saude Publica do Estado da Paraiba (LACEN-PB) | Laboratory of Respiratory Viruses and Measles, Oswaldo Cruz Institute, FIOCRUZ | Alice Sampaio Rocha; Ana Carolina Mendonca; Anna Carolina Paixao; Dalane Loudal Florentino Teixeira; Elisa Cavalcante Pereira; Fernando Motta; Joao Felipe Bezerra; Luciana Appolinario; Marilda Siqueira on behalf of the Fiocruz COVID-19 Genomic Surveillance Network; Paola Resende; Renata Serrano Lopes; Taina Venas |
| EPI_ISL_2274053, EPI_ISL_2274055, EPI_ISL_2274056, EPI_ISL_2274067, | Laboratorio Central de Saude Publica do Estado de Alagoas (LACEN/AL) | Laboratory of Respiratory Viruses and Measles, Oswaldo Cruz Institute, FIOCRUZ | Alice Sampaio Rocha; Ana Carolina Mendonca; Anderson Brandao Leite; Anna Carolina Paixao; Elisa Cavalcante Pereira; Fernando Motta; Luciana Appolinario; Marilda Siqueira on behalf of the Fiocruz COVID-19 Genomic Surveillance Network; Paola Resende; Renata Serrano Lopes; Taina Venas |

| | | | |
|---|---|---|---|
| EPI_ISL_2274068 | | | |
| EPI_ISL_2983196, EPI_ISL_2983197, EPI_ISL_2983198, EPI_ISL_2983199, EPI_ISL_2983200, EPI_ISL_2983201, EPI_ISL_2983202, EPI_ISL_2983280, EPI_ISL_2983281 | | | |
| see above | Laboratorio Central de Saude Publica do Estado do Maranhao (LACEN-MA) | Laboratory of Respiratory Viruses and Measles, Oswaldo Cruz Institute, FIOCRUZ | Agatha Cristinne Prudencio; Alice Sampaio Rocha; Ana Carolina Mendonca; Anna Carolina Paixao; Elisa Cavalcante Pereira; Fernando Motta; Ighor Leonardo Arantes Gomes; Lidio Gonçalves Lima Neto; Luciana Appolinario; Marilda Siqueira on behalf of the Fiocruz COVID-19 Genomic Surveillance Network; Paola Resende; Renata Serrano Lopes; Taina Moreira Venas; Tainá Venas |
| EPI_ISL_2274108, EPI_ISL_2274109, EPI_ISL_2274113, EPI_ISL_2274118, EPI_ISL_2274125, EPI_ISL_2274129 | Laboratorio Central de Saude Publica do Estado do Rio Grande do Sul (LACEN-RS) | Laboratory of Respiratory Viruses and Measles, Oswaldo Cruz Institute, FIOCRUZ | Alice Sampaio Rocha; Ana Carolina Mendonca; Anna Carolina Paixao; Elisa Cavalcante Pereira; Fernando Motta; Luciana Appolinario; Marilda Siqueira on behalf of the Fiocruz COVID-19 Genomic Surveillance Network; Paola Resende; Renata Serrano Lopes; Richard Salvato; Taina Venas; Tatiana Schaffer Gregianini |
| EPI_ISL_2196189, EPI_ISL_2196197, EPI_ISL_2196198, EPI_ISL_2196199, EPI_ISL_2196200, EPI_ISL_2196202, EPI_ISL_2196204, EPI_ISL_2196205, EPI_ISL_2196206, EPI_ISL_2196236 | | | |
| see above | Laboratorio Central de Saude Publica do Estado do Rio de Janeiro (LACEN-RJ) | Laboratory of Respiratory Viruses and Measles, Oswaldo Cruz Institute, FIOCRUZ | Alice Sampaio Rocha; Ana Carolina Mendonca; Andrea Cony Cavalcanti; Anna Carolina Paixao; Elisa Cavalcante Pereira; Fernando Motta; Luciana Appolinario; Marilda Siqueira on behalf of the Fiocruz COVID-19 Genomic Surveillance Network; Paola Resende; Renata Serrano Lopes; Taina Venas |
| EPI_ISL_3997110, EPI_ISL_3997228, EPI_ISL_3997230, EPI_ISL_3997231, EPI_ISL_3997240, EPI_ISL_3997739, EPI_ISL_3997833, EPI_ISL_3997877, EPI_ISL_3998018, EPI_ISL_3998051, EPI_ISL_3998169, EPI_ISL_3999620, EPI_ISL_4000067, EPI_ISL_4000229, EPI_ISL_4000270, EPI_ISL_4000384, EPI_ISL_4000695, EPI_ISL_4000717, EPI_ISL_4000718, EPI_ISL_4000719, EPI_ISL_4000801, EPI_ISL_4000802, EPI_ISL_4000803, EPI_ISL_4000804, EPI_ISL_4001402, EPI_ISL_4001403, EPI_ISL_4001404, EPI_ISL_4001405, EPI_ISL_4001562, EPI_ISL_4001708, EPI_ISL_4009398 | | | |
| see above | Laboratorio Central do Estado do Parana | Laboratorio Central do Estado do Parana | Guilherme Nardi Becker; Irina Nastassja Riediger; Maria do Carmo Debur Rossa; Mayra Presibella Giacomini; Nelson Quallio Marques |
| EPI_ISL_7716422, EPI_ISL_7716481 | Laboratorio Estatal de Salud Publica de Nuevo Leon | Laboratorio Estatal de Salud Publica de Nuevo Leon | Consuelo Treviño-Garza; Eduardo Isaac de la Rosa-Moreno; Else del Carmen Garcia-Garcia; Gloria Alejandra Jasso-de la Peña; Manuel Enrique de la O-Cavazos; Yadira Hilerio Garcia |
| EPI_ISL_5799817, EPI_ISL_5799818, EPI_ISL_5799819, EPI_ISL_5799853, EPI_ISL_5800194 | Laboratorio Municipal De Piracicaba | Instituto Butantan | Antonio Jorge Martins; Claudia Renata dos Santos Barros; David Schlesinger; Debora Botequio Moretti; Dimas Tadeu Covas; Elaine Cristina Marqueze; Elaine Vieira Santos; Evandra Strazza Rodrigues; Heidge Fukumasu; Jayme Augusto de Souza-Neto; José Salvatore Leister Patané; Luiz Alcantara; Luiz Lehmann Coutinho; Maria Carolina Elias; Maurício Lacerda Nogueira; Rafael dos Santos Bezerra; Raul Machado Neto; Rejane Maria Tommasini Grotto; Ricardo Haddad; Sandra Coccuzzo Sampaio Vessoni; Simone Kashima; Svetoslav Nanev Slavov; Vincent Louis Viala |
| EPI_ISL_3307443 | Laboratorio SYNLAB Colombia | Corporación para Investigaciones Biológicas-CIB | Jeanneth Mosquera Rendon; Jenny Santiago Cuesta; Marcela Mercado Reyes; Uriel A. Hurtado Paez |
| EPI_ISL_2427512, EPI_ISL_2427536, EPI_ISL_2427544, EPI_ISL_2427665, EPI_ISL_2427673, EPI_ISL_2427695, EPI_ISL_2427711, EPI_ISL_2427756, EPI_ISL_2427757, EPI_ISL_2427763, EPI_ISL_2427764, EPI_ISL_2427768, EPI_ISL_2427771, EPI_ISL_2427772, EPI_ISL_2427779 | | | |
| see above | Laboratorio de Biología Molecular Médica Uruguaya | Departments of Pathology and Medicine, New York University School of Medicine | Adriana Heguy; Cecilia Sorhouet; Christian Marier; Dacia Dimartino; Gonzalo Manrique; Maria Cristina Mogdasy; Maria Noel Zubillaga; Maria Victoria Elizondo; Paul Zappile |
| EPI_ISL_2777431, EPI_ISL_2777432, EPI_ISL_2777492, EPI_ISL_4030333 | Laboratorio de Ecologia de Doencas Transmissiveis na Amazonia, Instituto Leonidas e Maria Deane - Fiocruz Amazonia | Laboratorio de Ecologia de Doencas Transmissiveis na Amazonia, Instituto Leonidas e Maria Deane - Fiocruz Amazonia | Alex Martins; André Corado; Debora Duarte; Fabiola Mendonça da Silva Chui; Felipe Naveca; Fernanda Nascimento; Fernando Fonseca de Almeida Val; George Silva; Gisely Cardoso de Melo; Karina Pessoa; Luciana Gonçalves; Marcus Vinicius Guimarães de Lacerda; Maria Júlia Brandão; Maria Paula Gomes Mourão; Mariana Xavier Simão; Matilde Mejía; Michele Jesus; Valdinete Nascimento; Vanderson de Souza Sampaio; Victor Souza; Ágatha Costa |
| EPI_ISL_2728535 | Laboratorio de Infectologia y Virologia Molecular | Laboratory of Molecular Virology, School of Medicine, Pontificia Universidad Catolica de Chile | Alejandro Bhrun; Ana Maria Contreras; Andres E. Munoz-Marcos; Carlos Palma; Catalina Pardo-Roa; Constanza Maldonado; Constanza Martinez-Valdevenito; Eileen Serrano; Erick Salinas; Estefany Poblete; Francisco Melo; Jennifer Angulo; Jorge Levican; Leonardo I. Almonacid; M. Belen Leyton; Magdalena Vera; Marcela Ferres; Maria Jose Avendano; Rafael A. Medina; Tamara Garcia-Salum |
| EPI_ISL_3375951, EPI_ISL_3375955, EPI_ISL_3375956, EPI_ISL_3375980, EPI_ISL_3376183 | Laboratorio de Referencial Nacional de Virus Respiratorios | Laboratorio de Referencial Nacional de Virus Respiratorios | Carlos Padilla Rojas; Henri Bailon Calderon; Iris Silva Molina; Joseph Huayra Niquen; Lely Solari Zerpa; Luis Barcena Flores; Marco Galarza Perez; Nancy Rojas Serrano; Omar Caceres Rey; Orson Mestanza Millones; Priscila Lope Pari; Sandra Morales Ruiz; Steve Acedo Lazo; Veronica Hurtado Vela |
| EPI_ISL_1626619 | Laboratorio de Salud Pública Bogota | Gencore - Universidad de los Andes | Alejandro Gomez; Ana Maria Palacio; David Gonzalez; Gabriela Delgado; Johana Hernandez; Luisa Sacristan; Marcela Guevara; Silvia Restrepo |
| EPI_ISL_1673282, EPI_ISL_1673290, EPI_ISL_1673294, EPI_ISL_1673303, EPI_ISL_1673305 | Laboratorio de Virología HUCA | Laboratorio de Virología HUCA | Abreu F; Alvarez-Arguelles ME; Boga JA; Castelló C; Costales I; Coto E; Gómez de Oña J; Martín-Rodríguez G; Melón S; Perez-Martínez Z; Rojo S; Sandoval M |
| EPI_ISL_4405445 | Laboratorio del Hospital Interzonal General de Agudos " Evita " | Área de Secuenciación del Laboratorio de Virología del Hospital de Niños Dr. Ricardo Gutierrez on behalf of 'Proyecto Argentino Interinstitucional de genomica de SARS-CoV-2' (PAIS Consortium) | A; Acuña; D; Desimone; E; Goya; Grossi; I; L; LE; Luczac; Lusso; MI; MS; Musto; Nabaes Jodar; Natale; O; S; Serrano; Valinotto; Viegas, M. |
| EPI_ISL_1626571 | Laboratorio di Genetica Medica Ospedale Belcolle | INMI Lazzaro Spallanzani IRCCS | A Di Caro; B Bartolini; CEM Gruber; E Giombini; F Messina; F Natoni; F Santini; G Bonfiglio; G Pessina; M Rueca; MR Capobianchi; O Butera |
| EPI_ISL_1585338, EPI_ISL_1585340 | Laboratorio di Microbiologia | Laboratorio di Microbiologia | Martinetti Lucchini Gladys; Valeria Spina |
| EPI_ISL_1610369, EPI_ISL_1610445, EPI_ISL_1610454, EPI_ISL_1610462, EPI_ISL_1610537, EPI_ISL_1610720, EPI_ISL_1610775, EPI_ISL_1610817, EPI_ISL_1610849, EPI_ISL_1611033, EPI_ISL_1611062, EPI_ISL_1611072, EPI_ISL_1611104, EPI_ISL_1611109, EPI_ISL_1611110, EPI_ISL_1611133, EPI_ISL_1611135, EPI_ISL_1611139, EPI_ISL_1611144, EPI_ISL_1611146, EPI_ISL_1611147, EPI_ISL_1611149, EPI_ISL_1611150, EPI_ISL_1611151, EPI_ISL_1611159, EPI_ISL_1611170, EPI_ISL_1611182, EPI_ISL_1611186, EPI_ISL_1611192, EPI_ISL_1611247, EPI_ISL_1611271, EPI_ISL_1611288, EPI_ISL_1611337, EPI_ISL_1611346, EPI_ISL_1611400, EPI_ISL_1611423, EPI_ISL_1611441, EPI_ISL_1611445, EPI_ISL_1611490, EPI_ISL_1611495, EPI_ISL_1611497, EPI_ISL_1611502, EPI_ISL_1611503, EPI_ISL_1611515, EPI_ISL_1611522, EPI_ISL_1611540, EPI_ISL_1611560, EPI_ISL_1611568, EPI_ISL_1611609, EPI_ISL_1611686, EPI_ISL_1611708, EPI_ISL_1611827, EPI_ISL_1611927, EPI_ISL_1611946, EPI_ISL_1612018, EPI_ISL_1612032, EPI_ISL_1612108, EPI_ISL_1612109, EPI_ISL_1612122, EPI_ISL_1612160, EPI_ISL_1612200, EPI_ISL_1612462, EPI_ISL_1612485, EPI_ISL_1612509, EPI_ISL_1612607, EPI_ISL_1612616, EPI_ISL_1612742, EPI_ISL_1612797, EPI_ISL_1680471, EPI_ISL_1680503, EPI_ISL_1680520, EPI_ISL_1680522, EPI_ISL_1680556, EPI_ISL_1680558, EPI_ISL_1680823, EPI_ISL_1681710, EPI_ISL_1681735, EPI_ISL_1681784, EPI_ISL_1681789, EPI_ISL_1681847, EPI_ISL_1681853, EPI_ISL_1681914, EPI_ISL_1681921, EPI_ISL_1681924, EPI_ISL_1681925, EPI_ISL_1681948, EPI_ISL_1681959, EPI_ISL_1681962, EPI_ISL_1681974, EPI_ISL_1682001, EPI_ISL_1682002, EPI_ISL_1682011, EPI_ISL_1682013, EPI_ISL_1682020, EPI_ISL_1682064, EPI_ISL_1682084, EPI_ISL_1682086, EPI_ISL_1682102, EPI_ISL_1682103, EPI_ISL_1682104, EPI_ISL_1682112, EPI_ISL_1682138, EPI_ISL_1682144, EPI_ISL_1682150, EPI_ISL_1682154, EPI_ISL_1682169, EPI_ISL_1682200, EPI_ISL_1682237, EPI_ISL_1682247, EPI_ISL_1682262, EPI_ISL_1682268, EPI_ISL_1682272, EPI_ISL_1682275, EPI_ISL_1682277, EPI_ISL_1682309, EPI_ISL_1682315, EPI_ISL_1682389, EPI_ISL_1682405, EPI_ISL_1682415, EPI_ISL_1682418, EPI_ISL_1682423, EPI_ISL_1682438, EPI_ISL_1682441, EPI_ISL_1682464, EPI_ISL_1682484, EPI_ISL_1682512, EPI_ISL_1682605, EPI_ISL_1682614, EPI_ISL_1682646, EPI_ISL_1682658, EPI_ISL_1682667, EPI_ISL_1682675, EPI_ISL_1682684, EPI_ISL_1682714, EPI_ISL_1682720, EPI_ISL_1682724, EPI_ISL_1682730, EPI_ISL_1682732, EPI_ISL_1682751, EPI_ISL_1682772, EPI_ISL_1682778, EPI_ISL_1682987, EPI_ISL_1683019, EPI_ISL_1683151, EPI_ISL_1683160, EPI_ISL_1683184, EPI_ISL_1683195, EPI_ISL_1683216, EPI_ISL_1683219, EPI_ISL_1683257, EPI_ISL_1683269, EPI_ISL_1683284, EPI_ISL_1683293, EPI_ISL_1683306, EPI_ISL_1683314, EPI_ISL_1683318, EPI_ISL_1683474, EPI_ISL_1684345, EPI_ISL_1684367, EPI_ISL_1684376, EPI_ISL_1684427, EPI_ISL_1684639, EPI_ISL_1684670, EPI_ISL_1798435, EPI_ISL_4374996 | | | |
| see above | Laboratory Corporation of America | Centers for Disease Control and Prevention Division of Viral Diseases, Pathogen Discovery | Adrian Paskey; Amanda Douglas; Amanda Suchanek; Andrea Throop; Ayla Burns; Benjamin Rambo-Martin; Bobbi Croy; Brian Krueger; Brian Norvell; Christopher Gulvick; Christos Petropoulos; Clinton Paden; Clinton R. Paden; Craig Lukasik; Dakota Howard; Darlene Wagner; Debbie Boles; Dhwani Batra; Duncan MacCannell; Eyad Almasri; Goran Stevovic; Howard Engler; Hrushikesh Deshmukh; Jake Humphrey; Jana Schroth; Jason Caravas; Joe Voshell; John Pruitt; Jonathan Meltzer; Jonathan Williams; Kara Moser; Kimberly Wagner; Kristine Lacek; Lax Iyer; Lisa Pfefferle; Lyndon Tilson; Manoj Jain; Marcia Eisenberg; Mary Ann Cristobal; Mary Cristobal; Mary Williamson; Matthew Robinson; Matthew Schmerer; Michael Levandoski; Mike Sapeta; Mindy Nye; Minoo Agarwal; Mohan Kolli; Nuthawin Charoensri; Oren Cohen; Peter Cook; Peter W. Cook; Prashant Gupta; Qian Zeng; Rama Ghatti; Scott Parker; Scott Ryan; Scott Sammons; Shatavia Morrison; Stanley Letovsky; Steven Ragan; Suresh Babu Selvaraju; Suresh Selvaraju; Susan Countryman; Susan Hicks; Suzanne Dale; Thomas Urban; Tim Kuphal; Tricia Zwiefelhofer; Tymeckia Kendall; Victoria Caban Figueroa; Vincent Drouillon; Yvette Unoarumhi |
| EPI_ISL_2472918 | Laboratory of Infectious Diseases, Department of Biomedical and Clinical Sciences L. Sacco, University of Milan | Laboratory of Infectious Diseases, Department of Biomedical and Clinical Sciences L. Sacco, University of Milan | Alessia Lai; Annalisa Bergna; Carla Della Ventura; Claudia Balotta; Gianguglielmo Zehender on behalf of SARS-CoV-2 ITALIAN RESEARCH ENTERPRISE-(SCIRE) Collaborative Group; Massimo Galli |
| EPI_ISL_2443592 | Laboratory of Respiratory Viruses and Measles, Oswaldo Cruz Institute, FIOCRUZ | Laboratory of Respiratory Viruses and Measles, Oswaldo Cruz Institute, FIOCRUZ | Alice Sampaio Rocha; Ana Carolina Mendonca; Anna Carolina Paixao; Elisa Cavalcante Pereira; Fernando Motta; Luciana Appolinario; Marilda Siqueira on behalf of the Fiocruz COVID-19 Genomic Surveillance Network; Paola Resende; Renata Serrano Lopes; Taina Venas |
| EPI_ISL_2157524 | Laboratório Central de Saude Publica do Estado do Rio de Janeiro (LACEN/RJ) | Laboratory of Respiratory Viruses and Measles, Oswaldo Cruz Institute, FIOCRUZ | Alice Sampaio Rocha; Ana Carolina Mendonca; Andrea Cony Cavalcanti; Anna Carolina Paixao; Elisa Cavalcante Pereira; Fernando Motta; Luciana Appolinario; Marilda Siqueira on behalf of the Fiocruz COVID-19 Genomic Surveillance Network; Paola Resende; Renata Serrano Lopes; Taina Venas |
| EPI_ISL_4600563 | Laboratório Central de Saúde Pública Noel Nutels | Coordenação Geral de Laboratórios de Saúde Pública (CGLAB/DAEVS/SVS/MS) | Vagner Fonseca; et al. |
| EPI_ISL_2308416, EPI_ISL_4600511, EPI_ISL_4600529 | Laboratório Central de Saúde Pública de Pernambuco | Coordenação Geral de Laboratórios de Saúde Pública (CGLAB/DAEVS/SVS/MS) | Vagner Fonseca; et al. |
| EPI_ISL_4600569, EPI_ISL_4600578, EPI_ISL_4600580 | Laboratório Central de Saúde Pública de Santa Catarina | Coordenação Geral de Laboratórios de Saúde Pública (CGLAB/DAEVS/SVS/MS) | Vagner Fonseca; et al. |
| EPI_ISL_2777548, EPI_ISL_2777563, EPI_ISL_2777574, EPI_ISL_2777588, EPI_ISL_2777609, EPI_ISL_2777655, EPI_ISL_2777660, EPI_ISL_2777662, EPI_ISL_2777696, EPI_ISL_2777722, EPI_ISL_2777741, EPI_ISL_2777803, EPI_ISL_2777848, EPI_ISL_2777851 | | | |
| see above | Laboratório Central de Saúde Pública do Amazonas - LACEN- | Laboratorio de Ecologia de Doencas Transmissiveis na Amazonia, Instituto | André Corado; Debora Duarte; Felipe Naveca; Fernanda Nascimento; George Silva; Karina Pessoa; Luciana Gonçalves; Maria Júlia Brandão; Matilde Mejía; Michele Jesus; Valdinete Nascimento; Victor Souza; Ágatha Costa |

| | | | |
|---|---|---|---|
| | AM | Leonidas e Maria Deane - Fiocruz Amazonia | |
| EPI_ISL_3155987 | Laboratório de Biologia Integrativa | Laboratório de Biologia Integrativa | Alessandro Clayton de Souza Ferreira; Aline Brito de Lima; Carolina Moreira Voloch; Daniel Costa Queiroz; Danielle Alves Gomes Zauli; Diego Menezes Bonfim; Filipe Romero Rebello Moreira; Frederico Scott Varella Malta; Joice do Prado Silva; Lucyene Miguita Luiz; Nuno Rodrigues Faria; Paula Luize Camargos Fonseca; Rafael Marques de Souza; Renan Pedra de Souza; Renato Santana Aguiar; Rennan Garcias Moreira; Victor Cavalcanti Pardini; Victor Emmanuel Viana Geddes |
| EPI_ISL_6508482, EPI_ISL_6508484, EPI_ISL_6508550, EPI_ISL_6508556 | Laboratório de Biologia Integrativa/ UFMG | Laboratório de Biologia Integrativa/ UFMG | Adriana Aparecida Ribeiro; Alana Vitor Barbosa Costa; Alessandro Luís Gonçalves; Aline de Brito Lima; Ana Paula De Battisti Ribeiro; Ana Paula Salles Moura Fernandes; Andre Luiz Menezes; Bruna Walker Ferreira; Carolina Senra Alves de Souza; Cristiane P. T. Brito Mendonça; Daniel Costa Queiroz; Danielle Alves Gomes Zauli; Diego Menezes; Eneida Santos de Oliveira; Eva Lídia Arcoverde Medeiros; Felipe Campos de Melo Iani; Fernanda Gil de Souza; Fernanda Santos Mendes; Filipe Romero Rebello Moreira; Flávio Guimarães da Fonseca; Frederico Scott Varella Malta; Hugo Itaru Sato; Hugo José Alves; Igor Pereira Godinho; Jaqueline Silva de Oliveira; Joice do Prado Silva; José Nélio Januario; Juliana Wilke Saliba; Karine Lima Lourenço; Lucyene Miguita; Luige Biciati Alvim; Nara Oliveira Carvalho; Natiely Pereira Silva; Natália Rocha Guimarães; Paula Luize Camargos Fonseca; Pedro Henrique Barbosa de Paula Mendes; Rafael Marques de Souza; Renan Pedra de Souza; Renata Barbosa Peixoto Peixoto; Renato Santana de Aguiar; Rennan Garcias Moreira; Rillery Calixto Dias; Rubens Daniel Miserani Magalhães; Santuza Maria Ribeiro Teixeira; Talita Emile Ribeiro Adelino; Victor Emmanuel Viana Geddes; Walyson Coelho Costa |
| EPI_ISL_1578453 | Laboratório de Biologia Molecular do Hospital das Clínicas da Faculdade de Medicina de Botucatu/SP | Laboratórios Genômica Funcional (FCA/UNESP) e Biologia Molecular (FMB-HC/UNESP) - Rede de Vigilância Genômica (Vigenômica)/UNESP | Bianca Cechetto Carlos; Felipe Allan da Silva da Costa; Flavia Hebner Barbosa Trovão; Guilherme Targino Valente; Jayme A. Souza-Neto.; Patrícia Akemi Assato; Rejane Maria Tommasini Grotto |
| EPI_ISL_1578740, EPI_ISL_1579258, EPI_ISL_1580502 | Laboratório de Biologia Molecular do Hospital das Clínicas da Faculdade de Medicina de Botucatu/SP | Laboratórios de Genômica Funcional (FCA/UNESP) e Biologia Molecular (FMB-HC/UNESP) - Rede de Vigilância Genômica (Vigenômica)/UNESP | Bianca Cechetto Carlos; Felipe Allan da Silva da Costa; Flavia Hebner Barbosa Trovão; Guilherme Targino Valente; Jayme A. Souza-Neto.; Patrícia Akemi Assato; Rejane Maria Tommasini Grotto |
| EPI_ISL_2629790, EPI_ISL_2629791, EPI_ISL_2629792, EPI_ISL_2629794, EPI_ISL_2629795, EPI_ISL_2629796, EPI_ISL_2629797, EPI_ISL_2629798, EPI_ISL_2629799, EPI_ISL_2629800, EPI_ISL_2629801, EPI_ISL_2629802, EPI_ISL_2629803 | | | |
| see above | Laboratório de Virologia Molecular - Universidade Federal do Rio de Janeiro | Laboratório de Virologia Molecular - Universidade Federal do Rio de Janeiro | ; Alice Laschuk Herlinger; Amilcar Tanuri; André Felipe Andrade dos Santos; Carolina Moreira Voloch; Cássia Cristina Alves Gonçalves; Diana Mariani; Débora Souza Faffe; Filipe Romero Rebello Moreira; Francine Bittencourt Schiffler; Isabela de Carvalho Leitão; Marcelo Calado de Paula Tôrres; Matheus Augusto Calvano Cosentino; Mirela D'arc; Orlando da Costa Ferreira Junior; Rafael Mello Galliez; Raíssa Mirella dos Santos Cunha da Costa; Renato Santana de Aguiar; Terezinha Marta Pereira Pinto Castineiras; Thamiris dos Santos Miranda; Átila Duque Rossi |
| EPI_ISL_2196247, EPI_ISL_2196248, EPI_ISL_2196335, EPI_ISL_2196336, EPI_ISL_2196337, EPI_ISL_2196338, EPI_ISL_2196339, EPI_ISL_2196340, EPI_ISL_2196341, EPI_ISL_2196342, EPI_ISL_2196343, EPI_ISL_2196344, EPI_ISL_2196346, EPI_ISL_2196348, EPI_ISL_2196349, EPI_ISL_2196353, EPI_ISL_2196354, EPI_ISL_2443552, EPI_ISL_2536262 | | | |
| see above | Labortorio Central de Saude Publica do Estado de Santa Catarina (LACEN/SC) | Laboratory of Respiratory Viruses and Measles, Oswaldo Cruz Institute, FIOCRUZ | Alice Sampaio Rocha; Ana Carolina Mendonca; Anna Carolina Paixao; Darcita Buerger Rovaris; Elisa Cavalcante Pereira; Fernando Motta; Luciana Appolinario; Marilda Siqueira on behalf of the Fiocruz COVID-19 Genomic Surveillance Network; Paola Resende; Renata Serrano Lopes; Sandra Bianchini Fernandes; Taina Venas |
| EPI_ISL_2614376, EPI_ISL_2614377, EPI_ISL_2614378, EPI_ISL_2614379, EPI_ISL_2614380 | Labortorio Central de Saude Publica do Estado do Rio de Janeiro (LACEN/RJ) | Laboratory of Respiratory Viruses and Measles, Oswaldo Cruz Institute, FIOCRUZ | Alice Sampaio Rocha; Ana Carolina Mendonca; Andrea Cony Cavalcanti; Anna Carolina Paixao; Elisa Cavalcante Pereira; Fernando Motta; Luciana Appolinario; Marilda Siqueira on behalf of the Fiocruz COVID-19 Genomic Surveillance Network; Paola Resende; Renata Serrano Lopes; Taina Venas |
| EPI_ISL_2110771 | Landesgesundheitsamt Baden-Wurttemberg | Robert Koch Institute | |
| EPI_ISL_2196252, EPI_ISL_2196307, EPI_ISL_2196308, EPI_ISL_2196309, EPI_ISL_2196310, EPI_ISL_2196311, EPI_ISL_2443532, EPI_ISL_2443534, EPI_ISL_2443565, EPI_ISL_2603429, EPI_ISL_2603431, EPI_ISL_2603469 | | | |
| see above | Lboratorio Central de Saude Publica do Estado do Parana (LACEN/PR) | Laboratory of Respiratory Viruses and Measles, Oswaldo Cruz Institute, FIOCRUZ | Alice Sampaio Rocha; Ana Carolina Mendonca; Anna Carolina Paixao; Elisa Cavalcante Pereira; Fernando Motta; Irina Riediger; Luciana Appolinario; Marilda Siqueira on behalf of the Fiocruz COVID-19 Genomic Surveillance Network; Paola Resende; Renata Serrano Lopes; Taina Venas |
| EPI_ISL_1620769, EPI_ISL_1620793 | Libramont | Plateforme de testing Namuroise | Céline Maschietto; Degosserie Jonathan; Denis Olivier; Mullier François; Otto Gaetan |
| EPI_ISL_1518006, EPI_ISL_1519280 | Lighthouse Lab in Cambridge | Wellcome Sanger Institute for the COVID-19 Genomics UK (COG-UK) Consortium | Cordelia Langford; David K. Jackson; Dominic Kwiatkowski; Ewan Harrison; Ian Johnston; Jeffrey Barrett; John Sillitoe on behalf of the Wellcome Sanger Institute COVID-19 Surveillance Team; Rob Howes; Roberto Amato; Sonia Goncalves; The Lighthouse Lab in Cambridge and Alex Alderton |
| EPI_ISL_5778158 | Louisville Metro Public Health and Wellness Department/Louisville Metro Department of Corrections | University of Louisville Sequencing Technology Center | Elizabeth Hudson; Eric C. Rouchka; Julia H. Chariker; Leslie A. Wolf; Melissa L. Smith; William Lauer |
| EPI_ISL_1704738, EPI_ISL_2009369, EPI_ISL_2009371 | Lurie Children's Hospital of Chicago | Northwestern University - Ozer Lab | Egon A. Ozer; Judd F. Hultquist; Lacy M. Simons; Larry K. Kociolek; Michael G. Ison; Ramon Lorenzo-Redondo; Taylor J. Dean; William J. Muller; Xiaotian; Zheng |
| EPI_ISL_1531684 | M Health Fairview | Minnesota Department of Health, Public Health Laboratory | Alexandra Lorentz; Jacob Garfin; Matt Plumb; and Xiong Wang |
| EPI_ISL_2801328, EPI_ISL_2801329 | MATERNIDADE ESCOLA ASSIS CHATEAUBRIAND | Analytical Competence Molecular Epidemiology Lab/ACME, Oswaldo Cruz Foundation, Ceara (FIOCRUZ CE) | Cleber Furtado Aksenen e Suzana Porto Almeida; Fabio Miyajima; Fernando Braga Stehling; Francisco Eder de Moura Lopes; Jamille Maria Mendes Bezerra; Joaquim César do Nascimento Sousa Junior; Pedro Miguel Carneiro Jeronimo; Thais Ferreira de Oliveira; Thais de Oliveira Costa; Ticiane Cavalcante de Souza; Veridiana Pessoa Miyajima |
| EPI_ISL_3102263 | MATERNIDADE QUITERIA DE LIMA | Analytical Competence Molecular Epidemiology Lab/ACME, Oswaldo Cruz Foundation, Ceara (FIOCRUZ CE) | Cleber Furtado Aksenen; Fabio Miyajima; Fernando Braga Stehling; Francisco Eder de Moura Lopes; Jamille Maria Mendes Bezerra; Joaquim César do Nascimento Sousa Junior; Pedro Miguel Carneiro Jeronimo; Suzana Porto Almeida e Lucas Delerino; Thais Ferreira de Oliveira; Thais de Oliveira Costa; Ticiane Cavalcante de Souza; Veridiana Pessoa Miyajima |
| EPI_ISL_1594071 | MB-Cadham Provincial laboratory | National Microbiology Laboratory (NML) | Anna Majer; Anneliese Landgraff; CanCOGeN's metadata curation team; Darian Hole; David Alexander; Elsie Grudeski; Gary Van Domselaar; Grace Seo; Jared Bullard; Jennifer Tanner; Kerry Dust; Kirsten Biggar; Madison Chapel; Morag Graham; Natalie Knox; Nathalie Bastien; Paul Van Caeseele; Philip Mabon; Public Health Agency of Canada CanCOGeN team; Rhiannon Huzarewich; Russell Mandes; Shari Tyson; Timothy Booth; Yan Li |
| EPI_ISL_1759395 | ME Health and Environmental Testing Laboratory | Pathogen Discovery, Respiratory Viruses Branch, Division of Viral Diseases, Centers for Disease Control and Prevention | Adam Retchless; Anna Kelleher; Anna Montmayeur; Anna Uehara; Brian Lynch; Clinton R. Paden; Haibin Wang; Han Jia Justin Ng; Jing Zhang; Justin Lee; Krista Queen; Mark Burroughs; Peter Cook; Rachel Marine; Suxiang Tong; Yan Li; Ying Tao |
| EPI_ISL_1694877 | MEPHI, Aix Marseille University | MEPHI, Aix Marseille University | Anthony LEVASSEUR |
| EPI_ISL_4572673 | MS PHL | MS PHL | Beata Karolewicz; Daphne Ware; Jin Yiyao |
| EPI_ISL_2689885 | MS Public Health Laboratory | Centers for Disease Control and Prevention Division of Viral Diseases, Pathogen Discovery | Alison Laufer Halpin; Ben L. Rambo-Martin; Clinton R. Paden; Dakota Howard; Darlene Wagner; Dave Wentworth; Dhwani Batra; Jasmine Padilla; Justin Lee; Katie Dillon; Krista Queen; Kristen Knipe; Kristine Lacek; Mark Burroughs; Matthew Schmerer; Mili Sheth; Peter Cook; Sam Shepard; Sarah Nobles; Shoshona Le; Suxiang Tong; Vivien Dugan; Yvette Unoarumhi |
| EPI_ISL_1709863, EPI_ISL_5253371 | MSHS Clinical Microbiology Laboratories | MSHS Pathogen Surveillance Program | Adolfo García-Sastre; Adriana van de Guchte; Ajay Obla; Alberto Paniz-Mondolfi; Ana S. Gonzalez-Reiche; Angela Amoako; Ashley Salimbangon; Betsaida Salom Melo; Bremy Alburquerque; Brianne Ciferri; Charles Gleason; Daniel Floda; Deena R. Altman; Denise Jurczyszak; Emilia Mia Sordillo; Gintaras Deikus; Giulio Kleiner; Gopi Patel; Hala Alshammary; Harm van Bakel; Irina Oussenko; Jayeeta Dutta; Juan Soto; Julia Matthews; Katherine Beach; Kathryn Twyman; Kayla Russo; Komal Srivastava; Levy Sominsky; Mahmoud Awawda; Marta Luksza; Matthew M. Hernandez; Melissa Gitman; Michael D. Nowak; Mitchell J. Sullivan; Nancy Francoeur; Robert Sebra; Sarah Schaefer; Shelcie Fabre; Shwetha Hara Sridhar; Viviana Simon; Ying-Chih Wang; Zenab Khan |
| EPI_ISL_1638367 | MVZ Labor Dortmund | Robert Koch Institute | |
| EPI_ISL_1640682, EPI_ISL_1640683, EPI_ISL_1640692, EPI_ISL_1640733, EPI_ISL_1640738 | MVZ Labor Krone GbR | Robert Koch Institute | |
| EPI_ISL_1642343 | MVZ fÃ¼r Laboratoriumsmedizin und Mikrobiologie Koblenz-Mittelrhein (Labor Koblenz) | Robert Koch Institute | |
| EPI_ISL_1911102 | Maine Health and Environmental Testing Laboratory | Maine Health and Environmental Testing Laboratory | Barter, M.; Dewey, H.; Grieser, H.; H. and Tewhey, R.; Lynch, R.; Martha, J.; Matluk, N.; Meak, S.; Munger |
| EPI_ISL_3544226, EPI_ISL_3544238, EPI_ISL_3544240 | Maine Health and Environmental Testing Laboratory | Tewhey Lab, The Jackson Laboratory | Barter, M.; Dewey, H.; H. and Tewhey, R.; Iosue, F.; Lynch, R.; Matluk, N.; Munger |
| EPI_ISL_1696405 | Maryland Genomics, Institute for Genome Sciences, University of Maryland School of Medicine | Maryland Genomics, Institute for Genome Sciences, University of Maryland School of Medicine | Aditya; Claire M; Fraser; Holly; Humphrys; Jacques; Kranthi; Lisa D; Luke J; Mehta; Mike; Ott; Ravel; Roussey; Sadzewicz; Sandra; Tallon; Vavikolanu |
| EPI_ISL_1528105, EPI_ISL_1528106, EPI_ISL_1528149, EPI_ISL_1528150, EPI_ISL_1624535, EPI_ISL_1624686, EPI_ISL_2008330 | | | |
| see above | Massachusetts State Public Health Laboratory | Massachusetts State Public Health Laboratory | Andrew Lang; Glen Gallagher; Sandra Smole; Timelia Fink |
| EPI_ISL_1906667, EPI_ISL_2374680 | Michigan Department of Health and Human Services, Bureau of | Michigan Department of Health and Human Services, Bureau of | Blankenship HM; Riner D; Soehnlen MK |

| | | | |
|---|---|---|---|
| | Laboratories | Laboratories | |
| EPI_ISL_2365838 | Microbiologia e Virologia | Istituto Zooprofilattico Sperimentale delle Venezie | Adelaide Milani; Alessia Schivo; Alice Fusaro; Ambra Pastori; Annalisa Salviato; Antonia Ricci; Calogero Terregino; Edoardo Giussani; Elisa Palumbo; Erika Giorgia Quaranta; Isabella Monne; Luca Tassoni |
| EPI_ISL_1893011, EPI_ISL_1893034 | Microbiologia CATLAB | Can Ruti SARS-CoV-2 Sequencing Hub (HUGTiP/IrsiCaixa/IGTP) | Alba Sánchez; Anna Not; Antoni E Bordoy; Bonaventura Clotet; Cristina Casañ; Cristina Esteban; Francesc Catala-Moll; Gemma Clara; Ignacio Blanco; Marc Noguera-Julian; Maria Casadellà; Mariona Parera; Mercedes Guerrero; Montserrat Giménez; Pere-Joan Cardona; Pilar Armengol; Roger Paredes; Verónica Saludes; and Elisa Martró on behalf of the Can Ruti SARS-CoV-2 Sequencing Hub. |
| EPI_ISL_2140526, EPI_ISL_2140613, EPI_ISL_2140636 | Microbiology Department, Laboratori Clínic Metropolitana Nord. Hospital Universitari Germans Trias i Pujol | Can Ruti SARS-CoV-2 Sequencing Hub (HUGTiP/IrsiCaixa/IGTP) | Alba Sánchez; Anna Not; Antoni E Bordoy; Bonaventura Clotet; Cristina Casañ; Cristina Esteban; Francesc Catala-Moll; Gemma Clara; Ignacio Blanco; Marc Noguera-Julian; Maria Casadellà; Mariona Parera; Mercedes Guerrero; Montserrat Giménez; Pere-Joan Cardona; Pilar Armengol; Roger Paredes; Verónica Saludes; and Elisa Martró on behalf of the Can Ruti SARS-CoV-2 Sequencing Hub. |
| EPI_ISL_1547724, EPI_ISL_1661909, EPI_ISL_1661922, EPI_ISL_1661933, EPI_ISL_1661943, EPI_ISL_1661956, EPI_ISL_1661967, EPI_ISL_1661968, EPI_ISL_2035851 | | | |
| see above | Microbiology Department, Laboratori Clínic Metropolitana Nord. Hospital Universitari Germans Trias i Pujol. | Can Ruti SARS-CoV-2 Sequencing Hub (HUGTiP/IrsiCaixa/IGTP) | Alba Sánchez; Anna Not; Antoni E Bordoy; Bonaventura Clotet; Cristina Casañ; Cristina Esteban; Francesc Catala-Moll; Gemma Clara; Ignacio Blanco; Marc Noguera-Julian; Maria Casadellà; Mariona Parera; Mercedes Guerrero; Montserrat Giménez; Pere-Joan Cardona; Pilar Armengol; Roger Paredes; Verónica Saludes; and Elisa Martró on behalf of the Can Ruti SARS-CoV-2 Sequencing Hub. |
| EPI_ISL_1595725, EPI_ISL_1595727, EPI_ISL_2086123, EPI_ISL_2086124 | Microbiology Department. Complexo Hospitalario Universitario de Vigo | Microbiology Department. Complexo Hospitalario Universitario de Vigo | Alfaya N; Alonso I; Alvarez M; Cabrera JJ; Carballo R; Cores O; Cortizo S; Martinez L; Mediero G; Perez S; Potel C; Regueiro B; Rey S; Vasallo FJ; del-Campo V |
| EPI_ISL_1661436, EPI_ISL_1661454 | Microvida | Microvida | Jaco Verweij; Joep Stöhr; Suzan D. Pas |
| EPI_ISL_1534481, EPI_ISL_1534518, EPI_ISL_2936139, EPI_ISL_2936164, EPI_ISL_2936169, EPI_ISL_3032711, EPI_ISL_3063482, EPI_ISL_3063486 | | | |
| see above | Ministry of Health Turkey | Ministry of Health Turkey | Fatma Bayrakdar; Gulay Korukluoglu; Suleyman Yalcin; Yasemin Cosgun |
| EPI_ISL_1791376 | Missouri State Public Health Laboratory | Missouri State Public Health Laboratory | Ashley New; Joshua Barry; Matthew Sinn |
| EPI_ISL_1678371 | NH Dept. of Health and Human Services Public Health Labs | Centers for Disease Control and Prevention Division of Viral Diseases, Pathogen Discovery | Alison Laufer Halpin; Ben L. Rambo-Martin; Clinton R. Paden; Dakota Howard; Darlene Wagner; Dave Wentworth; Dhwani Batra; Jasmine Padilla; Justin Lee; Katie Dillon; Krista Queen; Kristen Knipe; Kristine Lacek; Mark Burroughs; Matthew Schmerer; Mili Sheth; Peter Cook; Sam Shepard; Sarah Nobles; Shoshona Le; Suxiang Tong; Vivien Dugan; Yvette Unoarumhi |
| EPI_ISL_1559352, EPI_ISL_1559353, EPI_ISL_1559356, EPI_ISL_1559357, EPI_ISL_1559361, EPI_ISL_1625884, EPI_ISL_1625907, EPI_ISL_1648108, EPI_ISL_1892590 | | | |
| see above | NJDOH, Public Health and Environmental Laboratories | NJ_PHEL | Byeong Jeong; Dana Woell; Jacquelyn Deverell; Lindsey Bodnar; Matthew Scarnati; Mohammad M Ali; Shiv K. Verma |
| EPI_ISL_1582291, EPI_ISL_1671640 | NORTHWELL HEALTH LABORATORIES | Wadsworth Center, New York State Department of Health | Alexis Russell; Catharine Prussing; Daryl M. Lamson; Erasmus Schneider; Erica Lasek-Nesselquist; John Kelly; Jonathan Plitnick; Kirsten St. George; Matthew Shudt; Melissa A Leisner; Navjot Singh |
| EPI_ISL_1966456, EPI_ISL_1966458 | NUCLEO DE SAUDE VILA FALCAO DE BAURU | Instituto Butantan / Mendelics | Antonio Jorge Martins; Bianca Cechetto Carlos. Mendelics: Bibiana Santos; Claudia Renata dos Santos Barros; Cíntia Bittar; David Schlesinger. Hemocentro Ribeirão Preto: Simone Kashima; Debora Botequio Moretti; Elaine Cristina Marqueze; Elaine Vieira dos Santos; Elisangela Chicaroni Mattos; Erika Freitas; Evandra Strazza Rodrigues; Felipe Allan da Silva da Costa; Flavia Aburjaile; Fábio Sossai Possebon; Guilherme Campos; Guilherme Targino Valente; Heidge Fukumasu. USP-Botucatu: Rejane Maria Tommasini Grotto; Helena Lage Ferreira; Instituto Butantan: Dimas Tadeu Covas; Jardelina de Souza Todao Bernardino; Jayme A. Souza-Neto; Jessika Cristina Chagas Lesbon; Jorge A. Petroli Marchesi; José Salvatore Leister Patané; João Paulo Kitajima; João Pessoa Araújo Jr.; Leila Sabrina Ullmann; Loyze Paola Oliveira de Lima; Luiz Aurelio de Campos Crispin. Centro de Genômica Funcional da ESALQ: Luiz Lehmann Coutinho; Luiz Carlos Junior de Alcantara; Lívia Sacchetto; Maisa C. Pereira Parra; Maria Carolina Elias; Marta Giovanetti; Marília Moraes; Maurício Lacerda Nogueira. Prefeitura de Sao Paulo: Melissa Palmieri.; Patricia Akemi Assato; Paula Rahal; Paulo Inacio da Costa; Rafael dos Santos Bezerra; Raquel de Lello Rocha Campos Cassano. NGS Soluções Genômicas: Pilar Drummond Sampaio Corrêa Mariani. FZEA-USP Pirassununga: Mirele Daiana Poleti; Raul Machado Neto; Ricardo Augusto Brassaloti; Ricardo Haddad; Rodrigo Tocantins Calado. FAMERP-SJRP: Cecília Artico Banho; Sandra Coccuzzo Sampaio; Svetoslav Nanev Slavov; Vagner Fonseca; Vincent Louis Viala |
| EPI_ISL_1532765 | NYU Langone Health | Departments of Pathology and Medicine, New York University School of Medicine | Adriana Heguy; Christian Marier; Dacia Dimartino; Emily Guzman; Gael Westby; Guiqing Wang; Paolo Cotzia; Paul Zappile; Peter Meyn; Sitharam Ramaswami; Yutong Zhang |
| EPI_ISL_1661782 | National Food and Veterinary Risk Assessment Institute | Vilnius University Hospital Santaros Klinikos, Center of Laboratory Medicine | Daniel Naumovas; Dovile Ezerskyte; Gytis Dudas; Ingrida Olendraite; Laimonas Griskevicius; Ligita Raugaite; Mindaugas Stoskus; Monika Katenaite; Rimvydas Norvilas |
| EPI_ISL_1657093 | National Institute of Infectious Diseases-Prof. Dr. Matei Bals Molecular Diagnostics Laboratory | National Institute of Infectious Diseases-Prof. Dr. Matei Bals Molecular Diagnostics Laboratory | Andreea Tudor; Corina Casangiu; Dan Otelea; Leontina Banica; Marius Surleac; Ovidiu Vlaicu; Simona Paraschiv |
| EPI_ISL_1524773, EPI_ISL_1623674, EPI_ISL_1623682, EPI_ISL_1665200, EPI_ISL_1719853, EPI_ISL_1801745 | National Platform bis UMONS/Jolimont | National Platform bis UMONS/Jolimont | Florian Juszczak; François Dufrasne; Gautier Detry; Guillaume Bayon-Vicente; Ruddy Wattiez |
| EPI_ISL_1444685, EPI_ISL_1626543, EPI_ISL_2030453 | Nebraska Public Health Laboratory | NPHL COVID-19 Response Team | NPHL COVID-19 Response Team |
| EPI_ISL_1620692 | North Dakota Department of Health, Public Health Laboratory | North Dakota Department of Health, Public Health Laboratory | Lisa Wingerter |
| EPI_ISL_1704688 | Northwestern Memorial Hospital | Northwestern University - Ozer Lab | Chad J. Achenbach; Chao Qi; Egon A. Ozer; Judd F. Hultquist; Lacy M. Simons; Lawrence J. Jennings; Michael G. Ison; Ramon Lorenzo-Redondo; Taylor J. Dean |
| EPI_ISL_1897570 | Norwegian Institute of Public Health, Department of Virology | Norwegian Institute of Public Health, Department of Virology | Atiya R Ali; Debech Nadia; Engebretsen Serina Beate; Garcia Llorente Ignacio; Hilde Elshaug; Hilde Vollan; Jon Bråte; Kamilla Heddeland Instefjord; Karoline Bragstad; Kathrine Stene-Johansen; Marie Paulsen Madsen; Olav Hungnes; Pedersen Benedikte Nevjen; Rasmus Riis Kopperud |
| EPI_ISL_5799815, EPI_ISL_5799816 | Nucleo De Saude Vila Falcao De Bauru | Instituto Butantan | Antonio Jorge Martins; Claudia Renata dos Santos Barros; David Schlesinger; Debora Botequio Moretti; Dimas Tadeu Covas; Elaine Cristina Marqueze; Elaine Vieira Santos; Evandra Strazza Rodrigues; Heidge Fukumasu; Jayme Augusto de Souza-Neto; José Salvatore Leister Patané; Luiz Alcantara; Luiz Lehmann Coutinho; Maria Carolina Elias; Maurício Lacerda Nogueira; Rafael dos Santos Bezerra; Raul Machado Neto; Rejane Maria Tommasini Grotto; Ricardo Haddad; Sandra Coccuzzo Sampaio Vessoni; Simone Kashima; Svetoslav Nanev Slavov; Vincent Louis Viala |
| EPI_ISL_1701786, EPI_ISL_1701787, EPI_ISL_1701788 | OHSU Lab Services Molecular Microbiology Lab | Oregon SARS-CoV-2 Genome Sequencing Center | Alec J. Hirsch; Andrew C. Adey; Benjamin N. Bimber; Brendan L. O'Connell; Brian J. O'Roak; Cierra LaBlanc; Daniel N. Streblow; Donna Hansel; Guang Fan; Kayla Carter; Ruth V. Nichols; Sally Grindstaff; Sonia Acharya; William B. Messer; Xuan Qin |
| EPI_ISL_1759374 | OK Public Health Laboratory, Oklahoma State DOH | Pathogen Discovery, Respiratory Viruses Branch, Division of Viral Diseases, Centers for Disease Control and Prevention | Adam Retchless; Anna Kelleher; Anna Montmayeur; Anna Uehara; Brian Lynch; Clinton R. Paden; Haibin Wang; Han Jia Justin Ng; Jing Zhang; Justin Lee; Krista Queen; Mark Burroughs; Peter Cook; Rachel Marine; Suxiang Tong; Yan Li; Ying Tao |
| EPI_ISL_1620231 | OLVZ Aalst | OLVZ Aalst | Astrid Holderbeke |
| EPI_ISL_1701833 | OR State PHL-Virology/Immunology Section | Centers for Disease Control and Prevention Division of Viral Diseases, Pathogen Discovery | Alison Laufer Halpin; Ben L. Rambo-Martin; Clinton R. Paden; Dakota Howard; Darlene Wagner; Dave Wentworth; Dhwani Batra; Jasmine Padilla; Justin Lee; Katie Dillon; Krista Queen; Kristen Knipe; Kristine Lacek; Mark Burroughs; Matthew Schmerer; Mili Sheth; Peter Cook; Sam Shepard; Sarah Nobles; Shoshona Le; Suxiang Tong; Vivien Dugan; Yvette Unoarumhi |
| EPI_ISL_1578011, EPI_ISL_1578016 | OSPEDALE SAN SALVATORE - MEDICINA DI LABORATORIO L'AQUILA(L'AQUILA) | Istituto Zooprofilattico Sperimentale dell'Abruzzo e Molise "G. Caporale" | Ancora M; Calistri P; Cammà C; Curini V; Di Domenico M; Di Pasquale A; Lorusso A; Mangone I; Marcacci M; Puglia I; Rinaldi A; Savini G; Scialabba S |
| EPI_ISL_1686350 | Ohio Department of Health Laboratory | Ohio Department of Health Laboratory | Allison Black; Brent Lee; Caitlin McDonnell; Eric Brandt; Erica Leasure; Glen McGillivary; Heather Blankenship; Holmes; Jade Mowery; Jennifer; Kelsey Florek; Keoni Omura; Kirtana Ramadugu; Quanta Brown; Stephanie Mccracken; Tyler Payne; and Tammy Bannerman |
| EPI_ISL_7275325, EPI_ISL_7275335, EPI_ISL_7275393, EPI_ISL_7275450 | Ontario's COVID-19 Genomics Rapid Response Coalition | McMaster University | Ahmed Draia; Allison McGeer; Andrew G. McArthur; Angel Li; Emily Panousis; Hooman Derakhshani; Jalees Nasir; Kuganya Nirmalarajah; Michael Surette; Patryk Aftanas; Samira Mubareka; Sheridan Baker |
| EPI_ISL_1511642, EPI_ISL_1585676 | Oregon State Public Health Laboratory | Oregon State Public Health Laboratory | Eugene Yeboah; John Fontana and Shane Sevey; Laura Tsaknaridis; Rafia Razzaque; Vanda Makris |
| EPI_ISL_1759354 | PA Department of Health, Bureau of Laboratories | Pathogen Discovery, Respiratory Viruses Branch, Division of Viral Diseases, Centers for Disease Control and Prevention | Adam Retchless; Anna Kelleher; Anna Montmayeur; Anna Uehara; Brian Lynch; Clinton R. Paden; Haibin Wang; Han Jia Justin Ng; Jing Zhang; Justin Lee; Krista Queen; Mark Burroughs; Peter Cook; Rachel Marine; Suxiang Tong; Yan Li; Ying Tao |
| EPI_ISL_1966467 | POLICLINICA COVID 19 ITAPETININGA | Instituto Butantan / Mendelics | Antonio Jorge Martins; Bianca Cechetto Carlos. Mendelics: Bibiana Santos; Claudia Renata dos Santos Barros; Cíntia Bittar; David Schlesinger. Hemocentro Ribeirão Preto: Simone Kashima; Debora Botequio Moretti; Elaine Cristina Marqueze; Elaine Vieira dos Santos; Elisangela Chicaroni Mattos; Erika Freitas; Evandra Strazza Rodrigues; Felipe Allan da Silva da Costa; Flavia Aburjaile; Fábio Sossai Possebon; Guilherme Campos; Guilherme Targino Valente; Heidge Fukumasu. USP-Botucatu: Rejane Maria Tommasini Grotto; Helena Lage Ferreira; Instituto Butantan: Dimas Tadeu Covas; Jardelina de Souza Todao Bernardino; Jayme A. Souza-Neto; Jessika Cristina Chagas Lesbon; Jorge A. Petroli Marchesi; José Salvatore Leister Patané; João Paulo Kitajima; João Pessoa Araújo Jr.; Leila Sabrina Ullmann; Loyze Paola Oliveira de Lima; Luiz Aurelio de Campos Crispin. Centro de Genômica Funcional da ESALQ: Luiz Lehmann |

| | | | |
|---|---|---|---|
| | | | Coutinho; Luiz Carlos Junior de Alcantara; Lívia Sacchetto; Maisa C. Pereira Parra; Maria Carolina Elias; Marta Giovanetti; Marília Moraes; Maurício Lacerda Nogueira. Prefeitura de Sao Paulo: Melissa Palmieri.; Patricia Akemi Assato; Paula Rahal; Paulo Inacio da Costa; Rafael dos Santos Bezerra; Raquel de Lello Rocha Campos Cassano. NGS Soluções Genômicas: Pilar Drummond Sampaio Corrêa Mariani. FZEA-USP Pirassununga: Mirele Daiana Poleti; Raul Machado Neto; Ricardo Augusto Brassaloti; Ricardo Haddad; Rodrigo Tocantins Calado. FAMERP-SJRP: Cecília Artico Banho; Sandra Coccuzzo Sampaio; Svetoslav Nanev Slavov; Vagner Fonseca; Vincent Louis Viala |
| EPI_ISL_5530149 | POSTO DE SAUDE DE QUIXERE | Analytical Competence Molecular Epidemiology Lab/ACME, Oswaldo Cruz Foundation, Ceara (FIOCRUZ CE) | Carlos Leonardo de Aragao Araujo; Cecilia Leite Costa & Eduardo Ruback dos Santos on behalf of COVID-19 FIOCRUZ Genomic Network; Cleber Furtado Aksenen; Fabio Miyajima; Fernando Braga Stehling; Francisco Eder de Moura Lopes; Igor Oliveira Duarte; Jamille Maria Mendes Bezerra; Joaquim Cesar do Nascimento Sousa Junior; Pedro Miguel Carneiro Jeronimo; Suzana Porto Almeida; Thais Ferreira de Oliveira; Thais de Oliveira Costa; Ticiane Cavalcante de Souza; Veridiana Pessoa Miyajima |
| EPI_ISL_1711685 | PR Public Health Lab | Centers for Disease Control and Prevention Division of Viral Diseases, Pathogen Discovery | Alison Laufer Halpin; Ben L. Rambo-Martin; Clinton R. Paden; Dakota Howard; Darlene Wagner; Dave Wentworth; Dhwani Batra; Jasmine Padilla; Justin Lee; Katie Dillon; Krista Queen; Kristen Knipe; Kristine Lacek; Mark Burroughs; Matthew Schmerer; Mili Sheth; Peter Cook; Sam Shepard; Sarah Nobles; Shoshona Le; Suxiang Tong; Vivien Dugan; Yvette Unoarumhi |
| EPI_ISL_2209408 | PRONTO ATENDIMENTO MUNICIPAL ALUMINIO | Instituto Butantan | Antonio Jorge Martins; Claudia Renata dos Santos Barros; David Schlesinger; Debora Botequio Moretti; Dimas Tadeu Covas; Elaine Cristina Marqueze; Elaine Vieira Santos; Evandra Strazza Rodrigues; Heidge Fukumasu; Jayme Augusto de Souza-Neto; José Salvatore Leister Patané; Luiz Alcantara; Luiz Lehmann Coutinho; Maria Carolina Elias; Maurício Lacerda Nogueira; Rafael dos Santos Bezerra; Raul Machado Neto; Rejane Maria Tommasini Grotto; Ricardo Haddad; Sandra Coccuzzo Sampaio Vessoni; Simone Kashima; Svetoslav Nanev Slavov; Vincent Louis Viala. |
| EPI_ISL_2344548 | PRONTO ATENDIMENTO MUNICIPAL ALUMINIO | Instituto Butantan / UNESP-Botucatu | Antonio Jorge Martins; Claudia Renata dos Santos Barros; David Schlesinger; Debora Botequio Moretti; Dimas Tadeu Covas; Elaine Cristina Marqueze; Elaine Vieira Santos; Evandra Strazza Rodrigues; Heidge Fukumasu; Jayme Augusto de Souza-Neto; José Salvatore Leister Patané; Luiz Alcantara; Luiz Lehmann Coutinho; Maria Carolina Elias; Maurício Lacerda Nogueira; Rafael dos Santos Bezerra; Raul Machado Neto; Rejane Maria Tommasini Grotto; Ricardo Haddad; Sandra Coccuzzo Sampaio Vessoni; Simone Kashima; Svetoslav Nanev Slavov; Vincent Louis Viala |
| EPI_ISL_1966508 | PRONTO SOCORRO MUNICIPAL DE SEVERINIA | Instituto Butantan / Mendelics | Antonio Jorge Martins; Bianca Cechetto Carlos. Mendelics: Bibiana Santos; Claudia Renata dos Santos Barros; Cíntia Bittar; David Schlesinger. Hemocentro Ribeirão Preto: Simone Kashima; Debora Botequio Moretti; Elaine Cristina Marqueze; Elaine Vieira dos Santos; Elisangela Chicaroni Mattos; Erika Freitas; Evandra Strazza Rodrigues; Felipe Allan da Silva da Costa; Flavia Aburjaile; Fábio Sossai Possebon; Guilherme Campos; Guilherme Targino Valente; Heidge Fukumasu. USP-Botucatu: Rejane Maria Tommasini Grotto; Helena Lage Ferreira; Instituto Butantan: Dimas Tadeu Covas; Jardelina de Souza Todao Bernardino; Jayme A. Souza-Neto; Jessika Cristina Chagas Lesbon; Jorge A. Petroli Marchesi; José Salvatore Leister Patané; João Paulo Kitajima; João Pessoa Araújo Jr.; Leila Sabrina Ullmann; Loyze Paola Oliveira de Lima; Luiz Aurelio de Campos Crispin. Centro de Genômica Funcional da ESALQ: Luiz Lehmann Coutinho; Luiz Carlos Junior de Alcantara; Lívia Sacchetto; Maisa C. Pereira Parra; Maria Carolina Elias; Marta Giovanetti; Marília Moraes; Maurício Lacerda Nogueira. Prefeitura de Sao Paulo: Melissa Palmieri.; Patricia Akemi Assato; Paula Rahal; Paulo Inacio da Costa; Rafael dos Santos Bezerra; Raquel de Lello Rocha Campos Cassano. NGS Soluções Genômicas: Pilar Drummond Sampaio Corrêa Mariani. FZEA-USP Pirassununga: Mirele Daiana Poleti; Raul Machado Neto; Ricardo Augusto Brassaloti; Ricardo Haddad; Rodrigo Tocantins Calado. FAMERP-SJRP: Cecília Artico Banho; Sandra Coccuzzo Sampaio; Svetoslav Nanev Slavov; Vagner Fonseca; Vincent Louis Viala |
| EPI_ISL_5529873 | PSF DE VILA ESPERANCA | Analytical Competence Molecular Epidemiology Lab/ACME, Oswaldo Cruz Foundation, Ceara (FIOCRUZ CE) | Carlos Leonardo de Aragao Araujo; Cecilia Leite Costa & Eduardo Ruback dos Santos on behalf of COVID-19 FIOCRUZ Genomic Network; Cleber Furtado Aksenen; Fabio Miyajima; Fernando Braga Stehling; Francisco Eder de Moura Lopes; Igor Oliveira Duarte; Jamille Maria Mendes Bezerra; Joaquim Cesar do Nascimento Sousa Junior; Pedro Miguel Carneiro Jeronimo; Suzana Porto Almeida; Thais Ferreira de Oliveira; Thais de Oliveira Costa; Ticiane Cavalcante de Souza; Veridiana Pessoa Miyajima |
| EPI_ISL_2209425 | PSF DR ANTONIO PIRES DE ALMEIDA PORTO FELIZ | Instituto Butantan | Antonio Jorge Martins; Claudia Renata dos Santos Barros; David Schlesinger; Debora Botequio Moretti; Dimas Tadeu Covas; Elaine Cristina Marqueze; Elaine Vieira Santos; Evandra Strazza Rodrigues; Heidge Fukumasu; Jayme Augusto de Souza-Neto; José Salvatore Leister Patané; Luiz Alcantara; Luiz Lehmann Coutinho; Maria Carolina Elias; Maurício Lacerda Nogueira; Rafael dos Santos Bezerra; Raul Machado Neto; Rejane Maria Tommasini Grotto; Ricardo Haddad; Sandra Coccuzzo Sampaio Vessoni; Simone Kashima; Svetoslav Nanev Slavov; Vincent Louis Viala. |
| EPI_ISL_2344584 | PSF DR ANTONIO PIRES DE ALMEIDA PORTO FELIZ | Instituto Butantan / UNESP-Botucatu | Antonio Jorge Martins; Claudia Renata dos Santos Barros; David Schlesinger; Debora Botequio Moretti; Dimas Tadeu Covas; Elaine Cristina Marqueze; Elaine Vieira Santos; Evandra Strazza Rodrigues; Heidge Fukumasu; Jayme Augusto de Souza-Neto; José Salvatore Leister Patané; Luiz Alcantara; Luiz Lehmann Coutinho; Maria Carolina Elias; Maurício Lacerda Nogueira; Rafael dos Santos Bezerra; Raul Machado Neto; Rejane Maria Tommasini Grotto; Ricardo Haddad; Sandra Coccuzzo Sampaio Vessoni; Simone Kashima; Svetoslav Nanev Slavov; Vincent Louis Viala |
| EPI_ISL_2209429, EPI_ISL_2209430, EPI_ISL_2209433 | PSF MARIA JOSE SALTO DE PIRAPORA | Instituto Butantan | Antonio Jorge Martins; Claudia Renata dos Santos Barros; David Schlesinger; Debora Botequio Moretti; Dimas Tadeu Covas; Elaine Cristina Marqueze; Elaine Vieira Santos; Evandra Strazza Rodrigues; Heidge Fukumasu; Jayme Augusto de Souza-Neto; José Salvatore Leister Patané; Luiz Alcantara; Luiz Lehmann Coutinho; Maria Carolina Elias; Maurício Lacerda Nogueira; Rafael dos Santos Bezerra; Raul Machado Neto; Rejane Maria Tommasini Grotto; Ricardo Haddad; Sandra Coccuzzo Sampaio Vessoni; Simone Kashima; Svetoslav Nanev Slavov; Vincent Louis Viala. |
| EPI_ISL_2344543, EPI_ISL_2344659, EPI_ISL_2345307 | PSF MARIA JOSE SALTO DE PIRAPORA | Instituto Butantan / UNESP-Botucatu | Antonio Jorge Martins; Claudia Renata dos Santos Barros; David Schlesinger; Debora Botequio Moretti; Dimas Tadeu Covas; Elaine Cristina Marqueze; Elaine Vieira Santos; Evandra Strazza Rodrigues; Heidge Fukumasu; Jayme Augusto de Souza-Neto; José Salvatore Leister Patané; Luiz Alcantara; Luiz Lehmann Coutinho; Maria Carolina Elias; Maurício Lacerda Nogueira; Rafael dos Santos Bezerra; Raul Machado Neto; Rejane Maria Tommasini Grotto; Ricardo Haddad; Sandra Coccuzzo Sampaio Vessoni; Simone Kashima; Svetoslav Nanev Slavov; Vincent Louis Viala |
| EPI_ISL_1542120, EPI_ISL_1542186, EPI_ISL_1542302, EPI_ISL_1542411, EPI_ISL_1542412, EPI_ISL_1542512, EPI_ISL_1542585, EPI_ISL_1542651, EPI_ISL_1542686, EPI_ISL_1542694, EPI_ISL_1542753, EPI_ISL_1542830, EPI_ISL_1542885, EPI_ISL_1542943, EPI_ISL_1543027, EPI_ISL_1543058 | | | |
| see above | Pandemic Response Lab - NYC | Pandemic Response Lab, R&D | Cybill del Castillo; Dylan Law; Haiping Hao; Henry Lee; Jon Laurent; Katharine Nelson; Melissa Hopkins; Michael Hammerling; Pradeep Bugga; Shinyoung Clair Kang; Sol Rey; William Ward |
| EPI_ISL_3982751, EPI_ISL_3982761, EPI_ISL_3982769, EPI_ISL_3982771 | Parana | LACEN PR | Guilherme Nardi Becker; Irina Nastassja Riediger; Maria do Carmo Debur Rossa; Mayra Presibella Giacomini; Nelson Quallio Marques |
| EPI_ISL_3982759, EPI_ISL_3982768 | Parana | Lacen-PR | Guilherme Nardi Becker; Irina Nastassja Riediger; Maria do Carmo Debur Rossa; Mayra Presibella Giacomini; Nelson Quallio Marques |
| EPI_ISL_3982749 | Parana | Lacen/PR | Guilherme Nardi Becker; Irina Nastassja Riediger; Maria do Carmo Debur Rossa; Mayra Presibella Giacomini; Nelson Quallio Marques |
| EPI_ISL_3982742, EPI_ISL_3982745, EPI_ISL_3982763 | Parana | Parana | Guilherme Nardi Becker; Irina Nastassja Riediger; Maria do Carmo Debur Rossa; Mayra Presibella Giacomini; Nelson Quallio Marques |
| EPI_ISL_2328297 | Pathogen Genomics Center, National Institute of Infectious Diseases | Pathogen Genomics Center, National Institute of Infectious Diseases | Kentaro Itokawa; Makoto Kuroda; Masanori Hashino; Rina Tanaka; Tsuyoshi Sekizuka |
| EPI_ISL_3046887 | Pennsylvania Department of Health Bureau of Laboratories | Pennsylvania Department of Health Bureau of Laboratories | Dongxiang Xia |
| EPI_ISL_7040720, EPI_ISL_7040753 | Pesaro | Microbiology University Politecnica delle Marche | Anna Valenza; Carla Acciarri; Katia Marinelli; Monica Lucia Ferreri; Patrizia Bagnarelli; Roberta Longo; Sara Caucci; Stefano Menzo |
| EPI_ISL_2663308 | Plataforma de Vigilancia Molecular (PVM) - FIOCRUZ/BA | Plataforma de Vigilancia Molecular (PVM) - FIOCRUZ/BA | Bruno Bezerril Andrade; Camila I. de Oliveira on behalf of the Fiocruz COVID-19 Genomic Surveillance Network.; Clarissa Araújo Gurgel; Leonardo Paiva Farias; Marina Cucco; Ricardo Khouri; Tiago Graf |
| EPI_ISL_1620203, EPI_ISL_1620218, EPI_ISL_1620222, EPI_ISL_1620773, EPI_ISL_1620797, EPI_ISL_1621199 | Plateforme de testing Namuroise | Plateforme de testing Namuroise | Céline Maschietto; Degosserie Jonathan; Denis Olivier; Mullier François; Otto Gaetan |
| EPI_ISL_1498361, EPI_ISL_1498362, EPI_ISL_1498366, EPI_ISL_1498371, EPI_ISL_1498372 | Platform BIS UZA/UAntwerpen | Labo Klinische Biologie, UZA | Basil Britto Xavier; Christine Lammens; Herman Goossens; Jasmine Coppens; Marie Le Mercier; Veerle Matheeussen |
| EPI_ISL_1558649 | Platform BIS UZA/UAntwerpen | UAntwerp, Laboratory of Medical Microbiology | Basil Britto Xavier; Christine Lammens; Herman Goossens; Jasmine Coppens; Marie Le Mercier; Veerle Matheeussen |
| EPI_ISL_5802191 | Policlinica Covid 19 Itapetininga | Instituto Butantan | Antonio Jorge Martins; Claudia Renata dos Santos Barros; David Schlesinger; Debora Botequio Moretti; Dimas Tadeu Covas; Elaine Cristina Marqueze; Elaine Vieira Santos; Evandra Strazza Rodrigues; Heidge Fukumasu; Jayme Augusto de Souza-Neto; José Salvatore Leister Patané; Luiz Alcantara; Luiz Lehmann Coutinho; Maria Carolina Elias; Maurício Lacerda Nogueira; Rafael dos Santos Bezerra; Raul Machado Neto; Rejane Maria Tommasini Grotto; Ricardo Haddad; Sandra Coccuzzo Sampaio Vessoni; Simone Kashima; Svetoslav Nanev Slavov; Vincent Louis Viala |
| EPI_ISL_5802153 | Pronto Socorro Municipal De Severinia | Instituto Butantan | Antonio Jorge Martins; Claudia Renata dos Santos Barros; David Schlesinger; Debora Botequio Moretti; Dimas Tadeu Covas; Elaine Cristina Marqueze; Elaine Vieira Santos; Evandra Strazza Rodrigues; Heidge Fukumasu; Jayme Augusto de Souza-Neto; José Salvatore Leister Patané; Luiz Alcantara; Luiz Lehmann Coutinho; Maria Carolina Elias; Maurício Lacerda Nogueira; Rafael dos Santos Bezerra; Raul Machado Neto; Rejane Maria Tommasini Grotto; Ricardo Haddad; Sandra Coccuzzo Sampaio Vessoni; Simone Kashima; Svetoslav Nanev Slavov; Vincent Louis Viala |
| EPI_ISL_5802183, EPI_ISL_5802184, EPI_ISL_5802185 | Psf Maria Jose Salto De Pirapora | Instituto Butantan | Antonio Jorge Martins; Claudia Renata dos Santos Barros; David Schlesinger; Debora Botequio Moretti; Dimas Tadeu Covas; Elaine Cristina Marqueze; Elaine Vieira Santos; Evandra Strazza Rodrigues; Heidge Fukumasu; Jayme Augusto de Souza-Neto; José Salvatore Leister Patané; Luiz Alcantara; Luiz Lehmann Coutinho; Maria Carolina Elias; Maurício Lacerda Nogueira; Rafael dos Santos Bezerra; Raul Machado Neto; Rejane Maria Tommasini Grotto; Ricardo Haddad; Sandra Coccuzzo Sampaio Vessoni; Simone Kashima; Svetoslav Nanev Slavov; Vincent Louis Viala |
| EPI_ISL_1581866, EPI_ISL_1581888, EPI_ISL_1582001, EPI_ISL_1582010, EPI_ISL_1582020, EPI_ISL_1582150, EPI_ISL_1695038, EPI_ISL_1695039, EPI_ISL_1695040, EPI_ISL_1695041, EPI_ISL_1695042, EPI_ISL_1695050, EPI_ISL_1695051, EPI_ISL_1695052, EPI_ISL_1695053, EPI_ISL_1695054, EPI_ISL_1695055, EPI_ISL_1695056, EPI_ISL_1695057, EPI_ISL_1695058, EPI_ISL_1695059, EPI_ISL_1695060, EPI_ISL_1695061, EPI_ISL_1695062, EPI_ISL_1695063, EPI_ISL_1695064, EPI_ISL_1695065, EPI_ISL_1695066, EPI_ISL_1695067, EPI_ISL_1695068, EPI_ISL_1695069, EPI_ISL_1695070, EPI_ISL_1695071, EPI_ISL_1695072, EPI_ISL_1695073, EPI_ISL_1695074, EPI_ISL_1695075, EPI_ISL_1695077, EPI_ISL_1695078, EPI_ISL_1695079, EPI_ISL_1695080, EPI_ISL_1695081, EPI_ISL_1695082, EPI_ISL_1695083, EPI_ISL_1695084, EPI_ISL_1695085, EPI_ISL_1695086, EPI_ISL_1695087, EPI_ISL_1695088, EPI_ISL_1695089, EPI_ISL_1695090, EPI_ISL_1695091, EPI_ISL_1695092, EPI_ISL_1695093, EPI_ISL_1695094, EPI_ISL_1695095, EPI_ISL_1695096, EPI_ISL_1695097, EPI_ISL_1695098, EPI_ISL_1695099, EPI_ISL_1695100, EPI_ISL_1695106, EPI_ISL_1695110, EPI_ISL_1695637, EPI_ISL_1695662, EPI_ISL_1695699, EPI_ISL_1695705, EPI_ISL_1695711, EPI_ISL_1695747, EPI_ISL_1695769, EPI_ISL_1702164, EPI_ISL_1702165, EPI_ISL_1702166, EPI_ISL_1702167, EPI_ISL_1702168, EPI_ISL_1702169, EPI_ISL_1702170, EPI_ISL_1702171, EPI_ISL_1702172, EPI_ISL_1702173, EPI_ISL_1702176, EPI_ISL_1702177, EPI_ISL_1702178, EPI_ISL_1753631, EPI_ISL_1753635, EPI_ISL_4366887, EPI_ISL_4370873 | | | |
| see above | Quest Diagnostics Incorporated | Centers for Disease Control and Prevention Division of Viral Diseases, Pathogen Discovery | A. Gerasimova; A. Perez; Adrian Paskey; B. Anderson; Benjamin Rambo-Martin; Christopher Gulvick; Clinton Paden; Clinton R. Paden; Dakota Howard; Darlene Wagner; Dhwani Batra; Duncan MacCannell; Erisa Sula; F. Lacbawan; I. A. Shlyakhter; I. Shlyakhter; Jason Caravas; K. Livingston; K.E. Livingston; Kara Moser; Kristine Lacek; L. Bernstein; L.E. Bernstein; M. Hua; Matthew Schmerer; P. Tanpaiboon; Peter Cook; Peter W. Cook; R. Kagan; R. M. Kagan; R. Owen; R. Rolando; R. V. Rolando; S. H. Rosenthal; S. Rosenthal; Scott Sammons; Shatavia Morrison; Tymeckia Kendall; Victoria Caban Figueroa; Y. Liu; Yvette Unoarumhi |
| EPI_ISL_1798708, EPI_ISL_1798713, EPI_ISL_1798732, EPI_ISL_1798741 | RI State Health Laboratories | Centers for Disease Control and Prevention Division of Viral Diseases, Pathogen Discovery | Alison Laufer Halpin; Ben L. Rambo-Martin; Clinton R. Paden; Dakota Howard; Darlene Wagner; Dave Wentworth; Dhwani Batra; Jasmine Padilla; Justin Lee; Katie Dillon; Krista Queen; Kristen Knipe; Kristine Lacek; Mark Burroughs; Matthew Schmerer; Mili Sheth; Peter Cook; Sam Shepard; Sarah Nobles; Shoshona Le; Suxiang Tong; Vivien Dugan; Yvette Unoarumhi |
| EPI_ISL_2628506 | Rady Children's Hospital - San Diego | Andersen lab at Scripps Research | Christina Clarke; David Dimmock; Denise Malicki; Kathryn Bouic; Linda Luo; SEARCH Alliance San Diego with Nanda Radamchar; Teresa Mueller |
| EPI_ISL_2249921, EPI_ISL_2249922 | Reditus Laboratories | Reditus Laboratories | Alexa Eichelberger; Cassy Philips; Joshua J. Geltz; M.S.; Ph.D.; Robert M. Sgambelluri |
| EPI_ISL_7156279, EPI_ISL_7156281, EPI_ISL_7156282, EPI_ISL_7156298, EPI_ISL_7156301, EPI_ISL_7156302, EPI_ISL_7156303, EPI_ISL_7156304, EPI_ISL_7156305, EPI_ISL_7156848, EPI_ISL_7156887 | | | |
| see above | Research Education in Disease Diagnosis and Intervention (REDDI) Lab, Clemson University | Research Education in Disease Diagnosis and Intervention (REDDI) Lab, Clemson University | Adib Shafi; Brian Krueger; Chloe Emerson; Christopher Parkinson; Christopher Saski; Congyue Peng; Delphine Dean; John Pruitt; Justin Napolitano; Kaitlyn Williams; Keegan Sell; Kylie King; Lax Iyer; Rachel Dango; Rachel Ham; Scott Parker; Stevin Wilson; Sujata Srikanth |
| EPI_ISL_2749808, EPI_ISL_2749813, | Respiratory Virus Unit, Microbiology Services | COVID-19 Genomics UK (COG-UK) Consortium | PHE Covid Sequencing Team |

| | | | |
|---|---|---|---|
| EPI_ISL_2749815, EPI_ISL_2749816 | Colindale, Public Health England | | |
| EPI_ISL_1710191, EPI_ISL_1710230, EPI_ISL_1710233 | Rhode Island Department of Health | Infectious Disease Program, Broad Institute of Harvard and MIT | Adams, G.; Azevedo, K.; B.L.; B.W.; Bauer, M.; Birren; Carter, A.; Chaluvadi, S.; D.J.; DeRuff, K.; Gladden-Young, A.; Huard, R.; J.E.; K.J.; King, E.; Lagerborg, K.; Lemieux; Loreth, C.; Miller, A.; Normandin, E.; P.C.; Park; Pearlman, L.; Reilly, S.; Rudy, M.; Sabeti; Siddle; Tomkins-Tinch, C.; and MacInnis |
| EPI_ISL_2799391 | Riga East University Hospital, National Microbiology Reference Laboratory | Riga East University Hospital, National Microbiology Reference Laboratory; Eurofins Genomics Europe Sequencing GmbH | Arzu Algulieva; Diāna Dušacka; Dārta Pūpola; Ilva Pole; Jevgenijs Bodrenko; Jūlija Čevere; Reinis Vangravs; Reinis Zeltmatis; Sergejs Ņikišins; Ģirts Šķenders |
| EPI_ISL_1966518 | SAE SERVICO DE ATENDIMENTO ESPECIALIZADO | Instituto Butantan / Mendelics | Antonio Jorge Martins; Bianca Cechetto Carlos. Mendelics: Bibiana Santos; Claudia Renata dos Santos Barros; Cíntia Bittar; David Schlesinger. Hemocentro Ribeirão Preto: Simone Kashima; Debora Botequio Moretti; Elaine Cristina Marqueze; Elaine Vieira dos Santos; Elisangela Chicaroni Mattos; Erika Freitas; Evandra Strazza Rodrigues; Felipe Allan da Silva da Costa; Flavia Aburjaile; Fábio Sossai Possebon; Guilherme Campos; Guilherme Targino Valente; Heidge Fukumasu. USP-Botucatu: Rejane Maria Tommasini Grotto; Helena Lage Ferreira; Instituto Butantan: Dimas Tadeu Covas; Jardelina de Souza Todao Bernardino; Jayme A. Souza-Neto; Jessika Cristina Chagas Lesbon; Jorge A. Petroli Marchesi; José Salvatore Leister Patané; João Paulo Kitajima; João Pessoa Araújo Jr.; Leila Sabrina Ullmann; Loyze Paola Oliveira de Lima; Luiz Aurelio de Campos Crispin. Centro de Genômica Funcional da ESALQ: Luiz Lehmann Coutinho; Luiz Carlos Junior de Alcantara; Lívia Sacchetto; Maisa C. Pereira Parra; Maria Carolina Elias; Marta Giovanetti; Marília Moraes; Maurício Lacerda Nogueira. Prefeitura de Sao Paulo: Melissa Palmieri.; Patricia Akemi Assato; Paula Rahal; Paulo Inacio da Costa; Rafael dos Santos Bezerra; Raquel de Lello Rocha Campos Cassano. NGS Soluções Genômicas: Pilar Drummond Sampaio Corrêa Mariani. FZEA-USP Pirassununga: Mirele Daiana Poleti; Raul Machado Neto; Ricardo Augusto Brassaloti; Ricardo Haddad; Rodrigo Tocantins Calado. FAMERP-SJRP: Cecília Artico Banho; Sandra Coccuzzo Sampaio; Svetoslav Nanev Slavov; Vagner Fonseca; Vincent Louis Viala |
| EPI_ISL_2209475, EPI_ISL_2209477, EPI_ISL_2209480 | SANTA CASA DE MISERICORDIA DE TIETE | Instituto Butantan | Antonio Jorge Martins; Claudia Renata dos Santos Barros; David Schlesinger; Debora Botequio Moretti; Dimas Tadeu Covas; Elaine Cristina Marqueze; Elaine Vieira Santos; Evandra Strazza Rodrigues; Heidge Fukumasu; Jayme Augusto de Souza-Neto; José Salvatore Leister Patané; Luiz Alcantara; Luiz Lehmann Coutinho; Maria Carolina Elias; Maurício Lacerda Nogueira; Rafael dos Santos Bezerra; Raul Machado Neto; Rejane Maria Tommasini Grotto; Ricardo Haddad; Sandra Coccuzzo Sampaio Vessoni; Simone Kashima; Svetoslav Nanev Slavov; Vincent Louis Viala. |
| EPI_ISL_2344656, EPI_ISL_2344681, EPI_ISL_2345308 | SANTA CASA DE MISERICORDIA DE TIETE | Instituto Butantan / UNESP-Botucatu | Antonio Jorge Martins; Claudia Renata dos Santos Barros; David Schlesinger; Debora Botequio Moretti; Dimas Tadeu Covas; Elaine Cristina Marqueze; Elaine Vieira Santos; Evandra Strazza Rodrigues; Heidge Fukumasu; Jayme Augusto de Souza-Neto; José Salvatore Leister Patané; Luiz Alcantara; Luiz Lehmann Coutinho; Maria Carolina Elias; Maurício Lacerda Nogueira; Rafael dos Santos Bezerra; Raul Machado Neto; Rejane Maria Tommasini Grotto; Ricardo Haddad; Sandra Coccuzzo Sampaio Vessoni; Simone Kashima; Svetoslav Nanev Slavov; Vincent Louis Viala |
| EPI_ISL_2209487 | SANTA CASA DE PORTO FELIZ | Instituto Butantan | Antonio Jorge Martins; Claudia Renata dos Santos Barros; David Schlesinger; Debora Botequio Moretti; Dimas Tadeu Covas; Elaine Cristina Marqueze; Elaine Vieira Santos; Evandra Strazza Rodrigues; Heidge Fukumasu; Jayme Augusto de Souza-Neto; José Salvatore Leister Patané; Luiz Alcantara; Luiz Lehmann Coutinho; Maria Carolina Elias; Maurício Lacerda Nogueira; Rafael dos Santos Bezerra; Raul Machado Neto; Rejane Maria Tommasini Grotto; Ricardo Haddad; Sandra Coccuzzo Sampaio Vessoni; Simone Kashima; Svetoslav Nanev Slavov; Vincent Louis Viala. |
| EPI_ISL_2344588 | SANTA CASA DE PORTO FELIZ | Instituto Butantan / UNESP-Botucatu | Antonio Jorge Martins; Claudia Renata dos Santos Barros; David Schlesinger; Debora Botequio Moretti; Dimas Tadeu Covas; Elaine Cristina Marqueze; Elaine Vieira Santos; Evandra Strazza Rodrigues; Heidge Fukumasu; Jayme Augusto de Souza-Neto; José Salvatore Leister Patané; Luiz Alcantara; Luiz Lehmann Coutinho; Maria Carolina Elias; Maurício Lacerda Nogueira; Rafael dos Santos Bezerra; Raul Machado Neto; Rejane Maria Tommasini Grotto; Ricardo Haddad; Sandra Coccuzzo Sampaio Vessoni; Simone Kashima; Svetoslav Nanev Slavov; Vincent Louis Viala |
| EPI_ISL_2648633 | SARS-CoV-2 Sequencing Castilla y Leon-Spain Consortium | SARS-CoV-2 Sequencing Castilla y Leon-Spain Consortium | Antonio Orduña-Domingo; Carlos Fuster Foz; Carmen Aldea-Mansilla; Carmen Gimeno Crespo; David Abad; Gregoria Megías Lobón; Jose María Eiros Bouza; Laura Sánchez de Prada; M. Isabel Fernandez-Natal; Marta Dominguez-Gil; Marta Hernandez; María Antonia García Castro; Mª Fe Brezmes-Valdivieso; Noelia Arenal Andrés; Silvia Rojo; Sonsoles Garcinuño Pérez |
| EPI_ISL_1927238, EPI_ISL_1927239 | SARS-CoV-2 testing team, National Institute of Infectious Diseases | Pathogen Genomics Center, National Institute of Infectious Diseases | Hazuka Y Furihata; Kentaro Itokawa; Makoto Kuroda; Masanori Hashino; Masumichi Saito; Naomi Nojiri; Nozomu Hanaoka; Rina Tanaka; Sana Uchikoba; Tsuguto Fujimoto; Tsuyoshi Sekizuka |
| EPI_ISL_2209490 | SAUDE COLETIVA CAPAO BONITO | Instituto Butantan | Antonio Jorge Martins; Claudia Renata dos Santos Barros; David Schlesinger; Debora Botequio Moretti; Dimas Tadeu Covas; Elaine Cristina Marqueze; Elaine Vieira Santos; Evandra Strazza Rodrigues; Heidge Fukumasu; Jayme Augusto de Souza-Neto; José Salvatore Leister Patané; Luiz Alcantara; Luiz Lehmann Coutinho; Maria Carolina Elias; Maurício Lacerda Nogueira; Rafael dos Santos Bezerra; Raul Machado Neto; Rejane Maria Tommasini Grotto; Ricardo Haddad; Sandra Coccuzzo Sampaio Vessoni; Simone Kashima; Svetoslav Nanev Slavov; Vincent Louis Viala. |
| EPI_ISL_2344657 | SAUDE COLETIVA CAPAO BONITO | Instituto Butantan / UNESP-Botucatu | Antonio Jorge Martins; Claudia Renata dos Santos Barros; David Schlesinger; Debora Botequio Moretti; Dimas Tadeu Covas; Elaine Cristina Marqueze; Elaine Vieira Santos; Evandra Strazza Rodrigues; Heidge Fukumasu; Jayme Augusto de Souza-Neto; José Salvatore Leister Patané; Luiz Alcantara; Luiz Lehmann Coutinho; Maria Carolina Elias; Maurício Lacerda Nogueira; Rafael dos Santos Bezerra; Raul Machado Neto; Rejane Maria Tommasini Grotto; Ricardo Haddad; Sandra Coccuzzo Sampaio Vessoni; Simone Kashima; Svetoslav Nanev Slavov; Vincent Louis Viala |
| EPI_ISL_1966484, EPI_ISL_1966487, EPI_ISL_1966491 | SECAO CENTRO DE DIAGNOSTICO SECEDI | Instituto Butantan / Mendelics | Antonio Jorge Martins; Bianca Cechetto Carlos. Mendelics: Bibiana Santos; Claudia Renata dos Santos Barros; Cíntia Bittar; David Schlesinger. Hemocentro Ribeirão Preto: Simone Kashima; Debora Botequio Moretti; Elaine Cristina Marqueze; Elaine Vieira dos Santos; Elisangela Chicaroni Mattos; Erika Freitas; Evandra Strazza Rodrigues; Felipe Allan da Silva da Costa; Flavia Aburjaile; Fábio Sossai Possebon; Guilherme Campos; Guilherme Targino Valente; Heidge Fukumasu. USP-Botucatu: Rejane Maria Tommasini Grotto; Helena Lage Ferreira; Instituto Butantan: Dimas Tadeu Covas; Jardelina de Souza Todao Bernardino; Jayme A. Souza-Neto; Jessika Cristina Chagas Lesbon; Jorge A. Petroli Marchesi; José Salvatore Leister Patané; João Paulo Kitajima; João Pessoa Araújo Jr.; Leila Sabrina Ullmann; Loyze Paola Oliveira de Lima; Luiz Aurelio de Campos Crispin. Centro de Genômica Funcional da ESALQ: Luiz Lehmann Coutinho; Luiz Carlos Junior de Alcantara; Lívia Sacchetto; Maisa C. Pereira Parra; Maria Carolina Elias; Marta Giovanetti; Marília Moraes; Maurício Lacerda Nogueira. Prefeitura de Sao Paulo: Melissa Palmieri.; Patricia Akemi Assato; Paula Rahal; Paulo Inacio da Costa; Rafael dos Santos Bezerra; Raquel de Lello Rocha Campos Cassano. NGS Soluções Genômicas: Pilar Drummond Sampaio Corrêa Mariani. FZEA-USP Pirassununga: Mirele Daiana Poleti; Raul Machado Neto; Ricardo Augusto Brassaloti; Ricardo Haddad; Rodrigo Tocantins Calado. FAMERP-SJRP: Cecília Artico Banho; Sandra Coccuzzo Sampaio; Svetoslav Nanev Slavov; Vagner Fonseca; Vincent Louis Viala |
| EPI_ISL_5530048 | SECRETARIA MUNICIPAL DA SAUDE DE ITAPIUNA | Analytical Competence Molecular Epidemiology Lab/ACME, Oswaldo Cruz Foundation, Ceara (FIOCRUZ CE) | Carlos Leonardo de Aragao Araujo; Cecilia Leite Costa & Eduardo Ruback dos Santos on behalf of COVID-19 FIOCRUZ Genomic Network; Cleber Furtado Aksenen; Fabio Miyajima; Fernando Braga Stehling; Francisco Eder de Moura Lopes; Igor Oliveira Duarte; Jamille Maria Mendes Bezerra; Joaquim Cesar do Nascimento Sousa Junior; Pedro Miguel Carneiro Jeronimo; Suzana Porto Almeida; Thais Ferreira de Oliveira; Thais de Oliveira Costa; Ticiane Cavalcante de Souza; Veridiana Pessoa Miyajima |
| EPI_ISL_1966551 | SECRETARIA MUNICIPAL DE SAUDE | Instituto Butantan / FZEA-USP (Pirassununga) | Antonio Jorge Martins; Bianca Cechetto Carlos. Mendelics: Bibiana Santos; Claudia Renata dos Santos Barros; Cíntia Bittar; David Schlesinger. Hemocentro Ribeirão Preto: Simone Kashima; Debora Botequio Moretti; Elaine Cristina Marqueze; Elaine Vieira dos Santos; Elisangela Chicaroni Mattos; Erika Freitas; Evandra Strazza Rodrigues; Felipe Allan da Silva da Costa; Flavia Aburjaile; Fábio Sossai Possebon; Guilherme Campos; Guilherme Targino Valente; Heidge Fukumasu. USP-Botucatu: Rejane Maria Tommasini Grotto; Helena Lage Ferreira; Instituto Butantan: Dimas Tadeu Covas; Jardelina de Souza Todao Bernardino; Jayme A. Souza-Neto; Jessika Cristina Chagas Lesbon; Jorge A. Petroli Marchesi; José Salvatore Leister Patané; João Paulo Kitajima; João Pessoa Araújo Jr.; Leila Sabrina Ullmann; Loyze Paola Oliveira de Lima; Luiz Aurelio de Campos Crispin. Centro de Genômica Funcional da ESALQ: Luiz Lehmann Coutinho; Luiz Carlos Junior de Alcantara; Lívia Sacchetto; Maisa C. Pereira Parra; Maria Carolina Elias; Marta Giovanetti; Marília Moraes; Maurício Lacerda Nogueira. Prefeitura de Sao Paulo: Melissa Palmieri.; Patricia Akemi Assato; Paula Rahal; Paulo Inacio da Costa; Rafael dos Santos Bezerra; Raquel de Lello Rocha Campos Cassano. NGS Soluções Genômicas: Pilar Drummond Sampaio Corrêa Mariani. FZEA-USP Pirassununga: Mirele Daiana Poleti; Raul Machado Neto; Ricardo Augusto Brassaloti; Ricardo Haddad; Rodrigo Tocantins Calado. FAMERP-SJRP: Cecília Artico Banho; Sandra Coccuzzo Sampaio; Svetoslav Nanev Slavov; Vagner Fonseca; Vincent Louis Viala |
| EPI_ISL_5530049 | SECRETARIA MUNICIPAL DE SAUDE DE JAGUARIBE | Analytical Competence Molecular Epidemiology Lab/ACME, Oswaldo Cruz Foundation, Ceara (FIOCRUZ CE) | Carlos Leonardo de Aragao Araujo; Cecilia Leite Costa & Eduardo Ruback dos Santos on behalf of COVID-19 FIOCRUZ Genomic Network; Cleber Furtado Aksenen; Fabio Miyajima; Fernando Braga Stehling; Francisco Eder de Moura Lopes; Igor Oliveira Duarte; Jamille Maria Mendes Bezerra; Joaquim Cesar do Nascimento Sousa Junior; Pedro Miguel Carneiro Jeronimo; Suzana Porto Almeida; Thais Ferreira de Oliveira; Thais de Oliveira Costa; Ticiane Cavalcante de Souza; Veridiana Pessoa Miyajima |
| EPI_ISL_3102297 | SECRETARIA MUNICIPAL DE SAUDE DE TIANGUA | Analytical Competence Molecular Epidemiology Lab/ACME, Oswaldo Cruz Foundation, Ceara (FIOCRUZ CE) | Cleber Furtado Aksenen; Fabio Miyajima; Fernando Braga Stehling; Francisco Eder de Moura Lopes; Jamille Maria Mendes Bezerra; Joaquim César do Nascimento Sousa Junior; Pedro Miguel Carneiro Jeronimo; Suzana Porto Almeida e Lucas Delerino; Thais Ferreira de Oliveira; Thais de Oliveira Costa; Ticiane Cavalcante de Souza; Veridiana Pessoa Miyajima |
| EPI_ISL_2209632, EPI_ISL_2209641, EPI_ISL_2209647 | SMS IPERO | Instituto Butantan | Antonio Jorge Martins; Claudia Renata dos Santos Barros; David Schlesinger; Debora Botequio Moretti; Dimas Tadeu Covas; Elaine Cristina Marqueze; Elaine Vieira Santos; Evandra Strazza Rodrigues; Heidge Fukumasu; Jayme Augusto de Souza-Neto; José Salvatore Leister Patané; Luiz Alcantara; Luiz Lehmann Coutinho; Maria Carolina Elias; Maurício Lacerda Nogueira; Rafael dos Santos Bezerra; Raul Machado Neto; Rejane Maria Tommasini Grotto; Ricardo Haddad; Sandra Coccuzzo Sampaio Vessoni; Simone Kashima; Svetoslav Nanev Slavov; Vincent Louis Viala. |
| EPI_ISL_2344541, EPI_ISL_2344586, EPI_ISL_2345642 | SMS IPERO | Instituto Butantan / UNESP-Botucatu | Antonio Jorge Martins; Claudia Renata dos Santos Barros; David Schlesinger; Debora Botequio Moretti; Dimas Tadeu Covas; Elaine Cristina Marqueze; Elaine Vieira Santos; Evandra Strazza Rodrigues; Heidge Fukumasu; Jayme Augusto de Souza-Neto; José Salvatore Leister Patané; Luiz Alcantara; Luiz Lehmann Coutinho; Maria Carolina Elias; Maurício Lacerda Nogueira; Rafael dos Santos Bezerra; Raul Machado Neto; Rejane Maria Tommasini Grotto; Ricardo Haddad; Sandra Coccuzzo Sampaio Vessoni; Simone Kashima; Svetoslav Nanev Slavov; Vincent Louis Viala |
| EPI_ISL_2209653, EPI_ISL_2209661 | SMS SECRETARIA MUNICIPAL DE SAUDE DE BOITUVA | Instituto Butantan | Antonio Jorge Martins; Claudia Renata dos Santos Barros; David Schlesinger; Debora Botequio Moretti; Dimas Tadeu Covas; Elaine Cristina Marqueze; Elaine Vieira Santos; Evandra Strazza Rodrigues; Heidge Fukumasu; Jayme Augusto de Souza-Neto; José Salvatore Leister Patané; Luiz Alcantara; Luiz Lehmann Coutinho; Maria Carolina Elias; Maurício Lacerda Nogueira; Rafael dos Santos Bezerra; Raul Machado Neto; Rejane Maria Tommasini Grotto; Ricardo Haddad; Sandra Coccuzzo Sampaio Vessoni; Simone Kashima; Svetoslav Nanev Slavov; Vincent Louis Viala. |
| EPI_ISL_2344591, EPI_ISL_2345306 | SMS SECRETARIA MUNICIPAL DE SAUDE DE BOITUVA | Instituto Butantan / UNESP-Botucatu | Antonio Jorge Martins; Claudia Renata dos Santos Barros; David Schlesinger; Debora Botequio Moretti; Dimas Tadeu Covas; Elaine Cristina Marqueze; Elaine Vieira Santos; Evandra Strazza Rodrigues; Heidge Fukumasu; Jayme Augusto de Souza-Neto; José Salvatore Leister Patané; Luiz Alcantara; Luiz Lehmann Coutinho; Maria Carolina Elias; Maurício Lacerda Nogueira; Rafael dos Santos Bezerra; Raul Machado Neto; Rejane Maria Tommasini Grotto; Ricardo Haddad; Sandra Coccuzzo Sampaio Vessoni; Simone Kashima; Svetoslav Nanev Slavov; Vincent Louis Viala |
| EPI_ISL_1671329 | STONY BROOK UNIVERSITY HOSPITAL | Wadsworth Center, New York State Department of Health | Alexis Russell; Catharine Prussing; Daryl M. Lamson; Erasmus Schneider; Erica Lasek-Nesselquist; John Kelly; Jonathan Plitnick; Kirsten St. George; Matthew Shudt; Melissa A Leisner; Navjot Singh |
| EPI_ISL_6512569 | SURA | Laboratorio Departamental de Salud Publica de Antioquia | Ana Victoria Valencia Duarte; Cristian Arbey Velarde Hoyos; Gloria Isabel Escobar; Idabely Betancur Ortiz; Juan P. Hernandez-Ortiz; Juan Pablo Isaza Agudelo; Maria Stella López |
| EPI_ISL_1820908 | SURA | Universidad Nacional de Colombia - Laboratorio Genómico One Health | Andres F. Cardona-Rios; Carlos Franco-Muñoz; Daniel O. Maldonado-Perez; Diego A. Álvarez-Díaz; Hector Alejandro Ruiz-Moreno; Idabely Betancur Ortiz; Jorge E. Osorio; Juan P. Hernandez-Ortiz; Karl A Ciuoderis; Katherine Laiton-Donato; Laura Silvana Perez; Lina M. Hurtado; Marcela Mercado-Reyes; Maria Angélica Maya; Maria Stella López; Rita Almanza Payares; Sandra Ines Cano; Simón Villegas Velásquez |
| EPI_ISL_1595678, EPI_ISL_1660649, EPI_ISL_1660653, EPI_ISL_1660654, EPI_ISL_1660682, EPI_ISL_1660699, EPI_ISL_1671953, EPI_ISL_1671960, EPI_ISL_1672227, EPI_ISL_1672232, EPI_ISL_1672237, EPI_ISL_1672238, EPI_ISL_1672239, EPI_ISL_1672240, EPI_ISL_1672249 | | | |
| see above | SYNLAB | GIGA Medical Genomics | Bouchra Boujemla; Cécile Meex; Keith Durkin; Maria Artesi; Marie-Pierre Hayette; Nathalie Renotte; Pierrette Melin; Raphaël Boreux; Sébastien Bontems; Vincent Bours |
| EPI_ISL_1571685 | SYNLAB Labor MÃ¼nchen Zentrum LMZ | Robert Koch Institute | |
| EPI_ISL_1574661 | SYNLAB MVZ Trier | Robert Koch Institute | |
| EPI_ISL_5802134 | Sae Servico De Atendimento Especializado | Instituto Butantan | Antonio Jorge Martins; Claudia Renata dos Santos Barros; David Schlesinger; Debora Botequio Moretti; Dimas Tadeu Covas; Elaine Cristina Marqueze; Elaine Vieira Santos; Evandra Strazza Rodrigues; Heidge Fukumasu; Jayme Augusto de Souza-Neto; José Salvatore Leister Patané; Luiz Alcantara; Luiz Lehmann Coutinho; Maria Carolina Elias; Maurício Lacerda Nogueira; Rafael dos Santos Bezerra; Raul Machado Neto; Rejane Maria Tommasini Grotto; Ricardo Haddad; Sandra Coccuzzo Sampaio Vessoni; Simone Kashima; Svetoslav Nanev Slavov; Vincent Louis Viala |
| EPI_ISL_1794617, EPI_ISL_1794626, EPI_ISL_1794631, EPI_ISL_1794644, EPI_ISL_1794953, EPI_ISL_1794979, EPI_ISL_1795017, EPI_ISL_1969410 | | | |
| see above | San Diego County Public Health Laboratory | Andersen lab at Scripps Research | Brett Austin; Jovan Shephard; SEARCH Alliance San Diego with Ashleigh Murphy; SEARCH Alliance San Diego with Tracy Basler |
| EPI_ISL_5802188, EPI_ISL_5802190, EPI_ISL_5802194 | Santa Casa De Misericordia De Tiete | Instituto Butantan | Antonio Jorge Martins; Claudia Renata dos Santos Barros; David Schlesinger; Debora Botequio Moretti; Dimas Tadeu Covas; Elaine Cristina Marqueze; Elaine Vieira Santos; Evandra Strazza Rodrigues; Heidge Fukumasu; Jayme Augusto de Souza-Neto; José Salvatore Leister Patané; Luiz Alcantara; Luiz Lehmann Coutinho; Maria Carolina Elias; Maurício Lacerda Nogueira; Rafael dos Santos Bezerra; Raul Machado Neto; Rejane Maria Tommasini Grotto; Ricardo Haddad; Sandra Coccuzzo Sampaio Vessoni; Simone Kashima; Svetoslav Nanev Slavov; Vincent Louis Viala |
| EPI_ISL_1508964, EPI_ISL_1508965 | Santa Clara County Public Health Laboratory | Santa Clara County Public Health Laboratory | Santa Clara County Public Health Department |
| EPI_ISL_1664279, EPI_ISL_1664286, EPI_ISL_1664290 | Santa Clara Valley Medical Center | Santa Clara County Public Health Laboratory | Santa Clara County Public Health Department |
| EPI_ISL_1794727, EPI_ISL_1794732, EPI_ISL_1794737, EPI_ISL_1794740 | Scripps Medical Laboratory | Andersen lab at Scripps Research | Ellen Stefanski; Ian Mchardy; SEARCH Alliance San Diego with Michael Quigley |
| EPI_ISL_2536651, | Seattle Flu Study | Seattle Flu Study | Amanda Adler; Barry R. Lutz; Benjamin Pelle; Caitlin R. Wolf; Chris D. Frazar; Deborah A. Nickerson; Elisabeth Brandstetter; Erica Ryke; Helen Y. Chu; Janet A. Englund; Jay Shendure; Jover Lee; Kairsten Fay; Kirsten Lacombe; Lea M. Starita; Mark J. Rieder; Matthew Richardson; Matthew Thompson; Melissa |

| | | | |
|---|---|---|---|
| EPI_ISL_2536654, EPI_ISL_2536686, EPI_ISL_2536692, EPI_ISL_2736350 | | | Truong; Michael Boeckh; Michael Famulare; Misja Ilcisin; Peter D. Han; Thomas R. Sibley; Trevor Bedford |
| EPI_ISL_5802198, EPI_ISL_5802204, EPI_ISL_5802216 | Secao Centro De Diagnostico Secedi | Instituto Butantan | Antonio Jorge Martins; Claudia Renata dos Santos Barros; David Schlesinger; Debora Botequio Moretti; Dimas Tadeu Covas; Elaine Cristina Marqueze; Elaine Vieira Santos; Evandra Strazza Rodrigues; Heidge Fukumasu; Jayme Augusto de Souza-Neto; José Salvatore Leister Patané; Luiz Alcantara; Luiz Lehmann Coutinho; Maria Carolina Elias; Maurício Lacerda Nogueira; Rafael dos Santos Bezerra; Raul Machado Neto; Rejane Maria Tommasini Grotto; Ricardo Haddad; Sandra Coccuzzo Sampaio Vessoni; Simone Kashima; Svetoslav Nanev Slavov; Vincent Louis Viala |
| EPI_ISL_5802145 | Secretaria Municipal De Saude | Instituto Butantan | Antonio Jorge Martins; Claudia Renata dos Santos Barros; David Schlesinger; Debora Botequio Moretti; Dimas Tadeu Covas; Elaine Cristina Marqueze; Elaine Vieira Santos; Evandra Strazza Rodrigues; Heidge Fukumasu; Jayme Augusto de Souza-Neto; José Salvatore Leister Patané; Luiz Alcantara; Luiz Lehmann Coutinho; Maria Carolina Elias; Maurício Lacerda Nogueira; Rafael dos Santos Bezerra; Raul Machado Neto; Rejane Maria Tommasini Grotto; Ricardo Haddad; Sandra Coccuzzo Sampaio Vessoni; Simone Kashima; Svetoslav Nanev Slavov; Vincent Louis Viala |
| EPI_ISL_2135335, EPI_ISL_2135683, EPI_ISL_2135685, EPI_ISL_2135692, EPI_ISL_2135694, EPI_ISL_2135703, EPI_ISL_2135706, EPI_ISL_2135709, EPI_ISL_2135713, EPI_ISL_2135715, EPI_ISL_2135989, EPI_ISL_2135990, EPI_ISL_2135991, EPI_ISL_2136090, EPI_ISL_2136094, EPI_ISL_6694437 | | | |
| see above | Servicio Virosis Respiratorias-Departamento Virología-INEI | Instituto Nacional Enfermedades Infecciosas C.G.Malbran | Avaro M.; Baumeister E.; Benedetti E.; Campos J.; Cisterna D.; Dattero ME; De Belder D.; Haim MS.; Lorenzo F.; Molina V.; Perandones C.; Poklepovich T.; Pontoriero A.; Russo M.; Sanchez Loria J.; Tuduri E. |
| EPI_ISL_3149732 | Servicio de Inmunología del Hospital Julio Perrando | Laboratorio de Biología Molecular, Instituto de Medicina Regional on behalf of 'Proyecto Argentino Interinstitucional de genomica de SARS-CoV-2' (PAIS Consortium) | Bettina Brusés; Florencia Vallejos Schulze; Griselda Oria; Horacio Lucero.; Javier Mussin; Laura Formichelli; María Delia Foussal; Melina Lorenzini Campos; Raúl Maximiliano Acevedo |
| EPI_ISL_1665127, EPI_ISL_1665136 | Servicio de Microbiología Hospital Ramón y Cajal | Servicio de Microbiología Hospital Ramón y Cajal | JC Galán; JM González-Alba; Laura Martínez. Melanie Abreu; Manuel Ponce |
| EPI_ISL_2016680 | Servicio de Microbiología. Hospital Universitario Doctor Peset | SeqCOVID-SPAIN consortium/IBV(CSIC) | José Miguel Nogueira Coito and SeqCOVID-SPAIN consortium; Juan Alberola Enguídanos; Juan José Camarena Miñana; Rosa González Pellicer |
| EPI_ISL_1657403 | Servizo de Microbioloxía. Hospital Lucus Augusti. Lugo. | Servizo de Microbioloxía. Complexo Hospitalario de Santiago de Compostela | Amparo Coira; Ana Rodríguez Macias; Antonio Aguilera; Antonio Moreno Flores; Carlos García_Riestra; Daniel Navarro; Gema Barbeito; Iria Roca; Javier Alba Domínguez; José Llovo; Julia Pita Carretero; Laura Millán; Laura Sante Fernández; Manuela Hernández; María Luisa Pérez_del_Molino; Mercedes Treviño; Mª José Gude González; Patricia Capón González; Rocío Trastoy; Teresa Lopez_Valiño; Xosé Costa |
| EPI_ISL_5316475, EPI_ISL_5316476, EPI_ISL_5316478, EPI_ISL_5316479, EPI_ISL_5316480, EPI_ISL_5316481, EPI_ISL_5316482, EPI_ISL_5316483, EPI_ISL_5316485, EPI_ISL_5316486, EPI_ISL_5316487, EPI_ISL_5316488, EPI_ISL_5316489, EPI_ISL_5316490, EPI_ISL_5316491, EPI_ISL_5316493, EPI_ISL_5316495, EPI_ISL_5316496, EPI_ISL_5316497, EPI_ISL_5316500, EPI_ISL_5316501, EPI_ISL_5316502, EPI_ISL_5316503, EPI_ISL_5316504, EPI_ISL_5316505, EPI_ISL_5316506, EPI_ISL_5316507, EPI_ISL_5316512, EPI_ISL_5316513, EPI_ISL_5316514, EPI_ISL_5316518, EPI_ISL_5316520, EPI_ISL_5316523, EPI_ISL_5316524, EPI_ISL_5316525, EPI_ISL_5316528, EPI_ISL_5316530, EPI_ISL_5316531, EPI_ISL_5316535, EPI_ISL_5316573, EPI_ISL_5316574, EPI_ISL_5316576, EPI_ISL_5316577, EPI_ISL_5316578, EPI_ISL_5316580, EPI_ISL_5316581, EPI_ISL_5316582, EPI_ISL_5316587, EPI_ISL_5316588, EPI_ISL_5316589, EPI_ISL_5316590, EPI_ISL_5316591, EPI_ISL_5316593, EPI_ISL_5316594, EPI_ISL_5316595, EPI_ISL_5316597, EPI_ISL_5316600, EPI_ISL_5316602, EPI_ISL_5316604, EPI_ISL_5316605, EPI_ISL_5316613, EPI_ISL_5316615, EPI_ISL_5316616, EPI_ISL_5316617, EPI_ISL_5316618, EPI_ISL_5316647, EPI_ISL_5316648, EPI_ISL_5316650, EPI_ISL_5316652, EPI_ISL_5316653, EPI_ISL_5316655, EPI_ISL_5316660, EPI_ISL_5316661, EPI_ISL_5316662, EPI_ISL_5316664, EPI_ISL_5316665, EPI_ISL_5316666, EPI_ISL_5316667, EPI_ISL_5316699, EPI_ISL_5316702, EPI_ISL_5316709, EPI_ISL_5316710, EPI_ISL_5316711, EPI_ISL_5316712, EPI_ISL_5316713, EPI_ISL_5316717, EPI_ISL_5316718, EPI_ISL_5316720, EPI_ISL_5316721, EPI_ISL_5316722, EPI_ISL_5316726, EPI_ISL_5316727, EPI_ISL_5316728, EPI_ISL_5316729, EPI_ISL_5316731, EPI_ISL_5316732, EPI_ISL_5316733, EPI_ISL_5316735 | | | |
| see above | Shared Hospital Laboratory | Shared Hospital Laboratory | Christie Vermeiren; Finlay Maguire; Kevin Katz; Patryk Aftanas; Robert Kozak; Samira Mubareka |
| EPI_ISL_1794755, EPI_ISL_1794766 | Sharp HealthCare Laboratory | Andersen lab at Scripps Research | Art Mendoza; Cathy Woerle; Jacquelyn Berumen; Liam McGinnis; Omid Bakhtar; SEARCH Alliance San Diego with Aaron Harding |
| EPI_ISL_2080163, EPI_ISL_2080165, EPI_ISL_2080174, EPI_ISL_2080175, EPI_ISL_2080189, EPI_ISL_2295927, EPI_ISL_2295932 | | | |
| see above | Simple Laboratories | Gagnon Lab, Southern Illinois University | Keith Gagnon |
| EPI_ISL_5802176, EPI_ISL_5802181, EPI_ISL_5802182 | Sms Ipero | Instituto Butantan | Antonio Jorge Martins; Claudia Renata dos Santos Barros; David Schlesinger; Debora Botequio Moretti; Dimas Tadeu Covas; Elaine Cristina Marqueze; Elaine Vieira Santos; Evandra Strazza Rodrigues; Heidge Fukumasu; Jayme Augusto de Souza-Neto; José Salvatore Leister Patané; Luiz Alcantara; Luiz Lehmann Coutinho; Maria Carolina Elias; Maurício Lacerda Nogueira; Rafael dos Santos Bezerra; Raul Machado Neto; Rejane Maria Tommasini Grotto; Ricardo Haddad; Sandra Coccuzzo Sampaio Vessoni; Simone Kashima; Svetoslav Nanev Slavov; Vincent Louis Viala |
| EPI_ISL_5802180, EPI_ISL_5802208 | Sms Secretaria Municipal De Saude De Boituva | Instituto Butantan | Antonio Jorge Martins; Claudia Renata dos Santos Barros; David Schlesinger; Debora Botequio Moretti; Dimas Tadeu Covas; Elaine Cristina Marqueze; Elaine Vieira Santos; Evandra Strazza Rodrigues; Heidge Fukumasu; Jayme Augusto de Souza-Neto; José Salvatore Leister Patané; Luiz Alcantara; Luiz Lehmann Coutinho; Maria Carolina Elias; Maurício Lacerda Nogueira; Rafael dos Santos Bezerra; Raul Machado Neto; Rejane Maria Tommasini Grotto; Ricardo Haddad; Sandra Coccuzzo Sampaio Vessoni; Simone Kashima; Svetoslav Nanev Slavov; Vincent Louis Viala |
| EPI_ISL_1688708, EPI_ISL_1909379, EPI_ISL_1909390, EPI_ISL_1909546 | Sonora Quest Laboratories | TGen North | "Jolene Bowers; Ashlyn Pfeiffer; Chris French; Darrin Lemmer; Dave Engelthaler; Hayley Yaglom; Heather Centner; Jolene Bowers; The Arizona COVID Genomics Union (ACGU); The Arizona COVID Genomics Union (ACGU)" |
| EPI_ISL_1833823 | St George's University Hospitals NHS Foundation Trust | COVID-19 Genomics UK (COG-UK) Consortium | Adam Witney; Cassie Pope; Irene Monahan; Joshua Taylor; Ken Laing; NgeeKeong Tan |
| EPI_ISL_3722147, EPI_ISL_3722148, EPI_ISL_3722159 | Stanford Health Care | Stanford University School of Medicine, Clinical Virology Laboratory | Becky Jiang; Bernadette Troung; Daniel Solis; James Zehnder; Malaya K. Sahoo; Mamdouh Sibai; Nathan Hammond; and Benjamin A. Pinsky |
| EPI_ISL_1770983, EPI_ISL_1771095 | State Laboratories Division, Hawaii State Department of Health | State Laboratories Division, Hawaii State Department of Health | Ayana Garnet; Drew Kuwazaki; Edward Desmond; Pamela O'Brien; Razvan Sultana |
| EPI_ISL_6782047, EPI_ISL_6782053 | State of New Hampshire Public Health Laboratories | State of New Hampshire Public Health Laboratories | Caitlin Mercier; Chris Benton; Jinfeng Li; Juan Bolanos; Xinglu Zhang |
| EPI_ISL_1808675, EPI_ISL_1902110, EPI_ISL_2257212, EPI_ISL_2257214, EPI_ISL_2257515, EPI_ISL_2257516, EPI_ISL_2416023 | | | |
| see above | Swedish national genomic surveillance program of SARS-CoV-2 | The Public Health Agency of Sweden | Alma Brolund; Maria Lind Karlberg; Maximilian Riess; Swedish national genomic surveillance program of SARS-CoV-2 |
| EPI_ISL_1591823, EPI_ISL_2000958 | TGen North | TGen North | "Jolene Bowers; Ashlyn Pfeiffer; Chris French; Darrin Lemmer; Dave Engelthaler; Hayley Yaglom; Heather Centner; Jolene Bowers; The Arizona COVID Genomics Union (ACGU); The Arizona COVID Genomics Union (ACGU)" |
| EPI_ISL_1710724, EPI_ISL_1754154, EPI_ISL_1754156, EPI_ISL_1754158, EPI_ISL_1754160, EPI_ISL_1754161, EPI_ISL_1754163, EPI_ISL_1754164, EPI_ISL_1754174, EPI_ISL_2081360 | | | |
| see above | Tampa General Hospital Esoteric Lab | Tampa General Hospital Esoteric Research & Development Lab | Amorce Lima; Deanna Becker; Dominic Uy; Grant Vestal; Suzane Silbert; Vicki Healer |
| EPI_ISL_1807253, EPI_ISL_1807288, EPI_ISL_1807289 | Texas Department of State Health Services (TXDSHS) | Texas Department of State Health Services (TXDSHS) | Anita Pokharel; Bonnie Oh; Chun Wang; Grace Kubin; Jenny Zhang; Lorraine Rodriguez; Maliha Rahman; Mayela Pedrueza; Myong Koag; Rachel Lee; Rashmi Tuladhar |
| EPI_ISL_1540817, EPI_ISL_1540831 | The Jackson Laboratory | The Jackson Laboratory | Adams M; Bergeron D; Kelly K; Li L; Long J; Omerza G; Renzette N |
| EPI_ISL_3930405 | The National University Hospital of Iceland | deCODE genetics | Agnar Helgason; Alma Moller; Arna B Agustsdottir; Arnaldur Gylfason; Asgeir Sigurdsson; Aslaug Jonasdottir; Berglind Eiriksdottir; Bjarni Thorbjornsson; Brynjar O Jensson; Daniel F Gudbjartsson; Droplaug N Magnusdottir; Elisabet E Gardarsdottir; Emil A Thorarensen; Gardar Sveinbjornsson; Gisli Masson; Gudmundur Georgsson; Gudmundur L Norddahl; Gudrun Sigmundsdottir; Hakon Jonsson; Hannes Eggertsson; Hilma Holm; Ingileif Jonsdottir; Jona Saemundsdottir; Kamilla S Josefsdottir; Kari Stefansson; Karl G Kristinsson; Kjartan R Gudmundsson; Kristin E Sveinsdottir; Kristjan E Hjorleifsson; Louise le Roux; Maney Sveinsdottir; Olafia S Gretarsdottir; Olafur T Magnusson; Pall Melsted; Patrick Sulem; Run Fridriksdottir; Solvi Rognvaldsson; Thora R Gunnarsdottir; Thordur Kristjansson; Thorolfur Gudnason; Unnur Thorsteinsdottir |
| EPI_ISL_2153744, EPI_ISL_2153745, EPI_ISL_2153761, EPI_ISL_2153767, EPI_ISL_2153781, EPI_ISL_2153784, EPI_ISL_2153789, EPI_ISL_2153794, EPI_ISL_2153800, EPI_ISL_2200124, EPI_ISL_2200133, EPI_ISL_2200143 | | | |
| see above | The Ohio State University Applied Microbiology Services Laboratory | The Ohio State University Applied Microbiology Services Laboratory | Seth A. Faith PhD |
| EPI_ISL_5530029, EPI_ISL_5530031, EPI_ISL_5530032, EPI_ISL_5530033 | UAPS AIDA SANTOS | Analytical Competence Molecular Epidemiology Lab/ACME, Oswaldo Cruz Foundation, Ceara (FIOCRUZ CE) | Carlos Leonardo de Aragao Araujo; Cecilia Leite Costa & Eduardo Ruback dos Santos on behalf of COVID-19 FIOCRUZ Genomic Network; Cleber Furtado Aksenen; Fabio Miyajima; Fernando Braga Stehling; Francisco Eder de Moura Lopes; Igor Oliveira Duarte; Jamille Maria Mendes Bezerra; Joaquim Cesar do Nascimento Sousa Junior; Pedro Miguel Carneiro Jeronimo; Suzana Porto Almeida; Thais Ferreira de Oliveira; Thais de Oliveira Costa; Ticiane Cavalcante de Souza; Veridiana Pessoa Miyajima |
| EPI_ISL_5529883 | UAPS ANASTACIO MAGALHAES | Analytical Competence Molecular Epidemiology Lab/ACME, Oswaldo Cruz Foundation, Ceara (FIOCRUZ CE) | Carlos Leonardo de Aragao Araujo; Cecilia Leite Costa & Eduardo Ruback dos Santos on behalf of COVID-19 FIOCRUZ Genomic Network; Cleber Furtado Aksenen; Fabio Miyajima; Fernando Braga Stehling; Francisco Eder de Moura Lopes; Igor Oliveira Duarte; Jamille Maria Mendes Bezerra; Joaquim Cesar do Nascimento Sousa Junior; Pedro Miguel Carneiro Jeronimo; Suzana Porto Almeida; Thais Ferreira de Oliveira; Thais de Oliveira Costa; Ticiane Cavalcante de Souza; Veridiana Pessoa Miyajima |
| EPI_ISL_5530036, EPI_ISL_5530037 | UAPS CARLOS RIBEIRO | Analytical Competence Molecular Epidemiology Lab/ACME, Oswaldo Cruz Foundation, Ceara (FIOCRUZ CE) | Carlos Leonardo de Aragao Araujo; Cecilia Leite Costa & Eduardo Ruback dos Santos on behalf of COVID-19 FIOCRUZ Genomic Network; Cleber Furtado Aksenen; Fabio Miyajima; Fernando Braga Stehling; Francisco Eder de Moura Lopes; Igor Oliveira Duarte; Jamille Maria Mendes Bezerra; Joaquim Cesar do Nascimento Sousa Junior; Pedro Miguel Carneiro Jeronimo; Suzana Porto Almeida; Thais Ferreira de Oliveira; Thais de Oliveira Costa; Ticiane Cavalcante de Souza; Veridiana Pessoa Miyajima |
| EPI_ISL_5530027 | UAPS HELIO GOES | Analytical Competence Molecular Epidemiology Lab/ACME, Oswaldo Cruz Foundation, Ceara (FIOCRUZ CE) | Carlos Leonardo de Aragao Araujo; Cecilia Leite Costa & Eduardo Ruback dos Santos on behalf of COVID-19 FIOCRUZ Genomic Network; Cleber Furtado Aksenen; Fabio Miyajima; Fernando Braga Stehling; Francisco Eder de Moura Lopes; Igor Oliveira Duarte; Jamille Maria Mendes Bezerra; Joaquim Cesar do Nascimento Sousa Junior; Pedro Miguel Carneiro Jeronimo; Suzana Porto Almeida; Thais Ferreira de Oliveira; Thais de Oliveira Costa; Ticiane Cavalcante de Souza; Veridiana Pessoa Miyajima |
| EPI_ISL_5530030 | UAPS RECAMONDE CAPELO | Analytical Competence Molecular Epidemiology Lab/ACME, Oswaldo Cruz Foundation, Ceara (FIOCRUZ CE) | Carlos Leonardo de Aragao Araujo; Cecilia Leite Costa & Eduardo Ruback dos Santos on behalf of COVID-19 FIOCRUZ Genomic Network; Cleber Furtado Aksenen; Fabio Miyajima; Fernando Braga Stehling; Francisco Eder de Moura Lopes; Igor Oliveira Duarte; Jamille Maria Mendes Bezerra; Joaquim Cesar do Nascimento Sousa Junior; Pedro Miguel Carneiro Jeronimo; Suzana Porto Almeida; Thais Ferreira de Oliveira; Thais de Oliveira Costa; Ticiane Cavalcante de Souza; Veridiana Pessoa Miyajima |
| EPI_ISL_5529885 | UAPS VALDEVINO DE CARVALHO | Analytical Competence Molecular Epidemiology Lab/ACME, Oswaldo | Carlos Leonardo de Aragao Araujo; Cecilia Leite Costa & Eduardo Ruback dos Santos on behalf of COVID-19 FIOCRUZ Genomic Network; Cleber Furtado Aksenen; Fabio Miyajima; Fernando Braga Stehling; Francisco Eder de Moura Lopes; Igor Oliveira Duarte; Jamille Maria Mendes Bezerra; Joaquim Cesar do Nascimento Sousa Junior; Pedro Miguel Carneiro Jeronimo; Suzana Porto Almeida; Thais Ferreira de Oliveira; Thais de Oliveira Costa; Ticiane Cavalcante de Souza; Veridiana Pessoa Miyajima |

| | | | |
|---|---|---|---|
| | | Cruz Foundation, Ceara (FIOCRUZ CE) | |
| EPI_ISL_1966543, EPI_ISL_1966544 | UBS DR ABELARDO PINHEIRO GUIMARAES SCRPARDO | Instituto Butantan / FZEA-USP (Pirassununga) | Antonio Jorge Martins; Bianca Cechetto Carlos. Mendelics: Bibiana Santos; Claudia Renata dos Santos Barros; Cíntia Bittar; David Schlesinger. Hemocentro Ribeirão Preto: Simone Kashima; Debora Botequio Moretti; Elaine Cristina Marqueze; Elaine Vieira dos Santos; Elisangela Chicaroni Mattos; Erika Freitas; Evandra Strazza Rodrigues; Felipe Allan da Silva da Costa; Flavia Aburjaile; Fábio Sossai Possebon; Guilherme Campos; Guilherme Targino Valente; Heidge Fukumasu. USP-Botucatu: Rejane Maria Tommasini Grotto; Helena Lage Ferreira; Instituto Butantan: Dimas Tadeu Covas; Jardelina de Souza Todao Bernardino; Jayme A. Souza-Neto; Jessika Cristina Chagas Lesbon; Jorge A. Petroli Marchesi; José Salvatore Leister Patané; João Paulo Kitajima; João Pessoa Araújo Jr.; Leila Sabrina Ullmann; Loyze Paola Oliveira de Lima; Luiz Aurelio de Campos Crispin. Centro de Genômica Funcional da ESALQ: Luiz Lehmann Coutinho; Luiz Carlos Junior de Alcantara; Lívia Sacchetto; Maisa C. Pereira Parra; Maria Carolina Elias; Marta Giovanetti; Marília Moraes; Maurício Lacerda Nogueira. Prefeitura de Sao Paulo: Melissa Palmieri.; Patricia Akemi Assato; Paula Rahal; Paulo Inacio da Costa; Rafael dos Santos Bezerra; Raquel de Lello Rocha Campos Cassano. NGS Soluções Genômicas: Pilar Drummond Sampaio Corrêa Mariani. FZEA-USP Pirassununga: Mirele Daiana Poleti; Raul Machado Neto; Ricardo Augusto Brassaloti; Ricardo Haddad; Rodrigo Tocantins Calado. FAMERP-SJRP: Cecília Artico Banho; Sandra Coccuzzo Sampaio; Svetoslav Nanev Slavov; Vagner Fonseca; Vincent Louis Viala |
| EPI_ISL_1966534 | UBS DR HELIO MIGLIARI | Instituto Butantan / FZEA-USP (Pirassununga) | Antonio Jorge Martins; Bianca Cechetto Carlos. Mendelics: Bibiana Santos; Claudia Renata dos Santos Barros; Cíntia Bittar; David Schlesinger. Hemocentro Ribeirão Preto: Simone Kashima; Debora Botequio Moretti; Elaine Cristina Marqueze; Elaine Vieira dos Santos; Elisangela Chicaroni Mattos; Erika Freitas; Evandra Strazza Rodrigues; Felipe Allan da Silva da Costa; Flavia Aburjaile; Fábio Sossai Possebon; Guilherme Campos; Guilherme Targino Valente; Heidge Fukumasu. USP-Botucatu: Rejane Maria Tommasini Grotto; Helena Lage Ferreira; Instituto Butantan: Dimas Tadeu Covas; Jardelina de Souza Todao Bernardino; Jayme A. Souza-Neto; Jessika Cristina Chagas Lesbon; Jorge A. Petroli Marchesi; José Salvatore Leister Patané; João Paulo Kitajima; João Pessoa Araújo Jr.; Leila Sabrina Ullmann; Loyze Paola Oliveira de Lima; Luiz Aurelio de Campos Crispin. Centro de Genômica Funcional da ESALQ: Luiz Lehmann Coutinho; Luiz Carlos Junior de Alcantara; Lívia Sacchetto; Maisa C. Pereira Parra; Maria Carolina Elias; Marta Giovanetti; Marília Moraes; Maurício Lacerda Nogueira. Prefeitura de Sao Paulo: Melissa Palmieri.; Patricia Akemi Assato; Paula Rahal; Paulo Inacio da Costa; Rafael dos Santos Bezerra; Raquel de Lello Rocha Campos Cassano. NGS Soluções Genômicas: Pilar Drummond Sampaio Corrêa Mariani. FZEA-USP Pirassununga: Mirele Daiana Poleti; Raul Machado Neto; Ricardo Augusto Brassaloti; Ricardo Haddad; Rodrigo Tocantins Calado. FAMERP-SJRP: Cecília Artico Banho; Sandra Coccuzzo Sampaio; Svetoslav Nanev Slavov; Vagner Fonseca; Vincent Louis Viala |
| EPI_ISL_1966546 | UBS DR WALDOMIRO FERREIRA NEVES SCRPARDO | Instituto Butantan / FZEA-USP (Pirassununga) | Antonio Jorge Martins; Bianca Cechetto Carlos. Mendelics: Bibiana Santos; Claudia Renata dos Santos Barros; Cíntia Bittar; David Schlesinger. Hemocentro Ribeirão Preto: Simone Kashima; Debora Botequio Moretti; Elaine Cristina Marqueze; Elaine Vieira dos Santos; Elisangela Chicaroni Mattos; Erika Freitas; Evandra Strazza Rodrigues; Felipe Allan da Silva da Costa; Flavia Aburjaile; Fábio Sossai Possebon; Guilherme Campos; Guilherme Targino Valente; Heidge Fukumasu. USP-Botucatu: Rejane Maria Tommasini Grotto; Helena Lage Ferreira; Instituto Butantan: Dimas Tadeu Covas; Jardelina de Souza Todao Bernardino; Jayme A. Souza-Neto; Jessika Cristina Chagas Lesbon; Jorge A. Petroli Marchesi; José Salvatore Leister Patané; João Paulo Kitajima; João Pessoa Araújo Jr.; Leila Sabrina Ullmann; Loyze Paola Oliveira de Lima; Luiz Aurelio de Campos Crispin. Centro de Genômica Funcional da ESALQ: Luiz Lehmann Coutinho; Luiz Carlos Junior de Alcantara; Lívia Sacchetto; Maisa C. Pereira Parra; Maria Carolina Elias; Marta Giovanetti; Marília Moraes; Maurício Lacerda Nogueira. Prefeitura de Sao Paulo: Melissa Palmieri.; Patricia Akemi Assato; Paula Rahal; Paulo Inacio da Costa; Rafael dos Santos Bezerra; Raquel de Lello Rocha Campos Cassano. NGS Soluções Genômicas: Pilar Drummond Sampaio Corrêa Mariani. FZEA-USP Pirassununga: Mirele Daiana Poleti; Raul Machado Neto; Ricardo Augusto Brassaloti; Ricardo Haddad; Rodrigo Tocantins Calado. FAMERP-SJRP: Cecília Artico Banho; Sandra Coccuzzo Sampaio; Svetoslav Nanev Slavov; Vagner Fonseca; Vincent Louis Viala |
| EPI_ISL_2209680 | UBS II DE TANABI MILTON MARTINS PERCHES | Instituto Butantan | Antonio Jorge Martins; Claudia Renata dos Santos Barros; David Schlesinger; Debora Botequio Moretti; Dimas Tadeu Covas; Elaine Cristina Marqueze; Elaine Vieira Santos; Evandra Strazza Rodrigues; Heidge Fukumasu; Jayme Augusto de Souza-Neto; José Salvatore Leister Patané; Luiz Alcantara; Luiz Lehmann Coutinho; Maria Carolina Elias; Maurício Lacerda Nogueira; Rafael dos Santos Bezerra; Raul Machado Neto; Rejane Maria Tommasini Grotto; Ricardo Haddad; Sandra Coccuzzo Sampaio Vessoni; Simone Kashima; Svetoslav Nanev Slavov; Vincent Louis Viala. |
| EPI_ISL_2344590 | UBS II DE TANABI MILTON MARTINS PERCHES | Instituto Butantan / UNESP-Botucatu | Antonio Jorge Martins; Claudia Renata dos Santos Barros; David Schlesinger; Debora Botequio Moretti; Dimas Tadeu Covas; Elaine Cristina Marqueze; Elaine Vieira Santos; Evandra Strazza Rodrigues; Heidge Fukumasu; Jayme Augusto de Souza-Neto; José Salvatore Leister Patané; Luiz Alcantara; Luiz Lehmann Coutinho; Maria Carolina Elias; Maurício Lacerda Nogueira; Rafael dos Santos Bezerra; Raul Machado Neto; Rejane Maria Tommasini Grotto; Ricardo Haddad; Sandra Coccuzzo Sampaio Vessoni; Simone Kashima; Svetoslav Nanev Slavov; Vincent Louis Viala |
| EPI_ISL_1966548 | UBS II DOUTOR ORLANDO BERTOLLI | Instituto Butantan / FZEA-USP (Pirassununga) | Antonio Jorge Martins; Bianca Cechetto Carlos. Mendelics: Bibiana Santos; Claudia Renata dos Santos Barros; Cíntia Bittar; David Schlesinger. Hemocentro Ribeirão Preto: Simone Kashima; Debora Botequio Moretti; Elaine Cristina Marqueze; Elaine Vieira dos Santos; Elisangela Chicaroni Mattos; Erika Freitas; Evandra Strazza Rodrigues; Felipe Allan da Silva da Costa; Flavia Aburjaile; Fábio Sossai Possebon; Guilherme Campos; Guilherme Targino Valente; Heidge Fukumasu. USP-Botucatu: Rejane Maria Tommasini Grotto; Helena Lage Ferreira; Instituto Butantan: Dimas Tadeu Covas; Jardelina de Souza Todao Bernardino; Jayme A. Souza-Neto; Jessika Cristina Chagas Lesbon; Jorge A. Petroli Marchesi; José Salvatore Leister Patané; João Paulo Kitajima; João Pessoa Araújo Jr.; Leila Sabrina Ullmann; Loyze Paola Oliveira de Lima; Luiz Aurelio de Campos Crispin. Centro de Genômica Funcional da ESALQ: Luiz Lehmann Coutinho; Luiz Carlos Junior de Alcantara; Lívia Sacchetto; Maisa C. Pereira Parra; Maria Carolina Elias; Marta Giovanetti; Marília Moraes; Maurício Lacerda Nogueira. Prefeitura de Sao Paulo: Melissa Palmieri.; Patricia Akemi Assato; Paula Rahal; Paulo Inacio da Costa; Rafael dos Santos Bezerra; Raquel de Lello Rocha Campos Cassano. NGS Soluções Genômicas: Pilar Drummond Sampaio Corrêa Mariani. FZEA-USP Pirassununga: Mirele Daiana Poleti; Raul Machado Neto; Ricardo Augusto Brassaloti; Ricardo Haddad; Rodrigo Tocantins Calado. FAMERP-SJRP: Cecília Artico Banho; Sandra Coccuzzo Sampaio; Svetoslav Nanev Slavov; Vagner Fonseca; Vincent Louis Viala |
| EPI_ISL_1966535 | UBS REGIAO OESTE DE OURINHOS | Instituto Butantan / FZEA-USP (Pirassununga) | Antonio Jorge Martins; Bianca Cechetto Carlos. Mendelics: Bibiana Santos; Claudia Renata dos Santos Barros; Cíntia Bittar; David Schlesinger. Hemocentro Ribeirão Preto: Simone Kashima; Debora Botequio Moretti; Elaine Cristina Marqueze; Elaine Vieira dos Santos; Elisangela Chicaroni Mattos; Erika Freitas; Evandra Strazza Rodrigues; Felipe Allan da Silva da Costa; Flavia Aburjaile; Fábio Sossai Possebon; Guilherme Campos; Guilherme Targino Valente; Heidge Fukumasu. USP-Botucatu: Rejane Maria Tommasini Grotto; Helena Lage Ferreira; Instituto Butantan: Dimas Tadeu Covas; Jardelina de Souza Todao Bernardino; Jayme A. Souza-Neto; Jessika Cristina Chagas Lesbon; Jorge A. Petroli Marchesi; José Salvatore Leister Patané; João Paulo Kitajima; João Pessoa Araújo Jr.; Leila Sabrina Ullmann; Loyze Paola Oliveira de Lima; Luiz Aurelio de Campos Crispin. Centro de Genômica Funcional da ESALQ: Luiz Lehmann Coutinho; Luiz Carlos Junior de Alcantara; Lívia Sacchetto; Maisa C. Pereira Parra; Maria Carolina Elias; Marta Giovanetti; Marília Moraes; Maurício Lacerda Nogueira. Prefeitura de Sao Paulo: Melissa Palmieri.; Patricia Akemi Assato; Paula Rahal; Paulo Inacio da Costa; Rafael dos Santos Bezerra; Raquel de Lello Rocha Campos Cassano. NGS Soluções Genômicas: Pilar Drummond Sampaio Corrêa Mariani. FZEA-USP Pirassununga: Mirele Daiana Poleti; Raul Machado Neto; Ricardo Augusto Brassaloti; Ricardo Haddad; Rodrigo Tocantins Calado. FAMERP-SJRP: Cecília Artico Banho; Sandra Coccuzzo Sampaio; Svetoslav Nanev Slavov; Vagner Fonseca; Vincent Louis Viala |
| EPI_ISL_3419550 | UC Davis Genome Center and Healthy Davis Together | UC Davis Genome Center | UC Davis |
| EPI_ISL_1707051 | UCLA Clinical Micro Lab | Los Angeles County Public Health Laboratories | P. Hemarajata et al. |
| EPI_ISL_2399017 | ULSS 5 Polesana | Istituto Zooprofilattico Sperimentale delle Venezie | Adelaide Milani; Alessia Schivo; Alice Fusaro; Ambra Pastori; Annalisa Salviato; Antonia Ricci; Calogero Terregino; Edoardo Giussani; Elisa Palumbo; Erika Giorgia Quaranta; Isabella Monne; Luca Tassoni |
| EPI_ISL_2365899 | ULSS 6 Euganea | Istituto Zooprofilattico Sperimentale delle Venezie | Adelaide Milani; Alessia Schivo; Alice Fusaro; Ambra Pastori; Annalisa Salviato; Antonia Ricci; Calogero Terregino; Edoardo Giussani; Elisa Palumbo; Erika Giorgia Quaranta; Isabella Monne; Luca Tassoni |
| EPI_ISL_3409101, EPI_ISL_3409102, EPI_ISL_3409263, EPI_ISL_3409833, EPI_ISL_3410078 | UMass Memorial Medical Center | Center for Microbiome Research | Doyle V. Ward; Karl J. Simon; Richard T. Ellison III |
| EPI_ISL_1966515 | UNIDADE BASICA DE SAUDE DE TAQUARAL CAETANO PITELLI | Instituto Butantan / Mendelics | Antonio Jorge Martins; Bianca Cechetto Carlos. Mendelics: Bibiana Santos; Claudia Renata dos Santos Barros; Cíntia Bittar; David Schlesinger. Hemocentro Ribeirão Preto: Simone Kashima; Debora Botequio Moretti; Elaine Cristina Marqueze; Elaine Vieira dos Santos; Elisangela Chicaroni Mattos; Erika Freitas; Evandra Strazza Rodrigues; Felipe Allan da Silva da Costa; Flavia Aburjaile; Fábio Sossai Possebon; Guilherme Campos; Guilherme Targino Valente; Heidge Fukumasu. USP-Botucatu: Rejane Maria Tommasini Grotto; Helena Lage Ferreira; Instituto Butantan: Dimas Tadeu Covas; Jardelina de Souza Todao Bernardino; Jayme A. Souza-Neto; Jessika Cristina Chagas Lesbon; Jorge A. Petroli Marchesi; José Salvatore Leister Patané; João Paulo Kitajima; João Pessoa Araújo Jr.; Leila Sabrina Ullmann; Loyze Paola Oliveira de Lima; Luiz Aurelio de Campos Crispin. Centro de Genômica Funcional da ESALQ: Luiz Lehmann Coutinho; Luiz Carlos Junior de Alcantara; Lívia Sacchetto; Maisa C. Pereira Parra; Maria Carolina Elias; Marta Giovanetti; Marília Moraes; Maurício Lacerda Nogueira. Prefeitura de Sao Paulo: Melissa Palmieri.; Patricia Akemi Assato; Paula Rahal; Paulo Inacio da Costa; Rafael dos Santos Bezerra; Raquel de Lello Rocha Campos Cassano. NGS Soluções Genômicas: Pilar Drummond Sampaio Corrêa Mariani. FZEA-USP Pirassununga: Mirele Daiana Poleti; Raul Machado Neto; Ricardo Augusto Brassaloti; Ricardo Haddad; Rodrigo Tocantins Calado. FAMERP-SJRP: Cecília Artico Banho; Sandra Coccuzzo Sampaio; Svetoslav Nanev Slavov; Vagner Fonseca; Vincent Louis Viala |
| EPI_ISL_3102311 | UNIDADE BASICA DE SAUDE DR FCO MARCIO FERNANDES | Analytical Competence Molecular Epidemiology Lab/ACME, Oswaldo Cruz Foundation, Ceara (FIOCRUZ CE) | Cleber Furtado Aksenen; Fabio Miyajima; Fernando Braga Stehling; Francisco Eder de Moura Lopes; Jamille Maria Mendes Bezerra; Joaquim César do Nascimento Sousa Junior; Pedro Miguel Carneiro Jeronimo; Suzana Porto Almeida e Lucas Delerino; Thais Ferreira de Oliveira; Thais de Oliveira Costa; Ticiane Cavalcante de Souza; Veridiana Pessoa Miyajima |
| EPI_ISL_1966555 | UNIDADE BASICA DE SAUDE II DE TARABAI | Instituto Butantan / FZEA-USP (Pirassununga) | Antonio Jorge Martins; Bianca Cechetto Carlos. Mendelics: Bibiana Santos; Claudia Renata dos Santos Barros; Cíntia Bittar; David Schlesinger. Hemocentro Ribeirão Preto: Simone Kashima; Debora Botequio Moretti; Elaine Cristina Marqueze; Elaine Vieira dos Santos; Elisangela Chicaroni Mattos; Erika Freitas; Evandra Strazza Rodrigues; Felipe Allan da Silva da Costa; Flavia Aburjaile; Fábio Sossai Possebon; Guilherme Campos; Guilherme Targino Valente; Heidge Fukumasu. USP-Botucatu: Rejane Maria Tommasini Grotto; Helena Lage Ferreira; Instituto Butantan: Dimas Tadeu Covas; Jardelina de Souza Todao Bernardino; Jayme A. Souza-Neto; Jessika Cristina Chagas Lesbon; Jorge A. Petroli Marchesi; José Salvatore Leister Patané; João Paulo Kitajima; João Pessoa Araújo Jr.; Leila Sabrina Ullmann; Loyze Paola Oliveira de Lima; Luiz Aurelio de Campos Crispin. Centro de Genômica Funcional da ESALQ: Luiz Lehmann Coutinho; Luiz Carlos Junior de Alcantara; Lívia Sacchetto; Maisa C. Pereira Parra; Maria Carolina Elias; Marta Giovanetti; Marília Moraes; Maurício Lacerda Nogueira. Prefeitura de Sao Paulo: Melissa Palmieri.; Patricia Akemi Assato; Paula Rahal; Paulo Inacio da Costa; Rafael dos Santos Bezerra; Raquel de Lello Rocha Campos Cassano. NGS Soluções Genômicas: Pilar Drummond Sampaio Corrêa Mariani. FZEA-USP Pirassununga: Mirele Daiana Poleti; Raul Machado Neto; Ricardo Augusto Brassaloti; Ricardo Haddad; Rodrigo Tocantins Calado. FAMERP-SJRP: Cecília Artico Banho; Sandra Coccuzzo Sampaio; Svetoslav Nanev Slavov; Vagner Fonseca; Vincent Louis Viala |
| EPI_ISL_1966549, EPI_ISL_1966550 | UNIDADE DE VIGILANCIA EM SAUDE | Instituto Butantan / FZEA-USP (Pirassununga) | Antonio Jorge Martins; Bianca Cechetto Carlos. Mendelics: Bibiana Santos; Claudia Renata dos Santos Barros; Cíntia Bittar; David Schlesinger. Hemocentro Ribeirão Preto: Simone Kashima; Debora Botequio Moretti; Elaine Cristina Marqueze; Elaine Vieira dos Santos; Elisangela Chicaroni Mattos; Erika Freitas; Evandra Strazza Rodrigues; Felipe Allan da Silva da Costa; Flavia Aburjaile; Fábio Sossai Possebon; Guilherme Campos; Guilherme Targino Valente; Heidge Fukumasu. USP-Botucatu: Rejane Maria Tommasini Grotto; Helena Lage Ferreira; Instituto Butantan: Dimas Tadeu Covas; Jardelina de Souza Todao Bernardino; Jayme A. Souza-Neto; Jessika Cristina Chagas Lesbon; Jorge A. Petroli Marchesi; José Salvatore Leister Patané; João Paulo Kitajima; João Pessoa Araújo Jr.; Leila Sabrina Ullmann; Loyze Paola Oliveira de Lima; Luiz Aurelio de Campos Crispin. Centro de Genômica Funcional da ESALQ: Luiz Lehmann Coutinho; Luiz Carlos Junior de Alcantara; Lívia Sacchetto; Maisa C. Pereira Parra; Maria Carolina Elias; Marta Giovanetti; Marília Moraes; Maurício Lacerda Nogueira. Prefeitura de Sao Paulo: Melissa Palmieri.; Patricia Akemi Assato; Paula Rahal; Paulo Inacio da Costa; Rafael dos Santos Bezerra; Raquel de Lello Rocha Campos Cassano. NGS Soluções Genômicas: Pilar Drummond Sampaio Corrêa Mariani. FZEA-USP Pirassununga: Mirele Daiana Poleti; Raul Machado Neto; Ricardo Augusto Brassaloti; Ricardo Haddad; Rodrigo Tocantins Calado. FAMERP-SJRP: Cecília Artico Banho; Sandra Coccuzzo Sampaio; Svetoslav Nanev Slavov; Vagner Fonseca; Vincent Louis Viala |
| EPI_ISL_2344688 | UNIDADE DE VIGILANCIA EPIDEMIOLOGICA | Instituto Butantan / FZEA-USP-Pirassununga | Antonio Jorge Martins; Claudia Renata dos Santos Barros; David Schlesinger; Debora Botequio Moretti; Dimas Tadeu Covas; Elaine Cristina Marqueze; Elaine Vieira Santos; Evandra Strazza Rodrigues; Heidge Fukumasu; Jayme Augusto de Souza-Neto; José Salvatore Leister Patané; Luiz Alcantara; Luiz Lehmann Coutinho; Maria Carolina Elias; Maurício Lacerda Nogueira; Rafael dos Santos Bezerra; Raul Machado Neto; Rejane Maria Tommasini Grotto; Ricardo Haddad; Sandra Coccuzzo Sampaio Vessoni; Simone Kashima; Svetoslav Nanev Slavov; Vincent Louis Viala |
| EPI_ISL_5647105 | UNIDADE MISTA DE TAIACU | Instituto Butantan | Antonio Jorge Martins; Claudia Renata dos Santos Barros; David Schlesinger; Debora Botequio Moretti; Dimas Tadeu Covas; Elaine Cristina Marqueze; Elaine Vieira Santos; Evandra Strazza Rodrigues; Heidge Fukumasu; Jayme Augusto de Souza-Neto; José Salvatore Leister Patané; Luiz Alcantara; Luiz Lehmann Coutinho; Maria Carolina Elias; Maurício Lacerda Nogueira; Rafael dos Santos Bezerra; Raul Machado Neto; Rejane Maria Tommasini Grotto; Ricardo Haddad; Sandra Coccuzzo Sampaio Vessoni; Simone Kashima; Svetoslav Nanev Slavov; Vincent Louis Viala |
| EPI_ISL_3102266 | UNIDADE PRONTO ATENDIMENTO CONJUNTO CEARA | Analytical Competence Molecular Epidemiology Lab/ACME, Oswaldo Cruz Foundation, Ceara (FIOCRUZ CE) | Cleber Furtado Aksenen; Fabio Miyajima; Fernando Braga Stehling; Francisco Eder de Moura Lopes; Jamille Maria Mendes Bezerra; Joaquim César do Nascimento Sousa Junior; Pedro Miguel Carneiro Jeronimo; Suzana Porto Almeida e Lucas Delerino; Thais Ferreira de Oliveira; Thais de Oliveira Costa; Ticiane Cavalcante de Souza; Veridiana Pessoa Miyajima |
| EPI_ISL_5530038 | UNIDADE PRONTO ATENDIMENTO JOSE WALTER | Analytical Competence Molecular Epidemiology Lab/ACME, Oswaldo Cruz Foundation, Ceara (FIOCRUZ CE) | Carlos Leonardo de Aragao Araujo; Cecilia Leite Costa & Eduardo Ruback dos Santos on behalf of COVID-19 FIOCRUZ Genomic Network; Cleber Furtado Aksenen; Fabio Miyajima; Fernando Braga Stehling; Francisco Eder de Moura Lopes; Igor Oliveira Duarte; Jamille Maria Mendes Bezerra; Joaquim Cesar do Nascimento Sousa Junior; Pedro Miguel Carneiro Jeronimo; Suzana Porto Almeida; Thais Ferreira de Oliveira; Thais de Oliveira Costa; Ticiane Cavalcante de Souza; Veridiana Pessoa Miyajima |
| EPI_ISL_5529878, EPI_ISL_5529970, EPI_ISL_5529971 | UNIDADE REFERENCIA COVID 19 | Analytical Competence Molecular Epidemiology Lab/ACME, Oswaldo Cruz Foundation, Ceara (FIOCRUZ CE) | Carlos Leonardo de Aragao Araujo; Cecilia Leite Costa & Eduardo Ruback dos Santos on behalf of COVID-19 FIOCRUZ Genomic Network; Cleber Furtado Aksenen; Fabio Miyajima; Fernando Braga Stehling; Francisco Eder de Moura Lopes; Igor Oliveira Duarte; Jamille Maria Mendes Bezerra; Joaquim Cesar do Nascimento Sousa Junior; Pedro Miguel Carneiro Jeronimo; Suzana Porto Almeida; Thais Ferreira de Oliveira; Thais de Oliveira Costa; Ticiane Cavalcante de Souza; Veridiana Pessoa Miyajima |
| EPI_ISL_3266999 | UNIGEM | Universidad Nacional de Colombia - Laboratorio Genómico One Health | Andres F. Cardona-Rios; Carlos Franco-Muñoz; Carolina Muñoz-Arango; Celeny Ortiz; Daniel O. Maldonado-Perez; Diego A. Álvarez-Díaz; Hector Alejandro Ruiz-Moreno; Idabely Betancur Ortiz; Jorge E. Osorio; Juan P. Hernandez-Ortiz; Karl A Ciuoderis; Katherine Laiton-Donato; Laura Silvana Perez; Lina M. Hurtado; Marcela Mercado-Reyes; Maria Angélica Maya; Maria Stella López; Rita Almanza Payares; Sandra Ines Cano; Simón Villegas Velásquez |
| EPI_ISL_1966436 | UPA DR AKIRA TADA | Instituto Butantan / Mendelics | Antonio Jorge Martins; Bianca Cechetto Carlos. Mendelics: Bibiana Santos; Claudia Renata dos Santos Barros; Cíntia Bittar; David Schlesinger. Hemocentro Ribeirão Preto: Simone Kashima; Debora Botequio Moretti; Elaine Cristina Marqueze; Elaine Vieira dos Santos; Elisangela Chicaroni Mattos; Erika Freitas; Evandra Strazza Rodrigues; Felipe Allan da Silva da Costa; Flavia Aburjaile; Fábio Sossai Possebon; Guilherme Campos; Guilherme Targino Valente; Heidge Fukumasu. USP-Botucatu: Rejane Maria Tommasini Grotto; Helena Lage Ferreira; Instituto Butantan: Dimas Tadeu Covas; Jardelina de Souza Todao Bernardino; Jayme A. Souza-Neto; Jessika Cristina Chagas Lesbon; Jorge A. Petroli Marchesi; José Salvatore Leister Patané; João Paulo Kitajima; João Pessoa Araújo Jr.; Leila Sabrina Ullmann; Loyze Paola Oliveira de Lima; Luiz Aurelio de Campos Crispin. Centro de Genômica Funcional da ESALQ: Luiz Lehmann Coutinho; Luiz Carlos Junior de Alcantara; Lívia Sacchetto; Maisa C. Pereira Parra; Maria Carolina Elias; Marta Giovanetti; Marília Moraes; Maurício Lacerda Nogueira. Prefeitura de Sao Paulo: Melissa Palmieri.; Patricia Akemi Assato; Paula Rahal; Paulo Inacio da Costa; Rafael dos Santos Bezerra; Raquel de Lello Rocha Campos Cassano. NGS Soluções Genômicas: Pilar Drummond Sampaio Corrêa Mariani. FZEA-USP Pirassununga: Mirele Daiana Poleti; Raul Machado Neto; Ricardo Augusto Brassaloti; Ricardo Haddad; Rodrigo Tocantins Calado. FAMERP-SJRP: Cecília Artico Banho; Sandra Coccuzzo Sampaio; Svetoslav Nanev Slavov; Vagner Fonseca; Vincent Louis Viala |
| EPI_ISL_1920125 | URMC LABS | Wadsworth Center, New York State Department of Health | Alexis Russell; Catharine Prussing; Daryl M. Lamson; Erasmus Schneider; Erica Lasek-Nesselquist; John Kelly; Jonathan Plitnick; Kirsten St. George; Matthew Shudt; Melissa A Leisner; Navjot Singh |
| EPI_ISL_1711121, EPI_ISL_1711128 | US Air Force School of Aerospace Medicine | US Air Force School of Aerospace Medicine | Amanda Javorina; Anthony Fries; Clarise Starr; Elizabeth Macias; Jennifer Meyer; Sarah Purves; William Buggele; William Gruner |
| EPI_ISL_1707080 | USC Clinical Lab | Los Angeles County Public Health Laboratories | P. Hemarajata et al. |
| EPI_ISL_1580549 | USCA Avezzano AVEZZANO(L'AQUILA) | Istituto Zooprofilattico Sperimentale dell'Abruzzo e Molise "G. Caporale" | Ancora M; Calistri P; Cammà C; Caporale M; Curini V; Delli Compagni E; Di Domenico M; Di Lollo Valeria; Di Pasquale A; Lorusso A; Mangone I; Marcacci M; Puglia I; Rinaldi A; Savini G; Scialabba S |

| | | | |
|---|---|---|---|
| EPI_ISL_1580604 | USCA Tagliacozzo TAGLIACOZZO(L'AQUILA) | Istituto Zooprofilattico Sperimentale dell'Abruzzo e Molise "G. Caporale" | Ancora M; Calistri P; Cammà C; Caporale M; Curini V; Delli Compagni E; Di Domenico M; Di Lollo Valeria; Di Pasquale A; Lorusso A; Mangone I; Marcacci M; Puglia I; Rinaldi A; Savini G; Scialabba S |
| EPI_ISL_1966542 | USF DR APARECIDO RODRIGUES MOUCO SCRPARDO | Instituto Butantan / FZEA-USP (Pirassununga) | Antonio Jorge Martins; Bianca Cechetto Carlos. Mendelics: Bibiana Santos; Claudia Renata dos Santos Barros; Cíntia Bittar; David Schlesinger. Hemocentro Ribeirão Preto: Simone Kashima; Debora Botequio Moretti; Elaine Cristina Marqueze; Elaine Vieira dos Santos; Elisangela Chicaroni Mattos; Erika Freitas; Evandra Strazza Rodrigues; Felipe Allan da Silva da Costa; Flavia Aburjaile; Fábio Sossai Possebon; Guilherme Campos; Guilherme Targino Valente; Heidge Fukumasu. USP-Botucatu: Rejane Maria Tommasini Grotto; Helena Lage Ferreira; Instituto Butantan: Dimas Tadeu Covas; Jardelina de Souza Todao Bernardino; Jayme A. Souza-Neto; Jessika Cristina Chagas Lesbon; Jorge A. Petroli Marchesi; José Salvatore Leister Patané; João Paulo Kitajima; João Pessoa Araújo Jr.; Leila Sabrina Ullmann; Loyze Paola Oliveira de Lima; Luiz Aurelio de Campos Crispin. Centro de Genômica Funcional da ESALQ: Luiz Lehmann Coutinho; Luiz Carlos Junior de Alcantara; Lívia Sacchetto; Maisa C. Pereira Parra; Maria Carolina Elias; Marta Giovanetti; Marília Moraes; Maurício Lacerda Nogueira. Prefeitura de Sao Paulo: Melissa Palmieri.; Patricia Akemi Assato; Paula Rahal; Paulo Inacio da Costa; Rafael dos Santos Bezerra; Raquel de Lello Rocha Campos Cassano. NGS Soluções Genômicas: Pilar Drummond Sampaio Corrêa Mariani. FZEA-USP Pirassununga: Mirele Daiana Poleti; Raul Machado Neto; Ricardo Augusto Brassaloti; Ricardo Haddad; Rodrigo Tocantins Calado. FAMERP-SJRP: Cecília Artico Banho; Sandra Coccuzzo Sampaio; Svetoslav Nanev Slavov; Vagner Fonseca; Vincent Louis Viala |
| EPI_ISL_2210010 | USF PAULISTA FERNANDOPOLIS ANTONIO PIVATO | Instituto Butantan | Antonio Jorge Martins; Claudia Renata dos Santos Barros; David Schlesinger; Debora Botequio Moretti; Dimas Tadeu Covas; Elaine Cristina Marqueze; Elaine Vieira Santos; Evandra Strazza Rodrigues; Heidge Fukumasu; Jayme Augusto de Souza-Neto; José Salvatore Leister Patané; Luiz Alcantara; Luiz Lehmann Coutinho; Maria Carolina Elias; Maurício Lacerda Nogueira; Rafael dos Santos Bezerra; Raul Machado Neto; Rejane Maria Tommasini Grotto; Ricardo Haddad; Sandra Coccuzzo Sampaio Vessoni; Simone Kashima; Svetoslav Nanev Slavov; Vincent Louis Viala. |
| EPI_ISL_1966526, EPI_ISL_1966527, EPI_ISL_1966528, EPI_ISL_1966529 | USF PAULISTA FERNANDOPOLIS ANTONIO PIVATO | Instituto Butantan / FZEA-USP (Pirassununga) | Antonio Jorge Martins; Bianca Cechetto Carlos. Mendelics: Bibiana Santos; Claudia Renata dos Santos Barros; Cíntia Bittar; David Schlesinger. Hemocentro Ribeirão Preto: Simone Kashima; Debora Botequio Moretti; Elaine Cristina Marqueze; Elaine Vieira dos Santos; Elisangela Chicaroni Mattos; Erika Freitas; Evandra Strazza Rodrigues; Felipe Allan da Silva da Costa; Flavia Aburjaile; Fábio Sossai Possebon; Guilherme Campos; Guilherme Targino Valente; Heidge Fukumasu. USP-Botucatu: Rejane Maria Tommasini Grotto; Helena Lage Ferreira; Instituto Butantan: Dimas Tadeu Covas; Jardelina de Souza Todao Bernardino; Jayme A. Souza-Neto; Jessika Cristina Chagas Lesbon; Jorge A. Petroli Marchesi; José Salvatore Leister Patané; João Paulo Kitajima; João Pessoa Araújo Jr.; Leila Sabrina Ullmann; Loyze Paola Oliveira de Lima; Luiz Aurelio de Campos Crispin. Centro de Genômica Funcional da ESALQ: Luiz Lehmann Coutinho; Luiz Carlos Junior de Alcantara; Lívia Sacchetto; Maisa C. Pereira Parra; Maria Carolina Elias; Marta Giovanetti; Marília Moraes; Maurício Lacerda Nogueira. Prefeitura de Sao Paulo: Melissa Palmieri.; Patricia Akemi Assato; Paula Rahal; Paulo Inacio da Costa; Rafael dos Santos Bezerra; Raquel de Lello Rocha Campos Cassano. NGS Soluções Genômicas: Pilar Drummond Sampaio Corrêa Mariani. FZEA-USP Pirassununga: Mirele Daiana Poleti; Raul Machado Neto; Ricardo Augusto Brassaloti; Ricardo Haddad; Rodrigo Tocantins Calado. FAMERP-SJRP: Cecília Artico Banho; Sandra Coccuzzo Sampaio; Svetoslav Nanev Slavov; Vagner Fonseca; Vincent Louis Viala |
| EPI_ISL_2344540 | USF PAULISTA FERNANDOPOLIS | Instituto Butantan / UNESP-Botucatu | Antonio Jorge Martins; Claudia Renata dos Santos Barros; David Schlesinger; Debora Botequio Moretti; Dimas Tadeu Covas; Elaine Cristina Marqueze; Elaine Vieira Santos; Evandra Strazza Rodrigues; Heidge Fukumasu; Jayme Augusto de Souza-Neto; José Salvatore Leister Patané; Luiz Alcantara; Luiz Lehmann Coutinho; Maria Carolina Elias; Maurício Lacerda Nogueira; Rafael dos Santos Bezerra; Raul Machado Neto; Rejane Maria Tommasini Grotto; Ricardo Haddad; Sandra Coccuzzo Sampaio Vessoni; Simone Kashima; Svetoslav Nanev Slavov; Vincent Louis Viala |
| EPI_ISL_1678288 | UT-Unified State Labs: Public Health Utah DOH | Centers for Disease Control and Prevention Division of Viral Diseases, Pathogen Discovery | Alison Laufer Halpin; Ben L. Rambo-Martin; Clinton R. Paden; Dakota Howard; Darlene Wagner; Dave Wentworth; Dhwani Batra; Jasmine Padilla; Justin Lee; Katie Dillon; Krista Queen; Kristen Knipe; Kristine Lacek; Mark Burroughs; Matthew Schmerer; Mili Sheth; Peter Cook; Sam Shepard; Sarah Nobles; Shoshona Le; Suxiang Tong; Vivien Dugan; Yvette Unoarumhi |
| EPI_ISL_1601467, EPI_ISL_1601519, EPI_ISL_1601522, EPI_ISL_1601541, EPI_ISL_1616631, EPI_ISL_1616632, EPI_ISL_1616637, EPI_ISL_1616644, EPI_ISL_1616645, EPI_ISL_1616653, EPI_ISL_1616654, EPI_ISL_1616659, EPI_ISL_1616663, EPI_ISL_1616666, EPI_ISL_1616727, EPI_ISL_1616742, EPI_ISL_1616748, EPI_ISL_1616758, EPI_ISL_1616825, EPI_ISL_1620820, EPI_ISL_1620954, EPI_ISL_1620955, EPI_ISL_1620956, EPI_ISL_1620957, EPI_ISL_1620958, EPI_ISL_1620959, EPI_ISL_1620960, EPI_ISL_1620961, EPI_ISL_1620962, EPI_ISL_1620963, EPI_ISL_1620964, EPI_ISL_1620965, EPI_ISL_1620966, EPI_ISL_1620967, EPI_ISL_1620968, EPI_ISL_1620969, EPI_ISL_1620970, EPI_ISL_1620971, EPI_ISL_1620972, EPI_ISL_1620973, EPI_ISL_1620974, EPI_ISL_1628163, EPI_ISL_1628164, EPI_ISL_1628165, EPI_ISL_1628166, EPI_ISL_1628167, EPI_ISL_1628168, EPI_ISL_1628181, EPI_ISL_1633577, EPI_ISL_1633590, EPI_ISL_1633597, EPI_ISL_1633615 | | | |
| see above | UW Virology Lab | UW Virology Lab | Alexander Greninger; Hong Xie; Keith R Jerome; Lasata Shrestha; Meei-Li Huang; Michelle Lin; Noah R. Baker; Pavitra Roychoudhury; Saraswathi Sathees; Sean Ellis; Shah Mohamed Bakhash |
| EPI_ISL_5802059, EPI_ISL_5802060 | Ubs Dr Abelardo Pinheiro Guimaraes Scrpardo | Instituto Butantan | Antonio Jorge Martins; Claudia Renata dos Santos Barros; David Schlesinger; Debora Botequio Moretti; Dimas Tadeu Covas; Elaine Cristina Marqueze; Elaine Vieira Santos; Evandra Strazza Rodrigues; Heidge Fukumasu; Jayme Augusto de Souza-Neto; José Salvatore Leister Patané; Luiz Alcantara; Luiz Lehmann Coutinho; Maria Carolina Elias; Maurício Lacerda Nogueira; Rafael dos Santos Bezerra; Raul Machado Neto; Rejane Maria Tommasini Grotto; Ricardo Haddad; Sandra Coccuzzo Sampaio Vessoni; Simone Kashima; Svetoslav Nanev Slavov; Vincent Louis Viala |
| EPI_ISL_5802055 | Ubs Dr Helio Migliari | Instituto Butantan | Antonio Jorge Martins; Claudia Renata dos Santos Barros; David Schlesinger; Debora Botequio Moretti; Dimas Tadeu Covas; Elaine Cristina Marqueze; Elaine Vieira Santos; Evandra Strazza Rodrigues; Heidge Fukumasu; Jayme Augusto de Souza-Neto; José Salvatore Leister Patané; Luiz Alcantara; Luiz Lehmann Coutinho; Maria Carolina Elias; Maurício Lacerda Nogueira; Rafael dos Santos Bezerra; Raul Machado Neto; Rejane Maria Tommasini Grotto; Ricardo Haddad; Sandra Coccuzzo Sampaio Vessoni; Simone Kashima; Svetoslav Nanev Slavov; Vincent Louis Viala |
| EPI_ISL_5802062 | Ubs Dr Waldomiro Ferreira Neves Scrpardo | Instituto Butantan | Antonio Jorge Martins; Claudia Renata dos Santos Barros; David Schlesinger; Debora Botequio Moretti; Dimas Tadeu Covas; Elaine Cristina Marqueze; Elaine Vieira Santos; Evandra Strazza Rodrigues; Heidge Fukumasu; Jayme Augusto de Souza-Neto; José Salvatore Leister Patané; Luiz Alcantara; Luiz Lehmann Coutinho; Maria Carolina Elias; Maurício Lacerda Nogueira; Rafael dos Santos Bezerra; Raul Machado Neto; Rejane Maria Tommasini Grotto; Ricardo Haddad; Sandra Coccuzzo Sampaio Vessoni; Simone Kashima; Svetoslav Nanev Slavov; Vincent Louis Viala |
| EPI_ISL_5802171 | Ubs Ii De Tanabi Milton Martins Perches | Instituto Butantan | Antonio Jorge Martins; Claudia Renata dos Santos Barros; David Schlesinger; Debora Botequio Moretti; Dimas Tadeu Covas; Elaine Cristina Marqueze; Elaine Vieira Santos; Evandra Strazza Rodrigues; Heidge Fukumasu; Jayme Augusto de Souza-Neto; José Salvatore Leister Patané; Luiz Alcantara; Luiz Lehmann Coutinho; Maria Carolina Elias; Maurício Lacerda Nogueira; Rafael dos Santos Bezerra; Raul Machado Neto; Rejane Maria Tommasini Grotto; Ricardo Haddad; Sandra Coccuzzo Sampaio Vessoni; Simone Kashima; Svetoslav Nanev Slavov; Vincent Louis Viala |
| EPI_ISL_5802083 | Ubs Ii Doutor Orlando Bertolli | Instituto Butantan | Antonio Jorge Martins; Claudia Renata dos Santos Barros; David Schlesinger; Debora Botequio Moretti; Dimas Tadeu Covas; Elaine Cristina Marqueze; Elaine Vieira Santos; Evandra Strazza Rodrigues; Heidge Fukumasu; Jayme Augusto de Souza-Neto; José Salvatore Leister Patané; Luiz Alcantara; Luiz Lehmann Coutinho; Maria Carolina Elias; Maurício Lacerda Nogueira; Rafael dos Santos Bezerra; Raul Machado Neto; Rejane Maria Tommasini Grotto; Ricardo Haddad; Sandra Coccuzzo Sampaio Vessoni; Simone Kashima; Svetoslav Nanev Slavov; Vincent Louis Viala |
| EPI_ISL_5802057 | Ubs Regiao Oeste De Ourinhos | Instituto Butantan | Antonio Jorge Martins; Claudia Renata dos Santos Barros; David Schlesinger; Debora Botequio Moretti; Dimas Tadeu Covas; Elaine Cristina Marqueze; Elaine Vieira Santos; Evandra Strazza Rodrigues; Heidge Fukumasu; Jayme Augusto de Souza-Neto; José Salvatore Leister Patané; Luiz Alcantara; Luiz Lehmann Coutinho; Maria Carolina Elias; Maurício Lacerda Nogueira; Rafael dos Santos Bezerra; Raul Machado Neto; Rejane Maria Tommasini Grotto; Ricardo Haddad; Sandra Coccuzzo Sampaio Vessoni; Simone Kashima; Svetoslav Nanev Slavov; Vincent Louis Viala |
| EPI_ISL_2391578, EPI_ISL_2391733 | Unidad de Investigación Médica de Yucatán (UIMY) | Instituto de Biotecnología de la UNAM | ; Alejandra Garcia-Gasca; Alejandra Hernández-Terán; Alejandro Sanchez-Flores; Alfredo Herrera-Estrella; Alicia Ocaña-Mondragón; Andreu Comas-García; Angel Gustavo Salas-Lais; Antonio Loza Román; Bernardo Martínez-Miguel; Blanca Taboada; Brenda Irasema Maldonado-Meza; Bruno Gomez-Gil; Carla Ivón Herrera-Najera; Carlos F. Arias; Celia Boukadida; Clara Esperanza Santacruz-Tinoco; Concepción Grajales-Muñiz; Consorcio Mexicano de Vigilancia Genómica (CoViGen-Mex). Authors (in alphabetical order): Julio Elias Alvarado-Yaah; Cristóbal Cháidez-Quiróz; Célida Duque Molina; Célida Martínez-Rodríguez; Daniel Fregoso-Rueda; Daniel Lira Morales; Eduardo Becerril-Vargas; Fernando Fontove-Herrera; Fidencio Mejía-Nepomuceno; Francisco Pulido; Gloria Elena Espinosa-Ayala; Gloria María Molina-Salinas; Gloria Vazquez; Hector Esteban Paz-Juárez; Hector Montoya-Fuentes; Helen Haydee Fernanda Ramírez-Plascencia; Irvin González-López; Jean Pierre González; Joel Armando Vázquez-Pérez.; Jorge Salas-Hernández; José Antonio Enciso-Moreno; José Arturo Martínez-Orozco; José Esteban Muñoz-Medina; José de Jesús Nuñez-Contreras; Juan Bautista Chale-Dzul; Julissa Enciso-Ibarra; Luis Alberto Ochoa-Carrera; Margarita Matías-Florentino; Mario Mújica-Sánchez; María Guadalupe Santiago-Mauricio; María Guadalupe de Jesús Mireles-Rivera; Nelly Sélem-Mojica; Pavel Isa; Ricardo Ciria Merce; Ricardo Grande; Rosa María Gutierrez Rios; Santiago Ávila-Ríos; Selene Zárate; Susana Lopez; Victor Eduardo Garcia-Arias; Víctor Hugo Borja-Aburto |
| EPI_ISL_5802151 | Unidade Basica De Saude De Taquaral Caetano Pitelli | Instituto Butantan | Antonio Jorge Martins; Claudia Renata dos Santos Barros; David Schlesinger; Debora Botequio Moretti; Dimas Tadeu Covas; Elaine Cristina Marqueze; Elaine Vieira Santos; Evandra Strazza Rodrigues; Heidge Fukumasu; Jayme Augusto de Souza-Neto; José Salvatore Leister Patané; Luiz Alcantara; Luiz Lehmann Coutinho; Maria Carolina Elias; Maurício Lacerda Nogueira; Rafael dos Santos Bezerra; Raul Machado Neto; Rejane Maria Tommasini Grotto; Ricardo Haddad; Sandra Coccuzzo Sampaio Vessoni; Simone Kashima; Svetoslav Nanev Slavov; Vincent Louis Viala |
| EPI_ISL_5802084 | Unidade Basica De Saude Ii De Tarabai | Instituto Butantan | Antonio Jorge Martins; Claudia Renata dos Santos Barros; David Schlesinger; Debora Botequio Moretti; Dimas Tadeu Covas; Elaine Cristina Marqueze; Elaine Vieira Santos; Evandra Strazza Rodrigues; Heidge Fukumasu; Jayme Augusto de Souza-Neto; José Salvatore Leister Patané; Luiz Alcantara; Luiz Lehmann Coutinho; Maria Carolina Elias; Maurício Lacerda Nogueira; Rafael dos Santos Bezerra; Raul Machado Neto; Rejane Maria Tommasini Grotto; Ricardo Haddad; Sandra Coccuzzo Sampaio Vessoni; Simone Kashima; Svetoslav Nanev Slavov; Vincent Louis Viala |
| EPI_ISL_5802144, EPI_ISL_5802146 | Unidade De Vigilancia Em Saude | Instituto Butantan | Antonio Jorge Martins; Claudia Renata dos Santos Barros; David Schlesinger; Debora Botequio Moretti; Dimas Tadeu Covas; Elaine Cristina Marqueze; Elaine Vieira Santos; Evandra Strazza Rodrigues; Heidge Fukumasu; Jayme Augusto de Souza-Neto; José Salvatore Leister Patané; Luiz Alcantara; Luiz Lehmann Coutinho; Maria Carolina Elias; Maurício Lacerda Nogueira; Rafael dos Santos Bezerra; Raul Machado Neto; Rejane Maria Tommasini Grotto; Ricardo Haddad; Sandra Coccuzzo Sampaio Vessoni; Simone Kashima; Svetoslav Nanev Slavov; Vincent Louis Viala |
| EPI_ISL_5782681 | Unidade De Vigilancia Epidemiologica | Instituto Butantan | Antonio Jorge Martins; Claudia Renata dos Santos Barros; David Schlesinger; Debora Botequio Moretti; Dimas Tadeu Covas; Elaine Cristina Marqueze; Elaine Vieira Santos; Evandra Strazza Rodrigues; Heidge Fukumasu; Jayme Augusto de Souza-Neto; José Salvatore Leister Patané; Luiz Alcantara; Luiz Lehmann Coutinho; Maria Carolina Elias; Maurício Lacerda Nogueira; Rafael dos Santos Bezerra; Raul Machado Neto; Rejane Maria Tommasini Grotto; Ricardo Haddad; Sandra Coccuzzo Sampaio Vessoni; Simone Kashima; Svetoslav Nanev Slavov; Vincent Louis Viala |
| EPI_ISL_1675334 | Universidad Nacional de Colombia - Laboratorio Genómico One Health | Universidad Nacional de Colombia - Laboratorio Genómico One Health | Andres F. Cardona-Rios; Carlos Franco-Muñoz; Daniel O. Maldonado-Perez; Diego A. Álvarez-Díaz; Hector Alejandro Ruiz-Moreno; Idabely Betancur Ortiz; Jorge E. Osorio; Juan P. Hernandez-Ortiz; Karl A Ciuoderis; Katherine Laiton-Donato; Laura Silvana Perez; Lina M. Hurtado; Marcela Mercado-Reyes; Maria Angélica Maya; Maria Stella López; Rita Almanza Payares; Sandra Ines Cano; Simón Villegas Velásquez |
| EPI_ISL_1498373 | University Hospital Antwerp (UZA), Drie Eikenstraat 655, 2650 Edegem, Belgium | Labo Klinische Biologie, UZA | Basil Britto Xavier; Christine Lammens; Herman Goossens; Jasmine Coppens; Marie Le Mercier; Veerle Matheeussen |
| EPI_ISL_1626570 | University Hospital Sant'Andrea-Sapienza | INMI Lazzaro Spallanzani IRCCS | A Di Caro; B Bartolini; CEM Gruber; E Giombini; F Messina; F Santini; G Bonfiglio; I Santino; M Rueca; M Simmaco; MR Capobianchi; O Butera |
| EPI_ISL_1533942, EPI_ISL_1533954, EPI_ISL_1660757, EPI_ISL_1660761 | University of Liège COVID-19 testing center | GIGA Medical Genomics | Bouchra Boujemla; Cécile Meex; Keith Durkin; Maria Artesi; Marie-Pierre Hayette; Nathalie Renotte; Pierrette Melin; Raphaël Boreux; Sébastien Bontems; Vincent Bours |
| EPI_ISL_1580367, EPI_ISL_1580372, EPI_ISL_1580379, EPI_ISL_1580393, EPI_ISL_1651560, EPI_ISL_1651561, EPI_ISL_1744527, EPI_ISL_2598264 | | | |
| see above | University of Michigan Clinical Microbiology Laboratory | Lauring Lab, University of Michigan, Department of Microbiology and Immunology | Valesano |
| EPI_ISL_2018923, EPI_ISL_2018927 | University of Rome Tor Vergata: Departm Experim Medicine Chair of Virology | University of Rome Tor Vergata: Departm Experim Medicine Chair of Virology | Francesca Ceccherini-Silberstein; Loredana Sarmati; Lorenzo Piermatteo; Luca Carioti; Marco Iannetta; Maria Botticelli; Maria Concetta Bellocchi; Massimo Andreoni; Rossana Scutari |
| EPI_ISL_1582475, EPI_ISL_1582481, EPI_ISL_1582594 | University of Wisconsin-Madison AIDS Vaccine Research Laboratories | University of Wisconsin-Madison AIDS Vaccine Research Laboratories | Gage Moreno; Katarina Braun; et al. AIDS Vaccine Research Laboratories |
| EPI_ISL_2020614, EPI_ISL_2020639, EPI_ISL_2455733, EPI_ISL_2465250, EPI_ISL_3014374 | Università Federico II - Dipartimento di scienze mediche traslazionali - Napoli | TIGEM | Antonio Grimaldi Patrizia Annunziata Francesco Panariello Teresa Giuliano Michele Cennamo Valentina Bouche Chiara Colantuono Lucio Di Filippo Mariano Fiorenza Anna Manfredi Marcello Salvi Giuseppe Portella Andrea Ballabio Davide Cacchiarelli |
| EPI_ISL_3270765 | Università Federico II - Dipartimento di scienze mediche traslazionali - Napoli | Telethon Institute of Genetics and Medicine (TIGEM) | Antonio Grimaldi Patrizia Annunziata Francesco Panariello Teresa Giuliano Michele Cennamo Valentina Bouche Chiara Colantuono Lucio Di Filippo Mariano Fiorenza Anna Manfredi Marcello Salvi Giuseppe Portella Andrea Ballabio Davide Cacchiarelli |
| EPI_ISL_1669943, EPI_ISL_1669945, EPI_ISL_1669946, EPI_ISL_1670647 | Università degli Studi di Perugia | Istituto Zooprofilattico Sperimentale dell'Abruzzo e Molise "G. Caporale" | Ancora M; Calistri P; Camilloni B; Cammà C; Caporale M; Curini V; Delli Compagni E; Di Domenico M; Di Lollo Valeria; Di Pasquale A; Lorusso A; Mangone I; Marcacci M; Mencacci A; Puglia I; Rinaldi A; Savini G; Scialabba S |
| EPI_ISL_5802280 | Upa Dr Akira Tada | Instituto Butantan | Antonio Jorge Martins; Claudia Renata dos Santos Barros; David Schlesinger; Debora Botequio Moretti; Dimas Tadeu Covas; Elaine Cristina Marqueze; Elaine Vieira Santos; Evandra Strazza Rodrigues; Heidge Fukumasu; Jayme Augusto de Souza-Neto; José Salvatore Leister Patané; Luiz Alcantara; Luiz Lehmann Coutinho; Maria Carolina Elias; Maurício Lacerda Nogueira; Rafael dos Santos Bezerra; Raul Machado Neto; Rejane Maria Tommasini Grotto; Ricardo Haddad; Sandra Coccuzzo Sampaio Vessoni; Simone Kashima; Svetoslav Nanev Slavov; Vincent Louis Viala |
| EPI_ISL_7040782, EPI_ISL_7040802 | Urbino | Microbiology University Politecnica delle Marche | Anna Valenza; Carla Acciarri; Katia Marinelli; Monica Lucia Ferreri; Patrizia Bagnarelli; Roberta Longo; Sara Caucci; Stefano Menzo |
| EPI_ISL_1972945 | Usansolo-Galdakao University Hospital | Cruces University Hospital | Ana Belén de la Hoz; Ana Gual-de-Torrella; Izaskun Alejo-Cancho; Mikel Gallego |
| EPI_ISL_5802058 | Usf Dr Aparecido Rodrigues Mouco Scrpardo | Instituto Butantan | Antonio Jorge Martins; Claudia Renata dos Santos Barros; David Schlesinger; Debora Botequio Moretti; Dimas Tadeu Covas; Elaine Cristina Marqueze; Elaine Vieira Santos; Evandra Strazza Rodrigues; Heidge Fukumasu; Jayme Augusto de Souza-Neto; José Salvatore Leister Patané; Luiz Alcantara; Luiz Lehmann Coutinho; Maria Carolina Elias; Maurício Lacerda Nogueira; Rafael dos Santos Bezerra; Raul Machado Neto; Rejane Maria Tommasini Grotto; Ricardo Haddad; Sandra Coccuzzo Sampaio Vessoni; Simone Kashima; Svetoslav Nanev Slavov; Vincent Louis Viala |
| EPI_ISL_5802149, | Usf Paulista Fernandopolis | Instituto Butantan | Antonio Jorge Martins; Claudia Renata dos Santos Barros; David Schlesinger; Debora Botequio Moretti; Dimas Tadeu Covas; Elaine Cristina Marqueze; Elaine Vieira Santos; Evandra Strazza Rodrigues; Heidge Fukumasu; Jayme Augusto de Souza-Neto; José Salvatore Leister Patané; Luiz Alcantara; Luiz Lehmann |

| | | | |
|---|---|---|---|
| EPI_ISL_5802150, EPI_ISL_5802154, EPI_ISL_5802160, EPI_ISL_5802162 | Antonio Pivato | | Coutinho; Maria Carolina Elias; Maurício Lacerda Nogueira; Rafael dos Santos Bezerra; Raul Machado Neto; Rejane Maria Tommasini Grotto; Ricardo Haddad; Sandra Coccuzzo Sampaio Vessoni; Simone Kashima; Svetoslav Nanev Slavov; Vincent Louis Viala |
| EPI_ISL_2081799, EPI_ISL_2467060, EPI_ISL_2677862, EPI_ISL_2679011, EPI_ISL_2679026 | Utah Public Health Laboratory | Utah Public Health Laboratory | Erin L. Young; Kelly F. Oakeson; Tara Gallagher |
| EPI_ISL_2283908 | VA Connecticut Healthcare System | Yale Center for Genomic Analysis | Brooke Sullivan; Curt Scharfe; Irina Tikhonova; Kaya Bilguvar; Shrikant Mane |
| EPI_ISL_3045958 | VIDOH (Virgin Islands Department of Health) | Grubaugh Lab - Yale School of Public Health | Anderson Brito; Annie Watkins; Brett Ellis; Chaney Kalinich; Chantal Vogels; Esther Ellis; Isabel Ott; Jendai Richards; Jessica Rothman; Joseph Fauver; Kendall Billig; Mallery Breban; Marlon Lawrence; Mary Petrone; Nathan Grubaugh; TaLesa Aderohunmu; Tara Alpert; Tobias Koch |
| EPI_ISL_1966525 | VIGILANCIA EM SAUDE | Instituto Butantan / FZEA-USP (Pirassununga) | Antonio Jorge Martins; Bianca Cechetto Carlos. Mendelics: Bibiana Santos; Claudia Renata dos Santos Barros; Cíntia Bittar; David Schlesinger. Hemocentro Ribeirão Preto: Simone Kashima; Debora Botequio Moretti; Elaine Cristina Marqueze; Elaine Vieira dos Santos; Elisangela Chicaroni Mattos; Erika Freitas; Evandra Strazza Rodrigues; Felipe Allan da Silva da Costa; Flavia Aburjaile; Fábio Sossai Possebon; Guilherme Campos; Guilherme Targino Valente; Heidge Fukumasu. USP-Botucatu: Rejane Maria Tommasini Grotto; Helena Lage Ferreira; Instituto Butantan: Dimas Tadeu Covas; Jardelina de Souza Todao Bernardino; Jayme A. Souza-Neto; Jessika Cristina Chagas Lesbon; Jorge A. Petroli Marchesi; José Salvatore Leister Patané; João Paulo Kitajima; João Pessoa Araújo Jr.; Leila Sabrina Ullmann; Loyze Paola Oliveira de Lima; Luiz Aurelio de Campos Crispin. Centro de Genômica Funcional da ESALQ: Luiz Lehmann Coutinho; Luiz Carlos Junior de Alcantara; Lívia Sacchetto; Maisa C. Pereira Parra; Maria Carolina Elias; Marta Giovanetti; Marília Moraes; Maurício Lacerda Nogueira. Prefeitura de Sao Paulo: Melissa Palmieri.; Patricia Akemi Assato; Paula Rahal; Paulo Inacio da Costa; Rafael dos Santos Bezerra; Raquel de Lello Rocha Campos Cassano. NGS Soluções Genômicas: Pilar Drummond Sampaio Corrêa Mariani. FZEA-USP Pirassununga: Mirele Daiana Poleti; Raul Machado Neto; Ricardo Augusto Brassaloti; Ricardo Haddad; Rodrigo Tocantins Calado. FAMERP-SJRP: Cecília Artico Banho; Sandra Coccuzzo Sampaio; Svetoslav Nanev Slavov; Vagner Fonseca; Vincent Louis Viala |
| EPI_ISL_2344694 | VIGILANCIA EPIDEMIOLOGICA DE LEME | Instituto Butantan / FZEA-USP-Pirassununga | Antonio Jorge Martins; Claudia Renata dos Santos Barros; David Schlesinger; Debora Botequio Moretti; Dimas Tadeu Covas; Elaine Cristina Marqueze; Elaine Vieira Santos; Evandra Strazza Rodrigues; Heidge Fukumasu; Jayme Augusto de Souza-Neto; José Salvatore Leister Patané; Luiz Alcantara; Luiz Lehmann Coutinho; Maria Carolina Elias; Maurício Lacerda Nogueira; Rafael dos Santos Bezerra; Raul Machado Neto; Rejane Maria Tommasini Grotto; Ricardo Haddad; Sandra Coccuzzo Sampaio Vessoni; Simone Kashima; Svetoslav Nanev Slavov; Vincent Louis Viala |
| EPI_ISL_2344692, EPI_ISL_2344695 | VIGILANCIA EPIDEMIOLOGICA E CONTROLE DE VETORES PIRASSUNUN | Instituto Butantan / FZEA-USP-Pirassununga | Antonio Jorge Martins; Claudia Renata dos Santos Barros; David Schlesinger; Debora Botequio Moretti; Dimas Tadeu Covas; Elaine Cristina Marqueze; Elaine Vieira Santos; Evandra Strazza Rodrigues; Heidge Fukumasu; Jayme Augusto de Souza-Neto; José Salvatore Leister Patané; Luiz Alcantara; Luiz Lehmann Coutinho; Maria Carolina Elias; Maurício Lacerda Nogueira; Rafael dos Santos Bezerra; Raul Machado Neto; Rejane Maria Tommasini Grotto; Ricardo Haddad; Sandra Coccuzzo Sampaio Vessoni; Simone Kashima; Svetoslav Nanev Slavov; Vincent Louis Viala |
| EPI_ISL_5780266, EPI_ISL_5780267 | VUMC Molecular Infectious Diseases Laboratory (MIDL) | Dr. Suman Das Lab - Vanderbilt University Medical Center (VUMC) | Bookyung Park; Grant Vestal; Helen Boone; Hunter Brown; Jonathan E. Schmitz; Meghan Shilts; Seesandra Rajagopala; Suman B. Pakala; Suman Das |
| EPI_ISL_5802109 | Vigilancia Em Saude | Instituto Butantan | Antonio Jorge Martins; Claudia Renata dos Santos Barros; David Schlesinger; Debora Botequio Moretti; Dimas Tadeu Covas; Elaine Cristina Marqueze; Elaine Vieira Santos; Evandra Strazza Rodrigues; Heidge Fukumasu; Jayme Augusto de Souza-Neto; José Salvatore Leister Patané; Luiz Alcantara; Luiz Lehmann Coutinho; Maria Carolina Elias; Maurício Lacerda Nogueira; Rafael dos Santos Bezerra; Raul Machado Neto; Rejane Maria Tommasini Grotto; Ricardo Haddad; Sandra Coccuzzo Sampaio Vessoni; Simone Kashima; Svetoslav Nanev Slavov; Vincent Louis Viala |
| EPI_ISL_5799689 | Vigilancia Epidemiologica De Leme | Instituto Butantan | Antonio Jorge Martins; Claudia Renata dos Santos Barros; David Schlesinger; Debora Botequio Moretti; Dimas Tadeu Covas; Elaine Cristina Marqueze; Elaine Vieira Santos; Evandra Strazza Rodrigues; Heidge Fukumasu; Jayme Augusto de Souza-Neto; José Salvatore Leister Patané; Luiz Alcantara; Luiz Lehmann Coutinho; Maria Carolina Elias; Maurício Lacerda Nogueira; Rafael dos Santos Bezerra; Raul Machado Neto; Rejane Maria Tommasini Grotto; Ricardo Haddad; Sandra Coccuzzo Sampaio Vessoni; Simone Kashima; Svetoslav Nanev Slavov; Vincent Louis Viala |
| EPI_ISL_5799687, EPI_ISL_5799690 | Vigilancia Epidemiologica E Controle De Vetores Pirassununga | Instituto Butantan | Antonio Jorge Martins; Claudia Renata dos Santos Barros; David Schlesinger; Debora Botequio Moretti; Dimas Tadeu Covas; Elaine Cristina Marqueze; Elaine Vieira Santos; Evandra Strazza Rodrigues; Heidge Fukumasu; Jayme Augusto de Souza-Neto; José Salvatore Leister Patané; Luiz Alcantara; Luiz Lehmann Coutinho; Maria Carolina Elias; Maurício Lacerda Nogueira; Rafael dos Santos Bezerra; Raul Machado Neto; Rejane Maria Tommasini Grotto; Ricardo Haddad; Sandra Coccuzzo Sampaio Vessoni; Simone Kashima; Svetoslav Nanev Slavov; Vincent Louis Viala |
| EPI_ISL_1683076 | Viollier AG | Department of Biosystems Science and Engineering, ETH Zürich | Andrea Patrignani; Andreia Cabral de Gouvea; Catharine Aquino; Chaoran Chen; Christiane Beckmann; Christoph Noppen; David Dreifuss; Doris Popovic; Griffin White; Ivan Topolsky; Jay Tracy; Katharina Jahn; Lara Fuhrmann; Laura Neff; Lennart Opitz; Maria Domenica Moccia; Maurice Redondo; Niko Beerenwinkel; Noemie Santamaria de Souza; Olivier Kobel; Philipp Jablonski; Ralph Schlapbach; Sarah Nadeau; Simon Grüter; Sophie Seidel; Tanja Stadler; Timothy Sykes |
| EPI_ISL_1760375 | Virginia Division of Consolidated Laboratory Services | Virginia Division of Consolidated Laboratory Services | Virginia DCLS |
| EPI_ISL_1636703 | Virology Department, Royal Infirmary of Edinburgh, NHS Lothian / School of Biological Sciences, University of Edinburgh | COVID-19 Genomics UK (COG-UK) Consortium | Colquhoun R; Cotton S; Dewar R; Hill V; Jackson B; McCrone JT; McHugh M; O'Toole Á; Rambaut A; Rooke S; Scher E; Templeton K; Yu X |
| EPI_ISL_1653847, EPI_ISL_1653868, EPI_ISL_1653877 | Washington State Department of Health Public Health Laboratories | Washington State Department of Health Public Health Laboratories | Avi Singh; Darren Lucas; Denny Russell; Drew MacKellar; Geoff Melly; Hannah Gray; Joenice Gonzalez; JohnAric Peterson; Philip Dykema; Rebecca Cao; Vanessa De Los Santos |
| EPI_ISL_2454549 | Wexner Medical Center | The Ohio State University College of Medicine | Koenig, S.; Seminetta, J. |
| EPI_ISL_1793861, EPI_ISL_1794124, EPI_ISL_1794316 | Wisconsin State Laboratory of Hygiene Communicable Disease Division | Wisconsin State Laboratory of Hygiene Communicable Disease Division | Abigail C. Shockey; Alicia J. Mooney; Kelsey R. Florek; Sara Wagner |
| EPI_ISL_1711271 | Wyoming Public Health Laboratory | Wyoming Public Health Laboratory | Ashley Norberg; Brian Dominguez; Brittany Oher; Cari Sloma; Channing Weber; Chayse Rowley; Elliot Thomasson; Jim Mildenberger; Marley Goetz; Sam Britz; Taylor Fearing; and Rob Christensen |
| EPI_ISL_1793506 | Yale Clinical Virology Lab | Yale Center for Genomic Analysis | Brooke Sullivan; Christopher Castaldi; Curt Scharfe; Irina Tikhonova; Kaya Bilguvar; Shrikant Mane |
| EPI_ISL_2613707 | cerballiance-IDF | Cerba lab | Aude Lessenne; Bénédicte Roquebert; Emmanuel Lecorche; Kader Merah; Laura Verdurme; Patrice Herisson; Sabine Trombert-Paolantoni; Stéphanie Haïm-Boukobza; Thierry Collin |
| EPI_ISL_1738993 | unknown | Instituto Nacional de Saude (INSA) | Borges et al |
