## Supplement 3 for "Rapid and Accurate Identification of SARS-CoV-2 Omicron Variants Using Droplet Digital PCR (RT-ddPCR)": All_0405-0411.pdf

We gratefully acknowledge the following Authors from the Originating laboratories responsible for obtaining the specimens, as well as the Submitting laboratories where the genome data were generated and shared via GISAID, on which this research is based.

All Submitters of data may be contacted directly via www.gisaid.org

Authors are sorted alphabetically.

| Accession ID | Originating Laboratory | Submitting Laboratory | Authors |
|---|---|---|---|
| EPI_ISL_1711723 | "NM Dept. Health, Scientific Laboratory Division " | Centers for Disease Control and Prevention Division of Viral Diseases, Pathogen Discovery | Alison Laufer Halpin; Ben L. Rambo-Martin; Clinton R. Paden; Dakota Howard; Darlene Wagner; Dave Wentworth; Dhwani Batra; Jasmine Padilla; Justin Lee; Katie Dillon; Krista Queen; Kristen Knipe; Kristine Lacek; Mark Burroughs; Matthew Schmerer; Mili Sheth; Peter Cook; Sam Shepard; Sarah Nobles; Shoshona Le; Suxiang Tong; Vivien Dugan; Yvette Unoarumhi |
| EPI_ISL_3347182 | ADILAB | Laboratorio Departamental de Salud Publica de Antioquia | Andres F. Cardona-Rios; Gloria Isabel Escobar; Idabely Betancur Ortiz; Juan P. Hernandez-Ortiz; Maria Stella López |
| EPI_ISL_1756002, EPI_ISL_1756003 | ALBANY MEDICAL CENTER | Wadsworth Center, New York State Department of Health | Alexis Russell; Catharine Prussing; Daryl M. Lamson; Erasmus Schneider; Erica Lasek-Nesselquist; John Kelly; Jonathan Plitnick; Kirsten St. George; Matthew Shudt; Melissa A Leisner; Navjot Singh |
| EPI_ISL_5915117, EPI_ISL_5915127, EPI_ISL_5915163, EPI_ISL_5915222, EPI_ISL_5915237, EPI_ISL_5915292, EPI_ISL_5915299, EPI_ISL_5915326, EPI_ISL_6573829, EPI_ISL_6573830, EPI_ISL_6573867, EPI_ISL_6573919 | | | |
| see above | AMALAB/FACISA/UFRN | WallauLab on behalf of Fiocruz COVID-19 Genomic Surveillance Network | Alexandre Freitas da Silva; Allan Roberto Dias Nunes; Antonio Marinho da Silva Neto; Cassia Docena; Constância Flávia Junqueira Ayres; Filipe Zimmer Dezordi; Gabriel Luz Wallau; Gustavo Barbosa de Lima; Joana Cristina Medeiros Tavares Marques; Katya Anaya Jacinto; Lais Ceschini Machado; Lilian Caroliny Amorim Silva; Marcelo Henrique dos Santos Paiva; Mariane dos Santos Duarte; Matheus Filgueira Bezerra; Sinval Pinto Brandão Filho |
| EPI_ISL_1967028 | AMBULATORIO DE MOLESTIAS INFECCIOSAS DE SALTO | Instituto Butantan / ESALQ-USP (Piracicaba) | Antonio Jorge Martins; Bianca Cechetto Carlos. Mendelics: Bibiana Santos; Claudia Renata dos Santos Barros; Cíntia Bittar; David Schlesinger. Hemocentro Ribeirão Preto: Simone Kashima; Debora Botequio Moretti; Elaine Cristina Marqueze; Elaine Vieira dos Santos; Elisangela Chicaroni Mattos; Erika Freitas; Evandra Strazza Rodrigues; Felipe Allan da Silva da Costa; Flavia Aburjaile; Fábio Sossai Possebon; Guilherme Campos; Guilherme Targino Valente; Heidge Fukumasu. USP-Botucatu: Rejane Maria Tommasini Grotto; Helena Lage Ferreira; Instituto Butantan: Dimas Tadeu Covas; Jardelina de Souza Todao Bernardino; Jayme A. Souza-Neto; Jessika Cristina Chagas Lesbon; Jorge A. Petroli Marchesi; José Salvatore Leister Patané; João Paulo Kitajima; João Pessoa Araújo Jr.; Leila Sabrina Ullmann; Loyze Paola Oliveira de Lima; Luiz Aurelio de Campos Crispin. Centro de Genômica Funcional da ESALQ: Luiz Lehmann Coutinho; Luiz Carlos Junior de Alcantara; Lívia Sacchetto; Maisa C. Pereira Parra; Maria Carolina Elias; Marta Giovanetti; Marília Moraes; Maurício Lacerda Nogueira. Prefeitura de Sao Paulo: Melissa Palmieri.; Patricia Akemi Assato; Paula Rahal; Paulo Inacio da Costa; Rafael dos Santos Bezerra; Raquel de Lello Rocha Campos Cassano. NGS Soluções Genômicas: Pilar Drummond Sampaio Corrêa Mariani. FZEA-USP Pirassununga: Mirele Daiana Poleti; Raul Machado Neto; Ricardo Augusto Brassaloti; Ricardo Haddad; Rodrigo Tocantins Calado. FAMERP-SJRP: Cecília Artico Banho; Sandra Coccuzzo Sampaio; Svetoslav Nanev Slavov; Vagner Fonseca; Vincent Louis Viala |
| EPI_ISL_1940188, EPI_ISL_1940249, EPI_ISL_1940277, EPI_ISL_1940278, EPI_ISL_1940300, EPI_ISL_1940304, EPI_ISL_1940370, EPI_ISL_1940434, EPI_ISL_1940504, EPI_ISL_1940520, EPI_ISL_1940569, EPI_ISL_1940680 | | | |
| see above | ASL Napoli 1 Centro | AMES Centro Polidiagnostico Strumentale S.r.l. | Antonella Di Carlo; Antonio Fico; Eloisa Evangelista; Giovanni Savarese; Luigi D'Amore; Luisa Circelli; Maurizio D'Amora; Monica Ianniello; Nadia Petrillo; Raffaella Ruggiero; Roberto Sirica |
| EPI_ISL_1599550 | AZ Klina | AZ KLina | Carl Vael - Lynsey Berckmans |
| EPI_ISL_1599551 | AZ Klina | AZ Klina | Carl Vael - Lynsey Berckmans |
| EPI_ISL_1972237 | AZ St.Jan Brugge-Oostende | AZ SINT-JAN BRUGGE | Jorn Hellemans; Laurien Hoornaert; Marijke Reynders; Patrick Descheemaeker; Thomas Van Landschoot |
| EPI_ISL_1904359, EPI_ISL_1904360 | AZDelta | AZDelta | Dieter De Smet; Geert Martens |
| EPI_ISL_1667433, EPI_ISL_1667501, EPI_ISL_1667573, EPI_ISL_1667626, EPI_ISL_1667669, EPI_ISL_1685735, EPI_ISL_1689880, EPI_ISL_1689902, EPI_ISL_1689906, EPI_ISL_1690148, EPI_ISL_1702134, EPI_ISL_1736358, EPI_ISL_1736359, EPI_ISL_1736389, EPI_ISL_1736423, EPI_ISL_1736430, EPI_ISL_1736432, EPI_ISL_1736463, EPI_ISL_1736478, EPI_ISL_1736522, EPI_ISL_1736543, EPI_ISL_1736552, EPI_ISL_1736594, EPI_ISL_1736597, EPI_ISL_1736598, EPI_ISL_1736604, EPI_ISL_1736629, EPI_ISL_1736638, EPI_ISL_1736639, EPI_ISL_1736657, EPI_ISL_1736712, EPI_ISL_1736755, EPI_ISL_1736757, EPI_ISL_1736761, EPI_ISL_1736762, EPI_ISL_1736801, EPI_ISL_1736807, EPI_ISL_1736812, EPI_ISL_1736832, EPI_ISL_1736861, EPI_ISL_1736870, EPI_ISL_1736876, EPI_ISL_1736879, EPI_ISL_1736948, EPI_ISL_1736963, EPI_ISL_1736973, EPI_ISL_1737113, EPI_ISL_1737599, EPI_ISL_1737616, EPI_ISL_1737640, EPI_ISL_1737662, EPI_ISL_1737673, EPI_ISL_1737680, EPI_ISL_1737816, EPI_ISL_1753545, EPI_ISL_1753558, EPI_ISL_1834462, EPI_ISL_1834524, EPI_ISL_1834532, EPI_ISL_1834533, EPI_ISL_1834580, EPI_ISL_1834597, EPI_ISL_1834629, EPI_ISL_1834688, EPI_ISL_1835366, EPI_ISL_1835388, EPI_ISL_1835395, EPI_ISL_1835396, EPI_ISL_1835413, EPI_ISL_1835419, EPI_ISL_1835420, EPI_ISL_1835421, EPI_ISL_1835423, EPI_ISL_1835424, EPI_ISL_1835425, EPI_ISL_1835427, EPI_ISL_1835430, EPI_ISL_1835432, EPI_ISL_1835440, EPI_ISL_1835445, EPI_ISL_1835462, EPI_ISL_1835466, EPI_ISL_1835500, EPI_ISL_1835513, EPI_ISL_1835534, EPI_ISL_1835567, EPI_ISL_1835602, EPI_ISL_1835614, EPI_ISL_1835640, EPI_ISL_1835674, EPI_ISL_1835679, EPI_ISL_1835765, EPI_ISL_1835826, EPI_ISL_1835857, EPI_ISL_1835894, EPI_ISL_1835914, EPI_ISL_1835915, EPI_ISL_1835927, EPI_ISL_1835936, EPI_ISL_1835982, EPI_ISL_1835992, EPI_ISL_1836021, EPI_ISL_1836083, EPI_ISL_1836086, EPI_ISL_1836115, EPI_ISL_1836116, EPI_ISL_1836203, EPI_ISL_1836213, EPI_ISL_1836264, EPI_ISL_1836270, EPI_ISL_1836279, EPI_ISL_1836317, EPI_ISL_1836323, EPI_ISL_1836326, EPI_ISL_1836333, EPI_ISL_1836334, EPI_ISL_1836464, EPI_ISL_1836536, EPI_ISL_1836649, EPI_ISL_1836670, EPI_ISL_1836709, EPI_ISL_1836731, EPI_ISL_1836735, EPI_ISL_1836839, EPI_ISL_1836841, EPI_ISL_1836879, EPI_ISL_1836880, EPI_ISL_1836897, EPI_ISL_1836906, EPI_ISL_1836930, EPI_ISL_1836955, EPI_ISL_1836973, EPI_ISL_1837069, EPI_ISL_1837107, EPI_ISL_1837138, EPI_ISL_1837172, EPI_ISL_1837218, EPI_ISL_1837308, EPI_ISL_1837311, EPI_ISL_1837327, EPI_ISL_1837343, EPI_ISL_1837673, EPI_ISL_1837677, EPI_ISL_1837710, EPI_ISL_1837722, EPI_ISL_1837729, EPI_ISL_1837747, EPI_ISL_1837794, EPI_ISL_1837806, EPI_ISL_1837819, EPI_ISL_1837820, EPI_ISL_1837822, EPI_ISL_1837929, EPI_ISL_1837948, EPI_ISL_1838020, EPI_ISL_1838108, EPI_ISL_1838146, EPI_ISL_1838177, EPI_ISL_1838181, EPI_ISL_1838240, EPI_ISL_1838300, EPI_ISL_1838394, EPI_ISL_1900196, EPI_ISL_1900212, EPI_ISL_1900345, EPI_ISL_1900367, EPI_ISL_1900379, EPI_ISL_1900384, EPI_ISL_1900387, EPI_ISL_1900389, EPI_ISL_1900415, EPI_ISL_1900417, EPI_ISL_1923120, EPI_ISL_1994977, EPI_ISL_1994999, EPI_ISL_1995021, EPI_ISL_1995041, EPI_ISL_1995077, EPI_ISL_1995094, EPI_ISL_1995141, EPI_ISL_1995155, EPI_ISL_1995255, EPI_ISL_1995362, EPI_ISL_1995407, EPI_ISL_1995521, EPI_ISL_1995533, EPI_ISL_1995550, EPI_ISL_1995561, EPI_ISL_1995565, EPI_ISL_1995587, EPI_ISL_1995588, EPI_ISL_1995610, EPI_ISL_1995612, EPI_ISL_1995615, EPI_ISL_1995635, EPI_ISL_1995664, EPI_ISL_1995683, EPI_ISL_1995695, EPI_ISL_1995699, EPI_ISL_1995758, EPI_ISL_1995765, EPI_ISL_1995811, EPI_ISL_1996196, EPI_ISL_1996269, EPI_ISL_1996295, EPI_ISL_1996301, EPI_ISL_1996340, EPI_ISL_1996351, EPI_ISL_1996361, EPI_ISL_1996373, EPI_ISL_1996378, EPI_ISL_1996379, EPI_ISL_1996454, EPI_ISL_1996455, EPI_ISL_1996469, EPI_ISL_1996480, EPI_ISL_1996496, EPI_ISL_1996524, EPI_ISL_1996553, EPI_ISL_1996610, EPI_ISL_1996624, EPI_ISL_1996683, EPI_ISL_1996685, EPI_ISL_1996686, EPI_ISL_1996688, EPI_ISL_1996689, EPI_ISL_1996690, EPI_ISL_1996701, EPI_ISL_1996714, EPI_ISL_1996772, EPI_ISL_1996773, EPI_ISL_1996785, EPI_ISL_1996834, EPI_ISL_1996835, EPI_ISL_1996836, EPI_ISL_1996858, EPI_ISL_1996860, EPI_ISL_1996872, EPI_ISL_1996874, EPI_ISL_1996881, EPI_ISL_1996887, EPI_ISL_1996934, EPI_ISL_1996942, EPI_ISL_1996976, EPI_ISL_1997051, EPI_ISL_1997167, EPI_ISL_1997168, EPI_ISL_1997252, EPI_ISL_1997289, EPI_ISL_1997344, EPI_ISL_1997345, EPI_ISL_1997351, EPI_ISL_1997360, EPI_ISL_1997384, EPI_ISL_1997769, EPI_ISL_1997791, EPI_ISL_1998219, EPI_ISL_1998229, EPI_ISL_1998241, EPI_ISL_1998563, EPI_ISL_1998955, EPI_ISL_1999879, EPI_ISL_2035053, EPI_ISL_2035114, EPI_ISL_2035153, EPI_ISL_2035156, EPI_ISL_2035159, EPI_ISL_2421418, EPI_ISL_2421437, EPI_ISL_4361517, EPI_ISL_4362153, EPI_ISL_4362946, EPI_ISL_4363024, EPI_ISL_4363546, EPI_ISL_4364118, EPI_ISL_4364410 | | | |
| see above | Aegis Sciences Corporation | Centers for Disease Control and Prevention Division of Viral Diseases, Pathogen Discovery | Adrian Paskey; Alec Vest; Benjamin Rambo-Martin; Christopher Gulvick; Clinton Paden; Clinton R. Paden; Cyndi Clark; Dakota Howard; Darlene Wagner; Dhwani Batra; Dillon Nall; Duncan MacCannell; Erisa Sula; Ethan Sanders; Holly Houdeshell; Jason Caravas; Kara Moser; Kristine Lacek; Matthew Hardison; Matthew Schmerer; Ola Kvalvaag; Patrick Campbell; Peter Cook; Peter W. Cook; Rob Case; Scott Sammons; Shatavia Morrison; Shaun Westlund; Tymeckia Kendall; Victoria Caban Figueroa; Vikramsinha Ghorpade; Yvette Unoarumhi |
| EPI_ISL_2169548, EPI_ISL_2169554, EPI_ISL_2169555, EPI_ISL_2169562, EPI_ISL_2169563, EPI_ISL_2169564, EPI_ISL_2169565, EPI_ISL_2169644, EPI_ISL_2169646, EPI_ISL_2169647, EPI_ISL_2169650, EPI_ISL_2169651, EPI_ISL_2169658, EPI_ISL_2169659, EPI_ISL_2169660, EPI_ISL_2169669, EPI_ISL_2169674, EPI_ISL_2169675, EPI_ISL_2169679, EPI_ISL_2169687, EPI_ISL_2169689, EPI_ISL_2169693, EPI_ISL_2169695, EPI_ISL_2169701, EPI_ISL_2169703, EPI_ISL_2169704, EPI_ISL_2169706, EPI_ISL_2169710, EPI_ISL_2169716, EPI_ISL_2169717, EPI_ISL_2169718, EPI_ISL_2169722, EPI_ISL_2169723, EPI_ISL_2169734, EPI_ISL_2169738, EPI_ISL_2169741, EPI_ISL_2169745, EPI_ISL_2169750, EPI_ISL_2169763, EPI_ISL_2169765, EPI_ISL_2169766, EPI_ISL_2169768, EPI_ISL_2169769, EPI_ISL_2169857, EPI_ISL_2169858, EPI_ISL_2169859, EPI_ISL_2169866, EPI_ISL_2169868, EPI_ISL_2169878, EPI_ISL_2169883, EPI_ISL_2169887, EPI_ISL_2169891, EPI_ISL_2169893, EPI_ISL_2169895, EPI_ISL_2169896, EPI_ISL_2169898, EPI_ISL_2169904, EPI_ISL_2169905, EPI_ISL_2169906, EPI_ISL_2169912, EPI_ISL_2169919, EPI_ISL_2169921, EPI_ISL_2169938, EPI_ISL_2169941, EPI_ISL_2170200, EPI_ISL_2170202, EPI_ISL_2170203, EPI_ISL_2170204, EPI_ISL_2170206, EPI_ISL_2170212, EPI_ISL_2170213, EPI_ISL_2170214, EPI_ISL_2170223, EPI_ISL_2170224, EPI_ISL_2170225, EPI_ISL_2170226, EPI_ISL_2170228, EPI_ISL_2170229, EPI_ISL_2170231, EPI_ISL_2170232, EPI_ISL_2170233, EPI_ISL_2170234, EPI_ISL_2170235, EPI_ISL_2170236, EPI_ISL_2170237, EPI_ISL_2170238, EPI_ISL_2170241, EPI_ISL_2170243, EPI_ISL_2170244, EPI_ISL_2170245, EPI_ISL_2170248, EPI_ISL_2170250, EPI_ISL_2170252, EPI_ISL_2170253, EPI_ISL_2170254, EPI_ISL_2170255, EPI_ISL_2170256, EPI_ISL_2170258, EPI_ISL_2170261, EPI_ISL_2170264, EPI_ISL_2170265, EPI_ISL_2170266, EPI_ISL_2170267, EPI_ISL_2170268, EPI_ISL_2170269, EPI_ISL_2170270, EPI_ISL_2170271, EPI_ISL_2170272, EPI_ISL_2170273, EPI_ISL_2170274, EPI_ISL_2170275, EPI_ISL_2170276, EPI_ISL_2170277, EPI_ISL_2170278, EPI_ISL_2170279, EPI_ISL_2170280, EPI_ISL_2170281, EPI_ISL_2170282, EPI_ISL_2170283, EPI_ISL_2170284, EPI_ISL_2170285, EPI_ISL_2170286, EPI_ISL_2170287, EPI_ISL_2170288, EPI_ISL_2170289, EPI_ISL_2170290, EPI_ISL_2170291, EPI_ISL_2170292, EPI_ISL_2170293, EPI_ISL_2170294, EPI_ISL_2170295, EPI_ISL_2170296, EPI_ISL_2170297, EPI_ISL_2170298, EPI_ISL_2170299, EPI_ISL_2170300, EPI_ISL_2170301, EPI_ISL_2170302, EPI_ISL_2170303, EPI_ISL_2170304, EPI_ISL_2170305, EPI_ISL_2170306, EPI_ISL_2170307, EPI_ISL_2170308, EPI_ISL_2170309, EPI_ISL_2170310, EPI_ISL_2170312, EPI_ISL_2170313, EPI_ISL_2170314, EPI_ISL_2170315, EPI_ISL_2170317, EPI_ISL_2170318, EPI_ISL_2170319, EPI_ISL_2170320, EPI_ISL_2170321, EPI_ISL_2170324, EPI_ISL_2170326, EPI_ISL_2170327, EPI_ISL_2170328, EPI_ISL_2170329, EPI_ISL_2170330, EPI_ISL_2170331, EPI_ISL_2170332, EPI_ISL_2170393, EPI_ISL_2170398, EPI_ISL_2170400, EPI_ISL_2170470, EPI_ISL_2170473, EPI_ISL_2170481, EPI_ISL_2170482 | | | |
| see above | Alberta Precision Labs (APL) | Public Health Agency of Canada (PHAC) National Microbiology Laboratory | Buss; Croxen M; Deo A; Dieu P; E; Ferrato C; Gill K; Khan F; Koleva P; Li V; Lloyd C; Lynch T; Ma R; Murphy S; Pabbaraju K; Shokoples S; Thayer J; Tipples G; Whitehouse M; Wong A; Yu C; Zelyas N |
| EPI_ISL_1822797, EPI_ISL_1822827, EPI_ISL_1822998, EPI_ISL_1823049, EPI_ISL_1823069, EPI_ISL_1823077, EPI_ISL_1926109, EPI_ISL_1926116, EPI_ISL_1926127, EPI_ISL_1926140, EPI_ISL_1926154, EPI_ISL_1926162, EPI_ISL_1926173, EPI_ISL_1926174, EPI_ISL_1926190 | | | |
| see above | Altius Institute | Seattle Flu Study | Alex Nguyen; Amanda Adler; Andrew Meuser; Barry R. Lutz; Benjamin Pelle; Caitlin R. Wolf; Chris D. Frazar; Clem Green; Daniel Bates; Deborah A. Nickerson; Elisabeth Brandstetter; Erica Ryke; Hannah Petersen; Helen Y. Chu; Jacob Rodriguez; Janet A. Englund; Jay Shendure; Jessica Halow; John Stamatoyannopoulos; Joshua Richards; Jover Lee; Julia Wald; Kairsten Fay; Kirsten Lacombe; Kneshay Harper; Lea M. Starita; Mark J. Rieder; Matt Hartman; Matthew Richardson; Matthew Thompson; Melissa Truong; Michael Boeckh; Michael Famulare; Misja Ilcisin; Muhammad Halimun; Olivia Waltner; Peter D. Han; Rebecca Bruders; Ryan Alexander; Sadie Patraw; Sofia Olsson; Stephanie DeBaun; Thomas R. Sibley; Tobias Ragoczy; Trevor Bedford; Truong Nguyen |
| EPI_ISL_5800210 | Ambulatorio De Molestias Infecciosas De Salto | Instituto Butantan | Antonio Jorge Martins; Claudia Renata dos Santos Barros; David Schlesinger; Debora Botequio Moretti; Dimas Tadeu Covas; Elaine Cristina Marqueze; Elaine Vieira dos Santos; Evandra Strazza Rodrigues; Heidge Fukumasu; Jayme Augusto de Souza-Neto; José Salvatore Leister Patané; Luiz Alcantara; Luiz Lehmann Coutinho; Maria Carolina Elias; Maurício Lacerda Nogueira; Rafael dos Santos Bezerra; Raul Machado Neto; Rejane Maria Tommasini Grotto; Ricardo Haddad; Sandra Coccuzzo Sampaio Vessoni; Simone Kashima; Svetoslav Nanev Slavov; Vincent Louis Viala |
| EPI_ISL_1516747 | Area of Virology, Serology and Virology Division (SAViD), New South Wales Health Pathology Randwick | Virology Research Laboratory; Area of Virology, Serology and Virology Division (SAViD), New South Wales Health Pathology Randwick | Au, J.; Bull, R.; Deveson, I.; Foster, C.; Rawlinson, W.; Ruiz Silva, M.; Van Hal, S. |
| EPI_ISL_2499835 | Arizona State Public Health Laboratory | Arizona State Public Health Laboratory | Jessica Escobar; Katherine Fullerton; Linda Getsinger; Nobuko Fukushima; Stacy White; Trung Huynh; Victor Waddell |
| EPI_ISL_1743682, EPI_ISL_1743683, EPI_ISL_1743684, EPI_ISL_1743685 | Arizona State University | Arizona State University | Efrem S. Lim; Joshua LaBaer; Joy M. Blain; LaRinda A. Holland; Nicholas J. Mellor; Peter T. Skidmore; Rabia Maqsood; Valerie Harris; Vel Murugan |
| EPI_ISL_1670611 | Arlon | Plateforme de testing Namuroise | Céline Maschietto; Degosserie Jonathan; Denis Olivier; Mullier François; Otto Gaetan |
| EPI_ISL_1822787, EPI_ISL_1822817, EPI_ISL_1822857, EPI_ISL_1822912, EPI_ISL_1822914, EPI_ISL_1823093 | Atlas Genomics | Seattle Flu Study | Amanda Adler; Barry R. Lutz; Benjamin Pelle; Brian Hiatt; Caitlin R. Wolf; Chris D. Frazar; Deborah A. Nickerson; Elisabeth Brandstetter; Erica Ryke; Geoff Melly; Helen Y. Chu; Janet A. Englund; Jay Shendure; Jover Lee; Kairsten Fay; Kirsten Lacombe; Lea M. Starita; Mark J. Rieder; Matthew Richardson; Matthew Thompson; Melissa Truong; Michael Boeckh; Michael Famulare; Misja Ilcisin; Peter D. Han; Philip Dykema; Romesh Gautom; Scott Lindquist; Thomas R. Sibley; Trevor Bedford |
| EPI_ISL_1854228, EPI_ISL_1854244, EPI_ISL_1854245 | Avelab | Instituto Nacional de Saude (INSA) and BioSystems & Integrative Sciences Institute (BioISI) Genomics Unit, FCUL | Borges et al |
| EPI_ISL_3266994, EPI_ISL_3266998, EPI_ISL_3267004, EPI_ISL_3267020, EPI_ISL_3267023, EPI_ISL_3267030, EPI_ISL_3267034, EPI_ISL_3267040, EPI_ISL_3267042, EPI_ISL_3276606, EPI_ISL_3276610, EPI_ISL_3276614, EPI_ISL_3276624 | | | |
| see above | Ayudas Diagnosticas Sura | Universidad Nacional de Colombia - Laboratorio Genómico One Health | Andres F. Cardona-Rios; Carlos Franco-Muñoz; Carolina Muñoz-Arango; Celeny Ortiz; Daniel O. Maldonado-Perez; Diego A. Álvarez-Díaz; Hector Alejandro Ruiz-Moreno; Idabely Betancur Ortiz; Jorge E. Osorio; Juan P. Hernandez-Ortiz; Karl A Ciuoderis; Katherine Laiton-Donato; Laura Silvana Perez; Lina M. Hurtado; Marcela Mercado-Reyes; Maria Angélica Maya; Maria Stella López; Rita Almanza Payares; Sandra Ines Cano; Simón Villegas Velásquez |
| EPI_ISL_3341942 | Azienda Ospedaliera Regionale San Carlo Potenza | Laboratorio di patologia clinica e microbiologia A.O.R - San Carlo Potenza | Angela Menchise; Anna Curci; Antonio Picerno; Chiara Autilio; Sara Pascale; Teresa Lopizzo |

| | | | |
|---|---|---|---|
| EPI_ISL_1941565, EPI_ISL_1941574, EPI_ISL_2007319, EPI_ISL_2007321 | Azienda Sanitaria dell'Alto Adige Laboratorio Aziendale di Microbiologia e Virologia | Istituto di Genomica Applicata | Davide Scaglione; Eleonora Paparelli; Elisa Masi; Elisabetta Giacobazzi; Elisabetta Pagani; Gabriele Magris; Irena Jurman; Irene Bianconi; Michele Morgante; Stefanie Wieser; Vera Vendramin |
| EPI_ISL_2653653, EPI_ISL_2653666, EPI_ISL_2653667, EPI_ISL_2653670, EPI_ISL_2653675, EPI_ISL_2653677, EPI_ISL_2653678, EPI_ISL_2653679, EPI_ISL_2653680, EPI_ISL_2653710, EPI_ISL_2653716, EPI_ISL_2653717, EPI_ISL_2653727, EPI_ISL_2653730, EPI_ISL_2653732, EPI_ISL_2653760, EPI_ISL_2653762, EPI_ISL_2653764, EPI_ISL_2653765, EPI_ISL_2653767, EPI_ISL_2653769, EPI_ISL_2653771, EPI_ISL_2653773, EPI_ISL_2653774, EPI_ISL_2653777, EPI_ISL_2653778, EPI_ISL_2653843, EPI_ISL_2653844, EPI_ISL_2653852, EPI_ISL_2653880, EPI_ISL_2653881, EPI_ISL_2653882, EPI_ISL_2653883, EPI_ISL_2653897, EPI_ISL_2653899, EPI_ISL_2653913, EPI_ISL_2653917, EPI_ISL_2653919, EPI_ISL_2653927, EPI_ISL_2653928, EPI_ISL_2653930, EPI_ISL_2653931, EPI_ISL_2653933, EPI_ISL_2653936, EPI_ISL_2653937, EPI_ISL_2653938, EPI_ISL_2653979, EPI_ISL_2653981, EPI_ISL_2653990, EPI_ISL_2653993, EPI_ISL_2653995, EPI_ISL_2653998, EPI_ISL_2654002, EPI_ISL_2654003, EPI_ISL_2654013, EPI_ISL_2654019, EPI_ISL_2654082, EPI_ISL_2654089, EPI_ISL_2654090, EPI_ISL_2654091, EPI_ISL_2654096, EPI_ISL_2654098, EPI_ISL_2654099, EPI_ISL_2654101, EPI_ISL_2654103, EPI_ISL_2654112, EPI_ISL_2654115, EPI_ISL_2654116, EPI_ISL_2654122, EPI_ISL_2654126, EPI_ISL_2654127, EPI_ISL_2654130, EPI_ISL_2654137, EPI_ISL_2654140, EPI_ISL_2654142, EPI_ISL_2654143, EPI_ISL_2654144, EPI_ISL_2654149, EPI_ISL_2654150, EPI_ISL_2654157, EPI_ISL_2654158, EPI_ISL_2654160, EPI_ISL_2654161, EPI_ISL_2654165, EPI_ISL_2654167, EPI_ISL_2654168, EPI_ISL_2654170, EPI_ISL_2654171, EPI_ISL_2654174, EPI_ISL_2654175, EPI_ISL_2654178, EPI_ISL_2654180, EPI_ISL_2654185, EPI_ISL_2654186, EPI_ISL_2654187, EPI_ISL_2654190, EPI_ISL_2654197, EPI_ISL_2654199, EPI_ISL_2654200, EPI_ISL_2654201, EPI_ISL_2654208, EPI_ISL_2654209, EPI_ISL_2654213, EPI_ISL_2654214, EPI_ISL_2654215, EPI_ISL_2654216, EPI_ISL_2654232, EPI_ISL_2654242, EPI_ISL_2654243, EPI_ISL_2654255, EPI_ISL_2654259, EPI_ISL_2654260, EPI_ISL_2654264, EPI_ISL_2654268, EPI_ISL_2654271, EPI_ISL_2654272, EPI_ISL_2654273, EPI_ISL_2654275, EPI_ISL_2654278, EPI_ISL_2654281, EPI_ISL_2654282, EPI_ISL_2654284, EPI_ISL_2654285, EPI_ISL_2654286, EPI_ISL_2654287, EPI_ISL_2654288, EPI_ISL_2654290, EPI_ISL_2654292, EPI_ISL_2654297, EPI_ISL_2654300, EPI_ISL_2654301, EPI_ISL_2654302, EPI_ISL_2654306, EPI_ISL_2654309, EPI_ISL_2654311, EPI_ISL_2654312, EPI_ISL_2654319, EPI_ISL_2654322, EPI_ISL_2654329, EPI_ISL_2654346, EPI_ISL_2654351, EPI_ISL_2654352, EPI_ISL_2654365, EPI_ISL_2654368, EPI_ISL_2654369, EPI_ISL_2654370, EPI_ISL_2654371, EPI_ISL_2654372, EPI_ISL_2654375, EPI_ISL_2654376, EPI_ISL_2654377, EPI_ISL_2654437, EPI_ISL_2654438, EPI_ISL_2654439, EPI_ISL_2654499, EPI_ISL_2654500, EPI_ISL_2654502, EPI_ISL_2654504, EPI_ISL_2654507, EPI_ISL_2654508, EPI_ISL_2654509, EPI_ISL_2654513, EPI_ISL_2654514, EPI_ISL_2654574, EPI_ISL_2654576, EPI_ISL_2654577, EPI_ISL_2654580, EPI_ISL_2654582, EPI_ISL_2654583, EPI_ISL_2654584, EPI_ISL_2654585, EPI_ISL_2654587, EPI_ISL_2654588, EPI_ISL_2654589, EPI_ISL_2654593, EPI_ISL_2654597, EPI_ISL_2654599, EPI_ISL_2654600, EPI_ISL_2654601, EPI_ISL_2654602, EPI_ISL_2654604, EPI_ISL_2654605, EPI_ISL_2654607, EPI_ISL_2654608, EPI_ISL_2654612, EPI_ISL_2654614, EPI_ISL_2654615, EPI_ISL_2654617, EPI_ISL_2654684, EPI_ISL_2654703, EPI_ISL_2654704, EPI_ISL_2654711, EPI_ISL_2654712, EPI_ISL_2654714, EPI_ISL_2654715, EPI_ISL_2654716, EPI_ISL_2654717, EPI_ISL_2654718, EPI_ISL_2654720, EPI_ISL_2654721, EPI_ISL_2654722, EPI_ISL_2654724, EPI_ISL_2654726, EPI_ISL_2654728, EPI_ISL_2654729, EPI_ISL_2654731, EPI_ISL_2654732, EPI_ISL_2654735, EPI_ISL_2654737, EPI_ISL_2654773, EPI_ISL_2654784, EPI_ISL_2654785, EPI_ISL_2654786, EPI_ISL_2654789, EPI_ISL_2654793, EPI_ISL_2654794, EPI_ISL_2654943, EPI_ISL_2654945, EPI_ISL_2654946, EPI_ISL_2654947, EPI_ISL_2654948, EPI_ISL_2654950, EPI_ISL_2654952, EPI_ISL_2654953, EPI_ISL_2654954, EPI_ISL_2654955, EPI_ISL_2654956, EPI_ISL_2654957, EPI_ISL_2654958, EPI_ISL_2654959, EPI_ISL_2654960, EPI_ISL_2654961, EPI_ISL_2654962, EPI_ISL_2654963, EPI_ISL_2654964, EPI_ISL_2654965, EPI_ISL_2654966, EPI_ISL_2654967, EPI_ISL_2654968, EPI_ISL_2654969, EPI_ISL_2654970, EPI_ISL_2654971, EPI_ISL_2654972, EPI_ISL_2654973, EPI_ISL_2654974, EPI_ISL_2654975, EPI_ISL_2654976, EPI_ISL_2654977, EPI_ISL_2654978, EPI_ISL_2654979, EPI_ISL_2654980, EPI_ISL_2654981, EPI_ISL_2654982, EPI_ISL_2654983, EPI_ISL_2654984, EPI_ISL_2654985, EPI_ISL_2654986, EPI_ISL_2654987, EPI_ISL_2654988, EPI_ISL_2654989, EPI_ISL_2654993, EPI_ISL_2655003, EPI_ISL_2655012, EPI_ISL_2655013, EPI_ISL_2655014, EPI_ISL_2655647, EPI_ISL_2655701, EPI_ISL_2655713, EPI_ISL_2656153, EPI_ISL_2656157, EPI_ISL_2718739, EPI_ISL_2718745, EPI_ISL_2718750, EPI_ISL_2718758, EPI_ISL_2718853, EPI_ISL_2718890, EPI_ISL_2718895, EPI_ISL_2718932, EPI_ISL_2718936, EPI_ISL_2718942, EPI_ISL_2718944, EPI_ISL_2718945, EPI_ISL_2718951, EPI_ISL_2718952, EPI_ISL_2718958, EPI_ISL_2718964, EPI_ISL_2718968, EPI_ISL_2719290, EPI_ISL_2720220, EPI_ISL_2720237, EPI_ISL_2720263, EPI_ISL_2720266, EPI_ISL_2720552, EPI_ISL_2721057, EPI_ISL_2721062, EPI_ISL_2721115, EPI_ISL_2721168, EPI_ISL_2721174, EPI_ISL_2721187, EPI_ISL_2721190, EPI_ISL_2721194, EPI_ISL_2721196, EPI_ISL_2721201, EPI_ISL_2721210, EPI_ISL_2721213, EPI_ISL_2721231, EPI_ISL_2721249, EPI_ISL_2721265, EPI_ISL_2721267, EPI_ISL_2721270, EPI_ISL_2721273, EPI_ISL_2721275, EPI_ISL_2721287, EPI_ISL_2721292, EPI_ISL_2721294, EPI_ISL_2721349, EPI_ISL_2721350, EPI_ISL_2721356, EPI_ISL_2721357 | | | |
| see above | BCCDC Public Health Laboratory | BCCDC Public Health Laboratory | Ana Pacagnella; Corrinne Ng; Dan Fornika; John Tyson; Kim Macdonald; Kimia Kamelian; Linda Hoang; Loretta Janz; Mel Krajden; Prystajecky Natalie; Robert Azana; Shannon Russell |
| EPI_ISL_1756356 | BIO-REFERENCE LABORATORIES | Wadsworth Center, New York State Department of Health | Alexis Russell; Catharine Prussing; Daryl M. Lamson; Erasmus Schneider; Erica Lasek-Nesselquist; John Kelly; Jonathan Plitnick; Kirsten St. George; Matthew Shudt; Melissa A Leisner; Navjot Singh |
| EPI_ISL_2544494 | Baker lab at Loyola Medical Center | Baker lab at Loyola Medical Center | Amanda Harrington; Amornrat O'Brien; BS; Cynthia Rowe; Gail E. Reid; MD; Monika Evdokimova; Nina M. Clark; PhD; Susan Uprichard; Xufang Deng; and Susan C. Baker |
| EPI_ISL_1739331 | Basurto University Hospital | Biocruces Bizkaia | Ana de la Hoz; Estibaliz Ugalde Zarraga; Jose Luis Díaz de Tuesta del Arco; Mikel Gallego Rodrigo; Mikel Urrutikoetxea-Gutiérrez; Mº Carmen Nieto Toboso |
| EPI_ISL_1706360 | Basurto University Hospital Clinical Microbiology Lab | Biocruces Bizkaia | Ana de la Hoz; Estibaliz Ugalde Zarraga; José Luis Díaz de Tuesta; Mikel Gallego Rodrigo; Mikel Urrutikoetxea-Gutierrez; Mª Carmen Nieto Toboso |
| EPI_ISL_1706277 | Basurto University Hospital Clinical Microbiology Laboratory | Biocruces Bizkaia | Ana de la Hoz; Estibaliz Ugalde Zarraga; José Luis Díaz de Tuesta del Arco.; Mikel Gallego Rodrigo; Mikel Urrutikoetxea-Gutierrez; Mª Carmen Nieto Toboso |
| EPI_ISL_2037414, EPI_ISL_2037443 | Basurto University Hospital: Clinical Microbiology Laboratory | Biocruces Bizkaia | Ana de la Hoz; Estibaliz Ugalde Zarraga; José Luis Díaz de Tuesta del Arco; Mikel Gallego Rodrigo; Mikel Urrutikoetxea-Gutiérrez; Mª Carmen Nieto Toboso |
| EPI_ISL_1627453, EPI_ISL_1627454, EPI_ISL_1733036 | Baylor Scott & White-Temple | Baylor Scott & White-Temple | Ari Rao; Caitlin Maloney; Kimberly Walker; Linden Morales; Marcus Volz; Shelby Hendrickson |
| EPI_ISL_1628519 | Berkeley Medical Center | WVU and Marshall University Combined Genomics Core Facilities | James Denvir; Peter Perrotta; Peter Stoilov; Ryan Percifield; Wesley Kimble |
| EPI_ISL_6667244 | Biological Threats Identification and Countermeasure Centre | Biological Threats Identification and Countermeasure Centre | Lepionka Tomasz; Rutyna Paweł; Skuza Katarzyna; Wójcicka Patrycja |
| EPI_ISL_3494304 | Boston University CTL | Boston University/National Emerging Infectious Disease Laboratories | Catherine Klapperich; Jacquelyn Turcinovic; John H. Connor; Lena Landeverde; Lynn Doucette-Stamm |
| EPI_ISL_3478918, EPI_ISL_3478919, EPI_ISL_3478921, EPI_ISL_3478922, EPI_ISL_3478923, EPI_ISL_3478924, EPI_ISL_3478926, EPI_ISL_3478930, EPI_ISL_3478935, EPI_ISL_3478936, EPI_ISL_3479446, EPI_ISL_3479451, EPI_ISL_3479455, EPI_ISL_3479472, EPI_ISL_3479474, EPI_ISL_3479477, EPI_ISL_3479478, EPI_ISL_3479479, EPI_ISL_3479481, EPI_ISL_3479485, EPI_ISL_3479489, EPI_ISL_3479490, EPI_ISL_3479491, EPI_ISL_3479492, EPI_ISL_3479493, EPI_ISL_3479496, EPI_ISL_3479497, EPI_ISL_3479498, EPI_ISL_3479499, EPI_ISL_3479505, EPI_ISL_3479506, EPI_ISL_3479507, EPI_ISL_3479508, EPI_ISL_3479509, EPI_ISL_3479510, EPI_ISL_3479511, EPI_ISL_3479513, EPI_ISL_3479514, EPI_ISL_3479515, EPI_ISL_3479516, EPI_ISL_3479518, EPI_ISL_3479519, EPI_ISL_3479521, EPI_ISL_3479522, EPI_ISL_3479523, EPI_ISL_3479524, EPI_ISL_3479525, EPI_ISL_3479526, EPI_ISL_3479527, EPI_ISL_3479530, EPI_ISL_3479531, EPI_ISL_3479532, EPI_ISL_3479536, EPI_ISL_3479537, EPI_ISL_3479538, EPI_ISL_3479539, EPI_ISL_3479540, EPI_ISL_3479541, EPI_ISL_3479545, EPI_ISL_3479546, EPI_ISL_3479547, EPI_ISL_3479559, EPI_ISL_3479560, EPI_ISL_3479561, EPI_ISL_3479565, EPI_ISL_3479567, EPI_ISL_3479570, EPI_ISL_3479573, EPI_ISL_3479574, EPI_ISL_3479576, EPI_ISL_3479577, EPI_ISL_3479578, EPI_ISL_3479581, EPI_ISL_3479583, EPI_ISL_3479584, EPI_ISL_3479594, EPI_ISL_3479595, EPI_ISL_3479596, EPI_ISL_3479597, EPI_ISL_3479598, EPI_ISL_3479599, EPI_ISL_3479600, EPI_ISL_3479601, EPI_ISL_3479602, EPI_ISL_3479603, EPI_ISL_3479605, EPI_ISL_3479606, EPI_ISL_3479607, EPI_ISL_3479609, EPI_ISL_3479610, EPI_ISL_3479613, EPI_ISL_3479615, EPI_ISL_3479616, EPI_ISL_3479617, EPI_ISL_3479618, EPI_ISL_3479619, EPI_ISL_3479622, EPI_ISL_3479623, EPI_ISL_3479637, EPI_ISL_3479640, EPI_ISL_3479644, EPI_ISL_3479645, EPI_ISL_3479647, EPI_ISL_3479650, EPI_ISL_3479651, EPI_ISL_3479652, EPI_ISL_3479653, EPI_ISL_3479654, EPI_ISL_3479655, EPI_ISL_3479657, EPI_ISL_3479658, EPI_ISL_3479659, EPI_ISL_3479662, EPI_ISL_3479663, EPI_ISL_3479669, EPI_ISL_3479671, EPI_ISL_3479672, EPI_ISL_3479674, EPI_ISL_3479675, EPI_ISL_3479676, EPI_ISL_3479677, EPI_ISL_3479678, EPI_ISL_3479680, EPI_ISL_3479682, EPI_ISL_3479683, EPI_ISL_3479684, EPI_ISL_3479685, EPI_ISL_3479686, EPI_ISL_3479688, EPI_ISL_3479689, EPI_ISL_3479690, EPI_ISL_3479692, EPI_ISL_3479693, EPI_ISL_3479694, EPI_ISL_3479695, EPI_ISL_3479696, EPI_ISL_3479697, EPI_ISL_3479698, EPI_ISL_3479700, EPI_ISL_3479701, EPI_ISL_3479702, EPI_ISL_3479706, EPI_ISL_3479707, EPI_ISL_3479708, EPI_ISL_3479713, EPI_ISL_3479725, EPI_ISL_3479737, EPI_ISL_3479745, EPI_ISL_3479746, EPI_ISL_3479747, EPI_ISL_3479749, EPI_ISL_3479751, EPI_ISL_3479752, EPI_ISL_3479753, EPI_ISL_3479755, EPI_ISL_3479756, EPI_ISL_3479757, EPI_ISL_3479758, EPI_ISL_3479761, EPI_ISL_3479762, EPI_ISL_3479763, EPI_ISL_3479765, EPI_ISL_3479769, EPI_ISL_3479773, EPI_ISL_3479777, EPI_ISL_3479778, EPI_ISL_3479779, EPI_ISL_3479781, EPI_ISL_3479784, EPI_ISL_3479785, EPI_ISL_3479786, EPI_ISL_3479787, EPI_ISL_3479788, EPI_ISL_3479791, EPI_ISL_3479793, EPI_ISL_3479794, EPI_ISL_3479795, EPI_ISL_3479796, EPI_ISL_3479799, EPI_ISL_3479800, EPI_ISL_3479801, EPI_ISL_3479803, EPI_ISL_3479805, EPI_ISL_3479808, EPI_ISL_3479809, EPI_ISL_3479817, EPI_ISL_3479818, EPI_ISL_3479820, EPI_ISL_3479821, EPI_ISL_3479822, EPI_ISL_3479823, EPI_ISL_3479824, EPI_ISL_3479825, EPI_ISL_3479826, EPI_ISL_3479827, EPI_ISL_3479828, EPI_ISL_3479829, EPI_ISL_3479831, EPI_ISL_3479832, EPI_ISL_3479833, EPI_ISL_3479834, EPI_ISL_3479835, EPI_ISL_3479836, EPI_ISL_3479837, EPI_ISL_3479839, EPI_ISL_3479840, EPI_ISL_3479841, EPI_ISL_3479842, EPI_ISL_3479844, EPI_ISL_3479845, EPI_ISL_3479846, EPI_ISL_3479847, EPI_ISL_3479850, EPI_ISL_3479851, EPI_ISL_3479858, EPI_ISL_3479859, EPI_ISL_3479861, EPI_ISL_3479863, EPI_ISL_3479864, EPI_ISL_3479865, EPI_ISL_3479868, EPI_ISL_3479874, EPI_ISL_3479875, EPI_ISL_3479876, EPI_ISL_3479877, EPI_ISL_3479879, EPI_ISL_3479880, EPI_ISL_3479881, EPI_ISL_3479882, EPI_ISL_3479884, EPI_ISL_3479885, EPI_ISL_3479886, EPI_ISL_3479887, EPI_ISL_3479889, EPI_ISL_3479890, EPI_ISL_3479896, EPI_ISL_3479898, EPI_ISL_3479899, EPI_ISL_3479901, EPI_ISL_3479902, EPI_ISL_3479903, EPI_ISL_3479904, EPI_ISL_3479905, EPI_ISL_3479907, EPI_ISL_3479908, EPI_ISL_3479909, EPI_ISL_3479910, EPI_ISL_3479911, EPI_ISL_3479913, EPI_ISL_3479914, EPI_ISL_3479915, EPI_ISL_3479916, EPI_ISL_3479917, EPI_ISL_3479918, EPI_ISL_3479920, EPI_ISL_3479921, EPI_ISL_3479923, EPI_ISL_3479924, EPI_ISL_3479927, EPI_ISL_3479929, EPI_ISL_3479930, EPI_ISL_3479932, EPI_ISL_3479933, EPI_ISL_3479934, EPI_ISL_3479938, EPI_ISL_3479939, EPI_ISL_3479941, EPI_ISL_3479947, EPI_ISL_3479948, EPI_ISL_3479950, EPI_ISL_3479952, EPI_ISL_3479955, EPI_ISL_3479959, EPI_ISL_3479960, EPI_ISL_3479963, EPI_ISL_3479964, EPI_ISL_3479965, EPI_ISL_3479966, EPI_ISL_3479967, EPI_ISL_3479968, EPI_ISL_3479969, EPI_ISL_3479970, EPI_ISL_3479974, EPI_ISL_3479975, EPI_ISL_3479977, EPI_ISL_3479980, EPI_ISL_3479981, EPI_ISL_3479983, EPI_ISL_3479986, EPI_ISL_3479989, EPI_ISL_3479990, EPI_ISL_3479993, EPI_ISL_3479996, EPI_ISL_3479998, EPI_ISL_3480000, EPI_ISL_3480004, EPI_ISL_3480006, EPI_ISL_3480007, EPI_ISL_3480008, EPI_ISL_3480009, EPI_ISL_3480010, EPI_ISL_3480011, EPI_ISL_3480012, EPI_ISL_3480013, EPI_ISL_3480014, EPI_ISL_3480015, EPI_ISL_3480016, EPI_ISL_3480017, EPI_ISL_3480019, EPI_ISL_3480020, EPI_ISL_3480022, EPI_ISL_3480023, EPI_ISL_3480041, EPI_ISL_3480052, EPI_ISL_3480053, EPI_ISL_3480068, EPI_ISL_3480079, EPI_ISL_3480112, EPI_ISL_3480113, EPI_ISL_3480119, EPI_ISL_3480127, EPI_ISL_3480152, EPI_ISL_3480153, EPI_ISL_3480155, EPI_ISL_3480211, EPI_ISL_3480212, EPI_ISL_3480213, EPI_ISL_3480215, EPI_ISL_3480226, EPI_ISL_3480231, EPI_ISL_3480232, EPI_ISL_3480233, EPI_ISL_3480234, EPI_ISL_3480237, EPI_ISL_3480238, EPI_ISL_3480240, EPI_ISL_3480241, EPI_ISL_3480242, EPI_ISL_3480244, EPI_ISL_3480245, EPI_ISL_3480246, EPI_ISL_3480247, EPI_ISL_3480248, EPI_ISL_3480249, EPI_ISL_3480250, EPI_ISL_3480251, EPI_ISL_3480252, EPI_ISL_3480253, EPI_ISL_3480254, EPI_ISL_3480255, EPI_ISL_3480256, EPI_ISL_3480257, EPI_ISL_3480259, EPI_ISL_3480260, EPI_ISL_3480261, EPI_ISL_3480262, EPI_ISL_3480263, EPI_ISL_3480266, EPI_ISL_3480267, EPI_ISL_3480268, EPI_ISL_3480269, EPI_ISL_3480270, EPI_ISL_3480271, EPI_ISL_3480272, EPI_ISL_3480273, EPI_ISL_3480274, EPI_ISL_3480278, EPI_ISL_3480280, EPI_ISL_3480282, EPI_ISL_3480284, EPI_ISL_3480285, EPI_ISL_3480287, EPI_ISL_3480289, EPI_ISL_3480290, EPI_ISL_3480291, EPI_ISL_3480297, EPI_ISL_3480298, EPI_ISL_3480299, EPI_ISL_3480301, EPI_ISL_3480302, EPI_ISL_3480312, EPI_ISL_3480369, EPI_ISL_3480376, EPI_ISL_3480383, EPI_ISL_3486462, EPI_ISL_3596344, EPI_ISL_3598071, EPI_ISL_3598072, EPI_ISL_3598080, EPI_ISL_3598086, EPI_ISL_3598090, EPI_ISL_3598092, EPI_ISL_3598097, EPI_ISL_3598101, EPI_ISL_3598103, EPI_ISL_3598105, EPI_ISL_3598109, EPI_ISL_3598110, EPI_ISL_3598115, EPI_ISL_3598116, EPI_ISL_3598123, EPI_ISL_3598124, EPI_ISL_3598127, EPI_ISL_3598132, EPI_ISL_3598136, EPI_ISL_3598137, EPI_ISL_3598148, EPI_ISL_3598149, EPI_ISL_3598150, EPI_ISL_3598151, EPI_ISL_3598155, EPI_ISL_3598156, EPI_ISL_3598164, EPI_ISL_3598165, EPI_ISL_3598167, EPI_ISL_3598179, EPI_ISL_3598181, EPI_ISL_3598184, EPI_ISL_3598189, EPI_ISL_3598193, EPI_ISL_3598196, EPI_ISL_3598198, EPI_ISL_3598201, EPI_ISL_3598203, EPI_ISL_3598213, EPI_ISL_3598215, EPI_ISL_3598220, EPI_ISL_3598226, EPI_ISL_3598227, EPI_ISL_3598231, EPI_ISL_3598232, EPI_ISL_3598234, EPI_ISL_3598239, EPI_ISL_3598244, EPI_ISL_3598245, EPI_ISL_3598247, EPI_ISL_3598253, EPI_ISL_3598255, EPI_ISL_3598256, EPI_ISL_3598257, EPI_ISL_3598261, EPI_ISL_3598263, EPI_ISL_3598267, EPI_ISL_3598270, EPI_ISL_3598280, EPI_ISL_3598281, EPI_ISL_3598283, EPI_ISL_3598288, EPI_ISL_3598289, EPI_ISL_3598301, EPI_ISL_3598305, EPI_ISL_3598316, EPI_ISL_3598320, EPI_ISL_3598324, EPI_ISL_3598325, EPI_ISL_3598326, EPI_ISL_3598329, EPI_ISL_3598332, EPI_ISL_3598346, EPI_ISL_3598347, EPI_ISL_3598355, EPI_ISL_3598356, EPI_ISL_3598359, EPI_ISL_3598387, EPI_ISL_3598388, EPI_ISL_3598406, EPI_ISL_3602401, EPI_ISL_4828391 | | | |
| see above | British Columbia Centre For Disease Control | BCCDC Public Health Laboratory | Ana Pacagnella; Corrinne Ng; Dan Fornika; John Tyson; Kim Macdonald; Kimia Kamelian; Linda Hoang; Loretta Janz; Mel Krajden; Prystajecky Natalie; Robert Azana; Shannon Russell |
| EPI_ISL_1757185, EPI_ISL_1757196, EPI_ISL_1757215, EPI_ISL_1757235, EPI_ISL_1757243, EPI_ISL_1757250, EPI_ISL_1757282, EPI_ISL_1757288, EPI_ISL_1757301 | | | |
| see above | Broad Institute Clinical Research Sequencing Platform | Infectious Disease Program, Broad Institute of Harvard and MIT | Adams, G.; B.L.; B.W.; Bauer, M.; Birren; Blumenstiel, B.; Brown, C.; Carter, A.; Chaluvadi, S.; D.J.; DeFelice, M.; DeRuff, K.; Dodge, S.; Gabriel, S.; Gallagher, G.; Gladden-Young, A.; Granger, B.; J.E.; K.J.; Lagerborg, K.; Larkin, K.; Lee, M.; Lemieux; Lennon, N.; Loreth, C.; Madoff, L.; McGovern, S.; Meldrim, J.; Normandin, E.; P.C.; Park; Pearlman, L.; Reilly, S.; Rudy, M.; Sabeti; Siddle; Smole, S.; Tomkins-Tinch, C.; Vicente, G.; and MacInnis |
| EPI_ISL_1661165, EPI_ISL_1661168, EPI_ISL_1661170, EPI_ISL_1661182, EPI_ISL_1661184 | Brussels Academic Hospital Lab (LHUB-ULB) | Labo Klinische Biologie, UZA | Basil Britto Xavier; Christine Lammens; Herman Goossens; Jasmine Coppens; Marie Le Mercier; Veerle Matheeussen |
| EPI_ISL_5529947 | CASA DE SAUDE ADILIA MARIA | Analytical Competence Molecular Epidemiology Lab/ACME, Oswaldo Cruz Foundation, Ceara (FIOCRUZ CE) | Carlos Leonardo de Aragao Araujo; Cecilia Leite Costa & Eduardo Ruback dos Santos on behalf of COVID-19 FIOCRUZ Genomic Network; Cleber Furtado Aksenen; Fabio Miyajima; Fernando Braga Stehling; Francisco Eder de Moura Lopes; Igor Oliveira Duarte; Jamille Maria Mendes Bezerra; Joaquim Cesar do Nascimento Sousa Junior; Pedro Miguel Carneiro Jeronimo; Suzana Porto Almeida; Thais Ferreira de Oliveira; Thais de Oliveira Costa; Ticiane Cavalcante de Souza; Veridiana Pessoa Miyajima |
| EPI_ISL_5530106 | CASA DE SAUDE MATERNIDADE CELESTINA COLARES | Analytical Competence Molecular Epidemiology Lab/ACME, Oswaldo Cruz Foundation, Ceara (FIOCRUZ CE) | Carlos Leonardo de Aragao Araujo; Cecilia Leite Costa & Eduardo Ruback dos Santos on behalf of COVID-19 FIOCRUZ Genomic Network; Cleber Furtado Aksenen; Fabio Miyajima; Fernando Braga Stehling; Francisco Eder de Moura Lopes; Igor Oliveira Duarte; Jamille Maria Mendes Bezerra; Joaquim Cesar do Nascimento Sousa Junior; Pedro Miguel Carneiro Jeronimo; Suzana Porto Almeida; Thais Ferreira de Oliveira; Thais de Oliveira Costa; Ticiane Cavalcante de Souza; Veridiana Pessoa Miyajima |
| EPI_ISL_2825817, EPI_ISL_2858320, EPI_ISL_2858351, EPI_ISL_2858453, EPI_ISL_2882791, EPI_ISL_2882822, EPI_ISL_2882842, EPI_ISL_2882843, EPI_ISL_2882895, EPI_ISL_2882953, EPI_ISL_2883018, EPI_ISL_2883022, EPI_ISL_2883038, EPI_ISL_2883042, EPI_ISL_5696543, EPI_ISL_5696545, EPI_ISL_5696546, EPI_ISL_5696553, EPI_ISL_5696554, EPI_ISL_5696556 | | | |
| see above | CDPH VBL | California Department of Public Health | CDPH-COVIDNet; Emily Smith on behalf of CDPH-COVIDNet and Priva Path Labs Inc.; UCLA Technology Center for Genomics & Bioinformatics |
| EPI_ISL_2018315 | CDPH, Viral and Rickettsial Disease Laboratory | Centers for Disease Control and Prevention Division of Viral Diseases, Pathogen Discovery | Alison Laufer Halpin; Ben L. Rambo-Martin; Clinton R. Paden; Dakota Howard; Darlene Wagner; Dave Wentworth; Dhwani Batra; Jasmine Padilla; Justin Lee; Katie Dillon; Krista Queen; Kristen Knipe; Kristine Lacek; Mark Burroughs; Matthew Schmerer; Mili Sheth; Peter Cook; Sam Shepard; Sarah Nobles; Shoshona Le; Suxiang Tong; Vivien Dugan; Yvette Unoarumhi |
| EPI_ISL_1966895, EPI_ISL_1966896, EPI_ISL_1966897, EPI_ISL_1966898, EPI_ISL_1966899, EPI_ISL_1966900, EPI_ISL_1966902, EPI_ISL_1966903, EPI_ISL_1966904 | | | |
| see above | CENTRO DE APOIO EPIDEMIOLOGICO | Instituto Butantan / ESALQ-USP (Piracicaba) | Antonio Jorge Martins; Bianca Cechetto Carlos. Mendelics: Bibiana Santos; Claudia Renata dos Santos Barros; Cíntia Bittar; David Schlesinger. Hemocentro Ribeirão Preto: Simone Kashima; Debora Botequio Moretti; Elaine Cristina Marqueze; Elaine Vieira dos Santos; Elisangela Chicaroni Mattos; Erika Freitas; Evandra Strazza Rodrigues; Felipe Allan da Silva da Costa; Flavia Aburjaile; Fábio Sossai Possebon; Guilherme Campos; Guilherme Targino Valente; Heidge Fukumasu. USP-Botucatu: Rejane Maria Tommasini Grotto; Helena Lage Ferreira; Instituto Butantan: Dimas Tadeu Covas; Jardelina de Souza Todao Bernardino; Jayme A. Souza-Neto; Jessika Cristina Chagas Lesbon; Jorge A. Petroli Marchesi; José Salvatore Leister Patané; João Paulo Kitajima; João Pessoa Araújo Jr.; Leila Sabrina Ullmann; Loyze Paola Oliveira de Lima; Luiz Aurelio de Campos Crispin. Centro de Genômica Funcional da ESALQ: Luiz Lehmann Coutinho; Luiz Carlos Junior de Alcantara; Lívia Sacchetto; Maisa C. Pereira Parra; Maria Carolina Elias; Marta Giovanetti; Marília Moraes; Maurício Lacerda Nogueira. Prefeitura de Sao Paulo: Melissa Palmieri.; Patricia Akemi Assato; Paula Rahal; Paulo Inacio da Costa; Rafael dos Santos Bezerra; Raquel de Lello Rocha Campos Cassano. NGS Soluções Genômicas: Pilar Drummond Sampaio Corrêa Mariani. FZEA-USP Pirassununga: Mirele Daiana Poleti; Raul Machado Neto; Ricardo Augusto Brassaloti; Ricardo Haddad; Rodrigo Tocantins Calado. FAMERP-SJRP: Cecília Artico Banho; Sandra Coccuzzo Sampaio; Svetoslav Nanev Slavov; Vagner Fonseca; Vincent |

| | | | |
|---|---|---|---|
| | | | Louis Viala |
| EPI_ISL_5530192 | CENTRO DE ATENCAO ESPECIALIZADA | Analytical Competence Molecular Epidemiology Lab/ACME, Oswaldo Cruz Foundation, Ceara (FIOCRUZ CE) | Carlos Leonardo de Aragao Araujo; Cecilia Leite Costa & Eduardo Ruback dos Santos on behalf of COVID-19 FIOCRUZ Genomic Network; Cleber Furtado Aksenen; Fabio Miyajima; Fernando Braga Stehling; Francisco Eder de Moura Lopes; Igor Oliveira Duarte; Jamille Maria Mendes Bezerra; Joaquim Cesar do Nascimento Sousa Junior; Pedro Miguel Carneiro Jeronimo; Suzana Porto Almeida; Thais Ferreira de Oliveira; Thais de Oliveira Costa; Ticiane Cavalcante de Souza; Veridiana Pessoa Miyajima |
| EPI_ISL_1966957, EPI_ISL_1966959, EPI_ISL_1966960 | CENTRO DE SAUDE DE MARACAI | Instituto Butantan / ESALQ-USP (Piracicaba) | Antonio Jorge Martins; Bianca Cechetto Carlos. Mendelics: Bibiana Santos; Claudia Renata dos Santos Barros; Cíntia Bittar; David Schlesinger. Hemocentro Ribeirão Preto: Simone Kashima; Debora Botequio Moretti; Elaine Cristina Marqueze; Elaine Vieira dos Santos; Elisangela Chicaroni Mattos; Erika Freitas; Evandra Strazza Rodrigues; Felipe Allan da Silva da Costa; Flavia Aburjaile; Fábio Sossai Possebon; Guilherme Campos; Guilherme Targino Valente; Heidge Fukumasu. USP-Botucatu: Rejane Maria Tommasini Grotto; Helena Lage Ferreira; Instituto Butantan: Dimas Tadeu Covas; Jardelina de Souza Todao Bernardino; Jayme A. Souza-Neto; Jessika Cristina Chagas Lesbon; Jorge A. Petroli Marchesi; José Salvatore Leister Patané; João Paulo Kitajima; João Pessoa Araújo Jr.; Leila Sabrina Ullmann; Loyze Paola Oliveira de Lima; Luiz Aurelio de Campos Crispin. Centro de Genômica Funcional da ESALQ: Luiz Lehmann Coutinho; Luiz Carlos Junior de Alcantara; Lívia Sacchetto; Maisa C. Pereira Parra; Maria Carolina Elias; Marta Giovanetti; Marília Moraes; Maurício Lacerda Nogueira. Prefeitura de Sao Paulo: Melissa Palmieri.; Patricia Akemi Assato; Paula Rahal; Paulo Inacio da Costa; Rafael dos Santos Bezerra; Raquel de Lello Rocha Campos Cassano. NGS Soluções Genômicas: Pilar Drummond Sampaio Corrêa Mariani. FZEA-USP Pirassununga: Mirele Daiana Poleti; Raul Machado Neto; Ricardo Augusto Brassaloti; Ricardo Haddad; Rodrigo Tocantins Calado. FAMERP-SJRP: Cecília Artico Banho; Sandra Coccuzzo Sampaio; Svetoslav Nanev Slavov; Vagner Fonseca; Vincent Louis Viala |
| EPI_ISL_2209265 | CENTRO DE SAUDE DR MARIO DIAS DE AGUIAR CAPIVARI | Instituto Butantan / FZEA-USP-Pirassununga | Antonio Jorge Martins; Claudia Renata dos Santos Barros; David Schlesinger; Debora Botequio Moretti; Dimas Tadeu Covas; Elaine Cristina Marqueze; Elaine Vieira Santos; Evandra Strazza Rodrigues; Heidge Fukumasu; Jayme Augusto de Souza-Neto; José Salvatore Leister Patané; Luiz Alcantara; Luiz Lehmann Coutinho; Maria Carolina Elias; Maurício Lacerda Nogueira; Rafael dos Santos Bezerra; Raul Machado Neto; Rejane Maria Tommasini Grotto; Ricardo Haddad; Sandra Coccuzzo Sampaio Vessoni; Simone Kashima; Svetoslav Nanev Slavov; Vincent Louis Viala |
| EPI_ISL_1966824, EPI_ISL_1966825, EPI_ISL_1966826, EPI_ISL_1966827, EPI_ISL_1966828, EPI_ISL_1966829 | CENTRO DE SAUDE DR RENATO DE CARVALHO RIBEIRO ANGATUBA | Instituto Butantan / ESALQ-USP (Piracicaba) | Antonio Jorge Martins; Bianca Cechetto Carlos. Mendelics: Bibiana Santos; Claudia Renata dos Santos Barros; Cíntia Bittar; David Schlesinger. Hemocentro Ribeirão Preto: Simone Kashima; Debora Botequio Moretti; Elaine Cristina Marqueze; Elaine Vieira dos Santos; Elisangela Chicaroni Mattos; Erika Freitas; Evandra Strazza Rodrigues; Felipe Allan da Silva da Costa; Flavia Aburjaile; Fábio Sossai Possebon; Guilherme Campos; Guilherme Targino Valente; Heidge Fukumasu. USP-Botucatu: Rejane Maria Tommasini Grotto; Helena Lage Ferreira; Instituto Butantan: Dimas Tadeu Covas; Jardelina de Souza Todao Bernardino; Jayme A. Souza-Neto; Jessika Cristina Chagas Lesbon; Jorge A. Petroli Marchesi; José Salvatore Leister Patané; João Paulo Kitajima; João Pessoa Araújo Jr.; Leila Sabrina Ullmann; Loyze Paola Oliveira de Lima; Luiz Aurelio de Campos Crispin. Centro de Genômica Funcional da ESALQ: Luiz Lehmann Coutinho; Luiz Carlos Junior de Alcantara; Lívia Sacchetto; Maisa C. Pereira Parra; Maria Carolina Elias; Marta Giovanetti; Marília Moraes; Maurício Lacerda Nogueira. Prefeitura de Sao Paulo: Melissa Palmieri.; Patricia Akemi Assato; Paula Rahal; Paulo Inacio da Costa; Rafael dos Santos Bezerra; Raquel de Lello Rocha Campos Cassano. NGS Soluções Genômicas: Pilar Drummond Sampaio Corrêa Mariani. FZEA-USP Pirassununga: Mirele Daiana Poleti; Raul Machado Neto; Ricardo Augusto Brassaloti; Ricardo Haddad; Rodrigo Tocantins Calado. FAMERP-SJRP: Cecília Artico Banho; Sandra Coccuzzo Sampaio; Svetoslav Nanev Slavov; Vagner Fonseca; Vincent Louis Viala |
| EPI_ISL_1967058, EPI_ISL_1967059 | CENTRO DE SAUDE ESF IV ZONA RURAL DOMINGOS DE S SJRIOPARDO | Instituto Butantan / ESALQ-USP (Piracicaba) | Antonio Jorge Martins; Bianca Cechetto Carlos. Mendelics: Bibiana Santos; Claudia Renata dos Santos Barros; Cíntia Bittar; David Schlesinger. Hemocentro Ribeirão Preto: Simone Kashima; Debora Botequio Moretti; Elaine Cristina Marqueze; Elaine Vieira dos Santos; Elisangela Chicaroni Mattos; Erika Freitas; Evandra Strazza Rodrigues; Felipe Allan da Silva da Costa; Flavia Aburjaile; Fábio Sossai Possebon; Guilherme Campos; Guilherme Targino Valente; Heidge Fukumasu. USP-Botucatu: Rejane Maria Tommasini Grotto; Helena Lage Ferreira; Instituto Butantan: Dimas Tadeu Covas; Jardelina de Souza Todao Bernardino; Jayme A. Souza-Neto; Jessika Cristina Chagas Lesbon; Jorge A. Petroli Marchesi; José Salvatore Leister Patané; João Paulo Kitajima; João Pessoa Araújo Jr.; Leila Sabrina Ullmann; Loyze Paola Oliveira de Lima; Luiz Aurelio de Campos Crispin. Centro de Genômica Funcional da ESALQ: Luiz Lehmann Coutinho; Luiz Carlos Junior de Alcantara; Lívia Sacchetto; Maisa C. Pereira Parra; Maria Carolina Elias; Marta Giovanetti; Marília Moraes; Maurício Lacerda Nogueira. Prefeitura de Sao Paulo: Melissa Palmieri.; Patricia Akemi Assato; Paula Rahal; Paulo Inacio da Costa; Rafael dos Santos Bezerra; Raquel de Lello Rocha Campos Cassano. NGS Soluções Genômicas: Pilar Drummond Sampaio Corrêa Mariani. FZEA-USP Pirassununga: Mirele Daiana Poleti; Raul Machado Neto; Ricardo Augusto Brassaloti; Ricardo Haddad; Rodrigo Tocantins Calado. FAMERP-SJRP: Cecília Artico Banho; Sandra Coccuzzo Sampaio; Svetoslav Nanev Slavov; Vagner Fonseca; Vincent Louis Viala |
| EPI_ISL_1966833 | CENTRO DE SAUDE I AVARE | Instituto Butantan / ESALQ-USP (Piracicaba) | Antonio Jorge Martins; Bianca Cechetto Carlos. Mendelics: Bibiana Santos; Claudia Renata dos Santos Barros; Cíntia Bittar; David Schlesinger. Hemocentro Ribeirão Preto: Simone Kashima; Debora Botequio Moretti; Elaine Cristina Marqueze; Elaine Vieira dos Santos; Elisangela Chicaroni Mattos; Erika Freitas; Evandra Strazza Rodrigues; Felipe Allan da Silva da Costa; Flavia Aburjaile; Fábio Sossai Possebon; Guilherme Campos; Guilherme Targino Valente; Heidge Fukumasu. USP-Botucatu: Rejane Maria Tommasini Grotto; Helena Lage Ferreira; Instituto Butantan: Dimas Tadeu Covas; Jardelina de Souza Todao Bernardino; Jayme A. Souza-Neto; Jessika Cristina Chagas Lesbon; Jorge A. Petroli Marchesi; José Salvatore Leister Patané; João Paulo Kitajima; João Pessoa Araújo Jr.; Leila Sabrina Ullmann; Loyze Paola Oliveira de Lima; Luiz Aurelio de Campos Crispin. Centro de Genômica Funcional da ESALQ: Luiz Lehmann Coutinho; Luiz Carlos Junior de Alcantara; Lívia Sacchetto; Maisa C. Pereira Parra; Maria Carolina Elias; Marta Giovanetti; Marília Moraes; Maurício Lacerda Nogueira. Prefeitura de Sao Paulo: Melissa Palmieri.; Patricia Akemi Assato; Paula Rahal; Paulo Inacio da Costa; Rafael dos Santos Bezerra; Raquel de Lello Rocha Campos Cassano. NGS Soluções Genômicas: Pilar Drummond Sampaio Corrêa Mariani. FZEA-USP Pirassununga: Mirele Daiana Poleti; Raul Machado Neto; Ricardo Augusto Brassaloti; Ricardo Haddad; Rodrigo Tocantins Calado. FAMERP-SJRP: Cecília Artico Banho; Sandra Coccuzzo Sampaio; Svetoslav Nanev Slavov; Vagner Fonseca; Vincent Louis Viala |
| EPI_ISL_1966912 | CENTRO DE SAUDE II DR ARCY BANDEIRA IBIUNA SP | Instituto Butantan / ESALQ-USP (Piracicaba) | Antonio Jorge Martins; Bianca Cechetto Carlos. Mendelics: Bibiana Santos; Claudia Renata dos Santos Barros; Cíntia Bittar; David Schlesinger. Hemocentro Ribeirão Preto: Simone Kashima; Debora Botequio Moretti; Elaine Cristina Marqueze; Elaine Vieira dos Santos; Elisangela Chicaroni Mattos; Erika Freitas; Evandra Strazza Rodrigues; Felipe Allan da Silva da Costa; Flavia Aburjaile; Fábio Sossai Possebon; Guilherme Campos; Guilherme Targino Valente; Heidge Fukumasu. USP-Botucatu: Rejane Maria Tommasini Grotto; Helena Lage Ferreira; Instituto Butantan: Dimas Tadeu Covas; Jardelina de Souza Todao Bernardino; Jayme A. Souza-Neto; Jessika Cristina Chagas Lesbon; Jorge A. Petroli Marchesi; José Salvatore Leister Patané; João Paulo Kitajima; João Pessoa Araújo Jr.; Leila Sabrina Ullmann; Loyze Paola Oliveira de Lima; Luiz Aurelio de Campos Crispin. Centro de Genômica Funcional da ESALQ: Luiz Lehmann Coutinho; Luiz Carlos Junior de Alcantara; Lívia Sacchetto; Maisa C. Pereira Parra; Maria Carolina Elias; Marta Giovanetti; Marília Moraes; Maurício Lacerda Nogueira. Prefeitura de Sao Paulo: Melissa Palmieri.; Patricia Akemi Assato; Paula Rahal; Paulo Inacio da Costa; Rafael dos Santos Bezerra; Raquel de Lello Rocha Campos Cassano. NGS Soluções Genômicas: Pilar Drummond Sampaio Corrêa Mariani. FZEA-USP Pirassununga: Mirele Daiana Poleti; Raul Machado Neto; Ricardo Augusto Brassaloti; Ricardo Haddad; Rodrigo Tocantins Calado. FAMERP-SJRP: Cecília Artico Banho; Sandra Coccuzzo Sampaio; Svetoslav Nanev Slavov; Vagner Fonseca; Vincent Louis Viala |
| EPI_ISL_1966955, EPI_ISL_1966956 | CENTRO DE SAUDE II MAIRINQUE MAIRINQUE | Instituto Butantan / ESALQ-USP (Piracicaba) | Antonio Jorge Martins; Bianca Cechetto Carlos. Mendelics: Bibiana Santos; Claudia Renata dos Santos Barros; Cíntia Bittar; David Schlesinger. Hemocentro Ribeirão Preto: Simone Kashima; Debora Botequio Moretti; Elaine Cristina Marqueze; Elaine Vieira dos Santos; Elisangela Chicaroni Mattos; Erika Freitas; Evandra Strazza Rodrigues; Felipe Allan da Silva da Costa; Flavia Aburjaile; Fábio Sossai Possebon; Guilherme Campos; Guilherme Targino Valente; Heidge Fukumasu. USP-Botucatu: Rejane Maria Tommasini Grotto; Helena Lage Ferreira; Instituto Butantan: Dimas Tadeu Covas; Jardelina de Souza Todao Bernardino; Jayme A. Souza-Neto; Jessika Cristina Chagas Lesbon; Jorge A. Petroli Marchesi; José Salvatore Leister Patané; João Paulo Kitajima; João Pessoa Araújo Jr.; Leila Sabrina Ullmann; Loyze Paola Oliveira de Lima; Luiz Aurelio de Campos Crispin. Centro de Genômica Funcional da ESALQ: Luiz Lehmann Coutinho; Luiz Carlos Junior de Alcantara; Lívia Sacchetto; Maisa C. Pereira Parra; Maria Carolina Elias; Marta Giovanetti; Marília Moraes; Maurício Lacerda Nogueira. Prefeitura de Sao Paulo: Melissa Palmieri.; Patricia Akemi Assato; Paula Rahal; Paulo Inacio da Costa; Rafael dos Santos Bezerra; Raquel de Lello Rocha Campos Cassano. NGS Soluções Genômicas: Pilar Drummond Sampaio Corrêa Mariani. FZEA-USP Pirassununga: Mirele Daiana Poleti; Raul Machado Neto; Ricardo Augusto Brassaloti; Ricardo Haddad; Rodrigo Tocantins Calado. FAMERP-SJRP: Cecília Artico Banho; Sandra Coccuzzo Sampaio; Svetoslav Nanev Slavov; Vagner Fonseca; Vincent Louis Viala |
| EPI_ISL_1967062, EPI_ISL_1967063, EPI_ISL_1967064 | CENTRO DE SAUDE II SAO MIGUEL ARCANJO | Instituto Butantan / ESALQ-USP (Piracicaba) | Antonio Jorge Martins; Bianca Cechetto Carlos. Mendelics: Bibiana Santos; Claudia Renata dos Santos Barros; Cíntia Bittar; David Schlesinger. Hemocentro Ribeirão Preto: Simone Kashima; Debora Botequio Moretti; Elaine Cristina Marqueze; Elaine Vieira dos Santos; Elisangela Chicaroni Mattos; Erika Freitas; Evandra Strazza Rodrigues; Felipe Allan da Silva da Costa; Flavia Aburjaile; Fábio Sossai Possebon; Guilherme Campos; Guilherme Targino Valente; Heidge Fukumasu. USP-Botucatu: Rejane Maria Tommasini Grotto; Helena Lage Ferreira; Instituto Butantan: Dimas Tadeu Covas; Jardelina de Souza Todao Bernardino; Jayme A. Souza-Neto; Jessika Cristina Chagas Lesbon; Jorge A. Petroli Marchesi; José Salvatore Leister Patané; João Paulo Kitajima; João Pessoa Araújo Jr.; Leila Sabrina Ullmann; Loyze Paola Oliveira de Lima; Luiz Aurelio de Campos Crispin. Centro de Genômica Funcional da ESALQ: Luiz Lehmann Coutinho; Luiz Carlos Junior de Alcantara; Lívia Sacchetto; Maisa C. Pereira Parra; Maria Carolina Elias; Marta Giovanetti; Marília Moraes; Maurício Lacerda Nogueira. Prefeitura de Sao Paulo: Melissa Palmieri.; Patricia Akemi Assato; Paula Rahal; Paulo Inacio da Costa; Rafael dos Santos Bezerra; Raquel de Lello Rocha Campos Cassano. NGS Soluções Genômicas: Pilar Drummond Sampaio Corrêa Mariani. FZEA-USP Pirassununga: Mirele Daiana Poleti; Raul Machado Neto; Ricardo Augusto Brassaloti; Ricardo Haddad; Rodrigo Tocantins Calado. FAMERP-SJRP: Cecília Artico Banho; Sandra Coccuzzo Sampaio; Svetoslav Nanev Slavov; Vagner Fonseca; Vincent Louis Viala |
| EPI_ISL_1967096, EPI_ISL_1967097 | CENTRO DE SAUDE SAO ROQUE DR JOSE CARVALHO BRITO | Instituto Butantan / ESALQ-USP (Piracicaba) | Antonio Jorge Martins; Bianca Cechetto Carlos. Mendelics: Bibiana Santos; Claudia Renata dos Santos Barros; Cíntia Bittar; David Schlesinger. Hemocentro Ribeirão Preto: Simone Kashima; Debora Botequio Moretti; Elaine Cristina Marqueze; Elaine Vieira dos Santos; Elisangela Chicaroni Mattos; Erika Freitas; Evandra Strazza Rodrigues; Felipe Allan da Silva da Costa; Flavia Aburjaile; Fábio Sossai Possebon; Guilherme Campos; Guilherme Targino Valente; Heidge Fukumasu. USP-Botucatu: Rejane Maria Tommasini Grotto; Helena Lage Ferreira; Instituto Butantan: Dimas Tadeu Covas; Jardelina de Souza Todao Bernardino; Jayme A. Souza-Neto; Jessika Cristina Chagas Lesbon; Jorge A. Petroli Marchesi; José Salvatore Leister Patané; João Paulo Kitajima; João Pessoa Araújo Jr.; Leila Sabrina Ullmann; Loyze Paola Oliveira de Lima; Luiz Aurelio de Campos Crispin. Centro de Genômica Funcional da ESALQ: Luiz Lehmann Coutinho; Luiz Carlos Junior de Alcantara; Lívia Sacchetto; Maisa C. Pereira Parra; Maria Carolina Elias; Marta Giovanetti; Marília Moraes; Maurício Lacerda Nogueira. Prefeitura de Sao Paulo: Melissa Palmieri.; Patricia Akemi Assato; Paula Rahal; Paulo Inacio da Costa; Rafael dos Santos Bezerra; Raquel de Lello Rocha Campos Cassano. NGS Soluções Genômicas: Pilar Drummond Sampaio Corrêa Mariani. FZEA-USP Pirassununga: Mirele Daiana Poleti; Raul Machado Neto; Ricardo Augusto Brassaloti; Ricardo Haddad; Rodrigo Tocantins Calado. FAMERP-SJRP: Cecília Artico Banho; Sandra Coccuzzo Sampaio; Svetoslav Nanev Slavov; Vagner Fonseca; Vincent Louis Viala |
| EPI_ISL_2344699, EPI_ISL_2344700 | CENTRO MEDICO DR NELSON SALOME DE CONCHAL | Instituto Butantan / FZEA-USP-Pirassununga | Antonio Jorge Martins; Claudia Renata dos Santos Barros; David Schlesinger; Debora Botequio Moretti; Dimas Tadeu Covas; Elaine Cristina Marqueze; Elaine Vieira Santos; Evandra Strazza Rodrigues; Heidge Fukumasu; Jayme Augusto de Souza-Neto; José Salvatore Leister Patané; Luiz Alcantara; Luiz Lehmann Coutinho; Maria Carolina Elias; Maurício Lacerda Nogueira; Rafael dos Santos Bezerra; Raul Machado Neto; Rejane Maria Tommasini Grotto; Ricardo Haddad; Sandra Coccuzzo Sampaio Vessoni; Simone Kashima; Svetoslav Nanev Slavov; Vincent Louis Viala |
| EPI_ISL_1628339, EPI_ISL_1628340, EPI_ISL_1628342 | CH de l'Ouest Guyanais | Institut Pasteur de la Guyane | A. Salmier; Anne Lavergne; Dominique Rousset |
| EPI_ISL_1756129, EPI_ISL_1756160, EPI_ISL_1756278 | CH.INTERCOMMUNAL DE CRETEIL | Department of Virology, Henri Mondor University Hospital, Assistance Publique Hôpitaux de Paris, Université Paris-Est Créteil, INSERM U955 | Alexandre Soulier; Christophe Rodriguez; Elisabeth Trawinski; Guillaume Gricourt; Jean-Michel Pawlotsky; Melissa N'Debi; Slim Fourati; Vanessa Demontant |
| EPI_ISL_1627097, EPI_ISL_1627102, EPI_ISL_1627104, EPI_ISL_1627105, EPI_ISL_1628334, EPI_ISL_1628335 | CHC Andrée Rosemon | Institut Pasteur de la Guyane | A. Salmier; Anne Lavergne; Dominique Rousset |
| EPI_ISL_1739049 | CHLC | Instituto Nacional de Saude (INSA) | Borges et al |
| EPI_ISL_2004178 | CHU Sao Joao, Porto | Instituto Nacional de Saude (INSA) and i3S - Instituto de Investigação e Inovação em Saúde | Borges et al |
| EPI_ISL_1854234, EPI_ISL_1854235, EPI_ISL_1854236 | CHULN - H Santa Maria | Instituto Nacional de Saude (INSA) | Borges et al |
| EPI_ISL_1655960 | CHWAPI - SITE NOTRE DAME | Institut de Pathologie et Genetique (IPG) | Jérémie Gras; Pascale Hilbert |
| EPI_ISL_1627099, EPI_ISL_1627100, EPI_ISL_1627101 | CNR Institut Pasteur de la Guyane | Institut Pasteur de la Guyane | Anne Lavergne; Dominique Rousset |
| EPI_ISL_1695421 | CNR Virus des Infections Respiratoires - France SUD | CNR Virus des Infections Respiratoires - France SUD | Antonin Bal; Bruno Lina; Bruno Simon; Gregory Destras; Gwendolyne Burfin; Hadrien Regue; Laurence Josset; Martine Valette; Quentin Semanas |
| EPI_ISL_2345961 | COMPLEXO HOSPITALAR OURO VERDE DE CAMPINAS | Instituto Butantan | Antonio Jorge Martins; Claudia Renata dos Santos Barros; David Schlesinger; Debora Botequio Moretti; Dimas Tadeu Covas; Elaine Cristina Marqueze; Elaine Vieira Santos; Evandra Strazza Rodrigues; Heidge Fukumasu; Jayme Augusto de Souza-Neto; José Salvatore Leister Patané; Luiz Alcantara; Luiz Lehmann Coutinho; Maria Carolina Elias; Maurício Lacerda Nogueira; Rafael dos Santos Bezerra; Raul Machado Neto; Rejane Maria Tommasini Grotto; Ricardo Haddad; Sandra Coccuzzo Sampaio Vessoni; Simone Kashima; Svetoslav Nanev Slavov; Vincent Louis Viala |
| EPI_ISL_2344702 | COORDENADORIA MUNICIPAL DE SAUDE DE IRACEMAPOLIS | Instituto Butantan / FZEA-USP-Pirassununga | Antonio Jorge Martins; Claudia Renata dos Santos Barros; David Schlesinger; Debora Botequio Moretti; Dimas Tadeu Covas; Elaine Cristina Marqueze; Elaine Vieira Santos; Evandra Strazza Rodrigues; Heidge Fukumasu; Jayme Augusto de Souza-Neto; José Salvatore Leister Patané; Luiz Alcantara; Luiz Lehmann Coutinho; Maria Carolina Elias; Maurício Lacerda Nogueira; Rafael dos Santos Bezerra; Raul Machado Neto; Rejane Maria Tommasini Grotto; Ricardo Haddad; Sandra Coccuzzo Sampaio Vessoni; Simone Kashima; Svetoslav Nanev Slavov; Vincent Louis Viala |
| EPI_ISL_1804964 | CQRC_QUALITY CONTROL CHEMICAL BIOLOGICAL RISK_AOOR Villa Sofia Cervello Palermo | CQRC_QUALITY CONTROL CHEMICAL BIOLOGICAL RISK_AOOR Villa Sofia Cervello Palermo | ; Brunacci, G.; Contino, F.; Di Gaudio, F. |
| EPI_ISL_1966834 | CS DE BALSAMO | Instituto Butantan / ESALQ-USP | Antonio Jorge Martins; Bianca Cechetto Carlos. Mendelics: Bibiana Santos; Claudia Renata dos Santos Barros; Cíntia Bittar; David Schlesinger. Hemocentro Ribeirão Preto: Simone Kashima; Debora Botequio Moretti; Elaine Cristina Marqueze; Elaine Vieira dos Santos; Elisangela Chicaroni Mattos; Erika Freitas; Evandra |

| | | | |
|---|---|---|---|
| | | (Piracicaba) | Strazza Rodrigues; Felipe Allan da Silva da Costa; Flavia Aburjaile; Fábio Sossai Possebon; Guilherme Campos; Guilherme Targino Valente; Heidge Fukumasu. USP-Botucatu: Rejane Maria Tommasini Grotto; Helena Lage Ferreira; Instituto Butantan: Dimas Tadeu Covas; Jardelina de Souza Todao Bernardino; Jayme A. Souza-Neto; Jessika Cristina Chagas Lesbon; Jorge A. Petroli Marchesi; José Salvatore Leister Patané; João Paulo Kitajima; João Pessoa Araújo Jr.; Leila Sabrina Ullmann; Loyze Paola Oliveira de Lima; Luiz Aurelio de Campos Crispin. Centro de Genômica Funcional da ESALQ: Luiz Lehmann Coutinho; Luiz Carlos Junior de Alcantara; Lívia Sacchetto; Maisa C. Pereira Parra; Maria Carolina Elias; Marta Giovanetti; Marília Moraes; Maurício Lacerda Nogueira. Prefeitura de Sao Paulo: Melissa Palmieri.; Patricia Akemi Assato; Paula Rahal; Paulo Inacio da Costa; Rafael dos Santos Bezerra; Raquel de Lello Rocha Campos Cassano. NGS Soluções Genômicas: Pilar Drummond Sampaio Corrêa Mariani. FZEA-USP Pirassununga: Mirele Daiana Poleti; Raul Machado Neto; Ricardo Augusto Brassaloti; Ricardo Haddad; Rodrigo Tocantins Calado. FAMERP-SJRP: Cecília Artico Banho; Sandra Coccuzzo Sampaio; Svetoslav Nanev Slavov; Vagner Fonseca; Vincent Louis Viala |
| EPI_ISL_1966868, EPI_ISL_1966869 | CS DE CATIGUA | Instituto Butantan / ESALQ-USP (Piracicaba) | Antonio Jorge Martins; Bianca Cechetto Carlos. Mendelics: Bibiana Santos; Claudia Renata dos Santos Barros; Cíntia Bittar; David Schlesinger. Hemocentro Ribeirão Preto: Simone Kashima; Debora Botequio Moretti; Elaine Cristina Marqueze; Elaine Vieira dos Santos; Elisangela Chicaroni Mattos; Erika Freitas; Evandra Strazza Rodrigues; Felipe Allan da Silva da Costa; Flavia Aburjaile; Fábio Sossai Possebon; Guilherme Campos; Guilherme Targino Valente; Heidge Fukumasu. USP-Botucatu: Rejane Maria Tommasini Grotto; Helena Lage Ferreira; Instituto Butantan: Dimas Tadeu Covas; Jardelina de Souza Todao Bernardino; Jayme A. Souza-Neto; Jessika Cristina Chagas Lesbon; Jorge A. Petroli Marchesi; José Salvatore Leister Patané; João Paulo Kitajima; João Pessoa Araújo Jr.; Leila Sabrina Ullmann; Loyze Paola Oliveira de Lima; Luiz Aurelio de Campos Crispin. Centro de Genômica Funcional da ESALQ: Luiz Lehmann Coutinho; Luiz Carlos Junior de Alcantara; Lívia Sacchetto; Maisa C. Pereira Parra; Maria Carolina Elias; Marta Giovanetti; Marília Moraes; Maurício Lacerda Nogueira. Prefeitura de Sao Paulo: Melissa Palmieri.; Patricia Akemi Assato; Paula Rahal; Paulo Inacio da Costa; Rafael dos Santos Bezerra; Raquel de Lello Rocha Campos Cassano. NGS Soluções Genômicas: Pilar Drummond Sampaio Corrêa Mariani. FZEA-USP Pirassununga: Mirele Daiana Poleti; Raul Machado Neto; Ricardo Augusto Brassaloti; Ricardo Haddad; Rodrigo Tocantins Calado. FAMERP-SJRP: Cecília Artico Banho; Sandra Coccuzzo Sampaio; Svetoslav Nanev Slavov; Vagner Fonseca; Vincent Louis Viala |
| EPI_ISL_2170895, EPI_ISL_2170896 | CS DE MONTE APRAZIVEL | Instituto Butantan / ESALQ-Piracicaba | Antonio Jorge Martins; Bianca Cechetto Carlos. Mendelics: Bibiana Santos; Claudia Renata dos Santos Barros; Cíntia Bittar; David Schlesinger. Hemocentro Ribeirão Preto: Simone Kashima; Debora Botequio Moretti; Elaine Cristina Marqueze; Elaine Vieira dos Santos; Elisangela Chicaroni Mattos; Erika Freitas; Evandra Strazza Rodrigues; Felipe Allan da Silva da Costa; Flavia Aburjaile; Fábio Sossai Possebon; Guilherme Campos; Guilherme Targino Valente; Heidge Fukumasu. USP-Botucatu: Rejane Maria Tommasini Grotto; Helena Lage Ferreira; Instituto Butantan: Dimas Tadeu Covas; Jardelina de Souza Todao Bernardino; Jayme A. Souza-Neto; Jessika Cristina Chagas Lesbon; Jorge A. Petroli Marchesi; José Salvatore Leister Patané; João Paulo Kitajima; João Pessoa Araújo Jr.; Leila Sabrina Ullmann; Loyze Paola Oliveira de Lima; Luiz Aurelio de Campos Crispin. Centro de Genômica Funcional da ESALQ: Luiz Lehmann Coutinho; Luiz Carlos Junior de Alcantara; Lívia Sacchetto; Maisa C. Pereira Parra; Maria Carolina Elias; Marta Giovanetti; Marília Moraes; Maurício Lacerda Nogueira. Prefeitura de Sao Paulo: Melissa Palmieri.; Patricia Akemi Assato; Paula Rahal; Paulo Inacio da Costa; Rafael dos Santos Bezerra; Raquel de Lello Rocha Campos Cassano. NGS Soluções Genômicas: Pilar Drummond Sampaio Corrêa Mariani. FZEA-USP Pirassununga: Mirele Daiana Poleti; Raul Machado Neto; Ricardo Augusto Brassaloti; Ricardo Haddad; Rodrigo Tocantins Calado. FAMERP-SJRP: Cecília Artico Banho; Sandra Coccuzzo Sampaio; Svetoslav Nanev Slavov; Vagner Fonseca; Vincent Louis Viala |
| EPI_ISL_1966961, EPI_ISL_1966962, EPI_ISL_1966963, EPI_ISL_1966964 | CS DE MONTE APRAZIVEL | Instituto Butantan / ESALQ-USP (Piracicaba) | Antonio Jorge Martins; Bianca Cechetto Carlos. Mendelics: Bibiana Santos; Claudia Renata dos Santos Barros; Cíntia Bittar; David Schlesinger. Hemocentro Ribeirão Preto: Simone Kashima; Debora Botequio Moretti; Elaine Cristina Marqueze; Elaine Vieira dos Santos; Elisangela Chicaroni Mattos; Erika Freitas; Evandra Strazza Rodrigues; Felipe Allan da Silva da Costa; Flavia Aburjaile; Fábio Sossai Possebon; Guilherme Campos; Guilherme Targino Valente; Heidge Fukumasu. USP-Botucatu: Rejane Maria Tommasini Grotto; Helena Lage Ferreira; Instituto Butantan: Dimas Tadeu Covas; Jardelina de Souza Todao Bernardino; Jayme A. Souza-Neto; Jessika Cristina Chagas Lesbon; Jorge A. Petroli Marchesi; José Salvatore Leister Patané; João Paulo Kitajima; João Pessoa Araújo Jr.; Leila Sabrina Ullmann; Loyze Paola Oliveira de Lima; Luiz Aurelio de Campos Crispin. Centro de Genômica Funcional da ESALQ: Luiz Lehmann Coutinho; Luiz Carlos Junior de Alcantara; Lívia Sacchetto; Maisa C. Pereira Parra; Maria Carolina Elias; Marta Giovanetti; Marília Moraes; Maurício Lacerda Nogueira. Prefeitura de Sao Paulo: Melissa Palmieri.; Patricia Akemi Assato; Paula Rahal; Paulo Inacio da Costa; Rafael dos Santos Bezerra; Raquel de Lello Rocha Campos Cassano. NGS Soluções Genômicas: Pilar Drummond Sampaio Corrêa Mariani. FZEA-USP Pirassununga: Mirele Daiana Poleti; Raul Machado Neto; Ricardo Augusto Brassaloti; Ricardo Haddad; Rodrigo Tocantins Calado. FAMERP-SJRP: Cecília Artico Banho; Sandra Coccuzzo Sampaio; Svetoslav Nanev Slavov; Vagner Fonseca; Vincent Louis Viala |
| EPI_ISL_1966984, EPI_ISL_1966985 | CS DE NEVES PAULISTA | Instituto Butantan / ESALQ-USP (Piracicaba) | Antonio Jorge Martins; Bianca Cechetto Carlos. Mendelics: Bibiana Santos; Claudia Renata dos Santos Barros; Cíntia Bittar; David Schlesinger. Hemocentro Ribeirão Preto: Simone Kashima; Debora Botequio Moretti; Elaine Cristina Marqueze; Elaine Vieira dos Santos; Elisangela Chicaroni Mattos; Erika Freitas; Evandra Strazza Rodrigues; Felipe Allan da Silva da Costa; Flavia Aburjaile; Fábio Sossai Possebon; Guilherme Campos; Guilherme Targino Valente; Heidge Fukumasu. USP-Botucatu: Rejane Maria Tommasini Grotto; Helena Lage Ferreira; Instituto Butantan: Dimas Tadeu Covas; Jardelina de Souza Todao Bernardino; Jayme A. Souza-Neto; Jessika Cristina Chagas Lesbon; Jorge A. Petroli Marchesi; José Salvatore Leister Patané; João Paulo Kitajima; João Pessoa Araújo Jr.; Leila Sabrina Ullmann; Loyze Paola Oliveira de Lima; Luiz Aurelio de Campos Crispin. Centro de Genômica Funcional da ESALQ: Luiz Lehmann Coutinho; Luiz Carlos Junior de Alcantara; Lívia Sacchetto; Maisa C. Pereira Parra; Maria Carolina Elias; Marta Giovanetti; Marília Moraes; Maurício Lacerda Nogueira. Prefeitura de Sao Paulo: Melissa Palmieri.; Patricia Akemi Assato; Paula Rahal; Paulo Inacio da Costa; Rafael dos Santos Bezerra; Raquel de Lello Rocha Campos Cassano. NGS Soluções Genômicas: Pilar Drummond Sampaio Corrêa Mariani. FZEA-USP Pirassununga: Mirele Daiana Poleti; Raul Machado Neto; Ricardo Augusto Brassaloti; Ricardo Haddad; Rodrigo Tocantins Calado. FAMERP-SJRP: Cecília Artico Banho; Sandra Coccuzzo Sampaio; Svetoslav Nanev Slavov; Vagner Fonseca; Vincent Louis Viala |
| EPI_ISL_1966988 | CS DE ORINDIUVA | Instituto Butantan / ESALQ-USP (Piracicaba) | Antonio Jorge Martins; Bianca Cechetto Carlos. Mendelics: Bibiana Santos; Claudia Renata dos Santos Barros; Cíntia Bittar; David Schlesinger. Hemocentro Ribeirão Preto: Simone Kashima; Debora Botequio Moretti; Elaine Cristina Marqueze; Elaine Vieira dos Santos; Elisangela Chicaroni Mattos; Erika Freitas; Evandra Strazza Rodrigues; Felipe Allan da Silva da Costa; Flavia Aburjaile; Fábio Sossai Possebon; Guilherme Campos; Guilherme Targino Valente; Heidge Fukumasu. USP-Botucatu: Rejane Maria Tommasini Grotto; Helena Lage Ferreira; Instituto Butantan: Dimas Tadeu Covas; Jardelina de Souza Todao Bernardino; Jayme A. Souza-Neto; Jessika Cristina Chagas Lesbon; Jorge A. Petroli Marchesi; José Salvatore Leister Patané; João Paulo Kitajima; João Pessoa Araújo Jr.; Leila Sabrina Ullmann; Loyze Paola Oliveira de Lima; Luiz Aurelio de Campos Crispin. Centro de Genômica Funcional da ESALQ: Luiz Lehmann Coutinho; Luiz Carlos Junior de Alcantara; Lívia Sacchetto; Maisa C. Pereira Parra; Maria Carolina Elias; Marta Giovanetti; Marília Moraes; Maurício Lacerda Nogueira. Prefeitura de Sao Paulo: Melissa Palmieri.; Patricia Akemi Assato; Paula Rahal; Paulo Inacio da Costa; Rafael dos Santos Bezerra; Raquel de Lello Rocha Campos Cassano. NGS Soluções Genômicas: Pilar Drummond Sampaio Corrêa Mariani. FZEA-USP Pirassununga: Mirele Daiana Poleti; Raul Machado Neto; Ricardo Augusto Brassaloti; Ricardo Haddad; Rodrigo Tocantins Calado. FAMERP-SJRP: Cecília Artico Banho; Sandra Coccuzzo Sampaio; Svetoslav Nanev Slavov; Vagner Fonseca; Vincent Louis Viala |
| EPI_ISL_1967050 | CS DE SANTA RITA DOESTE | Instituto Butantan / ESALQ-USP (Piracicaba) | Antonio Jorge Martins; Bianca Cechetto Carlos. Mendelics: Bibiana Santos; Claudia Renata dos Santos Barros; Cíntia Bittar; David Schlesinger. Hemocentro Ribeirão Preto: Simone Kashima; Debora Botequio Moretti; Elaine Cristina Marqueze; Elaine Vieira dos Santos; Elisangela Chicaroni Mattos; Erika Freitas; Evandra Strazza Rodrigues; Felipe Allan da Silva da Costa; Flavia Aburjaile; Fábio Sossai Possebon; Guilherme Campos; Guilherme Targino Valente; Heidge Fukumasu. USP-Botucatu: Rejane Maria Tommasini Grotto; Helena Lage Ferreira; Instituto Butantan: Dimas Tadeu Covas; Jardelina de Souza Todao Bernardino; Jayme A. Souza-Neto; Jessika Cristina Chagas Lesbon; Jorge A. Petroli Marchesi; José Salvatore Leister Patané; João Paulo Kitajima; João Pessoa Araújo Jr.; Leila Sabrina Ullmann; Loyze Paola Oliveira de Lima; Luiz Aurelio de Campos Crispin. Centro de Genômica Funcional da ESALQ: Luiz Lehmann Coutinho; Luiz Carlos Junior de Alcantara; Lívia Sacchetto; Maisa C. Pereira Parra; Maria Carolina Elias; Marta Giovanetti; Marília Moraes; Maurício Lacerda Nogueira. Prefeitura de Sao Paulo: Melissa Palmieri.; Patricia Akemi Assato; Paula Rahal; Paulo Inacio da Costa; Rafael dos Santos Bezerra; Raquel de Lello Rocha Campos Cassano. NGS Soluções Genômicas: Pilar Drummond Sampaio Corrêa Mariani. FZEA-USP Pirassununga: Mirele Daiana Poleti; Raul Machado Neto; Ricardo Augusto Brassaloti; Ricardo Haddad; Rodrigo Tocantins Calado. FAMERP-SJRP: Cecília Artico Banho; Sandra Coccuzzo Sampaio; Svetoslav Nanev Slavov; Vagner Fonseca; Vincent Louis Viala |
| EPI_ISL_1966992, EPI_ISL_1966993 | CS I DE OURINHOS | Instituto Butantan / ESALQ-USP (Piracicaba) | Antonio Jorge Martins; Bianca Cechetto Carlos. Mendelics: Bibiana Santos; Claudia Renata dos Santos Barros; Cíntia Bittar; David Schlesinger. Hemocentro Ribeirão Preto: Simone Kashima; Debora Botequio Moretti; Elaine Cristina Marqueze; Elaine Vieira dos Santos; Elisangela Chicaroni Mattos; Erika Freitas; Evandra Strazza Rodrigues; Felipe Allan da Silva da Costa; Flavia Aburjaile; Fábio Sossai Possebon; Guilherme Campos; Guilherme Targino Valente; Heidge Fukumasu. USP-Botucatu: Rejane Maria Tommasini Grotto; Helena Lage Ferreira; Instituto Butantan: Dimas Tadeu Covas; Jardelina de Souza Todao Bernardino; Jayme A. Souza-Neto; Jessika Cristina Chagas Lesbon; Jorge A. Petroli Marchesi; José Salvatore Leister Patané; João Paulo Kitajima; João Pessoa Araújo Jr.; Leila Sabrina Ullmann; Loyze Paola Oliveira de Lima; Luiz Aurelio de Campos Crispin. Centro de Genômica Funcional da ESALQ: Luiz Lehmann Coutinho; Luiz Carlos Junior de Alcantara; Lívia Sacchetto; Maisa C. Pereira Parra; Maria Carolina Elias; Marta Giovanetti; Marília Moraes; Maurício Lacerda Nogueira. Prefeitura de Sao Paulo: Melissa Palmieri.; Patricia Akemi Assato; Paula Rahal; Paulo Inacio da Costa; Rafael dos Santos Bezerra; Raquel de Lello Rocha Campos Cassano. NGS Soluções Genômicas: Pilar Drummond Sampaio Corrêa Mariani. FZEA-USP Pirassununga: Mirele Daiana Poleti; Raul Machado Neto; Ricardo Augusto Brassaloti; Ricardo Haddad; Rodrigo Tocantins Calado. FAMERP-SJRP: Cecília Artico Banho; Sandra Coccuzzo Sampaio; Svetoslav Nanev Slavov; Vagner Fonseca; Vincent Louis Viala |
| EPI_ISL_1966909 | CS II DR JAHYR DE PAULA RIBEIRO GUARA | Instituto Butantan / ESALQ-USP (Piracicaba) | Antonio Jorge Martins; Bianca Cechetto Carlos. Mendelics: Bibiana Santos; Claudia Renata dos Santos Barros; Cíntia Bittar; David Schlesinger. Hemocentro Ribeirão Preto: Simone Kashima; Debora Botequio Moretti; Elaine Cristina Marqueze; Elaine Vieira dos Santos; Elisangela Chicaroni Mattos; Erika Freitas; Evandra Strazza Rodrigues; Felipe Allan da Silva da Costa; Flavia Aburjaile; Fábio Sossai Possebon; Guilherme Campos; Guilherme Targino Valente; Heidge Fukumasu. USP-Botucatu: Rejane Maria Tommasini Grotto; Helena Lage Ferreira; Instituto Butantan: Dimas Tadeu Covas; Jardelina de Souza Todao Bernardino; Jayme A. Souza-Neto; Jessika Cristina Chagas Lesbon; Jorge A. Petroli Marchesi; José Salvatore Leister Patané; João Paulo Kitajima; João Pessoa Araújo Jr.; Leila Sabrina Ullmann; Loyze Paola Oliveira de Lima; Luiz Aurelio de Campos Crispin. Centro de Genômica Funcional da ESALQ: Luiz Lehmann Coutinho; Luiz Carlos Junior de Alcantara; Lívia Sacchetto; Maisa C. Pereira Parra; Maria Carolina Elias; Marta Giovanetti; Marília Moraes; Maurício Lacerda Nogueira. Prefeitura de Sao Paulo: Melissa Palmieri.; Patricia Akemi Assato; Paula Rahal; Paulo Inacio da Costa; Rafael dos Santos Bezerra; Raquel de Lello Rocha Campos Cassano. NGS Soluções Genômicas: Pilar Drummond Sampaio Corrêa Mariani. FZEA-USP Pirassununga: Mirele Daiana Poleti; Raul Machado Neto; Ricardo Augusto Brassaloti; Ricardo Haddad; Rodrigo Tocantins Calado. FAMERP-SJRP: Cecília Artico Banho; Sandra Coccuzzo Sampaio; Svetoslav Nanev Slavov; Vagner Fonseca; Vincent Louis Viala |
| EPI_ISL_1966989 | CS II DR MIGUEL VITALIANO ORLANDIA | Instituto Butantan / ESALQ-USP (Piracicaba) | Antonio Jorge Martins; Bianca Cechetto Carlos. Mendelics: Bibiana Santos; Claudia Renata dos Santos Barros; Cíntia Bittar; David Schlesinger. Hemocentro Ribeirão Preto: Simone Kashima; Debora Botequio Moretti; Elaine Cristina Marqueze; Elaine Vieira dos Santos; Elisangela Chicaroni Mattos; Erika Freitas; Evandra Strazza Rodrigues; Felipe Allan da Silva da Costa; Flavia Aburjaile; Fábio Sossai Possebon; Guilherme Campos; Guilherme Targino Valente; Heidge Fukumasu. USP-Botucatu: Rejane Maria Tommasini Grotto; Helena Lage Ferreira; Instituto Butantan: Dimas Tadeu Covas; Jardelina de Souza Todao Bernardino; Jayme A. Souza-Neto; Jessika Cristina Chagas Lesbon; Jorge A. Petroli Marchesi; José Salvatore Leister Patané; João Paulo Kitajima; João Pessoa Araújo Jr.; Leila Sabrina Ullmann; Loyze Paola Oliveira de Lima; Luiz Aurelio de Campos Crispin. Centro de Genômica Funcional da ESALQ: Luiz Lehmann Coutinho; Luiz Carlos Junior de Alcantara; Lívia Sacchetto; Maisa C. Pereira Parra; Maria Carolina Elias; Marta Giovanetti; Marília Moraes; Maurício Lacerda Nogueira. Prefeitura de Sao Paulo: Melissa Palmieri.; Patricia Akemi Assato; Paula Rahal; Paulo Inacio da Costa; Rafael dos Santos Bezerra; Raquel de Lello Rocha Campos Cassano. NGS Soluções Genômicas: Pilar Drummond Sampaio Corrêa Mariani. FZEA-USP Pirassununga: Mirele Daiana Poleti; Raul Machado Neto; Ricardo Augusto Brassaloti; Ricardo Haddad; Rodrigo Tocantins Calado. FAMERP-SJRP: Cecília Artico Banho; Sandra Coccuzzo Sampaio; Svetoslav Nanev Slavov; Vagner Fonseca; Vincent Louis Viala |
| EPI_ISL_2170897 | CS II EGIDIO BRUNHARA MORRO AGUDO | Instituto Butantan / ESALQ-Piracicaba | Antonio Jorge Martins; Bianca Cechetto Carlos. Mendelics: Bibiana Santos; Claudia Renata dos Santos Barros; Cíntia Bittar; David Schlesinger. Hemocentro Ribeirão Preto: Simone Kashima; Debora Botequio Moretti; Elaine Cristina Marqueze; Elaine Vieira dos Santos; Elisangela Chicaroni Mattos; Erika Freitas; Evandra Strazza Rodrigues; Felipe Allan da Silva da Costa; Flavia Aburjaile; Fábio Sossai Possebon; Guilherme Campos; Guilherme Targino Valente; Heidge Fukumasu. USP-Botucatu: Rejane Maria Tommasini Grotto; Helena Lage Ferreira; Instituto Butantan: Dimas Tadeu Covas; Jardelina de Souza Todao Bernardino; Jayme A. Souza-Neto; Jessika Cristina Chagas Lesbon; Jorge A. Petroli Marchesi; José Salvatore Leister Patané; João Paulo Kitajima; João Pessoa Araújo Jr.; Leila Sabrina Ullmann; Loyze Paola Oliveira de Lima; Luiz Aurelio de Campos Crispin. Centro de Genômica Funcional da ESALQ: Luiz Lehmann Coutinho; Luiz Carlos Junior de Alcantara; Lívia Sacchetto; Maisa C. Pereira Parra; Maria Carolina Elias; Marta Giovanetti; Marília Moraes; Maurício Lacerda Nogueira. Prefeitura de Sao Paulo: Melissa Palmieri.; Patricia Akemi Assato; Paula Rahal; Paulo Inacio da Costa; Rafael dos Santos Bezerra; Raquel de Lello Rocha Campos Cassano. NGS Soluções Genômicas: Pilar Drummond Sampaio Corrêa Mariani. FZEA-USP Pirassununga: Mirele Daiana Poleti; Raul Machado Neto; Ricardo Augusto Brassaloti; Ricardo Haddad; Rodrigo Tocantins Calado. FAMERP-SJRP: Cecília Artico Banho; Sandra Coccuzzo Sampaio; Svetoslav Nanev Slavov; Vagner Fonseca; Vincent Louis Viala |
| EPI_ISL_1966965, EPI_ISL_1966966, EPI_ISL_1966967, EPI_ISL_1966968, EPI_ISL_1966970, EPI_ISL_1966971, EPI_ISL_1966972, EPI_ISL_1966973, EPI_ISL_1966974, EPI_ISL_1966975, EPI_ISL_1966976, EPI_ISL_1966977, EPI_ISL_1966978, EPI_ISL_1966979, EPI_ISL_1966980, EPI_ISL_1966981, EPI_ISL_1966982, EPI_ISL_1966983 | | | |
| see above | CS II EGIDIO BRUNHARA MORRO AGUDO | Instituto Butantan / ESALQ-USP (Piracicaba) | Antonio Jorge Martins; Bianca Cechetto Carlos. Mendelics: Bibiana Santos; Claudia Renata dos Santos Barros; Cíntia Bittar; David Schlesinger. Hemocentro Ribeirão Preto: Simone Kashima; Debora Botequio Moretti; Elaine Cristina Marqueze; Elaine Vieira dos Santos; Elisangela Chicaroni Mattos; Erika Freitas; Evandra Strazza Rodrigues; Felipe Allan da Silva da Costa; Flavia Aburjaile; Fábio Sossai Possebon; Guilherme Campos; Guilherme Targino Valente; Heidge Fukumasu. USP-Botucatu: Rejane Maria Tommasini Grotto; Helena Lage Ferreira; Instituto Butantan: Dimas Tadeu Covas; Jardelina de Souza Todao Bernardino; Jayme A. Souza-Neto; Jessika Cristina Chagas Lesbon; Jorge A. Petroli Marchesi; José Salvatore Leister Patané; João Paulo Kitajima; João Pessoa Araújo Jr.; Leila Sabrina Ullmann; Loyze Paola Oliveira de Lima; Luiz Aurelio de Campos Crispin. Centro de Genômica Funcional da ESALQ: Luiz Lehmann Coutinho; Luiz Carlos Junior de Alcantara; Lívia Sacchetto; Maisa C. Pereira Parra; Maria Carolina Elias; Marta Giovanetti; Marília Moraes; Maurício Lacerda Nogueira. Prefeitura de Sao Paulo: Melissa Palmieri.; Patricia Akemi Assato; Paula Rahal; Paulo Inacio da Costa; Rafael dos Santos Bezerra; Raquel de Lello Rocha Campos Cassano. NGS Soluções Genômicas: Pilar Drummond Sampaio Corrêa Mariani. FZEA-USP Pirassununga: Mirele Daiana Poleti; Raul Machado Neto; Ricardo Augusto Brassaloti; Ricardo Haddad; Rodrigo Tocantins Calado. FAMERP-SJRP: Cecília Artico Banho; Sandra Coccuzzo Sampaio; Svetoslav Nanev Slavov; Vagner Fonseca; Vincent Louis Viala |
| EPI_ISL_1967004, EPI_ISL_1967006, EPI_ISL_1967007, EPI_ISL_1967008, EPI_ISL_1967010, EPI_ISL_1967011, EPI_ISL_1967012, EPI_ISL_1967013 | | | |
| see above | CSII DR WASHINGTON LUIS M RODRIGUES DA SILVA PITANGUEIRAS | Instituto Butantan / ESALQ-USP (Piracicaba) | Antonio Jorge Martins; Bianca Cechetto Carlos. Mendelics: Bibiana Santos; Claudia Renata dos Santos Barros; Cíntia Bittar; David Schlesinger. Hemocentro Ribeirão Preto: Simone Kashima; Debora Botequio Moretti; Elaine Cristina Marqueze; Elaine Vieira dos Santos; Elisangela Chicaroni Mattos; Erika Freitas; Evandra Strazza Rodrigues; Felipe Allan da Silva da Costa; Flavia Aburjaile; Fábio Sossai Possebon; Guilherme Campos; Guilherme Targino Valente; Heidge Fukumasu. USP-Botucatu: Rejane Maria Tommasini Grotto; Helena Lage Ferreira; Instituto Butantan: Dimas Tadeu Covas; Jardelina de Souza Todao Bernardino; Jayme A. Souza-Neto; Jessika Cristina Chagas Lesbon; Jorge A. Petroli Marchesi; José Salvatore Leister Patané; João Paulo Kitajima; João Pessoa Araújo Jr.; Leila Sabrina Ullmann; Loyze Paola Oliveira de Lima; Luiz Aurelio de Campos Crispin. Centro de Genômica Funcional da ESALQ: Luiz Lehmann Coutinho; Luiz Carlos Junior de Alcantara; Lívia Sacchetto; Maisa C. Pereira Parra; Maria Carolina Elias; Marta Giovanetti; Marília Moraes; Maurício Lacerda Nogueira. Prefeitura de Sao Paulo: Melissa Palmieri.; Patricia Akemi Assato; Paula Rahal; Paulo Inacio da Costa; Rafael dos Santos Bezerra; Raquel de Lello Rocha Campos Cassano. NGS Soluções Genômicas: Pilar Drummond Sampaio Corrêa Mariani. FZEA-USP Pirassununga: Mirele Daiana Poleti; Raul Machado Neto; Ricardo Augusto Brassaloti; Ricardo Haddad; Rodrigo Tocantins Calado. FAMERP-SJRP: Cecília Artico Banho; Sandra Coccuzzo Sampaio; Svetoslav Nanev Slavov; Vagner Fonseca; Vincent Louis Viala |
| EPI_ISL_2697916, EPI_ISL_2697917, EPI_ISL_2697918, EPI_ISL_2697956, EPI_ISL_2697957, EPI_ISL_2697958, EPI_ISL_2697959, EPI_ISL_2697960, EPI_ISL_2697975, EPI_ISL_2697981, EPI_ISL_2697982 | | | |
| see above | CTvacinas | CTvacinas | A.P.; B.L.; Coelho; D.B.; Dorlass; Durigon; E.G.; E.L.; F.G.; Fernandes; Fiorini, A.; Fonseca; G.P.; H.P.; K.L.; L.M.; Lourenco; Magalhaes; Oliveira; Ometto, T.; Peixoto, R.; R.D.; Sato, H.; Scagion; Teixeira, S.; Telezynski; Thomazelli |
| EPI_ISL_2234349, EPI_ISL_3536736, EPI_ISL_3536936 | California Department of Public Health | California Department of Public Health | CDPH IDLB COVIDNet et al |
| EPI_ISL_2837322, EPI_ISL_2837377, EPI_ISL_2837417, EPI_ISL_2837553, EPI_ISL_2837555, EPI_ISL_2837726, EPI_ISL_2837733, EPI_ISL_2837737, EPI_ISL_2837789, EPI_ISL_2837860, EPI_ISL_2837896, EPI_ISL_2837898, EPI_ISL_2837908, EPI_ISL_2837909 | | | |
| see above | California Department of Public Health Valencia Branch Laboratory (CDPH VBL) | California Department of Public Health | CDPH-COVIDNet |
| EPI_ISL_1904855 | Canterbury Health Laboratories | Institute of Environmental Science and Research (ESR) | Anja Werno; Antje van der Linden; Arlo Upton; Chris Mansell; David Hammer; Dragana Drinkovic; Erasmus Smit; Gary McAuliffe; Hana Sofia Andersson; Hermes Perez; James Ussher; Jill Sherwood; Jing Wang; Joep de Ligt; Josh Freeman; Julia Howard; Juliet Elvy; Lauren Jelly; Mary DeAlmeida; Matt Blakiston; Matt Storey; Matthew Rogers; Max Bloomfield; Michael Addidle; Michelle Balm; Muhammad Faisal; Nikki Freed; Olin Silander; Olivia Stroeven; Rachel Boyle; Sally Roberts; SallyAnn Harbison; Sarah Jefferies; Sharmini Muttaiyah; Susan Morpeth; Susan Taylor; Timothy Blackmore; Vani Sathyendran; Veronica Playle; Virginia Hope; Xiaoyun Ren |
| EPI_ISL_1752638 | Casa de Caridade Sao Vicente de Paulo Cajuru | Instituto Adolfo Lutz, Interdiciplinary Procedures Center, Strategic Laboratory | Caio Vinicius Dias Lopes; Claudia Regina Gonçalves; Claudio Tavares Sacchi; Erica Valessa Ramos Gomes; Karoline Rodrigues Campos; Katia Correa de Oliveira Santos; Leonardo Jose Tadeu de Araujo |
| EPI_ISL_7507915 | Cedars-Sinai Medical Center, Molecular Pathology Laboratory of Department of Pathology & Laboratory Medicine and Genomic Core | Cedars-Sinai Medical Center, Molecular Pathology Laboratory of Department of Pathology & Laboratory Medicine and Genomic Core | Brian Davis; Eric Vail; Jasmine T Plummer; Jorge Mario Sincuir Martinez; Stephanie Chen; Wenjuan Zhang |

| | | | |
|---|---|---|---|
| EPI_ISL_1843831 | Centogene; Dr. Bauer Laboratoriums GmbH | Robert Koch Institute | |
| EPI_ISL_2502414, EPI_ISL_2502457, EPI_ISL_2502458, EPI_ISL_2502459, EPI_ISL_2502460, EPI_ISL_2502461, EPI_ISL_2502462 | | | |
| see above | Central Laboratory, Bureau of Public Health (BOG) and Academic Hospital Paramaribo | Erasmus Medical Center | Bas B Oude Munnink; Cherise Beek; Consuella Partowidjojo; Dion Gajadin; Ed PF IJzerman; Emmanuelle Munger; Gary Gummels; Ingrid SK Krishnadath; Lycke Woittiez; Marion PG Koopmans; Mireille Van de Veer; Phyllis Pinas; Princes Wongsowidjojo; Radjesh Ori; Ranisha Doerbalie; Rohma Banwari; Soeradj Harkisoen; Stephen Vreden; Tilotmadebie Ramlal; Verne Nanhoe |
| EPI_ISL_3266078, EPI_ISL_3266081, EPI_ISL_3266089, EPI_ISL_3266090, EPI_ISL_3266092, EPI_ISL_3266094, EPI_ISL_3266106 | | | |
| see above | Central Public Health Laboratory - LACEN -Bahia, Salvador, Brazil | Central Public Health Laboratory - LACEN -Bahia, Salvador, Brazil | Arabela Leal; Breno Dominguez; Felicidade Pereira; Jaqueline Gomes; Luciana Oliveira; Luiz Alcantara; Marcela Gómez; Marta Giovanetti; Patrícia Cajado; Stephane Tosta; Vagner Fonseca; Vanessa Nardy |
| EPI_ISL_5800366, EPI_ISL_5800367, EPI_ISL_5800369, EPI_ISL_5800378, EPI_ISL_5800382, EPI_ISL_5800387, EPI_ISL_5800389, EPI_ISL_5800390, EPI_ISL_5800391 | | | |
| see above | Centro De Apoio Epidemiologico | Instituto Butantan | Antonio Jorge Martins; Claudia Renata dos Santos Barros; David Schlesinger; Debora Botequio Moretti; Dimas Tadeu Covas; Elaine Cristina Marqueze; Elaine Vieira Santos; Evandra Strazza Rodrigues; Heidge Fukumasu; Jayme Augusto de Souza-Neto; José Salvatore Leister Patané; Luiz Alcantara; Luiz Lehmann Coutinho; Maria Carolina Elias; Maurício Lacerda Nogueira; Rafael dos Santos Bezerra; Raul Machado Neto; Rejane Maria Tommasini Grotto; Ricardo Haddad; Sandra Coccuzzo Sampaio Vessoni; Simone Kashima; Svetoslav Nanev Slavov; Vincent Louis Viala |
| EPI_ISL_5800133, EPI_ISL_5800139, EPI_ISL_5800141 | Centro De Saude De Maracai | Instituto Butantan | Antonio Jorge Martins; Claudia Renata dos Santos Barros; David Schlesinger; Debora Botequio Moretti; Dimas Tadeu Covas; Elaine Cristina Marqueze; Elaine Vieira Santos; Evandra Strazza Rodrigues; Heidge Fukumasu; Jayme Augusto de Souza-Neto; José Salvatore Leister Patané; Luiz Alcantara; Luiz Lehmann Coutinho; Maria Carolina Elias; Maurício Lacerda Nogueira; Rafael dos Santos Bezerra; Raul Machado Neto; Rejane Maria Tommasini Grotto; Ricardo Haddad; Sandra Coccuzzo Sampaio Vessoni; Simone Kashima; Svetoslav Nanev Slavov; Vincent Louis Viala |
| EPI_ISL_5799710 | Centro De Saude Dr Mario Dias De Aguiar Capivari | Instituto Butantan | Antonio Jorge Martins; Claudia Renata dos Santos Barros; David Schlesinger; Debora Botequio Moretti; Dimas Tadeu Covas; Elaine Cristina Marqueze; Elaine Vieira Santos; Evandra Strazza Rodrigues; Heidge Fukumasu; Jayme Augusto de Souza-Neto; José Salvatore Leister Patané; Luiz Alcantara; Luiz Lehmann Coutinho; Maria Carolina Elias; Maurício Lacerda Nogueira; Rafael dos Santos Bezerra; Raul Machado Neto; Rejane Maria Tommasini Grotto; Ricardo Haddad; Sandra Coccuzzo Sampaio Vessoni; Simone Kashima; Svetoslav Nanev Slavov; Vincent Louis Viala |
| EPI_ISL_5800184, EPI_ISL_5800208, EPI_ISL_5800242, EPI_ISL_5800250, EPI_ISL_5800252, EPI_ISL_5800278 | Centro De Saude Dr Renato De Carvalho Ribeiro Angatuba | Instituto Butantan | Antonio Jorge Martins; Claudia Renata dos Santos Barros; David Schlesinger; Debora Botequio Moretti; Dimas Tadeu Covas; Elaine Cristina Marqueze; Elaine Vieira Santos; Evandra Strazza Rodrigues; Heidge Fukumasu; Jayme Augusto de Souza-Neto; José Salvatore Leister Patané; Luiz Alcantara; Luiz Lehmann Coutinho; Maria Carolina Elias; Maurício Lacerda Nogueira; Rafael dos Santos Bezerra; Raul Machado Neto; Rejane Maria Tommasini Grotto; Ricardo Haddad; Sandra Coccuzzo Sampaio Vessoni; Simone Kashima; Svetoslav Nanev Slavov; Vincent Louis Viala |
| EPI_ISL_5799966, EPI_ISL_5800006 | Centro De Saude Esf Iv Zona Rural Domingos De S Sjriopardo | Instituto Butantan | Antonio Jorge Martins; Claudia Renata dos Santos Barros; David Schlesinger; Debora Botequio Moretti; Dimas Tadeu Covas; Elaine Cristina Marqueze; Elaine Vieira Santos; Evandra Strazza Rodrigues; Heidge Fukumasu; Jayme Augusto de Souza-Neto; José Salvatore Leister Patané; Luiz Alcantara; Luiz Lehmann Coutinho; Maria Carolina Elias; Maurício Lacerda Nogueira; Rafael dos Santos Bezerra; Raul Machado Neto; Rejane Maria Tommasini Grotto; Ricardo Haddad; Sandra Coccuzzo Sampaio Vessoni; Simone Kashima; Svetoslav Nanev Slavov; Vincent Louis Viala |
| EPI_ISL_5800341 | Centro De Saude I Avare | Instituto Butantan | Antonio Jorge Martins; Claudia Renata dos Santos Barros; David Schlesinger; Debora Botequio Moretti; Dimas Tadeu Covas; Elaine Cristina Marqueze; Elaine Vieira Santos; Evandra Strazza Rodrigues; Heidge Fukumasu; Jayme Augusto de Souza-Neto; José Salvatore Leister Patané; Luiz Alcantara; Luiz Lehmann Coutinho; Maria Carolina Elias; Maurício Lacerda Nogueira; Rafael dos Santos Bezerra; Raul Machado Neto; Rejane Maria Tommasini Grotto; Ricardo Haddad; Sandra Coccuzzo Sampaio Vessoni; Simone Kashima; Svetoslav Nanev Slavov; Vincent Louis Viala |
| EPI_ISL_5800350 | Centro De Saude Ii Dr Arcy Bandeira Ibiuna Sp | Instituto Butantan | Antonio Jorge Martins; Claudia Renata dos Santos Barros; David Schlesinger; Debora Botequio Moretti; Dimas Tadeu Covas; Elaine Cristina Marqueze; Elaine Vieira Santos; Evandra Strazza Rodrigues; Heidge Fukumasu; Jayme Augusto de Souza-Neto; José Salvatore Leister Patané; Luiz Alcantara; Luiz Lehmann Coutinho; Maria Carolina Elias; Maurício Lacerda Nogueira; Rafael dos Santos Bezerra; Raul Machado Neto; Rejane Maria Tommasini Grotto; Ricardo Haddad; Sandra Coccuzzo Sampaio Vessoni; Simone Kashima; Svetoslav Nanev Slavov; Vincent Louis Viala |
| EPI_ISL_5800339, EPI_ISL_5800346 | Centro De Saude Ii Mairinque Mairinque | Instituto Butantan | Antonio Jorge Martins; Claudia Renata dos Santos Barros; David Schlesinger; Debora Botequio Moretti; Dimas Tadeu Covas; Elaine Cristina Marqueze; Elaine Vieira Santos; Evandra Strazza Rodrigues; Heidge Fukumasu; Jayme Augusto de Souza-Neto; José Salvatore Leister Patané; Luiz Alcantara; Luiz Lehmann Coutinho; Maria Carolina Elias; Maurício Lacerda Nogueira; Rafael dos Santos Bezerra; Raul Machado Neto; Rejane Maria Tommasini Grotto; Ricardo Haddad; Sandra Coccuzzo Sampaio Vessoni; Simone Kashima; Svetoslav Nanev Slavov; Vincent Louis Viala |
| EPI_ISL_5800335, EPI_ISL_5800344, EPI_ISL_5800345 | Centro De Saude Ii Sao Miguel Arcanjo | Instituto Butantan | Antonio Jorge Martins; Claudia Renata dos Santos Barros; David Schlesinger; Debora Botequio Moretti; Dimas Tadeu Covas; Elaine Cristina Marqueze; Elaine Vieira Santos; Evandra Strazza Rodrigues; Heidge Fukumasu; Jayme Augusto de Souza-Neto; José Salvatore Leister Patané; Luiz Alcantara; Luiz Lehmann Coutinho; Maria Carolina Elias; Maurício Lacerda Nogueira; Rafael dos Santos Bezerra; Raul Machado Neto; Rejane Maria Tommasini Grotto; Ricardo Haddad; Sandra Coccuzzo Sampaio Vessoni; Simone Kashima; Svetoslav Nanev Slavov; Vincent Louis Viala |
| EPI_ISL_5800307, EPI_ISL_5800340 | Centro De Saude Sao Roque Dr Jose Carvalho Brito | Instituto Butantan | Antonio Jorge Martins; Claudia Renata dos Santos Barros; David Schlesinger; Debora Botequio Moretti; Dimas Tadeu Covas; Elaine Cristina Marqueze; Elaine Vieira Santos; Evandra Strazza Rodrigues; Heidge Fukumasu; Jayme Augusto de Souza-Neto; José Salvatore Leister Patané; Luiz Alcantara; Luiz Lehmann Coutinho; Maria Carolina Elias; Maurício Lacerda Nogueira; Rafael dos Santos Bezerra; Raul Machado Neto; Rejane Maria Tommasini Grotto; Ricardo Haddad; Sandra Coccuzzo Sampaio Vessoni; Simone Kashima; Svetoslav Nanev Slavov; Vincent Louis Viala |
| EPI_ISL_5799693, EPI_ISL_5799694 | Centro Medico Dr Nelson Salome De Conchal | Instituto Butantan | Antonio Jorge Martins; Claudia Renata dos Santos Barros; David Schlesinger; Debora Botequio Moretti; Dimas Tadeu Covas; Elaine Cristina Marqueze; Elaine Vieira Santos; Evandra Strazza Rodrigues; Heidge Fukumasu; Jayme Augusto de Souza-Neto; José Salvatore Leister Patané; Luiz Alcantara; Luiz Lehmann Coutinho; Maria Carolina Elias; Maurício Lacerda Nogueira; Rafael dos Santos Bezerra; Raul Machado Neto; Rejane Maria Tommasini Grotto; Ricardo Haddad; Sandra Coccuzzo Sampaio Vessoni; Simone Kashima; Svetoslav Nanev Slavov; Vincent Louis Viala |
| EPI_ISL_2612341, EPI_ISL_2612342, EPI_ISL_2612344, EPI_ISL_5926837, EPI_ISL_5926838, EPI_ISL_5926847, EPI_ISL_5926848, EPI_ISL_5926849, EPI_ISL_5926850, EPI_ISL_5926851, EPI_ISL_5926852, EPI_ISL_5926853, EPI_ISL_5926854, EPI_ISL_5926855, EPI_ISL_5926856, EPI_ISL_5926857, EPI_ISL_5926858, EPI_ISL_5926859, EPI_ISL_5926860 | | | |
| see above | Centro de Infectologia Charles Mérieux/ Laboratório Rodolphe Mérieux, FUNDHACRE | Bioinformatics Laboratory / LNCC | Alessandra P Lamarca; Alexandra L Gerber; Aline de Freitas Souza; Ana Paula de C Guimarães; Ana Tereza R Vasconcelos; Andreas Stocker; Cirley Maria de Oliveira Lobato; Douglas Terra Machado; Janaína Mazaro; Janete Taynã Nascimento Rodrigues; Luiz Fellype Alves de Souza; Luiz G P de Almeida; Ronaldo da Silva F Jr; Rutilene B. Souza |
| EPI_ISL_3242632 | Clinica INDISA | "Facultad de Ciencias de la Vida, UNAB" | "Claudio Meneses; Ariel Orellana"; Claudio Olmos; Daniel Leon; Dayan Sanhueza; Eduardo Castro; Gonzalo Campaña; Macarena Bastias; Paola Pidal; Ricardo Yusta; Sebastian Wolter; Susana Saez; Víctor Monreal; Waldo Diaz |
| EPI_ISL_1965735, EPI_ISL_2500799 | Clinical Microbiology Laboratory, NewYork Presbyterian Hospital/Columbia University Irving Medical Center | Uhlemann Laboratory, Columbia University Irving Medical Center | Anne Kelley; Anne-Catrin Uhlemann; Maya Tagliavia; Medini K. Annavajhala |
| EPI_ISL_1664592 | Clinical Molecular Microbiology Laboratory, UNC Hospitals | Jeremy Wang | Alexander Rubinsteyn; Colleen Rice; Corbin Jones; Jason Smedberg; Jeremy Wang; Melissa Miller; Robert Hagan; Shawn Hawken |
| EPI_ISL_4300676 | Color Genomics | Chiu Laboratory, University of California, San Francisco | Alicia Sotomayor-Gonzalez; Alicia Zhou; Baolin Wang; Charles Chiu; Daniel Wong; Erika Torres; Madyson Wanner; Noah Brazer; Scott Topper; Venice Servellita; Yueyuan Zhang |
| EPI_ISL_1665871, EPI_ISL_1665875, EPI_ISL_1665881, EPI_ISL_2277163, EPI_ISL_6211359 | Colorado Department of Public Health and Environment | Colorado Department of Public Health and Environment | Alexandria Rossheim; Diana Ir; Emily A. Travanty; Laura Bankers; Mandy Waters; Michael Martin; Molly C. Hetherington-Rauth; Sarah Elizabeth Totten; Shannon Ely; Shannon R. Matzinger |
| EPI_ISL_1755861, EPI_ISL_1855286 | Columbia University Irving Medical Center | Wadsworth Center, New York State Department of Health | Alexis Russell; Catharine Prussing; Daryl M. Lamson; Erasmus Schneider; Erica Lasek-Nesselquist; John Kelly; Jonathan Plitnick; Kirsten St. George; Matthew Shudt; Melissa A Leisner; Navjot Singh |
| EPI_ISL_5802294 | Complexo Hospitalar Ouro Verde De Campinas | Instituto Butantan | Antonio Jorge Martins; Claudia Renata dos Santos Barros; David Schlesinger; Debora Botequio Moretti; Dimas Tadeu Covas; Elaine Cristina Marqueze; Elaine Vieira Santos; Evandra Strazza Rodrigues; Heidge Fukumasu; Jayme Augusto de Souza-Neto; José Salvatore Leister Patané; Luiz Alcantara; Luiz Lehmann Coutinho; Maria Carolina Elias; Maurício Lacerda Nogueira; Rafael dos Santos Bezerra; Raul Machado Neto; Rejane Maria Tommasini Grotto; Ricardo Haddad; Sandra Coccuzzo Sampaio Vessoni; Simone Kashima; Svetoslav Nanev Slavov; Vincent Louis Viala |
| EPI_ISL_2034753 | Connecticut DPH | Yale Center for Genomic Analysis | Anderson Brito; Annie Watkins; Anthony Muyombwe; Brooke Sullivan; Chaney Kalinich; Chantal Vogels; Curt Scharfe; Irina Tikhonova; Isabel Ott; Jafar Razeq; Jessica Rothman; Joseph Fauver; Kaya Bilguvar; Mallery Breban; Mary Petrone; Nathan Grubaugh; Randy Downing; Shrikant Mane; Stephen M. Bart; Tara Alpert |
| EPI_ISL_2497667 | Contra Costa County Public Health Lab | Chan-Zuckerberg Biohub | CZB Cliahub Consortium |
| EPI_ISL_5799696 | Coordenadoria Municipal De Saude De Iracemapolis | Instituto Butantan | Antonio Jorge Martins; Claudia Renata dos Santos Barros; David Schlesinger; Debora Botequio Moretti; Dimas Tadeu Covas; Elaine Cristina Marqueze; Elaine Vieira Santos; Evandra Strazza Rodrigues; Heidge Fukumasu; Jayme Augusto de Souza-Neto; José Salvatore Leister Patané; Luiz Alcantara; Luiz Lehmann Coutinho; Maria Carolina Elias; Maurício Lacerda Nogueira; Rafael dos Santos Bezerra; Raul Machado Neto; Rejane Maria Tommasini Grotto; Ricardo Haddad; Sandra Coccuzzo Sampaio Vessoni; Simone Kashima; Svetoslav Nanev Slavov; Vincent Louis Viala |
| EPI_ISL_2102747, EPI_ISL_2102830, EPI_ISL_2102890, EPI_ISL_2102893 | County of San Luis Obispo Public Health Laboratory | Chan-Zuckerberg Biohub | CZB Cliahub Consortium |
| EPI_ISL_5800200 | Cs De Balsamo | Instituto Butantan | Antonio Jorge Martins; Claudia Renata dos Santos Barros; David Schlesinger; Debora Botequio Moretti; Dimas Tadeu Covas; Elaine Cristina Marqueze; Elaine Vieira Santos; Evandra Strazza Rodrigues; Heidge Fukumasu; Jayme Augusto de Souza-Neto; José Salvatore Leister Patané; Luiz Alcantara; Luiz Lehmann Coutinho; Maria Carolina Elias; Maurício Lacerda Nogueira; Rafael dos Santos Bezerra; Raul Machado Neto; Rejane Maria Tommasini Grotto; Ricardo Haddad; Sandra Coccuzzo Sampaio Vessoni; Simone Kashima; Svetoslav Nanev Slavov; Vincent Louis Viala |
| EPI_ISL_5800195, EPI_ISL_5800196 | Cs De Catigua | Instituto Butantan | Antonio Jorge Martins; Claudia Renata dos Santos Barros; David Schlesinger; Debora Botequio Moretti; Dimas Tadeu Covas; Elaine Cristina Marqueze; Elaine Vieira Santos; Evandra Strazza Rodrigues; Heidge Fukumasu; Jayme Augusto de Souza-Neto; José Salvatore Leister Patané; Luiz Alcantara; Luiz Lehmann Coutinho; Maria Carolina Elias; Maurício Lacerda Nogueira; Rafael dos Santos Bezerra; Raul Machado Neto; Rejane Maria Tommasini Grotto; Ricardo Haddad; Sandra Coccuzzo Sampaio Vessoni; Simone Kashima; Svetoslav Nanev Slavov; Vincent Louis Viala |
| EPI_ISL_5800185, EPI_ISL_5800186, EPI_ISL_5800188, EPI_ISL_5800193, EPI_ISL_5800198, EPI_ISL_5800199 | Cs De Monte Aprazivel | Instituto Butantan | Antonio Jorge Martins; Claudia Renata dos Santos Barros; David Schlesinger; Debora Botequio Moretti; Dimas Tadeu Covas; Elaine Cristina Marqueze; Elaine Vieira Santos; Evandra Strazza Rodrigues; Heidge Fukumasu; Jayme Augusto de Souza-Neto; José Salvatore Leister Patané; Luiz Alcantara; Luiz Lehmann Coutinho; Maria Carolina Elias; Maurício Lacerda Nogueira; Rafael dos Santos Bezerra; Raul Machado Neto; Rejane Maria Tommasini Grotto; Ricardo Haddad; Sandra Coccuzzo Sampaio Vessoni; Simone Kashima; Svetoslav Nanev Slavov; Vincent Louis Viala |
| EPI_ISL_5800190, EPI_ISL_5800191 | Cs De Neves Paulista | Instituto Butantan | Antonio Jorge Martins; Claudia Renata dos Santos Barros; David Schlesinger; Debora Botequio Moretti; Dimas Tadeu Covas; Elaine Cristina Marqueze; Elaine Vieira Santos; Evandra Strazza Rodrigues; Heidge Fukumasu; Jayme Augusto de Souza-Neto; José Salvatore Leister Patané; Luiz Alcantara; Luiz Lehmann Coutinho; Maria Carolina Elias; Maurício Lacerda Nogueira; Rafael dos Santos Bezerra; Raul Machado Neto; Rejane Maria Tommasini Grotto; Ricardo Haddad; Sandra Coccuzzo Sampaio Vessoni; Simone Kashima; Svetoslav Nanev Slavov; Vincent Louis Viala |
| EPI_ISL_5800178 | Cs De Orindiuva | Instituto Butantan | Antonio Jorge Martins; Claudia Renata dos Santos Barros; David Schlesinger; Debora Botequio Moretti; Dimas Tadeu Covas; Elaine Cristina Marqueze; Elaine Vieira Santos; Evandra Strazza Rodrigues; Heidge Fukumasu; Jayme Augusto de Souza-Neto; José Salvatore Leister Patané; Luiz Alcantara; Luiz Lehmann Coutinho; Maria Carolina Elias; Maurício Lacerda Nogueira; Rafael dos Santos Bezerra; Raul Machado Neto; Rejane Maria Tommasini Grotto; Ricardo Haddad; Sandra Coccuzzo Sampaio Vessoni; Simone Kashima; Svetoslav Nanev Slavov; Vincent Louis Viala |
| EPI_ISL_5800189 | Cs De Santa Rita Doeste | Instituto Butantan | Antonio Jorge Martins; Claudia Renata dos Santos Barros; David Schlesinger; Debora Botequio Moretti; Dimas Tadeu Covas; Elaine Cristina Marqueze; Elaine Vieira Santos; Evandra Strazza Rodrigues; Heidge Fukumasu; Jayme Augusto de Souza-Neto; José Salvatore Leister Patané; Luiz Alcantara; Luiz Lehmann Coutinho; Maria Carolina Elias; Maurício Lacerda Nogueira; Rafael dos Santos Bezerra; Raul Machado Neto; Rejane Maria Tommasini Grotto; Ricardo Haddad; Sandra Coccuzzo Sampaio Vessoni; Simone Kashima; Svetoslav Nanev Slavov; Vincent Louis Viala |
| EPI_ISL_5799923, | Cs I De Ourinhos | Instituto Butantan | Antonio Jorge Martins; Claudia Renata dos Santos Barros; David Schlesinger; Debora Botequio Moretti; Dimas Tadeu Covas; Elaine Cristina Marqueze; Elaine Vieira Santos; Evandra Strazza Rodrigues; Heidge Fukumasu; Jayme Augusto de Souza-Neto; José Salvatore Leister Patané; Luiz Alcantara; Luiz Lehmann Coutinho; |

| | | | |
|---|---|---|---|
| EPI_ISL_5799931 | | | Maria Carolina Elias; Maurício Lacerda Nogueira; Rafael dos Santos Bezerra; Raul Machado Neto; Rejane Maria Tommasini Grotto; Ricardo Haddad; Sandra Coccuzzo Sampaio Vessoni; Simone Kashima; Svetoslav Nanev Slavov; Vincent Louis Viala |
| EPI_ISL_5800151 | Cs Ii Dr Jahyr De Paula Ribeiro Guara | Instituto Butantan | Antonio Jorge Martins; Claudia Renata dos Santos Barros; David Schlesinger; Debora Botequio Moretti; Dimas Tadeu Covas; Elaine Cristina Marqueze; Elaine Vieira Santos; Evandra Strazza Rodrigues; Heidge Fukumasu; Jayme Augusto de Souza-Neto; José Salvatore Leister Patané; Luiz Alcantara; Luiz Lehmann Coutinho; Maria Carolina Elias; Maurício Lacerda Nogueira; Rafael dos Santos Bezerra; Raul Machado Neto; Rejane Maria Tommasini Grotto; Ricardo Haddad; Sandra Coccuzzo Sampaio Vessoni; Simone Kashima; Svetoslav Nanev Slavov; Vincent Louis Viala |
| EPI_ISL_5799868 | Cs Ii Dr Miguel Vitaliano Orlandia | Instituto Butantan | Antonio Jorge Martins; Claudia Renata dos Santos Barros; David Schlesinger; Debora Botequio Moretti; Dimas Tadeu Covas; Elaine Cristina Marqueze; Elaine Vieira Santos; Evandra Strazza Rodrigues; Heidge Fukumasu; Jayme Augusto de Souza-Neto; José Salvatore Leister Patané; Luiz Alcantara; Luiz Lehmann Coutinho; Maria Carolina Elias; Maurício Lacerda Nogueira; Rafael dos Santos Bezerra; Raul Machado Neto; Rejane Maria Tommasini Grotto; Ricardo Haddad; Sandra Coccuzzo Sampaio Vessoni; Simone Kashima; Svetoslav Nanev Slavov; Vincent Louis Viala |
| EPI_ISL_5799892, EPI_ISL_5799897, EPI_ISL_5799906, EPI_ISL_5799918, EPI_ISL_5799925, EPI_ISL_5799943, EPI_ISL_5799945, EPI_ISL_5800132, EPI_ISL_5800135, EPI_ISL_5800137, EPI_ISL_5800142, EPI_ISL_5800143, EPI_ISL_5800144, EPI_ISL_5800145, EPI_ISL_5800146, EPI_ISL_5800147, EPI_ISL_5800148, EPI_ISL_5800149, EPI_ISL_5800150 | | | |
| see above | Cs Ii Egidio Brunhara Morro Agudo | Instituto Butantan | Antonio Jorge Martins; Claudia Renata dos Santos Barros; David Schlesinger; Debora Botequio Moretti; Dimas Tadeu Covas; Elaine Cristina Marqueze; Elaine Vieira Santos; Evandra Strazza Rodrigues; Heidge Fukumasu; Jayme Augusto de Souza-Neto; José Salvatore Leister Patané; Luiz Alcantara; Luiz Lehmann Coutinho; Maria Carolina Elias; Maurício Lacerda Nogueira; Rafael dos Santos Bezerra; Raul Machado Neto; Rejane Maria Tommasini Grotto; Ricardo Haddad; Sandra Coccuzzo Sampaio Vessoni; Simone Kashima; Svetoslav Nanev Slavov; Vincent Louis Viala |
| EPI_ISL_5800161, EPI_ISL_5800164, EPI_ISL_5800165, EPI_ISL_5800169, EPI_ISL_5800172, EPI_ISL_5800173, EPI_ISL_5800174, EPI_ISL_5800175 | | | |
| see above | Csii Dr Washington Luis M Rodrigues Da Silva Pitangueiras | Instituto Butantan | Antonio Jorge Martins; Claudia Renata dos Santos Barros; David Schlesinger; Debora Botequio Moretti; Dimas Tadeu Covas; Elaine Cristina Marqueze; Elaine Vieira Santos; Evandra Strazza Rodrigues; Heidge Fukumasu; Jayme Augusto de Souza-Neto; José Salvatore Leister Patané; Luiz Alcantara; Luiz Lehmann Coutinho; Maria Carolina Elias; Maurício Lacerda Nogueira; Rafael dos Santos Bezerra; Raul Machado Neto; Rejane Maria Tommasini Grotto; Ricardo Haddad; Sandra Coccuzzo Sampaio Vessoni; Simone Kashima; Svetoslav Nanev Slavov; Vincent Louis Viala |
| EPI_ISL_2507253, EPI_ISL_2755136, EPI_ISL_2755138, EPI_ISL_2755559, EPI_ISL_2755579, EPI_ISL_2755580, EPI_ISL_2755584 | | | |
| see above | Curative Labs | Curative Labs | Elias L. Salfati; Eugenia Khorosheva; George Way; J.Cesar Ignacio-Espinoza; Janet Chen; Mikhail Hanewich-Hollatz; Nabjot Sandhu; Sophia Quasem; Vladimir Slepnev; Zhiyi Xie |
| EPI_ISL_3452406 | Curative Labs | New Mexico Department of Health Scientific Laboratory | Anastacia Griego-Fisher; D'eldra Malone; Ellie Johnson; Jennifer Benoit; Justin Griego; Linda Salzar; Michael Sam; Monica Manginell |
| EPI_ISL_1798744, EPI_ISL_1798759, EPI_ISL_1798762 | DE Public Health Laboratory | Centers for Disease Control and Prevention Division of Viral Diseases, Pathogen Discovery | Alison Laufer Halpin; Ben L. Rambo-Martin; Clinton R. Paden; Dakota Howard; Darlene Wagner; Dave Wentworth; Dhwani Batra; Jasmine Padilla; Justin Lee; Katie Dillon; Krista Queen; Kristen Knipe; Kristine Lacek; Mark Burroughs; Matthew Schmerer; Mili Sheth; Peter Cook; Sam Shepard; Sarah Nobles; Shoshona Le; Suxiang Tong; Vivien Dugan; Yvette Unoarumhi |
| EPI_ISL_1967197 | DEPARTAMENTO DE SAUDE COLETIVA | Instituto Butantan / ESALQ-USP (Piracicaba) | Antonio Jorge Martins; Bianca Cechetto Carlos. Mendelics: Bibiana Santos; Claudia Renata dos Santos Barros; Cíntia Bittar; David Schlesinger. Hemocentro Ribeirão Preto: Simone Kashima; Debora Botequio Moretti; Elaine Cristina Marqueze; Elaine Vieira dos Santos; Elisangela Chicaroni Mattos; Erika Freitas; Evandra Strazza Rodrigues; Felipe Allan da Silva da Costa; Flavia Aburjaile; Fábio Sossai Possebon; Guilherme Campos; Guilherme Targino Valente; Heidge Fukumasu. USP-Botucatu: Rejane Maria Tommasini Grotto; Helena Lage Ferreira; Instituto Butantan: Dimas Tadeu Covas; Jardelina de Souza Todao Bernardino; Jayme A. Souza-Neto; Jessika Cristina Chagas Lesbon; Jorge A. Petroli Marchesi; José Salvatore Leister Patané; João Paulo Kitajima; João Pessoa Araújo Jr.; Leila Sabrina Ullmann; Loyze Paola Oliveira de Lima; Luiz Aurelio de Campos Crispin. Centro de Genômica Funcional da ESALQ: Luiz Lehmann Coutinho; Luiz Carlos Junior de Alcantara; Lívia Sacchetto; Maisa C. Pereira Parra; Maria Carolina Elias; Marta Giovanetti; Marília Moraes; Maurício Lacerda Nogueira. Prefeitura de Sao Paulo: Melissa Palmieri.; Patricia Akemi Assato; Paula Rahal; Paulo Inacio da Costa; Rafael dos Santos Bezerra; Raquel de Lello Rocha Campos Cassano. NGS Soluções Genômicas: Pilar Drummond Sampaio Corrêa Mariani. FZEA-USP Pirassununga: Mirele Daiana Poleti; Raul Machado Neto; Ricardo Augusto Brassaloti; Ricardo Haddad; Rodrigo Tocantins Calado. FAMERP-SJRP: Cecília Artico Banho; Sandra Coccuzzo Sampaio; Svetoslav Nanev Slavov; Vagner Fonseca; Vincent Louis Viala |
| EPI_ISL_1647119, EPI_ISL_1647164, EPI_ISL_1647169, EPI_ISL_1707558 | DIP. PREV. AVEZZANO SERVIZIO DI IGIENE EPIDEMIOLOGIA E SANITA' PUBBLICA | Istituto Zooprofilattico Sperimentale dell'Abruzzo e Molise "G. Caporale" | Ancora M; Calistri P; Cammà C; Caporale M; Curini V; Delli Compagni E; Di Domenico M; Di Lollo Valeria; Di Pasquale A; Lorusso A; Mangone I; Marcacci M; Puglia I; Rinaldi A; Savini G; Scialabba S |
| EPI_ISL_2362237 | DIRETORIA MUNICIPAL DE SAUDE DE ENGENHEIRO COELHO | Instituto Butantan / FZEA-USP-Pirassununga | Antonio Jorge Martins; Claudia Renata dos Santos Barros; David Schlesinger; Debora Botequio Moretti; Dimas Tadeu Covas; Elaine Cristina Marqueze; Elaine Vieira Santos; Evandra Strazza Rodrigues; Heidge Fukumasu; Jayme Augusto de Souza-Neto; José Salvatore Leister Patané; Luiz Alcantara; Luiz Lehmann Coutinho; Maria Carolina Elias; Maurício Lacerda Nogueira; Rafael dos Santos Bezerra; Raul Machado Neto; Rejane Maria Tommasini Grotto; Ricardo Haddad; Sandra Coccuzzo Sampaio Vessoni; Simone Kashima; Svetoslav Nanev Slavov; Vincent Louis Viala |
| EPI_ISL_5104491, EPI_ISL_5104493, EPI_ISL_5104500, EPI_ISL_5104504, EPI_ISL_5104506 | DNAGYN | LGBio (Laboratorio de Genetica & Biodiversidade) | Alex Honda Bernardes; Amanda Alves de Melo; Aparecido Divino da Cruz; Cintia Pelegrineti Targueta de Azevedo Brito; Daniela de Melo e Silva; Elisangela de Paula Silveira Lacerda; Juliana Santana de Curcio; Luiz Augusto Pereira; Marc Alexandre Duarte Gigonzac; Mariana Pires de Campos Telles; Ramilla dos Santos Braga; Renata de Oliveira Dias; Rhewter Nunes; Thais Cidália Vieira Gigonzac; Thais Guimarães Castro; Thays Millena Alves Pedroso |
| EPI_ISL_1804944, EPI_ISL_2262606 | DOHMH PHL | New York City Public Health Laboratory | Jade Wang; et al. |
| EPI_ISL_1909020, EPI_ISL_1909076 | DPHL | Delaware Public Health Lab | Rebecca Savage |
| EPI_ISL_1662688, EPI_ISL_1662689, EPI_ISL_1662690 | Delaware Public Health Lab | Delaware Public Health Lab | Rebecca Savage |
| EPI_ISL_5800282 | Departamento De Saude Coletiva | Instituto Butantan | Antonio Jorge Martins; Claudia Renata dos Santos Barros; David Schlesinger; Debora Botequio Moretti; Dimas Tadeu Covas; Elaine Cristina Marqueze; Elaine Vieira Santos; Evandra Strazza Rodrigues; Heidge Fukumasu; Jayme Augusto de Souza-Neto; José Salvatore Leister Patané; Luiz Alcantara; Luiz Lehmann Coutinho; Maria Carolina Elias; Maurício Lacerda Nogueira; Rafael dos Santos Bezerra; Raul Machado Neto; Rejane Maria Tommasini Grotto; Ricardo Haddad; Sandra Coccuzzo Sampaio Vessoni; Simone Kashima; Svetoslav Nanev Slavov; Vincent Louis Viala |
| EPI_ISL_1671954, EPI_ISL_1672226, EPI_ISL_1672241, EPI_ISL_1672242, EPI_ISL_1672243, EPI_ISL_1672247 | Department of Clinical Microbiology | GIGA Medical Genomics | Bouchra Boujemla; Cécile Meex; Keith Durkin; Maria Artesi; Marie-Pierre Hayette; Nathalie Renotte; Pierrette Melin; Raphaël Boreux; Sébastien Bontems; Vincent Bours |
| EPI_ISL_1795670 | Department of Public Health Microbiology Ljubljana, National Laboratory for Health, Environment and Food | Department for Public Health Microbiology Ljubljana, National Laboratory for Health, Environment and Food | José Gonçalves; Katarina Prosenc; Martin Bosilj; Metka Paragi; Natasa Berginc; Tom Koritnik |
| EPI_ISL_1669737, EPI_ISL_1669744, EPI_ISL_1669746, EPI_ISL_1669747, EPI_ISL_2757944, EPI_ISL_2757945 | Department of Virology, Istituto Zooprofilattico Sperimentale del Lazio e della Toscana (IZSLT) | Department of General Diagnostics; Department of Virology; Istituto Zooprofilattico Sperimentale del Lazio e della Toscana (IZSLT) | Alessia Franco; Antonella Cersini; Antonio Battisti.; Elena L. Diaconu; Fabiola Feltrin; Giuseppe Manna; Patricia Alba; Raffaela Conti; Teresa Scicluna; Virginia Carfora |
| EPI_ISL_3570075 | Dr. Risch Ostschweiz AG | Microbiology, Dr. Risch | Dominique Fabien Hilti; Faina Wehrli; Lorenz Risch; Martin Risch; Nadia Wohlwend; Sinem Kas; Thomas Bodmer |
| EPI_ISL_2145418, EPI_ISL_2145472, EPI_ISL_2145473, EPI_ISL_2145475, EPI_ISL_2145483, EPI_ISL_2145589 | Dutch COVID-19 response team | Erasmus Medical Center | Anne van der Linden; Annemiek van der Eijk; Bas Oude Munnink; Corine GeurtsvanKessel; David Nieuwenhuijse; Emmanuelle Munger; Irina Chestakova; Marion Koopmans; Marjan Boter; Reina Sikkema; Richard Molenkamp; on behalf of the Dutch national COVID-19 respo |
| EPI_ISL_1704983, EPI_ISL_1705117, EPI_ISL_1705273, EPI_ISL_1705388, EPI_ISL_1705485, EPI_ISL_1705536, EPI_ISL_1705556, EPI_ISL_1705566, EPI_ISL_1705665, EPI_ISL_1705734, EPI_ISL_1705827, EPI_ISL_1705853, EPI_ISL_1705864, EPI_ISL_1705867, EPI_ISL_1705954, EPI_ISL_1705962, EPI_ISL_1706015, EPI_ISL_1792431, EPI_ISL_1792432, EPI_ISL_1792434, EPI_ISL_1792435, EPI_ISL_1792436, EPI_ISL_1792445, EPI_ISL_1792447, EPI_ISL_1962268, EPI_ISL_1962269 | | | |
| see above | Dutch COVID-19 response team | National Institute for Public Health and the Environment (RIVM) | Adam Meijer; AnneMarie van den Brandt; Annelies Kroneman; Bas van der Veer; Chantal Reusken; Dennis Schmitz; Dirk Eggink; Euníce Then; Florian Zwagemaker; Harry Vennema; James Groot; Jeroen Cremer; Jolienke Hardeman; Karim Hajji; Kim Freriks; Linda van de Nes; Lisa Wijsman; Lynn Aarts; Melissa van Tuil; Robert Kohl; Ryanne Jaarsma; Sanne Bos; Sharon van den Brink; Sjoerd Kuiling; on behalf of the national COVID-19 response team |
| EPI_ISL_1966853 | ESF ROSA ROSSI MAIA | Instituto Butantan / ESALQ-USP (Piracicaba) | Antonio Jorge Martins; Bianca Cechetto Carlos. Mendelics: Bibiana Santos; Claudia Renata dos Santos Barros; Cíntia Bittar; David Schlesinger. Hemocentro Ribeirão Preto: Simone Kashima; Debora Botequio Moretti; Elaine Cristina Marqueze; Elaine Vieira dos Santos; Elisangela Chicaroni Mattos; Erika Freitas; Evandra Strazza Rodrigues; Felipe Allan da Silva da Costa; Flavia Aburjaile; Fábio Sossai Possebon; Guilherme Campos; Guilherme Targino Valente; Heidge Fukumasu. USP-Botucatu: Rejane Maria Tommasini Grotto; Helena Lage Ferreira; Instituto Butantan: Dimas Tadeu Covas; Jardelina de Souza Todao Bernardino; Jayme A. Souza-Neto; Jessika Cristina Chagas Lesbon; Jorge A. Petroli Marchesi; José Salvatore Leister Patané; João Paulo Kitajima; João Pessoa Araújo Jr.; Leila Sabrina Ullmann; Loyze Paola Oliveira de Lima; Luiz Aurelio de Campos Crispin. Centro de Genômica Funcional da ESALQ: Luiz Lehmann Coutinho; Luiz Carlos Junior de Alcantara; Lívia Sacchetto; Maisa C. Pereira Parra; Maria Carolina Elias; Marta Giovanetti; Marília Moraes; Maurício Lacerda Nogueira. Prefeitura de Sao Paulo: Melissa Palmieri.; Patricia Akemi Assato; Paula Rahal; Paulo Inacio da Costa; Rafael dos Santos Bezerra; Raquel de Lello Rocha Campos Cassano. NGS Soluções Genômicas: Pilar Drummond Sampaio Corrêa Mariani. FZEA-USP Pirassununga: Mirele Daiana Poleti; Raul Machado Neto; Ricardo Augusto Brassaloti; Ricardo Haddad; Rodrigo Tocantins Calado. FAMERP-SJRP: Cecília Artico Banho; Sandra Coccuzzo Sampaio; Svetoslav Nanev Slavov; Vagner Fonseca; Vincent Louis Viala |
| EPI_ISL_3215491, EPI_ISL_3215492, EPI_ISL_3215494, EPI_ISL_3215496, EPI_ISL_3215497, EPI_ISL_3215740 | EXCITE Lab | Andersen lab at Scripps Research | Abigail Schnapper; Angela Scioscia; Chip Schooley; Christine Mitchell + SEARCH; David Pride; Fred Wu; Helena Tubb; Natasha Martin Cheryl Anderson; Sawyer Farmer; Sharon Reed; Tommy Valles + SEARCH |
| EPI_ISL_2477643, EPI_ISL_2478296, EPI_ISL_2478297, EPI_ISL_2478298, EPI_ISL_2478299, EPI_ISL_2478302, EPI_ISL_2478305, EPI_ISL_2478310, EPI_ISL_2478317, EPI_ISL_2478319, EPI_ISL_3909805, EPI_ISL_3909807, EPI_ISL_3909808, EPI_ISL_3909810, EPI_ISL_3910025, EPI_ISL_3910040 | | | |
| see above | Edmonton Provincial Lab | Public Health Agency of Canada (PHAC) National Microbiology Laboratory | Buss; Croxen M; Deo A; Dieu P; E; Ferrato C; Gill K; Khan F; Koleva P; Li V; Lloyd C; Lynch T; Ma R; Murphy S; Pabbaraju K; Shokoples S; Thayer J; Tipples G; Whitehouse M; Wong A; Yu C; Zelyas N |
| EPI_ISL_5800124 | Esf Rosa Rossi Maia | Instituto Butantan | Antonio Jorge Martins; Claudia Renata dos Santos Barros; David Schlesinger; Debora Botequio Moretti; Dimas Tadeu Covas; Elaine Cristina Marqueze; Elaine Vieira Santos; Evandra Strazza Rodrigues; Heidge Fukumasu; Jayme Augusto de Souza-Neto; José Salvatore Leister Patané; Luiz Alcantara; Luiz Lehmann Coutinho; Maria Carolina Elias; Maurício Lacerda Nogueira; Rafael dos Santos Bezerra; Raul Machado Neto; Rejane Maria Tommasini Grotto; Ricardo Haddad; Sandra Coccuzzo Sampaio Vessoni; Simone Kashima; Svetoslav Nanev Slavov; Vincent Louis Viala |
| EPI_ISL_1845977, EPI_ISL_1845986, EPI_ISL_1845988, EPI_ISL_1845999, | Eurofins LifeCodexx GmbH | Robert Koch Institute | |

| | | | |
|---|---|---|---|
| EPI_ISL_1846031, EPI_ISL_1846187 | | | |
| EPI_ISL_2008454, EPI_ISL_2008459, EPI_ISL_2008460, EPI_ISL_2008498, EPI_ISL_2008507, EPI_ISL_2008511, EPI_ISL_2008515, EPI_ISL_2008583, EPI_ISL_2008742, EPI_ISL_2836037, EPI_ISL_2836038, EPI_ISL_2836039, EPI_ISL_2836043, EPI_ISL_2836047, EPI_ISL_2836370, EPI_ISL_2836376, EPI_ISL_2836377, EPI_ISL_4219004, EPI_ISL_5866487, EPI_ISL_5925188, EPI_ISL_6049963, EPI_ISL_6050023, EPI_ISL_6050024, EPI_ISL_6050025, EPI_ISL_6050026, EPI_ISL_6050027, EPI_ISL_6050041, EPI_ISL_6050607, EPI_ISL_6366961, EPI_ISL_6366962, EPI_ISL_6367472, EPI_ISL_6367531, EPI_ISL_6565963 | | | |
| see above | Florida Bureau of Public Health Laboratories | Florida Bureau of Public Health Laboratories | Jason Blanton; Jiaqi Li; Namratha Tarigopula; Sarah Schmedes; Tiffany Splatt |
| EPI_ISL_1918991 | Flow Health | Infectious Disease Program, Broad Institute of Harvard and MIT | Adams, G.; B.L.; B.W.; Bauer, M.; Birren; Carter, A.; Chaluvadi, S.; D.J.; DeRuff, K.; Gallagher, G.; Gladden-Young, A.; J.E.; K.J.; Lagerborg, K.; Lemieux; Loreth, C.; MacInnis; Normandin, E.; P.C.; Park; Reilly, S.; Rudy, M.; Siddle; Smole, S.; Tomkins-Tinch, C.; and Sabeti |
| EPI_ISL_2620907, EPI_ISL_2620908 | Fondazione IRCCS Ca' Granda Ospedale Maggiore Policlinico | Fondazione IRCCS Ca' Granda Ospedale Maggiore Policlinico | Ferruccio Ceriotti; Sara Uceda Renteria |
| EPI_ISL_1614589, EPI_ISL_1614614, EPI_ISL_1614667, EPI_ISL_1614680, EPI_ISL_1614698, EPI_ISL_1614707, EPI_ISL_1614734, EPI_ISL_1614747, EPI_ISL_1614751, EPI_ISL_1614753, EPI_ISL_1614755, EPI_ISL_1614780, EPI_ISL_1614787, EPI_ISL_1614801, EPI_ISL_1614806, EPI_ISL_1614841, EPI_ISL_1614847, EPI_ISL_1614867, EPI_ISL_1665945, EPI_ISL_1665958, EPI_ISL_1665984, EPI_ISL_1665991, EPI_ISL_1665992, EPI_ISL_1666057, EPI_ISL_1666081, EPI_ISL_1666085, EPI_ISL_1666088, EPI_ISL_1666100, EPI_ISL_1666101, EPI_ISL_1666102, EPI_ISL_1666112, EPI_ISL_1666117, EPI_ISL_1666224, EPI_ISL_1666240, EPI_ISL_1666274, EPI_ISL_1666309, EPI_ISL_1666320, EPI_ISL_1666322, EPI_ISL_1666323, EPI_ISL_1666324, EPI_ISL_1666325, EPI_ISL_1666326, EPI_ISL_1666329, EPI_ISL_1667118, EPI_ISL_1667119, EPI_ISL_1667176, EPI_ISL_1667177, EPI_ISL_1667185, EPI_ISL_1667186, EPI_ISL_1667199, EPI_ISL_1667202, EPI_ISL_1667203, EPI_ISL_1667221, EPI_ISL_1667226, EPI_ISL_1667242, EPI_ISL_1667245, EPI_ISL_1667277, EPI_ISL_1667284, EPI_ISL_1667289, EPI_ISL_1667290, EPI_ISL_1667293, EPI_ISL_1667313, EPI_ISL_1667338, EPI_ISL_1667360, EPI_ISL_1734004, EPI_ISL_1734014, EPI_ISL_1734073, EPI_ISL_1734076, EPI_ISL_1734093, EPI_ISL_1734124, EPI_ISL_1734135, EPI_ISL_1734136, EPI_ISL_1734152, EPI_ISL_1734154, EPI_ISL_1734169, EPI_ISL_1734193, EPI_ISL_1734216, EPI_ISL_1734217, EPI_ISL_1734219, EPI_ISL_1734267, EPI_ISL_1734276, EPI_ISL_1734279, EPI_ISL_1734447, EPI_ISL_1734481, EPI_ISL_1734501, EPI_ISL_1734512, EPI_ISL_1734534, EPI_ISL_1734540, EPI_ISL_1802677, EPI_ISL_1802694, EPI_ISL_1802695, EPI_ISL_1802699, EPI_ISL_2037730, EPI_ISL_2037760, EPI_ISL_2037765, EPI_ISL_2037773, EPI_ISL_2037774, EPI_ISL_2037775, EPI_ISL_2037776, EPI_ISL_2037784, EPI_ISL_2037786, EPI_ISL_2037801, EPI_ISL_2037803, EPI_ISL_2037815, EPI_ISL_2037823, EPI_ISL_2037824, EPI_ISL_2037831, EPI_ISL_2037834, EPI_ISL_2037856, EPI_ISL_2037875, EPI_ISL_2037880, EPI_ISL_2037882, EPI_ISL_2037885, EPI_ISL_2037916, EPI_ISL_2037921, EPI_ISL_2037947, EPI_ISL_2037951, EPI_ISL_2037956, EPI_ISL_2037963, EPI_ISL_2037966, EPI_ISL_2037998, EPI_ISL_2037999, EPI_ISL_2038005, EPI_ISL_2038010, EPI_ISL_2038021, EPI_ISL_2038022, EPI_ISL_2038145, EPI_ISL_2038209, EPI_ISL_2038214, EPI_ISL_2038232, EPI_ISL_2038235, EPI_ISL_2038251, EPI_ISL_2038255, EPI_ISL_2038364, EPI_ISL_2038372, EPI_ISL_2038397, EPI_ISL_2038398, EPI_ISL_2038399, EPI_ISL_2038400, EPI_ISL_2038434, EPI_ISL_2038440, EPI_ISL_2038456, EPI_ISL_4367703, EPI_ISL_4369599, EPI_ISL_4370575, EPI_ISL_4372349 | | | |
| see above | Fulgent Genetics | Centers for Disease Control and Prevention Division of Viral Diseases, Pathogen Discovery | Adrian Paskey; Becky Tsai; Benafsh Sapra; Benjamin Rambo-Martin; Christopher Gulvick; Clinton Paden; Clinton R. Paden; Dakota Howard; Darlene Wagner; Dhwani Batra; Doreen Ng; Duncan MacCannell; Erisa Sula; Harry Gao; James Xie; Jason Caravas; John Gao; Joseph Fierro; Kara Moser; Kristine Lacek; Matthew Schmerer; Mickey Li; Peter Cook; Peter W. Cook; Scott Sammons; Shatavia Morrison; Tymeckia Kendall; Victoria Caban Figueroa; Yan Meng; Yvette Unoarumhi |
| EPI_ISL_1937070, EPI_ISL_1937097, EPI_ISL_1937161, EPI_ISL_1937220, EPI_ISL_1937397, EPI_ISL_1937438 | Fulgent Genetics | Fulgent Genetics | Becky Tsai; Benafsh Sapra; Doreen Ng; Harry Gao; James Xie; John Gao; Joseph Fierro; Mickey Li; Yan Meng |
| EPI_ISL_3553531, EPI_ISL_3553532, EPI_ISL_3553535, EPI_ISL_3553538, EPI_ISL_3553540, EPI_ISL_3553541, EPI_ISL_3553549, EPI_ISL_3553551, EPI_ISL_3553552, EPI_ISL_3553556, EPI_ISL_3553558, EPI_ISL_3553560, EPI_ISL_3553563, EPI_ISL_3553564, EPI_ISL_3553565, EPI_ISL_3553571, EPI_ISL_3553572, EPI_ISL_3553573, EPI_ISL_3553574, EPI_ISL_3553578, EPI_ISL_3553579, EPI_ISL_3553580, EPI_ISL_3553642 | | | |
| see above | Fundação Ezequiel Dias (FUNED) | Fundação Ezequiel Dias | Andre Leal; Cynthia Vazquez; Elaine Cristina; Felipe Iani; Flavia Aburjaile; Gislene Garcia de Castro Lichs; Glauco Carvalho; Hegger Fritsch; Joilson Xavier; Luiz Alcantara.; Luiz Henrique Ferraz Demarchi; Luiz Takao Watanabe; Marina Castilhos Souza Umaki Zardin; Marta Giovanetti; Natalia Guimaraes; Raquel da Silva Ferreira; Talita Adelino; Vagner Fonseca; de Oliveira |
| EPI_ISL_2551523, EPI_ISL_2551524, EPI_ISL_2551530 | Fundação Ezequiel Dias - FUNED | Laboratório de Virologia Clínica e Molecular | Alex Fiorini; Ana Paula Salles Fernandes; Bruna Larotonda Telezynski; Danielle Bruna Leal Oliveira; Edison Luiz Durigon; Erick Gustavo Dorlass; Flavio Fonseca e Santuza Teixeira; Guilherme Pereira Scagion; Helena Perez Coelho; Hugo Sato; Karine Lima Lourenço; Luciano Matsumiya Thomazelli; Renata Peixoto; Rubens Daniel Miserani Magalhães; Tatiana Ometto |
| EPI_ISL_1756229, EPI_ISL_1756508 | GH A.CHENEVIER-H.MONDOR | Department of Virology, Henri Mondor University Hospital, Assistance Publique Hôpitaux de Paris, Université Paris-Est Créteil, INSERM U955 | Alexandre Soulier; Christophe Rodriguez; Elisabeth Trawinski; Guillaume Gricourt; Jean-Michel Pawlotsky; Melissa N'Debi; Slim Fourati; Vanessa Demontant |
| EPI_ISL_1757849, EPI_ISL_1757853, EPI_ISL_1757857, EPI_ISL_1757859 | GH de l'Est Francilien | Department of Virology, Henri Mondor University Hospital, Assistance Publique Hôpitaux de Paris, Université Paris-Est Créteil, INSERM U955 | Alexandre Soulier; Christophe Rodriguez; Elisabeth Trawinski; Guillaume Gricourt; Jean-Michel Pawlotsky; Melissa N'Debi; Slim Fourati; Vanessa Demontant |
| EPI_ISL_1756291 | GROUPE HOSPITALIER SUD ILE DE FRANCE | Department of Virology, Henri Mondor University Hospital, Assistance Publique Hôpitaux de Paris, Université Paris-Est Créteil, INSERM U955 | Alexandre Soulier; Christophe Rodriguez; Elisabeth Trawinski; Guillaume Gricourt; Jean-Michel Pawlotsky; Melissa N'Debi; Slim Fourati; Vanessa Demontant |
| EPI_ISL_2157195, EPI_ISL_2157196, EPI_ISL_2157197, EPI_ISL_2157198, EPI_ISL_2157201 | Gencore - Universidad de los Andes | Gencore - Universidad de los Andes | Ana Maria Palacio; Cristian Barrera; David Gonzalez; Gabriela Ariza; Luisa Sacristan; Marcela Guevara; Silvia Restrepo |
| EPI_ISL_1541055, EPI_ISL_1541058, EPI_ISL_1633360, EPI_ISL_1633365, EPI_ISL_1633367, EPI_ISL_1633368, EPI_ISL_1633379, EPI_ISL_1633387, EPI_ISL_1633388, EPI_ISL_1633392, EPI_ISL_1633399, EPI_ISL_1633430, EPI_ISL_1633432, EPI_ISL_1633433, EPI_ISL_1633524, EPI_ISL_1633526, EPI_ISL_1633540, EPI_ISL_1633541, EPI_ISL_1633544, EPI_ISL_1633549, EPI_ISL_1633550, EPI_ISL_1712324, EPI_ISL_1712339, EPI_ISL_1712340, EPI_ISL_1712341, EPI_ISL_1712346, EPI_ISL_2009196, EPI_ISL_2009236, EPI_ISL_2009239, EPI_ISL_2009291, EPI_ISL_2009304 | | | |
| see above | Genetica Molecular and Subdepartamento de Virologia ISP Chile | Instituto de Salud Publica de Chile | Andres Castillo; Barbara Parra; Gisselle Barra; Jaime Lagos; Javier Tognarelli; Jorge Fernandez; Karen Orostica; Loredana Arata; Patricia Bustos; Rodrigo Fasce; Soledad Ulloa |
| EPI_ISL_1739105, EPI_ISL_1739106, EPI_ISL_1739107, EPI_ISL_1739112, EPI_ISL_1739113, EPI_ISL_1739135, EPI_ISL_1739136 | | | |
| see above | Germano de Sousa | Instituto Nacional de Saude (INSA) | Borges et al |
| EPI_ISL_1854229 | Germano de Sousa | Instituto Nacional de Saude (INSA) and Instituto Gulbenkian de Ciencia (IGC) | Borges et al |
| EPI_ISL_5196284, EPI_ISL_5196285, EPI_ISL_5196296, EPI_ISL_5196297, EPI_ISL_5196308, EPI_ISL_5196348, EPI_ISL_5196352 | | | |
| see above | Gorgas Memorial Institute of Health Studies | Gorgas Memorial Institute of Health Studies | Castillo Jorge; Chen Maria; Franco Danilo; Gonzalez Claudia; Jessica Gondola; Leyda Abrego; Lopez-Verges Sandra; Marlenne Castillo; Martinez Alexander; Menacho Abdiel; Moreno Ambar; Moreno Brechla; Oris Chavarria; Ortiz Alma; Salazar Jacqueline |
| EPI_ISL_2601796 | Gravity Diagnostics, LLC | Gravity Diagnostics, LLC | Gravity Diagnostics |
| EPI_ISL_2339835, EPI_ISL_2603133, EPI_ISL_2603134, EPI_ISL_2603135, EPI_ISL_2603138, EPI_ISL_2603147, EPI_ISL_2603149, EPI_ISL_2603150, EPI_ISL_2603151, EPI_ISL_2603152, EPI_ISL_2603153, EPI_ISL_2603155, EPI_ISL_2603156, EPI_ISL_2603159, EPI_ISL_2603161, EPI_ISL_2603165, EPI_ISL_2603166, EPI_ISL_2603167, EPI_ISL_2603168, EPI_ISL_2603169, EPI_ISL_2603170, EPI_ISL_2603171, EPI_ISL_2603172, EPI_ISL_2603173, EPI_ISL_2603175 | | | |
| see above | Grupo de Investigación en Enfermedades Tropicales del Ejército (GINETEJ), Laboratorio de Referencia e Investigación, Dirección de Sanidad Ejército, Bogotá, Colombia | Centro de Investigaciones en Microbiología y Biotecnología-UR (CIMBIUR), Facultad de Ciencias Naturales, Universidad del Rosario, Bogotá, Colombia | Camilo A. Correa-Cárdenas; Carolina Oliveros; Claudia Méndez; Elizabeth K. Márquez; Frank de los Santos Ortiz; Juan David Ramírez; Julie Pérez; Lorena Albarracín; Luz H. Patiño; Maria Clara Duque; Marina Muñoz; María Teresa Alvarado; Nathalia Ballesteros; Sergio Castañeda; Sergio Gutíerrez-Riveros; Yanira Romero; Zulma Cucunubá |
| EPI_ISL_1632709, EPI_ISL_1632710 | Gundersen Clinical Microbiology Laboratory | Kabara Cancer Research Institute | Craig S. Richmond; Paraic A. Kenny |
| EPI_ISL_2402755, EPI_ISL_2402756, EPI_ISL_2402769 | Gyncentrum | Gyncentrum | Adam Pudełko; Agnieszka Polak; Aleksandra Skubis-Sikora; Celina Kruszniewska-Rajs; Emilia Morawiec; Jolanta Bartosiewicz- Wąsik; Magdalena Samul; Maria Maklasińska-Majdanik; Paweł Czerwiński; Robert Wojtyczka; Tomasz Wąsik |
| EPI_ISL_3912028, EPI_ISL_3912159 | H J M A HOSPITAL JOSE MARTINIANO DE ALENCAR | Analytical Competence Molecular Epidemiology Lab/ACME, Oswaldo Cruz Foundation, Ceara (FIOCRUZ CE) | Cleber Furtado Aksenen; Fabio Miyajima; Fernando Braga Stehling; Francisco Eder de Moura Lopes; Jamille Maria Mendes Bezerra; Joaquim Cesar do Nascimento Sousa Junior; Pedro Miguel Carneiro Jeronimo; Suzana Porto Almeida & Lucas Delerino on behalf of COVID-19 FIOCRUZ Genomic Network; Thais Ferreira de Oliveira; Thais de Oliveira Costa; Ticiane Cavalcante de Souza; Veridiana Pessoa Miyajima |
| EPI_ISL_1854249 | H Vila Franca Xira | Instituto Nacional de Saude (INSA) and Institute of Biomedicine (iBiMed), Universidade de Aveiro | Borges et al |
| EPI_ISL_2801365 | HEMOCE CENTRO DE HEMATOLOGIA E HEMOTERAPIA DO CEARA | Analytical Competence Molecular Epidemiology Lab/ACME, Oswaldo Cruz Foundation, Ceara (FIOCRUZ CE) | Cleber Furtado Aksenen e Suzana Porto Almeida; Fabio Miyajima; Fernando Braga Stehling; Francisco Eder de Moura Lopes; Jamille Maria Mendes Bezerra; Joaquim César do Nascimento Sousa Junior; Pedro Miguel Carneiro Jeronimo; Thais Ferreira de Oliveira; Thais de Oliveira Costa; Ticiane Cavalcante de Souza; Veridiana Pessoa Miyajima |
| EPI_ISL_3912111, | HGCC HOSPITAL GERAL DR | Analytical Competence Molecular | Carlos Leonardo de Aragao Araujo; Cleber Furtado Aksenen; Fabio Miyajima; Fernando Braga Stehling; Francisco Eder de Moura Lopes; Jamille Maria Mendes Bezerra; Joaquim Cesar do Nascimento Sousa Junior; Pedro Miguel Carneiro Jeronimo; Suzana Porto Almeida & Igor Oliveira Duarte on behalf of COVID-19 FIOCRUZ |

| | | | |
|---|---|---|---|
| EPI_ISL_3912124, EPI_ISL_3912163, EPI_ISL_3912164, EPI_ISL_3912165, EPI_ISL_5529974 | CESAR CALS | Epidemiology Lab/ACME, Oswaldo Cruz Foundation, Ceara (FIOCRUZ CE) | Genomic Network; Suzana Porto Almeida & Lucas Delerino on behalf of COVID-19 FIOCRUZ Genomic Network; Thais Ferreira de Oliveira; Thais de Oliveira Costa; Ticiane Cavalcante de Souza; Veridiana Pessoa Miyajima |
| EPI_ISL_3912064, EPI_ISL_3912068, EPI_ISL_3912119 | HIAS HOSPITAL INFANTIL ALBERT SABIN | Analytical Competence Molecular Epidemiology Lab/ACME, Oswaldo Cruz Foundation, Ceara (FIOCRUZ CE) | Cleber Furtado Aksenen; Fabio Miyajima; Fernando Braga Stehling; Francisco Eder de Moura Lopes; Jamille Maria Mendes Bezerra; Joaquim Cesar do Nascimento Sousa Junior; Pedro Miguel Carneiro Jeronimo; Suzana Porto Almeida & Lucas Delerino on behalf of COVID-19 FIOCRUZ Genomic Network; Thais Ferreira de Oliveira; Thais de Oliveira Costa; Ticiane Cavalcante de Souza; Veridiana Pessoa Miyajima |
| EPI_ISL_1805649 | HIC | Instituto Nacional de Salud- Dirección de Investigación en Salud Pública | Carlos Franco-Muñoz; Carmen Osorio; Diana Malo; Diego A. Álvarez-Díaz; Diego Andrés Prada; Gerardo Santamaría; Hector Alejandro Ruiz-Moreno; Jhonnatan Reales-González; Jorge Rivera; Juan Camilo Martinez; Julian Naizaque; Katherine Laiton-Donato; Lisseth Pardo; Magdalena Wiesner; Marcela Mercado-Reyes; Maria T. Herrera-Sepúlveda; Marta Lopez Blanco; Martha Lucia Ospina Martinez; Paola Rojas; Sergio Gomez; Sheryll Corchuelo; Ángela Alarcon Cruz |
| EPI_ISL_2017346, EPI_ISL_2017347, EPI_ISL_2017348, EPI_ISL_2017349, EPI_ISL_2017350, EPI_ISL_2017351, EPI_ISL_2017352, EPI_ISL_2017353, EPI_ISL_2017354, EPI_ISL_2017355, EPI_ISL_2017356, EPI_ISL_2017357, EPI_ISL_2017358, EPI_ISL_2017359, EPI_ISL_2017360, EPI_ISL_2017362, EPI_ISL_2017363, EPI_ISL_2017367, EPI_ISL_2017368, EPI_ISL_2017369, EPI_ISL_2187947, EPI_ISL_2187948, EPI_ISL_2187950, EPI_ISL_2187952, EPI_ISL_2187954, EPI_ISL_2187955, EPI_ISL_2187956, EPI_ISL_2187957, EPI_ISL_2187958, EPI_ISL_2187960, EPI_ISL_2187961, EPI_ISL_2187962, EPI_ISL_2187963, EPI_ISL_2187964, EPI_ISL_2187966, EPI_ISL_2187967, EPI_ISL_2187968, EPI_ISL_2187971, EPI_ISL_2187973, EPI_ISL_2187975, EPI_ISL_2187976, EPI_ISL_2187977, EPI_ISL_2187978, EPI_ISL_2187979, EPI_ISL_2187980, EPI_ISL_2187981, EPI_ISL_3254682 | | | |
| see above | HLAGYN - Laboratorio de Imunologia de Transplantes de Goias | HLAGYN - Laboratorio de Imunologia de Transplantes de Goias | Alessandro Leonardo Alvares Magalhaes; Daniel Ferreira de Sousa; Danielle de Paiva Rezende; Erika Lopes Rocha Batista; Fernando Antonio Vinhal dos Santos; Frederico Rodrigues Vinhal; Kamila Oliveira Reis De Freitas.; Lucas Carlos Gomes Pereira; Paola Cristina Resende Silva; Raphael Bessa Parmigiane; Sabrina Sara Moreira Duarte |
| EPI_ISL_3102235, EPI_ISL_3912031, EPI_ISL_3912100, EPI_ISL_3912123, EPI_ISL_3912178, EPI_ISL_3912179, EPI_ISL_3912180, EPI_ISL_3912181, EPI_ISL_3912182, EPI_ISL_3912183, EPI_ISL_3912184, EPI_ISL_3912185, EPI_ISL_3912186, EPI_ISL_3912187 | | | |
| see above | HM HOSPITAL DE MESSEJANA DR CARLOS ALBERTO STUDART GOMES | Analytical Competence Molecular Epidemiology Lab/ACME, Oswaldo Cruz Foundation, Ceara (FIOCRUZ CE) | Cleber Furtado Aksenen; Fabio Miyajima; Fernando Braga Stehling; Francisco Eder de Moura Lopes; Jamille Maria Mendes Bezerra; Joaquim Cesar do Nascimento Sousa Junior; Joaquim César do Nascimento Sousa Junior; Pedro Miguel Carneiro Jeronimo; Suzana Porto Almeida & Lucas Delerino on behalf of COVID-19 FIOCRUZ Genomic Network; Suzana Porto Almeida e Lucas Delerino; Thais Ferreira de Oliveira; Thais de Oliveira Costa; Ticiane Cavalcante de Souza; Veridiana Pessoa Miyajima |
| EPI_ISL_2345972 | HOSP DO SERV PUB ESTFCO MORATO DE OLIVEIRA SAO PAULO | Instituto Butantan | Antonio Jorge Martins; Claudia Renata dos Santos Barros; David Schlesinger; Debora Botequio Moretti; Dimas Tadeu Covas; Elaine Cristina Marqueze; Elaine Vieira Santos; Evandra Strazza Rodrigues; Heidge Fukumasu; Jayme Augusto de Souza-Neto; José Salvatore Leister Patané; Luiz Alcantara; Luiz Lehmann Coutinho; Maria Carolina Elias; Maurício Lacerda Nogueira; Rafael dos Santos Bezerra; Raul Machado Neto; Rejane Maria Tommasini Grotto; Ricardo Haddad; Sandra Coccuzzo Sampaio Vessoni; Simone Kashima; Svetoslav Nanev Slavov; Vincent Louis Viala |
| EPI_ISL_1967092, EPI_ISL_1967093, EPI_ISL_1967094, EPI_ISL_1967095 | HOSP DO SERV PUB ESTFCO MORATO DE OLIVEIRA SAO PAULO | Instituto Butantan / ESALQ-USP (Piracicaba) | Antonio Jorge Martins; Bianca Cechetto Carlos. Mendelics: Bibiana Santos; Claudia Renata dos Santos Barros; Cíntia Bittar; David Schlesinger. Hemocentro Ribeirão Preto: Simone Kashima; Debora Botequio Moretti; Elaine Cristina Marqueze; Elaine Vieira dos Santos; Elisangela Chicaroni Mattos; Erika Freitas; Evandra Strazza Rodrigues; Felipe Allan da Silva da Costa; Flavia Aburjaile; Fábio Sossai Possebon; Guilherme Campos; Guilherme Targino Valente; Heidge Fukumasu. USP-Botucatu: Rejane Maria Tommasini Grotto; Helena Lage Ferreira; Instituto Butantan: Dimas Tadeu Covas; Jardelina de Souza Todao Bernardino; Jayme A. Souza-Neto; Jessika Cristina Chagas Lesbon; Jorge A. Petroli Marchesi; José Salvatore Leister Patané; João Paulo Kitajima; João Pessoa Araújo Jr.; Leila Sabrina Ullmann; Loyze Paola Oliveira de Lima; Luiz Aurelio de Campos Crispin. Centro de Genômica Funcional da ESALQ: Luiz Lehmann Coutinho; Luiz Carlos Junior de Alcantara; Lívia Sacchetto; Maisa C. Pereira Parra; Maria Carolina Elias; Marta Giovanetti; Marília Moraes; Maurício Lacerda Nogueira. Prefeitura de Sao Paulo: Melissa Palmieri.; Patricia Akemi Assato; Paula Rahal; Paulo Inacio da Costa; Rafael dos Santos Bezerra; Raquel de Lello Rocha Campos Cassano. NGS Soluções Genômicas: Pilar Drummond Sampaio Corrêa Mariani. FZEA-USP Pirassununga: Mirele Daiana Poleti; Raul Machado Neto; Ricardo Augusto Brassaloti; Ricardo Haddad; Rodrigo Tocantins Calado. FAMERP-SJRP: Cecília Artico Banho; Sandra Coccuzzo Sampaio; Svetoslav Nanev Slavov; Vagner Fonseca; Vincent Louis Viala |
| EPI_ISL_2345882, EPI_ISL_2345886 | HOSP DO SERV PUB ESTFCO MORATO DE OLIVEIRA SAO PAULO | Instituto Butantan / FZEA-USP-Pirassununga | Antonio Jorge Martins; Claudia Renata dos Santos Barros; David Schlesinger; Debora Botequio Moretti; Dimas Tadeu Covas; Elaine Cristina Marqueze; Elaine Vieira Santos; Evandra Strazza Rodrigues; Heidge Fukumasu; Jayme Augusto de Souza-Neto; José Salvatore Leister Patané; Luiz Alcantara; Luiz Lehmann Coutinho; Maria Carolina Elias; Maurício Lacerda Nogueira; Rafael dos Santos Bezerra; Raul Machado Neto; Rejane Maria Tommasini Grotto; Ricardo Haddad; Sandra Coccuzzo Sampaio Vessoni; Simone Kashima; Svetoslav Nanev Slavov; Vincent Louis Viala |
| EPI_ISL_5530151, EPI_ISL_5530161, EPI_ISL_5530176 | HOSP DR THADEU DE PAULO BRITO | Analytical Competence Molecular Epidemiology Lab/ACME, Oswaldo Cruz Foundation, Ceara (FIOCRUZ CE) | Carlos Leonardo de Aragao Araujo; Cecilia Leite Costa & Eduardo Ruback dos Santos on behalf of COVID-19 FIOCRUZ Genomic Network; Cleber Furtado Aksenen; Fabio Miyajima; Fernando Braga Stehling; Francisco Eder de Moura Lopes; Igor Oliveira Duarte; Jamille Maria Mendes Bezerra; Joaquim Cesar do Nascimento Sousa Junior; Pedro Miguel Carneiro Jeronimo; Suzana Porto Almeida; Thais Ferreira de Oliveira; Thais de Oliveira Costa; Ticiane Cavalcante de Souza; Veridiana Pessoa Miyajima |
| EPI_ISL_1967029, EPI_ISL_1967030, EPI_ISL_1967031, EPI_ISL_1967032 | HOSP E MATERNIDADE MUNICIPAL N SRA MONTE SERRAT | Instituto Butantan / ESALQ-USP (Piracicaba) | Antonio Jorge Martins; Bianca Cechetto Carlos. Mendelics: Bibiana Santos; Claudia Renata dos Santos Barros; Cíntia Bittar; David Schlesinger. Hemocentro Ribeirão Preto: Simone Kashima; Debora Botequio Moretti; Elaine Cristina Marqueze; Elaine Vieira dos Santos; Elisangela Chicaroni Mattos; Erika Freitas; Evandra Strazza Rodrigues; Felipe Allan da Silva da Costa; Flavia Aburjaile; Fábio Sossai Possebon; Guilherme Campos; Guilherme Targino Valente; Heidge Fukumasu. USP-Botucatu: Rejane Maria Tommasini Grotto; Helena Lage Ferreira; Instituto Butantan: Dimas Tadeu Covas; Jardelina de Souza Todao Bernardino; Jayme A. Souza-Neto; Jessika Cristina Chagas Lesbon; Jorge A. Petroli Marchesi; José Salvatore Leister Patané; João Paulo Kitajima; João Pessoa Araújo Jr.; Leila Sabrina Ullmann; Loyze Paola Oliveira de Lima; Luiz Aurelio de Campos Crispin. Centro de Genômica Funcional da ESALQ: Luiz Lehmann Coutinho; Luiz Carlos Junior de Alcantara; Lívia Sacchetto; Maisa C. Pereira Parra; Maria Carolina Elias; Marta Giovanetti; Marília Moraes; Maurício Lacerda Nogueira. Prefeitura de Sao Paulo: Melissa Palmieri.; Patricia Akemi Assato; Paula Rahal; Paulo Inacio da Costa; Rafael dos Santos Bezerra; Raquel de Lello Rocha Campos Cassano. NGS Soluções Genômicas: Pilar Drummond Sampaio Corrêa Mariani. FZEA-USP Pirassununga: Mirele Daiana Poleti; Raul Machado Neto; Ricardo Augusto Brassaloti; Ricardo Haddad; Rodrigo Tocantins Calado. FAMERP-SJRP: Cecília Artico Banho; Sandra Coccuzzo Sampaio; Svetoslav Nanev Slavov; Vagner Fonseca; Vincent Louis Viala |
| EPI_ISL_5529887, EPI_ISL_5530068 | HOSP MATERN F FARIAS LEITAO | Analytical Competence Molecular Epidemiology Lab/ACME, Oswaldo Cruz Foundation, Ceara (FIOCRUZ CE) | Carlos Leonardo de Aragao Araujo; Cecilia Leite Costa & Eduardo Ruback dos Santos on behalf of COVID-19 FIOCRUZ Genomic Network; Cleber Furtado Aksenen; Fabio Miyajima; Fernando Braga Stehling; Francisco Eder de Moura Lopes; Igor Oliveira Duarte; Jamille Maria Mendes Bezerra; Joaquim Cesar do Nascimento Sousa Junior; Pedro Miguel Carneiro Jeronimo; Suzana Porto Almeida; Thais Ferreira de Oliveira; Thais de Oliveira Costa; Ticiane Cavalcante de Souza; Veridiana Pessoa Miyajima |
| EPI_ISL_5530091 | HOSP MATERN PAULO SARASATE | Analytical Competence Molecular Epidemiology Lab/ACME, Oswaldo Cruz Foundation, Ceara (FIOCRUZ CE) | Carlos Leonardo de Aragao Araujo; Cecilia Leite Costa & Eduardo Ruback dos Santos on behalf of COVID-19 FIOCRUZ Genomic Network; Cleber Furtado Aksenen; Fabio Miyajima; Fernando Braga Stehling; Francisco Eder de Moura Lopes; Igor Oliveira Duarte; Jamille Maria Mendes Bezerra; Joaquim Cesar do Nascimento Sousa Junior; Pedro Miguel Carneiro Jeronimo; Suzana Porto Almeida; Thais Ferreira de Oliveira; Thais de Oliveira Costa; Ticiane Cavalcante de Souza; Veridiana Pessoa Miyajima |
| EPI_ISL_5529946 | HOSP MATERN SANTA IZABEL ARACOIABA | Analytical Competence Molecular Epidemiology Lab/ACME, Oswaldo Cruz Foundation, Ceara (FIOCRUZ CE) | Carlos Leonardo de Aragao Araujo; Cecilia Leite Costa & Eduardo Ruback dos Santos on behalf of COVID-19 FIOCRUZ Genomic Network; Cleber Furtado Aksenen; Fabio Miyajima; Fernando Braga Stehling; Francisco Eder de Moura Lopes; Igor Oliveira Duarte; Jamille Maria Mendes Bezerra; Joaquim Cesar do Nascimento Sousa Junior; Pedro Miguel Carneiro Jeronimo; Suzana Porto Almeida; Thais Ferreira de Oliveira; Thais de Oliveira Costa; Ticiane Cavalcante de Souza; Veridiana Pessoa Miyajima |
| EPI_ISL_2345997, EPI_ISL_2346003, EPI_ISL_2346004 | HOSP MUN CIDADE TIRADENTES CARMEN PRUDENTE | Instituto Butantan | Antonio Jorge Martins; Claudia Renata dos Santos Barros; David Schlesinger; Debora Botequio Moretti; Dimas Tadeu Covas; Elaine Cristina Marqueze; Elaine Vieira Santos; Evandra Strazza Rodrigues; Heidge Fukumasu; Jayme Augusto de Souza-Neto; José Salvatore Leister Patané; Luiz Alcantara; Luiz Lehmann Coutinho; Maria Carolina Elias; Maurício Lacerda Nogueira; Rafael dos Santos Bezerra; Raul Machado Neto; Rejane Maria Tommasini Grotto; Ricardo Haddad; Sandra Coccuzzo Sampaio Vessoni; Simone Kashima; Svetoslav Nanev Slavov; Vincent Louis Viala |
| EPI_ISL_1967091 | HOSP MUN CIDADE TIRADENTES CARMEN PRUDENTE | Instituto Butantan / ESALQ-USP (Piracicaba) | Antonio Jorge Martins; Bianca Cechetto Carlos. Mendelics: Bibiana Santos; Claudia Renata dos Santos Barros; Cíntia Bittar; David Schlesinger. Hemocentro Ribeirão Preto: Simone Kashima; Debora Botequio Moretti; Elaine Cristina Marqueze; Elaine Vieira dos Santos; Elisangela Chicaroni Mattos; Erika Freitas; Evandra Strazza Rodrigues; Felipe Allan da Silva da Costa; Flavia Aburjaile; Fábio Sossai Possebon; Guilherme Campos; Guilherme Targino Valente; Heidge Fukumasu. USP-Botucatu: Rejane Maria Tommasini Grotto; Helena Lage Ferreira; Instituto Butantan: Dimas Tadeu Covas; Jardelina de Souza Todao Bernardino; Jayme A. Souza-Neto; Jessika Cristina Chagas Lesbon; Jorge A. Petroli Marchesi; José Salvatore Leister Patané; João Paulo Kitajima; João Pessoa Araújo Jr.; Leila Sabrina Ullmann; Loyze Paola Oliveira de Lima; Luiz Aurelio de Campos Crispin. Centro de Genômica Funcional da ESALQ: Luiz Lehmann Coutinho; Luiz Carlos Junior de Alcantara; Lívia Sacchetto; Maisa C. Pereira Parra; Maria Carolina Elias; Marta Giovanetti; Marília Moraes; Maurício Lacerda Nogueira. Prefeitura de Sao Paulo: Melissa Palmieri.; Patricia Akemi Assato; Paula Rahal; Paulo Inacio da Costa; Rafael dos Santos Bezerra; Raquel de Lello Rocha Campos Cassano. NGS Soluções Genômicas: Pilar Drummond Sampaio Corrêa Mariani. FZEA-USP Pirassununga: Mirele Daiana Poleti; Raul Machado Neto; Ricardo Augusto Brassaloti; Ricardo Haddad; Rodrigo Tocantins Calado. FAMERP-SJRP: Cecília Artico Banho; Sandra Coccuzzo Sampaio; Svetoslav Nanev Slavov; Vagner Fonseca; Vincent Louis Viala |
| EPI_ISL_2346022, EPI_ISL_2346024 | HOSP MUN JOSANIAS CASTANHA BRAGA | Instituto Butantan | Antonio Jorge Martins; Claudia Renata dos Santos Barros; David Schlesinger; Debora Botequio Moretti; Dimas Tadeu Covas; Elaine Cristina Marqueze; Elaine Vieira Santos; Evandra Strazza Rodrigues; Heidge Fukumasu; Jayme Augusto de Souza-Neto; José Salvatore Leister Patané; Luiz Alcantara; Luiz Lehmann Coutinho; Maria Carolina Elias; Maurício Lacerda Nogueira; Rafael dos Santos Bezerra; Raul Machado Neto; Rejane Maria Tommasini Grotto; Ricardo Haddad; Sandra Coccuzzo Sampaio Vessoni; Simone Kashima; Svetoslav Nanev Slavov; Vincent Louis Viala |
| EPI_ISL_2020126 | HOSPITAL CAN MISSES | HOSPITAL UNIVERSITARIO SON ESPASES | Antonio Oliver; Carla López-Causapé; Pablo Fraile-Ribot; SeqCovid |
| EPI_ISL_2346002 | HOSPITAL DE CAMPANHA COVID 19 CAIEIRAS | Instituto Butantan | Antonio Jorge Martins; Claudia Renata dos Santos Barros; David Schlesinger; Debora Botequio Moretti; Dimas Tadeu Covas; Elaine Cristina Marqueze; Elaine Vieira Santos; Evandra Strazza Rodrigues; Heidge Fukumasu; Jayme Augusto de Souza-Neto; José Salvatore Leister Patané; Luiz Alcantara; Luiz Lehmann Coutinho; Maria Carolina Elias; Maurício Lacerda Nogueira; Rafael dos Santos Bezerra; Raul Machado Neto; Rejane Maria Tommasini Grotto; Ricardo Haddad; Sandra Coccuzzo Sampaio Vessoni; Simone Kashima; Svetoslav Nanev Slavov; Vincent Louis Viala |
| EPI_ISL_1967165, EPI_ISL_1967166, EPI_ISL_1967167, EPI_ISL_1967168, EPI_ISL_1967169, EPI_ISL_1967170, EPI_ISL_1967171, EPI_ISL_1967172, EPI_ISL_1967173, EPI_ISL_1967174, EPI_ISL_1967175, EPI_ISL_1967176, EPI_ISL_1967177, EPI_ISL_1967178, EPI_ISL_1967179, EPI_ISL_1967180, EPI_ISL_1967181, EPI_ISL_1967182, EPI_ISL_1967183, EPI_ISL_1967184, EPI_ISL_1967185, EPI_ISL_1967186, EPI_ISL_1967187, EPI_ISL_1967188, EPI_ISL_1967189, EPI_ISL_1967190, EPI_ISL_1967191 | | | |
| see above | HOSPITAL DE CAMPANHA COVID 19 MUNICIPIO DE TAUBATE | Instituto Butantan / ESALQ-USP (Piracicaba) | Antonio Jorge Martins; Bianca Cechetto Carlos. Mendelics: Bibiana Santos; Claudia Renata dos Santos Barros; Cíntia Bittar; David Schlesinger. Hemocentro Ribeirão Preto: Simone Kashima; Debora Botequio Moretti; Elaine Cristina Marqueze; Elaine Vieira dos Santos; Elisangela Chicaroni Mattos; Erika Freitas; Evandra Strazza Rodrigues; Felipe Allan da Silva da Costa; Flavia Aburjaile; Fábio Sossai Possebon; Guilherme Campos; Guilherme Targino Valente; Heidge Fukumasu. USP-Botucatu: Rejane Maria Tommasini Grotto; Helena Lage Ferreira; Instituto Butantan: Dimas Tadeu Covas; Jardelina de Souza Todao Bernardino; Jayme A. Souza-Neto; Jessika Cristina Chagas Lesbon; Jorge A. Petroli Marchesi; José Salvatore Leister Patané; João Paulo Kitajima; João Pessoa Araújo Jr.; Leila Sabrina Ullmann; Loyze Paola Oliveira de Lima; Luiz Aurelio de Campos Crispin. Centro de Genômica Funcional da ESALQ: Luiz Lehmann Coutinho; Luiz Carlos Junior de Alcantara; Lívia Sacchetto; Maisa C. Pereira Parra; Maria Carolina Elias; Marta Giovanetti; Marília Moraes; Maurício Lacerda Nogueira. Prefeitura de Sao Paulo: Melissa Palmieri.; Patricia Akemi Assato; Paula Rahal; Paulo Inacio da Costa; Rafael dos Santos Bezerra; Raquel de Lello Rocha Campos Cassano. NGS Soluções Genômicas: Pilar Drummond Sampaio Corrêa Mariani. FZEA-USP Pirassununga: Mirele Daiana Poleti; Raul Machado Neto; Ricardo Augusto Brassaloti; Ricardo Haddad; Rodrigo Tocantins Calado. FAMERP-SJRP: Cecília Artico Banho; Sandra Coccuzzo Sampaio; Svetoslav Nanev Slavov; Vagner Fonseca; Vincent Louis Viala |
| EPI_ISL_1795106, EPI_ISL_2344556 | HOSPITAL DOS FORNCEDORES | Instituto Butantan / ESALQ-Piracicaba | Antonio Jorge Martins; Bianca Cechetto Carlos. Mendelics: Bibiana Santos; Claudia Renata dos Santos Barros; David Schlesinger; David Schlesinger. Hemocentro Ribeirão Preto: Simone Kashima; Debora Botequio Moretti; Debora Botequio Moretti. Centro de Genômica Funcional da ESALQ: Luiz Lehmann Coutinho; Dimas Tadeu Covas; Elaine Cristina Marqueze; Elaine Vieira Santos; Elaine Vieira dos Santos; Elisangela Chicaroni Mattos; Erika Freitas; Evandra Strazza Rodrigues; Felipe Allan da Silva da Costa; Flavia Aburjaile; Guilherme Targino Valente; Heidge Fukumasu; Heidge Fukumasu. USP-Botucatu: Rejane Maria Tommasini Grotto; Instituto Butantan: Alexander Roberto Precioso; Jayme A. Souza-Neto; Jayme Augusto de Souza-Neto; Jessika Cristina Chagas Lesbon; José Salvatore Leister Patané; João Paulo Kitajima; Luiz Alcantara; Luiz Carlos Junior de Alcantara; Luiz Lehmann Coutinho; Maria Carolina Elias; Marta Giovanetti; Maurício Lacerda Nogueira; Patricia Akemi Assato; Rafael dos Santos Bezerra; Raquel de Lello Rocha Campos Cassano. NGS Soluções Genômicas: Pilar Drummond Sampaio Corrêa Mariani. FZEA-USP Pirassununga: Mirele Daiana Poleti; Raul Machado Neto; Rejane Maria Tommasini Grotto; Ricardo Augusto Brassaloti; Ricardo Haddad; Rodrigo Tocantins Calado.; Sandra Coccuzzo Sampaio; Sandra Coccuzzo Sampaio Vessoni; Simone Kashima; Svetoslav Nanev Slavov; Vagner Fonseca; Vincent Louis Viala |
| EPI_ISL_3912022, EPI_ISL_3912054, EPI_ISL_3912196, EPI_ISL_3912197 | HOSPITAL E MATERNIDADE DRA ZILDA ARNS NEUMANN | Analytical Competence Molecular Epidemiology Lab/ACME, Oswaldo Cruz Foundation, Ceara (FIOCRUZ CE) | Cleber Furtado Aksenen; Fabio Miyajima; Fernando Braga Stehling; Francisco Eder de Moura Lopes; Jamille Maria Mendes Bezerra; Joaquim Cesar do Nascimento Sousa Junior; Pedro Miguel Carneiro Jeronimo; Suzana Porto Almeida & Lucas Delerino on behalf of COVID-19 FIOCRUZ Genomic Network; Thais Ferreira de Oliveira; Thais de Oliveira Costa; Ticiane Cavalcante de Souza; Veridiana Pessoa Miyajima |
| EPI_ISL_3102535, EPI_ISL_3912023, EPI_ISL_3912029, EPI_ISL_3912106, EPI_ISL_3912114, EPI_ISL_3912121, EPI_ISL_3912128, EPI_ISL_3912208 | | | |
| see above | HOSPITAL ESTADUAL LEONARDO DA VINCI | Analytical Competence Molecular Epidemiology Lab/ACME, Oswaldo Cruz Foundation, Ceara (FIOCRUZ CE) | Cleber Furtado Aksenen; Fabio Miyajima; Fernando Braga Stehling; Francisco Eder de Moura Lopes; Jamille Maria Mendes Bezerra; Joaquim Cesar do Nascimento Sousa Junior; Joaquim César do Nascimento Sousa Junior; Pedro Miguel Carneiro Jeronimo; Suzana Porto Almeida & Lucas Delerino on behalf of COVID-19 FIOCRUZ Genomic Network; Suzana Porto Almeida e Lucas Delerino; Thais Ferreira de Oliveira; Thais de Oliveira Costa; Ticiane Cavalcante de Souza; Veridiana Pessoa Miyajima |
| EPI_ISL_2150639 | HOSPITAL GENERAL VIRGEN DE LA LUZ | Instituto de Salud Carlos III | A. Monzón; F. Casas; I. Jiménez; I.RODRIGUEZ ESCUDERO; Iglesias-Caballero; M. Sandonís; MARIA JOSE; P. Zaballos; S. Camarero; S. Cuesta; S. Pozo; S. Varona; V. Vázquez-Morón |
| EPI_ISL_1967077 | HOSPITAL GERAL JESUS TEIXEIRA DA COSTA | Instituto Butantan / ESALQ-USP (Piracicaba) | Antonio Jorge Martins; Bianca Cechetto Carlos. Mendelics: Bibiana Santos; Claudia Renata dos Santos Barros; Cíntia Bittar; David Schlesinger. Hemocentro Ribeirão Preto: Simone Kashima; Debora Botequio Moretti; Elaine Cristina Marqueze; Elaine Vieira dos Santos; Elisangela Chicaroni Mattos; Erika Freitas; Evandra Strazza Rodrigues; Felipe Allan da Silva da Costa; Flavia Aburjaile; Fábio Sossai Possebon; Guilherme Campos; Guilherme Targino Valente; Heidge Fukumasu. USP-Botucatu: Rejane Maria Tommasini Grotto; Helena Lage Ferreira; Instituto Butantan: Dimas Tadeu Covas; Jardelina de Souza Todao Bernardino; Jayme A. |

| | | | |
|---|---|---|---|
| | GUAIANASES SAO PAULO | | Souza-Neto; Jessika Cristina Chagas Lesbon; Jorge A. Petroli Marchesi; José Salvatore Leister Patané; João Paulo Kitajima; João Pessoa Araújo Jr.; Leila Sabrina Ullmann; Loyze Paola Oliveira de Lima; Luiz Aurelio de Campos Crispin. Centro de Genômica Funcional da ESALQ: Luiz Lehmann Coutinho; Luiz Carlos Junior de Alcantara; Lívia Sacchetto; Maisa C. Pereira Parra; Maria Carolina Elias; Marta Giovanetti; Marília Moraes; Maurício Lacerda Nogueira. Prefeitura de Sao Paulo: Melissa Palmieri.; Patricia Akemi Assato; Paula Rahal; Paulo Inacio da Costa; Rafael dos Santos Bezerra; Raquel de Lello Rocha Campos Cassano. NGS Soluções Genômicas: Pilar Drummond Sampaio Corrêa Mariani. FZEA-USP Pirassununga: Mirele Daiana Poleti; Raul Machado Neto; Ricardo Augusto Brassaloti; Ricardo Haddad; Rodrigo Tocantins Calado. FAMERP-SJRP: Cecília Artico Banho; Sandra Coccuzzo Sampaio; Svetoslav Nanev Slavov; Vagner Fonseca; Vincent Louis Viala |
| EPI_ISL_2346037 | HOSPITAL MUN DR GUIDO GUIDA | Instituto Butantan | Antonio Jorge Martins; Claudia Renata dos Santos Barros; David Schlesinger; Debora Botequio Moretti; Dimas Tadeu Covas; Elaine Cristina Marqueze; Elaine Vieira Santos; Evandra Strazza Rodrigues; Heidge Fukumasu; Jayme Augusto de Souza-Neto; José Salvatore Leister Patané; Luiz Alcantara; Luiz Lehmann Coutinho; Maria Carolina Elias; Maurício Lacerda Nogueira; Rafael dos Santos Bezerra; Raul Machado Neto; Rejane Maria Tommasini Grotto; Ricardo Haddad; Sandra Coccuzzo Sampaio Vessoni; Simone Kashima; Svetoslav Nanev Slavov; Vincent Louis Viala |
| EPI_ISL_1966913, EPI_ISL_1966914 | HOSPITAL MUNICIPAL DE IBIUNA IBIUNA SP | Instituto Butantan / ESALQ-USP (Piracicaba) | Antonio Jorge Martins; Bianca Cechetto Carlos. Mendelics: Bibiana Santos; Claudia Renata dos Santos Barros; Cíntia Bittar; David Schlesinger. Hemocentro Ribeirão Preto: Simone Kashima; Debora Botequio Moretti; Elaine Cristina Marqueze; Elaine Vieira dos Santos; Elisangela Chicaroni Mattos; Erika Freitas; Evandra Strazza Rodrigues; Felipe Allan da Silva da Costa; Flavia Aburjaile; Fábio Sossai Possebon; Guilherme Campos; Guilherme Targino Valente; Heidge Fukumasu. USP-Botucatu: Rejane Maria Tommasini Grotto; Helena Lage Ferreira; Instituto Butantan: Dimas Tadeu Covas; Jardelina de Souza Todao Bernardino; Jayme A. Souza-Neto; Jessika Cristina Chagas Lesbon; Jorge A. Petroli Marchesi; José Salvatore Leister Patané; João Paulo Kitajima; João Pessoa Araújo Jr.; Leila Sabrina Ullmann; Loyze Paola Oliveira de Lima; Luiz Aurelio de Campos Crispin. Centro de Genômica Funcional da ESALQ: Luiz Lehmann Coutinho; Luiz Carlos Junior de Alcantara; Lívia Sacchetto; Maisa C. Pereira Parra; Maria Carolina Elias; Marta Giovanetti; Marília Moraes; Maurício Lacerda Nogueira. Prefeitura de Sao Paulo: Melissa Palmieri.; Patricia Akemi Assato; Paula Rahal; Paulo Inacio da Costa; Rafael dos Santos Bezerra; Raquel de Lello Rocha Campos Cassano. NGS Soluções Genômicas: Pilar Drummond Sampaio Corrêa Mariani. FZEA-USP Pirassununga: Mirele Daiana Poleti; Raul Machado Neto; Ricardo Augusto Brassaloti; Ricardo Haddad; Rodrigo Tocantins Calado. FAMERP-SJRP: Cecília Artico Banho; Sandra Coccuzzo Sampaio; Svetoslav Nanev Slavov; Vagner Fonseca; Vincent Louis Viala |
| EPI_ISL_1966919, EPI_ISL_1966920, EPI_ISL_1966921 | HOSPITAL MUNICIPAL DE ITABERA | Instituto Butantan / ESALQ-USP (Piracicaba) | Antonio Jorge Martins; Bianca Cechetto Carlos. Mendelics: Bibiana Santos; Claudia Renata dos Santos Barros; Cíntia Bittar; David Schlesinger. Hemocentro Ribeirão Preto: Simone Kashima; Debora Botequio Moretti; Elaine Cristina Marqueze; Elaine Vieira dos Santos; Elisangela Chicaroni Mattos; Erika Freitas; Evandra Strazza Rodrigues; Felipe Allan da Silva da Costa; Flavia Aburjaile; Fábio Sossai Possebon; Guilherme Campos; Guilherme Targino Valente; Heidge Fukumasu. USP-Botucatu: Rejane Maria Tommasini Grotto; Helena Lage Ferreira; Instituto Butantan: Dimas Tadeu Covas; Jardelina de Souza Todao Bernardino; Jayme A. Souza-Neto; Jessika Cristina Chagas Lesbon; Jorge A. Petroli Marchesi; José Salvatore Leister Patané; João Paulo Kitajima; João Pessoa Araújo Jr.; Leila Sabrina Ullmann; Loyze Paola Oliveira de Lima; Luiz Aurelio de Campos Crispin. Centro de Genômica Funcional da ESALQ: Luiz Lehmann Coutinho; Luiz Carlos Junior de Alcantara; Lívia Sacchetto; Maisa C. Pereira Parra; Maria Carolina Elias; Marta Giovanetti; Marília Moraes; Maurício Lacerda Nogueira. Prefeitura de Sao Paulo: Melissa Palmieri.; Patricia Akemi Assato; Paula Rahal; Paulo Inacio da Costa; Rafael dos Santos Bezerra; Raquel de Lello Rocha Campos Cassano. NGS Soluções Genômicas: Pilar Drummond Sampaio Corrêa Mariani. FZEA-USP Pirassununga: Mirele Daiana Poleti; Raul Machado Neto; Ricardo Augusto Brassaloti; Ricardo Haddad; Rodrigo Tocantins Calado. FAMERP-SJRP: Cecília Artico Banho; Sandra Coccuzzo Sampaio; Svetoslav Nanev Slavov; Vagner Fonseca; Vincent Louis Viala |
| EPI_ISL_1966942, EPI_ISL_1966945, EPI_ISL_1966946, EPI_ISL_1966947, EPI_ISL_1966949 | HOSPITAL MUNICIPAL DE ITAPIRA | Instituto Butantan / ESALQ-USP (Piracicaba) | Antonio Jorge Martins; Bianca Cechetto Carlos. Mendelics: Bibiana Santos; Claudia Renata dos Santos Barros; Cíntia Bittar; David Schlesinger. Hemocentro Ribeirão Preto: Simone Kashima; Debora Botequio Moretti; Elaine Cristina Marqueze; Elaine Vieira dos Santos; Elisangela Chicaroni Mattos; Erika Freitas; Evandra Strazza Rodrigues; Felipe Allan da Silva da Costa; Flavia Aburjaile; Fábio Sossai Possebon; Guilherme Campos; Guilherme Targino Valente; Heidge Fukumasu. USP-Botucatu: Rejane Maria Tommasini Grotto; Helena Lage Ferreira; Instituto Butantan: Dimas Tadeu Covas; Jardelina de Souza Todao Bernardino; Jayme A. Souza-Neto; Jessika Cristina Chagas Lesbon; Jorge A. Petroli Marchesi; José Salvatore Leister Patané; João Paulo Kitajima; João Pessoa Araújo Jr.; Leila Sabrina Ullmann; Loyze Paola Oliveira de Lima; Luiz Aurelio de Campos Crispin. Centro de Genômica Funcional da ESALQ: Luiz Lehmann Coutinho; Luiz Carlos Junior de Alcantara; Lívia Sacchetto; Maisa C. Pereira Parra; Maria Carolina Elias; Marta Giovanetti; Marília Moraes; Maurício Lacerda Nogueira. Prefeitura de Sao Paulo: Melissa Palmieri.; Patricia Akemi Assato; Paula Rahal; Paulo Inacio da Costa; Rafael dos Santos Bezerra; Raquel de Lello Rocha Campos Cassano. NGS Soluções Genômicas: Pilar Drummond Sampaio Corrêa Mariani. FZEA-USP Pirassununga: Mirele Daiana Poleti; Raul Machado Neto; Ricardo Augusto Brassaloti; Ricardo Haddad; Rodrigo Tocantins Calado. FAMERP-SJRP: Cecília Artico Banho; Sandra Coccuzzo Sampaio; Svetoslav Nanev Slavov; Vagner Fonseca; Vincent Louis Viala |
| EPI_ISL_5603215 | HOSPITAL MUNICIPAL DRA ZILDA SRNS NEUMAN | Analytical Competence Molecular Epidemiology Lab/ACME, Oswaldo Cruz Foundation, Ceara (FIOCRUZ CE) | Carlos Leonardo de Aragao Araujo; Cleber Furtado Aksenen; Fabio Miyajima; Fernando Braga Stehling; Francisco Eder de Moura Lopes; Jamille Maria Mendes Bezerra; Joaquim Cesar do Nascimento Sousa Junior; Pedro Miguel Carneiro Jeronimo; Suzana Porto Almeida & Igor Oliveira Duarte on behalf of COVID-19 FIOCRUZ Genomic Network; Thais Ferreira de Oliveira; Thais de Oliveira Costa; Ticiane Cavalcante de Souza; Veridiana Pessoa Miyajima |
| EPI_ISL_5530158 | HOSPITAL MUNICIPAL JOSE GONCALVES ROSA | Analytical Competence Molecular Epidemiology Lab/ACME, Oswaldo Cruz Foundation, Ceara (FIOCRUZ CE) | Carlos Leonardo de Aragao Araujo; Cecilia Leite Costa & Eduardo Ruback dos Santos on behalf of COVID-19 FIOCRUZ Genomic Network; Cleber Furtado Aksenen; Fabio Miyajima; Fernando Braga Stehling; Francisco Eder de Moura Lopes; Igor Oliveira Duarte; Jamille Maria Mendes Bezerra; Joaquim Cesar do Nascimento Sousa Junior; Pedro Miguel Carneiro Jeronimo; Suzana Porto Almeida; Thais Ferreira de Oliveira; Thais de Oliveira Costa; Ticiane Cavalcante de Souza; Veridiana Pessoa Miyajima |
| EPI_ISL_1967025, EPI_ISL_1967026 | HOSPITAL MUNICIPAL MARIA ROSA CARDOSO | Instituto Butantan / ESALQ-USP (Piracicaba) | Antonio Jorge Martins; Bianca Cechetto Carlos. Mendelics: Bibiana Santos; Claudia Renata dos Santos Barros; Cíntia Bittar; David Schlesinger. Hemocentro Ribeirão Preto: Simone Kashima; Debora Botequio Moretti; Elaine Cristina Marqueze; Elaine Vieira dos Santos; Elisangela Chicaroni Mattos; Erika Freitas; Evandra Strazza Rodrigues; Felipe Allan da Silva da Costa; Flavia Aburjaile; Fábio Sossai Possebon; Guilherme Campos; Guilherme Targino Valente; Heidge Fukumasu. USP-Botucatu: Rejane Maria Tommasini Grotto; Helena Lage Ferreira; Instituto Butantan: Dimas Tadeu Covas; Jardelina de Souza Todao Bernardino; Jayme A. Souza-Neto; Jessika Cristina Chagas Lesbon; Jorge A. Petroli Marchesi; José Salvatore Leister Patané; João Paulo Kitajima; João Pessoa Araújo Jr.; Leila Sabrina Ullmann; Loyze Paola Oliveira de Lima; Luiz Aurelio de Campos Crispin. Centro de Genômica Funcional da ESALQ: Luiz Lehmann Coutinho; Luiz Carlos Junior de Alcantara; Lívia Sacchetto; Maisa C. Pereira Parra; Maria Carolina Elias; Marta Giovanetti; Marília Moraes; Maurício Lacerda Nogueira. Prefeitura de Sao Paulo: Melissa Palmieri.; Patricia Akemi Assato; Paula Rahal; Paulo Inacio da Costa; Rafael dos Santos Bezerra; Raquel de Lello Rocha Campos Cassano. NGS Soluções Genômicas: Pilar Drummond Sampaio Corrêa Mariani. FZEA-USP Pirassununga: Mirele Daiana Poleti; Raul Machado Neto; Ricardo Augusto Brassaloti; Ricardo Haddad; Rodrigo Tocantins Calado. FAMERP-SJRP: Cecília Artico Banho; Sandra Coccuzzo Sampaio; Svetoslav Nanev Slavov; Vagner Fonseca; Vincent Louis Viala |
| EPI_ISL_1966846 | HOSPITAL MUNICIPAL REYNALDO GUERRA CAJATI | Instituto Butantan / ESALQ-USP (Piracicaba) | Antonio Jorge Martins; Bianca Cechetto Carlos. Mendelics: Bibiana Santos; Claudia Renata dos Santos Barros; Cíntia Bittar; David Schlesinger. Hemocentro Ribeirão Preto: Simone Kashima; Debora Botequio Moretti; Elaine Cristina Marqueze; Elaine Vieira dos Santos; Elisangela Chicaroni Mattos; Erika Freitas; Evandra Strazza Rodrigues; Felipe Allan da Silva da Costa; Flavia Aburjaile; Fábio Sossai Possebon; Guilherme Campos; Guilherme Targino Valente; Heidge Fukumasu. USP-Botucatu: Rejane Maria Tommasini Grotto; Helena Lage Ferreira; Instituto Butantan: Dimas Tadeu Covas; Jardelina de Souza Todao Bernardino; Jayme A. Souza-Neto; Jessika Cristina Chagas Lesbon; Jorge A. Petroli Marchesi; José Salvatore Leister Patané; João Paulo Kitajima; João Pessoa Araújo Jr.; Leila Sabrina Ullmann; Loyze Paola Oliveira de Lima; Luiz Aurelio de Campos Crispin. Centro de Genômica Funcional da ESALQ: Luiz Lehmann Coutinho; Luiz Carlos Junior de Alcantara; Lívia Sacchetto; Maisa C. Pereira Parra; Maria Carolina Elias; Marta Giovanetti; Marília Moraes; Maurício Lacerda Nogueira. Prefeitura de Sao Paulo: Melissa Palmieri.; Patricia Akemi Assato; Paula Rahal; Paulo Inacio da Costa; Rafael dos Santos Bezerra; Raquel de Lello Rocha Campos Cassano. NGS Soluções Genômicas: Pilar Drummond Sampaio Corrêa Mariani. FZEA-USP Pirassununga: Mirele Daiana Poleti; Raul Machado Neto; Ricardo Augusto Brassaloti; Ricardo Haddad; Rodrigo Tocantins Calado. FAMERP-SJRP: Cecília Artico Banho; Sandra Coccuzzo Sampaio; Svetoslav Nanev Slavov; Vagner Fonseca; Vincent Louis Viala |
| EPI_ISL_3502681 | HOSPITAL PRINCIPE DE ASTURIAS | HOSPITAL UNIVERSITARIO 12 DE OCTUBRE | Carmen Martín-Higuera; Esther Viedma; Irene Múñoz-Gallego; M.ª Dolores Folgueira; Mar Aguilera; Noelia Moral; Rafael Delgado; Sagrario Zurita |
| EPI_ISL_1966923, EPI_ISL_1966924, EPI_ISL_1966925, EPI_ISL_1966926 | HOSPITAL REGIONAL DE ITAPETININGA | Instituto Butantan / ESALQ-USP (Piracicaba) | Antonio Jorge Martins; Bianca Cechetto Carlos. Mendelics: Bibiana Santos; Claudia Renata dos Santos Barros; Cíntia Bittar; David Schlesinger. Hemocentro Ribeirão Preto: Simone Kashima; Debora Botequio Moretti; Elaine Cristina Marqueze; Elaine Vieira dos Santos; Elisangela Chicaroni Mattos; Erika Freitas; Evandra Strazza Rodrigues; Felipe Allan da Silva da Costa; Flavia Aburjaile; Fábio Sossai Possebon; Guilherme Campos; Guilherme Targino Valente; Heidge Fukumasu. USP-Botucatu: Rejane Maria Tommasini Grotto; Helena Lage Ferreira; Instituto Butantan: Dimas Tadeu Covas; Jardelina de Souza Todao Bernardino; Jayme A. Souza-Neto; Jessika Cristina Chagas Lesbon; Jorge A. Petroli Marchesi; José Salvatore Leister Patané; João Paulo Kitajima; João Pessoa Araújo Jr.; Leila Sabrina Ullmann; Loyze Paola Oliveira de Lima; Luiz Aurelio de Campos Crispin. Centro de Genômica Funcional da ESALQ: Luiz Lehmann Coutinho; Luiz Carlos Junior de Alcantara; Lívia Sacchetto; Maisa C. Pereira Parra; Maria Carolina Elias; Marta Giovanetti; Marília Moraes; Maurício Lacerda Nogueira. Prefeitura de Sao Paulo: Melissa Palmieri.; Patricia Akemi Assato; Paula Rahal; Paulo Inacio da Costa; Rafael dos Santos Bezerra; Raquel de Lello Rocha Campos Cassano. NGS Soluções Genômicas: Pilar Drummond Sampaio Corrêa Mariani. FZEA-USP Pirassununga: Mirele Daiana Poleti; Raul Machado Neto; Ricardo Augusto Brassaloti; Ricardo Haddad; Rodrigo Tocantins Calado. FAMERP-SJRP: Cecília Artico Banho; Sandra Coccuzzo Sampaio; Svetoslav Nanev Slavov; Vagner Fonseca; Vincent Louis Viala |
| EPI_ISL_1966954 | HOSPITAL SANTO ANTONIO DE JUQUIA JUQUIA | Instituto Butantan / ESALQ-USP (Piracicaba) | Antonio Jorge Martins; Bianca Cechetto Carlos. Mendelics: Bibiana Santos; Claudia Renata dos Santos Barros; Cíntia Bittar; David Schlesinger. Hemocentro Ribeirão Preto: Simone Kashima; Debora Botequio Moretti; Elaine Cristina Marqueze; Elaine Vieira dos Santos; Elisangela Chicaroni Mattos; Erika Freitas; Evandra Strazza Rodrigues; Felipe Allan da Silva da Costa; Flavia Aburjaile; Fábio Sossai Possebon; Guilherme Campos; Guilherme Targino Valente; Heidge Fukumasu. USP-Botucatu: Rejane Maria Tommasini Grotto; Helena Lage Ferreira; Instituto Butantan: Dimas Tadeu Covas; Jardelina de Souza Todao Bernardino; Jayme A. Souza-Neto; Jessika Cristina Chagas Lesbon; Jorge A. Petroli Marchesi; José Salvatore Leister Patané; João Paulo Kitajima; João Pessoa Araújo Jr.; Leila Sabrina Ullmann; Loyze Paola Oliveira de Lima; Luiz Aurelio de Campos Crispin. Centro de Genômica Funcional da ESALQ: Luiz Lehmann Coutinho; Luiz Carlos Junior de Alcantara; Lívia Sacchetto; Maisa C. Pereira Parra; Maria Carolina Elias; Marta Giovanetti; Marília Moraes; Maurício Lacerda Nogueira. Prefeitura de Sao Paulo: Melissa Palmieri.; Patricia Akemi Assato; Paula Rahal; Paulo Inacio da Costa; Rafael dos Santos Bezerra; Raquel de Lello Rocha Campos Cassano. NGS Soluções Genômicas: Pilar Drummond Sampaio Corrêa Mariani. FZEA-USP Pirassununga: Mirele Daiana Poleti; Raul Machado Neto; Ricardo Augusto Brassaloti; Ricardo Haddad; Rodrigo Tocantins Calado. FAMERP-SJRP: Cecília Artico Banho; Sandra Coccuzzo Sampaio; Svetoslav Nanev Slavov; Vagner Fonseca; Vincent Louis Viala |
| EPI_ISL_3912098, EPI_ISL_3912131 | HOSPITAL SAO JOSE DE DOENCAS INFECCIOSAS | Analytical Competence Molecular Epidemiology Lab/ACME, Oswaldo Cruz Foundation, Ceara (FIOCRUZ CE) | Cleber Furtado Aksenen; Fabio Miyajima; Fernando Braga Stehling; Francisco Eder de Moura Lopes; Jamille Maria Mendes Bezerra; Joaquim Cesar do Nascimento Sousa Junior; Pedro Miguel Carneiro Jeronimo; Suzana Porto Almeida & Lucas Delerino on behalf of COVID-19 FIOCRUZ Genomic Network; Thais Ferreira de Oliveira; Thais de Oliveira Costa; Ticiane Cavalcante de Souza; Veridiana Pessoa Miyajima |
| EPI_ISL_2209319 | HOSPITAL SAO JOSE ITIRAPINA | Instituto Butantan | Antonio Jorge Martins; Claudia Renata dos Santos Barros; David Schlesinger; Debora Botequio Moretti; Dimas Tadeu Covas; Elaine Cristina Marqueze; Elaine Vieira Santos; Evandra Strazza Rodrigues; Heidge Fukumasu; Jayme Augusto de Souza-Neto; José Salvatore Leister Patané; Luiz Alcantara; Luiz Lehmann Coutinho; Maria Carolina Elias; Maurício Lacerda Nogueira; Rafael dos Santos Bezerra; Raul Machado Neto; Rejane Maria Tommasini Grotto; Ricardo Haddad; Sandra Coccuzzo Sampaio Vessoni; Simone Kashima; Svetoslav Nanev Slavov; Vincent Louis Viala. |
| EPI_ISL_2210091 | HOSPITAL SAO JOSE ITIRAPINA | Instituto Butantan / FZEA-USP-Pirassununga | Antonio Jorge Martins; Claudia Renata dos Santos Barros; David Schlesinger; Debora Botequio Moretti; Dimas Tadeu Covas; Elaine Cristina Marqueze; Elaine Vieira Santos; Evandra Strazza Rodrigues; Heidge Fukumasu; Jayme Augusto de Souza-Neto; José Salvatore Leister Patané; Luiz Alcantara; Luiz Lehmann Coutinho; Maria Carolina Elias; Maurício Lacerda Nogueira; Rafael dos Santos Bezerra; Raul Machado Neto; Rejane Maria Tommasini Grotto; Ricardo Haddad; Sandra Coccuzzo Sampaio Vessoni; Simone Kashima; Svetoslav Nanev Slavov; Vincent Louis Viala |
| EPI_ISL_1967065, EPI_ISL_1967066 | HOSPITAL SAO MIGUEL ARCANJO | Instituto Butantan / ESALQ-USP (Piracicaba) | Antonio Jorge Martins; Bianca Cechetto Carlos. Mendelics: Bibiana Santos; Claudia Renata dos Santos Barros; Cíntia Bittar; David Schlesinger. Hemocentro Ribeirão Preto: Simone Kashima; Debora Botequio Moretti; Elaine Cristina Marqueze; Elaine Vieira dos Santos; Elisangela Chicaroni Mattos; Erika Freitas; Evandra Strazza Rodrigues; Felipe Allan da Silva da Costa; Flavia Aburjaile; Fábio Sossai Possebon; Guilherme Campos; Guilherme Targino Valente; Heidge Fukumasu. USP-Botucatu: Rejane Maria Tommasini Grotto; Helena Lage Ferreira; Instituto Butantan: Dimas Tadeu Covas; Jardelina de Souza Todao Bernardino; Jayme A. Souza-Neto; Jessika Cristina Chagas Lesbon; Jorge A. Petroli Marchesi; José Salvatore Leister Patané; João Paulo Kitajima; João Pessoa Araújo Jr.; Leila Sabrina Ullmann; Loyze Paola Oliveira de Lima; Luiz Aurelio de Campos Crispin. Centro de Genômica Funcional da ESALQ: Luiz Lehmann Coutinho; Luiz Carlos Junior de Alcantara; Lívia Sacchetto; Maisa C. Pereira Parra; Maria Carolina Elias; Marta Giovanetti; Marília Moraes; Maurício Lacerda Nogueira. Prefeitura de Sao Paulo: Melissa Palmieri.; Patricia Akemi Assato; Paula Rahal; Paulo Inacio da Costa; Rafael dos Santos Bezerra; Raquel de Lello Rocha Campos Cassano. NGS Soluções Genômicas: Pilar Drummond Sampaio Corrêa Mariani. FZEA-USP Pirassununga: Mirele Daiana Poleti; Raul Machado Neto; Ricardo Augusto Brassaloti; Ricardo Haddad; Rodrigo Tocantins Calado. FAMERP-SJRP: Cecília Artico Banho; Sandra Coccuzzo Sampaio; Svetoslav Nanev Slavov; Vagner Fonseca; Vincent Louis Viala |
| EPI_ISL_3502682, EPI_ISL_3502694, EPI_ISL_3502707, EPI_ISL_3502712, EPI_ISL_3502764, EPI_ISL_3532482 | HOSPITAL UNIVERSITARIO 12 DE OCTUBRE | HOSPITAL UNIVERSITARIO 12 DE OCTUBRE | Carmen Martín-Higuera; Esther Viedma; Irene Múñoz-Gallego; M.ª Dolores Folgueira; Mar Aguilera; Noelia Moral; Rafael Delgado; Sagrario Zurita |
| EPI_ISL_3385829 | HOSPITAL UNIVERSITARIO HERMANO MONCALEANO | Instituto Nacional de Salud-Dirección de Investigación en Salud Pública | Carlos Franco-Muñoz; Carmen Osorio; Diana Malo; Diego A. Álvarez-Díaz; Diego Andrés Prada; Gerardo Santamaría; Hector Alejandro Ruiz-Moreno; Jhonnatan Reales-González; Jorge Rivera; Juan Camilo Martinez; Julian Naizaque; Katherine Laiton-Donato; Lisseth Pardo; Magdalena Wiesner; Marcela Mercado-Reyes; Maria T. Herrera-Sepúlveda; Marta Lopez Blanco; Martha Lucia Ospina Martinez; Paola Rojas; Sergio Gomez; Sheryll Corchuelo; Ángela Alarcon Cruz |
| EPI_ISL_2020090, EPI_ISL_2020097, EPI_ISL_2020105 | HOSPITAL UNIVERSITARIO SON ESPASES | HOSPITAL UNIVERSITARIO SON ESPASES | Antonio Oliver; Carla López-Causapé; Pablo Fraile-Ribot; SeqCovid |
| EPI_ISL_7744025 | HU | Laboratório de Bioinformática - Universidade Federal de Santa Catarina | "Aline Daina Schlindwein"; "Ana Paula Christoff"; "Antuani Baptistela"; "Carolina Leite Martins"; "Darcita Buerger Rovaris"; "Dayane Azevedo Padilha"; "Doris Sobral Marques SouzaSobral"; "Edmundo Carlos Grisard"; "Eric Kazuo Kawagoe"; "Fernanda Luiza Ferrari"; "Fernanda Roesene Melo"; "Fernando Hartmann Barazzetti"; "Gislaine Fongaro"; "Glauber Wagner"; "Guilherme Augusto Maia"; "Guilherme Razzera"; "Guilherme Toledo e Silva"; "Julia Kinetz Wachter"; "Luiz Felipe de Oliveira"; "Marcos André Schörne"; "Marcus Vinícius Duarte Rodrigues"; "Maria Luiza Bazzo"; "Marlei Pickler Debiasi dos Anjos"; "Milene Moehr de Moraes"; "Nestor Wendt"; "Patrícia Hermes Stoco"; "Paula Sacchet"; "Renato Simões Moreira"; "Rodrigo de Paula Baptista"; "Tamela Zamboni Madaloz"; "Tatiany Aparecida Teixeira Soratto"; "Vilmar Benetti Filho" |
| EPI_ISL_2081100, EPI_ISL_2081106, EPI_ISL_2483366, EPI_ISL_2483379, EPI_ISL_2483424, EPI_ISL_2483432, EPI_ISL_2483443 | | | |
| see above | Hackensack Medical Center | New York Genome Center | Andre Corvelo; Barry Kreiswirth; David Perlin; Dayna M. Oschwald; Jose Mediavilla; Kaelea Composto; Kar Chow; Liang Chen; Marcus Cunningham; Michael Zody; Samantha Fennessey; Tom Maniatis |
| EPI_ISL_1679207, EPI_ISL_1679260, EPI_ISL_1679452, EPI_ISL_1734583, EPI_ISL_1734594, EPI_ISL_1734618, EPI_ISL_1734631, EPI_ISL_1734658, EPI_ISL_1734697, EPI_ISL_1734717, EPI_ISL_1734718, EPI_ISL_1734724, EPI_ISL_1734793, EPI_ISL_1734953, EPI_ISL_1734976, EPI_ISL_1734992, EPI_ISL_1735009, EPI_ISL_1735076, EPI_ISL_1735114, EPI_ISL_1735219, EPI_ISL_1735223, EPI_ISL_1735236, EPI_ISL_1735267, EPI_ISL_1735276, EPI_ISL_1735289, EPI_ISL_1735318, EPI_ISL_1735388, EPI_ISL_1735431, EPI_ISL_1735447, EPI_ISL_1735449, EPI_ISL_1735480, EPI_ISL_1735492, EPI_ISL_1735536, EPI_ISL_1735575, EPI_ISL_1735592, EPI_ISL_1735628, EPI_ISL_1735647, EPI_ISL_1735676, EPI_ISL_1753511, EPI_ISL_1796142, EPI_ISL_1796148, EPI_ISL_1796182, EPI_ISL_1796196, EPI_ISL_1796210, EPI_ISL_1796276, EPI_ISL_1796288, EPI_ISL_1796293, EPI_ISL_1796318, EPI_ISL_1796337, EPI_ISL_1796371, EPI_ISL_1796386, EPI_ISL_1796407, EPI_ISL_1796438, EPI_ISL_1796446, EPI_ISL_1796455, EPI_ISL_1796488, EPI_ISL_1796491, EPI_ISL_1796498, EPI_ISL_1796516, EPI_ISL_1796524, EPI_ISL_1796530, EPI_ISL_1796547, EPI_ISL_1796608, EPI_ISL_1796655, EPI_ISL_1796677, EPI_ISL_1796678, EPI_ISL_1796769, EPI_ISL_1796776, EPI_ISL_1796789, EPI_ISL_1796801, EPI_ISL_1796817, EPI_ISL_1796839, EPI_ISL_1796856, EPI_ISL_1796866, EPI_ISL_1796942, EPI_ISL_1796971, EPI_ISL_1796997, EPI_ISL_1797023, EPI_ISL_1797033, EPI_ISL_1797065, EPI_ISL_1797099, EPI_ISL_1797131, EPI_ISL_1797138, EPI_ISL_1797191, EPI_ISL_1797209, EPI_ISL_1797219, EPI_ISL_1797237, EPI_ISL_1797238, EPI_ISL_1797243, EPI_ISL_1797262, EPI_ISL_1797381, EPI_ISL_1797398, EPI_ISL_1797409, EPI_ISL_1803984, EPI_ISL_1804030, EPI_ISL_1804324, EPI_ISL_1804339, EPI_ISL_1804350, EPI_ISL_1804360, EPI_ISL_1804399, EPI_ISL_1804419, EPI_ISL_1804505, EPI_ISL_1804573, EPI_ISL_1804581, EPI_ISL_1804584, EPI_ISL_1804594, EPI_ISL_1804607, EPI_ISL_1804613, EPI_ISL_1804615, EPI_ISL_1804649, EPI_ISL_1804667, EPI_ISL_1804678, EPI_ISL_1804721, EPI_ISL_1817999, EPI_ISL_1818014, EPI_ISL_1818024, EPI_ISL_1818041, EPI_ISL_1818042, EPI_ISL_1818045, EPI_ISL_4346392, EPI_ISL_4347493, EPI_ISL_4347687 | | | |
| see above | Helix/Illumina | Centers for Disease Control and Prevention Division of Viral Diseases, Pathogen Discovery | Adrian Paskey; Alexandre Bolze; Ary Ascencio; Benjamin Rambo-Martin; Brad Sickler; Charlotte Rivera-Garcia; Christine Tran; Christopher Gulvick; Chrstine Tran; Clinton Paden; Clinton R. Paden; Dakota Howard; Darlene Wagner; David Becker; Dhwani Batra; Duncan MacCannell; Efren Sandoval; Eileen De Feo; Eileen de Feo; Elizabeth Cirulli; Eric Allen; Geraint Levan; James Lu; Jan Antico; Jason Caravas; Jason Nguyen; Jimmy Ramirez; Jingtao Liu; Kara Moser; Kelly Barrett; Kelly Schiabor Barrett; Kim Gietzen; Kristine Lacek; Magnus Isaksson; Marc Laurent; Matthew Schmerer; Matthew Tolentino; Nicole L. Washington; Nicole Washington; Peter Cook; Peter W. Cook; Phil Febbo; Ryan Cho; Scott Sammons; Shannon Wickline; Shatavia Morrison; Sherry Wang; Simon White; Tyler Cassens; William Lee; Yvette Unoarumhi |
| EPI_ISL_1811228 | Horquetas Rio Frio | Inciensa, Instituto Costarricense de Investigación y Enseñanza en | Pérez-Corrales C & Montiel-Ulloa G |

| | | | |
|---|---|---|---|
| | | Nutrición y Salud | |
| EPI_ISL_5800102, EPI_ISL_5800104, EPI_ISL_5800107, EPI_ISL_5800108, EPI_ISL_5802304, EPI_ISL_5802464, EPI_ISL_5802468 | | | |
| see above | Hosp Do Serv Pub Estfco Morato De Oliveira Sao Paulo | Instituto Butantan | Antonio Jorge Martins; Claudia Renata dos Santos Barros; David Schlesinger; Debora Botequio Moretti; Dimas Tadeu Covas; Elaine Cristina Marqueze; Elaine Vieira Santos; Evandra Strazza Rodrigues; Heidge Fukumasu; Jayme Augusto de Souza-Neto; José Salvatore Leister Patané; Luiz Alcantara; Luiz Lehmann Coutinho; Maria Carolina Elias; Maurício Lacerda Nogueira; Rafael dos Santos Bezerra; Raul Machado Neto; Rejane Maria Tommasini Grotto; Ricardo Haddad; Sandra Coccuzzo Sampaio Vessoni; Simone Kashima; Svetoslav Nanev Slavov; Vincent Louis Viala |
| EPI_ISL_5800212, EPI_ISL_5800229, EPI_ISL_5800231, EPI_ISL_5800233 | Hosp E Maternidade Municipal N Sra Monte Serrat | Instituto Butantan | Antonio Jorge Martins; Claudia Renata dos Santos Barros; David Schlesinger; Debora Botequio Moretti; Dimas Tadeu Covas; Elaine Cristina Marqueze; Elaine Vieira Santos; Evandra Strazza Rodrigues; Heidge Fukumasu; Jayme Augusto de Souza-Neto; José Salvatore Leister Patané; Luiz Alcantara; Luiz Lehmann Coutinho; Maria Carolina Elias; Maurício Lacerda Nogueira; Rafael dos Santos Bezerra; Raul Machado Neto; Rejane Maria Tommasini Grotto; Ricardo Haddad; Sandra Coccuzzo Sampaio Vessoni; Simone Kashima; Svetoslav Nanev Slavov; Vincent Louis Viala |
| EPI_ISL_5800088, EPI_ISL_5802330, EPI_ISL_5802335, EPI_ISL_5802336 | Hosp Mun Cidade Tiradentes Carmen Prudente | Instituto Butantan | Antonio Jorge Martins; Claudia Renata dos Santos Barros; David Schlesinger; Debora Botequio Moretti; Dimas Tadeu Covas; Elaine Cristina Marqueze; Elaine Vieira Santos; Evandra Strazza Rodrigues; Heidge Fukumasu; Jayme Augusto de Souza-Neto; José Salvatore Leister Patané; Luiz Alcantara; Luiz Lehmann Coutinho; Maria Carolina Elias; Maurício Lacerda Nogueira; Rafael dos Santos Bezerra; Raul Machado Neto; Rejane Maria Tommasini Grotto; Ricardo Haddad; Sandra Coccuzzo Sampaio Vessoni; Simone Kashima; Svetoslav Nanev Slavov; Vincent Louis Viala |
| EPI_ISL_5802352, EPI_ISL_5802354 | Hosp Mun Josanias Castanha Braga | Instituto Butantan | Antonio Jorge Martins; Claudia Renata dos Santos Barros; David Schlesinger; Debora Botequio Moretti; Dimas Tadeu Covas; Elaine Cristina Marqueze; Elaine Vieira Santos; Evandra Strazza Rodrigues; Heidge Fukumasu; Jayme Augusto de Souza-Neto; José Salvatore Leister Patané; Luiz Alcantara; Luiz Lehmann Coutinho; Maria Carolina Elias; Maurício Lacerda Nogueira; Rafael dos Santos Bezerra; Raul Machado Neto; Rejane Maria Tommasini Grotto; Ricardo Haddad; Sandra Coccuzzo Sampaio Vessoni; Simone Kashima; Svetoslav Nanev Slavov; Vincent Louis Viala |
| EPI_ISL_1700617, EPI_ISL_2030088 | Hospital | National Reference Center for Viruses of Respiratory Infections, Institut Pasteur, Paris | Amaury Vaysse; Angela Brisebarre; Camille Capel; Christophe Malabat; Corinne Maufrais; CéLine Ramanantsoa; Damien Mornico; Etienne Simon-Lorière; Frédéric Lemoine; Louise Lefrançois; Marianne Burgard; Marion Barbet; Maud Vanpeene; Méline Bizard; Sylvie Behillil; Sylvie Van der Werf; Vincent Enouf |
| EPI_ISL_1917315, EPI_ISL_1917317 | Hospital Center Emile Mayrisch | Laboratoire national de sante, Microbiology, Microbial Genomics Platform | Anke Wienecke-Baldacchino; Catherine Ragimbeau; Cynthia Oxacelay; Fatu Djabi; Jessica Tapp; Lise Pignon; Raoul Salmon; Tamir Abdelrahman |
| EPI_ISL_1917402 | Hospital Center Luxembourg | Laboratoire national de sante, Microbiology, Microbial Genomics Platform | Anke Wienecke-Baldacchino; Catherine Ragimbeau; Fatu Djabi; Jean-Hugues Francois; Jessica Tapp; Lise Pignon; Michel Kohnen; Raoul Salmon; Tamir Abdelrahman |
| EPI_ISL_5802334 | Hospital De Campanha Covid 19 Caieiras | Instituto Butantan | Antonio Jorge Martins; Claudia Renata dos Santos Barros; David Schlesinger; Debora Botequio Moretti; Dimas Tadeu Covas; Elaine Cristina Marqueze; Elaine Vieira Santos; Evandra Strazza Rodrigues; Heidge Fukumasu; Jayme Augusto de Souza-Neto; José Salvatore Leister Patané; Luiz Alcantara; Luiz Lehmann Coutinho; Maria Carolina Elias; Maurício Lacerda Nogueira; Rafael dos Santos Bezerra; Raul Machado Neto; Rejane Maria Tommasini Grotto; Ricardo Haddad; Sandra Coccuzzo Sampaio Vessoni; Simone Kashima; Svetoslav Nanev Slavov; Vincent Louis Viala |
| EPI_ISL_5800364, EPI_ISL_5800365, EPI_ISL_5800368, EPI_ISL_5800370, EPI_ISL_5800371, EPI_ISL_5800372, EPI_ISL_5800373, EPI_ISL_5800374, EPI_ISL_5800375, EPI_ISL_5800376, EPI_ISL_5800377, EPI_ISL_5800379, EPI_ISL_5800380, EPI_ISL_5800381, EPI_ISL_5800383, EPI_ISL_5800384, EPI_ISL_5800385, EPI_ISL_5800386, EPI_ISL_5800392, EPI_ISL_5800393, EPI_ISL_5800394, EPI_ISL_5800395, EPI_ISL_5800396, EPI_ISL_5800397, EPI_ISL_5800398, EPI_ISL_5800399, EPI_ISL_5800400 | | | |
| see above | Hospital De Campanha Covid 19 Municipio De Taubate | Instituto Butantan | Antonio Jorge Martins; Claudia Renata dos Santos Barros; David Schlesinger; Debora Botequio Moretti; Dimas Tadeu Covas; Elaine Cristina Marqueze; Elaine Vieira Santos; Evandra Strazza Rodrigues; Heidge Fukumasu; Jayme Augusto de Souza-Neto; José Salvatore Leister Patané; Luiz Alcantara; Luiz Lehmann Coutinho; Maria Carolina Elias; Maurício Lacerda Nogueira; Rafael dos Santos Bezerra; Raul Machado Neto; Rejane Maria Tommasini Grotto; Ricardo Haddad; Sandra Coccuzzo Sampaio Vessoni; Simone Kashima; Svetoslav Nanev Slavov; Vincent Louis Viala |
| EPI_ISL_1647783, EPI_ISL_1647841, EPI_ISL_1647844, EPI_ISL_1647845, EPI_ISL_1647851, EPI_ISL_1647852, EPI_ISL_1647854, EPI_ISL_1647855, EPI_ISL_1647858, EPI_ISL_1647859, EPI_ISL_1647944, EPI_ISL_1647945, EPI_ISL_1647946 | | | |
| see above | Hospital General Universitario Gregorio Marañón | Hospital General Universitario Gregorio Marañón | Cristina Rodriguez-Grande; Darío García de Viedma; Laura Pérez-Lago; Patricia Muñoz; Pedro Sola Campoy; Pilar Catalán; Sergio Buenestado Serrano |
| EPI_ISL_2271473, EPI_ISL_2271474, EPI_ISL_2271475 | Hospital General Universitario de Alicante - Instituto de Investigación Sanitaria y Biomédica de Alicante | SeqCOVID-SPAIN consortium/IBV(CSIC) | Carmen Molina Pardines and SeqCOVID-SPAIN consortium; Maripaz Ventero Martín |
| EPI_ISL_5800005 | Hospital Geral Jesus Teixeira Da Costa Guaianases Sao Paulo | Instituto Butantan | Antonio Jorge Martins; Claudia Renata dos Santos Barros; David Schlesinger; Debora Botequio Moretti; Dimas Tadeu Covas; Elaine Cristina Marqueze; Elaine Vieira Santos; Evandra Strazza Rodrigues; Heidge Fukumasu; Jayme Augusto de Souza-Neto; José Salvatore Leister Patané; Luiz Alcantara; Luiz Lehmann Coutinho; Maria Carolina Elias; Maurício Lacerda Nogueira; Rafael dos Santos Bezerra; Raul Machado Neto; Rejane Maria Tommasini Grotto; Ricardo Haddad; Sandra Coccuzzo Sampaio Vessoni; Simone Kashima; Svetoslav Nanev Slavov; Vincent Louis Viala |
| EPI_ISL_2551531 | Hospital Infantil Menino Jesus | Laboratório de Virologia Clínica e Molecular | Alex Fiorini; Ana Paula Salles Fernandes; Bruna Larotonda Telezynski; Danielle Bruna Leal Oliveira; Edison Luiz Durigon; Erick Gustavo Dorlass; Flavio Fonseca e Santuza Teixeira; Guilherme Pereira Scagion; Helena Perez Coelho; Hugo Sato; Karine Lima Lourenço; Luciano Matsumiya Thomazelli; Renata Peixoto; Rubens Daniel Miserani Magalhães; Tatiana Ometto |
| EPI_ISL_2363555, EPI_ISL_2363563, EPI_ISL_2363564 | Hospital Jaime Ferre - SAMCO Rafaela | Grupo de Genómica y Bioinformática del Instituto de Investigación de la Cadena Láctea CONICET-INTA on behalf of 'Proyecto Argentino Interinstitucional de genomica de SARS-CoV-2' (PAIS Consortium) | AF; Amadio; C; Eberhardt; Irazoqui; Isaia; JF; JM; MF; Pandolfi; Quaranta; V |
| EPI_ISL_7711732 | Hospital Monseñor Sanabria | Universidad de Costa Rica e Instituto Costarricense de Investigacion y Ensenanza en Nutricion y Salud | Brenes Hebleen; Camacho Erwin; Campos-Sanchez Rebeca; Ceballos Ana; Cordero Estela; Cristancho Marco; Duarte Francisco; Fernandez-Do Porto Dario; Herrera-Estrella Alfredo; Jimenez-Moraila Beatriz; Kreuze Jan; Mireles-Rivera Guadalupe; Molina-Mora Jose; Munoz-Medina Jose Esteban; Negri Tatiana; Nunes Gisele; Oliveira Guilherme; Oliveira Renato; Reales-Gonzalez Jhonnatan; Remes-Lenicov Federico; Reyes Alejandro; Sosa Ezequiel; Soto Claudio; Tsukayama Pablo; Turjanski Adrian; and Zayat Jonathan |
| EPI_ISL_5802366 | Hospital Mun Dr Guido Guida | Instituto Butantan | Antonio Jorge Martins; Claudia Renata dos Santos Barros; David Schlesinger; Debora Botequio Moretti; Dimas Tadeu Covas; Elaine Cristina Marqueze; Elaine Vieira Santos; Evandra Strazza Rodrigues; Heidge Fukumasu; Jayme Augusto de Souza-Neto; José Salvatore Leister Patané; Luiz Alcantara; Luiz Lehmann Coutinho; Maria Carolina Elias; Maurício Lacerda Nogueira; Rafael dos Santos Bezerra; Raul Machado Neto; Rejane Maria Tommasini Grotto; Ricardo Haddad; Sandra Coccuzzo Sampaio Vessoni; Simone Kashima; Svetoslav Nanev Slavov; Vincent Louis Viala |
| EPI_ISL_5800353, EPI_ISL_5800354 | Hospital Municipal De Ibiuna Ibiuna Sp | Instituto Butantan | Antonio Jorge Martins; Claudia Renata dos Santos Barros; David Schlesinger; Debora Botequio Moretti; Dimas Tadeu Covas; Elaine Cristina Marqueze; Elaine Vieira Santos; Evandra Strazza Rodrigues; Heidge Fukumasu; Jayme Augusto de Souza-Neto; José Salvatore Leister Patané; Luiz Alcantara; Luiz Lehmann Coutinho; Maria Carolina Elias; Maurício Lacerda Nogueira; Rafael dos Santos Bezerra; Raul Machado Neto; Rejane Maria Tommasini Grotto; Ricardo Haddad; Sandra Coccuzzo Sampaio Vessoni; Simone Kashima; Svetoslav Nanev Slavov; Vincent Louis Viala |
| EPI_ISL_5800310, EPI_ISL_5800313, EPI_ISL_5800315 | Hospital Municipal De Itabera | Instituto Butantan | Antonio Jorge Martins; Claudia Renata dos Santos Barros; David Schlesinger; Debora Botequio Moretti; Dimas Tadeu Covas; Elaine Cristina Marqueze; Elaine Vieira Santos; Evandra Strazza Rodrigues; Heidge Fukumasu; Jayme Augusto de Souza-Neto; José Salvatore Leister Patané; Luiz Alcantara; Luiz Lehmann Coutinho; Maria Carolina Elias; Maurício Lacerda Nogueira; Rafael dos Santos Bezerra; Raul Machado Neto; Rejane Maria Tommasini Grotto; Ricardo Haddad; Sandra Coccuzzo Sampaio Vessoni; Simone Kashima; Svetoslav Nanev Slavov; Vincent Louis Viala |
| EPI_ISL_5799972, EPI_ISL_5799995, EPI_ISL_5799996, EPI_ISL_5799997, EPI_ISL_5800027 | Hospital Municipal De Itapira | Instituto Butantan | Antonio Jorge Martins; Claudia Renata dos Santos Barros; David Schlesinger; Debora Botequio Moretti; Dimas Tadeu Covas; Elaine Cristina Marqueze; Elaine Vieira Santos; Evandra Strazza Rodrigues; Heidge Fukumasu; Jayme Augusto de Souza-Neto; José Salvatore Leister Patané; Luiz Alcantara; Luiz Lehmann Coutinho; Maria Carolina Elias; Maurício Lacerda Nogueira; Rafael dos Santos Bezerra; Raul Machado Neto; Rejane Maria Tommasini Grotto; Ricardo Haddad; Sandra Coccuzzo Sampaio Vessoni; Simone Kashima; Svetoslav Nanev Slavov; Vincent Louis Viala |
| EPI_ISL_1752640 | Hospital Municipal Dr Mario Gatti Campinas | Instituto Adolfo Lutz, Interdiciplinary Procedures Center, Strategic Laboratory | Caio Vinicius Dias Lopes; Claudia Regina Gonçalves; Claudio Tavares Sacchi; Erica Valessa Ramos Gomes; Karoline Rodrigues Campos; Katia Correa de Oliveira Santos; Leonardo Jose Tadeu de Araujo |
| EPI_ISL_5800255, EPI_ISL_5800265 | Hospital Municipal Maria Rosa Cardoso | Instituto Butantan | Antonio Jorge Martins; Claudia Renata dos Santos Barros; David Schlesinger; Debora Botequio Moretti; Dimas Tadeu Covas; Elaine Cristina Marqueze; Elaine Vieira Santos; Evandra Strazza Rodrigues; Heidge Fukumasu; Jayme Augusto de Souza-Neto; José Salvatore Leister Patané; Luiz Alcantara; Luiz Lehmann Coutinho; Maria Carolina Elias; Maurício Lacerda Nogueira; Rafael dos Santos Bezerra; Raul Machado Neto; Rejane Maria Tommasini Grotto; Ricardo Haddad; Sandra Coccuzzo Sampaio Vessoni; Simone Kashima; Svetoslav Nanev Slavov; Vincent Louis Viala |
| EPI_ISL_5800213 | Hospital Municipal Reynaldo Guerra Cajati | Instituto Butantan | Antonio Jorge Martins; Claudia Renata dos Santos Barros; David Schlesinger; Debora Botequio Moretti; Dimas Tadeu Covas; Elaine Cristina Marqueze; Elaine Vieira Santos; Evandra Strazza Rodrigues; Heidge Fukumasu; Jayme Augusto de Souza-Neto; José Salvatore Leister Patané; Luiz Alcantara; Luiz Lehmann Coutinho; Maria Carolina Elias; Maurício Lacerda Nogueira; Rafael dos Santos Bezerra; Raul Machado Neto; Rejane Maria Tommasini Grotto; Ricardo Haddad; Sandra Coccuzzo Sampaio Vessoni; Simone Kashima; Svetoslav Nanev Slavov; Vincent Louis Viala |
| EPI_ISL_1675328, EPI_ISL_1675329 | Hospital Pablo Tobón Uribe | Universidad Nacional de Colombia - Laboratorio Genómico One Health | Andres F. Cardona-Rios; Carlos Franco-Muñoz; Daniel O. Maldonado-Perez; Diego A. Álvarez-Díaz; Hector Alejandro Ruiz-Moreno; Idabely Betancur Ortiz; Jorge E. Osorio; Juan P. Hernandez-Ortiz; Karl A Ciuoderis; Katherine Laiton-Donato; Laura Silvana Perez; Lina M. Hurtado; Marcela Mercado-Reyes; Maria Angélica Maya; Maria Stella López; Rita Almanza Payares; Sandra Ines Cano; Simón Villegas Velásquez |
| EPI_ISL_5800239, EPI_ISL_5800241, EPI_ISL_5800243, EPI_ISL_5800248 | Hospital Regional De Itapetininga | Instituto Butantan | Antonio Jorge Martins; Claudia Renata dos Santos Barros; David Schlesinger; Debora Botequio Moretti; Dimas Tadeu Covas; Elaine Cristina Marqueze; Elaine Vieira Santos; Evandra Strazza Rodrigues; Heidge Fukumasu; Jayme Augusto de Souza-Neto; José Salvatore Leister Patané; Luiz Alcantara; Luiz Lehmann Coutinho; Maria Carolina Elias; Maurício Lacerda Nogueira; Rafael dos Santos Bezerra; Raul Machado Neto; Rejane Maria Tommasini Grotto; Ricardo Haddad; Sandra Coccuzzo Sampaio Vessoni; Simone Kashima; Svetoslav Nanev Slavov; Vincent Louis Viala |
| EPI_ISL_2614570 | Hospital Sagrada Família-Unidade Maua | Instituto Adolfo Lutz, Interdiciplinary Procedures Center, Strategic Laboratory | Caio Vinicius Dias Lopes; Claudia Regina Gonçalves; Claudio Tavares Sacchi; Erica Valessa Ramos Gomes; Karoline Rodrigues Campos; Leonardo Jose Tadeu de Araujo |
| EPI_ISL_7711733 | Hospital San Juan De Dios | Universidad de Costa Rica e Instituto Costarricense de Investigacion y Ensenanza en Nutricion y Salud | Brenes Hebleen; Camacho Erwin; Campos-Sanchez Rebeca; Ceballos Ana; Cordero Estela; Cristancho Marco; Duarte Francisco; Fernandez-Do Porto Dario; Herrera-Estrella Alfredo; Jimenez-Moraila Beatriz; Kreuze Jan; Mireles-Rivera Guadalupe; Molina-Mora Jose; Munoz-Medina Jose Esteban; Negri Tatiana; Nunes Gisele; Oliveira Guilherme; Oliveira Renato; Reales-Gonzalez Jhonnatan; Remes-Lenicov Federico; Reyes Alejandro; Sosa Ezequiel; Soto Claudio; Tsukayama Pablo; Turjanski Adrian; and Zayat Jonathan |
| EPI_ISL_5800157 | Hospital Santo Antonio De Juquia Juquia | Instituto Butantan | Antonio Jorge Martins; Claudia Renata dos Santos Barros; David Schlesinger; Debora Botequio Moretti; Dimas Tadeu Covas; Elaine Cristina Marqueze; Elaine Vieira Santos; Evandra Strazza Rodrigues; Heidge Fukumasu; Jayme Augusto de Souza-Neto; José Salvatore Leister Patané; Luiz Alcantara; Luiz Lehmann Coutinho; Maria Carolina Elias; Maurício Lacerda Nogueira; Rafael dos Santos Bezerra; Raul Machado Neto; Rejane Maria Tommasini Grotto; Ricardo Haddad; Sandra Coccuzzo Sampaio Vessoni; Simone Kashima; Svetoslav Nanev Slavov; Vincent Louis Viala |
| EPI_ISL_5799703 | Hospital Sao Jose Itirapina | Instituto Butantan | Antonio Jorge Martins; Claudia Renata dos Santos Barros; David Schlesinger; Debora Botequio Moretti; Dimas Tadeu Covas; Elaine Cristina Marqueze; Elaine Vieira Santos; Evandra Strazza Rodrigues; Heidge Fukumasu; Jayme Augusto de Souza-Neto; José Salvatore Leister Patané; Luiz Alcantara; Luiz Lehmann Coutinho; Maria Carolina Elias; Maurício Lacerda Nogueira; Rafael dos Santos Bezerra; Raul Machado Neto; Rejane Maria Tommasini Grotto; Ricardo Haddad; Sandra Coccuzzo Sampaio Vessoni; Simone Kashima; Svetoslav Nanev Slavov; Vincent Louis Viala |
| EPI_ISL_5800349, EPI_ISL_5800352 | Hospital Sao Miguel Arcanjo | Instituto Butantan | Antonio Jorge Martins; Claudia Renata dos Santos Barros; David Schlesinger; Debora Botequio Moretti; Dimas Tadeu Covas; Elaine Cristina Marqueze; Elaine Vieira Santos; Evandra Strazza Rodrigues; Heidge Fukumasu; Jayme Augusto de Souza-Neto; José Salvatore Leister Patané; Luiz Alcantara; Luiz Lehmann Coutinho; Maria Carolina Elias; Maurício Lacerda Nogueira; Rafael dos Santos Bezerra; Raul Machado Neto; Rejane Maria Tommasini Grotto; Ricardo Haddad; Sandra Coccuzzo Sampaio Vessoni; Simone Kashima; Svetoslav Nanev Slavov; Vincent Louis Viala |

| | | | |
|---|---|---|---|
| EPI_ISL_1669398 | Hospital Universitari Vall d'Hebron - Vall d'Hebron Institut de Recerca | Hospital Universitari Vall d'Hebron - Vall d'Hebron Institut de Recerca | Andrés Antón; Ariadna Rando; Carla Castillo; Cristina Andrés; Damir Garcia-Cehic; Josep Quer; Juliana Esperalba; Maria Carmen Martin; Maria Gema Codina; Maria Piñana; Tomàs Pumarola |
| EPI_ISL_2614569 | Hospital Universitário da USP | Instituto Adolfo Lutz, Interdiciplinary Procedures Center, Strategic Laboratory | Caio Vinicius Dias Lopes; Claudia Regina Gonçalves; Claudio Tavares Sacchi; Erica Valessa Ramos Gomes; Karoline Rodrigues Campos; Leonardo Jose Tadeu de Araujo |
| EPI_ISL_4196722 | Hospital Zonal Dr. Ramón Carrillo | Laboratorio Central Mg. Luis Alfredo Pianciola on behalf of 'Proyecto Argentino Interinstitucional de genomica de SARS-CoV-2' (PAIS Consortium) | C Pintos; C Rastellini; C Ziehm; J Ousset; L Pianciola; M Fernandez; M L Álvarez; M Mazzeo; P V Blanco.; Y Espasandin |
| EPI_ISL_3412032 | Hospital of the University of Pennsylvania Molecular Pathology Lab | Bushman | Andrew D. Marques; Aoife M. Roche; Arupa Ganguly; Ayannah S. Fitzgerald; Brendan Kelly; Jevon Graham-Wooten; John K. Everett; Kyle Rodino; Layla A. Khatib; Mike Feldman; Ronald G. Collman and Frederic Bushan; Samantha A. Whiteside; Scott Sherrill-Mix; Shantan Reddy; Young Hwang |
| EPI_ISL_2653034, EPI_ISL_3022685, EPI_ISL_3022748, EPI_ISL_3022756, EPI_ISL_3022760 | Hospital of the University of Pennsylvania Molecular Pathology Lab | Bushman Lab - University of Pennsylvania | Aoife M. Roche; Arupa Ganguly; Ayannah S. Fitzgerald; Brendan Kelly; Jevon Graham-Wooten; John K. Everett; Kyle Rodino; Layla A. Khatib; Mike Feldman; Ronald G. Collman and Frederic Bushan; Samantha A. Whiteside; Scott Sherrill-Mix; Shantan Reddy; Young Hwang |
| EPI_ISL_2211977, EPI_ISL_2212003, EPI_ISL_2213652, EPI_ISL_2213726, EPI_ISL_2216497, EPI_ISL_2216599, EPI_ISL_2216645 | | | |
| see above | Houston Methodist Hospital | Houston Methodist Hospital | Ilya J. Finkelstein; James J. Davis; Jessica Cambric; Jimmy Gollihar; Kristina Reppond; Layne Pruitt; Madison N. Shyer; Marcus Nguyen; Matthew Ojeda Saavedra; Paul A. Christensen; Prasanti Yerramilli; Randall J. Olsen; Robert Olson; Ryan Gadd; S. Wesley Long; Sishir Subedi; and James M. Musser |
| EPI_ISL_2467734 | Hôpital Bichat Claude Bernard, Laboratoire de Virologie | IAME UMR1137 Inserm, Université de Paris, Hôpital Bichat | Alexandre Storto; Amélie Recoing; Antoine Bridier-Nahmias; Benoit Visseaux; Charlotte Charpentier; Diane Descamps; Gilles Collin; Lena Daniel; Mélanie Bertine; Nadhira Houhou-Fidouh; Quentin Le Hingrat; Siham Hamri |
| EPI_ISL_1757581, EPI_ISL_1757582 | Hôpital Cochin | Department of Virology, Henri Mondor University Hospital, Assistance Publique Hôpitaux de Paris, Université Paris-Est Créteil, INSERM U955 | Alexandre Soulier; Christophe Rodriguez; Elisabeth Trawinski; Guillaume Gricourt; Jean-Michel Pawlotsky; Melissa N'Debi; Slim Fourati; Vanessa Demontant |
| EPI_ISL_1675331 | ICMT | Universidad Nacional de Colombia - Laboratorio Genómico One Health | Andres F. Cardona-Rios; Carlos Franco-Muñoz; Daniel O. Maldonado-Perez; Diego A. Álvarez-Díaz; Hector Alejandro Ruiz-Moreno; Idabely Betancur Ortiz; Jorge E. Osorio; Juan P. Hernandez-Ortiz; Karl A Ciuoderis; Katherine Laiton-Donato; Laura Silvana Perez; Lina M. Hurtado; Marcela Mercado-Reyes; Maria Angélica Maya; Maria Stella López; Rita Almanza Payares; Sandra Ines Cano; Simón Villegas Velásquez |
| EPI_ISL_1960074 | ICMT-APARTADO | Universidad Nacional de Colombia - Laboratorio Genómico One Health | Andres F. Cardona-Rios; Carlos Franco-Muñoz; Carolina Muñoz-Arango; Celeny Ortiz; Daniel O. Maldonado-Perez; Diego A. Álvarez-Díaz; Hector Alejandro Ruiz-Moreno; Idabely Betancur Ortiz; Jorge E. Osorio; Juan P. Hernandez-Ortiz; Karl A Ciuoderis; Katherine Laiton-Donato; Laura Silvana Perez; Lina M. Hurtado; Marcela Mercado-Reyes; Maria Angélica Maya; Maria Stella López; Rita Almanza Payares; Sandra Ines Cano; Simón Villegas Velásquez |
| EPI_ISL_3464693 | IEC- Instituto Evandro Chagas | ITV-Vale Institute of Technology | Amanda Vidal; Guilherme Oliveira; Mirleide Cordeiro dos Santos; Tatianne Costa Negri |
| EPI_ISL_2234905, EPI_ISL_4071893 | IICS-UNA | IICS-UNA | Adriana Valenzuela; Alejandra Rojas; Chyntia Diaz; Eva Nara; Fatima Cardozo; Florencia del Puerto; Joel Ortiz; Jonas Fernandez; Laura Franco; Laura Mendoza; Leticia Rojas; Magaly Martinez; Maria Eugenia Galeano. |
| EPI_ISL_1823782, EPI_ISL_1823788 | IL Department of Public Health Chicago Laboratory | Centers for Disease Control and Prevention Division of Viral Diseases, Pathogen Discovery | Alison Laufer Halpin; Ben L. Rambo-Martin; Clinton R. Paden; Dakota Howard; Darlene Wagner; Dave Wentworth; Dhwani Batra; Jasmine Padilla; Justin Lee; Katie Dillon; Krista Queen; Kristen Knipe; Kristine Lacek; Mark Burroughs; Matthew Schmerer; Mili Sheth; Peter Cook; Sam Shepard; Sarah Nobles; Shoshona Le; Suxiang Tong; Vivien Dugan; Yvette Unoarumhi |
| EPI_ISL_1798561, EPI_ISL_1798583 | IL Dept. of Public Health Springfield Laboratory | Centers for Disease Control and Prevention Division of Viral Diseases, Pathogen Discovery | Alison Laufer Halpin; Ben L. Rambo-Martin; Clinton R. Paden; Dakota Howard; Darlene Wagner; Dave Wentworth; Dhwani Batra; Jasmine Padilla; Justin Lee; Katie Dillon; Krista Queen; Kristen Knipe; Kristine Lacek; Mark Burroughs; Matthew Schmerer; Mili Sheth; Peter Cook; Sam Shepard; Sarah Nobles; Shoshona Le; Suxiang Tong; Vivien Dugan; Yvette Unoarumhi |
| EPI_ISL_2155041 | IMAT | Instituto Nacional de Salud-Dirección de Investigación en Salud Pública | Carlos Franco-Muñoz; Carmen Osorio; Diana Malo; Diego A. Álvarez-Díaz; Diego Andrés Prada; Gerardo Santamaría; Hector Alejandro Ruiz-Moreno; Jhonnatan Reales-González; Jorge Rivera; Juan Camilo Martinez; Julian Naizaque; Katherine Laiton-Donato; Lisseth Pardo; Magdalena Wiesner; Marcela Mercado-Reyes; Maria T. Herrera-Sepúlveda; Marta Lopez Blanco; Martha Lucia Ospina Martinez; Paola Rojas; Sergio Gomez; Sheryll Corchuelo; Ángela Alarcon Cruz |
| EPI_ISL_1722204, EPI_ISL_1722214, EPI_ISL_1722223, EPI_ISL_1844591 | IMD - Medizinisches Labor Rostock | Robert Koch Institute | |
| EPI_ISL_1824049 | IN State Department of Health Laboratory Services | Centers for Disease Control and Prevention Division of Viral Diseases, Pathogen Discovery | Alison Laufer Halpin; Ben L. Rambo-Martin; Clinton R. Paden; Dakota Howard; Darlene Wagner; Dave Wentworth; Dhwani Batra; Jasmine Padilla; Justin Lee; Katie Dillon; Krista Queen; Kristen Knipe; Kristine Lacek; Mark Burroughs; Matthew Schmerer; Mili Sheth; Peter Cook; Sam Shepard; Sarah Nobles; Shoshona Le; Suxiang Tong; Vivien Dugan; Yvette Unoarumhi |
| EPI_ISL_1648047, EPI_ISL_1648072, EPI_ISL_1648085, EPI_ISL_1710870, EPI_ISL_1710873, EPI_ISL_1818776 | IN State Department of Health Laboratory Services | IN State Department of Health Laboratory Services | Ankita Kashikar; Brian Pope; Cassandra Campion; Jamie Yeadon; Kyle Brownlee; Lixia Liu; Mark Glazier; Melissa Hindenlang |
| EPI_ISL_1656467, EPI_ISL_1656471, EPI_ISL_1697380, EPI_ISL_2483722 | INMI Lazzaro Spallanzani IRCCS | INMI Lazzaro Spallanzani IRCCS | A Di Caro; B Bartolini; CEM Gruber; E Giombini; F Messina; F Santini; G Bonfiglio; M Rueca; MR Capobianchi; O Butera |
| EPI_ISL_2665401 | INMI Lazzaro Spallanzani IRCCS | National Institute for Infectious Diseases (INMI) L. Spallanzani I.R.C.C.S | A Di Caro; B Bartolini; CEM Gruber; E Giombini; F Messina; F Santini; G Bonfiglio; M Rueca; MR Capobianchi; O Butera |
| EPI_ISL_2576989 | IRCCS San Gallicano Dermatological Institute | IRCCS Regina Elena National Cancer Institute | Aldo Morrone; Fabrizio Ensoli; Francesca De Nicola; Francesca Maione; Fulvia Pimpinelli; Gennaro Ciliberto; Giovanni Blandino; Giulia Orlandi; Matteo Pallocca; Maurizio Fanciulli |
| EPI_ISL_2628301 | IVIC | Laboratorio de Virología Molecular | Carmen L Loureiro; CoViVen Group; Domingo J Garzaro; Esmeralda Vizzi; Flor H Pujol; Héctor R Rangel; José Luis Zambrano; Lieska Rodríguez; Mariana Hidalgo; Pierina D´Angelo; Rossana C Jaspe; Víctor Alarcón; Yoneira Sulbaran; Zoila Moros |
| EPI_ISL_2427312, EPI_ISL_2500414 | IVY3 Central Lab, Vanderbilt University Medical Center | Lauring Lab, University of Michigan, Department of Microbiology and Immunology | Gilbert; Valesano |
| EPI_ISL_3191043 | Idaho Bureau of Laboratories | Idaho Bureau of Laboratories | "R. Beukelman; Aimee Ceniseros; Christian Loera; Christopher Ball"; Matthew Charles Burns; Robert L. Voermans |
| EPI_ISL_1663124, EPI_ISL_1663129, EPI_ISL_1663138, EPI_ISL_1663139, EPI_ISL_1742374, EPI_ISL_1742380, EPI_ISL_1742389, EPI_ISL_1809503, EPI_ISL_1809515, EPI_ISL_1809644, EPI_ISL_1809701, EPI_ISL_1809732, EPI_ISL_2800759, EPI_ISL_2800847 | | | |
| see above | Illinois Department of Public Health | Illinois Department of Public Health - Chicago Lab | Ira Heimler; Joel Price; Vineet K. Dhiman |
| EPI_ISL_1595524, EPI_ISL_1595527, EPI_ISL_1595529, EPI_ISL_1595530, EPI_ISL_1595531, EPI_ISL_1595533, EPI_ISL_1595535, EPI_ISL_1595536, EPI_ISL_1595539, EPI_ISL_1595546, EPI_ISL_1595547, EPI_ISL_1595553, EPI_ISL_1595554, EPI_ISL_1595555, EPI_ISL_1595556, EPI_ISL_1595557, EPI_ISL_1595562, EPI_ISL_1595569, EPI_ISL_1595578, EPI_ISL_1595579, EPI_ISL_1595580, EPI_ISL_1595582, EPI_ISL_1595583, EPI_ISL_1595584, EPI_ISL_1595585, EPI_ISL_1595586, EPI_ISL_1595592, EPI_ISL_1624091, EPI_ISL_1624095, EPI_ISL_1624101, EPI_ISL_1624107, EPI_ISL_1624109, EPI_ISL_1624110, EPI_ISL_1624113, EPI_ISL_1624119, EPI_ISL_1624127, EPI_ISL_1624129, EPI_ISL_1624141, EPI_ISL_1624150, EPI_ISL_1624158, EPI_ISL_1624160, EPI_ISL_1624171, EPI_ISL_1624174, EPI_ISL_1708236, EPI_ISL_1708240, EPI_ISL_1708247, EPI_ISL_1708248, EPI_ISL_1708261, EPI_ISL_1708262, EPI_ISL_1708314, EPI_ISL_1732590, EPI_ISL_1732600, EPI_ISL_1732603, EPI_ISL_1732608, EPI_ISL_1732610, EPI_ISL_1732612, EPI_ISL_1732615, EPI_ISL_1732617, EPI_ISL_1732618, EPI_ISL_1789153, EPI_ISL_1825692, EPI_ISL_1825693, EPI_ISL_1825695, EPI_ISL_1825696, EPI_ISL_1825700, EPI_ISL_1825701, EPI_ISL_1825704, EPI_ISL_1825705, EPI_ISL_1825707, EPI_ISL_1825751, EPI_ISL_1825752, EPI_ISL_1825777, EPI_ISL_1825778, EPI_ISL_1938496, EPI_ISL_1938501, EPI_ISL_1938523 | | | |
| see above | Illinois Department of Public Health - Springfield Lab | Illinois Department of Public Health - Springfield Lab | Bryan Sim; Gordon McCall |
| EPI_ISL_2136470 | Incyte Diagnostics Spokane | Seattle Flu Study | Amanda Adler; Barry R. Lutz; Benjamin Pelle; Brian Hiatt; Caitlin R. Wolf; Chris D. Frazar; Deborah A. Nickerson; Elisabeth Brandstetter; Erica Ryke; Geoff Melly; Helen Y. Chu; Janet A. Englund; Jay Shendure; Jover Lee; Kairsten Fay; Kirsten Lacombe; Lea M. Starita; Mark J. Rieder; Matthew Richardson; Matthew Thompson; Melissa Truong; Michael Boeckh; Michael Famulare; Misja Ilcisin; Peter D. Han; Philip Dykema; Romesh Gautom; Scott Lindquist; Thomas R. Sibley; Trevor Bedford |
| EPI_ISL_1823557, EPI_ISL_1823559, EPI_ISL_1823564 | Indiana Animal Disease Diagnostic Laboratory | Carpi Laboratory - Purdue University | Abebe A Fola; G Kenitra Hendrix; Giovanna Carpi; Ilinca I Ciubotariu; Jack Dorman; Jobin J Kattoor; Lev Gorenstein; Nicole M Perry; Rebecca P Wilkes |
| EPI_ISL_1733089, EPI_ISL_1795446, EPI_ISL_1795450, EPI_ISL_1795455, EPI_ISL_1795478, EPI_ISL_1795548, EPI_ISL_1795787, EPI_ISL_1802742, EPI_ISL_1802760, EPI_ISL_1802932, EPI_ISL_1803059, EPI_ISL_1803119, EPI_ISL_1803155, EPI_ISL_1803332, EPI_ISL_1803434, EPI_ISL_1817989, EPI_ISL_2013183, EPI_ISL_2013564, EPI_ISL_2013570, EPI_ISL_2013694, EPI_ISL_2013705, EPI_ISL_2047287, EPI_ISL_4386303, EPI_ISL_4390379 | | | |
| see above | Infinity Biologix | Centers for Disease Control and Prevention Division of Viral Diseases, Pathogen Discovery | Adrian Paskey; Benjamin Rambo-Martin; Chirayu Goswami; Christian Bixby; Christopher Gulvick; Clinton Paden; Clinton R. Paden; Dakota Howard; Darlene Wagner; Dhwani Batra; Duncan MacCannell; Erisa Sula; Jason Caravas; Jonathan Schultz; Kara Moser; Kristine Lacek; Matthew Schmerer; Peter Cook; Peter W. Cook; Robin Grimwood; Russ Hager; Scott Sammons; Shatavia Morrison; Tymeckia Kendall; Victoria Caban Figueroa; Yihe Wang; Yvette Unoarumhi |
| EPI_ISL_2193351 | Institut de Virologie du CHU | Institut de Virologie du CHU de | Fafi-Kremer Samira; Gallais Floriane; Gantner Pierre; Laugel Elodie; Solis Morgane; Velay Aurélie; Wendling Marie-Josée |

| | | | |
|---|---|---|---|
| | de Strasbourg | Strasbourg | |
| EPI_ISL_2100427 | Institute of Microbiology, Universidad San Francisco de Quito | Institute of Microbiology, Universidad San Francisco de Quito | Belén Prado-Vivar; Bernardo Gutiérrez; Gabriel Trueba; Guzmán Bernabéu Lorenzo; Juan José Guadalupe; Michelle Grunauer; Monica Becerra-Wong; Patricio Rojas-Silva; Paúl Cárdenas; Sully Márquez; Verónica Barragán |
| EPI_ISL_1752641, EPI_ISL_1752642, EPI_ISL_1752643 | Instituto Adolfo Lutz - Regional de Aracatuba | Instituto Adolfo Lutz, Interdiciplinary Procedures Center, Strategic Laboratory | Caio Vinicius Dias Lopes; Claudia Regina Gonçalves; Claudio Tavares Sacchi; Erica Valessa Ramos Gomes; Karoline Rodrigues Campos; Katia Correa de Oliveira Santos; Leonardo Jose Tadeu de Araujo |
| EPI_ISL_2919238 | Instituto Adolfo Lutz - Regional de Bauru | Instituto Adolfo Lutz, Interdiciplinary Procedures Center, Strategic Laboratory | Caio Vinicius Dias Lopes; Claudia Regina Gonçalves; Claudio Tavares Sacchi; Erica Valessa Ramos Gomes; Karoline Rodrigues Campos |
| EPI_ISL_2828687, EPI_ISL_2828688, EPI_ISL_2828689 | Instituto Adolfo Lutz - Regional de Campinas | Instituto Adolfo Lutz, Interdiciplinary Procedures Center, Strategic Laboratory | Caio Vinicius Dias Lopes; Claudia Regina Gonçalves; Claudio Tavares Sacchi; Erica Valessa Ramos Gomes; Karoline Rodrigues Campos |
| EPI_ISL_2614614, EPI_ISL_2614615, EPI_ISL_2614616 | Instituto Adolfo Lutz - Regional de Presidente Prudente | Instituto Adolfo Lutz, Interdiciplinary Procedures Center, Strategic Laboratory | Caio Vinicius Dias Lopes; Claudia Regina Gonçalves; Claudio Tavares Sacchi; Erica Valessa Ramos Gomes; Karoline Rodrigues Campos; Leonardo Jose Tadeu de Araujo |
| EPI_ISL_1752645 | Instituto Adolfo Lutz - Regional de Ribeirao Preto | Instituto Adolfo Lutz, Interdiciplinary Procedures Center, Strategic Laboratory | Caio Vinicius Dias Lopes; Claudia Regina Gonçalves; Claudio Tavares Sacchi; Erica Valessa Ramos Gomes; Karoline Rodrigues Campos; Katia Correa de Oliveira Santos; Leonardo Jose Tadeu de Araujo |
| EPI_ISL_2958841, EPI_ISL_2958842 | Instituto Adolfo Lutz - Regional de Sorocaba | Instituto Adolfo Lutz, Interdiciplinary Procedures Center, Strategic Laboratory | Caio Vinicius Dias Lopes; Claudia Regina Gonçalves; Claudio Tavares Sacchi; Erica Valessa Ramos Gomes; Karoline Rodrigues Campos |
| EPI_ISL_1715154, EPI_ISL_1715155, EPI_ISL_1715156, EPI_ISL_1715157, EPI_ISL_1715158, EPI_ISL_1731589, EPI_ISL_1731590, EPI_ISL_1731591, EPI_ISL_1731592, EPI_ISL_1731594, EPI_ISL_1731595, EPI_ISL_1731596, EPI_ISL_1731597, EPI_ISL_1752646, EPI_ISL_1752647, EPI_ISL_2003132, EPI_ISL_2756433, EPI_ISL_2756434, EPI_ISL_2756460, EPI_ISL_2756469, EPI_ISL_2756470, EPI_ISL_2756471 | | | |
| see above | Instituto Adolfo Lutz Central | Instituto Adolfo Lutz, Interdiciplinary Procedures Center, Strategic Laboratory | Caio Vinicius Dias Lopes; Claudia Regina Gonçalves; Claudio Tavares Sacchi; Erica Valessa Ramos Gomes; Karoline Rodrigues Campos; Katia Correa de Oliveira Santos; Leonardo Jose Tadeu de Araujo |
| EPI_ISL_4104722 | Instituto Butantan | Instituto Butantan | Antonio Jorge Martins; Claudia Renata dos Santos Barros; David Schlesinger; Debora Botequio Moretti; Dimas Tadeu Covas; Elaine Cristina Marqueze; Elaine Vieira Santos; Evandra Strazza Rodrigues; Heidge Fukumasu; Jayme Augusto de Souza-Neto; José Salvatore Leister Patané; Luiz Alcantara; Luiz Lehmann Coutinho; Maria Carolina Elias; Maurício Lacerda Nogueira; Rafael dos Santos Bezerra; Raul Machado Neto; Rejane Maria Tommasini Grotto; Ricardo Haddad; Sandra Coccuzzo Sampaio Vessoni; Simone Kashima; Svetoslav Nanev Slavov; Vincent Louis Viala |
| EPI_ISL_1824485, EPI_ISL_1824488 | Instituto Nacional de Medicina Genomica | Centro de Investigación en Enfermedades Infecciosas (CIENI), Instituto Nacional de Enfermedades Respiratorias (INER) | Arriaga-Canon C; Avila-Rios S; Boukadida C; Cedro-Tanda A; Herrera-Montalvo LA; Hidalgo-Miranda A; Matias-Florentino M; Mendoza-Vargas A; Perez-Garcia M; Reyes-Grajeda JP; Reyes-Teran G |
| EPI_ISL_2500927 | Instituto Nacional de Salud | Instituto Nacional de Salud-Dirección de Investigación en Salud Pública | Carlos Franco-Muñoz; Carmen Osorio; Diana Malo; Diego A. Álvarez-Díaz; Diego Andrés Prada; Gerardo Santamaría; Hector Alejandro Ruiz-Moreno; Jhonnatan Reales-González; Jorge Rivera; Juan Camilo Martinez; Julian Naizaque; Katherine Laiton-Donato; Lisseth Pardo; Magdalena Wiesner; Marcela Mercado-Reyes; Maria T. Herrera-Sepúlveda; Marta Lopez Blanco; Martha Lucia Ospina Martinez; Paola Rojas; Sergio Gomez; Sheryll Corchuelo; Ángela Alarcon Cruz |
| EPI_ISL_1854230, EPI_ISL_1854231, EPI_ISL_1854232, EPI_ISL_1854233 | Instituto Nacional de Saude (INSA) and Instituto Gulbenkian de Ciencia (IGC) | Instituto Nacional de Saude (INSA) and Instituto Gulbenkian de Ciencia (IGC) | Borges et al |
| EPI_ISL_2758988, EPI_ISL_2758992, EPI_ISL_2758994, EPI_ISL_2758995, EPI_ISL_2759001, EPI_ISL_2759002, EPI_ISL_2759004, EPI_ISL_2759006, EPI_ISL_2759012, EPI_ISL_2759016, EPI_ISL_2759017, EPI_ISL_2759020, EPI_ISL_2759021, EPI_ISL_2759022, EPI_ISL_2759024, EPI_ISL_2759026, EPI_ISL_2759028, EPI_ISL_2759035, EPI_ISL_2759041, EPI_ISL_2759045, EPI_ISL_2759046, EPI_ISL_2759047, EPI_ISL_2759050, EPI_ISL_2759051, EPI_ISL_2759052, EPI_ISL_2759057, EPI_ISL_2759059, EPI_ISL_2759061, EPI_ISL_2759063, EPI_ISL_2759066, EPI_ISL_2759068, EPI_ISL_2775421 | | | |
| see above | Instituto de Biologia Molecular do Paraná (LAC) | Instituto Carlos Chagas - Fiocruz | Alessandra De Melo Aguiar; Andreia Akemi Suzukawa; Andréa Rodrigues Ávila; Bruno Dallagiovanna; Dalila Zanette; Eduardo Balsanelli; Emanuel Maltempi de Souza; Fabio Passetti; Fabricio Klerynton Marchini; Fábio de Oliveira Pedrosa; Guilherme Becker; Helisson Faoro; Hellen Geremias dos Santos; Irina Nastassja Riediger; Letusa Albrecht; Lucas Blanes; Luis Gustavo Morello; Lysangela Ronalte Alves; Maria do Carmo Debur; Mauro de Medeiros Oliveira; Michelle Orane Schemberger; Paola Cristina Resende; Sheila Cristina Nardeli; Tiago Gräf; Valter Antônio de Baura |
| EPI_ISL_1734852, EPI_ISL_1734858, EPI_ISL_1734866, EPI_ISL_1734868, EPI_ISL_1734872, EPI_ISL_1734874, EPI_ISL_1734884, EPI_ISL_1911960, EPI_ISL_1911961, EPI_ISL_1911962, EPI_ISL_1911964, EPI_ISL_2223441, EPI_ISL_2223443, EPI_ISL_2223446, EPI_ISL_2223449, EPI_ISL_2223452, EPI_ISL_2295395, EPI_ISL_2295396, EPI_ISL_2295397, EPI_ISL_2295398, EPI_ISL_2295399 | | | |
| see above | Instituto de Biotecnologia - UNESP-Botucatu-SP | Instituto de Biotecnologia - UNESP-Botucatu-SP | Cecília Artico Banho; Cíntia Bittar; Fábio Sossai Possebon; Guilherme Campos; Helena Lage Ferreira; Jorge A. Petroli Marchesi; João Pessoa Araújo Jr.; Leila Sabrina Ullmann; Lívia Sacchetto; Maisa C. Pereira Parra; Marília Moraes; Maurício L. Nogueira; Paula Rahal; Paulo Inacio da Costa |
| EPI_ISL_2776178, EPI_ISL_2776179, EPI_ISL_2776181 | Instituto de Medicina Tropical & Salud Global (IMTSAG) | Grubaugh Lab - Yale School of Public Health | Alejandaro Vallejo Degaudenzi; Anderson Brito; Annie Watkins; Chaney Kalinich; Chantal Vogels; Elisa Contreras; Esperanza Mendoza; Isabel Ott; Jessica Rothman; Joseph Fauver; Kendall Billig; Mallery Breban; Mary Petrone; Nathan Grubaugh; Robert Paulino-Ramirez; Tara Alpert; Tobias Koch; Victor Virgilio Calderon |
| EPI_ISL_2691150 | Intituto Adolfo Lutz - Regional de Sorocaba | Instituto Adolfo Lutz, Interdiciplinary Procedures Center, Strategic Laboratory | Caio Vinicius Dias Lopes; Claudia Regina Gonçalves; Claudio Tavares Sacchi; Erica Valessa Ramos Gomes; Karoline Rodrigues Campos; Leonardo Jose Tadeu de Araujo |
| EPI_ISL_1714839, EPI_ISL_1714841, EPI_ISL_1714844, EPI_ISL_1715025, EPI_ISL_1793099, EPI_ISL_1793234, EPI_ISL_1793283, EPI_ISL_1793354, EPI_ISL_1793355, EPI_ISL_1793360, EPI_ISL_2029476, EPI_ISL_2029498 | | | |
| see above | Istituto Zooprofilattico Sperimentale del Mezzogiorno | TIGEM | Antonio Grimaldi Patrizia Annunziata Francesco Panariello Biancamaria Pierri Claudia Tiberio Teresa Giuliano Valentina Bouche Chiara Colantuono Maria Concetta Cuomo Denise Di Concilio Lucio Di Filippo Anna Manfredi Marcello Salvi Antonio Limone Luigi Atripaldi Pellegrino Cerino Andrea Ballabio Davide Cacchiarelli |
| EPI_ISL_1692673, EPI_ISL_1692689, EPI_ISL_1692864, EPI_ISL_1692959, EPI_ISL_1692980 | Istituto Zooprofilattico Sperimentale del Mezzogiorno | Telethon Institute of Genetics and Medicine (TIGEM) | Antonio Grimaldi Patrizia Annunziata Francesco Panariello Biancamaria Pierri Claudia Tiberio Teresa Giuliano Valentina Bouche Chiara Colantuono Maria Concetta Cuomo Denise Di Concilio Lucio Di Filippo Anna Manfredi Marcello Salvi Antonio Limone Luigi Atripaldi Pellegrino Cerino Andrea Ballabio Davide Cacchiarelli |
| EPI_ISL_1759558, EPI_ISL_1759559, EPI_ISL_1759560, EPI_ISL_1759562, EPI_ISL_1759564 | Istituto Zooprofilattico Sperimentale del Mezzogiorno - Azienda Ospedaliera "Pugliese Ciaccio" di Catanzaro | TIGEM | Antonio Grimaldi Patrizia Annunziata Francesco Panariello Biancamaria Pierri Claudia Tiberio Teresa Giuliano Valentina Bouche Chiara Colantuono Maria Concetta Cuomo Denise Di Concilio Lucio Di Filippo Anna Manfredi Pasquale Minchella Marcello Salvi Antonio Limone Luigi Atripaldi Pellegrino Cerino Andrea Ballabio Davide Cacchiarelli |
| EPI_ISL_1577770, EPI_ISL_1577796 | Jessa | Jessa | Cruys et al. on behalf of the Jessa_cmdLab |
| EPI_ISL_1739011, EPI_ISL_1739012, EPI_ISL_1739013, EPI_ISL_1739041 | Joaquim Chaves Saude | Instituto Nacional de Saude (INSA) and Institute of Biomedicine (iBiMed), Universidade de Aveiro | Borges et al |
| EPI_ISL_2246721 | John H Stroger Jr Hospital of Cook County | RIPHL at Rush University Medical Center | Ah-Young Sung; Felix Araujo Perez; Kevin Kunstman; Laura Furtado; Marieta Hyde; Max Kolton; Sharon Welbel; Stefan Green |
| EPI_ISL_1662968, EPI_ISL_1973409, EPI_ISL_1973458, EPI_ISL_2455319 | Johns Hopkins Hospital Department of Pathology | Johns Hopkins Hospital Department of Pathology | Adannaya Amadi; C. Paul Morris; Chun Huai Luo; Heba H. Mostafa; Matthew Schwartz; Nicholas Gallagher |
| EPI_ISL_2424322, EPI_ISL_2424326, EPI_ISL_2424339, EPI_ISL_2424343, EPI_ISL_2424363, EPI_ISL_2424401, EPI_ISL_2424406, EPI_ISL_2424407, EPI_ISL_2424408, EPI_ISL_2424409, EPI_ISL_2424449, EPI_ISL_2424451, EPI_ISL_2424462, EPI_ISL_2425113, EPI_ISL_2425145, EPI_ISL_2425149, EPI_ISL_2425153, EPI_ISL_2425157, EPI_ISL_2425158, EPI_ISL_2425159, EPI_ISL_2425162, EPI_ISL_2425163, EPI_ISL_2425164, EPI_ISL_2425165, EPI_ISL_2425172, EPI_ISL_2425177, EPI_ISL_2425178, EPI_ISL_2425179, EPI_ISL_2425186, EPI_ISL_2425189, EPI_ISL_2425191, EPI_ISL_2425192, EPI_ISL_2425193, EPI_ISL_2425198, EPI_ISL_2425206, EPI_ISL_2425207, EPI_ISL_2425208, EPI_ISL_2425209, EPI_ISL_2425210, EPI_ISL_2425211, EPI_ISL_2425214, EPI_ISL_2425215, EPI_ISL_2425216, EPI_ISL_2425224, EPI_ISL_2425226, EPI_ISL_2425227, EPI_ISL_2425229, EPI_ISL_2425231, EPI_ISL_2425232, EPI_ISL_2425324, EPI_ISL_2832962, EPI_ISL_2840570, EPI_ISL_2964732 | | | |
| see above | KU Leuven, Rega Institute, Clinical and Epidemiological Virology | KU Leuven, Rega Institute, Clinical and Epidemiological Virology | Bert Vanmechelen; Joan Marti-Carerras; Piet Maes; Tony Wawina-Bokalanga |
| EPI_ISL_2229749, EPI_ISL_2662925 | Kaleida Health Laboratories | University at Buffalo Genomics and Bioinformatics Core | Alyssa Pohlman; Amanda Boccolucci; Brandon Marzullo; Donald Yergeau; Jennifer Surtees; Jonathan Bard; Natalie Lamb; Norma Nowak |
| EPI_ISL_6375147, EPI_ISL_6375165, EPI_ISL_6375183 | Karolinska University Hospital Solna | Karolinska University Hospital | Annelie Bjerkner; Isak Sylvin; Jan Albert; Karolina Ininbergs; Lina Guerra Blomqvist; Lynda Eneh; Martin Ekman; Martina Wahlund; Robert Dyrdak; Sandra Broddesson; Tanja Normark; Tobias Allander; Valtteri Wirta; Zhibing Yun |
| EPI_ISL_1843745 | Klinisch Laboratorium ZNA | Klinisch Laboratorium ZNA | Verstrepen et al. |
| EPI_ISL_5529955 | LAB DE A CLIN MUNICIPAL DE CASCAVEL | Analytical Competence Molecular Epidemiology Lab/ACME, Oswaldo Cruz Foundation, Ceara (FIOCRUZ CE) | Carlos Leonardo de Aragao Araujo; Cecilia Leite Costa & Eduardo Ruback dos Santos on behalf of COVID-19 FIOCRUZ Genomic Network; Cleber Furtado Aksenen; Fabio Miyajima; Fernando Braga Stehling; Francisco Eder de Moura Lopes; Igor Oliveira Duarte; Jamille Maria Mendes Bezerra; Joaquim Cesar do Nascimento Sousa Junior; Pedro Miguel Carneiro Jeronimo; Suzana Porto Almeida; Thais Ferreira de Oliveira; Thais de Oliveira Costa; Ticiane Cavalcante de Souza; Veridiana Pessoa Miyajima |

| | | | |
|---|---|---|---|
| EPI_ISL_1788359, EPI_ISL_1788360, EPI_ISL_1788362 | LABORATOIRE DE BIOLOGIE MEDICALE | CNR Virus des Infections Respiratoires - France SUD | Antonin Bal; Bruno Lina; Bruno Simon; Gregory Destras; Gwendolyne Burfin; Hadrien Regue; Laurence Josset; Martine Valette; Quentin Semanas |
| EPI_ISL_2155045 | LABORATORIO CLINICO MASVIDA DE LA COSTA | Instituto Nacional de Salud-Dirección de Investigación en Salud Pública | Carlos Franco-Muñoz; Carmen Osorio; Diana Malo; Diego A. Álvarez-Díaz; Diego Andrés Prada; Gerardo Santamaría; Hector Alejandro Ruiz-Moreno; Jhonnatan Reales-González; Jorge Rivera; Juan Camilo Martinez; Julian Naizaque; Katherine Laiton-Donato; Lisseth Pardo; Magdalena Wiesner; Marcela Mercado-Reyes; Maria T. Herrera-Sepúlveda; Marta Lopez Blanco; Martha Lucia Ospina Martinez; Paola Rojas; Sergio Gomez; Sheryll Corchuelo; Ángela Alarcon Cruz |
| EPI_ISL_3355469 | LABORATORIO DE BIOLOGÍA MOLECULAR Y BIOTECNOLOGIA | Corporacion CorpoGen-Universidad de los Andes-Universidad Central | Christian Romero; Jorge Duitama; Juan Manuel Anzola; Laura González; Maryam Chaib De Mares; María Mercedes Zambrano; Nelly Díaz; Patricia Del Portillo; Silvia Restrepo |
| EPI_ISL_1966905 | LABORATORIO DE FRANCA | Instituto Butantan / ESALQ-USP (Piracicaba) | Antonio Jorge Martins; Bianca Cechetto Carlos. Mendelics: Bibiana Santos; Claudia Renata dos Santos Barros; Cíntia Bittar; David Schlesinger. Hemocentro Ribeirão Preto: Simone Kashima; Debora Botequio Moretti; Elaine Cristina Marqueze; Elaine Vieira dos Santos; Elisangela Chicaroni Mattos; Erika Freitas; Evandra Strazza Rodrigues; Felipe Allan da Silva da Costa; Flavia Aburjaile; Fábio Sossai Possebon; Guilherme Campos; Guilherme Targino Valente; Heidge Fukumasu. USP-Botucatu: Rejane Maria Tommasini Grotto; Helena Lage Ferreira; Instituto Butantan: Dimas Tadeu Covas; Jardelina de Souza Todao Bernardino; Jayme A. Souza-Neto; Jessika Cristina Chagas Lesbon; Jorge A. Petroli Marchesi; José Salvatore Leister Patané; João Paulo Kitajima; João Pessoa Araújo Jr.; Leila Sabrina Ullmann; Loyze Paola Oliveira de Lima; Luiz Aurelio de Campos Crispin. Centro de Genômica Funcional da ESALQ: Luiz Lehmann Coutinho; Luiz Carlos Junior de Alcantara; Lívia Sacchetto; Maisa C. Pereira Parra; Maria Carolina Elias; Marta Giovanetti; Marília Moraes; Maurício Lacerda Nogueira. Prefeitura de Sao Paulo: Melissa Palmieri.; Patricia Akemi Assato; Paula Rahal; Paulo Inacio da Costa; Rafael dos Santos Bezerra; Raquel de Lello Rocha Campos Cassano. NGS Soluções Genômicas: Pilar Drummond Sampaio Corrêa Mariani. FZEA-USP Pirassununga: Mirele Daiana Poleti; Raul Machado Neto; Ricardo Augusto Brassaloti; Ricardo Haddad; Rodrigo Tocantins Calado. FAMERP-SJRP: Cecília Artico Banho; Sandra Coccuzzo Sampaio; Svetoslav Nanev Slavov; Vagner Fonseca; Vincent Louis Viala |
| EPI_ISL_3275680 | LABORATORIO LAS AMERICAS | Universidad Nacional de Colombia - Laboratorio Genómico One Health | Andres F. Cardona-Rios; Carlos Franco-Muñoz; Carolina Muñoz-Arango; Celeny Ortiz; Daniel O. Maldonado-Perez; Diego A. Álvarez-Díaz; Hector Alejandro Ruiz-Moreno; Idabely Betancur Ortiz; Jorge E. Osorio; Juan P. Hernandez-Ortiz; Karl A Ciuoderis; Katherine Laiton-Donato; Laura Silvana Perez; Lina M. Hurtado; Marcela Mercado-Reyes; Maria Angélica Maya; Maria Stella López; Rita Almanza Payares; Sandra Ines Cano; Simón Villegas Velásquez |
| EPI_ISL_3102543 | LABORATORIO MUN ANALISES CLINICAS | Analytical Competence Molecular Epidemiology Lab/ACME, Oswaldo Cruz Foundation, Ceara (FIOCRUZ CE) | Cleber Furtado Aksenen; Fabio Miyajima; Fernando Braga Stehling; Francisco Eder de Moura Lopes; Jamille Maria Mendes Bezerra; Joaquim César do Nascimento Sousa Junior; Pedro Miguel Carneiro Jeronimo; Suzana Porto Almeida e Lucas Delerino; Thais Ferreira de Oliveira; Thais de Oliveira Costa; Ticiane Cavalcante de Souza; Veridiana Pessoa Miyajima |
| EPI_ISL_1966609, EPI_ISL_1966611 | LABORATORIO MUNICIPAL DE ANALISES CLINICAS DE ITANHAEM | Instituto Butantan / Mendelics | Antonio Jorge Martins; Bianca Cechetto Carlos. Mendelics: Bibiana Santos; Claudia Renata dos Santos Barros; Cíntia Bittar; David Schlesinger. Hemocentro Ribeirão Preto: Simone Kashima; Debora Botequio Moretti; Elaine Cristina Marqueze; Elaine Vieira dos Santos; Elisangela Chicaroni Mattos; Erika Freitas; Evandra Strazza Rodrigues; Felipe Allan da Silva da Costa; Flavia Aburjaile; Fábio Sossai Possebon; Guilherme Campos; Guilherme Targino Valente; Heidge Fukumasu. USP-Botucatu: Rejane Maria Tommasini Grotto; Helena Lage Ferreira; Instituto Butantan: Dimas Tadeu Covas; Jardelina de Souza Todao Bernardino; Jayme A. Souza-Neto; Jessika Cristina Chagas Lesbon; Jorge A. Petroli Marchesi; José Salvatore Leister Patané; João Paulo Kitajima; João Pessoa Araújo Jr.; Leila Sabrina Ullmann; Loyze Paola Oliveira de Lima; Luiz Aurelio de Campos Crispin. Centro de Genômica Funcional da ESALQ: Luiz Lehmann Coutinho; Luiz Carlos Junior de Alcantara; Lívia Sacchetto; Maisa C. Pereira Parra; Maria Carolina Elias; Marta Giovanetti; Marília Moraes; Maurício Lacerda Nogueira. Prefeitura de Sao Paulo: Melissa Palmieri.; Patricia Akemi Assato; Paula Rahal; Paulo Inacio da Costa; Rafael dos Santos Bezerra; Raquel de Lello Rocha Campos Cassano. NGS Soluções Genômicas: Pilar Drummond Sampaio Corrêa Mariani. FZEA-USP Pirassununga: Mirele Daiana Poleti; Raul Machado Neto; Ricardo Augusto Brassaloti; Ricardo Haddad; Rodrigo Tocantins Calado. FAMERP-SJRP: Cecília Artico Banho; Sandra Coccuzzo Sampaio; Svetoslav Nanev Slavov; Vagner Fonseca; Vincent Louis Viala |
| EPI_ISL_2209360 | LABORATORIO MUNICIPAL DE ANALISES CLINICAS DE RIO CLARO | Instituto Butantan | Antonio Jorge Martins; Claudia Renata dos Santos Barros; David Schlesinger; Debora Botequio Moretti; Dimas Tadeu Covas; Elaine Cristina Marqueze; Elaine Vieira Santos; Evandra Strazza Rodrigues; Heidge Fukumasu; Jayme Augusto de Souza-Neto; José Salvatore Leister Patané; Luiz Alcantara; Luiz Lehmann Coutinho; Maria Carolina Elias; Maurício Lacerda Nogueira; Rafael dos Santos Bezerra; Raul Machado Neto; Rejane Maria Tommasini Grotto; Ricardo Haddad; Sandra Coccuzzo Sampaio Vessoni; Simone Kashima; Svetoslav Nanev Slavov; Vincent Louis Viala. |
| EPI_ISL_2209122, EPI_ISL_2209252 | LABORATORIO MUNICIPAL DE ANALISES CLINICAS DE RIO CLARO | Instituto Butantan / FZEA-USP-Pirassununga | Antonio Jorge Martins; Claudia Renata dos Santos Barros; David Schlesinger; Debora Botequio Moretti; Dimas Tadeu Covas; Elaine Cristina Marqueze; Elaine Vieira Santos; Evandra Strazza Rodrigues; Heidge Fukumasu; Jayme Augusto de Souza-Neto; José Salvatore Leister Patané; Luiz Alcantara; Luiz Lehmann Coutinho; Maria Carolina Elias; Maurício Lacerda Nogueira; Rafael dos Santos Bezerra; Raul Machado Neto; Rejane Maria Tommasini Grotto; Ricardo Haddad; Sandra Coccuzzo Sampaio Vessoni; Simone Kashima; Svetoslav Nanev Slavov; Vincent Louis Viala |
| EPI_ISL_1795104, EPI_ISL_2344553 | LABORATORIO MUNICIPAL DE PIRACICABA | Instituto Butantan / ESALQ-Piracicaba | Antonio Jorge Martins; Bianca Cechetto Carlos. Mendelics: Bibiana Santos; Claudia Renata dos Santos Barros; David Schlesinger; David Schlesinger. Hemocentro Ribeirão Preto: Simone Kashima; Debora Botequio Moretti; Debora Botequio Moretti. Centro de Genômica Funcional da ESALQ: Luiz Lehmann Coutinho; Dimas Tadeu Covas; Elaine Cristina Marqueze; Elaine Vieira Santos; Elaine Vieira dos Santos; Elisangela Chicaroni Mattos; Erika Freitas; Evandra Strazza Rodrigues; Felipe Allan da Silva da Costa; Flavia Aburjaile; Guilherme Targino Valente; Heidge Fukumasu; Heidge Fukumasu. USP-Botucatu: Rejane Maria Tommasini Grotto; Instituto Butantan: Alexander Roberto Precioso; Jayme A. Souza-Neto; Jayme Augusto de Souza-Neto; Jessika Cristina Chagas Lesbon; José Salvatore Leister Patané; João Paulo Kitajima; Luiz Alcantara; Luiz Carlos Junior de Alcantara; Luiz Lehmann Coutinho; Maria Carolina Elias; Marta Giovanetti; Maurício Lacerda Nogueira; Patricia Akemi Assato; Rafael dos Santos Bezerra; Raquel de Lello Rocha Campos Cassano. NGS Soluções Genômicas: Pilar Drummond Sampaio Corrêa Mariani. FZEA-USP Pirassununga: Mirele Daiana Poleti; Raul Machado Neto; Rejane Maria Tommasini Grotto; Ricardo Augusto Brassaloti; Ricardo Haddad; Rodrigo Tocantins Calado.; Sandra Coccuzzo Sampaio; Sandra Coccuzzo Sampaio Vessoni; Simone Kashima; Svetoslav Nanev Slavov; Vagner Fonseca; Vincent Louis Viala |
| EPI_ISL_1966999, EPI_ISL_1967000, EPI_ISL_1967001, EPI_ISL_1967002, EPI_ISL_1967003 | LABORATORIO MUNICIPAL DE PIRACICABA | Instituto Butantan / ESALQ-USP (Piracicaba) | Antonio Jorge Martins; Bianca Cechetto Carlos. Mendelics: Bibiana Santos; Claudia Renata dos Santos Barros; Cíntia Bittar; David Schlesinger. Hemocentro Ribeirão Preto: Simone Kashima; Debora Botequio Moretti; Elaine Cristina Marqueze; Elaine Vieira dos Santos; Elisangela Chicaroni Mattos; Erika Freitas; Evandra Strazza Rodrigues; Felipe Allan da Silva da Costa; Flavia Aburjaile; Fábio Sossai Possebon; Guilherme Campos; Guilherme Targino Valente; Heidge Fukumasu. USP-Botucatu: Rejane Maria Tommasini Grotto; Helena Lage Ferreira; Instituto Butantan: Dimas Tadeu Covas; Jardelina de Souza Todao Bernardino; Jayme A. Souza-Neto; Jessika Cristina Chagas Lesbon; Jorge A. Petroli Marchesi; José Salvatore Leister Patané; João Paulo Kitajima; João Pessoa Araújo Jr.; Leila Sabrina Ullmann; Loyze Paola Oliveira de Lima; Luiz Aurelio de Campos Crispin. Centro de Genômica Funcional da ESALQ: Luiz Lehmann Coutinho; Luiz Carlos Junior de Alcantara; Lívia Sacchetto; Maisa C. Pereira Parra; Maria Carolina Elias; Marta Giovanetti; Marília Moraes; Maurício Lacerda Nogueira. Prefeitura de Sao Paulo: Melissa Palmieri.; Patricia Akemi Assato; Paula Rahal; Paulo Inacio da Costa; Rafael dos Santos Bezerra; Raquel de Lello Rocha Campos Cassano. NGS Soluções Genômicas: Pilar Drummond Sampaio Corrêa Mariani. FZEA-USP Pirassununga: Mirele Daiana Poleti; Raul Machado Neto; Ricardo Augusto Brassaloti; Ricardo Haddad; Rodrigo Tocantins Calado. FAMERP-SJRP: Cecília Artico Banho; Sandra Coccuzzo Sampaio; Svetoslav Nanev Slavov; Vagner Fonseca; Vincent Louis Viala |
| EPI_ISL_1967148, EPI_ISL_1967149, EPI_ISL_1967150, EPI_ISL_1967151, EPI_ISL_1967152, EPI_ISL_1967153, EPI_ISL_1967154, EPI_ISL_1967155, EPI_ISL_1967156, EPI_ISL_1967158 | | | |
| see above | LABORATORIO MUNICIPAL DE SUZANO | Instituto Butantan / ESALQ-USP (Piracicaba) | Antonio Jorge Martins; Bianca Cechetto Carlos. Mendelics: Bibiana Santos; Claudia Renata dos Santos Barros; Cíntia Bittar; David Schlesinger. Hemocentro Ribeirão Preto: Simone Kashima; Debora Botequio Moretti; Elaine Cristina Marqueze; Elaine Vieira dos Santos; Elisangela Chicaroni Mattos; Erika Freitas; Evandra Strazza Rodrigues; Felipe Allan da Silva da Costa; Flavia Aburjaile; Fábio Sossai Possebon; Guilherme Campos; Guilherme Targino Valente; Heidge Fukumasu. USP-Botucatu: Rejane Maria Tommasini Grotto; Helena Lage Ferreira; Instituto Butantan: Dimas Tadeu Covas; Jardelina de Souza Todao Bernardino; Jayme A. Souza-Neto; Jessika Cristina Chagas Lesbon; Jorge A. Petroli Marchesi; José Salvatore Leister Patané; João Paulo Kitajima; João Pessoa Araújo Jr.; Leila Sabrina Ullmann; Loyze Paola Oliveira de Lima; Luiz Aurelio de Campos Crispin. Centro de Genômica Funcional da ESALQ: Luiz Lehmann Coutinho; Luiz Carlos Junior de Alcantara; Lívia Sacchetto; Maisa C. Pereira Parra; Maria Carolina Elias; Marta Giovanetti; Marília Moraes; Maurício Lacerda Nogueira. Prefeitura de Sao Paulo: Melissa Palmieri.; Patricia Akemi Assato; Paula Rahal; Paulo Inacio da Costa; Rafael dos Santos Bezerra; Raquel de Lello Rocha Campos Cassano. NGS Soluções Genômicas: Pilar Drummond Sampaio Corrêa Mariani. FZEA-USP Pirassununga: Mirele Daiana Poleti; Raul Machado Neto; Ricardo Augusto Brassaloti; Ricardo Haddad; Rodrigo Tocantins Calado. FAMERP-SJRP: Cecília Artico Banho; Sandra Coccuzzo Sampaio; Svetoslav Nanev Slavov; Vagner Fonseca; Vincent Louis Viala |
| EPI_ISL_7744039, EPI_ISL_7744049, EPI_ISL_7744068, EPI_ISL_7744080, EPI_ISL_7744095, EPI_ISL_7744096 | LACEN | Laboratório de Bioinformática - Universidade Federal de Santa Catarina | "Aline Daina Schlindwein"; "Ana Paula Christoff"; "Antuani Baptistela"; "Carolina Leite Martins"; "Darcita Buerger Rovaris"; "Dayane Azevedo Padilha"; "Doris Sobral Marques SouzaSobral"; "Edmundo Carlos Grisard"; "Eric Kazuo Kawagoe"; "Fernanda Luiza Ferrari"; "Fernanda Roesene Melo"; "Fernando Hartmann Barazzetti"; "Gislaine Fongaro"; "Glauber Wagner"; "Guilherme Augusto Maia"; "Guilherme Razzera"; "Guilherme Toledo e Silva"; "Julia Kinetz Wachter"; "Luiz Felipe de Oliveira"; "Marcos André Schörne"; "Marcus Vinícius Duarte Rodrigues"; "Maria Luiza Bazzo"; "Marlei Pickler Debiasi dos Anjos"; "Milene Moehr de Moraes"; "Nestor Wendt"; "Patrícia Hermes Stoco"; "Paula Sacchet"; "Renato Simões Moreira"; "Rodrigo de Paula Baptista"; "Tamela Zamboni Madaloz"; "Tatiany Aparecida Teixeira Soratto"; "Vilmar Benetti Filho" |
| EPI_ISL_3316194, EPI_ISL_3316195 | LACEN do Estado de Mato Grosso | Instituto Adolfo Lutz, Interdiciplinary Procedures Center, Strategic Laboratory | Caio Vinicius Dias Lopes; Claudia Regina Gonçalves; Claudio Tavares Sacchi; Karoline Rodrigues Campos; Leonardo Tadeu de Araujo; Marlon Benedito Nascomento Santos |
| EPI_ISL_3691385 | LACEN do Estado do Mato Grosso do Sul | Instituto Adolfo Lutz, Interdiciplinary Procedures Center, Strategic Laboratory | Caio Vinicius Dias Lopes; Claudia Regina Gonçalves; Claudio Tavares Sacchi; Karoline Rodrigues Campos; Leonardo Tadeu de Araujo; Marlon Benedito Nascomento Santos |
| EPI_ISL_3134631 | LACEN/PE | WallauLab on behalf of Fiocruz COVID-19 Genomic Surveillance Network | Alexandre Freitas da Silva; Cassia Docena; Constância Flávia Junqueira Ayres; Filipe Zimmer Dezordi; Gabriel Luz Wallau; Gustavo Barbosa de Lima; Lais Ceschini Machado; Lilian Caroliny Amorim Silva; Marcelo Henrique dos Santos Paiva; Matheus Filgueira Bezerra; Sinval Pinto Brandão Filho |
| EPI_ISL_1960075 | LDSP | Universidad Nacional de Colombia - Laboratorio Genómico One Health | Andres F. Cardona-Rios; Carlos Franco-Muñoz; Carolina Muñoz-Arango; Celeny Ortiz; Daniel O. Maldonado-Perez; Diego A. Álvarez-Díaz; Hector Alejandro Ruiz-Moreno; Idabely Betancur Ortiz; Jorge E. Osorio; Juan P. Hernandez-Ortiz; Karl A Ciuoderis; Katherine Laiton-Donato; Laura Silvana Perez; Lina M. Hurtado; Marcela Mercado-Reyes; Maria Angélica Maya; Maria Stella López; Rita Almanza Payares; Sandra Ines Cano; Simón Villegas Velásquez |
| EPI_ISL_1805635, EPI_ISL_1805636, EPI_ISL_1805637, EPI_ISL_1805638, EPI_ISL_1805639, EPI_ISL_1805742 | LDSP FRONTERIZO | Instituto Nacional de Salud-Dirección de Investigación en Salud Pública | Carlos Franco-Muñoz; Carmen Osorio; Diana Malo; Diego A. Álvarez-Díaz; Diego Andrés Prada; Gerardo Santamaría; Hector Alejandro Ruiz-Moreno; Jhonnatan Reales-González; Jorge Rivera; Juan Camilo Martinez; Julian Naizaque; Katherine Laiton-Donato; Lisseth Pardo; Magdalena Wiesner; Marcela Mercado-Reyes; Maria T. Herrera-Sepúlveda; Marta Lopez Blanco; Martha Lucia Ospina Martinez; Paola Rojas; Sergio Gomez; Sheryll Corchuelo; Ángela Alarcon Cruz |
| EPI_ISL_1805498 | LESP Guanajuato | Instituto de Diagnostico y Referencia Epidemiologicos (INDRE) | Abril Rodriguez-Maldonado; Ariadna Medina-Benitez; Claudia Wong-Arambula; Ernesto Ramirez-Gonzalez.; Gisela Barrera-Badillo; Irma Lopez-Martinez; Joaquin Quiroz-Mercado; Lucia Hernandez-Rivas; Natividad Cruz-Ortiz; Sergio Rangel-Guerrero; Tatiana Nunez-Garcia; Vanessa Rivero-Arredondo |
| EPI_ISL_1805499, EPI_ISL_1805500 | LESP Nuevo Leon | Instituto de Diagnostico y Referencia Epidemiologicos (INDRE) | Abril Rodriguez-Maldonado; Ariadna Medina-Benitez; Claudia Wong-Arambula; Ernesto Ramirez-Gonzalez.; Gisela Barrera-Badillo; Irma Lopez-Martinez; Joaquin Quiroz-Mercado; Lucia Hernandez-Rivas; Natividad Cruz-Ortiz; Sergio Rangel-Guerrero; Tatiana Nunez-Garcia; Vanessa Rivero-Arredondo |
| EPI_ISL_2296088 | LESP Yucatan | Instituto de Diagnostico y Referencia Epidemiologicos (INDRE) | Abril Rodriguez-Maldonado; Ariadna Medina-Benitez; Claudia Wong-Arambula; Ernesto Ramirez-Gonzalez.; Gisela Barrera-Badillo; Irma Lopez-Martinez; Joaquin Quiroz-Mercado; Lucia Hernandez-Rivas; Natividad Cruz-Ortiz; Sergio Rangel-Guerrero; Tatiana Nunez-Garcia; Vanessa Rivero-Arredondo |
| EPI_ISL_1745177, EPI_ISL_1745193, EPI_ISL_1745196 | LHUB-ULB | Labo Klinische Biologie, UZA | Basil Britto Xavier; Christine Lammens; Herman Goossens; Jasmine Coppens; Marie Le Mercier; Veerle Matheeussen |
| EPI_ISL_3342149 | LIME | Laboratorio Departamental de Salud Publica de Antioquia | Andres F. Cardona-Rios; Gloria Isabel Escobar; Idabely Betancur Ortiz; Juan P. Hernandez-Ortiz; Maria Stella López |
| EPI_ISL_4196898, EPI_ISL_4212182 | LSP DISTRITAL BOGOTA | Gencore - Universidad de los Andes | Alejandro Gomez; Cristian Barrera; Gabriela Ariza; Gabriela Delgado; Johana Hernandez; Luisa Sacristan; Marcela Guevara; Marcela Mercado; Silvia Restrepo |
| EPI_ISL_1827184, EPI_ISL_1827196, EPI_ISL_1827220, EPI_ISL_1827221, EPI_ISL_1827222, EPI_ISL_1827223, EPI_ISL_1827231, EPI_ISL_1827242, EPI_ISL_1827246, EPI_ISL_1827316, EPI_ISL_1827320, EPI_ISL_1827340, EPI_ISL_1827358, EPI_ISL_1827459, EPI_ISL_1827460, EPI_ISL_2191967 | | | |
| see above | Lab voor klinische biologie | Lab voor klinische biologie | Bruno Verhasselt; Hannelore Hamerlinck; Marija Janevska |
| EPI_ISL_1738825, EPI_ISL_1738827, EPI_ISL_1739172, EPI_ISL_1739173, | Labeto - CAB - Leiria | Instituto Nacional de Saude (INSA) | Borges et al |

| | | | |
|---|---|---|---|
| EPI_ISL_1739174 | | | |
| EPI_ISL_1788821, EPI_ISL_1788822, EPI_ISL_1788823, EPI_ISL_1788824, EPI_ISL_1788825, EPI_ISL_1788826, EPI_ISL_1788827, EPI_ISL_1788828, EPI_ISL_1788829, EPI_ISL_1788830, EPI_ISL_1788831, EPI_ISL_1788832, EPI_ISL_1788833, EPI_ISL_1788834, EPI_ISL_1788835, EPI_ISL_1788836, EPI_ISL_1788837, EPI_ISL_1788838, EPI_ISL_1821634, EPI_ISL_1821635, EPI_ISL_1821636, EPI_ISL_1821637, EPI_ISL_1821638, EPI_ISL_1821639, EPI_ISL_1821640, EPI_ISL_1821641, EPI_ISL_1821642, EPI_ISL_1821653, EPI_ISL_1821654, EPI_ISL_1821657 | | | |
| see above | Labo Analyses Med | National Reference Center for Viruses of Respiratory Infections, Institut Pasteur, Paris | Amaury Vaysse; Angela Brisebarre; Camille Capel; Catherine Coignard; Christophe Malabat; Corinne Maufrais; Dominique Rousset; Emmanuelle Permal; Etienne Simon-Lorière; Frédéric Lemoine; Louise Lefrançois; Marion Barbet; Maud Vanpeene; Méline Bizard; Sylvie Behillil; Sylvie Van der Werf; Vincent Enouf |
| EPI_ISL_1643418, EPI_ISL_1643420, EPI_ISL_1643421, EPI_ISL_1643473, EPI_ISL_1720979 | Labor LÃ¼beck bzw. LaborÃ¤rztliche Gemeinschaftspraxis LÃ¼beck | Robert Koch Institute | |
| EPI_ISL_1627107, EPI_ISL_1628330, EPI_ISL_1628331, EPI_ISL_1628333, EPI_ISL_2227919, EPI_ISL_2227920, EPI_ISL_2227921, EPI_ISL_2227922 | | | |
| see above | Laboratoire Carage | Institut Pasteur de la Guyane | A. Salmier; Anne Lavergne; Dominique Rousset |
| EPI_ISL_2374086 | Laboratoire Virologie Saint Louis APHP | Laboratoire Virologie Saint Louis APHP | Constance Delauguerre; Jérome Le Goff; Linda Feghoul; Marie Laure Chaix; Marie Laure Néré; Maud Salmona; Severine Mercier Delarue; Sophia Achaibou |
| EPI_ISL_3458373, EPI_ISL_3458376, EPI_ISL_3458378, EPI_ISL_3458384, EPI_ISL_3458419, EPI_ISL_3458421, EPI_ISL_3458422, EPI_ISL_3458423, EPI_ISL_3458424, EPI_ISL_3458425, EPI_ISL_3508441, EPI_ISL_5872740, EPI_ISL_5872751, EPI_ISL_5872756, EPI_ISL_5872757, EPI_ISL_5872760, EPI_ISL_5872771, EPI_ISL_5872778, EPI_ISL_5873230, EPI_ISL_5887871, EPI_ISL_5887873, EPI_ISL_5887874, EPI_ISL_5887884, EPI_ISL_5891533, EPI_ISL_5891544, EPI_ISL_5891600, EPI_ISL_5891601, EPI_ISL_7713415, EPI_ISL_7713418, EPI_ISL_7713425, EPI_ISL_7713439, EPI_ISL_7713441, EPI_ISL_7713442, EPI_ISL_7713444, EPI_ISL_7713445, EPI_ISL_7713449, EPI_ISL_7713450, EPI_ISL_7713453, EPI_ISL_7713455, EPI_ISL_7713464, EPI_ISL_7713490, EPI_ISL_7713513, EPI_ISL_7713555, EPI_ISL_7713576 | | | |
| see above | Laboratoire de santé publique du Québec | Laboratoire de santé publique du Québec | Guillaume Bourque; Ioannis Ragoussis; Jesse Shapiro; Mark Lathrop and Judith Fafard on behalf of the CoVSeQ research group; Mark Lathrop and Michel Roger on behalf of the CoVSeQ research group; Sandrine Moreira |
| EPI_ISL_1918240, EPI_ISL_1918361 | Laboratoire national de sante, Microbiology, Virology | Laboratoire national de sante, Microbiology, Microbial Genomics Platform | Anke Wienecke-Baldacchino; Catherine Ragimbeau; Fatu Djabi; Jessica Tapp; Lise Pignon; Raoul Salmon; Tamir Abdelrahman; Trung Nguyen Nguyen |
| EPI_ISL_1917951 | Laboratoires Reunis | Laboratoire national de sante, Microbiology, Microbial Genomics Platform | Anke Wienecke-Baldacchino; Bernard Weber; Catherine Ragimbeau; Fatu Djabi; Jessica Tapp; Lise Pignon; Raoul Salmon; Tamir Abdelrahman |
| EPI_ISL_1917700 | Laboratoires d'analyses medicales - Ketterthill | Laboratoire national de sante, Microbiology, Microbial Genomics Platform | Anke Wienecke-Baldacchino; Caroline Scheiber; Catherine Ragimbeau; Fatu Djabi; Jessica Tapp; Lise Pignon; Raoul Salmon; Serge Vedy; Tamir Abdelrahman |
| EPI_ISL_1719835 | Laboratori Clinic Territorial de Girona | Can Ruti SARS-CoV-2 Sequencing Hub (HUGTiP/IrsiCaixa/IGTP) | Alba Sanchez; Anna Not; Antoni E Bordoy; Bonaventura Clotet; Cristina Casan; Cristina Esteban; Francesc Catala-Moll; Gemma Clara; Ignacio Blanco; Marc Noguera-Julian; Maria Casadella; Mariona Parera; Mercedes Guerrero; Montserrat Gimenez; Pere-Joan Cardona; Pilar Armengol; Roger Paredes; Veronica Saludes; and Elisa Martro on behalf of the Can Ruti SARS-CoV-2 Sequencing Hub. |
| EPI_ISL_2557356 | Laboratorio Central de Saude Publica do Estado de Minas Gerais (LACEN/MG) | Laboratory of Respiratory Viruses and Measles, Oswaldo Cruz Institute, FIOCRUZ | Alice Sampaio Rocha; Ana Carolina Mendonca; Andre Felipe Leal Bernardes; Anna Carolina Paixao; Elisa Cavalcante Pereira; Fernando Motta; Luciana Appolinario; Marilda Siqueira on behalf of the Fiocruz COVID-19 Genomic Surveillance Network; Paola Resende; Renata Serrano Lopes; Taina Venas |
| EPI_ISL_2196287, EPI_ISL_2196289, EPI_ISL_2196290, EPI_ISL_2196291, EPI_ISL_2196292, EPI_ISL_2196293, EPI_ISL_2196294, EPI_ISL_2196295, EPI_ISL_2196296, EPI_ISL_2196297, EPI_ISL_2196298, EPI_ISL_2196299, EPI_ISL_2196300, EPI_ISL_2196301, EPI_ISL_2443557, EPI_ISL_2443558, EPI_ISL_2443559 | | | |
| see above | Laboratorio Central de Saude Publica do Estado do Para (LACEN/PA) | Laboratory of Respiratory Viruses and Measles, Oswaldo Cruz Institute, FIOCRUZ | Alice Sampaio Rocha; Ana Carolina Mendonca; Anna Carolina Paixao; Elisa Cavalcante Pereira; Fernando Motta; Luciana Appolinario; Marilda Siqueira on behalf of the Fiocruz COVID-19 Genomic Surveillance Network; Paola Resende; Renata Serrano Lopes; Taina Venas; Valnete Andrade |
| EPI_ISL_1919605, EPI_ISL_1919606, EPI_ISL_1919608, EPI_ISL_1919609, EPI_ISL_1919610, EPI_ISL_1919611 | Laboratorio Analisi Osp. Città di Castello - Azienda USL Umbria1 | Istituto Zooprofilattico Sperimentale dell'Abruzzo e Molise "G. Caporale" | Ancora M; Calistri P; Cammà C; Caporale M; Curini V; Delli Compagni E; Di Domenico M; Di Lollo Valeria; Di Pasquale A; Lorusso A; Malagigi V; Mangone I; Marcacci M; Puglia I; Rinaldi A; Savini G; Scialabba S; Tacconi P |
| EPI_ISL_1858269, EPI_ISL_1858271, EPI_ISL_1858273, EPI_ISL_1858275, EPI_ISL_1858277, EPI_ISL_1858279, EPI_ISL_1858282, EPI_ISL_1858284, EPI_ISL_1858286, EPI_ISL_1858292, EPI_ISL_1858294, EPI_ISL_1858295, EPI_ISL_1858297, EPI_ISL_1858299, EPI_ISL_1858301, EPI_ISL_1858304, EPI_ISL_1858306, EPI_ISL_1858308, EPI_ISL_1858310, EPI_ISL_1858312, EPI_ISL_1858314, EPI_ISL_1858315, EPI_ISL_1858317, EPI_ISL_1858319, EPI_ISL_1858321, EPI_ISL_1858323, EPI_ISL_1858325, EPI_ISL_1858327, EPI_ISL_1858329, EPI_ISL_1858331, EPI_ISL_1858333, EPI_ISL_1858334, EPI_ISL_1858336, EPI_ISL_1858338, EPI_ISL_1858340, EPI_ISL_1858342, EPI_ISL_1858344, EPI_ISL_1858346, EPI_ISL_1858348, EPI_ISL_1858352, EPI_ISL_1858354, EPI_ISL_1858356, EPI_ISL_1858359, EPI_ISL_1858361, EPI_ISL_1858363, EPI_ISL_1858365, EPI_ISL_1858367, EPI_ISL_1858369, EPI_ISL_1858371, EPI_ISL_1858373, EPI_ISL_1858375, EPI_ISL_1858377, EPI_ISL_1858379, EPI_ISL_1858380, EPI_ISL_1858383, EPI_ISL_1858384, EPI_ISL_1858387, EPI_ISL_1858389, EPI_ISL_1858391, EPI_ISL_1858392, EPI_ISL_1858395, EPI_ISL_1858397, EPI_ISL_1858399, EPI_ISL_1858401, EPI_ISL_1858403, EPI_ISL_1858405, EPI_ISL_1858409, EPI_ISL_1858411, EPI_ISL_1858414, EPI_ISL_1858416, EPI_ISL_1858418, EPI_ISL_1858420, EPI_ISL_1858422, EPI_ISL_1858423, EPI_ISL_1858426, EPI_ISL_1858428, EPI_ISL_1858430, EPI_ISL_1858432, EPI_ISL_1858434, EPI_ISL_1858436, EPI_ISL_1858438, EPI_ISL_1858440, EPI_ISL_1858442, EPI_ISL_1858444, EPI_ISL_1858446, EPI_ISL_1858449, EPI_ISL_1858451, EPI_ISL_1858453, EPI_ISL_1858454, EPI_ISL_1858456, EPI_ISL_1858458, EPI_ISL_1858460, EPI_ISL_1858462, EPI_ISL_1858464, EPI_ISL_1858466, EPI_ISL_1858468, EPI_ISL_1858470, EPI_ISL_1858473, EPI_ISL_1858475, EPI_ISL_1858477, EPI_ISL_1858479, EPI_ISL_1858514, EPI_ISL_1858520, EPI_ISL_1858524, EPI_ISL_1858542, EPI_ISL_1858546, EPI_ISL_1858548, EPI_ISL_1858549, EPI_ISL_1858551, EPI_ISL_1858554, EPI_ISL_1858555, EPI_ISL_1858557, EPI_ISL_1858559, EPI_ISL_1858734, EPI_ISL_2837041, EPI_ISL_2837178 | | | |
| see above | Laboratorio Central Noel Nutels | Bioinformatics Laboratory / LNCC | Alessandra P Lamarca; Alexandra L Gerber; Amilcar Tanuri; Ana Paula de C Guimaraes; Ana Paula de C Guimarães; Ana Tereza R Vasconcelos; Andrea Cony Cavalcanti; Andréa Cony Cavalcanti; Caio Luiz Pereira Ribeiro; Cassia Alves; Cintia Policarpo; Claudia Maria Braga de Mello; Cristiane Gomes da Silva; Diana Mariani; Douglas Terra Machado; Flavio Dias da Silva; Flávio Dias da Silva; Gleidson da Silva de Oliveira; Leandro Magalhaes de Souza; Leandro Magalhães de Souza; Liliane Cavalcante; Luiz G P de Almeida; Marcio Henrique de Oliveira Garcia; Mario Sergio Ribeiro; Ronaldo da Silva F Jr; Silvia Carvalho; Thais Felix Cruz |
| EPI_ISL_3091094 | Laboratorio Central de Epidemiología (LCE) | Instituto de Biotecnología de la UNAM | ; Alejandra Garcia-Gasca; Alejandra Hernández-Terán; Alejandro Sanchez-Flores; Alfredo Herrera-Estrella; Alicia Ocaña-Mondragón; Andreu Comas-García; Angel Gustavo Salas-Lais; Antonio Loza Román; Bernardo Martínez-Miguel; Blanca Taboada; Brenda Irasema Maldonado-Meza; Bruno Gomez-Gil; Carla Ivón Herrera-Najera; Carlos F. Arias; Celia Boukadida; Clara Esperanza Santacruz-Tinoco; Concepción Grajales-Muñiz; Consorcio Mexicano de Vigilancia Genómica (CoViGen-Mex). Authors (in alphabetical order): Julio Elias Alvarado-Yaah; Cristóbal Cháidez-Quiróz; Célida Duque Molina; Célida Martínez- Rodríguez; Daniel Fregoso-Rueda; Daniel Lira Morales; Eduardo Becerril-Vargas; Fernando Fontove-Herrera; Fidencio Mejía-Nepomuceno; Francisco Pulido; Gloria Elena Espinosa-Ayala; Gloria María Molina-Salinas; Gloria Vazquez; Hector Esteban Paz-Juárez; Hector Montoya-Fuentes; Helen Haydee Fernanda Ramírez-Plascencia; Irvin González-López; Jean Pierre González; Joel Armando Vázquez-Pérez.; Jorge Salas-Hernández; José Antonio Enciso-Moreno; José Arturo Martínez-Orozco; José Esteban Muñoz-Medina; José de Jesús Nuñez-Contreras; Juan Bautista Chale-Dzul; Julissa Enciso-Ibarra; Luis Alberto Ochoa-Carrera; Margarita Matías-Florentino; Mario Mújica-Sánchez; María Guadalupe Santiago-Mauricio; María Guadalupe de Jesús Mireles-Rivera; Nelly Sélem-Mojica; Pavel Isa; Ricardo Ciria Merce; Ricardo Grande; Rosa María Gutierrez Rios; Santiago Ávila-Ríos; Selene Zárate; Susana Lopez; Victor Eduardo Garcia-Arias; Víctor Hugo Borja-Aburto |
| EPI_ISL_4518754 | Laboratorio Central de Salud Publica de Paraguay | Fundação Ezequiel Dias | Andre Leal; Andrea Gómez de la Fuente; Cynthia Vazquez; Elaine Cristina; Felipe Iani; Flavia Aburjaile; Gislene Garcia de Castro Lichs; Glauco Carvalho; Hegger Fritsch; Joilson Xavier; Juan Torales; Luiz Alcantara.; Luiz Henrique Ferraz Demarchi; Luiz Takao Watanabe; Marina Castilhos Souza Umaki Zardin; Marta Giovanetti; María José Ortega; María Liz Gamarra; Natalia Guimaraes; Raquel da Silva Ferreira; Shirley Villalba; Talita Adelino; Vagner Fonseca; de Oliveira |
| EPI_ISL_2157531 | Laboratorio Central de Saude Publica do Estado da Bahia (LACEN/BA) | Laboratory of Respiratory Viruses and Measles, Oswaldo Cruz Institute, FIOCRUZ | Alice Sampaio Rocha; Ana Carolina Mendonca; Anna Carolina Paixao; Elisa Cavalcante Pereira; Felicidade Pereira; Fernando Motta; Luciana Appolinario; Marilda Siqueira on behalf of the Fiocruz COVID-19 Genomic Surveillance Network; Paola Resende; Renata Serrano Lopes; Taina Venas |
| EPI_ISL_2536334, EPI_ISL_2536335, EPI_ISL_2536342, EPI_ISL_2536345, EPI_ISL_3061885, EPI_ISL_3061886, EPI_ISL_3061887 | | | |
| see above | Laboratorio Central de Saude Publica do Estado da Paraiba (LACEN-PB) | Laboratory of Respiratory Viruses and Measles, Oswaldo Cruz Institute, FIOCRUZ | Alice Sampaio Rocha; Ana Carolina Mendonca; Anna Carolina Paixao; Dalane Loudal Florentino Teixeira; Elisa Cavalcante Pereira; Fernando Motta; Joao Felipe Bezerra; Luciana Appolinario; Marilda Siqueira on behalf of the Fiocruz COVID-19 Genomic Surveillance Network; Paola Resende; Renata Serrano Lopes; Taina Venas |
| EPI_ISL_2274041, EPI_ISL_2274045, EPI_ISL_2274046, EPI_ISL_2274054, EPI_ISL_2274059, EPI_ISL_2274061, EPI_ISL_2274063 | | | |
| see above | Laboratorio Central de Saude Publica do Estado de Alagoas (LACEN/AL) | Laboratory of Respiratory Viruses and Measles, Oswaldo Cruz Institute, FIOCRUZ | Alice Sampaio Rocha; Ana Carolina Mendonca; Anderson Brandao Leite; Anna Carolina Paixao; Elisa Cavalcante Pereira; Fernando Motta; Luciana Appolinario; Marilda Siqueira on behalf of the Fiocruz COVID-19 Genomic Surveillance Network; Paola Resende; Renata Serrano Lopes; Taina Venas |
| EPI_ISL_2196351 | Laboratorio Central de Saude Publica do Estado de Sergipe (LACEN/SE) | Laboratory of Respiratory Viruses and Measles, Oswaldo Cruz Institute, FIOCRUZ | Alice Sampaio Rocha; Ana Carolina Mendonca; Anna Carolina Paixao; Cliomar Alves dos Santos; Elisa Cavalcante Pereira; Fernando Motta; Luciana Appolinario; Marilda Siqueira on behalf of the Fiocruz COVID-19 Genomic Surveillance Network; Paola Resende; Renata Serrano Lopes; Tainá Moreira Martins Venas |
| EPI_ISL_3045428, EPI_ISL_3045429, EPI_ISL_3045435, EPI_ISL_3045436, EPI_ISL_3045438, EPI_ISL_3045439, EPI_ISL_3045442, EPI_ISL_3045444, EPI_ISL_3045447, EPI_ISL_3045451, EPI_ISL_3045453, EPI_ISL_3045454, EPI_ISL_3045455, EPI_ISL_3045456, EPI_ISL_3045459 | | | |
| see above | Laboratorio Central de Saude Publica do Estado do Espirito Santo (LACEN/ES) | Laboratory of Respiratory Viruses and Measles, Oswaldo Cruz Institute, FIOCRUZ | Alice Sampaio Rocha; Ana Carolina Mendonca; Anna Carolina Paixao; Elisa Cavalcante Pereira; Fernando Motta; Luciana Appolinario; Marilda Siqueira on behalf of the Fiocruz COVID-19 Genomic Surveillance Network; Paola Resende; Renata Serrano Lopes; Rodrigo Ribeiro Rodrigues; Taina Venas |
| EPI_ISL_2983203, EPI_ISL_2983204, EPI_ISL_2983205, EPI_ISL_2983206, EPI_ISL_2983207, EPI_ISL_2983208, EPI_ISL_2983209, EPI_ISL_2983210, EPI_ISL_2983211, EPI_ISL_2983212 | | | |
| see above | Laboratorio Central de Saude Publica do Estado do Maranhao (LACEN-MA) | Laboratory of Respiratory Viruses and Measles, Oswaldo Cruz Institute, FIOCRUZ | Agatha Cristinne Prudencio; Alice Sampaio Rocha; Ana Carolina Mendonca; Anna Carolina Paixao; Elisa Cavalcante Pereira; Fernando Motta; Ighor Leonardo Arantes Gomes; Lidio Gonçalves Lima Neto; Luciana Appolinario; Marilda Siqueira on behalf of the Fiocruz COVID-19 Genomic Surveillance Network; Paola Resende; Renata Serrano Lopes; Tainá Venas |
| EPI_ISL_2758989, EPI_ISL_2758991, EPI_ISL_2758993, EPI_ISL_2758996, EPI_ISL_2758998, EPI_ISL_2758999, EPI_ISL_2759000, EPI_ISL_2759003, EPI_ISL_2759007, EPI_ISL_2759009, EPI_ISL_2759010, EPI_ISL_2759013, EPI_ISL_2759015, EPI_ISL_2759018, EPI_ISL_2759019, EPI_ISL_2759023, EPI_ISL_2759027, EPI_ISL_2759029, EPI_ISL_2759031, EPI_ISL_2759032, EPI_ISL_2759034, EPI_ISL_2759036, EPI_ISL_2759038, EPI_ISL_2759039, EPI_ISL_2759042, EPI_ISL_2759043, EPI_ISL_2759044, EPI_ISL_2759053, EPI_ISL_2759055, EPI_ISL_2759058, EPI_ISL_2759060, EPI_ISL_2759064, EPI_ISL_2759067, EPI_ISL_2759069, EPI_ISL_2775422 | | | |
| see above | Laboratorio Central de Saude Publica do Estado do Parana (Instituto de Biologia Molecular do Paraná (LAC)EN-PR) | Instituto Carlos Chagas - Fiocruz | Alessandra De Melo Aguiar; Andreia Akemi Suzukawa; Andréa Rodrigues Ávila; Bruno Dallagiovanna; Dalila Zanette; Eduardo Balsanelli; Emanuel Maltempi de Souza; Fabio Passetti; Fabricio Klerynton Marchini; Fábio de Oliveira Pedrosa; Guilherme Becker; Helisson Faoro; Hellen Geremias dos Santos; Irina Nastassja Riediger; Letusa Albrecht; Lucas Blanes; Luis Gustavo Morello; Lysangela Ronalte Alves; Maria do Carmo Debur; Mauro de Medeiros Oliveira; Michelle Orane Schemberger; Paola Cristina Resende; Sheila Cristina Nardeli; Tiago Gräf; Valter Antônio de Baura |
| EPI_ISL_2274115, EPI_ISL_2274117, EPI_ISL_2274123, EPI_ISL_2274127, EPI_ISL_2274128, EPI_ISL_2466126, EPI_ISL_2466138 | | | |
| see above | Laboratorio Central de Saude Publica do Estado do Rio Grande do Sul (LACEN-RS) | Laboratory of Respiratory Viruses and Measles, Oswaldo Cruz Institute, FIOCRUZ | Alice Sampaio Rocha; Ana Carolina Mendonca; Anna Carolina Paixao; Elisa Cavalcante Pereira; Fernando Motta; Luciana Appolinario; Marilda Siqueira on behalf of the Fiocruz COVID-19 Genomic Surveillance Network; Paola Resende; Renata Serrano Lopes; Richard Salvato; Taina Venas; Tatiana Schaffer Gregianini |
| EPI_ISL_3048791, EPI_ISL_3048792, EPI_ISL_3048793, | Laboratorio Central de Saude Publica do Estado do Rio Grande do Sul (LACEN- | Laboratório de Biologia Molecular da Universidade Federal de Ciências da Saúde de Porto | Adriana Seixas; Ana B. G. Veiga; Ana Paula Muterle Varela; Fabiana Quoos Mayer; Fernando Hayashi Sant'Anna; Janira Prichula; Letícia Garay Martins; Richard Steiner Salvato; Tatiana Schäffer Gregianini |

| | | | |
|---|---|---|---|
| EPI_ISL_3048794 | RS) | Alegre | |
| EPI_ISL_2196207, EPI_ISL_2196208, EPI_ISL_2196209 | Laboratorio Central de Saude Publica do Estado do Rio de Janeiro (LACEN-RJ) | Laboratory of Respiratory Viruses and Measles, Oswaldo Cruz Institute, FIOCRUZ | Alice Sampaio Rocha; Ana Carolina Mendonca; Andrea Cony Cavalcanti; Anna Carolina Paixao; Elisa Cavalcante Pereira; Fernando Motta; Luciana Appolinario; Marilda Siqueira on behalf of the Fiocruz COVID-19 Genomic Surveillance Network; Paola Resende; Renata Serrano Lopes; Taina Venas |
| EPI_ISL_2363559, EPI_ISL_2363560, EPI_ISL_2363568, EPI_ISL_2363569, EPI_ISL_2363570, EPI_ISL_2363573 | Laboratorio Central de la Ciudad de Santa Fe | Grupo de Genómica y Bioinformática del Instituto de Investigación de la Cadena Láctea CONICET-INTA on behalf of 'Proyecto Argentino Interinstitucional de genomica de SARS-CoV-2' (PAIS Consortium) | AF; Amadio; C; Eberhardt; G; Irazoqui; JM; MF; Mugna; Ojeda; Pastor; Rompato; V |
| EPI_ISL_3924651, EPI_ISL_3924929 | Laboratorio Clinico Toledo | Sequencing and Genomics facility (SGF) | Camille Zenon; Carlos Sariol; Filipa Godoy; Israel Candelaria Velez; Jose Carlos Velez; Jose Rodriguez Orengo; Limari Huertas; Marcia Cruz; Riccardo Papa; Silvia Planas; Steven Van Belleghem; Yadira Ortiz |
| EPI_ISL_4197233 | Laboratorio Clínico Analizar | Gencore - Universidad de los Andes | Cristian Barrera; David González; Gabriela Ariza; Luisa Sacristan; Marcela Guevara; Marcela Mercado; Silvia Restrepo |
| EPI_ISL_5800129 | Laboratorio De Franca | Instituto Butantan | Antonio Jorge Martins; Claudia Renata dos Santos Barros; David Schlesinger; Debora Botequio Moretti; Dimas Tadeu Covas; Elaine Cristina Marqueze; Elaine Vieira Santos; Evandra Strazza Rodrigues; Heidge Fukumasu; Jayme Augusto de Souza-Neto; José Salvatore Leister Patané; Luiz Alcantara; Luiz Lehmann Coutinho; Maria Carolina Elias; Maurício Lacerda Nogueira; Rafael dos Santos Bezerra; Raul Machado Neto; Rejane Maria Tommasini Grotto; Ricardo Haddad; Sandra Coccuzzo Sampaio Vessoni; Simone Kashima; Svetoslav Nanev Slavov; Vincent Louis Viala |
| EPI_ISL_5800022, EPI_ISL_5800025 | Laboratorio Municipal De Analises Clinicas De Itanhaem | Instituto Butantan | Antonio Jorge Martins; Claudia Renata dos Santos Barros; David Schlesinger; Debora Botequio Moretti; Dimas Tadeu Covas; Elaine Cristina Marqueze; Elaine Vieira Santos; Evandra Strazza Rodrigues; Heidge Fukumasu; Jayme Augusto de Souza-Neto; José Salvatore Leister Patané; Luiz Alcantara; Luiz Lehmann Coutinho; Maria Carolina Elias; Maurício Lacerda Nogueira; Rafael dos Santos Bezerra; Raul Machado Neto; Rejane Maria Tommasini Grotto; Ricardo Haddad; Sandra Coccuzzo Sampaio Vessoni; Simone Kashima; Svetoslav Nanev Slavov; Vincent Louis Viala |
| EPI_ISL_5799704, EPI_ISL_5799705 | Laboratorio Municipal De Analises Clinicas De Rio Claro | Instituto Butantan | Antonio Jorge Martins; Claudia Renata dos Santos Barros; David Schlesinger; Debora Botequio Moretti; Dimas Tadeu Covas; Elaine Cristina Marqueze; Elaine Vieira Santos; Evandra Strazza Rodrigues; Heidge Fukumasu; Jayme Augusto de Souza-Neto; José Salvatore Leister Patané; Luiz Alcantara; Luiz Lehmann Coutinho; Maria Carolina Elias; Maurício Lacerda Nogueira; Rafael dos Santos Bezerra; Raul Machado Neto; Rejane Maria Tommasini Grotto; Ricardo Haddad; Sandra Coccuzzo Sampaio Vessoni; Simone Kashima; Svetoslav Nanev Slavov; Vincent Louis Viala |
| EPI_ISL_5800158, EPI_ISL_5800167, EPI_ISL_5800262, EPI_ISL_5800401, EPI_ISL_5800402, EPI_ISL_5800405 | Laboratorio Municipal De Piracicaba | Instituto Butantan | Antonio Jorge Martins; Claudia Renata dos Santos Barros; David Schlesinger; Debora Botequio Moretti; Dimas Tadeu Covas; Elaine Cristina Marqueze; Elaine Vieira Santos; Evandra Strazza Rodrigues; Heidge Fukumasu; Jayme Augusto de Souza-Neto; José Salvatore Leister Patané; Luiz Alcantara; Luiz Lehmann Coutinho; Maria Carolina Elias; Maurício Lacerda Nogueira; Rafael dos Santos Bezerra; Raul Machado Neto; Rejane Maria Tommasini Grotto; Ricardo Haddad; Sandra Coccuzzo Sampaio Vessoni; Simone Kashima; Svetoslav Nanev Slavov; Vincent Louis Viala |
| EPI_ISL_2614585 | Laboratorio Municipal De Suzano | Instituto Adolfo Lutz, Interdiciplinary Procedures Center, Strategic Laboratory | Caio Vinicius Dias Lopes; Claudia Regina Gonçalves; Claudio Tavares Sacchi; Erica Valessa Ramos Gomes; Karoline Rodrigues Campos; Leonardo Jose Tadeu de Araujo |
| EPI_ISL_5799914, EPI_ISL_5799916, EPI_ISL_5799921, EPI_ISL_5799926, EPI_ISL_5799935, EPI_ISL_5799937, EPI_ISL_5799988, EPI_ISL_5799989, EPI_ISL_5799990, EPI_ISL_5799993 | | | |
| see above | Laboratorio Municipal De Suzano | Instituto Butantan | Antonio Jorge Martins; Claudia Renata dos Santos Barros; David Schlesinger; Debora Botequio Moretti; Dimas Tadeu Covas; Elaine Cristina Marqueze; Elaine Vieira Santos; Evandra Strazza Rodrigues; Heidge Fukumasu; Jayme Augusto de Souza-Neto; José Salvatore Leister Patané; Luiz Alcantara; Luiz Lehmann Coutinho; Maria Carolina Elias; Maurício Lacerda Nogueira; Rafael dos Santos Bezerra; Raul Machado Neto; Rejane Maria Tommasini Grotto; Ricardo Haddad; Sandra Coccuzzo Sampaio Vessoni; Simone Kashima; Svetoslav Nanev Slavov; Vincent Louis Viala |
| EPI_ISL_3307442, EPI_ISL_3307444, EPI_ISL_3307445 | Laboratorio SYNLAB Colombia | Corporación para Investigaciones Biológicas-CIB | Jeanneth Mosquera Rendon; Jenny Santiago Cuesta; Marcela Mercado Reyes; Uriel A. Hurtado Paez |
| EPI_ISL_2427505, EPI_ISL_2427506, EPI_ISL_2427514, EPI_ISL_2427521, EPI_ISL_2427522, EPI_ISL_2427530, EPI_ISL_2427537, EPI_ISL_2427545, EPI_ISL_2427561, EPI_ISL_2427567, EPI_ISL_2427573, EPI_ISL_2427574, EPI_ISL_2427581, EPI_ISL_2427582, EPI_ISL_2427583, EPI_ISL_2427590, EPI_ISL_2427693, EPI_ISL_2427694, EPI_ISL_2427697, EPI_ISL_2427700, EPI_ISL_2427701, EPI_ISL_2427702, EPI_ISL_2427704, EPI_ISL_2427708, EPI_ISL_2427709, EPI_ISL_2427712, EPI_ISL_2427717, EPI_ISL_2427719, EPI_ISL_2427724, EPI_ISL_2427725, EPI_ISL_2427727, EPI_ISL_2427728, EPI_ISL_2427731, EPI_ISL_2427732, EPI_ISL_2427733, EPI_ISL_2427734, EPI_ISL_2427738, EPI_ISL_2427739, EPI_ISL_2427740, EPI_ISL_2427742, EPI_ISL_2427746, EPI_ISL_2427747, EPI_ISL_2427749, EPI_ISL_2427750, EPI_ISL_2427754, EPI_ISL_2427755, EPI_ISL_2427758, EPI_ISL_2427762, EPI_ISL_2427765, EPI_ISL_2427776, EPI_ISL_2427777 | | | |
| see above | Laboratorio de Biología Molecular Médica Uruguaya | Departments of Pathology and Medicine, New York University School of Medicine | Adriana Heguy; Cecilia Sorhouet; Christian Marier; Dacia Dimartino; Gonzalo Manrique; Maria Cristina Mogdasy; Maria Noel Zubillaga; Maria Victoria Elizondo; Paul Zappile |
| EPI_ISL_4030308, EPI_ISL_4030309, EPI_ISL_4030310 | Laboratorio de Ecologia de Doencas Transmissiveis na Amazonia, Instituto Leonidas e Maria Deane - Fiocruz Amazonia | Laboratorio de Ecologia de Doencas Transmissiveis na Amazonia, Instituto Leonidas e Maria Deane - Fiocruz Amazonia | Alex Martins; André Corado; Debora Duarte; Fabiola Mendonça da Silva Chui; Felipe Naveca; Fernanda Nascimento; Fernando Fonseca de Almeida Val; George Silva; Gisely Cardoso de Melo; Karina Pessoa; Luciana Gonçalves; Marcus Vinicius Guimarães de Lacerda; Maria Júlia Brandão; Maria Paula Gomes Mourão; Mariana Xavier Simão; Matilde Mejía; Michele Jesus; Valdinete Nascimento; Vanderson de Souza Sampaio; Victor Souza; Ágatha Costa |
| EPI_ISL_3321104, EPI_ISL_3321107, EPI_ISL_3332891 | Laboratorio de Infectologia y Virologia Molecular | Laboratory of Molecular Virology, School of Medicine, Pontificia Universidad Catolica de Chile | Alejandro Bhrun; Ana Maria Contreras; Andres E. Munoz-Marcos; Carlos Palma; Catalina Pardo-Roa; Constanza Maldonado; Constanza Martinez-Valdevenito; Eileen Serrano; Erick Salinas; Estefany Poblete; Francisco Melo; Jennifer Angulo; Jorge Levican; Leonardo I. Almonacid; M. Belen Leyton; Magdalena Vera; Marcela Ferres; Maria Jose Avendano; Rafael A. Medina; Tamara Garcia-Salum |
| EPI_ISL_3023502, EPI_ISL_3023593, EPI_ISL_3375874, EPI_ISL_3375876, EPI_ISL_3375880, EPI_ISL_3375886, EPI_ISL_3375887, EPI_ISL_3375888, EPI_ISL_3375889, EPI_ISL_3375891, EPI_ISL_3375896, EPI_ISL_3375901, EPI_ISL_3375937, EPI_ISL_3375942, EPI_ISL_3375944, EPI_ISL_3375945, EPI_ISL_3375950, EPI_ISL_3375952, EPI_ISL_3375954, EPI_ISL_3375957, EPI_ISL_3375959, EPI_ISL_3376002, EPI_ISL_3376014, EPI_ISL_3376184, EPI_ISL_3376375, EPI_ISL_3376380, EPI_ISL_3376400, EPI_ISL_3376407, EPI_ISL_3376408 | | | |
| see above | Laboratorio de Referencial Nacional de Virus Respiratorios | Laboratorio de Referencial Nacional de Virus Respiratorios | Carlos Padilla Rojas; Henri Bailon Calderon; Iris Silva Molina; Joseph Huayra Niquen; Lely Solari Zerpa; Luis Barcena Flores; Marco Galarza Perez; Nancy Rojas Serrano; Omar Caceres Rey; Orson Mestanza Millones; Priscila Lope Pari; Sandra Morales Ruiz; Steve Acedo Lazo; Veronica Hurtado Vela |
| EPI_ISL_2157202, EPI_ISL_2157206, EPI_ISL_2157209 | Laboratorio de Salud Pública | Gencore - Universidad de los Andes | Alejandro Gomez; David Gonzalez; Gabriela Delgado; Johana Hernandez; Luisa Sacristan; Marcela Guevara; Silvia Restrepo |
| EPI_ISL_1673667, EPI_ISL_1673685, EPI_ISL_1701312, EPI_ISL_1701313, EPI_ISL_1701314, EPI_ISL_1913063, EPI_ISL_1913066 | | | |
| see above | Laboratorio de Virología HUCA | Laboratorio de Virología HUCA | Abreu F; Alvarez-Arguelles ME; Boga JA; Castelló C; Costales I; Coto E; Gómez de Oña J; Martín-Rodríguez G; Melón S; Perez-Martínez Z; Rojo S; Sandoval M |
| EPI_ISL_4405034, EPI_ISL_4405037, EPI_ISL_4405062 | Laboratorio de Virología del Hospital de Niños Dr. Ricardo Gutierrez | Área de Secuenciación del Laboratorio de Virología del Hospital de Niños Dr. Ricardo Gutierrez on behalf of 'Proyecto Argentino Interinstitucional de genomica de SARS-CoV-2' (PAIS Consortium) | AS; Acevedo; Acuña; Alexay; Alvarez Lopez; C; D; E; Goya; Gravis; Jacques; LE; Lusso; ME; MI; MS; Mistchenko; Nabaes Jodar; Natale; O; S; Valinotto; Viegas, M. |
| EPI_ISL_1680707, EPI_ISL_1680738, EPI_ISL_1680779, EPI_ISL_1680787, EPI_ISL_1680810, EPI_ISL_1680829, EPI_ISL_1680851, EPI_ISL_1680873, EPI_ISL_1680874, EPI_ISL_1680892, EPI_ISL_1680899, EPI_ISL_1680900, EPI_ISL_1680909, EPI_ISL_1680942, EPI_ISL_1680982, EPI_ISL_1680995, EPI_ISL_1681026, EPI_ISL_1681058, EPI_ISL_1681154, EPI_ISL_1681202, EPI_ISL_1681210, EPI_ISL_1681217, EPI_ISL_1681223, EPI_ISL_1681230, EPI_ISL_1681231, EPI_ISL_1681290, EPI_ISL_1681293, EPI_ISL_1681294, EPI_ISL_1681343, EPI_ISL_1681354, EPI_ISL_1681380, EPI_ISL_1681381, EPI_ISL_1681396, EPI_ISL_1681441, EPI_ISL_1681469, EPI_ISL_1681488, EPI_ISL_1681500, EPI_ISL_1681507, EPI_ISL_1681513, EPI_ISL_1681518, EPI_ISL_1681522, EPI_ISL_1681529, EPI_ISL_1681537, EPI_ISL_1681541, EPI_ISL_1681556, EPI_ISL_1681568, EPI_ISL_1683357, EPI_ISL_1683359, EPI_ISL_1683411, EPI_ISL_1683422, EPI_ISL_1683447, EPI_ISL_1683449, EPI_ISL_1683453, EPI_ISL_1683475, EPI_ISL_1683480, EPI_ISL_1683494, EPI_ISL_1683496, EPI_ISL_1683521, EPI_ISL_1683523, EPI_ISL_1683557, EPI_ISL_1683559, EPI_ISL_1683621, EPI_ISL_1683658, EPI_ISL_1683666, EPI_ISL_1683667, EPI_ISL_1683694, EPI_ISL_1683783, EPI_ISL_1683815, EPI_ISL_1683904, EPI_ISL_1683941, EPI_ISL_1683990, EPI_ISL_1684084, EPI_ISL_1684153, EPI_ISL_1684208, EPI_ISL_1684213, EPI_ISL_1684226, EPI_ISL_1684230, EPI_ISL_1684250, EPI_ISL_1684292, EPI_ISL_1684568, EPI_ISL_1684599, EPI_ISL_1684701, EPI_ISL_1684735, EPI_ISL_1684762, EPI_ISL_1684856, EPI_ISL_1684883, EPI_ISL_1685013, EPI_ISL_1685145, EPI_ISL_1685172, EPI_ISL_1685224, EPI_ISL_1798247, EPI_ISL_1798260, EPI_ISL_1798273, EPI_ISL_1798275, EPI_ISL_1798587, EPI_ISL_1798630, EPI_ISL_1798905, EPI_ISL_1798951, EPI_ISL_1798959, EPI_ISL_1799148, EPI_ISL_1799188, EPI_ISL_1799665, EPI_ISL_1799678, EPI_ISL_1799679, EPI_ISL_1799705, EPI_ISL_1799706, EPI_ISL_1799707, EPI_ISL_1799714, EPI_ISL_1799719, EPI_ISL_1799724, EPI_ISL_1799730, EPI_ISL_1799731, EPI_ISL_1799732, EPI_ISL_1799785, EPI_ISL_1799809, EPI_ISL_1799822, EPI_ISL_1799831, EPI_ISL_1799836, EPI_ISL_1799857, EPI_ISL_1799925, EPI_ISL_1799926, EPI_ISL_1799961, EPI_ISL_1799962, EPI_ISL_1799963, EPI_ISL_1799966, EPI_ISL_1800003, EPI_ISL_1800010, EPI_ISL_1800020, EPI_ISL_1800039, EPI_ISL_1800042, EPI_ISL_1800043, EPI_ISL_1800053, EPI_ISL_1800056, EPI_ISL_1800057, EPI_ISL_1800061, EPI_ISL_1800063, EPI_ISL_1800065, EPI_ISL_1800067, EPI_ISL_1800098, EPI_ISL_1800109, EPI_ISL_1800116, EPI_ISL_1800119, EPI_ISL_1800129, EPI_ISL_1800130, EPI_ISL_1800132, EPI_ISL_1800135, EPI_ISL_1800136, EPI_ISL_1800137, EPI_ISL_1800181, EPI_ISL_1800201, EPI_ISL_1800240, EPI_ISL_1800264, EPI_ISL_1800272, EPI_ISL_1800273, EPI_ISL_1800274, EPI_ISL_1800275, EPI_ISL_1800560, EPI_ISL_4374870 | | | |
| see above | Laboratory Corporation of America | Centers for Disease Control and Prevention Division of Viral Diseases, Pathogen Discovery | Adrian Paskey; Amanda Douglas; Amanda Suchanek; Andrea Throop; Ayla Burns; Benjamin Rambo-Martin; Bobbi Croy; Brian Krueger; Brian Norvell; Christopher Gulvick; Christos Petropoulos; Clinton Paden; Clinton R. Paden; Craig Lukasik; Dakota Howard; Darlene Wagner; Debbie Boles; Dhwani Batra; Duncan MacCannell; Eyad Almasri; Goran Stevovic; Howard Engler; Hrushikesh Deshmukh; Jake Humphrey; Jana Schroth; Jason Caravas; Joe Voshell; John Pruitt; Jonathan Meltzer; Jonathan Williams; Kara Moser; Kimberly Wagner; Kristine Lacek; Lax Iyer; Lisa Pfefferle; Lyndon Tilson; Manoj Jain; Marcia Eisenberg; Mary Ann Cristobal; Mary Cristobal; Mary Williamson; Matthew Robinson; Matthew Schmerer; Michael Levandoski; Mike Sapeta; Mindy Nye; Minoo Agarwal; Mohan Kolli; Nuthawin Charoensri; Oren Cohen; Peter Cook; Peter W. Cook; Prashant Gupta; Qian Zeng; Rama Ghatti; Scott Parker; Scott Ryan; Scott Sammons; Shatavia Morrison; Stanley Letovsky; Steven Ragan; Suresh Babu Selvaraju; Suresh Selvaraju; Susan Countryman; Susan Hicks; Suzanne Dale; Thomas Urban; Tim Kuphal; Tricia Zwiefelhofer; Tymeckia Kendall; Victoria Caban Figueroa; Vincent Drouillon; Yvette Unoarumhi |
| EPI_ISL_2443593, EPI_ISL_2443594, EPI_ISL_2536272, EPI_ISL_2536273, EPI_ISL_2536274, EPI_ISL_2557393, EPI_ISL_3190279, EPI_ISL_3832399, EPI_ISL_6943968, EPI_ISL_6943969, EPI_ISL_6943970 | | | |
| see above | Laboratory of Respiratory Viruses and Measles, Oswaldo Cruz Institute, FIOCRUZ | Laboratory of Respiratory Viruses and Measles, Oswaldo Cruz Institute, FIOCRUZ | Agatha Cristinne Prudencio; Agatha Soares; Alice Sampaio Rocha; Ana Carolina Mendonca; Anna Carolina Paixao; Bruna Mendonça da Silva; Elisa Cavalcante Pereira; Fernando Motta; Ighor Arantes; Ighor Leonardo Arantes Gomes; Jéssica Graça Macedo de Carvalho; Larissa Macedo Pinto; Luciana Appolinario; Marilda Siqueira on behalf of the Fiocruz COVID-19 Genomic Surveillance Network; Paola Resende; Patricia Brasil; Renata Serrano Lopes; Taina Venas; Victor Guimaraes |
| EPI_ISL_2696542 | Laboratory, Gravity Diagnostics | Laboratory, Gravity Diagnostics | Diagnostics, G. |
| EPI_ISL_2157532, EPI_ISL_2157533, EPI_ISL_2157534, EPI_ISL_2157535 | Laboratório Central de Saude Publica do Estado da Bahia (LACEN/BA) | Laboratory of Respiratory Viruses and Measles, Oswaldo Cruz Institute, FIOCRUZ | Alice Sampaio Rocha; Ana Carolina Mendonca; Anna Carolina Paixao; Elisa Cavalcante Pereira; Felicidade Pereira; Fernando Motta; Luciana Appolinario; Marilda Siqueira on behalf of the Fiocruz COVID-19 Genomic Surveillance Network; Paola Resende; Renata Serrano Lopes; Taina Venas |
| EPI_ISL_2274134 | Laboratório Central de Saude Publica do Estado de Santa Catarina (LACEN/SC) | Laboratory of Respiratory Viruses and Measles, Oswaldo Cruz Institute, FIOCRUZ | Alice Sampaio Rocha; Ana Carolina Mendonca; Anna Carolina Paixao; Darcita Buerger Rovaris; Elisa Cavalcante Pereira; Fernando Motta; Luciana Appolinario; Marilda Siqueira on behalf of the Fiocruz COVID-19 Genomic Surveillance Network; Paola Resende; Renata Serrano Lopes; Sandra Bianchini Fernandes; Taina Venas |

| | | | |
|---|---|---|---|
| EPI_ISL_2157525, EPI_ISL_2157526, EPI_ISL_2157528, EPI_ISL_2157529, EPI_ISL_2157530 | Laboratório Central de Saude Publica do Estado do Rio de Janeiro (LACEN/RJ) | Laboratory of Respiratory Viruses and Measles, Oswaldo Cruz Institute, FIOCRUZ | Alice Sampaio Rocha; Ana Carolina Mendonca; Andrea Cony Cavalcanti; Anna Carolina Paixao; Elisa Cavalcante Pereira; Fernando Motta; Luciana Appolinario; Marilda Siqueira on behalf of the Fiocruz COVID-19 Genomic Surveillance Network; Paola Resende; Renata Serrano Lopes; Taina Venas |
| EPI_ISL_4600552 | Laboratório Central de Saúde Pública Dr. Costa Alvarenga | Coordenação Geral de Laboratórios de Saúde Pública (CGLAB/DAEVS/SVS/MS) | Vagner Fonseca; et al. |
| EPI_ISL_4600561 | Laboratório Central de Saúde Pública Noel Nutels | Coordenação Geral de Laboratórios de Saúde Pública (CGLAB/DAEVS/SVS/MS) | Vagner Fonseca; et al. |
| EPI_ISL_4600526 | Laboratório Central de Saúde Pública de Pernambuco | Coordenação Geral de Laboratórios de Saúde Pública (CGLAB/DAEVS/SVS/MS) | Vagner Fonseca; et al. |
| EPI_ISL_4600570, EPI_ISL_4675985 | Laboratório Central de Saúde Pública de Santa Catarina | Coordenação Geral de Laboratórios de Saúde Pública (CGLAB/DAEVS/SVS/MS) | Vagner Fonseca; et al. |
| EPI_ISL_2777239, EPI_ISL_2777240, EPI_ISL_2777276, EPI_ISL_2777518, EPI_ISL_2777571, EPI_ISL_2777572, EPI_ISL_2777666, EPI_ISL_2777668, EPI_ISL_2777670, EPI_ISL_2777699, EPI_ISL_2777725, EPI_ISL_2777726, EPI_ISL_2777727, EPI_ISL_2777729, EPI_ISL_2777733, EPI_ISL_2777751, EPI_ISL_2777807, EPI_ISL_2777808, EPI_ISL_2777809, EPI_ISL_2777813, EPI_ISL_2777854, EPI_ISL_2777855, EPI_ISL_2777857, EPI_ISL_2777858, EPI_ISL_2777859, EPI_ISL_2777860, EPI_ISL_2777861, EPI_ISL_2777862, EPI_ISL_2777864, EPI_ISL_2777899 | | | |
| see above | Laboratório Central de Saúde Pública do Amazonas - LACEN-AM | Laboratorio de Ecologia de Doencas Transmissiveis na Amazonia, Instituto Leonidas e Maria Deane - Fiocruz Amazonia | André Corado; Debora Duarte; Felipe Naveca; Fernanda Nascimento; George Silva; Karina Pessoa; Luciana Gonçalves; Maria Júlia Brandão; Matilde Mejía; Michele Jesus; Valdinete Nascimento; Victor Souza; Ágatha Costa |
| EPI_ISL_3155984 | Laboratório de Biologia Integrativa | Laboratório de Biologia Integrativa | Alessandro Clayton de Souza Ferreira; Aline Brito de Lima; Carolina Moreira Voloch; Daniel Costa Queiroz; Danielle Alves Gomes Zauli; Diego Menezes Bonfim; Filipe Romero Rebello Moreira; Frederico Scott Varella Malta; Joice do Prado Silva; Lucyene Miguita Luiz; Nuno Rodrigues Faria; Paula Luize Camargos Fonseca; Rafael Marques de Souza; Renan Pedra de Souza; Renato Santana Aguiar; Rennan Garcias Moreira; Victor Cavalcanti Pardini; Victor Emmanuel Viana Geddes |
| EPI_ISL_6508515 | Laboratório de Biologia Integrativa/ UFMG | Laboratório de Biologia Integrativa/ UFMG | Adriana Aparecida Ribeiro; Alana Vitor Barbosa Costa; Alessandro Luís Gonçalves; Aline de Brito Lima; Ana Paula De Battisti Ribeiro; Ana Paula Salles Moura Fernandes; Andre Luiz Menezes; Bruna Walker Ferreira; Carolina Senra Alves de Souza; Cristiane P. T. Brito Mendonça; Daniel Costa Queiroz; Danielle Alves Gomes Zauli; Diego Menezes; Eneida Santos de Oliveira; Eva Lídia Arcoverde Medeiros; Felipe Campos de Melo Iani; Fernanda Gil de Souza; Fernanda Santos Mendes; Filipe Romero Rebello Moreira; Flávio Guimarães da Fonseca; Frederico Scott Varella Malta; Hugo Itaru Sato; Hugo José Alves; Igor Pereira Godinho; Jaqueline Silva de Oliveira; Joice do Prado Silva; José Nélio Januario; Juliana Wilke Saliba; Karine Lima Lourenço; Lucyene Miguita; Luige Biciati Alvim; Nara Oliveira Carvalho; Natiely Pereira Silva; Natália Rocha Guimarães; Paula Luize Camargos Fonseca; Pedro Henrique Barbosa de Paula Mendes; Rafael Marques de Souza; Renan Pedra de Souza; Renata Barbosa Peixoto Peixoto; Renato Santana de Aguiar; Rennan Garcias Moreira; Rillery Calixto Dias; Rubens Daniel Miserani Magalhães; Santuza Maria Ribeiro Teixeira; Talita Emile Ribeiro Adelino; Victor Emmanuel Viana Geddes; Walyson Coelho Costa |
| EPI_ISL_2466200, EPI_ISL_2466201, EPI_ISL_2466202, EPI_ISL_2466203, EPI_ISL_2466205, EPI_ISL_2466206 | Laboratório de Biologia Molecular de Doenças Infecciosas e do Câncer (LADIC - UFRN) | Laboratory of Respiratory Viruses and Measles, Oswaldo Cruz Institute, FIOCRUZ | Alice Sampaio Rocha; Ana Carolina Mendonca; Anna Carolina Paixao; Elisa Cavalcante Pereira; Fernando Motta; Josélio Araújo; Luciana Appolinario; Marilda Siqueira on behalf of the Fiocruz COVID-19 Genomic Surveillance Network; Paola Resende; Renata Serrano Lopes; Taina Venas |
| EPI_ISL_2629804, EPI_ISL_2629805, EPI_ISL_2629806, EPI_ISL_2629807, EPI_ISL_2629808, EPI_ISL_2629809, EPI_ISL_2629810, EPI_ISL_2629811 | | | |
| see above | Laboratório de Virologia Molecular - Universidade Federal do Rio de Janeiro | Laboratório de Virologia Molecular - Universidade Federal do Rio de Janeiro | ; Alice Laschuk Herlinger; Amilcar Tanuri; André Felipe Andrade dos Santos; Carolina Moreira Voloch; Cássia Cristina Alves Gonçalves; Diana Mariani; Débora Souza Faffe; Filipe Romero Rebello Moreira; Francine Bittencourt Schiffler; Isabela de Carvalho Leitão; Marcelo Calado de Paula Tôrres; Matheus Augusto Calvano Cosentino; Mirela D'arc; Orlando da Costa Ferreira Junior; Rafael Mello Galliez; Raíssa Mirella dos Santos Cunha da Costa; Renato Santana de Aguiar; Terezinha Marta Pereira Pinto Castineiras; Thamiris dos Santos Miranda; Átila Duque Rossi |
| EPI_ISL_2196249, EPI_ISL_2443551, EPI_ISL_2443685, EPI_ISL_2443687, EPI_ISL_2443688, EPI_ISL_2443690, EPI_ISL_2443691, EPI_ISL_2443692, EPI_ISL_2443694, EPI_ISL_2443699, EPI_ISL_2443701, EPI_ISL_2443702, EPI_ISL_2443703, EPI_ISL_2443705, EPI_ISL_2443709, EPI_ISL_2443712 | | | |
| see above | Labortorio Central de Saude Publica do Estado de Santa Catarina (LACEN/SC) | Laboratory of Respiratory Viruses and Measles, Oswaldo Cruz Institute, FIOCRUZ | Alice Sampaio Rocha; Ana Carolina Mendonca; Anna Carolina Paixao; Darcita Buerger Rovaris; Elisa Cavalcante Pereira; Fernando Motta; Luciana Appolinario; Marilda Siqueira on behalf of the Fiocruz COVID-19 Genomic Surveillance Network; Paola Resende; Renata Serrano Lopes; Sandra Bianchini Fernandes; Taina Venas |
| EPI_ISL_2196245, EPI_ISL_2196318, EPI_ISL_2196319, EPI_ISL_2196320, EPI_ISL_2196321, EPI_ISL_2196322 | Labortorio Central de Saude Publica do Estado do Rio de Janeiro (LACEN/RJ) | Laboratory of Respiratory Viruses and Measles, Oswaldo Cruz Institute, FIOCRUZ | Alice Sampaio Rocha; Ana Carolina Mendonca; Andrea Cony Cavalcanti; Anna Carolina Paixao; Elisa Cavalcante Pereira; Fernando Motta; Luciana Appolinario; Marilda Siqueira on behalf of the Fiocruz COVID-19 Genomic Surveillance Network; Paola Resende; Renata Serrano Lopes; Taina Venas |
| EPI_ISL_2157527, EPI_ISL_2196312, EPI_ISL_2274096, EPI_ISL_2443537, EPI_ISL_2443538, EPI_ISL_2443566, EPI_ISL_2443567, EPI_ISL_2603427, EPI_ISL_2603438, EPI_ISL_2603440, EPI_ISL_2603452, EPI_ISL_2603458, EPI_ISL_2603543, EPI_ISL_2603545 | | | |
| see above | Lboratorio Central de Saude Publica do Estado do Parana (LACEN/PR) | Laboratory of Respiratory Viruses and Measles, Oswaldo Cruz Institute, FIOCRUZ | Alice Sampaio Rocha; Ana Carolina Mendonca; Anna Carolina Paixao; Elisa Cavalcante Pereira; Fernando Motta; Irina Riediger; Luciana Appolinario; Marilda Siqueira on behalf of the Fiocruz COVID-19 Genomic Surveillance Network; Paola Resende; Renata Serrano Lopes; Taina Venas |
| EPI_ISL_1653472, EPI_ISL_2199984 | Lighthouse Lab in Cambridge | Wellcome Sanger Institute for the COVID-19 Genomics UK (COG-UK) Consortium | Cordelia Langford; David K. Jackson; Dominic Kwiatkowski; Ewan Harrison; Ian Johnston; Jeffrey Barrett; John Sillitoe on behalf of the Wellcome Sanger Institute COVID-19 Surveillance Team; Rob Howes; Roberto Amato; Sonia Goncalves; The Lighthouse Lab in Cambridge and Alex Alderton |
| EPI_ISL_1634819, EPI_ISL_1634918 | Lighthouse Lab in Glasgow | Wellcome Sanger Institute for the COVID-19 Genomics UK (COG-UK) Consortium | Anna Dominiczak and Alex Alderton; Carol Clugston; Cordelia Langford; David Gray; David K. Jackson; Dominic Kwiatkowski; Ewan Harrison; Harper VanSteenhouse; Ian Johnston; Jeffrey Barrett; John Sillitoe on behalf of the Wellcome Sanger Institute COVID-19 Surveillance Team; Roberto Amato; Sonia Goncalves; Yumi Kasai |
| EPI_ISL_1759891, EPI_ISL_2005677 | Lighthouse Lab in Milton Keynes | Wellcome Sanger Institute for the COVID-19 Genomics UK (COG-UK) Consortium | Cordelia Langford; David K. Jackson; Dominic Kwiatkowski; Ewan Harrison; Ian Johnston; Jeffrey Barrett; John Sillitoe on behalf of the Wellcome Sanger Institute COVID-19 Surveillance Team; Roberto Amato; Sonia Goncalves; The Lighthouse Lab in Milton Keynes and Alex Alderton |
| EPI_ISL_1704744, EPI_ISL_1704751, EPI_ISL_1704756 | Lurie Children's Hospital of Chicago | Northwestern University - Ozer Lab | Egon A. Ozer; Judd F. Hultquist; Lacy M. Simons; Larry K. Kociolek; Michael G. Ison; Ramon Lorenzo-Redondo; Taylor J. Dean; William J. Muller; Xiaotian; Zheng |
| EPI_ISL_1789611, EPI_ISL_1805300, EPI_ISL_1857182 | M Health Fairview | Minnesota Department of Health, Public Health Laboratory | Alexandra Lorentz; Jacob Garfin; Matt Plumb; and Xiong Wang |
| EPI_ISL_2801330 | MATERNIDADE ESCOLA ASSIS CHATEAUBRIAND | Analytical Competence Molecular Epidemiology Lab/ACME, Oswaldo Cruz Foundation, Ceara (FIOCRUZ CE) | Cleber Furtado Aksenen e Suzana Porto Almeida; Fabio Miyajima; Fernando Braga Stehling; Francisco Eder de Moura Lopes; Jamille Maria Mendes Bezerra; Joaquim César do Nascimento Sousa Junior; Pedro Miguel Carneiro Jeronimo; Thais Ferreira de Oliveira; Thais de Oliveira Costa; Ticiane Cavalcante de Souza; Veridiana Pessoa Miyajima |
| EPI_ISL_2495418, EPI_ISL_3356735, EPI_ISL_3356742, EPI_ISL_3356744 | MB-Cadham Provincial laboratory | National Microbiology Laboratory (NML) | Anna Majer; Anneliese Landgraff; CanCOGeN's metadata curation team; Darian Hole; David Alexander; Elsie Grudeski; Gary Van Domselaar; Grace Seo; Jared Bullard; Jennifer Tanner; Kerry Dust; Kirsten Biggar; Madison Chapel; Morag Graham; Natalie Knox; Nathalie Bastien; Paul Van Caeseele; Philip Mabon; Public Health Agency of Canada CanCOGeN team; Rhiannon Huzarewich; Russell Mandes; Shari Tyson; Timothy Booth; Yan Li |
| EPI_ISL_1731432 | MD PHL | MD PHL | Maryland Department of Health Laboratories Administration |
| EPI_ISL_1656547, EPI_ISL_1694908, EPI_ISL_1817105, EPI_ISL_2287041, EPI_ISL_2453376, EPI_ISL_2453389, EPI_ISL_2659662 | | | |
| see above | MEPHI, Aix Marseille University | MEPHI, Aix Marseille University | Anthony LEVASSEUR |
| EPI_ISL_2689876 | MS Public Health Laboratory | Centers for Disease Control and Prevention Division of Viral Diseases, Pathogen Discovery | Alison Laufer Halpin; Ben L. Rambo-Martin; Clinton R. Paden; Dakota Howard; Darlene Wagner; Dave Wentworth; Dhwani Batra; Jasmine Padilla; Justin Lee; Katie Dillon; Krista Queen; Kristen Knipe; Kristine Lacek; Mark Burroughs; Matthew Schmerer; Mili Sheth; Peter Cook; Sam Shepard; Sarah Nobles; Shoshona Le; Suxiang Tong; Vivien Dugan; Yvette Unoarumhi |
| EPI_ISL_1709940, EPI_ISL_1709956 | MSHS Clinical Microbiology Laboratories | MSHS Pathogen Surveillance Program | Adolfo García-Sastre; Adriana van de Guchte; Ajay Obla; Alberto Paniz-Mondolfi; Ana S. Gonzalez-Reiche; Angela Amoako; Ashley Salimbangon; Betsaida Salom Melo; Bremy Alburquerque; Brianne Ciferri; Charles Gleason; Daniel Floda; Deena R. Altman; Denise Jurczyszak; Emilia Mia Sordillo; Gintaras Deikus; Giulio Kleiner; Gopi Patel; Hala Alshammary; Harm van Bakel; Irina Oussenko; Jayeeta Dutta; Juan Soto; Julia Matthews; Katherine Beach; Kathryn Twyman; Kayla Russo; Komal Srivastava; Levy Sominsky; Mahmoud Awawda; Marta Luksza; Matthew M. Hernandez; Melissa Gitman; Michael D. Nowak; Mitchell J. Sullivan; Nancy Francoeur; Robert Sebra; Sarah Schaefer; Shelcie Fabre; Shwetha Hara Sridhar; Viviana Simon; Ying-Chih Wang; Zenab Khan |
| EPI_ISL_2009160 | MSK Microbiology Lab | MSK Microbiology Lab | Esther Babady; Krupa Jani; Tracy McMillen |
| EPI_ISL_1643478 | MVZ Dr. Eberhard & Partner Dortmund | Robert Koch Institute | |
| EPI_ISL_1644947, EPI_ISL_1722005 | MVZ Labor Krone GbR | Robert Koch Institute | |
| EPI_ISL_1725214, EPI_ISL_1725612 | MVZ fÃ¼r Laboratoriumsmedizin und Mikrobiologie Koblenz-Mittelrhein (Labor Koblenz) | Robert Koch Institute | |
| EPI_ISL_2802277 | Maine Health and | Tewhey Lab, The Jackson | Barter, M.; Dewey, H.; H. and Tewhey, R.; Iosue, F.; Lynch, R.; Matluk, N.; Munger |

| | | | |
|---|---|---|---|
| | Environmental Testing Laboratory | Laboratory | |
| EPI_ISL_1922569, EPI_ISL_1922577, EPI_ISL_1967502, EPI_ISL_1967511, EPI_ISL_1967512, EPI_ISL_1967513, EPI_ISL_1967518, EPI_ISL_1967539, EPI_ISL_1967579, EPI_ISL_2102332, EPI_ISL_2102479 | | | |
| see above | Maryland Genomics, Institute for Genome Sciences, University of Maryland School of Medicine | Maryland Genomics, Institute for Genome Sciences, University of Maryland School of Medicine | Aditya; Claire M; Fraser; Holly; Humphrys; Jacques; Kranthi; Lisa D; Luke J; Mehta; Mike; Ott; Ravel; Roussey; Sadzewicz; Sandra; Tallon; Vavikolanu |
| EPI_ISL_1624489, EPI_ISL_1624505, EPI_ISL_1624687, EPI_ISL_1624688, EPI_ISL_1624689, EPI_ISL_1624697, EPI_ISL_1710599, EPI_ISL_1710600, EPI_ISL_1710601, EPI_ISL_1710605, EPI_ISL_1710609, EPI_ISL_1710611, EPI_ISL_2008133, EPI_ISL_2008186 | | | |
| see above | Massachusetts State Public Health Laboratory | Massachusetts State Public Health Laboratory | Andrew Lang; Glen Gallagher; Sandra Smole; Timelia Fink |
| EPI_ISL_1640115 | Medizinisch-Diagnostisches Labor Kempten allgÃ¤ulab | Robert Koch Institute | |
| EPI_ISL_1850411 | Medizinisches Labor Wahl LÃ¼denscheid | Robert Koch Institute | |
| EPI_ISL_2246490, EPI_ISL_2246537 | Michigan Department of Health and Human Services, Bureau of Laboratories | Michigan Department of Health and Human Services, Bureau of Laboratories | Blankenship HM; Riner D; Soehnlen MK |
| EPI_ISL_1761495, EPI_ISL_1761507, EPI_ISL_1761541 | Microbiologia CATLAB | Can Ruti SARS-CoV-2 Sequencing Hub (HUGTiP/IrsiCaixa/IGTP) | Alba Sánchez; Anna Not; Antoni E Bordoy; Bonaventura Clotet; Cristina Casañ; Cristina Esteban; Francesc Catala-Moll; Gemma Clara; Ignacio Blanco; Marc Noguera-Julian; Maria Casadellà; Mariona Parera; Mercedes Guerrero; Montserrat Giménez; Pere-Joan Cardona; Pilar Armengol; Roger Paredes; Verónica Saludes; and Elisa Martró on behalf of the Can Ruti SARS-CoV-2 Sequencing Hub. |
| EPI_ISL_2140535, EPI_ISL_2140538 | Microbiology Department, Laboratori Clínic Metropolitana Nord. Hospital Universitari Germans Trias i Pujol | Can Ruti SARS-CoV-2 Sequencing Hub (HUGTiP/IrsiCaixa/IGTP) | Alba Sánchez; Anna Not; Antoni E Bordoy; Bonaventura Clotet; Cristina Casañ; Cristina Esteban; Francesc Catala-Moll; Gemma Clara; Ignacio Blanco; Marc Noguera-Julian; Maria Casadellà; Mariona Parera; Mercedes Guerrero; Montserrat Giménez; Pere-Joan Cardona; Pilar Armengol; Roger Paredes; Verónica Saludes; and Elisa Martró on behalf of the Can Ruti SARS-CoV-2 Sequencing Hub. |
| EPI_ISL_1636453, EPI_ISL_1636478, EPI_ISL_2351308 | Microbiology Department, Laboratori Clínic Metropolitana Nord. Hospital Universitari Germans Trias i Pujol. | Can Ruti SARS-CoV-2 Sequencing Hub (HUGTiP/IrsiCaixa/IGTP) | Alba Sánchez; Anna Not; Antoni E Bordoy; Bonaventura Clotet; Cristina Casañ; Cristina Esteban; Francesc Catala-Moll; Gemma Clara; Ignacio Blanco; Marc Noguera-Julian; Maria Casadellà; Mariona Parera; Mercedes Guerrero; Montserrat Giménez; Pere-Joan Cardona; Pilar Armengol; Roger Paredes; Verónica Saludes; and Elisa Martró on behalf of the Can Ruti SARS-CoV-2 Sequencing Hub. |
| EPI_ISL_2086126, EPI_ISL_2086127, EPI_ISL_2086136, EPI_ISL_2086138, EPI_ISL_2086167, EPI_ISL_2516968, EPI_ISL_2516969, EPI_ISL_2516970, EPI_ISL_2516972, EPI_ISL_2516973 | | | |
| see above | Microbiology Department. Complexo Hospitalario Universitario de Vigo | Microbiology Department. Complexo Hospitalario Universitario de Vigo | Alfaya N; Alonso I; Alvarez M; Cabrera JJ; Carballo R; Cores O; Cortizo S; Martinez L; Mediero G; Perez S; Potel C; Regueiro B; Rey S; Vasallo FJ; del-Campo V |
| EPI_ISL_1820845, EPI_ISL_1820848, EPI_ISL_1820849 | Microvida | Microvida | Jaco Verweij; Joep Stöhr; Suzan D. Pas |
| EPI_ISL_1710932, EPI_ISL_1760681 | Minnesota Department of Health, Public Health Laboratory | Minnesota Department of Health, Public Health Laboratory | Alexandra Lorentz; Jacob Garfin; Matt Plumb; and Xiong Wang |
| EPI_ISL_2443060 | Mirassol | Instituto Butantan / FAMERP | Antonio Jorge Martins; Claudia Renata dos Santos Barros; David Schlesinger; Debora Botequio Moretti; Dimas Tadeu Covas; Elaine Cristina Marqueze; Elaine Vieira Santos; Evandra Strazza Rodrigues; Heidge Fukumasu; Jayme Augusto de Souza-Neto; José Salvatore Leister Patané; Luiz Alcantara; Luiz Lehmann Coutinho; Maria Carolina Elias; Maurício Lacerda Nogueira; Rafael dos Santos Bezerra; Raul Machado Neto; Rejane Maria Tommasini Grotto; Ricardo Haddad; Sandra Coccuzzo Sampaio Vessoni; Simone Kashima; Svetoslav Nanev Slavov; Vincent Louis Viala |
| EPI_ISL_2035553 | Missouri State Public Health Laboratory | Missouri State Public Health Laboratory | Ashley New; Joshua Barry; Matthew Sinn |
| EPI_ISL_3142932 | Molecular & Genomic Pathology Laboratory, Thomas Jefferson University Hospital | Molecular & Genomic Pathology Laboratory, Thomas Jefferson University Hospital | Nitika Badjatia; Run Jin; Shannon Mahoney; Zi-xuan Wang |
| EPI_ISL_2978742 | Molecular Pathology Laboratory, Univ. of Illinois at Chicago | Igenbio, Inc | Benjamin Vaisvil; Daniel P. Schmitt; Joshua Bland; Renee Taylor; Shrihari S. Kadkol; Vinayak Kapatral |
| EPI_ISL_1648100, EPI_ISL_1648104, EPI_ISL_1960018 | NJDOH, Public Health and Environmental Laboratories | NJ_PHEL | Byeong Jeong; Dana Woell; Jacquelyn Deverell; Lindsey Bodnar; Matthew Scarnati; Shiv K. Verma |
| EPI_ISL_1671641 | NORTHWELL HEALTH LABORATORIES | Wadsworth Center, New York State Department of Health | Alexis Russell; Catharine Prussing; Daryl M. Lamson; Erasmus Schneider; Erica Lasek-Nesselquist; John Kelly; Jonathan Plitnick; Kirsten St. George; Matthew Shudt; Melissa A Leisner; Navjot Singh |
| EPI_ISL_2162038 | NS-QEII Health Sciences Centre | National Microbiology Laboratory (NML) | Anna Majer; Anneliese Landgraff; CanCOGeN's metadata curation team; Dan Gaston; Darian Hole; Elsie Grudeski; Gary Van Domselaar; Grace Seo; Janice Pettipas; Jason LeBlanc; Jennifer Tanner; Kirsten Biggar; Madison Chapel; Morag Graham; Natalie Knox; Nathalie Bastien; Philip Mabon; Public Health Agency of Canada CanCOGeN team; Rhiannon Huzarewich; Russell Mandes; Shari Tyson; Timothy Booth; Todd Hatchette; Yan Li |
| EPI_ISL_1966567 | NUCLEO DE SAUDE VILA FALCAO DE BAURU | Instituto Butantan / Mendelics | Antonio Jorge Martins; Bianca Cechetto Carlos. Mendelics: Bibiana Santos; Claudia Renata dos Santos Barros; Cíntia Bittar; David Schlesinger. Hemocentro Ribeirão Preto: Simone Kashima; Debora Botequio Moretti; Elaine Cristina Marqueze; Elaine Vieira dos Santos; Elisangela Chicaroni Mattos; Erika Freitas; Evandra Strazza Rodrigues; Felipe Allan da Silva da Costa; Flavia Aburjaile; Fábio Sossai Possebon; Guilherme Campos; Guilherme Targino Valente; Heidge Fukumasu. USP-Botucatu: Rejane Maria Tommasini Grotto; Helena Lage Ferreira; Instituto Butantan: Dimas Tadeu Covas; Jardelina de Souza Todao Bernardino; Jayme A. Souza-Neto; Jessika Cristina Chagas Lesbon; Jorge A. Petroli Marchesi; José Salvatore Leister Patané; João Paulo Kitajima; João Pessoa Araújo Jr.; Leila Sabrina Ullmann; Loyze Paola Oliveira de Lima; Luiz Aurelio de Campos Crispin. Centro de Genômica Funcional da ESALQ: Luiz Lehmann Coutinho; Luiz Carlos Junior de Alcantara; Lívia Sacchetto; Maisa C. Pereira Parra; Maria Carolina Elias; Marta Giovanetti; Marília Moraes; Maurício Lacerda Nogueira. Prefeitura de Sao Paulo: Melissa Palmieri.; Patricia Akemi Assato; Paula Rahal; Paulo Inacio da Costa; Rafael dos Santos Bezerra; Raquel de Lello Rocha Campos Cassano. NGS Soluções Genômicas: Pilar Drummond Sampaio Corrêa Mariani. FZEA-USP Pirassununga: Mirele Daiana Poleti; Raul Machado Neto; Ricardo Augusto Brassaloti; Ricardo Haddad; Rodrigo Tocantins Calado. FAMERP-SJRP: Cecília Artico Banho; Sandra Coccuzzo Sampaio; Svetoslav Nanev Slavov; Vagner Fonseca; Vincent Louis Viala |
| EPI_ISL_1654617, EPI_ISL_1654627, EPI_ISL_1789913, EPI_ISL_1789918 | NYU Langone Health | Departments of Pathology and Medicine, New York University School of Medicine | Adriana Heguy; Christian Marier; Dacia Dimartino; Emily Guzman; Gael Westby; Guiqing Wang; Paolo Cotzia; Paul Zappile; Peter Meyn; Sitharam Ramaswami; Yutong Zhang |
| EPI_ISL_1623659, EPI_ISL_1665199, EPI_ISL_1665211, EPI_ISL_1665215, EPI_ISL_1673662 | National Platform bis UMONS/Jolimont | National Platform bis UMONS/Jolimont | François Dufrasne; Gautier Detry; Guillaume Bayon-Vicente; Ruddy Wattiez |
| EPI_ISL_1543939, EPI_ISL_1620144 | National Public Health Laboratory, National Centre for Infectious Diseases | National Public Health Laboratory, National Centre for Infectious Diseases | Grace Jie Yin Ngan; Lin Cui; Raymond Tzer Pin Lin; Royce Ang; Tze Minn Mak; Zhenyang Zhou |
| EPI_ISL_1623005, EPI_ISL_1623006 | National Virus Reference Laboratory | National Virus Reference Laboratory | Charlene Bennett; Cillian F De Gascun; Gabriel Gonzalez; Jonathan Dean; Michael Carr; Zoe Yandle |
| EPI_ISL_1621157, EPI_ISL_1626556, EPI_ISL_2229828, EPI_ISL_2229831 | Nebraska Public Health Laboratory | NPHL COVID-19 Response Team | NPHL COVID-19 Response Team |
| EPI_ISL_1920038 | Northwell Health Laboratories | Wadsworth Center, New York State Department of Health | Alexis Russell; Catharine Prussing; Daryl M. Lamson; Erasmus Schneider; Erica Lasek-Nesselquist; John Kelly; Jonathan Plitnick; Kirsten St. George; Matthew Shudt; Melissa A Leisner; Navjot Singh |
| EPI_ISL_1822801, EPI_ISL_1822826, EPI_ISL_1822870 | Northwest Laboratories | Seattle Flu Study | Amanda Adler; Barry R. Lutz; Benjamin Pelle; Brian Hiatt; Caitlin R. Wolf; Chris D. Frazar; Deborah A. Nickerson; Elisabeth Brandstetter; Erica Ryke; Geoff Melly; Helen Y. Chu; Janet A. Englund; Jay Shendure; Jover Lee; Kairsten Fay; Kirsten Lacombe; Lea M. Starita; Mark J. Rieder; Matthew Richardson; Matthew Thompson; Melissa Truong; Michael Boeckh; Michael Famulare; Misja Ilcisin; Peter D. Han; Philip Dykema; Romesh Gautom; Scott Lindquist; Thomas R. Sibley; Trevor Bedford |
| EPI_ISL_1704698, EPI_ISL_1704705, EPI_ISL_1704714 | Northwestern Memorial Hospital | Northwestern University - Ozer Lab | Chad J. Achenbach; Chao Qi; Egon A. Ozer; Judd F. Hultquist; Lacy M. Simons; Lawrence J. Jennings; Michael G. Ison; Ramon Lorenzo-Redondo; Taylor J. Dean |
| EPI_ISL_2090677, EPI_ISL_2090678, EPI_ISL_2090679, EPI_ISL_2090683 | Northwestern Memorial Hospital | RIPHL at Rush University Medical Center | Felix Araujo Perez; Kevin Kunstman; Laura Furtado; Marieta Hyde; Max Kolton; Stefan Green |

| | | | |
|---|---|---|---|
| EPI_ISL_2443063 | Nova Aliança | Instituto Butantan / FAMERP | Antonio Jorge Martins; Claudia Renata dos Santos Barros; David Schlesinger; Debora Botequio Moretti; Dimas Tadeu Covas; Elaine Cristina Marqueze; Elaine Vieira Santos; Evandra Strazza Rodrigues; Heidge Fukumasu; Jayme Augusto de Souza-Neto; José Salvatore Leister Patané; Luiz Alcantara; Luiz Lehmann Coutinho; Maria Carolina Elias; Maurício Lacerda Nogueira; Rafael dos Santos Bezerra; Raul Machado Neto; Rejane Maria Tommasini Grotto; Ricardo Haddad; Sandra Coccuzzo Sampaio Vessoni; Simone Kashima; Svetoslav Nanev Slavov; Vincent Louis Viala |
| EPI_ISL_5799927 | Nucleo De Saude Vila Falcao De Bauru | Instituto Butantan | Antonio Jorge Martins; Claudia Renata dos Santos Barros; David Schlesinger; Debora Botequio Moretti; Dimas Tadeu Covas; Elaine Cristina Marqueze; Elaine Vieira Santos; Evandra Strazza Rodrigues; Heidge Fukumasu; Jayme Augusto de Souza-Neto; José Salvatore Leister Patané; Luiz Alcantara; Luiz Lehmann Coutinho; Maria Carolina Elias; Maurício Lacerda Nogueira; Rafael dos Santos Bezerra; Raul Machado Neto; Rejane Maria Tommasini Grotto; Ricardo Haddad; Sandra Coccuzzo Sampaio Vessoni; Simone Kashima; Svetoslav Nanev Slavov; Vincent Louis Viala |
| EPI_ISL_2452209 | OH Department of Health Laboratory | Centers for Disease Control and Prevention Division of Viral Diseases, Pathogen Discovery | Alison Laufer Halpin; Ben L. Rambo-Martin; Clinton R. Paden; Dakota Howard; Darlene Wagner; Dave Wentworth; Dhwani Batra; Jasmine Padilla; Justin Lee; Katie Dillon; Krista Queen; Kristen Knipe; Kristine Lacek; Mark Burroughs; Matthew Schmerer; Mili Sheth; Peter Cook; Sam Shepard; Sarah Nobles; Shoshona Le; Suxiang Tong; Vivien Dugan; Yvette Unoarumhi |
| EPI_ISL_2227666, EPI_ISL_2227667, EPI_ISL_2227675 | OHSU Lab Services Molecular Microbiology Lab | Oregon SARS-CoV-2 Genome Sequencing Center | Alec J. Hirsch; Andrew C. Adey; Benjamin N. Bimber; Brendan L. O'Connell; Brian J. O'Roak; Cierra LaBlanc; Daniel N. Streblow; Donna Hansel; Guang Fan; Kayla Carter; Ruth V. Nichols; Sally Grindstaff; Sonia Acharya; William B. Messer; Xuan Qin |
| EPI_ISL_1711792 | OR State PHL-Virology/Immunology Section | Centers for Disease Control and Prevention Division of Viral Diseases, Pathogen Discovery | Alison Laufer Halpin; Ben L. Rambo-Martin; Clinton R. Paden; Dakota Howard; Darlene Wagner; Dave Wentworth; Dhwani Batra; Jasmine Padilla; Justin Lee; Katie Dillon; Krista Queen; Kristen Knipe; Kristine Lacek; Mark Burroughs; Matthew Schmerer; Mili Sheth; Peter Cook; Sam Shepard; Sarah Nobles; Shoshona Le; Suxiang Tong; Vivien Dugan; Yvette Unoarumhi |
| EPI_ISL_1968980 | Omics Sciences Laboratory | Omics Sciences Laboratory | Darlyn Amaya; Derly Andrade Molina; Gabriel Morey León; Juan Carlos Fernández Cadena; Katheryn Sacheri Viteri; Rubén Armas González |
| EPI_ISL_2321651 | One Health | Universidad Nacional de Colombia - Laboratorio Genómico One Health | Andres F. Cardona-Rios; Carlos Franco-Muñoz; Daniel O. Maldonado-Perez; Diego A. Álvarez-Díaz; Hector Alejandro Ruiz-Moreno; Jorge E. Osorio; Juan P. Hernandez-Ortiz; Karl A Ciuoderis; Katherine Laiton-Donato; Laura Silvana Perez; Lina M. Hurtado; Marcela Mercado-Reyes; Maria Stella López; Monica Palma-Cuero; Simón Villegas Velásquez |
| EPI_ISL_7275888, EPI_ISL_7275904, EPI_ISL_7276299, EPI_ISL_7276317, EPI_ISL_7276321, EPI_ISL_7276347 | Ontario's COVID-19 Genomics Rapid Response Coalition | McMaster University | Ahmed Draia; Allison McGeer; Andrew G. McArthur; Angel Li; Emily Panousis; Hooman Derakhshani; Jalees Nasir; Kuganya Nirmalarajah; Michael Surette; Patryk Aftanas; Samira Mubareka; Sheridan Baker |
| EPI_ISL_1623696 | Oregon State Public Health Laboratory | Oregon State Public Health Laboratory | Eugene Yeboah; John Fontana and Shane Sevey; Laura Tsaknaridis; Rafia Razzaque; Vanda Makris |
| EPI_ISL_1656459, EPI_ISL_1656462 | Ospedale di Genzano - ASL RM 6 | INMI Lazzaro Spallanzani IRCCS | A Di Caro; B Bartolini; CEM Gruber; E Conti; E Giombini; F Messina; F Santini; G Bonfiglio; G Tramini; M Rueca; MR Capobianchi; O Butera |
| EPI_ISL_1856984, EPI_ISL_1856985, EPI_ISL_1856993 | Outre mer | National Reference Center for Viruses of Respiratory Infections, Institut Pasteur, Paris | Angela Brisebarre; Camille Capel; Christophe Malabat; Corinne Maufrais; Dominique Rousset; Etienne Simon-Lorière; Frédéric Lemoine; Gael Millot; Louise Lefrançois; Marion Barbet; Maud Vanpeene; Méline Bizard; Pierre Lechat; Sylvie Behillil; Sylvie Van der Werf; Vincent Enouf |
| EPI_ISL_1967005 | PA DE IBITIUVA DR OTAVIO BENETTI PITANGUEIRAS | Instituto Butantan / ESALQ-USP (Piracicaba) | Antonio Jorge Martins; Bianca Cechetto Carlos. Mendelics: Bibiana Santos; Claudia Renata dos Santos Barros; Cíntia Bittar; David Schlesinger. Hemocentro Ribeirão Preto: Simone Kashima; Debora Botequio Moretti; Elaine Cristina Marqueze; Elaine Vieira dos Santos; Elisangela Chicaroni Mattos; Erika Freitas; Evandra Strazza Rodrigues; Felipe Allan da Silva da Costa; Flavia Aburjaile; Fábio Sossai Possebon; Guilherme Campos; Guilherme Targino Valente; Heidge Fukumasu. USP-Botucatu: Rejane Maria Tommasini Grotto; Helena Lage Ferreira; Instituto Butantan: Dimas Tadeu Covas; Jardelina de Souza Todao Bernardino; Jayme A. Souza-Neto; Jessika Cristina Chagas Lesbon; Jorge A. Petroli Marchesi; José Salvatore Leister Patané; João Paulo Kitajima; João Pessoa Araújo Jr.; Leila Sabrina Ullmann; Loyze Paola Oliveira de Lima; Luiz Aurelio de Campos Crispin. Centro de Genômica Funcional da ESALQ: Luiz Lehmann Coutinho; Luiz Carlos Junior de Alcantara; Lívia Sacchetto; Maisa C. Pereira Parra; Maria Carolina Elias; Marta Giovanetti; Marília Moraes; Maurício Lacerda Nogueira. Prefeitura de Sao Paulo: Melissa Palmieri.; Patricia Akemi Assato; Paula Rahal; Paulo Inacio da Costa; Rafael dos Santos Bezerra; Raquel de Lello Rocha Campos Cassano. NGS Soluções Genômicas: Pilar Drummond Sampaio Corrêa Mariani. FZEA-USP Pirassununga: Mirele Daiana Poleti; Raul Machado Neto; Ricardo Augusto Brassaloti; Ricardo Haddad; Rodrigo Tocantins Calado. FAMERP-SJRP: Cecília Artico Banho; Sandra Coccuzzo Sampaio; Svetoslav Nanev Slavov; Vagner Fonseca; Vincent Louis Viala |
| EPI_ISL_1798808 | PA Department of Health, Bureau of Laboratories | Centers for Disease Control and Prevention Division of Viral Diseases, Pathogen Discovery | Alison Laufer Halpin; Ben L. Rambo-Martin; Clinton R. Paden; Dakota Howard; Darlene Wagner; Dave Wentworth; Dhwani Batra; Jasmine Padilla; Justin Lee; Katie Dillon; Krista Queen; Kristen Knipe; Kristine Lacek; Mark Burroughs; Matthew Schmerer; Mili Sheth; Peter Cook; Sam Shepard; Sarah Nobles; Shoshona Le; Suxiang Tong; Vivien Dugan; Yvette Unoarumhi |
| EPI_ISL_2467902 | PA Department of Health, Bureau of Laboratories | Genomics and Discovery, Respiratory Viruses Branch, Division of Viral Diseases, Centers for Disease Control and Prevention | Adam Retchless; Anna Kelleher; Anna Uehara; Brian Lynch; Clinton R. Paden; Dhwani Batra; Haibin Wang; Han Jia Justin Ng; Jasmine Padilla; Jing Zhang; Justin Lee; Krista Queen; Mark Burroughs; Mili Sheth; Morgan Davis; Peter Cook; Rachel Marine; Sarah Nobles; Suxiang Tong; Tara Coalter; Yan Li; Ying Tao |
| EPI_ISL_2345965, EPI_ISL_2345966 | PA NOVO OSASCO | Instituto Butantan | Antonio Jorge Martins; Claudia Renata dos Santos Barros; David Schlesinger; Debora Botequio Moretti; Dimas Tadeu Covas; Elaine Cristina Marqueze; Elaine Vieira Santos; Evandra Strazza Rodrigues; Heidge Fukumasu; Jayme Augusto de Souza-Neto; José Salvatore Leister Patané; Luiz Alcantara; Luiz Lehmann Coutinho; Maria Carolina Elias; Maurício Lacerda Nogueira; Rafael dos Santos Bezerra; Raul Machado Neto; Rejane Maria Tommasini Grotto; Ricardo Haddad; Sandra Coccuzzo Sampaio Vessoni; Simone Kashima; Svetoslav Nanev Slavov; Vincent Louis Viala |
| EPI_ISL_1960076 | PECET | Universidad Nacional de Colombia - Laboratorio Genómico One Health | Andres F. Cardona-Rios; Carlos Franco-Muñoz; Carolina Muñoz-Arango; Celeny Ortiz; Daniel O. Maldonado-Perez; Diego A. Álvarez-Díaz; Hector Alejandro Ruiz-Moreno; Idabely Betancur Ortiz; Jorge E. Osorio; Juan P. Hernandez-Ortiz; Karl A Ciuoderis; Katherine Laiton-Donato; Laura Silvana Perez; Lina M. Hurtado; Marcela Mercado-Reyes; Maria Angélica Maya; Maria Stella López; Rita Almanza Payares; Sandra Ines Cano; Simón Villegas Velásquez |
| EPI_ISL_1966916 | PENITENCIARIA ODON RAMOS MARANHAO IPERO | Instituto Butantan / ESALQ-USP (Piracicaba) | Antonio Jorge Martins; Bianca Cechetto Carlos. Mendelics: Bibiana Santos; Claudia Renata dos Santos Barros; Cíntia Bittar; David Schlesinger. Hemocentro Ribeirão Preto: Simone Kashima; Debora Botequio Moretti; Elaine Cristina Marqueze; Elaine Vieira dos Santos; Elisangela Chicaroni Mattos; Erika Freitas; Evandra Strazza Rodrigues; Felipe Allan da Silva da Costa; Flavia Aburjaile; Fábio Sossai Possebon; Guilherme Campos; Guilherme Targino Valente; Heidge Fukumasu. USP-Botucatu: Rejane Maria Tommasini Grotto; Helena Lage Ferreira; Instituto Butantan: Dimas Tadeu Covas; Jardelina de Souza Todao Bernardino; Jayme A. Souza-Neto; Jessika Cristina Chagas Lesbon; Jorge A. Petroli Marchesi; José Salvatore Leister Patané; João Paulo Kitajima; João Pessoa Araújo Jr.; Leila Sabrina Ullmann; Loyze Paola Oliveira de Lima; Luiz Aurelio de Campos Crispin. Centro de Genômica Funcional da ESALQ: Luiz Lehmann Coutinho; Luiz Carlos Junior de Alcantara; Lívia Sacchetto; Maisa C. Pereira Parra; Maria Carolina Elias; Marta Giovanetti; Marília Moraes; Maurício Lacerda Nogueira. Prefeitura de Sao Paulo: Melissa Palmieri.; Patricia Akemi Assato; Paula Rahal; Paulo Inacio da Costa; Rafael dos Santos Bezerra; Raquel de Lello Rocha Campos Cassano. NGS Soluções Genômicas: Pilar Drummond Sampaio Corrêa Mariani. FZEA-USP Pirassununga: Mirele Daiana Poleti; Raul Machado Neto; Ricardo Augusto Brassaloti; Ricardo Haddad; Rodrigo Tocantins Calado. FAMERP-SJRP: Cecília Artico Banho; Sandra Coccuzzo Sampaio; Svetoslav Nanev Slavov; Vagner Fonseca; Vincent Louis Viala |
| EPI_ISL_1966927, EPI_ISL_1966928, EPI_ISL_1966929, EPI_ISL_1966930, EPI_ISL_1966931, EPI_ISL_1966932, EPI_ISL_1966933, EPI_ISL_1966934 | | | |
| see above | POLICLINICA COVID 19 ITAPETININGA | Instituto Butantan / ESALQ-USP (Piracicaba) | Antonio Jorge Martins; Bianca Cechetto Carlos. Mendelics: Bibiana Santos; Claudia Renata dos Santos Barros; Cíntia Bittar; David Schlesinger. Hemocentro Ribeirão Preto: Simone Kashima; Debora Botequio Moretti; Elaine Cristina Marqueze; Elaine Vieira dos Santos; Elisangela Chicaroni Mattos; Erika Freitas; Evandra Strazza Rodrigues; Felipe Allan da Silva da Costa; Flavia Aburjaile; Fábio Sossai Possebon; Guilherme Campos; Guilherme Targino Valente; Heidge Fukumasu. USP-Botucatu: Rejane Maria Tommasini Grotto; Helena Lage Ferreira; Instituto Butantan: Dimas Tadeu Covas; Jardelina de Souza Todao Bernardino; Jayme A. Souza-Neto; Jessika Cristina Chagas Lesbon; Jorge A. Petroli Marchesi; José Salvatore Leister Patané; João Paulo Kitajima; João Pessoa Araújo Jr.; Leila Sabrina Ullmann; Loyze Paola Oliveira de Lima; Luiz Aurelio de Campos Crispin. Centro de Genômica Funcional da ESALQ: Luiz Lehmann Coutinho; Luiz Carlos Junior de Alcantara; Lívia Sacchetto; Maisa C. Pereira Parra; Maria Carolina Elias; Marta Giovanetti; Marília Moraes; Maurício Lacerda Nogueira. Prefeitura de Sao Paulo: Melissa Palmieri.; Patricia Akemi Assato; Paula Rahal; Paulo Inacio da Costa; Rafael dos Santos Bezerra; Raquel de Lello Rocha Campos Cassano. NGS Soluções Genômicas: Pilar Drummond Sampaio Corrêa Mariani. FZEA-USP Pirassununga: Mirele Daiana Poleti; Raul Machado Neto; Ricardo Augusto Brassaloti; Ricardo Haddad; Rodrigo Tocantins Calado. FAMERP-SJRP: Cecília Artico Banho; Sandra Coccuzzo Sampaio; Svetoslav Nanev Slavov; Vagner Fonseca; Vincent Louis Viala |
| EPI_ISL_2345978, EPI_ISL_2345980 | POLICLINICA JARDIM PARAISO | Instituto Butantan | Antonio Jorge Martins; Claudia Renata dos Santos Barros; David Schlesinger; Debora Botequio Moretti; Dimas Tadeu Covas; Elaine Cristina Marqueze; Elaine Vieira Santos; Evandra Strazza Rodrigues; Heidge Fukumasu; Jayme Augusto de Souza-Neto; José Salvatore Leister Patané; Luiz Alcantara; Luiz Lehmann Coutinho; Maria Carolina Elias; Maurício Lacerda Nogueira; Rafael dos Santos Bezerra; Raul Machado Neto; Rejane Maria Tommasini Grotto; Ricardo Haddad; Sandra Coccuzzo Sampaio Vessoni; Simone Kashima; Svetoslav Nanev Slavov; Vincent Louis Viala |
| EPI_ISL_2080492 | POOLE MICROBIOLOGY LABORATORY | COVID-19 Genomics UK (COG-UK) Consortium | PHE Covid Sequencing Team |
| EPI_ISL_5530190 | POSTO DE SAUDE DE LAGOINHA III | Analytical Competence Molecular Epidemiology Lab/ACME, Oswaldo Cruz Foundation, Ceara (FIOCRUZ CE) | Carlos Leonardo de Aragao Araujo; Cecilia Leite Costa & Eduardo Ruback dos Santos on behalf of COVID-19 FIOCRUZ Genomic Network; Cleber Furtado Aksenen; Fabio Miyajima; Fernando Braga Stehling; Francisco Eder de Moura Lopes; Igor Oliveira Duarte; Jamille Maria Mendes Bezerra; Joaquim Cesar do Nascimento Sousa Junior; Pedro Miguel Carneiro Jeronimo; Suzana Porto Almeida; Thais Ferreira de Oliveira; Thais de Oliveira Costa; Ticiane Cavalcante de Souza; Veridiana Pessoa Miyajima |
| EPI_ISL_3102533 | POSTO DE SAUDE MARIA DE LOURDES MAGALHAES MAIA | Analytical Competence Molecular Epidemiology Lab/ACME, Oswaldo Cruz Foundation, Ceara (FIOCRUZ CE) | Cleber Furtado Aksenen; Fabio Miyajima; Fernando Braga Stehling; Francisco Eder de Moura Lopes; Jamille Maria Mendes Bezerra; Joaquim César do Nascimento Sousa Junior; Pedro Miguel Carneiro Jeronimo; Suzana Porto Almeida e Lucas Delerino; Thais Ferreira de Oliveira; Thais de Oliveira Costa; Ticiane Cavalcante de Souza; Veridiana Pessoa Miyajima |
| EPI_ISL_1966910, EPI_ISL_1966911 | PR S DA FAMILIA UNIDADE DE SAUDE ADALBERTO ROCHA GUAREI | Instituto Butantan / ESALQ-USP (Piracicaba) | Antonio Jorge Martins; Bianca Cechetto Carlos. Mendelics: Bibiana Santos; Claudia Renata dos Santos Barros; Cíntia Bittar; David Schlesinger. Hemocentro Ribeirão Preto: Simone Kashima; Debora Botequio Moretti; Elaine Cristina Marqueze; Elaine Vieira dos Santos; Elisangela Chicaroni Mattos; Erika Freitas; Evandra Strazza Rodrigues; Felipe Allan da Silva da Costa; Flavia Aburjaile; Fábio Sossai Possebon; Guilherme Campos; Guilherme Targino Valente; Heidge Fukumasu. USP-Botucatu: Rejane Maria Tommasini Grotto; Helena Lage Ferreira; Instituto Butantan: Dimas Tadeu Covas; Jardelina de Souza Todao Bernardino; Jayme A. Souza-Neto; Jessika Cristina Chagas Lesbon; Jorge A. Petroli Marchesi; José Salvatore Leister Patané; João Paulo Kitajima; João Pessoa Araújo Jr.; Leila Sabrina Ullmann; Loyze Paola Oliveira de Lima; Luiz Aurelio de Campos Crispin. Centro de Genômica Funcional da ESALQ: Luiz Lehmann Coutinho; Luiz Carlos Junior de Alcantara; Lívia Sacchetto; Maisa C. Pereira Parra; Maria Carolina Elias; Marta Giovanetti; Marília Moraes; Maurício Lacerda Nogueira. Prefeitura de Sao Paulo: Melissa Palmieri.; Patricia Akemi Assato; Paula Rahal; Paulo Inacio da Costa; Rafael dos Santos Bezerra; Raquel de Lello Rocha Campos Cassano. NGS Soluções Genômicas: Pilar Drummond Sampaio Corrêa Mariani. FZEA-USP Pirassununga: Mirele Daiana Poleti; Raul Machado Neto; Ricardo Augusto Brassaloti; Ricardo Haddad; Rodrigo Tocantins Calado. FAMERP-SJRP: Cecília Artico Banho; Sandra Coccuzzo Sampaio; Svetoslav Nanev Slavov; Vagner Fonseca; Vincent Louis Viala |
| EPI_ISL_3275683, EPI_ISL_3275690 | PRIME DIAGNOSTICS | Universidad Nacional de Colombia - Laboratorio Genómico One Health | Andres F. Cardona-Rios; Carlos Franco-Muñoz; Carolina Muñoz-Arango; Celeny Ortiz; Daniel O. Maldonado-Perez; Diego A. Álvarez-Díaz; Hector Alejandro Ruiz-Moreno; Idabely Betancur Ortiz; Jorge E. Osorio; Juan P. Hernandez-Ortiz; Karl A Ciuoderis; Katherine Laiton-Donato; Laura Silvana Perez; Lina M. Hurtado; Marcela Mercado-Reyes; Maria Angélica Maya; Maria Stella López; Rita Almanza Payares; Sandra Ines Cano; Simón Villegas Velásquez |
| EPI_ISL_2345991, EPI_ISL_2345992, EPI_ISL_2346013, EPI_ISL_2346014 | PRONTO ATENDIMENTO DE VARGEM GRANDE PAULISTA | Instituto Butantan | Antonio Jorge Martins; Claudia Renata dos Santos Barros; David Schlesinger; Debora Botequio Moretti; Dimas Tadeu Covas; Elaine Cristina Marqueze; Elaine Vieira Santos; Evandra Strazza Rodrigues; Heidge Fukumasu; Jayme Augusto de Souza-Neto; José Salvatore Leister Patané; Luiz Alcantara; Luiz Lehmann Coutinho; Maria Carolina Elias; Maurício Lacerda Nogueira; Rafael dos Santos Bezerra; Raul Machado Neto; Rejane Maria Tommasini Grotto; Ricardo Haddad; Sandra Coccuzzo Sampaio Vessoni; Simone Kashima; Svetoslav Nanev Slavov; Vincent Louis Viala |
| EPI_ISL_2345962, EPI_ISL_2345963 | PRONTO ATENDIMENTO DR SERGIO AROUCA CAMPO GRANDE | Instituto Butantan | Antonio Jorge Martins; Claudia Renata dos Santos Barros; David Schlesinger; Debora Botequio Moretti; Dimas Tadeu Covas; Elaine Cristina Marqueze; Elaine Vieira Santos; Evandra Strazza Rodrigues; Heidge Fukumasu; Jayme Augusto de Souza-Neto; José Salvatore Leister Patané; Luiz Alcantara; Luiz Lehmann Coutinho; Maria Carolina Elias; Maurício Lacerda Nogueira; Rafael dos Santos Bezerra; Raul Machado Neto; Rejane Maria Tommasini Grotto; Ricardo Haddad; Sandra Coccuzzo Sampaio Vessoni; Simone Kashima; Svetoslav Nanev Slavov; Vincent Louis Viala |
| EPI_ISL_1966823 | PRONTO ATENDIMENTO MUNICIPAL ALUMINIO | Instituto Butantan / ESALQ-USP (Piracicaba) | Antonio Jorge Martins; Bianca Cechetto Carlos. Mendelics: Bibiana Santos; Claudia Renata dos Santos Barros; Cíntia Bittar; David Schlesinger. Hemocentro Ribeirão Preto: Simone Kashima; Debora Botequio Moretti; Elaine Cristina Marqueze; Elaine Vieira dos Santos; Elisangela Chicaroni Mattos; Erika Freitas; Evandra Strazza Rodrigues; Felipe Allan da Silva da Costa; Flavia Aburjaile; Fábio Sossai Possebon; Guilherme Campos; Guilherme Targino Valente; Heidge Fukumasu. USP-Botucatu: Rejane Maria Tommasini Grotto; Helena Lage Ferreira; Instituto Butantan: Dimas Tadeu Covas; Jardelina de Souza Todao Bernardino; Jayme A. Souza-Neto; Jessika Cristina Chagas Lesbon; Jorge A. Petroli Marchesi; José Salvatore Leister Patané; João Paulo Kitajima; João Pessoa Araújo Jr.; Leila Sabrina Ullmann; Loyze Paola Oliveira de Lima; Luiz Aurelio de Campos Crispin. Centro de Genômica Funcional da ESALQ: Luiz Lehmann Coutinho; Luiz Carlos Junior de Alcantara; Lívia Sacchetto; Maisa C. Pereira Parra; Maria Carolina Elias; Marta Giovanetti; Marília Moraes; Maurício Lacerda Nogueira. Prefeitura de Sao Paulo: Melissa Palmieri.; Patricia Akemi Assato; Paula Rahal; Paulo Inacio da Costa; Rafael dos Santos Bezerra; Raquel de Lello Rocha Campos Cassano. NGS Soluções Genômicas: Pilar Drummond Sampaio Corrêa Mariani. FZEA-USP Pirassununga: Mirele Daiana Poleti; Raul Machado Neto; Ricardo Augusto Brassaloti; Ricardo Haddad; Rodrigo Tocantins Calado. FAMERP-SJRP: Cecília Artico Banho; Sandra Coccuzzo Sampaio; Svetoslav Nanev Slavov; Vagner Fonseca; Vincent Louis Viala |
| EPI_ISL_1966952, EPI_ISL_1966953 | PRONTO ATENDIMENTO MUNICIPAL DE | Instituto Butantan / ESALQ-USP (Piracicaba) | Antonio Jorge Martins; Bianca Cechetto Carlos. Mendelics: Bibiana Santos; Claudia Renata dos Santos Barros; Cíntia Bittar; David Schlesinger. Hemocentro Ribeirão Preto: Simone Kashima; Debora Botequio Moretti; Elaine Cristina Marqueze; Elaine Vieira dos Santos; Elisangela Chicaroni Mattos; Erika Freitas; Evandra Strazza Rodrigues; Felipe Allan da Silva da Costa; Flavia Aburjaile; Fábio Sossai Possebon; Guilherme Campos; Guilherme Targino Valente; Heidge Fukumasu. USP-Botucatu: Rejane Maria Tommasini Grotto; Helena Lage Ferreira; Instituto Butantan: Dimas Tadeu Covas; Jardelina de Souza Todao Bernardino; Jayme A. |

| | | | |
|---|---|---|---|
| | JACUPIRANGA | | Souza-Neto; Jessika Cristina Chagas Lesbon; Jorge A. Petroli Marchesi; José Salvatore Leister Patané; João Paulo Kitajima; João Pessoa Araújo Jr.; Leila Sabrina Ullmann; Loyze Paola Oliveira de Lima; Luiz Aurelio de Campos Crispin. Centro de Genômica Funcional da ESALQ: Luiz Lehmann Coutinho; Luiz Carlos Junior de Alcantara; Lívia Sacchetto; Maisa C. Pereira Parra; Maria Carolina Elias; Marta Giovanetti; Marília Moraes; Maurício Lacerda Nogueira. Prefeitura de Sao Paulo: Melissa Palmieri.; Patricia Akemi Assato; Paula Rahal; Paulo Inacio da Costa; Rafael dos Santos Bezerra; Raquel de Lello Rocha Campos Cassano. NGS Soluções Genômicas: Pilar Drummond Sampaio Corrêa Mariani. FZEA-USP Pirassununga: Mirele Daiana Poleti; Raul Machado Neto; Ricardo Augusto Brassaloti; Ricardo Haddad; Rodrigo Tocantins Calado. FAMERP-SJRP: Cecília Artico Banho; Sandra Coccuzzo Sampaio; Svetoslav Nanev Slavov; Vagner Fonseca; Vincent Louis Viala |
| EPI_ISL_1966845 | PRONTO ATENDIMENTO MUNICIPAL ITALO SANTUCCI | Instituto Butantan / ESALQ-USP (Piracicaba) | Antonio Jorge Martins; Bianca Cechetto Carlos. Mendelics: Bibiana Santos; Claudia Renata dos Santos Barros; Cíntia Bittar; David Schlesinger. Hemocentro Ribeirão Preto: Simone Kashima; Debora Botequio Moretti; Elaine Cristina Marqueze; Elaine Vieira dos Santos; Elisangela Chicaroni Mattos; Erika Freitas; Evandra Strazza Rodrigues; Felipe Allan da Silva da Costa; Flavia Aburjaile; Fábio Sossai Possebon; Guilherme Campos; Guilherme Targino Valente; Heidge Fukumasu. USP-Botucatu: Rejane Maria Tommasini Grotto; Helena Lage Ferreira; Instituto Butantan: Dimas Tadeu Covas; Jardelina de Souza Todao Bernardino; Jayme A. Souza-Neto; Jessika Cristina Chagas Lesbon; Jorge A. Petroli Marchesi; José Salvatore Leister Patané; João Paulo Kitajima; João Pessoa Araújo Jr.; Leila Sabrina Ullmann; Loyze Paola Oliveira de Lima; Luiz Aurelio de Campos Crispin. Centro de Genômica Funcional da ESALQ: Luiz Lehmann Coutinho; Luiz Carlos Junior de Alcantara; Lívia Sacchetto; Maisa C. Pereira Parra; Maria Carolina Elias; Marta Giovanetti; Marília Moraes; Maurício Lacerda Nogueira. Prefeitura de Sao Paulo: Melissa Palmieri.; Patricia Akemi Assato; Paula Rahal; Paulo Inacio da Costa; Rafael dos Santos Bezerra; Raquel de Lello Rocha Campos Cassano. NGS Soluções Genômicas: Pilar Drummond Sampaio Corrêa Mariani. FZEA-USP Pirassununga: Mirele Daiana Poleti; Raul Machado Neto; Ricardo Augusto Brassaloti; Ricardo Haddad; Rodrigo Tocantins Calado. FAMERP-SJRP: Cecília Artico Banho; Sandra Coccuzzo Sampaio; Svetoslav Nanev Slavov; Vagner Fonseca; Vincent Louis Viala |
| EPI_ISL_1966876 | PRONTO SOCORRO CAUCAIA DO ALTO | Instituto Butantan / ESALQ-USP (Piracicaba) | Antonio Jorge Martins; Bianca Cechetto Carlos. Mendelics: Bibiana Santos; Claudia Renata dos Santos Barros; Cíntia Bittar; David Schlesinger. Hemocentro Ribeirão Preto: Simone Kashima; Debora Botequio Moretti; Elaine Cristina Marqueze; Elaine Vieira dos Santos; Elisangela Chicaroni Mattos; Erika Freitas; Evandra Strazza Rodrigues; Felipe Allan da Silva da Costa; Flavia Aburjaile; Fábio Sossai Possebon; Guilherme Campos; Guilherme Targino Valente; Heidge Fukumasu. USP-Botucatu: Rejane Maria Tommasini Grotto; Helena Lage Ferreira; Instituto Butantan: Dimas Tadeu Covas; Jardelina de Souza Todao Bernardino; Jayme A. Souza-Neto; Jessika Cristina Chagas Lesbon; Jorge A. Petroli Marchesi; José Salvatore Leister Patané; João Paulo Kitajima; João Pessoa Araújo Jr.; Leila Sabrina Ullmann; Loyze Paola Oliveira de Lima; Luiz Aurelio de Campos Crispin. Centro de Genômica Funcional da ESALQ: Luiz Lehmann Coutinho; Luiz Carlos Junior de Alcantara; Lívia Sacchetto; Maisa C. Pereira Parra; Maria Carolina Elias; Marta Giovanetti; Marília Moraes; Maurício Lacerda Nogueira. Prefeitura de Sao Paulo: Melissa Palmieri.; Patricia Akemi Assato; Paula Rahal; Paulo Inacio da Costa; Rafael dos Santos Bezerra; Raquel de Lello Rocha Campos Cassano. NGS Soluções Genômicas: Pilar Drummond Sampaio Corrêa Mariani. FZEA-USP Pirassununga: Mirele Daiana Poleti; Raul Machado Neto; Ricardo Augusto Brassaloti; Ricardo Haddad; Rodrigo Tocantins Calado. FAMERP-SJRP: Cecília Artico Banho; Sandra Coccuzzo Sampaio; Svetoslav Nanev Slavov; Vagner Fonseca; Vincent Louis Viala |
| EPI_ISL_2614568 | PS Jose Ibrahin | Instituto Adolfo Lutz, Interdiciplinary Procedures Center, Strategic Laboratory | Caio Vinicius Dias Lopes; Claudia Regina Gonçalves; Claudio Tavares Sacchi; Erica Valessa Ramos Gomes; Karoline Rodrigues Campos; Leonardo Jose Tadeu de Araujo |
| EPI_ISL_1967020, EPI_ISL_1967021, EPI_ISL_1967022, EPI_ISL_1967023, EPI_ISL_1967024 | PSF DR ANTONIO PIRES DE ALMEIDA PORTO FELIZ | Instituto Butantan / ESALQ-USP (Piracicaba) | Antonio Jorge Martins; Bianca Cechetto Carlos. Mendelics: Bibiana Santos; Claudia Renata dos Santos Barros; Cíntia Bittar; David Schlesinger. Hemocentro Ribeirão Preto: Simone Kashima; Debora Botequio Moretti; Elaine Cristina Marqueze; Elaine Vieira dos Santos; Elisangela Chicaroni Mattos; Erika Freitas; Evandra Strazza Rodrigues; Felipe Allan da Silva da Costa; Flavia Aburjaile; Fábio Sossai Possebon; Guilherme Campos; Guilherme Targino Valente; Heidge Fukumasu. USP-Botucatu: Rejane Maria Tommasini Grotto; Helena Lage Ferreira; Instituto Butantan: Dimas Tadeu Covas; Jardelina de Souza Todao Bernardino; Jayme A. Souza-Neto; Jessika Cristina Chagas Lesbon; Jorge A. Petroli Marchesi; José Salvatore Leister Patané; João Paulo Kitajima; João Pessoa Araújo Jr.; Leila Sabrina Ullmann; Loyze Paola Oliveira de Lima; Luiz Aurelio de Campos Crispin. Centro de Genômica Funcional da ESALQ: Luiz Lehmann Coutinho; Luiz Carlos Junior de Alcantara; Lívia Sacchetto; Maisa C. Pereira Parra; Maria Carolina Elias; Marta Giovanetti; Marília Moraes; Maurício Lacerda Nogueira. Prefeitura de Sao Paulo: Melissa Palmieri.; Patricia Akemi Assato; Paula Rahal; Paulo Inacio da Costa; Rafael dos Santos Bezerra; Raquel de Lello Rocha Campos Cassano. NGS Soluções Genômicas: Pilar Drummond Sampaio Corrêa Mariani. FZEA-USP Pirassununga: Mirele Daiana Poleti; Raul Machado Neto; Ricardo Augusto Brassaloti; Ricardo Haddad; Rodrigo Tocantins Calado. FAMERP-SJRP: Cecília Artico Banho; Sandra Coccuzzo Sampaio; Svetoslav Nanev Slavov; Vagner Fonseca; Vincent Louis Viala |
| EPI_ISL_1967033, EPI_ISL_1967034, EPI_ISL_1967035, EPI_ISL_1967036, EPI_ISL_1967037, EPI_ISL_1967038, EPI_ISL_1967039, EPI_ISL_1967040, EPI_ISL_1967041 | | | |
| see above | PSF MARIA JOSE SALTO DE PIRAPORA | Instituto Butantan / ESALQ-USP (Piracicaba) | Antonio Jorge Martins; Bianca Cechetto Carlos. Mendelics: Bibiana Santos; Claudia Renata dos Santos Barros; Cíntia Bittar; David Schlesinger. Hemocentro Ribeirão Preto: Simone Kashima; Debora Botequio Moretti; Elaine Cristina Marqueze; Elaine Vieira dos Santos; Elisangela Chicaroni Mattos; Erika Freitas; Evandra Strazza Rodrigues; Felipe Allan da Silva da Costa; Flavia Aburjaile; Fábio Sossai Possebon; Guilherme Campos; Guilherme Targino Valente; Heidge Fukumasu. USP-Botucatu: Rejane Maria Tommasini Grotto; Helena Lage Ferreira; Instituto Butantan: Dimas Tadeu Covas; Jardelina de Souza Todao Bernardino; Jayme A. Souza-Neto; Jessika Cristina Chagas Lesbon; Jorge A. Petroli Marchesi; José Salvatore Leister Patané; João Paulo Kitajima; João Pessoa Araújo Jr.; Leila Sabrina Ullmann; Loyze Paola Oliveira de Lima; Luiz Aurelio de Campos Crispin. Centro de Genômica Funcional da ESALQ: Luiz Lehmann Coutinho; Luiz Carlos Junior de Alcantara; Lívia Sacchetto; Maisa C. Pereira Parra; Maria Carolina Elias; Marta Giovanetti; Marília Moraes; Maurício Lacerda Nogueira. Prefeitura de Sao Paulo: Melissa Palmieri.; Patricia Akemi Assato; Paula Rahal; Paulo Inacio da Costa; Rafael dos Santos Bezerra; Raquel de Lello Rocha Campos Cassano. NGS Soluções Genômicas: Pilar Drummond Sampaio Corrêa Mariani. FZEA-USP Pirassununga: Mirele Daiana Poleti; Raul Machado Neto; Ricardo Augusto Brassaloti; Ricardo Haddad; Rodrigo Tocantins Calado. FAMERP-SJRP: Cecília Artico Banho; Sandra Coccuzzo Sampaio; Svetoslav Nanev Slavov; Vagner Fonseca; Vincent Louis Viala |
| EPI_ISL_2345960, EPI_ISL_2346010, EPI_ISL_2346021, EPI_ISL_2346032, EPI_ISL_2346043 | PSF PARQUE EMILIA | Instituto Butantan | Antonio Jorge Martins; Claudia Renata dos Santos Barros; David Schlesinger; Debora Botequio Moretti; Dimas Tadeu Covas; Elaine Cristina Marqueze; Elaine Vieira Santos; Evandra Strazza Rodrigues; Heidge Fukumasu; Jayme Augusto de Souza-Neto; José Salvatore Leister Patané; Luiz Alcantara; Luiz Lehmann Coutinho; Maria Carolina Elias; Maurício Lacerda Nogueira; Rafael dos Santos Bezerra; Raul Machado Neto; Rejane Maria Tommasini Grotto; Ricardo Haddad; Sandra Coccuzzo Sampaio Vessoni; Simone Kashima; Svetoslav Nanev Slavov; Vincent Louis Viala |
| EPI_ISL_5800162 | Pa De Ibitiuva Dr Otavio Benetti Pitangueiras | Instituto Butantan | Antonio Jorge Martins; Claudia Renata dos Santos Barros; David Schlesinger; Debora Botequio Moretti; Dimas Tadeu Covas; Elaine Cristina Marqueze; Elaine Vieira Santos; Evandra Strazza Rodrigues; Heidge Fukumasu; Jayme Augusto de Souza-Neto; José Salvatore Leister Patané; Luiz Alcantara; Luiz Lehmann Coutinho; Maria Carolina Elias; Maurício Lacerda Nogueira; Rafael dos Santos Bezerra; Raul Machado Neto; Rejane Maria Tommasini Grotto; Ricardo Haddad; Sandra Coccuzzo Sampaio Vessoni; Simone Kashima; Svetoslav Nanev Slavov; Vincent Louis Viala |
| EPI_ISL_5802298, EPI_ISL_5802299 | Pa Novo Osasco | Instituto Butantan | Antonio Jorge Martins; Claudia Renata dos Santos Barros; David Schlesinger; Debora Botequio Moretti; Dimas Tadeu Covas; Elaine Cristina Marqueze; Elaine Vieira Santos; Evandra Strazza Rodrigues; Heidge Fukumasu; Jayme Augusto de Souza-Neto; José Salvatore Leister Patané; Luiz Alcantara; Luiz Lehmann Coutinho; Maria Carolina Elias; Maurício Lacerda Nogueira; Rafael dos Santos Bezerra; Raul Machado Neto; Rejane Maria Tommasini Grotto; Ricardo Haddad; Sandra Coccuzzo Sampaio Vessoni; Simone Kashima; Svetoslav Nanev Slavov; Vincent Louis Viala |
| EPI_ISL_1542380, EPI_ISL_1542458, EPI_ISL_1542647, EPI_ISL_1542899, EPI_ISL_1543123, EPI_ISL_1543185, EPI_ISL_1543195, EPI_ISL_1543218, EPI_ISL_1543223, EPI_ISL_1543243, EPI_ISL_1543331, EPI_ISL_1543353, EPI_ISL_1543366, EPI_ISL_1543367, EPI_ISL_1543416, EPI_ISL_1543423, EPI_ISL_1543462, EPI_ISL_1543480, EPI_ISL_1543533, EPI_ISL_1543579, EPI_ISL_1543615, EPI_ISL_1543618, EPI_ISL_1543808, EPI_ISL_1543832, EPI_ISL_1543858, EPI_ISL_1543885, EPI_ISL_1543907, EPI_ISL_1635529, EPI_ISL_1635576, EPI_ISL_1635585, EPI_ISL_1635603, EPI_ISL_1635632, EPI_ISL_1635643, EPI_ISL_1635713, EPI_ISL_1635732, EPI_ISL_1635858, EPI_ISL_1635884, EPI_ISL_1635893, EPI_ISL_1635902, EPI_ISL_1636022, EPI_ISL_1636113, EPI_ISL_1636121, EPI_ISL_1636161, EPI_ISL_1636172, EPI_ISL_1636218, EPI_ISL_1636223 | | | |
| see above | Pandemic Response Lab - NYC | Pandemic Response Lab, R&D | Cybill del Castillo; Dylan Law; Haiping Hao; Henry Lee; Jon Laurent; Katharine Nelson; Melissa Hopkins; Michael Hammerling; Pradeep Bugga; Shinyoung Clair Kang; Sol Rey; William Ward |
| EPI_ISL_5800230 | Penitenciaria Odon Ramos Maranhao Ipero | Instituto Butantan | Antonio Jorge Martins; Claudia Renata dos Santos Barros; David Schlesinger; Debora Botequio Moretti; Dimas Tadeu Covas; Elaine Cristina Marqueze; Elaine Vieira Santos; Evandra Strazza Rodrigues; Heidge Fukumasu; Jayme Augusto de Souza-Neto; José Salvatore Leister Patané; Luiz Alcantara; Luiz Lehmann Coutinho; Maria Carolina Elias; Maurício Lacerda Nogueira; Rafael dos Santos Bezerra; Raul Machado Neto; Rejane Maria Tommasini Grotto; Ricardo Haddad; Sandra Coccuzzo Sampaio Vessoni; Simone Kashima; Svetoslav Nanev Slavov; Vincent Louis Viala |
| EPI_ISL_5521537 | Pennsylvania Department of Health Bureau of Laboratories | Pennsylvania Department of Health Bureau of Laboratories | Dongxiang Xia |
| EPI_ISL_7025335, EPI_ISL_7040614 | Pesaro | Microbiology University Politecnica delle Marche | Anna Valenza; Carla Acciarri; Katia Marinelli; Monica Lucia Ferreri; Patrizia Bagnarelli; Roberta Longo; Sara Caucci; Stefano Menzo |
| EPI_ISL_6907347, EPI_ISL_6907352, EPI_ISL_6907360, EPI_ISL_6907838, EPI_ISL_6907839, EPI_ISL_6908387, EPI_ISL_6908395, EPI_ISL_6908396, EPI_ISL_6908404, EPI_ISL_6908714, EPI_ISL_6908717, EPI_ISL_6908718, EPI_ISL_6908726, EPI_ISL_6908736, EPI_ISL_6908740, EPI_ISL_6908741, EPI_ISL_6908746 | | | |
| see above | Plataforma de Vigilancia Molecular (PVM) - FIOCRUZ/BA | Plataforma de Vigilancia Molecular (PVM) - FIOCRUZ/BA | Bruno Bezerril Andrade; Camila I. de Oliveira; Clarissa Araújo Gurgel; Eduardo Oyama; Francys A Rangel; Laila A da Silva; Laise de Moraes; Leonardo Paiva Farias; Marina Cucco; Pedro Silveira; Ricardo Khouri on behalf of the Fiocruz COVID-19 Genomic Surveillance Network.; Tiago Graf |
| EPI_ISL_1788097, EPI_ISL_1788110, EPI_ISL_1788120 | Plateforme de testing Namuroise | Plateforme de testing Namuroise | Céline Maschietto; Degosserie Jonathan; Denis Olivier; Mullier François; Otto Gaetan |
| EPI_ISL_1591150, EPI_ISL_1591151, EPI_ISL_1591156, EPI_ISL_1591168 | Platform BIS UZA/UAntwerpen | Labo Klinische Biologie, UZA | Basil Britto Xavier; Christine Lammens; Herman Goossens; Jasmine Coppens; Marie Le Mercier; Veerle Matheeussen |
| EPI_ISL_1558663 | Platform BIS UZA/UAntwerpen | UAntwerp, Laboratory of Medical Microbiology | Basil Britto Xavier; Christine Lammens; Herman Goossens; Jasmine Coppens; Marie Le Mercier; Veerle Matheeussen |
| EPI_ISL_5800263, EPI_ISL_5800267, EPI_ISL_5800276, EPI_ISL_5800285, EPI_ISL_5800287, EPI_ISL_5800289, EPI_ISL_5800291, EPI_ISL_5800298 | | | |
| see above | Policlinica Covid 19 Itapetininga | Instituto Butantan | Antonio Jorge Martins; Claudia Renata dos Santos Barros; David Schlesinger; Debora Botequio Moretti; Dimas Tadeu Covas; Elaine Cristina Marqueze; Elaine Vieira Santos; Evandra Strazza Rodrigues; Heidge Fukumasu; Jayme Augusto de Souza-Neto; José Salvatore Leister Patané; Luiz Alcantara; Luiz Lehmann Coutinho; Maria Carolina Elias; Maurício Lacerda Nogueira; Rafael dos Santos Bezerra; Raul Machado Neto; Rejane Maria Tommasini Grotto; Ricardo Haddad; Sandra Coccuzzo Sampaio Vessoni; Simone Kashima; Svetoslav Nanev Slavov; Vincent Louis Viala |
| EPI_ISL_5802310, EPI_ISL_5802311 | Policlinica Jardim Paraiso | Instituto Butantan | Antonio Jorge Martins; Claudia Renata dos Santos Barros; David Schlesinger; Debora Botequio Moretti; Dimas Tadeu Covas; Elaine Cristina Marqueze; Elaine Vieira Santos; Evandra Strazza Rodrigues; Heidge Fukumasu; Jayme Augusto de Souza-Neto; José Salvatore Leister Patané; Luiz Alcantara; Luiz Lehmann Coutinho; Maria Carolina Elias; Maurício Lacerda Nogueira; Rafael dos Santos Bezerra; Raul Machado Neto; Rejane Maria Tommasini Grotto; Ricardo Haddad; Sandra Coccuzzo Sampaio Vessoni; Simone Kashima; Svetoslav Nanev Slavov; Vincent Louis Viala |
| EPI_ISL_5800317, EPI_ISL_5800325 | Pr S Da Familia Unidade De Saude Adalberto Rocha Guarei | Instituto Butantan | Antonio Jorge Martins; Claudia Renata dos Santos Barros; David Schlesinger; Debora Botequio Moretti; Dimas Tadeu Covas; Elaine Cristina Marqueze; Elaine Vieira Santos; Evandra Strazza Rodrigues; Heidge Fukumasu; Jayme Augusto de Souza-Neto; José Salvatore Leister Patané; Luiz Alcantara; Luiz Lehmann Coutinho; Maria Carolina Elias; Maurício Lacerda Nogueira; Rafael dos Santos Bezerra; Raul Machado Neto; Rejane Maria Tommasini Grotto; Ricardo Haddad; Sandra Coccuzzo Sampaio Vessoni; Simone Kashima; Svetoslav Nanev Slavov; Vincent Louis Viala |
| EPI_ISL_5802324, EPI_ISL_5802325, EPI_ISL_5802344, EPI_ISL_5802345 | Pronto Atendimento De Vargem Grande Paulista | Instituto Butantan | Antonio Jorge Martins; Claudia Renata dos Santos Barros; David Schlesinger; Debora Botequio Moretti; Dimas Tadeu Covas; Elaine Cristina Marqueze; Elaine Vieira Santos; Evandra Strazza Rodrigues; Heidge Fukumasu; Jayme Augusto de Souza-Neto; José Salvatore Leister Patané; Luiz Alcantara; Luiz Lehmann Coutinho; Maria Carolina Elias; Maurício Lacerda Nogueira; Rafael dos Santos Bezerra; Raul Machado Neto; Rejane Maria Tommasini Grotto; Ricardo Haddad; Sandra Coccuzzo Sampaio Vessoni; Simone Kashima; Svetoslav Nanev Slavov; Vincent Louis Viala |
| EPI_ISL_5802295, EPI_ISL_5802296 | Pronto Atendimento Dr Sergio Arouca Campo Grande | Instituto Butantan | Antonio Jorge Martins; Claudia Renata dos Santos Barros; David Schlesinger; Debora Botequio Moretti; Dimas Tadeu Covas; Elaine Cristina Marqueze; Elaine Vieira Santos; Evandra Strazza Rodrigues; Heidge Fukumasu; Jayme Augusto de Souza-Neto; José Salvatore Leister Patané; Luiz Alcantara; Luiz Lehmann Coutinho; Maria Carolina Elias; Maurício Lacerda Nogueira; Rafael dos Santos Bezerra; Raul Machado Neto; Rejane Maria Tommasini Grotto; Ricardo Haddad; Sandra Coccuzzo Sampaio Vessoni; Simone Kashima; Svetoslav Nanev Slavov; Vincent Louis Viala |
| EPI_ISL_5800355 | Pronto Atendimento Municipal Aluminio | Instituto Butantan | Antonio Jorge Martins; Claudia Renata dos Santos Barros; David Schlesinger; Debora Botequio Moretti; Dimas Tadeu Covas; Elaine Cristina Marqueze; Elaine Vieira Santos; Evandra Strazza Rodrigues; Heidge Fukumasu; Jayme Augusto de Souza-Neto; José Salvatore Leister Patané; Luiz Alcantara; Luiz Lehmann Coutinho; Maria Carolina Elias; Maurício Lacerda Nogueira; Rafael dos Santos Bezerra; Raul Machado Neto; Rejane Maria Tommasini Grotto; Ricardo Haddad; Sandra Coccuzzo Sampaio Vessoni; Simone Kashima; Svetoslav Nanev Slavov; Vincent Louis Viala |
| EPI_ISL_5799884, EPI_ISL_5800171 | Pronto Atendimento Municipal De Jacupiranga | Instituto Butantan | Antonio Jorge Martins; Claudia Renata dos Santos Barros; David Schlesinger; Debora Botequio Moretti; Dimas Tadeu Covas; Elaine Cristina Marqueze; Elaine Vieira Santos; Evandra Strazza Rodrigues; Heidge Fukumasu; Jayme Augusto de Souza-Neto; José Salvatore Leister Patané; Luiz Alcantara; Luiz Lehmann Coutinho; Maria Carolina Elias; Maurício Lacerda Nogueira; Rafael dos Santos Bezerra; Raul Machado Neto; Rejane Maria Tommasini Grotto; Ricardo Haddad; Sandra Coccuzzo Sampaio Vessoni; Simone Kashima; Svetoslav Nanev Slavov; Vincent Louis Viala |
| EPI_ISL_5800254 | Pronto Atendimento Municipal Italo Santucci | Instituto Butantan | Antonio Jorge Martins; Claudia Renata dos Santos Barros; David Schlesinger; Debora Botequio Moretti; Dimas Tadeu Covas; Elaine Cristina Marqueze; Elaine Vieira Santos; Evandra Strazza Rodrigues; Heidge Fukumasu; Jayme Augusto de Souza-Neto; José Salvatore Leister Patané; Luiz Alcantara; Luiz Lehmann Coutinho; Maria Carolina Elias; Maurício Lacerda Nogueira; Rafael dos Santos Bezerra; Raul Machado Neto; Rejane Maria Tommasini Grotto; Ricardo Haddad; Sandra Coccuzzo Sampaio Vessoni; Simone Kashima; Svetoslav Nanev Slavov; Vincent Louis Viala |
| EPI_ISL_5799850 | Pronto Socorro Caucaia Do Alto | Instituto Butantan | Antonio Jorge Martins; Claudia Renata dos Santos Barros; David Schlesinger; Debora Botequio Moretti; Dimas Tadeu Covas; Elaine Cristina Marqueze; Elaine Vieira Santos; Evandra Strazza Rodrigues; Heidge Fukumasu; Jayme Augusto de Souza-Neto; José Salvatore Leister Patané; Luiz Alcantara; Luiz Lehmann Coutinho; Maria Carolina Elias; Maurício Lacerda Nogueira; Rafael dos Santos Bezerra; Raul Machado Neto; Rejane Maria Tommasini Grotto; Ricardo Haddad; Sandra Coccuzzo Sampaio Vessoni; Simone Kashima; Svetoslav Nanev Slavov; Vincent Louis Viala |
| EPI_ISL_2614571 | Pronto-Socorro Municipal Dr. Lauro Ribas Braga | Instituto Adolfo Lutz, Interdiciplinary Procedures Center, Strategic Laboratory | Caio Vinicius Dias Lopes; Claudia Regina Gonçalves; Claudio Tavares Sacchi; Erica Valessa Ramos Gomes; Karoline Rodrigues Campos; Leonardo Jose Tadeu de Araujo |
| EPI_ISL_2551285 | Providence Regional Medical Center Everett | Providence St. Joseph Health Molecular Genomics Laboratory | Alexa K Dowdell; Brian D Piening; Carlo B Bifulco; Fred L Robinson; Mary Campbell; Rogan Rattray |
| EPI_ISL_5800234, | Psf Dr Antonio Pires De | Instituto Butantan | Antonio Jorge Martins; Claudia Renata dos Santos Barros; David Schlesinger; Debora Botequio Moretti; Dimas Tadeu Covas; Elaine Cristina Marqueze; Elaine Vieira Santos; Evandra Strazza Rodrigues; Heidge Fukumasu; Jayme Augusto de Souza-Neto; José Salvatore Leister Patané; Luiz Alcantara; Luiz Lehmann Coutinho; |

| | | | |
|---|---|---|---|
| EPI_ISL_5800235, EPI_ISL_5800236, EPI_ISL_5800237, EPI_ISL_5800238 | Almeida Porto Feliz | | Maria Carolina Elias; Maurício Lacerda Nogueira; Rafael dos Santos Bezerra; Raul Machado Neto; Rejane Maria Tommasini Grotto; Ricardo Haddad; Sandra Coccuzzo Sampaio Vessoni; Simone Kashima; Svetoslav Nanev Slavov; Vincent Louis Viala |
| EPI_ISL_5800203, EPI_ISL_5800204, EPI_ISL_5800205, EPI_ISL_5800209, EPI_ISL_5800215, EPI_ISL_5800217, EPI_ISL_5800240, EPI_ISL_5800244, EPI_ISL_5800245 | | | |
| see above | Psf Maria Jose Salto De Pirapora | Instituto Butantan | Antonio Jorge Martins; Claudia Renata dos Santos Barros; David Schlesinger; Debora Botequio Moretti; Dimas Tadeu Covas; Elaine Cristina Marqueze; Elaine Vieira Santos; Evandra Strazza Rodrigues; Heidge Fukumasu; Jayme Augusto de Souza-Neto; José Salvatore Leister Patané; Luiz Alcantara; Luiz Lehmann Coutinho; Maria Carolina Elias; Maurício Lacerda Nogueira; Rafael dos Santos Bezerra; Raul Machado Neto; Rejane Maria Tommasini Grotto; Ricardo Haddad; Sandra Coccuzzo Sampaio Vessoni; Simone Kashima; Svetoslav Nanev Slavov; Vincent Louis Viala |
| EPI_ISL_5802289, EPI_ISL_5802290, EPI_ISL_5802291, EPI_ISL_5802292, EPI_ISL_5802293 | Psf Parque Emilia | Instituto Butantan | Antonio Jorge Martins; Claudia Renata dos Santos Barros; David Schlesinger; Debora Botequio Moretti; Dimas Tadeu Covas; Elaine Cristina Marqueze; Elaine Vieira Santos; Evandra Strazza Rodrigues; Heidge Fukumasu; Jayme Augusto de Souza-Neto; José Salvatore Leister Patané; Luiz Alcantara; Luiz Lehmann Coutinho; Maria Carolina Elias; Maurício Lacerda Nogueira; Rafael dos Santos Bezerra; Raul Machado Neto; Rejane Maria Tommasini Grotto; Ricardo Haddad; Sandra Coccuzzo Sampaio Vessoni; Simone Kashima; Svetoslav Nanev Slavov; Vincent Louis Viala |
| EPI_ISL_2688283 | Puerto Rico Department of Health | Centers for Disease Control and Prevention, Dengue Branch | Betzabel Flores; Gabriela Paz-Bailey; Gilberto A. Santiago; Glenda Gonzalez; Jorge L. Munoz-Jordan; Keyla Charriez |
| EPI_ISL_1695619, EPI_ISL_1695625, EPI_ISL_1695628, EPI_ISL_1695631, EPI_ISL_1695640, EPI_ISL_1695643, EPI_ISL_1695646, EPI_ISL_1695650, EPI_ISL_1695653, EPI_ISL_1695656, EPI_ISL_1695659, EPI_ISL_1695665, EPI_ISL_1695668, EPI_ISL_1695671, EPI_ISL_1695674, EPI_ISL_1695677, EPI_ISL_1695684, EPI_ISL_1695687, EPI_ISL_1695693, EPI_ISL_1695696, EPI_ISL_1695702, EPI_ISL_1695714, EPI_ISL_1695717, EPI_ISL_1695721, EPI_ISL_1695724, EPI_ISL_1695727, EPI_ISL_1695729, EPI_ISL_1711970, EPI_ISL_1711971, EPI_ISL_1711972, EPI_ISL_1711973, EPI_ISL_1753613, EPI_ISL_1753614, EPI_ISL_1753615, EPI_ISL_1753616, EPI_ISL_1753617, EPI_ISL_1753618, EPI_ISL_1753620, EPI_ISL_1753621, EPI_ISL_1753622, EPI_ISL_1753623, EPI_ISL_1753624, EPI_ISL_1753626, EPI_ISL_1753627, EPI_ISL_1753628, EPI_ISL_1753629, EPI_ISL_1753630, EPI_ISL_1753632, EPI_ISL_1753633, EPI_ISL_1753634, EPI_ISL_1753636, EPI_ISL_1753637, EPI_ISL_1753638, EPI_ISL_1753639, EPI_ISL_1753640, EPI_ISL_1753641, EPI_ISL_1753642, EPI_ISL_1753643, EPI_ISL_1753644, EPI_ISL_1753645, EPI_ISL_1753647, EPI_ISL_1753648, EPI_ISL_1753649, EPI_ISL_1753650, EPI_ISL_1753651, EPI_ISL_1753652, EPI_ISL_1753653, EPI_ISL_1753654, EPI_ISL_1753655, EPI_ISL_1753656, EPI_ISL_1753657, EPI_ISL_1753658, EPI_ISL_1753659, EPI_ISL_1753660, EPI_ISL_1753661, EPI_ISL_1761876, EPI_ISL_1761878, EPI_ISL_1761879, EPI_ISL_1761881, EPI_ISL_1761882, EPI_ISL_1761883, EPI_ISL_1761884, EPI_ISL_1761885, EPI_ISL_1761886, EPI_ISL_1761887, EPI_ISL_1761889, EPI_ISL_1761890, EPI_ISL_1761891, EPI_ISL_1761892, EPI_ISL_1761893, EPI_ISL_1761894, EPI_ISL_1761895, EPI_ISL_1761896, EPI_ISL_1761897, EPI_ISL_1804813, EPI_ISL_1804814, EPI_ISL_1804818, EPI_ISL_1804820, EPI_ISL_1818064, EPI_ISL_1818066, EPI_ISL_1818067, EPI_ISL_1818068, EPI_ISL_1818071, EPI_ISL_1900077, EPI_ISL_1900084, EPI_ISL_1900086, EPI_ISL_1900097, EPI_ISL_1900121, EPI_ISL_1900132, EPI_ISL_1900149, EPI_ISL_1900458, EPI_ISL_1935560, EPI_ISL_1935566, EPI_ISL_1935569, EPI_ISL_1935574, EPI_ISL_1935578, EPI_ISL_1935581, EPI_ISL_1935583, EPI_ISL_4372937 | | | |
| see above | Quest Diagnostics Incorporated | Centers for Disease Control and Prevention Division of Viral Diseases, Pathogen Discovery | A. Gerasimova; A. Perez; Adrian Paskey; B. Anderson; Benjamin Rambo-Martin; Christopher Gulvick; Clinton Paden; Clinton R. Paden; Dakota Howard; Darlene Wagner; Dhwani Batra; Duncan MacCannell; Erisa Sula; F. Lacbawan; I. A. Shlyakhter; I. Shlyakhter; Jason Caravas; K. Livingston; K.E. Livingston; Kara Moser; Kristine Lacek; L. Bernstein; L.E. Bernstein; M. Hua; Matthew Schmerer; P. Tanpaiboon; Peter Cook; Peter W. Cook; R. Kagan; R. M. Kagan; R. Owen; R. Rolando; R. V. Rolando; S. H. Rosenthal; S. Rosenthal; Scott Sammons; Shatavia Morrison; Tymeckia Kendall; Victoria Caban Figueroa; Y. Liu; Yvette Unoarumhi |
| EPI_ISL_1631841, EPI_ISL_1631888, EPI_ISL_1632206, EPI_ISL_1700431 | Randox Laboratories | Wellcome Sanger Institute for the COVID-19 Genomics UK (COG-UK) Consortium | Cordelia Langford; David K. Jackson; Dominic Kwiatkowski; Ewan Harrison; Ian Johnston; Jeffrey Barrett; John Sillitoe on behalf of the Wellcome Sanger Institute COVID-19 Surveillance Team; Randox Laboratories and Alex Alderton; Roberto Amato; Sonia Goncalves |
| EPI_ISL_2249818, EPI_ISL_2249918, EPI_ISL_2249919, EPI_ISL_2249920, EPI_ISL_2249923, EPI_ISL_2249924, EPI_ISL_2249925, EPI_ISL_2249926, EPI_ISL_2249927, EPI_ISL_2249928, EPI_ISL_2249929, EPI_ISL_2249930, EPI_ISL_2249931, EPI_ISL_2249932, EPI_ISL_2249933, EPI_ISL_2249934, EPI_ISL_2249935, EPI_ISL_2249936, EPI_ISL_2249937, EPI_ISL_2249938, EPI_ISL_2249939, EPI_ISL_2249940, EPI_ISL_2249941, EPI_ISL_2249942, EPI_ISL_2249943, EPI_ISL_2249944, EPI_ISL_2249945, EPI_ISL_2249946, EPI_ISL_2249947, EPI_ISL_2249948, EPI_ISL_2249949, EPI_ISL_2249950, EPI_ISL_2249951, EPI_ISL_2249952, EPI_ISL_2249953, EPI_ISL_2249954, EPI_ISL_2249955, EPI_ISL_2249956, EPI_ISL_2249958, EPI_ISL_2249959, EPI_ISL_2249960, EPI_ISL_2249961, EPI_ISL_2249962, EPI_ISL_2249963, EPI_ISL_2249965, EPI_ISL_2249966, EPI_ISL_2249967, EPI_ISL_2249968, EPI_ISL_2249969, EPI_ISL_2249970, EPI_ISL_2249971, EPI_ISL_2249972, EPI_ISL_2249973, EPI_ISL_2249974, EPI_ISL_2249975, EPI_ISL_2249976, EPI_ISL_2249977, EPI_ISL_2249978, EPI_ISL_2249979, EPI_ISL_2249980, EPI_ISL_2249981, EPI_ISL_2249982, EPI_ISL_2319629, EPI_ISL_2319631, EPI_ISL_2319632, EPI_ISL_2319633, EPI_ISL_2319634, EPI_ISL_2319635, EPI_ISL_2319636, EPI_ISL_2319637, EPI_ISL_2319640, EPI_ISL_2319646, EPI_ISL_2319647, EPI_ISL_2319648, EPI_ISL_2319651, EPI_ISL_2319654, EPI_ISL_2319657, EPI_ISL_2319658, EPI_ISL_2319660, EPI_ISL_2319662, EPI_ISL_2319663, EPI_ISL_2319664, EPI_ISL_2319666, EPI_ISL_2319670, EPI_ISL_2319671, EPI_ISL_2319674, EPI_ISL_2319676, EPI_ISL_2319678, EPI_ISL_2319679, EPI_ISL_2319682, EPI_ISL_2319683, EPI_ISL_2319684, EPI_ISL_2319685, EPI_ISL_2319686, EPI_ISL_2319687, EPI_ISL_2319691, EPI_ISL_2319693, EPI_ISL_2319694, EPI_ISL_2319696, EPI_ISL_2319698, EPI_ISL_2319701, EPI_ISL_2319702, EPI_ISL_2319704, EPI_ISL_2319705, EPI_ISL_2319706, EPI_ISL_2319707, EPI_ISL_2319709, EPI_ISL_2319715, EPI_ISL_2319723, EPI_ISL_2319738, EPI_ISL_2319752, EPI_ISL_2319753, EPI_ISL_2319755, EPI_ISL_2319756, EPI_ISL_2319757, EPI_ISL_2319758, EPI_ISL_2319759, EPI_ISL_2319760, EPI_ISL_2319761, EPI_ISL_2319763, EPI_ISL_2319765, EPI_ISL_2319766, EPI_ISL_2319767, EPI_ISL_2319769, EPI_ISL_2319770, EPI_ISL_2319771, EPI_ISL_2319772, EPI_ISL_2319774, EPI_ISL_2319775, EPI_ISL_2319776, EPI_ISL_2319777, EPI_ISL_2319778, EPI_ISL_2319786, EPI_ISL_2319800, EPI_ISL_2319801, EPI_ISL_2319804, EPI_ISL_2319805, EPI_ISL_2319807, EPI_ISL_2319808, EPI_ISL_2319810, EPI_ISL_2319812, EPI_ISL_2319982, EPI_ISL_2473882, EPI_ISL_2473893, EPI_ISL_2473909, EPI_ISL_2474088 | | | |
| see above | Reditus Laboratories | Reditus Laboratories | Alexa Eichelberger; Cassy Philips; Joshua J. Geltz; M.S.; Ph.D.; Rex Dyer; Robert M. Sgambelluri |
| EPI_ISL_7156306, EPI_ISL_7156307, EPI_ISL_7156308, EPI_ISL_7156309 | Research Education in Disease Diagnosis and Intervention (REDDI) Lab, Clemson University | Research Education in Disease Diagnosis and Intervention (REDDI) Lab, Clemson University | Adib Shafi; Brian Krueger; Chloe Emerson; Christopher Parkinson; Christopher Saski; Congyue Peng; Delphine Dean; John Pruitt; Justin Napolitano; Kaitlyn Williams; Keegan Sell; Kylie King; Lax Iyer; Rachel Dango; Rachel Ham; Scott Parker; Stevin Wilson; Sujata Srikanth |
| EPI_ISL_2749811 | Respiratory Virus Unit, Microbiology Services Colindale, Public Health England | COVID-19 Genomics UK (COG-UK) Consortium | PHE Covid Sequencing Team |
| EPI_ISL_1919165, EPI_ISL_1919167, EPI_ISL_2133632, EPI_ISL_2133678 | Rhode Island Department of Health | Infectious Disease Program, Broad Institute of Harvard and MIT | Adams, G.; Azevedo, K.; B.L.; B.W.; Bauer, M.; Birren; Carter, A.; Chaluvadi, S.; D.J.; DeRuff, K.; Gladden-Young, A.; Huard, R.; J.E.; K.J.; King, E.; Lagerborg, K.; Lemieux; Loreth, C.; Miller, A.; Normandin, E.; P.C.; Park; Pearlman, L.; Reilly, S.; Rudy, M.; Sabeti; Siddle; Tomkins-Tinch, C.; and MacInnis |
| EPI_ISL_2603870, EPI_ISL_2603897 | Rhode Island State Health Laboratory | Rhode Island State Health Laboratory | Ewa King; Kristin Carpenter-Azevedo; Richard C. Huard |
| EPI_ISL_3982694, EPI_ISL_3982698, EPI_ISL_3982700, EPI_ISL_3982704, EPI_ISL_3982708, EPI_ISL_3982712, EPI_ISL_3982715 | | | |
| see above | Ribeirao Preto | Laboratorio de Biologia Molecular Hemocentro de Ribeirao Preto | Antonio Jorge Martins; Claudia Renata dos Santos Barros; David Schlesinger; Debora Botequio Moretti; Dimas Tadeu Covas; Elaine Cristina Marqueze; Elaine Vieira Santos; Evandra Strazza Rodrigues; Heidge Fukumasu; Jayme Augusto de Souza-Neto; José Salvatore Leister Patané; Luiz Alcantara; Luiz Lehmann Coutinho; Maria Carolina Elias; Maurício Lacerda Nogueira; Rafael dos Santos Bezerra; Raul Machado Neto; Rejane Maria Tommasini Grotto; Ricardo Haddad; Sandra Coccuzzo Sampaio Vessoni; Simone Kashima; Svetoslav Nanev Slavov; Vincent Louis Viala |
| EPI_ISL_3982686 | Ribeirao Preto SP | Laboratorio de Biologia Molecular Hemocentro de Ribeirao Preto SP | Antonio Jorge Martins; Claudia Renata dos Santos Barros; David Schlesinger; Debora Botequio Moretti; Dimas Tadeu Covas; Elaine Cristina Marqueze; Elaine Vieira Santos; Evandra Strazza Rodrigues; Heidge Fukumasu; Jayme Augusto de Souza-Neto; José Salvatore Leister Patané; Luiz Alcantara; Luiz Lehmann Coutinho; Maria Carolina Elias; Maurício Lacerda Nogueira; Rafael dos Santos Bezerra; Raul Machado Neto; Rejane Maria Tommasini Grotto; Ricardo Haddad; Sandra Coccuzzo Sampaio Vessoni; Simone Kashima; Svetoslav Nanev Slavov; Vincent Louis Viala |
| EPI_ISL_3982689 | Ribeirao Preto SP | Laboratorio de Biologia Molecular Hemocentro de Ribeirao Preto | Antonio Jorge Martins; Claudia Renata dos Santos Barros; David Schlesinger; Debora Botequio Moretti; Dimas Tadeu Covas; Elaine Cristina Marqueze; Elaine Vieira Santos; Evandra Strazza Rodrigues; Heidge Fukumasu; Jayme Augusto de Souza-Neto; José Salvatore Leister Patané; Luiz Alcantara; Luiz Lehmann Coutinho; Maria Carolina Elias; Maurício Lacerda Nogueira; Rafael dos Santos Bezerra; Raul Machado Neto; Rejane Maria Tommasini Grotto; Ricardo Haddad; Sandra Coccuzzo Sampaio Vessoni; Simone Kashima; Svetoslav Nanev Slavov; Vincent Louis Viala |
| EPI_ISL_2346001 | SANTA CASA DE GUARAREMA | Instituto Butantan | Antonio Jorge Martins; Claudia Renata dos Santos Barros; David Schlesinger; Debora Botequio Moretti; Dimas Tadeu Covas; Elaine Cristina Marqueze; Elaine Vieira Santos; Evandra Strazza Rodrigues; Heidge Fukumasu; Jayme Augusto de Souza-Neto; José Salvatore Leister Patané; Luiz Alcantara; Luiz Lehmann Coutinho; Maria Carolina Elias; Maurício Lacerda Nogueira; Rafael dos Santos Bezerra; Raul Machado Neto; Rejane Maria Tommasini Grotto; Ricardo Haddad; Sandra Coccuzzo Sampaio Vessoni; Simone Kashima; Svetoslav Nanev Slavov; Vincent Louis Viala |
| EPI_ISL_1967192, EPI_ISL_1967193, EPI_ISL_1967194, EPI_ISL_1967195 | SANTA CASA DE MISERICORDIA DE TIETE | Instituto Butantan / ESALQ-USP (Piracicaba) | Antonio Jorge Martins; Bianca Cechetto Carlos. Mendelics: Bibiana Santos; Claudia Renata dos Santos Barros; Cíntia Bittar; David Schlesinger. Hemocentro Ribeirão Preto: Simone Kashima; Debora Botequio Moretti; Elaine Cristina Marqueze; Elaine Vieira dos Santos; Elisangela Chicaroni Mattos; Erika Freitas; Evandra Strazza Rodrigues; Felipe Allan da Silva da Costa; Flavia Aburjaile; Fábio Sossai Possebon; Guilherme Campos; Guilherme Targino Valente; Heidge Fukumasu. USP-Botucatu: Rejane Maria Tommasini Grotto; Helena Lage Ferreira; Instituto Butantan: Dimas Tadeu Covas; Jardelina de Souza Todao Bernardino; Jayme A. Souza-Neto; Jessika Cristina Chagas Lesbon; Jorge A. Petroli Marchesi; José Salvatore Leister Patané; João Paulo Kitajima; João Pessoa Araújo Jr.; Leila Sabrina Ullmann; Loyze Paola Oliveira de Lima; Luiz Aurelio de Campos Crispin. Centro de Genômica Funcional da ESALQ: Luiz Lehmann Coutinho; Luiz Carlos Junior de Alcantara; Lívia Sacchetto; Maisa C. Pereira Parra; Maria Carolina Elias; Marta Giovanetti; Marília Moraes; Maurício Lacerda Nogueira. Prefeitura de Sao Paulo: Melissa Palmieri.; Patricia Akemi Assato; Paula Rahal; Paulo Inacio da Costa; Rafael dos Santos Bezerra; Raquel de Lello Rocha Campos Cassano. NGS Soluções Genômicas: Pilar Drummond Sampaio Corrêa Mariani. FZEA-USP Pirassununga: Mirele Daiana Poleti; Raul Machado Neto; Ricardo Augusto Brassaloti; Ricardo Haddad; Rodrigo Tocantins Calado. FAMERP-SJRP: Cecília Artico Banho; Sandra Coccuzzo Sampaio; Svetoslav Nanev Slavov; Vagner Fonseca; Vincent Louis Viala |
| EPI_ISL_1967014, EPI_ISL_1967015, EPI_ISL_1967016, EPI_ISL_1967017, EPI_ISL_1967018, EPI_ISL_1967019 | SANTA CASA DE PORTO FELIZ | Instituto Butantan / ESALQ-USP (Piracicaba) | Antonio Jorge Martins; Bianca Cechetto Carlos. Mendelics: Bibiana Santos; Claudia Renata dos Santos Barros; Cíntia Bittar; David Schlesinger. Hemocentro Ribeirão Preto: Simone Kashima; Debora Botequio Moretti; Elaine Cristina Marqueze; Elaine Vieira dos Santos; Elisangela Chicaroni Mattos; Erika Freitas; Evandra Strazza Rodrigues; Felipe Allan da Silva da Costa; Flavia Aburjaile; Fábio Sossai Possebon; Guilherme Campos; Guilherme Targino Valente; Heidge Fukumasu. USP-Botucatu: Rejane Maria Tommasini Grotto; Helena Lage Ferreira; Instituto Butantan: Dimas Tadeu Covas; Jardelina de Souza Todao Bernardino; Jayme A. Souza-Neto; Jessika Cristina Chagas Lesbon; Jorge A. Petroli Marchesi; José Salvatore Leister Patané; João Paulo Kitajima; João Pessoa Araújo Jr.; Leila Sabrina Ullmann; Loyze Paola Oliveira de Lima; Luiz Aurelio de Campos Crispin. Centro de Genômica Funcional da ESALQ: Luiz Lehmann Coutinho; Luiz Carlos Junior de Alcantara; Lívia Sacchetto; Maisa C. Pereira Parra; Maria Carolina Elias; Marta Giovanetti; Marília Moraes; Maurício Lacerda Nogueira. Prefeitura de Sao Paulo: Melissa Palmieri.; Patricia Akemi Assato; Paula Rahal; Paulo Inacio da Costa; Rafael dos Santos Bezerra; Raquel de Lello Rocha Campos Cassano. NGS Soluções Genômicas: Pilar Drummond Sampaio Corrêa Mariani. FZEA-USP Pirassununga: Mirele Daiana Poleti; Raul Machado Neto; Ricardo Augusto Brassaloti; Ricardo Haddad; Rodrigo Tocantins Calado. FAMERP-SJRP: Cecília Artico Banho; Sandra Coccuzzo Sampaio; Svetoslav Nanev Slavov; Vagner Fonseca; Vincent Louis Viala |
| EPI_ISL_3102463 | SAO CARLOS DIAGNOSTICO POR IMAGEM | Analytical Competence Molecular Epidemiology Lab/ACME, Oswaldo Cruz Foundation, Ceara (FIOCRUZ CE) | Cleber Furtado Aksenen; Fabio Miyajima; Fernando Braga Stehling; Francisco Eder de Moura Lopes; Jamille Maria Mendes Bezerra; Joaquim César do Nascimento Sousa Junior; Pedro Miguel Carneiro Jeronimo; Suzana Porto Almeida e Lucas Delerino; Thais Ferreira de Oliveira; Thais de Oliveira Costa; Ticiane Cavalcante de Souza; Veridiana Pessoa Miyajima |
| EPI_ISL_2648769 | SARS-CoV-2 Sequencing Castilla y Leon-Spain Consortium | SARS-CoV-2 Sequencing Castilla y Leon-Spain Consortium | Antonio Orduña-Domingo; Carlos Fuster Foz; Carmen Aldea-Mansilla; Carmen Gimeno Crespo; David Abad; Gregoria Megías Lobón; Jose María Eiros Bouza; Laura Sánchez de Prada; M. Isabel Fernandez-Natal; Marta Dominguez-Gil; Marta Hernandez; María Antonia García Castro; Mª Fe Brezmes-Valdivieso; Noelia Arenal Andrés; Silvia Rojo; Sonsoles Garcinuño Pérez |
| EPI_ISL_2325346, EPI_ISL_2326595, EPI_ISL_2326596, EPI_ISL_2326597, EPI_ISL_2326602, EPI_ISL_2326610 | SARS-CoV-2 testing team, National Institute of Infectious Diseases | Pathogen Genomics Center, National Institute of Infectious Diseases | Hazuka Y Furihata; Kentaro Itokawa; Makoto Kuroda; Masanori Hashino; Masumichi Saito; Naomi Nojiri; Nozomu Hanaoka; Rina Tanaka; Sana Uchikoba; Tsuguto Fujimoto; Tsuyoshi Sekizuka |
| EPI_ISL_1711927 | SC Dept of Health and Env. Control-Bureau of Laboratories | Centers for Disease Control and Prevention Division of Viral Diseases, Pathogen Discovery | Alison Laufer Halpin; Ben L. Rambo-Martin; Clinton R. Paden; Dakota Howard; Darlene Wagner; Dave Wentworth; Dhwani Batra; Jasmine Padilla; Justin Lee; Katie Dillon; Krista Queen; Kristen Knipe; Kristine Lacek; Mark Burroughs; Matthew Schmerer; Mili Sheth; Peter Cook; Sam Shepard; Sarah Nobles; Shoshona Le; Suxiang Tong; Vivien Dugan; Yvette Unoarumhi |
| EPI_ISL_1966630, EPI_ISL_1966631 | SECAO CENTRO DE DIAGNOSTICO SECEDI | Instituto Butantan / Mendelics | Antonio Jorge Martins; Bianca Cechetto Carlos. Mendelics: Bibiana Santos; Claudia Renata dos Santos Barros; Cíntia Bittar; David Schlesinger. Hemocentro Ribeirão Preto: Simone Kashima; Debora Botequio Moretti; Elaine Cristina Marqueze; Elaine Vieira dos Santos; Elisangela Chicaroni Mattos; Erika Freitas; Evandra Strazza Rodrigues; Felipe Allan da Silva da Costa; Flavia Aburjaile; Fábio Sossai Possebon; Guilherme Campos; Guilherme Targino Valente; Heidge Fukumasu. USP-Botucatu: Rejane Maria Tommasini Grotto; Helena Lage Ferreira; Instituto Butantan: Dimas Tadeu Covas; Jardelina de Souza Todao Bernardino; Jayme A. Souza-Neto; Jessika Cristina Chagas Lesbon; Jorge A. Petroli Marchesi; José Salvatore Leister Patané; João Paulo Kitajima; João Pessoa Araújo Jr.; Leila Sabrina Ullmann; Loyze Paola Oliveira de Lima; Luiz Aurelio de Campos Crispin. Centro de Genômica Funcional da ESALQ: Luiz Lehmann Coutinho; Luiz Carlos Junior de Alcantara; Lívia Sacchetto; Maisa C. Pereira Parra; Maria Carolina Elias; Marta Giovanetti; Marília Moraes; Maurício Lacerda Nogueira. Prefeitura de Sao Paulo: Melissa Palmieri.; Patricia Akemi Assato; Paula Rahal; Paulo Inacio da Costa; Rafael dos Santos Bezerra; Raquel de Lello Rocha Campos Cassano. NGS Soluções Genômicas: Pilar Drummond Sampaio Corrêa Mariani. FZEA-USP Pirassununga: Mirele Daiana Poleti; Raul Machado Neto; Ricardo Augusto Brassaloti; Ricardo Haddad; Rodrigo Tocantins Calado. FAMERP-SJRP: Cecília Artico Banho; Sandra Coccuzzo Sampaio; Svetoslav Nanev Slavov; Vagner Fonseca; Vincent Louis Viala |
| EPI_ISL_2209505 | SECRETARIA DE SAUDE DE SANTA GERTRUDES | Instituto Butantan | Antonio Jorge Martins; Claudia Renata dos Santos Barros; David Schlesinger; Debora Botequio Moretti; Dimas Tadeu Covas; Elaine Cristina Marqueze; Elaine Vieira Santos; Evandra Strazza Rodrigues; Heidge Fukumasu; Jayme Augusto de Souza-Neto; José Salvatore Leister Patané; Luiz Alcantara; Luiz Lehmann Coutinho; Maria Carolina Elias; Maurício Lacerda Nogueira; Rafael dos Santos Bezerra; Raul Machado Neto; Rejane Maria Tommasini Grotto; Ricardo Haddad; Sandra Coccuzzo Sampaio Vessoni; Simone Kashima; Svetoslav Nanev Slavov; Vincent Louis Viala. |
| EPI_ISL_2209255, EPI_ISL_2344703 | SECRETARIA DE SAUDE DE SANTA GERTRUDES | Instituto Butantan / FZEA-USP-Pirassununga | Antonio Jorge Martins; Claudia Renata dos Santos Barros; David Schlesinger; Debora Botequio Moretti; Dimas Tadeu Covas; Elaine Cristina Marqueze; Elaine Vieira Santos; Evandra Strazza Rodrigues; Heidge Fukumasu; Jayme Augusto de Souza-Neto; José Salvatore Leister Patané; Luiz Alcantara; Luiz Lehmann Coutinho; Maria Carolina Elias; Maurício Lacerda Nogueira; Rafael dos Santos Bezerra; Raul Machado Neto; Rejane Maria Tommasini Grotto; Ricardo Haddad; Sandra Coccuzzo Sampaio Vessoni; Simone Kashima; Svetoslav Nanev Slavov; Vincent Louis Viala |
| EPI_ISL_2209259 | SECRETARIA DE SAUDE DE SAO PEDRO | Instituto Butantan / FZEA-USP-Pirassununga | Antonio Jorge Martins; Claudia Renata dos Santos Barros; David Schlesinger; Debora Botequio Moretti; Dimas Tadeu Covas; Elaine Cristina Marqueze; Elaine Vieira Santos; Evandra Strazza Rodrigues; Heidge Fukumasu; Jayme Augusto de Souza-Neto; José Salvatore Leister Patané; Luiz Alcantara; Luiz Lehmann Coutinho; Maria Carolina Elias; Maurício Lacerda Nogueira; Rafael dos Santos Bezerra; Raul Machado Neto; Rejane Maria Tommasini Grotto; Ricardo Haddad; Sandra Coccuzzo Sampaio Vessoni; Simone Kashima; Svetoslav Nanev Slavov; Vincent Louis Viala |

| | | | |
|---|---|---|---|
| EPI_ISL_5530113 | SECRETARIA DE SAUDE DO ESTADO DE PERNAMBUCO | Analytical Competence Molecular Epidemiology Lab/ACME, Oswaldo Cruz Foundation, Ceara (FIOCRUZ CE) | Carlos Leonardo de Aragao Araujo; Cecilia Leite Costa & Eduardo Ruback dos Santos on behalf of COVID-19 FIOCRUZ Genomic Network; Cleber Furtado Aksenen; Fabio Miyajima; Fernando Braga Stehling; Francisco Eder de Moura Lopes; Igor Oliveira Duarte; Jamille Maria Mendes Bezerra; Joaquim Cesar do Nascimento Sousa Junior; Pedro Miguel Carneiro Jeronimo; Suzana Porto Almeida; Thais Ferreira de Oliveira; Thaís de Oliveira Costa; Ticiane Cavalcante de Souza; Veridiana Pessoa Miyajima |
| EPI_ISL_1966950, EPI_ISL_1966951 | SECRETARIA MUNICIPAL DA SAUDE DE ITARARE | Instituto Butantan / ESALQ-USP (Piracicaba) | Antonio Jorge Martins; Bianca Cechetto Carlos. Mendelics: Bibiana Santos; Claudia Renata dos Santos Barros; Cíntia Bittar; David Schlesinger. Hemocentro Ribeirão Preto: Simone Kashima; Debora Botequio Moretti; Elaine Cristina Marqueze; Elaine Vieira dos Santos; Elisangela Chicaroni Mattos; Erika Freitas; Evandra Strazza Rodrigues; Felipe Allan da Silva da Costa; Flavia Aburjaile; Fábio Sossai Possebon; Guilherme Campos; Guilherme Targino Valente; Heidge Fukumasu. USP-Botucatu: Rejane Maria Tommasini Grotto; Helena Lage Ferreira; Instituto Butantan: Dimas Tadeu Covas; Jardelina de Souza Todao Bernardino; Jayme A. Souza-Neto; Jessika Cristina Chagas Lesbon; Jorge A. Petroli Marchesi; José Salvatore Leister Patané; João Paulo Kitajima; João Pessoa Araújo Jr.; Leila Sabrina Ullmann; Loyze Paola Oliveira de Lima; Luiz Aurelio de Campos Crispin. Centro de Genômica Funcional da ESALQ: Luiz Lehmann Coutinho; Luiz Carlos Junior de Alcantara; Lívia Sacchetto; Maisa C. Pereira Parra; Maria Carolina Elias; Marta Giovanetti; Marília Moraes; Maurício Lacerda Nogueira. Prefeitura de Sao Paulo: Melissa Palmieri.; Patricia Akemi Assato; Paula Rahal; Paulo Inacio da Costa; Rafael dos Santos Bezerra; Raquel de Lello Rocha Campos Cassano. NGS Soluções Genômicas: Pilar Drummond Sampaio Corrêa Mariani. FZEA-USP Pirassununga: Mirele Daiana Poleti; Raul Machado Neto; Ricardo Augusto Brassaloti; Ricardo Haddad; Rodrigo Tocantins Calado. FAMERP-SJRP: Cecília Artico Banho; Sandra Coccuzzo Sampaio; Svetoslav Nanev Slavov; Vagner Fonseca; Vincent Louis Viala |
| EPI_ISL_2209610 | SECRETARIA MUNICIPAL DE SAUDE DE CORDEIROPOLIS | Instituto Butantan | Antonio Jorge Martins; Claudia Renata dos Santos Barros; David Schlesinger; Debora Botequio Moretti; Dimas Tadeu Covas; Elaine Cristina Marqueze; Elaine Vieira Santos; Evandra Strazza Rodrigues; Heidge Fukumasu; Jayme Augusto de Souza-Neto; José Salvatore Leister Patané; Luiz Alcantara; Luiz Lehmann Coutinho; Maria Carolina Elias; Maurício Lacerda Nogueira; Rafael dos Santos Bezerra; Raul Machado Neto; Rejane Maria Tommasini Grotto; Ricardo Haddad; Sandra Coccuzzo Sampaio Vessoni; Simone Kashima; Svetoslav Nanev Slavov; Vincent Louis Viala. |
| EPI_ISL_2209137, EPI_ISL_2209145, EPI_ISL_2209271 | SECRETARIA MUNICIPAL DE SAUDE DE CORDEIROPOLIS | Instituto Butantan / FZEA-USP-Pirassununga | Antonio Jorge Martins; Claudia Renata dos Santos Barros; David Schlesinger; Debora Botequio Moretti; Dimas Tadeu Covas; Elaine Cristina Marqueze; Elaine Vieira Santos; Evandra Strazza Rodrigues; Heidge Fukumasu; Jayme Augusto de Souza-Neto; José Salvatore Leister Patané; Luiz Alcantara; Luiz Lehmann Coutinho; Maria Carolina Elias; Maurício Lacerda Nogueira; Rafael dos Santos Bezerra; Raul Machado Neto; Rejane Maria Tommasini Grotto; Ricardo Haddad; Sandra Coccuzzo Sampaio Vessoni; Simone Kashima; Svetoslav Nanev Slavov; Vincent Louis Viala |
| EPI_ISL_3102534, EPI_ISL_5530107, EPI_ISL_5530108, EPI_ISL_5530109 | SECRETARIA MUNICIPAL DE SAUDE DE TIANGUA | Analytical Competence Molecular Epidemiology Lab/ACME, Oswaldo Cruz Foundation, Ceara (FIOCRUZ CE) | Carlos Leonardo de Aragao Araujo; Cecilia Leite Costa & Eduardo Ruback dos Santos on behalf of COVID-19 FIOCRUZ Genomic Network; Cleber Furtado Aksenen; Fabio Miyajima; Fernando Braga Stehling; Francisco Eder de Moura Lopes; Igor Oliveira Duarte; Jamille Maria Mendes Bezerra; Joaquim Cesar do Nascimento Sousa Junior; Joaquim César do Nascimento Sousa Junior; Pedro Miguel Carneiro Jeronimo; Suzana Porto Almeida; Suzana Porto Almeida e Lucas Delerino; Thais Ferreira de Oliveira; Thaís de Oliveira Costa; Ticiane Cavalcante de Souza; Veridiana Pessoa Miyajima |
| EPI_ISL_1967098, EPI_ISL_1967099, EPI_ISL_1967100, EPI_ISL_1967101, EPI_ISL_1967102, EPI_ISL_1967103, EPI_ISL_1967104, EPI_ISL_1967105, EPI_ISL_1967106, EPI_ISL_1967107, EPI_ISL_1967108, EPI_ISL_1967109, EPI_ISL_1967110, EPI_ISL_1967111, EPI_ISL_1967112, EPI_ISL_1967113, EPI_ISL_1967114, EPI_ISL_1967115, EPI_ISL_1967116, EPI_ISL_1967117, EPI_ISL_1967118, EPI_ISL_1967119, EPI_ISL_1967120, EPI_ISL_1967121, EPI_ISL_1967122, EPI_ISL_1967123, EPI_ISL_1967124, EPI_ISL_1967125, EPI_ISL_1967126, EPI_ISL_1967127, EPI_ISL_1967128, EPI_ISL_1967129, EPI_ISL_1967130, EPI_ISL_1967131, EPI_ISL_1967132, EPI_ISL_1967133, EPI_ISL_1967134, EPI_ISL_1967135, EPI_ISL_1967136, EPI_ISL_1967137, EPI_ISL_1967138, EPI_ISL_1967139, EPI_ISL_1967140, EPI_ISL_1967141, EPI_ISL_1967142, EPI_ISL_1967143, EPI_ISL_1967144, EPI_ISL_1967145, EPI_ISL_1967146, EPI_ISL_1967147 | | | |
| see above | SECRETARIA MUNICIPAL DE SAUDE SOROCABA | Instituto Butantan / ESALQ-USP (Piracicaba) | Antonio Jorge Martins; Bianca Cechetto Carlos. Mendelics: Bibiana Santos; Claudia Renata dos Santos Barros; Cíntia Bittar; David Schlesinger. Hemocentro Ribeirão Preto: Simone Kashima; Debora Botequio Moretti; Elaine Cristina Marqueze; Elaine Vieira dos Santos; Elisangela Chicaroni Mattos; Erika Freitas; Evandra Strazza Rodrigues; Felipe Allan da Silva da Costa; Flavia Aburjaile; Fábio Sossai Possebon; Guilherme Campos; Guilherme Targino Valente; Heidge Fukumasu. USP-Botucatu: Rejane Maria Tommasini Grotto; Helena Lage Ferreira; Instituto Butantan: Dimas Tadeu Covas; Jardelina de Souza Todao Bernardino; Jayme A. Souza-Neto; Jessika Cristina Chagas Lesbon; Jorge A. Petroli Marchesi; José Salvatore Leister Patané; João Paulo Kitajima; João Pessoa Araújo Jr.; Leila Sabrina Ullmann; Loyze Paola Oliveira de Lima; Luiz Aurelio de Campos Crispin. Centro de Genômica Funcional da ESALQ: Luiz Lehmann Coutinho; Luiz Carlos Junior de Alcantara; Lívia Sacchetto; Maisa C. Pereira Parra; Maria Carolina Elias; Marta Giovanetti; Marília Moraes; Maurício Lacerda Nogueira. Prefeitura de Sao Paulo: Melissa Palmieri.; Patricia Akemi Assato; Paula Rahal; Paulo Inacio da Costa; Rafael dos Santos Bezerra; Raquel de Lello Rocha Campos Cassano. NGS Soluções Genômicas: Pilar Drummond Sampaio Corrêa Mariani. FZEA-USP Pirassununga: Mirele Daiana Poleti; Raul Machado Neto; Ricardo Augusto Brassaloti; Ricardo Haddad; Rodrigo Tocantins Calado. FAMERP-SJRP: Cecília Artico Banho; Sandra Coccuzzo Sampaio; Svetoslav Nanev Slavov; Vagner Fonseca; Vincent Louis Viala |
| EPI_ISL_2564553, EPI_ISL_2564555, EPI_ISL_2564557, EPI_ISL_2564587, EPI_ISL_2564589, EPI_ISL_2564866, EPI_ISL_2564876 | | | |
| see above | SK-Roy Romanow Provincial Laboratory | National Microbiology Laboratory (NML) | Amanda Lang; Anna Majer; Anneliese Landgraff; CanCOGeN's metadata curation team; Darian Hole; Elsie Grudeski; Gary Van Domselaar; Grace Seo; Jennifer Tanner; Jessica Minion; Kirsten Biggar; Madison Chapel; Morag Graham; Natalie Knox; Nathalie Bastien; Philip Mabon; Public Health Agency of Canada CanCOGeN team; Rachel DePaulo; Rhiannon Huzarewich; Russell Mandes; Ryan McDonald; Shari Tyson; Timothy Booth; Yan Li |
| EPI_ISL_2893533, EPI_ISL_2893534, EPI_ISL_5843947, EPI_ISL_5844001, EPI_ISL_5863934 | SK-Roy Romanow Provincial Laboratory | Saskatchewan - Roy Romanow Provincial Laboratory (RRPL) | Alanna Senecal; Amanda Lang; Anna Majer; Anneliese Landgraff; CanCOGeN's metadata curation team; Darian Hole; Elsie Grudeski; Gary Van Domselaar; Grace Seo; Jennifer Tanner; Jessica Minion; Kara Loos; Keith MacKenzie; Kirsten Biggar; Madison Chapel; Meredith Faires; Morag Graham; Natalie Knox; Nathalie Bastien; Philip Mabon; Public Health Agency of Canada CanCOGeN team; Rachel DePaulo; Rhiannon Huzarewich; Roy Romanow; Roy Romanow Provincial Laboratory - Molecular Diagnostics; Russell Mandes; Ryan McDonald; Shari Tyson; Timothy Booth; Yan Li |
| EPI_ISL_1966840, EPI_ISL_1966841, EPI_ISL_1966842, EPI_ISL_1966843, EPI_ISL_1966844 | SMS SECRETARIA MUNICIPAL DE SAUDE DE BOITUVA | Instituto Butantan / ESALQ-USP (Piracicaba) | Antonio Jorge Martins; Bianca Cechetto Carlos. Mendelics: Bibiana Santos; Claudia Renata dos Santos Barros; Cíntia Bittar; David Schlesinger. Hemocentro Ribeirão Preto: Simone Kashima; Debora Botequio Moretti; Elaine Cristina Marqueze; Elaine Vieira dos Santos; Elisangela Chicaroni Mattos; Erika Freitas; Evandra Strazza Rodrigues; Felipe Allan da Silva da Costa; Flavia Aburjaile; Fábio Sossai Possebon; Guilherme Campos; Guilherme Targino Valente; Heidge Fukumasu. USP-Botucatu: Rejane Maria Tommasini Grotto; Helena Lage Ferreira; Instituto Butantan: Dimas Tadeu Covas; Jardelina de Souza Todao Bernardino; Jayme A. Souza-Neto; Jessika Cristina Chagas Lesbon; Jorge A. Petroli Marchesi; José Salvatore Leister Patané; João Paulo Kitajima; João Pessoa Araújo Jr.; Leila Sabrina Ullmann; Loyze Paola Oliveira de Lima; Luiz Aurelio de Campos Crispin. Centro de Genômica Funcional da ESALQ: Luiz Lehmann Coutinho; Luiz Carlos Junior de Alcantara; Lívia Sacchetto; Maisa C. Pereira Parra; Maria Carolina Elias; Marta Giovanetti; Marília Moraes; Maurício Lacerda Nogueira. Prefeitura de Sao Paulo: Melissa Palmieri.; Patricia Akemi Assato; Paula Rahal; Paulo Inacio da Costa; Rafael dos Santos Bezerra; Raquel de Lello Rocha Campos Cassano. NGS Soluções Genômicas: Pilar Drummond Sampaio Corrêa Mariani. FZEA-USP Pirassununga: Mirele Daiana Poleti; Raul Machado Neto; Ricardo Augusto Brassaloti; Ricardo Haddad; Rodrigo Tocantins Calado. FAMERP-SJRP: Cecília Artico Banho; Sandra Coccuzzo Sampaio; Svetoslav Nanev Slavov; Vagner Fonseca; Vincent Louis Viala |
| EPI_ISL_1960049 | SOMER | Universidad Nacional de Colombia - Laboratorio Genómico One Health | Andres F. Cardona-Rios; Carlos Franco-Muñoz; Carolina Muñoz-Arango; Celeny Ortiz; Daniel O. Maldonado-Perez; Diego A. Álvarez-Díaz; Hector Alejandro Ruiz-Moreno; Idabely Betancur Ortiz; Jorge E. Osorio; Juan P. Hernandez-Ortiz; Karl A Ciuoderis; Katherine Laiton-Donato; Laura Silvana Perez; Lina M. Hurtado; Marcela Mercado-Reyes; Maria Angélica Maya; Maria Stella López; Rita Almanza Payares; Sandra Ines Cano; Simón Villegas Velásquez |
| EPI_ISL_6512553 | SURA | Laboratorio Departamental de Salud Publica de Antioquia | Ana Victoria Valencia Duarte; Cristian Arbey Velarde Hoyos; Gloria Isabel Escobar; Idabely Betancur Ortiz; Juan P. Hernandez-Ortiz; Juan Pablo Isaza Agudelo; Maria Stella López |
| EPI_ISL_1960040 | SURA | Universidad Nacional de Colombia - Laboratorio Genómico One Health | Andres F. Cardona-Rios; Carlos Franco-Muñoz; Carolina Muñoz-Arango; Celeny Ortiz; Daniel O. Maldonado-Perez; Diego A. Álvarez-Díaz; Hector Alejandro Ruiz-Moreno; Idabely Betancur Ortiz; Jorge E. Osorio; Juan P. Hernandez-Ortiz; Karl A Ciuoderis; Katherine Laiton-Donato; Laura Silvana Perez; Lina M. Hurtado; Marcela Mercado-Reyes; Maria Angélica Maya; Maria Stella López; Rita Almanza Payares; Sandra Ines Cano; Simón Villegas Velásquez |
| EPI_ISL_1671955, EPI_ISL_1671956, EPI_ISL_1671961, EPI_ISL_1671963, EPI_ISL_1672228, EPI_ISL_1672229, EPI_ISL_1672230, EPI_ISL_1672231, EPI_ISL_1672233, EPI_ISL_1672234, EPI_ISL_1672244, EPI_ISL_1672245, EPI_ISL_1672246, EPI_ISL_1672248, EPI_ISL_1793668 | | | |
| see above | SYNLAB | GIGA Medical Genomics | Bouchra Boujemla; Cécile Meex; Keith Durkin; Maria Artesi; Marie-Pierre Hayette; Nathalie Renotte; Pierrette Melin; Raphaël Boreux; Sébastien Bontems; Vincent Bours |
| EPI_ISL_1854238 | SYNLAB | Instituto Nacional de Saude (INSA) | Borges et al |
| EPI_ISL_1960048 | SYNLAB | Universidad Nacional de Colombia - Laboratorio Genómico One Health | Andres F. Cardona-Rios; Carlos Franco-Muñoz; Carolina Muñoz-Arango; Celeny Ortiz; Daniel O. Maldonado-Perez; Diego A. Álvarez-Díaz; Hector Alejandro Ruiz-Moreno; Idabely Betancur Ortiz; Jorge E. Osorio; Juan P. Hernandez-Ortiz; Karl A Ciuoderis; Katherine Laiton-Donato; Laura Silvana Perez; Lina M. Hurtado; Marcela Mercado-Reyes; Maria Angélica Maya; Maria Stella López; Rita Almanza Payares; Sandra Ines Cano; Simón Villegas Velásquez |
| EPI_ISL_3355506, EPI_ISL_3369942 | SYNLAB - ANGEL DIAGNOSTICA | Corporacion CorpoGen-Universidad de los Andes-Universidad Central | Christian Romero; Jorge Duitama; Juan Manuel Anzola; Laura González; Maryam Chaib De Mares; María Mercedes Zambrano; Nelly Díaz; Patricia Del Portillo; Silvia Restrepo |
| EPI_ISL_2155037 | SYNLAB -ANGEL DIAGNOSTICA | Instituto Nacional de Salud-Dirección de Investigación en Salud Pública | Carlos Franco-Muñoz; Carmen Osorio; Diana Malo; Diego A. Álvarez-Díaz; Diego Andrés Prada; Gerardo Santamaría; Hector Alejandro Ruiz-Moreno; Jhonnatan Reales-González; Jorge Rivera; Juan Camilo Martinez; Julian Naizaque; Katherine Laiton-Donato; Lisseth Pardo; Magdalena Wiesner; Marcela Mercado-Reyes; Maria T. Herrera-Sepúlveda; Marta Lopez Blanco; Martha Lucia Ospina Martinez; Paola Rojas; Sergio Gomez; Sheryll Corchuelo; Ángela Alarcon Cruz |
| EPI_ISL_1642947 | SYNLAB Jena Oncoscreen | Robert Koch Institute | |
| EPI_ISL_1641696 | SYNLAB MVZ Trier | Robert Koch Institute | |
| EPI_ISL_1791455, EPI_ISL_1794942, EPI_ISL_1794944, EPI_ISL_1795022, EPI_ISL_1969382, EPI_ISL_1969417, EPI_ISL_1969442, EPI_ISL_1969443, EPI_ISL_1969444, EPI_ISL_1969445, EPI_ISL_1969446, EPI_ISL_1969447, EPI_ISL_2423010 | | | |
| see above | San Diego County Public Health Laboratory | Andersen lab at Scripps Research | Brett Austin; Jovan Shephard; SEARCH Alliance San Diego with Ashleigh Murphy; SEARCH Alliance San Diego with Tracy Basler |
| EPI_ISL_1805403, EPI_ISL_1805404 | San Juan Regional Medical Center | New Mexico Department of Health Scientific Laboratory | Anastacia Griego-Fisher; D'eldra Malone; Ellie Johnson; Jennifer Benoit; Linda Salazar; Ratheesh Rajan |
| EPI_ISL_5802333 | Santa Casa De Guararema | Instituto Butantan | Antonio Jorge Martins; Claudia Renata dos Santos Barros; David Schlesinger; Debora Botequio Moretti; Dimas Tadeu Covas; Elaine Cristina Marqueze; Elaine Vieira Santos; Evandra Strazza Rodrigues; Heidge Fukumasu; Jayme Augusto de Souza-Neto; José Salvatore Leister Patané; Luiz Alcantara; Luiz Lehmann Coutinho; Maria Carolina Elias; Maurício Lacerda Nogueira; Rafael dos Santos Bezerra; Raul Machado Neto; Rejane Maria Tommasini Grotto; Ricardo Haddad; Sandra Coccuzzo Sampaio Vessoni; Simone Kashima; Svetoslav Nanev Slavov; Vincent Louis Viala |
| EPI_ISL_5800192, EPI_ISL_5800247, EPI_ISL_5800337, EPI_ISL_5800338 | Santa Casa De Misericordia De Tiete | Instituto Butantan | Antonio Jorge Martins; Claudia Renata dos Santos Barros; David Schlesinger; Debora Botequio Moretti; Dimas Tadeu Covas; Elaine Cristina Marqueze; Elaine Vieira Santos; Evandra Strazza Rodrigues; Heidge Fukumasu; Jayme Augusto de Souza-Neto; José Salvatore Leister Patané; Luiz Alcantara; Luiz Lehmann Coutinho; Maria Carolina Elias; Maurício Lacerda Nogueira; Rafael dos Santos Bezerra; Raul Machado Neto; Rejane Maria Tommasini Grotto; Ricardo Haddad; Sandra Coccuzzo Sampaio Vessoni; Simone Kashima; Svetoslav Nanev Slavov; Vincent Louis Viala |
| EPI_ISL_5800218, EPI_ISL_5800219, EPI_ISL_5800220, EPI_ISL_5800222, EPI_ISL_5800227, EPI_ISL_5800228 | Santa Casa De Porto Feliz | Instituto Butantan | Antonio Jorge Martins; Claudia Renata dos Santos Barros; David Schlesinger; Debora Botequio Moretti; Dimas Tadeu Covas; Elaine Cristina Marqueze; Elaine Vieira Santos; Evandra Strazza Rodrigues; Heidge Fukumasu; Jayme Augusto de Souza-Neto; José Salvatore Leister Patané; Luiz Alcantara; Luiz Lehmann Coutinho; Maria Carolina Elias; Maurício Lacerda Nogueira; Rafael dos Santos Bezerra; Raul Machado Neto; Rejane Maria Tommasini Grotto; Ricardo Haddad; Sandra Coccuzzo Sampaio Vessoni; Simone Kashima; Svetoslav Nanev Slavov; Vincent Louis Viala |
| EPI_ISL_1664266 | Santa Clara County Public Health Laboratory | Santa Clara County Public Health Laboratory | Santa Clara County Public Health Department |
| EPI_ISL_1913235, EPI_ISL_1913238, EPI_ISL_1913240, EPI_ISL_2691257 | Santa Clara Valley Medical Center | Santa Clara County Public Health Laboratory | Santa Clara County Public Health Department |
| EPI_ISL_1711429, EPI_ISL_1711465, EPI_ISL_1711589, EPI_ISL_2536671, EPI_ISL_2536693 | Seattle Flu Study | Seattle Flu Study | Amanda Adler; Barry R. Lutz; Benjamin Pelle; Caitlin R. Wolf; Chris D. Frazar; Deborah A. Nickerson; Elisabeth Brandstetter; Erica Ryke; Helen Y. Chu; Janet A. Englund; Jay Shendure; Jeff Duchin; Jover Lee; Kairsten Fay; Karen Cowgill; Kirsten Lacombe; Lea M. Starita; Mark J. Rieder; Matthew Richardson; Matthew Thompson; Melissa Truong; Michael Boeckh; Michael Famulare; Misja Ilcisin; Peter D. Han; Stephanie Schrag; Thomas R. Sibley; Trevor Bedford |
| EPI_ISL_5800062, EPI_ISL_5800073 | Secao Centro De Diagnostico Secedi | Instituto Butantan | Antonio Jorge Martins; Claudia Renata dos Santos Barros; David Schlesinger; Debora Botequio Moretti; Dimas Tadeu Covas; Elaine Cristina Marqueze; Elaine Vieira Santos; Evandra Strazza Rodrigues; Heidge Fukumasu; Jayme Augusto de Souza-Neto; José Salvatore Leister Patané; Luiz Alcantara; Luiz Lehmann Coutinho; Maria Carolina Elias; Maurício Lacerda Nogueira; Rafael dos Santos Bezerra; Raul Machado Neto; Rejane Maria Tommasini Grotto; Ricardo Haddad; Sandra Coccuzzo Sampaio Vessoni; Simone Kashima; Svetoslav Nanev Slavov; Vincent Louis Viala |

| | | | |
|---|---|---|---|
| EPI_ISL_5799697, EPI_ISL_5799706 | Secretaria De Saude De Santa Gertrudes | Instituto Butantan | Antonio Jorge Martins; Claudia Renata dos Santos Barros; David Schlesinger; Debora Botequio Moretti; Dimas Tadeu Covas; Elaine Cristina Marqueze; Elaine Vieira Santos; Evandra Strazza Rodrigues; Heidge Fukumasu; Jayme Augusto de Souza-Neto; José Salvatore Leister Patané; Luiz Alcantara; Luiz Lehmann Coutinho; Maria Carolina Elias; Maurício Lacerda Nogueira; Rafael dos Santos Bezerra; Raul Machado Neto; Rejane Maria Tommasini Grotto; Ricardo Haddad; Sandra Coccuzzo Sampaio Vessoni; Simone Kashima; Svetoslav Nanev Slavov; Vincent Louis Viala |
| EPI_ISL_5799707 | Secretaria De Saude De Sao Pedro | Instituto Butantan | Antonio Jorge Martins; Claudia Renata dos Santos Barros; David Schlesinger; Debora Botequio Moretti; Dimas Tadeu Covas; Elaine Cristina Marqueze; Elaine Vieira Santos; Evandra Strazza Rodrigues; Heidge Fukumasu; Jayme Augusto de Souza-Neto; José Salvatore Leister Patané; Luiz Alcantara; Luiz Lehmann Coutinho; Maria Carolina Elias; Maurício Lacerda Nogueira; Rafael dos Santos Bezerra; Raul Machado Neto; Rejane Maria Tommasini Grotto; Ricardo Haddad; Sandra Coccuzzo Sampaio Vessoni; Simone Kashima; Svetoslav Nanev Slavov; Vincent Louis Viala |
| EPI_ISL_5800246, EPI_ISL_5800251 | Secretaria Municipal Da Saude De Itarare | Instituto Butantan | Antonio Jorge Martins; Claudia Renata dos Santos Barros; David Schlesinger; Debora Botequio Moretti; Dimas Tadeu Covas; Elaine Cristina Marqueze; Elaine Vieira Santos; Evandra Strazza Rodrigues; Heidge Fukumasu; Jayme Augusto de Souza-Neto; José Salvatore Leister Patané; Luiz Alcantara; Luiz Lehmann Coutinho; Maria Carolina Elias; Maurício Lacerda Nogueira; Rafael dos Santos Bezerra; Raul Machado Neto; Rejane Maria Tommasini Grotto; Ricardo Haddad; Sandra Coccuzzo Sampaio Vessoni; Simone Kashima; Svetoslav Nanev Slavov; Vincent Louis Viala |
| EPI_ISL_5799711, EPI_ISL_5799712, EPI_ISL_5799713 | Secretaria Municipal De Saude De Cordeiropolis | Instituto Butantan | Antonio Jorge Martins; Claudia Renata dos Santos Barros; David Schlesinger; Debora Botequio Moretti; Dimas Tadeu Covas; Elaine Cristina Marqueze; Elaine Vieira Santos; Evandra Strazza Rodrigues; Heidge Fukumasu; Jayme Augusto de Souza-Neto; José Salvatore Leister Patané; Luiz Alcantara; Luiz Lehmann Coutinho; Maria Carolina Elias; Maurício Lacerda Nogueira; Rafael dos Santos Bezerra; Raul Machado Neto; Rejane Maria Tommasini Grotto; Ricardo Haddad; Sandra Coccuzzo Sampaio Vessoni; Simone Kashima; Svetoslav Nanev Slavov; Vincent Louis Viala |
| EPI_ISL_5800197, EPI_ISL_5800260, EPI_ISL_5800261, EPI_ISL_5800264, EPI_ISL_5800271, EPI_ISL_5800272, EPI_ISL_5800273, EPI_ISL_5800277, EPI_ISL_5800279, EPI_ISL_5800280, EPI_ISL_5800281, EPI_ISL_5800283, EPI_ISL_5800284, EPI_ISL_5800286, EPI_ISL_5800288, EPI_ISL_5800290, EPI_ISL_5800292, EPI_ISL_5800293, EPI_ISL_5800294, EPI_ISL_5800295, EPI_ISL_5800296, EPI_ISL_5800297, EPI_ISL_5800299, EPI_ISL_5800300, EPI_ISL_5800301, EPI_ISL_5800302, EPI_ISL_5800303, EPI_ISL_5800304, EPI_ISL_5800305, EPI_ISL_5800306, EPI_ISL_5800308, EPI_ISL_5800309, EPI_ISL_5800311, EPI_ISL_5800312, EPI_ISL_5800314, EPI_ISL_5800316, EPI_ISL_5800319, EPI_ISL_5800320, EPI_ISL_5800321, EPI_ISL_5800322, EPI_ISL_5800323, EPI_ISL_5800324, EPI_ISL_5800326, EPI_ISL_5800327, EPI_ISL_5800328, EPI_ISL_5800329, EPI_ISL_5800330, EPI_ISL_5800331, EPI_ISL_5800332, EPI_ISL_5800333 | | | |
| see above | Secretaria Municipal De Saude Sorocaba | Instituto Butantan | Antonio Jorge Martins; Claudia Renata dos Santos Barros; David Schlesinger; Debora Botequio Moretti; Dimas Tadeu Covas; Elaine Cristina Marqueze; Elaine Vieira Santos; Evandra Strazza Rodrigues; Heidge Fukumasu; Jayme Augusto de Souza-Neto; José Salvatore Leister Patané; Luiz Alcantara; Luiz Lehmann Coutinho; Maria Carolina Elias; Maurício Lacerda Nogueira; Rafael dos Santos Bezerra; Raul Machado Neto; Rejane Maria Tommasini Grotto; Ricardo Haddad; Sandra Coccuzzo Sampaio Vessoni; Simone Kashima; Svetoslav Nanev Slavov; Vincent Louis Viala |
| EPI_ISL_2179722, EPI_ISL_2179723, EPI_ISL_2179724, EPI_ISL_2179725, EPI_ISL_2179756 | Servicio Microbiología Hospital La Paz | Servicio Microbiología Hospital La Paz | Elie Dahdouh; Fernando Lázaro; Jesús Mingorance Cruz; Rubén Cáceres |
| EPI_ISL_2135339, EPI_ISL_2135688, EPI_ISL_2135693, EPI_ISL_2135716, EPI_ISL_2135717, EPI_ISL_2135718, EPI_ISL_2135719, EPI_ISL_2135720, EPI_ISL_2135726, EPI_ISL_2135988, EPI_ISL_2135992, EPI_ISL_2135993, EPI_ISL_2135994, EPI_ISL_2135999, EPI_ISL_2136000, EPI_ISL_2136011, EPI_ISL_2136117, EPI_ISL_2136168, EPI_ISL_2136175, EPI_ISL_2136176, EPI_ISL_2136177, EPI_ISL_2136182, EPI_ISL_6694434 | | | |
| see above | Servicio Virosis Respiratorias-Departamento Virología-INEI | Instituto Nacional Enfermedades Infecciosas C.G.Malbran | Avaro M.; Baumeister E.; Benedetti E.; Campos J.; Cisterna D.; Dattero ME; De Belder D.; Haim MS.; Lorenzo F.; Molina V.; Perandones C.; Poklepovich T.; Pontoriero A.; Russo M.; Sanchez Loria J.; Tuduri E. |
| EPI_ISL_2104415 | Servicio de Microbiologia Hospital Ramon y Cajal | Servicio de Microbiologia Hospital Ramon y Cajal | Galan JC; Martinez L. Abreu M; Ponce M; y Gonzalez-Alba JM |
| EPI_ISL_1665139, EPI_ISL_1665194, EPI_ISL_1665198, EPI_ISL_1665246 | Servicio de Microbiología Hospital Ramón y Cajal | Servicio de Microbiología Hospital Ramón y Cajal | JC Galán; JM González-Alba; Laura Martínez. Melanie Abreu; Manuel Ponce |
| EPI_ISL_1970050, EPI_ISL_1970054, EPI_ISL_1970055, EPI_ISL_1970061, EPI_ISL_1970076 | Servicio de Microbiología. Consorcio Hospital General Universitario de Valencia | SeqCOVID-SPAIN consortium/IBV(CSIC) | Begoña Fuster Escrivá; Carme Salvador García; Concepción Gimeno Cardona and SeqCOVID-SPAIN consortium; María Dolores Ocete; Rafael Medina González |
| EPI_ISL_2016681 | Servicio de Microbiología. Hospital Universitario Doctor Peset | SeqCOVID-SPAIN consortium/IBV(CSIC) | José Miguel Nogueira Coito and SeqCOVID-SPAIN consortium; Juan Alberola Enguídanos; Juan José Camarena Miñana; Rosa González Pellicer |
| EPI_ISL_2438680, EPI_ISL_2600575, EPI_ISL_2600576 | Servizo de Microbioloxía. Complexo Hospitalario de Santiago de Compostela | Servizo de Microbioloxía. Complexo Hospitalario de Santiago de Compostela | Amparo Coira; Antonio Aguilera; Carlos García_Riestra; Daniel Navarro; Gema Barbeito; Iria Roca; José Llovo; Laura Millán; Manuela Hernández; María Luisa Pérez_del_Molino; Mercedes Treviño; Rocío trastoy; Teresa Lopez_Valiño; Xosé Costa |
| EPI_ISL_5315354, EPI_ISL_5315359, EPI_ISL_5315360, EPI_ISL_5315361, EPI_ISL_5315362, EPI_ISL_5315368, EPI_ISL_5315370, EPI_ISL_5315371, EPI_ISL_5315380, EPI_ISL_5315387, EPI_ISL_5315388, EPI_ISL_5315389, EPI_ISL_5315390, EPI_ISL_5315391, EPI_ISL_5315393, EPI_ISL_5315395, EPI_ISL_5315396, EPI_ISL_5315397, EPI_ISL_5315398, EPI_ISL_5315399, EPI_ISL_5315400, EPI_ISL_5315402, EPI_ISL_5315406, EPI_ISL_5315410, EPI_ISL_5315411, EPI_ISL_5315413, EPI_ISL_5315415, EPI_ISL_5315416, EPI_ISL_5315417, EPI_ISL_5315420, EPI_ISL_5315423, EPI_ISL_5315424, EPI_ISL_5315427, EPI_ISL_5315428, EPI_ISL_5315430, EPI_ISL_5315431, EPI_ISL_5315432, EPI_ISL_5315433, EPI_ISL_5315434, EPI_ISL_5315436, EPI_ISL_5315437, EPI_ISL_5315439, EPI_ISL_5315440, EPI_ISL_5315441, EPI_ISL_5315442, EPI_ISL_5315457, EPI_ISL_5315458, EPI_ISL_5315463, EPI_ISL_5315465, EPI_ISL_5315466, EPI_ISL_5315471, EPI_ISL_5316432 | | | |
| see above | Shared Hospital Laboratory | Shared Hospital Laboratory | Christie Vermeiren; Finlay Maguire; Kevin Katz; Patryk Aftanas; Robert Kozak; Samira Mubareka |
| EPI_ISL_1794889, EPI_ISL_1794903 | Sharp HealthCare Laboratory | Andersen lab at Scripps Research | Art Mendoza; Cathy Woerle; Jacquelyn Berumen; Liam McGinnis; Omid Bakhtar; SEARCH Alliance San Diego with Aaron Harding |
| EPI_ISL_2080239, EPI_ISL_2080248 | Simple Laboratories | Gagnon Lab, Southern Illinois University | Keith Gagnon |
| EPI_ISL_5800216, EPI_ISL_5800249, EPI_ISL_5800256, EPI_ISL_5800342, EPI_ISL_5800343 | Sms Secretaria Municipal De Saude De Boituva | Instituto Butantan | Antonio Jorge Martins; Claudia Renata dos Santos Barros; David Schlesinger; Debora Botequio Moretti; Dimas Tadeu Covas; Elaine Cristina Marqueze; Elaine Vieira Santos; Evandra Strazza Rodrigues; Heidge Fukumasu; Jayme Augusto de Souza-Neto; José Salvatore Leister Patané; Luiz Alcantara; Luiz Lehmann Coutinho; Maria Carolina Elias; Maurício Lacerda Nogueira; Rafael dos Santos Bezerra; Raul Machado Neto; Rejane Maria Tommasini Grotto; Ricardo Haddad; Sandra Coccuzzo Sampaio Vessoni; Simone Kashima; Svetoslav Nanev Slavov; Vincent Louis Viala |
| EPI_ISL_1909276, EPI_ISL_1909336, EPI_ISL_1909524, EPI_ISL_1909530, EPI_ISL_1909536, EPI_ISL_1909596, EPI_ISL_1909752 | | | |
| see above | Sonora Quest Laboratories | TGen North | "Jolene Bowers; Ashlyn Pfeiffer; Chris French; Darrin Lemmer; Dave Engelthaler; Hayley Yaglom; Heather Centner; The Arizona COVID Genomics Union (ACGU)" |
| EPI_ISL_2691698 | Spital Männedorf AG | Institute of Medical Virology | Alexandra Trkola; Annette Audigé; Cyril Shah; Gabriela Ziltener; Guido Bloemberg; Jon Huder; Jürg Böni; Kevin Steiner; Maria Grünberg; Maryam Zaheri; Michael Huber; Riccarda Capaul; Stefan Schmutz; Verena Kufner |
| EPI_ISL_1924638, EPI_ISL_1924639 | Stanford Clinical Virology Laboratory | Santa Clara County Public Health Laboratory | Santa Clara County Public Health Department |
| EPI_ISL_3916216, EPI_ISL_3916217, EPI_ISL_3916231, EPI_ISL_3916254 | Stanford Health Care | Stanford University School of Medicine, Clinical Virology Laboratory | Becky Jiang; Fumiko Yamamoto; James Zehnder; Malaya K. Sahoo; Nathan Hammond; and Benjamin A. Pinsky |
| EPI_ISL_1771219, EPI_ISL_1771292, EPI_ISL_1771614, EPI_ISL_2007840, EPI_ISL_2007851 | State Laboratories Division, Hawaii State Department of Health | State Laboratories Division, Hawaii State Department of Health | Ayana Garnet; Drew Kuwazaki; Edward Desmond; Pamela O'Brien; Razvan Sultana |
| EPI_ISL_6782131, EPI_ISL_6782217 | State of New Hampshire Public Health Laboratories | State of New Hampshire Public Health Laboratories | Caitlin Mercier; Chris Benton; Jinfeng Li; Juan Bolanos; Xinglu Zhang |
| EPI_ISL_2208312, EPI_ISL_2416020 | Swedish national genomic surveillance program of SARS-CoV-2 | The Public Health Agency of Sweden | Alma Brolund; Maria Lind Karlberg; Maximilian Riess; Swedish national genomic surveillance program of SARS-CoV-2 |
| EPI_ISL_2621852, EPI_ISL_2621853, EPI_ISL_2621854, EPI_ISL_2621855 | Synlab | Universidad Nacional de Colombia - Laboratorio Genómico One Health | Andres F. Cardona-Rios; Carlos Franco-Muñoz; Carolina Muñoz-Arango; Celeny Ortiz; Daniel O. Maldonado-Perez; Diego A. Álvarez-Díaz; Hector Alejandro Ruiz-Moreno; Idabely Betancur Ortiz; Jorge E. Osorio; Juan P. Hernandez-Ortiz; Karl A Ciuoderis; Katherine Laiton-Donato; Laura Silvana Perez; Lina M. Hurtado; Marcela Mercado-Reyes; Maria Angélica Maya; Maria Stella López; Rita Almanza Payares; Sandra Ines Cano; Simón Villegas Velásquez |
| EPI_ISL_2443058, EPI_ISL_2443059, EPI_ISL_2443062, EPI_ISL_2443064, EPI_ISL_2443065 | São Jose do Rio Preto | Instituto Butantan / FAMERP | Antonio Jorge Martins; Claudia Renata dos Santos Barros; David Schlesinger; Debora Botequio Moretti; Dimas Tadeu Covas; Elaine Cristina Marqueze; Elaine Vieira Santos; Evandra Strazza Rodrigues; Heidge Fukumasu; Jayme Augusto de Souza-Neto; José Salvatore Leister Patané; Luiz Alcantara; Luiz Lehmann Coutinho; Maria Carolina Elias; Maurício Lacerda Nogueira; Rafael dos Santos Bezerra; Raul Machado Neto; Rejane Maria Tommasini Grotto; Ricardo Haddad; Sandra Coccuzzo Sampaio Vessoni; Simone Kashima; Svetoslav Nanev Slavov; Vincent Louis Viala |
| EPI_ISL_2000897 | TGen North | TGen North | "Jolene Bowers; Ashlyn Pfeiffer; Chris French; Darrin Lemmer; Dave Engelthaler; Hayley Yaglom; Heather Centner; The Arizona COVID Genomics Union (ACGU)" |
| EPI_ISL_2042410 | TN Division of Laboratory Services | Pathogen Discovery, Respiratory Viruses Branch, Division of Viral Diseases, Centers for Disease Control and Prevention | Adam Retchless; Anna Kelleher; Anna Uehara; Brian Lynch; Clinton R. Paden; Dhwani Batra; Haibin Wang; Han Jia Justin Ng; Jasmine Padilla; Jing Zhang; Justin Lee; Krista Queen; Mark Burroughs; Mili Sheth; Morgan Davis; Peter Cook; Rachel Marine; Sarah Nobles; Suxiang Tong; Tara Coalter; Yan Li; Ying Tao |
| EPI_ISL_1760561, EPI_ISL_1760594, EPI_ISL_1961099 | TXDSHS | TXDSHS | Anita Pokharel; Bonnie Oh; Chun Wang; Grace Kubin; Jenny Zhang; Lorraine Rodriguez; Maliha Rahman; Mayela Pedrueza; Myong Koag; Rachel Lee; Rashmi Tuladhar |
| EPI_ISL_2081379, EPI_ISL_2081382, EPI_ISL_2383082, EPI_ISL_2383084, EPI_ISL_2383090, EPI_ISL_2383092, EPI_ISL_2930433, EPI_ISL_2930435, EPI_ISL_2930438, EPI_ISL_2988592 | | | |
| see above | Tampa General Hospital Esoteric Lab | Tampa General Hospital Esoteric Research & Development Lab | Amorce Lima; Deanna Becker; Dominic Uy; Elaine Vendrone; Grant Vestal; Jon Faughn; Jorge Mecias-Frias; Suzane Silbert; Vicki Healer |
| EPI_ISL_1660605, EPI_ISL_1660608, EPI_ISL_1660609, EPI_ISL_1660611, EPI_ISL_1660612, EPI_ISL_1807248, EPI_ISL_1807275, EPI_ISL_1856932 | | | |
| see above | Texas Department of State | Texas Department of State Health | Anita Pokharel; Bonnie Oh; Chun Wang; Grace Kubin; Jenny Zhang; Lorraine Rodriguez; Maliha Rahman; Mayela Pedrueza; Myong Koag; Rachel Lee; Rashmi Tuladhar |

| | | | |
|---|---|---|---|
| | Health Services (TXDSHS) | Services (TXDSHS) | |
| EPI_ISL_1708575 | Thai Red Cross Emerging Infectious Diseases Health Science Centre, Chulalongkorn Hospital, Faculty of Medicine, Chulalongkorn University | Thai Red Cross Emerging Infectious Diseases Center and Faculty of Medicine, Chulalongkorn University | Apaporn Rodpan; Opass Putcharoen; Rome Buathong; Sininat Petcharat; Sopon Iamsirithaworn; Supaporn Wacharapluesadee; Thiravat Hemachudha; Weenassarin Ampoot; Wichai Thanasopon; Yutthana Joyjinda |
| EPI_ISL_1625680, EPI_ISL_1625681 | The Jackson Laboratory | The Jackson Laboratory | Adams M; Bergeron D; Kelly K; Li L; Long J; Omerza G; Renzette N |
| EPI_ISL_2006987, EPI_ISL_2341359, EPI_ISL_2341362, EPI_ISL_2341378, EPI_ISL_2341380, EPI_ISL_2341385, EPI_ISL_2341386, EPI_ISL_2341395 | | | |
| see above | The Ohio State University Applied Microbiology Services Laboratory | The Ohio State University Applied Microbiology Services Laboratory | Seth A. Faith PhD |
| EPI_ISL_2228215 | TriCore Reference Laboratories | Center for Global Health, University of New Mexico Health Sciences Center | Cecilia Thompson; Darrell Dinwiddie; Daryl Domman; Karissa Culbreath; Kendra Pesko; Kurt Schwalm; Valerie Morley |
| EPI_ISL_5530042 | UAPS DOM ALOISIO LORSCHEIDER | Analytical Competence Molecular Epidemiology Lab/ACME, Oswaldo Cruz Foundation, Ceara (FIOCRUZ CE) | Carlos Leonardo de Aragao Araujo; Cecilia Leite Costa & Eduardo Ruback dos Santos on behalf of COVID-19 FIOCRUZ Genomic Network; Cleber Furtado Aksenen; Fabio Miyajima; Fernando Braga Stehling; Francisco Eder de Moura Lopes; Igor Oliveira Duarte; Jamille Maria Mendes Bezerra; Joaquim Cesar do Nascimento Sousa Junior; Pedro Miguel Carneiro Jeronimo; Suzana Porto Almeida; Thais Ferreira de Oliveira; Thais de Oliveira Costa; Ticiane Cavalcante de Souza; Veridiana Pessoa Miyajima |
| EPI_ISL_5530041, EPI_ISL_5530140 | UAPS FRANCISCO MONTEIRO | Analytical Competence Molecular Epidemiology Lab/ACME, Oswaldo Cruz Foundation, Ceara (FIOCRUZ CE) | Carlos Leonardo de Aragao Araujo; Cecilia Leite Costa & Eduardo Ruback dos Santos on behalf of COVID-19 FIOCRUZ Genomic Network; Cleber Furtado Aksenen; Fabio Miyajima; Fernando Braga Stehling; Francisco Eder de Moura Lopes; Igor Oliveira Duarte; Jamille Maria Mendes Bezerra; Joaquim Cesar do Nascimento Sousa Junior; Pedro Miguel Carneiro Jeronimo; Suzana Porto Almeida; Thais Ferreira de Oliveira; Thais de Oliveira Costa; Ticiane Cavalcante de Souza; Veridiana Pessoa Miyajima |
| EPI_ISL_5530040 | UAPS PEDRO SAMPAIO | Analytical Competence Molecular Epidemiology Lab/ACME, Oswaldo Cruz Foundation, Ceara (FIOCRUZ CE) | Carlos Leonardo de Aragao Araujo; Cecilia Leite Costa & Eduardo Ruback dos Santos on behalf of COVID-19 FIOCRUZ Genomic Network; Cleber Furtado Aksenen; Fabio Miyajima; Fernando Braga Stehling; Francisco Eder de Moura Lopes; Igor Oliveira Duarte; Jamille Maria Mendes Bezerra; Joaquim Cesar do Nascimento Sousa Junior; Pedro Miguel Carneiro Jeronimo; Suzana Porto Almeida; Thais Ferreira de Oliveira; Thais de Oliveira Costa; Ticiane Cavalcante de Souza; Veridiana Pessoa Miyajima |
| EPI_ISL_1966877, EPI_ISL_1966878, EPI_ISL_1966879, EPI_ISL_1966880, EPI_ISL_1966881, EPI_ISL_1966882, EPI_ISL_1966883, EPI_ISL_1966884, EPI_ISL_1966885, EPI_ISL_1966886, EPI_ISL_1966887, EPI_ISL_1966888, EPI_ISL_1966890 | | | |
| see above | UBS ATALAIA DR MARIO AGUIAR FILHO | Instituto Butantan / ESALQ-USP (Piracicaba) | Antonio Jorge Martins; Bianca Cechetto Carlos. Mendelics: Bibiana Santos; Claudia Renata dos Santos Barros; Cíntia Bittar; David Schlesinger. Hemocentro Ribeirão Preto: Simone Kashima; Debora Botequio Moretti; Elaine Cristina Marqueze; Elaine Vieira dos Santos; Elisangela Chicaroni Mattos; Erika Freitas; Evandra Strazza Rodrigues; Felipe Allan da Silva da Costa; Flavia Aburjaile; Fábio Sossai Possebon; Guilherme Campos; Guilherme Targino Valente; Heidge Fukumasu. USP-Botucatu: Rejane Maria Tommasini Grotto; Helena Lage Ferreira; Instituto Butantan: Dimas Tadeu Covas; Jardelina de Souza Todao Bernardino; Jayme A. Souza-Neto; Jessika Cristina Chagas Lesbon; Jorge A. Petroli Marchesi; José Salvatore Leister Patané; João Paulo Kitajima; João Pessoa Araújo Jr.; Leila Sabrina Ullmann; Loyze Paola Oliveira de Lima; Luiz Aurelio de Campos Crispin. Centro de Genômica Funcional da ESALQ: Luiz Lehmann Coutinho; Luiz Carlos Junior de Alcantara; Lívia Sacchetto; Maisa C. Pereira Parra; Maria Carolina Elias; Marta Giovanetti; Marília Moraes; Maurício Lacerda Nogueira. Prefeitura de Sao Paulo: Melissa Palmieri.; Patricia Akemi Assato; Paula Rahal; Paulo Inacio da Costa; Rafael dos Santos Bezerra; Raquel de Lello Rocha Campos Cassano. NGS Soluções Genômicas: Pilar Drummond Sampaio Corrêa Mariani. FZEA-USP Pirassununga: Mirele Daiana Poleti; Raul Machado Neto; Ricardo Augusto Brassaloti; Ricardo Haddad; Rodrigo Tocantins Calado. FAMERP-SJRP: Cecília Artico Banho; Sandra Coccuzzo Sampaio; Svetoslav Nanev Slavov; Vagner Fonseca; Vincent Louis Viala |
| EPI_ISL_1967159, EPI_ISL_1967160, EPI_ISL_1967161, EPI_ISL_1967162 | UBS DE TABAPUA | Instituto Butantan / ESALQ-USP (Piracicaba) | Antonio Jorge Martins; Bianca Cechetto Carlos. Mendelics: Bibiana Santos; Claudia Renata dos Santos Barros; Cíntia Bittar; David Schlesinger. Hemocentro Ribeirão Preto: Simone Kashima; Debora Botequio Moretti; Elaine Cristina Marqueze; Elaine Vieira dos Santos; Elisangela Chicaroni Mattos; Erika Freitas; Evandra Strazza Rodrigues; Felipe Allan da Silva da Costa; Flavia Aburjaile; Fábio Sossai Possebon; Guilherme Campos; Guilherme Targino Valente; Heidge Fukumasu. USP-Botucatu: Rejane Maria Tommasini Grotto; Helena Lage Ferreira; Instituto Butantan: Dimas Tadeu Covas; Jardelina de Souza Todao Bernardino; Jayme A. Souza-Neto; Jessika Cristina Chagas Lesbon; Jorge A. Petroli Marchesi; José Salvatore Leister Patané; João Paulo Kitajima; João Pessoa Araújo Jr.; Leila Sabrina Ullmann; Loyze Paola Oliveira de Lima; Luiz Aurelio de Campos Crispin. Centro de Genômica Funcional da ESALQ: Luiz Lehmann Coutinho; Luiz Carlos Junior de Alcantara; Lívia Sacchetto; Maisa C. Pereira Parra; Maria Carolina Elias; Marta Giovanetti; Marília Moraes; Maurício Lacerda Nogueira. Prefeitura de Sao Paulo: Melissa Palmieri.; Patricia Akemi Assato; Paula Rahal; Paulo Inacio da Costa; Rafael dos Santos Bezerra; Raquel de Lello Rocha Campos Cassano. NGS Soluções Genômicas: Pilar Drummond Sampaio Corrêa Mariani. FZEA-USP Pirassununga: Mirele Daiana Poleti; Raul Machado Neto; Ricardo Augusto Brassaloti; Ricardo Haddad; Rodrigo Tocantins Calado. FAMERP-SJRP: Cecília Artico Banho; Sandra Coccuzzo Sampaio; Svetoslav Nanev Slavov; Vagner Fonseca; Vincent Louis Viala |
| EPI_ISL_1966990, EPI_ISL_1966994 | UBS DR HELIO MIGLIARI | Instituto Butantan / ESALQ-USP (Piracicaba) | Antonio Jorge Martins; Bianca Cechetto Carlos. Mendelics: Bibiana Santos; Claudia Renata dos Santos Barros; Cíntia Bittar; David Schlesinger. Hemocentro Ribeirão Preto: Simone Kashima; Debora Botequio Moretti; Elaine Cristina Marqueze; Elaine Vieira dos Santos; Elisangela Chicaroni Mattos; Erika Freitas; Evandra Strazza Rodrigues; Felipe Allan da Silva da Costa; Flavia Aburjaile; Fábio Sossai Possebon; Guilherme Campos; Guilherme Targino Valente; Heidge Fukumasu. USP-Botucatu: Rejane Maria Tommasini Grotto; Helena Lage Ferreira; Instituto Butantan: Dimas Tadeu Covas; Jardelina de Souza Todao Bernardino; Jayme A. Souza-Neto; Jessika Cristina Chagas Lesbon; Jorge A. Petroli Marchesi; José Salvatore Leister Patané; João Paulo Kitajima; João Pessoa Araújo Jr.; Leila Sabrina Ullmann; Loyze Paola Oliveira de Lima; Luiz Aurelio de Campos Crispin. Centro de Genômica Funcional da ESALQ: Luiz Lehmann Coutinho; Luiz Carlos Junior de Alcantara; Lívia Sacchetto; Maisa C. Pereira Parra; Maria Carolina Elias; Marta Giovanetti; Marília Moraes; Maurício Lacerda Nogueira. Prefeitura de Sao Paulo: Melissa Palmieri.; Patricia Akemi Assato; Paula Rahal; Paulo Inacio da Costa; Rafael dos Santos Bezerra; Raquel de Lello Rocha Campos Cassano. NGS Soluções Genômicas: Pilar Drummond Sampaio Corrêa Mariani. FZEA-USP Pirassununga: Mirele Daiana Poleti; Raul Machado Neto; Ricardo Augusto Brassaloti; Ricardo Haddad; Rodrigo Tocantins Calado. FAMERP-SJRP: Cecília Artico Banho; Sandra Coccuzzo Sampaio; Svetoslav Nanev Slavov; Vagner Fonseca; Vincent Louis Viala |
| EPI_ISL_1967163 | UBS ENRIQUE GUILHERME ERNESTO MARZEUSKI | Instituto Butantan / ESALQ-USP (Piracicaba) | Antonio Jorge Martins; Bianca Cechetto Carlos. Mendelics: Bibiana Santos; Claudia Renata dos Santos Barros; Cíntia Bittar; David Schlesinger. Hemocentro Ribeirão Preto: Simone Kashima; Debora Botequio Moretti; Elaine Cristina Marqueze; Elaine Vieira dos Santos; Elisangela Chicaroni Mattos; Erika Freitas; Evandra Strazza Rodrigues; Felipe Allan da Silva da Costa; Flavia Aburjaile; Fábio Sossai Possebon; Guilherme Campos; Guilherme Targino Valente; Heidge Fukumasu. USP-Botucatu: Rejane Maria Tommasini Grotto; Helena Lage Ferreira; Instituto Butantan: Dimas Tadeu Covas; Jardelina de Souza Todao Bernardino; Jayme A. Souza-Neto; Jessika Cristina Chagas Lesbon; Jorge A. Petroli Marchesi; José Salvatore Leister Patané; João Paulo Kitajima; João Pessoa Araújo Jr.; Leila Sabrina Ullmann; Loyze Paola Oliveira de Lima; Luiz Aurelio de Campos Crispin. Centro de Genômica Funcional da ESALQ: Luiz Lehmann Coutinho; Luiz Carlos Junior de Alcantara; Lívia Sacchetto; Maisa C. Pereira Parra; Maria Carolina Elias; Marta Giovanetti; Marília Moraes; Maurício Lacerda Nogueira. Prefeitura de Sao Paulo: Melissa Palmieri.; Patricia Akemi Assato; Paula Rahal; Paulo Inacio da Costa; Rafael dos Santos Bezerra; Raquel de Lello Rocha Campos Cassano. NGS Soluções Genômicas: Pilar Drummond Sampaio Corrêa Mariani. FZEA-USP Pirassununga: Mirele Daiana Poleti; Raul Machado Neto; Ricardo Augusto Brassaloti; Ricardo Haddad; Rodrigo Tocantins Calado. FAMERP-SJRP: Cecília Artico Banho; Sandra Coccuzzo Sampaio; Svetoslav Nanev Slavov; Vagner Fonseca; Vincent Louis Viala |
| EPI_ISL_1966995 | UBS III DE PARIQUERA ACU PARIQUERA ACU | Instituto Butantan / ESALQ-USP (Piracicaba) | Antonio Jorge Martins; Bianca Cechetto Carlos. Mendelics: Bibiana Santos; Claudia Renata dos Santos Barros; Cíntia Bittar; David Schlesinger. Hemocentro Ribeirão Preto: Simone Kashima; Debora Botequio Moretti; Elaine Cristina Marqueze; Elaine Vieira dos Santos; Elisangela Chicaroni Mattos; Erika Freitas; Evandra Strazza Rodrigues; Felipe Allan da Silva da Costa; Flavia Aburjaile; Fábio Sossai Possebon; Guilherme Campos; Guilherme Targino Valente; Heidge Fukumasu. USP-Botucatu: Rejane Maria Tommasini Grotto; Helena Lage Ferreira; Instituto Butantan: Dimas Tadeu Covas; Jardelina de Souza Todao Bernardino; Jayme A. Souza-Neto; Jessika Cristina Chagas Lesbon; Jorge A. Petroli Marchesi; José Salvatore Leister Patané; João Paulo Kitajima; João Pessoa Araújo Jr.; Leila Sabrina Ullmann; Loyze Paola Oliveira de Lima; Luiz Aurelio de Campos Crispin. Centro de Genômica Funcional da ESALQ: Luiz Lehmann Coutinho; Luiz Carlos Junior de Alcantara; Lívia Sacchetto; Maisa C. Pereira Parra; Maria Carolina Elias; Marta Giovanetti; Marília Moraes; Maurício Lacerda Nogueira. Prefeitura de Sao Paulo: Melissa Palmieri.; Patricia Akemi Assato; Paula Rahal; Paulo Inacio da Costa; Rafael dos Santos Bezerra; Raquel de Lello Rocha Campos Cassano. NGS Soluções Genômicas: Pilar Drummond Sampaio Corrêa Mariani. FZEA-USP Pirassununga: Mirele Daiana Poleti; Raul Machado Neto; Ricardo Augusto Brassaloti; Ricardo Haddad; Rodrigo Tocantins Calado. FAMERP-SJRP: Cecília Artico Banho; Sandra Coccuzzo Sampaio; Svetoslav Nanev Slavov; Vagner Fonseca; Vincent Louis Viala |
| EPI_ISL_2345970 | UBS IRMA AGUEDA MARIA JAIME | Instituto Butantan | Antonio Jorge Martins; Claudia Renata dos Santos Barros; David Schlesinger; Debora Botequio Moretti; Dimas Tadeu Covas; Elaine Cristina Marqueze; Elaine Vieira Santos; Evandra Strazza Rodrigues; Heidge Fukumasu; Jayme Augusto de Souza-Neto; José Salvatore Leister Patané; Luiz Alcantara; Luiz Lehmann Coutinho; Maria Carolina Elias; Maurício Lacerda Nogueira; Rafael dos Santos Bezerra; Raul Machado Neto; Rejane Maria Tommasini Grotto; Ricardo Haddad; Sandra Coccuzzo Sampaio Vessoni; Simone Kashima; Svetoslav Nanev Slavov; Vincent Louis Viala |
| EPI_ISL_2345982 | UBS JARDIM ACACIO | Instituto Butantan | Antonio Jorge Martins; Claudia Renata dos Santos Barros; David Schlesinger; Debora Botequio Moretti; Dimas Tadeu Covas; Elaine Cristina Marqueze; Elaine Vieira Santos; Evandra Strazza Rodrigues; Heidge Fukumasu; Jayme Augusto de Souza-Neto; José Salvatore Leister Patané; Luiz Alcantara; Luiz Lehmann Coutinho; Maria Carolina Elias; Maurício Lacerda Nogueira; Rafael dos Santos Bezerra; Raul Machado Neto; Rejane Maria Tommasini Grotto; Ricardo Haddad; Sandra Coccuzzo Sampaio Vessoni; Simone Kashima; Svetoslav Nanev Slavov; Vincent Louis Viala |
| EPI_ISL_2346025, EPI_ISL_2346026, EPI_ISL_2346031, EPI_ISL_2346034, EPI_ISL_2346039, EPI_ISL_2346042, EPI_ISL_2346044, EPI_ISL_2346047 | | | |
| see above | UBS JARDIM CAIUBY | Instituto Butantan | Antonio Jorge Martins; Claudia Renata dos Santos Barros; David Schlesinger; Debora Botequio Moretti; Dimas Tadeu Covas; Elaine Cristina Marqueze; Elaine Vieira Santos; Evandra Strazza Rodrigues; Heidge Fukumasu; Jayme Augusto de Souza-Neto; José Salvatore Leister Patané; Luiz Alcantara; Luiz Lehmann Coutinho; Maria Carolina Elias; Maurício Lacerda Nogueira; Rafael dos Santos Bezerra; Raul Machado Neto; Rejane Maria Tommasini Grotto; Ricardo Haddad; Sandra Coccuzzo Sampaio Vessoni; Simone Kashima; Svetoslav Nanev Slavov; Vincent Louis Viala |
| EPI_ISL_2346017, EPI_ISL_2346018, EPI_ISL_2346030, EPI_ISL_2346041 | UBS JARDIM MARAGOGIPE | Instituto Butantan | Antonio Jorge Martins; Claudia Renata dos Santos Barros; David Schlesinger; Debora Botequio Moretti; Dimas Tadeu Covas; Elaine Cristina Marqueze; Elaine Vieira Santos; Evandra Strazza Rodrigues; Heidge Fukumasu; Jayme Augusto de Souza-Neto; José Salvatore Leister Patané; Luiz Alcantara; Luiz Lehmann Coutinho; Maria Carolina Elias; Maurício Lacerda Nogueira; Rafael dos Santos Bezerra; Raul Machado Neto; Rejane Maria Tommasini Grotto; Ricardo Haddad; Sandra Coccuzzo Sampaio Vessoni; Simone Kashima; Svetoslav Nanev Slavov; Vincent Louis Viala |
| EPI_ISL_2345985, EPI_ISL_2345986, EPI_ISL_2345987, EPI_ISL_2345989, EPI_ISL_2345994 | UBS JARDIM ODETE | Instituto Butantan | Antonio Jorge Martins; Claudia Renata dos Santos Barros; David Schlesinger; Debora Botequio Moretti; Dimas Tadeu Covas; Elaine Cristina Marqueze; Elaine Vieira Santos; Evandra Strazza Rodrigues; Heidge Fukumasu; Jayme Augusto de Souza-Neto; José Salvatore Leister Patané; Luiz Alcantara; Luiz Lehmann Coutinho; Maria Carolina Elias; Maurício Lacerda Nogueira; Rafael dos Santos Bezerra; Raul Machado Neto; Rejane Maria Tommasini Grotto; Ricardo Haddad; Sandra Coccuzzo Sampaio Vessoni; Simone Kashima; Svetoslav Nanev Slavov; Vincent Louis Viala |
| EPI_ISL_2345993, EPI_ISL_2346023 | UBS JARDIM PAINEIRA | Instituto Butantan | Antonio Jorge Martins; Claudia Renata dos Santos Barros; David Schlesinger; Debora Botequio Moretti; Dimas Tadeu Covas; Elaine Cristina Marqueze; Elaine Vieira Santos; Evandra Strazza Rodrigues; Heidge Fukumasu; Jayme Augusto de Souza-Neto; José Salvatore Leister Patané; Luiz Alcantara; Luiz Lehmann Coutinho; Maria Carolina Elias; Maurício Lacerda Nogueira; Rafael dos Santos Bezerra; Raul Machado Neto; Rejane Maria Tommasini Grotto; Ricardo Haddad; Sandra Coccuzzo Sampaio Vessoni; Simone Kashima; Svetoslav Nanev Slavov; Vincent Louis Viala |
| EPI_ISL_2345976 | UBS JARDIM PRIMAVERA | Instituto Butantan | Antonio Jorge Martins; Claudia Renata dos Santos Barros; David Schlesinger; Debora Botequio Moretti; Dimas Tadeu Covas; Elaine Cristina Marqueze; Elaine Vieira Santos; Evandra Strazza Rodrigues; Heidge Fukumasu; Jayme Augusto de Souza-Neto; José Salvatore Leister Patané; Luiz Alcantara; Luiz Lehmann Coutinho; Maria Carolina Elias; Maurício Lacerda Nogueira; Rafael dos Santos Bezerra; Raul Machado Neto; Rejane Maria Tommasini Grotto; Ricardo Haddad; Sandra Coccuzzo Sampaio Vessoni; Simone Kashima; Svetoslav Nanev Slavov; Vincent Louis Viala |
| EPI_ISL_1966918 | UBS JARDIM VITORINO IPERO | Instituto Butantan / ESALQ-USP (Piracicaba) | Antonio Jorge Martins; Bianca Cechetto Carlos. Mendelics: Bibiana Santos; Claudia Renata dos Santos Barros; Cíntia Bittar; David Schlesinger. Hemocentro Ribeirão Preto: Simone Kashima; Debora Botequio Moretti; Elaine Cristina Marqueze; Elaine Vieira dos Santos; Elisangela Chicaroni Mattos; Erika Freitas; Evandra Strazza Rodrigues; Felipe Allan da Silva da Costa; Flavia Aburjaile; Fábio Sossai Possebon; Guilherme Campos; Guilherme Targino Valente; Heidge Fukumasu. USP-Botucatu: Rejane Maria Tommasini Grotto; Helena Lage Ferreira; Instituto Butantan: Dimas Tadeu Covas; Jardelina de Souza Todao Bernardino; Jayme A. Souza-Neto; Jessika Cristina Chagas Lesbon; Jorge A. Petroli Marchesi; José Salvatore Leister Patané; João Paulo Kitajima; João Pessoa Araújo Jr.; Leila Sabrina Ullmann; Loyze Paola Oliveira de Lima; Luiz Aurelio de Campos Crispin. Centro de Genômica Funcional da ESALQ: Luiz Lehmann Coutinho; Luiz Carlos Junior de Alcantara; Lívia Sacchetto; Maisa C. Pereira Parra; Maria Carolina Elias; Marta Giovanetti; Marília Moraes; Maurício Lacerda Nogueira. Prefeitura de Sao Paulo: Melissa Palmieri.; Patricia Akemi Assato; Paula Rahal; Paulo Inacio da Costa; Rafael dos Santos Bezerra; Raquel de Lello Rocha Campos Cassano. NGS Soluções Genômicas: Pilar Drummond Sampaio Corrêa Mariani. FZEA-USP Pirassununga: Mirele Daiana Poleti; Raul Machado Neto; Ricardo Augusto Brassaloti; Ricardo Haddad; Rodrigo Tocantins Calado. FAMERP-SJRP: Cecília Artico Banho; Sandra Coccuzzo Sampaio; Svetoslav Nanev Slavov; Vagner Fonseca; Vincent Louis Viala |
| EPI_ISL_2345981 | UBS JD PRESIDENTE DUTRA | Instituto Butantan | Antonio Jorge Martins; Claudia Renata dos Santos Barros; David Schlesinger; Debora Botequio Moretti; Dimas Tadeu Covas; Elaine Cristina Marqueze; Elaine Vieira Santos; Evandra Strazza Rodrigues; Heidge Fukumasu; Jayme Augusto de Souza-Neto; José Salvatore Leister Patané; Luiz Alcantara; Luiz Lehmann Coutinho; Maria Carolina Elias; Maurício Lacerda Nogueira; Rafael dos Santos Bezerra; Raul Machado Neto; Rejane Maria Tommasini Grotto; Ricardo Haddad; Sandra Coccuzzo Sampaio Vessoni; Simone Kashima; Svetoslav Nanev Slavov; Vincent Louis Viala |
| EPI_ISL_2614584 | UBS Jardim Jovaia | Instituto Adolfo Lutz, Interdiciplinary Procedures Center, Strategic Laboratory | Caio Vinicius Dias Lopes; Claudia Regina Gonçalves; Claudio Tavares Sacchi; Erica Valessa Ramos Gomes; Karoline Rodrigues Campos; Leonardo Jose Tadeu de Araujo |
| EPI_ISL_2345983, EPI_ISL_2346048 | UBS MARIA PIA DE OLIVEIRA | Instituto Butantan | Antonio Jorge Martins; Claudia Renata dos Santos Barros; David Schlesinger; Debora Botequio Moretti; Dimas Tadeu Covas; Elaine Cristina Marqueze; Elaine Vieira Santos; Evandra Strazza Rodrigues; Heidge Fukumasu; Jayme Augusto de Souza-Neto; José Salvatore Leister Patané; Luiz Alcantara; Luiz Lehmann Coutinho; Maria Carolina Elias; Maurício Lacerda Nogueira; Rafael dos Santos Bezerra; Raul Machado Neto; Rejane Maria Tommasini Grotto; Ricardo Haddad; Sandra Coccuzzo Sampaio Vessoni; Simone Kashima; Svetoslav Nanev Slavov; Vincent Louis Viala |
| EPI_ISL_2345977 | UBS MARINOPOLIS | Instituto Butantan | Antonio Jorge Martins; Claudia Renata dos Santos Barros; David Schlesinger; Debora Botequio Moretti; Dimas Tadeu Covas; Elaine Cristina Marqueze; Elaine Vieira Santos; Evandra Strazza Rodrigues; Heidge Fukumasu; Jayme Augusto de Souza-Neto; José Salvatore Leister Patané; Luiz Alcantara; Luiz Lehmann Coutinho; Maria Carolina Elias; Maurício Lacerda Nogueira; Rafael dos Santos Bezerra; Raul Machado Neto; Rejane Maria Tommasini Grotto; Ricardo Haddad; Sandra Coccuzzo Sampaio Vessoni; Simone Kashima; Svetoslav Nanev Slavov; Vincent Louis Viala |
| EPI_ISL_2346028, EPI_ISL_2346040 | UBS MORRO BRANCO | Instituto Butantan | Antonio Jorge Martins; Claudia Renata dos Santos Barros; David Schlesinger; Debora Botequio Moretti; Dimas Tadeu Covas; Elaine Cristina Marqueze; Elaine Vieira Santos; Evandra Strazza Rodrigues; Heidge Fukumasu; Jayme Augusto de Souza-Neto; José Salvatore Leister Patané; Luiz Alcantara; Luiz Lehmann Coutinho; Maria Carolina Elias; Maurício Lacerda Nogueira; Rafael dos Santos Bezerra; Raul Machado Neto; Rejane Maria Tommasini Grotto; Ricardo Haddad; Sandra Coccuzzo Sampaio Vessoni; Simone Kashima; Svetoslav Nanev Slavov; Vincent Louis Viala |
| EPI_ISL_2346027, | UBS PARQUE MARENGO | Instituto Butantan | Antonio Jorge Martins; Claudia Renata dos Santos Barros; David Schlesinger; Debora Botequio Moretti; Dimas Tadeu Covas; Elaine Cristina Marqueze; Elaine Vieira Santos; Evandra Strazza Rodrigues; Heidge Fukumasu; Jayme Augusto de Souza-Neto; José Salvatore Leister Patané; Luiz Alcantara; Luiz Lehmann Coutinho; |

| | | | |
|---|---|---|---|
| EPI_ISL_2346033 | | | Maria Carolina Elias; Maurício Lacerda Nogueira; Rafael dos Santos Bezerra; Raul Machado Neto; Rejane Maria Tommasini Grotto; Ricardo Haddad; Sandra Coccuzzo Sampaio Vessoni; Simone Kashima; Svetoslav Nanev Slavov; Vincent Louis Viala |
| EPI_ISL_2346015 | UBS RECANTO MONICA | Instituto Butantan | Antonio Jorge Martins; Claudia Renata dos Santos Barros; David Schlesinger; Debora Botequio Moretti; Dimas Tadeu Covas; Elaine Cristina Marqueze; Elaine Vieira Santos; Evandra Strazza Rodrigues; Heidge Fukumasu; Jayme Augusto de Souza-Neto; José Salvatore Leister Patané; Luiz Alcantara; Luiz Lehmann Coutinho; Maria Carolina Elias; Maurício Lacerda Nogueira; Rafael dos Santos Bezerra; Raul Machado Neto; Rejane Maria Tommasini Grotto; Ricardo Haddad; Sandra Coccuzzo Sampaio Vessoni; Simone Kashima; Svetoslav Nanev Slavov; Vincent Louis Viala |
| EPI_ISL_2345967, EPI_ISL_2345969, EPI_ISL_2345971 | UBS SANTA MARIA | Instituto Butantan | Antonio Jorge Martins; Claudia Renata dos Santos Barros; David Schlesinger; Debora Botequio Moretti; Dimas Tadeu Covas; Elaine Cristina Marqueze; Elaine Vieira Santos; Evandra Strazza Rodrigues; Heidge Fukumasu; Jayme Augusto de Souza-Neto; José Salvatore Leister Patané; Luiz Alcantara; Luiz Lehmann Coutinho; Maria Carolina Elias; Maurício Lacerda Nogueira; Rafael dos Santos Bezerra; Raul Machado Neto; Rejane Maria Tommasini Grotto; Ricardo Haddad; Sandra Coccuzzo Sampaio Vessoni; Simone Kashima; Svetoslav Nanev Slavov; Vincent Louis Viala |
| EPI_ISL_1966847, EPI_ISL_1966848, EPI_ISL_1966849, EPI_ISL_1966850, EPI_ISL_1966851, EPI_ISL_1966852 | UBS SAO JUDAS | Instituto Butantan / ESALQ-USP (Piracicaba) | Antonio Jorge Martins; Bianca Cechetto Carlos. Mendelics: Bibiana Santos; Claudia Renata dos Santos Barros; Cíntia Bittar; David Schlesinger. Hemocentro Ribeirão Preto: Simone Kashima; Debora Botequio Moretti; Elaine Cristina Marqueze; Elaine Vieira dos Santos; Elisangela Chicaroni Mattos; Erika Freitas; Evandra Strazza Rodrigues; Felipe Allan da Silva da Costa; Flavia Aburjaile; Fábio Sossai Possebon; Guilherme Campos; Guilherme Targino Valente; Heidge Fukumasu. USP-Botucatu: Rejane Maria Tommasini Grotto; Helena Lage Ferreira; Instituto Butantan: Dimas Tadeu Covas; Jardelina de Souza Todao Bernardino; Jayme A. Souza-Neto; Jessika Cristina Chagas Lesbon; Jorge A. Petroli Marchesi; José Salvatore Leister Patané; João Paulo Kitajima; João Pessoa Araújo Jr.; Leila Sabrina Ullmann; Loyze Paola Oliveira de Lima; Luiz Aurelio de Campos Crispin. Centro de Genômica Funcional da ESALQ: Luiz Lehmann Coutinho; Luiz Carlos Junior de Alcantara; Lívia Sacchetto; Maisa C. Pereira Parra; Maria Carolina Elias; Marta Giovanetti; Marília Moraes; Maurício Lacerda Nogueira. Prefeitura de Sao Paulo: Melissa Palmieri.; Patricia Akemi Assato; Paula Rahal; Paulo Inacio da Costa; Rafael dos Santos Bezerra; Raquel de Lello Rocha Campos Cassano. NGS Soluções Genômicas: Pilar Drummond Sampaio Corrêa Mariani. FZEA-USP Pirassununga: Mirele Daiana Poleti; Raul Machado Neto; Ricardo Augusto Brassaloti; Ricardo Haddad; Rodrigo Tocantins Calado. FAMERP-SJRP: Cecília Artico Banho; Sandra Coccuzzo Sampaio; Svetoslav Nanev Slavov; Vagner Fonseca; Vincent Louis Viala |
| EPI_ISL_3419561, EPI_ISL_3419562, EPI_ISL_3419566, EPI_ISL_3419569 | UC Davis Genome Center and Healthy Davis Together | UC Davis Genome Center | UC Davis |
| EPI_ISL_1675323, EPI_ISL_1675324, EPI_ISL_1675325 | UDEA | Universidad Nacional de Colombia - Laboratorio Genómico One Health | Andres F. Cardona-Rios; Carlos Franco-Muñoz; Daniel O. Maldonado-Perez; Diego A. Álvarez-Díaz; Hector Alejandro Ruiz-Moreno; Idabely Betancur Ortiz; Jorge E. Osorio; Juan P. Hernandez-Ortiz; Karl A Ciuoderis; Katherine Laiton-Donato; Laura Silvana Perez; Lina M. Hurtado; Marcela Mercado-Reyes; Maria Angélica Maya; Maria Stella López; Rita Almanza Payares; Sandra Ines Cano; Simón Villegas Velásquez |
| EPI_ISL_2365867 | ULSS 3 Venezia | Istituto Zooprofilattico Sperimentale delle Venezie | Adelaide Milani; Alessia Schivo; Alice Fusaro; Ambra Pastori; Annalisa Salviato; Antonia Ricci; Calogero Terregino; Edoardo Giussani; Elisa Palumbo; Erika Giorgia Quaranta; Isabella Monne; Luca Tassoni |
| EPI_ISL_2448413 | ULSS 6 Euganea | Istituto Zooprofilattico Sperimentale delle Venezie | Adelaide Milani; Alessia Schivo; Alice Fusaro; Ambra Pastori; Annalisa Salviato; Antonia Ricci; Calogero Terregino; Edoardo Giussani; Elisa Palumbo; Erika Giorgia Quaranta; Isabella Monne; Luca Tassoni |
| EPI_ISL_1712601, EPI_ISL_1712613 | UMC Groningen, Clinical Virology, Department of Medical Microbiology and Infection Prevention | UMC Groningen, Clinical Virology, Department of Medical Microbiology and Infection Prevention | Alexander Friedrich; Coretta Van Leer-Buter; Erley Lizarazo-Forero; Hubert Niesters; Lilli Gard; Marjolein Knoester; Monika Fliss; Sigrid Rosema; Xuewei Zhou |
| EPI_ISL_3409095, EPI_ISL_3409142, EPI_ISL_3409287, EPI_ISL_3409300, EPI_ISL_3409305, EPI_ISL_3409369, EPI_ISL_3409370, EPI_ISL_3409407, EPI_ISL_3409408, EPI_ISL_3409789, EPI_ISL_3410077, EPI_ISL_3410083 | | | |
| see above | UMass Memorial Medical Center | Center for Microbiome Research | Doyle V. Ward; Karl J. Simon; Richard T. Ellison III |
| EPI_ISL_1938251, EPI_ISL_1938256 | UNC Charlotte COVID-19 Testing Lab | UNC Charlotte Environmental Monitoring Laboratory | Angelica Martins; Cynthia Gibas; Jannatul Ferdous; Jessica Schlueter; Kevin Lambirth; Visva Barua |
| EPI_ISL_1854880, EPI_ISL_1854884 | UNC-CH COVID Surveillance Lab | Jeremy Wang | Alexander Rubinsteyn; Amir Barzin; Amy James Loftis; Corbin Jones; Jeremy Wang; Melissa Miller; Susan Fiscus |
| EPI_ISL_5530089 | UNIDADE BASICA DE SAUDE AFONSO HENRIQUE DE A MACHADO | Analytical Competence Molecular Epidemiology Lab/ACME, Oswaldo Cruz Foundation, Ceara (FIOCRUZ CE) | Carlos Leonardo de Aragao Araujo; Cecilia Leite Costa & Eduardo Ruback dos Santos on behalf of COVID-19 FIOCRUZ Genomic Network; Cleber Furtado Aksenen; Fabio Miyajima; Fernando Braga Stehling; Francisco Eder de Moura Lopes; Igor Oliveira Duarte; Jamille Maria Mendes Bezerra; Joaquim Cesar do Nascimento Sousa Junior; Pedro Miguel Carneiro Jeronimo; Suzana Porto Almeida; Thais Ferreira de Oliveira; Thais de Oliveira Costa; Ticiane Cavalcante de Souza; Veridiana Pessoa Miyajima |
| EPI_ISL_5530152 | UNIDADE BASICA DE SAUDE DO CENTRO | Analytical Competence Molecular Epidemiology Lab/ACME, Oswaldo Cruz Foundation, Ceara (FIOCRUZ CE) | Carlos Leonardo de Aragao Araujo; Cecilia Leite Costa & Eduardo Ruback dos Santos on behalf of COVID-19 FIOCRUZ Genomic Network; Cleber Furtado Aksenen; Fabio Miyajima; Fernando Braga Stehling; Francisco Eder de Moura Lopes; Igor Oliveira Duarte; Jamille Maria Mendes Bezerra; Joaquim Cesar do Nascimento Sousa Junior; Pedro Miguel Carneiro Jeronimo; Suzana Porto Almeida; Thais Ferreira de Oliveira; Thais de Oliveira Costa; Ticiane Cavalcante de Souza; Veridiana Pessoa Miyajima |
| EPI_ISL_5530090, EPI_ISL_5603287 | UNIDADE BASICA DE SAUDE LUIS PRATA GIRAO | Analytical Competence Molecular Epidemiology Lab/ACME, Oswaldo Cruz Foundation, Ceara (FIOCRUZ CE) | Carlos Leonardo de Aragao Araujo; Cecilia Leite Costa & Eduardo Ruback dos Santos on behalf of COVID-19 FIOCRUZ Genomic Network; Cleber Furtado Aksenen; Fabio Miyajima; Fernando Braga Stehling; Francisco Eder de Moura Lopes; Igor Oliveira Duarte; Jamille Maria Mendes Bezerra; Joaquim Cesar do Nascimento Sousa Junior; Pedro Miguel Carneiro Jeronimo; Suzana Porto Almeida; Thais Ferreira de Oliveira; Thais de Oliveira Costa; Ticiane Cavalcante de Souza; Veridiana Pessoa Miyajima |
| EPI_ISL_2345988 | UNIDADE DE PRONTO ATENDIMENTO DE VARZEA PAULISTA UPA II | Instituto Butantan | Antonio Jorge Martins; Claudia Renata dos Santos Barros; David Schlesinger; Debora Botequio Moretti; Dimas Tadeu Covas; Elaine Cristina Marqueze; Elaine Vieira Santos; Evandra Strazza Rodrigues; Heidge Fukumasu; Jayme Augusto de Souza-Neto; José Salvatore Leister Patané; Luiz Alcantara; Luiz Lehmann Coutinho; Maria Carolina Elias; Maurício Lacerda Nogueira; Rafael dos Santos Bezerra; Raul Machado Neto; Rejane Maria Tommasini Grotto; Ricardo Haddad; Sandra Coccuzzo Sampaio Vessoni; Simone Kashima; Svetoslav Nanev Slavov; Vincent Louis Viala |
| EPI_ISL_1966721 | UNIDADE DE PRONTO ATENDIMENTO DE VARZEA PAULISTA UPA II | Instituto Butantan / Mendelics | Antonio Jorge Martins; Bianca Cechetto Carlos. Mendelics: Bibiana Santos; Claudia Renata dos Santos Barros; Cíntia Bittar; David Schlesinger. Hemocentro Ribeirão Preto: Simone Kashima; Debora Botequio Moretti; Elaine Cristina Marqueze; Elaine Vieira dos Santos; Elisangela Chicaroni Mattos; Erika Freitas; Evandra Strazza Rodrigues; Felipe Allan da Silva da Costa; Flavia Aburjaile; Fábio Sossai Possebon; Guilherme Campos; Guilherme Targino Valente; Heidge Fukumasu. USP-Botucatu: Rejane Maria Tommasini Grotto; Helena Lage Ferreira; Instituto Butantan: Dimas Tadeu Covas; Jardelina de Souza Todao Bernardino; Jayme A. Souza-Neto; Jessika Cristina Chagas Lesbon; Jorge A. Petroli Marchesi; José Salvatore Leister Patané; João Paulo Kitajima; João Pessoa Araújo Jr.; Leila Sabrina Ullmann; Loyze Paola Oliveira de Lima; Luiz Aurelio de Campos Crispin. Centro de Genômica Funcional da ESALQ: Luiz Lehmann Coutinho; Luiz Carlos Junior de Alcantara; Lívia Sacchetto; Maisa C. Pereira Parra; Maria Carolina Elias; Marta Giovanetti; Marília Moraes; Maurício Lacerda Nogueira. Prefeitura de Sao Paulo: Melissa Palmieri.; Patricia Akemi Assato; Paula Rahal; Paulo Inacio da Costa; Rafael dos Santos Bezerra; Raquel de Lello Rocha Campos Cassano. NGS Soluções Genômicas: Pilar Drummond Sampaio Corrêa Mariani. FZEA-USP Pirassununga: Mirele Daiana Poleti; Raul Machado Neto; Ricardo Augusto Brassaloti; Ricardo Haddad; Rodrigo Tocantins Calado. FAMERP-SJRP: Cecília Artico Banho; Sandra Coccuzzo Sampaio; Svetoslav Nanev Slavov; Vagner Fonseca; Vincent Louis Viala |
| EPI_ISL_1966708 | UNIDADE DE PRONTO ATENDIMENTO UPA | Instituto Butantan / Mendelics | Antonio Jorge Martins; Bianca Cechetto Carlos. Mendelics: Bibiana Santos; Claudia Renata dos Santos Barros; Cíntia Bittar; David Schlesinger. Hemocentro Ribeirão Preto: Simone Kashima; Debora Botequio Moretti; Elaine Cristina Marqueze; Elaine Vieira dos Santos; Elisangela Chicaroni Mattos; Erika Freitas; Evandra Strazza Rodrigues; Felipe Allan da Silva da Costa; Flavia Aburjaile; Fábio Sossai Possebon; Guilherme Campos; Guilherme Targino Valente; Heidge Fukumasu. USP-Botucatu: Rejane Maria Tommasini Grotto; Helena Lage Ferreira; Instituto Butantan: Dimas Tadeu Covas; Jardelina de Souza Todao Bernardino; Jayme A. Souza-Neto; Jessika Cristina Chagas Lesbon; Jorge A. Petroli Marchesi; José Salvatore Leister Patané; João Paulo Kitajima; João Pessoa Araújo Jr.; Leila Sabrina Ullmann; Loyze Paola Oliveira de Lima; Luiz Aurelio de Campos Crispin. Centro de Genômica Funcional da ESALQ: Luiz Lehmann Coutinho; Luiz Carlos Junior de Alcantara; Lívia Sacchetto; Maisa C. Pereira Parra; Maria Carolina Elias; Marta Giovanetti; Marília Moraes; Maurício Lacerda Nogueira. Prefeitura de Sao Paulo: Melissa Palmieri.; Patricia Akemi Assato; Paula Rahal; Paulo Inacio da Costa; Rafael dos Santos Bezerra; Raquel de Lello Rocha Campos Cassano. NGS Soluções Genômicas: Pilar Drummond Sampaio Corrêa Mariani. FZEA-USP Pirassununga: Mirele Daiana Poleti; Raul Machado Neto; Ricardo Augusto Brassaloti; Ricardo Haddad; Rodrigo Tocantins Calado. FAMERP-SJRP: Cecília Artico Banho; Sandra Coccuzzo Sampaio; Svetoslav Nanev Slavov; Vagner Fonseca; Vincent Louis Viala |
| EPI_ISL_1967052, EPI_ISL_1967053, EPI_ISL_1967054, EPI_ISL_1967056, EPI_ISL_1967057 | UNIDADE DE VIGILANCIA EM SAUDE | Instituto Butantan / ESALQ-USP (Piracicaba) | Antonio Jorge Martins; Bianca Cechetto Carlos. Mendelics: Bibiana Santos; Claudia Renata dos Santos Barros; Cíntia Bittar; David Schlesinger. Hemocentro Ribeirão Preto: Simone Kashima; Debora Botequio Moretti; Elaine Cristina Marqueze; Elaine Vieira dos Santos; Elisangela Chicaroni Mattos; Erika Freitas; Evandra Strazza Rodrigues; Felipe Allan da Silva da Costa; Flavia Aburjaile; Fábio Sossai Possebon; Guilherme Campos; Guilherme Targino Valente; Heidge Fukumasu. USP-Botucatu: Rejane Maria Tommasini Grotto; Helena Lage Ferreira; Instituto Butantan: Dimas Tadeu Covas; Jardelina de Souza Todao Bernardino; Jayme A. Souza-Neto; Jessika Cristina Chagas Lesbon; Jorge A. Petroli Marchesi; José Salvatore Leister Patané; João Paulo Kitajima; João Pessoa Araújo Jr.; Leila Sabrina Ullmann; Loyze Paola Oliveira de Lima; Luiz Aurelio de Campos Crispin. Centro de Genômica Funcional da ESALQ: Luiz Lehmann Coutinho; Luiz Carlos Junior de Alcantara; Lívia Sacchetto; Maisa C. Pereira Parra; Maria Carolina Elias; Marta Giovanetti; Marília Moraes; Maurício Lacerda Nogueira. Prefeitura de Sao Paulo: Melissa Palmieri.; Patricia Akemi Assato; Paula Rahal; Paulo Inacio da Costa; Rafael dos Santos Bezerra; Raquel de Lello Rocha Campos Cassano. NGS Soluções Genômicas: Pilar Drummond Sampaio Corrêa Mariani. FZEA-USP Pirassununga: Mirele Daiana Poleti; Raul Machado Neto; Ricardo Augusto Brassaloti; Ricardo Haddad; Rodrigo Tocantins Calado. FAMERP-SJRP: Cecília Artico Banho; Sandra Coccuzzo Sampaio; Svetoslav Nanev Slavov; Vagner Fonseca; Vincent Louis Viala |
| EPI_ISL_2209856 | UNIDADE DE VIGILANCIA EPIDEMIOLOGICA | Instituto Butantan | Antonio Jorge Martins; Claudia Renata dos Santos Barros; David Schlesinger; Debora Botequio Moretti; Dimas Tadeu Covas; Elaine Cristina Marqueze; Elaine Vieira Santos; Evandra Strazza Rodrigues; Heidge Fukumasu; Jayme Augusto de Souza-Neto; José Salvatore Leister Patané; Luiz Alcantara; Luiz Lehmann Coutinho; Maria Carolina Elias; Maurício Lacerda Nogueira; Rafael dos Santos Bezerra; Raul Machado Neto; Rejane Maria Tommasini Grotto; Ricardo Haddad; Sandra Coccuzzo Sampaio Vessoni; Simone Kashima; Svetoslav Nanev Slavov; Vincent Louis Viala. |
| EPI_ISL_2209129, EPI_ISL_2209261 | UNIDADE DE VIGILANCIA EPIDEMIOLOGICA | Instituto Butantan / FZEA-USP-Pirassununga | Antonio Jorge Martins; Claudia Renata dos Santos Barros; David Schlesinger; Debora Botequio Moretti; Dimas Tadeu Covas; Elaine Cristina Marqueze; Elaine Vieira Santos; Evandra Strazza Rodrigues; Heidge Fukumasu; Jayme Augusto de Souza-Neto; José Salvatore Leister Patané; Luiz Alcantara; Luiz Lehmann Coutinho; Maria Carolina Elias; Maurício Lacerda Nogueira; Rafael dos Santos Bezerra; Raul Machado Neto; Rejane Maria Tommasini Grotto; Ricardo Haddad; Sandra Coccuzzo Sampaio Vessoni; Simone Kashima; Svetoslav Nanev Slavov; Vincent Louis Viala |
| EPI_ISL_1966915 | UNIDADE MISTA DE IGUAPE | Instituto Butantan / ESALQ-USP (Piracicaba) | Antonio Jorge Martins; Bianca Cechetto Carlos. Mendelics: Bibiana Santos; Claudia Renata dos Santos Barros; Cíntia Bittar; David Schlesinger. Hemocentro Ribeirão Preto: Simone Kashima; Debora Botequio Moretti; Elaine Cristina Marqueze; Elaine Vieira dos Santos; Elisangela Chicaroni Mattos; Erika Freitas; Evandra Strazza Rodrigues; Felipe Allan da Silva da Costa; Flavia Aburjaile; Fábio Sossai Possebon; Guilherme Campos; Guilherme Targino Valente; Heidge Fukumasu. USP-Botucatu: Rejane Maria Tommasini Grotto; Helena Lage Ferreira; Instituto Butantan: Dimas Tadeu Covas; Jardelina de Souza Todao Bernardino; Jayme A. Souza-Neto; Jessika Cristina Chagas Lesbon; Jorge A. Petroli Marchesi; José Salvatore Leister Patané; João Paulo Kitajima; João Pessoa Araújo Jr.; Leila Sabrina Ullmann; Loyze Paola Oliveira de Lima; Luiz Aurelio de Campos Crispin. Centro de Genômica Funcional da ESALQ: Luiz Lehmann Coutinho; Luiz Carlos Junior de Alcantara; Lívia Sacchetto; Maisa C. Pereira Parra; Maria Carolina Elias; Marta Giovanetti; Marília Moraes; Maurício Lacerda Nogueira. Prefeitura de Sao Paulo: Melissa Palmieri.; Patricia Akemi Assato; Paula Rahal; Paulo Inacio da Costa; Rafael dos Santos Bezerra; Raquel de Lello Rocha Campos Cassano. NGS Soluções Genômicas: Pilar Drummond Sampaio Corrêa Mariani. FZEA-USP Pirassununga: Mirele Daiana Poleti; Raul Machado Neto; Ricardo Augusto Brassaloti; Ricardo Haddad; Rodrigo Tocantins Calado. FAMERP-SJRP: Cecília Artico Banho; Sandra Coccuzzo Sampaio; Svetoslav Nanev Slavov; Vagner Fonseca; Vincent Louis Viala |
| EPI_ISL_1966863, EPI_ISL_1966864, EPI_ISL_1966865, EPI_ISL_1966866, EPI_ISL_1966867 | UNIDADE MISTA DE SAUDE CAPELA DO ALTO | Instituto Butantan / ESALQ-USP (Piracicaba) | Antonio Jorge Martins; Bianca Cechetto Carlos. Mendelics: Bibiana Santos; Claudia Renata dos Santos Barros; Cíntia Bittar; David Schlesinger. Hemocentro Ribeirão Preto: Simone Kashima; Debora Botequio Moretti; Elaine Cristina Marqueze; Elaine Vieira dos Santos; Elisangela Chicaroni Mattos; Erika Freitas; Evandra Strazza Rodrigues; Felipe Allan da Silva da Costa; Flavia Aburjaile; Fábio Sossai Possebon; Guilherme Campos; Guilherme Targino Valente; Heidge Fukumasu. USP-Botucatu: Rejane Maria Tommasini Grotto; Helena Lage Ferreira; Instituto Butantan: Dimas Tadeu Covas; Jardelina de Souza Todao Bernardino; Jayme A. Souza-Neto; Jessika Cristina Chagas Lesbon; Jorge A. Petroli Marchesi; José Salvatore Leister Patané; João Paulo Kitajima; João Pessoa Araújo Jr.; Leila Sabrina Ullmann; Loyze Paola Oliveira de Lima; Luiz Aurelio de Campos Crispin. Centro de Genômica Funcional da ESALQ: Luiz Lehmann Coutinho; Luiz Carlos Junior de Alcantara; Lívia Sacchetto; Maisa C. Pereira Parra; Maria Carolina Elias; Marta Giovanetti; Marília Moraes; Maurício Lacerda Nogueira. Prefeitura de Sao Paulo: Melissa Palmieri.; Patricia Akemi Assato; Paula Rahal; Paulo Inacio da Costa; Rafael dos Santos Bezerra; Raquel de Lello Rocha Campos Cassano. NGS Soluções Genômicas: Pilar Drummond Sampaio Corrêa Mariani. FZEA-USP Pirassununga: Mirele Daiana Poleti; Raul Machado Neto; Ricardo Augusto Brassaloti; Ricardo Haddad; Rodrigo Tocantins Calado. FAMERP-SJRP: Cecília Artico Banho; Sandra Coccuzzo Sampaio; Svetoslav Nanev Slavov; Vagner Fonseca; Vincent Louis Viala |
| EPI_ISL_1966986, EPI_ISL_1966987 | UNIDADE MISTA DE SAUDE DE ORIENTE | Instituto Butantan / ESALQ-USP (Piracicaba) | Antonio Jorge Martins; Bianca Cechetto Carlos. Mendelics: Bibiana Santos; Claudia Renata dos Santos Barros; Cíntia Bittar; David Schlesinger. Hemocentro Ribeirão Preto: Simone Kashima; Debora Botequio Moretti; Elaine Cristina Marqueze; Elaine Vieira dos Santos; Elisangela Chicaroni Mattos; Erika Freitas; Evandra Strazza Rodrigues; Felipe Allan da Silva da Costa; Flavia Aburjaile; Fábio Sossai Possebon; Guilherme Campos; Guilherme Targino Valente; Heidge Fukumasu. USP-Botucatu: Rejane Maria Tommasini Grotto; Helena Lage Ferreira; Instituto Butantan: Dimas Tadeu Covas; Jardelina de Souza Todao Bernardino; Jayme A. Souza-Neto; Jessika Cristina Chagas Lesbon; Jorge A. Petroli Marchesi; José Salvatore Leister Patané; João Paulo Kitajima; João Pessoa Araújo Jr.; Leila Sabrina Ullmann; Loyze Paola Oliveira de Lima; Luiz Aurelio de Campos Crispin. Centro de Genômica Funcional da ESALQ: Luiz Lehmann Coutinho; Luiz Carlos Junior de Alcantara; Lívia Sacchetto; Maisa C. Pereira Parra; Maria Carolina Elias; Marta Giovanetti; Marília Moraes; Maurício Lacerda Nogueira. Prefeitura de Sao Paulo: Melissa Palmieri.; Patricia Akemi Assato; Paula Rahal; Paulo Inacio da Costa; Rafael dos Santos Bezerra; Raquel de Lello Rocha Campos Cassano. NGS Soluções Genômicas: Pilar Drummond Sampaio Corrêa Mariani. FZEA-USP Pirassununga: Mirele Daiana Poleti; Raul Machado Neto; Ricardo Augusto Brassaloti; Ricardo Haddad; Rodrigo Tocantins Calado. FAMERP-SJRP: Cecília Artico Banho; Sandra Coccuzzo Sampaio; Svetoslav Nanev Slavov; Vagner Fonseca; Vincent Louis Viala |
| EPI_ISL_2344701 | UNIDADE MISTA OSVALDO P CAMARGO ELIAS FAUSTO | Instituto Butantan / FZEA-USP-Pirassununga | Antonio Jorge Martins; Claudia Renata dos Santos Barros; David Schlesinger; Debora Botequio Moretti; Dimas Tadeu Covas; Elaine Cristina Marqueze; Elaine Vieira Santos; Evandra Strazza Rodrigues; Heidge Fukumasu; Jayme Augusto de Souza-Neto; José Salvatore Leister Patané; Luiz Alcantara; Luiz Lehmann Coutinho; Maria Carolina Elias; Maurício Lacerda Nogueira; Rafael dos Santos Bezerra; Raul Machado Neto; Rejane Maria Tommasini Grotto; Ricardo Haddad; Sandra Coccuzzo Sampaio Vessoni; Simone Kashima; Svetoslav Nanev Slavov; Vincent Louis Viala |
| EPI_ISL_5529972 | UNIDADE REFERENCIA COVID 19 | Analytical Competence Molecular Epidemiology Lab/ACME, Oswaldo Cruz Foundation, Ceara (FIOCRUZ CE) | Carlos Leonardo de Aragao Araujo; Cecilia Leite Costa & Eduardo Ruback dos Santos on behalf of COVID-19 FIOCRUZ Genomic Network; Cleber Furtado Aksenen; Fabio Miyajima; Fernando Braga Stehling; Francisco Eder de Moura Lopes; Igor Oliveira Duarte; Jamille Maria Mendes Bezerra; Joaquim Cesar do Nascimento Sousa Junior; Pedro Miguel Carneiro Jeronimo; Suzana Porto Almeida; Thais Ferreira de Oliveira; Thais de Oliveira Costa; Ticiane Cavalcante de Souza; Veridiana Pessoa Miyajima |
| EPI_ISL_1966854, EPI_ISL_1966855, EPI_ISL_1966856, | UNIDADE SENTINELA COVID19 | Instituto Butantan / ESALQ-USP (Piracicaba) | Antonio Jorge Martins; Bianca Cechetto Carlos. Mendelics: Bibiana Santos; Claudia Renata dos Santos Barros; Cíntia Bittar; David Schlesinger. Hemocentro Ribeirão Preto: Simone Kashima; Debora Botequio Moretti; Elaine Cristina Marqueze; Elaine Vieira dos Santos; Elisangela Chicaroni Mattos; Erika Freitas; Evandra Strazza Rodrigues; Felipe Allan da Silva da Costa; Flavia Aburjaile; Fábio Sossai Possebon; Guilherme Campos; Guilherme Targino Valente; Heidge Fukumasu. USP-Botucatu: Rejane Maria Tommasini Grotto; Helena Lage Ferreira; Instituto Butantan: Dimas Tadeu Covas; Jardelina de Souza Todao Bernardino; Jayme A. Souza-Neto; Jessika Cristina Chagas Lesbon; Jorge A. Petroli Marchesi; José Salvatore Leister Patané; João Paulo Kitajima; João Pessoa Araújo Jr.; Leila Sabrina Ullmann; Loyze Paola Oliveira de Lima; Luiz Aurelio de Campos Crispin. Centro de Genômica Funcional da ESALQ: Luiz Lehmann Coutinho; Luiz Carlos Junior de |

| | | | |
|---|---|---|---|
| EPI_ISL_1966857, EPI_ISL_1966861 | | | Alcantara; Lívia Sacchetto; Maisa C. Pereira Parra; Maria Carolina Elias; Marta Giovanetti; Marília Moraes; Maurício Lacerda Nogueira. Prefeitura de Sao Paulo: Melissa Palmieri.; Patricia Akemi Assato; Paula Rahal; Paulo Inacio da Costa; Rafael dos Santos Bezerra; Raquel de Lello Rocha Campos Cassano. NGS Soluções Genômicas: Pilar Drummond Sampaio Corrêa Mariani. FZEA-USP Pirassununga: Mirele Daiana Poleti; Raul Machado Neto; Ricardo Augusto Brassaloti; Ricardo Haddad; Rodrigo Tocantins Calado. FAMERP-SJRP: Cecília Artico Banho; Sandra Coccuzzo Sampaio; Svetoslav Nanev Slavov; Vagner Fonseca; Vincent Louis Viala |
| EPI_ISL_1966859, EPI_ISL_1966860 | UNIDADE SENTINELA VILA APARECIDA | Instituto Butantan / ESALQ-USP (Piracicaba) | Antonio Jorge Martins; Bianca Cechetto Carlos. Mendelics: Bibiana Santos; Claudia Renata dos Santos Barros; Cíntia Bittar; David Schlesinger. Hemocentro Ribeirão Preto: Simone Kashima; Debora Botequio Moretti; Elaine Cristina Marqueze; Elaine Vieira dos Santos; Elisangela Chicaroni Mattos; Erika Freitas; Evandra Strazza Rodrigues; Felipe Allan da Silva da Costa; Flavia Aburjaile; Fábio Sossai Possebon; Guilherme Campos; Guilherme Targino Valente; Heidge Fukumasu. USP-Botucatu: Rejane Maria Tommasini Grotto; Helena Lage Ferreira; Instituto Butantan: Dimas Tadeu Covas; Jardelina de Souza Todao Bernardino; Jayme A. Souza-Neto; Jessika Cristina Chagas Lesbon; Jorge A. Petroli Marchesi; José Salvatore Leister Patané; João Paulo Kitajima; João Pessoa Araújo Jr.; Leila Sabrina Ullmann; Loyze Paola Oliveira de Lima; Luiz Aurelio de Campos Crispin. Centro de Genômica Funcional da ESALQ: Luiz Lehmann Coutinho; Luiz Carlos Junior de Alcantara; Lívia Sacchetto; Maisa C. Pereira Parra; Maria Carolina Elias; Marta Giovanetti; Marília Moraes; Maurício Lacerda Nogueira. Prefeitura de Sao Paulo: Melissa Palmieri.; Patricia Akemi Assato; Paula Rahal; Paulo Inacio da Costa; Rafael dos Santos Bezerra; Raquel de Lello Rocha Campos Cassano. NGS Soluções Genômicas: Pilar Drummond Sampaio Corrêa Mariani. FZEA-USP Pirassununga: Mirele Daiana Poleti; Raul Machado Neto; Ricardo Augusto Brassaloti; Ricardo Haddad; Rodrigo Tocantins Calado. FAMERP-SJRP: Cecília Artico Banho; Sandra Coccuzzo Sampaio; Svetoslav Nanev Slavov; Vagner Fonseca; Vincent Louis Viala |
| EPI_ISL_1960038, EPI_ISL_2136281, EPI_ISL_2136282, EPI_ISL_2136283, EPI_ISL_2136284, EPI_ISL_2136289, EPI_ISL_2136293, EPI_ISL_2136295, EPI_ISL_2136298, EPI_ISL_2136299, EPI_ISL_2136300, EPI_ISL_2136302, EPI_ISL_2136303, EPI_ISL_2136304, EPI_ISL_2136310, EPI_ISL_2136311, EPI_ISL_2136312 | | | |
| see above | UNIGEM | Universidad Nacional de Colombia - Laboratorio Genómico One Health | Andres F. Cardona-Rios; Carlos Franco-Muñoz; Carolina Muñoz-Arango; Celeny Ortiz; Daniel O. Maldonado-Perez; Diego A. Álvarez-Díaz; Hector Alejandro Ruiz-Moreno; Idabely Betancur Ortiz; Jorge E. Osorio; Juan P. Hernandez-Ortiz; Karl A Ciuoderis; Katherine Laiton-Donato; Laura Silvana Perez; Lina M. Hurtado; Marcela Mercado-Reyes; Maria Angélica Maya; Maria Stella López; Rita Almanza Payares; Sandra Ines Cano; Simón Villegas Velásquez |
| EPI_ISL_1738859, EPI_ISL_1738876, EPI_ISL_1738897, EPI_ISL_1738922, EPI_ISL_1738936, EPI_ISL_1738949, EPI_ISL_1738952, EPI_ISL_1738962, EPI_ISL_1738970, EPI_ISL_1738971, EPI_ISL_1738978, EPI_ISL_1738983, EPI_ISL_1739224, EPI_ISL_1744331 | | | |
| see above | UNILABS | Instituto Nacional de Saude (INSA) | Borges et al |
| EPI_ISL_1966889, EPI_ISL_1966891, EPI_ISL_1966892, EPI_ISL_1966893, EPI_ISL_1966894 | UPA ATALAIA DRA RITA DE CASSIA SORIO | Instituto Butantan / ESALQ-USP (Piracicaba) | Antonio Jorge Martins; Bianca Cechetto Carlos. Mendelics: Bibiana Santos; Claudia Renata dos Santos Barros; Cíntia Bittar; David Schlesinger. Hemocentro Ribeirão Preto: Simone Kashima; Debora Botequio Moretti; Elaine Cristina Marqueze; Elaine Vieira dos Santos; Elisangela Chicaroni Mattos; Erika Freitas; Evandra Strazza Rodrigues; Felipe Allan da Silva da Costa; Flavia Aburjaile; Fábio Sossai Possebon; Guilherme Campos; Guilherme Targino Valente; Heidge Fukumasu. USP-Botucatu: Rejane Maria Tommasini Grotto; Helena Lage Ferreira; Instituto Butantan: Dimas Tadeu Covas; Jardelina de Souza Todao Bernardino; Jayme A. Souza-Neto; Jessika Cristina Chagas Lesbon; Jorge A. Petroli Marchesi; José Salvatore Leister Patané; João Paulo Kitajima; João Pessoa Araújo Jr.; Leila Sabrina Ullmann; Loyze Paola Oliveira de Lima; Luiz Aurelio de Campos Crispin. Centro de Genômica Funcional da ESALQ: Luiz Lehmann Coutinho; Luiz Carlos Junior de Alcantara; Lívia Sacchetto; Maisa C. Pereira Parra; Maria Carolina Elias; Marta Giovanetti; Marília Moraes; Maurício Lacerda Nogueira. Prefeitura de Sao Paulo: Melissa Palmieri.; Patricia Akemi Assato; Paula Rahal; Paulo Inacio da Costa; Rafael dos Santos Bezerra; Raquel de Lello Rocha Campos Cassano. NGS Soluções Genômicas: Pilar Drummond Sampaio Corrêa Mariani. FZEA-USP Pirassununga: Mirele Daiana Poleti; Raul Machado Neto; Ricardo Augusto Brassaloti; Ricardo Haddad; Rodrigo Tocantins Calado. FAMERP-SJRP: Cecília Artico Banho; Sandra Coccuzzo Sampaio; Svetoslav Nanev Slavov; Vagner Fonseca; Vincent Louis Viala |
| EPI_ISL_2345996, EPI_ISL_2345998, EPI_ISL_2346000, EPI_ISL_2346005, EPI_ISL_2346007, EPI_ISL_2346008, EPI_ISL_2346009, EPI_ISL_2346011, EPI_ISL_2346016, EPI_ISL_2346019, EPI_ISL_2346020, EPI_ISL_2346029, EPI_ISL_2346036, EPI_ISL_2346038 | | | |
| see above | UPA DR FRANCO DA ROCHA | Instituto Butantan | Antonio Jorge Martins; Claudia Renata dos Santos Barros; David Schlesinger; Debora Botequio Moretti; Dimas Tadeu Covas; Elaine Cristina Marqueze; Elaine Vieira Santos; Evandra Strazza Rodrigues; Heidge Fukumasu; Jayme Augusto de Souza-Neto; José Salvatore Leister Patané; Luiz Alcantara; Luiz Lehmann Coutinho; Maria Carolina Elias; Maurício Lacerda Nogueira; Rafael dos Santos Bezerra; Raul Machado Neto; Rejane Maria Tommasini Grotto; Ricardo Haddad; Sandra Coccuzzo Sampaio Vessoni; Simone Kashima; Svetoslav Nanev Slavov; Vincent Louis Viala |
| EPI_ISL_1752639 | UPA Dr Luis Atilio Losi Viana Ribeirao Preto | Instituto Adolfo Lutz, Interdiciplinary Procedures Center, Strategic Laboratory | Caio Vinicius Dias Lopes; Claudia Regina Gonçalves; Claudio Tavares Sacchi; Erica Valessa Ramos Gomes; Karoline Rodrigues Campos; Katia Correa de Oliveira Santos; Leonardo Jose Tadeu de Araujo |
| EPI_ISL_2346049 | UPA SADAKO SEDOGUTI | Instituto Butantan | Antonio Jorge Martins; Claudia Renata dos Santos Barros; David Schlesinger; Debora Botequio Moretti; Dimas Tadeu Covas; Elaine Cristina Marqueze; Elaine Vieira Santos; Evandra Strazza Rodrigues; Heidge Fukumasu; Jayme Augusto de Souza-Neto; José Salvatore Leister Patané; Luiz Alcantara; Luiz Lehmann Coutinho; Maria Carolina Elias; Maurício Lacerda Nogueira; Rafael dos Santos Bezerra; Raul Machado Neto; Rejane Maria Tommasini Grotto; Ricardo Haddad; Sandra Coccuzzo Sampaio Vessoni; Simone Kashima; Svetoslav Nanev Slavov; Vincent Louis Viala |
| EPI_ISL_1966650, EPI_ISL_1966661, EPI_ISL_2170946, EPI_ISL_2170947 | UPA UNIDADE DE PRONTO ATENDIMENTO 24 HORAS BOM JESUS | Instituto Butantan / Mendelics | Antonio Jorge Martins; Bianca Cechetto Carlos. Mendelics: Bibiana Santos; Claudia Renata dos Santos Barros; Cíntia Bittar; David Schlesinger. Hemocentro Ribeirão Preto: Simone Kashima; Debora Botequio Moretti; Elaine Cristina Marqueze; Elaine Vieira dos Santos; Elisangela Chicaroni Mattos; Erika Freitas; Evandra Strazza Rodrigues; Felipe Allan da Silva da Costa; Flavia Aburjaile; Fábio Sossai Possebon; Guilherme Campos; Guilherme Targino Valente; Heidge Fukumasu. USP-Botucatu: Rejane Maria Tommasini Grotto; Helena Lage Ferreira; Instituto Butantan: Dimas Tadeu Covas; Jardelina de Souza Todao Bernardino; Jayme A. Souza-Neto; Jessika Cristina Chagas Lesbon; Jorge A. Petroli Marchesi; José Salvatore Leister Patané; João Paulo Kitajima; João Pessoa Araújo Jr.; Leila Sabrina Ullmann; Loyze Paola Oliveira de Lima; Luiz Aurelio de Campos Crispin. Centro de Genômica Funcional da ESALQ: Luiz Lehmann Coutinho; Luiz Carlos Junior de Alcantara; Lívia Sacchetto; Maisa C. Pereira Parra; Maria Carolina Elias; Marta Giovanetti; Marília Moraes; Maurício Lacerda Nogueira. Prefeitura de Sao Paulo: Melissa Palmieri.; Patricia Akemi Assato; Paula Rahal; Paulo Inacio da Costa; Rafael dos Santos Bezerra; Raquel de Lello Rocha Campos Cassano. NGS Soluções Genômicas: Pilar Drummond Sampaio Corrêa Mariani. FZEA-USP Pirassununga: Mirele Daiana Poleti; Raul Machado Neto; Ricardo Augusto Brassaloti; Ricardo Haddad; Rodrigo Tocantins Calado. FAMERP-SJRP: Cecília Artico Banho; Sandra Coccuzzo Sampaio; Svetoslav Nanev Slavov; Vagner Fonseca; Vincent Louis Viala |
| EPI_ISL_2614587, EPI_ISL_2614588 | UPA Vila Santa Catarina | Instituto Adolfo Lutz, Interdiciplinary Procedures Center, Strategic Laboratory | Caio Vinicius Dias Lopes; Claudia Regina Gonçalves; Claudio Tavares Sacchi; Erica Valessa Ramos Gomes; Karoline Rodrigues Campos; Leonardo Jose Tadeu de Araujo |
| EPI_ISL_2614567 | UPA de Jandira | Instituto Adolfo Lutz, Interdiciplinary Procedures Center, Strategic Laboratory | Caio Vinicius Dias Lopes; Claudia Regina Gonçalves; Claudio Tavares Sacchi; Erica Valessa Ramos Gomes; Karoline Rodrigues Campos; Leonardo Jose Tadeu de Araujo |
| EPI_ISL_1855230 | URMC LABS | Wadsworth Center, New York State Department of Health | Alexis Russell; Catharine Prussing; Daryl M. Lamson; Erasmus Schneider; Erica Lasek-Nesselquist; John Kelly; Jonathan Plitnick; Kirsten St. George; Matthew Shudt; Melissa A Leisner; Navjot Singh |
| EPI_ISL_2620770 | USC Clinical Lab | Los Angeles County Public Health Laboratories | P. Hemarajata et al. |
| EPI_ISL_1647126, EPI_ISL_1647157 | USCA Avezzano | Istituto Zooprofilattico Sperimentale dell'Abruzzo e Molise "G. Caporale" | Ancora M; Calistri P; Cammà C; Caporale M; Curini V; Delli Compagni E; Di Domenico M; Di Lollo Valeria; Di Pasquale A; Lorusso A; Mangone I; Marcacci M; Puglia I; Rinaldi A; Savini G; Scialabba S |
| EPI_ISL_1707574 | USCA Pescina | Istituto Zooprofilattico Sperimentale dell'Abruzzo e Molise "G. Caporale" | Ancora M; Calistri P; Cammà C; Caporale M; Curini V; Delli Compagni E; Di Domenico M; Di Lollo Valeria; Di Pasquale A; Lorusso A; Mangone I; Marcacci M; Puglia I; Rinaldi A; Savini G; Scialabba S |
| EPI_ISL_1707580, EPI_ISL_1707581 | USCA Tagliacozzo | Istituto Zooprofilattico Sperimentale dell'Abruzzo e Molise "G. Caporale" | Ancora M; Calistri P; Cammà C; Caporale M; Curini V; Delli Compagni E; Di Domenico M; Di Lollo Valeria; Di Pasquale A; Lorusso A; Mangone I; Marcacci M; Puglia I; Rinaldi A; Savini G; Scialabba S |
| EPI_ISL_1966907, EPI_ISL_1966908 | USF I DE GUAIMBE | Instituto Butantan / ESALQ-USP (Piracicaba) | Antonio Jorge Martins; Bianca Cechetto Carlos. Mendelics: Bibiana Santos; Claudia Renata dos Santos Barros; Cíntia Bittar; David Schlesinger. Hemocentro Ribeirão Preto: Simone Kashima; Debora Botequio Moretti; Elaine Cristina Marqueze; Elaine Vieira dos Santos; Elisangela Chicaroni Mattos; Erika Freitas; Evandra Strazza Rodrigues; Felipe Allan da Silva da Costa; Flavia Aburjaile; Fábio Sossai Possebon; Guilherme Campos; Guilherme Targino Valente; Heidge Fukumasu. USP-Botucatu: Rejane Maria Tommasini Grotto; Helena Lage Ferreira; Instituto Butantan: Dimas Tadeu Covas; Jardelina de Souza Todao Bernardino; Jayme A. Souza-Neto; Jessika Cristina Chagas Lesbon; Jorge A. Petroli Marchesi; José Salvatore Leister Patané; João Paulo Kitajima; João Pessoa Araújo Jr.; Leila Sabrina Ullmann; Loyze Paola Oliveira de Lima; Luiz Aurelio de Campos Crispin. Centro de Genômica Funcional da ESALQ: Luiz Lehmann Coutinho; Luiz Carlos Junior de Alcantara; Lívia Sacchetto; Maisa C. Pereira Parra; Maria Carolina Elias; Marta Giovanetti; Marília Moraes; Maurício Lacerda Nogueira. Prefeitura de Sao Paulo: Melissa Palmieri.; Patricia Akemi Assato; Paula Rahal; Paulo Inacio da Costa; Rafael dos Santos Bezerra; Raquel de Lello Rocha Campos Cassano. NGS Soluções Genômicas: Pilar Drummond Sampaio Corrêa Mariani. FZEA-USP Pirassununga: Mirele Daiana Poleti; Raul Machado Neto; Ricardo Augusto Brassaloti; Ricardo Haddad; Rodrigo Tocantins Calado. FAMERP-SJRP: Cecília Artico Banho; Sandra Coccuzzo Sampaio; Svetoslav Nanev Slavov; Vagner Fonseca; Vincent Louis Viala |
| EPI_ISL_2345959 | USF JARDIM DO LAGO ARTUR NOGUEIRA | Instituto Butantan | Antonio Jorge Martins; Claudia Renata dos Santos Barros; David Schlesinger; Debora Botequio Moretti; Dimas Tadeu Covas; Elaine Cristina Marqueze; Elaine Vieira Santos; Evandra Strazza Rodrigues; Heidge Fukumasu; Jayme Augusto de Souza-Neto; José Salvatore Leister Patané; Luiz Alcantara; Luiz Lehmann Coutinho; Maria Carolina Elias; Maurício Lacerda Nogueira; Rafael dos Santos Bezerra; Raul Machado Neto; Rejane Maria Tommasini Grotto; Ricardo Haddad; Sandra Coccuzzo Sampaio Vessoni; Simone Kashima; Svetoslav Nanev Slavov; Vincent Louis Viala |
| EPI_ISL_2345984 | USF JARDIM PIRATININGA ITAQUAQUECETUBA | Instituto Butantan | Antonio Jorge Martins; Claudia Renata dos Santos Barros; David Schlesinger; Debora Botequio Moretti; Dimas Tadeu Covas; Elaine Cristina Marqueze; Elaine Vieira Santos; Evandra Strazza Rodrigues; Heidge Fukumasu; Jayme Augusto de Souza-Neto; José Salvatore Leister Patané; Luiz Alcantara; Luiz Lehmann Coutinho; Maria Carolina Elias; Maurício Lacerda Nogueira; Rafael dos Santos Bezerra; Raul Machado Neto; Rejane Maria Tommasini Grotto; Ricardo Haddad; Sandra Coccuzzo Sampaio Vessoni; Simone Kashima; Svetoslav Nanev Slavov; Vincent Louis Viala |
| EPI_ISL_2346046 | USF PEQUENO CORACAO | Instituto Butantan | Antonio Jorge Martins; Claudia Renata dos Santos Barros; David Schlesinger; Debora Botequio Moretti; Dimas Tadeu Covas; Elaine Cristina Marqueze; Elaine Vieira Santos; Evandra Strazza Rodrigues; Heidge Fukumasu; Jayme Augusto de Souza-Neto; José Salvatore Leister Patané; Luiz Alcantara; Luiz Lehmann Coutinho; Maria Carolina Elias; Maurício Lacerda Nogueira; Rafael dos Santos Bezerra; Raul Machado Neto; Rejane Maria Tommasini Grotto; Ricardo Haddad; Sandra Coccuzzo Sampaio Vessoni; Simone Kashima; Svetoslav Nanev Slavov; Vincent Louis Viala |
| EPI_ISL_2345999 | USF PQ DOS TRABALHADORES ARTUR NOGUEIRA | Instituto Butantan | Antonio Jorge Martins; Claudia Renata dos Santos Barros; David Schlesinger; Debora Botequio Moretti; Dimas Tadeu Covas; Elaine Cristina Marqueze; Elaine Vieira Santos; Evandra Strazza Rodrigues; Heidge Fukumasu; Jayme Augusto de Souza-Neto; José Salvatore Leister Patané; Luiz Alcantara; Luiz Lehmann Coutinho; Maria Carolina Elias; Maurício Lacerda Nogueira; Rafael dos Santos Bezerra; Raul Machado Neto; Rejane Maria Tommasini Grotto; Ricardo Haddad; Sandra Coccuzzo Sampaio Vessoni; Simone Kashima; Svetoslav Nanev Slavov; Vincent Louis Viala |
| EPI_ISL_2345979 | USF SACILOTTO ARTUR NOGUEIRA | Instituto Butantan | Antonio Jorge Martins; Claudia Renata dos Santos Barros; David Schlesinger; Debora Botequio Moretti; Dimas Tadeu Covas; Elaine Cristina Marqueze; Elaine Vieira Santos; Evandra Strazza Rodrigues; Heidge Fukumasu; Jayme Augusto de Souza-Neto; José Salvatore Leister Patané; Luiz Alcantara; Luiz Lehmann Coutinho; Maria Carolina Elias; Maurício Lacerda Nogueira; Rafael dos Santos Bezerra; Raul Machado Neto; Rejane Maria Tommasini Grotto; Ricardo Haddad; Sandra Coccuzzo Sampaio Vessoni; Simone Kashima; Svetoslav Nanev Slavov; Vincent Louis Viala |
| EPI_ISL_1818072 | UT-Unified State Labs: Public Health Utah DOH | Centers for Disease Control and Prevention Division of Viral Diseases, Pathogen Discovery | Alison Laufer Halpin; Ben L. Rambo-Martin; Clinton R. Paden; Dakota Howard; Darlene Wagner; Dave Wentworth; Dhwani Batra; Jasmine Padilla; Justin Lee; Katie Dillon; Krista Queen; Kristen Knipe; Kristine Lacek; Mark Burroughs; Matthew Schmerer; Mili Sheth; Peter Cook; Sam Shepard; Sarah Nobles; Shoshona Le; Suxiang Tong; Vivien Dugan; Yvette Unoarumhi |
| EPI_ISL_1821703 | UTMG Pathology LLC | UTMG Pathology LLC | Abdallah Azouz; Kelsey Matande; Parasakthy Kumaravelu; Tiara Hymon; Timothy Hodge; Vickie Baselski |
| EPI_ISL_2295545 | UTMG Pathology LLC, University Clinical Health | UTMG Pathology LLC, University Clinical Health | Azouz, A.; Baselski, V.; Hodge, T.; Hymon, T.; Kumaravelu, P.; Matande, K. |
| EPI_ISL_1628162, EPI_ISL_1628170, EPI_ISL_1628171, EPI_ISL_1628172, EPI_ISL_1628173, EPI_ISL_1628174, EPI_ISL_1628175, EPI_ISL_1628176, EPI_ISL_1628178, EPI_ISL_1628179, EPI_ISL_1670795, EPI_ISL_1670796, EPI_ISL_1670797, EPI_ISL_1670798, EPI_ISL_1670799, EPI_ISL_1670800, EPI_ISL_1670801, EPI_ISL_1670802, EPI_ISL_1670803, EPI_ISL_1670804, EPI_ISL_1670805, EPI_ISL_1670806, EPI_ISL_1670807, EPI_ISL_1670808, EPI_ISL_1670809, EPI_ISL_1670810, EPI_ISL_1670811, EPI_ISL_1670813, EPI_ISL_1670814, EPI_ISL_1679827, EPI_ISL_1679830, EPI_ISL_1679836, EPI_ISL_1679837, EPI_ISL_1679848, EPI_ISL_1679892, EPI_ISL_1679901, EPI_ISL_1679907, EPI_ISL_1679910, EPI_ISL_1679913, EPI_ISL_1679916, EPI_ISL_1679981, EPI_ISL_1679991, EPI_ISL_1680056, EPI_ISL_1680170, EPI_ISL_1680187, EPI_ISL_1680202, EPI_ISL_1680204, EPI_ISL_1680259, EPI_ISL_1680265, EPI_ISL_1680343, EPI_ISL_1680344, EPI_ISL_1680350, EPI_ISL_1680357, EPI_ISL_1680358, EPI_ISL_1680359, EPI_ISL_1680372, EPI_ISL_1680391, EPI_ISL_1680395, EPI_ISL_1680397, EPI_ISL_1680399, EPI_ISL_1680404, EPI_ISL_1680412, EPI_ISL_1680413, EPI_ISL_1680448, EPI_ISL_1715445, EPI_ISL_1715466, EPI_ISL_1715494, EPI_ISL_1715503, EPI_ISL_1715538, EPI_ISL_1715540, EPI_ISL_1715555, EPI_ISL_1715557, EPI_ISL_1715572, EPI_ISL_1715575, EPI_ISL_1715577, EPI_ISL_1715597, EPI_ISL_1715601, EPI_ISL_1715604, EPI_ISL_1715640, EPI_ISL_1715641, EPI_ISL_1715647, EPI_ISL_1715650, EPI_ISL_1715656, EPI_ISL_1785635, EPI_ISL_1785688, EPI_ISL_1785708, EPI_ISL_1785731, EPI_ISL_1789217, EPI_ISL_1789221, EPI_ISL_1789247, EPI_ISL_1789248, EPI_ISL_1789288, EPI_ISL_1789289, EPI_ISL_1789302, EPI_ISL_1789314, EPI_ISL_1789318, EPI_ISL_1789328, EPI_ISL_1789364, EPI_ISL_1789365, EPI_ISL_1789368, EPI_ISL_1789369, EPI_ISL_1789391, EPI_ISL_1789392, EPI_ISL_2002510, EPI_ISL_2002516 | | | |
| see above | UW Virology Lab | UW Virology Lab | Alexander Greninger; Hong Xie; Keith R Jerome; Lasata Shrestha; Meei-Li Huang; Michelle Lin; Noah R. Baker; Pavitra Roychoudhury; Sean Ellis; Shah Mohamed Bakhash; Tien V. Nguyen |
| EPI_ISL_5799865, EPI_ISL_5799909, EPI_ISL_5799940, EPI_ISL_5799944, EPI_ISL_5799946, EPI_ISL_5799947, EPI_ISL_5799949, EPI_ISL_5799960, EPI_ISL_5799968, EPI_ISL_5800011, EPI_ISL_5800019, EPI_ISL_5800021, EPI_ISL_5800031 | | | |
| see above | Ubs Atalaia Dr Mario Aguiar Filho | Instituto Butantan | Antonio Jorge Martins; Claudia Renata dos Santos Barros; David Schlesinger; Debora Botequio Moretti; Dimas Tadeu Covas; Elaine Cristina Marqueze; Elaine Vieira Santos; Evandra Strazza Rodrigues; Heidge Fukumasu; Jayme Augusto de Souza-Neto; José Salvatore Leister Patané; Luiz Alcantara; Luiz Lehmann Coutinho; Maria Carolina Elias; Maurício Lacerda Nogueira; Rafael dos Santos Bezerra; Raul Machado Neto; Rejane Maria Tommasini Grotto; Ricardo Haddad; Sandra Coccuzzo Sampaio Vessoni; Simone Kashima; Svetoslav Nanev Slavov; Vincent Louis Viala |
| EPI_ISL_5800168, EPI_ISL_5800176, EPI_ISL_5800177, EPI_ISL_5800187 | Ubs De Tabapua | Instituto Butantan | Antonio Jorge Martins; Claudia Renata dos Santos Barros; David Schlesinger; Debora Botequio Moretti; Dimas Tadeu Covas; Elaine Cristina Marqueze; Elaine Vieira Santos; Evandra Strazza Rodrigues; Heidge Fukumasu; Jayme Augusto de Souza-Neto; José Salvatore Leister Patané; Luiz Alcantara; Luiz Lehmann Coutinho; Maria Carolina Elias; Maurício Lacerda Nogueira; Rafael dos Santos Bezerra; Raul Machado Neto; Rejane Maria Tommasini Grotto; Ricardo Haddad; Sandra Coccuzzo Sampaio Vessoni; Simone Kashima; Svetoslav Nanev Slavov; Vincent Louis Viala |
| EPI_ISL_5799877, EPI_ISL_5800000 | Ubs Dr Helio Migliari | Instituto Butantan | Antonio Jorge Martins; Claudia Renata dos Santos Barros; David Schlesinger; Debora Botequio Moretti; Dimas Tadeu Covas; Elaine Cristina Marqueze; Elaine Vieira Santos; Evandra Strazza Rodrigues; Heidge Fukumasu; Jayme Augusto de Souza-Neto; José Salvatore Leister Patané; Luiz Alcantara; Luiz Lehmann Coutinho; Maria Carolina Elias; Maurício Lacerda Nogueira; Rafael dos Santos Bezerra; Raul Machado Neto; Rejane Maria Tommasini Grotto; Ricardo Haddad; Sandra Coccuzzo Sampaio Vessoni; Simone Kashima; Svetoslav Nanev Slavov; Vincent Louis Viala |
| EPI_ISL_5800347 | Ubs Enrique Guilherme Ernesto Marzeuski | Instituto Butantan | Antonio Jorge Martins; Claudia Renata dos Santos Barros; David Schlesinger; Debora Botequio Moretti; Dimas Tadeu Covas; Elaine Cristina Marqueze; Elaine Vieira Santos; Evandra Strazza Rodrigues; Heidge Fukumasu; Jayme Augusto de Souza-Neto; José Salvatore Leister Patané; Luiz Alcantara; Luiz Lehmann Coutinho; Maria Carolina Elias; Maurício Lacerda Nogueira; Rafael dos Santos Bezerra; Raul Machado Neto; Rejane Maria Tommasini Grotto; Ricardo Haddad; Sandra Coccuzzo Sampaio Vessoni; Simone Kashima; Svetoslav Nanev Slavov; Vincent Louis Viala |

| | | | |
|---|---|---|---|
| EPI_ISL_5800126 | Ubs Iii De Pariquera Acu Pariquera Acu | Instituto Butantan | Antonio Jorge Martins; Claudia Renata dos Santos Barros; David Schlesinger; Debora Botequio Moretti; Dimas Tadeu Covas; Elaine Cristina Marqueze; Elaine Vieira Santos; Evandra Strazza Rodrigues; Heidge Fukumasu; Jayme Augusto de Souza-Neto; José Salvatore Leister Patané; Luiz Alcantara; Luiz Lehmann Coutinho; Maria Carolina Elias; Maurício Lacerda Nogueira; Rafael dos Santos Bezerra; Raul Machado Neto; Rejane Maria Tommasini Grotto; Ricardo Haddad; Sandra Coccuzzo Sampaio Vessoni; Simone Kashima; Svetoslav Nanev Slavov; Vincent Louis Viala |
| EPI_ISL_5802302 | Ubs Irma Agueda Maria Jaime | Instituto Butantan | Antonio Jorge Martins; Claudia Renata dos Santos Barros; David Schlesinger; Debora Botequio Moretti; Dimas Tadeu Covas; Elaine Cristina Marqueze; Elaine Vieira Santos; Evandra Strazza Rodrigues; Heidge Fukumasu; Jayme Augusto de Souza-Neto; José Salvatore Leister Patané; Luiz Alcantara; Luiz Lehmann Coutinho; Maria Carolina Elias; Maurício Lacerda Nogueira; Rafael dos Santos Bezerra; Raul Machado Neto; Rejane Maria Tommasini Grotto; Ricardo Haddad; Sandra Coccuzzo Sampaio Vessoni; Simone Kashima; Svetoslav Nanev Slavov; Vincent Louis Viala |
| EPI_ISL_5802314 | Ubs Jardim Acacio | Instituto Butantan | Antonio Jorge Martins; Claudia Renata dos Santos Barros; David Schlesinger; Debora Botequio Moretti; Dimas Tadeu Covas; Elaine Cristina Marqueze; Elaine Vieira Santos; Evandra Strazza Rodrigues; Heidge Fukumasu; Jayme Augusto de Souza-Neto; José Salvatore Leister Patané; Luiz Alcantara; Luiz Lehmann Coutinho; Maria Carolina Elias; Maurício Lacerda Nogueira; Rafael dos Santos Bezerra; Raul Machado Neto; Rejane Maria Tommasini Grotto; Ricardo Haddad; Sandra Coccuzzo Sampaio Vessoni; Simone Kashima; Svetoslav Nanev Slavov; Vincent Louis Viala |
| EPI_ISL_5802355, EPI_ISL_5802356, EPI_ISL_5802361, EPI_ISL_5802363, EPI_ISL_5802368, EPI_ISL_5802371, EPI_ISL_5802372, EPI_ISL_5802379 | | | |
| see above | Ubs Jardim Caiuby | Instituto Butantan | Antonio Jorge Martins; Claudia Renata dos Santos Barros; David Schlesinger; Debora Botequio Moretti; Dimas Tadeu Covas; Elaine Cristina Marqueze; Elaine Vieira Santos; Evandra Strazza Rodrigues; Heidge Fukumasu; Jayme Augusto de Souza-Neto; José Salvatore Leister Patané; Luiz Alcantara; Luiz Lehmann Coutinho; Maria Carolina Elias; Maurício Lacerda Nogueira; Rafael dos Santos Bezerra; Raul Machado Neto; Rejane Maria Tommasini Grotto; Ricardo Haddad; Sandra Coccuzzo Sampaio Vessoni; Simone Kashima; Svetoslav Nanev Slavov; Vincent Louis Viala |
| EPI_ISL_5802348, EPI_ISL_5802349, EPI_ISL_5802360, EPI_ISL_5802370 | Ubs Jardim Maragogipe | Instituto Butantan | Antonio Jorge Martins; Claudia Renata dos Santos Barros; David Schlesinger; Debora Botequio Moretti; Dimas Tadeu Covas; Elaine Cristina Marqueze; Elaine Vieira Santos; Evandra Strazza Rodrigues; Heidge Fukumasu; Jayme Augusto de Souza-Neto; José Salvatore Leister Patané; Luiz Alcantara; Luiz Lehmann Coutinho; Maria Carolina Elias; Maurício Lacerda Nogueira; Rafael dos Santos Bezerra; Raul Machado Neto; Rejane Maria Tommasini Grotto; Ricardo Haddad; Sandra Coccuzzo Sampaio Vessoni; Simone Kashima; Svetoslav Nanev Slavov; Vincent Louis Viala |
| EPI_ISL_5802319, EPI_ISL_5802320, EPI_ISL_5802321, EPI_ISL_5802322, EPI_ISL_5802327 | Ubs Jardim Odete | Instituto Butantan | Antonio Jorge Martins; Claudia Renata dos Santos Barros; David Schlesinger; Debora Botequio Moretti; Dimas Tadeu Covas; Elaine Cristina Marqueze; Elaine Vieira Santos; Evandra Strazza Rodrigues; Heidge Fukumasu; Jayme Augusto de Souza-Neto; José Salvatore Leister Patané; Luiz Alcantara; Luiz Lehmann Coutinho; Maria Carolina Elias; Maurício Lacerda Nogueira; Rafael dos Santos Bezerra; Raul Machado Neto; Rejane Maria Tommasini Grotto; Ricardo Haddad; Sandra Coccuzzo Sampaio Vessoni; Simone Kashima; Svetoslav Nanev Slavov; Vincent Louis Viala |
| EPI_ISL_5802326, EPI_ISL_5802353 | Ubs Jardim Paineira | Instituto Butantan | Antonio Jorge Martins; Claudia Renata dos Santos Barros; David Schlesinger; Debora Botequio Moretti; Dimas Tadeu Covas; Elaine Cristina Marqueze; Elaine Vieira Santos; Evandra Strazza Rodrigues; Heidge Fukumasu; Jayme Augusto de Souza-Neto; José Salvatore Leister Patané; Luiz Alcantara; Luiz Lehmann Coutinho; Maria Carolina Elias; Maurício Lacerda Nogueira; Rafael dos Santos Bezerra; Raul Machado Neto; Rejane Maria Tommasini Grotto; Ricardo Haddad; Sandra Coccuzzo Sampaio Vessoni; Simone Kashima; Svetoslav Nanev Slavov; Vincent Louis Viala |
| EPI_ISL_5802308 | Ubs Jardim Primavera | Instituto Butantan | Antonio Jorge Martins; Claudia Renata dos Santos Barros; David Schlesinger; Debora Botequio Moretti; Dimas Tadeu Covas; Elaine Cristina Marqueze; Elaine Vieira Santos; Evandra Strazza Rodrigues; Heidge Fukumasu; Jayme Augusto de Souza-Neto; José Salvatore Leister Patané; Luiz Alcantara; Luiz Lehmann Coutinho; Maria Carolina Elias; Maurício Lacerda Nogueira; Rafael dos Santos Bezerra; Raul Machado Neto; Rejane Maria Tommasini Grotto; Ricardo Haddad; Sandra Coccuzzo Sampaio Vessoni; Simone Kashima; Svetoslav Nanev Slavov; Vincent Louis Viala |
| EPI_ISL_5800253 | Ubs Jardim Vitorino Ipero | Instituto Butantan | Antonio Jorge Martins; Claudia Renata dos Santos Barros; David Schlesinger; Debora Botequio Moretti; Dimas Tadeu Covas; Elaine Cristina Marqueze; Elaine Vieira Santos; Evandra Strazza Rodrigues; Heidge Fukumasu; Jayme Augusto de Souza-Neto; José Salvatore Leister Patané; Luiz Alcantara; Luiz Lehmann Coutinho; Maria Carolina Elias; Maurício Lacerda Nogueira; Rafael dos Santos Bezerra; Raul Machado Neto; Rejane Maria Tommasini Grotto; Ricardo Haddad; Sandra Coccuzzo Sampaio Vessoni; Simone Kashima; Svetoslav Nanev Slavov; Vincent Louis Viala |
| EPI_ISL_5802312 | Ubs Jd Presidente Dutra | Instituto Butantan | Antonio Jorge Martins; Claudia Renata dos Santos Barros; David Schlesinger; Debora Botequio Moretti; Dimas Tadeu Covas; Elaine Cristina Marqueze; Elaine Vieira Santos; Evandra Strazza Rodrigues; Heidge Fukumasu; Jayme Augusto de Souza-Neto; José Salvatore Leister Patané; Luiz Alcantara; Luiz Lehmann Coutinho; Maria Carolina Elias; Maurício Lacerda Nogueira; Rafael dos Santos Bezerra; Raul Machado Neto; Rejane Maria Tommasini Grotto; Ricardo Haddad; Sandra Coccuzzo Sampaio Vessoni; Simone Kashima; Svetoslav Nanev Slavov; Vincent Louis Viala |
| EPI_ISL_5802315, EPI_ISL_5802380 | Ubs Maria Pia De Oliveira | Instituto Butantan | Antonio Jorge Martins; Claudia Renata dos Santos Barros; David Schlesinger; Debora Botequio Moretti; Dimas Tadeu Covas; Elaine Cristina Marqueze; Elaine Vieira Santos; Evandra Strazza Rodrigues; Heidge Fukumasu; Jayme Augusto de Souza-Neto; José Salvatore Leister Patané; Luiz Alcantara; Luiz Lehmann Coutinho; Maria Carolina Elias; Maurício Lacerda Nogueira; Rafael dos Santos Bezerra; Raul Machado Neto; Rejane Maria Tommasini Grotto; Ricardo Haddad; Sandra Coccuzzo Sampaio Vessoni; Simone Kashima; Svetoslav Nanev Slavov; Vincent Louis Viala |
| EPI_ISL_5802309 | Ubs Marinopolis | Instituto Butantan | Antonio Jorge Martins; Claudia Renata dos Santos Barros; David Schlesinger; Debora Botequio Moretti; Dimas Tadeu Covas; Elaine Cristina Marqueze; Elaine Vieira Santos; Evandra Strazza Rodrigues; Heidge Fukumasu; Jayme Augusto de Souza-Neto; José Salvatore Leister Patané; Luiz Alcantara; Luiz Lehmann Coutinho; Maria Carolina Elias; Maurício Lacerda Nogueira; Rafael dos Santos Bezerra; Raul Machado Neto; Rejane Maria Tommasini Grotto; Ricardo Haddad; Sandra Coccuzzo Sampaio Vessoni; Simone Kashima; Svetoslav Nanev Slavov; Vincent Louis Viala |
| EPI_ISL_5802358, EPI_ISL_5802369 | Ubs Morro Branco | Instituto Butantan | Antonio Jorge Martins; Claudia Renata dos Santos Barros; David Schlesinger; Debora Botequio Moretti; Dimas Tadeu Covas; Elaine Cristina Marqueze; Elaine Vieira Santos; Evandra Strazza Rodrigues; Heidge Fukumasu; Jayme Augusto de Souza-Neto; José Salvatore Leister Patané; Luiz Alcantara; Luiz Lehmann Coutinho; Maria Carolina Elias; Maurício Lacerda Nogueira; Rafael dos Santos Bezerra; Raul Machado Neto; Rejane Maria Tommasini Grotto; Ricardo Haddad; Sandra Coccuzzo Sampaio Vessoni; Simone Kashima; Svetoslav Nanev Slavov; Vincent Louis Viala |
| EPI_ISL_5802357, EPI_ISL_5802362 | Ubs Parque Marengo | Instituto Butantan | Antonio Jorge Martins; Claudia Renata dos Santos Barros; David Schlesinger; Debora Botequio Moretti; Dimas Tadeu Covas; Elaine Cristina Marqueze; Elaine Vieira Santos; Evandra Strazza Rodrigues; Heidge Fukumasu; Jayme Augusto de Souza-Neto; José Salvatore Leister Patané; Luiz Alcantara; Luiz Lehmann Coutinho; Maria Carolina Elias; Maurício Lacerda Nogueira; Rafael dos Santos Bezerra; Raul Machado Neto; Rejane Maria Tommasini Grotto; Ricardo Haddad; Sandra Coccuzzo Sampaio Vessoni; Simone Kashima; Svetoslav Nanev Slavov; Vincent Louis Viala |
| EPI_ISL_5802346 | Ubs Recanto Monica | Instituto Butantan | Antonio Jorge Martins; Claudia Renata dos Santos Barros; David Schlesinger; Debora Botequio Moretti; Dimas Tadeu Covas; Elaine Cristina Marqueze; Elaine Vieira Santos; Evandra Strazza Rodrigues; Heidge Fukumasu; Jayme Augusto de Souza-Neto; José Salvatore Leister Patané; Luiz Alcantara; Luiz Lehmann Coutinho; Maria Carolina Elias; Maurício Lacerda Nogueira; Rafael dos Santos Bezerra; Raul Machado Neto; Rejane Maria Tommasini Grotto; Ricardo Haddad; Sandra Coccuzzo Sampaio Vessoni; Simone Kashima; Svetoslav Nanev Slavov; Vincent Louis Viala |
| EPI_ISL_5802300, EPI_ISL_5802301, EPI_ISL_5802303 | Ubs Santa Maria | Instituto Butantan | Antonio Jorge Martins; Claudia Renata dos Santos Barros; David Schlesinger; Debora Botequio Moretti; Dimas Tadeu Covas; Elaine Cristina Marqueze; Elaine Vieira Santos; Evandra Strazza Rodrigues; Heidge Fukumasu; Jayme Augusto de Souza-Neto; José Salvatore Leister Patané; Luiz Alcantara; Luiz Lehmann Coutinho; Maria Carolina Elias; Maurício Lacerda Nogueira; Rafael dos Santos Bezerra; Raul Machado Neto; Rejane Maria Tommasini Grotto; Ricardo Haddad; Sandra Coccuzzo Sampaio Vessoni; Simone Kashima; Svetoslav Nanev Slavov; Vincent Louis Viala |
| EPI_ISL_5800106, EPI_ISL_5800109, EPI_ISL_5800111, EPI_ISL_5800114, EPI_ISL_5800120, EPI_ISL_5800122 | Ubs Sao Judas | Instituto Butantan | Antonio Jorge Martins; Claudia Renata dos Santos Barros; David Schlesinger; Debora Botequio Moretti; Dimas Tadeu Covas; Elaine Cristina Marqueze; Elaine Vieira Santos; Evandra Strazza Rodrigues; Heidge Fukumasu; Jayme Augusto de Souza-Neto; José Salvatore Leister Patané; Luiz Alcantara; Luiz Lehmann Coutinho; Maria Carolina Elias; Maurício Lacerda Nogueira; Rafael dos Santos Bezerra; Raul Machado Neto; Rejane Maria Tommasini Grotto; Ricardo Haddad; Sandra Coccuzzo Sampaio Vessoni; Simone Kashima; Svetoslav Nanev Slavov; Vincent Louis Viala |
| EPI_ISL_5800103, EPI_ISL_5802287 | Unidade De Pronto Atendimento De Varzea Paulista Upa Ii | Instituto Butantan | Antonio Jorge Martins; Claudia Renata dos Santos Barros; David Schlesinger; Debora Botequio Moretti; Dimas Tadeu Covas; Elaine Cristina Marqueze; Elaine Vieira Santos; Evandra Strazza Rodrigues; Heidge Fukumasu; Jayme Augusto de Souza-Neto; José Salvatore Leister Patané; Luiz Alcantara; Luiz Lehmann Coutinho; Maria Carolina Elias; Maurício Lacerda Nogueira; Rafael dos Santos Bezerra; Raul Machado Neto; Rejane Maria Tommasini Grotto; Ricardo Haddad; Sandra Coccuzzo Sampaio Vessoni; Simone Kashima; Svetoslav Nanev Slavov; Vincent Louis Viala |
| EPI_ISL_5800221 | Unidade De Pronto Atendimento Upa | Instituto Butantan | Antonio Jorge Martins; Claudia Renata dos Santos Barros; David Schlesinger; Debora Botequio Moretti; Dimas Tadeu Covas; Elaine Cristina Marqueze; Elaine Vieira Santos; Evandra Strazza Rodrigues; Heidge Fukumasu; Jayme Augusto de Souza-Neto; José Salvatore Leister Patané; Luiz Alcantara; Luiz Lehmann Coutinho; Maria Carolina Elias; Maurício Lacerda Nogueira; Rafael dos Santos Bezerra; Raul Machado Neto; Rejane Maria Tommasini Grotto; Ricardo Haddad; Sandra Coccuzzo Sampaio Vessoni; Simone Kashima; Svetoslav Nanev Slavov; Vincent Louis Viala |
| EPI_ISL_5800152, EPI_ISL_5800153, EPI_ISL_5800154, EPI_ISL_5800159, EPI_ISL_5800163 | Unidade De Vigilancia Em Saude | Instituto Butantan | Antonio Jorge Martins; Claudia Renata dos Santos Barros; David Schlesinger; Debora Botequio Moretti; Dimas Tadeu Covas; Elaine Cristina Marqueze; Elaine Vieira Santos; Evandra Strazza Rodrigues; Heidge Fukumasu; Jayme Augusto de Souza-Neto; José Salvatore Leister Patané; Luiz Alcantara; Luiz Lehmann Coutinho; Maria Carolina Elias; Maurício Lacerda Nogueira; Rafael dos Santos Bezerra; Raul Machado Neto; Rejane Maria Tommasini Grotto; Ricardo Haddad; Sandra Coccuzzo Sampaio Vessoni; Simone Kashima; Svetoslav Nanev Slavov; Vincent Louis Viala |
| EPI_ISL_5799708, EPI_ISL_5799709 | Unidade De Vigilancia Epidemiologica | Instituto Butantan | Antonio Jorge Martins; Claudia Renata dos Santos Barros; David Schlesinger; Debora Botequio Moretti; Dimas Tadeu Covas; Elaine Cristina Marqueze; Elaine Vieira Santos; Evandra Strazza Rodrigues; Heidge Fukumasu; Jayme Augusto de Souza-Neto; José Salvatore Leister Patané; Luiz Alcantara; Luiz Lehmann Coutinho; Maria Carolina Elias; Maurício Lacerda Nogueira; Rafael dos Santos Bezerra; Raul Machado Neto; Rejane Maria Tommasini Grotto; Ricardo Haddad; Sandra Coccuzzo Sampaio Vessoni; Simone Kashima; Svetoslav Nanev Slavov; Vincent Louis Viala |
| EPI_ISL_5800207 | Unidade Mista De Iguape | Instituto Butantan | Antonio Jorge Martins; Claudia Renata dos Santos Barros; David Schlesinger; Debora Botequio Moretti; Dimas Tadeu Covas; Elaine Cristina Marqueze; Elaine Vieira Santos; Evandra Strazza Rodrigues; Heidge Fukumasu; Jayme Augusto de Souza-Neto; José Salvatore Leister Patané; Luiz Alcantara; Luiz Lehmann Coutinho; Maria Carolina Elias; Maurício Lacerda Nogueira; Rafael dos Santos Bezerra; Raul Machado Neto; Rejane Maria Tommasini Grotto; Ricardo Haddad; Sandra Coccuzzo Sampaio Vessoni; Simone Kashima; Svetoslav Nanev Slavov; Vincent Louis Viala |
| EPI_ISL_5800356, EPI_ISL_5800357, EPI_ISL_5800358, EPI_ISL_5800360, EPI_ISL_5800361 | Unidade Mista De Saude Capela Do Alto | Instituto Butantan | Antonio Jorge Martins; Claudia Renata dos Santos Barros; David Schlesinger; Debora Botequio Moretti; Dimas Tadeu Covas; Elaine Cristina Marqueze; Elaine Vieira Santos; Evandra Strazza Rodrigues; Heidge Fukumasu; Jayme Augusto de Souza-Neto; José Salvatore Leister Patané; Luiz Alcantara; Luiz Lehmann Coutinho; Maria Carolina Elias; Maurício Lacerda Nogueira; Rafael dos Santos Bezerra; Raul Machado Neto; Rejane Maria Tommasini Grotto; Ricardo Haddad; Sandra Coccuzzo Sampaio Vessoni; Simone Kashima; Svetoslav Nanev Slavov; Vincent Louis Viala |
| EPI_ISL_5800121, EPI_ISL_5800123 | Unidade Mista De Saude De Oriente | Instituto Butantan | Antonio Jorge Martins; Claudia Renata dos Santos Barros; David Schlesinger; Debora Botequio Moretti; Dimas Tadeu Covas; Elaine Cristina Marqueze; Elaine Vieira Santos; Evandra Strazza Rodrigues; Heidge Fukumasu; Jayme Augusto de Souza-Neto; José Salvatore Leister Patané; Luiz Alcantara; Luiz Lehmann Coutinho; Maria Carolina Elias; Maurício Lacerda Nogueira; Rafael dos Santos Bezerra; Raul Machado Neto; Rejane Maria Tommasini Grotto; Ricardo Haddad; Sandra Coccuzzo Sampaio Vessoni; Simone Kashima; Svetoslav Nanev Slavov; Vincent Louis Viala |
| EPI_ISL_5799695 | Unidade Mista Osvaldo P Camargo Elias Fausto | Instituto Butantan | Antonio Jorge Martins; Claudia Renata dos Santos Barros; David Schlesinger; Debora Botequio Moretti; Dimas Tadeu Covas; Elaine Cristina Marqueze; Elaine Vieira Santos; Evandra Strazza Rodrigues; Heidge Fukumasu; Jayme Augusto de Souza-Neto; José Salvatore Leister Patané; Luiz Alcantara; Luiz Lehmann Coutinho; Maria Carolina Elias; Maurício Lacerda Nogueira; Rafael dos Santos Bezerra; Raul Machado Neto; Rejane Maria Tommasini Grotto; Ricardo Haddad; Sandra Coccuzzo Sampaio Vessoni; Simone Kashima; Svetoslav Nanev Slavov; Vincent Louis Viala |
| EPI_ISL_5800223, EPI_ISL_5800224, EPI_ISL_5800225, EPI_ISL_5800226, EPI_ISL_5800274 | Unidade Sentinela Covid19 | Instituto Butantan | Antonio Jorge Martins; Claudia Renata dos Santos Barros; David Schlesinger; Debora Botequio Moretti; Dimas Tadeu Covas; Elaine Cristina Marqueze; Elaine Vieira Santos; Evandra Strazza Rodrigues; Heidge Fukumasu; Jayme Augusto de Souza-Neto; José Salvatore Leister Patané; Luiz Alcantara; Luiz Lehmann Coutinho; Maria Carolina Elias; Maurício Lacerda Nogueira; Rafael dos Santos Bezerra; Raul Machado Neto; Rejane Maria Tommasini Grotto; Ricardo Haddad; Sandra Coccuzzo Sampaio Vessoni; Simone Kashima; Svetoslav Nanev Slavov; Vincent Louis Viala |
| EPI_ISL_5800258, EPI_ISL_5800266 | Unidade Sentinela Vila Aparecida | Instituto Butantan | Antonio Jorge Martins; Claudia Renata dos Santos Barros; David Schlesinger; Debora Botequio Moretti; Dimas Tadeu Covas; Elaine Cristina Marqueze; Elaine Vieira Santos; Evandra Strazza Rodrigues; Heidge Fukumasu; Jayme Augusto de Souza-Neto; José Salvatore Leister Patané; Luiz Alcantara; Luiz Lehmann Coutinho; Maria Carolina Elias; Maurício Lacerda Nogueira; Rafael dos Santos Bezerra; Raul Machado Neto; Rejane Maria Tommasini Grotto; Ricardo Haddad; Sandra Coccuzzo Sampaio Vessoni; Simone Kashima; Svetoslav Nanev Slavov; Vincent Louis Viala |
| EPI_ISL_2101386, EPI_ISL_2101407, EPI_ISL_2101430, EPI_ISL_2101566, EPI_ISL_2101567, EPI_ISL_2101568, EPI_ISL_2101569, EPI_ISL_2101570, EPI_ISL_2101571, EPI_ISL_2101572, EPI_ISL_2101638, EPI_ISL_2101639, EPI_ISL_2101640, EPI_ISL_2101641, EPI_ISL_2101642, EPI_ISL_2101643, EPI_ISL_2101660, EPI_ISL_2101661, EPI_ISL_2101662, EPI_ISL_2101663, EPI_ISL_2101664, EPI_ISL_2101722, EPI_ISL_2101731 | | | |
| see above | Unidade de apoio ao diagnóstico da COVID – UNADIG | Bioinformatics Laboratory / LNCC | Alessandra P Lamarca; Alexandra L Gerber; Amilcar Tanuri; Ana Paula de C Guimaraes; Ana Tereza R Vasconcelos; Andrea Cony Cavalcanti; Caio Luiz Pereira Ribeiro; Cassia Alves; Cintia Policarpo; Claudia Maria Braga de Mello; Cristiane Gomes da Silva; Diana Mariani; Douglas Terra Machado; Flavio Dias da Silva; Gleidson da Silva de Oliveira; Leandro Magalhaes de Souza; Liliane Cavalcante; Luiz G P de Almeida; Marcio Henrique de Oliveira Garcia; Mario Sergio Ribeiro; Ronaldo da Silva F Jr; Silvia Carvalho |
| EPI_ISL_3231187 | Universidad Autonoma de Yucatan | New York Genome Center | Amy Baldwin; Andre Corvelo; Aviles-Gomez E; Ayora-Talavera G; Chan-Gasca M; Dayna M. Oschwald; Flores-Quintal F; Granja-Perez P; Lopez-Coral L.; Michael Zody; Samantha Fennessey; Tom Maniatis; Villanueva-Jorge S; Yam-Pool E |
| EPI_ISL_1540707, EPI_ISL_1656146 | Universidad Tecnica Particular de Loja | Institute of Microbiology, Universidad San Francisco de Quito | Belén Prado-Vivar; Bernardo Gutiérrez; David Zuñiga; Fernando Serrano; Gabriel Trueba; Juan José Guadalupe; Katherine Ojeda; Luis Flores; Melissa Ortega; Michelle Grunauer; Monica Becerra-Wong; Paola Dalgo; Patricio Rojas-Silva; Paúl Cárdenas; Raiza Briceño; Raiza Briceño; Sully Márquez; Verónica Barragán |
| EPI_ISL_1714760 | University Hospitals Translational Laboratory (UHTL), University Hospitals | University Hospitals Translational Laboratory (UHTL), University Hospitals | Alouani, D.; Sadri, N.; Song, X. |
| EPI_ISL_2365939 | University of Bari Biomedical Sciences and Human Oncology, Policlinico | University of Bari Biomedical Sciences and Human Oncology | Anna Sallustio; Daniela Loconsole; Maria Chironna; Marisa Accogli |
| EPI_ISL_2246622, EPI_ISL_2246667, EPI_ISL_2692017, | University of Chicago Medicine | RIPHL at Rush University Medical Center | Cindy Bethel; Felix Araujo Perez; Kevin Kunstman; Laura Furtado; Lauren Megger; Marieta Hyde; Max Kolton; Stefan Green |

| | | | |
|---|---|---|---|
| EPI_ISL_2931111 | | | |
| EPI_ISL_1660774, EPI_ISL_1660775, EPI_ISL_1660782, EPI_ISL_1671951, EPI_ISL_1671952, EPI_ISL_1671959, EPI_ISL_1671962, EPI_ISL_1672235, EPI_ISL_1672236 | | | |
| see above | University of Liège COVID-19 testing center | GIGA Medical Genomics | Bouchra Boujemla; Cécile Meex; Keith Durkin; Maria Artesi; Marie-Pierre Hayette; Nathalie Renotte; Pierrette Melin; Raphaël Boreux; Sébastien Bontems; Vincent Bours |
| EPI_ISL_1651690 | University of Michigan Clinical Microbiology Laboratory | Lauring Lab, University of Michigan, Department of Microbiology and Immunology | Valesano |
| EPI_ISL_2018932, EPI_ISL_2018934, EPI_ISL_2018937, EPI_ISL_2018943 | University of Rome Tor Vergata: Departm Experim Medicine Chair of Virology | University of Rome Tor Vergata: Departm Experim Medicine Chair of Virology | Francesca Ceccherini-Silberstein; Loredana Sarmati; Lorenzo Piermatteo; Luca Carioti; Marco Iannetta; Maria Botticelli; Maria Concetta Bellocchi; Massimo Andreoni; Rossana Scutari |
| EPI_ISL_1822014 | University of Wisconsin-Madison AIDS Vaccine Research Laboratories | University of Wisconsin-Madison AIDS Vaccine Research Laboratories | Gage Moreno; Katarina Braun; et al. AIDS Vaccine Research Laboratories |
| EPI_ISL_2020636, EPI_ISL_2455693, EPI_ISL_2465270 | Università Federico II - Dipartimento di scienze mediche traslazionali - Napoli | TIGEM | Antonio Grimaldi Patrizia Annunziata Francesco Panariello Teresa Giuliano Michele Cennamo Valentina Bouche Chiara Colantuono Lucio Di Filippo Mariano Fiorenza Anna Manfredi Marcello Salvi Giuseppe Portella Andrea Ballabio Davide Cacchiarelli |
| EPI_ISL_1670655, EPI_ISL_1670656, EPI_ISL_1670657, EPI_ISL_1670658, EPI_ISL_1670659, EPI_ISL_1919640, EPI_ISL_1919643 | | | |
| see above | Università degli Studi di Perugia | Istituto Zooprofilattico Sperimentale dell'Abruzzo e Molise "G. Caporale" | Ancora M; Calistri P; Camilloni B; Cammà C; Caporale M; Curini V; Delli Compagni E; Di Domenico M; Di Lollo Valeria; Di Pasquale A; Lorusso A; Mangone I; Marcacci M; Mencacci A; Puglia I; Rinaldi A; Savini G; Scialabba S |
| EPI_ISL_5800030, EPI_ISL_5800078, EPI_ISL_5800080, EPI_ISL_5800083, EPI_ISL_5800087 | Upa Atalaia Dra Rita De Cassia Sorio | Instituto Butantan | Antonio Jorge Martins; Claudia Renata dos Santos Barros; David Schlesinger; Debora Botequio Moretti; Dimas Tadeu Covas; Elaine Cristina Marqueze; Elaine Vieira Santos; Evandra Strazza Rodrigues; Heidge Fukumasu; Jayme Augusto de Souza-Neto; José Salvatore Leister Patané; Luiz Alcantara; Luiz Lehmann Coutinho; Maria Carolina Elias; Maurício Lacerda Nogueira; Rafael dos Santos Bezerra; Raul Machado Neto; Rejane Maria Tommasini Grotto; Ricardo Haddad; Sandra Coccuzzo Sampaio Vessoni; Simone Kashima; Svetoslav Nanev Slavov; Vincent Louis Viala |
| EPI_ISL_5802329, EPI_ISL_5802331, EPI_ISL_5802332, EPI_ISL_5802337, EPI_ISL_5802339, EPI_ISL_5802340, EPI_ISL_5802341, EPI_ISL_5802342, EPI_ISL_5802347, EPI_ISL_5802350, EPI_ISL_5802351, EPI_ISL_5802359, EPI_ISL_5802365, EPI_ISL_5802367 | | | |
| see above | Upa Dr Franco Da Rocha | Instituto Butantan | Antonio Jorge Martins; Claudia Renata dos Santos Barros; David Schlesinger; Debora Botequio Moretti; Dimas Tadeu Covas; Elaine Cristina Marqueze; Elaine Vieira Santos; Evandra Strazza Rodrigues; Heidge Fukumasu; Jayme Augusto de Souza-Neto; José Salvatore Leister Patané; Luiz Alcantara; Luiz Lehmann Coutinho; Maria Carolina Elias; Maurício Lacerda Nogueira; Rafael dos Santos Bezerra; Raul Machado Neto; Rejane Maria Tommasini Grotto; Ricardo Haddad; Sandra Coccuzzo Sampaio Vessoni; Simone Kashima; Svetoslav Nanev Slavov; Vincent Louis Viala |
| EPI_ISL_5802381 | Upa Sadako Sedoguti | Instituto Butantan | Antonio Jorge Martins; Claudia Renata dos Santos Barros; David Schlesinger; Debora Botequio Moretti; Dimas Tadeu Covas; Elaine Cristina Marqueze; Elaine Vieira Santos; Evandra Strazza Rodrigues; Heidge Fukumasu; Jayme Augusto de Souza-Neto; José Salvatore Leister Patané; Luiz Alcantara; Luiz Lehmann Coutinho; Maria Carolina Elias; Maurício Lacerda Nogueira; Rafael dos Santos Bezerra; Raul Machado Neto; Rejane Maria Tommasini Grotto; Ricardo Haddad; Sandra Coccuzzo Sampaio Vessoni; Simone Kashima; Svetoslav Nanev Slavov; Vincent Louis Viala |
| EPI_ISL_5800052, EPI_ISL_5800060, EPI_ISL_5800077, EPI_ISL_5800085 | Upa Unidade De Pronto Atendimento 24 Horas Bom Jesus | Instituto Butantan | Antonio Jorge Martins; Claudia Renata dos Santos Barros; David Schlesinger; Debora Botequio Moretti; Dimas Tadeu Covas; Elaine Cristina Marqueze; Elaine Vieira Santos; Evandra Strazza Rodrigues; Heidge Fukumasu; Jayme Augusto de Souza-Neto; José Salvatore Leister Patané; Luiz Alcantara; Luiz Lehmann Coutinho; Maria Carolina Elias; Maurício Lacerda Nogueira; Rafael dos Santos Bezerra; Raul Machado Neto; Rejane Maria Tommasini Grotto; Ricardo Haddad; Sandra Coccuzzo Sampaio Vessoni; Simone Kashima; Svetoslav Nanev Slavov; Vincent Louis Viala |
| EPI_ISL_7019011, EPI_ISL_7019012, EPI_ISL_7040630, EPI_ISL_7040641, EPI_ISL_7040645, EPI_ISL_7040655, EPI_ISL_7040657 | | | |
| see above | Urbino | Microbiology University Politecnica delle Marche | Anna Valenza; Carla Acciarri; Katia Marinelli; Monica Lucia Ferreri; Patrizia Bagnarelli; Roberta Longo; Sara Caucci; Stefano Menzo |
| EPI_ISL_1972943, EPI_ISL_1972944, EPI_ISL_1972946, EPI_ISL_1972947 | Usansolo-Galdakao University Hospital | Cruces University Hospital | Ana Belén de la Hoz; Ana Gual-de-Torrella; Izaskun Alejo-Cancho; Mikel Gallego |
| EPI_ISL_5800094, EPI_ISL_5800125 | Usf I De Guaimbe | Instituto Butantan | Antonio Jorge Martins; Claudia Renata dos Santos Barros; David Schlesinger; Debora Botequio Moretti; Dimas Tadeu Covas; Elaine Cristina Marqueze; Elaine Vieira Santos; Evandra Strazza Rodrigues; Heidge Fukumasu; Jayme Augusto de Souza-Neto; José Salvatore Leister Patané; Luiz Alcantara; Luiz Lehmann Coutinho; Maria Carolina Elias; Maurício Lacerda Nogueira; Rafael dos Santos Bezerra; Raul Machado Neto; Rejane Maria Tommasini Grotto; Ricardo Haddad; Sandra Coccuzzo Sampaio Vessoni; Simone Kashima; Svetoslav Nanev Slavov; Vincent Louis Viala |
| EPI_ISL_5802284 | Usf Jardim Do Lago Artur Nogueira | Instituto Butantan | Antonio Jorge Martins; Claudia Renata dos Santos Barros; David Schlesinger; Debora Botequio Moretti; Dimas Tadeu Covas; Elaine Cristina Marqueze; Elaine Vieira Santos; Evandra Strazza Rodrigues; Heidge Fukumasu; Jayme Augusto de Souza-Neto; José Salvatore Leister Patané; Luiz Alcantara; Luiz Lehmann Coutinho; Maria Carolina Elias; Maurício Lacerda Nogueira; Rafael dos Santos Bezerra; Raul Machado Neto; Rejane Maria Tommasini Grotto; Ricardo Haddad; Sandra Coccuzzo Sampaio Vessoni; Simone Kashima; Svetoslav Nanev Slavov; Vincent Louis Viala |
| EPI_ISL_5802316, EPI_ISL_5802318 | Usf Jardim Piratininga Itaquaquecetuba | Instituto Butantan | Antonio Jorge Martins; Claudia Renata dos Santos Barros; David Schlesinger; Debora Botequio Moretti; Dimas Tadeu Covas; Elaine Cristina Marqueze; Elaine Vieira Santos; Evandra Strazza Rodrigues; Heidge Fukumasu; Jayme Augusto de Souza-Neto; José Salvatore Leister Patané; Luiz Alcantara; Luiz Lehmann Coutinho; Maria Carolina Elias; Maurício Lacerda Nogueira; Rafael dos Santos Bezerra; Raul Machado Neto; Rejane Maria Tommasini Grotto; Ricardo Haddad; Sandra Coccuzzo Sampaio Vessoni; Simone Kashima; Svetoslav Nanev Slavov; Vincent Louis Viala |
| EPI_ISL_5802378 | Usf Pequeno Coracao | Instituto Butantan | Antonio Jorge Martins; Claudia Renata dos Santos Barros; David Schlesinger; Debora Botequio Moretti; Dimas Tadeu Covas; Elaine Cristina Marqueze; Elaine Vieira Santos; Evandra Strazza Rodrigues; Heidge Fukumasu; Jayme Augusto de Souza-Neto; José Salvatore Leister Patané; Luiz Alcantara; Luiz Lehmann Coutinho; Maria Carolina Elias; Maurício Lacerda Nogueira; Rafael dos Santos Bezerra; Raul Machado Neto; Rejane Maria Tommasini Grotto; Ricardo Haddad; Sandra Coccuzzo Sampaio Vessoni; Simone Kashima; Svetoslav Nanev Slavov; Vincent Louis Viala |
| EPI_ISL_5802288 | Usf Pq Dos Trabalhadores Artur Nogueira | Instituto Butantan | Antonio Jorge Martins; Claudia Renata dos Santos Barros; David Schlesinger; Debora Botequio Moretti; Dimas Tadeu Covas; Elaine Cristina Marqueze; Elaine Vieira Santos; Evandra Strazza Rodrigues; Heidge Fukumasu; Jayme Augusto de Souza-Neto; José Salvatore Leister Patané; Luiz Alcantara; Luiz Lehmann Coutinho; Maria Carolina Elias; Maurício Lacerda Nogueira; Rafael dos Santos Bezerra; Raul Machado Neto; Rejane Maria Tommasini Grotto; Ricardo Haddad; Sandra Coccuzzo Sampaio Vessoni; Simone Kashima; Svetoslav Nanev Slavov; Vincent Louis Viala |
| EPI_ISL_5802286 | Usf Sacilotto Artur Nogueira | Instituto Butantan | Antonio Jorge Martins; Claudia Renata dos Santos Barros; David Schlesinger; Debora Botequio Moretti; Dimas Tadeu Covas; Elaine Cristina Marqueze; Elaine Vieira Santos; Evandra Strazza Rodrigues; Heidge Fukumasu; Jayme Augusto de Souza-Neto; José Salvatore Leister Patané; Luiz Alcantara; Luiz Lehmann Coutinho; Maria Carolina Elias; Maurício Lacerda Nogueira; Rafael dos Santos Bezerra; Raul Machado Neto; Rejane Maria Tommasini Grotto; Ricardo Haddad; Sandra Coccuzzo Sampaio Vessoni; Simone Kashima; Svetoslav Nanev Slavov; Vincent Louis Viala |
| EPI_ISL_2081801, EPI_ISL_2677893, EPI_ISL_2677894 | Utah Public Health Laboratory | Utah Public Health Laboratory | Erin L. Young; Kelly F. Oakeson; Tara Gallagher |
| EPI_ISL_2283946, EPI_ISL_2283952 | VA Connecticut Healthcare System | Yale Center for Genomic Analysis | Brooke Sullivan; Curt Scharfe; Irina Tikhonova; Kaya Bilguvar; Shrikant Mane |
| EPI_ISL_1966556, EPI_ISL_1966560 | VIGILANCIA EM SAUDE SAE VIGILANCIA EPIDEMIOLOGICA | Instituto Butantan / Mendelics | Antonio Jorge Martins; Bianca Cechetto Carlos. Mendelics: Bibiana Santos; Claudia Renata dos Santos Barros; Cíntia Bittar; David Schlesinger; David Schlesinger. Hemocentro Ribeirão Preto: Simone Kashima; Debora Botequio Moretti; Dimas Tadeu Covas; Elaine Cristina Marqueze; Elaine Vieira Santos; Elaine Vieira dos Santos; Elisangela Chicaroni Mattos; Erika Freitas; Evandra Strazza Rodrigues; Felipe Allan da Silva da Costa; Flavia Aburjaile; Fábio Sossai Possebon; Guilherme Campos; Guilherme Targino Valente; Heidge Fukumasu; Heidge Fukumasu. USP-Botucatu: Rejane Maria Tommasini Grotto; Helena Lage Ferreira; Instituto Butantan: Dimas Tadeu Covas; Jardelina de Souza Todao Bernardino; Jayme A. Souza-Neto; Jayme Augusto de Souza-Neto; Jessika Cristina Chagas Lesbon; Jorge A. Petroli Marchesi; José Salvatore Leister Patané; João Paulo Kitajima; João Pessoa Araújo Jr.; Leila Sabrina Ullmann; Loyze Paola Oliveira de Lima; Luiz Alcantara; Luiz Aurelio de Campos Crispin. Centro de Genômica Funcional da ESALQ: Luiz Lehmann Coutinho; Luiz Carlos Junior de Alcantara; Luiz Lehmann Coutinho; Lívia Sacchetto; Maisa C. Pereira Parra; Maria Carolina Elias; Marta Giovanetti; Marília Moraes; Maurício Lacerda Nogueira; Maurício Lacerda Nogueira. Prefeitura de Sao Paulo: Melissa Palmieri.; Patricia Akemi Assato; Paula Rahal; Paulo Inacio da Costa; Rafael dos Santos Bezerra; Raquel de Lello Rocha Campos Cassano. NGS Soluções Genômicas: Pilar Drummond Sampaio Corrêa Mariani. FZEA-USP Pirassununga: Mirele Daiana Poleti; Raul Machado Neto; Rejane Maria Tommasini Grotto; Ricardo Augusto Brassaloti; Ricardo Haddad; Rodrigo Tocantins Calado. FAMERP-SJRP: Cecília Artico Banho; Sandra Coccuzzo Sampaio; Sandra Coccuzzo Sampaio Vessoni; Simone Kashima; Svetoslav Nanev Slavov; Vagner Fonseca; Vincent Louis Viala |
| EPI_ISL_2210231, EPI_ISL_2210243 | VIGILANCIA EPIDEMIOLOGICA E CONTROLE DE VETORES PIRASSUNUN | Instituto Butantan | Antonio Jorge Martins; Claudia Renata dos Santos Barros; David Schlesinger; Debora Botequio Moretti; Dimas Tadeu Covas; Elaine Cristina Marqueze; Elaine Vieira Santos; Evandra Strazza Rodrigues; Heidge Fukumasu; Jayme Augusto de Souza-Neto; José Salvatore Leister Patané; Luiz Alcantara; Luiz Lehmann Coutinho; Maria Carolina Elias; Maurício Lacerda Nogueira; Rafael dos Santos Bezerra; Raul Machado Neto; Rejane Maria Tommasini Grotto; Ricardo Haddad; Sandra Coccuzzo Sampaio Vessoni; Simone Kashima; Svetoslav Nanev Slavov; Vincent Louis Viala. |
| EPI_ISL_2209443, EPI_ISL_2209448, EPI_ISL_2344697, EPI_ISL_2344698, EPI_ISL_2344704, EPI_ISL_2344706 | VIGILANCIA EPIDEMIOLOGICA E CONTROLE DE VETORES PIRASSUNUN | Instituto Butantan / FZEA-USP-Pirassununga | Antonio Jorge Martins; Claudia Renata dos Santos Barros; David Schlesinger; Debora Botequio Moretti; Dimas Tadeu Covas; Elaine Cristina Marqueze; Elaine Vieira Santos; Evandra Strazza Rodrigues; Heidge Fukumasu; Jayme Augusto de Souza-Neto; José Salvatore Leister Patané; Luiz Alcantara; Luiz Lehmann Coutinho; Maria Carolina Elias; Maurício Lacerda Nogueira; Rafael dos Santos Bezerra; Raul Machado Neto; Rejane Maria Tommasini Grotto; Ricardo Haddad; Sandra Coccuzzo Sampaio Vessoni; Simone Kashima; Svetoslav Nanev Slavov; Vincent Louis Viala |
| EPI_ISL_1966830, EPI_ISL_1966831 | VIGILANCIA EPIDIMIOLOGICA DE ARACARIGUAMA | Instituto Butantan / ESALQ-USP (Piracicaba) | Antonio Jorge Martins; Bianca Cechetto Carlos. Mendelics: Bibiana Santos; Claudia Renata dos Santos Barros; Cíntia Bittar; David Schlesinger. Hemocentro Ribeirão Preto: Simone Kashima; Debora Botequio Moretti; Elaine Cristina Marqueze; Elaine Vieira dos Santos; Elisangela Chicaroni Mattos; Erika Freitas; Evandra Strazza Rodrigues; Felipe Allan da Silva da Costa; Flavia Aburjaile; Fábio Sossai Possebon; Guilherme Campos; Guilherme Targino Valente; Heidge Fukumasu. USP-Botucatu: Rejane Maria Tommasini Grotto; Helena Lage Ferreira; Instituto Butantan: Dimas Tadeu Covas; Jardelina de Souza Todao Bernardino; Jayme A. Souza-Neto; Jessika Cristina Chagas Lesbon; Jorge A. Petroli Marchesi; José Salvatore Leister Patané; João Paulo Kitajima; João Pessoa Araújo Jr.; Leila Sabrina Ullmann; Loyze Paola Oliveira de Lima; Luiz Aurelio de Campos Crispin. Centro de Genômica Funcional da ESALQ: Luiz Lehmann Coutinho; Luiz Carlos Junior de Alcantara; Lívia Sacchetto; Maisa C. Pereira Parra; Maria Carolina Elias; Marta Giovanetti; Marília Moraes; Maurício Lacerda Nogueira. Prefeitura de Sao Paulo: Melissa Palmieri.; Patricia Akemi Assato; Paula Rahal; Paulo Inacio da Costa; Rafael dos Santos Bezerra; Raquel de Lello Rocha Campos Cassano. NGS Soluções Genômicas: Pilar Drummond Sampaio Corrêa Mariani. FZEA-USP Pirassununga: Mirele Daiana Poleti; Raul Machado Neto; Ricardo Augusto Brassaloti; Ricardo Haddad; Rodrigo Tocantins Calado. FAMERP-SJRP: Cecília Artico Banho; Sandra Coccuzzo Sampaio; Svetoslav Nanev Slavov; Vagner Fonseca; Vincent Louis Viala |
| EPI_ISL_2345973, EPI_ISL_2345974, EPI_ISL_2345975, EPI_ISL_2346012 | VIGILANCIA SANITARIA E VIG EPIDEMIOLOGICA DE ITAPEVI | Instituto Butantan | Antonio Jorge Martins; Claudia Renata dos Santos Barros; David Schlesinger; Debora Botequio Moretti; Dimas Tadeu Covas; Elaine Cristina Marqueze; Elaine Vieira Santos; Evandra Strazza Rodrigues; Heidge Fukumasu; Jayme Augusto de Souza-Neto; José Salvatore Leister Patané; Luiz Alcantara; Luiz Lehmann Coutinho; Maria Carolina Elias; Maurício Lacerda Nogueira; Rafael dos Santos Bezerra; Raul Machado Neto; Rejane Maria Tommasini Grotto; Ricardo Haddad; Sandra Coccuzzo Sampaio Vessoni; Simone Kashima; Svetoslav Nanev Slavov; Vincent Louis Viala |
| EPI_ISL_1966935, EPI_ISL_1966937, EPI_ISL_1966938, EPI_ISL_1966941 | VIGILANCIA SANITARIA E VIG EPIDEMIOLOGICA DE ITAPEVI | Instituto Butantan / ESALQ-USP (Piracicaba) | Antonio Jorge Martins; Bianca Cechetto Carlos. Mendelics: Bibiana Santos; Claudia Renata dos Santos Barros; Cíntia Bittar; David Schlesinger. Hemocentro Ribeirão Preto: Simone Kashima; Debora Botequio Moretti; Elaine Cristina Marqueze; Elaine Vieira dos Santos; Elisangela Chicaroni Mattos; Erika Freitas; Evandra Strazza Rodrigues; Felipe Allan da Silva da Costa; Flavia Aburjaile; Fábio Sossai Possebon; Guilherme Campos; Guilherme Targino Valente; Heidge Fukumasu. USP-Botucatu: Rejane Maria Tommasini Grotto; Helena Lage Ferreira; Instituto Butantan: Dimas Tadeu Covas; Jardelina de Souza Todao Bernardino; Jayme A. Souza-Neto; Jessika Cristina Chagas Lesbon; Jorge A. Petroli Marchesi; José Salvatore Leister Patané; João Paulo Kitajima; João Pessoa Araújo Jr.; Leila Sabrina Ullmann; Loyze Paola Oliveira de Lima; Luiz Aurelio de Campos Crispin. Centro de Genômica Funcional da ESALQ: Luiz Lehmann Coutinho; Luiz Carlos Junior de Alcantara; Lívia Sacchetto; Maisa C. Pereira Parra; Maria Carolina Elias; Marta Giovanetti; Marília Moraes; Maurício Lacerda Nogueira. Prefeitura de Sao Paulo: Melissa Palmieri.; Patricia Akemi Assato; Paula Rahal; Paulo Inacio da Costa; Rafael dos Santos Bezerra; Raquel de Lello Rocha Campos Cassano. NGS Soluções Genômicas: Pilar Drummond Sampaio Corrêa Mariani. FZEA-USP Pirassununga: Mirele Daiana Poleti; Raul Machado Neto; Ricardo Augusto Brassaloti; Ricardo Haddad; Rodrigo Tocantins Calado. FAMERP-SJRP: Cecília Artico Banho; Sandra Coccuzzo Sampaio; Svetoslav Nanev Slavov; Vagner Fonseca; Vincent Louis Viala |
| EPI_ISL_3667131 | VIROLOGY LABORATORY-CHU NICE | VIROLOGY LABORATORY-CHU NICE | Aicha El Yakine; Geraldine Gonfrier; Jean Machowiak; Sebastien Vitale; Valerie Giordanengo; Virginie Flahou |
| EPI_ISL_5780265 | VUMC Molecular Infectious | Dr. Suman Das Lab - Vanderbilt | Bookyung Park; Grant Vestal; Helen Boone; Hunter Brown; Jonathan E. Schmitz; Meghan Shilts; Seesandra Rajagopala; Suman B. Pakala; Suman Das |

| | | | |
|---|---|---|---|
| | Diseases Laboratory (MIDL) | University Medical Center (VUMC) | |
| EPI_ISL_5799958, EPI_ISL_5799965 | Vigilancia Em Saude Sae Vigilancia Epidemiologica | Instituto Butantan | Antonio Jorge Martins; Claudia Renata dos Santos Barros; David Schlesinger; Debora Botequio Moretti; Dimas Tadeu Covas; Elaine Cristina Marqueze; Elaine Vieira Santos; Evandra Strazza Rodrigues; Heidge Fukumasu; Jayme Augusto de Souza-Neto; José Salvatore Leister Patané; Luiz Alcantara; Luiz Lehmann Coutinho; Maria Carolina Elias; Maurício Lacerda Nogueira; Rafael dos Santos Bezerra; Raul Machado Neto; Rejane Maria Tommasini Grotto; Ricardo Haddad; Sandra Coccuzzo Sampaio Vessoni; Simone Kashima; Svetoslav Nanev Slavov; Vincent Louis Viala |
| EPI_ISL_5799691, EPI_ISL_5799692, EPI_ISL_5799698, EPI_ISL_5799700, EPI_ISL_5799701, EPI_ISL_5799702 | Vigilancia Epidemiologica E Controle De Vetores Pirassununga | Instituto Butantan | Antonio Jorge Martins; Claudia Renata dos Santos Barros; David Schlesinger; Debora Botequio Moretti; Dimas Tadeu Covas; Elaine Cristina Marqueze; Elaine Vieira Santos; Evandra Strazza Rodrigues; Heidge Fukumasu; Jayme Augusto de Souza-Neto; José Salvatore Leister Patané; Luiz Alcantara; Luiz Lehmann Coutinho; Maria Carolina Elias; Maurício Lacerda Nogueira; Rafael dos Santos Bezerra; Raul Machado Neto; Rejane Maria Tommasini Grotto; Ricardo Haddad; Sandra Coccuzzo Sampaio Vessoni; Simone Kashima; Svetoslav Nanev Slavov; Vincent Louis Viala |
| EPI_ISL_5800336, EPI_ISL_5800348 | Vigilancia Epidimiologica De Aracariguama | Instituto Butantan | Antonio Jorge Martins; Claudia Renata dos Santos Barros; David Schlesinger; Debora Botequio Moretti; Dimas Tadeu Covas; Elaine Cristina Marqueze; Elaine Vieira Santos; Evandra Strazza Rodrigues; Heidge Fukumasu; Jayme Augusto de Souza-Neto; José Salvatore Leister Patané; Luiz Alcantara; Luiz Lehmann Coutinho; Maria Carolina Elias; Maurício Lacerda Nogueira; Rafael dos Santos Bezerra; Raul Machado Neto; Rejane Maria Tommasini Grotto; Ricardo Haddad; Sandra Coccuzzo Sampaio Vessoni; Simone Kashima; Svetoslav Nanev Slavov; Vincent Louis Viala |
| EPI_ISL_5799861, EPI_ISL_5799869, EPI_ISL_5799887, EPI_ISL_5799900, EPI_ISL_5802305, EPI_ISL_5802306, EPI_ISL_5802307, EPI_ISL_5802343 | | | |
| see above | Vigilancia Sanitaria E Vig Epidemiologica De Itapevi | Instituto Butantan | Antonio Jorge Martins; Claudia Renata dos Santos Barros; David Schlesinger; Debora Botequio Moretti; Dimas Tadeu Covas; Elaine Cristina Marqueze; Elaine Vieira Santos; Evandra Strazza Rodrigues; Heidge Fukumasu; Jayme Augusto de Souza-Neto; José Salvatore Leister Patané; Luiz Alcantara; Luiz Lehmann Coutinho; Maria Carolina Elias; Maurício Lacerda Nogueira; Rafael dos Santos Bezerra; Raul Machado Neto; Rejane Maria Tommasini Grotto; Ricardo Haddad; Sandra Coccuzzo Sampaio Vessoni; Simone Kashima; Svetoslav Nanev Slavov; Vincent Louis Viala |
| EPI_ISL_2614586 | Vigilância Sanitaria E Epidemiologica | Instituto Adolfo Lutz, Interdiciplinary Procedures Center, Strategic Laboratory | Caio Vinicius Dias Lopes; Claudia Regina Gonçalves; Claudio Tavares Sacchi; Erica Valessa Ramos Gomes; Karoline Rodrigues Campos; Leonardo Jose Tadeu de Araujo |
| EPI_ISL_1659165, EPI_ISL_1659173 | Viollier AG | Department of Biosystems Science and Engineering, ETH Zurich | Andrea Patrignani; Andreia Cabral de Gouvea; Catharine Aquino; Chaoran Chen; Christiane Beckmann; Christoph Noppen; David Dreifuss; Deborah Penet; Doris Popovic; Emmanouil Dermitzakis; Griffin White; Henri Pegeot; Ioannis Xenarios; Ivan Topolsky; Jay Tracy; Katharina Jahn; Keith Harshman; Lara Fuhrmann; Laura Neff; Lennart Opitz; Lorenzo Cerutti; Maria Domenica Moccia; Maurice Redondo; Niko Beerenwinkel; Noemie Santamaria de Souza; Olivier Kobel; Philipp Jablonski; Ralph Schlapbach; Sarah Nadeau; Simon Gruter; Sophie Seidel; Tanja Stadler; Timothy Sykes |
| EPI_ISL_1822095 | Virginia Division of Consolidated Laboratory Services | Virginia Division of Consolidated Laboratory Services | Virginia DCLS |
| EPI_ISL_2434091 | Virology Laboratory, Scientific Department, Army Medical Center | Virology Laboratory, Scientific Department, Army Medical Center | Anella Monte; Anna Anselmo; Antonella Fortunato; Filippo Molinari; Florigio Lista.; Francesco Giordani; Giancarlo Petralito; Giandomenico Cerreto; Giulia Campoli; Lucia Nicosia; Marzia Cavalli; Riccardo De Sanctis; Rossella Brandi; Silvia Fillo; Vanessa Vera Fain |
| EPI_ISL_1671475 | WESTCHESTER MEDICAL CENTER | Wadsworth Center, New York State Department of Health | Alexis Russell; Catharine Prussing; Daryl M. Lamson; Erasmus Schneider; Erica Lasek-Nesselquist; John Kelly; Jonathan Plitnick; Kirsten St. George; Matthew Shudt; Melissa A Leisner; Navjot Singh |
| EPI_ISL_2295379 | WLA VAMC | Genomics and Discovery, Respiratory Viruses Branch, Division of Viral Diseases, Centers for Disease Control and Prevention | Adam Retchless; Anna Kelleher; Anna Uehara; Brian Lynch; Clinton R. Paden; Dhwani Batra; Haibin Wang; Han Jia Justin Ng; Jasmine Padilla; Jing Zhang; Justin Lee; Krista Queen; Mark Burroughs; Mili Sheth; Morgan Davis; Peter Cook; Rachel Marine; Sarah Nobles; Suxiang Tong; Tara Coalter; Yan Li; Ying Tao |
| EPI_ISL_1747421, EPI_ISL_1747437 | Washington State Department of Health Public Health Laboratories | Washington State Department of Health Public Health Laboratories | Avi Singh; Darren Lucas; Denny Russell; Drew MacKellar; Geoff Melly; Hannah Gray; Joenice Gonzalez; JohnAric Peterson; Philip Dykema; Rebecca Cao; Vanessa De Los Santos |
| EPI_ISL_2442283, EPI_ISL_2454540 | Wexner Medical Center | The Ohio State University College of Medicine | Koenig, S.; Seminetta, J. |
| EPI_ISL_1575419 | Wichita State University - Molecular Diagnostics Lab | Kansas Health and Environmental Lab | Ben Olsen; Jonathan Barnell; Mike Grose; and Phil Adam |
| EPI_ISL_1794087, EPI_ISL_1794115, EPI_ISL_1794239, EPI_ISL_2018546, EPI_ISL_2018614, EPI_ISL_2018675, EPI_ISL_2018676, EPI_ISL_2018692 | | | |
| see above | Wisconsin State Laboratory of Hygiene Communicable Disease Division | Wisconsin State Laboratory of Hygiene Communicable Disease Division | Abigail C. Shockey; Alicia J. Mooney; Erika M. Hanson; Kelsey R. Florek; Richard Griesser; Sara Wagner; Tonya Danz |
| EPI_ISL_3722611 | WolfTRACS Surveillance Lab | Harris/Rasmussen Lab | Angela Harris; David Rasmussen; Gina Chaput; Megan Jacob; Temi Ibitoye |
| EPI_ISL_1674660, EPI_ISL_1792971, EPI_ISL_1793058 | Yale Clinical Virology Lab | Grubaugh Lab - Yale School of Public Health | Anderson Brito; Annie Watkins; Chaney Kalinich; Chantal Vogels; Isabel Ott; Jessica Rothman; Joseph Fauver; Mallery Breban; Marie L. Landry; Mary Petrone; Nathan Grubaugh; Tara Alpert |
