## Supplement 3 for "Rapid and Accurate Identification of SARS-CoV-2 Omicron Variants Using Droplet Digital PCR (RT-ddPCR)": All_0412-0418.pdf

We gratefully acknowledge the following Authors from the Originating laboratories responsible for obtaining the specimens, as well as the Submitting laboratories where the genome data were generated and shared via GISAID, on which this research is based.

All Submitters of data may be contacted directly via www.gisaid.org

Authors are sorted alphabetically.

| Accession ID | Originating Laboratory | Submitting Laboratory | Authors |
|---|---|---|---|
| EPI_ISL_2645299, EPI_ISL_2645300, EPI_ISL_2645302, EPI_ISL_2645305 | "CO Dept. of Public Health and Environment, Lab Services Division" | Centers for Disease Control and Prevention Division of Viral Diseases, Pathogen Discovery | Alison Laufer Halpin; Ben L. Rambo-Martin; Clinton R. Paden; Dakota Howard; Darlene Wagner; Dave Wentworth; Dhwani Batra; Jasmine Padilla; Justin Lee; Katie Dillon; Krista Queen; Kristen Knipe; Kristine Lacek; Mark Burroughs; Matthew Schmerer; Mili Sheth; Peter Cook; Sam Shepard; Sarah Nobles; Shoshona Le; Suxiang Tong; Vivien Dugan; Yvette Unoarumhi |
| EPI_ISL_5899624 | A. Krumbholz, Labor Dr. Krause und Kollegen MVZ GmbH, Kiel | Charité Universitätsmedizin Berlin, Institut für Virologie | Barbara Mühlemann; Christian Drosten; Julia Schneider; Julia Tesch; Jörn Beheim-Schwarzbach; Talitha Veith; Terry Jones; Tobias Bleicker; Victor M Corman |
| EPI_ISL_2720406 | A.S.L. VERCELLI | Fondazione del Piemonte per l'Oncologia IRCCS | Antonino Sottile; Giorgio Giardina; Paola Marino; Silvia Brossa |
| EPI_ISL_3347191, EPI_ISL_3347192 | ADILAB | Laboratorio Departamental de Salud Publica de Antioquia | Andres F. Cardona-Rios; Gloria Isabel Escobar; Idabely Betancur Ortiz; Juan P. Hernandez-Ortiz; Maria Stella López |
| EPI_ISL_1960045 | ADILAB | Universidad Nacional de Colombia - Laboratorio Genómico One Health | Andres F. Cardona-Rios; Carlos Franco-Muñoz; Carolina Muñoz-Arango; Celeny Ortiz; Daniel O. Maldonado-Perez; Diego A. Álvarez-Díaz; Hector Alejandro Ruiz-Moreno; Idabely Betancur Ortiz; Jorge E. Osorio; Juan P. Hernandez-Ortiz; Karl A Ciuoderis; Katherine Laiton-Donato; Laura Silvana Perez; Lina M. Hurtado; Marcela Mercado-Reyes; Maria Angélica Maya; Maria Stella López; Rita Almanza Payares; Sandra Ines Cano; Simón Villegas Velásquez |
| EPI_ISL_1756015 | ALBANY MEDICAL CENTER | Wadsworth Center, New York State Department of Health | Alexis Russell; Catharine Prussing; Daryl M. Lamson; Erasmus Schneider; Erica Lasek-Nesselquist; John Kelly; Jonathan Plitnick; Kirsten St. George; Matthew Shudt; Melissa A Leisner; Navjot Singh |
| EPI_ISL_5915132, EPI_ISL_5915177, EPI_ISL_5915205, EPI_ISL_5915223, EPI_ISL_5915249, EPI_ISL_5915304, EPI_ISL_5915309, EPI_ISL_5915310, EPI_ISL_6573792, EPI_ISL_6573826, EPI_ISL_6573827, EPI_ISL_6573828, EPI_ISL_6573961 | | | |
| see above | AMALAB/FACISA/UFRN | WallauLab on behalf of Fiocruz COVID-19 Genomic Surveillance Network | Alexandre Freitas da Silva; Allan Roberto Dias Nunes; Antonio Marinho da Silva Neto; Cassia Docena; Constância Flávia Junqueira Ayres; Filipe Zimmer Dezordi; Gabriel Luz Wallau; Gustavo Barbosa de Lima; Joana Cristina Medeiros Tavares Marques; Katya Anaya Jacinto; Lais Ceschini Machado; Lilian Caroliny Amorim Silva; Marcelo Henrique dos Santos Paiva; Mariane dos Santos Duarte; Matheus Filgueira Bezerra; Sinval Pinto Brandão Filho |
| EPI_ISL_2344558, EPI_ISL_2344563 | AMBULATORIO DE ESPECIALIDADE V E MOGI MIRIM | Instituto Butantan / FZEA-USP-Pirassununga | Antonio Jorge Martins; Claudia Renata dos Santos Barros; David Schlesinger; Debora Botequio Moretti; Dimas Tadeu Covas; Elaine Cristina Marqueze; Elaine Vieira Santos; Evandra Strazza Rodrigues; Heidge Fukumasu; Jayme Augusto de Souza-Neto; José Salvatore Leister Patané; Luiz Alcantara; Luiz Lehmann Coutinho; Maria Carolina Elias; Maurício Lacerda Nogueira; Rafael dos Santos Bezerra; Raul Machado Neto; Rejane Maria Tommasini Grotto; Ricardo Haddad; Sandra Coccuzzo Sampaio Vessoni; Simone Kashima; Svetoslav Nanev Slavov; Vincent Louis Viala |
| EPI_ISL_2170964 | AMBULATORIO DR JOAO LANIA CENTRO JOSE BONIFACIO | Instituto Butantan / Mendelecis | Antonio Jorge Martins; Bianca Cechetto Carlos. Mendelics: Bibiana Santos; Claudia Renata dos Santos Barros; Cíntia Bittar; David Schlesinger. Hemocentro Ribeirão Preto: Simone Kashima; Debora Botequio Moretti; Elaine Cristina Marqueze; Elaine Vieira dos Santos; Elisangela Chicaroni Mattos; Erika Freitas; Evandra Strazza Rodrigues; Felipe Allan da Silva da Costa; Flavia Aburjaile; Fábio Sossai Possebon; Guilherme Campos; Guilherme Targino Valente; Heidge Fukumasu. USP-Botucatu: Rejane Maria Tommasini Grotto; Helena Lage Ferreira; Instituto Butantan: Dimas Tadeu Covas; Jardelina de Souza Todao Bernardino; Jayme A. Souza-Neto; Jessika Cristina Chagas Lesbon; Jorge A. Petroli Marchesi; José Salvatore Leister Patané; João Paulo Kitajima; João Pessoa Araújo Jr.; Leila Sabrina Ullmann; Loyze Paola Oliveira de Lima; Luiz Aurelio de Campos Crispin. Centro de Genômica Funcional da ESALQ: Luiz Lehmann Coutinho; Luiz Carlos Junior de Alcantara; Lívia Sacchetto; Maisa C. Pereira Parra; Maria Carolina Elias; Marta Giovanetti; Marília Moraes; Maurício Lacerda Nogueira. Prefeitura de Sao Paulo: Melissa Palmieri.; Patricia Akemi Assato; Paula Rahal; Paulo Inacio da Costa; Rafael dos Santos Bezerra; Raquel de Lello Rocha Campos Cassano. NGS Soluções Genômicas: Pilar Drummond Sampaio Corrêa Mariani. FZEA-USP Pirassununga: Mirele Daiana Poleti; Raul Machado Neto; Ricardo Augusto Brassaloti; Ricardo Haddad; Rodrigo Tocantins Calado. FAMERP-SJRP: Cecília Artico Banho; Sandra Coccuzzo Sampaio; Svetoslav Nanev Slavov; Vagner Fonseca; Vincent Louis Viala |
| EPI_ISL_2272981 | AREA DE SALUD CARRILLO | Inciensa, Instituto Costarricense de Investigación y Enseñanza en Nutrición y Salud | Adriana Godínez; Claudio Soto-Garita; Estela Cordero; Francisco Duarte; Hebleen Porras; Joselyn Prado & Adriana Bemúdez Espinoza; José Luis Vargas; Mariela Gutiérrez; Melany Calderón |
| EPI_ISL_1811234 | AS Santa Barbara | Inciensa, Instituto Costarricense de Investigación y Enseñanza en Nutrición y Salud | Pérez-Corrales C & Aguilar-Monge R |
| EPI_ISL_1940196, EPI_ISL_1940215, EPI_ISL_1940225, EPI_ISL_1940266, EPI_ISL_1940314, EPI_ISL_1940360, EPI_ISL_1940552 | | | |
| see above | ASL Napoli 1 Centro | AMES Centro Polidiagnostico Strumentale S.r.l. | Antonella Di Carlo; Antonio Fico; Eloisa Evangelista; Giovanni Savarese; Luigi D'Amore; Luisa Circelli; Maurizio D'Amora; Monica Ianniello; Nadia Petrillo; Raffaella Ruggiero; Roberto Sirica |
| EPI_ISL_1824600 | AZ Klina | AZ Klina | Carl Vael - Lynsey Berckmans |
| EPI_ISL_1904333 | AZDelta | AZDelta | Dieter De Smet; Geert Martens |
| EPI_ISL_1667693, EPI_ISL_1667694, EPI_ISL_1667703, EPI_ISL_1667718, EPI_ISL_1687502, EPI_ISL_1695521, EPI_ISL_1837427, EPI_ISL_1837442, EPI_ISL_1837454, EPI_ISL_1837486, EPI_ISL_1837492, EPI_ISL_1837525, EPI_ISL_1837526, EPI_ISL_1837537, EPI_ISL_1900402, EPI_ISL_1922902, EPI_ISL_1922930, EPI_ISL_1922950, EPI_ISL_1922958, EPI_ISL_1922959, EPI_ISL_1922970, EPI_ISL_1922976, EPI_ISL_1922977, EPI_ISL_1923016, EPI_ISL_1923092, EPI_ISL_1923146, EPI_ISL_1923148, EPI_ISL_1995348, EPI_ISL_1995349, EPI_ISL_1995418, EPI_ISL_1995434, EPI_ISL_1995438, EPI_ISL_1995485, EPI_ISL_1995937, EPI_ISL_1995939, EPI_ISL_1995986, EPI_ISL_1996152, EPI_ISL_1996153, EPI_ISL_1996222, EPI_ISL_1997461, EPI_ISL_1997492, EPI_ISL_1997494, EPI_ISL_1997517, EPI_ISL_1997602, EPI_ISL_1997607, EPI_ISL_1997621, EPI_ISL_1997738, EPI_ISL_1997838, EPI_ISL_1997887, EPI_ISL_1997889, EPI_ISL_1997913, EPI_ISL_1997996, EPI_ISL_1998008, EPI_ISL_1998010, EPI_ISL_1998025, EPI_ISL_1998026, EPI_ISL_1998042, EPI_ISL_1998098, EPI_ISL_1998179, EPI_ISL_1998188, EPI_ISL_1998194, EPI_ISL_1998195, EPI_ISL_1998290, EPI_ISL_1998302, EPI_ISL_1998314, EPI_ISL_1998346, EPI_ISL_1998350, EPI_ISL_1998374, EPI_ISL_1998395, EPI_ISL_1998418, EPI_ISL_1998431, EPI_ISL_1998438, EPI_ISL_1998441, EPI_ISL_1998518, EPI_ISL_1998522, EPI_ISL_1998544, EPI_ISL_1998555, EPI_ISL_1998561, EPI_ISL_1998564, EPI_ISL_1998573, EPI_ISL_1998612, EPI_ISL_1998636, EPI_ISL_1998676, EPI_ISL_1998679, EPI_ISL_1998713, EPI_ISL_1998735, EPI_ISL_1998751, EPI_ISL_1998754, EPI_ISL_1998770, EPI_ISL_1998787, EPI_ISL_1998795, EPI_ISL_1998802, EPI_ISL_1998808, EPI_ISL_1998810, EPI_ISL_1998855, EPI_ISL_1998868, EPI_ISL_1999015, EPI_ISL_1999034, EPI_ISL_1999057, EPI_ISL_1999085, EPI_ISL_1999086, EPI_ISL_1999161, EPI_ISL_1999177, EPI_ISL_1999178, EPI_ISL_1999217, EPI_ISL_1999239, EPI_ISL_1999252, EPI_ISL_1999322, EPI_ISL_1999334, EPI_ISL_1999346, EPI_ISL_1999375, EPI_ISL_1999389, EPI_ISL_1999465, EPI_ISL_1999488, EPI_ISL_1999574, EPI_ISL_1999582, EPI_ISL_1999604, EPI_ISL_1999607, EPI_ISL_1999619, EPI_ISL_1999663, EPI_ISL_1999668, EPI_ISL_1999749, EPI_ISL_1999949, EPI_ISL_1999959, EPI_ISL_1999961, EPI_ISL_1999966, EPI_ISL_1999977, EPI_ISL_2000154, EPI_ISL_2000155, EPI_ISL_2000167, EPI_ISL_2000179, EPI_ISL_2000195, EPI_ISL_2000208, EPI_ISL_2000266, EPI_ISL_2000270, EPI_ISL_2000271, EPI_ISL_2000296, EPI_ISL_2000335, EPI_ISL_2000336, EPI_ISL_2000345, EPI_ISL_2000349, EPI_ISL_2000370, EPI_ISL_2000373, EPI_ISL_2000413, EPI_ISL_2000425, EPI_ISL_2000427, EPI_ISL_2000458, EPI_ISL_2000489, EPI_ISL_2000496, EPI_ISL_2000509, EPI_ISL_2000512, EPI_ISL_2000525, EPI_ISL_2000539, EPI_ISL_2035209, EPI_ISL_2035213, EPI_ISL_2035219, EPI_ISL_2035225, EPI_ISL_2035227, EPI_ISL_2035235, EPI_ISL_2039244, EPI_ISL_2039316, EPI_ISL_2039404, EPI_ISL_2039551, EPI_ISL_2039579, EPI_ISL_2039617, EPI_ISL_2039716, EPI_ISL_2039719, EPI_ISL_2039727, EPI_ISL_2039733, EPI_ISL_2039842, EPI_ISL_2039974, EPI_ISL_2039976, EPI_ISL_2040012, EPI_ISL_2040080, EPI_ISL_2040085, EPI_ISL_2040108, EPI_ISL_2040423, EPI_ISL_2040505, EPI_ISL_2040513, EPI_ISL_2040523, EPI_ISL_2040689, EPI_ISL_2040725, EPI_ISL_2040736, EPI_ISL_2040794, EPI_ISL_2040851, EPI_ISL_2040885, EPI_ISL_2040899, EPI_ISL_2040900, EPI_ISL_2041010, EPI_ISL_2041012, EPI_ISL_2041018, EPI_ISL_2041031, EPI_ISL_2041046, EPI_ISL_2041212, EPI_ISL_2041215, EPI_ISL_2041216, EPI_ISL_2041243, EPI_ISL_2041730, EPI_ISL_2041980, EPI_ISL_2041984, EPI_ISL_2041996, EPI_ISL_2042007, EPI_ISL_2042009, EPI_ISL_2042128, EPI_ISL_2042129, EPI_ISL_2042150, EPI_ISL_2042350, EPI_ISL_2042451, EPI_ISL_2042482, EPI_ISL_2149527, EPI_ISL_2201145, EPI_ISL_2201168, EPI_ISL_2201173, EPI_ISL_2201195, EPI_ISL_2201198, EPI_ISL_2201201, EPI_ISL_2201231, EPI_ISL_2201369, EPI_ISL_2201428, EPI_ISL_2201449, EPI_ISL_2201486, EPI_ISL_2201547, EPI_ISL_2201562, EPI_ISL_2201628, EPI_ISL_2201638, EPI_ISL_2201761, EPI_ISL_2201774, EPI_ISL_2201815, EPI_ISL_2201864, EPI_ISL_2201884, EPI_ISL_2201886, EPI_ISL_2201920, EPI_ISL_2201925, EPI_ISL_2201942, EPI_ISL_2202002, EPI_ISL_2202004, EPI_ISL_2202005, EPI_ISL_2202011, EPI_ISL_2202051, EPI_ISL_2202058, EPI_ISL_2202110, EPI_ISL_2202112, EPI_ISL_2202114, EPI_ISL_2202115, EPI_ISL_2202126, EPI_ISL_2202134, EPI_ISL_2202161, EPI_ISL_2202258, EPI_ISL_2202265, EPI_ISL_2202272, EPI_ISL_2202284, EPI_ISL_2202321, EPI_ISL_2202410, EPI_ISL_2202518, EPI_ISL_2202522, EPI_ISL_2202565, EPI_ISL_2202609, EPI_ISL_2202618, EPI_ISL_2202699, EPI_ISL_2202760, EPI_ISL_2202821, EPI_ISL_2202840, EPI_ISL_2267993, EPI_ISL_2268032, EPI_ISL_2268035, EPI_ISL_2268040, EPI_ISL_2268045, EPI_ISL_2600207, EPI_ISL_2600227, EPI_ISL_2649914, EPI_ISL_4360623, EPI_ISL_4361138, EPI_ISL_4362596, EPI_ISL_4362679, EPI_ISL_4365447, EPI_ISL_4365481 | | | |
| see above | Aegis Sciences Corporation | Centers for Disease Control and Prevention Division of Viral Diseases, Pathogen Discovery | Adrian Paskey; Alec Vest; Benjamin Rambo-Martin; Christopher Gulvick; Clinton Paden; Clinton R. Paden; Cyndi Clark; Dakota Howard; Darlene Wagner; Dhwani Batra; Dillon Nall; Duncan MacCannell; Erisa Sula; Ethan Sanders; Holly Houdeshell; Jason Caravas; Kara Moser; Kristine Lacek; Matthew Hardison; Matthew Schmerer; Ola Kvalvaag; Patrick Campbell; Peter Cook; Peter W. Cook; Rob Case; Scott Sammons; Shatavia Morrison; Shaun Westlund; Tymeckia Kendall; Victoria Caban Figueroa; Vikramsinha Ghorpade; Yvette Unoarumhi |
| EPI_ISL_1739043 | Affidea | Instituto Nacional de Saude (INSA) | Borges et al |
| EPI_ISL_1854251 | Affidea | Instituto Nacional de Saude (INSA) and Institute of Biomedicine (iBiMed), Universidade de Aveiro | Borges et al |
| EPI_ISL_2945632 | Ages, MA15 Lieferdienst Veloce_Europe/Austria/Vienna | Elling and Cochella laboratories, IMBA/IMP | Alexander Stark; Ezgi Oezkan; Luisa Cochella and Ulrich Elling; Marcus Strobl; Olga Frank; Ramesh Yelagandula; Tanino Albanese |
| EPI_ISL_1920057, EPI_ISL_1921022, EPI_ISL_1921030 | Albany Medical Center | Wadsworth Center, New York State Department of Health | Alexis Russell; Catharine Prussing; Daryl M. Lamson; Erasmus Schneider; Erica Lasek-Nesselquist; John Kelly; Jonathan Plitnick; Kirsten St. George; Matthew Shudt; Melissa A Leisner; Navjot Singh |
| EPI_ISL_2169840, EPI_ISL_2169841, EPI_ISL_2169844, EPI_ISL_2169845, EPI_ISL_2169846, EPI_ISL_2169847, EPI_ISL_2169848, EPI_ISL_2169849, EPI_ISL_2169850, EPI_ISL_2169851, EPI_ISL_2170370, EPI_ISL_2170371, EPI_ISL_2170401, EPI_ISL_2170404, EPI_ISL_2170405, EPI_ISL_2170407, EPI_ISL_2170410, EPI_ISL_2170413, EPI_ISL_2170414, EPI_ISL_2170418, EPI_ISL_2170419, EPI_ISL_2170424, EPI_ISL_2170430, EPI_ISL_2170439, EPI_ISL_2170440, EPI_ISL_2170441, EPI_ISL_2170442, EPI_ISL_2170444, EPI_ISL_2170445, EPI_ISL_2170446, EPI_ISL_2170447, EPI_ISL_2170451, EPI_ISL_2170452, EPI_ISL_2170455, EPI_ISL_2170456, EPI_ISL_2170460, EPI_ISL_2170467, EPI_ISL_2170544, EPI_ISL_2170545, EPI_ISL_2170546 | | | |
| see above | Alberta Precision Labs (APL) | Public Health Agency of Canada (PHAC) National Microbiology Laboratory | Buss; Croxen M; Deo A; Dieu P; E; Ferrato C; Gill K; Khan F; Koleva P; Li V; Lloyd C; Lynch T; Ma R; Murphy S; Pabbaraju K; Shokoples S; Thayer J; Tipples G; Whitehouse M; Wong A; Yu C; Zelyas N |
| EPI_ISL_1697364 | Algemeen Medisch Labo | Labo Klinische Biologie, UZA | Basil Britto Xavier; Christine Lammens; Herman Goossens; Jasmine Coppens; Marie Le Mercier; Veerle Matheeussen |
| EPI_ISL_1926224, EPI_ISL_1926226 | Altius Institute | Seattle Flu Study | Alex Nguyen; Amanda Adler; Andrew Meuser; Barry R. Lutz; Benjamin Pelle; Caitlin R. Wolf; Chris D. Frazar; Clem Green; Daniel Bates; Deborah A. Nickerson; Elisabeth Brandstetter; Erica Ryke; Hannah Petersen; Helen Y. Chu; Jacob Rodriguez; Janet A. Englund; Jay Shendure; Jessica Halow; John Stamatoyannopoulos; Joshua Richards; Jover Lee; Julia Wald; Kairsten Fay; Kirsten Lacombe; Kneshay Harper; Lea M. Starita; Mark J. Rieder; Matt Hartman; Matthew Richardson; Matthew Thompson; Melissa Truong; Michael Boeckh; Michael Famulare; Misja Ilcisin; Muhammad Halimun; Olivia Waltner; Peter D. Han; Rebecca Bruders; Ryan Alexander; Sadie Patraw; Sofia Olsson; Stephanie DeBaun; Thomas R. Sibley; Tobias Ragoczy; Trevor Bedford; Truong Nguyen |
| EPI_ISL_5782466, EPI_ISL_5782471 | Ambulatorio De Especialidade V E Mogi Mirim | Instituto Butantan | Antonio Jorge Martins; Claudia Renata dos Santos Barros; David Schlesinger; Debora Botequio Moretti; Dimas Tadeu Covas; Elaine Cristina Marqueze; Elaine Vieira Santos; Evandra Strazza Rodrigues; Heidge Fukumasu; Jayme Augusto de Souza-Neto; José Salvatore Leister Patané; Luiz Alcantara; Luiz Lehmann Coutinho; Maria Carolina Elias; Maurício Lacerda Nogueira; Rafael dos Santos Bezerra; Raul Machado Neto; Rejane Maria Tommasini Grotto; Ricardo Haddad; Sandra Coccuzzo Sampaio Vessoni; Simone Kashima; Svetoslav Nanev Slavov; Vincent Louis Viala |
| EPI_ISL_5800467 | Ambulatorio Dr Joao Lania Centro Jose Bonifacio | Instituto Butantan | Antonio Jorge Martins; Claudia Renata dos Santos Barros; David Schlesinger; Debora Botequio Moretti; Dimas Tadeu Covas; Elaine Cristina Marqueze; Elaine Vieira Santos; Evandra Strazza Rodrigues; Heidge Fukumasu; Jayme Augusto de Souza-Neto; José Salvatore Leister Patané; Luiz Alcantara; Luiz Lehmann Coutinho; Maria Carolina Elias; Maurício Lacerda Nogueira; Rafael dos Santos Bezerra; Raul Machado Neto; Rejane Maria Tommasini Grotto; Ricardo Haddad; Sandra Coccuzzo Sampaio Vessoni; Simone Kashima; Svetoslav Nanev Slavov; Vincent Louis Viala |
| EPI_ISL_2086297 | Arizona State Public Health Laboratory | Arizona State Public Health Laboratory | Jessica Escobar; Katherine Fullerton; Linda Getsinger; Nobuko Fukushima; Stacy White; Trung Huynh; Victor Waddell |
| EPI_ISL_1743686, EPI_ISL_1910763, EPI_ISL_1910773, EPI_ISL_1910806, EPI_ISL_1910821, EPI_ISL_1910825, EPI_ISL_2007179, EPI_ISL_2153662 | | | |
| see above | Arizona State University | Arizona State University | Efrem S. Lim; Joshua LaBaer; Joy M. Blain; LaRinda A. Holland; Matthew F. Smith; Nicholas J. Mellor; Peter T. Skidmore; Rabia Maqsood; Valerie Harris; Vel Murugan |
| EPI_ISL_2423529 | Atlanta VA Medical Center | Genomics and Discovery, Respiratory Viruses Branch, Division of Viral Diseases, Centers for Disease Control and Prevention | Adam Retchless; Anna Kelleher; Anna Uehara; Brian Lynch; Clinton R. Paden; Dhwani Batra; Haibin Wang; Han Jia Justin Ng; Jasmine Padilla; Jing Zhang; Justin Lee; Krista Queen; Mark Burroughs; Mili Sheth; Morgan Davis; Peter Cook; Rachel Marine; Sarah Nobles; Suxiang Tong; Tara Coalter; Yan Li; Ying Tao |
| EPI_ISL_2011367, EPI_ISL_2011372, EPI_ISL_2011374, EPI_ISL_2011376, EPI_ISL_2011417, EPI_ISL_2136667, EPI_ISL_2136670, EPI_ISL_2136673 | | | |

| | | | |
|---|---|---|---|
| see above | Atlas Genomics | Seattle Flu Study | Amanda Adler; Barry R. Lutz; Benjamin Pelle; Brian Hiatt; Caitlin R. Wolf; Chris D. Frazar; Deborah A. Nickerson; Elisabeth Brandstetter; Erica Ryke; Geoff Melly; Helen Y. Chu; Janet A. Englund; Jay Shendure; Jover Lee; Kairsten Fay; Kirsten Lacombe; Lea M. Starita; Mark J. Rieder; Matthew Richardson; Matthew Thompson; Melissa Truong; Michael Boeckh; Michael Famulare; Misja Ilcisin; Peter D. Han; Philip Dykema; Romesh Gautom; Scott Lindquist; Thomas R. Sibley; Trevor Bedford |
| EPI_ISL_1854225, EPI_ISL_1854246, EPI_ISL_1854247, EPI_ISL_1854248 | Avelab | Instituto Nacional de Saude (INSA) and BioSystems & Integrative Sciences Institute (BioISI) Genomics Unit, FCUL | Borges et al |
| EPI_ISL_3267001, EPI_ISL_3267003, EPI_ISL_3267005, EPI_ISL_3267009, EPI_ISL_3267014, EPI_ISL_3267019, EPI_ISL_3267021, EPI_ISL_3267022, EPI_ISL_3267025, EPI_ISL_3267029, EPI_ISL_3267031, EPI_ISL_3267036, EPI_ISL_3276574, EPI_ISL_3276598, EPI_ISL_3276604, EPI_ISL_3276608, EPI_ISL_3276616, EPI_ISL_3276622, EPI_ISL_3276623, EPI_ISL_3276626, EPI_ISL_3276627 | | | |
| see above | Ayudas Diagnosticas Sura | Universidad Nacional de Colombia - Laboratorio Genómico One Health | Andres F. Cardona-Rios; Carlos Franco-Muñoz; Carolina Muñoz-Arango; Celeny Ortiz; Daniel O. Maldonado-Perez; Diego A. Álvarez-Díaz; Hector Alejandro Ruiz-Moreno; Idabely Betancur Ortiz; Jorge E. Osorio; Juan P. Hernandez-Ortiz; Karl A Ciuoderis; Katherine Laiton-Donato; Laura Silvana Perez; Lina M. Hurtado; Marcela Mercado-Reyes; Maria Angélica Maya; Maria Stella López; Rita Almanza Payares; Sandra Ines Cano; Simón Villegas Velásquez |
| EPI_ISL_1745250, EPI_ISL_1745318, EPI_ISL_1745320, EPI_ISL_1840710 | Azienda Sanitaria dell'Alto Adige Laboratorio Aziendale di Microbiologia e Virologia | Istituto di Genomica Applicata | Davide Scaglione; Eleonora Paparelli; Elisa Masi; Elisabetta Giacobazzi; Elisabetta Pagani; Gabriele Magris; Irena Jurman; Irene Bianconi; Michele Morgante; Stefanie Wieser; Vera Vendramin |
| EPI_ISL_2654402, EPI_ISL_2654408, EPI_ISL_2654412, EPI_ISL_2654421, EPI_ISL_2654424, EPI_ISL_2654431, EPI_ISL_2654511, EPI_ISL_2654517, EPI_ISL_2654527, EPI_ISL_2654536, EPI_ISL_2654539, EPI_ISL_2654540, EPI_ISL_2654542, EPI_ISL_2654559, EPI_ISL_2654560, EPI_ISL_2654618, EPI_ISL_2654621, EPI_ISL_2654628, EPI_ISL_2654631, EPI_ISL_2654632, EPI_ISL_2654633, EPI_ISL_2654634, EPI_ISL_2654640, EPI_ISL_2654641, EPI_ISL_2654642, EPI_ISL_2654643, EPI_ISL_2654645, EPI_ISL_2654646, EPI_ISL_2654647, EPI_ISL_2654652, EPI_ISL_2654657, EPI_ISL_2654659, EPI_ISL_2654660, EPI_ISL_2654661, EPI_ISL_2654666, EPI_ISL_2654667, EPI_ISL_2654668, EPI_ISL_2654686, EPI_ISL_2654739, EPI_ISL_2654741, EPI_ISL_2654742, EPI_ISL_2654745, EPI_ISL_2654760, EPI_ISL_2654761, EPI_ISL_2654762, EPI_ISL_2654764, EPI_ISL_2654765, EPI_ISL_2654766, EPI_ISL_2654863, EPI_ISL_2654887, EPI_ISL_2654888, EPI_ISL_2654889, EPI_ISL_2654941, EPI_ISL_2654942, EPI_ISL_2654991, EPI_ISL_2654992, EPI_ISL_2654995, EPI_ISL_2654999, EPI_ISL_2655008, EPI_ISL_2655009, EPI_ISL_2655010, EPI_ISL_2655015, EPI_ISL_2655019, EPI_ISL_2655022, EPI_ISL_2655027, EPI_ISL_2655034, EPI_ISL_2655038, EPI_ISL_2655043, EPI_ISL_2655044, EPI_ISL_2655045, EPI_ISL_2655046, EPI_ISL_2655047, EPI_ISL_2655052, EPI_ISL_2655053, EPI_ISL_2655055, EPI_ISL_2655058, EPI_ISL_2655060, EPI_ISL_2655061, EPI_ISL_2655064, EPI_ISL_2655065, EPI_ISL_2655073, EPI_ISL_2655081, EPI_ISL_2655083, EPI_ISL_2655085, EPI_ISL_2655086, EPI_ISL_2655093, EPI_ISL_2655094, EPI_ISL_2655102, EPI_ISL_2655103, EPI_ISL_2655107, EPI_ISL_2655112, EPI_ISL_2655113, EPI_ISL_2655114, EPI_ISL_2655116, EPI_ISL_2655119, EPI_ISL_2655123, EPI_ISL_2655124, EPI_ISL_2655126, EPI_ISL_2655128, EPI_ISL_2655134, EPI_ISL_2655136, EPI_ISL_2655137, EPI_ISL_2655141, EPI_ISL_2655144, EPI_ISL_2655146, EPI_ISL_2655153, EPI_ISL_2655156, EPI_ISL_2655162, EPI_ISL_2655163, EPI_ISL_2655164, EPI_ISL_2655166, EPI_ISL_2655170, EPI_ISL_2655171, EPI_ISL_2655172, EPI_ISL_2655427, EPI_ISL_2655431, EPI_ISL_2655433, EPI_ISL_2655435, EPI_ISL_2655438, EPI_ISL_2655440, EPI_ISL_2655441, EPI_ISL_2655444, EPI_ISL_2655445, EPI_ISL_2655449, EPI_ISL_2655455, EPI_ISL_2655456, EPI_ISL_2655459, EPI_ISL_2655460, EPI_ISL_2655461, EPI_ISL_2655462, EPI_ISL_2655550, EPI_ISL_2655551, EPI_ISL_2655552, EPI_ISL_2655553, EPI_ISL_2655554, EPI_ISL_2655555, EPI_ISL_2655556, EPI_ISL_2655557, EPI_ISL_2655559, EPI_ISL_2655560, EPI_ISL_2655561, EPI_ISL_2655610, EPI_ISL_2655612, EPI_ISL_2655614, EPI_ISL_2655622, EPI_ISL_2655624, EPI_ISL_2655631, EPI_ISL_2655633, EPI_ISL_2655634, EPI_ISL_2655636, EPI_ISL_2655648, EPI_ISL_2655718, EPI_ISL_2655719, EPI_ISL_2655720, EPI_ISL_2655721, EPI_ISL_2655722, EPI_ISL_2655724, EPI_ISL_2655725, EPI_ISL_2655728, EPI_ISL_2655734, EPI_ISL_2655736, EPI_ISL_2655740, EPI_ISL_2655741, EPI_ISL_2655743, EPI_ISL_2655745, EPI_ISL_2655750, EPI_ISL_2655754, EPI_ISL_2655756, EPI_ISL_2655759, EPI_ISL_2655760, EPI_ISL_2655761, EPI_ISL_2655764, EPI_ISL_2655765, EPI_ISL_2655766, EPI_ISL_2655767, EPI_ISL_2655768, EPI_ISL_2655769, EPI_ISL_2655771, EPI_ISL_2655774, EPI_ISL_2655777, EPI_ISL_2655778, EPI_ISL_2655781, EPI_ISL_2655783, EPI_ISL_2655784, EPI_ISL_2655785, EPI_ISL_2655793, EPI_ISL_2655795, EPI_ISL_2655797, EPI_ISL_2655798, EPI_ISL_2655800, EPI_ISL_2655802, EPI_ISL_2655806, EPI_ISL_2655809, EPI_ISL_2655811, EPI_ISL_2655812, EPI_ISL_2655815, EPI_ISL_2655816, EPI_ISL_2655818, EPI_ISL_2655820, EPI_ISL_2655821, EPI_ISL_2655823, EPI_ISL_2655830, EPI_ISL_2655831, EPI_ISL_2655832, EPI_ISL_2655834, EPI_ISL_2655835, EPI_ISL_2655836, EPI_ISL_2655839, EPI_ISL_2655840, EPI_ISL_2655841, EPI_ISL_2655842, EPI_ISL_2655843, EPI_ISL_2655845, EPI_ISL_2655848, EPI_ISL_2655850, EPI_ISL_2655851, EPI_ISL_2655852, EPI_ISL_2655854, EPI_ISL_2655855, EPI_ISL_2655856, EPI_ISL_2655857, EPI_ISL_2655859, EPI_ISL_2655860, EPI_ISL_2655864, EPI_ISL_2655865, EPI_ISL_2655866, EPI_ISL_2655868, EPI_ISL_2655869, EPI_ISL_2655871, EPI_ISL_2655872, EPI_ISL_2655874, EPI_ISL_2655875, EPI_ISL_2655877, EPI_ISL_2655881, EPI_ISL_2655882, EPI_ISL_2655885, EPI_ISL_2655886, EPI_ISL_2655887, EPI_ISL_2655888, EPI_ISL_2655889, EPI_ISL_2655891, EPI_ISL_2656162, EPI_ISL_2656165, EPI_ISL_2656167, EPI_ISL_2656169, EPI_ISL_2656170, EPI_ISL_2656176, EPI_ISL_2656180, EPI_ISL_2656184, EPI_ISL_2656189, EPI_ISL_2656193, EPI_ISL_2656194, EPI_ISL_2656197, EPI_ISL_2656201, EPI_ISL_2656204, EPI_ISL_2656206, EPI_ISL_2656207, EPI_ISL_2656209, EPI_ISL_2656211, EPI_ISL_2656214, EPI_ISL_2656217, EPI_ISL_2656220, EPI_ISL_2656221, EPI_ISL_2656224, EPI_ISL_2656230, EPI_ISL_2656231, EPI_ISL_2656233, EPI_ISL_2656234, EPI_ISL_2656236, EPI_ISL_2656239, EPI_ISL_2656244, EPI_ISL_2656245, EPI_ISL_2656246, EPI_ISL_2656248, EPI_ISL_2656249, EPI_ISL_2656250, EPI_ISL_2656251, EPI_ISL_2656253, EPI_ISL_2656255, EPI_ISL_2656256, EPI_ISL_2656257, EPI_ISL_2656259, EPI_ISL_2656262, EPI_ISL_2656266, EPI_ISL_2656271, EPI_ISL_2656273, EPI_ISL_2656276, EPI_ISL_2656277, EPI_ISL_2656278, EPI_ISL_2656279, EPI_ISL_2656280, EPI_ISL_2656283, EPI_ISL_2656287, EPI_ISL_2656288, EPI_ISL_2656290, EPI_ISL_2656294, EPI_ISL_2656296, EPI_ISL_2656304, EPI_ISL_2656307, EPI_ISL_2656308, EPI_ISL_2656309, EPI_ISL_2656313, EPI_ISL_2656315, EPI_ISL_2656317, EPI_ISL_2656320, EPI_ISL_2656323, EPI_ISL_2656325, EPI_ISL_2720248, EPI_ISL_2720264, EPI_ISL_2720272, EPI_ISL_2720281, EPI_ISL_2720556, EPI_ISL_2720557, EPI_ISL_2720558, EPI_ISL_2720561, EPI_ISL_2720562, EPI_ISL_2720567, EPI_ISL_2720568, EPI_ISL_2721070, EPI_ISL_2721125, EPI_ISL_2721132, EPI_ISL_2721133, EPI_ISL_2721337, EPI_ISL_2721341, EPI_ISL_2721342, EPI_ISL_2721364 | | | |
| see above | BCCDC Public Health Laboratory | BCCDC Public Health Laboratory | Ana Pacagnella; Corrinne Ng; Dan Fornika; John Tyson; Kim Macdonald; Kimia Kamelian; Linda Hoang; Loretta Janz; Mel Krajden; Prystajecky Natalie; Robert Azana; Shannon Russell |
| EPI_ISL_2544499 | Baker lab at Loyola Medical Center | Baker lab at Loyola Medical Center | Amanda Harrington; Amornrat O'Brien; BS; Cynthia Rowe; Gail E. Reid; MD; Monika Evdokimova; Nina M. Clark; PhD; Susan Uprichard; Xufang Deng; and Susan C. Baker |
| EPI_ISL_7672079, EPI_ISL_7672080, EPI_ISL_7672082, EPI_ISL_7672087, EPI_ISL_7672406, EPI_ISL_7672408 | Bambino Gesù Pediatric Hospital | Microbiology and Immunology Diagnosis Bambino Gesù Pediatric Hospital | Carlo Federico Perno; Claudia Alteri; Luna Colagrossi; Rossana Scutari; Valentino Costabile |
| EPI_ISL_2111394, EPI_ISL_2111406 | Bayerisches Landesamt fur Gesundheit und Lebensmittelsicherheit (LGL) | Robert Koch Institute | |
| EPI_ISL_2260469 | Bayerisches Landesamt für Gesundheit und Lebensmittelsicherheit (LGL) | Robert Koch Institute | |
| EPI_ISL_1840693 | Baylor Scott & White-Temple | Baylor Scott & White-Temple | Ari Rao; Kimberly Walker; Linden Morales; Marcus Volz; Shelby Hendrickson |
| EPI_ISL_5802503, EPI_ISL_5802504, EPI_ISL_5802559, EPI_ISL_5802560, EPI_ISL_5802561, EPI_ISL_5802562, EPI_ISL_5802563, EPI_ISL_5802564, EPI_ISL_5802579, EPI_ISL_5802603, EPI_ISL_5802604 | | | |
| see above | Biofast | Instituto Butantan | Antonio Jorge Martins; Claudia Renata dos Santos Barros; David Schlesinger; Debora Botequio Moretti; Dimas Tadeu Covas; Elaine Cristina Marqueze; Elaine Vieira Santos; Evandra Strazza Rodrigues; Heidge Fukumasu; Jayme Augusto de Souza-Neto; José Salvatore Leister Patané; Luiz Alcantara; Luiz Lehmann Coutinho; Maria Carolina Elias; Maurício Lacerda Nogueira; Rafael dos Santos Bezerra; Raul Machado Neto; Rejane Maria Tommasini Grotto; Ricardo Haddad; Sandra Coccuzzo Sampaio Vessoni; Simone Kashima; Svetoslav Nanev Slavov; Vincent Louis Viala |
| EPI_ISL_1756620, EPI_ISL_1757461 | Biogroup Bio Lam-LCD Saint-Denis | Department of Virology, Henri Mondor University Hospital, Assistance Publique Hôpitaux de Paris, Université Paris-Est Créteil, INSERM U955 | Alexandre Soulier; Christophe Rodriguez; Elisabeth Trawinski; Guillaume Gricourt; Jean-Michel Pawlotsky; Melissa N'Debi; Slim Fourati; Vanessa Demontant |
| EPI_ISL_1833931 | Bioinformatics and Biostatistics Lab, Advanced Sequencing Facility | COVID-19 Genomics UK (COG-UK) Consortium | Aengus Stewart; Chelsea Sawyer; Harshil Patel; Jerome Nicod; Laura Cubitt; Margaret Crawford |
| EPI_ISL_1916815 | BioneXt Lab | Laboratoire national de sante, Microbiology, Microbial Genomics Platform | Anke Wienecke-Baldacchino; Catherine Ragimbeau; Fatu Djabi; Jessica Tapp; Lise Pignon; Raoul Salmon; Tamir Abdelrahman; Thibault Ferrandon |
| EPI_ISL_3478941, EPI_ISL_3478945, EPI_ISL_3478949, EPI_ISL_3478953, EPI_ISL_3478954, EPI_ISL_3478955, EPI_ISL_3478958, EPI_ISL_3478960, EPI_ISL_3478961, EPI_ISL_3478969, EPI_ISL_3478970, EPI_ISL_3478974, EPI_ISL_3478975, EPI_ISL_3478976, EPI_ISL_3478977, EPI_ISL_3480088, EPI_ISL_3480089, EPI_ISL_3480090, EPI_ISL_3480091, EPI_ISL_3480092, EPI_ISL_3480095, EPI_ISL_3480096, EPI_ISL_3480098, EPI_ISL_3480099, EPI_ISL_3480100, EPI_ISL_3480101, EPI_ISL_3480103, EPI_ISL_3480104, EPI_ISL_3480105, EPI_ISL_3480106, EPI_ISL_3480116, EPI_ISL_3480134, EPI_ISL_3480154, EPI_ISL_3480156, EPI_ISL_3480158, EPI_ISL_3480160, EPI_ISL_3480164, EPI_ISL_3480166, EPI_ISL_3480167, EPI_ISL_3480168, EPI_ISL_3480169, EPI_ISL_3480170, EPI_ISL_3480172, EPI_ISL_3480173, EPI_ISL_3480177, EPI_ISL_3480179, EPI_ISL_3480180, EPI_ISL_3480185, EPI_ISL_3480187, EPI_ISL_3480189, EPI_ISL_3480198, EPI_ISL_3480199, EPI_ISL_3480202, EPI_ISL_3480204, EPI_ISL_3480209, EPI_ISL_3480216, EPI_ISL_3480223, EPI_ISL_3480224, EPI_ISL_3480225, EPI_ISL_3480230, EPI_ISL_3480288, EPI_ISL_3480292, EPI_ISL_3480295, EPI_ISL_3480296, EPI_ISL_3480306, EPI_ISL_3480307, EPI_ISL_3480309, EPI_ISL_3480310, EPI_ISL_3480323, EPI_ISL_3480330, EPI_ISL_3480332, EPI_ISL_3480337, EPI_ISL_3480339, EPI_ISL_3480345, EPI_ISL_3480346, EPI_ISL_3480348, EPI_ISL_3480355, EPI_ISL_3480386, EPI_ISL_3480391, EPI_ISL_3480393, EPI_ISL_3480394, EPI_ISL_3480396, EPI_ISL_3480398, EPI_ISL_3480400, EPI_ISL_3480413, EPI_ISL_3480417, EPI_ISL_3480422, EPI_ISL_3480426, EPI_ISL_3480428, EPI_ISL_3480429, EPI_ISL_3480430, EPI_ISL_3480431, EPI_ISL_3480433, EPI_ISL_3480434, EPI_ISL_3480436, EPI_ISL_3480437, EPI_ISL_3480438, EPI_ISL_3480439, EPI_ISL_3480447, EPI_ISL_3480448, EPI_ISL_3480449, EPI_ISL_3480453, EPI_ISL_3480454, EPI_ISL_3480456, EPI_ISL_3480457, EPI_ISL_3480459, EPI_ISL_3480460, EPI_ISL_3480462, EPI_ISL_3480463, EPI_ISL_3480464, EPI_ISL_3480466, EPI_ISL_3480467, EPI_ISL_3480469, EPI_ISL_3480472, EPI_ISL_3480473, EPI_ISL_3480474, EPI_ISL_3480482, EPI_ISL_3480484, EPI_ISL_3480488, EPI_ISL_3480490, EPI_ISL_3480501, EPI_ISL_3480503, EPI_ISL_3480509, EPI_ISL_3480510, EPI_ISL_3480511, EPI_ISL_3480512, EPI_ISL_3480514, EPI_ISL_3480515, EPI_ISL_3480516, EPI_ISL_3480521, EPI_ISL_3480523, EPI_ISL_3480524, EPI_ISL_3480528, EPI_ISL_3480534, EPI_ISL_3480535, EPI_ISL_3480536, EPI_ISL_3480538, EPI_ISL_3480539, EPI_ISL_3480540, EPI_ISL_3480541, EPI_ISL_3480543, EPI_ISL_3480544, EPI_ISL_3480545, EPI_ISL_3480547, EPI_ISL_3480551, EPI_ISL_3480552, EPI_ISL_3480554, EPI_ISL_3480560, EPI_ISL_3480571, EPI_ISL_3480572, EPI_ISL_3480576, EPI_ISL_3480578, EPI_ISL_3480583, EPI_ISL_3480586, EPI_ISL_3480587, EPI_ISL_3480591, EPI_ISL_3480594, EPI_ISL_3480595, EPI_ISL_3480596, EPI_ISL_3480597, EPI_ISL_3480600, EPI_ISL_3480601, EPI_ISL_3480604, EPI_ISL_3480605, EPI_ISL_3480606, EPI_ISL_3480608, EPI_ISL_3480609, EPI_ISL_3480611, EPI_ISL_3480612, EPI_ISL_3480613, EPI_ISL_3480614, EPI_ISL_3480616, EPI_ISL_3480618, EPI_ISL_3480619, EPI_ISL_3480620, EPI_ISL_3480621, EPI_ISL_3480622, EPI_ISL_3480623, EPI_ISL_3480624, EPI_ISL_3480626, EPI_ISL_3480627, EPI_ISL_3480631, EPI_ISL_3480633, EPI_ISL_3480634, EPI_ISL_3480635, EPI_ISL_3480636, EPI_ISL_3480638, EPI_ISL_3480639, EPI_ISL_3480642, EPI_ISL_3480649, EPI_ISL_3480650, EPI_ISL_3480654, EPI_ISL_3480655, EPI_ISL_3480656, EPI_ISL_3480657, EPI_ISL_3480660, EPI_ISL_3480661, EPI_ISL_3480665, EPI_ISL_3480668, EPI_ISL_3480672, EPI_ISL_3480676, EPI_ISL_3480677, EPI_ISL_3480681, EPI_ISL_3480689, EPI_ISL_3480694, EPI_ISL_3480700, EPI_ISL_3480701, EPI_ISL_3480702, EPI_ISL_3480704, EPI_ISL_3480706, EPI_ISL_3480708, EPI_ISL_3480734, EPI_ISL_3480735, EPI_ISL_3480736, EPI_ISL_3480737, EPI_ISL_3480738, EPI_ISL_3480740, EPI_ISL_3480741, EPI_ISL_3480745, EPI_ISL_3480747, EPI_ISL_3480748, EPI_ISL_3480750, EPI_ISL_3480751, EPI_ISL_3480752, EPI_ISL_3480754, EPI_ISL_3480755, EPI_ISL_3480757, EPI_ISL_3480758, EPI_ISL_3480760, EPI_ISL_3480761, EPI_ISL_3480767, EPI_ISL_3480776, EPI_ISL_3480779, EPI_ISL_3480782, EPI_ISL_3480783, EPI_ISL_3480785, EPI_ISL_3480787, EPI_ISL_3480791, EPI_ISL_3480792, EPI_ISL_3480803, EPI_ISL_3480805, EPI_ISL_3480806, EPI_ISL_3480807, EPI_ISL_3480808, EPI_ISL_3480810, EPI_ISL_3480811, EPI_ISL_3480812, EPI_ISL_3480813, EPI_ISL_3480815, EPI_ISL_3480816, EPI_ISL_3480819, EPI_ISL_3480826, EPI_ISL_3480842, EPI_ISL_3480843, EPI_ISL_3480851, EPI_ISL_3480852, EPI_ISL_3480853, EPI_ISL_3480992, EPI_ISL_3596347, EPI_ISL_3598429, EPI_ISL_3598431, EPI_ISL_3598438, EPI_ISL_3598442, EPI_ISL_3598443, EPI_ISL_3598444, EPI_ISL_3598458, EPI_ISL_3598461, EPI_ISL_3598462, EPI_ISL_3598466, EPI_ISL_3598467, EPI_ISL_3598470, EPI_ISL_3598480, EPI_ISL_3598489, EPI_ISL_3598491, EPI_ISL_3598494, EPI_ISL_3598495, EPI_ISL_3598500, EPI_ISL_3598502, EPI_ISL_3598510, EPI_ISL_3598511, EPI_ISL_3598512, EPI_ISL_3598514, EPI_ISL_3598521, EPI_ISL_3598522, EPI_ISL_3598523, EPI_ISL_3598524, EPI_ISL_3598528, EPI_ISL_3598529, EPI_ISL_3598534, EPI_ISL_3598541, EPI_ISL_3598545, EPI_ISL_3598553, EPI_ISL_3598560, EPI_ISL_3598566, EPI_ISL_3598567, EPI_ISL_3598569, EPI_ISL_3598571, EPI_ISL_3598579, EPI_ISL_3598580, EPI_ISL_3598581, EPI_ISL_3598584, EPI_ISL_3598589, EPI_ISL_3598593, EPI_ISL_3598609, EPI_ISL_3598618, EPI_ISL_3598627, EPI_ISL_3598639, EPI_ISL_3598665, EPI_ISL_3598668, EPI_ISL_3598671, EPI_ISL_3598682, EPI_ISL_3598712, EPI_ISL_3598725, EPI_ISL_3598740, EPI_ISL_3598799, EPI_ISL_3598802, EPI_ISL_3598805, EPI_ISL_3598811, EPI_ISL_3598876, EPI_ISL_4828386, EPI_ISL_4828387 | | | |
| see above | British Columbia Centre For Disease Control | BCCDC Public Health Laboratory | Ana Pacagnella; Corrinne Ng; Dan Fornika; John Tyson; Kim Macdonald; Kimia Kamelian; Linda Hoang; Loretta Janz; Mel Krajden; Prystajecky Natalie; Robert Azana; Shannon Russell |
| EPI_ISL_1743728, EPI_ISL_1743742, EPI_ISL_1743747, EPI_ISL_1743769, EPI_ISL_1743775, EPI_ISL_1743788, EPI_ISL_1743810, EPI_ISL_1743817, EPI_ISL_1743838, EPI_ISL_1743862, EPI_ISL_1743868, EPI_ISL_1743877, EPI_ISL_1743896, EPI_ISL_1743898, EPI_ISL_1743899, EPI_ISL_1743901, EPI_ISL_1743903, EPI_ISL_1743912, EPI_ISL_1743915, EPI_ISL_1743922, EPI_ISL_1743926, EPI_ISL_1743951, EPI_ISL_1743959, EPI_ISL_1743977, EPI_ISL_1743993, EPI_ISL_1743996, EPI_ISL_1744004, EPI_ISL_1744009, EPI_ISL_1744012, EPI_ISL_1744016, EPI_ISL_1744018, EPI_ISL_1744029, EPI_ISL_1744043, EPI_ISL_1744049, EPI_ISL_1744072, EPI_ISL_1744073, EPI_ISL_1744084, EPI_ISL_1744093, EPI_ISL_1744104, EPI_ISL_1744108, EPI_ISL_1744132, EPI_ISL_1744139, EPI_ISL_1744140, EPI_ISL_1744144, EPI_ISL_1744146, EPI_ISL_1744166, EPI_ISL_1744177, EPI_ISL_1744198, EPI_ISL_1744206, EPI_ISL_1744209, EPI_ISL_1744219, EPI_ISL_1744237, EPI_ISL_1744260, EPI_ISL_1744262, EPI_ISL_1744267, EPI_ISL_1744269, EPI_ISL_1744278, EPI_ISL_1744279, EPI_ISL_1744285, EPI_ISL_1744288, EPI_ISL_1744294, EPI_ISL_1744299, EPI_ISL_1786741, EPI_ISL_1786747, EPI_ISL_1786748, EPI_ISL_1786752, EPI_ISL_1786754, EPI_ISL_1786796, EPI_ISL_1786802, EPI_ISL_1786806, EPI_ISL_1786807, EPI_ISL_1786822, EPI_ISL_1786832, EPI_ISL_1786849, EPI_ISL_1786860, EPI_ISL_1786891, EPI_ISL_1786915, EPI_ISL_1786930, EPI_ISL_1786937, EPI_ISL_1786940, EPI_ISL_1786960, EPI_ISL_1786961, EPI_ISL_1786966, EPI_ISL_1786979, EPI_ISL_1786982, EPI_ISL_1786991, EPI_ISL_1786993, EPI_ISL_1786996, EPI_ISL_1787008, EPI_ISL_1787013, EPI_ISL_1787020, EPI_ISL_1787022, EPI_ISL_1787033, EPI_ISL_1787037, EPI_ISL_1787042, EPI_ISL_1787050, EPI_ISL_1787060, EPI_ISL_1787080, EPI_ISL_1787083, EPI_ISL_1787091, EPI_ISL_1787102, EPI_ISL_1787115, EPI_ISL_1787117, EPI_ISL_1787121, EPI_ISL_1787150, EPI_ISL_1787193, EPI_ISL_1787202, EPI_ISL_1787203, EPI_ISL_1787207, EPI_ISL_1787210, EPI_ISL_1787213, EPI_ISL_1787227, EPI_ISL_1918677, EPI_ISL_1919016, EPI_ISL_1919096, EPI_ISL_2016392 | | | |
| see above | Broad Institute Clinical Research Sequencing Platform | Infectious Disease Program, Broad Institute of Harvard and MIT | Adams, G.; B.L.; B.W.; Bauer, M.; Birren; Blumenstiel, B.; Brown, C.; Carter, A.; Chaluvadi, S.; D.J.; DeFelice, M.; DeRuff, K.; Dodge, S.; Gabriel, S.; Gallagher, G.; Gladden-Young, A.; Granger, B.; J.E.; K.J.; Lagerborg, K.; Larkin, K.; Lee, M.; Lemieux; Lennon, N.; Loreth, C.; Madoff, L.; McGovern, S.; Meldrim, J.; Normandin, E.; P.C.; Park; Pearlman, L.; Reilly, S.; Rudy, M.; Sabeti; Siddle; Smole, S.; Tomkins-Tinch, C.; Vicente, G.; and MacInnis |
| EPI_ISL_2103132, EPI_ISL_2103137 | CA DPH Viral and Rickettsial Disease Laboratory | Chan-Zuckerberg Biohub | CZB Cliahub Consortium |
| EPI_ISL_5530188, EPI_ISL_5603226, EPI_ISL_5603232, EPI_ISL_5603241, EPI_ISL_5603246, EPI_ISL_5603251 | CASA DE SAUDE ADILIA MARIA | Analytical Competence Molecular Epidemiology Lab/ACME, Oswaldo Cruz Foundation, Ceara (FIOCRUZ CE) | Carlos Leonardo de Aragao Araujo; Cecilia Leite Costa & Eduardo Ruback dos Santos on behalf of COVID-19 FIOCRUZ Genomic Network; Cleber Furtado Aksenen; Fabio Miyajima; Fernando Braga Stehling; Francisco Eder de Moura Lopes; Igor Oliveira Duarte; Jamille Maria Mendes Bezerra; Joaquim Cesar do Nascimento Sousa Junior; Pedro Miguel Carneiro Jeronimo; Suzana Porto Almeida; Thais Ferreira de Oliveira; Thais de Oliveira Costa; Ticiane Cavalcante de Souza; Veridiana Pessoa Miyajima |
| EPI_ISL_2825836, EPI_ISL_2825838, EPI_ISL_2825846, EPI_ISL_2825854, EPI_ISL_2825862, EPI_ISL_2882189, EPI_ISL_2890356, EPI_ISL_2890358, EPI_ISL_2890362, EPI_ISL_2891028, EPI_ISL_2891029, EPI_ISL_7335686 | | | |
| see above | CDPH VBL | California Department of Public Health | CDPH-COVIDNet; Emily Smith on behalf of CDPH-COVIDNet; UCLA Technology Center for Genomics & Bioinformatics |
| EPI_ISL_3102538 | CENTRO DE ATENDIMENTO PARA ENFRENTAMENTO AO COVID 19 | Analytical Competence Molecular Epidemiology Lab/ACME, Oswaldo Cruz Foundation, Ceara (FIOCRUZ CE) | Cleber Furtado Aksenen; Fabio Miyajima; Fernando Braga Stehling; Francisco Eder de Moura Lopes; Jamille Maria Mendes Bezerra; Joaquim César do Nascimento Sousa Junior; Pedro Miguel Carneiro Jeronimo; Suzana Porto Almeida e Lucas Delerino; Thais Ferreira de Oliveira; Thais de Oliveira Costa; Ticiane Cavalcante de Souza; Veridiana Pessoa Miyajima |
| EPI_ISL_5650286, EPI_ISL_5650287, EPI_ISL_5650288, | CENTRO DE REFERENCIA DO IDOSO DR HUMBERTO MENDES DE CARVALHO | Instituto Butantan | Antonio Jorge Martins; Claudia Renata dos Santos Barros; David Schlesinger; Debora Botequio Moretti; Dimas Tadeu Covas; Elaine Cristina Marqueze; Elaine Vieira Santos; Evandra Strazza Rodrigues; Heidge Fukumasu; Jayme Augusto de Souza-Neto; José Salvatore Leister Patané; Luiz Alcantara; Luiz Lehmann Coutinho; Maria Carolina Elias; Maurício Lacerda Nogueira; Rafael dos Santos Bezerra; Raul Machado Neto; Rejane Maria Tommasini Grotto; Ricardo Haddad; Sandra Coccuzzo Sampaio Vessoni; Simone Kashima; Svetoslav Nanev Slavov; Vincent Louis Viala |

| | | | |
|---|---|---|---|
| EPI_ISL_5650289, EPI_ISL_5650290, EPI_ISL_5650412 | | | |
| EPI_ISL_2209445 | CENTRO DE SAUDE DE ANALANDIA | Instituto Butantan / FZEA-USP-Pirassununga | Antonio Jorge Martins; Claudia Renata dos Santos Barros; David Schlesinger; Debora Botequio Moretti; Dimas Tadeu Covas; Elaine Cristina Marqueze; Elaine Vieira Santos; Evandra Strazza Rodrigues; Heidge Fukumasu; Jayme Augusto de Souza-Neto; José Salvatore Leister Patané; Luiz Alcantara; Luiz Lehmann Coutinho; Maria Carolina Elias; Maurício Lacerda Nogueira; Rafael dos Santos Bezerra; Raul Machado Neto; Rejane Maria Tommasini Grotto; Ricardo Haddad; Sandra Coccuzzo Sampaio Vessoni; Simone Kashima; Svetoslav Nanev Slavov; Vincent Louis Viala |
| EPI_ISL_2344710 | CENTRO DE SAUDE DR MARIO DIAS DE AGUIAR CAPIVARI | Instituto Butantan / FZEA-USP-Pirassununga | Antonio Jorge Martins; Claudia Renata dos Santos Barros; David Schlesinger; Debora Botequio Moretti; Dimas Tadeu Covas; Elaine Cristina Marqueze; Elaine Vieira Santos; Evandra Strazza Rodrigues; Heidge Fukumasu; Jayme Augusto de Souza-Neto; José Salvatore Leister Patané; Luiz Alcantara; Luiz Lehmann Coutinho; Maria Carolina Elias; Maurício Lacerda Nogueira; Rafael dos Santos Bezerra; Raul Machado Neto; Rejane Maria Tommasini Grotto; Ricardo Haddad; Sandra Coccuzzo Sampaio Vessoni; Simone Kashima; Svetoslav Nanev Slavov; Vincent Louis Viala |
| EPI_ISL_2344560 | CENTRO DE SAUDE II DR GABRIEL MESQUITA VARGEM GDE DO SUL | Instituto Butantan / FZEA-USP-Pirassununga | Antonio Jorge Martins; Claudia Renata dos Santos Barros; David Schlesinger; Debora Botequio Moretti; Dimas Tadeu Covas; Elaine Cristina Marqueze; Elaine Vieira Santos; Evandra Strazza Rodrigues; Heidge Fukumasu; Jayme Augusto de Souza-Neto; José Salvatore Leister Patané; Luiz Alcantara; Luiz Lehmann Coutinho; Maria Carolina Elias; Maurício Lacerda Nogueira; Rafael dos Santos Bezerra; Raul Machado Neto; Rejane Maria Tommasini Grotto; Ricardo Haddad; Sandra Coccuzzo Sampaio Vessoni; Simone Kashima; Svetoslav Nanev Slavov; Vincent Louis Viala |
| EPI_ISL_2208987, EPI_ISL_5650313, EPI_ISL_5650390, EPI_ISL_5650394 | CENTRO DE SAUDE III AFFONSO LUZZI SANTA CRUZ DAS PALMEIRAS | Instituto Butantan | Antonio Jorge Martins; Claudia Renata dos Santos Barros; David Schlesinger; Debora Botequio Moretti; Dimas Tadeu Covas; Elaine Cristina Marqueze; Elaine Vieira Santos; Evandra Strazza Rodrigues; Heidge Fukumasu; Jayme Augusto de Souza-Neto; José Salvatore Leister Patané; Luiz Alcantara; Luiz Lehmann Coutinho; Maria Carolina Elias; Maurício Lacerda Nogueira; Rafael dos Santos Bezerra; Raul Machado Neto; Rejane Maria Tommasini Grotto; Ricardo Haddad; Sandra Coccuzzo Sampaio Vessoni; Simone Kashima; Svetoslav Nanev Slavov; Vincent Louis Viala; Vincent Louis Viala. |
| EPI_ISL_2344567, EPI_ISL_2344570 | CENTRO DE SAUDE III AFFONSO LUZZI SANTA CRUZ DAS PALMEIRAS | Instituto Butantan / FZEA-USP-Pirassununga | Antonio Jorge Martins; Claudia Renata dos Santos Barros; David Schlesinger; Debora Botequio Moretti; Dimas Tadeu Covas; Elaine Cristina Marqueze; Elaine Vieira Santos; Evandra Strazza Rodrigues; Heidge Fukumasu; Jayme Augusto de Souza-Neto; José Salvatore Leister Patané; Luiz Alcantara; Luiz Lehmann Coutinho; Maria Carolina Elias; Maurício Lacerda Nogueira; Rafael dos Santos Bezerra; Raul Machado Neto; Rejane Maria Tommasini Grotto; Ricardo Haddad; Sandra Coccuzzo Sampaio Vessoni; Simone Kashima; Svetoslav Nanev Slavov; Vincent Louis Viala |
| EPI_ISL_5650260, EPI_ISL_5650261 | CENTRO DE SAUDE III BORBOREMA | Instituto Butantan | Antonio Jorge Martins; Claudia Renata dos Santos Barros; David Schlesinger; Debora Botequio Moretti; Dimas Tadeu Covas; Elaine Cristina Marqueze; Elaine Vieira Santos; Evandra Strazza Rodrigues; Heidge Fukumasu; Jayme Augusto de Souza-Neto; José Salvatore Leister Patané; Luiz Alcantara; Luiz Lehmann Coutinho; Maria Carolina Elias; Maurício Lacerda Nogueira; Rafael dos Santos Bezerra; Raul Machado Neto; Rejane Maria Tommasini Grotto; Ricardo Haddad; Sandra Coccuzzo Sampaio Vessoni; Simone Kashima; Svetoslav Nanev Slavov; Vincent Louis Viala |
| EPI_ISL_1966796 | CENTRO DE SAUDE TEREZINHA DE MORAES ARSILLA | Instituto Butantan / Mendelics | Antonio Jorge Martins; Bianca Cechetto Carlos. Mendelics: Bibiana Santos; Claudia Renata dos Santos Barros; Cíntia Bittar; David Schlesinger. Hemocentro Ribeirão Preto: Simone Kashima; Debora Botequio Moretti; Elaine Cristina Marqueze; Elaine Vieira dos Santos; Elisangela Chicaroni Mattos; Erika Freitas; Evandra Strazza Rodrigues; Felipe Allan da Silva da Costa; Flavia Aburjaile; Fábio Sossai Possebon; Guilherme Campos; Guilherme Targino Valente; Heidge Fukumasu. USP-Botucatu: Rejane Maria Tommasini Grotto; Helena Lage Ferreira; Instituto Butantan: Dimas Tadeu Covas; Jardelina de Souza Todao Bernardino; Jayme A. Souza-Neto; Jessika Cristina Chagas Lesbon; Jorge A. Petroli Marchesi; José Salvatore Leister Patané; João Paulo Kitajima; João Pessoa Araújo Jr.; Leila Sabrina Ullmann; Loyze Paola Oliveira de Lima; Luiz Aurelio de Campos Crispin. Centro de Genômica Funcional da ESALQ: Luiz Lehmann Coutinho; Luiz Carlos Junior de Alcantara; Lívia Sacchetto; Maisa C. Pereira Parra; Maria Carolina Elias; Marta Giovanetti; Marília Moraes; Maurício Lacerda Nogueira. Prefeitura de Sao Paulo: Melissa Palmieri.; Patricia Akemi Assato; Paula Rahal; Paulo Inacio da Costa; Rafael dos Santos Bezerra; Raquel de Lello Rocha Campos Cassano. NGS Soluções Genômicas: Pilar Drummond Sampaio Corrêa Mariani. FZEA-USP Pirassununga: Mirele Daiana Poleti; Raul Machado Neto; Ricardo Augusto Brassaloti; Ricardo Haddad; Rodrigo Tocantins Calado. FAMERP-SJRP: Cecília Artico Banho; Sandra Coccuzzo Sampaio; Svetoslav Nanev Slavov; Vagner Fonseca; Vincent Louis Viala |
| EPI_ISL_2209059 | CENTRO MEDICO DR NELSON SALOME DE CONCHAL | Instituto Butantan | Antonio Jorge Martins; Claudia Renata dos Santos Barros; David Schlesinger; Debora Botequio Moretti; Dimas Tadeu Covas; Elaine Cristina Marqueze; Elaine Vieira Santos; Evandra Strazza Rodrigues; Heidge Fukumasu; Jayme Augusto de Souza-Neto; José Salvatore Leister Patané; Luiz Alcantara; Luiz Lehmann Coutinho; Maria Carolina Elias; Maurício Lacerda Nogueira; Rafael dos Santos Bezerra; Raul Machado Neto; Rejane Maria Tommasini Grotto; Ricardo Haddad; Sandra Coccuzzo Sampaio Vessoni; Simone Kashima; Svetoslav Nanev Slavov; Vincent Louis Viala. |
| EPI_ISL_2344711 | CENTRO MEDICO DR NELSON SALOME DE CONCHAL | Instituto Butantan / FZEA-USP-Pirassununga | Antonio Jorge Martins; Claudia Renata dos Santos Barros; David Schlesinger; Debora Botequio Moretti; Dimas Tadeu Covas; Elaine Cristina Marqueze; Elaine Vieira Santos; Evandra Strazza Rodrigues; Heidge Fukumasu; Jayme Augusto de Souza-Neto; José Salvatore Leister Patané; Luiz Alcantara; Luiz Lehmann Coutinho; Maria Carolina Elias; Maurício Lacerda Nogueira; Rafael dos Santos Bezerra; Raul Machado Neto; Rejane Maria Tommasini Grotto; Ricardo Haddad; Sandra Coccuzzo Sampaio Vessoni; Simone Kashima; Svetoslav Nanev Slavov; Vincent Louis Viala |
| EPI_ISL_2209074 | CENTRO MUNICIPAL DE EPIDEMIOLOGIA E IMUNIZACOES LIMEIRA | Instituto Butantan | Antonio Jorge Martins; Claudia Renata dos Santos Barros; David Schlesinger; Debora Botequio Moretti; Dimas Tadeu Covas; Elaine Cristina Marqueze; Elaine Vieira Santos; Evandra Strazza Rodrigues; Heidge Fukumasu; Jayme Augusto de Souza-Neto; José Salvatore Leister Patané; Luiz Alcantara; Luiz Lehmann Coutinho; Maria Carolina Elias; Maurício Lacerda Nogueira; Rafael dos Santos Bezerra; Raul Machado Neto; Rejane Maria Tommasini Grotto; Ricardo Haddad; Sandra Coccuzzo Sampaio Vessoni; Simone Kashima; Svetoslav Nanev Slavov; Vincent Louis Viala. |
| EPI_ISL_2209982 | CENTRO MUNICIPAL DE EPIDEMIOLOGIA E IMUNIZACOES LIMEIRA | Instituto Butantan / FZEA-USP-Pirassununga | Antonio Jorge Martins; Claudia Renata dos Santos Barros; David Schlesinger; Debora Botequio Moretti; Dimas Tadeu Covas; Elaine Cristina Marqueze; Elaine Vieira Santos; Evandra Strazza Rodrigues; Heidge Fukumasu; Jayme Augusto de Souza-Neto; José Salvatore Leister Patané; Luiz Alcantara; Luiz Lehmann Coutinho; Maria Carolina Elias; Maurício Lacerda Nogueira; Rafael dos Santos Bezerra; Raul Machado Neto; Rejane Maria Tommasini Grotto; Ricardo Haddad; Sandra Coccuzzo Sampaio Vessoni; Simone Kashima; Svetoslav Nanev Slavov; Vincent Louis Viala |
| EPI_ISL_2562029, EPI_ISL_2562030 | CH Franck Joly | Cerba lab | Aude Lessenne; Bénédicte Roquebert; Emmanuel Lecorche; Kader Merah; Laura Verdurme; Patrice Herisson; Sabine Trombert-Paolantoni; Stéphanie Haïm-Boukobza; Thierry Collin |
| EPI_ISL_1757912 | CH. BEGIN | Department of Virology, Henri Mondor University Hospital, Assistance Publique Hôpitaux de Paris, Université Paris-Est Créteil, INSERM U955 | Alexandre Soulier; Christophe Rodriguez; Elisabeth Trawinski; Guillaume Gricourt; Jean-Michel Pawlotsky; Melissa N'Debi; Slim Fourati; Vanessa Demontant |
| EPI_ISL_1757696 | CH.INTERCOMMUNAL DE CRETEIL | Department of Virology, Henri Mondor University Hospital, Assistance Publique Hôpitaux de Paris, Université Paris-Est Créteil, INSERM U955 | Alexandre Soulier; Christophe Rodriguez; Elisabeth Trawinski; Guillaume Gricourt; Jean-Michel Pawlotsky; Melissa N'Debi; Slim Fourati; Vanessa Demontant |
| EPI_ISL_1628343, EPI_ISL_2227910 | CHC Andrée Rosemon | Institut Pasteur de la Guyane | A. Salmier; Anne Lavergne; Dominique Rousset |
| EPI_ISL_1854224 | CHLC | Instituto Nacional de Saude (INSA) | Borges et al |
| EPI_ISL_1739066 | CHMT | Instituto Nacional de Saude (INSA) | Borges et al |
| EPI_ISL_1826572 | CHR de La Citadelle | GIGA Medical Genomics | Bouchra Boujemla; Cécile Meex; Keith Durkin; Maria Artesi; Marie-Pierre Hayette; Nathalie Renotte; Pierrette Melin; Raphaël Boreux; Sébastien Bontems; Vincent Bours |
| EPI_ISL_2194032 | CHU Nantes Virology | CHU Nantes Virology | Audrey Rodallec; Berthe-Marie Imbert-Marcille; Celine Bressollette-Bodin; Thomas Drumel |
| EPI_ISL_1824622 | CHU Pontchaillou | CHU Pontchaillou | DE TAYRAC Marie; DENOUAL Florent; ETCHEVERRY Amandine; GALIBERT Marie Dominique; GROLHIER Claire; JAGLINE Steven; PRONIER Charlotte; SASSI Mohamed; THIBAULT Vincent |
| EPI_ISL_1678870, EPI_ISL_1678871, EPI_ISL_1678872, EPI_ISL_1678873 | CHWAPI - SITE NOTRE DAME | Institut de Pathologie et Genetique (IPG) | Jérémie Gras; Pascale Hilbert |
| EPI_ISL_1963928, EPI_ISL_1963929, EPI_ISL_1963930, EPI_ISL_1963933, EPI_ISL_1963936 | CLILAB | Microbiology Department | Aida Gonzalez-Diaz; Carmen Ardanuy; Jordi Camara; Jordi Niubó; Laura Calatayud; M Angeles Domínguez; Miguel Fernandez-Huerta; Sara Marti |
| EPI_ISL_1628336, EPI_ISL_1628337, EPI_ISL_1628338, EPI_ISL_2227911, EPI_ISL_2227912, EPI_ISL_2227913, EPI_ISL_2227914, EPI_ISL_2227915, EPI_ISL_2227917, EPI_ISL_2227918 | | | |
| see above | CNR Institut Pasteur de la Guyane | Institut Pasteur de la Guyane | A. Salmier; Anne Lavergne; Dominique Rousset |
| EPI_ISL_2383912 | CO Dept. of Public Health and Environment, Lab Services Division | Centers for Disease Control and Prevention Division of Viral Diseases, Pathogen Discovery | Alison Laufer Halpin; Ben L. Rambo-Martin; Clinton R. Paden; Dakota Howard; Darlene Wagner; Dave Wentworth; Dhwani Batra; Jasmine Padilla; Justin Lee; Katie Dillon; Krista Queen; Kristen Knipe; Kristine Lacek; Mark Burroughs; Matthew Schmerer; Mili Sheth; Peter Cook; Sam Shepard; Sarah Nobles; Shoshona Le; Suxiang Tong; Vivien Dugan; Yvette Unoarumhi |
| EPI_ISL_2209275, EPI_ISL_2344723 | COORDENADORIA MUNICIPAL DE SAUDE DE IRACEMAPOLIS | Instituto Butantan / FZEA-USP-Pirassununga | Antonio Jorge Martins; Claudia Renata dos Santos Barros; David Schlesinger; Debora Botequio Moretti; Dimas Tadeu Covas; Elaine Cristina Marqueze; Elaine Vieira Santos; Evandra Strazza Rodrigues; Heidge Fukumasu; Jayme Augusto de Souza-Neto; José Salvatore Leister Patané; Luiz Alcantara; Luiz Lehmann Coutinho; Maria Carolina Elias; Maurício Lacerda Nogueira; Rafael dos Santos Bezerra; Raul Machado Neto; Rejane Maria Tommasini Grotto; Ricardo Haddad; Sandra Coccuzzo Sampaio Vessoni; Simone Kashima; Svetoslav Nanev Slavov; Vincent Louis Viala |
| EPI_ISL_1804968, EPI_ISL_1804969, EPI_ISL_1804971, EPI_ISL_1804972, EPI_ISL_1804973 | CQRC_QUALITY CONTROL CHEMICAL BIOLOGICAL RISK_AOOR Villa Sofia Cervello Palermo | CQRC_QUALITY CONTROL CHEMICAL BIOLOGICAL RISK_AOOR Villa Sofia Cervello Palermo | ; Brunacci, G.; Contino, F.; Di Gaudio, F. |
| EPI_ISL_1966785 | CS DE MIRASSOLANDIA | Instituto Butantan / Mendelics | Antonio Jorge Martins; Bianca Cechetto Carlos. Mendelics: Bibiana Santos; Claudia Renata dos Santos Barros; Cíntia Bittar; David Schlesinger. Hemocentro Ribeirão Preto: Simone Kashima; Debora Botequio Moretti; Elaine Cristina Marqueze; Elaine Vieira dos Santos; Elisangela Chicaroni Mattos; Erika Freitas; Evandra Strazza Rodrigues; Felipe Allan da Silva da Costa; Flavia Aburjaile; Fábio Sossai Possebon; Guilherme Campos; Guilherme Targino Valente; Heidge Fukumasu. USP-Botucatu: Rejane Maria Tommasini Grotto; Helena Lage Ferreira; Instituto Butantan: Dimas Tadeu Covas; Jardelina de Souza Todao Bernardino; Jayme A. Souza-Neto; Jessika Cristina Chagas Lesbon; Jorge A. Petroli Marchesi; José Salvatore Leister Patané; João Paulo Kitajima; João Pessoa Araújo Jr.; Leila Sabrina Ullmann; Loyze Paola Oliveira de Lima; Luiz Aurelio de Campos Crispin. Centro de Genômica Funcional da ESALQ: Luiz Lehmann Coutinho; Luiz Carlos Junior de Alcantara; Lívia Sacchetto; Maisa C. Pereira Parra; Maria Carolina Elias; Marta Giovanetti; Marília Moraes; Maurício Lacerda Nogueira. Prefeitura de Sao Paulo: Melissa Palmieri.; Patricia Akemi Assato; Paula Rahal; Paulo Inacio da Costa; Rafael dos Santos Bezerra; Raquel de Lello Rocha Campos Cassano. NGS Soluções Genômicas: Pilar Drummond Sampaio Corrêa Mariani. FZEA-USP Pirassununga: Mirele Daiana Poleti; Raul Machado Neto; Ricardo Augusto Brassaloti; Ricardo Haddad; Rodrigo Tocantins Calado. FAMERP-SJRP: Cecília Artico Banho; Sandra Coccuzzo Sampaio; Svetoslav Nanev Slavov; Vagner Fonseca; Vincent Louis Viala |
| EPI_ISL_2697961, EPI_ISL_2697962, EPI_ISL_2697963, EPI_ISL_2697964, EPI_ISL_2697965, EPI_ISL_2697987 | CTvacinas | CTvacinas | A.P.; B.L.; Coelho; D.B.; Dorlass; Durigon; E.G.; E.L.; F.G.; Fernandes; Fiorini, A.; Fonseca; G.P.; H.P.; K.L.; L.M.; Lourenco; Magalhaes; Oliveira; Ometto, T.; Peixoto, R.; R.D.; Sato, H.; Scagion; Teixeira, S.; Telezynski; Thomazelli |
| EPI_ISL_2537462, EPI_ISL_2537474, EPI_ISL_2537477, EPI_ISL_3536710, EPI_ISL_3536731 | California Department of Public Health | California Department of Public Health | CDPH IDLB COVIDNet et al |
| EPI_ISL_4279958 | California Department of Public Health | Chiu Laboratory, University of California, San Francisco | Baolin Wang; Carl Hanson; Charles Chiu; Debra Wadford; Mary Kate Morris; Venice Servellita |
| EPI_ISL_2837352, EPI_ISL_2837489, EPI_ISL_2837490, EPI_ISL_2837494, EPI_ISL_2837516, EPI_ISL_2837612, EPI_ISL_2837627, EPI_ISL_2837628, EPI_ISL_2837629, EPI_ISL_2837688, EPI_ISL_2837767, EPI_ISL_2837770, EPI_ISL_2837795, EPI_ISL_2837807, EPI_ISL_2837810, EPI_ISL_2837811, EPI_ISL_2837817, EPI_ISL_2837820, EPI_ISL_2837863, EPI_ISL_2837971 | | | |
| see above | California Department of Public | California Department of Public Health | CDPH-COVIDNet |

| | | | |
|---|---|---|---|
| | Health Valencia Branch Laboratory (CDPH VBL) | | |
| EPI_ISL_7507397, EPI_ISL_7507414 | Cedars-Sinai Medical Center, Molecular Pathology Laboratory of Department of Pathology & Laboratory Medicine and Genomic Core | Cedars-Sinai Medical Center, Molecular Pathology Laboratory of Department of Pathology & Laboratory Medicine and Genomic Core | Brian Davis; Eric Vail; Jasmine T Plummer; Jorge Mario Sincuir Martinez; Stephanie Chen; Wenjuan Zhang |
| EPI_ISL_2502378, EPI_ISL_2502387, EPI_ISL_2502394, EPI_ISL_2502395, EPI_ISL_2502397, EPI_ISL_2502398, EPI_ISL_2502463, EPI_ISL_2502464, EPI_ISL_2502465, EPI_ISL_2502466, EPI_ISL_2502467, EPI_ISL_2502468, EPI_ISL_2502469, EPI_ISL_2502470, EPI_ISL_2502471, EPI_ISL_2502472, EPI_ISL_2502473 | | | |
| see above | Central Laboratory, Bureau of Public Health (BOG) and Academic Hospital Paramaribo | Erasmus Medical Center | Bas B Oude Munnink; Cherise Beek; Consuella Partowidjojo; Dion Gajadin; Ed PF IJzerman; Emmanuelle Munger; Gary Gummels; Ingrid SK Krishnadath; Lycke Woittiez; Marion PG Koopmans; Mireille Van de Veer; Phyllis Pinas; Princes Wongsowidjojo; Radjesh Ori; Ranisha Doerbalie; Rohma Banwari; Soeradj Harkisoen; Stephen Vreden; Tilotmadebie Ramlal; Verne Nanhoe |
| EPI_ISL_3255055, EPI_ISL_3255057, EPI_ISL_3255061, EPI_ISL_3255064, EPI_ISL_3255075, EPI_ISL_3255078, EPI_ISL_3255186, EPI_ISL_3266120, EPI_ISL_3944576 | | | |
| see above | Central Public Health Laboratory - LACEN -Bahia, Salvador, Brazil | Central Public Health Laboratory - LACEN -Bahia, Salvador, Brazil | Arabela Leal; Breno Dominguez; Felicidade Pereira; Gabriela Menezes; Jaqueline Gomes; Lenisa Dandara; Luciana Oliveira; Luiz Alcantara; Marcela Gómez; Marta Giovanetti; Patrícia Cajado; Stephane Tosta; Vagner Fonseca; Vanessa Nardy |
| EPI_ISL_5799742 | Centro De Saude De Analandia | Instituto Butantan | Antonio Jorge Martins; Claudia Renata dos Santos Barros; David Schlesinger; Debora Botequio Moretti; Dimas Tadeu Covas; Elaine Cristina Marqueze; Elaine Vieira Santos; Evandra Strazza Rodrigues; Heidge Fukumasu; Jayme Augusto de Souza-Neto; José Salvatore Leister Patané; Luiz Alcantara; Luiz Lehmann Coutinho; Maria Carolina Elias; Maurício Lacerda Nogueira; Rafael dos Santos Bezerra; Raul Machado Neto; Rejane Maria Tommasini Grotto; Ricardo Haddad; Sandra Coccuzzo Sampaio Vessoni; Simone Kashima; Svetoslav Nanev Slavov; Vincent Louis Viala |
| EPI_ISL_5799752 | Centro De Saude Dr Mario Dias De Aguiar Capivari | Instituto Butantan | Antonio Jorge Martins; Claudia Renata dos Santos Barros; David Schlesinger; Debora Botequio Moretti; Dimas Tadeu Covas; Elaine Cristina Marqueze; Elaine Vieira Santos; Evandra Strazza Rodrigues; Heidge Fukumasu; Jayme Augusto de Souza-Neto; José Salvatore Leister Patané; Luiz Alcantara; Luiz Lehmann Coutinho; Maria Carolina Elias; Maurício Lacerda Nogueira; Rafael dos Santos Bezerra; Raul Machado Neto; Rejane Maria Tommasini Grotto; Ricardo Haddad; Sandra Coccuzzo Sampaio Vessoni; Simone Kashima; Svetoslav Nanev Slavov; Vincent Louis Viala |
| EPI_ISL_5782468 | Centro De Saude Ii Dr Gabriel Mesquita Vargem Gde Do Sul | Instituto Butantan | Antonio Jorge Martins; Claudia Renata dos Santos Barros; David Schlesinger; Debora Botequio Moretti; Dimas Tadeu Covas; Elaine Cristina Marqueze; Elaine Vieira Santos; Evandra Strazza Rodrigues; Heidge Fukumasu; Jayme Augusto de Souza-Neto; José Salvatore Leister Patané; Luiz Alcantara; Luiz Lehmann Coutinho; Maria Carolina Elias; Maurício Lacerda Nogueira; Rafael dos Santos Bezerra; Raul Machado Neto; Rejane Maria Tommasini Grotto; Ricardo Haddad; Sandra Coccuzzo Sampaio Vessoni; Simone Kashima; Svetoslav Nanev Slavov; Vincent Louis Viala |
| EPI_ISL_5782475, EPI_ISL_5782478 | Centro De Saude Iii Affonso Luzzi Santa Cruz Das Palmeiras | Instituto Butantan | Antonio Jorge Martins; Claudia Renata dos Santos Barros; David Schlesinger; Debora Botequio Moretti; Dimas Tadeu Covas; Elaine Cristina Marqueze; Elaine Vieira Santos; Evandra Strazza Rodrigues; Heidge Fukumasu; Jayme Augusto de Souza-Neto; José Salvatore Leister Patané; Luiz Alcantara; Luiz Lehmann Coutinho; Maria Carolina Elias; Maurício Lacerda Nogueira; Rafael dos Santos Bezerra; Raul Machado Neto; Rejane Maria Tommasini Grotto; Ricardo Haddad; Sandra Coccuzzo Sampaio Vessoni; Simone Kashima; Svetoslav Nanev Slavov; Vincent Louis Viala |
| EPI_ISL_5800495 | Centro De Saude Terezinha De Moraes Arsilla | Instituto Butantan | Antonio Jorge Martins; Claudia Renata dos Santos Barros; David Schlesinger; Debora Botequio Moretti; Dimas Tadeu Covas; Elaine Cristina Marqueze; Elaine Vieira Santos; Evandra Strazza Rodrigues; Heidge Fukumasu; Jayme Augusto de Souza-Neto; José Salvatore Leister Patané; Luiz Alcantara; Luiz Lehmann Coutinho; Maria Carolina Elias; Maurício Lacerda Nogueira; Rafael dos Santos Bezerra; Raul Machado Neto; Rejane Maria Tommasini Grotto; Ricardo Haddad; Sandra Coccuzzo Sampaio Vessoni; Simone Kashima; Svetoslav Nanev Slavov; Vincent Louis Viala |
| EPI_ISL_5799753, EPI_ISL_5799754 | Centro Medico Dr Nelson Salome De Conchal | Instituto Butantan | Antonio Jorge Martins; Claudia Renata dos Santos Barros; David Schlesinger; Debora Botequio Moretti; Dimas Tadeu Covas; Elaine Cristina Marqueze; Elaine Vieira Santos; Evandra Strazza Rodrigues; Heidge Fukumasu; Jayme Augusto de Souza-Neto; José Salvatore Leister Patané; Luiz Alcantara; Luiz Lehmann Coutinho; Maria Carolina Elias; Maurício Lacerda Nogueira; Rafael dos Santos Bezerra; Raul Machado Neto; Rejane Maria Tommasini Grotto; Ricardo Haddad; Sandra Coccuzzo Sampaio Vessoni; Simone Kashima; Svetoslav Nanev Slavov; Vincent Louis Viala |
| EPI_ISL_5799727 | Centro Municipal De Epidemiologia E Imunizacoes Limeira | Instituto Butantan | Antonio Jorge Martins; Claudia Renata dos Santos Barros; David Schlesinger; Debora Botequio Moretti; Dimas Tadeu Covas; Elaine Cristina Marqueze; Elaine Vieira Santos; Evandra Strazza Rodrigues; Heidge Fukumasu; Jayme Augusto de Souza-Neto; José Salvatore Leister Patané; Luiz Alcantara; Luiz Lehmann Coutinho; Maria Carolina Elias; Maurício Lacerda Nogueira; Rafael dos Santos Bezerra; Raul Machado Neto; Rejane Maria Tommasini Grotto; Ricardo Haddad; Sandra Coccuzzo Sampaio Vessoni; Simone Kashima; Svetoslav Nanev Slavov; Vincent Louis Viala |
| EPI_ISL_4578883 | Centro Provincial de Referencia VIH/SIDA y Hepatitis Virales – Instituto Biológico Dr Tomás Perón | Área de Secuenciación del Laboratorio de Virología del Hospital de Niños Dr. Ricardo Gutierrez on behalf of 'Proyecto Argentino Interinstitucional de genomica de SARS-CoV-2' (PAIS Consortium) | A; Acuña; C; Carlos; Corazza; D; Dehaut; Dolcini; F; Fresina; Gatti; Gimenez; González; Goya; Gutierrez; K; L; LE; Lusso; M; MI; MS; Nabaes Jodar; Nardone; Natale; Nogueira Laspuir; Pagliari; R; Real; Romanko; S; Sanchez; Suarez; Szymanowski; Talavera; Ten Huver; V; Valinotto; Valle; Viegas, M. |
| EPI_ISL_2612390, EPI_ISL_2612391, EPI_ISL_2612392, EPI_ISL_5926804, EPI_ISL_5926805, EPI_ISL_5926806, EPI_ISL_5926807, EPI_ISL_5926808, EPI_ISL_5926833, EPI_ISL_5926839, EPI_ISL_5926842, EPI_ISL_5926843, EPI_ISL_5926844, EPI_ISL_5926845, EPI_ISL_5926846 | | | |
| see above | Centro de Infectologia Charles Mérieux/ Laboratório Rodolphe Mérieux, FUNDHACRE | Bioinformatics Laboratory / LNCC | Alessandra P Lamarca; Alexandra L Gerber; Aline de Freitas Souza; Ana Paula de C Guimarães; Ana Tereza R Vasconcelos; Andreas Stocker; Cirley Maria de Oliveira Lobato; Douglas Terra Machado; Janaína Mazaro; Janete Taynã Nascimento Rodrigues; Luiz Fellype Alves de Souza; Luiz G P de Almeida; Ronaldo da Silva F Jr; Rutilene B. Souza |
| EPI_ISL_2091312, EPI_ISL_2091317 | Centro de Investigación Biomédica de Occidente (CIBO) | Centro de Investigación en Enfermedades Infecciosas (CIENI), Instituto Nacional de Enfermedades Respiratorias (INER) | Alejandro Sanchez-Flores; Alfredo Herrera-Estrella; Alicia Ocaña-Mondragón; Angel Gustavo Salas-Lais; Bernardo Martínez-Miguel; Blanca Taboada; Brenda Irasema Maldonado-Meza; Carla Ivón Herrera-Najera; Carlos F. Arias; Celia Boukadida; Clara Esperanza Santacruz-Tinoco; Concepción Grajales-Muñiz; Consorcio Mexicano de Vigilancia Genómica (CoViGen-Mex). Authors (in alphabetical order): Julio Elias Alvarado-Yaah; Célida Duque Molina; Fernando Fontove-Herrera; Francisco Pulido; Gloria Elena Espinosa-Ayala; Gloria María Molina-Salinas; Gloria Vazquez; Hector Esteban Paz-Juárez; Hector Montoya-Fuentes; Helen Haydee Fernanda Ramírez-Plascencia; José Antonio Enciso-Moreno; José Esteban Muñoz-Medina; José de Jesús Nuñez-Contreras; Juan Bautista Chale-Dzul; Luis Alberto Ochoa-Carrera; Margarita Matías-Florentino; María Guadalupe Santiago-Mauricio; María Guadalupe de Jesús Mireles-Rivera; Nelly Sélem-Mojica; Pavel Isa; Ricardo Grande; Santiago Ávila-Ríos; Víctor Eduardo Garcia-Arias; Víctor Hugo Borja-Aburto |
| EPI_ISL_1854239 | Cintramedica | Instituto Nacional de Saude (INSA) | Borges et al |
| EPI_ISL_1805633 | Clinica los Nogales | Instituto Nacional de Salud- Dirección de Investigación en Salud Pública | Carlos Franco-Muñoz; Carmen Osorio; Diana Malo; Diego A. Álvarez-Díaz; Diego Andrés Prada; Gerardo Santamaría; Hector Alejandro Ruiz-Moreno; Jhonnatan Reales-González; Jorge Rivera; Juan Camilo Martinez; Julian Naizaque; Katherine Laiton-Donato; Lisseth Pardo; Magdalena Wiesner; Marcela Mercado-Reyes; Maria T. Herrera-Sepúlveda; Marta Lopez Blanco; Martha Lucia Ospina Martinez; Paola Rojas; Sergio Gomez; Sheryll Corchuelo; Ángela Alarcon Cruz |
| EPI_ISL_2500819 | Clinical Microbiology Laboratory, NewYork Presbyterian Hospital/Columbia University Irving Medical Center | Uhlemann Laboratory, Columbia University Irving Medical Center | Anne Kelley; Anne-Catrin Uhlemann; Maya Tagliavia; Medini K. Annavajhala |
| EPI_ISL_4300530 | Color Genomics | Chiu Laboratory, University of California, San Francisco | Alicia Sotomayor-Gonzalez; Alicia Zhou; Baolin Wang; Charles Chiu; Daniel Wong; Erika Torres; Madyson Wanner; Noah Brazer; Scott Topper; Venice Servellita; Yueyuan Zhang |
| EPI_ISL_2276629, EPI_ISL_2276861, EPI_ISL_6211358 | Colorado Department of Public Health and Environment | Colorado Department of Public Health and Environment | Alexandria Rossheim; Diana Ir; Emily A. Travanty; Laura Bankers; Mandy Waters; Michael Martin; Molly C. Hetherington-Rauth; Sarah Elizabeth Totten; Shannon R. Matzinger |
| EPI_ISL_2034747, EPI_ISL_3105469 | Connecticut DPH | Yale Center for Genomic Analysis | Anderson Brito; Annie Watkins; Anthony Muyombwe; Brooke Sullivan; Chaney Kalinich; Chantal Vogels; Curt Scharfe; Irina Tikhonova; Isabel Ott; Jafar Razeq; Jessica Rothman; Joseph Fauver; Kaya Bilguvar; Mallery Breban; Mary Petrone; Nathan Grubaugh; Randy Downing; Shrikant Mane; Stephen M. Bart; Tara Alpert |
| EPI_ISL_5799714, EPI_ISL_5799765 | Coordenadoria Municipal De Saude De Iracemapolis | Instituto Butantan | Antonio Jorge Martins; Claudia Renata dos Santos Barros; David Schlesinger; Debora Botequio Moretti; Dimas Tadeu Covas; Elaine Cristina Marqueze; Elaine Vieira Santos; Evandra Strazza Rodrigues; Heidge Fukumasu; Jayme Augusto de Souza-Neto; José Salvatore Leister Patané; Luiz Alcantara; Luiz Lehmann Coutinho; Maria Carolina Elias; Maurício Lacerda Nogueira; Rafael dos Santos Bezerra; Raul Machado Neto; Rejane Maria Tommasini Grotto; Ricardo Haddad; Sandra Coccuzzo Sampaio Vessoni; Simone Kashima; Svetoslav Nanev Slavov; Vincent Louis Viala |
| EPI_ISL_2103130 | County of San Luis Obispo Public Health Laboratory | Chan-Zuckerberg Biohub | CZB Cliahub Consortium |
| EPI_ISL_2658458 | Croatian Institute of Public Health | Croatian Institute of Public Health | Irena Tabain; Ivana Ferenčak |
| EPI_ISL_5800483 | Cs De Mirassolandia | Instituto Butantan | Antonio Jorge Martins; Claudia Renata dos Santos Barros; David Schlesinger; Debora Botequio Moretti; Dimas Tadeu Covas; Elaine Cristina Marqueze; Elaine Vieira Santos; Evandra Strazza Rodrigues; Heidge Fukumasu; Jayme Augusto de Souza-Neto; José Salvatore Leister Patané; Luiz Alcantara; Luiz Lehmann Coutinho; Maria Carolina Elias; Maurício Lacerda Nogueira; Rafael dos Santos Bezerra; Raul Machado Neto; Rejane Maria Tommasini Grotto; Ricardo Haddad; Sandra Coccuzzo Sampaio Vessoni; Simone Kashima; Svetoslav Nanev Slavov; Vincent Louis Viala |
| EPI_ISL_2754966, EPI_ISL_2754974, EPI_ISL_2754982, EPI_ISL_2754998, EPI_ISL_2754999, EPI_ISL_2755004, EPI_ISL_2755481, EPI_ISL_2755491, EPI_ISL_2755749, EPI_ISL_2755756, EPI_ISL_2755761, EPI_ISL_2756022 | | | |
| see above | Curative Labs | Curative Labs | Elias L. Salfati; Eugenia Khorosheva; George Way; J.Cesar Ignacio-Espinoza; Janet Chen; Mikhail Hanewich-Hollatz; Nabjot Sandhu; Sophia Quasem; Vladimir Slepnev; Zhiyi Xie |
| EPI_ISL_2725472 | DC Public Health Lab/ Dept. of Forensic Sciences | DC Public Health Lab/ Dept. of Forensic Sciences | Brittany Hamilton; Connie Maza; David Payne; Elizabeth Zelaya; Janis Doss; Jocelyn Hauser; Monica Mann; Sarah Scott; Scott Nguyen |
| EPI_ISL_5650243, EPI_ISL_5650246 | DEPARTAMENTO MUNICIPAL DE SAUDE DE BURITAMA | Instituto Butantan | Antonio Jorge Martins; Claudia Renata dos Santos Barros; David Schlesinger; Debora Botequio Moretti; Dimas Tadeu Covas; Elaine Cristina Marqueze; Elaine Vieira Santos; Evandra Strazza Rodrigues; Heidge Fukumasu; Jayme Augusto de Souza-Neto; José Salvatore Leister Patané; Luiz Alcantara; Luiz Lehmann Coutinho; Maria Carolina Elias; Maurício Lacerda Nogueira; Rafael dos Santos Bezerra; Raul Machado Neto; Rejane Maria Tommasini Grotto; Ricardo Haddad; Sandra Coccuzzo Sampaio Vessoni; Simone Kashima; Svetoslav Nanev Slavov; Vincent Louis Viala |
| EPI_ISL_1707601, EPI_ISL_1707605, EPI_ISL_1707607, EPI_ISL_1707656, EPI_ISL_1707660, EPI_ISL_1785155, EPI_ISL_1788750, EPI_ISL_1841214 | | | |
| see above | DIP. PREV. AVEZZANO SERVIZIO DI IGIENE EPIDEMIOLOGIA E SANITA' PUBBLICA | Istituto Zooprofilattico Sperimentale dell'Abruzzo e Molise "G. Caporale" | Ancora M; Calistri P; Cammà C; Caporale M; Curini V; Delli Compagni E; Di Domenico M; Di Lollo Valeria; Di Pasquale A; Lorusso A; Mangone I; Marcacci M; Puglia I; Rinaldi A; Savini G; Scialabba S |
| EPI_ISL_5104492, EPI_ISL_5104495, EPI_ISL_5104497, EPI_ISL_5104502, EPI_ISL_5104505, EPI_ISL_5104507, EPI_ISL_5104508 | | | |
| see above | DNAGYN | LGBio (Laboratorio de Genetica & Biodiversidade) | Alex Honda Bernardes; Amanda Alves de Melo; Aparecido Divino da Cruz; Cintia Pelegrineti Targueta de Azevedo Brito; Daniela de Melo e Silva; Elisangela de Paula Silveira Lacerda; Juliana Santana de Curcio; Luiz Augusto Pereira; Marc Alexandre Duarte Gigonzac; Mariana Pires de Campos Telles; Ramilla dos Santos Braga; Renata de Oliveira Dias; Rhewter Nunes; Thais Cidália Vieira Gigonzac; Thais Guimarães Castro; Thays Millena Alves Pedroso |
| EPI_ISL_1804954 | DOHMH Central Harlem | New York City Public Health Laboratory | Jade Wang; et al. |
| EPI_ISL_2262659 | DOHMH Morrisania | New York City Public Health Laboratory | Jade Wang; et al. |
| EPI_ISL_1694742, EPI_ISL_1694749 | DOHMH PHL | New York City Public Health Laboratory | Jade Wang; et al. |
| EPI_ISL_1908950, EPI_ISL_1908973 | DPHL | Delaware Public Health Lab | Rebecca Savage |
| EPI_ISL_4405169 | Departamento de Biología y genética molecular, IACA Laboratorios. | Área de Secuenciación del Laboratorio de Virología del Hospital de Niños Dr. Ricardo Gutierrez on behalf of 'Proyecto Argentino Interinstitucional de genomica de SARS-CoV-2' (PAIS | ; A; Acuña; D; Goya; LE; Lusso; MI; MS; Masciovecchio MV; Nabaes Jodar; Natale; S; Streitenberger ER; Suárez; Valinotto; Viegas, M. |

| | | | |
|---|---|---|---|
| | | Consortium) | |
| EPI_ISL_1708596, EPI_ISL_1708597, EPI_ISL_1793672, EPI_ISL_1826586, EPI_ISL_1826587, EPI_ISL_1826589, EPI_ISL_1826590 | | | |
| see above | Department of Clinical Microbiology | GIGA Medical Genomics | Bouchra Boujemla; Cécile Meex; Keith Durkin; Maria Artesi; Marie-Pierre Hayette; Nathalie Renotte; Pierrette Melin; Raphaël Boreux; Sébastien Bontems; Vincent Bours |
| EPI_ISL_1789529, EPI_ISL_1789530, EPI_ISL_1789531, EPI_ISL_1789536, EPI_ISL_2096061 | Department of Virology, Istituto Zooprofilattico Sperimentale del Lazio e della Toscana (IZSLT) | Department of General Diagnostics; Department of Virology; Istituto Zooprofilattico Sperimentale del Lazio e della Toscana (IZSLT) | Alessia Franco; Antonella Cersini; Antonio Battisti.; Elena L. Diaconu; Fabiola Feltrin; Giuseppe Manna; Patricia Alba; Raffaela Conti; Teresa Scicluna; Virginia Carfora |
| EPI_ISL_2449986 | Dutch COVID-19 response team | Erasmus Medical Center | Anne van der Linden; Annemiek van der Eijk; Bas Oude Munnink; Corine GeurtsvanKessel; David Nieuwenhuijse; Emmanuelle Munger; Irina Chestakova; Marion Koopmans; Marjan Boter; Reina Sikkema; Richard Molenkamp; on behalf of the Dutch national COVID-19 respo |
| EPI_ISL_1792428, EPI_ISL_1792437, EPI_ISL_1792439, EPI_ISL_1792440, EPI_ISL_1792441, EPI_ISL_1792442, EPI_ISL_1792443, EPI_ISL_1792444, EPI_ISL_1792446, EPI_ISL_1792448, EPI_ISL_1792449, EPI_ISL_1792450, EPI_ISL_1792451, EPI_ISL_1792452, EPI_ISL_1792453, EPI_ISL_1961137, EPI_ISL_1962253, EPI_ISL_1962257, EPI_ISL_1962258, EPI_ISL_1962260, EPI_ISL_1962261, EPI_ISL_1962272, EPI_ISL_1962275, EPI_ISL_2093852, EPI_ISL_2094397, EPI_ISL_3917324 | | | |
| see above | Dutch COVID-19 response team | National Institute for Public Health and the Environment (RIVM) | Adam Meijer; AnneMarie van den Brandt; Annelies Kroneman; Bas van der Veer; Chantal Reusken; Dennis Schmitz; Dirk Eggink; Euníce Then; Florian Zwagemaker; Harry Vennema; Ivo van Walle; James Groot; Jeroen Cremer; Karim Hajji; Kim Freriks; Linda van de Nes; Lisa Wijsman; Lynn Aarts; Melissa van Tuil; Robert Kohl; Ryanne Jaarsma; Sanne Bos; Sharon van den Brink; Sjoerd Kuiling; Stijn van Rossum; on behalf of the national COVID-19 response team |
| EPI_ISL_4060096, EPI_ISL_4060097, EPI_ISL_4060098 | Dynacare Manitoba | Manitoba Cadham Provincial Laboratory | David Alexander; Hilary Racher; Hongbin Li; Melissa Desaulnier; Tintu Abraham |
| EPI_ISL_3215466, EPI_ISL_3215487, EPI_ISL_3215493, EPI_ISL_3215601, EPI_ISL_3215639, EPI_ISL_3215673, EPI_ISL_3215693, EPI_ISL_3215697, EPI_ISL_3215727, EPI_ISL_3508990, EPI_ISL_3508991, EPI_ISL_3508992 | | | |
| see above | EXCITE Lab | Andersen lab at Scripps Research | Abigail Schnapper; Angela Scioscia; Cheryl Anderson; Chip Schooley; Christine Mitchell + SEARCH; David Pride; Fred Wu; Greg Humphrey; Helena Tubb; Natasha Martin; Natasha Martin Cheryl Anderson; Sawyer Farmer; Sharon Reed; Smruthi Karthikeyan; Sydney Morgan + Marni Jacobs + Celestine Magallanes + Holly Valentine + Louise C. Laurent + SEARCH; Tommy Valles + SEARCH; Tommy Valles + SEARCH |
| EPI_ISL_2477506, EPI_ISL_2477507, EPI_ISL_2477511, EPI_ISL_2477556, EPI_ISL_2477557, EPI_ISL_2477560, EPI_ISL_2477561, EPI_ISL_2477562, EPI_ISL_2477564, EPI_ISL_2477565, EPI_ISL_2477566, EPI_ISL_2477567, EPI_ISL_2477569, EPI_ISL_2477572, EPI_ISL_2477573, EPI_ISL_2477574, EPI_ISL_2477575, EPI_ISL_2477576, EPI_ISL_2477577, EPI_ISL_2477578, EPI_ISL_2477579, EPI_ISL_2477580, EPI_ISL_2477581, EPI_ISL_2477582, EPI_ISL_2477583, EPI_ISL_2477584, EPI_ISL_2477585, EPI_ISL_2477586, EPI_ISL_2477587, EPI_ISL_2477591, EPI_ISL_2477592, EPI_ISL_2477596, EPI_ISL_2477598, EPI_ISL_2477599, EPI_ISL_2477600, EPI_ISL_2477601, EPI_ISL_2477602, EPI_ISL_2477604, EPI_ISL_2477606, EPI_ISL_2477608, EPI_ISL_2477611, EPI_ISL_2477612, EPI_ISL_2477616, EPI_ISL_2477617, EPI_ISL_2477618, EPI_ISL_2477620, EPI_ISL_2477624, EPI_ISL_2477626, EPI_ISL_2477628, EPI_ISL_2477629, EPI_ISL_2477631, EPI_ISL_2477633, EPI_ISL_2477634, EPI_ISL_2477637, EPI_ISL_2477638, EPI_ISL_2477639, EPI_ISL_2477640, EPI_ISL_2477646, EPI_ISL_2477654, EPI_ISL_2477655, EPI_ISL_2477660, EPI_ISL_2477663, EPI_ISL_2477665, EPI_ISL_2477669, EPI_ISL_2477683, EPI_ISL_2477685, EPI_ISL_2477688, EPI_ISL_2477689, EPI_ISL_2477690, EPI_ISL_2477691, EPI_ISL_2477692, EPI_ISL_2477693, EPI_ISL_2477694, EPI_ISL_2477695, EPI_ISL_2477696, EPI_ISL_2477698, EPI_ISL_2477699, EPI_ISL_2477701, EPI_ISL_2477702, EPI_ISL_2477703, EPI_ISL_2477705, EPI_ISL_2477706, EPI_ISL_2477708, EPI_ISL_2477709, EPI_ISL_2477710, EPI_ISL_2477713, EPI_ISL_2477714, EPI_ISL_2477720, EPI_ISL_2477721, EPI_ISL_2477722, EPI_ISL_2477723, EPI_ISL_2477726, EPI_ISL_2477728, EPI_ISL_2477729, EPI_ISL_2477731, EPI_ISL_2477732, EPI_ISL_2477733, EPI_ISL_2477734, EPI_ISL_2477737, EPI_ISL_2477738, EPI_ISL_2477739, EPI_ISL_2477740, EPI_ISL_2477742, EPI_ISL_2477744, EPI_ISL_2477745, EPI_ISL_2477746, EPI_ISL_2477748, EPI_ISL_2477749, EPI_ISL_2477752, EPI_ISL_2477753, EPI_ISL_2477754, EPI_ISL_2477755, EPI_ISL_2477757, EPI_ISL_2477758, EPI_ISL_2477759, EPI_ISL_2477760, EPI_ISL_2477761, EPI_ISL_2477762, EPI_ISL_2477763, EPI_ISL_2477764, EPI_ISL_2477765, EPI_ISL_2477767, EPI_ISL_2477768, EPI_ISL_2477769, EPI_ISL_2477770, EPI_ISL_2477771, EPI_ISL_2477772, EPI_ISL_2477773, EPI_ISL_2477774, EPI_ISL_2477775, EPI_ISL_2477776, EPI_ISL_2477777, EPI_ISL_2477778, EPI_ISL_2477779, EPI_ISL_2477780, EPI_ISL_2477781, EPI_ISL_2477782, EPI_ISL_2477783, EPI_ISL_2477784, EPI_ISL_2477785, EPI_ISL_2477786, EPI_ISL_2477789, EPI_ISL_2477790, EPI_ISL_2477791, EPI_ISL_2477792, EPI_ISL_2477793, EPI_ISL_2477794, EPI_ISL_2477795, EPI_ISL_2477796, EPI_ISL_2477797, EPI_ISL_2477798, EPI_ISL_2477799, EPI_ISL_2477801, EPI_ISL_2477835, EPI_ISL_2477836, EPI_ISL_2477838, EPI_ISL_2477839, EPI_ISL_2477843, EPI_ISL_2477844, EPI_ISL_2477845, EPI_ISL_2477846, EPI_ISL_2477850, EPI_ISL_2477851, EPI_ISL_2477853, EPI_ISL_2477854, EPI_ISL_2477855, EPI_ISL_2477856, EPI_ISL_2477864, EPI_ISL_2477866, EPI_ISL_2477874, EPI_ISL_2477877, EPI_ISL_2477878, EPI_ISL_2477879, EPI_ISL_2477880, EPI_ISL_2477881, EPI_ISL_2477973, EPI_ISL_2477974, EPI_ISL_2477975, EPI_ISL_2478036, EPI_ISL_2478044, EPI_ISL_2478113, EPI_ISL_2478114, EPI_ISL_2478879, EPI_ISL_2478910, EPI_ISL_2479215, EPI_ISL_2479216, EPI_ISL_2479232, EPI_ISL_2479373, EPI_ISL_2479375, EPI_ISL_2479379, EPI_ISL_2479381, EPI_ISL_2479382, EPI_ISL_2479384, EPI_ISL_2479385, EPI_ISL_2479386, EPI_ISL_2479387, EPI_ISL_2479388, EPI_ISL_2479392, EPI_ISL_2479393, EPI_ISL_2479394, EPI_ISL_2479401, EPI_ISL_2479403, EPI_ISL_2479404, EPI_ISL_2479412, EPI_ISL_2479413, EPI_ISL_2479414, EPI_ISL_2479415, EPI_ISL_2479416, EPI_ISL_2479419, EPI_ISL_2479420, EPI_ISL_2479422, EPI_ISL_2479424, EPI_ISL_2479425, EPI_ISL_2479430, EPI_ISL_2479432, EPI_ISL_2479437, EPI_ISL_2479439, EPI_ISL_2479440, EPI_ISL_2941238, EPI_ISL_3909787, EPI_ISL_3909788, EPI_ISL_3909793, EPI_ISL_3909811, EPI_ISL_3909813, EPI_ISL_3909814, EPI_ISL_3909816, EPI_ISL_3909817, EPI_ISL_3909819, EPI_ISL_3909831, EPI_ISL_3910034, EPI_ISL_3910045, EPI_ISL_3910054, EPI_ISL_3910057, EPI_ISL_3910063 | | | |
| see above | Edmonton Provincial Lab | Public Health Agency of Canada (PHAC) National Microbiology Laboratory | Buss; Croxen M; Deo A; Dieu P; E; Ferrato C; Gill K; Khan F; Koleva P; Li V; Lloyd C; Lynch T; Ma R; Murphy S; Pabbaraju K; Shokoples S; Thayer J; Tipples G; Whitehouse M; Wong A; Yu C; Zelyas N |
| EPI_ISL_2107414, EPI_ISL_2107415 | Emory Molecular Diagnostics Laboratory, Emory Healthcare | Piantadosi Lab, Emory Department of Pathology | Ahmed Babiker; Anne Piantadosi |
| EPI_ISL_2110509, EPI_ISL_2110518, EPI_ISL_2110521, EPI_ISL_2110523, EPI_ISL_2110530, EPI_ISL_2110534, EPI_ISL_2110542, EPI_ISL_2110571, EPI_ISL_2110576, EPI_ISL_2110718 | | | |
| see above | Eurofins LifeCodexx GmbH | Robert Koch Institute | |
| EPI_ISL_2018301 | FL Bureau of Public Health Laboratories-Tampa | Centers for Disease Control and Prevention Division of Viral Diseases, Pathogen Discovery | Alison Laufer Halpin; Ben L. Rambo-Martin; Clinton R. Paden; Dakota Howard; Darlene Wagner; Dave Wentworth; Dhwani Batra; Jasmine Padilla; Justin Lee; Katie Dillon; Krista Queen; Kristen Knipe; Kristine Lacek; Mark Burroughs; Matthew Schmerer; Mili Sheth; Peter Cook; Sam Shepard; Sarah Nobles; Shoshona Le; Suxiang Tong; Vivien Dugan; Yvette Unoarumhi |
| EPI_ISL_2008503, EPI_ISL_2008599, EPI_ISL_2008616, EPI_ISL_2008645, EPI_ISL_2008659, EPI_ISL_2008709, EPI_ISL_2008711, EPI_ISL_2008729, EPI_ISL_2008768, EPI_ISL_2008775, EPI_ISL_2008813, EPI_ISL_2008823, EPI_ISL_2229989, EPI_ISL_2277955, EPI_ISL_2442656, EPI_ISL_6049961, EPI_ISL_6049970, EPI_ISL_6049974, EPI_ISL_6049976, EPI_ISL_6050123, EPI_ISL_6050148, EPI_ISL_6050608, EPI_ISL_6050756 | | | |
| see above | Florida Bureau of Public Health Laboratories | Florida Bureau of Public Health Laboratories | Jason Blanton; Namratha Tarigopula; Sarah Schmedes; Tiffany Splatt |
| EPI_ISL_2620911 | Fondazione IRCCS Ca' Granda Ospedale Maggiore Policlinico | Fondazione IRCCS Ca' Granda Ospedale Maggiore Policlinico | Ferruccio Ceriotti; Sara Uceda Renteria |
| EPI_ISL_1666131, EPI_ISL_1666132, EPI_ISL_1666144, EPI_ISL_1666154, EPI_ISL_1666160, EPI_ISL_1666161, EPI_ISL_1666165, EPI_ISL_1666176, EPI_ISL_1666193, EPI_ISL_1666195, EPI_ISL_1666346, EPI_ISL_1666376, EPI_ISL_1666383, EPI_ISL_1666389, EPI_ISL_1666391, EPI_ISL_1666395, EPI_ISL_1666396, EPI_ISL_1666432, EPI_ISL_1666443, EPI_ISL_1666455, EPI_ISL_1666471, EPI_ISL_1666480, EPI_ISL_1666492, EPI_ISL_1666504, EPI_ISL_1666511, EPI_ISL_1666513, EPI_ISL_1666515, EPI_ISL_1666542, EPI_ISL_1733866, EPI_ISL_1733868, EPI_ISL_1733874, EPI_ISL_1733877, EPI_ISL_1733879, EPI_ISL_1733887, EPI_ISL_1733897, EPI_ISL_1733903, EPI_ISL_1733915, EPI_ISL_1733935, EPI_ISL_1733936, EPI_ISL_1733952, EPI_ISL_1733954, EPI_ISL_1733969, EPI_ISL_1733971, EPI_ISL_1733973, EPI_ISL_1734419, EPI_ISL_1839064, EPI_ISL_1839088, EPI_ISL_1839089, EPI_ISL_1839198, EPI_ISL_1839201, EPI_ISL_1839280, EPI_ISL_1839284, EPI_ISL_1839290, EPI_ISL_1839300, EPI_ISL_1839301, EPI_ISL_1839302, EPI_ISL_1839305, EPI_ISL_1839313, EPI_ISL_1839320, EPI_ISL_1839328, EPI_ISL_1839333, EPI_ISL_1839379, EPI_ISL_1839414, EPI_ISL_1839437, EPI_ISL_1839513, EPI_ISL_1839521, EPI_ISL_1839522, EPI_ISL_1839525, EPI_ISL_1839529, EPI_ISL_1839530, EPI_ISL_1839545, EPI_ISL_1839636, EPI_ISL_1839656, EPI_ISL_1839662, EPI_ISL_1839669, EPI_ISL_1839672, EPI_ISL_1839682, EPI_ISL_1839684, EPI_ISL_1839690, EPI_ISL_1839693, EPI_ISL_1839707, EPI_ISL_1839708, EPI_ISL_1839713, EPI_ISL_1839729, EPI_ISL_1839736, EPI_ISL_1839743, EPI_ISL_1839773, EPI_ISL_1839782, EPI_ISL_1839905, EPI_ISL_1839910, EPI_ISL_1839917, EPI_ISL_1839930, EPI_ISL_1839937, EPI_ISL_1839938, EPI_ISL_1992239, EPI_ISL_1992469, EPI_ISL_1992472, EPI_ISL_1992489, EPI_ISL_1992502, EPI_ISL_1992503, EPI_ISL_1992528, EPI_ISL_1992529, EPI_ISL_1992534, EPI_ISL_2038036, EPI_ISL_2038042, EPI_ISL_2038059, EPI_ISL_2038062, EPI_ISL_2038080, EPI_ISL_2038083, EPI_ISL_2038087, EPI_ISL_2038096, EPI_ISL_2038100, EPI_ISL_2038103, EPI_ISL_2038121, EPI_ISL_2038132, EPI_ISL_2038133, EPI_ISL_2038141, EPI_ISL_2038155, EPI_ISL_2038161, EPI_ISL_2038174, EPI_ISL_2038185, EPI_ISL_2038190, EPI_ISL_2038196, EPI_ISL_2038199, EPI_ISL_2038200, EPI_ISL_2038268, EPI_ISL_2038297, EPI_ISL_2038309, EPI_ISL_2038310, EPI_ISL_2038321, EPI_ISL_2038470, EPI_ISL_2038473, EPI_ISL_2038491, EPI_ISL_2038515, EPI_ISL_2038537, EPI_ISL_2038541, EPI_ISL_2038549, EPI_ISL_2038555, EPI_ISL_2038572, EPI_ISL_2038576, EPI_ISL_2038600, EPI_ISL_2038644, EPI_ISL_2038645, EPI_ISL_2038649, EPI_ISL_2038664, EPI_ISL_2038666, EPI_ISL_2038683, EPI_ISL_2038695, EPI_ISL_2038723, EPI_ISL_4368448, EPI_ISL_4368570 | | | |
| see above | Fulgent Genetics | Centers for Disease Control and Prevention Division of Viral Diseases, Pathogen Discovery | Adrian Paskey; Becky Tsai; Benafsh Sapra; Benjamin Rambo-Martin; Christopher Gulvick; Clinton Paden; Clinton R. Paden; Dakota Howard; Darlene Wagner; Dhwani Batra; Doreen Ng; Duncan MacCannell; Erisa Sula; Harry Gao; James Xie; Jason Caravas; John Gao; Joseph Fierro; Kara Moser; Kristine Lacek; Matthew Schmerer; Mickey Li; Peter Cook; Peter W. Cook; Scott Sammons; Shatavia Morrison; Tymeckia Kendall; Victoria Caban Figueroa; Yan Meng; Yvette Unoarumhi |
| EPI_ISL_1937261, EPI_ISL_1937291, EPI_ISL_1937359, EPI_ISL_1937612, EPI_ISL_1937614 | Fulgent Genetics | Fulgent Genetics | Becky Tsai; Benafsh Sapra; Doreen Ng; Harry Gao; James Xie; John Gao; Joseph Fierro; Mickey Li; Yan Meng |
| EPI_ISL_3553534, EPI_ISL_3553539, EPI_ISL_3553542, EPI_ISL_3553545, EPI_ISL_3553548, EPI_ISL_3553553, EPI_ISL_3553554, EPI_ISL_3553557, EPI_ISL_3553561, EPI_ISL_3553562, EPI_ISL_3553583, EPI_ISL_3553590, EPI_ISL_3553598, EPI_ISL_3553601, EPI_ISL_3553605, EPI_ISL_3553621, EPI_ISL_3553623, EPI_ISL_3553624, EPI_ISL_3553636, EPI_ISL_3553643, EPI_ISL_3553644, EPI_ISL_3553645 | | | |
| see above | Fundação Ezequiel Dias (FUNED) | Fundação Ezequiel Dias | Andre Leal; Cynthia Vazquez; Elaine Cristina; Felipe Iani; Flavia Aburjaile; Gislene Garcia de Castro Lichs; Glauco Carvalho; Hegger Fritsch; Joilson Xavier; Luiz Alcantara.; Luiz Henrique Ferraz Demarchi; Luiz Takao Watanabe; Marina Castilhos Souza Umaki Zardin; Marta Giovanetti; Natalia Guimaraes; Raquel da Silva Ferreira; Talita Adelino; Vagner Fonseca; de Oliveira |
| EPI_ISL_1919420 | Furst Medical Laboratory | Norwegian Institute of Public Health, Department of Virology | Atiya R Ali; Debech Nadia; Engebretsen Serina Beate; Garcia Llorente Ignacio; Hilde Elshaug; Hilde Vollan; Jon Bråte; Kamilla Heddeland Instefjord; Karoline Bragstad; Kathrine Stene-Johansen; Marie Paulsen Madsen; Olav Hungnes; Pedersen Benedikte Nevjen; Rasmus Riis Kopperud |
| EPI_ISL_1937673 | GA Department of Public Health Laboratory | Centers for Disease Control and Prevention Division of Viral Diseases, Pathogen Discovery | Alison Laufer Halpin; Ben L. Rambo-Martin; Clinton R. Paden; Dakota Howard; Darlene Wagner; Dave Wentworth; Dhwani Batra; Jasmine Padilla; Justin Lee; Katie Dillon; Krista Queen; Kristen Knipe; Kristine Lacek; Mark Burroughs; Matthew Schmerer; Mili Sheth; Peter Cook; Sam Shepard; Sarah Nobles; Shoshona Le; Suxiang Tong; Vivien Dugan; Yvette Unoarumhi |
| EPI_ISL_1757348, EPI_ISL_1757405 | GH A.CHENEVIER-H.MONDOR | Department of Virology, Henri Mondor University Hospital, Assistance Publique Hôpitaux de Paris, Université Paris-Est Créteil, INSERM U955 | Alexandre Soulier; Christophe Rodriguez; Elisabeth Trawinski; Guillaume Gricourt; Jean-Michel Pawlotsky; Melissa N'Debi; Slim Fourati; Vanessa Demontant |
| EPI_ISL_1757685, EPI_ISL_1757708 | GROUPE HOSPITALIER SUD ILE DE FRANCE | Department of Virology, Henri Mondor University Hospital, Assistance Publique Hôpitaux de Paris, Université Paris-Est Créteil, INSERM U955 | Alexandre Soulier; Christophe Rodriguez; Elisabeth Trawinski; Guillaume Gricourt; Jean-Michel Pawlotsky; Melissa N'Debi; Slim Fourati; Vanessa Demontant |
| EPI_ISL_1966724, EPI_ISL_1966725, EPI_ISL_1966726 | GRUPO TECNICO DE VIGILANCIA SANITARIA E EPIDEMIOLOGICA | Instituto Butantan | Antonio Jorge Martins; Bianca Cechetto Carlos. Mendelics: Bibiana Santos; Claudia Renata dos Santos Barros; Cíntia Bittar; David Schlesinger. Hemocentro Ribeirão Preto: Simone Kashima; Debora Botequio Moretti; Elaine Cristina Marqueze; Elaine Vieira dos Santos; Elisangela Chicaroni Mattos; Erika Freitas; Evandra Strazza Rodrigues; Felipe Allan da Silva da Costa; Flavia Aburjaile; Fábio Sossai Possebon; Guilherme Campos; Guilherme Targino Valente; Heidge Fukumasu. USP-Botucatu: Rejane Maria Tommasini Grotto; Helena Lage Ferreira; Instituto Butantan: Dimas Tadeu Covas; Jardelina de Souza Todao Bernardino; Jayme A. Souza-Neto; Jessika Cristina Chagas Lesbon; Jorge A. Petroli Marchesi; José Salvatore Leister Patané; João Paulo Kitajima; João Pessoa Araújo Jr.; Leila Sabrina Ullmann; Loyze Paola Oliveira de Lima; Luiz Aurelio de Campos Crispin. Centro de Genômica Funcional da ESALQ: Luiz Lehmann Coutinho; Luiz Carlos Junior de Alcantara; Lívia Sacchetto; Maisa C. Pereira Parra; Maria Carolina Elias; Marta Giovanetti; Marília Moraes; Maurício Lacerda Nogueira. Prefeitura de Sao Paulo: Melissa Palmieri.; Patricia Akemi Assato; Paula Rahal; Paulo Inacio da Costa; Rafael dos Santos Bezerra; Raquel de Lello Rocha Campos Cassano. NGS Soluções Genômicas: Pilar Drummond Sampaio Corrêa Mariani. FZEA-USP Pirassununga: Mirele Daiana Poleti; Raul Machado Neto; Ricardo Augusto Brassaloti; Ricardo Haddad; Rodrigo Tocantins Calado. FAMERP-SJRP: Cecília Artico Banho; Sandra Coccuzzo Sampaio; Svetoslav Nanev Slavov; Vagner Fonseca; Vincent Louis Viala |
| EPI_ISL_2345835, EPI_ISL_2362247, EPI_ISL_2362271 | GRUPO TECNICO DE VIGILANCIA SANITARIA E EPIDEMIOLOGICA | Instituto Butantan / Mendelics | Antonio Jorge Martins; Claudia Renata dos Santos Barros; David Schlesinger; Debora Botequio Moretti; Dimas Tadeu Covas; Elaine Cristina Marqueze; Elaine Vieira Santos; Evandra Strazza Rodrigues; Heidge Fukumasu; Jayme Augusto de Souza-Neto; José Salvatore Leister Patané; Luiz Alcantara; Luiz Lehmann Coutinho; Maria Carolina Elias; Maurício Lacerda Nogueira; Rafael dos Santos Bezerra; Raul Machado Neto; Rejane Maria Tommasini Grotto; Ricardo Haddad; Sandra Coccuzzo Sampaio Vessoni; Simone Kashima; Svetoslav Nanev Slavov; Vincent Louis Viala |
| EPI_ISL_1712326, EPI_ISL_1712329, EPI_ISL_1712335, EPI_ISL_1712343, EPI_ISL_1712344, EPI_ISL_1712364, EPI_ISL_1712367, EPI_ISL_1712368, EPI_ISL_1712370, EPI_ISL_1712372, EPI_ISL_2009215, EPI_ISL_2009231, EPI_ISL_2009245, EPI_ISL_2009255, EPI_ISL_2009271, EPI_ISL_2009274, EPI_ISL_2009284, EPI_ISL_2009287, EPI_ISL_2009292, EPI_ISL_2009295, EPI_ISL_2009303, EPI_ISL_2009597, EPI_ISL_2009611, EPI_ISL_2009612, EPI_ISL_2009616, EPI_ISL_2009618, EPI_ISL_2009619, EPI_ISL_2009621, EPI_ISL_2597236, EPI_ISL_2597248, EPI_ISL_2597297, EPI_ISL_2597298, EPI_ISL_2597299, EPI_ISL_2597301 | | | |
| see above | Genetica Molecular and Subdepartamento de Virologia ISP Chile | Instituto de Salud Publica de Chile | Andres Castillo; Barbara Parra; Constanza Campano; Gisselle Barra; Javier Tognarelli; Jorge Fernandez; Karen Orostica; Loredana Arata; Patricia Bustos; Rodrigo Fasce; Soledad Ulloa |
| EPI_ISL_5196282 | Gorgas Memorial Institute of Health Studies | Gorgas Memorial Institute of Health Studies | Castillo Jorge; Chen Maria; Franco Danilo; Gonzalez Claudia; Jessica Gondola; Leyda Abrego; Lopez-Verges Sandra; Marlenne Castillo; Martinez Alexander; Menacho Abdiel; Moreno Ambar; Moreno Brechla; Oris Chavarria; Ortiz Alma; Salazar Jacqueline |
| EPI_ISL_2550797 | Grady Memorial Hospital, Clinical Microbiology Laboratory | Piantadosi Lab, Emory Department of Pathology | Anne Piantadosi; Dara Khosravi; Yun F (Wayne) Wang |

| | | | |
|---|---|---|---|
| EPI_ISL_2601789 | Gravity Diagnostics, LLC | Gravity Diagnostics, LLC | Gravity Diagnostics |
| EPI_ISL_5800468, EPI_ISL_5800469, EPI_ISL_5800470, EPI_ISL_5802404 | Grupo Tecnico De Vigilancia Sanitaria E Epidemiologica | Instituto Butantan | Antonio Jorge Martins; Claudia Renata dos Santos Barros; David Schlesinger; Debora Botequio Moretti; Dimas Tadeu Covas; Elaine Cristina Marqueze; Elaine Vieira Santos; Evandra Strazza Rodrigues; Heidge Fukumasu; Jayme Augusto de Souza-Neto; José Salvatore Leister Patané; Luiz Alcantara; Luiz Lehmann Coutinho; Maria Carolina Elias; Maurício Lacerda Nogueira; Rafael dos Santos Bezerra; Raul Machado Neto; Rejane Maria Tommasini Grotto; Ricardo Haddad; Sandra Coccuzzo Sampaio Vessoni; Simone Kashima; Svetoslav Nanev Slavov; Vincent Louis Viala |
| EPI_ISL_2339821, EPI_ISL_2339822, EPI_ISL_2339841 | Grupo de Investigación en Enfermedades Tropicales del Ejército (GINETEJ), Laboratorio de Referencia e Investigación, Dirección de Sanidad Ejército, Bogotá, Colombia | Centro de Investigaciones en Microbiología y Biotecnología-UR (CIMBIUR), Facultad de Ciencias Naturales, Universidad del Rosario, Bogotá, Colombia | Camilo A. Correa-Cárdenas; Carolina Oliveros; Claudia Méndez; Elizabeth K. Márquez; Frank de los Santos Ortiz; Juan David Ramírez; Julie Pérez; Lorena Albarracín; Luz H. Patiño; Maria Clara Duque; Marina Muñoz; María Teresa Alvarado; Nathalia Ballesteros; Sergio Castañeda; Sergio Gutiérrez-Riveros; Yanira Romero; Zulma Cucunubá |
| EPI_ISL_1969691, EPI_ISL_1969692 | Guangdong Provincial Center for Disease Control and Prevention | National Institute for Viral Disease Control and Prevention, China CDC | Baisheng; Min Kang; Xiang Zhao; Xiaoling Deng; Yang Song; Yao Hu; Zhencui Li |
| EPI_ISL_1854241 | H Garcia de Orta | Instituto Nacional de Saude (INSA) | Borges et al |
| EPI_ISL_1854250 | H Vila Franca Xira | Instituto Nacional de Saude (INSA) and Institute of Biomedicine (iBiMed), Universidade de Aveiro | Borges et al |
| EPI_ISL_3912025 | HGCC HOSPITAL GERAL DR CESAR CALS | Analytical Competence Molecular Epidemiology Lab/ACME, Oswaldo Cruz Foundation, Ceara (FIOCRUZ CE) | Cleber Furtado Aksenen; Fabio Miyajima; Fernando Braga Stehling; Francisco Eder de Moura Lopes; Jamille Maria Mendes Bezerra; Joaquim Cesar do Nascimento Sousa Junior; Pedro Miguel Carneiro Jeronimo; Suzana Porto Almeida & Lucas Delerino on behalf of COVID-19 FIOCRUZ Genomic Network; Thais Ferreira de Oliveira; Thais de Oliveira Costa; Ticiane Cavalcante de Souza; Veridiana Pessoa Miyajima |
| EPI_ISL_5603284 | HGF HOSPITAL GERAL DE FORTALEZA | Analytical Competence Molecular Epidemiology Lab/ACME, Oswaldo Cruz Foundation, Ceara (FIOCRUZ CE) | Carlos Leonardo de Aragao Araujo; Cecilia Leite Costa & Eduardo Ruback dos Santos on behalf of COVID-19 FIOCRUZ Genomic Network; Cleber Furtado Aksenen; Fabio Miyajima; Fernando Braga Stehling; Francisco Eder de Moura Lopes; Igor Oliveira Duarte; Jamille Maria Mendes Bezerra; Joaquim Cesar do Nascimento Sousa Junior; Pedro Miguel Carneiro Jeronimo; Suzana Porto Almeida; Thais Ferreira de Oliveira; Thais de Oliveira Costa; Ticiane Cavalcante de Souza; Veridiana Pessoa Miyajima |
| EPI_ISL_2384091 | HI Dept. of Health, State Laboratories Division | Centers for Disease Control and Prevention Division of Viral Diseases, Pathogen Discovery | Alison Laufer Halpin; Ben L. Rambo-Martin; Clinton R. Paden; Dakota Howard; Darlene Wagner; Dave Wentworth; Dhwani Batra; Jasmine Padilla; Justin Lee; Katie Dillon; Krista Queen; Kristen Knipe; Kristine Lacek; Mark Burroughs; Matthew Schmerer; Mili Sheth; Peter Cook; Sam Shepard; Sarah Nobles; Shoshona Le; Suxiang Tong; Vivien Dugan; Yvette Unoarumhi |
| EPI_ISL_3912060, EPI_ISL_3912065, EPI_ISL_3912102, EPI_ISL_3912118 | HIAS HOSPITAL INFANTIL ALBERT SABIN | Analytical Competence Molecular Epidemiology Lab/ACME, Oswaldo Cruz Foundation, Ceara (FIOCRUZ CE) | Cleber Furtado Aksenen; Fabio Miyajima; Fernando Braga Stehling; Francisco Eder de Moura Lopes; Jamille Maria Mendes Bezerra; Joaquim Cesar do Nascimento Sousa Junior; Pedro Miguel Carneiro Jeronimo; Suzana Porto Almeida & Lucas Delerino on behalf of COVID-19 FIOCRUZ Genomic Network; Thais Ferreira de Oliveira; Thais de Oliveira Costa; Ticiane Cavalcante de Souza; Veridiana Pessoa Miyajima |
| EPI_ISL_2017325, EPI_ISL_2017326, EPI_ISL_2017327, EPI_ISL_2017328, EPI_ISL_2017329, EPI_ISL_2017330, EPI_ISL_2017331, EPI_ISL_2017332, EPI_ISL_2017333, EPI_ISL_2017334, EPI_ISL_2017335, EPI_ISL_2017336, EPI_ISL_2017337, EPI_ISL_2017338, EPI_ISL_2017339, EPI_ISL_2017340, EPI_ISL_2017341, EPI_ISL_2017342, EPI_ISL_2017343, EPI_ISL_2017344, EPI_ISL_2017345, EPI_ISL_2017361, EPI_ISL_2017364, EPI_ISL_2017365, EPI_ISL_2017366, EPI_ISL_2017376, EPI_ISL_2017410, EPI_ISL_2017446, EPI_ISL_2017448, EPI_ISL_2017462, EPI_ISL_2017463, EPI_ISL_2017464, EPI_ISL_2017484, EPI_ISL_2102521, EPI_ISL_2187982, EPI_ISL_2187983, EPI_ISL_2187984 | | | |
| see above | HLAGYN - Laboratorio de Imunologia de Transplantes de Goias | HLAGYN - Laboratorio de Imunologia de Transplantes de Goias | Alessandro Leonardo Alvares Magalhaes; Daniel Ferreira de Sousa; Danielle de Paiva Rezende; Erika Lopes Rocha Batista; Fernando Antonio Vinhal dos Santos; Frederico Rodrigues Vinhal; Lucas Carlos Gomes Pereira; Paola Cristina Resende Silva; Raphael Bessa Parmigiane; Sabrina Sara Moreira Duarte |
| EPI_ISL_3102236, EPI_ISL_3912019, EPI_ISL_3912032, EPI_ISL_3912174, EPI_ISL_3912175, EPI_ISL_3912176 | HM HOSPITAL DE MESSEJANA DR CARLOS ALBERTO STUDART GOMES | Analytical Competence Molecular Epidemiology Lab/ACME, Oswaldo Cruz Foundation, Ceara (FIOCRUZ CE) | Cleber Furtado Aksenen; Fabio Miyajima; Fernando Braga Stehling; Francisco Eder de Moura Lopes; Jamille Maria Mendes Bezerra; Joaquim Cesar do Nascimento Sousa Junior; Joaquim César do Nascimento Sousa Junior; Pedro Miguel Carneiro Jeronimo; Suzana Porto Almeida & Lucas Delerino on behalf of COVID-19 FIOCRUZ Genomic Network; Suzana Porto Almeida e Lucas Delerino; Thais Ferreira de Oliveira; Thais de Oliveira Costa; Ticiane Cavalcante de Souza; Veridiana Pessoa Miyajima |
| EPI_ISL_1811236 | HNN | Inciensa, Instituto Costarricense de Investigación y Enseñanza en Nutrición y Salud | Pérez-Corrales C & Catalán-Marín F |
| EPI_ISL_2142666 | HOPITAL SAINT ANDRE | CNR Virus des Infections Respiratoires - France SUD | Antonin Bal; Bruno Lina; Bruno Simon; Gregory Destras; Gwendolyne Burfin; Hadrien Regue; Laurence Josset; Martine Valette; Quentin Semanas |
| EPI_ISL_5603233 | HOSP MATERN JOAO FERREIRA GOMES | Analytical Competence Molecular Epidemiology Lab/ACME, Oswaldo Cruz Foundation, Ceara (FIOCRUZ CE) | Carlos Leonardo de Aragao Araujo; Cecilia Leite Costa & Eduardo Ruback dos Santos on behalf of COVID-19 FIOCRUZ Genomic Network; Cleber Furtado Aksenen; Fabio Miyajima; Fernando Braga Stehling; Francisco Eder de Moura Lopes; Igor Oliveira Duarte; Jamille Maria Mendes Bezerra; Joaquim Cesar do Nascimento Sousa Junior; Pedro Miguel Carneiro Jeronimo; Suzana Porto Almeida; Thais Ferreira de Oliveira; Thais de Oliveira Costa; Ticiane Cavalcante de Souza; Veridiana Pessoa Miyajima |
| EPI_ISL_5603234, EPI_ISL_5603244, EPI_ISL_5603245, EPI_ISL_5603255, EPI_ISL_5603256, EPI_ISL_5603257 | HOSP MATERN PAULO SARASATE | Analytical Competence Molecular Epidemiology Lab/ACME, Oswaldo Cruz Foundation, Ceara (FIOCRUZ CE) | Carlos Leonardo de Aragao Araujo; Cecilia Leite Costa & Eduardo Ruback dos Santos on behalf of COVID-19 FIOCRUZ Genomic Network; Cleber Furtado Aksenen; Fabio Miyajima; Fernando Braga Stehling; Francisco Eder de Moura Lopes; Igor Oliveira Duarte; Jamille Maria Mendes Bezerra; Joaquim Cesar do Nascimento Sousa Junior; Pedro Miguel Carneiro Jeronimo; Suzana Porto Almeida; Thais Ferreira de Oliveira; Thais de Oliveira Costa; Ticiane Cavalcante de Souza; Veridiana Pessoa Miyajima |
| EPI_ISL_5603273 | HOSP MUN ABELARDO GADELHA DA ROCHA | Analytical Competence Molecular Epidemiology Lab/ACME, Oswaldo Cruz Foundation, Ceara (FIOCRUZ CE) | Carlos Leonardo de Aragao Araujo; Cecilia Leite Costa & Eduardo Ruback dos Santos on behalf of COVID-19 FIOCRUZ Genomic Network; Cleber Furtado Aksenen; Fabio Miyajima; Fernando Braga Stehling; Francisco Eder de Moura Lopes; Igor Oliveira Duarte; Jamille Maria Mendes Bezerra; Joaquim Cesar do Nascimento Sousa Junior; Pedro Miguel Carneiro Jeronimo; Suzana Porto Almeida; Thais Ferreira de Oliveira; Thais de Oliveira Costa; Ticiane Cavalcante de Souza; Veridiana Pessoa Miyajima |
| EPI_ISL_1966737 | HOSP MUN JOSANIAS CASTANHA BRAGA | Instituto Butantan | Antonio Jorge Martins; Bianca Cechetto Carlos. Mendelics: Bibiana Santos; Claudia Renata dos Santos Barros; Cíntia Bittar; David Schlesinger. Hemocentro Ribeirão Preto: Simone Kashima; Debora Botequio Moretti; Elaine Cristina Marqueze; Elaine Vieira dos Santos; Elisangela Chicaroni Mattos; Erika Freitas; Evandra Strazza Rodrigues; Felipe Allan da Silva da Costa; Flavia Aburjaile; Fábio Sossai Possebon; Guilherme Campos; Guilherme Targino Valente; Heidge Fukumasu. USP-Botucatu: Rejane Maria Tommasini Grotto; Helena Lage Ferreira; Instituto Butantan: Dimas Tadeu Covas; Jardelina de Souza Todao Bernardino; Jayme A. Souza-Neto; Jessika Cristina Chagas Lesbon; Jorge A. Petroli Marchesi; José Salvatore Leister Patané; João Paulo Kitajima; João Pessoa Araújo Jr.; Leila Sabrina Ullmann; Loyze Paola Oliveira de Lima; Luiz Aurelio de Campos Crispin. Centro de Genômica Funcional da ESALQ: Luiz Lehmann Coutinho; Luiz Carlos Junior de Alcantara; Lívia Sacchetto; Maisa C. Pereira Parra; Maria Carolina Elias; Marta Giovanetti; Marília Moraes; Maurício Lacerda Nogueira. Prefeitura de Sao Paulo: Melissa Palmieri.; Patricia Akemi Assato; Paula Rahal; Paulo Inacio da Costa; Rafael dos Santos Bezerra; Raquel de Lello Rocha Campos Cassano. NGS Soluções Genômicas: Pilar Drummond Sampaio Corrêa Mariani. FZEA-USP Pirassununga: Mirele Daiana Poleti; Raul Machado Neto; Ricardo Augusto Brassaloti; Ricardo Haddad; Rodrigo Tocantins Calado. FAMERP-SJRP: Cecília Artico Banho; Sandra Coccuzzo Sampaio; Svetoslav Nanev Slavov; Vagner Fonseca; Vincent Louis Viala |
| EPI_ISL_2272959 | HOSPITAL DR. ENRIQUE BALTODANO BRICEÑO | Inciensa, Instituto Costarricense de Investigación y Enseñanza en Nutrición y Salud | Adriana Godínez; Claudio Soto-Garita; Estela Cordero; Francisco Duarte; Hebleen Porras; Joselyn Prado & Arnoldo Zamora-Villegas; José Luis Vargas; Mariela Gutiérrez; Melany Calderón |
| EPI_ISL_5529978 | HOSPITAL E MATERNIDADE DRA ZILDA ARNS NEUMANN | Analytical Competence Molecular Epidemiology Lab/ACME, Oswaldo Cruz Foundation, Ceara (FIOCRUZ CE) | Carlos Leonardo de Aragao Araujo; Cleber Furtado Aksenen; Fabio Miyajima; Fernando Braga Stehling; Francisco Eder de Moura Lopes; Jamille Maria Mendes Bezerra; Joaquim Cesar do Nascimento Sousa Junior; Pedro Miguel Carneiro Jeronimo; Suzana Porto Almeida & Igor Oliveira Duarte on behalf of COVID-19 FIOCRUZ Genomic Network; Thais Ferreira de Oliveira; Thais de Oliveira Costa; Ticiane Cavalcante de Souza; Veridiana Pessoa Miyajima |
| EPI_ISL_1966738 | HOSPITAL GERAL DE VILA NOVA CACHOEIRINHA SAO PAULO | Instituto Butantan | Antonio Jorge Martins; Bianca Cechetto Carlos. Mendelics: Bibiana Santos; Claudia Renata dos Santos Barros; Cíntia Bittar; David Schlesinger. Hemocentro Ribeirão Preto: Simone Kashima; Debora Botequio Moretti; Elaine Cristina Marqueze; Elaine Vieira dos Santos; Elisangela Chicaroni Mattos; Erika Freitas; Evandra Strazza Rodrigues; Felipe Allan da Silva da Costa; Flavia Aburjaile; Fábio Sossai Possebon; Guilherme Campos; Guilherme Targino Valente; Heidge Fukumasu. USP-Botucatu: Rejane Maria Tommasini Grotto; Helena Lage Ferreira; Instituto Butantan: Dimas Tadeu Covas; Jardelina de Souza Todao Bernardino; Jayme A. Souza-Neto; Jessika Cristina Chagas Lesbon; Jorge A. Petroli Marchesi; José Salvatore Leister Patané; João Paulo Kitajima; João Pessoa Araújo Jr.; Leila Sabrina Ullmann; Loyze Paola Oliveira de Lima; Luiz Aurelio de Campos Crispin. Centro de Genômica Funcional da ESALQ: Luiz Lehmann Coutinho; Luiz Carlos Junior de Alcantara; Lívia Sacchetto; Maisa C. Pereira Parra; Maria Carolina Elias; Marta Giovanetti; Marília Moraes; Maurício Lacerda Nogueira. Prefeitura de Sao Paulo: Melissa Palmieri.; Patricia Akemi Assato; Paula Rahal; Paulo Inacio da Costa; Rafael dos Santos Bezerra; Raquel de Lello Rocha Campos Cassano. NGS Soluções Genômicas: Pilar Drummond Sampaio Corrêa Mariani. FZEA-USP Pirassununga: Mirele Daiana Poleti; Raul Machado Neto; Ricardo Augusto Brassaloti; Ricardo Haddad; Rodrigo Tocantins Calado. FAMERP-SJRP: Cecília Artico Banho; Sandra Coccuzzo Sampaio; Svetoslav Nanev Slavov; Vagner Fonseca; Vincent Louis Viala |
| EPI_ISL_4601286, EPI_ISL_4601287 | HOSPITAL INFANTA CRISTINA (BADAJOZ) | Instituto de Salud Carlos III | A. Monzón; F. Casas; I. Jiménez; I.MARTIN CORDERO; Iglesias-Caballero; M. Sandonís; P. Zaballos; PALOMA; S. Camarero; S. Cuesta; S. Pozo; S. Varona; V. Vázquez-Morón |
| EPI_ISL_2020122 | HOSPITAL MANACOR | HOSPITAL UNIVERSITARIO SON ESPASES | Antonio Oliver; Carla López-Causapé; Pablo Fraile-Ribot; SeqCovid |
| EPI_ISL_5603225, EPI_ISL_5603231, EPI_ISL_5603240, EPI_ISL_5603253 | HOSPITAL MUNICIPAL DE GUARAMIRANGA | Analytical Competence Molecular Epidemiology Lab/ACME, Oswaldo Cruz Foundation, Ceara (FIOCRUZ CE) | Carlos Leonardo de Aragao Araujo; Cecilia Leite Costa & Eduardo Ruback dos Santos on behalf of COVID-19 FIOCRUZ Genomic Network; Cleber Furtado Aksenen; Fabio Miyajima; Fernando Braga Stehling; Francisco Eder de Moura Lopes; Igor Oliveira Duarte; Jamille Maria Mendes Bezerra; Joaquim Cesar do Nascimento Sousa Junior; Pedro Miguel Carneiro Jeronimo; Suzana Porto Almeida; Thais Ferreira de Oliveira; Thais de Oliveira Costa; Ticiane Cavalcante de Souza; Veridiana Pessoa Miyajima |
| EPI_ISL_2209298, EPI_ISL_5650237, EPI_ISL_5650315 | HOSPITAL MUNICIPAL DE ITAPIRA | Instituto Butantan | Antonio Jorge Martins; Claudia Renata dos Santos Barros; David Schlesinger; Debora Botequio Moretti; Dimas Tadeu Covas; Elaine Cristina Marqueze; Elaine Vieira Santos; Evandra Strazza Rodrigues; Heidge Fukumasu; Jayme Augusto de Souza-Neto; José Salvatore Leister Patané; Luiz Alcantara; Luiz Lehmann Coutinho; Maria Carolina Elias; Maurício Lacerda Nogueira; Rafael dos Santos Bezerra; Raul Machado Neto; Rejane Maria Tommasini Grotto; Ricardo Haddad; Sandra Coccuzzo Sampaio Vessoni; Simone Kashima; Svetoslav Nanev Slavov; Vincent Louis Viala; Vincent Louis Viala. |
| EPI_ISL_2344569, EPI_ISL_2344571, EPI_ISL_2344572, EPI_ISL_2344573, EPI_ISL_2344574, EPI_ISL_2344578, EPI_ISL_2344579 | | | |
| see above | HOSPITAL MUNICIPAL DE ITAPIRA | Instituto Butantan / FZEA-USP-Pirassununga | Antonio Jorge Martins; Claudia Renata dos Santos Barros; David Schlesinger; Debora Botequio Moretti; Dimas Tadeu Covas; Elaine Cristina Marqueze; Elaine Vieira Santos; Evandra Strazza Rodrigues; Heidge Fukumasu; Jayme Augusto de Souza-Neto; José Salvatore Leister Patané; Luiz Alcantara; Luiz Lehmann Coutinho; Maria Carolina Elias; Maurício Lacerda Nogueira; Rafael dos Santos Bezerra; Raul Machado Neto; Rejane Maria Tommasini Grotto; Ricardo Haddad; Sandra Coccuzzo Sampaio Vessoni; Simone Kashima; Svetoslav Nanev Slavov; Vincent Louis Viala |
| EPI_ISL_5603224 | HOSPITAL MUNICIPAL HUMBERTO DE QUEIROZ | Analytical Competence Molecular Epidemiology Lab/ACME, Oswaldo Cruz Foundation, Ceara (FIOCRUZ CE) | Carlos Leonardo de Aragao Araujo; Cecilia Leite Costa & Eduardo Ruback dos Santos on behalf of COVID-19 FIOCRUZ Genomic Network; Cleber Furtado Aksenen; Fabio Miyajima; Fernando Braga Stehling; Francisco Eder de Moura Lopes; Igor Oliveira Duarte; Jamille Maria Mendes Bezerra; Joaquim Cesar do Nascimento Sousa Junior; Pedro Miguel Carneiro Jeronimo; Suzana Porto Almeida; Thais Ferreira de Oliveira; Thais de Oliveira Costa; Ticiane Cavalcante de Souza; Veridiana Pessoa Miyajima |
| EPI_ISL_2272973 | HOSPITAL SAN JUAN DE DIOS | Inciensa, Instituto Costarricense de Investigación y Enseñanza en Nutrición y Salud | Adriana Godínez; Claudio Soto-Garita; Estela Cordero; Francisco Duarte; Hebleen Porras; Joselyn Prado & Andony Cordero-Jiménez; José Luis Vargas; Mariela Gutiérrez; Melany Calderón |

| | | | |
|---|---|---|---|
| EPI_ISL_2272969 | HOSPITAL SAN VICENTE DE PAUL | Inciensa, Instituto Costarricense de Investigación y Enseñanza en Nutrición y Salud | Adriana Godínez; Claudio Soto-Garita; Estela Cordero; Francisco Duarte; Hebleen Porras; Joselyn Prado & Silvia Sáenz-García; José Luis Vargas; Mariela Gutiérrez; Melany Calderón |
| EPI_ISL_2209975, EPI_ISL_2344724 | HOSPITAL SAO JOSE ITIRAPINA | Instituto Butantan / FZEA-USP-Pirassununga | Antonio Jorge Martins; Claudia Renata dos Santos Barros; David Schlesinger; Debora Botequio Moretti; Dimas Tadeu Covas; Elaine Cristina Marqueze; Elaine Vieira Santos; Evandra Strazza Rodrigues; Heidge Fukumasu; Jayme Augusto de Souza-Neto; José Salvatore Leister Patané; Luiz Alcantara; Luiz Lehmann Coutinho; Maria Carolina Elias; Maurício Lacerda Nogueira; Rafael dos Santos Bezerra; Raul Machado Neto; Rejane Maria Tommasini Grotto; Ricardo Haddad; Sandra Coccuzzo Sampaio Vessoni; Simone Kashima; Svetoslav Nanev Slavov; Vincent Louis Viala |
| EPI_ISL_3102539 | HOSPITAL SAO MATEUS | Analytical Competence Molecular Epidemiology Lab/ACME, Oswaldo Cruz Foundation, Ceara (FIOCRUZ CE) | Cleber Furtado Aksenen; Fabio Miyajima; Fernando Braga Stehling; Francisco Eder de Moura Lopes; Jamille Maria Mendes Bezerra; Joaquim César do Nascimento Sousa Junior; Pedro Miguel Carneiro Jeronimo; Suzana Porto Almeida e Lucas Delerino; Thais Ferreira de Oliveira; Thais de Oliveira Costa; Ticiane Cavalcante de Souza; Veridiana Pessoa Miyajima |
| EPI_ISL_3502589, EPI_ISL_3502590, EPI_ISL_3502683, EPI_ISL_3502703 | HOSPITAL UNIVERSITARIO 12 DE OCTUBRE | HOSPITAL UNIVERSITARIO 12 DE OCTUBRE | Carmen Martín-Higuera; Esther Viedma; Irene Múñoz-Gallego; M.ª Dolores Folgueira; Mar Aguilera; Noelia Moral; Rafael Delgado; Sagrario Zurita |
| EPI_ISL_2020104, EPI_ISL_2020154 | HOSPITAL UNIVERSITARIO SON ESPASES | HOSPITAL UNIVERSITARIO SON ESPASES | Antonio Oliver; Carla López-Causapé; Pablo Fraile-Ribot; SeqCovid |
| EPI_ISL_2150641 | HOSPITAL UNIVERSITARIO VIRGEN DE LA ARRIXACA | Instituto de Salud Carlos III | A. Monzón; F. Casas; I. Jiménez; I.MORENO PARRADO; Iglesias-Caballero; LAURA; M. Sandonís; P. Zaballos; S. Camarero; S. Cuesta; S. Pozo; S. Varona; V. Vázquez-Morón |
| EPI_ISL_3912112 | HOSPITAL UNIVERSITARIO WALTER CANTIDIO | Analytical Competence Molecular Epidemiology Lab/ACME, Oswaldo Cruz Foundation, Ceara (FIOCRUZ CE) | Cleber Furtado Aksenen; Fabio Miyajima; Fernando Braga Stehling; Francisco Eder de Moura Lopes; Jamille Maria Mendes Bezerra; Joaquim Cesar do Nascimento Sousa Junior; Pedro Miguel Carneiro Jeronimo; Suzana Porto Almeida & Lucas Delerino on behalf of COVID-19 FIOCRUZ Genomic Network; Thais Ferreira de Oliveira; Thais de Oliveira Costa; Ticiane Cavalcante de Souza; Veridiana Pessoa Miyajima |
| EPI_ISL_1960043, EPI_ISL_1960044 | HPTU | Universidad Nacional de Colombia - Laboratorio Genómico One Health | Andres F. Cardona-Rios; Carlos Franco-Muñoz; Carolina Muñoz-Arango; Celeny Ortiz; Daniel O. Maldonado-Perez; Diego A. Álvarez-Díaz; Hector Alejandro Ruiz-Moreno; Idabely Betancur Ortiz; Jorge E. Osorio; Juan P. Hernandez-Ortiz; Karl A Ciuoderis; Katherine Laiton-Donato; Laura Silvana Perez; Lina M. Hurtado; Marcela Mercado-Reyes; Maria Angélica Maya; Maria Stella López; Rita Almanza Payares; Sandra Ines Cano; Simón Villegas Velásquez |
| EPI_ISL_7744048 | HU | Laboratório de Bioinformática - Universidade Federal de Santa Catarina | "Aline Daina Schlindwein"; "Ana Paula Christoff"; "Antuani Baptistela"; "Carolina Leite Martins"; "Darcita Buerger Rovaris"; "Dayane Azevedo Padilha"; "Doris Sobral Marques SouzaSobral"; "Edmundo Carlos Grisard"; "Eric Kazuo Kawagoe"; "Fernanda Luiza Ferrari"; "Fernanda Roesene Melo"; "Fernando Hartmann Barazzetti"; "Gislaine Fongaro"; "Glauber Wagner"; "Guilherme Augusto Maia"; "Guilherme Razzera"; "Guilherme Toledo e Silva"; "Julia Kinetz Wachter"; "Luiz Felipe de Oliveira"; "Marcos André Schörne"; "Marcus Vinícius Duarte Rodrigues"; "Maria Luiza Bazzo"; "Marlei Pickler Debiasi dos Anjos"; "Milene Moehr de Moraes"; "Nestor Wendt"; "Patrícia Hermes Stoco"; "Paula Sacchet"; "Renato Simões Moreira"; "Rodrigo de Paula Baptista"; "Tamela Zamboni Madaloz"; "Tatiany Aparecida Teixeira Soratto"; "Vilmar Benetti Filho" |
| EPI_ISL_2483425, EPI_ISL_2483426, EPI_ISL_2483428, EPI_ISL_2738475 | Hackensack Medical Center | New York Genome Center | Andre Corvelo; Barry Kreiswirth; David Perlin; Dayna M. Oschwald; Jose Mediavilla; Kaelea Composto; Kar Chow; Liang Chen; Marcus Cunningham; Michael Zody; Samantha Fennessey; Tom Maniatis |
| EPI_ISL_1803509, EPI_ISL_1803531, EPI_ISL_1803536, EPI_ISL_1803537, EPI_ISL_1803553, EPI_ISL_1803563, EPI_ISL_1803568, EPI_ISL_1803580, EPI_ISL_1803584, EPI_ISL_1803585, EPI_ISL_1803597, EPI_ISL_1803603, EPI_ISL_1803624, EPI_ISL_1803625, EPI_ISL_1803630, EPI_ISL_1803642, EPI_ISL_1803656, EPI_ISL_1803673, EPI_ISL_1803679, EPI_ISL_1803720, EPI_ISL_1803733, EPI_ISL_1803777, EPI_ISL_1803800, EPI_ISL_1803802, EPI_ISL_1803816, EPI_ISL_1803855, EPI_ISL_1803881, EPI_ISL_1803888, EPI_ISL_1803899, EPI_ISL_1803946, EPI_ISL_1803953, EPI_ISL_1804110, EPI_ISL_1804112, EPI_ISL_1804120, EPI_ISL_1804146, EPI_ISL_1804524, EPI_ISL_1804770, EPI_ISL_1804776, EPI_ISL_1906887, EPI_ISL_1906925, EPI_ISL_1907002, EPI_ISL_1907008, EPI_ISL_1907016, EPI_ISL_1907090, EPI_ISL_1907096, EPI_ISL_1907118, EPI_ISL_1907124, EPI_ISL_1907151, EPI_ISL_1907164, EPI_ISL_1907165, EPI_ISL_1907181, EPI_ISL_1907197, EPI_ISL_1907205, EPI_ISL_1907235, EPI_ISL_1907252, EPI_ISL_1907255, EPI_ISL_1907265, EPI_ISL_1907293, EPI_ISL_1907351, EPI_ISL_1907355, EPI_ISL_1907364, EPI_ISL_1907373, EPI_ISL_1907406, EPI_ISL_1907411, EPI_ISL_1907412, EPI_ISL_1907420, EPI_ISL_1907450, EPI_ISL_1907508, EPI_ISL_1907514, EPI_ISL_1907517, EPI_ISL_1907538, EPI_ISL_1907561, EPI_ISL_1907579, EPI_ISL_1907596, EPI_ISL_1907627, EPI_ISL_1907660, EPI_ISL_1907667, EPI_ISL_1907705, EPI_ISL_1907729, EPI_ISL_1907743, EPI_ISL_1907806, EPI_ISL_1907808, EPI_ISL_1907826, EPI_ISL_1907829, EPI_ISL_1907851, EPI_ISL_1907867, EPI_ISL_1907876, EPI_ISL_1907884, EPI_ISL_1907889, EPI_ISL_1907892, EPI_ISL_1907940, EPI_ISL_1907966, EPI_ISL_1907985, EPI_ISL_1908010, EPI_ISL_1908017, EPI_ISL_1908023, EPI_ISL_1908044, EPI_ISL_1908048, EPI_ISL_1908053, EPI_ISL_1908055, EPI_ISL_1908071, EPI_ISL_1908076, EPI_ISL_1908092, EPI_ISL_1908116, EPI_ISL_1908137, EPI_ISL_1923190, EPI_ISL_1923204, EPI_ISL_1923216, EPI_ISL_1923261, EPI_ISL_1923297, EPI_ISL_1923376, EPI_ISL_1923383, EPI_ISL_1923392, EPI_ISL_1923474, EPI_ISL_1923487, EPI_ISL_1923608, EPI_ISL_1923629, EPI_ISL_1923634, EPI_ISL_1923652, EPI_ISL_1923683, EPI_ISL_1923703, EPI_ISL_1923747, EPI_ISL_1923749, EPI_ISL_1923758, EPI_ISL_1923771, EPI_ISL_1923775, EPI_ISL_1923784, EPI_ISL_1923804, EPI_ISL_1923815, EPI_ISL_1923816, EPI_ISL_1923818, EPI_ISL_1923844, EPI_ISL_1923875, EPI_ISL_1923886, EPI_ISL_1923893, EPI_ISL_1923894, EPI_ISL_1923917, EPI_ISL_1923929, EPI_ISL_1923930, EPI_ISL_1923939, EPI_ISL_1923967, EPI_ISL_1923985, EPI_ISL_1923989, EPI_ISL_1923994, EPI_ISL_1924003, EPI_ISL_1924042, EPI_ISL_1924048, EPI_ISL_1924053, EPI_ISL_1924079, EPI_ISL_1924102, EPI_ISL_1924114, EPI_ISL_1924135, EPI_ISL_1924145, EPI_ISL_1924148, EPI_ISL_1924156, EPI_ISL_1924167, EPI_ISL_1924198, EPI_ISL_1924212, EPI_ISL_1924246, EPI_ISL_1924255, EPI_ISL_1924258, EPI_ISL_1924272, EPI_ISL_1935587, EPI_ISL_1935588, EPI_ISL_1935596, EPI_ISL_1942151, EPI_ISL_1942160, EPI_ISL_1942168, EPI_ISL_1990933, EPI_ISL_1990973, EPI_ISL_1991156, EPI_ISL_1991232, EPI_ISL_1991234, EPI_ISL_1991243, EPI_ISL_1991833, EPI_ISL_4346293, EPI_ISL_4346311, EPI_ISL_4347475 | | | |
| see above | Helix/Illumina | Centers for Disease Control and Prevention Division of Viral Diseases, Pathogen Discovery | Adrian Paskey; Alexandre Bolze; Ary Ascencio; Benjamin Rambo-Martin; Brad Sickler; Charlotte Rivera-Garcia; Christine Tran; Christopher Gulvick; Chrstine Tran; Clinton Paden; Clinton R. Paden; Dakota Howard; Darlene Wagner; David Becker; Dhwani Batra; Duncan MacCannell; Efren Sandoval; Eileen De Feo; Eileen de Feo; Elizabeth Cirulli; Eric Allen; Geraint Levan; James Lu; Jan Antico; Jason Caravas; Jason Nguyen; Jimmy Ramirez; Jingtao Liu; Kara Moser; Kelly Barrett; Kelly Schiabor Barrett; Kim Gietzen; Kristine Lacek; Magnus Isaksson; Marc Laurent; Matthew Schmerer; Matthew Tolentino; Nicole L. Washington; Nicole Washington; Peter Cook; Peter W. Cook; Phil Febbo; Ryan Cho; Scott Sammons; Shannon Wickline; Shatavia Morrison; Sherry Wang; Simon White; Tyler Cassens; William Lee; Yvette Unoarumhi |
| EPI_ISL_2501101 | Hennepin County Medical Center | Minnesota Department of Health, Public Health Laboratory | Alexandra Lorentz; Jacob Garfin; Matt Plumb; and Xiong Wang |
| EPI_ISL_5800462 | Hosp Mun Josanias Castanha Braga | Instituto Butantan | Antonio Jorge Martins; Claudia Renata dos Santos Barros; David Schlesinger; Debora Botequio Moretti; Dimas Tadeu Covas; Elaine Cristina Marqueze; Elaine Vieira Santos; Evandra Strazza Rodrigues; Heidge Fukumasu; Jayme Augusto de Souza-Neto; José Salvatore Leister Patané; Luiz Alcantara; Luiz Lehmann Coutinho; Maria Carolina Elias; Maurício Lacerda Nogueira; Rafael dos Santos Bezerra; Raul Machado Neto; Rejane Maria Tommasini Grotto; Ricardo Haddad; Sandra Coccuzzo Sampaio Vessoni; Simone Kashima; Svetoslav Nanev Slavov; Vincent Louis Viala |
| EPI_ISL_1700647, EPI_ISL_1700648, EPI_ISL_1700650, EPI_ISL_1821591 | Hospital | National Reference Center for Viruses of Respiratory Infections, Institut Pasteur, Paris | Adrien Pain; Angela Brisebarre; Camille Capel; Christophe Malabat; Corinne Maufrais; Damien Mornico; Etienne Simon-Lorière; Frédéric Lemoine; Laurence Louvet; Louise Lefrançois; Marion Barbet; Maud Vanpeene; Méline Bizard; Pascale Martres; Sylvie Behillil; Sylvie Van der Werf; Vincent Enouf |
| EPI_ISL_1917362, EPI_ISL_1917365 | Hospital Center Emile Mayrisch | Laboratoire national de sante, Microbiology, Microbial Genomics Platform | Anke Wienecke-Baldacchino; Catherine Ragimbeau; Cynthia Oxacelay; Fatu Djabi; Jessica Tapp; Lise Pignon; Raoul Salmon; Tamir Abdelrahman |
| EPI_ISL_2016620 | Hospital Francesc de Borja (Gandia) | SeqCOVID-SPAIN consortium/IBV(CSIC) | Inmaculada Vidal Catalá; Isabel García Nava and SeqCOVID-SPAIN consortium; Mª Rosa Navarro Cots; Nieves Orta Mira |
| EPI_ISL_1914361, EPI_ISL_1914362, EPI_ISL_1914363, EPI_ISL_1914364, EPI_ISL_1914365, EPI_ISL_1914366, EPI_ISL_1914367, EPI_ISL_1914368 | | | |
| see above | Hospital General Universitario Gregorio Marañón | Hospital General Universitario Gregorio Marañón | Cristina Rodriguez-Grande; Darío García de Viedma; Laura Pérez-Lago; Patricia Muñoz; Pedro Sola Campoy; Pilar Catalán; Sergio Buenestado Serrano |
| EPI_ISL_2271505 | Hospital General Universitario de Alicante - Instituto de Investigación Sanitaria y Biomédica de Alicante | SeqCOVID-SPAIN consortium/IBV(CSIC) | Carmen Molina Pardines and SeqCOVID-SPAIN consortium; Maripaz Ventero Martín |
| EPI_ISL_5800463 | Hospital Geral De Vila Nova Cachoeirinha Sao Paulo | Instituto Butantan | Antonio Jorge Martins; Claudia Renata dos Santos Barros; David Schlesinger; Debora Botequio Moretti; Dimas Tadeu Covas; Elaine Cristina Marqueze; Elaine Vieira Santos; Evandra Strazza Rodrigues; Heidge Fukumasu; Jayme Augusto de Souza-Neto; José Salvatore Leister Patané; Luiz Alcantara; Luiz Lehmann Coutinho; Maria Carolina Elias; Maurício Lacerda Nogueira; Rafael dos Santos Bezerra; Raul Machado Neto; Rejane Maria Tommasini Grotto; Ricardo Haddad; Sandra Coccuzzo Sampaio Vessoni; Simone Kashima; Svetoslav Nanev Slavov; Vincent Louis Viala |
| EPI_ISL_2614600 | Hospital Geral de Guaianazes | Instituto Adolfo Lutz, Interdiciplinary Procedures Center, Strategic Laboratory | Caio Vinicius Dias Lopes; Claudia Regina Gonçalves; Claudio Tavares Sacchi; Erica Valessa Ramos Gomes; Karoline Rodrigues Campos; Leonardo Jose Tadeu de Araujo |
| EPI_ISL_2614580 | Hospital Geral de Pedreira | Instituto Adolfo Lutz, Interdiciplinary Procedures Center, Strategic Laboratory | Caio Vinicius Dias Lopes; Claudia Regina Gonçalves; Claudio Tavares Sacchi; Erica Valessa Ramos Gomes; Karoline Rodrigues Campos; Leonardo Jose Tadeu de Araujo |
| EPI_ISL_2363558, EPI_ISL_2363565 | Hospital Jaime Ferre - SAMCO Rafaela | Grupo de Genómica y Bioinformática del Instituto de Investigación de la Cadena Láctea CONICET-INTA on behalf of 'Proyecto Argentino Interinstitucional de genomica de SARS-CoV-2' (PAIS Consortium) | AF; Amadio; C; Eberhardt; Irazoqui; Isaia; JF; JM; MF; Pandolfi; Quaranta; V |
| EPI_ISL_2614577 | Hospital Municipal Antonio Giglio | Instituto Adolfo Lutz, Interdiciplinary Procedures Center, Strategic Laboratory | Caio Vinicius Dias Lopes; Claudia Regina Gonçalves; Claudio Tavares Sacchi; Erica Valessa Ramos Gomes; Karoline Rodrigues Campos; Leonardo Jose Tadeu de Araujo |
| EPI_ISL_5782477, EPI_ISL_5782479, EPI_ISL_5782480, EPI_ISL_5782481, EPI_ISL_5782482, EPI_ISL_5782486, EPI_ISL_5782487 | | | |
| see above | Hospital Municipal De Itapira | Instituto Butantan | Antonio Jorge Martins; Claudia Renata dos Santos Barros; David Schlesinger; Debora Botequio Moretti; Dimas Tadeu Covas; Elaine Cristina Marqueze; Elaine Vieira Santos; Evandra Strazza Rodrigues; Heidge Fukumasu; Jayme Augusto de Souza-Neto; José Salvatore Leister Patané; Luiz Alcantara; Luiz Lehmann Coutinho; Maria Carolina Elias; Maurício Lacerda Nogueira; Rafael dos Santos Bezerra; Raul Machado Neto; Rejane Maria Tommasini Grotto; Ricardo Haddad; Sandra Coccuzzo Sampaio Vessoni; Simone Kashima; Svetoslav Nanev Slavov; Vincent Louis Viala |
| EPI_ISL_2614590 | Hospital Municipal Dr. Guido Guida | Instituto Adolfo Lutz, Interdiciplinary Procedures Center, Strategic Laboratory | Caio Vinicius Dias Lopes; Claudia Regina Gonçalves; Claudio Tavares Sacchi; Erica Valessa Ramos Gomes; Karoline Rodrigues Campos; Leonardo Jose Tadeu de Araujo |
| EPI_ISL_2614597 | Hospital Municipal de Mogi das Cruzes | Instituto Adolfo Lutz, Interdiciplinary Procedures Center, Strategic Laboratory | Caio Vinicius Dias Lopes; Claudia Regina Gonçalves; Claudio Tavares Sacchi; Erica Valessa Ramos Gomes; Karoline Rodrigues Campos; Leonardo Jose Tadeu de Araujo |
| EPI_ISL_2614594 | Hospital Regional de Cotia | Instituto Adolfo Lutz, Interdiciplinary Procedures Center, Strategic Laboratory | Caio Vinicius Dias Lopes; Claudia Regina Gonçalves; Claudio Tavares Sacchi; Erica Valessa Ramos Gomes; Karoline Rodrigues Campos; Leonardo Jose Tadeu de Araujo |
| EPI_ISL_5799726, EPI_ISL_5799766 | Hospital Sao Jose Itirapina | Instituto Butantan | Antonio Jorge Martins; Claudia Renata dos Santos Barros; David Schlesinger; Debora Botequio Moretti; Dimas Tadeu Covas; Elaine Cristina Marqueze; Elaine Vieira Santos; Evandra Strazza Rodrigues; Heidge Fukumasu; Jayme Augusto de Souza-Neto; José Salvatore Leister Patané; Luiz Alcantara; Luiz Lehmann Coutinho; Maria Carolina Elias; Maurício Lacerda Nogueira; Rafael dos Santos Bezerra; Raul Machado Neto; Rejane Maria Tommasini Grotto; Ricardo Haddad; Sandra Coccuzzo Sampaio Vessoni; Simone Kashima; Svetoslav Nanev Slavov; Vincent Louis Viala |
| EPI_ISL_3860415 | Hospital Señor del Milagro | Instituto de Patologia Vegetal (CIAP-INTA) on behalf of 'Proyecto Argentino Interinstitucional de genomica de SARS-CoV-2' (PAIS Consortium) | Debat; FD; Fernanda Agüero; Fernández; HJ; Lavaque Esteban; Marquez, N.; Pamela Cajal; Raskovsky Viviana; Veronica Lesser |
| EPI_ISL_6513196 | Hospital da Clinicas - UNICAMP | Laboratorio de Estudos de Virus | Bruno B. Benites; Daniel A. Toledo-Teixeira; Dewton Moraes Vasconcelos; Ester C. Sabino; Fabiana Granja; Fernando R. Spilki; Janine Schincariol Sabino; José Luiz Proenca-Modena; Julia Forato; Lia Furlaneto Marega; Luciana Souto Mofatto; Maria Marluce dos Santos Vilela; Mariene Ribeiro Amorim; Nuno R. |

| | | | |
|---|---|---|---|
| | | Emergentes | Faria; Raquel Silveira Bello Stucchi; William M. de Souza |
| EPI_ISL_2614575 | Hospital das Clinicas Luzia de Pinho Melo | Instituto Adolfo Lutz, Interdiciplinary Procedures Center, Strategic Laboratory | Caio Vinicius Dias Lopes; Claudia Regina Gonçalves; Claudio Tavares Sacchi; Erica Valessa Ramos Gomes; Karoline Rodrigues Campos; Leonardo Jose Tadeu de Araujo |
| EPI_ISL_1914379 | Hospital del Oriente de Asturias | Laboratorio de Virología HUCA | Abreu F; Alvarez-Arguelles ME; Boga JA; Castelló C; Costales I; Coto E; Gómez de Oña J; Martín-Rodríguez G; Melón S; Perez-Martínez Z; Rojo S; Sandoval M |
| EPI_ISL_2597572, EPI_ISL_2597577, EPI_ISL_2653036, EPI_ISL_3022828, EPI_ISL_3022860, EPI_ISL_3022861, EPI_ISL_3022865, EPI_ISL_3022874, EPI_ISL_3022886, EPI_ISL_3023231 | | | |
| see above | Hospital of the University of Pennsylvania Molecular Pathology Lab | Bushman Lab - University of Pennsylvania | Aoife M. Roche; Arupa Ganguly; Ayannah S. Fitzgerald; Brendan Kelly; Jevon Graham-Wooten; John K. Everett; Kyle Rodino; Layla A. Khatib; Mike Feldman; Ronald G. Collman and Frederic Bushan; Samantha A. Whiteside; Scott Sherrill-Mix; Shantan Reddy; Young Hwang |
| EPI_ISL_1672871 | Houston Health Dept. | Houston Health Dept. | Adolpho Lara; Pamela Brown; Ryker Penn; Yanlai Lai |
| EPI_ISL_2216933, EPI_ISL_2217100, EPI_ISL_2221466, EPI_ISL_2221525, EPI_ISL_2222823, EPI_ISL_2222845, EPI_ISL_2222893 | | | |
| see above | Houston Methodist Hospital | Houston Methodist Hospital | Ilya J. Finkelstein; James J. Davis; Jessica Cambric; Jimmy Gollihar; Kristina Reppond; Layne Pruitt; Madison N. Shyer; Marcus Nguyen; Matthew Ojeda Saavedra; Paul A. Christensen; Prasanti Yerramilli; Randall J. Olsen; Robert Olson; Ryan Gadd; S. Wesley Long; Sishir Subedi; and James M. Musser |
| EPI_ISL_1756322 | Hôpital Ambroise-Paré | Department of Virology, Henri Mondor University Hospital, Assistance Publique Hôpitaux de Paris, Université Paris-Est Créteil, INSERM U955 | Alexandre Soulier; Christophe Rodriguez; Elisabeth Trawinski; Guillaume Gricourt; Jean-Michel Pawlotsky; Melissa N'Debi; Slim Fourati; Vanessa Demontant |
| EPI_ISL_1757566, EPI_ISL_1757573, EPI_ISL_1757613 | Hôpital Avicenne | Department of Virology, Henri Mondor University Hospital, Assistance Publique Hôpitaux de Paris, Université Paris-Est Créteil, INSERM U955 | Alexandre Soulier; Christophe Rodriguez; Elisabeth Trawinski; Guillaume Gricourt; Jean-Michel Pawlotsky; Melissa N'Debi; Slim Fourati; Vanessa Demontant |
| EPI_ISL_2467748 | Hôpital Bichat Claude Bernard, Laboratoire de Virologie | IAME UMR1137 Inserm, Université de Paris, Hôpital Bichat | Alexandre Storto; Amélie Recoing; Antoine Bridier-Nahmias; Benoit Visseaux; Charlotte Charpentier; Diane Descamps; Gilles Collin; Lena Daniel; Mélanie Bertine; Nadhira Houhou-Fidouh; Quentin Le Hingrat; Siham Hamri |
| EPI_ISL_2444831, EPI_ISL_2444834, EPI_ISL_2444837, EPI_ISL_4071899, EPI_ISL_4071900 | IICS-UNA | IICS-UNA | Adriana Valenzuela; Alejandra Rojas; Chyntia Diaz; Eva Nara; Fatima Cardozo; Florencia del Puerto; Joel Ortiz; Jonas Fernandez; Laura Franco; Laura Mendoza; Leticia Rojas; Magaly Martinez; Maria Eugenia Galeano. |
| EPI_ISL_1937780 | IL Dept. of Public Health Springfield Laboratory | Centers for Disease Control and Prevention Division of Viral Diseases, Pathogen Discovery | Alison Laufer Halpin; Ben L. Rambo-Martin; Clinton R. Paden; Dakota Howard; Darlene Wagner; Dave Wentworth; Dhwani Batra; Jasmine Padilla; Justin Lee; Katie Dillon; Krista Queen; Kristen Knipe; Kristine Lacek; Mark Burroughs; Matthew Schmerer; Mili Sheth; Peter Cook; Sam Shepard; Sarah Nobles; Shoshona Le; Suxiang Tong; Vivien Dugan; Yvette Unoarumhi |
| EPI_ISL_1841026, EPI_ISL_1841032, EPI_ISL_1841035, EPI_ISL_1941628, EPI_ISL_1941643 | IN State Department of Health Laboratory Services | IN State Department of Health Laboratory Services | Brian Pope; Cassandra Campion; Jamie Yeadon; Kyle Brownlee; Lixia Liu; Mark Glazier; Melissa Hindenlang |
| EPI_ISL_2483725 | INMI Lazzaro Spallanzani IRCCS | INMI Lazzaro Spallanzani IRCCS | A Di Caro; B Bartolini; CEM Gruber; E Giombini; F Messina; F Santini; G Bonfiglio; M Rueca; MR Capobianchi; O Butera |
| EPI_ISL_1854217, EPI_ISL_1854220 | IPVC | Instituto Nacional de Saude (INSA) | Borges et al |
| EPI_ISL_2576998, EPI_ISL_2577001 | IRCCS San Gallicano Dermatological Institute | IRCCS Regina Elena National Cancer Institute | Aldo Morrone; Eleonora Sperandio; Fabrizio Ensoli; Francesca Sivori; Frauke Goeman; Fulvia Pimpinelli; Gennaro Ciliberto; Giovanni Blandino; Giulia Orlandi; Ludovica Ciuffreda; Martina Pontone; Maurizio Fanciulli; Sara Donzelli |
| EPI_ISL_2628302 | IVIC | Laboratorio de Virología Molecular | Carmen L Loureiro; CoViVen Group; Domingo J Garzaro; Esmeralda Vizzi; Flor H Pujol; Héctor R Rangel; José Luis Zambrano; Lieska Rodríguez; Mariana Hidalgo; Pierina D´Angelo; Rossana C Jaspe; Víctor Alarcón; Yoneira Sulbaran; Zoila Moros |
| EPI_ISL_2335136, EPI_ISL_2427270 | IVY3 Central Lab, Vanderbilt University Medical Center | Lauring Lab, University of Michigan, Department of Microbiology and Immunology | Gilbert |
| EPI_ISL_1742340, EPI_ISL_1742386, EPI_ISL_1742388, EPI_ISL_1742396, EPI_ISL_1742420, EPI_ISL_1809453, EPI_ISL_1809465, EPI_ISL_1809548, EPI_ISL_1809549, EPI_ISL_1809551, EPI_ISL_1809566, EPI_ISL_1809572, EPI_ISL_1809580, EPI_ISL_1809621, EPI_ISL_1809625, EPI_ISL_1809627, EPI_ISL_1809629, EPI_ISL_1809635, EPI_ISL_1809643, EPI_ISL_1809658, EPI_ISL_1809730, EPI_ISL_1964996, EPI_ISL_1965078 | | | |
| see above | Illinois Department of Public Health | Illinois Department of Public Health - Chicago Lab | Ira Heimler; Joel Price; Vineet K. Dhiman |
| EPI_ISL_1624148, EPI_ISL_1708241, EPI_ISL_1708243, EPI_ISL_1708244, EPI_ISL_1708254, EPI_ISL_1708258, EPI_ISL_1708304, EPI_ISL_1708305, EPI_ISL_1708309, EPI_ISL_1708312, EPI_ISL_1708313, EPI_ISL_1732552, EPI_ISL_1732553, EPI_ISL_1732556, EPI_ISL_1732558, EPI_ISL_1732559, EPI_ISL_1732562, EPI_ISL_1732570, EPI_ISL_1732575, EPI_ISL_1732578, EPI_ISL_1732579, EPI_ISL_1732627, EPI_ISL_1732628, EPI_ISL_1732629, EPI_ISL_1789122, EPI_ISL_1789123, EPI_ISL_1789129, EPI_ISL_1789132, EPI_ISL_1789141, EPI_ISL_1789142, EPI_ISL_1789143, EPI_ISL_1789146, EPI_ISL_1789148, EPI_ISL_1789174, EPI_ISL_1789175, EPI_ISL_1789176, EPI_ISL_1789177, EPI_ISL_1789178, EPI_ISL_1789194, EPI_ISL_1789196, EPI_ISL_1789199, EPI_ISL_1789201, EPI_ISL_1825708, EPI_ISL_1825711, EPI_ISL_1825713, EPI_ISL_1825714, EPI_ISL_1825715, EPI_ISL_1825716, EPI_ISL_1825720, EPI_ISL_1825722, EPI_ISL_1825723, EPI_ISL_1825725, EPI_ISL_1825726, EPI_ISL_1825731, EPI_ISL_1825732, EPI_ISL_1825736, EPI_ISL_1825738, EPI_ISL_1825739, EPI_ISL_1825740, EPI_ISL_1825742, EPI_ISL_1825743, EPI_ISL_1825744, EPI_ISL_1825745, EPI_ISL_1825748, EPI_ISL_1825758, EPI_ISL_1825776, EPI_ISL_1825779, EPI_ISL_1825781, EPI_ISL_1825783, EPI_ISL_1825784, EPI_ISL_1825786, EPI_ISL_1825787, EPI_ISL_1825788, EPI_ISL_1825821, EPI_ISL_1825863, EPI_ISL_1825864, EPI_ISL_1825866, EPI_ISL_1825868, EPI_ISL_1825873, EPI_ISL_1825874, EPI_ISL_1825876, EPI_ISL_1938485, EPI_ISL_1938487, EPI_ISL_1938491, EPI_ISL_1938495, EPI_ISL_1938498, EPI_ISL_1938499, EPI_ISL_1938520, EPI_ISL_1938522, EPI_ISL_1938596, EPI_ISL_2000783, EPI_ISL_2086537, EPI_ISL_2086538, EPI_ISL_2203534 | | | |
| see above | Illinois Department of Public Health - Springfield Lab | Illinois Department of Public Health - Springfield Lab | Bryan Sim; Gordon McCall |
| EPI_ISL_1708737 | Imeda Hospital | Imeda Hospital | Dagmar Obbels; Hanne Valgaeren; Johan Frans |
| EPI_ISL_1810847, EPI_ISL_1810883, EPI_ISL_1810903, EPI_ISL_1810904 | InDRE | Instituto Nacional de Medicina Genomica | Cedro-Tanda A; Garcia-Cardenas FJ; Gisela Barrera-Badillo; Gonzalez-Barrera D; Gonzalez-Woge MA; Herrera-Montalvo LA.; Hidalgo-Miranda A; Irma Lopez-Martinez; Jose Ernesto Ramirez González; Mendoza-Vargas A; Munguia-Garza P; Rangel-DeLeon D; Reyes-Grajeda JP |
| EPI_ISL_2011473, EPI_ISL_2011479, EPI_ISL_2011504, EPI_ISL_2011514, EPI_ISL_2136502, EPI_ISL_2136504, EPI_ISL_2136512 | | | |
| see above | Incyte Diagnostics Spokane | Seattle Flu Study | Amanda Adler; Barry R. Lutz; Benjamin Pelle; Brian Hiatt; Caitlin R. Wolf; Chris D. Frazar; Deborah A. Nickerson; Elisabeth Brandstetter; Erica Ryke; Geoff Melly; Helen Y. Chu; Janet A. Englund; Jay Shendure; Jover Lee; Kairsten Fay; Kirsten Lacombe; Lea M. Starita; Mark J. Rieder; Matthew Richardson; Matthew Thompson; Melissa Truong; Michael Boeckh; Michael Famulare; Misja Ilcisin; Peter D. Han; Philip Dykema; Romesh Gautom; Scott Lindquist; Thomas R. Sibley; Trevor Bedford |
| EPI_ISL_2304367 | Indiana Animal Disease Diagnostic Laboratory | Carpi Laboratory - Purdue University | Abebe A Fola; G Kenitra Hendrix; Giovanna Carpi; Ilinca I Ciubotariu; Jack Dorman; Jobin J Kattoor; Lev Gorenstein; Nicole M Perry; Rebecca P Wilkes |
| EPI_ISL_1795595, EPI_ISL_1803442, EPI_ISL_1817991, EPI_ISL_2013828, EPI_ISL_2013856, EPI_ISL_2013937, EPI_ISL_2014055, EPI_ISL_2014069, EPI_ISL_2014084, EPI_ISL_2014221, EPI_ISL_2014277, EPI_ISL_2014424, EPI_ISL_2014479, EPI_ISL_2014485, EPI_ISL_2014486, EPI_ISL_2014539, EPI_ISL_2014563, EPI_ISL_2014734, EPI_ISL_2047488, EPI_ISL_2186392, EPI_ISL_2186397, EPI_ISL_2283051, EPI_ISL_2283183, EPI_ISL_2283304, EPI_ISL_2283321, EPI_ISL_2283335, EPI_ISL_2283447, EPI_ISL_2283464, EPI_ISL_2283526, EPI_ISL_2283546, EPI_ISL_2283571, EPI_ISL_4383282, EPI_ISL_4385585, EPI_ISL_4385984, EPI_ISL_4386898, EPI_ISL_4387066, EPI_ISL_4388063, EPI_ISL_4389473, EPI_ISL_4389967 | | | |
| see above | Infinity Biologix | Centers for Disease Control and Prevention Division of Viral Diseases, Pathogen Discovery | Adrian Paskey; Benjamin Rambo-Martin; Chirayu Goswami; Christian Bixby; Christopher Gulvick; Clinton Paden; Clinton R. Paden; Dakota Howard; Darlene Wagner; Dhwani Batra; Duncan MacCannell; Erisa Sula; Jason Caravas; Jonathan Schultz; Kara Moser; Kristine Lacek; Matthew Schmerer; Peter Cook; Peter W. Cook; Robin Grimwood; Russ Hager; Scott Sammons; Shatavia Morrison; Tymeckia Kendall; Victoria Caban Figueroa; Yihe Wang; Yvette Unoarumhi |
| EPI_ISL_5782550, EPI_ISL_5782551, EPI_ISL_5782552, EPI_ISL_5782553, EPI_ISL_5782554, EPI_ISL_5782555, EPI_ISL_5782556, EPI_ISL_5782557, EPI_ISL_5782563, EPI_ISL_5782564, EPI_ISL_5782565, EPI_ISL_5782566, EPI_ISL_5782568, EPI_ISL_5782569, EPI_ISL_5782570, EPI_ISL_5782573, EPI_ISL_5782574, EPI_ISL_5782579, EPI_ISL_5782580, EPI_ISL_5782583, EPI_ISL_5782585, EPI_ISL_5782587, EPI_ISL_5782588, EPI_ISL_5782589, EPI_ISL_5782590, EPI_ISL_5782593, EPI_ISL_5782595, EPI_ISL_5782596, EPI_ISL_5782598, EPI_ISL_5782602, EPI_ISL_5782606, EPI_ISL_5782609, EPI_ISL_5782610, EPI_ISL_5782613, EPI_ISL_5782615, EPI_ISL_5782616, EPI_ISL_5782617, EPI_ISL_5782619, EPI_ISL_5782621, EPI_ISL_5782622, EPI_ISL_5782623, EPI_ISL_5782624, EPI_ISL_5782625, EPI_ISL_5782627, EPI_ISL_5782629, EPI_ISL_5782631, EPI_ISL_5782633, EPI_ISL_5782634, EPI_ISL_5782635, EPI_ISL_5782637, EPI_ISL_5782640, EPI_ISL_5782641, EPI_ISL_5782644, EPI_ISL_5782645, EPI_ISL_5782646, EPI_ISL_5782647, EPI_ISL_5782649, EPI_ISL_5782651, EPI_ISL_5782652, EPI_ISL_5782653, EPI_ISL_5782659 | | | |
| see above | Inside Diagnosticos | Instituto Butantan | Antonio Jorge Martins; Claudia Renata dos Santos Barros; David Schlesinger; Debora Botequio Moretti; Dimas Tadeu Covas; Elaine Cristina Marqueze; Elaine Vieira Santos; Evandra Strazza Rodrigues; Heidge Fukumasu; Jayme Augusto de Souza-Neto; José Salvatore Leister Patané; Luiz Alcantara; Luiz Lehmann Coutinho; Maria Carolina Elias; Maurício Lacerda Nogueira; Rafael dos Santos Bezerra; Raul Machado Neto; Rejane Maria Tommasini Grotto; Ricardo Haddad; Sandra Coccuzzo Sampaio Vessoni; Simone Kashima; Svetoslav Nanev Slavov; Vincent Louis Viala |
| EPI_ISL_1738807, EPI_ISL_2100428 | Institute of Microbiology, Universidad San Francisco de Quito | Institute of Microbiology, Universidad San Francisco de Quito | Belén Prado-Vivar; Bernardo Gutiérrez; Gabriel Trueba; Guzmán Bernabéu Lorenzo; Juan José Guadalupe; Manuel Jaramillo; Michelle Grunauer; Monica Becerra-Wong; Patricio Rojas-Silva; Paúl Cárdenas; Sully Márquez; Verónica Barragán |
| EPI_ISL_2691109, EPI_ISL_2919239 | Instituto Adolfo Lutz - Regional de Bauru | Instituto Adolfo Lutz, Interdiciplinary Procedures Center, Strategic Laboratory | Caio Vinicius Dias Lopes; Claudia Regina Gonçalves; Claudio Tavares Sacchi; Erica Valessa Ramos Gomes; Karoline Rodrigues Campos; Leonardo Jose Tadeu de Araujo |
| EPI_ISL_2958843 | Instituto Adolfo Lutz - Regional de Sorocaba | Instituto Adolfo Lutz, Interdiciplinary Procedures Center, Strategic Laboratory | Caio Vinicius Dias Lopes; Claudia Regina Gonçalves; Claudio Tavares Sacchi; Erica Valessa Ramos Gomes; Karoline Rodrigues Campos |
| EPI_ISL_2756435 | Instituto Adolfo Lutz Central | Instituto Adolfo Lutz, Interdiciplinary Procedures Center, Strategic Laboratory | Caio Vinicius Dias Lopes; Claudia Regina Gonçalves; Claudio Tavares Sacchi; Erica Valessa Ramos Gomes; Karoline Rodrigues Campos; Leonardo Jose Tadeu de Araujo |
| EPI_ISL_4104678, EPI_ISL_4104688, EPI_ISL_4104689, EPI_ISL_4104694, EPI_ISL_4104699, EPI_ISL_4104703, EPI_ISL_4104718, EPI_ISL_4104728, EPI_ISL_4104743, EPI_ISL_4104744 | | | |
| see above | Instituto Butantan | Instituto Butantan | Antonio Jorge Martins; Claudia Renata dos Santos Barros; David Schlesinger; Debora Botequio Moretti; Dimas Tadeu Covas; Elaine Cristina Marqueze; Elaine Vieira Santos; Evandra Strazza Rodrigues; Heidge Fukumasu; Jayme Augusto de Souza-Neto; José Salvatore Leister Patané; Luiz Alcantara; Luiz Lehmann Coutinho; Maria Carolina Elias; Maurício Lacerda Nogueira; Rafael dos Santos Bezerra; Raul Machado Neto; Rejane Maria Tommasini Grotto; Ricardo Haddad; Sandra Coccuzzo Sampaio Vessoni; Simone Kashima; Svetoslav Nanev Slavov; Vincent Louis Viala |
| EPI_ISL_1711293 | Instituto Nacional de Medicina Genomica | Instituto Nacional de Medicina Genomica | Alcaraz N; Canseco Mendez JC; Cedro-Tanda A; Garcia-Cardenas FJ; Gonzalez-Barrera D; Gonzalez-Woge MA; Herrera-Montalvo LA; Hidalgo-Miranda A; Mendoza-Vargas A; Miranda-Ortiz H; Munguia-Garza P; Ramirez-Vega O; Rangel-DeLeon D; Reyes-Grajeda JP; Rosas-Escobar P |
| EPI_ISL_2004134, | Instituto Nacional de Saude | Instituto Nacional de Saude (INSA) and | Borges et al |

| | | | |
|---|---|---|---|
| EPI_ISL_2004192 | (INSA) and i3S - Instituto de Investigação e Inovação em Saúde | i3S - Instituto de Investigação e Inovação em Saúde | |
| EPI_ISL_2295400, EPI_ISL_2295402, EPI_ISL_2295403, EPI_ISL_2295404, EPI_ISL_2295405, EPI_ISL_2295406, EPI_ISL_2295407, EPI_ISL_2295408, EPI_ISL_2295409, EPI_ISL_2295410, EPI_ISL_2295413, EPI_ISL_2295414, EPI_ISL_2295415, EPI_ISL_2295416 | | | |
| see above | Instituto de Biotecnologia - UNESP-Botucatu-SP | Instituto de Biotecnologia - UNESP-Botucatu-SP | Cecília Artico Banho; Cíntia Bittar; Fábio Sossai Possebon; Guilherme Campos; Helena Lage Ferreira; Jorge A. Petroli Marchesi; João Pessoa Araújo Jr.; Leila Sabrina Ullmann; Lívia Sacchetto; Maisa C. Pereira Parra; Marília Moraes; Maurício L. Nogueira; Paula Rahal; Paulo Inacio da Costa |
| EPI_ISL_2776171, EPI_ISL_2776172, EPI_ISL_2776174 | Instituto de Medicina Tropical & Salud Global (IMTSAG) | Grubaugh Lab - Yale School of Public Health | Alejandaro Vallejo Degaudenzi; Anderson Brito; Annie Watkins; Chaney Kalinich; Chantal Vogels; Elisa Contreras; Esperanza Mendoza; Isabel Ott; Jessica Rothman; Joseph Fauver; Kendall Billig; Mallery Breban; Mary Petrone; Nathan Grubaugh; Robert Paulino-Ramirez; Tara Alpert; Tobias Koch; Victor Virgilio Calderon |
| EPI_ISL_1714902, EPI_ISL_1714907, EPI_ISL_2020466, EPI_ISL_2020482, EPI_ISL_2020487, EPI_ISL_2020491, EPI_ISL_2020514, EPI_ISL_2029523, EPI_ISL_2029534, EPI_ISL_2137555 | | | |
| see above | Istituto Zooprofilattico Sperimentale del Mezzogiorno | TIGEM | Antonio Grimaldi Patrizia Annunziata Francesco Panariello Biancamaria Pierri Claudia Tiberio Teresa Giuliano Valentina Bouche Chiara Colantuono Maria Concetta Cuomo Denise Di Concilio Lucio Di Filippo Anna Manfredi Marcello Salvi Antonio Limone Luigi Atripaldi Pellegrino Cerino Andrea Ballabio Davide Cacchiarelli |
| EPI_ISL_2143796 | Istituto Zooprofilattico Sperimentale del Mezzogiorno | Telethon Institute of Genetics and Medicine (TIGEM) | Antonio Grimaldi Patrizia Annunziata Francesco Panariello Biancamaria Pierri Claudia Tiberio Teresa Giuliano Valentina Bouche Chiara Colantuono Maria Concetta Cuomo Denise Di Concilio Lucio Di Filippo Anna Manfredi Marcello Salvi Antonio Limone Luigi Atripaldi Pellegrino Cerino Andrea Ballabio Davide Cacchiarelli |
| EPI_ISL_1854242, EPI_ISL_1854243 | Joaquim Chaves Saude | Instituto Nacional de Saude (INSA) | Borges et al |
| EPI_ISL_1662944 | Johns Hopkins Hospital Department of Pathology | Johns Hopkins Hospital Department of Pathology | Adannaya Amadi; C. Paul Morris; Chun Huai Luo; Heba H. Mostafa; Matthew Schwartz |
| EPI_ISL_2424345, EPI_ISL_2424396, EPI_ISL_2424458, EPI_ISL_2424477, EPI_ISL_2425150, EPI_ISL_2425154, EPI_ISL_2425156, EPI_ISL_2425168, EPI_ISL_2425169, EPI_ISL_2425174, EPI_ISL_2425176, EPI_ISL_2425187, EPI_ISL_2425200, EPI_ISL_2425201, EPI_ISL_2425202, EPI_ISL_2425219, EPI_ISL_2425221, EPI_ISL_2425225, EPI_ISL_2425233, EPI_ISL_2425234, EPI_ISL_2425235, EPI_ISL_2425243, EPI_ISL_2425244, EPI_ISL_2425248, EPI_ISL_2425249, EPI_ISL_2425250, EPI_ISL_2832285, EPI_ISL_2832287, EPI_ISL_2832368, EPI_ISL_2832379, EPI_ISL_2832639, EPI_ISL_2832650, EPI_ISL_2832652, EPI_ISL_2832665, EPI_ISL_2832702, EPI_ISL_2832857, EPI_ISL_2832864, EPI_ISL_2832880, EPI_ISL_2832900, EPI_ISL_2832912, EPI_ISL_2832915, EPI_ISL_2832921, EPI_ISL_2832923, EPI_ISL_2832925, EPI_ISL_2832950, EPI_ISL_2833106, EPI_ISL_2834604 | | | |
| see above | KU Leuven, Rega Institute, Clinical and Epidemiological Virology | KU Leuven, Rega Institute, Clinical and Epidemiological Virology | Bert Vanmechelen; Joan Marti-Carerras; Piet Maes; Tony Wawina-Bokalanga |
| EPI_ISL_1823809 | LA Office of Public Health Laboratories | Centers for Disease Control and Prevention Division of Viral Diseases, Pathogen Discovery | Alison Laufer Halpin; Ben L. Rambo-Martin; Clinton R. Paden; Dakota Howard; Darlene Wagner; Dave Wentworth; Dhwani Batra; Jasmine Padilla; Justin Lee; Katie Dillon; Krista Queen; Kristen Knipe; Kristine Lacek; Mark Burroughs; Matthew Schmerer; Mili Sheth; Peter Cook; Sam Shepard; Sarah Nobles; Shoshona Le; Suxiang Tong; Vivien Dugan; Yvette Unoarumhi |
| EPI_ISL_1904980 | LABORATOIRE ALPHABIO - HOPITAL BEAUREGARD | CNR Virus des Infections Respiratoires - France SUD | Antonin Bal; Bruno Lina; Bruno Simon; Gregory Destras; Gwendolyne Burfin; Hadrien Regue; Laurence Josset; Martine Valette; Quentin Semanas |
| EPI_ISL_1934945, EPI_ISL_2142453 | LABORATOIRE DE BIOLOGIE MEDICALE | CNR Virus des Infections Respiratoires - France SUD | Antonin Bal; Bruno Lina; Bruno Simon; Gregory Destras; Gwendolyne Burfin; Hadrien Regue; Laurence Josset; Martine Valette; Quentin Semanas |
| EPI_ISL_2834731 | LABORATORIO CLINICO HIGUERA ESCALANTE | Instituto Nacional de Salud- Dirección de Investigación en Salud Pública | Carlos Franco-Muñoz; Carmen Osorio; Diana Malo; Diego A. Álvarez-Díaz; Diego Andrés Prada; Gerardo Santamaría; Hector Alejandro Ruiz-Moreno; Jhonnatan Reales-González; Jorge Rivera; Juan Camilo Martinez; Julian Naizaque; Katherine Laiton-Donato; Lisseth Pardo; Magdalena Wiesner; Marcela Mercado-Reyes; Maria T. Herrera-Sepúlveda; Marta Lopez Blanco; Martha Lucia Ospina Martinez; Paola Rojas; Sergio Gomez; Sheryll Corchuelo; Ángela Alarcon Cruz |
| EPI_ISL_3355470 | LABORATORIO DE BIOLOGÍA MOLECULAR Y BIOTECNOLOGIA | Corporacion CorpoGen-Universidad de los Andes-Universidad Central | Christian Romero; Jorge Duitama; Juan Manuel Anzola; Laura González; Maryam Chaib De Mares; María Mercedes Zambrano; Nelly Díaz; Patricia Del Portillo; Silvia Restrepo |
| EPI_ISL_2344583 | LABORATORIO DE FRANCA | Instituto Butantan / FZEA-USP-Pirassununga | Antonio Jorge Martins; Claudia Renata dos Santos Barros; David Schlesinger; Debora Botequio Moretti; Dimas Tadeu Covas; Elaine Cristina Marqueze; Elaine Vieira Santos; Evandra Strazza Rodrigues; Heidge Fukumasu; Jayme Augusto de Souza-Neto; José Salvatore Leister Patané; Luiz Alcantara; Luiz Lehmann Coutinho; Maria Carolina Elias; Maurício Lacerda Nogueira; Rafael dos Santos Bezerra; Raul Machado Neto; Rejane Maria Tommasini Grotto; Ricardo Haddad; Sandra Coccuzzo Sampaio Vessoni; Simone Kashima; Svetoslav Nanev Slavov; Vincent Louis Viala |
| EPI_ISL_2345658, EPI_ISL_2345698, EPI_ISL_2362241, EPI_ISL_2362242, EPI_ISL_2362253, EPI_ISL_2362264, EPI_ISL_2362274 | | | |
| see above | LABORATORIO DR PAULO EMILIO DALESSANDRO PINDAMONHANGABA | Instituto Butantan / Mendelics | Antonio Jorge Martins; Claudia Renata dos Santos Barros; David Schlesinger; Debora Botequio Moretti; Dimas Tadeu Covas; Elaine Cristina Marqueze; Elaine Vieira Santos; Evandra Strazza Rodrigues; Heidge Fukumasu; Jayme Augusto de Souza-Neto; José Salvatore Leister Patané; Luiz Alcantara; Luiz Lehmann Coutinho; Maria Carolina Elias; Maurício Lacerda Nogueira; Rafael dos Santos Bezerra; Raul Machado Neto; Rejane Maria Tommasini Grotto; Ricardo Haddad; Sandra Coccuzzo Sampaio Vessoni; Simone Kashima; Svetoslav Nanev Slavov; Vincent Louis Viala |
| EPI_ISL_3275681, EPI_ISL_3275682 | LABORATORIO LAS AMERICAS | Universidad Nacional de Colombia - Laboratorio Genómico One Health | Andres F. Cardona-Rios; Carlos Franco-Muñoz; Carolina Muñoz-Arango; Celeny Ortiz; Daniel O. Maldonado-Perez; Diego A. Álvarez-Díaz; Hector Alejandro Ruiz-Moreno; Idabely Betancur Ortiz; Jorge E. Osorio; Juan P. Hernandez-Ortiz; Karl A Ciuoderis; Katherine Laiton-Donato; Laura Silvana Perez; Lina M. Hurtado; Marcela Mercado-Reyes; Maria Angélica Maya; Maria Stella López; Rita Almanza Payares; Sandra Ines Cano; Simón Villegas Velásquez |
| EPI_ISL_2210088, EPI_ISL_2344716, EPI_ISL_2344728, EPI_ISL_2344729 | LABORATORIO MUNICIPAL DE ANALISES CLINICAS DE RIO CLARO | Instituto Butantan / FZEA-USP-Pirassununga | Antonio Jorge Martins; Claudia Renata dos Santos Barros; David Schlesinger; Debora Botequio Moretti; Dimas Tadeu Covas; Elaine Cristina Marqueze; Elaine Vieira Santos; Evandra Strazza Rodrigues; Heidge Fukumasu; Jayme Augusto de Souza-Neto; José Salvatore Leister Patané; Luiz Alcantara; Luiz Lehmann Coutinho; Maria Carolina Elias; Maurício Lacerda Nogueira; Rafael dos Santos Bezerra; Raul Machado Neto; Rejane Maria Tommasini Grotto; Ricardo Haddad; Sandra Coccuzzo Sampaio Vessoni; Simone Kashima; Svetoslav Nanev Slavov; Vincent Louis Viala |
| EPI_ISL_2344734 | LABORATORIO MUNICIPAL DE PIRACICABA | Instituto Butantan / ESALQ-Piracicaba | Antonio Jorge Martins; Claudia Renata dos Santos Barros; David Schlesinger; Debora Botequio Moretti; Dimas Tadeu Covas; Elaine Cristina Marqueze; Elaine Vieira Santos; Evandra Strazza Rodrigues; Heidge Fukumasu; Jayme Augusto de Souza-Neto; José Salvatore Leister Patané; Luiz Alcantara; Luiz Lehmann Coutinho; Maria Carolina Elias; Maurício Lacerda Nogueira; Rafael dos Santos Bezerra; Raul Machado Neto; Rejane Maria Tommasini Grotto; Ricardo Haddad; Sandra Coccuzzo Sampaio Vessoni; Simone Kashima; Svetoslav Nanev Slavov; Vincent Louis Viala |
| EPI_ISL_4877301 | LACEN do Estado de Goias | Instituto Adolfo Lutz, Interdiciplinary Procedures Center, Strategic Laboratory | Claudia Regina Gonçalves; Claudio Tavares Sacchi; Karoline Rodrigues Campos; Marlon Benedito Nascimento Santos |
| EPI_ISL_3316168, EPI_ISL_3316215 | LACEN do Estado de Mato Grosso | Instituto Adolfo Lutz, Interdiciplinary Procedures Center, Strategic Laboratory | Caio Vinicius Dias Lopes; Claudia Regina Gonçalves; Claudio Tavares Sacchi; Karoline Rodrigues Campos; Leonardo Tadeu de Araujo; Marlon Benedito Nascomento Santos |
| EPI_ISL_2919216, EPI_ISL_2919217, EPI_ISL_2919220, EPI_ISL_2919223, EPI_ISL_2919224 | LACEN do Estado de Tocantins | Instituto Adolfo Lutz, Interdiciplinary Procedures Center, Strategic Laboratory | Caio Vinicius Dias Lopes; Claudia Regina Gonçalves; Claudio Tavares Sacchi; Erica Valessa Ramos Gomes; Karoline Rodrigues Campos |
| EPI_ISL_3691382, EPI_ISL_3691383, EPI_ISL_3691384 | LACEN do Estado do Mato Grosso do Sul | Instituto Adolfo Lutz, Interdiciplinary Procedures Center, Strategic Laboratory | Caio Vinicius Dias Lopes; Claudia Regina Gonçalves; Claudio Tavares Sacchi; Karoline Rodrigues Campos; Leonardo Tadeu de Araujo; Marlon Benedito Nascomento Santos |
| EPI_ISL_3691681, EPI_ISL_3691682, EPI_ISL_3691683, EPI_ISL_3691684 | LACEN do Estado do Piaui | Instituto Adolfo Lutz, Interdiciplinary Procedures Center, Strategic Laboratory | Caio Vinicius Dias Lopes; Claudia Regina Gonçalves; Claudio Tavares Sacchi; Karoline Rodrigues Campos; Leonardo Tadeu de Araujo; Marlon Benedito Nascomento Santos |
| EPI_ISL_2958856 | LACEN do Mato Grosso do Sul | Instituto Adolfo Lutz, Interdiciplinary Procedures Center, Strategic Laboratory | Caio Vinicius Dias Lopes; Claudia Regina Gonçalves; Claudio Tavares Sacchi; Erica Valessa Ramos Gomes; Karoline Rodrigues Campos |
| EPI_ISL_1608128, EPI_ISL_1608129, EPI_ISL_1608130, EPI_ISL_1608131, EPI_ISL_1608132, EPI_ISL_1608133, EPI_ISL_1608134, EPI_ISL_1608135, EPI_ISL_1608136, EPI_ISL_1608137, EPI_ISL_1608138, EPI_ISL_1608139 | | | |
| see above | LBM ALPHABIO, Marseille | LBM ALPHABIO, Marseille | Vincent GARCIA |
| EPI_ISL_1934940 | LBM UNIBIO VALENTIN | CNR Virus des Infections Respiratoires - France SUD | Antonin Bal; Bruno Lina; Bruno Simon; Gregory Destras; Gwendolyne Burfin; Hadrien Regue; Laurence Josset; Martine Valette; Quentin Semanas |
| EPI_ISL_1960052, EPI_ISL_1960077 | LDSP | Universidad Nacional de Colombia - Laboratorio Genómico One Health | Andres F. Cardona-Rios; Carlos Franco-Muñoz; Carolina Muñoz-Arango; Celeny Ortiz; Daniel O. Maldonado-Perez; Diego A. Álvarez-Díaz; Hector Alejandro Ruiz-Moreno; Idabely Betancur Ortiz; Jorge E. Osorio; Juan P. Hernandez-Ortiz; Karl A Ciuoderis; Katherine Laiton-Donato; Laura Silvana Perez; Lina M. Hurtado; Marcela Mercado-Reyes; Maria Angélica Maya; Maria Stella López; Rita Almanza Payares; Sandra Ines Cano; Simón Villegas Velásquez |
| EPI_ISL_2155050 | LDSP AMAZONAS | Instituto Nacional de Salud- Dirección de Investigación en Salud Pública | Carlos Franco-Muñoz; Carmen Osorio; Diana Malo; Diego A. Álvarez-Díaz; Diego Andrés Prada; Gerardo Santamaría; Hector Alejandro Ruiz-Moreno; Jhonnatan Reales-González; Jorge Rivera; Juan Camilo Martinez; Julian Naizaque; Katherine Laiton-Donato; Lisseth Pardo; Magdalena Wiesner; Marcela Mercado-Reyes; Maria T. Herrera-Sepúlveda; Marta Lopez Blanco; Martha Lucia Ospina Martinez; Paola Rojas; Sergio Gomez; Sheryll Corchuelo; Ángela Alarcon Cruz |
| EPI_ISL_2155048, EPI_ISL_2155049 | LDSP CALDAS | Instituto Nacional de Salud- Dirección de Investigación en Salud Pública | Carlos Franco-Muñoz; Carmen Osorio; Diana Malo; Diego A. Álvarez-Díaz; Diego Andrés Prada; Gerardo Santamaría; Hector Alejandro Ruiz-Moreno; Jhonnatan Reales-González; Jorge Rivera; Juan Camilo Martinez; Julian Naizaque; Katherine Laiton-Donato; Lisseth Pardo; Magdalena Wiesner; Marcela Mercado-Reyes; Maria T. Herrera-Sepúlveda; Marta Lopez Blanco; Martha Lucia Ospina Martinez; Paola Rojas; Sergio Gomez; Sheryll Corchuelo; Ángela Alarcon Cruz |
| EPI_ISL_2296079, EPI_ISL_2296080, EPI_ISL_2296081, EPI_ISL_2342997 | LESP Ciudad de Mexico | Instituto de Diagnostico y Referencia Epidemiologicos (INDRE) | Abril Rodriguez-Maldonado; Ariadna Medina-Benitez; Claudia Wong-Arambula; Ernesto Ramirez-Gonzalez.; Gisela Barrera-Badillo; Irma Lopez-Martinez; Joaquin Quiroz-Mercado; Lucia Hernandez-Rivas; Natividad Cruz-Ortiz; Sergio Rangel-Guerrero; Tatiana Nunez-Garcia; Vanessa Rivero-Arredondo |
| EPI_ISL_2296082 | LESP Estado de Mexico | Instituto de Diagnostico y Referencia Epidemiologicos (INDRE) | Abril Rodriguez-Maldonado; Ariadna Medina-Benitez; Claudia Wong-Arambula; Ernesto Ramirez-Gonzalez.; Gisela Barrera-Badillo; Irma Lopez-Martinez; Joaquin Quiroz-Mercado; Lucia Hernandez-Rivas; Natividad Cruz-Ortiz; Sergio Rangel-Guerrero; Tatiana Nunez-Garcia; Vanessa Rivero-Arredondo |
| EPI_ISL_2105689, EPI_ISL_2105691 | LESP Quintana Roo | Instituto de Diagnostico y Referencia Epidemiologicos (INDRE) | Abril Rodriguez-Maldonado; Ariadna Medina-Benitez; Claudia Wong-Arambula; Ernesto Ramirez-Gonzalez.; Gisela Barrera-Badillo; Irma Lopez-Martinez; Joaquin Quiroz-Mercado; Lucia Hernandez-Rivas; Natividad Cruz-Ortiz; Sergio Rangel-Guerrero; Tatiana Nunez-Garcia; Vanessa Rivero-Arredondo |
| EPI_ISL_2674717 | LESP Sonora | Instituto de Diagnostico y Referencia Epidemiologicos (INDRE) | Abril Rodriguez-Maldonado; Ariadna Medina-Benitez; Claudia Wong-Arambula; Ernesto Ramirez-Gonzalez.; Gisela Barrera-Badillo; Irma Lopez-Martinez; Joaquin Quiroz-Mercado; Lucia Hernandez-Rivas; Natividad Cruz-Ortiz; Sergio Rangel-Guerrero; Tatiana Nunez-Garcia; Vanessa Rivero-Arredondo |
| EPI_ISL_2296084 | LESP Tamaulipas | Instituto de Diagnostico y Referencia Epidemiologicos (INDRE) | Abril Rodriguez-Maldonado; Ariadna Medina-Benitez; Claudia Wong-Arambula; Ernesto Ramirez-Gonzalez.; Gisela Barrera-Badillo; Irma Lopez-Martinez; Joaquin Quiroz-Mercado; Lucia Hernandez-Rivas; Natividad Cruz-Ortiz; Sergio Rangel-Guerrero; Tatiana Nunez-Garcia; Vanessa Rivero-Arredondo |

| | | | |
|---|---|---|---|
| EPI_ISL_1805501, EPI_ISL_1805502, EPI_ISL_2296089 | LESP Yucatan | Instituto de Diagnostico y Referencia Epidemiologicos (INDRE) | Abril Rodriguez-Maldonado; Ariadna Medina-Benitez; Claudia Wong-Arambula; Ernesto Ramirez-Gonzalez.; Gisela Barrera-Badillo; Irma Lopez-Martinez; Joaquin Quiroz-Mercado; Lucia Hernandez-Rivas; Natividad Cruz-Ortiz; Sergio Rangel-Guerrero; Tatiana Nunez-Garcia; Vanessa Rivero-Arredondo |
| EPI_ISL_1745061, EPI_ISL_1745176, EPI_ISL_1745186, EPI_ISL_1745187, EPI_ISL_1745188, EPI_ISL_1745189, EPI_ISL_1745190, EPI_ISL_1745195 | | | |
| see above | LHUB-ULB | Labo Klinische Biologie, UZA | Basil Britto Xavier; Christine Lammens; Herman Goossens; Jasmine Coppens; Marie Le Mercier; Veerle Matheeussen |
| EPI_ISL_3342150 | LIME | Laboratorio Departamental de Salud Publica de Antioquia | Andres F. Cardona-Rios; Gloria Isabel Escobar; Idabely Betancur Ortiz; Juan P. Hernandez-Ortiz; Maria Stella López |
| EPI_ISL_4196893, EPI_ISL_4196906 | LSP DISTRITAL BOGOTA | Gencore - Universidad de los Andes | Alejandro Gomez; Cristian Barrera; Gabriela Ariza; Gabriela Delgado; Johana Hernandez; Luisa Sacristan; Marcela Guevara; Silvia Restrepo |
| EPI_ISL_1827371, EPI_ISL_1827417, EPI_ISL_1827419, EPI_ISL_1827424, EPI_ISL_1827434, EPI_ISL_1827462, EPI_ISL_2191912, EPI_ISL_2191913, EPI_ISL_2191915, EPI_ISL_2191920, EPI_ISL_2191921, EPI_ISL_2191956, EPI_ISL_2192062 | | | |
| see above | Lab voor klinische biologie | Lab voor klinische biologie | Bruno Verhasselt; Hannelore Hamerlinck; Marija Janevska |
| EPI_ISL_1821643, EPI_ISL_1821644, EPI_ISL_1821645, EPI_ISL_1821646, EPI_ISL_1821647, EPI_ISL_1821648, EPI_ISL_1821649, EPI_ISL_1821650, EPI_ISL_1821651, EPI_ISL_1821652, EPI_ISL_1821655, EPI_ISL_1821656 | | | |
| see above | Labo Analyses Med | National Reference Center for Viruses of Respiratory Infections, Institut Pasteur, Paris | Amaury Vaysse; Angela Brisebarre; Camille Capel; Christophe Malabat; Corinne Maufrais; Dominique Rousset; Emmanuelle Permal; Etienne Simon-Lorière; Frédéric Lemoine; Louise Lefrançois; Marion Barbet; Maud Vanpeene; Méline Bizard; Sylvie Behillil; Sylvie Van der Werf; Vincent Enouf |
| EPI_ISL_2316525 | Labor Becker & Kollegen (Standort München) | Robert Koch Institute | |
| EPI_ISL_1727096 | Labor Dr. Heidrich & Kollegen MVZ GmbH Hamburg | Robert Koch Institute | |
| EPI_ISL_2125120 | Labor Dr. Spranger | Robert Koch Institute | |
| EPI_ISL_2227923, EPI_ISL_2227924, EPI_ISL_2227925, EPI_ISL_2227926 | Laboratoire Carage | Institut Pasteur de la Guyane | A. Salmier; Anne Lavergne; Dominique Rousset |
| EPI_ISL_2970546, EPI_ISL_3208003, EPI_ISL_3208004, EPI_ISL_3208005, EPI_ISL_3458365, EPI_ISL_3458366, EPI_ISL_3458367, EPI_ISL_3458368, EPI_ISL_3458369, EPI_ISL_3458370, EPI_ISL_3458383, EPI_ISL_3458385, EPI_ISL_3458418, EPI_ISL_3458434, EPI_ISL_3458436, EPI_ISL_3458467, EPI_ISL_3458472, EPI_ISL_3458474, EPI_ISL_3458477, EPI_ISL_3458625, EPI_ISL_3508444, EPI_ISL_3508445, EPI_ISL_3508446, EPI_ISL_3508447, EPI_ISL_3508448, EPI_ISL_5872427, EPI_ISL_5872444, EPI_ISL_5872469, EPI_ISL_5872479, EPI_ISL_5872786, EPI_ISL_5872804, EPI_ISL_5872818, EPI_ISL_5872819, EPI_ISL_5872837, EPI_ISL_5872845, EPI_ISL_5872856, EPI_ISL_5872864, EPI_ISL_5872878, EPI_ISL_5873258, EPI_ISL_5873302, EPI_ISL_5873440, EPI_ISL_5891603, EPI_ISL_5891604, EPI_ISL_5891605, EPI_ISL_5891606, EPI_ISL_5891607, EPI_ISL_5891608, EPI_ISL_5891609, EPI_ISL_5891610, EPI_ISL_7713459, EPI_ISL_7713486, EPI_ISL_7713488, EPI_ISL_7713498, EPI_ISL_7713499, EPI_ISL_7713500, EPI_ISL_7713508, EPI_ISL_7713511, EPI_ISL_7713520, EPI_ISL_7713548, EPI_ISL_7713549, EPI_ISL_7713552, EPI_ISL_7713574, EPI_ISL_7713582, EPI_ISL_7713620, EPI_ISL_7713656, EPI_ISL_7713657, EPI_ISL_7713659 | | | |
| see above | Laboratoire de santé publique du Québec | Laboratoire de santé publique du Québec | Guillaume Bourque; Ioannis Ragoussis; Jesse Shapiro; Mark Lathrop and Judith Fafard on behalf of the CoVSeQ research group; Mark Lathrop and Michel Roger on behalf of the CoVSeQ research group; Sandrine Moreira |
| EPI_ISL_1917732 | Laboratoires d'analyses medicales - Ketterthill | Laboratoire national de sante, Microbiology, Microbial Genomics Platform | Anke Wienecke-Baldacchino; Caroline Scheiber; Catherine Ragimbeau; Fatu Djabi; Jessica Tapp; Lise Pignon; Raoul Salmon; Serge Vedy; Tamir Abdelrahman |
| EPI_ISL_2308280 | Laboratori de Referencia de Catalunya | Laboratori de Referencia de Catalunya | Bellosillo B.; Canal M.; Hernandez JJ.; Padilla E.; Ramirez A.; Vilas A. |
| EPI_ISL_2196243, EPI_ISL_2196302, EPI_ISL_2196303, EPI_ISL_2443560 | Laboratorio Central de Saude Publica do Estado do Para (LACEN/PA) | Laboratory of Respiratory Viruses and Measles, Oswaldo Cruz Institute, FIOCRUZ | Alice Sampaio Rocha; Ana Carolina Mendonca; Anna Carolina Paixao; Elisa Cavalcante Pereira; Fernando Motta; Luciana Appolinario; Marilda Siqueira on behalf of the Fiocruz COVID-19 Genomic Surveillance Network; Paola Resende; Renata Serrano Lopes; Taina Venas; Valnete Andrade |
| EPI_ISL_1919607 | Laboratorio Analisi Osp. Città di Castello - Azienda USL Umbria1 | Istituto Zooprofilattico Sperimentale dell'Abruzzo e Molise "G. Caporale" | Ancora M; Calistri P; Cammà C; Caporale M; Curini V; Delli Compagni E; Di Domenico M; Di Lollo Valeria; Di Pasquale A; Lorusso A; Malagigi V; Mangone I; Marcacci M; Puglia I; Rinaldi A; Savini G; Scialabba S; Tacconi P |
| EPI_ISL_1804917 | Laboratorio Aziendale di Microbiologia e Virologia, Azienda Sanitaria dell'Alto Adige | Laboratorio Aziendale di Microbiologia e Virologia, Azienda Sanitaria dell'Alto Adige | Anne Picard; Bartolomeo Mobilio Rodriguez; Chiara Cantaloni; Claudia Volpato; Elisa Masi; Elisabetta Giacobazzi; Elisabetta Pagani; Irene Bianconi; Stefanie Wieser |
| EPI_ISL_1858350, EPI_ISL_1858486, EPI_ISL_1858488, EPI_ISL_1858490, EPI_ISL_1858492, EPI_ISL_1858494, EPI_ISL_1858496, EPI_ISL_1858498, EPI_ISL_1858500, EPI_ISL_1858504, EPI_ISL_1858506, EPI_ISL_1858510, EPI_ISL_1858526, EPI_ISL_1858528, EPI_ISL_1858530, EPI_ISL_1858532, EPI_ISL_1858534, EPI_ISL_1858536, EPI_ISL_1858538, EPI_ISL_1858540, EPI_ISL_1858561, EPI_ISL_1858563, EPI_ISL_1858565, EPI_ISL_1858567, EPI_ISL_1858569, EPI_ISL_1858571, EPI_ISL_1858572, EPI_ISL_1858575, EPI_ISL_1858577, EPI_ISL_1858578, EPI_ISL_1858580, EPI_ISL_1858582, EPI_ISL_1858584, EPI_ISL_1858586, EPI_ISL_1858590, EPI_ISL_1858592, EPI_ISL_1858594, EPI_ISL_1858596, EPI_ISL_1858598, EPI_ISL_1858599, EPI_ISL_1858602, EPI_ISL_1858603, EPI_ISL_1858606, EPI_ISL_1858607, EPI_ISL_1858610, EPI_ISL_1858611, EPI_ISL_1858613, EPI_ISL_1858615, EPI_ISL_1858617, EPI_ISL_1858619, EPI_ISL_1858621, EPI_ISL_1858623, EPI_ISL_1858625, EPI_ISL_1858627, EPI_ISL_1858629, EPI_ISL_1858631, EPI_ISL_1858633, EPI_ISL_1858636, EPI_ISL_1858637, EPI_ISL_1858639, EPI_ISL_1858641, EPI_ISL_1858643, EPI_ISL_1858645, EPI_ISL_1858648, EPI_ISL_1858650, EPI_ISL_1858651, EPI_ISL_1858653, EPI_ISL_1858655, EPI_ISL_1858656, EPI_ISL_1858658, EPI_ISL_1858660, EPI_ISL_1858662, EPI_ISL_1858664, EPI_ISL_1858666, EPI_ISL_1858668, EPI_ISL_1858670, EPI_ISL_1858672, EPI_ISL_1858674, EPI_ISL_1858675, EPI_ISL_1858677, EPI_ISL_1858680, EPI_ISL_1858682, EPI_ISL_1858684, EPI_ISL_1858686, EPI_ISL_1858688, EPI_ISL_1858691, EPI_ISL_1858693, EPI_ISL_1858694, EPI_ISL_1858697, EPI_ISL_1858699, EPI_ISL_1858701, EPI_ISL_1858703, EPI_ISL_1858705, EPI_ISL_1858707, EPI_ISL_1858709, EPI_ISL_1858711, EPI_ISL_1858713, EPI_ISL_1858715, EPI_ISL_1858717, EPI_ISL_1858719, EPI_ISL_1858721, EPI_ISL_1858722, EPI_ISL_1858724, EPI_ISL_1858728, EPI_ISL_1858730, EPI_ISL_1858732, EPI_ISL_1858736, EPI_ISL_1858738, EPI_ISL_1858740, EPI_ISL_1858741, EPI_ISL_1858743, EPI_ISL_1858745, EPI_ISL_1858747, EPI_ISL_1858749, EPI_ISL_1858751, EPI_ISL_1858752, EPI_ISL_1858754, EPI_ISL_1858756, EPI_ISL_2101721 | | | |
| see above | Laboratorio Central Noel Nutels | Bioinformatics Laboratory / LNCC | Alessandra P Lamarca; Alexandra L Gerber; Amilcar Tanuri; Ana Paula de C Guimaraes; Ana Paula de C Guimarães; Ana Tereza R Vasconcelos; Andrea Cony Cavalcanti; Andréa Cony Cavalcanti; Caio Luiz Pereira Ribeiro; Cassia Alves; Cintia Policarpo; Claudia Maria Braga de Mello; Cristiane Gomes da Silva; Diana Mariani; Douglas Terra Machado; Flavio Dias da Silva; Flávio Dias da Silva; Gleidson da Silva de Oliveira; Leandro Magalhaes de Souza; Leandro Magalhães de Souza; Liliane Cavalcante; Luiz G P de Almeida; Marcio Henrique de Oliveira Garcia; Mario Sergio Ribeiro; Ronaldo da Silva F Jr; Silvia Carvalho; Thais Felix Cruz |
| EPI_ISL_2091410 | Laboratorio Central de Epidemiología (LCE) | Centro de Investigación en Enfermedades Infecciosas (CIENI), Instituto Nacional de Enfermedades Respiratorias (INER) | Alejandro Sanchez-Flores; Alfredo Herrera-Estrella; Alicia Ocaña-Mondragón; Angel Gustavo Salas-Lais; Bernardo Martínez-Miguel; Blanca Taboada; Brenda Irasema Maldonado-Meza; Carla Ivón Herrera-Najera; Carlos F. Arias; Celia Boukadida; Clara Esperanza Santacruz-Tinoco; Concepción Grajales-Muñiz; Consorcio Mexicano de Vigilancia Genómica (CoViGen-Mex). Authors (in alphabetical order): Julio Elias Alvarado-Yaah; Célida Duque Molina; Fernando Fontove-Herrera; Francisco Pulido; Gloria Elena Espinosa-Ayala; Gloria María Molina-Salinas; Gloria Vazquez; Hector Esteban Paz-Juárez; Hector Montoya-Fuentes; Helen Haydee Fernanda Ramírez-Plascencia; José Antonio Enciso-Moreno; José Esteban Muñoz-Medina; José de Jesús Nuñez-Contreras; Juan Bautista Chale-Dzul; Luis Alberto Ochoa-Carrera; Margarita Matías-Florentino; María Guadalupe Santiago-Mauricio; María Guadalupe de Jesús Mireles-Rivera; Nelly Sélem-Mojica; Pavel Isa; Ricardo Grande; Santiago Ávila-Ríos; Victor Eduardo Garcia-Arias; Víctor Hugo Borja-Aburto |
| EPI_ISL_6944475, EPI_ISL_6944476, EPI_ISL_6944477, EPI_ISL_6944478, EPI_ISL_6944479, EPI_ISL_6944480, EPI_ISL_6944481, EPI_ISL_6944482, EPI_ISL_6944483, EPI_ISL_6944484 | | | |
| see above | Laboratorio Central de Salud Publica | Viral Special Pathogens Branch, Centers for Disease Control and Prevention | Andrea Gómez de la Fuente; Cynthia Vázquez; Emir Talundzic; Joel Montgomery; John Klena; Juan Torales; Justin Lee; María José Ortega; María Liz Gamarra; Shannon Whitmer; Shirley Villalba |
| EPI_ISL_4518741, EPI_ISL_4518748 | Laboratorio Central de Salud Publica de Paraguay | Fundação Ezequiel Dias | Andre Leal; Andrea Gómez de la Fuente; Cynthia Vazquez; Elaine Cristina; Felipe Iani; Flavia Aburjaile; Gislene Garcia de Castro Lichs; Glauco Carvalho; Hegger Fritsch; Joilson Xavier; Juan Torales; Luiz Alcantara.; Luiz Henrique Ferraz Demarchi; Luiz Takao Watanabe; Marina Castilhos Souza Umaki Zardin; Marta Giovanetti; María José Ortega; María Liz Gamarra; Natalia Guimaraes; Raquel da Silva Ferreira; Shirley Villalba; Talita Adelino; Vagner Fonseca; de Oliveira |
| EPI_ISL_2157541 | Laboratorio Central de Saude Publica do Esatado de Alagoas (LACEN/AL) | Laboratory of Respiratory Viruses and Measles, Oswaldo Cruz Institute, FIOCRUZ | Alice Sampaio Rocha; Ana Carolina Mendonca; Anderson Brandao Leite; Anna Carolina Paixao; Elisa Cavalcante Pereira; Fernando Motta; Luciana Appolinario; Marilda Siqueira on behalf of the Fiocruz COVID-19 Genomic Surveillance Network; Paola Resende; Renata Serrano Lopes; Taina Venas |
| EPI_ISL_2536325, EPI_ISL_2536339, EPI_ISL_2536340, EPI_ISL_3061880, EPI_ISL_3061881 | Laboratorio Central de Saude Publica do Estado da Paraiba (LACEN-PB) | Laboratory of Respiratory Viruses and Measles, Oswaldo Cruz Institute, FIOCRUZ | Alice Sampaio Rocha; Ana Carolina Mendonca; Anna Carolina Paixao; Dalane Loudal Florentino Teixeira; Elisa Cavalcante Pereira; Fernando Motta; Joao Felipe Bezerra; Luciana Appolinario; Marilda Siqueira on behalf of the Fiocruz COVID-19 Genomic Surveillance Network; Paola Resende; Renata Serrano Lopes; Taina Venas |
| EPI_ISL_2274042, EPI_ISL_2274043, EPI_ISL_2274047, EPI_ISL_2274049 | Laboratorio Central de Saude Publica do Estado de Alagoas (LACEN/AL) | Laboratory of Respiratory Viruses and Measles, Oswaldo Cruz Institute, FIOCRUZ | Alice Sampaio Rocha; Ana Carolina Mendonca; Anderson Brandao Leite; Anna Carolina Paixao; Elisa Cavalcante Pereira; Fernando Motta; Luciana Appolinario; Marilda Siqueira on behalf of the Fiocruz COVID-19 Genomic Surveillance Network; Paola Resende; Renata Serrano Lopes; Taina Venas |
| EPI_ISL_3045430, EPI_ISL_3045431, EPI_ISL_3045445, EPI_ISL_3045450, EPI_ISL_3045452, EPI_ISL_3045458 | Laboratorio Central de Saude Publica do Estado do Espirito Santo (LACEN/ES) | Laboratory of Respiratory Viruses and Measles, Oswaldo Cruz Institute, FIOCRUZ | Alice Sampaio Rocha; Ana Carolina Mendonca; Anna Carolina Paixao; Elisa Cavalcante Pereira; Fernando Motta; Luciana Appolinario; Marilda Siqueira on behalf of the Fiocruz COVID-19 Genomic Surveillance Network; Paola Resende; Renata Serrano Lopes; Rodrigo Ribeiro Rodrigues; Taina Venas |
| EPI_ISL_2983260, EPI_ISL_2983261, EPI_ISL_2983262, EPI_ISL_2983263, EPI_ISL_2983264, EPI_ISL_2983265, EPI_ISL_2983267 | | | |
| see above | Laboratorio Central de Saude Publica do Estado do Maranhao (LACEN-MA) | Laboratory of Respiratory Viruses and Measles, Oswaldo Cruz Institute, FIOCRUZ | Agatha Cristinne Prudencio; Alice Sampaio Rocha; Ana Carolina Mendonca; Anna Carolina Paixao; Elisa Cavalcante Pereira; Fernando Motta; Ighor Leonardo Arantes Gomes; Lidio Gonçalves Lima Neto; Luciana Appolinario; Marilda Siqueira on behalf of the Fiocruz COVID-19 Genomic Surveillance Network; Paola Resende; Renata Serrano Lopes; Taina Moreira Venas |
| EPI_ISL_2274131, EPI_ISL_2466129, EPI_ISL_2466131, EPI_ISL_2466132, EPI_ISL_2466133, EPI_ISL_2466134, EPI_ISL_2466135, EPI_ISL_2466269, EPI_ISL_2603435, EPI_ISL_2603580 | | | |
| see above | Laboratorio Central de Saude Publica do Estado do Rio Grande do Sul (LACEN-RS) | Laboratory of Respiratory Viruses and Measles, Oswaldo Cruz Institute, FIOCRUZ | Alice Sampaio Rocha; Ana Carolina Mendonca; Anderson Brandao Leite; Anna Carolina Paixao; Elisa Cavalcante Pereira; Fernando Motta; Luciana Appolinario; Marilda Siqueira on behalf of the Fiocruz COVID-19 Genomic Surveillance Network; Paola Resende; Renata Serrano Lopes; Richard Salvato; Taina Venas; Tatiana Schaffer Gregianini |
| EPI_ISL_3048796, EPI_ISL_3048798, EPI_ISL_3048800, EPI_ISL_3048801 | Laboratorio Central de Saude Publica do Estado do Rio Grande do Sul (LACEN-RS) | Laboratório de Biologia Molecular da Universidade Federal de Ciências da Saúde de Porto Alegre | Adriana Seixas; Ana B. G. Veiga; Ana Paula Muterle Varela; Fabiana Quoos Mayer; Fernando Hayashi Sant'Anna; Janira Prichula; Letícia Garay Martins; Richard Steiner Salvato; Tatiana Schäffer Gregianini |
| EPI_ISL_2363561, | Laboratorio Central de la | Grupo de Genómica y Bioinformática | AF; Amadio; C; Eberhardt; G; Irazoqui; JM; MF; Mugna; Ojeda; Pastor; Rompato; V |

| | | | |
|---|---|---|---|
| EPI_ISL_3033032 | Ciudad de Santa Fe | del Instituto de Investigación de la Cadena Láctea CONICET-INTA on behalf of 'Proyecto Argentino Interinstitucional de genomica de SARS-CoV-2' (PAIS Consortium) | |
| EPI_ISL_4197238, EPI_ISL_4197242, EPI_ISL_4197247, EPI_ISL_4197249 | Laboratorio Clínico Analizar | Gencore - Universidad de los Andes | Cristian Barrera; David González; Gabriela Ariza; Luisa Sacristan; Marcela Guevara; Marcela Mercado; Silvia Restrepo |
| EPI_ISL_5782491 | Laboratorio De Franca | Instituto Butantan | Antonio Jorge Martins; Claudia Renata dos Santos Barros; David Schlesinger; Debora Botequio Moretti; Dimas Tadeu Covas; Elaine Cristina Marqueze; Elaine Vieira Santos; Evandra Strazza Rodrigues; Heidge Fukumasu; Jayme Augusto de Souza-Neto; José Salvatore Leister Patané; Luiz Alcantara; Luiz Lehmann Coutinho; Maria Carolina Elias; Maurício Lacerda Nogueira; Rafael dos Santos Bezerra; Raul Machado Neto; Rejane Maria Tommasini Grotto; Ricardo Haddad; Sandra Coccuzzo Sampaio Vessoni; Simone Kashima; Svetoslav Nanev Slavov; Vincent Louis Viala |
| EPI_ISL_5802437, EPI_ISL_5802440, EPI_ISL_5802447, EPI_ISL_5802449, EPI_ISL_5802450, EPI_ISL_5802451, EPI_ISL_5802452, EPI_ISL_5802455, EPI_ISL_5802457, EPI_ISL_5802458 | | | |
| see above | Laboratorio Dr Paulo Emilio Dalessandro Pindamonhangaba | Instituto Butantan | Antonio Jorge Martins; Claudia Renata dos Santos Barros; David Schlesinger; Debora Botequio Moretti; Dimas Tadeu Covas; Elaine Cristina Marqueze; Elaine Vieira Santos; Evandra Strazza Rodrigues; Heidge Fukumasu; Jayme Augusto de Souza-Neto; José Salvatore Leister Patané; Luiz Alcantara; Luiz Lehmann Coutinho; Maria Carolina Elias; Maurício Lacerda Nogueira; Rafael dos Santos Bezerra; Raul Machado Neto; Rejane Maria Tommasini Grotto; Ricardo Haddad; Sandra Coccuzzo Sampaio Vessoni; Simone Kashima; Svetoslav Nanev Slavov; Vincent Louis Viala |
| EPI_ISL_5799748, EPI_ISL_5799759, EPI_ISL_5799770, EPI_ISL_5799771 | Laboratorio Municipal De Analises Clinicas De Rio Claro | Instituto Butantan | Antonio Jorge Martins; Claudia Renata dos Santos Barros; David Schlesinger; Debora Botequio Moretti; Dimas Tadeu Covas; Elaine Cristina Marqueze; Elaine Vieira Santos; Evandra Strazza Rodrigues; Heidge Fukumasu; Jayme Augusto de Souza-Neto; José Salvatore Leister Patané; Luiz Alcantara; Luiz Lehmann Coutinho; Maria Carolina Elias; Maurício Lacerda Nogueira; Rafael dos Santos Bezerra; Raul Machado Neto; Rejane Maria Tommasini Grotto; Ricardo Haddad; Sandra Coccuzzo Sampaio Vessoni; Simone Kashima; Svetoslav Nanev Slavov; Vincent Louis Viala |
| EPI_ISL_5800518 | Laboratorio Municipal De Piracicaba | Instituto Butantan | Antonio Jorge Martins; Claudia Renata dos Santos Barros; David Schlesinger; Debora Botequio Moretti; Dimas Tadeu Covas; Elaine Cristina Marqueze; Elaine Vieira Santos; Evandra Strazza Rodrigues; Heidge Fukumasu; Jayme Augusto de Souza-Neto; José Salvatore Leister Patané; Luiz Alcantara; Luiz Lehmann Coutinho; Maria Carolina Elias; Maurício Lacerda Nogueira; Rafael dos Santos Bezerra; Raul Machado Neto; Rejane Maria Tommasini Grotto; Ricardo Haddad; Sandra Coccuzzo Sampaio Vessoni; Simone Kashima; Svetoslav Nanev Slavov; Vincent Louis Viala |
| EPI_ISL_2614583, EPI_ISL_2614589 | Laboratorio Municipal De Suzano | Instituto Adolfo Lutz, Interdiciplinary Procedures Center, Strategic Laboratory | Caio Vinicius Dias Lopes; Claudia Regina Gonçalves; Claudio Tavares Sacchi; Erica Valessa Ramos Gomes; Karoline Rodrigues Campos; Leonardo Jose Tadeu de Araujo |
| EPI_ISL_3307446, EPI_ISL_3307447, EPI_ISL_3307448, EPI_ISL_3307449, EPI_ISL_3307450, EPI_ISL_3307452, EPI_ISL_3307453, EPI_ISL_3307454, EPI_ISL_3332692 | | | |
| see above | Laboratorio SYNLAB Colombia | Corporación para Investigaciones Biológicas-CIB | Jeanneth Mosquera Rendon; Jenny Santiago Cuesta; Marcela Mercado Reyes; Uriel A. Hurtado Paez |
| EPI_ISL_1909236 | Laboratorio Sondrio, ASST Valtellina e dell'Alto Lario | Laboratory of Clinical Microbiology, Virology and Bioemergencies, ASST Fatebenefratelli Sacco - Sacco Hospital | Alberto Rizzo; Alessandro Mancon; Fiorenza Bracchitta; Luca Rizzuto; Maria Rita Gismondo; Valeria Micheli |
| EPI_ISL_2427659, EPI_ISL_2427666, EPI_ISL_2427674, EPI_ISL_2427698, EPI_ISL_2427706, EPI_ISL_2427714, EPI_ISL_2427721, EPI_ISL_2427729, EPI_ISL_2427735, EPI_ISL_2427736, EPI_ISL_2427743, EPI_ISL_2427744, EPI_ISL_2427751, EPI_ISL_2427759, EPI_ISL_2427760, EPI_ISL_2427766, EPI_ISL_2427767 | | | |
| see above | Laboratorio de Biología Molecular Médica Uruguaya | Departments of Pathology and Medicine, New York University School of Medicine | Adriana Heguy; Cecilia Sorhouet; Christian Marier; Dacia Dimartino; Gonzalo Manrique; Maria Cristina Mogdasy; Maria Noel Zubillaga; Maria Victoria Elizondo; Paul Zappile |
| EPI_ISL_2777493 | Laboratorio de Ecologia de Doencas Transmissiveis na Amazonia, Instituto Leonidas e Maria Deane - Fiocruz Amazonia | Laboratorio de Ecologia de Doencas Transmissiveis na Amazonia, Instituto Leonidas e Maria Deane - Fiocruz Amazonia | André Corado; Debora Duarte; Felipe Naveca; Fernanda Nascimento; George Silva; Karina Pessoa; Luciana Gonçalves; Maria Júlia Brandão; Matilde Mejía; Michele Jesus; Valdinete Nascimento; Victor Souza; Ágatha Costa |
| EPI_ISL_2728536, EPI_ISL_2728538, EPI_ISL_2728544, EPI_ISL_2728548, EPI_ISL_2728550 | Laboratorio de Infectologia y Virologia Molecular | Laboratory of Molecular Virology, School of Medicine, Pontificia Universidad Catolica de Chile | Alejandro Bhrun; Ana Maria Contreras; Andres E. Munoz-Marcos; Carlos Palma; Catalina Pardo-Roa; Constanza Maldonado; Constanza Martinez-Valdevenito; Eileen Serrano; Erick Salinas; Estefany Poblete; Francisco Melo; Jennifer Angulo; Jorge Levican; Leonardo I. Almonacid; M. Belen Leyton; Magdalena Vera; Marcela Ferres; Maria Jose Avendano; Rafael A. Medina; Tamara Garcia-Salum |
| EPI_ISL_3048954, EPI_ISL_3048955 | Laboratorio de Pesquisa em Virologia, FAMERP, SJRP | Laboratorio de Pesquisa em Virologia, FAMERP, SJRP | Cecília Artico Banho; Cíntia Bittar; Fábio Sossai Possebon; Guilherme Campos; Helena Lage Ferreira; Jorge A. Petroli Marchesi; João Pessoa Araújo Jr.; Leila Sabrina Ullmann; Lívia Sacchetto; Maisa C. Pereira Parra; Marília Moraes; Maurício L. Nogueira.; Paula Rahal; Paulo Inacio da Costa |
| EPI_ISL_3023585, EPI_ISL_3375873, EPI_ISL_3375879, EPI_ISL_3375881, EPI_ISL_3375883, EPI_ISL_3375925, EPI_ISL_3375929, EPI_ISL_3375930, EPI_ISL_3375931, EPI_ISL_3375941, EPI_ISL_3375947, EPI_ISL_3375949, EPI_ISL_3375996, EPI_ISL_3376011, EPI_ISL_3376018, EPI_ISL_3376019, EPI_ISL_3376027 | | | |
| see above | Laboratorio de Referencial Nacional de Virus Respiratorios | Laboratorio de Referencial Nacional de Virus Respiratorios | Carlos Padilla Rojas; Henri Bailon Calderon; Iris Silva Molina; Joseph Huayra Niquen; Lely Solari Zerpa; Luis Barcena Flores; Marco Galarza Perez; Nancy Rojas Serrano; Omar Caceres Rey; Orson Mestanza Millones; Priscila Lope Pari; Sandra Morales Ruiz; Steve Acedo Lazo; Veronica Hurtado Vela |
| EPI_ISL_2157208 | Laboratorio de Salud Pública | Gencore - Universidad de los Andes | Alejandro Gomez; David Gonzalez; Gabriela Delgado; Johana Hernandez; Luisa Sacristan; Marcela Guevara; Silvia Restrepo |
| EPI_ISL_1914233, EPI_ISL_1914275 | Laboratorio de Virología HUCA | Laboratorio de Virología HUCA | Abreu F; Alvarez-Arguelles ME; Boga JA; Castelló C; Costales I; Coto E; Gómez de Oña J; Martín-Rodríguez G; Melón S; Perez-Martínez Z; Rojo S; Sandoval M |
| EPI_ISL_4405452 | Laboratorio del Hospital Interzonal General de Agudos " Evita " | Área de Secuenciación del Laboratorio de Virología del Hospital de Niños Dr. Ricardo Gutierrez on behalf of 'Proyecto Argentino Interinstitucional de genomica de SARS-CoV-2' (PAIS Consortium) | A; Acuña; D; Desimone; E; Goya; Grossi; I; L; LE; Luczac; Lusso; MI; MS; Musto; Nabaes Jodar; Natale; O; S; Serrano; Valinotto; Viegas, M. |
| EPI_ISL_1681686, EPI_ISL_1798359, EPI_ISL_1798403, EPI_ISL_1798683, EPI_ISL_1798978, EPI_ISL_1799020, EPI_ISL_1799040, EPI_ISL_1799051, EPI_ISL_1799079, EPI_ISL_1799089, EPI_ISL_1799217, EPI_ISL_1799271, EPI_ISL_1799371, EPI_ISL_1799490, EPI_ISL_1799525, EPI_ISL_1799546, EPI_ISL_1799561, EPI_ISL_1799614, EPI_ISL_1799622, EPI_ISL_1800290, EPI_ISL_1800372, EPI_ISL_1800374, EPI_ISL_1800378, EPI_ISL_1800390, EPI_ISL_1800427, EPI_ISL_1800428, EPI_ISL_1800432, EPI_ISL_1800519, EPI_ISL_1800550, EPI_ISL_1800553, EPI_ISL_1800568, EPI_ISL_1800579, EPI_ISL_1800588, EPI_ISL_1800589, EPI_ISL_1800597, EPI_ISL_1800598, EPI_ISL_1800599, EPI_ISL_1800600, EPI_ISL_1800607, EPI_ISL_1800642, EPI_ISL_1800658, EPI_ISL_1800698, EPI_ISL_1800754, EPI_ISL_1800776, EPI_ISL_1800811, EPI_ISL_1800824, EPI_ISL_1800838, EPI_ISL_1800888, EPI_ISL_1800973, EPI_ISL_1800974, EPI_ISL_1801014, EPI_ISL_1801042, EPI_ISL_1801043, EPI_ISL_1801080, EPI_ISL_1801110, EPI_ISL_1801125, EPI_ISL_1801220, EPI_ISL_1801234, EPI_ISL_1801251, EPI_ISL_1801281, EPI_ISL_1801284, EPI_ISL_1801294, EPI_ISL_1801303, EPI_ISL_1801316, EPI_ISL_1801319, EPI_ISL_1801325, EPI_ISL_1801332, EPI_ISL_1801333, EPI_ISL_1801383, EPI_ISL_1801415, EPI_ISL_1801440, EPI_ISL_1801475, EPI_ISL_1801485, EPI_ISL_1801516, EPI_ISL_1801559, EPI_ISL_1801564, EPI_ISL_1801605, EPI_ISL_1801637, EPI_ISL_1801639, EPI_ISL_1801642, EPI_ISL_1801648, EPI_ISL_1801652, EPI_ISL_1801676, EPI_ISL_1801682, EPI_ISL_1801684, EPI_ISL_1801686, EPI_ISL_1801687, EPI_ISL_1801694, EPI_ISL_1801697, EPI_ISL_1801700, EPI_ISL_1801701, EPI_ISL_1801702, EPI_ISL_1801715, EPI_ISL_1801725, EPI_ISL_1801736, EPI_ISL_1801752, EPI_ISL_1801754, EPI_ISL_1801810, EPI_ISL_1801811, EPI_ISL_1801831, EPI_ISL_1801832, EPI_ISL_1801836, EPI_ISL_1801842, EPI_ISL_1801871, EPI_ISL_1801898, EPI_ISL_1801913, EPI_ISL_1801944, EPI_ISL_1801945, EPI_ISL_1802040, EPI_ISL_1802041, EPI_ISL_1802048, EPI_ISL_1802049, EPI_ISL_1802051, EPI_ISL_1802062, EPI_ISL_1802063, EPI_ISL_1802065, EPI_ISL_1802066, EPI_ISL_1802068, EPI_ISL_1802069, EPI_ISL_1802073, EPI_ISL_1802092, EPI_ISL_1802095, EPI_ISL_1802127, EPI_ISL_1802140, EPI_ISL_1802141, EPI_ISL_1802147, EPI_ISL_1802149, EPI_ISL_1802153, EPI_ISL_1802154, EPI_ISL_1802157, EPI_ISL_1802159, EPI_ISL_1802165, EPI_ISL_1802166, EPI_ISL_1802168, EPI_ISL_1802169, EPI_ISL_1802196, EPI_ISL_1802199, EPI_ISL_1802201, EPI_ISL_1802202, EPI_ISL_1802203, EPI_ISL_1802205, EPI_ISL_1802210, EPI_ISL_1802216, EPI_ISL_1802218, EPI_ISL_1802221, EPI_ISL_1802223, EPI_ISL_1802226, EPI_ISL_1802227, EPI_ISL_1802230, EPI_ISL_1802232, EPI_ISL_1802236, EPI_ISL_1802271, EPI_ISL_1802274, EPI_ISL_1802276, EPI_ISL_1802285, EPI_ISL_1802334, EPI_ISL_1802346, EPI_ISL_1802353, EPI_ISL_1802356, EPI_ISL_1802366, EPI_ISL_1802374, EPI_ISL_1802377, EPI_ISL_1802380, EPI_ISL_1802382, EPI_ISL_1802383, EPI_ISL_1802384, EPI_ISL_1802385, EPI_ISL_1802397, EPI_ISL_1802398, EPI_ISL_1802399, EPI_ISL_1802401, EPI_ISL_1802404, EPI_ISL_1802414, EPI_ISL_1802463, EPI_ISL_1802469, EPI_ISL_1802472, EPI_ISL_1802477, EPI_ISL_1802498, EPI_ISL_1802499, EPI_ISL_1802504, EPI_ISL_1802513, EPI_ISL_1802517, EPI_ISL_1802551, EPI_ISL_1802561, EPI_ISL_1802562, EPI_ISL_1802568, EPI_ISL_1802588, EPI_ISL_1802616, EPI_ISL_1802617, EPI_ISL_1802640, EPI_ISL_1802649, EPI_ISL_1802658, EPI_ISL_1925674, EPI_ISL_1926336, EPI_ISL_1930656, EPI_ISL_2046441, EPI_ISL_4372762, EPI_ISL_4373536, EPI_ISL_4378996, EPI_ISL_4379055, EPI_ISL_4380279 | | | |
| see above | Laboratory Corporation of America | Centers for Disease Control and Prevention Division of Viral Diseases, Pathogen Discovery | Adrian Paskey; Amanda Douglas; Amanda Suchanek; Andrea Throop; Ayla Burns; Benjamin Rambo-Martin; Bobbi Croy; Brian Krueger; Brian Norvell; Christopher Gulvick; Christos Petropoulos; Clinton Paden; Clinton R. Paden; Craig Lukasik; Dakota Howard; Darlene Wagner; Debbie Boles; Dhwani Batra; Duncan MacCannell; Eyad Almasri; Goran Stevovic; Howard Engler; Hrushikesh Deshmukh; Jake Humphrey; Jana Schroth; Jason Caravas; Joe Voshell; John Pruitt; Jonathan Meltzer; Jonathan Williams; Kara Moser; Kimberly Wagner; Kristine Lacek; Lax Iyer; Lisa Pfefferle; Lyndon Tilson; Manoj Jain; Marcia Eisenberg; Mary Ann Cristobal; Mary Cristobal; Mary Williamson; Matthew Robinson; Matthew Schmerer; Michael Levandoski; Mike Sapeta; Mindy Nye; Minoo Agarwal; Mohan Kolli; Nuthawin Charoensri; Oren Cohen; Peter Cook; Peter W. Cook; Prashant Gupta; Qian Zeng; Rama Ghatti; Scott Parker; Scott Ryan; Scott Sammons; Shatavia Morrison; Stanley Letovsky; Steven Ragan; Suresh Babu Selvaraju; Suresh Selvaraju; Susan Countryman; Susan Hicks; Suzanne Dale; Thomas Urban; Tim Kuphal; Tricia Zwiefelhofer; Tymeckia Kendall; Victoria Caban Figueroa; Vincent Drouillon; Yvette Unoarumhi |
| EPI_ISL_2157540, EPI_ISL_2443596, EPI_ISL_2443597, EPI_ISL_2443598, EPI_ISL_6943983, EPI_ISL_6943984 | Laboratory of Respiratory Viruses and Measles, Oswaldo Cruz Institute, FIOCRUZ | Laboratory of Respiratory Viruses and Measles, Oswaldo Cruz Institute, FIOCRUZ | Alice Sampaio Rocha; Ana Carolina Mendonca; Andrea Cony Cavalcanti; Anna Carolina Paixao; Bruna Mendonça da Silva; Elisa Cavalcante Pereira; Fernando Motta; Ighor Arantes; Jéssica Graça Macedo de Carvalho; Larissa Macedo Pinto; Luciana Appolinario; Marilda Siqueira on behalf of the Fiocruz COVID-19 Genomic Surveillance Network; Paola Resende; Renata Serrano Lopes; Taina Venas; Victor Guimaraes |
| EPI_ISL_2696536 | Laboratory, Gravity Diagnostics | Laboratory, Gravity Diagnostics | Diagnostics, G. |
| EPI_ISL_2157536, EPI_ISL_2157537, EPI_ISL_2157538, EPI_ISL_2157539, EPI_ISL_2157549 | Laboratório Central de Saude Publica do Estado do Rio de Janeiro (LACEN/RJ) | Laboratory of Respiratory Viruses and Measles, Oswaldo Cruz Institute, FIOCRUZ | Alice Sampaio Rocha; Ana Carolina Mendonca; Andrea Cony Cavalcanti; Anna Carolina Paixao; Elisa Cavalcante Pereira; Fernando Motta; Luciana Appolinario; Marilda Siqueira on behalf of the Fiocruz COVID-19 Genomic Surveillance Network; Paola Resende; Renata Serrano Lopes; Taina Venas |
| EPI_ISL_4600507, EPI_ISL_4600515, EPI_ISL_4600516, EPI_ISL_4600523 | Laboratório Central de Saúde Pública de Pernambuco | Coordenação Geral de Laboratórios de Saúde Pública (CGLAB/DAEVS/SVS/MS) | Vagner Fonseca; et al. |
| EPI_ISL_4600568, EPI_ISL_4600579, EPI_ISL_4600581 | Laboratório Central de Saúde Pública de Santa Catarina | Coordenação Geral de Laboratórios de Saúde Pública (CGLAB/DAEVS/SVS/MS) | Vagner Fonseca; et al. |
| EPI_ISL_2777241, EPI_ISL_2777277, EPI_ISL_2777278, EPI_ISL_2777279, EPI_ISL_2777290, EPI_ISL_2777329, EPI_ISL_2777330, EPI_ISL_2777331, EPI_ISL_2777332, EPI_ISL_2777576, EPI_ISL_2777579, EPI_ISL_2777581, EPI_ISL_2777676, EPI_ISL_2777772, EPI_ISL_2777815, EPI_ISL_2777816, EPI_ISL_2777817, EPI_ISL_2777868, EPI_ISL_2777869, EPI_ISL_2777870, EPI_ISL_2777871, EPI_ISL_2777872, EPI_ISL_2777969 | | | |
| see above | Laboratório Central de Saúde Pública do Amazonas - LACEN-AM | Laboratorio de Ecologia de Doencas Transmissiveis na Amazonia, Instituto Leonidas e Maria Deane - Fiocruz Amazonia | André Corado; Debora Duarte; Felipe Naveca; Fernanda Nascimento; George Silva; Karina Pessoa; Luciana Gonçalves; Maria Júlia Brandão; Matilde Mejía; Michele Jesus; Valdinete Nascimento; Victor Souza; Ágatha Costa |

| | | | |
|---|---|---|---|
| EPI_ISL_4423935 | Laboratório de Baculovirus, Universidade de Brasília (UnB), Instituto de Ciências Biológicas (IB) | Laboratório de Virologia, Faculdade de Medicina, Universidade Federal de Mato Grosso (UFMT) | Bergman Morais Ribeiro; Fernando Lucas Melo; Francisco Scoffoni Kennedy de Azevedo; Gessica Fernanda Colnago de Lima; Renata Dezengrini Slhessarenko; Thais Campos Cruz |
| EPI_ISL_2375503, EPI_ISL_2375507, EPI_ISL_2375508, EPI_ISL_2375510, EPI_ISL_2375511, EPI_ISL_2376027, EPI_ISL_2376028, EPI_ISL_2376029, EPI_ISL_2376030, EPI_ISL_2376031, EPI_ISL_2382540, EPI_ISL_2382727, EPI_ISL_4051957 | | | |
| see above | Laboratório de Microbiologia Molecular - Universidade FEEVALE | Molecular Microbiology Laboratory | Alana Witt Hansen; Fernando Rosado Spilki; Flávio Silveira; Fágner Henrique Heldt; Juliana Schons Gularte; Juliane Deise Fleck; Mariana Soares da Silva; Matheus Nunes Weber; Meriane Demoliner; Micheli Filippi; Micheli Filippi.; Paula Rodrigues de Almeida; Vyctoria Malayhka de Abreu Góes Pereira. |
| EPI_ISL_2629812, EPI_ISL_2629813, EPI_ISL_2629814, EPI_ISL_2629815, EPI_ISL_2629816, EPI_ISL_2629817, EPI_ISL_2629818, EPI_ISL_2629819, EPI_ISL_2629820 | | | |
| see above | Laboratório de Virologia Molecular - Universidade Federal do Rio de Janeiro | Laboratório de Virologia Molecular - Universidade Federal do Rio de Janeiro | ; Alice Laschuk Herlinger; Amilcar Tanuri; André Felipe Andrade dos Santos; Carolina Moreira Voloch; Cássia Cristina Alves Gonçalves; Diana Mariani; Débora Souza Faffe; Filipe Romero Rebello Moreira; Francine Bittencourt Schiffler; Isabela de Carvalho Leitão; Marcelo Calado de Paula Tôrres; Matheus Augusto Calvano Cosentino; Mirela D'arc; Orlando da Costa Ferreira Junior; Rafael Mello Galliez; Raíssa Mirella dos Santos Cunha da Costa; Renato Santana de Aguiar; Terezinha Marta Pereira Pinto Castineiras; Thamiris dos Santos Miranda; Átila Duque Rossi |
| EPI_ISL_1843311 | Labormedizin Darmstadt | Robert Koch Institute | |
| EPI_ISL_2443686, EPI_ISL_2443689, EPI_ISL_2443693, EPI_ISL_2443695, EPI_ISL_2443696, EPI_ISL_2443697, EPI_ISL_2443698, EPI_ISL_2443700, EPI_ISL_2443704, EPI_ISL_2443706, EPI_ISL_2443707 | | | |
| see above | Labortorio Central de Saude Publica do Estado de Santa Catarina (LACEN/SC) | Laboratory of Respiratory Viruses and Measles, Oswaldo Cruz Institute, FIOCRUZ | Alice Sampaio Rocha; Ana Carolina Mendonca; Anna Carolina Paixao; Darcita Buerger Rovaris; Elisa Cavalcante Pereira; Fernando Motta; Luciana Appolinario; Marilda Siqueira on behalf of the Fiocruz COVID-19 Genomic Surveillance Network; Paola Resende; Renata Serrano Lopes; Sandra Bianchini Fernandes; Taina Venas |
| EPI_ISL_2196313, EPI_ISL_2196314, EPI_ISL_2196315, EPI_ISL_2196316, EPI_ISL_2196317, EPI_ISL_2196324, EPI_ISL_2196325, EPI_ISL_2196326, EPI_ISL_2196327, EPI_ISL_2196328, EPI_ISL_2196329, EPI_ISL_2196330, EPI_ISL_2196331, EPI_ISL_2196332, EPI_ISL_2196356, EPI_ISL_2274099, EPI_ISL_2443615, EPI_ISL_2443616, EPI_ISL_2443617, EPI_ISL_2443618, EPI_ISL_2443619, EPI_ISL_2443620, EPI_ISL_2443621, EPI_ISL_2443622, EPI_ISL_2443626, EPI_ISL_2443627, EPI_ISL_2443628, EPI_ISL_2443629, EPI_ISL_2443632 | | | |
| see above | Labortorio Central de Saude Publica do Estado do Rio de Janeiro (LACEN/RJ) | Laboratory of Respiratory Viruses and Measles, Oswaldo Cruz Institute, FIOCRUZ | Alice Sampaio Rocha; Ana Carolina Mendonca; Andrea Cony Cavalcanti; Anna Carolina Paixao; Elisa Cavalcante Pereira; Fernando Motta; Luciana Appolinario; Marilda Siqueira on behalf of the Fiocruz COVID-19 Genomic Surveillance Network; Paola Resende; Renata Serrano Lopes; Taina Venas |
| EPI_ISL_2443539, EPI_ISL_2443540, EPI_ISL_2443553, EPI_ISL_2443568, EPI_ISL_2443569, EPI_ISL_2443570, EPI_ISL_2603425, EPI_ISL_2603426, EPI_ISL_2603437, EPI_ISL_2603459, EPI_ISL_2603465, EPI_ISL_2603544, EPI_ISL_2603546, EPI_ISL_2613816, EPI_ISL_2613817, EPI_ISL_2613818, EPI_ISL_2613819, EPI_ISL_2613820, EPI_ISL_2613821, EPI_ISL_2613822, EPI_ISL_2613823, EPI_ISL_2613824, EPI_ISL_2645488, EPI_ISL_2645489, EPI_ISL_2645490, EPI_ISL_2863890 | | | |
| see above | Lboratorio Central de Saude Publica do Estado do Parana (LACEN/PR) | Laboratory of Respiratory Viruses and Measles, Oswaldo Cruz Institute, FIOCRUZ | Agatha Cristinne Prudencio Soares; Alice Sampaio Rocha; Ana Carolina Mendonca; Anna Carolina Paixao; Elisa Cavalcante Pereira; Fernando Motta; Ighor Leonardo Arantes Gomes; Irina Riediger; Luciana Appolinario; Marilda Siqueira on behalf of the Fiocruz COVID-19 Genomic Surveillance Network; Paola Resende; Renata Serrano Lopes; Taina Venas |
| EPI_ISL_1718434 | Lighthouse Lab in Alderley Park | Wellcome Sanger Institute for the COVID-19 Genomics UK (COG-UK) Consortium | Cordelia Langford; David K. Jackson; Dominic Kwiatkowski; Ewan Harrison; Ian Johnston; Jacquelyn Wynn; Jeffrey Barrett; John Sillitoe on behalf of the Wellcome Sanger Institute COVID-19 Surveillance Team; Mairead Hyland; Roberto Amato; Sonia Goncalves; The Lighthouse Lab in Alderley Park and Alex Alderton |
| EPI_ISL_1653315, EPI_ISL_1697704, EPI_ISL_1698366, EPI_ISL_1719527, EPI_ISL_2199983 | Lighthouse Lab in Cambridge | Wellcome Sanger Institute for the COVID-19 Genomics UK (COG-UK) Consortium | Cordelia Langford; David K. Jackson; Dominic Kwiatkowski; Ewan Harrison; Ian Johnston; Jeffrey Barrett; John Sillitoe on behalf of the Wellcome Sanger Institute COVID-19 Surveillance Team; Rob Howes; Roberto Amato; Sonia Goncalves; The Lighthouse Lab in Cambridge and Alex Alderton |
| EPI_ISL_1673030 | Lighthouse Lab in Glasgow | Wellcome Sanger Institute for the COVID-19 Genomics UK (COG-UK) Consortium | Anna Dominiczak and Alex Alderton; Carol Clugston; Cordelia Langford; David Gray; David K. Jackson; Dominic Kwiatkowski; Ewan Harrison; Harper VanSteenhouse; Ian Johnston; Jeffrey Barrett; John Sillitoe on behalf of the Wellcome Sanger Institute COVID-19 Surveillance Team; Roberto Amato; Sonia Goncalves; Yumi Kasai |
| EPI_ISL_1697974, EPI_ISL_1698023, EPI_ISL_1698047, EPI_ISL_1698072, EPI_ISL_1698105, EPI_ISL_1699787, EPI_ISL_1718902, EPI_ISL_1718906, EPI_ISL_1719025, EPI_ISL_1830142 | | | |
| see above | Lighthouse Lab in Milton Keynes | Wellcome Sanger Institute for the COVID-19 Genomics UK (COG-UK) Consortium | Cordelia Langford; David K. Jackson; Dominic Kwiatkowski; Ewan Harrison; Ian Johnston; Jeffrey Barrett; John Sillitoe on behalf of the Wellcome Sanger Institute COVID-19 Surveillance Team; Roberto Amato; Sonia Goncalves; The Lighthouse Lab in Milton Keynes and Alex Alderton |
| EPI_ISL_2009307, EPI_ISL_2009389, EPI_ISL_2009394, EPI_ISL_2009395 | Lurie Children's Hospital of Chicago | Northwestern University - Ozer Lab | Egon A. Ozer; Judd F. Hultquist; Lacy M. Simons; Larry K. Kociolek; Michael G. Ison; Ramon Lorenzo-Redondo; Taylor J. Dean; William J. Muller; Xiaotian; Zheng |
| EPI_ISL_2036066 | M Health Fairview | Minnesota Department of Health, Public Health Laboratory | Alexandra Lorentz; Jacob Garfin; Matt Plumb; and Xiong Wang |
| EPI_ISL_5650247, EPI_ISL_5650250 | MACRO II CENTRO DE SAUDE DE PENAPOLIS | Instituto Butantan | Antonio Jorge Martins; Claudia Renata dos Santos Barros; David Schlesinger; Debora Botequio Moretti; Dimas Tadeu Covas; Elaine Cristina Marqueze; Elaine Vieira Santos; Evandra Strazza Rodrigues; Heidge Fukumasu; Jayme Augusto de Souza-Neto; José Salvatore Leister Patané; Luiz Alcantara; Luiz Lehmann Coutinho; Maria Carolina Elias; Maurício Lacerda Nogueira; Rafael dos Santos Bezerra; Raul Machado Neto; Rejane Maria Tommasini Grotto; Ricardo Haddad; Sandra Coccuzzo Sampaio Vessoni; Simone Kashima; Svetoslav Nanev Slavov; Vincent Louis Viala |
| EPI_ISL_1672366 | MD PHL | MD PHL | Maryland Department of Health Laboratories Administration |
| EPI_ISL_1847198 | MDI Limbach Berlin GmbH; MVZ Labor Berlin | Robert Koch Institute | |
| EPI_ISL_1817057, EPI_ISL_1964718, EPI_ISL_1964719, EPI_ISL_2036370, EPI_ISL_2036582, EPI_ISL_2597089, EPI_ISL_2597136, EPI_ISL_2597196 | | | |
| see above | MEPHI, Aix Marseille University | MEPHI, Aix Marseille University | Anthony LEVASSEUR |
| EPI_ISL_1855324 | MONTEFIORE MEDICAL CENTER LABORATORIES | Wadsworth Center, New York State Department of Health | Alexis Russell; Catharine Prussing; Daryl M. Lamson; Erasmus Schneider; Erica Lasek-Nesselquist; John Kelly; Jonathan Plitnick; Kirsten St. George; Matthew Shudt; Melissa A Leisner; Navjot Singh |
| EPI_ISL_5253372, EPI_ISL_6492422, EPI_ISL_6492447, EPI_ISL_6492452, EPI_ISL_6492471, EPI_ISL_6492479, EPI_ISL_6492505 | | | |
| see above | MSHS Clinical Microbiology Laboratories | MSHS Pathogen Surveillance Program | Adolfo García-Sastre; Adriana van de Guchte; Ajay Obla; Alberto Paniz-Mondolfi; Ana S. Gonzalez-Reiche; Angela Amoako; Ashley Salimbangon; Betsaida Salom Melo; Bremy Alburquerque; Brianne Ciferri; Charles Gleason; Daniel Floda; Deena R. Altman; Denise Jurczyszak; Emilia Mia Sordillo; Gintaras Deikus; Giulio Kleiner; Gopi Patel; Hala Alshammary; Harm van Bakel; Irina Oussenko; Jayeeta Dutta; Juan Soto; Julia Matthews; Katherine Beach; Kathryn Twyman; Kayla Russo; Komal Srivastava; Levy Sominsky; Mahmoud Awawda; Marta Luksza; Matthew M. Hernandez; Melissa Gitman; Michael D. Nowak; Mitchell J. Sullivan; Nancy Francoeur; Robert Sebra; Sarah Schaefer; Shelcie Fabre; Shwetha Hara Sridhar; Viviana Simon; Ying-Chih Wang; Zenab Khan |
| EPI_ISL_2690685 | MSK Microbiology Lab | MSK Microbiology Lab | Esther Babady; Krupa Jani; Tracy McMillen |
| EPI_ISL_1847231 | MVZ fÃ¼r Laboratoriumsmedizin und Mikrobiologie Koblenz-Mittelrhein (Labor Koblenz) | Robert Koch Institute | |
| EPI_ISL_1821048, EPI_ISL_1821053 | Maine HETL | Health and Environmental Testing Laboratory | Barter, M.; Dewey, H.; H. and Tewhey, R.; Lynch, R.; Martha, J.; Matluk, N.; Munger |
| EPI_ISL_1826325, EPI_ISL_1826326, EPI_ISL_1826327, EPI_ISL_1826328, EPI_ISL_1826329, EPI_ISL_1973041, EPI_ISL_1973054, EPI_ISL_1973056, EPI_ISL_1973064, EPI_ISL_1973065, EPI_ISL_1973066, EPI_ISL_2802302, EPI_ISL_2802331, EPI_ISL_5366447, EPI_ISL_5366448, EPI_ISL_5366449, EPI_ISL_5366451, EPI_ISL_5366452 | | | |
| see above | Maine Health and Environmental Testing Laboratory | Tewhey Lab, The Jackson Laboratory | Barter, M.; Dewey, H.; H. and Tewhey, R.; Iosue, F.; Lynch, R.; Matluk, N.; Munger |
| EPI_ISL_3011674, EPI_ISL_3011712 | Maryland Genomics, Institute for Genome Sciences, University of Maryland School of Medicine | Maryland Genomics, Institute for Genome Sciences, University of Maryland School of Medicine | Claire M; Fraser; Hazen; Holly; Humphrys; Ivette; Jacques; Jonathan; Kranthi; Lim; Lisa D; Luke J; Mike; Ott; Ravel; Roussey; Sadzewicz; Sandra; Santana-Cruz; Tallon; Tracy; Vavikolanu |
| EPI_ISL_2008152, EPI_ISL_2008160, EPI_ISL_2008172, EPI_ISL_2008315, EPI_ISL_2008318, EPI_ISL_2008329 | Massachusetts State Public Health Laboratory | Massachusetts State Public Health Laboratory | Andrew Lang; Glen Gallagher; Sandra Smole; Timelia Fink |
| EPI_ISL_2374706, EPI_ISL_3129091 | Michigan Department of Health and Human Services, Bureau of Laboratories | Michigan Department of Health and Human Services, Bureau of Laboratories | Blankenship HM; Riner D; Soehnlen MK |
| EPI_ISL_1892866, EPI_ISL_1892891, EPI_ISL_1892915, EPI_ISL_1892940 | Microbiologia CATLAB | Can Ruti SARS-CoV-2 Sequencing Hub (HUGTiP/IrsiCaixa/IGTP) | Alba Sánchez; Anna Not; Antoni E Bordoy; Bonaventura Clotet; Cristina Casañ; Cristina Esteban; Francesc Catala-Moll; Gemma Clara; Ignacio Blanco; Marc Noguera-Julian; Maria Casadellà; Mariona Parera; Mercedes Guerrero; Montserrat Giménez; Pere-Joan Cardona; Pilar Armengol; Roger Paredes; Verónica Saludes; and Elisa Martró on behalf of the Can Ruti SARS-CoV-2 Sequencing Hub. |
| EPI_ISL_1719794, EPI_ISL_1719796, EPI_ISL_1719797, EPI_ISL_1719805, EPI_ISL_1719819, EPI_ISL_1719827, EPI_ISL_1719841, EPI_ISL_1719843, EPI_ISL_1761474, EPI_ISL_1761547 | | | |
| see above | Microbiology Department, Laboratori Clínic Metropolitana Nord. Hospital Universitari Germans Trias i Pujol. | Can Ruti SARS-CoV-2 Sequencing Hub (HUGTiP/IrsiCaixa/IGTP) | Alba Sánchez; Anna Not; Antoni E Bordoy; Bonaventura Clotet; Cristina Casañ; Cristina Esteban; Francesc Catala-Moll; Gemma Clara; Ignacio Blanco; Marc Noguera-Julian; Maria Casadellà; Mariona Parera; Mercedes Guerrero; Montserrat Giménez; Pere-Joan Cardona; Pilar Armengol; Roger Paredes; Verónica Saludes; and Elisa Martró on behalf of the Can Ruti SARS-CoV-2 Sequencing Hub. |

| | | | |
|---|---|---|---|
| EPI_ISL_2086140, EPI_ISL_2086141, EPI_ISL_2086144, EPI_ISL_2086145, EPI_ISL_2086146, EPI_ISL_2086147, EPI_ISL_2086148, EPI_ISL_2086149, EPI_ISL_2086151, EPI_ISL_2086152, EPI_ISL_2086154, EPI_ISL_2516974, EPI_ISL_2516976 | | | |
| see above | Microbiology Department. Complexo Hospitalario Universitario de Vigo | Microbiology Department. Complexo Hospitalario Universitario de Vigo | Alfaya N; Alonso I; Alvarez M; Cabrera JJ; Carballo R; Cores O; Cortizo S; Martinez L; Mediero G; Perez S; Potel C; Regueiro B; Rey S; Vasallo FJ; del-Campo V |
| EPI_ISL_2106148, EPI_ISL_2289147 | Microbiology Division, SC DHEC | Microbiology Division, SC DHEC | Flores, H.; Freeman, J. |
| EPI_ISL_2362677 | Microbiology and Virology Unit, Florence Careggi University Hospital | Microbiology and Virology Unit, Florence Careggi University Hospital | Alberto Antonelli; Emanuele Gori; Fabio Morecchiato; Gian Maria Rossolini; Ilaria Baccani; Marco Coppi; Nicla Giovacchini; Noemi Aiezza; Vincenzo Di Pilato |
| EPI_ISL_1820824 | Microvida | Microvida | Jaco Verweij; Joep Stöhr; Suzan D. Pas |
| EPI_ISL_1711038, EPI_ISL_1733517, EPI_ISL_1733533, EPI_ISL_1760757, EPI_ISL_1857496, EPI_ISL_1857497, EPI_ISL_1857559, EPI_ISL_2015211, EPI_ISL_2038809 | | | |
| see above | Minnesota Department of Health, Public Health Laboratory | Minnesota Department of Health, Public Health Laboratory | Alexandra Lorentz; Jacob Garfin; Matt Plumb; and Xiong Wang |
| EPI_ISL_2035524, EPI_ISL_2035549, EPI_ISL_2035550 | Missouri State Public Health Laboratory | Missouri State Public Health Laboratory | Ashley New; Joshua Barry; Matthew Sinn |
| EPI_ISL_5598876 | Molecular Biology Laboratory, Vidant Medical Center | Brody Integrative Genomics Core, East Carolina University | Changhong Yin; Heather Duncan; John T. Fallon; Kimberly P. Briley; Weihua Huang |
| EPI_ISL_1913001, EPI_ISL_2943188 | Montana Public Health Laboratory | Montana Public Health Laboratory | Carrie Biskupiak; Deborah Gibson; Joy Ritter; Michael Dills; Michelle Mozer |
| EPI_ISL_1920004 | Montefiore Medical Center Laboratories | Wadsworth Center, New York State Department of Health | Alexis Russell; Catharine Prussing; Daryl M. Lamson; Erasmus Schneider; Erica Lasek-Nesselquist; John Kelly; Jonathan Plitnick; Kirsten St. George; Matthew Shudt; Melissa A Leisner; Navjot Singh |
| EPI_ISL_4772366 | NAMRU-6 | NAMRU-6 | Cristhopher Cruz; Eugenio Abente; Gilda Troncos; Greg Rice; Luz Cedano Del Aguila; Marita Silva; Paul Graf.; Sonia Ampuero; Victoria Espejo; Yeny Tinoco |
| EPI_ISL_1823999 | NH Dept. of Health and Human Services Public Health Labs | Centers for Disease Control and Prevention Division of Viral Diseases, Pathogen Discovery | Alison Laufer Halpin; Ben L. Rambo-Martin; Clinton R. Paden; Dakota Howard; Darlene Wagner; Dave Wentworth; Dhwani Batra; Jasmine Padilla; Justin Lee; Katie Dillon; Krista Queen; Kristen Knipe; Kristine Lacek; Mark Burroughs; Matthew Schmerer; Mili Sheth; Peter Cook; Sam Shepard; Sarah Nobles; Shoshona Le; Suxiang Tong; Vivien Dugan; Yvette Unoarumhi |
| EPI_ISL_1732139, EPI_ISL_1960009 | NJDOH, Public Health and Environmental Laboratories | NJ_PHEL | Byeong Jeong; Dana Woell; Jacquelyn Deverell; Lindsey Bodnar; Matthew Scarnati; Shiv K. Verma |
| EPI_ISL_2161792 | NL-Dr. Leonard A. Miller Centre for Health Services | National Microbiology Laboratory (NML) | Adel Malek; Anna Majer; Anneliese Landgraff; CanCOGeN's metadata curation team; Darian Hole; Elsie Grudeski; Gary Van Domselaar; George Zahariadis; Grace Seo; Jennifer Tanner; Kerri Smith; Kirsten Biggar; Laura Gilbert; Madison Chapel; Morag Graham; Natalie Knox; Nathalie Bastien; Philip Mabon; Public Health Agency of Canada CanCOGeN team; Rhiannon Huzarewich; Robert Needle; Russell Mandes; Shari Tyson; Timothy Booth; Yan Li; Yang Yu |
| EPI_ISL_5650291, EPI_ISL_5650292, EPI_ISL_5650293, EPI_ISL_5650294, EPI_ISL_5650295, EPI_ISL_5650296, EPI_ISL_5650297, EPI_ISL_5650298, EPI_ISL_5650299, EPI_ISL_5650300, EPI_ISL_5650301, EPI_ISL_5650302, EPI_ISL_5650303, EPI_ISL_5650304, EPI_ISL_5650305, EPI_ISL_5650306, EPI_ISL_5650307, EPI_ISL_5650308, EPI_ISL_5650309, EPI_ISL_5650310, EPI_ISL_5650311 | | | |
| see above | NUCLEO DE SAUDE VILA FALCAO DE BAURU | Instituto Butantan | Antonio Jorge Martins; Claudia Renata dos Santos Barros; David Schlesinger; Debora Botequio Moretti; Dimas Tadeu Covas; Elaine Cristina Marqueze; Elaine Vieira Santos; Evandra Strazza Rodrigues; Heidge Fukumasu; Jayme Augusto de Souza-Neto; José Salvatore Leister Patané; Luiz Alcantara; Luiz Lehmann Coutinho; Maria Carolina Elias; Maurício Lacerda Nogueira; Rafael dos Santos Bezerra; Raul Machado Neto; Rejane Maria Tommasini Grotto; Ricardo Haddad; Sandra Coccuzzo Sampaio Vessoni; Simone Kashima; Svetoslav Nanev Slavov; Vincent Louis Viala |
| EPI_ISL_1790047 | NYU Langone Health | Departments of Pathology and Medicine, New York University School of Medicine | Adriana Heguy; Christian Marier; Dacia Dimartino; Emily Guzman; Gael Westby; Guiqing Wang; Paolo Cotzia; Paul Zappile; Peter Meyn; Sitharam Ramaswami; Yutong Zhang |
| EPI_ISL_1969931 | National Institute of Infectious Diseases-Prof. Dr. Matei Bals Molecular Diagnostics Laboratory | National Institute of Infectious Diseases-Prof. Dr. Matei Bals Molecular Diagnostics Laboratory | Andreea Tudor; Corina Casangiu; Dan Otelea; Leontina Banica; Marius Surleac; Ovidiu Vlaicu; Simona Paraschiv |
| EPI_ISL_1665203, EPI_ISL_1665222, EPI_ISL_1665223, EPI_ISL_1665233, EPI_ISL_1665234, EPI_ISL_1801772, EPI_ISL_1810942 | | | |
| see above | National Platform bis UMONS/Jolimont | National Platform bis UMONS/Jolimont | Florian Juszczak; François Dufrasne; Gautier Detry; Guillaume Bayon-Vicente; Ruddy Wattiez |
| EPI_ISL_1914659, EPI_ISL_1914660 | National Public Health Laboratory, National Centre for Infectious Diseases | National Public Health Laboratory, National Centre for Infectious Diseases | Grace Jie Yin Ngan; Lin Cui; Raymond Tzer Pin Lin; Royce Ang; Tze Minn Mak; Zhenyang Zhou |
| EPI_ISL_1785271 | National Virus Reference Laboratory | National Virus Reference Laboratory | Charlene Bennett; Cillian F De Gascun; Gabriel Gonzalez; Jonathan Dean; Michael Carr; Zoe Yandle |
| EPI_ISL_1738803, EPI_ISL_6340244 | Nebraska Public Health Laboratory | NPHL COVID-19 Response Team | NPHL COVID-19 Response Team |
| EPI_ISL_1939412 | North Dakota Department of Health, Public Health Laboratory | North Dakota Department of Health, Public Health Laboratory | Lisa Wingerter |
| EPI_ISL_1822833, EPI_ISL_1822838, EPI_ISL_1822873, EPI_ISL_1822965, EPI_ISL_1823021, EPI_ISL_1823048, EPI_ISL_1823087, EPI_ISL_1925978, EPI_ISL_1926003, EPI_ISL_1926006, EPI_ISL_1926041, EPI_ISL_1926046, EPI_ISL_1926057, EPI_ISL_1926065, EPI_ISL_1926085 | | | |
| see above | Northwest Laboratories | Seattle Flu Study | Amanda Adler; Barry R. Lutz; Benjamin Pelle; Brian Hiatt; Caitlin R. Wolf; Chris D. Frazar; Deborah A. Nickerson; Elisabeth Brandstetter; Erica Ryke; Geoff Melly; Helen Y. Chu; Janet A. Englund; Jay Shendure; Jover Lee; Kairsten Fay; Kirsten Lacombe; Lea M. Starita; Mark J. Rieder; Matthew Richardson; Matthew Thompson; Melissa Truong; Michael Boeckh; Michael Famulare; Misja Ilcisin; Peter D. Han; Philip Dykema; Romesh Gautom; Scott Lindquist; Thomas R. Sibley; Trevor Bedford |
| EPI_ISL_2009315, EPI_ISL_2009316, EPI_ISL_2009318, EPI_ISL_2009330 | Northwestern Memorial Hospital | Northwestern University - Ozer Lab | Chad J. Achenbach; Chao Qi; Egon A. Ozer; Judd F. Hultquist; Lacy M. Simons; Lawrence J. Jennings; Michael G. Ison; Ramon Lorenzo-Redondo; Taylor J. Dean |
| EPI_ISL_2080748, EPI_ISL_2080749, EPI_ISL_2080750, EPI_ISL_2080751, EPI_ISL_2080752, EPI_ISL_2080753, EPI_ISL_2080754, EPI_ISL_2080755, EPI_ISL_2080756, EPI_ISL_2080757, EPI_ISL_2080758, EPI_ISL_2080759, EPI_ISL_2080760, EPI_ISL_2080761, EPI_ISL_2080762, EPI_ISL_2227676, EPI_ISL_2227677, EPI_ISL_2227678, EPI_ISL_2227679, EPI_ISL_2227680, EPI_ISL_2227681 | | | |
| see above | OHSU Lab Services Molecular Microbiology Lab | Oregon SARS-CoV-2 Genome Sequencing Center | Alec J. Hirsch; Andrew C. Adey; Benjamin N. Bimber; Brendan L. O'Connell; Brian J. O'Roak; Cierra LaBlanc; Daniel N. Streblow; Donna Hansel; Guang Fan; Kayla Carter; Ruth V. Nichols; Sally Grindstaff; Sonia Acharya; William B. Messer; Xuan Qin |
| EPI_ISL_1841221 | OSPEDALE CIVILE SANT'OMERO - PRONTO SOCCORSO | Istituto Zooprofilattico Sperimentale dell'Abruzzo e Molise "G. Caporale" | Ancora M; Calistri P; Cammà C; Caporale M; Curini V; Delli Compagni E; Di Domenico M; Di Lollo Valeria; Di Pasquale A; Lorusso A; Mangone I; Marcacci M; Puglia I; Rinaldi A; Savini G; Scialabba S |
| EPI_ISL_2321652, EPI_ISL_2321653 | One Health | Universidad Nacional de Colombia - Laboratorio Genómico One Health | Andres F. Cardona-Rios; Carlos Franco-Muñoz; Daniel O. Maldonado-Perez; Diego A. Álvarez-Díaz; Hector Alejandro Ruiz-Moreno; Jorge E. Osorio; Juan P. Hernandez-Ortiz; Karl A Ciuoderis; Katherine Laiton-Donato; Laura Silvana Perez; Lina M. Hurtado; Marcela Mercado-Reyes; Maria Stella López; Monica Palma-Cuero; Simón Villegas Velásquez |
| EPI_ISL_7276572, EPI_ISL_7276630, EPI_ISL_7276664, EPI_ISL_7276743, EPI_ISL_7276841 | Ontario's COVID-19 Genomics Rapid Response Coalition | McMaster University | Ahmed Draia; Allison McGeer; Andrew G. McArthur; Angel Li; Emily Panousis; Hooman Derakhshani; Jalees Nasir; Kuganya Nirmalarajah; Michael Surette; Patryk Aftanas; Samira Mubareka; Sheridan Baker |
| EPI_ISL_1964166, EPI_ISL_1964167 | Oregon State Public Health Laboratory | Oregon State Public Health Laboratory | Eugene Yeboah; John Fontana and Shane Sevey; Laura Tsaknaridis; Rafia Razzaque; Vanda Makris |
| EPI_ISL_1730802 | Originating lab: Wales Specialist Virology Centre Sequencing lab: Pathogen Genomics Unit | Public Health Wales Microbiology Cardiff Wales Specialist Virology Centre | Alec Birchley; Alexander Adams; Amy Gaskin; Angela Marchbank; Bree Gatica-Wilcox; Catherine Moore; Jason Coombes; Joanne Watkins; Joel Southgate; Johnathan Evans; Laura Gifford; Lauren Gilbert; Lee Graham; Malorie Perry; Matthew Bull; Nicole Pacchiarini; Sally Corden; Sara Kumziene-Summerhayes; Sara Rey; Sarah Taylor; Simon Cottrell; Sophie Jones; Tom Connor |
| EPI_ISL_1856986, EPI_ISL_1856987, EPI_ISL_1856989, EPI_ISL_1856990, EPI_ISL_2293328, EPI_ISL_2293329, EPI_ISL_2293330, EPI_ISL_2293332, EPI_ISL_2293334, EPI_ISL_2293335, EPI_ISL_2293336, EPI_ISL_2293337, EPI_ISL_2293338, EPI_ISL_2293340, EPI_ISL_2293341, EPI_ISL_2293342, EPI_ISL_2293343, EPI_ISL_2293345, EPI_ISL_2293346, EPI_ISL_2293347, EPI_ISL_2293348, EPI_ISL_2293352, EPI_ISL_2293354, EPI_ISL_2293355 | | | |
| see above | Outre mer | National Reference Center for Viruses of Respiratory Infections, Institut Pasteur, Paris | Angela Brisebarre; Camille Capel; Christophe Malabat; Corinne Maufrais; Dominique Rousset; Etienne Simon-Lorière; Frédéric Lemoine; Louise Lefrançois; Marion Barbet; Maud Vanpeene; Méline Bizard; Pierre Lechat; Sylvie Behillil; Sylvie Van der Werf; Vincent Enouf |
| EPI_ISL_5603260 | POSTO DE SAUDE DE QUIXERE | Analytical Competence Molecular Epidemiology Lab/ACME, Oswaldo Cruz Foundation, Ceara (FIOCRUZ CE) | Carlos Leonardo de Aragao Araujo; Cecilia Leite Costa & Eduardo Ruback dos Santos on behalf of COVID-19 FIOCRUZ Genomic Network; Cleber Furtado Aksenen; Fabio Miyajima; Fernando Braga Stehling; Francisco Eder de Moura Lopes; Igor Oliveira Duarte; Jamille Maria Mendes Bezerra; Joaquim Cesar do Nascimento Sousa Junior; Pedro Miguel Carneiro Jeronimo; Suzana Porto Almeida; Thais Ferreira de Oliveira; Thais de Oliveira Costa; Ticiane Cavalcante de Souza; Veridiana Pessoa Miyajima |
| EPI_ISL_2344557 | POSTO MEDICO NATAL DIEGUES DE ESTIVA GERBI | Instituto Butantan / FZEA-USP-Pirassununga | Antonio Jorge Martins; Claudia Renata dos Santos Barros; David Schlesinger; Debora Botequio Moretti; Dimas Tadeu Covas; Elaine Cristina Marqueze; Elaine Vieira Santos; Evandra Strazza Rodrigues; Heidge Fukumasu; Jayme Augusto de Souza-Neto; José Salvatore Leister Patané; Luiz Alcantara; Luiz Lehmann Coutinho; Maria Carolina Elias; Maurício Lacerda Nogueira; Rafael dos Santos Bezerra; Raul Machado Neto; Rejane Maria Tommasini Grotto; Ricardo Haddad; Sandra Coccuzzo Sampaio Vessoni; Simone Kashima; Svetoslav Nanev Slavov; Vincent Louis Viala |
| EPI_ISL_1966727 | PPA MARCIO VALDEVINO BATISTA | Instituto Butantan | Antonio Jorge Martins; Bianca Cechetto Carlos. Mendelics: Bibiana Santos; Claudia Renata dos Santos Barros; Cíntia Bittar; David Schlesinger. Hemocentro Ribeirão Preto: Simone Kashima; Debora Botequio Moretti; Elaine Cristina Marqueze; Elaine Vieira dos Santos; Elisangela Chicaroni Mattos; Erika Freitas; Evandra Strazza Rodrigues; Felipe Allan da Silva da Costa; Flavia Aburjaile; Fábio Sossai Possebon; Guilherme Campos; Guilherme Targino Valente; Heidge Fukumasu. USP-Botucatu: Rejane Maria Tommasini Grotto; Helena Lage Ferreira; Instituto Butantan: Dimas Tadeu Covas; Jardelina de Souza Todao Bernardino; Jayme A. Souza-Neto; Jessika Cristina Chagas Lesbon; Jorge A. Petroli Marchesi; José Salvatore Leister Patané; João Paulo Kitajima; João Pessoa Araújo Jr.; Leila Sabrina Ullmann; Loyze Paola Oliveira de Lima; Luiz Aurelio de Campos Crispin. Centro de Genômica Funcional da ESALQ: Luiz Lehmann Coutinho; Luiz Carlos Junior de Alcantara; Lívia Sacchetto; Maisa C. Pereira Parra; Maria Carolina Elias; Marta Giovanetti; Marília Moraes; Maurício Lacerda Nogueira. Prefeitura de Sao Paulo: Melissa Palmieri.; Patricia Akemi Assato; Paula Rahal; Paulo Inacio da Costa; Rafael dos Santos Bezerra; Raquel de Lello |

| | | | |
|---|---|---|---|
| | | | Rocha Campos Cassano. NGS Soluções Genômicas: Pilar Drummond Sampaio Corrêa Mariani. FZEA-USP Pirassununga: Mirele Daiana Poleti; Raul Machado Neto; Ricardo Augusto Brassaloti; Ricardo Haddad; Rodrigo Tocantins Calado. FAMERP-SJRP: Cecília Artico Banho; Sandra Coccuzzo Sampaio; Svetoslav Nanev Slavov; Vagner Fonseca; Vincent Louis Viala |
| EPI_ISL_3354801, EPI_ISL_3354802 | PR Public Health Lab | Centers for Disease Control and Prevention Division of Viral Diseases, Pathogen Discovery | Alex Burgin; Ben Rambo-Martin; Clinton Paden; Dakota Howard; Dave Wentworth; Dhwani Batra; Jasmine Padilla; Justin Lee; Krista Queen; Kristen Knipe; Kristine Lacek; Mark Burroughs; Matthew Schmerer; Meghan Bentz; Mili Sheth; Peter Cook; Sam Shepard; Sarah Nobles; Suxiang Tong; Vivien Dugan; Yvette Unoarumhi |
| EPI_ISL_2345742 | PRONTO ATENDIMENTO MUNICIPAL ITALO SANTUCCI | Instituto Butantan / Mendelics | Antonio Jorge Martins; Claudia Renata dos Santos Barros; David Schlesinger; Debora Botequio Moretti; Dimas Tadeu Covas; Elaine Cristina Marqueze; Elaine Vieira Santos; Evandra Strazza Rodrigues; Heidge Fukumasu; Jayme Augusto de Souza-Neto; José Salvatore Leister Patané; Luiz Alcantara; Luiz Lehmann Coutinho; Maria Carolina Elias; Maurício Lacerda Nogueira; Rafael dos Santos Bezerra; Raul Machado Neto; Rejane Maria Tommasini Grotto; Ricardo Haddad; Sandra Coccuzzo Sampaio Vessoni; Simone Kashima; Svetoslav Nanev Slavov; Vincent Louis Viala |
| EPI_ISL_1966735, EPI_ISL_1966736 | PRONTO SOCORRO PESTANA P S ANDRE SACCO | Instituto Butantan | Antonio Jorge Martins; Bianca Cechetto Carlos. Mendelics: Bibiana Santos; Claudia Renata dos Santos Barros; Cíntia Bittar; David Schlesinger. Hemocentro Ribeirão Preto: Simone Kashima; Debora Botequio Moretti; Elaine Cristina Marqueze; Elaine Vieira dos Santos; Elisangela Chicaroni Mattos; Erika Freitas; Evandra Strazza Rodrigues; Felipe Allan da Silva da Costa; Flavia Aburjaile; Fábio Sossai Possebon; Guilherme Campos; Guilherme Targino Valente; Heidge Fukumasu. USP-Botucatu: Rejane Maria Tommasini Grotto; Helena Lage Ferreira; Instituto Butantan: Dimas Tadeu Covas; Jardelina de Souza Todao Bernardino; Jayme A. Souza-Neto; Jessika Cristina Chagas Lesbon; Jorge A. Petroli Marchesi; José Salvatore Leister Patané; João Paulo Kitajima; João Pessoa Araújo Jr.; Leila Sabrina Ullmann; Loyze Paola Oliveira de Lima; Luiz Aurelio de Campos Crispin. Centro de Genômica Funcional da ESALQ: Luiz Lehmann Coutinho; Luiz Carlos Junior de Alcantara; Lívia Sacchetto; Maisa C. Pereira Parra; Maria Carolina Elias; Marta Giovanetti; Marília Moraes; Maurício Lacerda Nogueira. Prefeitura de Sao Paulo: Melissa Palmieri.; Patricia Akemi Assato; Paula Rahal; Paulo Inacio da Costa; Rafael dos Santos Bezerra; Raquel de Lello Rocha Campos Cassano. NGS Soluções Genômicas: Pilar Drummond Sampaio Corrêa Mariani. FZEA-USP Pirassununga: Mirele Daiana Poleti; Raul Machado Neto; Ricardo Augusto Brassaloti; Ricardo Haddad; Rodrigo Tocantins Calado. FAMERP-SJRP: Cecília Artico Banho; Sandra Coccuzzo Sampaio; Svetoslav Nanev Slavov; Vagner Fonseca; Vincent Louis Viala |
| EPI_ISL_2614579 | PS Municipal Dr Alceu Lot | Instituto Adolfo Lutz, Interdiciplinary Procedures Center, Strategic Laboratory | Caio Vinicius Dias Lopes; Claudia Regina Gonçalves; Claudio Tavares Sacchi; Erica Valessa Ramos Gomes; Karoline Rodrigues Campos; Leonardo Jose Tadeu de Araujo |
| EPI_ISL_5650258, EPI_ISL_5650259, EPI_ISL_5650262 | PSF NOVA EUROPA | Instituto Butantan | Antonio Jorge Martins; Claudia Renata dos Santos Barros; David Schlesinger; Debora Botequio Moretti; Dimas Tadeu Covas; Elaine Cristina Marqueze; Elaine Vieira Santos; Evandra Strazza Rodrigues; Heidge Fukumasu; Jayme Augusto de Souza-Neto; José Salvatore Leister Patané; Luiz Alcantara; Luiz Lehmann Coutinho; Maria Carolina Elias; Maurício Lacerda Nogueira; Rafael dos Santos Bezerra; Raul Machado Neto; Rejane Maria Tommasini Grotto; Ricardo Haddad; Sandra Coccuzzo Sampaio Vessoni; Simone Kashima; Svetoslav Nanev Slavov; Vincent Louis Viala |
| EPI_ISL_1633723, EPI_ISL_1633730, EPI_ISL_1633778, EPI_ISL_1633810, EPI_ISL_1633834, EPI_ISL_1633841, EPI_ISL_1633842, EPI_ISL_1633860, EPI_ISL_1633867, EPI_ISL_1633869, EPI_ISL_1633877, EPI_ISL_1633901, EPI_ISL_1633981, EPI_ISL_1634062, EPI_ISL_1634087, EPI_ISL_1634242, EPI_ISL_1634254, EPI_ISL_1634290, EPI_ISL_1634309, EPI_ISL_1634325, EPI_ISL_1635917, EPI_ISL_1635954, EPI_ISL_1636080, EPI_ISL_1636248, EPI_ISL_1636344, EPI_ISL_1636349, EPI_ISL_1636382, EPI_ISL_1636384, EPI_ISL_1717111, EPI_ISL_1717112, EPI_ISL_1717136, EPI_ISL_1717153, EPI_ISL_1717159, EPI_ISL_1717196, EPI_ISL_1717222, EPI_ISL_1717244, EPI_ISL_1717300, EPI_ISL_1717309, EPI_ISL_1717399, EPI_ISL_1717452, EPI_ISL_1717467, EPI_ISL_1717478, EPI_ISL_1717482, EPI_ISL_1717500, EPI_ISL_1717530, EPI_ISL_1717644, EPI_ISL_1717672 | | | |
| see above | Pandemic Response Lab - NYC | Pandemic Response Lab, R&D | Cybill del Castillo; Dylan Law; Haiping Hao; Henry Lee; Jon Laurent; Katharine Nelson; Melissa Hopkins; Michael Hammerling; Pradeep Bugga; Shinyoung Clair Kang; Sol Rey; William Ward |
| EPI_ISL_2993480 | Pathology, UAB | Pathology, UAB | E.J.; Hendrickson; Hernandez, J.; Leal; Lefkowitz; Moates, D.; R.C.; S.M. |
| EPI_ISL_3030296 | Pesaro | Microbiology University Politecnica delle Marche | Anna Valenza; Carla Acciarri; Katia Marinelli; Monica Lucia Ferreri; Patrizia Bagnarelli; Roberta Longo; Sara Caucci; Stefano Menzo |
| EPI_ISL_2663309, EPI_ISL_2663310, EPI_ISL_6907362, EPI_ISL_6907848, EPI_ISL_6907853, EPI_ISL_6907858, EPI_ISL_6907864, EPI_ISL_6907870, EPI_ISL_6907881, EPI_ISL_6907882, EPI_ISL_6907887, EPI_ISL_6908419, EPI_ISL_6908420, EPI_ISL_6908427, EPI_ISL_6908431, EPI_ISL_6908752, EPI_ISL_6908753, EPI_ISL_6908757, EPI_ISL_6908760, EPI_ISL_6908766, EPI_ISL_6908767, EPI_ISL_6908783 | | | |
| see above | Plataforma de Vigilancia Molecular (PVM) - FIOCRUZ/BA | Plataforma de Vigilancia Molecular (PVM) - FIOCRUZ/BA | Bruno Bezerril Andrade; Camila I. de Oliveira; Camila I. de Oliveira on behalf of the Fiocruz COVID-19 Genomic Surveillance Network.; Clarissa Araújo Gurgel; Eduardo Oyama; Francys A Rangel; Laila A da Silva; Laise de Moraes; Leonardo Paiva Farias; Marina Cucco; Pedro Silveira; Ricardo Khouri; Ricardo Khouri on behalf of the Fiocruz COVID-19 Genomic Surveillance Network.; Tiago Graf |
| EPI_ISL_2629157, EPI_ISL_2629160, EPI_ISL_2629161, EPI_ISL_2629162 | Plateforme Covid IDF, site Broussais | HEGP - Laboratoire de Virologie | David Veyer; Hélène Péré; Julien Puech; Maxime Wack; Nabil Gastli; Nicolas Robillard; Sandrine Imbeaud |
| EPI_ISL_1670617, EPI_ISL_1670621, EPI_ISL_1670623 | Plateforme de testing Namuroise | Plateforme de testing Namuroise | Céline Maschietto; Degosserie Jonathan; Denis Olivier; Mullier François; Otto Gaetan |
| EPI_ISL_1839887 | Plateforme de testing namuroise | Plateforme de testing namuroise | Céline Maschietto; Degosserie Jonathan; Denis Olivier; Mullier François; Otto Gaetan |
| EPI_ISL_1661192, EPI_ISL_1661211, EPI_ISL_1661222, EPI_ISL_1661235, EPI_ISL_1661247, EPI_ISL_1697302, EPI_ISL_1697312, EPI_ISL_1697338, EPI_ISL_1745063 | | | |
| see above | Platform BIS UZA/UAntwerpen | Labo Klinische Biologie, UZA | Basil Britto Xavier; Christine Lammens; Herman Goossens; Jasmine Coppens; Marie Le Mercier; Veerle Matheeussen |
| EPI_ISL_2259232 | Pomona Valley Hospital Medical Center | Los Angeles County Public Health Laboratories | P. Hemarajata et al. |
| EPI_ISL_5782465 | Posto Medico Natal Diegues De Estiva Gerbi | Instituto Butantan | Antonio Jorge Martins; Claudia Renata dos Santos Barros; David Schlesinger; Debora Botequio Moretti; Dimas Tadeu Covas; Elaine Cristina Marqueze; Elaine Vieira Santos; Evandra Strazza Rodrigues; Heidge Fukumasu; Jayme Augusto de Souza-Neto; José Salvatore Leister Patané; Luiz Alcantara; Luiz Lehmann Coutinho; Maria Carolina Elias; Maurício Lacerda Nogueira; Rafael dos Santos Bezerra; Raul Machado Neto; Rejane Maria Tommasini Grotto; Ricardo Haddad; Sandra Coccuzzo Sampaio Vessoni; Simone Kashima; Svetoslav Nanev Slavov; Vincent Louis Viala |
| EPI_ISL_5800413 | Ppa Marcio Valdevino Batista | Instituto Butantan | Antonio Jorge Martins; Claudia Renata dos Santos Barros; David Schlesinger; Debora Botequio Moretti; Dimas Tadeu Covas; Elaine Cristina Marqueze; Elaine Vieira Santos; Evandra Strazza Rodrigues; Heidge Fukumasu; Jayme Augusto de Souza-Neto; José Salvatore Leister Patané; Luiz Alcantara; Luiz Lehmann Coutinho; Maria Carolina Elias; Maurício Lacerda Nogueira; Rafael dos Santos Bezerra; Raul Machado Neto; Rejane Maria Tommasini Grotto; Ricardo Haddad; Sandra Coccuzzo Sampaio Vessoni; Simone Kashima; Svetoslav Nanev Slavov; Vincent Louis Viala |
| EPI_ISL_2344593, EPI_ISL_2344594, EPI_ISL_2344595, EPI_ISL_2344596, EPI_ISL_2344597, EPI_ISL_2344598, EPI_ISL_2344599, EPI_ISL_2344600, EPI_ISL_2344601, EPI_ISL_2344602, EPI_ISL_2344603, EPI_ISL_2344604, EPI_ISL_2344605, EPI_ISL_2344606, EPI_ISL_2344607, EPI_ISL_2344608, EPI_ISL_2344609, EPI_ISL_2344610, EPI_ISL_2344611, EPI_ISL_2344612, EPI_ISL_2344613, EPI_ISL_2344614, EPI_ISL_2344615, EPI_ISL_2344616, EPI_ISL_2344617, EPI_ISL_2344618, EPI_ISL_2344619, EPI_ISL_2344620, EPI_ISL_2344622, EPI_ISL_2344623, EPI_ISL_2344625, EPI_ISL_2344626, EPI_ISL_2344627, EPI_ISL_2344628, EPI_ISL_2344629, EPI_ISL_2344630, EPI_ISL_2344631, EPI_ISL_2344632, EPI_ISL_2344633, EPI_ISL_2344634, EPI_ISL_2344635, EPI_ISL_2344636, EPI_ISL_2344637, EPI_ISL_2344638, EPI_ISL_2344639, EPI_ISL_2344640, EPI_ISL_2344641, EPI_ISL_2344642, EPI_ISL_2344643, EPI_ISL_2344644, EPI_ISL_2344645, EPI_ISL_2344646, EPI_ISL_2344647, EPI_ISL_2344648, EPI_ISL_2344649, EPI_ISL_2344650, EPI_ISL_2344651, EPI_ISL_2344652, EPI_ISL_2344653, EPI_ISL_2344654, EPI_ISL_2344655, EPI_ISL_2346083, EPI_ISL_2346084, EPI_ISL_2346085, EPI_ISL_2346086, EPI_ISL_2346087, EPI_ISL_2346088, EPI_ISL_2346089, EPI_ISL_2346090, EPI_ISL_2346091, EPI_ISL_2346092, EPI_ISL_2346093 | | | |
| see above | Prefeirtura de SP | Instituto Butantan | Antonio Jorge Martins; Claudia Renata dos Santos Barros; David Schlesinger; Debora Botequio Moretti; Dimas Tadeu Covas; Elaine Cristina Marqueze; Elaine Vieira Santos; Evandra Strazza Rodrigues; Heidge Fukumasu; Jayme Augusto de Souza-Neto; José Salvatore Leister Patané; Luiz Alcantara; Luiz Lehmann Coutinho; Maria Carolina Elias; Maurício Lacerda Nogueira; Rafael dos Santos Bezerra; Raul Machado Neto; Rejane Maria Tommasini Grotto; Ricardo Haddad; Sandra Coccuzzo Sampaio Vessoni; Simone Kashima; Svetoslav Nanev Slavov; Vincent Louis Viala |
| EPI_ISL_2483729 | Presidio Ospedaliero G. B. Grassi ASL RM3 | INMI Lazzaro Spallanzani IRCCS | A Di Caro; B Bartolini; CEM Gruber; E Giombini; E Ristori; F Messina; F Santini; F Tabacco; G Bonfiglio; M Rueca; MR Capobianchi; O Butera |
| EPI_ISL_2437996 | Prime Diagnostics | Universidad Nacional de Colombia - Laboratorio Genómico One Health | Andres F. Cardona-Rios; Carlos Franco-Muñoz; Carolina Muñoz-Arango; Celeny Ortiz; Daniel O. Maldonado-Perez; Diego A. Álvarez-Díaz; Hector Alejandro Ruiz-Moreno; Idabely Betancur Ortiz; Jorge E. Osorio; Juan P. Hernandez-Ortiz; Karl A Ciuoderis; Katherine Laiton-Donato; Laura Silvana Perez; Lina M. Hurtado; Marcela Mercado-Reyes; Maria Angélica Maya; Maria Stella López; Rita Almanza Payares; Sandra Ines Cano; Simón Villegas Velásquez |
| EPI_ISL_7406413 | Programa Laboratorio de Salud Pública "Dr Dalmiro Pérez Laborda" | Área de Secuenciación del Laboratorio de Virología del Hospital de Niños Dr. Ricardo Gutierrez on behalf of 'Proyecto Argentino Interinstitucional de genomica de SARS-CoV-2' (PAIS Consortium) | A; AM; Acuña; Aguilera; BH; Bohn; Cabral Bombardieri; Campos; Carrizo; D; E; Esteves; F; G; Goya; J; JM; JP; Jofré; L; LE; Lacaze; Lusso; M; MA; MI; MS; Mastrodonato; Molina; Nabaes Jodar; Natale; Olivera; Perez Diaz; Peñalva; Quijano; Rivero; Rosales; S; Talia; Valinotto; Viegas, M. |
| EPI_ISL_5802427 | Pronto Atendimento Municipal Italo Santucci | Instituto Butantan | Antonio Jorge Martins; Claudia Renata dos Santos Barros; David Schlesinger; Debora Botequio Moretti; Dimas Tadeu Covas; Elaine Cristina Marqueze; Elaine Vieira Santos; Evandra Strazza Rodrigues; Heidge Fukumasu; Jayme Augusto de Souza-Neto; José Salvatore Leister Patané; Luiz Alcantara; Luiz Lehmann Coutinho; Maria Carolina Elias; Maurício Lacerda Nogueira; Rafael dos Santos Bezerra; Raul Machado Neto; Rejane Maria Tommasini Grotto; Ricardo Haddad; Sandra Coccuzzo Sampaio Vessoni; Simone Kashima; Svetoslav Nanev Slavov; Vincent Louis Viala |
| EPI_ISL_2841606 | Pronto Atendimento de Vargem Grande Paulista | Instituto Adolfo Lutz, Interdiciplinary Procedures Center, Strategic Laboratory | Caio Vinicius Dias Lopes; Claudia Regina Gonçalves; Claudio Tavares Sacchi; Erica Valessa Ramos Gomes; Karoline Rodrigues Campos |
| EPI_ISL_2614573 | Pronto Socorro Andre Sacco - Pestana | Instituto Adolfo Lutz, Interdiciplinary Procedures Center, Strategic Laboratory | Caio Vinicius Dias Lopes; Claudia Regina Gonçalves; Claudio Tavares Sacchi; Erica Valessa Ramos Gomes; Karoline Rodrigues Campos; Leonardo Jose Tadeu de Araujo |
| EPI_ISL_5802407 | Pronto Socorro Caucaia Do Alto | Instituto Butantan | Antonio Jorge Martins; Claudia Renata dos Santos Barros; David Schlesinger; Debora Botequio Moretti; Dimas Tadeu Covas; Elaine Cristina Marqueze; Elaine Vieira Santos; Evandra Strazza Rodrigues; Heidge Fukumasu; Jayme Augusto de Souza-Neto; José Salvatore Leister Patané; Luiz Alcantara; Luiz Lehmann Coutinho; Maria Carolina Elias; Maurício Lacerda Nogueira; Rafael dos Santos Bezerra; Raul Machado Neto; Rejane Maria Tommasini Grotto; Ricardo Haddad; Sandra Coccuzzo Sampaio Vessoni; Simone Kashima; Svetoslav Nanev Slavov; Vincent Louis Viala |
| EPI_ISL_5800449, EPI_ISL_5800461 | Pronto Socorro Pestana P S Andre Sacco | Instituto Butantan | Antonio Jorge Martins; Claudia Renata dos Santos Barros; David Schlesinger; Debora Botequio Moretti; Dimas Tadeu Covas; Elaine Cristina Marqueze; Elaine Vieira Santos; Evandra Strazza Rodrigues; Heidge Fukumasu; Jayme Augusto de Souza-Neto; José Salvatore Leister Patané; Luiz Alcantara; Luiz Lehmann Coutinho; Maria Carolina Elias; Maurício Lacerda Nogueira; Rafael dos Santos Bezerra; Raul Machado Neto; Rejane Maria Tommasini Grotto; Ricardo Haddad; Sandra Coccuzzo Sampaio Vessoni; Simone Kashima; Svetoslav Nanev Slavov; Vincent Louis Viala |
| EPI_ISL_2551396, EPI_ISL_2551410, EPI_ISL_2551412 | Providence Regional Medical Center Everett | Providence St. Joseph Health Molecular Genomics Laboratory | Alexa K Dowdell; Brian D Piening; Carlo B Bifulco; Fred L Robinson; Mary Campbell; Rogan Rattray |
| EPI_ISL_2295677, EPI_ISL_2295734, EPI_ISL_2698785, EPI_ISL_2698795 | Public Health Laboratory, Minnesota Department of Health | University of Minnesota Genomics Center | Corbin Dirkx; Daryl M. Gohl; Jaquelyn Kuriger-Laber; John Garbe; and Sean Wang |
| EPI_ISL_1804815, EPI_ISL_1804816, EPI_ISL_1818051, EPI_ISL_1818052, EPI_ISL_1818053, EPI_ISL_1818054, EPI_ISL_1818056, EPI_ISL_1818059, EPI_ISL_1818062, EPI_ISL_1818065, EPI_ISL_1818070, EPI_ISL_1900043, EPI_ISL_1900048, EPI_ISL_1900059, EPI_ISL_1900062, EPI_ISL_1900070, EPI_ISL_1900074, EPI_ISL_1900082, EPI_ISL_1900092, EPI_ISL_1900100, EPI_ISL_1900102, EPI_ISL_1900105, EPI_ISL_1900128, EPI_ISL_1900134, EPI_ISL_1900142, EPI_ISL_1900144, EPI_ISL_1900152, EPI_ISL_1900155, EPI_ISL_1900425, EPI_ISL_1900426, EPI_ISL_1900427, EPI_ISL_1900428, EPI_ISL_1900430, EPI_ISL_1900432, EPI_ISL_1900433, EPI_ISL_1900434, EPI_ISL_1900435, EPI_ISL_1900437, EPI_ISL_1900438, EPI_ISL_1900439, EPI_ISL_1900440, EPI_ISL_1900441, EPI_ISL_1900442, EPI_ISL_1900443, EPI_ISL_1900444, EPI_ISL_1900445, EPI_ISL_1900446, EPI_ISL_1900447, EPI_ISL_1900448, EPI_ISL_1900449, EPI_ISL_1900450, EPI_ISL_1900451, EPI_ISL_1900452, EPI_ISL_1900453, EPI_ISL_1900454, EPI_ISL_1900455, EPI_ISL_1900456, EPI_ISL_1900457, EPI_ISL_1900459, EPI_ISL_1900460, EPI_ISL_1900461, EPI_ISL_1900462, EPI_ISL_1900463, EPI_ISL_1900465, EPI_ISL_1935556, EPI_ISL_1935557, EPI_ISL_1935558, EPI_ISL_1935559, EPI_ISL_1935561, EPI_ISL_1935562, EPI_ISL_1935563, EPI_ISL_1935564, EPI_ISL_1935565, EPI_ISL_1935567, EPI_ISL_1935568, EPI_ISL_1935570, EPI_ISL_1935571, EPI_ISL_1935573, EPI_ISL_1935576, EPI_ISL_1935582, EPI_ISL_1942169, EPI_ISL_1942170, EPI_ISL_1942171, EPI_ISL_1942172, EPI_ISL_1942173, EPI_ISL_1942174, EPI_ISL_1942176, EPI_ISL_1942177, EPI_ISL_1942178, EPI_ISL_1942179, EPI_ISL_1942181, EPI_ISL_1942182, EPI_ISL_1942183, EPI_ISL_1942190, EPI_ISL_1942193, EPI_ISL_1942195, EPI_ISL_1942196, EPI_ISL_1942197, EPI_ISL_1942200, EPI_ISL_1942203, EPI_ISL_1942204, EPI_ISL_1942205, EPI_ISL_1942206, EPI_ISL_1942207, EPI_ISL_1942208, EPI_ISL_1942213, EPI_ISL_1942215, EPI_ISL_1942216, EPI_ISL_1990052, EPI_ISL_2034716, EPI_ISL_2034751, EPI_ISL_2034811, EPI_ISL_2034817, EPI_ISL_2035308, EPI_ISL_2035327, EPI_ISL_2035333, EPI_ISL_2035339, EPI_ISL_2035341, EPI_ISL_2140759, EPI_ISL_2140761, EPI_ISL_2140762, EPI_ISL_2140764, EPI_ISL_2140765, EPI_ISL_2140770, EPI_ISL_2140771, EPI_ISL_2140772, EPI_ISL_2140773, EPI_ISL_2140776, EPI_ISL_2140793, EPI_ISL_2140794, EPI_ISL_4367959 | | | |
| see above | Quest Diagnostics Incorporated | Centers for Disease Control and Prevention Division of Viral Diseases, Pathogen Discovery | A. Gerasimova; A. Perez; Adrian Paskey; B. Anderson; Benjamin Rambo-Martin; Christopher Gulvick; Clinton Paden; Clinton R. Paden; Dakota Howard; Darlene Wagner; Dhwani Batra; Duncan MacCannell; Erisa Sula; F. Lacbawan; I. A. Shlyakhter; I. Shlyakhter; Jason Caravas; K. Livingston; K.E. Livingston; Kara Moser; Kristine Lacek; L. Bernstein; L.E. Bernstein; M. Hua; Matthew Schmerer; P. Tanpaiboon; Peter Cook; Peter W. Cook; R. Kagan; R. M. Kagan; R. Owen; R. Rolando; R. V. Rolando; S. H. Rosenthal; S. Rosenthal; Scott Sammons; Shatavia Morrison; Tymeckia Kendall; Victoria Caban Figueroa; Y. Liu; Yvette Unoarumhi |
| EPI_ISL_2018331, EPI_ISL_2018343 | RI State Health Laboratories | Centers for Disease Control and Prevention Division of Viral Diseases, Pathogen Discovery | Alison Laufer Halpin; Ben L. Rambo-Martin; Clinton R. Paden; Dakota Howard; Darlene Wagner; Dave Wentworth; Dhwani Batra; Jasmine Padilla; Justin Lee; Katie Dillon; Krista Queen; Kristen Knipe; Kristine Lacek; Mark Burroughs; Matthew Schmerer; Mili Sheth; Peter Cook; Sam Shepard; Sarah Nobles; Shoshona Le; Suxiang Tong; Vivien Dugan; Yvette Unoarumhi |
| EPI_ISL_2628503 | Rady Children's Hospital - San | Andersen lab at Scripps Research | Christina Clarke; David Dimmock; Denise Malicki; Kathryn Bouic; Linda Luo; SEARCH Alliance San Diego with Nanda Radamchar; Teresa Mueller |

| | | | |
|---|---|---|---|
| | Diego | | |
| EPI_ISL_2249655, EPI_ISL_2249983, EPI_ISL_2249984, EPI_ISL_2249985, EPI_ISL_2249986, EPI_ISL_2249987, EPI_ISL_2249988, EPI_ISL_2249989, EPI_ISL_2249990, EPI_ISL_2249991, EPI_ISL_2249992, EPI_ISL_2249993, EPI_ISL_2249994, EPI_ISL_2249995, EPI_ISL_2249996, EPI_ISL_2249997, EPI_ISL_2249998, EPI_ISL_2249999, EPI_ISL_2250000, EPI_ISL_2250001, EPI_ISL_2250002, EPI_ISL_2250003, EPI_ISL_2250004, EPI_ISL_2250005, EPI_ISL_2250006, EPI_ISL_2250007, EPI_ISL_2250008, EPI_ISL_2250009, EPI_ISL_2250010, EPI_ISL_2250011, EPI_ISL_2250012, EPI_ISL_2250013, EPI_ISL_2250014, EPI_ISL_2250015, EPI_ISL_2250016, EPI_ISL_2319816, EPI_ISL_2319818, EPI_ISL_2319833, EPI_ISL_2319834, EPI_ISL_2319835, EPI_ISL_2473840, EPI_ISL_2473863, EPI_ISL_2473868, EPI_ISL_2473872, EPI_ISL_2473874, EPI_ISL_2473875, EPI_ISL_2473889, EPI_ISL_2473944, EPI_ISL_2473977, EPI_ISL_2474075, EPI_ISL_2474142, EPI_ISL_2662115, EPI_ISL_2662116, EPI_ISL_2662117, EPI_ISL_2662118, EPI_ISL_2662119, EPI_ISL_2662120, EPI_ISL_2662121, EPI_ISL_2662122, EPI_ISL_2662123, EPI_ISL_2662124, EPI_ISL_2662125, EPI_ISL_2662126, EPI_ISL_2662127, EPI_ISL_2662128, EPI_ISL_2662129, EPI_ISL_2662130, EPI_ISL_2662131, EPI_ISL_2662132, EPI_ISL_2662133, EPI_ISL_2662134, EPI_ISL_2662135, EPI_ISL_2662136, EPI_ISL_2662137, EPI_ISL_2662138, EPI_ISL_2662139, EPI_ISL_2662140, EPI_ISL_2662141, EPI_ISL_2662142, EPI_ISL_2662143, EPI_ISL_2662144, EPI_ISL_2662145, EPI_ISL_2662146, EPI_ISL_2662147, EPI_ISL_2662148, EPI_ISL_2662149, EPI_ISL_2662150, EPI_ISL_2662151, EPI_ISL_2662152, EPI_ISL_2662153, EPI_ISL_2662154, EPI_ISL_2662155, EPI_ISL_2662156, EPI_ISL_2662157, EPI_ISL_2662158, EPI_ISL_2662159, EPI_ISL_2662160, EPI_ISL_2662161, EPI_ISL_2662162, EPI_ISL_2662163, EPI_ISL_2662164, EPI_ISL_2662165, EPI_ISL_2662166, EPI_ISL_2662167, EPI_ISL_2662168, EPI_ISL_2662169, EPI_ISL_2662170 | | | |
| see above | Reditus Laboratories | Reditus Laboratories | Alexa Eichelberger; Cassy Philips; Joshua J. Geltz; M.S.; Ph.D.; Rex Dyer; Robert M. Sgambelluri |
| EPI_ISL_2253163, EPI_ISL_2254416, EPI_ISL_2254417, EPI_ISL_2254419, EPI_ISL_2254469, EPI_ISL_2254473, EPI_ISL_2254475 | | | |
| see above | Respiratory Viruses Branch, Centers for Disease Control and Prevention | Respiratory Viruses Branch, Centers for Disease Control and Prevention | Anderson, B.; Batra, D.; Bernstein, L.; C.R.; Campbell, P.; Caravas, J.; Case, R.; Clark, C.; Cook; Gerasimova, A.; Ghorpade, V.; Gulvick, C.; Hardison, M.; Houdeshell, H.; Howard, D.; Hua, M.; I.A.; Kagan; Kvalvaag, O.; Lacbawan, F.; Liu, Y.; Livingston, K.; MacCannell, D.; Morrison, S.; Moser, K.; Nall, D.; Owen, R.; P.W.; Paden; Paskey, A.; Perez, A.; R.M.; R.V.; Rambo-Martin, B.; Rolando; Rosenthal; S.H.; Sammons, S.; Sanders, E.; Schmerer, M.; Shlyakhter; Tanpaiboon, P.; Unoarumhi, Y.; Vest, A.; Wagner, D.; Westlund, S. |
| EPI_ISL_1919142, EPI_ISL_1919143, EPI_ISL_1919149, EPI_ISL_1919151, EPI_ISL_1919186, EPI_ISL_1919194, EPI_ISL_1919224 | | | |
| see above | Rhode Island Department of Health | Infectious Disease Program, Broad Institute of Harvard and MIT | Adams, G.; Azevedo, K.; B.L.; B.W.; Bauer, M.; Birren; Carter, A.; Chaluvadi, S.; D.J.; DeRuff, K.; Gladden-Young, A.; Huard, R.; J.E.; K.J.; King, E.; Lagerborg, K.; Lemieux; Loreth, C.; Miller, A.; Normandin, E.; P.C.; Park; Pearlman, L.; Reilly, S.; Rudy, M.; Sabeti; Siddle; Tomkins-Tinch, C.; and MacInnis |
| EPI_ISL_2604833 | Rhode Island State Health Laboratories | Rhode Island State Health Laboratories | Ewa King; Kristin Carpenter-Azevedo; Richard C. Huard |
| EPI_ISL_2502625 | S.C. Patologia Clinica, Ospedale Sant'Andrea, ASL 5 | U.O. Igiene, Ospedale Policlinico San Martino | Battolla Enrico; Bruzzone Bianca; Caligiuri Patrizia; De Pace Vanessa; Domnich Alexander; Icardi Giancarlo; Orsi Andrea; Ricucci Valentina |
| EPI_ISL_2321587 | SAN VICENTE FUNDACION | Universidad Nacional de Colombia - Laboratorio Genómico One Health | Andres F. Cardona-Rios; Carlos Franco-Muñoz; Carolina Muñoz-Arango; Celeny Ortiz; Daniel O. Maldonado-Perez; Diego A. Álvarez-Díaz; Hector Alejandro Ruiz-Moreno; Idabely Betancur Ortiz; Jorge E. Osorio; Juan P. Hernandez-Ortiz; Karl A Ciuoderis; Katherine Laiton-Donato; Laura Silvana Perez; Lina M. Hurtado; Marcela Mercado-Reyes; Maria Angélica Maya; Maria Stella López; Rita Almanza Payares; Sandra Ines Cano; Simón Villegas Velásquez |
| EPI_ISL_5650245 | SANTA CASA DE BIRIGUI | Instituto Butantan | Antonio Jorge Martins; Claudia Renata dos Santos Barros; David Schlesinger; Debora Botequio Moretti; Dimas Tadeu Covas; Elaine Cristina Marqueze; Elaine Vieira Santos; Evandra Strazza Rodrigues; Heidge Fukumasu; Jayme Augusto de Souza-Neto; José Salvatore Leister Patané; Luiz Alcantara; Luiz Lehmann Coutinho; Maria Carolina Elias; Maurício Lacerda Nogueira; Rafael dos Santos Bezerra; Raul Machado Neto; Rejane Maria Tommasini Grotto; Ricardo Haddad; Sandra Coccuzzo Sampaio Vessoni; Simone Kashima; Svetoslav Nanev Slavov; Vincent Louis Viala |
| EPI_ISL_2345648 | SANTA CASA DE MISERICORDIA SAO JOSE | Instituto Butantan / Mendelics | Antonio Jorge Martins; Claudia Renata dos Santos Barros; David Schlesinger; Debora Botequio Moretti; Dimas Tadeu Covas; Elaine Cristina Marqueze; Elaine Vieira Santos; Evandra Strazza Rodrigues; Heidge Fukumasu; Jayme Augusto de Souza-Neto; José Salvatore Leister Patané; Luiz Alcantara; Luiz Lehmann Coutinho; Maria Carolina Elias; Maurício Lacerda Nogueira; Rafael dos Santos Bezerra; Raul Machado Neto; Rejane Maria Tommasini Grotto; Ricardo Haddad; Sandra Coccuzzo Sampaio Vessoni; Simone Kashima; Svetoslav Nanev Slavov; Vincent Louis Viala |
| EPI_ISL_2325490, EPI_ISL_2325492, EPI_ISL_2335174 | SARS-CoV-2 testing team, National Institute of Infectious Diseases | Pathogen Genomics Center, National Institute of Infectious Diseases | Hazuka Y Furihata; Kentaro Itokawa; Makoto Kuroda; Masanori Hashino; Masumichi Saito; Naomi Nojiri; Nozomu Hanaoka; Rina Tanaka; Sana Uchikoba; Tsuguto Fujimoto; Tsuyoshi Sekizuka |
| EPI_ISL_5650239, EPI_ISL_5650265, EPI_ISL_5650266, EPI_ISL_5650267, EPI_ISL_5650268, EPI_ISL_5650269, EPI_ISL_5650271, EPI_ISL_5650272, EPI_ISL_5650273, EPI_ISL_5650275, EPI_ISL_5650278, EPI_ISL_5650281, EPI_ISL_5650282 | | | |
| see above | SECAO CENTRO DE DIAGNOSTICO SECEDI | Instituto Butantan | Antonio Jorge Martins; Claudia Renata dos Santos Barros; David Schlesinger; Debora Botequio Moretti; Dimas Tadeu Covas; Elaine Cristina Marqueze; Elaine Vieira Santos; Evandra Strazza Rodrigues; Heidge Fukumasu; Jayme Augusto de Souza-Neto; José Salvatore Leister Patané; Luiz Alcantara; Luiz Lehmann Coutinho; Maria Carolina Elias; Maurício Lacerda Nogueira; Rafael dos Santos Bezerra; Raul Machado Neto; Rejane Maria Tommasini Grotto; Ricardo Haddad; Sandra Coccuzzo Sampaio Vessoni; Simone Kashima; Svetoslav Nanev Slavov; Vincent Louis Viala |
| EPI_ISL_2345750 | SECRETARIA DE SAUDE | Instituto Butantan / Mendelics | Antonio Jorge Martins; Claudia Renata dos Santos Barros; David Schlesinger; Debora Botequio Moretti; Dimas Tadeu Covas; Elaine Cristina Marqueze; Elaine Vieira Santos; Evandra Strazza Rodrigues; Heidge Fukumasu; Jayme Augusto de Souza-Neto; José Salvatore Leister Patané; Luiz Alcantara; Luiz Lehmann Coutinho; Maria Carolina Elias; Maurício Lacerda Nogueira; Rafael dos Santos Bezerra; Raul Machado Neto; Rejane Maria Tommasini Grotto; Ricardo Haddad; Sandra Coccuzzo Sampaio Vessoni; Simone Kashima; Svetoslav Nanev Slavov; Vincent Louis Viala |
| EPI_ISL_2209496 | SECRETARIA DE SAUDE DE RAFARD | Instituto Butantan | Antonio Jorge Martins; Claudia Renata dos Santos Barros; David Schlesinger; Debora Botequio Moretti; Dimas Tadeu Covas; Elaine Cristina Marqueze; Elaine Vieira Santos; Evandra Strazza Rodrigues; Heidge Fukumasu; Jayme Augusto de Souza-Neto; José Salvatore Leister Patané; Luiz Alcantara; Luiz Lehmann Coutinho; Maria Carolina Elias; Maurício Lacerda Nogueira; Rafael dos Santos Bezerra; Raul Machado Neto; Rejane Maria Tommasini Grotto; Ricardo Haddad; Sandra Coccuzzo Sampaio Vessoni; Simone Kashima; Svetoslav Nanev Slavov; Vincent Louis Viala. |
| EPI_ISL_2344733 | SECRETARIA DE SAUDE DE RAFARD | Instituto Butantan / ESALQ-Piracicaba | Antonio Jorge Martins; Claudia Renata dos Santos Barros; David Schlesinger; Debora Botequio Moretti; Dimas Tadeu Covas; Elaine Cristina Marqueze; Elaine Vieira Santos; Evandra Strazza Rodrigues; Heidge Fukumasu; Jayme Augusto de Souza-Neto; José Salvatore Leister Patané; Luiz Alcantara; Luiz Lehmann Coutinho; Maria Carolina Elias; Maurício Lacerda Nogueira; Rafael dos Santos Bezerra; Raul Machado Neto; Rejane Maria Tommasini Grotto; Ricardo Haddad; Sandra Coccuzzo Sampaio Vessoni; Simone Kashima; Svetoslav Nanev Slavov; Vincent Louis Viala |
| EPI_ISL_2344726, EPI_ISL_2344727 | SECRETARIA DE SAUDE DE RAFARD | Instituto Butantan / FZEA-USP-Pirassununga | Antonio Jorge Martins; Claudia Renata dos Santos Barros; David Schlesinger; Debora Botequio Moretti; Dimas Tadeu Covas; Elaine Cristina Marqueze; Elaine Vieira Santos; Evandra Strazza Rodrigues; Heidge Fukumasu; Jayme Augusto de Souza-Neto; José Salvatore Leister Patané; Luiz Alcantara; Luiz Lehmann Coutinho; Maria Carolina Elias; Maurício Lacerda Nogueira; Rafael dos Santos Bezerra; Raul Machado Neto; Rejane Maria Tommasini Grotto; Ricardo Haddad; Sandra Coccuzzo Sampaio Vessoni; Simone Kashima; Svetoslav Nanev Slavov; Vincent Louis Viala |
| EPI_ISL_2209522 | SECRETARIA DE SAUDE DE SANTA GERTRUDES | Instituto Butantan | Antonio Jorge Martins; Claudia Renata dos Santos Barros; David Schlesinger; Debora Botequio Moretti; Dimas Tadeu Covas; Elaine Cristina Marqueze; Elaine Vieira Santos; Evandra Strazza Rodrigues; Heidge Fukumasu; Jayme Augusto de Souza-Neto; José Salvatore Leister Patané; Luiz Alcantara; Luiz Lehmann Coutinho; Maria Carolina Elias; Maurício Lacerda Nogueira; Rafael dos Santos Bezerra; Raul Machado Neto; Rejane Maria Tommasini Grotto; Ricardo Haddad; Sandra Coccuzzo Sampaio Vessoni; Simone Kashima; Svetoslav Nanev Slavov; Vincent Louis Viala. |
| EPI_ISL_2209937, EPI_ISL_2209947, EPI_ISL_2344730 | SECRETARIA DE SAUDE DE SANTA GERTRUDES | Instituto Butantan / FZEA-USP-Pirassununga | Antonio Jorge Martins; Claudia Renata dos Santos Barros; David Schlesinger; Debora Botequio Moretti; Dimas Tadeu Covas; Elaine Cristina Marqueze; Elaine Vieira Santos; Evandra Strazza Rodrigues; Heidge Fukumasu; Jayme Augusto de Souza-Neto; José Salvatore Leister Patané; Luiz Alcantara; Luiz Lehmann Coutinho; Maria Carolina Elias; Maurício Lacerda Nogueira; Rafael dos Santos Bezerra; Raul Machado Neto; Rejane Maria Tommasini Grotto; Ricardo Haddad; Sandra Coccuzzo Sampaio Vessoni; Simone Kashima; Svetoslav Nanev Slavov; Vincent Louis Viala |
| EPI_ISL_2209537 | SECRETARIA DE SAUDE DE SAO PEDRO | Instituto Butantan | Antonio Jorge Martins; Claudia Renata dos Santos Barros; David Schlesinger; Debora Botequio Moretti; Dimas Tadeu Covas; Elaine Cristina Marqueze; Elaine Vieira Santos; Evandra Strazza Rodrigues; Heidge Fukumasu; Jayme Augusto de Souza-Neto; José Salvatore Leister Patané; Luiz Alcantara; Luiz Lehmann Coutinho; Maria Carolina Elias; Maurício Lacerda Nogueira; Rafael dos Santos Bezerra; Raul Machado Neto; Rejane Maria Tommasini Grotto; Ricardo Haddad; Sandra Coccuzzo Sampaio Vessoni; Simone Kashima; Svetoslav Nanev Slavov; Vincent Louis Viala. |
| EPI_ISL_2209212, EPI_ISL_2209955, EPI_ISL_2344731 | SECRETARIA DE SAUDE DE SAO PEDRO | Instituto Butantan / FZEA-USP-Pirassununga | Antonio Jorge Martins; Claudia Renata dos Santos Barros; David Schlesinger; Debora Botequio Moretti; Dimas Tadeu Covas; Elaine Cristina Marqueze; Elaine Vieira Santos; Evandra Strazza Rodrigues; Heidge Fukumasu; Jayme Augusto de Souza-Neto; José Salvatore Leister Patané; Luiz Alcantara; Luiz Lehmann Coutinho; Maria Carolina Elias; Maurício Lacerda Nogueira; Rafael dos Santos Bezerra; Raul Machado Neto; Rejane Maria Tommasini Grotto; Ricardo Haddad; Sandra Coccuzzo Sampaio Vessoni; Simone Kashima; Svetoslav Nanev Slavov; Vincent Louis Viala |
| EPI_ISL_5650253 | SECRETARIA MUNICIPAL DE SAUDE | Instituto Butantan | Antonio Jorge Martins; Claudia Renata dos Santos Barros; David Schlesinger; Debora Botequio Moretti; Dimas Tadeu Covas; Elaine Cristina Marqueze; Elaine Vieira Santos; Evandra Strazza Rodrigues; Heidge Fukumasu; Jayme Augusto de Souza-Neto; José Salvatore Leister Patané; Luiz Alcantara; Luiz Lehmann Coutinho; Maria Carolina Elias; Maurício Lacerda Nogueira; Rafael dos Santos Bezerra; Raul Machado Neto; Rejane Maria Tommasini Grotto; Ricardo Haddad; Sandra Coccuzzo Sampaio Vessoni; Simone Kashima; Svetoslav Nanev Slavov; Vincent Louis Viala |
| EPI_ISL_2344559, EPI_ISL_2344564 | SECRETARIA MUNICIPAL DE SAUDE | Instituto Butantan / FZEA-USP-Pirassununga | Antonio Jorge Martins; Claudia Renata dos Santos Barros; David Schlesinger; Debora Botequio Moretti; Dimas Tadeu Covas; Elaine Cristina Marqueze; Elaine Vieira Santos; Evandra Strazza Rodrigues; Heidge Fukumasu; Jayme Augusto de Souza-Neto; José Salvatore Leister Patané; Luiz Alcantara; Luiz Lehmann Coutinho; Maria Carolina Elias; Maurício Lacerda Nogueira; Rafael dos Santos Bezerra; Raul Machado Neto; Rejane Maria Tommasini Grotto; Ricardo Haddad; Sandra Coccuzzo Sampaio Vessoni; Simone Kashima; Svetoslav Nanev Slavov; Vincent Louis Viala |
| EPI_ISL_2344737 | SECRETARIA MUNICIPAL DE SAUDE DE CORDEIROPOLIS | Instituto Butantan / ESALQ-Piracicaba | Antonio Jorge Martins; Claudia Renata dos Santos Barros; David Schlesinger; Debora Botequio Moretti; Dimas Tadeu Covas; Elaine Cristina Marqueze; Elaine Vieira Santos; Evandra Strazza Rodrigues; Heidge Fukumasu; Jayme Augusto de Souza-Neto; José Salvatore Leister Patané; Luiz Alcantara; Luiz Lehmann Coutinho; Maria Carolina Elias; Maurício Lacerda Nogueira; Rafael dos Santos Bezerra; Raul Machado Neto; Rejane Maria Tommasini Grotto; Ricardo Haddad; Sandra Coccuzzo Sampaio Vessoni; Simone Kashima; Svetoslav Nanev Slavov; Vincent Louis Viala |
| EPI_ISL_2209457, EPI_ISL_2209462, EPI_ISL_2210037, EPI_ISL_2210082 | SECRETARIA MUNICIPAL DE SAUDE DE CORDEIROPOLIS | Instituto Butantan / FZEA-USP-Pirassununga | Antonio Jorge Martins; Claudia Renata dos Santos Barros; David Schlesinger; Debora Botequio Moretti; Dimas Tadeu Covas; Elaine Cristina Marqueze; Elaine Vieira Santos; Evandra Strazza Rodrigues; Heidge Fukumasu; Jayme Augusto de Souza-Neto; José Salvatore Leister Patané; Luiz Alcantara; Luiz Lehmann Coutinho; Maria Carolina Elias; Maurício Lacerda Nogueira; Rafael dos Santos Bezerra; Raul Machado Neto; Rejane Maria Tommasini Grotto; Ricardo Haddad; Sandra Coccuzzo Sampaio Vessoni; Simone Kashima; Svetoslav Nanev Slavov; Vincent Louis Viala |
| EPI_ISL_5603259 | SECRETARIA MUNICIPAL DE SAUDE DE SAO JOAO DO JAGUARIBE | Analytical Competence Molecular Epidemiology Lab/ACME, Oswaldo Cruz Foundation, Ceara (FIOCRUZ CE) | Carlos Leonardo de Aragao Araujo; Cecilia Leite Costa & Eduardo Ruback dos Santos on behalf of COVID-19 FIOCRUZ Genomic Network; Cleber Furtado Aksenen; Fabio Miyajima; Fernando Braga Stehling; Francisco Eder de Moura Lopes; Igor Oliveira Duarte; Jamille Maria Mendes Bezerra; Joaquim Cesar do Nascimento Sousa Junior; Pedro Miguel Carneiro Jeronimo; Suzana Porto Almeida; Thais Ferreira de Oliveira; Thais de Oliveira Costa; Ticiane Cavalcante de Souza; Veridiana Pessoa Miyajima |
| EPI_ISL_3102537 | SECRETARIA MUNICIPAL DE SAUDE DE TIANGUA | Analytical Competence Molecular Epidemiology Lab/ACME, Oswaldo Cruz Foundation, Ceara (FIOCRUZ CE) | Cleber Furtado Aksenen; Fabio Miyajima; Fernando Braga Stehling; Francisco Eder de Moura Lopes; Jamille Maria Mendes Bezerra; Joaquim César do Nascimento Sousa Junior; Pedro Miguel Carneiro Jeronimo; Suzana Porto Almeida e Lucas Delerino; Thais Ferreira de Oliveira; Thais de Oliveira Costa; Ticiane Cavalcante de Souza; Veridiana Pessoa Miyajima |
| EPI_ISL_2151747 | SELAS MEDILYS | Department of Virology, Henri Mondor University Hospital, Assistance Publique Hôpitaux de Paris, Université Paris-Est Créteil, INSERM U955 | Alexandre Soulier; Christophe Rodriguez; Elisabeth Trawinski; Guillaume Gricourt; Jean-Michel Pawlotsky; Melissa N'Debi; Slim Fourati; Vanessa Demontant |
| EPI_ISL_1707673 | SIESP CHIETI - DRIVE IN ORTONA | Istituto Zooprofilattico Sperimentale dell'Abruzzo e Molise "G. Caporale" | Ancora M; Calistri P; Cammà C; Caporale M; Curini V; Delli Compagni E; Di Domenico M; Di Lollo Valeria; Di Pasquale A; Lorusso A; Mangone I; Marcacci M; Puglia I; Rinaldi A; Savini G; Scialabba S |
| EPI_ISL_1841217 | SIESP DIPARTIMENTO DI PREVENZIONE TERAMO | Istituto Zooprofilattico Sperimentale dell'Abruzzo e Molise "G. Caporale" | Ancora M; Calistri P; Cammà C; Caporale M; Curini V; Delli Compagni E; Di Domenico M; Di Lollo Valeria; Di Pasquale A; Lorusso A; Mangone I; Marcacci M; Puglia I; Rinaldi A; Savini G; Scialabba S |
| EPI_ISL_2564457, EPI_ISL_2564458, EPI_ISL_2564915, EPI_ISL_2564947, EPI_ISL_2564959, EPI_ISL_2586056, EPI_ISL_2586076, EPI_ISL_2586132, EPI_ISL_2586185 | | | |
| see above | SK-Roy Romanow Provincial Laboratory | National Microbiology Laboratory (NML) | Amanda Lang; Anna Majer; Anneliese Landgraff; CanCOGeN's metadata curation team; Darian Hole; Elsie Grudeski; Gary Van Domselaar; Grace Seo; Jennifer Tanner; Jessica Minion; Kirsten Biggar; Madison Chapel; Morag Graham; Natalie Knox; Nathalie Bastien; Philip Mabon; Public Health Agency of Canada CanCOGeN team; Rachel DePaulo; Rhiannon Huzarewich; Russell Mandes; Ryan McDonald; Shari Tyson; Timothy Booth; Yan Li |
| EPI_ISL_2893541, EPI_ISL_2893542, EPI_ISL_2893543, EPI_ISL_2893544, EPI_ISL_2893545, EPI_ISL_2893546, EPI_ISL_2893547, EPI_ISL_2893548, EPI_ISL_2893549, EPI_ISL_2893550, EPI_ISL_5844113, EPI_ISL_5844170, EPI_ISL_5844225, EPI_ISL_5844276, EPI_ISL_5844281, EPI_ISL_5844283, EPI_ISL_5844284, EPI_ISL_5844287, EPI_ISL_5844288, EPI_ISL_5844290, EPI_ISL_5844292, EPI_ISL_5844326, EPI_ISL_5844333, EPI_ISL_5844336, EPI_ISL_5844344, EPI_ISL_5844346, EPI_ISL_5844351 | | | |
| see above | SK-Roy Romanow Provincial Laboratory | Saskatchewan - Roy Romanow Provincial Laboratory (RRPL) | Alanna Senecal; Amanda Lang; Anna Majer; Anneliese Landgraff; CanCOGeN's metadata curation team; Darian Hole; Elsie Grudeski; Gary Van Domselaar; Grace Seo; Jennifer Tanner; Jessica Minion; Kara Loos; Keith MacKenzie; Kirsten Biggar; Madison Chapel; Meredith Faires; Morag Graham; Natalie Knox; Nathalie Bastien; Philip Mabon; Public Health Agency of Canada CanCOGeN team; Rachel DePaulo; Rhiannon Huzarewich; Roy Romanow Provincial Laboratory - Molecular Diagnostics; Russell Mandes; Ryan McDonald; Shari Tyson; Timothy Booth; Yan Li |
| EPI_ISL_2321643, EPI_ISL_2321645, EPI_ISL_3276558 | SOMER | Universidad Nacional de Colombia - Laboratorio Genómico One Health | Andres F. Cardona-Rios; Carlos Franco-Muñoz; Carolina Muñoz-Arango; Celeny Ortiz; Daniel O. Maldonado-Perez; Diego A. Álvarez-Díaz; Hector Alejandro Ruiz-Moreno; Idabely Betancur Ortiz; Jorge E. Osorio; Juan P. Hernandez-Ortiz; Karl A Ciuoderis; Katherine Laiton-Donato; Laura Silvana Perez; Lina M. Hurtado; Marcela Mercado-Reyes; Maria Angélica Maya; Maria Stella López; Rita Almanza Payares; Sandra Ines Cano; Simón Villegas Velásquez |
| EPI_ISL_1960039, EPI_ISL_1960041, EPI_ISL_1960053 | SURA | Universidad Nacional de Colombia - Laboratorio Genómico One Health | Andres F. Cardona-Rios; Carlos Franco-Muñoz; Carolina Muñoz-Arango; Celeny Ortiz; Daniel O. Maldonado-Perez; Diego A. Álvarez-Díaz; Hector Alejandro Ruiz-Moreno; Idabely Betancur Ortiz; Jorge E. Osorio; Juan P. Hernandez-Ortiz; Karl A Ciuoderis; Katherine Laiton-Donato; Laura Silvana Perez; Lina M. Hurtado; Marcela Mercado-Reyes; Maria Angélica Maya; Maria Stella López; Rita Almanza Payares; Sandra Ines Cano; Simón Villegas Velásquez |
| EPI_ISL_1793669, EPI_ISL_1793670, EPI_ISL_1793673, EPI_ISL_1793779, EPI_ISL_1826558, EPI_ISL_1826598, EPI_ISL_1826600, EPI_ISL_1826619, EPI_ISL_1826635, EPI_ISL_1826647, EPI_ISL_1826670, EPI_ISL_1826683, EPI_ISL_1826684, EPI_ISL_1826690, EPI_ISL_1826692, EPI_ISL_1826699, EPI_ISL_1826705, EPI_ISL_1826708, EPI_ISL_1826713, EPI_ISL_1826724, EPI_ISL_1970806, EPI_ISL_1970809, EPI_ISL_2003364, EPI_ISL_2003366 | | | |
| see above | SYNLAB | GIGA Medical Genomics | Bouchra Boujemla; Cécile Meex; Keith Durkin; Maria Artesi; Marie-Pierre Hayette; Nathalie Renotte; Pierrette Melin; Raphaël Boreux; Sébastien Bontems; Vincent Bours |
| EPI_ISL_2004325 | SYNLAB | Instituto Nacional de Saude (INSA) | Borges et al |
| EPI_ISL_3355507 | SYNLAB - ANGEL DIAGNOSTICA | Corporacion CorpoGen-Universidad de los Andes-Universidad Central | Christian Romero; Jorge Duitama; Juan Manuel Anzola; Laura González; Maryam Chaib De Mares; María Mercedes Zambrano; Nelly Díaz; Patricia Del Portillo; Silvia Restrepo |

| | | | |
|---|---|---|---|
| EPI_ISL_2155039 | SYNLAB -ANGEL DIAGNOSTICA | Instituto Nacional de Salud- Dirección de Investigación en Salud Pública | Carlos Franco-Muñoz; Carmen Osorio; Diana Malo; Diego A. Álvarez-Díaz; Diego Andrés Prada; Gerardo Santamaría; Hector Alejandro Ruiz-Moreno; Jhonnatan Reales-González; Jorge Rivera; Juan Camilo Martinez; Julian Naizaque; Katherine Laiton-Donato; Lisseth Pardo; Magdalena Wiesner; Marcela Mercado-Reyes; Maria T. Herrera-Sepúlveda; Marta Lopez Blanco; Martha Lucia Ospina Martinez; Paola Rojas; Sergio Gomez; Sheryll Corchuelo; Ángela Alarcon Cruz |
| EPI_ISL_2009059 | SYNLAB ANGEL DIAGNOSTICA | Instituto Nacional de Salud- Dirección de Investigación en Salud Pública | Carlos Franco-Muñoz; Carmen Osorio; Diana Malo; Diego A. Álvarez-Díaz; Diego Andrés Prada; Gerardo Santamaría; Hector Alejandro Ruiz-Moreno; Jhonnatan Reales-González; Jorge Rivera; Juan Camilo Martinez; Julian Naizaque; Katherine Laiton-Donato; Lisseth Pardo; Magdalena Wiesner; Marcela Mercado-Reyes; Maria T. Herrera-Sepúlveda; Marta Lopez Blanco; Martha Lucia Ospina Martinez; Paola Rojas; Sergio Gomez; Sheryll Corchuelo; Ángela Alarcon Cruz |
| EPI_ISL_1725040, EPI_ISL_1727806 | SYNLAB MVZ Trier | Robert Koch Institute | |
| EPI_ISL_2329538, EPI_ISL_2329542, EPI_ISL_2329543, EPI_ISL_2329547, EPI_ISL_2329567, EPI_ISL_2329573, EPI_ISL_2329584, EPI_ISL_2329602 | | | |
| see above | San Diego County Public Health Laboratory | Andersen lab at Scripps Research | Brett Austin; Jovan Shephard; SEARCH Alliance San Diego with Ashleigh Murphy |
| EPI_ISL_1970923, EPI_ISL_1970952 | San Juan Regional Medical Center | New Mexico Department of Health Scientific Laboratory | Anastacia Griego-Fisher; D'eldra Malone; Ellie Johnson; Jennifer Benoit; Linda Salazar; Mark Willmon; Ratheesh Rajan |
| EPI_ISL_5802431, EPI_ISL_5802432 | Santa Casa De Misericordia Sao Jose | Instituto Butantan | Antonio Jorge Martins; Claudia Renata dos Santos Barros; David Schlesinger; Debora Botequio Moretti; Dimas Tadeu Covas; Elaine Cristina Marqueze; Elaine Vieira Santos; Evandra Strazza Rodrigues; Heidge Fukumasu; Jayme Augusto de Souza-Neto; José Salvatore Leister Patané; Luiz Alcantara; Luiz Lehmann Coutinho; Maria Carolina Elias; Maurício Lacerda Nogueira; Rafael dos Santos Bezerra; Raul Machado Neto; Rejane Maria Tommasini Grotto; Ricardo Haddad; Sandra Coccuzzo Sampaio Vessoni; Simone Kashima; Svetoslav Nanev Slavov; Vincent Louis Viala |
| EPI_ISL_2500207 | Santa Clara County Public Health Laboratory | Chan-Zuckerberg Biohub | CZB Cliahub Consortium |
| EPI_ISL_1969535, EPI_ISL_1969536, EPI_ISL_1969544, EPI_ISL_1969548 | Scripps Medical Laboratory | Andersen lab at Scripps Research | Ellen Stefanski; Ian Mchardy; SEARCH Alliance San Diego with Michael Quigley |
| EPI_ISL_2011269, EPI_ISL_2011272, EPI_ISL_2011275, EPI_ISL_2011322, EPI_ISL_2011327, EPI_ISL_2011328, EPI_ISL_2011438, EPI_ISL_2536657, EPI_ISL_2536679, EPI_ISL_2692507 | | | |
| see above | Seattle Flu Study | Seattle Flu Study | Amanda Adler; Barry R. Lutz; Benjamin Pelle; Caitlin R. Wolf; Chris D. Frazar; Deborah A. Nickerson; Elisabeth Brandstetter; Erica Ryke; Helen Y. Chu; Janet A. Englund; Jay Shendure; Jeff Duchin; Jover Lee; Kairsten Fay; Karen Cowgill; Kirsten Lacombe; Lea M. Starita; Mark J. Rieder; Matthew Richardson; Matthew Thompson; Melissa Truong; Michael Boeckh; Michael Famulare; Misja Ilcisin; Peter D. Han; Stephanie Schrag; Thomas R. Sibley; Trevor Bedford |
| EPI_ISL_5802441 | Secretaria De Saude | Instituto Butantan | Antonio Jorge Martins; Claudia Renata dos Santos Barros; David Schlesinger; Debora Botequio Moretti; Dimas Tadeu Covas; Elaine Cristina Marqueze; Elaine Vieira Santos; Evandra Strazza Rodrigues; Heidge Fukumasu; Jayme Augusto de Souza-Neto; José Salvatore Leister Patané; Luiz Alcantara; Luiz Lehmann Coutinho; Maria Carolina Elias; Maurício Lacerda Nogueira; Rafael dos Santos Bezerra; Raul Machado Neto; Rejane Maria Tommasini Grotto; Ricardo Haddad; Sandra Coccuzzo Sampaio Vessoni; Simone Kashima; Svetoslav Nanev Slavov; Vincent Louis Viala |
| EPI_ISL_5799768, EPI_ISL_5799769, EPI_ISL_5800517 | Secretaria De Saude De Rafard | Instituto Butantan | Antonio Jorge Martins; Claudia Renata dos Santos Barros; David Schlesinger; Debora Botequio Moretti; Dimas Tadeu Covas; Elaine Cristina Marqueze; Elaine Vieira Santos; Evandra Strazza Rodrigues; Heidge Fukumasu; Jayme Augusto de Souza-Neto; José Salvatore Leister Patané; Luiz Alcantara; Luiz Lehmann Coutinho; Maria Carolina Elias; Maurício Lacerda Nogueira; Rafael dos Santos Bezerra; Raul Machado Neto; Rejane Maria Tommasini Grotto; Ricardo Haddad; Sandra Coccuzzo Sampaio Vessoni; Simone Kashima; Svetoslav Nanev Slavov; Vincent Louis Viala |
| EPI_ISL_5799731, EPI_ISL_5799732, EPI_ISL_5799772 | Secretaria De Saude De Santa Gertrudes | Instituto Butantan | Antonio Jorge Martins; Claudia Renata dos Santos Barros; David Schlesinger; Debora Botequio Moretti; Dimas Tadeu Covas; Elaine Cristina Marqueze; Elaine Vieira Santos; Evandra Strazza Rodrigues; Heidge Fukumasu; Jayme Augusto de Souza-Neto; José Salvatore Leister Patané; Luiz Alcantara; Luiz Lehmann Coutinho; Maria Carolina Elias; Maurício Lacerda Nogueira; Rafael dos Santos Bezerra; Raul Machado Neto; Rejane Maria Tommasini Grotto; Ricardo Haddad; Sandra Coccuzzo Sampaio Vessoni; Simone Kashima; Svetoslav Nanev Slavov; Vincent Louis Viala |
| EPI_ISL_5799733, EPI_ISL_5799734, EPI_ISL_5799773 | Secretaria De Saude De Sao Pedro | Instituto Butantan | Antonio Jorge Martins; Claudia Renata dos Santos Barros; David Schlesinger; Debora Botequio Moretti; Dimas Tadeu Covas; Elaine Cristina Marqueze; Elaine Vieira Santos; Evandra Strazza Rodrigues; Heidge Fukumasu; Jayme Augusto de Souza-Neto; José Salvatore Leister Patané; Luiz Alcantara; Luiz Lehmann Coutinho; Maria Carolina Elias; Maurício Lacerda Nogueira; Rafael dos Santos Bezerra; Raul Machado Neto; Rejane Maria Tommasini Grotto; Ricardo Haddad; Sandra Coccuzzo Sampaio Vessoni; Simone Kashima; Svetoslav Nanev Slavov; Vincent Louis Viala |
| EPI_ISL_5782467, EPI_ISL_5782472 | Secretaria Municipal De Saude | Instituto Butantan | Antonio Jorge Martins; Claudia Renata dos Santos Barros; David Schlesinger; Debora Botequio Moretti; Dimas Tadeu Covas; Elaine Cristina Marqueze; Elaine Vieira Santos; Evandra Strazza Rodrigues; Heidge Fukumasu; Jayme Augusto de Souza-Neto; José Salvatore Leister Patané; Luiz Alcantara; Luiz Lehmann Coutinho; Maria Carolina Elias; Maurício Lacerda Nogueira; Rafael dos Santos Bezerra; Raul Machado Neto; Rejane Maria Tommasini Grotto; Ricardo Haddad; Sandra Coccuzzo Sampaio Vessoni; Simone Kashima; Svetoslav Nanev Slavov; Vincent Louis Viala |
| EPI_ISL_5799720, EPI_ISL_5799745, EPI_ISL_5799746, EPI_ISL_5799747, EPI_ISL_5800546 | Secretaria Municipal De Saude De Cordeiropolis | Instituto Butantan | Antonio Jorge Martins; Claudia Renata dos Santos Barros; David Schlesinger; Debora Botequio Moretti; Dimas Tadeu Covas; Elaine Cristina Marqueze; Elaine Vieira Santos; Evandra Strazza Rodrigues; Heidge Fukumasu; Jayme Augusto de Souza-Neto; José Salvatore Leister Patané; Luiz Alcantara; Luiz Lehmann Coutinho; Maria Carolina Elias; Maurício Lacerda Nogueira; Rafael dos Santos Bezerra; Raul Machado Neto; Rejane Maria Tommasini Grotto; Ricardo Haddad; Sandra Coccuzzo Sampaio Vessoni; Simone Kashima; Svetoslav Nanev Slavov; Vincent Louis Viala |
| EPI_ISL_3030284, EPI_ISL_3030347 | Senigallia | Microbiology University Politecnica delle Marche | Anna Valenza; Carla Acciarri; Katia Marinelli; Monica Lucia Ferreri; Patrizia Bagnarelli; Roberta Longo; Sara Caucci; Stefano Menzo |
| EPI_ISL_2179675, EPI_ISL_2179677, EPI_ISL_2179678, EPI_ISL_2179707, EPI_ISL_2179721, EPI_ISL_2179726, EPI_ISL_2179727 | | | |
| see above | Servicio Microbiología Hospital La Paz | Servicio Microbiología Hospital La Paz | Elie Dahdouh; Fernando Lázaro; Jesús Mingorance Cruz; Rubén Cáceres |
| EPI_ISL_2136006, EPI_ISL_2136012, EPI_ISL_2136015, EPI_ISL_2136029, EPI_ISL_2136059, EPI_ISL_2136060, EPI_ISL_2136061, EPI_ISL_2136063, EPI_ISL_2136100, EPI_ISL_2136103, EPI_ISL_2136105, EPI_ISL_2136107, EPI_ISL_2136112, EPI_ISL_2136113, EPI_ISL_2136114, EPI_ISL_2136115, EPI_ISL_2136116, EPI_ISL_2136119, EPI_ISL_2136124, EPI_ISL_2136141, EPI_ISL_2136143, EPI_ISL_2136153, EPI_ISL_2136155, EPI_ISL_2136160, EPI_ISL_2140066, EPI_ISL_2140109, EPI_ISL_2140112, EPI_ISL_2158695, EPI_ISL_2158796, EPI_ISL_2158813, EPI_ISL_2158828, EPI_ISL_4082784 | | | |
| see above | Servicio Virosis Respiratorias-Departamento Virología-INEI | Instituto Nacional Enfermedades Infecciosas C.G.Malbran | Avaro M.; Baumeister E.; Benedetti E.; Campos J.; Cisterna D.; Dattero ME; De Belder D.; Haim MS.; Lorenzo F.; Molina V.; Perandones C.; Poklepovich T.; Pontoriero A.; Russo M.; Sanchez Loria J.; Tuduri E. |
| EPI_ISL_1912296, EPI_ISL_1912302, EPI_ISL_2104441 | Servicio de Microbiologia Hospital Ramon y Cajal | Servicio de Microbiologia Hospital Ramon y Cajal | Galan JC; Martinez L. Abreu M; Ponce M; y Gonzalez-Alba JM |
| EPI_ISL_1970073, EPI_ISL_1970075, EPI_ISL_1970078, EPI_ISL_1970079, EPI_ISL_2271616, EPI_ISL_2271617, EPI_ISL_2271618, EPI_ISL_2271619, EPI_ISL_2271620, EPI_ISL_2271625, EPI_ISL_2271626, EPI_ISL_2271638 | | | |
| see above | Servicio de Microbiología. Consorcio Hospital General Universitario de Valencia | SeqCOVID-SPAIN consortium/IBV(CSIC) | Begoña Fuster Escrivá; Carme Salvador García; Concepción Gimeno Cardona and SeqCOVID-SPAIN consortium; María Dolores Ocete; Rafael Medina González |
| EPI_ISL_1916260 | Servicio de Microbiología. Hospital General Universitario de Castellón | SeqCOVID-SPAIN consortium/IBV(CSIC) | María Dolores Tirado Balaguer and SeqCOVID-SPAIN consortium; Rosario Moreno Muñoz |
| EPI_ISL_2323415 | Servicio de Microbiología. Hospital Universitario Doctor Peset | SeqCOVID-SPAIN consortium/IBV(CSIC) | José Miguel Nogueira Coito and SeqCOVID-SPAIN consortium; Juan Alberola Enguídanos; Juan José Camarena Miñana; Rosa González Pellicer |
| EPI_ISL_2600567 | Servizo de Microbioloxía. Complexo Hospitalario de Santiago de Compostela | Servizo de Microbioloxía. Complexo Hospitalario de Santiago de Compostela | Amparo Coira; Antonio Aguilera; Carlos García_Riestra; Daniel Navarro; Gema Barbeito; Iria Roca; José Llovo; Laura Millán; Manuela Hernández; María Luisa Pérez_del_Molino; Mercedes Treviño; Rocío trastoy; Teresa Lopez_Valiño; Xosé Costa |
| EPI_ISL_5316363, EPI_ISL_5316364, EPI_ISL_5316365, EPI_ISL_5316366, EPI_ISL_5316368, EPI_ISL_5316369, EPI_ISL_5316370, EPI_ISL_5316377, EPI_ISL_5316381, EPI_ISL_5316385, EPI_ISL_5316389, EPI_ISL_5316390, EPI_ISL_5316392, EPI_ISL_5316393, EPI_ISL_5316394, EPI_ISL_5316395, EPI_ISL_5316396, EPI_ISL_5316397, EPI_ISL_5316404, EPI_ISL_5316405, EPI_ISL_5316407, EPI_ISL_5316408, EPI_ISL_5316409, EPI_ISL_5316410, EPI_ISL_5316411, EPI_ISL_5316417, EPI_ISL_5316419, EPI_ISL_5316422, EPI_ISL_5316423, EPI_ISL_5316424, EPI_ISL_5316425, EPI_ISL_5316428, EPI_ISL_5316430 | | | |
| see above | Shared Hospital Laboratory | Shared Hospital Laboratory | Christie Vermeiren; Finlay Maguire; Kevin Katz; Patryk Aftanas; Robert Kozak; Samira Mubareka |
| EPI_ISL_1969590, EPI_ISL_1969594, EPI_ISL_1969598 | Sharp HealthCare Laboratory | Andersen lab at Scripps Research | Art Mendoza; Cathy Woerle; Jacquelyn Berumen; Liam McGinnis; Omid Bakhtar; SEARCH Alliance San Diego with Aaron Harding |
| EPI_ISL_2295973, EPI_ISL_2295981, EPI_ISL_2295986, EPI_ISL_2295988, EPI_ISL_2295989, EPI_ISL_2295992, EPI_ISL_2295994 | | | |
| see above | Simple Laboratories | Gagnon Lab, Southern Illinois University | Keith Gagnon |
| EPI_ISL_3394022 | Sonic Reference Laboratory | Sonic Reference Laboratory | "Robert Moller; Bijaya Dhakal; Cora Lahey; Ellen Pizarek"; John Woody; Krishna Maddula |
| EPI_ISL_1909329, EPI_ISL_1909569 | Sonora Quest Laboratories | TGen North | "Jolene Bowers; Ashlyn Pfeiffer; Chris French; Darrin Lemmer; Dave Engelthaler; Hayley Yaglom; Heather Centner; The Arizona COVID Genomics Union (ACGU)" |
| EPI_ISL_2462410 | South Eastern Area Laboratory Services (SEALS) | NSW Health Pathology - Institute of Clinical Pathology and Medical Research; Westmead Hospital; University of Sydney | CIDM-PH et al. |
| EPI_ISL_1739286, EPI_ISL_1739287, EPI_ISL_1739288 | Southern Nevada Public Health Laboratory | Southern Nevada Public Health Laboratory | Michael Picker |
| EPI_ISL_1826573 | St. Nikolaus-Hospital Eupen | GIGA Medical Genomics | Bouchra Boujemla; Cécile Meex; Keith Durkin; Maria Artesi; Marie-Pierre Hayette; Nathalie Renotte; Pierrette Melin; Raphaël Boreux; Sébastien Bontems; Vincent Bours |
| EPI_ISL_4496737, EPI_ISL_4496753, EPI_ISL_4496770 | Stanford Health Care | Stanford University School of Medicine, Clinical Virology Laboratory | Becky Jiang; Bernadette Troung; Hannah Wang; James Zehnder; Malaya K. Sahoo; and Benjamin A. Pinsky |
| EPI_ISL_1771593, EPI_ISL_1771623, EPI_ISL_1771627, EPI_ISL_1771632, EPI_ISL_1771636, EPI_ISL_1771641, EPI_ISL_2007865, EPI_ISL_2007910, EPI_ISL_2007933 | | | |
| see above | State Laboratories Division, Hawaii State Department of Health | State Laboratories Division, Hawaii State Department of Health | Ayana Garnet; Drew Kuwazaki; Edward Desmond; Pamela O'Brien; Razvan Sultana |

| | | | |
|---|---|---|---|
| EPI_ISL_2378131, EPI_ISL_2378207, EPI_ISL_2378210, EPI_ISL_2378213, EPI_ISL_2378221, EPI_ISL_2378222, EPI_ISL_2378225, EPI_ISL_2378228, EPI_ISL_2378234, EPI_ISL_2378238, EPI_ISL_2378248, EPI_ISL_2378253, EPI_ISL_2378274, EPI_ISL_2378291, EPI_ISL_2378297, EPI_ISL_2378302, EPI_ISL_2378329, EPI_ISL_2378347, EPI_ISL_2378350, EPI_ISL_2378356, EPI_ISL_2378357, EPI_ISL_2378494, EPI_ISL_2378495, EPI_ISL_2378504, EPI_ISL_2378516, EPI_ISL_2378523, EPI_ISL_2378526, EPI_ISL_2378531, EPI_ISL_2378539, EPI_ISL_2473052, EPI_ISL_2800561, EPI_ISL_2800562, EPI_ISL_2800563, EPI_ISL_2800564, EPI_ISL_2800565 | | | |
| see above | State Testing Facility | Altius Institute for Biomedical Research | Alex Isner; Alex Nguyen; Amanda Gale; Audra Johnson; Clem Green; Daniel Bates; Eric Thorland; Jacob Rodriguez; Jean Robinston; Jemma Nelson; Jessica Kunder; John Stamatoyannopoulos; Joshua Richards; Julia Wald; Kneshay Harper; Lauren Mitchell; Mark Frerker; Matt Hartman; Michael Buckley; Muhammad Halimun; Rebecca Bruders; Sadie Patraw; Sofia Olsson; Tobias Ragoczy |
| EPI_ISL_2198321, EPI_ISL_2208428, EPI_ISL_3781496 | Swedish national genomic surveillance program of SARS-CoV-2 | The Public Health Agency of Sweden | Alma Brolund; Maria Lind Karlberg; Maximilian Riess; Swedish national genomic surveillance program of SARS-CoV-2 |
| EPI_ISL_2621857, EPI_ISL_2621858 | Synlab | Universidad Nacional de Colombia - Laboratorio Genómico One Health | Andres F. Cardona-Rios; Carlos Franco-Muñoz; Carolina Muñoz-Arango; Celeny Ortiz; Daniel O. Maldonado-Perez; Diego A. Álvarez-Díaz; Hector Alejandro Ruiz-Moreno; Idabely Betancur Ortiz; Jorge E. Osorio; Juan P. Hernandez-Ortiz; Karl A Ciuoderis; Katherine Laiton-Donato; Laura Silvana Perez; Lina M. Hurtado; Marcela Mercado-Reyes; Maria Angélica Maya; Maria Stella López; Rita Almanza Payares; Sandra Ines Cano; Simón Villegas Velásquez |
| EPI_ISL_1725712, EPI_ISL_1729627, EPI_ISL_1729640 | Synlab MVZ Augsburg | Robert Koch Institute | |
| EPI_ISL_1855153, EPI_ISL_1935613 | TEMPUS LABS INC | Wadsworth Center, New York State Department of Health | Alexis Russell; Catharine Prussing; Daryl M. Lamson; Erasmus Schneider; Erica Lasek-Nesselquist; John Kelly; Jonathan Plitnick; Kirsten St. George; Matthew Shudt; Melissa A Leisner; Navjot Singh |
| EPI_ISL_1707371, EPI_ISL_2000899, EPI_ISL_2000974 | TGen North | TGen North | "Jolene Bowers; Ashlyn Pfeiffer; Chris French; Darrin Lemmer; Dave Engelthaler; Hayley Yaglom; Heather Centner; The Arizona COVID Genomics Union (ACGU)" |
| EPI_ISL_1760558, EPI_ISL_1961073, EPI_ISL_1961119, EPI_ISL_1961120, EPI_ISL_1971410, EPI_ISL_1971414, EPI_ISL_1971451, EPI_ISL_1971459, EPI_ISL_2017909, EPI_ISL_2141450 | | | |
| see above | TXDSHS | TXDSHS | Anita Pokharel; Bonnie Oh; Chun Wang; Grace Kubin; Jenny Zhang; Karen Bobier; Lorraine Rodriguez; Maliha Rahman; Mayela Pedrueza; Myong Koag; Rachel Lee; Rashmi Tuladhar |
| EPI_ISL_2484171, EPI_ISL_2484202, EPI_ISL_2626628, EPI_ISL_2626631, EPI_ISL_2727550 | Tampa General Hospital Esoteric Lab | Tampa General Hospital Esoteric Research & Development Lab | Amorce Lima; Deanna Becker; Dominic Uy; Elaine Vendrone; Jon Faughn; Jorge Mecias-Frias; Suzane Silbert; Vicki Healer |
| EPI_ISL_1760070, EPI_ISL_1760107, EPI_ISL_1760109, EPI_ISL_1760110, EPI_ISL_1760111, EPI_ISL_1760112 | Texas Department of State Health Services (TXDSHS) | Texas Department of State Health Services (TXDSHS) | Anita Pokharel; Bonnie Oh; Chun Wang; Grace Kubin; Jenny Zhang; Lorraine Rodriguez; Maliha Rahman; Mayela Pedrueza; Myong Koag; Rachel Lee; Rashmi Tuladhar |
| EPI_ISL_1739426, EPI_ISL_1739478 | The Jackson Laboratory | The Jackson Laboratory | Adams M; Bergeron D; Kelly K; Li L; Long J; Omerza G; Renzette N |
| EPI_ISL_1824149, EPI_ISL_2006936, EPI_ISL_2006942, EPI_ISL_2006945, EPI_ISL_2006996, EPI_ISL_2006997 | The Ohio State University Applied Microbiology Services Laboratory | The Ohio State University Applied Microbiology Services Laboratory | Seth A. Faith PhD |
| EPI_ISL_3568741, EPI_ISL_3568742, EPI_ISL_3568744, EPI_ISL_3668142, EPI_ISL_3668143, EPI_ISL_3668145 | The University of Alabama | Synergy Laboratories | Megan Cornwell |
| EPI_ISL_1965489 | Tyrolpath Obrist Brunhuber GmbH | Bergthaler laboratory, CeMM Research Center for Molecular Medicine of the Austrian Academy of Sciences | Andreas Bergthaler; Anna Schedl; Bekir Erguner; Benedikt Agerer; Christoph Bock; Fabian Amman; Jan Laine; Lukas Endler; Maelle Le Moing; Martin Senekowitsch; Michael Schuster; Petr Triska; Thomas Penz |
| EPI_ISL_3184481 | U.O. Microbiologia Laboratorio Unico Centro Servizi - AUSL della Romagna | U.O. Microbiologia, Laboratorio Unico Centro Servizi - AUSL della Romagna | Giorgio Dirani |
| EPI_ISL_1960604 | UAB Diagnostikos laboratorija | National Public Health Surveillance Laboratory | Ana Steponkiene; Danas Baksa; Jelena Razmuk; Lukas Vasionis; Lukas Zemaitis; Migle Gabrielaite; Svajune Muralyte |
| EPI_ISL_5529937 | UAPS GRASIELA BARROSO | Analytical Competence Molecular Epidemiology Lab/ACME, Oswaldo Cruz Foundation, Ceara (FIOCRUZ CE) | Carlos Leonardo de Aragao Araujo; Cecilia Leite Costa & Eduardo Ruback dos Santos on behalf of COVID-19 FIOCRUZ Genomic Network; Cleber Furtado Aksenen; Fabio Miyajima; Fernando Braga Stehling; Francisco Eder de Moura Lopes; Igor Oliveira Duarte; Jamille Maria Mendes Bezerra; Joaquim Cesar do Nascimento Sousa Junior; Pedro Miguel Carneiro Jeronimo; Suzana Porto Almeida; Thais Ferreira de Oliveira; Thais de Oliveira Costa; Ticiane Cavalcante de Souza; Veridiana Pessoa Miyajima |
| EPI_ISL_5650414 | UBS 01 VILA BANDEIRANTES BIRIGUI | Instituto Butantan | Antonio Jorge Martins; Claudia Renata dos Santos Barros; David Schlesinger; Debora Botequio Moretti; Dimas Tadeu Covas; Elaine Cristina Marqueze; Elaine Vieira Santos; Evandra Strazza Rodrigues; Heidge Fukumasu; Jayme Augusto de Souza-Neto; José Salvatore Leister Patané; Luiz Alcantara; Luiz Lehmann Coutinho; Maria Carolina Elias; Maurício Lacerda Nogueira; Rafael dos Santos Bezerra; Raul Machado Neto; Rejane Maria Tommasini Grotto; Ricardo Haddad; Sandra Coccuzzo Sampaio Vessoni; Simone Kashima; Svetoslav Nanev Slavov; Vincent Louis Viala |
| EPI_ISL_5650244, EPI_ISL_5650248 | UBS 06 PARQUE DAS NACOES BIRIGUI | Instituto Butantan | Antonio Jorge Martins; Claudia Renata dos Santos Barros; David Schlesinger; Debora Botequio Moretti; Dimas Tadeu Covas; Elaine Cristina Marqueze; Elaine Vieira Santos; Evandra Strazza Rodrigues; Heidge Fukumasu; Jayme Augusto de Souza-Neto; José Salvatore Leister Patané; Luiz Alcantara; Luiz Lehmann Coutinho; Maria Carolina Elias; Maurício Lacerda Nogueira; Rafael dos Santos Bezerra; Raul Machado Neto; Rejane Maria Tommasini Grotto; Ricardo Haddad; Sandra Coccuzzo Sampaio Vessoni; Simone Kashima; Svetoslav Nanev Slavov; Vincent Louis Viala |
| EPI_ISL_5650236, EPI_ISL_5650249 | UBS 11 DR SERGIO AUGUSTO CLARK XAVIER SOARES PORTAL II | Instituto Butantan | Antonio Jorge Martins; Claudia Renata dos Santos Barros; David Schlesinger; Debora Botequio Moretti; Dimas Tadeu Covas; Elaine Cristina Marqueze; Elaine Vieira Santos; Evandra Strazza Rodrigues; Heidge Fukumasu; Jayme Augusto de Souza-Neto; José Salvatore Leister Patané; Luiz Alcantara; Luiz Lehmann Coutinho; Maria Carolina Elias; Maurício Lacerda Nogueira; Rafael dos Santos Bezerra; Raul Machado Neto; Rejane Maria Tommasini Grotto; Ricardo Haddad; Sandra Coccuzzo Sampaio Vessoni; Simone Kashima; Svetoslav Nanev Slavov; Vincent Louis Viala |
| EPI_ISL_2614596 | UBS Atalaia | Instituto Adolfo Lutz, Interdiciplinary Procedures Center, Strategic Laboratory | Caio Vinicius Dias Lopes; Claudia Regina Gonçalves; Claudio Tavares Sacchi; Erica Valessa Ramos Gomes; Karoline Rodrigues Campos; Leonardo Jose Tadeu de Araujo |
| EPI_ISL_5650251 | UBS DR ALFREDO DANTAS DE SOUZA UMUARAMA | Instituto Butantan | Antonio Jorge Martins; Claudia Renata dos Santos Barros; David Schlesinger; Debora Botequio Moretti; Dimas Tadeu Covas; Elaine Cristina Marqueze; Elaine Vieira Santos; Evandra Strazza Rodrigues; Heidge Fukumasu; Jayme Augusto de Souza-Neto; José Salvatore Leister Patané; Luiz Alcantara; Luiz Lehmann Coutinho; Maria Carolina Elias; Maurício Lacerda Nogueira; Rafael dos Santos Bezerra; Raul Machado Neto; Rejane Maria Tommasini Grotto; Ricardo Haddad; Sandra Coccuzzo Sampaio Vessoni; Simone Kashima; Svetoslav Nanev Slavov; Vincent Louis Viala |
| EPI_ISL_2362270 | UBS GUILHERMINA DE NOBREGA ABREU | Instituto Butantan / Mendelics | Antonio Jorge Martins; Claudia Renata dos Santos Barros; David Schlesinger; Debora Botequio Moretti; Dimas Tadeu Covas; Elaine Cristina Marqueze; Elaine Vieira Santos; Evandra Strazza Rodrigues; Heidge Fukumasu; Jayme Augusto de Souza-Neto; José Salvatore Leister Patané; Luiz Alcantara; Luiz Lehmann Coutinho; Maria Carolina Elias; Maurício Lacerda Nogueira; Rafael dos Santos Bezerra; Raul Machado Neto; Rejane Maria Tommasini Grotto; Ricardo Haddad; Sandra Coccuzzo Sampaio Vessoni; Simone Kashima; Svetoslav Nanev Slavov; Vincent Louis Viala |
| EPI_ISL_5650417 | UBS III DE BENTO DE ABREU | Instituto Butantan | Antonio Jorge Martins; Claudia Renata dos Santos Barros; David Schlesinger; Debora Botequio Moretti; Dimas Tadeu Covas; Elaine Cristina Marqueze; Elaine Vieira Santos; Evandra Strazza Rodrigues; Heidge Fukumasu; Jayme Augusto de Souza-Neto; José Salvatore Leister Patané; Luiz Alcantara; Luiz Lehmann Coutinho; Maria Carolina Elias; Maurício Lacerda Nogueira; Rafael dos Santos Bezerra; Raul Machado Neto; Rejane Maria Tommasini Grotto; Ricardo Haddad; Sandra Coccuzzo Sampaio Vessoni; Simone Kashima; Svetoslav Nanev Slavov; Vincent Louis Viala |
| EPI_ISL_1966728, EPI_ISL_1966730 | UBS JOSE SABINO FERREIRA | Instituto Butantan | Antonio Jorge Martins; Bianca Cechetto Carlos. Mendelics: Bibiana Santos; Claudia Renata dos Santos Barros; Cíntia Bittar; David Schlesinger. Hemocentro Ribeirão Preto: Simone Kashima; Debora Botequio Moretti; Elaine Cristina Marqueze; Elaine Vieira dos Santos; Elisangela Chicaroni Mattos; Erika Freitas; Evandra Strazza Rodrigues; Felipe Allan da Silva da Costa; Flavia Aburjaile; Fábio Sossai Possebon; Guilherme Campos; Guilherme Targino Valente; Heidge Fukumasu. USP-Botucatu: Rejane Maria Tommasini Grotto; Helena Lage Ferreira; Instituto Butantan: Dimas Tadeu Covas; Jardelina de Souza Todao Bernardino; Jayme A. Souza-Neto; Jessika Cristina Chagas Lesbon; Jorge A. Petroli Marchesi; José Salvatore Leister Patané; João Paulo Kitajima; João Pessoa Araújo Jr.; Leila Sabrina Ullmann; Loyze Paola Oliveira de Lima; Luiz Aurelio de Campos Crispin. Centro de Genômica Funcional da ESALQ: Luiz Lehmann Coutinho; Luiz Carlos Junior de Alcantara; Lívia Sacchetto; Maisa C. Pereira Parra; Maria Carolina Elias; Marta Giovanetti; Marília Moraes; Maurício Lacerda Nogueira. Prefeitura de Sao Paulo: Melissa Palmieri.; Patricia Akemi Assato; Paula Rahal; Paulo Inacio da Costa; Rafael dos Santos Bezerra; Raquel de Lello Rocha Campos Cassano. NGS Soluções Genômicas: Pilar Drummond Sampaio Corrêa Mariani. FZEA-USP Pirassununga: Mirele Daiana Poleti; Raul Machado Neto; Ricardo Augusto Brassaloti; Ricardo Haddad; Rodrigo Tocantins Calado. FAMERP-SJRP: Cecília Artico Banho; Sandra Coccuzzo Sampaio; Svetoslav Nanev Slavov; Vagner Fonseca; Vincent Louis Viala |
| EPI_ISL_2614593 | UBS Maria Pia de Oliveira | Instituto Adolfo Lutz, Interdiciplinary Procedures Center, Strategic Laboratory | Caio Vinicius Dias Lopes; Claudia Regina Gonçalves; Claudio Tavares Sacchi; Erica Valessa Ramos Gomes; Karoline Rodrigues Campos; Leonardo Jose Tadeu de Araujo |
| EPI_ISL_1966731, EPI_ISL_1966732, EPI_ISL_1966733, EPI_ISL_1966734 | UBS RAIMUNDA CAVALCANTE DE SOUZA | Instituto Butantan | Antonio Jorge Martins; Bianca Cechetto Carlos. Mendelics: Bibiana Santos; Claudia Renata dos Santos Barros; Cíntia Bittar; David Schlesinger. Hemocentro Ribeirão Preto: Simone Kashima; Debora Botequio Moretti; Elaine Cristina Marqueze; Elaine Vieira dos Santos; Elisangela Chicaroni Mattos; Erika Freitas; Evandra Strazza Rodrigues; Felipe Allan da Silva da Costa; Flavia Aburjaile; Fábio Sossai Possebon; Guilherme Campos; Guilherme Targino Valente; Heidge Fukumasu. USP-Botucatu: Rejane Maria Tommasini Grotto; Helena Lage Ferreira; Instituto Butantan: Dimas Tadeu Covas; Jardelina de Souza Todao Bernardino; Jayme A. Souza-Neto; Jessika Cristina Chagas Lesbon; Jorge A. Petroli Marchesi; José Salvatore Leister Patané; João Paulo Kitajima; João Pessoa Araújo Jr.; Leila Sabrina Ullmann; Loyze Paola Oliveira de Lima; Luiz Aurelio de Campos Crispin. Centro de Genômica Funcional da ESALQ: Luiz Lehmann Coutinho; Luiz Carlos Junior de Alcantara; Lívia Sacchetto; Maisa C. Pereira Parra; Maria Carolina Elias; Marta Giovanetti; Marília Moraes; Maurício Lacerda Nogueira. Prefeitura de Sao Paulo: Melissa Palmieri.; Patricia Akemi Assato; Paula Rahal; Paulo Inacio da Costa; Rafael dos Santos Bezerra; Raquel de Lello Rocha Campos Cassano. NGS Soluções Genômicas: Pilar Drummond Sampaio Corrêa Mariani. FZEA-USP Pirassununga: Mirele Daiana Poleti; Raul Machado Neto; Ricardo Augusto Brassaloti; Ricardo Haddad; Rodrigo Tocantins Calado. FAMERP-SJRP: Cecília Artico Banho; Sandra Coccuzzo Sampaio; Svetoslav Nanev Slavov; Vagner Fonseca; Vincent Louis Viala |
| EPI_ISL_2210207 | UBS RAIMUNDA CAVALCANTE DE SOUZA | Instituto Butantan / Mendelics | Antonio Jorge Martins; Claudia Renata dos Santos Barros; David Schlesinger; Debora Botequio Moretti; Dimas Tadeu Covas; Elaine Cristina Marqueze; Elaine Vieira Santos; Evandra Strazza Rodrigues; Heidge Fukumasu; Jayme Augusto de Souza-Neto; José Salvatore Leister Patané; Luiz Alcantara; Luiz Lehmann Coutinho; Maria Carolina Elias; Maurício Lacerda Nogueira; Rafael dos Santos Bezerra; Raul Machado Neto; Rejane Maria Tommasini Grotto; Ricardo Haddad; Sandra Coccuzzo Sampaio Vessoni; Simone Kashima; Svetoslav Nanev Slavov; Vincent Louis Viala |
| EPI_ISL_2614592 | UBS Silvio Joao Luiz de Lucia | Instituto Adolfo Lutz, Interdiciplinary Procedures Center, Strategic Laboratory | Caio Vinicius Dias Lopes; Claudia Regina Gonçalves; Claudio Tavares Sacchi; Erica Valessa Ramos Gomes; Karoline Rodrigues Campos; Leonardo Jose Tadeu de Araujo |
| EPI_ISL_2614578 | UBS São Lourenco da Serra | Instituto Adolfo Lutz, Interdiciplinary Procedures Center, Strategic Laboratory | Caio Vinicius Dias Lopes; Claudia Regina Gonçalves; Claudio Tavares Sacchi; Erica Valessa Ramos Gomes; Karoline Rodrigues Campos; Leonardo Jose Tadeu de Araujo |
| EPI_ISL_3419574, EPI_ISL_3419575 | UC Davis Genome Center and Healthy Davis Together | UC Davis Genome Center | UC Davis |

| | | | |
|---|---|---|---|
| EPI_ISL_2620767 | UCLA Clinical Micro Lab | Los Angeles County Public Health Laboratories | P. Hemarajata et al. |
| EPI_ISL_1789504, EPI_ISL_1789512, EPI_ISL_1789523 | UMC Groningen, Clinical Virology, Department of Medical Microbiology and Infection Prevention | UMC Groningen, Clinical Virology, Department of Medical Microbiology and Infection Prevention | Alexander Friedrich; Coretta Van Leer-Buter; Erley Lizarazo-Forero; Hubert Niesters; Lilli Gard; Marjolein Knoester; Monika Fliss; Sigrid Rosema; Xuewei Zhou |
| EPI_ISL_3409113, EPI_ISL_3409126, EPI_ISL_3409157, EPI_ISL_3409168, EPI_ISL_3409192, EPI_ISL_3409202, EPI_ISL_3409227, EPI_ISL_3409315, EPI_ISL_3409409, EPI_ISL_3409807, EPI_ISL_3409942, EPI_ISL_3409963, EPI_ISL_3409989, EPI_ISL_3410034, EPI_ISL_3410079, EPI_ISL_3410082, EPI_ISL_3410903 | | | |
| see above | UMass Memorial Medical Center | Center for Microbiome Research | Doyle V. Ward; Karl J. Simon; Richard T. Ellison III |
| EPI_ISL_2344562 | UNIDADE BASICA DE SAUDE ALBERTO CARLOS BOINTONI | Instituto Butantan / FZEA-USP-Pirassununga | Antonio Jorge Martins; Claudia Renata dos Santos Barros; David Schlesinger; Debora Botequio Moretti; Dimas Tadeu Covas; Elaine Cristina Marqueze; Elaine Vieira Santos; Evandra Strazza Rodrigues; Heidge Fukumasu; Jayme Augusto de Souza-Neto; José Salvatore Leister Patané; Luiz Alcantara; Luiz Lehmann Coutinho; Maria Carolina Elias; Maurício Lacerda Nogueira; Rafael dos Santos Bezerra; Raul Machado Neto; Rejane Maria Tommasini Grotto; Ricardo Haddad; Sandra Coccuzzo Sampaio Vessoni; Simone Kashima; Svetoslav Nanev Slavov; Vincent Louis Viala |
| EPI_ISL_2209724 | UNIDADE BASICA DE SAUDE DE IPEUNA | Instituto Butantan | Antonio Jorge Martins; Claudia Renata dos Santos Barros; David Schlesinger; Debora Botequio Moretti; Dimas Tadeu Covas; Elaine Cristina Marqueze; Elaine Vieira Santos; Evandra Strazza Rodrigues; Heidge Fukumasu; Jayme Augusto de Souza-Neto; José Salvatore Leister Patané; Luiz Alcantara; Luiz Lehmann Coutinho; Maria Carolina Elias; Maurício Lacerda Nogueira; Rafael dos Santos Bezerra; Raul Machado Neto; Rejane Maria Tommasini Grotto; Ricardo Haddad; Sandra Coccuzzo Sampaio Vessoni; Simone Kashima; Svetoslav Nanev Slavov; Vincent Louis Viala. |
| EPI_ISL_2210056 | UNIDADE BASICA DE SAUDE DE IPEUNA | Instituto Butantan / FZEA-USP-Pirassununga | Antonio Jorge Martins; Claudia Renata dos Santos Barros; David Schlesinger; Debora Botequio Moretti; Dimas Tadeu Covas; Elaine Cristina Marqueze; Elaine Vieira Santos; Evandra Strazza Rodrigues; Heidge Fukumasu; Jayme Augusto de Souza-Neto; José Salvatore Leister Patané; Luiz Alcantara; Luiz Lehmann Coutinho; Maria Carolina Elias; Maurício Lacerda Nogueira; Rafael dos Santos Bezerra; Raul Machado Neto; Rejane Maria Tommasini Grotto; Ricardo Haddad; Sandra Coccuzzo Sampaio Vessoni; Simone Kashima; Svetoslav Nanev Slavov; Vincent Louis Viala |
| EPI_ISL_2209769 | UNIDADE BASICA DE SAUDE DR MATHEUS GABRIEL BONASSA | Instituto Butantan | Antonio Jorge Martins; Claudia Renata dos Santos Barros; David Schlesinger; Debora Botequio Moretti; Dimas Tadeu Covas; Elaine Cristina Marqueze; Elaine Vieira Santos; Evandra Strazza Rodrigues; Heidge Fukumasu; Jayme Augusto de Souza-Neto; José Salvatore Leister Patané; Luiz Alcantara; Luiz Lehmann Coutinho; Maria Carolina Elias; Maurício Lacerda Nogueira; Rafael dos Santos Bezerra; Raul Machado Neto; Rejane Maria Tommasini Grotto; Ricardo Haddad; Sandra Coccuzzo Sampaio Vessoni; Simone Kashima; Svetoslav Nanev Slavov; Vincent Louis Viala. |
| EPI_ISL_2209183, EPI_ISL_2209996, EPI_ISL_2344708 | UNIDADE BASICA DE SAUDE DR MATHEUS GABRIEL BONASSA | Instituto Butantan / FZEA-USP-Pirassununga | Antonio Jorge Martins; Claudia Renata dos Santos Barros; David Schlesinger; Debora Botequio Moretti; Dimas Tadeu Covas; Elaine Cristina Marqueze; Elaine Vieira Santos; Evandra Strazza Rodrigues; Heidge Fukumasu; Jayme Augusto de Souza-Neto; José Salvatore Leister Patané; Luiz Alcantara; Luiz Lehmann Coutinho; Maria Carolina Elias; Maurício Lacerda Nogueira; Rafael dos Santos Bezerra; Raul Machado Neto; Rejane Maria Tommasini Grotto; Ricardo Haddad; Sandra Coccuzzo Sampaio Vessoni; Simone Kashima; Svetoslav Nanev Slavov; Vincent Louis Viala |
| EPI_ISL_2345790 | UNIDADE DE PRONTO ATENDIMENTO CENTRAL DE CARAGUATATUBA | Instituto Butantan / Mendelics | Antonio Jorge Martins; Claudia Renata dos Santos Barros; David Schlesinger; Debora Botequio Moretti; Dimas Tadeu Covas; Elaine Cristina Marqueze; Elaine Vieira Santos; Evandra Strazza Rodrigues; Heidge Fukumasu; Jayme Augusto de Souza-Neto; José Salvatore Leister Patané; Luiz Alcantara; Luiz Lehmann Coutinho; Maria Carolina Elias; Maurício Lacerda Nogueira; Rafael dos Santos Bezerra; Raul Machado Neto; Rejane Maria Tommasini Grotto; Ricardo Haddad; Sandra Coccuzzo Sampaio Vessoni; Simone Kashima; Svetoslav Nanev Slavov; Vincent Louis Viala |
| EPI_ISL_2209816 | UNIDADE DE PRONTO ATENDIMENTO JD AMANDA | Instituto Butantan | Antonio Jorge Martins; Claudia Renata dos Santos Barros; David Schlesinger; Debora Botequio Moretti; Dimas Tadeu Covas; Elaine Cristina Marqueze; Elaine Vieira Santos; Evandra Strazza Rodrigues; Heidge Fukumasu; Jayme Augusto de Souza-Neto; José Salvatore Leister Patané; Luiz Alcantara; Luiz Lehmann Coutinho; Maria Carolina Elias; Maurício Lacerda Nogueira; Rafael dos Santos Bezerra; Raul Machado Neto; Rejane Maria Tommasini Grotto; Ricardo Haddad; Sandra Coccuzzo Sampaio Vessoni; Simone Kashima; Svetoslav Nanev Slavov; Vincent Louis Viala. |
| EPI_ISL_2344577 | UNIDADE DE PRONTO ATENDIMENTO JD AMANDA | Instituto Butantan / FZEA-USP-Pirassununga | Antonio Jorge Martins; Claudia Renata dos Santos Barros; David Schlesinger; Debora Botequio Moretti; Dimas Tadeu Covas; Elaine Cristina Marqueze; Elaine Vieira Santos; Evandra Strazza Rodrigues; Heidge Fukumasu; Jayme Augusto de Souza-Neto; José Salvatore Leister Patané; Luiz Alcantara; Luiz Lehmann Coutinho; Maria Carolina Elias; Maurício Lacerda Nogueira; Rafael dos Santos Bezerra; Raul Machado Neto; Rejane Maria Tommasini Grotto; Ricardo Haddad; Sandra Coccuzzo Sampaio Vessoni; Simone Kashima; Svetoslav Nanev Slavov; Vincent Louis Viala |
| EPI_ISL_2209222, EPI_ISL_2209237 | UNIDADE DE VIGILANCIA EPIDEMIOLOGICA | Instituto Butantan / FZEA-USP-Pirassununga | Antonio Jorge Martins; Claudia Renata dos Santos Barros; David Schlesinger; Debora Botequio Moretti; Dimas Tadeu Covas; Elaine Cristina Marqueze; Elaine Vieira Santos; Evandra Strazza Rodrigues; Heidge Fukumasu; Jayme Augusto de Souza-Neto; José Salvatore Leister Patané; Luiz Alcantara; Luiz Lehmann Coutinho; Maria Carolina Elias; Maurício Lacerda Nogueira; Rafael dos Santos Bezerra; Raul Machado Neto; Rejane Maria Tommasini Grotto; Ricardo Haddad; Sandra Coccuzzo Sampaio Vessoni; Simone Kashima; Svetoslav Nanev Slavov; Vincent Louis Viala |
| EPI_ISL_2209900 | UNIDADE MISTA DE SAUDE MOMBUCA | Instituto Butantan | Antonio Jorge Martins; Claudia Renata dos Santos Barros; David Schlesinger; Debora Botequio Moretti; Dimas Tadeu Covas; Elaine Cristina Marqueze; Elaine Vieira Santos; Evandra Strazza Rodrigues; Heidge Fukumasu; Jayme Augusto de Souza-Neto; José Salvatore Leister Patané; Luiz Alcantara; Luiz Lehmann Coutinho; Maria Carolina Elias; Maurício Lacerda Nogueira; Rafael dos Santos Bezerra; Raul Machado Neto; Rejane Maria Tommasini Grotto; Ricardo Haddad; Sandra Coccuzzo Sampaio Vessoni; Simone Kashima; Svetoslav Nanev Slavov; Vincent Louis Viala. |
| EPI_ISL_2209990, EPI_ISL_2344725 | UNIDADE MISTA DE SAUDE MOMBUCA | Instituto Butantan / FZEA-USP-Pirassununga | Antonio Jorge Martins; Claudia Renata dos Santos Barros; David Schlesinger; Debora Botequio Moretti; Dimas Tadeu Covas; Elaine Cristina Marqueze; Elaine Vieira Santos; Evandra Strazza Rodrigues; Heidge Fukumasu; Jayme Augusto de Souza-Neto; José Salvatore Leister Patané; Luiz Alcantara; Luiz Lehmann Coutinho; Maria Carolina Elias; Maurício Lacerda Nogueira; Rafael dos Santos Bezerra; Raul Machado Neto; Rejane Maria Tommasini Grotto; Ricardo Haddad; Sandra Coccuzzo Sampaio Vessoni; Simone Kashima; Svetoslav Nanev Slavov; Vincent Louis Viala |
| EPI_ISL_2209908 | UNIDADE MISTA OSVALDO P CAMARGO ELIAS FAUSTO | Instituto Butantan | Antonio Jorge Martins; Claudia Renata dos Santos Barros; David Schlesinger; Debora Botequio Moretti; Dimas Tadeu Covas; Elaine Cristina Marqueze; Elaine Vieira Santos; Evandra Strazza Rodrigues; Heidge Fukumasu; Jayme Augusto de Souza-Neto; José Salvatore Leister Patané; Luiz Alcantara; Luiz Lehmann Coutinho; Maria Carolina Elias; Maurício Lacerda Nogueira; Rafael dos Santos Bezerra; Raul Machado Neto; Rejane Maria Tommasini Grotto; Ricardo Haddad; Sandra Coccuzzo Sampaio Vessoni; Simone Kashima; Svetoslav Nanev Slavov; Vincent Louis Viala. |
| EPI_ISL_2209246, EPI_ISL_2209248, EPI_ISL_2362239 | UNIDADE MISTA OSVALDO P CAMARGO ELIAS FAUSTO | Instituto Butantan / FZEA-USP-Pirassununga | Antonio Jorge Martins; Claudia Renata dos Santos Barros; David Schlesinger; Debora Botequio Moretti; Dimas Tadeu Covas; Elaine Cristina Marqueze; Elaine Vieira Santos; Evandra Strazza Rodrigues; Heidge Fukumasu; Jayme Augusto de Souza-Neto; José Salvatore Leister Patané; Luiz Alcantara; Luiz Lehmann Coutinho; Maria Carolina Elias; Maurício Lacerda Nogueira; Rafael dos Santos Bezerra; Raul Machado Neto; Rejane Maria Tommasini Grotto; Ricardo Haddad; Sandra Coccuzzo Sampaio Vessoni; Simone Kashima; Svetoslav Nanev Slavov; Vincent Louis Viala |
| EPI_ISL_2209916, EPI_ISL_2209931 | UNIDADE RESPIRATORIA NOVA HORTOLANDIA | Instituto Butantan | Antonio Jorge Martins; Claudia Renata dos Santos Barros; David Schlesinger; Debora Botequio Moretti; Dimas Tadeu Covas; Elaine Cristina Marqueze; Elaine Vieira Santos; Evandra Strazza Rodrigues; Heidge Fukumasu; Jayme Augusto de Souza-Neto; José Salvatore Leister Patané; Luiz Alcantara; Luiz Lehmann Coutinho; Maria Carolina Elias; Maurício Lacerda Nogueira; Rafael dos Santos Bezerra; Raul Machado Neto; Rejane Maria Tommasini Grotto; Ricardo Haddad; Sandra Coccuzzo Sampaio Vessoni; Simone Kashima; Svetoslav Nanev Slavov; Vincent Louis Viala. |
| EPI_ISL_2344565, EPI_ISL_2344580, EPI_ISL_2344581, EPI_ISL_2344582 | UNIDADE RESPIRATORIA NOVA HORTOLANDIA | Instituto Butantan / FZEA-USP-Pirassununga | Antonio Jorge Martins; Claudia Renata dos Santos Barros; David Schlesinger; Debora Botequio Moretti; Dimas Tadeu Covas; Elaine Cristina Marqueze; Elaine Vieira Santos; Evandra Strazza Rodrigues; Heidge Fukumasu; Jayme Augusto de Souza-Neto; José Salvatore Leister Patané; Luiz Alcantara; Luiz Lehmann Coutinho; Maria Carolina Elias; Maurício Lacerda Nogueira; Rafael dos Santos Bezerra; Raul Machado Neto; Rejane Maria Tommasini Grotto; Ricardo Haddad; Sandra Coccuzzo Sampaio Vessoni; Simone Kashima; Svetoslav Nanev Slavov; Vincent Louis Viala |
| EPI_ISL_2136314, EPI_ISL_2136324, EPI_ISL_2136325, EPI_ISL_2136328, EPI_ISL_2136329, EPI_ISL_2136331, EPI_ISL_2136334, EPI_ISL_2321592, EPI_ISL_2321593, EPI_ISL_2321601, EPI_ISL_2321602, EPI_ISL_2321603, EPI_ISL_2321606, EPI_ISL_2321607, EPI_ISL_2321608, EPI_ISL_2321612, EPI_ISL_2321613, EPI_ISL_2321614, EPI_ISL_2321615, EPI_ISL_2321616, EPI_ISL_2321617, EPI_ISL_2321618, EPI_ISL_2321619, EPI_ISL_2321622, EPI_ISL_2321625, EPI_ISL_2321626, EPI_ISL_3267000 | | | |
| see above | UNIGEM | Universidad Nacional de Colombia - Laboratorio Genómico One Health | Andres F. Cardona-Rios; Carlos Franco-Muñoz; Carolina Muñoz-Arango; Celeny Ortiz; Daniel O. Maldonado-Perez; Diego A. Álvarez-Díaz; Hector Alejandro Ruiz-Moreno; Idabely Betancur Ortiz; Jorge E. Osorio; Juan P. Hernandez-Ortiz; Karl A Ciuoderis; Katherine Laiton-Donato; Laura Silvana Perez; Lina M. Hurtado; Marcela Mercado-Reyes; Maria Angélica Maya; Maria Stella López; Rita Almanza Payares; Sandra Ines Cano; Simón Villegas Velásquez |
| EPI_ISL_2614574 | UPA Akira Tada | Instituto Adolfo Lutz, Interdiciplinary Procedures Center, Strategic Laboratory | Caio Vinicius Dias Lopes; Claudia Regina Gonçalves; Claudio Tavares Sacchi; Erica Valessa Ramos Gomes; Karoline Rodrigues Campos; Leonardo Jose Tadeu de Araujo |
| EPI_ISL_2209040 | UPA DR AKIRA TADA | Instituto Butantan / Mendelics | Antonio Jorge Martins; Claudia Renata dos Santos Barros; David Schlesinger; Debora Botequio Moretti; Dimas Tadeu Covas; Elaine Cristina Marqueze; Elaine Vieira Santos; Evandra Strazza Rodrigues; Heidge Fukumasu; Jayme Augusto de Souza-Neto; José Salvatore Leister Patané; Luiz Alcantara; Luiz Lehmann Coutinho; Maria Carolina Elias; Maurício Lacerda Nogueira; Rafael dos Santos Bezerra; Raul Machado Neto; Rejane Maria Tommasini Grotto; Ricardo Haddad; Sandra Coccuzzo Sampaio Vessoni; Simone Kashima; Svetoslav Nanev Slavov; Vincent Louis Viala |
| EPI_ISL_2208904, EPI_ISL_2362256 | UPA DR FRANCO DA ROCHA | Instituto Butantan / Mendelics | Antonio Jorge Martins; Claudia Renata dos Santos Barros; David Schlesinger; Debora Botequio Moretti; Dimas Tadeu Covas; Elaine Cristina Marqueze; Elaine Vieira Santos; Evandra Strazza Rodrigues; Heidge Fukumasu; Jayme Augusto de Souza-Neto; José Salvatore Leister Patané; Luiz Alcantara; Luiz Lehmann Coutinho; Maria Carolina Elias; Maurício Lacerda Nogueira; Rafael dos Santos Bezerra; Raul Machado Neto; Rejane Maria Tommasini Grotto; Ricardo Haddad; Sandra Coccuzzo Sampaio Vessoni; Simone Kashima; Svetoslav Nanev Slavov; Vincent Louis Viala |
| EPI_ISL_1966729 | UPA JOSE CAMPOS BARRETO ZEQUINHA BARRETO | Instituto Butantan | Antonio Jorge Martins; Bianca Cechetto Carlos. Mendelics: Bibiana Santos; Claudia Renata dos Santos Barros; Cíntia Bittar; David Schlesinger. Hemocentro Ribeirão Preto: Simone Kashima; Debora Botequio Moretti; Elaine Cristina Marqueze; Elaine Vieira dos Santos; Elisangela Chicaroni Mattos; Erika Freitas; Evandra Strazza Rodrigues; Felipe Allan da Silva da Costa; Flavia Aburjaile; Fábio Sossai Possebon; Guilherme Campos; Guilherme Targino Valente; Heidge Fukumasu. USP-Botucatu: Rejane Maria Tommasini Grotto; Helena Lage Ferreira; Instituto Butantan: Dimas Tadeu Covas; Jardelina de Souza Todao Bernardino; Jayme A. Souza-Neto; Jessika Cristina Chagas Lesbon; Jorge A. Petroli Marchesi; José Salvatore Leister Patané; João Paulo Kitajima; João Pessoa Araújo Jr.; Leila Sabrina Ullmann; Loyze Paola Oliveira de Lima; Luiz Aurelio de Campos Crispin. Centro de Genômica Funcional da ESALQ: Luiz Lehmann Coutinho; Luiz Carlos Junior de Alcantara; Lívia Sacchetto; Maisa C. Pereira Parra; Maria Carolina Elias; Marta Giovanetti; Marília Moraes; Maurício Lacerda Nogueira. Prefeitura de Sao Paulo: Melissa Palmieri.; Patricia Akemi Assato; Paula Rahal; Paulo Inacio da Costa; Rafael dos Santos Bezerra; Raquel de Lello Rocha Campos Cassano. NGS Soluções Genômicas: Pilar Drummond Sampaio Corrêa Mariani. FZEA-USP Pirassununga: Mirele Daiana Poleti; Raul Machado Neto; Ricardo Augusto Brassaloti; Ricardo Haddad; Rodrigo Tocantins Calado. FAMERP-SJRP: Cecília Artico Banho; Sandra Coccuzzo Sampaio; Svetoslav Nanev Slavov; Vagner Fonseca; Vincent Louis Viala |
| EPI_ISL_2344575, EPI_ISL_2344576 | UPA UNIDADE DE PRONTO ATENDIMENTO 24 HORAS BOM JESUS | Instituto Butantan / FZEA-USP-Pirassununga | Antonio Jorge Martins; Claudia Renata dos Santos Barros; David Schlesinger; Debora Botequio Moretti; Dimas Tadeu Covas; Elaine Cristina Marqueze; Elaine Vieira Santos; Evandra Strazza Rodrigues; Heidge Fukumasu; Jayme Augusto de Souza-Neto; José Salvatore Leister Patané; Luiz Alcantara; Luiz Lehmann Coutinho; Maria Carolina Elias; Maurício Lacerda Nogueira; Rafael dos Santos Bezerra; Raul Machado Neto; Rejane Maria Tommasini Grotto; Ricardo Haddad; Sandra Coccuzzo Sampaio Vessoni; Simone Kashima; Svetoslav Nanev Slavov; Vincent Louis Viala |
| EPI_ISL_2614595 | UPA Vila Santa Catarina | Instituto Adolfo Lutz, Interdiciplinary Procedures Center, Strategic Laboratory | Caio Vinicius Dias Lopes; Claudia Regina Gonçalves; Claudio Tavares Sacchi; Erica Valessa Ramos Gomes; Karoline Rodrigues Campos; Leonardo Jose Tadeu de Araujo |
| EPI_ISL_2260375, EPI_ISL_2260390 | USC Clinical Lab | Los Angeles County Public Health Laboratories | P. Hemarajata et al. |
| EPI_ISL_1707666, EPI_ISL_1841218, EPI_ISL_1841219 | USCA Avezzano | Istituto Zooprofilattico Sperimentale dell'Abruzzo e Molise "G. Caporale" | Ancora M; Calistri P; Cammà C; Caporale M; Curini V; Delli Compagni E; Di Domenico M; Di Lollo Valeria; Di Pasquale A; Lorusso A; Mangone I; Marcacci M; Puglia I; Rinaldi A; Savini G; Scialabba S |
| EPI_ISL_1785196 | USCA Tagliacozzo | Istituto Zooprofilattico Sperimentale dell'Abruzzo e Molise "G. Caporale" | Ancora M; Calistri P; Cammà C; Caporale M; Curini V; Delli Compagni E; Di Domenico M; Di Lollo Valeria; Di Pasquale A; Lorusso A; Mangone I; Marcacci M; Puglia I; Rinaldi A; Savini G; Scialabba S |
| EPI_ISL_1715490, EPI_ISL_1715506, EPI_ISL_1715531, EPI_ISL_1715709, EPI_ISL_1715711, EPI_ISL_1715738, EPI_ISL_1715739, EPI_ISL_1715747, EPI_ISL_1715752, EPI_ISL_1715753, EPI_ISL_1715754, EPI_ISL_1715760, EPI_ISL_1715762, EPI_ISL_1715765, EPI_ISL_1785623, EPI_ISL_1785641, EPI_ISL_1785646, EPI_ISL_1785657, EPI_ISL_1785660, EPI_ISL_1785673, EPI_ISL_1785674, EPI_ISL_1785779, EPI_ISL_1785797, EPI_ISL_1785831, EPI_ISL_1785839, EPI_ISL_1785842, EPI_ISL_1785855, EPI_ISL_1785858, EPI_ISL_1785864, EPI_ISL_1785920, EPI_ISL_1785924, EPI_ISL_1785927, EPI_ISL_1785928, EPI_ISL_1785934, EPI_ISL_1785936, EPI_ISL_1785941, EPI_ISL_1785949, EPI_ISL_1785951, EPI_ISL_1785957, EPI_ISL_1789429, EPI_ISL_1789452, EPI_ISL_1789457, EPI_ISL_1789459, EPI_ISL_1789461, EPI_ISL_1825889, EPI_ISL_1825893, EPI_ISL_1825895, EPI_ISL_1825921, EPI_ISL_1825925, EPI_ISL_1825927, EPI_ISL_1826086, EPI_ISL_1826087, EPI_ISL_1826088, EPI_ISL_1826089, EPI_ISL_1826090, EPI_ISL_1826091, EPI_ISL_1826092, EPI_ISL_1826093, EPI_ISL_1826094, EPI_ISL_1826095, EPI_ISL_1826096, EPI_ISL_1826097, EPI_ISL_1826098, EPI_ISL_1826099, EPI_ISL_1826100, EPI_ISL_1826101, EPI_ISL_1826102, EPI_ISL_1826103, EPI_ISL_1826104, EPI_ISL_1826105, EPI_ISL_1826106, EPI_ISL_1826107, EPI_ISL_1826108, EPI_ISL_1826109, EPI_ISL_1826110, EPI_ISL_1826111, EPI_ISL_1826112, EPI_ISL_1826113, EPI_ISL_1826114, EPI_ISL_1826115, EPI_ISL_1826116, EPI_ISL_1826117, EPI_ISL_1826118, EPI_ISL_1826119, EPI_ISL_1826120, EPI_ISL_1826121, EPI_ISL_1826122, EPI_ISL_1826123, EPI_ISL_1826124, EPI_ISL_1826125, EPI_ISL_1849063, EPI_ISL_1849074, EPI_ISL_1849242, EPI_ISL_1849262, EPI_ISL_1849267, EPI_ISL_1849277, EPI_ISL_1849342, EPI_ISL_1849408, EPI_ISL_1849429, EPI_ISL_1849438, EPI_ISL_1849458, EPI_ISL_1849468, EPI_ISL_1849502, EPI_ISL_1849526, EPI_ISL_1849582, EPI_ISL_1849620, EPI_ISL_1849635, EPI_ISL_1849659, EPI_ISL_1849723, EPI_ISL_1849733, EPI_ISL_1849737, EPI_ISL_1849739, EPI_ISL_1849777, EPI_ISL_1849801, EPI_ISL_1849827, EPI_ISL_1849833, EPI_ISL_1849871, EPI_ISL_1849889, EPI_ISL_1849902, EPI_ISL_1850064, EPI_ISL_1850095, EPI_ISL_1850126, EPI_ISL_1850135, EPI_ISL_1850166, EPI_ISL_1850251, EPI_ISL_1850296, EPI_ISL_1850444, EPI_ISL_1850485, EPI_ISL_1850491, EPI_ISL_2002140, EPI_ISL_2002160, EPI_ISL_2002168, EPI_ISL_2002188, EPI_ISL_2002229, EPI_ISL_2002235, EPI_ISL_2002237, EPI_ISL_2002239, EPI_ISL_2002244, EPI_ISL_2002271, EPI_ISL_2002272, EPI_ISL_2002273, EPI_ISL_2002274, EPI_ISL_2002287, EPI_ISL_2002521, EPI_ISL_2002524, EPI_ISL_2002536, EPI_ISL_2002548, EPI_ISL_2002563, EPI_ISL_2002575, EPI_ISL_2002936, EPI_ISL_2002937, EPI_ISL_2002938, EPI_ISL_2002939, EPI_ISL_2002940, EPI_ISL_2172433, EPI_ISL_2172468, EPI_ISL_2172470, EPI_ISL_2172471, EPI_ISL_2172472, EPI_ISL_4395955 | | | |
| see above | UW Virology Lab | UW Virology Lab | Alexander Greninger; Hong Xie; Keith R Jerome; Lasata Shrestha; Maria Lukes; Meei-Li Huang; Michelle Lin; Noah R. Baker; Pavitra Roychoudhury; Ricardo Perez; Savanna S. Carmack; Sean Ellis; Shah Mohamed Bakhash; Tien V. Nguyen |
| EPI_ISL_5802417 | Ubs Getulino Jose Dias | Instituto Butantan | Antonio Jorge Martins; Claudia Renata dos Santos Barros; David Schlesinger; Debora Botequio Moretti; Dimas Tadeu Covas; Elaine Cristina Marqueze; Elaine Vieira Santos; Evandra Strazza Rodrigues; Heidge Fukumasu; Jayme Augusto de Souza-Neto; José Salvatore Leister Patané; Luiz Alcantara; Luiz Lehmann Coutinho; Maria Carolina Elias; Maurício Lacerda Nogueira; Rafael dos Santos Bezerra; Raul Machado Neto; Rejane Maria Tommasini Grotto; Ricardo Haddad; Sandra Coccuzzo Sampaio Vessoni; Simone Kashima; Svetoslav Nanev Slavov; Vincent Louis Viala |
| EPI_ISL_5802419 | Ubs Guilhermina De Nobrega Abreu | Instituto Butantan | Antonio Jorge Martins; Claudia Renata dos Santos Barros; David Schlesinger; Debora Botequio Moretti; Dimas Tadeu Covas; Elaine Cristina Marqueze; Elaine Vieira Santos; Evandra Strazza Rodrigues; Heidge Fukumasu; Jayme Augusto de Souza-Neto; José Salvatore Leister Patané; Luiz Alcantara; Luiz Lehmann Coutinho; Maria Carolina Elias; Maurício Lacerda Nogueira; Rafael dos Santos Bezerra; Raul Machado Neto; Rejane Maria Tommasini Grotto; Ricardo Haddad; Sandra Coccuzzo Sampaio Vessoni; Simone Kashima; Svetoslav Nanev Slavov; Vincent Louis Viala |
| EPI_ISL_5800417, EPI_ISL_5800430 | Ubs Jose Sabino Ferreira | Instituto Butantan | Antonio Jorge Martins; Claudia Renata dos Santos Barros; David Schlesinger; Debora Botequio Moretti; Dimas Tadeu Covas; Elaine Cristina Marqueze; Elaine Vieira Santos; Evandra Strazza Rodrigues; Heidge Fukumasu; Jayme Augusto de Souza-Neto; José Salvatore Leister Patané; Luiz Alcantara; Luiz Lehmann Coutinho; Maria Carolina Elias; Maurício Lacerda Nogueira; Rafael dos Santos Bezerra; Raul Machado Neto; Rejane Maria Tommasini Grotto; Ricardo Haddad; Sandra Coccuzzo Sampaio Vessoni; Simone Kashima; Svetoslav Nanev Slavov; Vincent Louis Viala |
| EPI_ISL_5800432, EPI_ISL_5800433, EPI_ISL_5800436, EPI_ISL_5800439, EPI_ISL_5802410 | Ubs Raimunda Cavalcante De Souza | Instituto Butantan | Antonio Jorge Martins; Claudia Renata dos Santos Barros; David Schlesinger; Debora Botequio Moretti; Dimas Tadeu Covas; Elaine Cristina Marqueze; Elaine Vieira Santos; Evandra Strazza Rodrigues; Heidge Fukumasu; Jayme Augusto de Souza-Neto; José Salvatore Leister Patané; Luiz Alcantara; Luiz Lehmann Coutinho; Maria Carolina Elias; Maurício Lacerda Nogueira; Rafael dos Santos Bezerra; Raul Machado Neto; Rejane Maria Tommasini Grotto; Ricardo Haddad; Sandra Coccuzzo Sampaio Vessoni; Simone Kashima; Svetoslav Nanev Slavov; Vincent Louis Viala |
| EPI_ISL_2402020 | Unidad de Investigación Médica | Unidad de Genomica Avanzada | Alejandro Sanchez-Flores; Alfredo Herrera-Estrella; Alicia Ocana-Mondragon; Angel Gustavo Salas-Lais; Bernardo Martinez-Miguel; Blanca Taboada; Brenda Irasema Maldonado-Meza; Carla Ivon Herrera-Najera; Carlos F. Arias; Celia Boukadida; Clara Esperanza Santacruz-Tinoco; Concepcion Grajales-Muniz; |

| | | | |
|---|---|---|---|
| | de Yucatán (UIMY) | | Consorcio Mexicano de Vigilancia Genomica (CoViGen-Mex). Authors (in alphabetical order): Julio Elias Alvarado-Yaah; Fernando Fontove-Herrera; Francisco Pulido; Gloria Elena Espinoza-Ayala; Gloria Maria Molina-Salinas; Gloria Vazquez; Hector Esteban Paz-Juarez; Hector Montoya-Fuentes; Helen Haydee Fernanda Ramirez-Plascencia; Jorge Ivan Salinal-Nevarez; Jose Antonio Enciso-Moreno; Jose Esteban Munoz-Medina; Jose de Jesus Nunez-Contreras; Juan Bautista Chale-Dzul; Luis Alberto Ochoa-Carrera; Margarita Matias-Florentino; Maria Guadalupe Santiago-Mauricio; Maria Guadalupe de Jesus Mireles-Rivera; Nelly Selem-Mojica; Pavel Isa; Ricardo Grande; Santiago Avila-Rios; Victor Hugo Borja-Aburto |
| EPI_ISL_5782470 | Unidade Basica De Saude Alberto Carlos Bointoni | Instituto Butantan | Antonio Jorge Martins; Claudia Renata dos Santos Barros; David Schlesinger; Debora Botequio Moretti; Dimas Tadeu Covas; Elaine Cristina Marqueze; Elaine Vieira Santos; Evandra Strazza Rodrigues; Heidge Fukumasu; Jayme Augusto de Souza-Neto; José Salvatore Leister Patané; Luiz Alcantara; Luiz Lehmann Coutinho; Maria Carolina Elias; Maurício Lacerda Nogueira; Rafael dos Santos Bezerra; Raul Machado Neto; Rejane Maria Tommasini Grotto; Ricardo Haddad; Sandra Coccuzzo Sampaio Vessoni; Simone Kashima; Svetoslav Nanev Slavov; Vincent Louis Viala |
| EPI_ISL_5799724 | Unidade Basica De Saude De Ipeuna | Instituto Butantan | Antonio Jorge Martins; Claudia Renata dos Santos Barros; David Schlesinger; Debora Botequio Moretti; Dimas Tadeu Covas; Elaine Cristina Marqueze; Elaine Vieira Santos; Evandra Strazza Rodrigues; Heidge Fukumasu; Jayme Augusto de Souza-Neto; José Salvatore Leister Patané; Luiz Alcantara; Luiz Lehmann Coutinho; Maria Carolina Elias; Maurício Lacerda Nogueira; Rafael dos Santos Bezerra; Raul Machado Neto; Rejane Maria Tommasini Grotto; Ricardo Haddad; Sandra Coccuzzo Sampaio Vessoni; Simone Kashima; Svetoslav Nanev Slavov; Vincent Louis Viala |
| EPI_ISL_5799729, EPI_ISL_5799730, EPI_ISL_5799749 | Unidade Basica De Saude Dr Matheus Gabriel Bonassa | Instituto Butantan | Antonio Jorge Martins; Claudia Renata dos Santos Barros; David Schlesinger; Debora Botequio Moretti; Dimas Tadeu Covas; Elaine Cristina Marqueze; Elaine Vieira Santos; Evandra Strazza Rodrigues; Heidge Fukumasu; Jayme Augusto de Souza-Neto; José Salvatore Leister Patané; Luiz Alcantara; Luiz Lehmann Coutinho; Maria Carolina Elias; Maurício Lacerda Nogueira; Rafael dos Santos Bezerra; Raul Machado Neto; Rejane Maria Tommasini Grotto; Ricardo Haddad; Sandra Coccuzzo Sampaio Vessoni; Simone Kashima; Svetoslav Nanev Slavov; Vincent Louis Viala |
| EPI_ISL_5802434 | Unidade De Pronto Atendimento Central De Caraguatatuba | Instituto Butantan | Antonio Jorge Martins; Claudia Renata dos Santos Barros; David Schlesinger; Debora Botequio Moretti; Dimas Tadeu Covas; Elaine Cristina Marqueze; Elaine Vieira Santos; Evandra Strazza Rodrigues; Heidge Fukumasu; Jayme Augusto de Souza-Neto; José Salvatore Leister Patané; Luiz Alcantara; Luiz Lehmann Coutinho; Maria Carolina Elias; Maurício Lacerda Nogueira; Rafael dos Santos Bezerra; Raul Machado Neto; Rejane Maria Tommasini Grotto; Ricardo Haddad; Sandra Coccuzzo Sampaio Vessoni; Simone Kashima; Svetoslav Nanev Slavov; Vincent Louis Viala |
| EPI_ISL_5782485 | Unidade De Pronto Atendimento Jd Amanda | Instituto Butantan | Antonio Jorge Martins; Claudia Renata dos Santos Barros; David Schlesinger; Debora Botequio Moretti; Dimas Tadeu Covas; Elaine Cristina Marqueze; Elaine Vieira Santos; Evandra Strazza Rodrigues; Heidge Fukumasu; Jayme Augusto de Souza-Neto; José Salvatore Leister Patané; Luiz Alcantara; Luiz Lehmann Coutinho; Maria Carolina Elias; Maurício Lacerda Nogueira; Rafael dos Santos Bezerra; Raul Machado Neto; Rejane Maria Tommasini Grotto; Ricardo Haddad; Sandra Coccuzzo Sampaio Vessoni; Simone Kashima; Svetoslav Nanev Slavov; Vincent Louis Viala |
| EPI_ISL_5799735, EPI_ISL_5799737 | Unidade De Vigilancia Epidemiologica | Instituto Butantan | Antonio Jorge Martins; Claudia Renata dos Santos Barros; David Schlesinger; Debora Botequio Moretti; Dimas Tadeu Covas; Elaine Cristina Marqueze; Elaine Vieira Santos; Evandra Strazza Rodrigues; Heidge Fukumasu; Jayme Augusto de Souza-Neto; José Salvatore Leister Patané; Luiz Alcantara; Luiz Lehmann Coutinho; Maria Carolina Elias; Maurício Lacerda Nogueira; Rafael dos Santos Bezerra; Raul Machado Neto; Rejane Maria Tommasini Grotto; Ricardo Haddad; Sandra Coccuzzo Sampaio Vessoni; Simone Kashima; Svetoslav Nanev Slavov; Vincent Louis Viala |
| EPI_ISL_5799728, EPI_ISL_5799767 | Unidade Mista De Saude Mombuca | Instituto Butantan | Antonio Jorge Martins; Claudia Renata dos Santos Barros; David Schlesinger; Debora Botequio Moretti; Dimas Tadeu Covas; Elaine Cristina Marqueze; Elaine Vieira Santos; Evandra Strazza Rodrigues; Heidge Fukumasu; Jayme Augusto de Souza-Neto; José Salvatore Leister Patané; Luiz Alcantara; Luiz Lehmann Coutinho; Maria Carolina Elias; Maurício Lacerda Nogueira; Rafael dos Santos Bezerra; Raul Machado Neto; Rejane Maria Tommasini Grotto; Ricardo Haddad; Sandra Coccuzzo Sampaio Vessoni; Simone Kashima; Svetoslav Nanev Slavov; Vincent Louis Viala |
| EPI_ISL_5799739, EPI_ISL_5799740, EPI_ISL_5799741 | Unidade Mista Osvaldo P Camargo Elias Fausto | Instituto Butantan | Antonio Jorge Martins; Claudia Renata dos Santos Barros; David Schlesinger; Debora Botequio Moretti; Dimas Tadeu Covas; Elaine Cristina Marqueze; Elaine Vieira Santos; Evandra Strazza Rodrigues; Heidge Fukumasu; Jayme Augusto de Souza-Neto; José Salvatore Leister Patané; Luiz Alcantara; Luiz Lehmann Coutinho; Maria Carolina Elias; Maurício Lacerda Nogueira; Rafael dos Santos Bezerra; Raul Machado Neto; Rejane Maria Tommasini Grotto; Ricardo Haddad; Sandra Coccuzzo Sampaio Vessoni; Simone Kashima; Svetoslav Nanev Slavov; Vincent Louis Viala |
| EPI_ISL_5782473, EPI_ISL_5782488, EPI_ISL_5782489, EPI_ISL_5782490 | Unidade Respiratoria Nova Hortolandia | Instituto Butantan | Antonio Jorge Martins; Claudia Renata dos Santos Barros; David Schlesinger; Debora Botequio Moretti; Dimas Tadeu Covas; Elaine Cristina Marqueze; Elaine Vieira Santos; Evandra Strazza Rodrigues; Heidge Fukumasu; Jayme Augusto de Souza-Neto; José Salvatore Leister Patané; Luiz Alcantara; Luiz Lehmann Coutinho; Maria Carolina Elias; Maurício Lacerda Nogueira; Rafael dos Santos Bezerra; Raul Machado Neto; Rejane Maria Tommasini Grotto; Ricardo Haddad; Sandra Coccuzzo Sampaio Vessoni; Simone Kashima; Svetoslav Nanev Slavov; Vincent Louis Viala |
| EPI_ISL_2101387, EPI_ISL_2101391, EPI_ISL_2101431, EPI_ISL_2101432, EPI_ISL_2101433, EPI_ISL_2101455, EPI_ISL_2101456, EPI_ISL_2101457, EPI_ISL_2101458, EPI_ISL_2101459, EPI_ISL_2101460, EPI_ISL_2101461, EPI_ISL_2101462, EPI_ISL_2101463, EPI_ISL_2101470, EPI_ISL_2101495, EPI_ISL_2101505, EPI_ISL_2101506, EPI_ISL_2101522, EPI_ISL_2101524, EPI_ISL_2101525, EPI_ISL_2101526, EPI_ISL_2101527, EPI_ISL_2101612, EPI_ISL_2101644, EPI_ISL_2101645, EPI_ISL_2101646, EPI_ISL_2101647, EPI_ISL_2101648, EPI_ISL_2101649, EPI_ISL_2101650, EPI_ISL_2101651, EPI_ISL_2101652, EPI_ISL_2101653, EPI_ISL_2101654, EPI_ISL_2101655, EPI_ISL_2101656, EPI_ISL_2101657, EPI_ISL_2101658, EPI_ISL_2101659, EPI_ISL_2101665, EPI_ISL_2101666, EPI_ISL_2101667, EPI_ISL_2101668, EPI_ISL_2101669, EPI_ISL_2101670, EPI_ISL_2101676, EPI_ISL_2101677, EPI_ISL_2101678, EPI_ISL_2101679, EPI_ISL_2101680, EPI_ISL_2101715, EPI_ISL_2101724, EPI_ISL_2101727, EPI_ISL_2101740, EPI_ISL_2101741, EPI_ISL_2101744 | | | |
| see above | Unidade de apoio ao diagnóstico da COVID – UNADIG | Bioinformatics Laboratory / LNCC | Alessandra P Lamarca; Alexandra L Gerber; Amilcar Tanuri; Ana Paula de C Guimaraes; Ana Tereza R Vasconcelos; Andrea Cony Cavalcanti; Caio Luiz Pereira Ribeiro; Cassia Alves; Cintia Policarpo; Claudia Maria Braga de Mello; Cristiane Gomes da Silva; Diana Mariani; Douglas Terra Machado; Flavio Dias da Silva; Gleidson da Silva de Oliveira; Leandro Magalhaes de Souza; Liliane Cavalcante; Luiz G P de Almeida; Marcio Henrique de Oliveira Garcia; Mario Sergio Ribeiro; Ronaldo da Silva F Jr; Silvia Carvalho |
| EPI_ISL_1911626 | Unilabs | Institute of Medical Virology | Alexandra Trkola; Annette AudigŽ; Cyril Shah; Gabriela Ziltener; Guido Bloemberg; Jon Huder; JŸrg Bšni; Kevin Steiner; Maria GrŸnberg; Maryam Zaheri; Michael Huber; Riccarda Capaul; Stefan Schmutz; Verena Kufner |
| EPI_ISL_3671011 | Universidad Nacional sede Medellin Laboratorio One Health | Universidad Nacional de Colombia - Laboratorio Genómico One Health | Andres F. Cardona-Rios; Carlos Franco-Muñoz; Carolina Muñoz-Arango; Celeny Ortiz; Daniel O. Maldonado-Perez; Diego A. Álvarez-Díaz; Hector Alejandro Ruiz-Moreno; Idabely Betancur Ortiz; Jorge E. Osorio; Juan P. Hernandez-Ortiz; Karl A Ciuoderis; Katherine Laiton-Donato; Laura Silvana Perez; Lina M. Hurtado; Marcela Mercado-Reyes; Maria Angélica Maya; Maria Stella López; Rita Almanza Payares; Sandra Ines Cano; Simón Villegas Velásquez |
| EPI_ISL_3671022 | Universidad de Antioquia Laboratorio LIME | Universidad Nacional de Colombia - Laboratorio Genómico One Health | Andres F. Cardona-Rios; Carlos Franco-Muñoz; Carolina Muñoz-Arango; Celeny Ortiz; Daniel O. Maldonado-Perez; Diego A. Álvarez-Díaz; Hector Alejandro Ruiz-Moreno; Idabely Betancur Ortiz; Jorge E. Osorio; Juan P. Hernandez-Ortiz; Karl A Ciuoderis; Katherine Laiton-Donato; Laura Silvana Perez; Lina M. Hurtado; Marcela Mercado-Reyes; Maria Angélica Maya; Maria Stella López; Rita Almanza Payares; Sandra Ines Cano; Simón Villegas Velásquez |
| EPI_ISL_1714761, EPI_ISL_1714762, EPI_ISL_1714763, EPI_ISL_1714764, EPI_ISL_1714765, EPI_ISL_1714766, EPI_ISL_1714767, EPI_ISL_1714768, EPI_ISL_1714769 | | | |
| see above | University Hospitals Translational Laboratory (UHTL), University Hospitals | University Hospitals Translational Laboratory (UHTL), University Hospitals | Alouani, D.; Sadri, N.; Song, X. |
| EPI_ISL_1708132 | University Hospitals of Geneva, Laboratory of Virology | HUG, Laboratory of Virology and the Health2030 Genome Center | Ana Rita Goncalves; Deborah Penet; Emmanouil Dermitzakis; Henri Pegeot; Ioannis Xenarios; Keith Harshman; Laurent Kaiser; Lorenzo Cerutti; Melyssa Elies; Samuel Cordey |
| EPI_ISL_2939098, EPI_ISL_2939100 | University of Arkansas for Medical Sciences | Center for Global Health, University of New Mexico Health Sciences Center | Catherine Kirkpatrick; Darrell Dinwiddie; Daryl Domman; David Ussery; Jennifer Laudadio; Joshua L. Kennedy; Kurt Schwalm; Susan Harley; Uday Chalwadi; Valerie Morley |
| EPI_ISL_2246581, EPI_ISL_2246585, EPI_ISL_2246590, EPI_ISL_2246605, EPI_ISL_2692016 | University of Chicago Medicine | RIPHL at Rush University Medical Center | Cindy Bethel; Felix Araujo Perez; Kevin Kunstman; Laura Furtado; Lauren Megger; Marieta Hyde; Max Kolton; Stefan Green |
| EPI_ISL_1793671, EPI_ISL_1793674, EPI_ISL_1793675, EPI_ISL_1793676, EPI_ISL_1793679, EPI_ISL_1793680, EPI_ISL_1793681, EPI_ISL_1793778, EPI_ISL_1793780, EPI_ISL_2003348, EPI_ISL_2003349 | | | |
| see above | University of Liège COVID-19 testing center | GIGA Medical Genomics | Bouchra Boujemla; Cécile Meex; Keith Durkin; Maria Artesi; Marie-Pierre Hayette; Nathalie Renotte; Pierrette Melin; Raphaël Boreux; Sébastien Bontems; Vincent Bours |
| EPI_ISL_1919868, EPI_ISL_1919933 | University of Michigan Clinical Microbiology Laboratory | Lauring Lab, University of Michigan, Department of Microbiology and Immunology | Valesano |
| EPI_ISL_1791529 | University of Oregon COVID-19 MAP Laboratory | University of Oregon Genomics and Cell Characterization Core Facility (GC3F) | Ariana White; Douglas Turnbull; Jason Carriere; Jeff Bishop |
| EPI_ISL_2018944, EPI_ISL_2018946 | University of Rome Tor Vergata: Departm Experim Medicine Chair of Virology | University of Rome Tor Vergata: Departm Experim Medicine Chair of Virology | Francesca Ceccherini-Silberstein; Loredana Sarmati; Lorenzo Piermatteo; Luca Carioti; Marco Iannetta; Maria Botticelli; Maria Concetta Bellocchi; Massimo Andreoni; Rossana Scutari |
| EPI_ISL_1855380, EPI_ISL_1855383 | University of Wisconsin-Madison AIDS Vaccine Research Laboratories | University of Wisconsin-Madison AIDS Vaccine Research Laboratories | Gage Moreno; Katarina Braun; et al. AIDS Vaccine Research Laboratories |
| EPI_ISL_2137426, EPI_ISL_2137449, EPI_ISL_2227027, EPI_ISL_2455590, EPI_ISL_2465269, EPI_ISL_2465284 | Università Federico II - Dipartimento di scienze mediche traslazionali - Napoli | TIGEM | Antonio Grimaldi Patrizia Annunziata Francesco Panariello Teresa Giuliano Michele Cennamo Valentina Bouche Chiara Colantuono Lucio Di Filippo Mariano Fiorenza Anna Manfredi Marcello Salvi Giuseppe Portella Andrea Ballabio Davide Cacchiarelli |
| EPI_ISL_3270766 | Università Federico II - Dipartimento di scienze mediche traslazionali - Napoli | Telethon Institute of Genetics and Medicine (TIGEM) | Antonio Grimaldi Patrizia Annunziata Francesco Panariello Teresa Giuliano Michele Cennamo Valentina Bouche Chiara Colantuono Lucio Di Filippo Mariano Fiorenza Anna Manfredi Marcello Salvi Giuseppe Portella Andrea Ballabio Davide Cacchiarelli |
| EPI_ISL_1919603, EPI_ISL_1919641 | Università degli Studi di Perugia | Istituto Zooprofilattico Sperimentale dell'Abruzzo e Molise "G. Caporale" | Ancora M; Calistri P; Camilloni B; Cammà C; Curini V; Delli Compagni E; Di Domenico M; Di Pasquale A; Lorusso A; Mangone I; Marcacci M; Mencacci A; Puglia I; Rinaldi A; Savini G; Scialabba S |
| EPI_ISL_5802414, EPI_ISL_5802415 | Upa Dr Akira Tada | Instituto Butantan | Antonio Jorge Martins; Claudia Renata dos Santos Barros; David Schlesinger; Debora Botequio Moretti; Dimas Tadeu Covas; Elaine Cristina Marqueze; Elaine Vieira Santos; Evandra Strazza Rodrigues; Heidge Fukumasu; Jayme Augusto de Souza-Neto; José Salvatore Leister Patané; Luiz Alcantara; Luiz Lehmann Coutinho; Maria Carolina Elias; Maurício Lacerda Nogueira; Rafael dos Santos Bezerra; Raul Machado Neto; Rejane Maria Tommasini Grotto; Ricardo Haddad; Sandra Coccuzzo Sampaio Vessoni; Simone Kashima; Svetoslav Nanev Slavov; Vincent Louis Viala |
| EPI_ISL_5802387, EPI_ISL_5802388, EPI_ISL_5802392, EPI_ISL_5802393 | Upa Dr Franco Da Rocha | Instituto Butantan | Antonio Jorge Martins; Claudia Renata dos Santos Barros; David Schlesinger; Debora Botequio Moretti; Dimas Tadeu Covas; Elaine Cristina Marqueze; Elaine Vieira Santos; Evandra Strazza Rodrigues; Heidge Fukumasu; Jayme Augusto de Souza-Neto; José Salvatore Leister Patané; Luiz Alcantara; Luiz Lehmann Coutinho; Maria Carolina Elias; Maurício Lacerda Nogueira; Rafael dos Santos Bezerra; Raul Machado Neto; Rejane Maria Tommasini Grotto; Ricardo Haddad; Sandra Coccuzzo Sampaio Vessoni; Simone Kashima; Svetoslav Nanev Slavov; Vincent Louis Viala |
| EPI_ISL_5800421 | Upa Jose Campos Barreto Zequinha Barreto | Instituto Butantan | Antonio Jorge Martins; Claudia Renata dos Santos Barros; David Schlesinger; Debora Botequio Moretti; Dimas Tadeu Covas; Elaine Cristina Marqueze; Elaine Vieira Santos; Evandra Strazza Rodrigues; Heidge Fukumasu; Jayme Augusto de Souza-Neto; José Salvatore Leister Patané; Luiz Alcantara; Luiz Lehmann Coutinho; Maria Carolina Elias; Maurício Lacerda Nogueira; Rafael dos Santos Bezerra; Raul Machado Neto; Rejane Maria Tommasini Grotto; Ricardo Haddad; Sandra Coccuzzo Sampaio Vessoni; Simone Kashima; Svetoslav Nanev Slavov; Vincent Louis Viala |
| EPI_ISL_5782483, EPI_ISL_5782484 | Upa Unidade De Pronto Atendimento 24 Horas Bom Jesus | Instituto Butantan | Antonio Jorge Martins; Claudia Renata dos Santos Barros; David Schlesinger; Debora Botequio Moretti; Dimas Tadeu Covas; Elaine Cristina Marqueze; Elaine Vieira Santos; Evandra Strazza Rodrigues; Heidge Fukumasu; Jayme Augusto de Souza-Neto; José Salvatore Leister Patané; Luiz Alcantara; Luiz Lehmann Coutinho; Maria Carolina Elias; Maurício Lacerda Nogueira; Rafael dos Santos Bezerra; Raul Machado Neto; Rejane Maria Tommasini Grotto; Ricardo Haddad; Sandra Coccuzzo Sampaio Vessoni; Simone Kashima; Svetoslav Nanev Slavov; Vincent Louis Viala |
| EPI_ISL_1972948 | Usansolo-Galdakao University Hospital | Cruces University Hospital | Ana Belén de la Hoz; Ana Gual-de-Torrella; Izaskun Alejo-Cancho; Mikel Gallego |
| EPI_ISL_2291787, EPI_ISL_2458289, EPI_ISL_2467052, EPI_ISL_2496699, EPI_ISL_2496714, EPI_ISL_2679231, EPI_ISL_2679572, EPI_ISL_3269709 | | | |
| see above | Utah Public Health Laboratory | Utah Public Health Laboratory | Erin L. Young; Kelly F. Oakeson; Olinto Linares-Perdomo; Tara Gallagher |
| EPI_ISL_5650263 | VIGILANCIA EM SAUDE SAE VIGILANCIA EPIDEMIOLOGICA | Instituto Butantan | Antonio Jorge Martins; Claudia Renata dos Santos Barros; David Schlesinger; Debora Botequio Moretti; Dimas Tadeu Covas; Elaine Cristina Marqueze; Elaine Vieira Santos; Evandra Strazza Rodrigues; Heidge Fukumasu; Jayme Augusto de Souza-Neto; José Salvatore Leister Patané; Luiz Alcantara; Luiz Lehmann Coutinho; Maria Carolina Elias; Maurício Lacerda Nogueira; Rafael dos Santos Bezerra; Raul Machado Neto; Rejane Maria Tommasini Grotto; Ricardo Haddad; Sandra Coccuzzo Sampaio Vessoni; Simone Kashima; Svetoslav Nanev Slavov; Vincent Louis Viala |
| EPI_ISL_2210106 | VIGILANCIA EPIDEMIOLOGICA | Instituto Butantan | Antonio Jorge Martins; Claudia Renata dos Santos Barros; David Schlesinger; Debora Botequio Moretti; Dimas Tadeu Covas; Elaine Cristina Marqueze; Elaine Vieira Santos; Evandra Strazza Rodrigues; Heidge Fukumasu; Jayme Augusto de Souza-Neto; José Salvatore Leister Patané; Luiz Alcantara; Luiz Lehmann |

| | | | |
|---|---|---|---|
| | DE LEME | | Coutinho; Maria Carolina Elias; Maurício Lacerda Nogueira; Rafael dos Santos Bezerra; Raul Machado Neto; Rejane Maria Tommasini Grotto; Ricardo Haddad; Sandra Coccuzzo Sampaio Vessoni; Simone Kashima; Svetoslav Nanev Slavov; Vincent Louis Viala. |
| EPI_ISL_2344746 | VIGILANCIA EPIDEMIOLOGICA DE LEME | Instituto Butantan / ESALQ-Piracicaba | Antonio Jorge Martins; Claudia Renata dos Santos Barros; David Schlesinger; Debora Botequio Moretti; Dimas Tadeu Covas; Elaine Cristina Marqueze; Elaine Vieira Santos; Evandra Strazza Rodrigues; Heidge Fukumasu; Jayme Augusto de Souza-Neto; José Salvatore Leister Patané; Luiz Alcantara; Luiz Lehmann Coutinho; Maria Carolina Elias; Maurício Lacerda Nogueira; Rafael dos Santos Bezerra; Raul Machado Neto; Rejane Maria Tommasini Grotto; Ricardo Haddad; Sandra Coccuzzo Sampaio Vessoni; Simone Kashima; Svetoslav Nanev Slavov; Vincent Louis Viala |
| EPI_ISL_2210041, EPI_ISL_2210045, EPI_ISL_2210048, EPI_ISL_2344713, EPI_ISL_2344714, EPI_ISL_2344715, EPI_ISL_2344720 | | | |
| see above | VIGILANCIA EPIDEMIOLOGICA DE LEME | Instituto Butantan / FZEA-USP-Pirassununga | Antonio Jorge Martins; Claudia Renata dos Santos Barros; David Schlesinger; Debora Botequio Moretti; Dimas Tadeu Covas; Elaine Cristina Marqueze; Elaine Vieira Santos; Evandra Strazza Rodrigues; Heidge Fukumasu; Jayme Augusto de Souza-Neto; José Salvatore Leister Patané; Luiz Alcantara; Luiz Lehmann Coutinho; Maria Carolina Elias; Maurício Lacerda Nogueira; Rafael dos Santos Bezerra; Raul Machado Neto; Rejane Maria Tommasini Grotto; Ricardo Haddad; Sandra Coccuzzo Sampaio Vessoni; Simone Kashima; Svetoslav Nanev Slavov; Vincent Louis Viala |
| EPI_ISL_2210257, EPI_ISL_2210281 | VIGILANCIA EPIDEMIOLOGICA E CONTROLE DE VETORES PIRASSUNUN | Instituto Butantan | Antonio Jorge Martins; Claudia Renata dos Santos Barros; David Schlesinger; Debora Botequio Moretti; Dimas Tadeu Covas; Elaine Cristina Marqueze; Elaine Vieira Santos; Evandra Strazza Rodrigues; Heidge Fukumasu; Jayme Augusto de Souza-Neto; José Salvatore Leister Patané; Luiz Alcantara; Luiz Lehmann Coutinho; Maria Carolina Elias; Maurício Lacerda Nogueira; Rafael dos Santos Bezerra; Raul Machado Neto; Rejane Maria Tommasini Grotto; Ricardo Haddad; Sandra Coccuzzo Sampaio Vessoni; Simone Kashima; Svetoslav Nanev Slavov; Vincent Louis Viala. |
| EPI_ISL_2209717, EPI_ISL_2210019, EPI_ISL_2210030, EPI_ISL_2344719 | VIGILANCIA EPIDEMIOLOGICA E CONTROLE DE VETORES PIRASSUNUN | Instituto Butantan / FZEA-USP-Pirassununga | Antonio Jorge Martins; Claudia Renata dos Santos Barros; David Schlesinger; Debora Botequio Moretti; Dimas Tadeu Covas; Elaine Cristina Marqueze; Elaine Vieira Santos; Evandra Strazza Rodrigues; Heidge Fukumasu; Jayme Augusto de Souza-Neto; José Salvatore Leister Patané; Luiz Alcantara; Luiz Lehmann Coutinho; Maria Carolina Elias; Maurício Lacerda Nogueira; Rafael dos Santos Bezerra; Raul Machado Neto; Rejane Maria Tommasini Grotto; Ricardo Haddad; Sandra Coccuzzo Sampaio Vessoni; Simone Kashima; Svetoslav Nanev Slavov; Vincent Louis Viala |
| EPI_ISL_3667127 | VIROLOGY LABORATORY-CHU NICE | VIROLOGY LABORATORY-CHU NICE | Aicha El Yakine; Geraldine Gonfrier; Jean Machowiak; Sebastien Vitale; Valerie Giordanengo; Virginie Flahou |
| EPI_ISL_5799721, EPI_ISL_5799722, EPI_ISL_5799723, EPI_ISL_5799756, EPI_ISL_5799757, EPI_ISL_5799758, EPI_ISL_5799762, EPI_ISL_5800578 | | | |
| see above | Vigilancia Epidemiologica De Leme | Instituto Butantan | Antonio Jorge Martins; Claudia Renata dos Santos Barros; David Schlesinger; Debora Botequio Moretti; Dimas Tadeu Covas; Elaine Cristina Marqueze; Elaine Vieira Santos; Evandra Strazza Rodrigues; Heidge Fukumasu; Jayme Augusto de Souza-Neto; José Salvatore Leister Patané; Luiz Alcantara; Luiz Lehmann Coutinho; Maria Carolina Elias; Maurício Lacerda Nogueira; Rafael dos Santos Bezerra; Raul Machado Neto; Rejane Maria Tommasini Grotto; Ricardo Haddad; Sandra Coccuzzo Sampaio Vessoni; Simone Kashima; Svetoslav Nanev Slavov; Vincent Louis Viala |
| EPI_ISL_5799761 | Vigilancia Epidemiologica E Controle De Vetores Pirassunun | Instituto Butantan | Antonio Jorge Martins; Claudia Renata dos Santos Barros; David Schlesinger; Debora Botequio Moretti; Dimas Tadeu Covas; Elaine Cristina Marqueze; Elaine Vieira Santos; Evandra Strazza Rodrigues; Heidge Fukumasu; Jayme Augusto de Souza-Neto; José Salvatore Leister Patané; Luiz Alcantara; Luiz Lehmann Coutinho; Maria Carolina Elias; Maurício Lacerda Nogueira; Rafael dos Santos Bezerra; Raul Machado Neto; Rejane Maria Tommasini Grotto; Ricardo Haddad; Sandra Coccuzzo Sampaio Vessoni; Simone Kashima; Svetoslav Nanev Slavov; Vincent Louis Viala |
| EPI_ISL_5799717, EPI_ISL_5799718, EPI_ISL_5799719 | Vigilancia Epidemiologica E Controle De Vetores Pirassununga | Instituto Butantan | Antonio Jorge Martins; Claudia Renata dos Santos Barros; David Schlesinger; Debora Botequio Moretti; Dimas Tadeu Covas; Elaine Cristina Marqueze; Elaine Vieira Santos; Evandra Strazza Rodrigues; Heidge Fukumasu; Jayme Augusto de Souza-Neto; José Salvatore Leister Patané; Luiz Alcantara; Luiz Lehmann Coutinho; Maria Carolina Elias; Maurício Lacerda Nogueira; Rafael dos Santos Bezerra; Raul Machado Neto; Rejane Maria Tommasini Grotto; Ricardo Haddad; Sandra Coccuzzo Sampaio Vessoni; Simone Kashima; Svetoslav Nanev Slavov; Vincent Louis Viala |
| EPI_ISL_1914208 | Viollier AG | Department of Biosystems Science and Engineering, ETH Zurich | Chaoran Chen; Christiane Beckmann; Christoph Noppen; David Dreifuss; Deborah Penet; Emmanouil Dermitzakis; Henri Pegeot; Ioannis Xenarios; Ivan Topolsky; Katharina Jahn; Keith Harshman; Lara Fuhrmann; Lorenzo Cerutti; Maurice Redondo; Niko Beerenwinkel; Noemie Santamaria de Souza; Olivier Kobel; Philipp Jablonski; Sarah Nadeau; Sophie Seidel; Tanja Stadler |
| EPI_ISL_1750646 | Viollier AG | Department of Biosystems Science and Engineering, ETH Zürich | Chaoran Chen; Christian Beisel; Christiane Beckmann; Christoph Noppen; David Dreifuss; Elodie Burcklen; Ina Nissen; Ivan Topolsky; Katharina Jahn; Lara Fuhrmann; Maurice Redondo; Mirjam Feldkamp; Natascha Santacroce; Niko Beerenwinkel; Noemie Santamaria de Souza; Olivier Kobel; Philipp Jablonski; Rebecca Denes; Sarah Nadeau; Sophie Seidel; Tanja Stadler |
| EPI_ISL_2081447, EPI_ISL_2362006 | Virginia Division of Consolidated Laboratory Services | Virginia Division of Consolidated Laboratory Services | Virginia DCLS |
| EPI_ISL_2434099, EPI_ISL_2884082, EPI_ISL_2884083, EPI_ISL_2884090, EPI_ISL_2884091 | Virology Laboratory, Scientific Department, Army Medical Center | Virology Laboratory, Scientific Department, Army Medical Center | Anella Monte; Anna Anselmo; Antonella Fortunato; Filippo Molinari; Florigio Lista; Florigio Lista.; Francesco Giordani; Giancarlo Petralito; Giandomenico Cerreto; Giulia Campoli; Lucia Nicosia; Marzia Cavalli; Riccardo De Sanctis; Rossella Brandi; Silvia Fillo; Vanessa Vera Fain |
| EPI_ISL_1823830 | WA State Department of Health | Centers for Disease Control and Prevention Division of Viral Diseases, Pathogen Discovery | Alison Laufer Halpin; Ben L. Rambo-Martin; Clinton R. Paden; Dakota Howard; Darlene Wagner; Dave Wentworth; Dhwani Batra; Jasmine Padilla; Justin Lee; Katie Dillon; Krista Queen; Kristen Knipe; Kristine Lacek; Mark Burroughs; Matthew Schmerer; Mili Sheth; Peter Cook; Sam Shepard; Sarah Nobles; Shoshona Le; Suxiang Tong; Vivien Dugan; Yvette Unoarumhi |
| EPI_ISL_1920121 | Wadsworth Center, New York State Department of Health | Wadsworth Center, New York State Department of Health | Alexis Russell; Catharine Prussing; Daryl M. Lamson; Erasmus Schneider; Erica Lasek-Nesselquist; John Kelly; Jonathan Plitnick; Kirsten St. George; Matthew Shudt; Melissa A Leisner; Navjot Singh |
| EPI_ISL_1653885, EPI_ISL_1747415, EPI_ISL_1747435, EPI_ISL_1747442, EPI_ISL_1747444, EPI_ISL_1747450, EPI_ISL_1897378, EPI_ISL_1897379, EPI_ISL_1897380, EPI_ISL_1897420, EPI_ISL_1897431 | | | |
| see above | Washington State Department of Health Public Health Laboratories | Washington State Department of Health Public Health Laboratories | Avi Singh; Darren Lucas; Denny Russell; Drew MacKellar; Geoff Melly; Hannah Gray; Joenice Gonzalez; JohnAric Peterson; Philip Dykema; Rebecca Cao; Vanessa De Los Santos |
| EPI_ISL_2081152 | Weill Cornell Medicine | New York Genome Center | Andre Corvelo; Arryn Craney; Chris Mason; Dayna M. Oschwald; Hanna Rennert; Lars F Westblade; Margaret Elizabeth Ross; Melissa Cushing; Michael Zody; Olivier Elemento; Priya Velu; Samantha Fennessey; Tom Maniatis |
| EPI_ISL_2453332, EPI_ISL_2454296 | Wexner Medical Center | The Ohio State University College of Medicine | Koenig, S.; Seminetta, J. |
| EPI_ISL_1652085, EPI_ISL_1652086, EPI_ISL_1652087, EPI_ISL_1652089, EPI_ISL_1760185, EPI_ISL_1760189, EPI_ISL_1760190, EPI_ISL_1760197, EPI_ISL_1760199, EPI_ISL_1805460 | | | |
| see above | Wichita State University - Molecular Diagnostics Lab | Kansas Health and Environmental Lab | Ben Olsen; Jonathan Barnell; Mike Grose; and Phil Adam |
| EPI_ISL_1794445, EPI_ISL_1794530, EPI_ISL_1794549, EPI_ISL_2018581, EPI_ISL_2018802, EPI_ISL_2018804 | Wisconsin State Laboratory of Hygiene Communicable Disease Division | Wisconsin State Laboratory of Hygiene Communicable Disease Division | Abigail C. Shockey; Alicia J. Mooney; Erika M. Hanson; Kelsey R. Florek; Richard Griesser; Sara Wagner; Tonya Danz |
| EPI_ISL_1793089, EPI_ISL_1963744 | Yale Clinical Virology Lab | Grubaugh Lab - Yale School of Public Health | Anderson Brito; Annie Watkins; Chaney Kalinich; Chantal Vogels; Isabel Ott; Jessica Rothman; Joseph Fauver; Mallery Breban; Marie L. Landry; Mary Petrone; Nathan Grubaugh; Tara Alpert |
| EPI_ISL_2034918, EPI_ISL_2034998 | Yale Clinical Virology Lab | Yale Center for Genomic Analysis | Brooke Sullivan; Curt Scharfe; Irina Tikhonova; Kaya Bilguvar; Shrikant Mane |
