## Supplement 3 for "Rapid and Accurate Identification of SARS-CoV-2 Omicron Variants Using Droplet Digital PCR (RT-ddPCR)": All_0419-0425.pdf

We gratefully acknowledge the following Authors from the Originating laboratories responsible for obtaining the specimens, as well as the Submitting laboratories where the genome data were generated and shared via GISAID, on which this research is based.

All Submitters of data may be contacted directly via www.gisaid.org

Authors are sorted alphabetically.

| Accession ID | Originating Laboratory | Submitting Laboratory | Authors |
|---|---|---|---|
| EPI_ISL_2650645 | A.S.L. ASTI - PRESIDIO OSP. CARDINAL G. MASSAIA - POLIAMBULATORIO OSPEDALIERO - | Fondazione del Piemonte per l'Oncologia IRCCS | Antonino Sottile; Giorgio Giardina; Paola Marino; Silvia Brossa |
| EPI_ISL_2650606, EPI_ISL_2650611 | A.S.L. BIELLA - OSPEDALE DEGLI INFERMI - SC LABORATORIO ANALISI - | Fondazione del Piemonte per l'Oncologia IRCCS | Antonino Sottile; Giorgio Giardina; Paola Marino; Silvia Brossa |
| EPI_ISL_2650612 | A.S.L. CUNEO 1 - OSPEDALE DI SAVIGLIANO - S.S. LABORATORIO ANALISI CN1 - | Fondazione del Piemonte per l'Oncologia IRCCS | Antonino Sottile; Giorgio Giardina; Paola Marino; Silvia Brossa |
| EPI_ISL_2650703 | A.S.L. NOVARA - ISTITUTO S.S. TRINITA' - BORGOMANERO - S.S. LABORATORIO ANALISI CHIMICO CLINICHE - | Fondazione del Piemonte per l'Oncologia IRCCS | Antonino Sottile; Giorgio Giardina; Paola Marino; Silvia Brossa |
| EPI_ISL_3347178, EPI_ISL_3347180, EPI_ISL_3347193, EPI_ISL_3347194, EPI_ISL_3347195, EPI_ISL_3347202 | ADILAB | Laboratorio Departamental de Salud Publica de Antioquia | Andres F. Cardona-Rios; Gloria Isabel Escobar; Idabely Betancur Ortiz; Juan P. Hernandez-Ortiz; Maria Stella López |
| EPI_ISL_3276549 | ADILAB | Universidad Nacional de Colombia - Laboratorio Genómico One Health | Andres F. Cardona-Rios; Carlos Franco-Muñoz; Carolina Muñoz-Arango; Celeny Ortiz; Daniel O. Maldonado-Perez; Diego A. Álvarez-Díaz; Hector Alejandro Ruiz-Moreno; Idabely Betancur Ortiz; Jorge E. Osorio; Juan P. Hernandez-Ortiz; Karl A Ciuoderis; Katherine Laiton-Donato; Laura Silvana Perez; Lina M. Hurtado; Marcela Mercado-Reyes; Maria Angélica Maya; Maria Stella López; Rita Almanza Payares; Sandra Ines Cano; Simón Villegas Velásquez |
| EPI_ISL_5802489, EPI_ISL_5802490, EPI_ISL_5802491, EPI_ISL_5802496 | AFIP | Instituto Butantan | Antonio Jorge Martins; Claudia Renata dos Santos Barros; David Schlesinger; Debora Botequio Moretti; Dimas Tadeu Covas; Elaine Cristina Marqueze; Elaine Vieira Santos; Evandra Strazza Rodrigues; Heidge Fukumasu; Jayme Augusto de Souza-Neto; José Salvatore Leister Patané; Luiz Alcantara; Luiz Lehmann Coutinho; Maria Carolina Elias; Maurício Lacerda Nogueira; Rafael dos Santos Bezerra; Raul Machado Neto; Rejane Maria Tommasini Grotto; Ricardo Haddad; Sandra Coccuzzo Sampaio Vessoni; Simone Kashima; Svetoslav Nanev Slavov; Vincent Louis Viala |
| EPI_ISL_5915123, EPI_ISL_5915206, EPI_ISL_5915207, EPI_ISL_5915238, EPI_ISL_6573824, EPI_ISL_6573825 | AMALAB/FACISA/UFRN | WallauLab on behalf of Fiocruz COVID-19 Genomic Surveillance Network | Alexandre Freitas da Silva; Allan Roberto Dias Nunes; Antonio Marinho da Silva Neto; Cassia Docena; Constância Flávia Junqueira Ayres; Filipe Zimmer Dezordi; Gabriel Luz Wallau; Gustavo Barbosa de Lima; Joana Cristina Medeiros Tavares Marques; Katya Anaya Jacinto; Lais Ceschini Machado; Lilian Caroliny Amorim Silva; Marcelo Henrique dos Santos Paiva; Mariane dos Santos Duarte; Matheus Filgueira Bezerra; Sinval Pinto Brandão Filho |
| EPI_ISL_2208874 | AMB MEDICO DR FRANCISCO FELIX DE MENDONCA SAUDADE | Instituto Butantan / ESALQ-Piracicaba | Antonio Jorge Martins; Claudia Renata dos Santos Barros; David Schlesinger; Debora Botequio Moretti; Dimas Tadeu Covas; Elaine Cristina Marqueze; Elaine Vieira Santos; Evandra Strazza Rodrigues; Heidge Fukumasu; Jayme Augusto de Souza-Neto; José Salvatore Leister Patané; Luiz Alcantara; Luiz Lehmann Coutinho; Maria Carolina Elias; Maurício Lacerda Nogueira; Rafael dos Santos Bezerra; Raul Machado Neto; Rejane Maria Tommasini Grotto; Ricardo Haddad; Sandra Coccuzzo Sampaio Vessoni; Simone Kashima; Svetoslav Nanev Slavov; Vincent Louis Viala |
| EPI_ISL_2344807 | AMBULATORIO DE ESPECIALIDADE V E MOGI MIRIM | Instituto Butantan / ESALQ-Piracicaba | Antonio Jorge Martins; Claudia Renata dos Santos Barros; David Schlesinger; Debora Botequio Moretti; Dimas Tadeu Covas; Elaine Cristina Marqueze; Elaine Vieira Santos; Evandra Strazza Rodrigues; Heidge Fukumasu; Jayme Augusto de Souza-Neto; José Salvatore Leister Patané; Luiz Alcantara; Luiz Lehmann Coutinho; Maria Carolina Elias; Maurício Lacerda Nogueira; Rafael dos Santos Bezerra; Raul Machado Neto; Rejane Maria Tommasini Grotto; Ricardo Haddad; Sandra Coccuzzo Sampaio Vessoni; Simone Kashima; Svetoslav Nanev Slavov; Vincent Louis Viala |
| EPI_ISL_2674309 | ANALIZAR LABORATORIO CLINICO AUTOMATIZADO - SYNLAB | Instituto Nacional de Salud-Dirección de Investigación en Salud Pública | Carlos Franco-Muñoz; Carmen Osorio; Diana Malo; Diego A. Álvarez-Díaz; Diego Andrés Prada; Gerardo Santamaría; Hector Alejandro Ruiz-Moreno; Jhonnatan Reales-González; Jorge Rivera; Juan Camilo Martinez; Julian Naizaque; Katherine Laiton-Donato; Lisseth Pardo; Magdalena Wiesner; Marcela Mercado-Reyes; Maria T. Herrera-Sepúlveda; Marta Lopez Blanco; Martha Lucia Ospina Martinez; Paola Rojas; Sergio Gomez; Sheryll Corchuelo; Ángela Alarcon Cruz |
| EPI_ISL_2018198, EPI_ISL_2018203 | AR Dept. of Health-PHL, Molecular Diagnostics | Centers for Disease Control and Prevention Division of Viral Diseases, Pathogen Discovery | Alison Laufer Halpin; Ben L. Rambo-Martin; Clinton R. Paden; Dakota Howard; Darlene Wagner; Dave Wentworth; Dhwani Batra; Jasmine Padilla; Justin Lee; Katie Dillon; Krista Queen; Kristen Knipe; Kristine Lacek; Mark Burroughs; Matthew Schmerer; Mili Sheth; Peter Cook; Sam Shepard; Sarah Nobles; Shoshona Le; Suxiang Tong; Vivien Dugan; Yvette Unoarumhi |
| EPI_ISL_2273001 | AREA DE SALUD CORONADO | Inciensa, Instituto Costarricense de Investigación y Enseñanza en Nutrición y Salud | Adriana Godínez; Claudio Soto-Garita; Estela Cordero; Francisco Duarte; Hebleen Porras; Joselyn Prado & Mariel López; José Luis Vargas; Mariela Gutiérrez; Melany Calderón |
| EPI_ISL_2272983 | AREA DE SALUD GOICOECHEA 2 - CLINICA DR. JIMENEZ NUÑEZ | Inciensa, Instituto Costarricense de Investigación y Enseñanza en Nutrición y Salud | Adriana Godínez; Claudio Soto-Garita; Estela Cordero; Francisco Duarte; Hebleen Porras; Joselyn Prado & Juan Carlos Cartes Parra; José Luis Vargas; Mariela Gutiérrez; Melany Calderón |
| EPI_ISL_2272988 | AREA DE SALUD MORAVIA | Inciensa, Instituto Costarricense de Investigación y Enseñanza en Nutrición y Salud | Adriana Godínez; Claudio Soto-Garita; Estela Cordero; Francisco Duarte; Hebleen Porras; Joselyn Prado & Mariel López; José Luis Vargas; Mariela Gutiérrez; Melany Calderón |
| EPI_ISL_1940377, EPI_ISL_1940426, EPI_ISL_1940453, EPI_ISL_1940587, EPI_ISL_1940609, EPI_ISL_1940627, EPI_ISL_1940638, EPI_ISL_1940678 | | | |
| see above | ASL Napoli 1 Centro | AMES Centro Polidiagnostico Strumentale S.r.l. | Antonella Di Carlo; Antonio Fico; Eloisa Evangelista; Giovanni Savarese; Luigi D'Amore; Luisa Circelli; Maurizio D'Amora; Monica Ianniello; Nadia Petrillo; Raffaella Ruggiero; Roberto Sirica |
| EPI_ISL_1824639, EPI_ISL_1824679 | AZ Klina | AZ Klina | Carl Vael - Lynsey Berckmans |
| EPI_ISL_1905193, EPI_ISL_1905201, EPI_ISL_1905214, EPI_ISL_1972236 | AZ St.Jan Brugge-Oostende | AZ SINT-JAN BRUGGE | Jorn Hellemans; Laurien Hoornaert; Marijke Reynders; Patrick Descheemaeker; Thomas Van Landschoot |
| EPI_ISL_1999796, EPI_ISL_1999817, EPI_ISL_1999835, EPI_ISL_1999841, EPI_ISL_1999873, EPI_ISL_2000052, EPI_ISL_2000056, EPI_ISL_2000082, EPI_ISL_2000093, EPI_ISL_2000100, EPI_ISL_2000104, EPI_ISL_2000107, EPI_ISL_2000116, EPI_ISL_2000119, EPI_ISL_2035245, EPI_ISL_2038996, EPI_ISL_2039001, EPI_ISL_2039022, EPI_ISL_2039060, EPI_ISL_2039064, EPI_ISL_2039073, EPI_ISL_2039083, EPI_ISL_2039096, EPI_ISL_2039416, EPI_ISL_2039460, EPI_ISL_2039469, EPI_ISL_2039470, EPI_ISL_2039485, EPI_ISL_2039527, EPI_ISL_2039541, EPI_ISL_2039765, EPI_ISL_2039771, EPI_ISL_2039779, EPI_ISL_2039783, EPI_ISL_2039823, EPI_ISL_2039843, EPI_ISL_2039845, EPI_ISL_2039850, EPI_ISL_2039895, EPI_ISL_2039897, EPI_ISL_2039919, EPI_ISL_2040165, EPI_ISL_2040201, EPI_ISL_2040252, EPI_ISL_2040267, EPI_ISL_2040294, EPI_ISL_2040299, EPI_ISL_2040306, EPI_ISL_2040340, EPI_ISL_2040348, EPI_ISL_2040368, EPI_ISL_2040376, EPI_ISL_2040377, EPI_ISL_2040378, EPI_ISL_2040536, EPI_ISL_2040620, EPI_ISL_2040629, EPI_ISL_2040657, EPI_ISL_2041076, EPI_ISL_2041078, EPI_ISL_2041123, EPI_ISL_2041155, EPI_ISL_2041164, EPI_ISL_2041165, EPI_ISL_2041179, EPI_ISL_2041192, EPI_ISL_2041280, EPI_ISL_2041303, EPI_ISL_2041349, EPI_ISL_2041362, EPI_ISL_2041404, EPI_ISL_2041405, EPI_ISL_2041407, EPI_ISL_2041423, EPI_ISL_2041424, EPI_ISL_2041432, EPI_ISL_2041456, EPI_ISL_2041471, EPI_ISL_2041500, EPI_ISL_2041517, EPI_ISL_2041519, EPI_ISL_2041521, EPI_ISL_2041527, EPI_ISL_2041564, EPI_ISL_2041590, EPI_ISL_2041601, EPI_ISL_2041633, EPI_ISL_2041642, EPI_ISL_2041721, EPI_ISL_2041723, EPI_ISL_2041739, EPI_ISL_2041807, EPI_ISL_2041816, EPI_ISL_2041820, EPI_ISL_2041837, EPI_ISL_2041843, EPI_ISL_2041845, EPI_ISL_2042059, EPI_ISL_2042103, EPI_ISL_2042104, EPI_ISL_2042393, EPI_ISL_2043517, EPI_ISL_2089591, EPI_ISL_2089605, EPI_ISL_2089608, EPI_ISL_2089609, EPI_ISL_2089626, EPI_ISL_2089627, EPI_ISL_2089703, EPI_ISL_2089735, EPI_ISL_2089844, EPI_ISL_2089857, EPI_ISL_2089866, EPI_ISL_2089912, EPI_ISL_2089959, EPI_ISL_2089987, EPI_ISL_2089992, EPI_ISL_2089993, EPI_ISL_2089995, EPI_ISL_2089996, EPI_ISL_2090010, EPI_ISL_2090030, EPI_ISL_2090102, EPI_ISL_2090257, EPI_ISL_2090266, EPI_ISL_2090271, EPI_ISL_2090343, EPI_ISL_2090352, EPI_ISL_2090354, EPI_ISL_2090357, EPI_ISL_2090361, EPI_ISL_2090365, EPI_ISL_2090368, EPI_ISL_2090390, EPI_ISL_2090395, EPI_ISL_2090398, EPI_ISL_2090402, EPI_ISL_2090403, EPI_ISL_2090455, EPI_ISL_2090456, EPI_ISL_2132545, EPI_ISL_2149216, EPI_ISL_2149217, EPI_ISL_2149302, EPI_ISL_2149311, EPI_ISL_2149318, EPI_ISL_2149322, EPI_ISL_2149333, EPI_ISL_2149340, EPI_ISL_2149356, EPI_ISL_2149385, EPI_ISL_2149391, EPI_ISL_2149393, EPI_ISL_2149418, EPI_ISL_2149499, EPI_ISL_2149528, EPI_ISL_2149536, EPI_ISL_2149538, EPI_ISL_2149539, EPI_ISL_2149541, EPI_ISL_2149542, EPI_ISL_2149543, EPI_ISL_2149544, EPI_ISL_2149549, EPI_ISL_2149565, EPI_ISL_2149587, EPI_ISL_2149589, EPI_ISL_2149594, EPI_ISL_2149595, EPI_ISL_2149611, EPI_ISL_2149632, EPI_ISL_2149655, EPI_ISL_2149656, EPI_ISL_2149697, EPI_ISL_2149707, EPI_ISL_2149728, EPI_ISL_2149745, EPI_ISL_2149765, EPI_ISL_2149774, EPI_ISL_2149817, EPI_ISL_2149829, EPI_ISL_2149847, EPI_ISL_2149861, EPI_ISL_2149919, EPI_ISL_2149922, EPI_ISL_2149926, EPI_ISL_2149939, EPI_ISL_2149971, EPI_ISL_2149972, EPI_ISL_2149992, EPI_ISL_2150067, EPI_ISL_2150079, EPI_ISL_2150082, EPI_ISL_2150085, EPI_ISL_2150099, EPI_ISL_2150117, EPI_ISL_2150191, EPI_ISL_2150192, EPI_ISL_2150195, EPI_ISL_2150218, EPI_ISL_2150222, EPI_ISL_2150312, EPI_ISL_2150315, EPI_ISL_2150324, EPI_ISL_2150325, EPI_ISL_2150330, EPI_ISL_2150361, EPI_ISL_2150373, EPI_ISL_2150378, EPI_ISL_2150384, EPI_ISL_2150433, EPI_ISL_2150462, EPI_ISL_2150465, EPI_ISL_2150466, EPI_ISL_2150470, EPI_ISL_2150481, EPI_ISL_2150562, EPI_ISL_2150567, EPI_ISL_2150673, EPI_ISL_2150685, EPI_ISL_2150686, EPI_ISL_2150734, EPI_ISL_2150786, EPI_ISL_2150792, EPI_ISL_2150805, EPI_ISL_2150852, EPI_ISL_2150854, EPI_ISL_2150898, EPI_ISL_2150902, EPI_ISL_2150906, EPI_ISL_2150907, EPI_ISL_2150937, EPI_ISL_2150960, EPI_ISL_2150964, EPI_ISL_2150971, EPI_ISL_2187534, EPI_ISL_2187545, EPI_ISL_2187551, EPI_ISL_2187556, EPI_ISL_2187558, EPI_ISL_2187566, EPI_ISL_2202961, EPI_ISL_2202991, EPI_ISL_2203009, EPI_ISL_2203094, EPI_ISL_2203219, EPI_ISL_2203221, EPI_ISL_2203229, EPI_ISL_2203257, EPI_ISL_2268115, EPI_ISL_2268128, EPI_ISL_2370268, EPI_ISL_2370278, EPI_ISL_2370336, EPI_ISL_2372445, EPI_ISL_2372452, EPI_ISL_2421841, EPI_ISL_2421843, EPI_ISL_2421882, EPI_ISL_2421885, EPI_ISL_2421886, EPI_ISL_2421896, EPI_ISL_2421934, EPI_ISL_2489875, EPI_ISL_2600264, EPI_ISL_2600265, EPI_ISL_2600266, EPI_ISL_2600269, EPI_ISL_2600270, EPI_ISL_2600278, EPI_ISL_2600290, EPI_ISL_4360862, EPI_ISL_4361856, EPI_ISL_4362137, EPI_ISL_4362163, EPI_ISL_4362682, EPI_ISL_4362862, EPI_ISL_4365350 | | | |
| see above | Aegis Sciences Corporation | Centers for Disease Control and Prevention Division of Viral Diseases, Pathogen Discovery | Adrian Paskey; Alec Vest; Benjamin Rambo-Martin; Christopher Gulvick; Clinton Paden; Clinton R. Paden; Cyndi Clark; Dakota Howard; Darlene Wagner; Dhwani Batra; Dillon Nall; Duncan MacCannell; Erisa Sula; Ethan Sanders; Holly Houdeshell; Jason Caravas; Kara Moser; Kristine Lacek; Matthew Hardison; Matthew Schmerer; Ola Kvalvaag; Patrick Campbell; Peter Cook; Peter W. Cook; Rob Case; Scott Sammons; Shatavia Morrison; Shaun Westlund; Tymeckia Kendall; Victoria Caban Figueroa; Vikramsinha Ghorpade; Yvette Unoarumhi |
| EPI_ISL_1920059 | Albany Medical Center | Wadsworth Center, New York State Department of Health | Alexis Russell; Catharine Prussing; Daryl M. Lamson; Erasmus Schneider; Erica Lasek-Nesselquist; John Kelly; Jonathan Plitnick; Kirsten St. George; Matthew Shudt; Melissa A Leisner; Navjot Singh |
| EPI_ISL_5800816 | Amb Medico Dr Francisco Felix De Mendonca Saudade | Instituto Butantan | Antonio Jorge Martins; Claudia Renata dos Santos Barros; David Schlesinger; Debora Botequio Moretti; Dimas Tadeu Covas; Elaine Cristina Marqueze; Elaine Vieira Santos; Evandra Strazza Rodrigues; Heidge Fukumasu; Jayme Augusto de Souza-Neto; José Salvatore Leister Patané; Luiz Alcantara; Luiz Lehmann Coutinho; Maria Carolina Elias; Maurício Lacerda Nogueira; Rafael dos Santos Bezerra; Raul Machado Neto; Rejane Maria Tommasini Grotto; Ricardo Haddad; Sandra Coccuzzo Sampaio Vessoni; Simone Kashima; Svetoslav Nanev Slavov; Vincent Louis Viala |
| EPI_ISL_5800734 | Ambulatorio De Especialidade V E Mogi Mirim | Instituto Butantan | Antonio Jorge Martins; Claudia Renata dos Santos Barros; David Schlesinger; Debora Botequio Moretti; Dimas Tadeu Covas; Elaine Cristina Marqueze; Elaine Vieira Santos; Evandra Strazza Rodrigues; Heidge Fukumasu; Jayme Augusto de Souza-Neto; José Salvatore Leister Patané; Luiz Alcantara; Luiz Lehmann Coutinho; Maria Carolina Elias; Maurício Lacerda Nogueira; Rafael dos Santos Bezerra; Raul Machado Neto; Rejane Maria Tommasini Grotto; Ricardo Haddad; Sandra Coccuzzo Sampaio Vessoni; Simone Kashima; Svetoslav Nanev Slavov; Vincent Louis Viala |
| EPI_ISL_1911183 | Area of Virology, Serology and Virology Division (SAViD), New South Wales Health Pathology Randwick | Virology Research Laboratory; Area of Virology, Serology and Virology Division (SAViD), New South Wales Health Pathology Randwick | Au, J.; Bull, R.; Deveson, I.; Foster, C.; Rawlinson, W.; Ruiz Silva, M.; Van Hal, S. |

| | | | |
|---|---|---|---|
| EPI_ISL_2007151, EPI_ISL_2007164, EPI_ISL_2007173, EPI_ISL_2007174, EPI_ISL_2015335, EPI_ISL_2015344, EPI_ISL_2015346, EPI_ISL_2153562, EPI_ISL_2153565 | | | |
| see above | Arizona State University | Arizona State University | Efrem S. Lim; Joshua LaBaer; Joy M. Blain; LaRinda A. Holland; Matthew F. Smith; Nicholas J. Mellor; Peter T. Skidmore; Rabia Maqsood; Valerie Harris; Vel Murugan |
| EPI_ISL_2136678, EPI_ISL_2136688, EPI_ISL_2136703, EPI_ISL_2136707, EPI_ISL_2136712, EPI_ISL_2136727 | Atlas Genomics | Seattle Flu Study | Amanda Adler; Barry R. Lutz; Benjamin Pelle; Brian Hiatt; Caitlin R. Wolf; Chris D. Frazar; Deborah A. Nickerson; Elisabeth Brandstetter; Erica Ryke; Geoff Melly; Helen Y. Chu; Janet A. Englund; Jay Shendure; Jover Lee; Kairsten Fay; Kirsten Lacombe; Lea M. Starita; Mark J. Rieder; Matthew Richardson; Matthew Thompson; Melissa Truong; Michael Boeckh; Michael Famulare; Misja Ilcisin; Peter D. Han; Philip Dykema; Romesh Gautom; Scott Lindquist; Thomas R. Sibley; Trevor Bedford |
| EPI_ISL_3276566, EPI_ISL_3276571, EPI_ISL_3276573, EPI_ISL_3276579, EPI_ISL_3276580, EPI_ISL_3276581, EPI_ISL_3276583, EPI_ISL_3276596, EPI_ISL_3276609, EPI_ISL_3671026 | | | |
| see above | Ayudas Diagnosticas Sura | Universidad Nacional de Colombia - Laboratorio Genómico One Health | Andres F. Cardona-Rios; Carlos Franco-Muñoz; Carolina Muñoz-Arango; Celeny Ortiz; Daniel O. Maldonado-Perez; Diego A. Álvarez-Díaz; Hector Alejandro Ruiz-Moreno; Idabely Betancur Ortiz; Jorge E. Osorio; Juan P. Hernandez-Ortiz; Karl A Ciuoderis; Katherine Laiton-Donato; Laura Silvana Perez; Lina M. Hurtado; Marcela Mercado-Reyes; Maria Angélica Maya; Maria Stella López; Rita Almanza Payares; Sandra Ines Cano; Simón Villegas Velásquez |
| EPI_ISL_1785240 | Azienda Ospedaliera Terni | Istituto Zooprofilattico Sperimentale dell'Abruzzo e Molise "G. Caporale" | Ancora M; Calistri P; Cammà C; Caporale M; Curini V; Delli Compagni E; Di Domenico M; Di Lollo Valeria; Di Pasquale A; Lorusso A; Mangone I; Marcacci M; Palumbo M; Puglia I; Rinaldi A; Savini G; Scaccetti A; Scialabba S |
| EPI_ISL_1840728, EPI_ISL_1840731, EPI_ISL_1840732, EPI_ISL_1840750, EPI_ISL_1840758, EPI_ISL_1840770, EPI_ISL_1840781 | | | |
| see above | Azienda Sanitaria dell'Alto Adige Laboratorio Aziendale di Microbiologia e Virologia | Istituto di Genomica Applicata | Davide Scaglione; Eleonora Paparelli; Elisa Masi; Elisabetta Giacobazzi; Elisabetta Pagani; Gabriele Magris; Irena Jurman; Irene Bianconi; Michele Morgante; Stefanie Wieser; Vera Vendramin |
| EPI_ISL_1727239 | Azienda USL Valle d'Aosta | U.O. Genomics, S.S. Genetics and Advanced Omics Techniques, Istituto Zooprofilattico Sperimentale del Piemonte, Liguria e Valle d'Aosta | Chiara Beltramo; Elena Bozzetta; Massimo Di Benedetto; Patrizia Agnese Falcone; Pier Luigi Acutis; Simone Peletto |
| EPI_ISL_2655238, EPI_ISL_2655264, EPI_ISL_2655288, EPI_ISL_2655335, EPI_ISL_2655342, EPI_ISL_2655347, EPI_ISL_2655350, EPI_ISL_2655352, EPI_ISL_2655355, EPI_ISL_2655375, EPI_ISL_2655381, EPI_ISL_2655382, EPI_ISL_2655399, EPI_ISL_2655400, EPI_ISL_2655403, EPI_ISL_2655404, EPI_ISL_2655405, EPI_ISL_2655407, EPI_ISL_2655465, EPI_ISL_2655471, EPI_ISL_2655475, EPI_ISL_2655476, EPI_ISL_2655482, EPI_ISL_2655483, EPI_ISL_2655487, EPI_ISL_2655489, EPI_ISL_2655493, EPI_ISL_2655494, EPI_ISL_2655495, EPI_ISL_2655503, EPI_ISL_2655504, EPI_ISL_2655507, EPI_ISL_2655510, EPI_ISL_2655512, EPI_ISL_2655516, EPI_ISL_2655517, EPI_ISL_2655521, EPI_ISL_2655531, EPI_ISL_2655534, EPI_ISL_2655535, EPI_ISL_2655539, EPI_ISL_2655568, EPI_ISL_2655569, EPI_ISL_2655570, EPI_ISL_2655571, EPI_ISL_2655576, EPI_ISL_2655577, EPI_ISL_2655581, EPI_ISL_2655586, EPI_ISL_2655587, EPI_ISL_2655588, EPI_ISL_2655593, EPI_ISL_2655595, EPI_ISL_2655596, EPI_ISL_2655598, EPI_ISL_2655599, EPI_ISL_2655601, EPI_ISL_2655607, EPI_ISL_2655927, EPI_ISL_2655928, EPI_ISL_2655929, EPI_ISL_2655930, EPI_ISL_2655931, EPI_ISL_2655935, EPI_ISL_2655936, EPI_ISL_2655937, EPI_ISL_2655939, EPI_ISL_2655941, EPI_ISL_2655942, EPI_ISL_2655943, EPI_ISL_2655944, EPI_ISL_2655945, EPI_ISL_2655950, EPI_ISL_2655953, EPI_ISL_2655954, EPI_ISL_2655959, EPI_ISL_2655961, EPI_ISL_2655963, EPI_ISL_2655967, EPI_ISL_2655970, EPI_ISL_2655973, EPI_ISL_2655975, EPI_ISL_2655976, EPI_ISL_2655979, EPI_ISL_2655980, EPI_ISL_2655983, EPI_ISL_2655986, EPI_ISL_2655988, EPI_ISL_2655989, EPI_ISL_2655990, EPI_ISL_2655991, EPI_ISL_2655993, EPI_ISL_2655994, EPI_ISL_2655999, EPI_ISL_2656005, EPI_ISL_2656011, EPI_ISL_2656014, EPI_ISL_2656018, EPI_ISL_2656022, EPI_ISL_2656026, EPI_ISL_2656027, EPI_ISL_2656033, EPI_ISL_2656035, EPI_ISL_2656037, EPI_ISL_2656040, EPI_ISL_2656042, EPI_ISL_2656046, EPI_ISL_2656048, EPI_ISL_2656052, EPI_ISL_2656054, EPI_ISL_2656055, EPI_ISL_2656056, EPI_ISL_2656058, EPI_ISL_2656059, EPI_ISL_2656060, EPI_ISL_2656061, EPI_ISL_2656063, EPI_ISL_2656064, EPI_ISL_2656071, EPI_ISL_2656072, EPI_ISL_2656074, EPI_ISL_2656075, EPI_ISL_2656078, EPI_ISL_2656080, EPI_ISL_2656081, EPI_ISL_2656082, EPI_ISL_2656086, EPI_ISL_2656367, EPI_ISL_2656368, EPI_ISL_2656369, EPI_ISL_2656370, EPI_ISL_2656371, EPI_ISL_2656372, EPI_ISL_2656373, EPI_ISL_2656374, EPI_ISL_2656375, EPI_ISL_2656376, EPI_ISL_2656377, EPI_ISL_2656379, EPI_ISL_2656381, EPI_ISL_2656382, EPI_ISL_2656384, EPI_ISL_2656386, EPI_ISL_2656387, EPI_ISL_2656388, EPI_ISL_2656389, EPI_ISL_2656391, EPI_ISL_2656392, EPI_ISL_2656394, EPI_ISL_2656396, EPI_ISL_2656404, EPI_ISL_2656406, EPI_ISL_2656409, EPI_ISL_2656410, EPI_ISL_2656411, EPI_ISL_2656412, EPI_ISL_2656413, EPI_ISL_2656415, EPI_ISL_2656417, EPI_ISL_2656418, EPI_ISL_2656422, EPI_ISL_2656426, EPI_ISL_2656428, EPI_ISL_2656430, EPI_ISL_2656431, EPI_ISL_2656433, EPI_ISL_2656439, EPI_ISL_2656445, EPI_ISL_2656446, EPI_ISL_2656447, EPI_ISL_2656451, EPI_ISL_2656464, EPI_ISL_2656466, EPI_ISL_2656467, EPI_ISL_2656469, EPI_ISL_2656471, EPI_ISL_2656473, EPI_ISL_2656480, EPI_ISL_2656484, EPI_ISL_2656485, EPI_ISL_2656486, EPI_ISL_2656489, EPI_ISL_2656490, EPI_ISL_2656498, EPI_ISL_2656499, EPI_ISL_2656500, EPI_ISL_2656501, EPI_ISL_2656504, EPI_ISL_2656506, EPI_ISL_2656508, EPI_ISL_2656510, EPI_ISL_2656511, EPI_ISL_2656513, EPI_ISL_2656519, EPI_ISL_2656520, EPI_ISL_2656527, EPI_ISL_2656532, EPI_ISL_2656533, EPI_ISL_2656536, EPI_ISL_2656710, EPI_ISL_2656713, EPI_ISL_2656779, EPI_ISL_2656801, EPI_ISL_2656817, EPI_ISL_2656887, EPI_ISL_2656927, EPI_ISL_2656931, EPI_ISL_2656943, EPI_ISL_2656946, EPI_ISL_2656968, EPI_ISL_2656998, EPI_ISL_2657008, EPI_ISL_2657044, EPI_ISL_2657051, EPI_ISL_2657065, EPI_ISL_2657093, EPI_ISL_2657102, EPI_ISL_2657103, EPI_ISL_2657120, EPI_ISL_2718978, EPI_ISL_2718979, EPI_ISL_2718980, EPI_ISL_2718981, EPI_ISL_2718982, EPI_ISL_2718983, EPI_ISL_2718984, EPI_ISL_2718986, EPI_ISL_2718987, EPI_ISL_2718988, EPI_ISL_2718992, EPI_ISL_2718994, EPI_ISL_2719004, EPI_ISL_2719008, EPI_ISL_2719010, EPI_ISL_2719012, EPI_ISL_2719013, EPI_ISL_2719015, EPI_ISL_2719017, EPI_ISL_2719020, EPI_ISL_2719022, EPI_ISL_2719026, EPI_ISL_2719035, EPI_ISL_2719038, EPI_ISL_2719039, EPI_ISL_2719040, EPI_ISL_2719049, EPI_ISL_2719051, EPI_ISL_2719052, EPI_ISL_2719053, EPI_ISL_2719056, EPI_ISL_2719057, EPI_ISL_2719058, EPI_ISL_2719059, EPI_ISL_2719060, EPI_ISL_2719061, EPI_ISL_2719062, EPI_ISL_2719063, EPI_ISL_2719064, EPI_ISL_2719065, EPI_ISL_2719066, EPI_ISL_2719067, EPI_ISL_2719069, EPI_ISL_2719074, EPI_ISL_2719077, EPI_ISL_2719078, EPI_ISL_2719079, EPI_ISL_2719083, EPI_ISL_2719084, EPI_ISL_2719085, EPI_ISL_2719087, EPI_ISL_2719091, EPI_ISL_2719092, EPI_ISL_2719314, EPI_ISL_2719319, EPI_ISL_2719328, EPI_ISL_2719331, EPI_ISL_2719334, EPI_ISL_2719335, EPI_ISL_2719336, EPI_ISL_2719338, EPI_ISL_2719340, EPI_ISL_2719341, EPI_ISL_2719342, EPI_ISL_2719343, EPI_ISL_2719344, EPI_ISL_2719349, EPI_ISL_2719350, EPI_ISL_2719354, EPI_ISL_2719356, EPI_ISL_2719358, EPI_ISL_2719359, EPI_ISL_2719362, EPI_ISL_2719363, EPI_ISL_2719364, EPI_ISL_2719366, EPI_ISL_2719367, EPI_ISL_2719370, EPI_ISL_2719371, EPI_ISL_2719377, EPI_ISL_2719378, EPI_ISL_2719380, EPI_ISL_2719384, EPI_ISL_2719405, EPI_ISL_2719412, EPI_ISL_2719413, EPI_ISL_2719415, EPI_ISL_2719416, EPI_ISL_2719417, EPI_ISL_2719420, EPI_ISL_2719425, EPI_ISL_2719428, EPI_ISL_2719429, EPI_ISL_2719430, EPI_ISL_2719431, EPI_ISL_2719435, EPI_ISL_2719436, EPI_ISL_2719438, EPI_ISL_2719441, EPI_ISL_2719442, EPI_ISL_2719445, EPI_ISL_2719447, EPI_ISL_2719674, EPI_ISL_2720056, EPI_ISL_2720057, EPI_ISL_2720058, EPI_ISL_2720063, EPI_ISL_2720064, EPI_ISL_2720065, EPI_ISL_2720066, EPI_ISL_2720067, EPI_ISL_2720068, EPI_ISL_2720069, EPI_ISL_2720070, EPI_ISL_2720073, EPI_ISL_2720074, EPI_ISL_2720075, EPI_ISL_2720076, EPI_ISL_2720077, EPI_ISL_2720078, EPI_ISL_2720080, EPI_ISL_2720081, EPI_ISL_2720082, EPI_ISL_2720084, EPI_ISL_2720085, EPI_ISL_2720086, EPI_ISL_2720087, EPI_ISL_2720088, EPI_ISL_2720092, EPI_ISL_2720093, EPI_ISL_2720094, EPI_ISL_2720096, EPI_ISL_2720097, EPI_ISL_2720098, EPI_ISL_2720100, EPI_ISL_2720101, EPI_ISL_2720102, EPI_ISL_2720106, EPI_ISL_2720107, EPI_ISL_2720148, EPI_ISL_2720149, EPI_ISL_2720150, EPI_ISL_2720151, EPI_ISL_2720152, EPI_ISL_2720159, EPI_ISL_2720160, EPI_ISL_2720161, EPI_ISL_2720165, EPI_ISL_2720166, EPI_ISL_2720167, EPI_ISL_2720169, EPI_ISL_2720171, EPI_ISL_2720172, EPI_ISL_2720174, EPI_ISL_2720177, EPI_ISL_2720178, EPI_ISL_2720179, EPI_ISL_2720180, EPI_ISL_2720183, EPI_ISL_2720185, EPI_ISL_2720186, EPI_ISL_2720187, EPI_ISL_2720189, EPI_ISL_2720190, EPI_ISL_2720191, EPI_ISL_2720192, EPI_ISL_2720193, EPI_ISL_2720194, EPI_ISL_2720195, EPI_ISL_2720197, EPI_ISL_2720225, EPI_ISL_2720227, EPI_ISL_2720251, EPI_ISL_2720254, EPI_ISL_2720259, EPI_ISL_2720572, EPI_ISL_2720790, EPI_ISL_2720793, EPI_ISL_2720794, EPI_ISL_2720795, EPI_ISL_2720796, EPI_ISL_2720800, EPI_ISL_2720801, EPI_ISL_2720803, EPI_ISL_2720805, EPI_ISL_2720812, EPI_ISL_2720813, EPI_ISL_2720815, EPI_ISL_2720816, EPI_ISL_2720819, EPI_ISL_2720820, EPI_ISL_2720881, EPI_ISL_2720882, EPI_ISL_2720884, EPI_ISL_2720885, EPI_ISL_2720892, EPI_ISL_2720896, EPI_ISL_2720897, EPI_ISL_2720898, EPI_ISL_2720912, EPI_ISL_2720913, EPI_ISL_2720914, EPI_ISL_2720915, EPI_ISL_2720918, EPI_ISL_2720919, EPI_ISL_2721969, EPI_ISL_2722040 | | | |
| see above | BCCDC Public Health Laboratory | BCCDC Public Health Laboratory | Ana Pacagnella; Corrinne Ng; Dan Fornika; John Tyson; Kim Macdonald; Kimia Kamelian; Linda Hoang; Loretta Janz; Mel Krajden; Prystajecky Natalie; Robert Azana; Shannon Russell |
| EPI_ISL_2097241 | BOSTON HEART DIAGNOSTICS CORP | Wadsworth Center, New York State Department of Health | Alexis Russell; Catharine Prussing; Daryl M. Lamson; Erasmus Schneider; Erica Lasek-Nesselquist; John Kelly; Jonathan Plitnick; Kirsten St. George; Matthew Shudt; Melissa A Leisner; Navjot Singh |
| EPI_ISL_7672115, EPI_ISL_7672116 | Bambino Gesù Pediatric Hospital | Microbiology and Immunology Diagnosis Bambino Gesù Pediatric Hospital | Carlo Federico Perno; Claudia Alteri; Luna Colagrossi; Rossana Scutari; Valentino Costabile |
| EPI_ISL_2260452, EPI_ISL_2262706 | Bayerisches Landesamt für Gesundheit und Lebensmittelsicherheit (LGL) | Robert Koch Institute | |
| EPI_ISL_1855894, EPI_ISL_1972275 | Baylor Scott & White-Temple | Baylor Scott & White-Temple | Ari Rao; Kimberly Walker; Linden Morales; Marcus Volz; Shelby Hendrickson |
| EPI_ISL_5802497, EPI_ISL_5802498, EPI_ISL_5802505, EPI_ISL_5802506, EPI_ISL_5802507, EPI_ISL_5802508, EPI_ISL_5802509, EPI_ISL_5802510, EPI_ISL_5802511, EPI_ISL_5802512, EPI_ISL_5802513, EPI_ISL_5802515, EPI_ISL_5802516, EPI_ISL_5802517, EPI_ISL_5802518, EPI_ISL_5802519, EPI_ISL_5802520, EPI_ISL_5802521, EPI_ISL_5802522, EPI_ISL_5802523, EPI_ISL_5802524, EPI_ISL_5802525, EPI_ISL_5802526, EPI_ISL_5802527, EPI_ISL_5802528, EPI_ISL_5802529, EPI_ISL_5802530, EPI_ISL_5802565, EPI_ISL_5802566, EPI_ISL_5802567, EPI_ISL_5802568, EPI_ISL_5802569, EPI_ISL_5802570, EPI_ISL_5802571, EPI_ISL_5802572, EPI_ISL_5802573, EPI_ISL_5802584, EPI_ISL_5802585, EPI_ISL_5802586, EPI_ISL_5802587, EPI_ISL_5802588, EPI_ISL_5802589, EPI_ISL_5802590, EPI_ISL_5802591, EPI_ISL_5802592, EPI_ISL_5802593, EPI_ISL_5802594 | | | |
| see above | Biofast | Instituto Butantan | Antonio Jorge Martins; Claudia Renata dos Santos Barros; David Schlesinger; Debora Botequio Moretti; Dimas Tadeu Covas; Elaine Cristina Marqueze; Elaine Vieira Santos; Evandra Strazza Rodrigues; Heidge Fukumasu; Jayme Augusto de Souza-Neto; José Salvatore Leister Patané; Luiz Alcantara; Luiz Lehmann Coutinho; Maria Carolina Elias; Maurício Lacerda Nogueira; Rafael dos Santos Bezerra; Raul Machado Neto; Rejane Maria Tommasini Grotto; Ricardo Haddad; Sandra Coccuzzo Sampaio Vessoni; Simone Kashima; Svetoslav Nanev Slavov; Vincent Louis Viala |
| EPI_ISL_1916588, EPI_ISL_1917064, EPI_ISL_1917098, EPI_ISL_1917111, EPI_ISL_1917129, EPI_ISL_3145466 | BioneXt Lab | Laboratoire national de sante, Microbiology, Microbial Genomics Platform | Anke Wienecke-Baldacchino; Catherine Ragimbeau; Elodie Solarino; Fatu Djabi; Jessica Tapp; Lise Pignon; Raoul Salmon; Tamir Abdelrahman; Thibault Ferrandon; Virginie Jover |
| EPI_ISL_3494426, EPI_ISL_3494429, EPI_ISL_3494433 | Boston University CTL | Boston University/National Emerging Infectious Disease Laboratories | Catherine Klapperich; Jacquelyn Turcinovic; John H. Connor; Lena Landeverde; Lynn Doucette-Stamm |
| EPI_ISL_3478991, EPI_ISL_3478992, EPI_ISL_3478995, EPI_ISL_3478996, EPI_ISL_3478997, EPI_ISL_3480876, EPI_ISL_3480877, EPI_ISL_3480881, EPI_ISL_3480882, EPI_ISL_3480883, EPI_ISL_3480884, EPI_ISL_3480886, EPI_ISL_3480887, EPI_ISL_3480890, EPI_ISL_3480892, EPI_ISL_3480893, EPI_ISL_3480896, EPI_ISL_3480907, EPI_ISL_3480908, EPI_ISL_3480914, EPI_ISL_3480915, EPI_ISL_3480922, EPI_ISL_3480923, EPI_ISL_3480924, EPI_ISL_3480928, EPI_ISL_3480929, EPI_ISL_3480932, EPI_ISL_3480933, EPI_ISL_3480934, EPI_ISL_3480935, EPI_ISL_3480936, EPI_ISL_3480937, EPI_ISL_3480939, EPI_ISL_3480945, EPI_ISL_3480947, EPI_ISL_3480948, EPI_ISL_3480950, EPI_ISL_3480955, EPI_ISL_3480958, EPI_ISL_3480962, EPI_ISL_3480964, EPI_ISL_3480965, EPI_ISL_3480966, EPI_ISL_3480967, EPI_ISL_3480970, EPI_ISL_3480971, EPI_ISL_3480973, EPI_ISL_3480974, EPI_ISL_3480977, EPI_ISL_3480978, EPI_ISL_3480979, EPI_ISL_3480980, EPI_ISL_3480981, EPI_ISL_3480985, EPI_ISL_3480988, EPI_ISL_3480990, EPI_ISL_3480991, EPI_ISL_3480993, EPI_ISL_3480994, EPI_ISL_3480995, EPI_ISL_3480996, EPI_ISL_3481002, EPI_ISL_3481005, EPI_ISL_3481009, EPI_ISL_3481011, EPI_ISL_3481016, EPI_ISL_3481018, EPI_ISL_3481021, EPI_ISL_3481036, EPI_ISL_3481038, EPI_ISL_3481039, EPI_ISL_3481040, EPI_ISL_3481041, EPI_ISL_3481043, EPI_ISL_3481045, EPI_ISL_3481046, EPI_ISL_3481049, EPI_ISL_3481050, EPI_ISL_3481058, EPI_ISL_3481059, EPI_ISL_3481062, EPI_ISL_3481063, EPI_ISL_3481066, EPI_ISL_3481068, EPI_ISL_3481076, EPI_ISL_3481077, EPI_ISL_3481078, EPI_ISL_3481079, EPI_ISL_3481080, EPI_ISL_3481081, EPI_ISL_3481086, EPI_ISL_3481087, EPI_ISL_3481089, EPI_ISL_3481090, EPI_ISL_3481093, EPI_ISL_3481094, EPI_ISL_3481095, EPI_ISL_3481096, EPI_ISL_3481099, EPI_ISL_3481101, EPI_ISL_3481102, EPI_ISL_3481103, EPI_ISL_3481104, EPI_ISL_3481105, EPI_ISL_3481106, EPI_ISL_3481107, EPI_ISL_3481109, EPI_ISL_3481110, EPI_ISL_3481119, EPI_ISL_3481127, EPI_ISL_3481130, EPI_ISL_3481136, EPI_ISL_3481137, EPI_ISL_3481139, EPI_ISL_3481141, EPI_ISL_3481144, EPI_ISL_3481149, EPI_ISL_3481150, EPI_ISL_3481151, EPI_ISL_3481152, EPI_ISL_3481154, EPI_ISL_3481155, EPI_ISL_3481156, EPI_ISL_3481159, EPI_ISL_3481161, EPI_ISL_3481162, EPI_ISL_3481163, EPI_ISL_3481164, EPI_ISL_3481165, EPI_ISL_3481166, EPI_ISL_3481167, EPI_ISL_3481168, EPI_ISL_3481169, EPI_ISL_3481179, EPI_ISL_3481187, EPI_ISL_3481188, EPI_ISL_3481192, EPI_ISL_3481194, EPI_ISL_3481195, EPI_ISL_3481196, EPI_ISL_3481202, EPI_ISL_3481203, EPI_ISL_3481206, EPI_ISL_3481212, EPI_ISL_3481213, EPI_ISL_3481214, EPI_ISL_3481216, EPI_ISL_3481218, EPI_ISL_3481220, EPI_ISL_3481221, EPI_ISL_3481225, EPI_ISL_3481226, EPI_ISL_3481228, EPI_ISL_3481230, EPI_ISL_3481231, EPI_ISL_3481232, EPI_ISL_3481233, EPI_ISL_3481236, EPI_ISL_3481237, EPI_ISL_3481241, EPI_ISL_3481242, EPI_ISL_3481243, EPI_ISL_3481247, EPI_ISL_3481250, EPI_ISL_3481497, EPI_ISL_3598895, EPI_ISL_3598911, EPI_ISL_3598919, EPI_ISL_3598920, EPI_ISL_3598921, EPI_ISL_3598929, EPI_ISL_3598934, EPI_ISL_3598936, EPI_ISL_3598938, EPI_ISL_3598943, EPI_ISL_3598947, EPI_ISL_3598953, EPI_ISL_3598954, EPI_ISL_3598955, EPI_ISL_3598958, EPI_ISL_3598960, EPI_ISL_3598962, EPI_ISL_3598964, EPI_ISL_3598970, EPI_ISL_3598980, EPI_ISL_3598984, EPI_ISL_3599004, EPI_ISL_3599005, EPI_ISL_3599011, EPI_ISL_3599016, EPI_ISL_3599017, EPI_ISL_3599026, EPI_ISL_3599029, EPI_ISL_3599030, EPI_ISL_3599033, EPI_ISL_3599041, EPI_ISL_4423489, EPI_ISL_4423491, EPI_ISL_4423494, EPI_ISL_4423503, EPI_ISL_4423507, EPI_ISL_4423511, EPI_ISL_4423517, EPI_ISL_4423524, EPI_ISL_4423537, EPI_ISL_4423558, EPI_ISL_4423570, EPI_ISL_4423578, EPI_ISL_4423579, EPI_ISL_4423583, EPI_ISL_4423592, EPI_ISL_4423593, EPI_ISL_4423595, EPI_ISL_4423640 | | | |
| see above | British Columbia Centre For Disease Control | BCCDC Public Health Laboratory | Ana Pacagnella; Corrinne Ng; Dan Fornika; John Tyson; Kim Macdonald; Kimia Kamelian; Linda Hoang; Loretta Janz; Mel Krajden; Prystajecky Natalie; Robert Azana; Shannon Russell |
| EPI_ISL_1786788, EPI_ISL_1786853, EPI_ISL_1786870, EPI_ISL_1786964, EPI_ISL_1787055, EPI_ISL_1787119, EPI_ISL_1787142, EPI_ISL_1825156, EPI_ISL_1825161, EPI_ISL_1825167, EPI_ISL_1825180, EPI_ISL_1825186, EPI_ISL_1825202, EPI_ISL_1825231, EPI_ISL_1825261, EPI_ISL_1825273, EPI_ISL_1825358, EPI_ISL_1825377, EPI_ISL_1825378, EPI_ISL_1825380, EPI_ISL_1825386, EPI_ISL_1825387, EPI_ISL_1825392, EPI_ISL_1825407, EPI_ISL_1825409, EPI_ISL_1825417, EPI_ISL_1825419, EPI_ISL_1825421, EPI_ISL_1825426, EPI_ISL_1825430, EPI_ISL_1825438, EPI_ISL_1825442, EPI_ISL_1825474, EPI_ISL_1825526, EPI_ISL_1825585, EPI_ISL_1825591, EPI_ISL_1825597, EPI_ISL_1825607, EPI_ISL_1825610, EPI_ISL_1825611, EPI_ISL_1825617, EPI_ISL_1825638, EPI_ISL_1825641, EPI_ISL_1825649, EPI_ISL_1825650, EPI_ISL_1825651, EPI_ISL_1825657, EPI_ISL_1825659, EPI_ISL_1825674, EPI_ISL_1825675, EPI_ISL_1918417, EPI_ISL_1918422, EPI_ISL_1918423, EPI_ISL_1918433, EPI_ISL_1918441, EPI_ISL_1918443, EPI_ISL_1918445, EPI_ISL_1918446, EPI_ISL_1918451, EPI_ISL_1918454, EPI_ISL_1918463, EPI_ISL_1918475, EPI_ISL_1918479, EPI_ISL_1918486, EPI_ISL_1918488, EPI_ISL_1918492, EPI_ISL_1918493, EPI_ISL_1918495, EPI_ISL_1918496, EPI_ISL_1918500, EPI_ISL_1918509, EPI_ISL_1918511, EPI_ISL_1918516, EPI_ISL_1918518, EPI_ISL_1918520, EPI_ISL_1918531, EPI_ISL_1918539, EPI_ISL_1918542, EPI_ISL_1918547, EPI_ISL_1918554, EPI_ISL_1918555, EPI_ISL_1918559, EPI_ISL_1918562, EPI_ISL_1918568, EPI_ISL_1918569, EPI_ISL_1918571, EPI_ISL_1918590, EPI_ISL_1918596, EPI_ISL_1918600, EPI_ISL_1918603, EPI_ISL_1918604, EPI_ISL_1918608, EPI_ISL_1918609, EPI_ISL_1918617, EPI_ISL_1918626, EPI_ISL_1918644, EPI_ISL_1918656, EPI_ISL_1918659, EPI_ISL_1918661, EPI_ISL_1918669, EPI_ISL_1918676, EPI_ISL_1918694, EPI_ISL_1918699, EPI_ISL_1918701, EPI_ISL_1918704, EPI_ISL_1918706, EPI_ISL_1918708, EPI_ISL_1918710, EPI_ISL_1918719, EPI_ISL_1918720, EPI_ISL_1918742, EPI_ISL_1918747, EPI_ISL_1918759, EPI_ISL_1918762, EPI_ISL_1918764, EPI_ISL_1918767, EPI_ISL_1918787, EPI_ISL_1918796, EPI_ISL_1918798, EPI_ISL_1918802, EPI_ISL_1918807, EPI_ISL_1918808, EPI_ISL_1918814, EPI_ISL_1918818, EPI_ISL_1918819, EPI_ISL_1918841, EPI_ISL_1918849, EPI_ISL_1918854, EPI_ISL_1918857, EPI_ISL_1918868, EPI_ISL_1918878, EPI_ISL_1918883, EPI_ISL_1918894, EPI_ISL_1918896, EPI_ISL_1918899, EPI_ISL_1918902, EPI_ISL_1918905, EPI_ISL_1918910, EPI_ISL_1918920, EPI_ISL_1918924, EPI_ISL_1918926, EPI_ISL_1918928, EPI_ISL_1918931, EPI_ISL_1918937, EPI_ISL_1967972, EPI_ISL_2016413, EPI_ISL_2016415 | | | |
| see above | Broad Institute Clinical Research Sequencing Platform | Infectious Disease Program, Broad Institute of Harvard and MIT | Adams, G.; B.L.; B.W.; Bauer, M.; Birren; Blumenstiel, B.; Brown, C.; Carter, A.; Chaluvadi, S.; D.J.; DeFelice, M.; DeRuff, K.; Dodge, S.; Gabriel, S.; Gallagher, G.; Gladden-Young, A.; Granger, B.; J.E.; K.J.; Lagerborg, K.; Larkin, K.; Lee, M.; Lemieux; Lennon, N.; Loreth, C.; Madoff, L.; McGovern, S.; Meldrim, J.; Normandin, E.; P.C.; Park; Pearlman, L.; Reilly, S.; Rudy, M.; Sabeti; Siddle; Smole, S.; Tomkins-Tinch, C.; Vicente, G.; and MacInnis |
| EPI_ISL_2341419, EPI_ISL_2341425, EPI_ISL_2341513, EPI_ISL_2341568, EPI_ISL_2341647 | CA DPH Viral and Rickettsial Disease Laboratory | Chan-Zuckerberg Biohub | CZB Cliahub Consortium |
| EPI_ISL_5603235, EPI_ISL_5603306 | CASA DE SAUDE MATERNIDADE CELESTINA COLARES | Analytical Competence Molecular Epidemiology Lab/ACME, Oswaldo Cruz Foundation, Ceara (FIOCRUZ CE) | Carlos Leonardo de Aragao Araujo; Cecilia Leite Costa & Eduardo Ruback dos Santos on behalf of COVID-19 FIOCRUZ Genomic Network; Cleber Furtado Aksenen; Fabio Miyajima; Fernando Braga Stehling; Francisco Eder de Moura Lopes; Igor Oliveira Duarte; Jamille Maria Mendes Bezerra; Joaquim Cesar do Nascimento Sousa Junior; Pedro Miguel Carneiro Jeronimo; Suzana Porto Almeida; Thais Ferreira de Oliveira; Thais de Oliveira Costa; Ticiane Cavalcante de Souza; Veridiana Pessoa Miyajima |
| EPI_ISL_2825870, EPI_ISL_2825874, EPI_ISL_2882162, EPI_ISL_2882282, EPI_ISL_2882291, EPI_ISL_2882483, EPI_ISL_2882484, EPI_ISL_2882635, EPI_ISL_2890084, EPI_ISL_2891886, EPI_ISL_2891913, EPI_ISL_2892041, EPI_ISL_2892374, EPI_ISL_2921765, EPI_ISL_2921766, EPI_ISL_2921767, EPI_ISL_2921839, EPI_ISL_2922138, EPI_ISL_2922158, EPI_ISL_2922244, EPI_ISL_5696570, EPI_ISL_5696576, | | | |

| | | | |
|---|---|---|---|
| EPI_ISL_7335595, EPI_ISL_7335654 | | | |
| see above | CDPH VBL | California Department of Public Health | CDPH-COVIDNet; Emily Smith on behalf of CDPH-COVIDNet; Emily Smith on behalf of CDPH-COVIDNet and Priva Path Labs Inc.; UCLA Technology Center for Genomics & Bioinformatics |
| EPI_ISL_2097252 | CENTERS LAB NJ, LLC (DBA MEDLABS DIAGNOSTICS) | Wadsworth Center, New York State Department of Health | Alexis Russell; Catharine Prussing; Daryl M. Lamson; Erasmus Schneider; Erica Lasek-Nesselquist; John Kelly; Jonathan Plitnick; Kirsten St. George; Matthew Shudt; Melissa A Leisner; Navjot Singh |
| EPI_ISL_2344961, EPI_ISL_2344963, EPI_ISL_2344967, EPI_ISL_2344974, EPI_ISL_2344981, EPI_ISL_2344984, EPI_ISL_2344986, EPI_ISL_2344990, EPI_ISL_2344991, EPI_ISL_2344992, EPI_ISL_2344993, EPI_ISL_2344994, EPI_ISL_2344996, EPI_ISL_2344998 | | | |
| see above | CENTRO DE APOIO EPIDEMIOLOGICO | Instituto Butantan / ESALQ-Piracicaba | Antonio Jorge Martins; Claudia Renata dos Santos Barros; David Schlesinger; Debora Botequio Moretti; Dimas Tadeu Covas; Elaine Cristina Marqueze; Elaine Vieira Santos; Evandra Strazza Rodrigues; Heidge Fukumasu; Jayme Augusto de Souza-Neto; José Salvatore Leister Patané; Luiz Alcantara; Luiz Lehmann Coutinho; Maria Carolina Elias; Maurício Lacerda Nogueira; Rafael dos Santos Bezerra; Raul Machado Neto; Rejane Maria Tommasini Grotto; Ricardo Haddad; Sandra Coccuzzo Sampaio Vessoni; Simone Kashima; Svetoslav Nanev Slavov; Vincent Louis Viala |
| EPI_ISL_5603218 | CENTRO DE ATENDIMENTO AO ENFRENTAMENTO A COVID 19 | Analytical Competence Molecular Epidemiology Lab/ACME, Oswaldo Cruz Foundation, Ceara (FIOCRUZ CE) | Carlos Leonardo de Aragao Araujo; Cecilia Leite Costa & Eduardo Ruback dos Santos on behalf of COVID-19 FIOCRUZ Genomic Network; Cleber Furtado Aksenen; Fabio Miyajima; Fernando Braga Stehling; Francisco Eder de Moura Lopes; Igor Oliveira Duarte; Jamille Maria Mendes Bezerra; Joaquim Cesar do Nascimento Sousa Junior; Pedro Miguel Carneiro Jeronimo; Suzana Porto Almeida; Thais Ferreira de Oliveira; Thais de Oliveira Costa; Ticiane Cavalcante de Souza; Veridiana Pessoa Miyajima |
| EPI_ISL_3102511, EPI_ISL_3102512 | CENTRO DE ATENDIMENTO PARA ENFRENTAMENTO A COVID 19 | Analytical Competence Molecular Epidemiology Lab/ACME, Oswaldo Cruz Foundation, Ceara (FIOCRUZ CE) | Cleber Furtado Aksenen; Fabio Miyajima; Fernando Braga Stehling; Francisco Eder de Moura Lopes; Jamille Maria Mendes Bezerra; Joaquim César do Nascimento Sousa Junior; Pedro Miguel Carneiro Jeronimo; Suzana Porto Almeida e Lucas Delerino; Thais Ferreira de Oliveira; Thais de Oliveira Costa; Ticiane Cavalcante de Souza; Veridiana Pessoa Miyajima |
| EPI_ISL_1967251 | CENTRO DE DETENCAO PROVISORIA DE NOVA INDEPENDENCIA | Instituto Butantan / Mendelics | Antonio Jorge Martins; Bianca Cechetto Carlos. Mendelics: Bibiana Santos; Claudia Renata dos Santos Barros; Cíntia Bittar; David Schlesinger. Hemocentro Ribeirão Preto: Simone Kashima; Debora Botequio Moretti; Elaine Cristina Marqueze; Elaine Vieira dos Santos; Elisangela Chicaroni Mattos; Erika Freitas; Evandra Strazza Rodrigues; Felipe Allan da Silva da Costa; Flavia Aburjaile; Fábio Sossai Possebon; Guilherme Campos; Guilherme Targino Valente; Heidge Fukumasu. USP-Botucatu: Rejane Maria Tommasini Grotto; Helena Lage Ferreira; Instituto Butantan: Dimas Tadeu Covas; Jardelina de Souza Todao Bernardino; Jayme A. Souza-Neto; Jessika Cristina Chagas Lesbon; Jorge A. Petroli Marchesi; José Salvatore Leister Patané; João Paulo Kitajima; João Pessoa Araújo Jr.; Leila Sabrina Ullmann; Loyze Paola Oliveira de Lima; Luiz Aurelio de Campos Crispin. Centro de Genômica Funcional da ESALQ: Luiz Lehmann Coutinho; Luiz Carlos Junior de Alcantara; Lívia Sacchetto; Maisa C. Pereira Parra; Maria Carolina Elias; Marta Giovanetti; Marília Moraes; Maurício Lacerda Nogueira. Prefeitura de Sao Paulo: Melissa Palmieri.; Patricia Akemi Assato; Paula Rahal; Paulo Inacio da Costa; Rafael dos Santos Bezerra; Raquel de Lello Rocha Campos Cassano. NGS Soluções Genômicas: Pilar Drummond Sampaio Corrêa Mariani. FZEA-USP Pirassununga: Mirele Daiana Poleti; Raul Machado Neto; Ricardo Augusto Brassaloti; Ricardo Haddad; Rodrigo Tocantins Calado. FAMERP-SJRP: Cecília Artico Banho; Sandra Coccuzzo Sampaio; Svetoslav Nanev Slavov; Vagner Fonseca; Vincent Louis Viala |
| EPI_ISL_2344792 | CENTRO DE ESPECIALIDADES DE AGUAI AGUAI | Instituto Butantan / ESALQ-Piracicaba | Antonio Jorge Martins; Claudia Renata dos Santos Barros; David Schlesinger; Debora Botequio Moretti; Dimas Tadeu Covas; Elaine Cristina Marqueze; Elaine Vieira Santos; Evandra Strazza Rodrigues; Heidge Fukumasu; Jayme Augusto de Souza-Neto; José Salvatore Leister Patané; Luiz Alcantara; Luiz Lehmann Coutinho; Maria Carolina Elias; Maurício Lacerda Nogueira; Rafael dos Santos Bezerra; Raul Machado Neto; Rejane Maria Tommasini Grotto; Ricardo Haddad; Sandra Coccuzzo Sampaio Vessoni; Simone Kashima; Svetoslav Nanev Slavov; Vincent Louis Viala |
| EPI_ISL_5603228 | CENTRO DE SAUDE DA FAMILIA DR MARLON FERNANDES | Analytical Competence Molecular Epidemiology Lab/ACME, Oswaldo Cruz Foundation, Ceara (FIOCRUZ CE) | Carlos Leonardo de Aragao Araujo; Cecilia Leite Costa & Eduardo Ruback dos Santos on behalf of COVID-19 FIOCRUZ Genomic Network; Cleber Furtado Aksenen; Fabio Miyajima; Fernando Braga Stehling; Francisco Eder de Moura Lopes; Igor Oliveira Duarte; Jamille Maria Mendes Bezerra; Joaquim Cesar do Nascimento Sousa Junior; Pedro Miguel Carneiro Jeronimo; Suzana Porto Almeida; Thais Ferreira de Oliveira; Thais de Oliveira Costa; Ticiane Cavalcante de Souza; Veridiana Pessoa Miyajima |
| EPI_ISL_2344852 | CENTRO DE SAUDE DE ANALANDIA | Instituto Butantan / ESALQ-Piracicaba | Antonio Jorge Martins; Claudia Renata dos Santos Barros; David Schlesinger; Debora Botequio Moretti; Dimas Tadeu Covas; Elaine Cristina Marqueze; Elaine Vieira Santos; Evandra Strazza Rodrigues; Heidge Fukumasu; Jayme Augusto de Souza-Neto; José Salvatore Leister Patané; Luiz Alcantara; Luiz Lehmann Coutinho; Maria Carolina Elias; Maurício Lacerda Nogueira; Rafael dos Santos Bezerra; Raul Machado Neto; Rejane Maria Tommasini Grotto; Ricardo Haddad; Sandra Coccuzzo Sampaio Vessoni; Simone Kashima; Svetoslav Nanev Slavov; Vincent Louis Viala |
| EPI_ISL_1967212 | CENTRO DE SAUDE DE BARBOSA | Instituto Butantan / Mendelics | Antonio Jorge Martins; Bianca Cechetto Carlos. Mendelics: Bibiana Santos; Claudia Renata dos Santos Barros; Cíntia Bittar; David Schlesinger. Hemocentro Ribeirão Preto: Simone Kashima; Debora Botequio Moretti; Elaine Cristina Marqueze; Elaine Vieira dos Santos; Elisangela Chicaroni Mattos; Erika Freitas; Evandra Strazza Rodrigues; Felipe Allan da Silva da Costa; Flavia Aburjaile; Fábio Sossai Possebon; Guilherme Campos; Guilherme Targino Valente; Heidge Fukumasu. USP-Botucatu: Rejane Maria Tommasini Grotto; Helena Lage Ferreira; Instituto Butantan: Dimas Tadeu Covas; Jardelina de Souza Todao Bernardino; Jayme A. Souza-Neto; Jessika Cristina Chagas Lesbon; Jorge A. Petroli Marchesi; José Salvatore Leister Patané; João Paulo Kitajima; João Pessoa Araújo Jr.; Leila Sabrina Ullmann; Loyze Paola Oliveira de Lima; Luiz Aurelio de Campos Crispin. Centro de Genômica Funcional da ESALQ: Luiz Lehmann Coutinho; Luiz Carlos Junior de Alcantara; Lívia Sacchetto; Maisa C. Pereira Parra; Maria Carolina Elias; Marta Giovanetti; Marília Moraes; Maurício Lacerda Nogueira. Prefeitura de Sao Paulo: Melissa Palmieri.; Patricia Akemi Assato; Paula Rahal; Paulo Inacio da Costa; Rafael dos Santos Bezerra; Raquel de Lello Rocha Campos Cassano. NGS Soluções Genômicas: Pilar Drummond Sampaio Corrêa Mariani. FZEA-USP Pirassununga: Mirele Daiana Poleti; Raul Machado Neto; Ricardo Augusto Brassaloti; Ricardo Haddad; Rodrigo Tocantins Calado. FAMERP-SJRP: Cecília Artico Banho; Sandra Coccuzzo Sampaio; Svetoslav Nanev Slavov; Vagner Fonseca; Vincent Louis Viala |
| EPI_ISL_2344774, EPI_ISL_2344777 | CENTRO DE SAUDE DE CHAVANTES | Instituto Butantan / ESALQ-Piracicaba | Antonio Jorge Martins; Claudia Renata dos Santos Barros; David Schlesinger; Debora Botequio Moretti; Dimas Tadeu Covas; Elaine Cristina Marqueze; Elaine Vieira Santos; Evandra Strazza Rodrigues; Heidge Fukumasu; Jayme Augusto de Souza-Neto; José Salvatore Leister Patané; Luiz Alcantara; Luiz Lehmann Coutinho; Maria Carolina Elias; Maurício Lacerda Nogueira; Rafael dos Santos Bezerra; Raul Machado Neto; Rejane Maria Tommasini Grotto; Ricardo Haddad; Sandra Coccuzzo Sampaio Vessoni; Simone Kashima; Svetoslav Nanev Slavov; Vincent Louis Viala |
| EPI_ISL_2209396 | CENTRO DE SAUDE DE IBIRAREMA | Instituto Butantan / ESALQ-Piracicaba | Antonio Jorge Martins; Claudia Renata dos Santos Barros; David Schlesinger; Debora Botequio Moretti; Dimas Tadeu Covas; Elaine Cristina Marqueze; Elaine Vieira Santos; Evandra Strazza Rodrigues; Heidge Fukumasu; Jayme Augusto de Souza-Neto; José Salvatore Leister Patané; Luiz Alcantara; Luiz Lehmann Coutinho; Maria Carolina Elias; Maurício Lacerda Nogueira; Rafael dos Santos Bezerra; Raul Machado Neto; Rejane Maria Tommasini Grotto; Ricardo Haddad; Sandra Coccuzzo Sampaio Vessoni; Simone Kashima; Svetoslav Nanev Slavov; Vincent Louis Viala |
| EPI_ISL_5603267, EPI_ISL_5603268, EPI_ISL_5603278 | CENTRO DE SAUDE DE IPAPORANGA | Analytical Competence Molecular Epidemiology Lab/ACME, Oswaldo Cruz Foundation, Ceara (FIOCRUZ CE) | Carlos Leonardo de Aragao Araujo; Cecilia Leite Costa & Eduardo Ruback dos Santos on behalf of COVID-19 FIOCRUZ Genomic Network; Cleber Furtado Aksenen; Fabio Miyajima; Fernando Braga Stehling; Francisco Eder de Moura Lopes; Igor Oliveira Duarte; Jamille Maria Mendes Bezerra; Joaquim Cesar do Nascimento Sousa Junior; Pedro Miguel Carneiro Jeronimo; Suzana Porto Almeida; Thais Ferreira de Oliveira; Thais de Oliveira Costa; Ticiane Cavalcante de Souza; Veridiana Pessoa Miyajima |
| EPI_ISL_2344787 | CENTRO DE SAUDE DE LUTECIA | Instituto Butantan / ESALQ-Piracicaba | Antonio Jorge Martins; Claudia Renata dos Santos Barros; David Schlesinger; Debora Botequio Moretti; Dimas Tadeu Covas; Elaine Cristina Marqueze; Elaine Vieira Santos; Evandra Strazza Rodrigues; Heidge Fukumasu; Jayme Augusto de Souza-Neto; José Salvatore Leister Patané; Luiz Alcantara; Luiz Lehmann Coutinho; Maria Carolina Elias; Maurício Lacerda Nogueira; Rafael dos Santos Bezerra; Raul Machado Neto; Rejane Maria Tommasini Grotto; Ricardo Haddad; Sandra Coccuzzo Sampaio Vessoni; Simone Kashima; Svetoslav Nanev Slavov; Vincent Louis Viala |
| EPI_ISL_2344785 | CENTRO DE SAUDE DE OCAUCU | Instituto Butantan / ESALQ-Piracicaba | Antonio Jorge Martins; Claudia Renata dos Santos Barros; David Schlesinger; Debora Botequio Moretti; Dimas Tadeu Covas; Elaine Cristina Marqueze; Elaine Vieira Santos; Evandra Strazza Rodrigues; Heidge Fukumasu; Jayme Augusto de Souza-Neto; José Salvatore Leister Patané; Luiz Alcantara; Luiz Lehmann Coutinho; Maria Carolina Elias; Maurício Lacerda Nogueira; Rafael dos Santos Bezerra; Raul Machado Neto; Rejane Maria Tommasini Grotto; Ricardo Haddad; Sandra Coccuzzo Sampaio Vessoni; Simone Kashima; Svetoslav Nanev Slavov; Vincent Louis Viala |
| EPI_ISL_2344791 | CENTRO DE SAUDE DE SPEDRO DO TURVO | Instituto Butantan / ESALQ-Piracicaba | Antonio Jorge Martins; Claudia Renata dos Santos Barros; David Schlesinger; Debora Botequio Moretti; Dimas Tadeu Covas; Elaine Cristina Marqueze; Elaine Vieira Santos; Evandra Strazza Rodrigues; Heidge Fukumasu; Jayme Augusto de Souza-Neto; José Salvatore Leister Patané; Luiz Alcantara; Luiz Lehmann Coutinho; Maria Carolina Elias; Maurício Lacerda Nogueira; Rafael dos Santos Bezerra; Raul Machado Neto; Rejane Maria Tommasini Grotto; Ricardo Haddad; Sandra Coccuzzo Sampaio Vessoni; Simone Kashima; Svetoslav Nanev Slavov; Vincent Louis Viala |
| EPI_ISL_2344744 | CENTRO DE SAUDE DO INDAIA DO AGUAPEI FLORIDA PAULISTA | Instituto Butantan / ESALQ-Piracicaba | Antonio Jorge Martins; Claudia Renata dos Santos Barros; David Schlesinger; Debora Botequio Moretti; Dimas Tadeu Covas; Elaine Cristina Marqueze; Elaine Vieira Santos; Evandra Strazza Rodrigues; Heidge Fukumasu; Jayme Augusto de Souza-Neto; José Salvatore Leister Patané; Luiz Alcantara; Luiz Lehmann Coutinho; Maria Carolina Elias; Maurício Lacerda Nogueira; Rafael dos Santos Bezerra; Raul Machado Neto; Rejane Maria Tommasini Grotto; Ricardo Haddad; Sandra Coccuzzo Sampaio Vessoni; Simone Kashima; Svetoslav Nanev Slavov; Vincent Louis Viala |
| EPI_ISL_2344781 | CENTRO DE SAUDE DR MARIO DIAS DE AGUIAR CAPIVARI | Instituto Butantan / ESALQ-Piracicaba | Antonio Jorge Martins; Claudia Renata dos Santos Barros; David Schlesinger; Debora Botequio Moretti; Dimas Tadeu Covas; Elaine Cristina Marqueze; Elaine Vieira Santos; Evandra Strazza Rodrigues; Heidge Fukumasu; Jayme Augusto de Souza-Neto; José Salvatore Leister Patané; Luiz Alcantara; Luiz Lehmann Coutinho; Maria Carolina Elias; Maurício Lacerda Nogueira; Rafael dos Santos Bezerra; Raul Machado Neto; Rejane Maria Tommasini Grotto; Ricardo Haddad; Sandra Coccuzzo Sampaio Vessoni; Simone Kashima; Svetoslav Nanev Slavov; Vincent Louis Viala |
| EPI_ISL_5603212, EPI_ISL_5603274, EPI_ISL_5603281, EPI_ISL_5603282 | CENTRO DE SAUDE DR MIRANDA TAVARES | Analytical Competence Molecular Epidemiology Lab/ACME, Oswaldo Cruz Foundation, Ceara (FIOCRUZ CE) | Carlos Leonardo de Aragao Araujo; Cecilia Leite Costa & Eduardo Ruback dos Santos on behalf of COVID-19 FIOCRUZ Genomic Network; Cleber Furtado Aksenen; Fabio Miyajima; Fernando Braga Stehling; Francisco Eder de Moura Lopes; Igor Oliveira Duarte; Jamille Maria Mendes Bezerra; Joaquim Cesar do Nascimento Sousa Junior; Pedro Miguel Carneiro Jeronimo; Suzana Porto Almeida; Thais Ferreira de Oliveira; Thais de Oliveira Costa; Ticiane Cavalcante de Souza; Veridiana Pessoa Miyajima |
| EPI_ISL_2344797, EPI_ISL_2344798 | CENTRO DE SAUDE DR NELSON DA CUNHA BASTOS | Instituto Butantan / ESALQ-Piracicaba | Antonio Jorge Martins; Claudia Renata dos Santos Barros; David Schlesinger; Debora Botequio Moretti; Dimas Tadeu Covas; Elaine Cristina Marqueze; Elaine Vieira Santos; Evandra Strazza Rodrigues; Heidge Fukumasu; Jayme Augusto de Souza-Neto; José Salvatore Leister Patané; Luiz Alcantara; Luiz Lehmann Coutinho; Maria Carolina Elias; Maurício Lacerda Nogueira; Rafael dos Santos Bezerra; Raul Machado Neto; Rejane Maria Tommasini Grotto; Ricardo Haddad; Sandra Coccuzzo Sampaio Vessoni; Simone Kashima; Svetoslav Nanev Slavov; Vincent Louis Viala |
| EPI_ISL_2344790 | CENTRO DE SAUDE DR PLINIO ALBERS DE GALIA | Instituto Butantan / ESALQ-Piracicaba | Antonio Jorge Martins; Claudia Renata dos Santos Barros; David Schlesinger; Debora Botequio Moretti; Dimas Tadeu Covas; Elaine Cristina Marqueze; Elaine Vieira Santos; Evandra Strazza Rodrigues; Heidge Fukumasu; Jayme Augusto de Souza-Neto; José Salvatore Leister Patané; Luiz Alcantara; Luiz Lehmann Coutinho; Maria Carolina Elias; Maurício Lacerda Nogueira; Rafael dos Santos Bezerra; Raul Machado Neto; Rejane Maria Tommasini Grotto; Ricardo Haddad; Sandra Coccuzzo Sampaio Vessoni; Simone Kashima; Svetoslav Nanev Slavov; Vincent Louis Viala |
| EPI_ISL_2344832, EPI_ISL_2344873, EPI_ISL_2344904, EPI_ISL_2344905, EPI_ISL_2344907 | CENTRO DE SAUDE DR RENATO DE CARVALHO RIBEIRO ANGATUBA | Instituto Butantan / ESALQ-Piracicaba | Antonio Jorge Martins; Claudia Renata dos Santos Barros; David Schlesinger; Debora Botequio Moretti; Dimas Tadeu Covas; Elaine Cristina Marqueze; Elaine Vieira Santos; Evandra Strazza Rodrigues; Heidge Fukumasu; Jayme Augusto de Souza-Neto; José Salvatore Leister Patané; Luiz Alcantara; Luiz Lehmann Coutinho; Maria Carolina Elias; Maurício Lacerda Nogueira; Rafael dos Santos Bezerra; Raul Machado Neto; Rejane Maria Tommasini Grotto; Ricardo Haddad; Sandra Coccuzzo Sampaio Vessoni; Simone Kashima; Svetoslav Nanev Slavov; Vincent Louis Viala |
| EPI_ISL_2344779, EPI_ISL_2344783, EPI_ISL_2344784 | CENTRO DE SAUDE ESF IV ZONA RURAL DOMINGOS DE S SJRIOPARDO | Instituto Butantan / ESALQ-Piracicaba | Antonio Jorge Martins; Claudia Renata dos Santos Barros; David Schlesinger; Debora Botequio Moretti; Dimas Tadeu Covas; Elaine Cristina Marqueze; Elaine Vieira Santos; Evandra Strazza Rodrigues; Heidge Fukumasu; Jayme Augusto de Souza-Neto; José Salvatore Leister Patané; Luiz Alcantara; Luiz Lehmann Coutinho; Maria Carolina Elias; Maurício Lacerda Nogueira; Rafael dos Santos Bezerra; Raul Machado Neto; Rejane Maria Tommasini Grotto; Ricardo Haddad; Sandra Coccuzzo Sampaio Vessoni; Simone Kashima; Svetoslav Nanev Slavov; Vincent Louis Viala |
| EPI_ISL_2209436 | CENTRO DE SAUDE II MAIRINQUE MAIRINQUE | Instituto Butantan / ESALQ-Piracicaba | Antonio Jorge Martins; Claudia Renata dos Santos Barros; David Schlesinger; Debora Botequio Moretti; Dimas Tadeu Covas; Elaine Cristina Marqueze; Elaine Vieira Santos; Evandra Strazza Rodrigues; Heidge Fukumasu; Jayme Augusto de Souza-Neto; José Salvatore Leister Patané; Luiz Alcantara; Luiz Lehmann Coutinho; Maria Carolina Elias; Maurício Lacerda Nogueira; Rafael dos Santos Bezerra; Raul Machado Neto; Rejane Maria Tommasini Grotto; Ricardo Haddad; Sandra Coccuzzo Sampaio Vessoni; Simone Kashima; Svetoslav Nanev Slavov; Vincent Louis Viala |
| EPI_ISL_2209032, EPI_ISL_2344908 | CENTRO DE SAUDE TEREZINHA DE MORAES ARSILLA | Instituto Butantan / ESALQ-Piracicaba | Antonio Jorge Martins; Claudia Renata dos Santos Barros; David Schlesinger; Debora Botequio Moretti; Dimas Tadeu Covas; Elaine Cristina Marqueze; Elaine Vieira Santos; Evandra Strazza Rodrigues; Heidge Fukumasu; Jayme Augusto de Souza-Neto; José Salvatore Leister Patané; Luiz Alcantara; Luiz Lehmann Coutinho; Maria Carolina Elias; Maurício Lacerda Nogueira; Rafael dos Santos Bezerra; Raul Machado Neto; Rejane Maria Tommasini Grotto; Ricardo Haddad; Sandra Coccuzzo Sampaio Vessoni; Simone Kashima; Svetoslav Nanev Slavov; Vincent Louis Viala |
| EPI_ISL_2344749, EPI_ISL_2344752, EPI_ISL_2344757, EPI_ISL_2344759 | CENTRO DE TRIAGEM COVID 19 | Instituto Butantan / ESALQ-Piracicaba | Antonio Jorge Martins; Claudia Renata dos Santos Barros; David Schlesinger; Debora Botequio Moretti; Dimas Tadeu Covas; Elaine Cristina Marqueze; Elaine Vieira Santos; Evandra Strazza Rodrigues; Heidge Fukumasu; Jayme Augusto de Souza-Neto; José Salvatore Leister Patané; Luiz Alcantara; Luiz Lehmann Coutinho; Maria Carolina Elias; Maurício Lacerda Nogueira; Rafael dos Santos Bezerra; Raul Machado Neto; Rejane Maria Tommasini Grotto; Ricardo Haddad; Sandra Coccuzzo Sampaio Vessoni; Simone Kashima; Svetoslav Nanev Slavov; Vincent Louis Viala |
| EPI_ISL_1994018 | CH Roubaix | CHU Lille | AIT YAHYA Emilie; ALIDJINOU Enagnon Kazali; BOCKET Laurence; CREPIN Michel; DEMAY Christophe; ENGELMANN Ilka; GEFFROY Sandrine; GUIGON Aurélie; LAMBERT Valérie; LAZREK Mouna; NOBILLIAUX Florian; PREVOST Brigitte; THUILLIER Caroline; TINEZ Claire |
| EPI_ISL_2226343, EPI_ISL_2226354, EPI_ISL_2226358 | CH SAINTONGE | CHU Poitiers | Agnes BEBY-DEFAUX; Birama N'DIAYE; Caroline MICHAUD; Magali GARCIA; Manon PRAT; Maxime PICHON; Nicolas LEVEQUE; Valentin BON-BARET |
| EPI_ISL_1994005, EPI_ISL_1994006 | CH St Omer | CHU Lille | AIT YAHYA Emilie; ALIDJINOU Enagnon Kazali; BOCKET Laurence; CREPIN Michel; DEMAY Christophe; ENGELMANN Ilka; GEFFROY Sandrine; GUIGON Aurélie; LAMBERT Valérie; LAZREK Mouna; NOBILLIAUX Florian; PREVOST Brigitte; THUILLIER Caroline; TINEZ Claire |
| EPI_ISL_1757879, EPI_ISL_1757887 | CH.INTERCOMMUNAL DE CRETEIL | Department of Virology, Henri Mondor University Hospital, Assistance Publique Hôpitaux de Paris, Université Paris-Est Créteil, INSERM U955 | Alexandre Soulier; Christophe Rodriguez; Elisabeth Trawinski; Guillaume Gricourt; Jean-Michel Pawlotsky; Melissa N'Debi; Slim Fourati; Vanessa Demontant |
| EPI_ISL_1919717 | CHR Verviers | GIGA Medical Genomics | Bouchra Boujemla; Cécile Meex; Keith Durkin; Maria Artesi; Marie-Pierre Hayette; Nathalie Renotte; Pierrette Melin; Raphaël Boreux; Sébastien Bontems; Vincent Bours |
| EPI_ISL_1920214 | CHU Lille | CHU Lille | AIT YAHYA Emilie; ALIDJINOU Enagnon Kazali; BOCKET Laurence; CREPIN Michel; DEMAY Christophe; ENGELMANN Ilka; GEFFROY Sandrine; GUIGON Aurélie; LAMBERT Valérie; LAZREK Mouna; NOBILLIAUX Florian; PREVOST Brigitte; THUILLIER Caroline; TINEZ Claire |
| EPI_ISL_2017661, EPI_ISL_2017665, | CLILAB | Microbiology Department | Aida Gonzalez-Diaz; Carmen Ardanuy; Jordi Camara; Jordi Niubó; Laura Calatayud; M Angeles Domínguez; Miguel Fernandez-Huerta; Sara Marti |

| | | | |
|---|---|---|---|
| EPI_ISL_2017668 | | | |
| EPI_ISL_1797447, EPI_ISL_1970689 | CLINIQUE N-D DE GRACE | Institut de Pathologie et Genetique (IPG) | Jérémie Gras; Pascale Hilbert |
| EPI_ISL_1804974, EPI_ISL_1804975, EPI_ISL_1804977, EPI_ISL_1804984, EPI_ISL_1804988, EPI_ISL_1804989, EPI_ISL_1804990, EPI_ISL_1804991, EPI_ISL_1804992 | | | |
| see above | CQRC_QUALITY CONTROL CHEMICAL BIOLOGICAL RISK_AOOR Villa Sofia Cervello Palermo | CQRC_QUALITY CONTROL CHEMICAL BIOLOGICAL RISK_AOOR Villa Sofia Cervello Palermo | ; Bonvissuto, M.; Brunacci, G.; Buffa, V.; Contino, F.; Di Chiara, V.; Di Gaudio, F.; Di Sclafani, A.; Lungari, A.; Orlando, V.; Seidita, G.; Todaro, I. |
| EPI_ISL_2344863 | CS DE BALSAMO | Instituto Butantan / ESALQ-Piracicaba | Antonio Jorge Martins; Claudia Renata dos Santos Barros; David Schlesinger; Debora Botequio Moretti; Dimas Tadeu Covas; Elaine Cristina Marqueze; Elaine Vieira Santos; Evandra Strazza Rodrigues; Heidge Fukumasu; Jayme Augusto de Souza-Neto; José Salvatore Leister Patané; Luiz Alcantara; Luiz Lehmann Coutinho; Maria Carolina Elias; Maurício Lacerda Nogueira; Rafael dos Santos Bezerra; Raul Machado Neto; Rejane Maria Tommasini Grotto; Ricardo Haddad; Sandra Coccuzzo Sampaio Vessoni; Simone Kashima; Svetoslav Nanev Slavov; Vincent Louis Viala |
| EPI_ISL_2344823, EPI_ISL_2344824, EPI_ISL_2344857, EPI_ISL_2344860 | CS DE CATIGUA | Instituto Butantan / ESALQ-Piracicaba | Antonio Jorge Martins; Claudia Renata dos Santos Barros; David Schlesinger; Debora Botequio Moretti; Dimas Tadeu Covas; Elaine Cristina Marqueze; Elaine Vieira Santos; Evandra Strazza Rodrigues; Heidge Fukumasu; Jayme Augusto de Souza-Neto; José Salvatore Leister Patané; Luiz Alcantara; Luiz Lehmann Coutinho; Maria Carolina Elias; Maurício Lacerda Nogueira; Rafael dos Santos Bezerra; Raul Machado Neto; Rejane Maria Tommasini Grotto; Ricardo Haddad; Sandra Coccuzzo Sampaio Vessoni; Simone Kashima; Svetoslav Nanev Slavov; Vincent Louis Viala |
| EPI_ISL_2209177, EPI_ISL_2209397, EPI_ISL_2344871, EPI_ISL_2344874 | CS DE COSMORAMA | Instituto Butantan / ESALQ-Piracicaba | Antonio Jorge Martins; Claudia Renata dos Santos Barros; David Schlesinger; Debora Botequio Moretti; Dimas Tadeu Covas; Elaine Cristina Marqueze; Elaine Vieira Santos; Evandra Strazza Rodrigues; Heidge Fukumasu; Jayme Augusto de Souza-Neto; José Salvatore Leister Patané; Luiz Alcantara; Luiz Lehmann Coutinho; Maria Carolina Elias; Maurício Lacerda Nogueira; Rafael dos Santos Bezerra; Raul Machado Neto; Rejane Maria Tommasini Grotto; Ricardo Haddad; Sandra Coccuzzo Sampaio Vessoni; Simone Kashima; Svetoslav Nanev Slavov; Vincent Louis Viala |
| EPI_ISL_2344879, EPI_ISL_2344881 | CS DE FLOREAL | Instituto Butantan / ESALQ-Piracicaba | Antonio Jorge Martins; Claudia Renata dos Santos Barros; David Schlesinger; Debora Botequio Moretti; Dimas Tadeu Covas; Elaine Cristina Marqueze; Elaine Vieira Santos; Evandra Strazza Rodrigues; Heidge Fukumasu; Jayme Augusto de Souza-Neto; José Salvatore Leister Patané; Luiz Alcantara; Luiz Lehmann Coutinho; Maria Carolina Elias; Maurício Lacerda Nogueira; Rafael dos Santos Bezerra; Raul Machado Neto; Rejane Maria Tommasini Grotto; Ricardo Haddad; Sandra Coccuzzo Sampaio Vessoni; Simone Kashima; Svetoslav Nanev Slavov; Vincent Louis Viala |
| EPI_ISL_2344882 | CS DE MARAPOAMA | Instituto Butantan / ESALQ-Piracicaba | Antonio Jorge Martins; Claudia Renata dos Santos Barros; David Schlesinger; Debora Botequio Moretti; Dimas Tadeu Covas; Elaine Cristina Marqueze; Elaine Vieira Santos; Evandra Strazza Rodrigues; Heidge Fukumasu; Jayme Augusto de Souza-Neto; José Salvatore Leister Patané; Luiz Alcantara; Luiz Lehmann Coutinho; Maria Carolina Elias; Maurício Lacerda Nogueira; Rafael dos Santos Bezerra; Raul Machado Neto; Rejane Maria Tommasini Grotto; Ricardo Haddad; Sandra Coccuzzo Sampaio Vessoni; Simone Kashima; Svetoslav Nanev Slavov; Vincent Louis Viala |
| EPI_ISL_2208881, EPI_ISL_2344872, EPI_ISL_2344875, EPI_ISL_2344887, EPI_ISL_2344889 | CS DE ONDA VERDE | Instituto Butantan / ESALQ-Piracicaba | Antonio Jorge Martins; Claudia Renata dos Santos Barros; David Schlesinger; Debora Botequio Moretti; Dimas Tadeu Covas; Elaine Cristina Marqueze; Elaine Vieira Santos; Evandra Strazza Rodrigues; Heidge Fukumasu; Jayme Augusto de Souza-Neto; José Salvatore Leister Patané; Luiz Alcantara; Luiz Lehmann Coutinho; Maria Carolina Elias; Maurício Lacerda Nogueira; Rafael dos Santos Bezerra; Raul Machado Neto; Rejane Maria Tommasini Grotto; Ricardo Haddad; Sandra Coccuzzo Sampaio Vessoni; Simone Kashima; Svetoslav Nanev Slavov; Vincent Louis Viala |
| EPI_ISL_2209164, EPI_ISL_2344859 | CS DE PALMARES PAULISTA | Instituto Butantan / ESALQ-Piracicaba | Antonio Jorge Martins; Claudia Renata dos Santos Barros; David Schlesinger; Debora Botequio Moretti; Dimas Tadeu Covas; Elaine Cristina Marqueze; Elaine Vieira Santos; Evandra Strazza Rodrigues; Heidge Fukumasu; Jayme Augusto de Souza-Neto; José Salvatore Leister Patané; Luiz Alcantara; Luiz Lehmann Coutinho; Maria Carolina Elias; Maurício Lacerda Nogueira; Rafael dos Santos Bezerra; Raul Machado Neto; Rejane Maria Tommasini Grotto; Ricardo Haddad; Sandra Coccuzzo Sampaio Vessoni; Simone Kashima; Svetoslav Nanev Slavov; Vincent Louis Viala |
| EPI_ISL_2344769, EPI_ISL_2344772 | CS DE RINOPOLIS | Instituto Butantan / ESALQ-Piracicaba | Antonio Jorge Martins; Claudia Renata dos Santos Barros; David Schlesinger; Debora Botequio Moretti; Dimas Tadeu Covas; Elaine Cristina Marqueze; Elaine Vieira Santos; Evandra Strazza Rodrigues; Heidge Fukumasu; Jayme Augusto de Souza-Neto; José Salvatore Leister Patané; Luiz Alcantara; Luiz Lehmann Coutinho; Maria Carolina Elias; Maurício Lacerda Nogueira; Rafael dos Santos Bezerra; Raul Machado Neto; Rejane Maria Tommasini Grotto; Ricardo Haddad; Sandra Coccuzzo Sampaio Vessoni; Simone Kashima; Svetoslav Nanev Slavov; Vincent Louis Viala |
| EPI_ISL_2344854, EPI_ISL_2344855, EPI_ISL_2344856 | CS DE SANTA ALBERTINA | Instituto Butantan / ESALQ-Piracicaba | Antonio Jorge Martins; Claudia Renata dos Santos Barros; David Schlesinger; Debora Botequio Moretti; Dimas Tadeu Covas; Elaine Cristina Marqueze; Elaine Vieira Santos; Evandra Strazza Rodrigues; Heidge Fukumasu; Jayme Augusto de Souza-Neto; José Salvatore Leister Patané; Luiz Alcantara; Luiz Lehmann Coutinho; Maria Carolina Elias; Maurício Lacerda Nogueira; Rafael dos Santos Bezerra; Raul Machado Neto; Rejane Maria Tommasini Grotto; Ricardo Haddad; Sandra Coccuzzo Sampaio Vessoni; Simone Kashima; Svetoslav Nanev Slavov; Vincent Louis Viala |
| EPI_ISL_2344862 | CS DE VALENTIM GENTIL | Instituto Butantan / ESALQ-Piracicaba | Antonio Jorge Martins; Claudia Renata dos Santos Barros; David Schlesinger; Debora Botequio Moretti; Dimas Tadeu Covas; Elaine Cristina Marqueze; Elaine Vieira Santos; Evandra Strazza Rodrigues; Heidge Fukumasu; Jayme Augusto de Souza-Neto; José Salvatore Leister Patané; Luiz Alcantara; Luiz Lehmann Coutinho; Maria Carolina Elias; Maurício Lacerda Nogueira; Rafael dos Santos Bezerra; Raul Machado Neto; Rejane Maria Tommasini Grotto; Ricardo Haddad; Sandra Coccuzzo Sampaio Vessoni; Simone Kashima; Svetoslav Nanev Slavov; Vincent Louis Viala |
| EPI_ISL_2344778 | CS IACRI | Instituto Butantan / ESALQ-Piracicaba | Antonio Jorge Martins; Claudia Renata dos Santos Barros; David Schlesinger; Debora Botequio Moretti; Dimas Tadeu Covas; Elaine Cristina Marqueze; Elaine Vieira Santos; Evandra Strazza Rodrigues; Heidge Fukumasu; Jayme Augusto de Souza-Neto; José Salvatore Leister Patané; Luiz Alcantara; Luiz Lehmann Coutinho; Maria Carolina Elias; Maurício Lacerda Nogueira; Rafael dos Santos Bezerra; Raul Machado Neto; Rejane Maria Tommasini Grotto; Ricardo Haddad; Sandra Coccuzzo Sampaio Vessoni; Simone Kashima; Svetoslav Nanev Slavov; Vincent Louis Viala |
| EPI_ISL_2344831 | CSA COMPLEXO DE SAUDE ANHANGUERA LTDA | Instituto Butantan | Antonio Jorge Martins; Claudia Renata dos Santos Barros; David Schlesinger; Debora Botequio Moretti; Dimas Tadeu Covas; Elaine Cristina Marqueze; Elaine Vieira Santos; Evandra Strazza Rodrigues; Heidge Fukumasu; Jayme Augusto de Souza-Neto; José Salvatore Leister Patané; Luiz Alcantara; Luiz Lehmann Coutinho; Maria Carolina Elias; Maurício Lacerda Nogueira; Rafael dos Santos Bezerra; Raul Machado Neto; Rejane Maria Tommasini Grotto; Ricardo Haddad; Sandra Coccuzzo Sampaio Vessoni; Simone Kashima; Svetoslav Nanev Slavov; Vincent Louis Viala |
| EPI_ISL_2344906 | CSA COMPLEXO DE SAUDE ANHANGUERA TUCURUVI | Instituto Butantan | Antonio Jorge Martins; Claudia Renata dos Santos Barros; David Schlesinger; Debora Botequio Moretti; Dimas Tadeu Covas; Elaine Cristina Marqueze; Elaine Vieira Santos; Evandra Strazza Rodrigues; Heidge Fukumasu; Jayme Augusto de Souza-Neto; José Salvatore Leister Patané; Luiz Alcantara; Luiz Lehmann Coutinho; Maria Carolina Elias; Maurício Lacerda Nogueira; Rafael dos Santos Bezerra; Raul Machado Neto; Rejane Maria Tommasini Grotto; Ricardo Haddad; Sandra Coccuzzo Sampaio Vessoni; Simone Kashima; Svetoslav Nanev Slavov; Vincent Louis Viala |
| EPI_ISL_2344925, EPI_ISL_2344943 | CSA COMPLEXO DE SAUDE ANHANGUERA VILA JAGUARA | Instituto Butantan | Antonio Jorge Martins; Claudia Renata dos Santos Barros; David Schlesinger; Debora Botequio Moretti; Dimas Tadeu Covas; Elaine Cristina Marqueze; Elaine Vieira Santos; Evandra Strazza Rodrigues; Heidge Fukumasu; Jayme Augusto de Souza-Neto; José Salvatore Leister Patané; Luiz Alcantara; Luiz Lehmann Coutinho; Maria Carolina Elias; Maurício Lacerda Nogueira; Rafael dos Santos Bezerra; Raul Machado Neto; Rejane Maria Tommasini Grotto; Ricardo Haddad; Sandra Coccuzzo Sampaio Vessoni; Simone Kashima; Svetoslav Nanev Slavov; Vincent Louis Viala |
| EPI_ISL_2500422, EPI_ISL_2500423, EPI_ISL_2500428, EPI_ISL_2501533 | CT Department of Public Health | CT Department of Public Health | Claire_Pearson; Tu_N_Nguyen |
| EPI_ISL_2105160 | CT-Dr. Katherine A. Kelley State Public Health Lab | Centers for Disease Control and Prevention Division of Viral Diseases, Pathogen Discovery | Alison Laufer Halpin; Ben L. Rambo-Martin; Clinton R. Paden; Dakota Howard; Darlene Wagner; Dave Wentworth; Dhwani Batra; Jasmine Padilla; Justin Lee; Katie Dillon; Krista Queen; Kristen Knipe; Kristine Lacek; Mark Burroughs; Matthew Schmerer; Mili Sheth; Peter Cook; Sam Shepard; Sarah Nobles; Shoshona Le; Suxiang Tong; Vivien Dugan; Yvette Unoarumhi |
| EPI_ISL_2698082 | CTvacinas | CTvacinas | A.P.; B.L.; Coelho; D.B.; Dorlass; Durigon; E.G.; E.L.; F.G.; Fernandes; Fiorini, A.; Fonseca; G.P.; H.P.; K.L.; L.M.; Lourenco; Magalhaes; Oliveira; Ometto, T.; Peixoto, R.; R.D.; Sato, H.; Scagion; Teixeira, S.; Telezynski; Thomazelli |
| EPI_ISL_2537460, EPI_ISL_2537463, EPI_ISL_2537464 | California Department of Public Health | California Department of Public Health | CDPH IDLB COVIDNet et al |
| EPI_ISL_2778847, EPI_ISL_2837498 | California Department of Public Health Valencia Branch Laboratory (CDPH VBL) | California Department of Public Health | CDPH-COVIDNet; UCLA Technology Center for Genomics & Bioinformatics |
| EPI_ISL_7507981, EPI_ISL_7508040, EPI_ISL_7508086 | Cedars-Sinai Medical Center, Molecular Pathology Laboratory of Department of Pathology & Laboratory Medicine and Genomic Core | Cedars-Sinai Medical Center, Molecular Pathology Laboratory of Department of Pathology & Laboratory Medicine and Genomic Core | Brian Davis; Eric Vail; Jasmine T Plummer; Jorge Mario Sincuir Martinez; Stephanie Chen; Wenjuan Zhang |
| EPI_ISL_2502386, EPI_ISL_2502439, EPI_ISL_2502475, EPI_ISL_2502476, EPI_ISL_2502477 | Central Laboratory, Bureau of Public Health (BOG) and Academic Hospital Paramaribo | Erasmus Medical Center | Bas B Oude Munnink; Cherise Beek; Consuella Partowidjojo; Dion Gajadin; Ed PF IJzerman; Emmanuelle Munger; Gary Gummels; Ingrid SK Krishnadath; Lycke Woittiez; Marion PG Koopmans; Mireille Van de Veer; Phyllis Pinas; Princes Wongsowidjojo; Radjesh Ori; Ranisha Doerbalie; Rohma Banwari; Soeradj Harkisoen; Stephen Vreden; Tilotmadebie Ramlal; Verne Nanhoe |
| EPI_ISL_3255136, EPI_ISL_3255172 | Central Public Health Laboratory - LACEN -Bahia, Salvador, Brazil | Central Public Health Laboratory - LACEN -Bahia, Salvador, Brazil | Arabela Leal; Breno Dominguez; Felicidade Pereira; Jaqueline Gomes; Luciana Oliveira; Luiz Alcantara; Marcela Gómez; Marta Giovanetti; Patrícia Cajado; Stephane Tosta; Vagner Fonseca; Vanessa Nardy |
| EPI_ISL_1853299, EPI_ISL_1853348 | Centre Hospitalier Universitaire Clermont-Ferrand | CHU Clermont-Ferrand, service de virologie | Bisseux Maxime; Combes Patricia; Henquell Cécile; Mirand Audrey |
| EPI_ISL_5800955, EPI_ISL_5800959, EPI_ISL_5800963, EPI_ISL_5800970, EPI_ISL_5800978, EPI_ISL_5800982, EPI_ISL_5800984, EPI_ISL_5800989, EPI_ISL_5800993, EPI_ISL_5800994, EPI_ISL_5800995, EPI_ISL_5800996, EPI_ISL_5800997, EPI_ISL_5800999, EPI_ISL_5801001 | | | |
| see above | Centro De Apoio Epidemiologico | Instituto Butantan | Antonio Jorge Martins; Claudia Renata dos Santos Barros; David Schlesinger; Debora Botequio Moretti; Dimas Tadeu Covas; Elaine Cristina Marqueze; Elaine Vieira Santos; Evandra Strazza Rodrigues; Heidge Fukumasu; Jayme Augusto de Souza-Neto; José Salvatore Leister Patané; Luiz Alcantara; Luiz Lehmann Coutinho; Maria Carolina Elias; Maurício Lacerda Nogueira; Rafael dos Santos Bezerra; Raul Machado Neto; Rejane Maria Tommasini Grotto; Ricardo Haddad; Sandra Coccuzzo Sampaio Vessoni; Simone Kashima; Svetoslav Nanev Slavov; Vincent Louis Viala |
| EPI_ISL_5800855 | Centro De Detencao Provisoria De Nova Independencia | Instituto Butantan | Antonio Jorge Martins; Claudia Renata dos Santos Barros; David Schlesinger; Debora Botequio Moretti; Dimas Tadeu Covas; Elaine Cristina Marqueze; Elaine Vieira Santos; Evandra Strazza Rodrigues; Heidge Fukumasu; Jayme Augusto de Souza-Neto; José Salvatore Leister Patané; Luiz Alcantara; Luiz Lehmann Coutinho; Maria Carolina Elias; Maurício Lacerda Nogueira; Rafael dos Santos Bezerra; Raul Machado Neto; Rejane Maria Tommasini Grotto; Ricardo Haddad; Sandra Coccuzzo Sampaio Vessoni; Simone Kashima; Svetoslav Nanev Slavov; Vincent Louis Viala |
| EPI_ISL_5800694 | Centro De Especialidades De Aguai Aguai | Instituto Butantan | Antonio Jorge Martins; Claudia Renata dos Santos Barros; David Schlesinger; Debora Botequio Moretti; Dimas Tadeu Covas; Elaine Cristina Marqueze; Elaine Vieira Santos; Evandra Strazza Rodrigues; Heidge Fukumasu; Jayme Augusto de Souza-Neto; José Salvatore Leister Patané; Luiz Alcantara; Luiz Lehmann Coutinho; Maria Carolina Elias; Maurício Lacerda Nogueira; Rafael dos Santos Bezerra; Raul Machado Neto; Rejane Maria Tommasini Grotto; Ricardo Haddad; Sandra Coccuzzo Sampaio Vessoni; Simone Kashima; Svetoslav Nanev Slavov; Vincent Louis Viala |
| EPI_ISL_5802641, EPI_ISL_5802642, EPI_ISL_5802643, EPI_ISL_5802644, EPI_ISL_5802647, EPI_ISL_5802648 | Centro De Genomas | Instituto Butantan | Antonio Jorge Martins; Claudia Renata dos Santos Barros; David Schlesinger; Debora Botequio Moretti; Dimas Tadeu Covas; Elaine Cristina Marqueze; Elaine Vieira Santos; Evandra Strazza Rodrigues; Heidge Fukumasu; Jayme Augusto de Souza-Neto; José Salvatore Leister Patané; Luiz Alcantara; Luiz Lehmann Coutinho; Maria Carolina Elias; Maurício Lacerda Nogueira; Rafael dos Santos Bezerra; Raul Machado Neto; Rejane Maria Tommasini Grotto; Ricardo Haddad; Sandra Coccuzzo Sampaio Vessoni; Simone Kashima; Svetoslav Nanev Slavov; Vincent Louis Viala |
| EPI_ISL_5800800 | Centro De Saude De Analandia | Instituto Butantan | Antonio Jorge Martins; Claudia Renata dos Santos Barros; David Schlesinger; Debora Botequio Moretti; Dimas Tadeu Covas; Elaine Cristina Marqueze; Elaine Vieira Santos; Evandra Strazza Rodrigues; Heidge Fukumasu; Jayme Augusto de Souza-Neto; José Salvatore Leister Patané; Luiz Alcantara; Luiz Lehmann Coutinho; Maria Carolina Elias; Maurício Lacerda Nogueira; Rafael dos Santos Bezerra; Raul Machado Neto; Rejane Maria Tommasini Grotto; Ricardo Haddad; Sandra Coccuzzo Sampaio Vessoni; Simone Kashima; Svetoslav Nanev Slavov; Vincent Louis Viala |
| EPI_ISL_5800817 | Centro De Saude De Barbosa | Instituto Butantan | Antonio Jorge Martins; Claudia Renata dos Santos Barros; David Schlesinger; Debora Botequio Moretti; Dimas Tadeu Covas; Elaine Cristina Marqueze; Elaine Vieira Santos; Evandra Strazza Rodrigues; Heidge Fukumasu; Jayme Augusto de Souza-Neto; José Salvatore Leister Patané; Luiz Alcantara; Luiz Lehmann Coutinho; Maria Carolina Elias; Maurício Lacerda Nogueira; Rafael dos Santos Bezerra; Raul Machado Neto; Rejane Maria Tommasini Grotto; Ricardo Haddad; Sandra Coccuzzo Sampaio Vessoni; Simone Kashima; Svetoslav Nanev Slavov; Vincent Louis Viala |
| EPI_ISL_5800642, EPI_ISL_5800662 | Centro De Saude De Chavantes | Instituto Butantan | Antonio Jorge Martins; Claudia Renata dos Santos Barros; David Schlesinger; Debora Botequio Moretti; Dimas Tadeu Covas; Elaine Cristina Marqueze; Elaine Vieira Santos; Evandra Strazza Rodrigues; Heidge Fukumasu; Jayme Augusto de Souza-Neto; José Salvatore Leister Patané; Luiz Alcantara; Luiz Lehmann Coutinho; Maria Carolina Elias; Maurício Lacerda Nogueira; Rafael dos Santos Bezerra; Raul Machado Neto; Rejane Maria Tommasini Grotto; Ricardo Haddad; Sandra Coccuzzo Sampaio Vessoni; Simone Kashima; Svetoslav Nanev Slavov; Vincent Louis Viala |

| | | | |
|---|---|---|---|
| EPI_ISL_5800630, EPI_ISL_5800737 | Centro De Saude De Ibirarema | Instituto Butantan | Antonio Jorge Martins; Claudia Renata dos Santos Barros; David Schlesinger; Debora Botequio Moretti; Dimas Tadeu Covas; Elaine Cristina Marqueze; Elaine Vieira Santos; Evandra Strazza Rodrigues; Heidge Fukumasu; Jayme Augusto de Souza-Neto; José Salvatore Leister Patané; Luiz Alcantara; Luiz Lehmann Coutinho; Maria Carolina Elias; Maurício Lacerda Nogueira; Rafael dos Santos Bezerra; Raul Machado Neto; Rejane Maria Tommasini Grotto; Ricardo Haddad; Sandra Coccuzzo Sampaio Vessoni; Simone Kashima; Svetoslav Nanev Slavov; Vincent Louis Viala |
| EPI_ISL_5800688 | Centro De Saude De Lutecia | Instituto Butantan | Antonio Jorge Martins; Claudia Renata dos Santos Barros; David Schlesinger; Debora Botequio Moretti; Dimas Tadeu Covas; Elaine Cristina Marqueze; Elaine Vieira Santos; Evandra Strazza Rodrigues; Heidge Fukumasu; Jayme Augusto de Souza-Neto; José Salvatore Leister Patané; Luiz Alcantara; Luiz Lehmann Coutinho; Maria Carolina Elias; Maurício Lacerda Nogueira; Rafael dos Santos Bezerra; Raul Machado Neto; Rejane Maria Tommasini Grotto; Ricardo Haddad; Sandra Coccuzzo Sampaio Vessoni; Simone Kashima; Svetoslav Nanev Slavov; Vincent Louis Viala |
| EPI_ISL_5800680 | Centro De Saude De Ocaucu | Instituto Butantan | Antonio Jorge Martins; Claudia Renata dos Santos Barros; David Schlesinger; Debora Botequio Moretti; Dimas Tadeu Covas; Elaine Cristina Marqueze; Elaine Vieira Santos; Evandra Strazza Rodrigues; Heidge Fukumasu; Jayme Augusto de Souza-Neto; José Salvatore Leister Patané; Luiz Alcantara; Luiz Lehmann Coutinho; Maria Carolina Elias; Maurício Lacerda Nogueira; Rafael dos Santos Bezerra; Raul Machado Neto; Rejane Maria Tommasini Grotto; Ricardo Haddad; Sandra Coccuzzo Sampaio Vessoni; Simone Kashima; Svetoslav Nanev Slavov; Vincent Louis Viala |
| EPI_ISL_5800693 | Centro De Saude De Spedro Do Turvo | Instituto Butantan | Antonio Jorge Martins; Claudia Renata dos Santos Barros; David Schlesinger; Debora Botequio Moretti; Dimas Tadeu Covas; Elaine Cristina Marqueze; Elaine Vieira Santos; Evandra Strazza Rodrigues; Heidge Fukumasu; Jayme Augusto de Souza-Neto; José Salvatore Leister Patané; Luiz Alcantara; Luiz Lehmann Coutinho; Maria Carolina Elias; Maurício Lacerda Nogueira; Rafael dos Santos Bezerra; Raul Machado Neto; Rejane Maria Tommasini Grotto; Ricardo Haddad; Sandra Coccuzzo Sampaio Vessoni; Simone Kashima; Svetoslav Nanev Slavov; Vincent Louis Viala |
| EPI_ISL_5800575 | Centro De Saude Do Indaia Do Aguapei Florida Paulista | Instituto Butantan | Antonio Jorge Martins; Claudia Renata dos Santos Barros; David Schlesinger; Debora Botequio Moretti; Dimas Tadeu Covas; Elaine Cristina Marqueze; Elaine Vieira Santos; Evandra Strazza Rodrigues; Heidge Fukumasu; Jayme Augusto de Souza-Neto; José Salvatore Leister Patané; Luiz Alcantara; Luiz Lehmann Coutinho; Maria Carolina Elias; Maurício Lacerda Nogueira; Rafael dos Santos Bezerra; Raul Machado Neto; Rejane Maria Tommasini Grotto; Ricardo Haddad; Sandra Coccuzzo Sampaio Vessoni; Simone Kashima; Svetoslav Nanev Slavov; Vincent Louis Viala |
| EPI_ISL_5800672 | Centro De Saude Dr Mario Dias De Aguiar Capivari | Instituto Butantan | Antonio Jorge Martins; Claudia Renata dos Santos Barros; David Schlesinger; Debora Botequio Moretti; Dimas Tadeu Covas; Elaine Cristina Marqueze; Elaine Vieira Santos; Evandra Strazza Rodrigues; Heidge Fukumasu; Jayme Augusto de Souza-Neto; José Salvatore Leister Patané; Luiz Alcantara; Luiz Lehmann Coutinho; Maria Carolina Elias; Maurício Lacerda Nogueira; Rafael dos Santos Bezerra; Raul Machado Neto; Rejane Maria Tommasini Grotto; Ricardo Haddad; Sandra Coccuzzo Sampaio Vessoni; Simone Kashima; Svetoslav Nanev Slavov; Vincent Louis Viala |
| EPI_ISL_5800720, EPI_ISL_5800721 | Centro De Saude Dr Nelson Da Cunha Bastos | Instituto Butantan | Antonio Jorge Martins; Claudia Renata dos Santos Barros; David Schlesinger; Debora Botequio Moretti; Dimas Tadeu Covas; Elaine Cristina Marqueze; Elaine Vieira Santos; Evandra Strazza Rodrigues; Heidge Fukumasu; Jayme Augusto de Souza-Neto; José Salvatore Leister Patané; Luiz Alcantara; Luiz Lehmann Coutinho; Maria Carolina Elias; Maurício Lacerda Nogueira; Rafael dos Santos Bezerra; Raul Machado Neto; Rejane Maria Tommasini Grotto; Ricardo Haddad; Sandra Coccuzzo Sampaio Vessoni; Simone Kashima; Svetoslav Nanev Slavov; Vincent Louis Viala |
| EPI_ISL_5800691 | Centro De Saude Dr Plinio Albers De Galia | Instituto Butantan | Antonio Jorge Martins; Claudia Renata dos Santos Barros; David Schlesinger; Debora Botequio Moretti; Dimas Tadeu Covas; Elaine Cristina Marqueze; Elaine Vieira Santos; Evandra Strazza Rodrigues; Heidge Fukumasu; Jayme Augusto de Souza-Neto; José Salvatore Leister Patané; Luiz Alcantara; Luiz Lehmann Coutinho; Maria Carolina Elias; Maurício Lacerda Nogueira; Rafael dos Santos Bezerra; Raul Machado Neto; Rejane Maria Tommasini Grotto; Ricardo Haddad; Sandra Coccuzzo Sampaio Vessoni; Simone Kashima; Svetoslav Nanev Slavov; Vincent Louis Viala |
| EPI_ISL_5800773, EPI_ISL_5800832, EPI_ISL_5800881, EPI_ISL_5800884, EPI_ISL_5800888 | Centro De Saude Dr Renato De Carvalho Ribeiro Angatuba | Instituto Butantan | Antonio Jorge Martins; Claudia Renata dos Santos Barros; David Schlesinger; Debora Botequio Moretti; Dimas Tadeu Covas; Elaine Cristina Marqueze; Elaine Vieira Santos; Evandra Strazza Rodrigues; Heidge Fukumasu; Jayme Augusto de Souza-Neto; José Salvatore Leister Patané; Luiz Alcantara; Luiz Lehmann Coutinho; Maria Carolina Elias; Maurício Lacerda Nogueira; Rafael dos Santos Bezerra; Raul Machado Neto; Rejane Maria Tommasini Grotto; Ricardo Haddad; Sandra Coccuzzo Sampaio Vessoni; Simone Kashima; Svetoslav Nanev Slavov; Vincent Louis Viala |
| EPI_ISL_5800665, EPI_ISL_5800677, EPI_ISL_5800678 | Centro De Saude Esf Iv Zona Rural Domingos De S Sjriopardo | Instituto Butantan | Antonio Jorge Martins; Claudia Renata dos Santos Barros; David Schlesinger; Debora Botequio Moretti; Dimas Tadeu Covas; Elaine Cristina Marqueze; Elaine Vieira Santos; Evandra Strazza Rodrigues; Heidge Fukumasu; Jayme Augusto de Souza-Neto; José Salvatore Leister Patané; Luiz Alcantara; Luiz Lehmann Coutinho; Maria Carolina Elias; Maurício Lacerda Nogueira; Rafael dos Santos Bezerra; Raul Machado Neto; Rejane Maria Tommasini Grotto; Ricardo Haddad; Sandra Coccuzzo Sampaio Vessoni; Simone Kashima; Svetoslav Nanev Slavov; Vincent Louis Viala |
| EPI_ISL_5800770 | Centro De Saude Ii Mairinque Mairinque | Instituto Butantan | Antonio Jorge Martins; Claudia Renata dos Santos Barros; David Schlesinger; Debora Botequio Moretti; Dimas Tadeu Covas; Elaine Cristina Marqueze; Elaine Vieira Santos; Evandra Strazza Rodrigues; Heidge Fukumasu; Jayme Augusto de Souza-Neto; José Salvatore Leister Patané; Luiz Alcantara; Luiz Lehmann Coutinho; Maria Carolina Elias; Maurício Lacerda Nogueira; Rafael dos Santos Bezerra; Raul Machado Neto; Rejane Maria Tommasini Grotto; Ricardo Haddad; Sandra Coccuzzo Sampaio Vessoni; Simone Kashima; Svetoslav Nanev Slavov; Vincent Louis Viala |
| EPI_ISL_5800885, EPI_ISL_5800889 | Centro De Saude Terezinha De Moraes Arsilla | Instituto Butantan | Antonio Jorge Martins; Claudia Renata dos Santos Barros; David Schlesinger; Debora Botequio Moretti; Dimas Tadeu Covas; Elaine Cristina Marqueze; Elaine Vieira Santos; Evandra Strazza Rodrigues; Heidge Fukumasu; Jayme Augusto de Souza-Neto; José Salvatore Leister Patané; Luiz Alcantara; Luiz Lehmann Coutinho; Maria Carolina Elias; Maurício Lacerda Nogueira; Rafael dos Santos Bezerra; Raul Machado Neto; Rejane Maria Tommasini Grotto; Ricardo Haddad; Sandra Coccuzzo Sampaio Vessoni; Simone Kashima; Svetoslav Nanev Slavov; Vincent Louis Viala |
| EPI_ISL_5800601, EPI_ISL_5800608, EPI_ISL_5800614, EPI_ISL_5800617 | Centro De Triagem Covid 19 | Instituto Butantan | Antonio Jorge Martins; Claudia Renata dos Santos Barros; David Schlesinger; Debora Botequio Moretti; Dimas Tadeu Covas; Elaine Cristina Marqueze; Elaine Vieira Santos; Evandra Strazza Rodrigues; Heidge Fukumasu; Jayme Augusto de Souza-Neto; José Salvatore Leister Patané; Luiz Alcantara; Luiz Lehmann Coutinho; Maria Carolina Elias; Maurício Lacerda Nogueira; Rafael dos Santos Bezerra; Raul Machado Neto; Rejane Maria Tommasini Grotto; Ricardo Haddad; Sandra Coccuzzo Sampaio Vessoni; Simone Kashima; Svetoslav Nanev Slavov; Vincent Louis Viala |
| EPI_ISL_5926861, EPI_ISL_5926862 | Centro de Infectologia Charles Mérieux/ Laboratório Rodolphe Mérieux, FUNDHACRE | Bioinformatics Laboratory / LNCC | Alessandra P Lamarca; Alexandra L Gerber; Aline de Freitas Souza; Ana Paula de C Guimarães; Ana Tereza R Vasconcelos; Andreas Stocker; Cirley Maria de Oliveira Lobato; Douglas Terra Machado; Janaína Mazaro; Janete Taynã Nascimento Rodrigues; Luiz Fellype Alves de Souza; Luiz G P de Almeida; Ronaldo da Silva F Jr; Rutilene B. Souza |
| EPI_ISL_2402153 | Centro de Investigación Biomédica de Occidente (CIBO) | Unidad de Genomica Avanzada | Alejandro Sanchez-Flores; Alfredo Herrera-Estrella; Alicia Ocana-Mondragon; Angel Gustavo Salas-Lais; Bernardo Martinez-Miguel; Blanca Taboada; Brenda Irasema Maldonado-Meza; Carla Ivon Herrera-Najera; Carlos F. Arias; Celia Boukadida; Clara Esperanza Santacruz-Tinoco; Concepcion Grajales-Muniz; Consorcio Mexicano de Vigilancia Genomica (CoViGen-Mex). Authors (in alphabetical order): Julio Elias Alvarado-Yaah; Fernando Fontove-Herrera; Francisco Pulido; Gloria Elena Espinoza-Ayala; Gloria Maria Molina-Salinas; Gloria Vazquez; Hector Esteban Paz-Juarez; Hector Montoya-Fuentes; Helen Haydee Fernanda Ramirez-Plascencia; Jorge Ivan Salinal-Nevarez; Jose Antonio Enciso-Moreno; Jose Esteban Munoz-Medina; Jose de Jesus Nunez-Contreras; Juan Bautista Chale-Dzul; Luis Alberto Ochoa-Carrera; Margarita Matias-Florentino; Maria Guadalupe Santiago-Mauricio; Maria Guadalupe de Jesus Mireles-Rivera; Nelly Selem-Mojica; Pavel Isa; Ricardo Grande; Santiago Avila-Rios; Victor Hugo Borja-Aburto |
| EPI_ISL_2401944, EPI_ISL_2401953, EPI_ISL_2401966 | Centro de Investigación Biomédica del Noreste (CIBIN) | Unidad de Genomica Avanzada | Alejandro Sanchez-Flores; Alfredo Herrera-Estrella; Alicia Ocana-Mondragon; Angel Gustavo Salas-Lais; Bernardo Martinez-Miguel; Blanca Taboada; Brenda Irasema Maldonado-Meza; Carla Ivon Herrera-Najera; Carlos F. Arias; Celia Boukadida; Clara Esperanza Santacruz-Tinoco; Concepcion Grajales-Muniz; Consorcio Mexicano de Vigilancia Genomica (CoViGen-Mex). Authors (in alphabetical order): Julio Elias Alvarado-Yaah; Fernando Fontove-Herrera; Francisco Pulido; Gloria Elena Espinoza-Ayala; Gloria Maria Molina-Salinas; Gloria Vazquez; Hector Esteban Paz-Juarez; Hector Montoya-Fuentes; Helen Haydee Fernanda Ramirez-Plascencia; Jorge Ivan Salinal-Nevarez; Jose Antonio Enciso-Moreno; Jose Esteban Munoz-Medina; Jose de Jesus Nunez-Contreras; Juan Bautista Chale-Dzul; Luis Alberto Ochoa-Carrera; Margarita Matias-Florentino; Maria Guadalupe Santiago-Mauricio; Maria Guadalupe de Jesus Mireles-Rivera; Nelly Selem-Mojica; Pavel Isa; Ricardo Grande; Santiago Avila-Rios; Victor Hugo Borja-Aburto |
| EPI_ISL_2142004 | Centrum Medycyny Klinicznej dr n.med. Karol Majewski - DiMedical | 1. National Institute of Public Health - National Institute of Hygiene; 2. Eurofins Genomics Europe Sequencing GmbH | ECDC COVID-19 WGS support team; Eurofins Genomics Europe Sequencing Team; Gierczyński Rafał; Sadkowska-Todys Małgorzata; Wołkowicz Tomasz; Zacharczuk Katarzyna |
| EPI_ISL_4498203 | Children’s Hospital of Philadelphia | Planet | Ahmed M. Moustafa; Alex Arvanitis; Azad Ahmed; Brandy Neide; Colleen Bianco; Josh Chang Mell; Lidiya Denu; Paul J. Planet; Rebecca M. Harris; Susan Coffin; Swetha Rajagopal |
| EPI_ISL_2004300, EPI_ISL_2004301 | Cintramedica | Instituto Nacional de Saude (INSA) | Borges et al |
| EPI_ISL_2467089, EPI_ISL_2467090, EPI_ISL_2467091, EPI_ISL_2467103, EPI_ISL_2467113, EPI_ISL_2467130, EPI_ISL_2467133, EPI_ISL_2467135, EPI_ISL_2467139, EPI_ISL_2467140, EPI_ISL_2467142, EPI_ISL_2467343, EPI_ISL_2490296, EPI_ISL_2490298, EPI_ISL_2490305 | | | |
| see above | Clinica INDISA | "Facultad de Ciencias de la Vida, UNAB" | "Claudio Meneses; Ariel Orellana"; Claudio Olmos; Daniel Leon; Dayan Sanhueza; Eduardo Castro; Gonzalo Campaña; Macarena Bastias; Paola Pidal; Ricardo Yusta; Sebastian Wolter; Susana Saez; Víctor Monreal; Waldo Diaz |
| EPI_ISL_1854946, EPI_ISL_1854952, EPI_ISL_2098778, EPI_ISL_2098779 | Clinical Molecular Microbiology Laboratory, UNC Hospitals | Jeremy Wang | Alexander Rubinsteyn; Brianna de la Houssaye; Colleen Rice; Corbin Jones; Jason Smedberg; Jeremy Wang; Jose Luis Torres Castillo; Melissa Miller; Robert Hagan; Shawn Hawken |
| EPI_ISL_1805786 | Clinical Reference Laboratory | Kansas Health and Environmental Lab | Ben Olsen; Jonathan Barnell; Mike Grose; and Phil Adam |
| EPI_ISL_4300610 | Color Genomics | Chiu Laboratory, University of California, San Francisco | Alicia Sotomayor-Gonzalez; Alicia Zhou; Baolin Wang; Charles Chiu; Daniel Wong; Erika Torres; Madyson Wanner; Noah Brazer; Scott Topper; Venice Servellita; Yueyuan Zhang |
| EPI_ISL_2277552, EPI_ISL_2277564, EPI_ISL_2277565, EPI_ISL_2309060, EPI_ISL_2309124, EPI_ISL_6211190 | Colorado Department of Public Health and Environment | Colorado Department of Public Health and Environment | Alexandria Rossheim; Diana Ir; Emily A. Travanty; Laura Bankers; Mandy Waters; Michael Martin; Molly C. Hetherington-Rauth; Sarah Elizabeth Totten; Shannon R. Matzinger |
| EPI_ISL_2496904, EPI_ISL_2496910, EPI_ISL_2496912, EPI_ISL_2496914 | Contra Costa County Public Health Lab | Chan-Zuckerberg Biohub | CZB Cliahub Consortium |
| EPI_ISL_2103008 | County of San Luis Obispo Public Health Laboratory | Chan-Zuckerberg Biohub | CZB Cliahub Consortium |
| EPI_ISL_7727192 | Covid Laboratory Biogem | Covid Laboratory Biogem | Alessandra Fucci and Michele Caraglia; Alessia Maria Cossu; Cinzia Miarelli; Clara Iannarone; Egildo Luca D'andrea; Federica Melisi; Giovambattista Capasso; Marco Bocchetti; Marianna Scrima; Michele Ceccarelli; Piera Grisolia; Teresa Maria Rosaria Noviello; Ylenia Abruzzese |
| EPI_ISL_2658466 | Croatian Institute of Public Health | Croatian Institute of Public Health | Irena Tabain; Ivana Ferenčak |
| EPI_ISL_5800814 | Cs De Balsamo | Instituto Butantan | Antonio Jorge Martins; Claudia Renata dos Santos Barros; David Schlesinger; Debora Botequio Moretti; Dimas Tadeu Covas; Elaine Cristina Marqueze; Elaine Vieira Santos; Evandra Strazza Rodrigues; Heidge Fukumasu; Jayme Augusto de Souza-Neto; José Salvatore Leister Patané; Luiz Alcantara; Luiz Lehmann Coutinho; Maria Carolina Elias; Maurício Lacerda Nogueira; Rafael dos Santos Bezerra; Raul Machado Neto; Rejane Maria Tommasini Grotto; Ricardo Haddad; Sandra Coccuzzo Sampaio Vessoni; Simone Kashima; Svetoslav Nanev Slavov; Vincent Louis Viala |
| EPI_ISL_5800759, EPI_ISL_5800760, EPI_ISL_5800805, EPI_ISL_5800811 | Cs De Catigua | Instituto Butantan | Antonio Jorge Martins; Claudia Renata dos Santos Barros; David Schlesinger; Debora Botequio Moretti; Dimas Tadeu Covas; Elaine Cristina Marqueze; Elaine Vieira Santos; Evandra Strazza Rodrigues; Heidge Fukumasu; Jayme Augusto de Souza-Neto; José Salvatore Leister Patané; Luiz Alcantara; Luiz Lehmann Coutinho; Maria Carolina Elias; Maurício Lacerda Nogueira; Rafael dos Santos Bezerra; Raul Machado Neto; Rejane Maria Tommasini Grotto; Ricardo Haddad; Sandra Coccuzzo Sampaio Vessoni; Simone Kashima; Svetoslav Nanev Slavov; Vincent Louis Viala |
| EPI_ISL_5800824, EPI_ISL_5800825, EPI_ISL_5800830, EPI_ISL_5800833 | Cs De Cosmorama | Instituto Butantan | Antonio Jorge Martins; Claudia Renata dos Santos Barros; David Schlesinger; Debora Botequio Moretti; Dimas Tadeu Covas; Elaine Cristina Marqueze; Elaine Vieira Santos; Evandra Strazza Rodrigues; Heidge Fukumasu; Jayme Augusto de Souza-Neto; José Salvatore Leister Patané; Luiz Alcantara; Luiz Lehmann Coutinho; Maria Carolina Elias; Maurício Lacerda Nogueira; Rafael dos Santos Bezerra; Raul Machado Neto; Rejane Maria Tommasini Grotto; Ricardo Haddad; Sandra Coccuzzo Sampaio Vessoni; Simone Kashima; Svetoslav Nanev Slavov; Vincent Louis Viala |
| EPI_ISL_5800838, EPI_ISL_5800841 | Cs De Floreal | Instituto Butantan | Antonio Jorge Martins; Claudia Renata dos Santos Barros; David Schlesinger; Debora Botequio Moretti; Dimas Tadeu Covas; Elaine Cristina Marqueze; Elaine Vieira Santos; Evandra Strazza Rodrigues; Heidge Fukumasu; Jayme Augusto de Souza-Neto; José Salvatore Leister Patané; Luiz Alcantara; Luiz Lehmann Coutinho; Maria Carolina Elias; Maurício Lacerda Nogueira; Rafael dos Santos Bezerra; Raul Machado Neto; Rejane Maria Tommasini Grotto; Ricardo Haddad; Sandra Coccuzzo Sampaio Vessoni; Simone Kashima; Svetoslav Nanev Slavov; Vincent Louis Viala |
| EPI_ISL_5800842 | Cs De Marapoama | Instituto Butantan | Antonio Jorge Martins; Claudia Renata dos Santos Barros; David Schlesinger; Debora Botequio Moretti; Dimas Tadeu Covas; Elaine Cristina Marqueze; Elaine Vieira Santos; Evandra Strazza Rodrigues; Heidge Fukumasu; Jayme Augusto de Souza-Neto; José Salvatore Leister Patané; Luiz Alcantara; Luiz Lehmann Coutinho; Maria Carolina Elias; Maurício Lacerda Nogueira; Rafael dos Santos Bezerra; Raul Machado Neto; Rejane Maria Tommasini Grotto; Ricardo Haddad; Sandra Coccuzzo Sampaio Vessoni; Simone Kashima; Svetoslav Nanev Slavov; Vincent Louis Viala |
| EPI_ISL_5800826, EPI_ISL_5800831, EPI_ISL_5800834, | Cs De Onda Verde | Instituto Butantan | Antonio Jorge Martins; Claudia Renata dos Santos Barros; David Schlesinger; Debora Botequio Moretti; Dimas Tadeu Covas; Elaine Cristina Marqueze; Elaine Vieira Santos; Evandra Strazza Rodrigues; Heidge Fukumasu; Jayme Augusto de Souza-Neto; José Salvatore Leister Patané; Luiz Alcantara; Luiz Lehmann Coutinho; Maria Carolina Elias; Maurício Lacerda Nogueira; Rafael dos Santos Bezerra; Raul Machado Neto; Rejane Maria Tommasini Grotto; Ricardo Haddad; Sandra Coccuzzo Sampaio Vessoni; Simone Kashima; Svetoslav Nanev Slavov; Vincent Louis Viala |

| | | | |
|---|---|---|---|
| EPI_ISL_5800850, EPI_ISL_5800853 | | | |
| EPI_ISL_5800809, EPI_ISL_5800810 | Cs De Palmares Paulista | Instituto Butantan | Antonio Jorge Martins; Claudia Renata dos Santos Barros; David Schlesinger; Debora Botequio Moretti; Dimas Tadeu Covas; Elaine Cristina Marqueze; Elaine Vieira Santos; Evandra Strazza Rodrigues; Heidge Fukumasu; Jayme Augusto de Souza-Neto; José Salvatore Leister Patané; Luiz Alcantara; Luiz Lehmann Coutinho; Maria Carolina Elias; Maurício Lacerda Nogueira; Rafael dos Santos Bezerra; Raul Machado Neto; Rejane Maria Tommasini Grotto; Ricardo Haddad; Sandra Coccuzzo Sampaio Vessoni; Simone Kashima; Svetoslav Nanev Slavov; Vincent Louis Viala |
| EPI_ISL_5800634, EPI_ISL_5800637 | Cs De Rinopolis | Instituto Butantan | Antonio Jorge Martins; Claudia Renata dos Santos Barros; David Schlesinger; Debora Botequio Moretti; Dimas Tadeu Covas; Elaine Cristina Marqueze; Elaine Vieira Santos; Evandra Strazza Rodrigues; Heidge Fukumasu; Jayme Augusto de Souza-Neto; José Salvatore Leister Patané; Luiz Alcantara; Luiz Lehmann Coutinho; Maria Carolina Elias; Maurício Lacerda Nogueira; Rafael dos Santos Bezerra; Raul Machado Neto; Rejane Maria Tommasini Grotto; Ricardo Haddad; Sandra Coccuzzo Sampaio Vessoni; Simone Kashima; Svetoslav Nanev Slavov; Vincent Louis Viala |
| EPI_ISL_5800802, EPI_ISL_5800803, EPI_ISL_5800804 | Cs De Santa Albertina | Instituto Butantan | Antonio Jorge Martins; Claudia Renata dos Santos Barros; David Schlesinger; Debora Botequio Moretti; Dimas Tadeu Covas; Elaine Cristina Marqueze; Elaine Vieira Santos; Evandra Strazza Rodrigues; Heidge Fukumasu; Jayme Augusto de Souza-Neto; José Salvatore Leister Patané; Luiz Alcantara; Luiz Lehmann Coutinho; Maria Carolina Elias; Maurício Lacerda Nogueira; Rafael dos Santos Bezerra; Raul Machado Neto; Rejane Maria Tommasini Grotto; Ricardo Haddad; Sandra Coccuzzo Sampaio Vessoni; Simone Kashima; Svetoslav Nanev Slavov; Vincent Louis Viala |
| EPI_ISL_5800813 | Cs De Valentim Gentil | Instituto Butantan | Antonio Jorge Martins; Claudia Renata dos Santos Barros; David Schlesinger; Debora Botequio Moretti; Dimas Tadeu Covas; Elaine Cristina Marqueze; Elaine Vieira Santos; Evandra Strazza Rodrigues; Heidge Fukumasu; Jayme Augusto de Souza-Neto; José Salvatore Leister Patané; Luiz Alcantara; Luiz Lehmann Coutinho; Maria Carolina Elias; Maurício Lacerda Nogueira; Rafael dos Santos Bezerra; Raul Machado Neto; Rejane Maria Tommasini Grotto; Ricardo Haddad; Sandra Coccuzzo Sampaio Vessoni; Simone Kashima; Svetoslav Nanev Slavov; Vincent Louis Viala |
| EPI_ISL_5800663 | Cs Iacri | Instituto Butantan | Antonio Jorge Martins; Claudia Renata dos Santos Barros; David Schlesinger; Debora Botequio Moretti; Dimas Tadeu Covas; Elaine Cristina Marqueze; Elaine Vieira Santos; Evandra Strazza Rodrigues; Heidge Fukumasu; Jayme Augusto de Souza-Neto; José Salvatore Leister Patané; Luiz Alcantara; Luiz Lehmann Coutinho; Maria Carolina Elias; Maurício Lacerda Nogueira; Rafael dos Santos Bezerra; Raul Machado Neto; Rejane Maria Tommasini Grotto; Ricardo Haddad; Sandra Coccuzzo Sampaio Vessoni; Simone Kashima; Svetoslav Nanev Slavov; Vincent Louis Viala |
| EPI_ISL_5800768 | Csa Complexo De Saude Anhanguera Ltda | Instituto Butantan | Antonio Jorge Martins; Claudia Renata dos Santos Barros; David Schlesinger; Debora Botequio Moretti; Dimas Tadeu Covas; Elaine Cristina Marqueze; Elaine Vieira Santos; Evandra Strazza Rodrigues; Heidge Fukumasu; Jayme Augusto de Souza-Neto; José Salvatore Leister Patané; Luiz Alcantara; Luiz Lehmann Coutinho; Maria Carolina Elias; Maurício Lacerda Nogueira; Rafael dos Santos Bezerra; Raul Machado Neto; Rejane Maria Tommasini Grotto; Ricardo Haddad; Sandra Coccuzzo Sampaio Vessoni; Simone Kashima; Svetoslav Nanev Slavov; Vincent Louis Viala |
| EPI_ISL_5800887 | Csa Complexo De Saude Anhanguera Tucuruvi | Instituto Butantan | Antonio Jorge Martins; Claudia Renata dos Santos Barros; David Schlesinger; Debora Botequio Moretti; Dimas Tadeu Covas; Elaine Cristina Marqueze; Elaine Vieira Santos; Evandra Strazza Rodrigues; Heidge Fukumasu; Jayme Augusto de Souza-Neto; José Salvatore Leister Patané; Luiz Alcantara; Luiz Lehmann Coutinho; Maria Carolina Elias; Maurício Lacerda Nogueira; Rafael dos Santos Bezerra; Raul Machado Neto; Rejane Maria Tommasini Grotto; Ricardo Haddad; Sandra Coccuzzo Sampaio Vessoni; Simone Kashima; Svetoslav Nanev Slavov; Vincent Louis Viala |
| EPI_ISL_5800914, EPI_ISL_5800933 | Csa Complexo De Saude Anhanguera Vila Jaguara | Instituto Butantan | Antonio Jorge Martins; Claudia Renata dos Santos Barros; David Schlesinger; Debora Botequio Moretti; Dimas Tadeu Covas; Elaine Cristina Marqueze; Elaine Vieira Santos; Evandra Strazza Rodrigues; Heidge Fukumasu; Jayme Augusto de Souza-Neto; José Salvatore Leister Patané; Luiz Alcantara; Luiz Lehmann Coutinho; Maria Carolina Elias; Maurício Lacerda Nogueira; Rafael dos Santos Bezerra; Raul Machado Neto; Rejane Maria Tommasini Grotto; Ricardo Haddad; Sandra Coccuzzo Sampaio Vessoni; Simone Kashima; Svetoslav Nanev Slavov; Vincent Louis Viala |
| EPI_ISL_2755045, EPI_ISL_2755050, EPI_ISL_2755061, EPI_ISL_2755080, EPI_ISL_2755124, EPI_ISL_2755340, EPI_ISL_2755484, EPI_ISL_2755509, EPI_ISL_2755525, EPI_ISL_2755551, EPI_ISL_2755564, EPI_ISL_2755570, EPI_ISL_2755770, EPI_ISL_2755775, EPI_ISL_2755782, EPI_ISL_2755803 | | | |
| see above | Curative Labs | Curative Labs | Elias L. Salfati; Eugenia Khorosheva; George Way; J.Cesar Ignacio-Espinoza; Janet Chen; Mikhail Hanewich-Hollatz; Nabjot Sandhu; Sophia Quasem; Vladimir Slepnev; Zhiyi Xie |
| EPI_ISL_2229246 | DC Public Health Lab/ Dept. of Forensic Sciences | Centers for Disease Control and Prevention Division of Viral Diseases, Pathogen Discovery | Alison Laufer Halpin; Ben L. Rambo-Martin; Clinton R. Paden; Dakota Howard; Darlene Wagner; Dave Wentworth; Dhwani Batra; Jasmine Padilla; Justin Lee; Katie Dillon; Krista Queen; Kristen Knipe; Kristine Lacek; Mark Burroughs; Matthew Schmerer; Mili Sheth; Peter Cook; Sam Shepard; Sarah Nobles; Shoshona Le; Suxiang Tong; Vivien Dugan; Yvette Unoarumhi |
| EPI_ISL_2100159 | DC Public Health Lab/ Dept. of Forensic Sciences | DC Public Health Lab/ Dept. of Forensic Sciences | Brittany Hamilton; Connie Maza; David Payne; Elizabeth Zelaya; Janis Doss; Jocelyn Hauser; Monica Mann; Sarah Scott; Scott Nguyen |
| EPI_ISL_1785127, EPI_ISL_1841216, EPI_ISL_1841220 | DIP. PREV. AVEZZANO SERVIZIO DI IGIENE EPIDEMIOLOGIA E SANITA' PUBBLICA | Istituto Zooprofilattico Sperimentale dell'Abruzzo e Molise "G. Caporale" | Ancora M; Calistri P; Cammà C; Caporale M; Curini V; Delli Compagni E; Di Domenico M; Di Lollo Valeria; Di Pasquale A; Lorusso A; Mangone I; Marcacci M; Puglia I; Rinaldi A; Savini G; Scialabba S |
| EPI_ISL_5104494, EPI_ISL_5104496 | DNAGYN | LGBio (Laboratorio de Genetica & Biodiversidade) | Alex Honda Bernardes; Amanda Alves de Melo; Aparecido Divino da Cruz; Cintia Pelegrineti Targueta de Azevedo Brito; Daniela de Melo e Silva; Elisangela de Paula Silveira Lacerda; Juliana Santana de Curcio; Luiz Augusto Pereira; Marc Alexandre Duarte Gigonzac; Mariana Pires de Campos Telles; Ramilla dos Santos Braga; Renata de Oliveira Dias; Rhewter Nunes; Thais Cidália Vieira Gigonzac; Thais Guimarães Castro; Thays Millena Alves Pedroso |
| EPI_ISL_1804958 | DOHMH Jamaica | New York City Public Health Laboratory | Jade Wang; et al. |
| EPI_ISL_2383674 | DPHL | Delaware Public Health Lab | Rebecca Savage |
| EPI_ISL_2107619 | Delaware Public Health Lab | Delaware Public Health Lab | Rebecca Savage |
| EPI_ISL_1826801, EPI_ISL_1826828 | Department of Clinical Microbiology | GIGA Medical Genomics | Bouchra Boujemla; Cécile Meex; Keith Durkin; Maria Artesi; Marie-Pierre Hayette; Nathalie Renotte; Pierrette Melin; Raphaël Boreux; Sébastien Bontems; Vincent Bours |
| EPI_ISL_5428913 | Dr. Risch Ostschweiz AG | Microbiology | Dominique Fabien Hilti; Faina Wehrli; Lorenz Risch; Martin Risch; Nadia Wohlwend; Sinem Kas; Thomas Bodmer |
| EPI_ISL_3798365, EPI_ISL_4210137 | Dr. Risch Ostschweiz AG | Microbiology, Dr. Risch | Dominique Fabien Hilti; Faina Wehrli; Lorenz Risch; Martin Risch; Nadia Wohlwend; Sinem Kas; Thomas Bodmer |
| EPI_ISL_2145686 | Dutch COVID-19 response team | Erasmus Medical Center | Anne van der Linden; Annemiek van der Eijk; Bas Oude Munnink; Corine GeurtsvanKessel; David Nieuwenhuijse; Emmanuelle Munger; Irina Chestakova; Marion Koopmans; Marjan Boter; Reina Sikkema; Richard Molenkamp; on behalf of the Dutch national COVID-19 respo |
| EPI_ISL_2030790 | Dutch COVID-19 response team | Medical Microbiology, Maastricht University Medical Centre | Brian van der Veer*; Carmen Reumkens; Christian Hoebe; Erik Beuken; Jozef Dingemans*; Lieke van Alphen; Paul Savelkoul |
| EPI_ISL_1962254, EPI_ISL_1962255, EPI_ISL_1962256, EPI_ISL_1962259, EPI_ISL_1962262, EPI_ISL_1962263, EPI_ISL_1962264, EPI_ISL_1962265, EPI_ISL_1962266, EPI_ISL_1962267, EPI_ISL_1962271, EPI_ISL_1962276, EPI_ISL_1962277, EPI_ISL_1962278, EPI_ISL_1962279, EPI_ISL_1962280, EPI_ISL_1962281, EPI_ISL_2093854, EPI_ISL_2093856, EPI_ISL_2093858, EPI_ISL_2093864, EPI_ISL_2093865, EPI_ISL_2093866, EPI_ISL_2093867, EPI_ISL_2093868, EPI_ISL_2093870, EPI_ISL_2093871, EPI_ISL_2093874, EPI_ISL_2093875, EPI_ISL_2093892, EPI_ISL_2093893, EPI_ISL_2219018, EPI_ISL_2405922, EPI_ISL_6230263 | | | |
| see above | Dutch COVID-19 response team | National Institute for Public Health and the Environment (RIVM) | Adam Meijer; AnneMarie van den Brandt; Annelies Kroneman; Bas van der Veer; Chantal Reusken; Dennis Schmitz; Dirk Eggink; Euníce Then; Florian Zwagemaker; Harry Vennema; Ivo van Walle; James Groot; Jeroen Cremer; Karim Hajji; Kim Freriks; Linda van de Nes; Lisa Wijsman; Lynn Aarts; Melissa van Tuil; Ryanne Jaarsma; Sanne Bos; Sharon van den Brink; Sjoerd Kuiling; Stijn van Rossum; on behalf of the national COVID-19 response team |
| EPI_ISL_4060099, EPI_ISL_4060100, EPI_ISL_4060102, EPI_ISL_4060104, EPI_ISL_4060105, EPI_ISL_4060108 | Dynacare Manitoba | Manitoba Cadham Provincial Laboratory | David Alexander; Hilary Racher; Hongbin Li; Melissa Desaulnier; Tintu Abraham |
| EPI_ISL_2344869 | E S F DR EDSON GIANETTO JD CARLOS CASSETARI | Instituto Butantan / ESALQ-Piracicaba | Antonio Jorge Martins; Claudia Renata dos Santos Barros; David Schlesinger; Debora Botequio Moretti; Dimas Tadeu Covas; Elaine Cristina Marqueze; Elaine Vieira Santos; Evandra Strazza Rodrigues; Heidge Fukumasu; Jayme Augusto de Souza-Neto; José Salvatore Leister Patané; Luiz Alcantara; Luiz Lehmann Coutinho; Maria Carolina Elias; Maurício Lacerda Nogueira; Rafael dos Santos Bezerra; Raul Machado Neto; Rejane Maria Tommasini Grotto; Ricardo Haddad; Sandra Coccuzzo Sampaio Vessoni; Simone Kashima; Svetoslav Nanev Slavov; Vincent Louis Viala |
| EPI_ISL_5800823 | E S F Dr Edson Gianetto Jd Carlos Cassetari | Instituto Butantan | Antonio Jorge Martins; Claudia Renata dos Santos Barros; David Schlesinger; Debora Botequio Moretti; Dimas Tadeu Covas; Elaine Cristina Marqueze; Elaine Vieira Santos; Evandra Strazza Rodrigues; Heidge Fukumasu; Jayme Augusto de Souza-Neto; José Salvatore Leister Patané; Luiz Alcantara; Luiz Lehmann Coutinho; Maria Carolina Elias; Maurício Lacerda Nogueira; Rafael dos Santos Bezerra; Raul Machado Neto; Rejane Maria Tommasini Grotto; Ricardo Haddad; Sandra Coccuzzo Sampaio Vessoni; Simone Kashima; Svetoslav Nanev Slavov; Vincent Louis Viala |
| EPI_ISL_2344877 | E S F WALDO DSTEFANO JD PRIMAVERA JOSE BONIFACIO | Instituto Butantan / ESALQ-Piracicaba | Antonio Jorge Martins; Claudia Renata dos Santos Barros; David Schlesinger; Debora Botequio Moretti; Dimas Tadeu Covas; Elaine Cristina Marqueze; Elaine Vieira Santos; Evandra Strazza Rodrigues; Heidge Fukumasu; Jayme Augusto de Souza-Neto; José Salvatore Leister Patané; Luiz Alcantara; Luiz Lehmann Coutinho; Maria Carolina Elias; Maurício Lacerda Nogueira; Rafael dos Santos Bezerra; Raul Machado Neto; Rejane Maria Tommasini Grotto; Ricardo Haddad; Sandra Coccuzzo Sampaio Vessoni; Simone Kashima; Svetoslav Nanev Slavov; Vincent Louis Viala |
| EPI_ISL_5800836 | E S F Waldo Dstefano Jd Primavera Jose Bonifacio | Instituto Butantan | Antonio Jorge Martins; Claudia Renata dos Santos Barros; David Schlesinger; Debora Botequio Moretti; Dimas Tadeu Covas; Elaine Cristina Marqueze; Elaine Vieira Santos; Evandra Strazza Rodrigues; Heidge Fukumasu; Jayme Augusto de Souza-Neto; José Salvatore Leister Patané; Luiz Alcantara; Luiz Lehmann Coutinho; Maria Carolina Elias; Maurício Lacerda Nogueira; Rafael dos Santos Bezerra; Raul Machado Neto; Rejane Maria Tommasini Grotto; Ricardo Haddad; Sandra Coccuzzo Sampaio Vessoni; Simone Kashima; Svetoslav Nanev Slavov; Vincent Louis Viala |
| EPI_ISL_1941841 | EHNV | Laboratory of genomics and metagenomics | Claire Bertelli; Damien Jacot; Gilbert Greub; Sebastien Aeby; Trestan Pillonel |
| EPI_ISL_2344834, EPI_ISL_2344835, EPI_ISL_2344845, EPI_ISL_2344861 | EMERGENCIA RESPIRATORIA DE NOVA GRANADA | Instituto Butantan / ESALQ-Piracicaba | Antonio Jorge Martins; Claudia Renata dos Santos Barros; David Schlesinger; Debora Botequio Moretti; Dimas Tadeu Covas; Elaine Cristina Marqueze; Elaine Vieira Santos; Evandra Strazza Rodrigues; Heidge Fukumasu; Jayme Augusto de Souza-Neto; José Salvatore Leister Patané; Luiz Alcantara; Luiz Lehmann Coutinho; Maria Carolina Elias; Maurício Lacerda Nogueira; Rafael dos Santos Bezerra; Raul Machado Neto; Rejane Maria Tommasini Grotto; Ricardo Haddad; Sandra Coccuzzo Sampaio Vessoni; Simone Kashima; Svetoslav Nanev Slavov; Vincent Louis Viala |
| EPI_ISL_2209628 | ESF M1 UNI AMERICA DE JALES | Instituto Butantan / ESALQ-Piracicaba | Antonio Jorge Martins; Claudia Renata dos Santos Barros; David Schlesinger; Debora Botequio Moretti; Dimas Tadeu Covas; Elaine Cristina Marqueze; Elaine Vieira Santos; Evandra Strazza Rodrigues; Heidge Fukumasu; Jayme Augusto de Souza-Neto; José Salvatore Leister Patané; Luiz Alcantara; Luiz Lehmann Coutinho; Maria Carolina Elias; Maurício Lacerda Nogueira; Rafael dos Santos Bezerra; Raul Machado Neto; Rejane Maria Tommasini Grotto; Ricardo Haddad; Sandra Coccuzzo Sampaio Vessoni; Simone Kashima; Svetoslav Nanev Slavov; Vincent Louis Viala |
| EPI_ISL_2344870 | ESF RURAL DR JOSE CICERO FONTES XAVIER | Instituto Butantan / ESALQ-Piracicaba | Antonio Jorge Martins; Claudia Renata dos Santos Barros; David Schlesinger; Debora Botequio Moretti; Dimas Tadeu Covas; Elaine Cristina Marqueze; Elaine Vieira Santos; Evandra Strazza Rodrigues; Heidge Fukumasu; Jayme Augusto de Souza-Neto; José Salvatore Leister Patané; Luiz Alcantara; Luiz Lehmann Coutinho; Maria Carolina Elias; Maurício Lacerda Nogueira; Rafael dos Santos Bezerra; Raul Machado Neto; Rejane Maria Tommasini Grotto; Ricardo Haddad; Sandra Coccuzzo Sampaio Vessoni; Simone Kashima; Svetoslav Nanev Slavov; Vincent Louis Viala |
| EPI_ISL_3215604 | EXCITE Lab | Andersen lab at Scripps Research | Abigail Schnapper; Angela Scioscia; Chip Schooley; David Pride; Helena Tubb; Natasha Martin Cheryl Anderson; Sawyer Farmer; Sharon Reed; Tommy Valles + SEARCH |
| EPI_ISL_2477517, EPI_ISL_2477518, EPI_ISL_2477520, EPI_ISL_2477522, EPI_ISL_2477524, EPI_ISL_2477976, EPI_ISL_2477979, EPI_ISL_2477980, EPI_ISL_2477982, EPI_ISL_2477983, EPI_ISL_2477984, EPI_ISL_2477985, EPI_ISL_2477986, EPI_ISL_2477987, EPI_ISL_2477988, EPI_ISL_2477989, EPI_ISL_2477990, EPI_ISL_2477991, EPI_ISL_2478105, EPI_ISL_2478229, EPI_ISL_2478241, EPI_ISL_2478266, EPI_ISL_2478271, EPI_ISL_2478291, EPI_ISL_2478292, EPI_ISL_2478293, EPI_ISL_2478294, EPI_ISL_2478295, EPI_ISL_2478421, EPI_ISL_2478422, EPI_ISL_2478423, EPI_ISL_2478424, EPI_ISL_2478680, EPI_ISL_2478681, EPI_ISL_2478912, EPI_ISL_2478913, EPI_ISL_2478914, EPI_ISL_2478915, EPI_ISL_2478919, EPI_ISL_2478920, EPI_ISL_2478921, EPI_ISL_2478922, EPI_ISL_2479197, EPI_ISL_2479200, EPI_ISL_2479202, EPI_ISL_2479209, EPI_ISL_2479211, EPI_ISL_2479219, EPI_ISL_2479220, EPI_ISL_2479222, EPI_ISL_2479223, EPI_ISL_2479234, EPI_ISL_2479235, EPI_ISL_2479236, EPI_ISL_2479240, EPI_ISL_2479243, EPI_ISL_2479248, EPI_ISL_2479249, EPI_ISL_2479250, EPI_ISL_2479251, EPI_ISL_2479252, EPI_ISL_2479256, EPI_ISL_2479261, EPI_ISL_2479389, EPI_ISL_2479390, EPI_ISL_2479579, EPI_ISL_2479797, EPI_ISL_2940418, EPI_ISL_2940475, EPI_ISL_2941298, EPI_ISL_2941330, EPI_ISL_2941334, EPI_ISL_2941349, EPI_ISL_2941379, EPI_ISL_2941425, EPI_ISL_2941427, EPI_ISL_2941502, EPI_ISL_3909748, EPI_ISL_3909749, EPI_ISL_3909750, EPI_ISL_3909753, EPI_ISL_3909754, EPI_ISL_3909755, EPI_ISL_3909756, EPI_ISL_3909757, EPI_ISL_3909758, EPI_ISL_3909759, EPI_ISL_3909760, EPI_ISL_3909764, EPI_ISL_3909823, EPI_ISL_3909825, EPI_ISL_3910028, EPI_ISL_3910031, EPI_ISL_3910271 | | | |
| see above | Edmonton Provincial Lab | Public Health Agency of Canada (PHAC) National Microbiology Laboratory | Buss; Croxen M; Deo A; Dieu P; E; Ferrato C; Gill K; Khan F; Koleva P; Li V; Lloyd C; Lynch T; Ma R; Murphy S; Pabbaraju K; Shokoples S; Thayer J; Tipples G; Whitehouse M; Wong A; Yu C; Zelyas N |
| EPI_ISL_5800779, EPI_ISL_5800780, EPI_ISL_5800791, EPI_ISL_5800812 | Emergencia Respiratoria De Nova Granada | Instituto Butantan | Antonio Jorge Martins; Claudia Renata dos Santos Barros; David Schlesinger; Debora Botequio Moretti; Dimas Tadeu Covas; Elaine Cristina Marqueze; Elaine Vieira Santos; Evandra Strazza Rodrigues; Heidge Fukumasu; Jayme Augusto de Souza-Neto; José Salvatore Leister Patané; Luiz Alcantara; Luiz Lehmann Coutinho; Maria Carolina Elias; Maurício Lacerda Nogueira; Rafael dos Santos Bezerra; Raul Machado Neto; Rejane Maria Tommasini Grotto; Ricardo Haddad; Sandra Coccuzzo Sampaio Vessoni; Simone Kashima; Svetoslav Nanev Slavov; Vincent Louis Viala |
| EPI_ISL_2346154 | Emory Molecular Diagnostics Laboratory, Emory Healthcare | Piantadosi Lab, Emory Department of Pathology | Ahmed Babiker; Anne Piantadosi |
| EPI_ISL_5800827 | Esf M1 Uni America De Jales | Instituto Butantan | Antonio Jorge Martins; Claudia Renata dos Santos Barros; David Schlesinger; Debora Botequio Moretti; Dimas Tadeu Covas; Elaine Cristina Marqueze; Elaine Vieira Santos; Evandra Strazza Rodrigues; Heidge Fukumasu; Jayme Augusto de Souza-Neto; José Salvatore Leister Patané; Luiz Alcantara; Luiz Lehmann Coutinho; Maria Carolina Elias; Maurício Lacerda Nogueira; Rafael dos Santos Bezerra; Raul Machado Neto; Rejane Maria Tommasini Grotto; Ricardo Haddad; Sandra Coccuzzo Sampaio Vessoni; Simone Kashima; Svetoslav Nanev Slavov; Vincent Louis Viala |
| EPI_ISL_5800829 | Esf Rural Dr Jose Cicero Fontes Xavier | Instituto Butantan | Antonio Jorge Martins; Claudia Renata dos Santos Barros; David Schlesinger; Debora Botequio Moretti; Dimas Tadeu Covas; Elaine Cristina Marqueze; Elaine Vieira Santos; Evandra Strazza Rodrigues; Heidge Fukumasu; Jayme Augusto de Souza-Neto; José Salvatore Leister Patané; Luiz Alcantara; Luiz Lehmann Coutinho; Maria Carolina Elias; Maurício Lacerda Nogueira; Rafael dos Santos Bezerra; Raul Machado Neto; Rejane Maria Tommasini Grotto; Ricardo Haddad; Sandra Coccuzzo Sampaio Vessoni; Simone Kashima; Svetoslav Nanev Slavov; Vincent Louis Viala |

| | | | |
|---|---|---|---|
| EPI_ISL_2123732, EPI_ISL_2123737, EPI_ISL_2123803, EPI_ISL_2123804 | Eurofins LifeCodexx GmbH | Robert Koch Institute | |
| EPI_ISL_2018238 | FL Bur. of Public Health Laboratories-Jacksonville | Centers for Disease Control and Prevention Division of Viral Diseases, Pathogen Discovery | Alison Laufer Halpin; Ben L. Rambo-Martin; Clinton R. Paden; Dakota Howard; Darlene Wagner; Dave Wentworth; Dhwani Batra; Jasmine Padilla; Justin Lee; Katie Dillon; Krista Queen; Kristen Knipe; Kristine Lacek; Mark Burroughs; Matthew Schmerer; Mili Sheth; Peter Cook; Sam Shepard; Sarah Nobles; Shoshona Le; Suxiang Tong; Vivien Dugan; Yvette Unoarumhi |
| EPI_ISL_2018168 | FL Bureau of Public Health Laboratories-Miami | Centers for Disease Control and Prevention Division of Viral Diseases, Pathogen Discovery | Alison Laufer Halpin; Ben L. Rambo-Martin; Clinton R. Paden; Dakota Howard; Darlene Wagner; Dave Wentworth; Dhwani Batra; Jasmine Padilla; Justin Lee; Katie Dillon; Krista Queen; Kristen Knipe; Kristine Lacek; Mark Burroughs; Matthew Schmerer; Mili Sheth; Peter Cook; Sam Shepard; Sarah Nobles; Shoshona Le; Suxiang Tong; Vivien Dugan; Yvette Unoarumhi |
| EPI_ISL_3355485 | FUNDACIÓN HOSPITAL SAN PEDRO | Corporacion CorpoGen-Universidad de los Andes-Universidad Central | Christian Romero; Jorge Duitama; Juan Manuel Anzola; Laura González; Maryam Chaib De Mares; María Mercedes Zambrano; Nelly Díaz; Patricia Del Portillo; Silvia Restrepo |
| EPI_ISL_5802683, EPI_ISL_5802684, EPI_ISL_5802685, EPI_ISL_5802686, EPI_ISL_5802687, EPI_ISL_5802688, EPI_ISL_5802689 | | | |
| see above | Faculdade De Medicina Abc | Instituto Butantan | Antonio Jorge Martins; Claudia Renata dos Santos Barros; David Schlesinger; Debora Botequio Moretti; Dimas Tadeu Covas; Elaine Cristina Marqueze; Elaine Vieira Santos; Evandra Strazza Rodrigues; Heidge Fukumasu; Jayme Augusto de Souza-Neto; José Salvatore Leister Patané; Luiz Alcantara; Luiz Lehmann Coutinho; Maria Carolina Elias; Maurício Lacerda Nogueira; Rafael dos Santos Bezerra; Raul Machado Neto; Rejane Maria Tommasini Grotto; Ricardo Haddad; Sandra Coccuzzo Sampaio Vessoni; Simone Kashima; Svetoslav Nanev Slavov; Vincent Louis Viala |
| EPI_ISL_2008736, EPI_ISL_2008744, EPI_ISL_2008745, EPI_ISL_2008827, EPI_ISL_2008832, EPI_ISL_2008902, EPI_ISL_2008905, EPI_ISL_2229992, EPI_ISL_2277654, EPI_ISL_2277919, EPI_ISL_2277958, EPI_ISL_2277959, EPI_ISL_2277960, EPI_ISL_2277961, EPI_ISL_2277962, EPI_ISL_2277963, EPI_ISL_2442669, EPI_ISL_2442719, EPI_ISL_2836748, EPI_ISL_2836770, EPI_ISL_2836771, EPI_ISL_4218610, EPI_ISL_4218614, EPI_ISL_4218686, EPI_ISL_4218906, EPI_ISL_4218925, EPI_ISL_4218954, EPI_ISL_4218955, EPI_ISL_4218975, EPI_ISL_4218982, EPI_ISL_4218983, EPI_ISL_4218988, EPI_ISL_5866159, EPI_ISL_5924992, EPI_ISL_5925025, EPI_ISL_5925174, EPI_ISL_5925175, EPI_ISL_5925176, EPI_ISL_6049935, EPI_ISL_6049968, EPI_ISL_6049972, EPI_ISL_6049979, EPI_ISL_6049981, EPI_ISL_6049983, EPI_ISL_6049994, EPI_ISL_6050072, EPI_ISL_6050077, EPI_ISL_6050079, EPI_ISL_6050094, EPI_ISL_6050120, EPI_ISL_6050147, EPI_ISL_6050172, EPI_ISL_6050199, EPI_ISL_6050223, EPI_ISL_6050624, EPI_ISL_6050677, EPI_ISL_6050750 | | | |
| see above | Florida Bureau of Public Health Laboratories | Florida Bureau of Public Health Laboratories | Jason Blanton; Jiaqi Li; Namratha Tarigopula; Sarah Schmedes; Tiffany Splatt |
| EPI_ISL_2620939, EPI_ISL_2620950, EPI_ISL_2620952 | Fondazione IRCCS Ca' Granda Ospedale Maggiore Policlinico | Fondazione IRCCS Ca' Granda Ospedale Maggiore Policlinico | Ferruccio Ceriotti; Sara Uceda Renteria |
| EPI_ISL_1839111, EPI_ISL_1839119, EPI_ISL_1839128, EPI_ISL_1839140, EPI_ISL_1839142, EPI_ISL_1839145, EPI_ISL_1839146, EPI_ISL_1839163, EPI_ISL_1839212, EPI_ISL_1839228, EPI_ISL_1839229, EPI_ISL_1839238, EPI_ISL_1839250, EPI_ISL_1839261, EPI_ISL_1839269, EPI_ISL_1839384, EPI_ISL_1839386, EPI_ISL_1839392, EPI_ISL_1839393, EPI_ISL_1839451, EPI_ISL_1839485, EPI_ISL_1839489, EPI_ISL_1839491, EPI_ISL_1839532, EPI_ISL_1839538, EPI_ISL_1839596, EPI_ISL_1839600, EPI_ISL_1839608, EPI_ISL_1839613, EPI_ISL_1839614, EPI_ISL_1839850, EPI_ISL_1839855, EPI_ISL_1839857, EPI_ISL_1839870, EPI_ISL_1839871, EPI_ISL_1839889, EPI_ISL_1839893, EPI_ISL_1839949, EPI_ISL_1839952, EPI_ISL_1839953, EPI_ISL_1839971, EPI_ISL_1839977, EPI_ISL_1839980, EPI_ISL_1839982, EPI_ISL_1839997, EPI_ISL_1840001, EPI_ISL_1840014, EPI_ISL_1840019, EPI_ISL_1840063, EPI_ISL_1840074, EPI_ISL_1840076, EPI_ISL_1840083, EPI_ISL_1840092, EPI_ISL_1840093, EPI_ISL_1840116, EPI_ISL_1840122, EPI_ISL_1840124, EPI_ISL_1840125, EPI_ISL_1992173, EPI_ISL_1992281, EPI_ISL_1992284, EPI_ISL_1992287, EPI_ISL_1992293, EPI_ISL_1992317, EPI_ISL_1992397, EPI_ISL_1992412, EPI_ISL_1992427, EPI_ISL_1992430, EPI_ISL_1992441, EPI_ISL_1992448, EPI_ISL_1992449, EPI_ISL_1992457, EPI_ISL_1992484, EPI_ISL_1992524, EPI_ISL_1992542, EPI_ISL_1992569, EPI_ISL_1992587, EPI_ISL_1992588, EPI_ISL_1992632, EPI_ISL_1992634, EPI_ISL_1992643, EPI_ISL_1992646, EPI_ISL_1992648, EPI_ISL_1992660, EPI_ISL_1992666, EPI_ISL_1992679, EPI_ISL_1992705, EPI_ISL_1992713, EPI_ISL_1992719, EPI_ISL_1992723, EPI_ISL_1992746, EPI_ISL_1992760, EPI_ISL_1992762, EPI_ISL_1992764, EPI_ISL_1992800, EPI_ISL_1992809, EPI_ISL_1992813, EPI_ISL_2038681, EPI_ISL_2038718, EPI_ISL_2038734, EPI_ISL_2038741, EPI_ISL_2038749, EPI_ISL_2038757, EPI_ISL_2038761, EPI_ISL_4367333, EPI_ISL_4370171, EPI_ISL_4371027 | | | |
| see above | Fulgent Genetics | Centers for Disease Control and Prevention Division of Viral Diseases, Pathogen Discovery | Adrian Paskey; Becky Tsai; Benafsh Sapra; Benjamin Rambo-Martin; Christopher Gulvick; Clinton Paden; Clinton R. Paden; Dakota Howard; Darlene Wagner; Dhwani Batra; Doreen Ng; Duncan MacCannell; Erisa Sula; Harry Gao; James Xie; Jason Caravas; John Gao; Joseph Fierro; Kara Moser; Kristine Lacek; Matthew Schmerer; Mickey Li; Peter Cook; Peter W. Cook; Scott Sammons; Shatavia Morrison; Tymeckia Kendall; Victoria Caban Figueroa; Yan Meng; Yvette Unoarumhi |
| EPI_ISL_2229306 | GA Department of Public Health Laboratory | Centers for Disease Control and Prevention Division of Viral Diseases, Pathogen Discovery | Alison Laufer Halpin; Ben L. Rambo-Martin; Clinton R. Paden; Dakota Howard; Darlene Wagner; Dave Wentworth; Dhwani Batra; Jasmine Padilla; Justin Lee; Katie Dillon; Krista Queen; Kristen Knipe; Kristine Lacek; Mark Burroughs; Matthew Schmerer; Mili Sheth; Peter Cook; Sam Shepard; Sarah Nobles; Shoshona Le; Suxiang Tong; Vivien Dugan; Yvette Unoarumhi |
| EPI_ISL_1855145, EPI_ISL_2421160 | GLENS FALLS HOSPITAL LABORATORY | Wadsworth Center, New York State Department of Health | Alexis Russell; Catharine Prussing; Daryl M. Lamson; Erasmus Schneider; Erica Lasek-Nesselquist; John Kelly; Jonathan Plitnick; Kirsten St. George; Matthew Shudt; Melissa A Leisner; Navjot Singh |
| EPI_ISL_2009537, EPI_ISL_2009538, EPI_ISL_2009539, EPI_ISL_2009540, EPI_ISL_2009546, EPI_ISL_2009549, EPI_ISL_2009552, EPI_ISL_2009554, EPI_ISL_2009555, EPI_ISL_2009600, EPI_ISL_2009601, EPI_ISL_2009603, EPI_ISL_2009604, EPI_ISL_2009614, EPI_ISL_2009624, EPI_ISL_2009625, EPI_ISL_2391013, EPI_ISL_2391040, EPI_ISL_2597253, EPI_ISL_2597255, EPI_ISL_2597256, EPI_ISL_2597260, EPI_ISL_2597263, EPI_ISL_2597265, EPI_ISL_2597267, EPI_ISL_2597268, EPI_ISL_2597271, EPI_ISL_2597272, EPI_ISL_2597273, EPI_ISL_2597274, EPI_ISL_2597278, EPI_ISL_2597279, EPI_ISL_2597280, EPI_ISL_2597281, EPI_ISL_2597283, EPI_ISL_2597284, EPI_ISL_2597285, EPI_ISL_2597286, EPI_ISL_2597287, EPI_ISL_2597288, EPI_ISL_2597305 | | | |
| see above | Genetica Molecular and Subdepartamento de Virologia ISP Chile | Instituto de Salud Publica de Chile | Andres Castillo; Barbara Parra; Constanza Campano; Gisselle Barra; Javier Tognarelli; Jorge Fernandez; Karen Orostica; Loredana Arata; Patricia Bustos; Rodrigo Fasce; Soledad Ulloa |
| EPI_ISL_5196287, EPI_ISL_5196335, EPI_ISL_5196354 | Gorgas Memorial Institute of Health Studies | Gorgas Memorial Institute of Health Studies | Castillo Jorge; Chen Maria; Franco Danilo; Gonzalez Claudia; Jessica Gondola; Leyda Abrego; Lopez-Verges Sandra; Marlenne Castillo; Martinez Alexander; Menacho Abdiel; Moreno Ambar; Moreno Brechla; Oris Chavarria; Ortiz Alma; Salazar Jacqueline |
| EPI_ISL_2346153 | Grady Memorial Hospital, Clinical Microbiology Laboratory | Piantadosi Lab, Emory Department of Pathology | Anne Piantadosi; Yun F (Wayne) Wang |
| EPI_ISL_2601869, EPI_ISL_2601908, EPI_ISL_2601911 | Gravity Diagnostics, LLC | Gravity Diagnostics, LLC | Gravity Diagnostics |
| EPI_ISL_2371255 | Greek Genome Center, Biomedical Research Foundation of the Academy of Athens (BRFAA) | Greek Genome Center, Biomedical Research Foundation of the Academy of Athens (BRFAA) | Dimitrios Thanos; Emmanouil Athanasiadis; Giannis Vatsellas; Katerina Zoi; Theodoros Loupis |
| EPI_ISL_1745232 | Gundersen Clinical Microbiology Laboratory | Kabara Cancer Research Institute | Craig S. Richmond; Paraic A. Kenny |
| EPI_ISL_1854240 | H Evora | Instituto Nacional de Saude (INSA) | Borges et al |
| EPI_ISL_2945006, EPI_ISL_2945032 | HG Pharma | Elling and Cochella laboratories, IMBA/IMP | Alexander Stark; Ezgi Oezkan; Luisa Cochella and Ulrich Elling; Marcus Strobl; Olga Frank; Ramesh Yelagandula; Tanino Albanese |
| EPI_ISL_3102335 | HGCC HOSPITAL GERAL DR CESAR CALS | Analytical Competence Molecular Epidemiology Lab/ACME, Oswaldo Cruz Foundation, Ceara (FIOCRUZ CE) | Cleber Furtado Aksenen; Fabio Miyajima; Fernando Braga Stehling; Francisco Eder de Moura Lopes; Jamille Maria Mendes Bezerra; Joaquim César do Nascimento Sousa Junior; Pedro Miguel Carneiro Jeronimo; Suzana Porto Almeida e Lucas Delerino; Thais Ferreira de Oliveira; Thais de Oliveira Costa; Ticiane Cavalcante de Souza; Veridiana Pessoa Miyajima |
| EPI_ISL_5603219, EPI_ISL_5603266 | HGF HOSPITAL GERAL DE FORTALEZA | Analytical Competence Molecular Epidemiology Lab/ACME, Oswaldo Cruz Foundation, Ceara (FIOCRUZ CE) | Carlos Leonardo de Aragao Araujo; Cecilia Leite Costa & Eduardo Ruback dos Santos on behalf of COVID-19 FIOCRUZ Genomic Network; Cleber Furtado Aksenen; Fabio Miyajima; Fernando Braga Stehling; Francisco Eder de Moura Lopes; Igor Oliveira Duarte; Jamille Maria Mendes Bezerra; Joaquim Cesar do Nascimento Sousa Junior; Pedro Miguel Carneiro Jeronimo; Suzana Porto Almeida; Thais Ferreira de Oliveira; Thais de Oliveira Costa; Ticiane Cavalcante de Souza; Veridiana Pessoa Miyajima |
| EPI_ISL_1941834 | HIB | Laboratory of genomics and metagenomics | Claire Bertelli; Damien Jacot; Gilbert Greub; Sebastien Aeby; Trestan Pillonel |
| EPI_ISL_2017370, EPI_ISL_2017371, EPI_ISL_2017372, EPI_ISL_2017373, EPI_ISL_2017374, EPI_ISL_2017375, EPI_ISL_2017377, EPI_ISL_2017378, EPI_ISL_2017379, EPI_ISL_2017380, EPI_ISL_2017381, EPI_ISL_2017382, EPI_ISL_2017383, EPI_ISL_2017384, EPI_ISL_2017385, EPI_ISL_2017386, EPI_ISL_2017387, EPI_ISL_2017388, EPI_ISL_2017389, EPI_ISL_2017390, EPI_ISL_2017482, EPI_ISL_2187985, EPI_ISL_2187986, EPI_ISL_2187987, EPI_ISL_4061646 | | | |
| see above | HLAGYN - Laboratorio de Imunologia de Transplantes de Goias | HLAGYN - Laboratorio de Imunologia de Transplantes de Goias | Alessandro Leonardo Alvares Magalhaes; Daniel Ferreira de Sousa; Danielle de Paiva Rezende; Erika Lopes Rocha Batista; Fernando Antonio Vinhal dos Santos; Frederico Rodrigues Vinhal; Kamila Oliveira Reis De Freitas.; Lucas Carlos Gomes Pereira; Paola Cristina Resende Silva; Sabrina Sara Moreira Duarte |
| EPI_ISL_2345003 | HOSP DE TRANSPLANT DO EST DE SP EURYCLIDES DE JESUS ZERBINI | Instituto Butantan | Antonio Jorge Martins; Claudia Renata dos Santos Barros; David Schlesinger; Debora Botequio Moretti; Dimas Tadeu Covas; Elaine Cristina Marqueze; Elaine Vieira Santos; Evandra Strazza Rodrigues; Heidge Fukumasu; Jayme Augusto de Souza-Neto; José Salvatore Leister Patané; Luiz Alcantara; Luiz Lehmann Coutinho; Maria Carolina Elias; Maurício Lacerda Nogueira; Rafael dos Santos Bezerra; Raul Machado Neto; Rejane Maria Tommasini Grotto; Ricardo Haddad; Sandra Coccuzzo Sampaio Vessoni; Simone Kashima; Svetoslav Nanev Slavov; Vincent Louis Viala |
| EPI_ISL_2209313, EPI_ISL_2344909, EPI_ISL_2344914, EPI_ISL_2344947, EPI_ISL_2344948 | HOSP E MATERNIDADE MUNICIPAL N SRA MONTE SERRAT | Instituto Butantan / ESALQ-Piracicaba | Antonio Jorge Martins; Claudia Renata dos Santos Barros; David Schlesinger; Debora Botequio Moretti; Dimas Tadeu Covas; Elaine Cristina Marqueze; Elaine Vieira Santos; Evandra Strazza Rodrigues; Heidge Fukumasu; Jayme Augusto de Souza-Neto; José Salvatore Leister Patané; Luiz Alcantara; Luiz Lehmann Coutinho; Maria Carolina Elias; Maurício Lacerda Nogueira; Rafael dos Santos Bezerra; Raul Machado Neto; Rejane Maria Tommasini Grotto; Ricardo Haddad; Sandra Coccuzzo Sampaio Vessoni; Simone Kashima; Svetoslav Nanev Slavov; Vincent Louis Viala |
| EPI_ISL_5603252, EPI_ISL_5603275 | HOSP MATERN F FARIAS LEITAO | Analytical Competence Molecular Epidemiology Lab/ACME, Oswaldo Cruz Foundation, Ceara (FIOCRUZ CE) | Carlos Leonardo de Aragao Araujo; Cecilia Leite Costa & Eduardo Ruback dos Santos on behalf of COVID-19 FIOCRUZ Genomic Network; Cleber Furtado Aksenen; Fabio Miyajima; Fernando Braga Stehling; Francisco Eder de Moura Lopes; Igor Oliveira Duarte; Jamille Maria Mendes Bezerra; Joaquim Cesar do Nascimento Sousa Junior; Pedro Miguel Carneiro Jeronimo; Suzana Porto Almeida; Thais Ferreira de Oliveira; Thais de Oliveira Costa; Ticiane Cavalcante de Souza; Veridiana Pessoa Miyajima |
| EPI_ISL_5603270, EPI_ISL_5603310 | HOSP MATERN PAULO SARASATE | Analytical Competence Molecular Epidemiology Lab/ACME, Oswaldo Cruz Foundation, Ceara (FIOCRUZ CE) | Carlos Leonardo de Aragao Araujo; Cecilia Leite Costa & Eduardo Ruback dos Santos on behalf of COVID-19 FIOCRUZ Genomic Network; Cleber Furtado Aksenen; Fabio Miyajima; Fernando Braga Stehling; Francisco Eder de Moura Lopes; Igor Oliveira Duarte; Jamille Maria Mendes Bezerra; Joaquim Cesar do Nascimento Sousa Junior; Pedro Miguel Carneiro Jeronimo; Suzana Porto Almeida; Thais Ferreira de Oliveira; Thais de Oliveira Costa; Ticiane Cavalcante de Souza; Veridiana Pessoa Miyajima |
| EPI_ISL_2209243, EPI_ISL_2345019, EPI_ISL_2345020, EPI_ISL_2345022, EPI_ISL_2345023, EPI_ISL_2345024, EPI_ISL_2345025, EPI_ISL_2345026, EPI_ISL_2345027 | | | |
| see above | HOSP MUN JOSANIAS CASTANHA BRAGA | Instituto Butantan | Antonio Jorge Martins; Claudia Renata dos Santos Barros; David Schlesinger; Debora Botequio Moretti; Dimas Tadeu Covas; Elaine Cristina Marqueze; Elaine Vieira Santos; Evandra Strazza Rodrigues; Heidge Fukumasu; Jayme Augusto de Souza-Neto; José Salvatore Leister Patané; Luiz Alcantara; Luiz Lehmann Coutinho; Maria Carolina Elias; Maurício Lacerda Nogueira; Rafael dos Santos Bezerra; Raul Machado Neto; Rejane Maria Tommasini Grotto; Ricardo Haddad; Sandra Coccuzzo Sampaio Vessoni; Simone Kashima; Svetoslav Nanev Slavov; Vincent Louis Viala; Vincent Louis Viala. |
| EPI_ISL_2344953, EPI_ISL_2344954, EPI_ISL_2344955, EPI_ISL_2344956, EPI_ISL_2344957, EPI_ISL_2344973, EPI_ISL_2344975, EPI_ISL_2344976, EPI_ISL_2344977, EPI_ISL_2344978, EPI_ISL_2344980, EPI_ISL_2344987, EPI_ISL_2344989, EPI_ISL_2344997, EPI_ISL_2345005, EPI_ISL_2345008, EPI_ISL_2345011, EPI_ISL_2345018, EPI_ISL_2345021 | | | |
| see above | HOSPITAL DE CAMPANHA | Instituto Butantan / ESALQ- | Antonio Jorge Martins; Claudia Renata dos Santos Barros; David Schlesinger; Debora Botequio Moretti; Dimas Tadeu Covas; Elaine Cristina Marqueze; Elaine Vieira Santos; Evandra Strazza Rodrigues; Heidge Fukumasu; Jayme Augusto de Souza-Neto; José Salvatore Leister Patané; Luiz Alcantara; Luiz Lehmann |

| | | | |
|---|---|---|---|
| | COVID 19 MUNICIPIO DE TAUBATE | Piracicaba | Coutinho; Maria Carolina Elias; Maurício Lacerda Nogueira; Rafael dos Santos Bezerra; Raul Machado Neto; Rejane Maria Tommasini Grotto; Ricardo Haddad; Sandra Coccuzzo Sampaio Vessoni; Simone Kashima; Svetoslav Nanev Slavov; Vincent Louis Viala |
| EPI_ISL_2344848, EPI_ISL_2344849 | HOSPITAL DE MISERICORDIA ALTINOPOLIS | Instituto Butantan / ESALQ-Piracicaba | Antonio Jorge Martins; Claudia Renata dos Santos Barros; David Schlesinger; Debora Botequio Moretti; Dimas Tadeu Covas; Elaine Cristina Marqueze; Elaine Vieira Santos; Evandra Strazza Rodrigues; Heidge Fukumasu; Jayme Augusto de Souza-Neto; José Salvatore Leister Patané; Luiz Alcantara; Luiz Lehmann Coutinho; Maria Carolina Elias; Maurício Lacerda Nogueira; Rafael dos Santos Bezerra; Raul Machado Neto; Rejane Maria Tommasini Grotto; Ricardo Haddad; Sandra Coccuzzo Sampaio Vessoni; Simone Kashima; Svetoslav Nanev Slavov; Vincent Louis Viala |
| EPI_ISL_2344982 | HOSPITAL E MATERNIDADE NOSSA SENHORA DA AJUDA | Instituto Butantan / ESALQ-Piracicaba | Antonio Jorge Martins; Claudia Renata dos Santos Barros; David Schlesinger; Debora Botequio Moretti; Dimas Tadeu Covas; Elaine Cristina Marqueze; Elaine Vieira Santos; Evandra Strazza Rodrigues; Heidge Fukumasu; Jayme Augusto de Souza-Neto; José Salvatore Leister Patané; Luiz Alcantara; Luiz Lehmann Coutinho; Maria Carolina Elias; Maurício Lacerda Nogueira; Rafael dos Santos Bezerra; Raul Machado Neto; Rejane Maria Tommasini Grotto; Ricardo Haddad; Sandra Coccuzzo Sampaio Vessoni; Simone Kashima; Svetoslav Nanev Slavov; Vincent Louis Viala |
| EPI_ISL_2209004, EPI_ISL_2209269, EPI_ISL_2209315, EPI_ISL_2344899, EPI_ISL_2344900, EPI_ISL_2344901, EPI_ISL_2344911, EPI_ISL_2344942, EPI_ISL_2344944, EPI_ISL_2344945, EPI_ISL_2344946 | | | |
| see above | HOSPITAL E PRONTO SOCORRO PORTINARI | Instituto Butantan | Antonio Jorge Martins; Claudia Renata dos Santos Barros; David Schlesinger; Debora Botequio Moretti; Dimas Tadeu Covas; Elaine Cristina Marqueze; Elaine Vieira Santos; Evandra Strazza Rodrigues; Heidge Fukumasu; Jayme Augusto de Souza-Neto; José Salvatore Leister Patané; Luiz Alcantara; Luiz Lehmann Coutinho; Maria Carolina Elias; Maurício Lacerda Nogueira; Rafael dos Santos Bezerra; Raul Machado Neto; Rejane Maria Tommasini Grotto; Ricardo Haddad; Sandra Coccuzzo Sampaio Vessoni; Simone Kashima; Svetoslav Nanev Slavov; Vincent Louis Viala. |
| EPI_ISL_1967299 | HOSPITAL E PRONTO SOCORRO PORTINARI | Instituto Butantan / Mendelics | Antonio Jorge Martins; Bianca Cechetto Carlos. Mendelics: Bibiana Santos; Claudia Renata dos Santos Barros; Cíntia Bittar; David Schlesinger. Hemocentro Ribeirão Preto: Simone Kashima; Debora Botequio Moretti; Elaine Cristina Marqueze; Elaine Vieira dos Santos; Elisangela Chicaroni Mattos; Erika Freitas; Evandra Strazza Rodrigues; Felipe Allan da Silva da Costa; Flavia Aburjaile; Fábio Sossai Possebon; Guilherme Campos; Guilherme Targino Valente; Heidge Fukumasu. USP-Botucatu: Rejane Maria Tommasini Grotto; Helena Lage Ferreira; Instituto Butantan: Dimas Tadeu Covas; Jardelina de Souza Todao Bernardino; Jayme A. Souza-Neto; Jessika Cristina Chagas Lesbon; Jorge A. Petroli Marchesi; José Salvatore Leister Patané; João Paulo Kitajima; João Pessoa Araújo Jr.; Leila Sabrina Ullmann; Loyze Paola Oliveira de Lima; Luiz Aurelio de Campos Crispin. Centro de Genômica Funcional da ESALQ: Luiz Lehmann Coutinho; Luiz Carlos Junior de Alcantara; Lívia Sacchetto; Maisa C. Pereira Parra; Maria Carolina Elias; Marta Giovanetti; Marília Moraes; Maurício Lacerda Nogueira. Prefeitura de Sao Paulo: Melissa Palmieri.; Patricia Akemi Assato; Paula Rahal; Paulo Inacio da Costa; Rafael dos Santos Bezerra; Raquel de Lello Rocha Campos Cassano. NGS Soluções Genômicas: Pilar Drummond Sampaio Corrêa Mariani. FZEA-USP Pirassununga: Mirele Daiana Poleti; Raul Machado Neto; Ricardo Augusto Brassaloti; Ricardo Haddad; Rodrigo Tocantins Calado. FAMERP-SJRP: Cecília Artico Banho; Sandra Coccuzzo Sampaio; Svetoslav Nanev Slavov; Vagner Fonseca; Vincent Louis Viala |
| EPI_ISL_2345029 | HOSPITAL ESCOLA DA UNIVERSIDADE DE TAUBATE | Instituto Butantan / ESALQ-Piracicaba | Antonio Jorge Martins; Claudia Renata dos Santos Barros; David Schlesinger; Debora Botequio Moretti; Dimas Tadeu Covas; Elaine Cristina Marqueze; Elaine Vieira Santos; Evandra Strazza Rodrigues; Heidge Fukumasu; Jayme Augusto de Souza-Neto; José Salvatore Leister Patané; Luiz Alcantara; Luiz Lehmann Coutinho; Maria Carolina Elias; Maurício Lacerda Nogueira; Rafael dos Santos Bezerra; Raul Machado Neto; Rejane Maria Tommasini Grotto; Ricardo Haddad; Sandra Coccuzzo Sampaio Vessoni; Simone Kashima; Svetoslav Nanev Slavov; Vincent Louis Viala |
| EPI_ISL_3102514 | HOSPITAL ESTADUAL LEONARDO DA VINCI | Analytical Competence Molecular Epidemiology Lab/ACME, Oswaldo Cruz Foundation, Ceara (FIOCRUZ CE) | Cleber Furtado Aksenen; Fabio Miyajima; Fernando Braga Stehling; Francisco Eder de Moura Lopes; Jamille Maria Mendes Bezerra; Joaquim César do Nascimento Sousa Junior; Pedro Miguel Carneiro Jeronimo; Suzana Porto Almeida e Lucas Delerino; Thais Ferreira de Oliveira; Thais de Oliveira Costa; Ticiane Cavalcante de Souza; Veridiana Pessoa Miyajima |
| EPI_ISL_4601277 | HOSPITAL INFANTA CRISTINA (BADAJOZ) | Instituto de Salud Carlos III | A. Monzón; F. Casas; I. Jiménez; I.MARTIN CORDERO; Iglesias-Caballero; M. Sandonís; P. Zaballos; PALOMA; S. Camarero; S. Cuesta; S. Pozo; S. Varona; V. Vázquez-Morón |
| EPI_ISL_5603254, EPI_ISL_5603258, EPI_ISL_5603263, EPI_ISL_5603304 | HOSPITAL JOSE MARIA PHILOMENO GOMES | Analytical Competence Molecular Epidemiology Lab/ACME, Oswaldo Cruz Foundation, Ceara (FIOCRUZ CE) | Carlos Leonardo de Aragao Araujo; Cecilia Leite Costa & Eduardo Ruback dos Santos on behalf of COVID-19 FIOCRUZ Genomic Network; Cleber Furtado Aksenen; Fabio Miyajima; Fernando Braga Stehling; Francisco Eder de Moura Lopes; Igor Oliveira Duarte; Jamille Maria Mendes Bezerra; Joaquim Cesar do Nascimento Sousa Junior; Pedro Miguel Carneiro Jeronimo; Suzana Porto Almeida; Thais Ferreira de Oliveira; Thais de Oliveira Costa; Ticiane Cavalcante de Souza; Veridiana Pessoa Miyajima |
| EPI_ISL_2344767, EPI_ISL_2344788 | HOSPITAL MUNICIPAL DE ITAPIRA | Instituto Butantan / ESALQ-Piracicaba | Antonio Jorge Martins; Claudia Renata dos Santos Barros; David Schlesinger; Debora Botequio Moretti; Dimas Tadeu Covas; Elaine Cristina Marqueze; Elaine Vieira Santos; Evandra Strazza Rodrigues; Heidge Fukumasu; Jayme Augusto de Souza-Neto; José Salvatore Leister Patané; Luiz Alcantara; Luiz Lehmann Coutinho; Maria Carolina Elias; Maurício Lacerda Nogueira; Rafael dos Santos Bezerra; Raul Machado Neto; Rejane Maria Tommasini Grotto; Ricardo Haddad; Sandra Coccuzzo Sampaio Vessoni; Simone Kashima; Svetoslav Nanev Slavov; Vincent Louis Viala |
| EPI_ISL_2344800 | HOSPITAL MUNICIPAL DR TABAJARA RAMOS | Instituto Butantan / ESALQ-Piracicaba | Antonio Jorge Martins; Claudia Renata dos Santos Barros; David Schlesinger; Debora Botequio Moretti; Dimas Tadeu Covas; Elaine Cristina Marqueze; Elaine Vieira Santos; Evandra Strazza Rodrigues; Heidge Fukumasu; Jayme Augusto de Souza-Neto; José Salvatore Leister Patané; Luiz Alcantara; Luiz Lehmann Coutinho; Maria Carolina Elias; Maurício Lacerda Nogueira; Rafael dos Santos Bezerra; Raul Machado Neto; Rejane Maria Tommasini Grotto; Ricardo Haddad; Sandra Coccuzzo Sampaio Vessoni; Simone Kashima; Svetoslav Nanev Slavov; Vincent Louis Viala |
| EPI_ISL_2344960, EPI_ISL_2344969 | HOSPITAL MUNICIPAL REYNALDO GUERRA CAJATI | Instituto Butantan / ESALQ-Piracicaba | Antonio Jorge Martins; Claudia Renata dos Santos Barros; David Schlesinger; Debora Botequio Moretti; Dimas Tadeu Covas; Elaine Cristina Marqueze; Elaine Vieira Santos; Evandra Strazza Rodrigues; Heidge Fukumasu; Jayme Augusto de Souza-Neto; José Salvatore Leister Patané; Luiz Alcantara; Luiz Lehmann Coutinho; Maria Carolina Elias; Maurício Lacerda Nogueira; Rafael dos Santos Bezerra; Raul Machado Neto; Rejane Maria Tommasini Grotto; Ricardo Haddad; Sandra Coccuzzo Sampaio Vessoni; Simone Kashima; Svetoslav Nanev Slavov; Vincent Louis Viala |
| EPI_ISL_3102513 | HOSPITAL OTOCLINICA | Analytical Competence Molecular Epidemiology Lab/ACME, Oswaldo Cruz Foundation, Ceara (FIOCRUZ CE) | Cleber Furtado Aksenen; Fabio Miyajima; Fernando Braga Stehling; Francisco Eder de Moura Lopes; Jamille Maria Mendes Bezerra; Joaquim César do Nascimento Sousa Junior; Pedro Miguel Carneiro Jeronimo; Suzana Porto Almeida e Lucas Delerino; Thais Ferreira de Oliveira; Thais de Oliveira Costa; Ticiane Cavalcante de Souza; Veridiana Pessoa Miyajima |
| EPI_ISL_2344898 | HOSPITAL REGIONAL DE ITAPETININGA | Instituto Butantan / ESALQ-Piracicaba | Antonio Jorge Martins; Claudia Renata dos Santos Barros; David Schlesinger; Debora Botequio Moretti; Dimas Tadeu Covas; Elaine Cristina Marqueze; Elaine Vieira Santos; Evandra Strazza Rodrigues; Heidge Fukumasu; Jayme Augusto de Souza-Neto; José Salvatore Leister Patané; Luiz Alcantara; Luiz Lehmann Coutinho; Maria Carolina Elias; Maurício Lacerda Nogueira; Rafael dos Santos Bezerra; Raul Machado Neto; Rejane Maria Tommasini Grotto; Ricardo Haddad; Sandra Coccuzzo Sampaio Vessoni; Simone Kashima; Svetoslav Nanev Slavov; Vincent Louis Viala |
| EPI_ISL_5603265 | HOSPITAL REGIONAL NORTE | Analytical Competence Molecular Epidemiology Lab/ACME, Oswaldo Cruz Foundation, Ceara (FIOCRUZ CE) | Carlos Leonardo de Aragao Araujo; Cecilia Leite Costa & Eduardo Ruback dos Santos on behalf of COVID-19 FIOCRUZ Genomic Network; Cleber Furtado Aksenen; Fabio Miyajima; Fernando Braga Stehling; Francisco Eder de Moura Lopes; Igor Oliveira Duarte; Jamille Maria Mendes Bezerra; Joaquim Cesar do Nascimento Sousa Junior; Pedro Miguel Carneiro Jeronimo; Suzana Porto Almeida; Thais Ferreira de Oliveira; Thais de Oliveira Costa; Ticiane Cavalcante de Souza; Veridiana Pessoa Miyajima |
| EPI_ISL_2502719 | HOSPITAL SAN VICENTE DE PAUL | Inciensa, Instituto Costarricense de InvestigaciOn y Enseñanza en NutriciOn y Salud | Adriana Godínez; Claudio Soto-Garita; Estela Cordero; Francisco Duarte; Hebleen Porras; Jose Luis Vargas; Joselyn Prado & Silvia Saenz-Garcia; Mariela Gutierrez; Melany CalderOn |
| EPI_ISL_2170983, EPI_ISL_2170984 | HOSPITAL UNIVERSITARIO SAO FRANCISCO BRAGANCA PAULISTA | Instituto Butantan / Mendelics | Antonio Jorge Martins; Bianca Cechetto Carlos. Mendelics: Bibiana Santos; Claudia Renata dos Santos Barros; Cíntia Bittar; David Schlesinger. Hemocentro Ribeirão Preto: Simone Kashima; Debora Botequio Moretti; Elaine Cristina Marqueze; Elaine Vieira dos Santos; Elisangela Chicaroni Mattos; Erika Freitas; Evandra Strazza Rodrigues; Felipe Allan da Silva da Costa; Flavia Aburjaile; Fábio Sossai Possebon; Guilherme Campos; Guilherme Targino Valente; Heidge Fukumasu. USP-Botucatu: Rejane Maria Tommasini Grotto; Helena Lage Ferreira; Instituto Butantan: Dimas Tadeu Covas; Jardelina de Souza Todao Bernardino; Jayme A. Souza-Neto; Jessika Cristina Chagas Lesbon; Jorge A. Petroli Marchesi; José Salvatore Leister Patané; João Paulo Kitajima; João Pessoa Araújo Jr.; Leila Sabrina Ullmann; Loyze Paola Oliveira de Lima; Luiz Aurelio de Campos Crispin. Centro de Genômica Funcional da ESALQ: Luiz Lehmann Coutinho; Luiz Carlos Junior de Alcantara; Lívia Sacchetto; Maisa C. Pereira Parra; Maria Carolina Elias; Marta Giovanetti; Marília Moraes; Maurício Lacerda Nogueira. Prefeitura de Sao Paulo: Melissa Palmieri.; Patricia Akemi Assato; Paula Rahal; Paulo Inacio da Costa; Rafael dos Santos Bezerra; Raquel de Lello Rocha Campos Cassano. NGS Soluções Genômicas: Pilar Drummond Sampaio Corrêa Mariani. FZEA-USP Pirassununga: Mirele Daiana Poleti; Raul Machado Neto; Ricardo Augusto Brassaloti; Ricardo Haddad; Rodrigo Tocantins Calado. FAMERP-SJRP: Cecília Artico Banho; Sandra Coccuzzo Sampaio; Svetoslav Nanev Slavov; Vagner Fonseca; Vincent Louis Viala |
| EPI_ISL_2100328, EPI_ISL_2100340 | HOSPITAL UNIVERSITARIO SON ESPASES | HOSPITAL UNIVERSITARIO SON ESPASES | Antonio Oliver; Carla López-Causapé; Pablo Fraile-Ribot; SeqCovid |
| EPI_ISL_2621859 | HPTU | Universidad Nacional de Colombia - Laboratorio Genómico One Health | Andres F. Cardona-Rios; Carlos Franco-Muñoz; Carolina Muñoz-Arango; Celeny Ortiz; Daniel O. Maldonado-Perez; Diego A. Álvarez-Díaz; Hector Alejandro Ruiz-Moreno; Idabely Betancur Ortiz; Jorge E. Osorio; Juan P. Hernandez-Ortiz; Karl A Ciuoderis; Katherine Laiton-Donato; Laura Silvana Perez; Lina M. Hurtado; Marcela Mercado-Reyes; Maria Angélica Maya; Maria Stella López; Rita Almanza Payares; Sandra Ines Cano; Simón Villegas Velásquez |
| EPI_ISL_2081119 | Hackensack Medical Center | New York Genome Center | Andre Corvelo; Barry Kreiswirth; David Perlin; Dayna M. Oschwald; Jose Mediavilla; Kaelea Composto; Kar Chow; Liang Chen; Marcus Cunningham; Michael Zody; Samantha Fennessey; Tom Maniatis |
| EPI_ISL_2006656 | Health Services Laboratories | Wellcome Sanger Institute for the COVID-19 Genomics UK (COG-UK) Consortium | Cordelia Langford; David K. Jackson; Dominic Kwiatkowski; Ewan Harrison; Health Services Laboratories and Alex Alderton; Ian Johnston; Jeffrey Barrett; John Sillitoe on behalf of the Wellcome Sanger Institute COVID-19 Surveillance Team; Roberto Amato; Sonia Goncalves |
| EPI_ISL_2376215 | Heilig hart Lier | Imeda Hospital | Dagmar Obbels; Hanne Valgaeren; Johan Frans |
| EPI_ISL_1990577, EPI_ISL_1990589, EPI_ISL_1990612, EPI_ISL_1990624, EPI_ISL_1990644, EPI_ISL_1990668, EPI_ISL_1990707, EPI_ISL_1990717, EPI_ISL_1990732, EPI_ISL_1990737, EPI_ISL_1990757, EPI_ISL_1990781, EPI_ISL_1990804, EPI_ISL_1990814, EPI_ISL_1990827, EPI_ISL_1990883, EPI_ISL_1990887, EPI_ISL_1990901, EPI_ISL_1990903, EPI_ISL_1990916, EPI_ISL_1990920, EPI_ISL_1990959, EPI_ISL_1990990, EPI_ISL_1990993, EPI_ISL_1991018, EPI_ISL_1991027, EPI_ISL_1991034, EPI_ISL_1991048, EPI_ISL_1991058, EPI_ISL_1991170, EPI_ISL_1991374, EPI_ISL_1991382, EPI_ISL_1991389, EPI_ISL_1991394, EPI_ISL_1991406, EPI_ISL_1991421, EPI_ISL_1991422, EPI_ISL_1991467, EPI_ISL_1991471, EPI_ISL_1991480, EPI_ISL_1991493, EPI_ISL_1991524, EPI_ISL_1991526, EPI_ISL_1991540, EPI_ISL_1991563, EPI_ISL_1991564, EPI_ISL_1991580, EPI_ISL_1991601, EPI_ISL_1991605, EPI_ISL_1991667, EPI_ISL_1991675, EPI_ISL_1991677, EPI_ISL_1991702, EPI_ISL_1991709, EPI_ISL_1991711, EPI_ISL_1991721, EPI_ISL_1991725, EPI_ISL_1991729, EPI_ISL_1991803, EPI_ISL_2009667, EPI_ISL_2009725, EPI_ISL_2009729, EPI_ISL_2009737, EPI_ISL_2009745, EPI_ISL_2009776, EPI_ISL_2009777, EPI_ISL_2009807, EPI_ISL_2009818, EPI_ISL_2009828, EPI_ISL_2009832, EPI_ISL_2009875, EPI_ISL_2009880, EPI_ISL_2009889, EPI_ISL_2009898, EPI_ISL_2009900, EPI_ISL_2009914, EPI_ISL_2009921, EPI_ISL_2009929, EPI_ISL_2009932, EPI_ISL_2009951, EPI_ISL_2009959, EPI_ISL_2010001, EPI_ISL_2010002, EPI_ISL_2010010, EPI_ISL_2010013, EPI_ISL_2010031, EPI_ISL_2010037, EPI_ISL_2010051, EPI_ISL_2010081, EPI_ISL_2010084, EPI_ISL_2010087, EPI_ISL_2010096, EPI_ISL_2010097, EPI_ISL_2010115, EPI_ISL_2010122, EPI_ISL_2010130, EPI_ISL_2010132, EPI_ISL_2010149, EPI_ISL_2010185, EPI_ISL_2010206, EPI_ISL_2010250, EPI_ISL_2010274, EPI_ISL_2010282, EPI_ISL_2010312, EPI_ISL_2010351, EPI_ISL_2010395, EPI_ISL_2010400, EPI_ISL_2010420, EPI_ISL_2010422, EPI_ISL_2010428, EPI_ISL_2010442, EPI_ISL_2010454, EPI_ISL_2010462, EPI_ISL_2010485, EPI_ISL_2010502, EPI_ISL_2010515, EPI_ISL_2010521, EPI_ISL_2010534, EPI_ISL_2010580, EPI_ISL_2010584, EPI_ISL_2010620, EPI_ISL_2010622, EPI_ISL_2010637, EPI_ISL_2010646, EPI_ISL_2010648, EPI_ISL_2010653, EPI_ISL_2010671, EPI_ISL_2010741, EPI_ISL_2010753, EPI_ISL_2010754, EPI_ISL_2010760, EPI_ISL_2010807, EPI_ISL_2010868, EPI_ISL_2034881, EPI_ISL_2034956, EPI_ISL_2035267, EPI_ISL_2035280, EPI_ISL_2035282, EPI_ISL_2035296, EPI_ISL_2035298, EPI_ISL_2097415, EPI_ISL_2097433, EPI_ISL_2097540, EPI_ISL_2097544, EPI_ISL_2097549, EPI_ISL_2097573, EPI_ISL_2097593, EPI_ISL_2097594, EPI_ISL_2097708, EPI_ISL_2097747, EPI_ISL_2097754, EPI_ISL_2097755, EPI_ISL_2097757, EPI_ISL_2097759, EPI_ISL_2097761, EPI_ISL_2097762, EPI_ISL_2097779, EPI_ISL_2097834, EPI_ISL_2097850, EPI_ISL_2097862, EPI_ISL_2097874, EPI_ISL_2097886, EPI_ISL_2097895, EPI_ISL_2097903, EPI_ISL_2097924, EPI_ISL_2097950, EPI_ISL_2097957, EPI_ISL_2097969, EPI_ISL_2097973, EPI_ISL_2098014, EPI_ISL_2098016, EPI_ISL_2098037, EPI_ISL_2098070, EPI_ISL_2098075, EPI_ISL_2098086, EPI_ISL_2098097, EPI_ISL_2098105, EPI_ISL_2098148, EPI_ISL_2098205, EPI_ISL_2132610, EPI_ISL_2132611, EPI_ISL_4346073, EPI_ISL_4346790, EPI_ISL_4347005 | | | |
| see above | Helix/Illumina | Centers for Disease Control and Prevention Division of Viral Diseases, Pathogen Discovery | Adrian Paskey; Alexandre Bolze; Ary Ascencio; Benjamin Rambo-Martin; Brad Sickler; Charlotte Rivera-Garcia; Christine Tran; Christopher Gulvick; Chrstine Tran; Clinton Paden; Clinton R. Paden; Dakota Howard; Darlene Wagner; David Becker; Dhwani Batra; Duncan MacCannell; Efren Sandoval; Eileen De Feo; Eileen de Feo; Elizabeth Cirulli; Eric Allen; Geraint Levan; James Lu; Jan Antico; Jason Caravas; Jason Nguyen; Jimmy Ramirez; Jingtao Liu; Kara Moser; Kelly Barrett; Kelly Schiabor Barrett; Kim Gietzen; Kristine Lacek; Magnus Isaksson; Marc Laurent; Matthew Schmerer; Matthew Tolentino; Nicole L. Washington; Nicole Washington; Peter Cook; Peter W. Cook; Phil Febbo; Ryan Cho; Scott Sammons; Shannon Wickline; Shatavia Morrison; Sherry Wang; Simon White; Tyler Cassens; William Lee; Yvette Unoarumhi |
| EPI_ISL_5801008 | Hosp De Transplant Do Est De Sp Euryclides De Jesus Zerbini | Instituto Butantan | Antonio Jorge Martins; Claudia Renata dos Santos Barros; David Schlesinger; Debora Botequio Moretti; Dimas Tadeu Covas; Elaine Cristina Marqueze; Elaine Vieira Santos; Evandra Strazza Rodrigues; Heidge Fukumasu; Jayme Augusto de Souza-Neto; José Salvatore Leister Patané; Luiz Alcantara; Luiz Lehmann Coutinho; Maria Carolina Elias; Maurício Lacerda Nogueira; Rafael dos Santos Bezerra; Raul Machado Neto; Rejane Maria Tommasini Grotto; Ricardo Haddad; Sandra Coccuzzo Sampaio Vessoni; Simone Kashima; Svetoslav Nanev Slavov; Vincent Louis Viala |
| EPI_ISL_5800886, EPI_ISL_5800890, EPI_ISL_5800896, EPI_ISL_5800937, EPI_ISL_5800938 | Hosp E Maternidade Municipal N Sra Monte Serrat | Instituto Butantan | Antonio Jorge Martins; Claudia Renata dos Santos Barros; David Schlesinger; Debora Botequio Moretti; Dimas Tadeu Covas; Elaine Cristina Marqueze; Elaine Vieira Santos; Evandra Strazza Rodrigues; Heidge Fukumasu; Jayme Augusto de Souza-Neto; José Salvatore Leister Patané; Luiz Alcantara; Luiz Lehmann Coutinho; Maria Carolina Elias; Maurício Lacerda Nogueira; Rafael dos Santos Bezerra; Raul Machado Neto; Rejane Maria Tommasini Grotto; Ricardo Haddad; Sandra Coccuzzo Sampaio Vessoni; Simone Kashima; Svetoslav Nanev Slavov; Vincent Louis Viala |
| EPI_ISL_5801028, EPI_ISL_5801029, EPI_ISL_5801031, EPI_ISL_5801032, EPI_ISL_5801033, EPI_ISL_5801034, EPI_ISL_5801035, EPI_ISL_5801036 | | | |
| see above | Hosp Mun Josanias Castanha Braga | Instituto Butantan | Antonio Jorge Martins; Claudia Renata dos Santos Barros; David Schlesinger; Debora Botequio Moretti; Dimas Tadeu Covas; Elaine Cristina Marqueze; Elaine Vieira Santos; Evandra Strazza Rodrigues; Heidge Fukumasu; Jayme Augusto de Souza-Neto; José Salvatore Leister Patané; Luiz Alcantara; Luiz Lehmann Coutinho; Maria Carolina Elias; Maurício Lacerda Nogueira; Rafael dos Santos Bezerra; Raul Machado Neto; Rejane Maria Tommasini Grotto; Ricardo Haddad; Sandra Coccuzzo Sampaio Vessoni; Simone Kashima; Svetoslav Nanev Slavov; Vincent Louis Viala |
| EPI_ISL_3149730 | Hosp. 4 de Junio Dr. Ramón Carrillo | Laboratorio de Biología Molecular, Instituto de Medicina Regional on behalf of 'Proyecto Argentino Interinstitucional de genomica de SARS-CoV-2' (PAIS Consortium) | Andrea Carolina Piedrabuena; Bettina Brusés; Diana Pérez Quintana; Gabriela Jurasek; Griselda Oria; Horacio Lucero.; Javier Mussin; Laura Formichelli; Mayra Mendoza; Melina Lorenzini Campos; Raúl Maximiliano Acevedo |
| EPI_ISL_2029664, EPI_ISL_2029772, EPI_ISL_2029808, EPI_ISL_2029930 | Hospital | National Reference Center for Viruses of Respiratory Infections, Institut Pasteur, Paris | Angela Brisebarre; Camille Capel; Christophe Malabat; Corinne Maufrais; Etienne Simon-Lorière; Frédéric Lemoine; Louise Lefrançois; Marion Barbet; Maud Vanpeene; Méline Bizard; Nabil Gastli; Sylvie Behillil; Sylvie Van der Werf; Vincent Enouf |
| EPI_ISL_5800947, EPI_ISL_5800948, EPI_ISL_5800949, EPI_ISL_5800950, EPI_ISL_5800951, EPI_ISL_5800969, EPI_ISL_5800971, EPI_ISL_5800972, EPI_ISL_5800974, EPI_ISL_5800975, EPI_ISL_5800977, EPI_ISL_5800990, EPI_ISL_5800992, EPI_ISL_5801000, EPI_ISL_5801010, EPI_ISL_5801013, EPI_ISL_5801016, EPI_ISL_5801027, EPI_ISL_5801030 | | | |
| see above | Hospital De Campanha Covid 19 Municipio De Taubate | Instituto Butantan | Antonio Jorge Martins; Claudia Renata dos Santos Barros; David Schlesinger; Debora Botequio Moretti; Dimas Tadeu Covas; Elaine Cristina Marqueze; Elaine Vieira Santos; Evandra Strazza Rodrigues; Heidge Fukumasu; Jayme Augusto de Souza-Neto; José Salvatore Leister Patané; Luiz Alcantara; Luiz Lehmann Coutinho; Maria Carolina Elias; Maurício Lacerda Nogueira; Rafael dos Santos Bezerra; Raul Machado Neto; Rejane Maria Tommasini Grotto; Ricardo Haddad; Sandra Coccuzzo Sampaio Vessoni; Simone Kashima; Svetoslav Nanev Slavov; Vincent Louis Viala |

| | | | |
|---|---|---|---|
| EPI_ISL_5800795, EPI_ISL_5800796 | Hospital De Misericordia Altinopolis | Instituto Butantan | Antonio Jorge Martins; Claudia Renata dos Santos Barros; David Schlesinger; Debora Botequio Moretti; Dimas Tadeu Covas; Elaine Cristina Marqueze; Elaine Vieira Santos; Evandra Strazza Rodrigues; Heidge Fukumasu; Jayme Augusto de Souza-Neto; José Salvatore Leister Patané; Luiz Alcantara; Luiz Lehmann Coutinho; Maria Carolina Elias; Maurício Lacerda Nogueira; Rafael dos Santos Bezerra; Raul Machado Neto; Rejane Maria Tommasini Grotto; Ricardo Haddad; Sandra Coccuzzo Sampaio Vessoni; Simone Kashima; Svetoslav Nanev Slavov; Vincent Louis Viala |
| EPI_ISL_5800980 | Hospital E Maternidade Nossa Senhora Da Ajuda | Instituto Butantan | Antonio Jorge Martins; Claudia Renata dos Santos Barros; David Schlesinger; Debora Botequio Moretti; Dimas Tadeu Covas; Elaine Cristina Marqueze; Elaine Vieira Santos; Evandra Strazza Rodrigues; Heidge Fukumasu; Jayme Augusto de Souza-Neto; José Salvatore Leister Patané; Luiz Alcantara; Luiz Lehmann Coutinho; Maria Carolina Elias; Maurício Lacerda Nogueira; Rafael dos Santos Bezerra; Raul Machado Neto; Rejane Maria Tommasini Grotto; Ricardo Haddad; Sandra Coccuzzo Sampaio Vessoni; Simone Kashima; Svetoslav Nanev Slavov; Vincent Louis Viala |
| EPI_ISL_5800874, EPI_ISL_5800875, EPI_ISL_5800876, EPI_ISL_5800877, EPI_ISL_5800892, EPI_ISL_5800899, EPI_ISL_5800932, EPI_ISL_5800934, EPI_ISL_5800935, EPI_ISL_5800936, EPI_ISL_5800958 | | | |
| see above | Hospital E Pronto Socorro Portinari | Instituto Butantan | Antonio Jorge Martins; Claudia Renata dos Santos Barros; David Schlesinger; Debora Botequio Moretti; Dimas Tadeu Covas; Elaine Cristina Marqueze; Elaine Vieira Santos; Evandra Strazza Rodrigues; Heidge Fukumasu; Jayme Augusto de Souza-Neto; José Salvatore Leister Patané; Luiz Alcantara; Luiz Lehmann Coutinho; Maria Carolina Elias; Maurício Lacerda Nogueira; Rafael dos Santos Bezerra; Raul Machado Neto; Rejane Maria Tommasini Grotto; Ricardo Haddad; Sandra Coccuzzo Sampaio Vessoni; Simone Kashima; Svetoslav Nanev Slavov; Vincent Louis Viala |
| EPI_ISL_5801038 | Hospital Escola Da Universidade De Taubate | Instituto Butantan | Antonio Jorge Martins; Claudia Renata dos Santos Barros; David Schlesinger; Debora Botequio Moretti; Dimas Tadeu Covas; Elaine Cristina Marqueze; Elaine Vieira Santos; Evandra Strazza Rodrigues; Heidge Fukumasu; Jayme Augusto de Souza-Neto; José Salvatore Leister Patané; Luiz Alcantara; Luiz Lehmann Coutinho; Maria Carolina Elias; Maurício Lacerda Nogueira; Rafael dos Santos Bezerra; Raul Machado Neto; Rejane Maria Tommasini Grotto; Ricardo Haddad; Sandra Coccuzzo Sampaio Vessoni; Simone Kashima; Svetoslav Nanev Slavov; Vincent Louis Viala |
| EPI_ISL_2016621, EPI_ISL_2016622, EPI_ISL_2016623 | Hospital Francesc de Borja (Gandia) | SeqCOVID-SPAIN consortium/IBV(CSIC) | Inmaculada Vidal Catalá; Isabel García Nava and SeqCOVID-SPAIN consortium; Mª Rosa Navarro Cots; Nieves Orta Mira |
| EPI_ISL_1914327, EPI_ISL_3505904, EPI_ISL_3505905, EPI_ISL_3505906, EPI_ISL_3505907, EPI_ISL_3505908, EPI_ISL_3505909, EPI_ISL_3505911, EPI_ISL_3505912, EPI_ISL_3505913, EPI_ISL_3505914, EPI_ISL_3505915 | | | |
| see above | Hospital General Universitario Gregorio Marañón | Hospital General Universitario Gregorio Marañón | Cristina Rodriguez-Grande; Darío García de Viedma; Julia Suárez; Laura Pérez-Lago; Marta Herranz Martin; Patricia Muñoz; Pedro Sola Campoy; Pilar Catalán; Sergio Buenestado Serrano; Victor Manuel de la Cueva |
| EPI_ISL_2271517, EPI_ISL_2374104 | Hospital General Universitario de Alicante - Instituto de Investigación Sanitaria y Biomédica de Alicante | SeqCOVID-SPAIN consortium/IBV(CSIC) | Carmen Molina Pardines and SeqCOVID-SPAIN consortium; Maripaz Ventero Martín |
| EPI_ISL_4405067 | Hospital General de Agudos Dr. Cosme Argerich | Área de Secuenciación del Laboratorio de Virología del Hospital de Niños Dr. Ricardo Gutierrez on behalf of 'Proyecto Argentino Interinstitucional de genomica de SARS-CoV-2' (PAIS Consortium) | Acuña; Andrea Fernández; D; Florencia Funez; Florencia Rodríguez; Goya; Jésica Galeano; Karina Polanski; LE; Lusso; MI; MS; Marcia Pozzati; Nabaes Jodar; Natale; S; Valinotto; Viegas, M. |
| EPI_ISL_2614581 | Hospital Geral do Grajau | Instituto Adolfo Lutz, Interdiciplinary Procedures Center, Strategic Laboratory | Caio Vinicius Dias Lopes; Claudia Regina Gonçalves; Claudio Tavares Sacchi; Erica Valessa Ramos Gomes; Karoline Rodrigues Campos; Leonardo Jose Tadeu de Araujo |
| EPI_ISL_2551532 | Hospital Infantnil Darcy Vargas | Laboratório de Virologia Clínica e Molecular | Alex Fiorini; Ana Paula Salles Fernandes; Bruna Larotonda Telezynski; Danielle Bruna Leal Oliveira; Edison Luiz Durigon; Erick Gustavo Dorlass; Flavio Fonseca e Santuza Teixeira; Guilherme Pereira Scagion; Helena Perez Coelho; Hugo Sato; Karine Lima Lourenço; Luciano Matsumiya Thomazelli; Renata Peixoto; Rubens Daniel Miserani Magalhães; Tatiana Ometto |
| EPI_ISL_3033050, EPI_ISL_3033051 | Hospital Jaime Ferre - SAMCO Rafaela | Grupo de Genómica y Bioinformática del Instituto de Investigación de la Cadena Láctea CONICET-INTA on behalf of 'Proyecto Argentino Interinstitucional de genomica de SARS-CoV-2' (PAIS Consortium) | AF; Amadio; C; Eberhardt; Irazoqui; Isaia; JF; JM; MF; Pandolfi; Quaranta; V |
| EPI_ISL_5800631, EPI_ISL_5800689 | Hospital Municipal De Itapira | Instituto Butantan | Antonio Jorge Martins; Claudia Renata dos Santos Barros; David Schlesinger; Debora Botequio Moretti; Dimas Tadeu Covas; Elaine Cristina Marqueze; Elaine Vieira Santos; Evandra Strazza Rodrigues; Heidge Fukumasu; Jayme Augusto de Souza-Neto; José Salvatore Leister Patané; Luiz Alcantara; Luiz Lehmann Coutinho; Maria Carolina Elias; Maurício Lacerda Nogueira; Rafael dos Santos Bezerra; Raul Machado Neto; Rejane Maria Tommasini Grotto; Ricardo Haddad; Sandra Coccuzzo Sampaio Vessoni; Simone Kashima; Svetoslav Nanev Slavov; Vincent Louis Viala |
| EPI_ISL_5800725 | Hospital Municipal Dr Tabajara Ramos | Instituto Butantan | Antonio Jorge Martins; Claudia Renata dos Santos Barros; David Schlesinger; Debora Botequio Moretti; Dimas Tadeu Covas; Elaine Cristina Marqueze; Elaine Vieira Santos; Evandra Strazza Rodrigues; Heidge Fukumasu; Jayme Augusto de Souza-Neto; José Salvatore Leister Patané; Luiz Alcantara; Luiz Lehmann Coutinho; Maria Carolina Elias; Maurício Lacerda Nogueira; Rafael dos Santos Bezerra; Raul Machado Neto; Rejane Maria Tommasini Grotto; Ricardo Haddad; Sandra Coccuzzo Sampaio Vessoni; Simone Kashima; Svetoslav Nanev Slavov; Vincent Louis Viala |
| EPI_ISL_5800954, EPI_ISL_5800965 | Hospital Municipal Reynaldo Guerra Cajati | Instituto Butantan | Antonio Jorge Martins; Claudia Renata dos Santos Barros; David Schlesinger; Debora Botequio Moretti; Dimas Tadeu Covas; Elaine Cristina Marqueze; Elaine Vieira Santos; Evandra Strazza Rodrigues; Heidge Fukumasu; Jayme Augusto de Souza-Neto; José Salvatore Leister Patané; Luiz Alcantara; Luiz Lehmann Coutinho; Maria Carolina Elias; Maurício Lacerda Nogueira; Rafael dos Santos Bezerra; Raul Machado Neto; Rejane Maria Tommasini Grotto; Ricardo Haddad; Sandra Coccuzzo Sampaio Vessoni; Simone Kashima; Svetoslav Nanev Slavov; Vincent Louis Viala |
| EPI_ISL_5800870 | Hospital Regional De Itapetininga | Instituto Butantan | Antonio Jorge Martins; Claudia Renata dos Santos Barros; David Schlesinger; Debora Botequio Moretti; Dimas Tadeu Covas; Elaine Cristina Marqueze; Elaine Vieira Santos; Evandra Strazza Rodrigues; Heidge Fukumasu; Jayme Augusto de Souza-Neto; José Salvatore Leister Patané; Luiz Alcantara; Luiz Lehmann Coutinho; Maria Carolina Elias; Maurício Lacerda Nogueira; Rafael dos Santos Bezerra; Raul Machado Neto; Rejane Maria Tommasini Grotto; Ricardo Haddad; Sandra Coccuzzo Sampaio Vessoni; Simone Kashima; Svetoslav Nanev Slavov; Vincent Louis Viala |
| EPI_ISL_3230021 | Hospital Señor del Milagro | Instituto de Patologia Vegetal (CIAP-INTA) on behalf of 'Proyecto Argentino Interinstitucional de genomica de SARS-CoV-2' (PAIS Consortium) | Debat, HJ.; FD; Fernanda Aguero; Fernández; Lavaque Esteban; Marquez, N.; Pamela Cajal; Raskovsky Viviana; Veronica Lesser |
| EPI_ISL_5800564, EPI_ISL_5800583 | Hospital Universitario Sao Francisco Braganca Paulista | Instituto Butantan | Antonio Jorge Martins; Claudia Renata dos Santos Barros; David Schlesinger; Debora Botequio Moretti; Dimas Tadeu Covas; Elaine Cristina Marqueze; Elaine Vieira Santos; Evandra Strazza Rodrigues; Heidge Fukumasu; Jayme Augusto de Souza-Neto; José Salvatore Leister Patané; Luiz Alcantara; Luiz Lehmann Coutinho; Maria Carolina Elias; Maurício Lacerda Nogueira; Rafael dos Santos Bezerra; Raul Machado Neto; Rejane Maria Tommasini Grotto; Ricardo Haddad; Sandra Coccuzzo Sampaio Vessoni; Simone Kashima; Svetoslav Nanev Slavov; Vincent Louis Viala |
| EPI_ISL_2614565 | Hospital de Base de Sao Jose do Rio Preto | Instituto Adolfo Lutz, Interdiciplinary Procedures Center, Strategic Laboratory | Caio Vinicius Dias Lopes; Claudia Regina Gonçalves; Claudio Tavares Sacchi; Erica Valessa Ramos Gomes; Karoline Rodrigues Campos; Leonardo Jose Tadeu de Araujo |
| EPI_ISL_2614598 | Hospital de Campanha Covid-19 Mairipora | Instituto Adolfo Lutz, Interdiciplinary Procedures Center, Strategic Laboratory | Caio Vinicius Dias Lopes; Claudia Regina Gonçalves; Claudio Tavares Sacchi; Erica Valessa Ramos Gomes; Karoline Rodrigues Campos; Leonardo Jose Tadeu de Araujo |
| EPI_ISL_2597608, EPI_ISL_2597615, EPI_ISL_3022946, EPI_ISL_3022959, EPI_ISL_3023115, EPI_ISL_3023120 | Hospital of the University of Pennsylvania Molecular Pathology Lab | Bushman Lab - University of Pennsylvania | Aoife M. Roche; Arupa Ganguly; Ayannah S. Fitzgerald; Brendan Kelly; Jevon Graham-Wooten; John K. Everett; Kyle Rodino; Layla A. Khatib; Mike Feldman; Ronald G. Collman and Frederic Bushan; Samantha A. Whiteside; Scott Sherrill-Mix; Shantan Reddy; Young Hwang |
| EPI_ISL_2223003, EPI_ISL_2223017, EPI_ISL_2224440, EPI_ISL_2224487, EPI_ISL_2224488, EPI_ISL_2225125 | Houston Methodist Hospital | Houston Methodist Hospital | Ilya J. Finkelstein; James J. Davis; Jessica Cambric; Jimmy Gollihar; Kristina Reppond; Layne Pruitt; Madison N. Shyer; Marcus Nguyen; Matthew Ojeda Saavedra; Paul A. Christensen; Prasanti Yerramilli; Randall J. Olsen; Robert Olson; Ryan Gadd; S. Wesley Long; Sishir Subedi; and James M. Musser |
| EPI_ISL_2151548, EPI_ISL_2151562, EPI_ISL_2151583 | Hôpital Avicenne | Department of Virology, Henri Mondor University Hospital, Assistance Publique Hôpitaux de Paris, Université Paris-Est Créteil, INSERM U955 | Alexandre Soulier; Christophe Rodriguez; Elisabeth Trawinski; Guillaume Gricourt; Jean-Michel Pawlotsky; Melissa N'Debi; Slim Fourati; Vanessa Demontant |
| EPI_ISL_1937747 | IA State Hygienic Laboratory | Centers for Disease Control and Prevention Division of Viral Diseases, Pathogen Discovery | Alison Laufer Halpin; Ben L. Rambo-Martin; Clinton R. Paden; Dakota Howard; Darlene Wagner; Dave Wentworth; Dhwani Batra; Jasmine Padilla; Justin Lee; Katie Dillon; Krista Queen; Kristen Knipe; Kristine Lacek; Mark Burroughs; Matthew Schmerer; Mili Sheth; Peter Cook; Sam Shepard; Sarah Nobles; Shoshona Le; Suxiang Tong; Vivien Dugan; Yvette Unoarumhi |
| EPI_ISL_4198475, EPI_ISL_4198476, EPI_ISL_4198477 | IDIME | Gencore - Universidad de los Andes | Cristian Barrera; David González; Gabriela Ariza; Luisa Sacristan; Marcela Guevara; Marcela Mercado; Silvia Restrepo |
| EPI_ISL_2444813, EPI_ISL_2444828, EPI_ISL_2444829, EPI_ISL_4071898 | IICS-UNA | IICS-UNA | Adriana Valenzuela; Alejandra Rojas; Chyntia Diaz; Eva Nara; Fatima Cardozo; Florencia del Puerto; Joel Ortiz; Jonas Fernandez; Laura Franco; Laura Mendoza; Leticia Rojas; Magaly Martinez; Maria Eugenia Galeano. |
| EPI_ISL_1937768, EPI_ISL_1937771, EPI_ISL_1937777, EPI_ISL_1937781, EPI_ISL_1989977 | IL Dept. of Public Health Springfield Laboratory | Centers for Disease Control and Prevention Division of Viral Diseases, Pathogen Discovery | Alison Laufer Halpin; Ben L. Rambo-Martin; Clinton R. Paden; Dakota Howard; Darlene Wagner; Dave Wentworth; Dhwani Batra; Jasmine Padilla; Justin Lee; Katie Dillon; Krista Queen; Kristen Knipe; Kristine Lacek; Mark Burroughs; Matthew Schmerer; Mili Sheth; Peter Cook; Sam Shepard; Sarah Nobles; Shoshona Le; Suxiang Tong; Vivien Dugan; Yvette Unoarumhi |
| EPI_ISL_1841017, EPI_ISL_1941597, EPI_ISL_1941605, EPI_ISL_1994874, EPI_ISL_1994876, EPI_ISL_1994879, EPI_ISL_1994881, EPI_ISL_1994904, EPI_ISL_1994906, EPI_ISL_1994907, EPI_ISL_1994908, EPI_ISL_1994909, EPI_ISL_1994920, EPI_ISL_2097176 | | | |
| see above | IN State Department of Health Laboratory Services | IN State Department of Health Laboratory Services | Brian Pope; Cassandra Campion; Jamie Yeadon; Kyle Brownlee; Lixia Liu; Mark Glazier; Melissa Hindenlang |
| EPI_ISL_2483720 | INMI Lazzaro Spallanzani IRCCS | INMI Lazzaro Spallanzani IRCCS | A Di Caro; B Bartolini; CEM Gruber; E Giombini; F Messina; F Santini; G Bonfiglio; M Rueca; MR Capobianchi; O Butera |
| EPI_ISL_1701391 | INT Fondazione Pascale | INT Fondazione Pascale | INT Fondazione Pascale |

| | | | |
|---|---|---|---|
| EPI_ISL_2545033, EPI_ISL_2577012, EPI_ISL_2577015, EPI_ISL_2577016, EPI_ISL_2577017, EPI_ISL_2577018, EPI_ISL_2577024, EPI_ISL_2716745, EPI_ISL_2716750, EPI_ISL_2716752 | | | |
| see above | IRCCS San Gallicano Dermatological Institute | IRCCS Regina Elena National Cancer Institute | Aldo Morrone; Alice Massacci; Andrea Cazzani; Arianna Mastrofrancesco; Eleonora Sperandio; Fabrizio Ensoli; Francesca De Nicola; Francesca Sivori; Francisco Obregon; Frauke Goeman; Fulvia Pimpinelli; Gennaro Ciliberto; Giovanni Blandino; Giulia Orlandi; Grazia Prignano; Ilaria Cavallo; Ilaria Celesti; Ludovica Ciuffreda; Matteo Pallocca; Maurizio Fanciulli; Sabrina Strano; Sara Donzelli |
| EPI_ISL_2628303 | IVIC | Laboratorio de Virología Molecular | Carmen L Loureiro; CoViVen Group; Domingo J Garzaro; Esmeralda Vizzi; Flor H Pujol; Héctor R Rangel; José Luis Zambrano; Lieska Rodríguez; Mariana Hidalgo; Pierina D´Angelo; Rossana C Jaspe; Víctor Alarcón; Yoneira Sulbaran; Zoila Moros |
| EPI_ISL_2427309, EPI_ISL_2427310, EPI_ISL_2427311, EPI_ISL_2598284 | IVY3 Central Lab, Vanderbilt University Medical Center | Lauring Lab, University of Michigan, Department of Microbiology and Immunology | Gilbert; Valesano |
| EPI_ISL_2786785, EPI_ISL_2786786 | Ibaraki Prefectural Institute of Public Health | Ibaraki Prefectural Institute of Public Health | Keiko Goto; Kentaro Itokawa; Makoto Kuroda; Masanori Hashino; Rina Tanaka; Tsuyoshi Sekizuka |
| EPI_ISL_2295870, EPI_ISL_2295874 | Illinois Department of Public Health | Gagnon Lab, Southern Illinois University | Keith Gagnon |
| EPI_ISL_1809660, EPI_ISL_1809677, EPI_ISL_1964979, EPI_ISL_1964984, EPI_ISL_1964987, EPI_ISL_1964998, EPI_ISL_1965031, EPI_ISL_1965051, EPI_ISL_1965054, EPI_ISL_1965065, EPI_ISL_1965068, EPI_ISL_1965071, EPI_ISL_1965072, EPI_ISL_1965073, EPI_ISL_1965081, EPI_ISL_1965090, EPI_ISL_1965092, EPI_ISL_1965100, EPI_ISL_1965116, EPI_ISL_2140027, EPI_ISL_2140031 | | | |
| see above | Illinois Department of Public Health | Illinois Department of Public Health - Chicago Lab | Ira Heimler; Joel Price; Vineet K. Dhiman |
| EPI_ISL_1825749, EPI_ISL_1825750, EPI_ISL_1825766, EPI_ISL_1825767, EPI_ISL_1825771, EPI_ISL_1825791, EPI_ISL_1825792, EPI_ISL_1825795, EPI_ISL_1825796, EPI_ISL_1825797, EPI_ISL_1825800, EPI_ISL_1825802, EPI_ISL_1825804, EPI_ISL_1825806, EPI_ISL_1825807, EPI_ISL_1825809, EPI_ISL_1825810, EPI_ISL_1825811, EPI_ISL_1825812, EPI_ISL_1825813, EPI_ISL_1825837, EPI_ISL_1825839, EPI_ISL_1825842, EPI_ISL_1825844, EPI_ISL_1825846, EPI_ISL_1825847, EPI_ISL_1825850, EPI_ISL_1825851, EPI_ISL_1825852, EPI_ISL_1825854, EPI_ISL_1825861, EPI_ISL_1825865, EPI_ISL_1938507, EPI_ISL_1938510, EPI_ISL_1938540, EPI_ISL_1938549, EPI_ISL_1938554, EPI_ISL_1938558, EPI_ISL_1938559, EPI_ISL_1938562, EPI_ISL_1938567, EPI_ISL_1938571, EPI_ISL_1938572, EPI_ISL_1938573, EPI_ISL_1938575, EPI_ISL_1938576, EPI_ISL_1938577, EPI_ISL_1938578, EPI_ISL_1938579, EPI_ISL_1938581, EPI_ISL_1938582, EPI_ISL_1938583, EPI_ISL_1938584, EPI_ISL_1938585, EPI_ISL_1938586, EPI_ISL_1938590, EPI_ISL_1938591, EPI_ISL_1938606, EPI_ISL_1938611, EPI_ISL_1938636, EPI_ISL_1938637, EPI_ISL_2000784, EPI_ISL_2000785, EPI_ISL_2000805, EPI_ISL_2086540, EPI_ISL_2086543, EPI_ISL_2086545, EPI_ISL_2086549, EPI_ISL_2086560, EPI_ISL_2086561, EPI_ISL_2086562, EPI_ISL_2135058, EPI_ISL_2135065, EPI_ISL_2135070, EPI_ISL_2135073, EPI_ISL_2135075, EPI_ISL_2135118, EPI_ISL_2135119, EPI_ISL_2203505, EPI_ISL_2203506, EPI_ISL_2203511, EPI_ISL_2203512, EPI_ISL_2203513, EPI_ISL_2203518, EPI_ISL_2203519, EPI_ISL_2203521, EPI_ISL_2203524, EPI_ISL_2203525, EPI_ISL_2203528, EPI_ISL_2203530, EPI_ISL_2203531, EPI_ISL_2203533, EPI_ISL_2339393 | | | |
| see above | Illinois Department of Public Health - Springfield Lab | Illinois Department of Public Health - Springfield Lab | Bryan Sim; Gordon McCall |
| EPI_ISL_2011492, EPI_ISL_2011496, EPI_ISL_2011497, EPI_ISL_2011499, EPI_ISL_2011500, EPI_ISL_2011524, EPI_ISL_2011527, EPI_ISL_2011535, EPI_ISL_2011542, EPI_ISL_2011543, EPI_ISL_2011546, EPI_ISL_2011553, EPI_ISL_2011557, EPI_ISL_2011560, EPI_ISL_2011561, EPI_ISL_2011564, EPI_ISL_2011567, EPI_ISL_2011623, EPI_ISL_2011624, EPI_ISL_2011625, EPI_ISL_2011633, EPI_ISL_2011635, EPI_ISL_2011636, EPI_ISL_2011637, EPI_ISL_2011639, EPI_ISL_2011640, EPI_ISL_2536379 | | | |
| see above | Incyte Diagnostics Spokane | Seattle Flu Study | Amanda Adler; Barry R. Lutz; Benjamin Pelle; Brian Hiatt; Caitlin R. Wolf; Chris D. Frazar; Deborah A. Nickerson; Elisabeth Brandstetter; Erica Ryke; Geoff Melly; Helen Y. Chu; Janet A. Englund; Jay Shendure; Jover Lee; Kairsten Fay; Kirsten Lacombe; Lea M. Starita; Mark J. Rieder; Matthew Richardson; Matthew Thompson; Melissa Truong; Michael Boeckh; Michael Famulare; Misja Ilcisin; Peter D. Han; Philip Dykema; Romesh Gautom; Scott Lindquist; Thomas R. Sibley; Trevor Bedford |
| EPI_ISL_2304410 | Indiana Animal Disease Diagnostic Laboratory | Carpi Laboratory - Purdue University | Abebe A Fola; G Kenitra Hendrix; Giovanna Carpi; Ilinca I Ciubotariu; Jack Dorman; Jobin J Kattoor; Lev Gorenstein; Nicole M Perry; Rebecca P Wilkes |
| EPI_ISL_2444235, EPI_ISL_2444242 | Indiana State Department of Health | Quantigen Biosciences | Paul J. Chidress |
| EPI_ISL_2087169, EPI_ISL_2087178, EPI_ISL_2087180, EPI_ISL_2087204, EPI_ISL_2087239, EPI_ISL_2087245, EPI_ISL_2087284, EPI_ISL_2087351, EPI_ISL_2087374, EPI_ISL_2087463, EPI_ISL_2087476, EPI_ISL_2087567, EPI_ISL_2087622, EPI_ISL_2087707, EPI_ISL_2087827, EPI_ISL_2087900, EPI_ISL_2087935, EPI_ISL_2088221, EPI_ISL_2088241, EPI_ISL_2088250, EPI_ISL_2088395, EPI_ISL_2132374, EPI_ISL_2283322, EPI_ISL_2283380, EPI_ISL_2283441, EPI_ISL_4382863, EPI_ISL_4383824, EPI_ISL_4384406, EPI_ISL_4384408, EPI_ISL_4385231, EPI_ISL_4386378, EPI_ISL_4386637, EPI_ISL_4388420, EPI_ISL_4389906, EPI_ISL_4390652 | | | |
| see above | Infinity Biologix | Centers for Disease Control and Prevention Division of Viral Diseases, Pathogen Discovery | Adrian Paskey; Benjamin Rambo-Martin; Chirayu Goswami; Christian Bixby; Christopher Gulvick; Clinton Paden; Clinton R. Paden; Dakota Howard; Darlene Wagner; Dhwani Batra; Duncan MacCannell; Erisa Sula; Jason Caravas; Jonathan Schultz; Kara Moser; Kristine Lacek; Matthew Schmerer; Peter Cook; Peter W. Cook; Robin Grimwood; Russ Hager; Scott Sammons; Shatavia Morrison; Tymeckia Kendall; Victoria Caban Figueroa; Yihe Wang; Yvette Unoarumhi |
| EPI_ISL_5802580, EPI_ISL_5802582, EPI_ISL_5802583, EPI_ISL_5802605, EPI_ISL_5802606, EPI_ISL_5802607, EPI_ISL_5802608, EPI_ISL_5802609, EPI_ISL_5802610, EPI_ISL_5802611, EPI_ISL_5802612, EPI_ISL_5802613, EPI_ISL_5802614, EPI_ISL_5802615, EPI_ISL_5802616, EPI_ISL_5802617, EPI_ISL_5802618, EPI_ISL_5802619, EPI_ISL_5802620, EPI_ISL_5802622, EPI_ISL_5802623, EPI_ISL_5802624, EPI_ISL_5802625, EPI_ISL_5802626, EPI_ISL_5802627, EPI_ISL_5802628, EPI_ISL_5802629, EPI_ISL_5802630, EPI_ISL_5802631, EPI_ISL_5802632, EPI_ISL_5802633, EPI_ISL_5802634, EPI_ISL_5802635, EPI_ISL_5802636, EPI_ISL_5802637, EPI_ISL_5802638, EPI_ISL_5802639, EPI_ISL_5802640 | | | |
| see above | Inside | Instituto Butantan | Antonio Jorge Martins; Claudia Renata dos Santos Barros; David Schlesinger; Debora Botequio Moretti; Dimas Tadeu Covas; Elaine Cristina Marqueze; Elaine Vieira Santos; Evandra Strazza Rodrigues; Heidge Fukumasu; Jayme Augusto de Souza-Neto; José Salvatore Leister Patané; Luiz Alcantara; Luiz Lehmann Coutinho; Maria Carolina Elias; Maurício Lacerda Nogueira; Rafael dos Santos Bezerra; Raul Machado Neto; Rejane Maria Tommasini Grotto; Ricardo Haddad; Sandra Coccuzzo Sampaio Vessoni; Simone Kashima; Svetoslav Nanev Slavov; Vincent Louis Viala |
| EPI_ISL_5782548, EPI_ISL_5782549, EPI_ISL_5782560, EPI_ISL_5782581, EPI_ISL_5782582, EPI_ISL_5782592, EPI_ISL_5782599, EPI_ISL_5782600, EPI_ISL_5782601, EPI_ISL_5782603, EPI_ISL_5782607, EPI_ISL_5782612, EPI_ISL_5782626, EPI_ISL_5782628, EPI_ISL_5782636, EPI_ISL_5782638, EPI_ISL_5782643, EPI_ISL_5782660, EPI_ISL_5802649, EPI_ISL_5802650, EPI_ISL_5802651, EPI_ISL_5802652, EPI_ISL_5802653, EPI_ISL_5802654, EPI_ISL_5802655, EPI_ISL_5802656, EPI_ISL_5802657, EPI_ISL_5802658, EPI_ISL_5802659, EPI_ISL_5802660, EPI_ISL_5802661, EPI_ISL_5802662, EPI_ISL_5802663, EPI_ISL_5802664, EPI_ISL_5802665, EPI_ISL_5802666, EPI_ISL_5802667, EPI_ISL_5802668, EPI_ISL_5802669, EPI_ISL_5802670, EPI_ISL_5802671, EPI_ISL_5802672, EPI_ISL_5802673, EPI_ISL_5802674, EPI_ISL_5802675, EPI_ISL_5802676, EPI_ISL_5802677, EPI_ISL_5802678, EPI_ISL_5802679, EPI_ISL_5802681, EPI_ISL_5802682 | | | |
| see above | Inside Diagnosticos | Instituto Butantan | Antonio Jorge Martins; Claudia Renata dos Santos Barros; David Schlesinger; Debora Botequio Moretti; Dimas Tadeu Covas; Elaine Cristina Marqueze; Elaine Vieira Santos; Evandra Strazza Rodrigues; Heidge Fukumasu; Jayme Augusto de Souza-Neto; José Salvatore Leister Patané; Luiz Alcantara; Luiz Lehmann Coutinho; Maria Carolina Elias; Maurício Lacerda Nogueira; Rafael dos Santos Bezerra; Raul Machado Neto; Rejane Maria Tommasini Grotto; Ricardo Haddad; Sandra Coccuzzo Sampaio Vessoni; Simone Kashima; Svetoslav Nanev Slavov; Vincent Louis Viala |
| EPI_ISL_2691157, EPI_ISL_2691158 | Instituo Adolfo Lutz Central | Instituto Adolfo Lutz, Interdiciplinary Procedures Center, Strategic Laboratory | Caio Vinicius Dias Lopes; Claudia Regina Gonçalves; Claudio Tavares Sacchi; Erica Valessa Ramos Gomes; Karoline Rodrigues Campos; Leonardo Jose Tadeu de Araujo |
| EPI_ISL_1848371 | Institut fÃ¼r Immunologie und Genetik Kaiserslautern: Medizinisches Labor Dr. med. B. Thiele | Robert Koch Institute | |
| EPI_ISL_1916517 | Institute for Infectious Diseases, University of Bern, Switzerland | Institute for Infectious Diseases, University of Bern, Switzerland | Alban Ramette; Christian Baumann; Cora Sägesser; Franziska Suter-Riniker; Miguel A Terrazos Miani; Pascal Bittel; Peter Keller; Stefan Neuenschwander; Stephen L Leib |
| EPI_ISL_1739244 | Institute of Microbiology, Universidad San Francisco de Quito | Institute of Microbiology, Universidad San Francisco de Quito | Belén Prado-Vivar; Bernardo Gutiérrez; Gabriel Trueba; Gabriela Zambrano; Juan José Guadalupe; Michelle Grunauer; Monica Becerra-Wong; Patricio Rojas-Silva; Paúl Cárdenas; Sully Márquez; Verónica Barragán |
| EPI_ISL_2691115, EPI_ISL_2691116, EPI_ISL_2691122, EPI_ISL_2919247, EPI_ISL_2919258 | Instituto Adolfo Lutz - Regional de Bauru | Instituto Adolfo Lutz, Interdiciplinary Procedures Center, Strategic Laboratory | Caio Vinicius Dias Lopes; Claudia Regina Gonçalves; Claudio Tavares Sacchi; Erica Valessa Ramos Gomes; Karoline Rodrigues Campos; Leonardo Jose Tadeu de Araujo |
| EPI_ISL_2828686, EPI_ISL_2828691 | Instituto Adolfo Lutz - Regional de Campinas | Instituto Adolfo Lutz, Interdiciplinary Procedures Center, Strategic Laboratory | Caio Vinicius Dias Lopes; Claudia Regina Gonçalves; Claudio Tavares Sacchi; Erica Valessa Ramos Gomes; Karoline Rodrigues Campos |
| EPI_ISL_2614626, EPI_ISL_2614627, EPI_ISL_2614628, EPI_ISL_2691089, EPI_ISL_2691090 | Instituto Adolfo Lutz - Regional de Marilia | Instituto Adolfo Lutz, Interdiciplinary Procedures Center, Strategic Laboratory | Caio Vinicius Dias Lopes; Claudia Regina Gonçalves; Claudio Tavares Sacchi; Erica Valessa Ramos Gomes; Karoline Rodrigues Campos; Leonardo Jose Tadeu de Araujo |
| EPI_ISL_2958834, EPI_ISL_2958835 | Instituto Adolfo Lutz - Regional de Sao Jose do Rio Preto | Instituto Adolfo Lutz, Interdiciplinary Procedures Center, Strategic Laboratory | Caio Vinicius Dias Lopes; Claudia Regina Gonçalves; Claudio Tavares Sacchi; Erica Valessa Ramos Gomes; Karoline Rodrigues Campos |
| EPI_ISL_2958845, EPI_ISL_2958846 | Instituto Adolfo Lutz - Regional de Sorocaba | Instituto Adolfo Lutz, Interdiciplinary Procedures Center, Strategic Laboratory | Caio Vinicius Dias Lopes; Claudia Regina Gonçalves; Claudio Tavares Sacchi; Erica Valessa Ramos Gomes; Karoline Rodrigues Campos |
| EPI_ISL_4104716, EPI_ISL_4104720, EPI_ISL_4104724, EPI_ISL_4104729 | Instituto Butantan | Instituto Butantan | Antonio Jorge Martins; Claudia Renata dos Santos Barros; David Schlesinger; Debora Botequio Moretti; Dimas Tadeu Covas; Elaine Cristina Marqueze; Elaine Vieira Santos; Evandra Strazza Rodrigues; Heidge Fukumasu; Jayme Augusto de Souza-Neto; José Salvatore Leister Patané; Luiz Alcantara; Luiz Lehmann Coutinho; Maria Carolina Elias; Maurício Lacerda Nogueira; Rafael dos Santos Bezerra; Raul Machado Neto; Rejane Maria Tommasini Grotto; Ricardo Haddad; Sandra Coccuzzo Sampaio Vessoni; Simone Kashima; Svetoslav Nanev Slavov; Vincent Louis Viala |
| EPI_ISL_2160668 | Instituto Nacional de Medicina Genomica | Centro de Investigación en Enfermedades Infecciosas (CIENI), Instituto Nacional de Enfermedades Respiratorias (INER) | Arriaga-Canon C; Avila-Rios S; Boukadida C; Cedro-Tanda A; Herrera-Montalvo LA; Hidalgo-Miranda A; Matias-Florentino M; Mendoza-Vargas A; Perez-Garcia M; Reyes-Grajeda JP; Reyes-Teran G |
| EPI_ISL_1807358, EPI_ISL_2080635 | Instituto Nacional de Medicina Genomica | Instituto Nacional de Medicina Genomica | Alcaraz N; Cedro-Tanda A; Escobar-Arrazola; Garcia-Cardenas FJ; Gonzalez-Barrera D; Gonzalez-Woge MA; Herrera-Montalvo LA; Herrera-Montalvo LA.; Hidalgo-Miranda A; M; Mendoza-Vargas A; Munguia-Garza P; Ramirez-Vega O; Rangel-DeLeon D; Reyes-Grajeda JP |
| EPI_ISL_2155036, EPI_ISL_2500946, EPI_ISL_2500950 | Instituto Nacional de Salud | Instituto Nacional de Salud- Dirección de Investigación en Salud Pública | Carlos Franco-Muñoz; Carmen Osorio; Diana Malo; Diego A. Álvarez-Díaz; Diego Andrés Prada; Gerardo Santamaría; Hector Alejandro Ruiz-Moreno; Jhonnatan Reales-González; Jorge Rivera; Juan Camilo Martinez; Julian Naizaque; Katherine Laiton-Donato; Lisseth Pardo; Magdalena Wiesner; Marcela Mercado-Reyes; Maria T. Herrera-Sepúlveda; Marta Lopez Blanco; Martha Lucia Ospina Martinez; Paola Rojas; Sergio Gomez; Sheryll Corchuelo; Ángela Alarcon Cruz |
| EPI_ISL_2295417, EPI_ISL_2295418, EPI_ISL_2295419, EPI_ISL_2295420, EPI_ISL_2295422, EPI_ISL_2295423, EPI_ISL_2295425, EPI_ISL_2295426 | | | |
| see above | Instituto de Biotecnologia - UNESP-Botucatu-SP | Instituto de Biotecnologia - UNESP-Botucatu-SP | Cecília Artico Banho; Cíntia Bittar; Fábio Sossai Possebon; Guilherme Campos; Helena Lage Ferreira; Jorge A. Petroli Marchesi; João Pessoa Araújo Jr.; Leila Sabrina Ullmann; Lívia Sacchetto; Maisa C. Pereira Parra; Marília Moraes; Maurício L. Nogueira; Paula Rahal; Paulo Inacio da Costa |
| EPI_ISL_2756556 | Instituto de Investigaciones Biomédicas en Retrovirus y SIDA (INBIRS), Universidad de | INEI - ANLIS. Instituto Nacional de Enfermedades Infecciosas, Administración Nacional de. | Ana Ceballos; Augusto Varese; Federico Remes Lenicov; Ignacio Mazzitelli; Jorge Geffner.; Valeria Ochoa |

| | | | |
|---|---|---|---|
| | Buenos Aires (UBA)-CONICET, Buenos Aires, Argentina. | Laboratorios e institutos de Salud "Dr. Carlos G. Malbran". | |
| EPI_ISL_2756604 | Instituto de Investigaciones Biomédicas en Retrovirus y SIDA (INBIRS), Universidad de Buenos Aires (UBA)-CONICET, Buenos Aires, Argentina. | INEI - ANLIS. Instituto Nacional de Enfermedades Infecciosas, Administración Nacional de. Laboratorios e institutos de Salud "Dr. Carlos G. Malbran"./INBIRS. | Ana Ceballos; Augusto Varese; Federico Remes Lenicov; Ignacio Mazzitelli; Jorge Geffner.; Valeria Ochoa |
| EPI_ISL_2756521 | Instituto de Investigaciones Biomédicas en Retrovirus y SIDA (INBIRS), Universidad de Buenos Aires (UBA)-CONICET, Buenos Aires, Argentina. | INEI - ANLIS. Instituto Nacional de Enfermedades Infecciosas, Administración Nacional de. Laboratorios e institutos de Salud Dr. Carlos G. Malbran | Ana Ceballos; Augusto Varese; Federico Remes Lenicov; Ignacio Mazzitelli; Jorge Geffner; Valeria Ochoa |
| EPI_ISL_2691146 | Intituto Adolfo Lutz - Regional de Sorocaba | Instituto Adolfo Lutz, Interdiciplinary Procedures Center, Strategic Laboratory | Caio Vinicius Dias Lopes; Claudia Regina Gonçalves; Claudio Tavares Sacchi; Erica Valessa Ramos Gomes; Karoline Rodrigues Campos; Leonardo Jose Tadeu de Araujo |
| EPI_ISL_1759413, EPI_ISL_1759455, EPI_ISL_1759458, EPI_ISL_1759459, EPI_ISL_1759483, EPI_ISL_1759632, EPI_ISL_1759634, EPI_ISL_1759674, EPI_ISL_2137671, EPI_ISL_2137699, EPI_ISL_2137713, EPI_ISL_2137714, EPI_ISL_2153253, EPI_ISL_2171301, EPI_ISL_2171328, EPI_ISL_2171365, EPI_ISL_2226904 | | | |
| see above | Istituto Zooprofilattico Sperimentale del Mezzogiorno | TIGEM | Antonio Grimaldi Patrizia Annunziata Francesco Panariello Biancamaria Pierri Claudia Tiberio Teresa Giuliano Valentina Bouche Chiara Colantuono Maria Concetta Cuomo Denise Di Concilio Lucio Di Filippo Anna Manfredi Marcello Salvi Antonio Limone Luigi Atripaldi Pellegrino Cerino Andrea Ballabio Davide Cacchiarelli |
| EPI_ISL_2143876, EPI_ISL_2143954, EPI_ISL_2143955, EPI_ISL_2143957, EPI_ISL_2143960, EPI_ISL_2143987, EPI_ISL_2144029 | | | |
| see above | Istituto Zooprofilattico Sperimentale del Mezzogiorno | Telethon Institute of Genetics and Medicine (TIGEM) | Antonio Grimaldi Patrizia Annunziata Francesco Panariello Biancamaria Pierri Claudia Tiberio Teresa Giuliano Valentina Bouche Chiara Colantuono Maria Concetta Cuomo Denise Di Concilio Lucio Di Filippo Anna Manfredi Marcello Salvi Antonio Limone Luigi Atripaldi Pellegrino Cerino Andrea Ballabio Davide Cacchiarelli |
| EPI_ISL_2246725, EPI_ISL_2246726, EPI_ISL_2246728 | John H Stroger Jr Hospital of Cook County | RIPHL at Rush University Medical Center | Ah-Young Sung; Felix Araujo Perez; Kevin Kunstman; Laura Furtado; Marieta Hyde; Max Kolton; Sharon Welbel; Stefan Green |
| EPI_ISL_2331670 | Johns Hopkins Hospital Department of Pathology | Johns Hopkins Hospital Department of Pathology | Adannaya Amadi; C. Paul Morris; Chun Huai Luo; Heba H. Mostafa; Matthew Schwartz; Nicholas Gallagher |
| EPI_ISL_2424268, EPI_ISL_2424275, EPI_ISL_2424434, EPI_ISL_2424469, EPI_ISL_2425106, EPI_ISL_2425107, EPI_ISL_2425155, EPI_ISL_2425180, EPI_ISL_2425196, EPI_ISL_2425203, EPI_ISL_2425217, EPI_ISL_2425218, EPI_ISL_2425222, EPI_ISL_2832252, EPI_ISL_2832277, EPI_ISL_2832321, EPI_ISL_2832435, EPI_ISL_2832640, EPI_ISL_2832723, EPI_ISL_2832942, EPI_ISL_2832943, EPI_ISL_2832944, EPI_ISL_2832945, EPI_ISL_2832946, EPI_ISL_2832947, EPI_ISL_2832948, EPI_ISL_2833059, EPI_ISL_2833062, EPI_ISL_2833063, EPI_ISL_2833067, EPI_ISL_2833069, EPI_ISL_2833071, EPI_ISL_2833074, EPI_ISL_2833076, EPI_ISL_2833084, EPI_ISL_2833094, EPI_ISL_2833124, EPI_ISL_2833140, EPI_ISL_2833186, EPI_ISL_2833190, EPI_ISL_2833194, EPI_ISL_2833197, EPI_ISL_2833460, EPI_ISL_2833711, EPI_ISL_2833727, EPI_ISL_2834567 | | | |
| see above | KU Leuven, Rega Institute, Clinical and Epidemiological Virology | KU Leuven, Rega Institute, Clinical and Epidemiological Virology | Bert Vanmechelen; Joan Marti-Carerras; Piet Maes; Tony Wawina-Bokalanga |
| EPI_ISL_3737713, EPI_ISL_3737714, EPI_ISL_3737715 | Kentucky State Public Health Lab | Kentucky State Public Health Lab | Anjayya Tummala; Joshua Tobias; Karim George; Matthew Johnson; Rachel Zinner; Rhonda Lucas; Vaneet Arora; William Grooms |
| EPI_ISL_1843772 | Klinisch Laboratorium ZNA | Klinisch Laboratorium ZNA | Verstrepen et al. |
| EPI_ISL_5603250 | LAB DE A CLIN MUNICIPAL DE CASCAVEL | Analytical Competence Molecular Epidemiology Lab/ACME, Oswaldo Cruz Foundation, Ceara (FIOCRUZ CE) | Carlos Leonardo de Aragao Araujo; Cecilia Leite Costa & Eduardo Ruback dos Santos on behalf of COVID-19 FIOCRUZ Genomic Network; Cleber Furtado Aksenen; Fabio Miyajima; Fernando Braga Stehling; Francisco Eder de Moura Lopes; Igor Oliveira Duarte; Jamille Maria Mendes Bezerra; Joaquim Cesar do Nascimento Sousa Junior; Pedro Miguel Carneiro Jeronimo; Suzana Porto Almeida; Thais Ferreira de Oliveira; Thais de Oliveira Costa; Ticiane Cavalcante de Souza; Veridiana Pessoa Miyajima |
| EPI_ISL_2142664 | LABORATOIRE DE BIOLOGIE MEDICALE | CNR Virus des Infections Respiratoires - France SUD | Antonin Bal; Bruno Lina; Bruno Simon; Gregory Destras; Gwendolyne Burfin; Hadrien Regue; Laurence Josset; Martine Valette; Quentin Semanas |
| EPI_ISL_2834727 | LABORATORIO CLINICO HIGUERA ESCALANTE | Instituto Nacional de Salud- Dirección de Investigación en Salud Pública | Carlos Franco-Muñoz; Carmen Osorio; Diana Malo; Diego A. Álvarez-Díaz; Diego Andrés Prada; Gerardo Santamaría; Hector Alejandro Ruiz-Moreno; Jhonnatan Reales-González; Jorge Rivera; Juan Camilo Martinez; Julian Naizaque; Katherine Laiton-Donato; Lisseth Pardo; Magdalena Wiesner; Marcela Mercado-Reyes; Maria T. Herrera-Sepúlveda; Marta Lopez Blanco; Martha Lucia Ospina Martinez; Paola Rojas; Sergio Gomez; Sheryll Corchuelo; Ángela Alarcon Cruz |
| EPI_ISL_2344827, EPI_ISL_2344828, EPI_ISL_2344836, EPI_ISL_2344837, EPI_ISL_2344839, EPI_ISL_2344840, EPI_ISL_2344841, EPI_ISL_2344842, EPI_ISL_2344843, EPI_ISL_2344844 | | | |
| see above | LABORATORIO DE FRANCA | Instituto Butantan / ESALQ-Piracicaba | Antonio Jorge Martins; Claudia Renata dos Santos Barros; David Schlesinger; Debora Botequio Moretti; Dimas Tadeu Covas; Elaine Cristina Marqueze; Elaine Vieira Santos; Evandra Strazza Rodrigues; Heidge Fukumasu; Jayme Augusto de Souza-Neto; José Salvatore Leister Patané; Luiz Alcantara; Luiz Lehmann Coutinho; Maria Carolina Elias; Maurício Lacerda Nogueira; Rafael dos Santos Bezerra; Raul Machado Neto; Rejane Maria Tommasini Grotto; Ricardo Haddad; Sandra Coccuzzo Sampaio Vessoni; Simone Kashima; Svetoslav Nanev Slavov; Vincent Louis Viala |
| EPI_ISL_2344801, EPI_ISL_2344826, EPI_ISL_2344838, EPI_ISL_2344851, EPI_ISL_2344864, EPI_ISL_2344884, EPI_ISL_2344915, EPI_ISL_2344917, EPI_ISL_2344971, EPI_ISL_2344972, EPI_ISL_2344979, EPI_ISL_2344983, EPI_ISL_2344985, EPI_ISL_2344988, EPI_ISL_2345028, EPI_ISL_2345033, EPI_ISL_2345034, EPI_ISL_2345118, EPI_ISL_2345169 | | | |
| see above | LABORATORIO MUNICIPAL DE PIRACICABA | Instituto Butantan / ESALQ-Piracicaba | Antonio Jorge Martins; Claudia Renata dos Santos Barros; David Schlesinger; Debora Botequio Moretti; Dimas Tadeu Covas; Elaine Cristina Marqueze; Elaine Vieira Santos; Evandra Strazza Rodrigues; Heidge Fukumasu; Jayme Augusto de Souza-Neto; José Salvatore Leister Patané; Luiz Alcantara; Luiz Lehmann Coutinho; Maria Carolina Elias; Maurício Lacerda Nogueira; Rafael dos Santos Bezerra; Raul Machado Neto; Rejane Maria Tommasini Grotto; Ricardo Haddad; Sandra Coccuzzo Sampaio Vessoni; Simone Kashima; Svetoslav Nanev Slavov; Vincent Louis Viala |
| EPI_ISL_7744026, EPI_ISL_7744027, EPI_ISL_7744040, EPI_ISL_7744100, EPI_ISL_7744121 | LACEN | Laboratório de Bioinformática - Universidade Federal de Santa Catarina | "Aline Daina Schlindwein"; "Ana Paula Christoff"; "Antuani Baptistela"; "Carolina Leite Martins"; "Darcita Buerger Rovaris"; "Dayane Azevedo Padilha"; "Doris Sobral Marques SouzaSobral"; "Edmundo Carlos Grisard"; "Eric Kazuo Kawagoe"; "Fernanda Luiza Ferrari"; "Fernanda Roesene Melo"; "Fernando Hartmann Barazzetti"; "Gislaine Fongaro"; "Glauber Wagner"; "Guilherme Augusto Maia"; "Guilherme Razzera"; "Guilherme Toledo e Silva"; "Julia Kinetz Wachter"; "Luiz Felipe de Oliveira"; "Marcos André Schörne"; "Marcus Vinícius Duarte Rodrigues"; "Maria Luiza Bazzo"; "Marlei Pickler Debiasi dos Anjos"; "Milene Moehr de Moraes"; "Nestor Wendt"; "Patrícia Hermes Stoco"; "Paula Sacchet"; "Renato Simões Moreira"; "Rodrigo de Paula Baptista"; "Tamela Zamboni Madaloz"; "Tatiany Aparecida Teixeira Soratto"; "Vilmar Benetti Filho" |
| EPI_ISL_2691208, EPI_ISL_2691216, EPI_ISL_2691217 | LACEN do Estado de Goias | Instituto Adolfo Lutz, Interdiciplinary Procedures Center, Strategic Laboratory | Caio Vinicius Dias Lopes; Claudia Regina Gonçalves; Claudio Tavares Sacchi; Erica Valessa Ramos Gomes; Karoline Rodrigues Campos; Leonardo Jose Tadeu de Araujo |
| EPI_ISL_3316153 | LACEN do Estado de Mato Grosso | Instituto Adolfo Lutz, Interdiciplinary Procedures Center, Strategic Laboratory | Caio Vinicius Dias Lopes; Claudia Regina Gonçalves; Claudio Tavares Sacchi; Karoline Rodrigues Campos; Leonardo Tadeu de Araujo; Marlon Benedito Nascomento Santos |
| EPI_ISL_2919218, EPI_ISL_2919219, EPI_ISL_2919225, EPI_ISL_2919226 | LACEN do Estado de Tocantins | Instituto Adolfo Lutz, Interdiciplinary Procedures Center, Strategic Laboratory | Caio Vinicius Dias Lopes; Claudia Regina Gonçalves; Claudio Tavares Sacchi; Erica Valessa Ramos Gomes; Karoline Rodrigues Campos |
| EPI_ISL_3691377, EPI_ISL_3691378, EPI_ISL_3691379, EPI_ISL_3691380, EPI_ISL_3691381 | LACEN do Estado do Mato Grosso do Sul | Instituto Adolfo Lutz, Interdiciplinary Procedures Center, Strategic Laboratory | Caio Vinicius Dias Lopes; Claudia Regina Gonçalves; Claudio Tavares Sacchi; Karoline Rodrigues Campos; Leonardo Tadeu de Araujo; Marlon Benedito Nascomento Santos |
| EPI_ISL_2958858, EPI_ISL_2958859, EPI_ISL_2958860, EPI_ISL_2958861, EPI_ISL_2958863, EPI_ISL_2958864, EPI_ISL_2958865, EPI_ISL_2958866, EPI_ISL_2958867, EPI_ISL_2958868, EPI_ISL_2958869, EPI_ISL_2958870, EPI_ISL_2958871 | | | |
| see above | LACEN do Mato Grosso do Sul | Instituto Adolfo Lutz, Interdiciplinary Procedures Center, Strategic Laboratory | Caio Vinicius Dias Lopes; Claudia Regina Gonçalves; Claudio Tavares Sacchi; Erica Valessa Ramos Gomes; Karoline Rodrigues Campos |
| EPI_ISL_3134550, EPI_ISL_3134551, EPI_ISL_3134552, EPI_ISL_3134554, EPI_ISL_3134555, EPI_ISL_3134556, EPI_ISL_3703158, EPI_ISL_3703169, EPI_ISL_3703170, EPI_ISL_3703171, EPI_ISL_3703172, EPI_ISL_3703173, EPI_ISL_3703174, EPI_ISL_3703175, EPI_ISL_3703176, EPI_ISL_3703177, EPI_ISL_3703178, EPI_ISL_3703179, EPI_ISL_3703180, EPI_ISL_3703181, EPI_ISL_3703182, EPI_ISL_3703183, EPI_ISL_3703184, EPI_ISL_3703185, EPI_ISL_3703188, EPI_ISL_3703197, EPI_ISL_3703207, EPI_ISL_3703216, EPI_ISL_3703226, EPI_ISL_3703236, EPI_ISL_3703246, EPI_ISL_3703255, EPI_ISL_3703265, EPI_ISL_3703275, EPI_ISL_3703285, EPI_ISL_3703295, EPI_ISL_3703306, EPI_ISL_3703334, EPI_ISL_3703344, EPI_ISL_3703370 | | | |
| see above | LACEN/PE | WallauLab on behalf of Fiocruz COVID-19 Genomic Surveillance Network | Alexandre Freitas da Silva; Cassia Docena; Constância Flávia Junqueira Ayres; Filipe Zimmer Dezordi; Gabriel Luz Wallau; Gustavo Barbosa de Lima; Lais Ceschini Machado; Lilian Caroliny Amorim Silva; Marcelo Henrique dos Santos Paiva; Matheus Filgueira Bezerra; Sinval Pinto Brandão Filho |
| EPI_ISL_4487022, EPI_ISL_4488035, EPI_ISL_4488038, EPI_ISL_4488041 | LACEN/PI | WallauLab on behalf of Fiocruz COVID-19 Genomic Surveillance Network | Adelino Soares Lima Neto; Alexandre Freitas da Silva; Antônio Marinho; Cassia Docena; Constância Flávia Junqueira Ayres; Filipe Zimmer Dezordi; Gabriel Luz Wallau; Gustavo Barbosa de Lima; Hellen de Oliveira Amaral; Jacenir Reis dos Santos Mallet; Joana Carolina Viana Lima; Lais Ceschini Machado; Leandro de Mattos; Lilian Caroliny Amorim Silva; Marcela de Lacerda Valença Queiroz; Marcelo Adriano da Cunha e Silva Vieira; Marcelo Henrique dos Santos Paiva; Matheus Filgueira Bezerra; Sinval Pinto Brandão Filho; Túlio de Lima Campos; Vladimir Costa Silva; Walterlene de Carvalho Goçalves |
| EPI_ISL_2296086 | LESP Baja California | Instituto de Diagnostico y Referencia Epidemiologicos (INDRE) | Abril Rodriguez-Maldonado; Ariadna Medina-Benitez; Claudia Wong-Arambula; Ernesto Ramirez-Gonzalez.; Gisela Barrera-Badillo; Irma Lopez-Martinez; Joaquin Quiroz-Mercado; Lucia Hernandez-Rivas; Natividad Cruz-Ortiz; Sergio Rangel-Guerrero; Tatiana Nunez-Garcia; Vanessa Rivero-Arredondo |
| EPI_ISL_2296085 | LESP Chiapas | Instituto de Diagnostico y Referencia Epidemiologicos (INDRE) | Abril Rodriguez-Maldonado; Ariadna Medina-Benitez; Claudia Wong-Arambula; Ernesto Ramirez-Gonzalez.; Gisela Barrera-Badillo; Irma Lopez-Martinez; Joaquin Quiroz-Mercado; Lucia Hernandez-Rivas; Natividad Cruz-Ortiz; Sergio Rangel-Guerrero; Tatiana Nunez-Garcia; Vanessa Rivero-Arredondo |
| EPI_ISL_2105694, EPI_ISL_2105716, EPI_ISL_2105717 | LESP Ciudad de Mexico | Instituto de Diagnostico y Referencia Epidemiologicos (INDRE) | Abril Rodriguez-Maldonado; Ariadna Medina-Benitez; Claudia Wong-Arambula; Ernesto Ramirez-Gonzalez.; Gisela Barrera-Badillo; Irma Lopez-Martinez; Joaquin Quiroz-Mercado; Lucia Hernandez-Rivas; Natividad Cruz-Ortiz; Sergio Rangel-Guerrero; Tatiana Nunez-Garcia; Vanessa Rivero-Arredondo |
| EPI_ISL_2105682, EPI_ISL_2105683, EPI_ISL_2105686, EPI_ISL_2105687, | LESP Guanajuato | Instituto de Diagnostico y Referencia Epidemiologicos (INDRE) | Abril Rodriguez-Maldonado; Ariadna Medina-Benitez; Claudia Wong-Arambula; Ernesto Ramirez-Gonzalez.; Gisela Barrera-Badillo; Irma Lopez-Martinez; Joaquin Quiroz-Mercado; Lucia Hernandez-Rivas; Natividad Cruz-Ortiz; Sergio Rangel-Guerrero; Tatiana Nunez-Garcia; Vanessa Rivero-Arredondo |

| | | | |
|---|---|---|---|
| EPI_ISL_2296069 | | | |
| EPI_ISL_2296070 | LESP Jalisco | Instituto de Diagnostico y Referencia Epidemiologicos (INDRE) | Abril Rodriguez-Maldonado; Ariadna Medina-Benitez; Claudia Wong-Arambula; Ernesto Ramirez-Gonzalez.; Gisela Barrera-Badillo; Irma Lopez-Martinez; Joaquin Quiroz-Mercado; Lucia Hernandez-Rivas; Natividad Cruz-Ortiz; Sergio Rangel-Guerrero; Tatiana Nunez-Garcia; Vanessa Rivero-Arredondo |
| EPI_ISL_2105680, EPI_ISL_2296068 | LESP Nuevo Leon | Instituto de Diagnostico y Referencia Epidemiologicos (INDRE) | Abril Rodriguez-Maldonado; Ariadna Medina-Benitez; Claudia Wong-Arambula; Ernesto Ramirez-Gonzalez.; Gisela Barrera-Badillo; Irma Lopez-Martinez; Joaquin Quiroz-Mercado; Lucia Hernandez-Rivas; Natividad Cruz-Ortiz; Sergio Rangel-Guerrero; Tatiana Nunez-Garcia; Vanessa Rivero-Arredondo |
| EPI_ISL_2296087 | LESP Quintana Roo | Instituto de Diagnostico y Referencia Epidemiologicos (INDRE) | Abril Rodriguez-Maldonado; Ariadna Medina-Benitez; Claudia Wong-Arambula; Ernesto Ramirez-Gonzalez.; Gisela Barrera-Badillo; Irma Lopez-Martinez; Joaquin Quiroz-Mercado; Lucia Hernandez-Rivas; Natividad Cruz-Ortiz; Sergio Rangel-Guerrero; Tatiana Nunez-Garcia; Vanessa Rivero-Arredondo |
| EPI_ISL_2296090, EPI_ISL_2296091 | LESP Yucatan | Instituto de Diagnostico y Referencia Epidemiologicos (INDRE) | Abril Rodriguez-Maldonado; Ariadna Medina-Benitez; Claudia Wong-Arambula; Ernesto Ramirez-Gonzalez.; Gisela Barrera-Badillo; Irma Lopez-Martinez; Joaquin Quiroz-Mercado; Lucia Hernandez-Rivas; Natividad Cruz-Ortiz; Sergio Rangel-Guerrero; Tatiana Nunez-Garcia; Vanessa Rivero-Arredondo |
| EPI_ISL_1963334, EPI_ISL_1963338, EPI_ISL_1963448 | LHUB-ULB | Labo Klinische Biologie, UZA | Basil Britto Xavier; Christine Lammens; Herman Goossens; Jasmine Coppens; Marie Le Mercier; Veerle Matheeussen |
| EPI_ISL_4212136, EPI_ISL_4212142, EPI_ISL_4212148 | LSP DISTRITAL BOGOTA | Gencore - Universidad de los Andes | Alejandro Gomez; Cristian Barrera; Gabriela Ariza; Gabriela Delgado; Johana Hernandez; Luisa Sacristan; Marcela Guevara; Marcela Mercado; Silvia Restrepo |
| EPI_ISL_1913065 | LUM Laboratorio Unico Metropolitano - AUSL BOLOGNA | U.O. Microbiologia, Laboratorio Unico Centro Servizi - AUSL della Romagna | Francesca Taddei; Giorgio Dirani; Giulia Gatti; Ilaria Poggianti; Rita Mancini; Silvia Zannoli; Vittorio Sambri |
| EPI_ISL_2191972, EPI_ISL_2191983, EPI_ISL_2191985, EPI_ISL_2191999, EPI_ISL_2192001, EPI_ISL_2192019, EPI_ISL_2192041, EPI_ISL_2192043, EPI_ISL_2192046, EPI_ISL_2192060, EPI_ISL_2192067, EPI_ISL_2192107, EPI_ISL_2192133, EPI_ISL_2192139, EPI_ISL_2192161, EPI_ISL_2192170, EPI_ISL_2192173, EPI_ISL_2192193, EPI_ISL_2192239, EPI_ISL_2192337 | | | |
| see above | Lab voor klinische biologie | Lab voor klinische biologie | Bruno Verhasselt; Hannelore Hamerlinck; Marija Janevska |
| EPI_ISL_2386799 | Labor Becker & Kollegen (Standort München) | Robert Koch Institute | |
| EPI_ISL_1840806, EPI_ISL_1840823, EPI_ISL_1840826, EPI_ISL_1840857, EPI_ISL_2107861, EPI_ISL_2107882, EPI_ISL_2107885 | | | |
| see above | Labor Berlin Charité Vivantes GmbH / Institut für Virologie | Charité Universitätsmedizin Berlin, Institut für Virologie/Labor Berlin | Barbara Mühlemann; Christian Drosten; Christine Stephan; Peter Menzel; Rolf Schwarzer; Terry Jones; Victor M Corman |
| EPI_ISL_2125124 | Labor Dr. Spranger | Robert Koch Institute | |
| EPI_ISL_2108352, EPI_ISL_2108406 | Labor Lubeck bzw. Laborarztliche Gemeinschaftspraxis Lubeck | Robert Koch Institute | |
| EPI_ISL_2111585 | Labor ZOTZ\|KLIMAS; MVZ Dusseldorf-Centrum | Robert Koch Institute | |
| EPI_ISL_3021525, EPI_ISL_3021532, EPI_ISL_3458428, EPI_ISL_3458442, EPI_ISL_3458444, EPI_ISL_3458445, EPI_ISL_3458456, EPI_ISL_3458457, EPI_ISL_3458458, EPI_ISL_3458464, EPI_ISL_3458465, EPI_ISL_3458478, EPI_ISL_3458483, EPI_ISL_3458484, EPI_ISL_3458488, EPI_ISL_3458492, EPI_ISL_3458493, EPI_ISL_3458495, EPI_ISL_3458502, EPI_ISL_3458507, EPI_ISL_3458523, EPI_ISL_3458524, EPI_ISL_3458623, EPI_ISL_3458624, EPI_ISL_3458626, EPI_ISL_3458627, EPI_ISL_3458628, EPI_ISL_3458651, EPI_ISL_3458652, EPI_ISL_3458653, EPI_ISL_3458654, EPI_ISL_3458655, EPI_ISL_3458656, EPI_ISL_3508451, EPI_ISL_3508452, EPI_ISL_3508453, EPI_ISL_3508454, EPI_ISL_3508455, EPI_ISL_3508456, EPI_ISL_5872626, EPI_ISL_5872631, EPI_ISL_5872640, EPI_ISL_5872643, EPI_ISL_5872673, EPI_ISL_5872898, EPI_ISL_5872901, EPI_ISL_5872908, EPI_ISL_5872909, EPI_ISL_5872921, EPI_ISL_5872926, EPI_ISL_7686765, EPI_ISL_7713564, EPI_ISL_7713573, EPI_ISL_7713654 | | | |
| see above | Laboratoire de santé publique du Québec | Laboratoire de santé publique du Québec | Guillaume Bourque; Ioannis Ragoussis; Jesse Shapiro; Mark Lathrop and Judith Fafard on behalf of the CoVSeQ research group; Mark Lathrop and Michel Roger on behalf of the CoVSeQ research group; Sandrine Moreira |
| EPI_ISL_1918150 | Laboratoire national de sante, Microbiology, Virology | Laboratoire national de sante, Microbiology, Microbial Genomics Platform | Anke Wienecke-Baldacchino; Catherine Ragimbeau; Fatu Djabi; Jessica Tapp; Lise Pignon; Raoul Salmon; Tamir Abdelrahman; Trung Nguyen Nguyen |
| EPI_ISL_1917858 | Laboratoires d'analyses medicales - Ketterthill | Laboratoire national de sante, Microbiology, Microbial Genomics Platform | Anke Wienecke-Baldacchino; Caroline Scheiber; Catherine Ragimbeau; Fatu Djabi; Jessica Tapp; Lise Pignon; Raoul Salmon; Serge Vedy; Tamir Abdelrahman |
| EPI_ISL_2196304, EPI_ISL_2196305, EPI_ISL_2443561, EPI_ISL_2443562, EPI_ISL_2645422 | Laboratorio Central de Saude Publica do Estado do Para (LACEN/PA) | Laboratory of Respiratory Viruses and Measles, Oswaldo Cruz Institute, FIOCRUZ | Alice Sampaio Rocha; Ana Carolina Mendonca; Anna Carolina Paixao; Elisa Cavalcante Pereira; Fernando Motta; Luciana Appolinario; Marilda Siqueira on behalf of the Fiocruz COVID-19 Genomic Surveillance Network; Paola Resende; Renata Serrano Lopes; Taina Venas; Valnete Andrade |
| EPI_ISL_1785214, EPI_ISL_1785224, EPI_ISL_1785228, EPI_ISL_1785229, EPI_ISL_1785245, EPI_ISL_1785251, EPI_ISL_2001448, EPI_ISL_2001449 | | | |
| see above | Laboratorio Analisi Osp. Città di Castello - Azienda USL Umbria1 | Istituto Zooprofilattico Sperimentale dell'Abruzzo e Molise "G. Caporale" | Ancora M; Calistri P; Cammà C; Caporale M; Curini V; Delli Compagni E; Di Domenico M; Di Lollo Valeria; Di Pasquale A; Lorusso A; Malagigi V; Mangone I; Marcacci M; Puglia I; Rinaldi A; Savini G; Scialabba S; Tacconi P |
| EPI_ISL_1923562 | Laboratorio Aziendale di Microbiologia e Virologia, Azienda Sanitaria dell'Alto Adige | Laboratorio Aziendale di Microbiologia e Virologia, Azienda Sanitaria dell'Alto Adige | Anne Picard; Bartolomeo Mobilio Rodriguez; Chiara Cantaloni; Claudia Volpato; Elisa Masi; Elisabetta Giacobazzi; Elisabetta Pagani; Irene Bianconi; Stefanie Wieser |
| EPI_ISL_4196759 | Laboratorio Central Mg. Luis Alfredo Pianciola | Laboratorio Central Mg. Luis Alfredo Pianciola on behalf of 'Proyecto Argentino Interinstitucional de genomica de SARS-CoV-2' (PAIS Consortium) | C Pintos; C Rastellini.; C Ziehm; J Ousset; L Pianciola; M Fernandez; M Mazzeo |
| EPI_ISL_2101389, EPI_ISL_2101403, EPI_ISL_2101404, EPI_ISL_2101406, EPI_ISL_2101408, EPI_ISL_2101409, EPI_ISL_2101410, EPI_ISL_2101411, EPI_ISL_2101412, EPI_ISL_2101413, EPI_ISL_2101414, EPI_ISL_2101415, EPI_ISL_2101416, EPI_ISL_2101417, EPI_ISL_2101418, EPI_ISL_2101419, EPI_ISL_2101420, EPI_ISL_2101421, EPI_ISL_2101422, EPI_ISL_2101423, EPI_ISL_2101424, EPI_ISL_2101425, EPI_ISL_2101426, EPI_ISL_2101434, EPI_ISL_2101435, EPI_ISL_2101436, EPI_ISL_2101437, EPI_ISL_2101438, EPI_ISL_2101439, EPI_ISL_2101440, EPI_ISL_2101441, EPI_ISL_2101442, EPI_ISL_2101444, EPI_ISL_2101445, EPI_ISL_2101446, EPI_ISL_2101447, EPI_ISL_2101448, EPI_ISL_2101449, EPI_ISL_2101450, EPI_ISL_2101451, EPI_ISL_2101452, EPI_ISL_2101453, EPI_ISL_2101454, EPI_ISL_2101464, EPI_ISL_2101465, EPI_ISL_2101466, EPI_ISL_2101467, EPI_ISL_2101469, EPI_ISL_2101474, EPI_ISL_2101476, EPI_ISL_2101477, EPI_ISL_2101478, EPI_ISL_2101479, EPI_ISL_2101480, EPI_ISL_2101481, EPI_ISL_2101486, EPI_ISL_2101487, EPI_ISL_2101488, EPI_ISL_2101489, EPI_ISL_2101490, EPI_ISL_2101491, EPI_ISL_2101492, EPI_ISL_2101493, EPI_ISL_2101496, EPI_ISL_2101497, EPI_ISL_2101498, EPI_ISL_2101499, EPI_ISL_2101500, EPI_ISL_2101501, EPI_ISL_2101502, EPI_ISL_2101503, EPI_ISL_2101504, EPI_ISL_2101507, EPI_ISL_2101514, EPI_ISL_2101515, EPI_ISL_2101516, EPI_ISL_2101517, EPI_ISL_2101518, EPI_ISL_2101519, EPI_ISL_2101528, EPI_ISL_2101529, EPI_ISL_2101530, EPI_ISL_2101531, EPI_ISL_2101532, EPI_ISL_2101533, EPI_ISL_2101534, EPI_ISL_2101535, EPI_ISL_2101536, EPI_ISL_2101542, EPI_ISL_2101543, EPI_ISL_2101544, EPI_ISL_2101545, EPI_ISL_2101546, EPI_ISL_2101547, EPI_ISL_2101548, EPI_ISL_2101549, EPI_ISL_2101550, EPI_ISL_2101551, EPI_ISL_2101552, EPI_ISL_2101553, EPI_ISL_2101554, EPI_ISL_2101555, EPI_ISL_2101556, EPI_ISL_2101557, EPI_ISL_2101558, EPI_ISL_2101559, EPI_ISL_2101560, EPI_ISL_2101561, EPI_ISL_2101562, EPI_ISL_2101563, EPI_ISL_2101564, EPI_ISL_2101565, EPI_ISL_2101573, EPI_ISL_2101574, EPI_ISL_2101575, EPI_ISL_2101576, EPI_ISL_2101577, EPI_ISL_2101578, EPI_ISL_2101579, EPI_ISL_2101580, EPI_ISL_2101582, EPI_ISL_2101583, EPI_ISL_2101584, EPI_ISL_2101586, EPI_ISL_2101587, EPI_ISL_2101588, EPI_ISL_2101589, EPI_ISL_2101590, EPI_ISL_2101591, EPI_ISL_2101592, EPI_ISL_2101593, EPI_ISL_2101594, EPI_ISL_2101595, EPI_ISL_2101596, EPI_ISL_2101597, EPI_ISL_2101598, EPI_ISL_2101599, EPI_ISL_2101600, EPI_ISL_2101601, EPI_ISL_2101602, EPI_ISL_2101603, EPI_ISL_2101604, EPI_ISL_2101605, EPI_ISL_2101606, EPI_ISL_2101607, EPI_ISL_2101608, EPI_ISL_2101609, EPI_ISL_2101610, EPI_ISL_2101611, EPI_ISL_2101613, EPI_ISL_2101615, EPI_ISL_2101616, EPI_ISL_2101617, EPI_ISL_2101618, EPI_ISL_2101622, EPI_ISL_2101623, EPI_ISL_2101624, EPI_ISL_2101625, EPI_ISL_2101626, EPI_ISL_2101627, EPI_ISL_2101628, EPI_ISL_2101629, EPI_ISL_2101630, EPI_ISL_2101631, EPI_ISL_2101632, EPI_ISL_2101633, EPI_ISL_2101634, EPI_ISL_2101635, EPI_ISL_2101636, EPI_ISL_2101637, EPI_ISL_2101672, EPI_ISL_2101673, EPI_ISL_2101674, EPI_ISL_2101675, EPI_ISL_2101691, EPI_ISL_2101697, EPI_ISL_2101698, EPI_ISL_2101699, EPI_ISL_2101700, EPI_ISL_2101701, EPI_ISL_2101702, EPI_ISL_2101703, EPI_ISL_2101704, EPI_ISL_2101705, EPI_ISL_2101706, EPI_ISL_2101707, EPI_ISL_2101716, EPI_ISL_2101717, EPI_ISL_2101719, EPI_ISL_2101720, EPI_ISL_2101723, EPI_ISL_2101725, EPI_ISL_2101728, EPI_ISL_2101729, EPI_ISL_2101730, EPI_ISL_2101735, EPI_ISL_2101736, EPI_ISL_2101737, EPI_ISL_2101738, EPI_ISL_2101739, EPI_ISL_2101745 | | | |
| see above | Laboratorio Central Noel Nutels | Bioinformatics Laboratory / LNCC | Alessandra P Lamarca; Alexandra L Gerber; Amilcar Tanuri; Ana Paula de C Guimaraes; Ana Tereza R Vasconcelos; Andrea Cony Cavalcanti; Caio Luiz Pereira Ribeiro; Cassia Alves; Cintia Policarpo; Claudia Maria Braga de Mello; Cristiane Gomes da Silva; Diana Mariani; Douglas Terra Machado; Flavio Dias da Silva; Gleidson da Silva de Oliveira; Leandro Magalhaes de Souza; Liliane Cavalcante; Luiz G P de Almeida; Marcio Henrique de Oliveira Garcia; Mario Sergio Ribeiro; Ronaldo da Silva F Jr; Silvia Carvalho |
| EPI_ISL_4196736 | Laboratorio Central Sección Virología, Hospital Regional Artemides Zatti | Laboratorio Central Mg. Luis Alfredo Pianciola on behalf of 'Proyecto Argentino Interinstitucional de genomica de SARS-CoV-2' (PAIS Consortium) | A De Fino.; C Pintos; C Rastellini; C Ziehm; D F di Prátula; J Ousset; L Pianciola; M Fernandez; M Mazzeo |
| EPI_ISL_2402216 | Laboratorio Central de Epidemiología (LCE) | Unidad de Genomica Avanzada | Alejandro Sanchez-Flores; Alfredo Herrera-Estrella; Alicia Ocana-Mondragon; Angel Gustavo Salas-Lais; Bernardo Martinez-Miguel; Blanca Taboada; Brenda Irasema Maldonado-Meza; Carla Ivon Herrera-Najera; Carlos F. Arias; Celia Boukadida; Clara Esperanza Santacruz-Tinoco; Concepcion Grajales-Muniz; Consorcio Mexicano de Vigilancia Genomica (CoViGen-Mex). Authors (in alphabetical order): Julio Elias Alvarado-Yaah; Fernando Fontove-Herrera; Francisco Pulido; Gloria Elena Espinoza-Ayala; Gloria Maria Molina-Salinas; Gloria Vazquez; Hector Esteban Paz-Juarez; Hector Montoya-Fuentes; Helen Haydee Fernanda Ramirez-Plascencia; Jorge Ivan Salinal-Nevarez; Jose Antonio Enciso-Moreno; Jose Esteban Munoz-Medina; Jose de Jesus Nunez-Contreras; Juan Bautista Chale-Dzul; Luis Alberto Ochoa-Carrera; Margarita Matias-Florentino; Maria Guadalupe Santiago-Mauricio; Maria Guadalupe de Jesus Mireles-Rivera; Nelly Selem-Mojica; Pavel Isa; Ricardo Grande; Santiago Avila-Rios; Victor Hugo Borja-Aburto |
| EPI_ISL_6944355, EPI_ISL_6944488, EPI_ISL_6944489 | Laboratorio Central de Salud Publica | Viral Special Pathogens Branch, Centers for Disease Control and Prevention | Andrea Gómez de la Fuente; Cynthia Vázquez; Emir Talundzic; Joel Montgomery; John Klena; Juan Torales; Justin Lee; María José Ortega; María Liz Gamarra; Shannon Whitmer; Shirley Villalba |
| EPI_ISL_2536338, EPI_ISL_3061882, EPI_ISL_3061889, EPI_ISL_3061890 | Laboratorio Central de Saude Publica do Estado da Paraiba (LACEN-PB) | Laboratory of Respiratory Viruses and Measles, Oswaldo Cruz Institute, FIOCRUZ | Alice Sampaio Rocha; Ana Carolina Mendonca; Anna Carolina Paixao; Dalane Loudal Florentino Teixeira; Elisa Cavalcante Pereira; Fernando Motta; Joao Felipe Bezerra; Luciana Appolinario; Marilda Siqueira on behalf of the Fiocruz COVID-19 Genomic Surveillance Network; Paola Resende; Renata Serrano Lopes; Taina Venas |
| EPI_ISL_2274044, EPI_ISL_2274065, EPI_ISL_2274069, EPI_ISL_2274070, EPI_ISL_2274071, EPI_ISL_2466246, EPI_ISL_2466247, EPI_ISL_2466248, EPI_ISL_2466249, EPI_ISL_2466250, EPI_ISL_2466251, EPI_ISL_2466252, EPI_ISL_2466253, EPI_ISL_2466254, EPI_ISL_2466255, EPI_ISL_2466256, EPI_ISL_2466257, EPI_ISL_2466258, EPI_ISL_2466259, EPI_ISL_2557335 | | | |
| see above | Laboratorio Central de Saude Publica do Estado de Alagoas (LACEN/AL) | Laboratory of Respiratory Viruses and Measles, Oswaldo Cruz Institute, FIOCRUZ | Alice Sampaio Rocha; Ana Carolina Mendonca; Anderson Brandao Leite; Anna Carolina Paixao; Elisa Cavalcante Pereira; Fernando Motta; Luciana Appolinario; Marilda Siqueira on behalf of the Fiocruz COVID-19 Genomic Surveillance Network; Paola Resende; Renata Serrano Lopes; Taina Venas |
| EPI_ISL_3045427, EPI_ISL_3045432, EPI_ISL_3045433, | Laboratorio Central de Saude Publica do Estado do Espirito Santo (LACEN/ES) | Laboratory of Respiratory Viruses and Measles, Oswaldo Cruz Institute, FIOCRUZ | Alice Sampaio Rocha; Ana Carolina Mendonca; Anna Carolina Paixao; Elisa Cavalcante Pereira; Fernando Motta; Luciana Appolinario; Marilda Siqueira on behalf of the Fiocruz COVID-19 Genomic Surveillance Network; Paola Resende; Renata Serrano Lopes; Rodrigo Ribeiro Rodrigues; Taina Venas |

| | | | |
|---|---|---|---|
| EPI_ISL_3045448, EPI_ISL_3190238 | | | |
| EPI_ISL_2983253, EPI_ISL_2983266, EPI_ISL_2983268, EPI_ISL_2983269, EPI_ISL_2983270, EPI_ISL_2983271, EPI_ISL_2983272, EPI_ISL_2983273, EPI_ISL_2983274, EPI_ISL_2983275, EPI_ISL_2983276, EPI_ISL_2983277, EPI_ISL_2983278, EPI_ISL_2983279, EPI_ISL_2983282, EPI_ISL_2983283, EPI_ISL_2983284, EPI_ISL_2983304, EPI_ISL_2983325 | | | |
| see above | Laboratorio Central de Saude Publica do Estado do Maranhao (LACEN-MA) | Laboratory of Respiratory Viruses and Measles, Oswaldo Cruz Institute, FIOCRUZ | Agatha Cristinne Prudencio; Alice Sampaio Rocha; Ana Carolina Mendonca; Anna Carolina Paixao; Elisa Cavalcante Pereira; Fernando Motta; Ighor Leonardo Arantes Gomes; Lidio Gonçalves Lima Neto; Luciana Appolinario; Marilda Siqueira on behalf of the Fiocruz COVID-19 Genomic Surveillance Network; Paola Resende; Renata Serrano Lopes; Taina Moreira Venas |
| EPI_ISL_2536354 | Laboratorio Central de Saude Publica do Estado do Para (LACEN/PA) | Laboratory of Respiratory Viruses and Measles, Oswaldo Cruz Institute, FIOCRUZ | Alice Sampaio Rocha; Ana Carolina Mendonca; Anna Carolina Paixao; Elisa Cavalcante Pereira; Fernando Motta; Luciana Appolinario; Marilda Siqueira on behalf of the Fiocruz COVID-19 Genomic Surveillance Network; Paola Resende; Renata Serrano Lopes; Taina Venas; Valnete Andrade |
| EPI_ISL_2603449, EPI_ISL_2603450, EPI_ISL_2603581, EPI_ISL_2603582, EPI_ISL_2603583, EPI_ISL_2603584, EPI_ISL_2603585, EPI_ISL_2603586, EPI_ISL_2603587, EPI_ISL_2603589, EPI_ISL_2603590 | | | |
| see above | Laboratorio Central de Saude Publica do Estado do Rio Grande do Sul (LACEN-RS) | Laboratory of Respiratory Viruses and Measles, Oswaldo Cruz Institute, FIOCRUZ | Alice Sampaio Rocha; Ana Carolina Mendonca; Anderson Brandao Leite; Anna Carolina Paixao; Elisa Cavalcante Pereira; Fernando Motta; Luciana Appolinario; Marilda Siqueira on behalf of the Fiocruz COVID-19 Genomic Surveillance Network; Paola Resende; Renata Serrano Lopes; Taina Venas |
| EPI_ISL_3048802, EPI_ISL_3048803, EPI_ISL_3048805, EPI_ISL_3048807 | Laboratorio Central de Saude Publica do Estado do Rio Grande do Sul (LACEN-RS) | Laboratório de Biologia Molecular da Universidade Federal de Ciências da Saúde de Porto Alegre | Adriana Seixas; Ana B. G. Veiga; Ana Paula Muterle Varela; Fabiana Quoos Mayer; Fernando Hayashi Sant'Anna; Janira Prichula; Letícia Garay Martins; Richard Steiner Salvato; Tatiana Schäffer Gregianini |
| EPI_ISL_3033035, EPI_ISL_3033037, EPI_ISL_3033040 | Laboratorio Central de la Ciudad de Santa Fe | Grupo de Genómica y Bioinformática del Instituto de Investigación de la Cadena Láctea CONICET-INTA on behalf of 'Proyecto Argentino Interinstitucional de genomica de SARS-CoV-2' (PAIS Consortium) | AF; Amadio; C; Eberhardt; G; Irazoqui; JM; MF; Mugna; Ojeda; Pastor; Rompato; V |
| EPI_ISL_4197261, EPI_ISL_4197262 | Laboratorio Clínico Analizar | Gencore - Universidad de los Andes | Cristian Barrera; David González; Gabriela Ariza; Luisa Sacristan; Marcela Guevara; Marcela Mercado; Silvia Restrepo |
| EPI_ISL_5800763, EPI_ISL_5800764, EPI_ISL_5800781, EPI_ISL_5800783, EPI_ISL_5800785, EPI_ISL_5800786, EPI_ISL_5800787, EPI_ISL_5800788, EPI_ISL_5800789, EPI_ISL_5800790 | | | |
| see above | Laboratorio De Franca | Instituto Butantan | Antonio Jorge Martins; Claudia Renata dos Santos Barros; David Schlesinger; Debora Botequio Moretti; Dimas Tadeu Covas; Elaine Cristina Marqueze; Elaine Vieira Santos; Evandra Strazza Rodrigues; Heidge Fukumasu; Jayme Augusto de Souza-Neto; José Salvatore Leister Patané; Luiz Alcantara; Luiz Lehmann Coutinho; Maria Carolina Elias; Maurício Lacerda Nogueira; Rafael dos Santos Bezerra; Raul Machado Neto; Rejane Maria Tommasini Grotto; Ricardo Haddad; Sandra Coccuzzo Sampaio Vessoni; Simone Kashima; Svetoslav Nanev Slavov; Vincent Louis Viala |
| EPI_ISL_5199340 | Laboratorio Echavarria | Instituto Nacional de Salud-Dirección de Investigación en Salud Pública | Beatriz de Arco; Carlos Franco-Muñoz; Carmen Osorio; Diana Malo; Diego A. Álvarez-Díaz; Diego Andrés Prada; Gerardo Santamaría; Hector Alejandro Ruiz-Moreno; Jhonnatan Reales-González; Jorge Rivera; Juan Camilo Martinez; Julian Naizaque; Katherine Laiton-Donato; Lisseth Pardo; Magdalena Wiesner; Marcela Mercado-Reyes; Maria T. Herrera-Sepúlveda; Marta Lopez Blanco; Martha Lucia Ospina Martinez; Paola Rojas; Sergio Gomez; Sheryll Corchuelo; Tatiana Cobos; Ángela Alarcon Cruz |
| EPI_ISL_5800726, EPI_ISL_5800762, EPI_ISL_5800784, EPI_ISL_5800798, EPI_ISL_5800815, EPI_ISL_5800846, EPI_ISL_5800898, EPI_ISL_5800901, EPI_ISL_5800967, EPI_ISL_5800968, EPI_ISL_5800976, EPI_ISL_5800981, EPI_ISL_5800983, EPI_ISL_5800991, EPI_ISL_5801037, EPI_ISL_5801044, EPI_ISL_5801045, EPI_ISL_5801157, EPI_ISL_5801237 | | | |
| see above | Laboratorio Municipal De Piracicaba | Instituto Butantan | Antonio Jorge Martins; Claudia Renata dos Santos Barros; David Schlesinger; Debora Botequio Moretti; Dimas Tadeu Covas; Elaine Cristina Marqueze; Elaine Vieira Santos; Evandra Strazza Rodrigues; Heidge Fukumasu; Jayme Augusto de Souza-Neto; José Salvatore Leister Patané; Luiz Alcantara; Luiz Lehmann Coutinho; Maria Carolina Elias; Maurício Lacerda Nogueira; Rafael dos Santos Bezerra; Raul Machado Neto; Rejane Maria Tommasini Grotto; Ricardo Haddad; Sandra Coccuzzo Sampaio Vessoni; Simone Kashima; Svetoslav Nanev Slavov; Vincent Louis Viala |
| EPI_ISL_3307455, EPI_ISL_3307456, EPI_ISL_3307457, EPI_ISL_3307458 | Laboratorio SYNLAB Colombia | Corporación para Investigaciones Biológicas-CIB | Jeanneth Mosquera Rendon; Jenny Santiago Cuesta; Marcela Mercado Reyes; Uriel A. Hurtado Paez |
| EPI_ISL_2777494, EPI_ISL_4030327, EPI_ISL_4030335, EPI_ISL_4030340 | Laboratorio de Ecologia de Doencas Transmissiveis na Amazonia, Instituto Leonidas e Maria Deane - Fiocruz Amazonia | Laboratorio de Ecologia de Doencas Transmissiveis na Amazonia, Instituto Leonidas e Maria Deane - Fiocruz Amazonia | Alex Martins; André Corado; Debora Duarte; Fabiola Mendonça da Silva Chui; Felipe Naveca; Fernanda Nascimento; Fernando Fonseca de Almeida Val; George Silva; Gisely Cardoso de Melo; Karina Pessoa; Luciana Gonçalves; Marcus Vinicius Guimarães de Lacerda; Maria Júlia Brandão; Maria Paula Gomes Mourão; Mariana Xavier Simão; Matilde Mejía; Michele Jesus; Valdinete Nascimento; Vanderson de Souza Sampaio; Victor Souza; Ágatha Costa |
| EPI_ISL_2728552, EPI_ISL_2728564, EPI_ISL_2728565, EPI_ISL_2728571, EPI_ISL_3944410 | Laboratorio de Infectologia y Virologia Molecular | Laboratory of Molecular Virology, School of Medicine, Pontificia Universidad Catolica de Chile | Alejandro Bhrun; Ana Maria Contreras; Andres E. Munoz-Marcos; Carlos Palma; Catalina Pardo-Roa; Constanza Maldonado; Constanza Martinez-Valdevenito; Eileen Serrano; Erick Salinas; Estefany Poblete; Francisco Melo; Jennifer Angulo; Jorge Levican; Leonardo I. Almonacid; M. Belen Leyton; Magdalena Vera; Marcela Ferres; Maria Jose Avendano; Rafael A. Medina; Tamara Garcia-Salum |
| EPI_ISL_2854131 | Laboratorio de Medicina Molecular, Universidad de Magallanes | Centro Asistencial Docente y de Investigacion, Universidad de Magallanes | Constanza Ceroni; Diego Alvarez; Jacqueline Aldridge; Jorge González; Marcelo Navarrete; Roberto Uribe-Paredes |
| EPI_ISL_2008946, EPI_ISL_2008947, EPI_ISL_2008949, EPI_ISL_2008950, EPI_ISL_2008952, EPI_ISL_2008953, EPI_ISL_2008954, EPI_ISL_2008955, EPI_ISL_2008957, EPI_ISL_2008958, EPI_ISL_2008959, EPI_ISL_2008960, EPI_ISL_2008963, EPI_ISL_2008964, EPI_ISL_2008967, EPI_ISL_3048956, EPI_ISL_3048957, EPI_ISL_3048958, EPI_ISL_3761525, EPI_ISL_3761749 | | | |
| see above | Laboratorio de Pesquisa em Virologia, FAMERP, SJRP | Laboratorio de Pesquisa em Virologia, FAMERP, SJRP | Beatriz de Carvalho Marques; Cecília Artico Banho; Cíntia Bittar; Fábio Sossai Possebon; Guilherme Campos; Helena Lage Ferreira; Jorge A. Petroli Marchesi; João Pessoa Araújo Jr.; Leila Sabrina Ullmann; Lívia Sacchetto; Maisa C. Pereira Parra; Marília Moraes; Maurício L. Nogueira.; Paula Rahal; Paulo Inacio da Costa |
| EPI_ISL_3401523 | Laboratorio de Referencia Nacional de Virus Respiratorios. Centro Nacional de Salud Publica. Instituto Nacional de Salud Peru. | Laboratorio de Referencia Nacional de Virus Respiratorios. Centro Nacional de Salud Publica. Instituto Nacional de Salud Peru. | Carlos Padilla Rojas; Henri Bailon Calderon; Iris Silva Molina; Joseph Huayra Niquen; Lely Solari Zerpa; Luis Barcena Flores; Marco Galarza Perez; Nancy Rojas Serrano; Nieves Sevilla Castañeda; Omar Caceres Rey; Orson Mestanza Millones; Princesa Medrano Alhuay; Priscila Lope Pari; Sandra Morales Ruiz; Sara Gordillo Vilchez; Steve Acedo Lazo; Veronica Hurtado Vela; Victor Jimenez Vasquez; Wendy Lizarraga Olivares |
| EPI_ISL_3023587, EPI_ISL_3023599, EPI_ISL_3375928, EPI_ISL_3376004, EPI_ISL_3376005, EPI_ISL_3376007, EPI_ISL_3376013, EPI_ISL_3376020, EPI_ISL_3376022, EPI_ISL_3376029, EPI_ISL_3376030 | | | |
| see above | Laboratorio de Referencial Nacional de Virus Respiratorios | Laboratorio de Referencial Nacional de Virus Respiratorios | Carlos Padilla Rojas; Henri Bailon Calderon; Iris Silva Molina; Joseph Huayra Niquen; Lely Solari Zerpa; Luis Barcena Flores; Marco Galarza Perez; Nancy Rojas Serrano; Omar Caceres Rey; Orson Mestanza Millones; Priscila Lope Pari; Sandra Morales Ruiz; Steve Acedo Lazo; Veronica Hurtado Vela |
| EPI_ISL_4405065, EPI_ISL_4405070, EPI_ISL_4405073 | Laboratorio de Virología del Hospital de Niños Dr. Ricardo Gutierrez | Área de Secuenciación del Laboratorio de Virología del Hospital de Niños Dr. Ricardo Gutierrez on behalf of 'Proyecto Argentino Interinstitucional de genomica de SARS-CoV-2' (PAIS Consortium) | AS; Acevedo; Acuña; Alexay; Alvarez Lopez; C; D; E; Goya; Gravis; Jacques; LE; Lusso; ME; MI; MS; Mistchenko; Nabaes Jodar; Natale; O; S; Valinotto; Viegas, M. |
| EPI_ISL_4197199, EPI_ISL_4197214, EPI_ISL_4212170, EPI_ISL_4212172 | Laboratorio de la Clínica Los Nogales | Gencore - Universidad de los Andes | Cristian Barrera; David González; Gabriela Ariza; Luisa Sacristan; Marcela Guevara; Marcela Mercado; Silvia Restrepo |
| EPI_ISL_4405378 | Laboratorio de salud pública, Facultad de Ciencias Exactas, Universidad Nacional de La Plata | Área de Secuenciación del Laboratorio de Virología del Hospital de Niños Dr. Ricardo Gutierrez on behalf of 'Proyecto Argentino Interinstitucional de genomica de SARS-CoV-2' (PAIS Consortium) | A; Acuña; Angeletti; Cordero; D; Goya; LE; Lusso; MI; MS; Nabaes Jodar; Nadalich; Natale; R; S; Toro; V; Valinotto; Viegas, M. |
| EPI_ISL_4405468 | Laboratorio del Hospital Interzonal General de Agudos " Evita " | Área de Secuenciación del Laboratorio de Virología del Hospital de Niños Dr. Ricardo Gutierrez on behalf of 'Proyecto Argentino Interinstitucional de genomica de SARS-CoV-2' (PAIS Consortium) | A; Acuña; D; Desimone; E; Goya; Grossi; I; L; LE; Luczac; Lusso; MI; MS; Musto; Nabaes Jodar; Natale; O; S; Serrano; Valinotto; Viegas, M. |
| EPI_ISL_1840646 | Laboratorio di Microbiologia | Laboratorio di Microbiologia | Martinetti Lucchini Gladys; Valeria Spina |
| EPI_ISL_1925323 | Laboratorio di Microbiologia e Virologia, Università Vita-Salute San Raffaele, Milano | Laboratorio di Microbiologia e Virologia, Università Vita-Salute San Raffaele, Milano | Elena Criscuolo; Enzo Boeri; Massimo Clementi; Matteo Castelli; Michela Sampaolo; Nicasio Mancini; Nicola Clementi; Roberta Antonia Diotti; Roberto Ferrarese |
| EPI_ISL_2450848, EPI_ISL_2450849, EPI_ISL_2450850, EPI_ISL_2450851 | Laboratorio di Patologia Clinica, Ospedale San Paolo in Valloria, ASL 2 Liguria | U.O. Igiene, Ospedale Policlinico San Martino | Bruzzone Bianca; Caligiuri Patrizia; De Pace Vanessa; Domnich Alexander; Icardi Giancarlo on behalf of SARS-CoV-2 ITALIAN RESEARCH ENTERPRISE-(SCIRE) Collaborative Group; Lillo Flavia; Orsi Andrea; Ricucci Valentina |
| EPI_ISL_2657857 | Laboratorios del Valle SAS | Instituto Nacional de Salud-Dirección de Investigación en Salud Pública | Carlos Franco-Muñoz; Carmen Osorio; Diana Malo; Diego A. Álvarez-Díaz; Diego Andrés Prada; Gerardo Santamaría; Hector Alejandro Ruiz-Moreno; Jhonnatan Reales-González; Jorge Rivera; Juan Camilo Martinez; Julian Naizaque; Katherine Laiton-Donato; Lisseth Pardo; Magdalena Wiesner; Marcela Mercado-Reyes; Maria T. Herrera-Sepúlveda; Marta Lopez Blanco; Martha Lucia Ospina Martinez; Paola Rojas; Sergio Gomez; Sheryll Corchuelo; Ángela Alarcon Cruz |

| | | | |
|---|---|---|---|
| EPI_ISL_2015713 | Laboratorium Analiz Lekarskich Alab Rzeszów | 1. National Institute of Public Health - National Institute of Hygiene, Warsaw, Poland 2. Biobank Lab, University of Lodz 3. Laboratory of Respiratory Viruses, Teaching and Clinical Center of the Medical University of Lodz | Dominik Strapagiel; Izabela Dróżdż; Jakub Lach; Katarzyna Zacharczuk; Klaudyna Królikowska; Maciej Borowiec; Magdalena Nowakowska; Magdalena Traczyk-Borszyńska; Marcin Słomka; Marta Sobalska-Kwapis; Małgorzata Sadkowska-Todys; Tomasz Płoszaj; Tomasz Wołkowicz |
| EPI_ISL_1801991, EPI_ISL_1802005, EPI_ISL_1802007, EPI_ISL_1802033, EPI_ISL_1925716, EPI_ISL_1925717, EPI_ISL_1925718, EPI_ISL_1925723, EPI_ISL_1925724, EPI_ISL_1925726, EPI_ISL_1925731, EPI_ISL_1925735, EPI_ISL_1925739, EPI_ISL_1925743, EPI_ISL_1925788, EPI_ISL_1925837, EPI_ISL_1926111, EPI_ISL_1926122, EPI_ISL_1926181, EPI_ISL_1926185, EPI_ISL_1926188, EPI_ISL_1926200, EPI_ISL_1926237, EPI_ISL_1926238, EPI_ISL_1926281, EPI_ISL_1926284, EPI_ISL_1926294, EPI_ISL_1926298, EPI_ISL_1926306, EPI_ISL_1926314, EPI_ISL_1926315, EPI_ISL_1926316, EPI_ISL_1926383, EPI_ISL_1926386, EPI_ISL_1926411, EPI_ISL_1926424, EPI_ISL_1926441, EPI_ISL_1926483, EPI_ISL_1926496, EPI_ISL_1926516, EPI_ISL_1926545, EPI_ISL_1926589, EPI_ISL_1926612, EPI_ISL_1926615, EPI_ISL_1926618, EPI_ISL_1926620, EPI_ISL_1926621, EPI_ISL_1926626, EPI_ISL_1926647, EPI_ISL_1926649, EPI_ISL_1926667, EPI_ISL_1926675, EPI_ISL_1926678, EPI_ISL_1926681, EPI_ISL_1926685, EPI_ISL_1926690, EPI_ISL_1926692, EPI_ISL_1926715, EPI_ISL_1926748, EPI_ISL_1926813, EPI_ISL_1926829, EPI_ISL_1926830, EPI_ISL_1926832, EPI_ISL_1926835, EPI_ISL_1926836, EPI_ISL_1926838, EPI_ISL_1926839, EPI_ISL_1926977, EPI_ISL_1926980, EPI_ISL_1926989, EPI_ISL_1926998, EPI_ISL_1927003, EPI_ISL_1927006, EPI_ISL_1927009, EPI_ISL_1927018, EPI_ISL_1927033, EPI_ISL_1927036, EPI_ISL_1927049, EPI_ISL_1927055, EPI_ISL_1927057, EPI_ISL_1927072, EPI_ISL_1927085, EPI_ISL_1927103, EPI_ISL_1927134, EPI_ISL_1927140, EPI_ISL_1927142, EPI_ISL_1927250, EPI_ISL_1927259, EPI_ISL_1927272, EPI_ISL_1927277, EPI_ISL_1927281, EPI_ISL_1927307, EPI_ISL_1927470, EPI_ISL_1927600, EPI_ISL_1927619, EPI_ISL_1927625, EPI_ISL_1927856, EPI_ISL_1927859, EPI_ISL_1927877, EPI_ISL_1927983, EPI_ISL_1928034, EPI_ISL_1928118, EPI_ISL_1928418, EPI_ISL_1928628, EPI_ISL_1928660, EPI_ISL_1928720, EPI_ISL_1928723, EPI_ISL_1928728, EPI_ISL_1928730, EPI_ISL_1928731, EPI_ISL_1928733, EPI_ISL_1928734, EPI_ISL_1928741, EPI_ISL_1928742, EPI_ISL_1928754, EPI_ISL_1928771, EPI_ISL_1928787, EPI_ISL_1928800, EPI_ISL_1928803, EPI_ISL_1928804, EPI_ISL_1928805, EPI_ISL_1928807, EPI_ISL_1928809, EPI_ISL_1928810, EPI_ISL_1928811, EPI_ISL_1928812, EPI_ISL_1928814, EPI_ISL_1928818, EPI_ISL_1928828, EPI_ISL_1928829, EPI_ISL_1928837, EPI_ISL_1928838, EPI_ISL_1928839, EPI_ISL_1928879, EPI_ISL_1928885, EPI_ISL_1928897, EPI_ISL_1928901, EPI_ISL_1928906, EPI_ISL_1928907, EPI_ISL_1928908, EPI_ISL_1928909, EPI_ISL_1928912, EPI_ISL_1928914, EPI_ISL_1928915, EPI_ISL_1928916, EPI_ISL_1928928, EPI_ISL_1928929, EPI_ISL_1928932, EPI_ISL_1928934, EPI_ISL_1928935, EPI_ISL_1928937, EPI_ISL_1928980, EPI_ISL_1928982, EPI_ISL_1928986, EPI_ISL_1928987, EPI_ISL_1929039, EPI_ISL_1929048, EPI_ISL_1929066, EPI_ISL_1929111, EPI_ISL_1929137, EPI_ISL_1929161, EPI_ISL_1929198, EPI_ISL_1929553, EPI_ISL_1929562, EPI_ISL_1929565, EPI_ISL_1929706, EPI_ISL_1929733, EPI_ISL_1929736, EPI_ISL_1929786, EPI_ISL_1929867, EPI_ISL_1929882, EPI_ISL_1929905, EPI_ISL_1929984, EPI_ISL_1930079, EPI_ISL_1930081, EPI_ISL_1930084, EPI_ISL_1930095, EPI_ISL_1930106, EPI_ISL_1930112, EPI_ISL_1930140, EPI_ISL_1930150, EPI_ISL_1930153, EPI_ISL_1930162, EPI_ISL_1930176, EPI_ISL_1930234, EPI_ISL_1930237, EPI_ISL_1930239, EPI_ISL_1930270, EPI_ISL_1930276, EPI_ISL_1930310, EPI_ISL_1930313, EPI_ISL_1930391, EPI_ISL_1930765, EPI_ISL_1930769, EPI_ISL_1930775, EPI_ISL_1930805, EPI_ISL_1930831, EPI_ISL_1930883, EPI_ISL_1931193, EPI_ISL_1931258, EPI_ISL_1931384, EPI_ISL_1931585, EPI_ISL_1931703, EPI_ISL_1931714, EPI_ISL_1931848, EPI_ISL_2044060, EPI_ISL_2044063, EPI_ISL_4375968, EPI_ISL_4376642, EPI_ISL_4378998 | | | |
| see above | Laboratory Corporation of America | Centers for Disease Control and Prevention Division of Viral Diseases, Pathogen Discovery | Adrian Paskey; Amanda Douglas; Amanda Suchanek; Andrea Throop; Ayla Burns; Benjamin Rambo-Martin; Bobbi Croy; Brian Krueger; Brian Norvell; Christopher Gulvick; Christos Petropoulos; Clinton Paden; Clinton R. Paden; Craig Lukasik; Dakota Howard; Darlene Wagner; Debbie Boles; Dhwani Batra; Duncan MacCannell; Eyad Almasri; Goran Stevovic; Howard Engler; Hrushikesh Deshmukh; Jake Humphrey; Jana Schroth; Jason Caravas; Joe Voshell; John Pruitt; Jonathan Meltzer; Jonathan Williams; Kara Moser; Kimberly Wagner; Kristine Lacek; Lax Iyer; Lisa Pfefferle; Lyndon Tilson; Manoj Jain; Marcia Eisenberg; Mary Ann Cristobal; Mary Cristobal; Mary Williamson; Matthew Robinson; Matthew Schmerer; Michael Levandoski; Mike Sapeta; Mindy Nye; Minoo Agarwal; Mohan Kolli; Nuthawin Charoensri; Oren Cohen; Peter Cook; Peter W. Cook; Prashant Gupta; Qian Zeng; Rama Ghatti; Scott Parker; Scott Ryan; Scott Sammons; Shatavia Morrison; Stanley Letovsky; Steven Ragan; Suresh Babu Selvaraju; Suresh Selvaraju; Susan Countryman; Susan Hicks; Suzanne Dale; Thomas Urban; Tim Kuphal; Tricia Zwiefelhofer; Tymeckia Kendall; Victoria Caban Figueroa; Vincent Drouillon; Yvette Unoarumhi |
| EPI_ISL_2835679 | Laboratory Medicine, University of Washington | Laboratory Medicine, University of Washington | Castor; J.L. |
| EPI_ISL_1797466 | Laboratory of Clinical Microbiology, Virology and Bioemergencies, ASST Fatebenefratelli Sacco - Sacco Hospital | Laboratory of Clinical Microbiology, Virology and Bioemergencies, ASST Fatebenefratelli Sacco - Sacco Hospital | Alberto Rizzo; Alessandro Mancon; Fiorenza Bracchitta; Luca Rizzuto; Maria Rita Gismondo; Valeria Micheli |
| EPI_ISL_2157545, EPI_ISL_2443601, EPI_ISL_3434727 | Laboratory of Respiratory Viruses and Measles, Oswaldo Cruz Institute, FIOCRUZ | Laboratory of Respiratory Viruses and Measles, Oswaldo Cruz Institute, FIOCRUZ | Agatha Cristinne Prudencio Soares; Alice Sampaio Rocha; Ana Carolina Mendonca; Andrea Cony Cavalcanti; Anna Carolina Paixao; Elisa Cavalcante Pereira; Fernando Motta; Ighor Leonardo Arantes Gomes; Luciana Appolinario; Marilda Siqueira on behalf of the Fiocruz COVID-19 Genomic Surveillance Network; Paola Resende; Renata Serrano Lopes; Taina Venas |
| EPI_ISL_2696612, EPI_ISL_2696651, EPI_ISL_2696654 | Laboratory, Gravity Diagnostics | Laboratory, Gravity Diagnostics | Diagnostics, G. |
| EPI_ISL_2157542, EPI_ISL_2157543, EPI_ISL_2157544 | Laboratório Central de Saude Publica do Estado da Bahia (LACEN/BA) | Laboratory of Respiratory Viruses and Measles, Oswaldo Cruz Institute, FIOCRUZ | Alice Sampaio Rocha; Ana Carolina Mendonca; Anna Carolina Paixao; Elisa Cavalcante Pereira; Felicidade Pereira; Fernando Motta; Luciana Appolinario; Marilda Siqueira on behalf of the Fiocruz COVID-19 Genomic Surveillance Network; Paola Resende; Renata Serrano Lopes; Taina Venas |
| EPI_ISL_4600530 | Laboratório Central de Saúde Pública de Pernambuco | Coordenação Geral de Laboratórios de Saúde Pública (CGLAB/DAEVS/SVS/MS) | Vagner Fonseca; et al. |
| EPI_ISL_2777235, EPI_ISL_2777280, EPI_ISL_2777311, EPI_ISL_2777313, EPI_ISL_2777314, EPI_ISL_2777333, EPI_ISL_2777529, EPI_ISL_2777550, EPI_ISL_2777600, EPI_ISL_2777622, EPI_ISL_2777753, EPI_ISL_2777820, EPI_ISL_2777821, EPI_ISL_2777823, EPI_ISL_2777874, EPI_ISL_2777875, EPI_ISL_2777876, EPI_ISL_2777877, EPI_ISL_2777878, EPI_ISL_2777972, EPI_ISL_2777973, EPI_ISL_2777974, EPI_ISL_2777975, EPI_ISL_2777976, EPI_ISL_2778008 | | | |
| see above | Laboratório Central de Saúde Pública do Amazonas - LACEN-AM | Laboratorio de Ecologia de Doencas Transmissiveis na Amazonia, Instituto Leonidas e Maria Deane - Fiocruz Amazonia | André Corado; Debora Duarte; Felipe Naveca; Fernanda Nascimento; George Silva; Karina Pessoa; Luciana Gonçalves; Maria Júlia Brandão; Matilde Mejía; Michele Jesus; Valdinete Nascimento; Victor Souza; Ágatha Costa |
| EPI_ISL_2375842, EPI_ISL_2375843, EPI_ISL_2375844, EPI_ISL_2375845, EPI_ISL_2375847, EPI_ISL_2376034, EPI_ISL_2376138, EPI_ISL_2382535, EPI_ISL_2382536, EPI_ISL_2382543, EPI_ISL_3873644, EPI_ISL_3873646, EPI_ISL_3873670 | | | |
| see above | Laboratório de Microbiologia Molecular - Universidade FEEVALE | Molecular Microbiology Laboratory | Alana Witt Hansen; Fernando Rosado Spilki; Flávio Silveira; Fágner Henrique Heldt; Juliana Schons Gularte; Juliane Deise Fleck; Mariana Soares da Silva; Matheus Nunes Weber; Meriane Demoliner; Micheli Filippi; Micheli Filippi.; Paula Rodrigues de Almeida; Vyctoria Malayhka de Abreu Góes Pereira. |
| EPI_ISL_2629821, EPI_ISL_2629823, EPI_ISL_2629824, EPI_ISL_2629825, EPI_ISL_2629826, EPI_ISL_2629827, EPI_ISL_2629828 | | | |
| see above | Laboratório de Virologia Molecular - Universidade Federal do Rio de Janeiro | Laboratório de Virologia Molecular - Universidade Federal do Rio de Janeiro | ; Alice Laschuk Herlinger; Amilcar Tanuri; André Felipe Andrade dos Santos; Carolina Moreira Voloch; Cássia Cristina Alves Gonçalves; Diana Mariani; Débora Souza Faffe; Filipe Romero Rebello Moreira; Francine Bittencourt Schiffler; Isabela de Carvalho Leitão; Marcelo Calado de Paula Tôrres; Matheus Augusto Calvano Cosentino; Mirela D'arc; Orlando da Costa Ferreira Junior; Rafael Mello Galliez; Raíssa Mirella dos Santos Cunha da Costa; Renato Santana de Aguiar; Terezinha Marta Pereira Pinto Castineiras; Thamiris dos Santos Miranda; Átila Duque Rossi |
| EPI_ISL_4520590 | Laboratório de Virologia e Cultivo Celular - LABVICC | Laboratório Baculovírus | Aline Belmok; Bergmann Morais Ribeiro; Clayton Luiz Borges; Déborah Anjos; Elen Regozino Muniz; Fabiola Souza Fiaccadori; Fernando L Melo; Gabriela Silvério Bazilio; Juliana Alves Parente Rocha; Keila Correia de Alcântara; Menira Souza; Nathânia Dábilla; Valéria Christina de Rezende Feres |
| EPI_ISL_2108265, EPI_ISL_2112830 | Labormedizin Darmstadt | Robert Koch Institute | |
| EPI_ISL_2443529, EPI_ISL_2443675, EPI_ISL_2443676, EPI_ISL_2443677, EPI_ISL_2443678, EPI_ISL_2443679, EPI_ISL_2443680, EPI_ISL_2443681, EPI_ISL_2443682, EPI_ISL_2443683, EPI_ISL_2443684, EPI_ISL_2443708 | | | |
| see above | Labortorio Central de Saude Publica do Estado de Santa Catarina (LACEN/SC) | Laboratory of Respiratory Viruses and Measles, Oswaldo Cruz Institute, FIOCRUZ | Alice Sampaio Rocha; Ana Carolina Mendonca; Anna Carolina Paixao; Darcita Buerger Rovaris; Elisa Cavalcante Pereira; Fernando Motta; Luciana Appolinario; Marilda Siqueira on behalf of the Fiocruz COVID-19 Genomic Surveillance Network; Paola Resende; Renata Serrano Lopes; Sandra Bianchini Fernandes; Taina Venas |
| EPI_ISL_2196333, EPI_ISL_2196334, EPI_ISL_2274098, EPI_ISL_2443637, EPI_ISL_2443638, EPI_ISL_2443639, EPI_ISL_2443640, EPI_ISL_2443641, EPI_ISL_2443648, EPI_ISL_2443651, EPI_ISL_2557376, EPI_ISL_2557377, EPI_ISL_2557378, EPI_ISL_2557384 | | | |
| see above | Labortorio Central de Saude Publica do Estado do Rio de Janeiro (LACEN/RJ) | Laboratory of Respiratory Viruses and Measles, Oswaldo Cruz Institute, FIOCRUZ | Alice Sampaio Rocha; Ana Carolina Mendonca; Andrea Cony Cavalcanti; Anna Carolina Paixao; Elisa Cavalcante Pereira; Fernando Motta; Luciana Appolinario; Marilda Siqueira on behalf of the Fiocruz COVID-19 Genomic Surveillance Network; Paola Resende; Renata Serrano Lopes; Taina Venas |
| EPI_ISL_2443528, EPI_ISL_2443542, EPI_ISL_2443543, EPI_ISL_2443544, EPI_ISL_2443549, EPI_ISL_2443550, EPI_ISL_2443572, EPI_ISL_2443573, EPI_ISL_2443574, EPI_ISL_2603460, EPI_ISL_2603461, EPI_ISL_2603466, EPI_ISL_2863855, EPI_ISL_2863856, EPI_ISL_2863857, EPI_ISL_2863858, EPI_ISL_2863859, EPI_ISL_2863860, EPI_ISL_2863861, EPI_ISL_2863862, EPI_ISL_2863863, EPI_ISL_2863864, EPI_ISL_2863867, EPI_ISL_2863895, EPI_ISL_2863896 | | | |
| see above | Lboratorio Central de Saude Publica do Estado do Parana (LACEN/PR) | Laboratory of Respiratory Viruses and Measles, Oswaldo Cruz Institute, FIOCRUZ | Agatha Cristinne Prudencio Soares; Alice Sampaio Rocha; Ana Carolina Mendonca; Anna Carolina Paixao; Elisa Cavalcante Pereira; Fernando Motta; Ighor Leonardo Arantes Gomes; Irina Riediger; Luciana Appolinario; Marilda Siqueira on behalf of the Fiocruz COVID-19 Genomic Surveillance Network; Paola Resende; Renata Serrano Lopes; Taina Venas |
| EPI_ISL_1790765, EPI_ISL_1830458 | Lighthouse Lab in Alderley Park | Wellcome Sanger Institute for the COVID-19 Genomics UK (COG-UK) Consortium | Cordelia Langford; David K. Jackson; Dominic Kwiatkowski; Ewan Harrison; Ian Johnston; Jacquelyn Wynn; Jeffrey Barrett; John Sillitoe on behalf of the Wellcome Sanger Institute COVID-19 Surveillance Team; Mairead Hyland; Roberto Amato; Sonia Goncalves; The Lighthouse Lab in Alderley Park and Alex Alderton |
| EPI_ISL_1740541, EPI_ISL_1740689 | Lighthouse Lab in Cambridge | Wellcome Sanger Institute for the COVID-19 Genomics UK (COG-UK) Consortium | Cordelia Langford; David K. Jackson; Dominic Kwiatkowski; Ewan Harrison; Ian Johnston; Jeffrey Barrett; John Sillitoe on behalf of the Wellcome Sanger Institute COVID-19 Surveillance Team; Rob Howes; Roberto Amato; Sonia Goncalves; The Lighthouse Lab in Cambridge and Alex Alderton |
| EPI_ISL_1790811 | Lighthouse Lab in Glasgow | Wellcome Sanger Institute for the COVID-19 Genomics UK (COG-UK) Consortium | Anna Dominiczak and Alex Alderton; Carol Clugston; Cordelia Langford; David Gray; David K. Jackson; Dominic Kwiatkowski; Ewan Harrison; Harper VanSteenhouse; Ian Johnston; Jeffrey Barrett; John Sillitoe on behalf of the Wellcome Sanger Institute COVID-19 Surveillance Team; Roberto Amato; Sonia Goncalves; Yumi Kasai |
| EPI_ISL_1759260, EPI_ISL_1759893, EPI_ISL_1790999, EPI_ISL_1831282, EPI_ISL_1986931, EPI_ISL_1987187, EPI_ISL_2172155, EPI_ISL_2608287 | | | |
| see above | Lighthouse Lab in Milton Keynes | Wellcome Sanger Institute for the COVID-19 Genomics UK (COG-UK) Consortium | Cordelia Langford; David K. Jackson; Dominic Kwiatkowski; Ewan Harrison; Ian Johnston; Jeffrey Barrett; John Sillitoe on behalf of the Wellcome Sanger Institute COVID-19 Surveillance Team; Roberto Amato; Sonia Goncalves; The Lighthouse Lab in Milton Keynes and Alex Alderton |
| EPI_ISL_1853241 | Limbach - MVZ Humangenetik Ulm | Robert Koch Institute | |
| EPI_ISL_2259243, EPI_ISL_2259248 | Los Angeles County Public Health Laboratories | Los Angeles County Public Health Laboratories | P. Hemarajata et al. |
| EPI_ISL_2373055, EPI_ISL_2373057, EPI_ISL_2373061, EPI_ISL_2373062, EPI_ISL_2373064 | Lurie Children's Hospital of Chicago | Northwestern University - Ozer Lab | Egon A. Ozer; Judd F. Hultquist; Lacy M. Simons; Larry K. Kociolek; Michael G. Ison; Ramon Lorenzo-Redondo; Taylor J. Dean; William J. Muller; Xiaotian; Zheng |
| EPI_ISL_2036067 | M Health Fairview | Minnesota Department of Health, Public Health Laboratory | Alexandra Lorentz; Jacob Garfin; Matt Plumb; and Xiong Wang |
| EPI_ISL_2801331 | MATERNIDADE ESCOLA ASSIS | Analytical Competence Molecular | Cleber Furtado Aksenen e Suzana Porto Almeida; Fabio Miyajima; Fernando Braga Stehling; Francisco Eder de Moura Lopes; Jamille Maria Mendes Bezerra; Joaquim César do Nascimento Sousa Junior; Pedro Miguel Carneiro Jeronimo; Thais Ferreira de Oliveira; Thais de Oliveira Costa; Ticiane Cavalcante de Souza; |

| | | | |
|---|---|---|---|
| | CHATEAUBRIAND | Epidemiology Lab/ACME, Oswaldo Cruz Foundation, Ceara (FIOCRUZ CE) | Veridiana Pessoa Miyajima |
| EPI_ISL_1811581 | MD PHL | MD PHL | Maryland Department of Health Laboratories Administration |
| EPI_ISL_1964592, EPI_ISL_1964628, EPI_ISL_2036337, EPI_ISL_2036451, EPI_ISL_2036606, EPI_ISL_2036673, EPI_ISL_2036719, EPI_ISL_2036722, EPI_ISL_2286019, EPI_ISL_2286283, EPI_ISL_2288733, EPI_ISL_2288734, EPI_ISL_2288735 | | | |
| see above | MEPHI, Aix Marseille University | MEPHI, Aix Marseille University | Anthony LEVASSEUR |
| EPI_ISL_5253373, EPI_ISL_6492531, EPI_ISL_6492561 | MSHS Clinical Microbiology Laboratories | MSHS Pathogen Surveillance Program | Adolfo García-Sastre; Adriana van de Guchte; Ajay Obla; Alberto Paniz-Mondolfi; Ana S. Gonzalez-Reiche; Angela Amoako; Ashley Salimbangon; Betsaida Salom Melo; Bremy Alburquerque; Brianne Ciferri; Charles Gleason; Daniel Floda; Deena R. Altman; Denise Jurczyszak; Emilia Mia Sordillo; Gintaras Deikus; Giulio Kleiner; Gopi Patel; Hala Alshammary; Harm van Bakel; Irina Oussenko; Jayeeta Dutta; Juan Soto; Julia Matthews; Katherine Beach; Kathryn Twyman; Kayla Russo; Komal Srivastava; Levy Sominsky; Mahmoud Awawda; Marta Luksza; Matthew M. Hernandez; Melissa Gitman; Michael D. Nowak; Mitchell J. Sullivan; Nancy Francoeur; Robert Sebra; Sarah Schaefer; Shelcie Fabre; Shwetha Hara Sridhar; Viviana Simon; Ying-Chih Wang; Zenab Khan |
| EPI_ISL_1937562, EPI_ISL_2018362 | MT Public Health Laboratory | Centers for Disease Control and Prevention Division of Viral Diseases, Pathogen Discovery | Alison Laufer Halpin; Ben L. Rambo-Martin; Clinton R. Paden; Dakota Howard; Darlene Wagner; Dave Wentworth; Dhwani Batra; Jasmine Padilla; Justin Lee; Katie Dillon; Krista Queen; Kristen Knipe; Kristine Lacek; Mark Burroughs; Matthew Schmerer; Mili Sheth; Peter Cook; Sam Shepard; Sarah Nobles; Shoshona Le; Suxiang Tong; Vivien Dugan; Yvette Unoarumhi |
| EPI_ISL_2261363 | MVZ Labor Dr. Quade & Kollegen GmbH | Robert Koch Institute | |
| EPI_ISL_1849414, EPI_ISL_1849483 | MVZ Labor Krone GbR | Robert Koch Institute | |
| EPI_ISL_2100987, EPI_ISL_2101010, EPI_ISL_2101014, EPI_ISL_2501159, EPI_ISL_2501190 | Maine HETL | Health and Environmental Testing Laboratory | Barter, M.; Dewey, H.; Grieser, H.; H. and Tewhey, R.; Lynch, R.; Martha, J.; Matluk, N.; Meak, S.; Munger |
| EPI_ISL_1826244, EPI_ISL_1826249, EPI_ISL_1826330, EPI_ISL_1826331, EPI_ISL_1826332, EPI_ISL_1826333, EPI_ISL_1972976, EPI_ISL_2245616, EPI_ISL_4083775 | | | |
| see above | Maine Health and Environmental Testing Laboratory | Tewhey Lab, The Jackson Laboratory | Barter, M.; Dewey, H.; H. and Tewhey, R.; Iosue, F.; Lynch, R.; Matluk, N.; Munger |
| EPI_ISL_2161617, EPI_ISL_2377654, EPI_ISL_2377655, EPI_ISL_3011745, EPI_ISL_3011837, EPI_ISL_3011863, EPI_ISL_3011865 | | | |
| see above | Maryland Genomics, Institute for Genome Sciences, University of Maryland School of Medicine | Maryland Genomics, Institute for Genome Sciences, University of Maryland School of Medicine | Aditya; Claire M; Fraser; Hazen; Holly; Humphrys; Ivette; Jacques; Jonathan; Kranthi; Lim; Lisa D; Luke J; Mehta; Mike; Ott; Ravel; Roussey; Sadzewicz; Sandra; Santana-Cruz; Tallon; Tracy; Vavikolanu |
| EPI_ISL_6680586 | Mass General Brigham | Mass General Brigham | A.E.; Adams, G.; Anahtar, M.; B.L.; B.W.; Bauer, M.; Birren; Branda, J.; Carter, A.; Cerrato, F.; Chaluvadi, S.; Chapman; Cusick, C.; D.J.; DeRuff, K.; E. and Sabeti; Flowers, K.; Gallagher, G.; Gladden-Young, A.; Gnirke, A.; Harris, J.; J.E.; K.J.; LaRocque, R.; Lagerborg, K.; Lemieux; Lin; Loreth, C.; MacInnis; Neumann, A.; Normandin, E.; P.C.; Park; Pierce, V.; Reilly, S.; Rosenberg; Rudy, M.; Ryan, E.; S.B.; Shaw, B.; Siddle; Slater, D.; Smole, S.; Tomkins-Tinch, C.; Turbett, S. |
| EPI_ISL_2016473 | Massachusetts General Hospital | Infectious Disease Program, Broad Institute of Harvard and MIT | A.E.; Adams, G.; Anahtar, M.; B.L.; B.W.; Bauer, M.; Birren; Branda, J.; Carter, A.; Cerrato, F.; Chaluvadi, S.; Chapman; Cusick, C.; D.J.; DeRuff, K.; E. and Sabeti; Flowers, K.; Gallagher, G.; Gladden-Young, A.; Gnirke, A.; Harris, J.; J.E.; K.J.; LaRocque, R.; Lagerborg, K.; Lemieux; Lin; Loreth, C.; MacInnis; Neumann, A.; Normandin, E.; P.C.; Park; Pierce, V.; Reilly, S.; Rosenberg; Rudy, M.; Ryan, E.; S.B.; Shaw, B.; Siddle; Slater, D.; Smole, S.; Tomkins-Tinch, C.; Turbett, S. |
| EPI_ISL_2008127, EPI_ISL_2008128, EPI_ISL_2008129, EPI_ISL_2008132, EPI_ISL_2008148, EPI_ISL_2008153, EPI_ISL_2008195, EPI_ISL_2008326, EPI_ISL_2008341, EPI_ISL_2008344, EPI_ISL_2229528, EPI_ISL_2483775 | | | |
| see above | Massachusetts State Public Health Laboratory | Massachusetts State Public Health Laboratory | Andrew Lang; Glen Gallagher; Sandra Smole; Timelia Fink |
| EPI_ISL_2095175, EPI_ISL_2095176 | Max von Pettenkofer Institute, Virology, National Reference Center for Retroviruses, LMU Munich | Laboratory for Functional Genome Analysis; Dept. Genomics; Gene Center of the LMU Munich | Alexander Graf; Helmut Blum; Max Muenchhoff; Oliver Keppler; Stefan Krebs |
| EPI_ISL_1999665, EPI_ISL_1999698, EPI_ISL_1999701, EPI_ISL_1999734, EPI_ISL_1999766, EPI_ISL_1999797, EPI_ISL_1999828, EPI_ISL_1999831, EPI_ISL_1999848 | | | |
| see above | Microbiologia CATLAB | Can Ruti SARS-CoV-2 Sequencing Hub (HUGTiP/IrsiCaixa/IGTP) | Alba Sánchez; Anna Not; Antoni E Bordoy; Bonaventura Clotet; Cristina Casañ; Cristina Esteban; Francesc Catala-Moll; Gemma Clara; Ignacio Blanco; Marc Noguera-Julian; Maria Casadellà; Mariona Parera; Mercedes Guerrero; Montserrat Giménez; Pere-Joan Cardona; Pilar Armengol; Roger Paredes; Verónica Saludes; and Elisa Martró on behalf of the Can Ruti SARS-CoV-2 Sequencing Hub. |
| EPI_ISL_1792926 | Microbiologia e Virologia Cotugno | Microbiologia e Virologia Cotugno | Antonio Grimaldi Patrizia Annunziata Francesco Panariello Biancamaria Pierri Claudia Tiberio Teresa Giuliano Anna Perfetti Valentina Bouche Chiara Colantuono Maria Concetta Cuomo Denise Di Concilio Lucio Di Filippo Anna Manfredi Marcello Salvi Antonio Limone Luigi Atripaldi Pellegrino Cerino Andrea Ballabio Davide Cacchiarelli |
| EPI_ISL_1892991, EPI_ISL_2000011, EPI_ISL_2000122, EPI_ISL_2136876, EPI_ISL_2136877, EPI_ISL_2136888, EPI_ISL_2136901, EPI_ISL_2136912 | | | |
| see above | Microbiologia CATLAB | Can Ruti SARS-CoV-2 Sequencing Hub (HUGTiP/IrsiCaixa/IGTP) | Alba Sánchez; Anna Not; Antoni E Bordoy; Bonaventura Clotet; Cristina Casañ; Cristina Esteban; Francesc Catala-Moll; Gemma Clara; Ignacio Blanco; Marc Noguera-Julian; Maria Casadellà; Mariona Parera; Mercedes Guerrero; Montserrat Giménez; Pere-Joan Cardona; Pilar Armengol; Roger Paredes; Verónica Saludes; and Elisa Martró on behalf of the Can Ruti SARS-CoV-2 Sequencing Hub. |
| EPI_ISL_2372690 | Microbiology Department, Laboratori Clínic Metropolitana Nord. Hospital Universitari Germans Trias i Pujol | Can Ruti SARS-CoV-2 Sequencing Hub (HUGTiP/IrsiCaixa/IGTP) | Anna Not; Antoni E Bordoy; Bonaventura Clotet; Cristina Casañ; Cristina Esteban; Francesc Catala-Moll; Gemma Clara; Ignacio Blanco; Marc Noguera-Julian; Maria Casadellà; Mercedes Guerrero; Montserrat Giménez; Pere-Joan Cardona; Pilar Armengol; Roger Pared |
| EPI_ISL_1761504, EPI_ISL_1761538, EPI_ISL_1831601, EPI_ISL_1831612, EPI_ISL_1831615, EPI_ISL_1831662, EPI_ISL_1892968, EPI_ISL_1893046 | | | |
| see above | Microbiology Department, Laboratori Clínic Metropolitana Nord. Hospital Universitari Germans Trias i Pujol. | Can Ruti SARS-CoV-2 Sequencing Hub (HUGTiP/IrsiCaixa/IGTP) | Alba Sánchez; Anna Not; Antoni E Bordoy; Bonaventura Clotet; Cristina Casañ; Cristina Esteban; Francesc Catala-Moll; Gemma Clara; Ignacio Blanco; Marc Noguera-Julian; Maria Casadellà; Mariona Parera; Mercedes Guerrero; Montserrat Giménez; Pere-Joan Cardona; Pilar Armengol; Roger Paredes; Verónica Saludes; and Elisa Martró on behalf of the Can Ruti SARS-CoV-2 Sequencing Hub. |
| EPI_ISL_2086155, EPI_ISL_2086156, EPI_ISL_2086162, EPI_ISL_2086163, EPI_ISL_2086165, EPI_ISL_2086168, EPI_ISL_2086169, EPI_ISL_2086170, EPI_ISL_2086171, EPI_ISL_2086172, EPI_ISL_2086173 | | | |
| see above | Microbiology Department. Complexo Hospitalario Universitario de Vigo | Microbiology Department. Complexo Hospitalario Universitario de Vigo | Alfaya N; Alonso I; Alvarez M; Cabrera JJ; Carballo R; Cores O; Cortizo S; Martinez L; Mediero G; Perez S; Potel C; Regueiro B; Rey S; Vasallo FJ; del-Campo V |
| EPI_ISL_2293686, EPI_ISL_2312976 | Microbiology Division, SC DHEC | Microbiology Division, SC DHEC | Flores, H.; Freeman, J. |
| EPI_ISL_1820837, EPI_ISL_1820843, EPI_ISL_2029101 | Microvida | Microvida | Jaco Verweij; Joep Stöhr; Suzan D. Pas |
| EPI_ISL_2080566 | Minnesota Department of Health, Public Health Laboratory | Minnesota Department of Health, Public Health Laboratory | Alexandra Lorentz; Jacob Garfin; Matt Plumb; and Xiong Wang |
| EPI_ISL_2035540, EPI_ISL_2035556, EPI_ISL_2035591, EPI_ISL_2035592, EPI_ISL_2035594, EPI_ISL_2035596, EPI_ISL_2158679, EPI_ISL_2158681 | | | |
| see above | Missouri State Public Health Laboratory | Missouri State Public Health Laboratory | Ashley New; Joshua Barry; Matthew Sinn |
| EPI_ISL_3668146 | Mobile County Health Department | Synergy Laboratories | Megan Cornwell |
| EPI_ISL_5598977 | Molecular Biology Laboratory, Vidant Medical Center | Brody Integrative Genomics Core, East Carolina University | Changhong Yin; Heather Duncan; John T. Fallon; Kimberly P. Briley; Weihua Huang |
| EPI_ISL_2102124 | Molecular Diagnostics Pathology Department Mater Dei Hospital Malta | Molecular Diagnostics Pathology Department Mater Dei Hospital Malta | C Cilia; G Zahra; L Grech; R Borg |
| EPI_ISL_1913000, EPI_ISL_1913002, EPI_ISL_2941753, EPI_ISL_2941759, EPI_ISL_2941771 | Montana Public Health Laboratory | Montana Public Health Laboratory | Carrie Biskupiak; Deborah Gibson; Joy Ritter; Michael Dills; Michelle Mozer |
| EPI_ISL_4772372 | NAMRU-6 | NAMRU-6 | Cristhopher Cruz; Eugenio Abente; Gilda Troncos; Greg Rice; Luz Cedano Del Aguila; Marita Silva; Paul Graf.; Sonia Ampuero; Victoria Espejo; Yeny Tinoco |
| EPI_ISL_4772380 | NAMRU-6 | NAMRU-7 | Cristhopher Cruz; Eugenio Abente; Gilda Troncos; Greg Rice; Luz Cedano Del Aguila; Marita Silva; Paul Graf.; Sonia Ampuero; Victoria Espejo; Yeny Tinoco |
| EPI_ISL_1969002, EPI_ISL_1969075, | NJDOH, Public Health and Environmental Laboratories | NJ_PHEL | Byeong Jeong; Dana Woell; Jacquelyn Deverell; Lindsey Bodnar; Matthew Scarnati; Shiv K. Verma |

| | | | |
|---|---|---|---|
| EPI_ISL_2185580, EPI_ISL_2185581 | | | |
| EPI_ISL_2097266, EPI_ISL_2097277, EPI_ISL_2097289 | NORTHWELL HEALTH LABORATORIES | Wadsworth Center, New York State Department of Health | Alexis Russell; Catharine Prussing; Daryl M. Lamson; Erasmus Schneider; Erica Lasek-Nesselquist; John Kelly; Jonathan Plitnick; Kirsten St. George; Matthew Shudt; Melissa A Leisner; Navjot Singh |
| EPI_ISL_3399764 | NS-QEII Health Sciences Centre | National Microbiology Laboratory (NML) | Anna Majer; Anneliese Landgraff; Dan Gaston; Elsie Grudeski; Gary Van Domselaar; Grace Seo; Janice Pettipas; Jason LeBlanc; Jennifer Tanner; Morag Graham; Natalie Knox; Nathal; Philip Mabon; Rhiannon Huzarewich; Russell Mandes; Shari Tyson; Todd Hatchette |
| EPI_ISL_2098603, EPI_ISL_2098618 | NYC Pandemic Response Lab | Wadsworth Center, New York State Department of Health | Alexis Russell; Catharine Prussing; Daryl M. Lamson; Erasmus Schneider; Erica Lasek-Nesselquist; John Kelly; Jonathan Plitnick; Kirsten St. George; Matthew Shudt; Melissa A Leisner; Navjot Singh |
| EPI_ISL_2018244 | NYSDOH Wadsworth Center, Virology Lab | Centers for Disease Control and Prevention Division of Viral Diseases, Pathogen Discovery | Alison Laufer Halpin; Ben L. Rambo-Martin; Clinton R. Paden; Dakota Howard; Darlene Wagner; Dave Wentworth; Dhwani Batra; Jasmine Padilla; Justin Lee; Katie Dillon; Krista Queen; Kristen Knipe; Kristine Lacek; Mark Burroughs; Matthew Schmerer; Mili Sheth; Peter Cook; Sam Shepard; Sarah Nobles; Shoshona Le; Suxiang Tong; Vivien Dugan; Yvette Unoarumhi |
| EPI_ISL_2428621 | Nacionalinis maisto ir veterinarijos rizikos vertinimo institutas | Vilnius University Hospital Santaros Klinikos, Center of Laboratory Medicine | Daniel Naumovas; Dovile Ezerskyte; Gytis Dudas; Ingrida Olendraite; Ligita Raugaite; Monika Katenaite; Rimvydas Norvilas |
| EPI_ISL_1720614, EPI_ISL_1801756, EPI_ISL_1801787, EPI_ISL_1801797, EPI_ISL_1971472, EPI_ISL_1971482, EPI_ISL_1971488, EPI_ISL_1971492 | | | |
| see above | National Platform bis UMONS/Jolimont | National Platform bis UMONS/Jolimont | Florian Juszczak; François Dufrasne; Gautier Detry; Guillaume Bayon-Vicente; Ruddy Wattiez |
| EPI_ISL_1821289 | Nebraska Public Health Laboratory | NPHL COVID-19 Response Team | NPHL COVID-19 Response Team |
| EPI_ISL_1970970, EPI_ISL_2322770 | New Mexico Department of Health Scientific Laboratory | New Mexico Department of Health Scientific Laboratory | Anastacia Griego-Fisher; D'eldra Malone; Ellie Johnson; Jennifer Benoit; Linda Salazar; Mark Willmon; Ratheesh Rajan |
| EPI_ISL_3245679 | North Dakota Department of Health, Public Health Laboratory | North Dakota Department of Health, Public Health Laboratory | Lisa Wingerter |
| EPI_ISL_1920010, EPI_ISL_1920014 | Northwell Health Laboratories | Wadsworth Center, New York State Department of Health | Alexis Russell; Catharine Prussing; Daryl M. Lamson; Erasmus Schneider; Erica Lasek-Nesselquist; John Kelly; Jonathan Plitnick; Kirsten St. George; Matthew Shudt; Melissa A Leisner; Navjot Singh |
| EPI_ISL_2136525, EPI_ISL_2136540, EPI_ISL_2136541, EPI_ISL_2136545, EPI_ISL_2136546, EPI_ISL_2136554, EPI_ISL_2136557, EPI_ISL_2136564, EPI_ISL_2136566, EPI_ISL_2136575, EPI_ISL_2136588, EPI_ISL_2136595, EPI_ISL_2136602, EPI_ISL_2136609, EPI_ISL_2136615, EPI_ISL_2136616, EPI_ISL_2136622, EPI_ISL_2136627, EPI_ISL_2136635, EPI_ISL_2136643 | | | |
| see above | Northwest Laboratory | Seattle Flu Study | Amanda Adler; Barry R. Lutz; Benjamin Pelle; Brian Hiatt; Caitlin R. Wolf; Chris D. Frazar; Deborah A. Nickerson; Elisabeth Brandstetter; Erica Ryke; Geoff Melly; Helen Y. Chu; Janet A. Englund; Jay Shendure; Jover Lee; Kairsten Fay; Kirsten Lacombe; Lea M. Starita; Mark J. Rieder; Matthew Richardson; Matthew Thompson; Melissa Truong; Michael Boeckh; Michael Famulare; Misja Ilcisin; Peter D. Han; Philip Dykema; Romesh Gautom; Scott Lindquist; Thomas R. Sibley; Trevor Bedford |
| EPI_ISL_2009333, EPI_ISL_2009339 | Northwestern Memorial Hospital | Northwestern University - Ozer Lab | Chad J. Achenbach; Chao Qi; Egon A. Ozer; Judd F. Hultquist; Lacy M. Simons; Lawrence J. Jennings; Michael G. Ison; Ramon Lorenzo-Redondo; Taylor J. Dean |
| EPI_ISL_3388717, EPI_ISL_3388722 | Northwestern Memorial Hospital | RIPHL at Rush University Medical Center | Amber Kimble; Chao Qi; Edith Perez; Felix Araujo Perez; Kevin Kunstman; Laura Furtado; Marieta Hyde; Max Kolton; Stefan Green |
| EPI_ISL_2105172, EPI_ISL_2105214 | OH Department of Health Laboratory | Centers for Disease Control and Prevention Division of Viral Diseases, Pathogen Discovery | Alison Laufer Halpin; Ben L. Rambo-Martin; Clinton R. Paden; Dakota Howard; Darlene Wagner; Dave Wentworth; Dhwani Batra; Jasmine Padilla; Justin Lee; Katie Dillon; Krista Queen; Kristen Knipe; Kristine Lacek; Mark Burroughs; Matthew Schmerer; Mili Sheth; Peter Cook; Sam Shepard; Sarah Nobles; Shoshona Le; Suxiang Tong; Vivien Dugan; Yvette Unoarumhi |
| EPI_ISL_2227683, EPI_ISL_2227684, EPI_ISL_2227685, EPI_ISL_2227686, EPI_ISL_2227687, EPI_ISL_2227688, EPI_ISL_2227689, EPI_ISL_2227690, EPI_ISL_2227691, EPI_ISL_2227692, EPI_ISL_2227693, EPI_ISL_2378900, EPI_ISL_2378901, EPI_ISL_2378904, EPI_ISL_2378908, EPI_ISL_2378909, EPI_ISL_2378914, EPI_ISL_2378916, EPI_ISL_2378926, EPI_ISL_2378932, EPI_ISL_2378969, EPI_ISL_2378977, EPI_ISL_2378981, EPI_ISL_2462076, EPI_ISL_2462090, EPI_ISL_2462091, EPI_ISL_2462103, EPI_ISL_2462105, EPI_ISL_2462117 | | | |
| see above | OHSU Lab Services Molecular Microbiology Lab | Oregon SARS-CoV-2 Genome Sequencing Center | Alec J. Hirsch; Andrew C. Adey; Benjamin N. Bimber; Brendan L. O'Connell; Brian J. O'Roak; Cierra LaBlanc; Daniel N. Streblow; Donna Hansel; Guang Fan; Kayla Carter; Ruth V. Nichols; Sally Grindstaff; Sonia Acharya; William B. Messer; Xuan Qin |
| EPI_ISL_3065800, EPI_ISL_3065805, EPI_ISL_3065811 | OHSU-MM Lab | OHSU-MM Lab | Rebecca O'Gara |
| EPI_ISL_1904866 | OLVZ Aalst | OLVZ Aalst | Anne Vankeerberghen |
| EPI_ISL_3354886, EPI_ISL_3354911 | OR State PHL-Virology/Immunology Section | Centers for Disease Control and Prevention Division of Viral Diseases, Pathogen Discovery | Alex Burgin; Ben Rambo-Martin; Clinton Paden; Dakota Howard; Dave Wentworth; Dhwani Batra; Jasmine Padilla; Justin Lee; Krista Queen; Kristen Knipe; Kristine Lacek; Mark Burroughs; Matthew Schmerer; Meghan Bentz; Mili Sheth; Peter Cook; Sam Shepard; Sarah Nobles; Suxiang Tong; Vivien Dugan; Yvette Unoarumhi |
| EPI_ISL_1897447, EPI_ISL_1897467, EPI_ISL_1967717, EPI_ISL_1967718, EPI_ISL_2404739 | Oregon State Public Health Laboratory | Oregon State Public Health Laboratory | Eugene Yeboah; John Fontana and Shane Sevey; Laura Tsaknaridis; Rafia Razzaque; Vanda Makris |
| EPI_ISL_1825089, EPI_ISL_1825094, EPI_ISL_1825097 | Ospedale "F. Spaziani" Frosinone | INMI Lazzaro Spallanzani IRCCS | B Bartolini; C Sias; CEM Gruber; E Giombini; E Sperandio; F Messina; G Brocco; G Orlandi; GF Inger; M Pallocca; M Rueca; O Butera; R Pulselli |
| EPI_ISL_1819245 | Ospedale Casa Sollievo della Sofferenza | Istituto Zooprofilattico Sperimentale della Puglia e della Basilicata | Bianco A.; Capozzi L.; Del Sambro L.; Difato L.; Giannico A; Labonia M.; Parisi A.; Ridolfi D.; Simone D. |
| EPI_ISL_1824592, EPI_ISL_1824594 | Ospedale San Giovanni Evangelista | INMI Lazzaro Spallanzani IRCCS | A Massacci; B Bartolini; CEM Gruber; D Cerini; D Di Fusco; E Sperandio; F De Nicola; F Messina; F Santini; G Bonfiglio; GF Inger; M Rueca; O Butera |
| EPI_ISL_1824571, EPI_ISL_1824573, EPI_ISL_1825099, EPI_ISL_1825101, EPI_ISL_1825106 | Ospedale Sandro Pertini ASL Roma2 | INMI Lazzaro Spallanzani IRCCS | A Massacci; B Bartolini; CEM Gruber; E Giombini; E Sperandio; F Messina; F Santini; G Bonfiglio; G Cappiello; G Orlandi; GF Inger; L Ciuffreda; M Pallocca; M Rueca; MC Cava; O Butera; R Longo; S Donzelli; S Romano; V Michela |
| EPI_ISL_2371853 | Ospedale di Genzano - ASL RM 6 | INMI Lazzaro Spallanzani IRCCS | A Di Caro; B Bartolini; CEM Gruber; E Conti; E Giombini; F Messina; F Santini; G Bonfiglio; G Tramini; M Rueca; MR Capobianchi; O Butera |
| EPI_ISL_2293349, EPI_ISL_2293350, EPI_ISL_2293351, EPI_ISL_2293356, EPI_ISL_2293357, EPI_ISL_2293358, EPI_ISL_2293360, EPI_ISL_2293363, EPI_ISL_2293365, EPI_ISL_2293367, EPI_ISL_2293368, EPI_ISL_2293369, EPI_ISL_2293371 | | | |
| see above | Outre mer | National Reference Center for Viruses of Respiratory Infections, Institut Pasteur, Paris | Angela Brisebarre; Camille Capel; Christophe Malabat; Corinne Maufrais; Dominique Rousset; Etienne Simon-Lorière; Frédéric Lemoine; Louise Lefrançois; Marion Barbet; Maud Vanpeene; Méline Bizard; Pierre Lechat; Sylvie Behillil; Sylvie Van der Werf; Vincent Enouf |
| EPI_ISL_1960047 | PECET | Universidad Nacional de Colombia - Laboratorio Genómico One Health | Andres F. Cardona-Rios; Carlos Franco-Muñoz; Carolina Muñoz-Arango; Celeny Ortiz; Daniel O. Maldonado-Perez; Diego A. Álvarez-Díaz; Hector Alejandro Ruiz-Moreno; Idabely Betancur Ortiz; Jorge E. Osorio; Juan P. Hernandez-Ortiz; Karl A Ciuoderis; Katherine Laiton-Donato; Laura Silvana Perez; Lina M. Hurtado; Marcela Mercado-Reyes; Maria Angélica Maya; Maria Stella López; Rita Almanza Payares; Sandra Ines Cano; Simón Villegas Velásquez |
| EPI_ISL_2344927, EPI_ISL_2344941 | POLICLINICA COVID 19 ITAPETININGA | Instituto Butantan / ESALQ-Piracicaba | Antonio Jorge Martins; Claudia Renata dos Santos Barros; David Schlesinger; Debora Botequio Moretti; Dimas Tadeu Covas; Elaine Cristina Marqueze; Elaine Vieira Santos; Evandra Strazza Rodrigues; Heidge Fukumasu; Jayme Augusto de Souza-Neto; José Salvatore Leister Patané; Luiz Alcantara; Luiz Lehmann Coutinho; Maria Carolina Elias; Maurício Lacerda Nogueira; Rafael dos Santos Bezerra; Raul Machado Neto; Rejane Maria Tommasini Grotto; Ricardo Haddad; Sandra Coccuzzo Sampaio Vessoni; Simone Kashima; Svetoslav Nanev Slavov; Vincent Louis Viala |
| EPI_ISL_5603279 | POSTO DE SAUDE DO JUA | Analytical Competence Molecular Epidemiology Lab/ACME, Oswaldo Cruz Foundation, Ceara (FIOCRUZ CE) | Carlos Leonardo de Aragao Araujo; Cecilia Leite Costa & Eduardo Ruback dos Santos on behalf of COVID-19 FIOCRUZ Genomic Network; Cleber Furtado Aksenen; Fabio Miyajima; Fernando Braga Stehling; Francisco Eder de Moura Lopes; Igor Oliveira Duarte; Jamille Maria Mendes Bezerra; Joaquim Cesar do Nascimento Sousa Junior; Pedro Miguel Carneiro Jeronimo; Suzana Porto Almeida; Thais Ferreira de Oliveira; Thais de Oliveira Costa; Ticiane Cavalcante de Souza; Veridiana Pessoa Miyajima |
| EPI_ISL_5603271 | POSTO DE SAUDE FRANCISCA ROMANA DE OLIVEIRA | Analytical Competence Molecular Epidemiology Lab/ACME, Oswaldo Cruz Foundation, Ceara (FIOCRUZ CE) | Carlos Leonardo de Aragao Araujo; Cecilia Leite Costa & Eduardo Ruback dos Santos on behalf of COVID-19 FIOCRUZ Genomic Network; Cleber Furtado Aksenen; Fabio Miyajima; Fernando Braga Stehling; Francisco Eder de Moura Lopes; Igor Oliveira Duarte; Jamille Maria Mendes Bezerra; Joaquim Cesar do Nascimento Sousa Junior; Pedro Miguel Carneiro Jeronimo; Suzana Porto Almeida; Thais Ferreira de Oliveira; Thais de Oliveira Costa; Ticiane Cavalcante de Souza; Veridiana Pessoa Miyajima |
| EPI_ISL_2344789 | PRONTO ATENDIMENTO MUNICIPAL DE JACUPIRANGA | Instituto Butantan / ESALQ-Piracicaba | Antonio Jorge Martins; Claudia Renata dos Santos Barros; David Schlesinger; Debora Botequio Moretti; Dimas Tadeu Covas; Elaine Cristina Marqueze; Elaine Vieira Santos; Evandra Strazza Rodrigues; Heidge Fukumasu; Jayme Augusto de Souza-Neto; José Salvatore Leister Patané; Luiz Alcantara; Luiz Lehmann Coutinho; Maria Carolina Elias; Maurício Lacerda Nogueira; Rafael dos Santos Bezerra; Raul Machado Neto; Rejane Maria Tommasini Grotto; Ricardo Haddad; Sandra Coccuzzo Sampaio Vessoni; Simone Kashima; Svetoslav Nanev Slavov; Vincent Louis Viala |
| EPI_ISL_2344910 | PSF MARIA JOSE SALTO DE PIRAPORA | Instituto Butantan / ESALQ-Piracicaba | Antonio Jorge Martins; Claudia Renata dos Santos Barros; David Schlesinger; Debora Botequio Moretti; Dimas Tadeu Covas; Elaine Cristina Marqueze; Elaine Vieira Santos; Evandra Strazza Rodrigues; Heidge Fukumasu; Jayme Augusto de Souza-Neto; José Salvatore Leister Patané; Luiz Alcantara; Luiz Lehmann Coutinho; Maria Carolina Elias; Maurício Lacerda Nogueira; Rafael dos Santos Bezerra; Raul Machado Neto; Rejane Maria Tommasini Grotto; Ricardo Haddad; Sandra Coccuzzo Sampaio Vessoni; Simone Kashima; Svetoslav Nanev Slavov; Vincent Louis Viala |
| EPI_ISL_2344918 | PSF PALMITALZINHO APIAI | Instituto Butantan / ESALQ-Piracicaba | Antonio Jorge Martins; Claudia Renata dos Santos Barros; David Schlesinger; Debora Botequio Moretti; Dimas Tadeu Covas; Elaine Cristina Marqueze; Elaine Vieira Santos; Evandra Strazza Rodrigues; Heidge Fukumasu; Jayme Augusto de Souza-Neto; José Salvatore Leister Patané; Luiz Alcantara; Luiz Lehmann Coutinho; Maria Carolina Elias; Maurício Lacerda Nogueira; Rafael dos Santos Bezerra; Raul Machado Neto; Rejane Maria Tommasini Grotto; Ricardo Haddad; Sandra Coccuzzo Sampaio Vessoni; Simone Kashima; Svetoslav Nanev Slavov; Vincent Louis Viala |
| EPI_ISL_1717679, EPI_ISL_1717818, EPI_ISL_1717823, EPI_ISL_1717859, EPI_ISL_1717903, EPI_ISL_1717950, EPI_ISL_1717972, EPI_ISL_1717978, EPI_ISL_1717995, EPI_ISL_1717997, EPI_ISL_1718013, EPI_ISL_1718046, EPI_ISL_1718064, EPI_ISL_1718159, EPI_ISL_1718186, EPI_ISL_1718233, EPI_ISL_1828065, EPI_ISL_1828107, EPI_ISL_1828114, EPI_ISL_1828173, EPI_ISL_1828185, EPI_ISL_1828191, EPI_ISL_1828238, EPI_ISL_1828240, EPI_ISL_1828297, EPI_ISL_1828354, EPI_ISL_2965366, EPI_ISL_4096583 | | | |
| see above | Pandemic Response Lab - NYC | Pandemic Response Lab, R&D | Cybill del Castillo; Dylan Law; Haiping Hao; Henry Lee; Isabel Fernandez Escapa; Jon Laurent; Katharine Nelson; Melissa Hopkins; Michael Hammerling; Pradeep Bugga; Shinyoung Clair Kang; Sol Rey; William Ward |

| | | | |
|---|---|---|---|
| EPI_ISL_3187420, EPI_ISL_3193312 | Pathogen Genomics Center, National Institute of Infectious Diseases | Pathogen Genomics Center, National Institute of Infectious Diseases | Kentaro Itokawa; Makoto Kuroda; Masanori Hashino; Rina Tanaka; Tsuyoshi Sekizuka |
| EPI_ISL_3015186 | Pesaro | Microbiology University Politecnica delle Marche | Anna Valenza; Carla Acciarri; Katia Marinelli; Monica Lucia Ferreri; Patrizia Bagnarelli; Roberta Longo; Sara Caucci; Stefano Menzo |
| EPI_ISL_4405576 | Plataforma de Servicios Biotecnólogicos: UTTIPP/PSB , Universidad Nacional de Quilmes. | Área de Secuenciación del Laboratorio de Virología del Hospital de Niños Dr. Ricardo Gutierrez on behalf of 'Proyecto Argentino Interinstitucional de genomica de SARS-CoV-2' (PAIS Consortium) | A; Acuña; Cardama; Castello; D; Farina; G; Goya; Goñi; H; LE; Lusso; MI; MS; Nabaes Jodar; Natale; S; Valinotto; Viegas, M. |
| EPI_ISL_6907895, EPI_ISL_6908432, EPI_ISL_6908433, EPI_ISL_6908434, EPI_ISL_6908435, EPI_ISL_6908774, EPI_ISL_6908782, EPI_ISL_6908786, EPI_ISL_6908791, EPI_ISL_6908794, EPI_ISL_6908795, EPI_ISL_6908799, EPI_ISL_6908806, EPI_ISL_6908807, EPI_ISL_6908808, EPI_ISL_6908809, EPI_ISL_6908810, EPI_ISL_6908811, EPI_ISL_6908812, EPI_ISL_6908813, EPI_ISL_6908820 | | | |
| see above | Plataforma de Vigilancia Molecular (PVM) - FIOCRUZ/BA | Plataforma de Vigilancia Molecular (PVM) - FIOCRUZ/BA | Bruno Bezerril Andrade; Camila I. de Oliveira; Clarissa Araújo Gurgel; Eduardo Oyama; Francys A Rangel; Laila A da Silva; Laise de Moraes; Leonardo Paiva Farias; Marina Cucco; Pedro Silveira; Ricardo Khouri on behalf of the Fiocruz COVID-19 Genomic Surveillance Network.; Tiago Graf |
| EPI_ISL_1804869, EPI_ISL_1804876, EPI_ISL_1804884 | Plateforme de testing Namuroise | Plateforme de testing Namuroise | Céline Maschietto; Degosserie Jonathan; Denis Olivier; Mullier François; Otto Gaetan |
| EPI_ISL_1745062, EPI_ISL_1745178, EPI_ISL_1745179, EPI_ISL_1745180, EPI_ISL_1745181, EPI_ISL_1745182, EPI_ISL_1745183, EPI_ISL_1745184, EPI_ISL_1745185, EPI_ISL_1745191, EPI_ISL_1745192, EPI_ISL_1841416 | | | |
| see above | Platform BIS UZA/UAntwerpen | Labo Klinische Biologie, UZA | Basil Britto Xavier; Christine Lammens; Herman Goossens; Jasmine Coppens; Marie Le Mercier; Veerle Matheeussen |
| EPI_ISL_5800916, EPI_ISL_5800931 | Policlinica Covid 19 Itapetininga | Instituto Butantan | Antonio Jorge Martins; Claudia Renata dos Santos Barros; David Schlesinger; Debora Botequio Moretti; Dimas Tadeu Covas; Elaine Cristina Marqueze; Elaine Vieira Santos; Evandra Strazza Rodrigues; Heidge Fukumasu; Jayme Augusto de Souza-Neto; José Salvatore Leister Patané; Luiz Alcantara; Luiz Lehmann Coutinho; Maria Carolina Elias; Maurício Lacerda Nogueira; Rafael dos Santos Bezerra; Raul Machado Neto; Rejane Maria Tommasini Grotto; Ricardo Haddad; Sandra Coccuzzo Sampaio Vessoni; Simone Kashima; Svetoslav Nanev Slavov; Vincent Louis Viala |
| EPI_ISL_2259233, EPI_ISL_2259235 | Pomona Valley Hospital Medical Center | Los Angeles County Public Health Laboratories | P. Hemarajata et al. |
| EPI_ISL_2345906, EPI_ISL_2345907, EPI_ISL_2345908, EPI_ISL_2345909, EPI_ISL_2345910, EPI_ISL_2345911, EPI_ISL_2345912, EPI_ISL_2345913, EPI_ISL_2345914, EPI_ISL_2345915, EPI_ISL_2345916, EPI_ISL_2345917, EPI_ISL_2345918, EPI_ISL_2345919, EPI_ISL_2345920, EPI_ISL_2345921, EPI_ISL_2345922, EPI_ISL_2345923, EPI_ISL_2345924, EPI_ISL_2345925, EPI_ISL_2345926, EPI_ISL_2345927, EPI_ISL_2345928, EPI_ISL_2345929, EPI_ISL_2345930, EPI_ISL_2345931, EPI_ISL_2345932, EPI_ISL_2345933, EPI_ISL_2345934, EPI_ISL_2345935, EPI_ISL_2345937, EPI_ISL_2345938, EPI_ISL_2346094, EPI_ISL_2346095, EPI_ISL_2346096, EPI_ISL_2346097, EPI_ISL_2346098, EPI_ISL_2346099 | | | |
| see above | Prefeirtura de SP | Instituto Butantan | Antonio Jorge Martins; Claudia Renata dos Santos Barros; David Schlesinger; Debora Botequio Moretti; Dimas Tadeu Covas; Elaine Cristina Marqueze; Elaine Vieira Santos; Evandra Strazza Rodrigues; Heidge Fukumasu; Jayme Augusto de Souza-Neto; José Salvatore Leister Patané; Luiz Alcantara; Luiz Lehmann Coutinho; Maria Carolina Elias; Maurício Lacerda Nogueira; Rafael dos Santos Bezerra; Raul Machado Neto; Rejane Maria Tommasini Grotto; Ricardo Haddad; Sandra Coccuzzo Sampaio Vessoni; Simone Kashima; Svetoslav Nanev Slavov; Vincent Louis Viala |
| EPI_ISL_7406388, EPI_ISL_7406412 | Programa Laboratorio de Salud Pública "Dr Dalmiro Pérez Laborda" | Área de Secuenciación del Laboratorio de Virología del Hospital de Niños Dr. Ricardo Gutierrez on behalf of 'Proyecto Argentino Interinstitucional de genomica de SARS-CoV-2' (PAIS Consortium) | A; AM; Acuña; Aguilera; BH; Bohn; Cabral Bombardieri; Campos; Carrizo; D; E; Esteves; F; G; Goya; J; JM; JP; Jofré; L; LE; Lacaze; Lusso; M; MA; MI; MS; Mastrodonato; Molina; Nabaes Jodar; Natale; Olivera; Perez Diaz; Peñalva; Quijano; Rivero; Rosales; S; Talia; Valinotto; Viegas, M. |
| EPI_ISL_5800690 | Pronto Atendimento Municipal De Jacupiranga | Instituto Butantan | Antonio Jorge Martins; Claudia Renata dos Santos Barros; David Schlesinger; Debora Botequio Moretti; Dimas Tadeu Covas; Elaine Cristina Marqueze; Elaine Vieira Santos; Evandra Strazza Rodrigues; Heidge Fukumasu; Jayme Augusto de Souza-Neto; José Salvatore Leister Patané; Luiz Alcantara; Luiz Lehmann Coutinho; Maria Carolina Elias; Maurício Lacerda Nogueira; Rafael dos Santos Bezerra; Raul Machado Neto; Rejane Maria Tommasini Grotto; Ricardo Haddad; Sandra Coccuzzo Sampaio Vessoni; Simone Kashima; Svetoslav Nanev Slavov; Vincent Louis Viala |
| EPI_ISL_2551494, EPI_ISL_2551497 | Providence Oregon Regional Laboratories | Providence St. Joseph Health Molecular Genomics Laboratory | Alexa K Dowdell; Brian D Piening; Carlo B Bifulco; Fred L Robinson; Mary Campbell; Rogan Rattray |
| EPI_ISL_2551430 | Providence Regional Medical Center Everett | Providence St. Joseph Health Molecular Genomics Laboratory | Alexa K Dowdell; Brian D Piening; Carlo B Bifulco; Fred L Robinson; Mary Campbell; Rogan Rattray |
| EPI_ISL_5800891 | Psf Maria Jose Salto De Pirapora | Instituto Butantan | Antonio Jorge Martins; Claudia Renata dos Santos Barros; David Schlesinger; Debora Botequio Moretti; Dimas Tadeu Covas; Elaine Cristina Marqueze; Elaine Vieira Santos; Evandra Strazza Rodrigues; Heidge Fukumasu; Jayme Augusto de Souza-Neto; José Salvatore Leister Patané; Luiz Alcantara; Luiz Lehmann Coutinho; Maria Carolina Elias; Maurício Lacerda Nogueira; Rafael dos Santos Bezerra; Raul Machado Neto; Rejane Maria Tommasini Grotto; Ricardo Haddad; Sandra Coccuzzo Sampaio Vessoni; Simone Kashima; Svetoslav Nanev Slavov; Vincent Louis Viala |
| EPI_ISL_5800903 | Psf Palmitalzinho Apiai | Instituto Butantan | Antonio Jorge Martins; Claudia Renata dos Santos Barros; David Schlesinger; Debora Botequio Moretti; Dimas Tadeu Covas; Elaine Cristina Marqueze; Elaine Vieira Santos; Evandra Strazza Rodrigues; Heidge Fukumasu; Jayme Augusto de Souza-Neto; José Salvatore Leister Patané; Luiz Alcantara; Luiz Lehmann Coutinho; Maria Carolina Elias; Maurício Lacerda Nogueira; Rafael dos Santos Bezerra; Raul Machado Neto; Rejane Maria Tommasini Grotto; Ricardo Haddad; Sandra Coccuzzo Sampaio Vessoni; Simone Kashima; Svetoslav Nanev Slavov; Vincent Louis Viala |
| EPI_ISL_2698949, EPI_ISL_2699007, EPI_ISL_2699253 | Public Health Laboratory, Minnesota Department of Health | University of Minnesota Genomics Center | Corbin Dirkx; Daryl M. Gohl; Jaquelyn Kuriger-Laber; John Garbe; and Sean Wang |
| EPI_ISL_1942175, EPI_ISL_1942186, EPI_ISL_1942187, EPI_ISL_1942188, EPI_ISL_1942189, EPI_ISL_1942191, EPI_ISL_1942198, EPI_ISL_1942199, EPI_ISL_1942202, EPI_ISL_1942209, EPI_ISL_1942210, EPI_ISL_1942211, EPI_ISL_1989980, EPI_ISL_1989981, EPI_ISL_1989982, EPI_ISL_1989983, EPI_ISL_1989985, EPI_ISL_1989986, EPI_ISL_1989988, EPI_ISL_1989990, EPI_ISL_1989991, EPI_ISL_1989993, EPI_ISL_1989994, EPI_ISL_1989995, EPI_ISL_1989998, EPI_ISL_1989999, EPI_ISL_1990000, EPI_ISL_2034712, EPI_ISL_2034714, EPI_ISL_2034718, EPI_ISL_2034720, EPI_ISL_2034722, EPI_ISL_2034724, EPI_ISL_2034732, EPI_ISL_2034736, EPI_ISL_2034739, EPI_ISL_2034740, EPI_ISL_2034742, EPI_ISL_2034744, EPI_ISL_2034746, EPI_ISL_2034755, EPI_ISL_2034758, EPI_ISL_2034762, EPI_ISL_2034769, EPI_ISL_2034773, EPI_ISL_2034776, EPI_ISL_2034782, EPI_ISL_2034785, EPI_ISL_2034793, EPI_ISL_2034796, EPI_ISL_2034799, EPI_ISL_2034808, EPI_ISL_2034815, EPI_ISL_2034822, EPI_ISL_2034829, EPI_ISL_2034833, EPI_ISL_2034840, EPI_ISL_2034844, EPI_ISL_2035302, EPI_ISL_2035303, EPI_ISL_2035304, EPI_ISL_2035305, EPI_ISL_2035306, EPI_ISL_2035307, EPI_ISL_2035309, EPI_ISL_2035310, EPI_ISL_2035311, EPI_ISL_2035312, EPI_ISL_2035313, EPI_ISL_2035314, EPI_ISL_2035315, EPI_ISL_2035316, EPI_ISL_2035317, EPI_ISL_2035318, EPI_ISL_2035319, EPI_ISL_2035320, EPI_ISL_2035321, EPI_ISL_2035322, EPI_ISL_2035323, EPI_ISL_2035325, EPI_ISL_2035326, EPI_ISL_2035328, EPI_ISL_2035329, EPI_ISL_2035330, EPI_ISL_2035331, EPI_ISL_2035332, EPI_ISL_2035335, EPI_ISL_2035336, EPI_ISL_2035337, EPI_ISL_2035338, EPI_ISL_2035340, EPI_ISL_2035342, EPI_ISL_2035343, EPI_ISL_2035345, EPI_ISL_2035346, EPI_ISL_2035347, EPI_ISL_2035348, EPI_ISL_2035349, EPI_ISL_2035350, EPI_ISL_2035351, EPI_ISL_2035352, EPI_ISL_2035353, EPI_ISL_2035354, EPI_ISL_2035355, EPI_ISL_2035356, EPI_ISL_2035357, EPI_ISL_2035358, EPI_ISL_2035359, EPI_ISL_2035360, EPI_ISL_2035361, EPI_ISL_2035362, EPI_ISL_2035363, EPI_ISL_2035364, EPI_ISL_2132574, EPI_ISL_2132575, EPI_ISL_2132577, EPI_ISL_2132578, EPI_ISL_2132579, EPI_ISL_2132580, EPI_ISL_2132581, EPI_ISL_2132582, EPI_ISL_2132583, EPI_ISL_2132584, EPI_ISL_2132585, EPI_ISL_2132586, EPI_ISL_2132619, EPI_ISL_2132621, EPI_ISL_2132623, EPI_ISL_2132624, EPI_ISL_2132627, EPI_ISL_2132629, EPI_ISL_2140732, EPI_ISL_2140739, EPI_ISL_2140760, EPI_ISL_2140763, EPI_ISL_2140767, EPI_ISL_2140769, EPI_ISL_2140791, EPI_ISL_2140792, EPI_ISL_2187307, EPI_ISL_2187313, EPI_ISL_2241642, EPI_ISL_4372618 | | | |
| see above | Quest Diagnostics Incorporated | Centers for Disease Control and Prevention Division of Viral Diseases, Pathogen Discovery | A. Gerasimova; A. Perez; Adrian Paskey; B. Anderson; Benjamin Rambo-Martin; Christopher Gulvick; Clinton Paden; Clinton R. Paden; Dakota Howard; Darlene Wagner; Dhwani Batra; Duncan MacCannell; Erisa Sula; F. Lacbawan; I. A. Shlyakhter; I. Shlyakhter; Jason Caravas; K. Livingston; K.E. Livingston; Kara Moser; Kristine Lacek; L. Bernstein; L.E. Bernstein; M. Hua; Matthew Schmerer; P. Tanpaiboon; Peter Cook; Peter W. Cook; R. Kagan; R. M. Kagan; R. Owen; R. Rolando; R. V. Rolando; S. H. Rosenthal; S. Rosenthal; Scott Sammons; Shatavia Morrison; Tymeckia Kendall; Victoria Caban Figueroa; Y. Liu; Yvette Unoarumhi |
| EPI_ISL_2105113, EPI_ISL_2105129, EPI_ISL_2105133, EPI_ISL_2105144 | RI State Health Laboratories | Centers for Disease Control and Prevention Division of Viral Diseases, Pathogen Discovery | Alison Laufer Halpin; Ben L. Rambo-Martin; Clinton R. Paden; Dakota Howard; Darlene Wagner; Dave Wentworth; Dhwani Batra; Jasmine Padilla; Justin Lee; Katie Dillon; Krista Queen; Kristen Knipe; Kristine Lacek; Mark Burroughs; Matthew Schmerer; Mili Sheth; Peter Cook; Sam Shepard; Sarah Nobles; Shoshona Le; Suxiang Tong; Vivien Dugan; Yvette Unoarumhi |
| EPI_ISL_2005712 | Randox Laboratories | Wellcome Sanger Institute for the COVID-19 Genomics UK (COG-UK) Consortium | Cordelia Langford; David K. Jackson; Dominic Kwiatkowski; Ewan Harrison; Ian Johnston; Jeffrey Barrett; John Sillitoe on behalf of the Wellcome Sanger Institute COVID-19 Surveillance Team; Randox Laboratories and Alex Alderton; Roberto Amato; Sonia Goncalves |
| EPI_ISL_2319838 | Reditus Laboratories | Reditus Laboratories | Alexa Eichelberger; Cassy Philips; Joshua J. Geltz; M.S.; Ph.D.; Rex Dyer; Robert M. Sgambelluri |
| EPI_ISL_7156471 | Research Education in Disease Diagnosis and Intervention (REDDI) Lab, Clemson University | Research Education in Disease Diagnosis and Intervention (REDDI) Lab, Clemson University | Christopher Parkinson; Christopher Saski; Congyue Peng; Delphine Dean; Justin Napolitano; Kaitlyn Williams; Keegan Sell; Kylie King; Lior Rennert; Rachel Ham; Stevin Wilson; Sujata Srikanth |
| EPI_ISL_2741730 | Respiratory Virus Unit, Microbiology Services Colindale, Public Health England | COVID-19 Genomics UK (COG-UK) Consortium | PHE Covid Sequencing Team |
| EPI_ISL_2106070, EPI_ISL_2106073, EPI_ISL_2106079, EPI_ISL_2106086, EPI_ISL_2106100, EPI_ISL_2106103, EPI_ISL_2106117, EPI_ISL_2253128, EPI_ISL_2253134, EPI_ISL_2253139, EPI_ISL_2253143, EPI_ISL_2253144, EPI_ISL_2253145, EPI_ISL_2253146, EPI_ISL_2253147, EPI_ISL_2253148, EPI_ISL_2253150, EPI_ISL_2253151, EPI_ISL_2253153, EPI_ISL_2253154, EPI_ISL_2253156, EPI_ISL_2253157, EPI_ISL_2253158, EPI_ISL_2253159, EPI_ISL_2253161, EPI_ISL_2253164, EPI_ISL_2253165, EPI_ISL_2254335, EPI_ISL_2254358, EPI_ISL_2254370, EPI_ISL_2254375, EPI_ISL_2254376, EPI_ISL_2254377, EPI_ISL_2254391, EPI_ISL_2254399, EPI_ISL_2254438, EPI_ISL_2254457 | | | |
| see above | Respiratory Viruses Branch, Centers for Disease Control and Prevention | Respiratory Viruses Branch, Centers for Disease Control and Prevention | Anderson, B.; Baker; Bakhash; Batra, D.; Bernstein, L.; Breit, N.; C.R.; Campbell, P.; Caravas, J.; Case, R.; Clark, C.; Cook; Ellis, S.; Gerasimova, A.; Ghorpade, V.; Greninger, A.; Gulvick, C.; Hardison, M.; Houdeshell, H.; Howard, D.; Hua, M.; Huang; I.A.; Jerome; K.R.; Kagan; Kvalvaag, O.; Lacbawan, F.; Lin, M.; Liu, Y.; Livingston; Livingston, K.; M.-L.; M.S.; MacCannell, D.; Mathias, P.; Morrison, S.; Moser, K.; N.R.; Nall, D.; Nguyen; Owen, R.; P.W.; Paden; Paskey, A.; Perez, A.; Perez, R.; R.J.; R.M.; R.V.; Rambo-Martin, B.; Rolando; Rosenthal; Roychoudhury; S.H.; S.M.; Sammons, S.; Sanders, E.; Schmerer, M.; Shlyakhter; Shrestha, L.; T.V.; Tanpaiboon, P.; Unoarumhi, Y.; Vest, A.; Wagner, D.; Westlund, S.; Xie, H. |
| EPI_ISL_2016490, EPI_ISL_2016491, EPI_ISL_2016492, EPI_ISL_2016504, EPI_ISL_2016505, EPI_ISL_2016507, EPI_ISL_2016515, EPI_ISL_2016516, EPI_ISL_2016523, EPI_ISL_2016528, EPI_ISL_2016554, EPI_ISL_2016556, EPI_ISL_2016574, EPI_ISL_2016581, EPI_ISL_2016594, EPI_ISL_2016597, EPI_ISL_2016600, EPI_ISL_2016608 | | | |
| see above | Rhode Island Department of Health | Infectious Disease Program, Broad Institute of Harvard and MIT | Adams, G.; Azevedo, K.; B.L.; B.W.; Bauer, M.; Birren; Carter, A.; Chaluvadi, S.; D.J.; DeRuff, K.; Gladden-Young, A.; Huard, R.; J.E.; K.J.; King, E.; Lagerborg, K.; Lemieux; Loreth, C.; Miller, A.; Normandin, E.; P.C.; Park; Pearlman, L.; Reilly, S.; Rudy, M.; Sabeti; Siddle; Tomkins-Tinch, C.; and MacInnis |
| EPI_ISL_2604861 | Rhode Island State Health Laboratories | Rhode Island State Health Laboratories | Ewa King; Kristin Carpenter-Azevedo; Richard C. Huard |
| EPI_ISL_2450815, EPI_ISL_2450817, EPI_ISL_2450819, EPI_ISL_2450820, EPI_ISL_2450821, EPI_ISL_2450822 | S.C. Patologia Clinica, Ospedale Sant'Andrea, ASL 5 | U.O. Igiene, Ospedale Policlinico San Martino | Battolla Enrico; Bruzzone Bianca; Caligiuri Patrizia; De Pace Vanessa; Domnich Alexander; Icardi Giancarlo on behalf of SARS-CoV-2 ITALIAN RESEARCH ENTERPRISE-(SCIRE) Collaborative Group; Orsi Andrea; Ricucci Valentina |
| EPI_ISL_2344739, EPI_ISL_2344740, EPI_ISL_2344741, EPI_ISL_2344742, EPI_ISL_2344745, EPI_ISL_2344747, EPI_ISL_2344802, EPI_ISL_2344803, EPI_ISL_2344804, EPI_ISL_2344805, EPI_ISL_2344808, EPI_ISL_2344810, EPI_ISL_2344811, EPI_ISL_2344812, EPI_ISL_2344813, EPI_ISL_2344815, EPI_ISL_2344816, EPI_ISL_2344817, EPI_ISL_2344819 | | | |
| see above | SAE CTA SERV ATEND ESPEC TESTAGEM E ACONSELHAMENTO | Instituto Butantan / ESALQ-Piracicaba | Antonio Jorge Martins; Claudia Renata dos Santos Barros; David Schlesinger; Debora Botequio Moretti; Dimas Tadeu Covas; Elaine Cristina Marqueze; Elaine Vieira Santos; Evandra Strazza Rodrigues; Heidge Fukumasu; Jayme Augusto de Souza-Neto; José Salvatore Leister Patané; Luiz Alcantara; Luiz Lehmann Coutinho; Maria Carolina Elias; Maurício Lacerda Nogueira; Rafael dos Santos Bezerra; Raul Machado Neto; Rejane Maria Tommasini Grotto; Ricardo Haddad; Sandra Coccuzzo Sampaio Vessoni; Simone Kashima; Svetoslav Nanev Slavov; Vincent Louis Viala |
| EPI_ISL_2209543, EPI_ISL_2345035, | SANTA CASA DE GUARAREMA | Instituto Butantan | Antonio Jorge Martins; Claudia Renata dos Santos Barros; David Schlesinger; Debora Botequio Moretti; Dimas Tadeu Covas; Elaine Cristina Marqueze; Elaine Vieira Santos; Evandra Strazza Rodrigues; Heidge Fukumasu; Jayme Augusto de Souza-Neto; José Salvatore Leister Patané; Luiz Alcantara; Luiz Lehmann Coutinho; Maria Carolina Elias; Maurício Lacerda Nogueira; Rafael dos Santos Bezerra; Raul Machado Neto; Rejane Maria Tommasini Grotto; Ricardo Haddad; Sandra Coccuzzo Sampaio Vessoni; Simone Kashima; Svetoslav Nanev Slavov; Vincent Louis Viala |

| | | | |
|---|---|---|---|
| EPI_ISL_2345036, EPI_ISL_2345037, EPI_ISL_2345038 | | | |
| EPI_ISL_2170997 | SANTA CASA DE GUARAREMA | Instituto Butantan / Mendelics | Antonio Jorge Martins; Bianca Cechetto Carlos. Mendelics: Bibiana Santos; Claudia Renata dos Santos Barros; Cíntia Bittar; David Schlesinger. Hemocentro Ribeirão Preto: Simone Kashima; Debora Botequio Moretti; Elaine Cristina Marqueze; Elaine Vieira dos Santos; Elisangela Chicaroni Mattos; Erika Freitas; Evandra Strazza Rodrigues; Felipe Allan da Silva da Costa; Flavia Aburjaile; Fábio Sossai Possebon; Guilherme Campos; Guilherme Targino Valente; Heidge Fukumasu. USP-Botucatu: Rejane Maria Tommasini Grotto; Helena Lage Ferreira; Instituto Butantan: Dimas Tadeu Covas; Jardelina de Souza Todao Bernardino; Jayme A. Souza-Neto; Jessika Cristina Chagas Lesbon; Jorge A. Petroli Marchesi; José Salvatore Leister Patané; João Paulo Kitajima; João Pessoa Araújo Jr.; Leila Sabrina Ullmann; Loyze Paola Oliveira de Lima; Luiz Aurelio de Campos Crispin. Centro de Genômica Funcional da ESALQ: Luiz Lehmann Coutinho; Luiz Carlos Junior de Alcantara; Lívia Sacchetto; Maisa C. Pereira Parra; Maria Carolina Elias; Marta Giovanetti; Marília Moraes; Maurício Lacerda Nogueira. Prefeitura de Sao Paulo: Melissa Palmieri.; Patricia Akemi Assato; Paula Rahal; Paulo Inacio da Costa; Rafael dos Santos Bezerra; Raquel de Lello Rocha Campos Cassano. NGS Soluções Genômicas: Pilar Drummond Sampaio Corrêa Mariani. FZEA-USP Pirassununga: Mirele Daiana Poleti; Raul Machado Neto; Ricardo Augusto Brassaloti; Ricardo Haddad; Rodrigo Tocantins Calado. FAMERP-SJRP: Cecília Artico Banho; Sandra Coccuzzo Sampaio; Svetoslav Nanev Slavov; Vagner Fonseca; Vincent Louis Viala |
| EPI_ISL_2209171, EPI_ISL_2209630, EPI_ISL_2344865, EPI_ISL_2344876, EPI_ISL_2344878, EPI_ISL_2344880, EPI_ISL_2344883 | | | |
| see above | SANTA CASA DE JOSE BONIFACIO | Instituto Butantan / ESALQ-Piracicaba | Antonio Jorge Martins; Claudia Renata dos Santos Barros; David Schlesinger; Debora Botequio Moretti; Dimas Tadeu Covas; Elaine Cristina Marqueze; Elaine Vieira Santos; Evandra Strazza Rodrigues; Heidge Fukumasu; Jayme Augusto de Souza-Neto; José Salvatore Leister Patané; Luiz Alcantara; Luiz Lehmann Coutinho; Maria Carolina Elias; Maurício Lacerda Nogueira; Rafael dos Santos Bezerra; Raul Machado Neto; Rejane Maria Tommasini Grotto; Ricardo Haddad; Sandra Coccuzzo Sampaio Vessoni; Simone Kashima; Svetoslav Nanev Slavov; Vincent Louis Viala |
| EPI_ISL_2209108, EPI_ISL_2345004, EPI_ISL_2345007, EPI_ISL_2345009, EPI_ISL_2345010, EPI_ISL_2345012, EPI_ISL_2345013 | | | |
| see above | SANTA CASA DE MISERICORDIA SAO JOSE | Instituto Butantan / ESALQ-Piracicaba | Antonio Jorge Martins; Claudia Renata dos Santos Barros; David Schlesinger; Debora Botequio Moretti; Dimas Tadeu Covas; Elaine Cristina Marqueze; Elaine Vieira Santos; Evandra Strazza Rodrigues; Heidge Fukumasu; Jayme Augusto de Souza-Neto; José Salvatore Leister Patané; Luiz Alcantara; Luiz Lehmann Coutinho; Maria Carolina Elias; Maurício Lacerda Nogueira; Rafael dos Santos Bezerra; Raul Machado Neto; Rejane Maria Tommasini Grotto; Ricardo Haddad; Sandra Coccuzzo Sampaio Vessoni; Simone Kashima; Svetoslav Nanev Slavov; Vincent Louis Viala |
| EPI_ISL_2344890, EPI_ISL_2344891, EPI_ISL_2344892, EPI_ISL_2344894 | SANTA CASA DE PORTO FELIZ | Instituto Butantan / ESALQ-Piracicaba | Antonio Jorge Martins; Claudia Renata dos Santos Barros; David Schlesinger; Debora Botequio Moretti; Dimas Tadeu Covas; Elaine Cristina Marqueze; Elaine Vieira Santos; Evandra Strazza Rodrigues; Heidge Fukumasu; Jayme Augusto de Souza-Neto; José Salvatore Leister Patané; Luiz Alcantara; Luiz Lehmann Coutinho; Maria Carolina Elias; Maurício Lacerda Nogueira; Rafael dos Santos Bezerra; Raul Machado Neto; Rejane Maria Tommasini Grotto; Ricardo Haddad; Sandra Coccuzzo Sampaio Vessoni; Simone Kashima; Svetoslav Nanev Slavov; Vincent Louis Viala |
| EPI_ISL_2649245, EPI_ISL_2791723 | SARS-CoV-2 Sequencing Castilla y Leon-Spain Consortium | SARS-CoV-2 Sequencing Castilla y Leon-Spain Consortium | Antonio Orduña-Domingo; Carlos Fuster Foz; Carmen Aldea-Mansilla; Carmen Gimeno Crespo; David Abad; Gregoria Megías Lobón; Jose María Eiros Bouza; Laura Sánchez de Prada; M. Isabel Fernandez-Natal; Marta Dominguez-Gil; Marta Hernandez; María Antonia García Castro; Mª Fe Brezmes-Valdivieso; Noelia Arenal Andrés; Silvia Rojo; Sonsoles Garcinuño Pérez |
| EPI_ISL_2131662, EPI_ISL_2336130, EPI_ISL_2336133, EPI_ISL_2337801 | SARS-CoV-2 testing team, National Institute of Infectious Diseases | Pathogen Genomics Center, National Institute of Infectious Diseases | Hazuka Y Furihata; Kentaro Itokawa; Makoto Kuroda; Masanori Hashino; Masumichi Saito; Naomi Nojiri; Nozomu Hanaoka; Rina Tanaka; Sana Uchikoba; Tsuguto Fujimoto; Tsuyoshi Sekizuka |
| EPI_ISL_2018303 | SC Dept of Health and Env. Control-Bureau of Laboratories | Centers for Disease Control and Prevention Division of Viral Diseases, Pathogen Discovery | Alison Laufer Halpin; Ben L. Rambo-Martin; Clinton R. Paden; Dakota Howard; Darlene Wagner; Dave Wentworth; Dhwani Batra; Jasmine Padilla; Justin Lee; Katie Dillon; Krista Queen; Kristen Knipe; Kristine Lacek; Mark Burroughs; Matthew Schmerer; Mili Sheth; Peter Cook; Sam Shepard; Sarah Nobles; Shoshona Le; Suxiang Tong; Vivien Dugan; Yvette Unoarumhi |
| EPI_ISL_2344820, EPI_ISL_2344822 | SECRETARIA DE SAUDE DE SAO PEDRO | Instituto Butantan / ESALQ-Piracicaba | Antonio Jorge Martins; Claudia Renata dos Santos Barros; David Schlesinger; Debora Botequio Moretti; Dimas Tadeu Covas; Elaine Cristina Marqueze; Elaine Vieira Santos; Evandra Strazza Rodrigues; Heidge Fukumasu; Jayme Augusto de Souza-Neto; José Salvatore Leister Patané; Luiz Alcantara; Luiz Lehmann Coutinho; Maria Carolina Elias; Maurício Lacerda Nogueira; Rafael dos Santos Bezerra; Raul Machado Neto; Rejane Maria Tommasini Grotto; Ricardo Haddad; Sandra Coccuzzo Sampaio Vessoni; Simone Kashima; Svetoslav Nanev Slavov; Vincent Louis Viala |
| EPI_ISL_2344895, EPI_ISL_2344896 | SECRETARIA MUNICIPAL DA SAUDE DE ITAPEVA SP | Instituto Butantan / ESALQ-Piracicaba | Antonio Jorge Martins; Claudia Renata dos Santos Barros; David Schlesinger; Debora Botequio Moretti; Dimas Tadeu Covas; Elaine Cristina Marqueze; Elaine Vieira Santos; Evandra Strazza Rodrigues; Heidge Fukumasu; Jayme Augusto de Souza-Neto; José Salvatore Leister Patané; Luiz Alcantara; Luiz Lehmann Coutinho; Maria Carolina Elias; Maurício Lacerda Nogueira; Rafael dos Santos Bezerra; Raul Machado Neto; Rejane Maria Tommasini Grotto; Ricardo Haddad; Sandra Coccuzzo Sampaio Vessoni; Simone Kashima; Svetoslav Nanev Slavov; Vincent Louis Viala |
| EPI_ISL_2344903, EPI_ISL_2344912 | SECRETARIA MUNICIPAL DA SAUDE DE ITARARE | Instituto Butantan / ESALQ-Piracicaba | Antonio Jorge Martins; Claudia Renata dos Santos Barros; David Schlesinger; Debora Botequio Moretti; Dimas Tadeu Covas; Elaine Cristina Marqueze; Elaine Vieira Santos; Evandra Strazza Rodrigues; Heidge Fukumasu; Jayme Augusto de Souza-Neto; José Salvatore Leister Patané; Luiz Alcantara; Luiz Lehmann Coutinho; Maria Carolina Elias; Maurício Lacerda Nogueira; Rafael dos Santos Bezerra; Raul Machado Neto; Rejane Maria Tommasini Grotto; Ricardo Haddad; Sandra Coccuzzo Sampaio Vessoni; Simone Kashima; Svetoslav Nanev Slavov; Vincent Louis Viala |
| EPI_ISL_2344858 | SECRETARIA MUNICIPAL DE SAUDE DE CORDEIROPOLIS | Instituto Butantan / ESALQ-Piracicaba | Antonio Jorge Martins; Claudia Renata dos Santos Barros; David Schlesinger; Debora Botequio Moretti; Dimas Tadeu Covas; Elaine Cristina Marqueze; Elaine Vieira Santos; Evandra Strazza Rodrigues; Heidge Fukumasu; Jayme Augusto de Souza-Neto; José Salvatore Leister Patané; Luiz Alcantara; Luiz Lehmann Coutinho; Maria Carolina Elias; Maurício Lacerda Nogueira; Rafael dos Santos Bezerra; Raul Machado Neto; Rejane Maria Tommasini Grotto; Ricardo Haddad; Sandra Coccuzzo Sampaio Vessoni; Simone Kashima; Svetoslav Nanev Slavov; Vincent Louis Viala |
| EPI_ISL_5603261, EPI_ISL_5603262 | SECRETARIA MUNICIPAL DE SAUDE DE JAGUARIBE | Analytical Competence Molecular Epidemiology Lab/ACME, Oswaldo Cruz Foundation, Ceara (FIOCRUZ CE) | Carlos Leonardo de Aragao Araujo; Cecilia Leite Costa & Eduardo Ruback dos Santos on behalf of COVID-19 FIOCRUZ Genomic Network; Cleber Furtado Aksenen; Fabio Miyajima; Fernando Braga Stehling; Francisco Eder de Moura Lopes; Igor Oliveira Duarte; Jamille Maria Mendes Bezerra; Joaquim Cesar do Nascimento Sousa Junior; Pedro Miguel Carneiro Jeronimo; Suzana Porto Almeida; Thais Ferreira de Oliveira; Thais de Oliveira Costa; Ticiane Cavalcante de Souza; Veridiana Pessoa Miyajima |
| EPI_ISL_3102541 | SECRETARIA MUNICIPAL DE SAUDE DE TIANGUA | Analytical Competence Molecular Epidemiology Lab/ACME, Oswaldo Cruz Foundation, Ceara (FIOCRUZ CE) | Cleber Furtado Aksenen; Fabio Miyajima; Fernando Braga Stehling; Francisco Eder de Moura Lopes; Jamille Maria Mendes Bezerra; Joaquim César do Nascimento Sousa Junior; Pedro Miguel Carneiro Jeronimo; Suzana Porto Almeida e Lucas Delerino; Thais Ferreira de Oliveira; Thais de Oliveira Costa; Ticiane Cavalcante de Souza; Veridiana Pessoa Miyajima |
| EPI_ISL_1967253, EPI_ISL_1967255, EPI_ISL_1967256, EPI_ISL_1967257 | SECRETARIA MUNICIPAL DE SAUDE PORTO FERREIRA | Instituto Butantan / Mendelics | Antonio Jorge Martins; Bianca Cechetto Carlos. Mendelics: Bibiana Santos; Claudia Renata dos Santos Barros; Cíntia Bittar; David Schlesinger. Hemocentro Ribeirão Preto: Simone Kashima; Debora Botequio Moretti; Elaine Cristina Marqueze; Elaine Vieira dos Santos; Elisangela Chicaroni Mattos; Erika Freitas; Evandra Strazza Rodrigues; Felipe Allan da Silva da Costa; Flavia Aburjaile; Fábio Sossai Possebon; Guilherme Campos; Guilherme Targino Valente; Heidge Fukumasu. USP-Botucatu: Rejane Maria Tommasini Grotto; Helena Lage Ferreira; Instituto Butantan: Dimas Tadeu Covas; Jardelina de Souza Todao Bernardino; Jayme A. Souza-Neto; Jessika Cristina Chagas Lesbon; Jorge A. Petroli Marchesi; José Salvatore Leister Patané; João Paulo Kitajima; João Pessoa Araújo Jr.; Leila Sabrina Ullmann; Loyze Paola Oliveira de Lima; Luiz Aurelio de Campos Crispin. Centro de Genômica Funcional da ESALQ: Luiz Lehmann Coutinho; Luiz Carlos Junior de Alcantara; Lívia Sacchetto; Maisa C. Pereira Parra; Maria Carolina Elias; Marta Giovanetti; Marília Moraes; Maurício Lacerda Nogueira. Prefeitura de Sao Paulo: Melissa Palmieri.; Patricia Akemi Assato; Paula Rahal; Paulo Inacio da Costa; Rafael dos Santos Bezerra; Raquel de Lello Rocha Campos Cassano. NGS Soluções Genômicas: Pilar Drummond Sampaio Corrêa Mariani. FZEA-USP Pirassununga: Mirele Daiana Poleti; Raul Machado Neto; Ricardo Augusto Brassaloti; Ricardo Haddad; Rodrigo Tocantins Calado. FAMERP-SJRP: Cecília Artico Banho; Sandra Coccuzzo Sampaio; Svetoslav Nanev Slavov; Vagner Fonseca; Vincent Louis Viala |
| EPI_ISL_2209285, EPI_ISL_2344867, EPI_ISL_2344897, EPI_ISL_2344919, EPI_ISL_2344920, EPI_ISL_2344921, EPI_ISL_2344922, EPI_ISL_2344923, EPI_ISL_2344924, EPI_ISL_2344926, EPI_ISL_2344928, EPI_ISL_2344929, EPI_ISL_2344930, EPI_ISL_2344931, EPI_ISL_2344932, EPI_ISL_2344933, EPI_ISL_2344934, EPI_ISL_2344935, EPI_ISL_2344936, EPI_ISL_2344937, EPI_ISL_2344938, EPI_ISL_2344939 | | | |
| see above | SECRETARIA MUNICIPAL DE SAUDE SOROCABA | Instituto Butantan / ESALQ-Piracicaba | Antonio Jorge Martins; Claudia Renata dos Santos Barros; David Schlesinger; Debora Botequio Moretti; Dimas Tadeu Covas; Elaine Cristina Marqueze; Elaine Vieira Santos; Evandra Strazza Rodrigues; Heidge Fukumasu; Jayme Augusto de Souza-Neto; José Salvatore Leister Patané; Luiz Alcantara; Luiz Lehmann Coutinho; Maria Carolina Elias; Maurício Lacerda Nogueira; Rafael dos Santos Bezerra; Raul Machado Neto; Rejane Maria Tommasini Grotto; Ricardo Haddad; Sandra Coccuzzo Sampaio Vessoni; Simone Kashima; Svetoslav Nanev Slavov; Vincent Louis Viala |
| EPI_ISL_1785097, EPI_ISL_1785098 | SIESP DIPARTIMENTO DI PREVENZIONE TERAMO | Istituto Zooprofilattico Sperimentale dell'Abruzzo e Molise "G. Caporale" | Ancora M; Calistri P; Cammà C; Caporale M; Curini V; Delli Compagni E; Di Domenico M; Di Lollo Valeria; Di Pasquale A; Lorusso A; Mangone I; Marcacci M; Puglia I; Rinaldi A; Savini G; Scialabba S |
| EPI_ISL_1785119, EPI_ISL_1917529, EPI_ISL_1917531 | SIESP SULMONA | Istituto Zooprofilattico Sperimentale dell'Abruzzo e Molise "G. Caporale" | Ancora M; Calistri P; Cammà C; Caporale M; Curini V; Delli Compagni E; Di Domenico M; Di Lollo Valeria; Di Pasquale A; Lorusso A; Mangone I; Marcacci M; Puglia I; Rinaldi A; Savini G; Scialabba S |
| EPI_ISL_2564796, EPI_ISL_2564797, EPI_ISL_2564798, EPI_ISL_2565021, EPI_ISL_2585453, EPI_ISL_2585811, EPI_ISL_2585813, EPI_ISL_2585816, EPI_ISL_2586125, EPI_ISL_2586639, EPI_ISL_2586657, EPI_ISL_2586694, EPI_ISL_2586698, EPI_ISL_2586701, EPI_ISL_2586713 | | | |
| see above | SK-Roy Romanow Provincial Laboratory | National Microbiology Laboratory (NML) | Amanda Lang; Anna Majer; Anneliese Landgraff; CanCOGeN's metadata curation team; Darian Hole; Elsie Grudeski; Gary Van Domselaar; Grace Seo; Jennifer Tanner; Jessica Minion; Kirsten Biggar; Madison Chapel; Morag Graham; Natalie Knox; Nathalie Bastien; Philip Mabon; Public Health Agency of Canada CanCOGeN team; Rachel DePaulo; Rhiannon Huzarewich; Russell Mandes; Ryan McDonald; Shari Tyson; Timothy Booth; Yan Li |
| EPI_ISL_5844360, EPI_ISL_5844391, EPI_ISL_6124670, EPI_ISL_6124677 | SK-Roy Romanow Provincial Laboratory | Saskatchewan - Roy Romanow Provincial Laboratory (RRPL) | Alanna Senecal; Amanda Lang; Jessica Minion; Kara Loos; Keith MacKenzie; Meredith Faires; Rachel DePaulo; Roy Romanow Provincial Laboratory - Molecular Diagnostics; Ryan McDonald |
| EPI_ISL_2344940 | SMS IPERO | Instituto Butantan / ESALQ-Piracicaba | Antonio Jorge Martins; Claudia Renata dos Santos Barros; David Schlesinger; Debora Botequio Moretti; Dimas Tadeu Covas; Elaine Cristina Marqueze; Elaine Vieira Santos; Evandra Strazza Rodrigues; Heidge Fukumasu; Jayme Augusto de Souza-Neto; José Salvatore Leister Patané; Luiz Alcantara; Luiz Lehmann Coutinho; Maria Carolina Elias; Maurício Lacerda Nogueira; Rafael dos Santos Bezerra; Raul Machado Neto; Rejane Maria Tommasini Grotto; Ricardo Haddad; Sandra Coccuzzo Sampaio Vessoni; Simone Kashima; Svetoslav Nanev Slavov; Vincent Louis Viala |
| EPI_ISL_2344888, EPI_ISL_2344913 | SMS SECRETARIA MUNICIPAL DE SAUDE DE BOITUVA | Instituto Butantan / ESALQ-Piracicaba | Antonio Jorge Martins; Claudia Renata dos Santos Barros; David Schlesinger; Debora Botequio Moretti; Dimas Tadeu Covas; Elaine Cristina Marqueze; Elaine Vieira Santos; Evandra Strazza Rodrigues; Heidge Fukumasu; Jayme Augusto de Souza-Neto; José Salvatore Leister Patané; Luiz Alcantara; Luiz Lehmann Coutinho; Maria Carolina Elias; Maurício Lacerda Nogueira; Rafael dos Santos Bezerra; Raul Machado Neto; Rejane Maria Tommasini Grotto; Ricardo Haddad; Sandra Coccuzzo Sampaio Vessoni; Simone Kashima; Svetoslav Nanev Slavov; Vincent Louis Viala |
| EPI_ISL_2097297 | STONY BROOK UNIVERSITY HOSPITAL | Wadsworth Center, New York State Department of Health | Alexis Russell; Catharine Prussing; Daryl M. Lamson; Erasmus Schneider; Erica Lasek-Nesselquist; John Kelly; Jonathan Plitnick; Kirsten St. George; Matthew Shudt; Melissa A Leisner; Navjot Singh |
| EPI_ISL_5155577, EPI_ISL_5155579, EPI_ISL_5155741, EPI_ISL_5155824, EPI_ISL_5155963, EPI_ISL_5155970, EPI_ISL_5156533, EPI_ISL_5156583, EPI_ISL_5156584, EPI_ISL_5156585, EPI_ISL_5156586 | | | |
| see above | SUNY Upstate Medical University | 505 Irving Avenue, Syracuse, NY 13210 - Frank Middleton Lab | Brian Pavlovitz; Frank Middleton |
| EPI_ISL_4666450 | SURA | Laboratorio Departamental de Salud Publica de Antioquia | Andres F. Cardona-Rios; Cristian Arbey Velarde Hoyos; Gloria Isabel Escobar; Idabely Betancur Ortiz; Juan P. Hernandez-Ortiz; Maria Stella López |
| EPI_ISL_4878291 | SURA Bogota | Instituto Nacional de Salud-Dirección de Investigación en Salud Pública | Carlos Franco-Muñoz; Carmen Osorio; Diana Malo; Diego A. Álvarez-Díaz; Diego Andrés Prada; Gerardo Santamaría; Hector Alejandro Ruiz-Moreno; Jhonnatan Reales-González; Jorge Rivera; Juan Camilo Martinez; Julian Naizaque; Katherine Laiton-Donato; Lisseth Pardo; Magdalena Wiesner; Marcela Mercado-Reyes; Maria T. Herrera-Sepúlveda; Marta Lopez Blanco; Martha Lucia Ospina Martinez; Paola Rojas; Sergio Gomez; Sheryll Corchuelo; Ángela Alarcon Cruz |
| EPI_ISL_1826671, EPI_ISL_1826691, EPI_ISL_1826702, EPI_ISL_1826709, EPI_ISL_1826710, EPI_ISL_1826761, EPI_ISL_1826792, EPI_ISL_1919658, EPI_ISL_1919659, EPI_ISL_1919677, EPI_ISL_1919705, EPI_ISL_1919706, EPI_ISL_1919718, EPI_ISL_1919719, EPI_ISL_1919727, EPI_ISL_1919734, EPI_ISL_1919738, EPI_ISL_1970770, EPI_ISL_1970771, EPI_ISL_1970780, EPI_ISL_2003211 | | | |
| see above | SYNLAB | GIGA Medical Genomics | Bouchra Boujemla; Cécile Meex; Keith Durkin; Maria Artesi; Marie-Pierre Hayette; Nathalie Renotte; Pierrette Melin; Raphaël Boreux; Sébastien Bontems; Vincent Bours |
| EPI_ISL_2004327, EPI_ISL_2004328, EPI_ISL_2004329, EPI_ISL_2004332 | SYNLAB | Instituto Nacional de Saude (INSA) | Borges et al |
| EPI_ISL_1848074, EPI_ISL_2110055 | SYNLAB Jena Oncoscreen | Robert Koch Institute | |
| EPI_ISL_1851692 | SYNLAB MVZ Weiden | Robert Koch Institute | |
| EPI_ISL_5800569, EPI_ISL_5800570, EPI_ISL_5800571, EPI_ISL_5800573, EPI_ISL_5800576, EPI_ISL_5800580, EPI_ISL_5800729, EPI_ISL_5800730, EPI_ISL_5800731, EPI_ISL_5800732, EPI_ISL_5800735, EPI_ISL_5800738, EPI_ISL_5800739, EPI_ISL_5800740, EPI_ISL_5800741, EPI_ISL_5800743, EPI_ISL_5800745, EPI_ISL_5800746, EPI_ISL_5800751 | | | |
| see above | Sae Cta Serv Atend Espec Testagem E Aconselhamento | Instituto Butantan | Antonio Jorge Martins; Claudia Renata dos Santos Barros; David Schlesinger; Debora Botequio Moretti; Dimas Tadeu Covas; Elaine Cristina Marqueze; Elaine Vieira Santos; Evandra Strazza Rodrigues; Heidge Fukumasu; Jayme Augusto de Souza-Neto; José Salvatore Leister Patané; Luiz Alcantara; Luiz Lehmann Coutinho; Maria Carolina Elias; Maurício Lacerda Nogueira; Rafael dos Santos Bezerra; Raul Machado Neto; Rejane Maria Tommasini Grotto; Ricardo Haddad; Sandra Coccuzzo Sampaio Vessoni; Simone Kashima; Svetoslav Nanev Slavov; Vincent Louis Viala |

| | | | |
|---|---|---|---|
| EPI_ISL_2231178, EPI_ISL_2329589, EPI_ISL_2329595, EPI_ISL_2329601, EPI_ISL_2329608, EPI_ISL_2422984, EPI_ISL_2422993, EPI_ISL_2628627, EPI_ISL_2628630 | | | |
| see above | San Diego County Public Health Laboratory | Andersen lab at Scripps Research | Brett Austin; Jovan Shephard; SEARCH Alliance San Diego with Ashleigh Murphy |
| EPI_ISL_1824063, EPI_ISL_1824066, EPI_ISL_1824069, EPI_ISL_1824070, EPI_ISL_1824075, EPI_ISL_1824079, EPI_ISL_1824122 | | | |
| see above | San Gallicano Dermatological Institute I.F.O. | INMI Lazzaro Spallanzani IRCCS | A Massacci; B Bartolini; CEM Gruber; E Giombini; E Sperandio; F De Nicola; F Messina; F Santini; G Bonfiglio; G Orlandi; GF Inger; L Ciuffreda; M Pallocca; M Rueca; O Butera |
| EPI_ISL_1970975, EPI_ISL_1970978 | San Juan Regional Medical Center | New Mexico Department of Health Scientific Laboratory | Anastacia Griego-Fisher; D'eldra Malone; Ellie Johnson; Jennifer Benoit; Linda Salazar; Mark Willmon; Ratheesh Rajan |
| EPI_ISL_5801041, EPI_ISL_5801042, EPI_ISL_5801046, EPI_ISL_5801047, EPI_ISL_5801048, EPI_ISL_5801049 | Santa Casa De Guararema | Instituto Butantan | Antonio Jorge Martins; Claudia Renata dos Santos Barros; David Schlesinger; Debora Botequio Moretti; Dimas Tadeu Covas; Elaine Cristina Marqueze; Elaine Vieira Santos; Evandra Strazza Rodrigues; Heidge Fukumasu; Jayme Augusto de Souza-Neto; José Salvatore Leister Patané; Luiz Alcantara; Luiz Lehmann Coutinho; Maria Carolina Elias; Maurício Lacerda Nogueira; Rafael dos Santos Bezerra; Raul Machado Neto; Rejane Maria Tommasini Grotto; Ricardo Haddad; Sandra Coccuzzo Sampaio Vessoni; Simone Kashima; Svetoslav Nanev Slavov; Vincent Louis Viala |
| EPI_ISL_5800818, EPI_ISL_5800821, EPI_ISL_5800828, EPI_ISL_5800835, EPI_ISL_5800837, EPI_ISL_5800839, EPI_ISL_5800844 | | | |
| see above | Santa Casa De Jose Bonifacio | Instituto Butantan | Antonio Jorge Martins; Claudia Renata dos Santos Barros; David Schlesinger; Debora Botequio Moretti; Dimas Tadeu Covas; Elaine Cristina Marqueze; Elaine Vieira Santos; Evandra Strazza Rodrigues; Heidge Fukumasu; Jayme Augusto de Souza-Neto; José Salvatore Leister Patané; Luiz Alcantara; Luiz Lehmann Coutinho; Maria Carolina Elias; Maurício Lacerda Nogueira; Rafael dos Santos Bezerra; Raul Machado Neto; Rejane Maria Tommasini Grotto; Ricardo Haddad; Sandra Coccuzzo Sampaio Vessoni; Simone Kashima; Svetoslav Nanev Slavov; Vincent Louis Viala |
| EPI_ISL_5801009, EPI_ISL_5801012, EPI_ISL_5801014, EPI_ISL_5801015, EPI_ISL_5801017, EPI_ISL_5801018, EPI_ISL_5801019 | | | |
| see above | Santa Casa De Misericordia Sao Jose | Instituto Butantan | Antonio Jorge Martins; Claudia Renata dos Santos Barros; David Schlesinger; Debora Botequio Moretti; Dimas Tadeu Covas; Elaine Cristina Marqueze; Elaine Vieira Santos; Evandra Strazza Rodrigues; Heidge Fukumasu; Jayme Augusto de Souza-Neto; José Salvatore Leister Patané; Luiz Alcantara; Luiz Lehmann Coutinho; Maria Carolina Elias; Maurício Lacerda Nogueira; Rafael dos Santos Bezerra; Raul Machado Neto; Rejane Maria Tommasini Grotto; Ricardo Haddad; Sandra Coccuzzo Sampaio Vessoni; Simone Kashima; Svetoslav Nanev Slavov; Vincent Louis Viala |
| EPI_ISL_5800856, EPI_ISL_5800857, EPI_ISL_5800859, EPI_ISL_5800863 | Santa Casa De Porto Feliz | Instituto Butantan | Antonio Jorge Martins; Claudia Renata dos Santos Barros; David Schlesinger; Debora Botequio Moretti; Dimas Tadeu Covas; Elaine Cristina Marqueze; Elaine Vieira Santos; Evandra Strazza Rodrigues; Heidge Fukumasu; Jayme Augusto de Souza-Neto; José Salvatore Leister Patané; Luiz Alcantara; Luiz Lehmann Coutinho; Maria Carolina Elias; Maurício Lacerda Nogueira; Rafael dos Santos Bezerra; Raul Machado Neto; Rejane Maria Tommasini Grotto; Ricardo Haddad; Sandra Coccuzzo Sampaio Vessoni; Simone Kashima; Svetoslav Nanev Slavov; Vincent Louis Viala |
| EPI_ISL_1824109, EPI_ISL_2659047, EPI_ISL_2659075, EPI_ISL_2659082 | Santa Clara Valley Medical Center | Santa Clara County Public Health Laboratory | Santa Clara County Public Health Department |
| EPI_ISL_1969513, EPI_ISL_1969518, EPI_ISL_1969519, EPI_ISL_1969532 | Scripps Medical Laboratory | Andersen lab at Scripps Research | Ellen Stefanski; Ian Mchardy; SEARCH Alliance San Diego with Michael Quigley |
| EPI_ISL_2489977, EPI_ISL_2489983, EPI_ISL_2489989, EPI_ISL_3105055, EPI_ISL_3372972 | Seattle Flu Study | Seattle Flu Study | Amanda Adler; Barry R. Lutz; Benjamin Pelle; Caitlin R. Wolf; Chris D. Frazar; Deborah A. Nickerson; Elisabeth Brandstetter; Erica Ryke; Helen Y. Chu; Janet A. Englund; Jay Shendure; Jeff Duchin; Jover Lee; Kairsten Fay; Karen Cowgill; Kirsten Lacombe; Lea M. Starita; Mark J. Rieder; Matthew Richardson; Matthew Thompson; Melissa Truong; Michael Boeckh; Michael Famulare; Misja Ilcisin; Peter D. Han; Stephanie Schrag; Thomas R. Sibley; Trevor Bedford |
| EPI_ISL_5800756, EPI_ISL_5800758 | Secretaria De Saude De Sao Pedro | Instituto Butantan | Antonio Jorge Martins; Claudia Renata dos Santos Barros; David Schlesinger; Debora Botequio Moretti; Dimas Tadeu Covas; Elaine Cristina Marqueze; Elaine Vieira Santos; Evandra Strazza Rodrigues; Heidge Fukumasu; Jayme Augusto de Souza-Neto; José Salvatore Leister Patané; Luiz Alcantara; Luiz Lehmann Coutinho; Maria Carolina Elias; Maurício Lacerda Nogueira; Rafael dos Santos Bezerra; Raul Machado Neto; Rejane Maria Tommasini Grotto; Ricardo Haddad; Sandra Coccuzzo Sampaio Vessoni; Simone Kashima; Svetoslav Nanev Slavov; Vincent Louis Viala |
| EPI_ISL_5800866, EPI_ISL_5800867 | Secretaria Municipal Da Saude De Itapeva Sp | Instituto Butantan | Antonio Jorge Martins; Claudia Renata dos Santos Barros; David Schlesinger; Debora Botequio Moretti; Dimas Tadeu Covas; Elaine Cristina Marqueze; Elaine Vieira Santos; Evandra Strazza Rodrigues; Heidge Fukumasu; Jayme Augusto de Souza-Neto; José Salvatore Leister Patané; Luiz Alcantara; Luiz Lehmann Coutinho; Maria Carolina Elias; Maurício Lacerda Nogueira; Rafael dos Santos Bezerra; Raul Machado Neto; Rejane Maria Tommasini Grotto; Ricardo Haddad; Sandra Coccuzzo Sampaio Vessoni; Simone Kashima; Svetoslav Nanev Slavov; Vincent Louis Viala |
| EPI_ISL_5800880, EPI_ISL_5800893 | Secretaria Municipal Da Saude De Itarare | Instituto Butantan | Antonio Jorge Martins; Claudia Renata dos Santos Barros; David Schlesinger; Debora Botequio Moretti; Dimas Tadeu Covas; Elaine Cristina Marqueze; Elaine Vieira Santos; Evandra Strazza Rodrigues; Heidge Fukumasu; Jayme Augusto de Souza-Neto; José Salvatore Leister Patané; Luiz Alcantara; Luiz Lehmann Coutinho; Maria Carolina Elias; Maurício Lacerda Nogueira; Rafael dos Santos Bezerra; Raul Machado Neto; Rejane Maria Tommasini Grotto; Ricardo Haddad; Sandra Coccuzzo Sampaio Vessoni; Simone Kashima; Svetoslav Nanev Slavov; Vincent Louis Viala |
| EPI_ISL_5800808 | Secretaria Municipal De Saude De Cordeiropolis | Instituto Butantan | Antonio Jorge Martins; Claudia Renata dos Santos Barros; David Schlesinger; Debora Botequio Moretti; Dimas Tadeu Covas; Elaine Cristina Marqueze; Elaine Vieira Santos; Evandra Strazza Rodrigues; Heidge Fukumasu; Jayme Augusto de Souza-Neto; José Salvatore Leister Patané; Luiz Alcantara; Luiz Lehmann Coutinho; Maria Carolina Elias; Maurício Lacerda Nogueira; Rafael dos Santos Bezerra; Raul Machado Neto; Rejane Maria Tommasini Grotto; Ricardo Haddad; Sandra Coccuzzo Sampaio Vessoni; Simone Kashima; Svetoslav Nanev Slavov; Vincent Louis Viala |
| EPI_ISL_5800647, EPI_ISL_5800656, EPI_ISL_5800667, EPI_ISL_5800673 | Secretaria Municipal De Saude Porto Ferreira | Instituto Butantan | Antonio Jorge Martins; Claudia Renata dos Santos Barros; David Schlesinger; Debora Botequio Moretti; Dimas Tadeu Covas; Elaine Cristina Marqueze; Elaine Vieira Santos; Evandra Strazza Rodrigues; Heidge Fukumasu; Jayme Augusto de Souza-Neto; José Salvatore Leister Patané; Luiz Alcantara; Luiz Lehmann Coutinho; Maria Carolina Elias; Maurício Lacerda Nogueira; Rafael dos Santos Bezerra; Raul Machado Neto; Rejane Maria Tommasini Grotto; Ricardo Haddad; Sandra Coccuzzo Sampaio Vessoni; Simone Kashima; Svetoslav Nanev Slavov; Vincent Louis Viala |
| EPI_ISL_5800820, EPI_ISL_5800868, EPI_ISL_5800905, EPI_ISL_5800908, EPI_ISL_5800909, EPI_ISL_5800911, EPI_ISL_5800912, EPI_ISL_5800913, EPI_ISL_5800915, EPI_ISL_5800917, EPI_ISL_5800918, EPI_ISL_5800919, EPI_ISL_5800920, EPI_ISL_5800921, EPI_ISL_5800922, EPI_ISL_5800923, EPI_ISL_5800924, EPI_ISL_5800925, EPI_ISL_5800926, EPI_ISL_5800927, EPI_ISL_5800928, EPI_ISL_5800929 | | | |
| see above | Secretaria Municipal De Saude Sorocaba | Instituto Butantan | Antonio Jorge Martins; Claudia Renata dos Santos Barros; David Schlesinger; Debora Botequio Moretti; Dimas Tadeu Covas; Elaine Cristina Marqueze; Elaine Vieira Santos; Evandra Strazza Rodrigues; Heidge Fukumasu; Jayme Augusto de Souza-Neto; José Salvatore Leister Patané; Luiz Alcantara; Luiz Lehmann Coutinho; Maria Carolina Elias; Maurício Lacerda Nogueira; Rafael dos Santos Bezerra; Raul Machado Neto; Rejane Maria Tommasini Grotto; Ricardo Haddad; Sandra Coccuzzo Sampaio Vessoni; Simone Kashima; Svetoslav Nanev Slavov; Vincent Louis Viala |
| EPI_ISL_2179729, EPI_ISL_2179732, EPI_ISL_2179733, EPI_ISL_2179752, EPI_ISL_2179753, EPI_ISL_2179754, EPI_ISL_2179755, EPI_ISL_2179757, EPI_ISL_2179758, EPI_ISL_2179794 | | | |
| see above | Servicio Microbiología Hospital La Paz | Servicio Microbiología Hospital La Paz | Elie Dahdouh; Fernando Lázaro; Jesús Mingorance Cruz; Rubén Cáceres |
| EPI_ISL_2136043, EPI_ISL_2136051, EPI_ISL_2136097, EPI_ISL_2136099, EPI_ISL_2136110, EPI_ISL_2136120, EPI_ISL_2136122, EPI_ISL_2136125, EPI_ISL_2136126, EPI_ISL_2136127, EPI_ISL_2136128, EPI_ISL_2136130, EPI_ISL_2136132, EPI_ISL_2136133, EPI_ISL_2136134, EPI_ISL_2140039, EPI_ISL_2140040, EPI_ISL_2140041, EPI_ISL_2140042, EPI_ISL_2140044, EPI_ISL_2140045, EPI_ISL_2140046, EPI_ISL_2140047, EPI_ISL_2140052, EPI_ISL_2140054, EPI_ISL_2140055, EPI_ISL_2140057, EPI_ISL_2140060, EPI_ISL_2140069, EPI_ISL_2140075, EPI_ISL_2140077, EPI_ISL_2140078, EPI_ISL_2140080, EPI_ISL_2140082, EPI_ISL_2140083, EPI_ISL_2140084, EPI_ISL_2140085, EPI_ISL_2140086, EPI_ISL_2140087, EPI_ISL_2140093, EPI_ISL_2140096, EPI_ISL_2140097, EPI_ISL_2140098, EPI_ISL_2140101, EPI_ISL_2140113, EPI_ISL_2140116, EPI_ISL_2140135, EPI_ISL_2140136, EPI_ISL_2158701, EPI_ISL_2158709, EPI_ISL_2158710, EPI_ISL_2158720, EPI_ISL_2158730, EPI_ISL_2158756, EPI_ISL_2158782, EPI_ISL_2158797, EPI_ISL_4082219, EPI_ISL_4082447, EPI_ISL_4082449, EPI_ISL_4082823, EPI_ISL_4082825 | | | |
| see above | Servicio Virosis Respiratorias-Departamento Virología-INEI | Instituto Nacional Enfermedades Infecciosas C.G.Malbran | Avaro M.; Baumeister E.; Benedetti E.; Campos J.; Cisterna D.; Dattero ME; De Belder D.; Haim MS.; Lorenzo F.; Molina V.; Perandones C.; Poklepovich T.; Pontoriero A.; Russo M.; Sanchez Loria J.; Tuduri E. |
| EPI_ISL_2104421, EPI_ISL_2104437, EPI_ISL_2426653 | Servicio de Microbiologia Hospital Ramon y Cajal | Servicio de Microbiologia Hospital Ramon y Cajal | Galan JC; Martinez L. Abreu M; Ponce M; y Gonzalez-Alba JM |
| EPI_ISL_1922315, EPI_ISL_1922316, EPI_ISL_1922318, EPI_ISL_1922321, EPI_ISL_1922479 | Servicio de Microbiología Clínica (Complejo Hospitalario de Navarra, Pamplona) | Centro de Secuenciación NASERTIC | Ana Miqueleiz; Ana Navascués; Carmen Ezpeleta Baquedano |
| EPI_ISL_2545376 | Servicio de Microbiología, Laboratori Clínic Metropolitana Nord. Hospital Universitari Germans Trias i Pujol. Institut d'Investigació en Ciències de la Salut Germans Trias i Pujol (IGTP) | SeqCOVID-SPAIN consortium/IBV(CSIC) | Adrián Antuori; Anabel Fernández; Anna Not; Antoni E. Bordoy; Cristina Casañ and SeqCOVID-SPAIN consortium; Elisa Martró; Nona Romaní; Verónica Saludes |
| EPI_ISL_2271593, EPI_ISL_2271594, EPI_ISL_2271599, EPI_ISL_2271600, EPI_ISL_2271601, EPI_ISL_2271606, EPI_ISL_2271610, EPI_ISL_2271613 | | | |
| see above | Servicio de Microbiología. Consorcio Hospital General Universitario de Valencia | SeqCOVID-SPAIN consortium/IBV(CSIC) | Begoña Fuster Escrivá; Carme Salvador García; Concepción Gimeno Cardona and SeqCOVID-SPAIN consortium; María Dolores Ocete; Rafael Medina González |
| EPI_ISL_2016629 | Servicio de Microbiología. Hospital Clínico Universitario de Valencia | SeqCOVID-SPAIN consortium/IBV(CSIC) | David Navarro Ortega; Eliseo Albert Vicent; Ignacio Torres and SeqCOVID-SPAIN consortium |
| EPI_ISL_2323444, EPI_ISL_2323450 | Servicio de Microbiología. Hospital Universitario Doctor Peset | SeqCOVID-SPAIN consortium/IBV(CSIC) | José Miguel Nogueira Coito and SeqCOVID-SPAIN consortium; Juan Alberola Enguídanos; Juan José Camarena Miñana; Rosa González Pellicer |
| EPI_ISL_5316135, EPI_ISL_5316137, EPI_ISL_5316138, EPI_ISL_5316139, EPI_ISL_5316141, EPI_ISL_5316142, EPI_ISL_5316143, EPI_ISL_5316148, EPI_ISL_5316152, EPI_ISL_5316157, EPI_ISL_5316158, EPI_ISL_5316162, EPI_ISL_5316164, EPI_ISL_5316167, EPI_ISL_5316168, EPI_ISL_5316169, EPI_ISL_5316170, EPI_ISL_5316171, EPI_ISL_5316175, EPI_ISL_5316176, EPI_ISL_5316177, EPI_ISL_5316181, EPI_ISL_5316182, EPI_ISL_5316183, EPI_ISL_5316184, EPI_ISL_5316185, EPI_ISL_5316186, EPI_ISL_5316188, EPI_ISL_5316190, EPI_ISL_5316191, EPI_ISL_5316194, EPI_ISL_5316197, EPI_ISL_5316198, EPI_ISL_5316204, EPI_ISL_5316205, EPI_ISL_5316206, EPI_ISL_5316209, EPI_ISL_5316210, EPI_ISL_5316211, EPI_ISL_5316212, EPI_ISL_5316213, EPI_ISL_5316214, EPI_ISL_5316215, EPI_ISL_5316217, EPI_ISL_5316218, EPI_ISL_5316219, EPI_ISL_5316220, EPI_ISL_5316221, EPI_ISL_5316222, EPI_ISL_5316224, EPI_ISL_5316225, EPI_ISL_5316226, EPI_ISL_5316228, EPI_ISL_5316230, EPI_ISL_5316231, EPI_ISL_5316238, EPI_ISL_5316239, EPI_ISL_5316240, EPI_ISL_5316242, EPI_ISL_5316244, EPI_ISL_5316246, EPI_ISL_5316247, EPI_ISL_5316252, EPI_ISL_5316254, EPI_ISL_5316257, EPI_ISL_5316259, EPI_ISL_5316260, EPI_ISL_5316261, EPI_ISL_5316262, EPI_ISL_5316263, EPI_ISL_5316264, EPI_ISL_5316269, EPI_ISL_5316270, EPI_ISL_5316273, EPI_ISL_5316274, EPI_ISL_5316275, EPI_ISL_5316276, EPI_ISL_5316278, EPI_ISL_5316279, EPI_ISL_5316280, EPI_ISL_5316288, EPI_ISL_5316289, EPI_ISL_5316290, EPI_ISL_5316295, EPI_ISL_5316296, EPI_ISL_5316297, EPI_ISL_5316298, EPI_ISL_5316299, EPI_ISL_5316300, EPI_ISL_5316301, EPI_ISL_5316302, EPI_ISL_5316304, EPI_ISL_5316305, EPI_ISL_5316310, EPI_ISL_5316311, EPI_ISL_5316313, EPI_ISL_5316315, EPI_ISL_5316319, EPI_ISL_5316320, EPI_ISL_5316321, EPI_ISL_5316325, EPI_ISL_5316330, EPI_ISL_5316334, EPI_ISL_5316335 | | | |
| see above | Shared Hospital Laboratory | Shared Hospital Laboratory | Christie Vermeiren; Finlay Maguire; Kevin Katz; Patryk Aftanas; Robert Kozak; Samira Mubareka |
| EPI_ISL_2231176, EPI_ISL_2231177 | Sharp HealthCare Laboratory | Andersen lab at Scripps Research | Art Mendoza; Cathy Woerle; Jacquelyn Berumen; Liam McGinnis; Omid Bakhtar; SEARCH Alliance San Diego with Aaron Harding |
| EPI_ISL_2296008, EPI_ISL_2296014, | Simple Laboratories | Gagnon Lab, Southern Illinois University | Keith Gagnon |

| | | | |
|---|---|---|---|
| EPI_ISL_2296019 | | | |
| EPI_ISL_5800930 | Sms Ipero | Instituto Butantan | Antonio Jorge Martins; Claudia Renata dos Santos Barros; David Schlesinger; Debora Botequio Moretti; Dimas Tadeu Covas; Elaine Cristina Marqueze; Elaine Vieira Santos; Evandra Strazza Rodrigues; Heidge Fukumasu; Jayme Augusto de Souza-Neto; José Salvatore Leister Patané; Luiz Alcantara; Luiz Lehmann Coutinho; Maria Carolina Elias; Maurício Lacerda Nogueira; Rafael dos Santos Bezerra; Raul Machado Neto; Rejane Maria Tommasini Grotto; Ricardo Haddad; Sandra Coccuzzo Sampaio Vessoni; Simone Kashima; Svetoslav Nanev Slavov; Vincent Louis Viala |
| EPI_ISL_5800852, EPI_ISL_5800894 | Sms Secretaria Municipal De Saude De Boituva | Instituto Butantan | Antonio Jorge Martins; Claudia Renata dos Santos Barros; David Schlesinger; Debora Botequio Moretti; Dimas Tadeu Covas; Elaine Cristina Marqueze; Elaine Vieira Santos; Evandra Strazza Rodrigues; Heidge Fukumasu; Jayme Augusto de Souza-Neto; José Salvatore Leister Patané; Luiz Alcantara; Luiz Lehmann Coutinho; Maria Carolina Elias; Maurício Lacerda Nogueira; Rafael dos Santos Bezerra; Raul Machado Neto; Rejane Maria Tommasini Grotto; Ricardo Haddad; Sandra Coccuzzo Sampaio Vessoni; Simone Kashima; Svetoslav Nanev Slavov; Vincent Louis Viala |
| EPI_ISL_2006896 | South Dakota Public Health Laboratory | South Dakota Public Health Laboratory | Jacob Garfin and Chris Carlson |
| EPI_ISL_2102287 | Spital Limmattal | Institute of Medical Virology | Alexandra Trkola; Annette AudigŽ; Catharine Aquino; Cyril Shah; Daniel Ehrsam; Gabriela Ziltener; Guido Bloemberg; Hubert Rehrauer; Isabel StŸrmer; Joel Wirz; Jon Huder; JŸrg Bšni; Kevin Steiner; Maria GrŸnberg; Maryam Zaheri; Michael Huber; Riccarda Capaul; Stefan Schmutz; Verena Kufner; Weihong Qi |
| EPI_ISL_1826790, EPI_ISL_1919660, EPI_ISL_1919668, EPI_ISL_1919669, EPI_ISL_1970768, EPI_ISL_1970773, EPI_ISL_1970775, EPI_ISL_2003319 | | | |
| see above | St. Nikolaus-Hospital Eupen | GIGA Medical Genomics | Bouchra Boujemla; Cécile Meex; Keith Durkin; Maria Artesi; Marie-Pierre Hayette; Nathalie Renotte; Pierrette Melin; Raphaël Boreux; Sébastien Bontems; Vincent Bours |
| EPI_ISL_2659061 | Stanford Clinical Virology Laboratory | Santa Clara County Public Health Laboratory | Santa Clara County Public Health Department |
| EPI_ISL_4496781, EPI_ISL_4496782, EPI_ISL_4496783 | Stanford Health Care | Stanford University School of Medicine, Clinical Virology Laboratory | Becky Jiang; Bernadette Troung; Hannah Wang; James Zehnder; Malaya K. Sahoo; and Benjamin A. Pinsky |
| EPI_ISL_2007994, EPI_ISL_2007997, EPI_ISL_2008007, EPI_ISL_2008014, EPI_ISL_2724559 | State Laboratories Division, Hawaii State Department of Health | State Laboratories Division, Hawaii State Department of Health | Ayana Garnet; Drew Kuwazaki; Edward Desmond; Pamela O'Brien; Razvan Sultana |
| EPI_ISL_2377909, EPI_ISL_2377929, EPI_ISL_2377939, EPI_ISL_2377942, EPI_ISL_2378010, EPI_ISL_2378027, EPI_ISL_2378374, EPI_ISL_2378397, EPI_ISL_2378439, EPI_ISL_2472999, EPI_ISL_2473064, EPI_ISL_2473087, EPI_ISL_2800560, EPI_ISL_2800566 | | | |
| see above | State Testing Facility | Altius Institute for Biomedical Research | Alex Isner; Alex Nguyen; Amanda Gale; Audra Johnson; Clem Green; Daniel Bates; Eric Thorland; Jacob Rodriguez; Jean Robinston; Jemma Nelson; Jessica Kunder; John Stamatoyannopoulos; Joshua Richards; Julia Wald; Kneshay Harper; Lauren Mitchell; Mark Frerker; Matt Hartman; Michael Buckley; Muhammad Halimun; Rebecca Bruders; Sadie Patraw; Sofia Olsson; Tobias Ragoczy |
| EPI_ISL_2033275, EPI_ISL_2033311, EPI_ISL_2257985 | Swedish national genomic surveillance program of SARS-CoV-2 | The Public Health Agency of Sweden | Alma Brolund; Maria Lind Karlberg; Maximilian Riess; Swedish national genomic surveillance program of SARS-CoV-2 |
| EPI_ISL_1850250, EPI_ISL_1851649, EPI_ISL_1851706, EPI_ISL_1852551, EPI_ISL_1852796, EPI_ISL_1852803 | Synlab MVZ Augsburg | Robert Koch Institute | |
| EPI_ISL_2000871, EPI_ISL_2281466, EPI_ISL_2281501, EPI_ISL_2281516, EPI_ISL_2281617, EPI_ISL_2281675, EPI_ISL_2281736, EPI_ISL_2307138 | | | |
| see above | TGen North | TGen North | "Jolene Bowers; Ashlyn Pfeiffer; Chris French; Darrin Lemmer; Dave Engelthaler; Hayley Yaglom; Heather Centner; The Arizona COVID Genomics Union (ACGU)" |
| EPI_ISL_2017899, EPI_ISL_2017911, EPI_ISL_2017919, EPI_ISL_2177435, EPI_ISL_2177492, EPI_ISL_2280073, EPI_ISL_2280074, EPI_ISL_2280075, EPI_ISL_2280077, EPI_ISL_2280078 | | | |
| see above | TXDSHS | TXDSHS | Anita Pokharel; Bonnie Oh; Chun Wang; Grace Kubin; Jenny Zhang; Karen Bobier; Lorraine Rodriguez; Maliha Rahman; Mayela Pedrueza; Myong Koag; Rachel Lee; Rashmi Tuladhar |
| EPI_ISL_2649802, EPI_ISL_2649803, EPI_ISL_2649809 | Tampa General Hospital Esoteric Lab | Tampa General Hospital Esoteric Research & Development Lab | Amorce Lima; Deanna Becker; Dominic Uy; Jon Faughn; Suzane Silbert; Vicki Healer |
| EPI_ISL_2134998 | Texas Children's Hospital | Texas Children's Microbiome Center | Ila Singh; James Dunn; James Versalovic; Jennifer K. Spinler; Ruth Ann Luna |
| EPI_ISL_2389200, EPI_ISL_2389205, EPI_ISL_2389207, EPI_ISL_2545186, EPI_ISL_2545188, EPI_ISL_2545189, EPI_ISL_2545190 | | | |
| see above | The Caribbean Public Health Agency | Carrington Lab, Department of PreClinical Sciences, Faculty of Medical Sciences, The University of the West Indies | Adesh Ramsubhag; Anushka Ramjag; Arianne Brown-Jordan; Avery Hinds; Chinna Chinnadurai; Christine V. F. Carrington; Christopher Oura; Gabriel Escobar; Jaya Jayaraman; Jerome Foster; Karla Georges; Marsha Ivey; Naresh Nandram; Nikita S. D. Sahadeo; Nuno Faria; Oliver Pybus; Rahul Naidu; Rajini Haraksingh; Risha Singh; Roshan Parasram; Sarah Hill; Stanley Giddings; SueMin Nathaniel; Vernie Ramkissoon |
| EPI_ISL_2658392, EPI_ISL_2658397, EPI_ISL_2658400, EPI_ISL_2658401 | The Caribbean Public Health Agency | Carrington Lab, Department of Preclinical Sciences, Faculty of Medical Sciences, The University of the West Indies, St Augustine Campus | Anushka Ramjag; Arianne Brown-Jordan; Avery Hinds; Christine V. F. Carrington; Christopher Oura; Gabriel Escobar; Karla Georges; Naresh Nandram; Nikita S. D. Sahadeo; Nuno Faria; Oliver Pybus; Risha Singh; Roshan Parasram; Sarah Hill; SueMin Nathaniel; Vernie Ramkissoon |
| EPI_ISL_1826486, EPI_ISL_1826494, EPI_ISL_1826530, EPI_ISL_1826533 | The Jackson Laboratory | The Jackson Laboratory | Adams M; Bergeron D; Kelly K; Li L; Long J; Omerza G; Renzette N |
| EPI_ISL_1911088 | The Ohio State University Applied Microbiology Services Laboratory | The Ohio State University Applied Microbiology Services Laboratory | Seth A. Faith PhD |
| EPI_ISL_2227507, EPI_ISL_2227542 | TriCore Reference Laboratories | Center for Global Health, University of New Mexico Health Sciences Center | Cecilia Thompson; Darrell Dinwiddie; Daryl Domman; Karissa Culbreath; Kendra Pesko; Kurt Schwalm; Valerie Morley |
| EPI_ISL_2389190, EPI_ISL_2389192 | Trinidad Public Health Laboratory | Carrington Lab, Department of PreClinical Sciences, Faculty of Medical Sciences, The University of the West Indies | Adesh Ramsubhag; Arianne Brown-Jordan; Avery Hinds; Chinna Chinnadurai; Christine V. F. Carrington; Christopher Oura; Gabriel Escobar; Jaya Jayaraman; Jerome Foster; Karla Georges; Marsha Ivey; Naresh Nandram; Nikita S. D. Sahadeo; Nuno Faria; Oliver Pybus; Rahul Naidu; Rajini Haraksingh; Risha Singh; Roshan Parasram; Sarah Hill; Stanley Giddings; SueMin Nathaniel; Vernie Ramkissoon |
| EPI_ISL_3184594, EPI_ISL_3184622 | U.O. Microbiologia Laboratorio Unico Centro Servizi - AUSL della Romagna | U.O. Microbiologia, Laboratorio Unico Centro Servizi - AUSL della Romagna | Giorgio Dirani |
| EPI_ISL_5603264 | UAPS CASEMIRO FILHO | Analytical Competence Molecular Epidemiology Lab/ACME, Oswaldo Cruz Foundation, Ceara (FIOCRUZ CE) | Carlos Leonardo de Aragao Araujo; Cecilia Leite Costa & Eduardo Ruback dos Santos on behalf of COVID-19 FIOCRUZ Genomic Network; Cleber Furtado Aksenen; Fabio Miyajima; Fernando Braga Stehling; Francisco Eder de Moura Lopes; Igor Oliveira Duarte; Jamille Maria Mendes Bezerra; Joaquim Cesar do Nascimento Sousa Junior; Pedro Miguel Carneiro Jeronimo; Suzana Porto Almeida; Thais Ferreira de Oliveira; Thais de Oliveira Costa; Ticiane Cavalcante de Souza; Veridiana Pessoa Miyajima |
| EPI_ISL_5603220 | UBS CENTRO DE SAUDE I | Analytical Competence Molecular Epidemiology Lab/ACME, Oswaldo Cruz Foundation, Ceara (FIOCRUZ CE) | Carlos Leonardo de Aragao Araujo; Cecilia Leite Costa & Eduardo Ruback dos Santos on behalf of COVID-19 FIOCRUZ Genomic Network; Cleber Furtado Aksenen; Fabio Miyajima; Fernando Braga Stehling; Francisco Eder de Moura Lopes; Igor Oliveira Duarte; Jamille Maria Mendes Bezerra; Joaquim Cesar do Nascimento Sousa Junior; Pedro Miguel Carneiro Jeronimo; Suzana Porto Almeida; Thais Ferreira de Oliveira; Thais de Oliveira Costa; Ticiane Cavalcante de Souza; Veridiana Pessoa Miyajima |
| EPI_ISL_2344847 | UBS DE PEDRANOPOLIS | Instituto Butantan / ESALQ-Piracicaba | Antonio Jorge Martins; Claudia Renata dos Santos Barros; David Schlesinger; Debora Botequio Moretti; Dimas Tadeu Covas; Elaine Cristina Marqueze; Elaine Vieira Santos; Evandra Strazza Rodrigues; Heidge Fukumasu; Jayme Augusto de Souza-Neto; José Salvatore Leister Patané; Luiz Alcantara; Luiz Lehmann Coutinho; Maria Carolina Elias; Maurício Lacerda Nogueira; Rafael dos Santos Bezerra; Raul Machado Neto; Rejane Maria Tommasini Grotto; Ricardo Haddad; Sandra Coccuzzo Sampaio Vessoni; Simone Kashima; Svetoslav Nanev Slavov; Vincent Louis Viala |
| EPI_ISL_2344850 | UBS DE SAO JOAO DAS DUAS PONTES | Instituto Butantan / ESALQ-Piracicaba | Antonio Jorge Martins; Claudia Renata dos Santos Barros; David Schlesinger; Debora Botequio Moretti; Dimas Tadeu Covas; Elaine Cristina Marqueze; Elaine Vieira Santos; Evandra Strazza Rodrigues; Heidge Fukumasu; Jayme Augusto de Souza-Neto; José Salvatore Leister Patané; Luiz Alcantara; Luiz Lehmann Coutinho; Maria Carolina Elias; Maurício Lacerda Nogueira; Rafael dos Santos Bezerra; Raul Machado Neto; Rejane Maria Tommasini Grotto; Ricardo Haddad; Sandra Coccuzzo Sampaio Vessoni; Simone Kashima; Svetoslav Nanev Slavov; Vincent Louis Viala |
| EPI_ISL_1967205, EPI_ISL_1967206, EPI_ISL_1967207, EPI_ISL_1967208, EPI_ISL_1967209 | UBS DR ALFREDO DANTAS DE SOUZA UMUARAMA | Instituto Butantan / Mendelics | Antonio Jorge Martins; Bianca Cechetto Carlos. Mendelics: Bibiana Santos; Claudia Renata dos Santos Barros; Cíntia Bittar; David Schlesinger. Hemocentro Ribeirão Preto: Simone Kashima; Debora Botequio Moretti; Elaine Cristina Marqueze; Elaine Vieira dos Santos; Elisangela Chicaroni Mattos; Erika Freitas; Evandra Strazza Rodrigues; Felipe Allan da Silva da Costa; Flavia Aburjaile; Fábio Sossai Possebon; Guilherme Campos; Guilherme Targino Valente; Heidge Fukumasu. USP-Botucatu: Rejane Maria Tommasini Grotto; Helena Lage Ferreira; Instituto Butantan: Dimas Tadeu Covas; Jardelina de Souza Todao Bernardino; Jayme A. Souza-Neto; Jessika Cristina Chagas Lesbon; Jorge A. Petroli Marchesi; José Salvatore Leister Patané; João Paulo Kitajima; João Pessoa Araújo Jr.; Leila Sabrina Ullmann; Loyze Paola Oliveira de Lima; Luiz Aurelio de Campos Crispin. Centro de Genômica Funcional da ESALQ: Luiz Lehmann Coutinho; Luiz Carlos Junior de Alcantara; Lívia Sacchetto; Maisa C. Pereira Parra; Maria Carolina Elias; Marta Giovanetti; Marília Moraes; Maurício Lacerda Nogueira. Prefeitura de Sao Paulo: Melissa Palmieri.; Patricia Akemi Assato; Paula Rahal; Paulo Inacio da Costa; Rafael dos Santos Bezerra; Raquel de Lello Rocha Campos Cassano. NGS Soluções Genômicas: Pilar Drummond Sampaio Corrêa Mariani. FZEA-USP Pirassununga: Mirele Daiana Poleti; Raul Machado Neto; Ricardo Augusto Brassaloti; Ricardo Haddad; Rodrigo Tocantins Calado. FAMERP-SJRP: Cecília Artico Banho; Sandra Coccuzzo Sampaio; Svetoslav Nanev Slavov; Vagner Fonseca; Vincent Louis Viala |
| EPI_ISL_2344949 | UBS ENRIQUE GUILHERME ERNESTO MARZEUSKI | Instituto Butantan / ESALQ-Piracicaba | Antonio Jorge Martins; Claudia Renata dos Santos Barros; David Schlesinger; Debora Botequio Moretti; Dimas Tadeu Covas; Elaine Cristina Marqueze; Elaine Vieira Santos; Evandra Strazza Rodrigues; Heidge Fukumasu; Jayme Augusto de Souza-Neto; José Salvatore Leister Patané; Luiz Alcantara; Luiz Lehmann Coutinho; Maria Carolina Elias; Maurício Lacerda Nogueira; Rafael dos Santos Bezerra; Raul Machado Neto; Rejane Maria Tommasini Grotto; Ricardo Haddad; Sandra Coccuzzo Sampaio Vessoni; Simone Kashima; Svetoslav Nanev Slavov; Vincent Louis Viala |
| EPI_ISL_2344753, EPI_ISL_2344761 | UBS II DE PIRAPOZINHO C SERVICO DE EACS E ESF | Instituto Butantan / ESALQ-Piracicaba | Antonio Jorge Martins; Claudia Renata dos Santos Barros; David Schlesinger; Debora Botequio Moretti; Dimas Tadeu Covas; Elaine Cristina Marqueze; Elaine Vieira Santos; Evandra Strazza Rodrigues; Heidge Fukumasu; Jayme Augusto de Souza-Neto; José Salvatore Leister Patané; Luiz Alcantara; Luiz Lehmann Coutinho; Maria Carolina Elias; Maurício Lacerda Nogueira; Rafael dos Santos Bezerra; Raul Machado Neto; Rejane Maria Tommasini Grotto; Ricardo Haddad; Sandra Coccuzzo Sampaio Vessoni; Simone Kashima; Svetoslav Nanev Slavov; Vincent Louis Viala |
| EPI_ISL_2344751 | UBS II DE TACIBA | Instituto Butantan / ESALQ-Piracicaba | Antonio Jorge Martins; Claudia Renata dos Santos Barros; David Schlesinger; Debora Botequio Moretti; Dimas Tadeu Covas; Elaine Cristina Marqueze; Elaine Vieira Santos; Evandra Strazza Rodrigues; Heidge Fukumasu; Jayme Augusto de Souza-Neto; José Salvatore Leister Patané; Luiz Alcantara; Luiz Lehmann Coutinho; Maria Carolina Elias; Maurício Lacerda Nogueira; Rafael dos Santos Bezerra; Raul Machado Neto; Rejane Maria Tommasini Grotto; Ricardo Haddad; Sandra Coccuzzo Sampaio Vessoni; Simone Kashima; Svetoslav Nanev Slavov; Vincent Louis Viala |
| EPI_ISL_1967285, EPI_ISL_1967286 | UBS JARDIM CABUCU | Instituto Butantan / Mendelics | Antonio Jorge Martins; Bianca Cechetto Carlos. Mendelics: Bibiana Santos; Claudia Renata dos Santos Barros; Cíntia Bittar; David Schlesinger. Hemocentro Ribeirão Preto: Simone Kashima; Debora Botequio Moretti; Elaine Cristina Marqueze; Elaine Vieira dos Santos; Elisangela Chicaroni Mattos; Erika Freitas; Evandra Strazza Rodrigues; Felipe Allan da Silva da Costa; Flavia Aburjaile; Fábio Sossai Possebon; Guilherme Campos; Guilherme Targino Valente; Heidge Fukumasu. USP-Botucatu: Rejane Maria Tommasini Grotto; Helena Lage Ferreira; Instituto Butantan: Dimas Tadeu Covas; Jardelina de Souza Todao Bernardino; Jayme A. Souza-Neto; Jessika Cristina Chagas Lesbon; Jorge A. Petroli Marchesi; José Salvatore Leister Patané; João Paulo Kitajima; João Pessoa Araújo Jr.; Leila Sabrina Ullmann; Loyze Paola Oliveira de Lima; Luiz Aurelio de Campos Crispin. Centro de Genômica Funcional da ESALQ: Luiz Lehmann Coutinho; Luiz Carlos Junior |

| | | | |
|---|---|---|---|
| | | | de Alcantara; Lívia Sacchetto; Maisa C. Pereira Parra; Maria Carolina Elias; Marta Giovanetti; Marília Moraes; Maurício Lacerda Nogueira. Prefeitura de Sao Paulo: Melissa Palmieri.; Patricia Akemi Assato; Paula Rahal; Paulo Inacio da Costa; Rafael dos Santos Bezerra; Raquel de Lello Rocha Campos Cassano. NGS Soluções Genômicas: Pilar Drummond Sampaio Corrêa Mariani. FZEA-USP Pirassununga: Mirele Daiana Poleti; Raul Machado Neto; Ricardo Augusto Brassaloti; Ricardo Haddad; Rodrigo Tocantins Calado. FAMERP-SJRP: Cecília Artico Banho; Sandra Coccuzzo Sampaio; Svetoslav Nanev Slavov; Vagner Fonseca; Vincent Louis Viala |
| EPI_ISL_2171000 | UBS JARDIM MUNHOZ | Instituto Butantan / Mendelics | Antonio Jorge Martins; Bianca Cechetto Carlos. Mendelics: Bibiana Santos; Claudia Renata dos Santos Barros; Cíntia Bittar; David Schlesinger. Hemocentro Ribeirão Preto: Simone Kashima; Debora Botequio Moretti; Elaine Cristina Marqueze; Elaine Vieira dos Santos; Elisangela Chicaroni Mattos; Erika Freitas; Evandra Strazza Rodrigues; Felipe Allan da Silva da Costa; Flavia Aburjaile; Fábio Sossai Possebon; Guilherme Campos; Guilherme Targino Valente; Heidge Fukumasu. USP-Botucatu: Rejane Maria Tommasini Grotto; Helena Lage Ferreira; Instituto Butantan: Dimas Tadeu Covas; Jardelina de Souza Todao Bernardino; Jayme A. Souza-Neto; Jessika Cristina Chagas Lesbon; Jorge A. Petroli Marchesi; José Salvatore Leister Patané; João Paulo Kitajima; João Pessoa Araújo Jr.; Leila Sabrina Ullmann; Loyze Paola Oliveira de Lima; Luiz Aurelio de Campos Crispin. Centro de Genômica Funcional da ESALQ: Luiz Lehmann Coutinho; Luiz Carlos Junior de Alcantara; Lívia Sacchetto; Maisa C. Pereira Parra; Maria Carolina Elias; Marta Giovanetti; Marília Moraes; Maurício Lacerda Nogueira. Prefeitura de Sao Paulo: Melissa Palmieri.; Patricia Akemi Assato; Paula Rahal; Paulo Inacio da Costa; Rafael dos Santos Bezerra; Raquel de Lello Rocha Campos Cassano. NGS Soluções Genômicas: Pilar Drummond Sampaio Corrêa Mariani. FZEA-USP Pirassununga: Mirele Daiana Poleti; Raul Machado Neto; Ricardo Augusto Brassaloti; Ricardo Haddad; Rodrigo Tocantins Calado. FAMERP-SJRP: Cecília Artico Banho; Sandra Coccuzzo Sampaio; Svetoslav Nanev Slavov; Vagner Fonseca; Vincent Louis Viala |
| EPI_ISL_2344902 | UBS JARDIM VITORINO IPERO | Instituto Butantan / ESALQ-Piracicaba | Antonio Jorge Martins; Claudia Renata dos Santos Barros; David Schlesinger; Debora Botequio Moretti; Dimas Tadeu Covas; Elaine Cristina Marqueze; Elaine Vieira Santos; Evandra Strazza Rodrigues; Heidge Fukumasu; Jayme Augusto de Souza-Neto; José Salvatore Leister Patané; Luiz Alcantara; Luiz Lehmann Coutinho; Maria Carolina Elias; Maurício Lacerda Nogueira; Rafael dos Santos Bezerra; Raul Machado Neto; Rejane Maria Tommasini Grotto; Ricardo Haddad; Sandra Coccuzzo Sampaio Vessoni; Simone Kashima; Svetoslav Nanev Slavov; Vincent Louis Viala |
| EPI_ISL_2344825 | UBS JD SALTENSE | Instituto Butantan / ESALQ-Piracicaba | Antonio Jorge Martins; Claudia Renata dos Santos Barros; David Schlesinger; Debora Botequio Moretti; Dimas Tadeu Covas; Elaine Cristina Marqueze; Elaine Vieira Santos; Evandra Strazza Rodrigues; Heidge Fukumasu; Jayme Augusto de Souza-Neto; José Salvatore Leister Patané; Luiz Alcantara; Luiz Lehmann Coutinho; Maria Carolina Elias; Maurício Lacerda Nogueira; Rafael dos Santos Bezerra; Raul Machado Neto; Rejane Maria Tommasini Grotto; Ricardo Haddad; Sandra Coccuzzo Sampaio Vessoni; Simone Kashima; Svetoslav Nanev Slavov; Vincent Louis Viala |
| EPI_ISL_5603221, EPI_ISL_5603227, EPI_ISL_5603229, EPI_ISL_5603309 | UBS MARATOAN | Analytical Competence Molecular Epidemiology Lab/ACME, Oswaldo Cruz Foundation, Ceara (FIOCRUZ CE) | Carlos Leonardo de Aragao Araujo; Cecilia Leite Costa & Eduardo Ruback dos Santos on behalf of COVID-19 FIOCRUZ Genomic Network; Cleber Furtado Aksenen; Fabio Miyajima; Fernando Braga Stehling; Francisco Eder de Moura Lopes; Igor Oliveira Duarte; Jamille Maria Mendes Bezerra; Joaquim Cesar do Nascimento Sousa Junior; Pedro Miguel Carneiro Jeronimo; Suzana Porto Almeida; Thais Ferreira de Oliveira; Thais de Oliveira Costa; Ticiane Cavalcante de Souza; Veridiana Pessoa Miyajima |
| EPI_ISL_2170998 | UBS RECREIO SAO JORGE | Instituto Butantan / Mendelics | Antonio Jorge Martins; Bianca Cechetto Carlos. Mendelics: Bibiana Santos; Claudia Renata dos Santos Barros; Cíntia Bittar; David Schlesinger. Hemocentro Ribeirão Preto: Simone Kashima; Debora Botequio Moretti; Elaine Cristina Marqueze; Elaine Vieira dos Santos; Elisangela Chicaroni Mattos; Erika Freitas; Evandra Strazza Rodrigues; Felipe Allan da Silva da Costa; Flavia Aburjaile; Fábio Sossai Possebon; Guilherme Campos; Guilherme Targino Valente; Heidge Fukumasu. USP-Botucatu: Rejane Maria Tommasini Grotto; Helena Lage Ferreira; Instituto Butantan: Dimas Tadeu Covas; Jardelina de Souza Todao Bernardino; Jayme A. Souza-Neto; Jessika Cristina Chagas Lesbon; Jorge A. Petroli Marchesi; José Salvatore Leister Patané; João Paulo Kitajima; João Pessoa Araújo Jr.; Leila Sabrina Ullmann; Loyze Paola Oliveira de Lima; Luiz Aurelio de Campos Crispin. Centro de Genômica Funcional da ESALQ: Luiz Lehmann Coutinho; Luiz Carlos Junior de Alcantara; Lívia Sacchetto; Maisa C. Pereira Parra; Maria Carolina Elias; Marta Giovanetti; Marília Moraes; Maurício Lacerda Nogueira. Prefeitura de Sao Paulo: Melissa Palmieri.; Patricia Akemi Assato; Paula Rahal; Paulo Inacio da Costa; Rafael dos Santos Bezerra; Raquel de Lello Rocha Campos Cassano. NGS Soluções Genômicas: Pilar Drummond Sampaio Corrêa Mariani. FZEA-USP Pirassununga: Mirele Daiana Poleti; Raul Machado Neto; Ricardo Augusto Brassaloti; Ricardo Haddad; Rodrigo Tocantins Calado. FAMERP-SJRP: Cecília Artico Banho; Sandra Coccuzzo Sampaio; Svetoslav Nanev Slavov; Vagner Fonseca; Vincent Louis Viala |
| EPI_ISL_2841605 | UBS Silvio Joao Luiz de Lucia | Instituto Adolfo Lutz, Interdiciplinary Procedures Center, Strategic Laboratory | Caio Vinicius Dias Lopes; Claudia Regina Gonçalves; Claudio Tavares Sacchi; Erica Valessa Ramos Gomes; Karoline Rodrigues Campos |
| EPI_ISL_3419578, EPI_ISL_3419579, EPI_ISL_3419580, EPI_ISL_3419581, EPI_ISL_3419582, EPI_ISL_3419583 | UC Davis Genome Center and Healthy Davis Together | UC Davis Genome Center | UC Davis |
| EPI_ISL_3321293, EPI_ISL_3321294, EPI_ISL_3321295, EPI_ISL_3321296, EPI_ISL_3321297, EPI_ISL_3321298, EPI_ISL_3321299 | | | |
| see above | UC-Christus Clinical Hospital Laboratory | Laboratory of Molecular Virology, School of Medicine, Pontificia Universidad Catolica de Chile | Ana Maria Guzman; Andres E. Munoz-Marcos; Catalina Pardo-Roa; Eileen Serrano; Erick Salinas; Estefany Poblete; Francisco Melo; Jorge Levican; Leonardo I. Almonacid; Maria Jose Avendano; Maria Patricia Vega; Rafael A. Medina; Ricardo Enrique de la Barra; Tamara Garcia-Salum |
| EPI_ISL_2392171, EPI_ISL_2620751 | UCLA Clinical Micro Lab | Los Angeles County Public Health Laboratories | P. Hemarajata et al. |
| EPI_ISL_1960046, EPI_ISL_1960063, EPI_ISL_2438086 | UDEA | Universidad Nacional de Colombia - Laboratorio Genómico One Health | Andres F. Cardona-Rios; Carlos Franco-Muñoz; Carolina Muñoz-Arango; Celeny Ortiz; Daniel O. Maldonado-Perez; Diego A. Álvarez-Díaz; Hector Alejandro Ruiz-Moreno; Idabely Betancur Ortiz; Jorge E. Osorio; Juan P. Hernandez-Ortiz; Karl A Ciuoderis; Katherine Laiton-Donato; Laura Silvana Perez; Lina M. Hurtado; Marcela Mercado-Reyes; Maria Angélica Maya; Maria Stella López; Rita Almanza Payares; Sandra Ines Cano; Simón Villegas Velásquez |
| EPI_ISL_2448438 | ULSS 2 Marca Trevigiana | Istituto Zooprofilattico Sperimentale delle Venezie | Adelaide Milani; Alessia Schivo; Alice Fusaro; Ambra Pastori; Annalisa Salviato; Antonia Ricci; Calogero Terregino; Edoardo Giussani; Elisa Palumbo; Erika Giorgia Quaranta; Isabella Monne; Luca Tassoni |
| EPI_ISL_2448402 | ULSS 3 Venezia | Istituto Zooprofilattico Sperimentale delle Venezie | Adelaide Milani; Alessia Schivo; Alice Fusaro; Ambra Pastori; Annalisa Salviato; Antonia Ricci; Calogero Terregino; Edoardo Giussani; Elisa Palumbo; Erika Giorgia Quaranta; Isabella Monne; Luca Tassoni |
| EPI_ISL_3152116 | UMC Groningen, Clinical Virology, Department of Medical Microbiology and Infection Prevention | UMC Groningen, Clinical Virology, Department of Medical Microbiology and Infection Prevention | Alexander Friedrich; Coretta Van Leer-Buter; Erley Lizarazo-Forero; Hubert Niesters; Lilli Gard; Marjolein Knoester; Monika Fliss; Sigrid Rosema; Xuewei Zhou |
| EPI_ISL_2344952 | UMS PIRAPORA DO BOM JESUS | Instituto Butantan | Antonio Jorge Martins; Claudia Renata dos Santos Barros; David Schlesinger; Debora Botequio Moretti; Dimas Tadeu Covas; Elaine Cristina Marqueze; Elaine Vieira Santos; Evandra Strazza Rodrigues; Heidge Fukumasu; Jayme Augusto de Souza-Neto; José Salvatore Leister Patané; Luiz Alcantara; Luiz Lehmann Coutinho; Maria Carolina Elias; Maurício Lacerda Nogueira; Rafael dos Santos Bezerra; Raul Machado Neto; Rejane Maria Tommasini Grotto; Ricardo Haddad; Sandra Coccuzzo Sampaio Vessoni; Simone Kashima; Svetoslav Nanev Slavov; Vincent Louis Viala |
| EPI_ISL_3409099, EPI_ISL_3409116, EPI_ISL_3409128, EPI_ISL_3409135, EPI_ISL_3409200, EPI_ISL_3409216, EPI_ISL_3409288, EPI_ISL_3409608, EPI_ISL_3409777, EPI_ISL_3410080, EPI_ISL_3410084 | | | |
| see above | UMass Memorial Medical Center | Center for Microbiome Research | Doyle V. Ward; Karl J. Simon; Richard T. Ellison III |
| EPI_ISL_5603222 | UNID MISTA DE MORAUJO | Analytical Competence Molecular Epidemiology Lab/ACME, Oswaldo Cruz Foundation, Ceara (FIOCRUZ CE) | Carlos Leonardo de Aragao Araujo; Cecilia Leite Costa & Eduardo Ruback dos Santos on behalf of COVID-19 FIOCRUZ Genomic Network; Cleber Furtado Aksenen; Fabio Miyajima; Fernando Braga Stehling; Francisco Eder de Moura Lopes; Igor Oliveira Duarte; Jamille Maria Mendes Bezerra; Joaquim Cesar do Nascimento Sousa Junior; Pedro Miguel Carneiro Jeronimo; Suzana Porto Almeida; Thais Ferreira de Oliveira; Thais de Oliveira Costa; Ticiane Cavalcante de Souza; Veridiana Pessoa Miyajima |
| EPI_ISL_2344755, EPI_ISL_2344760 | UNIDADE BASICA DE SAUDE III DE ALFREDO MARCONDES | Instituto Butantan / ESALQ-Piracicaba | Antonio Jorge Martins; Claudia Renata dos Santos Barros; David Schlesinger; Debora Botequio Moretti; Dimas Tadeu Covas; Elaine Cristina Marqueze; Elaine Vieira Santos; Evandra Strazza Rodrigues; Heidge Fukumasu; Jayme Augusto de Souza-Neto; José Salvatore Leister Patané; Luiz Alcantara; Luiz Lehmann Coutinho; Maria Carolina Elias; Maurício Lacerda Nogueira; Rafael dos Santos Bezerra; Raul Machado Neto; Rejane Maria Tommasini Grotto; Ricardo Haddad; Sandra Coccuzzo Sampaio Vessoni; Simone Kashima; Svetoslav Nanev Slavov; Vincent Louis Viala |
| EPI_ISL_2344885, EPI_ISL_2344886 | UNIDADE DE PRONTO ATENDIMENTO DE JALES | Instituto Butantan / ESALQ-Piracicaba | Antonio Jorge Martins; Claudia Renata dos Santos Barros; David Schlesinger; Debora Botequio Moretti; Dimas Tadeu Covas; Elaine Cristina Marqueze; Elaine Vieira Santos; Evandra Strazza Rodrigues; Heidge Fukumasu; Jayme Augusto de Souza-Neto; José Salvatore Leister Patané; Luiz Alcantara; Luiz Lehmann Coutinho; Maria Carolina Elias; Maurício Lacerda Nogueira; Rafael dos Santos Bezerra; Raul Machado Neto; Rejane Maria Tommasini Grotto; Ricardo Haddad; Sandra Coccuzzo Sampaio Vessoni; Simone Kashima; Svetoslav Nanev Slavov; Vincent Louis Viala |
| EPI_ISL_2209018 | UNIDADE DE PRONTO ATENDIMENTO UPA | Instituto Butantan / ESALQ-Piracicaba | Antonio Jorge Martins; Claudia Renata dos Santos Barros; David Schlesinger; Debora Botequio Moretti; Dimas Tadeu Covas; Elaine Cristina Marqueze; Elaine Vieira Santos; Evandra Strazza Rodrigues; Heidge Fukumasu; Jayme Augusto de Souza-Neto; José Salvatore Leister Patané; Luiz Alcantara; Luiz Lehmann Coutinho; Maria Carolina Elias; Maurício Lacerda Nogueira; Rafael dos Santos Bezerra; Raul Machado Neto; Rejane Maria Tommasini Grotto; Ricardo Haddad; Sandra Coccuzzo Sampaio Vessoni; Simone Kashima; Svetoslav Nanev Slavov; Vincent Louis Viala |
| EPI_ISL_2344818 | UNIDADE MISTA DE IGUAPE | Instituto Butantan / ESALQ-Piracicaba | Antonio Jorge Martins; Claudia Renata dos Santos Barros; David Schlesinger; Debora Botequio Moretti; Dimas Tadeu Covas; Elaine Cristina Marqueze; Elaine Vieira Santos; Evandra Strazza Rodrigues; Heidge Fukumasu; Jayme Augusto de Souza-Neto; José Salvatore Leister Patané; Luiz Alcantara; Luiz Lehmann Coutinho; Maria Carolina Elias; Maurício Lacerda Nogueira; Rafael dos Santos Bezerra; Raul Machado Neto; Rejane Maria Tommasini Grotto; Ricardo Haddad; Sandra Coccuzzo Sampaio Vessoni; Simone Kashima; Svetoslav Nanev Slavov; Vincent Louis Viala |
| EPI_ISL_2344950 | UNIDADE MISTA DE SAUDE CAPELA DO ALTO | Instituto Butantan / ESALQ-Piracicaba | Antonio Jorge Martins; Claudia Renata dos Santos Barros; David Schlesinger; Debora Botequio Moretti; Dimas Tadeu Covas; Elaine Cristina Marqueze; Elaine Vieira Santos; Evandra Strazza Rodrigues; Heidge Fukumasu; Jayme Augusto de Souza-Neto; José Salvatore Leister Patané; Luiz Alcantara; Luiz Lehmann Coutinho; Maria Carolina Elias; Maurício Lacerda Nogueira; Rafael dos Santos Bezerra; Raul Machado Neto; Rejane Maria Tommasini Grotto; Ricardo Haddad; Sandra Coccuzzo Sampaio Vessoni; Simone Kashima; Svetoslav Nanev Slavov; Vincent Louis Viala |
| EPI_ISL_3102542, EPI_ISL_5603243 | UNIDADE MISTA DE SAUDE DE MISSAO VELHA | Analytical Competence Molecular Epidemiology Lab/ACME, Oswaldo Cruz Foundation, Ceara (FIOCRUZ CE) | Carlos Leonardo de Aragao Araujo; Cecilia Leite Costa & Eduardo Ruback dos Santos on behalf of COVID-19 FIOCRUZ Genomic Network; Cleber Furtado Aksenen; Fabio Miyajima; Fernando Braga Stehling; Francisco Eder de Moura Lopes; Igor Oliveira Duarte; Jamille Maria Mendes Bezerra; Joaquim Cesar do Nascimento Sousa Junior; Joaquim César do Nascimento Sousa Junior; Pedro Miguel Carneiro Jeronimo; Suzana Porto Almeida; Suzana Porto Almeida e Lucas Delerino; Thais Ferreira de Oliveira; Thais de Oliveira Costa; Ticiane Cavalcante de Souza; Veridiana Pessoa Miyajima |
| EPI_ISL_2344893 | UNIDADE MISTA DE SAUDE MOMBUCA | Instituto Butantan / ESALQ-Piracicaba | Antonio Jorge Martins; Claudia Renata dos Santos Barros; David Schlesinger; Debora Botequio Moretti; Dimas Tadeu Covas; Elaine Cristina Marqueze; Elaine Vieira Santos; Evandra Strazza Rodrigues; Heidge Fukumasu; Jayme Augusto de Souza-Neto; José Salvatore Leister Patané; Luiz Alcantara; Luiz Lehmann Coutinho; Maria Carolina Elias; Maurício Lacerda Nogueira; Rafael dos Santos Bezerra; Raul Machado Neto; Rejane Maria Tommasini Grotto; Ricardo Haddad; Sandra Coccuzzo Sampaio Vessoni; Simone Kashima; Svetoslav Nanev Slavov; Vincent Louis Viala |
| EPI_ISL_2344750 | UNIDADE MISTA OSVALDO P CAMARGO ELIAS FAUSTO | Instituto Butantan / ESALQ-Piracicaba | Antonio Jorge Martins; Claudia Renata dos Santos Barros; David Schlesinger; Debora Botequio Moretti; Dimas Tadeu Covas; Elaine Cristina Marqueze; Elaine Vieira Santos; Evandra Strazza Rodrigues; Heidge Fukumasu; Jayme Augusto de Souza-Neto; José Salvatore Leister Patané; Luiz Alcantara; Luiz Lehmann Coutinho; Maria Carolina Elias; Maurício Lacerda Nogueira; Rafael dos Santos Bezerra; Raul Machado Neto; Rejane Maria Tommasini Grotto; Ricardo Haddad; Sandra Coccuzzo Sampaio Vessoni; Simone Kashima; Svetoslav Nanev Slavov; Vincent Louis Viala |
| EPI_ISL_2344916 | UNIDADE PSF PINHEIROS APIAI | Instituto Butantan / ESALQ-Piracicaba | Antonio Jorge Martins; Claudia Renata dos Santos Barros; David Schlesinger; Debora Botequio Moretti; Dimas Tadeu Covas; Elaine Cristina Marqueze; Elaine Vieira Santos; Evandra Strazza Rodrigues; Heidge Fukumasu; Jayme Augusto de Souza-Neto; José Salvatore Leister Patané; Luiz Alcantara; Luiz Lehmann Coutinho; Maria Carolina Elias; Maurício Lacerda Nogueira; Rafael dos Santos Bezerra; Raul Machado Neto; Rejane Maria Tommasini Grotto; Ricardo Haddad; Sandra Coccuzzo Sampaio Vessoni; Simone Kashima; Svetoslav Nanev Slavov; Vincent Louis Viala |
| EPI_ISL_2136323, EPI_ISL_2321594 | UNIGEM | Universidad Nacional de Colombia - Laboratorio Genómico One Health | Andres F. Cardona-Rios; Carlos Franco-Muñoz; Carolina Muñoz-Arango; Celeny Ortiz; Daniel O. Maldonado-Perez; Diego A. Álvarez-Díaz; Hector Alejandro Ruiz-Moreno; Idabely Betancur Ortiz; Jorge E. Osorio; Juan P. Hernandez-Ortiz; Karl A Ciuoderis; Katherine Laiton-Donato; Laura Silvana Perez; Lina M. Hurtado; Marcela Mercado-Reyes; Maria Angélica Maya; Maria Stella López; Rita Almanza Payares; Sandra Ines Cano; Simón Villegas Velásquez |
| EPI_ISL_2209401, EPI_ISL_2209404, EPI_ISL_2344754, EPI_ISL_2344764, EPI_ISL_2344768, EPI_ISL_2344770, EPI_ISL_2344773 | | | |
| see above | UPA 24 HORAS ANA JACINTA | Instituto Butantan / ESALQ-Piracicaba | Antonio Jorge Martins; Claudia Renata dos Santos Barros; David Schlesinger; Debora Botequio Moretti; Dimas Tadeu Covas; Elaine Cristina Marqueze; Elaine Vieira Santos; Evandra Strazza Rodrigues; Heidge Fukumasu; Jayme Augusto de Souza-Neto; José Salvatore Leister Patané; Luiz Alcantara; Luiz Lehmann Coutinho; Maria Carolina Elias; Maurício Lacerda Nogueira; Rafael dos Santos Bezerra; Raul Machado Neto; Rejane Maria Tommasini Grotto; Ricardo Haddad; Sandra Coccuzzo Sampaio Vessoni; Simone Kashima; Svetoslav Nanev Slavov; Vincent Louis Viala |
| EPI_ISL_2209379, EPI_ISL_2344758, EPI_ISL_2344762, EPI_ISL_2344765, EPI_ISL_2344766 | UPA 24 HORAS DR ALOISIO MUNIZ DE ANDRADE | Instituto Butantan / ESALQ-Piracicaba | Antonio Jorge Martins; Claudia Renata dos Santos Barros; David Schlesinger; Debora Botequio Moretti; Dimas Tadeu Covas; Elaine Cristina Marqueze; Elaine Vieira Santos; Evandra Strazza Rodrigues; Heidge Fukumasu; Jayme Augusto de Souza-Neto; José Salvatore Leister Patané; Luiz Alcantara; Luiz Lehmann Coutinho; Maria Carolina Elias; Maurício Lacerda Nogueira; Rafael dos Santos Bezerra; Raul Machado Neto; Rejane Maria Tommasini Grotto; Ricardo Haddad; Sandra Coccuzzo Sampaio Vessoni; Simone Kashima; Svetoslav Nanev Slavov; Vincent Louis Viala |
| EPI_ISL_1967223, EPI_ISL_1967225, EPI_ISL_1967226, EPI_ISL_2170972, EPI_ISL_2170973 | UPA DE BEBEDOURO | Instituto Butantan / Mendelics | Antonio Jorge Martins; Bianca Cechetto Carlos. Mendelics: Bibiana Santos; Claudia Renata dos Santos Barros; Cíntia Bittar; David Schlesinger. Hemocentro Ribeirão Preto: Simone Kashima; Debora Botequio Moretti; Elaine Cristina Marqueze; Elaine Vieira dos Santos; Elisangela Chicaroni Mattos; Erika Freitas; Evandra Strazza Rodrigues; Felipe Allan da Silva da Costa; Flavia Aburjaile; Fábio Sossai Possebon; Guilherme Campos; Guilherme Targino Valente; Heidge Fukumasu. USP-Botucatu: Rejane Maria Tommasini Grotto; Helena Lage Ferreira; Instituto Butantan: Dimas Tadeu Covas; Jardelina de Souza Todao Bernardino; Jayme A. Souza-Neto; Jessika Cristina Chagas Lesbon; Jorge A. Petroli Marchesi; José Salvatore Leister Patané; João Paulo Kitajima; João Pessoa Araújo Jr.; Leila Sabrina Ullmann; Loyze Paola Oliveira de Lima; Luiz Aurelio de Campos Crispin. Centro de Genômica Funcional da ESALQ: Luiz Lehmann Coutinho; Luiz Carlos Junior de Alcantara; Lívia Sacchetto; Maisa C. Pereira Parra; Maria Carolina Elias; Marta Giovanetti; Marília Moraes; Maurício Lacerda Nogueira. Prefeitura de Sao Paulo: Melissa Palmieri.; Patricia Akemi Assato; Paula Rahal; Paulo Inacio da Costa; Rafael dos Santos Bezerra; Raquel de Lello Rocha Campos Cassano. NGS Soluções Genômicas: Pilar Drummond Sampaio Corrêa Mariani. FZEA-USP Pirassununga: Mirele Daiana Poleti; Raul Machado Neto; Ricardo Augusto Brassaloti; Ricardo Haddad; Rodrigo Tocantins Calado. FAMERP-SJRP: Cecília Artico Banho; Sandra Coccuzzo Sampaio; Svetoslav Nanev Slavov; Vagner Fonseca; Vincent Louis Viala |
| EPI_ISL_1967306, EPI_ISL_2171025, | UPA DR AKIRA TADA | Instituto Butantan / Mendelics | Antonio Jorge Martins; Bianca Cechetto Carlos. Mendelics: Bibiana Santos; Claudia Renata dos Santos Barros; Cíntia Bittar; David Schlesinger. Hemocentro Ribeirão Preto: Simone Kashima; Debora Botequio Moretti; Elaine Cristina Marqueze; Elaine Vieira dos Santos; Elisangela Chicaroni Mattos; Erika Freitas; Evandra Strazza Rodrigues; Felipe Allan da Silva da Costa; Flavia Aburjaile; Fábio Sossai Possebon; Guilherme Campos; Guilherme Targino Valente; Heidge Fukumasu. USP-Botucatu: Rejane Maria Tommasini Grotto; Helena Lage Ferreira; Instituto Butantan: Dimas Tadeu Covas; Jardelina de Souza Todao Bernardino; Jayme A. |

| | | | |
|---|---|---|---|
| EPI_ISL_2171026, EPI_ISL_2171028 | | | Souza-Neto; Jessika Cristina Chagas Lesbon; Jorge A. Petroli Marchesi; José Salvatore Leister Patané; João Paulo Kitajima; João Pessoa Araújo Jr.; Leila Sabrina Ullmann; Loyze Paola Oliveira de Lima; Luiz Aurelio de Campos Crispin. Centro de Genômica Funcional da ESALQ: Luiz Lehmann Coutinho; Luiz Carlos Junior de Alcantara; Lívia Sacchetto; Maisa C. Pereira Parra; Maria Carolina Elias; Marta Giovanetti; Marília Moraes; Maurício Lacerda Nogueira. Prefeitura de Sao Paulo: Melissa Palmieri.; Patricia Akemi Assato; Paula Rahal; Paulo Inacio da Costa; Rafael dos Santos Bezerra; Raquel de Lello Rocha Campos Cassano. NGS Soluções Genômicas: Pilar Drummond Sampaio Corrêa Mariani. FZEA-USP Pirassununga: Mirele Daiana Poleti; Raul Machado Neto; Ricardo Augusto Brassaloti; Ricardo Haddad; Rodrigo Tocantins Calado. FAMERP-SJRP: Cecília Artico Banho; Sandra Coccuzzo Sampaio; Svetoslav Nanev Slavov; Vagner Fonseca; Vincent Louis Viala |
| EPI_ISL_2345014, EPI_ISL_2345016, EPI_ISL_2345017 | UPA DR FRANCO DA ROCHA | Instituto Butantan | Antonio Jorge Martins; Claudia Renata dos Santos Barros; David Schlesinger; Debora Botequio Moretti; Dimas Tadeu Covas; Elaine Cristina Marqueze; Elaine Vieira Santos; Evandra Strazza Rodrigues; Heidge Fukumasu; Jayme Augusto de Souza-Neto; José Salvatore Leister Patané; Luiz Alcantara; Luiz Lehmann Coutinho; Maria Carolina Elias; Maurício Lacerda Nogueira; Rafael dos Santos Bezerra; Rejane Maria Tommasini Grotto; Ricardo Haddad; Sandra Coccuzzo Sampaio Vessoni; Simone Kashima; Svetoslav Nanev Slavov; Vincent Louis Viala |
| EPI_ISL_2170996 | UPA DR FRANCO DA ROCHA | Instituto Butantan / Mendelics | Antonio Jorge Martins; Bianca Cechetto Carlos. Mendelics: Bibiana Santos; Claudia Renata dos Santos Barros; Cíntia Bittar; David Schlesinger. Hemocentro Ribeirão Preto: Simone Kashima; Debora Botequio Moretti; Elaine Cristina Marqueze; Elaine Vieira dos Santos; Elisangela Chicaroni Mattos; Erika Freitas; Evandra Strazza Rodrigues; Felipe Allan da Silva da Costa; Flavia Aburjaile; Fábio Sossai Possebon; Guilherme Campos; Guilherme Targino Valente; Heidge Fukumasu. USP-Botucatu: Rejane Maria Tommasini Grotto; Helena Lage Ferreira; Instituto Butantan: Dimas Tadeu Covas; Jardelina de Souza Todao Bernardino; Jayme A. Souza-Neto; Jessika Cristina Chagas Lesbon; Jorge A. Petroli Marchesi; José Salvatore Leister Patané; João Paulo Kitajima; João Pessoa Araújo Jr.; Leila Sabrina Ullmann; Loyze Paola Oliveira de Lima; Luiz Aurelio de Campos Crispin. Centro de Genômica Funcional da ESALQ: Luiz Lehmann Coutinho; Luiz Carlos Junior de Alcantara; Lívia Sacchetto; Maisa C. Pereira Parra; Maria Carolina Elias; Marta Giovanetti; Marília Moraes; Maurício Lacerda Nogueira. Prefeitura de Sao Paulo: Melissa Palmieri.; Patricia Akemi Assato; Paula Rahal; Paulo Inacio da Costa; Rafael dos Santos Bezerra; Raquel de Lello Rocha Campos Cassano. NGS Soluções Genômicas: Pilar Drummond Sampaio Corrêa Mariani. FZEA-USP Pirassununga: Mirele Daiana Poleti; Raul Machado Neto; Ricardo Augusto Brassaloti; Ricardo Haddad; Rodrigo Tocantins Calado. FAMERP-SJRP: Cecília Artico Banho; Sandra Coccuzzo Sampaio; Svetoslav Nanev Slavov; Vagner Fonseca; Vincent Louis Viala |
| EPI_ISL_2210003, EPI_ISL_2344999, EPI_ISL_2345000, EPI_ISL_2345001, EPI_ISL_2345002, EPI_ISL_2345030 | UPA VILA SANTA CATARINA | Instituto Butantan | Antonio Jorge Martins; Claudia Renata dos Santos Barros; David Schlesinger; Debora Botequio Moretti; Dimas Tadeu Covas; Elaine Cristina Marqueze; Elaine Vieira Santos; Evandra Strazza Rodrigues; Heidge Fukumasu; Jayme Augusto de Souza-Neto; José Salvatore Leister Patané; Luiz Alcantara; Luiz Lehmann Coutinho; Maria Carolina Elias; Maurício Lacerda Nogueira; Rafael dos Santos Bezerra; Raul Machado Neto; Rejane Maria Tommasini Grotto; Ricardo Haddad; Sandra Coccuzzo Sampaio Vessoni; Simone Kashima; Svetoslav Nanev Slavov; Vincent Louis Viala; Vincent Louis Viala. |
| EPI_ISL_1967298, EPI_ISL_2171020, EPI_ISL_2171022 | UPA VILA SANTA CATARINA | Instituto Butantan / Mendelics | Antonio Jorge Martins; Bianca Cechetto Carlos. Mendelics: Bibiana Santos; Claudia Renata dos Santos Barros; Cíntia Bittar; David Schlesinger. Hemocentro Ribeirão Preto: Simone Kashima; Debora Botequio Moretti; Elaine Cristina Marqueze; Elaine Vieira dos Santos; Elisangela Chicaroni Mattos; Erika Freitas; Evandra Strazza Rodrigues; Felipe Allan da Silva da Costa; Flavia Aburjaile; Fábio Sossai Possebon; Guilherme Campos; Guilherme Targino Valente; Heidge Fukumasu. USP-Botucatu: Rejane Maria Tommasini Grotto; Helena Lage Ferreira; Instituto Butantan: Dimas Tadeu Covas; Jardelina de Souza Todao Bernardino; Jayme A. Souza-Neto; Jessika Cristina Chagas Lesbon; Jorge A. Petroli Marchesi; José Salvatore Leister Patané; João Paulo Kitajima; João Pessoa Araújo Jr.; Leila Sabrina Ullmann; Loyze Paola Oliveira de Lima; Luiz Aurelio de Campos Crispin. Centro de Genômica Funcional da ESALQ: Luiz Lehmann Coutinho; Luiz Carlos Junior de Alcantara; Lívia Sacchetto; Maisa C. Pereira Parra; Maria Carolina Elias; Marta Giovanetti; Marília Moraes; Maurício Lacerda Nogueira. Prefeitura de Sao Paulo: Melissa Palmieri.; Patricia Akemi Assato; Paula Rahal; Paulo Inacio da Costa; Rafael dos Santos Bezerra; Raquel de Lello Rocha Campos Cassano. NGS Soluções Genômicas: Pilar Drummond Sampaio Corrêa Mariani. FZEA-USP Pirassununga: Mirele Daiana Poleti; Raul Machado Neto; Ricardo Augusto Brassaloti; Ricardo Haddad; Rodrigo Tocantins Calado. FAMERP-SJRP: Cecília Artico Banho; Sandra Coccuzzo Sampaio; Svetoslav Nanev Slavov; Vagner Fonseca; Vincent Louis Viala |
| EPI_ISL_2402238, EPI_ISL_2402252 | US Air Force School of Aerospace Medicine | US Air Force School of Aerospace Medicine | Amanda Javorina; Anthony Fries; Carol Garrett; Clarise Starr; Elizabeth Macias; Jennifer Meyer; Sarah Purves; William Gruner |
| EPI_ISL_2260377, EPI_ISL_2260381 | USC Clinical Lab | Los Angeles County Public Health Laboratories | P. Hemarajata et al. |
| EPI_ISL_1917530 | USCA Avezzano | Istituto Zooprofilattico Sperimentale dell'Abruzzo e Molise "G. Caporale" | Ancora M; Calistri P; Cammà C; Caporale M; Curini V; Delli Compagni E; Di Domenico M; Di Lollo Valeria; Di Pasquale A; Lorusso A; Mangone I; Marcacci M; Puglia I; Rinaldi A; Savini G; Scialabba S |
| EPI_ISL_1841215 | USCA Pescina | Istituto Zooprofilattico Sperimentale dell'Abruzzo e Molise "G. Caporale" | Ancora M; Calistri P; Cammà C; Caporale M; Curini V; Delli Compagni E; Di Domenico M; Di Lollo Valeria; Di Pasquale A; Lorusso A; Mangone I; Marcacci M; Puglia I; Rinaldi A; Savini G; Scialabba S |
| EPI_ISL_2344763 | USF II ALADIA ARAN RODRIGUES | Instituto Butantan / ESALQ-Piracicaba | Antonio Jorge Martins; Claudia Renata dos Santos Barros; David Schlesinger; Debora Botequio Moretti; Dimas Tadeu Covas; Elaine Cristina Marqueze; Elaine Vieira Santos; Evandra Strazza Rodrigues; Heidge Fukumasu; Jayme Augusto de Souza-Neto; José Salvatore Leister Patané; Luiz Alcantara; Luiz Lehmann Coutinho; Maria Carolina Elias; Maurício Lacerda Nogueira; Rafael dos Santos Bezerra; Raul Machado Neto; Rejane Maria Tommasini Grotto; Ricardo Haddad; Sandra Coccuzzo Sampaio Vessoni; Simone Kashima; Svetoslav Nanev Slavov; Vincent Louis Viala |
| EPI_ISL_1827719, EPI_ISL_1827722, EPI_ISL_2002204, EPI_ISL_2002207, EPI_ISL_2002213, EPI_ISL_2002221, EPI_ISL_2002295, EPI_ISL_2002299, EPI_ISL_2002312, EPI_ISL_2002313, EPI_ISL_2002347, EPI_ISL_2002375, EPI_ISL_2002400, EPI_ISL_2002407, EPI_ISL_2002420, EPI_ISL_2002433, EPI_ISL_2002435, EPI_ISL_2002444, EPI_ISL_2002471, EPI_ISL_2002481, EPI_ISL_2002718, EPI_ISL_2002739, EPI_ISL_2002741, EPI_ISL_2002748, EPI_ISL_2002753, EPI_ISL_2002755, EPI_ISL_2002765, EPI_ISL_2002776, EPI_ISL_2002789, EPI_ISL_2002816, EPI_ISL_2002831, EPI_ISL_2002838, EPI_ISL_2002839, EPI_ISL_2002847, EPI_ISL_2002848, EPI_ISL_2002859, EPI_ISL_2002875, EPI_ISL_2002876, EPI_ISL_2002881, EPI_ISL_2002899, EPI_ISL_2002906, EPI_ISL_2002907, EPI_ISL_2002911, EPI_ISL_2002915, EPI_ISL_2002918, EPI_ISL_2031284, EPI_ISL_2031292, EPI_ISL_2031316, EPI_ISL_2031480, EPI_ISL_2031481, EPI_ISL_2031482, EPI_ISL_2031485, EPI_ISL_2031486, EPI_ISL_2031487, EPI_ISL_2031488, EPI_ISL_2031489, EPI_ISL_2031490, EPI_ISL_2031491, EPI_ISL_2031492, EPI_ISL_2031493, EPI_ISL_2031494, EPI_ISL_2031495, EPI_ISL_2031496, EPI_ISL_2031497, EPI_ISL_2031498, EPI_ISL_2031499, EPI_ISL_2037449, EPI_ISL_2037454, EPI_ISL_2037458, EPI_ISL_2037465, EPI_ISL_2037469, EPI_ISL_2037472, EPI_ISL_2037474, EPI_ISL_2037483, EPI_ISL_2037485, EPI_ISL_2037489, EPI_ISL_2037492, EPI_ISL_2037493, EPI_ISL_2037496, EPI_ISL_2037545, EPI_ISL_2037550, EPI_ISL_2037551, EPI_ISL_2037556, EPI_ISL_2037574, EPI_ISL_2037579, EPI_ISL_2037582, EPI_ISL_2037584, EPI_ISL_2037586, EPI_ISL_2037587, EPI_ISL_2037592, EPI_ISL_2037597, EPI_ISL_2037601, EPI_ISL_2037608, EPI_ISL_2037612, EPI_ISL_2037630, EPI_ISL_2037642, EPI_ISL_2037652, EPI_ISL_2037654, EPI_ISL_2037656, EPI_ISL_2037664, EPI_ISL_2037666, EPI_ISL_2037667, EPI_ISL_2037680, EPI_ISL_2037682, EPI_ISL_2037684, EPI_ISL_2082483, EPI_ISL_2082486, EPI_ISL_2082513, EPI_ISL_2082514, EPI_ISL_2082517, EPI_ISL_2082518, EPI_ISL_2082519, EPI_ISL_2082520, EPI_ISL_2082528, EPI_ISL_2082534, EPI_ISL_2082542, EPI_ISL_2082561, EPI_ISL_2082568, EPI_ISL_2082569, EPI_ISL_2082570, EPI_ISL_2082572, EPI_ISL_2082573, EPI_ISL_2082592, EPI_ISL_2082595, EPI_ISL_2082599, EPI_ISL_2082632, EPI_ISL_2082638, EPI_ISL_2082639, EPI_ISL_2082641, EPI_ISL_2082650, EPI_ISL_2082657, EPI_ISL_2082668, EPI_ISL_2082671, EPI_ISL_2082675, EPI_ISL_2082677, EPI_ISL_2082681 | | | |
| see above | UW Virology Lab | UW Virology Lab | Alexander Greninger; Hong Xie; Keith R Jerome; Lasata Shrestha; Meei-Li Huang; Michelle Lin; Nathan Breit; Noah R. Baker; Patrick Mathias; Pavitra Roychoudhury; Ricardo Perez; Robert J. Livingston; Sean Ellis; Shah Mohamed Bakhash; Tien V. Nguyen |
| EPI_ISL_5800794 | Ubs De Pedranopolis | Instituto Butantan | Antonio Jorge Martins; Claudia Renata dos Santos Barros; David Schlesinger; Debora Botequio Moretti; Dimas Tadeu Covas; Elaine Cristina Marqueze; Elaine Vieira Santos; Evandra Strazza Rodrigues; Heidge Fukumasu; Jayme Augusto de Souza-Neto; José Salvatore Leister Patané; Luiz Alcantara; Luiz Lehmann Coutinho; Maria Carolina Elias; Maurício Lacerda Nogueira; Rafael dos Santos Bezerra; Raul Machado Neto; Rejane Maria Tommasini Grotto; Ricardo Haddad; Sandra Coccuzzo Sampaio Vessoni; Simone Kashima; Svetoslav Nanev Slavov; Vincent Louis Viala |
| EPI_ISL_5800797 | Ubs De Sao Joao Das Duas Pontes | Instituto Butantan | Antonio Jorge Martins; Claudia Renata dos Santos Barros; David Schlesinger; Debora Botequio Moretti; Dimas Tadeu Covas; Elaine Cristina Marqueze; Elaine Vieira Santos; Evandra Strazza Rodrigues; Heidge Fukumasu; Jayme Augusto de Souza-Neto; José Salvatore Leister Patané; Luiz Alcantara; Luiz Lehmann Coutinho; Maria Carolina Elias; Maurício Lacerda Nogueira; Rafael dos Santos Bezerra; Raul Machado Neto; Rejane Maria Tommasini Grotto; Ricardo Haddad; Sandra Coccuzzo Sampaio Vessoni; Simone Kashima; Svetoslav Nanev Slavov; Vincent Louis Viala |
| EPI_ISL_5800747, EPI_ISL_5800749, EPI_ISL_5800752, EPI_ISL_5800753, EPI_ISL_5800754 | Ubs Dr Alfredo Dantas De Souza Umuarama | Instituto Butantan | Antonio Jorge Martins; Claudia Renata dos Santos Barros; David Schlesinger; Debora Botequio Moretti; Dimas Tadeu Covas; Elaine Cristina Marqueze; Elaine Vieira Santos; Evandra Strazza Rodrigues; Heidge Fukumasu; Jayme Augusto de Souza-Neto; José Salvatore Leister Patané; Luiz Alcantara; Luiz Lehmann Coutinho; Maria Carolina Elias; Maurício Lacerda Nogueira; Rafael dos Santos Bezerra; Raul Machado Neto; Rejane Maria Tommasini Grotto; Ricardo Haddad; Sandra Coccuzzo Sampaio Vessoni; Simone Kashima; Svetoslav Nanev Slavov; Vincent Louis Viala |
| EPI_ISL_5800939 | Ubs Enrique Guilherme Ernesto Marzeuski | Instituto Butantan | Antonio Jorge Martins; Claudia Renata dos Santos Barros; David Schlesinger; Debora Botequio Moretti; Dimas Tadeu Covas; Elaine Cristina Marqueze; Elaine Vieira Santos; Evandra Strazza Rodrigues; Heidge Fukumasu; Jayme Augusto de Souza-Neto; José Salvatore Leister Patané; Luiz Alcantara; Luiz Lehmann Coutinho; Maria Carolina Elias; Maurício Lacerda Nogueira; Rafael dos Santos Bezerra; Raul Machado Neto; Rejane Maria Tommasini Grotto; Ricardo Haddad; Sandra Coccuzzo Sampaio Vessoni; Simone Kashima; Svetoslav Nanev Slavov; Vincent Louis Viala |
| EPI_ISL_5800609, EPI_ISL_5800619 | Ubs Ii De Pirapozinho C Servico De Eacs E Esf | Instituto Butantan | Antonio Jorge Martins; Claudia Renata dos Santos Barros; David Schlesinger; Debora Botequio Moretti; Dimas Tadeu Covas; Elaine Cristina Marqueze; Elaine Vieira Santos; Evandra Strazza Rodrigues; Heidge Fukumasu; Jayme Augusto de Souza-Neto; José Salvatore Leister Patané; Luiz Alcantara; Luiz Lehmann Coutinho; Maria Carolina Elias; Maurício Lacerda Nogueira; Rafael dos Santos Bezerra; Raul Machado Neto; Rejane Maria Tommasini Grotto; Ricardo Haddad; Sandra Coccuzzo Sampaio Vessoni; Simone Kashima; Svetoslav Nanev Slavov; Vincent Louis Viala |
| EPI_ISL_5800607 | Ubs Ii De Taciba | Instituto Butantan | Antonio Jorge Martins; Claudia Renata dos Santos Barros; David Schlesinger; Debora Botequio Moretti; Dimas Tadeu Covas; Elaine Cristina Marqueze; Elaine Vieira Santos; Evandra Strazza Rodrigues; Heidge Fukumasu; Jayme Augusto de Souza-Neto; José Salvatore Leister Patané; Luiz Alcantara; Luiz Lehmann Coutinho; Maria Carolina Elias; Maurício Lacerda Nogueira; Rafael dos Santos Bezerra; Raul Machado Neto; Rejane Maria Tommasini Grotto; Ricardo Haddad; Sandra Coccuzzo Sampaio Vessoni; Simone Kashima; Svetoslav Nanev Slavov; Vincent Louis Viala |
| EPI_ISL_5800771, EPI_ISL_5800772 | Ubs Jardim Cabucu | Instituto Butantan | Antonio Jorge Martins; Claudia Renata dos Santos Barros; David Schlesinger; Debora Botequio Moretti; Dimas Tadeu Covas; Elaine Cristina Marqueze; Elaine Vieira Santos; Evandra Strazza Rodrigues; Heidge Fukumasu; Jayme Augusto de Souza-Neto; José Salvatore Leister Patané; Luiz Alcantara; Luiz Lehmann Coutinho; Maria Carolina Elias; Maurício Lacerda Nogueira; Rafael dos Santos Bezerra; Raul Machado Neto; Rejane Maria Tommasini Grotto; Ricardo Haddad; Sandra Coccuzzo Sampaio Vessoni; Simone Kashima; Svetoslav Nanev Slavov; Vincent Louis Viala |
| EPI_ISL_5800840 | Ubs Jardim Munhoz | Instituto Butantan | Antonio Jorge Martins; Claudia Renata dos Santos Barros; David Schlesinger; Debora Botequio Moretti; Dimas Tadeu Covas; Elaine Cristina Marqueze; Elaine Vieira Santos; Evandra Strazza Rodrigues; Heidge Fukumasu; Jayme Augusto de Souza-Neto; José Salvatore Leister Patané; Luiz Alcantara; Luiz Lehmann Coutinho; Maria Carolina Elias; Maurício Lacerda Nogueira; Rafael dos Santos Bezerra; Raul Machado Neto; Rejane Maria Tommasini Grotto; Ricardo Haddad; Sandra Coccuzzo Sampaio Vessoni; Simone Kashima; Svetoslav Nanev Slavov; Vincent Louis Viala |
| EPI_ISL_5800878 | Ubs Jardim Vitorino Ipero | Instituto Butantan | Antonio Jorge Martins; Claudia Renata dos Santos Barros; David Schlesinger; Debora Botequio Moretti; Dimas Tadeu Covas; Elaine Cristina Marqueze; Elaine Vieira Santos; Evandra Strazza Rodrigues; Heidge Fukumasu; Jayme Augusto de Souza-Neto; José Salvatore Leister Patané; Luiz Alcantara; Luiz Lehmann Coutinho; Maria Carolina Elias; Maurício Lacerda Nogueira; Rafael dos Santos Bezerra; Raul Machado Neto; Rejane Maria Tommasini Grotto; Ricardo Haddad; Sandra Coccuzzo Sampaio Vessoni; Simone Kashima; Svetoslav Nanev Slavov; Vincent Louis Viala |
| EPI_ISL_5800761 | Ubs Jd Saltense | Instituto Butantan | Antonio Jorge Martins; Claudia Renata dos Santos Barros; David Schlesinger; Debora Botequio Moretti; Dimas Tadeu Covas; Elaine Cristina Marqueze; Elaine Vieira Santos; Evandra Strazza Rodrigues; Heidge Fukumasu; Jayme Augusto de Souza-Neto; José Salvatore Leister Patané; Luiz Alcantara; Luiz Lehmann Coutinho; Maria Carolina Elias; Maurício Lacerda Nogueira; Rafael dos Santos Bezerra; Raul Machado Neto; Rejane Maria Tommasini Grotto; Ricardo Haddad; Sandra Coccuzzo Sampaio Vessoni; Simone Kashima; Svetoslav Nanev Slavov; Vincent Louis Viala |
| EPI_ISL_5800744 | Ubs Recreio Sao Jorge | Instituto Butantan | Antonio Jorge Martins; Claudia Renata dos Santos Barros; David Schlesinger; Debora Botequio Moretti; Dimas Tadeu Covas; Elaine Cristina Marqueze; Elaine Vieira Santos; Evandra Strazza Rodrigues; Heidge Fukumasu; Jayme Augusto de Souza-Neto; José Salvatore Leister Patané; Luiz Alcantara; Luiz Lehmann Coutinho; Maria Carolina Elias; Maurício Lacerda Nogueira; Rafael dos Santos Bezerra; Raul Machado Neto; Rejane Maria Tommasini Grotto; Ricardo Haddad; Sandra Coccuzzo Sampaio Vessoni; Simone Kashima; Svetoslav Nanev Slavov; Vincent Louis Viala |
| EPI_ISL_2382297 | Uconn Health Clinical Microbiology | Brenton Graveley Lab, UConn Health | Brenton Graveley; Christina Caron; Kimberly Hayes; Lori Avery; Michael Senh; Sara Olson; Stacey Ferraro; Xintao Wei |
| EPI_ISL_5800946 | Ums Pirapora Do Bom Jesus | Instituto Butantan | Antonio Jorge Martins; Claudia Renata dos Santos Barros; David Schlesinger; Debora Botequio Moretti; Dimas Tadeu Covas; Elaine Cristina Marqueze; Elaine Vieira Santos; Evandra Strazza Rodrigues; Heidge Fukumasu; Jayme Augusto de Souza-Neto; José Salvatore Leister Patané; Luiz Alcantara; Luiz Lehmann Coutinho; Maria Carolina Elias; Maurício Lacerda Nogueira; Rafael dos Santos Bezerra; Raul Machado Neto; Rejane Maria Tommasini Grotto; Ricardo Haddad; Sandra Coccuzzo Sampaio Vessoni; Simone Kashima; Svetoslav Nanev Slavov; Vincent Louis Viala |
| EPI_ISL_2402076 | Unidad de Investigación Biomédica de Zacatecas (UIBZ) | Unidad de Genomica Avanzada | Alejandro Sanchez-Flores; Alfredo Herrera-Estrella; Alicia Ocana-Mondragon; Angel Gustavo Salas-Lais; Bernardo Martinez-Miguel; Blanca Taboada; Brenda Irasema Maldonado-Meza; Carla Ivon Herrera-Najera; Carlos F. Arias; Celia Boukadida; Clara Esperanza Santacruz-Tinoco; Concepcion Grajales-Muniz; Consorcio Mexicano de Vigilancia Genomica (CoViGen-Mex). Authors (in alphabetical order): Julio Elias Alvarado-Yaah; Fernando Fontove-Herrera; Francisco Pulido; Gloria Elena Espinoza-Ayala; Gloria Maria Molina-Salinas; Gloria Vazquez; Hector Esteban Paz-Juarez; Hector Montoya-Fuentes; Helen Haydee Fernanda Ramirez-Plascencia; Jorge Ivan Salinal-Nevarez; Jose Antonio Enciso-Moreno; Jose Esteban Munoz-Medina; Jose de Jesus Nunez-Contreras; Juan Bautista Chale-Dzul; Luis Alberto Ochoa-Carrera; Margarita Matias-Florentino; Maria Guadalupe Santiago-Mauricio; Maria Guadalupe de Jesus Mireles-Rivera; Nelly Selem-Mojica; Pavel Isa; Ricardo Grande; Santiago Avila-Rios; Victor Hugo Borja-Aburto |
| EPI_ISL_2402043 | Unidad de Investigación Médica de Yucatán (UIMY) | Unidad de Genomica Avanzada | Alejandro Sanchez-Flores; Alfredo Herrera-Estrella; Alicia Ocana-Mondragon; Angel Gustavo Salas-Lais; Bernardo Martinez-Miguel; Blanca Taboada; Brenda Irasema Maldonado-Meza; Carla Ivon Herrera-Najera; Carlos F. Arias; Celia Boukadida; Clara Esperanza Santacruz-Tinoco; Concepcion Grajales-Muniz; Consorcio Mexicano de Vigilancia Genomica (CoViGen-Mex). Authors (in alphabetical order): Julio Elias Alvarado-Yaah; Fernando Fontove-Herrera; Francisco Pulido; Gloria Elena Espinoza-Ayala; Gloria Maria Molina-Salinas; Gloria Vazquez; Hector Esteban Paz-Juarez; Hector Montoya-Fuentes; Helen Haydee Fernanda Ramirez-Plascencia; Jorge Ivan Salinal-Nevarez; Jose Antonio Enciso-Moreno; Jose Esteban Munoz-Medina; Jose de Jesus Nunez-Contreras; Juan Bautista Chale-Dzul; Luis Alberto Ochoa-Carrera; Margarita Matias-Florentino; Maria Guadalupe Santiago-Mauricio; Maria Guadalupe de Jesus Mireles-Rivera; Nelly Selem-Mojica; Pavel Isa; Ricardo Grande; Santiago Avila-Rios; Victor Hugo Borja-Aburto |
| EPI_ISL_5800612, EPI_ISL_5800618 | Unidade Basica De Saude Iii De Alfredo Marcondes | Instituto Butantan | Antonio Jorge Martins; Claudia Renata dos Santos Barros; David Schlesinger; Debora Botequio Moretti; Dimas Tadeu Covas; Elaine Cristina Marqueze; Elaine Vieira Santos; Evandra Strazza Rodrigues; Heidge Fukumasu; Jayme Augusto de Souza-Neto; José Salvatore Leister Patané; Luiz Alcantara; Luiz Lehmann Coutinho; Maria Carolina Elias; Maurício Lacerda Nogueira; Rafael dos Santos Bezerra; Raul Machado Neto; Rejane Maria Tommasini Grotto; Ricardo Haddad; Sandra Coccuzzo Sampaio Vessoni; Simone Kashima; Svetoslav Nanev Slavov; Vincent Louis Viala |
| EPI_ISL_5800847, EPI_ISL_5800849 | Unidade De Pronto Atendimento De Jales | Instituto Butantan | Antonio Jorge Martins; Claudia Renata dos Santos Barros; David Schlesinger; Debora Botequio Moretti; Dimas Tadeu Covas; Elaine Cristina Marqueze; Elaine Vieira Santos; Evandra Strazza Rodrigues; Heidge Fukumasu; Jayme Augusto de Souza-Neto; José Salvatore Leister Patané; Luiz Alcantara; Luiz Lehmann Coutinho; Maria Carolina Elias; Maurício Lacerda Nogueira; Rafael dos Santos Bezerra; Raul Machado Neto; Rejane Maria Tommasini Grotto; Ricardo Haddad; Sandra Coccuzzo Sampaio Vessoni; Simone Kashima; Svetoslav Nanev Slavov; Vincent Louis Viala |
| EPI_ISL_5800882 | Unidade De Pronto Atendimento Upa | Instituto Butantan | Antonio Jorge Martins; Claudia Renata dos Santos Barros; David Schlesinger; Debora Botequio Moretti; Dimas Tadeu Covas; Elaine Cristina Marqueze; Elaine Vieira Santos; Evandra Strazza Rodrigues; Heidge Fukumasu; Jayme Augusto de Souza-Neto; José Salvatore Leister Patané; Luiz Alcantara; Luiz Lehmann Coutinho; Maria Carolina Elias; Maurício Lacerda Nogueira; Rafael dos Santos Bezerra; Raul Machado Neto; Rejane Maria Tommasini Grotto; Ricardo Haddad; Sandra Coccuzzo Sampaio Vessoni; Simone Kashima; Svetoslav Nanev Slavov; Vincent Louis Viala |
| EPI_ISL_5800750 | Unidade Mista De Iguape | Instituto Butantan | Antonio Jorge Martins; Claudia Renata dos Santos Barros; David Schlesinger; Debora Botequio Moretti; Dimas Tadeu Covas; Elaine Cristina Marqueze; Elaine Vieira Santos; Evandra Strazza Rodrigues; Heidge Fukumasu; Jayme Augusto de Souza-Neto; José Salvatore Leister Patané; Luiz Alcantara; Luiz Lehmann Coutinho; Maria Carolina Elias; Maurício Lacerda Nogueira; Rafael dos Santos Bezerra; Raul Machado Neto; Rejane Maria Tommasini Grotto; Ricardo Haddad; Sandra Coccuzzo Sampaio Vessoni; Simone Kashima; Svetoslav Nanev Slavov; Vincent Louis Viala |
| EPI_ISL_5800940 | Unidade Mista De Saude Capela Do Alto | Instituto Butantan | Antonio Jorge Martins; Claudia Renata dos Santos Barros; David Schlesinger; Debora Botequio Moretti; Dimas Tadeu Covas; Elaine Cristina Marqueze; Elaine Vieira Santos; Evandra Strazza Rodrigues; Heidge Fukumasu; Jayme Augusto de Souza-Neto; José Salvatore Leister Patané; Luiz Alcantara; Luiz Lehmann Coutinho; Maria Carolina Elias; Maurício Lacerda Nogueira; Rafael dos Santos Bezerra; Raul Machado Neto; Rejane Maria Tommasini Grotto; Ricardo Haddad; Sandra Coccuzzo Sampaio Vessoni; Simone Kashima; Svetoslav Nanev Slavov; Vincent Louis Viala |
| EPI_ISL_5800860 | Unidade Mista De Saude Mombuca | Instituto Butantan | Antonio Jorge Martins; Claudia Renata dos Santos Barros; David Schlesinger; Debora Botequio Moretti; Dimas Tadeu Covas; Elaine Cristina Marqueze; Elaine Vieira Santos; Evandra Strazza Rodrigues; Heidge Fukumasu; Jayme Augusto de Souza-Neto; José Salvatore Leister Patané; Luiz Alcantara; Luiz Lehmann Coutinho; Maria Carolina Elias; Maurício Lacerda Nogueira; Rafael dos Santos Bezerra; Raul Machado Neto; Rejane Maria Tommasini Grotto; Ricardo Haddad; Sandra Coccuzzo Sampaio Vessoni; Simone Kashima; Svetoslav Nanev Slavov; Vincent Louis Viala |
| EPI_ISL_5800603 | Unidade Mista Osvaldo P Camargo Elias Fausto | Instituto Butantan | Antonio Jorge Martins; Claudia Renata dos Santos Barros; David Schlesinger; Debora Botequio Moretti; Dimas Tadeu Covas; Elaine Cristina Marqueze; Elaine Vieira Santos; Evandra Strazza Rodrigues; Heidge Fukumasu; Jayme Augusto de Souza-Neto; José Salvatore Leister Patané; Luiz Alcantara; Luiz Lehmann Coutinho; Maria Carolina Elias; Maurício Lacerda Nogueira; Rafael dos Santos Bezerra; Raul Machado Neto; Rejane Maria Tommasini Grotto; Ricardo Haddad; Sandra Coccuzzo Sampaio Vessoni; Simone Kashima; Svetoslav Nanev Slavov; Vincent Louis Viala |
| EPI_ISL_5800900 | Unidade Psf Pinheiros Apiai | Instituto Butantan | Antonio Jorge Martins; Claudia Renata dos Santos Barros; David Schlesinger; Debora Botequio Moretti; Dimas Tadeu Covas; Elaine Cristina Marqueze; Elaine Vieira Santos; Evandra Strazza Rodrigues; Heidge Fukumasu; Jayme Augusto de Souza-Neto; José Salvatore Leister Patané; Luiz Alcantara; Luiz Lehmann |

| | | | |
|---|---|---|---|
| | | | Coutinho; Maria Carolina Elias; Maurício Lacerda Nogueira; Rafael dos Santos Bezerra; Raul Machado Neto; Rejane Maria Tommasini Grotto; Ricardo Haddad; Sandra Coccuzzo Sampaio Vessoni; Simone Kashima; Svetoslav Nanev Slavov; Vincent Louis Viala |
| EPI_ISL_2101390, EPI_ISL_2101471, EPI_ISL_2101472, EPI_ISL_2101473, EPI_ISL_2101475, EPI_ISL_2101483, EPI_ISL_2101484, EPI_ISL_2101508, EPI_ISL_2101509, EPI_ISL_2101510, EPI_ISL_2101511, EPI_ISL_2101512, EPI_ISL_2101513, EPI_ISL_2101671, EPI_ISL_2101681, EPI_ISL_2101682, EPI_ISL_2101683, EPI_ISL_2101684, EPI_ISL_2101685, EPI_ISL_2101686, EPI_ISL_2101687, EPI_ISL_2101688, EPI_ISL_2101692, EPI_ISL_2101693, EPI_ISL_2101694, EPI_ISL_2101695, EPI_ISL_2101696, EPI_ISL_2101714, EPI_ISL_2101734 | | | |
| see above | Unidade de apoio ao diagnóstico da COVID – UNADIG | Bioinformatics Laboratory / LNCC | Alessandra P Lamarca; Alexandra L Gerber; Amilcar Tanuri; Ana Paula de C Guimaraes; Ana Tereza R Vasconcelos; Andrea Cony Cavalcanti; Caio Luiz Pereira Ribeiro; Cassia Alves; Cintia Policarpo; Claudia Maria Braga de Mello; Cristiane Gomes da Silva; Diana Mariani; Douglas Terra Machado; Flavio Dias da Silva; Gleidson da Silva de Oliveira; Leandro Magalhaes de Souza; Liliane Cavalcante; Luiz G P de Almeida; Marcio Henrique de Oliveira Garcia; Mario Sergio Ribeiro; Ronaldo da Silva F Jr; Silvia Carvalho |
| EPI_ISL_3671020 | Universidad Nacional sede Medellin Laboratorio One Health | Universidad Nacional de Colombia - Laboratorio Genómico One Health | Andres F. Cardona-Rios; Carlos Franco-Muñoz; Carolina Muñoz-Arango; Celeny Ortiz; Daniel O. Maldonado-Perez; Diego A. Álvarez-Díaz; Hector Alejandro Ruiz-Moreno; Idabely Betancur Ortiz; Jorge E. Osorio; Juan P. Hernandez-Ortiz; Karl A Ciuoderis; Katherine Laiton-Donato; Laura Silvana Perez; Lina M. Hurtado; Marcela Mercado-Reyes; Maria Angélica Maya; Maria Stella López; Rita Almanza Payares; Sandra Ines Cano; Simón Villegas Velásquez |
| EPI_ISL_1963271, EPI_ISL_1963272 | University Hospitals of Geneva, Laboratory of Virology | HUG, Laboratory of Virology and the Health2030 Genome Center | Ana Rita Goncalves; Deborah Penet; Emmanouil Dermitzakis; Henri Pegeot; Ioannis Xenarios; Keith Harshman; Laurent Kaiser; Lorenzo Cerutti; Melyssa Elies; Samuel Cordey |
| EPI_ISL_2365938 | University of Bari Biomedical Sciences and Human Oncology, Policlinico | University of Bari Biomedical Sciences and Human Oncology | Maria Chironna |
| EPI_ISL_2246607, EPI_ISL_2692004, EPI_ISL_2692007 | University of Chicago Medicine | RIPHL at Rush University Medical Center | Cindy Bethel; Felix Araujo Perez; Kevin Kunstman; Laura Furtado; Lauren Megger; Marieta Hyde; Max Kolton; Stefan Green |
| EPI_ISL_1970738, EPI_ISL_1970747, EPI_ISL_1970759 | University of Liège COVID-19 testing center | GIGA Medical Genomics | Bouchra Boujemla; Cécile Meex; Keith Durkin; Maria Artesi; Marie-Pierre Hayette; Nathalie Renotte; Pierrette Melin; Raphaël Boreux; Sébastien Bontems; Vincent Bours |
| EPI_ISL_2001220, EPI_ISL_2001240, EPI_ISL_2001258 | University of Michigan Clinical Microbiology Laboratory | Lauring Lab, University of Michigan, Department of Microbiology and Immunology | Valesano |
| EPI_ISL_2007825, EPI_ISL_2007829 | University of Oregon COVID-19 MAP Laboratory | University of Oregon Genomics and Cell Characterization Core Facility (GC3F) | Ariana White; Douglas Turnbull; Jason Carriere; Jason Sydes; Jeff Bishop; Peter Batzel |
| EPI_ISL_2018960, EPI_ISL_2018961, EPI_ISL_2285343, EPI_ISL_2285347 | University of Rome Tor Vergata: Departm Experim Medicine Chair of Virology | University of Rome Tor Vergata: Departm Experim Medicine Chair of Virology | Francesca Ceccherini-Silberstein; Loredana Sarmati; Lorenzo Piermatteo; Luca Carioti; Marco Iannetta; Maria Botticelli; Maria Concetta Bellocchi; Massimo Andreoni; Rossana Scutari |
| EPI_ISL_1822054, EPI_ISL_1822070, EPI_ISL_1822084, EPI_ISL_2225535 | University of Wisconsin-Madison AIDS Vaccine Research Laboratories | University of Wisconsin-Madison AIDS Vaccine Research Laboratories | Gage Moreno; Katarina Braun; et al. AIDS Vaccine Research Laboratories |
| EPI_ISL_1850441 | UniversitÃ¤tsklinikum MÃ¼nster Institut fÃ¼r Virologie | Robert Koch Institute | |
| EPI_ISL_2137468, EPI_ISL_2137484, EPI_ISL_2227047, EPI_ISL_2227054, EPI_ISL_2455598, EPI_ISL_2455618, EPI_ISL_2455623, EPI_ISL_2455626, EPI_ISL_2455660, EPI_ISL_3014220, EPI_ISL_3014221 | | | |
| see above | Università Federico II - Dipartimento di scienze mediche traslazionali - Napoli | TIGEM | Antonio Grimaldi Patrizia Annunziata Francesco Panariello Teresa Giuliano Michele Cennamo Valentina Bouche Chiara Colantuono Lucio Di Filippo Mariano Fiorenza Anna Manfredi Marcello Salvi Giuseppe Portella Andrea Ballabio Davide Cacchiarelli |
| EPI_ISL_3270773 | Università Federico II - Dipartimento di scienze mediche traslazionali - Napoli | Telethon Institute of Genetics and Medicine (TIGEM) | Antonio Grimaldi Patrizia Annunziata Francesco Panariello Teresa Giuliano Michele Cennamo Valentina Bouche Chiara Colantuono Lucio Di Filippo Mariano Fiorenza Anna Manfredi Marcello Salvi Giuseppe Portella Andrea Ballabio Davide Cacchiarelli |
| EPI_ISL_1785209, EPI_ISL_1785210, EPI_ISL_1785221 | Università degli Studi di Perugia | Istituto Zooprofilattico Sperimentale dell'Abruzzo e Molise "G. Caporale" | Ancora M; Calistri P; Camilloni B; Cammà C; Caporale M; Curini V; Delli Compagni E; Di Domenico M; Di Lollo Valeria; Di Pasquale A; Lorusso A; Mangone I; Marcacci M; Mencacci A; Puglia I; Rinaldi A; Savini G; Scialabba S |
| EPI_ISL_5800610, EPI_ISL_5800622, EPI_ISL_5800629, EPI_ISL_5800632, EPI_ISL_5800633, EPI_ISL_5800635, EPI_ISL_5800638 | | | |
| see above | Upa 24 Horas Ana Jacinta | Instituto Butantan | Antonio Jorge Martins; Claudia Renata dos Santos Barros; David Schlesinger; Debora Botequio Moretti; Dimas Tadeu Covas; Elaine Cristina Marqueze; Elaine Vieira Santos; Evandra Strazza Rodrigues; Heidge Fukumasu; Jayme Augusto de Souza-Neto; José Salvatore Leister Patané; Luiz Alcantara; Luiz Lehmann Coutinho; Maria Carolina Elias; Maurício Lacerda Nogueira; Rafael dos Santos Bezerra; Raul Machado Neto; Rejane Maria Tommasini Grotto; Ricardo Haddad; Sandra Coccuzzo Sampaio Vessoni; Simone Kashima; Svetoslav Nanev Slavov; Vincent Louis Viala |
| EPI_ISL_5800615, EPI_ISL_5800616, EPI_ISL_5800620, EPI_ISL_5800624, EPI_ISL_5800626, EPI_ISL_5800628 | Upa 24 Horas Dr Aloisio Muniz De Andrade | Instituto Butantan | Antonio Jorge Martins; Claudia Renata dos Santos Barros; David Schlesinger; Debora Botequio Moretti; Dimas Tadeu Covas; Elaine Cristina Marqueze; Elaine Vieira Santos; Evandra Strazza Rodrigues; Heidge Fukumasu; Jayme Augusto de Souza-Neto; José Salvatore Leister Patané; Luiz Alcantara; Luiz Lehmann Coutinho; Maria Carolina Elias; Maurício Lacerda Nogueira; Rafael dos Santos Bezerra; Raul Machado Neto; Rejane Maria Tommasini Grotto; Ricardo Haddad; Sandra Coccuzzo Sampaio Vessoni; Simone Kashima; Svetoslav Nanev Slavov; Vincent Louis Viala |
| EPI_ISL_5800623, EPI_ISL_5800625, EPI_ISL_5800640, EPI_ISL_5800650, EPI_ISL_5800659 | Upa De Bebedouro | Instituto Butantan | Antonio Jorge Martins; Claudia Renata dos Santos Barros; David Schlesinger; Debora Botequio Moretti; Dimas Tadeu Covas; Elaine Cristina Marqueze; Elaine Vieira Santos; Evandra Strazza Rodrigues; Heidge Fukumasu; Jayme Augusto de Souza-Neto; José Salvatore Leister Patané; Luiz Alcantara; Luiz Lehmann Coutinho; Maria Carolina Elias; Maurício Lacerda Nogueira; Rafael dos Santos Bezerra; Raul Machado Neto; Rejane Maria Tommasini Grotto; Ricardo Haddad; Sandra Coccuzzo Sampaio Vessoni; Simone Kashima; Svetoslav Nanev Slavov; Vincent Louis Viala |
| EPI_ISL_5800879, EPI_ISL_5800897, EPI_ISL_5800907, EPI_ISL_5800957 | Upa Dr Akira Tada | Instituto Butantan | Antonio Jorge Martins; Claudia Renata dos Santos Barros; David Schlesinger; Debora Botequio Moretti; Dimas Tadeu Covas; Elaine Cristina Marqueze; Elaine Vieira Santos; Evandra Strazza Rodrigues; Heidge Fukumasu; Jayme Augusto de Souza-Neto; José Salvatore Leister Patané; Luiz Alcantara; Luiz Lehmann Coutinho; Maria Carolina Elias; Maurício Lacerda Nogueira; Rafael dos Santos Bezerra; Raul Machado Neto; Rejane Maria Tommasini Grotto; Ricardo Haddad; Sandra Coccuzzo Sampaio Vessoni; Simone Kashima; Svetoslav Nanev Slavov; Vincent Louis Viala |
| EPI_ISL_5801020, EPI_ISL_5801021, EPI_ISL_5801023, EPI_ISL_5801024 | Upa Dr Franco Da Rocha | Instituto Butantan | Antonio Jorge Martins; Claudia Renata dos Santos Barros; David Schlesinger; Debora Botequio Moretti; Dimas Tadeu Covas; Elaine Cristina Marqueze; Elaine Vieira Santos; Evandra Strazza Rodrigues; Heidge Fukumasu; Jayme Augusto de Souza-Neto; José Salvatore Leister Patané; Luiz Alcantara; Luiz Lehmann Coutinho; Maria Carolina Elias; Maurício Lacerda Nogueira; Rafael dos Santos Bezerra; Raul Machado Neto; Rejane Maria Tommasini Grotto; Ricardo Haddad; Sandra Coccuzzo Sampaio Vessoni; Simone Kashima; Svetoslav Nanev Slavov; Vincent Louis Viala |
| EPI_ISL_5801002, EPI_ISL_5801003, EPI_ISL_5801004, EPI_ISL_5801005, EPI_ISL_5801006, EPI_ISL_5801007, EPI_ISL_5801025, EPI_ISL_5801039 | | | |
| see above | Upa Vila Santa Catarina | Instituto Butantan | Antonio Jorge Martins; Claudia Renata dos Santos Barros; David Schlesinger; Debora Botequio Moretti; Dimas Tadeu Covas; Elaine Cristina Marqueze; Elaine Vieira Santos; Evandra Strazza Rodrigues; Heidge Fukumasu; Jayme Augusto de Souza-Neto; José Salvatore Leister Patané; Luiz Alcantara; Luiz Lehmann Coutinho; Maria Carolina Elias; Maurício Lacerda Nogueira; Rafael dos Santos Bezerra; Raul Machado Neto; Rejane Maria Tommasini Grotto; Ricardo Haddad; Sandra Coccuzzo Sampaio Vessoni; Simone Kashima; Svetoslav Nanev Slavov; Vincent Louis Viala |
| EPI_ISL_3015187, EPI_ISL_3015194 | Urbino | Microbiology University Politecnica delle Marche | Anna Valenza; Carla Acciarri; Katia Marinelli; Monica Lucia Ferreri; Patrizia Bagnarelli; Roberta Longo; Sara Caucci; Stefano Menzo |
| EPI_ISL_5800621 | Usf Ii Aladia Aran Rodrigues | Instituto Butantan | Antonio Jorge Martins; Claudia Renata dos Santos Barros; David Schlesinger; Debora Botequio Moretti; Dimas Tadeu Covas; Elaine Cristina Marqueze; Elaine Vieira Santos; Evandra Strazza Rodrigues; Heidge Fukumasu; Jayme Augusto de Souza-Neto; José Salvatore Leister Patané; Luiz Alcantara; Luiz Lehmann Coutinho; Maria Carolina Elias; Maurício Lacerda Nogueira; Rafael dos Santos Bezerra; Raul Machado Neto; Rejane Maria Tommasini Grotto; Ricardo Haddad; Sandra Coccuzzo Sampaio Vessoni; Simone Kashima; Svetoslav Nanev Slavov; Vincent Louis Viala |
| EPI_ISL_2290853, EPI_ISL_2292139, EPI_ISL_2467046, EPI_ISL_2679747, EPI_ISL_3269714 | Utah Public Health Laboratory | Utah Public Health Laboratory | Erin L. Young; Kelly F. Oakeson; Olinto Linares-Perdomo; Tara Gallagher |
| EPI_ISL_2210083, EPI_ISL_2344951, EPI_ISL_2344958, EPI_ISL_2344959, EPI_ISL_2344962, EPI_ISL_2344964, EPI_ISL_2344965, EPI_ISL_2344966, EPI_ISL_2344968, EPI_ISL_2344970, EPI_ISL_2345039, EPI_ISL_2345040, EPI_ISL_2345041, EPI_ISL_2479926 | | | |
| see above | VIGILANCIA EM SAUDE DE FCO MORATO | Instituto Butantan | Antonio Jorge Martins; Claudia Renata dos Santos Barros; David Schlesinger; Debora Botequio Moretti; Dimas Tadeu Covas; Elaine Cristina Marqueze; Elaine Vieira Santos; Evandra Strazza Rodrigues; Heidge Fukumasu; Jayme Augusto de Souza-Neto; José Salvatore Leister Patané; Luiz Alcantara; Luiz Lehmann Coutinho; Maria Carolina Elias; Maurício Lacerda Nogueira; Rafael dos Santos Bezerra; Raul Machado Neto; Rejane Maria Tommasini Grotto; Ricardo Haddad; Sandra Coccuzzo Sampaio Vessoni; Simone Kashima; Svetoslav Nanev Slavov; Vincent Louis Viala; Vincent Louis Viala. |
| EPI_ISL_2170986, EPI_ISL_2170989, EPI_ISL_2170990, EPI_ISL_2170992, EPI_ISL_2170993, EPI_ISL_2170994 | VIGILANCIA EM SAUDE DE FCO MORATO | Instituto Butantan / Mendelics | Antonio Jorge Martins; Bianca Cechetto Carlos. Mendelics: Bibiana Santos; Claudia Renata dos Santos Barros; Cíntia Bittar; David Schlesinger; David Schlesinger. Hemocentro Ribeirão Preto: Simone Kashima; Debora Botequio Moretti; Dimas Tadeu Covas; Elaine Cristina Marqueze; Elaine Vieira Santos; Elaine Vieira dos Santos; Elisangela Chicaroni Mattos; Erika Freitas; Evandra Strazza Rodrigues; Felipe Allan da Silva da Costa; Flavia Aburjaile; Fábio Sossai Possebon; Guilherme Campos; Guilherme Targino Valente; Heidge Fukumasu; Heidge Fukumasu. USP-Botucatu: Rejane Maria Tommasini Grotto; Helena Lage Ferreira; Instituto Butantan: Dimas Tadeu Covas; Jardelina de Souza Todao Bernardino; Jayme A. Souza-Neto; Jayme Augusto de Souza-Neto; Jessika Cristina Chagas Lesbon; Jorge A. Petroli Marchesi; José Salvatore Leister Patané; João Paulo Kitajima; João Pessoa Araújo Jr.; Leila Sabrina Ullmann; Loyze Paola Oliveira de Lima; Luiz Alcantara; Luiz Aurelio de Campos Crispin. Centro de Genômica Funcional da ESALQ: Luiz Lehmann Coutinho; Luiz Carlos Junior de Alcantara; Luiz Lehmann Coutinho; Lívia Sacchetto; Maisa C. Pereira Parra; Maria Carolina Elias; Marta Giovanetti; Marília Moraes; Maurício Lacerda Nogueira; Maurício Lacerda Nogueira. Prefeitura de Sao Paulo: Melissa Palmieri.; Patricia Akemi Assato; Paula Rahal; Paulo Inacio da Costa; Rafael dos Santos Bezerra; Raquel de Lello Rocha Campos Cassano. NGS Soluções Genômicas: Pilar Drummond Sampaio Corrêa Mariani. FZEA-USP Pirassununga: Mirele Daiana Poleti; Raul Machado Neto; Rejane Maria Tommasini Grotto; Ricardo Augusto Brassaloti; Ricardo Haddad; Rodrigo Tocantins Calado. FAMERP-SJRP: Cecília Artico Banho; Sandra Coccuzzo Sampaio; Sandra Coccuzzo Sampaio Vessoni; Simone Kashima; Svetoslav Nanev Slavov; Vagner Fonseca; Vincent Louis Viala |
| EPI_ISL_1967199, EPI_ISL_1967200, EPI_ISL_1967201, EPI_ISL_1967202 | VIGILANCIA EM SAUDE SAE VIGILANCIA EPIDEMIOLOGICA | Instituto Butantan / Mendelics | Antonio Jorge Martins; Bianca Cechetto Carlos. Mendelics: Bibiana Santos; Claudia Renata dos Santos Barros; Cíntia Bittar; David Schlesinger. Hemocentro Ribeirão Preto: Simone Kashima; Debora Botequio Moretti; Elaine Cristina Marqueze; Elaine Vieira dos Santos; Elisangela Chicaroni Mattos; Erika Freitas; Evandra Strazza Rodrigues; Felipe Allan da Silva da Costa; Flavia Aburjaile; Fábio Sossai Possebon; Guilherme Campos; Guilherme Targino Valente; Heidge Fukumasu. USP-Botucatu: Rejane Maria Tommasini Grotto; Helena Lage Ferreira; Instituto Butantan: Dimas Tadeu Covas; Jardelina de Souza Todao Bernardino; Jayme A. Souza-Neto; Jessika Cristina Chagas Lesbon; Jorge A. Petroli Marchesi; José Salvatore Leister Patané; João Paulo Kitajima; João Pessoa Araújo Jr.; Leila Sabrina Ullmann; Loyze Paola Oliveira de Lima; Luiz Aurelio de Campos Crispin. Centro de Genômica Funcional da ESALQ: Luiz Lehmann Coutinho; Luiz Carlos Junior de Alcantara; Lívia Sacchetto; Maisa C. Pereira Parra; Maria Carolina Elias; Marta Giovanetti; Marília Moraes; Maurício Lacerda Nogueira. Prefeitura de Sao Paulo: Melissa Palmieri.; Patricia Akemi Assato; Paula Rahal; Paulo Inacio da Costa; Rafael dos Santos Bezerra; Raquel de Lello Rocha Campos Cassano. NGS Soluções Genômicas: Pilar Drummond Sampaio Corrêa Mariani. FZEA-USP Pirassununga: Mirele Daiana Poleti; Raul Machado Neto; Ricardo Augusto Brassaloti; Ricardo Haddad; Rodrigo Tocantins Calado. FAMERP-SJRP: Cecília Artico Banho; Sandra Coccuzzo Sampaio; Svetoslav Nanev Slavov; Vagner Fonseca; |

| | | | |
|---|---|---|---|
| | | | Vincent Louis Viala |
| EPI_ISL_2344809 | VIGILANCIA EPIDEMIOLOGICA DE LEME | Instituto Butantan / ESALQ-Piracicaba | Antonio Jorge Martins; Claudia Renata dos Santos Barros; David Schlesinger; Debora Botequio Moretti; Dimas Tadeu Covas; Elaine Cristina Marqueze; Elaine Vieira Santos; Evandra Strazza Rodrigues; Heidge Fukumasu; Jayme Augusto de Souza-Neto; José Salvatore Leister Patané; Luiz Alcantara; Luiz Lehmann Coutinho; Maria Carolina Elias; Maurício Lacerda Nogueira; Rafael dos Santos Bezerra; Raul Machado Neto; Rejane Maria Tommasini Grotto; Ricardo Haddad; Sandra Coccuzzo Sampaio Vessoni; Simone Kashima; Svetoslav Nanev Slavov; Vincent Louis Viala |
| EPI_ISL_2344771, EPI_ISL_2344830 | VIGILANCIA EPIDEMIOLOGICA E CONTROLE DE VETORES PIRASSUNUN | Instituto Butantan / ESALQ-Piracicaba | Antonio Jorge Martins; Claudia Renata dos Santos Barros; David Schlesinger; Debora Botequio Moretti; Dimas Tadeu Covas; Elaine Cristina Marqueze; Elaine Vieira Santos; Evandra Strazza Rodrigues; Heidge Fukumasu; Jayme Augusto de Souza-Neto; José Salvatore Leister Patané; Luiz Alcantara; Luiz Lehmann Coutinho; Maria Carolina Elias; Maurício Lacerda Nogueira; Rafael dos Santos Bezerra; Raul Machado Neto; Rejane Maria Tommasini Grotto; Ricardo Haddad; Sandra Coccuzzo Sampaio Vessoni; Simone Kashima; Svetoslav Nanev Slavov; Vincent Louis Viala |
| EPI_ISL_2109687, EPI_ISL_2109691 | VIROLOGY LABORATORY-CHU NICE | VIROLOGY LABORATORY-CHU NICE | Aicha El Yakine; Geraldine Gonfrier; Jean Machowiak; Sebastien Vitale; Valerie Giordanengo; Virginie Flahou |
| EPI_ISL_3058896, EPI_ISL_3059940 | Vienna Bio Center | Elling and Cochella laboratories, IMBA/IMP | Alexander Stark; Ezgi Oezkan; Luisa Cochella and Ulrich Elling; Marcus Strobl; Olga Frank; Ramesh Yelagandula; Tanino Albanese |
| EPI_ISL_5800799, EPI_ISL_5800807, EPI_ISL_5800861, EPI_ISL_5800941, EPI_ISL_5800943, EPI_ISL_5800944, EPI_ISL_5800945, EPI_ISL_5800952, EPI_ISL_5800953, EPI_ISL_5800956, EPI_ISL_5800960, EPI_ISL_5800961, EPI_ISL_5800962, EPI_ISL_5800964, EPI_ISL_5800966, EPI_ISL_5801050, EPI_ISL_5801051, EPI_ISL_5801052 | | | |
| see above | Vigilancia Em Saude De Fco Morato | Instituto Butantan | Antonio Jorge Martins; Claudia Renata dos Santos Barros; David Schlesinger; Debora Botequio Moretti; Dimas Tadeu Covas; Elaine Cristina Marqueze; Elaine Vieira Santos; Evandra Strazza Rodrigues; Heidge Fukumasu; Jayme Augusto de Souza-Neto; José Salvatore Leister Patané; Luiz Alcantara; Luiz Lehmann Coutinho; Maria Carolina Elias; Maurício Lacerda Nogueira; Rafael dos Santos Bezerra; Raul Machado Neto; Rejane Maria Tommasini Grotto; Ricardo Haddad; Sandra Coccuzzo Sampaio Vessoni; Simone Kashima; Svetoslav Nanev Slavov; Vincent Louis Viala |
| EPI_ISL_5800649, EPI_ISL_5800654, EPI_ISL_5800655, EPI_ISL_5800723 | Vigilancia Em Saude Sae Vigilancia Epidemiologica | Instituto Butantan | Antonio Jorge Martins; Claudia Renata dos Santos Barros; David Schlesinger; Debora Botequio Moretti; Dimas Tadeu Covas; Elaine Cristina Marqueze; Elaine Vieira Santos; Evandra Strazza Rodrigues; Heidge Fukumasu; Jayme Augusto de Souza-Neto; José Salvatore Leister Patané; Luiz Alcantara; Luiz Lehmann Coutinho; Maria Carolina Elias; Maurício Lacerda Nogueira; Rafael dos Santos Bezerra; Raul Machado Neto; Rejane Maria Tommasini Grotto; Ricardo Haddad; Sandra Coccuzzo Sampaio Vessoni; Simone Kashima; Svetoslav Nanev Slavov; Vincent Louis Viala |
| EPI_ISL_5800736, EPI_ISL_5802691 | Vigilancia Epidemiologica De Leme | Instituto Butantan | Antonio Jorge Martins; Claudia Renata dos Santos Barros; David Schlesinger; Debora Botequio Moretti; Dimas Tadeu Covas; Elaine Cristina Marqueze; Elaine Vieira Santos; Evandra Strazza Rodrigues; Heidge Fukumasu; Jayme Augusto de Souza-Neto; José Salvatore Leister Patané; Luiz Alcantara; Luiz Lehmann Coutinho; Maria Carolina Elias; Maurício Lacerda Nogueira; Rafael dos Santos Bezerra; Raul Machado Neto; Rejane Maria Tommasini Grotto; Ricardo Haddad; Sandra Coccuzzo Sampaio Vessoni; Simone Kashima; Svetoslav Nanev Slavov; Vincent Louis Viala |
| EPI_ISL_5800636, EPI_ISL_5800767 | Vigilancia Epidemiologica E Controle De Vetores Pirassunun | Instituto Butantan | Antonio Jorge Martins; Claudia Renata dos Santos Barros; David Schlesinger; Debora Botequio Moretti; Dimas Tadeu Covas; Elaine Cristina Marqueze; Elaine Vieira Santos; Evandra Strazza Rodrigues; Heidge Fukumasu; Jayme Augusto de Souza-Neto; José Salvatore Leister Patané; Luiz Alcantara; Luiz Lehmann Coutinho; Maria Carolina Elias; Maurício Lacerda Nogueira; Rafael dos Santos Bezerra; Raul Machado Neto; Rejane Maria Tommasini Grotto; Ricardo Haddad; Sandra Coccuzzo Sampaio Vessoni; Simone Kashima; Svetoslav Nanev Slavov; Vincent Louis Viala |
| EPI_ISL_1914207, EPI_ISL_1914209 | Viollier AG | Department of Biosystems Science and Engineering, ETH Zurich | Andrea Patrignani; Andreia Cabral de Gouvea; Catharine Aquino; Chaoran Chen; Christian Beisel; Christiane Beckmann; Christoph Noppen; David Dreifuss; Doris Popovic; Elodie Burcklen; Griffin White; Ina Nissen; Ivan Topolsky; Jay Tracy; Katharina Jahn; Lara Fuhrmann; Laura Neff; Lennart Opitz; Maria Domenica Moccia; Maurice Redondo; Mirjam Feldkamp; Natascha Santacroce; Niko Beerenwinkel; Noemie Santamaria de Souza; Olivier Kobel; Philipp Jablonski; Ralph Schlapbach; Rebecca Denes; Sarah Nadeau; Simon Gruter; Sophie Seidel; Tanja Stadler; Timothy Sykes |
| EPI_ISL_2017206, EPI_ISL_2017207 | Viollier AG | Department of Biosystems Science and Engineering, ETH Zürich | Chaoran Chen; Christian Beisel; Christiane Beckmann; Christoph Noppen; David Dreifuss; Elodie Burcklen; Ina Nissen; Ivan Topolsky; Katharina Jahn; Lara Fuhrmann; Maurice Redondo; Mirjam Feldkamp; Natascha Santacroce; Niko Beerenwinkel; Noemie Santamaria de Souza; Olivier Kobel; Philipp Jablonski; Rebecca Denes; Sarah Nadeau; Sophie Seidel; Tanja Stadler |
| EPI_ISL_2003686, EPI_ISL_2101940, EPI_ISL_2344505 | Virginia Division of Consolidated Laboratory Services | Virginia Division of Consolidated Laboratory Services | Virginia DCLS |
| EPI_ISL_2884098, EPI_ISL_2884101, EPI_ISL_2884108, EPI_ISL_2884213 | Virology Laboratory, Scientific Department, Army Medical Center | Virology Laboratory, Scientific Department, Army Medical Center | Anella Monte; Anna Anselmo; Antonella Fortunato; Filippo Molinari; Florigio Lista; Francesco Giordani; Giancarlo Petralito; Giandomenico Cerreto; Giulia Campoli; Lucia Nicosia; Marzia Cavalli; Riccardo De Sanctis; Rossella Brandi; Silvia Fillo; Vanessa Vera Fain |
| EPI_ISL_2097308 | WESTCHESTER MEDICAL CENTER | Wadsworth Center, New York State Department of Health | Alexis Russell; Catharine Prussing; Daryl M. Lamson; Erasmus Schneider; Erica Lasek-Nesselquist; John Kelly; Jonathan Plitnick; Kirsten St. George; Matthew Shudt; Melissa A Leisner; Navjot Singh |
| EPI_ISL_2015657 | WSSE w Krakowie | 1. National Institute of Public Health - National Institute of Hygiene, Warsaw, Poland 2. Biobank Lab, University of Lodz 3. Laboratory of Respiratory Viruses, Teaching and Clinical Center of the Medical University of Lodz | Dominik Strapagiel; Izabela Dróżdż; Jakub Lach; Katarzyna Zacharczuk; Klaudyna Królikowska; Maciej Borowiec; Magdalena Nowakowska; Magdalena Traczyk-Borszyńska; Marcin Słomka; Marta Sobalska-Kwapis; Małgorzata Sadkowska-Todys; Tomasz Płoszaj; Tomasz Wołkowicz |
| EPI_ISL_1747455, EPI_ISL_1897377, EPI_ISL_1897403, EPI_ISL_1897404, EPI_ISL_1897405, EPI_ISL_1897406, EPI_ISL_1897407, EPI_ISL_1897408, EPI_ISL_1897409, EPI_ISL_1897416, EPI_ISL_2096104, EPI_ISL_2096105, EPI_ISL_2096106, EPI_ISL_2096107, EPI_ISL_2096108, EPI_ISL_2096113, EPI_ISL_2336164, EPI_ISL_2336225, EPI_ISL_2336226, EPI_ISL_2336230, EPI_ISL_2336231, EPI_ISL_2336232, EPI_ISL_2336233, EPI_ISL_2336234, EPI_ISL_2378824, EPI_ISL_2378825 | | | |
| see above | Washington State Department of Health Public Health Laboratories | Washington State Department of Health Public Health Laboratories | Avi Singh; Darren Lucas; Denny Russell; Drew MacKellar; Geoff Melly; Hannah Gray; Joenice Gonzalez; JohnAric Peterson; Philip Dykema; Rebecca Cao; Vanessa De Los Santos |
| EPI_ISL_2399525, EPI_ISL_2399759, EPI_ISL_2400232 | Wexner Medical Center | The Ohio State University College of Medicine | Koenig, S.; Seminetta, J. |
| EPI_ISL_1700771, EPI_ISL_1760181, EPI_ISL_1760184, EPI_ISL_1760191, EPI_ISL_1760192, EPI_ISL_1760195, EPI_ISL_1760196, EPI_ISL_1760198, EPI_ISL_1805458, EPI_ISL_1805780, EPI_ISL_1807191, EPI_ISL_1807192, EPI_ISL_1807195, EPI_ISL_1807196, EPI_ISL_1807198, EPI_ISL_1807199, EPI_ISL_1807202, EPI_ISL_1819283, EPI_ISL_1819291, EPI_ISL_1819292, EPI_ISL_1819295, EPI_ISL_1819296 | | | |
| see above | Wichita State University - Molecular Diagnostics Lab | Kansas Health and Environmental Lab | Ben Olsen; Jonathan Barnell; Mike Grose; and Phil Adam |
| EPI_ISL_2102723 | Willis-Knighton Medical Center Hospital Laboratory | LSUHS Emerging Viral Threat Laboratory | Alexander Mijalis; Andrew D. Yurochko; Christopher G. Kevil; Gregory L. Ware; Jennifer L. Carroll; Jeremy P. Kamil; John A. Vanchiere; Maarten Van Diest; Rona S. Scott |
| EPI_ISL_2018713, EPI_ISL_2278999, EPI_ISL_2279001, EPI_ISL_2279053, EPI_ISL_2279054, EPI_ISL_2279062, EPI_ISL_2279087, EPI_ISL_2279180 | | | |
| see above | Wisconsin State Laboratory of Hygiene Communicable Disease Division | Wisconsin State Laboratory of Hygiene Communicable Disease Division | Abigail C. Shockey; Alicia J. Mooney; Erika M. Hanson; Kelsey R. Florek; Richard Griesser; Sara Wagner; Tonya Danz |
| EPI_ISL_1913010, EPI_ISL_1913011, EPI_ISL_1913012 | Wyoming Public Health Laboratory | Wyoming Public Health Laboratory | Ashley Norberg; Brian Dominguez; Cari Sloma; Channing Weber; Chayse Rowley; Elliot Thomasson; Jim Mildenberger; Lynette Gumbleton; Marley Goetz; Noah Hull; Sam Britz; Taylor Fearing; Wanda Manley; and Rob Christensen |
| EPI_ISL_1963807, EPI_ISL_2023095, EPI_ISL_2023097, EPI_ISL_2023126, EPI_ISL_2023143, EPI_ISL_2159667, EPI_ISL_2598696 | | | |
| see above | Yale Clinical Virology Lab | Grubaugh Lab - Yale School of Public Health | Anderson Brito; Anne Wyllie; Annie Watkins; Chaney Kalinich; Chantal Vogels; Isabel Ott; Jessica Rothman; Joseph Fauver; Mallery Breban; Marie L. Landry; Mary Petrone; Nathan Grubaugh; Tara Alpert |
| EPI_ISL_2020846, EPI_ISL_2035072, EPI_ISL_2035093 | Yale Clinical Virology Lab | Yale Center for Genomic Analysis | Brooke Sullivan; Curt Scharfe; Irina Tikhonova; Kaya Bilguvar; Shrikant Mane |
