## Supplement 3 for "Rapid and Accurate Identification of SARS-CoV-2 Omicron Variants Using Droplet Digital PCR (RT-ddPCR)": All_0426-0502.pdf

We gratefully acknowledge the following Authors from the Originating laboratories responsible for obtaining the specimens, as well as the Submitting laboratories where the genome data were generated and shared via GISAID, on which this research is based.

All Submitters of data may be contacted directly via www.gisaid.org

Authors are sorted alphabetically.

| Accession ID | Originating Laboratory | Submitting Laboratory | Authors |
|---|---|---|---|
| EPI_ISL_3354922 | "AR Dept. of Health-PHL, Molecular Diagnostics" | Centers for Disease Control and Prevention Division of Viral Diseases, Pathogen Discovery | Alex Burgin; Ben Rambo-Martin; Clinton Paden; Dakota Howard; Dave Wentworth; Dhwani Batra; Jasmine Padilla; Justin Lee; Krista Queen; Kristen Knipe; Kristine Lacek; Mark Burroughs; Matthew Schmerer; Meghan Bentz; Mili Sheth; Peter Cook; Sam Shepard; Sarah Nobles; Suxiang Tong; Vivien Dugan; Yvette Unoarumhi |
| EPI_ISL_5899625, EPI_ISL_5899626, EPI_ISL_5899627, EPI_ISL_5899628 | A. Krumbholz, Labor Dr. Krause und Kollegen MVZ GmbH, Kiel | Charité Universitätsmedizin Berlin, Institut für Virologie | Barbara Mühlemann; Christian Drosten; Julia Schneider; Julia Tesch; Jörn Beheim-Schwarzbach; Talitha Veith; Terry Jones; Tobias Bleicker; Victor M Corman |
| EPI_ISL_3347189, EPI_ISL_3347190, EPI_ISL_3347196, EPI_ISL_3347197, EPI_ISL_3347198, EPI_ISL_3347199, EPI_ISL_3347200, EPI_ISL_3347201 | | | |
| see above | ADILAB | Laboratorio Departamental de Salud Publica de Antioquia | Andres F. Cardona-Rios; Gloria Isabel Escobar; Idabely Betancur Ortiz; Juan P. Hernandez-Ortiz; Maria Stella López |
| EPI_ISL_5782505, EPI_ISL_5782506, EPI_ISL_5782507, EPI_ISL_5782508, EPI_ISL_5782509, EPI_ISL_5782510, EPI_ISL_5782511, EPI_ISL_5782512, EPI_ISL_5782513, EPI_ISL_5782514, EPI_ISL_5782515, EPI_ISL_5782516, EPI_ISL_5782517, EPI_ISL_5782518, EPI_ISL_5782519, EPI_ISL_5782520, EPI_ISL_5782521, EPI_ISL_5782522, EPI_ISL_5782523, EPI_ISL_5782524, EPI_ISL_5782525, EPI_ISL_5782526, EPI_ISL_5782527, EPI_ISL_5782528, EPI_ISL_5782529, EPI_ISL_5782530, EPI_ISL_5782531, EPI_ISL_5782532, EPI_ISL_5782533, EPI_ISL_5782534, EPI_ISL_5782535, EPI_ISL_5782536, EPI_ISL_5782537, EPI_ISL_5782538, EPI_ISL_5782539, EPI_ISL_5782540, EPI_ISL_5782541 | | | |
| see above | AFIP | Instituto Butantan | Antonio Jorge Martins; Claudia Renata dos Santos Barros; David Schlesinger; Debora Botequio Moretti; Dimas Tadeu Covas; Elaine Cristina Marqueze; Elaine Vieira Santos; Evandra Strazza Rodrigues; Heidge Fukumasu; Jayme Augusto de Souza-Neto; José Salvatore Leister Patané; Luiz Alcantara; Luiz Lehmann Coutinho; Maria Carolina Elias; Maurício Lacerda Nogueira; Rafael dos Santos Bezerra; Raul Machado Neto; Rejane Maria Tommasini Grotto; Ricardo Haddad; Sandra Coccuzzo Sampaio Vessoni; Simone Kashima; Svetoslav Nanev Slavov; Vincent Louis Viala |
| EPI_ISL_2229280, EPI_ISL_2229281, EPI_ISL_2229282 | AL Dept. of Public Health Bureau of Clinical Laboratories | Centers for Disease Control and Prevention Division of Viral Diseases, Pathogen Discovery | Alison Laufer Halpin; Ben L. Rambo-Martin; Clinton R. Paden; Dakota Howard; Darlene Wagner; Dave Wentworth; Dhwani Batra; Jasmine Padilla; Justin Lee; Katie Dillon; Krista Queen; Kristen Knipe; Kristine Lacek; Mark Burroughs; Matthew Schmerer; Mili Sheth; Peter Cook; Sam Shepard; Sarah Nobles; Shoshona Le; Suxiang Tong; Vivien Dugan; Yvette Unoarumhi |
| EPI_ISL_2098500, EPI_ISL_2274622, EPI_ISL_2274623 | ALBANY MEDICAL CENTER | Wadsworth Center, New York State Department of Health | Alexis Russell; Catharine Prussing; Daryl M. Lamson; Erasmus Schneider; Erica Lasek-Nesselquist; John Kelly; Jonathan Plitnick; Kirsten St. George; Matthew Shudt; Melissa A Leisner; Navjot Singh |
| EPI_ISL_5915135, EPI_ISL_5915151, EPI_ISL_5915224, EPI_ISL_5915251, EPI_ISL_5915252, EPI_ISL_5915253, EPI_ISL_5915327, EPI_ISL_6573822, EPI_ISL_6573823, EPI_ISL_6573862, EPI_ISL_6573866, EPI_ISL_6573879, EPI_ISL_6573957 | | | |
| see above | AMALAB/FACISA/UFRN | WallauLab on behalf of Fiocruz COVID-19 Genomic Surveillance Network | Alexandre Freitas da Silva; Allan Roberto Dias Nunes; Antonio Marinho da Silva Neto; Cassia Docena; Constância Flávia Junqueira Ayres; Filipe Zimmer Dezordi; Gabriel Luz Wallau; Gustavo Barbosa de Lima; Joana Cristina Medeiros Tavares Marques; Katya Anaya Jacinto; Lais Ceschini Machado; Lilian Caroliny Amorim Silva; Marcelo Henrique dos Santos Paiva; Mariane dos Santos Duarte; Matheus Filgueira Bezerra; Sinval Pinto Brandão Filho |
| EPI_ISL_2155825 | AR Dept. of Health-PHL, Molecular Diagnostics | Pathogen Discovery, Respiratory Viruses Branch, Division of Viral Diseases, Centers for Disease Control and Prevention | Adam Retchless; Anna Kelleher; Anna Uehara; Brian Lynch; Clinton R. Paden; Dhwani Batra; Haibin Wang; Han Jia Justin Ng; Jasmine Padilla; Jing Zhang; Justin Lee; Krista Queen; Mark Burroughs; Mili Sheth; Morgan Davis; Peter Cook; Rachel Marine; Sarah Nobles; Suxiang Tong; Tara Coalter; Yan Li; Ying Tao |
| EPI_ISL_2502728 | AREA DE SALUD CARRILLO | Inciensa, Instituto Costarricense de InvestigaciOn y Enseñanza en NutriciOn y Salud | Adriana Godínez; Claudio Soto-Garita; Estela Cordero; Francisco Duarte; Hebleen Porras; Jose Luis Vargas; Joselyn Prado & Ivanna Krize-Morún; Mariela Gutierrez; Melany CalderOn |
| EPI_ISL_2502725 | AREA DE SALUD CURRIDABAT 2 | Inciensa, Instituto Costarricense de InvestigaciOn y Enseñanza en NutriciOn y Salud | Adriana Godínez; Claudio Soto-Garita; Estela Cordero; Francisco Duarte; Hebleen Porras; Jose Luis Vargas; Joselyn Prado & Mariel LOpez; Mariela Gutierrez; Melany CalderOn |
| EPI_ISL_2273004 | AREA DE SALUD SAN JUAN-SAN DIEGO-CONCEPCION 2 | Inciensa, Instituto Costarricense de Investigación y Enseñanza en Nutrición y Salud | Adriana Godínez; Claudio Soto-Garita; Estela Cordero; Francisco Duarte; Hebleen Porras; Joselyn Prado & Mariel López; José Luis Vargas; Mariela Gutiérrez; Melany Calderón |
| EPI_ISL_2658269 | AREA DE SALUD ZAPOTE-CATEDRAL - CLINICA DR. CARLOS DURAN | Inciensa, Instituto Costarricense de Investigación y Enseñanza en Nutrición y Salud | Adriana Godínez; Claudio Soto-Garita; Estela Cordero; Francisco Duarte; Hebleen Porras; Joselyn Prado & Andony Cordero-Jiménez; José Luis Vargas; Mariela Gutiérrez; Melany Calderón |
| EPI_ISL_4850555 | AYUDAS DIAGNOSTICAS SURA | Instituto Nacional de Salud- Dirección de Investigación en Salud Pública | Carlos Franco-Muñoz; Carmen Osorio; Diana Malo; Diego A. Álvarez-Díaz; Diego Andrés Prada; Gerardo Santamaría; Hector Alejandro Ruiz-Moreno; Jhonnatan Reales-González; Jorge Rivera; Juan Camilo Martinez; Julian Naizaque; Katherine Laiton-Donato; Lisseth Pardo; Magdalena Wiesner; Marcela Mercado-Reyes; Maria T. Herrera-Sepúlveda; Marta Lopez Blanco; Martha Lucia Ospina Martinez; Paola Rojas; Sergio Gomez; Sheryll Corchuelo; Ángela Alarcon Cruz |
| EPI_ISL_2137045, EPI_ISL_2137047, EPI_ISL_2137380 | AZ Klina | AZ Klina | Carl Vael - Lynsey Berckmans |
| EPI_ISL_2036101, EPI_ISL_2036102, EPI_ISL_2036113 | AZDelta | AZ Delta Medical Laboratories in Roeselare, Belgium | Dieter De Smet; Geert Martens; Merijn Vanhee; on behalf of AZ Delta COVID-19 Genomics core (member of Genomic surveillance of SARS-CoV-2 in Belgium network) |
| EPI_ISL_2438003 | Adilab | Universidad Nacional de Colombia - Laboratorio Genómico One Health | Andres F. Cardona-Rios; Carlos Franco-Muñoz; Carolina Muñoz-Arango; Celeny Ortiz; Daniel O. Maldonado-Perez; Diego A. Álvarez-Díaz; Hector Alejandro Ruiz-Moreno; Idabely Betancur Ortiz; Jorge E. Osorio; Juan P. Hernandez-Ortiz; Karl A Ciuoderis; Katherine Laiton-Donato; Laura Silvana Perez; Lina M. Hurtado; Marcela Mercado-Reyes; Maria Angélica Maya; Maria Stella López; Rita Almanza Payares; Sandra Ines Cano; Simón Villegas Velásquez |
| EPI_ISL_2039124, EPI_ISL_2039125, EPI_ISL_2039127, EPI_ISL_2039135, EPI_ISL_2039144, EPI_ISL_2039149, EPI_ISL_2039153, EPI_ISL_2039158, EPI_ISL_2039163, EPI_ISL_2039390, EPI_ISL_2039622, EPI_ISL_2039650, EPI_ISL_2039666, EPI_ISL_2039672, EPI_ISL_2041658, EPI_ISL_2041681, EPI_ISL_2041705, EPI_ISL_2041892, EPI_ISL_2041893, EPI_ISL_2041921, EPI_ISL_2041936, EPI_ISL_2041942, EPI_ISL_2041961, EPI_ISL_2041969, EPI_ISL_2042220, EPI_ISL_2042228, EPI_ISL_2042229, EPI_ISL_2042248, EPI_ISL_2042494, EPI_ISL_2042505, EPI_ISL_2042550, EPI_ISL_2042605, EPI_ISL_2042611, EPI_ISL_2042629, EPI_ISL_2042646, EPI_ISL_2042650, EPI_ISL_2042675, EPI_ISL_2042711, EPI_ISL_2042721, EPI_ISL_2042729, EPI_ISL_2042745, EPI_ISL_2042780, EPI_ISL_2042794, EPI_ISL_2042813, EPI_ISL_2042822, EPI_ISL_2042824, EPI_ISL_2042844, EPI_ISL_2042853, EPI_ISL_2042896, EPI_ISL_2042900, EPI_ISL_2042917, EPI_ISL_2042951, EPI_ISL_2042953, EPI_ISL_2042969, EPI_ISL_2042980, EPI_ISL_2043034, EPI_ISL_2043041, EPI_ISL_2043044, EPI_ISL_2043050, EPI_ISL_2043100, EPI_ISL_2043105, EPI_ISL_2043116, EPI_ISL_2043117, EPI_ISL_2043171, EPI_ISL_2043172, EPI_ISL_2043175, EPI_ISL_2043177, EPI_ISL_2043178, EPI_ISL_2043181, EPI_ISL_2043196, EPI_ISL_2043197, EPI_ISL_2043213, EPI_ISL_2043216, EPI_ISL_2043220, EPI_ISL_2043238, EPI_ISL_2043246, EPI_ISL_2043249, EPI_ISL_2043253, EPI_ISL_2043259, EPI_ISL_2043278, EPI_ISL_2043318, EPI_ISL_2043358, EPI_ISL_2043361, EPI_ISL_2043365, EPI_ISL_2043398, EPI_ISL_2043430, EPI_ISL_2043432, EPI_ISL_2043447, EPI_ISL_2043466, EPI_ISL_2043489, EPI_ISL_2043495, EPI_ISL_2043499, EPI_ISL_2043503, EPI_ISL_2043505, EPI_ISL_2043511, EPI_ISL_2043523, EPI_ISL_2043527, EPI_ISL_2043535, EPI_ISL_2043545, EPI_ISL_2043549, EPI_ISL_2089455, EPI_ISL_2089479, EPI_ISL_2089502, EPI_ISL_2089506, EPI_ISL_2089512, EPI_ISL_2089516, EPI_ISL_2089527, EPI_ISL_2089660, EPI_ISL_2089680, EPI_ISL_2089686, EPI_ISL_2089707, EPI_ISL_2089739, EPI_ISL_2089742, EPI_ISL_2089782, EPI_ISL_2089785, EPI_ISL_2089786, EPI_ISL_2089832, EPI_ISL_2089842, EPI_ISL_2089849, EPI_ISL_2089852, EPI_ISL_2089953, EPI_ISL_2090063, EPI_ISL_2090311, EPI_ISL_2090317, EPI_ISL_2090329, EPI_ISL_2145803, EPI_ISL_2145804, EPI_ISL_2145808, EPI_ISL_2145821, EPI_ISL_2145823, EPI_ISL_2145837, EPI_ISL_2145841, EPI_ISL_2145845, EPI_ISL_2145851, EPI_ISL_2145866, EPI_ISL_2145869, EPI_ISL_2145870, EPI_ISL_2145884, EPI_ISL_2145891, EPI_ISL_2145899, EPI_ISL_2145903, EPI_ISL_2145911, EPI_ISL_2145912, EPI_ISL_2145914, EPI_ISL_2145925, EPI_ISL_2145965, EPI_ISL_2145972, EPI_ISL_2145979, EPI_ISL_2146010, EPI_ISL_2147667, EPI_ISL_2147671, EPI_ISL_2147755, EPI_ISL_2147815, EPI_ISL_2147852, EPI_ISL_2147920, EPI_ISL_2147921, EPI_ISL_2148006, EPI_ISL_2148008, EPI_ISL_2148024, EPI_ISL_2148030, EPI_ISL_2148036, EPI_ISL_2148037, EPI_ISL_2148257, EPI_ISL_2148258, EPI_ISL_2148264, EPI_ISL_2148267, EPI_ISL_2148277, EPI_ISL_2148292, EPI_ISL_2148312, EPI_ISL_2148322, EPI_ISL_2148362, EPI_ISL_2148373, EPI_ISL_2148378, EPI_ISL_2148403, EPI_ISL_2148406, EPI_ISL_2148415, EPI_ISL_2148433, EPI_ISL_2148451, EPI_ISL_2148505, EPI_ISL_2148507, EPI_ISL_2148521, EPI_ISL_2148527, EPI_ISL_2148541, EPI_ISL_2148570, EPI_ISL_2148576, EPI_ISL_2148592, EPI_ISL_2148619, EPI_ISL_2148632, EPI_ISL_2148635, EPI_ISL_2148638, EPI_ISL_2148644, EPI_ISL_2148651, EPI_ISL_2148668, EPI_ISL_2148703, EPI_ISL_2148710, EPI_ISL_2148728, EPI_ISL_2148762, EPI_ISL_2148763, EPI_ISL_2148766, EPI_ISL_2148767, EPI_ISL_2148768, EPI_ISL_2148778, EPI_ISL_2148782, EPI_ISL_2148925, EPI_ISL_2149437, EPI_ISL_2149442, EPI_ISL_2149448, EPI_ISL_2149449, EPI_ISL_2150138, EPI_ISL_2150139, EPI_ISL_2150140, EPI_ISL_2150151, EPI_ISL_2150159, EPI_ISL_2150172, EPI_ISL_2150178, EPI_ISL_2150295, EPI_ISL_2150494, EPI_ISL_2150501, EPI_ISL_2150507, EPI_ISL_2150522, EPI_ISL_2150535, EPI_ISL_2150539, EPI_ISL_2150542, EPI_ISL_2150544, EPI_ISL_2150554, EPI_ISL_2159209, EPI_ISL_2159215, EPI_ISL_2159220, EPI_ISL_2159250, EPI_ISL_2159252, EPI_ISL_2159263, EPI_ISL_2159269, EPI_ISL_2159430, EPI_ISL_2159431, EPI_ISL_2187359, EPI_ISL_2187363, EPI_ISL_2187490, EPI_ISL_2187492, EPI_ISL_2187496, EPI_ISL_2187505, EPI_ISL_2187513, EPI_ISL_2187582, EPI_ISL_2187608, EPI_ISL_2187614, EPI_ISL_2489829, EPI_ISL_2489837, EPI_ISL_2489843, EPI_ISL_2489846, EPI_ISL_2489877, EPI_ISL_2489883, EPI_ISL_2489884, EPI_ISL_2489887, EPI_ISL_4360241, EPI_ISL_4360581, EPI_ISL_4361184, EPI_ISL_4362196, EPI_ISL_4363398, EPI_ISL_4363791, EPI_ISL_4364813, EPI_ISL_4364917, EPI_ISL_4365865, EPI_ISL_4365909 | | | |
| see above | Aegis Sciences Corporation | Centers for Disease Control and Prevention Division of Viral Diseases, Pathogen Discovery | Adrian Paskey; Alec Vest; Benjamin Rambo-Martin; Christopher Gulvick; Clinton Paden; Clinton R. Paden; Cyndi Clark; Dakota Howard; Darlene Wagner; Dhwani Batra; Dillon Nall; Duncan MacCannell; Erisa Sula; Ethan Sanders; Holly Houdeshell; Jason Caravas; Kara Moser; Kristine Lacek; Matthew Hardison; Matthew Schmerer; Ola Kvalvaag; Patrick Campbell; Peter Cook; Peter W. Cook; Rob Case; Scott Sammons; Shatavia Morrison; Shaun Westlund; Tymeckia Kendall; Victoria Caban Figueroa; Vikramsinha Ghorpade; Yvette Unoarumhi |
| EPI_ISL_2946646 | Ages,Labor Dr. Mustafa_S | Elling and Cochella laboratories, IMBA/IMP | Alexander Stark; Ezgi Oezkan; Luisa Cochella and Ulrich Elling; Marcus Strobl; Olga Frank; Ramesh Yelagandula; Tanino Albanese |
| EPI_ISL_1967940, EPI_ISL_2008991, EPI_ISL_2429085, EPI_ISL_2429091, EPI_ISL_3118609 | Alaska State Virology Laboratory | Alaska State Virology Laboratory | Elva House; Jack Chen; Jacob Zidek; Lisa Smith; Ph.D.; Stephanie DeRonde |
| EPI_ISL_2137028 | Althaia. Xarxa Assistencial Universitària de Manresa | IrsiCaixa | Bonaventura Clotet; Bonaventura Clotet Gloria Trujillo; Carolina Gonzalez Fernandez; Eulalia Grau; Francesc Catala-Moll; Jaume Trape Pujol; Marc Noguera-Julian; Maria Casadellà; Mariona Parera; Miquel Micó; Pilar Armengol; Rafel Perez Vidal; Roger Paredes |
| EPI_ISL_5782492, EPI_ISL_5782493, EPI_ISL_5782494, EPI_ISL_5782495, EPI_ISL_5782496, EPI_ISL_5782497, EPI_ISL_5782498, EPI_ISL_5782499, EPI_ISL_5782500, EPI_ISL_5782501 | | | |
| see above | Ambulatorio Ib | Instituto Butantan | Antonio Jorge Martins; Claudia Renata dos Santos Barros; David Schlesinger; Debora Botequio Moretti; Dimas Tadeu Covas; Elaine Cristina Marqueze; Elaine Vieira Santos; Evandra Strazza Rodrigues; Heidge Fukumasu; Jayme Augusto de Souza-Neto; José Salvatore Leister Patané; Luiz Alcantara; Luiz Lehmann Coutinho; Maria Carolina Elias; Maurício Lacerda Nogueira; Rafael dos Santos Bezerra; Raul Machado Neto; Rejane Maria Tommasini Grotto; Ricardo Haddad; Sandra Coccuzzo Sampaio Vessoni; Simone Kashima; Svetoslav Nanev Slavov; Vincent Louis Viala |
| EPI_ISL_3015180 | Ancona | Microbiology University Politecnica delle Marche | Anna Valenza; Carla Acciarri; Katia Marinelli; Monica Lucia Ferreri; Patrizia Bagnarelli; Roberta Longo; Sara Caucci; Stefano Menzo |
| EPI_ISL_2082479 | Arizona State Public Health Laboratory | Arizona State Public Health Laboratory | Jessica Escobar; Katherine Fullerton; Linda Getsinger; Nobuko Fukushima; Stacy White; Trung Huynh; Victor Waddell |
| EPI_ISL_2007150, EPI_ISL_2007165, EPI_ISL_2007175, EPI_ISL_2007176, EPI_ISL_2007177, EPI_ISL_2007178, EPI_ISL_2153617, EPI_ISL_2153659, EPI_ISL_2153660, EPI_ISL_2153661, EPI_ISL_2153663, EPI_ISL_2153664, EPI_ISL_2250100, EPI_ISL_2250102, EPI_ISL_2250111, EPI_ISL_2250113, EPI_ISL_2250119, EPI_ISL_2250123, EPI_ISL_2250130, EPI_ISL_2250132, EPI_ISL_2250147 | | | |
| see above | Arizona State University | Arizona State University | Efrem S. Lim; Joshua LaBaer; Joy M. Blain; LaRinda A. Holland; Matthew F. Smith; Nicholas J. Mellor; Peter T. Skidmore; Rabia Maqsood; Valerie Harris; Vel Murugan |
| EPI_ISL_2484741 | Arkansas Children's Hospital | Center for Global Health, University of New Mexico Health Sciences Center | Amanda Novack; Darrell Dinwiddie; Daryl Domman; Dirk Haselow; Joshua L. Kennedy; Kurt Schwalm; Valerie Morley |

| | | | |
|---|---|---|---|
| EPI_ISL_2484740, EPI_ISL_2939042 | Arkansas Department of Health | Center for Global Health, University of New Mexico Health Sciences Center | Atul Kothari; Darrell Dinwiddie; Daryl Domman; Joshua L. Kennedy; Kurt Schwalm; Valerie Morley |
| EPI_ISL_3015177 | Ascoli Piceno | Microbiology University Politecnica delle Marche | Anna Valenza; Carla Acciarri; Katia Marinelli; Monica Lucia Ferreri; Patrizia Bagnarelli; Roberta Longo; Sara Caucci; Stefano Menzo |
| EPI_ISL_3276578, EPI_ISL_3276588, EPI_ISL_3276589, EPI_ISL_3276591, EPI_ISL_3276592, EPI_ISL_3276593, EPI_ISL_3276594, EPI_ISL_3276595, EPI_ISL_3276619, EPI_ISL_3721597 | | | |
| see above | Ayudas Diagnosticas Sura | Universidad Nacional de Colombia - Laboratorio Genómico One Health | Andres F. Cardona-Rios; Carlos Franco-Muñoz; Carolina Muñoz-Arango; Celeny Ortiz; Daniel O. Maldonado-Perez; Diego A. Álvarez-Díaz; Hector Alejandro Ruiz-Moreno; Idabely Betancur Ortiz; Jorge E. Osorio; Juan P. Hernandez-Ortiz; Karl A Ciuoderis; Katherine Laiton-Donato; Laura Silvana Perez; Lina M. Hurtado; Marcela Mercado-Reyes; Maria Angélica Maya; Maria Stella López; Rita Almanza Payares; Sandra Ines Cano; Simón Villegas Velásquez |
| EPI_ISL_2080258, EPI_ISL_2080274, EPI_ISL_2080282, EPI_ISL_2080294, EPI_ISL_2080295, EPI_ISL_2080297 | Azienda Sanitaria dell'Alto Adige Laboratorio Aziendale di Microbiologia e Virologia | Istituto di Genomica Applicata | Davide Scaglione; Eleonora Paparelli; Elisa Masi; Elisabetta Giacobazzi; Elisabetta Pagani; Gabriele Magris; Irena Jurman; Irene Bianconi; Michele Morgante; Stefanie Wieser; Vera Vendramin |
| EPI_ISL_2444173 | BADY BASSITT | Instituto Butantan / FAMERP | Antonio Jorge Martins; Claudia Renata dos Santos Barros; David Schlesinger; Debora Botequio Moretti; Dimas Tadeu Covas; Elaine Cristina Marqueze; Elaine Vieira Santos; Evandra Strazza Rodrigues; Heidge Fukumasu; Jayme Augusto de Souza-Neto; José Salvatore Leister Patané; Luiz Alcantara; Luiz Lehmann Coutinho; Maria Carolina Elias; Maurício Lacerda Nogueira; Rafael dos Santos Bezerra; Raul Machado Neto; Rejane Maria Tommasini Grotto; Ricardo Haddad; Sandra Coccuzzo Sampaio Vessoni; Simone Kashima; Svetoslav Nanev Slavov; Vincent Louis Viala |
| EPI_ISL_2656709, EPI_ISL_2656770, EPI_ISL_2656821, EPI_ISL_2656892, EPI_ISL_2656920, EPI_ISL_2656949, EPI_ISL_2656981, EPI_ISL_2657020, EPI_ISL_2657027, EPI_ISL_2657068, EPI_ISL_2657072, EPI_ISL_2657075, EPI_ISL_2657090, EPI_ISL_2657105, EPI_ISL_2719127, EPI_ISL_2719128, EPI_ISL_2719129, EPI_ISL_2719131, EPI_ISL_2719132, EPI_ISL_2719142, EPI_ISL_2719143, EPI_ISL_2719149, EPI_ISL_2719158, EPI_ISL_2719159, EPI_ISL_2719165, EPI_ISL_2719169, EPI_ISL_2719170, EPI_ISL_2719172, EPI_ISL_2719176, EPI_ISL_2719177, EPI_ISL_2719179, EPI_ISL_2719182, EPI_ISL_2719186, EPI_ISL_2719189, EPI_ISL_2719191, EPI_ISL_2719194, EPI_ISL_2719196, EPI_ISL_2719198, EPI_ISL_2719199, EPI_ISL_2719200, EPI_ISL_2719203, EPI_ISL_2719204, EPI_ISL_2719206, EPI_ISL_2719207, EPI_ISL_2719211, EPI_ISL_2719212, EPI_ISL_2719213, EPI_ISL_2719214, EPI_ISL_2719215, EPI_ISL_2719216, EPI_ISL_2719225, EPI_ISL_2719226, EPI_ISL_2719227, EPI_ISL_2719229, EPI_ISL_2719234, EPI_ISL_2719238, EPI_ISL_2719247, EPI_ISL_2719248, EPI_ISL_2719249, EPI_ISL_2719250, EPI_ISL_2719254, EPI_ISL_2719257, EPI_ISL_2719259, EPI_ISL_2719263, EPI_ISL_2719269, EPI_ISL_2719271, EPI_ISL_2719272, EPI_ISL_2719273, EPI_ISL_2719274, EPI_ISL_2719475, EPI_ISL_2719479, EPI_ISL_2719482, EPI_ISL_2719486, EPI_ISL_2719487, EPI_ISL_2719489, EPI_ISL_2719491, EPI_ISL_2719492, EPI_ISL_2719494, EPI_ISL_2719497, EPI_ISL_2719498, EPI_ISL_2719499, EPI_ISL_2719501, EPI_ISL_2719502, EPI_ISL_2719504, EPI_ISL_2719505, EPI_ISL_2719508, EPI_ISL_2719512, EPI_ISL_2719514, EPI_ISL_2719515, EPI_ISL_2719519, EPI_ISL_2719521, EPI_ISL_2719523, EPI_ISL_2719525, EPI_ISL_2719526, EPI_ISL_2719528, EPI_ISL_2719533, EPI_ISL_2719534, EPI_ISL_2719536, EPI_ISL_2719540, EPI_ISL_2719543, EPI_ISL_2719544, EPI_ISL_2719546, EPI_ISL_2719547, EPI_ISL_2719553, EPI_ISL_2719556, EPI_ISL_2719560, EPI_ISL_2719565, EPI_ISL_2719566, EPI_ISL_2719567, EPI_ISL_2719569, EPI_ISL_2719573, EPI_ISL_2719574, EPI_ISL_2719575, EPI_ISL_2719576, EPI_ISL_2719579, EPI_ISL_2719581, EPI_ISL_2719582, EPI_ISL_2719583, EPI_ISL_2719584, EPI_ISL_2719585, EPI_ISL_2719587, EPI_ISL_2719589, EPI_ISL_2719591, EPI_ISL_2719593, EPI_ISL_2719595, EPI_ISL_2719598, EPI_ISL_2719601, EPI_ISL_2719610, EPI_ISL_2719612, EPI_ISL_2719618, EPI_ISL_2719621, EPI_ISL_2719624, EPI_ISL_2719626, EPI_ISL_2719629, EPI_ISL_2719630, EPI_ISL_2719632, EPI_ISL_2719635, EPI_ISL_2719636, EPI_ISL_2719639, EPI_ISL_2719644, EPI_ISL_2719648, EPI_ISL_2719651, EPI_ISL_2719653, EPI_ISL_2719661, EPI_ISL_2719666, EPI_ISL_2719668, EPI_ISL_2719672, EPI_ISL_2719679, EPI_ISL_2719680, EPI_ISL_2719681, EPI_ISL_2719682, EPI_ISL_2719689, EPI_ISL_2719691, EPI_ISL_2719697, EPI_ISL_2719700, EPI_ISL_2719708, EPI_ISL_2719710, EPI_ISL_2719711, EPI_ISL_2719725, EPI_ISL_2719727, EPI_ISL_2719728, EPI_ISL_2719737, EPI_ISL_2719738, EPI_ISL_2719740, EPI_ISL_2719742, EPI_ISL_2719743, EPI_ISL_2719749, EPI_ISL_2719754, EPI_ISL_2719755, EPI_ISL_2719758, EPI_ISL_2719760, EPI_ISL_2719765, EPI_ISL_2719768, EPI_ISL_2719770, EPI_ISL_2719773, EPI_ISL_2719774, EPI_ISL_2719777, EPI_ISL_2719780, EPI_ISL_2719783, EPI_ISL_2719785, EPI_ISL_2719787, EPI_ISL_2719788, EPI_ISL_2719790, EPI_ISL_2719791, EPI_ISL_2719793, EPI_ISL_2719794, EPI_ISL_2719798, EPI_ISL_2719799, EPI_ISL_2719800, EPI_ISL_2719803, EPI_ISL_2719845, EPI_ISL_2719846, EPI_ISL_2719847, EPI_ISL_2719849, EPI_ISL_2719851, EPI_ISL_2719865, EPI_ISL_2719870, EPI_ISL_2719871, EPI_ISL_2719874, EPI_ISL_2719876, EPI_ISL_2719877, EPI_ISL_2719881, EPI_ISL_2719886, EPI_ISL_2719892, EPI_ISL_2719899, EPI_ISL_2719900, EPI_ISL_2719908, EPI_ISL_2719911, EPI_ISL_2719913, EPI_ISL_2719914, EPI_ISL_2719918, EPI_ISL_2719919, EPI_ISL_2719921, EPI_ISL_2719924, EPI_ISL_2719926, EPI_ISL_2719930, EPI_ISL_2719936, EPI_ISL_2719937, EPI_ISL_2719942, EPI_ISL_2719944, EPI_ISL_2719946, EPI_ISL_2719947, EPI_ISL_2719948, EPI_ISL_2719950, EPI_ISL_2719951, EPI_ISL_2719953, EPI_ISL_2719954, EPI_ISL_2719956, EPI_ISL_2719957, EPI_ISL_2719958, EPI_ISL_2719964, EPI_ISL_2719965, EPI_ISL_2719966, EPI_ISL_2719967, EPI_ISL_2719969, EPI_ISL_2719974, EPI_ISL_2719975, EPI_ISL_2719977, EPI_ISL_2719978, EPI_ISL_2719979, EPI_ISL_2719981, EPI_ISL_2719985, EPI_ISL_2719986, EPI_ISL_2720027, EPI_ISL_2720028, EPI_ISL_2720029, EPI_ISL_2720030, EPI_ISL_2720031, EPI_ISL_2720032, EPI_ISL_2720033, EPI_ISL_2720034, EPI_ISL_2720035, EPI_ISL_2720036, EPI_ISL_2720037, EPI_ISL_2720038, EPI_ISL_2720127, EPI_ISL_2720130, EPI_ISL_2720131, EPI_ISL_2720132, EPI_ISL_2720133, EPI_ISL_2720214, EPI_ISL_2720215, EPI_ISL_2720216, EPI_ISL_2720304, EPI_ISL_2720305, EPI_ISL_2720306, EPI_ISL_2720310, EPI_ISL_2720313, EPI_ISL_2720314, EPI_ISL_2720317, EPI_ISL_2720319, EPI_ISL_2720324, EPI_ISL_2720325, EPI_ISL_2720328, EPI_ISL_2720592, EPI_ISL_2720593, EPI_ISL_2720598, EPI_ISL_2720599, EPI_ISL_2720602, EPI_ISL_2720607, EPI_ISL_2720617, EPI_ISL_2720622, EPI_ISL_2720626, EPI_ISL_2720627, EPI_ISL_2720783, EPI_ISL_2720786, EPI_ISL_2720789, EPI_ISL_2720832, EPI_ISL_2720833, EPI_ISL_2720834, EPI_ISL_2720835, EPI_ISL_2720836, EPI_ISL_2720837, EPI_ISL_2720842, EPI_ISL_2720847, EPI_ISL_2720848, EPI_ISL_2720849, EPI_ISL_2720850, EPI_ISL_2720852, EPI_ISL_2720853, EPI_ISL_2720854, EPI_ISL_2720855, EPI_ISL_2720860, EPI_ISL_2720861, EPI_ISL_2720865, EPI_ISL_2720866, EPI_ISL_2720867, EPI_ISL_2720868, EPI_ISL_2720869, EPI_ISL_2720870, EPI_ISL_2720871, EPI_ISL_2720872, EPI_ISL_2720873, EPI_ISL_2720874, EPI_ISL_2720875, EPI_ISL_2720876, EPI_ISL_2720945, EPI_ISL_2720946, EPI_ISL_2720947, EPI_ISL_2720948, EPI_ISL_2720949, EPI_ISL_2720957, EPI_ISL_2720958, EPI_ISL_2720959, EPI_ISL_2720960, EPI_ISL_2720961, EPI_ISL_2720962, EPI_ISL_2720963, EPI_ISL_2720964, EPI_ISL_2720965, EPI_ISL_2720970, EPI_ISL_2720971, EPI_ISL_2720972, EPI_ISL_2720977, EPI_ISL_2720978, EPI_ISL_2720979, EPI_ISL_2720980, EPI_ISL_2720981, EPI_ISL_2720982, EPI_ISL_2720983, EPI_ISL_2720988, EPI_ISL_2720990, EPI_ISL_2720998, EPI_ISL_2720999, EPI_ISL_2721000, EPI_ISL_2721006, EPI_ISL_2721007, EPI_ISL_2721008, EPI_ISL_2721009, EPI_ISL_2721010, EPI_ISL_2721011, EPI_ISL_2721012, EPI_ISL_2721013, EPI_ISL_2721014, EPI_ISL_2721023, EPI_ISL_2721365, EPI_ISL_2721389, EPI_ISL_2721402, EPI_ISL_2721406, EPI_ISL_2721407, EPI_ISL_2721516, EPI_ISL_2721517, EPI_ISL_2721554, EPI_ISL_2721666, EPI_ISL_2721672, EPI_ISL_2721677, EPI_ISL_2721750, EPI_ISL_2721774, EPI_ISL_2721776, EPI_ISL_2721793, EPI_ISL_2721797, EPI_ISL_2721802, EPI_ISL_2721807, EPI_ISL_2721817, EPI_ISL_2721839, EPI_ISL_2721859, EPI_ISL_2721879, EPI_ISL_2721888, EPI_ISL_2721890, EPI_ISL_2721942, EPI_ISL_2721962, EPI_ISL_2721964 | | | |
| see above | BCCDC Public Health Laboratory | BCCDC Public Health Laboratory | Ana Pacagnella; Corrinne Ng; Dan Fornika; John Tyson; Kim Macdonald; Kimia Kamelian; Linda Hoang; Loretta Janz; Mel Krajden; Prystajecky Natalie; Robert Azana; Shannon Russell |
| EPI_ISL_2152465 | BIOMNIS EUROFINS IVRY | Department of Virology, Henri Mondor University Hospital, Assistance Publique Hôpitaux de Paris, Université Paris-Est Créteil, INSERM U955 | Alexandre Soulier; Christophe Rodriguez; Elisabeth Trawinski; Guillaume Gricourt; Jean-Michel Pawlotsky; Melissa N'Debi; Slim Fourati; Vanessa Demontant |
| EPI_ISL_2544513 | Baker lab at Loyola Medical Center | Baker lab at Loyola Medical Center | Amanda Harrington; Amornrat O'Brien; BS; Cynthia Rowe; Gail E. Reid; MD; Monika Evdokimova; Nina M. Clark; PhD; Susan Uprichard; Xufang Deng; and Susan C. Baker |
| EPI_ISL_7672122 | Bambino Gesù Pediatric Hospital | Microbiology and Immunology Diagnosis Bambino Gesù Pediatric Hospital | Carlo Federico Perno; Claudia Alteri; Luna Colagrossi; Rossana Scutari; Valentino Costabile |
| EPI_ISL_2260422, EPI_ISL_2262754, EPI_ISL_2262815, EPI_ISL_2390488 | Bayerisches Landesamt für Gesundheit und Lebensmittelsicherheit (LGL) | Robert Koch Institute | |
| EPI_ISL_1972301, EPI_ISL_2193229, EPI_ISL_2193230 | Baylor Scott & White-Temple | Baylor Scott & White-Temple | Ari Rao; Kimberly Walker; Linden Morales; Marcus Volz; Shelby Hendrickson |
| EPI_ISL_2204446 | Berkeley Medical Center | WVU and Marshall University Combined Genomics Core Facilities | James Denvir; Peter Perrotta; Peter Stoilov; Ryan Percifield; Wesley Kimble |
| EPI_ISL_5802500, EPI_ISL_5802501, EPI_ISL_5802502, EPI_ISL_5802532, EPI_ISL_5802533, EPI_ISL_5802534, EPI_ISL_5802536, EPI_ISL_5802537, EPI_ISL_5802541, EPI_ISL_5802542, EPI_ISL_5802543, EPI_ISL_5802544, EPI_ISL_5802545, EPI_ISL_5802546, EPI_ISL_5802547, EPI_ISL_5802548, EPI_ISL_5802549, EPI_ISL_5802550, EPI_ISL_5802551, EPI_ISL_5802553, EPI_ISL_5802554, EPI_ISL_5802555, EPI_ISL_5802558, EPI_ISL_5802574, EPI_ISL_5802575, EPI_ISL_5802577, EPI_ISL_5802578, EPI_ISL_5802595, EPI_ISL_5802598, EPI_ISL_5802599, EPI_ISL_5802600, EPI_ISL_5802601, EPI_ISL_5802602 | | | |
| see above | Biofast | Instituto Butantan | Antonio Jorge Martins; Claudia Renata dos Santos Barros; David Schlesinger; Debora Botequio Moretti; Dimas Tadeu Covas; Elaine Cristina Marqueze; Elaine Vieira Santos; Evandra Strazza Rodrigues; Heidge Fukumasu; Jayme Augusto de Souza-Neto; José Salvatore Leister Patané; Luiz Alcantara; Luiz Lehmann Coutinho; Maria Carolina Elias; Maurício Lacerda Nogueira; Rafael dos Santos Bezerra; Raul Machado Neto; Rejane Maria Tommasini Grotto; Ricardo Haddad; Sandra Coccuzzo Sampaio Vessoni; Simone Kashima; Svetoslav Nanev Slavov; Vincent Louis Viala |
| EPI_ISL_2151882 | Biogroup Bio Lam-LCD Saint-Denis | Department of Virology, Henri Mondor University Hospital, Assistance Publique Hôpitaux de Paris, Université Paris-Est Créteil, INSERM U955 | Alexandre Soulier; Christophe Rodriguez; Elisabeth Trawinski; Guillaume Gricourt; Jean-Michel Pawlotsky; Melissa N'Debi; Slim Fourati; Vanessa Demontant |
| EPI_ISL_2105670, EPI_ISL_2105671 | Biolab Diagnostic Laboratories | Biolab Diagnostic Laboratories | Ahmad Tibi; Amid Abdelnour; Badia Saddedin; Eiad Atwa; Issa Abu-Dayyeh; Lama Hussein; Shayma Ali |
| EPI_ISL_2698103, EPI_ISL_2698104 | Biology, UFLA, Universidade Federal de Lavras | Biology, UFLA | Barcante, J.; Cherem, J.; Fernandes, G.; Luciano, P.; Melo, D.; Pylro, V. |
| EPI_ISL_2401290, EPI_ISL_2401348, EPI_ISL_2401349, EPI_ISL_3145467, EPI_ISL_3145468, EPI_ISL_3145469, EPI_ISL_3145470, EPI_ISL_3145471 | | | |
| see above | BioneXt Lab | Laboratoire national de sante, Microbiology, Microbial Genomics Platform | Anke Wienecke-Baldacchino; Catherine Ragimbeau; Elodie Solarino; Fatu Djabi; Jessica Tapp; Lise Pignon; Raoul Salmon; Tamir Abdelrahman; Thibault Ferrandon; Virginie Jover |
| EPI_ISL_7570530, EPI_ISL_7570534, EPI_ISL_7592538 | Bluewater Diagnostic Laboratory, LLC | University of Louisville Sequencing Technology Center | Alonzo Shepherd; Aryan Neupane; Elizabeth Hudson; Eric C. Rouchka; Erik Korte; Julia H. Chariker; Melissa L. Smith; William Lauer |
| EPI_ISL_3494458, EPI_ISL_3494466, EPI_ISL_3494467 | Boston University CTL | Boston University/National Emerging Infectious Disease Laboratories | Catherine Klapperich; Jacquelyn Turcinovic; John H. Connor; Lena Landeverde; Lynn Doucette-Stamm |
| EPI_ISL_3478998, EPI_ISL_3479000, EPI_ISL_3479001, EPI_ISL_3479003, EPI_ISL_3479004, EPI_ISL_3479005, EPI_ISL_3479006, EPI_ISL_3481397, EPI_ISL_3481398, EPI_ISL_3481399, EPI_ISL_3481401, EPI_ISL_3481406, EPI_ISL_3481407, EPI_ISL_3481408, EPI_ISL_3481411, EPI_ISL_3481422, EPI_ISL_3481424, EPI_ISL_3481429, EPI_ISL_3481431, EPI_ISL_3481434, EPI_ISL_3481438, EPI_ISL_3481441, EPI_ISL_3481443, EPI_ISL_3481447, EPI_ISL_3481449, EPI_ISL_3481450, EPI_ISL_3481458, EPI_ISL_3481460, EPI_ISL_3481464, EPI_ISL_3481469, EPI_ISL_3481471, EPI_ISL_3481476, EPI_ISL_3481481, EPI_ISL_3481484, EPI_ISL_3481498, EPI_ISL_3481500, EPI_ISL_3481505, EPI_ISL_3481509, EPI_ISL_3481511, EPI_ISL_3481516, EPI_ISL_3481518, EPI_ISL_3481520, EPI_ISL_3481521, EPI_ISL_3481523, EPI_ISL_3481527, EPI_ISL_3481537, EPI_ISL_3481538, EPI_ISL_3481541, EPI_ISL_3481543, EPI_ISL_3481545, EPI_ISL_3481547, EPI_ISL_3481552, EPI_ISL_3481556, EPI_ISL_3481559, EPI_ISL_3481561, EPI_ISL_3481574, EPI_ISL_3481579, EPI_ISL_3481582, EPI_ISL_3481584, EPI_ISL_3481587, EPI_ISL_3481590, EPI_ISL_3481594, EPI_ISL_3481597, EPI_ISL_3481599, EPI_ISL_3481603, EPI_ISL_3481605, EPI_ISL_3481608, EPI_ISL_3481616, EPI_ISL_3481620, EPI_ISL_3481623, EPI_ISL_3481624, EPI_ISL_3481626, EPI_ISL_3481628, EPI_ISL_3481629, EPI_ISL_3481631, EPI_ISL_3481633, EPI_ISL_3481635, EPI_ISL_3481642, EPI_ISL_3481643, EPI_ISL_3481660, EPI_ISL_3481662, EPI_ISL_3481666, EPI_ISL_3481669, EPI_ISL_3481671, EPI_ISL_3481673, EPI_ISL_3481674, EPI_ISL_3481676, EPI_ISL_3481678, EPI_ISL_3481682, EPI_ISL_3481684, EPI_ISL_3481685, EPI_ISL_3481687, EPI_ISL_3481689, EPI_ISL_3481691, EPI_ISL_3481692, EPI_ISL_3481700, EPI_ISL_3481701, EPI_ISL_3481703, EPI_ISL_3481705, EPI_ISL_3481707, EPI_ISL_3481709, EPI_ISL_3481710, EPI_ISL_3481712, EPI_ISL_3481714, EPI_ISL_3481715, EPI_ISL_3481721, EPI_ISL_3481724, EPI_ISL_3481725, EPI_ISL_3481727, EPI_ISL_3481728, EPI_ISL_3481736, EPI_ISL_3481739, EPI_ISL_3481741, EPI_ISL_3481743, EPI_ISL_3481745, EPI_ISL_3481748, EPI_ISL_3481750, EPI_ISL_3481756, EPI_ISL_3481757, EPI_ISL_3481761, EPI_ISL_3481762, EPI_ISL_3481764, EPI_ISL_3481771, EPI_ISL_3481773, EPI_ISL_3481774, EPI_ISL_3481775, EPI_ISL_3481789, EPI_ISL_3481791, EPI_ISL_3481798, EPI_ISL_3481800, EPI_ISL_3481803, EPI_ISL_3481804, EPI_ISL_3481806, EPI_ISL_3481809, EPI_ISL_3481811, EPI_ISL_3481812, EPI_ISL_3481813, EPI_ISL_3481814, EPI_ISL_3481815, EPI_ISL_3481817, EPI_ISL_3481818, EPI_ISL_3481822, EPI_ISL_3481829, EPI_ISL_3481831, EPI_ISL_3481832, EPI_ISL_3481833, EPI_ISL_3481834, EPI_ISL_3481836, EPI_ISL_3481837, EPI_ISL_3481839, EPI_ISL_3481840, EPI_ISL_3481841, EPI_ISL_3481843, EPI_ISL_3481844, EPI_ISL_3481845, EPI_ISL_3481847, EPI_ISL_3481848, EPI_ISL_3481862, EPI_ISL_3481863, EPI_ISL_3481865, EPI_ISL_3481866, EPI_ISL_3481867, EPI_ISL_3481869, EPI_ISL_3481870, EPI_ISL_3481876, EPI_ISL_3481878, EPI_ISL_3481879, EPI_ISL_3481880, EPI_ISL_3481883, EPI_ISL_3481884, EPI_ISL_3481885, EPI_ISL_3481886, EPI_ISL_3481888, EPI_ISL_3481889, EPI_ISL_3481891, EPI_ISL_3481892, EPI_ISL_3481893, EPI_ISL_3481894, EPI_ISL_3481895, EPI_ISL_3481898, EPI_ISL_3481899, EPI_ISL_3481900, EPI_ISL_3481902, EPI_ISL_3481904, EPI_ISL_3481905, EPI_ISL_3481906, EPI_ISL_3481907, EPI_ISL_3481911, EPI_ISL_3481912, EPI_ISL_3481917, EPI_ISL_3481921, EPI_ISL_3481922, EPI_ISL_3481923, EPI_ISL_3481924, EPI_ISL_3481925, EPI_ISL_3481928, EPI_ISL_3481929, EPI_ISL_3481930, EPI_ISL_3481931, EPI_ISL_3481932, EPI_ISL_3481934, EPI_ISL_3481935, EPI_ISL_3481936, EPI_ISL_3481939, EPI_ISL_3481941, EPI_ISL_3481942, EPI_ISL_3481943, EPI_ISL_3481944, EPI_ISL_3481947, EPI_ISL_3481948, EPI_ISL_3481952, EPI_ISL_3481954, EPI_ISL_3481955, EPI_ISL_3481960, EPI_ISL_3481962, EPI_ISL_3481964, EPI_ISL_3481966, EPI_ISL_3481968, EPI_ISL_3481969, EPI_ISL_3481970, EPI_ISL_3481971, EPI_ISL_3481973, EPI_ISL_3481976, EPI_ISL_3481981, EPI_ISL_3481983, EPI_ISL_3481991, EPI_ISL_3481993, EPI_ISL_3481999, EPI_ISL_3482001, EPI_ISL_3482004, EPI_ISL_3482006, EPI_ISL_3482010, EPI_ISL_3482021, EPI_ISL_3482026, EPI_ISL_3482029, EPI_ISL_3482030, EPI_ISL_3482031, EPI_ISL_3482032, EPI_ISL_3482033, EPI_ISL_3482035, EPI_ISL_3482037, EPI_ISL_3482041, EPI_ISL_3482047, EPI_ISL_3482054, EPI_ISL_3482056, EPI_ISL_3482059, EPI_ISL_3482061, EPI_ISL_3482068, EPI_ISL_3482079, EPI_ISL_3482081, EPI_ISL_3482083, EPI_ISL_3482087, EPI_ISL_3482089, EPI_ISL_3482091, EPI_ISL_3482092, EPI_ISL_3482093, EPI_ISL_3482094, EPI_ISL_3482095, EPI_ISL_3482097, EPI_ISL_3482099, EPI_ISL_3482100, EPI_ISL_3482101, EPI_ISL_3482102, EPI_ISL_3482105, EPI_ISL_3482106, EPI_ISL_3482107, EPI_ISL_3482109, EPI_ISL_3482111, EPI_ISL_3482112, EPI_ISL_3482113, EPI_ISL_3482116, EPI_ISL_3482117, EPI_ISL_3482119, EPI_ISL_3482122, EPI_ISL_3482124, EPI_ISL_3482125, EPI_ISL_3482126, EPI_ISL_3482129, EPI_ISL_3482131, EPI_ISL_3482132, EPI_ISL_3482133, EPI_ISL_3482134, EPI_ISL_3482135, EPI_ISL_3482136, EPI_ISL_3482137, EPI_ISL_3482146, EPI_ISL_3482147, EPI_ISL_3482149, EPI_ISL_3482150, EPI_ISL_3482151, EPI_ISL_3482152, EPI_ISL_3482156, EPI_ISL_3482157, EPI_ISL_3482158, EPI_ISL_3482159, EPI_ISL_3482160, EPI_ISL_3482164, EPI_ISL_3482166, EPI_ISL_3482167, EPI_ISL_3482168, EPI_ISL_3482169, EPI_ISL_3482171, EPI_ISL_3482173, EPI_ISL_3482176, EPI_ISL_3482177, EPI_ISL_3482178, EPI_ISL_3482179, EPI_ISL_3482180, EPI_ISL_3482181, EPI_ISL_3482183, EPI_ISL_3482184, EPI_ISL_3482185, EPI_ISL_3482186, EPI_ISL_3482189, EPI_ISL_3482190, EPI_ISL_3482192, EPI_ISL_3482193, EPI_ISL_3482194, EPI_ISL_3482195, EPI_ISL_3482197, EPI_ISL_3482198, EPI_ISL_3482200, EPI_ISL_3482202, EPI_ISL_3482203, EPI_ISL_3482204, EPI_ISL_3482206, EPI_ISL_3482207, EPI_ISL_3482208, EPI_ISL_3482209, EPI_ISL_3482211, EPI_ISL_3482212, EPI_ISL_3482216, EPI_ISL_3482217, EPI_ISL_3482218, EPI_ISL_3482221, EPI_ISL_3482222, EPI_ISL_3482224, EPI_ISL_3482227, EPI_ISL_3482229, EPI_ISL_3482230, EPI_ISL_3482231, EPI_ISL_3482233, EPI_ISL_3482234, EPI_ISL_3482235, EPI_ISL_3482236, EPI_ISL_3482238, EPI_ISL_3482239, EPI_ISL_3482240, EPI_ISL_3482241, EPI_ISL_3482242, EPI_ISL_3482243, EPI_ISL_3482247, EPI_ISL_3482248, EPI_ISL_3482251, EPI_ISL_3482254, EPI_ISL_3482255, EPI_ISL_3482256, EPI_ISL_3482257, EPI_ISL_3482258, EPI_ISL_3482259, EPI_ISL_3482260, EPI_ISL_3482261, EPI_ISL_3482262, EPI_ISL_3482263, EPI_ISL_3482264, EPI_ISL_3482265, EPI_ISL_3482266, EPI_ISL_3482269, EPI_ISL_3482270, EPI_ISL_3482271, EPI_ISL_3482272, EPI_ISL_3482275, EPI_ISL_3482276, EPI_ISL_3482278, EPI_ISL_3482281, EPI_ISL_3482282, EPI_ISL_3482285, EPI_ISL_3482287, EPI_ISL_3482289, EPI_ISL_3482290, EPI_ISL_3482291, EPI_ISL_3482293, EPI_ISL_3482294, EPI_ISL_3482296, EPI_ISL_3482297, EPI_ISL_3482298, EPI_ISL_3482299, EPI_ISL_3482300, EPI_ISL_3482301, EPI_ISL_3482302, EPI_ISL_3482303, EPI_ISL_3482306, EPI_ISL_3482307, EPI_ISL_3482308, EPI_ISL_3482309, EPI_ISL_3482310, EPI_ISL_3482311, EPI_ISL_3482312, EPI_ISL_3482313, EPI_ISL_3482314, EPI_ISL_3482315, EPI_ISL_3482317, EPI_ISL_3482318, EPI_ISL_3482319, EPI_ISL_3482320, EPI_ISL_3482321, EPI_ISL_3482322, EPI_ISL_3482323, EPI_ISL_3482324, EPI_ISL_3482327, EPI_ISL_3482328, EPI_ISL_3482329, EPI_ISL_3482332, EPI_ISL_3482333, EPI_ISL_3482335, EPI_ISL_3482336, EPI_ISL_3482338, EPI_ISL_3482341, EPI_ISL_3482342, EPI_ISL_3482343, EPI_ISL_3482344, EPI_ISL_3482345, EPI_ISL_3482352, EPI_ISL_3482353, EPI_ISL_3482355, EPI_ISL_3482357, EPI_ISL_3482360, EPI_ISL_3482361, EPI_ISL_3482363, EPI_ISL_3482364, EPI_ISL_3482365, EPI_ISL_3482366, EPI_ISL_3482371, EPI_ISL_3482374, EPI_ISL_3482375, EPI_ISL_3482376, EPI_ISL_3482377, EPI_ISL_3482378, EPI_ISL_3482379, EPI_ISL_3482380, EPI_ISL_3482382, EPI_ISL_3482383, EPI_ISL_3482385, EPI_ISL_3482387, EPI_ISL_3482390, EPI_ISL_3482391, EPI_ISL_3482392, EPI_ISL_3482396, EPI_ISL_3482398, EPI_ISL_3482400, EPI_ISL_3482401, EPI_ISL_3482402, EPI_ISL_3482403, EPI_ISL_3482404, EPI_ISL_3482406, EPI_ISL_3482407, EPI_ISL_3482409, EPI_ISL_3482411, EPI_ISL_3482412, EPI_ISL_3482413, EPI_ISL_3482414, EPI_ISL_3482416, EPI_ISL_3482417, EPI_ISL_3482419, EPI_ISL_3482420, EPI_ISL_3482421, EPI_ISL_3482423, EPI_ISL_3482424, EPI_ISL_3482426, EPI_ISL_3482429, EPI_ISL_3482430, EPI_ISL_3482431, EPI_ISL_3482432, EPI_ISL_3482433, EPI_ISL_3482434, EPI_ISL_3482436, EPI_ISL_3482439, EPI_ISL_3482442, EPI_ISL_3482443, EPI_ISL_3482448, EPI_ISL_3482449, EPI_ISL_3482451, EPI_ISL_3482452, EPI_ISL_3482455, EPI_ISL_3482458, EPI_ISL_3482459, EPI_ISL_3482461, EPI_ISL_3482462, EPI_ISL_3482465, EPI_ISL_3482466, EPI_ISL_3482467, EPI_ISL_3482468, EPI_ISL_3482475, EPI_ISL_3482476, EPI_ISL_3482477, EPI_ISL_3482479, EPI_ISL_3482481, EPI_ISL_3482483, EPI_ISL_3482486, EPI_ISL_3482488, EPI_ISL_3482491, EPI_ISL_3482492, EPI_ISL_3482496, EPI_ISL_3482497, EPI_ISL_3482498, EPI_ISL_3482501, | | | |

| | | | |
|---|---|---|---|
| EPI_ISL_3482502, EPI_ISL_3482503, EPI_ISL_3482504, EPI_ISL_3482508, EPI_ISL_3482509, EPI_ISL_3482510, EPI_ISL_3482515, EPI_ISL_3482527, EPI_ISL_3482532, EPI_ISL_3482535, EPI_ISL_3482537, EPI_ISL_3482540, EPI_ISL_3482545, EPI_ISL_3482548, EPI_ISL_3482550, EPI_ISL_3482553, EPI_ISL_3482570, EPI_ISL_3482573, EPI_ISL_3482576, EPI_ISL_3482584, EPI_ISL_3482589, EPI_ISL_3482591, EPI_ISL_3482594, EPI_ISL_3482599, EPI_ISL_3482612, EPI_ISL_3482621, EPI_ISL_3482622, EPI_ISL_3482626, EPI_ISL_3482627, EPI_ISL_3482640, EPI_ISL_3482641, EPI_ISL_3482642, EPI_ISL_3482645, EPI_ISL_3482646, EPI_ISL_3482647, EPI_ISL_3482648, EPI_ISL_3482650, EPI_ISL_3482652, EPI_ISL_3482654, EPI_ISL_3482655, EPI_ISL_3482656, EPI_ISL_3482658, EPI_ISL_3482659, EPI_ISL_3482660, EPI_ISL_3482661, EPI_ISL_3482667, EPI_ISL_3482669, EPI_ISL_3482672, EPI_ISL_3482674, EPI_ISL_3482679, EPI_ISL_3482680, EPI_ISL_3482718, EPI_ISL_3482721, EPI_ISL_3482724, EPI_ISL_3482941, EPI_ISL_3483081, EPI_ISL_3483084, EPI_ISL_3491224, EPI_ISL_3596356, EPI_ISL_3596359, EPI_ISL_3596365, EPI_ISL_3596368, EPI_ISL_3596382, EPI_ISL_3596385, EPI_ISL_3599156, EPI_ISL_3599174, EPI_ISL_3599175, EPI_ISL_3599182, EPI_ISL_3599210, EPI_ISL_3599211, EPI_ISL_3599218, EPI_ISL_3599263, EPI_ISL_3599288, EPI_ISL_3599294, EPI_ISL_3599296, EPI_ISL_3599303, EPI_ISL_3599308, EPI_ISL_3599325, EPI_ISL_3599327, EPI_ISL_3599334, EPI_ISL_3599345, EPI_ISL_3599352, EPI_ISL_3599364, EPI_ISL_3599366, EPI_ISL_3599367, EPI_ISL_3599369, EPI_ISL_3599385, EPI_ISL_3599387, EPI_ISL_3599393, EPI_ISL_3599396, EPI_ISL_3599408, EPI_ISL_3599411, EPI_ISL_3599413, EPI_ISL_3599416, EPI_ISL_3599447, EPI_ISL_3599460, EPI_ISL_3599462, EPI_ISL_3599463, EPI_ISL_3599464, EPI_ISL_3599469, EPI_ISL_3599473, EPI_ISL_3599486, EPI_ISL_3599500, EPI_ISL_3599501, EPI_ISL_3599503, EPI_ISL_3599508, EPI_ISL_3599521, EPI_ISL_3599527, EPI_ISL_3599536, EPI_ISL_3599540, EPI_ISL_3599551, EPI_ISL_3599556, EPI_ISL_3599560, EPI_ISL_3599561, EPI_ISL_3599562, EPI_ISL_3599566, EPI_ISL_3599567, EPI_ISL_3599568, EPI_ISL_3599582, EPI_ISL_3599593, EPI_ISL_3599594, EPI_ISL_3599600, EPI_ISL_3599619, EPI_ISL_3599625, EPI_ISL_3599629, EPI_ISL_3599659, EPI_ISL_3599660, EPI_ISL_3599661, EPI_ISL_3599679, EPI_ISL_3599685, EPI_ISL_3599686, EPI_ISL_3599706, EPI_ISL_3599711, EPI_ISL_3599712, EPI_ISL_3599713, EPI_ISL_3599724, EPI_ISL_3599728, EPI_ISL_3599732, EPI_ISL_3599749, EPI_ISL_3599762, EPI_ISL_3599768, EPI_ISL_3599772, EPI_ISL_3599804, EPI_ISL_3599810, EPI_ISL_3599812, EPI_ISL_3599840, EPI_ISL_3599841, EPI_ISL_3599850, EPI_ISL_3599852, EPI_ISL_3599854, EPI_ISL_3599856, EPI_ISL_3599871, EPI_ISL_3599877, EPI_ISL_3599881, EPI_ISL_3599886, EPI_ISL_3599891, EPI_ISL_3599897, EPI_ISL_3602426, EPI_ISL_4420711, EPI_ISL_4420712, EPI_ISL_4423606, EPI_ISL_4423637, EPI_ISL_4423642, EPI_ISL_4424916, EPI_ISL_4424923, EPI_ISL_4424927, EPI_ISL_4424932, EPI_ISL_4424934, EPI_ISL_4424936, EPI_ISL_4424938, EPI_ISL_4424941, EPI_ISL_4424946, EPI_ISL_4424948, EPI_ISL_4424951, EPI_ISL_4424956, EPI_ISL_4424960, EPI_ISL_4424963, EPI_ISL_4424970, EPI_ISL_4424971, EPI_ISL_4424972, EPI_ISL_4424973, EPI_ISL_4424974, EPI_ISL_4424975, EPI_ISL_4424977, EPI_ISL_4424979, EPI_ISL_4424980, EPI_ISL_4424982, EPI_ISL_4424983, EPI_ISL_4424984, EPI_ISL_4424991, EPI_ISL_4424992, EPI_ISL_4424993, EPI_ISL_4424994, EPI_ISL_4424995, EPI_ISL_4424996, EPI_ISL_4424997, EPI_ISL_4424998, EPI_ISL_4425005, EPI_ISL_4425006, EPI_ISL_4425007, EPI_ISL_4425008, EPI_ISL_4425009, EPI_ISL_4425011, EPI_ISL_4425012, EPI_ISL_4425014, EPI_ISL_4425017, EPI_ISL_4425020, EPI_ISL_4425021, EPI_ISL_4425022, EPI_ISL_4425023, EPI_ISL_4425024, EPI_ISL_4425025, EPI_ISL_4425026, EPI_ISL_4425029, EPI_ISL_4425030, EPI_ISL_4425031, EPI_ISL_4425032, EPI_ISL_4425035, EPI_ISL_4425036, EPI_ISL_4425037, EPI_ISL_4425039, EPI_ISL_4425040, EPI_ISL_4425043, EPI_ISL_4425044, EPI_ISL_4425045, EPI_ISL_4425059, EPI_ISL_4425061, EPI_ISL_4425062, EPI_ISL_4425065, EPI_ISL_4425067, EPI_ISL_4425178, EPI_ISL_4425295, EPI_ISL_4425350 | | | |
| see above | British Columbia Centre For Disease Control | BCCDC Public Health Laboratory | Ana Pacagnella; Corrinne Ng; Dan Fornika; John Tyson; Kim Macdonald; Kimia Kamelian; Linda Hoang; Loretta Janz; Mel Krajden; Prystajecky Natalie; Robert Azana; Shannon Russell |
| EPI_ISL_1967964, EPI_ISL_1967973, EPI_ISL_1967983, EPI_ISL_1968005, EPI_ISL_1968009, EPI_ISL_1968013, EPI_ISL_1968016, EPI_ISL_1968020, EPI_ISL_1968025, EPI_ISL_1968036, EPI_ISL_1968048, EPI_ISL_1968050, EPI_ISL_1968052, EPI_ISL_1968058, EPI_ISL_1968060, EPI_ISL_1968062, EPI_ISL_1968075, EPI_ISL_1968084, EPI_ISL_1968103, EPI_ISL_1968120, EPI_ISL_1968124, EPI_ISL_1968134, EPI_ISL_1968137, EPI_ISL_1968138, EPI_ISL_1968156, EPI_ISL_1968158, EPI_ISL_1968169, EPI_ISL_1968173, EPI_ISL_1968175, EPI_ISL_1968199, EPI_ISL_1968200, EPI_ISL_1968205, EPI_ISL_1968207, EPI_ISL_1968238, EPI_ISL_1968241, EPI_ISL_1968246, EPI_ISL_1968248, EPI_ISL_1968249, EPI_ISL_1968257, EPI_ISL_1968269, EPI_ISL_1968271, EPI_ISL_1968288, EPI_ISL_1968289, EPI_ISL_1968290, EPI_ISL_1968292, EPI_ISL_1968304, EPI_ISL_1968310, EPI_ISL_1968314, EPI_ISL_1968318, EPI_ISL_1968320, EPI_ISL_1968328, EPI_ISL_1968332, EPI_ISL_1968337, EPI_ISL_1968340, EPI_ISL_1968341, EPI_ISL_1968344, EPI_ISL_1968345, EPI_ISL_1968350, EPI_ISL_1968354, EPI_ISL_1968360, EPI_ISL_1968362, EPI_ISL_1968364, EPI_ISL_1968374, EPI_ISL_1968386, EPI_ISL_1968395, EPI_ISL_1968400, EPI_ISL_1968401, EPI_ISL_1968403, EPI_ISL_1968405, EPI_ISL_1968408, EPI_ISL_1968412, EPI_ISL_1968413, EPI_ISL_1968418, EPI_ISL_1968420, EPI_ISL_1968437, EPI_ISL_1968443, EPI_ISL_1968451, EPI_ISL_1968453, EPI_ISL_1968461, EPI_ISL_1968479, EPI_ISL_1968528, EPI_ISL_1971534, EPI_ISL_1971538, EPI_ISL_1971550, EPI_ISL_1971558, EPI_ISL_1971560, EPI_ISL_1971564, EPI_ISL_1971568, EPI_ISL_1971572, EPI_ISL_1971578, EPI_ISL_1971579, EPI_ISL_1971584, EPI_ISL_1971594, EPI_ISL_1971600, EPI_ISL_1971609, EPI_ISL_1971611, EPI_ISL_1971630, EPI_ISL_1971633, EPI_ISL_1971637, EPI_ISL_1971642, EPI_ISL_1971651, EPI_ISL_1971660, EPI_ISL_1971662, EPI_ISL_1971672, EPI_ISL_1971673, EPI_ISL_1971675, EPI_ISL_1971682, EPI_ISL_1971684, EPI_ISL_1971689, EPI_ISL_1971693, EPI_ISL_1971694, EPI_ISL_1971701, EPI_ISL_1971706, EPI_ISL_1971712, EPI_ISL_1971720, EPI_ISL_1971722, EPI_ISL_1971730, EPI_ISL_1971734, EPI_ISL_1971736, EPI_ISL_1971740, EPI_ISL_1971741, EPI_ISL_1971742, EPI_ISL_1971749, EPI_ISL_1971751, EPI_ISL_1971756, EPI_ISL_1971757, EPI_ISL_1971759, EPI_ISL_1971760, EPI_ISL_1971765, EPI_ISL_1971772, EPI_ISL_1971776, EPI_ISL_1971780, EPI_ISL_1971784, EPI_ISL_1971787, EPI_ISL_1971794, EPI_ISL_1971796, EPI_ISL_1971798, EPI_ISL_1971812, EPI_ISL_1971815, EPI_ISL_1971818, EPI_ISL_1971823, EPI_ISL_1971825, EPI_ISL_1971841, EPI_ISL_1971848, EPI_ISL_1971849, EPI_ISL_1971860, EPI_ISL_1971864, EPI_ISL_1971867, EPI_ISL_1971876, EPI_ISL_1971879, EPI_ISL_1971882, EPI_ISL_1971887, EPI_ISL_1971889, EPI_ISL_1971891, EPI_ISL_1971894, EPI_ISL_1971903, EPI_ISL_1971907, EPI_ISL_1971910, EPI_ISL_1971913, EPI_ISL_1971914, EPI_ISL_1971916, EPI_ISL_1971924, EPI_ISL_1971928, EPI_ISL_1971934, EPI_ISL_1971943, EPI_ISL_1971944, EPI_ISL_1971954, EPI_ISL_1971956, EPI_ISL_1971957, EPI_ISL_1971962, EPI_ISL_1971967, EPI_ISL_1971974, EPI_ISL_1971977, EPI_ISL_1971978, EPI_ISL_1971999, EPI_ISL_1972003, EPI_ISL_1972009, EPI_ISL_1972012, EPI_ISL_1972031, EPI_ISL_1972069, EPI_ISL_2096276, EPI_ISL_2096278, EPI_ISL_2096322, EPI_ISL_2096425, EPI_ISL_2096464, EPI_ISL_2096484, EPI_ISL_2096523, EPI_ISL_2096529, EPI_ISL_2096537, EPI_ISL_2096604, EPI_ISL_2096619, EPI_ISL_2096642 | | | |
| see above | Broad Institute Clinical Research Sequencing Platform | Infectious Disease Program, Broad Institute of Harvard and MIT | Adams, G.; B.L.; B.W.; Bauer, M.; Birren; Blumenstiel, B.; Brown, C.; Carter, A.; Chaluvadi, S.; D.J.; DeFelice, M.; DeRuff, K.; Dodge, S.; Gabriel, S.; Gallagher, G.; Gladden-Young, A.; Granger, B.; J.E.; K.J.; Lagerborg, K.; Larkin, K.; Lee, M.; Lemieux; Lennon, N.; Loreth, C.; Madoff, L.; McGovern, S.; Meldrim, J.; Normandin, E.; P.C.; Park; Pearlman, L.; Reilly, S.; Rudy, M.; Sabeti; Siddle; Smole, S.; Tomkins-Tinch, C.; Vicente, G.; and MacInnis |
| EPI_ISL_2500044, EPI_ISL_2500067, EPI_ISL_2500083, EPI_ISL_2500084, EPI_ISL_2500102, EPI_ISL_2500120, EPI_ISL_2500149, EPI_ISL_2500164, EPI_ISL_2500170, EPI_ISL_2500186, EPI_ISL_2500210, EPI_ISL_2658581 | | | |
| see above | CA DPH Viral and Rickettsial Disease Laboratory | Chan-Zuckerberg Biohub | CZB Cliahub Consortium |
| EPI_ISL_2105135, EPI_ISL_2248730 | CA-Los Angeles County Public Health Laboratory | Centers for Disease Control and Prevention Division of Viral Diseases, Pathogen Discovery | Alison Laufer Halpin; Ben L. Rambo-Martin; Clinton R. Paden; Dakota Howard; Darlene Wagner; Dave Wentworth; Dhwani Batra; Jasmine Padilla; Justin Lee; Katie Dillon; Krista Queen; Kristen Knipe; Kristine Lacek; Mark Burroughs; Matthew Schmerer; Mili Sheth; Peter Cook; Sam Shepard; Sarah Nobles; Shoshona Le; Suxiang Tong; Vivien Dugan; Yvette Unoarumhi |
| EPI_ISL_6229698 | CADEIA PUBLICA DE JUAZEIRO DO NORTE TOURINHO | ACME Lab, Oswaldo Cruz Foundation, FIOCRUZ/CE | Carlos Leonardo de Aragao Araujo; Cecilia Leite Costa & Eduardo Ruback dos Santos on behalf of COVID-19 FIOCRUZ Genomic Network; Cleber Furtado Aksenen; Fabio Miyajima; Fernando Braga Stehling; Francisco Eder de Moura Lopes; Igor Oliveira Duarte; Jamille Maria Mendes Bezerra; Joaquim Cesar do Nascimento Sousa Junior; Pedro Miguel Carneiro Jeronimo; Suzana Porto Almeida; Thais Ferreira de Oliveira; Thais de Oliveira Costa; Ticiane Cavalcante de Souza; Veridiana Pessoa Miyajima |
| EPI_ISL_2825980, EPI_ISL_2825981, EPI_ISL_2825990, EPI_ISL_2826003, EPI_ISL_2826004, EPI_ISL_2826022, EPI_ISL_2891986, EPI_ISL_2891998, EPI_ISL_2892000, EPI_ISL_2892222, EPI_ISL_2892223, EPI_ISL_2892226, EPI_ISL_2892228, EPI_ISL_2892233, EPI_ISL_2892261, EPI_ISL_2892287, EPI_ISL_2892337, EPI_ISL_2892339, EPI_ISL_2892343, EPI_ISL_2892357, EPI_ISL_2892359, EPI_ISL_2892360, EPI_ISL_2921703, EPI_ISL_2921827, EPI_ISL_2921828, EPI_ISL_2921837, EPI_ISL_2921838, EPI_ISL_2921840, EPI_ISL_2921841, EPI_ISL_2921860, EPI_ISL_2921862, EPI_ISL_2921886, EPI_ISL_2921892, EPI_ISL_2921915, EPI_ISL_2921916, EPI_ISL_2921940, EPI_ISL_2921967, EPI_ISL_2921968, EPI_ISL_2921970, EPI_ISL_2922041, EPI_ISL_2922042, EPI_ISL_2922063, EPI_ISL_2922139, EPI_ISL_2922153, EPI_ISL_2922154, EPI_ISL_2922177, EPI_ISL_2922178, EPI_ISL_2922180, EPI_ISL_2922246, EPI_ISL_2922256, EPI_ISL_2922278, EPI_ISL_2922279, EPI_ISL_2922280, EPI_ISL_2923548, EPI_ISL_2923609, EPI_ISL_2923616, EPI_ISL_2923648, EPI_ISL_2976970, EPI_ISL_2976984, EPI_ISL_2976988, EPI_ISL_2977002, EPI_ISL_2977003, EPI_ISL_2977007, EPI_ISL_2977020, EPI_ISL_2977025, EPI_ISL_2977046, EPI_ISL_2977053, EPI_ISL_2977067, EPI_ISL_2977075, EPI_ISL_2977076, EPI_ISL_2977093, EPI_ISL_2977108, EPI_ISL_2977148, EPI_ISL_2977156, EPI_ISL_2977165, EPI_ISL_2977170, EPI_ISL_2978154, EPI_ISL_7335814, EPI_ISL_7335902 | | | |
| see above | CDPH VBL | California Department of Public Health | CDPH-COVIDNet; Emily Smith on behalf of CDPH-COVIDNet; UCLA Technology Center for Genomics & Bioinformatics; UCSD EXCITE lab |
| EPI_ISL_2209086, EPI_ISL_2209721, EPI_ISL_2345689, EPI_ISL_2345735, EPI_ISL_2345779, EPI_ISL_2345830, EPI_ISL_2345878 | | | |
| see above | CENTRO DE APOIO EPIDEMIOLOGICO | Instituto Butantan / Mendelics | Antonio Jorge Martins; Claudia Renata dos Santos Barros; David Schlesinger; Debora Botequio Moretti; Dimas Tadeu Covas; Elaine Cristina Marqueze; Elaine Vieira Santos; Evandra Strazza Rodrigues; Heidge Fukumasu; Jayme Augusto de Souza-Neto; José Salvatore Leister Patané; Luiz Alcantara; Luiz Lehmann Coutinho; Maria Carolina Elias; Maurício Lacerda Nogueira; Rafael dos Santos Bezerra; Raul Machado Neto; Rejane Maria Tommasini Grotto; Ricardo Haddad; Sandra Coccuzzo Sampaio Vessoni; Simone Kashima; Svetoslav Nanev Slavov; Vincent Louis Viala |
| EPI_ISL_3102309, EPI_ISL_3102329, EPI_ISL_3102515 | CENTRO DE ATENDIMENTO PARA ENFRENTAMENTO A COVID 19 | Analytical Competence Molecular Epidemiology Lab/ACME, Oswaldo Cruz Foundation, Ceara (FIOCRUZ CE) | Cleber Furtado Aksenen; Fabio Miyajima; Fernando Braga Stehling; Francisco Eder de Moura Lopes; Jamille Maria Mendes Bezerra; Joaquim César do Nascimento Sousa Junior; Pedro Miguel Carneiro Jeronimo; Suzana Porto Almeida e Lucas Delerino; Thais Ferreira de Oliveira; Thais de Oliveira Costa; Ticiane Cavalcante de Souza; Veridiana Pessoa Miyajima |
| EPI_ISL_2210219 | CENTRO DE ESPECIALIDADES DE PRIMAVERA | Instituto Butantan / Mendelics | Antonio Jorge Martins; Claudia Renata dos Santos Barros; David Schlesinger; Debora Botequio Moretti; Dimas Tadeu Covas; Elaine Cristina Marqueze; Elaine Vieira Santos; Evandra Strazza Rodrigues; Heidge Fukumasu; Jayme Augusto de Souza-Neto; José Salvatore Leister Patané; Luiz Alcantara; Luiz Lehmann Coutinho; Maria Carolina Elias; Maurício Lacerda Nogueira; Rafael dos Santos Bezerra; Raul Machado Neto; Rejane Maria Tommasini Grotto; Ricardo Haddad; Sandra Coccuzzo Sampaio Vessoni; Simone Kashima; Svetoslav Nanev Slavov; Vincent Louis Viala |
| EPI_ISL_2345213, EPI_ISL_2345214, EPI_ISL_2345220, EPI_ISL_2345221, EPI_ISL_2345222 | CENTRO DE SAUDE DE AVAI | Instituto Butantan / ESALQ-Piracicaba | Antonio Jorge Martins; Claudia Renata dos Santos Barros; David Schlesinger; Debora Botequio Moretti; Dimas Tadeu Covas; Elaine Cristina Marqueze; Elaine Vieira Santos; Evandra Strazza Rodrigues; Heidge Fukumasu; Jayme Augusto de Souza-Neto; José Salvatore Leister Patané; Luiz Alcantara; Luiz Lehmann Coutinho; Maria Carolina Elias; Maurício Lacerda Nogueira; Rafael dos Santos Bezerra; Raul Machado Neto; Rejane Maria Tommasini Grotto; Ricardo Haddad; Sandra Coccuzzo Sampaio Vessoni; Simone Kashima; Svetoslav Nanev Slavov; Vincent Louis Viala |
| EPI_ISL_2345153 | CENTRO DE SAUDE DE BARBOSA | Instituto Butantan / ESALQ-Piracicaba | Antonio Jorge Martins; Claudia Renata dos Santos Barros; David Schlesinger; Debora Botequio Moretti; Dimas Tadeu Covas; Elaine Cristina Marqueze; Elaine Vieira Santos; Evandra Strazza Rodrigues; Heidge Fukumasu; Jayme Augusto de Souza-Neto; José Salvatore Leister Patané; Luiz Alcantara; Luiz Lehmann Coutinho; Maria Carolina Elias; Maurício Lacerda Nogueira; Rafael dos Santos Bezerra; Raul Machado Neto; Rejane Maria Tommasini Grotto; Ricardo Haddad; Sandra Coccuzzo Sampaio Vessoni; Simone Kashima; Svetoslav Nanev Slavov; Vincent Louis Viala |
| EPI_ISL_2345236 | CENTRO DE SAUDE DE OCAUCU | Instituto Butantan / ESALQ-Piracicaba | Antonio Jorge Martins; Claudia Renata dos Santos Barros; David Schlesinger; Debora Botequio Moretti; Dimas Tadeu Covas; Elaine Cristina Marqueze; Elaine Vieira Santos; Evandra Strazza Rodrigues; Heidge Fukumasu; Jayme Augusto de Souza-Neto; José Salvatore Leister Patané; Luiz Alcantara; Luiz Lehmann Coutinho; Maria Carolina Elias; Maurício Lacerda Nogueira; Rafael dos Santos Bezerra; Raul Machado Neto; Rejane Maria Tommasini Grotto; Ricardo Haddad; Sandra Coccuzzo Sampaio Vessoni; Simone Kashima; Svetoslav Nanev Slavov; Vincent Louis Viala |
| EPI_ISL_2345192, EPI_ISL_2345201 | CENTRO DE SAUDE DE OSVALDO CRUZ | Instituto Butantan / ESALQ-Piracicaba | Antonio Jorge Martins; Claudia Renata dos Santos Barros; David Schlesinger; Debora Botequio Moretti; Dimas Tadeu Covas; Elaine Cristina Marqueze; Elaine Vieira Santos; Evandra Strazza Rodrigues; Heidge Fukumasu; Jayme Augusto de Souza-Neto; José Salvatore Leister Patané; Luiz Alcantara; Luiz Lehmann Coutinho; Maria Carolina Elias; Maurício Lacerda Nogueira; Rafael dos Santos Bezerra; Raul Machado Neto; Rejane Maria Tommasini Grotto; Ricardo Haddad; Sandra Coccuzzo Sampaio Vessoni; Simone Kashima; Svetoslav Nanev Slavov; Vincent Louis Viala |
| EPI_ISL_2345685, EPI_ISL_2362246 | CENTRO DE SAUDE DR JOAO JOAQUIM FERREIRA CRISTAIS PAULISTA | Instituto Butantan / Mendelics | Antonio Jorge Martins; Claudia Renata dos Santos Barros; David Schlesinger; Debora Botequio Moretti; Dimas Tadeu Covas; Elaine Cristina Marqueze; Elaine Vieira Santos; Evandra Strazza Rodrigues; Heidge Fukumasu; Jayme Augusto de Souza-Neto; José Salvatore Leister Patané; Luiz Alcantara; Luiz Lehmann Coutinho; Maria Carolina Elias; Maurício Lacerda Nogueira; Rafael dos Santos Bezerra; Raul Machado Neto; Rejane Maria Tommasini Grotto; Ricardo Haddad; Sandra Coccuzzo Sampaio Vessoni; Simone Kashima; Svetoslav Nanev Slavov; Vincent Louis Viala |
| EPI_ISL_2345189 | CENTRO DE SAUDE II DE BROTAS | Instituto Butantan / ESALQ-Piracicaba | Antonio Jorge Martins; Claudia Renata dos Santos Barros; David Schlesinger; Debora Botequio Moretti; Dimas Tadeu Covas; Elaine Cristina Marqueze; Elaine Vieira Santos; Evandra Strazza Rodrigues; Heidge Fukumasu; Jayme Augusto de Souza-Neto; José Salvatore Leister Patané; Luiz Alcantara; Luiz Lehmann Coutinho; Maria Carolina Elias; Maurício Lacerda Nogueira; Rafael dos Santos Bezerra; Raul Machado Neto; Rejane Maria Tommasini Grotto; Ricardo Haddad; Sandra Coccuzzo Sampaio Vessoni; Simone Kashima; Svetoslav Nanev Slavov; Vincent Louis Viala |
| EPI_ISL_2209189, EPI_ISL_2345650, EPI_ISL_2345695, EPI_ISL_2345844, EPI_ISL_2362262 | CENTRO DE SAUDE II DR GABRIEL MESQUITA VARGEM GDE DO SUL | Instituto Butantan / Mendelics | Antonio Jorge Martins; Claudia Renata dos Santos Barros; David Schlesinger; Debora Botequio Moretti; Dimas Tadeu Covas; Elaine Cristina Marqueze; Elaine Vieira Santos; Evandra Strazza Rodrigues; Heidge Fukumasu; Jayme Augusto de Souza-Neto; José Salvatore Leister Patané; Luiz Alcantara; Luiz Lehmann Coutinho; Maria Carolina Elias; Maurício Lacerda Nogueira; Rafael dos Santos Bezerra; Raul Machado Neto; Rejane Maria Tommasini Grotto; Ricardo Haddad; Sandra Coccuzzo Sampaio Vessoni; Simone Kashima; Svetoslav Nanev Slavov; Vincent Louis Viala |
| EPI_ISL_2209527, EPI_ISL_2345644, EPI_ISL_2345649, EPI_ISL_2345743, EPI_ISL_2345747, EPI_ISL_2345791 | CENTRO DE SAUDE II IBITINGA | Instituto Butantan / Mendelics | Antonio Jorge Martins; Claudia Renata dos Santos Barros; David Schlesinger; Debora Botequio Moretti; Dimas Tadeu Covas; Elaine Cristina Marqueze; Elaine Vieira Santos; Evandra Strazza Rodrigues; Heidge Fukumasu; Jayme Augusto de Souza-Neto; José Salvatore Leister Patané; Luiz Alcantara; Luiz Lehmann Coutinho; Maria Carolina Elias; Maurício Lacerda Nogueira; Rafael dos Santos Bezerra; Raul Machado Neto; Rejane Maria Tommasini Grotto; Ricardo Haddad; Sandra Coccuzzo Sampaio Vessoni; Simone Kashima; Svetoslav Nanev Slavov; Vincent Louis Viala |
| EPI_ISL_2345825 | CENTRO DE SAUDE II MATAO | Instituto Butantan / Mendelics | Antonio Jorge Martins; Claudia Renata dos Santos Barros; David Schlesinger; Debora Botequio Moretti; Dimas Tadeu Covas; Elaine Cristina Marqueze; Elaine Vieira Santos; Evandra Strazza Rodrigues; Heidge Fukumasu; Jayme Augusto de Souza-Neto; José Salvatore Leister Patané; Luiz Alcantara; Luiz Lehmann Coutinho; Maria Carolina Elias; Maurício Lacerda Nogueira; Rafael dos Santos Bezerra; Raul Machado Neto; Rejane Maria Tommasini Grotto; Ricardo Haddad; Sandra Coccuzzo Sampaio Vessoni; Simone Kashima; Svetoslav Nanev Slavov; Vincent Louis Viala |
| EPI_ISL_2209307, EPI_ISL_2209312, EPI_ISL_2345792, EPI_ISL_2362260 | CENTRO DE SAUDE III AFFONSO LUZZI SANTA CRUZ DAS PALMEIRAS | Instituto Butantan / Mendelics | Antonio Jorge Martins; Claudia Renata dos Santos Barros; David Schlesinger; Debora Botequio Moretti; Dimas Tadeu Covas; Elaine Cristina Marqueze; Elaine Vieira Santos; Evandra Strazza Rodrigues; Heidge Fukumasu; Jayme Augusto de Souza-Neto; José Salvatore Leister Patané; Luiz Alcantara; Luiz Lehmann Coutinho; Maria Carolina Elias; Maurício Lacerda Nogueira; Rafael dos Santos Bezerra; Raul Machado Neto; Rejane Maria Tommasini Grotto; Ricardo Haddad; Sandra Coccuzzo Sampaio Vessoni; Simone Kashima; Svetoslav Nanev Slavov; Vincent Louis Viala |
| EPI_ISL_2345195 | CENTRO DE SAUDE III ALTO ALEGRE | Instituto Butantan / ESALQ-Piracicaba | Antonio Jorge Martins; Claudia Renata dos Santos Barros; David Schlesinger; Debora Botequio Moretti; Dimas Tadeu Covas; Elaine Cristina Marqueze; Elaine Vieira Santos; Evandra Strazza Rodrigues; Heidge Fukumasu; Jayme Augusto de Souza-Neto; José Salvatore Leister Patané; Luiz Alcantara; Luiz Lehmann Coutinho; Maria Carolina Elias; Maurício Lacerda Nogueira; Rafael dos Santos Bezerra; Raul Machado Neto; Rejane Maria Tommasini Grotto; Ricardo Haddad; Sandra Coccuzzo Sampaio Vessoni; Simone Kashima; Svetoslav Nanev Slavov; Vincent Louis Viala |
| EPI_ISL_2345187, EPI_ISL_2345188 | CENTRO DE SAUDE III DE BOCAINA | Instituto Butantan / ESALQ-Piracicaba | Antonio Jorge Martins; Claudia Renata dos Santos Barros; David Schlesinger; Debora Botequio Moretti; Dimas Tadeu Covas; Elaine Cristina Marqueze; Elaine Vieira Santos; Evandra Strazza Rodrigues; Heidge Fukumasu; Jayme Augusto de Souza-Neto; José Salvatore Leister Patané; Luiz Alcantara; Luiz Lehmann Coutinho; Maria Carolina Elias; Maurício Lacerda Nogueira; Rafael dos Santos Bezerra; Raul Machado Neto; Rejane Maria Tommasini Grotto; Ricardo Haddad; Sandra Coccuzzo Sampaio Vessoni; Simone Kashima; Svetoslav Nanev Slavov; Vincent Louis Viala |
| EPI_ISL_2345206 | CENTRO DE SAUDE III DE IGARACU DO TIETE | Instituto Butantan / ESALQ-Piracicaba | Antonio Jorge Martins; Claudia Renata dos Santos Barros; David Schlesinger; Debora Botequio Moretti; Dimas Tadeu Covas; Elaine Cristina Marqueze; Elaine Vieira Santos; Evandra Strazza Rodrigues; Heidge Fukumasu; Jayme Augusto de Souza-Neto; José Salvatore Leister Patané; Luiz Alcantara; Luiz Lehmann Coutinho; Maria Carolina Elias; Maurício Lacerda Nogueira; Rafael dos Santos Bezerra; Raul Machado Neto; Rejane Maria Tommasini Grotto; Ricardo Haddad; Sandra Coccuzzo Sampaio Vessoni; Simone Kashima; Svetoslav Nanev Slavov; Vincent Louis Viala |
| EPI_ISL_2345129 | CENTRO MEDICO PMESP | Instituto Butantan / ESALQ-Piracicaba | Antonio Jorge Martins; Claudia Renata dos Santos Barros; David Schlesinger; Debora Botequio Moretti; Dimas Tadeu Covas; Elaine Cristina Marqueze; Elaine Vieira Santos; Evandra Strazza Rodrigues; Heidge Fukumasu; Jayme Augusto de Souza-Neto; José Salvatore Leister Patané; Luiz Alcantara; Luiz Lehmann Coutinho; Maria Carolina Elias; Maurício Lacerda Nogueira; Rafael dos Santos Bezerra; Raul Machado Neto; Rejane Maria Tommasini Grotto; Ricardo Haddad; Sandra Coccuzzo Sampaio Vessoni; Simone Kashima; Svetoslav Nanev Slavov; Vincent Louis Viala |
| EPI_ISL_2015908 | CH DE SAINTONGE | CNR Virus des Infections Respiratoires - France SUD | Antonin Bal; Bruno Lina; Bruno Simon; Gregory Destras; Gwendolyne Burfin; Hadrien Regue; Laurence Josset; Martine Valette; Quentin Semanas |
| EPI_ISL_2151720 | CHI VILLENEUVE ST GEORGES | Department of Virology, Henri Mondor University Hospital, Assistance Publique Hôpitaux de Paris, Université Paris-Est Créteil, INSERM U955 | Alexandre Soulier; Christophe Rodriguez; Elisabeth Trawinski; Guillaume Gricourt; Jean-Michel Pawlotsky; Melissa N'Debi; Slim Fourati; Vanessa Demontant |
| EPI_ISL_2004290, EPI_ISL_2004298, | CHLC | Instituto Nacional de Saude (INSA) | Borges et al |

| | | | |
|---|---|---|---|
| EPI_ISL_2004299 | | | |
| EPI_ISL_2249177, EPI_ISL_2249179 | CHULN - H Santa Maria | Instituto Nacional de Saude (INSA) and Centro de Investigaçao em Biodiversidade e Recursos Geneticos (CIBIO), Universidade do Porto | Borges et al |
| EPI_ISL_1941821 | CHUV | Laboratory of genomics and metagenomics | Claire Bertelli; Damien Jacot; Gilbert Greub; Sebastien Aeby; Trestan Pillonel |
| EPI_ISL_2351959, EPI_ISL_2351960, EPI_ISL_2351964, EPI_ISL_2351967, EPI_ISL_2351968, EPI_ISL_2351969, EPI_ISL_2351970, EPI_ISL_2351971 | | | |
| see above | CLILAB | Microbiology Department | Aida Gonzalez-Diaz; Carmen Ardanuy; Jordi Camara; Jordi Niubó; Laura Calatayud; M Angeles Domínguez; Miguel Fernandez-Huerta; Sara Marti |
| EPI_ISL_3355515 | CLINICA PUTUMAYO | Corporacion CorpoGen-Universidad de los Andes-Universidad Central | Christian Romero; Jorge Duitama; Juan Manuel Anzola; Laura González; Maryam Chaib De Mares; María Mercedes Zambrano; Nelly Díaz; Patricia Del Portillo; Silvia Restrepo |
| EPI_ISL_1970690 | CLINIQUE N-D DE GRACE | Institut de Pathologie et Genetique (IPG) | Jérémie Gras; Pascale Hilbert |
| EPI_ISL_2674315, EPI_ISL_2674316 | COMFANDI - SAN NICOLAS | Instituto Nacional de Salud- Dirección de Investigación en Salud Pública | Carlos Franco-Muñoz; Carmen Osorio; Diana Malo; Diego A. Álvarez-Díaz; Diego Andrés Prada; Gerardo Santamaría; Hector Alejandro Ruiz-Moreno; Jhonnatan Reales-González; Jorge Rivera; Juan Camilo Martinez; Julian Naizaque; Katherine Laiton-Donato; Lisseth Pardo; Magdalena Wiesner; Marcela Mercado-Reyes; Maria T. Herrera-Sepúlveda; Marta Lopez Blanco; Martha Lucia Ospina Martinez; Paola Rojas; Sergio Gomez; Sheryll Corchuelo; Ángela Alarcon Cruz |
| EPI_ISL_2674337 | CORPOMEDICA | Instituto Nacional de Salud- Dirección de Investigación en Salud Pública | Carlos Franco-Muñoz; Carmen Osorio; Diana Malo; Diego A. Álvarez-Díaz; Diego Andrés Prada; Gerardo Santamaría; Hector Alejandro Ruiz-Moreno; Jhonnatan Reales-González; Jorge Rivera; Juan Camilo Martinez; Julian Naizaque; Katherine Laiton-Donato; Lisseth Pardo; Magdalena Wiesner; Marcela Mercado-Reyes; Maria T. Herrera-Sepúlveda; Marta Lopez Blanco; Martha Lucia Ospina Martinez; Paola Rojas; Sergio Gomez; Sheryll Corchuelo; Ángela Alarcon Cruz |
| EPI_ISL_2345078, EPI_ISL_2345194, EPI_ISL_2345198, EPI_ISL_2345200 | CS II DR MIGUEL VITALIANO ORLANDIA | Instituto Butantan / ESALQ-Piracicaba | Antonio Jorge Martins; Claudia Renata dos Santos Barros; David Schlesinger; Debora Botequio Moretti; Dimas Tadeu Covas; Elaine Cristina Marqueze; Elaine Vieira Santos; Evandra Strazza Rodrigues; Heidge Fukumasu; Jayme Augusto de Souza-Neto; José Salvatore Leister Patané; Luiz Alcantara; Luiz Lehmann Coutinho; Maria Carolina Elias; Maurício Lacerda Nogueira; Rafael dos Santos Bezerra; Raul Machado Neto; Rejane Maria Tommasini Grotto; Ricardo Haddad; Sandra Coccuzzo Sampaio Vessoni; Simone Kashima; Svetoslav Nanev Slavov; Vincent Louis Viala |
| EPI_ISL_2209850 | CS II DR SEBASTIAO RIBEIRO DO VALLE | Instituto Butantan / Mendelics | Antonio Jorge Martins; Claudia Renata dos Santos Barros; David Schlesinger; Debora Botequio Moretti; Dimas Tadeu Covas; Elaine Cristina Marqueze; Elaine Vieira Santos; Evandra Strazza Rodrigues; Heidge Fukumasu; Jayme Augusto de Souza-Neto; José Salvatore Leister Patané; Luiz Alcantara; Luiz Lehmann Coutinho; Maria Carolina Elias; Maurício Lacerda Nogueira; Rafael dos Santos Bezerra; Raul Machado Neto; Rejane Maria Tommasini Grotto; Ricardo Haddad; Sandra Coccuzzo Sampaio Vessoni; Simone Kashima; Svetoslav Nanev Slavov; Vincent Louis Viala |
| EPI_ISL_2345273, EPI_ISL_2345274, EPI_ISL_2345278 | CS IOLANDA FLEURY DE CAMPOS GUEDES NUPORANGA | Instituto Butantan / ESALQ-Piracicaba | Antonio Jorge Martins; Claudia Renata dos Santos Barros; David Schlesinger; Debora Botequio Moretti; Dimas Tadeu Covas; Elaine Cristina Marqueze; Elaine Vieira Santos; Evandra Strazza Rodrigues; Heidge Fukumasu; Jayme Augusto de Souza-Neto; José Salvatore Leister Patané; Luiz Alcantara; Luiz Lehmann Coutinho; Maria Carolina Elias; Maurício Lacerda Nogueira; Rafael dos Santos Bezerra; Raul Machado Neto; Rejane Maria Tommasini Grotto; Ricardo Haddad; Sandra Coccuzzo Sampaio Vessoni; Simone Kashima; Svetoslav Nanev Slavov; Vincent Louis Viala |
| EPI_ISL_2345171, EPI_ISL_2345246 | CSII DR JOSE FERREIRA TELLES | Instituto Butantan / ESALQ-Piracicaba | Antonio Jorge Martins; Claudia Renata dos Santos Barros; David Schlesinger; Debora Botequio Moretti; Dimas Tadeu Covas; Elaine Cristina Marqueze; Elaine Vieira Santos; Evandra Strazza Rodrigues; Heidge Fukumasu; Jayme Augusto de Souza-Neto; José Salvatore Leister Patané; Luiz Alcantara; Luiz Lehmann Coutinho; Maria Carolina Elias; Maurício Lacerda Nogueira; Rafael dos Santos Bezerra; Raul Machado Neto; Rejane Maria Tommasini Grotto; Ricardo Haddad; Sandra Coccuzzo Sampaio Vessoni; Simone Kashima; Svetoslav Nanev Slavov; Vincent Louis Viala |
| EPI_ISL_2105892, EPI_ISL_2105895 | CSL, Maryland Department of Health | CSL, Maryland Department of Health | Keller, E.; Loomis, J.; Low, K. |
| EPI_ISL_2501536 | CT Department of Public Health | CT Department of Public Health | Claire_Pearson; Tu_N_Nguyen |
| EPI_ISL_2537469, EPI_ISL_2842810, EPI_ISL_3536727, EPI_ISL_3536925, EPI_ISL_3536926, EPI_ISL_3536927 | California Department of Public Health | California Department of Public Health | CDPH IDLB COVIDNet et al |
| EPI_ISL_2778870, EPI_ISL_2778895, EPI_ISL_2778918, EPI_ISL_2778922, EPI_ISL_2778935, EPI_ISL_2778953 | California Department of Public Health Valencia Branch Laboratory (CDPH VBL) | California Department of Public Health | CDPH-COVIDNet; UCLA Technology Center for Genomics & Bioinformatics |
| EPI_ISL_7508010 | Cedars-Sinai Medical Center, Molecular Pathology Laboratory of Department of Pathology & Laboratory Medicine and Genomic Core | Cedars-Sinai Medical Center, Molecular Pathology Laboratory of Department of Pathology & Laboratory Medicine and Genomic Core | Brian Davis; Eric Vail; Jasmine T Plummer; Jorge Mario Sincuir Martinez; Stephanie Chen; Wenjuan Zhang |
| EPI_ISL_1939271, EPI_ISL_1939272 | Center for Laboratory Medicine | Center for Laboratory Medicine | Yannick Gerth |
| EPI_ISL_2502377, EPI_ISL_2502383, EPI_ISL_2502392, EPI_ISL_2502406, EPI_ISL_2502430, EPI_ISL_2502435, EPI_ISL_2502478, EPI_ISL_2502479, EPI_ISL_2502480 | | | |
| see above | Central Laboratory, Bureau of Public Health (BOG) and Academic Hospital Paramaribo | Erasmus Medical Center | Bas B Oude Munnink; Cherise Beek; Consuella Partowidjojo; Dion Gajadin; Ed PF IJzerman; Emmanuelle Munger; Gary Gummels; Ingrid SK Krishnadath; Lycke Woittiez; Marion PG Koopmans; Mireille Van de Veer; Phyllis Pinas; Princes Wongsowidjojo; Radjesh Ori; Ranisha Doerbalie; Rohma Banwari; Soeradj Harkisoen; Stephen Vreden; Tilotmadebie Ramlal; Verne Nanhoe |
| EPI_ISL_3255073, EPI_ISL_3255074, EPI_ISL_3255076, EPI_ISL_3255139, EPI_ISL_3255194, EPI_ISL_3255256, EPI_ISL_5146189 | | | |
| see above | Central Public Health Laboratory - LACEN -Bahia, Salvador, Brazil | Central Public Health Laboratory - LACEN -Bahia, Salvador, Brazil | Arabela Leal; Breno Dominguez; Felicidade Pereira; Gabriela Menezes; Jaqueline Gomes; Lenisa Dandara; Luciana Oliveira; Luiz Alcantara; Marcela Gómez; Marta Giovanetti; Patrícia Cajado; Stephane Tosta; Vagner Fonseca; Vanessa Nardy |
| EPI_ISL_2002663, EPI_ISL_2002664 | Centre Hospitalier Universitaire Clermont-Ferrand | CHU Clermont-Ferrand, service de virologie | Bisseux Maxime; Combes Patricia; Henquell Cécile; Mirand Audrey |
| EPI_ISL_2401761 | Centre Hospitalier du Nord | Laboratoire national de sante, Microbiology, Microbial Genomics Platform | Anke Wienecke-Baldacchino; Catherine Ragimbeau; Fatiha Boulmerka; Fatu Djabi; Jacqueline Parmentier; Jessica Tapp; Lise Pignon; Raoul Salmon; Tamir Abdelrahman |
| EPI_ISL_5801496, EPI_ISL_5801498, EPI_ISL_5801499, EPI_ISL_5801500, EPI_ISL_5801501, EPI_ISL_5801502, EPI_ISL_5801503 | | | |
| see above | Centro De Apoio Epidemiologico | Instituto Butantan | Antonio Jorge Martins; Claudia Renata dos Santos Barros; David Schlesinger; Debora Botequio Moretti; Dimas Tadeu Covas; Elaine Cristina Marqueze; Elaine Vieira Santos; Evandra Strazza Rodrigues; Heidge Fukumasu; Jayme Augusto de Souza-Neto; José Salvatore Leister Patané; Luiz Alcantara; Luiz Lehmann Coutinho; Maria Carolina Elias; Maurício Lacerda Nogueira; Rafael dos Santos Bezerra; Raul Machado Neto; Rejane Maria Tommasini Grotto; Ricardo Haddad; Sandra Coccuzzo Sampaio Vessoni; Simone Kashima; Svetoslav Nanev Slavov; Vincent Louis Viala |
| EPI_ISL_5801461 | Centro De Especialidades De Primavera | Instituto Butantan | Antonio Jorge Martins; Claudia Renata dos Santos Barros; David Schlesinger; Debora Botequio Moretti; Dimas Tadeu Covas; Elaine Cristina Marqueze; Elaine Vieira Santos; Evandra Strazza Rodrigues; Heidge Fukumasu; Jayme Augusto de Souza-Neto; José Salvatore Leister Patané; Luiz Alcantara; Luiz Lehmann Coutinho; Maria Carolina Elias; Maurício Lacerda Nogueira; Rafael dos Santos Bezerra; Raul Machado Neto; Rejane Maria Tommasini Grotto; Ricardo Haddad; Sandra Coccuzzo Sampaio Vessoni; Simone Kashima; Svetoslav Nanev Slavov; Vincent Louis Viala |
| EPI_ISL_5782542, EPI_ISL_5782543, EPI_ISL_5782544, EPI_ISL_5782546 | Centro De Genomas | Instituto Butantan | Antonio Jorge Martins; Claudia Renata dos Santos Barros; David Schlesinger; Debora Botequio Moretti; Dimas Tadeu Covas; Elaine Cristina Marqueze; Elaine Vieira Santos; Evandra Strazza Rodrigues; Heidge Fukumasu; Jayme Augusto de Souza-Neto; José Salvatore Leister Patané; Luiz Alcantara; Luiz Lehmann Coutinho; Maria Carolina Elias; Maurício Lacerda Nogueira; Rafael dos Santos Bezerra; Raul Machado Neto; Rejane Maria Tommasini Grotto; Ricardo Haddad; Sandra Coccuzzo Sampaio Vessoni; Simone Kashima; Svetoslav Nanev Slavov; Vincent Louis Viala |
| EPI_ISL_5802711 | Centro De Saude De Analandia | Instituto Butantan | Antonio Jorge Martins; Claudia Renata dos Santos Barros; David Schlesinger; Debora Botequio Moretti; Dimas Tadeu Covas; Elaine Cristina Marqueze; Elaine Vieira Santos; Evandra Strazza Rodrigues; Heidge Fukumasu; Jayme Augusto de Souza-Neto; José Salvatore Leister Patané; Luiz Alcantara; Luiz Lehmann Coutinho; Maria Carolina Elias; Maurício Lacerda Nogueira; Rafael dos Santos Bezerra; Raul Machado Neto; Rejane Maria Tommasini Grotto; Ricardo Haddad; Sandra Coccuzzo Sampaio Vessoni; Simone Kashima; Svetoslav Nanev Slavov; Vincent Louis Viala |
| EPI_ISL_5801308, EPI_ISL_5801310, EPI_ISL_5801321, EPI_ISL_5801322, EPI_ISL_5801323 | Centro De Saude De Avai | Instituto Butantan | Antonio Jorge Martins; Claudia Renata dos Santos Barros; David Schlesinger; Debora Botequio Moretti; Dimas Tadeu Covas; Elaine Cristina Marqueze; Elaine Vieira Santos; Evandra Strazza Rodrigues; Heidge Fukumasu; Jayme Augusto de Souza-Neto; José Salvatore Leister Patané; Luiz Alcantara; Luiz Lehmann Coutinho; Maria Carolina Elias; Maurício Lacerda Nogueira; Rafael dos Santos Bezerra; Raul Machado Neto; Rejane Maria Tommasini Grotto; Ricardo Haddad; Sandra Coccuzzo Sampaio Vessoni; Simone Kashima; Svetoslav Nanev Slavov; Vincent Louis Viala |
| EPI_ISL_5801204 | Centro De Saude De Barbosa | Instituto Butantan | Antonio Jorge Martins; Claudia Renata dos Santos Barros; David Schlesinger; Debora Botequio Moretti; Dimas Tadeu Covas; Elaine Cristina Marqueze; Elaine Vieira Santos; Evandra Strazza Rodrigues; Heidge Fukumasu; Jayme Augusto de Souza-Neto; José Salvatore Leister Patané; Luiz Alcantara; Luiz Lehmann Coutinho; Maria Carolina Elias; Maurício Lacerda Nogueira; Rafael dos Santos Bezerra; Raul Machado Neto; Rejane Maria Tommasini Grotto; Ricardo Haddad; Sandra Coccuzzo Sampaio Vessoni; Simone Kashima; Svetoslav Nanev Slavov; Vincent Louis Viala |
| EPI_ISL_5801337 | Centro De Saude De Ocaucu | Instituto Butantan | Antonio Jorge Martins; Claudia Renata dos Santos Barros; David Schlesinger; Debora Botequio Moretti; Dimas Tadeu Covas; Elaine Cristina Marqueze; Elaine Vieira Santos; Evandra Strazza Rodrigues; Heidge Fukumasu; Jayme Augusto de Souza-Neto; José Salvatore Leister Patané; Luiz Alcantara; Luiz Lehmann Coutinho; Maria Carolina Elias; Maurício Lacerda Nogueira; Rafael dos Santos Bezerra; Raul Machado Neto; Rejane Maria Tommasini Grotto; Ricardo Haddad; Sandra Coccuzzo Sampaio Vessoni; Simone Kashima; Svetoslav Nanev Slavov; Vincent Louis Viala |
| EPI_ISL_5801277, EPI_ISL_5801291 | Centro De Saude De Osvaldo Cruz | Instituto Butantan | Antonio Jorge Martins; Claudia Renata dos Santos Barros; David Schlesinger; Debora Botequio Moretti; Dimas Tadeu Covas; Elaine Cristina Marqueze; Elaine Vieira Santos; Evandra Strazza Rodrigues; Heidge Fukumasu; Jayme Augusto de Souza-Neto; José Salvatore Leister Patané; Luiz Alcantara; Luiz Lehmann Coutinho; Maria Carolina Elias; Maurício Lacerda Nogueira; Rafael dos Santos Bezerra; Raul Machado Neto; Rejane Maria Tommasini Grotto; Ricardo Haddad; Sandra Coccuzzo Sampaio Vessoni; Simone Kashima; Svetoslav Nanev Slavov; Vincent Louis Viala |
| EPI_ISL_5801148, EPI_ISL_5801302 | Centro De Saude Dr Joao Joaquim Ferreira Cristais Paulista | Instituto Butantan | Antonio Jorge Martins; Claudia Renata dos Santos Barros; David Schlesinger; Debora Botequio Moretti; Dimas Tadeu Covas; Elaine Cristina Marqueze; Elaine Vieira Santos; Evandra Strazza Rodrigues; Heidge Fukumasu; Jayme Augusto de Souza-Neto; José Salvatore Leister Patané; Luiz Alcantara; Luiz Lehmann Coutinho; Maria Carolina Elias; Maurício Lacerda Nogueira; Rafael dos Santos Bezerra; Raul Machado Neto; Rejane Maria Tommasini Grotto; Ricardo Haddad; Sandra Coccuzzo Sampaio Vessoni; Simone Kashima; Svetoslav Nanev Slavov; Vincent Louis Viala |
| EPI_ISL_5802725 | Centro De Saude Dr Mario Dias De Aguiar Capivari | Instituto Butantan | Antonio Jorge Martins; Claudia Renata dos Santos Barros; David Schlesinger; Debora Botequio Moretti; Dimas Tadeu Covas; Elaine Cristina Marqueze; Elaine Vieira Santos; Evandra Strazza Rodrigues; Heidge Fukumasu; Jayme Augusto de Souza-Neto; José Salvatore Leister Patané; Luiz Alcantara; Luiz Lehmann Coutinho; Maria Carolina Elias; Maurício Lacerda Nogueira; Rafael dos Santos Bezerra; Raul Machado Neto; Rejane Maria Tommasini Grotto; Ricardo Haddad; Sandra Coccuzzo Sampaio Vessoni; Simone Kashima; Svetoslav Nanev Slavov; Vincent Louis Viala |
| EPI_ISL_5801274 | Centro De Saude Ii De Brotas | Instituto Butantan | Antonio Jorge Martins; Claudia Renata dos Santos Barros; David Schlesinger; Debora Botequio Moretti; Dimas Tadeu Covas; Elaine Cristina Marqueze; Elaine Vieira Santos; Evandra Strazza Rodrigues; Heidge Fukumasu; Jayme Augusto de Souza-Neto; José Salvatore Leister Patané; Luiz Alcantara; Luiz Lehmann Coutinho; Maria Carolina Elias; Maurício Lacerda Nogueira; Rafael dos Santos Bezerra; Raul Machado Neto; Rejane Maria Tommasini Grotto; Ricardo Haddad; Sandra Coccuzzo Sampaio Vessoni; Simone Kashima; Svetoslav Nanev Slavov; Vincent Louis Viala |
| EPI_ISL_5801201, EPI_ISL_5801208, EPI_ISL_5801338, EPI_ISL_5801339, | Centro De Saude Ii Dr Gabriel Mesquita Vargem Gde Do Sul | Instituto Butantan | Antonio Jorge Martins; Claudia Renata dos Santos Barros; David Schlesinger; Debora Botequio Moretti; Dimas Tadeu Covas; Elaine Cristina Marqueze; Elaine Vieira Santos; Evandra Strazza Rodrigues; Heidge Fukumasu; Jayme Augusto de Souza-Neto; José Salvatore Leister Patané; Luiz Alcantara; Luiz Lehmann Coutinho; Maria Carolina Elias; Maurício Lacerda Nogueira; Rafael dos Santos Bezerra; Raul Machado Neto; Rejane Maria Tommasini Grotto; Ricardo Haddad; Sandra Coccuzzo Sampaio Vessoni; Simone Kashima; Svetoslav Nanev Slavov; Vincent Louis Viala |

| | | | |
|---|---|---|---|
| EPI_ISL_5801340 | | | |
| EPI_ISL_5802698 | Centro De Saude Ii Dr Jose De Felipe E S Pinhal Sp | Instituto Butantan | Antonio Jorge Martins; Claudia Renata dos Santos Barros; David Schlesinger; Debora Botequio Moretti; Dimas Tadeu Covas; Elaine Cristina Marqueze; Elaine Vieira Santos; Evandra Strazza Rodrigues; Heidge Fukumasu; Jayme Augusto de Souza-Neto; José Salvatore Leister Patané; Luiz Alcantara; Luiz Lehmann Coutinho; Maria Carolina Elias; Maurício Lacerda Nogueira; Rafael dos Santos Bezerra; Raul Machado Neto; Rejane Maria Tommasini Grotto; Ricardo Haddad; Sandra Coccuzzo Sampaio Vessoni; Simone Kashima; Svetoslav Nanev Slavov; Vincent Louis Viala |
| EPI_ISL_5802727 | Centro De Saude Ii Dr Jose Paione Mococa | Instituto Butantan | Antonio Jorge Martins; Claudia Renata dos Santos Barros; David Schlesinger; Debora Botequio Moretti; Dimas Tadeu Covas; Elaine Cristina Marqueze; Elaine Vieira Santos; Evandra Strazza Rodrigues; Heidge Fukumasu; Jayme Augusto de Souza-Neto; José Salvatore Leister Patané; Luiz Alcantara; Luiz Lehmann Coutinho; Maria Carolina Elias; Maurício Lacerda Nogueira; Rafael dos Santos Bezerra; Raul Machado Neto; Rejane Maria Tommasini Grotto; Ricardo Haddad; Sandra Coccuzzo Sampaio Vessoni; Simone Kashima; Svetoslav Nanev Slavov; Vincent Louis Viala |
| EPI_ISL_5801213, EPI_ISL_5801214, EPI_ISL_5801217, EPI_ISL_5801219, EPI_ISL_5801224, EPI_ISL_5801225, EPI_ISL_5801227 | | | |
| see above | Centro De Saude Ii Ibitinga | Instituto Butantan | Antonio Jorge Martins; Claudia Renata dos Santos Barros; David Schlesinger; Debora Botequio Moretti; Dimas Tadeu Covas; Elaine Cristina Marqueze; Elaine Vieira Santos; Evandra Strazza Rodrigues; Heidge Fukumasu; Jayme Augusto de Souza-Neto; José Salvatore Leister Patané; Luiz Alcantara; Luiz Lehmann Coutinho; Maria Carolina Elias; Maurício Lacerda Nogueira; Rafael dos Santos Bezerra; Raul Machado Neto; Rejane Maria Tommasini Grotto; Ricardo Haddad; Sandra Coccuzzo Sampaio Vessoni; Simone Kashima; Svetoslav Nanev Slavov; Vincent Louis Viala |
| EPI_ISL_5801311 | Centro De Saude Ii Matao | Instituto Butantan | Antonio Jorge Martins; Claudia Renata dos Santos Barros; David Schlesinger; Debora Botequio Moretti; Dimas Tadeu Covas; Elaine Cristina Marqueze; Elaine Vieira Santos; Evandra Strazza Rodrigues; Heidge Fukumasu; Jayme Augusto de Souza-Neto; José Salvatore Leister Patané; Luiz Alcantara; Luiz Lehmann Coutinho; Maria Carolina Elias; Maurício Lacerda Nogueira; Rafael dos Santos Bezerra; Raul Machado Neto; Rejane Maria Tommasini Grotto; Ricardo Haddad; Sandra Coccuzzo Sampaio Vessoni; Simone Kashima; Svetoslav Nanev Slavov; Vincent Louis Viala |
| EPI_ISL_5801344, EPI_ISL_5801348, EPI_ISL_5801351, EPI_ISL_5801354, EPI_ISL_5801357 | Centro De Saude Iii Affonso Luzzi Santa Cruz Das Palmeiras | Instituto Butantan | Antonio Jorge Martins; Claudia Renata dos Santos Barros; David Schlesinger; Debora Botequio Moretti; Dimas Tadeu Covas; Elaine Cristina Marqueze; Elaine Vieira Santos; Evandra Strazza Rodrigues; Heidge Fukumasu; Jayme Augusto de Souza-Neto; José Salvatore Leister Patané; Luiz Alcantara; Luiz Lehmann Coutinho; Maria Carolina Elias; Maurício Lacerda Nogueira; Rafael dos Santos Bezerra; Raul Machado Neto; Rejane Maria Tommasini Grotto; Ricardo Haddad; Sandra Coccuzzo Sampaio Vessoni; Simone Kashima; Svetoslav Nanev Slavov; Vincent Louis Viala |
| EPI_ISL_5801280 | Centro De Saude Iii Alto Alegre | Instituto Butantan | Antonio Jorge Martins; Claudia Renata dos Santos Barros; David Schlesinger; Debora Botequio Moretti; Dimas Tadeu Covas; Elaine Cristina Marqueze; Elaine Vieira Santos; Evandra Strazza Rodrigues; Heidge Fukumasu; Jayme Augusto de Souza-Neto; José Salvatore Leister Patané; Luiz Alcantara; Luiz Lehmann Coutinho; Maria Carolina Elias; Maurício Lacerda Nogueira; Rafael dos Santos Bezerra; Raul Machado Neto; Rejane Maria Tommasini Grotto; Ricardo Haddad; Sandra Coccuzzo Sampaio Vessoni; Simone Kashima; Svetoslav Nanev Slavov; Vincent Louis Viala |
| EPI_ISL_5802709 | Centro De Saude Iii De Aguas De Sao Pedro | Instituto Butantan | Antonio Jorge Martins; Claudia Renata dos Santos Barros; David Schlesinger; Debora Botequio Moretti; Dimas Tadeu Covas; Elaine Cristina Marqueze; Elaine Vieira Santos; Evandra Strazza Rodrigues; Heidge Fukumasu; Jayme Augusto de Souza-Neto; José Salvatore Leister Patané; Luiz Alcantara; Luiz Lehmann Coutinho; Maria Carolina Elias; Maurício Lacerda Nogueira; Rafael dos Santos Bezerra; Raul Machado Neto; Rejane Maria Tommasini Grotto; Ricardo Haddad; Sandra Coccuzzo Sampaio Vessoni; Simone Kashima; Svetoslav Nanev Slavov; Vincent Louis Viala |
| EPI_ISL_5801272, EPI_ISL_5801273 | Centro De Saude Iii De Bocaina | Instituto Butantan | Antonio Jorge Martins; Claudia Renata dos Santos Barros; David Schlesinger; Debora Botequio Moretti; Dimas Tadeu Covas; Elaine Cristina Marqueze; Elaine Vieira Santos; Evandra Strazza Rodrigues; Heidge Fukumasu; Jayme Augusto de Souza-Neto; José Salvatore Leister Patané; Luiz Alcantara; Luiz Lehmann Coutinho; Maria Carolina Elias; Maurício Lacerda Nogueira; Rafael dos Santos Bezerra; Raul Machado Neto; Rejane Maria Tommasini Grotto; Ricardo Haddad; Sandra Coccuzzo Sampaio Vessoni; Simone Kashima; Svetoslav Nanev Slavov; Vincent Louis Viala |
| EPI_ISL_5802690 | Centro De Saude Iii De Divinolandia | Instituto Butantan | Antonio Jorge Martins; Claudia Renata dos Santos Barros; David Schlesinger; Debora Botequio Moretti; Dimas Tadeu Covas; Elaine Cristina Marqueze; Elaine Vieira Santos; Evandra Strazza Rodrigues; Heidge Fukumasu; Jayme Augusto de Souza-Neto; José Salvatore Leister Patané; Luiz Alcantara; Luiz Lehmann Coutinho; Maria Carolina Elias; Maurício Lacerda Nogueira; Rafael dos Santos Bezerra; Raul Machado Neto; Rejane Maria Tommasini Grotto; Ricardo Haddad; Sandra Coccuzzo Sampaio Vessoni; Simone Kashima; Svetoslav Nanev Slavov; Vincent Louis Viala |
| EPI_ISL_5801299 | Centro De Saude Iii De Igaracu Do Tiete | Instituto Butantan | Antonio Jorge Martins; Claudia Renata dos Santos Barros; David Schlesinger; Debora Botequio Moretti; Dimas Tadeu Covas; Elaine Cristina Marqueze; Elaine Vieira Santos; Evandra Strazza Rodrigues; Heidge Fukumasu; Jayme Augusto de Souza-Neto; José Salvatore Leister Patané; Luiz Alcantara; Luiz Lehmann Coutinho; Maria Carolina Elias; Maurício Lacerda Nogueira; Rafael dos Santos Bezerra; Raul Machado Neto; Rejane Maria Tommasini Grotto; Ricardo Haddad; Sandra Coccuzzo Sampaio Vessoni; Simone Kashima; Svetoslav Nanev Slavov; Vincent Louis Viala |
| EPI_ISL_5802710 | Centro Medico Dr Nelson Salome De Conchal | Instituto Butantan | Antonio Jorge Martins; Claudia Renata dos Santos Barros; David Schlesinger; Debora Botequio Moretti; Dimas Tadeu Covas; Elaine Cristina Marqueze; Elaine Vieira Santos; Evandra Strazza Rodrigues; Heidge Fukumasu; Jayme Augusto de Souza-Neto; José Salvatore Leister Patané; Luiz Alcantara; Luiz Lehmann Coutinho; Maria Carolina Elias; Maurício Lacerda Nogueira; Rafael dos Santos Bezerra; Raul Machado Neto; Rejane Maria Tommasini Grotto; Ricardo Haddad; Sandra Coccuzzo Sampaio Vessoni; Simone Kashima; Svetoslav Nanev Slavov; Vincent Louis Viala |
| EPI_ISL_5801172 | Centro Medico Pmesp | Instituto Butantan | Antonio Jorge Martins; Claudia Renata dos Santos Barros; David Schlesinger; Debora Botequio Moretti; Dimas Tadeu Covas; Elaine Cristina Marqueze; Elaine Vieira Santos; Evandra Strazza Rodrigues; Heidge Fukumasu; Jayme Augusto de Souza-Neto; José Salvatore Leister Patané; Luiz Alcantara; Luiz Lehmann Coutinho; Maria Carolina Elias; Maurício Lacerda Nogueira; Rafael dos Santos Bezerra; Raul Machado Neto; Rejane Maria Tommasini Grotto; Ricardo Haddad; Sandra Coccuzzo Sampaio Vessoni; Simone Kashima; Svetoslav Nanev Slavov; Vincent Louis Viala |
| EPI_ISL_4405261 | Centro Provincial de Referencia VIH/SIDA y Hepatitis Virales – Instituto Biológico Dr Tomás Perón | Área de Secuenciación del Laboratorio de Virología del Hospital de Niños Dr. Ricardo Gutierrez on behalf of 'Proyecto Argentino Interinstitucional de genomica de SARS-CoV-2' (PAIS Consortium) | A; Acuña; C; Carlos; Corazza; D; Dehaut; Dolcini; F; Fresina; Gatti; Gimenez; González; Goya; Gutierrez; K; L; LE; Lusso; M; MI; MS; Nabaes Jodar; Nardone; Natale; Nogueira Laspuir; Pagliari; R; Real; Romanko; S; Sanchez; Suarez; Szymanowski; Talavera; Ten Huver; V; Valinotto; Valle; Viegas, M. |
| EPI_ISL_4405195 | Centro de Investigaciones Básicas y Aplicadas, UNNOBA | Área de Secuenciación del Laboratorio de Virología del Hospital de Niños Dr. Ricardo Gutierrez on behalf of 'Proyecto Argentino Interinstitucional de genomica de SARS-CoV-2' (PAIS Consortium) | A; Acuña; Alaniz; Bagnis; Barbero; Bonadeo; Brandone; C; Cassarini; Castro; Chimento; Cristina; D; Demarchi; Español; F; Fernández; G; Goya; Gracia Balbi; Hernández del Pin; I; Icardi; L; LE; Lusso; M; MI; MS; Menite; Moroni; Morro; N; Nabaes Jodar; Natale; P; Palumbo; Pasquinelli; Perrone; R; Romano; Rosales; S; Sevic; Spinelli; V; Valinotto; Valla; Viegas, M.; Villafañe; Vitale |
| EPI_ISL_2402171 | Centro de Investigación Biomédica de Occidente (CIBO) | Unidad de Genomica Avanzada | Alejandro Sanchez-Flores; Alfredo Herrera-Estrella; Alicia Ocana-Mondragon; Angel Gustavo Salas-Lais; Bernardo Martinez-Miguel; Blanca Taboada; Brenda Irasema Maldonado-Meza; Carla Ivon Herrera-Najera; Carlos F. Arias; Celia Boukadida; Clara Esperanza Santacruz-Tinoco; Concepcion Grajales-Muniz; Consorcio Mexicano de Vigilancia Genomica (CoViGen-Mex). Authors (in alphabetical order): Julio Elias Alvarado-Yaah; Fernando Fontove-Herrera; Francisco Pulido; Gloria Elena Espinoza-Ayala; Gloria Maria Molina-Salinas; Gloria Vazquez; Hector Esteban Paz-Juarez; Hector Montoya-Fuentes; Helen Haydee Fernanda Ramirez-Plascencia; Jorge Ivan Salinal-Nevarez; Jose Antonio Enciso-Moreno; Jose Esteban Munoz-Medina; Jose de Jesus Nunez-Contreras; Juan Bautista Chale-Dzul; Luis Alberto Ochoa-Carrera; Margarita Matias-Florentino; Maria Guadalupe Santiago-Mauricio; Maria Guadalupe de Jesus Mireles-Rivera; Nelly Selem-Mojica; Pavel Isa; Ricardo Grande; Santiago Avila-Rios; Victor Hugo Borja-Aburto |
| EPI_ISL_2401970, EPI_ISL_2401975, EPI_ISL_2401979, EPI_ISL_2401991 | Centro de Investigación Biomédica del Noreste (CIBIN) | Unidad de Genomica Avanzada | Alejandro Sanchez-Flores; Alfredo Herrera-Estrella; Alicia Ocana-Mondragon; Angel Gustavo Salas-Lais; Bernardo Martinez-Miguel; Blanca Taboada; Brenda Irasema Maldonado-Meza; Carla Ivon Herrera-Najera; Carlos F. Arias; Celia Boukadida; Clara Esperanza Santacruz-Tinoco; Concepcion Grajales-Muniz; Consorcio Mexicano de Vigilancia Genomica (CoViGen-Mex). Authors (in alphabetical order): Julio Elias Alvarado-Yaah; Fernando Fontove-Herrera; Francisco Pulido; Gloria Elena Espinoza-Ayala; Gloria Maria Molina-Salinas; Gloria Vazquez; Hector Esteban Paz-Juarez; Hector Montoya-Fuentes; Helen Haydee Fernanda Ramirez-Plascencia; Jorge Ivan Salinal-Nevarez; Jose Antonio Enciso-Moreno; Jose Esteban Munoz-Medina; Jose de Jesus Nunez-Contreras; Juan Bautista Chale-Dzul; Luis Alberto Ochoa-Carrera; Margarita Matias-Florentino; Maria Guadalupe Santiago-Mauricio; Maria Guadalupe de Jesus Mireles-Rivera; Nelly Selem-Mojica; Pavel Isa; Ricardo Grande; Santiago Avila-Rios; Victor Hugo Borja-Aburto |
| EPI_ISL_4261865, EPI_ISL_4261866 | Chiu Laboratory, University of California, San Francisco | Chiu Laboratory, University of California, San Francisco | Alicia Sotomayor-Gonzalez; Baolin Wang; Candace Wang; Charles Chiu; Daniel Wong; Dustin Glasner; Venice Servellita; Xianding (Wayne) Deng; Yueyuan Zhang |
| EPI_ISL_2500223, EPI_ISL_2500224, EPI_ISL_2500226, EPI_ISL_2500235, EPI_ISL_2500246, EPI_ISL_2500247, EPI_ISL_2617558, EPI_ISL_2617565, EPI_ISL_2617566, EPI_ISL_2617570, EPI_ISL_2617582 | | | |
| see above | City of Milwaukee Health Department Laboratory | City of Milwaukee Health Department Laboratory | Amy Bauer; Jennifer Lentz; Manjeet Khubbar; Samantha Scott; Sanjib Bhattacharyya |
| EPI_ISL_2467156, EPI_ISL_2467159, EPI_ISL_2467170, EPI_ISL_2467171, EPI_ISL_2467173, EPI_ISL_2467174, EPI_ISL_2467179, EPI_ISL_2467181, EPI_ISL_2467182, EPI_ISL_2467185, EPI_ISL_2467188, EPI_ISL_2467200, EPI_ISL_2467203, EPI_ISL_2467208, EPI_ISL_2467210, EPI_ISL_2467214, EPI_ISL_2467215, EPI_ISL_2467216, EPI_ISL_2467217, EPI_ISL_2467218, EPI_ISL_2467219, EPI_ISL_2467220, EPI_ISL_2467352, EPI_ISL_3242638, EPI_ISL_3242639, EPI_ISL_3242640, EPI_ISL_3242645, EPI_ISL_3242649 | | | |
| see above | Clinica INDISA | "Facultad de Ciencias de la Vida, UNAB" | "Claudio Meneses; Ariel Orellana"; Claudio Olmos; Daniel Leon; Dayan Sanhueza; Eduardo Castro; Gonzalo Campaña; Macarena Bastias; Paola Pidal; Ricardo Yusta; Sebastian Wolter; Susana Saez; Víctor Monreal; Waldo Diaz |
| EPI_ISL_2098734 | Clinical Molecular Microbiology Laboratory, UNC Hospitals | Jeremy Wang | Alexander Rubinsteyn; Brianna de la Houssaye; Colleen Rice; Corbin Jones; Jason Smedberg; Jeremy Wang; Jose Luis Torres Castillo; Melissa Miller; Robert Hagan; Shawn Hawken |
| EPI_ISL_1935600 | Cliniques universitaires Saint-Luc | UCLouvain/IREC/MBLG | Benoit Kabamba Mukadi; Eleonore Ngyuvula; Jean Ruelle |
| EPI_ISL_4300518 | Color Genomics | Chiu Laboratory, University of California, San Francisco | Alicia Sotomayor-Gonzalez; Alicia Zhou; Baolin Wang; Charles Chiu; Daniel Wong; Erika Torres; Madyson Wanner; Noah Brazer; Scott Topper; Venice Servellita; Yueyuan Zhang |
| EPI_ISL_2309507, EPI_ISL_2309567, EPI_ISL_2309958, EPI_ISL_2309972, EPI_ISL_2310039, EPI_ISL_3160056, EPI_ISL_3988533, EPI_ISL_3988565 | | | |
| see above | Colorado Department of Public Health and Environment | Colorado Department of Public Health and Environment | Alexandria Rossheim; Diana Ir; Emily A. Travanty; Laura Bankers; Mandy Waters; Michael Martin; Molly C. Hetherington-Rauth; Sarah Elizabeth Totten; Shannon R. Matzinger |
| EPI_ISL_5802701 | Coordenadoria Municipal De Saude De Iracemapolis | Instituto Butantan | Antonio Jorge Martins; Claudia Renata dos Santos Barros; David Schlesinger; Debora Botequio Moretti; Dimas Tadeu Covas; Elaine Cristina Marqueze; Elaine Vieira Santos; Evandra Strazza Rodrigues; Heidge Fukumasu; Jayme Augusto de Souza-Neto; José Salvatore Leister Patané; Luiz Alcantara; Luiz Lehmann Coutinho; Maria Carolina Elias; Maurício Lacerda Nogueira; Rafael dos Santos Bezerra; Raul Machado Neto; Rejane Maria Tommasini Grotto; Ricardo Haddad; Sandra Coccuzzo Sampaio Vessoni; Simone Kashima; Svetoslav Nanev Slavov; Vincent Louis Viala |
| EPI_ISL_7727223 | Covid Laboratory Biogem | Covid Laboratory Biogem | Alessandra Fucci and Michele Caraglia; Alessia Maria Cossu; Cinzia Miarelli; Clara Iannarone; Egildo Luca D'andrea; Federica Melisi; Giovambattista Capasso; Marco Bocchetti; Marianna Scrima; Michele Ceccarelli; Piera Grisolia; Teresa Maria Rosaria Noviello; Ylenia Abruzzese |
| EPI_ISL_2562431 | Creighton University Medical Center | Creighton COVID Consortium | Cynthia Watson; Holly Stessman; Jake Siedlik; Michael Belshan; Richard Goering |
| EPI_ISL_5801097, EPI_ISL_5801279, EPI_ISL_5801288, EPI_ISL_5801290 | Cs Ii Dr Miguel Vitaliano Orlandia | Instituto Butantan | Antonio Jorge Martins; Claudia Renata dos Santos Barros; David Schlesinger; Debora Botequio Moretti; Dimas Tadeu Covas; Elaine Cristina Marqueze; Elaine Vieira Santos; Evandra Strazza Rodrigues; Heidge Fukumasu; Jayme Augusto de Souza-Neto; José Salvatore Leister Patané; Luiz Alcantara; Luiz Lehmann Coutinho; Maria Carolina Elias; Maurício Lacerda Nogueira; Rafael dos Santos Bezerra; Raul Machado Neto; Rejane Maria Tommasini Grotto; Ricardo Haddad; Sandra Coccuzzo Sampaio Vessoni; Simone Kashima; Svetoslav Nanev Slavov; Vincent Louis Viala |
| EPI_ISL_5801281 | Cs Ii Dr Sebastiao Ribeiro Do Valle | Instituto Butantan | Antonio Jorge Martins; Claudia Renata dos Santos Barros; David Schlesinger; Debora Botequio Moretti; Dimas Tadeu Covas; Elaine Cristina Marqueze; Elaine Vieira Santos; Evandra Strazza Rodrigues; Heidge Fukumasu; Jayme Augusto de Souza-Neto; José Salvatore Leister Patané; Luiz Alcantara; Luiz Lehmann Coutinho; Maria Carolina Elias; Maurício Lacerda Nogueira; Rafael dos Santos Bezerra; Raul Machado Neto; Rejane Maria Tommasini Grotto; Ricardo Haddad; Sandra Coccuzzo Sampaio Vessoni; Simone Kashima; Svetoslav Nanev Slavov; Vincent Louis Viala |
| EPI_ISL_5801384, EPI_ISL_5801385, EPI_ISL_5801389 | Cs Iolanda Fleury De Campos Guedes Nuporanga | Instituto Butantan | Antonio Jorge Martins; Claudia Renata dos Santos Barros; David Schlesinger; Debora Botequio Moretti; Dimas Tadeu Covas; Elaine Cristina Marqueze; Elaine Vieira Santos; Evandra Strazza Rodrigues; Heidge Fukumasu; Jayme Augusto de Souza-Neto; José Salvatore Leister Patané; Luiz Alcantara; Luiz Lehmann Coutinho; Maria Carolina Elias; Maurício Lacerda Nogueira; Rafael dos Santos Bezerra; Raul Machado Neto; Rejane Maria Tommasini Grotto; Ricardo Haddad; Sandra Coccuzzo Sampaio Vessoni; Simone Kashima; Svetoslav Nanev Slavov; Vincent Louis Viala |
| EPI_ISL_5801243, EPI_ISL_5801355 | Csii Dr Jose Ferreira Telles | Instituto Butantan | Antonio Jorge Martins; Claudia Renata dos Santos Barros; David Schlesinger; Debora Botequio Moretti; Dimas Tadeu Covas; Elaine Cristina Marqueze; Elaine Vieira Santos; Evandra Strazza Rodrigues; Heidge Fukumasu; Jayme Augusto de Souza-Neto; José Salvatore Leister Patané; Luiz Alcantara; Luiz Lehmann Coutinho; Maria Carolina Elias; Maurício Lacerda Nogueira; Rafael dos Santos Bezerra; Raul Machado Neto; Rejane Maria Tommasini Grotto; Ricardo Haddad; Sandra Coccuzzo Sampaio Vessoni; Simone Kashima; Svetoslav Nanev Slavov; Vincent Louis Viala |
| EPI_ISL_5802707 | Csiii Amalia Gothardo Manesco Santa Maria Da Serra | Instituto Butantan | Antonio Jorge Martins; Claudia Renata dos Santos Barros; David Schlesinger; Debora Botequio Moretti; Dimas Tadeu Covas; Elaine Cristina Marqueze; Elaine Vieira Santos; Evandra Strazza Rodrigues; Heidge Fukumasu; Jayme Augusto de Souza-Neto; José Salvatore Leister Patané; Luiz Alcantara; Luiz Lehmann Coutinho; Maria Carolina Elias; Maurício Lacerda Nogueira; Rafael dos Santos Bezerra; Raul Machado Neto; Rejane Maria Tommasini Grotto; Ricardo Haddad; Sandra Coccuzzo Sampaio Vessoni; Simone Kashima; Svetoslav Nanev Slavov; Vincent Louis Viala |
| EPI_ISL_2507235, EPI_ISL_2755068, EPI_ISL_2755070, EPI_ISL_2755087, EPI_ISL_2755104, EPI_ISL_2755128, EPI_ISL_2755150, EPI_ISL_2755206, EPI_ISL_2755234, EPI_ISL_2755266, EPI_ISL_2755515, EPI_ISL_2755575, EPI_ISL_2755576, EPI_ISL_2755577, EPI_ISL_2755589, EPI_ISL_2755608, EPI_ISL_2755609, EPI_ISL_2755650, EPI_ISL_2755651, EPI_ISL_2755654, EPI_ISL_2755743, EPI_ISL_2755827, EPI_ISL_2755828, EPI_ISL_2755834, EPI_ISL_2755840, EPI_ISL_2755847, EPI_ISL_2755856, EPI_ISL_2755858, EPI_ISL_2755859, EPI_ISL_2755866, EPI_ISL_2755867, EPI_ISL_2755880, EPI_ISL_2755881, EPI_ISL_2755895, EPI_ISL_2755918, EPI_ISL_2755934, EPI_ISL_2755995, EPI_ISL_2755996 | | | |
| see above | Curative Labs | Curative Labs | Elias L. Salfati; Eugenia Khorosheva; George Way; J.Cesar Ignacio-Espinoza; Janet Chen; Mikhail Hanewich-Hollatz; Nabjot Sandhu; Sophia Quasem; Vladimir Slepnev; Zhiyi Xie |
| EPI_ISL_3663910, EPI_ISL_3663921, EPI_ISL_3664000, EPI_ISL_3664021, EPI_ISL_3664024, EPI_ISL_3664025, EPI_ISL_3664036, EPI_ISL_3664045, EPI_ISL_3664067, EPI_ISL_3664098, EPI_ISL_4428139, EPI_ISL_4428141, EPI_ISL_4428144, EPI_ISL_4428147 | | | |
| see above | DASA | DASA | Adriano Bonaldi; Angelica Hristov; Annelise Lopes; Bianca Cota; Cristina Oliveira; Jose Levi; Lidia Yamamoto; Paulo Pierry; Rodrigo Guarischi; Rodrigo Salazar |
| EPI_ISL_2384000 | DC Public Health Lab/ Dept. of Forensic Sciences | Centers for Disease Control and Prevention Division of Viral Diseases, Pathogen Discovery | Alison Laufer Halpin; Ben L. Rambo-Martin; Clinton R. Paden; Dakota Howard; Darlene Wagner; Dave Wentworth; Dhwani Batra; Jasmine Padilla; Justin Lee; Katie Dillon; Krista Queen; Kristen Knipe; Kristine Lacek; Mark Burroughs; Matthew Schmerer; Mili Sheth; Peter Cook; Sam Shepard; Sarah Nobles; Shoshona Le; Suxiang Tong; Vivien Dugan; Yvette Unoarumhi |

| | | | |
|---|---|---|---|
| EPI_ISL_2345239, EPI_ISL_2345240, EPI_ISL_2345241, EPI_ISL_2345243, EPI_ISL_2345244, EPI_ISL_2345249, EPI_ISL_2345252, EPI_ISL_2345253, EPI_ISL_2345254, EPI_ISL_2345255, EPI_ISL_2345256, EPI_ISL_2345257, EPI_ISL_2345258, EPI_ISL_2345259, EPI_ISL_2345260 | | | |
| see above | DEPARTAMENTO DE SAUDE MUNICIPAL SOCORRO SP | Instituto Butantan / ESALQ-Piracicaba | Antonio Jorge Martins; Claudia Renata dos Santos Barros; David Schlesinger; Debora Botequio Moretti; Dimas Tadeu Covas; Elaine Cristina Marqueze; Elaine Vieira Santos; Evandra Strazza Rodrigues; Heidge Fukumasu; Jayme Augusto de Souza-Neto; José Salvatore Leister Patané; Luiz Alcantara; Luiz Lehmann Coutinho; Maria Carolina Elias; Maurício Lacerda Nogueira; Rafael dos Santos Bezerra; Raul Machado Neto; Rejane Maria Tommasini Grotto; Ricardo Haddad; Sandra Coccuzzo Sampaio Vessoni; Simone Kashima; Svetoslav Nanev Slavov; Vincent Louis Viala |
| EPI_ISL_2001353 | DIP. PREV. AVEZZANO SERVIZIO DI IGIENE EPIDEMIOLOGIA E SANITA' PUBBLICA | Istituto Zooprofilattico Sperimentale dell'Abruzzo e Molise "G. Caporale" | Ancora M; Calistri P; Cammà C; Caporale M; Curini V; Delli Compagni E; Di Domenico M; Di Lollo Valeria; Di Pasquale A; Lorusso A; Mangone I; Marcacci M; Puglia I; Rinaldi A; Savini G; Scialabba S |
| EPI_ISL_5104498, EPI_ISL_5104499, EPI_ISL_5104501, EPI_ISL_5104503, EPI_ISL_5104509 | DNAGYN | LGBio (Laboratorio de Genetica & Biodiversidade) | Alex Honda Bernardes; Amanda Alves de Melo; Aparecido Divino da Cruz; Cintia Pelegrineti Targueta de Azevedo Brito; Daniela de Melo e Silva; Elisangela de Paula Silveira Lacerda; Juliana Santana de Curcio; Luiz Augusto Pereira; Marc Alexandre Duarte Gigonzac; Mariana Pires de Campos Telles; Ramilla dos Santos Braga; Renata de Oliveira Dias; Rhewter Nunes; Thais Cidália Vieira Gigonzac; Thais Guimarães Castro; Thays Millena Alves Pedroso |
| EPI_ISL_2151376 | DOHMH Crown Heights | New York City Public Health Laboratory | Jade Wang; et al. |
| EPI_ISL_2151389 | DOHMH Jamaica | New York City Public Health Laboratory | Jade Wang; et al. |
| EPI_ISL_2151381 | DOHMH Morrisania | New York City Public Health Laboratory | Jade Wang; et al. |
| EPI_ISL_2151394 | DOHMH PHL | New York City Public Health Laboratory | Jade Wang; et al. |
| EPI_ISL_2383814, EPI_ISL_2383847, EPI_ISL_2383877, EPI_ISL_3692669 | DPHL | Delaware Public Health Lab | Rebecca Savage |
| EPI_ISL_5801345, EPI_ISL_5801346, EPI_ISL_5801347, EPI_ISL_5801350, EPI_ISL_5801352, EPI_ISL_5801359, EPI_ISL_5801362, EPI_ISL_5801363, EPI_ISL_5801364, EPI_ISL_5801365, EPI_ISL_5801366, EPI_ISL_5801367, EPI_ISL_5801368, EPI_ISL_5801369, EPI_ISL_5801370 | | | |
| see above | Departamento De Saude Municipal Socorro Sp | Instituto Butantan | Antonio Jorge Martins; Claudia Renata dos Santos Barros; David Schlesinger; Debora Botequio Moretti; Dimas Tadeu Covas; Elaine Cristina Marqueze; Elaine Vieira Santos; Evandra Strazza Rodrigues; Heidge Fukumasu; Jayme Augusto de Souza-Neto; José Salvatore Leister Patané; Luiz Alcantara; Luiz Lehmann Coutinho; Maria Carolina Elias; Maurício Lacerda Nogueira; Rafael dos Santos Bezerra; Raul Machado Neto; Rejane Maria Tommasini Grotto; Ricardo Haddad; Sandra Coccuzzo Sampaio Vessoni; Simone Kashima; Svetoslav Nanev Slavov; Vincent Louis Viala |
| EPI_ISL_2930633 | Department of Clinical Microbiology | GIGA Medical Genomics | Bouchra Boujemla; Claire Gourzonès; Cécile Meex; Keith Durkin; Laurent Gillet; Maria Artesi; Marie-Pierre Hayette; Nadine Cambisano; Nathalie Renotte; Olivier Ek; Sébastien Bontems; Vincent Bours |
| EPI_ISL_2099850, EPI_ISL_2099851 | Department of Public Health Bucharest | National Institute of Infectious Diseases-Prof. Dr. Matei Bals Molecular Diagnostics Laboratory | Andreea Tudor; Corina Casangiu; Dan Otelea; Leontina Banica; Marius Surleac; Ovidiu Vlaicu; Simona Paraschiv |
| EPI_ISL_2023036, EPI_ISL_2023037, EPI_ISL_2023038, EPI_ISL_2023040, EPI_ISL_2023041, EPI_ISL_2023045, EPI_ISL_2023046, EPI_ISL_2757950 | | | |
| see above | Department of Virology, Istituto Zooprofilattico Sperimentale del Lazio e della Toscana (IZSLT) | Department of General Diagnostics; Department of Virology; Istituto Zooprofilattico Sperimentale del Lazio e della Toscana (IZSLT) | Alessia Franco; Antonella Cersini; Antonio Battisti.; Elena L. Diaconu; Fabiola Feltrin; Giuseppe Manna; Patricia Alba; Raffaela Conti; Teresa Scicluna; Virginia Carfora |
| EPI_ISL_5069975 | Dr. Risch Ostschweiz AG | Microbiology | Dominique Fabien Hilti; Faina Wehrli; Lorenz Risch; Martin Risch; Nadia Wohlwend; Sinem Kas; Thomas Bodmer |
| EPI_ISL_3798148, EPI_ISL_3798153, EPI_ISL_3798164, EPI_ISL_3798184, EPI_ISL_3798210, EPI_ISL_4210139, EPI_ISL_4210144 | | | |
| see above | Dr. Risch Ostschweiz AG | Microbiology, Dr. Risch | Dominique Fabien Hilti; Faina Wehrli; Lorenz Risch; Martin Risch; Nadia Wohlwend; Sinem Kas; Thomas Bodmer |
| EPI_ISL_2093859, EPI_ISL_2093861, EPI_ISL_2093863, EPI_ISL_2093876, EPI_ISL_2093877, EPI_ISL_2093878, EPI_ISL_2093879, EPI_ISL_2093880, EPI_ISL_2093881, EPI_ISL_2093882, EPI_ISL_2093883, EPI_ISL_2093884, EPI_ISL_2093885, EPI_ISL_2093886, EPI_ISL_2093887, EPI_ISL_2093888, EPI_ISL_2093890, EPI_ISL_2093891, EPI_ISL_2094385, EPI_ISL_2219050, EPI_ISL_2219054, EPI_ISL_2219062, EPI_ISL_2219072, EPI_ISL_2219075, EPI_ISL_2219077, EPI_ISL_2219080, EPI_ISL_2219083, EPI_ISL_2219085, EPI_ISL_2219088, EPI_ISL_2475677 | | | |
| see above | Dutch COVID-19 response team | National Institute for Public Health and the Environment (RIVM) | Adam Meijer; AnneMarie van den Brandt; Annelies Kroneman; Bas van der Veer; Chantal Reusken; Dennis Schmitz; Dirk Eggink; Euníce Then; Florian Zwagemaker; Harry Vennema; James Groot; Jeroen Cremer; Karim Hajji; Kim Freriks; Linda van de Nes; Lisa Wijsman; Lynn Aarts; Melissa van Tuil; Ryanne Jaarsma; Sanne Bos; Sharon van den Brink; Sjoerd Kuiling; on behalf of the national COVID-19 response team |
| EPI_ISL_2345128 | ESF COHAB IV | Instituto Butantan / ESALQ-Piracicaba | Antonio Jorge Martins; Claudia Renata dos Santos Barros; David Schlesinger; Debora Botequio Moretti; Dimas Tadeu Covas; Elaine Cristina Marqueze; Elaine Vieira Santos; Evandra Strazza Rodrigues; Heidge Fukumasu; Jayme Augusto de Souza-Neto; José Salvatore Leister Patané; Luiz Alcantara; Luiz Lehmann Coutinho; Maria Carolina Elias; Maurício Lacerda Nogueira; Rafael dos Santos Bezerra; Raul Machado Neto; Rejane Maria Tommasini Grotto; Ricardo Haddad; Sandra Coccuzzo Sampaio Vessoni; Simone Kashima; Svetoslav Nanev Slavov; Vincent Louis Viala |
| EPI_ISL_3215593, EPI_ISL_3215602, EPI_ISL_3215610, EPI_ISL_3215669, EPI_ISL_3215692, EPI_ISL_3215710, EPI_ISL_3215725, EPI_ISL_3215733, EPI_ISL_3509000 | | | |
| see above | EXCITE Lab | Andersen lab at Scripps Research | Abigail Schnapper; Angela Scioscia; Chip Schooley; Christine Mitchell + SEARCH; David Pride; Fred Wu; Helena Tubb; Natasha Martin Cheryl Anderson; Sawyer Farmer; Sharon Reed; Sydney Morgan + Marni Jacobs + Celestine Magallanes + Holly Valentine + Louise C. Laurent + SEARCH; Tommy Valles + SEARCH |
| EPI_ISL_2478420, EPI_ISL_2478555, EPI_ISL_2478556, EPI_ISL_2478557, EPI_ISL_2478558, EPI_ISL_2478562, EPI_ISL_2478819, EPI_ISL_2478899, EPI_ISL_2478900, EPI_ISL_2478901, EPI_ISL_2478905, EPI_ISL_2478907, EPI_ISL_2478908, EPI_ISL_2479073, EPI_ISL_2479078, EPI_ISL_2479087, EPI_ISL_2479093, EPI_ISL_2479095, EPI_ISL_2479099, EPI_ISL_2479175, EPI_ISL_2479176, EPI_ISL_2479294, EPI_ISL_2479296, EPI_ISL_2479307, EPI_ISL_2479340, EPI_ISL_2479584, EPI_ISL_2479586, EPI_ISL_2479624, EPI_ISL_2479642, EPI_ISL_2479647, EPI_ISL_2479652, EPI_ISL_2479801, EPI_ISL_2940130, EPI_ISL_2940190, EPI_ISL_2941032, EPI_ISL_2941033, EPI_ISL_3117256, EPI_ISL_3909630, EPI_ISL_3909631, EPI_ISL_3909719, EPI_ISL_3909720, EPI_ISL_3909721 | | | |
| see above | Edmonton Provincial Lab | Public Health Agency of Canada (PHAC) National Microbiology Laboratory | Buss; Croxen M; Deo A; Dieu P; E; Ferrato C; Gill K; Khan F; Koleva P; Li V; Lloyd C; Lynch T; Ma R; Murphy S; Pabbaraju K; Shokoples S; Thayer J; Tipples G; Whitehouse M; Wong A; Yu C; Zelyas N |
| EPI_ISL_2346155 | Emory Molecular Diagnostics Laboratory, Emory Healthcare | Piantadosi Lab, Emory Department of Pathology | Ahmed Babiker; Anne Piantadosi |
| EPI_ISL_5801171 | Esf Cohab Iv | Instituto Butantan | Antonio Jorge Martins; Claudia Renata dos Santos Barros; David Schlesinger; Debora Botequio Moretti; Dimas Tadeu Covas; Elaine Cristina Marqueze; Elaine Vieira Santos; Evandra Strazza Rodrigues; Heidge Fukumasu; Jayme Augusto de Souza-Neto; José Salvatore Leister Patané; Luiz Alcantara; Luiz Lehmann Coutinho; Maria Carolina Elias; Maurício Lacerda Nogueira; Rafael dos Santos Bezerra; Raul Machado Neto; Rejane Maria Tommasini Grotto; Ricardo Haddad; Sandra Coccuzzo Sampaio Vessoni; Simone Kashima; Svetoslav Nanev Slavov; Vincent Louis Viala |
| EPI_ISL_2229055 | FL Bureau of Public Health Laboratories-Miami | Centers for Disease Control and Prevention Division of Viral Diseases, Pathogen Discovery | Alison Laufer Halpin; Ben L. Rambo-Martin; Clinton R. Paden; Dakota Howard; Darlene Wagner; Dave Wentworth; Dhwani Batra; Jasmine Padilla; Justin Lee; Katie Dillon; Krista Queen; Kristen Knipe; Kristine Lacek; Mark Burroughs; Matthew Schmerer; Mili Sheth; Peter Cook; Sam Shepard; Sarah Nobles; Shoshona Le; Suxiang Tong; Vivien Dugan; Yvette Unoarumhi |
| EPI_ISL_5782503 | Faculdade De Medicina Abc | Instituto Butantan | Antonio Jorge Martins; Claudia Renata dos Santos Barros; David Schlesinger; Debora Botequio Moretti; Dimas Tadeu Covas; Elaine Cristina Marqueze; Elaine Vieira Santos; Evandra Strazza Rodrigues; Heidge Fukumasu; Jayme Augusto de Souza-Neto; José Salvatore Leister Patané; Luiz Alcantara; Luiz Lehmann Coutinho; Maria Carolina Elias; Maurício Lacerda Nogueira; Rafael dos Santos Bezerra; Raul Machado Neto; Rejane Maria Tommasini Grotto; Ricardo Haddad; Sandra Coccuzzo Sampaio Vessoni; Simone Kashima; Svetoslav Nanev Slavov; Vincent Louis Viala |
| EPI_ISL_2230005, EPI_ISL_2230015, EPI_ISL_2277710, EPI_ISL_2277737, EPI_ISL_2277740, EPI_ISL_2277918, EPI_ISL_2277956, EPI_ISL_2277964, EPI_ISL_2277965, EPI_ISL_2277966, EPI_ISL_2277967, EPI_ISL_2277968, EPI_ISL_2277969, EPI_ISL_2442736, EPI_ISL_2442739, EPI_ISL_2442745, EPI_ISL_2442748, EPI_ISL_2442773, EPI_ISL_2442774, EPI_ISL_2442776, EPI_ISL_2442781, EPI_ISL_2442785, EPI_ISL_2442795, EPI_ISL_2442800, EPI_ISL_2442809, EPI_ISL_2442824, EPI_ISL_2442831, EPI_ISL_2442833, EPI_ISL_2567091, EPI_ISL_2567092, EPI_ISL_2567093, EPI_ISL_2567099, EPI_ISL_2567101, EPI_ISL_2836027, EPI_ISL_2836028, EPI_ISL_2836029, EPI_ISL_2836031, EPI_ISL_2836033, EPI_ISL_2836034, EPI_ISL_2836270, EPI_ISL_2836298, EPI_ISL_2836448, EPI_ISL_2836449, EPI_ISL_2836491, EPI_ISL_2836625, EPI_ISL_2836676, EPI_ISL_2836681, EPI_ISL_2836687, EPI_ISL_6049952, EPI_ISL_6049984, EPI_ISL_6049986, EPI_ISL_6049988, EPI_ISL_6049990, EPI_ISL_6049996, EPI_ISL_6049997, EPI_ISL_6049999, EPI_ISL_6050011, EPI_ISL_6050013, EPI_ISL_6050015, EPI_ISL_6050017, EPI_ISL_6050019, EPI_ISL_6050021, EPI_ISL_6050022, EPI_ISL_6050054, EPI_ISL_6050055, EPI_ISL_6050071, EPI_ISL_6050073, EPI_ISL_6050074, EPI_ISL_6050188, EPI_ISL_6050231, EPI_ISL_6050609, EPI_ISL_6050619, EPI_ISL_6050640, EPI_ISL_6050732 | | | |
| see above | Florida Bureau of Public Health Laboratories | Florida Bureau of Public Health Laboratories | Jason Blanton; Namratha Tarigopula; Sarah Schmedes; Tiffany Splatt |
| EPI_ISL_2405211 | Flow Diagnostics Lab | Los Angeles County Public Health Laboratories | P. Hemarajata et al. |
| EPI_ISL_2620954 | Fondazione IRCCS Ca' Granda Ospedale Maggiore Policlinico | Fondazione IRCCS Ca' Granda Ospedale Maggiore Policlinico | Ferruccio Ceriotti; Sara Uceda Renteria |
| EPI_ISL_4237489 | Fulgent Diagnostic Laboratory | Centers for Disease Control and Prevention Division of Viral Diseases, Pathogen Discovery | Alex Burgin; Ben Rambo-Martin; Clinton Paden; Dakota Howard; Dave Wentworth; Dhwani Batra; Jasmine Padilla; Justin Lee; Krista Queen; Kristen Knipe; Kristine Lacek; Mark Burroughs; Matthew Schmerer; Meghan Bentz; Mili Sheth; Peter Cook; Sam Shepard; Sarah Nobles; Suxiang Tong; Vivien Dugan; Yvette Unoarumhi |
| EPI_ISL_1992023, EPI_ISL_1992028, EPI_ISL_1992036, EPI_ISL_1992046, EPI_ISL_1992053, EPI_ISL_1992063, EPI_ISL_1992070, EPI_ISL_1992082, EPI_ISL_1992088, EPI_ISL_1992096, EPI_ISL_1992097, EPI_ISL_1992112, EPI_ISL_1992121, EPI_ISL_1992143, EPI_ISL_1992149, EPI_ISL_1992152, EPI_ISL_1992174, EPI_ISL_1992178, EPI_ISL_1992179, EPI_ISL_1992190, EPI_ISL_1992196, EPI_ISL_1992201, EPI_ISL_1992202, EPI_ISL_1992203, EPI_ISL_1992206, EPI_ISL_1992212, EPI_ISL_1992324, EPI_ISL_1992327, EPI_ISL_1992341, EPI_ISL_1992345, EPI_ISL_1992348, EPI_ISL_1992364, EPI_ISL_1992370, EPI_ISL_1992377, EPI_ISL_1992392, EPI_ISL_1992824, EPI_ISL_1992855, EPI_ISL_1992884, EPI_ISL_1992892, EPI_ISL_1992921, EPI_ISL_1992968, EPI_ISL_1992969, EPI_ISL_1992977, EPI_ISL_1992981, EPI_ISL_1992984, EPI_ISL_1992985, EPI_ISL_1993019, EPI_ISL_1993020, EPI_ISL_1993032, EPI_ISL_1993040, EPI_ISL_1993067, EPI_ISL_1993076, EPI_ISL_1993087, EPI_ISL_1993095, EPI_ISL_1993119, EPI_ISL_1993129, EPI_ISL_1993134, EPI_ISL_1993151, EPI_ISL_1993170, EPI_ISL_1993173, EPI_ISL_1993175, EPI_ISL_1993176, EPI_ISL_1993177, EPI_ISL_1993181, EPI_ISL_1993183, EPI_ISL_1993185, EPI_ISL_1993190, EPI_ISL_1993191, EPI_ISL_1993195, EPI_ISL_1993199, EPI_ISL_1993202, EPI_ISL_1993293, EPI_ISL_1993355, EPI_ISL_1993357, EPI_ISL_1993370, EPI_ISL_1993373, EPI_ISL_2043630, EPI_ISL_2043633, EPI_ISL_2043636, EPI_ISL_2043661, EPI_ISL_2043685, EPI_ISL_2043693, EPI_ISL_2043695, EPI_ISL_2043699, EPI_ISL_2043701, EPI_ISL_2043704, EPI_ISL_2043705, EPI_ISL_2043782, EPI_ISL_2043788, EPI_ISL_2043796, EPI_ISL_2043800, EPI_ISL_2043805, EPI_ISL_2043824, EPI_ISL_2043828, EPI_ISL_2043840, EPI_ISL_2043865, EPI_ISL_2043880, EPI_ISL_2043933, EPI_ISL_2134514, EPI_ISL_2134520, EPI_ISL_2134525, EPI_ISL_2134551, EPI_ISL_2134562, EPI_ISL_2134564, EPI_ISL_2134576, EPI_ISL_2134577, EPI_ISL_2179864, EPI_ISL_2179871, EPI_ISL_4370652 | | | |
| see above | Fulgent Genetics | Centers for Disease Control and Prevention Division of Viral Diseases, Pathogen Discovery | Adrian Paskey; Becky Tsai; Benafsh Sapra; Benjamin Rambo-Martin; Christopher Gulvick; Clinton Paden; Clinton R. Paden; Dakota Howard; Darlene Wagner; Dhwani Batra; Doreen Ng; Duncan MacCannell; Erisa Sula; Harry Gao; James Xie; Jason Caravas; John Gao; Joseph Fierro; Kara Moser; Kristine Lacek; Matthew Schmerer; Mickey Li; Peter Cook; Peter W. Cook; Scott Sammons; Shatavia Morrison; Tymeckia Kendall; Victoria Caban Figueroa; Yan Meng; Yvette Unoarumhi |
| EPI_ISL_6701386, EPI_ISL_6701387, EPI_ISL_6701388, EPI_ISL_6701389, EPI_ISL_6701415 | Fundação Ezequiel Dias | Instituto René Rachou / Fiocruz Minas | André Bernardes; Anna Salim; Caroline Penido; Felipe Iani; Flávio Araújo; Gabriel Fernandes; Pedro Alves; Rubens do Monte Neto; Thaís Silva |
| EPI_ISL_2311749, EPI_ISL_2422457 | GA Department of Public Health Laboratory | Genomics and Discovery, Respiratory Viruses Branch, Division of Viral Diseases, Centers for Disease Control and Prevention | Adam Retchless; Anna Kelleher; Anna Uehara; Brian Lynch; Clinton R. Paden; Dhwani Batra; Haibin Wang; Han Jia Justin Ng; Jasmine Padilla; Jing Zhang; Justin Lee; Krista Queen; Mark Burroughs; Mili Sheth; Morgan Davis; Peter Cook; Rachel Marine; Sarah Nobles; Suxiang Tong; Tara Coalter; Yan Li; Ying Tao |
| EPI_ISL_1915595 | GHDC - SITE ST JOSEPH | Institut de Pathologie et Genetique | Jérémie Gras; Pascale Hilbert |

| | | | |
|---|---|---|---|
| | | (IPG) | |
| EPI_ISL_2421158 | GLENS FALLS HOSPITAL LABORATORY | Wadsworth Center, New York State Department of Health | Alexis Russell; Catharine Prussing; Daryl M. Lamson; Erasmus Schneider; Erica Lasek-Nesselquist; John Kelly; Jonathan Plitnick; Kirsten St. George; Matthew Shudt; Melissa A Leisner; Navjot Singh |
| EPI_ISL_2009535, EPI_ISL_2009541, EPI_ISL_2009556, EPI_ISL_2391042, EPI_ISL_2391044, EPI_ISL_2391045, EPI_ISL_2391049, EPI_ISL_2391052, EPI_ISL_2391061, EPI_ISL_2391064, EPI_ISL_2391077, EPI_ISL_2391087, EPI_ISL_2391089, EPI_ISL_2391091, EPI_ISL_2391092, EPI_ISL_2391093, EPI_ISL_2391099, EPI_ISL_2391100, EPI_ISL_2391106, EPI_ISL_2391107, EPI_ISL_2391108, EPI_ISL_2391123, EPI_ISL_2391126, EPI_ISL_2391127, EPI_ISL_2391128, EPI_ISL_2391130, EPI_ISL_2391131, EPI_ISL_2391132, EPI_ISL_2391133, EPI_ISL_2391145, EPI_ISL_2391148, EPI_ISL_2391153, EPI_ISL_2391157, EPI_ISL_2391166, EPI_ISL_2391178, EPI_ISL_2391270, EPI_ISL_2391298, EPI_ISL_2391391 | | | |
| see above | Genetica Molecular and Subdepartamento de Virologia ISP Chile | Instituto de Salud Publica de Chile | Andres Castillo; Barbara Parra; Constanza Campano; Gisselle Barra; Javier Tognarelli; Jorge Fernandez; Karen Orostica; Loredana Arata; Patricia Bustos; Rodrigo Fasce; Soledad Ulloa |
| EPI_ISL_5196283 | Gorgas Memorial Institute of Health Studies | Gorgas Memorial Institute of Health Studies | Castillo Jorge; Chen Maria; Franco Danilo; Gonzalez Claudia; Jessica Gondola; Leyda Abrego; Lopez-Verges Sandra; Marlenne Castillo; Martinez Alexander; Menacho Abdiel; Moreno Ambar; Moreno Brechla; Oris Chavarria; Ortiz Alma; Salazar Jacqueline |
| EPI_ISL_2601808, EPI_ISL_2601814, EPI_ISL_2601821, EPI_ISL_2601994, EPI_ISL_2602027, EPI_ISL_2602040, EPI_ISL_2602075, EPI_ISL_2602076, EPI_ISL_2602079, EPI_ISL_2854812, EPI_ISL_2854839, EPI_ISL_2854858, EPI_ISL_2854860, EPI_ISL_2854877, EPI_ISL_2854906, EPI_ISL_2854913, EPI_ISL_2854917, EPI_ISL_2854931 | | | |
| see above | Gravity Diagnostics, LLC | Gravity Diagnostics, LLC | Gravity Diagnostics |
| EPI_ISL_4926393 | Grupo Inmunovirología, Universidad de Antioquia | Grupo Inmunovirología, Universidad de Antioquia | Francisco J. Diaz; Maria T. Rugeles; Wbeimar Aguilar-Jimenez |
| EPI_ISL_7307236 | Grupo de Investigación en Enfermedades Tropicales del Ejército (GINETEJ), Laboratorio de Referencia e Investigación, Dirección de Sanidad Ejército, Bogotá, Colombia | Centro de Investigaciones en Microbiología y Biotecnología-UR (CIMBIUR), Facultad de Ciencias Naturales, Universidad del Rosario, Bogotá, Colombia | Alberto Paniz-Mondolfi; Angie Ramírez; Beatriz Ariza; Camilo A. Correa-Cárdenas; Carlos Gómez-Restrepo; Claudia Cardozo-Romero; Claudia Méndez; David-Santiago Quevedo; Guido España; Hernando Díaz; Juan David Ramírez; Juliana Cuervo-Rojas; Julie Pérez; Luz H. Patiño; Manuel-Antonio Franco; Maria-Clara Duque; Marina Muñoz; Nathalia Ballesteros; Nicolas Luna; Sergio Castañeda; Zulma M. Cucunubá |
| EPI_ISL_2451655 | Guam Public Health Laboratory | Centers for Disease Control and Prevention Division of Viral Diseases, Pathogen Discovery | Alison Laufer Halpin; Ben L. Rambo-Martin; Clinton R. Paden; Dakota Howard; Darlene Wagner; Dave Wentworth; Dhwani Batra; Jasmine Padilla; Justin Lee; Katie Dillon; Krista Queen; Kristen Knipe; Kristine Lacek; Mark Burroughs; Matthew Schmerer; Mili Sheth; Peter Cook; Sam Shepard; Sarah Nobles; Shoshona Le; Suxiang Tong; Vivien Dugan; Yvette Unoarumhi |
| EPI_ISL_1960108, EPI_ISL_1960121 | Gundersen Clinical Microbiology Laboratory | Kabara Cancer Research Institute | Craig S. Richmond; Paraic A. Kenny |
| EPI_ISL_2945749, EPI_ISL_2945763, EPI_ISL_2945781, EPI_ISL_2945888, EPI_ISL_2945925, EPI_ISL_2978815, EPI_ISL_2978824 | | | |
| see above | HG Pharma | Elling and Cochella laboratories, IMBA/IMP | Alexander Stark; Ezgi Oezkan; Luisa Cochella and Ulrich Elling; Marcus Strobl; Olga Frank; Ramesh Yelagandula; Tanino Albanese |
| EPI_ISL_3102213 | HGCC HOSPITAL GERAL DR CESAR CALS | Analytical Competence Molecular Epidemiology Lab/ACME, Oswaldo Cruz Foundation, Ceara (FIOCRUZ CE) | Cleber Furtado Aksenen; Fabio Miyajima; Fernando Braga Stehling; Francisco Eder de Moura Lopes; Jamille Maria Mendes Bezerra; Joaquim César do Nascimento Sousa Junior; Pedro Miguel Carneiro Jeronimo; Suzana Porto Almeida e Lucas Delerino; Thais Ferreira de Oliveira; Thais de Oliveira Costa; Ticiane Cavalcante de Souza; Veridiana Pessoa Miyajima |
| EPI_ISL_2017431, EPI_ISL_2017432, EPI_ISL_2017433, EPI_ISL_2017434, EPI_ISL_2017435, EPI_ISL_2017436, EPI_ISL_2017437, EPI_ISL_2017438, EPI_ISL_2017439, EPI_ISL_2017440, EPI_ISL_2187988, EPI_ISL_2187990, EPI_ISL_2187991, EPI_ISL_2497437, EPI_ISL_2497438, EPI_ISL_2497439, EPI_ISL_4890125 | | | |
| see above | HLAGYN - Laboratorio de Imunologia de Transplantes de Goias | HLAGYN - Laboratorio de Imunologia de Transplantes de Goias | Alessandro Leonardo Alvares Magalhaes; Daniel Ferreira de Sousa; Danielle de Paiva Rezende; Erika Lopes Rocha Batista; Fernando Antonio Vinhal dos Santos; Frederico Rodrigues Vinhal; Kamila Oliveira Reis De Freitas.; Lucas Carlos Gomes Pereira; Paola Cristina Resende Silva; Sabrina Sara Moreira Duarte |
| EPI_ISL_2345764, EPI_ISL_2362254 | HOSP E MATERNIDADE MUNICIPAL N SRA MONTE SERRAT | Instituto Butantan / Mendelics | Antonio Jorge Martins; Claudia Renata dos Santos Barros; David Schlesinger; Debora Botequio Moretti; Dimas Tadeu Covas; Elaine Cristina Marqueze; Elaine Vieira Santos; Evandra Strazza Rodrigues; Heidge Fukumasu; Jayme Augusto de Souza-Neto; José Salvatore Leister Patané; Luiz Alcantara; Luiz Lehmann Coutinho; Maria Carolina Elias; Maurício Lacerda Nogueira; Rafael dos Santos Bezerra; Raul Machado Neto; Rejane Maria Tommasini Grotto; Ricardo Haddad; Sandra Coccuzzo Sampaio Vessoni; Simone Kashima; Svetoslav Nanev Slavov; Vincent Louis Viala |
| EPI_ISL_2345074, EPI_ISL_2345075 | HOSP MUN CIDADE TIRADENTES CARMEN PRUDENTE | Instituto Butantan / ESALQ-Piracicaba | Antonio Jorge Martins; Claudia Renata dos Santos Barros; David Schlesinger; Debora Botequio Moretti; Dimas Tadeu Covas; Elaine Cristina Marqueze; Elaine Vieira Santos; Evandra Strazza Rodrigues; Heidge Fukumasu; Jayme Augusto de Souza-Neto; José Salvatore Leister Patané; Luiz Alcantara; Luiz Lehmann Coutinho; Maria Carolina Elias; Maurício Lacerda Nogueira; Rafael dos Santos Bezerra; Raul Machado Neto; Rejane Maria Tommasini Grotto; Ricardo Haddad; Sandra Coccuzzo Sampaio Vessoni; Simone Kashima; Svetoslav Nanev Slavov; Vincent Louis Viala |
| EPI_ISL_2345068, EPI_ISL_2345071, EPI_ISL_2345072, EPI_ISL_2345073 | HOSP MUN DE MOGI DAS CRUZES PREF WALDEMAR COSTA FILHO | Instituto Butantan / ESALQ-Piracicaba | Antonio Jorge Martins; Claudia Renata dos Santos Barros; David Schlesinger; Debora Botequio Moretti; Dimas Tadeu Covas; Elaine Cristina Marqueze; Elaine Vieira Santos; Evandra Strazza Rodrigues; Heidge Fukumasu; Jayme Augusto de Souza-Neto; José Salvatore Leister Patané; Luiz Alcantara; Luiz Lehmann Coutinho; Maria Carolina Elias; Maurício Lacerda Nogueira; Rafael dos Santos Bezerra; Raul Machado Neto; Rejane Maria Tommasini Grotto; Ricardo Haddad; Sandra Coccuzzo Sampaio Vessoni; Simone Kashima; Svetoslav Nanev Slavov; Vincent Louis Viala |
| EPI_ISL_2100335, EPI_ISL_2100346 | HOSPITAL CAN MISSES | HOSPITAL UNIVERSITARIO SON ESPASES | Antonio Oliver; Carla López-Causapé; Pablo Fraile-Ribot; SeqCovid |
| EPI_ISL_2209101, EPI_ISL_2209363, EPI_ISL_2209883, EPI_ISL_2210101, EPI_ISL_2210236, EPI_ISL_2345676, EPI_ISL_2345722, EPI_ISL_2345731, EPI_ISL_2345819, EPI_ISL_2345826, EPI_ISL_2345870 | | | |
| see above | HOSPITAL DE CAMPANHA COVID 19 MUNICIPIO DE TAUBATE | Instituto Butantan / Mendelics | Antonio Jorge Martins; Claudia Renata dos Santos Barros; David Schlesinger; Debora Botequio Moretti; Dimas Tadeu Covas; Elaine Cristina Marqueze; Elaine Vieira Santos; Evandra Strazza Rodrigues; Heidge Fukumasu; Jayme Augusto de Souza-Neto; José Salvatore Leister Patané; Luiz Alcantara; Luiz Lehmann Coutinho; Maria Carolina Elias; Maurício Lacerda Nogueira; Rafael dos Santos Bezerra; Raul Machado Neto; Rejane Maria Tommasini Grotto; Ricardo Haddad; Sandra Coccuzzo Sampaio Vessoni; Simone Kashima; Svetoslav Nanev Slavov; Vincent Louis Viala |
| EPI_ISL_3502660 | HOSPITAL DE GETAFE | HOSPITAL UNIVERSITARIO 12 DE OCTUBRE | Carmen Martín-Higuera; Esther Viedma; Irene Múñoz-Gallego; M.ª Dolores Folgueira; Mar Aguilera; Noelia Moral; Rafael Delgado; Sagrario Zurita |
| EPI_ISL_2674336, EPI_ISL_2674338, EPI_ISL_2674340 | HOSPITAL DEPARTAMENTAL MARIA INMACULADA | Instituto Nacional de Salud- Dirección de Investigación en Salud Pública | Carlos Franco-Muñoz; Carmen Osorio; Diana Malo; Diego A. Álvarez-Díaz; Diego Andrés Prada; Gerardo Santamaría; Hector Alejandro Ruiz-Moreno; Jhonnatan Reales-González; Jorge Rivera; Juan Camilo Martinez; Julian Naizaque; Katherine Laiton-Donato; Lisseth Pardo; Magdalena Wiesner; Marcela Mercado-Reyes; Maria T. Herrera-Sepúlveda; Marta Lopez Blanco; Martha Lucia Ospina Martinez; Paola Rojas; Sergio Gomez; Sheryll Corchuelo; Ángela Alarcon Cruz |
| EPI_ISL_2345131 | HOSPITAL DR OSIRIS FLORINDO COELHO FERRAZ DE VASCONCELOS | Instituto Butantan / ESALQ-Piracicaba | Antonio Jorge Martins; Claudia Renata dos Santos Barros; David Schlesinger; Debora Botequio Moretti; Dimas Tadeu Covas; Elaine Cristina Marqueze; Elaine Vieira Santos; Evandra Strazza Rodrigues; Heidge Fukumasu; Jayme Augusto de Souza-Neto; José Salvatore Leister Patané; Luiz Alcantara; Luiz Lehmann Coutinho; Maria Carolina Elias; Maurício Lacerda Nogueira; Rafael dos Santos Bezerra; Raul Machado Neto; Rejane Maria Tommasini Grotto; Ricardo Haddad; Sandra Coccuzzo Sampaio Vessoni; Simone Kashima; Svetoslav Nanev Slavov; Vincent Louis Viala |
| EPI_ISL_2209292 | HOSPITAL E MATERNIDADE NOSSA SENHORA DA AJUDA | Instituto Butantan / Mendelics | Antonio Jorge Martins; Claudia Renata dos Santos Barros; David Schlesinger; Debora Botequio Moretti; Dimas Tadeu Covas; Elaine Cristina Marqueze; Elaine Vieira Santos; Evandra Strazza Rodrigues; Heidge Fukumasu; Jayme Augusto de Souza-Neto; José Salvatore Leister Patané; Luiz Alcantara; Luiz Lehmann Coutinho; Maria Carolina Elias; Maurício Lacerda Nogueira; Rafael dos Santos Bezerra; Raul Machado Neto; Rejane Maria Tommasini Grotto; Ricardo Haddad; Sandra Coccuzzo Sampaio Vessoni; Simone Kashima; Svetoslav Nanev Slavov; Vincent Louis Viala |
| EPI_ISL_2345077, EPI_ISL_2345086, EPI_ISL_2345143 | HOSPITAL GERAL JESUS TEIXEIRA DA COSTA GUAIANASES SAO PAULO | Instituto Butantan / ESALQ-Piracicaba | Antonio Jorge Martins; Claudia Renata dos Santos Barros; David Schlesinger; Debora Botequio Moretti; Dimas Tadeu Covas; Elaine Cristina Marqueze; Elaine Vieira Santos; Evandra Strazza Rodrigues; Heidge Fukumasu; Jayme Augusto de Souza-Neto; José Salvatore Leister Patané; Luiz Alcantara; Luiz Lehmann Coutinho; Maria Carolina Elias; Maurício Lacerda Nogueira; Rafael dos Santos Bezerra; Raul Machado Neto; Rejane Maria Tommasini Grotto; Ricardo Haddad; Sandra Coccuzzo Sampaio Vessoni; Simone Kashima; Svetoslav Nanev Slavov; Vincent Louis Viala |
| EPI_ISL_2208890 | HOSPITAL MUNICIPAL DE IBIUNA IBIUNA SP | Instituto Butantan / Mendelics | Antonio Jorge Martins; Claudia Renata dos Santos Barros; David Schlesinger; Debora Botequio Moretti; Dimas Tadeu Covas; Elaine Cristina Marqueze; Elaine Vieira Santos; Evandra Strazza Rodrigues; Heidge Fukumasu; Jayme Augusto de Souza-Neto; José Salvatore Leister Patané; Luiz Alcantara; Luiz Lehmann Coutinho; Maria Carolina Elias; Maurício Lacerda Nogueira; Rafael dos Santos Bezerra; Raul Machado Neto; Rejane Maria Tommasini Grotto; Ricardo Haddad; Sandra Coccuzzo Sampaio Vessoni; Simone Kashima; Svetoslav Nanev Slavov; Vincent Louis Viala |
| EPI_ISL_2801361, EPI_ISL_2801368, EPI_ISL_3102296 | HOSPITAL MUNICIPAL NOSSA SENHORA DOS MILAGRES | Analytical Competence Molecular Epidemiology Lab/ACME, Oswaldo Cruz Foundation, Ceara (FIOCRUZ CE) | Cleber Furtado Aksenen; Cleber Furtado Aksenen e Suzana Porto Almeida; Fabio Miyajima; Fernando Braga Stehling; Francisco Eder de Moura Lopes; Jamille Maria Mendes Bezerra; Joaquim César do Nascimento Sousa Junior; Pedro Miguel Carneiro Jeronimo; Suzana Porto Almeida e Lucas Delerino; Thais Ferreira de Oliveira; Thais de Oliveira Costa; Ticiane Cavalcante de Souza; Veridiana Pessoa Miyajima |
| EPI_ISL_3102267 | HOSPITAL REGIONAL NORTE | Analytical Competence Molecular Epidemiology Lab/ACME, Oswaldo Cruz Foundation, Ceara (FIOCRUZ CE) | Cleber Furtado Aksenen; Fabio Miyajima; Fernando Braga Stehling; Francisco Eder de Moura Lopes; Jamille Maria Mendes Bezerra; Joaquim César do Nascimento Sousa Junior; Pedro Miguel Carneiro Jeronimo; Suzana Porto Almeida e Lucas Delerino; Thais Ferreira de Oliveira; Thais de Oliveira Costa; Ticiane Cavalcante de Souza; Veridiana Pessoa Miyajima |
| EPI_ISL_2345202 | HOSPITAL SANTA THEREZINHA BROTAS | Instituto Butantan / ESALQ-Piracicaba | Antonio Jorge Martins; Claudia Renata dos Santos Barros; David Schlesinger; Debora Botequio Moretti; Dimas Tadeu Covas; Elaine Cristina Marqueze; Elaine Vieira Santos; Evandra Strazza Rodrigues; Heidge Fukumasu; Jayme Augusto de Souza-Neto; José Salvatore Leister Patané; Luiz Alcantara; Luiz Lehmann Coutinho; Maria Carolina Elias; Maurício Lacerda Nogueira; Rafael dos Santos Bezerra; Raul Machado Neto; Rejane Maria Tommasini Grotto; Ricardo Haddad; Sandra Coccuzzo Sampaio Vessoni; Simone Kashima; Svetoslav Nanev Slavov; Vincent Louis Viala |
| EPI_ISL_2801369 | HOSPITAL SAO MATEUS | Analytical Competence Molecular Epidemiology Lab/ACME, Oswaldo Cruz Foundation, Ceara (FIOCRUZ CE) | Cleber Furtado Aksenen e Suzana Porto Almeida; Fabio Miyajima; Fernando Braga Stehling; Francisco Eder de Moura Lopes; Jamille Maria Mendes Bezerra; Joaquim César do Nascimento Sousa Junior; Pedro Miguel Carneiro Jeronimo; Thais Ferreira de Oliveira; Thais de Oliveira Costa; Ticiane Cavalcante de Souza; Veridiana Pessoa Miyajima |
| EPI_ISL_3502741, EPI_ISL_3502744, EPI_ISL_3502746, EPI_ISL_3502750, EPI_ISL_3502754, EPI_ISL_3502760 | HOSPITAL UNIVERSITARIO 12 DE OCTUBRE | HOSPITAL UNIVERSITARIO 12 DE OCTUBRE | Carmen Martín-Higuera; Esther Viedma; Irene Múñoz-Gallego; M.ª Dolores Folgueira; Mar Aguilera; Noelia Moral; Rafael Delgado; Sagrario Zurita |
| EPI_ISL_2100300, EPI_ISL_2100312, EPI_ISL_2100342, EPI_ISL_2241449 | HOSPITAL UNIVERSITARIO SON ESPASES | HOSPITAL UNIVERSITARIO SON ESPASES | Antonio Oliver; Carla López-Causapé; Pablo Fraile-Ribot; SeqCovid |
| EPI_ISL_2005563, EPI_ISL_2022533, EPI_ISL_2080385 | Health Services Laboratories | Wellcome Sanger Institute for the COVID-19 Genomics UK (COG-UK) Consortium | Cordelia Langford; David K. Jackson; Dominic Kwiatkowski; Ewan Harrison; Health Services Laboratories and Alex Alderton; Ian Johnston; Jeffrey Barrett; John Sillitoe on behalf of the Wellcome Sanger Institute COVID-19 Surveillance Team; Roberto Amato; Sonia Goncalves |
| EPI_ISL_2097388, EPI_ISL_2097397, EPI_ISL_2097425, EPI_ISL_2097448, EPI_ISL_2097462, EPI_ISL_2097463, EPI_ISL_2097466, EPI_ISL_2097467, EPI_ISL_2097476, EPI_ISL_2097498, EPI_ISL_2097605, EPI_ISL_2097627, EPI_ISL_2097635, EPI_ISL_2097639, EPI_ISL_2097647, EPI_ISL_2097654, EPI_ISL_2097659, EPI_ISL_2097668, EPI_ISL_2097753, EPI_ISL_2098108, EPI_ISL_2144069, EPI_ISL_2144084, EPI_ISL_2144092, EPI_ISL_2144100, EPI_ISL_2144101, EPI_ISL_2144111, EPI_ISL_2144116, EPI_ISL_2144121, EPI_ISL_2144126, EPI_ISL_2144133, EPI_ISL_2144140, EPI_ISL_2144194, EPI_ISL_2144205, EPI_ISL_2144211, EPI_ISL_2144224, EPI_ISL_2144233, EPI_ISL_2144276, EPI_ISL_2144299, EPI_ISL_2144321, EPI_ISL_2144322, EPI_ISL_2144346, EPI_ISL_2144372, EPI_ISL_2144376, EPI_ISL_2144391, EPI_ISL_2144393, EPI_ISL_2144396, EPI_ISL_2144400, EPI_ISL_2144401, EPI_ISL_2144404, EPI_ISL_2144414, EPI_ISL_2144417, EPI_ISL_2144437, EPI_ISL_2144451, EPI_ISL_2144455, EPI_ISL_2144476, EPI_ISL_2144483, EPI_ISL_2144598, EPI_ISL_2144605, EPI_ISL_2144635, EPI_ISL_2144640, EPI_ISL_2144693, EPI_ISL_2144708, EPI_ISL_2144724, EPI_ISL_2144742, EPI_ISL_2144744, EPI_ISL_2144756, EPI_ISL_2144759, EPI_ISL_2144771, EPI_ISL_2144773, EPI_ISL_2144774, EPI_ISL_2144799, EPI_ISL_2144918, EPI_ISL_2144934, EPI_ISL_2144955, EPI_ISL_2144956, EPI_ISL_2144959, EPI_ISL_2144977, EPI_ISL_2144982, EPI_ISL_2144998, EPI_ISL_2145002, EPI_ISL_2145018, EPI_ISL_2145029, EPI_ISL_2145031, EPI_ISL_2145039, EPI_ISL_2145054, EPI_ISL_2145061, EPI_ISL_2145082, EPI_ISL_2145132, EPI_ISL_2145144, EPI_ISL_2145169, EPI_ISL_2145185, EPI_ISL_2145198, EPI_ISL_2145201, EPI_ISL_2145236, EPI_ISL_2145268, EPI_ISL_2145314, EPI_ISL_2145320, EPI_ISL_2145349, EPI_ISL_2145352, EPI_ISL_2145372, EPI_ISL_2145384, EPI_ISL_2145433, EPI_ISL_2145512, EPI_ISL_2145556, EPI_ISL_2145663, EPI_ISL_2145761, EPI_ISL_2159478, EPI_ISL_2159479, EPI_ISL_2159485, EPI_ISL_2159490, EPI_ISL_2159491, EPI_ISL_2159492, EPI_ISL_2159497, EPI_ISL_2159508, EPI_ISL_2159518, EPI_ISL_2159529, EPI_ISL_2159535, EPI_ISL_2159538, EPI_ISL_2159557, EPI_ISL_2159571, EPI_ISL_2159590, EPI_ISL_2159628, EPI_ISL_2159635, EPI_ISL_2159697, EPI_ISL_2159713, EPI_ISL_2159728, EPI_ISL_2159734, EPI_ISL_2159736, EPI_ISL_2159764, EPI_ISL_2159773, EPI_ISL_2159775, EPI_ISL_2159778, EPI_ISL_2159781, EPI_ISL_2159784, EPI_ISL_2159786, EPI_ISL_2159790, EPI_ISL_2159812, EPI_ISL_2159824, EPI_ISL_2159826, EPI_ISL_2159831, EPI_ISL_2159865, EPI_ISL_2159868, EPI_ISL_2159889, EPI_ISL_2159906, EPI_ISL_2159907, EPI_ISL_2159919, EPI_ISL_2159940, EPI_ISL_2159943, EPI_ISL_2159950, EPI_ISL_2159959, EPI_ISL_2159976, EPI_ISL_2159977, EPI_ISL_2159997, EPI_ISL_2160021, | | | |

| | | | |
|---|---|---|---|
| EPI_ISL_2160037, EPI_ISL_2160048, EPI_ISL_2160052, EPI_ISL_2160054, EPI_ISL_2160093, EPI_ISL_2160102, EPI_ISL_2160106, EPI_ISL_2160125, EPI_ISL_2160127, EPI_ISL_2160147, EPI_ISL_2160163, EPI_ISL_2160181, EPI_ISL_2160182, EPI_ISL_2160194, EPI_ISL_2160206, EPI_ISL_2160212, EPI_ISL_2160221, EPI_ISL_2160223, EPI_ISL_2160228, EPI_ISL_2160271, EPI_ISL_2160408, EPI_ISL_2160476, EPI_ISL_2160493, EPI_ISL_2160524, EPI_ISL_2160526, EPI_ISL_2160527, EPI_ISL_2160537, EPI_ISL_2160545, EPI_ISL_2160556, EPI_ISL_2160586, EPI_ISL_2160610, EPI_ISL_2160617, EPI_ISL_2160624, EPI_ISL_2160634, EPI_ISL_2160645, EPI_ISL_2160664, EPI_ISL_2160683, EPI_ISL_2160712, EPI_ISL_2160722, EPI_ISL_2160728, EPI_ISL_2187320, EPI_ISL_2187334, EPI_ISL_2187339, EPI_ISL_2187619, EPI_ISL_2187623, EPI_ISL_2187638, EPI_ISL_2247672, EPI_ISL_2247715, EPI_ISL_2247820, EPI_ISL_2247822, EPI_ISL_2247834, EPI_ISL_2247846, EPI_ISL_2247848, EPI_ISL_2247864, EPI_ISL_2247874, EPI_ISL_2247877, EPI_ISL_2247899, EPI_ISL_2248232, EPI_ISL_2248233, EPI_ISL_2248251, EPI_ISL_2248252, EPI_ISL_2248293, EPI_ISL_2248329, EPI_ISL_2248335, EPI_ISL_2248355, EPI_ISL_2248368, EPI_ISL_4345828 | | | |
| see above | Helix/Illumina | Centers for Disease Control and Prevention Division of Viral Diseases, Pathogen Discovery | Adrian Paskey; Alexandre Bolze; Ary Ascencio; Benjamin Rambo-Martin; Brad Sickler; Charlotte Rivera-Garcia; Christine Tran; Christopher Gulvick; Chrstine Tran; Clinton Paden; Clinton R. Paden; Dakota Howard; Darlene Wagner; David Becker; Dhwani Batra; Duncan MacCannell; Efren Sandoval; Eileen De Feo; Eileen de Feo; Elizabeth Cirulli; Eric Allen; Geraint Levan; James Lu; Jan Antico; Jason Caravas; Jason Nguyen; Jimmy Ramirez; Jingtao Liu; Kara Moser; Kelly Barrett; Kelly Schiabor Barrett; Kim Gietzen; Kristine Lacek; Magnus Isaksson; Marc Laurent; Matthew Schmerer; Matthew Tolentino; Nicole L. Washington; Nicole Washington; Peter Cook; Peter W. Cook; Phil Febbo; Ryan Cho; Scott Sammons; Shannon Wickline; Shatavia Morrison; Sherry Wang; Simon White; Tyler Cassens; William Lee; Yvette Unoarumhi |
| EPI_ISL_5801425, EPI_ISL_5801426 | Hosp E Maternidade Municipal N Sra Monte Serrat | Instituto Butantan | Antonio Jorge Martins; Claudia Renata dos Santos Barros; David Schlesinger; Debora Botequio Moretti; Dimas Tadeu Covas; Elaine Cristina Marqueze; Elaine Vieira Santos; Evandra Strazza Rodrigues; Heidge Fukumasu; Jayme Augusto de Souza-Neto; José Salvatore Leister Patané; Luiz Alcantara; Luiz Lehmann Coutinho; Maria Carolina Elias; Maurício Lacerda Nogueira; Rafael dos Santos Bezerra; Raul Machado Neto; Rejane Maria Tommasini Grotto; Ricardo Haddad; Sandra Coccuzzo Sampaio Vessoni; Simone Kashima; Svetoslav Nanev Slavov; Vincent Louis Viala |
| EPI_ISL_5801092, EPI_ISL_5801093 | Hosp Mun Cidade Tiradentes Carmen Prudente | Instituto Butantan | Antonio Jorge Martins; Claudia Renata dos Santos Barros; David Schlesinger; Debora Botequio Moretti; Dimas Tadeu Covas; Elaine Cristina Marqueze; Elaine Vieira Santos; Evandra Strazza Rodrigues; Heidge Fukumasu; Jayme Augusto de Souza-Neto; José Salvatore Leister Patané; Luiz Alcantara; Luiz Lehmann Coutinho; Maria Carolina Elias; Maurício Lacerda Nogueira; Rafael dos Santos Bezerra; Raul Machado Neto; Rejane Maria Tommasini Grotto; Ricardo Haddad; Sandra Coccuzzo Sampaio Vessoni; Simone Kashima; Svetoslav Nanev Slavov; Vincent Louis Viala |
| EPI_ISL_5801086, EPI_ISL_5801089, EPI_ISL_5801090, EPI_ISL_5801091 | Hosp Mun De Mogi Das Cruzes Pref Waldemar Costa Filho | Instituto Butantan | Antonio Jorge Martins; Claudia Renata dos Santos Barros; David Schlesinger; Debora Botequio Moretti; Dimas Tadeu Covas; Elaine Cristina Marqueze; Elaine Vieira Santos; Evandra Strazza Rodrigues; Heidge Fukumasu; Jayme Augusto de Souza-Neto; José Salvatore Leister Patané; Luiz Alcantara; Luiz Lehmann Coutinho; Maria Carolina Elias; Maurício Lacerda Nogueira; Rafael dos Santos Bezerra; Raul Machado Neto; Rejane Maria Tommasini Grotto; Ricardo Haddad; Sandra Coccuzzo Sampaio Vessoni; Simone Kashima; Svetoslav Nanev Slavov; Vincent Louis Viala |
| EPI_ISL_2030070, EPI_ISL_2188427, EPI_ISL_2279645 | Hospital | National Reference Center for Viruses of Respiratory Infections, Institut Pasteur, Paris | Alain Delepine; Amaury Vaysse; Angela Brisebarre; Camille Capel; Christophe Malabat; Corinne Maufrais; Etienne Simon-Lorière; Frédéric Lemoine; Louise Lefrançois; Marion Barbet; Maud Vanpeene; Méline Bizard; Pascale Martres; StéPhanie Marque Juillet; Sylvie Behillil; Sylvie Van der Werf; Vincent Enouf |
| EPI_ISL_5801480, EPI_ISL_5801483, EPI_ISL_5801484, EPI_ISL_5801485, EPI_ISL_5801486, EPI_ISL_5801488, EPI_ISL_5801490, EPI_ISL_5801492, EPI_ISL_5801494, EPI_ISL_5801509, EPI_ISL_5801511, EPI_ISL_5801512, EPI_ISL_5801513, EPI_ISL_5801515, EPI_ISL_5801516, EPI_ISL_5801517 | | | |
| see above | Hospital De Campanha Covid 19 Municipio De Taubate | Instituto Butantan | Antonio Jorge Martins; Claudia Renata dos Santos Barros; David Schlesinger; Debora Botequio Moretti; Dimas Tadeu Covas; Elaine Cristina Marqueze; Elaine Vieira Santos; Evandra Strazza Rodrigues; Heidge Fukumasu; Jayme Augusto de Souza-Neto; José Salvatore Leister Patané; Luiz Alcantara; Luiz Lehmann Coutinho; Maria Carolina Elias; Maurício Lacerda Nogueira; Rafael dos Santos Bezerra; Raul Machado Neto; Rejane Maria Tommasini Grotto; Ricardo Haddad; Sandra Coccuzzo Sampaio Vessoni; Simone Kashima; Svetoslav Nanev Slavov; Vincent Louis Viala |
| EPI_ISL_5801175 | Hospital Dr Osiris Florindo Coelho Ferraz De Vasconcelos | Instituto Butantan | Antonio Jorge Martins; Claudia Renata dos Santos Barros; David Schlesinger; Debora Botequio Moretti; Dimas Tadeu Covas; Elaine Cristina Marqueze; Elaine Vieira Santos; Evandra Strazza Rodrigues; Heidge Fukumasu; Jayme Augusto de Souza-Neto; José Salvatore Leister Patané; Luiz Alcantara; Luiz Lehmann Coutinho; Maria Carolina Elias; Maurício Lacerda Nogueira; Rafael dos Santos Bezerra; Raul Machado Neto; Rejane Maria Tommasini Grotto; Ricardo Haddad; Sandra Coccuzzo Sampaio Vessoni; Simone Kashima; Svetoslav Nanev Slavov; Vincent Louis Viala |
| EPI_ISL_5801481, EPI_ISL_5801482 | Hospital E Maternidade Nossa Senhora Da Ajuda | Instituto Butantan | Antonio Jorge Martins; Claudia Renata dos Santos Barros; David Schlesinger; Debora Botequio Moretti; Dimas Tadeu Covas; Elaine Cristina Marqueze; Elaine Vieira Santos; Evandra Strazza Rodrigues; Heidge Fukumasu; Jayme Augusto de Souza-Neto; José Salvatore Leister Patané; Luiz Alcantara; Luiz Lehmann Coutinho; Maria Carolina Elias; Maurício Lacerda Nogueira; Rafael dos Santos Bezerra; Raul Machado Neto; Rejane Maria Tommasini Grotto; Ricardo Haddad; Sandra Coccuzzo Sampaio Vessoni; Simone Kashima; Svetoslav Nanev Slavov; Vincent Louis Viala |
| EPI_ISL_1964180, EPI_ISL_1964181, EPI_ISL_1964216, EPI_ISL_3505917, EPI_ISL_3505921, EPI_ISL_3505922, EPI_ISL_3505923, EPI_ISL_3505927 | | | |
| see above | Hospital General Universitario Gregorio Marañón | Hospital General Universitario Gregorio Marañón | Cristina Rodriguez-Grande; Darío García de Viedma; Julia Suárez; Laura Pérez-Lago; Marta Herranz Martin; Patricia Muñoz; Pedro Sola Campoy; Pilar Catalán; Sergio Buenestado Serrano; Victor Manuel de la Cueva |
| EPI_ISL_2271588, EPI_ISL_2374111, EPI_ISL_2374120 | Hospital General Universitario de Alicante - Instituto de Investigación Sanitaria y Biomédica de Alicante | SeqCOVID-SPAIN consortium/IBV(CSIC) | Carmen Molina Pardines and SeqCOVID-SPAIN consortium; Maripaz Ventero Martín |
| EPI_ISL_5801096, EPI_ISL_5801109, EPI_ISL_5801190 | Hospital Geral Jesus Teixeira Da Costa Guaianases Sao Paulo | Instituto Butantan | Antonio Jorge Martins; Claudia Renata dos Santos Barros; David Schlesinger; Debora Botequio Moretti; Dimas Tadeu Covas; Elaine Cristina Marqueze; Elaine Vieira Santos; Evandra Strazza Rodrigues; Heidge Fukumasu; Jayme Augusto de Souza-Neto; José Salvatore Leister Patané; Luiz Alcantara; Luiz Lehmann Coutinho; Maria Carolina Elias; Maurício Lacerda Nogueira; Rafael dos Santos Bezerra; Raul Machado Neto; Rejane Maria Tommasini Grotto; Ricardo Haddad; Sandra Coccuzzo Sampaio Vessoni; Simone Kashima; Svetoslav Nanev Slavov; Vincent Louis Viala |
| EPI_ISL_3033049 | Hospital Jaime Ferre - SAMCO Rafaela | Grupo de Genómica y Bioinformática del Instituto de Investigación de la Cadena Láctea CONICET-INTA on behalf of 'Proyecto Argentino Interinstitucional de genomica de SARS-CoV-2' (PAIS Consortium) | AF; Amadio; C; Eberhardt; Irazoqui; Isaia; JF; JM; MF; Pandolfi; Quaranta; V |
| EPI_ISL_5801427 | Hospital Municipal De Ibiuna Ibiuna Sp | Instituto Butantan | Antonio Jorge Martins; Claudia Renata dos Santos Barros; David Schlesinger; Debora Botequio Moretti; Dimas Tadeu Covas; Elaine Cristina Marqueze; Elaine Vieira Santos; Evandra Strazza Rodrigues; Heidge Fukumasu; Jayme Augusto de Souza-Neto; José Salvatore Leister Patané; Luiz Alcantara; Luiz Lehmann Coutinho; Maria Carolina Elias; Maurício Lacerda Nogueira; Rafael dos Santos Bezerra; Raul Machado Neto; Rejane Maria Tommasini Grotto; Ricardo Haddad; Sandra Coccuzzo Sampaio Vessoni; Simone Kashima; Svetoslav Nanev Slavov; Vincent Louis Viala |
| EPI_ISL_5801294 | Hospital Municipal Dr Tabajara Ramos | Instituto Butantan | Antonio Jorge Martins; Claudia Renata dos Santos Barros; David Schlesinger; Debora Botequio Moretti; Dimas Tadeu Covas; Elaine Cristina Marqueze; Elaine Vieira Santos; Evandra Strazza Rodrigues; Heidge Fukumasu; Jayme Augusto de Souza-Neto; José Salvatore Leister Patané; Luiz Alcantara; Luiz Lehmann Coutinho; Maria Carolina Elias; Maurício Lacerda Nogueira; Rafael dos Santos Bezerra; Raul Machado Neto; Rejane Maria Tommasini Grotto; Ricardo Haddad; Sandra Coccuzzo Sampaio Vessoni; Simone Kashima; Svetoslav Nanev Slavov; Vincent Louis Viala |
| EPI_ISL_2365419 | Hospital San Agustín | Laboratorio de Virología HUCA | Abreu F; Alvarez-Arguelles ME; Boga JA; Castelló C; Costales I; Coto E; Gómez de Oña J; Martín-Rodríguez G; Melón S; Perez-Martínez Z; Rojo S; Sandoval M |
| EPI_ISL_7711734 | Hospital San Vicente De Paul | Universidad de Costa Rica e Instituto Costarricense de Investigacion y Ensenanza en Nutricion y Salud | Brenes Hebleen; Camacho Erwin; Campos-Sanchez Rebeca; Ceballos Ana; Cordero Estela; Cristancho Marco; Duarte Francisco; Fernandez-Do Porto Dario; Herrera-Estrella Alfredo; Jimenez-Moraila Beatriz; Kreuze Jan; Mireles-Rivera Guadalupe; Molina-Mora Jose; Munoz-Medina Jose Esteban; Negri Tatiana; Nunes Gisele; Oliveira Guilherme; Oliveira Renato; Reales-Gonzalez Jhonnatan; Remes-Lenicov Federico; Reyes Alejandro; Sosa Ezequiel; Soto Claudio; Tsukayama Pablo; Turjanski Adrian; and Zayat Jonathan |
| EPI_ISL_5801293 | Hospital Santa Therezinha Brotas | Instituto Butantan | Antonio Jorge Martins; Claudia Renata dos Santos Barros; David Schlesinger; Debora Botequio Moretti; Dimas Tadeu Covas; Elaine Cristina Marqueze; Elaine Vieira Santos; Evandra Strazza Rodrigues; Heidge Fukumasu; Jayme Augusto de Souza-Neto; José Salvatore Leister Patané; Luiz Alcantara; Luiz Lehmann Coutinho; Maria Carolina Elias; Maurício Lacerda Nogueira; Rafael dos Santos Bezerra; Raul Machado Neto; Rejane Maria Tommasini Grotto; Ricardo Haddad; Sandra Coccuzzo Sampaio Vessoni; Simone Kashima; Svetoslav Nanev Slavov; Vincent Louis Viala |
| EPI_ISL_5802716 | Hospital Sao Jose Itirapina | Instituto Butantan | Antonio Jorge Martins; Claudia Renata dos Santos Barros; David Schlesinger; Debora Botequio Moretti; Dimas Tadeu Covas; Elaine Cristina Marqueze; Elaine Vieira Santos; Evandra Strazza Rodrigues; Heidge Fukumasu; Jayme Augusto de Souza-Neto; José Salvatore Leister Patané; Luiz Alcantara; Luiz Lehmann Coutinho; Maria Carolina Elias; Maurício Lacerda Nogueira; Rafael dos Santos Bezerra; Raul Machado Neto; Rejane Maria Tommasini Grotto; Ricardo Haddad; Sandra Coccuzzo Sampaio Vessoni; Simone Kashima; Svetoslav Nanev Slavov; Vincent Louis Viala |
| EPI_ISL_2017673, EPI_ISL_2017674 | Hospital Universitari Bellvitge | Microbiology Department | Aida Gonzalez-Diaz; Carmen Ardanuy; Jordi Camara; Jordi Niubó; Laura Calatayud; M Angeles Domínguez; Miguel Fernandez-Huerta; Sara Marti |
| EPI_ISL_1908890, EPI_ISL_1908893 | Hospital Universitari Joan XXIII de Tarragona | Hospital Universitari Vall d'Hebron - Vall d'Hebron Institut de Recerca | Andrés Antón; Ariadna Rando; Carla Castillo; Cristina Andrés; Damir Garcia-Cehic; Josep Quer; Juliana Esperalba; Maria Carmen Martin; Maria Gema Codina; Maria Piñana; Tomàs Pumarola |
| EPI_ISL_2047711, EPI_ISL_2047712, EPI_ISL_2047721, EPI_ISL_2162247 | Hospital Universitari Vall d'Hebron - Vall d'Hebron Institut de Recerca | Hospital Universitari Vall d'Hebron - Vall d'Hebron Institut de Recerca | Alejandra González-Sánchez; Andrés Antón; Ariadna Rando; Carla Castillo; Cristina Andrés; Damir Garcia-Cehic; Josep Quer; Juliana Esperalba; Maria Carmen Martin; Maria Gema Codina; Maria Piñana; Tomàs Pumarola |
| EPI_ISL_2323396 | Hospital Universitari i Politècnic La Fe de València | SeqCOVID-SPAIN consortium/IBV(CSIC) | Ana Gil Brusola; Eva González Barberá; José Luis López Hontangas and SeqCOVID-SPAIN consortium; María Dolores Gómez Ruiz; Salvador Giner Almaraz |
| EPI_ISL_7307259, EPI_ISL_7307291 | Hospital Universitario San Ignacio | Centro de Investigaciones en Microbiología y Biotecnología-UR (CIMBIUR), Facultad de Ciencias Naturales, Universidad del Rosario, Bogotá, Colombia | Alberto Paniz-Mondolfi; Angie Ramírez; Beatriz Ariza; Camilo A. Correa-Cárdenas; Carlos Gómez-Restrepo; Claudia Cardozo-Romero; Claudia Méndez; David-Santiago Quevedo; Guido España; Hernando Díaz; Juan David Ramírez; Juliana Cuervo-Rojas; Julie Pérez; Luz H. Patiño; Manuel-Antonio Franco; Maria-Clara Duque; Marina Muñoz; Nathalia Ballesteros; Nicolas Luna; Sergio Castañeda; Zulma M. Cucunubá |
| EPI_ISL_2465946 | Hospital Universitario Virgen de las Nieves de Granada-SAS | SeqCOVID-SPAIN consortium/IBV(CSIC) | Irene Pedrosa Corral; José M. Navarro-Marí and SeqCOVID-SPAIN consortium; Sara Sanbonmatsu Gámez |
| EPI_ISL_2238703, EPI_ISL_2238704 | Hospital de Cabueñes | Laboratorio de Virología HUCA | Abreu F; Alvarez-Arguelles ME; Boga JA; Castelló C; Costales I; Coto E; Gómez de Oña J; Martín-Rodríguez G; Melón S; Perez-Martínez Z; Rojo S; Sandoval M |
| EPI_ISL_3022992, EPI_ISL_3023011, EPI_ISL_3023015, EPI_ISL_3023037, EPI_ISL_3023045, EPI_ISL_3023111, EPI_ISL_3023255 | | | |
| see above | Hospital of the University of Pennsylvania Molecular Pathology Lab | Bushman Lab - University of Pennsylvania | Aoife M. Roche; Arupa Ganguly; Ayannah S. Fitzgerald; Brendan Kelly; Jevon Graham-Wooten; John K. Everett; Kyle Rodino; Layla A. Khatib; Mike Feldman; Ronald G. Collman and Frederic Bushan; Samantha A. Whiteside; Scott Sherrill-Mix; Shantan Reddy; Young Hwang |
| EPI_ISL_4196730 | Hospital zonal Francisco Lopez Lima General Roca | Laboratorio Central Mg. Luis Alfredo Pianciola on behalf of 'Proyecto Argentino Interinstitucional de genomica de SARS-CoV-2' (PAIS Consortium) | A Montecinos.; C Pintos; C Rastellini; C Ziehm; D Durany; J Laher; J Ousset; L Pianciola; M Fernandez; M Mazzeo; T Paniceres |
| EPI_ISL_1961020 | Houston Health Dept. | Houston Health Dept. | Adolpho Lara; Pamela Brown; Ryker Penn; Yanlai Lai |
| EPI_ISL_2224718, EPI_ISL_2225228, EPI_ISL_2225268, EPI_ISL_2225269, EPI_ISL_2225308, EPI_ISL_2225324, EPI_ISL_2225349, EPI_ISL_2225382, EPI_ISL_2225387, EPI_ISL_2225419, EPI_ISL_2225425, EPI_ISL_2225468 | | | |
| see above | Houston Methodist Hospital | Houston Methodist Hospital | Ilya J. Finkelstein; James J. Davis; Jessica Cambric; Jimmy Gollihar; Kristina Reppond; Layne Pruitt; Madison N. Shyer; Marcus Nguyen; Matthew Ojeda Saavedra; Paul A. Christensen; Prasanti Yerramilli; Randall J. Olsen; Robert Olson; Ryan Gadd; S. Wesley Long; Sishir Subedi; and James M. Musser |
| EPI_ISL_2228289, EPI_ISL_2228314, EPI_ISL_2341509 | Humboldt County Public Health Laboratory | Chan-Zuckerberg Biohub | CZB Cliahub Consortium |
| EPI_ISL_2452079 | ID Bureau of Laboratories | Centers for Disease Control and Prevention Division of Viral Diseases, Pathogen Discovery | Alison Laufer Halpin; Ben L. Rambo-Martin; Clinton R. Paden; Dakota Howard; Darlene Wagner; Dave Wentworth; Dhwani Batra; Jasmine Padilla; Justin Lee; Katie Dillon; Krista Queen; Kristen Knipe; Kristine Lacek; Mark Burroughs; Matthew Schmerer; Mili Sheth; Peter Cook; Sam Shepard; Sarah Nobles; Shoshona Le; Suxiang Tong; Vivien Dugan; Yvette Unoarumhi |
| EPI_ISL_2444814, EPI_ISL_2444815, EPI_ISL_2444823, EPI_ISL_2444824, EPI_ISL_2444826, EPI_ISL_2444827, EPI_ISL_2444830 | | | |
| see above | IICS-UNA | IICS-UNA | Adriana Valenzuela; Alejandra Rojas; Chyntia Diaz; Eva Nara; Fatima Cardozo; Florencia del Puerto; Joel Ortiz; Jonas Fernandez; Laura Franco; Laura Mendoza; Leticia Rojas; Magaly Martinez; Maria Eugenia Galeano. |
| EPI_ISL_2115383 | IMD - Institut fur Medizinische | Robert Koch Institute | |

| | | | |
|---|---|---|---|
| | Diagnostik Berlin-Potsdam | | |
| EPI_ISL_2097206, EPI_ISL_2134587, EPI_ISL_2134592, EPI_ISL_2134625, EPI_ISL_2134626, EPI_ISL_2134628, EPI_ISL_2134629, EPI_ISL_2242527, EPI_ISL_2242529, EPI_ISL_2242531, EPI_ISL_2242536, EPI_ISL_2242542, EPI_ISL_2242547, EPI_ISL_2304496, EPI_ISL_2361368, EPI_ISL_2361369, EPI_ISL_2361371, EPI_ISL_2361380, EPI_ISL_2361425 | | | |
| see above | IN State Department of Health Laboratory Services | IN State Department of Health Laboratory Services | Brian Pope; Cassandra Campion; Jamie Yeadon; Kyle Brownlee; Lixia Liu; Mark Glazier; Melissa Hindenlang |
| EPI_ISL_2134579 | INSA | Instituto Nacional de Saude (INSA) | Borges et al |
| EPI_ISL_2444184 | IPIGUA | Instituto Butantan / FAMERP | Antonio Jorge Martins; Claudia Renata dos Santos Barros; David Schlesinger; Debora Botequio Moretti; Dimas Tadeu Covas; Elaine Cristina Marqueze; Elaine Vieira Santos; Evandra Strazza Rodrigues; Heidge Fukumasu; Jayme Augusto de Souza-Neto; José Salvatore Leister Patané; Luiz Alcantara; Luiz Lehmann Coutinho; Maria Carolina Elias; Maurício Lacerda Nogueira; Rafael dos Santos Bezerra; Raul Machado Neto; Rejane Maria Tommasini Grotto; Ricardo Haddad; Sandra Coccuzzo Sampaio Vessoni; Simone Kashima; Svetoslav Nanev Slavov; Vincent Louis Viala |
| EPI_ISL_2612491 | ISDH | Quantigen Biosciences | Paul J. Childress |
| EPI_ISL_2500373, EPI_ISL_2500407 | IVY3 Central Lab, Vanderbilt University Medical Center | Lauring Lab, University of Michigan, Department of Microbiology and Immunology | Valesano |
| EPI_ISL_2031620, EPI_ISL_2140150, EPI_ISL_3217242 | Idaho Bureau of Laboratories | Idaho Bureau of Laboratories | "R. Beukelman; Aimee Ceniseros; Christian Loera; Christopher Ball; Christopher Ball"; Matthew Charles Burns; R. Beukelman; Robert L. Voermans |
| EPI_ISL_2295877, EPI_ISL_2295878, EPI_ISL_2295879, EPI_ISL_2295882, EPI_ISL_3670779 | Illinois Department of Public Health | Gagnon Lab, Southern Illinois University | Keith Gagnon |
| EPI_ISL_1965085, EPI_ISL_2139923, EPI_ISL_2139926, EPI_ISL_2139928, EPI_ISL_2139929, EPI_ISL_2139930, EPI_ISL_2139936, EPI_ISL_2139954, EPI_ISL_2139956, EPI_ISL_2140016, EPI_ISL_2140019, EPI_ISL_2140025, EPI_ISL_2140026, EPI_ISL_2140028, EPI_ISL_2140029, EPI_ISL_2140030, EPI_ISL_2300494, EPI_ISL_2300525, EPI_ISL_2300626 | | | |
| see above | Illinois Department of Public Health | Illinois Department of Public Health - Chicago Lab | Ira Heimler; Joel Price; Vineet K. Dhiman |
| EPI_ISL_1938505, EPI_ISL_1938517, EPI_ISL_1938532, EPI_ISL_1938551, EPI_ISL_1938557, EPI_ISL_1938574, EPI_ISL_1938594, EPI_ISL_1938607, EPI_ISL_1938612, EPI_ISL_1938613, EPI_ISL_1938614, EPI_ISL_1938615, EPI_ISL_1938621, EPI_ISL_1938631, EPI_ISL_1938634, EPI_ISL_1938635, EPI_ISL_1938638, EPI_ISL_1938640, EPI_ISL_1938641, EPI_ISL_1938643, EPI_ISL_1938645, EPI_ISL_1938650, EPI_ISL_1938651, EPI_ISL_1938653, EPI_ISL_1938654, EPI_ISL_1938655, EPI_ISL_1938656, EPI_ISL_1938657, EPI_ISL_1938658, EPI_ISL_1938662, EPI_ISL_1938664, EPI_ISL_1938665, EPI_ISL_1938668, EPI_ISL_1938669, EPI_ISL_2000780, EPI_ISL_2000787, EPI_ISL_2000788, EPI_ISL_2000792, EPI_ISL_2000795, EPI_ISL_2000796, EPI_ISL_2000799, EPI_ISL_2000800, EPI_ISL_2000815, EPI_ISL_2000816, EPI_ISL_2000820, EPI_ISL_2000823, EPI_ISL_2000825, EPI_ISL_2000826, EPI_ISL_2000827, EPI_ISL_2000828, EPI_ISL_2000830, EPI_ISL_2000831, EPI_ISL_2000840, EPI_ISL_2000841, EPI_ISL_2000851, EPI_ISL_2000852, EPI_ISL_2000853, EPI_ISL_2000856, EPI_ISL_2086491, EPI_ISL_2086492, EPI_ISL_2086496, EPI_ISL_2086499, EPI_ISL_2086500, EPI_ISL_2086502, EPI_ISL_2086506, EPI_ISL_2086507, EPI_ISL_2086508, EPI_ISL_2086510, EPI_ISL_2086513, EPI_ISL_2086517, EPI_ISL_2086520, EPI_ISL_2086533, EPI_ISL_2135038, EPI_ISL_2135039, EPI_ISL_2135040, EPI_ISL_2135041, EPI_ISL_2135061, EPI_ISL_2135084, EPI_ISL_2135110, EPI_ISL_2135111, EPI_ISL_2135113, EPI_ISL_2135114, EPI_ISL_2135122, EPI_ISL_2135123, EPI_ISL_2135124, EPI_ISL_2135128, EPI_ISL_2135129, EPI_ISL_2135131, EPI_ISL_2203492, EPI_ISL_2203498, EPI_ISL_2203594, EPI_ISL_2203596, EPI_ISL_2203624, EPI_ISL_2203632, EPI_ISL_2203635, EPI_ISL_2203648, EPI_ISL_2339403, EPI_ISL_2403028, EPI_ISL_2403104, EPI_ISL_2403153 | | | |
| see above | Illinois Department of Public Health - Springfield Lab | Illinois Department of Public Health - Springfield Lab | Bryan Sim; Gordon McCall |
| EPI_ISL_2230846, EPI_ISL_2230855, EPI_ISL_2230887, EPI_ISL_2230897 | InDRE | Instituto Nacional de Medicina Genomica | Cedro-Tanda A; Garcia-Cardenas FJ; Gisela Barrera-Badillo; Gonzalez-Barrera D; Gonzalez-Woge MA; Herrera-Montalvo LA.; Hidalgo-Miranda A; Irma Lopez-Martinez; Jose Ernesto Ramirez González; Mendoza-Vargas A; Munguia-Garza P; Rangel-DeLeon D; Reyes-Grajeda JP |
| EPI_ISL_2136745, EPI_ISL_2536388, EPI_ISL_2536390, EPI_ISL_2536392, EPI_ISL_2536396, EPI_ISL_2536409, EPI_ISL_2536418, EPI_ISL_2536420, EPI_ISL_2536424, EPI_ISL_2536428, EPI_ISL_2536432, EPI_ISL_2536437, EPI_ISL_2536438 | | | |
| see above | Incyte Diagnostics Spokane | Seattle Flu Study | Amanda Adler; Barry R. Lutz; Benjamin Pelle; Brian Hiatt; Caitlin R. Wolf; Chris D. Frazar; Deborah A. Nickerson; Elisabeth Brandstetter; Erica Ryke; Geoff Melly; Helen Y. Chu; Janet A. Englund; Jay Shendure; Jover Lee; Kairsten Fay; Kirsten Lacombe; Lea M. Starita; Mark J. Rieder; Matthew Richardson; Matthew Thompson; Melissa Truong; Michael Boeckh; Michael Famulare; Misja Ilcisin; Peter D. Han; Philip Dykema; Romesh Gautom; Scott Lindquist; Thomas R. Sibley; Trevor Bedford |
| EPI_ISL_2304418, EPI_ISL_2304419, EPI_ISL_2304422, EPI_ISL_2304423, EPI_ISL_2304447, EPI_ISL_2304448, EPI_ISL_2304449 | | | |
| see above | Indiana Animal Disease Diagnostic Laboratory | Carpi Laboratory - Purdue University | Abebe A Fola; G Kenitra Hendrix; Giovanna Carpi; Ilinca I Ciubotariu; Jack Dorman; Jobin J Kattoor; Lev Gorenstein; Nicole M Perry; Rebecca P Wilkes |
| EPI_ISL_2087621, EPI_ISL_2087772, EPI_ISL_2088376, EPI_ISL_2088412, EPI_ISL_2088418, EPI_ISL_2088456, EPI_ISL_2088514, EPI_ISL_2088517, EPI_ISL_2088564, EPI_ISL_2088619, EPI_ISL_2088651, EPI_ISL_2088708, EPI_ISL_2088723, EPI_ISL_2088832, EPI_ISL_2088858, EPI_ISL_2088939, EPI_ISL_2088951, EPI_ISL_2089006, EPI_ISL_2089012, EPI_ISL_2089026, EPI_ISL_2089046, EPI_ISL_2089149, EPI_ISL_2089204, EPI_ISL_2089212, EPI_ISL_2089223, EPI_ISL_2089265, EPI_ISL_2089302, EPI_ISL_2089320, EPI_ISL_2089330, EPI_ISL_2089416, EPI_ISL_2089442, EPI_ISL_2132514, EPI_ISL_2186713, EPI_ISL_2186743, EPI_ISL_2186747, EPI_ISL_2186768, EPI_ISL_2186815, EPI_ISL_2186829, EPI_ISL_2186862, EPI_ISL_2186898, EPI_ISL_2186919, EPI_ISL_2186961, EPI_ISL_2186971, EPI_ISL_2186991, EPI_ISL_2187001, EPI_ISL_2187091, EPI_ISL_2187129, EPI_ISL_2187222, EPI_ISL_2187223, EPI_ISL_2323966, EPI_ISL_4388766 | | | |
| see above | Infinity Biologix | Centers for Disease Control and Prevention Division of Viral Diseases, Pathogen Discovery | Adrian Paskey; Benjamin Rambo-Martin; Chirayu Goswami; Christian Bixby; Christopher Gulvick; Clinton Paden; Clinton R. Paden; Dakota Howard; Darlene Wagner; Dhwani Batra; Duncan MacCannell; Erisa Sula; Jason Caravas; Jonathan Schultz; Kara Moser; Kristine Lacek; Matthew Schmerer; Peter Cook; Peter W. Cook; Robin Grimwood; Russ Hager; Scott Sammons; Shatavia Morrison; Tymeckia Kendall; Victoria Caban Figueroa; Yihe Wang; Yvette Unoarumhi |
| EPI_ISL_5782561, EPI_ISL_5782567, EPI_ISL_5782571, EPI_ISL_5782575, EPI_ISL_5782576, EPI_ISL_5782577, EPI_ISL_5782578, EPI_ISL_5782584, EPI_ISL_5782586, EPI_ISL_5782591, EPI_ISL_5782594, EPI_ISL_5782604, EPI_ISL_5782608, EPI_ISL_5782611, EPI_ISL_5782614, EPI_ISL_5782630, EPI_ISL_5782632, EPI_ISL_5782639, EPI_ISL_5782642, EPI_ISL_5782648, EPI_ISL_5782650, EPI_ISL_5782654, EPI_ISL_5782655, EPI_ISL_5782656, EPI_ISL_5782661 | | | |
| see above | Inside Diagnosticos | Instituto Butantan | Antonio Jorge Martins; Claudia Renata dos Santos Barros; David Schlesinger; Debora Botequio Moretti; Dimas Tadeu Covas; Elaine Cristina Marqueze; Elaine Vieira Santos; Evandra Strazza Rodrigues; Heidge Fukumasu; Jayme Augusto de Souza-Neto; José Salvatore Leister Patané; Luiz Alcantara; Luiz Lehmann Coutinho; Maria Carolina Elias; Maurício Lacerda Nogueira; Rafael dos Santos Bezerra; Raul Machado Neto; Rejane Maria Tommasini Grotto; Ricardo Haddad; Sandra Coccuzzo Sampaio Vessoni; Simone Kashima; Svetoslav Nanev Slavov; Vincent Louis Viala |
| EPI_ISL_2828666, EPI_ISL_2828667 | Instituo Adolfo Lutz - Reginal de Ribeirao Preto | Instituto Adolfo Lutz, Interdiciplinary Procedures Center, Strategic Laboratory | Caio Vinicius Dias Lopes; Claudia Regina Gonçalves; Claudio Tavares Sacchi; Erica Valessa Ramos Gomes; Karoline Rodrigues Campos |
| EPI_ISL_2352020, EPI_ISL_2367261 | Institut de Virologie du CHU de Strasbourg | Institut de Virologie du CHU de Strasbourg | Fafi-Kremer Samira; Gallais Floriane; Gantner Pierre; Laugel Elodie; Solis Morgane; Velay Aurélie; Wendling Marie-Josée |
| EPI_ISL_2691119, EPI_ISL_2919240, EPI_ISL_2919248, EPI_ISL_2919261 | Instituto Adolfo Lutz - Regional de Bauru | Instituto Adolfo Lutz, Interdiciplinary Procedures Center, Strategic Laboratory | Caio Vinicius Dias Lopes; Claudia Regina Gonçalves; Claudio Tavares Sacchi; Erica Valessa Ramos Gomes; Karoline Rodrigues Campos; Leonardo Jose Tadeu de Araujo |
| EPI_ISL_2828690, EPI_ISL_2828692, EPI_ISL_2828693, EPI_ISL_2828694, EPI_ISL_2828695 | Instituto Adolfo Lutz - Regional de Campinas | Instituto Adolfo Lutz, Interdiciplinary Procedures Center, Strategic Laboratory | Caio Vinicius Dias Lopes; Claudia Regina Gonçalves; Claudio Tavares Sacchi; Erica Valessa Ramos Gomes; Karoline Rodrigues Campos |
| EPI_ISL_2691091, EPI_ISL_2691092, EPI_ISL_2691093, EPI_ISL_2691094, EPI_ISL_2691095 | Instituto Adolfo Lutz - Regional de Marilia | Instituto Adolfo Lutz, Interdiciplinary Procedures Center, Strategic Laboratory | Caio Vinicius Dias Lopes; Claudia Regina Gonçalves; Claudio Tavares Sacchi; Erica Valessa Ramos Gomes; Karoline Rodrigues Campos; Leonardo Jose Tadeu de Araujo |
| EPI_ISL_2614618, EPI_ISL_2614622 | Instituto Adolfo Lutz - Regional de Presidente Prudente | Instituto Adolfo Lutz, Interdiciplinary Procedures Center, Strategic Laboratory | Caio Vinicius Dias Lopes; Claudia Regina Gonçalves; Claudio Tavares Sacchi; Erica Valessa Ramos Gomes; Karoline Rodrigues Campos; Leonardo Jose Tadeu de Araujo |
| EPI_ISL_2691123 | Instituto Adolfo Lutz - Regional de Ribeirao Preto | Instituto Adolfo Lutz, Interdiciplinary Procedures Center, Strategic Laboratory | Caio Vinicius Dias Lopes; Claudia Regina Gonçalves; Claudio Tavares Sacchi; Erica Valessa Ramos Gomes; Karoline Rodrigues Campos; Leonardo Jose Tadeu de Araujo |
| EPI_ISL_2828696, EPI_ISL_2828697, EPI_ISL_2828700, EPI_ISL_2828703 | Instituto Adolfo Lutz - Regional de Santos | Instituto Adolfo Lutz, Interdiciplinary Procedures Center, Strategic Laboratory | Caio Vinicius Dias Lopes; Claudia Regina Gonçalves; Claudio Tavares Sacchi; Erica Valessa Ramos Gomes; Karoline Rodrigues Campos |
| EPI_ISL_2919264, EPI_ISL_2958836, EPI_ISL_2958837, EPI_ISL_2958838, EPI_ISL_2958839 | Instituto Adolfo Lutz - Regional de Sao Jose do Rio Preto | Instituto Adolfo Lutz, Interdiciplinary Procedures Center, Strategic Laboratory | Caio Vinicius Dias Lopes; Claudia Regina Gonçalves; Claudio Tavares Sacchi; Erica Valessa Ramos Gomes; Karoline Rodrigues Campos |
| EPI_ISL_2958844 | Instituto Adolfo Lutz - Regional de Sorocaba | Instituto Adolfo Lutz, Interdiciplinary Procedures Center, Strategic Laboratory | Caio Vinicius Dias Lopes; Claudia Regina Gonçalves; Claudio Tavares Sacchi; Erica Valessa Ramos Gomes; Karoline Rodrigues Campos |
| EPI_ISL_2691129, EPI_ISL_2691130, EPI_ISL_2691133, EPI_ISL_2691138, EPI_ISL_2691140, EPI_ISL_2691141, EPI_ISL_2756427, EPI_ISL_2756429, EPI_ISL_2756430 | | | |
| see above | Instituto Adolfo Lutz Central | Instituto Adolfo Lutz, Interdiciplinary Procedures Center, Strategic Laboratory | Caio Vinicius Dias Lopes; Claudia Regina Gonçalves; Claudio Tavares Sacchi; Erica Valessa Ramos Gomes; Karoline Rodrigues Campos; Leonardo Jose Tadeu de Araujo |
| EPI_ISL_4104677, EPI_ISL_4104681, EPI_ISL_4104691, EPI_ISL_4104693, EPI_ISL_4104695, EPI_ISL_4104704, EPI_ISL_4104705, EPI_ISL_4104709, EPI_ISL_4104710, EPI_ISL_4104711, EPI_ISL_4104715, EPI_ISL_4104719, EPI_ISL_4104725, EPI_ISL_4104734, EPI_ISL_4104736 | | | |
| see above | Instituto Butantan | Instituto Butantan | Antonio Jorge Martins; Claudia Renata dos Santos Barros; David Schlesinger; Debora Botequio Moretti; Dimas Tadeu Covas; Elaine Cristina Marqueze; Elaine Vieira Santos; Evandra Strazza Rodrigues; Heidge Fukumasu; Jayme Augusto de Souza-Neto; José Salvatore Leister Patané; Luiz Alcantara; Luiz Lehmann Coutinho; Maria Carolina Elias; Maurício Lacerda Nogueira; Rafael dos Santos Bezerra; Raul Machado Neto; Rejane Maria Tommasini Grotto; Ricardo Haddad; Sandra Coccuzzo Sampaio Vessoni; Simone Kashima; Svetoslav Nanev Slavov; Vincent Louis Viala |
| EPI_ISL_2348790 | Instituto Nacional De | NIC-Instituto Nacional de | Alfredo Bruno; Domenica de Mora.; Jimmy Garcés; Johanna Laines; Lizbeth Patiño; Manuel Gonzalez; Maritza Olmedo; Michelle Páez |

| | | | |
|---|---|---|---|
| | Investigación En Salud Pública- Crn De Influenza Y Otros Virus Respiratorios | Investigación en Salud Pública | |
| EPI_ISL_2346467 | Instituto Nacional de Enfermedades Respiratorias (INER) | Centro de Investigación en Enfermedades Infecciosas (CIENI), Instituto Nacional de Enfermedades Respiratorias (INER) | Alejandra Hernández-Terán; Alejandro Sanchez-Flores; Alfredo Herrera-Estrella; Alicia Ocaña-Mondragón; Angel Gustavo Salas-Lais; Bernardo Martínez-Miguel; Blanca Taboada; Brenda Irasema Maldonado-Meza; Carla Ivón Herrera-Najera; Carlos F. Arias; Celia Boukadida; Clara Esperanza Santacruz-Tinoco; Concepción Grajales-Muñiz; Consorcio Mexicano de Vigilancia Genómica (CoViGen-Mex). Authors (in alphabetical order): Julio Elias Alvarado-Yaah; Célida Duque Molina; Eduardo Becerril-Vargas; Fernando Fontove-Herrera; Fidencio Mejía-Nepomuceno; Francisco Pulido; Gloria Elena Espinosa-Ayala; Gloria María Molina-Salinas; Gloria Vazquez; Hector Esteban Paz-Juárez; Hector Montoya-Fuentes; Helen Haydee Fernanda Ramírez-Plascencia; Joel Armando Vázquez-Pérez; Jorge Salas-Hernández; José Antonio Enciso-Moreno; José Arturo Martínez-Orozco; José Esteban Muñoz-Medina; José de Jesús Nuñez-Contreras; Juan Bautista Chale-Dzul; Luis Alberto Ochoa-Carrera; Margarita Matías-Florentino; Mario Mújica-Sánchez; María Guadalupe Santiago-Mauricio; María Guadalupe de Jesús Mireles-Rivera; Nelly Sélem-Mojica; Pavel Isa; Ricardo Grande; Santiago Ávila-Ríos; Victor Eduardo Garcia-Arias; Víctor Hugo Borja-Aburto |
| EPI_ISL_2080693, EPI_ISL_2230812, EPI_ISL_2230858 | Instituto Nacional de Medicina Genomica | Instituto Nacional de Medicina Genomica | Alcaraz N; Cedro-Tanda A; Cisneros- Villanueva M; Escobar-Arrazola; Gonzalez-Barrera D; Herrera-Montalvo LA.; Hidalgo-Miranda A; M; Mendoza-Vargas A; Munguia-Garza P; Ramirez-Vega O; Rangel-DeLeon D; Reyes-Grajeda JP |
| EPI_ISL_2500957 | Instituto Nacional de Salud | Instituto Nacional de Salud- Dirección de Investigación en Salud Pública | Carlos Franco-Muñoz; Carmen Osorio; Diana Malo; Diego A. Álvarez-Díaz; Diego Andrés Prada; Gerardo Santamaría; Hector Alejandro Ruiz-Moreno; Jhonnatan Reales-González; Jorge Rivera; Juan Camilo Martinez; Julian Naizaque; Katherine Laiton-Donato; Lisseth Pardo; Magdalena Wiesner; Marcela Mercado-Reyes; Maria T. Herrera-Sepúlveda; Marta Lopez Blanco; Martha Lucia Ospina Martinez; Paola Rojas; Sergio Gomez; Sheryll Corchuelo; Ángela Alarcon Cruz |
| EPI_ISL_2295427, EPI_ISL_2323121, EPI_ISL_2323126, EPI_ISL_2378640, EPI_ISL_2378641, EPI_ISL_2378642, EPI_ISL_2378643 | | | |
| see above | Instituto de Biotecnologia - UNESP-Botucatu-SP | Instituto de Biotecnologia - UNESP-Botucatu-SP | Cecília Artico Banho; Cíntia Bittar; Fábio Sossai Possebon; Guilherme Campos; Helena Lage Ferreira; Jorge A. Petroli Marchesi; João Pessoa Araújo Jr.; Leila Sabrina Ullmann; Lívia Sacchetto; Maisa C. Pereira Parra; Marília Moraes; Maurício L. Nogueira; Paula Rahal; Paulo Inacio da Costa |
| EPI_ISL_5147044, EPI_ISL_5147045 | Instituto de Medicina Tropical Alexander Von Humboldt, Sede Cusco | Laboratorio de Genómica Microbiana, Universidad Peruana Cayetano Heredia | Alejandra Dávila-Barclay; Diego Cuicapuza; Fátima Concha; Guillermo Salvatierra; Janet Huancachoque; Luis González; Maria Luisa Morales; Martha Fernández; Miguel Cabada; Pablo Tsukayama; Pedro E. Romero; Pool Marcos |
| EPI_ISL_2536524, EPI_ISL_2536526, EPI_ISL_2536533, EPI_ISL_2536536 | Interpath Laboratory | Seattle Flu Study | Amanda Adler; Barry R. Lutz; Benjamin Pelle; Brian Hiatt; Caitlin R. Wolf; Chris D. Frazar; Deborah A. Nickerson; Elisabeth Brandstetter; Erica Ryke; Geoff Melly; Helen Y. Chu; Janet A. Englund; Jay Shendure; Jover Lee; Kairsten Fay; Kirsten Lacombe; Lea M. Starita; Mark J. Rieder; Matthew Richardson; Matthew Thompson; Melissa Truong; Michael Boeckh; Michael Famulare; Misja Ilcisin; Peter D. Han; Philip Dykema; Romesh Gautom; Scott Lindquist; Thomas R. Sibley; Trevor Bedford |
| EPI_ISL_2691147, EPI_ISL_2691148 | Intituto Adolfo Lutz - Regional de Sorocaba | Instituto Adolfo Lutz, Interdiciplinary Procedures Center, Strategic Laboratory | Caio Vinicius Dias Lopes; Claudia Regina Gonçalves; Claudio Tavares Sacchi; Erica Valessa Ramos Gomes; Karoline Rodrigues Campos; Leonardo Jose Tadeu de Araujo |
| EPI_ISL_2183435 | Israel Central Virology laboratory | Israel National Consortium for SARS-CoV-2 sequencing | Dana Bar-Ilan; Efrat Dahan Bucris; Efrat Glick-Saar; Ella Mendelson; Gideon Rechavi; Michal Mandelboim; Miranda Geva; Neta Zuckerman; Netanel Abu; Omri Nayshool; Oran Erster; Orna Mor |
| EPI_ISL_2171436, EPI_ISL_2171498, EPI_ISL_2171540, EPI_ISL_2171588, EPI_ISL_2227083 | Istituto Zooprofilattico Sperimentale del Mezzogiorno | TIGEM | Antonio Grimaldi Patrizia Annunziata Francesco Panariello Biancamaria Pierri Claudia Tiberio Teresa Giuliano Valentina Bouche Chiara Colantuono Maria Concetta Cuomo Denise Di Concilio Lucio Di Filippo Anna Manfredi Marcello Salvi Antonio Limone Luigi Atripaldi Pellegrino Cerino Andrea Ballabio Davide Cacchiarelli |
| EPI_ISL_2444186 | JOSE BONIFACIO | Instituto Butantan / FAMERP | Antonio Jorge Martins; Claudia Renata dos Santos Barros; David Schlesinger; Debora Botequio Moretti; Dimas Tadeu Covas; Elaine Cristina Marqueze; Elaine Vieira Santos; Evandra Strazza Rodrigues; Heidge Fukumasu; Jayme Augusto de Souza-Neto; José Salvatore Leister Patané; Luiz Alcantara; Luiz Lehmann Coutinho; Maria Carolina Elias; Maurício Lacerda Nogueira; Rafael dos Santos Bezerra; Raul Machado Neto; Rejane Maria Tommasini Grotto; Ricardo Haddad; Sandra Coccuzzo Sampaio Vessoni; Simone Kashima; Svetoslav Nanev Slavov; Vincent Louis Viala |
| EPI_ISL_2007210 | Jessa | Jessa | Berden et al. on behalf of the Jessa_cmdLab |
| EPI_ISL_2476376, EPI_ISL_2476380, EPI_ISL_2476381 | John H Stroger Jr Hospital of Cook County | RIPHL at Rush University Medical Center | Ah-Young Sung; Felix Araujo Perez; Kevin Kunstman; Laura Furtado; Marieta Hyde; Max Kolton; Sharon Welbel; Stefan Green |
| EPI_ISL_1973505, EPI_ISL_2331506 | Johns Hopkins Hospital Department of Pathology | Johns Hopkins Hospital Department of Pathology | Adannaya Amadi; C. Paul Morris; Chun Huai Luo; Heba H. Mostafa; Matthew Schwartz; Nicholas Gallagher |
| EPI_ISL_2726406, EPI_ISL_2726407, EPI_ISL_2832425, EPI_ISL_2832426, EPI_ISL_2832427, EPI_ISL_2832428, EPI_ISL_2832430, EPI_ISL_2832431, EPI_ISL_2832432, EPI_ISL_2832452, EPI_ISL_2832608, EPI_ISL_2832618, EPI_ISL_2832696, EPI_ISL_2832698, EPI_ISL_2832704, EPI_ISL_2832712, EPI_ISL_2833150, EPI_ISL_2833261, EPI_ISL_2833276, EPI_ISL_2833343, EPI_ISL_2833344, EPI_ISL_2833379, EPI_ISL_2833419, EPI_ISL_2833421, EPI_ISL_2833425, EPI_ISL_2833426, EPI_ISL_2833427 | | | |
| see above | KU Leuven, Rega Institute, Clinical and Epidemiological Virology | KU Leuven, Rega Institute, Clinical and Epidemiological Virology | Bert Vanmechelen; Joan Marti-Carerras; Piet Maes; Tony Wawina-Bokalanga |
| EPI_ISL_2383979 | KY State Public Health Lab | Centers for Disease Control and Prevention Division of Viral Diseases, Pathogen Discovery | Alison Laufer Halpin; Ben L. Rambo-Martin; Clinton R. Paden; Dakota Howard; Darlene Wagner; Dave Wentworth; Dhwani Batra; Jasmine Padilla; Justin Lee; Katie Dillon; Krista Queen; Kristen Knipe; Kristine Lacek; Mark Burroughs; Matthew Schmerer; Mili Sheth; Peter Cook; Sam Shepard; Sarah Nobles; Shoshona Le; Suxiang Tong; Vivien Dugan; Yvette Unoarumhi |
| EPI_ISL_1805462, EPI_ISL_2271719 | Kansas Health and Environmental Lab | Kansas Health and Environmental Lab | Ben Olsen; Jonathan Barnell; Katherine Wiggins; Mike Grose; and Phil Adam |
| EPI_ISL_4080587 | Kentucky State Public Health Lab | Kentucky State Public Health Lab | Anjayya Tummala; Chitralipi Kattunga; Joshua Tobias; Karim George; Matthew Johnson; Rachel Zinner; Rhonda Lucas; Vaneet Arora; William Grooms |
| EPI_ISL_3354895 | LA Office of Public Health Laboratories | Centers for Disease Control and Prevention Division of Viral Diseases, Pathogen Discovery | Alex Burgin; Ben Rambo-Martin; Clinton Paden; Dakota Howard; Dave Wentworth; Dhwani Batra; Jasmine Padilla; Justin Lee; Krista Queen; Kristen Knipe; Kristine Lacek; Mark Burroughs; Matthew Schmerer; Meghan Bentz; Mili Sheth; Peter Cook; Sam Shepard; Sarah Nobles; Suxiang Tong; Vivien Dugan; Yvette Unoarumhi |
| EPI_ISL_3385825 | LABORATORIO CLINICO HIGUERA ESCALANTE | Instituto Nacional de Salud- Dirección de Investigación en Salud Pública | Carlos Franco-Muñoz; Carmen Osorio; Diana Malo; Diego A. Álvarez-Díaz; Diego Andrés Prada; Gerardo Santamaría; Hector Alejandro Ruiz-Moreno; Jhonnatan Reales-González; Jorge Rivera; Juan Camilo Martinez; Julian Naizaque; Katherine Laiton-Donato; Lisseth Pardo; Magdalena Wiesner; Marcela Mercado-Reyes; Maria T. Herrera-Sepúlveda; Marta Lopez Blanco; Martha Lucia Ospina Martinez; Paola Rojas; Sergio Gomez; Sheryll Corchuelo; Ángela Alarcon Cruz |
| EPI_ISL_2674327 | LABORATORIO DE INFECCION E INMUNIDAD | Instituto Nacional de Salud- Dirección de Investigación en Salud Pública | Carlos Franco-Muñoz; Carmen Osorio; Diana Malo; Diego A. Álvarez-Díaz; Diego Andrés Prada; Gerardo Santamaría; Hector Alejandro Ruiz-Moreno; Jhonnatan Reales-González; Jorge Rivera; Juan Camilo Martinez; Julian Naizaque; Katherine Laiton-Donato; Lisseth Pardo; Magdalena Wiesner; Marcela Mercado-Reyes; Maria T. Herrera-Sepúlveda; Marta Lopez Blanco; Martha Lucia Ospina Martinez; Paola Rojas; Sergio Gomez; Sheryll Corchuelo; Ángela Alarcon Cruz |
| EPI_ISL_2621840, EPI_ISL_2621841 | LABORATORIO ECHAVARRIA | Universidad Nacional de Colombia - Laboratorio Genómico One Health | Andres F. Cardona-Rios; Carlos Franco-Muñoz; Carolina Muñoz-Arango; Celeny Ortiz; Daniel O. Maldonado-Perez; Diego A. Álvarez-Díaz; Hector Alejandro Ruiz-Moreno; Idabely Betancur Ortiz; Jorge E. Osorio; Juan P. Hernandez-Ortiz; Karl A Ciuoderis; Katherine Laiton-Donato; Laura Silvana Perez; Lina M. Hurtado; Marcela Mercado-Reyes; Maria Angélica Maya; Maria Stella López; Rita Almanza Payares; Sandra Ines Cano; Simón Villegas Velásquez |
| EPI_ISL_2345141, EPI_ISL_2345142 | LABORATORIO LOCAL DE ITAPECERICA DA SERRA | Instituto Butantan / ESALQ-Piracicaba | Antonio Jorge Martins; Claudia Renata dos Santos Barros; David Schlesinger; Debora Botequio Moretti; Dimas Tadeu Covas; Elaine Cristina Marqueze; Elaine Vieira Santos; Evandra Strazza Rodrigues; Heidge Fukumasu; Jayme Augusto de Souza-Neto; José Salvatore Leister Patané; Luiz Alcantara; Luiz Lehmann Coutinho; Maria Carolina Elias; Maurício Lacerda Nogueira; Rafael dos Santos Bezerra; Raul Machado Neto; Rejane Maria Tommasini Grotto; Ricardo Haddad; Sandra Coccuzzo Sampaio Vessoni; Simone Kashima; Svetoslav Nanev Slavov; Vincent Louis Viala |
| EPI_ISL_2345186, EPI_ISL_2345248, EPI_ISL_2345250, EPI_ISL_2345251, EPI_ISL_2345267, EPI_ISL_2345271, EPI_ISL_2345288, EPI_ISL_2345289, EPI_ISL_2345290, EPI_ISL_2345291, EPI_ISL_2345292, EPI_ISL_2345293, EPI_ISL_2345295, EPI_ISL_2345296, EPI_ISL_2345297, EPI_ISL_2345298 | | | |
| see above | LABORATORIO MUNICIPAL DE PIRACICABA | Instituto Butantan / ESALQ-Piracicaba | Antonio Jorge Martins; Claudia Renata dos Santos Barros; David Schlesinger; Debora Botequio Moretti; Dimas Tadeu Covas; Elaine Cristina Marqueze; Elaine Vieira Santos; Evandra Strazza Rodrigues; Heidge Fukumasu; Jayme Augusto de Souza-Neto; José Salvatore Leister Patané; Luiz Alcantara; Luiz Lehmann Coutinho; Maria Carolina Elias; Maurício Lacerda Nogueira; Rafael dos Santos Bezerra; Raul Machado Neto; Rejane Maria Tommasini Grotto; Ricardo Haddad; Sandra Coccuzzo Sampaio Vessoni; Simone Kashima; Svetoslav Nanev Slavov; Vincent Louis Viala |
| EPI_ISL_3404848 | LACEN | LGBio (Laboratorio de Genetica & Biodiversidade) | Alex Honda Bernardes; Amanda Alves de Melo; Aparecido Divino da Cruz; Cintia Pelegrineti Targueta de Azevedo Brito; Daniela de Melo e Silva; Elisangela de Paula Silveira Lacerda; Juliana Santana de Curcio; Luiz Augusto Pereira; Marc Alexandre Duarte Gigonzac; Mariana Pires de Campos Telles; Ramilla dos Santos Braga; Renata de Oliveira Dias; Rhewter Nunes; Thais Cidália Vieira Gigonzac; Thais Guimarães Castro; Thays Millena Alves Pedroso |
| EPI_ISL_2691212, EPI_ISL_2691213 | LACEN do Estado de Goias | Instituto Adolfo Lutz, Interdiciplinary Procedures Center, Strategic Laboratory | Caio Vinicius Dias Lopes; Claudia Regina Gonçalves; Claudio Tavares Sacchi; Erica Valessa Ramos Gomes; Karoline Rodrigues Campos; Leonardo Jose Tadeu de Araujo |
| EPI_ISL_3316189 | LACEN do Estado de Mato Grosso | Instituto Adolfo Lutz, Interdiciplinary Procedures Center, Strategic Laboratory | Caio Vinicius Dias Lopes; Claudia Regina Gonçalves; Claudio Tavares Sacchi; Karoline Rodrigues Campos; Leonardo Tadeu de Araujo; Marlon Benedito Nascomento Santos |
| EPI_ISL_2919210 | LACEN do Estado de Tocantins | Instituto Adolfo Lutz, Interdiciplinary Procedures Center, Strategic Laboratory | Caio Vinicius Dias Lopes; Claudia Regina Gonçalves; Claudio Tavares Sacchi; Erica Valessa Ramos Gomes; Karoline Rodrigues Campos |
| EPI_ISL_3691376 | LACEN do Estado do Mato Grosso do Sul | Instituto Adolfo Lutz, Interdiciplinary Procedures Center, Strategic Laboratory | Caio Vinicius Dias Lopes; Claudia Regina Gonçalves; Claudio Tavares Sacchi; Karoline Rodrigues Campos; Leonardo Tadeu de Araujo; Marlon Benedito Nascomento Santos |
| EPI_ISL_2958874, EPI_ISL_2958875, EPI_ISL_2958876, EPI_ISL_2958877, EPI_ISL_2958878, EPI_ISL_2958879 | LACEN do Mato Grosso do Sul | Instituto Adolfo Lutz, Interdiciplinary Procedures Center, Strategic Laboratory | Caio Vinicius Dias Lopes; Claudia Regina Gonçalves; Claudio Tavares Sacchi; Erica Valessa Ramos Gomes; Karoline Rodrigues Campos |
| EPI_ISL_2821275, EPI_ISL_3134558, EPI_ISL_3134559, EPI_ISL_3134560, EPI_ISL_3134561, EPI_ISL_3134562, EPI_ISL_3134563, EPI_ISL_3134564, EPI_ISL_3134565, EPI_ISL_3134566, EPI_ISL_3134567, EPI_ISL_3134568, EPI_ISL_3134569, EPI_ISL_3134570, EPI_ISL_3134571, EPI_ISL_3134572, EPI_ISL_3134573, EPI_ISL_3134574, EPI_ISL_3134575, EPI_ISL_3134576, EPI_ISL_3134577, EPI_ISL_3134578, EPI_ISL_3134580, EPI_ISL_3134581, EPI_ISL_3134582, EPI_ISL_3134584, EPI_ISL_3134585, EPI_ISL_3134586, EPI_ISL_3134593, EPI_ISL_3134594, EPI_ISL_3134856, EPI_ISL_3447589, EPI_ISL_3703362, EPI_ISL_3703385, EPI_ISL_3703396, EPI_ISL_3703399, EPI_ISL_3703403, EPI_ISL_3703407, EPI_ISL_3703417, EPI_ISL_3703421, EPI_ISL_3703428, EPI_ISL_3703432, EPI_ISL_3703436, EPI_ISL_3703451, EPI_ISL_3703461, EPI_ISL_3703471, EPI_ISL_3703475, EPI_ISL_3703479, EPI_ISL_3703490, EPI_ISL_3703498, EPI_ISL_3703508, EPI_ISL_3703511, EPI_ISL_3703515, EPI_ISL_3703519, EPI_ISL_3703523, EPI_ISL_3703527, EPI_ISL_3703540, EPI_ISL_3703544, EPI_ISL_3703550, EPI_ISL_3703558, EPI_ISL_3703569, EPI_ISL_3703572, EPI_ISL_3703575, EPI_ISL_3703580, EPI_ISL_3703584, EPI_ISL_3703588, EPI_ISL_3703600, EPI_ISL_3703603, EPI_ISL_3703607, EPI_ISL_3703612, EPI_ISL_3703621, EPI_ISL_3703630, EPI_ISL_3703635, EPI_ISL_3703638, EPI_ISL_3703642, EPI_ISL_3703646, EPI_ISL_3703650, EPI_ISL_3703654, EPI_ISL_3703657, EPI_ISL_3703661, EPI_ISL_3703665, EPI_ISL_5254418 | | | |
| see above | LACEN/PE | WallauLab on behalf of Fiocruz COVID-19 Genomic Surveillance Network | Alexandre Freitas da Silva; Antonio Marinho da Silva Neto; Antonio Mauro Rezende; Cassia Docena; Constância Flávia Junqueira Ayres; Cássia Docena; Duschinka Ribeiro Duarte Guedes; Elisama Helvecio; Filipe Zimmer Dezordi; Gabriel Luz Wallau; Gustavo Barbosa de Lima; Lais Ceschini Machado; Larissa Krokovsky; Laís Ceschini Machado; Lilian Caroliny Amorim Silva; Marcelo Henrique dos Santos Paiva; Matheus Filgueira Bezerra; Sinval Pinto Brandão Filho |
| EPI_ISL_4488010, EPI_ISL_4488013, EPI_ISL_4488016, EPI_ISL_4488019, EPI_ISL_4488022, EPI_ISL_4488025, EPI_ISL_4488027, EPI_ISL_4488029, EPI_ISL_4488032, EPI_ISL_4488056, EPI_ISL_4488071 | | | |
| see above | LACEN/PI | WallauLab on behalf of Fiocruz | Adelino Soares Lima Neto; Alexandre Freitas da Silva; Antônio Marinho; Cassia Docena; Constância Flávia Junqueira Ayres; Filipe Zimmer Dezordi; Gabriel Luz Wallau; Gustavo Barbosa de Lima; Hellen de Oliveira Amaral; Jacenir Reis dos Santos Mallet; Joana Carolina Viana Lima; Lais Ceschini Machado; |

| | | | |
|---|---|---|---|
| | | COVID-19 Genomic Surveillance Network | Leandro de Mattos; Lilian Caroliny Amorim Silva; Marcela de Lacerda Valença Queiroz; Marcelo Adriano da Cunha e Silva Vieira; Marcelo Henrique dos Santos Paiva; Matheus Filgueira Bezerra; Sinval Pinto Brandão Filho; Túlio de Lima Campos; Vladimir Costa Silva; Walterlene de Carvalho Goçalves |
| EPI_ISL_2373664 | LBM UNIBIO VALENTIN | CNR Virus des Infections Respiratoires - France SUD | Antonin Bal; Bruno Lina; Bruno Simon; Gregory Destras; Gwendolyne Burfin; Hadrien Regue; Laurence Josset; Martine Valette; Quentin Semanas |
| EPI_ISL_3385852 | LDSP BOYACA | Instituto Nacional de Salud- Dirección de Investigación en Salud Pública | Carlos Franco-Muñoz; Carmen Osorio; Diana Malo; Diego A. Álvarez-Díaz; Diego Andrés Prada; Gerardo Santamaría; Hector Alejandro Ruiz-Moreno; Jhonnatan Reales-González; Jorge Rivera; Juan Camilo Martinez; Julian Naizaque; Katherine Laiton-Donato; Lisseth Pardo; Magdalena Wiesner; Marcela Mercado-Reyes; Maria T. Herrera-Sepúlveda; Marta Lopez Blanco; Martha Lucia Ospina Martinez; Paola Rojas; Sergio Gomez; Sheryll Corchuelo; Ángela Alarcon Cruz |
| EPI_ISL_2674289 | LDSP CUNDINAMARCA | Instituto Nacional de Salud- Dirección de Investigación en Salud Pública | Carlos Franco-Muñoz; Carmen Osorio; Diana Malo; Diego A. Álvarez-Díaz; Diego Andrés Prada; Gerardo Santamaría; Hector Alejandro Ruiz-Moreno; Jhonnatan Reales-González; Jorge Rivera; Juan Camilo Martinez; Julian Naizaque; Katherine Laiton-Donato; Lisseth Pardo; Magdalena Wiesner; Marcela Mercado-Reyes; Maria T. Herrera-Sepúlveda; Marta Lopez Blanco; Martha Lucia Ospina Martinez; Paola Rojas; Sergio Gomez; Sheryll Corchuelo; Ángela Alarcon Cruz |
| EPI_ISL_2674295 | LDSP LETICIA | Instituto Nacional de Salud- Dirección de Investigación en Salud Pública | Carlos Franco-Muñoz; Carmen Osorio; Diana Malo; Diego A. Álvarez-Díaz; Diego Andrés Prada; Gerardo Santamaría; Hector Alejandro Ruiz-Moreno; Jhonnatan Reales-González; Jorge Rivera; Juan Camilo Martinez; Julian Naizaque; Katherine Laiton-Donato; Lisseth Pardo; Magdalena Wiesner; Marcela Mercado-Reyes; Maria T. Herrera-Sepúlveda; Marta Lopez Blanco; Martha Lucia Ospina Martinez; Paola Rojas; Sergio Gomez; Sheryll Corchuelo; Ángela Alarcon Cruz |
| EPI_ISL_2674312 | LDSP NARIÑO | Instituto Nacional de Salud- Dirección de Investigación en Salud Pública | Carlos Franco-Muñoz; Carmen Osorio; Diana Malo; Diego A. Álvarez-Díaz; Diego Andrés Prada; Gerardo Santamaría; Hector Alejandro Ruiz-Moreno; Jhonnatan Reales-González; Jorge Rivera; Juan Camilo Martinez; Julian Naizaque; Katherine Laiton-Donato; Lisseth Pardo; Magdalena Wiesner; Marcela Mercado-Reyes; Maria T. Herrera-Sepúlveda; Marta Lopez Blanco; Martha Lucia Ospina Martinez; Paola Rojas; Sergio Gomez; Sheryll Corchuelo; Ángela Alarcon Cruz |
| EPI_ISL_2296092 | LESP Chihuahua | Instituto de Diagnostico y Referencia Epidemiologicos (INDRE) | Abril Rodriguez-Maldonado; Ariadna Medina-Benitez; Claudia Wong-Arambula; Ernesto Ramirez-Gonzalez.; Gisela Barrera-Badillo; Irma Lopez-Martinez; Joaquin Quiroz-Mercado; Lucia Hernandez-Rivas; Natividad Cruz-Ortiz; Sergio Rangel-Guerrero; Tatiana Nunez-Garcia; Vanessa Rivero-Arredondo |
| EPI_ISL_2105718, EPI_ISL_2296060 | LESP Ciudad de Mexico | Instituto de Diagnostico y Referencia Epidemiologicos (INDRE) | Abril Rodriguez-Maldonado; Ariadna Medina-Benitez; Claudia Wong-Arambula; Ernesto Ramirez-Gonzalez.; Gisela Barrera-Badillo; Irma Lopez-Martinez; Joaquin Quiroz-Mercado; Lucia Hernandez-Rivas; Natividad Cruz-Ortiz; Sergio Rangel-Guerrero; Tatiana Nunez-Garcia; Vanessa Rivero-Arredondo |
| EPI_ISL_2296093 | LESP Guanajuato | Instituto de Diagnostico y Referencia Epidemiologicos (INDRE) | Abril Rodriguez-Maldonado; Ariadna Medina-Benitez; Claudia Wong-Arambula; Ernesto Ramirez-Gonzalez.; Gisela Barrera-Badillo; Irma Lopez-Martinez; Joaquin Quiroz-Mercado; Lucia Hernandez-Rivas; Natividad Cruz-Ortiz; Sergio Rangel-Guerrero; Tatiana Nunez-Garcia; Vanessa Rivero-Arredondo |
| EPI_ISL_2105715 | LESP Jalisco | Instituto de Diagnostico y Referencia Epidemiologicos (INDRE) | Abril Rodriguez-Maldonado; Ariadna Medina-Benitez; Claudia Wong-Arambula; Ernesto Ramirez-Gonzalez.; Gisela Barrera-Badillo; Irma Lopez-Martinez; Joaquin Quiroz-Mercado; Lucia Hernandez-Rivas; Natividad Cruz-Ortiz; Sergio Rangel-Guerrero; Tatiana Nunez-Garcia; Vanessa Rivero-Arredondo |
| EPI_ISL_2296061 | LESP Quintana Roo | Instituto de Diagnostico y Referencia Epidemiologicos (INDRE) | Abril Rodriguez-Maldonado; Ariadna Medina-Benitez; Claudia Wong-Arambula; Ernesto Ramirez-Gonzalez.; Gisela Barrera-Badillo; Irma Lopez-Martinez; Joaquin Quiroz-Mercado; Lucia Hernandez-Rivas; Natividad Cruz-Ortiz; Sergio Rangel-Guerrero; Tatiana Nunez-Garcia; Vanessa Rivero-Arredondo |
| EPI_ISL_2296065 | LESP Tamaulipas | Instituto de Diagnostico y Referencia Epidemiologicos (INDRE) | Abril Rodriguez-Maldonado; Ariadna Medina-Benitez; Claudia Wong-Arambula; Ernesto Ramirez-Gonzalez.; Gisela Barrera-Badillo; Irma Lopez-Martinez; Joaquin Quiroz-Mercado; Lucia Hernandez-Rivas; Natividad Cruz-Ortiz; Sergio Rangel-Guerrero; Tatiana Nunez-Garcia; Vanessa Rivero-Arredondo |
| EPI_ISL_2016801, EPI_ISL_2016817, EPI_ISL_2016818, EPI_ISL_2016824 | LHUB-ULB | Labo Klinische Biologie, UZA | Basil Britto Xavier; Christine Lammens; Herman Goossens; Jasmine Coppens; Marie Le Mercier; Veerle Matheeussen |
| EPI_ISL_4196914, EPI_ISL_4212161, EPI_ISL_4212162, EPI_ISL_4212165 | LSP DISTRITAL BOGOTA | Gencore - Universidad de los Andes | Alejandro Gomez; Cristian Barrera; Gabriela Ariza; Gabriela Delgado; Johana Hernandez; Luisa Sacristan; Marcela Guevara; Marcela Mercado; Silvia Restrepo |
| EPI_ISL_2192206, EPI_ISL_2192347, EPI_ISL_2192351, EPI_ISL_2192354, EPI_ISL_2192391, EPI_ISL_2362701 | Lab voor klinische biologie | Lab voor klinische biologie | Bruno Verhasselt; Hannelore Hamerlinck; Marija Janevska |
| EPI_ISL_2029739 | Labo Analyses Med | National Reference Center for Viruses of Respiratory Infections, Institut Pasteur, Paris | Angela Brisebarre; Arditi Cocco; Camille Capel; Christophe Malabat; Corinne Maufrais; Etienne Simon-Lorière; Frédéric Lemoine; Louise Lefrançois; Marion Barbet; Maud Vanpeene; Méline Bizard; Pierre Lechat; Sylvie Behillil; Sylvie Van der Werf; Vincent Enouf |
| EPI_ISL_2279669 | Labo analyses med | National Reference Center for Viruses of Respiratory Infections, Institut Pasteur, Paris | Angela Brisebarre; Camille Capel; Christophe Malabat; Corinne Maufrais; Etienne Simon-Lorière; Frédéric Lemoine; Louise Lefrançois; Marion Barbet; Maud Vanpeene; Méline Bizard; OphéLie Said-Delattre; Sylvie Behillil; Sylvie Van der Werf; Victoire Baillet; Vincent Enouf |
| EPI_ISL_2389922, EPI_ISL_2389956 | Labor Becker & Kollegen (Standort München) | Robert Koch Institute | |
| EPI_ISL_2107829, EPI_ISL_2107855, EPI_ISL_2107858, EPI_ISL_2107862, EPI_ISL_2107866, EPI_ISL_2107867, EPI_ISL_2339439, EPI_ISL_2339447, EPI_ISL_2339448, EPI_ISL_2339451 | | | |
| see above | Labor Berlin Charité Vivantes GmbH / Institut für Virologie | Charité Universitätsmedizin Berlin, Institut für Virologie/Labor Berlin | Barbara Mühlemann; Christian Drosten; Christine Stephan; Peter Menzel; Rolf Schwarzer; Terry Jones; Victor M Corman |
| EPI_ISL_2384910 | Labor Dr. Fenner und Kollegen | Heinrich Pette Institute, Leibniz Institute for Experimental Virology | Adam Grundhoff; Alexis Robitaille; Johannes Knobloch; Martin Aepfelbacher; Nicole Fischer; Thomas Günther |
| EPI_ISL_2122353 | Labor Lobeck bzw. Laborarztliche Gemeinschaftspraxis Lobeck | Robert Koch Institute | |
| EPI_ISL_2115463 | Labor Lubeck bzw. Laborarztliche Gemeinschaftspraxis Lubeck | Robert Koch Institute | |
| EPI_ISL_2122404, EPI_ISL_2122501 | Laborarztpraxis Osnabruck | Robert Koch Institute | |
| EPI_ISL_2970548, EPI_ISL_2970550, EPI_ISL_2970551, EPI_ISL_2970553, EPI_ISL_2970555, EPI_ISL_2970556, EPI_ISL_2970557, EPI_ISL_2970559, EPI_ISL_3458527, EPI_ISL_3458529, EPI_ISL_3458541, EPI_ISL_3458543, EPI_ISL_3458545, EPI_ISL_3458546, EPI_ISL_3458547, EPI_ISL_3458548, EPI_ISL_3458552, EPI_ISL_3458559, EPI_ISL_3458718, EPI_ISL_3458719, EPI_ISL_3458720, EPI_ISL_3458724, EPI_ISL_3458726, EPI_ISL_3458730, EPI_ISL_3458736, EPI_ISL_3458737, EPI_ISL_5873496, EPI_ISL_5873509, EPI_ISL_5873523, EPI_ISL_5873528, EPI_ISL_5873535, EPI_ISL_5873541, EPI_ISL_7713622, EPI_ISL_7713623, EPI_ISL_7713642, EPI_ISL_7713643, EPI_ISL_7713644, EPI_ISL_7713645, EPI_ISL_7713646, EPI_ISL_7713647, EPI_ISL_7713650 | | | |
| see above | Laboratoire de santé publique du Québec | Laboratoire de santé publique du Québec | Guillaume Bourque; Ioannis Ragoussis; Jesse Shapiro; Mark Lathrop and Judith Fafard on behalf of the CoVSeQ research group; Mark Lathrop and Michel Roger on behalf of the CoVSeQ research group; Sandrine Moreira |
| EPI_ISL_2400867 | Laboratoires Reunis | Laboratoire national de sante, Microbiology, Microbial Genomics Platform | Anke Wienecke-Baldacchino; Bernard Weber; Catherine Ragimbeau; Fatu Djabi; Jessica Tapp; Lise Pignon; Raoul Salmon; Tamir Abdelrahman |
| EPI_ISL_2400614, EPI_ISL_2400615, EPI_ISL_2400618, EPI_ISL_2400619, EPI_ISL_2400631, EPI_ISL_2400637 | Laboratoires d'analyses medicales - Ketterthill | Laboratoire national de sante, Microbiology, Microbial Genomics Platform | Anke Wienecke-Baldacchino; Caroline Scheiber; Catherine Ragimbeau; Fatu Djabi; Jessica Tapp; Lise Pignon; Raoul Salmon; Serge Vedy; Tamir Abdelrahman |
| EPI_ISL_2136904 | Laboratori Clínic Territorial de Girona | Can Ruti SARS-CoV-2 Sequencing Hub (HUGTiP/IrsiCaixa/IGTP) | Alba Sánchez; Anna Not; Antoni E Bordoy; Bonaventura Clotet; Cristina Casañ; Cristina Esteban; Francesc Catala-Moll; Gemma Clara; Ignacio Blanco; Marc Noguera-Julian; Maria Casadellà; Mariona Parera; Mercedes Guerrero; Montserrat Giménez; Pere-Joan Cardona; Pilar Armengol; Roger Paredes; Verónica Saludes; and Elisa Martró on behalf of the Can Ruti SARS-CoV-2 Sequencing Hub. |
| EPI_ISL_4080767 | Laboratorio Central de Saude Publica do Estado do Para (LACEN/PA) | Laboratory of Respiratory Viruses and Measles, Oswaldo Cruz Institute, FIOCRUZ | Agatha Soares; Alice Sampaio Rocha; Ana Carolina Mendonca; Anna Carolina Paixao; Elisa Cavalcante Pereira; Fernando Motta; Ighor Arantes; Luciana Appolinario; Marilda Siqueira on behalf of the Fiocruz COVID-19 Genomic Surveillance Network; Paola Resende; Renata Serrano Lopes; Taina Venas; Valnete Andrade |
| EPI_ISL_2001450, EPI_ISL_2131454, EPI_ISL_2131455, EPI_ISL_2131456, EPI_ISL_2131457, EPI_ISL_2131458 | Laboratorio Analisi Osp. Città di Castello - Azienda USL Umbria1 | Istituto Zooprofilattico Sperimentale dell'Abruzzo e Molise "G. Caporale" | Ancora M; Calistri P; Cammà C; Curini V; Delli Compagni E; Di Domenico M; Di Pasquale A; Lorusso A; Malagigi V; Mangone I; Marcacci M; Puglia I; Rinaldi A; Savini G; Scialabba S; Tacconi P |
| EPI_ISL_2047598, EPI_ISL_2047600 | Laboratorio Aziendale di Microbiologia e Virologia, Azienda Sanitaria dell'Alto Adige | Laboratorio Aziendale di Microbiologia e Virologia, Azienda Sanitaria dell'Alto Adige | Anne Picard; Bartolomeo Mobilio Rodriguez; Chiara Cantaloni; Claudia Volpato; Elisa Masi; Elisabetta Giacobazzi; Elisabetta Pagani; Irene Bianconi; Stefanie Wieser |
| EPI_ISL_2101388, EPI_ISL_2101405, EPI_ISL_2101482, EPI_ISL_2101537, EPI_ISL_2101539, EPI_ISL_2101540, EPI_ISL_2101619, EPI_ISL_2101620, EPI_ISL_2101689, EPI_ISL_2101690, EPI_ISL_2101708, EPI_ISL_2101709, EPI_ISL_2101710, EPI_ISL_2101711, EPI_ISL_2101712, EPI_ISL_2101713, EPI_ISL_2101726, EPI_ISL_2101732, EPI_ISL_2101733, EPI_ISL_2385432, EPI_ISL_2385433, EPI_ISL_2385434, EPI_ISL_2385435, EPI_ISL_2385436, EPI_ISL_2385437, EPI_ISL_2385438, EPI_ISL_2385439, EPI_ISL_2385440, EPI_ISL_2385441, EPI_ISL_2385442, EPI_ISL_2385443, EPI_ISL_2385444, EPI_ISL_2385445, EPI_ISL_2385446, EPI_ISL_2385447, EPI_ISL_2385448, EPI_ISL_2385449, EPI_ISL_2385450, EPI_ISL_2385451, EPI_ISL_2385452, EPI_ISL_2385453, EPI_ISL_2385454, EPI_ISL_2385455, EPI_ISL_2385456, EPI_ISL_2385457, EPI_ISL_2385458, EPI_ISL_2385459, EPI_ISL_2385460, EPI_ISL_2385461, EPI_ISL_2385462, EPI_ISL_2385463, EPI_ISL_2385464, EPI_ISL_2385465, EPI_ISL_2385466, EPI_ISL_2385467, EPI_ISL_2385468, EPI_ISL_2385469, EPI_ISL_2385470, EPI_ISL_2385471, EPI_ISL_2385472, EPI_ISL_2385474, EPI_ISL_2385478, EPI_ISL_2385480, EPI_ISL_2385481, EPI_ISL_2385482, EPI_ISL_2385483, EPI_ISL_2385484, EPI_ISL_2385485, EPI_ISL_2385486, EPI_ISL_2385487, EPI_ISL_2385490, EPI_ISL_2385491, EPI_ISL_2385492, EPI_ISL_2385493, EPI_ISL_2385494, EPI_ISL_2385495, EPI_ISL_2385496, EPI_ISL_2385497, EPI_ISL_2385498, EPI_ISL_2385506, EPI_ISL_2385507, EPI_ISL_2385510, EPI_ISL_2385511, EPI_ISL_2385513, EPI_ISL_2385517, EPI_ISL_2385518, EPI_ISL_2385519, EPI_ISL_2385537, EPI_ISL_2385546 | | | |
| see above | Laboratorio Central Noel Nutels | Bioinformatics Laboratory / LNCC | Alessandra P Lamarca; Alexandra L Gerber; Amilcar Tanuri; Ana Paula de C Guimaraes; Ana Tereza R Vasconcelos; Andrea Cony Cavalcanti; Caio Luiz Pereira Ribeiro; Cassia Alves; Cintia Policarpo; Claudia Maria Braga de Mello; Cristiane Gomes da Silva; Diana Mariani; Douglas Terra Machado; Flavio Dias da Silva; Gleidson da Silva de Oliveira; Leandro Magalhaes de Souza; Liliane Cavalcante; Luiz G P de Almeida; Marcio Henrique de Oliveira Garcia; Mario Sergio Ribeiro; Ronaldo da Silva F Jr; Silvia Carvalho |
| EPI_ISL_2391526, EPI_ISL_2391632, EPI_ISL_2391711, EPI_ISL_3091097 | Laboratorio Central de Epidemiología (LCE) | Instituto de Biotecnología de la UNAM | ; Alejandra Garcia-Gasca; Alejandra Hernández-Terán; Alejandro Sanchez-Flores; Alfredo Herrera-Estrella; Alicia Ocaña-Mondragón; Andreu Comas-García; Angel Gustavo Salas-Lais; Antonio Loza Román; Bernardo Martínez-Miguel; Blanca Taboada; Brenda Irasema Maldonado-Meza; Bruno Gomez-Gil; Carla Ivón Herrera-Najera; Carlos F. Arias; Celia Boukadida; Clara Esperanza Santacruz-Tinoco; Concepción Grajales-Muñiz; Consorcio Mexicano de Vigilancia Genómica (CoViGen-Mex). Authors (in alphabetical order): Julio Elias Alvarado-Yaah; Cristóbal Cháidez-Quiróz; Célida Duque Molina; Célida Martínez-Rodríguez; Daniel Fregoso-Rueda; Daniel Lira Morales; Eduardo Becerril-Vargas; Fernando Fontove-Herrera; Fidencio Mejía-Nepomuceno; Francisco Pulido; Gloria Elena Espinosa-Ayala; Gloria María Molina-Salinas; Gloria Vazquez; Hector Esteban Paz-Juárez; Hector Montoya-Fuentes; Helen Haydee Fernanda Ramírez-Plascencia; Irvin González-López; Jean Pierre González; Joel Armando Vázquez-Pérez.; Jorge Salas-Hernández; José Antonio Enciso-Moreno; José Arturo Martínez-Orozco; José Esteban Muñoz-Medina; José de Jesús Nuñez-Contreras; Juan Bautista Chale-Dzul; Julissa Enciso-Ibarra; Luis Alberto Ochoa- |

| | | | |
|---|---|---|---|
| | | | Carrera; Margarita Matías-Florentino; Mario Mújica-Sánchez; María Guadalupe Santiago-Mauricio; María Guadalupe de Jesús Mireles-Rivera; Nelly Sélem-Mojica; Pavel Isa; Ricardo Ciria Merce; Ricardo Grande; Rosa María Gutierrez Rios; Santiago Ávila-Ríos; Selene Zárate; Susana Lopez; Victor Eduardo García-Arias; Víctor Hugo Borja-Aburto |
| EPI_ISL_2402225 | Laboratorio Central de Epidemiología (LCE) | Unidad de Genomica Avanzada | Alejandro Sanchez-Flores; Alfredo Herrera-Estrella; Alicia Ocana-Mondragon; Angel Gustavo Salas-Lais; Bernardo Martinez-Miguel; Blanca Taboada; Brenda Irasema Maldonado-Meza; Carla Ivon Herrera-Najera; Carlos F. Arias; Celia Boukadida; Clara Esperanza Santacruz-Tinoco; Concepcion Grajales-Muniz; Consorcio Mexicano de Vigilancia Genomica (CoViGen-Mex). Authors (in alphabetical order): Julio Elias Alvarado-Yaah; Fernando Fontove-Herrera; Francisco Pulido; Gloria Elena Espinoza-Ayala; Gloria Maria Molina-Salinas; Gloria Vazquez; Hector Esteban Paz-Juarez; Hector Montoya-Fuentes; Helen Haydee Fernanda Ramirez-Plascencia; Jorge Ivan Salinal-Nevarez; Jose Antonio Enciso-Moreno; Jose Esteban Munoz-Medina; Jose de Jesus Nunez-Contreras; Juan Bautista Chale-Dzul; Luis Alberto Ochoa-Carrera; Margarita Matias-Florentino; Maria Guadalupe Santiago-Mauricio; Maria Guadalupe de Jesus Mireles-Rivera; Nelly Selem-Mojica; Pavel Isa; Ricardo Grande; Santiago Avila-Rios; Victor Hugo Borja-Aburto |
| EPI_ISL_6944527 | Laboratorio Central de Salud Publica | Viral Special Pathogens Branch, Centers for Disease Control and Prevention | Andrea Gómez de la Fuente; Cynthia Vázquez; Emir Talundzic; Joel Montgomery; John Klena; Juan Torales; Justin Lee; María José Ortega; María Liz Gamarra; Shannon Whitmer; Shirley Villalba |
| EPI_ISL_2196253, EPI_ISL_2196260, EPI_ISL_2196261 | Laboratorio Central de Saude Publica do Estado da Bahia (LACEN/BA) | Laboratory of Respiratory Viruses and Measles, Oswaldo Cruz Institute, FIOCRUZ | Alice Sampaio Rocha; Ana Carolina Mendonca; Anna Carolina Paixao; Elisa Cavalcante Pereira; Felicidade Pereira; Fernando Motta; Luciana Appolinario; Marilda Siqueira on behalf of the Fiocruz COVID-19 Genomic Surveillance Network; Paola Resende; Renata Serrano Lopes; Taina Venas |
| EPI_ISL_2536344, EPI_ISL_3061891 | Laboratorio Central de Saude Publica do Estado da Paraiba (LACEN-PB) | Laboratory of Respiratory Viruses and Measles, Oswaldo Cruz Institute, FIOCRUZ | Alice Sampaio Rocha; Ana Carolina Mendonca; Anna Carolina Paixao; Dalane Loudal Florentino Teixeira; Elisa Cavalcante Pereira; Fernando Motta; Joao Felipe Bezerra; Luciana Appolinario; Marilda Siqueira on behalf of the Fiocruz COVID-19 Genomic Surveillance Network; Paola Resende; Renata Serrano Lopes; Taina Venas |
| EPI_ISL_2466232, EPI_ISL_2466233, EPI_ISL_2466234, EPI_ISL_2466235, EPI_ISL_2466236, EPI_ISL_2466237, EPI_ISL_2466238, EPI_ISL_2466239, EPI_ISL_2466240, EPI_ISL_2466241, EPI_ISL_2557310, EPI_ISL_2557311, EPI_ISL_2557316, EPI_ISL_2557317, EPI_ISL_2557318, EPI_ISL_2557319, EPI_ISL_2557332, EPI_ISL_2557336 | | | |
| see above | Laboratorio Central de Saude Publica do Estado de Alagoas (LACEN/AL) | Laboratory of Respiratory Viruses and Measles, Oswaldo Cruz Institute, FIOCRUZ | Alice Sampaio Rocha; Ana Carolina Mendonca; Anderson Brandao Leite; Anna Carolina Paixao; Elisa Cavalcante Pereira; Fernando Motta; Luciana Appolinario; Marilda Siqueira on behalf of the Fiocruz COVID-19 Genomic Surveillance Network; Paola Resende; Renata Serrano Lopes; Taina Venas |
| EPI_ISL_3832396, EPI_ISL_3832443, EPI_ISL_3832444, EPI_ISL_3832471, EPI_ISL_4061434, EPI_ISL_4061435, EPI_ISL_4061436 | | | |
| see above | Laboratorio Central de Saude Publica do Estado de Santa Catarina (LACEN/SC) | Laboratory of Respiratory Viruses and Measles, Oswaldo Cruz Institute, FIOCRUZ | Alice Sampaio Rocha; Ana Carolina Mendonca; Anna Carolina Paixao; Darcita Buerger Rovaris; Elisa Cavalcante Pereira; Fernando Motta; Luciana Appolinario; Marilda Siqueira on behalf of the Fiocruz COVID-19 Genomic Surveillance Network; Paola Resende; Renata Serrano Lopes; Sandra Bianchini Fernandes; Taina Venas |
| EPI_ISL_2196262, EPI_ISL_3190243 | Laboratorio Central de Saude Publica do Estado do Espirito Santo (LACEN/ES) | Laboratory of Respiratory Viruses and Measles, Oswaldo Cruz Institute, FIOCRUZ | Alice Sampaio Rocha; Ana Carolina Mendonca; Anna Carolina Paixao; Eilisa Cavalcante Pereira; Elisa Cavalcante Pereira; Fernando Motta; Luciana Appolinario; Marilda Siqueira on behalf of the Fiocruz COVID-19 Genomic Surveillance Network; Paola Resende; Renata Serrano Lopes; Rodrigo Ribeiro Rodrigues; Taina Venas |
| EPI_ISL_2983250, EPI_ISL_2983254, EPI_ISL_2983285, EPI_ISL_2983286, EPI_ISL_2983287, EPI_ISL_2983288, EPI_ISL_2983289, EPI_ISL_2983290, EPI_ISL_2983291, EPI_ISL_2983292, EPI_ISL_2983333, EPI_ISL_2983335 | | | |
| see above | Laboratorio Central de Saude Publica do Estado do Maranhao (LACEN-MA) | Laboratory of Respiratory Viruses and Measles, Oswaldo Cruz Institute, FIOCRUZ | Agatha Cristinne Prudencio; Alice Sampaio Rocha; Ana Carolina Mendonca; Anna Carolina Paixao; Elisa Cavalcante Pereira; Fernando Motta; Ighor Leonardo Arantes Gomes; Lidio Gonçalves Lima Neto; Luciana Appolinario; Marilda Siqueira on behalf of the Fiocruz COVID-19 Genomic Surveillance Network; Paola Resende; Renata Serrano Lopes; Taina Moreira Venas |
| EPI_ISL_4061296, EPI_ISL_4061297, EPI_ISL_4061300, EPI_ISL_4061302, EPI_ISL_4061303, EPI_ISL_4061304, EPI_ISL_4061305, EPI_ISL_4061306, EPI_ISL_4061307, EPI_ISL_4061309, EPI_ISL_4061310, EPI_ISL_4061311, EPI_ISL_4061315, EPI_ISL_4061317, EPI_ISL_4061318, EPI_ISL_4061320, EPI_ISL_4061324, EPI_ISL_4061325 | | | |
| see above | Laboratorio Central de Saude Publica do Estado do Parana (LACEN/PR) | Laboratory of Respiratory Viruses and Measles, Oswaldo Cruz Institute, FIOCRUZ | Agatha Soares; Alice Sampaio Rocha; Ana Carolina Mendonca; Anna Carolina Paixao; Elisa Cavalcante Pereira; Fernando Motta; Ighor Leonardo Arantes; Irina Riediger; Luciana Appolinario; Marilda Siqueira on behalf of the Fiocruz COVID-19 Genomic Surveillance Network; Paola Resende; Renata Serrano Lopes; Taina Venas |
| EPI_ISL_2603436, EPI_ISL_2603577, EPI_ISL_2603578, EPI_ISL_2603579, EPI_ISL_2603588 | Laboratorio Central de Saude Publica do Estado do Rio Grande do Sul (LACEN-RS) | Laboratory of Respiratory Viruses and Measles, Oswaldo Cruz Institute, FIOCRUZ | Alice Sampaio Rocha; Ana Carolina Mendonca; Anderson Brandao Leite; Anna Carolina Paixao; Elisa Cavalcante Pereira; Fernando Motta; Luciana Appolinario; Marilda Siqueira on behalf of the Fiocruz COVID-19 Genomic Surveillance Network; Paola Resende; Renata Serrano Lopes; Taina Venas |
| EPI_ISL_3048808 | Laboratorio Central de Saude Publica do Estado do Rio Grande do Sul (LACEN-RS) | Laboratório de Biologia Molecular da Universidade Federal de Ciências da Saúde de Porto Alegre | Adriana Seixas; Ana B. G. Veiga; Ana Paula Muterle Varela; Fabiana Quoos Mayer; Fernando Hayashi Sant'Anna; Janira Prichula; Letícia Garay Martins; Richard Steiner Salvato; Tatiana Schäffer Gregianini |
| EPI_ISL_3033043, EPI_ISL_3033044, EPI_ISL_3033045, EPI_ISL_3033052 | Laboratorio Central de la Ciudad de Santa Fe | Grupo de Genómica y Bioinformática del Instituto de Investigación de la Cadena Láctea CONICET-INTA on behalf of 'Proyecto Argentino Interinstitucional de genomica de SARS-CoV-2' (PAIS Consortium) | AF; Amadio; C; Eberhardt; G; Irazoqui; JM; MF; Mugna; Ojeda; Pastor; Rompato; V |
| EPI_ISL_2271688 | Laboratorio Central, Ministerio de Salud Cordoba | Instituto de Patologia Vegetal (CIAP-INTA) on behalf of 'Proyecto Argentino Interinstitucional de genomica de SARS-CoV-2' (PAIS Consortium) | Barbas, G.; Castro, G.; Debat, HJ.; FD; Fernandez; M; M.B.; Marquez, N.; Pisano; Re, V. |
| EPI_ISL_2674335 | Laboratorio Clínica Putumayo | Instituto Nacional de Salud- Dirección de Investigación en Salud Pública | Carlos Franco-Muñoz; Carmen Osorio; Diana Malo; Diego A. Álvarez-Díaz; Diego Andrés Prada; Gerardo Santamaría; Hector Alejandro Ruiz-Moreno; Jhonnatan Reales-González; Jorge Rivera; Juan Camilo Martinez; Julian Naizaque; Katherine Laiton-Donato; Lisseth Pardo; Magdalena Wiesner; Marcela Mercado-Reyes; Maria T. Herrera-Sepúlveda; Marta Lopez Blanco; Martha Lucia Ospina Martinez; Paola Rojas; Sergio Gomez; Sheryll Corchuelo; Ángela Alarcon Cruz |
| EPI_ISL_4197220 | Laboratorio Clínico Analizar | Gencore - Universidad de los Andes | Cristian Barrera; David González; Gabriela Ariza; Luisa Sacristan; Marcela Guevara; Marcela Mercado; Silvia Restrepo |
| EPI_ISL_3333565, EPI_ISL_3333566, EPI_ISL_3333567 | Laboratorio Clínico Gamma | MELISA Institute Genomics and Proteomics SpA | Cristian Vargas; Elard Koch; Francisco Alvarez; Guillermo Nourdin; Mauricio Hernandez |
| EPI_ISL_5801188, EPI_ISL_5801189 | Laboratorio Local De Itapecerica Da Serra | Instituto Butantan | Antonio Jorge Martins; Claudia Renata dos Santos Barros; David Schlesinger; Debora Botequio Moretti; Dimas Tadeu Covas; Elaine Cristina Marqueze; Elaine Vieira Santos; Evandra Strazza Rodrigues; Heidge Fukumasu; Jayme Augusto de Souza-Neto; José Salvatore Leister Patané; Luiz Alcantara; Luiz Lehmann Coutinho; Maria Carolina Elias; Maurício Lacerda Nogueira; Rafael dos Santos Bezerra; Raul Machado Neto; Rejane Maria Tommasini Grotto; Ricardo Haddad; Sandra Coccuzzo Sampaio Vessoni; Simone Kashima; Svetoslav Nanev Slavov; Vincent Louis Viala |
| EPI_ISL_5802703, EPI_ISL_5802718 | Laboratorio Municipal De Analises Clinicas De Rio Claro | Instituto Butantan | Antonio Jorge Martins; Claudia Renata dos Santos Barros; David Schlesinger; Debora Botequio Moretti; Dimas Tadeu Covas; Elaine Cristina Marqueze; Elaine Vieira Santos; Evandra Strazza Rodrigues; Heidge Fukumasu; Jayme Augusto de Souza-Neto; José Salvatore Leister Patané; Luiz Alcantara; Luiz Lehmann Coutinho; Maria Carolina Elias; Maurício Lacerda Nogueira; Rafael dos Santos Bezerra; Raul Machado Neto; Rejane Maria Tommasini Grotto; Ricardo Haddad; Sandra Coccuzzo Sampaio Vessoni; Simone Kashima; Svetoslav Nanev Slavov; Vincent Louis Viala |
| EPI_ISL_5801269, EPI_ISL_5801358, EPI_ISL_5801360, EPI_ISL_5801361, EPI_ISL_5801378, EPI_ISL_5801382, EPI_ISL_5801401, EPI_ISL_5801403, EPI_ISL_5801405, EPI_ISL_5801410, EPI_ISL_5801411, EPI_ISL_5801412, EPI_ISL_5801421, EPI_ISL_5801424, EPI_ISL_5801437, EPI_ISL_5801438, EPI_ISL_5801440 | | | |
| see above | Laboratorio Municipal De Piracicaba | Instituto Butantan | Antonio Jorge Martins; Claudia Renata dos Santos Barros; David Schlesinger; Debora Botequio Moretti; Dimas Tadeu Covas; Elaine Cristina Marqueze; Elaine Vieira Santos; Evandra Strazza Rodrigues; Heidge Fukumasu; Jayme Augusto de Souza-Neto; José Salvatore Leister Patané; Luiz Alcantara; Luiz Lehmann Coutinho; Maria Carolina Elias; Maurício Lacerda Nogueira; Rafael dos Santos Bezerra; Raul Machado Neto; Rejane Maria Tommasini Grotto; Ricardo Haddad; Sandra Coccuzzo Sampaio Vessoni; Simone Kashima; Svetoslav Nanev Slavov; Vincent Louis Viala |
| EPI_ISL_5158240 | Laboratorio Nacional de Salud Publica | Inciensa, Instituto Costarricense de Investigación y Enseñanza en Nutrición y Salud | Carlos Guerra; Denis Jovel; Laboratorio de Genómica INCIENSA; Ruth Vasquez |
| EPI_ISL_3275191 | Laboratorio Nacional de Salud, Ministerio de Salud Publica y Asistencia Social | Genomics and Proteomics Departament, Gorgas Memorial Institute For Health Studies | Alexander A Martinez; Ambar Moreno; Claudia Estrada; Claudia Gonzalez V; César Roberto Conde Pereira; Jessica Gondola; Leyda Abrego; Marlene Castillo; Oris Chavarria |
| EPI_ISL_5199348 | Laboratorio SYNLAB Colombia | Instituto Nacional de Salud- Dirección de Investigación en Salud Pública | Beatriz de Arco; Carlos Franco-Muñoz; Carmen Osorio; Diana Malo; Diego A. Álvarez-Díaz; Diego Andrés Prada; Gerardo Santamaría; Hector Alejandro Ruiz-Moreno; Jhonnatan Reales-González; Jorge Rivera; Juan Camilo Martinez; Julian Naizaque; Katherine Laiton-Donato; Lisseth Pardo; Magdalena Wiesner; Marcela Mercado-Reyes; Maria T. Herrera-Sepúlveda; Marta Lopez Blanco; Martha Lucia Ospina Martinez; Paola Rojas; Sergio Gomez; Sheryll Corchuelo; Tatiana Cobos; Ángela Alarcon Cruz |
| EPI_ISL_5771874 | Laboratorio Salud Publica Mendoza | Nodo de Secuenciación Tierra del Fuego - Hospital Regional Ushuaia - Centro Austral De Investigaciones Científicas - Universidad Nacional De Tierra Del Fuego | AE; CF; Ceballos; F; Gallego; Gramundi; ID; Nardi; Rojas; SG |
| EPI_ISL_6121561 | Laboratorio de Alta Complejidad (LACMI), Instituto de Genética Humana (IgeHM) Parque de la Salud de la Provincia de Misiones, Universidad Nacional de Misiones | Grupo de Genómica y Bioinformática del Instituto de Investigación de la Cadena Láctea CONICET-INTA on behalf of 'Proyecto Argentino Interinstitucional de genomica de SARS-CoV-2' (PAIS Consortium) | AF; Aguirre; Amadio; Benitez; C; D; Eberhardt; Fontana; G; Gamarra; Gonzales; Irazoqui; JM; K; Ludojoski; M; MF; R; Salvatierra; Silva; TA; Urquiza; Vogel |
| EPI_ISL_4405289 | Laboratorio de Biología Molecular Bolívar | Área de Secuenciación del Laboratorio de Virología del Hospital de Niños Dr. Ricardo Gutierrez on behalf of 'Proyecto Argentino Interinstitucional de genomica de SARS-CoV-2' (PAIS Consortium) | Acuña; Cabrerizo; Caviglia; D; Di Claudio; D'Andrea; F; Goya; Jofré; LE; Larreche; Lescano; Lusso; M; ME; MI; ML; MR; MS; Martínez; Nabaes Jodar; Natale; R; S; Ticeira; Valinotto; Viegas, M. |
| EPI_ISL_4030337, EPI_ISL_4030339, EPI_ISL_4030341, EPI_ISL_4030357, EPI_ISL_4030361 | Laboratorio de Ecologia de Doencas Transmissiveis na Amazonia, Instituto Leonidas e Maria Deane - Fiocruz Amazonia | Laboratorio de Ecologia de Doencas Transmissiveis na Amazonia, Instituto Leonidas e Maria Deane - Fiocruz Amazonia | Alex Martins; André Corado; Debora Duarte; Fabiola Mendonça da Silva Chui; Felipe Naveca; Fernanda Nascimento; Fernando Fonseca de Almeida Val; George Silva; Gisely Cardoso de Melo; Karina Pessoa; Luciana Gonçalves; Marcus Vinicius Guimarães de Lacerda; Maria Júlia Brandão; Maria Paula Gomes Mourão; Mariana Xavier Simão; Matilde Mejía; Michele Jesus; Valdinete Nascimento; Vanderson de Souza Sampaio; Victor Souza; Ágatha Costa |

| | | | |
|---|---|---|---|
| EPI_ISL_2728553, EPI_ISL_2728554, EPI_ISL_2728556, EPI_ISL_2728557, EPI_ISL_2728568, EPI_ISL_2728569 | Laboratorio de Infectologia y Virologia Molecular | Laboratory of Molecular Virology, School of Medicine, Pontificia Universidad Catolica de Chile | Alejandro Bhrun; Ana Maria Contreras; Andres E. Munoz-Marcos; Carlos Palma; Catalina Pardo-Roa; Constanza Maldonado; Constanza Martinez-Valdevenito; Eileen Serrano; Erick Salinas; Estefany Poblete; Francisco Melo; Jennifer Angulo; Jorge Levican; Leonardo I. Almonacid; M. Belen Leyton; Magdalena Vera; Marcela Ferres; Maria Jose Avendano; Rafael A. Medina; Tamara Garcia-Salum |
| EPI_ISL_2854130, EPI_ISL_2854134, EPI_ISL_2854137, EPI_ISL_2854140, EPI_ISL_2854142, EPI_ISL_2854143, EPI_ISL_2854144, EPI_ISL_2854145, EPI_ISL_2854148, EPI_ISL_2854150, EPI_ISL_2907515 | | | |
| see above | Laboratorio de Medicina Molecular, Universidad de Magallanes | Centro Asistencial Docente y de Investigacion, Universidad de Magallanes | Constanza Ceroni; Diego Alvarez; Inés Cid; Jacqueline Aldridge; Jorge González; Marcelo Navarrete; Roberto Uribe-Paredes |
| EPI_ISL_2008913, EPI_ISL_2008915, EPI_ISL_2008923, EPI_ISL_2008927, EPI_ISL_2008928, EPI_ISL_2008931, EPI_ISL_2008932, EPI_ISL_2107301, EPI_ISL_3048959, EPI_ISL_3048960 | | | |
| see above | Laboratorio de Pesquisa em Virologia, FAMERP, SJRP | Laboratorio de Pesquisa em Virologia, FAMERP, SJRP | Cecília Artico Banho; Cíntia Bittar; Fábio Sossai Possebon; Guilherme Campos; Helena Lage Ferreira; Jorge A. Petroli Marchesi; João Pessoa Araújo Jr.; Leila Sabrina Ullmann; Lívia Sacchetto; Maisa C. Pereira Parra; Marília Moraes; Maurício L. Nogueira.; Paula Rahal; Paulo Inacio da Costa |
| EPI_ISL_3401448, EPI_ISL_3401455, EPI_ISL_3401456, EPI_ISL_3401467 | Laboratorio de Referencia Nacional de Virus Respiratorios. Centro Nacional de Salud Publica. Instituto Nacional de Salud Peru. | Laboratorio de Referencia Nacional de Virus Respiratorios. Centro Nacional de Salud Publica. Instituto Nacional de Salud Peru. | Carlos Padilla Rojas; Henri Bailon Calderon; Iris Silva Molina; Joseph Huayra Niquen; Lely Solari Zerpa; Luis Barcena Flores; Marco Galarza Perez; Nancy Rojas Serrano; Nieves Sevilla Castañeda; Omar Caceres Rey; Orson Mestanza Millones; Princesa Medrano Alhuay; Priscila Lope Pari; Sandra Morales Ruiz; Sara Gordillo Vilchez; Steve Acedo Lazo; Veronica Hurtado Vela; Victor Jimenez Vasquez; Wendy Lizarraga Olivares |
| EPI_ISL_3023588, EPI_ISL_3375905, EPI_ISL_3375953, EPI_ISL_3376032, EPI_ISL_3376377, EPI_ISL_3376411 | Laboratorio de Referencial Nacional de Virus Respiratorios | Laboratorio de Referencial Nacional de Virus Respiratorios | Carlos Padilla Rojas; Henri Bailon Calderon; Iris Silva Molina; Joseph Huayra Niquen; Lely Solari Zerpa; Luis Barcena Flores; Marco Galarza Perez; Nancy Rojas Serrano; Omar Caceres Rey; Orson Mestanza Millones; Priscila Lope Pari; Sandra Morales Ruiz; Steve Acedo Lazo; Veronica Hurtado Vela |
| EPI_ISL_4405471 | Laboratorio del Hospital Interzonal General de Agudos " Evita " | Área de Secuenciación del Laboratorio de Virología del Hospital de Niños Dr. Ricardo Gutierrez on behalf of 'Proyecto Argentino Interinstitucional de genomica de SARS-CoV-2' (PAIS Consortium) | A; Acuña; D; Desimone; E; Goya; Grossi; I; L; LE; Luczac; Lusso; MI; MS; Musto; Nabaes Jodar; Natale; O; S; Serrano; Valinotto; Viegas, M. |
| EPI_ISL_1993809 | Laboratorio di Microbiologia | Laboratorio di Microbiologia | Martinetti Lucchini Gladys; Valeria Spina |
| EPI_ISL_1930508, EPI_ISL_1930531, EPI_ISL_1930579, EPI_ISL_1930587, EPI_ISL_1930606, EPI_ISL_2044009, EPI_ISL_2044017, EPI_ISL_2044024, EPI_ISL_2044030, EPI_ISL_2044041, EPI_ISL_2044047, EPI_ISL_2044051, EPI_ISL_2044061, EPI_ISL_2044092, EPI_ISL_2044098, EPI_ISL_2044125, EPI_ISL_2044127, EPI_ISL_2044131, EPI_ISL_2044183, EPI_ISL_2044197, EPI_ISL_2044205, EPI_ISL_2044214, EPI_ISL_2044267, EPI_ISL_2044271, EPI_ISL_2044278, EPI_ISL_2044281, EPI_ISL_2044295, EPI_ISL_2044300, EPI_ISL_2044427, EPI_ISL_2044447, EPI_ISL_2044456, EPI_ISL_2044457, EPI_ISL_2044471, EPI_ISL_2044472, EPI_ISL_2044480, EPI_ISL_2044487, EPI_ISL_2044489, EPI_ISL_2044490, EPI_ISL_2044496, EPI_ISL_2044499, EPI_ISL_2044501, EPI_ISL_2044505, EPI_ISL_2044506, EPI_ISL_2044532, EPI_ISL_2044555, EPI_ISL_2044582, EPI_ISL_2044594, EPI_ISL_2044602, EPI_ISL_2044629, EPI_ISL_2044671, EPI_ISL_2044674, EPI_ISL_2044683, EPI_ISL_2044697, EPI_ISL_2044698, EPI_ISL_2044702, EPI_ISL_2044710, EPI_ISL_2044720, EPI_ISL_2044723, EPI_ISL_2044815, EPI_ISL_2044853, EPI_ISL_2044871, EPI_ISL_2044910, EPI_ISL_2044937, EPI_ISL_2044967, EPI_ISL_2044969, EPI_ISL_2045023, EPI_ISL_2045038, EPI_ISL_2045040, EPI_ISL_2045108, EPI_ISL_2045121, EPI_ISL_2045127, EPI_ISL_2045128, EPI_ISL_2045129, EPI_ISL_2045132, EPI_ISL_2045134, EPI_ISL_2045145, EPI_ISL_2045155, EPI_ISL_2045159, EPI_ISL_2045162, EPI_ISL_2045164, EPI_ISL_2045172, EPI_ISL_2045183, EPI_ISL_2045187, EPI_ISL_2045196, EPI_ISL_2045200, EPI_ISL_2045201, EPI_ISL_2045202, EPI_ISL_2045204, EPI_ISL_2045208, EPI_ISL_2045209, EPI_ISL_2045212, EPI_ISL_2045215, EPI_ISL_2045225, EPI_ISL_2045231, EPI_ISL_2045238, EPI_ISL_2045239, EPI_ISL_2045247, EPI_ISL_2045250, EPI_ISL_2045253, EPI_ISL_2045257, EPI_ISL_2045265, EPI_ISL_2045266, EPI_ISL_2045275, EPI_ISL_2045307, EPI_ISL_2045311, EPI_ISL_2045314, EPI_ISL_2045318, EPI_ISL_2045321, EPI_ISL_2045322, EPI_ISL_2045323, EPI_ISL_2045324, EPI_ISL_2045326, EPI_ISL_2045334, EPI_ISL_2045335, EPI_ISL_2045337, EPI_ISL_2045380, EPI_ISL_2045404, EPI_ISL_2045405, EPI_ISL_2045409, EPI_ISL_2045410, EPI_ISL_2045412, EPI_ISL_2045416, EPI_ISL_2045420, EPI_ISL_2045456, EPI_ISL_2045458, EPI_ISL_2045461, EPI_ISL_2045470, EPI_ISL_2045504, EPI_ISL_2045537, EPI_ISL_2045556, EPI_ISL_2045581, EPI_ISL_2045585, EPI_ISL_2045586, EPI_ISL_2045588, EPI_ISL_2045630, EPI_ISL_2045631, EPI_ISL_2045634, EPI_ISL_2045637, EPI_ISL_2045639, EPI_ISL_2045669, EPI_ISL_2045670, EPI_ISL_2045687, EPI_ISL_2045714, EPI_ISL_2045730, EPI_ISL_2045740, EPI_ISL_2045751, EPI_ISL_2045759, EPI_ISL_2045763, EPI_ISL_2045764, EPI_ISL_2045767, EPI_ISL_2045771, EPI_ISL_2045774, EPI_ISL_2045776, EPI_ISL_2045779, EPI_ISL_2045781, EPI_ISL_2045784, EPI_ISL_2045820, EPI_ISL_2045822, EPI_ISL_2045824, EPI_ISL_2045857, EPI_ISL_2045858, EPI_ISL_2045864, EPI_ISL_2045872, EPI_ISL_2045877, EPI_ISL_2045926, EPI_ISL_2046013, EPI_ISL_2046076, EPI_ISL_2046190, EPI_ISL_2046216, EPI_ISL_2046220, EPI_ISL_2046240, EPI_ISL_2046269, EPI_ISL_2046300, EPI_ISL_2046328, EPI_ISL_2046342, EPI_ISL_2046372, EPI_ISL_2046385, EPI_ISL_2046410, EPI_ISL_2046411, EPI_ISL_2046425, EPI_ISL_2046426, EPI_ISL_2046478, EPI_ISL_2046488, EPI_ISL_2046521, EPI_ISL_2046524, EPI_ISL_2046527, EPI_ISL_2046536, EPI_ISL_2046540, EPI_ISL_2046554, EPI_ISL_2046564, EPI_ISL_2046566, EPI_ISL_2046569, EPI_ISL_2046571, EPI_ISL_2046592, EPI_ISL_2046603, EPI_ISL_2046616, EPI_ISL_2046617, EPI_ISL_2046622, EPI_ISL_2046623, EPI_ISL_2046649, EPI_ISL_2046691, EPI_ISL_2046702, EPI_ISL_2183480, EPI_ISL_2183486, EPI_ISL_2183491, EPI_ISL_2183493, EPI_ISL_2183509, EPI_ISL_2183512, EPI_ISL_2183537, EPI_ISL_2183540, EPI_ISL_2183550, EPI_ISL_2183603, EPI_ISL_2183623, EPI_ISL_2183626, EPI_ISL_2183868, EPI_ISL_2183883, EPI_ISL_2183903, EPI_ISL_2183915, EPI_ISL_2184056, EPI_ISL_4373563, EPI_ISL_4373969, EPI_ISL_4374005, EPI_ISL_4378206, EPI_ISL_4378935, EPI_ISL_4380406, EPI_ISL_4381522 | | | |
| see above | Laboratory Corporation of America | Centers for Disease Control and Prevention Division of Viral Diseases, Pathogen Discovery | Adrian Paskey; Amanda Douglas; Amanda Suchanek; Andrea Throop; Ayla Burns; Benjamin Rambo-Martin; Bobbi Croy; Brian Krueger; Brian Norvell; Christopher Gulvick; Christos Petropoulos; Clinton Paden; Clinton R. Paden; Craig Lukasik; Dakota Howard; Darlene Wagner; Debbie Boles; Dhwani Batra; Duncan MacCannell; Eyad Almasri; Goran Stevovic; Howard Engler; Hrushikesh Deshmukh; Jake Humphrey; Jana Schroth; Jason Caravas; Joe Voshell; John Pruitt; Jonathan Meltzer; Jonathan Williams; Kara Moser; Kimberly Wagner; Kristine Lacek; Lax Iyer; Lisa Pfefferle; Lyndon Tilson; Manoj Jain; Marcia Eisenberg; Mary Ann Cristobal; Mary Cristobal; Mary Williamson; Matthew Robinson; Matthew Schmerer; Michael Levandoski; Mike Sapeta; Mindy Nye; Minoo Agarwal; Mohan Kolli; Nuthawin Charoensri; Oren Cohen; Peter Cook; Peter W. Cook; Prashant Gupta; Qian Zeng; Rama Ghatti; Scott Parker; Scott Ryan; Scott Sammons; Shatavia Morrison; Stanley Letovsky; Steven Ragan; Suresh Babu Selvaraju; Suresh Selvaraju; Susan Countryman; Susan Hicks; Suzanne Dale; Thomas Urban; Tim Kuphal; Tricia Zwiefelhofer; Tymeckia Kendall; Victoria Caban Figueroa; Vincent Drouillon; Yvette Unoarumhi |
| EPI_ISL_2196251, EPI_ISL_2443605, EPI_ISL_2443606, EPI_ISL_2443607, EPI_ISL_2557403, EPI_ISL_3434728, EPI_ISL_5158269, EPI_ISL_6172740, EPI_ISL_6943985, EPI_ISL_6943986 | | | |
| see above | Laboratory of Respiratory Viruses and Measles, Oswaldo Cruz Institute, FIOCRUZ | Laboratory of Respiratory Viruses and Measles, Oswaldo Cruz Institute, FIOCRUZ | Agatha Cristinne Prudencio; Agatha Cristinne Prudencio Soares; Alice Sampaio Rocha; Ana Carolina Mendonca; Anna Carolina Paixao; Bruna Mendonça da Silva; Elisa Cavalcante Pereira; Fernando Motta; Ighor Arantes; Ighor Leonardo Arantes Gomes; Jéssica Graça Macedo de Carvalho; Larissa Macedo Pinto; Luciana Appolinario; Marilda Siqueira on behalf of the Fiocruz COVID-19 Genomic Surveillance Network; Paola Resende; Patricia Brasil; Renata Serrano Lopes; Taina Venas; Thais Aguiar; Victor Guimaraes; Vitor Guimaraes Ribeiro |
| EPI_ISL_2696312, EPI_ISL_2696345, EPI_ISL_2696358, EPI_ISL_2696393, EPI_ISL_2696394, EPI_ISL_2696396, EPI_ISL_2696555, EPI_ISL_2696561, EPI_ISL_2696567, EPI_ISL_3025474, EPI_ISL_3025502, EPI_ISL_3025522, EPI_ISL_3025524, EPI_ISL_3025541, EPI_ISL_3025572, EPI_ISL_3025580, EPI_ISL_3025584, EPI_ISL_3025597 | | | |
| see above | Laboratory, Gravity Diagnostics | Laboratory, Gravity Diagnostics | Diagnostics, G. |
| EPI_ISL_4600524, EPI_ISL_4600527, EPI_ISL_4600537 | Laboratório Central de Saúde Pública de Pernambuco | Coordenação Geral de Laboratórios de Saúde Pública (CGLAB/DAEVS/SVS/MS) | Vagner Fonseca; et al. |
| EPI_ISL_2777281, EPI_ISL_2777282, EPI_ISL_2777292, EPI_ISL_2777334, EPI_ISL_2777346, EPI_ISL_2777347, EPI_ISL_2777513, EPI_ISL_2777514, EPI_ISL_2777515, EPI_ISL_2777517, EPI_ISL_2777530, EPI_ISL_2777547, EPI_ISL_2777573, EPI_ISL_2777575, EPI_ISL_2777577, EPI_ISL_2777578, EPI_ISL_2777582, EPI_ISL_2777583, EPI_ISL_2777586, EPI_ISL_2777591, EPI_ISL_2777594, EPI_ISL_2777597, EPI_ISL_2777601, EPI_ISL_2777603, EPI_ISL_2777604, EPI_ISL_2777605, EPI_ISL_2777610, EPI_ISL_2777614, EPI_ISL_2777623, EPI_ISL_2777628, EPI_ISL_2777630, EPI_ISL_2777633, EPI_ISL_2777634, EPI_ISL_2777686, EPI_ISL_2777687, EPI_ISL_2777702, EPI_ISL_2777756, EPI_ISL_2777757, EPI_ISL_2777760, EPI_ISL_2777761, EPI_ISL_2777764, EPI_ISL_2777765, EPI_ISL_2777770, EPI_ISL_2777832, EPI_ISL_2777833, EPI_ISL_2777834, EPI_ISL_2777835, EPI_ISL_2777836, EPI_ISL_2777837, EPI_ISL_2777839, EPI_ISL_2777879, EPI_ISL_2777880, EPI_ISL_2777881, EPI_ISL_2777882, EPI_ISL_2777883, EPI_ISL_2777931, EPI_ISL_2777932, EPI_ISL_2777936, EPI_ISL_2777978, EPI_ISL_2777979, EPI_ISL_2777980, EPI_ISL_2777981, EPI_ISL_2777982, EPI_ISL_2777989, EPI_ISL_2777990, EPI_ISL_2777994 | | | |
| see above | Laboratório Central de Saúde Pública do Amazonas - LACEN-AM | Laboratorio de Ecologia de Doencas Transmissiveis na Amazonia, Instituto Leonidas e Maria Deane - Fiocruz Amazonia | André Corado; Debora Duarte; Felipe Naveca; Fernanda Nascimento; George Silva; Karina Pessoa; Luciana Gonçalves; Maria Júlia Brandão; Matilde Mejía; Michele Jesus; Valdinete Nascimento; Victor Souza; Ágatha Costa |
| EPI_ISL_4565991 | Laboratório de Agentes Infecciosos e Vetores -LAIVE-Bahia, Barreiras, Brazil | Laboratório Baculovírus | Aline Belmok; Bergmann M Ribeiro; Fernando L Melo; Jaime Amorim; Josilene Pinheiro |
| EPI_ISL_3873627 | Laboratório de Microbiologia Molecular - Universidade FEEVALE | Molecular Microbiology Laboratory | Alana Witt Hansen; Fernando Rosado Spilki; Fágner Henrique Heldt; Juliana Schons Gularte; Juliane Deise Fleck; Mariana Soares da Silva; Matheus Nunes Weber; Meriane Demoliner; Micheli Filippi; Paula Rodrigues de Almeida; Vyctoria Malayhka de Abreu Góes Pereira. |
| EPI_ISL_2629829, EPI_ISL_2629830, EPI_ISL_2629831, EPI_ISL_2629833, EPI_ISL_2629834, EPI_ISL_2629835 | Laboratório de Virologia Molecular - Universidade Federal do Rio de Janeiro | Laboratório de Virologia Molecular - Universidade Federal do Rio de Janeiro | ; Alice Laschuk Herlinger; Amilcar Tanuri; André Felipe Andrade dos Santos; Carolina Moreira Voloch; Cássia Cristina Alves Gonçalves; Diana Mariani; Débora Souza Faffe; Filipe Romero Rebello Moreira; Francine Bittencourt Schiffler; Isabela de Carvalho Leitão; Marcelo Calado de Paula Tôrres; Matheus Augusto Calvano Cosentino; Mirela D'arc; Orlando da Costa Ferreira Junior; Rafael Mello Galliez; Raíssa Mirella dos Santos Cunha da Costa; Renato Santana de Aguiar; Terezinha Marta Pereira Pinto Castineiras; Thamiris dos Santos Miranda; Átila Duque Rossi |
| EPI_ISL_4520591 | Laboratório de Virologia e Cultivo Celular - LABVICC | Laboratório Baculovírus | Aline Belmok; Bergmann Morais Ribeiro; Clayton Luiz Borges; Déborah Anjos; Elen Regozino Muniz; Fabiola Souza Fiaccadori; Fernando L Melo; Gabriela Silvério Bazilio; Juliana Alves Parente Rocha; Keila Correia de Alcântara; Menira Souza; Nathânia Dábilla; Valéria Christina de Rezende Feres |
| EPI_ISL_2443710, EPI_ISL_2466260, EPI_ISL_2536228, EPI_ISL_2536230, EPI_ISL_2536233, EPI_ISL_2536234, EPI_ISL_2536237, EPI_ISL_2536239, EPI_ISL_2536241, EPI_ISL_2536244, EPI_ISL_2536247, EPI_ISL_2536251, EPI_ISL_2536252, EPI_ISL_2536254, EPI_ISL_2536255, EPI_ISL_2536256, EPI_ISL_2536257, EPI_ISL_2536261, EPI_ISL_2536263 | | | |
| see above | Labortorio Central de Saude Publica do Estado de Santa Catarina (LACEN/SC) | Laboratory of Respiratory Viruses and Measles, Oswaldo Cruz Institute, FIOCRUZ | Alice Sampaio Rocha; Ana Carolina Mendonca; Anna Carolina Paixao; Darcita Buerger Rovaris; Elisa Cavalcante Pereira; Fernando Motta; Luciana Appolinario; Marilda Siqueira on behalf of the Fiocruz COVID-19 Genomic Surveillance Network; Paola Resende; Renata Serrano Lopes; Sandra Bianchini Fernandes; Taina Venas |
| EPI_ISL_2443653, EPI_ISL_2443654, EPI_ISL_2443655, EPI_ISL_2443656, EPI_ISL_2443657, EPI_ISL_2443658, EPI_ISL_2557379, EPI_ISL_2557404, EPI_ISL_3061894, EPI_ISL_3061895, EPI_ISL_3061897 | | | |
| see above | Labortorio Central de Saude Publica do Estado do Rio de Janeiro (LACEN/RJ) | Laboratory of Respiratory Viruses and Measles, Oswaldo Cruz Institute, FIOCRUZ | Alice Sampaio Rocha; Ana Carolina Mendonca; Andrea Cony Cavalcanti; Anna Carolina Paixao; Elisa Cavalcante Pereira; Fernando Motta; Luciana Appolinario; Marilda Siqueira on behalf of the Fiocruz COVID-19 Genomic Surveillance Network; Paola Resende; Renata Serrano Lopes; Taina Venas |
| EPI_ISL_2443547, EPI_ISL_2443548, EPI_ISL_2443554, EPI_ISL_2443576, EPI_ISL_2603430, EPI_ISL_2603453, EPI_ISL_2603456, EPI_ISL_2603462, EPI_ISL_2603463, EPI_ISL_2603464, EPI_ISL_2603470, EPI_ISL_2863865, EPI_ISL_2863866, EPI_ISL_2863868, EPI_ISL_2863869, EPI_ISL_2863870, EPI_ISL_2863871, EPI_ISL_2863872, EPI_ISL_2863873, EPI_ISL_2863874, EPI_ISL_2863875, EPI_ISL_2863876, EPI_ISL_2863877, EPI_ISL_2863878, EPI_ISL_2863879, EPI_ISL_2863880, EPI_ISL_2863881, EPI_ISL_2863882, EPI_ISL_2863883, EPI_ISL_2863884, EPI_ISL_2863885, EPI_ISL_2863886, EPI_ISL_2863893 | | | |
| see above | Lboratorio Central de Saude Publica do Estado do Parana (LACEN/PR) | Laboratory of Respiratory Viruses and Measles, Oswaldo Cruz Institute, FIOCRUZ | Agatha Cristinne Prudencio Soares; Alice Sampaio Rocha; Ana Carolina Mendonca; Anna Carolina Paixao; Elisa Cavalcante Pereira; Fernando Motta; Ighor Leonardo Arantes Gomes; Irina Riediger; Luciana Appolinario; Marilda Siqueira on behalf of the Fiocruz COVID-19 Genomic Surveillance Network; Paola Resende; Renata Serrano Lopes; Taina Venas |
| EPI_ISL_2692138, EPI_ISL_2692139, EPI_ISL_2692140 | Lifescan | RIPHL at Rush University Medical Center | Chicago Department of Public Health; Felix Araujo Perez; Kevin Kunstman; Laura Furtado; Marieta Hyde; Max Kolton; Stefan Green |
| EPI_ISL_1912739, EPI_ISL_2122825 | Lighthouse Lab in Glasgow | Wellcome Sanger Institute for the COVID-19 Genomics UK (COG-UK) Consortium | Anna Dominiczak and Alex Alderton; Carol Clugston; Cordelia Langford; David Gray; David K. Jackson; Dominic Kwiatkowski; Ewan Harrison; Harper VanSteenhouse; Ian Johnston; Jeffrey Barrett; John Sillitoe on behalf of the Wellcome Sanger Institute COVID-19 Surveillance Team; Roberto Amato; Sonia Goncalves; Yumi Kasai |
| EPI_ISL_1829361, EPI_ISL_1921451, EPI_ISL_1921551, EPI_ISL_1921677, EPI_ISL_2021041, EPI_ISL_2021062, EPI_ISL_2021081, EPI_ISL_2021666, EPI_ISL_2021696, EPI_ISL_2022037 | | | |
| see above | Lighthouse Lab in Milton Keynes | Wellcome Sanger Institute for the COVID-19 Genomics UK (COG-UK) | Cordelia Langford; David K. Jackson; Dominic Kwiatkowski; Ewan Harrison; Ian Johnston; Jeffrey Barrett; John Sillitoe on behalf of the Wellcome Sanger Institute COVID-19 Surveillance Team; Roberto Amato; Sonia Goncalves; The Lighthouse Lab in Milton Keynes and Alex Alderton |

| | | | |
|---|---|---|---|
| | | Consortium | |
| EPI_ISL_3086531, EPI_ISL_3086532 | Louisiana Office of Public Heath Laboratory | LSUHS Emerging Viral Threat Laboratory | Alexander Mijalis; Andrew D. Yurochko; Christopher G. Kevil; Gregory L. Ware; Jennifer L. Carroll; Jeremy P. Kamil; John A. Vanchiere; Maarten Van Diest; Rona S. Scott |
| EPI_ISL_2373069, EPI_ISL_2373076, EPI_ISL_2373079, EPI_ISL_2373081 | Lurie Children's Hospital of Chicago | Northwestern University - Ozer Lab | Egon A. Ozer; Judd F. Hultquist; Lacy M. Simons; Larry K. Kociolek; Michael G. Ison; Ramon Lorenzo-Redondo; Taylor J. Dean; William J. Muller; Xiaotian; Zheng |
| EPI_ISL_2249576 | M Health Fairview | Minnesota Department of Health, Public Health Laboratory | Alexandra Lorentz; Jacob Garfin; Matt Plumb; and Xiong Wang |
| EPI_ISL_2495419, EPI_ISL_2495425, EPI_ISL_2495426, EPI_ISL_2495427, EPI_ISL_2495610 | MB-Cadham Provincial laboratory | National Microbiology Laboratory (NML) | Anna Majer; Anneliese Landgraff; CanCOGeN's metadata curation team; Darian Hole; David Alexander; Elsie Grudeski; Gary Van Domselaar; Grace Seo; Jared Bullard; Jennifer Tanner; Kerry Dust; Kirsten Biggar; Madison Chapel; Morag Graham; Natalie Knox; Nathalie Bastien; Paul Van Caeseele; Philip Mabon; Public Health Agency of Canada CanCOGeN team; Rhiannon Huzarewich; Russell Mandes; Shari Tyson; Timothy Booth; Yan Li |
| EPI_ISL_2227429 | MD PHL | MD PHL | Maryland Department of Health Laboratories Administration |
| EPI_ISL_2036426, EPI_ISL_2036450, EPI_ISL_2286043, EPI_ISL_2286217, EPI_ISL_2286285, EPI_ISL_2286779, EPI_ISL_2286840, EPI_ISL_2288732, EPI_ISL_2288736, EPI_ISL_2288737, EPI_ISL_2288738, EPI_ISL_2288739, EPI_ISL_2288740, EPI_ISL_2454243, EPI_ISL_2843621 | | | |
| see above | MEPHI, Aix Marseille University | MEPHI, Aix Marseille University | Anthony LEVASSEUR |
| EPI_ISL_3354883 | MO State Public Health Laboratory | Centers for Disease Control and Prevention Division of Viral Diseases, Pathogen Discovery | Alex Burgin; Ben Rambo-Martin; Clinton Paden; Dakota Howard; Dave Wentworth; Dhwani Batra; Jasmine Padilla; Justin Lee; Krista Queen; Kristen Knipe; Kristine Lacek; Mark Burroughs; Matthew Schmerer; Meghan Bentz; Mili Sheth; Peter Cook; Sam Shepard; Sarah Nobles; Suxiang Tong; Vivien Dugan; Yvette Unoarumhi |
| EPI_ISL_2274608 | MONTEFIORE MEDICAL CENTER LABORATORIES | Wadsworth Center, New York State Department of Health | Alexis Russell; Catharine Prussing; Daryl M. Lamson; Erasmus Schneider; Erica Lasek-Nesselquist; John Kelly; Jonathan Plitnick; Kirsten St. George; Matthew Shudt; Melissa A Leisner; Navjot Singh |
| EPI_ISL_6492575, EPI_ISL_6492586, EPI_ISL_6492608, EPI_ISL_6492616, EPI_ISL_6492635 | MSHS Clinical Microbiology Laboratories | MSHS Pathogen Surveillance Program | Adolfo García-Sastre; Adriana van de Guchte; Ajay Obla; Alberto Paniz-Mondolfi; Ana S. Gonzalez-Reiche; Angela Amoako; Ashley Salimbangon; Betsaida Salom Melo; Bremy Alburquerque; Brianne Ciferri; Charles Gleason; Daniel Floda; Deena R. Altman; Denise Jurczyszak; Emilia Mia Sordillo; Gintaras Deikus; Giulio Kleiner; Gopi Patel; Hala Alshammary; Harm van Bakel; Irina Oussenko; Jayeeta Dutta; Juan Soto; Julia Matthews; Katherine Beach; Kathryn Twyman; Kayla Russo; Komal Srivastava; Levy Sominsky; Mahmoud Awawda; Marta Luksza; Matthew M. Hernandez; Melissa Gitman; Michael D. Nowak; Mitchell J. Sullivan; Nancy Francoeur; Robert Sebra; Sarah Schaefer; Shelcie Fabre; Shwetha Hara Sridhar; Viviana Simon; Ying-Chih Wang; Zenab Khan |
| EPI_ISL_2105154 | MT Public Health Laboratory | Centers for Disease Control and Prevention Division of Viral Diseases, Pathogen Discovery | Alison Laufer Halpin; Ben L. Rambo-Martin; Clinton R. Paden; Dakota Howard; Darlene Wagner; Dave Wentworth; Dhwani Batra; Jasmine Padilla; Justin Lee; Katie Dillon; Krista Queen; Kristen Knipe; Kristine Lacek; Mark Burroughs; Matthew Schmerer; Mili Sheth; Peter Cook; Sam Shepard; Sarah Nobles; Shoshona Le; Suxiang Tong; Vivien Dugan; Yvette Unoarumhi |
| EPI_ISL_2111276 | MVZ Labor Dr. Fenner und Kollegen (Standort Hamburg) | Robert Koch Institute | |
| EPI_ISL_2124016, EPI_ISL_2124020, EPI_ISL_2259757 | MVZ Labor Dr. Limbach & Kollegen GbR | Robert Koch Institute | |
| EPI_ISL_2802394, EPI_ISL_2802396, EPI_ISL_2802398, EPI_ISL_2802401, EPI_ISL_2802402, EPI_ISL_4083794, EPI_ISL_4083798, EPI_ISL_4083810 | | | |
| see above | Maine Health and Environmental Testing Laboratory | Tewhey Lab, The Jackson Laboratory | Barter, M.; Dewey, H.; H. and Tewhey, R.; Iosue, F.; Lynch, R.; Matluk, N.; Munger |
| EPI_ISL_3011907, EPI_ISL_3011921, EPI_ISL_3011940 | Maryland Genomics, Institute for Genome Sciences, University of Maryland School of Medicine | Maryland Genomics, Institute for Genome Sciences, University of Maryland School of Medicine | Claire M; Fraser; Hazen; Holly; Humphrys; Ivette; Jacques; Jonathan; Kranthi; Lim; Lisa D; Luke J; Mike; Ott; Ravel; Roussey; Sadzewicz; Sandra; Santana-Cruz; Tallon; Tracy; Vavikolanu |
| EPI_ISL_2016481, EPI_ISL_2016483, EPI_ISL_2362372 | Massachusetts General Hospital | Infectious Disease Program, Broad Institute of Harvard and MIT | A.E.; Adams, G.; Anahtar, M.; B.L.; B.W.; Bauer, M.; Birren; Branda, J.; Carter, A.; Cerrato, F.; Chaluvadi, S.; Chapman; Cusick, C.; D.J.; DeRuff, K.; E. and Sabeti; Flowers, K.; Gallagher, G.; Gladden-Young, A.; Gnirke, A.; Harris, J.; J.E.; K.J.; LaRocque, R.; Lagerborg, K.; Lemieux; Lin; Loreth, C.; MacInnis; Neumann, A.; Normandin, E.; P.C.; Park; Pierce, V.; Reilly, S.; Rosenberg; Rudy, M.; Ryan, E.; S.B.; Shaw, B.; Siddle; Slater, D.; Smole, S.; Tomkins-Tinch, C.; Turbett, S. |
| EPI_ISL_2008173, EPI_ISL_2008217, EPI_ISL_2008348, EPI_ISL_2229485, EPI_ISL_2229492, EPI_ISL_2229493, EPI_ISL_2229521, EPI_ISL_2229523, EPI_ISL_2229532, EPI_ISL_2229777, EPI_ISL_2400260 | | | |
| see above | Massachusetts State Public Health Laboratory | Massachusetts State Public Health Laboratory | Andrew Lang; Glen Gallagher; Sandra Smole; Timelia Fink |
| EPI_ISL_2095177, EPI_ISL_2095178 | Max von Pettenkofer Institute, Virology, National Reference Center for Retroviruses, LMU Munich | Laboratory for Functional Genome Analysis; Dept. Genomics; Gene Center of the LMU Munich | Alexander Graf; Helmut Blum; Max Muenchhoff; Oliver Keppler; Stefan Krebs |
| EPI_ISL_2136412 | Mayo Clinic & Mayo Clinic Laboratories | Minnesota Department of Health, Public Health Laboratory | Alexandra Lorentz; Jacob Garfin; Matt Plumb; and Xiong Wang |
| EPI_ISL_2136865, EPI_ISL_2136900, EPI_ISL_2136913, EPI_ISL_2136924, EPI_ISL_2136935, EPI_ISL_2136947 | Microbiologia CATLAB | Can Ruti SARS-CoV-2 Sequencing Hub (HUGTiP/IrsiCaixa/IGTP) | Alba Sánchez; Anna Not; Antoni E Bordoy; Bonaventura Clotet; Cristina Casañ; Cristina Esteban; Francesc Catala-Moll; Gemma Clara; Ignacio Blanco; Marc Noguera-Julian; Maria Casadellà; Mariona Parera; Mercedes Guerrero; Montserrat Giménez; Pere-Joan Cardona; Pilar Armengol; Roger Paredes; Verónica Saludes; and Elisa Martró on behalf of the Can Ruti SARS-CoV-2 Sequencing Hub. |
| EPI_ISL_2380035, EPI_ISL_2380046 | Microbiology Department, Laboratori Clínic Metropolitana Nord. Hospital Universitari Germans Trias i Pujol. | Can Ruti SARS-CoV-2 Sequencing Hub (HUGTiP/IrsiCaixa/IGTP) | Anna Not; Antoni E Bordoy; Bonaventura Clotet; Cristina Casañ; Cristina Esteban; Francesc Catala-Moll; Gemma Clara; Ignacio Blanco; Marc Noguera-Julian; Maria Casadellà; Mercedes Guerrero; Montserrat Giménez; Pere-Joan Cardona; Pilar Armengol; Roger Pared |
| EPI_ISL_2086157, EPI_ISL_2086158, EPI_ISL_2086159, EPI_ISL_2967793, EPI_ISL_2967849 | Microbiology Department. Complexo Hospitalario Universitario de Vigo | Microbiology Department. Complexo Hospitalario Universitario de Vigo | Alfaya N; Alonso I; Alvarez M; Cabrera JJ; Carballo R; Cores O; Cortizo S; Davina C; Martinez L; Mediero G; Perez S; Potel C; Regueiro B; Rey S; Vasallo FJ; del-Campo V |
| EPI_ISL_2029078, EPI_ISL_2029097, EPI_ISL_2029098, EPI_ISL_2029099, EPI_ISL_2029100, EPI_ISL_2029102, EPI_ISL_2029103, EPI_ISL_2259124, EPI_ISL_2259129, EPI_ISL_2259143, EPI_ISL_2259150, EPI_ISL_2259156, EPI_ISL_2259189 | | | |
| see above | Microvida | Microvida | Jaco Verweij; Joep Stohr; Joep Stöhr; Suzan D. Pas |
| EPI_ISL_2003439 | Minnesota Department of Health, Public Health Laboratory | Minnesota Department of Health, Public Health Laboratory | Alexandra Lorentz; Jacob Garfin; Matt Plumb; and Xiong Wang |
| EPI_ISL_2035571, EPI_ISL_2158668, EPI_ISL_2158678, EPI_ISL_2158682, EPI_ISL_2158684 | Missouri State Public Health Laboratory | Missouri State Public Health Laboratory | Ashley New; Joshua Barry; Matthew Sinn |
| EPI_ISL_3142933 | Molecular & Genomic Pathology Laboratory, Thomas Jefferson University Hospital | Molecular & Genomic Pathology Laboratory, Thomas Jefferson University Hospital | Nitika Badjatia; Run Jin; Shannon Mahoney; Zi-xuan Wang |
| EPI_ISL_5589636, EPI_ISL_5589735, EPI_ISL_5599060, EPI_ISL_5599064, EPI_ISL_5599067, EPI_ISL_5599198, EPI_ISL_5599267, EPI_ISL_5599270, EPI_ISL_5599274 | | | |
| see above | Molecular Biology Laboratory, Vidant Medical Center | Brody Integrative Genomics Core, East Carolina University | Changhong Yin; Heather Duncan; John T. Fallon; Kimberly P. Briley; Weihua Huang |
| EPI_ISL_2105942, EPI_ISL_2105943, EPI_ISL_2105944, EPI_ISL_2105945, EPI_ISL_2105946, EPI_ISL_2105947, EPI_ISL_2105948, EPI_ISL_2105949, EPI_ISL_2105950, EPI_ISL_2105951, EPI_ISL_2105952, EPI_ISL_2105953, EPI_ISL_2105954, EPI_ISL_2105956, EPI_ISL_2105957 | | | |
| see above | Molecular Diagnostics Pathology Department Mater Dei Hospital Malta | Molecular Diagnostics Pathology Department Mater Dei Hospital Malta | C Cilia; G Zahra; L Grech; R Borg |
| EPI_ISL_2834617 | Montana DPHHS Public Health Laboratory | Center for Global Health, University of New Mexico Health Sciences Center | Carrie Biskupiak; Darrell Dinwiddie; Daryl Domman; Deborah Gibson; Joy Ritter; Kurt Schwalm; Valerie Morley |
| EPI_ISL_2023375, EPI_ISL_2941729, EPI_ISL_2941746 | Montana Public Health Laboratory | Montana Public Health Laboratory | Carrie Biskupiak; Deborah Gibson; Joy Ritter; Michael Dills; Michelle Mozer |
| EPI_ISL_2308295 | Monterey County Public Health | Monterey County Public Health | Monterey County Public Health Laboratory |

| | | | |
|---|---|---|---|
| | Laboratory | Laboratory | |
| EPI_ISL_4025759 | NB-Hôpital Georges L. Dumont | New Brunswick - Vitalité Health Network | Allain E.; Chacko S.; Crapoulet N.; Desnoyers G.; Garceau R.; Lacroix J.; Shaw W. |
| EPI_ISL_2105180 | NC State Laboratory of Public Health | Centers for Disease Control and Prevention Division of Viral Diseases, Pathogen Discovery | Alison Laufer Halpin; Ben L. Rambo-Martin; Clinton R. Paden; Dakota Howard; Darlene Wagner; Dave Wentworth; Dhwani Batra; Jasmine Padilla; Justin Lee; Katie Dillon; Krista Queen; Kristen Knipe; Kristine Lacek; Mark Burroughs; Matthew Schmerer; Mili Sheth; Peter Cook; Sam Shepard; Sarah Nobles; Shoshona Le; Suxiang Tong; Vivien Dugan; Yvette Unoarumhi |
| EPI_ISL_2185587, EPI_ISL_2281856, EPI_ISL_3023749 | NJDOH, Public Health and Environmental Laboratories | NJ_PHEL | Allison Roder; Byeong Jeong; Chelsea San Filippo; Dana Woell; Jacquelyn Deverell; Lindsey Bodnar; Matthew Scarnati; Ryan Pachucki; Shiv K. Verma |
| EPI_ISL_2484584, EPI_ISL_2484587, EPI_ISL_2484588, EPI_ISL_2484591, EPI_ISL_2484618, EPI_ISL_2484619 | NL-Dr. Leonard A. Miller Centre for Health Services | National Microbiology Laboratory (NML) | Adel Malek; Anna Majer; Anneliese Landgraff; CanCOGeN's metadata curation team; Darian Hole; Elsie Grudeski; Gary Van Domselaar; George Zahariadis; Grace Seo; Jennifer Tanner; Kerri Smith; Kirsten Biggar; Laura Gilbert; Madison Chapel; Morag Graham; Natalie Knox; Nathalie Bastien; Philip Mabon; Public Health Agency of Canada CanCOGeN team; Rhiannon Huzarewich; Robert Needle; Russell Mandes; Shari Tyson; Timothy Booth; Yan Li; Yang Yu |
| EPI_ISL_4061014 | NL-Dr. Leonard A. Miller Centre for Health Services | Newfoundland and Labrador - Eastern Health | Adel Malek; Anna Majer; Anneliese Landgraff; Elsie Grudeski; Gary Van Domselaar; George Zahariad; Grace Seo; Jennifer Tanner; Laura Gilbert; Morag Graham; Natalie Knox; Philip Mabon; Rhiannon Huzarewich; Robert Needle; Russell Mandes; Shari Tyson; Yang Yu |
| EPI_ISL_2097278 | NORTHWELL HEALTH LABORATORIES | Wadsworth Center, New York State Department of Health | Alexis Russell; Catharine Prussing; Daryl M. Lamson; Erasmus Schneider; Erica Lasek-Nesselquist; John Kelly; Jonathan Plitnick; Kirsten St. George; Matthew Shudt; Melissa A Leisner; Navjot Singh |
| EPI_ISL_2345165, EPI_ISL_2345174, EPI_ISL_2345176, EPI_ISL_2345177 | NUCLEO DE ESPECIALIDADES EM SAUDE | Instituto Butantan / ESALQ-Piracicaba | Antonio Jorge Martins; Claudia Renata dos Santos Barros; David Schlesinger; Debora Botequio Moretti; Dimas Tadeu Covas; Elaine Cristina Marqueze; Elaine Vieira Santos; Evandra Strazza Rodrigues; Heidge Fukumasu; Jayme Augusto de Souza-Neto; José Salvatore Leister Patané; Luiz Alcantara; Luiz Lehmann Coutinho; Maria Carolina Elias; Maurício Lacerda Nogueira; Rafael dos Santos Bezerra; Raul Machado Neto; Rejane Maria Tommasini Grotto; Ricardo Haddad; Sandra Coccuzzo Sampaio Vessoni; Simone Kashima; Svetoslav Nanev Slavov; Vincent Louis Viala |
| EPI_ISL_2345205, EPI_ISL_2345207, EPI_ISL_2345208, EPI_ISL_2345215, EPI_ISL_2345216, EPI_ISL_2345217, EPI_ISL_2345218, EPI_ISL_2345226, EPI_ISL_2345242, EPI_ISL_2345245, EPI_ISL_2345261, EPI_ISL_2345263, EPI_ISL_2345276, EPI_ISL_2345280, EPI_ISL_2345282, EPI_ISL_2345283 | | | |
| see above | NUCLEO DE SAUDE VILA FALCAO DE BAURU | Instituto Butantan / ESALQ-Piracicaba | Antonio Jorge Martins; Claudia Renata dos Santos Barros; David Schlesinger; Debora Botequio Moretti; Dimas Tadeu Covas; Elaine Cristina Marqueze; Elaine Vieira Santos; Evandra Strazza Rodrigues; Heidge Fukumasu; Jayme Augusto de Souza-Neto; José Salvatore Leister Patané; Luiz Alcantara; Luiz Lehmann Coutinho; Maria Carolina Elias; Maurício Lacerda Nogueira; Rafael dos Santos Bezerra; Raul Machado Neto; Rejane Maria Tommasini Grotto; Ricardo Haddad; Sandra Coccuzzo Sampaio Vessoni; Simone Kashima; Svetoslav Nanev Slavov; Vincent Louis Viala |
| EPI_ISL_2345084, EPI_ISL_2345091, EPI_ISL_2345101, EPI_ISL_2345112, EPI_ISL_2345122, EPI_ISL_2345132, EPI_ISL_2345144, EPI_ISL_2345145, EPI_ISL_2345146, EPI_ISL_2345147, EPI_ISL_2345148, EPI_ISL_2345151, EPI_ISL_2345164, EPI_ISL_2345180, EPI_ISL_2345181, EPI_ISL_2345182 | | | |
| see above | NUCLEO SAUDE III N HABITACIONAL PRESIDENTE GEISEL BAURU | Instituto Butantan / ESALQ-Piracicaba | Antonio Jorge Martins; Claudia Renata dos Santos Barros; David Schlesinger; Debora Botequio Moretti; Dimas Tadeu Covas; Elaine Cristina Marqueze; Elaine Vieira Santos; Evandra Strazza Rodrigues; Heidge Fukumasu; Jayme Augusto de Souza-Neto; José Salvatore Leister Patané; Luiz Alcantara; Luiz Lehmann Coutinho; Maria Carolina Elias; Maurício Lacerda Nogueira; Rafael dos Santos Bezerra; Raul Machado Neto; Rejane Maria Tommasini Grotto; Ricardo Haddad; Sandra Coccuzzo Sampaio Vessoni; Simone Kashima; Svetoslav Nanev Slavov; Vincent Louis Viala |
| EPI_ISL_2161703, EPI_ISL_2161708, EPI_ISL_2161714, EPI_ISL_2161718, EPI_ISL_2161775 | NYU Langone Health | Departments of Pathology and Medicine, New York University School of Medicine | Adriana Heguy; Christian Marier; Dacia Dimartino; Emily Guzman; Gael Westby; Guiqing Wang; Paolo Cotzia; Paul Zappile; Peter Meyn; Sitharam Ramaswami; Yutong Zhang |
| EPI_ISL_2621315 | National Center of Infectious and Parasitic Diseases | National Center of Infectious and Parasitic Diseases | Alexiev et al |
| EPI_ISL_4602047 | National Influenza Centre | National Influenza Centre | ; Benjamiin B. Lindsey; Benjamin H. Foulkes; Dennis Laryea; Ernest Asiedu; Franklin Asiedu-Bekoe; Gordon Awandare; Ivy A. Asante; Joseph Oliver-Commey; Joyce Ngoi; Linda Boatemaa; Lorreta Kwasah; Mathew D. Parker; Michael Marks; Mildred Adusei-Poku; Sharon Hsu; Thushan I de Silva; William K. Ampofo |
| EPI_ISL_2562083, EPI_ISL_2562084 | National Institute of Public Health | State Veterinary Institute Prague | A; D; H; J; Jirincova; Nagy; Suri; T; Trnka; Vecerova |
| EPI_ISL_1971476, EPI_ISL_1971480, EPI_ISL_1971484, EPI_ISL_1971500, EPI_ISL_1971501, EPI_ISL_1971502, EPI_ISL_1971506, EPI_ISL_2131812, EPI_ISL_2131813, EPI_ISL_2131814, EPI_ISL_2131822, EPI_ISL_2136196, EPI_ISL_2136198, EPI_ISL_2136203, EPI_ISL_2136204, EPI_ISL_2136207, EPI_ISL_2136209, EPI_ISL_2179660, EPI_ISL_2271915, EPI_ISL_2271935, EPI_ISL_2271951 | | | |
| see above | National Platform bis UMONS/Jolimont | National Platform bis UMONS/Jolimont | Eric Tarantino; Florian Juszczak; François Dufrasne; Gautier Detry; Guillaume Bayon-Vicente; Ruddy Wattiez |
| EPI_ISL_2349709 | National Public Health Laboratory, National Centre for Infectious Diseases | National Public Health Laboratory, National Centre for Infectious Diseases | Grace Jie Yin Ngan; Lin Cui; Raymond Tzer Pin Lin; Royce Ang; Tze Minn Mak; Zhenyang Zhou |
| EPI_ISL_2023408, EPI_ISL_2023409, EPI_ISL_2689668 | Nebraska Public Health Laboratory | NPHL COVID-19 Response Team | NPHL COVID-19 Response Team |
| EPI_ISL_4877713, EPI_ISL_4877953 | Nevada State Public Health Laboratory | Nevada State Public Health Laboratory | Andrew Gorzalski; Mark Pandori |
| EPI_ISL_2322768, EPI_ISL_2322775, EPI_ISL_2322789, EPI_ISL_2322792, EPI_ISL_2322795 | New Mexico Department of Health Scientific Laboratory | New Mexico Department of Health Scientific Laboratory | Anastacia Griego-Fisher; D'eldra Malone; Ellie Johnson; Jennifer Benoit |
| EPI_ISL_2023285, EPI_ISL_2023286, EPI_ISL_2023287, EPI_ISL_2023288, EPI_ISL_2023289, EPI_ISL_2023290 | North Carolina State Laboratory of Public Health (NCSLPH) | North Carolina State Laboratory of Public Health (NCSLPH) | Chase K; Glover W; Greene S; Miller MC |
| EPI_ISL_2151476, EPI_ISL_2151478, EPI_ISL_2151479, EPI_ISL_3062288, EPI_ISL_3062290 | North Dakota Department of Health, Public Health Laboratory | North Dakota Department of Health, Public Health Laboratory | Lisa Wingerter |
| EPI_ISL_2136636, EPI_ISL_2136638, EPI_ISL_2490096, EPI_ISL_2490097, EPI_ISL_2490101, EPI_ISL_2490102, EPI_ISL_2490103, EPI_ISL_2490106, EPI_ISL_2490107, EPI_ISL_2490111, EPI_ISL_2490119, EPI_ISL_2490122, EPI_ISL_2490123, EPI_ISL_2490126, EPI_ISL_2490130, EPI_ISL_2490132, EPI_ISL_2490136, EPI_ISL_2490138, EPI_ISL_2490150, EPI_ISL_2490154, EPI_ISL_2490159, EPI_ISL_2490173, EPI_ISL_2490176, EPI_ISL_2490183, EPI_ISL_2490189, EPI_ISL_2490223 | | | |
| see above | Northwest Laboratory | Seattle Flu Study | Amanda Adler; Barry R. Lutz; Benjamin Pelle; Brian Hiatt; Caitlin R. Wolf; Chris D. Frazar; Deborah A. Nickerson; Elisabeth Brandstetter; Erica Ryke; Geoff Melly; Helen Y. Chu; Janet A. Englund; Jay Shendure; Jover Lee; Kairsten Fay; Kirsten Lacombe; Lea M. Starita; Mark J. Rieder; Matthew Richardson; Matthew Thompson; Melissa Truong; Michael Boeckh; Michael Famulare; Misja Ilcisin; Peter D. Han; Philip Dykema; Romesh Gautom; Scott Lindquist; Thomas R. Sibley; Trevor Bedford |
| EPI_ISL_2009347, EPI_ISL_2372954, EPI_ISL_2372956 | Northwestern Memorial Hospital | Northwestern University - Ozer Lab | Chad J. Achenbach; Chao Qi; Egon A. Ozer; Judd F. Hultquist; Lacy M. Simons; Lawrence J. Jennings; Michael G. Ison; Ramon Lorenzo-Redondo; Taylor J. Dean |
| EPI_ISL_3388716, EPI_ISL_3388718 | Northwestern Memorial Hospital | RIPHL at Rush University Medical Center | Amber Kimble; Chao Qi; Edith Perez; Felix Araujo Perez; Kevin Kunstman; Laura Furtado; Marieta Hyde; Max Kolton; Stefan Green |
| EPI_ISL_2346268, EPI_ISL_2400517 | Notre Dame COVID Response and Testing Laboratory | Notre Dame Genomics & Bioinformatics Core Facility | Joseph Sarro; Melissa Stephens; Michael Pfrender |
| EPI_ISL_5801233, EPI_ISL_5801250, EPI_ISL_5801254, EPI_ISL_5801256 | Nucleo De Especialidades Em Saude | Instituto Butantan | Antonio Jorge Martins; Claudia Renata dos Santos Barros; David Schlesinger; Debora Botequio Moretti; Dimas Tadeu Covas; Elaine Cristina Marqueze; Elaine Vieira Santos; Evandra Strazza Rodrigues; Heidge Fukumasu; Jayme Augusto de Souza-Neto; José Salvatore Leister Patané; Luiz Alcantara; Luiz Lehmann Coutinho; Maria Carolina Elias; Maurício Lacerda Nogueira; Rafael dos Santos Bezerra; Raul Machado Neto; Rejane Maria Tommasini Grotto; Ricardo Haddad; Sandra Coccuzzo Sampaio Vessoni; Simone Kashima; Svetoslav Nanev Slavov; Vincent Louis Viala |
| EPI_ISL_5801298, EPI_ISL_5801300, EPI_ISL_5801301, EPI_ISL_5801312, EPI_ISL_5801317, EPI_ISL_5801318, EPI_ISL_5801319, EPI_ISL_5801327, EPI_ISL_5801341, EPI_ISL_5801349, EPI_ISL_5801353, EPI_ISL_5801371, EPI_ISL_5801373, EPI_ISL_5801387, EPI_ISL_5801391, EPI_ISL_5801393, EPI_ISL_5801394 | | | |
| see above | Nucleo De Saude Vila Falcao De Bauru | Instituto Butantan | Antonio Jorge Martins; Claudia Renata dos Santos Barros; David Schlesinger; Debora Botequio Moretti; Dimas Tadeu Covas; Elaine Cristina Marqueze; Elaine Vieira Santos; Evandra Strazza Rodrigues; Heidge Fukumasu; Jayme Augusto de Souza-Neto; José Salvatore Leister Patané; Luiz Alcantara; Luiz Lehmann Coutinho; Maria Carolina Elias; Maurício Lacerda Nogueira; Rafael dos Santos Bezerra; Raul Machado Neto; Rejane Maria Tommasini Grotto; Ricardo Haddad; Sandra Coccuzzo Sampaio Vessoni; Simone Kashima; Svetoslav Nanev Slavov; Vincent Louis Viala |
| EPI_ISL_5801107, EPI_ISL_5801114, EPI_ISL_5801131, EPI_ISL_5801145, EPI_ISL_5801165, EPI_ISL_5801176, EPI_ISL_5801192, EPI_ISL_5801193, EPI_ISL_5801194, EPI_ISL_5801195, EPI_ISL_5801196, EPI_ISL_5801202, EPI_ISL_5801231, EPI_ISL_5801263, EPI_ISL_5801264, EPI_ISL_5801265 | | | |
| see above | Nucleo Saude Iii N Habitacional Presidente Geisel Bauru | Instituto Butantan | Antonio Jorge Martins; Claudia Renata dos Santos Barros; David Schlesinger; Debora Botequio Moretti; Dimas Tadeu Covas; Elaine Cristina Marqueze; Elaine Vieira Santos; Evandra Strazza Rodrigues; Heidge Fukumasu; Jayme Augusto de Souza-Neto; José Salvatore Leister Patané; Luiz Alcantara; Luiz Lehmann Coutinho; Maria Carolina Elias; Maurício Lacerda Nogueira; Rafael dos Santos Bezerra; Raul Machado Neto; Rejane Maria Tommasini Grotto; Ricardo Haddad; Sandra Coccuzzo Sampaio Vessoni; Simone Kashima; Svetoslav Nanev Slavov; Vincent Louis Viala |
| EPI_ISL_2378934, EPI_ISL_2378942, EPI_ISL_2378945, EPI_ISL_2462125, EPI_ISL_2462135, EPI_ISL_2462136, EPI_ISL_2462137, EPI_ISL_2462138, EPI_ISL_2462140, EPI_ISL_2462144, EPI_ISL_2462147, EPI_ISL_2462153, EPI_ISL_2462164, EPI_ISL_2462173, EPI_ISL_2462174, EPI_ISL_2462178, EPI_ISL_2462179, EPI_ISL_2462182, EPI_ISL_2462184, EPI_ISL_2462185, EPI_ISL_2462187, EPI_ISL_2462198, EPI_ISL_2462202, EPI_ISL_2462221, EPI_ISL_2462263, EPI_ISL_2462270, EPI_ISL_2462272, EPI_ISL_2462275, EPI_ISL_2462276, EPI_ISL_2462280, EPI_ISL_2462281, EPI_ISL_2462285, EPI_ISL_2462288, EPI_ISL_2462294, EPI_ISL_2462297, EPI_ISL_2462302, EPI_ISL_2462304, EPI_ISL_2462305, EPI_ISL_2462308, EPI_ISL_2663623, EPI_ISL_2663628 | | | |
| see above | OHSU Lab Services Molecular Microbiology Lab | Oregon SARS-CoV-2 Genome Sequencing Center | Alec J. Hirsch; Andrew C. Adey; Benjamin N. Bimber; Brendan L. O'Connell; Brian J. O'Roak; Cierra LaBlanc; Daniel N. Streblow; Donna Hansel; Guang Fan; Kayla Carter; Ruth V. Nichols; Sally Grindstaff; Sonia Acharya; William B. Messer; Xuan Qin |
| EPI_ISL_3354890, | OR State PHL- | Centers for Disease Control and | Alex Burgin; Ben Rambo-Martin; Clinton Paden; Dakota Howard; Dave Wentworth; Dhwani Batra; Jasmine Padilla; Justin Lee; Krista Queen; Kristen Knipe; Kristine Lacek; Mark Burroughs; Matthew Schmerer; Meghan Bentz; Mili Sheth; Peter Cook; Sam Shepard; Sarah Nobles; Suxiang Tong; Vivien Dugan; |

| | | | |
|---|---|---|---|
| EPI_ISL_3354936 | Virology/Immunology Section | Prevention Division of Viral Diseases, Pathogen Discovery | Yvette Unoarumhi |
| EPI_ISL_2858702, EPI_ISL_2858703 | Ohio Department of Health Laboratory | Ohio Department of Health Laboratory | Allison Black; Brent Lee; Caitlin McDonnell; Eric Brandt; Erica Leasure; Glen McGillivary; Heather Blankenship; Holmes; Jade Mowery; Jennifer; Kelsey Florek; Keoni Omura; Kirtana Ramadugu; Quanta Brown; Stephanie Mccracken; Tyler Payne; and Tammy Bannerman |
| EPI_ISL_2566361 | Operation Infectious Diseases, Naval Health Research Center | Operation Infectious Diseases, Naval Health Research Center | A.E.; Balansay-Ames; C.A.; Fierro, M.; Iniguez-Stevens, E.; Kriner, P.; M.S.; Moser, K.; Myers; Pan; Phippard; R.S.; R.W.; Underwood |
| EPI_ISL_2023470, EPI_ISL_2081513, EPI_ISL_2086696, EPI_ISL_2138892, EPI_ISL_2139551, EPI_ISL_2139560, EPI_ISL_2139565, EPI_ISL_2203067, EPI_ISL_2424247, EPI_ISL_2557921 | | | |
| see above | Oregon State Public Health Laboratory | Oregon State Public Health Laboratory | Eugene Yeboah; John Fontana and Shane Sevey; Laura Tsaknaridis; Rafia Razzaque; Vanda Makris |
| EPI_ISL_2420465 | Ospedale San Filippo Neri | INMI Lazzaro Spallanzani IRCCS | A Di Caro; A Tamburro; B Bartolini; CEM Gruber; E Giombini; F Messina; F Santini; G Bonfiglio; M Meledandri; M Rueca; ML Schiavone; MR Capobianchi; O Butera |
| EPI_ISL_2450369 | Ospedale Sandro Pertini ASL Roma 2 | INMI Lazzaro Spallanzani IRCCS | A Di Caro; B Bartolini; CEM Gruber; E Giombini; F Messina; F Santini; G Bonfiglio; G Cappiello; M Rueca; MC Cava; MR Capobianchi; O Butera; R Longo |
| EPI_ISL_2293362, EPI_ISL_2293364, EPI_ISL_2293370, EPI_ISL_2293373, EPI_ISL_2293374, EPI_ISL_2293375, EPI_ISL_2293380 | | | |
| see above | Outre mer | National Reference Center for Viruses of Respiratory Infections, Institut Pasteur, Paris | Angela Brisebarre; Camille Capel; Christophe Malabat; Corinne Maufrais; Dominique Rousset; Etienne Simon-Lorière; Frédéric Lemoine; Louise Lefrançois; Marion Barbet; Maud Vanpeene; Méline Bizard; Pierre Lechat; Sylvie Behillil; Sylvie Van der Werf; Vincent Enouf |
| EPI_ISL_2209348, EPI_ISL_2345675 | PAS JOAO ANTONIO DO NASCIMENTO | Instituto Butantan / Mendelics | Antonio Jorge Martins; Claudia Renata dos Santos Barros; David Schlesinger; Debora Botequio Moretti; Dimas Tadeu Covas; Elaine Cristina Marqueze; Elaine Vieira Santos; Evandra Strazza Rodrigues; Heidge Fukumasu; Jayme Augusto de Souza-Neto; José Salvatore Leister Patané; Luiz Alcantara; Luiz Lehmann Coutinho; Maria Carolina Elias; Maurício Lacerda Nogueira; Rafael dos Santos Bezerra; Raul Machado Neto; Rejane Maria Tommasini Grotto; Ricardo Haddad; Sandra Coccuzzo Sampaio Vessoni; Simone Kashima; Svetoslav Nanev Slavov; Vincent Louis Viala |
| EPI_ISL_2208911 | PENITENCIARIA DE PRESIDENTE PRUDENTE | Instituto Butantan / Mendelics | Antonio Jorge Martins; Claudia Renata dos Santos Barros; David Schlesinger; Debora Botequio Moretti; Dimas Tadeu Covas; Elaine Cristina Marqueze; Elaine Vieira Santos; Evandra Strazza Rodrigues; Heidge Fukumasu; Jayme Augusto de Souza-Neto; José Salvatore Leister Patané; Luiz Alcantara; Luiz Lehmann Coutinho; Maria Carolina Elias; Maurício Lacerda Nogueira; Rafael dos Santos Bezerra; Raul Machado Neto; Rejane Maria Tommasini Grotto; Ricardo Haddad; Sandra Coccuzzo Sampaio Vessoni; Simone Kashima; Svetoslav Nanev Slavov; Vincent Louis Viala |
| EPI_ISL_2345809, EPI_ISL_2345814, EPI_ISL_2345859 | POLICLINICA COVID 19 ITAPETININGA | Instituto Butantan / Mendelics | Antonio Jorge Martins; Claudia Renata dos Santos Barros; David Schlesinger; Debora Botequio Moretti; Dimas Tadeu Covas; Elaine Cristina Marqueze; Elaine Vieira Santos; Evandra Strazza Rodrigues; Heidge Fukumasu; Jayme Augusto de Souza-Neto; José Salvatore Leister Patané; Luiz Alcantara; Luiz Lehmann Coutinho; Maria Carolina Elias; Maurício Lacerda Nogueira; Rafael dos Santos Bezerra; Raul Machado Neto; Rejane Maria Tommasini Grotto; Ricardo Haddad; Sandra Coccuzzo Sampaio Vessoni; Simone Kashima; Svetoslav Nanev Slavov; Vincent Louis Viala |
| EPI_ISL_2345265 | POLICLINICA HORTOLANDIA | Instituto Butantan / ESALQ-Piracicaba | Antonio Jorge Martins; Claudia Renata dos Santos Barros; David Schlesinger; Debora Botequio Moretti; Dimas Tadeu Covas; Elaine Cristina Marqueze; Elaine Vieira Santos; Evandra Strazza Rodrigues; Heidge Fukumasu; Jayme Augusto de Souza-Neto; José Salvatore Leister Patané; Luiz Alcantara; Luiz Lehmann Coutinho; Maria Carolina Elias; Maurício Lacerda Nogueira; Rafael dos Santos Bezerra; Raul Machado Neto; Rejane Maria Tommasini Grotto; Ricardo Haddad; Sandra Coccuzzo Sampaio Vessoni; Simone Kashima; Svetoslav Nanev Slavov; Vincent Louis Viala |
| EPI_ISL_3354889 | PR Public Health Lab | Centers for Disease Control and Prevention Division of Viral Diseases, Pathogen Discovery | Alex Burgin; Ben Rambo-Martin; Clinton Paden; Dakota Howard; Dave Wentworth; Dhwani Batra; Jasmine Padilla; Justin Lee; Krista Queen; Kristen Knipe; Kristine Lacek; Mark Burroughs; Matthew Schmerer; Meghan Bentz; Mili Sheth; Peter Cook; Sam Shepard; Sarah Nobles; Suxiang Tong; Vivien Dugan; Yvette Unoarumhi |
| EPI_ISL_2345183, EPI_ISL_2345184, EPI_ISL_2345185, EPI_ISL_2345199 | PRONTO SOCORRO MUNICIPAL DE SEVERINIA | Instituto Butantan / ESALQ-Piracicaba | Antonio Jorge Martins; Claudia Renata dos Santos Barros; David Schlesinger; Debora Botequio Moretti; Dimas Tadeu Covas; Elaine Cristina Marqueze; Elaine Vieira Santos; Evandra Strazza Rodrigues; Heidge Fukumasu; Jayme Augusto de Souza-Neto; José Salvatore Leister Patané; Luiz Alcantara; Luiz Lehmann Coutinho; Maria Carolina Elias; Maurício Lacerda Nogueira; Rafael dos Santos Bezerra; Raul Machado Neto; Rejane Maria Tommasini Grotto; Ricardo Haddad; Sandra Coccuzzo Sampaio Vessoni; Simone Kashima; Svetoslav Nanev Slavov; Vincent Louis Viala |
| EPI_ISL_2209716 | PSF DO PARQUE DOS PINHEIROS | Instituto Butantan / Mendelics | Antonio Jorge Martins; Claudia Renata dos Santos Barros; David Schlesinger; Debora Botequio Moretti; Dimas Tadeu Covas; Elaine Cristina Marqueze; Elaine Vieira Santos; Evandra Strazza Rodrigues; Heidge Fukumasu; Jayme Augusto de Souza-Neto; José Salvatore Leister Patané; Luiz Alcantara; Luiz Lehmann Coutinho; Maria Carolina Elias; Maurício Lacerda Nogueira; Rafael dos Santos Bezerra; Raul Machado Neto; Rejane Maria Tommasini Grotto; Ricardo Haddad; Sandra Coccuzzo Sampaio Vessoni; Simone Kashima; Svetoslav Nanev Slavov; Vincent Louis Viala |
| EPI_ISL_2345759 | PSF DR ANTONIO PIRES DE ALMEIDA PORTO FELIZ | Instituto Butantan / Mendelics | Antonio Jorge Martins; Claudia Renata dos Santos Barros; David Schlesinger; Debora Botequio Moretti; Dimas Tadeu Covas; Elaine Cristina Marqueze; Elaine Vieira Santos; Evandra Strazza Rodrigues; Heidge Fukumasu; Jayme Augusto de Souza-Neto; José Salvatore Leister Patané; Luiz Alcantara; Luiz Lehmann Coutinho; Maria Carolina Elias; Maurício Lacerda Nogueira; Rafael dos Santos Bezerra; Raul Machado Neto; Rejane Maria Tommasini Grotto; Ricardo Haddad; Sandra Coccuzzo Sampaio Vessoni; Simone Kashima; Svetoslav Nanev Slavov; Vincent Louis Viala |
| EPI_ISL_2345672, EPI_ISL_2345796 | PSF MARIA JOSE SALTO DE PIRAPORA | Instituto Butantan / Mendelics | Antonio Jorge Martins; Claudia Renata dos Santos Barros; David Schlesinger; Debora Botequio Moretti; Dimas Tadeu Covas; Elaine Cristina Marqueze; Elaine Vieira Santos; Evandra Strazza Rodrigues; Heidge Fukumasu; Jayme Augusto de Souza-Neto; José Salvatore Leister Patané; Luiz Alcantara; Luiz Lehmann Coutinho; Maria Carolina Elias; Maurício Lacerda Nogueira; Rafael dos Santos Bezerra; Raul Machado Neto; Rejane Maria Tommasini Grotto; Ricardo Haddad; Sandra Coccuzzo Sampaio Vessoni; Simone Kashima; Svetoslav Nanev Slavov; Vincent Louis Viala |
| EPI_ISL_1828375, EPI_ISL_1828400, EPI_ISL_1828427, EPI_ISL_1828466, EPI_ISL_1828491, EPI_ISL_1828499, EPI_ISL_1828506, EPI_ISL_1828520, EPI_ISL_1828569, EPI_ISL_1828585, EPI_ISL_1828614, EPI_ISL_1828615, EPI_ISL_1828618, EPI_ISL_1828625, EPI_ISL_1828640, EPI_ISL_1828674, EPI_ISL_1828683, EPI_ISL_1828686, EPI_ISL_1828688, EPI_ISL_1828695, EPI_ISL_2036988, EPI_ISL_2037071, EPI_ISL_2037072, EPI_ISL_2037088, EPI_ISL_2037091, EPI_ISL_2037109, EPI_ISL_2037133, EPI_ISL_2037142, EPI_ISL_2037153, EPI_ISL_2037181, EPI_ISL_2037188, EPI_ISL_2037191, EPI_ISL_4096591 | | | |
| see above | Pandemic Response Lab - NYC | Pandemic Response Lab, R&D | Cybill del Castillo; Dylan Law; Haiping Hao; Henry Lee; Jon Laurent; Katharine Nelson; Melissa Hopkins; Michael Hammerling; Pradeep Bugga; Shinyoung Clair Kang; Sol Rey; William Ward |
| EPI_ISL_5801448, EPI_ISL_5801451 | Pas Joao Antonio Do Nascimento | Instituto Butantan | Antonio Jorge Martins; Claudia Renata dos Santos Barros; David Schlesinger; Debora Botequio Moretti; Dimas Tadeu Covas; Elaine Cristina Marqueze; Elaine Vieira Santos; Evandra Strazza Rodrigues; Heidge Fukumasu; Jayme Augusto de Souza-Neto; José Salvatore Leister Patané; Luiz Alcantara; Luiz Lehmann Coutinho; Maria Carolina Elias; Maurício Lacerda Nogueira; Rafael dos Santos Bezerra; Raul Machado Neto; Rejane Maria Tommasini Grotto; Ricardo Haddad; Sandra Coccuzzo Sampaio Vessoni; Simone Kashima; Svetoslav Nanev Slavov; Vincent Louis Viala |
| EPI_ISL_3194067 | Pathogen Genomics Center, National Institute of Infectious Diseases | Pathogen Genomics Center, National Institute of Infectious Diseases | Kentaro Itokawa; Makoto Kuroda; Masanori Hashino; Rina Tanaka; Tsuyoshi Sekizuka |
| EPI_ISL_5801439 | Penitenciaria De Presidente Prudente | Instituto Butantan | Antonio Jorge Martins; Claudia Renata dos Santos Barros; David Schlesinger; Debora Botequio Moretti; Dimas Tadeu Covas; Elaine Cristina Marqueze; Elaine Vieira Santos; Evandra Strazza Rodrigues; Heidge Fukumasu; Jayme Augusto de Souza-Neto; José Salvatore Leister Patané; Luiz Alcantara; Luiz Lehmann Coutinho; Maria Carolina Elias; Maurício Lacerda Nogueira; Rafael dos Santos Bezerra; Raul Machado Neto; Rejane Maria Tommasini Grotto; Ricardo Haddad; Sandra Coccuzzo Sampaio Vessoni; Simone Kashima; Svetoslav Nanev Slavov; Vincent Louis Viala |
| EPI_ISL_6907901, EPI_ISL_6907906, EPI_ISL_6908436, EPI_ISL_6908437, EPI_ISL_6908438, EPI_ISL_6908439, EPI_ISL_6908821, EPI_ISL_6908826, EPI_ISL_6908829, EPI_ISL_6908830, EPI_ISL_6908836, EPI_ISL_6908842, EPI_ISL_6908848, EPI_ISL_6908849 | | | |
| see above | Plataforma de Vigilancia Molecular (PVM) - FIOCRUZ/BA | Plataforma de Vigilancia Molecular (PVM) - FIOCRUZ/BA | Bruno Bezerril Andrade; Camila I. de Oliveira; Clarissa Araújo Gurgel; Eduardo Oyama; Francys A Rangel; Laila A da Silva; Laise de Moraes; Leonardo Paiva Farias; Marina Cucco; Pedro Silveira; Ricardo Khouri on behalf of the Fiocruz COVID-19 Genomic Surveillance Network.; Tiago Graf |
| EPI_ISL_1915903, EPI_ISL_1915905, EPI_ISL_1993652 | Plateforme de testing Namuroise | Plateforme de testing Namuroise | Céline Maschietto; Degosserie Jonathan; Denis Olivier; Lesly Nyinkeu Kemamen; Mullier François; Otto Gaetan |
| EPI_ISL_1963345, EPI_ISL_1963431, EPI_ISL_2047633, EPI_ISL_2047665, EPI_ISL_2047667, EPI_ISL_2292728 | Platform BIS UZA/UAntwerpen | Labo Klinische Biologie, UZA | Basil Britto Xavier; Christine Lammens; Herman Goossens; Jasmine Coppens; Marie Le Mercier; Veerle Matheeussen |
| EPI_ISL_5801413, EPI_ISL_5801418, EPI_ISL_5801420 | Policlinica Covid 19 Itapetininga | Instituto Butantan | Antonio Jorge Martins; Claudia Renata dos Santos Barros; David Schlesinger; Debora Botequio Moretti; Dimas Tadeu Covas; Elaine Cristina Marqueze; Elaine Vieira Santos; Evandra Strazza Rodrigues; Heidge Fukumasu; Jayme Augusto de Souza-Neto; José Salvatore Leister Patané; Luiz Alcantara; Luiz Lehmann Coutinho; Maria Carolina Elias; Maurício Lacerda Nogueira; Rafael dos Santos Bezerra; Raul Machado Neto; Rejane Maria Tommasini Grotto; Ricardo Haddad; Sandra Coccuzzo Sampaio Vessoni; Simone Kashima; Svetoslav Nanev Slavov; Vincent Louis Viala |
| EPI_ISL_5801376 | Policlinica Hortolandia | Instituto Butantan | Antonio Jorge Martins; Claudia Renata dos Santos Barros; David Schlesinger; Debora Botequio Moretti; Dimas Tadeu Covas; Elaine Cristina Marqueze; Elaine Vieira Santos; Evandra Strazza Rodrigues; Heidge Fukumasu; Jayme Augusto de Souza-Neto; José Salvatore Leister Patané; Luiz Alcantara; Luiz Lehmann Coutinho; Maria Carolina Elias; Maurício Lacerda Nogueira; Rafael dos Santos Bezerra; Raul Machado Neto; Rejane Maria Tommasini Grotto; Ricardo Haddad; Sandra Coccuzzo Sampaio Vessoni; Simone Kashima; Svetoslav Nanev Slavov; Vincent Louis Viala |
| EPI_ISL_2391750, EPI_ISL_2391780 | Pomona Valley Hospital Medical Center | Los Angeles County Public Health Laboratories | P. Hemarajata et al. |
| EPI_ISL_4052652, EPI_ISL_4052654 | Precision Genomics | MUSC Molecular Pathology Laboratory | Ana Olivo; Ana Vallari; Barbara Harris; Dariusz Pytel; Frederick S. Nolte; Gavin Cloherty; Jaclyn Dunne; Jeremy Stuart; Jim Madory; Julie W. Hirschhorn; Kristen Maurer; Mary Rodgers; Scott Curry; Todd Meyer; W. Bailey Glen Jr |
| EPI_ISL_5801266, EPI_ISL_5801267, EPI_ISL_5801268, EPI_ISL_5801289 | Pronto Socorro Municipal De Severinia | Instituto Butantan | Antonio Jorge Martins; Claudia Renata dos Santos Barros; David Schlesinger; Debora Botequio Moretti; Dimas Tadeu Covas; Elaine Cristina Marqueze; Elaine Vieira Santos; Evandra Strazza Rodrigues; Heidge Fukumasu; Jayme Augusto de Souza-Neto; José Salvatore Leister Patané; Luiz Alcantara; Luiz Lehmann Coutinho; Maria Carolina Elias; Maurício Lacerda Nogueira; Rafael dos Santos Bezerra; Raul Machado Neto; Rejane Maria Tommasini Grotto; Ricardo Haddad; Sandra Coccuzzo Sampaio Vessoni; Simone Kashima; Svetoslav Nanev Slavov; Vincent Louis Viala |
| EPI_ISL_2551470, EPI_ISL_2551473, EPI_ISL_2551484, EPI_ISL_2551488, EPI_ISL_2551496 | Providence Oregon Regional Laboratories | Providence St. Joseph Health Molecular Genomics Laboratory | Alexa K Dowdell; Brian D Piening; Carlo B Bifulco; Fred L Robinson; Mary Campbell; Rogan Rattray |
| EPI_ISL_2551446, EPI_ISL_2551453, EPI_ISL_2551454 | Providence Regional Medical Center Everett | Providence St. Joseph Health Molecular Genomics Laboratory | Alexa K Dowdell; Brian D Piening; Carlo B Bifulco; Fred L Robinson; Mary Campbell; Rogan Rattray |
| EPI_ISL_5801472 | Psf Do Parque Dos Pinheiros | Instituto Butantan | Antonio Jorge Martins; Claudia Renata dos Santos Barros; David Schlesinger; Debora Botequio Moretti; Dimas Tadeu Covas; Elaine Cristina Marqueze; Elaine Vieira Santos; Evandra Strazza Rodrigues; Heidge Fukumasu; Jayme Augusto de Souza-Neto; José Salvatore Leister Patané; Luiz Alcantara; Luiz Lehmann Coutinho; Maria Carolina Elias; Maurício Lacerda Nogueira; Rafael dos Santos Bezerra; Raul Machado Neto; Rejane Maria Tommasini Grotto; Ricardo Haddad; Sandra Coccuzzo Sampaio Vessoni; Simone Kashima; Svetoslav Nanev Slavov; Vincent Louis Viala |
| EPI_ISL_5801422 | Psf Dr Antonio Pires De Almeida Porto Feliz | Instituto Butantan | Antonio Jorge Martins; Claudia Renata dos Santos Barros; David Schlesinger; Debora Botequio Moretti; Dimas Tadeu Covas; Elaine Cristina Marqueze; Elaine Vieira Santos; Evandra Strazza Rodrigues; Heidge Fukumasu; Jayme Augusto de Souza-Neto; José Salvatore Leister Patané; Luiz Alcantara; Luiz Lehmann Coutinho; Maria Carolina Elias; Maurício Lacerda Nogueira; Rafael dos Santos Bezerra; Raul Machado Neto; Rejane Maria Tommasini Grotto; Ricardo Haddad; Sandra Coccuzzo Sampaio Vessoni; Simone Kashima; Svetoslav Nanev Slavov; Vincent Louis Viala |
| EPI_ISL_5801457, EPI_ISL_5801464 | Psf Maria Jose Salto De Pirapora | Instituto Butantan | Antonio Jorge Martins; Claudia Renata dos Santos Barros; David Schlesinger; Debora Botequio Moretti; Dimas Tadeu Covas; Elaine Cristina Marqueze; Elaine Vieira Santos; Evandra Strazza Rodrigues; Heidge Fukumasu; Jayme Augusto de Souza-Neto; José Salvatore Leister Patané; Luiz Alcantara; Luiz Lehmann Coutinho; Maria Carolina Elias; Maurício Lacerda Nogueira; Rafael dos Santos Bezerra; Raul Machado Neto; Rejane Maria Tommasini Grotto; Ricardo Haddad; Sandra Coccuzzo Sampaio Vessoni; Simone Kashima; Svetoslav Nanev Slavov; Vincent Louis Viala |
| EPI_ISL_2294568, EPI_ISL_2697716, EPI_ISL_2697779, EPI_ISL_2697800, EPI_ISL_2697856, EPI_ISL_2697864, EPI_ISL_2698363, EPI_ISL_2698387, EPI_ISL_2698394, EPI_ISL_2698429, EPI_ISL_2698728, EPI_ISL_2699183, EPI_ISL_2699308 | | | |
| see above | Public Health Laboratory, Minnesota Department of Health | University of Minnesota Genomics Center | Corbin Dirkx; Daryl M. Gohl; Jaquelyn Kuriger-Laber; John Garbe; and Sean Wang |
| EPI_ISL_2196082, EPI_ISL_2196150 | QLabs | WVU and Marshall University Combined Genomics Core Facilities | James Denvir; Peter Perrotta; Peter Stoilov; Ryan Percifield; Wesley Kimble |
| EPI_ISL_2132620, EPI_ISL_2132625, EPI_ISL_2132626, EPI_ISL_2132628, EPI_ISL_2132630, EPI_ISL_2132631, EPI_ISL_2140734, EPI_ISL_2140736, EPI_ISL_2140737, EPI_ISL_2140738, EPI_ISL_2140740, EPI_ISL_2140742, EPI_ISL_2140743, EPI_ISL_2140745, EPI_ISL_2140746, EPI_ISL_2140747, EPI_ISL_2140748, EPI_ISL_2140750, EPI_ISL_2140751, EPI_ISL_2140752, EPI_ISL_2140753, EPI_ISL_2140754, | | | |

| | | | |
|---|---|---|---|
| EPI_ISL_2140755, EPI_ISL_2140756, EPI_ISL_2140758, EPI_ISL_2140766, EPI_ISL_2140768, EPI_ISL_2140774, EPI_ISL_2140775, EPI_ISL_2140777, EPI_ISL_2140778, EPI_ISL_2140779, EPI_ISL_2140780, EPI_ISL_2140781, EPI_ISL_2140782, EPI_ISL_2140783, EPI_ISL_2140784, EPI_ISL_2140786, EPI_ISL_2140787, EPI_ISL_2140788, EPI_ISL_2140789, EPI_ISL_2140790, EPI_ISL_2140795, EPI_ISL_2140796, EPI_ISL_2140797, EPI_ISL_2140798, EPI_ISL_2143220, EPI_ISL_2187230, EPI_ISL_2187231, EPI_ISL_2187233, EPI_ISL_2187234, EPI_ISL_2187235, EPI_ISL_2187236, EPI_ISL_2187237, EPI_ISL_2187239, EPI_ISL_2187241, EPI_ISL_2187243, EPI_ISL_2187245, EPI_ISL_2187246, EPI_ISL_2187247, EPI_ISL_2187248, EPI_ISL_2187249, EPI_ISL_2187250, EPI_ISL_2187251, EPI_ISL_2187253, EPI_ISL_2187254, EPI_ISL_2187256, EPI_ISL_2187257, EPI_ISL_2187258, EPI_ISL_2187259, EPI_ISL_2187260, EPI_ISL_2187261, EPI_ISL_2187262, EPI_ISL_2187263, EPI_ISL_2187264, EPI_ISL_2187265, EPI_ISL_2187266, EPI_ISL_2187267, EPI_ISL_2187268, EPI_ISL_2187269, EPI_ISL_2187270, EPI_ISL_2187271, EPI_ISL_2187272, EPI_ISL_2187273, EPI_ISL_2187274, EPI_ISL_2187275, EPI_ISL_2187276, EPI_ISL_2187277, EPI_ISL_2187278, EPI_ISL_2187279, EPI_ISL_2187280, EPI_ISL_2187282, EPI_ISL_2187283, EPI_ISL_2187285, EPI_ISL_2187286, EPI_ISL_2187287, EPI_ISL_2187288, EPI_ISL_2187289, EPI_ISL_2187290, EPI_ISL_2187291, EPI_ISL_2187292, EPI_ISL_2187293, EPI_ISL_2187294, EPI_ISL_2187295, EPI_ISL_2187296, EPI_ISL_2187297, EPI_ISL_2187298, EPI_ISL_2187299, EPI_ISL_2187300, EPI_ISL_2187301, EPI_ISL_2187302, EPI_ISL_2187303, EPI_ISL_2187304, EPI_ISL_2187305, EPI_ISL_2187306, EPI_ISL_2187308, EPI_ISL_2187309, EPI_ISL_2187310, EPI_ISL_2187311, EPI_ISL_2187314, EPI_ISL_2187315, EPI_ISL_2187316, EPI_ISL_2187317, EPI_ISL_2241628, EPI_ISL_2241631, EPI_ISL_2241633, EPI_ISL_2241636, EPI_ISL_2241639, EPI_ISL_2268144, EPI_ISL_2268152, EPI_ISL_2268159, EPI_ISL_2268161, EPI_ISL_2268164, EPI_ISL_2268166, EPI_ISL_2268169, EPI_ISL_2268171, EPI_ISL_2268174, EPI_ISL_2268176, EPI_ISL_2268178, EPI_ISL_2268181, EPI_ISL_2268186, EPI_ISL_2268189, EPI_ISL_2268319, EPI_ISL_2268331, EPI_ISL_2268388, EPI_ISL_2280120, EPI_ISL_2280127, EPI_ISL_2280134, EPI_ISL_2280137, EPI_ISL_2280170, EPI_ISL_2280172, EPI_ISL_2307159, EPI_ISL_2371708, EPI_ISL_2371717, EPI_ISL_2372421, EPI_ISL_4371799, EPI_ISL_4372301 | | | |
| see above | Quest Diagnostics Incorporated | Centers for Disease Control and Prevention Division of Viral Diseases, Pathogen Discovery | A. Gerasimova; A. Perez; Adrian Paskey; B. Anderson; Benjamin Rambo-Martin; Christopher Gulvick; Clinton Paden; Clinton R. Paden; Dakota Howard; Darlene Wagner; Dhwani Batra; Duncan MacCannell; Erisa Sula; F. Lacbawan; I. A. Shlyakhter; I. Shlyakhter; Jason Caravas; K. Livingston; K.E. Livingston; Kara Moser; Kristine Lacek; L. Bernstein; L.E. Bernstein; M. Hua; Matthew Schmerer; P. Tanpaiboon; Peter Cook; Peter W. Cook; R. Kagan; R. M. Kagan; R. Owen; R. Rolando; R. V. Rolando; S. H. Rosenthal; S. Rosenthal; Scott Sammons; Shatavia Morrison; Tymeckia Kendall; Victoria Caban Figueroa; Y. Liu; Yvette Unoarumhi |
| EPI_ISL_1987296, EPI_ISL_2004447, EPI_ISL_2004611 | Randox Laboratories | Wellcome Sanger Institute for the COVID-19 Genomics UK (COG-UK) Consortium | Cordelia Langford; David K. Jackson; Dominic Kwiatkowski; Ewan Harrison; Ian Johnston; Jeffrey Barrett; John Sillitoe on behalf of the Wellcome Sanger Institute COVID-19 Surveillance Team; Randox Laboratories and Alex Alderton; Roberto Amato; Sonia Goncalves |
| EPI_ISL_2319840, EPI_ISL_2319841, EPI_ISL_2319842, EPI_ISL_2319844, EPI_ISL_2319845, EPI_ISL_2319848, EPI_ISL_2319857, EPI_ISL_2319865, EPI_ISL_2319867, EPI_ISL_2319869, EPI_ISL_2319870, EPI_ISL_2319877, EPI_ISL_2319885, EPI_ISL_2319893, EPI_ISL_2319895, EPI_ISL_2319906, EPI_ISL_2319907, EPI_ISL_2319908, EPI_ISL_2319909, EPI_ISL_2319910, EPI_ISL_2319911, EPI_ISL_2319912, EPI_ISL_2319914, EPI_ISL_2319915, EPI_ISL_2319918, EPI_ISL_2319919, EPI_ISL_2319920, EPI_ISL_2319921, EPI_ISL_2319923, EPI_ISL_2319924, EPI_ISL_2319925, EPI_ISL_2319926, EPI_ISL_2319928, EPI_ISL_2319935, EPI_ISL_2319941, EPI_ISL_2319942, EPI_ISL_2319943, EPI_ISL_2319944, EPI_ISL_2319946, EPI_ISL_2319948, EPI_ISL_2319953, EPI_ISL_2319956, EPI_ISL_2319957, EPI_ISL_2319958, EPI_ISL_2319959, EPI_ISL_2319960, EPI_ISL_2319961, EPI_ISL_2319963, EPI_ISL_2319964, EPI_ISL_2319965, EPI_ISL_2319967, EPI_ISL_2319968, EPI_ISL_2319969, EPI_ISL_2319972, EPI_ISL_2319975, EPI_ISL_2319976, EPI_ISL_2319977, EPI_ISL_2319980, EPI_ISL_2319981, EPI_ISL_2473827, EPI_ISL_2473830, EPI_ISL_2473835, EPI_ISL_2473837, EPI_ISL_2473838, EPI_ISL_2473839, EPI_ISL_2473842, EPI_ISL_2473845, EPI_ISL_2473851, EPI_ISL_2473853, EPI_ISL_2473854, EPI_ISL_2473855, EPI_ISL_2473856, EPI_ISL_2473857, EPI_ISL_2473860, EPI_ISL_2473861, EPI_ISL_2473862, EPI_ISL_2473865, EPI_ISL_2473867, EPI_ISL_2473869, EPI_ISL_2473870, EPI_ISL_2473871, EPI_ISL_2473873, EPI_ISL_2473878, EPI_ISL_2473884, EPI_ISL_2473885, EPI_ISL_2473886, EPI_ISL_2473887, EPI_ISL_2473890, EPI_ISL_2473895, EPI_ISL_2473904, EPI_ISL_2473906, EPI_ISL_2473908, EPI_ISL_2473910, EPI_ISL_2473911, EPI_ISL_2473915, EPI_ISL_2473916, EPI_ISL_2473919, EPI_ISL_2473920, EPI_ISL_2473921, EPI_ISL_2473924, EPI_ISL_2473933, EPI_ISL_2473942, EPI_ISL_2473943, EPI_ISL_2473946, EPI_ISL_2473947, EPI_ISL_2473961, EPI_ISL_2473963, EPI_ISL_2473965, EPI_ISL_2473966, EPI_ISL_2473968, EPI_ISL_2473969, EPI_ISL_2473975, EPI_ISL_2473978, EPI_ISL_2473985, EPI_ISL_2473986, EPI_ISL_2474032, EPI_ISL_2474053, EPI_ISL_2474061, EPI_ISL_2474069, EPI_ISL_2474070, EPI_ISL_2474074, EPI_ISL_2474076, EPI_ISL_2474078, EPI_ISL_2474083, EPI_ISL_2474096, EPI_ISL_2474104, EPI_ISL_2474136, EPI_ISL_2474151, EPI_ISL_2474169, EPI_ISL_2662171, EPI_ISL_2662172, EPI_ISL_2662173, EPI_ISL_2662174, EPI_ISL_2662175, EPI_ISL_2662176, EPI_ISL_2662177, EPI_ISL_2662178, EPI_ISL_2662179, EPI_ISL_2662180, EPI_ISL_2662181, EPI_ISL_2662182, EPI_ISL_2662183, EPI_ISL_2662184, EPI_ISL_2662185, EPI_ISL_2662186, EPI_ISL_2662187, EPI_ISL_2662188, EPI_ISL_2662189, EPI_ISL_2662190, EPI_ISL_2662191, EPI_ISL_2662192, EPI_ISL_2662193, EPI_ISL_2662194, EPI_ISL_2662195, EPI_ISL_2662196, EPI_ISL_2662197, EPI_ISL_2662198, EPI_ISL_2662199, EPI_ISL_2662200, EPI_ISL_2662201, EPI_ISL_2662202, EPI_ISL_2662203, EPI_ISL_2662204, EPI_ISL_2662205, EPI_ISL_2662206, EPI_ISL_2662207, EPI_ISL_2662208 | | | |
| see above | Reditus Laboratories | Reditus Laboratories | Alexa Eichelberger; Cassy Philips; Joshua J. Geltz; M.S.; Ph.D.; Rex Dyer; Robert M. Sgambelluri |
| EPI_ISL_2742286, EPI_ISL_3777139 | Respiratory Virus Unit, Microbiology Services Colindale, Public Health England | COVID-19 Genomics UK (COG-UK) Consortium | PHE Covid Sequencing Team |
| EPI_ISL_2106006, EPI_ISL_2106010, EPI_ISL_2106015, EPI_ISL_2106025, EPI_ISL_2106026, EPI_ISL_2106030, EPI_ISL_2106042, EPI_ISL_2106049, EPI_ISL_2106051, EPI_ISL_2253129, EPI_ISL_2253130, EPI_ISL_2253132, EPI_ISL_2253136, EPI_ISL_2253137, EPI_ISL_2253138, EPI_ISL_2253140, EPI_ISL_2253141, EPI_ISL_2253142, EPI_ISL_2253149, EPI_ISL_2254265, EPI_ISL_2254274, EPI_ISL_2254293, EPI_ISL_2254319, EPI_ISL_2254320, EPI_ISL_2254326, EPI_ISL_2254331, EPI_ISL_2254332, EPI_ISL_2254351, EPI_ISL_2254461, EPI_ISL_2254467, EPI_ISL_2254484 | | | |
| see above | Respiratory Viruses Branch, Centers for Disease Control and Prevention | Respiratory Viruses Branch, Centers for Disease Control and Prevention | Anderson, B.; Baker; Bakhash; Batra, D.; Bernstein, L.; Breit, N.; C.R.; Campbell, P.; Caravas, J.; Case, R.; Clark, C.; Cook; Ellis, S.; Gerasimova, A.; Ghorpade, V.; Greninger, A.; Gulvick, C.; Hardison, M.; Houdeshell, H.; Howard, D.; Hua, M.; Huang; I.A.; Jerome; K.R.; Kagan; Kvalvaag, O.; Lacbawan, F.; Lin, M.; Liu, Y.; Livingston; Livingston, K.; M.-L.; M.S.; MacCannell, D.; Mathias, P.; Morrison, S.; Moser, K.; N.R.; Nall, D.; Nguyen; Owen, R.; P.W.; Paden; Paskey, A.; Perez, A.; Perez, R.; R.J.; R.M.; R.V.; Rambo-Martin, B.; Rolando; Rosenthal; Roychoudhury; S.H.; S.M.; Sammons, S.; Sanders, E.; Schmerer, M.; Shlyakhter; Shrestha, L.; T.V.; Tanpaiboon, P.; Unoarumhi, Y.; Vest, A.; Wagner, D.; Westlund, S.; Xie, H. |
| EPI_ISL_2133713, EPI_ISL_2133759, EPI_ISL_2133763, EPI_ISL_2133808, EPI_ISL_2133812, EPI_ISL_2133814, EPI_ISL_2133829, EPI_ISL_2133866, EPI_ISL_2133872, EPI_ISL_2133873, EPI_ISL_2133874, EPI_ISL_2133879, EPI_ISL_2133886, EPI_ISL_2133888, EPI_ISL_2133897, EPI_ISL_2133900, EPI_ISL_2133910, EPI_ISL_2133911, EPI_ISL_2133917, EPI_ISL_2133926 | | | |
| see above | Rhode Island Department of Health | Infectious Disease Program, Broad Institute of Harvard and MIT | Adams, G.; Azevedo, K.; B.L.; B.W.; Bauer, M.; Birren; Carter, A.; Chaluvadi, S.; D.J.; DeRuff, K.; Gladden-Young, A.; Huard, R.; J.E.; K.J.; King, E.; Lagerborg, K.; Lemieux; Loreth, C.; Miller, A.; Normandin, E.; P.C.; Park; Pearlman, L.; Reilly, S.; Rudy, M.; Sabeti; Siddle; Tomkins-Tinch, C.; and MacInnis |
| EPI_ISL_2604789, EPI_ISL_2604798, EPI_ISL_2604800, EPI_ISL_2604802, EPI_ISL_2604813 | Rhode Island State Health Laboratories | Rhode Island State Health Laboratories | Ewa King; Kristin Carpenter-Azevedo; Richard C. Huard |
| EPI_ISL_2425522 | Rhode Island State Health Laboratory | Rhode Island State Health Laboratory | Ewa King; Kristin Carpenter-Azevedo; Richard C. Huard |
| EPI_ISL_2691919, EPI_ISL_2691926 | Rush University Medical Center | RIPHL at Rush University Medical Center | Diane Springer; Felix Araujo Perez; Joyce Houlihan; Kevin Kunstman; Laura Furtado; Marieta Hyde; Mary Hayden; Max Kolton; Stefan Green |
| EPI_ISL_2840616 | S.C. Laboratorio Analisi, ASL 3 Liguria | U.O. Igiene, Ospedale Policlinico San Martino | Bruzzone Bianca; Caligiuri Patrizia; De Pace Vanessa; Domnich Alexander; Icardi Giancarlo on behalf of SARS-CoV-2 ITALIAN RESEARCH ENTERPRISE-(SCIRE) Collaborative Group; Orsi Andrea; Ricucci Valentina; Spitaleri Antonino |
| EPI_ISL_2209872, EPI_ISL_2210133 | SANTA CASA DE MISERICORDIA DE UBATUBA | Instituto Butantan / Mendelics | Antonio Jorge Martins; Claudia Renata dos Santos Barros; David Schlesinger; Debora Botequio Moretti; Dimas Tadeu Covas; Elaine Cristina Marqueze; Elaine Vieira Santos; Evandra Strazza Rodrigues; Heidge Fukumasu; Jayme Augusto de Souza-Neto; José Salvatore Leister Patané; Luiz Alcantara; Luiz Lehmann Coutinho; Maria Carolina Elias; Maurício Lacerda Nogueira; Rafael dos Santos Bezerra; Raul Machado Neto; Rejane Maria Tommasini Grotto; Ricardo Haddad; Sandra Coccuzzo Sampaio Vessoni; Simone Kashima; Svetoslav Nanev Slavov; Vincent Louis Viala |
| EPI_ISL_2209407, EPI_ISL_2209888, EPI_ISL_2210250, EPI_ISL_2345682, EPI_ISL_2345727, EPI_ISL_2345739, EPI_ISL_2345783, EPI_ISL_2362269 | | | |
| see above | SANTA CASA DE MISERICORDIA SAO JOSE | Instituto Butantan / Mendelics | Antonio Jorge Martins; Claudia Renata dos Santos Barros; David Schlesinger; Debora Botequio Moretti; Dimas Tadeu Covas; Elaine Cristina Marqueze; Elaine Vieira Santos; Evandra Strazza Rodrigues; Heidge Fukumasu; Jayme Augusto de Souza-Neto; José Salvatore Leister Patané; Luiz Alcantara; Luiz Lehmann Coutinho; Maria Carolina Elias; Maurício Lacerda Nogueira; Rafael dos Santos Bezerra; Raul Machado Neto; Rejane Maria Tommasini Grotto; Ricardo Haddad; Sandra Coccuzzo Sampaio Vessoni; Simone Kashima; Svetoslav Nanev Slavov; Vincent Louis Viala |
| EPI_ISL_2444154, EPI_ISL_2444157, EPI_ISL_2444158, EPI_ISL_2444159, EPI_ISL_2444160, EPI_ISL_2444161, EPI_ISL_2444163, EPI_ISL_2444164, EPI_ISL_2444166, EPI_ISL_2444167, EPI_ISL_2444168, EPI_ISL_2444169, EPI_ISL_2444171, EPI_ISL_2444174, EPI_ISL_2444176, EPI_ISL_2444178, EPI_ISL_2444179, EPI_ISL_2444180, EPI_ISL_2444181, EPI_ISL_2444182, EPI_ISL_2444185, EPI_ISL_2444187, EPI_ISL_2444188, EPI_ISL_2444190, EPI_ISL_2455396 | | | |
| see above | SAO JOSE DO RIO PRETO | Instituto Butantan / FAMERP | Antonio Jorge Martins; Claudia Renata dos Santos Barros; David Schlesinger; Debora Botequio Moretti; Dimas Tadeu Covas; Elaine Cristina Marqueze; Elaine Vieira Santos; Evandra Strazza Rodrigues; Heidge Fukumasu; Jayme Augusto de Souza-Neto; José Salvatore Leister Patané; Luiz Alcantara; Luiz Lehmann Coutinho; Maria Carolina Elias; Maurício Lacerda Nogueira; Rafael dos Santos Bezerra; Raul Machado Neto; Rejane Maria Tommasini Grotto; Ricardo Haddad; Sandra Coccuzzo Sampaio Vessoni; Simone Kashima; Svetoslav Nanev Slavov; Vincent Louis Viala |
| EPI_ISL_2098626 | SARATOGA HOSPITAL LABORATORY | Wadsworth Center, New York State Department of Health | Alexis Russell; Catharine Prussing; Daryl M. Lamson; Erasmus Schneider; Erica Lasek-Nesselquist; John Kelly; Jonathan Plitnick; Kirsten St. George; Matthew Shudt; Melissa A Leisner; Navjot Singh |
| EPI_ISL_2649551, EPI_ISL_2791690, EPI_ISL_2791821 | SARS-CoV-2 Sequencing Castilla y Leon-Spain Consortium | SARS-CoV-2 Sequencing Castilla y Leon-Spain Consortium | Antonio Orduña-Domingo; Carlos Fuster Foz; Carmen Aldea-Mansilla; Carmen Gimeno Crespo; David Abad; Gregoria Megías Lobón; Jose María Eiros Bouza; Laura Sánchez de Prada; M. Isabel Fernandez-Natal; Marta Dominguez-Gil; Marta Hernandez; María Antonia García Castro; Mª Fe Brezmes-Valdivieso; Noelia Arenal Andrés; Silvia Rojo; Sonsoles Garcinuño Pérez |
| EPI_ISL_2131669, EPI_ISL_2338152, EPI_ISL_2338191, EPI_ISL_2338193, EPI_ISL_2338194, EPI_ISL_2338195, EPI_ISL_2338196 | | | |
| see above | SARS-CoV-2 testing team, National Institute of Infectious Diseases | Pathogen Genomics Center, National Institute of Infectious Diseases | Hazuka Y Furihata; Kentaro Itokawa; Makoto Kuroda; Masanori Hashino; Masumichi Saito; Naomi Nojiri; Nozomu Hanaoka; Rina Tanaka; Sana Uchikoba; Tsuguto Fujimoto; Tsuyoshi Sekizuka |
| EPI_ISL_2345855 | SECRETARIA MUNICIPAL DA SAUDE DE ITARARE | Instituto Butantan / Mendelics | Antonio Jorge Martins; Claudia Renata dos Santos Barros; David Schlesinger; Debora Botequio Moretti; Dimas Tadeu Covas; Elaine Cristina Marqueze; Elaine Vieira Santos; Evandra Strazza Rodrigues; Heidge Fukumasu; Jayme Augusto de Souza-Neto; José Salvatore Leister Patané; Luiz Alcantara; Luiz Lehmann Coutinho; Maria Carolina Elias; Maurício Lacerda Nogueira; Rafael dos Santos Bezerra; Raul Machado Neto; Rejane Maria Tommasini Grotto; Ricardo Haddad; Sandra Coccuzzo Sampaio Vessoni; Simone Kashima; Svetoslav Nanev Slavov; Vincent Louis Viala |
| EPI_ISL_2345237, EPI_ISL_2345238 | SECRETARIA MUNICIPAL DE SAUDE DE JARINU | Instituto Butantan / ESALQ-Piracicaba | Antonio Jorge Martins; Claudia Renata dos Santos Barros; David Schlesinger; Debora Botequio Moretti; Dimas Tadeu Covas; Elaine Cristina Marqueze; Elaine Vieira Santos; Evandra Strazza Rodrigues; Heidge Fukumasu; Jayme Augusto de Souza-Neto; José Salvatore Leister Patané; Luiz Alcantara; Luiz Lehmann Coutinho; Maria Carolina Elias; Maurício Lacerda Nogueira; Rafael dos Santos Bezerra; Raul Machado Neto; Rejane Maria Tommasini Grotto; Ricardo Haddad; Sandra Coccuzzo Sampaio Vessoni; Simone Kashima; Svetoslav Nanev Slavov; Vincent Louis Viala |
| EPI_ISL_2345209, EPI_ISL_2345211 | SECRETARIA MUNICIPAL DE SAUDE DE MACATUBA | Instituto Butantan / ESALQ-Piracicaba | Antonio Jorge Martins; Claudia Renata dos Santos Barros; David Schlesinger; Debora Botequio Moretti; Dimas Tadeu Covas; Elaine Cristina Marqueze; Elaine Vieira Santos; Evandra Strazza Rodrigues; Heidge Fukumasu; Jayme Augusto de Souza-Neto; José Salvatore Leister Patané; Luiz Alcantara; Luiz Lehmann Coutinho; Maria Carolina Elias; Maurício Lacerda Nogueira; Rafael dos Santos Bezerra; Raul Machado Neto; Rejane Maria Tommasini Grotto; Ricardo Haddad; Sandra Coccuzzo Sampaio Vessoni; Simone Kashima; Svetoslav Nanev Slavov; Vincent Louis Viala |
| EPI_ISL_2801356 | SECRETARIA MUNICIPAL DE SAUDE DE TIANGUA | Analytical Competence Molecular Epidemiology Lab/ACME, Oswaldo Cruz Foundation, Ceara (FIOCRUZ CE) | Cleber Furtado Aksenen e Suzana Porto Almeida; Fabio Miyajima; Fernando Braga Stehling; Francisco Eder de Moura Lopes; Jamille Maria Mendes Bezerra; Joaquim César do Nascimento Sousa Junior; Pedro Miguel Carneiro Jeronimo; Thais Ferreira de Oliveira; Thais de Oliveira Costa; Ticiane Cavalcante de Souza; Veridiana Pessoa Miyajima |
| EPI_ISL_2209068, EPI_ISL_2209197, EPI_ISL_2209511, EPI_ISL_2209664, EPI_ISL_2209878, EPI_ISL_2210183, EPI_ISL_2345655, EPI_ISL_2345660, EPI_ISL_2345664, EPI_ISL_2345700, EPI_ISL_2345751, EPI_ISL_2345755, EPI_ISL_2345805, EPI_ISL_2345848, EPI_ISL_2345852 | | | |
| see above | SECRETARIA MUNICIPAL DE SAUDE SOROCABA | Instituto Butantan / Mendelics | Antonio Jorge Martins; Claudia Renata dos Santos Barros; David Schlesinger; Debora Botequio Moretti; Dimas Tadeu Covas; Elaine Cristina Marqueze; Elaine Vieira Santos; Evandra Strazza Rodrigues; Heidge Fukumasu; Jayme Augusto de Souza-Neto; José Salvatore Leister Patané; Luiz Alcantara; Luiz Lehmann Coutinho; Maria Carolina Elias; Maurício Lacerda Nogueira; Rafael dos Santos Bezerra; Raul Machado Neto; Rejane Maria Tommasini Grotto; Ricardo Haddad; Sandra Coccuzzo Sampaio Vessoni; Simone Kashima; Svetoslav Nanev Slavov; Vincent Louis Viala |
| EPI_ISL_2001356, EPI_ISL_2001392, EPI_ISL_2131368, EPI_ISL_2131369, EPI_ISL_2131370 | SIESP DIPARTIMENTO DI PREVENZIONE TERAMO | Istituto Zooprofilattico Sperimentale dell'Abruzzo e Molise "G. Caporale" | Ancora M; Calistri P; Cammà C; Caporale M; Curini V; Delli Compagni E; Di Domenico M; Di Lollo Valeria; Di Pasquale A; Lorusso A; Mangone I; Marcacci M; Puglia I; Rinaldi A; Savini G; Scialabba S |
| EPI_ISL_2001342, EPI_ISL_2001343, EPI_ISL_2001344, EPI_ISL_2001406 | SIESP SULMONA | Istituto Zooprofilattico Sperimentale dell'Abruzzo e Molise "G. Caporale" | Ancora M; Calistri P; Cammà C; Caporale M; Curini V; Delli Compagni E; Di Domenico M; Di Lollo Valeria; Di Pasquale A; Lorusso A; Mangone I; Marcacci M; Puglia I; Rinaldi A; Savini G; Scialabba S |
| EPI_ISL_2585962, EPI_ISL_2585967, EPI_ISL_2586310, EPI_ISL_2586412, EPI_ISL_2586418, EPI_ISL_2586429 | SK-Roy Romanow Provincial Laboratory | National Microbiology Laboratory (NML) | Amanda Lang; Anna Majer; Anneliese Landgraff; CanCOGeN's metadata curation team; Darian Hole; Elsie Grudeski; Gary Van Domselaar; Grace Seo; Jennifer Tanner; Jessica Minion; Kirsten Biggar; Madison Chapel; Morag Graham; Natalie Knox; Nathalie Bastien; Philip Mabon; Public Health Agency of Canada CanCOGeN team; Rachel DePaulo; Rhiannon Huzarewich; Russell Mandes; Ryan McDonald; Shari Tyson; Timothy Booth; Yan Li |
| EPI_ISL_6124734, EPI_ISL_6124746, EPI_ISL_6124772 | SK-Roy Romanow Provincial Laboratory | Saskatchewan - Roy Romanow Provincial Laboratory (RRPL) | Alanna Senecal; Amanda Lang; Jessica Minion; Kara Loos; Keith MacKenzie; Meredith Faires; Rachel DePaulo; Roy Romanow Provincial Laboratory - Molecular Diagnostics; Ryan McDonald |
| EPI_ISL_2209419 | SMS IPERO | Instituto Butantan / Mendelics | Antonio Jorge Martins; Claudia Renata dos Santos Barros; David Schlesinger; Debora Botequio Moretti; Dimas Tadeu Covas; Elaine Cristina Marqueze; Elaine Vieira Santos; Evandra Strazza Rodrigues; Heidge Fukumasu; Jayme Augusto de Souza-Neto; José Salvatore Leister Patané; Luiz Alcantara; Luiz Lehmann Coutinho; Maria Carolina Elias; Maurício Lacerda Nogueira; Rafael dos Santos Bezerra; Raul Machado Neto; Rejane Maria Tommasini Grotto; Ricardo Haddad; Sandra Coccuzzo Sampaio Vessoni; Simone Kashima; Svetoslav Nanev Slavov; Vincent Louis Viala |
| EPI_ISL_2210223, | SMS SECRETARIA MUNICIPAL DE | Instituto Butantan / Mendelics | Antonio Jorge Martins; Claudia Renata dos Santos Barros; David Schlesinger; Debora Botequio Moretti; Dimas Tadeu Covas; Elaine Cristina Marqueze; Elaine Vieira Santos; Evandra Strazza Rodrigues; Heidge Fukumasu; Jayme Augusto de Souza-Neto; José Salvatore Leister Patané; Luiz Alcantara; Luiz |

| | | | |
|---|---|---|---|
| EPI_ISL_2345708, EPI_ISL_2345713, EPI_ISL_2345718 | SAUDE DE BOITUVA | | Lehmann Coutinho; Maria Carolina Elias; Maurício Lacerda Nogueira; Rafael dos Santos Bezerra; Raul Machado Neto; Rejane Maria Tommasini Grotto; Ricardo Haddad; Sandra Coccuzzo Sampaio Vessoni; Simone Kashima; Svetoslav Nanev Slavov; Vincent Louis Viala |
| EPI_ISL_2321632, EPI_ISL_2321649 | SOMER | Universidad Nacional de Colombia - Laboratorio Genómico One Health | Andres F. Cardona-Rios; Carlos Franco-Muñoz; Carolina Muñoz-Arango; Celeny Ortiz; Daniel O. Maldonado-Perez; Diego A. Álvarez-Díaz; Hector Alejandro Ruiz-Moreno; Idabely Betancur Ortiz; Jorge E. Osorio; Juan P. Hernandez-Ortiz; Karl A Ciuoderis; Katherine Laiton-Donato; Laura Silvana Perez; Lina M. Hurtado; Marcela Mercado-Reyes; Maria Angélica Maya; Maria Stella López; Rita Almanza Payares; Sandra Ines Cano; Simón Villegas Velásquez |
| EPI_ISL_2098138 | SUNY UPSTATE MEDICAL UNIVERSITY | Wadsworth Center, New York State Department of Health | Alexis Russell; Catharine Prussing; Daryl M. Lamson; Erasmus Schneider; Erica Lasek-Nesselquist; John Kelly; Jonathan Plitnick; Kirsten St. George; Matthew Shudt; Melissa A Leisner; Navjot Singh |
| EPI_ISL_5155780, EPI_ISL_5155972, EPI_ISL_5156587 | SUNY Upstate Medical University | 505 Irving Avenue, Syracuse, NY 13210 - Frank Middleton Lab | Brian Pavlovitz; Frank Middleton |
| EPI_ISL_4820603 | SURA | Laboratorio Departamental de Salud Publica de Antioquia | Cristian Arbey Velarde Hoyos; Gloria Isabel Escobar; Idabely Betancur Ortiz; Juan P. Hernandez-Ortiz; Maria Stella López |
| EPI_ISL_1919735, EPI_ISL_2003217, EPI_ISL_2003221, EPI_ISL_2003311, EPI_ISL_2003315, EPI_ISL_2003377, EPI_ISL_2003378, EPI_ISL_2003392, EPI_ISL_2003397, EPI_ISL_2003419, EPI_ISL_2003421, EPI_ISL_2139716, EPI_ISL_2139723, EPI_ISL_2139724, EPI_ISL_2139744 | | | |
| see above | SYNLAB | GIGA Medical Genomics | Bouchra Boujemla; Cécile Meex; Keith Durkin; Maria Artesi; Marie-Pierre Hayette; Nathalie Renotte; Pierrette Melin; Raphaël Boreux; Sébastien Bontems; Vincent Bours |
| EPI_ISL_2321576 | SYNLAB | Universidad Nacional de Colombia - Laboratorio Genómico One Health | Andres F. Cardona-Rios; Carlos Franco-Muñoz; Carolina Muñoz-Arango; Celeny Ortiz; Daniel O. Maldonado-Perez; Diego A. Álvarez-Díaz; Hector Alejandro Ruiz-Moreno; Idabely Betancur Ortiz; Jorge E. Osorio; Juan P. Hernandez-Ortiz; Karl A Ciuoderis; Katherine Laiton-Donato; Laura Silvana Perez; Lina M. Hurtado; Marcela Mercado-Reyes; Maria Angélica Maya; Maria Stella López; Rita Almanza Payares; Sandra Ines Cano; Simón Villegas Velásquez |
| EPI_ISL_2113048 | SYNLAB MVZ Berlin | Robert Koch Institute | |
| EPI_ISL_2261832, EPI_ISL_2262086 | SYNLAB MVZ Leinfelden-Echterdingen | Robert Koch Institute | |
| EPI_ISL_2111664, EPI_ISL_2111754, EPI_ISL_2111934, EPI_ISL_2113539 | SYNLAB MVZ Trier | Robert Koch Institute | |
| EPI_ISL_2105765, EPI_ISL_2105772, EPI_ISL_2105794, EPI_ISL_2105823, EPI_ISL_2105831, EPI_ISL_2105835, EPI_ISL_2230843, EPI_ISL_2230898, EPI_ISL_2230899, EPI_ISL_2230901, EPI_ISL_2230902, EPI_ISL_2230903 | | | |
| see above | Salud Digna | Instituto Nacional de Medicina Genomica | Abraham Campos-Romero; Cedro-Tanda A; Escobar-Arrazola; Gonzalez-Barrera D; Herrera-Montalvo LA.; Hidalgo-Miranda A; Luna-Ruiz Marco; M; Mendoza-Vargas A; Moreno-Camacho José Luis; Munguia-Garza P; Ramirez-Vega O; Rangel-DeLeon D; Reyes-Grajeda JP; Rodriguez-Gallegos Jorge |
| EPI_ISL_2231072, EPI_ISL_2231075, EPI_ISL_2423003, EPI_ISL_2423011, EPI_ISL_2628677 | San Diego County Public Health Laboratory | Andersen lab at Scripps Research | Brett Austin; Jovan Shephard; SEARCH Alliance San Diego with Ashleigh Murphy |
| EPI_ISL_2420723 | San Gallicano Dermatological Institute I.F.O. | INMI Lazzaro Spallanzani IRCCS | A Di Caro; B Bartolini; CEM Gruber; E Giombini; F Ensoli; F Messina; F Pimpinelli; F Santini; G Bonfiglio; I Cavallo; M Rueca; MR Capobianchi; O Butera |
| EPI_ISL_2211292 | San Juan Regional Medical Center | New Mexico Department of Health Scientific Laboratory | Anastacia Griego-Fisher; D'eldra Malone; Ellie Johnson; Jennifer Benoit; Linda Salazar; Ratheesh Rajan |
| EPI_ISL_5801479, EPI_ISL_5801495 | Santa Casa De Misericordia De Ubatuba | Instituto Butantan | Antonio Jorge Martins; Claudia Renata dos Santos Barros; David Schlesinger; Debora Botequio Moretti; Dimas Tadeu Covas; Elaine Cristina Marqueze; Elaine Vieira Santos; Evandra Strazza Rodrigues; Heidge Fukumasu; Jayme Augusto de Souza-Neto; José Salvatore Leister Patané; Luiz Alcantara; Luiz Lehmann Coutinho; Maria Carolina Elias; Maurício Lacerda Nogueira; Rafael dos Santos Bezerra; Raul Machado Neto; Rejane Maria Tommasini Grotto; Ricardo Haddad; Sandra Coccuzzo Sampaio Vessoni; Simone Kashima; Svetoslav Nanev Slavov; Vincent Louis Viala |
| EPI_ISL_5801408, EPI_ISL_5801489, EPI_ISL_5801493, EPI_ISL_5801504, EPI_ISL_5801505, EPI_ISL_5801506, EPI_ISL_5801508, EPI_ISL_5801510 | | | |
| see above | Santa Casa De Misericordia Sao Jose | Instituto Butantan | Antonio Jorge Martins; Claudia Renata dos Santos Barros; David Schlesinger; Debora Botequio Moretti; Dimas Tadeu Covas; Elaine Cristina Marqueze; Elaine Vieira Santos; Evandra Strazza Rodrigues; Heidge Fukumasu; Jayme Augusto de Souza-Neto; José Salvatore Leister Patané; Luiz Alcantara; Luiz Lehmann Coutinho; Maria Carolina Elias; Maurício Lacerda Nogueira; Rafael dos Santos Bezerra; Raul Machado Neto; Rejane Maria Tommasini Grotto; Ricardo Haddad; Sandra Coccuzzo Sampaio Vessoni; Simone Kashima; Svetoslav Nanev Slavov; Vincent Louis Viala |
| EPI_ISL_2659092, EPI_ISL_2659102, EPI_ISL_2659111, EPI_ISL_2659114 | Santa Clara Valley Medical Center | Santa Clara County Public Health Laboratory | Santa Clara County Public Health Department |
| EPI_ISL_3373746, EPI_ISL_3543807 | Scripps Medical Laboratory | Andersen lab at Scripps Research | Ellen Stefanski; Ian Mchardy; SEARCH Alliance San Diego with Michael Quigley |
| EPI_ISL_2490028, EPI_ISL_2490029, EPI_ISL_2490033, EPI_ISL_2490039 | Seattle Flu Study | Seattle Flu Study | Amanda Adler; Barry R. Lutz; Benjamin Pelle; Caitlin R. Wolf; Chris D. Frazar; Deborah A. Nickerson; Elisabeth Brandstetter; Erica Ryke; Helen Y. Chu; Janet A. Englund; Jay Shendure; Jeff Duchin; Jover Lee; Kairsten Fay; Karen Cowgill; Kirsten Lacombe; Lea M. Starita; Mark J. Rieder; Matthew Richardson; Matthew Thompson; Melissa Truong; Michael Boeckh; Michael Famulare; Misja Ilcisin; Peter D. Han; Stephanie Schrag; Thomas R. Sibley; Trevor Bedford |
| EPI_ISL_5802702 | Secretaria De Saude De Rafard | Instituto Butantan | Antonio Jorge Martins; Claudia Renata dos Santos Barros; David Schlesinger; Debora Botequio Moretti; Dimas Tadeu Covas; Elaine Cristina Marqueze; Elaine Vieira Santos; Evandra Strazza Rodrigues; Heidge Fukumasu; Jayme Augusto de Souza-Neto; José Salvatore Leister Patané; Luiz Alcantara; Luiz Lehmann Coutinho; Maria Carolina Elias; Maurício Lacerda Nogueira; Rafael dos Santos Bezerra; Raul Machado Neto; Rejane Maria Tommasini Grotto; Ricardo Haddad; Sandra Coccuzzo Sampaio Vessoni; Simone Kashima; Svetoslav Nanev Slavov; Vincent Louis Viala |
| EPI_ISL_5802706 | Secretaria De Saude De Santa Gertrudes | Instituto Butantan | Antonio Jorge Martins; Claudia Renata dos Santos Barros; David Schlesinger; Debora Botequio Moretti; Dimas Tadeu Covas; Elaine Cristina Marqueze; Elaine Vieira Santos; Evandra Strazza Rodrigues; Heidge Fukumasu; Jayme Augusto de Souza-Neto; José Salvatore Leister Patané; Luiz Alcantara; Luiz Lehmann Coutinho; Maria Carolina Elias; Maurício Lacerda Nogueira; Rafael dos Santos Bezerra; Raul Machado Neto; Rejane Maria Tommasini Grotto; Ricardo Haddad; Sandra Coccuzzo Sampaio Vessoni; Simone Kashima; Svetoslav Nanev Slavov; Vincent Louis Viala |
| EPI_ISL_5802696, EPI_ISL_5802697, EPI_ISL_5802719, EPI_ISL_5802724 | Secretaria De Saude De Sao Pedro | Instituto Butantan | Antonio Jorge Martins; Claudia Renata dos Santos Barros; David Schlesinger; Debora Botequio Moretti; Dimas Tadeu Covas; Elaine Cristina Marqueze; Elaine Vieira Santos; Evandra Strazza Rodrigues; Heidge Fukumasu; Jayme Augusto de Souza-Neto; José Salvatore Leister Patané; Luiz Alcantara; Luiz Lehmann Coutinho; Maria Carolina Elias; Maurício Lacerda Nogueira; Rafael dos Santos Bezerra; Raul Machado Neto; Rejane Maria Tommasini Grotto; Ricardo Haddad; Sandra Coccuzzo Sampaio Vessoni; Simone Kashima; Svetoslav Nanev Slavov; Vincent Louis Viala |
| EPI_ISL_5801436 | Secretaria Municipal Da Saude De Itarare | Instituto Butantan | Antonio Jorge Martins; Claudia Renata dos Santos Barros; David Schlesinger; Debora Botequio Moretti; Dimas Tadeu Covas; Elaine Cristina Marqueze; Elaine Vieira Santos; Evandra Strazza Rodrigues; Heidge Fukumasu; Jayme Augusto de Souza-Neto; José Salvatore Leister Patané; Luiz Alcantara; Luiz Lehmann Coutinho; Maria Carolina Elias; Maurício Lacerda Nogueira; Rafael dos Santos Bezerra; Raul Machado Neto; Rejane Maria Tommasini Grotto; Ricardo Haddad; Sandra Coccuzzo Sampaio Vessoni; Simone Kashima; Svetoslav Nanev Slavov; Vincent Louis Viala |
| EPI_ISL_5802712 | Secretaria Municipal De Saude De Cordeiropolis | Instituto Butantan | Antonio Jorge Martins; Claudia Renata dos Santos Barros; David Schlesinger; Debora Botequio Moretti; Dimas Tadeu Covas; Elaine Cristina Marqueze; Elaine Vieira Santos; Evandra Strazza Rodrigues; Heidge Fukumasu; Jayme Augusto de Souza-Neto; José Salvatore Leister Patané; Luiz Alcantara; Luiz Lehmann Coutinho; Maria Carolina Elias; Maurício Lacerda Nogueira; Rafael dos Santos Bezerra; Raul Machado Neto; Rejane Maria Tommasini Grotto; Ricardo Haddad; Sandra Coccuzzo Sampaio Vessoni; Simone Kashima; Svetoslav Nanev Slavov; Vincent Louis Viala |
| EPI_ISL_5801342, EPI_ISL_5801343 | Secretaria Municipal De Saude De Jarinu | Instituto Butantan | Antonio Jorge Martins; Claudia Renata dos Santos Barros; David Schlesinger; Debora Botequio Moretti; Dimas Tadeu Covas; Elaine Cristina Marqueze; Elaine Vieira Santos; Evandra Strazza Rodrigues; Heidge Fukumasu; Jayme Augusto de Souza-Neto; José Salvatore Leister Patané; Luiz Alcantara; Luiz Lehmann Coutinho; Maria Carolina Elias; Maurício Lacerda Nogueira; Rafael dos Santos Bezerra; Raul Machado Neto; Rejane Maria Tommasini Grotto; Ricardo Haddad; Sandra Coccuzzo Sampaio Vessoni; Simone Kashima; Svetoslav Nanev Slavov; Vincent Louis Viala |
| EPI_ISL_5801303, EPI_ISL_5801305 | Secretaria Municipal De Saude De Macatuba | Instituto Butantan | Antonio Jorge Martins; Claudia Renata dos Santos Barros; David Schlesinger; Debora Botequio Moretti; Dimas Tadeu Covas; Elaine Cristina Marqueze; Elaine Vieira Santos; Evandra Strazza Rodrigues; Heidge Fukumasu; Jayme Augusto de Souza-Neto; José Salvatore Leister Patané; Luiz Alcantara; Luiz Lehmann Coutinho; Maria Carolina Elias; Maurício Lacerda Nogueira; Rafael dos Santos Bezerra; Raul Machado Neto; Rejane Maria Tommasini Grotto; Ricardo Haddad; Sandra Coccuzzo Sampaio Vessoni; Simone Kashima; Svetoslav Nanev Slavov; Vincent Louis Viala |
| EPI_ISL_5801404, EPI_ISL_5801407, EPI_ISL_5801409, EPI_ISL_5801414, EPI_ISL_5801416, EPI_ISL_5801417, EPI_ISL_5801430, EPI_ISL_5801432, EPI_ISL_5801433, EPI_ISL_5801434, EPI_ISL_5801441, EPI_ISL_5801442, EPI_ISL_5801443, EPI_ISL_5801444, EPI_ISL_5801445, EPI_ISL_5801446 | | | |
| see above | Secretaria Municipal De Saude Sorocaba | Instituto Butantan | Antonio Jorge Martins; Claudia Renata dos Santos Barros; David Schlesinger; Debora Botequio Moretti; Dimas Tadeu Covas; Elaine Cristina Marqueze; Elaine Vieira Santos; Evandra Strazza Rodrigues; Heidge Fukumasu; Jayme Augusto de Souza-Neto; José Salvatore Leister Patané; Luiz Alcantara; Luiz Lehmann Coutinho; Maria Carolina Elias; Maurício Lacerda Nogueira; Rafael dos Santos Bezerra; Raul Machado Neto; Rejane Maria Tommasini Grotto; Ricardo Haddad; Sandra Coccuzzo Sampaio Vessoni; Simone Kashima; Svetoslav Nanev Slavov; Vincent Louis Viala |
| EPI_ISL_2179731, EPI_ISL_2179759, EPI_ISL_2179809 | Servicio Microbiología Hospital La Paz | Servicio Microbiología Hospital La Paz | Elie Dahdouh; Fernando Lázaro; Jesús Mingorance Cruz; Rubén Cáceres |
| EPI_ISL_2136136, EPI_ISL_2140106, EPI_ISL_2140108, EPI_ISL_2140143, EPI_ISL_2140144, EPI_ISL_2140145, EPI_ISL_2140146, EPI_ISL_2158697, EPI_ISL_2158700, EPI_ISL_2158703, EPI_ISL_2158704, EPI_ISL_2158705, EPI_ISL_2158706, EPI_ISL_2158734, EPI_ISL_2158735, EPI_ISL_2158737, EPI_ISL_2158738, EPI_ISL_2158740, EPI_ISL_2158764, EPI_ISL_2158767, EPI_ISL_2158781, EPI_ISL_2158783, EPI_ISL_2158798, EPI_ISL_2158799, EPI_ISL_2158814, EPI_ISL_2158818, EPI_ISL_2158824, EPI_ISL_2158827, EPI_ISL_2158829, EPI_ISL_2158831, EPI_ISL_2158834, EPI_ISL_4082199, EPI_ISL_6694832 | | | |
| see above | Servicio Virosis Respiratorias-Departamento Virología-INEI | Instituto Nacional Enfermedades Infecciosas C.G.Malbran | Avaro M.; Baumeister E.; Benedetti E.; Campos J.; Cisterna D.; Dattero ME; De Belder D.; Haim MS.; Lorenzo F.; Molina V.; Perandones C.; Poklepovich T.; Pontoriero A.; Russo M.; Sanchez Loria J.; Tuduri E. |
| EPI_ISL_2104452, EPI_ISL_2104454, EPI_ISL_2283668 | Servicio de Microbiologia Hospital Ramon y Cajal | Servicio de Microbiologia Hospital Ramon y Cajal | Galan JC; Martinez L. Abreu M; Ponce M; y Gonzalez-Alba JM |
| EPI_ISL_1922394, EPI_ISL_1922397, EPI_ISL_2133001, EPI_ISL_2133002, EPI_ISL_2133003, EPI_ISL_2133006, EPI_ISL_2510579, EPI_ISL_2516676 | | | |
| see above | Servicio de Microbiología Clínica (Complejo Hospitalario de Navarra, Pamplona) | Centro de Secuenciación NASERTIC | Ana Miqueleiz; Ana Navascués; Carmen Ezpeleta Baquedano |
| EPI_ISL_2545375, EPI_ISL_2545401, EPI_ISL_2545405 | Servicio de Microbiología, Laboratori Clínic Metropolitana Nord. Hospital Universitari Germans Trias i Pujol. Institut d'Investigació en Ciències de la Salut Germans Trias i Pujol (IGTP) | SeqCOVID-SPAIN consortium/IBV(CSIC) | Adrián Antuori; Anabel Fernández; Anna Not; Antoni E. Bordoy; Cristina Casañ and SeqCOVID-SPAIN consortium; Elisa Martró; Nona Romaní; Verónica Saludes |
| EPI_ISL_2271646, EPI_ISL_2271647, EPI_ISL_2271650, EPI_ISL_2362280 | Servicio de Microbiología. Consorcio Hospital General Universitario de Valencia | SeqCOVID-SPAIN consortium/IBV(CSIC) | Begoña Fuster Escrivá; Carme Salvador García; Concepción Gimeno Cardona and SeqCOVID-SPAIN consortium; María Dolores Ocete; Rafael Medina González |

| | | | |
|---|---|---|---|
| EPI_ISL_5315921, EPI_ISL_5315923, EPI_ISL_5315925, EPI_ISL_5315926, EPI_ISL_5315929, EPI_ISL_5315930, EPI_ISL_5315932, EPI_ISL_5315933, EPI_ISL_5315934, EPI_ISL_5315935, EPI_ISL_5315936, EPI_ISL_5315937, EPI_ISL_5315938, EPI_ISL_5315941, EPI_ISL_5315942, EPI_ISL_5315943, EPI_ISL_5315944, EPI_ISL_5315945, EPI_ISL_5315946, EPI_ISL_5315947, EPI_ISL_5315948, EPI_ISL_5315950, EPI_ISL_5315951, EPI_ISL_5315952, EPI_ISL_5315953, EPI_ISL_5315954, EPI_ISL_5315956, EPI_ISL_5315958, EPI_ISL_5315959, EPI_ISL_5315960, EPI_ISL_5315961, EPI_ISL_5315969, EPI_ISL_5315974, EPI_ISL_5315977, EPI_ISL_5315981, EPI_ISL_5315982, EPI_ISL_5315983, EPI_ISL_5315984, EPI_ISL_5315985, EPI_ISL_5315986, EPI_ISL_5315988, EPI_ISL_5315989, EPI_ISL_5315992, EPI_ISL_5315993, EPI_ISL_5315994, EPI_ISL_5315997, EPI_ISL_5316000, EPI_ISL_5316003, EPI_ISL_5316004, EPI_ISL_5316005, EPI_ISL_5316006, EPI_ISL_5316008, EPI_ISL_5316009, EPI_ISL_5316010, EPI_ISL_5316012, EPI_ISL_5316013, EPI_ISL_5316014, EPI_ISL_5316016, EPI_ISL_5316017, EPI_ISL_5316018, EPI_ISL_5316019, EPI_ISL_5316020, EPI_ISL_5316022, EPI_ISL_5316025, EPI_ISL_5316026, EPI_ISL_5316028, EPI_ISL_5316030, EPI_ISL_5316032, EPI_ISL_5316034, EPI_ISL_5316035, EPI_ISL_5316037, EPI_ISL_5316038, EPI_ISL_5316040, EPI_ISL_5316041, EPI_ISL_5316042, EPI_ISL_5316043, EPI_ISL_5316046, EPI_ISL_5316052, EPI_ISL_5316053, EPI_ISL_5316054, EPI_ISL_5316055, EPI_ISL_5316057, EPI_ISL_5316061, EPI_ISL_5316062, EPI_ISL_5316064, EPI_ISL_5316067, EPI_ISL_5316070, EPI_ISL_5316072, EPI_ISL_5316073, EPI_ISL_5316074, EPI_ISL_5316075, EPI_ISL_5316076, EPI_ISL_5316077, EPI_ISL_5316078, EPI_ISL_5316079, EPI_ISL_5316080, EPI_ISL_5316082, EPI_ISL_5316107, EPI_ISL_5316670, EPI_ISL_5316673, EPI_ISL_5316674, EPI_ISL_5316675, EPI_ISL_5316682, EPI_ISL_5316683, EPI_ISL_5316684, EPI_ISL_5316685, EPI_ISL_5316686, EPI_ISL_5316687, EPI_ISL_5316688, EPI_ISL_5316689, EPI_ISL_5316692, EPI_ISL_5316695, EPI_ISL_5316696 | | | |
| see above | Shared Hospital Laboratory | Shared Hospital Laboratory | Christie Vermeiren; Finlay Maguire; Kevin Katz; Patryk Aftanas; Robert Kozak; Samira Mubareka |
| EPI_ISL_2231179, EPI_ISL_2231180, EPI_ISL_2231181, EPI_ISL_2231182, EPI_ISL_2231183, EPI_ISL_2231184, EPI_ISL_2231185 | | | |
| see above | Sharp HealthCare Laboratory | Andersen lab at Scripps Research | Art Mendoza; Cathy Woerle; Jacquelyn Berumen; Liam McGinnis; Omid Bakhtar; SEARCH Alliance San Diego with Aaron Harding |
| EPI_ISL_2296024, EPI_ISL_2296034, EPI_ISL_2296040, EPI_ISL_2296044, EPI_ISL_2800969, EPI_ISL_2800995, EPI_ISL_2801029 | | | |
| see above | Simple Laboratories | Gagnon Lab, Southern Illinois University | Keith Gagnon |
| EPI_ISL_5801447 | Sms Ipero | Instituto Butantan | Antonio Jorge Martins; Claudia Renata dos Santos Barros; David Schlesinger; Debora Botequio Moretti; Dimas Tadeu Covas; Elaine Cristina Marqueze; Elaine Vieira Santos; Evandra Strazza Rodrigues; Heidge Fukumasu; Jayme Augusto de Souza-Neto; José Salvatore Leister Patané; Luiz Alcantara; Luiz Lehmann Coutinho; Maria Carolina Elias; Maurício Lacerda Nogueira; Rafael dos Santos Bezerra; Raul Machado Neto; Rejane Maria Tommasini Grotto; Ricardo Haddad; Sandra Coccuzzo Sampaio Vessoni; Simone Kashima; Svetoslav Nanev Slavov; Vincent Louis Viala |
| EPI_ISL_5801406, EPI_ISL_5801415, EPI_ISL_5801429, EPI_ISL_5801431 | Sms Secretaria Municipal De Saude De Boituva | Instituto Butantan | Antonio Jorge Martins; Claudia Renata dos Santos Barros; David Schlesinger; Debora Botequio Moretti; Dimas Tadeu Covas; Elaine Cristina Marqueze; Elaine Vieira Santos; Evandra Strazza Rodrigues; Heidge Fukumasu; Jayme Augusto de Souza-Neto; José Salvatore Leister Patané; Luiz Alcantara; Luiz Lehmann Coutinho; Maria Carolina Elias; Maurício Lacerda Nogueira; Rafael dos Santos Bezerra; Raul Machado Neto; Rejane Maria Tommasini Grotto; Ricardo Haddad; Sandra Coccuzzo Sampaio Vessoni; Simone Kashima; Svetoslav Nanev Slavov; Vincent Louis Viala |
| EPI_ISL_2438025 | Somer | Universidad Nacional de Colombia - Laboratorio Genómico One Health | Andres F. Cardona-Rios; Carlos Franco-Muñoz; Carolina Muñoz-Arango; Celeny Ortiz; Daniel O. Maldonado-Perez; Diego A. Álvarez-Díaz; Hector Alejandro Ruiz-Moreno; Idabely Betancur Ortiz; Jorge E. Osorio; Juan P. Hernandez-Ortiz; Karl A Ciuoderis; Katherine Laiton-Donato; Laura Silvana Perez; Lina M. Hurtado; Marcela Mercado-Reyes; Maria Angélica Maya; Maria Stella López; Rita Almanza Payares; Sandra Ines Cano; Simón Villegas Velásquez |
| EPI_ISL_2230717 | Southern Nevada Public Health Laboratory | Southern Nevada Public Health Laboratory | Michael Picker |
| EPI_ISL_4496811, EPI_ISL_4496812, EPI_ISL_4496813, EPI_ISL_4496815, EPI_ISL_4496816 | Stanford Health Care | Stanford University School of Medicine, Clinical Virology Laboratory | Becky Jiang; James Zehnder; Malaya K. Sahoo; Selamawit Bihon; and Benjamin A. Pinsky |
| EPI_ISL_3418888, EPI_ISL_3418895, EPI_ISL_3418896, EPI_ISL_3418957, EPI_ISL_3418973 | StarMed Healthcare | UNC Charlotte Environmental Monitoring Laboratory | Adam France; Cynthia Gibas; Erin Stiers; Jannatul Ferdous; Jessica Schlueter; Kevin Lambirth; Sam Kunkleman |
| EPI_ISL_2135474, EPI_ISL_2135509, EPI_ISL_2724517, EPI_ISL_2724550 | State Laboratories Division, Hawaii State Department of Health | State Laboratories Division, Hawaii State Department of Health | Ayana Garnet; Drew Kuwazaki; Edward Desmond; Pamela O'Brien; Razvan Sultana |
| EPI_ISL_2377960, EPI_ISL_2377968, EPI_ISL_2377981, EPI_ISL_2377993, EPI_ISL_2377995, EPI_ISL_2378045, EPI_ISL_2378054, EPI_ISL_2378163, EPI_ISL_2378167, EPI_ISL_2378181, EPI_ISL_2378182, EPI_ISL_2378400, EPI_ISL_2378401, EPI_ISL_2378415, EPI_ISL_2378430, EPI_ISL_2378432, EPI_ISL_2473013, EPI_ISL_2473079, EPI_ISL_2473102, EPI_ISL_2473109, EPI_ISL_2473124, EPI_ISL_2473136, EPI_ISL_2473147, EPI_ISL_2473153, EPI_ISL_2473179, EPI_ISL_2473183, EPI_ISL_2473217, EPI_ISL_2473219, EPI_ISL_2473251, EPI_ISL_2473255, EPI_ISL_2473270, EPI_ISL_2473281, EPI_ISL_2473283 | | | |
| see above | State Testing Facility | Altius Institute for Biomedical Research | Alex Isner; Alex Nguyen; Amanda Gale; Audra Johnson; Clem Green; Daniel Bates; Eric Thorland; Jacob Rodriguez; Jean Robinston; Jemma Nelson; Jessica Kunder; John Stamatoyannopoulos; Joshua Richards; Julia Wald; Kneshay Harper; Lauren Mitchell; Mark Frerker; Matt Hartman; Michael Buckley; Muhammad Halimun; Rebecca Bruders; Sadie Patraw; Sofia Olsson; Tobias Ragoczy |
| EPI_ISL_6794666, EPI_ISL_6794688 | State of New Hampshire Public Health Laboratories | State of New Hampshire Public Health Laboratories | Caitlin Mercier; Chris Benton; Jinfeng Li; Juan Bolanos; Xinglu Zhang |
| EPI_ISL_2438018 | Synlab | Universidad Nacional de Colombia - Laboratorio Genómico One Health | Andres F. Cardona-Rios; Carlos Franco-Muñoz; Carolina Muñoz-Arango; Celeny Ortiz; Daniel O. Maldonado-Perez; Diego A. Álvarez-Díaz; Hector Alejandro Ruiz-Moreno; Idabely Betancur Ortiz; Jorge E. Osorio; Juan P. Hernandez-Ortiz; Karl A Ciuoderis; Katherine Laiton-Donato; Laura Silvana Perez; Lina M. Hurtado; Marcela Mercado-Reyes; Maria Angélica Maya; Maria Stella López; Rita Almanza Payares; Sandra Ines Cano; Simón Villegas Velásquez |
| EPI_ISL_2281451, EPI_ISL_2281511, EPI_ISL_2281726, EPI_ISL_2423586, EPI_ISL_2423702, EPI_ISL_2424098, EPI_ISL_2454613 | | | |
| see above | TGen North | TGen North | "Jolene Bowers; Ashlyn Pfeiffer; Chris French; Darrin Lemmer; Dave Engelthaler; Hayley Yaglom; Heather Centner; Jolene Bowers; The Arizona COVID Genomics Union (ACGU); The Arizona COVID Genomics Union (ACGU)" |
| EPI_ISL_2141435, EPI_ISL_2141441, EPI_ISL_2177457, EPI_ISL_2177470, EPI_ISL_2177497, EPI_ISL_2177509, EPI_ISL_2280027 | | | |
| see above | TXDSHS | TXDSHS | Anita Pokharel; Bonnie Oh; Chun Wang; Grace Kubin; Jenny Zhang; Karen Bobier; Lorraine Rodriguez; Maliha Rahman; Mayela Pedrueza; Myong Koag; Rachel Lee; Rashmi Tuladhar |
| EPI_ISL_2727436 | Tampa General Hospital Esoteric Lab | Tampa General Hospital Esoteric Research & Development Lab | Amorce Lima; Deanna Becker; Jon Faughn; Suzane Silbert |
| EPI_ISL_2631408, EPI_ISL_2631410, EPI_ISL_2631411, EPI_ISL_2631412, EPI_ISL_2631414, EPI_ISL_2631416, EPI_ISL_2631421, EPI_ISL_2631423, EPI_ISL_2631426, EPI_ISL_2631427, EPI_ISL_2631428, EPI_ISL_2631432, EPI_ISL_2631433, EPI_ISL_2631435 | | | |
| see above | The Caribbean Public Health Agency | Carrington Lab, Department of PreClinical Sciences, Faculty of Medical Sciences, The University of the West Indies, St Augustine Campus | Anushka Ramjag; Arianne Brown-Jordan; Avery Hinds; Christine V. F. Carrington; Christopher Oura; Gabriel Escobar; Karla Georges; Naresh Nandram; Nikita S. D. Sahadeo; Nuno Faria; Oliver Pybus; Risha Singh; Roshan Parasram; Sarah Hill; SueMin Nathaniel; Vernie Ramkissoon |
| EPI_ISL_2649743, EPI_ISL_2649750, EPI_ISL_2649754, EPI_ISL_2649755, EPI_ISL_2649758, EPI_ISL_2649768, EPI_ISL_2649770, EPI_ISL_2649771, EPI_ISL_2649788, EPI_ISL_2649790, EPI_ISL_2658370, EPI_ISL_2658374, EPI_ISL_2658375, EPI_ISL_2658377, EPI_ISL_2658380, EPI_ISL_2658382, EPI_ISL_2658389, EPI_ISL_6944295, EPI_ISL_6944296, EPI_ISL_6944297, EPI_ISL_6944298, EPI_ISL_6944299, EPI_ISL_6944300 | | | |
| see above | The Caribbean Public Health Agency | Carrington Lab, Department of Preclinical Sciences, Faculty of Medical Sciences, The University of the West Indies, St Augustine Campus | Anushka Ramjag; Arianne Brown-Jordan; Avery Hinds; Christine V. F. Carrington; Christopher Oura; Gabriel Escobar; Karla Georges; Naresh Nandram; Nikita S. D. Sahadeo; Nuno Faria; Oliver Pybus; Risha Singh; Roshan Parasram; Sarah Hill; Soren Nicholls; SueMin Nathaniel; Vernie Ramkissoon |
| EPI_ISL_1970632, EPI_ISL_1970663, EPI_ISL_1970674 | The Jackson Laboratory | The Jackson Laboratory | Adams M; Bergeron D; Kelly K; Li L; Omerza G; Renzette N |
| EPI_ISL_2631429 | The Trinidad and Tobago Public Health Laboratory | Carrington Lab, Department of PreClinical Sciences, Faculty of Medical Sciences, The University of the West Indies, St Augustine Campus | Anushka Ramjag; Arianne Brown-Jordan; Avery Hinds; Christine V. F. Carrington; Christopher Oura; Gabriel Escobar; Karla Georges; Naresh Nandram; Nikita S. D. Sahadeo; Nuno Faria; Oliver Pybus; Risha Singh; Roshan Parasram; Sarah Hill; SueMin Nathaniel; Vernie Ramkissoon |
| EPI_ISL_2649760, EPI_ISL_2649761, EPI_ISL_2649762, EPI_ISL_2649763, EPI_ISL_2658408, EPI_ISL_2658409, EPI_ISL_2658410 | | | |
| see above | The Trinidad and Tobago Public Health Laboratory | Carrington Lab, Department of Preclinical Sciences, Faculty of Medical Sciences, The University of the West Indies, St Augustine Campus | Anushka Ramjag; Arianne Brown-Jordan; Avery Hinds; Christine V. F. Carrington; Christopher Oura; Gabriel Escobar; Karla Georges; Naresh Nandram; Nikita S. D. Sahadeo; Nuno Faria; Oliver Pybus; Risha Singh; Roshan Parasram; Sarah Hill; SueMin Nathaniel; Vernie Ramkissoon |
| EPI_ISL_2454361 | TriCore Reference Laboratories | Center for Global Health, University of New Mexico Health Sciences Center | Cecilia Thompson; Darrell Dinwiddie; Daryl Domman; Karissa Culbreath; Kendra Pesko; Kurt Schwalm; Valerie Morley |
| EPI_ISL_3184496, EPI_ISL_3184497, EPI_ISL_3184512, EPI_ISL_3184671, EPI_ISL_3184672, EPI_ISL_3184685, EPI_ISL_3184708, EPI_ISL_3184711, EPI_ISL_3184712, EPI_ISL_3184731, EPI_ISL_3184733 | | | |
| see above | U.O. Microbiologia Laboratorio Unico Centro Servizi - AUSL della Romagna | U.O. Microbiologia, Laboratorio Unico Centro Servizi - AUSL della Romagna | Giorgio Dirani |
| EPI_ISL_5603286, EPI_ISL_5603313 | UBASF DE PEDRA | Analytical Competence Molecular Epidemiology Lab/ACME, Oswaldo Cruz Foundation, Ceara (FIOCRUZ CE) | Carlos Leonardo de Aragao Araujo; Cecilia Leite Costa & Eduardo Ruback dos Santos on behalf of COVID-19 FIOCRUZ Genomic Network; Cleber Furtado Aksenen; Fabio Miyajima; Fernando Braga Stehling; Francisco Eder de Moura Lopes; Igor Oliveira Duarte; Jamille Maria Mendes Bezerra; Joaquim Cesar do Nascimento Sousa Junior; Pedro Miguel Carneiro Jeronimo; Suzana Porto Almeida; Thais Ferreira de Oliveira; Thais de Oliveira Costa; Ticiane Cavalcante de Souza; Veridiana Pessoa Miyajima |
| EPI_ISL_2208930 | UBS ALAMBARI | Instituto Butantan / Mendelics | Antonio Jorge Martins; Claudia Renata dos Santos Barros; David Schlesinger; Debora Botequio Moretti; Dimas Tadeu Covas; Elaine Cristina Marqueze; Elaine Vieira Santos; Evandra Strazza Rodrigues; Heidge Fukumasu; Jayme Augusto de Souza-Neto; José Salvatore Leister Patané; Luiz Alcantara; Luiz Lehmann Coutinho; Maria Carolina Elias; Maurício Lacerda Nogueira; Rafael dos Santos Bezerra; Raul Machado Neto; Rejane Maria Tommasini Grotto; Ricardo Haddad; Sandra Coccuzzo Sampaio Vessoni; Simone Kashima; Svetoslav Nanev Slavov; Vincent Louis Viala |
| EPI_ISL_2345154, EPI_ISL_2345155, EPI_ISL_2345157 | UBS BONFIM | Instituto Butantan / ESALQ-Piracicaba | Antonio Jorge Martins; Claudia Renata dos Santos Barros; David Schlesinger; Debora Botequio Moretti; Dimas Tadeu Covas; Elaine Cristina Marqueze; Elaine Vieira Santos; Evandra Strazza Rodrigues; Heidge Fukumasu; Jayme Augusto de Souza-Neto; José Salvatore Leister Patané; Luiz Alcantara; Luiz Lehmann Coutinho; Maria Carolina Elias; Maurício Lacerda Nogueira; Rafael dos Santos Bezerra; Raul Machado Neto; Rejane Maria Tommasini Grotto; Ricardo Haddad; Sandra Coccuzzo Sampaio Vessoni; Simone Kashima; Svetoslav Nanev Slavov; Vincent Louis Viala |
| EPI_ISL_2345190, EPI_ISL_2345191 | UBS DR ABELARDO PINHEIRO GUIMARAES SCRPARDO | Instituto Butantan / ESALQ-Piracicaba | Antonio Jorge Martins; Claudia Renata dos Santos Barros; David Schlesinger; Debora Botequio Moretti; Dimas Tadeu Covas; Elaine Cristina Marqueze; Elaine Vieira Santos; Evandra Strazza Rodrigues; Heidge Fukumasu; Jayme Augusto de Souza-Neto; José Salvatore Leister Patané; Luiz Alcantara; Luiz Lehmann Coutinho; Maria Carolina Elias; Maurício Lacerda Nogueira; Rafael dos Santos Bezerra; Raul Machado Neto; Rejane Maria Tommasini Grotto; Ricardo Haddad; Sandra Coccuzzo Sampaio Vessoni; Simone Kashima; Svetoslav Nanev Slavov; Vincent Louis Viala |

| | | | |
|---|---|---|---|
| EPI_ISL_2345156, EPI_ISL_2345158, EPI_ISL_2345159, EPI_ISL_2345162, EPI_ISL_2345163, EPI_ISL_2345166, EPI_ISL_2345167, EPI_ISL_2345168, EPI_ISL_2345173, EPI_ISL_2345175, EPI_ISL_2345179 | | | |
| see above | UBS DR ALFREDO DANTAS DE SOUZA UMUARAMA | Instituto Butantan / ESALQ-Piracicaba | Antonio Jorge Martins; Claudia Renata dos Santos Barros; David Schlesinger; Debora Botequio Moretti; Dimas Tadeu Covas; Elaine Cristina Marqueze; Elaine Vieira Santos; Evandra Strazza Rodrigues; Heidge Fukumasu; Jayme Augusto de Souza-Neto; José Salvatore Leister Patané; Luiz Alcantara; Luiz Lehmann Coutinho; Maria Carolina Elias; Maurício Lacerda Nogueira; Rafael dos Santos Bezerra; Raul Machado Neto; Rejane Maria Tommasini Grotto; Ricardo Haddad; Sandra Coccuzzo Sampaio Vessoni; Simone Kashima; Svetoslav Nanev Slavov; Vincent Louis Viala |
| EPI_ISL_2345178 | UBS GLENDA RODELLA DUMAS | Instituto Butantan / ESALQ-Piracicaba | Antonio Jorge Martins; Claudia Renata dos Santos Barros; David Schlesinger; Debora Botequio Moretti; Dimas Tadeu Covas; Elaine Cristina Marqueze; Elaine Vieira Santos; Evandra Strazza Rodrigues; Heidge Fukumasu; Jayme Augusto de Souza-Neto; José Salvatore Leister Patané; Luiz Alcantara; Luiz Lehmann Coutinho; Maria Carolina Elias; Maurício Lacerda Nogueira; Rafael dos Santos Bezerra; Raul Machado Neto; Rejane Maria Tommasini Grotto; Ricardo Haddad; Sandra Coccuzzo Sampaio Vessoni; Simone Kashima; Svetoslav Nanev Slavov; Vincent Louis Viala |
| EPI_ISL_2345822, EPI_ISL_2345858 | UBS II DE ALVARES MACHADO | Instituto Butantan / Mendelics | Antonio Jorge Martins; Claudia Renata dos Santos Barros; David Schlesinger; Debora Botequio Moretti; Dimas Tadeu Covas; Elaine Cristina Marqueze; Elaine Vieira Santos; Evandra Strazza Rodrigues; Heidge Fukumasu; Jayme Augusto de Souza-Neto; José Salvatore Leister Patané; Luiz Alcantara; Luiz Lehmann Coutinho; Maria Carolina Elias; Maurício Lacerda Nogueira; Rafael dos Santos Bezerra; Raul Machado Neto; Rejane Maria Tommasini Grotto; Ricardo Haddad; Sandra Coccuzzo Sampaio Vessoni; Simone Kashima; Svetoslav Nanev Slavov; Vincent Louis Viala |
| EPI_ISL_2209600, EPI_ISL_2210148 | UBS II DR JOSE CARLOS MACUCO JANINI | Instituto Butantan / Mendelics | Antonio Jorge Martins; Claudia Renata dos Santos Barros; David Schlesinger; Debora Botequio Moretti; Dimas Tadeu Covas; Elaine Cristina Marqueze; Elaine Vieira Santos; Evandra Strazza Rodrigues; Heidge Fukumasu; Jayme Augusto de Souza-Neto; José Salvatore Leister Patané; Luiz Alcantara; Luiz Lehmann Coutinho; Maria Carolina Elias; Maurício Lacerda Nogueira; Rafael dos Santos Bezerra; Raul Machado Neto; Rejane Maria Tommasini Grotto; Ricardo Haddad; Sandra Coccuzzo Sampaio Vessoni; Simone Kashima; Svetoslav Nanev Slavov; Vincent Louis Viala |
| EPI_ISL_2209494 | UBS III DE PARIQUERA ACU PARIQUERA ACU | Instituto Butantan / Mendelics | Antonio Jorge Martins; Claudia Renata dos Santos Barros; David Schlesinger; Debora Botequio Moretti; Dimas Tadeu Covas; Elaine Cristina Marqueze; Elaine Vieira Santos; Evandra Strazza Rodrigues; Heidge Fukumasu; Jayme Augusto de Souza-Neto; José Salvatore Leister Patané; Luiz Alcantara; Luiz Lehmann Coutinho; Maria Carolina Elias; Maurício Lacerda Nogueira; Rafael dos Santos Bezerra; Raul Machado Neto; Rejane Maria Tommasini Grotto; Ricardo Haddad; Sandra Coccuzzo Sampaio Vessoni; Simone Kashima; Svetoslav Nanev Slavov; Vincent Louis Viala |
| EPI_ISL_2345193, EPI_ISL_2345196 | UBS III MURUTINGA DO SUL | Instituto Butantan / ESALQ-Piracicaba | Antonio Jorge Martins; Claudia Renata dos Santos Barros; David Schlesinger; Debora Botequio Moretti; Dimas Tadeu Covas; Elaine Cristina Marqueze; Elaine Vieira Santos; Evandra Strazza Rodrigues; Heidge Fukumasu; Jayme Augusto de Souza-Neto; José Salvatore Leister Patané; Luiz Alcantara; Luiz Lehmann Coutinho; Maria Carolina Elias; Maurício Lacerda Nogueira; Rafael dos Santos Bezerra; Raul Machado Neto; Rejane Maria Tommasini Grotto; Ricardo Haddad; Sandra Coccuzzo Sampaio Vessoni; Simone Kashima; Svetoslav Nanev Slavov; Vincent Louis Viala |
| EPI_ISL_2345223, EPI_ISL_2345224, EPI_ISL_2345225, EPI_ISL_2345228, EPI_ISL_2345229, EPI_ISL_2345230, EPI_ISL_2345231, EPI_ISL_2345232, EPI_ISL_2345233, EPI_ISL_2345234, EPI_ISL_2345235 | | | |
| see above | UBS IV GUARARAPES | Instituto Butantan / ESALQ-Piracicaba | Antonio Jorge Martins; Claudia Renata dos Santos Barros; David Schlesinger; Debora Botequio Moretti; Dimas Tadeu Covas; Elaine Cristina Marqueze; Elaine Vieira Santos; Evandra Strazza Rodrigues; Heidge Fukumasu; Jayme Augusto de Souza-Neto; José Salvatore Leister Patané; Luiz Alcantara; Luiz Lehmann Coutinho; Maria Carolina Elias; Maurício Lacerda Nogueira; Rafael dos Santos Bezerra; Raul Machado Neto; Rejane Maria Tommasini Grotto; Ricardo Haddad; Sandra Coccuzzo Sampaio Vessoni; Simone Kashima; Svetoslav Nanev Slavov; Vincent Louis Viala |
| EPI_ISL_2345134 | UBS VILA RIBEIRO | Instituto Butantan / ESALQ-Piracicaba | Antonio Jorge Martins; Claudia Renata dos Santos Barros; David Schlesinger; Debora Botequio Moretti; Dimas Tadeu Covas; Elaine Cristina Marqueze; Elaine Vieira Santos; Evandra Strazza Rodrigues; Heidge Fukumasu; Jayme Augusto de Souza-Neto; José Salvatore Leister Patané; Luiz Alcantara; Luiz Lehmann Coutinho; Maria Carolina Elias; Maurício Lacerda Nogueira; Rafael dos Santos Bezerra; Raul Machado Neto; Rejane Maria Tommasini Grotto; Ricardo Haddad; Sandra Coccuzzo Sampaio Vessoni; Simone Kashima; Svetoslav Nanev Slavov; Vincent Louis Viala |
| EPI_ISL_3419589 | UC Davis Genome Center and Healthy Davis Together | UC Davis Genome Center | UC Davis |
| EPI_ISL_3321301, EPI_ISL_3321303, EPI_ISL_3535719 | UC-Christus Clinical Hospital Laboratory | Laboratory of Molecular Virology, School of Medicine, Pontificia Universidad Catolica de Chile | Ana Maira Guzman; Andres E. Munoz-Marcos; Catalina Pardo-Roa; Eileen Serrano; Erick Salinas; Estefany Poblete; Francisco Melo; Jorge Levican; Leonardo I. Almonacid; Maria Jose Avendano; Maria Patricia Vega; Rafael A. Medina; Ricardo Enrique de la Barra; Tamara Garcia-Salum |
| EPI_ISL_2424196 | UCLA Clinical Micro Lab | Los Angeles County Public Health Laboratories | P. Hemarajata et al. |
| EPI_ISL_2376401 | ULSS 2 Marca Trevigiana | Istituto Zooprofilattico Sperimentale delle Venezie | Adelaide Milani; Alessia Schivo; Alice Fusaro; Ambra Pastori; Annalisa Salviato; Antonia Ricci; Calogero Terregino; Edoardo Giussani; Elisa Palumbo; Erika Giorgia Quaranta; Isabella Monne; Luca Tassoni |
| EPI_ISL_2099504, EPI_ISL_2099524, EPI_ISL_2099528 | UMC Groningen, Clinical Virology, Department of Medical Microbiology and Infection Prevention | UMC Groningen, Clinical Virology, Department of Medical Microbiology and Infection Prevention | Alexander Friedrich; Coretta Van Leer-Buter; Erley Lizarazo-Forero; Hubert Niesters; Lilli Gard; Marjolein Knoester; Monika Fliss; Sigrid Rosema; Xuewei Zhou |
| EPI_ISL_3409285, EPI_ISL_3409293, EPI_ISL_3409318, EPI_ISL_3409852, EPI_ISL_3410081, EPI_ISL_3410932 | UMass Memorial Medical Center | Center for Microbiome Research | Doyle V. Ward; Karl J. Simon; Richard T. Ellison III |
| EPI_ISL_5603283, EPI_ISL_5603285 | UNIDADE BASICA DE SAUDE ALTO BRILHANTE | Analytical Competence Molecular Epidemiology Lab/ACME, Oswaldo Cruz Foundation, Ceara (FIOCRUZ CE) | Carlos Leonardo de Aragao Araujo; Cecilia Leite Costa & Eduardo Ruback dos Santos on behalf of COVID-19 FIOCRUZ Genomic Network; Cleber Furtado Aksenen; Fabio Miyajima; Fernando Braga Stehling; Francisco Eder de Moura Lopes; Igor Oliveira Duarte; Jamille Maria Mendes Bezerra; Joaquim Cesar do Nascimento Sousa Junior; Pedro Miguel Carneiro Jeronimo; Suzana Porto Almeida; Thais Ferreira de Oliveira; Thais de Oliveira Costa; Ticiane Cavalcante de Souza; Veridiana Pessoa Miyajima |
| EPI_ISL_5603288 | UNIDADE BASICA DE SAUDE ALTO NELANDIA | Analytical Competence Molecular Epidemiology Lab/ACME, Oswaldo Cruz Foundation, Ceara (FIOCRUZ CE) | Carlos Leonardo de Aragao Araujo; Cecilia Leite Costa & Eduardo Ruback dos Santos on behalf of COVID-19 FIOCRUZ Genomic Network; Cleber Furtado Aksenen; Fabio Miyajima; Fernando Braga Stehling; Francisco Eder de Moura Lopes; Igor Oliveira Duarte; Jamille Maria Mendes Bezerra; Joaquim Cesar do Nascimento Sousa Junior; Pedro Miguel Carneiro Jeronimo; Suzana Porto Almeida; Thais Ferreira de Oliveira; Thais de Oliveira Costa; Ticiane Cavalcante de Souza; Veridiana Pessoa Miyajima |
| EPI_ISL_5603242 | UNIDADE BASICA DE SAUDE CACIMBAS | Analytical Competence Molecular Epidemiology Lab/ACME, Oswaldo Cruz Foundation, Ceara (FIOCRUZ CE) | Carlos Leonardo de Aragao Araujo; Cecilia Leite Costa & Eduardo Ruback dos Santos on behalf of COVID-19 FIOCRUZ Genomic Network; Cleber Furtado Aksenen; Fabio Miyajima; Fernando Braga Stehling; Francisco Eder de Moura Lopes; Igor Oliveira Duarte; Jamille Maria Mendes Bezerra; Joaquim Cesar do Nascimento Sousa Junior; Pedro Miguel Carneiro Jeronimo; Suzana Porto Almeida; Thais Ferreira de Oliveira; Thais de Oliveira Costa; Ticiane Cavalcante de Souza; Veridiana Pessoa Miyajima |
| EPI_ISL_5603272, EPI_ISL_5603276 | UNIDADE BASICA DE SAUDE COLIBRIS | Analytical Competence Molecular Epidemiology Lab/ACME, Oswaldo Cruz Foundation, Ceara (FIOCRUZ CE) | Carlos Leonardo de Aragao Araujo; Cecilia Leite Costa & Eduardo Ruback dos Santos on behalf of COVID-19 FIOCRUZ Genomic Network; Cleber Furtado Aksenen; Fabio Miyajima; Fernando Braga Stehling; Francisco Eder de Moura Lopes; Igor Oliveira Duarte; Jamille Maria Mendes Bezerra; Joaquim Cesar do Nascimento Sousa Junior; Pedro Miguel Carneiro Jeronimo; Suzana Porto Almeida; Thais Ferreira de Oliveira; Thais de Oliveira Costa; Ticiane Cavalcante de Souza; Veridiana Pessoa Miyajima |
| EPI_ISL_5603239 | UNIDADE BASICA DE SAUDE DA GIRILANDIA | Analytical Competence Molecular Epidemiology Lab/ACME, Oswaldo Cruz Foundation, Ceara (FIOCRUZ CE) | Carlos Leonardo de Aragao Araujo; Cecilia Leite Costa & Eduardo Ruback dos Santos on behalf of COVID-19 FIOCRUZ Genomic Network; Cleber Furtado Aksenen; Fabio Miyajima; Fernando Braga Stehling; Francisco Eder de Moura Lopes; Igor Oliveira Duarte; Jamille Maria Mendes Bezerra; Joaquim Cesar do Nascimento Sousa Junior; Pedro Miguel Carneiro Jeronimo; Suzana Porto Almeida; Thais Ferreira de Oliveira; Thais de Oliveira Costa; Ticiane Cavalcante de Souza; Veridiana Pessoa Miyajima |
| EPI_ISL_5603230 | UNIDADE BASICA DE SAUDE DA SEDE | Analytical Competence Molecular Epidemiology Lab/ACME, Oswaldo Cruz Foundation, Ceara (FIOCRUZ CE) | Carlos Leonardo de Aragao Araujo; Cecilia Leite Costa & Eduardo Ruback dos Santos on behalf of COVID-19 FIOCRUZ Genomic Network; Cleber Furtado Aksenen; Fabio Miyajima; Fernando Braga Stehling; Francisco Eder de Moura Lopes; Igor Oliveira Duarte; Jamille Maria Mendes Bezerra; Joaquim Cesar do Nascimento Sousa Junior; Pedro Miguel Carneiro Jeronimo; Suzana Porto Almeida; Thais Ferreira de Oliveira; Thais de Oliveira Costa; Ticiane Cavalcante de Souza; Veridiana Pessoa Miyajima |
| EPI_ISL_2345203, EPI_ISL_2345204 | UNIDADE BASICA DE SAUDE DE TARUMA | Instituto Butantan / ESALQ-Piracicaba | Antonio Jorge Martins; Claudia Renata dos Santos Barros; David Schlesinger; Debora Botequio Moretti; Dimas Tadeu Covas; Elaine Cristina Marqueze; Elaine Vieira Santos; Evandra Strazza Rodrigues; Heidge Fukumasu; Jayme Augusto de Souza-Neto; José Salvatore Leister Patané; Luiz Alcantara; Luiz Lehmann Coutinho; Maria Carolina Elias; Maurício Lacerda Nogueira; Rafael dos Santos Bezerra; Raul Machado Neto; Rejane Maria Tommasini Grotto; Ricardo Haddad; Sandra Coccuzzo Sampaio Vessoni; Simone Kashima; Svetoslav Nanev Slavov; Vincent Louis Viala |
| EPI_ISL_5603237 | UNIDADE BASICA DE SAUDE DESERTO | Analytical Competence Molecular Epidemiology Lab/ACME, Oswaldo Cruz Foundation, Ceara (FIOCRUZ CE) | Carlos Leonardo de Aragao Araujo; Cecilia Leite Costa & Eduardo Ruback dos Santos on behalf of COVID-19 FIOCRUZ Genomic Network; Cleber Furtado Aksenen; Fabio Miyajima; Fernando Braga Stehling; Francisco Eder de Moura Lopes; Igor Oliveira Duarte; Jamille Maria Mendes Bezerra; Joaquim Cesar do Nascimento Sousa Junior; Pedro Miguel Carneiro Jeronimo; Suzana Porto Almeida; Thais Ferreira de Oliveira; Thais de Oliveira Costa; Ticiane Cavalcante de Souza; Veridiana Pessoa Miyajima |
| EPI_ISL_2209732, EPI_ISL_2210126, EPI_ISL_2345770 | UNIDADE BASICA DE SAUDE II ESTRELA DO NORTE | Instituto Butantan / Mendelics | Antonio Jorge Martins; Claudia Renata dos Santos Barros; David Schlesinger; Debora Botequio Moretti; Dimas Tadeu Covas; Elaine Cristina Marqueze; Elaine Vieira Santos; Evandra Strazza Rodrigues; Heidge Fukumasu; Jayme Augusto de Souza-Neto; José Salvatore Leister Patané; Luiz Alcantara; Luiz Lehmann Coutinho; Maria Carolina Elias; Maurício Lacerda Nogueira; Rafael dos Santos Bezerra; Raul Machado Neto; Rejane Maria Tommasini Grotto; Ricardo Haddad; Sandra Coccuzzo Sampaio Vessoni; Simone Kashima; Svetoslav Nanev Slavov; Vincent Louis Viala |
| EPI_ISL_5603277, EPI_ISL_5603280 | UNIDADE BASICA DE SAUDE JOSE OSIMO | Analytical Competence Molecular Epidemiology Lab/ACME, Oswaldo Cruz Foundation, Ceara (FIOCRUZ CE) | Carlos Leonardo de Aragao Araujo; Cecilia Leite Costa & Eduardo Ruback dos Santos on behalf of COVID-19 FIOCRUZ Genomic Network; Cleber Furtado Aksenen; Fabio Miyajima; Fernando Braga Stehling; Francisco Eder de Moura Lopes; Igor Oliveira Duarte; Jamille Maria Mendes Bezerra; Joaquim Cesar do Nascimento Sousa Junior; Pedro Miguel Carneiro Jeronimo; Suzana Porto Almeida; Thais Ferreira de Oliveira; Thais de Oliveira Costa; Ticiane Cavalcante de Souza; Veridiana Pessoa Miyajima |
| EPI_ISL_5603269, EPI_ISL_5603307 | UNIDADE BASICA DE SAUDE TAUAZINHO | Analytical Competence Molecular Epidemiology Lab/ACME, Oswaldo Cruz Foundation, Ceara (FIOCRUZ CE) | Carlos Leonardo de Aragao Araujo; Cecilia Leite Costa & Eduardo Ruback dos Santos on behalf of COVID-19 FIOCRUZ Genomic Network; Cleber Furtado Aksenen; Fabio Miyajima; Fernando Braga Stehling; Francisco Eder de Moura Lopes; Igor Oliveira Duarte; Jamille Maria Mendes Bezerra; Joaquim Cesar do Nascimento Sousa Junior; Pedro Miguel Carneiro Jeronimo; Suzana Porto Almeida; Thais Ferreira de Oliveira; Thais de Oliveira Costa; Ticiane Cavalcante de Souza; Veridiana Pessoa Miyajima |
| EPI_ISL_2345100 | UNIDADE DE PRONTO ATENDIMENTO UPA DRA ANA OLIVIA BENTIVOGLIO | Instituto Butantan / ESALQ-Piracicaba | Antonio Jorge Martins; Claudia Renata dos Santos Barros; David Schlesinger; Debora Botequio Moretti; Dimas Tadeu Covas; Elaine Cristina Marqueze; Elaine Vieira Santos; Evandra Strazza Rodrigues; Heidge Fukumasu; Jayme Augusto de Souza-Neto; José Salvatore Leister Patané; Luiz Alcantara; Luiz Lehmann Coutinho; Maria Carolina Elias; Maurício Lacerda Nogueira; Rafael dos Santos Bezerra; Raul Machado Neto; Rejane Maria Tommasini Grotto; Ricardo Haddad; Sandra Coccuzzo Sampaio Vessoni; Simone Kashima; Svetoslav Nanev Slavov; Vincent Louis Viala |
| EPI_ISL_2345210, EPI_ISL_2345212, EPI_ISL_2345219, EPI_ISL_2345227 | UNIDADE DE SAUDE DA FAMILIA NOVO HORIZONTE | Instituto Butantan / ESALQ-Piracicaba | Antonio Jorge Martins; Claudia Renata dos Santos Barros; David Schlesinger; Debora Botequio Moretti; Dimas Tadeu Covas; Elaine Cristina Marqueze; Elaine Vieira Santos; Evandra Strazza Rodrigues; Heidge Fukumasu; Jayme Augusto de Souza-Neto; José Salvatore Leister Patané; Luiz Alcantara; Luiz Lehmann Coutinho; Maria Carolina Elias; Maurício Lacerda Nogueira; Rafael dos Santos Bezerra; Raul Machado Neto; Rejane Maria Tommasini Grotto; Ricardo Haddad; Sandra Coccuzzo Sampaio Vessoni; Simone Kashima; Svetoslav Nanev Slavov; Vincent Louis Viala |
| EPI_ISL_2345269, EPI_ISL_2345270, EPI_ISL_2345272, EPI_ISL_2345275, EPI_ISL_2345277, EPI_ISL_2345279, EPI_ISL_2345281, EPI_ISL_2345284, EPI_ISL_2345285, EPI_ISL_2345286, EPI_ISL_2345287 | | | |
| see above | UNIDADE DE VIGILANCIA EM SAUDE | Instituto Butantan / ESALQ-Piracicaba | Antonio Jorge Martins; Claudia Renata dos Santos Barros; David Schlesinger; Debora Botequio Moretti; Dimas Tadeu Covas; Elaine Cristina Marqueze; Elaine Vieira Santos; Evandra Strazza Rodrigues; Heidge Fukumasu; Jayme Augusto de Souza-Neto; José Salvatore Leister Patané; Luiz Alcantara; Luiz Lehmann Coutinho; Maria Carolina Elias; Maurício Lacerda Nogueira; Rafael dos Santos Bezerra; Raul Machado Neto; Rejane Maria Tommasini Grotto; Ricardo Haddad; Sandra Coccuzzo Sampaio Vessoni; Simone Kashima; Svetoslav Nanev Slavov; Vincent Louis Viala |
| EPI_ISL_2345079, EPI_ISL_2345109, EPI_ISL_2345136, EPI_ISL_2345139, EPI_ISL_2345160 | UNIDADE REFERENCIAL SUDOESTE | Instituto Butantan / ESALQ-Piracicaba | Antonio Jorge Martins; Claudia Renata dos Santos Barros; David Schlesinger; Debora Botequio Moretti; Dimas Tadeu Covas; Elaine Cristina Marqueze; Elaine Vieira Santos; Evandra Strazza Rodrigues; Heidge Fukumasu; Jayme Augusto de Souza-Neto; José Salvatore Leister Patané; Luiz Alcantara; Luiz Lehmann Coutinho; Maria Carolina Elias; Maurício Lacerda Nogueira; Rafael dos Santos Bezerra; Raul Machado Neto; Rejane Maria Tommasini Grotto; Ricardo Haddad; Sandra Coccuzzo Sampaio Vessoni; Simone Kashima; Svetoslav Nanev Slavov; Vincent Louis Viala |
| EPI_ISL_2345247 | UNIDADE SAUDE FAMILIA DR PIERRE ANTONIO COPPIETERS AREALVA | Instituto Butantan / ESALQ-Piracicaba | Antonio Jorge Martins; Claudia Renata dos Santos Barros; David Schlesinger; Debora Botequio Moretti; Dimas Tadeu Covas; Elaine Cristina Marqueze; Elaine Vieira Santos; Evandra Strazza Rodrigues; Heidge Fukumasu; Jayme Augusto de Souza-Neto; José Salvatore Leister Patané; Luiz Alcantara; Luiz Lehmann Coutinho; Maria Carolina Elias; Maurício Lacerda Nogueira; Rafael dos Santos Bezerra; Raul Machado Neto; Rejane Maria Tommasini Grotto; Ricardo Haddad; Sandra Coccuzzo Sampaio Vessoni; Simone Kashima; Svetoslav Nanev Slavov; Vincent Louis Viala |
| EPI_ISL_2208916 | UNIDADE SENTINELA COVID19 | Instituto Butantan / Mendelics | Antonio Jorge Martins; Claudia Renata dos Santos Barros; David Schlesinger; Debora Botequio Moretti; Dimas Tadeu Covas; Elaine Cristina Marqueze; Elaine Vieira Santos; Evandra Strazza Rodrigues; Heidge Fukumasu; Jayme Augusto de Souza-Neto; José Salvatore Leister Patané; Luiz Alcantara; Luiz Lehmann Coutinho; Maria Carolina Elias; Maurício Lacerda Nogueira; Rafael dos Santos Bezerra; Raul Machado Neto; Rejane Maria Tommasini Grotto; Ricardo Haddad; Sandra Coccuzzo Sampaio Vessoni; Simone Kashima; Svetoslav Nanev Slavov; Vincent Louis Viala |
| EPI_ISL_3385823 | UNIVERSIDAD DE CALDAS | Instituto Nacional de Salud- Dirección de Investigación en Salud Pública | Carlos Franco-Muñoz; Carmen Osorio; Diana Malo; Diego A. Álvarez-Díaz; Diego Andrés Prada; Gerardo Santamaría; Hector Alejandro Ruiz-Moreno; Jhonnatan Reales-González; Jorge Rivera; Juan Camilo Martinez; Julian Naizaque; Katherine Laiton-Donato; Lisseth Pardo; Magdalena Wiesner; Marcela Mercado-Reyes; Maria T. Herrera-Sepúlveda; Marta Lopez Blanco; Martha Lucia Ospina Martinez; Paola Rojas; Sergio Gomez; Sheryll Corchuelo; Ángela Alarcon Cruz |
| EPI_ISL_2345268 | UPA IPIRANGA | Instituto Butantan / ESALQ-Piracicaba | Antonio Jorge Martins; Claudia Renata dos Santos Barros; David Schlesinger; Debora Botequio Moretti; Dimas Tadeu Covas; Elaine Cristina Marqueze; Elaine Vieira Santos; Evandra Strazza Rodrigues; Heidge Fukumasu; Jayme Augusto de Souza-Neto; José Salvatore Leister Patané; Luiz Alcantara; Luiz Lehmann Coutinho; Maria Carolina Elias; Maurício Lacerda Nogueira; Rafael dos Santos Bezerra; Raul Machado Neto; Rejane Maria Tommasini Grotto; Ricardo Haddad; Sandra Coccuzzo Sampaio Vessoni; Simone Kashima; Svetoslav Nanev Slavov; Vincent Louis Viala |
| EPI_ISL_2402291, | US Air Force School of Aerospace | US Air Force School of Aerospace | Amanda Javorina; Anthony Fries; Carol Garrett; Clarise Starr; Elizabeth Macias; Jennifer Meyer; Sarah Purves; William Gruner |

| | | | |
|---|---|---|---|
| EPI_ISL_2426709, EPI_ISL_2426773 | Medicine | Medicine | |
| EPI_ISL_2131372, EPI_ISL_2235325, EPI_ISL_2235336 | USCA Avezzano | Istituto Zooprofilattico Sperimentale dell'Abruzzo e Molise "G. Caporale" | Ancora M; Calistri P; Cammà C; Caporale M; Curini V; Delli Compagni E; Di Domenico M; Di Lollo Valeria; Di Pasquale A; Lorusso A; Mangone I; Marcacci M; Puglia I; Rinaldi A; Savini G; Scialabba S |
| EPI_ISL_2131378 | USCA Tagliacozzo | Istituto Zooprofilattico Sperimentale dell'Abruzzo e Molise "G. Caporale" | Ancora M; Calistri P; Cammà C; Caporale M; Curini V; Delli Compagni E; Di Domenico M; Di Lollo Valeria; Di Pasquale A; Lorusso A; Mangone I; Marcacci M; Puglia I; Rinaldi A; Savini G; Scialabba S |
| EPI_ISL_2020974, EPI_ISL_2086337, EPI_ISL_2086352, EPI_ISL_2086356, EPI_ISL_2086360, EPI_ISL_2086370, EPI_ISL_2086397, EPI_ISL_2086404, EPI_ISL_2086416, EPI_ISL_2086422, EPI_ISL_2104831, EPI_ISL_2104842, EPI_ISL_2104865, EPI_ISL_2104869, EPI_ISL_2104915, EPI_ISL_2104956, EPI_ISL_2104967, EPI_ISL_2104971, EPI_ISL_2104979, EPI_ISL_2104984, EPI_ISL_2104990, EPI_ISL_2104999, EPI_ISL_2105034, EPI_ISL_2105038, EPI_ISL_2105039, EPI_ISL_2105055, EPI_ISL_2105081, EPI_ISL_2233113, EPI_ISL_2233117, EPI_ISL_2233124, EPI_ISL_2233152, EPI_ISL_2233153, EPI_ISL_2233155, EPI_ISL_2233172, EPI_ISL_2233179, EPI_ISL_2233184, EPI_ISL_2233185, EPI_ISL_2233194, EPI_ISL_2233208, EPI_ISL_2233225, EPI_ISL_2502832, EPI_ISL_2502833, EPI_ISL_2502836, EPI_ISL_2502847, EPI_ISL_2502868, EPI_ISL_2502870, EPI_ISL_2502872, EPI_ISL_2502881, EPI_ISL_2502913, EPI_ISL_2502919, EPI_ISL_2502924, EPI_ISL_2502932, EPI_ISL_2502943, EPI_ISL_2502944, EPI_ISL_2502952, EPI_ISL_2502958, EPI_ISL_2502971, EPI_ISL_2503004, EPI_ISL_2503006, EPI_ISL_2503012, EPI_ISL_2503013, EPI_ISL_2503020, EPI_ISL_2503028, EPI_ISL_2503055, EPI_ISL_2503062, EPI_ISL_2503065, EPI_ISL_2503066, EPI_ISL_2503083, EPI_ISL_2503092, EPI_ISL_2503110, EPI_ISL_2503115, EPI_ISL_2503120, EPI_ISL_2503130, EPI_ISL_2503133, EPI_ISL_2618128, EPI_ISL_2618154, EPI_ISL_2618156, EPI_ISL_2618157, EPI_ISL_2618169, EPI_ISL_2618170, EPI_ISL_2618177, EPI_ISL_2618180, EPI_ISL_2618188, EPI_ISL_2618205, EPI_ISL_2618214, EPI_ISL_2618258, EPI_ISL_2618262, EPI_ISL_2618269, EPI_ISL_2618278, EPI_ISL_2618297, EPI_ISL_2618311, EPI_ISL_2618314, EPI_ISL_2618319, EPI_ISL_2618363, EPI_ISL_2618403, EPI_ISL_2618405, EPI_ISL_2618412, EPI_ISL_2618438, EPI_ISL_2618442, EPI_ISL_2618459, EPI_ISL_2618471, EPI_ISL_2618481, EPI_ISL_2618499, EPI_ISL_2618530, EPI_ISL_2618536, EPI_ISL_2618583, EPI_ISL_2618585, EPI_ISL_2618592, EPI_ISL_2618595, EPI_ISL_2618597, EPI_ISL_2618620, EPI_ISL_2618631, EPI_ISL_2618636, EPI_ISL_2618637, EPI_ISL_2618638, EPI_ISL_2618648, EPI_ISL_2618649, EPI_ISL_2618653, EPI_ISL_2618660, EPI_ISL_2618665 | | | |
| see above | UW Virology Lab | UW Virology Lab | Alexander Greninger; Hong Xie; Keith R Jerome; Lasata Shrestha; Meei-Li Huang; Michelle Lin; Nathan Breit; Noah R. Baker; Patrick Mathias; Pavitra Roychoudhury; Ricardo Perez; Robert J. Livingston; Sean Ellis; Shah Mohamed Bakhash; Tien V. Nguyen |
| EPI_ISL_5801452 | Ubs Alambari | Instituto Butantan | Antonio Jorge Martins; Claudia Renata dos Santos Barros; David Schlesinger; Debora Botequio Moretti; Dimas Tadeu Covas; Elaine Cristina Marqueze; Elaine Vieira Santos; Evandra Strazza Rodrigues; Heidge Fukumasu; Jayme Augusto de Souza-Neto; José Salvatore Leister Patané; Luiz Alcantara; Luiz Lehmann Coutinho; Maria Carolina Elias; Maurício Lacerda Nogueira; Rafael dos Santos Bezerra; Raul Machado Neto; Rejane Maria Tommasini Grotto; Ricardo Haddad; Sandra Coccuzzo Sampaio Vessoni; Simone Kashima; Svetoslav Nanev Slavov; Vincent Louis Viala |
| EPI_ISL_5801205, EPI_ISL_5801206, EPI_ISL_5801209 | Ubs Bonfim | Instituto Butantan | Antonio Jorge Martins; Claudia Renata dos Santos Barros; David Schlesinger; Debora Botequio Moretti; Dimas Tadeu Covas; Elaine Cristina Marqueze; Elaine Vieira Santos; Evandra Strazza Rodrigues; Heidge Fukumasu; Jayme Augusto de Souza-Neto; José Salvatore Leister Patané; Luiz Alcantara; Luiz Lehmann Coutinho; Maria Carolina Elias; Maurício Lacerda Nogueira; Rafael dos Santos Bezerra; Raul Machado Neto; Rejane Maria Tommasini Grotto; Ricardo Haddad; Sandra Coccuzzo Sampaio Vessoni; Simone Kashima; Svetoslav Nanev Slavov; Vincent Louis Viala |
| EPI_ISL_5801275, EPI_ISL_5801276 | Ubs Dr Abelardo Pinheiro Guimaraes Scrpardo | Instituto Butantan | Antonio Jorge Martins; Claudia Renata dos Santos Barros; David Schlesinger; Debora Botequio Moretti; Dimas Tadeu Covas; Elaine Cristina Marqueze; Elaine Vieira Santos; Evandra Strazza Rodrigues; Heidge Fukumasu; Jayme Augusto de Souza-Neto; José Salvatore Leister Patané; Luiz Alcantara; Luiz Lehmann Coutinho; Maria Carolina Elias; Maurício Lacerda Nogueira; Rafael dos Santos Bezerra; Raul Machado Neto; Rejane Maria Tommasini Grotto; Ricardo Haddad; Sandra Coccuzzo Sampaio Vessoni; Simone Kashima; Svetoslav Nanev Slavov; Vincent Louis Viala |
| EPI_ISL_5801207, EPI_ISL_5801220, EPI_ISL_5801222, EPI_ISL_5801229, EPI_ISL_5801230, EPI_ISL_5801234, EPI_ISL_5801235, EPI_ISL_5801236, EPI_ISL_5801248, EPI_ISL_5801252, EPI_ISL_5801262 | | | |
| see above | Ubs Dr Alfredo Dantas De Souza Umuarama | Instituto Butantan | Antonio Jorge Martins; Claudia Renata dos Santos Barros; David Schlesinger; Debora Botequio Moretti; Dimas Tadeu Covas; Elaine Cristina Marqueze; Elaine Vieira Santos; Evandra Strazza Rodrigues; Heidge Fukumasu; Jayme Augusto de Souza-Neto; José Salvatore Leister Patané; Luiz Alcantara; Luiz Lehmann Coutinho; Maria Carolina Elias; Maurício Lacerda Nogueira; Rafael dos Santos Bezerra; Raul Machado Neto; Rejane Maria Tommasini Grotto; Ricardo Haddad; Sandra Coccuzzo Sampaio Vessoni; Simone Kashima; Svetoslav Nanev Slavov; Vincent Louis Viala |
| EPI_ISL_5801261 | Ubs Glenda Rodella Dumas | Instituto Butantan | Antonio Jorge Martins; Claudia Renata dos Santos Barros; David Schlesinger; Debora Botequio Moretti; Dimas Tadeu Covas; Elaine Cristina Marqueze; Elaine Vieira Santos; Evandra Strazza Rodrigues; Heidge Fukumasu; Jayme Augusto de Souza-Neto; José Salvatore Leister Patané; Luiz Alcantara; Luiz Lehmann Coutinho; Maria Carolina Elias; Maurício Lacerda Nogueira; Rafael dos Santos Bezerra; Raul Machado Neto; Rejane Maria Tommasini Grotto; Ricardo Haddad; Sandra Coccuzzo Sampaio Vessoni; Simone Kashima; Svetoslav Nanev Slavov; Vincent Louis Viala |
| EPI_ISL_5801475, EPI_ISL_5801476 | Ubs Ii De Alvares Machado | Instituto Butantan | Antonio Jorge Martins; Claudia Renata dos Santos Barros; David Schlesinger; Debora Botequio Moretti; Dimas Tadeu Covas; Elaine Cristina Marqueze; Elaine Vieira Santos; Evandra Strazza Rodrigues; Heidge Fukumasu; Jayme Augusto de Souza-Neto; José Salvatore Leister Patané; Luiz Alcantara; Luiz Lehmann Coutinho; Maria Carolina Elias; Maurício Lacerda Nogueira; Rafael dos Santos Bezerra; Raul Machado Neto; Rejane Maria Tommasini Grotto; Ricardo Haddad; Sandra Coccuzzo Sampaio Vessoni; Simone Kashima; Svetoslav Nanev Slavov; Vincent Louis Viala |
| EPI_ISL_5801454, EPI_ISL_5801456 | Ubs Ii Dr Jose Carlos Macuco Janini | Instituto Butantan | Antonio Jorge Martins; Claudia Renata dos Santos Barros; David Schlesinger; Debora Botequio Moretti; Dimas Tadeu Covas; Elaine Cristina Marqueze; Elaine Vieira Santos; Evandra Strazza Rodrigues; Heidge Fukumasu; Jayme Augusto de Souza-Neto; José Salvatore Leister Patané; Luiz Alcantara; Luiz Lehmann Coutinho; Maria Carolina Elias; Maurício Lacerda Nogueira; Rafael dos Santos Bezerra; Raul Machado Neto; Rejane Maria Tommasini Grotto; Ricardo Haddad; Sandra Coccuzzo Sampaio Vessoni; Simone Kashima; Svetoslav Nanev Slavov; Vincent Louis Viala |
| EPI_ISL_5801147 | Ubs Iii De Pariquera Acu Pariquera Acu | Instituto Butantan | Antonio Jorge Martins; Claudia Renata dos Santos Barros; David Schlesinger; Debora Botequio Moretti; Dimas Tadeu Covas; Elaine Cristina Marqueze; Elaine Vieira Santos; Evandra Strazza Rodrigues; Heidge Fukumasu; Jayme Augusto de Souza-Neto; José Salvatore Leister Patané; Luiz Alcantara; Luiz Lehmann Coutinho; Maria Carolina Elias; Maurício Lacerda Nogueira; Rafael dos Santos Bezerra; Raul Machado Neto; Rejane Maria Tommasini Grotto; Ricardo Haddad; Sandra Coccuzzo Sampaio Vessoni; Simone Kashima; Svetoslav Nanev Slavov; Vincent Louis Viala |
| EPI_ISL_5801278, EPI_ISL_5801282 | Ubs Iii Murutinga Do Sul | Instituto Butantan | Antonio Jorge Martins; Claudia Renata dos Santos Barros; David Schlesinger; Debora Botequio Moretti; Dimas Tadeu Covas; Elaine Cristina Marqueze; Elaine Vieira Santos; Evandra Strazza Rodrigues; Heidge Fukumasu; Jayme Augusto de Souza-Neto; José Salvatore Leister Patané; Luiz Alcantara; Luiz Lehmann Coutinho; Maria Carolina Elias; Maurício Lacerda Nogueira; Rafael dos Santos Bezerra; Raul Machado Neto; Rejane Maria Tommasini Grotto; Ricardo Haddad; Sandra Coccuzzo Sampaio Vessoni; Simone Kashima; Svetoslav Nanev Slavov; Vincent Louis Viala |
| EPI_ISL_5801324, EPI_ISL_5801325, EPI_ISL_5801326, EPI_ISL_5801329, EPI_ISL_5801330, EPI_ISL_5801331, EPI_ISL_5801332, EPI_ISL_5801333, EPI_ISL_5801334, EPI_ISL_5801335, EPI_ISL_5801336 | | | |
| see above | Ubs Iv Guararapes | Instituto Butantan | Antonio Jorge Martins; Claudia Renata dos Santos Barros; David Schlesinger; Debora Botequio Moretti; Dimas Tadeu Covas; Elaine Cristina Marqueze; Elaine Vieira Santos; Evandra Strazza Rodrigues; Heidge Fukumasu; Jayme Augusto de Souza-Neto; José Salvatore Leister Patané; Luiz Alcantara; Luiz Lehmann Coutinho; Maria Carolina Elias; Maurício Lacerda Nogueira; Rafael dos Santos Bezerra; Raul Machado Neto; Rejane Maria Tommasini Grotto; Ricardo Haddad; Sandra Coccuzzo Sampaio Vessoni; Simone Kashima; Svetoslav Nanev Slavov; Vincent Louis Viala |
| EPI_ISL_5801178 | Ubs Vila Ribeiro | Instituto Butantan | Antonio Jorge Martins; Claudia Renata dos Santos Barros; David Schlesinger; Debora Botequio Moretti; Dimas Tadeu Covas; Elaine Cristina Marqueze; Elaine Vieira Santos; Evandra Strazza Rodrigues; Heidge Fukumasu; Jayme Augusto de Souza-Neto; José Salvatore Leister Patané; Luiz Alcantara; Luiz Lehmann Coutinho; Maria Carolina Elias; Maurício Lacerda Nogueira; Rafael dos Santos Bezerra; Raul Machado Neto; Rejane Maria Tommasini Grotto; Ricardo Haddad; Sandra Coccuzzo Sampaio Vessoni; Simone Kashima; Svetoslav Nanev Slavov; Vincent Louis Viala |
| EPI_ISL_2970075 | Unidad de Investigacion Medica de Yucatan (UIMY) | Unidad de Genomica Avanzada | ; Alejandra Garcia-Gasca; Alejandra Hernandez-Teran; Alejandro Sanchez-Flores; Alfredo Herrera-Estrella; Alicia Ocaña-Mondragon; Andreu Comas-Garcia; Angel Gustavo Salas-Lais; Antonio Loza Roman; Bernardo Martinez-Miguel; Blanca Taboada; Brenda Irasema Maldonado-Meza; Bruno Gomez-Gil; Carla Ivon Herrera-Najera; Carlos F. Arias; Celia Boukadida; Celida Duque Molina; Celida Martinez- Rodriguez; Clara Esperanza Santacruz-Tinoco; Concepcion Grajales-Muñiz; Consorcio Mexicano de Vigilancia Genomica (CoViGen-Mex). Authors (in alphabetical order): Julio Elias Alvarado-Yaah; Cristobal Chaidez-Quiroz; Daniel Fregoso-Rueda; Daniel Lira Morales; Eduardo Becerril-Vargas; Fernando Fontove-Herrera; Fidencio Mejia-Nepomuceno; Francisco Pulido; Gloria Elena Espinosa-Ayala; Gloria Maria Molina-Salinas; Gloria Vazquez; Hector Esteban Paz-Juarez; Hector Montoya-Fuentes; Helen Haydee Fernanda Ramirez-Plascencia; Irvin Gonzalez-Lopez; Jean Pierre Gonzalez; Jesus Hernandez; Joel Armando Vazquez-Perez.; Jorge Salas-Hernandez; Jose Antonio Enciso-Moreno; Jose Arturo Martinez-Orozco; Jose Esteban Muñoz-Medina; Jose de Jesus Nuñez-Contreras; Juan Bautista Chale-Dzul; Julissa Enciso-Ibarra; Luis Alberto Ochoa-Carrera; Margarita Matias-Florentino; Maria Guadalupe Santiago-Mauricio; Maria Guadalupe de Jesus Mireles-Rivera; Mario Mujica-Sanchez; Marissa Perez-Garcia; Nelly Selem-Mojica; Pavel Isa; Ricardo Ciria Merce; Ricardo Grande; Rosa Maria Gutierrez Rios; Santiago avila-Rios; Selene Zarate; Susana Lopez; Veronica Mata-Haro; Victor Eduardo Garcia-Arias; Victor Hugo Borja-Aburto |
| EPI_ISL_2490367, EPI_ISL_2490368, EPI_ISL_2490369 | Unidad de Investigación Médica de Yucatán (UIMY) | Centro de Investigación en Enfermedades Infecciosas (CIENI), Instituto Nacional de Enfermedades Respiratorias (INER) | ; Alejandra Garcia-Gasca; Alejandra Hernández-Terán; Alejandro Sanchez-Flores; Alfredo Herrera-Estrella; Alicia Ocaña-Mondragón; Andreu Comas-García; Angel Gustavo Salas-Lais; Antonio Loza Román; Bernardo Martínez-Miguel; Blanca Taboada; Brenda Irasema Maldonado-Meza; Bruno Gomez-Gil; Carla Ivón Herrera-Najera; Carlos F. Arias; Celia Boukadida; Clara Esperanza Santacruz-Tinoco; Concepción Grajales-Muñiz; Consorcio Mexicano de Vigilancia Genómica (CoViGen-Mex). Authors (in alphabetical order): Julio Elias Alvarado-Yaah; Cristóbal Cháidez-Quiróz; Célida Duque Molina; Célida Martínez-Rodríguez; Daniel Fregoso-Rueda; Daniel Lira Morales; Eduardo Becerril-Vargas; Fernando Fontove-Herrera; Fidencio Mejía-Nepomuceno; Francisco Pulido; Gloria Elena Espinosa-Ayala; Gloria María Molina-Salinas; Gloria Vazquez; Hector Esteban Paz-Juárez; Hector Montoya-Fuentes; Helen Haydee Fernanda Ramírez-Plascencia; Irvin González-López; Jean Pierre González; Joel Armando Vázquez-Pérez.; Jorge Salas-Hernández; José Antonio Enciso-Moreno; José Arturo Martínez-Orozco; José Esteban Muñoz-Medina; José de Jesús Nuñez-Contreras; Juan Bautista Chale-Dzul; Julissa Enciso-Ibarra; Luis Alberto Ochoa-Carrera; Margarita Matías-Florentino; Mario Mújica-Sánchez; Marissa Perez-Garcia; María Guadalupe Santiago-Mauricio; María Guadalupe de Jesús Mireles-Rivera; Nelly Sélem-Mojica; Pavel Isa; Ricardo Ciria Merce; Ricardo Grande; Rosa María Gutierrez Rios; Santiago Ávila-Ríos; Selene Zárate; Susana Lopez; Victor Eduardo Garcia-Arias; Víctor Hugo Borja-Aburto |
| EPI_ISL_2402062 | Unidad de Investigación Médica de Yucatán (UIMY) | Unidad de Genomica Avanzada | Alejandro Sanchez-Flores; Alfredo Herrera-Estrella; Alicia Ocana-Mondragon; Angel Gustavo Salas-Lais; Bernardo Martinez-Miguel; Blanca Taboada; Brenda Irasema Maldonado-Meza; Carla Ivon Herrera-Najera; Carlos F. Arias; Celia Boukadida; Clara Esperanza Santacruz-Tinoco; Concepcion Grajales-Muniz; Consorcio Mexicano de Vigilancia Genomica (CoViGen-Mex). Authors (in alphabetical order): Julio Elias Alvarado-Yaah; Fernando Fontove-Herrera; Francisco Pulido; Gloria Elena Espinoza-Ayala; Gloria Maria Molina-Salinas; Gloria Vazquez; Hector Esteban Paz-Juarez; Hector Montoya-Fuentes; Helen Haydee Fernanda Ramirez-Plascencia; Jorge Ivan Salinal-Nevarez; Jose Antonio Enciso-Moreno; Jose Esteban Munoz-Medina; Jose de Jesus Nunez-Contreras; Juan Bautista Chale-Dzul; Luis Alberto Ochoa-Carrera; Margarita Matias-Florentino; Maria Guadalupe Santiago-Mauricio; Maria Guadalupe de Jesus Mireles-Rivera; Nelly Selem-Mojica; Pavel Isa; Ricardo Grande; Santiago Avila-Rios; Victor Hugo Borja-Aburto |
| EPI_ISL_5802700, EPI_ISL_5802715 | Unidade Basica De Saude De Ipeuna | Instituto Butantan | Antonio Jorge Martins; Claudia Renata dos Santos Barros; David Schlesinger; Debora Botequio Moretti; Dimas Tadeu Covas; Elaine Cristina Marqueze; Elaine Vieira Santos; Evandra Strazza Rodrigues; Heidge Fukumasu; Jayme Augusto de Souza-Neto; José Salvatore Leister Patané; Luiz Alcantara; Luiz Lehmann Coutinho; Maria Carolina Elias; Maurício Lacerda Nogueira; Rafael dos Santos Bezerra; Raul Machado Neto; Rejane Maria Tommasini Grotto; Ricardo Haddad; Sandra Coccuzzo Sampaio Vessoni; Simone Kashima; Svetoslav Nanev Slavov; Vincent Louis Viala |
| EPI_ISL_5801296, EPI_ISL_5801297 | Unidade Basica De Saude De Taruma | Instituto Butantan | Antonio Jorge Martins; Claudia Renata dos Santos Barros; David Schlesinger; Debora Botequio Moretti; Dimas Tadeu Covas; Elaine Cristina Marqueze; Elaine Vieira Santos; Evandra Strazza Rodrigues; Heidge Fukumasu; Jayme Augusto de Souza-Neto; José Salvatore Leister Patané; Luiz Alcantara; Luiz Lehmann Coutinho; Maria Carolina Elias; Maurício Lacerda Nogueira; Rafael dos Santos Bezerra; Raul Machado Neto; Rejane Maria Tommasini Grotto; Ricardo Haddad; Sandra Coccuzzo Sampaio Vessoni; Simone Kashima; Svetoslav Nanev Slavov; Vincent Louis Viala |
| EPI_ISL_5802704, EPI_ISL_5802705 | Unidade Basica De Saude Dr Matheus Gabriel Bonassa | Instituto Butantan | Antonio Jorge Martins; Claudia Renata dos Santos Barros; David Schlesinger; Debora Botequio Moretti; Dimas Tadeu Covas; Elaine Cristina Marqueze; Elaine Vieira Santos; Evandra Strazza Rodrigues; Heidge Fukumasu; Jayme Augusto de Souza-Neto; José Salvatore Leister Patané; Luiz Alcantara; Luiz Lehmann Coutinho; Maria Carolina Elias; Maurício Lacerda Nogueira; Rafael dos Santos Bezerra; Raul Machado Neto; Rejane Maria Tommasini Grotto; Ricardo Haddad; Sandra Coccuzzo Sampaio Vessoni; Simone Kashima; Svetoslav Nanev Slavov; Vincent Louis Viala |
| EPI_ISL_5801453, EPI_ISL_5801458, EPI_ISL_5801463 | Unidade Basica De Saude Ii Estrela Do Norte | Instituto Butantan | Antonio Jorge Martins; Claudia Renata dos Santos Barros; David Schlesinger; Debora Botequio Moretti; Dimas Tadeu Covas; Elaine Cristina Marqueze; Elaine Vieira Santos; Evandra Strazza Rodrigues; Heidge Fukumasu; Jayme Augusto de Souza-Neto; José Salvatore Leister Patané; Luiz Alcantara; Luiz Lehmann Coutinho; Maria Carolina Elias; Maurício Lacerda Nogueira; Rafael dos Santos Bezerra; Raul Machado Neto; Rejane Maria Tommasini Grotto; Ricardo Haddad; Sandra Coccuzzo Sampaio Vessoni; Simone Kashima; Svetoslav Nanev Slavov; Vincent Louis Viala |
| EPI_ISL_5801125, EPI_ISL_5801128 | Unidade De Pronto Atendimento Upa Dra Ana Olivia Bentivoglio | Instituto Butantan | Antonio Jorge Martins; Claudia Renata dos Santos Barros; David Schlesinger; Debora Botequio Moretti; Dimas Tadeu Covas; Elaine Cristina Marqueze; Elaine Vieira Santos; Evandra Strazza Rodrigues; Heidge Fukumasu; Jayme Augusto de Souza-Neto; José Salvatore Leister Patané; Luiz Alcantara; Luiz Lehmann Coutinho; Maria Carolina Elias; Maurício Lacerda Nogueira; Rafael dos Santos Bezerra; Raul Machado Neto; Rejane Maria Tommasini Grotto; Ricardo Haddad; Sandra Coccuzzo Sampaio Vessoni; Simone Kashima; Svetoslav Nanev Slavov; Vincent Louis Viala |
| EPI_ISL_5801304, EPI_ISL_5801307, EPI_ISL_5801320, EPI_ISL_5801328 | Unidade De Saude Da Familia Novo Horizonte | Instituto Butantan | Antonio Jorge Martins; Claudia Renata dos Santos Barros; David Schlesinger; Debora Botequio Moretti; Dimas Tadeu Covas; Elaine Cristina Marqueze; Elaine Vieira Santos; Evandra Strazza Rodrigues; Heidge Fukumasu; Jayme Augusto de Souza-Neto; José Salvatore Leister Patané; Luiz Alcantara; Luiz Lehmann Coutinho; Maria Carolina Elias; Maurício Lacerda Nogueira; Rafael dos Santos Bezerra; Raul Machado Neto; Rejane Maria Tommasini Grotto; Ricardo Haddad; Sandra Coccuzzo Sampaio Vessoni; Simone Kashima; Svetoslav Nanev Slavov; Vincent Louis Viala |
| EPI_ISL_5801380, EPI_ISL_5801381, EPI_ISL_5801383, EPI_ISL_5801386, EPI_ISL_5801388, EPI_ISL_5801390, EPI_ISL_5801392, EPI_ISL_5801395, EPI_ISL_5801396, EPI_ISL_5801397, EPI_ISL_5801398 | | | |
| see above | Unidade De Vigilancia Em Saude | Instituto Butantan | Antonio Jorge Martins; Claudia Renata dos Santos Barros; David Schlesinger; Debora Botequio Moretti; Dimas Tadeu Covas; Elaine Cristina Marqueze; Elaine Vieira Santos; Evandra Strazza Rodrigues; Heidge Fukumasu; Jayme Augusto de Souza-Neto; José Salvatore Leister Patané; Luiz Alcantara; Luiz Lehmann Coutinho; Maria Carolina Elias; Maurício Lacerda Nogueira; Rafael dos Santos Bezerra; Raul Machado Neto; Rejane Maria Tommasini Grotto; Ricardo Haddad; Sandra Coccuzzo Sampaio Vessoni; Simone Kashima; Svetoslav Nanev Slavov; Vincent Louis Viala |
| EPI_ISL_5802708 | Unidade De Vigilancia Epidemiologica | Instituto Butantan | Antonio Jorge Martins; Claudia Renata dos Santos Barros; David Schlesinger; Debora Botequio Moretti; Dimas Tadeu Covas; Elaine Cristina Marqueze; Elaine Vieira Santos; Evandra Strazza Rodrigues; Heidge Fukumasu; Jayme Augusto de Souza-Neto; José Salvatore Leister Patané; Luiz Alcantara; Luiz Lehmann Coutinho; Maria Carolina Elias; Maurício Lacerda Nogueira; Rafael dos Santos Bezerra; Raul Machado Neto; Rejane Maria Tommasini Grotto; Ricardo Haddad; Sandra Coccuzzo Sampaio Vessoni; Simone Kashima; Svetoslav Nanev Slavov; Vincent Louis Viala |
| EPI_ISL_5802695, EPI_ISL_5802717 | Unidade Mista De Saude Mombuca | Instituto Butantan | Antonio Jorge Martins; Claudia Renata dos Santos Barros; David Schlesinger; Debora Botequio Moretti; Dimas Tadeu Covas; Elaine Cristina Marqueze; Elaine Vieira Santos; Evandra Strazza Rodrigues; Heidge Fukumasu; Jayme Augusto de Souza-Neto; José Salvatore Leister Patané; Luiz Alcantara; Luiz Lehmann Coutinho; Maria Carolina Elias; Maurício Lacerda Nogueira; Rafael dos Santos Bezerra; Raul Machado Neto; Rejane Maria Tommasini Grotto; Ricardo Haddad; Sandra Coccuzzo Sampaio Vessoni; Simone Kashima; Svetoslav Nanev Slavov; Vincent Louis Viala |
| EPI_ISL_5801102, EPI_ISL_5801142, EPI_ISL_5801180, EPI_ISL_5801185, EPI_ISL_5801226 | Unidade Referencial Sudoeste | Instituto Butantan | Antonio Jorge Martins; Claudia Renata dos Santos Barros; David Schlesinger; Debora Botequio Moretti; Dimas Tadeu Covas; Elaine Cristina Marqueze; Elaine Vieira Santos; Evandra Strazza Rodrigues; Heidge Fukumasu; Jayme Augusto de Souza-Neto; José Salvatore Leister Patané; Luiz Alcantara; Luiz Lehmann Coutinho; Maria Carolina Elias; Maurício Lacerda Nogueira; Rafael dos Santos Bezerra; Raul Machado Neto; Rejane Maria Tommasini Grotto; Ricardo Haddad; Sandra Coccuzzo Sampaio Vessoni; Simone Kashima; Svetoslav Nanev Slavov; Vincent Louis Viala |
| EPI_ISL_5801356 | Unidade Saude Familia Dr Pierre Antonio Coppieters Arealva | Instituto Butantan | Antonio Jorge Martins; Claudia Renata dos Santos Barros; David Schlesinger; Debora Botequio Moretti; Dimas Tadeu Covas; Elaine Cristina Marqueze; Elaine Vieira Santos; Evandra Strazza Rodrigues; Heidge Fukumasu; Jayme Augusto de Souza-Neto; José Salvatore Leister Patané; Luiz Alcantara; Luiz Lehmann Coutinho; Maria Carolina Elias; Maurício Lacerda Nogueira; Rafael dos Santos Bezerra; Raul Machado Neto; Rejane Maria Tommasini Grotto; Ricardo Haddad; Sandra Coccuzzo Sampaio Vessoni; Simone Kashima; Svetoslav Nanev Slavov; Vincent Louis Viala |
| EPI_ISL_5801423 | Unidade Sentinela Covid19 | Instituto Butantan | Antonio Jorge Martins; Claudia Renata dos Santos Barros; David Schlesinger; Debora Botequio Moretti; Dimas Tadeu Covas; Elaine Cristina Marqueze; Elaine Vieira Santos; Evandra Strazza Rodrigues; Heidge Fukumasu; Jayme Augusto de Souza-Neto; José Salvatore Leister Patané; Luiz Alcantara; Luiz Lehmann Coutinho; Maria Carolina Elias; Maurício Lacerda Nogueira; Rafael dos Santos Bezerra; Raul Machado Neto; Rejane Maria Tommasini Grotto; Ricardo Haddad; Sandra Coccuzzo Sampaio Vessoni; Simone Kashima; Svetoslav Nanev Slavov; Vincent Louis Viala |
| EPI_ISL_2691641 | Unidade de apoio ao diagnostico da COVID - UNADIG | Bioinformatics Laboratory / LNCC | Alessandra P Lamarca; Alexandra L Gerber; Amilcar Tanuri; Ana Paula de C Guimaraes; Ana Tereza R Vasconcelos; Andrea Cony Cavalcanti; Caio Luiz Pereira Ribeiro; Cassia Alves; Cintia Policarpo; Claudia Maria Braga de Mello; Cristiane Gomes da Silva; Diana Mariani; Douglas Terra Machado; Flavio Dias da Silva; Gleidson da Silva de Oliveira; Leandro Magalhaes de Souza; Liliane Cavalcante; Luiz G P de Almeida; Marcio Henrique de Oliveira Garcia; Mario Sergio Ribeiro; Ronaldo da Silva F Jr; Silvia Carvalho |
| EPI_ISL_2385669, EPI_ISL_2385670, EPI_ISL_2385671, EPI_ISL_2385672, EPI_ISL_2385673, EPI_ISL_2385674, EPI_ISL_2385675, EPI_ISL_2385676, EPI_ISL_2385677, EPI_ISL_2385678, EPI_ISL_2385679, EPI_ISL_2385680, EPI_ISL_2385681, EPI_ISL_2385682, EPI_ISL_2385683, EPI_ISL_2385684, EPI_ISL_2385685, EPI_ISL_2385686, EPI_ISL_2385687, EPI_ISL_2385689, EPI_ISL_2385697, EPI_ISL_2385700, EPI_ISL_2385701, EPI_ISL_2385702, EPI_ISL_2385703, EPI_ISL_2385704, EPI_ISL_2385705, EPI_ISL_2385706, EPI_ISL_2385707, EPI_ISL_2385708, EPI_ISL_2385709, EPI_ISL_2385710, EPI_ISL_2385711, EPI_ISL_2385713, EPI_ISL_2385715, EPI_ISL_2385716, EPI_ISL_2385717, EPI_ISL_2385718, EPI_ISL_2385719, EPI_ISL_2385720, EPI_ISL_2385721, EPI_ISL_2385724, EPI_ISL_2385725, EPI_ISL_2385726, EPI_ISL_2385727, EPI_ISL_2385728, EPI_ISL_2385729, EPI_ISL_2385731, EPI_ISL_2385732, EPI_ISL_2385733, EPI_ISL_2385734, EPI_ISL_2385735, EPI_ISL_2385736, EPI_ISL_2385737, EPI_ISL_2385738, EPI_ISL_2385739, EPI_ISL_2385740, EPI_ISL_2385741, EPI_ISL_2385742, EPI_ISL_2385743, EPI_ISL_2385744, EPI_ISL_2385745, EPI_ISL_2385746, EPI_ISL_2385747, EPI_ISL_2385748, EPI_ISL_2385749, EPI_ISL_2385750, EPI_ISL_2385751, EPI_ISL_2385752, EPI_ISL_2385753, EPI_ISL_2385754, EPI_ISL_2385755, EPI_ISL_2385757, EPI_ISL_2385758, EPI_ISL_2385759, EPI_ISL_2385760, EPI_ISL_2385761, EPI_ISL_2385762, EPI_ISL_2385763, EPI_ISL_2385764, EPI_ISL_2385765, EPI_ISL_2385766, EPI_ISL_2385767, EPI_ISL_2385768, EPI_ISL_2385769, EPI_ISL_2385770, EPI_ISL_2385771, EPI_ISL_2385772, EPI_ISL_2385773, EPI_ISL_2385774, EPI_ISL_2385775, EPI_ISL_2385776, EPI_ISL_2385777, EPI_ISL_2385778, EPI_ISL_2385779, EPI_ISL_2385780, EPI_ISL_2385781, EPI_ISL_2385782, EPI_ISL_2385783, EPI_ISL_2385784, EPI_ISL_2385785, EPI_ISL_2385786 | | | |
| see above | Unidade de apoio ao diagnóstico | Bioinformatics Laboratory / LNCC | Alessandra P Lamarca; Alexandra L Gerber; Amilcar Tanuri; Ana Paula de C Guimaraes; Ana Tereza R Vasconcelos; Andrea Cony Cavalcanti; Caio Luiz Pereira Ribeiro; Cassia Alves; Cintia Policarpo; Claudia Maria Braga de Mello; Cristiane Gomes da Silva; Diana Mariani; Douglas Terra Machado; Flavio Dias da |

| | | | |
|---|---|---|---|
| | da COVID – UNADIG | | Silva; Gleidson da Silva de Oliveira; Leandro Magalhaes de Souza; Liliane Cavalcante; Luiz G P de Almeida; Marcio Henrique de Oliveira Garcia; Mario Sergio Ribeiro; Ronaldo da Silva F Jr; Silvia Carvalho |
| EPI_ISL_1975177 | University College London, Great Ormond Street Hospital for Children NHS Foundation Trust, Imperial College Healthcare NHS Trust | COVID-19 Genomics UK (COG-UK) Consortium | Alison Holmes; Charlotte Williams; Helena Tutill; Jacqueline Findlay; James Price; Judith Breuer; Julianne Brown; Kathryn Harris; Leysa Forrest; Mark Kristiansen; Paola Niola; Paola Resende Silva; Patricia Dyal; Paul Randell; Rachel Williams; Samuel Weeks; Sergi Castellano; Sunando Roy; Tony Brooks; Yasmin Panchbhaya |
| EPI_ISL_2091058, EPI_ISL_2091059, EPI_ISL_2091060, EPI_ISL_2091061, EPI_ISL_2091062, EPI_ISL_2091063, EPI_ISL_2091064, EPI_ISL_2091109, EPI_ISL_2091112, EPI_ISL_2091113, EPI_ISL_2091114 | | | |
| see above | University Hospitals Translational Laboratory (UHTL), University Hospitals | University Hospitals Translational Laboratory (UHTL), University Hospitals | Alouani, D.; Sadri, N.; Song, X. |
| EPI_ISL_1963270 | University Hospitals of Geneva, Laboratory of Virology | HUG, Laboratory of Virology and the Health2030 Genome Center | Ana Rita Goncalves; Deborah Penet; Emmanouil Dermitzakis; Henri Pegeot; Ioannis Xenarios; Keith Harshman; Laurent Kaiser; Lorenzo Cerutti; Melyssa Elies; Samuel Cordey |
| EPI_ISL_2931009 | University of Arkansas for Medical Sciences | Center for Global Health, University of New Mexico Health Sciences Center | Catherine Kirkpatrick; Darrell Dinwiddie; Daryl Domman; David Ussery; Jennifer Laudadio; Joshua L. Kennedy; Kurt Schwalm; Susan Harley; Uday Chalwadi; Valerie Morley |
| EPI_ISL_2476350, EPI_ISL_2691960, EPI_ISL_2692011, EPI_ISL_2692013, EPI_ISL_2692015 | University of Chicago Medicine | RIPHL at Rush University Medical Center | Felix Araujo Perez; Kevin Kunstman; Laura Furtado; Lauren Megger; Marieta Hyde; Max Kolton; Stefan Green |
| EPI_ISL_2692093 | University of Illinois at Chicago | RIPHL at Rush University Medical Center | Felix Araujo Perez; Julie Bentea; Kevin Kunstman; Laura Furtado; Marieta Hyde; Max Kolton; Omar Perez; Stefan Green |
| EPI_ISL_2003251, EPI_ISL_2003255, EPI_ISL_2003257, EPI_ISL_2003267 | University of Liège COVID-19 testing center | GIGA Medical Genomics | Bouchra Boujemla; Cécile Meex; Keith Durkin; Maria Artesi; Marie-Pierre Hayette; Nathalie Renotte; Pierrette Melin; Raphaël Boreux; Sébastien Bontems; Vincent Bours |
| EPI_ISL_2100375, EPI_ISL_2100384 | University of Michigan Clinical Microbiology Laboratory | Lauring Lab, University of Michigan, Department of Microbiology and Immunology | Valesano |
| EPI_ISL_2285352, EPI_ISL_2285361 | University of Rome Tor Vergata: Departm Experim Medicine Chair of Virology | University of Rome Tor Vergata: Departm Experim Medicine Chair of Virology | Francesca Ceccherini-Silberstein; Loredana Sarmati; Lorenzo Piermatteo; Luca Carioti; Marco Iannetta; Maria Botticelli; Maria Concetta Bellocchi; Massimo Andreoni; Rossana Scutari |
| EPI_ISL_2225543, EPI_ISL_2225555, EPI_ISL_2225567, EPI_ISL_2225572 | University of Wisconsin-Madison AIDS Vaccine Research Laboratories | University of Wisconsin-Madison AIDS Vaccine Research Laboratories | Gage Moreno; Katarina Braun; et al. AIDS Vaccine Research Laboratories |
| EPI_ISL_2227060, EPI_ISL_2227072 | Università Federico II - Dipartimento di scienze mediche traslazionali - Napoli | TIGEM | Antonio Grimaldi Patrizia Annunziata Francesco Panariello Teresa Giuliano Michele Cennamo Valentina Bouche Chiara Colantuono Lucio Di Filippo Mariano Fiorenza Anna Manfredi Marcello Salvi Giuseppe Portella Andrea Ballabio Davide Cacchiarelli |
| EPI_ISL_3270791 | Università Federico II - Dipartimento di scienze mediche traslazionali - Napoli | Telethon Institute of Genetics and Medicine (TIGEM) | Antonio Grimaldi Patrizia Annunziata Francesco Panariello Teresa Giuliano Michele Cennamo Valentina Bouche Chiara Colantuono Lucio Di Filippo Mariano Fiorenza Anna Manfredi Marcello Salvi Giuseppe Portella Andrea Ballabio Davide Cacchiarelli |
| EPI_ISL_2001434, EPI_ISL_2001437, EPI_ISL_2001438, EPI_ISL_2001447 | Università degli Studi di Perugia | Istituto Zooprofilattico Sperimentale dell'Abruzzo e Molise "G. Caporale" | Ancora M; Calistri P; Camilloni B; Cammà C; Curini V; Di Domenico M; Di Pasquale A; Lorusso A; Mangone I; Marcacci M; Mencacci A; Puglia I; Rinaldi A; Savini G; Scialabba S |
| EPI_ISL_2086872 | UniversitŠtsSpital ZŸrich | Institute of Medical Virology | Alexandra Trkola; Annette AudigŽ; Catharine Aquino; Cyril Shah; Daniel Ehrsam; Gabriela Ziltener; Guido Bloemberg; Hubert Rehrauer; Isabel StŸrmer; Joel Wirz; Jon Huder; JŸrg Bšni; Kevin Steiner; Maria GrŸnberg; Maryam Zaheri; Michael Huber; Riccarda Capaul; Stefan Schmutz; Verena Kufner; Weihong Qi |
| EPI_ISL_2086800 | UniversitŠtsSpital ZŸrich 046 | Institute of Medical Virology | Alexandra Trkola; Annette AudigŽ; Catharine Aquino; Cyril Shah; Daniel Ehrsam; Gabriela Ziltener; Guido Bloemberg; Hubert Rehrauer; Isabel StŸrmer; Joel Wirz; Jon Huder; JŸrg Bšni; Kevin Steiner; Maria GrŸnberg; Maryam Zaheri; Michael Huber; Riccarda Capaul; Stefan Schmutz; Verena Kufner; Weihong Qi |
| EPI_ISL_5801379 | Upa Ipiranga | Instituto Butantan | Antonio Jorge Martins; Claudia Renata dos Santos Barros; David Schlesinger; Debora Botequio Moretti; Dimas Tadeu Covas; Elaine Cristina Marqueze; Elaine Vieira Santos; Evandra Strazza Rodrigues; Heidge Fukumasu; Jayme Augusto de Souza-Neto; José Salvatore Leister Patané; Luiz Alcantara; Luiz Lehmann Coutinho; Maria Carolina Elias; Maurício Lacerda Nogueira; Rafael dos Santos Bezerra; Raul Machado Neto; Rejane Maria Tommasini Grotto; Ricardo Haddad; Sandra Coccuzzo Sampaio Vessoni; Simone Kashima; Svetoslav Nanev Slavov; Vincent Louis Viala |
| EPI_ISL_3030219, EPI_ISL_3030310 | Urbino | Microbiology University Politecnica delle Marche | Anna Valenza; Carla Acciarri; Katia Marinelli; Monica Lucia Ferreri; Patrizia Bagnarelli; Roberta Longo; Sara Caucci; Stefano Menzo |
| EPI_ISL_2152435 | Usansolo-Galdakao University Hospital | Cruces University Hospital | Ana Belén de la Hoz; Ana Gual-de-Torrella; Izaskun Alejo-Cancho; Mikel Gallego |
| EPI_ISL_2284852, EPI_ISL_2292949, EPI_ISL_2857455, EPI_ISL_2857528, EPI_ISL_2857531, EPI_ISL_3263711, EPI_ISL_3263913 | | | |
| see above | Utah Public Health Laboratory | Utah Public Health Laboratory | Erin L. Young; Kelly F. Oakeson; Olinto Linares-Perdomo; Tara Gallagher |
| EPI_ISL_2209304, EPI_ISL_2209309 | VIGILANCIA EM SAUDE SAE VIGILANCIA EPIDEMIOLOGICA | Instituto Butantan / Mendelics | Antonio Jorge Martins; Claudia Renata dos Santos Barros; David Schlesinger; Debora Botequio Moretti; Dimas Tadeu Covas; Elaine Cristina Marqueze; Elaine Vieira Santos; Evandra Strazza Rodrigues; Heidge Fukumasu; Jayme Augusto de Souza-Neto; José Salvatore Leister Patané; Luiz Alcantara; Luiz Lehmann Coutinho; Maria Carolina Elias; Maurício Lacerda Nogueira; Rafael dos Santos Bezerra; Raul Machado Neto; Rejane Maria Tommasini Grotto; Ricardo Haddad; Sandra Coccuzzo Sampaio Vessoni; Simone Kashima; Svetoslav Nanev Slavov; Vincent Louis Viala |
| EPI_ISL_2209094, EPI_ISL_2345645, EPI_ISL_2345744, EPI_ISL_2345782, EPI_ISL_2345787, EPI_ISL_2345869 | VIGILANCIA EPIDEMIOLOGICA JARDINOPOLIS SP | Instituto Butantan / Mendelics | Antonio Jorge Martins; Claudia Renata dos Santos Barros; David Schlesinger; Debora Botequio Moretti; Dimas Tadeu Covas; Elaine Cristina Marqueze; Elaine Vieira Santos; Evandra Strazza Rodrigues; Heidge Fukumasu; Jayme Augusto de Souza-Neto; José Salvatore Leister Patané; Luiz Alcantara; Luiz Lehmann Coutinho; Maria Carolina Elias; Maurício Lacerda Nogueira; Rafael dos Santos Bezerra; Raul Machado Neto; Rejane Maria Tommasini Grotto; Ricardo Haddad; Sandra Coccuzzo Sampaio Vessoni; Simone Kashima; Svetoslav Nanev Slavov; Vincent Louis Viala |
| EPI_ISL_2209777, EPI_ISL_2345668 | VIGILANCIA EPIDIMIOLOGICA DE ARACARIGUAMA | Instituto Butantan / Mendelics | Antonio Jorge Martins; Claudia Renata dos Santos Barros; David Schlesinger; Debora Botequio Moretti; Dimas Tadeu Covas; Elaine Cristina Marqueze; Elaine Vieira Santos; Evandra Strazza Rodrigues; Heidge Fukumasu; Jayme Augusto de Souza-Neto; José Salvatore Leister Patané; Luiz Alcantara; Luiz Lehmann Coutinho; Maria Carolina Elias; Maurício Lacerda Nogueira; Rafael dos Santos Bezerra; Raul Machado Neto; Rejane Maria Tommasini Grotto; Ricardo Haddad; Sandra Coccuzzo Sampaio Vessoni; Simone Kashima; Svetoslav Nanev Slavov; Vincent Louis Viala |
| EPI_ISL_5780270, EPI_ISL_5780271 | VUMC Molecular Infectious Diseases Laboratory (MIDL) | Dr. Suman Das Lab - Vanderbilt University Medical Center (VUMC) | Bookyung Park; Grant Vestal; Helen Boone; Hunter Brown; Jonathan E. Schmitz; Meghan Shilts; Seesandra Rajagopala; Suman B. Pakala; Suman Das |
| EPI_ISL_5801212, EPI_ISL_5801271 | Vigilancia Em Saude Sae Vigilancia Epidemiologica | Instituto Butantan | Antonio Jorge Martins; Claudia Renata dos Santos Barros; David Schlesinger; Debora Botequio Moretti; Dimas Tadeu Covas; Elaine Cristina Marqueze; Elaine Vieira Santos; Evandra Strazza Rodrigues; Heidge Fukumasu; Jayme Augusto de Souza-Neto; José Salvatore Leister Patané; Luiz Alcantara; Luiz Lehmann Coutinho; Maria Carolina Elias; Maurício Lacerda Nogueira; Rafael dos Santos Bezerra; Raul Machado Neto; Rejane Maria Tommasini Grotto; Ricardo Haddad; Sandra Coccuzzo Sampaio Vessoni; Simone Kashima; Svetoslav Nanev Slavov; Vincent Louis Viala |
| EPI_ISL_5802699, EPI_ISL_5802714, EPI_ISL_5802729 | Vigilancia Epidemiologica De Leme | Instituto Butantan | Antonio Jorge Martins; Claudia Renata dos Santos Barros; David Schlesinger; Debora Botequio Moretti; Dimas Tadeu Covas; Elaine Cristina Marqueze; Elaine Vieira Santos; Evandra Strazza Rodrigues; Heidge Fukumasu; Jayme Augusto de Souza-Neto; José Salvatore Leister Patané; Luiz Alcantara; Luiz Lehmann Coutinho; Maria Carolina Elias; Maurício Lacerda Nogueira; Rafael dos Santos Bezerra; Raul Machado Neto; Rejane Maria Tommasini Grotto; Ricardo Haddad; Sandra Coccuzzo Sampaio Vessoni; Simone Kashima; Svetoslav Nanev Slavov; Vincent Louis Viala |
| EPI_ISL_5802694, EPI_ISL_5802731 | Vigilancia Epidemiologica E Controle De Vetores Pirassununga | Instituto Butantan | Antonio Jorge Martins; Claudia Renata dos Santos Barros; David Schlesinger; Debora Botequio Moretti; Dimas Tadeu Covas; Elaine Cristina Marqueze; Elaine Vieira Santos; Evandra Strazza Rodrigues; Heidge Fukumasu; Jayme Augusto de Souza-Neto; José Salvatore Leister Patané; Luiz Alcantara; Luiz Lehmann Coutinho; Maria Carolina Elias; Maurício Lacerda Nogueira; Rafael dos Santos Bezerra; Raul Machado Neto; Rejane Maria Tommasini Grotto; Ricardo Haddad; Sandra Coccuzzo Sampaio Vessoni; Simone Kashima; Svetoslav Nanev Slavov; Vincent Louis Viala |
| EPI_ISL_5801245, EPI_ISL_5801246, EPI_ISL_5801255, EPI_ISL_5801257, EPI_ISL_5801258, EPI_ISL_5801259, EPI_ISL_5801260 | | | |
| see above | Vigilancia Epidemiologica Jardinopolis Sp | Instituto Butantan | Antonio Jorge Martins; Claudia Renata dos Santos Barros; David Schlesinger; Debora Botequio Moretti; Dimas Tadeu Covas; Elaine Cristina Marqueze; Elaine Vieira Santos; Evandra Strazza Rodrigues; Heidge Fukumasu; Jayme Augusto de Souza-Neto; José Salvatore Leister Patané; Luiz Alcantara; Luiz Lehmann Coutinho; Maria Carolina Elias; Maurício Lacerda Nogueira; Rafael dos Santos Bezerra; Raul Machado Neto; Rejane Maria Tommasini Grotto; Ricardo Haddad; Sandra Coccuzzo Sampaio Vessoni; Simone Kashima; Svetoslav Nanev Slavov; Vincent Louis Viala |
| EPI_ISL_5801435, EPI_ISL_5801462, EPI_ISL_5801465 | Vigilancia Epidimiologica De Aracariguama | Instituto Butantan | Antonio Jorge Martins; Claudia Renata dos Santos Barros; David Schlesinger; Debora Botequio Moretti; Dimas Tadeu Covas; Elaine Cristina Marqueze; Elaine Vieira Santos; Evandra Strazza Rodrigues; Heidge Fukumasu; Jayme Augusto de Souza-Neto; José Salvatore Leister Patané; Luiz Alcantara; Luiz Lehmann Coutinho; Maria Carolina Elias; Maurício Lacerda Nogueira; Rafael dos Santos Bezerra; Raul Machado Neto; Rejane Maria Tommasini Grotto; Ricardo Haddad; Sandra Coccuzzo Sampaio Vessoni; Simone Kashima; Svetoslav Nanev Slavov; Vincent Louis Viala |
| EPI_ISL_2019017, EPI_ISL_2019183, EPI_ISL_2212759, EPI_ISL_2307663 | Viollier AG | Department of Biosystems Science and Engineering, ETH Zürich | Andrea Patrignani; Andreia Cabral de Gouvea; Catharine Aquino; Chaoran Chen; Christian Beisel; Christiane Beckmann; Christoph Noppen; Daniel Ehrsam; David Dreifuss; Doris Popovic; Elodie Burcklen; Griffin White; Ina Nissen; Isabel Stürmer; Ivan Topolsky; Jay Tracy; Katharina Jahn; Lara Fuhrmann; Laura Neff; Lennart Opitz; Maria Domenica Moccia; Maurice Redondo; Mirjam Feldkamp; Natascha Santacroce; Niko Beerenwinkel; Noemie Santamaria de Souza; Olivier Kobel; Philipp Jablonski; Ralph Schlapbach; Rebecca Denes; Sarah Nadeau; Simon Grüter; Sophie Seidel; Tanja Stadler; Timothy Sykes |
| EPI_ISL_2101937, EPI_ISL_2229478 | Virginia Division of Consolidated Laboratory Services | Virginia Division of Consolidated Laboratory Services | Virginia DCLS |
| EPI_ISL_2128476 | Virology Department, Sheffield Teaching Hospitals NHS Foundation Trust/Department of Infection, Immunity and Cardiovascular Disease, The Medical School, University of | COVID-19 Genomics UK (COG-UK) Consortium | Adri Angyal; Alex Keeley; Benjamin Lindsey; Cariad Evans; Danielle Groves; Dave Partridge; Katie Johnson; Laura Carrilero; Luke Green; Matthew Parker; Matthew Wyles; Mehmet Yavuz; Mohammad Raza; Nikki Smith; Paul Parsons; Rachel Tucker; Rebecca Brown; Thushan de Silva |

| | | | |
|---|---|---|---|
| | Sheffield | | |
| EPI_ISL_2883999, EPI_ISL_2884139, EPI_ISL_2884140, EPI_ISL_2884143, EPI_ISL_2884144, EPI_ISL_2884220, EPI_ISL_2884222 | | | |
| see above | Virology Laboratory, Scientific Department, Army Medical Center | Virology Laboratory, Scientific Department, Army Medical Center | Anella Monte; Anna Anselmo; Antonella Fortunato; Filippo Molinari; Florigio Lista; Francesco Giordani; Giancarlo Petralito; Giandomenico Cerreto; Giulia Campoli; Lucia Nicosia; Margherita De Santis; Maria Anna Spinelli; Marzia Cavalli; Riccardo De Sanctis; Rossella Brandi; Silvia Fillo; Vanessa Vera Fain |
| EPI_ISL_2105163 | WA State Department of Health | Centers for Disease Control and Prevention Division of Viral Diseases, Pathogen Discovery | Alison Laufer Halpin; Ben L. Rambo-Martin; Clinton R. Paden; Dakota Howard; Darlene Wagner; Dave Wentworth; Dhwani Batra; Jasmine Padilla; Justin Lee; Katie Dillon; Krista Queen; Kristen Knipe; Kristine Lacek; Mark Burroughs; Matthew Schmerer; Mili Sheth; Peter Cook; Sam Shepard; Sarah Nobles; Shoshona Le; Suxiang Tong; Vivien Dugan; Yvette Unoarumhi |
| EPI_ISL_2097314 | WESTCHESTER MEDICAL CENTER | Wadsworth Center, New York State Department of Health | Alexis Russell; Catharine Prussing; Daryl M. Lamson; Erasmus Schneider; Erica Lasek-Nesselquist; John Kelly; Jonathan Plitnick; Kirsten St. George; Matthew Shudt; Melissa A Leisner; Navjot Singh |
| EPI_ISL_2533758 | WLA VAMC | Genomics and Discovery, Respiratory Viruses Branch, Division of Viral Diseases, Centers for Disease Control and Prevention | Adam Retchless; Anna Kelleher; Anna Uehara; Brian Lynch; Clinton R. Paden; Dhwani Batra; Haibin Wang; Han Jia Justin Ng; Jasmine Padilla; Jing Zhang; Justin Lee; Krista Queen; Mark Burroughs; Mili Sheth; Morgan Davis; Peter Cook; Rachel Marine; Sarah Nobles; Suxiang Tong; Tara Coalter; Yan Li; Ying Tao |
| EPI_ISL_2127876 | Wales Specialist Virology Centre Sequencing lab: Pathogen Genomics Unit | Public Health Wales Microbiology Cardiff Wales Specialist Virology Centre | Alec Birchley; Alexander Adams; Amy Gaskin; Angela Marchbank; Bree Gatica-Wilcox; Catherine Moore; Jason Coombes; Joanne Watkins; Joel Southgate; Johnathan Evans; Laura Gifford; Lauren Gilbert; Lee Graham; Malorie Perry; Matthew Bull; Nicole Pacchiarini; Sally Corden; Sara Kumziene-Summerhayes; Sara Rey; Sarah Taylor; Simon Cottrell; Sophie Jones; Tom Connor |
| EPI_ISL_1897429, EPI_ISL_2096103, EPI_ISL_2336153, EPI_ISL_2336157, EPI_ISL_2336168, EPI_ISL_2336170, EPI_ISL_2336171, EPI_ISL_2336172, EPI_ISL_2336173, EPI_ISL_2336245, EPI_ISL_2336253 | | | |
| see above | Washington State Department of Health Public Health Laboratories | Washington State Department of Health Public Health Laboratories | Avi Singh; Darren Lucas; Denny Russell; Drew MacKellar; Geoff Melly; Hannah Gray; Joenice Gonzalez; JohnAric Peterson; Philip Dykema; Rebecca Cao; Vanessa De Los Santos |
| EPI_ISL_2376267, EPI_ISL_2376268, EPI_ISL_2376278, EPI_ISL_2376282, EPI_ISL_2376284, EPI_ISL_2377899, EPI_ISL_2382336, EPI_ISL_2382339 | | | |
| see above | Wexner Medical Center | The Ohio State University College of Medicine | Koenig, S.; Seminetta, J. |
| EPI_ISL_1823109, EPI_ISL_1823112, EPI_ISL_1823115, EPI_ISL_1823117, EPI_ISL_1823119, EPI_ISL_1823123, EPI_ISL_1854740, EPI_ISL_1854741, EPI_ISL_1854742, EPI_ISL_1854743, EPI_ISL_1854744, EPI_ISL_1854746, EPI_ISL_1854748, EPI_ISL_1854751, EPI_ISL_1857038, EPI_ISL_1857039, EPI_ISL_1857044, EPI_ISL_1857047, EPI_ISL_1857051, EPI_ISL_1857056, EPI_ISL_1857060, EPI_ISL_1857061, EPI_ISL_1857062, EPI_ISL_1857066, EPI_ISL_1857070, EPI_ISL_1857087, EPI_ISL_1857093, EPI_ISL_1921877, EPI_ISL_1922707, EPI_ISL_1940143, EPI_ISL_1940145, EPI_ISL_1940151, EPI_ISL_1961018 | | | |
| see above | Wichita State University - Molecular Diagnostics Lab | Kansas Health and Environmental Lab | Ben Olsen; Jonathan Barnell; Mike Grose; and Phil Adam |
| EPI_ISL_2279132, EPI_ISL_2279133, EPI_ISL_2279137, EPI_ISL_2279143, EPI_ISL_2279292, EPI_ISL_2279316, EPI_ISL_2279322, EPI_ISL_2279327, EPI_ISL_2279367, EPI_ISL_2279371, EPI_ISL_2279394, EPI_ISL_2279414, EPI_ISL_2279430, EPI_ISL_2279435, EPI_ISL_2279446, EPI_ISL_2279454, EPI_ISL_2279456, EPI_ISL_2279460, EPI_ISL_2279464, EPI_ISL_2279466, EPI_ISL_2279470, EPI_ISL_2279474, EPI_ISL_2279482 | | | |
| see above | Wisconsin State Laboratory of Hygiene Communicable Disease Division | Wisconsin State Laboratory of Hygiene Communicable Disease Division | Abigail C. Shockey; Alicia J. Mooney; Erika M. Hanson; Kelsey R. Florek; Richard Griesser; Sara Wagner; Tonya Danz |
| EPI_ISL_2015270, EPI_ISL_2226816, EPI_ISL_2402351, EPI_ISL_2444755 | Wyoming Public Health Laboratory | Wyoming Public Health Laboratory | Ashley Norberg; Brian Dominguez; Brittany Oher; Cari Sloma; Channing Weber; Chayse Rowley; Elliot Thomasson; Jim Mildenberger; Marley Goetz; Sam Britz; Taylor Fearing; and Rob Christensen |
| EPI_ISL_2023184 | Yale Clinical Virology Lab | Grubaugh Lab - Yale School of Public Health | Anderson Brito; Annie Watkins; Chaney Kalinich; Chantal Vogels; Isabel Ott; Jessica Rothman; Joseph Fauver; Mallery Breban; Marie L. Landry; Mary Petrone; Nathan Grubaugh; Tara Alpert |
| EPI_ISL_2159078 | Yale Clinical Virology Lab | Yale Center for Genomic Analysis | Brooke Sullivan; Curt Scharfe; Irina Tikhonova; Kaya Bilguvar; Shrikant Mane |
