## Supplement 3 for "Rapid and Accurate Identification of SARS-CoV-2 Omicron Variants Using Droplet Digital PCR (RT-ddPCR)": All_0503-0509.pdf

We gratefully acknowledge the following Authors from the Originating laboratories responsible for obtaining the specimens, as well as the Submitting laboratories where the genome data were generated and shared via GISAID, on which this research is based.

All Submitters of data may be contacted directly via www.gisaid.org

Authors are sorted alphabetically.

| Accession ID | Originating Laboratory | Submitting Laboratory | Authors |
|---|---|---|---|
| EPI_ISL_5899671 | A. Krumbholz, Labor Dr. Krause und Kollegen MVZ GmbH, Kiel | Charité Universitätsmedizin Berlin, Institut für Virologie | Barbara Mühlemann; Christian Drosten; Julia Schneider; Julia Tesch; Jörn Beheim-Schwarzbach; Talitha Veith; Terry Jones; Tobias Bleicker; Victor M Corman |
| EPI_ISL_5915138, EPI_ISL_5915152, EPI_ISL_5915194, EPI_ISL_5915202, EPI_ISL_5915225, EPI_ISL_5915254, EPI_ISL_5915255, EPI_ISL_5915256, EPI_ISL_6573820, EPI_ISL_6573821, EPI_ISL_6573873, EPI_ISL_6573930 | | | |
| see above | AMALAB/FACISA/UFRN | WallauLab on behalf of Fiocruz COVID-19 Genomic Surveillance Network | Alexandre Freitas da Silva; Allan Roberto Dias Nunes; Antonio Marinho da Silva Neto; Cassia Docena; Constância Flávia Junqueira Ayres; Filipe Zimmer Dezordi; Gabriel Luz Wallau; Gustavo Barbosa de Lima; Joana Cristina Medeiros Tavares Marques; Katya Anaya Jacinto; Lais Ceschini Machado; Lilian Caroliny Amorim Silva; Marcelo Henrique dos Santos Paiva; Mariane dos Santos Duarte; Matheus Filgueira Bezerra; Sinval Pinto Brandão Filho |
| EPI_ISL_2502727 | AREA DE SALUD SAN JUAN-SAN DIEGO-CONCEPCION 2 | Inciensa, Instituto Costarricense de InvestigaciOn y Enseñanza en NutriciOn y Salud | Adriana Godínez; Claudio Soto-Garita; Estela Cordero; Francisco Duarte; Hebleen Porras; Jose Luis Vargas; Joselyn Prado & Mariel LOpez; Mariela Gutierrez; Melany CalderOn |
| EPI_ISL_3495376 | ASST GOM NIGUARDA | ASST Grande ospedale Metropolitano Niguarda | Alice Nava |
| EPI_ISL_2137724, EPI_ISL_2137725, EPI_ISL_2137734 | AZ Klina | AZ Klina | Carl Vael - Lynsey Berckmans |
| EPI_ISL_2146068, EPI_ISL_2146070, EPI_ISL_2146074, EPI_ISL_2146087, EPI_ISL_2146088, EPI_ISL_2146089, EPI_ISL_2146112, EPI_ISL_2146137, EPI_ISL_2146138, EPI_ISL_2146167, EPI_ISL_2146186, EPI_ISL_2146237, EPI_ISL_2146239, EPI_ISL_2146257, EPI_ISL_2146258, EPI_ISL_2146280, EPI_ISL_2146297, EPI_ISL_2146301, EPI_ISL_2146313, EPI_ISL_2146319, EPI_ISL_2146322, EPI_ISL_2146325, EPI_ISL_2146326, EPI_ISL_2146334, EPI_ISL_2146337, EPI_ISL_2146354, EPI_ISL_2146365, EPI_ISL_2146387, EPI_ISL_2146399, EPI_ISL_2146401, EPI_ISL_2146405, EPI_ISL_2146448, EPI_ISL_2146466, EPI_ISL_2146467, EPI_ISL_2146468, EPI_ISL_2146493, EPI_ISL_2146521, EPI_ISL_2146522, EPI_ISL_2146528, EPI_ISL_2146529, EPI_ISL_2146541, EPI_ISL_2146544, EPI_ISL_2146551, EPI_ISL_2146562, EPI_ISL_2146568, EPI_ISL_2146581, EPI_ISL_2146583, EPI_ISL_2146611, EPI_ISL_2146628, EPI_ISL_2146644, EPI_ISL_2146653, EPI_ISL_2146657, EPI_ISL_2146684, EPI_ISL_2146694, EPI_ISL_2146706, EPI_ISL_2146709, EPI_ISL_2146717, EPI_ISL_2146719, EPI_ISL_2146722, EPI_ISL_2146729, EPI_ISL_2146736, EPI_ISL_2146775, EPI_ISL_2146780, EPI_ISL_2146784, EPI_ISL_2146795, EPI_ISL_2146824, EPI_ISL_2146833, EPI_ISL_2146858, EPI_ISL_2146877, EPI_ISL_2146886, EPI_ISL_2146894, EPI_ISL_2146909, EPI_ISL_2146921, EPI_ISL_2146948, EPI_ISL_2146950, EPI_ISL_2146951, EPI_ISL_2146957, EPI_ISL_2146959, EPI_ISL_2147010, EPI_ISL_2147040, EPI_ISL_2147045, EPI_ISL_2147046, EPI_ISL_2147071, EPI_ISL_2147101, EPI_ISL_2147103, EPI_ISL_2147109, EPI_ISL_2147115, EPI_ISL_2147117, EPI_ISL_2147126, EPI_ISL_2147131, EPI_ISL_2147132, EPI_ISL_2147133, EPI_ISL_2147147, EPI_ISL_2147159, EPI_ISL_2147188, EPI_ISL_2147240, EPI_ISL_2147244, EPI_ISL_2147246, EPI_ISL_2147252, EPI_ISL_2147270, EPI_ISL_2147398, EPI_ISL_2147400, EPI_ISL_2147403, EPI_ISL_2147405, EPI_ISL_2147411, EPI_ISL_2147419, EPI_ISL_2147421, EPI_ISL_2147454, EPI_ISL_2147461, EPI_ISL_2147467, EPI_ISL_2147478, EPI_ISL_2147484, EPI_ISL_2147503, EPI_ISL_2147520, EPI_ISL_2147522, EPI_ISL_2147537, EPI_ISL_2147542, EPI_ISL_2147547, EPI_ISL_2147548, EPI_ISL_2147564, EPI_ISL_2147572, EPI_ISL_2147580, EPI_ISL_2147594, EPI_ISL_2147603, EPI_ISL_2147604, EPI_ISL_2147605, EPI_ISL_2147606, EPI_ISL_2147616, EPI_ISL_2147809, EPI_ISL_2147873, EPI_ISL_2147887, EPI_ISL_2147890, EPI_ISL_2147891, EPI_ISL_2147929, EPI_ISL_2147943, EPI_ISL_2147949, EPI_ISL_2147954, EPI_ISL_2147956, EPI_ISL_2147963, EPI_ISL_2147967, EPI_ISL_2147968, EPI_ISL_2148142, EPI_ISL_2148147, EPI_ISL_2148190, EPI_ISL_2148195, EPI_ISL_2148200, EPI_ISL_2148211, EPI_ISL_2148239, EPI_ISL_2148245, EPI_ISL_2148248, EPI_ISL_2148853, EPI_ISL_2148895, EPI_ISL_2148902, EPI_ISL_2148905, EPI_ISL_2148913, EPI_ISL_2148914, EPI_ISL_2148939, EPI_ISL_2148947, EPI_ISL_2148948, EPI_ISL_2149145, EPI_ISL_2158851, EPI_ISL_2158858, EPI_ISL_2158874, EPI_ISL_2158883, EPI_ISL_2158900, EPI_ISL_2158911, EPI_ISL_2158919, EPI_ISL_2158936, EPI_ISL_2158961, EPI_ISL_2158962, EPI_ISL_2158973, EPI_ISL_2158982, EPI_ISL_2158985, EPI_ISL_2158996, EPI_ISL_2159049, EPI_ISL_2159125, EPI_ISL_2159128, EPI_ISL_2159135, EPI_ISL_2159158, EPI_ISL_2159281, EPI_ISL_2159297, EPI_ISL_2159298, EPI_ISL_2159304, EPI_ISL_2159306, EPI_ISL_2159307, EPI_ISL_2159346, EPI_ISL_2159351, EPI_ISL_2159359, EPI_ISL_2159366, EPI_ISL_2159367, EPI_ISL_2159463, EPI_ISL_2180537, EPI_ISL_2180582, EPI_ISL_2180586, EPI_ISL_2180656, EPI_ISL_2180666, EPI_ISL_2180723, EPI_ISL_2180739, EPI_ISL_2181174, EPI_ISL_2181180, EPI_ISL_2181251, EPI_ISL_2187377, EPI_ISL_2187386, EPI_ISL_2187389, EPI_ISL_2187397, EPI_ISL_2187402, EPI_ISL_2187403, EPI_ISL_2187404, EPI_ISL_2187407, EPI_ISL_2187408, EPI_ISL_2187409, EPI_ISL_2187417, EPI_ISL_2187432, EPI_ISL_2187442, EPI_ISL_2187455, EPI_ISL_2187486, EPI_ISL_2187487, EPI_ISL_2187609, EPI_ISL_2241643, EPI_ISL_2241655, EPI_ISL_4363109 | | | |
| see above | Aegis Sciences Corporation | Centers for Disease Control and Prevention Division of Viral Diseases, Pathogen Discovery | Adrian Paskey; Alec Vest; Benjamin Rambo-Martin; Christopher Gulvick; Clinton Paden; Clinton R. Paden; Cyndi Clark; Dakota Howard; Darlene Wagner; Dhwani Batra; Dillon Nall; Duncan MacCannell; Erisa Sula; Ethan Sanders; Holly Houdeshell; Jason Caravas; Kara Moser; Kristine Lacek; Matthew Hardison; Matthew Schmerer; Ola Kvalvaag; Patrick Campbell; Peter Cook; Peter W. Cook; Rob Case; Scott Sammons; Shatavia Morrison; Shaun Westlund; Tymeckia Kendall; Victoria Caban Figueroa; Vikramsinha Ghorpade; Yvette Unoarumhi |
| EPI_ISL_2249137 | Affidea | Instituto Nacional de Saude (INSA) and Centro de Investigaçao em Biodiversidade e Recursos Geneticos (CIBIO), Universidade do Porto | Borges et al |
| EPI_ISL_2949294 | Ages, Lifebrain COVID 19 Labor GmbH Klinik Penzing Pavillon 17_Europe/Austria/Vienna | Elling and Cochella laboratories, IMBA/IMP | Alexander Stark; Ezgi Oezkan; Luisa Cochella and Ulrich Elling; Marcus Strobl; Olga Frank; Ramesh Yelagandula; Tanino Albanese |
| EPI_ISL_2101759, EPI_ISL_2136442, EPI_ISL_2187659, EPI_ISL_2213196, EPI_ISL_2213199, EPI_ISL_2376069, EPI_ISL_2376070 | | | |
| see above | Alaska State Virology Laboratory | Alaska State Virology Laboratory | Elva House; Jack Chen; Jacob Zidek; Lisa Smith; Ph.D.; Stephanie DeRonde |
| EPI_ISL_2134634, EPI_ISL_2134636 | Algemeen Medisch Labo | Labo Klinische Biologie, UZA | Basil Britto Xavier; Christine Lammens; Herman Goossens; Jasmine Coppens; Marie Le Mercier; Veerle Matheeussen |
| EPI_ISL_3243025, EPI_ISL_3243028, EPI_ISL_3243029, EPI_ISL_3243030, EPI_ISL_3243031 | Applied DNA Sciences Inc | Applied DNA Sciences Inc | Huan Dong; Lucas Liu; Spencer William; Yuhua Sun |
| EPI_ISL_2227488, EPI_ISL_2227489 | Arizona State Public Health Laboratory | Arizona State Public Health Laboratory | Jessica Escobar; Katherine Fullerton; Linda Getsinger; Nobuko Fukushima; Stacy White; Trung Huynh; Victor Waddell |
| EPI_ISL_2250017, EPI_ISL_2250018, EPI_ISL_2250021, EPI_ISL_2250032, EPI_ISL_2250036, EPI_ISL_2250047, EPI_ISL_2250051, EPI_ISL_2250058, EPI_ISL_2250060, EPI_ISL_2250085, EPI_ISL_3386449 | | | |
| see above | Arizona State University | Arizona State University | Ajeet Bains; Efrem S. Lim; Joshua LaBaer; Joy M. Blain; LaRinda A. Holland; Matthew F. Smith; Nathaniel Johnson; Nicholas J. Mellor; Peter T. Skidmore; Rabia Maqsood; Valerie Harris; Vel Murugan |
| EPI_ISL_2484742 | Arkansas Children's Hospital | Center for Global Health, University of New Mexico Health Sciences Center | Amanda Novack; Darrell Dinwiddie; Daryl Domman; Dirk Haselow; Joshua L. Kennedy; Kurt Schwalm; Valerie Morley |
| EPI_ISL_2939077 | Arkansas Department of Health | Center for Global Health, University of New Mexico Health Sciences Center | Amanda Novack; Darrell Dinwiddie; Daryl Domman; Dirk Haselow; Joshua L. Kennedy; Kurt Schwalm; Valerie Morley |
| EPI_ISL_2597989 | Atlanta VA Medical Center | Genomics and Discovery, Respiratory Viruses Branch, Division of Viral Diseases, Centers for Disease Control and Prevention | Adam Retchless; Anna Kelleher; Anna Uehara; Brian Lynch; Clinton R. Paden; Dhwani Batra; Haibin Wang; Han Jia Justin Ng; Jasmine Padilla; Jing Zhang; Justin Lee; Krista Queen; Mark Burroughs; Mili Sheth; Morgan Davis; Peter Cook; Rachel Marine; Sarah Nobles; Suxiang Tong; Tara Coalter; Yan Li; Ying Tao |
| EPI_ISL_2692292, EPI_ISL_2692300, EPI_ISL_2692305, EPI_ISL_2692459 | Atlas Genomics | Seattle Flu Study | Amanda Adler; Barry R. Lutz; Benjamin Pelle; Brian Hiatt; Caitlin R. Wolf; Chris D. Frazar; Deborah A. Nickerson; Elisabeth Brandstetter; Erica Ryke; Geoff Melly; Helen Y. Chu; Janet A. Englund; Jay Shendure; Jover Lee; Kairsten Fay; Kirsten Lacombe; Lea M. Starita; Mark J. Rieder; Matthew Richardson; Matthew Thompson; Melissa Truong; Michael Boeckh; Michael Famulare; Misja Ilcisin; Peter D. Han; Philip Dykema; Romesh Gautom; Scott Lindquist; Thomas R. Sibley; Trevor Bedford |
| EPI_ISL_2249139 | Avelab | Instituto Nacional de Saude (INSA) and Centro de Investigaçao em Biodiversidade e Recursos Geneticos (CIBIO), Universidade do Porto | Borges et al |
| EPI_ISL_2396879 | Azienda Ospedaliera Pugliese Ciaccio di Catanzaro SOC Microbiologia e Virologia | Azienda Ospedaliera Pugliese Ciaccio di Catanzaro SOC Microbiologia e Virologia | Rossana Tallerico Federica Pasceri Marco De Fazio Ilenia Talotta Giuseppina Panduri Pasquale Minchella |
| EPI_ISL_2420737 | Azienda Ospedaliera San Camillo Forlanini | INMI Lazzaro Spallanzani IRCCS | A Di Caro; B Bartolini; CEM Gruber; D Gallone; E Giombini; F Basile; F Messina; F Santini; G Bonfiglio; G Parisi; M Rueca; MR Capobianchi; O Butera |
| EPI_ISL_2872695 | Azienda Ospedaliero Universitaria di Sassari | AMES Centro Polidiagnostico Strumentale S.r.l. | Anna Puggioni; Bianca Paglietti; Caterina Serra; Claudia Piu; Elena Rimini; Erica Mura; Flavia Angioj; Gabriele Ibba; Giulia Rocca; Laura Firino; Rosalba Govoni; Salvatore Rubino; Sergio Uzzau; Vincenzo Lai |
| EPI_ISL_2325127 | Azienda Sanitaria dell'Alto Adige - Laboratorio Aziendale di Microbiologia e Virologia | Azienda Sanitaria dell'Alto Adige | Irene Bianconi |
| EPI_ISL_2444212 | BARRETOS | Instituto Butantan / FAMERP | Antonio Jorge Martins; Claudia Renata dos Santos Barros; David Schlesinger; Debora Botequio Moretti; Dimas Tadeu Covas; Elaine Cristina Marqueze; Elaine Vieira Santos; Evandra Strazza Rodrigues; Heidge Fukumasu; Jayme Augusto de Souza-Neto; José Salvatore Leister Patané; Luiz Alcantara; Luiz Lehmann Coutinho; Maria Carolina Elias; Maurício Lacerda Nogueira; Rafael dos Santos Bezerra; Raul Machado Neto; Rejane Maria Tommasini Grotto; Ricardo Haddad; Sandra Coccuzzo Sampaio Vessoni; Simone Kashima; Svetoslav Nanev Slavov; Vincent Louis Viala |
| EPI_ISL_2656541, EPI_ISL_2656543, EPI_ISL_2656544, EPI_ISL_2656545, EPI_ISL_2656546, EPI_ISL_2656550, EPI_ISL_2656556, EPI_ISL_2656557, EPI_ISL_2656559, EPI_ISL_2656560, EPI_ISL_2656563, EPI_ISL_2656564, EPI_ISL_2656566, EPI_ISL_2656567, EPI_ISL_2656569, EPI_ISL_2656574, EPI_ISL_2656575, EPI_ISL_2656578, EPI_ISL_2656579, EPI_ISL_2656580, EPI_ISL_2656584, EPI_ISL_2656586, EPI_ISL_2656588, EPI_ISL_2656590, EPI_ISL_2656593, EPI_ISL_2656595, EPI_ISL_2656597, EPI_ISL_2656598, EPI_ISL_2656600, EPI_ISL_2656605, EPI_ISL_2656609, EPI_ISL_2656614, EPI_ISL_2656616, EPI_ISL_2656618, EPI_ISL_2656619, EPI_ISL_2656626, EPI_ISL_2656628, EPI_ISL_2656630, EPI_ISL_2656633, EPI_ISL_2656638, EPI_ISL_2656639, EPI_ISL_2656641, EPI_ISL_2656643, EPI_ISL_2656644, EPI_ISL_2656649, EPI_ISL_2656650, EPI_ISL_2656652, EPI_ISL_2656653, EPI_ISL_2656655, EPI_ISL_2656656, EPI_ISL_2656658, EPI_ISL_2656663, EPI_ISL_2656665, EPI_ISL_2656673, EPI_ISL_2656674, EPI_ISL_2656675, EPI_ISL_2656676, EPI_ISL_2656677, EPI_ISL_2656680, EPI_ISL_2656682, EPI_ISL_2656689, EPI_ISL_2656692, EPI_ISL_2656694, EPI_ISL_2656695, EPI_ISL_2656700, EPI_ISL_2656702, EPI_ISL_2656704, EPI_ISL_2656714, EPI_ISL_2656721, EPI_ISL_2656737, EPI_ISL_2656741, EPI_ISL_2656743, EPI_ISL_2656746, EPI_ISL_2656749, EPI_ISL_2656750, EPI_ISL_2656751, EPI_ISL_2656757, EPI_ISL_2656761, EPI_ISL_2656762, EPI_ISL_2656776, EPI_ISL_2656781, EPI_ISL_2656786, EPI_ISL_2656788, EPI_ISL_2656789, EPI_ISL_2656795, EPI_ISL_2656797, EPI_ISL_2656815, EPI_ISL_2656823, EPI_ISL_2656829, EPI_ISL_2656836, EPI_ISL_2656841, EPI_ISL_2656843, EPI_ISL_2656847, EPI_ISL_2656856, EPI_ISL_2656859, EPI_ISL_2656863, EPI_ISL_2656867, EPI_ISL_2656874, EPI_ISL_2656879, EPI_ISL_2656885, EPI_ISL_2656889, EPI_ISL_2656895, EPI_ISL_2656896, EPI_ISL_2656898, EPI_ISL_2656899, EPI_ISL_2656907, EPI_ISL_2656913, EPI_ISL_2656924, EPI_ISL_2656925, EPI_ISL_2656932, EPI_ISL_2656937, EPI_ISL_2656941, EPI_ISL_2656942, EPI_ISL_2656950, EPI_ISL_2656951, EPI_ISL_2656961, EPI_ISL_2656969, EPI_ISL_2656973, EPI_ISL_2656974, EPI_ISL_2657007, EPI_ISL_2657062, EPI_ISL_2657083, EPI_ISL_2657121, EPI_ISL_2720371, EPI_ISL_2720372, EPI_ISL_2720373, EPI_ISL_2720375, EPI_ISL_2720379, EPI_ISL_2720382, EPI_ISL_2720385, EPI_ISL_2720390, EPI_ISL_2720392, EPI_ISL_2720401, EPI_ISL_2720403, EPI_ISL_2720405, EPI_ISL_2720415, EPI_ISL_2720453, EPI_ISL_2720466, EPI_ISL_2720473, EPI_ISL_2720485, EPI_ISL_2720487, EPI_ISL_2720494, EPI_ISL_2720501, EPI_ISL_2720505, EPI_ISL_2720508, EPI_ISL_2720515, EPI_ISL_2720531, EPI_ISL_2720533, EPI_ISL_2720535, EPI_ISL_2720537, EPI_ISL_2720538, EPI_ISL_2720543, EPI_ISL_2720545, EPI_ISL_2720548, EPI_ISL_2720550, EPI_ISL_2720672, EPI_ISL_2720675, EPI_ISL_2720677, EPI_ISL_2720680, EPI_ISL_2720684, EPI_ISL_2720686, EPI_ISL_2720688, EPI_ISL_2720692, EPI_ISL_2720693, EPI_ISL_2720694, EPI_ISL_2720697, EPI_ISL_2720707, EPI_ISL_2720709, EPI_ISL_2720711, EPI_ISL_2720716, EPI_ISL_2720717, EPI_ISL_2720721, EPI_ISL_2720722, EPI_ISL_2720724, EPI_ISL_2720726, EPI_ISL_2720729, EPI_ISL_2720730, EPI_ISL_2720732, EPI_ISL_2720735, EPI_ISL_2720738, EPI_ISL_2720739, EPI_ISL_2720741, EPI_ISL_2720744, EPI_ISL_2720745, EPI_ISL_2720754, EPI_ISL_2720755, EPI_ISL_2720756, EPI_ISL_2720757, EPI_ISL_2720759, EPI_ISL_2720760, EPI_ISL_2720762, EPI_ISL_2720764, EPI_ISL_2720766, EPI_ISL_2720771, EPI_ISL_2720772, EPI_ISL_2720776, EPI_ISL_2720777, EPI_ISL_2720778, | | | |

| | | | |
|---|---|---|---|
| EPI_ISL_2720779, EPI_ISL_2720782, EPI_ISL_2720788, EPI_ISL_2721375, EPI_ISL_2721379, EPI_ISL_2721391, EPI_ISL_2721409, EPI_ISL_2721415, EPI_ISL_2721435, EPI_ISL_2721436, EPI_ISL_2721437, EPI_ISL_2721438, EPI_ISL_2721439, EPI_ISL_2721440, EPI_ISL_2721443, EPI_ISL_2721446, EPI_ISL_2721454, EPI_ISL_2721455, EPI_ISL_2721457, EPI_ISL_2721468, EPI_ISL_2721471, EPI_ISL_2721476, EPI_ISL_2721481, EPI_ISL_2721486, EPI_ISL_2721487, EPI_ISL_2721497, EPI_ISL_2721503, EPI_ISL_2721506, EPI_ISL_2721509, EPI_ISL_2721510, EPI_ISL_2721511, EPI_ISL_2721515, EPI_ISL_2721519, EPI_ISL_2721525, EPI_ISL_2721530, EPI_ISL_2721544, EPI_ISL_2721545, EPI_ISL_2721547, EPI_ISL_2721555, EPI_ISL_2721564, EPI_ISL_2721568, EPI_ISL_2721570, EPI_ISL_2721575, EPI_ISL_2721582, EPI_ISL_2721587, EPI_ISL_2721592, EPI_ISL_2721604, EPI_ISL_2721605, EPI_ISL_2721621, EPI_ISL_2721622, EPI_ISL_2721623, EPI_ISL_2721627, EPI_ISL_2721630, EPI_ISL_2721631, EPI_ISL_2721632, EPI_ISL_2721633, EPI_ISL_2721634, EPI_ISL_2721640, EPI_ISL_2721641, EPI_ISL_2721648, EPI_ISL_2721656, EPI_ISL_2721661, EPI_ISL_2721663, EPI_ISL_2721664, EPI_ISL_2721665, EPI_ISL_2721667, EPI_ISL_2721678, EPI_ISL_2721687, EPI_ISL_2721697, EPI_ISL_2721708, EPI_ISL_2721712, EPI_ISL_2721718, EPI_ISL_2721719, EPI_ISL_2721720, EPI_ISL_2721723, EPI_ISL_2721752, EPI_ISL_2721758, EPI_ISL_2721768, EPI_ISL_2721772, EPI_ISL_2721782, EPI_ISL_2721792, EPI_ISL_2721795, EPI_ISL_2721796, EPI_ISL_2721798, EPI_ISL_2721801, EPI_ISL_2721804, EPI_ISL_2721805, EPI_ISL_2721810, EPI_ISL_2721811, EPI_ISL_2721819, EPI_ISL_2721821, EPI_ISL_2721841, EPI_ISL_2721845, EPI_ISL_2721848, EPI_ISL_2721849, EPI_ISL_2721854, EPI_ISL_2721875, EPI_ISL_2721920, EPI_ISL_2721933, EPI_ISL_2721936, EPI_ISL_2721938, EPI_ISL_2721941, EPI_ISL_2721957, EPI_ISL_2721958, EPI_ISL_2721959 | | | |
| see above | BCCDC Public Health Laboratory | BCCDC Public Health Laboratory | Ana Pacagnella; Corrinne Ng; Dan Fornika; John Tyson; Kim Macdonald; Kimia Kamelian; Linda Hoang; Loretta Janz; Mel Krajden; Prystajecky Natalie; Robert Azana; Shannon Russell |
| EPI_ISL_2421233 | BOSTON HEART DIAGNOSTICS CORP | Wadsworth Center, New York State Department of Health | Alexis Russell; Catharine Prussing; Daryl M. Lamson; Erasmus Schneider; Erica Lasek-Nesselquist; John Kelly; Jonathan Plitnick; Kirsten St. George; Matthew Shudt; Melissa A Leisner; Navjot Singh |
| EPI_ISL_2484739 | Baptist Health Medical Center | Center for Global Health, University of New Mexico Health Sciences Center | Amanda Novack; Darrell Dinwiddie; Daryl Domman; Dirk Haselow; Joshua L. Kennedy; Kurt Schwalm; Valerie Morley |
| EPI_ISL_2083061, EPI_ISL_2083080, EPI_ISL_2193240 | Baylor Scott & White-Temple | Baylor Scott & White-Temple | Ari Rao; Kimberly Walker; Linden Morales; Marcus Volz; Shelby Hendrickson |
| EPI_ISL_2401375, EPI_ISL_2401448, EPI_ISL_3145472, EPI_ISL_3145473, EPI_ISL_3145474 | BioneXt Lab | Laboratoire national de sante, Microbiology, Microbial Genomics Platform | Anke Wienecke-Baldacchino; Catherine Ragimbeau; Elodie Solarino; Fatu Djabi; Jessica Tapp; Lise Pignon; Raoul Salmon; Tamir Abdelrahman; Thibault Ferrandon; Virginie Jover |
| EPI_ISL_3494472, EPI_ISL_3494480, EPI_ISL_3494483, EPI_ISL_3494485 | Boston University CTL | Boston University/National Emerging Infectious Disease Laboratories | Catherine Klapperich; Jacquelyn Turcinovic; John H. Connor; Lena Landeverde; Lynn Doucette-Stamm |
| EPI_ISL_3479009, EPI_ISL_3479011, EPI_ISL_3479012, EPI_ISL_3479013, EPI_ISL_3479014, EPI_ISL_3479017, EPI_ISL_3479018, EPI_ISL_3479019, EPI_ISL_3479020, EPI_ISL_3479021, EPI_ISL_3479023, EPI_ISL_3482745, EPI_ISL_3482750, EPI_ISL_3482751, EPI_ISL_3482753, EPI_ISL_3482757, EPI_ISL_3482759, EPI_ISL_3482760, EPI_ISL_3482761, EPI_ISL_3482804, EPI_ISL_3482807, EPI_ISL_3482810, EPI_ISL_3482812, EPI_ISL_3482813, EPI_ISL_3482819, EPI_ISL_3482820, EPI_ISL_3482822, EPI_ISL_3482823, EPI_ISL_3482824, EPI_ISL_3482829, EPI_ISL_3482844, EPI_ISL_3482845, EPI_ISL_3482850, EPI_ISL_3482851, EPI_ISL_3482852, EPI_ISL_3482857, EPI_ISL_3482862, EPI_ISL_3482878, EPI_ISL_3482880, EPI_ISL_3482884, EPI_ISL_3482891, EPI_ISL_3482919, EPI_ISL_3482927, EPI_ISL_3482929, EPI_ISL_3482994, EPI_ISL_3482995, EPI_ISL_3483000, EPI_ISL_3483006, EPI_ISL_3483009, EPI_ISL_3483010, EPI_ISL_3483011, EPI_ISL_3483013, EPI_ISL_3483016, EPI_ISL_3483017, EPI_ISL_3483019, EPI_ISL_3483021, EPI_ISL_3483025, EPI_ISL_3483026, EPI_ISL_3483027, EPI_ISL_3483031, EPI_ISL_3483039, EPI_ISL_3483041, EPI_ISL_3483042, EPI_ISL_3483053, EPI_ISL_3483054, EPI_ISL_3483055, EPI_ISL_3483064, EPI_ISL_3483070, EPI_ISL_3483072, EPI_ISL_3483092, EPI_ISL_3483093, EPI_ISL_3483094, EPI_ISL_3483095, EPI_ISL_3483096, EPI_ISL_3483097, EPI_ISL_3483099, EPI_ISL_3483100, EPI_ISL_3483101, EPI_ISL_3483103, EPI_ISL_3483106, EPI_ISL_3483107, EPI_ISL_3483109, EPI_ISL_3483113, EPI_ISL_3483114, EPI_ISL_3483116, EPI_ISL_3483117, EPI_ISL_3483119, EPI_ISL_3483121, EPI_ISL_3483123, EPI_ISL_3483124, EPI_ISL_3483125, EPI_ISL_3483126, EPI_ISL_3483129, EPI_ISL_3483130, EPI_ISL_3483131, EPI_ISL_3483132, EPI_ISL_3483134, EPI_ISL_3483139, EPI_ISL_3483146, EPI_ISL_3483148, EPI_ISL_3483152, EPI_ISL_3483154, EPI_ISL_3483155, EPI_ISL_3483157, EPI_ISL_3483159, EPI_ISL_3483161, EPI_ISL_3483162, EPI_ISL_3483167, EPI_ISL_3483168, EPI_ISL_3483169, EPI_ISL_3483176, EPI_ISL_3483178, EPI_ISL_3483179, EPI_ISL_3483183, EPI_ISL_3483194, EPI_ISL_3483199, EPI_ISL_3483202, EPI_ISL_3483204, EPI_ISL_3483205, EPI_ISL_3483215, EPI_ISL_3483216, EPI_ISL_3483218, EPI_ISL_3483221, EPI_ISL_3483223, EPI_ISL_3483228, EPI_ISL_3483229, EPI_ISL_3483230, EPI_ISL_3483231, EPI_ISL_3483232, EPI_ISL_3483236, EPI_ISL_3483244, EPI_ISL_3483250, EPI_ISL_3483257, EPI_ISL_3483260, EPI_ISL_3483265, EPI_ISL_3483266, EPI_ISL_3483267, EPI_ISL_3483269, EPI_ISL_3483272, EPI_ISL_3483277, EPI_ISL_3483281, EPI_ISL_3483282, EPI_ISL_3483292, EPI_ISL_3483308, EPI_ISL_3483311, EPI_ISL_3483312, EPI_ISL_3483318, EPI_ISL_3483321, EPI_ISL_3483322, EPI_ISL_3483323, EPI_ISL_3483325, EPI_ISL_3483326, EPI_ISL_3483327, EPI_ISL_3483328, EPI_ISL_3483334, EPI_ISL_3483336, EPI_ISL_3483337, EPI_ISL_3483338, EPI_ISL_3483339, EPI_ISL_3483340, EPI_ISL_3483341, EPI_ISL_3483343, EPI_ISL_3483346, EPI_ISL_3483348, EPI_ISL_3483349, EPI_ISL_3483350, EPI_ISL_3483353, EPI_ISL_3483355, EPI_ISL_3483361, EPI_ISL_3483362, EPI_ISL_3483363, EPI_ISL_3483364, EPI_ISL_3483367, EPI_ISL_3483368, EPI_ISL_3483369, EPI_ISL_3483385, EPI_ISL_3483387, EPI_ISL_3483389, EPI_ISL_3483391, EPI_ISL_3483393, EPI_ISL_3483397, EPI_ISL_3483398, EPI_ISL_3483401, EPI_ISL_3483402, EPI_ISL_3483404, EPI_ISL_3483406, EPI_ISL_3483408, EPI_ISL_3483409, EPI_ISL_3483411, EPI_ISL_3483412, EPI_ISL_3483413, EPI_ISL_3483414, EPI_ISL_3483415, EPI_ISL_3483417, EPI_ISL_3483419, EPI_ISL_3483420, EPI_ISL_3483425, EPI_ISL_3483428, EPI_ISL_3483435, EPI_ISL_3483437, EPI_ISL_3483438, EPI_ISL_3483440, EPI_ISL_3483441, EPI_ISL_3483442, EPI_ISL_3483443, EPI_ISL_3483444, EPI_ISL_3483445, EPI_ISL_3483449, EPI_ISL_3483453, EPI_ISL_3483455, EPI_ISL_3483456, EPI_ISL_3483457, EPI_ISL_3483459, EPI_ISL_3483463, EPI_ISL_3483464, EPI_ISL_3483470, EPI_ISL_3483473, EPI_ISL_3483475, EPI_ISL_3483480, EPI_ISL_3483493, EPI_ISL_3483498, EPI_ISL_3483504, EPI_ISL_3483508, EPI_ISL_3483511, EPI_ISL_3483514, EPI_ISL_3483519, EPI_ISL_3483521, EPI_ISL_3483528, EPI_ISL_3483530, EPI_ISL_3483533, EPI_ISL_3483535, EPI_ISL_3483537, EPI_ISL_3483540, EPI_ISL_3483542, EPI_ISL_3483545, EPI_ISL_3483550, EPI_ISL_3483552, EPI_ISL_3483557, EPI_ISL_3483559, EPI_ISL_3483560, EPI_ISL_3483561, EPI_ISL_3483562, EPI_ISL_3483563, EPI_ISL_3483564, EPI_ISL_3483565, EPI_ISL_3483567, EPI_ISL_3483568, EPI_ISL_3483569, EPI_ISL_3483572, EPI_ISL_3483573, EPI_ISL_3483574, EPI_ISL_3483577, EPI_ISL_3483578, EPI_ISL_3483580, EPI_ISL_3483582, EPI_ISL_3483586, EPI_ISL_3483587, EPI_ISL_3483588, EPI_ISL_3483589, EPI_ISL_3483610, EPI_ISL_3483611, EPI_ISL_3483613, EPI_ISL_3483617, EPI_ISL_3483619, EPI_ISL_3483622, EPI_ISL_3483623, EPI_ISL_3483631, EPI_ISL_3483645, EPI_ISL_3483648, EPI_ISL_3483650, EPI_ISL_3483651, EPI_ISL_3483652, EPI_ISL_3483653, EPI_ISL_3483655, EPI_ISL_3483656, EPI_ISL_3483701, EPI_ISL_3483988, EPI_ISL_3483990, EPI_ISL_3483992, EPI_ISL_3483994, EPI_ISL_3484007, EPI_ISL_3484011, EPI_ISL_3484045, EPI_ISL_3484060, EPI_ISL_3484062, EPI_ISL_3484183, EPI_ISL_3484192, EPI_ISL_3484194, EPI_ISL_3484196, EPI_ISL_3484198, EPI_ISL_3484201, EPI_ISL_3484203, EPI_ISL_3484219, EPI_ISL_3484401, EPI_ISL_3484403, EPI_ISL_3484406, EPI_ISL_3484462, EPI_ISL_3484463, EPI_ISL_3484465, EPI_ISL_3484467, EPI_ISL_3484470, EPI_ISL_3484471, EPI_ISL_3484472, EPI_ISL_3484473, EPI_ISL_3484476, EPI_ISL_3484478, EPI_ISL_3484486, EPI_ISL_3484490, EPI_ISL_3484494, EPI_ISL_3484506, EPI_ISL_3484513, EPI_ISL_3484516, EPI_ISL_3484521, EPI_ISL_3484522, EPI_ISL_3484524, EPI_ISL_3484530, EPI_ISL_3484549, EPI_ISL_3484551, EPI_ISL_3484552, EPI_ISL_3484553, EPI_ISL_3484557, EPI_ISL_3484561, EPI_ISL_3484562, EPI_ISL_3484565, EPI_ISL_3484574, EPI_ISL_3485074, EPI_ISL_3485188, EPI_ISL_3596396, EPI_ISL_3596409, EPI_ISL_3596412, EPI_ISL_3596415, EPI_ISL_3596418, EPI_ISL_3596447, EPI_ISL_3596450, EPI_ISL_3596453, EPI_ISL_3596459, EPI_ISL_3600014, EPI_ISL_3600015, EPI_ISL_3600039, EPI_ISL_3600041, EPI_ISL_3600043, EPI_ISL_3600057, EPI_ISL_3600082, EPI_ISL_3600096, EPI_ISL_3600100, EPI_ISL_3600106, EPI_ISL_3600109, EPI_ISL_3600130, EPI_ISL_3600131, EPI_ISL_3600147, EPI_ISL_3600154, EPI_ISL_3600157, EPI_ISL_3600165, EPI_ISL_3600181, EPI_ISL_3600183, EPI_ISL_3600193, EPI_ISL_3600194, EPI_ISL_3600197, EPI_ISL_3600199, EPI_ISL_3600201, EPI_ISL_3600202, EPI_ISL_3600209, EPI_ISL_3600211, EPI_ISL_3600212, EPI_ISL_3600215, EPI_ISL_3600219, EPI_ISL_3600224, EPI_ISL_3600254, EPI_ISL_3600260, EPI_ISL_3600261, EPI_ISL_3600265, EPI_ISL_3600267, EPI_ISL_3600288, EPI_ISL_3600298, EPI_ISL_3600299, EPI_ISL_3600306, EPI_ISL_3600307, EPI_ISL_3600309, EPI_ISL_3600310, EPI_ISL_3600318, EPI_ISL_3600319, EPI_ISL_3600324, EPI_ISL_3600333, EPI_ISL_3600340, EPI_ISL_3600346, EPI_ISL_3600351, EPI_ISL_3600355, EPI_ISL_3600358, EPI_ISL_3600362, EPI_ISL_3600365, EPI_ISL_3600372, EPI_ISL_3600377, EPI_ISL_3600378, EPI_ISL_3600380, EPI_ISL_3600382, EPI_ISL_3600385, EPI_ISL_3600408, EPI_ISL_3600417, EPI_ISL_3600425, EPI_ISL_3600428, EPI_ISL_3600433, EPI_ISL_3600439, EPI_ISL_3600444, EPI_ISL_3600445, EPI_ISL_3600446, EPI_ISL_3600447, EPI_ISL_3600449, EPI_ISL_3600452, EPI_ISL_3600460, EPI_ISL_3600461, EPI_ISL_3600465, EPI_ISL_3600466, EPI_ISL_3600467, EPI_ISL_3600470, EPI_ISL_3600473, EPI_ISL_3600474, EPI_ISL_3600476, EPI_ISL_3600477, EPI_ISL_3600478, EPI_ISL_3600480, EPI_ISL_3600483, EPI_ISL_3600484, EPI_ISL_3600485, EPI_ISL_3600487, EPI_ISL_3600492, EPI_ISL_3600493, EPI_ISL_3600494, EPI_ISL_3600498, EPI_ISL_3600505, EPI_ISL_3600514, EPI_ISL_3600516, EPI_ISL_3600517, EPI_ISL_3600518, EPI_ISL_3600522, EPI_ISL_3600551, EPI_ISL_3600554, EPI_ISL_3600567, EPI_ISL_3600571, EPI_ISL_3600575, EPI_ISL_3600578, EPI_ISL_3600579, EPI_ISL_3600580, EPI_ISL_3600594, EPI_ISL_3600600, EPI_ISL_3600610, EPI_ISL_3600614, EPI_ISL_3600620, EPI_ISL_3600621, EPI_ISL_3600623, EPI_ISL_3600624, EPI_ISL_3600633, EPI_ISL_3600636, EPI_ISL_3600638, EPI_ISL_3600647, EPI_ISL_3600649, EPI_ISL_3600662, EPI_ISL_3600691, EPI_ISL_3600702, EPI_ISL_3600712, EPI_ISL_3600713, EPI_ISL_3600719, EPI_ISL_3600723, EPI_ISL_3600732, EPI_ISL_3600746, EPI_ISL_3600752, EPI_ISL_3600755, EPI_ISL_3600757, EPI_ISL_3600758, EPI_ISL_3600762, EPI_ISL_3600766, EPI_ISL_3600770, EPI_ISL_3600787, EPI_ISL_3600788, EPI_ISL_3600790, EPI_ISL_3600801, EPI_ISL_3600807, EPI_ISL_3600809, EPI_ISL_3600821, EPI_ISL_3601112, EPI_ISL_4423643, EPI_ISL_4425188, EPI_ISL_4425195, EPI_ISL_4425200, EPI_ISL_4425257, EPI_ISL_4425264, EPI_ISL_4425269, EPI_ISL_4425274, EPI_ISL_4425288, EPI_ISL_4425330, EPI_ISL_7314600 | | | |
| see above | British Columbia Centre For Disease Control | BCCDC Public Health Laboratory | Ana Pacagnella; Corrinne Ng; Dan Fornika; John Tyson; Kim Macdonald; Kimia Kamelian; Linda Hoang; Loretta Janz; Mel Krajden; Prystajecky Natalie; Robert Azana; Shannon Russell |
| EPI_ISL_2096218, EPI_ISL_2096222, EPI_ISL_2096224, EPI_ISL_2096228, EPI_ISL_2096230, EPI_ISL_2096235, EPI_ISL_2096238, EPI_ISL_2096239, EPI_ISL_2096244, EPI_ISL_2096252, EPI_ISL_2096254, EPI_ISL_2096258, EPI_ISL_2096259, EPI_ISL_2096262, EPI_ISL_2096306, EPI_ISL_2096315, EPI_ISL_2096319, EPI_ISL_2096327, EPI_ISL_2096337, EPI_ISL_2096340, EPI_ISL_2096362, EPI_ISL_2096381, EPI_ISL_2096388, EPI_ISL_2096394, EPI_ISL_2096402, EPI_ISL_2096418, EPI_ISL_2096424, EPI_ISL_2096428, EPI_ISL_2096431, EPI_ISL_2096435, EPI_ISL_2096441, EPI_ISL_2096452, EPI_ISL_2096454, EPI_ISL_2096458, EPI_ISL_2096479, EPI_ISL_2096500, EPI_ISL_2096511, EPI_ISL_2096536, EPI_ISL_2096540, EPI_ISL_2096543, EPI_ISL_2096557, EPI_ISL_2096560, EPI_ISL_2096571, EPI_ISL_2096574, EPI_ISL_2096588, EPI_ISL_2096596, EPI_ISL_2096614, EPI_ISL_2096622, EPI_ISL_2096625, EPI_ISL_2096632, EPI_ISL_2096639, EPI_ISL_2096649, EPI_ISL_2096652, EPI_ISL_2096663, EPI_ISL_2096673, EPI_ISL_2096719, EPI_ISL_2096726, EPI_ISL_2103829, EPI_ISL_2103830, EPI_ISL_2103836, EPI_ISL_2103839, EPI_ISL_2103844, EPI_ISL_2103857, EPI_ISL_2103858, EPI_ISL_2103861, EPI_ISL_2103864, EPI_ISL_2103866, EPI_ISL_2103869, EPI_ISL_2103874, EPI_ISL_2103879, EPI_ISL_2103883, EPI_ISL_2103886, EPI_ISL_2103891, EPI_ISL_2103892, EPI_ISL_2103901, EPI_ISL_2103912, EPI_ISL_2103913, EPI_ISL_2103921, EPI_ISL_2103932, EPI_ISL_2103941, EPI_ISL_2103949, EPI_ISL_2103965, EPI_ISL_2103974, EPI_ISL_2103978, EPI_ISL_2103984, EPI_ISL_2103985, EPI_ISL_2103991, EPI_ISL_2103998, EPI_ISL_2104008, EPI_ISL_2104013, EPI_ISL_2104017, EPI_ISL_2104019, EPI_ISL_2104020, EPI_ISL_2104031, EPI_ISL_2104039, EPI_ISL_2104043, EPI_ISL_2104046, EPI_ISL_2104061, EPI_ISL_2104081, EPI_ISL_2104088, EPI_ISL_2104093, EPI_ISL_2104117, EPI_ISL_2104122, EPI_ISL_2104150, EPI_ISL_2104151, EPI_ISL_2104155, EPI_ISL_2104161, EPI_ISL_2104162, EPI_ISL_2104168, EPI_ISL_2104171, EPI_ISL_2104172, EPI_ISL_2104173, EPI_ISL_2104183, EPI_ISL_2104192, EPI_ISL_2104194, EPI_ISL_2104202, EPI_ISL_2104203, EPI_ISL_2104205, EPI_ISL_2104212, EPI_ISL_2104217, EPI_ISL_2104225, EPI_ISL_2104231, EPI_ISL_2104238, EPI_ISL_2104244, EPI_ISL_2104247, EPI_ISL_2104252, EPI_ISL_2104259, EPI_ISL_2104267, EPI_ISL_2104271, EPI_ISL_2104273, EPI_ISL_2104277, EPI_ISL_2104280, EPI_ISL_2104286, EPI_ISL_2104287, EPI_ISL_2104319, EPI_ISL_2104331, EPI_ISL_2104381 | | | |
| see above | Broad Institute Clinical Research Sequencing Platform | Infectious Disease Program, Broad Institute of Harvard and MIT | Adams, G.; B.L.; B.W.; Bauer, M.; Birren; Blumenstiel, B.; Brown, C.; Carter, A.; Chaluvadi, S.; D.J.; DeFelice, M.; DeRuff, K.; Dodge, S.; Gabriel, S.; Gallagher, G.; Gladden-Young, A.; Granger, B.; J.E.; K.J.; Lagerborg, K.; Larkin, K.; Lee, M.; Lemieux; Lennon, N.; Loreth, C.; Madoff, L.; McGovern, S.; Meldrim, J.; Normandin, E.; P.C.; Park; Pearlman, L.; Reilly, S.; Rudy, M.; Sabeti; Siddle; Smole, S.; Tomkins-Tinch, C.; Vicente, G.; and MacInnis |
| EPI_ISL_2496900, EPI_ISL_2496906, EPI_ISL_2496909, EPI_ISL_2497648, EPI_ISL_2497649, EPI_ISL_2497650, EPI_ISL_2497651, EPI_ISL_2497652, EPI_ISL_2497655, EPI_ISL_2497658, EPI_ISL_2497665, EPI_ISL_2497668 | | | |
| see above | CA DPH Viral and Rickettsial Disease Laboratory | Chan-Zuckerberg Biohub | CZB Cliahub Consortium |
| EPI_ISL_2383986, EPI_ISL_2383987 | CA-Los Angeles County Public Health Laboratory | Centers for Disease Control and Prevention Division of Viral Diseases, Pathogen Discovery | Alison Laufer Halpin; Ben L. Rambo-Martin; Clinton R. Paden; Dakota Howard; Darlene Wagner; Dave Wentworth; Dhwani Batra; Jasmine Padilla; Justin Lee; Katie Dillon; Krista Queen; Kristen Knipe; Kristine Lacek; Mark Burroughs; Matthew Schmerer; Mili Sheth; Peter Cook; Sam Shepard; Sarah Nobles; Shoshona Le; Suxiang Tong; Vivien Dugan; Yvette Unoarumhi |
| EPI_ISL_2891748, EPI_ISL_2891793, EPI_ISL_2891816, EPI_ISL_2891819, EPI_ISL_2891821, EPI_ISL_2891824, EPI_ISL_2891826, EPI_ISL_2891837, EPI_ISL_2891846, EPI_ISL_2891848, EPI_ISL_2891852, EPI_ISL_2891855, EPI_ISL_2891920, EPI_ISL_2891922, EPI_ISL_2891929, EPI_ISL_2891930, EPI_ISL_2891939, EPI_ISL_2891951, EPI_ISL_2891953, EPI_ISL_2892018, EPI_ISL_2892115, EPI_ISL_2892127, EPI_ISL_2892197, EPI_ISL_2892198, EPI_ISL_2892244, EPI_ISL_2892362, EPI_ISL_2892365, EPI_ISL_2921887, EPI_ISL_2921888, EPI_ISL_2921947, EPI_ISL_2921948, EPI_ISL_2921975, EPI_ISL_2921980, EPI_ISL_2977096, EPI_ISL_2977113, EPI_ISL_2977159, EPI_ISL_2977188, EPI_ISL_2977198, EPI_ISL_2977205, EPI_ISL_2977212, EPI_ISL_2977229, EPI_ISL_2977236, EPI_ISL_2977267, EPI_ISL_2977271, EPI_ISL_2977281, EPI_ISL_2977288, EPI_ISL_2977299, EPI_ISL_2977307, EPI_ISL_2977313, EPI_ISL_2977315, EPI_ISL_2977319, EPI_ISL_2977322, EPI_ISL_2977326, EPI_ISL_2977330, EPI_ISL_2977338, EPI_ISL_2977339, EPI_ISL_2977346, EPI_ISL_2977350, EPI_ISL_2977352, EPI_ISL_2977356, EPI_ISL_2977361, EPI_ISL_2977372, EPI_ISL_2977378, EPI_ISL_2977385, EPI_ISL_2977390, EPI_ISL_2977391, EPI_ISL_2977393, EPI_ISL_2977405, EPI_ISL_2977417, EPI_ISL_2977449, EPI_ISL_2978168, EPI_ISL_3243751, EPI_ISL_3243752, EPI_ISL_3243757, EPI_ISL_3243767, EPI_ISL_3243781, EPI_ISL_3243785, EPI_ISL_3243791, EPI_ISL_3243796, EPI_ISL_3243809, EPI_ISL_3243810, EPI_ISL_3243812, EPI_ISL_3243822, EPI_ISL_3243834, EPI_ISL_3243837, EPI_ISL_5696584, EPI_ISL_7335366, EPI_ISL_7335566 | | | |
| see above | CDPH VBL | California Department of Public Health | CDPH-COVIDNet; Emily Smith on behalf of CDPH-COVIDNet; Emily Smith on behalf of CDPH-COVIDNet and Priva Path Labs Inc.; Emily Smith on behalf of CDPH-COVIDNet and UCSD EXCITE lab; UCSD EXCITE lab |
| EPI_ISL_2260022 | CENTOGENE Frankfurt Laboratory: Niederlassung Industriepark Höchst | Robert Koch Institute | |
| EPI_ISL_6229699 | CENTRO DE ATENDIMENTO COVID 19 TEREZINHA NUNES DA ROCHA | ACME Lab, Oswaldo Cruz Foundation, FIOCRUZ/CE | Carlos Leonardo de Aragao Araujo; Cecilia Leite Costa & Eduardo Ruback dos Santos on behalf of COVID-19 FIOCRUZ Genomic Network; Cleber Furtado Aksenen; Fabio Miyajima; Fernando Braga Stehling; Francisco Eder de Moura Lopes; Igor Oliveira Duarte; Jamille Maria Mendes Bezerra; Joaquim Cesar do Nascimento Sousa Junior; Pedro Miguel Carneiro Jeronimo; Suzana Porto Almeida; Thais Ferreira de Oliveira; Thais de Oliveira Costa; Ticiane Cavalcante de Souza; Veridiana Pessoa Miyajima |
| EPI_ISL_2345715 | CENTRO DE ESPECIALIDADES DE PRIMAVERA | Instituto Butantan / Mendelics | Antonio Jorge Martins; Claudia Renata dos Santos Barros; David Schlesinger; Debora Botequio Moretti; Dimas Tadeu Covas; Elaine Cristina Marqueze; Elaine Vieira Santos; Evandra Strazza Rodrigues; Heidge Fukumasu; Jayme Augusto de Souza-Neto; José Salvatore Leister Patané; Luiz Alcantara; Luiz Lehmann Coutinho; Maria Carolina Elias; Maurício Lacerda Nogueira; Rafael dos Santos Bezerra; Raul Machado Neto; Rejane Maria Tommasini Grotto; Ricardo Haddad; Sandra Coccuzzo Sampaio Vessoni; Simone Kashima; Svetoslav Nanev Slavov; Vincent Louis Viala |
| EPI_ISL_5650324 | CENTRO DE ESPECIALIDDS MEDICAS IRMA LEOPOLDINA PIRASSUNUNGA | Instituto Butantan | Antonio Jorge Martins; Claudia Renata dos Santos Barros; David Schlesinger; Debora Botequio Moretti; Dimas Tadeu Covas; Elaine Cristina Marqueze; Elaine Vieira Santos; Evandra Strazza Rodrigues; Heidge Fukumasu; Jayme Augusto de Souza-Neto; José Salvatore Leister Patané; Luiz Alcantara; Luiz Lehmann Coutinho; Maria Carolina Elias; Maurício Lacerda Nogueira; Rafael dos Santos Bezerra; Raul Machado Neto; Rejane Maria Tommasini Grotto; Ricardo Haddad; Sandra Coccuzzo Sampaio Vessoni; Simone Kashima; Svetoslav Nanev Slavov; Vincent Louis Viala |
| EPI_ISL_2345815, EPI_ISL_2345860, EPI_ISL_2345865 | CENTRO DE REFERENCIA DO IDOSO DR HUMBERTO MENDES DE CARVALHO | Instituto Butantan / Mendelics | Antonio Jorge Martins; Claudia Renata dos Santos Barros; David Schlesinger; Debora Botequio Moretti; Dimas Tadeu Covas; Elaine Cristina Marqueze; Elaine Vieira Santos; Evandra Strazza Rodrigues; Heidge Fukumasu; Jayme Augusto de Souza-Neto; José Salvatore Leister Patané; Luiz Alcantara; Luiz Lehmann Coutinho; Maria Carolina Elias; Maurício Lacerda Nogueira; Rafael dos Santos Bezerra; Raul Machado Neto; Rejane Maria Tommasini Grotto; Ricardo Haddad; Sandra Coccuzzo Sampaio Vessoni; Simone Kashima; Svetoslav Nanev Slavov; Vincent Louis Viala |
| EPI_ISL_6229700, EPI_ISL_6229858, EPI_ISL_6229870 | CENTRO DE SAUDE DA FAMILIA DE MASSAPE SEDE IV | ACME Lab, Oswaldo Cruz Foundation, FIOCRUZ/CE | Carlos Leonardo de Aragao Araujo; Cecilia Leite Costa & Eduardo Ruback dos Santos on behalf of COVID-19 FIOCRUZ Genomic Network; Cleber Furtado Aksenen; Fabio Miyajima; Fernando Braga Stehling; Francisco Eder de Moura Lopes; Igor Oliveira Duarte; Jamille Maria Mendes Bezerra; Joaquim Cesar do Nascimento Sousa Junior; Pedro Miguel Carneiro Jeronimo; Suzana Porto Almeida; Thais Ferreira de Oliveira; Thais de Oliveira Costa; Ticiane Cavalcante de Souza; Veridiana Pessoa Miyajima |
| EPI_ISL_2209593, EPI_ISL_2345657, EPI_ISL_2345702, EPI_ISL_2345753 | CENTRO DE SAUDE DE MARACAI | Instituto Butantan / Mendelics | Antonio Jorge Martins; Claudia Renata dos Santos Barros; David Schlesinger; Debora Botequio Moretti; Dimas Tadeu Covas; Elaine Cristina Marqueze; Elaine Vieira Santos; Evandra Strazza Rodrigues; Heidge Fukumasu; Jayme Augusto de Souza-Neto; José Salvatore Leister Patané; Luiz Alcantara; Luiz Lehmann Coutinho; Maria Carolina Elias; Maurício Lacerda Nogueira; Rafael dos Santos Bezerra; Raul Machado Neto; Rejane Maria Tommasini Grotto; Ricardo Haddad; Sandra Coccuzzo Sampaio Vessoni; Simone Kashima; Svetoslav Nanev Slavov; Vincent Louis Viala |
| EPI_ISL_6229701, EPI_ISL_6229702, EPI_ISL_6229825, EPI_ISL_6229872 | CENTRO DE SAUDE DE PENTECOSTE | ACME Lab, Oswaldo Cruz Foundation, FIOCRUZ/CE | Carlos Leonardo de Aragao Araujo; Cecilia Leite Costa & Eduardo Ruback dos Santos on behalf of COVID-19 FIOCRUZ Genomic Network; Cleber Furtado Aksenen; Fabio Miyajima; Fernando Braga Stehling; Francisco Eder de Moura Lopes; Igor Oliveira Duarte; Jamille Maria Mendes Bezerra; Joaquim Cesar do Nascimento Sousa Junior; Pedro Miguel Carneiro Jeronimo; Suzana Porto Almeida; Thais Ferreira de Oliveira; Thais de Oliveira Costa; Ticiane Cavalcante de Souza; Veridiana Pessoa Miyajima |
| EPI_ISL_5603312 | CENTRO DE SAUDE DR MIRANDA TAVARES | Analytical Competence Molecular Epidemiology Lab/ACME, Oswaldo Cruz Foundation, Ceara (FIOCRUZ CE) | Carlos Leonardo de Aragao Araujo; Cecilia Leite Costa & Eduardo Ruback dos Santos on behalf of COVID-19 FIOCRUZ Genomic Network; Cleber Furtado Aksenen; Fabio Miyajima; Fernando Braga Stehling; Francisco Eder de Moura Lopes; Igor Oliveira Duarte; Jamille Maria Mendes Bezerra; Joaquim Cesar do Nascimento Sousa Junior; Pedro Miguel Carneiro Jeronimo; Suzana Porto Almeida; Thais Ferreira de Oliveira; Thais de Oliveira Costa; Ticiane Cavalcante de Souza; Veridiana Pessoa Miyajima |
| EPI_ISL_2208974, EPI_ISL_2209353, | CENTRO DE SAUDE II IBITINGA | Instituto Butantan / Mendelics | Antonio Jorge Martins; Claudia Renata dos Santos Barros; David Schlesinger; Debora Botequio Moretti; Dimas Tadeu Covas; Elaine Cristina Marqueze; Elaine Vieira Santos; Evandra Strazza Rodrigues; Heidge Fukumasu; Jayme Augusto de Souza-Neto; José Salvatore Leister Patané; Luiz Alcantara; Luiz Lehmann Coutinho; Maria Carolina Elias; Maurício Lacerda Nogueira; Rafael dos Santos Bezerra; Raul Machado Neto; Rejane Maria Tommasini Grotto; Ricardo Haddad; Sandra Coccuzzo Sampaio Vessoni; Simone Kashima; Svetoslav Nanev Slavov; Vincent Louis Viala |

| | | | |
|---|---|---|---|
| EPI_ISL_2209368, EPI_ISL_2345724, EPI_ISL_2345766, EPI_ISL_2345861 | | | |
| EPI_ISL_5650241, EPI_ISL_5650242 | CENTRO DE SAUDE III DE AGUAS DE SAO PEDRO | Instituto Butantan | Antonio Jorge Martins; Claudia Renata dos Santos Barros; David Schlesinger; Debora Botequio Moretti; Dimas Tadeu Covas; Elaine Cristina Marqueze; Elaine Vieira Santos; Evandra Strazza Rodrigues; Heidge Fukumasu; Jayme Augusto de Souza-Neto; José Salvatore Leister Patané; Luiz Alcantara; Luiz Lehmann Coutinho; Maria Carolina Elias; Maurício Lacerda Nogueira; Rafael dos Santos Bezerra; Raul Machado Neto; Rejane Maria Tommasini Grotto; Ricardo Haddad; Sandra Coccuzzo Sampaio Vessoni; Simone Kashima; Svetoslav Nanev Slavov; Vincent Louis Viala |
| EPI_ISL_2209702 | CENTRO DE SAUDE III NELCIDIO DA SILVEIRA BASTOS | Instituto Butantan / Mendelics | Antonio Jorge Martins; Claudia Renata dos Santos Barros; David Schlesinger; Debora Botequio Moretti; Dimas Tadeu Covas; Elaine Cristina Marqueze; Elaine Vieira Santos; Evandra Strazza Rodrigues; Heidge Fukumasu; Jayme Augusto de Souza-Neto; José Salvatore Leister Patané; Luiz Alcantara; Luiz Lehmann Coutinho; Maria Carolina Elias; Maurício Lacerda Nogueira; Rafael dos Santos Bezerra; Raul Machado Neto; Rejane Maria Tommasini Grotto; Ricardo Haddad; Sandra Coccuzzo Sampaio Vessoni; Simone Kashima; Svetoslav Nanev Slavov; Vincent Louis Viala |
| EPI_ISL_2208938, EPI_ISL_2209968, EPI_ISL_2210119, EPI_ISL_2345697, EPI_ISL_2345842 | CENTRO DE SAUDE III SALES OLIVEIRA | Instituto Butantan / Mendelics | Antonio Jorge Martins; Claudia Renata dos Santos Barros; David Schlesinger; Debora Botequio Moretti; Dimas Tadeu Covas; Elaine Cristina Marqueze; Elaine Vieira Santos; Evandra Strazza Rodrigues; Heidge Fukumasu; Jayme Augusto de Souza-Neto; José Salvatore Leister Patané; Luiz Alcantara; Luiz Lehmann Coutinho; Maria Carolina Elias; Maurício Lacerda Nogueira; Rafael dos Santos Bezerra; Raul Machado Neto; Rejane Maria Tommasini Grotto; Ricardo Haddad; Sandra Coccuzzo Sampaio Vessoni; Simone Kashima; Svetoslav Nanev Slavov; Vincent Louis Viala |
| EPI_ISL_5650321 | CENTRO MEDICO DR NELSON SALOME DE CONCHAL | Instituto Butantan | Antonio Jorge Martins; Claudia Renata dos Santos Barros; David Schlesinger; Debora Botequio Moretti; Dimas Tadeu Covas; Elaine Cristina Marqueze; Elaine Vieira Santos; Evandra Strazza Rodrigues; Heidge Fukumasu; Jayme Augusto de Souza-Neto; José Salvatore Leister Patané; Luiz Alcantara; Luiz Lehmann Coutinho; Maria Carolina Elias; Maurício Lacerda Nogueira; Rafael dos Santos Bezerra; Raul Machado Neto; Rejane Maria Tommasini Grotto; Ricardo Haddad; Sandra Coccuzzo Sampaio Vessoni; Simone Kashima; Svetoslav Nanev Slavov; Vincent Louis Viala |
| EPI_ISL_2248789, EPI_ISL_2248792, EPI_ISL_2248794 | CHEDV | Instituto Nacional de Saude (INSA) | Borges et al |
| EPI_ISL_2688692 | CHI Elbeuf | Centre Hospitalier Universitaire de Rouen Laboratoire de Virologie | Alice Moisan; Fabienne De Oliveira; Marie Leoz |
| EPI_ISL_2248827 | CHLC | Instituto Nacional de Saude (INSA) | Borges et al |
| EPI_ISL_2566671 | CHU NIMES | CHU NIMES | Agathe Boudet; Marie-josée Carles; Sophie Bravo; Stephan Robin |
| EPI_ISL_2533931 | CIAD-Hermosillo | Immunology Laboratory | ; Alejandra García-Gasca; Alejandra Hernández-Terán; Alejandro Sánchez-Flores; Alfredo Herrera-Estrella; Alicia Ocaña-Mondragón; Andreu Comas-García; Angel Gustavo Salas-Lais; Antonio Loza Román; Bernardo Martínez-Miguel; Blanca Taboada; Brenda Irasema Maldonado-Meza; Bruno Gómez-Gil; Carla Ivón Herrera-Najera; Carlos F. Arias; Celia Boukadida; Clara Esperanza Santacruz-Tinoco; Concepción Grajales-Muñiz; Consorcio Mexicano de Vigilancia Genómica (CoViGen-Mex). Authors (in alphabetical order): Julio Elias Alvarado-Yaah; Cristóbal Cháidez-Quiróz; Célida Duque Molina; Célida Martínez- Rodríguez; Daniel Fregoso-Rueda; Daniel Lira Morales; Eduardo Becerril-Vargas; Fernando Fontove-Herrera; Fidencio Mejía-Nepomuceno; Francisco Pulido; Gloria Elena Espinosa-Ayala; Gloria María Molina-Salinas; Gloria Vazquez; Hector Esteban Paz-Juárez; Hector Montoya-Fuentes; Helen Haydee Fernanda Ramírez-Plascencia; Irvin González-López; Jean Pierre González; Jesús Hernández; Joel Armando Vázquez-Pérez.; Jorge Salas-Hernández; José Antonio Enciso-Moreno; José Arturo Martínez-Orozco; José Esteban Muñoz-Medina; José de Jesús Nuñez-Contreras; Juan Bautista Chale-Dzul; Julissa Enciso-Ibarra; Luis Alberto Ochoa-Carrera; Margarita Matías-Florentino; Mario Mújica-Sánchez; Marissa Perez-Garcia; María Guadalupe Santiago-Mauricio; María Guadalupe de Jesús Mireles-Rivera; Nelly Sélem-Mojica; Pavel Isa; Ricardo Ciria Merce; Ricardo Grande; Rosa María Gutiérrez Rios; Santiago Ávila-Ríos; Selene Zárate; Susana Lopez; Verónica Mata-Haro; Victor Eduardo García-Arias; Víctor Hugo Borja-Aburto |
| EPI_ISL_2351955, EPI_ISL_2351957, EPI_ISL_2351958, EPI_ISL_2351974, EPI_ISL_2351978 | CLILAB | Microbiology Department | Aida Gonzalez-Diaz; Carmen Ardanuy; Jordi Camara; Jordi Niubó; Laura Calatayud; M Angeles Domínguez; Miguel Fernandez-Huerta; Sara Marti |
| EPI_ISL_6229708 | CSF DO PE DA SERRA MANOEL CLARINDO LOPES | ACME Lab, Oswaldo Cruz Foundation, FIOCRUZ/CE | Carlos Leonardo de Aragao Araujo; Cecilia Leite Costa & Eduardo Ruback dos Santos on behalf of COVID-19 FIOCRUZ Genomic Network; Cleber Furtado Aksenen; Fabio Miyajima; Fernando Braga Stehling; Francisco Eder de Moura Lopes; Igor Oliveira Duarte; Jamille Maria Mendes Bezerra; Joaquim Cesar do Nascimento Sousa Junior; Pedro Miguel Carneiro Jeronimo; Suzana Porto Almeida; Thais Ferreira de Oliveira; Thais de Oliveira Costa; Ticiane Cavalcante de Souza; Veridiana Pessoa Miyajima |
| EPI_ISL_6229709 | CSF DO TANGENTE CICERO JACINTO DE OLIVEIRA | ACME Lab, Oswaldo Cruz Foundation, FIOCRUZ/CE | Carlos Leonardo de Aragao Araujo; Cecilia Leite Costa & Eduardo Ruback dos Santos on behalf of COVID-19 FIOCRUZ Genomic Network; Cleber Furtado Aksenen; Fabio Miyajima; Fernando Braga Stehling; Francisco Eder de Moura Lopes; Igor Oliveira Duarte; Jamille Maria Mendes Bezerra; Joaquim Cesar do Nascimento Sousa Junior; Pedro Miguel Carneiro Jeronimo; Suzana Porto Almeida; Thais Ferreira de Oliveira; Thais de Oliveira Costa; Ticiane Cavalcante de Souza; Veridiana Pessoa Miyajima |
| EPI_ISL_6229710 | CSF JUNCO | ACME Lab, Oswaldo Cruz Foundation, FIOCRUZ/CE | Carlos Leonardo de Aragao Araujo; Cecilia Leite Costa & Eduardo Ruback dos Santos on behalf of COVID-19 FIOCRUZ Genomic Network; Cleber Furtado Aksenen; Fabio Miyajima; Fernando Braga Stehling; Francisco Eder de Moura Lopes; Igor Oliveira Duarte; Jamille Maria Mendes Bezerra; Joaquim Cesar do Nascimento Sousa Junior; Pedro Miguel Carneiro Jeronimo; Suzana Porto Almeida; Thais Ferreira de Oliveira; Thais de Oliveira Costa; Ticiane Cavalcante de Souza; Veridiana Pessoa Miyajima |
| EPI_ISL_2345868 | CSII DR WASHINGTON LUIS M RODRIGUES DA SILVA PITANGUEIRAS | Instituto Butantan / Mendelics | Antonio Jorge Martins; Claudia Renata dos Santos Barros; David Schlesinger; Debora Botequio Moretti; Dimas Tadeu Covas; Elaine Cristina Marqueze; Elaine Vieira Santos; Evandra Strazza Rodrigues; Heidge Fukumasu; Jayme Augusto de Souza-Neto; José Salvatore Leister Patané; Luiz Alcantara; Luiz Lehmann Coutinho; Maria Carolina Elias; Maurício Lacerda Nogueira; Rafael dos Santos Bezerra; Raul Machado Neto; Rejane Maria Tommasini Grotto; Ricardo Haddad; Sandra Coccuzzo Sampaio Vessoni; Simone Kashima; Svetoslav Nanev Slavov; Vincent Louis Viala |
| EPI_ISL_2350401 | CSL, Maryland Department of Health | CSL, Maryland Department of Health | Keller, E.; Washington, Y. |
| EPI_ISL_2892464, EPI_ISL_2892507, EPI_ISL_2892509, EPI_ISL_3536732, EPI_ISL_3536733, EPI_ISL_4005512, EPI_ISL_4005514, EPI_ISL_4005519, EPI_ISL_4005520, EPI_ISL_4005522 | | | |
| see above | California Department of Public Health | California Department of Public Health | CDPH IDLB COVIDNet et al |
| EPI_ISL_2502388, EPI_ISL_2502393, EPI_ISL_2502404, EPI_ISL_2502426, EPI_ISL_2502481, EPI_ISL_2502482 | Central Laboratory, Bureau of Public Health (BOG) and Academic Hospital Paramaribo | Erasmus Medical Center | Bas B Oude Munnink; Cherise Beek; Consuella Partowidjojo; Dion Gajadin; Ed PF IJzerman; Emmanuelle Munger; Gary Gummels; Ingrid SK Krishnadath; Lycke Woittiez; Marion PG Koopmans; Mireille Van de Veer; Phyllis Pinas; Princes Wongsowidjojo; Radjesh Ori; Ranisha Doerbalie; Rohma Banwari; Soeradj Harkisoen; Stephen Vreden; Tilotmadebie Ramlal; Verne Nanhoe |
| EPI_ISL_4296390 | Central Medical Laboratory | Baylor College of Medicine | Adrianna Maliga; Alexander Kneubehl; Allison Lino; Gerhaldine Morazan; Kristy Murray; Russell Manzanero; Sarah Gunter; Sarah Strobel; Shannon Ronca |
| EPI_ISL_3255065, EPI_ISL_3255066, EPI_ISL_3255067, EPI_ISL_3255068, EPI_ISL_3255069, EPI_ISL_3255071, EPI_ISL_3255072, EPI_ISL_3255081, EPI_ISL_3255083, EPI_ISL_3255143, EPI_ISL_3255197, EPI_ISL_3255207, EPI_ISL_3255223 | | | |
| see above | Central Public Health Laboratory - LACEN -Bahia, Salvador, Brazil | Central Public Health Laboratory - LACEN -Bahia, Salvador, Brazil | Arabela Leal; Breno Dominguez; Felicidade Pereira; Jaqueline Gomes; Luciana Oliveira; Luiz Alcantara; Marcela Gómez; Marta Giovanetti; Patrícia Cajado; Stephane Tosta; Vagner Fonseca; Vanessa Nardy |
| EPI_ISL_2134734 | Centre Hospitalier Universitaire Clermont-Ferrand | CHU Clermont-Ferrand, service de virologie | Bisseux Maxime; Combes Patricia; Henquell Cécile; Mirand Audrey |
| EPI_ISL_5801699 | Centro De Especialidades De Primavera | Instituto Butantan | Antonio Jorge Martins; Claudia Renata dos Santos Barros; David Schlesinger; Debora Botequio Moretti; Dimas Tadeu Covas; Elaine Cristina Marqueze; Elaine Vieira Santos; Evandra Strazza Rodrigues; Heidge Fukumasu; Jayme Augusto de Souza-Neto; José Salvatore Leister Patané; Luiz Alcantara; Luiz Lehmann Coutinho; Maria Carolina Elias; Maurício Lacerda Nogueira; Rafael dos Santos Bezerra; Raul Machado Neto; Rejane Maria Tommasini Grotto; Ricardo Haddad; Sandra Coccuzzo Sampaio Vessoni; Simone Kashima; Svetoslav Nanev Slavov; Vincent Louis Viala |
| EPI_ISL_5801566, EPI_ISL_5801567, EPI_ISL_5801683 | Centro De Referencia Do Idoso Dr Humberto Mendes De Carvalho | Instituto Butantan | Antonio Jorge Martins; Claudia Renata dos Santos Barros; David Schlesinger; Debora Botequio Moretti; Dimas Tadeu Covas; Elaine Cristina Marqueze; Elaine Vieira Santos; Evandra Strazza Rodrigues; Heidge Fukumasu; Jayme Augusto de Souza-Neto; José Salvatore Leister Patané; Luiz Alcantara; Luiz Lehmann Coutinho; Maria Carolina Elias; Maurício Lacerda Nogueira; Rafael dos Santos Bezerra; Raul Machado Neto; Rejane Maria Tommasini Grotto; Ricardo Haddad; Sandra Coccuzzo Sampaio Vessoni; Simone Kashima; Svetoslav Nanev Slavov; Vincent Louis Viala |
| EPI_ISL_5801580, EPI_ISL_5801585, EPI_ISL_5801593, EPI_ISL_5801597 | Centro De Saude De Maracai | Instituto Butantan | Antonio Jorge Martins; Claudia Renata dos Santos Barros; David Schlesinger; Debora Botequio Moretti; Dimas Tadeu Covas; Elaine Cristina Marqueze; Elaine Vieira Santos; Evandra Strazza Rodrigues; Heidge Fukumasu; Jayme Augusto de Souza-Neto; José Salvatore Leister Patané; Luiz Alcantara; Luiz Lehmann Coutinho; Maria Carolina Elias; Maurício Lacerda Nogueira; Rafael dos Santos Bezerra; Raul Machado Neto; Rejane Maria Tommasini Grotto; Ricardo Haddad; Sandra Coccuzzo Sampaio Vessoni; Simone Kashima; Svetoslav Nanev Slavov; Vincent Louis Viala |
| EPI_ISL_5801618, EPI_ISL_5801619, EPI_ISL_5801620, EPI_ISL_5801621, EPI_ISL_5801651, EPI_ISL_5801664, EPI_ISL_5801667 | | | |
| see above | Centro De Saude Ii Ibitinga | Instituto Butantan | Antonio Jorge Martins; Claudia Renata dos Santos Barros; David Schlesinger; Debora Botequio Moretti; Dimas Tadeu Covas; Elaine Cristina Marqueze; Elaine Vieira Santos; Evandra Strazza Rodrigues; Heidge Fukumasu; Jayme Augusto de Souza-Neto; José Salvatore Leister Patané; Luiz Alcantara; Luiz Lehmann Coutinho; Maria Carolina Elias; Maurício Lacerda Nogueira; Rafael dos Santos Bezerra; Raul Machado Neto; Rejane Maria Tommasini Grotto; Ricardo Haddad; Sandra Coccuzzo Sampaio Vessoni; Simone Kashima; Svetoslav Nanev Slavov; Vincent Louis Viala |
| EPI_ISL_5801703 | Centro De Saude Iii Nelcidio Da Silveira Bastos | Instituto Butantan | Antonio Jorge Martins; Claudia Renata dos Santos Barros; David Schlesinger; Debora Botequio Moretti; Dimas Tadeu Covas; Elaine Cristina Marqueze; Elaine Vieira Santos; Evandra Strazza Rodrigues; Heidge Fukumasu; Jayme Augusto de Souza-Neto; José Salvatore Leister Patané; Luiz Alcantara; Luiz Lehmann Coutinho; Maria Carolina Elias; Maurício Lacerda Nogueira; Rafael dos Santos Bezerra; Raul Machado Neto; Rejane Maria Tommasini Grotto; Ricardo Haddad; Sandra Coccuzzo Sampaio Vessoni; Simone Kashima; Svetoslav Nanev Slavov; Vincent Louis Viala |
| EPI_ISL_5801606, EPI_ISL_5801608, EPI_ISL_5801611, EPI_ISL_5801612, EPI_ISL_5801617 | Centro De Saude Iii Sales Oliveira | Instituto Butantan | Antonio Jorge Martins; Claudia Renata dos Santos Barros; David Schlesinger; Debora Botequio Moretti; Dimas Tadeu Covas; Elaine Cristina Marqueze; Elaine Vieira Santos; Evandra Strazza Rodrigues; Heidge Fukumasu; Jayme Augusto de Souza-Neto; José Salvatore Leister Patané; Luiz Alcantara; Luiz Lehmann Coutinho; Maria Carolina Elias; Maurício Lacerda Nogueira; Rafael dos Santos Bezerra; Raul Machado Neto; Rejane Maria Tommasini Grotto; Ricardo Haddad; Sandra Coccuzzo Sampaio Vessoni; Simone Kashima; Svetoslav Nanev Slavov; Vincent Louis Viala |
| EPI_ISL_2674352 | Centro de Atención y Daignóstico de Enfermedades Infecciosas-CDI | Instituto Nacional de Salud-Dirección de Investigación en Salud Pública | Carlos Franco-Muñoz; Carmen Osorio; Diana Malo; Diego A. Álvarez-Díaz; Diego Andrés Prada; Gerardo Santamaría; Hector Alejandro Ruiz-Moreno; Jhonnatan Reales-González; Jorge Rivera; Juan Camilo Martinez; Julian Naizaque; Katherine Laiton-Donato; Lisseth Pardo; Magdalena Wiesner; Marcela Mercado-Reyes; Maria T. Herrera-Sepúlveda; Marta Lopez Blanco; Martha Lucia Ospina Martinez; Paola Rojas; Sergio Gomez; Sheryll Corchuelo; Ángela Alarcon Cruz |
| EPI_ISL_5926864, EPI_ISL_5926872 | Centro de Infectologia Charles Mérieux/ Laboratório Rodolphe Mérieux, FUNDHACRE | Bioinformatics Laboratory / LNCC | Alessandra P Lamarca; Alexandra L Gerber; Aline de Freitas Souza; Ana Paula de C Guimarães; Ana Tereza R Vasconcelos; Andreas Stocker; Cirley Maria de Oliveira Lobato; Douglas Terra Machado; Janaína Mazaro; Janete Taynã Nascimento Rodrigues; Luiz Fellype Alves de Souza; Luiz G P de Almeida; Ronaldo da Silva F Jr; Rutilene B. Souza |
| EPI_ISL_2490359, EPI_ISL_2490360, EPI_ISL_2490361, EPI_ISL_2490362, EPI_ISL_2490363, EPI_ISL_2490364, EPI_ISL_2490365, EPI_ISL_2490366 | | | |
| see above | Centro de Investigación Biomédica del Noreste (CIBIN) | Centro de Investigación en Enfermedades Infecciosas (CIENI), Instituto Nacional de Enfermedades Respiratorias (INER) | ; Alejandra Garcia-Gasca; Alejandra Hernández-Terán; Alejandro Sanchez-Flores; Alfredo Herrera-Estrella; Alicia Ocaña-Mondragón; Andreu Comas-García; Angel Gustavo Salas-Lais; Antonio Loza Román; Bernardo Martínez-Miguel; Blanca Taboada; Brenda Irasema Maldonado-Meza; Bruno Gomez-Gil; Carla Ivón Herrera-Najera; Carlos F. Arias; Celia Boukadida; Clara Esperanza Santacruz-Tinoco; Concepción Grajales-Muñiz; Consorcio Mexicano de Vigilancia Genómica (CoViGen-Mex). Authors (in alphabetical order): Julio Elias Alvarado-Yaah; Cristóbal Cháidez-Quiróz; Célida Duque Molina; Célida Martínez- Rodríguez; Daniel Fregoso-Rueda; Daniel Lira Morales; Eduardo Becerril-Vargas; Fernando Fontove-Herrera; Fidencio Mejía-Nepomuceno; Francisco Pulido; Gloria Elena Espinosa-Ayala; Gloria María Molina-Salinas; Gloria Vazquez; Hector Esteban Paz-Juárez; Hector Montoya-Fuentes; Helen Haydee Fernanda Ramírez-Plascencia; Irvin González-López; Jean Pierre González; Joel Armando Vázquez-Pérez.; Jorge Salas-Hernández; José Antonio Enciso-Moreno; José Arturo Martínez-Orozco; José Esteban Muñoz-Medina; José de Jesús Nuñez-Contreras; Juan Bautista Chale-Dzul; Julissa Enciso-Ibarra; Luis Alberto Ochoa-Carrera; Margarita Matías-Florentino; Mario Mújica-Sánchez; Marissa Perez-Garcia; María Guadalupe Santiago-Mauricio; María Guadalupe de Jesús Mireles-Rivera; Nelly Sélem-Mojica; Pavel Isa; Ricardo Ciria Merce; Ricardo Grande; Rosa María Gutierrez Rios; Santiago Ávila-Ríos; Selene Zárate; Susana Lopez; Victor Eduardo Garcia-Arias; Víctor Hugo Borja-Aburto |
| EPI_ISL_4498213, EPI_ISL_4498430, EPI_ISL_4498435, EPI_ISL_4498549 | Children's Hospital of Philadelphia | Planet | Ahmed M. Moustafa; Alex Arvanitis; Azad Ahmed; Brandy Neide; Colleen Bianco; Josh Chang Mell; Lidiya Denu; Paul J. Planet; Rebecca M. Harris; Susan Coffin; Swetha Rajagopal |
| EPI_ISL_4261886 | Chiu Laboratory, University of California, San Francisco | Chiu Laboratory, University of California, San Francisco | Alicia Sotomayor-Gonzalez; Baolin Wang; Candace Wang; Charles Chiu; Daniel Wong; Dustin Glasner; Venice Servellita; Xianding (Wayne) Deng; Yueyuan Zhang |

| | | | |
|---|---|---|---|
| EPI_ISL_2258223, EPI_ISL_2258229, EPI_ISL_2258230, EPI_ISL_2258231, EPI_ISL_2258240, EPI_ISL_2258241, EPI_ISL_2258252, EPI_ISL_2273845, EPI_ISL_2273849, EPI_ISL_2273853, EPI_ISL_2273859 | | | |
| see above | City of Milwaukee Health Department Laboratory | City of Milwaukee Health Department Laboratory | Amy Bauer; Jennifer Lentz; Manjeet Khubbar; Samantha Scott; Sanjib Bhattacharyya |
| EPI_ISL_2467227, EPI_ISL_2467230, EPI_ISL_2467232, EPI_ISL_2467240, EPI_ISL_2467246, EPI_ISL_2467269, EPI_ISL_2467273, EPI_ISL_2467274, EPI_ISL_2467275, EPI_ISL_2467278, EPI_ISL_2467283, EPI_ISL_2467287, EPI_ISL_3242637, EPI_ISL_3242643, EPI_ISL_3242651, EPI_ISL_3242674, EPI_ISL_3242675 | | | |
| see above | Clinica INDISA | "Facultad de Ciencias de la Vida, UNAB" | "Claudio Meneses; Ariel Orellana"; Claudio Olmos; Daniel Leon; Dayan Sanhueza; Eduardo Castro; Gonzalo Campaña; Macarena Bastias; Paola Pidal; Ricardo Yusta; Sebastian Wolter; Susana Saez; Víctor Monreal; Waldo Diaz |
| EPI_ISL_2500847 | Clinical Microbiology Laboratory, NewYork Presbyterian Hospital/Columbia University Irving Medical Center | Uhlemann Laboratory, Columbia University Irving Medical Center | Anne Kelley; Anne-Catrin Uhlemann; Maya Tagliavia; Medini K. Annavajhala |
| EPI_ISL_5323440 | Color Genomics | Chiu Laboratory, University of California, San Francisco | Alicia Sotomayor-Gonzalez; Alicia Zhou; Baolin Wang; Charles Chiu; Daniel Wong; Erika Torres; Noah Brazer; Scott Topper; Venice Servellita |
| EPI_ISL_2309879, EPI_ISL_2310449, EPI_ISL_2310621, EPI_ISL_2311040, EPI_ISL_3696553, EPI_ISL_3842256, EPI_ISL_3842258, EPI_ISL_3842260, EPI_ISL_3842269, EPI_ISL_3842294, EPI_ISL_3842320, EPI_ISL_3842321, EPI_ISL_3842507, EPI_ISL_3842993, EPI_ISL_4749962, EPI_ISL_6211527 | | | |
| see above | Colorado Department of Public Health and Environment | Colorado Department of Public Health and Environment | Alexandria Rossheim; Diana Ir; Emily A. Travanty; Laura Bankers; Mandy Waters; Michael A. Martin; Michael Martin; Molly C. Hetherington-Rauth; Sarah Elizabeth Totten; Shannon R. Matzinger |
| EPI_ISL_7727227, EPI_ISL_7727234, EPI_ISL_7727235, EPI_ISL_7727239, EPI_ISL_7727243 | Covid Laboratory Biogem | Covid Laboratory Biogem | Alessandra Fucci and Michele Caraglia; Alessia Maria Cossu; Cinzia Miarelli; Clara Iannarone; Egildo Luca D'andrea; Federica Melisi; Giovambattista Capasso; Marco Bocchetti; Marianna Scrima; Michele Ceccarelli; Piera Grisolia; Teresa Maria Rosaria Noviello; Ylenia Abruzzese |
| EPI_ISL_5801689 | Csii Dr Washington Luis M Rodrigues Da Silva Pitangueiras | Instituto Butantan | Antonio Jorge Martins; Claudia Renata dos Santos Barros; David Schlesinger; Debora Botequio Moretti; Dimas Tadeu Covas; Elaine Cristina Marqueze; Elaine Vieira Santos; Evandra Strazza Rodrigues; Heidge Fukumasu; Jayme Augusto de Souza-Neto; José Salvatore Leister Patané; Luiz Alcantara; Luiz Lehmann Coutinho; Maria Carolina Elias; Maurício Lacerda Nogueira; Rafael dos Santos Bezerra; Raul Machado Neto; Rejane Maria Tommasini Grotto; Ricardo Haddad; Sandra Coccuzzo Sampaio Vessoni; Simone Kashima; Svetoslav Nanev Slavov; Vincent Louis Viala |
| EPI_ISL_2755153, EPI_ISL_2755157, EPI_ISL_2755191, EPI_ISL_2755221, EPI_ISL_2755222, EPI_ISL_2755243, EPI_ISL_2755251, EPI_ISL_2755259, EPI_ISL_2755273, EPI_ISL_2755274, EPI_ISL_2755283, EPI_ISL_2755310, EPI_ISL_2755422, EPI_ISL_2755424, EPI_ISL_2755586, EPI_ISL_2755592, EPI_ISL_2755593, EPI_ISL_2755599, EPI_ISL_2755601, EPI_ISL_2755614, EPI_ISL_2755615, EPI_ISL_2755616, EPI_ISL_2755624, EPI_ISL_2755636, EPI_ISL_2755659, EPI_ISL_2755675, EPI_ISL_2755677, EPI_ISL_2755852, EPI_ISL_2755876, EPI_ISL_2755908, EPI_ISL_2755929, EPI_ISL_2756040, EPI_ISL_2756041, EPI_ISL_3064598, EPI_ISL_3064620, EPI_ISL_3270248, EPI_ISL_3270257, EPI_ISL_3270404 | | | |
| see above | Curative Labs | Curative Labs | Elias L. Salfati; Eugenia Khorosheva; George Way; J.Cesar Ignacio-Espinoza; Janet Chen; Mikhail Hanewich-Hollatz; Nabjot Sandhu; Sophia Quasem; Vladimir Slepnev; Zhiyi Xie |
| EPI_ISL_3663780, EPI_ISL_3663781, EPI_ISL_3663787, EPI_ISL_3663792, EPI_ISL_3663795, EPI_ISL_3663796, EPI_ISL_3663797, EPI_ISL_3663799, EPI_ISL_3663806, EPI_ISL_3663809, EPI_ISL_3663810, EPI_ISL_3663813, EPI_ISL_3663815, EPI_ISL_3663817, EPI_ISL_3663821, EPI_ISL_3663823, EPI_ISL_3663825, EPI_ISL_3663829, EPI_ISL_3663836, EPI_ISL_3663839, EPI_ISL_3663842, EPI_ISL_3663843, EPI_ISL_3663845, EPI_ISL_3663846, EPI_ISL_3663848, EPI_ISL_3663849, EPI_ISL_3663851, EPI_ISL_3663852, EPI_ISL_3663854, EPI_ISL_3663855, EPI_ISL_3663856, EPI_ISL_3663858, EPI_ISL_3663861, EPI_ISL_3663863, EPI_ISL_3663867, EPI_ISL_3663868, EPI_ISL_3663869, EPI_ISL_3663871, EPI_ISL_3663872, EPI_ISL_3663873, EPI_ISL_3663875, EPI_ISL_3663879, EPI_ISL_3663882, EPI_ISL_3663883, EPI_ISL_3663885, EPI_ISL_3663887, EPI_ISL_3663888, EPI_ISL_3663892, EPI_ISL_3663893, EPI_ISL_3663897, EPI_ISL_3663899, EPI_ISL_3663900, EPI_ISL_3663903, EPI_ISL_3663906, EPI_ISL_3663907, EPI_ISL_3663908, EPI_ISL_3663909, EPI_ISL_3663911, EPI_ISL_3663914, EPI_ISL_3663917, EPI_ISL_3663918, EPI_ISL_3663919, EPI_ISL_3663920, EPI_ISL_3663925, EPI_ISL_3663926, EPI_ISL_3663927, EPI_ISL_3663928, EPI_ISL_3663929, EPI_ISL_3663931, EPI_ISL_3663932, EPI_ISL_3663933, EPI_ISL_3663935, EPI_ISL_3663936, EPI_ISL_3663937, EPI_ISL_3663938, EPI_ISL_3663939, EPI_ISL_3663940, EPI_ISL_3663942, EPI_ISL_3663943, EPI_ISL_3663944, EPI_ISL_3663945, EPI_ISL_3663946, EPI_ISL_3663947, EPI_ISL_3663948, EPI_ISL_3663950, EPI_ISL_3663951, EPI_ISL_3663952, EPI_ISL_3663953, EPI_ISL_3663955, EPI_ISL_3663956, EPI_ISL_3663957, EPI_ISL_3663958, EPI_ISL_3663959, EPI_ISL_3663961, EPI_ISL_3663962, EPI_ISL_3663964, EPI_ISL_3663965, EPI_ISL_3663967, EPI_ISL_3663968, EPI_ISL_3663971, EPI_ISL_3663972, EPI_ISL_3663976, EPI_ISL_3663977, EPI_ISL_3663978, EPI_ISL_3663982, EPI_ISL_3663983, EPI_ISL_3663984, EPI_ISL_3663987, EPI_ISL_3663990, EPI_ISL_3663991, EPI_ISL_3663992, EPI_ISL_3663993, EPI_ISL_3663995, EPI_ISL_3663997, EPI_ISL_3663998, EPI_ISL_3663999, EPI_ISL_3664001, EPI_ISL_3664002, EPI_ISL_3664003, EPI_ISL_3664004, EPI_ISL_3664005, EPI_ISL_3664006, EPI_ISL_3664007, EPI_ISL_3664008, EPI_ISL_3664009, EPI_ISL_3664010, EPI_ISL_3664015, EPI_ISL_3664016, EPI_ISL_3664017, EPI_ISL_3664018, EPI_ISL_3664019, EPI_ISL_3664023, EPI_ISL_3664027, EPI_ISL_3664028, EPI_ISL_3664030, EPI_ISL_3664033, EPI_ISL_3664035, EPI_ISL_3664038, EPI_ISL_3664041, EPI_ISL_3664046, EPI_ISL_3664048, EPI_ISL_3664049, EPI_ISL_3664051, EPI_ISL_3664052, EPI_ISL_3664053, EPI_ISL_3664057, EPI_ISL_3664058, EPI_ISL_3664060, EPI_ISL_3664063, EPI_ISL_3664064, EPI_ISL_3664065, EPI_ISL_3664066, EPI_ISL_3664068, EPI_ISL_3664071, EPI_ISL_3664072, EPI_ISL_3664073, EPI_ISL_3664076, EPI_ISL_3664077, EPI_ISL_3664080, EPI_ISL_3664082, EPI_ISL_3664083, EPI_ISL_3664084, EPI_ISL_3664085, EPI_ISL_3664087, EPI_ISL_3664089, EPI_ISL_3664091, EPI_ISL_3664095, EPI_ISL_3664097, EPI_ISL_3664102, EPI_ISL_3664119, EPI_ISL_3664124, EPI_ISL_3664125, EPI_ISL_3664128, EPI_ISL_3664129, EPI_ISL_3664130, EPI_ISL_3664138, EPI_ISL_3664142, EPI_ISL_3664144, EPI_ISL_3664160, EPI_ISL_3664162, EPI_ISL_3664163, EPI_ISL_3664164, EPI_ISL_3664165, EPI_ISL_3664166, EPI_ISL_3664168, EPI_ISL_3664169, EPI_ISL_3664171, EPI_ISL_3664175, EPI_ISL_3664176, EPI_ISL_3664177, EPI_ISL_3664179, EPI_ISL_3664182, EPI_ISL_3664188, EPI_ISL_3664190, EPI_ISL_3664192, EPI_ISL_3664194, EPI_ISL_3664195, EPI_ISL_3664197, EPI_ISL_3664208, EPI_ISL_3664212, EPI_ISL_3664213, EPI_ISL_3664216, EPI_ISL_3664218, EPI_ISL_3664219, EPI_ISL_3664220, EPI_ISL_3664222, EPI_ISL_3664223, EPI_ISL_3664226, EPI_ISL_3664230, EPI_ISL_3664233, EPI_ISL_3664235, EPI_ISL_3664236, EPI_ISL_3664237, EPI_ISL_3664239, EPI_ISL_3664242, EPI_ISL_3664247, EPI_ISL_3664248, EPI_ISL_3664249, EPI_ISL_3664251, EPI_ISL_3664252, EPI_ISL_3664253, EPI_ISL_3664260, EPI_ISL_3664261, EPI_ISL_3664263, EPI_ISL_3664264, EPI_ISL_3664265, EPI_ISL_3664267, EPI_ISL_3664269, EPI_ISL_3664270, EPI_ISL_3664273, EPI_ISL_3664276, EPI_ISL_3664277, EPI_ISL_3664278, EPI_ISL_4426669, EPI_ISL_4426697, EPI_ISL_4426700, EPI_ISL_4426712, EPI_ISL_4426746, EPI_ISL_4426751, EPI_ISL_4426849, EPI_ISL_4428008, EPI_ISL_4428011, EPI_ISL_4428015, EPI_ISL_4428018, EPI_ISL_4428022, EPI_ISL_4428025, EPI_ISL_4428027, EPI_ISL_4428028, EPI_ISL_4428031, EPI_ISL_4428034, EPI_ISL_4428037, EPI_ISL_4428041, EPI_ISL_4428044, EPI_ISL_4428047, EPI_ISL_4428050, EPI_ISL_4428054, EPI_ISL_4428057, EPI_ISL_4428060, EPI_ISL_4428064, EPI_ISL_4428067, EPI_ISL_4428070, EPI_ISL_4428074, EPI_ISL_4428077, EPI_ISL_4428080, EPI_ISL_4428083, EPI_ISL_4428086, EPI_ISL_4428089, EPI_ISL_4428092, EPI_ISL_4428096, EPI_ISL_4428098, EPI_ISL_4428101, EPI_ISL_4428104, EPI_ISL_4428107, EPI_ISL_4428109, EPI_ISL_4428112, EPI_ISL_4428115, EPI_ISL_4428118, EPI_ISL_4428121, EPI_ISL_4428123, EPI_ISL_4428126, EPI_ISL_4428129, EPI_ISL_4428132, EPI_ISL_4428136, EPI_ISL_4428176, EPI_ISL_4428186, EPI_ISL_4428229, EPI_ISL_4428269 | | | |
| see above | DASA | DASA | Adriano Bonaldi; Angelica Hristov; Annelise Lopes; Bianca Cota; Cristina Oliveira; Jose Levi; Lidia Yamamoto; Paulo Pierry; Rodrigo Guarischi; Rodrigo Salazar |
| EPI_ISL_2451775 | DC Public Health Lab/ Dept. of Forensic Sciences | Centers for Disease Control and Prevention Division of Viral Diseases, Pathogen Discovery | Alison Laufer Halpin; Ben L. Rambo-Martin; Clinton R. Paden; Dakota Howard; Darlene Wagner; Dave Wentworth; Dhwani Batra; Jasmine Padilla; Justin Lee; Katie Dillon; Krista Queen; Kristen Knipe; Kristine Lacek; Mark Burroughs; Matthew Schmerer; Mili Sheth; Peter Cook; Sam Shepard; Sarah Nobles; Shoshona Le; Suxiang Tong; Vivien Dugan; Yvette Unoarumhi |
| EPI_ISL_2228976 | DE Public Health Laboratory | Centers for Disease Control and Prevention Division of Viral Diseases, Pathogen Discovery | Alison Laufer Halpin; Ben L. Rambo-Martin; Clinton R. Paden; Dakota Howard; Darlene Wagner; Dave Wentworth; Dhwani Batra; Jasmine Padilla; Justin Lee; Katie Dillon; Krista Queen; Kristen Knipe; Kristine Lacek; Mark Burroughs; Matthew Schmerer; Mili Sheth; Peter Cook; Sam Shepard; Sarah Nobles; Shoshona Le; Suxiang Tong; Vivien Dugan; Yvette Unoarumhi |
| EPI_ISL_2345827, EPI_ISL_2345871 | DEPARTAMENTO DE SAUDE MUNICIPAL SOCORRO SP | Instituto Butantan / Mendelics | Antonio Jorge Martins; Claudia Renata dos Santos Barros; David Schlesinger; Debora Botequio Moretti; Dimas Tadeu Covas; Elaine Cristina Marqueze; Elaine Vieira Santos; Evandra Strazza Rodrigues; Heidge Fukumasu; Jayme Augusto de Souza-Neto; José Salvatore Leister Patané; Luiz Alcantara; Luiz Lehmann Coutinho; Maria Carolina Elias; Maurício Lacerda Nogueira; Rafael dos Santos Bezerra; Raul Machado Neto; Rejane Maria Tommasini Grotto; Ricardo Haddad; Sandra Coccuzzo Sampaio Vessoni; Simone Kashima; Svetoslav Nanev Slavov; Vincent Louis Viala |
| EPI_ISL_2235371 | DIP. PREV. AVEZZANO SERVIZIO DI IGIENE EPIDEMIOLOGIA E SANITA' PUBBLICA | Istituto Zooprofilattico Sperimentale dell'Abruzzo e Molise "G. Caporale" | Ancora M; Calistri P; Cammà C; Caporale M; Curini V; Delli Compagni E; Di Domenico M; Di Lollo Valeria; Di Pasquale A; Lorusso A; Mangone I; Marcacci M; Puglia I; Rinaldi A; Savini G; Scialabba S |
| EPI_ISL_2151413 | DOHMH Central Harlem | New York City Public Health Laboratory | Jade Wang; et al. |
| EPI_ISL_3692620, EPI_ISL_3692624 | DPHL | Delaware Public Health Lab | Rebecca Savage |
| EPI_ISL_5801638, EPI_ISL_5801640 | Departamento De Saude Municipal Socorro Sp | Instituto Butantan | Antonio Jorge Martins; Claudia Renata dos Santos Barros; David Schlesinger; Debora Botequio Moretti; Dimas Tadeu Covas; Elaine Cristina Marqueze; Elaine Vieira Santos; Evandra Strazza Rodrigues; Heidge Fukumasu; Jayme Augusto de Souza-Neto; José Salvatore Leister Patané; Luiz Alcantara; Luiz Lehmann Coutinho; Maria Carolina Elias; Maurício Lacerda Nogueira; Rafael dos Santos Bezerra; Raul Machado Neto; Rejane Maria Tommasini Grotto; Ricardo Haddad; Sandra Coccuzzo Sampaio Vessoni; Simone Kashima; Svetoslav Nanev Slavov; Vincent Louis Viala |
| EPI_ISL_2139880, EPI_ISL_2426619 | Department of Clinical Microbiology | GIGA Medical Genomics | Bouchra Boujemla; Cécile Meex; Keith Durkin; Maria Artesi; Marie-Pierre Hayette; Nathalie Renotte; Pierrette Melin; Raphaël Boreux; Sébastien Bontems; Vincent Bours |
| EPI_ISL_2146022 | Department of Virology, Istituto Zooprofilattico Sperimentale del Lazio e della Toscana (IZSLT) | Department of General Diagnostics; Department of Virology; Istituto Zooprofilattico Sperimentale del Lazio e della Toscana (IZSLT) | Alessia Franco; Antonella Cersini; Antonio Battisti.; Elena L. Diaconu; Fabiola Feltrin; Giuseppe Manna; Patricia Alba; Raffaela Conti; Teresa Scicluna; Virginia Carfora |
| EPI_ISL_3635749 | Dr. Risch Ostschweiz AG | Microbiology, Dr. Risch | Dominique Fabien Hilti; Faina Wehrli; Lorenz Risch; Martin Risch; Nadia Wohlwend; Sinem Kas; Thomas Bodmer |
| EPI_ISL_2380049 | Drive in Ohain | Plateforme Fédérale COVID ULB-IBC | A. Marchand; B. Haerlingen; C. Henin; M-L Delforge; R. De Mendonça |
| EPI_ISL_2145628 | Dutch COVID-19 response team | Erasmus Medical Center | Anne van der Linden; Annemiek van der Eijk; Bas Oude Munnink; Corine GeurtsvanKessel; David Nieuwenhuijse; Emmanuelle Munger; Irina Chestakova; Marion Koopmans; Marjan Boter; Reina Sikkema; Richard Molenkamp; on behalf of the Dutch national COVID-19 respo |
| EPI_ISL_2365493 | Dutch COVID-19 response team | Medical Microbiology, Maastricht University Medical Centre | Brian van der Veer*; Carmen Reumkens; Christian Hoebe; Erik Beuken; Jozef Dingemans*; Lieke van Alphen; Paul Savelkoul |
| EPI_ISL_2217848, EPI_ISL_2219009, EPI_ISL_2219010, EPI_ISL_2219013, EPI_ISL_2219015, EPI_ISL_2219019, EPI_ISL_2219024, EPI_ISL_2219027, EPI_ISL_2219030, EPI_ISL_2219033, EPI_ISL_2219038, EPI_ISL_2219041, EPI_ISL_2219043, EPI_ISL_2219046, EPI_ISL_2219048, EPI_ISL_2219057, EPI_ISL_2219091, EPI_ISL_2219094, EPI_ISL_2219095, EPI_ISL_2219098, EPI_ISL_2219100, EPI_ISL_2219102, EPI_ISL_2303352, EPI_ISL_2303353, EPI_ISL_2303354, EPI_ISL_2303356, EPI_ISL_2303357, EPI_ISL_2303358, EPI_ISL_2303360, EPI_ISL_2303366, EPI_ISL_2303377, EPI_ISL_2303378, EPI_ISL_2303388, EPI_ISL_2303389, EPI_ISL_2406433, EPI_ISL_2475710 | | | |
| see above | Dutch COVID-19 response team | National Institute for Public Health and the Environment (RIVM) | Adam Meijer; AnneMarie van den Brandt; Annelies Kroneman; Bas van der Veer; Chantal Reusken; Dennis Schmitz; Dirk Eggink; Euníce Then; Florian Zwagemaker; Harry Vennema; James Groot; Jeroen Cremer; Karim Hajji; Kim Freriks; Linda van de Nes; Lisa Wijsman; Lynn Aarts; Melissa van Tuil; Ryanne Jaarsma; Sanne Bos; Sharon van den Brink; Sjoerd Kuiling; on behalf of the national COVID-19 response team |
| EPI_ISL_3215650, EPI_ISL_3215668, EPI_ISL_3215674, EPI_ISL_3215686, EPI_ISL_3215743 | EXCITE Lab | Andersen lab at Scripps Research | Abigail Schnapper; Angela Scioscia; Chip Schooley; Christine Mitchell + SEARCH; David Pride; Fred Wu; Helena Tubb; Natasha Martin Cheryl Anderson; Sawyer Farmer; Sharon Reed; Tommy Valles + SEARCH |
| EPI_ISL_2478955, EPI_ISL_2478958, EPI_ISL_2478961, EPI_ISL_2479367, EPI_ISL_2479447, EPI_ISL_2479448, EPI_ISL_2479450, EPI_ISL_2479451, EPI_ISL_2479452, EPI_ISL_2479704, EPI_ISL_2479812, EPI_ISL_2940806, EPI_ISL_2940856, EPI_ISL_2940871, EPI_ISL_2940883, EPI_ISL_2940884, EPI_ISL_2940885, EPI_ISL_2940889, EPI_ISL_3116935, EPI_ISL_3116936, EPI_ISL_3116937, EPI_ISL_3116938, EPI_ISL_3116939, EPI_ISL_3117257, EPI_ISL_3117258, EPI_ISL_3117269 | | | |
| see above | Edmonton Provincial Lab | Public Health Agency of Canada (PHAC) National Microbiology Laboratory | Buss; Croxen M; Deo A; Dieu P; E; Ferrato C; Gill K; Khan F; Koleva P; Li V; Lloyd C; Lynch T; Ma R; Murphy S; Pabbaraju K; Shokoples S; Thayer J; Tipples G; Whitehouse M; Wong A; Yu C; Zelyas N |
| EPI_ISL_2315599, EPI_ISL_2316877, EPI_ISL_2316938, EPI_ISL_2316950, EPI_ISL_2316952, EPI_ISL_2316955, EPI_ISL_2316974, EPI_ISL_2316984, EPI_ISL_2317019, EPI_ISL_2317020, EPI_ISL_2317031 | | | |
| see above | Eurofins LifeCodexx GmbH | Robert Koch Institute | |
| EPI_ISL_3385801 | FUNDACION HOSPITAL SAN PEDRO DIOCESIS DE PASTO | Instituto Nacional de Salud-Dirección de Investigación en Salud Pública | Carlos Franco-Muñoz; Carmen Osorio; Diana Malo; Diego A. Álvarez-Díaz; Diego Andrés Prada; Gerardo Santamaría; Hector Alejandro Ruiz-Moreno; Jhonnatan Reales-González; Jorge Rivera; Juan Camilo Martinez; Julian Naizaque; Katherine Laiton-Donato; Lisseth Pardo; Magdalena Wiesner; Marcela Mercado-Reyes; Maria T. Herrera-Sepúlveda; Marta Lopez Blanco; Martha Lucia Ospina Martinez; Paola Rojas; Sergio Gomez; Sheryll Corchuelo; Ángela Alarcon Cruz |
| EPI_ISL_2277736, EPI_ISL_2277948, EPI_ISL_2277970, EPI_ISL_2277971, EPI_ISL_2277972, EPI_ISL_2277973, EPI_ISL_2277974, EPI_ISL_2277975, EPI_ISL_2442837, EPI_ISL_2442838, EPI_ISL_2442898, EPI_ISL_2442899, EPI_ISL_2442900, EPI_ISL_2442901, EPI_ISL_2442902, EPI_ISL_2442926, EPI_ISL_2442927, EPI_ISL_2567120, EPI_ISL_2567121, EPI_ISL_2567122, EPI_ISL_2567137, EPI_ISL_2567138, EPI_ISL_2836021, EPI_ISL_2836022, EPI_ISL_2836024, EPI_ISL_2836049, EPI_ISL_2836051, EPI_ISL_2836057, EPI_ISL_2836058, EPI_ISL_2836061, EPI_ISL_2836062, EPI_ISL_2836107, EPI_ISL_2836111, EPI_ISL_2836290, EPI_ISL_2836297, EPI_ISL_2836305, EPI_ISL_2836497, EPI_ISL_6049992, EPI_ISL_6050001, EPI_ISL_6050002, EPI_ISL_6050004, EPI_ISL_6050006, EPI_ISL_6050008, EPI_ISL_6050009, EPI_ISL_6050029, EPI_ISL_6050039, EPI_ISL_6050040, EPI_ISL_6050102 | | | |
| see above | Florida Bureau of Public Health Laboratories | Florida Bureau of Public Health Laboratories | Jason Blanton; Namratha Tarigopula; Sarah Schmedes; Tiffany Splatt |
| EPI_ISL_2620683, EPI_ISL_2620684, | Fondazione IRCCS Ca' Granda Ospedale Maggiore Policlinico | Fondazione IRCCS Ca' Granda Ospedale Maggiore Policlinico | Ferruccio Ceriotti; Sara Uceda Renteria |

| | | | |
|---|---|---|---|
| EPI_ISL_2620686, EPI_ISL_2620687, EPI_ISL_2620688, EPI_ISL_2620709 | | | |
| EPI_ISL_2662824 | Fondazione Policlinico Tor Vergata | National Institute for Infectious Diseases (INMI) L. Spallanzani I.R.C.C.S | A Di Caro; B Bartolini; CEM Gruber; E Giombini; F Messina; F Santini; G Bonfiglio; M Rueca; MR Capobianchi; O Butera |
| EPI_ISL_1993217, EPI_ISL_1993238, EPI_ISL_1993246, EPI_ISL_1993270, EPI_ISL_2043712, EPI_ISL_2043716, EPI_ISL_2043737, EPI_ISL_2043740, EPI_ISL_2043753, EPI_ISL_2043758, EPI_ISL_2043760, EPI_ISL_2043771, EPI_ISL_2043772, EPI_ISL_2043951, EPI_ISL_2043953, EPI_ISL_2043954, EPI_ISL_2043958, EPI_ISL_2043969, EPI_ISL_2043971, EPI_ISL_2043972, EPI_ISL_2043973, EPI_ISL_2043976, EPI_ISL_2043977, EPI_ISL_2043988, EPI_ISL_2043989, EPI_ISL_2043991, EPI_ISL_2133990, EPI_ISL_2133995, EPI_ISL_2134019, EPI_ISL_2134052, EPI_ISL_2134077, EPI_ISL_2134084, EPI_ISL_2134096, EPI_ISL_2134144, EPI_ISL_2134148, EPI_ISL_2134151, EPI_ISL_2134156, EPI_ISL_2134177, EPI_ISL_2134186, EPI_ISL_2134188, EPI_ISL_2134189, EPI_ISL_2134191, EPI_ISL_2134192, EPI_ISL_2134194, EPI_ISL_2134208, EPI_ISL_2134215, EPI_ISL_2134225, EPI_ISL_2134235, EPI_ISL_2134241, EPI_ISL_2134245, EPI_ISL_2134262, EPI_ISL_2134312, EPI_ISL_2134321, EPI_ISL_2134323, EPI_ISL_2134328, EPI_ISL_2134332, EPI_ISL_2134337, EPI_ISL_2134345, EPI_ISL_2134366, EPI_ISL_2134371, EPI_ISL_2134373, EPI_ISL_2134375, EPI_ISL_2134389, EPI_ISL_2134394, EPI_ISL_2134410, EPI_ISL_2134430, EPI_ISL_2134433, EPI_ISL_2134434, EPI_ISL_2134442, EPI_ISL_2134448, EPI_ISL_2134449, EPI_ISL_2134463, EPI_ISL_2134464, EPI_ISL_2134465, EPI_ISL_2134466, EPI_ISL_2134471, EPI_ISL_2134487, EPI_ISL_2134488, EPI_ISL_2134499, EPI_ISL_2134502, EPI_ISL_2134506, EPI_ISL_2134508, EPI_ISL_2160753, EPI_ISL_2160762, EPI_ISL_2160765, EPI_ISL_2160773, EPI_ISL_2160778, EPI_ISL_2160863, EPI_ISL_2160884, EPI_ISL_2160903, EPI_ISL_2160904, EPI_ISL_2160909, EPI_ISL_2179852, EPI_ISL_2179857, EPI_ISL_2179858, EPI_ISL_2179860, EPI_ISL_2179894, EPI_ISL_2179900, EPI_ISL_2269267, EPI_ISL_2269541, EPI_ISL_2269553, EPI_ISL_2269562, EPI_ISL_2269581, EPI_ISL_2269584, EPI_ISL_2269592, EPI_ISL_2269605, EPI_ISL_2269608, EPI_ISL_4370716 | | | |
| see above | Fulgent Genetics | Centers for Disease Control and Prevention Division of Viral Diseases, Pathogen Discovery | Adrian Paskey; Becky Tsai; Benafsh Sapra; Benjamin Rambo-Martin; Christopher Gulvick; Clinton Paden; Clinton R. Paden; Dakota Howard; Darlene Wagner; Dhwani Batra; Doreen Ng; Duncan MacCannell; Erisa Sula; Harry Gao; James Xie; Jason Caravas; John Gao; Joseph Fierro; Kara Moser; Kristine Lacek; Matthew Schmerer; Mickey Li; Peter Cook; Peter W. Cook; Scott Sammons; Shatavia Morrison; Tymeckia Kendall; Victoria Caban Figueroa; Yan Meng; Yvette Unoarumhi |
| EPI_ISL_6701390, EPI_ISL_6701392 | Fundação Ezequiel Dias | Instituto René Rachou / Fiocruz Minas | André Bernardes; Anna Salim; Caroline Penido; Felipe Iani; Flávio Araújo; Gabriel Fernandes; Pedro Alves; Rubens do Monte Neto; Thaís Silva |
| EPI_ISL_2345701, EPI_ISL_2345709, EPI_ISL_2345756 | GRIPARIO FALCAO | Instituto Butantan / Mendelics | Antonio Jorge Martins; Claudia Renata dos Santos Barros; David Schlesinger; Debora Botequio Moretti; Dimas Tadeu Covas; Elaine Cristina Marqueze; Elaine Vieira Santos; Evandra Strazza Rodrigues; Heidge Fukumasu; Jayme Augusto de Souza-Neto; José Salvatore Leister Patané; Luiz Alcantara; Luiz Lehmann Coutinho; Maria Carolina Elias; Maurício Lacerda Nogueira; Rafael dos Santos Bezerra; Raul Machado Neto; Rejane Maria Tommasini Grotto; Ricardo Haddad; Sandra Coccuzzo Sampaio Vessoni; Simone Kashima; Svetoslav Nanev Slavov; Vincent Louis Viala |
| EPI_ISL_2228704 | GROUPE HOSPITALIER SUD ILE DE FRANCE | Department of Virology, Henri Mondor University Hospital, Assistance Publique Hôpitaux de Paris, Université Paris-Est Créteil, INSERM U955 | Alexandre Soulier; Christophe Rodriguez; Elisabeth Trawinski; Guillaume Gricourt; Jean-Michel Pawlotsky; Melissa N'Debi; Slim Fourati; Vanessa Demontant |
| EPI_ISL_3491798 | Gencore - Universidad de los Andes | Gencore - Universidad de los Andes | Catherine Jaller; Cristian Barrera; David González; Luisa Sacristan; Marcela Guevara; Silvia Restrepo |
| EPI_ISL_2391141, EPI_ISL_2391173, EPI_ISL_2391180, EPI_ISL_2391182, EPI_ISL_2391185, EPI_ISL_2391187, EPI_ISL_2391190, EPI_ISL_2391196, EPI_ISL_2391228, EPI_ISL_2391229, EPI_ISL_2391230, EPI_ISL_2391238, EPI_ISL_2391239, EPI_ISL_2391241, EPI_ISL_2391246, EPI_ISL_2391247, EPI_ISL_2391248, EPI_ISL_2391250, EPI_ISL_2391252, EPI_ISL_2391253, EPI_ISL_2391254, EPI_ISL_2391255, EPI_ISL_2391258, EPI_ISL_2391259, EPI_ISL_2391260, EPI_ISL_2391261, EPI_ISL_2391263, EPI_ISL_2391265, EPI_ISL_2391266, EPI_ISL_2391268, EPI_ISL_2391289, EPI_ISL_2391291, EPI_ISL_2391293, EPI_ISL_2391296, EPI_ISL_2391297, EPI_ISL_2391299, EPI_ISL_2391300, EPI_ISL_2391316, EPI_ISL_2391317, EPI_ISL_2391325, EPI_ISL_2391326, EPI_ISL_2391328, EPI_ISL_2391330, EPI_ISL_2391331, EPI_ISL_2391332, EPI_ISL_2391341, EPI_ISL_2391355, EPI_ISL_2391356, EPI_ISL_2391359, EPI_ISL_2391398, EPI_ISL_2391400, EPI_ISL_2391402, EPI_ISL_2391403, EPI_ISL_2391407, EPI_ISL_2391408, EPI_ISL_2391411, EPI_ISL_2391412, EPI_ISL_2391413, EPI_ISL_2391414, EPI_ISL_2391415, EPI_ISL_2391416, EPI_ISL_2391417, EPI_ISL_2391418, EPI_ISL_2391419, EPI_ISL_2391420, EPI_ISL_2391433, EPI_ISL_2391437, EPI_ISL_2391439, EPI_ISL_2391440, EPI_ISL_2391442, EPI_ISL_2391444, EPI_ISL_2391445, EPI_ISL_2391446, EPI_ISL_2391448, EPI_ISL_2391449, EPI_ISL_2391450, EPI_ISL_2391452, EPI_ISL_2391453, EPI_ISL_2391454, EPI_ISL_2391455, EPI_ISL_2391456, EPI_ISL_2391461, EPI_ISL_2391467, EPI_ISL_2391468, EPI_ISL_2391470, EPI_ISL_2391471, EPI_ISL_2391472, EPI_ISL_2391475, EPI_ISL_2391476, EPI_ISL_2509133, EPI_ISL_2509140, EPI_ISL_2509141, EPI_ISL_2509324, EPI_ISL_2659190 | | | |
| see above | Genetica Molecular and Subdepartamento de Virologia ISP Chile | Instituto de Salud Publica de Chile | Andres Castillo; Barbara Parra; Constanza Campano; Gisselle Barra; Javier Tognarelli; Jorge Fernandez; Karen Orostica; Loredana Arata; Patricia Bustos; Rodrigo Fasce; Soledad Ulloa |
| EPI_ISL_2340148, EPI_ISL_2340184, EPI_ISL_2340199, EPI_ISL_2340221, EPI_ISL_2340225 | Germano de Sousa | Instituto Nacional de Saude (INSA) | Borges et al |
| EPI_ISL_2602055, EPI_ISL_2602060, EPI_ISL_2602062, EPI_ISL_2602080, EPI_ISL_2602099, EPI_ISL_2602102, EPI_ISL_2602108, EPI_ISL_2602123, EPI_ISL_2602132, EPI_ISL_2602141, EPI_ISL_2602147, EPI_ISL_2602149, EPI_ISL_2602151, EPI_ISL_2602165, EPI_ISL_2602248, EPI_ISL_2602250, EPI_ISL_2602251, EPI_ISL_2602258, EPI_ISL_2602273, EPI_ISL_2602275, EPI_ISL_2602278, EPI_ISL_2602286, EPI_ISL_2602287, EPI_ISL_2602288, EPI_ISL_2602301, EPI_ISL_2602309, EPI_ISL_2602313, EPI_ISL_2602361, EPI_ISL_2602362, EPI_ISL_2602377, EPI_ISL_2602381, EPI_ISL_2602395, EPI_ISL_2613840, EPI_ISL_2613841, EPI_ISL_2613860, EPI_ISL_2613863, EPI_ISL_2613873, EPI_ISL_2613874, EPI_ISL_2613877, EPI_ISL_2613908, EPI_ISL_2614011, EPI_ISL_2614012, EPI_ISL_2614024, EPI_ISL_2614026, EPI_ISL_2614027, EPI_ISL_2614063 | | | |
| see above | Gravity Diagnostics, LLC | Gravity Diagnostics, LLC | Gravity Diagnostics |
| EPI_ISL_5801577, EPI_ISL_5801582, EPI_ISL_5801590 | Gripario Falcao | Instituto Butantan | Antonio Jorge Martins; Claudia Renata dos Santos Barros; David Schlesinger; Debora Botequio Moretti; Dimas Tadeu Covas; Elaine Cristina Marqueze; Elaine Vieira Santos; Evandra Strazza Rodrigues; Heidge Fukumasu; Jayme Augusto de Souza-Neto; José Salvatore Leister Patané; Luiz Alcantara; Luiz Lehmann Coutinho; Maria Carolina Elias; Maurício Lacerda Nogueira; Rafael dos Santos Bezerra; Raul Machado Neto; Rejane Maria Tommasini Grotto; Ricardo Haddad; Sandra Coccuzzo Sampaio Vessoni; Simone Kashima; Svetoslav Nanev Slavov; Vincent Louis Viala |
| EPI_ISL_2157237, EPI_ISL_2157239, EPI_ISL_2157248 | Gundersen Clinical Microbiology Laboratory | Kabara Cancer Research Institute | Craig S. Richmond; Paraic A. Kenny |
| EPI_ISL_2187992, EPI_ISL_2187994, EPI_ISL_2187995, EPI_ISL_2187997, EPI_ISL_2187999, EPI_ISL_2188001, EPI_ISL_2188002, EPI_ISL_2188004, EPI_ISL_2188005, EPI_ISL_2188009, EPI_ISL_2188012, EPI_ISL_2188013, EPI_ISL_2188014, EPI_ISL_2188015, EPI_ISL_2188016, EPI_ISL_2188017, EPI_ISL_2188018, EPI_ISL_2188019, EPI_ISL_2188020, EPI_ISL_2188021, EPI_ISL_2188023, EPI_ISL_2188024, EPI_ISL_2188025, EPI_ISL_2188026, EPI_ISL_2188027, EPI_ISL_2222900, EPI_ISL_2222918, EPI_ISL_2227564, EPI_ISL_2348604, EPI_ISL_2348605, EPI_ISL_2348606, EPI_ISL_2348607, EPI_ISL_5438932, EPI_ISL_5438946 | | | |
| see above | HLAGYN - Laboratorio de Imunologia de Transplantes de Goias | HLAGYN - Laboratorio de Imunologia de Transplantes de Goias | Alessandro Leonardo Alvares Magalhaes; Daniel Ferreira de Sousa; Erika Lopes Rocha Batista; Fernando Antonio Vinhal dos Santos; Frederico Rodrigues Vinhal; Kamila Oliveira Reis De Freitas.; Lucas Carlos Gomes Pereira; Sabrina Sara Moreira Duarte |
| EPI_ISL_6229723 | HOSP MATERN PAULO SARASATE | ACME Lab, Oswaldo Cruz Foundation, FIOCRUZ/CE | Carlos Leonardo de Aragao Araujo; Cecilia Leite Costa & Eduardo Ruback dos Santos on behalf of COVID-19 FIOCRUZ Genomic Network; Cleber Furtado Aksenen; Fabio Miyajima; Fernando Braga Stehling; Francisco Eder de Moura Lopes; Igor Oliveira Duarte; Jamille Maria Mendes Bezerra; Joaquim Cesar do Nascimento Sousa Junior; Pedro Miguel Carneiro Jeronimo; Suzana Porto Almeida; Thais Ferreira de Oliveira; Thais de Oliveira Costa; Ticiane Cavalcante de Souza; Veridiana Pessoa Miyajima |
| EPI_ISL_6229724 | HOSP MATERN SANTA IZABEL ARACOIABA | ACME Lab, Oswaldo Cruz Foundation, FIOCRUZ/CE | Carlos Leonardo de Aragao Araujo; Cecilia Leite Costa & Eduardo Ruback dos Santos on behalf of COVID-19 FIOCRUZ Genomic Network; Cleber Furtado Aksenen; Fabio Miyajima; Fernando Braga Stehling; Francisco Eder de Moura Lopes; Igor Oliveira Duarte; Jamille Maria Mendes Bezerra; Joaquim Cesar do Nascimento Sousa Junior; Pedro Miguel Carneiro Jeronimo; Suzana Porto Almeida; Thais Ferreira de Oliveira; Thais de Oliveira Costa; Ticiane Cavalcante de Souza; Veridiana Pessoa Miyajima |
| EPI_ISL_3502579, EPI_ISL_3502666 | HOSPITAL DE GETAFE | HOSPITAL UNIVERSITARIO 12 DE OCTUBRE | Carmen Martín-Higuera; Esther Viedma; Irene Múñoz-Gallego; M.ª Dolores Folgueira; Mar Aguilera; Noelia Moral; Rafael Delgado; Sagrario Zurita |
| EPI_ISL_3102510, EPI_ISL_3102516 | HOSPITAL ESTADUAL LEONARDO DA VINCI | Analytical Competence Molecular Epidemiology Lab/ACME, Oswaldo Cruz Foundation, Ceara (FIOCRUZ CE) | Cleber Furtado Aksenen; Fabio Miyajima; Fernando Braga Stehling; Francisco Eder de Moura Lopes; Jamille Maria Mendes Bezerra; Joaquim César do Nascimento Sousa Junior; Pedro Miguel Carneiro Jeronimo; Suzana Porto Almeida e Lucas Delerino; Thais Ferreira de Oliveira; Thais de Oliveira Costa; Ticiane Cavalcante de Souza; Veridiana Pessoa Miyajima |
| EPI_ISL_4414777 | HOSPITAL GENERAL UNIVERSITARIO DE CIUDAD REAL | Instituto de Salud Carlos III | A. Monzón; F. Casas; I. Jiménez; I.ILLESCAS FERNANDEZ-BERMEJO; Iglesias-Caballero; M. SOLEDAD; M. Sandonís; P. Zaballos; S. Camarero; S. Cuesta; S. Pozo; S. Varona; V. Vázquez-Morón |
| EPI_ISL_2208870, EPI_ISL_2209499, EPI_ISL_2209603, EPI_ISL_2345774, EPI_ISL_2345833, EPI_ISL_2362248 | HOSPITAL MUNICIPAL DE ITAPIRA | Instituto Butantan / Mendelics | Antonio Jorge Martins; Claudia Renata dos Santos Barros; David Schlesinger; Debora Botequio Moretti; Dimas Tadeu Covas; Elaine Cristina Marqueze; Elaine Vieira Santos; Evandra Strazza Rodrigues; Heidge Fukumasu; Jayme Augusto de Souza-Neto; José Salvatore Leister Patané; Luiz Alcantara; Luiz Lehmann Coutinho; Maria Carolina Elias; Maurício Lacerda Nogueira; Rafael dos Santos Bezerra; Raul Machado Neto; Rejane Maria Tommasini Grotto; Ricardo Haddad; Sandra Coccuzzo Sampaio Vessoni; Simone Kashima; Svetoslav Nanev Slavov; Vincent Louis Viala |
| EPI_ISL_5603301 | HOSPITAL MUNICIPAL DE JIJOCA DE JERICOACOARA | Analytical Competence Molecular Epidemiology Lab/ACME, Oswaldo Cruz Foundation, Ceara (FIOCRUZ CE) | Carlos Leonardo de Aragao Araujo; Cecilia Leite Costa & Eduardo Ruback dos Santos on behalf of COVID-19 FIOCRUZ Genomic Network; Cleber Furtado Aksenen; Fabio Miyajima; Fernando Braga Stehling; Francisco Eder de Moura Lopes; Igor Oliveira Duarte; Jamille Maria Mendes Bezerra; Joaquim Cesar do Nascimento Sousa Junior; Pedro Miguel Carneiro Jeronimo; Suzana Porto Almeida; Thais Ferreira de Oliveira; Thais de Oliveira Costa; Ticiane Cavalcante de Souza; Veridiana Pessoa Miyajima |
| EPI_ISL_6229691 | HOSPITAL MUNICIPAL DR EUDASIO BARROSO HMEB | ACME Lab, Oswaldo Cruz Foundation, FIOCRUZ/CE | Carlos Leonardo de Aragao Araujo; Cecilia Leite Costa & Eduardo Ruback dos Santos on behalf of COVID-19 FIOCRUZ Genomic Network; Cleber Furtado Aksenen; Fabio Miyajima; Fernando Braga Stehling; Francisco Eder de Moura Lopes; Igor Oliveira Duarte; Jamille Maria Mendes Bezerra; Joaquim Cesar do Nascimento Sousa Junior; Pedro Miguel Carneiro Jeronimo; Suzana Porto Almeida; Thais Ferreira de Oliveira; Thais de Oliveira Costa; Ticiane Cavalcante de Souza; Veridiana Pessoa Miyajima |
| EPI_ISL_5825557, EPI_ISL_5825562 | HOSPITAL MUNICIPAL DR EUDASIO BARROSO HMEB | Analytical Competence Molecular Epidemiology Lab/ACME, Oswaldo Cruz Foundation, Ceara (FIOCRUZ CE) | Carlos Leonardo de Aragao Araujo; Cecilia Leite Costa & Eduardo Ruback dos Santos on behalf of COVID-19 FIOCRUZ Genomic Network; Cleber Furtado Aksenen; Fabio Miyajima; Fernando Braga Stehling; Francisco Eder de Moura Lopes; Igor Oliveira Duarte; Jamille Maria Mendes Bezerra; Joaquim Cesar do Nascimento Sousa Junior; Pedro Miguel Carneiro Jeronimo; Suzana Porto Almeida; Thais Ferreira de Oliveira; Thais de Oliveira Costa; Ticiane Cavalcante de Souza; Veridiana Pessoa Miyajima |
| EPI_ISL_2345693, EPI_ISL_2362259 | HOSPITAL MUNICIPAL DR WALDEMAR TEBALDI | Instituto Butantan / Mendelics | Antonio Jorge Martins; Claudia Renata dos Santos Barros; David Schlesinger; Debora Botequio Moretti; Dimas Tadeu Covas; Elaine Cristina Marqueze; Elaine Vieira Santos; Evandra Strazza Rodrigues; Heidge Fukumasu; Jayme Augusto de Souza-Neto; José Salvatore Leister Patané; Luiz Alcantara; Luiz Lehmann Coutinho; Maria Carolina Elias; Maurício Lacerda Nogueira; Rafael dos Santos Bezerra; Raul Machado Neto; Rejane Maria Tommasini Grotto; Ricardo Haddad; Sandra Coccuzzo Sampaio Vessoni; Simone Kashima; Svetoslav Nanev Slavov; Vincent Louis Viala |
| EPI_ISL_6229729 | HOSPITAL MUNICIPAL JOAO LEOPOLDO PINHEIRO LANDIM | ACME Lab, Oswaldo Cruz Foundation, FIOCRUZ/CE | Carlos Leonardo de Aragao Araujo; Cecilia Leite Costa & Eduardo Ruback dos Santos on behalf of COVID-19 FIOCRUZ Genomic Network; Cleber Furtado Aksenen; Fabio Miyajima; Fernando Braga Stehling; Francisco Eder de Moura Lopes; Igor Oliveira Duarte; Jamille Maria Mendes Bezerra; Joaquim Cesar do Nascimento Sousa Junior; Pedro Miguel Carneiro Jeronimo; Suzana Porto Almeida; Thais Ferreira de Oliveira; Thais de Oliveira Costa; Ticiane Cavalcante de Souza; Veridiana Pessoa Miyajima |
| EPI_ISL_2493441, EPI_ISL_5650323 | HOSPITAL MUNICIPAL JORGE CURY DE SANTA MARIA DA SERRA | Instituto Butantan | Antonio Jorge Martins; Claudia Renata dos Santos Barros; David Schlesinger; Debora Botequio Moretti; Dimas Tadeu Covas; Elaine Cristina Marqueze; Elaine Vieira Santos; Evandra Strazza Rodrigues; Heidge Fukumasu; Jayme Augusto de Souza-Neto; José Salvatore Leister Patané; Luiz Alcantara; Luiz Lehmann Coutinho; Maria Carolina Elias; Maurício Lacerda Nogueira; Rafael dos Santos Bezerra; Raul Machado Neto; Rejane Maria Tommasini Grotto; Ricardo Haddad; Sandra Coccuzzo Sampaio Vessoni; Simone Kashima; Svetoslav Nanev Slavov; Vincent Louis Viala |
| EPI_ISL_2801305, EPI_ISL_2801362 | HOSPITAL MUNICIPAL NOSSA SENHORA DOS MILAGRES | Analytical Competence Molecular Epidemiology Lab/ACME, Oswaldo Cruz Foundation, Ceara (FIOCRUZ CE) | Cleber Furtado Aksenen e Suzana Porto Almeida; Fabio Miyajima; Fernando Braga Stehling; Francisco Eder de Moura Lopes; Jamille Maria Mendes Bezerra; Joaquim César do Nascimento Sousa Junior; Pedro Miguel Carneiro Jeronimo; Thais Ferreira de Oliveira; Thais de Oliveira Costa; Ticiane Cavalcante de Souza; Veridiana Pessoa Miyajima |
| EPI_ISL_6229731, EPI_ISL_6229732, EPI_ISL_6229733, EPI_ISL_6229844 | HOSPITAL MUNICIPAL SENADOR CARLOS JEREISSATI | ACME Lab, Oswaldo Cruz Foundation, FIOCRUZ/CE | Carlos Leonardo de Aragao Araujo; Cecilia Leite Costa & Eduardo Ruback dos Santos on behalf of COVID-19 FIOCRUZ Genomic Network; Cleber Furtado Aksenen; Fabio Miyajima; Fernando Braga Stehling; Francisco Eder de Moura Lopes; Igor Oliveira Duarte; Jamille Maria Mendes Bezerra; Joaquim Cesar do Nascimento Sousa Junior; Pedro Miguel Carneiro Jeronimo; Suzana Porto Almeida; Thais Ferreira de Oliveira; Thais de Oliveira Costa; Ticiane Cavalcante de Souza; Veridiana Pessoa Miyajima |
| EPI_ISL_6229736, EPI_ISL_6229737 | HOSPITAL REGIONAL DE IGUATU | ACME Lab, Oswaldo Cruz Foundation, FIOCRUZ/CE | Carlos Leonardo de Aragao Araujo; Cecilia Leite Costa & Eduardo Ruback dos Santos on behalf of COVID-19 FIOCRUZ Genomic Network; Cleber Furtado Aksenen; Fabio Miyajima; Fernando Braga Stehling; Francisco Eder de Moura Lopes; Igor Oliveira Duarte; Jamille Maria Mendes Bezerra; Joaquim Cesar do Nascimento Sousa Junior; Pedro Miguel Carneiro Jeronimo; Suzana Porto Almeida; Thais Ferreira de Oliveira; Thais de Oliveira Costa; Ticiane Cavalcante de Souza; Veridiana Pessoa Miyajima |

| | | | |
|---|---|---|---|
| EPI_ISL_6229835 | HOSPITAL SENADOR OZIRES PONTES | ACME Lab, Oswaldo Cruz Foundation, FIOCRUZ/CE | Carlos Leonardo de Aragao Araujo; Cecilia Leite Costa & Eduardo Ruback dos Santos on behalf of COVID-19 FIOCRUZ Genomic Network; Cleber Furtado Aksenen; Fabio Miyajima; Fernando Braga Stehling; Francisco Eder de Moura Lopes; Igor Oliveira Duarte; Jamille Maria Mendes Bezerra; Joaquim Cesar do Nascimento Sousa Junior; Pedro Miguel Carneiro Jeronimo; Suzana Porto Almeida; Thais Ferreira de Oliveira; Thais de Oliveira Costa; Ticiane Cavalcante de Souza; Veridiana Pessoa Miyajima |
| EPI_ISL_3502597, EPI_ISL_3532489 | HOSPITAL UNIVERSITARIO 12 DE OCTUBRE | HOSPITAL UNIVERSITARIO 12 DE OCTUBRE | Carmen Martín-Higuera; Esther Viedma; Irene Múñoz-Gallego; M.ª Dolores Folgueira; Mar Aguilera; Noelia Moral; Rafael Delgado; Sagrario Zurita |
| EPI_ISL_4414802 | HOSPITAL UNIVERSITARIO DE CEUTA | Instituto de Salud Carlos III | A. Monzón; F. Casas; I. Jiménez; I.HIJANO VILLEGAS; M. Sandonís; P. Zaballos; S. Cuesta; S. Iglesias-Caballero; S. Pozo; S. Varona; SALOME; V. Camarero; Vázquez-Morón |
| EPI_ISL_2134884 | HOSPITAL UNIVERSITARIO SON ESPASES | HOSPITAL UNIVERSITARIO SON ESPASES | Antonio Oliver; Carla López-Causapé; Pablo Fraile-Ribot; SeqCovid |
| EPI_ISL_4414793 | HOSPITAL VIRGEN DE LA SALUD DE TOLEDO | Instituto de Salud Carlos III | A. Monzón; EVA; F. Casas; I. Jiménez; I.HEREDERO GALVEZ; Iglesias-Caballero; M. Sandonís; P. Zaballos; S. Camarero; S. Cuesta; S. Pozo; S. Varona; V. Vázquez-Morón |
| EPI_ISL_2621174 | HPTU | Universidad Nacional de Colombia - Laboratorio Genómico One Health | Andres F. Cardona-Rios; Carlos Franco-Muñoz; Carolina Muñoz-Arango; Celeny Ortiz; Daniel O. Maldonado-Perez; Diego A. Álvarez-Díaz; Hector Alejandro Ruiz-Moreno; Idabely Betancur Ortiz; Jorge E. Osorio; Juan P. Hernandez-Ortiz; Karl A Ciuoderis; Katherine Laiton-Donato; Laura Silvana Perez; Lina M. Hurtado; Marcela Mercado-Reyes; Maria Angélica Maya; Maria Stella López; Rita Almanza Payares; Sandra Ines Cano; Simón Villegas Velásquez |
| EPI_ISL_3235358 | Hackensack Medical Center | New York Genome Center | Amy Baldwin; Andre Corvelo; Barry Kreiswirth; David Perlin; Dayna M. Oschwald; Jose Mediavilla; Kaelea Composto; Kar Chow; Liang Chen; Marcus Cunningham; Michael Zody; Samantha Fennessey; Tom Maniatis |
| EPI_ISL_2080392 | Health Services Laboratories | Wellcome Sanger Institute for the COVID-19 Genomics UK (COG-UK) Consortium | Cordelia Langford; David K. Jackson; Dominic Kwiatkowski; Ewan Harrison; Health Services Laboratories and Alex Alderton; Ian Johnston; Jeffrey Barrett; John Sillitoe on behalf of the Wellcome Sanger Institute COVID-19 Surveillance Team; Roberto Amato; Sonia Goncalves |
| EPI_ISL_2160218, EPI_ISL_2160418, EPI_ISL_2247366, EPI_ISL_2247373, EPI_ISL_2247374, EPI_ISL_2247391, EPI_ISL_2247406, EPI_ISL_2247446, EPI_ISL_2247455, EPI_ISL_2247484, EPI_ISL_2247512, EPI_ISL_2247521, EPI_ISL_2247523, EPI_ISL_2247532, EPI_ISL_2247539, EPI_ISL_2247561, EPI_ISL_2247573, EPI_ISL_2247581, EPI_ISL_2247588, EPI_ISL_2247596, EPI_ISL_2247602, EPI_ISL_2247612, EPI_ISL_2247620, EPI_ISL_2247624, EPI_ISL_2247628, EPI_ISL_2247630, EPI_ISL_2247631, EPI_ISL_2247638, EPI_ISL_2247746, EPI_ISL_2247794, EPI_ISL_2247800, EPI_ISL_2247807, EPI_ISL_2247808, EPI_ISL_2247817, EPI_ISL_2247917, EPI_ISL_2247946, EPI_ISL_2248015, EPI_ISL_2248057, EPI_ISL_2248067, EPI_ISL_2248069, EPI_ISL_2248070, EPI_ISL_2248072, EPI_ISL_2248089, EPI_ISL_2248097, EPI_ISL_2248148, EPI_ISL_2248156, EPI_ISL_2248298, EPI_ISL_2248384, EPI_ISL_2248385, EPI_ISL_2248394, EPI_ISL_2248409, EPI_ISL_2248420, EPI_ISL_2248432, EPI_ISL_2248434, EPI_ISL_2268352, EPI_ISL_2268363, EPI_ISL_2269624, EPI_ISL_2269627, EPI_ISL_2269635, EPI_ISL_2269637, EPI_ISL_2269649, EPI_ISL_2269669, EPI_ISL_2269670, EPI_ISL_2269706, EPI_ISL_2269709, EPI_ISL_2269733, EPI_ISL_2269750, EPI_ISL_2269768, EPI_ISL_2269773, EPI_ISL_2269797, EPI_ISL_2269812, EPI_ISL_2269813, EPI_ISL_2269817, EPI_ISL_2269839, EPI_ISL_2269844, EPI_ISL_2269873, EPI_ISL_2269888, EPI_ISL_2269913, EPI_ISL_2269945, EPI_ISL_2269947, EPI_ISL_2269976, EPI_ISL_2269979, EPI_ISL_2269982, EPI_ISL_2269986, EPI_ISL_2269988, EPI_ISL_2270010, EPI_ISL_2270014, EPI_ISL_2270030, EPI_ISL_2270042, EPI_ISL_2270047, EPI_ISL_2270075, EPI_ISL_2270097, EPI_ISL_2270105, EPI_ISL_2270111, EPI_ISL_2270119, EPI_ISL_2270140, EPI_ISL_2270148, EPI_ISL_2270168, EPI_ISL_2270172, EPI_ISL_2270173, EPI_ISL_2270199, EPI_ISL_2270203, EPI_ISL_2270224, EPI_ISL_2270231, EPI_ISL_2270234, EPI_ISL_2270241, EPI_ISL_2270255, EPI_ISL_2270257, EPI_ISL_2270258, EPI_ISL_2270260, EPI_ISL_2270264, EPI_ISL_2270302, EPI_ISL_2270309, EPI_ISL_2270315, EPI_ISL_2270316, EPI_ISL_2270317, EPI_ISL_2270324, EPI_ISL_2270333, EPI_ISL_2270360, EPI_ISL_2270369, EPI_ISL_2270386, EPI_ISL_2270387, EPI_ISL_2270388, EPI_ISL_2270407, EPI_ISL_2270412, EPI_ISL_2270414, EPI_ISL_2270445, EPI_ISL_2270456, EPI_ISL_2270461, EPI_ISL_2270474, EPI_ISL_2270479, EPI_ISL_2270491, EPI_ISL_2270499, EPI_ISL_2270508, EPI_ISL_2270519, EPI_ISL_2270523, EPI_ISL_2270529, EPI_ISL_2270534, EPI_ISL_2270536, EPI_ISL_2270559, EPI_ISL_2270566, EPI_ISL_2270571, EPI_ISL_2270584, EPI_ISL_2270595, EPI_ISL_2270596, EPI_ISL_2270597, EPI_ISL_2270598, EPI_ISL_2270603, EPI_ISL_2270604, EPI_ISL_2270611, EPI_ISL_2270613, EPI_ISL_2270624, EPI_ISL_2270628, EPI_ISL_2270637, EPI_ISL_2270652, EPI_ISL_2270655, EPI_ISL_2270671, EPI_ISL_2270692, EPI_ISL_2270693, EPI_ISL_2270700, EPI_ISL_2270703, EPI_ISL_2270706, EPI_ISL_2270714, EPI_ISL_2270715, EPI_ISL_2270719, EPI_ISL_2270728, EPI_ISL_2270739, EPI_ISL_2270747, EPI_ISL_2270761, EPI_ISL_2270772, EPI_ISL_2270779, EPI_ISL_2270783, EPI_ISL_2270784, EPI_ISL_2270791, EPI_ISL_2270792, EPI_ISL_2270797, EPI_ISL_2270800, EPI_ISL_2270828, EPI_ISL_2270830, EPI_ISL_2270833, EPI_ISL_2280178, EPI_ISL_2280191, EPI_ISL_2280199, EPI_ISL_2280209, EPI_ISL_2280210, EPI_ISL_2280219, EPI_ISL_2320016, EPI_ISL_2320030, EPI_ISL_2320041, EPI_ISL_2320046, EPI_ISL_2320054, EPI_ISL_2320059, EPI_ISL_2320079, EPI_ISL_2320100, EPI_ISL_2320101, EPI_ISL_2320116, EPI_ISL_2320126, EPI_ISL_2320137, EPI_ISL_2320167, EPI_ISL_2320540, EPI_ISL_2320541, EPI_ISL_2320548, EPI_ISL_2320551, EPI_ISL_2320581, EPI_ISL_2320583, EPI_ISL_2320584, EPI_ISL_2320595, EPI_ISL_2320600, EPI_ISL_2320665, EPI_ISL_2320675, EPI_ISL_2320687, EPI_ISL_2320691, EPI_ISL_2320702, EPI_ISL_2320709, EPI_ISL_2320722, EPI_ISL_2320731, EPI_ISL_2320733, EPI_ISL_2320743, EPI_ISL_2320752, EPI_ISL_2320753, EPI_ISL_2320851, EPI_ISL_2320857, EPI_ISL_2320862, EPI_ISL_2320867, EPI_ISL_2320877, EPI_ISL_2320882, EPI_ISL_2320898, EPI_ISL_2320905, EPI_ISL_2320908, EPI_ISL_2320929, EPI_ISL_2320939, EPI_ISL_2320949, EPI_ISL_2320953, EPI_ISL_2320958, EPI_ISL_2320965, EPI_ISL_2320974, EPI_ISL_2320979, EPI_ISL_2320982, EPI_ISL_2320993, EPI_ISL_2321139, EPI_ISL_2367609, EPI_ISL_2367628, EPI_ISL_2367648, EPI_ISL_2440265, EPI_ISL_4345817, EPI_ISL_4345834, EPI_ISL_4346732, EPI_ISL_4346768, EPI_ISL_4347314, EPI_ISL_4347508 | | | |
| see above | Helix/Illumina | Centers for Disease Control and Prevention Division of Viral Diseases, Pathogen Discovery | Adrian Paskey; Alexandre Bolze; Ary Ascencio; Benjamin Rambo-Martin; Brad Sickler; Charlotte Rivera-Garcia; Christine Tran; Christopher Gulvick; Chrstine Tran; Clinton Paden; Clinton R. Paden; Dakota Howard; Darlene Wagner; David Becker; Dhwani Batra; Duncan MacCannell; Efren Sandoval; Eileen De Feo; Eileen de Feo; Elizabeth Cirulli; Eric Allen; Geraint Levan; James Lu; Jan Antico; Jason Caravas; Jason Nguyen; Jimmy Ramirez; Jingtao Liu; Kara Moser; Kelly Barrett; Kelly Schiabor Barrett; Kim Gietzen; Kristine Lacek; Magnus Isaksson; Marc Laurent; Matthew Schmerer; Matthew Tolentino; Nicole L. Washington; Nicole Washington; Peter Cook; Peter W. Cook; Phil Febbo; Ryan Cho; Scott Sammons; Shannon Wickline; Shatavia Morrison; Sherry Wang; Simon White; Tyler Cassens; William Lee; Yvette Unoarumhi |
| EPI_ISL_2401838 | Hospital Center Emile Mayrisch | Laboratoire national de sante, Microbiology, Microbial Genomics Platform | Anke Wienecke-Baldacchino; Catherine Ragimbeau; Cynthia Oxacelay; Fatu Djabi; Jessica Tapp; Lise Pignon; Raoul Salmon; Tamir Abdelrahman |
| EPI_ISL_2439984 | Hospital Francesc de Borja (Gandia) | SeqCOVID-SPAIN consortium/IBV(CSIC) | Inmaculada Vidal Catalá; Isabel García Nava and SeqCOVID-SPAIN consortium; Mª Rosa Navarro Cots; Nieves Orta Mira |
| EPI_ISL_2080886, EPI_ISL_2080922, EPI_ISL_2080923, EPI_ISL_3505935, EPI_ISL_3505937 | Hospital General Universitario Gregorio Marañón | Hospital General Universitario Gregorio Marañón | Cristina Rodriguez-Grande; Darío García de Viedma; Julia Suárez; Laura Pérez-Lago; Marta Herranz Martin; Patricia Muñoz; Pedro Sola Campoy; Pilar Catalán; Sergio Buenestado Serrano; Victor Manuel de la Cueva |
| EPI_ISL_4405512 | Hospital General de Agudos Dr. Cosme Argerich | Área de Secuenciación del Laboratorio de Virología del Hospital de Niños Dr. Ricardo Gutierrez on behalf of 'Proyecto Argentino Interinstitucional de genomica de SARS-CoV-2' (PAIS Consortium) | Acuña; Andrea Fernández; D; Florencia Funez; Florencia Rodríguez; Goya; Jésica Galeano; Karina Polanski; LE; Lusso; MI; MS; Marcia Pozzati; Nabaes Jodar; Natale; S; Valinotto; Viegas, M. |
| EPI_ISL_5801603, EPI_ISL_5801646, EPI_ISL_5801668, EPI_ISL_5801669, EPI_ISL_5801670, EPI_ISL_5801671 | Hospital Municipal De Itapira | Instituto Butantan | Antonio Jorge Martins; Claudia Renata dos Santos Barros; David Schlesinger; Debora Botequio Moretti; Dimas Tadeu Covas; Elaine Cristina Marqueze; Elaine Vieira Santos; Evandra Strazza Rodrigues; Heidge Fukumasu; Jayme Augusto de Souza-Neto; José Salvatore Leister Patané; Luiz Alcantara; Luiz Lehmann Coutinho; Maria Carolina Elias; Maurício Lacerda Nogueira; Rafael dos Santos Bezerra; Raul Machado Neto; Rejane Maria Tommasini Grotto; Ricardo Haddad; Sandra Coccuzzo Sampaio Vessoni; Simone Kashima; Svetoslav Nanev Slavov; Vincent Louis Viala |
| EPI_ISL_5801674, EPI_ISL_5801678 | Hospital Municipal Dr Waldemar Tebaldi | Instituto Butantan | Antonio Jorge Martins; Claudia Renata dos Santos Barros; David Schlesinger; Debora Botequio Moretti; Dimas Tadeu Covas; Elaine Cristina Marqueze; Elaine Vieira Santos; Evandra Strazza Rodrigues; Heidge Fukumasu; Jayme Augusto de Souza-Neto; José Salvatore Leister Patané; Luiz Alcantara; Luiz Lehmann Coutinho; Maria Carolina Elias; Maurício Lacerda Nogueira; Rafael dos Santos Bezerra; Raul Machado Neto; Rejane Maria Tommasini Grotto; Ricardo Haddad; Sandra Coccuzzo Sampaio Vessoni; Simone Kashima; Svetoslav Nanev Slavov; Vincent Louis Viala |
| EPI_ISL_2662822 | Hospital San Agustín | Laboratorio de Virología HUCA | Abreu F; Alvarez-Arguelles ME; Boga JA; Castelló C; Costales I; Coto E; Gómez de Oña J; Martín-Rodríguez G; Melón S; Perez-Martínez Z; Rojo S; Sandoval M |
| EPI_ISL_3230019 | Hospital Señor del Milagro | Instituto de Patologia Vegetal (CIAP-INTA) on behalf of 'Proyecto Argentino Interinstitucional de genomica de SARS-CoV-2' (PAIS Consortium) | Debat, HJ.; FD; Fernanda Aguero; Fernández; Lavaque Esteban; Marquez, N.; Pamela Cajal; Raskovsky Viviana; Veronica Lesser |
| EPI_ISL_2351935, EPI_ISL_2351989 | Hospital Universitari Bellvitge | Microbiology Department | Aida Gonzalez-Diaz; Carmen Ardanuy; Jordi Camara; Jordi Niubó; Laura Calatayud; M Angeles Domínguez; Miguel Fernandez-Huerta; Sara Marti |
| EPI_ISL_2047850 | Hospital Universitari Joan XXIII de Tarragona | Hospital Universitari Vall d'Hebron - Vall d'Hebron Institut de Recerca | Alejandra González-Sánchez; Andrés Antón; Ariadna Rando; Carla Castillo; Cristina Andrés; Damir Garcia-Cehic; Josep Quer; Juliana Esperalba; Maria Carmen Martin; Maria Gema Codina; Maria Piñana; Tomàs Pumarola |
| EPI_ISL_2162252, EPI_ISL_2162253, EPI_ISL_2200155 | Hospital Universitari Vall d'Hebron - Vall d'Hebron Institut de Recerca | Hospital Universitari Vall d'Hebron - Vall d'Hebron Institut de Recerca | Alejandra González-Sánchez; Andrés Antón; Ariadna Rando; Carla Castillo; Cristina Andrés; Damir Garcia-Cehic; Josep Quer; Juliana Esperalba; Maria Carmen Martin; Maria Gema Codina; Maria Piñana; Tomàs Pumarola |
| EPI_ISL_2383259 | Hospital Universitari i Politècnic La Fe de València | SeqCOVID-SPAIN consortium/IBV(CSIC) | Ana Gil Brusola; Eva González Barberá; José Luis López Hontangas and SeqCOVID-SPAIN consortium; María Dolores Gómez Ruiz; Salvador Giner Almaraz |
| EPI_ISL_6809428 | Hospital Universitario Fundación Alcorcón | Hospital Universitario Fundación Alcorcón | Ana Santos; Isabel Fradejas; Jorge do Campo; Marta Casas; Rosa Frutos |
| EPI_ISL_7307293, EPI_ISL_7307297 | Hospital Universitario San Ignacio | Centro de Investigaciones en Microbiología y Biotecnología-UR (CIMBIUR), Facultad de Ciencias Naturales, Universidad del Rosario, Bogotá, Colombia | Alberto Paniz-Mondolfi; Angie Ramírez; Beatriz Ariza; Camilo A. Correa-Cárdenas; Carlos Gómez-Restrepo; Claudia Cardozo-Romero; Claudia Méndez; David-Santiago Quevedo; Guido España; Hernando Díaz; Juan David Ramírez; Juliana Cuervo-Rojas; Julie Pérez; Luz H. Patiño; Manuel-Antonio Franco; Maria-Clara Duque; Marina Muñoz; Nathalia Ballesteros; Nicolas Luna; Sergio Castañeda; Zulma M. Cucunubá |
| EPI_ISL_4196725 | Hospital Zonal Dr. Ramón Carrillo | Laboratorio Central Mg. Luis Alfredo Pianciola on behalf of 'Proyecto Argentino Interinstitucional de genomica de SARS-CoV-2' (PAIS Consortium) | C Pintos; C Rastellini; C Ziehm; J Ousset; L Pianciola; M Fernandez; M L Álvarez; M Mazzeo; P V Blanco.; Y Espasandin |
| EPI_ISL_2238720 | Hospital de San Agustin | Laboratorio de Virología HUCA | Abreu F; Alvarez-Arguelles ME; Boga JA; Castelló C; Costales I; Coto E; Gómez de Oña J; Martín-Rodríguez G; Melón S; Perez-Martínez Z; Rojo S; Sandoval M |
| EPI_ISL_2407652 | Hospital de San Agustín | Laboratorio de Virología HUCA | Abreu F; Alvarez-Arguelles ME; Boga JA; Castelló C; Costales I; Coto E; Gómez de Oña J; Martín-Rodríguez G; Melón S; Perez-Martínez Z; Rojo S; Sandoval M |
| EPI_ISL_5070078, EPI_ISL_5070148, EPI_ISL_5070158, EPI_ISL_5070187, EPI_ISL_5070219, EPI_ISL_5070242, EPI_ISL_5070244, EPI_ISL_5070271, EPI_ISL_5070274, EPI_ISL_5070280, EPI_ISL_5070305, EPI_ISL_5070310, EPI_ISL_5070353 | | | |
| see above | Houston Methodist Hospital | Houston Methodist Hospital | Ilya J. Finkelstein; James J. Davis; Jessica Cambric; Jimmy Gollihar; Kristina Reppond; Layne Pruitt; Madison N. Shyer; Marcus Nguyen; Matthew Ojeda Saavedra; Paul A. Christensen; Prasanti Yerramilli; Randall J. Olsen; Robert Olson; Ryan Gadd; S. Wesley Long; Sishir Subedi; and James M. Musser |
| EPI_ISL_2341449, EPI_ISL_2500133 | Humboldt County Public Health Laboratory | Chan-Zuckerberg Biohub | CZB Cliahub Consortium |
| EPI_ISL_2621884, EPI_ISL_2651215, EPI_ISL_2651217, EPI_ISL_2651220 | ICMT-APARTADO | Universidad Nacional de Colombia - Laboratorio Genómico One Health | Andres F. Cardona-Rios; Carlos Franco-Muñoz; Carolina Muñoz-Arango; Celeny Ortiz; Daniel O. Maldonado-Perez; Diego A. Álvarez-Díaz; Hector Alejandro Ruiz-Moreno; Idabely Betancur Ortiz; Jorge E. Osorio; Juan P. Hernandez-Ortiz; Karl A Ciuoderis; Katherine Laiton-Donato; Laura Silvana Perez; Lina M. Hurtado; Marcela Mercado-Reyes; Maria Angélica Maya; Maria Stella López; Rita Almanza Payares; Sandra Ines Cano; Simón Villegas Velásquez |
| EPI_ISL_2621157, EPI_ISL_2621158, EPI_ISL_2621159 | ICMT-SABANETA | Universidad Nacional de Colombia - Laboratorio Genómico One Health | Andres F. Cardona-Rios; Carlos Franco-Muñoz; Carolina Muñoz-Arango; Celeny Ortiz; Daniel O. Maldonado-Perez; Diego A. Álvarez-Díaz; Hector Alejandro Ruiz-Moreno; Idabely Betancur Ortiz; Jorge E. Osorio; Juan P. Hernandez-Ortiz; Karl A Ciuoderis; Katherine Laiton-Donato; Laura Silvana Perez; Lina M. Hurtado; Marcela Mercado-Reyes; Maria Angélica Maya; Maria Stella López; Rita Almanza Payares; Sandra Ines Cano; Simón Villegas Velásquez |
| EPI_ISL_2444819, EPI_ISL_2444820, | IICS-UNA | IICS-UNA | Adriana Valenzuela; Alejandra Rojas; Chyntia Diaz; Eva Nara; Fatima Cardozo; Florencia del Puerto; Joel Ortiz; Jonas Fernandez; Laura Franco; Laura Mendoza; Leticia Rojas; Magaly Martinez; Maria Eugenia Galeano. |

| | | | |
|---|---|---|---|
| EPI_ISL_2444822 | | | |
| EPI_ISL_2229216, EPI_ISL_2229221, EPI_ISL_2229223, EPI_ISL_2229226, EPI_ISL_2229237, EPI_ISL_2229238, EPI_ISL_2229239, EPI_ISL_2229241 | | | |
| see above | IL Dept. of Public Health Springfield Laboratory | Centers for Disease Control and Prevention Division of Viral Diseases, Pathogen Discovery | Alison Laufer Halpin; Ben L. Rambo-Martin; Clinton R. Paden; Dakota Howard; Darlene Wagner; Dave Wentworth; Dhwani Batra; Jasmine Padilla; Justin Lee; Katie Dillon; Krista Queen; Kristen Knipe; Kristine Lacek; Mark Burroughs; Matthew Schmerer; Mili Sheth; Peter Cook; Sam Shepard; Sarah Nobles; Shoshona Le; Suxiang Tong; Vivien Dugan; Yvette Unoarumhi |
| EPI_ISL_2097192, EPI_ISL_2097213, EPI_ISL_2242546, EPI_ISL_2242581, EPI_ISL_2242591, EPI_ISL_2361384, EPI_ISL_2361389, EPI_ISL_2361411, EPI_ISL_2361414, EPI_ISL_2361423, EPI_ISL_2361428, EPI_ISL_2361434, EPI_ISL_2361437, EPI_ISL_2361449, EPI_ISL_2420155, EPI_ISL_3007516 | | | |
| see above | IN State Department of Health Laboratory Services | IN State Department of Health Laboratory Services | Brian Pope; Cassandra Campion; Jamie Yeadon; Kyle Brownlee; Lixia Liu; Mark Glazier; Melissa Hindenlang |
| EPI_ISL_2628297 | INHRR | Laboratorio de Virología Molecular | Carmen L Loureiro; CoViVen Group; Domingo J Garzaro; Esmeralda Vizzi; Flor H Pujol; Héctor R Rangel; José Luis Zambrano; Lieska Rodríguez; Mariana Hidalgo; Pierina D´Angelo; Rossana C Jaspe; Víctor Alarcón; Yoneira Sulbaran; Zoila Moros |
| EPI_ISL_2445050, EPI_ISL_2445066 | INSIDE DIAGNÓSTICOS NORTE BRASILÂNDIA/FREGUESIA DO Ó | Instituto Butantan | Antonio Jorge Martins; Claudia Renata dos Santos Barros; David Schlesinger; Debora Botequio Moretti; Dimas Tadeu Covas; Elaine Cristina Marqueze; Elaine Vieira Santos; Evandra Strazza Rodrigues; Heidge Fukumasu; Jayme Augusto de Souza-Neto; José Salvatore Leister Patané; Luiz Alcantara; Luiz Lehmann Coutinho; Maria Carolina Elias; Maurício Lacerda Nogueira; Rafael dos Santos Bezerra; Raul Machado Neto; Rejane Maria Tommasini Grotto; Ricardo Haddad; Sandra Coccuzzo Sampaio Vessoni; Simone Kashima; Svetoslav Nanev Slavov; Vincent Louis Viala |
| EPI_ISL_2445070 | INSIDE DIAGNÓSTICOS NORTE CACHOEIRINHA/CASA VERDE | Instituto Butantan | Antonio Jorge Martins; Claudia Renata dos Santos Barros; David Schlesinger; Debora Botequio Moretti; Dimas Tadeu Covas; Elaine Cristina Marqueze; Elaine Vieira Santos; Evandra Strazza Rodrigues; Heidge Fukumasu; Jayme Augusto de Souza-Neto; José Salvatore Leister Patané; Luiz Alcantara; Luiz Lehmann Coutinho; Maria Carolina Elias; Maurício Lacerda Nogueira; Rafael dos Santos Bezerra; Raul Machado Neto; Rejane Maria Tommasini Grotto; Ricardo Haddad; Sandra Coccuzzo Sampaio Vessoni; Simone Kashima; Svetoslav Nanev Slavov; Vincent Louis Viala |
| EPI_ISL_2445062, EPI_ISL_2445071 | INSIDE DIAGNÓSTICOS NORTE PERUS | Instituto Butantan | Antonio Jorge Martins; Claudia Renata dos Santos Barros; David Schlesinger; Debora Botequio Moretti; Dimas Tadeu Covas; Elaine Cristina Marqueze; Elaine Vieira Santos; Evandra Strazza Rodrigues; Heidge Fukumasu; Jayme Augusto de Souza-Neto; José Salvatore Leister Patané; Luiz Alcantara; Luiz Lehmann Coutinho; Maria Carolina Elias; Maurício Lacerda Nogueira; Rafael dos Santos Bezerra; Raul Machado Neto; Rejane Maria Tommasini Grotto; Ricardo Haddad; Sandra Coccuzzo Sampaio Vessoni; Simone Kashima; Svetoslav Nanev Slavov; Vincent Louis Viala |
| EPI_ISL_2445056 | INSIDE DIAGNÓSTICOS SUDESTE MOOCA/ARICANDUVA | Instituto Butantan | Antonio Jorge Martins; Claudia Renata dos Santos Barros; David Schlesinger; Debora Botequio Moretti; Dimas Tadeu Covas; Elaine Cristina Marqueze; Elaine Vieira Santos; Evandra Strazza Rodrigues; Heidge Fukumasu; Jayme Augusto de Souza-Neto; José Salvatore Leister Patané; Luiz Alcantara; Luiz Lehmann Coutinho; Maria Carolina Elias; Maurício Lacerda Nogueira; Rafael dos Santos Bezerra; Raul Machado Neto; Rejane Maria Tommasini Grotto; Ricardo Haddad; Sandra Coccuzzo Sampaio Vessoni; Simone Kashima; Svetoslav Nanev Slavov; Vincent Louis Viala |
| EPI_ISL_2445052 | INSIDE DIAGNÓSTICOS SUDESTE VILA MARIANA/JABAQUARA | Instituto Butantan | Antonio Jorge Martins; Claudia Renata dos Santos Barros; David Schlesinger; Debora Botequio Moretti; Dimas Tadeu Covas; Elaine Cristina Marqueze; Elaine Vieira Santos; Evandra Strazza Rodrigues; Heidge Fukumasu; Jayme Augusto de Souza-Neto; José Salvatore Leister Patané; Luiz Alcantara; Luiz Lehmann Coutinho; Maria Carolina Elias; Maurício Lacerda Nogueira; Rafael dos Santos Bezerra; Raul Machado Neto; Rejane Maria Tommasini Grotto; Ricardo Haddad; Sandra Coccuzzo Sampaio Vessoni; Simone Kashima; Svetoslav Nanev Slavov; Vincent Louis Viala |
| EPI_ISL_2445079 | INSIDE DIAGNÓSTICOS SUDESTE VILA PRUDENTE/SAPOPEMBA | Instituto Butantan | Antonio Jorge Martins; Claudia Renata dos Santos Barros; David Schlesinger; Debora Botequio Moretti; Dimas Tadeu Covas; Elaine Cristina Marqueze; Elaine Vieira Santos; Evandra Strazza Rodrigues; Heidge Fukumasu; Jayme Augusto de Souza-Neto; José Salvatore Leister Patané; Luiz Alcantara; Luiz Lehmann Coutinho; Maria Carolina Elias; Maurício Lacerda Nogueira; Rafael dos Santos Bezerra; Raul Machado Neto; Rejane Maria Tommasini Grotto; Ricardo Haddad; Sandra Coccuzzo Sampaio Vessoni; Simone Kashima; Svetoslav Nanev Slavov; Vincent Louis Viala |
| EPI_ISL_2458114 | INSPI-CRN DE INFLUENZA Y OTROS VIRUS RESPIRATORIOS | NIC-INSPI | Alfredo Bruno; Domenica de Mora.; Jimmy Garcés; Johanna Laines; Lizbeth Patiño; Manuel Gonzalez; Maritza Olmedo; Michelle Páez |
| EPI_ISL_2716746, EPI_ISL_2716747, EPI_ISL_2716748, EPI_ISL_2716825 | IRCCS San Gallicano Dermatological Institute | IRCCS Regina Elena National Cancer Institute | Aldo Morrone; Alice Massacci; Carla Mottini; Eleonora Sperandio; Fabrizio Ensoli; Francesca De Nicola; Frauke Goeman; Fulvia Pimpinelli; Gennaro Ciliberto; Giovanna D'agosto; Giovanni Blandino; Giulia Orlandi; Martina Pontone; Matteo Pallocca; Maurizio Fanciulli; Sabrina Strano; Sara Donzelli; Sara Petrolo |
| EPI_ISL_2500380, EPI_ISL_2500419, EPI_ISL_2500420, EPI_ISL_2598272 | IVY3 Central Lab, Vanderbilt University Medical Center | Lauring Lab, University of Michigan, Department of Microbiology and Immunology | Valesano |
| EPI_ISL_2933102, EPI_ISL_2933105 | IZSM | IZSM-U.O.C. Virologia | Antonio Limone; Claudio de Martinis; Esterina De Carlo; Giovanna Fusco; Lorena Cardillo; Maurizio Viscardi |
| EPI_ISL_2834650, EPI_ISL_2834653, EPI_ISL_2834656, EPI_ISL_2834676 | Idaho Bureau of Laboratories | Center for Global Health, University of New Mexico Health Sciences Center | Christopher Ball; Darrell Dinwiddie; Daryl Domman; Kurt Schwalm; Matthew Burns; Robert Voermans; Valerie Morley |
| EPI_ISL_2295917, EPI_ISL_2960185, EPI_ISL_2960186 | Illinois Department of Public Health | Gagnon Lab, Southern Illinois University | Keith Gagnon |
| EPI_ISL_2300286, EPI_ISL_2300328, EPI_ISL_2300353, EPI_ISL_2300387, EPI_ISL_2300407, EPI_ISL_2300414, EPI_ISL_2300416, EPI_ISL_2300425, EPI_ISL_2300531, EPI_ISL_2300536, EPI_ISL_2300582, EPI_ISL_2300631, EPI_ISL_2300634, EPI_ISL_2467401, EPI_ISL_2467403, EPI_ISL_2467446, EPI_ISL_2800860 | | | |
| see above | Illinois Department of Public Health | Illinois Department of Public Health - Chicago Lab | Ira Heimler; Joel Price; Vineet K. Dhiman |
| EPI_ISL_2000802, EPI_ISL_2000803, EPI_ISL_2086570, EPI_ISL_2086572, EPI_ISL_2086573, EPI_ISL_2135044, EPI_ISL_2135047, EPI_ISL_2135048, EPI_ISL_2135050, EPI_ISL_2135051, EPI_ISL_2135052, EPI_ISL_2135089, EPI_ISL_2135090, EPI_ISL_2135092, EPI_ISL_2135094, EPI_ISL_2135098, EPI_ISL_2135099, EPI_ISL_2135100, EPI_ISL_2135101, EPI_ISL_2135102, EPI_ISL_2135104, EPI_ISL_2135105, EPI_ISL_2135106, EPI_ISL_2135108, EPI_ISL_2203538, EPI_ISL_2203540, EPI_ISL_2203542, EPI_ISL_2203548, EPI_ISL_2203552, EPI_ISL_2203553, EPI_ISL_2203555, EPI_ISL_2203556, EPI_ISL_2203558, EPI_ISL_2203559, EPI_ISL_2203561, EPI_ISL_2203562, EPI_ISL_2203564, EPI_ISL_2203565, EPI_ISL_2203567, EPI_ISL_2203573, EPI_ISL_2203574, EPI_ISL_2203577, EPI_ISL_2203580, EPI_ISL_2203587, EPI_ISL_2203616, EPI_ISL_2203619, EPI_ISL_2203627, EPI_ISL_2203630, EPI_ISL_2203650, EPI_ISL_2203663, EPI_ISL_2203671, EPI_ISL_2203679, EPI_ISL_2203685, EPI_ISL_2203741, EPI_ISL_2339400, EPI_ISL_2339401, EPI_ISL_2403015, EPI_ISL_2403021, EPI_ISL_2403044, EPI_ISL_2403106, EPI_ISL_2403107 | | | |
| see above | Illinois Department of Public Health - Springfield Lab | Illinois Department of Public Health - Springfield Lab | Bryan Sim; Gordon McCall |
| EPI_ISL_2376230, EPI_ISL_2376231 | Imeda Hospital | Imeda Hospital | Dagmar Obbels; Hanne Valgaeren; Johan Frans |
| EPI_ISL_2987711, EPI_ISL_2987712 | Imágenes Diagnósticas de Mazatlán | Microbial Genomics Laboratory | ; Alejandra García-Gasca; Alejandra Hernández-Terán; Alejandro Sánchez-Flores; Alfredo Herrera-Estrella; Alicia Ocaña-Mondragón; Andreu Comas-García; Angel Gustavo Salas-Lais; Antonio Loza Román; Bernardo Martínez-Miguel; Blanca Taboada; Brenda Irasema Maldonado-Meza; Bruno Gómez-Gil; Carla Ivón Herrera-Najera; Carlos F. Arias; Celia Boukadida; Clara Esperanza Santacruz-Tinoco; Concepción Grajales-Muñiz; Consorcio Mexicano de Vigilancia Genómica (CoViGen-Mex). Authors (in alphabetical order): Julio Elias Alvarado-Yaah; Cristóbal Cháidez-Quiróz; Célida Duque Molina; Célida Martínez- Rodríguez; Daniel Fregoso-Rueda; Daniel Lira Morales; Eduardo Becerril-Vargas; Fernando Fontove-Herrera; Fidencio Mejía-Nepomuceno; Francisco Pulido; Gloria Elena Espinosa-Ayala; Gloria María Molina-Salinas; Gloria Vazquez; Hector Esteban Paz-Juárez; Hector Montoya-Fuentes; Helen Haydee Fernanda Ramírez-Plascencia; Irvin González-López; Jean Pierre González; Jesús Hernández; Joel Armando Vázquez-Pérez.; Jorge Salas-Hernández; José Antonio Enciso-Moreno; José Arturo Martínez-Orozco; José Esteban Muñoz-Medina; José de Jesús Nuñez-Contreras; Juan Bautista Chale-Dzul; Julissa Enciso-Ibarra; Luis Alberto Ochoa-Carrera; Margarita Matías-Florentino; Mario Mújica-Sánchez; Marissa Perez-Garcia; María Guadalupe Santiago-Mauricio; María Guadalupe de Jesús Mireles-Rivera; Nelly Sélem-Mojica; Pavel Isa; Ricardo Ciria Merce; Ricardo Grande; Rosa María Gutiérrez Rios; Santiago Ávila-Ríos; Selene Zárate; Susana Lopez; Verónica Mata-Haro; Victor Eduardo García-Arias; Víctor Hugo Borja-Aburto |
| EPI_ISL_2490282, EPI_ISL_2536459, EPI_ISL_2536466, EPI_ISL_2536469, EPI_ISL_2536488 | Incyte Diagnostics Spokane | Seattle Flu Study | Amanda Adler; Barry R. Lutz; Benjamin Pelle; Brian Hiatt; Caitlin R. Wolf; Chris D. Frazar; Deborah A. Nickerson; Elisabeth Brandstetter; Erica Ryke; Geoff Melly; Helen Y. Chu; Janet A. Englund; Jay Shendure; Jover Lee; Kairsten Fay; Kirsten Lacombe; Lea M. Starita; Mark J. Rieder; Matthew Richardson; Matthew Thompson; Melissa Truong; Michael Boeckh; Michael Famulare; Misja Ilcisin; Peter D. Han; Philip Dykema; Romesh Gautom; Scott Lindquist; Thomas R. Sibley; Trevor Bedford |
| EPI_ISL_2304452, EPI_ISL_2304454, EPI_ISL_2304455 | Indiana Animal Disease Diagnostic Laboratory | Carpi Laboratory - Purdue University | Abebe A Fola; G Kenitra Hendrix; Giovanna Carpi; Ilinca I Ciubotariu; Jack Dorman; Jobin J Kattoor; Lev Gorenstein; Nicole M Perry; Rebecca P Wilkes |
| EPI_ISL_2088742, EPI_ISL_2186443, EPI_ISL_2186446, EPI_ISL_2186466, EPI_ISL_2186534, EPI_ISL_2186561, EPI_ISL_2186568, EPI_ISL_2186620, EPI_ISL_2186668, EPI_ISL_2186793, EPI_ISL_2282269, EPI_ISL_2282286, EPI_ISL_2282306, EPI_ISL_2282321, EPI_ISL_2282322, EPI_ISL_2282331, EPI_ISL_2282388, EPI_ISL_2282415, EPI_ISL_2282417, EPI_ISL_2282428, EPI_ISL_2282465, EPI_ISL_2282561, EPI_ISL_2282575, EPI_ISL_2282596, EPI_ISL_2282601, EPI_ISL_2282608, EPI_ISL_2282639, EPI_ISL_2282640, EPI_ISL_2282655, EPI_ISL_2282693, EPI_ISL_2282714, EPI_ISL_2282727, EPI_ISL_2282768, EPI_ISL_2282787, EPI_ISL_2282796, EPI_ISL_2282799, EPI_ISL_2282803, EPI_ISL_2282862, EPI_ISL_2282869, EPI_ISL_2282871, EPI_ISL_2282899, EPI_ISL_2284205, EPI_ISL_2323698, EPI_ISL_2323705, EPI_ISL_2323970, EPI_ISL_2324024, EPI_ISL_2324039, EPI_ISL_2324096, EPI_ISL_2324124, EPI_ISL_4383378, EPI_ISL_4383441 | | | |
| see above | Infinity Biologix | Centers for Disease Control and Prevention Division of Viral Diseases, Pathogen Discovery | Adrian Paskey; Benjamin Rambo-Martin; Chirayu Goswami; Christian Bixby; Christopher Gulvick; Clinton Paden; Clinton R. Paden; Dakota Howard; Darlene Wagner; Dhwani Batra; Duncan MacCannell; Erisa Sula; Jason Caravas; Jonathan Schultz; Kara Moser; Kristine Lacek; Matthew Schmerer; Peter Cook; Peter W. Cook; Robin Grimwood; Russ Hager; Scott Sammons; Shatavia Morrison; Tymeckia Kendall; Victoria Caban Figueroa; Yihe Wang; Yvette Unoarumhi |
| EPI_ISL_2828662 | Instituo Adolfo Lutz - Reginal de Ribeirao Preto | Instituto Adolfo Lutz, Interdiciplinary Procedures Center, Strategic Laboratory | Caio Vinicius Dias Lopes; Claudia Regina Gonçalves; Claudio Tavares Sacchi; Erica Valessa Ramos Gomes; Karoline Rodrigues Campos |
| EPI_ISL_2828655, EPI_ISL_2828656 | Instituo Adolfo Lutz Central | Instituto Adolfo Lutz, Interdiciplinary Procedures Center, Strategic Laboratory | Caio Vinicius Dias Lopes; Claudia Regina Gonçalves; Claudio Tavares Sacchi; Erica Valessa Ramos Gomes; Karoline Rodrigues Campos |
| EPI_ISL_2357234, EPI_ISL_2357732 | Institut de Virologie du CHU de Strasbourg | Institut de Virologie du CHU de Strasbourg | Fafi-Kremer Samira; Gallais Floriane; Gantner Pierre; Laugel Elodie; Solis Morgane; Velay Aurélie; Wendling Marie-Josée |
| EPI_ISL_2948375, EPI_ISL_2948404, EPI_ISL_2948405, EPI_ISL_2948420, EPI_ISL_2948426, EPI_ISL_2948448, EPI_ISL_2948453, EPI_ISL_2948455, EPI_ISL_2948474, EPI_ISL_2948526, EPI_ISL_2948529, EPI_ISL_2948549, EPI_ISL_2948551, EPI_ISL_2948568, EPI_ISL_2948569, EPI_ISL_2948581, EPI_ISL_2948585, EPI_ISL_2948619, EPI_ISL_2948621, EPI_ISL_2948626, EPI_ISL_2948643 | | | |
| see above | Institut fr Virologie am Department fr Hygiene, Mikrobiologie und Publich Health der Medizinischen Universitt Innsbruck | Elling and Cochella laboratories, IMBA/IMP | Alexander Stark; Ezgi Oezkan; Luisa Cochella and Ulrich Elling; Marcus Strobl; Olga Frank; Ramesh Yelagandula; Tanino Albanese |
| EPI_ISL_2277682 | Institute for Infectious Diseases | Institute for Infectious Diseases | Alban Ramette; Christian Baumann; Cora Sägesser; Franziska Suter-Riniker; Miguel A Terrazos Miani; Pascal Bittel; Peter Keller; Stefan Neuenschwander; Stephen L Leib |
| EPI_ISL_2100435 | Institute of Microbiology, Universidad San Francisco de Quito | Institute of Microbiology, Universidad San Francisco de Quito | Belén Prado-Vivar; Bernardo Gutiérrez; Gabriel Trueba; Juan José Guadalupe; Luis Morales; Michelle Grunauer; Monica Becerra-Wong; Patricio Rojas-Silva; Paúl Cárdenas; Sully Márquez; Verónica Barragán |
| EPI_ISL_2691110, EPI_ISL_2691111, EPI_ISL_2691113, EPI_ISL_2919250, EPI_ISL_2919251, EPI_ISL_3316179 | Instituto Adolfo Lutz - Regional de Bauru | Instituto Adolfo Lutz, Interdiciplinary Procedures Center, Strategic Laboratory | Caio Vinicius Dias Lopes; Claudia Regina Gonçalves; Claudio Tavares Sacchi; Erica Valessa Ramos Gomes; Karoline Rodrigues Campos; Leonardo Jose Tadeu de Araujo; Leonardo Tadeu de Araujo; Marlon Benedito Nascomento Santos |
| EPI_ISL_3316200 | Instituto Adolfo Lutz - Regional de Campinas | Instituto Adolfo Lutz, Interdiciplinary Procedures Center, Strategic | Caio Vinicius Dias Lopes; Claudia Regina Gonçalves; Claudio Tavares Sacchi; Karoline Rodrigues Campos; Leonardo Tadeu de Araujo; Marlon Benedito Nascomento Santos |

| | | | |
|---|---|---|---|
| | | Laboratory | |
| EPI_ISL_2614620, EPI_ISL_2614621, EPI_ISL_2614625, EPI_ISL_3316156, EPI_ISL_3316234, EPI_ISL_3316238 | Instituto Adolfo Lutz - Regional de Presidente Prudente | Instituto Adolfo Lutz, Interdiciplinary Procedures Center, Strategic Laboratory | Caio Vinicius Dias Lopes; Claudia Regina Gonçalves; Claudio Tavares Sacchi; Erica Valessa Ramos Gomes; Karoline Rodrigues Campos; Leonardo Jose Tadeu de Araujo; Leonardo Tadeu de Araujo; Marlon Benedito Nascomento Santos |
| EPI_ISL_2691124 | Instituto Adolfo Lutz - Regional de Ribeirao Preto | Instituto Adolfo Lutz, Interdiciplinary Procedures Center, Strategic Laboratory | Caio Vinicius Dias Lopes; Claudia Regina Gonçalves; Claudio Tavares Sacchi; Erica Valessa Ramos Gomes; Karoline Rodrigues Campos; Leonardo Jose Tadeu de Araujo |
| EPI_ISL_2828698, EPI_ISL_2828699, EPI_ISL_2828702 | Instituto Adolfo Lutz - Regional de Santos | Instituto Adolfo Lutz, Interdiciplinary Procedures Center, Strategic Laboratory | Caio Vinicius Dias Lopes; Claudia Regina Gonçalves; Claudio Tavares Sacchi; Erica Valessa Ramos Gomes; Karoline Rodrigues Campos |
| EPI_ISL_2691126, EPI_ISL_2691127, EPI_ISL_2691132, EPI_ISL_2691134, EPI_ISL_2691136, EPI_ISL_2691139, EPI_ISL_2691142, EPI_ISL_2691143, EPI_ISL_2691144, EPI_ISL_2756426, EPI_ISL_5640226 | | | |
| see above | Instituto Adolfo Lutz Central | Instituto Adolfo Lutz, Interdiciplinary Procedures Center, Strategic Laboratory | Caio Vinicius Dias Lopes; Claudia Regina Gonçalves; Claudio Tavares Sacchi; Erica Valessa Ramos Gomes; Karoline Rodrigues Campos; Leonardo Jose Tadeu de Araujo; Leonardo Tadeu de Araujo; Marlon Benedito Nascimento Santos |
| EPI_ISL_4104672, EPI_ISL_4104727 | Instituto Butantan | Instituto Butantan | Antonio Jorge Martins; Claudia Renata dos Santos Barros; David Schlesinger; Debora Botequio Moretti; Dimas Tadeu Covas; Elaine Cristina Marqueze; Elaine Vieira Santos; Evandra Strazza Rodrigues; Heidge Fukumasu; Jayme Augusto de Souza-Neto; José Salvatore Leister Patané; Luiz Alcantara; Luiz Lehmann Coutinho; Maria Carolina Elias; Maurício Lacerda Nogueira; Rafael dos Santos Bezerra; Raul Machado Neto; Rejane Maria Tommasini Grotto; Ricardo Haddad; Sandra Coccuzzo Sampaio Vessoni; Simone Kashima; Svetoslav Nanev Slavov; Vincent Louis Viala |
| EPI_ISL_2490587 | Instituto Nacional de Enfermedades Respiratorias (INER) | Centro de Investigación en Enfermedades Infecciosas (CIENI), Instituto Nacional de Enfermedades Respiratorias (INER) | ; Alejandra Garcia-Gasca; Alejandra Hernández-Terán; Alejandro Sanchez-Flores; Alfredo Herrera-Estrella; Alicia Ocaña-Mondragón; Andreu Comas-García; Angel Gustavo Salas-Lais; Antonio Loza Román; Bernardo Martínez-Miguel; Blanca Taboada; Brenda Irasema Maldonado-Meza; Bruno Gomez-Gil; Carla Ivón Herrera-Najera; Carlos F. Arias; Celia Boukadida; Clara Esperanza Santacruz-Tinoco; Concepción Grajales-Muñiz; Consorcio Mexicano de Vigilancia Genómica (CoViGen-Mex). Authors (in alphabetical order): Julio Elias Alvarado-Yaah; Cristóbal Cháidez-Quiróz; Célida Duque Molina; Célida Martínez- Rodríguez; Daniel Fregoso-Rueda; Daniel Lira Morales; Eduardo Becerril-Vargas; Fernando Fontove-Herrera; Fidencio Mejía-Nepomuceno; Francisco Pulido; Gloria Elena Espinosa-Ayala; Gloria María Molina-Salinas; Gloria Vazquez; Hector Esteban Paz-Juárez; Hector Montoya-Fuentes; Helen Haydee Fernanda Ramírez-Plascencia; Irvin González-López; Jean Pierre González; Joel Armando Vázquez-Pérez.; Jorge Salas-Hernández; José Antonio Enciso-Moreno; José Arturo Martínez-Orozco; José Esteban Muñoz-Medina; José de Jesús Nuñez-Contreras; Juan Bautista Chale-Dzul; Julissa Enciso-Ibarra; Luis Alberto Ochoa-Carrera; Margarita Matías-Florentino; Mario Mújica-Sánchez; Marissa Perez-Garcia; María Guadalupe Santiago-Mauricio; María Guadalupe de Jesús Mireles-Rivera; Nelly Sélem-Mojica; Pavel Isa; Ricardo Ciria Merce; Ricardo Grande; Rosa María Gutierrez Rios; Santiago Ávila-Ríos; Selene Zárate; Susana Lopez; Victor Eduardo Garcia-Arias; Víctor Hugo Borja-Aburto |
| EPI_ISL_2230807, EPI_ISL_2230829, EPI_ISL_2230861 | Instituto Nacional de Medicina Genomica | Instituto Nacional de Medicina Genomica | Cedro-Tanda A; Cisneros- Villanueva M; Escobar-Arrazola; Gonzalez-Barrera D; Herrera-Montalvo LA.; Hidalgo-Miranda A; M; Mendoza-Vargas A; Munguia-Garza P; Ramirez-Vega O; Rangel-DeLeon D; Reyes-Grajeda JP |
| EPI_ISL_2323128, EPI_ISL_2323130, EPI_ISL_2323136, EPI_ISL_2323139, EPI_ISL_2336150, EPI_ISL_2378644, EPI_ISL_2378645, EPI_ISL_2378646, EPI_ISL_2378647, EPI_ISL_2378648, EPI_ISL_2378649, EPI_ISL_2378650, EPI_ISL_2378651, EPI_ISL_2378652, EPI_ISL_2378653, EPI_ISL_2378654, EPI_ISL_2378655, EPI_ISL_2378656, EPI_ISL_2378657, EPI_ISL_2378658, EPI_ISL_2378659 | | | |
| see above | Instituto de Biotecnologia - UNESP-Botucatu-SP | Instituto de Biotecnologia - UNESP-Botucatu-SP | Cecília Artico Banho; Cíntia Bittar; Fábio Sossai Possebon; Guilherme Campos; Helena Lage Ferreira; Jorge A. Petroli Marchesi; João Pessoa Araújo Jr.; Leila Sabrina Ullmann; Lívia Sacchetto; Maisa C. Pereira Parra; Marília Moraes; Maurício L. Nogueira; Paula Rahal; Paulo Inacio da Costa |
| EPI_ISL_2183556, EPI_ISL_2183660 | Israel Central Virology laboratory | Israel National Consortium for SARS-CoV-2 sequencing | Dana Bar-Ilan; Efrat Dahan Bucris; Efrat Glick-Saar; Ella Mendelson; Gideon Rechavi; Michal Mandelboim; Miranda Geva; Neta Zuckerman; Netanel Abu; Omri Nayshool; Oran Erster; Orna Mor |
| EPI_ISL_2194291, EPI_ISL_2194335, EPI_ISL_2194336, EPI_ISL_2194342, EPI_ISL_2194355, EPI_ISL_2194401, EPI_ISL_2194436, EPI_ISL_2194445, EPI_ISL_2194453, EPI_ISL_2194454, EPI_ISL_2194459, EPI_ISL_2194495, EPI_ISL_2194501, EPI_ISL_2194520, EPI_ISL_2194521, EPI_ISL_2194522, EPI_ISL_2194534, EPI_ISL_2194535, EPI_ISL_2194538, EPI_ISL_2226888, EPI_ISL_2281905, EPI_ISL_2281906, EPI_ISL_2356918, EPI_ISL_2356920, EPI_ISL_2357055, EPI_ISL_2357057 | | | |
| see above | Istituto Zooprofilattico Sperimentale del Mezzogiorno | TIGEM | Antonio Grimaldi Patrizia Annunziata Francesco Panariello Biancamaria Pierri Claudia Tiberio Teresa Giuliano Valentina Bouche Chiara Colantuono Maria Concetta Cuomo Denise Di Concilio Lucio Di Filippo Anna Manfredi Marcello Salvi Antonio Limone Luigi Atripaldi Pellegrino Cerino Andrea Ballabio Davide Cacchiarelli |
| EPI_ISL_2248906, EPI_ISL_2248907, EPI_ISL_2248910, EPI_ISL_2248911 | Joaquim Chaves Saude | Instituto Nacional de Saude (INSA) | Borges et al |
| EPI_ISL_2476384, EPI_ISL_2476386, EPI_ISL_2597978 | John H Stroger Jr Hospital of Cook County | RIPHL at Rush University Medical Center | Ah-Young Sung; Felix Araujo Perez; Kevin Kunstman; Laura Furtado; Marieta Hyde; Max Kolton; Sharon Welbel; Stefan Green |
| EPI_ISL_2331499, EPI_ISL_2331546 | Johns Hopkins Hospital Department of Pathology | Johns Hopkins Hospital Department of Pathology | Adannaya Amadi; C. Paul Morris; Chun Huai Luo; Heba H. Mostafa; Matthew Schwartz; Nicholas Gallagher |
| EPI_ISL_2384201 | KS Health and Environmental Laboratories | Centers for Disease Control and Prevention Division of Viral Diseases, Pathogen Discovery | Alison Laufer Halpin; Ben L. Rambo-Martin; Clinton R. Paden; Dakota Howard; Darlene Wagner; Dave Wentworth; Dhwani Batra; Jasmine Padilla; Justin Lee; Katie Dillon; Krista Queen; Kristen Knipe; Kristine Lacek; Mark Burroughs; Matthew Schmerer; Mili Sheth; Peter Cook; Sam Shepard; Sarah Nobles; Shoshona Le; Suxiang Tong; Vivien Dugan; Yvette Unoarumhi |
| EPI_ISL_2725607, EPI_ISL_2725646, EPI_ISL_2725647, EPI_ISL_2725648, EPI_ISL_2725649, EPI_ISL_2725671, EPI_ISL_2726412, EPI_ISL_2726415, EPI_ISL_2726416, EPI_ISL_2726417, EPI_ISL_2726418, EPI_ISL_2726419, EPI_ISL_2726420, EPI_ISL_2726421, EPI_ISL_2726422, EPI_ISL_2726423, EPI_ISL_2726424, EPI_ISL_2726425, EPI_ISL_2726426, EPI_ISL_2726427, EPI_ISL_2726428, EPI_ISL_2726429, EPI_ISL_2726430, EPI_ISL_2726431, EPI_ISL_2726434, EPI_ISL_2726435, EPI_ISL_2726444, EPI_ISL_2726462, EPI_ISL_2726507, EPI_ISL_4008001, EPI_ISL_4008003, EPI_ISL_4008006 | | | |
| see above | KU Leuven, Rega Institute, Clinical and Epidemiological Virology | KU Leuven, Rega Institute, Clinical and Epidemiological Virology | Bert Vanmechelen; Joan Marti-Carerras; Piet Maes; Tony Wawina-Bokalanga |
| EPI_ISL_3561359, EPI_ISL_3561554, EPI_ISL_3561832, EPI_ISL_3561959, EPI_ISL_3561968, EPI_ISL_3561990, EPI_ISL_3562087, EPI_ISL_4005465 | | | |
| see above | Kaiser Permanente NW Reginal Lab | OHSU MM Lab | Yun Wu |
| EPI_ISL_3504935, EPI_ISL_3504984, EPI_ISL_3504997, EPI_ISL_3505017, EPI_ISL_3505069, EPI_ISL_3505095, EPI_ISL_3505098, EPI_ISL_3505101, EPI_ISL_3505106, EPI_ISL_3505107, EPI_ISL_3505114, EPI_ISL_3505129 | | | |
| see above | Kaiser Permanente NW Reginal Lab | OHSU-MM Lab | Yun Wu |
| EPI_ISL_2934067, EPI_ISL_2934068 | Kaiser Regional | Santa Clara County Public Health Laboratory | Santa Clara County Public Health Department |
| EPI_ISL_2662973 | Kaleida Health Laboratories | University at Buffalo Genomics and Bioinformatics Core | Alyssa Pohlman; Amanda Boccolucci; Brandon Marzullo; Donald Yergeau; Jennifer Surtees; Jonathan Bard; Natalie Lamb; Norma Nowak |
| EPI_ISL_1964227, EPI_ISL_2037393, EPI_ISL_2092834 | Kansas Health and Environmental Lab | Kansas Health and Environmental Lab | Ben Olsen; Jonathan Barnell; Mike Grose; and Phil Adam |
| EPI_ISL_1964231, EPI_ISL_1964234, EPI_ISL_2091026, EPI_ISL_2091032, EPI_ISL_2091034, EPI_ISL_2091040, EPI_ISL_2091041, EPI_ISL_2091043 | | | |
| see above | Kansas Health and Environmental Lab | Wichita State University - Molecular Diagnostics Lab | Ben Olsen; Jonathan Barnell; Mike Grose; and Phil Adam |
| EPI_ISL_3742788, EPI_ISL_3758007, EPI_ISL_4080591 | Kentucky State Public Health Lab | Kentucky State Public Health Lab | Anjayya Tummala; Chitralipi Kattunga; Joshua Tobias; Karim George; Matthew Johnson; Rachel Zinner; Rhonda Lucas; Vaneet Arora; William Grooms |
| EPI_ISL_2386751 | Klinikum Osnabrück Medizinisches Labor | Robert Koch Institute | |
| EPI_ISL_2228057 | Klinisch Laboratorium ZNA | Klinisch Laboratorium ZNA | Verstrepen et al. |
| EPI_ISL_2976196 | Kootenai Health | Idaho Bureau of Laboratories | "R. Beukelman; Aimee Ceniseros; Christian Loera; Christopher Ball"; Matthew Charles Burns; Robert L. Voermans |
| EPI_ISL_2289224 | LABORATOIRE DE BIOLOGIE MEDICALE | CNR Virus des Infections Respiratoires - France SUD | Antonin Bal; Bruno Lina; Bruno Simon; Gregory Destras; Gwendolyne Burfin; Hadrien Regue; Laurence Josset; Martine Valette; Quentin Semanas |
| EPI_ISL_2674299 | LABORATORIO CARVAJAL | Instituto Nacional de Salud-Dirección de Investigación en Salud Pública | Carlos Franco-Muñoz; Carmen Osorio; Diana Malo; Diego A. Álvarez-Díaz; Diego Andrés Prada; Gerardo Santamaría; Hector Alejandro Ruiz-Moreno; Jhonnatan Reales-González; Jorge Rivera; Juan Camilo Martinez; Julian Naizaque; Katherine Laiton-Donato; Lisseth Pardo; Magdalena Wiesner; Marcela Mercado-Reyes; Maria T. Herrera-Sepúlveda; Marta Lopez Blanco; Martha Lucia Ospina Martinez; Paola Rojas; Sergio Gomez; Sheryll Corchuelo; Ángela Alarcon Cruz |
| EPI_ISL_2674326 | LABORATORIO DE INFECCION E INMUNIDAD | Instituto Nacional de Salud-Dirección de Investigación en Salud Pública | Carlos Franco-Muñoz; Carmen Osorio; Diana Malo; Diego A. Álvarez-Díaz; Diego Andrés Prada; Gerardo Santamaría; Hector Alejandro Ruiz-Moreno; Jhonnatan Reales-González; Jorge Rivera; Juan Camilo Martinez; Julian Naizaque; Katherine Laiton-Donato; Lisseth Pardo; Magdalena Wiesner; Marcela Mercado-Reyes; Maria T. Herrera-Sepúlveda; Marta Lopez Blanco; Martha Lucia Ospina Martinez; Paola Rojas; Sergio Gomez; Sheryll Corchuelo; Ángela Alarcon Cruz |
| EPI_ISL_2621847, EPI_ISL_2621848, EPI_ISL_2621862, EPI_ISL_2621863, EPI_ISL_2621864, EPI_ISL_2621867 | LABORATORIO ECHAVARRIA | Universidad Nacional de Colombia - Laboratorio Genómico One Health | Andres F. Cardona-Rios; Carlos Franco-Muñoz; Carolina Muñoz-Arango; Celeny Ortiz; Daniel O. Maldonado-Perez; Diego A. Álvarez-Díaz; Hector Alejandro Ruiz-Moreno; Idabely Betancur Ortiz; Jorge E. Osorio; Juan P. Hernandez-Ortiz; Karl A Ciuoderis; Katherine Laiton-Donato; Laura Silvana Perez; Lina M. Hurtado; Marcela Mercado-Reyes; Maria Angélica Maya; Maria Stella López; Rita Almanza Payares; Sandra Ines Cano; Simón Villegas Velásquez |
| EPI_ISL_5650395 | LABORATORIO MUNICIPAL DE ANALISES CLINICAS DE RIO CLARO | Instituto Butantan | Antonio Jorge Martins; Claudia Renata dos Santos Barros; David Schlesinger; Debora Botequio Moretti; Dimas Tadeu Covas; Elaine Cristina Marqueze; Elaine Vieira Santos; Evandra Strazza Rodrigues; Heidge Fukumasu; Jayme Augusto de Souza-Neto; José Salvatore Leister Patané; Luiz Alcantara; Luiz Lehmann Coutinho; Maria Carolina Elias; Maurício Lacerda Nogueira; Rafael dos Santos Bezerra; Raul Machado Neto; Rejane Maria Tommasini Grotto; Ricardo Haddad; Sandra Coccuzzo Sampaio Vessoni; Simone Kashima; Svetoslav Nanev Slavov; Vincent Louis Viala |
| EPI_ISL_2493449, EPI_ISL_5650325, EPI_ISL_5650387, EPI_ISL_5650413 | LABORATORIO MUNICIPAL DE PIRACICABA | Instituto Butantan | Antonio Jorge Martins; Claudia Renata dos Santos Barros; David Schlesinger; Debora Botequio Moretti; Dimas Tadeu Covas; Elaine Cristina Marqueze; Elaine Vieira Santos; Evandra Strazza Rodrigues; Heidge Fukumasu; Jayme Augusto de Souza-Neto; José Salvatore Leister Patané; Luiz Alcantara; Luiz Lehmann Coutinho; Maria Carolina Elias; Maurício Lacerda Nogueira; Rafael dos Santos Bezerra; Raul Machado Neto; Rejane Maria Tommasini Grotto; Ricardo Haddad; Sandra Coccuzzo Sampaio Vessoni; Simone Kashima; Svetoslav Nanev Slavov; Vincent Louis Viala |

| | | | |
|---|---|---|---|
| EPI_ISL_2210259, EPI_ISL_2345741 | LABORATORIO MUNICIPAL DE SUZANO | Instituto Butantan / Mendelics | Antonio Jorge Martins; Claudia Renata dos Santos Barros; David Schlesinger; Debora Botequio Moretti; Dimas Tadeu Covas; Elaine Cristina Marqueze; Elaine Vieira Santos; Evandra Strazza Rodrigues; Heidge Fukumasu; Jayme Augusto de Souza-Neto; José Salvatore Leister Patané; Luiz Alcantara; Luiz Lehmann Coutinho; Maria Carolina Elias; Maurício Lacerda Nogueira; Rafael dos Santos Bezerra; Raul Machado Neto; Rejane Maria Tommasini Grotto; Ricardo Haddad; Sandra Coccuzzo Sampaio Vessoni; Simone Kashima; Svetoslav Nanev Slavov; Vincent Louis Viala |
| EPI_ISL_6721791 | LACEN - Laboratório Central de Saúde Pública de Pernambuco | Evandro Chagas Institute | Delana Andreza Melo Bezerra; Dielle Monteiro Teixeira; Jedson Ferreira Cardoso; Jessylene de Almeida Ferreira; Kenny da Costa Pinheiro; Luana Soares Barbagelata; Luana da Silva Soares; Mirleide Cordeiro dos Santos; Patrícia dos Santos Lobo; Rayssa Layna da Silva Bedran; Sandro Patroca da Silva |
| EPI_ISL_3316312, EPI_ISL_3316313, EPI_ISL_3316314 | LACEN do Distrito Federal | Instituto Adolfo Lutz, Interdiciplinary Procedures Center, Strategic Laboratory | Caio Vinicius Dias Lopes; Claudia Regina Gonçalves; Claudio Tavares Sacchi; Karoline Rodrigues Campos; Leonardo Tadeu de Araujo; Marlon Benedito Nascomento Santos |
| EPI_ISL_2691206, EPI_ISL_2691207, EPI_ISL_2691211 | LACEN do Estado de Goias | Instituto Adolfo Lutz, Interdiciplinary Procedures Center, Strategic Laboratory | Caio Vinicius Dias Lopes; Claudia Regina Gonçalves; Claudio Tavares Sacchi; Erica Valessa Ramos Gomes; Karoline Rodrigues Campos; Leonardo Jose Tadeu de Araujo |
| EPI_ISL_3316166, EPI_ISL_3316212, EPI_ISL_3316223 | LACEN do Estado de Mato Grosso | Instituto Adolfo Lutz, Interdiciplinary Procedures Center, Strategic Laboratory | Caio Vinicius Dias Lopes; Claudia Regina Gonçalves; Claudio Tavares Sacchi; Karoline Rodrigues Campos; Leonardo Tadeu de Araujo; Marlon Benedito Nascomento Santos |
| EPI_ISL_2919212, EPI_ISL_2919214, EPI_ISL_2919215 | LACEN do Estado de Tocantins | Instituto Adolfo Lutz, Interdiciplinary Procedures Center, Strategic Laboratory | Caio Vinicius Dias Lopes; Claudia Regina Gonçalves; Claudio Tavares Sacchi; Erica Valessa Ramos Gomes; Karoline Rodrigues Campos |
| EPI_ISL_3691371, EPI_ISL_3691372, EPI_ISL_3691373, EPI_ISL_3691374, EPI_ISL_3691375 | LACEN do Estado do Mato Grosso do Sul | Instituto Adolfo Lutz, Interdiciplinary Procedures Center, Strategic Laboratory | Caio Vinicius Dias Lopes; Claudia Regina Gonçalves; Claudio Tavares Sacchi; Karoline Rodrigues Campos; Leonardo Tadeu de Araujo; Marlon Benedito Nascomento Santos |
| EPI_ISL_3691685, EPI_ISL_3691686, EPI_ISL_3691687, EPI_ISL_4206941, EPI_ISL_4206969 | LACEN do Estado do Piaui | Instituto Adolfo Lutz, Interdiciplinary Procedures Center, Strategic Laboratory | Caio Vinicius Dias Lopes; Claudia Regina Gonçalves; Claudio Tavares Sacchi; Karoline Rodrigues Campos; Leonardo Tadeu de Araujo; Marlon Benedito Nascimento Santos; Marlon Benedito Nascomento Santos |
| EPI_ISL_2821274, EPI_ISL_3134595, EPI_ISL_3134596, EPI_ISL_3134597, EPI_ISL_3134598, EPI_ISL_3134599, EPI_ISL_3134600, EPI_ISL_3134601, EPI_ISL_3134602, EPI_ISL_3134603, EPI_ISL_3134604, EPI_ISL_3134605, EPI_ISL_3134606, EPI_ISL_3134607, EPI_ISL_3134608, EPI_ISL_3134609, EPI_ISL_3134610, EPI_ISL_3134611, EPI_ISL_3134612, EPI_ISL_3134613, EPI_ISL_3134614, EPI_ISL_3134615, EPI_ISL_3134616, EPI_ISL_3134617, EPI_ISL_3134618, EPI_ISL_3134619, EPI_ISL_3134620, EPI_ISL_3134621, EPI_ISL_3134622, EPI_ISL_3134623, EPI_ISL_3134624, EPI_ISL_3134625, EPI_ISL_3134626, EPI_ISL_3134627, EPI_ISL_3134628, EPI_ISL_3134629, EPI_ISL_3134630, EPI_ISL_3134632, EPI_ISL_3134640, EPI_ISL_3134642, EPI_ISL_3134645, EPI_ISL_3134646, EPI_ISL_3134647, EPI_ISL_3134649, EPI_ISL_3134652, EPI_ISL_3134835, EPI_ISL_3134836, EPI_ISL_3134837, EPI_ISL_3134841, EPI_ISL_3152237, EPI_ISL_5254419 | | | |
| see above | LACEN/PE | WallauLab on behalf of Fiocruz COVID-19 Genomic Surveillance Network | Alexandre Freitas da Silva; Antonio Marinho da Silva Neto; Antonio Mauro Rezende; Cassia Docena; Constância Flávia Junqueira Ayres; Cássia Docena; Duschinka Ribeiro Duarte Guedes; Elisama Helvecio; Filipe Zimmer Dezordi; Gabriel Luz Wallau; Gustavo Barbosa de Lima; Lais Ceschini Machado; Larissa Krokovsky; Laís Ceschini Machado; Lilian Caroliny Amorim Silva; Marcelo Henrique dos Santos Paiva; Matheus Filgueira Bezerra; Sinval Pinto Brandão Filho |
| EPI_ISL_4487033, EPI_ISL_4487962, EPI_ISL_4487967, EPI_ISL_4487972, EPI_ISL_4487978, EPI_ISL_4487981, EPI_ISL_4487985, EPI_ISL_4487988, EPI_ISL_4487993, EPI_ISL_4488002, EPI_ISL_4488008 | | | |
| see above | LACEN/PI | WallauLab on behalf of Fiocruz COVID-19 Genomic Surveillance Network | Adelino Soares Lima Neto; Alexandre Freitas da Silva; Antônio Marinho; Cassia Docena; Constância Flávia Junqueira Ayres; Filipe Zimmer Dezordi; Gabriel Luz Wallau; Gustavo Barbosa de Lima; Hellen de Oliveira Amaral; Jacenir Reis dos Santos Mallet; Joana Carolina Viana Lima; Lais Ceschini Machado; Leandro de Mattos; Lilian Caroliny Amorim Silva; Marcela de Lacerda Valença Queiroz; Marcelo Adriano da Cunha e Silva Vieira; Marcelo Henrique dos Santos Paiva; Matheus Filgueira Bezerra; Sinval Pinto Brandão Filho; Túlio de Lima Campos; Vladimir Costa Silva; Walterlene de Carvalho Goçalves |
| EPI_ISL_5193463, EPI_ISL_5193570, EPI_ISL_5193571, EPI_ISL_5193572, EPI_ISL_5193573, EPI_ISL_5193574, EPI_ISL_5193575, EPI_ISL_5193576, EPI_ISL_5193577, EPI_ISL_5193578, EPI_ISL_5193601, EPI_ISL_5193661 | | | |
| see above | LACEN/RR | Laboratorio de Ecologia de Doencas Transmissiveis na Amazonia, Instituto Leonidas e Maria Deane - Fiocruz Amazonia | André Corado; Cátia Alexandra Ribeiro Meneses; Felipe Naveca; Fernanda Nascimento; George Silva; Ilma de Aguiar Antony; Janis Cassimiro Lopes; Luciana Gonçalves; Maria Júlia Brandão; Matilde Mejía; Pricila Carlos Veloso; Rodrigo Melo Maito; Valdinete Nascimento; Victor Souza |
| EPI_ISL_3276560 | LAS AMERICAS | Universidad Nacional de Colombia - Laboratorio Genómico One Health | Andres F. Cardona-Rios; Carlos Franco-Muñoz; Carolina Muñoz-Arango; Celeny Ortiz; Daniel O. Maldonado-Perez; Diego A. Álvarez-Díaz; Hector Alejandro Ruiz-Moreno; Idabely Betancur Ortiz; Jorge E. Osorio; Juan P. Hernandez-Ortiz; Karl A Ciuoderis; Katherine Laiton-Donato; Laura Silvana Perez; Lina M. Hurtado; Marcela Mercado-Reyes; Maria Angélica Maya; Maria Stella López; Rita Almanza Payares; Sandra Ines Cano; Simón Villegas Velásquez |
| EPI_ISL_3010780, EPI_ISL_3010813, EPI_ISL_3010855, EPI_ISL_3010922, EPI_ISL_3010930 | LATE - Laboratório de Técnicas Especiais - Hospital Israelita Albert Einstein | LATE - Laboratório de Técnicas Especiais - Hospital Israelita Albert Einstein | Alexandre Hideaki Takara; Ana Paula Moreira Salles; Anelisie da Silva Santos; Deyvid Amgarten; Erick Gustavo Dorlass; Fernanda de Mello Malta; João Renato Rebello Pinho; Marcio Anunciacao Menezes; Pedro Henrique Sebe Rodrigues; Raquel Riyuzo |
| EPI_ISL_2483064 | LBM CBM Le Chesnay | Eurofins Biomnis | Véronique Jacomo; Xavier Naudot |
| EPI_ISL_2674301, EPI_ISL_2674302, EPI_ISL_2674303, EPI_ISL_2674304 | LDSP HUILA | Instituto Nacional de Salud-Dirección de Investigación en Salud Pública | Carlos Franco-Muñoz; Carmen Osorio; Diana Malo; Diego A. Álvarez-Díaz; Diego Andrés Prada; Gerardo Santamaría; Hector Alejandro Ruiz-Moreno; Jhonnatan Reales-González; Jorge Rivera; Juan Camilo Martinez; Julian Naizaque; Katherine Laiton-Donato; Lisseth Pardo; Magdalena Wiesner; Marcela Mercado-Reyes; Maria T. Herrera-Sepúlveda; Marta Lopez Blanco; Martha Lucia Ospina Martinez; Paola Rojas; Sergio Gomez; Sheryll Corchuelo; Ángela Alarcon Cruz |
| EPI_ISL_2296072, EPI_ISL_2296074, EPI_ISL_2296075, EPI_ISL_2296078, EPI_ISL_2296083 | LESP Ciudad de Mexico | Instituto de Diagnostico y Referencia Epidemiologicos (INDRE) | Abril Rodriguez-Maldonado; Ariadna Medina-Benitez; Claudia Wong-Arambula; Ernesto Ramirez-Gonzalez.; Gisela Barrera-Badillo; Irma Lopez-Martinez; Joaquin Quiroz-Mercado; Lucia Hernandez-Rivas; Natividad Cruz-Ortiz; Sergio Rangel-Guerrero; Tatiana Nunez-Garcia; Vanessa Rivero-Arredondo |
| EPI_ISL_2479892 | LESP Colima | Instituto de Diagnostico y Referencia Epidemiologicos (INDRE) | Abril Rodriguez-Maldonado; Ariadna Medina-Benitez; Claudia Wong-Arambula; Ernesto Ramirez-Gonzalez.; Gisela Barrera-Badillo; Irma Lopez-Martinez; Joaquin Quiroz-Mercado; Lucia Hernandez-Rivas; Natividad Cruz-Ortiz; Sergio Rangel-Guerrero; Tatiana Nunez-Garcia; Vanessa Rivero-Arredondo |
| EPI_ISL_2296095, EPI_ISL_2296096, EPI_ISL_2476413, EPI_ISL_2476415, EPI_ISL_2476424, EPI_ISL_2476426, EPI_ISL_2674628, EPI_ISL_2674656, EPI_ISL_2674657 | | | |
| see above | LESP Guanajuato | Instituto de Diagnostico y Referencia Epidemiologicos (INDRE) | Abril Rodriguez-Maldonado; Ariadna Medina-Benitez; Claudia Wong-Arambula; Ernesto Ramirez-Gonzalez.; Gisela Barrera-Badillo; Irma Lopez-Martinez; Joaquin Quiroz-Mercado; Lucia Hernandez-Rivas; Natividad Cruz-Ortiz; Sergio Rangel-Guerrero; Tatiana Nunez-Garcia; Vanessa Rivero-Arredondo |
| EPI_ISL_2361362 | LESP Guanajuato | Instituto de diagnóstico y Referencia Epidemiologicos (INDRE) Departamento de Virología | Abril Rodriguez-Maldonado; Ariadna Medina-Benitez; Claudia Wong-Arambula; Ernesto Ramirez-Gonzalez.; Gisela Barrera-Badillo; Irma Lopez-Martinez; Joaquin Quiroz-Mercado; Lucia Hernandez-Rivas; Natividad Cruz-Ortiz; Sergio Rangel-Guerrero; Tatiana Nunez-Garcia; Vanessa Rivero-Arredondo |
| EPI_ISL_2479911 | LESP Hidalgo | Instituto de Diagnostico y Referencia Epidemiologicos (INDRE) | Abril Rodriguez-Maldonado; Ariadna Medina-Benitez; Claudia Wong-Arambula; Ernesto Ramirez-Gonzalez.; Gisela Barrera-Badillo; Irma Lopez-Martinez; Joaquin Quiroz-Mercado; Lucia Hernandez-Rivas; Natividad Cruz-Ortiz; Sergio Rangel-Guerrero; Tatiana Nunez-Garcia; Vanessa Rivero-Arredondo |
| EPI_ISL_2795657 | LESP Hidalgo | LESP Hidalgo GENE2LIFE | CASTILLO-ORTEGA LAURA SOFIA; GARCIA-GONZALEZ OCTAVIO; GARCIA-REYES MELITO; HERRERA-GOMEZ CONNIE ALEJANDRA; VIGUERAS-MORALES YAJAIRA SULIM; VILLAFUENTES-TELLEZ HECTOR ALBERTO |
| EPI_ISL_2476440 | LESP Morelos | Instituto de Diagnostico y Referencia Epidemiologicos (INDRE) | Abril Rodriguez-Maldonado; Ariadna Medina-Benitez; Claudia Wong-Arambula; Ernesto Ramirez-Gonzalez.; Gisela Barrera-Badillo; Irma Lopez-Martinez; Joaquin Quiroz-Mercado; Lucia Hernandez-Rivas; Natividad Cruz-Ortiz; Sergio Rangel-Guerrero; Tatiana Nunez-Garcia; Vanessa Rivero-Arredondo |
| EPI_ISL_2479902 | LESP Nuevo Leon | Instituto de Diagnostico y Referencia Epidemiologicos (INDRE) | Abril Rodriguez-Maldonado; Ariadna Medina-Benitez; Claudia Wong-Arambula; Ernesto Ramirez-Gonzalez.; Gisela Barrera-Badillo; Irma Lopez-Martinez; Joaquin Quiroz-Mercado; Lucia Hernandez-Rivas; Natividad Cruz-Ortiz; Sergio Rangel-Guerrero; Tatiana Nunez-Garcia; Vanessa Rivero-Arredondo |
| EPI_ISL_2545682, EPI_ISL_2545714 | LESP Queretaro | Instituto de Diagnostico y Referencia Epidemiologicos (INDRE) | Abril Rodriguez-Maldonado; Ariadna Medina-Benitez; Claudia Wong-Arambula; Ernesto Ramirez-Gonzalez.; Gisela Barrera-Badillo; Irma Lopez-Martinez; Joaquin Quiroz-Mercado; Lucia Hernandez-Rivas; Natividad Cruz-Ortiz; Sergio Rangel-Guerrero; Tatiana Nunez-Garcia; Vanessa Rivero-Arredondo |
| EPI_ISL_2479891 | LESP Quintana Roo | Instituto de Diagnostico y Referencia Epidemiologicos (INDRE) | Abril Rodriguez-Maldonado; Ariadna Medina-Benitez; Claudia Wong-Arambula; Ernesto Ramirez-Gonzalez.; Gisela Barrera-Badillo; Irma Lopez-Martinez; Joaquin Quiroz-Mercado; Lucia Hernandez-Rivas; Natividad Cruz-Ortiz; Sergio Rangel-Guerrero; Tatiana Nunez-Garcia; Vanessa Rivero-Arredondo |
| EPI_ISL_2476416, EPI_ISL_2476425, EPI_ISL_2476431, EPI_ISL_2476433, EPI_ISL_2476434, EPI_ISL_2476435, EPI_ISL_2545680, EPI_ISL_2545681 | | | |
| see above | LESP Tamaulipas | Instituto de Diagnostico y Referencia Epidemiologicos (INDRE) | Abril Rodriguez-Maldonado; Ariadna Medina-Benitez; Claudia Wong-Arambula; Ernesto Ramirez-Gonzalez.; Gisela Barrera-Badillo; Irma Lopez-Martinez; Joaquin Quiroz-Mercado; Lucia Hernandez-Rivas; Natividad Cruz-Ortiz; Sergio Rangel-Guerrero; Tatiana Nunez-Garcia; Vanessa Rivero-Arredondo |
| EPI_ISL_2016836, EPI_ISL_2131349, EPI_ISL_2131352, EPI_ISL_2131354, EPI_ISL_2131359, EPI_ISL_2131360, EPI_ISL_2131363, EPI_ISL_2131364 | | | |
| see above | LHUB-ULB | Labo Klinische Biologie, UZA | Basil Britto Xavier; Christine Lammens; Herman Goossens; Jasmine Coppens; Marie Le Mercier; Veerle Matheeussen |
| EPI_ISL_2192444, EPI_ISL_2192450, EPI_ISL_2192476, EPI_ISL_2192508, EPI_ISL_2192516, EPI_ISL_2192522, EPI_ISL_2192546, EPI_ISL_2192547, EPI_ISL_2192548, EPI_ISL_2362850 | | | |
| see above | Lab voor klinische biologie | Lab voor klinische biologie | Bruno Verhasselt; Hannelore Hamerlinck; Marija Janevska |
| EPI_ISL_2678400, EPI_ISL_2678404, EPI_ISL_2678406, EPI_ISL_2678409, EPI_ISL_2678413, EPI_ISL_2678425, EPI_ISL_2678428, EPI_ISL_2678432, EPI_ISL_2678434 | | | |
| see above | Labor Becker & Kollegen (Standort München) | Robert Koch Institute | |
| EPI_ISL_2227356, EPI_ISL_2339444 | Labor Berlin Charité Vivantes GmbH / Institut für Virologie | Charité Universitätsmedizin Berlin, Institut für Virologie/Labor Berlin | Barbara Mühlemann; Christian Drosten; Christine Stephan; Peter Menzel; Rolf Schwarzer; Terry Jones; Victor M Corman |
| EPI_ISL_2384917 | Labor Dr. Fenner und Kollegen | Heinrich Pette Institute, Leibniz Institute for Experimental Virology | Adam Grundhoff; Alexis Robitaille; Johannes Knobloch; Martin Aepfelbacher; Nicole Fischer; Thomas Günther |
| EPI_ISL_2129701 | Labor Dr. Heidrich & Kollegen MVZ GmbH Hamburg | Robert Koch Institute | |

| | | | |
|---|---|---|---|
| EPI_ISL_2129083 | Labor Monchengladbach MVZ Dr. Stein + Kollegen GbR | Robert Koch Institute | |
| EPI_ISL_2125029 | Labor Prof. Dr. G. Enders MVZ GbR | Robert Koch Institute | |
| EPI_ISL_2264860 | Labor ZOTZ\|KLIMAS; MVZ Düsseldorf-Centrum | Robert Koch Institute | |
| EPI_ISL_2227903 | Laboratoire Carage | Institut Pasteur de la Guyane | A. Enfissi; A. Salmier; Anne Lavergne; Dominique Rousset |
| EPI_ISL_2406505 | Laboratoire Virologie Saint Louis APHP | Laboratoire Virologie Saint Louis APHP | Constance Delauguerre; Jérome Le Goff; Linda Feghoul; Marie Laure Chaix; Marie Laure Néré; Maud Salmona; Severine Mercier Delarue; Sophia Achaibou |
| EPI_ISL_3208006, EPI_ISL_3458566, EPI_ISL_3458567, EPI_ISL_3458574, EPI_ISL_3458578, EPI_ISL_3458579, EPI_ISL_3458580, EPI_ISL_3458619, EPI_ISL_3458646, EPI_ISL_3458711, EPI_ISL_3458714, EPI_ISL_3458715, EPI_ISL_3458716, EPI_ISL_5867800, EPI_ISL_5873445, EPI_ISL_5873555, EPI_ISL_7713661 | | | |
| see above | Laboratoire de santé publique du Québec | Laboratoire de santé publique du Québec | Guillaume Bourque; Ioannis Ragoussis; Jesse Shapiro; Mark Lathrop and Judith Fafard on behalf of the CoVSeQ research group; Mark Lathrop and Michel Roger on behalf of the CoVSeQ research group; Sandrine Moreira |
| EPI_ISL_2401055, EPI_ISL_2401056, EPI_ISL_2401063, EPI_ISL_2401072, EPI_ISL_2401076, EPI_ISL_2401098, EPI_ISL_2401107 | | | |
| see above | Laboratoire national de sante, Microbiology, Virology | Laboratoire national de sante, Microbiology, Microbial Genomics Platform | Anke Wienecke-Baldacchino; Catherine Ragimbeau; Fatu Djabi; Jessica Tapp; Lise Pignon; Raoul Salmon; Tamir Abdelrahman; Trung Nguyen Nguyen |
| EPI_ISL_2400875, EPI_ISL_2400882 | Laboratoires Reunis | Laboratoire national de sante, Microbiology, Microbial Genomics Platform | Anke Wienecke-Baldacchino; Bernard Weber; Catherine Ragimbeau; Fatu Djabi; Jessica Tapp; Lise Pignon; Raoul Salmon; Tamir Abdelrahman |
| EPI_ISL_2400675, EPI_ISL_2400677, EPI_ISL_2400715, EPI_ISL_2400721, EPI_ISL_2400725, EPI_ISL_2400731, EPI_ISL_3148148, EPI_ISL_3148149 | | | |
| see above | Laboratoires d'analyses medicales - Ketterthill | Laboratoire national de sante, Microbiology, Microbial Genomics Platform | Anke Wienecke-Baldacchino; Caroline Scheiber; Catherine Ragimbeau; Elodie Solarino; Fatu Djabi; Jessica Tapp; Lise Pignon; Raoul Salmon; Serge Vedy; Tamir Abdelrahman; Virginie Jover |
| EPI_ISL_2645424, EPI_ISL_2645429, EPI_ISL_2645436, EPI_ISL_2645438 | Laboratorio Central de Saude Publica do Estado do Para (LACEN/PA) | Laboratory of Respiratory Viruses and Measles, Oswaldo Cruz Institute, FIOCRUZ | Alice Sampaio Rocha; Ana Carolina Mendonca; Anna Carolina Paixao; Elisa Cavalcante Pereira; Fernando Motta; Luciana Appolinario; Marilda Siqueira on behalf of the Fiocruz COVID-19 Genomic Surveillance Network; Paola Resende; Renata Serrano Lopes; Taina Venas; Valnete Andrade |
| EPI_ISL_2156063, EPI_ISL_2156074 | Laboratorio Aziendale di Microbiologia e Virologia, Azienda Sanitaria dell'Alto Adige | Laboratorio Aziendale di Microbiologia e Virologia, Azienda Sanitaria dell'Alto Adige | Anne Picard; Bartolomeo Mobilio Rodriguez; Chiara Cantaloni; Claudia Volpato; Elisa Masi; Elisabetta Giacobazzi; Elisabetta Pagani; Irene Bianconi; Stefanie Wieser |
| EPI_ISL_4196762, EPI_ISL_4196764, EPI_ISL_4196765, EPI_ISL_4196766 | Laboratorio Central Mg. Luis Alfredo Pianciola | Laboratorio Central Mg. Luis Alfredo Pianciola on behalf of 'Proyecto Argentino Interinstitucional de genomica de SARS-CoV-2' (PAIS Consortium) | C Pintos; C Rastellini.; C Ziehm; J Ousset; L Pianciola; M Fernandez; M Mazzeo |
| EPI_ISL_2385499, EPI_ISL_2385500, EPI_ISL_2385501, EPI_ISL_2385508, EPI_ISL_2385509, EPI_ISL_2385520, EPI_ISL_2385521, EPI_ISL_2385522, EPI_ISL_2385523, EPI_ISL_2385524, EPI_ISL_2385527, EPI_ISL_2385528, EPI_ISL_2385529, EPI_ISL_2385530, EPI_ISL_2385532, EPI_ISL_2385533, EPI_ISL_2385534, EPI_ISL_2385535, EPI_ISL_2385536, EPI_ISL_2385538, EPI_ISL_2385539, EPI_ISL_2385540, EPI_ISL_2385541, EPI_ISL_2385542, EPI_ISL_2385543, EPI_ISL_2385544, EPI_ISL_2385548, EPI_ISL_2385549, EPI_ISL_2385550, EPI_ISL_2385551, EPI_ISL_2385552, EPI_ISL_2385553, EPI_ISL_2385554, EPI_ISL_2385555, EPI_ISL_2385556, EPI_ISL_2385557, EPI_ISL_2385558, EPI_ISL_2385559, EPI_ISL_2385560, EPI_ISL_2385561, EPI_ISL_2385562, EPI_ISL_2385563, EPI_ISL_2385564, EPI_ISL_2385565, EPI_ISL_2385566, EPI_ISL_2385567, EPI_ISL_2385568, EPI_ISL_2385569, EPI_ISL_2385570, EPI_ISL_2385571, EPI_ISL_2385572, EPI_ISL_2385573, EPI_ISL_2385574, EPI_ISL_2385575, EPI_ISL_2385576, EPI_ISL_2385577, EPI_ISL_2385578, EPI_ISL_2385579, EPI_ISL_2385580, EPI_ISL_2385581, EPI_ISL_2385583, EPI_ISL_2385584, EPI_ISL_2385585, EPI_ISL_2385586, EPI_ISL_2385587, EPI_ISL_2385588, EPI_ISL_2385589, EPI_ISL_2385590, EPI_ISL_2385592, EPI_ISL_2385593, EPI_ISL_2385594, EPI_ISL_2385595, EPI_ISL_2385596, EPI_ISL_2385597, EPI_ISL_2385598, EPI_ISL_2385599, EPI_ISL_2385600, EPI_ISL_2385601, EPI_ISL_2385602, EPI_ISL_2385603, EPI_ISL_2385604, EPI_ISL_2385605, EPI_ISL_2385606, EPI_ISL_2385607, EPI_ISL_2385608, EPI_ISL_2385609, EPI_ISL_2385610, EPI_ISL_2385611, EPI_ISL_2385612, EPI_ISL_2385613, EPI_ISL_2385614, EPI_ISL_2385615, EPI_ISL_2385616, EPI_ISL_2385617, EPI_ISL_2385618, EPI_ISL_2385619, EPI_ISL_2385620, EPI_ISL_2385621, EPI_ISL_2385622, EPI_ISL_2385623, EPI_ISL_2385624, EPI_ISL_2385625, EPI_ISL_2385626, EPI_ISL_2385627, EPI_ISL_2385628, EPI_ISL_2385629, EPI_ISL_2385630, EPI_ISL_2385631, EPI_ISL_2385632, EPI_ISL_2385633, EPI_ISL_2385634, EPI_ISL_2385635, EPI_ISL_2385636, EPI_ISL_2385637, EPI_ISL_2385638, EPI_ISL_2385639, EPI_ISL_2385640, EPI_ISL_2385641, EPI_ISL_2385642, EPI_ISL_2385643, EPI_ISL_2385644, EPI_ISL_2385645, EPI_ISL_2385646, EPI_ISL_2385647, EPI_ISL_2385648, EPI_ISL_2385649, EPI_ISL_2385650, EPI_ISL_2385651, EPI_ISL_2385652, EPI_ISL_2385653, EPI_ISL_2385654, EPI_ISL_2385655, EPI_ISL_2385656, EPI_ISL_2385657, EPI_ISL_2385658, EPI_ISL_2385659, EPI_ISL_2385660, EPI_ISL_2385661, EPI_ISL_2385662, EPI_ISL_2385663, EPI_ISL_2385664, EPI_ISL_2385665, EPI_ISL_2385666, EPI_ISL_2385667, EPI_ISL_2385668 | | | |
| see above | Laboratorio Central Noel Nutels | Bioinformatics Laboratory / LNCC | Alessandra P Lamarca; Alexandra L Gerber; Amilcar Tanuri; Ana Paula de C Guimaraes; Ana Tereza R Vasconcelos; Andrea Cony Cavalcanti; Caio Luiz Pereira Ribeiro; Cassia Alves; Cintia Policarpo; Claudia Maria Braga de Mello; Cristiane Gomes da Silva; Diana Mariani; Douglas Terra Machado; Flavio Dias da Silva; Gleidson da Silva de Oliveira; Leandro Magalhaes de Souza; Liliane Cavalcante; Luiz G P de Almeida; Marcio Henrique de Oliveira Garcia; Mario Sergio Ribeiro; Ronaldo da Silva F Jr; Silvia Carvalho |
| EPI_ISL_2490387, EPI_ISL_2490389, EPI_ISL_2490393 | Laboratorio Central de Epidemiología (LCE) | Centro de Investigación en Enfermedades Infecciosas (CIENI), Instituto Nacional de Enfermedades Respiratorias (INER) | ; Alejandra Garcia-Gasca; Alejandra Hernández-Terán; Alejandro Sanchez-Flores; Alfredo Herrera-Estrella; Alicia Ocaña-Mondragón; Andreu Comas-García; Angel Gustavo Salas-Lais; Antonio Loza Román; Bernardo Martínez-Miguel; Blanca Taboada; Brenda Irasema Maldonado-Meza; Bruno Gomez-Gil; Carla Ivón Herrera-Najera; Carlos F. Arias; Celia Boukadida; Clara Esperanza Santacruz-Tinoco; Concepción Grajales-Muñiz; Consorcio Mexicano de Vigilancia Genómica (CoViGen-Mex). Authors (in alphabetical order): Julio Elias Alvarado-Yaah; Cristóbal Cháidez-Quiróz; Célida Duque Molina; Célida Martínez- Rodríguez; Daniel Fregoso-Rueda; Daniel Lira Morales; Eduardo Becerril-Vargas; Fernando Fontove-Herrera; Fidencio Mejía-Nepomuceno; Francisco Pulido; Gloria Elena Espinosa-Ayala; Gloria María Molina-Salinas; Gloria Vazquez; Hector Esteban Paz-Juárez; Hector Montoya-Fuentes; Helen Haydee Fernanda Ramírez-Plascencia; Irvin González-López; Jean Pierre González; Joel Armando Vázquez-Pérez.; Jorge Salas-Hernández; José Antonio Enciso-Moreno; José Arturo Martínez-Orozco; José Esteban Muñoz-Medina; José de Jesús Nuñez-Contreras; Juan Bautista Chale-Dzul; Julissa Enciso-Ibarra; Luis Alberto Ochoa-Carrera; Margarita Matías-Florentino; Mario Mújica-Sánchez; Marissa Perez-Garcia; María Guadalupe Santiago-Mauricio; María Guadalupe de Jesús Mireles-Rivera; Nelly Sélem-Mojica; Pavel Isa; Ricardo Ciria Merce; Ricardo Grande; Rosa María Gutierrez Rios; Santiago Ávila-Ríos; Selene Zárate; Susana Lopez; Víctor Eduardo Garcia-Arias; Víctor Hugo Borja-Aburto |
| EPI_ISL_2391520, EPI_ISL_2391549, EPI_ISL_2391609, EPI_ISL_2391681, EPI_ISL_2391716 | Laboratorio Central de Epidemiología (LCE) | Instituto de Biotecnología de la UNAM | ; Alejandra Garcia-Gasca; Alejandra Hernández-Terán; Alejandro Sanchez-Flores; Alfredo Herrera-Estrella; Alicia Ocaña-Mondragón; Andreu Comas-García; Angel Gustavo Salas-Lais; Antonio Loza Román; Bernardo Martínez-Miguel; Blanca Taboada; Brenda Irasema Maldonado-Meza; Bruno Gomez-Gil; Carla Ivón Herrera-Najera; Carlos F. Arias; Celia Boukadida; Clara Esperanza Santacruz-Tinoco; Concepción Grajales-Muñiz; Consorcio Mexicano de Vigilancia Genómica (CoViGen-Mex). Authors (in alphabetical order): Julio Elias Alvarado-Yaah; Cristóbal Cháidez-Quiróz; Célida Duque Molina; Célida Martínez- Rodríguez; Daniel Fregoso-Rueda; Daniel Lira Morales; Eduardo Becerril-Vargas; Fernando Fontove-Herrera; Fidencio Mejía-Nepomuceno; Francisco Pulido; Gloria Elena Espinosa-Ayala; Gloria María Molina-Salinas; Gloria Vazquez; Hector Esteban Paz-Juárez; Hector Montoya-Fuentes; Helen Haydee Fernanda Ramírez-Plascencia; Irvin González-López; Jean Pierre González; Joel Armando Vázquez-Pérez.; Jorge Salas-Hernández; José Antonio Enciso-Moreno; José Arturo Martínez-Orozco; José Esteban Muñoz-Medina; José de Jesús Nuñez-Contreras; Juan Bautista Chale-Dzul; Julissa Enciso-Ibarra; Luis Alberto Ochoa-Carrera; Margarita Matías-Florentino; Mario Mújica-Sánchez; María Guadalupe Santiago-Mauricio; María Guadalupe de Jesús Mireles-Rivera; Nelly Sélem-Mojica; Pavel Isa; Ricardo Ciria Merce; Ricardo Grande; Rosa María Gutierrez Rios; Santiago Ávila-Ríos; Selene Zárate; Susana Lopez; Víctor Eduardo Garcia-Arias; Víctor Hugo Borja-Aburto |
| EPI_ISL_3149735, EPI_ISL_3149736 | Laboratorio Central de Salud Publica | Laboratorio de Biología Molecular, Instituto de Medicina Regional on behalf of 'Proyecto Argentino Interinstitucional de genomica de SARS-CoV-2' (PAIS Consortium) | Andrea Ayala; Bettina Brusés; Erica Struss; Esteban Paredes; Griselda Oria; Horacio Lucero.; Javier Mussin; Laura Formichelli; Melina Lorenzini Campos; Raúl Maximiliano Acevedo; Verónica Gómez; Victoria Femenías |
| EPI_ISL_6944490, EPI_ISL_6944492, EPI_ISL_6944499, EPI_ISL_6944502, EPI_ISL_6944505, EPI_ISL_6944506, EPI_ISL_6944513, EPI_ISL_6944514, EPI_ISL_6944523, EPI_ISL_6944528, EPI_ISL_6944529, EPI_ISL_6944532, EPI_ISL_6944553 | | | |
| see above | Laboratorio Central de Salud Publica | Viral Special Pathogens Branch, Centers for Disease Control and Prevention | Andrea Gómez de la Fuente; Cynthia Vázquez; Emir Talundzic; Joel Montgomery; John Klena; Juan Torales; Justin Lee; María José Ortega; María Liz Gamarra; Shannon Whitmer; Shirley Villalba |
| EPI_ISL_2274073, EPI_ISL_2274074 | Laboratorio Central de Saude Publica do Estado da Bahia (LACEN/BA) | Laboratory of Respiratory Viruses and Measles, Oswaldo Cruz Institute, FIOCRUZ | Alice Sampaio Rocha; Ana Carolina Mendonca; Anderson Brandao Leite; Anna Carolina Paixao; Elisa Cavalcante Pereira; Fernando Motta; Luciana Appolinario; Marilda Siqueira on behalf of the Fiocruz COVID-19 Genomic Surveillance Network; Paola Resende; Renata Serrano Lopes; Taina Venas |
| EPI_ISL_3190189 | Laboratorio Central de Saude Publica do Estado da Paraiba (LACEN/PB) | Laboratory of Respiratory Viruses and Measles, Oswaldo Cruz Institute, FIOCRUZ | Alice Sampaio Rocha; Ana Carolina Mendonca; Anna Carolina Paixao; Dalane Loudal Florentino Teixeira; Elisa Cavalcante Pereira; Fernando Motta; Joao Felipe Bezerra; Luciana Appolinario; Marilda Siqueira on behalf of the Fiocruz COVID-19 Genomic Surveillance Network; Paola Resende; Renata Serrano Lopes; Taina Venas |
| EPI_ISL_2466231, EPI_ISL_2557313, EPI_ISL_2557314, EPI_ISL_2557315, EPI_ISL_2557320, EPI_ISL_2557321, EPI_ISL_2557322, EPI_ISL_2557323, EPI_ISL_2557324, EPI_ISL_2557325, EPI_ISL_2557326, EPI_ISL_2557327, EPI_ISL_2557328, EPI_ISL_2557329, EPI_ISL_2557330, EPI_ISL_2557331, EPI_ISL_2557333, EPI_ISL_2557334, EPI_ISL_2645388 | | | |
| see above | Laboratorio Central de Saude Publica do Estado de Alagoas (LACEN/AL) | Laboratory of Respiratory Viruses and Measles, Oswaldo Cruz Institute, FIOCRUZ | Alice Sampaio Rocha; Ana Carolina Mendonca; Anderson Brandao Leite; Anna Carolina Paixao; Elisa Cavalcante Pereira; Fernando Motta; Luciana Appolinario; Marilda Siqueira on behalf of the Fiocruz COVID-19 Genomic Surveillance Network; Paola Resende; Renata Serrano Lopes; Taina Venas |
| EPI_ISL_3235302 | Laboratorio Central de Saude Publica do Estado de Santa Catarina (LACEN/SC) | Laboratory of Respiratory Viruses and Measles, Oswaldo Cruz Institute, FIOCRUZ | Alice Sampaio Rocha; Ana Carolina Mendonca; Anna Carolina Paixao; Darcita Buerger Rovaris; Elisa Cavalcante Pereira; Fernando Motta; Luciana Appolinario; Marilda Siqueira on behalf of the Fiocruz COVID-19 Genomic Surveillance Network; Paola Resende; Renata Serrano Lopes; Sandra Bianchini Fernandes; Taina Venas |
| EPI_ISL_2863619, EPI_ISL_2863625, EPI_ISL_2863628, EPI_ISL_2863632, EPI_ISL_2863635 | Laboratorio Central de Saude Publica do Estado de Sergipe (LACEN/SE) | Laboratory of Respiratory Viruses and Measles, Oswaldo Cruz Institute, FIOCRUZ | Alice Sampaio Rocha; Ana Carolina Mendonca; Anna Carolina Paixao; Cliomar Alves dos Santos; Elisa Cavalcante Pereira; Fernando Motta; Luciana Appolinario; Marilda Siqueira on behalf of the Fiocruz COVID-19 Genomic Surveillance Network; Paola Resende; Renata Serrano Lopes; Tainá Moreira Martins Venas |
| EPI_ISL_3190240, EPI_ISL_3190242 | Laboratorio Central de Saude Publica do Estado do Espirito Santo (LACEN/ES) | Laboratory of Respiratory Viruses and Measles, Oswaldo Cruz Institute, FIOCRUZ | Alice Sampaio Rocha; Ana Carolina Mendonca; Anna Carolina Paixao; Elisa Cavalcante Pereira; Fernando Motta; Luciana Appolinario; Marilda Siqueira on behalf of the Fiocruz COVID-19 Genomic Surveillance Network; Paola Resende; Renata Serrano Lopes; Rodrigo Ribeiro Rodrigues; Taina Venas |
| EPI_ISL_2983238, EPI_ISL_2983239, EPI_ISL_2983251, EPI_ISL_2983293, EPI_ISL_2983294, EPI_ISL_2983295, EPI_ISL_2983296, EPI_ISL_2983297, EPI_ISL_2983298, EPI_ISL_2983334, EPI_ISL_2983336 | | | |
| see above | Laboratorio Central de Saude Publica do Estado do Maranhao (LACEN-MA) | Laboratory of Respiratory Viruses and Measles, Oswaldo Cruz Institute, FIOCRUZ | Agatha Cristinne Prudencio; Alice Sampaio Rocha; Ana Carolina Mendonca; Anna Carolina Paixao; Elisa Cavalcante Pereira; Fernando Motta; Ighor Leonardo Arantes Gomes; Lidio Gonçalves Lima Neto; Luciana Appolinario; Marilda Siqueira on behalf of the Fiocruz COVID-19 Genomic Surveillance Network; Paola Resende; Renata Serrano Lopes; Taina Moreira Venas |
| EPI_ISL_2443673, EPI_ISL_2603433, EPI_ISL_2603434, EPI_ISL_2603444, EPI_ISL_2603445, EPI_ISL_2603448, EPI_ISL_2603555, EPI_ISL_2603556, EPI_ISL_2603565, EPI_ISL_2603566, EPI_ISL_2603567, EPI_ISL_2603568, EPI_ISL_2603569, EPI_ISL_2603570, EPI_ISL_2603571, EPI_ISL_2603572, EPI_ISL_2603573, EPI_ISL_2603574, EPI_ISL_2603575, EPI_ISL_2603576 | | | |
| see above | Laboratorio Central de Saude Publica do Estado do Rio Grande do Sul (LACEN-RS) | Laboratory of Respiratory Viruses and Measles, Oswaldo Cruz Institute, FIOCRUZ | Alice Sampaio Rocha; Ana Carolina Mendonca; Anderson Brandao Leite; Anna Carolina Paixao; Elisa Cavalcante Pereira; Fernando Motta; Luciana Appolinario; Marilda Siqueira on behalf of the Fiocruz COVID-19 Genomic Surveillance Network; Paola Resende; Renata Serrano Lopes; Richard Salvato; Taina Venas; Tatiana Schaffer Gregianini |
| EPI_ISL_2983213 | Laboratorio Central de Saude Publica do Estado do Rio de Janeiro (LACEN-RJ) | Laboratory of Respiratory Viruses and Measles, Oswaldo Cruz Institute, FIOCRUZ | Agatha Cristinne Prudencio; Alice Sampaio Rocha; Ana Carolina Mendonca; Andrea Cony Cavalcanti; Anna Carolina Paixao; Elisa Cavalcante Pereira; Fernando Motta; Ighor Leonardo Arantes Gomes; Luciana Appolinario; Marilda Siqueira on behalf of the Fiocruz COVID-19 Genomic Surveillance Network; Paola Resende; Renata Serrano Lopes; Taina Venas |

| | | | |
|---|---|---|---|
| EPI_ISL_3033053, EPI_ISL_3033054, EPI_ISL_3033055, EPI_ISL_3033057 | Laboratorio Central de la Ciudad de Santa Fe | Grupo de Genómica y Bioinformática del Instituto de Investigación de la Cadena Láctea CONICET-INTA on behalf of 'Proyecto Argentino Interinstitucional de genomica de SARS-CoV-2' (PAIS Consortium) | AF; Amadio; C; Eberhardt; G; Irazoqui; JM; MF; Mugna; Ojeda; Pastor; Rompato; V |
| EPI_ISL_2271689, EPI_ISL_2271690 | Laboratorio Central, Ministerio de Salud Cordoba | Instituto de Patologia Vegetal (CIAP-INTA) on behalf of 'Proyecto Argentino Interinstitucional de genomica de SARS-CoV-2' (PAIS Consortium) | Barbas, G.; Castro, G.; Debat, HJ.; FD; Fernandez; M; M.B.; Marquez, N.; Pisano; Re, V. |
| EPI_ISL_4196714, EPI_ISL_4196718 | Laboratorio MICROBIOM | Laboratorio Central Mg. Luis Alfredo Pianciola on behalf of 'Proyecto Argentino Interinstitucional de genomica de SARS-CoV-2' (PAIS Consortium) | C Pintos; C Rastellini; C Ziehm; J Ousset; L Pianciola; M Fernandez; M Mazzeo; S Cecchi. |
| EPI_ISL_5801692, EPI_ISL_5801695 | Laboratorio Municipal De Suzano | Instituto Butantan | Antonio Jorge Martins; Claudia Renata dos Santos Barros; David Schlesinger; Debora Botequio Moretti; Dimas Tadeu Covas; Elaine Cristina Marqueze; Elaine Vieira Santos; Evandra Strazza Rodrigues; Heidge Fukumasu; Jayme Augusto de Souza-Neto; José Salvatore Leister Patané; Luiz Alcantara; Luiz Lehmann Coutinho; Maria Carolina Elias; Maurício Lacerda Nogueira; Rafael dos Santos Bezerra; Raul Machado Neto; Rejane Maria Tommasini Grotto; Ricardo Haddad; Sandra Coccuzzo Sampaio Vessoni; Simone Kashima; Svetoslav Nanev Slavov; Vincent Louis Viala |
| EPI_ISL_5199343, EPI_ISL_5199344 | Laboratorio SYNLAB Colombia | Instituto Nacional de Salud-Dirección de Investigación en Salud Pública | Beatriz de Arco; Carlos Franco-Muñoz; Carmen Osorio; Diana Malo; Diego A. Álvarez-Díaz; Diego Andrés Prada; Gerardo Santamaría; Hector Alejandro Ruiz-Moreno; Jhonnatan Reales-González; Jorge Rivera; Juan Camilo Martinez; Julian Naizaque; Katherine Laiton-Donato; Lisseth Pardo; Magdalena Wiesner; Marcela Mercado-Reyes; Maria T. Herrera-Sepúlveda; Marta Lopez Blanco; Martha Lucia Ospina Martinez; Paola Rojas; Sergio Gomez; Sheryll Corchuelo; Tatiana Cobos; Ángela Alarcon Cruz |
| EPI_ISL_2462062 | Laboratorio de Biologia Molecular, Hospital San Pedro Claver | Laboratorio de Biologia Molecular, Hospital San Pedro Claver | Aneth Vasquez Michel; Carmen Delgado Barrera; Oscar M. Rollano-Peñaloza; Sandra Miranda Sardon |
| EPI_ISL_4405292 | Laboratorio de Biología Molecular Bolívar | Área de Secuenciación del Laboratorio de Virología del Hospital de Niños Dr. Ricardo Gutierrez on behalf of 'Proyecto Argentino Interinstitucional de genomica de SARS-CoV-2' (PAIS Consortium) | Acuña; Cabrerizo; Caviglia; D; Di Claudio; D'Andrea; F; Goya; Jofré; LE; Larreche; Lescano; Lusso; M; ME; MI; ML; MR; MS; Martínez; Nabaes Jodar; Natale; R; S; Ticeira; Valinotto; Viegas, M. |
| EPI_ISL_2777433, EPI_ISL_2777434, EPI_ISL_4030346, EPI_ISL_4030349, EPI_ISL_4030364 | Laboratorio de Ecologia de Doencas Transmissiveis na Amazonia, Instituto Leonidas e Maria Deane - Fiocruz Amazonia | Laboratorio de Ecologia de Doencas Transmissiveis na Amazonia, Instituto Leonidas e Maria Deane - Fiocruz Amazonia | Alex Martins; André Corado; Debora Duarte; Fabiola Mendonça da Silva Chui; Felipe Naveca; Fernanda Nascimento; Fernando Fonseca de Almeida Val; George Silva; Gisely Cardoso de Melo; Karina Pessoa; Luciana Gonçalves; Marcus Vinicius Guimarães de Lacerda; Maria Júlia Brandão; Maria Paula Gomes Mourão; Mariana Xavier Simão; Matilde Mejía; Michele Jesus; Valdinete Nascimento; Vanderson de Souza Sampaio; Victor Souza; Ágatha Costa |
| EPI_ISL_2728572, EPI_ISL_2728573 | Laboratorio de Infectologia y Virologia Molecular | Laboratory of Molecular Virology, School of Medicine, Pontificia Universidad Catolica de Chile | Alejandro Bhrun; Ana Maria Contreras; Andres E. Munoz-Marcos; Carlos Palma; Catalina Pardo-Roa; Constanza Maldonado; Constanza Martinez-Valdevenito; Eileen Serrano; Erick Salinas; Estefany Poblete; Francisco Melo; Jennifer Angulo; Jorge Levican; Leonardo I. Almonacid; M. Belen Leyton; Magdalena Vera; Marcela Ferres; Maria Jose Avendano; Rafael A. Medina; Tamara Garcia-Salum |
| EPI_ISL_2854110, EPI_ISL_2854111, EPI_ISL_2854114, EPI_ISL_2854116, EPI_ISL_2854119, EPI_ISL_2854120, EPI_ISL_2854121, EPI_ISL_2907491, EPI_ISL_2907492, EPI_ISL_2907493, EPI_ISL_2907494, EPI_ISL_2907495, EPI_ISL_3134268 | | | |
| see above | Laboratorio de Medicina Molecular, Universidad de Magallanes | Centro Asistencial Docente y de Investigacion, Universidad de Magallanes | Carolina Pérez; Constanza Ceroni; Diego Alvarez; Inés Cid; Jacqueline Aldridge; Jorge González; Juan Ríos; Marcelo Navarrete; Roberto Uribe-Paredes |
| EPI_ISL_3048961, EPI_ISL_3048962, EPI_ISL_3048963, EPI_ISL_3048964, EPI_ISL_3048966, EPI_ISL_3048967, EPI_ISL_3761636, EPI_ISL_3761752, EPI_ISL_3761763 | | | |
| see above | Laboratorio de Pesquisa em Virologia, FAMERP, SJRP | Laboratorio de Pesquisa em Virologia, FAMERP, SJRP | Beatriz de Carvalho Marques; Cecília Artico Banho; Cíntia Bittar; Fábio Sossai Possebon; Guilherme Campos; Helena Lage Ferreira; Jorge A. Petroli Marchesi; João Pessoa Araújo Jr.; Leila Sabrina Ullmann; Lívia Sacchetto; Maisa C. Pereira Parra; Marília Moraes; Maurício L. Nogueira.; Paula Rahal; Paulo Inacio da Costa |
| EPI_ISL_3401458, EPI_ISL_3401459, EPI_ISL_3401464, EPI_ISL_3401469, EPI_ISL_3672413, EPI_ISL_3826392, EPI_ISL_3826393, EPI_ISL_3826394, EPI_ISL_3826397, EPI_ISL_3826398, EPI_ISL_3826399, EPI_ISL_3826400, EPI_ISL_3826413, EPI_ISL_3991421, EPI_ISL_3991675, EPI_ISL_4052715, EPI_ISL_4052716, EPI_ISL_4052717, EPI_ISL_4052718, EPI_ISL_4052730, EPI_ISL_4052737, EPI_ISL_4052741, EPI_ISL_5934954 | | | |
| see above | Laboratorio de Referencia Nacional de Virus Respiratorios. Centro Nacional de Salud Publica. Instituto Nacional de Salud Peru. | Laboratorio de Referencia Nacional de Virus Respiratorios. Centro Nacional de Salud Publica. Instituto Nacional de Salud Peru. | Carlos Padilla Rojas; Henri Bailon Calderon; Iris Silva Molina; Joseph Huayra Niquen; Lely Solari Zerpa; Luis Barcena Flores; Marco Galarza Perez; Nancy Rojas Serrano; Nieves Sevilla Castañeda; Omar Caceres Rey; Orson Mestanza Millones; Princesa Medrano Alhuay; Priscila Lope Pari; Sandra Morales Ruiz; Sara Gordillo Vilchez; Steve Acedo Lazo; Veronica Hurtado Vela; Victor Jimenez Vasquez; Wendy Lizarraga Olivares |
| EPI_ISL_3375938, EPI_ISL_3375960, EPI_ISL_3376382, EPI_ISL_3376383, EPI_ISL_3376403 | Laboratorio de Referencial Nacional de Virus Respiratorios | Laboratorio de Referencial Nacional de Virus Respiratorios | Carlos Padilla Rojas; Henri Bailon Calderon; Iris Silva Molina; Joseph Huayra Niquen; Lely Solari Zerpa; Luis Barcena Flores; Marco Galarza Perez; Nancy Rojas Serrano; Omar Caceres Rey; Orson Mestanza Millones; Priscila Lope Pari; Sandra Morales Ruiz; Steve Acedo Lazo; Veronica Hurtado Vela |
| EPI_ISL_4405088, EPI_ISL_4405097, EPI_ISL_4405101 | Laboratorio de Virología del Hospital de Niños Dr. Ricardo Gutierrez | Área de Secuenciación del Laboratorio de Virología del Hospital de Niños Dr. Ricardo Gutierrez on behalf of 'Proyecto Argentino Interinstitucional de genomica de SARS-CoV-2' (PAIS Consortium) | AS; Acevedo; Acuña; Alexay; Alvarez Lopez; C; D; E; Goya; Gravis; Jacques; LE; Lusso; ME; MI; MS; Mistchenko; Nabaes Jodar; Natale; O; S; Valinotto; Viegas, M. |
| EPI_ISL_4405217, EPI_ISL_4405222 | Laboratorio del Hospital Interzonal General de Agudos "Eva Perón" | Área de Secuenciación del Laboratorio de Virología del Hospital de Niños Dr. Ricardo Gutierrez on behalf of 'Proyecto Argentino Interinstitucional de genomica de SARS-CoV-2' (PAIS Consortium) | Acuña; C; Carulla; D; Goya; Kairiyama; LE; Lusso; M; MI; MS; Nabaes Jodar; Natale; Pengue; Piñeyro C; S; Valinotto; Viegas, M. |
| EPI_ISL_5771877 | Laboratorio del Hospital Regional Ushuaia Gdor. Ernesto Campos | Nodo de Secuenciación Tierra del Fuego - Hospital Regional Ushuaia - Centro Austral De Investigaciones Científicas - Universidad Nacional De Tierra Del Fuego | AE; Boutureira, MF.; CA; CB; CF; Castro; Ceballos; Cáceres; De Roccis; F; G; Gallego; Gramundi; ID; Nardi; Rojas; SB; SG; Yulan |
| EPI_ISL_7620344, EPI_ISL_7620345, EPI_ISL_7620346, EPI_ISL_7620349, EPI_ISL_7620373, EPI_ISL_7620374 | Laboratorio del Hospital Regional Ushuaia Gdor. Ernesto Campos | Nodo de Secuenciación Tierra del Fuego - Hospital Regional Ushuaia - Centro Austral De Investigaciones Científicas - Universidad Nacional De Tierra Del Fuego on behalf PAIS project and ANLIS-Malbran | AE; Boutureira, MF.; CA; CB; CF; Castro; Ceballos; Cáceres; De Roccis; F; G; Gallego; Gramundi; ID; Nardi; Rojas; SB; SG; Yulan |
| EPI_ISL_1993810 | Laboratorio di Microbiologia | Laboratorio di Microbiologia | Martinetti Lucchini Gladys; Valeria Spina |
| EPI_ISL_2046738, EPI_ISL_2046739, EPI_ISL_2046740, EPI_ISL_2046796, EPI_ISL_2046806, EPI_ISL_2181664, EPI_ISL_2181665, EPI_ISL_2181674, EPI_ISL_2181680, EPI_ISL_2181682, EPI_ISL_2181685, EPI_ISL_2181704, EPI_ISL_2181716, EPI_ISL_2181717, EPI_ISL_2181718, EPI_ISL_2181733, EPI_ISL_2181749, EPI_ISL_2181751, EPI_ISL_2181778, EPI_ISL_2181784, EPI_ISL_2181786, EPI_ISL_2181796, EPI_ISL_2181799, EPI_ISL_2181825, EPI_ISL_2181850, EPI_ISL_2181906, EPI_ISL_2181995, EPI_ISL_2182018, EPI_ISL_2182083, EPI_ISL_2182097, EPI_ISL_2182100, EPI_ISL_2182110, EPI_ISL_2182155, EPI_ISL_2182169, EPI_ISL_2182182, EPI_ISL_2182211, EPI_ISL_2182217, EPI_ISL_2182231, EPI_ISL_2182239, EPI_ISL_2182256, EPI_ISL_2182273, EPI_ISL_2182313, EPI_ISL_2182319, EPI_ISL_2182333, EPI_ISL_2182341, EPI_ISL_2182349, EPI_ISL_2182399, EPI_ISL_2182437, EPI_ISL_2182439, EPI_ISL_2182442, EPI_ISL_2182452, EPI_ISL_2182463, EPI_ISL_2182477, EPI_ISL_2182535, EPI_ISL_2182544, EPI_ISL_2182586, EPI_ISL_2182622, EPI_ISL_2182628, EPI_ISL_2182658, EPI_ISL_2182666, EPI_ISL_2182674, EPI_ISL_2182705, EPI_ISL_2182725, EPI_ISL_2182728, EPI_ISL_2182755, EPI_ISL_2182761, EPI_ISL_2182803, EPI_ISL_2182805, EPI_ISL_2182823, EPI_ISL_2182853, EPI_ISL_2182883, EPI_ISL_2182896, EPI_ISL_2182965, EPI_ISL_2182976, EPI_ISL_2183167, EPI_ISL_2183757, EPI_ISL_2183764, EPI_ISL_2183771, EPI_ISL_2183778, EPI_ISL_2183781, EPI_ISL_2183816, EPI_ISL_2183821, EPI_ISL_2183825, EPI_ISL_2183847, EPI_ISL_2183855, EPI_ISL_2183864, EPI_ISL_2183870, EPI_ISL_2183874, EPI_ISL_2183878, EPI_ISL_2183884, EPI_ISL_2183898, EPI_ISL_2183902, EPI_ISL_2183912, EPI_ISL_2183917, EPI_ISL_2183918, EPI_ISL_2183987, EPI_ISL_2183988, EPI_ISL_2183992, EPI_ISL_2184000, EPI_ISL_2184003, EPI_ISL_2184004, EPI_ISL_2184005, EPI_ISL_2184013, EPI_ISL_2184016, EPI_ISL_2184028, EPI_ISL_2184031, EPI_ISL_2184032, EPI_ISL_2184033, EPI_ISL_2184035, EPI_ISL_2184037, EPI_ISL_2184125, EPI_ISL_2184174, EPI_ISL_2184178, EPI_ISL_2184189, EPI_ISL_2184190, EPI_ISL_2184195, EPI_ISL_2184228, EPI_ISL_2184235, EPI_ISL_2184257, EPI_ISL_2184265, EPI_ISL_2184348, EPI_ISL_2184364, EPI_ISL_2184400, EPI_ISL_2184440, EPI_ISL_2184456, EPI_ISL_2184462, EPI_ISL_2184499, EPI_ISL_2184525, EPI_ISL_2184550, EPI_ISL_2184551, EPI_ISL_2184554, EPI_ISL_2184558, EPI_ISL_2184559, EPI_ISL_2184561, EPI_ISL_2184563, EPI_ISL_2184568, EPI_ISL_2184569, EPI_ISL_2184580, EPI_ISL_2184581, EPI_ISL_2184588, EPI_ISL_2184596, EPI_ISL_2184603, EPI_ISL_2184612, EPI_ISL_2184620, EPI_ISL_2184629, EPI_ISL_2184637, EPI_ISL_2184639, EPI_ISL_2184640, EPI_ISL_2184689, EPI_ISL_2184712, EPI_ISL_2184713, EPI_ISL_2184721, EPI_ISL_2184722, EPI_ISL_2184725, EPI_ISL_2184726, EPI_ISL_2184730, EPI_ISL_2184731, EPI_ISL_2184732, EPI_ISL_2184736, EPI_ISL_2184753, EPI_ISL_2184765, EPI_ISL_2184766, EPI_ISL_2184770, EPI_ISL_2184773, EPI_ISL_2184775, EPI_ISL_2184788, EPI_ISL_2184811, EPI_ISL_2184820, EPI_ISL_2184823, EPI_ISL_2184833, EPI_ISL_2184900, EPI_ISL_2184905, EPI_ISL_2184913, EPI_ISL_2184915, EPI_ISL_2184980, EPI_ISL_2185040, EPI_ISL_2185049, EPI_ISL_2185069, EPI_ISL_2185076, EPI_ISL_2185077, EPI_ISL_2185082, EPI_ISL_2185083, EPI_ISL_2185084, EPI_ISL_2185086, EPI_ISL_2185122, EPI_ISL_2185134, EPI_ISL_2185136, EPI_ISL_2185149, EPI_ISL_2185161, EPI_ISL_2185219, EPI_ISL_2185224, EPI_ISL_2185233, EPI_ISL_2185301, EPI_ISL_2185317, EPI_ISL_2185334, EPI_ISL_2185420, EPI_ISL_2185434, EPI_ISL_2185507, EPI_ISL_2305667, EPI_ISL_2305672, EPI_ISL_2305674, EPI_ISL_2305676, EPI_ISL_2305682, EPI_ISL_2305684, EPI_ISL_2305712, EPI_ISL_2305729, EPI_ISL_2305730, EPI_ISL_2305745, EPI_ISL_2305842, EPI_ISL_2305962 | | | |
| see above | Laboratory Corporation of America | Centers for Disease Control and Prevention Division of Viral Diseases, Pathogen Discovery | Adrian Paskey; Amanda Douglas; Amanda Suchanek; Andrea Throop; Ayla Burns; Benjamin Rambo-Martin; Bobbi Croy; Brian Krueger; Brian Norvell; Christopher Gulvick; Christos Petropoulos; Clinton R. Paden; Craig Lukasik; Dakota Howard; Darlene Wagner; Debbie Boles; Dhwani Batra; Duncan MacCannell; Eyad Almasri; Goran Stevovic; Howard Engler; Hrushikesh Deshmukh; Jake Humphrey; Jana Schroth; Jason Caravas; Joe Voshell; John Pruitt; Jonathan Meltzer; Jonathan Williams; Kara Moser; Kimberly Wagner; Lax Iyer; Lyndon Tilson; Manoj Jain; Marcia Eisenberg; Mary Ann Cristobal; Mary Williamson; Matthew Schmerer; Michael Levandoski; Mike Sapeta; Mindy Nye; Minoo Agarwal; Mohan Kolli; Nuthawin Charoensri; Oren Cohen; Peter W. Cook; Prashant Gupta; Qian Zeng; Rama Ghatti; Scott Parker; Scott Ryan; Scott Sammons; Shatavia Morrison; Stanley Letovsky; Steven Ragan; Suresh Babu Selvaraju; Susan Countryman; Susan Hicks; Suzanne Dale; Thomas Urban; Tim Kuphal; Tricia Zwiefelhofer; Vincent Drouillon; Yvette Unoarumhi |
| EPI_ISL_2811862 | Laboratory Medicine, University of Washington | Laboratory Medicine, University of Washington | Castor; J.L. |
| EPI_ISL_2443610, EPI_ISL_2443611, EPI_ISL_2443613, EPI_ISL_3045381, EPI_ISL_4237272, EPI_ISL_4237348, EPI_ISL_4237349, EPI_ISL_4237350, EPI_ISL_4237435, EPI_ISL_6172741, EPI_ISL_6172743, EPI_ISL_6172744, EPI_ISL_6172745, EPI_ISL_6172746, EPI_ISL_6970953, EPI_ISL_6970958, EPI_ISL_6970972, EPI_ISL_6970981 | | | |
| see above | Laboratory of Respiratory Viruses and Measles, Oswaldo Cruz Institute, FIOCRUZ | Laboratory of Respiratory Viruses and Measles, Oswaldo Cruz Institute, FIOCRUZ | Agatha Cristinne Prudencio; Alice Sampaio Rocha; Ana Carolina Mendonca; Anna Carolina Paixao; Bruna Mendonça da Silva; Elisa Cavalcante Pereira; Fernando Motta; Ighor Arantes; Ighor Leonardo Arantes Gomes; Jéssica Graça Macedo de Carvalho; Larissa Macedo Pinto; Luciana Appolinario; Marilda Siqueira on behalf of the Fiocruz COVID-19 Genomic Surveillance Network; Paola Resende; Renata Serrano Lopes; Taina Venas; Victor Guimaraes; Vitor Guimaraes Ribeiro |

| | | | |
|---|---|---|---|
| EPI_ISL_2696373, EPI_ISL_2696378, EPI_ISL_2696380, EPI_ISL_2696397, EPI_ISL_2696417, EPI_ISL_2696420, EPI_ISL_2696426, EPI_ISL_2696440, EPI_ISL_2696449, EPI_ISL_2696458, EPI_ISL_2696464, EPI_ISL_2696466, EPI_ISL_2696468, EPI_ISL_2696480, EPI_ISL_2696754, EPI_ISL_2696756, EPI_ISL_2696757, EPI_ISL_2696765, EPI_ISL_2696781, EPI_ISL_2696783, EPI_ISL_2696792, EPI_ISL_2696793, EPI_ISL_2696794, EPI_ISL_2696806, EPI_ISL_2696815, EPI_ISL_2696818, EPI_ISL_2696857, EPI_ISL_2696858, EPI_ISL_2696877, EPI_ISL_2696890 | | | |
| see above | Laboratory, Gravity Diagnostics | Laboratory, Gravity Diagnostics | Diagnostics, G. |
| EPI_ISL_4600517, EPI_ISL_4600519, EPI_ISL_4600531 | Laboratório Central de Saúde Pública de Pernambuco | Coordenação Geral de Laboratórios de Saúde Pública (CGLAB/DAEVS/SVS/MS) | Vagner Fonseca; et al. |
| EPI_ISL_2777242, EPI_ISL_2777243, EPI_ISL_2777246, EPI_ISL_2777283, EPI_ISL_2777288, EPI_ISL_2777295, EPI_ISL_2777335, EPI_ISL_2777336, EPI_ISL_2777337, EPI_ISL_2777348, EPI_ISL_2777527, EPI_ISL_2777531, EPI_ISL_2777532, EPI_ISL_2777553, EPI_ISL_2777559, EPI_ISL_2777580, EPI_ISL_2777593, EPI_ISL_2777606, EPI_ISL_2777612, EPI_ISL_2777617, EPI_ISL_2777619, EPI_ISL_2777629, EPI_ISL_2777632, EPI_ISL_2777635, EPI_ISL_2777636, EPI_ISL_2777643, EPI_ISL_2777693, EPI_ISL_2777773, EPI_ISL_2777774, EPI_ISL_2777844, EPI_ISL_2777845, EPI_ISL_2777846, EPI_ISL_2777849, EPI_ISL_2777850, EPI_ISL_2777884, EPI_ISL_2777885, EPI_ISL_2777886, EPI_ISL_2777887, EPI_ISL_2777888, EPI_ISL_2777889, EPI_ISL_2777900, EPI_ISL_2777907, EPI_ISL_2777913, EPI_ISL_2777914, EPI_ISL_2777918, EPI_ISL_2777919, EPI_ISL_2777920, EPI_ISL_2777922, EPI_ISL_2777927, EPI_ISL_2777937, EPI_ISL_2777943, EPI_ISL_2777950, EPI_ISL_2777954, EPI_ISL_2777962, EPI_ISL_2777963, EPI_ISL_2777983, EPI_ISL_2777984 | | | |
| see above | Laboratório Central de Saúde Pública do Amazonas - LACEN-AM | Laboratorio de Ecologia de Doencas Transmissiveis na Amazonia, Instituto Leonidas e Maria Deane - Fiocruz Amazonia | André Corado; Debora Duarte; Felipe Naveca; Fernanda Nascimento; George Silva; Karina Pessoa; Luciana Gonçalves; Maria Júlia Brandão; Matilde Mejía; Michele Jesus; Valdinete Nascimento; Victor Souza; Ágatha Costa |
| EPI_ISL_7260679 | Laboratório Central de Saúde Pública do Estado do Tocantins (LACEN/TO) | Bioinformatics and Biotechnology Laboratory (Labinftec/Federal University of Tocantins, Campus of Gurupi) | Fabrício Souza Campos; Jucimária Dantas Galvão; Raíssa Nunes dos Santos; Ueric José Borges de Souza |
| EPI_ISL_2466190, EPI_ISL_2466191, EPI_ISL_2466192, EPI_ISL_2466193, EPI_ISL_2466194, EPI_ISL_2466195, EPI_ISL_2466196, EPI_ISL_2466197, EPI_ISL_2466198, EPI_ISL_2466199 | | | |
| see above | Laboratório de Biologia Molecular de Doenças Infecciosas e do Câncer (LADIC - UFRN) | Laboratory of Respiratory Viruses and Measles, Oswaldo Cruz Institute, FIOCRUZ | Alice Sampaio Rocha; Ana Carolina Mendonca; Anna Carolina Paixao; Elisa Cavalcante Pereira; Fernando Motta; Josélio Araújo; Luciana Appolinario; Marilda Siqueira on behalf of the Fiocruz COVID-19 Genomic Surveillance Network; Paola Resende; Renata Serrano Lopes; Taina Venas |
| EPI_ISL_3873652, EPI_ISL_3873656, EPI_ISL_4051953, EPI_ISL_4051955 | Laboratório de Microbiologia Molecular - Universidade FEEVALE | Molecular Microbiology Laboratory | Alana Witt Hansen; Fernando Rosado Spilki; Fágner Henrique Heldt; Juliana Schons Gularte; Juliane Deise Fleck; Mariana Soares da Silva; Matheus Nunes Weber; Meriane Demoliner; Micheli Filippi; Paula Rodrigues de Almeida; Vyctoria Malayhka de Abreu Góes Pereira. |
| EPI_ISL_2629836, EPI_ISL_2629837, EPI_ISL_2629838 | Laboratório de Virologia Molecular - Universidade Federal do Rio de Janeiro | Laboratório de Virologia Molecular - Universidade Federal do Rio de Janeiro | ; Alice Laschuk Herlinger; Amilcar Tanuri; André Felipe Andrade dos Santos; Carolina Moreira Voloch; Cássia Cristina Alves Gonçalves; Diana Mariani; Débora Souza Faffe; Filipe Romero Rebello Moreira; Francine Bittencourt Schiffler; Isabela de Carvalho Leitão; Marcelo Calado de Paula Tôrres; Matheus Augusto Calvano Cosentino; Mirela D’arc; Orlando da Costa Ferreira Junior; Rafael Mello Galliez; Raíssa Mirella dos Santos Cunha da Costa; Renato Santana de Aguiar; Terezinha Marta Pereira Pinto Castineiras; Thamiris dos Santos Miranda; Átila Duque Rossi |
| EPI_ISL_4520592 | Laboratório de Virologia e Cultivo Celular - LABVICC | Laboratório Baculovírus | Aline Belmok; Bergmann Morais Ribeiro; Clayton Luiz Borges; Déborah Anjos; Elen Regozino Muniz; Fabiola Souza Fiaccadori; Fernando L Melo; Gabriela Silvério Bazilio; Juliana Alves Parente Rocha; Keila Correia de Alcântara; Menira Souza; Nathânia Dábilla; Valéria Christina de Rezende Feres |
| EPI_ISL_2536243, EPI_ISL_2536246, EPI_ISL_2536248, EPI_ISL_2536250, EPI_ISL_2536253, EPI_ISL_2536258, EPI_ISL_2536260, EPI_ISL_3061899 | | | |
| see above | Labortorio Central de Saude Publica do Estado de Santa Catarina (LACEN/SC) | Laboratory of Respiratory Viruses and Measles, Oswaldo Cruz Institute, FIOCRUZ | Alice Sampaio Rocha; Ana Carolina Mendonca; Anna Carolina Paixao; Darcita Buerger Rovaris; Elisa Cavalcante Pereira; Fernando Motta; Luciana Appolinario; Marilda Siqueira on behalf of the Fiocruz COVID-19 Genomic Surveillance Network; Paola Resende; Renata Serrano Lopes; Sandra Bianchini Fernandes; Taina Venas |
| EPI_ISL_2466230, EPI_ISL_2536275, EPI_ISL_2536276, EPI_ISL_2536299, EPI_ISL_2536300, EPI_ISL_2536301, EPI_ISL_2536302 | | | |
| see above | Labortorio Central de Saude Publica do Estado do Rio de Janeiro (LACEN/RJ) | Laboratory of Respiratory Viruses and Measles, Oswaldo Cruz Institute, FIOCRUZ | Alice Sampaio Rocha; Ana Carolina Mendonca; Andrea Cony Cavalcanti; Anna Carolina Paixao; Elisa Cavalcante Pereira; Fernando Motta; Luciana Appolinario; Marilda Siqueira on behalf of the Fiocruz COVID-19 Genomic Surveillance Network; Paola Resende; Renata Serrano Lopes; Taina Venas |
| EPI_ISL_2314852 | Landesgesundheitsamt Baden-Württemberg | Robert Koch Institute | |
| EPI_ISL_2557359, EPI_ISL_2557360, EPI_ISL_2557361, EPI_ISL_2557366, EPI_ISL_2557367, EPI_ISL_2557368, EPI_ISL_2557369, EPI_ISL_2557370, EPI_ISL_2557372, EPI_ISL_2557374, EPI_ISL_2603451, EPI_ISL_2603454, EPI_ISL_2603455, EPI_ISL_2645440, EPI_ISL_2645441, EPI_ISL_2645442, EPI_ISL_2645443, EPI_ISL_2863682, EPI_ISL_2863725, EPI_ISL_2863795, EPI_ISL_2863887, EPI_ISL_2863888 | | | |
| see above | Lboratorio Central de Saude Publica do Estado do Parana (LACEN/PR) | Laboratory of Respiratory Viruses and Measles, Oswaldo Cruz Institute, FIOCRUZ | Agatha Cristinne Prudencio; Agatha Cristinne Prudencio Soares; Alice Sampaio Rocha; Ana Carolina Mendonca; Anna Carolina Paixao; Elisa Cavalcante Pereira; Fernando Motta; Ighor Leonardo Arantes Gomes; Irina Riediger; Luciana Appolinario; Marilda Siqueira on behalf of the Fiocruz COVID-19 Genomic Surveillance Network; Paola Resende; Renata Serrano Lopes; Taina Venas |
| EPI_ISL_2692141 | Lifescan | RIPHL at Rush University Medical Center | Chicago Department of Public Health; Felix Araujo Perez; Kevin Kunstman; Laura Furtado; Marieta Hyde; Max Kolton; Stefan Green |
| EPI_ISL_2119734 | Lighthouse Lab in Alderley Park | Wellcome Sanger Institute for the COVID-19 Genomics UK (COG-UK) Consortium | Cordelia Langford; David K. Jackson; Dominic Kwiatkowski; Ewan Harrison; Ian Johnston; Jacquelyn Wynn; Jeffrey Barrett; John Sillitoe on behalf of the Wellcome Sanger Institute COVID-19 Surveillance Team; Mairead Hyland; Roberto Amato; Sonia Goncalves; The Lighthouse Lab in Alderley Park and Alex Alderton |
| EPI_ISL_2004797, EPI_ISL_2118938, EPI_ISL_2119450, EPI_ISL_2119812, EPI_ISL_2121617, EPI_ISL_2121861 | Lighthouse Lab in Milton Keynes | Wellcome Sanger Institute for the COVID-19 Genomics UK (COG-UK) Consortium | Cordelia Langford; David K. Jackson; Dominic Kwiatkowski; Ewan Harrison; Ian Johnston; Jeffrey Barrett; John Sillitoe on behalf of the Wellcome Sanger Institute COVID-19 Surveillance Team; Roberto Amato; Sonia Goncalves; The Lighthouse Lab in Milton Keynes and Alex Alderton |
| EPI_ISL_2120565 | Limbach - MVZ Labor Ludwigsburg | Robert Koch Institute | |
| EPI_ISL_2612550 | Lincoln Diagnostics | New York City Public Health Laboratory | Jade Wang; et al. |
| EPI_ISL_2373094 | Lurie Children's Hospital of Chicago | Northwestern University - Ozer Lab | Egon A. Ozer; Judd F. Hultquist; Lacy M. Simons; Larry K. Kociolek; Michael G. Ison; Ramon Lorenzo-Redondo; Taylor J. Dean; William J. Muller; Xiaotian; Zheng |
| EPI_ISL_2249573 | M Health Fairview | Minnesota Department of Health, Public Health Laboratory | Alexandra Lorentz; Jacob Garfin; Matt Plumb; and Xiong Wang |
| EPI_ISL_2451813, EPI_ISL_2451821 | MA State Public Health Laboratory | Centers for Disease Control and Prevention Division of Viral Diseases, Pathogen Discovery | Alison Laufer Halpin; Ben L. Rambo-Martin; Clinton R. Paden; Dakota Howard; Darlene Wagner; Dave Wentworth; Dhwani Batra; Jasmine Padilla; Justin Lee; Katie Dillon; Krista Queen; Kristen Knipe; Kristine Lacek; Mark Burroughs; Matthew Schmerer; Mili Sheth; Peter Cook; Sam Shepard; Sarah Nobles; Shoshona Le; Suxiang Tong; Vivien Dugan; Yvette Unoarumhi |
| EPI_ISL_2801332 | MATERNIDADE ESCOLA ASSIS CHATEAUBRIAND | Analytical Competence Molecular Epidemiology Lab/ACME, Oswaldo Cruz Foundation, Ceara (FIOCRUZ CE) | Cleber Furtado Aksenen e Suzana Porto Almeida; Fabio Miyajima; Fernando Braga Stehling; Francisco Eder de Moura Lopes; Jamille Maria Mendes Bezerra; Joaquim César do Nascimento Sousa Junior; Pedro Miguel Carneiro Jeronimo; Thais Ferreira de Oliveira; Thais de Oliveira Costa; Ticiane Cavalcante de Souza; Veridiana Pessoa Miyajima |
| EPI_ISL_3356779, EPI_ISL_3356780, EPI_ISL_3356781, EPI_ISL_3356793, EPI_ISL_3356796, EPI_ISL_3356798, EPI_ISL_3356800, EPI_ISL_3356801, EPI_ISL_3356803, EPI_ISL_3356805, EPI_ISL_3356806 | | | |
| see above | MB-Cadham Provincial laboratory | Manitoba Cadham Provincial Laboratory | Anna Majer; Anneliese Landgraff; CanCOGeN's metadata curation team; Darian Hole; David Alexander; Elsie Grudeski; Gary Van Domselaar; Grace Seo; Jared Bullard; Jennifer Tanner; Kerry Dust; Kirsten Biggar; Madison Chapel; Morag Graham; Natalie Knox; Nathalie Bastien; Paul Van Caeseele; Philip Mabon; Public Health Agency of Canada CanCOGeN team; Rhiannon Huzarewich; Russell Mandes; Shari Tyson; Timothy Booth; Yan Li |
| EPI_ISL_2495420, EPI_ISL_2495421, EPI_ISL_2495428, EPI_ISL_2495611, EPI_ISL_2495612, EPI_ISL_2495613, EPI_ISL_2495614, EPI_ISL_3356772, EPI_ISL_3356773 | | | |
| see above | MB-Cadham Provincial laboratory | National Microbiology Laboratory (NML) | Anna Majer; Anneliese Landgraff; CanCOGeN's metadata curation team; Darian Hole; David Alexander; Elsie Grudeski; Gary Van Domselaar; Grace Seo; Jared Bullard; Jennifer Tanner; Kerry Dust; Kirsten Biggar; Madison Chapel; Morag Graham; Natalie Knox; Nathalie Bastien; Paul Van Caeseele; Philip Mabon; Public Health Agency of Canada CanCOGeN team; Rhiannon Huzarewich; Russell Mandes; Shari Tyson; Timothy Booth; Yan Li |
| EPI_ISL_2112740, EPI_ISL_2314672 | MDI Limbach Berlin GmbH; MVZ Labor Berlin | Robert Koch Institute | |
| EPI_ISL_2286824, EPI_ISL_2454192 | MEPHI, Aix Marseille University | MEPHI, Aix Marseille University | Anthony LEVASSEUR |
| EPI_ISL_2229261 | MO State Public Health Laboratory | Centers for Disease Control and Prevention Division of Viral Diseases, Pathogen Discovery | Alison Laufer Halpin; Ben L. Rambo-Martin; Clinton R. Paden; Dakota Howard; Darlene Wagner; Dave Wentworth; Dhwani Batra; Jasmine Padilla; Justin Lee; Katie Dillon; Krista Queen; Kristen Knipe; Kristine Lacek; Mark Burroughs; Matthew Schmerer; Mili Sheth; Peter Cook; Sam Shepard; Sarah Nobles; Shoshona Le; Suxiang Tong; Vivien Dugan; Yvette Unoarumhi |
| EPI_ISL_2444210 | MONTE APRAZIVEL | Instituto Butantan / FAMERP | Antonio Jorge Martins; Claudia Renata dos Santos Barros; David Schlesinger; Debora Botequio Moretti; Dimas Tadeu Covas; Elaine Cristina Marqueze; Elaine Vieira Santos; Evandra Strazza Rodrigues; Heidge Fukumasu; Jayme Augusto de Souza-Neto; José Salvatore Leister Patané; Luiz Alcantara; Luiz Lehmann Coutinho; Maria Carolina Elias; Maurício Lacerda Nogueira; Rafael dos Santos Bezerra; Raul Machado Neto; Rejane Maria Tommasini Grotto; Ricardo Haddad; Sandra Coccuzzo Sampaio Vessoni; Simone Kashima; Svetoslav Nanev Slavov; Vincent Louis Viala |
| EPI_ISL_6492650, EPI_ISL_6492651, EPI_ISL_6492677 | MSHS Clinical Microbiology Laboratories | MSHS Pathogen Surveillance Program | Adolfo García-Sastre; Adriana van de Guchte; Ajay Obla; Alberto Paniz-Mondolfi; Ana S. Gonzalez-Reiche; Angela Amoako; Ashley Salimbangon; Betsaida Salom Melo; Bremy Alburquerque; Brianne Ciferri; Charles Gleason; Daniel Floda; Deena R. Altman; Denise Jurczyszak; Emilia Mia Sordillo; Gintaras Deikus; Giulio Kleiner; Gopi Patel; Hala Alshammary; Harm van Bakel; Irina Oussenko; Jayeeta Dutta; Juan Soto; Julia Matthews; Katherine Beach; Kathryn Twyman; Kayla Russo; Komal Srivastava; Levy Sominsky; Mahmoud Awawda; Marta Luksza; Matthew M. Hernandez; Melissa Gitman; Michael D. Nowak; Mitchell J. Sullivan; Nancy Francoeur; Robert Sebra; Sarah Schaefer; Shelcie Fabre; Shwetha Hara Sridhar; Viviana Simon; Ying-Chih Wang; Zenab Khan |
| EPI_ISL_2690695 | MSK Microbiology Lab | MSK Microbiology Lab | Esther Babady; Krupa Jani; Tracy McMillen |
| EPI_ISL_2264097, EPI_ISL_2264138 | MVZ Labor Dr. Fenner und Kollegen (Standort Hamburg) | Robert Koch Institute | |
| EPI_ISL_2259678, EPI_ISL_2259680, EPI_ISL_2259682, EPI_ISL_2259683, EPI_ISL_2259684, EPI_ISL_2259685, EPI_ISL_2259689, EPI_ISL_2259690, EPI_ISL_2259691, EPI_ISL_2259693, EPI_ISL_2259695, EPI_ISL_2313170, EPI_ISL_2313188 | | | |
| see above | MVZ Labor Dr. Limbach & Kollegen GbR | Robert Koch Institute | |
| EPI_ISL_2128958 | MVZ fur Laboratoriumsmedizin und | Robert Koch Institute | |

| | | | |
|---|---|---|---|
| | Mikrobiologie Koblenz-Mittelrhein (Labor Koblenz) | | |
| EPI_ISL_2260045, EPI_ISL_2264722 | MVZ für Laboratoriumsmedizin und Mikrobiologie Koblenz-Mittelrhein (Labor Koblenz) | Robert Koch Institute | |
| EPI_ISL_2245511, EPI_ISL_2245512, EPI_ISL_2245527, EPI_ISL_2802520, EPI_ISL_2802525, EPI_ISL_2802526, EPI_ISL_2802527, EPI_ISL_2802528, EPI_ISL_2802529, EPI_ISL_2802530, EPI_ISL_2802531, EPI_ISL_2802532, EPI_ISL_4083816, EPI_ISL_6532618, EPI_ISL_6532620, EPI_ISL_6532627, EPI_ISL_6532629, EPI_ISL_6532631, EPI_ISL_6532633, EPI_ISL_6532662, EPI_ISL_6532666 | | | |
| see above | Maine Health and Environmental Testing Laboratory | Tewhey Lab, The Jackson Laboratory | Barter, M.; Dewey, H.; H. and Tewhey, R.; Iosue, F.; Lynch, R.; Matluk, N.; Munger |
| EPI_ISL_2284996, EPI_ISL_2285048, EPI_ISL_2285093, EPI_ISL_2285103, EPI_ISL_2307182, EPI_ISL_2307184, EPI_ISL_2307204, EPI_ISL_2307272, EPI_ISL_2307289, EPI_ISL_2488894, EPI_ISL_2488959, EPI_ISL_2488979, EPI_ISL_2489020, EPI_ISL_2489032, EPI_ISL_2489040, EPI_ISL_2489042, EPI_ISL_2646386, EPI_ISL_2646418, EPI_ISL_2646421, EPI_ISL_2646444, EPI_ISL_2646447, EPI_ISL_2646459, EPI_ISL_2646463, EPI_ISL_2646464, EPI_ISL_2646474, EPI_ISL_2646505, EPI_ISL_2646507, EPI_ISL_2646512, EPI_ISL_2646516, EPI_ISL_2646523, EPI_ISL_4347951 | | | |
| see above | Mako Medical | Centers for Disease Control and Prevention Division of Viral Diseases, Pathogen Discovery | Adrian Paskey; Benjamin Rambo-Martin; Christopher Gulvick; Clinton Paden; Clinton R. Paden; Dakota Howard; Darlene Wagner; Dhwani Batra; Duncan MacCannell; Erisa Sula; Jason Caravas; Kara Moser; Kristine Lacek; Lauren Moon; Matthew Schmerer; Matthew Tugwell; Peter Cook; Peter W. Cook; Scott Sammons; Shatavia Morrison; Tymeckia Kendall; Victoria Caban Figueroa; Yvette Unoarumhi |
| EPI_ISL_2464619, EPI_ISL_2501758, EPI_ISL_3012064, EPI_ISL_3012073 | Maryland Genomics, Institute for Genome Sciences, University of Maryland School of Medicine | Maryland Genomics, Institute for Genome Sciences, University of Maryland School of Medicine | Aditya; Claire M; Fraser; Hazen; Holly; Humphrys; Ivette; Jacques; Jonathan; Kranthi; Lim; Lisa D; Luke J; Mehta; Mike; Ott; Ravel; Roussey; Sadzewicz; Sandra; Santana-Cruz; Tallon; Tracy; Vavikolanu |
| EPI_ISL_6666506, EPI_ISL_6666534, EPI_ISL_6666552, EPI_ISL_6680576, EPI_ISL_6680590, EPI_ISL_6680605, EPI_ISL_6680640, EPI_ISL_6680648, EPI_ISL_6680661, EPI_ISL_6680683, EPI_ISL_6680707, EPI_ISL_6680721, EPI_ISL_6680744, EPI_ISL_6680750, EPI_ISL_6680754, EPI_ISL_6680819, EPI_ISL_6680825, EPI_ISL_6680856, EPI_ISL_6680877, EPI_ISL_6680879, EPI_ISL_6680904, EPI_ISL_6680932, EPI_ISL_6680938, EPI_ISL_6680948, EPI_ISL_6680964, EPI_ISL_6680978, EPI_ISL_6680988, EPI_ISL_6681000, EPI_ISL_6681022, EPI_ISL_6681026, EPI_ISL_6681030, EPI_ISL_6681035, EPI_ISL_6681036 | | | |
| see above | Mass General Brigham | Mass General Brigham | A.E.; Adams, G.; Anahtar, M.; B.L.; B.W.; Bauer, M.; Birren; Branda, J.; Carter, A.; Cerrato, F.; Chaluvadi, S.; Chapman; Cusick, C.; D.J.; DeRuff, K.; E. and Sabeti; Flowers, K.; Gallagher, G.; Gladden-Young, A.; Gnirke, A.; Harris, J.; J.E.; K.J.; LaRocque, R.; Lagerborg, K.; Lemieux; Lin; Loreth, C.; MacInnis; Neumann, A.; Normandin, E.; P.C.; Park; Pierce, V.; Reilly, S.; Rosenberg; Rudy, M.; Ryan, E.; S.B.; Shaw, B.; Siddle; Slater, D.; Smole, S.; Tomkins-Tinch, C.; Turbett, S. |
| EPI_ISL_2229481, EPI_ISL_2229490, EPI_ISL_2229799, EPI_ISL_2483774, EPI_ISL_2483799, EPI_ISL_2483806, EPI_ISL_2483810 | | | |
| see above | Massachusetts State Public Health Laboratory | Massachusetts State Public Health Laboratory | Andrew Lang; Glen Gallagher; Sandra Smole; Timelia Fink |
| EPI_ISL_2450316 | Max von Pettenkofer Institute, Virology, National Reference Center for Retroviruses, LMU Munich | Laboratory for Functional Genome Analysis; Dept. Genomics; Gene Center of the LMU Munich | Alexander Graf; Helmut Blum; Max Muenchhoff; Oliver Keppler; Stefan Krebs |
| EPI_ISL_2140625 | Microbiology Department, Laboratori Clínic Metropolitana Nord. Hospital Universitari Germans Trias i Pujol | Can Ruti SARS-CoV-2 Sequencing Hub (HUGTiP/IrsiCaixa/IGTP) | Alba Sánchez; Anna Not; Antoni E Bordoy; Bonaventura Clotet; Cristina Casañ; Cristina Esteban; Francesc Catala-Moll; Gemma Clara; Ignacio Blanco; Marc Noguera-Julian; Maria Casadellà; Mariona Parera; Mercedes Guerrero; Montserrat Giménez; Pere-Joan Cardona; Pilar Armengol; Roger Paredes; Verónica Saludes; and Elisa Martró on behalf of the Can Ruti SARS-CoV-2 Sequencing Hub. |
| EPI_ISL_2380037 | Microbiology Department, Laboratori Clínic Metropolitana Nord. Hospital Universitari Germans Trias i Pujol. | Can Ruti SARS-CoV-2 Sequencing Hub (HUGTiP/IrsiCaixa/IGTP) | Anna Not; Antoni E Bordoy; Bonaventura Clotet; Cristina Casañ; Cristina Esteban; Francesc Catala-Moll; Gemma Clara; Ignacio Blanco; Marc Noguera-Julian; Maria Casadellà; Mercedes Guerrero; Montserrat Giménez; Pere-Joan Cardona; Pilar Armengol; Roger Pared |
| EPI_ISL_2036092 | Microbiology Department, University Hospital Donostia | Microbiology Department, University Hospital Donostia | Cilla G.; Gomez M; Marimon JM; Montes M; Piñeiro L; Sorarrain A |
| EPI_ISL_2285139, EPI_ISL_2967838, EPI_ISL_2967854 | Microbiology Department. Complexo Hospitalario Universitario de Vigo | Microbiology Department. Complexo Hospitalario Universitario de Vigo | Alfaya N; Alonso I; Alvarez M; Cabrera JJ; Carballo R; Cores O; Cortizo S; Davina C; Martinez L; Mediero G; Perez S; Potel C; Regueiro B; Rey S; Vasallo FJ; del-Campo V |
| EPI_ISL_2313038 | Microbiology Division, SC DHEC | Microbiology Division, SC DHEC | Flores, H.; Freeman, J. |
| EPI_ISL_2259151, EPI_ISL_2259160, EPI_ISL_2259191, EPI_ISL_2259192, EPI_ISL_2259193 | Microvida | Microvida | Jaco Verweij; Joep Stohr; Suzan D. Pas |
| EPI_ISL_2092803, EPI_ISL_2136384, EPI_ISL_2136395, EPI_ISL_2136397, EPI_ISL_2136426, EPI_ISL_2249519, EPI_ISL_2658537 | | | |
| see above | Minnesota Department of Health, Public Health Laboratory | Minnesota Department of Health, Public Health Laboratory | Alexandra Lorentz; Jacob Garfin; Matt Plumb; and Xiong Wang |
| EPI_ISL_2304242, EPI_ISL_2304250, EPI_ISL_2304253, EPI_ISL_2304254, EPI_ISL_2304260, EPI_ISL_2304268 | Missouri State Public Health Laboratory | Missouri State Public Health Laboratory | Ashley New; Joshua Barry; Matthew Sinn |
| EPI_ISL_2107812, EPI_ISL_2107827 | Molecular Diagnostics Pathology Department Mater Dei Hospital Malta | Molecular Diagnostics Pathology Department Mater Dei Hospital Malta | C Cilia; G Zahra; L Grech; R Borg |
| EPI_ISL_2646995 | Montana Public Health Laboratory | Montana Public Health Laboratory | Carrie Biskupiak; Deborah Gibson; Joy Ritter; Michael Dills; Michelle Mozer |
| EPI_ISL_3354969 | NH Dept. of Health and Human Services Public Health Labs | Centers for Disease Control and Prevention Division of Viral Diseases, Pathogen Discovery | Alex Burgin; Ben Rambo-Martin; Clinton Paden; Dakota Howard; Dave Wentworth; Dhwani Batra; Jasmine Padilla; Justin Lee; Krista Queen; Kristen Knipe; Kristine Lacek; Mark Burroughs; Matthew Schmerer; Meghan Bentz; Mili Sheth; Peter Cook; Sam Shepard; Sarah Nobles; Suxiang Tong; Vivien Dugan; Yvette Unoarumhi |
| EPI_ISL_2185595, EPI_ISL_2185610, EPI_ISL_2376137 | NJDOH, Public Health and Environmental Laboratories | NJ_PHEL | Allison Roder; Byeong Jeong; Dana Woell; Jacquelyn Deverell; Lindsey Bodnar; Matthew Scarnati; Shiv K. Verma |
| EPI_ISL_4061019, EPI_ISL_4061020, EPI_ISL_4061021, EPI_ISL_4061023, EPI_ISL_4061026, EPI_ISL_4061028 | NL-Dr. Leonard A. Miller Centre for Health Services | Newfoundland and Labrador - Eastern Health | Adel Malek; Anna Majer; Anneliese Landgraff; Elsie Grudeski; Gary Van Domselaar; George Zahariad; Grace Seo; Jennifer Tanner; Laura Gilbert; Morag Graham; Natalie Knox; Philip Mabon; Rhiannon Huzarewich; Robert Needle; Russell Mandes; Shari Tyson; Yang Yu |
| EPI_ISL_2345800 | NUCLEO DE SAUDE MARY DOTA DE BAURU | Instituto Butantan / Mendelics | Antonio Jorge Martins; Claudia Renata dos Santos Barros; David Schlesinger; Debora Botequio Moretti; Dimas Tadeu Covas; Elaine Cristina Marqueze; Elaine Vieira Santos; Evandra Strazza Rodrigues; Heidge Fukumasu; Jayme Augusto de Souza-Neto; José Salvatore Leister Patané; Luiz Alcantara; Luiz Lehmann Coutinho; Maria Carolina Elias; Maurício Lacerda Nogueira; Rafael dos Santos Bezerra; Raul Machado Neto; Rejane Maria Tommasini Grotto; Ricardo Haddad; Sandra Coccuzzo Sampaio Vessoni; Simone Kashima; Svetoslav Nanev Slavov; Vincent Louis Viala |
| EPI_ISL_2161747, EPI_ISL_2427406, EPI_ISL_2427408 | NYU Langone Health | Departments of Pathology and Medicine, New York University School of Medicine | Adriana Heguy; Christian Marier; Dacia Dimartino; Emily Guzman; Gael Westby; Guiqing Wang; Paolo Cotzia; Paul Zappile; Peter Meyn; Sitharam Ramaswami; Yutong Zhang |
| EPI_ISL_2562076, EPI_ISL_2562077, EPI_ISL_2562078, EPI_ISL_2562080 | National Institute of Public Health | State Veterinary Institute Prague | A; D; H; J; Jirincova; Nagy; Suri; T; Trnka; Vecerova |
| EPI_ISL_1971494, EPI_ISL_2131808, EPI_ISL_2131809, EPI_ISL_2131811, EPI_ISL_2132217, EPI_ISL_2132227 | National Platform bis UMONS/Jolimont | National Platform bis UMONS/Jolimont | Florian Juszczak; François Dufrasne; Gautier Detry; Guillaume Bayon-Vicente; Ruddy Wattiez |
| EPI_ISL_2400270, EPI_ISL_2400274 | Nebraska Public Health Laboratory | NPHL COVID-19 Response Team | NPHL COVID-19 Response Team |
| EPI_ISL_2381909 | New Mexico Department of Health Scientific Laboratory | New Mexico Department of Health Scientific Laboratory | Anastacia Griego-Fisher; D'eldra Malone; Ellie Johnson; Jennifer Benoit; Keila Gutierrez; Linda Salazar; Ratheesh Rajan |
| EPI_ISL_2239608 | Northumbria University / South Tees Hospitals NHS Foundation Trust / North Cumbria Integrated Care NHS Foundation Trust / North Tees and Hartlepool NHS Foundation Trust / Newcastle Hospitals NHS Foundation Trust | COVID-19 Genomics UK (COG-UK) Consortium | Andrew Nelson; Brendan Payne; Clive Graham; Darren L Smith; Debra Padgett; Edward Barton; Emma Swindells; Garren Scott; Gary Black; Gary Eltringham; Giles S Holt; Greg R Young; Jane Greenaway; Jennifer Collins; John Allan; Joshua Loh; Lynn Dover; Matthew Bashton; Mohammad A Tariq; Paul Baker; Sarah Essex; Steve Liggett; Wen C Yew; Yusri Taha |
| EPI_ISL_2490191, EPI_ISL_2490193, EPI_ISL_2490195, EPI_ISL_2490208, EPI_ISL_2490214, EPI_ISL_2490219, EPI_ISL_2490224, EPI_ISL_2490239, EPI_ISL_2490240, EPI_ISL_2490251, EPI_ISL_2536556 | | | |

| | | | |
|---|---|---|---|
| see above | Northwest Laboratory | Seattle Flu Study | Amanda Adler; Barry R. Lutz; Benjamin Pelle; Brian Hiatt; Caitlin R. Wolf; Chris D. Frazar; Deborah A. Nickerson; Elisabeth Brandstetter; Erica Ryke; Geoff Melly; Helen Y. Chu; Janet A. Englund; Jay Shendure; Jover Lee; Kairsten Fay; Kirsten Lacombe; Lea M. Starita; Mark J. Rieder; Matthew Richardson; Matthew Thompson; Melissa Truong; Michael Boeckh; Michael Famulare; Misja Ilcisin; Peter D. Han; Philip Dykema; Romesh Gautom; Scott Lindquist; Thomas R. Sibley; Trevor Bedford |
| EPI_ISL_2372974, EPI_ISL_2372978, EPI_ISL_2372979, EPI_ISL_2372981, EPI_ISL_2372982, EPI_ISL_2372984, EPI_ISL_2372992, EPI_ISL_2372993 | | | |
| see above | Northwestern Memorial Hospital | Northwestern University - Ozer Lab | Chad J. Achenbach; Chao Qi; Egon A. Ozer; Judd F. Hultquist; Lacy M. Simons; Lawrence J. Jennings; Michael G. Ison; Ramon Lorenzo-Redondo; Taylor J. Dean |
| EPI_ISL_2692045 | Northwestern Memorial Hospital | RIPHL at Rush University Medical Center | Amber Kimble; Chao Qi; Felix Araujo Perez; Kevin Kunstman; Laura Furtado; Marieta Hyde; Max Kolton; Stefan Green |
| EPI_ISL_5801535 | Nucleo De Saude Mary Dota De Bauru | Instituto Butantan | Antonio Jorge Martins; Claudia Renata dos Santos Barros; David Schlesinger; Debora Botequio Moretti; Dimas Tadeu Covas; Elaine Cristina Marqueze; Elaine Vieira Santos; Evandra Strazza Rodrigues; Heidge Fukumasu; Jayme Augusto de Souza-Neto; José Salvatore Leister Patané; Luiz Alcantara; Luiz Lehmann Coutinho; Maria Carolina Elias; Maurício Lacerda Nogueira; Rafael dos Santos Bezerra; Raul Machado Neto; Rejane Maria Tommasini Grotto; Ricardo Haddad; Sandra Coccuzzo Sampaio Vessoni; Simone Kashima; Svetoslav Nanev Slavov; Vincent Louis Viala |
| EPI_ISL_2462148, EPI_ISL_2462223, EPI_ISL_2462230, EPI_ISL_2462231, EPI_ISL_2462232, EPI_ISL_2462241, EPI_ISL_2462242, EPI_ISL_2663642, EPI_ISL_2663643, EPI_ISL_2663648, EPI_ISL_2663651, EPI_ISL_2663652, EPI_ISL_2663654, EPI_ISL_2663669, EPI_ISL_2663671, EPI_ISL_2663672, EPI_ISL_2663679, EPI_ISL_2663687, EPI_ISL_2663691, EPI_ISL_2663703, EPI_ISL_2663706, EPI_ISL_2663707, EPI_ISL_2663715, EPI_ISL_2663716, EPI_ISL_2663722, EPI_ISL_2663723 | | | |
| see above | OHSU Lab Services Molecular Microbiology Lab | Oregon SARS-CoV-2 Genome Sequencing Center | Alec J. Hirsch; Andrew C. Adey; Benjamin N. Bimber; Brendan L. O'Connell; Brian J. O'Roak; Cierra LaBlanc; Daniel N. Streblow; Donna Hansel; Guang Fan; Ruth V. Nichols; Sonia Acharya; William B. Messer; Xuan Qin |
| EPI_ISL_3065608, EPI_ISL_3065609, EPI_ISL_3065612, EPI_ISL_3065628, EPI_ISL_3065637, EPI_ISL_3065639, EPI_ISL_3065644, EPI_ISL_3065648, EPI_ISL_3065661, EPI_ISL_3065665, EPI_ISL_3065668, EPI_ISL_3065669, EPI_ISL_3065672, EPI_ISL_3065674, EPI_ISL_3065676, EPI_ISL_3065679, EPI_ISL_3065680, EPI_ISL_3065683, EPI_ISL_3065686, EPI_ISL_3065689, EPI_ISL_3065699, EPI_ISL_3065702, EPI_ISL_3065704, EPI_ISL_3065710, EPI_ISL_3065713, EPI_ISL_3065716, EPI_ISL_3065720, EPI_ISL_3065722, EPI_ISL_3065724, EPI_ISL_3065725, EPI_ISL_3065728, EPI_ISL_3065731, EPI_ISL_3065799, EPI_ISL_3065803 | | | |
| see above | OHSU-MM Lab | OHSU-MM Lab | Rebecca O'Gara |
| EPI_ISL_2311773 | OK Public Health Laboratory, Oklahoma State DOH | Genomics and Discovery, Respiratory Viruses Branch, Division of Viral Diseases, Centers for Disease Control and Prevention | Adam Retchless; Anna Kelleher; Anna Uehara; Brian Lynch; Clinton R. Paden; Dhwani Batra; Haibin Wang; Han Jia Justin Ng; Jasmine Padilla; Jing Zhang; Justin Lee; Krista Queen; Mark Burroughs; Mili Sheth; Morgan Davis; Peter Cook; Rachel Marine; Sarah Nobles; Suxiang Tong; Tara Coalter; Yan Li; Ying Tao |
| EPI_ISL_2131270, EPI_ISL_2131272, EPI_ISL_2131276, EPI_ISL_2131283 | OLVZ Aalst | OLVZ Aalst | Astrid Holderbeke |
| EPI_ISL_2399429 | OR State PHL-Virology/Immunology Section | Centers for Disease Control and Prevention Division of Viral Diseases, Pathogen Discovery | Alison Laufer Halpin; Ben L. Rambo-Martin; Clinton R. Paden; Dakota Howard; Darlene Wagner; Dave Wentworth; Dhwani Batra; Jasmine Padilla; Justin Lee; Katie Dillon; Krista Queen; Kristen Knipe; Kristine Lacek; Mark Burroughs; Matthew Schmerer; Mili Sheth; Peter Cook; Sam Shepard; Sarah Nobles; Shoshona Le; Suxiang Tong; Vivien Dugan; Yvette Unoarumhi |
| EPI_ISL_2753961, EPI_ISL_2753965, EPI_ISL_2754855, EPI_ISL_2755100, EPI_ISL_2756219, EPI_ISL_2768100 | Ohio Department of Health Laboratory | Ohio Department of Health Laboratory | Allison Black; Brent Lee; Caitlin McDonnell; Eric Brandt; Erica Leasure; Glen McGillivary; Heather Blankenship; Holmes; Jade Mowery; Jennifer; Kelsey Florek; Keoni Omura; Kirtana Ramadugu; Quanta Brown; Stephanie Mccracken; Tyler Payne; and Tammy Bannerman |
| EPI_ISL_2500059, EPI_ISL_2500104 | Orange County Public Health Lab | Chan-Zuckerberg Biohub | CZB Cliahub Consortium |
| EPI_ISL_2023471, EPI_ISL_2023472, EPI_ISL_2023473, EPI_ISL_2023477, EPI_ISL_2081522, EPI_ISL_2203066, EPI_ISL_2203070, EPI_ISL_2203071, EPI_ISL_2203095, EPI_ISL_2339326, EPI_ISL_2348927, EPI_ISL_2404748, EPI_ISL_2473671 | | | |
| see above | Oregon State Public Health Laboratory | Oregon State Public Health Laboratory | Eugene Yeboah; John Fontana and Shane Sevey; Laura Tsaknaridis; Rafia Razzaque; Vanda Makris |
| EPI_ISL_2375465 | Ospedale Sandro Pertini ASL Roma2 | INMI Lazzaro Spallanzani IRCCS | A Di Caro; B Bartolini; CEM Gruber; E Giombini; F Messina; F Santini; G Bonfiglio; M Rueca; MR Capobianchi; O Butera |
| EPI_ISL_3120101 | Ospedale Valtellina Valchiavenna | Laboratory of Clinical Microbiology, Virology and Bioemergencies, ASST Fatebenefratelli Sacco - Sacco Hospital | Valeria Micheli |
| EPI_ISL_2450374 | Ospedale di Genzano - ASL RM 6 | INMI Lazzaro Spallanzani IRCCS | A Di Caro; B Bartolini; CEM Gruber; E Conti; E Giombini; F Messina; F Santini; G Bonfiglio; G Tramini; M Rueca; MR Capobianchi; O Butera |
| EPI_ISL_2293376, EPI_ISL_2293377, EPI_ISL_2293378, EPI_ISL_2293379, EPI_ISL_2293381, EPI_ISL_2293382 | Outre mer | National Reference Center for Viruses of Respiratory Infections, Institut Pasteur, Paris | Angela Brisebarre; Camille Capel; Christophe Malabat; Corinne Maufrais; Dominique Rousset; Etienne Simon-Lorière; Frédéric Lemoine; Louise Lefrançois; Marion Barbet; Maud Vanpeene; Méline Bizard; Pierre Lechat; Sylvie Behillil; Sylvie Van der Werf; Vincent Enouf |
| EPI_ISL_2345714 | PRONTO ATENDIMENTO MUNICIPAL | Instituto Butantan / Mendelics | Antonio Jorge Martins; Claudia Renata dos Santos Barros; David Schlesinger; Debora Botequio Moretti; Dimas Tadeu Covas; Elaine Cristina Marqueze; Elaine Vieira Santos; Evandra Strazza Rodrigues; Heidge Fukumasu; Jayme Augusto de Souza-Neto; José Salvatore Leister Patané; Luiz Alcantara; Luiz Lehmann Coutinho; Maria Carolina Elias; Maurício Lacerda Nogueira; Rafael dos Santos Bezerra; Raul Machado Neto; Rejane Maria Tommasini Grotto; Ricardo Haddad; Sandra Coccuzzo Sampaio Vessoni; Simone Kashima; Svetoslav Nanev Slavov; Vincent Louis Viala |
| EPI_ISL_2345729 | PRONTO SOCORRO MUNICIPAL TAMBAU | Instituto Butantan / Mendelics | Antonio Jorge Martins; Claudia Renata dos Santos Barros; David Schlesinger; Debora Botequio Moretti; Dimas Tadeu Covas; Elaine Cristina Marqueze; Elaine Vieira Santos; Evandra Strazza Rodrigues; Heidge Fukumasu; Jayme Augusto de Souza-Neto; José Salvatore Leister Patané; Luiz Alcantara; Luiz Lehmann Coutinho; Maria Carolina Elias; Maurício Lacerda Nogueira; Rafael dos Santos Bezerra; Raul Machado Neto; Rejane Maria Tommasini Grotto; Ricardo Haddad; Sandra Coccuzzo Sampaio Vessoni; Simone Kashima; Svetoslav Nanev Slavov; Vincent Louis Viala |
| EPI_ISL_2037216, EPI_ISL_2037241, EPI_ISL_2037256, EPI_ISL_2037304, EPI_ISL_2037314, EPI_ISL_2037317, EPI_ISL_2037375, EPI_ISL_2106316, EPI_ISL_2106325, EPI_ISL_2106346, EPI_ISL_2106360, EPI_ISL_2106362, EPI_ISL_2106365, EPI_ISL_2106382, EPI_ISL_2106383, EPI_ISL_2106392, EPI_ISL_2106426, EPI_ISL_2106428, EPI_ISL_2106430, EPI_ISL_2106447, EPI_ISL_2106473, EPI_ISL_2106484, EPI_ISL_2106488, EPI_ISL_4096596 | | | |
| see above | Pandemic Response Lab - NYC | Pandemic Response Lab, R&D | Cybill del Castillo; Dylan Law; Haiping Hao; Henry Lee; Jon Laurent; Katharine Nelson; Melissa Hopkins; Michael Hammerling; Pradeep Bugga; Shinyoung Clair Kang; Sol Rey; William Ward |
| EPI_ISL_3194222 | Pathogen Genomics Center, National Institute of Infectious Diseases | Pathogen Genomics Center, National Institute of Infectious Diseases | Kentaro Itokawa; Makoto Kuroda; Masanori Hashino; Rina Tanaka; Tsuyoshi Sekizuka |
| EPI_ISL_3046970 | Pennsylvania Department of Health Bureau of Laboratories | Pennsylvania Department of Health Bureau of Laboratories | Dongxiang Xia |
| EPI_ISL_3030318 | Pesaro | Microbiology University Politecnica delle Marche | Anna Valenza; Carla Acciarri; Katia Marinelli; Monica Lucia Ferreri; Patrizia Bagnarelli; Roberta Longo; Sara Caucci; Stefano Menzo |
| EPI_ISL_6907911, EPI_ISL_6907917, EPI_ISL_6907921, EPI_ISL_6907922, EPI_ISL_6908440, EPI_ISL_6908441, EPI_ISL_6908451, EPI_ISL_6908455, EPI_ISL_6908456, EPI_ISL_6908462, EPI_ISL_6908855, EPI_ISL_6908860, EPI_ISL_6908862, EPI_ISL_6908865, EPI_ISL_6908870, EPI_ISL_6908874, EPI_ISL_6908878, EPI_ISL_6908879, EPI_ISL_6908885, EPI_ISL_6941438 | | | |
| see above | Plataforma de Vigilancia Molecular (PVM) - FIOCRUZ/BA | Plataforma de Vigilancia Molecular (PVM) - FIOCRUZ/BA | Bruno Bezerril Andrade; Camila I. de Oliveira; Clarissa Araújo Gurgel; Eduardo Oyama; Francys A Rangel; Laila A da Silva; Laise de Moraes; Leonardo Paiva Farias; Marina Cucco; Pedro Silveira; Ricardo Khouri on behalf of the Fiocruz COVID-19 Genomic Surveillance Network.; Tiago Graf |
| EPI_ISL_2015272, EPI_ISL_2015273, EPI_ISL_2037416, EPI_ISL_2037417, EPI_ISL_2037420, EPI_ISL_2037432, EPI_ISL_2037434, EPI_ISL_2037441, EPI_ISL_2151140, EPI_ISL_2151144, EPI_ISL_2151145, EPI_ISL_2151146, EPI_ISL_2151149 | | | |
| see above | Plateforme de testing Namuroise | Plateforme de testing Namuroise | Degosserie Jonathan; Denis Olivier; Lesly Nyinkeu Kemamen; Mullier François; Otto Gaetan |
| EPI_ISL_2016753, EPI_ISL_2016761, EPI_ISL_2016763, EPI_ISL_2131289, EPI_ISL_2272761 | Platform BIS UZA/UAntwerpen | Labo Klinische Biologie, UZA | Basil Britto Xavier; Christine Lammens; Herman Goossens; Jasmine Coppens; Marie Le Mercier; Veerle Matheeussen |
| EPI_ISL_2420727 | Policlinico Universitario Campus Bio-Medico | INMI Lazzaro Spallanzani IRCCS | A Di Caro; B Bartolini; CEM Gruber; E Giombini; E Riva; F Messina; F Santini; G Bonfiglio; M Fogolari; M Rueca; MR Capobianchi; O Butera; S Angeletti |
| EPI_ISL_2488760 | Policlinico Universitario Campus Bio-Medico, Rome, Italy | Policlinico Universitario Campus Bio-Medico, Rome, Italy | Elisabetta Riva; Francesca Antonelli; Lucia de Florio; Maria Francesconi; Marta Fogolori; Marta Giovanetti; Massimo Ciccozzi; Roberta Veralli; Silvia Angeletti |
| EPI_ISL_2232606 | Premier Medical Lab | Minnesota Department of Health, Public Health Laboratory | Alexandra Lorentz; Jacob Garfin; Matt Plumb; and Xiong Wang |
| EPI_ISL_5801551 | Pronto Atendimento Municipal | Instituto Butantan | Antonio Jorge Martins; Claudia Renata dos Santos Barros; David Schlesinger; Debora Botequio Moretti; Dimas Tadeu Covas; Elaine Cristina Marqueze; Elaine Vieira Santos; Evandra Strazza Rodrigues; Heidge Fukumasu; Jayme Augusto de Souza-Neto; José Salvatore Leister Patané; Luiz Alcantara; Luiz Lehmann Coutinho; Maria Carolina Elias; Maurício Lacerda Nogueira; Rafael dos Santos Bezerra; Raul Machado Neto; Rejane Maria Tommasini Grotto; Ricardo Haddad; Sandra Coccuzzo Sampaio Vessoni; Simone Kashima; Svetoslav Nanev Slavov; Vincent Louis Viala |
| EPI_ISL_5801613 | Pronto Socorro Municipal Tambau | Instituto Butantan | Antonio Jorge Martins; Claudia Renata dos Santos Barros; David Schlesinger; Debora Botequio Moretti; Dimas Tadeu Covas; Elaine Cristina Marqueze; Elaine Vieira Santos; Evandra Strazza Rodrigues; Heidge Fukumasu; Jayme Augusto de Souza-Neto; José Salvatore Leister Patané; Luiz Alcantara; Luiz Lehmann Coutinho; Maria Carolina Elias; Maurício Lacerda Nogueira; Rafael dos Santos Bezerra; Raul Machado Neto; Rejane Maria Tommasini Grotto; Ricardo Haddad; Sandra Coccuzzo Sampaio Vessoni; Simone Kashima; Svetoslav Nanev Slavov; Vincent Louis Viala |
| EPI_ISL_2551500, EPI_ISL_2551501 | Providence Oregon Regional Laboratories | Providence St. Joseph Health Molecular Genomics Laboratory | Alexa K Dowdell; Brian D Piening; Carlo B Bifulco; Fred L Robinson; Mary Campbell; Rogan Rattray |
| EPI_ISL_2551461 | Providence Regional Medical Center Everett | Providence St. Joseph Health Molecular Genomics Laboratory | Alexa K Dowdell; Brian D Piening; Carlo B Bifulco; Fred L Robinson; Mary Campbell; Rogan Rattray |
| EPI_ISL_2698135, EPI_ISL_2698150, EPI_ISL_2698216, EPI_ISL_2698232, EPI_ISL_2698234, EPI_ISL_2698247, EPI_ISL_2698317, EPI_ISL_2698423, EPI_ISL_2698546, EPI_ISL_2698550, EPI_ISL_2698568, EPI_ISL_2699298, EPI_ISL_2699319, EPI_ISL_2699331, EPI_ISL_2699399 | | | |
| see above | Public Health Laboratory, Minnesota Department of Health | University of Minnesota Genomics Center | Corbin Dirkx; Daryl M. Gohl; Jaquelyn Kuriger-Laber; John Garbe; and Sean Wang |
| EPI_ISL_2187312, EPI_ISL_2241627, EPI_ISL_2241629, EPI_ISL_2241630, EPI_ISL_2241634, EPI_ISL_2241635, EPI_ISL_2241637, EPI_ISL_2241638, EPI_ISL_2241640, EPI_ISL_2268154, EPI_ISL_2268156, EPI_ISL_2268271, EPI_ISL_2268273, EPI_ISL_2268275, EPI_ISL_2268278, EPI_ISL_2268280, EPI_ISL_2268282, EPI_ISL_2268287, EPI_ISL_2268291, EPI_ISL_2268293, EPI_ISL_2268295, EPI_ISL_2268297, EPI_ISL_2268299, EPI_ISL_2268302, EPI_ISL_2268304, EPI_ISL_2268307, EPI_ISL_2268309, EPI_ISL_2268311, EPI_ISL_2268314, EPI_ISL_2268325, EPI_ISL_2268328, EPI_ISL_2268333, EPI_ISL_2268335, EPI_ISL_2268336, EPI_ISL_2268366, EPI_ISL_2268367, EPI_ISL_2268368, EPI_ISL_2268369, EPI_ISL_2268370, EPI_ISL_2268371, EPI_ISL_2268373, EPI_ISL_2268374, EPI_ISL_2268375, EPI_ISL_2268376, EPI_ISL_2268377, EPI_ISL_2268378, EPI_ISL_2268379, EPI_ISL_2268380, EPI_ISL_2268381, EPI_ISL_2268382, EPI_ISL_2268384, EPI_ISL_2268385, EPI_ISL_2268386, EPI_ISL_2268389, EPI_ISL_2268390, EPI_ISL_2268391, EPI_ISL_2268393, EPI_ISL_2280080, EPI_ISL_2280081, EPI_ISL_2280082, EPI_ISL_2280084, EPI_ISL_2280085, EPI_ISL_2280086, EPI_ISL_2280087, EPI_ISL_2280088, EPI_ISL_2280089, | | | |

| | | | |
|---|---|---|---|
| EPI_ISL_2280090, EPI_ISL_2280091, EPI_ISL_2280092, EPI_ISL_2280096, EPI_ISL_2280100, EPI_ISL_2280102, EPI_ISL_2280104, EPI_ISL_2280105, EPI_ISL_2280106, EPI_ISL_2280107, EPI_ISL_2280108, EPI_ISL_2280109, EPI_ISL_2280110, EPI_ISL_2280111, EPI_ISL_2280112, EPI_ISL_2280113, EPI_ISL_2280114, EPI_ISL_2280115, EPI_ISL_2280116, EPI_ISL_2280117, EPI_ISL_2280118, EPI_ISL_2280119, EPI_ISL_2280121, EPI_ISL_2280123, EPI_ISL_2280124, EPI_ISL_2280125, EPI_ISL_2280126, EPI_ISL_2280128, EPI_ISL_2280129, EPI_ISL_2280130, EPI_ISL_2280131, EPI_ISL_2280133, EPI_ISL_2280135, EPI_ISL_2280136, EPI_ISL_2280138, EPI_ISL_2280139, EPI_ISL_2280140, EPI_ISL_2280141, EPI_ISL_2280142, EPI_ISL_2280143, EPI_ISL_2280144, EPI_ISL_2280145, EPI_ISL_2280146, EPI_ISL_2280147, EPI_ISL_2280148, EPI_ISL_2280149, EPI_ISL_2280150, EPI_ISL_2280151, EPI_ISL_2280152, EPI_ISL_2280153, EPI_ISL_2280154, EPI_ISL_2280155, EPI_ISL_2280156, EPI_ISL_2280157, EPI_ISL_2280158, EPI_ISL_2280159, EPI_ISL_2280160, EPI_ISL_2280161, EPI_ISL_2280162, EPI_ISL_2280164, EPI_ISL_2280166, EPI_ISL_2280167, EPI_ISL_2280169, EPI_ISL_2280171, EPI_ISL_2280173, EPI_ISL_2280175, EPI_ISL_2307141, EPI_ISL_2307143, EPI_ISL_2307144, EPI_ISL_2307149, EPI_ISL_2307154, EPI_ISL_2307157, EPI_ISL_2307158, EPI_ISL_2307160, EPI_ISL_2307161, EPI_ISL_2307162, EPI_ISL_2307164, EPI_ISL_2371692, EPI_ISL_2371698, EPI_ISL_2371705, EPI_ISL_2371711, EPI_ISL_2371719, EPI_ISL_2371723, EPI_ISL_2372410, EPI_ISL_2372411, EPI_ISL_2372413, EPI_ISL_2372414, EPI_ISL_2372415, EPI_ISL_2372416, EPI_ISL_2372417, EPI_ISL_2372418, EPI_ISL_2372419, EPI_ISL_2372422, EPI_ISL_4366573, EPI_ISL_4371700 | | | |
| see above | Quest Diagnostics Incorporated | Centers for Disease Control and Prevention Division of Viral Diseases, Pathogen Discovery | A. Gerasimova; A. Perez; Adrian Paskey; B. Anderson; Benjamin Rambo-Martin; Christopher Gulvick; Clinton Paden; Clinton R. Paden; Dakota Howard; Darlene Wagner; Dhwani Batra; Duncan MacCannell; Erisa Sula; F. Lacbawan; I. A. Shlyakhter; I. Shlyakhter; Jason Caravas; K. Livingston; K.E. Livingston; Kara Moser; Kristine Lacek; L. Bernstein; L.E. Bernstein; M. Hua; Matthew Schmerer; P. Tanpaiboon; Peter Cook; Peter W. Cook; R. Kagan; R. M. Kagan; R. Owen; R. Rolando; R. V. Rolando; S. H. Rosenthal; S. Rosenthal; Scott Sammons; Shatavia Morrison; Tymeckia Kendall; Victoria Caban Figueroa; Y. Liu; Yvette Unoarumhi |
| EPI_ISL_2384137, EPI_ISL_2384146, EPI_ISL_2384147, EPI_ISL_2384148, EPI_ISL_2384149, EPI_ISL_2384152, EPI_ISL_2384154 | | | |
| see above | RI State Health Laboratories | Centers for Disease Control and Prevention Division of Viral Diseases, Pathogen Discovery | Alison Laufer Halpin; Ben L. Rambo-Martin; Clinton R. Paden; Dakota Howard; Darlene Wagner; Dave Wentworth; Dhwani Batra; Jasmine Padilla; Justin Lee; Katie Dillon; Krista Queen; Kristen Knipe; Kristine Lacek; Mark Burroughs; Matthew Schmerer; Mili Sheth; Peter Cook; Sam Shepard; Sarah Nobles; Shoshona Le; Suxiang Tong; Vivien Dugan; Yvette Unoarumhi |
| EPI_ISL_2021751 | Randox Laboratories | Wellcome Sanger Institute for the COVID-19 Genomics UK (COG-UK) Consortium | Cordelia Langford; David K. Jackson; Dominic Kwiatkowski; Ewan Harrison; Ian Johnston; Jeffrey Barrett; John Sillitoe on behalf of the Wellcome Sanger Institute COVID-19 Surveillance Team; Randox Laboratories and Alex Alderton; Roberto Amato; Sonia Goncalves |
| EPI_ISL_2473834, EPI_ISL_2473836, EPI_ISL_2473841, EPI_ISL_2473843, EPI_ISL_2473844, EPI_ISL_2473846, EPI_ISL_2473847, EPI_ISL_2473850, EPI_ISL_2473852, EPI_ISL_2473858, EPI_ISL_2473859, EPI_ISL_2473866, EPI_ISL_2473876, EPI_ISL_2473879, EPI_ISL_2473880, EPI_ISL_2473881, EPI_ISL_2473883, EPI_ISL_2473891, EPI_ISL_2473894, EPI_ISL_2473896, EPI_ISL_2473897, EPI_ISL_2473898, EPI_ISL_2473899, EPI_ISL_2473900, EPI_ISL_2473901, EPI_ISL_2473902, EPI_ISL_2473905, EPI_ISL_2473907, EPI_ISL_2473917, EPI_ISL_2473922, EPI_ISL_2473927, EPI_ISL_2473930, EPI_ISL_2473932, EPI_ISL_2473948, EPI_ISL_2473949, EPI_ISL_2473970, EPI_ISL_2473971, EPI_ISL_2473979, EPI_ISL_2474005, EPI_ISL_2474006, EPI_ISL_2474033, EPI_ISL_2474044, EPI_ISL_2474046, EPI_ISL_2474049, EPI_ISL_2474062, EPI_ISL_2474063, EPI_ISL_2474072, EPI_ISL_2474077, EPI_ISL_2474090, EPI_ISL_2474092, EPI_ISL_2474106, EPI_ISL_2474107, EPI_ISL_2474125, EPI_ISL_2474126, EPI_ISL_2474172, EPI_ISL_2474173, EPI_ISL_2474176, EPI_ISL_2474177, EPI_ISL_2662209, EPI_ISL_2662210, EPI_ISL_2662211, EPI_ISL_2662212, EPI_ISL_2662213, EPI_ISL_2662214, EPI_ISL_2662215, EPI_ISL_2662216, EPI_ISL_2662217, EPI_ISL_2662218, EPI_ISL_2662219, EPI_ISL_2662220, EPI_ISL_2662221, EPI_ISL_2662222, EPI_ISL_2662223, EPI_ISL_2662224, EPI_ISL_2662225, EPI_ISL_2662226, EPI_ISL_2662227, EPI_ISL_2662228, EPI_ISL_2662229, EPI_ISL_2662230, EPI_ISL_2662231, EPI_ISL_2662232, EPI_ISL_2662233, EPI_ISL_2662234, EPI_ISL_2662235, EPI_ISL_2662236, EPI_ISL_2662237, EPI_ISL_2662238, EPI_ISL_2662239, EPI_ISL_2662240, EPI_ISL_2662241, EPI_ISL_2662242, EPI_ISL_2662243, EPI_ISL_2662244, EPI_ISL_2662245, EPI_ISL_2662246, EPI_ISL_2662247, EPI_ISL_2662248, EPI_ISL_2662249, EPI_ISL_2662250, EPI_ISL_2662251, EPI_ISL_2662252, EPI_ISL_2662253, EPI_ISL_2662254, EPI_ISL_2662255, EPI_ISL_2662256, EPI_ISL_2662257, EPI_ISL_2662258, EPI_ISL_2662259, EPI_ISL_2662260, EPI_ISL_2662261, EPI_ISL_2662262, EPI_ISL_2662263, EPI_ISL_2662264, EPI_ISL_2662265, EPI_ISL_2662266, EPI_ISL_2662267, EPI_ISL_2662268, EPI_ISL_2662269, EPI_ISL_2662270, EPI_ISL_2662271, EPI_ISL_2662272, EPI_ISL_2662273, EPI_ISL_2662274, EPI_ISL_2662275, EPI_ISL_2662276, EPI_ISL_2662277, EPI_ISL_2662278, EPI_ISL_2662279, EPI_ISL_2662280, EPI_ISL_2662281, EPI_ISL_2662282, EPI_ISL_2662283, EPI_ISL_2662284, EPI_ISL_2662285, EPI_ISL_2662286 | | | |
| see above | Reditus Laboratories | Reditus Laboratories | Alexa Eichelberger; Cassy Philips; Joshua J. Geltz; M.S.; Ph.D.; Rex Dyer; Robert M. Sgambelluri |
| EPI_ISL_2742343, EPI_ISL_2742380, EPI_ISL_3776071, EPI_ISL_3776073, EPI_ISL_3777148 | Respiratory Virus Unit, Microbiology Services Colindale, Public Health England | COVID-19 Genomics UK (COG-UK) Consortium | PHE Covid Sequencing Team |
| EPI_ISL_2232415, EPI_ISL_2232434, EPI_ISL_2232435, EPI_ISL_2232439, EPI_ISL_2232443, EPI_ISL_2232444, EPI_ISL_2232445, EPI_ISL_2232451, EPI_ISL_2232463, EPI_ISL_2232508, EPI_ISL_2232530, EPI_ISL_2232532, EPI_ISL_2232534, EPI_ISL_2232536, EPI_ISL_2232537, EPI_ISL_2232538 | | | |
| see above | Rhode Island Department of Health | Infectious Disease Program, Broad Institute of Harvard and MIT | Adams, G.; Azevedo, K.; B.L.; B.W.; Bauer, M.; Birren; Carter, A.; Chaluvadi, S.; D.J.; DeRuff, K.; Gladden-Young, A.; Huard, R.; J.E.; K.J.; King, E.; Lagerborg, K.; Lemieux; Loreth, C.; Miller, A.; Normandin, E.; P.C.; Park; Pearlman, L.; Reilly, S.; Rudy, M.; Sabeti; Siddle; Tomkins-Tinch, C.; and MacInnis |
| EPI_ISL_2425499, EPI_ISL_2425504, EPI_ISL_2425519, EPI_ISL_2425520, EPI_ISL_2426817, EPI_ISL_2725577 | Rhode Island State Health Laboratory | Rhode Island State Health Laboratory | Ewa King; Kristin Carpenter-Azevedo; Richard C. Huard |
| EPI_ISL_6604398 | Rush University Medical Center | RIPHL at Rush University Medical Center | Alyse Kittner; Diane Springer; Edith Perez; Felix Araujo Perez; Joyce Houlihan; Kevin Kunstman; Laura Furtado; Marieta Hyde; Mary Hayden; Sofiya Bobrovska; Stefan Green |
| EPI_ISL_2444211, EPI_ISL_2444213, EPI_ISL_2444214, EPI_ISL_2444215, EPI_ISL_2444216, EPI_ISL_2444218, EPI_ISL_2444219, EPI_ISL_2444220, EPI_ISL_2444221, EPI_ISL_2444222 | | | |
| see above | SAO JOSE DO RIO PRETO | Instituto Butantan / FAMERP | Antonio Jorge Martins; Claudia Renata dos Santos Barros; David Schlesinger; Debora Botequio Moretti; Dimas Tadeu Covas; Elaine Cristina Marqueze; Elaine Vieira Santos; Evandra Strazza Rodrigues; Heidge Fukumasu; Jayme Augusto de Souza-Neto; José Salvatore Leister Patané; Luiz Alcantara; Luiz Lehmann Coutinho; Maria Carolina Elias; Maurício Lacerda Nogueira; Rafael dos Santos Bezerra; Raul Machado Neto; Rejane Maria Tommasini Grotto; Ricardo Haddad; Sandra Coccuzzo Sampaio Vessoni; Simone Kashima; Svetoslav Nanev Slavov; Vincent Louis Viala |
| EPI_ISL_2791991, EPI_ISL_2792004, EPI_ISL_2792138, EPI_ISL_2792253, EPI_ISL_2957619 | SARS-CoV-2 Sequencing Castilla y Leon-Spain Consortium | SARS-CoV-2 Sequencing Castilla y Leon-Spain Consortium | Antonio Orduña-Domingo; Carlos Fuster Foz; Carmen Aldea-Mansilla; Carmen Gimeno Crespo; David Abad; Gregoria Megías Lobón; Jose María Eiros Bouza; Laura Sánchez de Prada; M. Isabel Fernandez-Natal; Marta Dominguez-Gil; Marta Hernandez; María Antonia García Castro; Mª Fe Brezmes-Valdivieso; Noelia Arenal Andrés; Silvia Rojo; Sonsoles Garcinuño Pérez |
| EPI_ISL_2131663, EPI_ISL_2131664, EPI_ISL_2131665, EPI_ISL_2131666, EPI_ISL_2338211, EPI_ISL_2338221 | SARS-CoV-2 testing team, National Institute of Infectious Diseases | Pathogen Genomics Center, National Institute of Infectious Diseases | Hazuka Y Furihata; Kentaro Itokawa; Makoto Kuroda; Masanori Hashino; Masumichi Saito; Naomi Nojiri; Nozomu Hanaoka; Rina Tanaka; Sana Uchikoba; Tsuguto Fujimoto; Tsuyoshi Sekizuka |
| EPI_ISL_2205561 | SC (UCO) Igiene e Sanità Pubblica, ASUGI, Trieste | ARGO Laboratorio Genomica ed Epigenomica | D'Agaro P; Dal Monego S; Degasperi M; Licastro D; Marcello A; Piscianz E; Segat L |
| EPI_ISL_2674350 | SECCION SERVICIOS INTEGRALES DE SALUD Y DESARROLLO PSICOSOCIAL DE BIENESTAR UNIVERSITARIO UIS | Instituto Nacional de Salud-Dirección de Investigación en Salud Pública | Carlos Franco-Muñoz; Carmen Osorio; Diana Malo; Diego A. Álvarez-Díaz; Diego Andrés Prada; Gerardo Santamaría; Hector Alejandro Ruiz-Moreno; Jhonnatan Reales-González; Jorge Rivera; Juan Camilo Martinez; Julian Naizaque; Katherine Laiton-Donato; Lisseth Pardo; Magdalena Wiesner; Marcela Mercado-Reyes; Maria T. Herrera-Sepúlveda; Marta Lopez Blanco; Martha Lucia Ospina Martinez; Paola Rojas; Sergio Gomez; Sheryll Corchuelo; Ángela Alarcon Cruz |
| EPI_ISL_5650322 | SECRETARIA DE SAUDE DE SANTA GERTRUDES | Instituto Butantan | Antonio Jorge Martins; Claudia Renata dos Santos Barros; David Schlesinger; Debora Botequio Moretti; Dimas Tadeu Covas; Elaine Cristina Marqueze; Elaine Vieira Santos; Evandra Strazza Rodrigues; Heidge Fukumasu; Jayme Augusto de Souza-Neto; José Salvatore Leister Patané; Luiz Alcantara; Luiz Lehmann Coutinho; Maria Carolina Elias; Maurício Lacerda Nogueira; Rafael dos Santos Bezerra; Raul Machado Neto; Rejane Maria Tommasini Grotto; Ricardo Haddad; Sandra Coccuzzo Sampaio Vessoni; Simone Kashima; Svetoslav Nanev Slavov; Vincent Louis Viala |
| EPI_ISL_5650240, EPI_ISL_5650319 | SECRETARIA DE SAUDE DE SAO PEDRO | Instituto Butantan | Antonio Jorge Martins; Claudia Renata dos Santos Barros; David Schlesinger; Debora Botequio Moretti; Dimas Tadeu Covas; Elaine Cristina Marqueze; Elaine Vieira Santos; Evandra Strazza Rodrigues; Heidge Fukumasu; Jayme Augusto de Souza-Neto; José Salvatore Leister Patané; Luiz Alcantara; Luiz Lehmann Coutinho; Maria Carolina Elias; Maurício Lacerda Nogueira; Rafael dos Santos Bezerra; Raul Machado Neto; Rejane Maria Tommasini Grotto; Ricardo Haddad; Sandra Coccuzzo Sampaio Vessoni; Simone Kashima; Svetoslav Nanev Slavov; Vincent Louis Viala |
| EPI_ISL_2209516, EPI_ISL_2209905, EPI_ISL_2210305, EPI_ISL_2345661, EPI_ISL_2345802, EPI_ISL_2345849 | SECRETARIA DE SAUDE PUBLICA DE PRAIA GRANDE | Instituto Butantan / Mendelics | Antonio Jorge Martins; Claudia Renata dos Santos Barros; David Schlesinger; Debora Botequio Moretti; Dimas Tadeu Covas; Elaine Cristina Marqueze; Elaine Vieira Santos; Evandra Strazza Rodrigues; Heidge Fukumasu; Jayme Augusto de Souza-Neto; José Salvatore Leister Patané; Luiz Alcantara; Luiz Lehmann Coutinho; Maria Carolina Elias; Maurício Lacerda Nogueira; Rafael dos Santos Bezerra; Raul Machado Neto; Rejane Maria Tommasini Grotto; Ricardo Haddad; Sandra Coccuzzo Sampaio Vessoni; Simone Kashima; Svetoslav Nanev Slavov; Vincent Louis Viala |
| EPI_ISL_2345875 | SECRETARIA MUNICIPAL DA SAUDE DE JOANOPOLIS | Instituto Butantan / Mendelics | Antonio Jorge Martins; Claudia Renata dos Santos Barros; David Schlesinger; Debora Botequio Moretti; Dimas Tadeu Covas; Elaine Cristina Marqueze; Elaine Vieira Santos; Evandra Strazza Rodrigues; Heidge Fukumasu; Jayme Augusto de Souza-Neto; José Salvatore Leister Patané; Luiz Alcantara; Luiz Lehmann Coutinho; Maria Carolina Elias; Maurício Lacerda Nogueira; Rafael dos Santos Bezerra; Raul Machado Neto; Rejane Maria Tommasini Grotto; Ricardo Haddad; Sandra Coccuzzo Sampaio Vessoni; Simone Kashima; Svetoslav Nanev Slavov; Vincent Louis Viala |
| EPI_ISL_5825559, EPI_ISL_5825560, EPI_ISL_5825561 | SECRETARIA MUNICIPAL DE SAUDE DE CEDRO CEARA | Analytical Competence Molecular Epidemiology Lab/ACME, Oswaldo Cruz Foundation, Ceara (FIOCRUZ CE) | Carlos Leonardo de Aragao Araujo; Cecilia Leite Costa & Eduardo Ruback dos Santos on behalf of COVID-19 FIOCRUZ Genomic Network; Cleber Furtado Aksenen; Fabio Miyajima; Fernando Braga Stehling; Francisco Eder de Moura Lopes; Igor Oliveira Duarte; Jamille Maria Mendes Bezerra; Joaquim Cesar do Nascimento Sousa Junior; Pedro Miguel Carneiro Jeronimo; Suzana Porto Almeida; Thais Ferreira de Oliveira; Thais de Oliveira Costa; Ticiane Cavalcante de Souza; Veridiana Pessoa Miyajima |
| EPI_ISL_5650318 | SECRETARIA MUNICIPAL DE SAUDE DE CORDEIROPOLIS | Instituto Butantan | Antonio Jorge Martins; Claudia Renata dos Santos Barros; David Schlesinger; Debora Botequio Moretti; Dimas Tadeu Covas; Elaine Cristina Marqueze; Elaine Vieira Santos; Evandra Strazza Rodrigues; Heidge Fukumasu; Jayme Augusto de Souza-Neto; José Salvatore Leister Patané; Luiz Alcantara; Luiz Lehmann Coutinho; Maria Carolina Elias; Maurício Lacerda Nogueira; Rafael dos Santos Bezerra; Raul Machado Neto; Rejane Maria Tommasini Grotto; Ricardo Haddad; Sandra Coccuzzo Sampaio Vessoni; Simone Kashima; Svetoslav Nanev Slavov; Vincent Louis Viala |
| EPI_ISL_6229770, EPI_ISL_6229771, EPI_ISL_6229772, EPI_ISL_6229773, EPI_ISL_6229774, EPI_ISL_6229775, EPI_ISL_6229847 | | | |
| see above | SECRETARIA MUNICIPAL DE SAUDE DE ITAREMA | ACME Lab, Oswaldo Cruz Foundation, FIOCRUZ/CE | Carlos Leonardo de Aragao Araujo; Cecilia Leite Costa & Eduardo Ruback dos Santos on behalf of COVID-19 FIOCRUZ Genomic Network; Cleber Furtado Aksenen; Fabio Miyajima; Fernando Braga Stehling; Francisco Eder de Moura Lopes; Igor Oliveira Duarte; Jamille Maria Mendes Bezerra; Joaquim Cesar do Nascimento Sousa Junior; Pedro Miguel Carneiro Jeronimo; Suzana Porto Almeida; Thais Ferreira de Oliveira; Thais de Oliveira Costa; Ticiane Cavalcante de Souza; Veridiana Pessoa Miyajima |
| EPI_ISL_2209341 | SERV DE VIG SANITARIA EPIDEMIO E CTRL DE ZOONOZES GUARUJA | Instituto Butantan / Mendelics | Antonio Jorge Martins; Claudia Renata dos Santos Barros; David Schlesinger; Debora Botequio Moretti; Dimas Tadeu Covas; Elaine Cristina Marqueze; Elaine Vieira Santos; Evandra Strazza Rodrigues; Heidge Fukumasu; Jayme Augusto de Souza-Neto; José Salvatore Leister Patané; Luiz Alcantara; Luiz Lehmann Coutinho; Maria Carolina Elias; Maurício Lacerda Nogueira; Rafael dos Santos Bezerra; Raul Machado Neto; Rejane Maria Tommasini Grotto; Ricardo Haddad; Sandra Coccuzzo Sampaio Vessoni; Simone Kashima; Svetoslav Nanev Slavov; Vincent Louis Viala |
| EPI_ISL_2131404 | SIESP CHIETI - DRIVE IN CHIETI | Istituto Zooprofilattico Sperimentale dell'Abruzzo e Molise "G. Caporale" | Ancora M; Calistri P; Cammà C; Caporale M; Curini V; Delli Compagni E; Di Domenico M; Di Lollo Valeria; Di Pasquale A; Lorusso A; Mangone I; Marcacci M; Puglia I; Rinaldi A; Savini G; Scialabba S |
| EPI_ISL_2235387, EPI_ISL_2235388 | SIESP DIPARTIMENTO DI PREVENZIONE CHIETI | Istituto Zooprofilattico Sperimentale dell'Abruzzo e Molise "G. Caporale" | Ancora M; Calistri P; Cammà C; Caporale M; Curini V; Delli Compagni E; Di Domenico M; Di Lollo Valeria; Di Pasquale A; Lorusso A; Mangone I; Marcacci M; Puglia I; Rinaldi A; Savini G; Scialabba S |
| EPI_ISL_2131412, EPI_ISL_2131421, EPI_ISL_2235355, EPI_ISL_2235361 | SIESP DIPARTIMENTO DI PREVENZIONE TERAMO | Istituto Zooprofilattico Sperimentale dell'Abruzzo e Molise "G. Caporale" | Ancora M; Calistri P; Cammà C; Caporale M; Curini V; Delli Compagni E; Di Domenico M; Di Lollo Valeria; Di Pasquale A; Lorusso A; Mangone I; Marcacci M; Puglia I; Rinaldi A; Savini G; Scialabba S |
| EPI_ISL_2587149, EPI_ISL_2587151, EPI_ISL_2587339, EPI_ISL_2587341, EPI_ISL_2587699 | SK-Roy Romanow Provincial Laboratory | National Microbiology Laboratory (NML) | Amanda Lang; Anna Majer; Anneliese Landgraff; CanCOGeN's metadata curation team; Darian Hole; Elsie Grudeski; Gary Van Domselaar; Grace Seo; Jennifer Tanner; Jessica Minion; Kirsten Biggar; Madison Chapel; Morag Graham; Natalie Knox; Nathalie Bastien; Philip Mabon; Public Health Agency of Canada CanCOGeN team; Rachel DePaulo; Rhiannon Huzarewich; Russell Mandes; Ryan McDonald; Shari Tyson; Timothy Booth; Yan Li |
| EPI_ISL_6124764, EPI_ISL_6124813, EPI_ISL_6124853, EPI_ISL_6124860, EPI_ISL_6124888, EPI_ISL_6124916, EPI_ISL_6124918, EPI_ISL_6124922, EPI_ISL_6124932, EPI_ISL_6124942, EPI_ISL_6124949, EPI_ISL_6124953, EPI_ISL_6124983, EPI_ISL_6125024 | | | |
| see above | SK-Roy Romanow Provincial Laboratory | Saskatchewan - Roy Romanow | Alanna Senecal; Amanda Lang; Jessica Minion; Kara Loos; Keith MacKenzie; Meredith Faires; Rachel DePaulo; Roy Romanow Provincial Laboratory - Molecular Diagnostics; Ryan McDonald |

| | | | |
|---|---|---|---|
| | | Provincial Laboratory (RRPL) | |
| EPI_ISL_6565014 | SRC VB "Vector", "Collection of microorganisms" Department. | SRC VB "Vector", "Collection of microorganisms" Department. | Alexander N. Shvalov; Anastasiya A. Nazarenko; Anastasiya M. Smirnova; Elena V. Gavrilova; Oleg V. Pyankov; Rinat A. Maksyutov; Sergey A. Bodnev; Tatyana V. Tregubchak |
| EPI_ISL_5155888, EPI_ISL_5155971, EPI_ISL_5155973 | SUNY Upstate Medical University | 505 Irving Avenue, Syracuse, NY 13210 - Frank Middleton Lab | Brian Pavlovitz; Frank Middleton |
| EPI_ISL_2674305 | SURA | Instituto Nacional de Salud-Dirección de Investigación en Salud Pública | Carlos Franco-Muñoz; Carmen Osorio; Diana Malo; Diego A. Álvarez-Díaz; Diego Andrés Prada; Gerardo Santamaría; Hector Alejandro Ruiz-Moreno; Jhonnatan Reales-González; Jorge Rivera; Juan Camilo Martinez; Julian Naizaque; Katherine Laiton-Donato; Lisseth Pardo; Magdalena Wiesner; Marcela Mercado-Reyes; Maria T. Herrera-Sepúlveda; Marta Lopez Blanco; Martha Lucia Ospina Martinez; Paola Rojas; Sergio Gomez; Sheryll Corchuelo; Ángela Alarcon Cruz |
| EPI_ISL_2139715, EPI_ISL_2139717, EPI_ISL_2139718, EPI_ISL_2139719, EPI_ISL_2139720, EPI_ISL_2139721, EPI_ISL_2139725, EPI_ISL_2139876, EPI_ISL_2139877, EPI_ISL_2139878, EPI_ISL_2139881, EPI_ISL_2139882, EPI_ISL_2245220, EPI_ISL_2245237, EPI_ISL_2245252, EPI_ISL_2245262, EPI_ISL_2245404, EPI_ISL_2245420, EPI_ISL_2245421, EPI_ISL_2245422, EPI_ISL_2245423, EPI_ISL_2245428, EPI_ISL_2245429, EPI_ISL_2245435, EPI_ISL_2245436 | | | |
| see above | SYNLAB | GIGA Medical Genomics | Bouchra Boujemla; Cécile Meex; Keith Durkin; Maria Artesi; Marie-Pierre Hayette; Nathalie Renotte; Pierrette Melin; Raphaël Boreux; Sébastien Bontems; Vincent Bours |
| EPI_ISL_3355473, EPI_ISL_3355475, EPI_ISL_3355476 | SYNLAB - ANGEL DIAGNOSTICA | Corporacion CorpoGen-Universidad de los Andes-Universidad Central | Christian Romero; Jorge Duitama; Juan Manuel Anzola; Laura González; Maryam Chaib De Mares; María Mercedes Zambrano; Nelly Díaz; Patricia Del Portillo; Silvia Restrepo |
| EPI_ISL_2124105 | SYNLAB Labor MÃ¼nchen Zentrum LMZ | Robert Koch Institute | |
| EPI_ISL_2124700 | SYNLAB MVZ Dachau | Robert Koch Institute | |
| EPI_ISL_2264253 | SYNLAB MVZ Leinfelden-Echterdingen | Robert Koch Institute | |
| EPI_ISL_2116332, EPI_ISL_2116720 | SYNLAB MVZ Leverkusen | Robert Koch Institute | |
| EPI_ISL_2118294 | SYNLAB MVZ Trier | Robert Koch Institute | |
| EPI_ISL_2105731, EPI_ISL_2105847, EPI_ISL_2105848, EPI_ISL_2105857, EPI_ISL_2105871, EPI_ISL_2230831, EPI_ISL_2230900, EPI_ISL_2319184, EPI_ISL_2319195, EPI_ISL_2319198, EPI_ISL_2319216, EPI_ISL_2319227, EPI_ISL_2319231, EPI_ISL_2319233, EPI_ISL_2319236, EPI_ISL_2319237, EPI_ISL_2319238, EPI_ISL_2319393, EPI_ISL_2319399 | | | |
| see above | Salud Digna | Instituto Nacional de Medicina Genomica | Abraham Campos-Romero; Cedro-Tanda A; Escobar-Arrazola; Gonzalez-Barrera D; Herrera-Montalvo LA.; Hidalgo-Miranda A; Luna-Ruiz Marco; M; Mendoza-Vargas A; Moreno-Camacho José Luis; Munguia-Garza P; Ramirez-Vega O; Rangel-DeLeon D; Reyes-Grajeda JP; Rodriguez-Gallegos Jorge |
| EPI_ISL_2329537, EPI_ISL_2628612, EPI_ISL_2628614, EPI_ISL_2628742, EPI_ISL_2628786 | San Diego County Public Health Laboratory | Andersen lab at Scripps Research | Brett Austin; Jovan Shephard; SEARCH Alliance San Diego with Ashleigh Murphy |
| EPI_ISL_2287924 | San Juan Regional Medical Center | New Mexico Department of Health Scientific Laboratory | Anastacia Griego-Fisher; D'eldra Malone; Ellie Johnson; Jennifer Benoit |
| EPI_ISL_2659064 | Santa Clara County Public Health Laboratory | Santa Clara County Public Health Laboratory | Santa Clara County Public Health Department |
| EPI_ISL_2490090, EPI_ISL_2490092, EPI_ISL_2692210, EPI_ISL_2692215, EPI_ISL_2692503, EPI_ISL_3372886, EPI_ISL_3372926 | | | |
| see above | Seattle Flu Study | Seattle Flu Study | Amanda Adler; Barry R. Lutz; Benjamin Pelle; Caitlin R. Wolf; Chris D. Frazar; Deborah A. Nickerson; Elisabeth Brandstetter; Erica Ryke; Helen Y. Chu; Janet A. Englund; Jay Shendure; Jeff Duchin; Jover Lee; Kairsten Fay; Karen Cowgill; Kirsten Lacombe; Lea M. Starita; Mark J. Rieder; Matthew Richardson; Matthew Thompson; Melissa Truong; Michael Boeckh; Michael Famulare; Misja Ilcisin; Peter D. Han; Stephanie Schrag; Thomas R. Sibley; Trevor Bedford |
| EPI_ISL_5801576, EPI_ISL_5801581, EPI_ISL_5801584, EPI_ISL_5801586, EPI_ISL_5801591, EPI_ISL_5801592, EPI_ISL_5801598 | | | |
| see above | Secretaria De Saude Publica De Praia Grande | Instituto Butantan | Antonio Jorge Martins; Claudia Renata dos Santos Barros; David Schlesinger; Debora Botequio Moretti; Dimas Tadeu Covas; Elaine Cristina Marqueze; Elaine Vieira Santos; Evandra Strazza Rodrigues; Heidge Fukumasu; Jayme Augusto de Souza-Neto; José Salvatore Leister Patané; Luiz Alcantara; Luiz Lehmann Coutinho; Maria Carolina Elias; Maurício Lacerda Nogueira; Rafael dos Santos Bezerra; Raul Machado Neto; Rejane Maria Tommasini Grotto; Ricardo Haddad; Sandra Coccuzzo Sampaio Vessoni; Simone Kashima; Svetoslav Nanev Slavov; Vincent Louis Viala |
| EPI_ISL_5801682 | Secretaria Municipal Da Saude De Joanopolis | Instituto Butantan | Antonio Jorge Martins; Claudia Renata dos Santos Barros; David Schlesinger; Debora Botequio Moretti; Dimas Tadeu Covas; Elaine Cristina Marqueze; Elaine Vieira Santos; Evandra Strazza Rodrigues; Heidge Fukumasu; Jayme Augusto de Souza-Neto; José Salvatore Leister Patané; Luiz Alcantara; Luiz Lehmann Coutinho; Maria Carolina Elias; Maurício Lacerda Nogueira; Rafael dos Santos Bezerra; Raul Machado Neto; Rejane Maria Tommasini Grotto; Ricardo Haddad; Sandra Coccuzzo Sampaio Vessoni; Simone Kashima; Svetoslav Nanev Slavov; Vincent Louis Viala |
| EPI_ISL_5801531 | Serv De Vig Sanitaria Epidemio E Ctrl De Zoonozes Guaruja | Instituto Butantan | Antonio Jorge Martins; Claudia Renata dos Santos Barros; David Schlesinger; Debora Botequio Moretti; Dimas Tadeu Covas; Elaine Cristina Marqueze; Elaine Vieira Santos; Evandra Strazza Rodrigues; Heidge Fukumasu; Jayme Augusto de Souza-Neto; José Salvatore Leister Patané; Luiz Alcantara; Luiz Lehmann Coutinho; Maria Carolina Elias; Maurício Lacerda Nogueira; Rafael dos Santos Bezerra; Raul Machado Neto; Rejane Maria Tommasini Grotto; Ricardo Haddad; Sandra Coccuzzo Sampaio Vessoni; Simone Kashima; Svetoslav Nanev Slavov; Vincent Louis Viala |
| EPI_ISL_2179784, EPI_ISL_2179802, EPI_ISL_2179805 | Servicio Microbiología Hospital La Paz | Servicio Microbiología Hospital La Paz | Elie Dahdouh; Fernando Lázaro; Jesús Mingorance Cruz; Rubén Cáceres |
| EPI_ISL_2158741, EPI_ISL_2158742, EPI_ISL_2158746, EPI_ISL_2158747, EPI_ISL_2158748, EPI_ISL_2158750, EPI_ISL_2158754, EPI_ISL_2158755, EPI_ISL_2158765, EPI_ISL_2158766, EPI_ISL_2158769, EPI_ISL_2158770, EPI_ISL_2158776, EPI_ISL_2158777, EPI_ISL_2158778, EPI_ISL_2158785, EPI_ISL_2158794, EPI_ISL_2158817, EPI_ISL_2158825, EPI_ISL_2158840, EPI_ISL_2158844, EPI_ISL_2158847, EPI_ISL_2158849, EPI_ISL_4082184, EPI_ISL_4082188, EPI_ISL_4082189, EPI_ISL_4082191, EPI_ISL_4082193, EPI_ISL_4082195, EPI_ISL_4082200, EPI_ISL_4082201, EPI_ISL_4082202, EPI_ISL_4082203, EPI_ISL_4082210, EPI_ISL_4082213, EPI_ISL_4082215, EPI_ISL_4082216, EPI_ISL_4082217, EPI_ISL_4082227, EPI_ISL_4082228, EPI_ISL_4082229, EPI_ISL_4082233, EPI_ISL_4082243, EPI_ISL_4082246, EPI_ISL_4082329, EPI_ISL_4082374, EPI_ISL_4082459, EPI_ISL_4083228, EPI_ISL_6694361 | | | |
| see above | Servicio Virosis Respiratorias-Departamento Virología-INEI | Instituto Nacional Enfermedades Infecciosas C.G.Malbran | Avaro M.; Baumeister E.; Benedetti E.; Campos J.; Cisterna D.; Dattero ME; De Belder D.; Haim MS.; Lorenzo F.; Molina V.; Perandones C.; Poklepovich T.; Pontoriero A.; Russo M.; Sanchez Loria J.; Tuduri E. |
| EPI_ISL_2283646, EPI_ISL_2283655, EPI_ISL_2283665, EPI_ISL_2283669, EPI_ISL_2283670, EPI_ISL_2861033 | Servicio de Microbiologia Hospital Ramon y Cajal | Servicio de Microbiologia Hospital Ramon y Cajal | Galan JC; Martinez L. Abreu M; Ponce M; y Gonzalez-Alba JM |
| EPI_ISL_2133015, EPI_ISL_2133016, EPI_ISL_2133017, EPI_ISL_2133019 | Servicio de Microbiología Clínica (Complejo Hospitalario de Navarra, Pamplona) | Centro de Secuenciación NASERTIC | Ana Miqueleiz; Ana Navascués; Carmen Ezpeleta Baquedano |
| EPI_ISL_2187013 | Servicio de Microbiología, Hospital General Universitario de Ciudad Real | Servicio de Microbiología, Hospital General Universitario de Ciudad Real | Jose Martinez Alarcon; Soledad Illescas |
| EPI_ISL_2362312, EPI_ISL_2362323, EPI_ISL_2362326 | Servicio de Microbiología. Consorcio Hospital General Universitario de Valencia | SeqCOVID-SPAIN consortium/IBV(CSIC) | Begoña Fuster Escrivá; Carme Salvador García; Concepción Gimeno Cardona and SeqCOVID-SPAIN consortium; María Dolores Ocete; Rafael Medina González |
| EPI_ISL_2323408 | Servicio de Microbiología. Hospital Clínico Universitario de Valencia | SeqCOVID-SPAIN consortium/IBV(CSIC) | David Navarro Ortega; Eliseo Albert Vicent; Ignacio Torres and SeqCOVID-SPAIN consortium |
| EPI_ISL_2162847, EPI_ISL_2162848 | Servizo de Microbioloxía. Complexo Hospitalario de Santiago de Compostela | Servizo de Microbioloxía. Complexo Hospitalario de Santiago de Compostela | Amparo Coira; Antonio Aguilera; Carlos García_Riestra; Daniel Navarro; Gema Barbeito; Iria Roca; José Llovo; Laura Millán; Manuela Hernández; María Luisa Pérez_del_Molino; Mercedes Treviño; Rocío trastoy; Teresa Lopez_Valiño; Xosé Costa |
| EPI_ISL_5315444, EPI_ISL_5315445, EPI_ISL_5315451, EPI_ISL_5315452, EPI_ISL_5315453, EPI_ISL_5315861, EPI_ISL_5315866, EPI_ISL_5315869, EPI_ISL_5315870, EPI_ISL_5315871, EPI_ISL_5315873, EPI_ISL_5315874, EPI_ISL_5315875, EPI_ISL_5315877, EPI_ISL_5315878, EPI_ISL_5315880, EPI_ISL_5315881, EPI_ISL_5315882, EPI_ISL_5315883, EPI_ISL_5315884, EPI_ISL_5315885, EPI_ISL_5315887, EPI_ISL_5315888, EPI_ISL_5315890, EPI_ISL_5315891, EPI_ISL_5315892, EPI_ISL_5315896, EPI_ISL_5315898, EPI_ISL_5315899, EPI_ISL_5315900, EPI_ISL_5315901, EPI_ISL_5315902, EPI_ISL_5315903, EPI_ISL_5315904, EPI_ISL_5315907, EPI_ISL_5315910, EPI_ISL_5315911, EPI_ISL_5315912, EPI_ISL_5315913, EPI_ISL_5315914, EPI_ISL_5315918, EPI_ISL_5315919, EPI_ISL_5316567, EPI_ISL_5316568, EPI_ISL_5316569, EPI_ISL_5316572 | | | |
| see above | Shared Hospital Laboratory | Shared Hospital Laboratory | Christie Vermeiren; Finlay Maguire; Kevin Katz; Patryk Aftanas; Robert Kozak; Samira Mubareka |
| EPI_ISL_2329427, EPI_ISL_2329445, EPI_ISL_2329450, EPI_ISL_2329452, EPI_ISL_2329453, EPI_ISL_2329459, EPI_ISL_2329462, EPI_ISL_2329463, EPI_ISL_2422827, EPI_ISL_2422833 | | | |
| see above | Sharp HealthCare Laboratory | Andersen lab at Scripps Research | Art Mendoza; Cathy Woerle; Jacquelyn Berumen; Liam McGinnis; Omid Bakhtar; SEARCH Alliance San Diego with Aaron Harding |
| EPI_ISL_2800958, EPI_ISL_2800961, EPI_ISL_2801115 | Simple Laboratories | Gagnon Lab, Southern Illinois University | Keith Gagnon |
| EPI_ISL_2438030, EPI_ISL_2438031, EPI_ISL_2438036, EPI_ISL_2621882, EPI_ISL_2651210 | Somer | Universidad Nacional de Colombia - Laboratorio Genómico One Health | Andres F. Cardona-Rios; Carlos Franco-Muñoz; Carolina Muñoz-Arango; Celeny Ortiz; Daniel O. Maldonado-Perez; Diego A. Álvarez-Díaz; Hector Alejandro Ruiz-Moreno; Idabely Betancur Ortiz; Jorge E. Osorio; Juan P. Hernandez-Ortiz; Karl A Ciuoderis; Katherine Laiton-Donato; Laura Silvana Perez; Lina M. Hurtado; Marcela Mercado-Reyes; Maria Angélica Maya; Maria Stella López; Rita Almanza Payares; Sandra Ines Cano; Simón Villegas Velásquez |
| EPI_ISL_3393976 | Sonic Reference Laboratory | Sonic Reference Laboratory | "Robert Moller; Bijaya Dhakal; Cora Lahey; Ellen Pizarek"; John Woody; Krishna Maddula |
| EPI_ISL_2230733, EPI_ISL_2230734, EPI_ISL_2230735, EPI_ISL_2230739 | Southern Nevada Public Health Laboratory | Southern Nevada Public Health Laboratory | Michael Picker |
| EPI_ISL_2137014 | Spital Limmattal | Institute of Medical Virology | Alexandra Trkola; Annette AudigŽ; Catharine Aquino; Cyril Shah; Daniel Ehrsam; Gabriela Ziltener; Guido Bloemberg; Hubert Rehrauer; Isabel StŸrmer; Joel Wirz; Jon Huder; JŸrg Bšni; Kevin Steiner; Maria GrŸnberg; Maryam Zaheri; Michael Huber; Riccarda Capaul; Stefan Schmutz; Verena Kufner; Weihong Qi |

| | | | |
|---|---|---|---|
| EPI_ISL_2136954, EPI_ISL_2136986, EPI_ISL_2136995, EPI_ISL_2137005 | Spital MŠnnedorf AG | Institute of Medical Virology | Alexandra Trkola; Annette AudigŽ; Catharine Aquino; Cyril Shah; Daniel Ehrsam; Gabriela Ziltener; Guido Bloemberg; Hubert Rehrauer; Isabel StŸrmer; Joel Wirz; Jon Huder; JŸrg Bšni; Kevin Steiner; Maria GrŸnberg; Maryam Zaheri; Michael Huber; Riccarda Capaul; Stefan Schmutz; Verena Kufner; Weihong Qi |
| EPI_ISL_4496856 | Stanford Health Care | Stanford University School of Medicine, Clinical Virology Laboratory | Becky Jiang; James Zehnder; Malaya K. Sahoo; Selamawit Bihon; and Benjamin A. Pinsky |
| EPI_ISL_2724555, EPI_ISL_2724574, EPI_ISL_2724590, EPI_ISL_2724602, EPI_ISL_2724611, EPI_ISL_2724624, EPI_ISL_2724739 | | | |
| see above | State Laboratories Division, Hawaii State Department of Health | State Laboratories Division, Hawaii State Department of Health | Ayana Garnet; Drew Kuwazaki; Edward Desmond; Pamela O'Brien; Razvan Sultana |
| EPI_ISL_2378546, EPI_ISL_2378547, EPI_ISL_2378548, EPI_ISL_2378552, EPI_ISL_2378553, EPI_ISL_2378555, EPI_ISL_2378559, EPI_ISL_2378560, EPI_ISL_2378574, EPI_ISL_2378578, EPI_ISL_2378580, EPI_ISL_2378588, EPI_ISL_2378592, EPI_ISL_2378601, EPI_ISL_2378603, EPI_ISL_2378615, EPI_ISL_2378616, EPI_ISL_2378625, EPI_ISL_2378627, EPI_ISL_2472919, EPI_ISL_2472922, EPI_ISL_2472930, EPI_ISL_2472941, EPI_ISL_2472942, EPI_ISL_2472954, EPI_ISL_2472989, EPI_ISL_2473038, EPI_ISL_2473061, EPI_ISL_2473570, EPI_ISL_2473579, EPI_ISL_2473601, EPI_ISL_2473604, EPI_ISL_2473608, EPI_ISL_2473613, EPI_ISL_2473614, EPI_ISL_2473624, EPI_ISL_2473631, EPI_ISL_2473638, EPI_ISL_2473649, EPI_ISL_2473650 | | | |
| see above | State Testing Facility | Altius Institute for Biomedical Research | Alex Isner; Alex Nguyen; Amanda Gale; Audra Johnson; Clem Green; Daniel Bates; Eric Thorland; Jacob Rodriguez; Jean Robinston; Jemma Nelson; Jessica Kunder; John Stamatoyannopoulos; Joshua Richards; Julia Wald; Kneshay Harper; Lauren Mitchell; Mark Frerker; Matt Hartman; Michael Buckley; Muhammad Halimun; Rebecca Bruders; Sadie Patraw; Sofia Olsson; Tobias Ragoczy |
| EPI_ISL_2391790, EPI_ISL_2391791 | Sun Clinical Laboratory | Los Angeles County Public Health Laboratories | P. Hemarajata et al. |
| EPI_ISL_2413875, EPI_ISL_2413880, EPI_ISL_2413956, EPI_ISL_2414220, EPI_ISL_2414270, EPI_ISL_2414279 | Swedish national genomic surveillance program of SARS-CoV-2 | The Public Health Agency of Sweden | Alma Brolund; Maria Lind Karlberg; Maximilian Riess; Swedish national genomic surveillance program of SARS-CoV-2 |
| EPI_ISL_2249001 | Synlab | Instituto Nacional de Saude (INSA) | Borges et al |
| EPI_ISL_2438048 | Synlab | Universidad Nacional de Colombia - Laboratorio Genómico One Health | Andres F. Cardona-Rios; Carlos Franco-Muñoz; Carolina Muñoz-Arango; Celeny Ortiz; Daniel O. Maldonado-Perez; Diego A. Álvarez-Díaz; Hector Alejandro Ruiz-Moreno; Idabely Betancur Ortiz; Jorge E. Osorio; Juan P. Hernandez-Ortiz; Karl A Ciuoderis; Katherine Laiton-Donato; Laura Silvana Perez; Lina M. Hurtado; Marcela Mercado-Reyes; Maria Angélica Maya; Maria Stella López; Rita Almanza Payares; Sandra Ines Cano; Simón Villegas Velásquez |
| EPI_ISL_2124435, EPI_ISL_2126492, EPI_ISL_2126806, EPI_ISL_2130003 | Synlab MVZ Augsburg | Robert Koch Institute | |
| EPI_ISL_2281719, EPI_ISL_2491085, EPI_ISL_2491212, EPI_ISL_2600392, EPI_ISL_2671981, EPI_ISL_2672023 | TGen North | TGen North | "Jolene Bowers; Ashlyn Pfeiffer; Chris French; Darrin Lemmer; Dave Engelthaler; Hayley Yaglom; Heather Centner; Jolene Bowers; The Arizona COVID Genomics Union (ACGU); The Arizona COVID Genomics Union (ACGU)" |
| EPI_ISL_2141477, EPI_ISL_2141478, EPI_ISL_2141479, EPI_ISL_2141480, EPI_ISL_2141481, EPI_ISL_2141482, EPI_ISL_2141483, EPI_ISL_2141484, EPI_ISL_2141485, EPI_ISL_2141486, EPI_ISL_2141487, EPI_ISL_2141488, EPI_ISL_2141489, EPI_ISL_2141490, EPI_ISL_2280002, EPI_ISL_2280038, EPI_ISL_2280040, EPI_ISL_2280041, EPI_ISL_2280054, EPI_ISL_2280055, EPI_ISL_2334068, EPI_ISL_2334077, EPI_ISL_2334092, EPI_ISL_2334093, EPI_ISL_2443305, EPI_ISL_2443309, EPI_ISL_2443311 | | | |
| see above | TXDSHS | TXDSHS | Anita Pokharel; Bonnie Oh; Chun Wang; Grace Kubin; Jenny Zhang; Karen Bobier; Lorraine Rodriguez; Maliha Rahman; Mayela Pedrueza; Myong Koag; Rachel Lee; Rashmi Tuladhar |
| EPI_ISL_2790479, EPI_ISL_2790508, EPI_ISL_2790509, EPI_ISL_2790544 | Tampa General Hospital Esoteric Lab | Tampa General Hospital Esoteric Research & Development Lab | Amorce Lima; Deanna Becker; Dominic Uy; Elaine Vendrone; Jon Faughn; Jorge Mecias-Frias; Suzane Silbert; Vicki Healer |
| EPI_ISL_2621688, EPI_ISL_2621689, EPI_ISL_2621690, EPI_ISL_2621692, EPI_ISL_2621693, EPI_ISL_2621695, EPI_ISL_2621701, EPI_ISL_2621702 | | | |
| see above | The Caribbean Public Health Agency | Carrington Lab, Department of Preclinical Sciences, Faculty of Medical Sciences, The University of the West Indies | Anushka Ramjag; Arianne Brown-Jordan; Avery Hinds; Christine V. F. Carrington; Christopher Oura; Gabriel Escobar; Karla Georges; Nikita S. D. Sahadeo; Nuno Faria; Oliver Pybus; Risha Singh; Roshan Parasram; Sarah Hill; SueMin Nathaniel; Vernie Ramkissoon |
| EPI_ISL_2678126, EPI_ISL_2678129, EPI_ISL_2678132, EPI_ISL_2678136, EPI_ISL_2678143, EPI_ISL_2678144, EPI_ISL_2678145, EPI_ISL_2678161, EPI_ISL_2678173, EPI_ISL_2678174, EPI_ISL_2678177, EPI_ISL_2678178, EPI_ISL_2678179, EPI_ISL_2678180, EPI_ISL_2678181, EPI_ISL_2678182, EPI_ISL_2678183, EPI_ISL_2678187, EPI_ISL_2678190, EPI_ISL_2678191, EPI_ISL_2716593, EPI_ISL_2716594, EPI_ISL_2716596, EPI_ISL_2716597, EPI_ISL_2716598, EPI_ISL_2756588, EPI_ISL_2756589, EPI_ISL_2756590, EPI_ISL_2756603, EPI_ISL_3634450, EPI_ISL_3634451, EPI_ISL_3634452, EPI_ISL_3634453, EPI_ISL_3634454, EPI_ISL_3634457, EPI_ISL_3634458, EPI_ISL_3634459, EPI_ISL_3634460, EPI_ISL_3634461, EPI_ISL_3634462, EPI_ISL_3634463, EPI_ISL_3634464, EPI_ISL_3634465, EPI_ISL_3634467, EPI_ISL_3634468, EPI_ISL_3634469, EPI_ISL_3634470, EPI_ISL_3634471, EPI_ISL_3634472, EPI_ISL_3634473, EPI_ISL_3634474, EPI_ISL_3634475 | | | |
| see above | The Caribbean Public Health Agency | Carrington Lab, Department of Preclinical Sciences, Faculty of Medical Sciences, The University of the West Indies, St Augustine Campus | Anushka Ramjag; Arianne Brown-Jordan; Avery Hinds; Christine V. F. Carrington; Christopher Oura; Gabriel Escobar; Nikita S. D. Sahadeo; Nuno Faria; Oliver Pybus; Risha Singh; Roshan Parasram; Sarah Hill; SueMin Nathaniel; Vernie Ramkissoon |
| EPI_ISL_2133353 | The Jackson Laboratory | The Jackson Laboratory | Adams M; Bergeron D; Kelly K; Li L; Long J; Omerza G; Renzette N |
| EPI_ISL_2598094, EPI_ISL_2598100, EPI_ISL_2598116, EPI_ISL_3274696 | TriCore Reference Laboratories | Center for Global Health, University of New Mexico Health Sciences Center | Cecilia Thompson; Darrell Dinwiddie; Daryl Domman; Karissa Culbreath; Kendra Pesko; Kurt Schwalm; Valerie Morley |
| EPI_ISL_2334732 | UAB Diagnostikos laboratorija | National Public Health Surveillance Laboratory | Ana Steponkiene; Danas Baksa; Jelena Razmuk; Lukas Vasionis; Lukas Zemaitis; Migle Gabrielaite; Svajune Muralyte |
| EPI_ISL_6229826 | UBASF PSF SEDE I | ACME Lab, Oswaldo Cruz Foundation, FIOCRUZ/CE | Carlos Leonardo de Aragao Araujo; Cecilia Leite Costa & Eduardo Ruback dos Santos on behalf of COVID-19 FIOCRUZ Genomic Network; Cleber Furtado Aksenen; Fabio Miyajima; Fernando Braga Stehling; Francisco Eder de Moura Lopes; Igor Oliveira Duarte; Jamille Maria Mendes Bezerra; Joaquim Cesar do Nascimento Sousa Junior; Pedro Miguel Carneiro Jeronimo; Suzana Porto Almeida; Thais Ferreira de Oliveira; Thais de Oliveira Costa; Ticiane Cavalcante de Souza; Veridiana Pessoa Miyajima |
| EPI_ISL_5650316 | UBS CUMBICA | Instituto Butantan | Antonio Jorge Martins; Claudia Renata dos Santos Barros; David Schlesinger; Debora Botequio Moretti; Dimas Tadeu Covas; Elaine Cristina Marqueze; Elaine Vieira Santos; Evandra Strazza Rodrigues; Heidge Fukumasu; Jayme Augusto de Souza-Neto; José Salvatore Leister Patané; Luiz Alcantara; Luiz Lehmann Coutinho; Maria Carolina Elias; Maurício Lacerda Nogueira; Rafael dos Santos Bezerra; Raul Machado Neto; Rejane Maria Tommasini Grotto; Ricardo Haddad; Sandra Coccuzzo Sampaio Vessoni; Simone Kashima; Svetoslav Nanev Slavov; Vincent Louis Viala |
| EPI_ISL_2345646, EPI_ISL_2345745 | UBS DR ALFREDO DANTAS DE SOUZA UMUARAMA | Instituto Butantan / Mendelics | Antonio Jorge Martins; Claudia Renata dos Santos Barros; David Schlesinger; Debora Botequio Moretti; Dimas Tadeu Covas; Elaine Cristina Marqueze; Elaine Vieira Santos; Evandra Strazza Rodrigues; Heidge Fukumasu; Jayme Augusto de Souza-Neto; José Salvatore Leister Patané; Luiz Alcantara; Luiz Lehmann Coutinho; Maria Carolina Elias; Maurício Lacerda Nogueira; Rafael dos Santos Bezerra; Raul Machado Neto; Rejane Maria Tommasini Grotto; Ricardo Haddad; Sandra Coccuzzo Sampaio Vessoni; Simone Kashima; Svetoslav Nanev Slavov; Vincent Louis Viala |
| EPI_ISL_2345803 | UBS DR ERNESTO GAION | Instituto Butantan / Mendelics | Antonio Jorge Martins; Claudia Renata dos Santos Barros; David Schlesinger; Debora Botequio Moretti; Dimas Tadeu Covas; Elaine Cristina Marqueze; Elaine Vieira Santos; Evandra Strazza Rodrigues; Heidge Fukumasu; Jayme Augusto de Souza-Neto; José Salvatore Leister Patané; Luiz Alcantara; Luiz Lehmann Coutinho; Maria Carolina Elias; Maurício Lacerda Nogueira; Rafael dos Santos Bezerra; Raul Machado Neto; Rejane Maria Tommasini Grotto; Ricardo Haddad; Sandra Coccuzzo Sampaio Vessoni; Simone Kashima; Svetoslav Nanev Slavov; Vincent Louis Viala |
| EPI_ISL_2345757 | UBS DR WALDOMIRO FERREIRA NEVES SCRPARDO | Instituto Butantan / Mendelics | Antonio Jorge Martins; Claudia Renata dos Santos Barros; David Schlesinger; Debora Botequio Moretti; Dimas Tadeu Covas; Elaine Cristina Marqueze; Elaine Vieira Santos; Evandra Strazza Rodrigues; Heidge Fukumasu; Jayme Augusto de Souza-Neto; José Salvatore Leister Patané; Luiz Alcantara; Luiz Lehmann Coutinho; Maria Carolina Elias; Maurício Lacerda Nogueira; Rafael dos Santos Bezerra; Raul Machado Neto; Rejane Maria Tommasini Grotto; Ricardo Haddad; Sandra Coccuzzo Sampaio Vessoni; Simone Kashima; Svetoslav Nanev Slavov; Vincent Louis Viala |
| EPI_ISL_2209618 | UBS II DE REGENTE FEIJO | Instituto Butantan / Mendelics | Antonio Jorge Martins; Claudia Renata dos Santos Barros; David Schlesinger; Debora Botequio Moretti; Dimas Tadeu Covas; Elaine Cristina Marqueze; Elaine Vieira Santos; Evandra Strazza Rodrigues; Heidge Fukumasu; Jayme Augusto de Souza-Neto; José Salvatore Leister Patané; Luiz Alcantara; Luiz Lehmann Coutinho; Maria Carolina Elias; Maurício Lacerda Nogueira; Rafael dos Santos Bezerra; Raul Machado Neto; Rejane Maria Tommasini Grotto; Ricardo Haddad; Sandra Coccuzzo Sampaio Vessoni; Simone Kashima; Svetoslav Nanev Slavov; Vincent Louis Viala |
| EPI_ISL_2345666, EPI_ISL_2345710, EPI_ISL_2362265 | UBS III DE RANCHARIA | Instituto Butantan / Mendelics | Antonio Jorge Martins; Claudia Renata dos Santos Barros; David Schlesinger; Debora Botequio Moretti; Dimas Tadeu Covas; Elaine Cristina Marqueze; Elaine Vieira Santos; Evandra Strazza Rodrigues; Heidge Fukumasu; Jayme Augusto de Souza-Neto; José Salvatore Leister Patané; Luiz Alcantara; Luiz Lehmann Coutinho; Maria Carolina Elias; Maurício Lacerda Nogueira; Rafael dos Santos Bezerra; Raul Machado Neto; Rejane Maria Tommasini Grotto; Ricardo Haddad; Sandra Coccuzzo Sampaio Vessoni; Simone Kashima; Svetoslav Nanev Slavov; Vincent Louis Viala |
| EPI_ISL_5977773 | UC Davis Genome Center | UC Davis Genome Center | Healthy Davis Together; UC Davis |
| EPI_ISL_3535720 | UC-Christus Clinical Hospital Laboratory | Laboratory of Molecular Virology, School of Medicine, Pontificia Universidad Catolica de Chile | Ana Maira Guzman; Andres E. Munoz-Marcos; Catalina Pardo-Roa; Eileen Serrano; Erick Salinas; Estefany Poblete; Francisco Melo; Jorge Levican; Leonardo I. Almonacid; Maria Jose Avendano; Maria Patricia Vega; Rafael A. Medina; Ricardo Enrique de la Barra; Tamara Garcia-Salum |
| EPI_ISL_3104695, EPI_ISL_3104696 | UCC (Universidad Central del Caribe) | Grubaugh Lab - Yale School of Public Health | Anderson Brito; Andrea Firpo; Annie Watkins; Chaney Kalinich; Chantal Vogels; David Rivera Aponte; Ernesto Vazquez; Fabiola Cruz-Lopez; Fabiola Fontanet; Isabel Ott; Jessica Rothman; Joseph Fauver; Karolane Gonzalez; Kendall Billig; Mallery Breban; Mary Petrone; Natalie Machargo; Nathan Grubaugh; Patricia Serrano; Tania Mitwalli; Tara Alpert; Tobias Koch; Vianca Aponte |
| EPI_ISL_2404857, EPI_ISL_2405167 | UCLA Clinical Micro Lab | Los Angeles County Public Health Laboratories | P. Hemarajata et al. |
| EPI_ISL_2448471 | ULSS 5 Polesana | Istituto Zooprofilattico Sperimentale delle Venezie | Adelaide Milani; Alessia Schivo; Alice Fusaro; Ambra Pastori; Annalisa Salviato; Antonia Ricci; Calogero Terregino; Edoardo Giussani; Elisa Palumbo; Erika Giorgia Quaranta; Isabella Monne; Luca Tassoni |
| EPI_ISL_3409164, EPI_ISL_3409229, EPI_ISL_3409298, EPI_ISL_3409341, EPI_ISL_3410085 | UMass Memorial Medical Center | Center for Microbiome Research | Doyle V. Ward; Karl J. Simon; Richard T. Ellison III |
| EPI_ISL_5825558 | UNIDADE BASICA DE SAUDE DA FAMILIA MARIA DE FATIMA MARQUES | Analytical Competence Molecular Epidemiology Lab/ACME, Oswaldo Cruz Foundation, Ceara (FIOCRUZ | Carlos Leonardo de Aragao Araujo; Cecilia Leite Costa & Eduardo Ruback dos Santos on behalf of COVID-19 FIOCRUZ Genomic Network; Cleber Furtado Aksenen; Fabio Miyajima; Fernando Braga Stehling; Francisco Eder de Moura Lopes; Igor Oliveira Duarte; Jamille Maria Mendes Bezerra; Joaquim Cesar do Nascimento Sousa Junior; Pedro Miguel Carneiro Jeronimo; Suzana Porto Almeida; Thais Ferreira de Oliveira; Thais de Oliveira Costa; Ticiane Cavalcante de Souza; Veridiana Pessoa Miyajima |

| | | | |
|---|---|---|---|
| | | CE) | |
| EPI_ISL_6229803, EPI_ISL_6229804 | UNIDADE BASICA DE SAUDE FLORES | ACME Lab, Oswaldo Cruz Foundation, FIOCRUZ/CE | Carlos Leonardo de Aragao Araujo; Cecilia Leite Costa & Eduardo Ruback dos Santos on behalf of COVID-19 FIOCRUZ Genomic Network; Cleber Furtado Aksenen; Fabio Miyajima; Fernando Braga Stehling; Francisco Eder de Moura Lopes; Igor Oliveira Duarte; Jamille Maria Mendes Bezerra; Joaquim Cesar do Nascimento Sousa Junior; Pedro Miguel Carneiro Jeronimo; Suzana Porto Almeida; Thais Ferreira de Oliveira; Thais de Oliveira Costa; Ticiane Cavalcante de Souza; Veridiana Pessoa Miyajima |
| EPI_ISL_2209351, EPI_ISL_2210141, EPI_ISL_2345673, EPI_ISL_2345719, EPI_ISL_2345723 | UNIDADE BASICA DE SAUDE JARDIM DO LAGO | Instituto Butantan / Mendelics | Antonio Jorge Martins; Claudia Renata dos Santos Barros; David Schlesinger; Debora Botequio Moretti; Dimas Tadeu Covas; Elaine Cristina Marqueze; Elaine Vieira Santos; Evandra Strazza Rodrigues; Heidge Fukumasu; Jayme Augusto de Souza-Neto; José Salvatore Leister Patané; Luiz Alcantara; Luiz Lehmann Coutinho; Maria Carolina Elias; Maurício Lacerda Nogueira; Rafael dos Santos Bezerra; Raul Machado Neto; Rejane Maria Tommasini Grotto; Ricardo Haddad; Sandra Coccuzzo Sampaio Vessoni; Simone Kashima; Svetoslav Nanev Slavov; Vincent Louis Viala |
| EPI_ISL_2345789 | UNIDADE BASICA SAUDE NATAL ABADIO LACERDA | Instituto Butantan / Mendelics | Antonio Jorge Martins; Claudia Renata dos Santos Barros; David Schlesinger; Debora Botequio Moretti; Dimas Tadeu Covas; Elaine Cristina Marqueze; Elaine Vieira Santos; Evandra Strazza Rodrigues; Heidge Fukumasu; Jayme Augusto de Souza-Neto; José Salvatore Leister Patané; Luiz Alcantara; Luiz Lehmann Coutinho; Maria Carolina Elias; Maurício Lacerda Nogueira; Rafael dos Santos Bezerra; Raul Machado Neto; Rejane Maria Tommasini Grotto; Ricardo Haddad; Sandra Coccuzzo Sampaio Vessoni; Simone Kashima; Svetoslav Nanev Slavov; Vincent Louis Viala |
| EPI_ISL_6229805 | UNIDADE DE PRONTO ATENDIMENTO 24H DR ANTONIO MAGALHAES UPA | ACME Lab, Oswaldo Cruz Foundation, FIOCRUZ/CE | Carlos Leonardo de Aragao Araujo; Cecilia Leite Costa & Eduardo Ruback dos Santos on behalf of COVID-19 FIOCRUZ Genomic Network; Cleber Furtado Aksenen; Fabio Miyajima; Fernando Braga Stehling; Francisco Eder de Moura Lopes; Igor Oliveira Duarte; Jamille Maria Mendes Bezerra; Joaquim Cesar do Nascimento Sousa Junior; Pedro Miguel Carneiro Jeronimo; Suzana Porto Almeida; Thais Ferreira de Oliveira; Thais de Oliveira Costa; Ticiane Cavalcante de Souza; Veridiana Pessoa Miyajima |
| EPI_ISL_6229845 | UNIDADE DE PRONTO ATENDIMENTO DE IGUATU UPA | ACME Lab, Oswaldo Cruz Foundation, FIOCRUZ/CE | Carlos Leonardo de Aragao Araujo; Cecilia Leite Costa & Eduardo Ruback dos Santos on behalf of COVID-19 FIOCRUZ Genomic Network; Cleber Furtado Aksenen; Fabio Miyajima; Fernando Braga Stehling; Francisco Eder de Moura Lopes; Igor Oliveira Duarte; Jamille Maria Mendes Bezerra; Joaquim Cesar do Nascimento Sousa Junior; Pedro Miguel Carneiro Jeronimo; Suzana Porto Almeida; Thais Ferreira de Oliveira; Thais de Oliveira Costa; Ticiane Cavalcante de Souza; Veridiana Pessoa Miyajima |
| EPI_ISL_2801357 | UNIDADE PRONTO ATENDIMENTO ITAPERI | Analytical Competence Molecular Epidemiology Lab/ACME, Oswaldo Cruz Foundation, Ceara (FIOCRUZ CE) | Cleber Furtado Aksenen e Suzana Porto Almeida; Fabio Miyajima; Fernando Braga Stehling; Francisco Eder de Moura Lopes; Jamille Maria Mendes Bezerra; Joaquim César do Nascimento Sousa Junior; Pedro Miguel Carneiro Jeronimo; Thais Ferreira de Oliveira; Thais de Oliveira Costa; Ticiane Cavalcante de Souza; Veridiana Pessoa Miyajima |
| EPI_ISL_6229812 | UNIDADE REFERENCIA COVID 19 | ACME Lab, Oswaldo Cruz Foundation, FIOCRUZ/CE | Carlos Leonardo de Aragao Araujo; Cecilia Leite Costa & Eduardo Ruback dos Santos on behalf of COVID-19 FIOCRUZ Genomic Network; Cleber Furtado Aksenen; Fabio Miyajima; Fernando Braga Stehling; Francisco Eder de Moura Lopes; Igor Oliveira Duarte; Jamille Maria Mendes Bezerra; Joaquim Cesar do Nascimento Sousa Junior; Pedro Miguel Carneiro Jeronimo; Suzana Porto Almeida; Thais Ferreira de Oliveira; Thais de Oliveira Costa; Ticiane Cavalcante de Souza; Veridiana Pessoa Miyajima |
| EPI_ISL_5825563, EPI_ISL_5825564 | UNIDADE REFERENCIA COVID 19 | Analytical Competence Molecular Epidemiology Lab/ACME, Oswaldo Cruz Foundation, Ceara (FIOCRUZ CE) | Carlos Leonardo de Aragao Araujo; Cecilia Leite Costa & Eduardo Ruback dos Santos on behalf of COVID-19 FIOCRUZ Genomic Network; Cleber Furtado Aksenen; Fabio Miyajima; Fernando Braga Stehling; Francisco Eder de Moura Lopes; Igor Oliveira Duarte; Jamille Maria Mendes Bezerra; Joaquim Cesar do Nascimento Sousa Junior; Pedro Miguel Carneiro Jeronimo; Suzana Porto Almeida; Thais Ferreira de Oliveira; Thais de Oliveira Costa; Ticiane Cavalcante de Souza; Veridiana Pessoa Miyajima |
| EPI_ISL_3355488 | UNIVERSIDAD DEL QUINDIO | Corporacion CorpoGen-Universidad de los Andes-Universidad Central | Christian Romero; Jorge Duitama; Juan Manuel Anzola; Laura González; Maryam Chaib De Mares; María Mercedes Zambrano; Nelly Díaz; Patricia Del Portillo; Silvia Restrepo |
| EPI_ISL_2820953, EPI_ISL_2820958 | UOC Microbiologia e Virologia, Azienda Ospedaliera Universitaria Senese, Siena, Italy | Dipartimento di Biotecnologie Mediche | Claudia Gandolfo; David Pinzauti; Francesco Santoro; Gabriele Anichini; Gianni Gori Savellini; Gianni Pozzi; Maria Grazia Cusi |
| EPI_ISL_2208944 | UPA DE BEBEDOURO | Instituto Butantan / Mendelics | Antonio Jorge Martins; Claudia Renata dos Santos Barros; David Schlesinger; Debora Botequio Moretti; Dimas Tadeu Covas; Elaine Cristina Marqueze; Elaine Vieira Santos; Evandra Strazza Rodrigues; Heidge Fukumasu; Jayme Augusto de Souza-Neto; José Salvatore Leister Patané; Luiz Alcantara; Luiz Lehmann Coutinho; Maria Carolina Elias; Maurício Lacerda Nogueira; Rafael dos Santos Bezerra; Raul Machado Neto; Rejane Maria Tommasini Grotto; Ricardo Haddad; Sandra Coccuzzo Sampaio Vessoni; Simone Kashima; Svetoslav Nanev Slavov; Vincent Louis Viala |
| EPI_ISL_2209746 | UPA MARY DOTA | Instituto Butantan / Mendelics | Antonio Jorge Martins; Claudia Renata dos Santos Barros; David Schlesinger; Debora Botequio Moretti; Dimas Tadeu Covas; Elaine Cristina Marqueze; Elaine Vieira Santos; Evandra Strazza Rodrigues; Heidge Fukumasu; Jayme Augusto de Souza-Neto; José Salvatore Leister Patané; Luiz Alcantara; Luiz Lehmann Coutinho; Maria Carolina Elias; Maurício Lacerda Nogueira; Rafael dos Santos Bezerra; Raul Machado Neto; Rejane Maria Tommasini Grotto; Ricardo Haddad; Sandra Coccuzzo Sampaio Vessoni; Simone Kashima; Svetoslav Nanev Slavov; Vincent Louis Viala |
| EPI_ISL_2345820, EPI_ISL_2362243 | UPA UNIDADE DE PRONTO ATENDIMENTO 24 HORAS BOM JESUS | Instituto Butantan / Mendelics | Antonio Jorge Martins; Claudia Renata dos Santos Barros; David Schlesinger; Debora Botequio Moretti; Dimas Tadeu Covas; Elaine Cristina Marqueze; Elaine Vieira Santos; Evandra Strazza Rodrigues; Heidge Fukumasu; Jayme Augusto de Souza-Neto; José Salvatore Leister Patané; Luiz Alcantara; Luiz Lehmann Coutinho; Maria Carolina Elias; Maurício Lacerda Nogueira; Rafael dos Santos Bezerra; Raul Machado Neto; Rejane Maria Tommasini Grotto; Ricardo Haddad; Sandra Coccuzzo Sampaio Vessoni; Simone Kashima; Svetoslav Nanev Slavov; Vincent Louis Viala |
| EPI_ISL_2426705, EPI_ISL_2426720, EPI_ISL_2426768, EPI_ISL_2426771, EPI_ISL_2426774, EPI_ISL_2426783, EPI_ISL_2450425 | | | |
| see above | US Air Force School of Aerospace Medicine | US Air Force School of Aerospace Medicine | Amanda Javorina; Anthony Fries; Carol Garrett; Clarise Starr; Cole Anderson; Elizabeth Macias; Fritz Castillo; Jennifer Meyer; Sarah Purves; William Gruner |
| EPI_ISL_2391784 | USC Clinical Lab | Los Angeles County Public Health Laboratories | P. Hemarajata et al. |
| EPI_ISL_2235429, EPI_ISL_2235430 | USCA Avezzano | Istituto Zooprofilattico Sperimentale dell'Abruzzo e Molise "G. Caporale" | Ancora M; Calistri P; Cammà C; Caporale M; Curini V; Delli Compagni E; Di Domenico M; Di Lollo Valeria; Di Pasquale A; Lorusso A; Mangone I; Marcacci M; Puglia I; Rinaldi A; Savini G; Scialabba S |
| EPI_ISL_2235332 | USCA Pescina | Istituto Zooprofilattico Sperimentale dell'Abruzzo e Molise "G. Caporale" | Ancora M; Calistri P; Cammà C; Caporale M; Curini V; Delli Compagni E; Di Domenico M; Di Lollo Valeria; Di Pasquale A; Lorusso A; Mangone I; Marcacci M; Puglia I; Rinaldi A; Savini G; Scialabba S |
| EPI_ISL_2345746 | USF ARCADAS | Instituto Butantan / Mendelics | Antonio Jorge Martins; Claudia Renata dos Santos Barros; David Schlesinger; Debora Botequio Moretti; Dimas Tadeu Covas; Elaine Cristina Marqueze; Elaine Vieira Santos; Evandra Strazza Rodrigues; Heidge Fukumasu; Jayme Augusto de Souza-Neto; José Salvatore Leister Patané; Luiz Alcantara; Luiz Lehmann Coutinho; Maria Carolina Elias; Maurício Lacerda Nogueira; Rafael dos Santos Bezerra; Raul Machado Neto; Rejane Maria Tommasini Grotto; Ricardo Haddad; Sandra Coccuzzo Sampaio Vessoni; Simone Kashima; Svetoslav Nanev Slavov; Vincent Louis Viala |
| EPI_ISL_2104837, EPI_ISL_2104840, EPI_ISL_2104855, EPI_ISL_2104871, EPI_ISL_2104885, EPI_ISL_2104889, EPI_ISL_2104898, EPI_ISL_2104924, EPI_ISL_2104987, EPI_ISL_2105057, EPI_ISL_2105063, EPI_ISL_2161831, EPI_ISL_2161833, EPI_ISL_2161842, EPI_ISL_2161859, EPI_ISL_2161881, EPI_ISL_2161903, EPI_ISL_2161957, EPI_ISL_2161985, EPI_ISL_2162011, EPI_ISL_2162027, EPI_ISL_2194045, EPI_ISL_2194047, EPI_ISL_2194049, EPI_ISL_2194050, EPI_ISL_2194051, EPI_ISL_2194076, EPI_ISL_2194086, EPI_ISL_2194087, EPI_ISL_2194089, EPI_ISL_2194098, EPI_ISL_2194108, EPI_ISL_2194109, EPI_ISL_2194130, EPI_ISL_2194133, EPI_ISL_2194138, EPI_ISL_2194142, EPI_ISL_2194174, EPI_ISL_2194213, EPI_ISL_2194216, EPI_ISL_2233144, EPI_ISL_2233257, EPI_ISL_2233293, EPI_ISL_2233305, EPI_ISL_2233320, EPI_ISL_2299432, EPI_ISL_2299441, EPI_ISL_2299451, EPI_ISL_2299454, EPI_ISL_2299456, EPI_ISL_2299488, EPI_ISL_2340696, EPI_ISL_2340768, EPI_ISL_2340799, EPI_ISL_2618309, EPI_ISL_2618326, EPI_ISL_2618367, EPI_ISL_2618370 | | | |
| see above | UW Virology Lab | UW Virology Lab | Alexander Greninger; Hong Xie; Keith R Jerome; Lasata Shrestha; Meei-Li Huang; Michelle Lin; Nathan Breit; Noah R. Baker; Patrick Mathias; Pavitra Roychoudhury; Ricardo Perez; Robert J. Livingston; Sean Ellis; Shah Mohamed Bakhash; Tien V. Nguyen |
| EPI_ISL_5801583, EPI_ISL_5801615 | Ubs Dr Alfredo Dantas De Souza Umuarama | Instituto Butantan | Antonio Jorge Martins; Claudia Renata dos Santos Barros; David Schlesinger; Debora Botequio Moretti; Dimas Tadeu Covas; Elaine Cristina Marqueze; Elaine Vieira Santos; Evandra Strazza Rodrigues; Heidge Fukumasu; Jayme Augusto de Souza-Neto; José Salvatore Leister Patané; Luiz Alcantara; Luiz Lehmann Coutinho; Maria Carolina Elias; Maurício Lacerda Nogueira; Rafael dos Santos Bezerra; Raul Machado Neto; Rejane Maria Tommasini Grotto; Ricardo Haddad; Sandra Coccuzzo Sampaio Vessoni; Simone Kashima; Svetoslav Nanev Slavov; Vincent Louis Viala |
| EPI_ISL_5801519 | Ubs Dr Ernesto Gaion | Instituto Butantan | Antonio Jorge Martins; Claudia Renata dos Santos Barros; David Schlesinger; Debora Botequio Moretti; Dimas Tadeu Covas; Elaine Cristina Marqueze; Elaine Vieira Santos; Evandra Strazza Rodrigues; Heidge Fukumasu; Jayme Augusto de Souza-Neto; José Salvatore Leister Patané; Luiz Alcantara; Luiz Lehmann Coutinho; Maria Carolina Elias; Maurício Lacerda Nogueira; Rafael dos Santos Bezerra; Raul Machado Neto; Rejane Maria Tommasini Grotto; Ricardo Haddad; Sandra Coccuzzo Sampaio Vessoni; Simone Kashima; Svetoslav Nanev Slavov; Vincent Louis Viala |
| EPI_ISL_5801574 | Ubs Dr Waldomiro Ferreira Neves Scrpardo | Instituto Butantan | Antonio Jorge Martins; Claudia Renata dos Santos Barros; David Schlesinger; Debora Botequio Moretti; Dimas Tadeu Covas; Elaine Cristina Marqueze; Elaine Vieira Santos; Evandra Strazza Rodrigues; Heidge Fukumasu; Jayme Augusto de Souza-Neto; José Salvatore Leister Patané; Luiz Alcantara; Luiz Lehmann Coutinho; Maria Carolina Elias; Maurício Lacerda Nogueira; Rafael dos Santos Bezerra; Raul Machado Neto; Rejane Maria Tommasini Grotto; Ricardo Haddad; Sandra Coccuzzo Sampaio Vessoni; Simone Kashima; Svetoslav Nanev Slavov; Vincent Louis Viala |
| EPI_ISL_5801698 | Ubs Ii De Regente Feijo | Instituto Butantan | Antonio Jorge Martins; Claudia Renata dos Santos Barros; David Schlesinger; Debora Botequio Moretti; Dimas Tadeu Covas; Elaine Cristina Marqueze; Elaine Vieira Santos; Evandra Strazza Rodrigues; Heidge Fukumasu; Jayme Augusto de Souza-Neto; José Salvatore Leister Patané; Luiz Alcantara; Luiz Lehmann Coutinho; Maria Carolina Elias; Maurício Lacerda Nogueira; Rafael dos Santos Bezerra; Raul Machado Neto; Rejane Maria Tommasini Grotto; Ricardo Haddad; Sandra Coccuzzo Sampaio Vessoni; Simone Kashima; Svetoslav Nanev Slavov; Vincent Louis Viala |
| EPI_ISL_5801705, EPI_ISL_5801712, EPI_ISL_5801714 | Ubs Iii De Rancharia | Instituto Butantan | Antonio Jorge Martins; Claudia Renata dos Santos Barros; David Schlesinger; Debora Botequio Moretti; Dimas Tadeu Covas; Elaine Cristina Marqueze; Elaine Vieira Santos; Evandra Strazza Rodrigues; Heidge Fukumasu; Jayme Augusto de Souza-Neto; José Salvatore Leister Patané; Luiz Alcantara; Luiz Lehmann Coutinho; Maria Carolina Elias; Maurício Lacerda Nogueira; Rafael dos Santos Bezerra; Raul Machado Neto; Rejane Maria Tommasini Grotto; Ricardo Haddad; Sandra Coccuzzo Sampaio Vessoni; Simone Kashima; Svetoslav Nanev Slavov; Vincent Louis Viala |
| EPI_ISL_2490385 | Unidad de Investigación Biomédica de Zacatecas (UIBZ) | Centro de Investigación en Enfermedades Infecciosas (CIENI), Instituto Nacional de Enfermedades Respiratorias (INER) | ; Alejandra Garcia-Gasca; Alejandra Hernández-Terán; Alejandro Sanchez-Flores; Alfredo Herrera-Estrella; Alicia Ocaña-Mondragón; Andreu Comas-García; Angel Gustavo Salas-Lais; Antonio Loza Román; Bernardo Martínez-Miguel; Blanca Taboada; Brenda Irasema Maldonado-Meza; Bruno Gomez-Gil; Carla Ivón Herrera-Najera; Carlos F. Arias; Celia Boukadida; Clara Esperanza Santacruz-Tinoco; Concepción Grajales-Muñiz; Consorcio Mexicano de Vigilancia Genómica (CoViGen-Mex). Authors (in alphabetical order): Julio Elias Alvarado-Yaah; Cristóbal Cháidez-Quiróz; Célida Duque Molina; Célida Martínez- Rodríguez; Daniel Fregoso-Rueda; Daniel Lira Morales; Eduardo Becerril-Vargas; Fernando Fontove-Herrera; Fidencio Mejía-Nepomuceno; Francisco Pulido; Gloria Elena Espinosa-Ayala; Gloria María Molina-Salinas; Gloria Vazquez; Hector Esteban Paz-Juárez; Hector Montoya-Fuentes; Helen Haydee Fernanda Ramírez-Plascencia; Irvin González-López; Jean Pierre González; Joel Armando Vázquez-Pérez.; Jorge Salas-Hernández; José Antonio Enciso-Moreno; José Arturo Martínez-Orozco; José Esteban Muñoz-Medina; José de Jesús Nuñez-Contreras; Juan Bautista Chale-Dzul; Julissa Enciso-Ibarra; Luis Alberto Ochoa-Carrera; Margarita Matías-Florentino; Mario Mújica-Sánchez; Marissa Perez-Garcia; María Guadalupe Santiago-Mauricio; María Guadalupe de Jesús Mireles-Rivera; Nelly Sélem-Mojica; Pavel Isa; Ricardo Ciria Merce; Ricardo Grande; Rosa María Gutierrez Rios; Santiago Ávila-Ríos; Selene Zárate; Susana Lopez; Victor Eduardo Garcia-Arias; Víctor Hugo Borja-Aburto |
| EPI_ISL_2490371, EPI_ISL_2490372, EPI_ISL_2490373, EPI_ISL_2490374, EPI_ISL_2490375, EPI_ISL_2490376, EPI_ISL_2490377 | | | |
| see above | Unidad de Investigación Médica de Yucatán (UIMY) | Centro de Investigación en Enfermedades Infecciosas (CIENI), Instituto Nacional de Enfermedades Respiratorias (INER) | ; Alejandra Garcia-Gasca; Alejandra Hernández-Terán; Alejandro Sanchez-Flores; Alfredo Herrera-Estrella; Alicia Ocaña-Mondragón; Andreu Comas-García; Angel Gustavo Salas-Lais; Antonio Loza Román; Bernardo Martínez-Miguel; Blanca Taboada; Brenda Irasema Maldonado-Meza; Bruno Gomez-Gil; Carla Ivón Herrera-Najera; Carlos F. Arias; Celia Boukadida; Clara Esperanza Santacruz-Tinoco; Concepción Grajales-Muñiz; Consorcio Mexicano de Vigilancia Genómica (CoViGen-Mex). Authors (in alphabetical order): Julio Elias Alvarado-Yaah; Cristóbal Cháidez-Quiróz; Célida Duque Molina; Célida Martínez- Rodríguez; Daniel Fregoso-Rueda; Daniel Lira Morales; Eduardo Becerril-Vargas; Fernando Fontove-Herrera; Fidencio Mejía-Nepomuceno; Francisco Pulido; Gloria Elena Espinosa-Ayala; Gloria María Molina-Salinas; Gloria Vazquez; Hector Esteban Paz-Juárez; Hector Montoya-Fuentes; Helen Haydee Fernanda Ramírez-Plascencia; Irvin González-López; Jean Pierre González; Joel Armando Vázquez-Pérez.; Jorge Salas-Hernández; José Antonio Enciso-Moreno; José Arturo Martínez-Orozco; José Esteban Muñoz-Medina; José de Jesús Nuñez-Contreras; Juan Bautista Chale-Dzul; Julissa Enciso-Ibarra; Luis Alberto Ochoa-Carrera; Margarita Matías-Florentino; Mario Mújica-Sánchez; Marissa Perez-Garcia; María Guadalupe Santiago-Mauricio; María Guadalupe de Jesús Mireles-Rivera; Nelly Sélem-Mojica; Pavel Isa; Ricardo Ciria Merce; Ricardo Grande; Rosa María Gutierrez Rios; Santiago Ávila-Ríos; Selene Zárate; Susana Lopez; Victor Eduardo Garcia-Arias; Víctor Hugo Borja-Aburto |
| EPI_ISL_5801626, EPI_ISL_5801630, EPI_ISL_5801639, EPI_ISL_5801650, EPI_ISL_5801658 | Unidade Basica De Saude Jardim Do Lago | Instituto Butantan | Antonio Jorge Martins; Claudia Renata dos Santos Barros; David Schlesinger; Debora Botequio Moretti; Dimas Tadeu Covas; Elaine Cristina Marqueze; Elaine Vieira Santos; Evandra Strazza Rodrigues; Heidge Fukumasu; Jayme Augusto de Souza-Neto; José Salvatore Leister Patané; Luiz Alcantara; Luiz Lehmann Coutinho; Maria Carolina Elias; Maurício Lacerda Nogueira; Rafael dos Santos Bezerra; Raul Machado Neto; Rejane Maria Tommasini Grotto; Ricardo Haddad; Sandra Coccuzzo Sampaio Vessoni; Simone Kashima; Svetoslav Nanev Slavov; Vincent Louis Viala |
| EPI_ISL_5801602 | Unidade Basica Saude Natal Abadio Lacerda | Instituto Butantan | Antonio Jorge Martins; Claudia Renata dos Santos Barros; David Schlesinger; Debora Botequio Moretti; Dimas Tadeu Covas; Elaine Cristina Marqueze; Elaine Vieira Santos; Evandra Strazza Rodrigues; Heidge Fukumasu; Jayme Augusto de Souza-Neto; José Salvatore Leister Patané; Luiz Alcantara; Luiz Lehmann Coutinho; Maria Carolina Elias; Maurício Lacerda Nogueira; Rafael dos Santos Bezerra; Raul Machado Neto; Rejane Maria Tommasini Grotto; Ricardo Haddad; Sandra Coccuzzo Sampaio Vessoni; Simone Kashima; Svetoslav Nanev Slavov; Vincent Louis Viala |
| EPI_ISL_2385698, EPI_ISL_2385714, EPI_ISL_2385722, EPI_ISL_2385723 | Unidade de apoio ao diagnóstico da COVID – UNADIG | Bioinformatics Laboratory / LNCC | Alessandra P Lamarca; Alexandra L Gerber; Amilcar Tanuri; Ana Paula de C Guimaraes; Ana Tereza R Vasconcelos; Andrea Cony Cavalcanti; Caio Luiz Pereira Ribeiro; Cassia Alves; Cintia Policarpo; Claudia Maria Braga de Mello; Cristiane Gomes da Silva; Diana Mariani; Douglas Terra Machado; Flavio Dias da Silva; Gleidson da Silva de Oliveira; Leandro Magalhaes de Souza; Liliane Cavalcante; Luiz G P de Almeida; Marcio Henrique de Oliveira Garcia; Mario Sergio Ribeiro; Ronaldo da Silva F Jr; Silvia Carvalho |
| EPI_ISL_3023942 | Universidad Autonoma de Yucatan | New York Genome Center | Amy Baldwin; Andre Corvelo; Aviles-Gomez E; Ayora-Talavera G; Chan-Gasca M; Dayna M. Oschwald; Flores-Quintal F; Granja-Perez P; Lopez-Coral L.; Michael Zody; Samantha Fennessey; Tom Maniatis; Villanueva-Jorge S; Yam-Pool E |
| EPI_ISL_2438094 | Universidad Nacional de Colombia - Laboratorio Genómico One Health | Universidad Nacional de Colombia - Laboratorio Genómico One Health | Andres F. Cardona-Rios; Carlos Franco-Muñoz; Carolina Muñoz-Arango; Celeny Ortiz; Daniel O. Maldonado-Perez; Diego A. Álvarez-Díaz; Hector Alejandro Ruiz-Moreno; Idabely Betancur Ortiz; Jorge E. Osorio; Juan P. Hernandez-Ortiz; Karl A Ciuoderis; Katherine Laiton-Donato; Laura Silvana Perez; Lina M. Hurtado; Marcela Mercado-Reyes; Maria Angélica Maya; Maria Stella López; Rita Almanza Payares; Sandra Ines Cano; Simón Villegas Velásquez |
| EPI_ISL_5199341 | Universidad de Caldas | Instituto Nacional de Salud-Dirección de Investigación en Salud Pública | Beatriz de Arco; Carlos Franco-Muñoz; Carmen Osorio; Diana Malo; Diego A. Álvarez-Díaz; Diego Andrés Prada; Gerardo Santamaría; Hector Alejandro Ruiz-Moreno; Jhonnatan Reales-González; Jorge Rivera; Juan Camilo Martinez; Julian Naizaque; Katherine Laiton-Donato; Lisseth Pardo; Magdalena Wiesner; Marcela Mercado-Reyes; Maria T. Herrera-Sepúlveda; Marta Lopez Blanco; Martha Lucia Ospina Martinez; Paola Rojas; Sergio Gomez; Sheryll Corchuelo; Tatiana Cobos; Ángela Alarcon Cruz |
| EPI_ISL_2091078, EPI_ISL_2091079, EPI_ISL_2091080, EPI_ISL_2091111, EPI_ISL_2193435, EPI_ISL_2193436, EPI_ISL_2193437, EPI_ISL_2193438, EPI_ISL_2193440 | | | |
| see above | University Hospitals Translational | University Hospitals Translational | Alouani, D.; Sadri, N.; Song, X. |

| | | | |
|---|---|---|---|
| | Laboratory (UHTL), University Hospitals | Laboratory (UHTL), University Hospitals | |
| EPI_ISL_2104674 | University Hospitals of Geneva, Laboratory of Virology | HUG, Laboratory of Virology and the Health2030 Genome Center | Ana Rita Goncalves; Deborah Penet; Emmanouil Dermitzakis; Henri Pegeot; Ioannis Xenarios; Keith Harshman; Laurent Kaiser; Lorenzo Cerutti; Melyssa Elies; Samuel Cordey |
| EPI_ISL_2408064, EPI_ISL_2408065, EPI_ISL_2408066, EPI_ISL_2408067 | University of Bari Biomedical Sciences and Human Oncology | University of Bari Biomedical Sciences and Human Oncology | Accogli M.; Chironna M.; Loconsole D.; Sallustio A. |
| EPI_ISL_2832102, EPI_ISL_2832103, EPI_ISL_2832104 | University of Bari Biomedical Sciences and Human Oncology, Policlinico | University of Bari Biomedical Sciences and Human Oncology | Accogli M.; Chironna M.; Loconsole D.; Sallustio A. |
| EPI_ISL_2476365, EPI_ISL_2476370, EPI_ISL_2476371, EPI_ISL_2476372, EPI_ISL_2691964, EPI_ISL_2691967, EPI_ISL_2692012 | | | |
| see above | University of Chicago Medicine | RIPHL at Rush University Medical Center | Felix Araujo Perez; Kevin Kunstman; Laura Furtado; Lauren Megger; Marieta Hyde; Max Kolton; Stefan Green |
| EPI_ISL_2476390, EPI_ISL_2476391, EPI_ISL_2476393, EPI_ISL_2476400, EPI_ISL_2692086 | University of Illinois at Chicago | RIPHL at Rush University Medical Center | Felix Araujo Perez; Julie Bentea; Kevin Kunstman; Laura Furtado; Marieta Hyde; Max Kolton; Omar Perez; Stefan Green |
| EPI_ISL_2139707, EPI_ISL_2139708, EPI_ISL_2139722, EPI_ISL_2139879 | University of Liège COVID-19 testing center | GIGA Medical Genomics | Bouchra Boujemla; Cécile Meex; Fabrice Bureau; Keith Durkin; Laurent Gillet; Maria Artesi; Marie-Pierre Hayette; Nathalie Renotte; Sébastien Bontems; Vincent Bours; Wouter Coppieters |
| EPI_ISL_2100396, EPI_ISL_2136244 | University of Michigan Clinical Microbiology Laboratory | Lauring Lab, University of Michigan, Department of Microbiology and Immunology | Valesano |
| EPI_ISL_2304022 | University of Oregon COVID-19 MAP Laboratory | University of Oregon Genomics and Cell Charactericzation Core Facility (GC3F) | Ariana White; Douglas Turnbull; Jason Carriere; Jason Sydes; Jeff Bishop; Peter Batzel |
| EPI_ISL_2285364, EPI_ISL_2285365, EPI_ISL_2285368, EPI_ISL_2285369, EPI_ISL_2285371, EPI_ISL_2285372, EPI_ISL_2285375 | | | |
| see above | University of Rome Tor Vergata: Departm Experim Medicine Chair of Virology | University of Rome Tor Vergata: Departm Experim Medicine Chair of Virology | Francesca Ceccherini-Silberstein; Loredana Sarmati; Lorenzo Piermatteo; Luca Carioti; Marco Iannetta; Maria Botticelli; Maria Concetta Bellocchi; Massimo Andreoni; Rossana Scutari |
| EPI_ISL_2225561, EPI_ISL_2230609 | University of Wisconsin-Madison AIDS Vaccine Research Laboratories | University of Wisconsin-Madison AIDS Vaccine Research Laboratories | Gage Moreno; Katarina Braun; et al. AIDS Vaccine Research Laboratories |
| EPI_ISL_3014394 | Università Federico II - Dipartimento di scienze mediche traslazionali - Napoli | TIGEM | Antonio Grimaldi Patrizia Annunziata Francesco Panariello Teresa Giuliano Michele Cennamo Valentina Bouche Chiara Colantuono Lucio Di Filippo Mariano Fiorenza Anna Manfredi Marcello Salvi Giuseppe Portella Andrea Ballabio Davide Cacchiarelli |
| EPI_ISL_3270814 | Università Federico II - Dipartimento di scienze mediche traslazionali - Napoli | Telethon Institute of Genetics and Medicine (TIGEM) | Antonio Grimaldi Patrizia Annunziata Francesco Panariello Teresa Giuliano Michele Cennamo Valentina Bouche Chiara Colantuono Lucio Di Filippo Mariano Fiorenza Anna Manfredi Marcello Salvi Giuseppe Portella Andrea Ballabio Davide Cacchiarelli |
| EPI_ISL_2131461, EPI_ISL_2131463, EPI_ISL_2131464, EPI_ISL_2131470, EPI_ISL_2131471, EPI_ISL_2131472, EPI_ISL_2131474, EPI_ISL_2131479, EPI_ISL_2235438, EPI_ISL_2235439, EPI_ISL_2235440 | | | |
| see above | Università degli Studi di Perugia | Istituto Zooprofilattico Sperimentale dell'Abruzzo e Molise "G. Caporale" | Ancora M; Calistri P; Camilloni B; Cammà C; Caporale M; Curini V; Delli Compagni E; Di Domenico M; Di Lollo Valeria; Di Pasquale A; Lorusso A; Mangone I; Marcacci M; Mencacci A; Puglia I; Rinaldi A; Savini G; Scialabba S |
| EPI_ISL_2137018 | UniversitŠtsSpital ZŸrich | Institute of Medical Virology | Alexandra Trkola; Annette AudigŽ; Catharine Aquino; Cyril Shah; Daniel Ehrsam; Gabriela Ziltener; Guido Bloemberg; Hubert Rehrauer; Isabel StŸrmer; Joel Wirz; Jon Huder; JŸrg Bšni; Kevin Steiner; Maria GrŸnberg; Maryam Zaheri; Michael Huber; Riccarda Capaul; Stefan Schmutz; Verena Kufner; Weihong Qi |
| EPI_ISL_5801569 | Upa De Bebedouro | Instituto Butantan | Antonio Jorge Martins; Claudia Renata dos Santos Barros; David Schlesinger; Debora Botequio Moretti; Dimas Tadeu Covas; Elaine Cristina Marqueze; Elaine Vieira Santos; Evandra Strazza Rodrigues; Heidge Fukumasu; Jayme Augusto de Souza-Neto; José Salvatore Leister Patané; Luiz Alcantara; Luiz Lehmann Coutinho; Maria Carolina Elias; Maurício Lacerda Nogueira; Rafael dos Santos Bezerra; Raul Machado Neto; Rejane Maria Tommasini Grotto; Ricardo Haddad; Sandra Coccuzzo Sampaio Vessoni; Simone Kashima; Svetoslav Nanev Slavov; Vincent Louis Viala |
| EPI_ISL_5801536 | Upa Mary Dota | Instituto Butantan | Antonio Jorge Martins; Claudia Renata dos Santos Barros; David Schlesinger; Debora Botequio Moretti; Dimas Tadeu Covas; Elaine Cristina Marqueze; Elaine Vieira Santos; Evandra Strazza Rodrigues; Heidge Fukumasu; Jayme Augusto de Souza-Neto; José Salvatore Leister Patané; Luiz Alcantara; Luiz Lehmann Coutinho; Maria Carolina Elias; Maurício Lacerda Nogueira; Rafael dos Santos Bezerra; Raul Machado Neto; Rejane Maria Tommasini Grotto; Ricardo Haddad; Sandra Coccuzzo Sampaio Vessoni; Simone Kashima; Svetoslav Nanev Slavov; Vincent Louis Viala |
| EPI_ISL_5801653, EPI_ISL_5801654 | Upa Unidade De Pronto Atendimento 24 Horas Bom Jesus | Instituto Butantan | Antonio Jorge Martins; Claudia Renata dos Santos Barros; David Schlesinger; Debora Botequio Moretti; Dimas Tadeu Covas; Elaine Cristina Marqueze; Elaine Vieira Santos; Evandra Strazza Rodrigues; Heidge Fukumasu; Jayme Augusto de Souza-Neto; José Salvatore Leister Patané; Luiz Alcantara; Luiz Lehmann Coutinho; Maria Carolina Elias; Maurício Lacerda Nogueira; Rafael dos Santos Bezerra; Raul Machado Neto; Rejane Maria Tommasini Grotto; Ricardo Haddad; Sandra Coccuzzo Sampaio Vessoni; Simone Kashima; Svetoslav Nanev Slavov; Vincent Louis Viala |
| EPI_ISL_3030381 | Urbino | Microbiology University Politecnica delle Marche | Anna Valenza; Carla Acciarri; Katia Marinelli; Monica Lucia Ferreri; Patrizia Bagnarelli; Roberta Longo; Sara Caucci; Stefano Menzo |
| EPI_ISL_2408062, EPI_ISL_2408063 | Usansolo-Galdakao University Hospital | Cruces University Hospital | Ana Belén de la Hoz; Ana Gual-de-Torrella; Izaskun Alejo-Cancho; Mikel Gallego |
| EPI_ISL_5801604 | Usf Arcadas | Instituto Butantan | Antonio Jorge Martins; Claudia Renata dos Santos Barros; David Schlesinger; Debora Botequio Moretti; Dimas Tadeu Covas; Elaine Cristina Marqueze; Elaine Vieira Santos; Evandra Strazza Rodrigues; Heidge Fukumasu; Jayme Augusto de Souza-Neto; José Salvatore Leister Patané; Luiz Alcantara; Luiz Lehmann Coutinho; Maria Carolina Elias; Maurício Lacerda Nogueira; Rafael dos Santos Bezerra; Raul Machado Neto; Rejane Maria Tommasini Grotto; Ricardo Haddad; Sandra Coccuzzo Sampaio Vessoni; Simone Kashima; Svetoslav Nanev Slavov; Vincent Louis Viala |
| EPI_ISL_2292566, EPI_ISL_2292801, EPI_ISL_2456350, EPI_ISL_2456508, EPI_ISL_2456649, EPI_ISL_2456735, EPI_ISL_2458489 | | | |
| see above | Utah Public Health Laboratory | Utah Public Health Laboratory | Erin L. Young; Kelly F. Oakeson; Tara Gallagher |
| EPI_ISL_2479921 | VIGILANCIA EM SAUDE SAE VIGILANCIA EPIDEMIOLOGICA | Instituto Butantan | Antonio Jorge Martins; Claudia Renata dos Santos Barros; David Schlesinger; Debora Botequio Moretti; Dimas Tadeu Covas; Elaine Cristina Marqueze; Elaine Vieira Santos; Evandra Strazza Rodrigues; Heidge Fukumasu; Jayme Augusto de Souza-Neto; José Salvatore Leister Patané; Luiz Alcantara; Luiz Lehmann Coutinho; Maria Carolina Elias; Maurício Lacerda Nogueira; Rafael dos Santos Bezerra; Raul Machado Neto; Rejane Maria Tommasini Grotto; Ricardo Haddad; Sandra Coccuzzo Sampaio Vessoni; Simone Kashima; Svetoslav Nanev Slavov; Vincent Louis Viala |
| EPI_ISL_2362250 | VIGILANCIA EM SAUDE SAE VIGILANCIA EPIDEMIOLOGICA | Instituto Butantan / Mendelics | Antonio Jorge Martins; Claudia Renata dos Santos Barros; David Schlesinger; Debora Botequio Moretti; Dimas Tadeu Covas; Elaine Cristina Marqueze; Elaine Vieira Santos; Evandra Strazza Rodrigues; Heidge Fukumasu; Jayme Augusto de Souza-Neto; José Salvatore Leister Patané; Luiz Alcantara; Luiz Lehmann Coutinho; Maria Carolina Elias; Maurício Lacerda Nogueira; Rafael dos Santos Bezerra; Raul Machado Neto; Rejane Maria Tommasini Grotto; Ricardo Haddad; Sandra Coccuzzo Sampaio Vessoni; Simone Kashima; Svetoslav Nanev Slavov; Vincent Louis Viala |
| EPI_ISL_5650320 | VIGILANCIA EPIDEMIOLOGICA DE LEME | Instituto Butantan | Antonio Jorge Martins; Claudia Renata dos Santos Barros; David Schlesinger; Debora Botequio Moretti; Dimas Tadeu Covas; Elaine Cristina Marqueze; Elaine Vieira Santos; Evandra Strazza Rodrigues; Heidge Fukumasu; Jayme Augusto de Souza-Neto; José Salvatore Leister Patané; Luiz Alcantara; Luiz Lehmann Coutinho; Maria Carolina Elias; Maurício Lacerda Nogueira; Rafael dos Santos Bezerra; Raul Machado Neto; Rejane Maria Tommasini Grotto; Ricardo Haddad; Sandra Coccuzzo Sampaio Vessoni; Simone Kashima; Svetoslav Nanev Slavov; Vincent Louis Viala |
| EPI_ISL_2404818, EPI_ISL_2404831, EPI_ISL_2404832, EPI_ISL_2404833, EPI_ISL_2404834, EPI_ISL_2404835 | VMC | Santa Clara County Public Health Laboratory | Santa Clara County Public Health Department |
| EPI_ISL_4551860 | Via Pansini 5 - Napoli - Italy | TIGEM | Antonio Grimaldi Patrizia Annunziata Francesco Panariello Teresa Giuliano Michele Cennamo Valentina Bouche Chiara Colantuono Lucio Di Filippo Mariano Fiorenza Anna Manfredi Marcello Salvi Giuseppe Portella Andrea Ballabio Davide Cacchiarelli |
| EPI_ISL_5801550 | Vigilancia Em Saude Sae Vigilancia Epidemiologica | Instituto Butantan | Antonio Jorge Martins; Claudia Renata dos Santos Barros; David Schlesinger; Debora Botequio Moretti; Dimas Tadeu Covas; Elaine Cristina Marqueze; Elaine Vieira Santos; Evandra Strazza Rodrigues; Heidge Fukumasu; Jayme Augusto de Souza-Neto; José Salvatore Leister Patané; Luiz Alcantara; Luiz Lehmann Coutinho; Maria Carolina Elias; Maurício Lacerda Nogueira; Rafael dos Santos Bezerra; Raul Machado Neto; Rejane Maria Tommasini Grotto; Ricardo Haddad; Sandra Coccuzzo Sampaio Vessoni; Simone Kashima; Svetoslav Nanev Slavov; Vincent Louis Viala |
| EPI_ISL_2151956, EPI_ISL_2151963, EPI_ISL_2151992, EPI_ISL_2307864 | Viollier AG | Department of Biosystems Science and Engineering, ETH Zürich | Andrea Patrignani; Andreia Cabral de Gouvea; Catharine Aquino; Chaoran Chen; Christian Beisel; Christiane Beckmann; Christoph Noppen; Daniel Ehrsam; David Dreifuss; Doris Popovic; Elodie Burcklen; Griffin White; Ina Nissen; Isabel Stürmer; Ivan Topolsky; Jay Tracy; Katharina Jahn; Lara Fuhrmann; Laura Neff; Lennart Opitz; Maria Domenica Moccia; Maurice Redondo; Mirjam Feldkamp; Natascha Santacroce; Niko Beerenwinkel; Noemie Santamaria de Souza; Olivier Kobel; Philipp Jablonski; Ralph Schlapbach; Rebecca Denes; Sarah Nadeau; Simon Grüter; Sophie Seidel; Tanja Stadler; Timothy Sykes |
| EPI_ISL_2229446, EPI_ISL_2229453, EPI_ISL_2344478, EPI_ISL_2344485, EPI_ISL_2344490, EPI_ISL_2603338 | Virginia Division of Consolidated Laboratory Services | Virginia Division of Consolidated Laboratory Services | Virginia DCLS |
| EPI_ISL_2884319, EPI_ISL_2884320, EPI_ISL_2884560 | Virology Laboratory, Scientific Department, Army Medical Center | Virology Laboratory, Scientific Department, Army Medical Center | Anella Monte; Anna Anselmo; Antonella Fortunato; Filippo Molinari; Florigio Lista; Francesco Giordani; Giancarlo Petralito; Giandomenico Cerreto; Giulia Campoli; Lucia Nicosia; Margherita De Santis; Maria Anna Spinelli; Marzia Cavalli; Riccardo De Sanctis; Rossella Brandi; Silvia Fillo; Vanessa Vera Fain |
| EPI_ISL_2274650 | WESTCHESTER MEDICAL CENTER | Wadsworth Center, New York State Department of Health | Alexis Russell; Catharine Prussing; Daryl M. Lamson; Erasmus Schneider; Erica Lasek-Nesselquist; John Kelly; Jonathan Plitnick; Kirsten St. George; Matthew Shudt; Melissa A Leisner; Navjot Singh |
| EPI_ISL_2248729 | WI State Laboratory of Hygiene | Centers for Disease Control and Prevention Division of Viral Diseases, Pathogen Discovery | Alison Laufer Halpin; Ben L. Rambo-Martin; Clinton R. Paden; Dakota Howard; Darlene Wagner; Dave Wentworth; Dhwani Batra; Jasmine Padilla; Justin Lee; Katie Dillon; Krista Queen; Kristen Knipe; Kristine Lacek; Mark Burroughs; Matthew Schmerer; Mili Sheth; Peter Cook; Sam Shepard; Sarah Nobles; Shoshona Le; Suxiang Tong; Vivien Dugan; Yvette Unoarumhi |
| EPI_ISL_2096109, EPI_ISL_2096110, EPI_ISL_2096111, EPI_ISL_2096112, EPI_ISL_2096114, EPI_ISL_2336177, EPI_ISL_2336191, EPI_ISL_2336192, EPI_ISL_2336209, EPI_ISL_2336224, EPI_ISL_2336242, EPI_ISL_2336246, EPI_ISL_2378840 | | | |
| see above | Washington State Department of Health | Washington State Department of | Avi Singh; Darren Lucas; Denny Russell; Drew MacKellar; Geoff Melly; Hannah Gray; Joenice Gonzalez; JohnAric Peterson; Philip Dykema; Rebecca Cao; Vanessa De Los Santos |

| | | | |
|---|---|---|---|
| | Public Health Laboratories | Health Public Health Laboratories | |
| EPI_ISL_2466523, EPI_ISL_2466837 | Wexner Medical Center | The Ohio State University College of Medicine | Koenig, S.; Seminetta, J. |
| EPI_ISL_2615914 | Wheeling Hospital | WVU and Marshall University Combined Genomics Core Facilities | James Denvir; Peter Perrotta; Peter Stoilov; Ryan Percifield; Wesley Kimble |
| EPI_ISL_1961015, EPI_ISL_2023319, EPI_ISL_2023321, EPI_ISL_2023333, EPI_ISL_2023334, EPI_ISL_2023336, EPI_ISL_2023341, EPI_ISL_2023343, EPI_ISL_2023347, EPI_ISL_2023354, EPI_ISL_2037405, EPI_ISL_2037410, EPI_ISL_2037411, EPI_ISL_2037413, EPI_ISL_2086424, EPI_ISL_2086425, EPI_ISL_2086426, EPI_ISL_2086433, EPI_ISL_2086435, EPI_ISL_2086439, EPI_ISL_2086440, EPI_ISL_2086442, EPI_ISL_2086451, EPI_ISL_2092837 | | | |
| see above | Wichita State University - Molecular Diagnostics Lab | Kansas Health and Environmental Lab | Ben Olsen; Jonathan Barnell; Mike Grose; and Phil Adam |
| EPI_ISL_2229382 | Willis-Knighton Medical Center Hospital Laboratory | LSUHS Emerging Viral Threat Laboratory | Alexander Mijalis; Andrew D. Yurochko; Christopher G. Kevil; Gregory L. Ware; Jennifer L. Carroll; Jeremy P. Kamil; John A. Vanchiere; Maarten Van Diest; Rona S. Scott |
| EPI_ISL_2279286, EPI_ISL_2279337, EPI_ISL_2279497, EPI_ISL_2558094, EPI_ISL_2558097, EPI_ISL_2558150, EPI_ISL_2558168, EPI_ISL_2558174, EPI_ISL_2558222, EPI_ISL_2558243 | | | |
| see above | Wisconsin State Laboratory of Hygiene Communicable Disease Division | Wisconsin State Laboratory of Hygiene Communicable Disease Division | Abigail C. Shockey; Alicia J. Mooney; Erika M. Hanson; Kelsey R. Florek; Richard Griesser; Sara Wagner; Tonya Danz |
| EPI_ISL_2152436, EPI_ISL_2226781 | Wyoming Public Health Laboratory | Wyoming Public Health Laboratory | Ashley Norberg; Brian Dominguez; Brittany Oher; Cari Sloma; Channing Weber; Chayse Rowley; Elliot Thomasson; Jim Mildenberger; Marley Goetz; Sam Britz; Taylor Fearing; and Rob Christensen |
| EPI_ISL_2159642, EPI_ISL_2296383, EPI_ISL_2296387, EPI_ISL_2296389 | Yale Clinical Virology Lab | Grubaugh Lab - Yale School of Public Health | Anderson Brito; Annie Watkins; Chaney Kalinich; Chantal Vogels; Isabel Ott; Jessica Rothman; Joseph Fauver; Mallery Breban; Marie L. Landry; Mary Petrone; Nathan Grubaugh; Tara Alpert |
| EPI_ISL_2381476 | ZOTZ KLIMAS MVZ Düsseldorf-Centrum GbR ÜBAG für Labormedizin, Genetik, Zytologie, Pathologie | Center of Medical Microbiology, Virology, and Hospital Hygiene, University of Duesseldorf | Alexander Dilthey; Andreas Walker; Daniel Strelow; Jessica Nicolai; Jörg Timm; Katrin Hoffmann; Klaus Pfeffer; Lisanna Hülse; Malte Kohns Vasconcelos; Maximilian Damagnez; Nadine Lübke; Patrick Finzer; Rainer Zotz; Tobias Wienemann; Torsten Houwaart |
