## Supplement 3 for "Rapid and Accurate Identification of SARS-CoV-2 Omicron Variants Using Droplet Digital PCR (RT-ddPCR)": All_0510-0516.pdf

We gratefully acknowledge the following Authors from the Originating laboratories responsible for obtaining the specimens, as well as the Submitting laboratories where the genome data were generated and shared via GISAID, on which this research is based.

All Submitters of data may be contacted directly via www.gisaid.org

Authors are sorted alphabetically.

| Accession ID | Originating Laboratory | Submitting Laboratory | Authors |
|---|---|---|---|
| EPI_ISL_2721253 | A.S.L. ALESSANDRIA | Fondazione del Piemonte per l'Oncologia IRCCS | Antonino Sottile; Giorgio Giardina; Paola Marino; Silvia Brossa |
| EPI_ISL_2383983 | AL Dept. of Public Health Bureau of Clinical Laboratories | Centers for Disease Control and Prevention Division of Viral Diseases, Pathogen Discovery | Alison Laufer Halpin; Ben L. Rambo-Martin; Clinton R. Paden; Dakota Howard; Darlene Wagner; Dave Wentworth; Dhwani Batra; Jasmine Padilla; Justin Lee; Katie Dillon; Krista Queen; Kristen Knipe; Kristine Lacek; Mark Burroughs; Matthew Schmerer; Mili Sheth; Peter Cook; Sam Shepard; Sarah Nobles; Shoshona Le; Suxiang Tong; Vivien Dugan; Yvette Unoarumhi |
| EPI_ISL_2279505 | ALBANY MEDICAL CENTER | Wadsworth Center, New York State Department of Health | Alexis Russell; Catharine Prussing; Daryl M. Lamson; Erasmus Schneider; Erica Lasek-Nesselquist; John Kelly; Jonathan Plitnick; Kirsten St. George; Matthew Shudt; Melissa A Leisner; Navjot Singh |
| EPI_ISL_5915128, EPI_ISL_5915129, EPI_ISL_5915140, EPI_ISL_5915169, EPI_ISL_5915209, EPI_ISL_5915210, EPI_ISL_5915233, EPI_ISL_5915257, EPI_ISL_5915258, EPI_ISL_5915306, EPI_ISL_5915318, EPI_ISL_6573791, EPI_ISL_6573875, EPI_ISL_6573929 | | | |
| see above | AMALAB/FACISA/UFRN | WallauLab on behalf of Fiocruz COVID-19 Genomic Surveillance Network | Alexandre Freitas da Silva; Allan Roberto Dias Nunes; Antonio Marinho da Silva Neto; Cassia Docena; Constância Flávia Junqueira Ayres; Filipe Zimmer Dezordi; Gabriel Luz Wallau; Gustavo Barbosa de Lima; Joana Cristina Medeiros Tavares Marques; Katya Anaya Jacinto; Lais Ceschini Machado; Lilian Caroliny Amorim Silva; Marcelo Henrique dos Santos Paiva; Mariane dos Santos Duarte; Matheus Filgueira Bezerra; Sinval Pinto Brandão Filho |
| EPI_ISL_2445419, EPI_ISL_2445420 | AMBULATORIO DE ESPECIALIDADE V E MOGI MIRIM | Instituto Butantan | Antonio Jorge Martins; Claudia Renata dos Santos Barros; David Schlesinger; Debora Botequio Moretti; Dimas Tadeu Covas; Elaine Cristina Marqueze; Elaine Vieira Santos; Evandra Strazza Rodrigues; Heidge Fukumasu; Jayme Augusto de Souza-Neto; José Salvatore Leister Patané; Luiz Alcantara; Luiz Lehmann Coutinho; Maria Carolina Elias; Maurício Lacerda Nogueira; Rafael dos Santos Bezerra; Raul Machado Neto; Rejane Maria Tommasini Grotto; Ricardo Haddad; Sandra Coccuzzo Sampaio Vessoni; Simone Kashima; Svetoslav Nanev Slavov; Vincent Louis Viala |
| EPI_ISL_2502747 | AREA DE SALUD PARAISO-CERVANTES | Inciensa, Instituto Costarricense de InvestigaciOn y Enseñanza en NutriciOn y Salud | Adriana Godínez; Claudio Soto-Garita; Estela Cordero; Francisco Duarte; Hebleen Porras; Jose Luis Vargas; Mariela Gutierrez & Joselyn Prado; Melany CalderOn |
| EPI_ISL_2382002 | AZ St.Jan campus Oostende | AZ Sint Jan | Jorn Hellemans; Laurien Hoornaert; Marie Madeleine Chabert-Consen; Marijke Reynders; Patrick Descheemaeker; Thomas Van Landschoot |
| EPI_ISL_2135658, EPI_ISL_2135666, EPI_ISL_2135861, EPI_ISL_2135873 | AZDelta | AZ Delta Medical Laboratories in Roeselare, Belgium | Dieter De Smet; Geert Martens; Merijn Vanhee; on behalf of AZ Delta COVID-19 Genomics core (member of Genomic surveillance of SARS-CoV-2 in Belgium network) |
| EPI_ISL_2180720, EPI_ISL_2180853, EPI_ISL_2180906, EPI_ISL_2180910, EPI_ISL_2180948, EPI_ISL_2180961, EPI_ISL_2181054, EPI_ISL_2181055, EPI_ISL_2181101, EPI_ISL_2181156, EPI_ISL_2181175, EPI_ISL_2181185, EPI_ISL_2181196, EPI_ISL_2181199, EPI_ISL_2181225, EPI_ISL_2181226, EPI_ISL_2181234, EPI_ISL_2181275, EPI_ISL_2181278, EPI_ISL_2181279, EPI_ISL_2181280, EPI_ISL_2181292, EPI_ISL_2181297, EPI_ISL_2181298, EPI_ISL_2181308, EPI_ISL_2181338, EPI_ISL_2181349, EPI_ISL_2181372, EPI_ISL_2181375, EPI_ISL_2181376, EPI_ISL_2181387, EPI_ISL_2181388, EPI_ISL_2181389, EPI_ISL_2181403, EPI_ISL_2181434, EPI_ISL_2181446, EPI_ISL_2181450, EPI_ISL_2181496, EPI_ISL_2181550, EPI_ISL_2181554, EPI_ISL_2181556, EPI_ISL_2181560, EPI_ISL_2181578, EPI_ISL_2181604, EPI_ISL_2181610, EPI_ISL_2181615, EPI_ISL_2181633, EPI_ISL_2181637, EPI_ISL_2181641, EPI_ISL_2181644, EPI_ISL_2204513, EPI_ISL_2204519, EPI_ISL_2204524, EPI_ISL_2204549, EPI_ISL_2204604, EPI_ISL_2204618, EPI_ISL_2204622, EPI_ISL_2204628, EPI_ISL_2204655, EPI_ISL_2204690, EPI_ISL_2204734, EPI_ISL_2204736, EPI_ISL_2204739, EPI_ISL_2204749, EPI_ISL_2204771, EPI_ISL_2204795, EPI_ISL_2204808, EPI_ISL_2204817, EPI_ISL_2204857, EPI_ISL_2204937, EPI_ISL_2204986, EPI_ISL_2241669, EPI_ISL_2241671, EPI_ISL_2241678, EPI_ISL_2241689, EPI_ISL_2268247, EPI_ISL_2280287, EPI_ISL_2280329, EPI_ISL_2280354, EPI_ISL_2280467, EPI_ISL_2280472, EPI_ISL_2280544, EPI_ISL_2280547, EPI_ISL_2280553, EPI_ISL_2280554, EPI_ISL_2280564, EPI_ISL_2280566, EPI_ISL_2280579, EPI_ISL_2280585, EPI_ISL_2280597, EPI_ISL_2280601, EPI_ISL_2280604, EPI_ISL_2280620, EPI_ISL_2280669, EPI_ISL_2280670, EPI_ISL_2280675, EPI_ISL_2280678, EPI_ISL_2280681, EPI_ISL_2280725, EPI_ISL_2280734, EPI_ISL_2280736, EPI_ISL_2280753, EPI_ISL_2280760, EPI_ISL_2280761, EPI_ISL_2280762, EPI_ISL_2280763, EPI_ISL_2280793, EPI_ISL_2280804, EPI_ISL_2280805, EPI_ISL_2280810, EPI_ISL_2280825, EPI_ISL_2280829, EPI_ISL_2280830, EPI_ISL_2280838, EPI_ISL_2280847, EPI_ISL_2280854, EPI_ISL_2280859, EPI_ISL_2280876, EPI_ISL_2280891, EPI_ISL_2280917, EPI_ISL_2280933, EPI_ISL_2280934, EPI_ISL_2280948, EPI_ISL_2280952, EPI_ISL_2280963, EPI_ISL_2280974, EPI_ISL_2280976, EPI_ISL_2280987, EPI_ISL_2280988, EPI_ISL_2281006, EPI_ISL_2283813, EPI_ISL_2283828, EPI_ISL_2283831, EPI_ISL_2283835, EPI_ISL_2283836, EPI_ISL_2283843, EPI_ISL_2283846, EPI_ISL_2283848, EPI_ISL_2283852, EPI_ISL_2283854, EPI_ISL_2283856, EPI_ISL_2283859, EPI_ISL_2283861, EPI_ISL_2321252, EPI_ISL_2321257, EPI_ISL_2321259, EPI_ISL_2321267, EPI_ISL_2321286, EPI_ISL_2321288, EPI_ISL_2321301, EPI_ISL_2321310, EPI_ISL_2321324, EPI_ISL_2321338, EPI_ISL_2321356, EPI_ISL_2321358, EPI_ISL_2321362, EPI_ISL_2321365, EPI_ISL_2321370, EPI_ISL_2367654, EPI_ISL_2367658, EPI_ISL_2367673, EPI_ISL_2372306 | | | |
| see above | Aegis Sciences Corporation | Centers for Disease Control and Prevention Division of Viral Diseases, Pathogen Discovery | Adrian Paskey; Alec Vest; Benjamin Rambo-Martin; Christopher Gulvick; Clinton R. Paden; Cyndi Clark; Dakota Howard; Darlene Wagner; Dhwani Batra; Dillon Nall; Duncan MacCannell; Ethan Sanders; Holly Houdeshell; Jason Caravas; Kara Moser; Matthew Hardison; Matthew Schmerer; Ola Kvalvaag; Patrick Campbell; Peter W. Cook; Rob Case; Scott Sammons; Shatavia Morrison; Shaun Westlund; Vikramsinha Ghorpade; Yvette Unoarumhi |
| EPI_ISL_2951275 | Ages,Lifebrain COVID 19 Labor GmbH Klinik Penzing Pavillon 17 | Elling and Cochella laboratories, IMBA/IMP | Alexander Stark; Ezgi Oezkan; Luisa Cochella and Ulrich Elling; Marcus Strobl; Olga Frank; Ramesh Yelagandula; Tanino Albanese |
| EPI_ISL_2948867 | Ages,Zentralinstitut fr Med. und Chem. Labordiagnostik LKH-Universittskliniken Innsbruck_Europe/Austria/Tirol | Elling and Cochella laboratories, IMBA/IMP | Alexander Stark; Ezgi Oezkan; Luisa Cochella and Ulrich Elling; Marcus Strobl; Olga Frank; Ramesh Yelagandula; Tanino Albanese |
| EPI_ISL_2154069, EPI_ISL_2187648, EPI_ISL_2272531, EPI_ISL_2272538, EPI_ISL_2308647, EPI_ISL_2376089, EPI_ISL_2376098 | | | |
| see above | Alaska State Virology Laboratory | Alaska State Virology Laboratory | Elva House; Jack Chen; Jacob Zidek; Lisa Smith; Ph.D.; Stephanie DeRonde |
| EPI_ISL_2272728, EPI_ISL_2272746 | Algemeen Medisch Labo | Labo Klinische Biologie, UZA | Basil Britto Xavier; Christine Lammens; Herman Goossens; Jasmine Coppens; Marie Le Mercier; Veerle Matheeussen |
| EPI_ISL_2227487, EPI_ISL_2325174 | Arizona State Public Health Laboratory | Arizona State Public Health Laboratory | Jessica Escobar; Katherine Fullerton; Linda Getsinger; Nobuko Fukushima; Stacy White; Trung Huynh; Victor Waddell |
| EPI_ISL_2250027, EPI_ISL_2250041, EPI_ISL_2250043, EPI_ISL_2250045, EPI_ISL_2250057, EPI_ISL_2250068, EPI_ISL_2250082, EPI_ISL_2339283, EPI_ISL_2339300, EPI_ISL_2484044 | | | |
| see above | Arizona State University | Arizona State University | Efrem S. Lim; Joshua LaBaer; Joy M. Blain; LaRinda A. Holland; Matthew F. Smith; Nicholas J. Mellor; Peter T. Skidmore; Rabia Maqsood; Valerie Harris; Vel Murugan |
| EPI_ISL_2931027, EPI_ISL_2931039, EPI_ISL_2931040 | Arkansas Children's Hospital | Center for Global Health, University of New Mexico Health Sciences Center | Amanda Novack; Darrell Dinwiddie; Daryl Domman; Dirk Haselow; Joshua L. Kennedy; Kurt Schwalm; Valerie Morley |
| EPI_ISL_2939081, EPI_ISL_2939084 | Arkansas Department of Health | Center for Global Health, University of New Mexico Health Sciences Center | Amanda Novack; Darrell Dinwiddie; Daryl Domman; Dirk Haselow; Joshua L. Kennedy; Kurt Schwalm; Valerie Morley |
| EPI_ISL_2692445, EPI_ISL_2692447, EPI_ISL_2692455, EPI_ISL_2692457, EPI_ISL_2692467, EPI_ISL_2692469, EPI_ISL_2692470, EPI_ISL_2692478 | | | |
| see above | Atlas Genomics | Seattle Flu Study | Amanda Adler; Barry R. Lutz; Benjamin Pelle; Brian Hiatt; Caitlin R. Wolf; Chris D. Frazar; Deborah A. Nickerson; Elisabeth Brandstetter; Erica Ryke; Geoff Melly; Helen Y. Chu; Janet A. Englund; Jay Shendure; Jover Lee; Kairsten Fay; Kirsten Lacombe; Lea M. Starita; Mark J. Rieder; Matthew Richardson; Matthew Thompson; Melissa Truong; Michael Boeckh; Michael Famulare; Misja Ilcisin; Peter D. Han; Philip Dykema; Romesh Gautom; Scott Lindquist; Thomas R. Sibley; Trevor Bedford |
| EPI_ISL_2427096, EPI_ISL_2427168, EPI_ISL_2427172, EPI_ISL_2485062, EPI_ISL_2485063 | Austrian Agency for Health and Food Safety (AGES) | Bergthaler laboratory, CeMM Research Center for Molecular Medicine of the Austrian Academy of Sciences | Andreas Bergthaler; Anna Schedl; Bekir Erguner; Benedikt Agerer; Christoph Bock; Fabian Amman; Jan Laine; Lukas Endler; Maelle Le Moing; Martin Senekowitsch; Michael Schuster; Petr Triska; Thomas Penz |
| EPI_ISL_2249140 | Avelab | Instituto Nacional de Saude (INSA) and Centro de Investigaçao em Biodiversidade e Recursos Geneticos (CIBIO), Universidade do Porto | Borges et al |
| EPI_ISL_2396877, EPI_ISL_2396880, EPI_ISL_2396881 | Azienda Ospedaliera Pugliese Ciaccio di Catanzaro SOC Microbiologia e Virologia | Azienda Ospedaliera Pugliese Ciaccio di Catanzaro SOC Microbiologia e Virologia | Rossana Tallerico Federica Pasceri Marco De Fazio Ilenia Talotta Giuseppina Panduri Pasquale Minchella |
| EPI_ISL_2450377 | Azienda Ospedaliera San Giovanni Addolorata | INMI Lazzaro Spallanzani IRCCS | A Di Caro; B Bartolini; CEM Gruber; E Giombini; F Messina; F Santini; G Bonfiglio; M Gaudio; M Rueca; MR Capobianchi; O Butera; PM Placanica |
| EPI_ISL_2308683 | Azienda Ospedaliera Terni | Istituto Zooprofilattico Sperimentale dell'Abruzzo e Molise "G. Caporale" | Ancora M; Calistri P; Cammà C; Curini V; Delli Compagni E; Di Domenico M; Di Pasquale A; Lorusso A; Mangone I; Marcacci M; Palumbo M; Puglia I; Rinaldi A; Savini G; Scaccetti A; Scialabba S |
| EPI_ISL_2872700, EPI_ISL_2872703, EPI_ISL_2872711 | Azienda Ospedaliero Universitaria di Sassari | AMES Centro Polidiagnostico Strumentale S.r.l. | Anna Puggioni; Bianca Paglietti; Caterina Serra; Claudia Piu; Elena Rimini; Erica Mura; Flavia Angioj; Gabriele Ibba; Giulia Rocca; Laura Firino; Rosalba Govoni; Salvatore Rubino; Sergio Uzzau; Vincenzo Lai |
| EPI_ISL_2318904, EPI_ISL_2318906, EPI_ISL_2318908, EPI_ISL_2318909, EPI_ISL_2318914, EPI_ISL_2318916, EPI_ISL_2318921, EPI_ISL_2318932, EPI_ISL_2318952, EPI_ISL_2318956, EPI_ISL_2318958, EPI_ISL_2318964, EPI_ISL_2318970, EPI_ISL_2318986, EPI_ISL_2318988 | | | |
| see above | Azienda Sanitaria dell'Alto Adige Laboratorio Aziendale di Microbiologia e Virologia | Istituto di Genomica Applicata | Davide Scaglione; Eleonora Paparelli; Elisa Masi; Elisabetta Giacobazzi; Elisabetta Pagani; Gabriele Magris; Irena Jurman; Irene Bianconi; Michele Morgante; Stefanie Wieser; Vera Vendramin |
| EPI_ISL_2308700, EPI_ISL_2308701 | Azienda USL Umbria 2 | Istituto Zooprofilattico Sperimentale dell'Abruzzo e Molise "G. Caporale" | Ancora M; Calistri P; Cammà C; Curini V; Delli Compagni E; Di Domenico M; Di Pasquale A; Lorusso A; Mangone I; Marcacci M; Pistoni E; Proietti A; Puglia I; Rinaldi A; Savini G; Scialabba S |
| EPI_ISL_2721366, EPI_ISL_2721380, EPI_ISL_2721388, EPI_ISL_2721392, EPI_ISL_2721394, EPI_ISL_2721397, EPI_ISL_2721408, EPI_ISL_2721416, EPI_ISL_2721423, EPI_ISL_2721427, EPI_ISL_2721448, EPI_ISL_2721450, EPI_ISL_2721463, EPI_ISL_2721469, EPI_ISL_2721477, EPI_ISL_2721478, EPI_ISL_2721482, EPI_ISL_2721484, EPI_ISL_2721491, EPI_ISL_2721499, EPI_ISL_2721508, EPI_ISL_2721535, EPI_ISL_2721536, EPI_ISL_2721539, EPI_ISL_2721542, EPI_ISL_2721546, EPI_ISL_2721550, EPI_ISL_2721561, EPI_ISL_2721565, EPI_ISL_2721573, EPI_ISL_2721585, EPI_ISL_2721590, EPI_ISL_2721591, EPI_ISL_2721597, EPI_ISL_2721601, EPI_ISL_2721608, EPI_ISL_2721613, EPI_ISL_2721614, EPI_ISL_2721625, EPI_ISL_2721646, EPI_ISL_2721660, EPI_ISL_2721670, EPI_ISL_2721675, EPI_ISL_2721676, EPI_ISL_2721681, EPI_ISL_2721685, EPI_ISL_2721686, EPI_ISL_2721690, EPI_ISL_2721695, EPI_ISL_2721699, EPI_ISL_2721705, EPI_ISL_2721710, EPI_ISL_2721729, EPI_ISL_2721732, EPI_ISL_2721736, EPI_ISL_2721756, EPI_ISL_2722002, EPI_ISL_2722038 | | | |
| see above | BCCDC Public Health Laboratory | BCCDC Public Health Laboratory | Ana Pacagnella; Corrinne Ng; Dan Fornika; John Tyson; Kim Macdonald; Kimia Kamelian; Linda Hoang; Loretta Janz; Mel Krajden; Prystajecky Natalie; Robert Azana; Shannon Russell |

| | | | |
|---|---|---|---|
| EPI_ISL_2445085 | BIOFAST CENTRO SANTA CECÍLIA | Instituto Butantan | Antonio Jorge Martins; Claudia Renata dos Santos Barros; David Schlesinger; Debora Botequio Moretti; Dimas Tadeu Covas; Elaine Cristina Marqueze; Elaine Vieira Santos; Evandra Strazza Rodrigues; Heidge Fukumasu; Jayme Augusto de Souza-Neto; José Salvatore Leister Patané; Luiz Alcantara; Luiz Lehmann Coutinho; Maria Carolina Elias; Maurício Lacerda Nogueira; Rafael dos Santos Bezerra; Raul Machado Neto; Rejane Maria Tommasini Grotto; Ricardo Haddad; Sandra Coccuzzo Sampaio Vessoni; Simone Kashima; Svetoslav Nanev Slavov; Vincent Louis Viala |
| EPI_ISL_2445086 | BIOFAST CENTRO SÉ | Instituto Butantan | Antonio Jorge Martins; Claudia Renata dos Santos Barros; David Schlesinger; Debora Botequio Moretti; Dimas Tadeu Covas; Elaine Cristina Marqueze; Elaine Vieira Santos; Evandra Strazza Rodrigues; Heidge Fukumasu; Jayme Augusto de Souza-Neto; José Salvatore Leister Patané; Luiz Alcantara; Luiz Lehmann Coutinho; Maria Carolina Elias; Maurício Lacerda Nogueira; Rafael dos Santos Bezerra; Raul Machado Neto; Rejane Maria Tommasini Grotto; Ricardo Haddad; Sandra Coccuzzo Sampaio Vessoni; Simone Kashima; Svetoslav Nanev Slavov; Vincent Louis Viala |
| EPI_ISL_2445093, EPI_ISL_2445094, EPI_ISL_2445095 | BIOFAST LESTE GUAIANASES | Instituto Butantan | Antonio Jorge Martins; Claudia Renata dos Santos Barros; David Schlesinger; Debora Botequio Moretti; Dimas Tadeu Covas; Elaine Cristina Marqueze; Elaine Vieira Santos; Evandra Strazza Rodrigues; Heidge Fukumasu; Jayme Augusto de Souza-Neto; José Salvatore Leister Patané; Luiz Alcantara; Luiz Lehmann Coutinho; Maria Carolina Elias; Maurício Lacerda Nogueira; Rafael dos Santos Bezerra; Raul Machado Neto; Rejane Maria Tommasini Grotto; Ricardo Haddad; Sandra Coccuzzo Sampaio Vessoni; Simone Kashima; Svetoslav Nanev Slavov; Vincent Louis Viala |
| EPI_ISL_2445099, EPI_ISL_2445100, EPI_ISL_2445101, EPI_ISL_2445102 | BIOFAST LESTE ITAIM PAULISTA | Instituto Butantan | Antonio Jorge Martins; Claudia Renata dos Santos Barros; David Schlesinger; Debora Botequio Moretti; Dimas Tadeu Covas; Elaine Cristina Marqueze; Elaine Vieira Santos; Evandra Strazza Rodrigues; Heidge Fukumasu; Jayme Augusto de Souza-Neto; José Salvatore Leister Patané; Luiz Alcantara; Luiz Lehmann Coutinho; Maria Carolina Elias; Maurício Lacerda Nogueira; Rafael dos Santos Bezerra; Raul Machado Neto; Rejane Maria Tommasini Grotto; Ricardo Haddad; Sandra Coccuzzo Sampaio Vessoni; Simone Kashima; Svetoslav Nanev Slavov; Vincent Louis Viala |
| EPI_ISL_2445087, EPI_ISL_2445088, EPI_ISL_2445090, EPI_ISL_2445091, EPI_ISL_2445092 | BIOFAST LESTE ITAQUERA | Instituto Butantan | Antonio Jorge Martins; Claudia Renata dos Santos Barros; David Schlesinger; Debora Botequio Moretti; Dimas Tadeu Covas; Elaine Cristina Marqueze; Elaine Vieira Santos; Evandra Strazza Rodrigues; Heidge Fukumasu; Jayme Augusto de Souza-Neto; José Salvatore Leister Patané; Luiz Alcantara; Luiz Lehmann Coutinho; Maria Carolina Elias; Maurício Lacerda Nogueira; Rafael dos Santos Bezerra; Raul Machado Neto; Rejane Maria Tommasini Grotto; Ricardo Haddad; Sandra Coccuzzo Sampaio Vessoni; Simone Kashima; Svetoslav Nanev Slavov; Vincent Louis Viala |
| EPI_ISL_2445096, EPI_ISL_2445097, EPI_ISL_2445098 | BIOFAST LESTE SÃO MIGUEL PAULISTA | Instituto Butantan | Antonio Jorge Martins; Claudia Renata dos Santos Barros; David Schlesinger; Debora Botequio Moretti; Dimas Tadeu Covas; Elaine Cristina Marqueze; Elaine Vieira Santos; Evandra Strazza Rodrigues; Heidge Fukumasu; Jayme Augusto de Souza-Neto; José Salvatore Leister Patané; Luiz Alcantara; Luiz Lehmann Coutinho; Maria Carolina Elias; Maurício Lacerda Nogueira; Rafael dos Santos Bezerra; Raul Machado Neto; Rejane Maria Tommasini Grotto; Ricardo Haddad; Sandra Coccuzzo Sampaio Vessoni; Simone Kashima; Svetoslav Nanev Slavov; Vincent Louis Viala |
| EPI_ISL_2445103, EPI_ISL_2445104 | BIOFAST LESTE TIRADENTES | Instituto Butantan | Antonio Jorge Martins; Claudia Renata dos Santos Barros; David Schlesinger; Debora Botequio Moretti; Dimas Tadeu Covas; Elaine Cristina Marqueze; Elaine Vieira Santos; Evandra Strazza Rodrigues; Heidge Fukumasu; Jayme Augusto de Souza-Neto; José Salvatore Leister Patané; Luiz Alcantara; Luiz Lehmann Coutinho; Maria Carolina Elias; Maurício Lacerda Nogueira; Rafael dos Santos Bezerra; Raul Machado Neto; Rejane Maria Tommasini Grotto; Ricardo Haddad; Sandra Coccuzzo Sampaio Vessoni; Simone Kashima; Svetoslav Nanev Slavov; Vincent Louis Viala |
| EPI_ISL_2445105, EPI_ISL_2445106, EPI_ISL_2445107, EPI_ISL_2473811 | BIOFAST NORTE SANTANA/JAÇANA/TUCURUVI | Instituto Butantan | Antonio Jorge Martins; Claudia Renata dos Santos Barros; David Schlesinger; Debora Botequio Moretti; Dimas Tadeu Covas; Elaine Cristina Marqueze; Elaine Vieira Santos; Evandra Strazza Rodrigues; Heidge Fukumasu; Jayme Augusto de Souza-Neto; José Salvatore Leister Patané; Luiz Alcantara; Luiz Lehmann Coutinho; Maria Carolina Elias; Maurício Lacerda Nogueira; Rafael dos Santos Bezerra; Raul Machado Neto; Rejane Maria Tommasini Grotto; Ricardo Haddad; Sandra Coccuzzo Sampaio Vessoni; Simone Kashima; Svetoslav Nanev Slavov; Vincent Louis Viala |
| EPI_ISL_2445108, EPI_ISL_2445109, EPI_ISL_2445110, EPI_ISL_2445111, EPI_ISL_2445112, EPI_ISL_2445113 | BIOFAST SUL SANTO AMARO/CIDADE ADEMAR | Instituto Butantan | Antonio Jorge Martins; Claudia Renata dos Santos Barros; David Schlesinger; Debora Botequio Moretti; Dimas Tadeu Covas; Elaine Cristina Marqueze; Elaine Vieira Santos; Evandra Strazza Rodrigues; Heidge Fukumasu; Jayme Augusto de Souza-Neto; José Salvatore Leister Patané; Luiz Alcantara; Luiz Lehmann Coutinho; Maria Carolina Elias; Maurício Lacerda Nogueira; Rafael dos Santos Bezerra; Raul Machado Neto; Rejane Maria Tommasini Grotto; Ricardo Haddad; Sandra Coccuzzo Sampaio Vessoni; Simone Kashima; Svetoslav Nanev Slavov; Vincent Louis Viala |
| EPI_ISL_2373844 | BIOMNIS LYON | CNR Virus des Infections Respiratoires - France SUD | Antonin Bal; Bruno Lina; Bruno Simon; Gregory Destras; Gwendolyne Burfin; Hadrien Regue; Laurence Josset; Martine Valette; Quentin Semanas |
| EPI_ISL_2193283, EPI_ISL_2443472, EPI_ISL_2443507 | Baylor Scott & White-Temple | Baylor Scott & White-Temple | Ari Rao; Kimberly Walker; Linden Morales; Marcus Volz; Shelby Hendrickson; Shelby Johnson |
| EPI_ISL_2401553, EPI_ISL_2401558, EPI_ISL_2401579 | BioneXt Lab | Laboratoire national de sante, Microbiology, Microbial Genomics Platform | Anke Wienecke-Baldacchino; Catherine Ragimbeau; Fatu Djabi; Jessica Tapp; Lise Pignon; Raoul Salmon; Tamir Abdelrahman; Thibault Ferrandon |
| EPI_ISL_3494491 | Boston University CTL | Boston University/National Emerging Infectious Disease Laboratories | Catherine Klapperich; Jacquelyn Turcinovic; John H. Connor; Lena Landeverde; Lynn Doucette-Stamm |
| EPI_ISL_3479026, EPI_ISL_3479027, EPI_ISL_3479028, EPI_ISL_3479030, EPI_ISL_3479032, EPI_ISL_3479034, EPI_ISL_3479037, EPI_ISL_3479038, EPI_ISL_3479041, EPI_ISL_3479042, EPI_ISL_3479043, EPI_ISL_3479044, EPI_ISL_3479045, EPI_ISL_3479052, EPI_ISL_3479056, EPI_ISL_3479057, EPI_ISL_3479062, EPI_ISL_3479063, EPI_ISL_3479066, EPI_ISL_3479067, EPI_ISL_3483816, EPI_ISL_3483819, EPI_ISL_3483835, EPI_ISL_3483837, EPI_ISL_3483843, EPI_ISL_3483845, EPI_ISL_3483856, EPI_ISL_3483860, EPI_ISL_3483871, EPI_ISL_3483874, EPI_ISL_3483879, EPI_ISL_3483881, EPI_ISL_3483960, EPI_ISL_3483962, EPI_ISL_3483964, EPI_ISL_3483966, EPI_ISL_3483978, EPI_ISL_3483984, EPI_ISL_3483985, EPI_ISL_3484076, EPI_ISL_3484097, EPI_ISL_3484099, EPI_ISL_3484122, EPI_ISL_3484124, EPI_ISL_3484137, EPI_ISL_3484144, EPI_ISL_3484146, EPI_ISL_3484150, EPI_ISL_3484152, EPI_ISL_3484161, EPI_ISL_3484170, EPI_ISL_3484173, EPI_ISL_3484175, EPI_ISL_3484180, EPI_ISL_3484214, EPI_ISL_3484237, EPI_ISL_3484242, EPI_ISL_3484244, EPI_ISL_3484247, EPI_ISL_3484258, EPI_ISL_3484262, EPI_ISL_3484264, EPI_ISL_3484267, EPI_ISL_3484270, EPI_ISL_3484276, EPI_ISL_3484280, EPI_ISL_3484295, EPI_ISL_3484296, EPI_ISL_3484301, EPI_ISL_3484303, EPI_ISL_3484304, EPI_ISL_3484311, EPI_ISL_3484313, EPI_ISL_3484315, EPI_ISL_3484320, EPI_ISL_3484322, EPI_ISL_3484324, EPI_ISL_3484337, EPI_ISL_3484340, EPI_ISL_3484362, EPI_ISL_3484366, EPI_ISL_3484373, EPI_ISL_3484374, EPI_ISL_3484377, EPI_ISL_3484379, EPI_ISL_3484381, EPI_ISL_3484383, EPI_ISL_3484384, EPI_ISL_3484385, EPI_ISL_3484388, EPI_ISL_3484393, EPI_ISL_3484394, EPI_ISL_3484395, EPI_ISL_3484408, EPI_ISL_3484410, EPI_ISL_3484411, EPI_ISL_3484415, EPI_ISL_3484417, EPI_ISL_3484418, EPI_ISL_3484424, EPI_ISL_3484426, EPI_ISL_3484432, EPI_ISL_3484440, EPI_ISL_3484441, EPI_ISL_3484445, EPI_ISL_3484449, EPI_ISL_3484450, EPI_ISL_3484479, EPI_ISL_3484481, EPI_ISL_3484482, EPI_ISL_3484483, EPI_ISL_3484484, EPI_ISL_3484485, EPI_ISL_3484532, EPI_ISL_3484576, EPI_ISL_3484594, EPI_ISL_3484597, EPI_ISL_3484598, EPI_ISL_3484602, EPI_ISL_3484603, EPI_ISL_3484605, EPI_ISL_3484609, EPI_ISL_3484613, EPI_ISL_3484615, EPI_ISL_3484619, EPI_ISL_3484620, EPI_ISL_3484621, EPI_ISL_3484623, EPI_ISL_3484626, EPI_ISL_3484628, EPI_ISL_3484629, EPI_ISL_3484631, EPI_ISL_3484637, EPI_ISL_3484638, EPI_ISL_3484642, EPI_ISL_3484655, EPI_ISL_3484657, EPI_ISL_3484660, EPI_ISL_3484661, EPI_ISL_3484662, EPI_ISL_3484668, EPI_ISL_3484675, EPI_ISL_3484676, EPI_ISL_3484677, EPI_ISL_3484678, EPI_ISL_3484679, EPI_ISL_3484680, EPI_ISL_3484688, EPI_ISL_3484690, EPI_ISL_3484693, EPI_ISL_3484701, EPI_ISL_3484703, EPI_ISL_3484704, EPI_ISL_3484709, EPI_ISL_3484710, EPI_ISL_3484711, EPI_ISL_3484713, EPI_ISL_3484714, EPI_ISL_3484715, EPI_ISL_3484716, EPI_ISL_3484717, EPI_ISL_3484724, EPI_ISL_3484725, EPI_ISL_3484730, EPI_ISL_3484731, EPI_ISL_3484735, EPI_ISL_3484737, EPI_ISL_3484739, EPI_ISL_3484742, EPI_ISL_3484744, EPI_ISL_3484748, EPI_ISL_3484749, EPI_ISL_3484751, EPI_ISL_3484752, EPI_ISL_3484753, EPI_ISL_3484755, EPI_ISL_3484761, EPI_ISL_3484763, EPI_ISL_3484772, EPI_ISL_3484773, EPI_ISL_3484774, EPI_ISL_3484778, EPI_ISL_3484779, EPI_ISL_3484780, EPI_ISL_3484781, EPI_ISL_3484783, EPI_ISL_3484786, EPI_ISL_3484787, EPI_ISL_3484789, EPI_ISL_3484792, EPI_ISL_3484793, EPI_ISL_3484797, EPI_ISL_3484798, EPI_ISL_3484800, EPI_ISL_3484801, EPI_ISL_3484802, EPI_ISL_3484803, EPI_ISL_3484804, EPI_ISL_3484805, EPI_ISL_3484808, EPI_ISL_3484816, EPI_ISL_3484819, EPI_ISL_3484820, EPI_ISL_3484821, EPI_ISL_3484822, EPI_ISL_3484825, EPI_ISL_3484826, EPI_ISL_3484829, EPI_ISL_3484832, EPI_ISL_3484834, EPI_ISL_3484844, EPI_ISL_3484846, EPI_ISL_3484847, EPI_ISL_3484848, EPI_ISL_3484849, EPI_ISL_3484851, EPI_ISL_3484852, EPI_ISL_3484856, EPI_ISL_3484857, EPI_ISL_3484858, EPI_ISL_3484860, EPI_ISL_3484861, EPI_ISL_3484862, EPI_ISL_3484863, EPI_ISL_3484864, EPI_ISL_3484865, EPI_ISL_3484866, EPI_ISL_3484868, EPI_ISL_3484870, EPI_ISL_3484871, EPI_ISL_3484872, EPI_ISL_3484873, EPI_ISL_3484874, EPI_ISL_3484875, EPI_ISL_3484887, EPI_ISL_3484888, EPI_ISL_3484890, EPI_ISL_3484891, EPI_ISL_3484892, EPI_ISL_3484893, EPI_ISL_3484894, EPI_ISL_3484895, EPI_ISL_3484898, EPI_ISL_3484899, EPI_ISL_3484900, EPI_ISL_3484901, EPI_ISL_3484903, EPI_ISL_3484904, EPI_ISL_3484905, EPI_ISL_3484909, EPI_ISL_3484910, EPI_ISL_3484911, EPI_ISL_3484914, EPI_ISL_3484915, EPI_ISL_3484917, EPI_ISL_3484918, EPI_ISL_3484921, EPI_ISL_3484923, EPI_ISL_3484930, EPI_ISL_3484932, EPI_ISL_3484933, EPI_ISL_3484935, EPI_ISL_3484936, EPI_ISL_3484937, EPI_ISL_3484939, EPI_ISL_3484941, EPI_ISL_3484943, EPI_ISL_3484944, EPI_ISL_3484946, EPI_ISL_3484947, EPI_ISL_3484949, EPI_ISL_3484954, EPI_ISL_3484955, EPI_ISL_3484969, EPI_ISL_3484979, EPI_ISL_3484980, EPI_ISL_3484985, EPI_ISL_3484988, EPI_ISL_3484990, EPI_ISL_3484991, EPI_ISL_3484993, EPI_ISL_3484994, EPI_ISL_3484998, EPI_ISL_3485001, EPI_ISL_3485002, EPI_ISL_3485004, EPI_ISL_3485007, EPI_ISL_3485009, EPI_ISL_3485011, EPI_ISL_3485012, EPI_ISL_3485018, EPI_ISL_3485020, EPI_ISL_3485033, EPI_ISL_3485034, EPI_ISL_3485038, EPI_ISL_3485041, EPI_ISL_3485042, EPI_ISL_3485053, EPI_ISL_3485058, EPI_ISL_3485077, EPI_ISL_3485079, EPI_ISL_3485082, EPI_ISL_3485086, EPI_ISL_3485093, EPI_ISL_3485094, EPI_ISL_3485096, EPI_ISL_3485101, EPI_ISL_3485103, EPI_ISL_3485111, EPI_ISL_3485114, EPI_ISL_3485119, EPI_ISL_3485125, EPI_ISL_3485127, EPI_ISL_3485130, EPI_ISL_3485134, EPI_ISL_3485135, EPI_ISL_3485139, EPI_ISL_3485153, EPI_ISL_3485154, EPI_ISL_3485156, EPI_ISL_3485158, EPI_ISL_3485161, EPI_ISL_3485164, EPI_ISL_3485169, EPI_ISL_3485186, EPI_ISL_3485197, EPI_ISL_3485200, EPI_ISL_3485208, EPI_ISL_3485210, EPI_ISL_3485212, EPI_ISL_3485214, EPI_ISL_3485227, EPI_ISL_3485232, EPI_ISL_3485233, EPI_ISL_3485235, EPI_ISL_3485238, EPI_ISL_3485239, EPI_ISL_3485241, EPI_ISL_3485243, EPI_ISL_3485244, EPI_ISL_3485246, EPI_ISL_3485248, EPI_ISL_3485250, EPI_ISL_3485255, EPI_ISL_3485262, EPI_ISL_3485264, EPI_ISL_3485266, EPI_ISL_3485271, EPI_ISL_3485279, EPI_ISL_3485284, EPI_ISL_3485286, EPI_ISL_3485288, EPI_ISL_3485290, EPI_ISL_3485300, EPI_ISL_3485302, EPI_ISL_3485304, EPI_ISL_3485305, EPI_ISL_3485313, EPI_ISL_3485315, EPI_ISL_3485323, EPI_ISL_3485336, EPI_ISL_3485343, EPI_ISL_3485348, EPI_ISL_3485349, EPI_ISL_3485351, EPI_ISL_3485353, EPI_ISL_3485354, EPI_ISL_3485356, EPI_ISL_3485357, EPI_ISL_3485360, EPI_ISL_3485361, EPI_ISL_3485364, EPI_ISL_3485365, EPI_ISL_3485366, EPI_ISL_3485367, EPI_ISL_3485368, EPI_ISL_3485378, EPI_ISL_3485380, EPI_ISL_3485381, EPI_ISL_3485383, EPI_ISL_3485384, EPI_ISL_3485389, EPI_ISL_3485390, EPI_ISL_3485400, EPI_ISL_3485401, EPI_ISL_3485402, EPI_ISL_3485404, EPI_ISL_3485407, EPI_ISL_3485410, EPI_ISL_3485411, EPI_ISL_3485412, EPI_ISL_3485413, EPI_ISL_3485414, EPI_ISL_3485416, EPI_ISL_3485419, EPI_ISL_3485420, EPI_ISL_3485421, EPI_ISL_3485422, EPI_ISL_3485426, EPI_ISL_3485428, EPI_ISL_3485429, EPI_ISL_3485430, EPI_ISL_3485432, EPI_ISL_3485434, EPI_ISL_3485441, EPI_ISL_3485442, EPI_ISL_3485445, EPI_ISL_3485448, EPI_ISL_3485449, EPI_ISL_3485451, EPI_ISL_3485452, EPI_ISL_3485459, EPI_ISL_3485460, EPI_ISL_3485461, EPI_ISL_3485466, EPI_ISL_3485467, EPI_ISL_3485468, EPI_ISL_3485473, EPI_ISL_3485474, EPI_ISL_3485488, EPI_ISL_3485490, EPI_ISL_3485491, EPI_ISL_3485493, EPI_ISL_3485505, EPI_ISL_3485506, EPI_ISL_3485511, EPI_ISL_3485512, EPI_ISL_3485513, EPI_ISL_3485515, EPI_ISL_3485517, EPI_ISL_3485518, EPI_ISL_3485519, EPI_ISL_3485521, EPI_ISL_3485527, EPI_ISL_3485528, EPI_ISL_3485530, EPI_ISL_3485531, EPI_ISL_3485538, EPI_ISL_3485542, EPI_ISL_3485544, EPI_ISL_3485545, EPI_ISL_3485546, EPI_ISL_3485552, EPI_ISL_3485553, EPI_ISL_3485556, EPI_ISL_3485561, EPI_ISL_3485563, EPI_ISL_3485565, EPI_ISL_3485566, EPI_ISL_3485570, EPI_ISL_3485571, EPI_ISL_3485574, EPI_ISL_3485575, EPI_ISL_3485576, EPI_ISL_3485578, EPI_ISL_3485581, EPI_ISL_3485583, EPI_ISL_3485584, EPI_ISL_3485587, EPI_ISL_3485588, EPI_ISL_3485592, EPI_ISL_3485636, EPI_ISL_3485639, EPI_ISL_3485678, EPI_ISL_3485748, EPI_ISL_3485749, EPI_ISL_3485750, EPI_ISL_3485752, EPI_ISL_3485753, EPI_ISL_3485754, EPI_ISL_3485755, EPI_ISL_3485756, EPI_ISL_3485757, EPI_ISL_3485760, EPI_ISL_3485785, EPI_ISL_3485788, EPI_ISL_3485791, EPI_ISL_3485796, EPI_ISL_3485799, EPI_ISL_3485800, EPI_ISL_3485802, EPI_ISL_3485804, EPI_ISL_3485805, EPI_ISL_3485807, EPI_ISL_3485810, EPI_ISL_3485812, EPI_ISL_3485813, EPI_ISL_3485816, EPI_ISL_3485818, EPI_ISL_3485819, EPI_ISL_3485821, EPI_ISL_3485822, EPI_ISL_3485824, EPI_ISL_3485826, EPI_ISL_3485830, EPI_ISL_3485843, EPI_ISL_3485848, EPI_ISL_3485850, EPI_ISL_3485851, EPI_ISL_3485853, EPI_ISL_3485855, EPI_ISL_3485856, EPI_ISL_3485858, EPI_ISL_3485859, EPI_ISL_3485861, EPI_ISL_3485868, EPI_ISL_3485878, EPI_ISL_3485883, EPI_ISL_3485898, EPI_ISL_3485901, EPI_ISL_3485906, EPI_ISL_3485909, EPI_ISL_3485911, EPI_ISL_3485923, EPI_ISL_3485925, EPI_ISL_3485957, EPI_ISL_3485958, EPI_ISL_3485960, EPI_ISL_3485964, EPI_ISL_3485965, EPI_ISL_3485992, EPI_ISL_3485993, EPI_ISL_3485994, EPI_ISL_3485995, EPI_ISL_3485996, EPI_ISL_3486107, EPI_ISL_3486111, EPI_ISL_3486112, EPI_ISL_3486118, EPI_ISL_3486120, EPI_ISL_3486135, EPI_ISL_3486138, EPI_ISL_3486140, EPI_ISL_3486142, EPI_ISL_3486143, EPI_ISL_3486161, EPI_ISL_3486162, EPI_ISL_3486163, EPI_ISL_3486166, EPI_ISL_3486167, EPI_ISL_3486172, EPI_ISL_3486178, EPI_ISL_3486179, EPI_ISL_3486180, EPI_ISL_3486182, EPI_ISL_3486192, EPI_ISL_3486272, EPI_ISL_3486277, EPI_ISL_3486280, EPI_ISL_3486282, EPI_ISL_3486287, EPI_ISL_3486288, EPI_ISL_3486304, EPI_ISL_3486305, EPI_ISL_3486307, EPI_ISL_3486310, EPI_ISL_3486311, EPI_ISL_3486313, EPI_ISL_3486326, EPI_ISL_3486340, EPI_ISL_3486360, EPI_ISL_3486378, EPI_ISL_3486394, EPI_ISL_3596489, EPI_ISL_3596491, EPI_ISL_3600875, EPI_ISL_3600876, EPI_ISL_3600877, EPI_ISL_3600888, EPI_ISL_3600894, EPI_ISL_3600895, EPI_ISL_3600897, EPI_ISL_3600903, EPI_ISL_3600912, EPI_ISL_3600926, EPI_ISL_3600927, EPI_ISL_3600930, EPI_ISL_3600939, EPI_ISL_3600946, EPI_ISL_3600949, EPI_ISL_3600952, EPI_ISL_3600976, EPI_ISL_3600977, EPI_ISL_3600981, EPI_ISL_3600983, EPI_ISL_3600986, EPI_ISL_3600987, EPI_ISL_3601000, EPI_ISL_3601001, EPI_ISL_3601002, EPI_ISL_3601006, EPI_ISL_3601017, EPI_ISL_3601032, EPI_ISL_3601034, EPI_ISL_3601035, EPI_ISL_3601037, EPI_ISL_3601040, EPI_ISL_3601045, EPI_ISL_3601048, EPI_ISL_3601053, EPI_ISL_3601054, EPI_ISL_3601064, EPI_ISL_3601091, EPI_ISL_3601098, EPI_ISL_3601136, EPI_ISL_3601137, EPI_ISL_3601138, EPI_ISL_3601139, EPI_ISL_3601140, EPI_ISL_3601147, EPI_ISL_3601153, EPI_ISL_3601160, EPI_ISL_3601161, EPI_ISL_3601167, EPI_ISL_3601169, EPI_ISL_3601171, EPI_ISL_3601172, EPI_ISL_3601174, EPI_ISL_3601175, EPI_ISL_3601179, EPI_ISL_3601186, EPI_ISL_3601187, EPI_ISL_3601189, EPI_ISL_3601190, EPI_ISL_3601191, EPI_ISL_3601192, EPI_ISL_3601194, EPI_ISL_3601201, EPI_ISL_3601205, EPI_ISL_3601206, EPI_ISL_3601207, EPI_ISL_3601208, EPI_ISL_3601209, EPI_ISL_3601211, EPI_ISL_3601214, EPI_ISL_3601219, EPI_ISL_3601221, EPI_ISL_3601227, EPI_ISL_3601234, EPI_ISL_3601239, EPI_ISL_3601249, EPI_ISL_3601265, EPI_ISL_3601268, EPI_ISL_3601271, EPI_ISL_3601272, EPI_ISL_3601273, EPI_ISL_3601274, EPI_ISL_3601275, EPI_ISL_3601279, EPI_ISL_3601283, EPI_ISL_3601285, EPI_ISL_3601286, EPI_ISL_3601293, EPI_ISL_3601294, EPI_ISL_3601297, EPI_ISL_3601300, EPI_ISL_3601301, EPI_ISL_3601302, EPI_ISL_3601303, EPI_ISL_3601309, EPI_ISL_3601317, EPI_ISL_3601324, EPI_ISL_3601327, EPI_ISL_3601330, EPI_ISL_3601336, EPI_ISL_3601338, EPI_ISL_3601353, EPI_ISL_3601355, EPI_ISL_3601356, EPI_ISL_3601361, EPI_ISL_3601363, EPI_ISL_3601364, EPI_ISL_3601366, EPI_ISL_3601367, EPI_ISL_3601368, EPI_ISL_3601371, EPI_ISL_3601389, EPI_ISL_3601390, EPI_ISL_3601391, EPI_ISL_3601392, EPI_ISL_3601401, EPI_ISL_3601404, EPI_ISL_3601462 | | | |
| see above | British Columbia Centre For Disease Control | BCCDC Public Health Laboratory | Ana Pacagnella; Corrinne Ng; Dan Fornika; John Tyson; Kim Macdonald; Kimia Kamelian; Linda Hoang; Loretta Janz; Mel Krajden; Prystajecky Natalie; Robert Azana; Shannon Russell |
| EPI_ISL_2192596, EPI_ISL_2192598, EPI_ISL_2192611, EPI_ISL_2192627, EPI_ISL_2192629, EPI_ISL_2192641, EPI_ISL_2192658, EPI_ISL_2192663, EPI_ISL_2192686, EPI_ISL_2192695, EPI_ISL_2192710, EPI_ISL_2192721, EPI_ISL_2192731, EPI_ISL_2192732, EPI_ISL_2192735, EPI_ISL_2192736, EPI_ISL_2192739, EPI_ISL_2192747, EPI_ISL_2192751, EPI_ISL_2192766, EPI_ISL_2192770, EPI_ISL_2192777, EPI_ISL_2192789, EPI_ISL_2192790, EPI_ISL_2192804, EPI_ISL_2192818, EPI_ISL_2192833, EPI_ISL_2192842, EPI_ISL_2192847, EPI_ISL_2192849, EPI_ISL_2192852, EPI_ISL_2192857, EPI_ISL_2192861, EPI_ISL_2192863, EPI_ISL_2192875, EPI_ISL_2192880, EPI_ISL_2192882, EPI_ISL_2192888, EPI_ISL_2192899, EPI_ISL_2192900, EPI_ISL_2192910, EPI_ISL_2192919, EPI_ISL_2192928, EPI_ISL_2192929, EPI_ISL_2192931, EPI_ISL_2192932, EPI_ISL_2192940, EPI_ISL_2192947, EPI_ISL_2192949, EPI_ISL_2192953, EPI_ISL_2192981, EPI_ISL_2192988, EPI_ISL_2193000, EPI_ISL_2193011, EPI_ISL_2193017, EPI_ISL_2193028, EPI_ISL_2193033, EPI_ISL_2193034, EPI_ISL_2193036, EPI_ISL_2193040, EPI_ISL_2193053, EPI_ISL_2193054, EPI_ISL_2193055, EPI_ISL_2193058, EPI_ISL_2193060, EPI_ISL_2193066, EPI_ISL_2193067, EPI_ISL_2193072, EPI_ISL_2193094, EPI_ISL_2193100, EPI_ISL_2193103, EPI_ISL_2193107, EPI_ISL_2193110, EPI_ISL_2193117, EPI_ISL_2193118, EPI_ISL_2193128, EPI_ISL_2193139, EPI_ISL_2193151, EPI_ISL_2193152, EPI_ISL_2193185, EPI_ISL_2311884, EPI_ISL_2311885, EPI_ISL_2311899, EPI_ISL_2311916, EPI_ISL_2311967, EPI_ISL_2311974, EPI_ISL_2312001, EPI_ISL_2312032, EPI_ISL_2312059, EPI_ISL_2312114, EPI_ISL_2312115, EPI_ISL_2312117, EPI_ISL_2312125, EPI_ISL_2312159, EPI_ISL_2312224, EPI_ISL_2312282, EPI_ISL_2312304, EPI_ISL_2312305, EPI_ISL_2312319, EPI_ISL_2312363, EPI_ISL_2312402, EPI_ISL_2312408, EPI_ISL_2312419 | | | |
| see above | Broad Institute Clinical Research Sequencing Platform | Infectious Disease Program, Broad Institute of Harvard and MIT | Adams, G.; B.L.; B.W.; Bauer, M.; Birren; Blumenstiel, B.; Brown, C.; Carter, A.; Chaluvadi, S.; D.J.; DeFelice, M.; DeRuff, K.; Dodge, S.; Gabriel, S.; Gallagher, G.; Gladden-Young, A.; Granger, B.; J.E.; K.J.; Lagerborg, K.; Larkin, K.; Lee, M.; Lemieux; Lennon, N.; Loreth, C.; Madoff, L.; McGovern, S.; Meldrim, J.; Normandin, E.; P.C.; Park; Pearlman, L.; Reilly, S.; Rudy, M.; Sabeti; Siddle; Smole, S.; Tomkins-Tinch, C.; Vicente, G.; and MacInnis |
| EPI_ISL_2496778, EPI_ISL_2496896, EPI_ISL_2496898, EPI_ISL_2496899, EPI_ISL_2496901, EPI_ISL_2496902, EPI_ISL_2496903, EPI_ISL_2496905, EPI_ISL_2496907, EPI_ISL_2496908, EPI_ISL_2496911, EPI_ISL_2496913 | | | |
| see above | CA DPH Viral and Rickettsial Disease Laboratory | Chan-Zuckerberg Biohub | CZB Cliahub Consortium |
| EPI_ISL_2445508, EPI_ISL_2445512, EPI_ISL_2445514, EPI_ISL_2473788 | CADIP VILA REGINA FERNANDOPOLIS | Instituto Butantan | Antonio Jorge Martins; Claudia Renata dos Santos Barros; David Schlesinger; Debora Botequio Moretti; Dimas Tadeu Covas; Elaine Cristina Marqueze; Elaine Vieira Santos; Evandra Strazza Rodrigues; Heidge Fukumasu; Jayme Augusto de Souza-Neto; José Salvatore Leister Patané; Luiz Alcantara; Luiz Lehmann Coutinho; Maria Carolina Elias; Maurício Lacerda Nogueira; Rafael dos Santos Bezerra; Raul Machado Neto; Rejane Maria Tommasini Grotto; Ricardo Haddad; Sandra Coccuzzo Sampaio Vessoni; Simone Kashima; Svetoslav Nanev Slavov; Vincent Louis Viala |
| EPI_ISL_2891723, EPI_ISL_2891724, EPI_ISL_2891725, EPI_ISL_2891769, EPI_ISL_2891770, EPI_ISL_2891839, EPI_ISL_2891896, EPI_ISL_2891915, EPI_ISL_2891918, EPI_ISL_2892323, EPI_ISL_2922205, EPI_ISL_2922206, EPI_ISL_2922208, EPI_ISL_2922225, EPI_ISL_2922250, EPI_ISL_2922251, EPI_ISL_2922295, EPI_ISL_2922296, EPI_ISL_3243836, EPI_ISL_3243847, EPI_ISL_3243855, EPI_ISL_3243856, EPI_ISL_3243857, EPI_ISL_3243875, EPI_ISL_3243878, EPI_ISL_3243879, EPI_ISL_3243902, EPI_ISL_3243909, EPI_ISL_3243912, EPI_ISL_3243914, EPI_ISL_3243921, EPI_ISL_3243932, EPI_ISL_3243936, EPI_ISL_3243941, EPI_ISL_3243945, EPI_ISL_3243956, EPI_ISL_3243960, EPI_ISL_3243994, EPI_ISL_3244010, EPI_ISL_3244011, EPI_ISL_3244013, EPI_ISL_3244022, EPI_ISL_3244026, EPI_ISL_3244053, EPI_ISL_3244057, EPI_ISL_3244061, EPI_ISL_3244066, EPI_ISL_3244071, EPI_ISL_3244074, EPI_ISL_3244081, EPI_ISL_3247435, EPI_ISL_5696586 | | | |
| see above | CDPH VBL | California Department of Public Health | CDPH-COVIDNet; Emily Smith on behalf of CDPH-COVIDNet and Priva Path Labs Inc.; Emily Smith on behalf of CDPH-COVIDNet and UCSD EXCITE lab |
| EPI_ISL_2267957, EPI_ISL_2268077, EPI_ISL_2268135 | CENTOGENE Frankfurt Laboratory: Niederlassung Industriepark Höchst | Robert Koch Institute | |
| EPI_ISL_3102270 | CENTRO DE ATENDIMENTO PARA ENFRENTAMENTO AO COVID 19 | Analytical Competence Molecular Epidemiology Lab/ACME, Oswaldo Cruz Foundation, Ceara (FIOCRUZ CE) | Cleber Furtado Aksenen; Fabio Miyajima; Fernando Braga Stehling; Francisco Eder de Moura Lopes; Jamille Maria Mendes Bezerra; Joaquim César do Nascimento Sousa Junior; Pedro Miguel Carneiro Jeronimo; Suzana Porto Almeida e Lucas Delerino; Thais Ferreira de Oliveira; Thais de Oliveira Costa; Ticiane Cavalcante de Souza; Veridiana Pessoa Miyajima |

| | | | |
|---|---|---|---|
| EPI_ISL_2445551, EPI_ISL_2445553 | CENTRO DE ESPECIALIDADES DE PRIMAVERA | Instituto Butantan | Antonio Jorge Martins; Claudia Renata dos Santos Barros; David Schlesinger; Debora Botequio Moretti; Dimas Tadeu Covas; Elaine Cristina Marqueze; Elaine Vieira Santos; Evandra Strazza Rodrigues; Heidge Fukumasu; Jayme Augusto de Souza-Neto; José Salvatore Leister Patané; Luiz Alcantara; Luiz Lehmann Coutinho; Maria Carolina Elias; Maurício Lacerda Nogueira; Rafael dos Santos Bezerra; Raul Machado Neto; Rejane Maria Tommasini Grotto; Ricardo Haddad; Sandra Coccuzzo Sampaio Vessoni; Simone Kashima; Svetoslav Nanev Slavov; Vincent Louis Viala |
| EPI_ISL_2445539 | CENTRO DE ESPECIALIDADES MARIAS DA GLORIA | Instituto Butantan | Antonio Jorge Martins; Claudia Renata dos Santos Barros; David Schlesinger; Debora Botequio Moretti; Dimas Tadeu Covas; Elaine Cristina Marqueze; Elaine Vieira Santos; Evandra Strazza Rodrigues; Heidge Fukumasu; Jayme Augusto de Souza-Neto; José Salvatore Leister Patané; Luiz Alcantara; Luiz Lehmann Coutinho; Maria Carolina Elias; Maurício Lacerda Nogueira; Rafael dos Santos Bezerra; Raul Machado Neto; Rejane Maria Tommasini Grotto; Ricardo Haddad; Sandra Coccuzzo Sampaio Vessoni; Simone Kashima; Svetoslav Nanev Slavov; Vincent Louis Viala |
| EPI_ISL_2445340 | CENTRO DE ESPECIALIDADES MUNICIPAL CESAR TORREZAN R BONITO | Instituto Butantan | Antonio Jorge Martins; Claudia Renata dos Santos Barros; David Schlesinger; Debora Botequio Moretti; Dimas Tadeu Covas; Elaine Cristina Marqueze; Elaine Vieira Santos; Evandra Strazza Rodrigues; Heidge Fukumasu; Jayme Augusto de Souza-Neto; José Salvatore Leister Patané; Luiz Alcantara; Luiz Lehmann Coutinho; Maria Carolina Elias; Maurício Lacerda Nogueira; Rafael dos Santos Bezerra; Raul Machado Neto; Rejane Maria Tommasini Grotto; Ricardo Haddad; Sandra Coccuzzo Sampaio Vessoni; Simone Kashima; Svetoslav Nanev Slavov; Vincent Louis Viala |
| EPI_ISL_2445615, EPI_ISL_2473809, EPI_ISL_2473810 | CENTRO DE GENOMAS | Instituto Butantan | Antonio Jorge Martins; Claudia Renata dos Santos Barros; David Schlesinger; Debora Botequio Moretti; Dimas Tadeu Covas; Elaine Cristina Marqueze; Elaine Vieira Santos; Evandra Strazza Rodrigues; Heidge Fukumasu; Jayme Augusto de Souza-Neto; José Salvatore Leister Patané; Luiz Alcantara; Luiz Lehmann Coutinho; Maria Carolina Elias; Maurício Lacerda Nogueira; Rafael dos Santos Bezerra; Raul Machado Neto; Rejane Maria Tommasini Grotto; Ricardo Haddad; Sandra Coccuzzo Sampaio Vessoni; Simone Kashima; Svetoslav Nanev Slavov; Vincent Louis Viala |
| EPI_ISL_2445428, EPI_ISL_2445429, EPI_ISL_2445430, EPI_ISL_2445431, EPI_ISL_2445433, EPI_ISL_2445434 | CENTRO DE SAUDE DE LUCELIA | Instituto Butantan | Antonio Jorge Martins; Claudia Renata dos Santos Barros; David Schlesinger; Debora Botequio Moretti; Dimas Tadeu Covas; Elaine Cristina Marqueze; Elaine Vieira Santos; Evandra Strazza Rodrigues; Heidge Fukumasu; Jayme Augusto de Souza-Neto; José Salvatore Leister Patané; Luiz Alcantara; Luiz Lehmann Coutinho; Maria Carolina Elias; Maurício Lacerda Nogueira; Rafael dos Santos Bezerra; Raul Machado Neto; Rejane Maria Tommasini Grotto; Ricardo Haddad; Sandra Coccuzzo Sampaio Vessoni; Simone Kashima; Svetoslav Nanev Slavov; Vincent Louis Viala |
| EPI_ISL_2445441, EPI_ISL_2445442, EPI_ISL_2445445 | CENTRO DE SAUDE DR NELSON DA CUNHA BASTOS | Instituto Butantan | Antonio Jorge Martins; Claudia Renata dos Santos Barros; David Schlesinger; Debora Botequio Moretti; Dimas Tadeu Covas; Elaine Cristina Marqueze; Elaine Vieira Santos; Evandra Strazza Rodrigues; Heidge Fukumasu; Jayme Augusto de Souza-Neto; José Salvatore Leister Patané; Luiz Alcantara; Luiz Lehmann Coutinho; Maria Carolina Elias; Maurício Lacerda Nogueira; Rafael dos Santos Bezerra; Raul Machado Neto; Rejane Maria Tommasini Grotto; Ricardo Haddad; Sandra Coccuzzo Sampaio Vessoni; Simone Kashima; Svetoslav Nanev Slavov; Vincent Louis Viala |
| EPI_ISL_2445440 | CENTRO DE SAUDE ECHAPORA | Instituto Butantan | Antonio Jorge Martins; Claudia Renata dos Santos Barros; David Schlesinger; Debora Botequio Moretti; Dimas Tadeu Covas; Elaine Cristina Marqueze; Elaine Vieira Santos; Evandra Strazza Rodrigues; Heidge Fukumasu; Jayme Augusto de Souza-Neto; José Salvatore Leister Patané; Luiz Alcantara; Luiz Lehmann Coutinho; Maria Carolina Elias; Maurício Lacerda Nogueira; Rafael dos Santos Bezerra; Raul Machado Neto; Rejane Maria Tommasini Grotto; Ricardo Haddad; Sandra Coccuzzo Sampaio Vessoni; Simone Kashima; Svetoslav Nanev Slavov; Vincent Louis Viala |
| EPI_ISL_2473787 | CENTRO DE SAUDE II CARDOSO | Instituto Butantan | Antonio Jorge Martins; Claudia Renata dos Santos Barros; David Schlesinger; Debora Botequio Moretti; Dimas Tadeu Covas; Elaine Cristina Marqueze; Elaine Vieira Santos; Evandra Strazza Rodrigues; Heidge Fukumasu; Jayme Augusto de Souza-Neto; José Salvatore Leister Patané; Luiz Alcantara; Luiz Lehmann Coutinho; Maria Carolina Elias; Maurício Lacerda Nogueira; Rafael dos Santos Bezerra; Raul Machado Neto; Rejane Maria Tommasini Grotto; Ricardo Haddad; Sandra Coccuzzo Sampaio Vessoni; Simone Kashima; Svetoslav Nanev Slavov; Vincent Louis Viala |
| EPI_ISL_2445529, EPI_ISL_2445531, EPI_ISL_2445533 | CENTRO DE SAUDE II DE POTIRENDABA CESARINO BENFATTI | Instituto Butantan | Antonio Jorge Martins; Claudia Renata dos Santos Barros; David Schlesinger; Debora Botequio Moretti; Dimas Tadeu Covas; Elaine Cristina Marqueze; Elaine Vieira Santos; Evandra Strazza Rodrigues; Heidge Fukumasu; Jayme Augusto de Souza-Neto; José Salvatore Leister Patané; Luiz Alcantara; Luiz Lehmann Coutinho; Maria Carolina Elias; Maurício Lacerda Nogueira; Rafael dos Santos Bezerra; Raul Machado Neto; Rejane Maria Tommasini Grotto; Ricardo Haddad; Sandra Coccuzzo Sampaio Vessoni; Simone Kashima; Svetoslav Nanev Slavov; Vincent Louis Viala |
| EPI_ISL_2445317 | CENTRO DE SAUDE II IBITINGA | Instituto Butantan | Antonio Jorge Martins; Claudia Renata dos Santos Barros; David Schlesinger; Debora Botequio Moretti; Dimas Tadeu Covas; Elaine Cristina Marqueze; Elaine Vieira Santos; Evandra Strazza Rodrigues; Heidge Fukumasu; Jayme Augusto de Souza-Neto; José Salvatore Leister Patané; Luiz Alcantara; Luiz Lehmann Coutinho; Maria Carolina Elias; Maurício Lacerda Nogueira; Rafael dos Santos Bezerra; Raul Machado Neto; Rejane Maria Tommasini Grotto; Ricardo Haddad; Sandra Coccuzzo Sampaio Vessoni; Simone Kashima; Svetoslav Nanev Slavov; Vincent Louis Viala |
| EPI_ISL_2445603 | CENTRO DE SAUDE II SAO MIGUEL ARCANJO | Instituto Butantan | Antonio Jorge Martins; Claudia Renata dos Santos Barros; David Schlesinger; Debora Botequio Moretti; Dimas Tadeu Covas; Elaine Cristina Marqueze; Elaine Vieira Santos; Evandra Strazza Rodrigues; Heidge Fukumasu; Jayme Augusto de Souza-Neto; José Salvatore Leister Patané; Luiz Alcantara; Luiz Lehmann Coutinho; Maria Carolina Elias; Maurício Lacerda Nogueira; Rafael dos Santos Bezerra; Raul Machado Neto; Rejane Maria Tommasini Grotto; Ricardo Haddad; Sandra Coccuzzo Sampaio Vessoni; Simone Kashima; Svetoslav Nanev Slavov; Vincent Louis Viala |
| EPI_ISL_2494282, EPI_ISL_2494284 | CENTRO DE SAUDE III DE AGUAS DE SAO PEDRO | Instituto Butantan | Antonio Jorge Martins; Claudia Renata dos Santos Barros; David Schlesinger; Debora Botequio Moretti; Dimas Tadeu Covas; Elaine Cristina Marqueze; Elaine Vieira Santos; Evandra Strazza Rodrigues; Heidge Fukumasu; Jayme Augusto de Souza-Neto; José Salvatore Leister Patané; Luiz Alcantara; Luiz Lehmann Coutinho; Maria Carolina Elias; Maurício Lacerda Nogueira; Rafael dos Santos Bezerra; Raul Machado Neto; Rejane Maria Tommasini Grotto; Ricardo Haddad; Sandra Coccuzzo Sampaio Vessoni; Simone Kashima; Svetoslav Nanev Slavov; Vincent Louis Viala |
| EPI_ISL_2445548, EPI_ISL_2445557 | CENTRO DE SAUDE III NELCIDIO DA SILVEIRA BASTOS | Instituto Butantan | Antonio Jorge Martins; Claudia Renata dos Santos Barros; David Schlesinger; Debora Botequio Moretti; Dimas Tadeu Covas; Elaine Cristina Marqueze; Elaine Vieira Santos; Evandra Strazza Rodrigues; Heidge Fukumasu; Jayme Augusto de Souza-Neto; José Salvatore Leister Patané; Luiz Alcantara; Luiz Lehmann Coutinho; Maria Carolina Elias; Maurício Lacerda Nogueira; Rafael dos Santos Bezerra; Raul Machado Neto; Rejane Maria Tommasini Grotto; Ricardo Haddad; Sandra Coccuzzo Sampaio Vessoni; Simone Kashima; Svetoslav Nanev Slavov; Vincent Louis Viala |
| EPI_ISL_2340842, EPI_ISL_2445348, EPI_ISL_2445358, EPI_ISL_2445360, EPI_ISL_2445446, EPI_ISL_2445447, EPI_ISL_2445448, EPI_ISL_2445449, EPI_ISL_2445451, EPI_ISL_2445452, EPI_ISL_2473697 | | | |
| see above | CENTRO INTEGRADO DE SAUDE | Instituto Butantan | Antonio Jorge Martins; Claudia Renata dos Santos Barros; David Schlesinger; Debora Botequio Moretti; Dimas Tadeu Covas; Elaine Cristina Marqueze; Elaine Vieira Santos; Evandra Strazza Rodrigues; Heidge Fukumasu; Jayme Augusto de Souza-Neto; José Salvatore Leister Patané; Luiz Alcantara; Luiz Lehmann Coutinho; Maria Carolina Elias; Maurício Lacerda Nogueira; Rafael dos Santos Bezerra; Raul Machado Neto; Rejane Maria Tommasini Grotto; Ricardo Haddad; Sandra Coccuzzo Sampaio Vessoni; Simone Kashima; Svetoslav Nanev Slavov; Vincent Louis Viala |
| EPI_ISL_2493445 | CENTRO SAUDE III DR JOSE TADEU MOURAO | Instituto Butantan | Antonio Jorge Martins; Claudia Renata dos Santos Barros; David Schlesinger; Debora Botequio Moretti; Dimas Tadeu Covas; Elaine Cristina Marqueze; Elaine Vieira Santos; Evandra Strazza Rodrigues; Heidge Fukumasu; Jayme Augusto de Souza-Neto; José Salvatore Leister Patané; Luiz Alcantara; Luiz Lehmann Coutinho; Maria Carolina Elias; Maurício Lacerda Nogueira; Rafael dos Santos Bezerra; Raul Machado Neto; Rejane Maria Tommasini Grotto; Ricardo Haddad; Sandra Coccuzzo Sampaio Vessoni; Simone Kashima; Svetoslav Nanev Slavov; Vincent Louis Viala |
| EPI_ISL_2621206 | CENTROLAB | Universidad Nacional de Colombia - Laboratorio Genómico One Health | Andres F. Cardona-Rios; Carlos Franco-Muñoz; Carolina Muñoz-Arango; Celeny Ortiz; Daniel O. Maldonado-Perez; Diego A. Álvarez-Díaz; Hector Alejandro Ruiz-Moreno; Idabely Betancur Ortiz; Jorge E. Osorio; Juan P. Hernandez-Ortiz; Karl A Ciuoderis; Katherine Laiton-Donato; Laura Silvana Perez; Lina M. Hurtado; Marcela Mercado-Reyes; Maria Angélica Maya; Maria Stella López; Rita Almanza Payares; Sandra Ines Cano; Simón Villegas Velásquez |
| EPI_ISL_2614611, EPI_ISL_2614612 | CETESB - Companhia Ambiental do Estado de Sao Paulo | Instituto Adolfo Lutz, Interdiciplinary Procedures Center, Strategic Laboratory | Caio Vinicius Dias Lopes; Claudia Regina Gonçalves; Claudio Tavares Sacchi; Erica Valessa Ramos Gomes; Karoline Rodrigues Campos; Leonardo Jose Tadeu de Araujo; Luisa Zanolli Moreno; Maria Ines Zanoli Sato; Mikaela R. F. Barbosa |
| EPI_ISL_2361524, EPI_ISL_2361526 | CH ROUBAIX | CHU Lille - Laboratoire de Virologie | AIT YAHYA Emilie; ALIDJINOU Enagnon Kazali; BOCKET Laurence; CREPIN Michel; DEMAY Christophe; ENGELMANN Ilka; GEFFROY Sandrine; GUIGON Aurélie; LAMBERT Valérie; LAZREK Mouna; NOBILLIAUX Florian; PREVOST Brigitte; TCHANTCHOU NJOSSE YANICK; THUILLIER Caroline; TINEZ Claire |
| EPI_ISL_2333715 | CH.INTERCOMMUNAL DE CRETEIL | Department of Virology, Henri Mondor University Hospital, Assistance Publique Hôpitaux de Paris, Université Paris-Est Créteil, INSERM U955 | Alexandre Soulier; Christophe Rodriguez; Elisabeth Trawinski; Guillaume Gricourt; Jean-Michel Pawlotsky; Melissa N'Debi; Slim Fourati; Vanessa Demontant |
| EPI_ISL_2227891 | CHC Andrée Rosemon | Institut Pasteur de la Guyane | A. Enfissi; A. Salmier; Anne Lavergne; Dominique Rousset |
| EPI_ISL_2688742 | CHI Elbeuf | Centre Hospitalier Universitaire de Rouen Laboratoire de Virologie | Alice Moisan; Fabienne De Oliveira; Marie Leoz |
| EPI_ISL_2510151 | CHUGA-IBP-laboratoire de Virologie | IBP-laboratoire de virologie | Alban Caporossi; Anne Signori -Schmuck; Anne-Karen Faure; Aurélie Truffot; Benjamin Nemoz; Hugo Jardin; Julien Andréani; Julien Lupo; Léa Ponderand; Pascal Poignard; Patrice Morand; Raphaële Germi; Sylvie Larrat |
| EPI_ISL_2516497, EPI_ISL_2516531 | CLILAB | Microbiology Department | Aida Gonzalez-Diaz; Carmen Ardanuy; Jordi Camara; Jordi Niubó; Laura Calatayud; M Angeles Domínguez; Miguel Fernandez-Huerta; Sara Marti |
| EPI_ISL_3355516 | CLINICA PUTUMAYO | Corporacion CorpoGen-Universidad de los Andes-Universidad Central | Christian Romero; Jorge Duitama; Juan Manuel Anzola; Laura González; Maryam Chaib De Mares; María Mercedes Zambrano; Nelly Díaz; Patricia Del Portillo; Silvia Restrepo |
| EPI_ISL_2227892, EPI_ISL_2227893, EPI_ISL_2227894, EPI_ISL_2227895, EPI_ISL_2227896, EPI_ISL_2227897, EPI_ISL_2227898, EPI_ISL_2227899, EPI_ISL_2227900, EPI_ISL_2227901 | | | |
| see above | CNR Institut Pasteur de la Guyane | Institut Pasteur de la Guyane | A. Enfissi; A. Salmier; Anne Lavergne; Dominique Rousset |
| EPI_ISL_2493446 | COORDENADORIA MUNICIPAL DE SAUDE DE IRACEMAPOLIS | Instituto Butantan | Antonio Jorge Martins; Claudia Renata dos Santos Barros; David Schlesinger; Debora Botequio Moretti; Dimas Tadeu Covas; Elaine Cristina Marqueze; Elaine Vieira Santos; Evandra Strazza Rodrigues; Heidge Fukumasu; Jayme Augusto de Souza-Neto; José Salvatore Leister Patané; Luiz Alcantara; Luiz Lehmann Coutinho; Maria Carolina Elias; Maurício Lacerda Nogueira; Rafael dos Santos Bezerra; Raul Machado Neto; Rejane Maria Tommasini Grotto; Ricardo Haddad; Sandra Coccuzzo Sampaio Vessoni; Simone Kashima; Svetoslav Nanev Slavov; Vincent Louis Viala |
| EPI_ISL_2445437, EPI_ISL_2445438, EPI_ISL_2445443 | CS DE QUINTANA | Instituto Butantan | Antonio Jorge Martins; Claudia Renata dos Santos Barros; David Schlesinger; Debora Botequio Moretti; Dimas Tadeu Covas; Elaine Cristina Marqueze; Elaine Vieira Santos; Evandra Strazza Rodrigues; Heidge Fukumasu; Jayme Augusto de Souza-Neto; José Salvatore Leister Patané; Luiz Alcantara; Luiz Lehmann Coutinho; Maria Carolina Elias; Maurício Lacerda Nogueira; Rafael dos Santos Bezerra; Raul Machado Neto; Rejane Maria Tommasini Grotto; Ricardo Haddad; Sandra Coccuzzo Sampaio Vessoni; Simone Kashima; Svetoslav Nanev Slavov; Vincent Louis Viala |
| EPI_ISL_2445455, EPI_ISL_2445458, EPI_ISL_2445459, EPI_ISL_2445461, EPI_ISL_2445462 | CS II DR JAHYR DE PAULA RIBEIRO GUARA | Instituto Butantan | Antonio Jorge Martins; Claudia Renata dos Santos Barros; David Schlesinger; Debora Botequio Moretti; Dimas Tadeu Covas; Elaine Cristina Marqueze; Elaine Vieira Santos; Evandra Strazza Rodrigues; Heidge Fukumasu; Jayme Augusto de Souza-Neto; José Salvatore Leister Patané; Luiz Alcantara; Luiz Lehmann Coutinho; Maria Carolina Elias; Maurício Lacerda Nogueira; Rafael dos Santos Bezerra; Raul Machado Neto; Rejane Maria Tommasini Grotto; Ricardo Haddad; Sandra Coccuzzo Sampaio Vessoni; Simone Kashima; Svetoslav Nanev Slavov; Vincent Louis Viala |
| EPI_ISL_2445406 | CS II DR MIGUEL VITALIANO ORLANDIA | Instituto Butantan | Antonio Jorge Martins; Claudia Renata dos Santos Barros; David Schlesinger; Debora Botequio Moretti; Dimas Tadeu Covas; Elaine Cristina Marqueze; Elaine Vieira Santos; Evandra Strazza Rodrigues; Heidge Fukumasu; Jayme Augusto de Souza-Neto; José Salvatore Leister Patané; Luiz Alcantara; Luiz Lehmann Coutinho; Maria Carolina Elias; Maurício Lacerda Nogueira; Rafael dos Santos Bezerra; Raul Machado Neto; Rejane Maria Tommasini Grotto; Ricardo Haddad; Sandra Coccuzzo Sampaio Vessoni; Simone Kashima; Svetoslav Nanev Slavov; Vincent Louis Viala |
| EPI_ISL_6229706 | CSF BILHEIRA | ACME Lab, Oswaldo Cruz Foundation, FIOCRUZ/CE | Carlos Leonardo de Aragao Araujo; Cecilia Leite Costa & Eduardo Ruback dos Santos on behalf of COVID-19 FIOCRUZ Genomic Network; Cleber Furtado Aksenen; Fabio Miyajima; Fernando Braga Stehling; Francisco Eder de Moura Lopes; Igor Oliveira Duarte; Jamille Maria Mendes Bezerra; Joaquim Cesar do Nascimento Sousa Junior; Pedro Miguel Carneiro Jeronimo; Suzana Porto Almeida; Thais Ferreira de Oliveira; Thais de Oliveira Costa; Ticiane Cavalcante de Souza; Veridiana Pessoa Miyajima |
| EPI_ISL_6229707 | CSF CONJUNTO SANTO ANTONIO | ACME Lab, Oswaldo Cruz Foundation, FIOCRUZ/CE | Carlos Leonardo de Aragao Araujo; Cecilia Leite Costa & Eduardo Ruback dos Santos on behalf of COVID-19 FIOCRUZ Genomic Network; Cleber Furtado Aksenen; Fabio Miyajima; Fernando Braga Stehling; Francisco Eder de Moura Lopes; Igor Oliveira Duarte; Jamille Maria Mendes Bezerra; Joaquim Cesar do Nascimento Sousa Junior; Pedro Miguel Carneiro Jeronimo; Suzana Porto Almeida; Thais Ferreira de Oliveira; Thais de Oliveira Costa; Ticiane Cavalcante de Souza; Veridiana Pessoa Miyajima |
| EPI_ISL_6229711 | CSF RAFAEL ARRUDA | ACME Lab, Oswaldo Cruz Foundation, FIOCRUZ/CE | Carlos Leonardo de Aragao Araujo; Cecilia Leite Costa & Eduardo Ruback dos Santos on behalf of COVID-19 FIOCRUZ Genomic Network; Cleber Furtado Aksenen; Fabio Miyajima; Fernando Braga Stehling; Francisco Eder de Moura Lopes; Igor Oliveira Duarte; Jamille Maria Mendes Bezerra; Joaquim Cesar do Nascimento Sousa Junior; Pedro Miguel Carneiro Jeronimo; Suzana Porto Almeida; Thais Ferreira de Oliveira; Thais de Oliveira Costa; Ticiane Cavalcante de Souza; Veridiana Pessoa Miyajima |
| EPI_ISL_6229712 | CSF TAPERUABA | ACME Lab, Oswaldo Cruz Foundation, FIOCRUZ/CE | Carlos Leonardo de Aragao Araujo; Cecilia Leite Costa & Eduardo Ruback dos Santos on behalf of COVID-19 FIOCRUZ Genomic Network; Cleber Furtado Aksenen; Fabio Miyajima; Fernando Braga Stehling; Francisco Eder de Moura Lopes; Igor Oliveira Duarte; Jamille Maria Mendes Bezerra; Joaquim Cesar do Nascimento Sousa Junior; Pedro Miguel Carneiro Jeronimo; Suzana Porto Almeida; Thais Ferreira de Oliveira; Thais de Oliveira Costa; Ticiane Cavalcante de Souza; Veridiana Pessoa Miyajima |
| EPI_ISL_2710379, EPI_ISL_2710381 | CT Department of Public Health | CT Department of Public Health | Claire Pearson; Tu N. Nguyen |
| EPI_ISL_2892489, EPI_ISL_3536707, EPI_ISL_3536928, EPI_ISL_3536929, EPI_ISL_3536930, EPI_ISL_4005534, EPI_ISL_4005535, EPI_ISL_4005537, EPI_ISL_4005542, EPI_ISL_4005543 | | | |
| see above | California Department of Public Health | California Department of Public Health | CDPH IDLB COVIDNet et al |
| EPI_ISL_7507603 | Cedars-Sinai Medical Center, Molecular Pathology Laboratory of Department of Pathology & Laboratory Medicine and Genomic Core | Cedars-Sinai Medical Center, Molecular Pathology Laboratory of Department of Pathology & Laboratory Medicine and Genomic Core | Brian Davis; Eric Vail; Jasmine T Plummer; Jorge Mario Sincuir Martinez; Stephanie Chen; Wenjuan Zhang |
| EPI_ISL_2296515, EPI_ISL_2296516 | Centracare Laboratory Services | Minnesota Department of Health, Public Health Laboratory | Alexandra Lorentz; Jacob Garfin; Matt Plumb; and Xiong Wang |

| | | | |
|---|---|---|---|
| EPI_ISL_2502376, EPI_ISL_2502380, EPI_ISL_2502396, EPI_ISL_2502441, EPI_ISL_2502483, EPI_ISL_2502484, EPI_ISL_2502485, EPI_ISL_2502486, EPI_ISL_2502487, EPI_ISL_2502488, EPI_ISL_2502489, EPI_ISL_2502490, EPI_ISL_2502491, EPI_ISL_2502492, EPI_ISL_2502493, EPI_ISL_2502494, EPI_ISL_2502495, EPI_ISL_2502496, EPI_ISL_2502497, EPI_ISL_2502498, EPI_ISL_2502508, EPI_ISL_3462622 | | | |
| see above | Central Laboratory, Bureau of Public Health (BOG) and Academic Hospital Paramaribo | Erasmus Medical Center | Bas B Oude Munnink; Cherise Beek; Consuella Partowidjojo; Dion Gajadin; Ed PF IJzerman; Emmanuelle Munger; Gary Gummels; Ingrid SK Krishnadath; Lycke Woittiez; Marion PG Koopmans; Mireille Van de Veer; Phyllis Pinas; Princes Wongsowidjojo; Radjesh Ori; Ranisha Doerbalie; Rohma Banwari; Soeradj Harkisoen; Stephen Vreden; Tilotmadebie Ramlal; Verne Nanhoe |
| EPI_ISL_3255059, EPI_ISL_3255062, EPI_ISL_3255080, EPI_ISL_3255133, EPI_ISL_3255150 | Central Public Health Laboratory - LACEN - Bahia, Salvador, Brazil | Central Public Health Laboratory - LACEN -Bahia, Salvador, Brazil | Arabela Leal; Breno Dominguez; Felicidade Pereira; Jaqueline Gomes; Luciana Oliveira; Luiz Alcantara; Marcela Gómez; Marta Giovanetti; Patrícia Cajado; Stephane Tosta; Vagner Fonseca; Vanessa Nardy |
| EPI_ISL_5926865 | Centro de Infectologia Charles Mérieux/ Laboratório Rodolphe Mérieux, FUNDHACRE | Bioinformatics Laboratory / LNCC | Alessandra P Lamarca; Alexandra L Gerber; Aline de Freitas Souza; Ana Paula de C Guimarães; Ana Tereza R Vasconcelos; Andreas Stocker; Cirley Maria de Oliveira Lobato; Douglas Terra Machado; Janaína Mazaro; Janete Taynã Nascimento Rodrigues; Luiz Fellype Alves de Souza; Luiz G P de Almeida; Ronaldo da Silva F Jr; Rutilene B. Souza |
| EPI_ISL_2490383, EPI_ISL_2490384 | Centro de Investigación Biomédica de Occidente (CIBO) | Centro de Investigación en Enfermedades Infecciosas (CIENI), Instituto Nacional de Enfermedades Respiratorias (INER) | ; Alejandra Garcia-Gasca; Alejandra Hernández-Terán; Alejandro Sanchez-Flores; Alfredo Herrera-Estrella; Alicia Ocaña-Mondragón; Andreu Comas-García; Angel Gustavo Salas-Lais; Antonio Loza Román; Bernardo Martínez-Miguel; Blanca Taboada; Brenda Irasema Maldonado-Meza; Bruno Gomez-Gil; Carla Ivón Herrera-Najera; Carlos F. Arias; Celia Boukadida; Clara Esperanza Santacruz-Tinoco; Concepción Grajales-Muñiz; Consorcio Mexicano de Vigilancia Genómica (CoViGen-Mex). Authors (in alphabetical order): Julio Elias Alvarado-Yaah; Cristóbal Cháidez-Quiróz; Célida Duque Molina; Célida Martínez- Rodríguez; Daniel Fregoso-Rueda; Daniel Lira Morales; Eduardo Becerril-Vargas; Fernando Fontove-Herrera; Fidencio Mejía-Nepomuceno; Francisco Pulido; Gloria Elena Espinosa-Ayala; Gloria María Molina-Salinas; Gloria Vazquez; Hector Esteban Paz-Juárez; Hector Montoya-Fuentes; Helen Haydee Fernanda Ramírez-Plascencia; Irvin González-López; Jean Pierre González; Joel Armando Vázquez-Pérez.; Jorge Salas-Hernández; José Antonio Enciso-Moreno; José Arturo Martínez-Orozco; José Esteban Muñoz-Medina; José de Jesús Nuñez-Contreras; Juan Bautista Chale-Dzul; Julissa Enciso-Ibarra; Luis Alberto Ochoa-Carrera; Margarita Matías-Florentino; Mario Mújica-Sánchez; Marissa Perez-Garcia; María Guadalupe Santiago-Mauricio; María Guadalupe de Jesús Mireles-Rivera; Nelly Sélem-Mojica; Pavel Isa; Ricardo Ciria Merce; Ricardo Grande; Rosa María Gutierrez Rios; Santiago Ávila-Ríos; Selene Zárate; Susana Lopez; Victor Eduardo Garcia-Arias; Víctor Hugo Borja-Aburto |
| EPI_ISL_4262058, EPI_ISL_5055016 | Chiu Laboratory, University of California, San Francisco | Chiu Laboratory, University of California, San Francisco | Alicia Sotomayor-Gonzalez; Baolin Wang; Candace Wang; Charles Chiu; Daniel Wong; Dustin Glasner; Venice Servellita; Xianding (Wayne) Deng; Yueyuan Zhang |
| EPI_ISL_3074070, EPI_ISL_3074075, EPI_ISL_3074080, EPI_ISL_3074085, EPI_ISL_3074100, EPI_ISL_3074103 | City of Milwaukee Health Department Laboratory | City of Milwaukee Health Department Laboratory | Amy Bauer; Jennifer Lentz; Manjeet Khubbar; Samantha Scott; Sanjib Bhattacharyya |
| EPI_ISL_2467294, EPI_ISL_2467298, EPI_ISL_3242485, EPI_ISL_3242487, EPI_ISL_3242494, EPI_ISL_3242511, EPI_ISL_3242519, EPI_ISL_3242548, EPI_ISL_3242646, EPI_ISL_3242657, EPI_ISL_3242658, EPI_ISL_3242673 | | | |
| see above | Clinica INDISA | "Facultad de Ciencias de la Vida, UNAB" | "Claudio Meneses; Ariel Orellana"; Claudio Olmos; Daniel Leon; Dayan Sanhueza; Eduardo Castro; Gonzalo Campaña; Macarena Bastias; Paola Pidal; Ricardo Yusta; Sebastian Wolter; Susana Saez; Víctor Monreal; Waldo Diaz |
| EPI_ISL_3232278, EPI_ISL_3232385 | Clinical Microbiology Laboratory, NewYork Presbyterian Hospital/Columbia University Irving Medical Center | Uhlemann Laboratory, Columbia University Irving Medical Center | Anne Kelley; Anne-Catrin Uhlemann; Maya Tagliavia; Medini K. Annavajhala |
| EPI_ISL_4635663 | Clinical Microbiology, University of Texas Medical Branch (UTMB), Galveston | ClickSeq Lab, University of Texas Medical Branch, UTMB, Galveston | Dong J.; Jaworski E.; Judy B.; Ksiazek T.; Langsjoen R.M.; Machado R.R; McConnell A.J.; Menachery V.D.; Miller A.L.; Mitchell B.M.; Morris V.G.; Newman P.; Plante J.A.; Plante K.S.; Pyles R.B.; Ren P.; Routh A.; Saada N.I.; Sotcheff S.L.; Swetnam D.; Thompson J.K.; Weaver S.C; Widen S.; Zhou Y. |
| EPI_ISL_2380845, EPI_ISL_2380856 | Clinical Molecular Microbiology Laboratory, UNC Hospitals | Jeremy Wang | Alexander Rubinsteyn; Colleen Rice; Corbin Jones; Jason Smedberg; Jeremy Wang; Melissa Miller; Robert Hagan; Shawn Hawken |
| EPI_ISL_2230784, EPI_ISL_2230785, EPI_ISL_2230787, EPI_ISL_2230788, EPI_ISL_2230789, EPI_ISL_2319619, EPI_ISL_2319620, EPI_ISL_2319621, EPI_ISL_2319622, EPI_ISL_2319623 | | | |
| see above | Cliniques universitaires Saint-Luc | UCLouvain/IREC/MBLG | Benoit Kabamba Mukadi; Jean Ruelle; Nicolas Pinte; Ophélie Simon |
| EPI_ISL_5260657, EPI_ISL_5323489 | Color Genomics | Chiu Laboratory, University of California, San Francisco | Alicia Sotomayor-Gonzalez; Alicia Zhou; Baolin Wang; Charles Chiu; Daniel Wong; Erika Torres; Noah Brazer; Scott Topper; Venice Servellita |
| EPI_ISL_2310521, EPI_ISL_2310553, EPI_ISL_2310601, EPI_ISL_2310633, EPI_ISL_2310643, EPI_ISL_2310644, EPI_ISL_2310692, EPI_ISL_2310745, EPI_ISL_2311034, EPI_ISL_2549913, EPI_ISL_2549925, EPI_ISL_2549960, EPI_ISL_2549993, EPI_ISL_2549998, EPI_ISL_2550187, EPI_ISL_2550260, EPI_ISL_3160136, EPI_ISL_3582757, EPI_ISL_3582921, EPI_ISL_3582979, EPI_ISL_3583599, EPI_ISL_3583715, EPI_ISL_3583782, EPI_ISL_3583797, EPI_ISL_3583804, EPI_ISL_3583882, EPI_ISL_3583885, EPI_ISL_6211421, EPI_ISL_6211424, EPI_ISL_6211425, EPI_ISL_6211427, EPI_ISL_6211428, EPI_ISL_6211429, EPI_ISL_6211430, EPI_ISL_6211431, EPI_ISL_6211432, EPI_ISL_6211438, EPI_ISL_6211511, EPI_ISL_6211524 | | | |
| see above | Colorado Department of Public Health and Environment | Colorado Department of Public Health and Environment | Alexandria Rossheim; Diana Ir; Emily A. Travanty; Laura Bankers; Mandy Waters; Michael Martin; Molly C. Hetherington-Rauth; Sarah Elizabeth Totten; Shannon R. Matzinger |
| EPI_ISL_2285125 | Complejo Hospitalario Universitario de Pontevedra | Microbiology Department. Complexo Hospitalario Universitario de Vigo | Campello M; Perez S; Regueiro B; Trigo M |
| EPI_ISL_2632126 | Corona-Testzentrum ifp Institut für Produktqualität GmbH | Robert Koch Institute | |
| EPI_ISL_7727231, EPI_ISL_7727246, EPI_ISL_7727250, EPI_ISL_7727254, EPI_ISL_7727257 | Covid Laboratory Biogem | Covid Laboratory Biogem | Alessandra Fucci and Michele Caraglia; Alessia Maria Cossu; Cinzia Miarelli; Clara Iannarone; Egildo Luca D'andrea; Federica Melisi; Giovambattista Capasso; Marco Bocchetti; Marianna Scrima; Michele Ceccarelli; Piera Grisolia; Teresa Maria Rosaria Noviello; Ylenia Abruzzese |
| EPI_ISL_2755195, EPI_ISL_2755229, EPI_ISL_2755230, EPI_ISL_2755260, EPI_ISL_2755318, EPI_ISL_2755475, EPI_ISL_2755602, EPI_ISL_2755606, EPI_ISL_2755607, EPI_ISL_2755628, EPI_ISL_2755630, EPI_ISL_2755646, EPI_ISL_2755647, EPI_ISL_2755649, EPI_ISL_2755665, EPI_ISL_2755685, EPI_ISL_2755707, EPI_ISL_2755721, EPI_ISL_2755894, EPI_ISL_2755997, EPI_ISL_2783516, EPI_ISL_2783519, EPI_ISL_2783521, EPI_ISL_2783522, EPI_ISL_2783524, EPI_ISL_2783667, EPI_ISL_2783678, EPI_ISL_2783679, EPI_ISL_2783681, EPI_ISL_2783684, EPI_ISL_2783796, EPI_ISL_2783797, EPI_ISL_2783798, EPI_ISL_2783799, EPI_ISL_2783887, EPI_ISL_2783988, EPI_ISL_2783992, EPI_ISL_2783997, EPI_ISL_3064605, EPI_ISL_3270135, EPI_ISL_3270380, EPI_ISL_3270394, EPI_ISL_3273332 | | | |
| see above | Curative Labs | Curative Labs | Elias L. Salfati; Eugenia Khorosheva; George Way; J.Cesar Ignacio-Espinoza; Janet Chen; Mikhail Hanewich-Hollatz; Nabjot Sandhu; Sophia Quasem; Vladimir Slepnev; Zhiyi Xie |
| EPI_ISL_3663778, EPI_ISL_3663779, EPI_ISL_3663782, EPI_ISL_3663783, EPI_ISL_3663784, EPI_ISL_3663785, EPI_ISL_3663786, EPI_ISL_3663788, EPI_ISL_3663789, EPI_ISL_3663790, EPI_ISL_3663791, EPI_ISL_3663793, EPI_ISL_3663798, EPI_ISL_3663800, EPI_ISL_3663801, EPI_ISL_3663802, EPI_ISL_3663803, EPI_ISL_3663804, EPI_ISL_3663805, EPI_ISL_3663807, EPI_ISL_3663811, EPI_ISL_3663812, EPI_ISL_3663814, EPI_ISL_3663816, EPI_ISL_3663818, EPI_ISL_3663819, EPI_ISL_3663820, EPI_ISL_3663822, EPI_ISL_3663824, EPI_ISL_3663826, EPI_ISL_3663828, EPI_ISL_3663830, EPI_ISL_3663831, EPI_ISL_3663832, EPI_ISL_3663833, EPI_ISL_3663834, EPI_ISL_3663835, EPI_ISL_3663837, EPI_ISL_3663838, EPI_ISL_3663840, EPI_ISL_3663841, EPI_ISL_3663844, EPI_ISL_3663853, EPI_ISL_3663857, EPI_ISL_3663862, EPI_ISL_3663864, EPI_ISL_3663865, EPI_ISL_3663866, EPI_ISL_3663870, EPI_ISL_3663874, EPI_ISL_3663876, EPI_ISL_3663877, EPI_ISL_3663878, EPI_ISL_3663880, EPI_ISL_3663881, EPI_ISL_3663884, EPI_ISL_3663886, EPI_ISL_3663889, EPI_ISL_3663890, EPI_ISL_3663891, EPI_ISL_3663894, EPI_ISL_3663895, EPI_ISL_3663896, EPI_ISL_3663898, EPI_ISL_3663902, EPI_ISL_3663904, EPI_ISL_3663905, EPI_ISL_3663912, EPI_ISL_3663913, EPI_ISL_3663915, EPI_ISL_3663916, EPI_ISL_3663922, EPI_ISL_3663923, EPI_ISL_3663924, EPI_ISL_3663930, EPI_ISL_3663934, EPI_ISL_3663949, EPI_ISL_3663954, EPI_ISL_3663960, EPI_ISL_3663963, EPI_ISL_3663966, EPI_ISL_3663969, EPI_ISL_3663970, EPI_ISL_3663973, EPI_ISL_3663974, EPI_ISL_3663975, EPI_ISL_3663979, EPI_ISL_3663980, EPI_ISL_3663981, EPI_ISL_3663985, EPI_ISL_3663986, EPI_ISL_3663988, EPI_ISL_3663989, EPI_ISL_3663994, EPI_ISL_3663996, EPI_ISL_3664011, EPI_ISL_3664012, EPI_ISL_3664013, EPI_ISL_3664014, EPI_ISL_3664020, EPI_ISL_3664022, EPI_ISL_3664026, EPI_ISL_3664029, EPI_ISL_3664031, EPI_ISL_3664032, EPI_ISL_3664034, EPI_ISL_3664037, EPI_ISL_3664039, EPI_ISL_3664042, EPI_ISL_3664043, EPI_ISL_3664047, EPI_ISL_3664050, EPI_ISL_3664054, EPI_ISL_3664055, EPI_ISL_3664056, EPI_ISL_3664059, EPI_ISL_3664061, EPI_ISL_3664062, EPI_ISL_3664069, EPI_ISL_3664070, EPI_ISL_3664074, EPI_ISL_3664075, EPI_ISL_3664078, EPI_ISL_3664079, EPI_ISL_3664081, EPI_ISL_3664086, EPI_ISL_3664088, EPI_ISL_3664090, EPI_ISL_3664092, EPI_ISL_3664093, EPI_ISL_3664094, EPI_ISL_3664096, EPI_ISL_3664099, EPI_ISL_3664100, EPI_ISL_3664101, EPI_ISL_3664103, EPI_ISL_3664104, EPI_ISL_3664106, EPI_ISL_3664107, EPI_ISL_3664108, EPI_ISL_3664109, EPI_ISL_3664110, EPI_ISL_3664111, EPI_ISL_3664112, EPI_ISL_3664113, EPI_ISL_3664114, EPI_ISL_3664115, EPI_ISL_3664116, EPI_ISL_3664117, EPI_ISL_3664126, EPI_ISL_3664127, EPI_ISL_3664131, EPI_ISL_3664132, EPI_ISL_3664133, EPI_ISL_3664134, EPI_ISL_3664135, EPI_ISL_3664136, EPI_ISL_3664139, EPI_ISL_3664140, EPI_ISL_3664141, EPI_ISL_3664143, EPI_ISL_3664145, EPI_ISL_3664146, EPI_ISL_3664147, EPI_ISL_3664148, EPI_ISL_3664149, EPI_ISL_3664150, EPI_ISL_3664151, EPI_ISL_3664152, EPI_ISL_3664153, EPI_ISL_3664154, EPI_ISL_3664155, EPI_ISL_3664156, EPI_ISL_3664157, EPI_ISL_3664158, EPI_ISL_3664159, EPI_ISL_3664161, EPI_ISL_3664167, EPI_ISL_3664170, EPI_ISL_3664172, EPI_ISL_3664173, EPI_ISL_3664174, EPI_ISL_3664178, EPI_ISL_3664180, EPI_ISL_3664181, EPI_ISL_3664183, EPI_ISL_3664184, EPI_ISL_3664185, EPI_ISL_3664187, EPI_ISL_3664189, EPI_ISL_3664191, EPI_ISL_3664193, EPI_ISL_3664196, EPI_ISL_3664199, EPI_ISL_3664200, EPI_ISL_3664201, EPI_ISL_3664202, EPI_ISL_3664203, EPI_ISL_3664204, EPI_ISL_3664205, EPI_ISL_3664207, EPI_ISL_3664209, EPI_ISL_3664210, EPI_ISL_3664211, EPI_ISL_3664214, EPI_ISL_3664215, EPI_ISL_3664217, EPI_ISL_3664221, EPI_ISL_3664224, EPI_ISL_3664225, EPI_ISL_3664227, EPI_ISL_3664228, EPI_ISL_3664229, EPI_ISL_3664231, EPI_ISL_3664232, EPI_ISL_3664234, EPI_ISL_3664238, EPI_ISL_3664240, EPI_ISL_3664241, EPI_ISL_3664244, EPI_ISL_3664245, EPI_ISL_3664246, EPI_ISL_3664250, EPI_ISL_3664256, EPI_ISL_3664257, EPI_ISL_3664258, EPI_ISL_3664259, EPI_ISL_3664262, EPI_ISL_3664266, EPI_ISL_3664268, EPI_ISL_3664271, EPI_ISL_4426665, EPI_ISL_4426692, EPI_ISL_4426706, EPI_ISL_4426734, EPI_ISL_4426737, EPI_ISL_4426777, EPI_ISL_4426785, EPI_ISL_4426787, EPI_ISL_4426810, EPI_ISL_4426816, EPI_ISL_4426828, EPI_ISL_4426831, EPI_ISL_4426840, EPI_ISL_4427883, EPI_ISL_4427885, EPI_ISL_4427887, EPI_ISL_4427889, EPI_ISL_4427891, EPI_ISL_4427893, EPI_ISL_4427895, EPI_ISL_4427896, EPI_ISL_4427900, EPI_ISL_4427903, EPI_ISL_4427906, EPI_ISL_4427909, EPI_ISL_4427912, EPI_ISL_4427915, EPI_ISL_4427918, EPI_ISL_4427921, EPI_ISL_4427925, EPI_ISL_4427928, EPI_ISL_4427931, EPI_ISL_4427934, EPI_ISL_4427937, EPI_ISL_4427940, EPI_ISL_4427944, EPI_ISL_4427947, EPI_ISL_4427950, EPI_ISL_4427953, EPI_ISL_4427956, EPI_ISL_4427959, EPI_ISL_4427962, EPI_ISL_4427965, EPI_ISL_4427968, EPI_ISL_4427971, EPI_ISL_4427974, EPI_ISL_4427978, EPI_ISL_4427981, EPI_ISL_4427984, EPI_ISL_4427986, EPI_ISL_4427989, EPI_ISL_4427992, EPI_ISL_4427995, EPI_ISL_4427999, EPI_ISL_4428002, EPI_ISL_4428005, EPI_ISL_4428207, EPI_ISL_4428232, EPI_ISL_4428245, EPI_ISL_4428317, EPI_ISL_4428320, EPI_ISL_4428323, EPI_ISL_6851416, EPI_ISL_6851420, EPI_ISL_7695039, EPI_ISL_7743918, EPI_ISL_7743919 | | | |
| see above | DASA | DASA | Adriano Bonaldi; Angelica Hristov; Annelise Lopes; Bianca Cota; Camila Romano; Cristina Oliveira; Jose Levi; Lidia Yamamoto; Luciane Sussuchi; Paulo Pierry; Rodrigo Guarischi; Rodrigo Salazar |
| EPI_ISL_3010122 | DC Public Health Lab/ Dept. of Forensic Sciences | DC Public Health Lab/ Dept. of Forensic Sciences | Brittany Hamilton; Connie Maza; David Payne; Elizabeth Zelaya; Janis Doss; Jocelyn Hauser; Monica Mann; Sarah Scott; Scott Nguyen |
| EPI_ISL_2452011 | DE Public Health Laboratory | Centers for Disease Control and Prevention Division of Viral Diseases, Pathogen Discovery | Alison Laufer Halpin; Ben L. Rambo-Martin; Clinton R. Paden; Dakota Howard; Darlene Wagner; Dave Wentworth; Dhwani Batra; Jasmine Padilla; Justin Lee; Katie Dillon; Krista Queen; Kristen Knipe; Kristine Lacek; Mark Burroughs; Matthew Schmerer; Mili Sheth; Peter Cook; Sam Shepard; Sarah Nobles; Shoshona Le; Suxiang Tong; Vivien Dugan; Yvette Unoarumhi |
| EPI_ISL_2270843, EPI_ISL_2270892, EPI_ISL_2270893, EPI_ISL_2270904 | DIP. PREV. AVEZZANO SERVIZIO DI IGIENE EPIDEMIOLOGIA E SANITA' PUBBLICA | Istituto Zooprofilattico Sperimentale dell'Abruzzo e Molise "G. Caporale" | Ancora M; Calistri P; Cammà C; Caporale M; Curini V; Delli Compagni E; Di Domenico M; Di Lollo Valeria; Di Pasquale A; Lorusso A; Mangone I; Marcacci M; Puglia I; Rinaldi A; Savini G; Scialabba S |
| EPI_ISL_2493374 | DIRETORIA MUNICIPAL DE SAUDE DE ENGENHEIRO COELHO | Instituto Butantan | Antonio Jorge Martins; Claudia Renata dos Santos Barros; David Schlesinger; Debora Botequio Moretti; Dimas Tadeu Covas; Elaine Cristina Marqueze; Elaine Vieira Santos; Evandra Strazza Rodrigues; Heidge Fukumasu; Jayme Augusto de Souza-Neto; José Salvatore Leister Patané; Luiz Alcantara; Luiz Lehmann Coutinho; Maria Carolina Elias; Maurício Lacerda Nogueira; Rafael dos Santos Bezerra; Raul Machado Neto; Rejane Maria Tommasini Grotto; Ricardo Haddad; Sandra Coccuzzo Sampaio Vessoni; Simone Kashima; Svetoslav Nanev Slavov; Vincent Louis Viala |
| EPI_ISL_2179185, EPI_ISL_2322907, EPI_ISL_2400430 | DOHMH Crown Heights | New York City Public Health Laboratory | Jade Wang; et al. |
| EPI_ISL_2179184 | DOHMH Morrisania | New York City Public Health Laboratory | Jade Wang; et al. |
| EPI_ISL_2322910 | DOHMH PHL | New York City Public Health Laboratory | Jade Wang; et al. |
| EPI_ISL_2383713 | DPHL | Delaware Public Health Lab | Rebecca Savage |
| EPI_ISL_2245249, EPI_ISL_2245250, EPI_ISL_2289044 | Department of Clinical Microbiology | GIGA Medical Genomics | Bouchra Boujemla; Cécile Meex; Keith Durkin; Maria Artesi; Marie-Pierre Hayette; Nathalie Renotte; Pierrette Melin; Raphaël Boreux; Sébastien Bontems; Vincent Bours |
| EPI_ISL_2246272, EPI_ISL_2246275, EPI_ISL_2420638, EPI_ISL_2420641 | Department of Virology, Istituto Zooprofilattico Sperimentale del Lazio e della Toscana (IZSLT) | Department of General Diagnostics; Department of Virology; Istituto Zooprofilattico Sperimentale del Lazio e della Toscana (IZSLT) | Alessia Franco; Antonella Cersini; Antonio Battisti.; Elena L. Diaconu; Fabiola Feltrin; Giuseppe Manna; Patricia Alba; Raffaela Conti; Teresa Scicluna; Virginia Carfora |

| | | | |
|---|---|---|---|
| EPI_ISL_2315005, EPI_ISL_2636434 | Diagnosticum - Labor Neukirchen | Robert Koch Institute | |
| EPI_ISL_3578231, EPI_ISL_3578252, EPI_ISL_3578253 | Dr. Risch Ostschweiz AG | Microbiology, Dr. Risch | Dominique Fabien Hilti; Faina Wehrli; Lorenz Risch; Martin Risch; Nadia Wohlwend; Sinem Kas; Thomas Bodmer |
| EPI_ISL_2483497 | Dutch COVID-19 response team | Erasmus Medical Center | Anne van der Linden; Annemiek van der Eijk; Bas Oude Munnink; Corine GeurtsvanKessel; David Nieuwenhuijse; Emmanuelle Munger; Irina Chestakova; Marion Koopmans; Marjan Boter; Reina Sikkema; Richard Molenkamp; on behalf of the Dutch national COVID-19 respo |
| EPI_ISL_2302188, EPI_ISL_2302189, EPI_ISL_2303359, EPI_ISL_2303361, EPI_ISL_2303362, EPI_ISL_2303363, EPI_ISL_2303364, EPI_ISL_2303365, EPI_ISL_2303368, EPI_ISL_2303369, EPI_ISL_2303370, EPI_ISL_2303371, EPI_ISL_2303372, EPI_ISL_2303373, EPI_ISL_2303374, EPI_ISL_2303376, EPI_ISL_2303379, EPI_ISL_2303380, EPI_ISL_2303381, EPI_ISL_2303382, EPI_ISL_2303383, EPI_ISL_2303384, EPI_ISL_2303385, EPI_ISL_2303386, EPI_ISL_2303387, EPI_ISL_2303390, EPI_ISL_2303391, EPI_ISL_2405430, EPI_ISL_2405451, EPI_ISL_2405534, EPI_ISL_2405753, EPI_ISL_2405786, EPI_ISL_2405788, EPI_ISL_2406150, EPI_ISL_2406440, EPI_ISL_2673276 | | | |
| see above | Dutch COVID-19 response team | National Institute for Public Health and the Environment (RIVM) | Adam Meijer; AnneMarie van den Brandt; Annelies Kroneman; Bas van der Veer; Chantal Reusken; Dennis Schmitz; Dirk Eggink; Euníce Then; Florian Zwagemaker; Harry Vennema; James Groot; Jeroen Cremer; Karim Hajji; Kim Freriks; Linda van de Nes; Lisa Wijsman; Lynn Aarts; Melissa van Tuil; Ryanne Jaarsma; Sanne Bos; Sharon van den Brink; Sjoerd Kuiling; on behalf of the national COVID-19 response team |
| EPI_ISL_2445535, EPI_ISL_2473790 | ESF I E II | Instituto Butantan | Antonio Jorge Martins; Claudia Renata dos Santos Barros; David Schlesinger; Debora Botequio Moretti; Dimas Tadeu Covas; Elaine Cristina Marqueze; Elaine Vieira Santos; Evandra Strazza Rodrigues; Heidge Fukumasu; Jayme Augusto de Souza-Neto; José Salvatore Leister Patané; Luiz Alcantara; Luiz Lehmann Coutinho; Maria Carolina Elias; Maurício Lacerda Nogueira; Rafael dos Santos Bezerra; Raul Machado Neto; Rejane Maria Tommasini Grotto; Ricardo Haddad; Sandra Coccuzzo Sampaio Vessoni; Simone Kashima; Svetoslav Nanev Slavov; Vincent Louis Viala |
| EPI_ISL_2445515, EPI_ISL_2473691 | ESF NOVA TANABI II | Instituto Butantan | Antonio Jorge Martins; Claudia Renata dos Santos Barros; David Schlesinger; Debora Botequio Moretti; Dimas Tadeu Covas; Elaine Cristina Marqueze; Elaine Vieira Santos; Evandra Strazza Rodrigues; Heidge Fukumasu; Jayme Augusto de Souza-Neto; José Salvatore Leister Patané; Luiz Alcantara; Luiz Lehmann Coutinho; Maria Carolina Elias; Maurício Lacerda Nogueira; Rafael dos Santos Bezerra; Raul Machado Neto; Rejane Maria Tommasini Grotto; Ricardo Haddad; Sandra Coccuzzo Sampaio Vessoni; Simone Kashima; Svetoslav Nanev Slavov; Vincent Louis Viala |
| EPI_ISL_3215691, EPI_ISL_3215700 | EXCITE Lab | Andersen lab at Scripps Research | Abigail Schnapper; Angela Scioscia; Cheryl Anderson; Chip Schooley; David Pride; Greg Humphrey; Helena Tubb; Natasha Martin; Natasha Martin Cheryl Anderson; Sawyer Farmer; Sharon Reed; Smruthi Karthikeyan; Tommy Valles + SEARCH; Tommy Valles + SEARCH |
| EPI_ISL_2940118, EPI_ISL_2940119, EPI_ISL_2940129, EPI_ISL_2940189, EPI_ISL_2940409, EPI_ISL_2940410, EPI_ISL_2940436, EPI_ISL_2940438, EPI_ISL_2940439, EPI_ISL_2940440, EPI_ISL_2940441, EPI_ISL_2940442, EPI_ISL_2940463, EPI_ISL_2940496, EPI_ISL_2940498, EPI_ISL_2940499, EPI_ISL_2940500, EPI_ISL_2940501, EPI_ISL_2940502, EPI_ISL_2940503, EPI_ISL_2940504, EPI_ISL_2940514, EPI_ISL_2940630, EPI_ISL_2940638, EPI_ISL_2940639, EPI_ISL_2940640, EPI_ISL_2940642, EPI_ISL_2940802, EPI_ISL_2940804, EPI_ISL_2941043, EPI_ISL_2941092, EPI_ISL_2941122, EPI_ISL_3116885, EPI_ISL_3116886, EPI_ISL_3117414 | | | |
| see above | Edmonton Provincial Lab | Public Health Agency of Canada (PHAC) National Microbiology Laboratory | Buss; Croxen M; Deo A; Dieu P; E; Ferrato C; Gill K; Khan F; Koleva P; Li V; Lloyd C; Lynch T; Ma R; Murphy S; Pabbaraju K; Shokoples S; Thayer J; Tipples G; Whitehouse M; Wong A; Yu C; Zelyas N |
| EPI_ISL_3758745 | Emory Molecular Diagnostics Laboratory, Emory Healthcare | Piantadosi Lab, Emory Department of Pathology | Ahmed Babiker; Anne Piantadosi; Dara Khosravi |
| EPI_ISL_5647439 | FACULDADE DE MEDICINA ABC | Instituto Butantan | Antonio Jorge Martins; Claudia Renata dos Santos Barros; David Schlesinger; Debora Botequio Moretti; Dimas Tadeu Covas; Elaine Cristina Marqueze; Elaine Vieira Santos; Evandra Strazza Rodrigues; Heidge Fukumasu; Jayme Augusto de Souza-Neto; José Salvatore Leister Patané; Luiz Alcantara; Luiz Lehmann Coutinho; Maria Carolina Elias; Maurício Lacerda Nogueira; Rafael dos Santos Bezerra; Raul Machado Neto; Rejane Maria Tommasini Grotto; Ricardo Haddad; Sandra Coccuzzo Sampaio Vessoni; Simone Kashima; Svetoslav Nanev Slavov; Vincent Louis Viala |
| EPI_ISL_2473694 | FUNDAÇÃO ABC | Instituto Butantan | Antonio Jorge Martins; Claudia Renata dos Santos Barros; David Schlesinger; Debora Botequio Moretti; Dimas Tadeu Covas; Elaine Cristina Marqueze; Elaine Vieira Santos; Evandra Strazza Rodrigues; Heidge Fukumasu; Jayme Augusto de Souza-Neto; José Salvatore Leister Patané; Luiz Alcantara; Luiz Lehmann Coutinho; Maria Carolina Elias; Maurício Lacerda Nogueira; Rafael dos Santos Bezerra; Raul Machado Neto; Rejane Maria Tommasini Grotto; Ricardo Haddad; Sandra Coccuzzo Sampaio Vessoni; Simone Kashima; Svetoslav Nanev Slavov; Vincent Louis Viala |
| EPI_ISL_2442930, EPI_ISL_2442946, EPI_ISL_2442947, EPI_ISL_2442959, EPI_ISL_2442960, EPI_ISL_2442965, EPI_ISL_2442967, EPI_ISL_2442995, EPI_ISL_2567147, EPI_ISL_2567151, EPI_ISL_2567156, EPI_ISL_2567162, EPI_ISL_2567164, EPI_ISL_2567165, EPI_ISL_2567170, EPI_ISL_2567178, EPI_ISL_2567181, EPI_ISL_2567185, EPI_ISL_2567186, EPI_ISL_2567187, EPI_ISL_2836066, EPI_ISL_2836105, EPI_ISL_2836141, EPI_ISL_2836158, EPI_ISL_2836454, EPI_ISL_2836469, EPI_ISL_2836470, EPI_ISL_2836816, EPI_ISL_6050030, EPI_ISL_6050031, EPI_ISL_6050032, EPI_ISL_6050050, EPI_ISL_6050051, EPI_ISL_6050052, EPI_ISL_6050053, EPI_ISL_6050119, EPI_ISL_6050127, EPI_ISL_6050164, EPI_ISL_6050258 | | | |
| see above | Florida Bureau of Public Health Laboratories | Florida Bureau of Public Health Laboratories | Jason Blanton; Namratha Tarigopula; Sarah Schmedes; Tiffany Splatt |
| EPI_ISL_2620701, EPI_ISL_2620703 | Fondazione IRCCS Ca' Granda Ospedale Maggiore Policlinico | Fondazione IRCCS Ca' Granda Ospedale Maggiore Policlinico | Ferruccio Ceriotti; Sara Uceda Renteria |
| EPI_ISL_2133941, EPI_ISL_2133948, EPI_ISL_2133988, EPI_ISL_2160821, EPI_ISL_2160825, EPI_ISL_2160827, EPI_ISL_2160829, EPI_ISL_2160834, EPI_ISL_2160836, EPI_ISL_2160837, EPI_ISL_2160845, EPI_ISL_2160846, EPI_ISL_2160847, EPI_ISL_2160935, EPI_ISL_2160984, EPI_ISL_2160986, EPI_ISL_2160988, EPI_ISL_2161001, EPI_ISL_2161010, EPI_ISL_2161019, EPI_ISL_2161020, EPI_ISL_2161024, EPI_ISL_2269221, EPI_ISL_2269251, EPI_ISL_2269256, EPI_ISL_2269257, EPI_ISL_2269264, EPI_ISL_2269277, EPI_ISL_2269401, EPI_ISL_2269436, EPI_ISL_2269455, EPI_ISL_2269486, EPI_ISL_2269489, EPI_ISL_2269502, EPI_ISL_2269503, EPI_ISL_2269509, EPI_ISL_2269515, EPI_ISL_2269524, EPI_ISL_2269525, EPI_ISL_2269530, EPI_ISL_2269554, EPI_ISL_2269599, EPI_ISL_2280263, EPI_ISL_2368085, EPI_ISL_2368115, EPI_ISL_2441831, EPI_ISL_2441834, EPI_ISL_2441846, EPI_ISL_2525439, EPI_ISL_2525441, EPI_ISL_4367085, EPI_ISL_4369959, EPI_ISL_4370784, EPI_ISL_4370791, EPI_ISL_4371236, EPI_ISL_4372328 | | | |
| see above | Fulgent Genetics | Centers for Disease Control and Prevention Division of Viral Diseases, Pathogen Discovery | Adrian Paskey; Becky Tsai; Benafsh Sapra; Benjamin Rambo-Martin; Christopher Gulvick; Clinton Paden; Clinton R. Paden; Dakota Howard; Darlene Wagner; Dhwani Batra; Doreen Ng; Duncan MacCannell; Erisa Sula; Harry Gao; James Xie; Jason Caravas; John Gao; Joseph Fierro; Kara Moser; Kristine Lacek; Matthew Schmerer; Mickey Li; Peter Cook; Peter W. Cook; Scott Sammons; Shatavia Morrison; Tymeckia Kendall; Victoria Caban Figueroa; Yan Meng; Yvette Unoarumhi |
| EPI_ISL_6701394, EPI_ISL_6701395, EPI_ISL_6701396, EPI_ISL_6701397, EPI_ISL_6701424, EPI_ISL_6701425 | Fundação Ezequiel Dias | Instituto René Rachou / Fiocruz Minas | André Bernardes; Anna Salim; Caroline Penido; Felipe Iani; Flávio Araújo; Gabriel Fernandes; Pedro Alves; Rubens do Monte Neto; Thaís Silva |
| EPI_ISL_2452229 | GA Department of Public Health Laboratory | Centers for Disease Control and Prevention Division of Viral Diseases, Pathogen Discovery | Alison Laufer Halpin; Ben L. Rambo-Martin; Clinton R. Paden; Dakota Howard; Darlene Wagner; Dave Wentworth; Dhwani Batra; Jasmine Padilla; Justin Lee; Katie Dillon; Krista Queen; Kristen Knipe; Kristine Lacek; Mark Burroughs; Matthew Schmerer; Mili Sheth; Peter Cook; Sam Shepard; Sarah Nobles; Shoshona Le; Suxiang Tong; Vivien Dugan; Yvette Unoarumhi |
| EPI_ISL_2422461 | GA Department of Public Health Laboratory | Genomics and Discovery, Respiratory Viruses Branch, Division of Viral Diseases, Centers for Disease Control and Prevention | Adam Retchless; Anna Kelleher; Anna Uehara; Brian Lynch; Clinton R. Paden; Dhwani Batra; Haibin Wang; Han Jia Justin Ng; Jasmine Padilla; Jing Zhang; Justin Lee; Krista Queen; Mark Burroughs; Mili Sheth; Morgan Davis; Peter Cook; Rachel Marine; Sarah Nobles; Suxiang Tong; Tara Coalter; Yan Li; Ying Tao |
| EPI_ISL_2228873, EPI_ISL_2333802 | GH A.CHENEVIER-H.MONDOR | Department of Virology, Henri Mondor University Hospital, Assistance Publique Hôpitaux de Paris, Université Paris-Est Créteil, INSERM U955 | Alexandre Soulier; Christophe Rodriguez; Elisabeth Trawinski; Guillaume Gricourt; Jean-Michel Pawlotsky; Melissa N'Debi; Slim Fourati; Vanessa Demontant |
| EPI_ISL_2333688 | GROUPE HOSPITALIER SUD ILE DE FRANCE | Department of Virology, Henri Mondor University Hospital, Assistance Publique Hôpitaux de Paris, Université Paris-Est Créteil, INSERM U955 | Alexandre Soulier; Christophe Rodriguez; Elisabeth Trawinski; Guillaume Gricourt; Jean-Michel Pawlotsky; Melissa N'Debi; Slim Fourati; Vanessa Demontant |
| EPI_ISL_2421228 | Garnet Health Medical Center Laboratory | Wadsworth Center, New York State Department of Health | Alexis Russell; Catharine Prussing; Daryl M. Lamson; Erasmus Schneider; Erica Lasek-Nesselquist; John Kelly; Jonathan Plitnick; Kirsten St. George; Matthew Shudt; Melissa A Leisner; Navjot Singh |
| EPI_ISL_3491799, EPI_ISL_3491800, EPI_ISL_3491803, EPI_ISL_3491805, EPI_ISL_3491810, EPI_ISL_3491820, EPI_ISL_3491822, EPI_ISL_3491824, EPI_ISL_3491825 | | | |
| see above | Gencore - Universidad de los Andes | Gencore - Universidad de los Andes | Catherine Jaller; Cristian Barrera; David González; Luisa Sacristan; Marcela Guevara; Silvia Restrepo |
| EPI_ISL_2391303, EPI_ISL_2391306, EPI_ISL_2391310, EPI_ISL_2391320, EPI_ISL_2391321, EPI_ISL_2391322, EPI_ISL_2391323, EPI_ISL_2391336, EPI_ISL_2391345, EPI_ISL_2391348, EPI_ISL_2391427, EPI_ISL_2391429, EPI_ISL_2391430, EPI_ISL_2508548, EPI_ISL_2508551, EPI_ISL_2508553, EPI_ISL_2508554, EPI_ISL_2508556, EPI_ISL_2508562, EPI_ISL_2508567, EPI_ISL_2508572, EPI_ISL_2508589, EPI_ISL_2508596, EPI_ISL_2508597, EPI_ISL_2508599, EPI_ISL_2508603, EPI_ISL_2508604, EPI_ISL_2508605, EPI_ISL_2508606, EPI_ISL_2508608, EPI_ISL_2508609, EPI_ISL_2508610, EPI_ISL_2508611, EPI_ISL_2508612, EPI_ISL_2508613, EPI_ISL_2508614, EPI_ISL_2508616, EPI_ISL_2509125, EPI_ISL_2509126, EPI_ISL_2509127, EPI_ISL_2509128, EPI_ISL_2509132, EPI_ISL_2509134, EPI_ISL_2509135, EPI_ISL_2509136, EPI_ISL_2509138, EPI_ISL_2509139, EPI_ISL_2509145, EPI_ISL_2509146, EPI_ISL_2509152, EPI_ISL_2509155, EPI_ISL_2509156, EPI_ISL_2509157, EPI_ISL_2509158, EPI_ISL_2509159, EPI_ISL_2509161, EPI_ISL_2509170, EPI_ISL_2509284, EPI_ISL_2543598, EPI_ISL_2566531, EPI_ISL_2566532, EPI_ISL_2566533, EPI_ISL_2566534, EPI_ISL_2566535, EPI_ISL_2566536, EPI_ISL_2566537, EPI_ISL_2892601 | | | |
| see above | Genetica Molecular and Subdepartamento de Virologia ISP Chile | Instituto de Salud Publica de Chile | Andres Castillo; Barbara Parra; Constanza Campano; Gisselle Barra; Javier Tognarelli; Jorge Fernandez; Karen Orostica; Loredana Arata; Patricia Bustos; Rodrigo Fasce; Soledad Ulloa |
| EPI_ISL_2248888 | Germano de Sousa | Instituto Nacional de Saude (INSA) | Borges et al |
| EPI_ISL_2340344 | Germano de Sousa | Instituto Nacional de Saude (INSA) and BioSystems & Integrative Sciences Institute (BioISI) Genomics Unit, FCUL | Borges et al |
| EPI_ISL_2360224, EPI_ISL_2360225, EPI_ISL_2360226, EPI_ISL_2360227 | Gesundheitsdirektion Zürich | Institute of Medical Virology | Alexandra Trkola; Annette Audigé; Cyril Shah; Gabriela Ziltener; Guido Bloemberg; Jon Huder; Jürg Böni; Kevin Steiner; Maria Grünberg; Maryam Zaheri; Michael Huber; Riccarda Capaul; Stefan Schmutz; Verena Kufner |
| EPI_ISL_2535843, EPI_ISL_2535845, EPI_ISL_2535855, EPI_ISL_2535863, EPI_ISL_2535873, EPI_ISL_2535878, EPI_ISL_2535884, EPI_ISL_2535885, EPI_ISL_2535887, EPI_ISL_2535894, EPI_ISL_2535895, EPI_ISL_2613881, EPI_ISL_2613885, EPI_ISL_2613889, EPI_ISL_2613892, EPI_ISL_2613952, EPI_ISL_2613974, EPI_ISL_2614039, EPI_ISL_2614059 | | | |
| see above | Gravity Diagnostics, LLC | Gravity Diagnostics, LLC | Gravity Diagnostics |
| EPI_ISL_7476656 | Grupo de Investigación en Enfermedades Tropicales del Ejército (GINETEJ), Laboratorio de Referencia e Investigación, Dirección de Sanidad Ejército, Bogotá, Colombia | Centro de Investigaciones en Microbiología y Biotecnología-UR (CIMBIUR), Facultad de Ciencias Naturales, Universidad del Rosario, Bogotá, Colombia | Alberto E. Paniz-Mondolfi; Angie L. Ramírez; Camilo Correa-Cardenas; Claudia Mendez; Enzo Guerrero-Araya; Juan David Ramírez; Julie Perez; Luz H. Patiño; Maria Clara Duque; Marina Muñoz; Nathalia Ballesteros; Nicolás Luna; Sergio Castañeda |
| EPI_ISL_2376238 | Gundersen Clinical Microbiology Laboratory | Kabara Cancer Research Institute | Craig S. Richmond; Paraic A. Kenny |
| EPI_ISL_6229832 | HGMAP HOSPITAL DE CAMPANHA COVID 19 | ACME Lab, Oswaldo Cruz Foundation, FIOCRUZ/CE | Carlos Leonardo de Aragao Araujo; Cecilia Leite Costa & Eduardo Ruback dos Santos on behalf of COVID-19 FIOCRUZ Genomic Network; Cleber Furtado Aksenen; Fabio Miyajima; Fernando Braga Stehling; Francisco Eder de Moura Lopes; Igor Oliveira Duarte; Jamille Maria Mendes Bezerra; Joaquim Cesar do Nascimento Sousa Junior; Pedro Miguel Carneiro Jeronimo; Suzana Porto Almeida; Thais Ferreira de Oliveira; Thais de Oliveira Costa; Ticiane Cavalcante de Souza; Veridiana Pessoa Miyajima |
| EPI_ISL_2452110, EPI_ISL_2452112, EPI_ISL_2452113, | HI Dept. of Health, State Laboratories Division | Centers for Disease Control and Prevention Division of Viral Diseases, Pathogen Discovery | Alison Laufer Halpin; Ben L. Rambo-Martin; Clinton R. Paden; Dakota Howard; Darlene Wagner; Dave Wentworth; Dhwani Batra; Jasmine Padilla; Justin Lee; Katie Dillon; Krista Queen; Kristen Knipe; Kristine Lacek; Mark Burroughs; Matthew Schmerer; Mili Sheth; Peter Cook; Sam Shepard; Sarah Nobles; Shoshona Le; Suxiang Tong; Vivien Dugan; Yvette Unoarumhi |

| | | | |
|---|---|---|---|
| EPI_ISL_2452117, EPI_ISL_2452130 | | | |
| EPI_ISL_2222902, EPI_ISL_2222905, EPI_ISL_2222908, EPI_ISL_2222911, EPI_ISL_2222913, EPI_ISL_2222916, EPI_ISL_2222917, EPI_ISL_2227565, EPI_ISL_2348608, EPI_ISL_2348609, EPI_ISL_2348610, EPI_ISL_2348611, EPI_ISL_2348612, EPI_ISL_2348613, EPI_ISL_2348614, EPI_ISL_2348615, EPI_ISL_2466446, EPI_ISL_2466464, EPI_ISL_2466465, EPI_ISL_2497440, EPI_ISL_2617584, EPI_ISL_2617585, EPI_ISL_3254465 | | | |
| see above | HLAGYN - Laboratorio de Imunologia de Transplantes de Goias | HLAGYN - Laboratorio de Imunologia de Transplantes de Goias | Alessandro Leonardo Alvares Magalhaes; Daniel Ferreira de Sousa; Erika Lopes Rocha Batista; Fernando Antonio Vinhal dos Santos; Frederico Rodrigues Vinhal; Kamila Oliveira Reis De Freitas.; Lucas Carlos Gomes Pereira; Sabrina Sara Moreira Duarte |
| EPI_ISL_6229719, EPI_ISL_6229720, EPI_ISL_6229721, EPI_ISL_6229722, EPI_ISL_6229868 | HOSP MATERN F FARIAS LEITAO | ACME Lab, Oswaldo Cruz Foundation, FIOCRUZ/CE | Carlos Leonardo de Aragao Araujo; Cecilia Leite Costa & Eduardo Ruback dos Santos on behalf of COVID-19 FIOCRUZ Genomic Network; Cleber Furtado Aksenen; Fabio Miyajima; Fernando Braga Stehling; Francisco Eder de Moura Lopes; Igor Oliveira Duarte; Jamille Maria Mendes Bezerra; Joaquim Cesar do Nascimento Sousa Junior; Pedro Miguel Carneiro Jeronimo; Suzana Porto Almeida; Thais Ferreira de Oliveira; Thais de Oliveira Costa; Ticiane Cavalcante de Souza; Veridiana Pessoa Miyajima |
| EPI_ISL_2445305 | HOSP PRESIDENTE | Instituto Butantan | Antonio Jorge Martins; Claudia Renata dos Santos Barros; David Schlesinger; Debora Botequio Moretti; Dimas Tadeu Covas; Elaine Cristina Marqueze; Elaine Vieira Santos; Evandra Strazza Rodrigues; Heidge Fukumasu; Jayme Augusto de Souza-Neto; José Salvatore Leister Patané; Luiz Alcantara; Luiz Lehmann Coutinho; Maria Carolina Elias; Maurício Lacerda Nogueira; Rafael dos Santos Bezerra; Raul Machado Neto; Rejane Maria Tommasini Grotto; Ricardo Haddad; Sandra Coccuzzo Sampaio Vessoni; Simone Kashima; Svetoslav Nanev Slavov; Vincent Louis Viala |
| EPI_ISL_3102518 | HOSPISTAL MATERNIDADE DR LUIS DE GONZAGA DA FONSECA MOTA | Analytical Competence Molecular Epidemiology Lab/ACME, Oswaldo Cruz Foundation, Ceara (FIOCRUZ CE) | Cleber Furtado Aksenen; Fabio Miyajima; Fernando Braga Stehling; Francisco Eder de Moura Lopes; Jamille Maria Mendes Bezerra; Joaquim César do Nascimento Sousa Junior; Pedro Miguel Carneiro Jeronimo; Suzana Porto Almeida e Lucas Delerino; Thais Ferreira de Oliveira; Thais de Oliveira Costa; Ticiane Cavalcante de Souza; Veridiana Pessoa Miyajima |
| EPI_ISL_2445572, EPI_ISL_2445577, EPI_ISL_2445579, EPI_ISL_2445583, EPI_ISL_2445591, EPI_ISL_2473800 | HOSPITAL DE CAMPANHA COVID 19 MUNICIPIO DE TAUBATE | Instituto Butantan | Antonio Jorge Martins; Claudia Renata dos Santos Barros; David Schlesinger; Debora Botequio Moretti; Dimas Tadeu Covas; Elaine Cristina Marqueze; Elaine Vieira Santos; Evandra Strazza Rodrigues; Heidge Fukumasu; Jayme Augusto de Souza-Neto; José Salvatore Leister Patané; Luiz Alcantara; Luiz Lehmann Coutinho; Maria Carolina Elias; Maurício Lacerda Nogueira; Rafael dos Santos Bezerra; Raul Machado Neto; Rejane Maria Tommasini Grotto; Ricardo Haddad; Sandra Coccuzzo Sampaio Vessoni; Simone Kashima; Svetoslav Nanev Slavov; Vincent Louis Viala |
| EPI_ISL_2445274 | HOSPITAL DR OSIRIS FLORINDO COELHO FERRAZ DE VASCONCELOS | Instituto Butantan | Antonio Jorge Martins; Claudia Renata dos Santos Barros; David Schlesinger; Debora Botequio Moretti; Dimas Tadeu Covas; Elaine Cristina Marqueze; Elaine Vieira Santos; Evandra Strazza Rodrigues; Heidge Fukumasu; Jayme Augusto de Souza-Neto; José Salvatore Leister Patané; Luiz Alcantara; Luiz Lehmann Coutinho; Maria Carolina Elias; Maurício Lacerda Nogueira; Rafael dos Santos Bezerra; Raul Machado Neto; Rejane Maria Tommasini Grotto; Ricardo Haddad; Sandra Coccuzzo Sampaio Vessoni; Simone Kashima; Svetoslav Nanev Slavov; Vincent Louis Viala |
| EPI_ISL_3102276, EPI_ISL_3102289 | HOSPITAL ESTADUAL LEONARDO DA VINCI | Analytical Competence Molecular Epidemiology Lab/ACME, Oswaldo Cruz Foundation, Ceara (FIOCRUZ CE) | Cleber Furtado Aksenen; Fabio Miyajima; Fernando Braga Stehling; Francisco Eder de Moura Lopes; Jamille Maria Mendes Bezerra; Joaquim César do Nascimento Sousa Junior; Pedro Miguel Carneiro Jeronimo; Suzana Porto Almeida e Lucas Delerino; Thais Ferreira de Oliveira; Thais de Oliveira Costa; Ticiane Cavalcante de Souza; Veridiana Pessoa Miyajima |
| EPI_ISL_3260824 | HOSPITAL GENERAL UNIVERSITARIO DE GUADALAJARA | Instituto de Salud Carlos III | A. Monzón; ALEJANDRO; F. Casas; I. Jiménez; I.GONZALEZ PRAETORIUS; M. Sandonís; P. Zaballos; S. Cuesta; S. Iglesias-Caballero; S. Pozo; S. Varona; V. Camarero; Vázquez-Morón |
| EPI_ISL_4414859, EPI_ISL_4414860, EPI_ISL_4414861, EPI_ISL_4414862 | HOSPITAL GENERAL UNIVERSITARIO REINA SOFIA | Instituto de Salud Carlos III | A. Monzón; F. Casas; I. Jiménez; I.NUÑEZ TRIGUEROS; Iglesias-Caballero; M. Sandonís; Mª LUZ; P. Zaballos; S. Camarero; S. Cuesta; S. Pozo; S. Varona; V. Vázquez-Morón |
| EPI_ISL_3102287, EPI_ISL_3102320 | HOSPITAL MATERNIDADE SANTA RITA | Analytical Competence Molecular Epidemiology Lab/ACME, Oswaldo Cruz Foundation, Ceara (FIOCRUZ CE) | Cleber Furtado Aksenen; Fabio Miyajima; Fernando Braga Stehling; Francisco Eder de Moura Lopes; Jamille Maria Mendes Bezerra; Joaquim César do Nascimento Sousa Junior; Pedro Miguel Carneiro Jeronimo; Suzana Porto Almeida e Lucas Delerino; Thais Ferreira de Oliveira; Thais de Oliveira Costa; Ticiane Cavalcante de Souza; Veridiana Pessoa Miyajima |
| EPI_ISL_2445605, EPI_ISL_2473698 | HOSPITAL MUNICIPAL DE IBIUNA IBIUNA SP | Instituto Butantan | Antonio Jorge Martins; Claudia Renata dos Santos Barros; David Schlesinger; Debora Botequio Moretti; Dimas Tadeu Covas; Elaine Cristina Marqueze; Elaine Vieira Santos; Evandra Strazza Rodrigues; Heidge Fukumasu; Jayme Augusto de Souza-Neto; José Salvatore Leister Patané; Luiz Alcantara; Luiz Lehmann Coutinho; Maria Carolina Elias; Maurício Lacerda Nogueira; Rafael dos Santos Bezerra; Raul Machado Neto; Rejane Maria Tommasini Grotto; Ricardo Haddad; Sandra Coccuzzo Sampaio Vessoni; Simone Kashima; Svetoslav Nanev Slavov; Vincent Louis Viala |
| EPI_ISL_2445500, EPI_ISL_2445506, EPI_ISL_2445507, EPI_ISL_2445509, EPI_ISL_2445511 | HOSPITAL MUNICIPAL DR TABAJARA RAMOS | Instituto Butantan | Antonio Jorge Martins; Claudia Renata dos Santos Barros; David Schlesinger; Debora Botequio Moretti; Dimas Tadeu Covas; Elaine Cristina Marqueze; Elaine Vieira Santos; Evandra Strazza Rodrigues; Heidge Fukumasu; Jayme Augusto de Souza-Neto; José Salvatore Leister Patané; Luiz Alcantara; Luiz Lehmann Coutinho; Maria Carolina Elias; Maurício Lacerda Nogueira; Rafael dos Santos Bezerra; Raul Machado Neto; Rejane Maria Tommasini Grotto; Ricardo Haddad; Sandra Coccuzzo Sampaio Vessoni; Simone Kashima; Svetoslav Nanev Slavov; Vincent Louis Viala |
| EPI_ISL_2801352, EPI_ISL_2801358, EPI_ISL_3102517 | HOSPITAL MUNICIPAL NOSSA SENHORA DOS MILAGRES | Analytical Competence Molecular Epidemiology Lab/ACME, Oswaldo Cruz Foundation, Ceara (FIOCRUZ CE) | Cleber Furtado Aksenen; Cleber Furtado Aksenen e Suzana Porto Almeida; Fabio Miyajima; Fernando Braga Stehling; Francisco Eder de Moura Lopes; Jamille Maria Mendes Bezerra; Joaquim César do Nascimento Sousa Junior; Pedro Miguel Carneiro Jeronimo; Suzana Porto Almeida e Lucas Delerino; Thais Ferreira de Oliveira; Thais de Oliveira Costa; Ticiane Cavalcante de Souza; Veridiana Pessoa Miyajima |
| EPI_ISL_2445223, EPI_ISL_2445228, EPI_ISL_2445229, EPI_ISL_2445230, EPI_ISL_2445232, EPI_ISL_2445235, EPI_ISL_2445239 | | | |
| see above | HOSPITAL MUNICIPAL REYNALDO GUERRA CAJATI | Instituto Butantan | Antonio Jorge Martins; Claudia Renata dos Santos Barros; David Schlesinger; Debora Botequio Moretti; Dimas Tadeu Covas; Elaine Cristina Marqueze; Elaine Vieira Santos; Evandra Strazza Rodrigues; Heidge Fukumasu; Jayme Augusto de Souza-Neto; José Salvatore Leister Patané; Luiz Alcantara; Luiz Lehmann Coutinho; Maria Carolina Elias; Maurício Lacerda Nogueira; Rafael dos Santos Bezerra; Raul Machado Neto; Rejane Maria Tommasini Grotto; Ricardo Haddad; Sandra Coccuzzo Sampaio Vessoni; Simone Kashima; Svetoslav Nanev Slavov; Vincent Louis Viala |
| EPI_ISL_2801367, EPI_ISL_3102319 | HOSPITAL OTOCLINICA | Analytical Competence Molecular Epidemiology Lab/ACME, Oswaldo Cruz Foundation, Ceara (FIOCRUZ CE) | Cleber Furtado Aksenen; Cleber Furtado Aksenen e Suzana Porto Almeida; Fabio Miyajima; Fernando Braga Stehling; Francisco Eder de Moura Lopes; Jamille Maria Mendes Bezerra; Joaquim César do Nascimento Sousa Junior; Pedro Miguel Carneiro Jeronimo; Suzana Porto Almeida e Lucas Delerino; Thais Ferreira de Oliveira; Thais de Oliveira Costa; Ticiane Cavalcante de Souza; Veridiana Pessoa Miyajima |
| EPI_ISL_6229734, EPI_ISL_6229735 | HOSPITAL REGIONAL DE IGUATU | ACME Lab, Oswaldo Cruz Foundation, FIOCRUZ/CE | Carlos Leonardo de Aragao Araujo; Cecilia Leite Costa & Eduardo Ruback dos Santos on behalf of COVID-19 FIOCRUZ Genomic Network; Cleber Furtado Aksenen; Fabio Miyajima; Fernando Braga Stehling; Francisco Eder de Moura Lopes; Igor Oliveira Duarte; Jamille Maria Mendes Bezerra; Joaquim Cesar do Nascimento Sousa Junior; Pedro Miguel Carneiro Jeronimo; Suzana Porto Almeida; Thais Ferreira de Oliveira; Thais de Oliveira Costa; Ticiane Cavalcante de Souza; Veridiana Pessoa Miyajima |
| EPI_ISL_2445550 | HOSPITAL REGIONAL DE ITAPETININGA | Instituto Butantan | Antonio Jorge Martins; Claudia Renata dos Santos Barros; David Schlesinger; Debora Botequio Moretti; Dimas Tadeu Covas; Elaine Cristina Marqueze; Elaine Vieira Santos; Evandra Strazza Rodrigues; Heidge Fukumasu; Jayme Augusto de Souza-Neto; José Salvatore Leister Patané; Luiz Alcantara; Luiz Lehmann Coutinho; Maria Carolina Elias; Maurício Lacerda Nogueira; Rafael dos Santos Bezerra; Raul Machado Neto; Rejane Maria Tommasini Grotto; Ricardo Haddad; Sandra Coccuzzo Sampaio Vessoni; Simone Kashima; Svetoslav Nanev Slavov; Vincent Louis Viala |
| EPI_ISL_2445540, EPI_ISL_2473690 | HOSPITAL REGIONAL DE REGISTRO REGISTRO | Instituto Butantan | Antonio Jorge Martins; Claudia Renata dos Santos Barros; David Schlesinger; Debora Botequio Moretti; Dimas Tadeu Covas; Elaine Cristina Marqueze; Elaine Vieira Santos; Evandra Strazza Rodrigues; Heidge Fukumasu; Jayme Augusto de Souza-Neto; José Salvatore Leister Patané; Luiz Alcantara; Luiz Lehmann Coutinho; Maria Carolina Elias; Maurício Lacerda Nogueira; Rafael dos Santos Bezerra; Raul Machado Neto; Rejane Maria Tommasini Grotto; Ricardo Haddad; Sandra Coccuzzo Sampaio Vessoni; Simone Kashima; Svetoslav Nanev Slavov; Vincent Louis Viala |
| EPI_ISL_2801334 | HOSPITAL REGIONAL DO CARIRI | Analytical Competence Molecular Epidemiology Lab/ACME, Oswaldo Cruz Foundation, Ceara (FIOCRUZ CE) | Cleber Furtado Aksenen e Suzana Porto Almeida; Fabio Miyajima; Fernando Braga Stehling; Francisco Eder de Moura Lopes; Jamille Maria Mendes Bezerra; Joaquim César do Nascimento Sousa Junior; Pedro Miguel Carneiro Jeronimo; Thais Ferreira de Oliveira; Thais de Oliveira Costa; Ticiane Cavalcante de Souza; Veridiana Pessoa Miyajima |
| EPI_ISL_6229747, EPI_ISL_6229748 | HOSPITAL REGIONAL DO SERTAO CENTRAL | ACME Lab, Oswaldo Cruz Foundation, FIOCRUZ/CE | Carlos Leonardo de Aragao Araujo; Cecilia Leite Costa & Eduardo Ruback dos Santos on behalf of COVID-19 FIOCRUZ Genomic Network; Cleber Furtado Aksenen; Fabio Miyajima; Fernando Braga Stehling; Francisco Eder de Moura Lopes; Igor Oliveira Duarte; Jamille Maria Mendes Bezerra; Joaquim Cesar do Nascimento Sousa Junior; Pedro Miguel Carneiro Jeronimo; Suzana Porto Almeida; Thais Ferreira de Oliveira; Thais de Oliveira Costa; Ticiane Cavalcante de Souza; Veridiana Pessoa Miyajima |
| EPI_ISL_5603302, EPI_ISL_5825569 | HOSPITAL REGIONAL DO SERTAO CENTRAL | Analytical Competence Molecular Epidemiology Lab/ACME, Oswaldo Cruz Foundation, Ceara (FIOCRUZ CE) | Carlos Leonardo de Aragao Araujo; Cecilia Leite Costa & Eduardo Ruback dos Santos on behalf of COVID-19 FIOCRUZ Genomic Network; Cleber Furtado Aksenen; Fabio Miyajima; Fernando Braga Stehling; Francisco Eder de Moura Lopes; Igor Oliveira Duarte; Jamille Maria Mendes Bezerra; Joaquim Cesar do Nascimento Sousa Junior; Pedro Miguel Carneiro Jeronimo; Suzana Porto Almeida; Thais Ferreira de Oliveira; Thais de Oliveira Costa; Ticiane Cavalcante de Souza; Veridiana Pessoa Miyajima |
| EPI_ISL_2445242 | HOSPITAL SAO FRANCISCO SC LTDA | Instituto Butantan | Antonio Jorge Martins; Claudia Renata dos Santos Barros; David Schlesinger; Debora Botequio Moretti; Dimas Tadeu Covas; Elaine Cristina Marqueze; Elaine Vieira Santos; Evandra Strazza Rodrigues; Heidge Fukumasu; Jayme Augusto de Souza-Neto; José Salvatore Leister Patané; Luiz Alcantara; Luiz Lehmann Coutinho; Maria Carolina Elias; Maurício Lacerda Nogueira; Rafael dos Santos Bezerra; Raul Machado Neto; Rejane Maria Tommasini Grotto; Ricardo Haddad; Sandra Coccuzzo Sampaio Vessoni; Simone Kashima; Svetoslav Nanev Slavov; Vincent Louis Viala |
| EPI_ISL_3502726, EPI_ISL_3502727, EPI_ISL_3502728, EPI_ISL_3502729, EPI_ISL_3532472 | HOSPITAL UNIVERSITARIO 12 DE OCTUBRE | HOSPITAL UNIVERSITARIO 12 DE OCTUBRE | Carmen Martín-Higuera; Esther Viedma; Irene Múñoz-Gallego; M.ª Dolores Folgueira; Mar Aguilera; Noelia Moral; Rafael Delgado; Sagrario Zurita |
| EPI_ISL_2241457 | HOSPITAL UNIVERSITARIO SON ESPASES | HOSPITAL UNIVERSITARIO SON ESPASES | Antonio Oliver; Carla López-Causapé; Pablo Fraile-Ribot; SeqCovid |
| EPI_ISL_2241452 | HOSPITAL UNIVERSITARIO SON LLATZER | HOSPITAL UNIVERSITARIO SON ESPASES | Antonio Oliver; Carla López-Causapé; Pablo Fraile-Ribot; SeqCovid |
| EPI_ISL_2621176, EPI_ISL_2651225, EPI_ISL_2651226 | HPTU | Universidad Nacional de Colombia - Laboratorio Genómico One Health | Andres F. Cardona-Rios; Carlos Franco-Muñoz; Carolina Muñoz-Arango; Celeny Ortiz; Daniel O. Maldonado-Perez; Diego A. Álvarez-Díaz; Hector Alejandro Ruiz-Moreno; Idabely Betancur Ortiz; Jorge E. Osorio; Juan P. Hernandez-Ortiz; Karl A Ciuoderis; Katherine Laiton-Donato; Laura Silvana Perez; Lina M. Hurtado; Marcela Mercado-Reyes; Maria Angélica Maya; Maria Stella López; Rita Almanza Payares; Sandra Ines Cano; Simón Villegas Velásquez |
| EPI_ISL_2738508, EPI_ISL_2738515, EPI_ISL_2738517, EPI_ISL_2738626, EPI_ISL_3235356 | Hackensack Medical Center | New York Genome Center | Amy Baldwin; Andre Corvelo; Barry Kreiswirth; David Perlin; Dayna M. Oschwald; Jose Mediavilla; Kaelea Composto; Kar Chow; Liang Chen; Marcus Cunningham; Michael Zody; Samantha Fennessey; Tom Maniatis |
| EPI_ISL_2727870 | HealthPartners Central Lab | Minnesota Department of Health, Public Health Laboratory | Alexandra Lorentz; Jacob Garfin; Matt Plumb; and Xiong Wang |
| EPI_ISL_2320150, EPI_ISL_2320151, EPI_ISL_2320152, EPI_ISL_2320171, EPI_ISL_2320174, EPI_ISL_2320175, EPI_ISL_2320182, EPI_ISL_2320184, EPI_ISL_2320199, EPI_ISL_2320218, EPI_ISL_2320223, EPI_ISL_2320258, EPI_ISL_2320278, EPI_ISL_2320279, EPI_ISL_2320293, EPI_ISL_2320298, EPI_ISL_2320330, EPI_ISL_2320338, EPI_ISL_2320340, EPI_ISL_2320351, EPI_ISL_2320354, EPI_ISL_2320378, EPI_ISL_2320387, EPI_ISL_2320388, EPI_ISL_2320394, EPI_ISL_2320414, EPI_ISL_2320427, EPI_ISL_2320438, EPI_ISL_2320459, EPI_ISL_2320469, EPI_ISL_2320479, EPI_ISL_2320481, EPI_ISL_2320506, EPI_ISL_2320519, EPI_ISL_2320694, EPI_ISL_2320772, EPI_ISL_2320784, EPI_ISL_2320792, EPI_ISL_2320799, EPI_ISL_2320804, EPI_ISL_2320838, EPI_ISL_2320952, EPI_ISL_2367611, EPI_ISL_2367614, | | | |

| | | | |
|---|---|---|---|
| EPI_ISL_2367617, EPI_ISL_2367638, EPI_ISL_2440059, EPI_ISL_2440065, EPI_ISL_2440073, EPI_ISL_2440075, EPI_ISL_2440091, EPI_ISL_2440106, EPI_ISL_2440116, EPI_ISL_2440130, EPI_ISL_2440131, EPI_ISL_2440132, EPI_ISL_2440144, EPI_ISL_2440162, EPI_ISL_2440167, EPI_ISL_2440172, EPI_ISL_2440176, EPI_ISL_2440183, EPI_ISL_2440187, EPI_ISL_2440192, EPI_ISL_2440195, EPI_ISL_2440198, EPI_ISL_2440203, EPI_ISL_2440215, EPI_ISL_2440225, EPI_ISL_2440227, EPI_ISL_2440236, EPI_ISL_2440242, EPI_ISL_2440245, EPI_ISL_2440266, EPI_ISL_2440267, EPI_ISL_2440284, EPI_ISL_2440309, EPI_ISL_2440344, EPI_ISL_2440350, EPI_ISL_2440351, EPI_ISL_2440352, EPI_ISL_2440358, EPI_ISL_2440367, EPI_ISL_2440372, EPI_ISL_2440382, EPI_ISL_2440384, EPI_ISL_2440390, EPI_ISL_2440395, EPI_ISL_2440399, EPI_ISL_2440406, EPI_ISL_2440412, EPI_ISL_2440413, EPI_ISL_2440416, EPI_ISL_2440429, EPI_ISL_2440437, EPI_ISL_2440461, EPI_ISL_2440464, EPI_ISL_2440466, EPI_ISL_2440467, EPI_ISL_2440469, EPI_ISL_2440483, EPI_ISL_2440484, EPI_ISL_2440489, EPI_ISL_2440516, EPI_ISL_2440517, EPI_ISL_2440519, EPI_ISL_2440523, EPI_ISL_2440526, EPI_ISL_2451495, EPI_ISL_2451500, EPI_ISL_2598845, EPI_ISL_2598855, EPI_ISL_2598856, EPI_ISL_2598858, EPI_ISL_2598860, EPI_ISL_2598880, EPI_ISL_2598886, EPI_ISL_2598893, EPI_ISL_2598904, EPI_ISL_2598907, EPI_ISL_2598912, EPI_ISL_2598914, EPI_ISL_2598936, EPI_ISL_2598955, EPI_ISL_2598960, EPI_ISL_2598962, EPI_ISL_2598964, EPI_ISL_2598968, EPI_ISL_2598975, EPI_ISL_2598977, EPI_ISL_2598988, EPI_ISL_2598990, EPI_ISL_2598997, EPI_ISL_2599002, EPI_ISL_2599010, EPI_ISL_2599019, EPI_ISL_2599023, EPI_ISL_2599035, EPI_ISL_2599039, EPI_ISL_2599057, EPI_ISL_2599060, EPI_ISL_2599077, EPI_ISL_2599083, EPI_ISL_2599098, EPI_ISL_2599100, EPI_ISL_2599120, EPI_ISL_2599123, EPI_ISL_2599165, EPI_ISL_2599239, EPI_ISL_2599240, EPI_ISL_2599249, EPI_ISL_2599260, EPI_ISL_2599262, EPI_ISL_2599264, EPI_ISL_2599280, EPI_ISL_2599289, EPI_ISL_2599294, EPI_ISL_2599296, EPI_ISL_2599298, EPI_ISL_2599304, EPI_ISL_2599320, EPI_ISL_2599342, EPI_ISL_2599345, EPI_ISL_2599347, EPI_ISL_2599353, EPI_ISL_2599361, EPI_ISL_2599369, EPI_ISL_2599393, EPI_ISL_2599401, EPI_ISL_2599402, EPI_ISL_2599416, EPI_ISL_2599417, EPI_ISL_2599431, EPI_ISL_2599441, EPI_ISL_2599443, EPI_ISL_2599450, EPI_ISL_2599452, EPI_ISL_2599485, EPI_ISL_2599494, EPI_ISL_2599496, EPI_ISL_2599516, EPI_ISL_2599519, EPI_ISL_2599538, EPI_ISL_2599549, EPI_ISL_2599560, EPI_ISL_2599563, EPI_ISL_2599611, EPI_ISL_2599623, EPI_ISL_2649916, EPI_ISL_2649917, EPI_ISL_2649926, EPI_ISL_2649927, EPI_ISL_2649936, EPI_ISL_4347583 | | | |
| see above | Helix/Illumina | Centers for Disease Control and Prevention Division of Viral Diseases, Pathogen Discovery | Adrian Paskey; Alexandre Bolze; Ary Ascencio; Benjamin Rambo-Martin; Brad Sickler; Charlotte Rivera-Garcia; Christine Tran; Christopher Gulvick; Chrstine Tran; Clinton Paden; Clinton R. Paden; Dakota Howard; Darlene Wagner; David Becker; Dhwani Batra; Duncan MacCannell; Efren Sandoval; Eileen De Feo; Eileen de Feo; Elizabeth Cirulli; Eric Allen; Geraint Levan; James Lu; Jan Antico; Jason Caravas; Jason Nguyen; Jimmy Ramirez; Jingtao Liu; Kara Moser; Kelly Barrett; Kelly Schiabor Barrett; Kim Gietzen; Kristine Lacek; Magnus Isaksson; Marc Laurent; Matthew Schmerer; Matthew Tolentino; Nicole L. Washington; Nicole Washington; Peter Cook; Peter W. Cook; Phil Febbo; Ryan Cho; Scott Sammons; Shannon Wickline; Shatavia Morrison; Sherry Wang; Simon White; Tyler Cassens; William Lee; Yvette Unoarumhi |
| EPI_ISL_2454299 | Hellenic National Blood Transfusion Center - EKEA | Greek Genome Center, Biomedical Research Foundation of the Academy of Athens (BRFAA) | Dimitrios Thanos; Efthimia Petinaki; Emmanouil Athanasiadis; Giannis Vatsellas; Katerina Zoi; Kostas Stamoulis; Theodoros Loupis |
| EPI_ISL_3149728 | Hosp. 4 de Junio Dr. Ramón Carrillo | Laboratorio de Biología Molecular, Instituto de Medicina Regional on behalf of 'Proyecto Argentino Interinstitucional de genomica de SARS-CoV-2' (PAIS Consortium) | Andrea Carolina Piedrabuena; Bettina Brusés; Diana Pérez Quintana; Gabriela Jurasek; Griselda Oria; Horacio Lucero.; Javier Mussin; Laura Formichelli; Mayra Mendoza; Melina Lorenzini Campos; Raúl Maximiliano Acevedo |
| EPI_ISL_2374380, EPI_ISL_2462941 | Hospital | National Reference Center for Viruses of Respiratory Infections, Institut Pasteur, Paris | Amaury Vaysse; Angela Brisebarre; Axelle Paquin; Camille Capel; Christophe Malabat; Corinne Maufrais; Emmanuelle Permal; Eric Farfour; Etienne Simon-Lorière; Frédéric Lemoine; Louise Lefrançois; Marion Barbet; Maud Vanpeene; Méline Bizard; Sylvie Behillil; Sylvie Van der Werf; Vincent Enouf |
| EPI_ISL_2178828, EPI_ISL_3505940, EPI_ISL_3505942, EPI_ISL_3505943, EPI_ISL_3505948, EPI_ISL_3505949, EPI_ISL_3505952, EPI_ISL_3505953 | | | |
| see above | Hospital General Universitario Gregorio Marañón | Hospital General Universitario Gregorio Marañón | Cristina Rodriguez-Grande; Darío García de Viedma; Julia Suárez; Laura Pérez-Lago; Marta Herranz Martin; Patricia Muñoz; Pedro Sola Campoy; Pilar Catalán; Sergio Buenestado Serrano; Victor Manuel de la Cueva; Victor de la Cueva |
| EPI_ISL_2567563 | Hospital General Universitario de Alicante - Instituto de Investigación Sanitaria y Biomédica de Alicante | SeqCOVID-SPAIN consortium/IBV(CSIC) | Carmen Molina Pardines and SeqCOVID-SPAIN consortium; Maripaz Ventero Martín |
| EPI_ISL_3230026 | Hospital Señor del Milagro | Instituto de Patologia Vegetal (CIAP-INTA) on behalf of 'Proyecto Argentino Interinstitucional de genomica de SARS-CoV-2' (PAIS Consortium) | Debat, HJ.; FD; Fernanda Aguero; Fernández; Lavaque Esteban; Marquez, N.; Pamela Cajal; Raskovsky Viviana; Veronica Lesser |
| EPI_ISL_2284955, EPI_ISL_2284956, EPI_ISL_2284957, EPI_ISL_2284958, EPI_ISL_2284968, EPI_ISL_2284969 | Hospital Universitari Arnau de Vilanova | Hospital Universitari Vall d'Hebron - Vall d'Hebron Institut de Recerca | Alejandra González-Sánchez; Andrés Antón; Ariadna Rando; Carla Castillo; Cristina Andrés; Damir Garcia-Cehic; Josep Quer; Juliana Esperalba; Maria Carmen Martin; Maria Gema Codina; Maria Piñana; Tomàs Pumarola |
| EPI_ISL_2516404, EPI_ISL_2516522, EPI_ISL_2516527 | Hospital Universitari Bellvitge | Microbiology Department | Aida Gonzalez-Diaz; Carmen Ardanuy; Jordi Camara; Jordi Niubó; Laura Calatayud; M Angeles Domínguez; Miguel Fernandez-Huerta; Sara Marti |
| EPI_ISL_2284932 | Hospital Universitari Vall d'Hebron - Vall d'Hebron Institut de Recerca | Hospital Universitari Vall d'Hebron - Vall d'Hebron Institut de Recerca | Alejandra González-Sánchez; Andrés Antón; Ariadna Rando; Carla Castillo; Cristina Andrés; Damir Garcia-Cehic; Josep Quer; Juliana Esperalba; Maria Carmen Martin; Maria Gema Codina; Maria Piñana; Tomàs Pumarola |
| EPI_ISL_2383275 | Hospital Universitari i Politècnic La Fe de València | SeqCOVID-SPAIN consortium/IBV(CSIC) | Ana Gil Brusola; Eva González Barberá; José Luis López Hontangas and SeqCOVID-SPAIN consortium; María Dolores Gómez Ruiz; Salvador Giner Almaraz |
| EPI_ISL_7307276, EPI_ISL_7307282 | Hospital Universitario San Ignacio | Centro de Investigaciones en Microbiología y Biotecnología-UR (CIMBIUR), Facultad de Ciencias Naturales, Universidad del Rosario, Bogotá, Colombia | Alberto Paniz-Mondolfi; Angie Ramírez; Beatriz Ariza; Camilo A. Correa-Cárdenas; Carlos Gómez-Restrepo; Claudia Cardozo-Romero; Claudia Méndez; David-Santiago Quevedo; Guido España; Hernando Díaz; Juan David Ramírez; Juliana Cuervo-Rojas; Julie Pérez; Luz H. Patiño; Manuel-Antonio Franco; Maria-Clara Duque; Marina Muñoz; Nathalia Ballesteros; Nicolas Luna; Sergio Castañeda; Zulma M. Cucunubá |
| EPI_ISL_4196726 | Hospital Zonal Dr. Ramón Carrillo | Laboratorio Central Mg. Luis Alfredo Pianciola on behalf of 'Proyecto Argentino Interinstitucional de genomica de SARS-CoV-2' (PAIS Consortium) | C Pintos; C Rastellini; C Ziehm; J Ousset; L Pianciola; M Fernandez; M L Álvarez; M Mazzeo; P V Blanco.; Y Espasandin |
| EPI_ISL_6513480 | Hospital da Clinicas - UNICAMP | Laboratorio de Estudos de Virus Emergentes | Bruno B. Benites; Daniel A. Toledo-Teixeira; Dewton Moraes Vasconcelos; Ester C. Sabino; Fabiana Granja; Fernando R. Spilki; Janine Schincariol Sabino; José Luiz Proenca-Modena; Julia Forato; Lia Furlaneto Marega; Luciana Souto Mofatto; Maria Marluce dos Santos Vilela; Mariene Ribeiro Amorim; Nuno R. Faria; Raquel Silveira Bello Stucchi; William M. de Souza |
| EPI_ISL_2597665, EPI_ISL_2597679, EPI_ISL_3023140 | Hospital of the University of Pennsylvania Molecular Pathology Lab | Bushman Lab - University of Pennsylvania | Aoife M. Roche; Arupa Ganguly; Ayannah S. Fitzgerald; Brendan Kelly; Jevon Graham-Wooten; John K. Everett; Kyle Rodino; Layla A. Khatib; Mike Feldman; Ronald G. Collman and Frederic Bushan; Samantha A. Whiteside; Scott Sherrill-Mix; Shantan Reddy; Young Hwang |
| EPI_ISL_5070421, EPI_ISL_5070428, EPI_ISL_5070527, EPI_ISL_5070562, EPI_ISL_5070566, EPI_ISL_5070612, EPI_ISL_5070719, EPI_ISL_5070730 | | | |
| see above | Houston Methodist Hospital | Houston Methodist Hospital | Ilya J. Finkelstein; James J. Davis; Jessica Cambric; Jimmy Gollihar; Kristina Reppond; Layne Pruitt; Madison N. Shyer; Marcus Nguyen; Matthew Ojeda Saavedra; Paul A. Christensen; Prasanti Yerramilli; Randall J. Olsen; Robert Olson; Ryan Gadd; S. Wesley Long; Sishir Subedi; and James M. Musser |
| EPI_ISL_2497657, EPI_ISL_2497662, EPI_ISL_2500025, EPI_ISL_2500061, EPI_ISL_2500157, EPI_ISL_2658738 | Humboldt County Public Health Laboratory | Chan-Zuckerberg Biohub | CZB Cliahub Consortium |
| EPI_ISL_2445617, EPI_ISL_2445620, EPI_ISL_2445621, EPI_ISL_2445623, EPI_ISL_2445625, EPI_ISL_2445626, EPI_ISL_2445630 | | | |
| see above | Humboldt County Public Health Laboratory | Humboldt County Public Health Laboratory | Jeremy Corrigan |
| EPI_ISL_2333773 | Hôpital Cochin | Department of Virology, Henri Mondor University Hospital, Assistance Publique Hôpitaux de Paris, Université Paris-Est Créteil, INSERM U955 | Alexandre Soulier; Christophe Rodriguez; Elisabeth Trawinski; Guillaume Gricourt; Jean-Michel Pawlotsky; Melissa N'Debi; Slim Fourati; Vanessa Demontant |
| EPI_ISL_2621169 | ICMT-APARTADO | Universidad Nacional de Colombia - Laboratorio Genómico One Health | Andres F. Cardona-Rios; Carlos Franco-Muñoz; Carolina Muñoz-Arango; Celeny Ortiz; Daniel O. Maldonado-Perez; Diego A. Álvarez-Díaz; Hector Alejandro Ruiz-Moreno; Idabely Betancur Ortiz; Jorge E. Osorio; Juan P. Hernandez-Ortiz; Karl A Ciuoderis; Katherine Laiton-Donato; Laura Silvana Perez; Lina M. Hurtado; Marcela Mercado-Reyes; Maria Angélica Maya; Maria Stella López; Rita Almanza Payares; Sandra Ines Cano; Simón Villegas Velásquez |
| EPI_ISL_2451791, EPI_ISL_2451803, EPI_ISL_2451809, EPI_ISL_2463002 | IL Dept. of Public Health Springfield Laboratory | Centers for Disease Control and Prevention Division of Viral Diseases, Pathogen Discovery | Alison Laufer Halpin; Ben L. Rambo-Martin; Clinton R. Paden; Dakota Howard; Darlene Wagner; Dave Wentworth; Dhwani Batra; Jasmine Padilla; Justin Lee; Katie Dillon; Krista Queen; Kristen Knipe; Kristine Lacek; Mark Burroughs; Matthew Schmerer; Mili Sheth; Peter Cook; Sam Shepard; Sarah Nobles; Shoshona Le; Suxiang Tong; Vivien Dugan; Yvette Unoarumhi |
| EPI_ISL_2266154, EPI_ISL_2266158 | IMD - Institut für Medizinische Diagnostik Berlin-Potsdam | Robert Koch Institute | |
| EPI_ISL_2420139, EPI_ISL_2420152, EPI_ISL_2420158, EPI_ISL_2420163, EPI_ISL_2420170, EPI_ISL_2420173, EPI_ISL_2420208, EPI_ISL_2420213, EPI_ISL_2420216, EPI_ISL_2420244, EPI_ISL_2439147, EPI_ISL_2439149, EPI_ISL_2439150, EPI_ISL_2439167, EPI_ISL_2439169, EPI_ISL_2544560, EPI_ISL_2544561, EPI_ISL_2544635, EPI_ISL_3007487 | | | |
| see above | IN State Department of Health Laboratory Services | IN State Department of Health Laboratory Services | Brian Pope; Cassandra Campion; Jamie Yeadon; Kyle Brownlee; Lixia Liu; Mark Glazier; Melissa Hindenlang |
| EPI_ISL_2319321, EPI_ISL_2319335, EPI_ISL_2319340, EPI_ISL_2319349, EPI_ISL_2319369, EPI_ISL_2319370, EPI_ISL_2319372, EPI_ISL_2319373, EPI_ISL_2319374, EPI_ISL_2319380 | | | |
| see above | INDRE | Instituto Nacional de Medicina Genomica | Cedro-Tanda A; Garcia-Cardenas FJ; Gisela Barrera-Badillo; Gonzalez-Barrera D; Gonzalez-Woge MA; Herrera-Montalvo LA.; Hidalgo-Miranda A; Irma Lopez-Martinez; Jose Ernesto Ramirez González; Mendoza-Vargas A; Munguia-Garza P; Rangel-DeLeon D; Reyes-Grajeda JP |
| EPI_ISL_2628307 | INHRR | Laboratorio de Virología Molecular | Carmen L Loureiro; CoViVen Group; Domingo J Garzaro; Esmeralda Vizzi; Flor H Pujol; Héctor R Rangel; José Luis Zambrano; Lieska Rodríguez; Mariana Hidalgo; Pierina D´Angelo; Rossana C Jaspe; Víctor Alarcón; Yoneira Sulbaran; Zoila Moros |
| EPI_ISL_2493162, EPI_ISL_2493247, EPI_ISL_2493715, EPI_ISL_2493721, EPI_ISL_2493723, | INSIDE DIAGNÓSTICOS | Instituto Butantan | Antonio Jorge Martins; Claudia Renata dos Santos Barros; David Schlesinger; Debora Botequio Moretti; Dimas Tadeu Covas; Elaine Cristina Marqueze; Elaine Vieira Santos; Evandra Strazza Rodrigues; Heidge Fukumasu; Jayme Augusto de Souza-Neto; José Salvatore Leister Patané; Luiz Alcantara; Luiz Lehmann Coutinho; Maria Carolina Elias; Maurício Lacerda Nogueira; Rafael dos Santos Bezerra; Raul Machado Neto; Rejane Maria Tommasini Grotto; Ricardo Haddad; Sandra Coccuzzo Sampaio Vessoni; Simone Kashima; Svetoslav Nanev Slavov; Vincent Louis Viala |

| | | | |
|---|---|---|---|
| EPI_ISL_2493726 | | | |
| EPI_ISL_2445061, EPI_ISL_2445081 | INSIDE DIAGNÓSTICOS NORTE BRASILÂNDIA/FREGUESIA DO Ó | Instituto Butantan | Antonio Jorge Martins; Claudia Renata dos Santos Barros; David Schlesinger; Debora Botequio Moretti; Dimas Tadeu Covas; Elaine Cristina Marqueze; Elaine Vieira Santos; Evandra Strazza Rodrigues; Heidge Fukumasu; Jayme Augusto de Souza-Neto; José Salvatore Leister Patané; Luiz Alcantara; Luiz Lehmann Coutinho; Maria Carolina Elias; Maurício Lacerda Nogueira; Rafael dos Santos Bezerra; Raul Machado Neto; Rejane Maria Tommasini Grotto; Ricardo Haddad; Sandra Coccuzzo Sampaio Vessoni; Simone Kashima; Svetoslav Nanev Slavov; Vincent Louis Viala |
| EPI_ISL_2445059, EPI_ISL_2445065 | INSIDE DIAGNÓSTICOS NORTE CACHOEIRINHA/CASA VERDE | Instituto Butantan | Antonio Jorge Martins; Claudia Renata dos Santos Barros; David Schlesinger; Debora Botequio Moretti; Dimas Tadeu Covas; Elaine Cristina Marqueze; Elaine Vieira Santos; Evandra Strazza Rodrigues; Heidge Fukumasu; Jayme Augusto de Souza-Neto; José Salvatore Leister Patané; Luiz Alcantara; Luiz Lehmann Coutinho; Maria Carolina Elias; Maurício Lacerda Nogueira; Rafael dos Santos Bezerra; Raul Machado Neto; Rejane Maria Tommasini Grotto; Ricardo Haddad; Sandra Coccuzzo Sampaio Vessoni; Simone Kashima; Svetoslav Nanev Slavov; Vincent Louis Viala |
| EPI_ISL_2445058 | INSIDE DIAGNÓSTICOS NORTE PIRITUBA | Instituto Butantan | Antonio Jorge Martins; Claudia Renata dos Santos Barros; David Schlesinger; Debora Botequio Moretti; Dimas Tadeu Covas; Elaine Cristina Marqueze; Elaine Vieira Santos; Evandra Strazza Rodrigues; Heidge Fukumasu; Jayme Augusto de Souza-Neto; José Salvatore Leister Patané; Luiz Alcantara; Luiz Lehmann Coutinho; Maria Carolina Elias; Maurício Lacerda Nogueira; Rafael dos Santos Bezerra; Raul Machado Neto; Rejane Maria Tommasini Grotto; Ricardo Haddad; Sandra Coccuzzo Sampaio Vessoni; Simone Kashima; Svetoslav Nanev Slavov; Vincent Louis Viala |
| EPI_ISL_2445047, EPI_ISL_2445060, EPI_ISL_2445064 | INSIDE DIAGNÓSTICOS NORTE VILA MARIA/VILA GUILHERME | Instituto Butantan | Antonio Jorge Martins; Claudia Renata dos Santos Barros; David Schlesinger; Debora Botequio Moretti; Dimas Tadeu Covas; Elaine Cristina Marqueze; Elaine Vieira Santos; Evandra Strazza Rodrigues; Heidge Fukumasu; Jayme Augusto de Souza-Neto; José Salvatore Leister Patané; Luiz Alcantara; Luiz Lehmann Coutinho; Maria Carolina Elias; Maurício Lacerda Nogueira; Rafael dos Santos Bezerra; Raul Machado Neto; Rejane Maria Tommasini Grotto; Ricardo Haddad; Sandra Coccuzzo Sampaio Vessoni; Simone Kashima; Svetoslav Nanev Slavov; Vincent Louis Viala |
| EPI_ISL_2445053, EPI_ISL_2445074, EPI_ISL_2445077 | INSIDE DIAGNÓSTICOS OESTE BUTANTÃ | Instituto Butantan | Antonio Jorge Martins; Claudia Renata dos Santos Barros; David Schlesinger; Debora Botequio Moretti; Dimas Tadeu Covas; Elaine Cristina Marqueze; Elaine Vieira Santos; Evandra Strazza Rodrigues; Heidge Fukumasu; Jayme Augusto de Souza-Neto; José Salvatore Leister Patané; Luiz Alcantara; Luiz Lehmann Coutinho; Maria Carolina Elias; Maurício Lacerda Nogueira; Rafael dos Santos Bezerra; Raul Machado Neto; Rejane Maria Tommasini Grotto; Ricardo Haddad; Sandra Coccuzzo Sampaio Vessoni; Simone Kashima; Svetoslav Nanev Slavov; Vincent Louis Viala |
| EPI_ISL_2445048, EPI_ISL_2445073 | INSIDE DIAGNÓSTICOS OESTE LAPA/PINHEIROS | Instituto Butantan | Antonio Jorge Martins; Claudia Renata dos Santos Barros; David Schlesinger; Debora Botequio Moretti; Dimas Tadeu Covas; Elaine Cristina Marqueze; Elaine Vieira Santos; Evandra Strazza Rodrigues; Heidge Fukumasu; Jayme Augusto de Souza-Neto; José Salvatore Leister Patané; Luiz Alcantara; Luiz Lehmann Coutinho; Maria Carolina Elias; Maurício Lacerda Nogueira; Rafael dos Santos Bezerra; Raul Machado Neto; Rejane Maria Tommasini Grotto; Ricardo Haddad; Sandra Coccuzzo Sampaio Vessoni; Simone Kashima; Svetoslav Nanev Slavov; Vincent Louis Viala |
| EPI_ISL_2445067, EPI_ISL_2445072, EPI_ISL_2445076, EPI_ISL_2445084 | INSIDE DIAGNÓSTICOS SUDESTE IPIRANGA | Instituto Butantan | Antonio Jorge Martins; Claudia Renata dos Santos Barros; David Schlesinger; Debora Botequio Moretti; Dimas Tadeu Covas; Elaine Cristina Marqueze; Elaine Vieira Santos; Evandra Strazza Rodrigues; Heidge Fukumasu; Jayme Augusto de Souza-Neto; José Salvatore Leister Patané; Luiz Alcantara; Luiz Lehmann Coutinho; Maria Carolina Elias; Maurício Lacerda Nogueira; Rafael dos Santos Bezerra; Raul Machado Neto; Rejane Maria Tommasini Grotto; Ricardo Haddad; Sandra Coccuzzo Sampaio Vessoni; Simone Kashima; Svetoslav Nanev Slavov; Vincent Louis Viala |
| EPI_ISL_2473815 | INSIDE DIAGNÓSTICOS SUDESTE VILA MARIANA/JABAQUARA | Instituto Butantan | Antonio Jorge Martins; Claudia Renata dos Santos Barros; David Schlesinger; Debora Botequio Moretti; Dimas Tadeu Covas; Elaine Cristina Marqueze; Elaine Vieira Santos; Evandra Strazza Rodrigues; Heidge Fukumasu; Jayme Augusto de Souza-Neto; José Salvatore Leister Patané; Luiz Alcantara; Luiz Lehmann Coutinho; Maria Carolina Elias; Maurício Lacerda Nogueira; Rafael dos Santos Bezerra; Raul Machado Neto; Rejane Maria Tommasini Grotto; Ricardo Haddad; Sandra Coccuzzo Sampaio Vessoni; Simone Kashima; Svetoslav Nanev Slavov; Vincent Louis Viala |
| EPI_ISL_2445063, EPI_ISL_2445082 | INSIDE DIAGNÓSTICOS SUDESTE VILA PRUDENTE/SAPOPEMBA | Instituto Butantan | Antonio Jorge Martins; Claudia Renata dos Santos Barros; David Schlesinger; Debora Botequio Moretti; Dimas Tadeu Covas; Elaine Cristina Marqueze; Elaine Vieira Santos; Evandra Strazza Rodrigues; Heidge Fukumasu; Jayme Augusto de Souza-Neto; José Salvatore Leister Patané; Luiz Alcantara; Luiz Lehmann Coutinho; Maria Carolina Elias; Maurício Lacerda Nogueira; Rafael dos Santos Bezerra; Raul Machado Neto; Rejane Maria Tommasini Grotto; Ricardo Haddad; Sandra Coccuzzo Sampaio Vessoni; Simone Kashima; Svetoslav Nanev Slavov; Vincent Louis Viala |
| EPI_ISL_2445054, EPI_ISL_2445055, EPI_ISL_2445057, EPI_ISL_2445069, EPI_ISL_2445080 | INSIDE DIAGNÓSTICOS SUL CAPELA DO SOCORRO | Instituto Butantan | Antonio Jorge Martins; Claudia Renata dos Santos Barros; David Schlesinger; Debora Botequio Moretti; Dimas Tadeu Covas; Elaine Cristina Marqueze; Elaine Vieira Santos; Evandra Strazza Rodrigues; Heidge Fukumasu; Jayme Augusto de Souza-Neto; José Salvatore Leister Patané; Luiz Alcantara; Luiz Lehmann Coutinho; Maria Carolina Elias; Maurício Lacerda Nogueira; Rafael dos Santos Bezerra; Raul Machado Neto; Rejane Maria Tommasini Grotto; Ricardo Haddad; Sandra Coccuzzo Sampaio Vessoni; Simone Kashima; Svetoslav Nanev Slavov; Vincent Louis Viala |
| EPI_ISL_2445049, EPI_ISL_2445051, EPI_ISL_2445068, EPI_ISL_2445075 | INSIDE DIAGNÓSTICOS SUL PARELHEIROS | Instituto Butantan | Antonio Jorge Martins; Claudia Renata dos Santos Barros; David Schlesinger; Debora Botequio Moretti; Dimas Tadeu Covas; Elaine Cristina Marqueze; Elaine Vieira Santos; Evandra Strazza Rodrigues; Heidge Fukumasu; Jayme Augusto de Souza-Neto; José Salvatore Leister Patané; Luiz Alcantara; Luiz Lehmann Coutinho; Maria Carolina Elias; Maurício Lacerda Nogueira; Rafael dos Santos Bezerra; Raul Machado Neto; Rejane Maria Tommasini Grotto; Ricardo Haddad; Sandra Coccuzzo Sampaio Vessoni; Simone Kashima; Svetoslav Nanev Slavov; Vincent Louis Viala |
| EPI_ISL_2445046 | INSIDE LABORATÓRIOS NORTE PIRITUBA | Instituto Butantan | Antonio Jorge Martins; Claudia Renata dos Santos Barros; David Schlesinger; Debora Botequio Moretti; Dimas Tadeu Covas; Elaine Cristina Marqueze; Elaine Vieira Santos; Evandra Strazza Rodrigues; Heidge Fukumasu; Jayme Augusto de Souza-Neto; José Salvatore Leister Patané; Luiz Alcantara; Luiz Lehmann Coutinho; Maria Carolina Elias; Maurício Lacerda Nogueira; Rafael dos Santos Bezerra; Raul Machado Neto; Rejane Maria Tommasini Grotto; Ricardo Haddad; Sandra Coccuzzo Sampaio Vessoni; Simone Kashima; Svetoslav Nanev Slavov; Vincent Louis Viala |
| EPI_ISL_2458115 | INSPI-CRN DE INFLUENZA Y OTROS VIRUS RESPIRATORIOS | NIC-INSPI | Alfredo Bruno; Domenica de Mora.; Jimmy Garcés; Johanna Laines; Lizbeth Patiño; Manuel Gonzalez; Maritza Olmedo; Michelle Páez |
| EPI_ISL_2340228 | IPO Lisboa | Instituto Nacional de Saude (INSA) | Borges et al |
| EPI_ISL_5476569 | IRCCS Sacro Cuore Don Calabria Hospital, Department of Infectious, Tropical Diseases & Microbiology | IRCCS Sacro Cuore Don Calabria Hospital, Department of Infectious, Tropical Diseases & Microbiology | ; Antonio Mori; Chiara Piubelli; Elena Pomari; Michela Deiana |
| EPI_ISL_2716786, EPI_ISL_2716789, EPI_ISL_2716793, EPI_ISL_2716796, EPI_ISL_2716797, EPI_ISL_2716801, EPI_ISL_2716810, EPI_ISL_2716816, EPI_ISL_2716818, EPI_ISL_2716819, EPI_ISL_2716820, EPI_ISL_2716821 | | | |
| see above | IRCCS San Gallicano Dermatological Institute | IRCCS Regina Elena National Cancer Institute | Aldo Morrone; Alessia Lauretti; Alice Massacci; Andrea Cazzani; Antonio Federico; Arianna Mastrofrancesco; Eleonora Sperandio; Fabrizio Ensoli; Francesca De Nicola; Frauke Goeman; Fulvia Fraticelli; Fulvia Pimpinelli; Gennaro Ciliberto; Giovanna D’agosto; Giovanni Blandino; Giulia Orlandi; Ilaria Celesti; Lorenzo Furzi; Ludovica Ciuffreda; Martina Diano; Matteo Pallocca; Maurizio Fanciulli; Sabrina Strano; Sara Donzelli; Serena Salvo; di Domenico Enea Gino |
| EPI_ISL_2712986 | ISDH | Quantigen Biosciences | Paul J. Childress |
| EPI_ISL_2628306 | IVIC | Laboratorio de Virología Molecular | Carmen L Loureiro; CoViVen Group; Domingo J Garzaro; Esmeralda Vizzi; Flor H Pujol; Héctor R Rangel; José Luis Zambrano; Lieska Rodríguez; Mariana Hidalgo; Pierina D´Angelo; Rossana C Jaspe; Víctor Alarcón; Yoneira Sulbaran; Zoila Moros |
| EPI_ISL_2789982, EPI_ISL_2801466 | IVY3 Central Lab, Vanderbilt University Medical Center | Lauring Lab, University of Michigan, Department of Microbiology and Immunology | Valesano |
| EPI_ISL_2933103, EPI_ISL_2933104, EPI_ISL_2933106 | IZSM | IZSM-U.O.C. Virologia | Antonio Limone; Claudio de Martinis; Esterina De Carlo; Giovanna Fusco; Lorena Cardillo; Maurizio Viscardi |
| EPI_ISL_2598130 | Idaho Bureau of Laboratories | Center for Global Health, University of New Mexico Health Sciences Center | Christopher Ball; Darrell Dinwiddie; Daryl Domman; Kurt Schwalm; Matthew Burns; Robert Voermans; Valerie Morley |
| EPI_ISL_2801135, EPI_ISL_2801136, EPI_ISL_2801138, EPI_ISL_2801139 | Illinois Department of Public Health | Gagnon Lab, Southern Illinois University | Keith Gagnon |
| EPI_ISL_2300283, EPI_ISL_2300298, EPI_ISL_2300396, EPI_ISL_2300432, EPI_ISL_2300467, EPI_ISL_2300470, EPI_ISL_2300486, EPI_ISL_2300519, EPI_ISL_2467398, EPI_ISL_2467402, EPI_ISL_2467406, EPI_ISL_2467407, EPI_ISL_2467409, EPI_ISL_2467410, EPI_ISL_2467414, EPI_ISL_2467415, EPI_ISL_2467444, EPI_ISL_2467452, EPI_ISL_2467453, EPI_ISL_2467455, EPI_ISL_2467461, EPI_ISL_2467467, EPI_ISL_2467470, EPI_ISL_2467472, EPI_ISL_2467474, EPI_ISL_2467477 | | | |
| see above | Illinois Department of Public Health | Illinois Department of Public Health - Chicago Lab | Ira Heimler; Joel Price; Vineet K. Dhiman |
| EPI_ISL_2135063, EPI_ISL_2135069, EPI_ISL_2135088, EPI_ISL_2135103, EPI_ISL_2203537, EPI_ISL_2203626, EPI_ISL_2203629, EPI_ISL_2203689, EPI_ISL_2203692, EPI_ISL_2203696, EPI_ISL_2203700, EPI_ISL_2203705, EPI_ISL_2203712, EPI_ISL_2203722, EPI_ISL_2203726, EPI_ISL_2203736, EPI_ISL_2203744, EPI_ISL_2203755, EPI_ISL_2203759, EPI_ISL_2203763, EPI_ISL_2339343, EPI_ISL_2339348, EPI_ISL_2339352, EPI_ISL_2339353, EPI_ISL_2339354, EPI_ISL_2339355, EPI_ISL_2339357, EPI_ISL_2339358, EPI_ISL_2339362, EPI_ISL_2339367, EPI_ISL_2339368, EPI_ISL_2339371, EPI_ISL_2339374, EPI_ISL_2339378, EPI_ISL_2339379, EPI_ISL_2339382, EPI_ISL_2339387, EPI_ISL_2339389, EPI_ISL_2339391, EPI_ISL_2339392, EPI_ISL_2339394, EPI_ISL_2339396, EPI_ISL_2339397, EPI_ISL_2339398, EPI_ISL_2339399, EPI_ISL_2339415, EPI_ISL_2403016, EPI_ISL_2403017, EPI_ISL_2403022, EPI_ISL_2403023, EPI_ISL_2403024, EPI_ISL_2403026, EPI_ISL_2403031, EPI_ISL_2403034, EPI_ISL_2403036, EPI_ISL_2403037, EPI_ISL_2403043, EPI_ISL_2403080, EPI_ISL_2403088, EPI_ISL_2403120, EPI_ISL_2403126, EPI_ISL_2403132, EPI_ISL_2403138, EPI_ISL_2403155, EPI_ISL_2598209 | | | |
| see above | Illinois Department of Public Health - Springfield Lab | Illinois Department of Public Health - Springfield Lab | Bryan Sim; Gordon McCall |
| EPI_ISL_2490285, EPI_ISL_2692422, EPI_ISL_2692425, EPI_ISL_2692433, EPI_ISL_2714922, EPI_ISL_2714927, EPI_ISL_2714937, EPI_ISL_2714938, EPI_ISL_2714941, EPI_ISL_2714942, EPI_ISL_2714943 | | | |
| see above | Incyte Diagnostics Spokane | Seattle Flu Study | Amanda Adler; Barry R. Lutz; Benjamin Pelle; Brian Hiatt; Caitlin R. Wolf; Chris D. Frazar; Deborah A. Nickerson; Elisabeth Brandstetter; Erica Ryke; Geoff Melly; Helen Y. Chu; Janet A. Englund; Jay Shendure; Jover Lee; Kairsten Fay; Kirsten Lacombe; Lea M. Starita; Mark J. Rieder; Matthew Richardson; Matthew Thompson; Melissa Truong; Michael Boeckh; Michael Famulare; Misja Ilcisin; Peter D. Han; Philip Dykema; Romesh Gautom; Scott Lindquist; Thomas R. Sibley; Trevor Bedford |
| EPI_ISL_2323476, EPI_ISL_2323479, EPI_ISL_2323487, EPI_ISL_2323491, EPI_ISL_2323521, EPI_ISL_2323648, EPI_ISL_2323660, EPI_ISL_2323673, EPI_ISL_2323677, EPI_ISL_2323678, EPI_ISL_2323701, EPI_ISL_2323732, EPI_ISL_2323758, EPI_ISL_2323812, EPI_ISL_2323828, EPI_ISL_2323904, EPI_ISL_2323915, EPI_ISL_2323919, EPI_ISL_2323958, EPI_ISL_2324037, EPI_ISL_2324060, EPI_ISL_2324075, EPI_ISL_2324078, EPI_ISL_2324088, EPI_ISL_2324094, EPI_ISL_2324110, EPI_ISL_2324158, EPI_ISL_2375598, EPI_ISL_2375684, EPI_ISL_2375781, EPI_ISL_2652715, EPI_ISL_2652896, EPI_ISL_4382968, EPI_ISL_4383185, EPI_ISL_4387002 | | | |
| see above | Infinity Biologix | Centers for Disease Control and Prevention Division of Viral Diseases, Pathogen Discovery | Adrian Paskey; Benjamin Rambo-Martin; Chirayu Goswami; Christian Bixby; Christopher Gulvick; Clinton Paden; Clinton R. Paden; Dakota Howard; Darlene Wagner; Dhwani Batra; Duncan MacCannell; Erisa Sula; Jason Caravas; Jonathan Schultz; Kara Moser; Kristine Lacek; Matthew Schmerer; Peter Cook; Peter W. Cook; Robin Grimwood; Russ Hager; Scott Sammons; Shatavia Morrison; Tymeckia Kendall; Victoria Caban Figueroa; Yihe Wang; Yvette Unoarumhi |
| EPI_ISL_2828654, EPI_ISL_2828657, EPI_ISL_2828659 | Instituo Adolfo Lutz Central | Instituto Adolfo Lutz, Interdiciplinary Procedures Center, Strategic Laboratory | Caio Vinicius Dias Lopes; Claudia Regina Gonçalves; Claudio Tavares Sacchi; Erica Valessa Ramos Gomes; Karoline Rodrigues Campos |
| EPI_ISL_2949758, EPI_ISL_2949771, EPI_ISL_2949804, EPI_ISL_2949828, EPI_ISL_2949832, EPI_ISL_2949838, EPI_ISL_2949839, EPI_ISL_2949848, EPI_ISL_2949860, EPI_ISL_2949903, EPI_ISL_2949927, EPI_ISL_2949933, EPI_ISL_2949956, EPI_ISL_2949960, EPI_ISL_2949986, EPI_ISL_2950001, EPI_ISL_2950005, EPI_ISL_2950008, EPI_ISL_2950020, EPI_ISL_2950034, EPI_ISL_2950035, EPI_ISL_2950085, EPI_ISL_2950103, EPI_ISL_2950181 | | | |
| see above | Institut fr Virologie am Department fr Hygiene, Mikrobiologie und Publich Health der Medizinischen Universitt Innsbruck | Elling and Cochella laboratories, IMBA/IMP | Alexander Stark; Ezgi Oezkan; Luisa Cochella and Ulrich Elling; Marcus Strobl; Olga Frank; Ramesh Yelagandula; Tanino Albanese |
| EPI_ISL_2153449 | Institute for Infectious Diseases, University of Bern | Institute for Infectious Diseases, University of Bern | Alban Ramette; Christian Baumann; Cora Sägesser; Franziska Suter-Riniker; Miguel A Terrazos Miani; Pascal Bittel; Peter Keller; Stefan Neuenschwander; Stephen L Leib |
| EPI_ISL_2382058 | Institute of Legal Medicine, Medical University of Innsbruck | Bergthaler laboratory, CeMM Research Center for Molecular Medicine of the Austrian Academy of Sciences | Andreas Bergthaler; Anna Schedl; Bekir Erguner; Benedikt Agerer; Christoph Bock; Fabian Amman; Jan Laine; Lukas Endler; Maelle Le Moing; Martin Senekowitsch; Michael Schuster; Petr Triska; Thomas Penz |
| EPI_ISL_2691171, EPI_ISL_2691172 | Instituto Adolfo Lutz - Reginal de Sao José do Rio Preto | Instituto Adolfo Lutz, Interdiciplinary Procedures Center, Strategic Laboratory | Caio Vinicius Dias Lopes; Claudia Regina Gonçalves; Claudio Tavares Sacchi; Erica Valessa Ramos Gomes; Karoline Rodrigues Campos; Leonardo Jose Tadeu de Araujo |
| EPI_ISL_2919241, | Instituto Adolfo Lutz - Regional de Bauru | Instituto Adolfo Lutz, Interdiciplinary | Caio Vinicius Dias Lopes; Claudia Regina Gonçalves; Claudio Tavares Sacchi; Erica Valessa Ramos Gomes; Karoline Rodrigues Campos |

| | | | |
|---|---|---|---|
| EPI_ISL_2919252, EPI_ISL_2919253, EPI_ISL_2919259, EPI_ISL_2919260 | | Procedures Center, Strategic Laboratory | |
| EPI_ISL_3316207, EPI_ISL_3316216 | Instituto Adolfo Lutz - Regional de Campinas | Instituto Adolfo Lutz, Interdiciplinary Procedures Center, Strategic Laboratory | Caio Vinicius Dias Lopes; Claudia Regina Gonçalves; Claudio Tavares Sacchi; Karoline Rodrigues Campos; Leonardo Tadeu de Araujo; Marlon Benedito Nascomento Santos |
| EPI_ISL_2614619, EPI_ISL_2614623, EPI_ISL_2614624, EPI_ISL_3316154, EPI_ISL_3316235 | Instituto Adolfo Lutz - Regional de Presidente Prudente | Instituto Adolfo Lutz, Interdiciplinary Procedures Center, Strategic Laboratory | Caio Vinicius Dias Lopes; Claudia Regina Gonçalves; Claudio Tavares Sacchi; Erica Valessa Ramos Gomes; Karoline Rodrigues Campos; Leonardo Jose Tadeu de Araujo; Leonardo Tadeu de Araujo; Marlon Benedito Nascomento Santos |
| EPI_ISL_2691145 | Instituto Adolfo Lutz - Regional de Sao Jose do Rio Preto | Instituto Adolfo Lutz, Interdiciplinary Procedures Center, Strategic Laboratory | Caio Vinicius Dias Lopes; Claudia Regina Gonçalves; Claudio Tavares Sacchi; Erica Valessa Ramos Gomes; Karoline Rodrigues Campos; Leonardo Jose Tadeu de Araujo |
| EPI_ISL_2241317, EPI_ISL_2241318, EPI_ISL_2241319, EPI_ISL_2241320, EPI_ISL_2241321, EPI_ISL_2241322, EPI_ISL_2241323, EPI_ISL_2241324, EPI_ISL_2241325, EPI_ISL_2241326 | | | |
| see above | Instituto Butantan | Instituto Butantan | Antonio Jorge Martins; Claudia Renata dos Santos Barros; David Schlesinger; Debora Botequio Moretti; Dimas Tadeu Covas; Elaine Cristina Marqueze; Elaine Vieira Santos; Evandra Strazza Rodrigues; Heidge Fukumasu; Jayme Augusto de Souza-Neto; José Salvatore Leister Patané; Luiz Alcantara; Luiz Lehmann Coutinho; Maria Carolina Elias; Maurício Lacerda Nogueira; Rafael dos Santos Bezerra; Raul Machado Neto; Rejane Maria Tommasini Grotto; Ricardo Haddad; Sandra Coccuzzo Sampaio Vessoni; Simone Kashima; Svetoslav Nanev Slavov; Vincent Louis Viala |
| EPI_ISL_2348761 | Instituto Nacional De Investigación En Salud Pública-Crn De Influenza Y Otros Virus Respiratorios | NIC-Instituto Nacional de Investigación en Salud Pública | Alfredo Bruno; Domenica de Mora.; Jimmy Garcés; Johanna Laines; Lizbeth Patiño; Manuel Gonzalez; Maritza Olmedo; Michelle Páez |
| EPI_ISL_4405348 | Instituto Nacional de Epidemiología "Dr. Jara" | Área de Secuenciación del Laboratorio de Virología del Hospital de Niños Dr. Ricardo Gutierrez on behalf of 'Proyecto Argentino Interinstitucional de genomica de SARS-CoV-2' (PAIS Consortium) | Acuña; CJ; Cimmino; D; Goya; I; LE; Lusso; MI; MS; Nabaes Jodar; Natale; O; Pagano; S; Uez; Valinotto; Viegas, M. |
| EPI_ISL_2230826, EPI_ISL_2319390, EPI_ISL_2350098, EPI_ISL_2350099, EPI_ISL_2881833 | Instituto Nacional de Medicina Genomica | Instituto Nacional de Medicina Genomica | Abraham Campos-Romero; Cedro-Tanda A; Cisneros- Villanueva M; Escobar-Arrazola; Gonzalez-Barrera D; Herrera-Montalvo LA.; Hidalgo-Miranda A; Luna-Ruiz Marco; M; Mendoza-Vargas A; Moreno-Camacho José Luis; Munguia-Garza P; Ramirez-Vega O; Rangel-DeLeon D; Reyes-Grajeda JP; Rodriguez-Gallegos Jorge |
| EPI_ISL_2295428, EPI_ISL_2295430, EPI_ISL_2295432, EPI_ISL_2295434, EPI_ISL_2295435, EPI_ISL_2295436, EPI_ISL_2295437, EPI_ISL_2295438, EPI_ISL_2295439, EPI_ISL_2295440, EPI_ISL_2295442, EPI_ISL_2295444, EPI_ISL_2295445, EPI_ISL_2295447, EPI_ISL_2295449, EPI_ISL_2295450, EPI_ISL_2295451, EPI_ISL_2295452, EPI_ISL_2378660, EPI_ISL_2378661, EPI_ISL_2378662, EPI_ISL_2378663, EPI_ISL_2378664, EPI_ISL_2378665, EPI_ISL_2378666, EPI_ISL_2378667, EPI_ISL_2378668, EPI_ISL_2378669, EPI_ISL_2378670, EPI_ISL_2378672, EPI_ISL_2378673, EPI_ISL_2378674, EPI_ISL_2378676, EPI_ISL_2378677, EPI_ISL_2378678, EPI_ISL_2378679, EPI_ISL_2378680, EPI_ISL_2378681, EPI_ISL_2378682, EPI_ISL_2378683, EPI_ISL_2378684, EPI_ISL_2382534 | | | |
| see above | Instituto de Biotecnologia - UNESP-Botucatu-SP | Instituto de Biotecnologia - UNESP-Botucatu-SP | Cecília Artico Banho; Cíntia Bittar; Fábio Sossai Possebon; Guilherme Campos; Helena Lage Ferreira; Jorge A. Petroli Marchesi; João Pessoa Araújo Jr.; Leila Sabrina Ullmann; Lívia Sacchetto; Maisa C. Pereira Parra; Marília Moraes; Maurício L. Nogueira; Paula Rahal; Paulo Inacio da Costa |
| EPI_ISL_3104701, EPI_ISL_3104706, EPI_ISL_3104707, EPI_ISL_3104708 | Instituto de Medicina Tropical & Salud Global Universidad Iberoamericana | Grubaugh Lab - Yale School of Public Health | Anderson Brito; Andrea Firpo; Annie Watkins; Chaney Kalinich; Chantal Vogels; David Rivera Aponte; Ernesto Vazquez; Fabiola Cruz-Lopez; Fabiola Fontanet; Isabel Ott; Jessica Rothman; Joseph Fauver; Karolane Gonzalez; Kendall Billig; Mallery Breban; Mary Petrone; Natalie Machargo; Nathan Grubaugh; Patricia Serrano; Tania Mitwalli; Tara Alpert; Tobias Koch; Vianca Aponte |
| EPI_ISL_5147024, EPI_ISL_5147060 | Instituto de Medicina Tropical Alexander Von Humboldt, Sede Cusco | Laboratorio de Genómica Microbiana, Universidad Peruana Cayetano Heredia | Alejandra Dávila-Barclay; Diego Cuicapuza; Fátima Concha; Guillermo Salvatierra; Janet Huancachoque; Luis González; Maria Luisa Morales; Martha Fernández; Miguel Cabada; Pablo Tsukayama; Pedro E. Romero; Pool Marcos |
| EPI_ISL_2714967, EPI_ISL_2714968, EPI_ISL_2714978, EPI_ISL_2715007 | Interpath Laboratory | Seattle Flu Study | Amanda Adler; Barry R. Lutz; Benjamin Pelle; Brian Hiatt; Caitlin R. Wolf; Chris D. Frazar; Deborah A. Nickerson; Elisabeth Brandstetter; Erica Ryke; Geoff Melly; Helen Y. Chu; Janet A. Englund; Jay Shendure; Jover Lee; Kairsten Fay; Kirsten Lacombe; Lea M. Starita; Mark J. Rieder; Matthew Richardson; Matthew Thompson; Melissa Truong; Michael Boeckh; Michael Famulare; Misja Ilcisin; Peter D. Han; Philip Dykema; Romesh Gautom; Scott Lindquist; Thomas R. Sibley; Trevor Bedford |
| EPI_ISL_2282005, EPI_ISL_2282006, EPI_ISL_2282012, EPI_ISL_2282039, EPI_ISL_2282090, EPI_ISL_2357095, EPI_ISL_2357105, EPI_ISL_2357106, EPI_ISL_2357113, EPI_ISL_2357122, EPI_ISL_2357123, EPI_ISL_2357124, EPI_ISL_2357127, EPI_ISL_2357130 | | | |
| see above | Istituto Zooprofilattico Sperimentale del Mezzogiorno | TIGEM | Antonio Grimaldi Patrizia Annunziata Francesco Panariello Biancamaria Pierri Claudia Tiberio Teresa Giuliano Valentina Bouche Chiara Colantuono Maria Concetta Cuomo Denise Di Concilio Lucio Di Filippo Anna Manfredi Marcello Salvi Antonio Limone Luigi Atripaldi Pellegrino Cerino Andrea Ballabio Davide Cacchiarelli |
| EPI_ISL_2615918, EPI_ISL_2615919 | J.W. Ruby Memorial Hospital | WVU and Marshall University Combined Genomics Core Facilities | James Denvir; Peter Perrotta; Peter Stoilov; Ryan Percifield; Wesley Kimble |
| EPI_ISL_2140209, EPI_ISL_2140210 | Jessa | Jessa | Berden et al. on behalf of the Jessa_cmdLab |
| EPI_ISL_2692063, EPI_ISL_2692064, EPI_ISL_2692068 | John H Stroger Jr Hospital of Cook County | RIPHL at Rush University Medical Center | Ah-Young Sung; Felix Araujo Perez; Kevin Kunstman; Laura Furtado; Marieta Hyde; Max Kolton; Sharon Welbel; Stefan Green |
| EPI_ISL_2384209 | KS Health and Environmental Laboratories | Centers for Disease Control and Prevention Division of Viral Diseases, Pathogen Discovery | Alison Laufer Halpin; Ben L. Rambo-Martin; Clinton R. Paden; Dakota Howard; Darlene Wagner; Dave Wentworth; Dhwani Batra; Jasmine Padilla; Justin Lee; Katie Dillon; Krista Queen; Kristen Knipe; Kristine Lacek; Mark Burroughs; Matthew Schmerer; Mili Sheth; Peter Cook; Sam Shepard; Sarah Nobles; Shoshona Le; Suxiang Tong; Vivien Dugan; Yvette Unoarumhi |
| EPI_ISL_2725605, EPI_ISL_2725642, EPI_ISL_2725650, EPI_ISL_2725655, EPI_ISL_2725672, EPI_ISL_2726436, EPI_ISL_2726437, EPI_ISL_2726438, EPI_ISL_2726439, EPI_ISL_2726440, EPI_ISL_2726441, EPI_ISL_2726442, EPI_ISL_2726443, EPI_ISL_2726445, EPI_ISL_2726447, EPI_ISL_2726448, EPI_ISL_2726449, EPI_ISL_2726450, EPI_ISL_2726451, EPI_ISL_2726452, EPI_ISL_2726453, EPI_ISL_2726454, EPI_ISL_2726455, EPI_ISL_2726456, EPI_ISL_2726457, EPI_ISL_2726458, EPI_ISL_2726460, EPI_ISL_2726461, EPI_ISL_2726463, EPI_ISL_2726464, EPI_ISL_2726465, EPI_ISL_2726467, EPI_ISL_2726468, EPI_ISL_2726469, EPI_ISL_2726470, EPI_ISL_2726471, EPI_ISL_2726472, EPI_ISL_2726473, EPI_ISL_2726497, EPI_ISL_2726498, EPI_ISL_2726499, EPI_ISL_2726510, EPI_ISL_2726511, EPI_ISL_2726515, EPI_ISL_2825042, EPI_ISL_2825048, EPI_ISL_2825051, EPI_ISL_2825060, EPI_ISL_2834779 | | | |
| see above | KU Leuven, Rega Institute, Clinical and Epidemiological Virology | KU Leuven, Rega Institute, Clinical and Epidemiological Virology | Bert Vanmechelen; Joan Marti-Carerras; Piet Maes; Tony Wawina-Bokalanga |
| EPI_ISL_3561053, EPI_ISL_3561071, EPI_ISL_3561093, EPI_ISL_3561110, EPI_ISL_3561142, EPI_ISL_3561152, EPI_ISL_3561180, EPI_ISL_3561198, EPI_ISL_3561208, EPI_ISL_3561231, EPI_ISL_3561261, EPI_ISL_3561266, EPI_ISL_3561270, EPI_ISL_3561280, EPI_ISL_3561285, EPI_ISL_3561295, EPI_ISL_3561309, EPI_ISL_3561318, EPI_ISL_3561322, EPI_ISL_3561372, EPI_ISL_3561382, EPI_ISL_3561478, EPI_ISL_3561486, EPI_ISL_3561491, EPI_ISL_3561509 | | | |
| see above | Kaiser Permanente NW Reginal Lab | OHSU MM Lab | Yun Wu |
| EPI_ISL_3504864, EPI_ISL_3504890, EPI_ISL_3504893, EPI_ISL_3504909, EPI_ISL_3504929, EPI_ISL_3504939, EPI_ISL_3504963, EPI_ISL_3504977, EPI_ISL_3505081, EPI_ISL_3505090, EPI_ISL_3505126 | | | |
| see above | Kaiser Permanente NW Reginal Lab | OHSU-MM Lab | Yun Wu |
| EPI_ISL_2092832, EPI_ISL_2092833, EPI_ISL_2101370, EPI_ISL_2101374, EPI_ISL_2156041 | Kansas Health and Environmental Lab | Kansas Health and Environmental Lab | Ben Olsen; Jonathan Barnell; Katherine Wiggins; Mike Grose; and Phil Adam |
| EPI_ISL_3742793, EPI_ISL_4080578, EPI_ISL_4080579, EPI_ISL_4080580, EPI_ISL_4080581, EPI_ISL_4080583, EPI_ISL_4080584, EPI_ISL_4080589, EPI_ISL_4080590 | | | |
| see above | Kentucky State Public Health Lab | Kentucky State Public Health Lab | Anjayya Tummala; Chitralipi Kattunga; Joshua Tobias; Karim George; Matthew Johnson; Rachel Zinner; Rhonda Lucas; Vaneet Arora; William Grooms |
| EPI_ISL_2400396 | Klinisch Laboratorium ZNA | Klinisch Laboratorium ZNA | Verstrepen et al. |
| EPI_ISL_3385805 | LABORATORIO CLINICO HIGUERA ESCALANTE | Instituto Nacional de Salud-Dirección de Investigación en Salud Pública | Carlos Franco-Muñoz; Carmen Osorio; Diana Malo; Diego A. Álvarez-Díaz; Diego Andrés Prada; Gerardo Santamaría; Hector Alejandro Ruiz-Moreno; Jhonnatan Reales-González; Jorge Rivera; Juan Camilo Martinez; Julian Naizaque; Katherine Laiton-Donato; Lisseth Pardo; Magdalena Wiesner; Marcela Mercado-Reyes; Maria T. Herrera-Sepúlveda; Marta Lopez Blanco; Martha Lucia Ospina Martinez; Paola Rojas; Sergio Gomez; Sheryll Corchuelo; Ángela Alarcon Cruz |
| EPI_ISL_2445495, EPI_ISL_2445497, EPI_ISL_2473783 | LABORATORIO DE FRANCA | Instituto Butantan | Antonio Jorge Martins; Claudia Renata dos Santos Barros; David Schlesinger; Debora Botequio Moretti; Dimas Tadeu Covas; Elaine Cristina Marqueze; Elaine Vieira Santos; Evandra Strazza Rodrigues; Heidge Fukumasu; Jayme Augusto de Souza-Neto; José Salvatore Leister Patané; Luiz Alcantara; Luiz Lehmann Coutinho; Maria Carolina Elias; Maurício Lacerda Nogueira; Rafael dos Santos Bezerra; Raul Machado Neto; Rejane Maria Tommasini Grotto; Ricardo Haddad; Sandra Coccuzzo Sampaio Vessoni; Simone Kashima; Svetoslav Nanev Slavov; Vincent Louis Viala |
| EPI_ISL_2445561, EPI_ISL_2445562 | LABORATORIO DR PAULO EMILIO DALESSANDRO PINDAMONHANGABA | Instituto Butantan | Antonio Jorge Martins; Claudia Renata dos Santos Barros; David Schlesinger; Debora Botequio Moretti; Dimas Tadeu Covas; Elaine Cristina Marqueze; Elaine Vieira Santos; Evandra Strazza Rodrigues; Heidge Fukumasu; Jayme Augusto de Souza-Neto; José Salvatore Leister Patané; Luiz Alcantara; Luiz Lehmann Coutinho; Maria Carolina Elias; Maurício Lacerda Nogueira; Rafael dos Santos Bezerra; Raul Machado Neto; Rejane Maria Tommasini Grotto; Ricardo Haddad; Sandra Coccuzzo Sampaio Vessoni; Simone Kashima; Svetoslav Nanev Slavov; Vincent Louis Viala |
| EPI_ISL_2621844, EPI_ISL_2621860, EPI_ISL_2621874 | LABORATORIO ECHAVARRIA | Universidad Nacional de Colombia - Laboratorio Genómico One Health | Andres F. Cardona-Rios; Carlos Franco-Muñoz; Carolina Muñoz-Arango; Celeny Ortiz; Daniel O. Maldonado-Perez; Diego A. Álvarez-Díaz; Hector Alejandro Ruiz-Moreno; Idabely Betancur Ortiz; Jorge E. Osorio; Juan P. Hernandez-Ortiz; Karl A Ciuoderis; Katherine Laiton-Donato; Laura Silvana Perez; Lina M. Hurtado; Marcela Mercado-Reyes; Maria Angélica Maya; Maria Stella López; Rita Almanza Payares; Sandra Ines Cano; Simón Villegas Velásquez |
| EPI_ISL_2445349, EPI_ISL_2473692 | LABORATORIO MUNICIPAL DE ANALISES CLINICAS DE ITANHAEM | Instituto Butantan | Antonio Jorge Martins; Claudia Renata dos Santos Barros; David Schlesinger; Debora Botequio Moretti; Dimas Tadeu Covas; Elaine Cristina Marqueze; Elaine Vieira Santos; Evandra Strazza Rodrigues; Heidge Fukumasu; Jayme Augusto de Souza-Neto; José Salvatore Leister Patané; Luiz Alcantara; Luiz Lehmann Coutinho; Maria Carolina Elias; Maurício Lacerda Nogueira; Rafael dos Santos Bezerra; Raul Machado Neto; Rejane Maria Tommasini Grotto; Ricardo Haddad; Sandra Coccuzzo Sampaio Vessoni; Simone Kashima; Svetoslav Nanev Slavov; Vincent Louis Viala |
| EPI_ISL_2493368 | LABORATORIO MUNICIPAL DE ANALISES CLINICAS DE RIO CLARO | Instituto Butantan | Antonio Jorge Martins; Claudia Renata dos Santos Barros; David Schlesinger; Debora Botequio Moretti; Dimas Tadeu Covas; Elaine Cristina Marqueze; Elaine Vieira Santos; Evandra Strazza Rodrigues; Heidge Fukumasu; Jayme Augusto de Souza-Neto; José Salvatore Leister Patané; Luiz Alcantara; Luiz Lehmann Coutinho; Maria Carolina Elias; Maurício Lacerda Nogueira; Rafael dos Santos Bezerra; Raul Machado Neto; Rejane Maria Tommasini Grotto; Ricardo Haddad; Sandra Coccuzzo Sampaio Vessoni; Simone Kashima; Svetoslav Nanev Slavov; Vincent Louis Viala |
| EPI_ISL_2383033, | LABORATORIO MUNICIPAL DE PIRACICABA | Instituto Butantan | Antonio Jorge Martins; Claudia Renata dos Santos Barros; David Schlesinger; Debora Botequio Moretti; Dimas Tadeu Covas; Elaine Cristina Marqueze; Elaine Vieira Santos; Evandra Strazza Rodrigues; Heidge Fukumasu; Jayme Augusto de Souza-Neto; José Salvatore Leister Patané; Luiz Alcantara; |

| | | | |
|---|---|---|---|
| EPI_ISL_2493453, EPI_ISL_2493454, EPI_ISL_2493455 | | | Luiz Lehmann Coutinho; Maria Carolina Elias; Maurício Lacerda Nogueira; Rafael dos Santos Bezerra; Raul Machado Neto; Rejane Maria Tommasini Grotto; Ricardo Haddad; Sandra Coccuzzo Sampaio Vessoni; Simone Kashima; Svetoslav Nanev Slavov; Vincent Louis Viala |
| EPI_ISL_3316315, EPI_ISL_3316316 | LACEN do Distrito Federal | Instituto Adolfo Lutz, Interdiciplinary Procedures Center, Strategic Laboratory | Caio Vinicius Dias Lopes; Claudia Regina Gonçalves; Claudio Tavares Sacchi; Karoline Rodrigues Campos; Leonardo Tadeu de Araujo; Marlon Benedito Nascomento Santos |
| EPI_ISL_2691209, EPI_ISL_2691210 | LACEN do Estado de Goias | Instituto Adolfo Lutz, Interdiciplinary Procedures Center, Strategic Laboratory | Caio Vinicius Dias Lopes; Claudia Regina Gonçalves; Claudio Tavares Sacchi; Erica Valessa Ramos Gomes; Karoline Rodrigues Campos; Leonardo Jose Tadeu de Araujo |
| EPI_ISL_3316182, EPI_ISL_3316185, EPI_ISL_3316190, EPI_ISL_3316203, EPI_ISL_3316211, EPI_ISL_3316226, EPI_ISL_3316229, EPI_ISL_3316231, EPI_ISL_3316233, EPI_ISL_3316241 | | | |
| see above | LACEN do Estado de Mato Grosso | Instituto Adolfo Lutz, Interdiciplinary Procedures Center, Strategic Laboratory | Caio Vinicius Dias Lopes; Claudia Regina Gonçalves; Claudio Tavares Sacchi; Karoline Rodrigues Campos; Leonardo Tadeu de Araujo; Marlon Benedito Nascomento Santos |
| EPI_ISL_2919207, EPI_ISL_2919211, EPI_ISL_3316155 | LACEN do Estado de Tocantins | Instituto Adolfo Lutz, Interdiciplinary Procedures Center, Strategic Laboratory | Caio Vinicius Dias Lopes; Claudia Regina Gonçalves; Claudio Tavares Sacchi; Erica Valessa Ramos Gomes; Karoline Rodrigues Campos; Leonardo Tadeu de Araujo; Marlon Benedito Nascomento Santos |
| EPI_ISL_3545810, EPI_ISL_3545811, EPI_ISL_3691368, EPI_ISL_3691369, EPI_ISL_3691370 | LACEN do Estado do Mato Grosso do Sul | Instituto Adolfo Lutz, Interdiciplinary Procedures Center, Strategic Laboratory | Caio Vinicius Dias Lopes; Claudia Regina Gonçalves; Claudio Tavares Sacchi; Karoline Rodrigues Campos; Leonardo Tadeu de Araujo; Marlon Benedito Nascomento Santos |
| EPI_ISL_3134654, EPI_ISL_3134655, EPI_ISL_3134656, EPI_ISL_3134657, EPI_ISL_3134658, EPI_ISL_3134659, EPI_ISL_3134660, EPI_ISL_3134661, EPI_ISL_3134662, EPI_ISL_3134663, EPI_ISL_3134838, EPI_ISL_3134840, EPI_ISL_3134842, EPI_ISL_3134843, EPI_ISL_3134844, EPI_ISL_3134845, EPI_ISL_3134846, EPI_ISL_3134847, EPI_ISL_3134848, EPI_ISL_3134849, EPI_ISL_3134850, EPI_ISL_3134851, EPI_ISL_3134852, EPI_ISL_3134857, EPI_ISL_3447435, EPI_ISL_3447591 | | | |
| see above | LACEN/PE | WallauLab on behalf of Fiocruz COVID-19 Genomic Surveillance Network | Alexandre Freitas da Silva; Cassia Docena; Constância Flávia Junqueira Ayres; Filipe Zimmer Dezordi; Gabriel Luz Wallau; Gustavo Barbosa de Lima; Lais Ceschini Machado; Lilian Caroliny Amorim Silva; Marcelo Henrique dos Santos Paiva; Matheus Filgueira Bezerra; Sinval Pinto Brandão Filho |
| EPI_ISL_4487897, EPI_ISL_4487908, EPI_ISL_4487915, EPI_ISL_4487922, EPI_ISL_4487931, EPI_ISL_4487936, EPI_ISL_4487943, EPI_ISL_4487948, EPI_ISL_4487955 | | | |
| see above | LACEN/PI | WallauLab on behalf of Fiocruz COVID-19 Genomic Surveillance Network | Adelino Soares Lima Neto; Alexandre Freitas da Silva; Antônio Marinho; Cassia Docena; Constância Flávia Junqueira Ayres; Filipe Zimmer Dezordi; Gabriel Luz Wallau; Gustavo Barbosa de Lima; Hellen de Oliveira Amaral; Jacenir Reis dos Santos Mallet; Joana Carolina Viana Lima; Lais Ceschini Machado; Leandro de Mattos; Lilian Caroliny Amorim Silva; Marcela de Lacerda Valença Queiroz; Marcelo Adriano da Cunha e Silva Vieira; Marcelo Henrique dos Santos Paiva; Matheus Filgueira Bezerra; Sinval Pinto Brandão Filho; Túlio de Lima Campos; Vladimir Costa Silva; Walterlene de Carvalho Goçalves |
| EPI_ISL_5193460, EPI_ISL_5193564, EPI_ISL_5193568, EPI_ISL_5193569, EPI_ISL_5193585 | LACEN/RR | Laboratorio de Ecologia de Doencas Transmissiveis na Amazonia, Instituto Leonidas e Maria Deane - Fiocruz Amazonia | André Corado; Cátia Alexandra Ribeiro Meneses; Felipe Naveca; Fernanda Nascimento; George Silva; Ilma de Aguiar Antony; Janis Cassimiro Lopes; Luciana Gonçalves; Maria Júlia Brandão; Matilde Mejía; Pricila Carlos Veloso; Rodrigo Melo Maito; Valdinete Nascimento; Victor Souza |
| EPI_ISL_3010766, EPI_ISL_3010771, EPI_ISL_3010779, EPI_ISL_3010789, EPI_ISL_3010797, EPI_ISL_3010804, EPI_ISL_3010811, EPI_ISL_3010812, EPI_ISL_3010822, EPI_ISL_3010852, EPI_ISL_3010858, EPI_ISL_3010873, EPI_ISL_3010882 | | | |
| see above | LATE - Laboratório de Técnicas Especiais - Hospital Israelita Albert Einstein | LATE - Laboratório de Técnicas Especiais - Hospital Israelita Albert Einstein | Alexandre Hideaki Takara; Ana Paula Moreira Salles; Anelisie da Silva Santos; Deyvid Amgarten; Erick Gustavo Dorlass; Fernanda de Mello Malta; João Renato Rebello Pinho; Marcio Anunciacao Menezes; Pedro Henrique Sebe Rodrigues; Raquel Riyuzo |
| EPI_ISL_2332620 | LBM ALPHABIO, Marseille | LBM ALPHABIO, Marseille | Vincent GARCIA |
| EPI_ISL_2476412, EPI_ISL_2476423, EPI_ISL_2476428, EPI_ISL_2476429, EPI_ISL_2545815 | LESP Ciudad de Mexico | Instituto de Diagnostico y Referencia Epidemiologicos (INDRE) | Abril Rodriguez-Maldonado; Ariadna Medina-Benitez; Claudia Wong-Arambula; Ernesto Ramirez-Gonzalez.; Gisela Barrera-Badillo; Irma Lopez-Martinez; Joaquin Quiroz-Mercado; Lucia Hernandez-Rivas; Natividad Cruz-Ortiz; Sergio Rangel-Guerrero; Tatiana Nunez-Garcia; Vanessa Rivero-Arredondo |
| EPI_ISL_2476438, EPI_ISL_2479910 | LESP Colima | Instituto de Diagnostico y Referencia Epidemiologicos (INDRE) | Abril Rodriguez-Maldonado; Ariadna Medina-Benitez; Claudia Wong-Arambula; Ernesto Ramirez-Gonzalez.; Gisela Barrera-Badillo; Irma Lopez-Martinez; Joaquin Quiroz-Mercado; Lucia Hernandez-Rivas; Natividad Cruz-Ortiz; Sergio Rangel-Guerrero; Tatiana Nunez-Garcia; Vanessa Rivero-Arredondo |
| EPI_ISL_2296073 | LESP Estado de Mexico | Instituto de Diagnostico y Referencia Epidemiologicos (INDRE) | Abril Rodriguez-Maldonado; Ariadna Medina-Benitez; Claudia Wong-Arambula; Ernesto Ramirez-Gonzalez.; Gisela Barrera-Badillo; Irma Lopez-Martinez; Joaquin Quiroz-Mercado; Lucia Hernandez-Rivas; Natividad Cruz-Ortiz; Sergio Rangel-Guerrero; Tatiana Nunez-Garcia; Vanessa Rivero-Arredondo |
| EPI_ISL_2476418, EPI_ISL_2479899 | LESP Guanajuato | Instituto de Diagnostico y Referencia Epidemiologicos (INDRE) | Abril Rodriguez-Maldonado; Ariadna Medina-Benitez; Claudia Wong-Arambula; Ernesto Ramirez-Gonzalez.; Gisela Barrera-Badillo; Irma Lopez-Martinez; Joaquin Quiroz-Mercado; Lucia Hernandez-Rivas; Natividad Cruz-Ortiz; Sergio Rangel-Guerrero; Tatiana Nunez-Garcia; Vanessa Rivero-Arredondo |
| EPI_ISL_2644834 | LESP HIDALGO | LESP HIDALGO GENE2LIFE | CASTILLO-ORTEGA LAURA SOFIA; GARCIA-GONZALEZ OCTAVIO; GARCIA-REYES MELITO; HERRERA-GOMEZ CONNIE ALEJANDRA; VIGUERAS-MORALES YAJAIRA SULIM; VILLAFUENTES-TELLEZ HECTOR ALBERTO |
| EPI_ISL_2479912 | LESP Hidalgo | Instituto de Diagnostico y Referencia Epidemiologicos (INDRE) | Abril Rodriguez-Maldonado; Ariadna Medina-Benitez; Claudia Wong-Arambula; Ernesto Ramirez-Gonzalez.; Gisela Barrera-Badillo; Irma Lopez-Martinez; Joaquin Quiroz-Mercado; Lucia Hernandez-Rivas; Natividad Cruz-Ortiz; Sergio Rangel-Guerrero; Tatiana Nunez-Garcia; Vanessa Rivero-Arredondo |
| EPI_ISL_2479880 | LESP Queretaro | Instituto de Diagnostico y Referencia Epidemiologicos (INDRE) | Abril Rodriguez-Maldonado; Ariadna Medina-Benitez; Claudia Wong-Arambula; Ernesto Ramirez-Gonzalez.; Gisela Barrera-Badillo; Irma Lopez-Martinez; Joaquin Quiroz-Mercado; Lucia Hernandez-Rivas; Natividad Cruz-Ortiz; Sergio Rangel-Guerrero; Tatiana Nunez-Garcia; Vanessa Rivero-Arredondo |
| EPI_ISL_2479889, EPI_ISL_2479890, EPI_ISL_2479915, EPI_ISL_2859053 | LESP Quintana Roo | Instituto de Diagnostico y Referencia Epidemiologicos (INDRE) | Abril Rodriguez-Maldonado; Ariadna Medina-Benitez; Claudia Wong-Arambula; Ernesto Ramirez-Gonzalez.; Gisela Barrera-Badillo; Irma Lopez-Martinez; Joaquin Quiroz-Mercado; Lucia Hernandez-Rivas; Maribel Gonzalez-Villa; Natividad Cruz-Ortiz; Sergio Rangel-Guerrero; Tatiana Nunez-Garcia; Vanessa Rivero-Arredondo |
| EPI_ISL_2545813, EPI_ISL_2545814, EPI_ISL_2545819 | LESP San Luis Potosi | Instituto de Diagnostico y Referencia Epidemiologicos (INDRE) | Abril Rodriguez-Maldonado; Ariadna Medina-Benitez; Claudia Wong-Arambula; Ernesto Ramirez-Gonzalez.; Gisela Barrera-Badillo; Irma Lopez-Martinez; Joaquin Quiroz-Mercado; Lucia Hernandez-Rivas; Natividad Cruz-Ortiz; Sergio Rangel-Guerrero; Tatiana Nunez-Garcia; Vanessa Rivero-Arredondo |
| EPI_ISL_2476432, EPI_ISL_2476436 | LESP Tamaulipas | Instituto de Diagnostico y Referencia Epidemiologicos (INDRE) | Abril Rodriguez-Maldonado; Ariadna Medina-Benitez; Claudia Wong-Arambula; Ernesto Ramirez-Gonzalez.; Gisela Barrera-Badillo; Irma Lopez-Martinez; Joaquin Quiroz-Mercado; Lucia Hernandez-Rivas; Natividad Cruz-Ortiz; Sergio Rangel-Guerrero; Tatiana Nunez-Garcia; Vanessa Rivero-Arredondo |
| EPI_ISL_2479886, EPI_ISL_2545733, EPI_ISL_2545734 | LESP Yucatan | Instituto de Diagnostico y Referencia Epidemiologicos (INDRE) | Abril Rodriguez-Maldonado; Ariadna Medina-Benitez; Claudia Wong-Arambula; Ernesto Ramirez-Gonzalez.; Gisela Barrera-Badillo; Irma Lopez-Martinez; Joaquin Quiroz-Mercado; Lucia Hernandez-Rivas; Natividad Cruz-Ortiz; Sergio Rangel-Guerrero; Tatiana Nunez-Garcia; Vanessa Rivero-Arredondo |
| EPI_ISL_2331861, EPI_ISL_2358236, EPI_ISL_2358237 | LHUB-ULB | Labo Klinische Biologie, UZA | Basil Britto Xavier; Christine Lammens; Herman Goossens; Jasmine Coppens; Marie Le Mercier; Veerle Matheeussen |
| EPI_ISL_4878288 | Lab Ayudas Diagnosticas SURA- sede Cali | Instituto Nacional de Salud- Dirección de Investigación en Salud Pública | Carlos Franco-Muñoz; Carmen Osorio; Diana Malo; Diego A. Álvarez-Díaz; Diego Andrés Prada; Gerardo Santamaría; Hector Alejandro Ruiz-Moreno; Jhonnatan Reales-González; Jorge Rivera; Juan Camilo Martinez; Julian Naizaque; Katherine Laiton-Donato; Lisseth Pardo; Magdalena Wiesner; Marcela Mercado-Reyes; Maria T. Herrera-Sepúlveda; Marta Lopez Blanco; Martha Lucia Ospina Martinez; Paola Rojas; Sergio Gomez; Sheryll Corchuelo; Ángela Alarcon Cruz |
| EPI_ISL_2362793, EPI_ISL_2362862, EPI_ISL_2362868, EPI_ISL_2362905, EPI_ISL_2362910 | Lab voor klinische biologie | Lab voor klinische biologie | Bruno Verhasselt; Hannelore Hamerlinck; Marija Janevska |
| EPI_ISL_2275868, EPI_ISL_2275869, EPI_ISL_2275871, EPI_ISL_2275872 | Lab. Microbiologia e Virologia Cotugno A.O. dei Colli - Istituto Zooprofilattico Sperimentale del Mezzogiorno | TIGEM | Antonio Grimaldi Patrizia Annunziata Francesco Panariello Biancamaria Pierri Claudia Tiberio Teresa Giuliano Valentina Bouche Chiara Colantuono Maria Concetta Cuomo Denise Di Concilio Lucio Di Filippo Anna Manfredi Marcello Salvi Antonio Limone Luigi Atripaldi Pellegrino Cerino Andrea Ballabio Davide Cacchiarelli |
| EPI_ISL_2374425 | Labo Analyses Med | National Reference Center for Viruses of Respiratory Infections, Institut Pasteur, Paris | Angela Brisebarre; Benedicte Baccouch; Camille Capel; Christophe Malabat; Corinne Maufrais; Emmanuelle Permal; Etienne Simon-Lorière; Frédéric Lemoine; Louise Lefrançois; Marion Barbet; Maud Vanpeene; Méline Bizard; Sylvie Behillil; Sylvie Van der Werf; Vincent Enouf |
| EPI_ISL_2635528, EPI_ISL_2635534, EPI_ISL_2635545 | Labor Becker & Kollegen (Standort München) | Robert Koch Institute | |
| EPI_ISL_2227904, EPI_ISL_2227905, EPI_ISL_2227907, EPI_ISL_2362533, EPI_ISL_2362534, EPI_ISL_2362536, EPI_ISL_2362537, EPI_ISL_2362538, EPI_ISL_2426980 | | | |
| see above | Laboratoire Carage | Institut Pasteur de la Guyane | A. Enfissi; A. Salmier; Anne Lavergne; Antoine Enfissi; Arielle Salmier; Dominique Rousset |
| EPI_ISL_2228963 | Laboratoire LENYS | Department of Virology, Henri Mondor University Hospital, Assistance Publique Hôpitaux de Paris, Université Paris-Est Créteil, | Alexandre Soulier; Christophe Rodriguez; Elisabeth Trawinski; Guillaume Gricourt; Jean-Michel Pawlotsky; Melissa N'Debi; Slim Fourati; Vanessa Demontant |

| | | | |
|---|---|---|---|
| | INSERM U955 | | |
| EPI_ISL_2492499, EPI_ISL_2492500, EPI_ISL_2492501, EPI_ISL_2492502, EPI_ISL_2492503, EPI_ISL_2492504, EPI_ISL_2492505, EPI_ISL_2492512, EPI_ISL_2492546 | | | |
| see above | Laboratoire National de Santé Publique – LNSP (HAITI - LNSP) | Laboratory of Respiratory Viruses and Measles, Oswaldo Cruz Institute, FIOCRUZ | Alice Sampaio Rocha; Ana Carolina Mendonca; Anna Carolina Paixao; Elisa Cavalcante Pereira; Fernando Motta; Ito Journel; Jaques Boncy; Luciana Appolinario; Marilda Siqueira on behalf of the Fiocruz COVID-19 Genomic Surveillance Network; Paola Resende; Patrick Delly; Renata Serrano Lopes; Taina Venas |
| EPI_ISL_3021517, EPI_ISL_3458630, EPI_ISL_3458647, EPI_ISL_3458648, EPI_ISL_3458649, EPI_ISL_5867801, EPI_ISL_5867802, EPI_ISL_5873460 | | | |
| see above | Laboratoire de santé publique du Québec | Laboratoire de santé publique du Québec | Guillaume Bourque; Ioannis Ragoussis; Jesse Shapiro; Mark Lathrop and Michel Roger on behalf of the CoVSeQ research group; Sandrine Moreira |
| EPI_ISL_2401128, EPI_ISL_2401143, EPI_ISL_2401150 | Laboratoire national de sante, Microbiology, Virology | Laboratoire national de sante, Microbiology, Microbial Genomics Platform | Anke Wienecke-Baldacchino; Catherine Ragimbeau; Fatu Djabi; Jessica Tapp; Lise Pignon; Raoul Salmon; Tamir Abdelrahman; Trung Nguyen Nguyen |
| EPI_ISL_2400913, EPI_ISL_2400928, EPI_ISL_2400929, EPI_ISL_2400933 | Laboratoires Reunis | Laboratoire national de sante, Microbiology, Microbial Genomics Platform | Anke Wienecke-Baldacchino; Bernard Weber; Catherine Ragimbeau; Fatu Djabi; Jessica Tapp; Lise Pignon; Raoul Salmon; Tamir Abdelrahman |
| EPI_ISL_2400750, EPI_ISL_2400775 | Laboratoires d'analyses medicales - Ketterthill | Laboratoire national de sante, Microbiology, Microbial Genomics Platform | Anke Wienecke-Baldacchino; Caroline Scheiber; Catherine Ragimbeau; Fatu Djabi; Jessica Tapp; Lise Pignon; Raoul Salmon; Serge Vedy; Tamir Abdelrahman |
| EPI_ISL_2497876, EPI_ISL_2497877, EPI_ISL_2626433 | Laboratori de Referencia de Catalunya | Laboratori de Referencia de Catalunya | Bellosillo B.; Canal M.; Hernandez JJ.; Padilla E.; Ramirez A.; Vilas A. |
| EPI_ISL_2645421, EPI_ISL_2645423, EPI_ISL_2645425, EPI_ISL_2645426, EPI_ISL_2645428, EPI_ISL_2645431, EPI_ISL_2645432, EPI_ISL_2645433, EPI_ISL_2645437, EPI_ISL_2645439 | | | |
| see above | Laboratorio Central de Saude Publica do Estado do Para (LACEN/PA) | Laboratory of Respiratory Viruses and Measles, Oswaldo Cruz Institute, FIOCRUZ | Alice Sampaio Rocha; Ana Carolina Mendonca; Anna Carolina Paixao; Elisa Cavalcante Pereira; Fernando Motta; Luciana Appolinario; Marilda Siqueira on behalf of the Fiocruz COVID-19 Genomic Surveillance Network; Paola Resende; Renata Serrano Lopes; Taina Venas; Valnete Andrade |
| EPI_ISL_2308719 | Laboratorio Analisi Osp. Città di Castello - Azienda USL Umbria10 | Istituto Zooprofilattico Sperimentale dell'Abruzzo e Molise "G. Caporale" | Ancora M; Calistri P; Cammà C; Curini V; Delli Compagni E; Di Domenico M; Di Pasquale A; Lorusso A; Malagigi V; Mangone I; Marcacci M; Puglia I; Rinaldi A; Savini G; Scialabba S; Tacconi P |
| EPI_ISL_2308720 | Laboratorio Analisi Osp. Città di Castello - Azienda USL Umbria11 | Istituto Zooprofilattico Sperimentale dell'Abruzzo e Molise "G. Caporale" | Ancora M; Calistri P; Cammà C; Curini V; Delli Compagni E; Di Domenico M; Di Pasquale A; Lorusso A; Malagigi V; Mangone I; Marcacci M; Puglia I; Rinaldi A; Savini G; Scialabba S; Tacconi P |
| EPI_ISL_2308713 | Laboratorio Analisi Osp. Città di Castello - Azienda USL Umbria4 | Istituto Zooprofilattico Sperimentale dell'Abruzzo e Molise "G. Caporale" | Ancora M; Calistri P; Cammà C; Curini V; Delli Compagni E; Di Domenico M; Di Pasquale A; Lorusso A; Malagigi V; Mangone I; Marcacci M; Puglia I; Rinaldi A; Savini G; Scialabba S; Tacconi P |
| EPI_ISL_2308714 | Laboratorio Analisi Osp. Città di Castello - Azienda USL Umbria5 | Istituto Zooprofilattico Sperimentale dell'Abruzzo e Molise "G. Caporale" | Ancora M; Calistri P; Cammà C; Curini V; Delli Compagni E; Di Domenico M; Di Pasquale A; Lorusso A; Malagigi V; Mangone I; Marcacci M; Puglia I; Rinaldi A; Savini G; Scialabba S; Tacconi P |
| EPI_ISL_2308715 | Laboratorio Analisi Osp. Città di Castello - Azienda USL Umbria6 | Istituto Zooprofilattico Sperimentale dell'Abruzzo e Molise "G. Caporale" | Ancora M; Calistri P; Cammà C; Curini V; Delli Compagni E; Di Domenico M; Di Pasquale A; Lorusso A; Malagigi V; Mangone I; Marcacci M; Puglia I; Rinaldi A; Savini G; Scialabba S; Tacconi P |
| EPI_ISL_2308716 | Laboratorio Analisi Osp. Città di Castello - Azienda USL Umbria7 | Istituto Zooprofilattico Sperimentale dell'Abruzzo e Molise "G. Caporale" | Ancora M; Calistri P; Cammà C; Curini V; Delli Compagni E; Di Domenico M; Di Pasquale A; Lorusso A; Malagigi V; Mangone I; Marcacci M; Puglia I; Rinaldi A; Savini G; Scialabba S; Tacconi P |
| EPI_ISL_2308717 | Laboratorio Analisi Osp. Città di Castello - Azienda USL Umbria8 | Istituto Zooprofilattico Sperimentale dell'Abruzzo e Molise "G. Caporale" | Ancora M; Calistri P; Cammà C; Curini V; Delli Compagni E; Di Domenico M; Di Pasquale A; Lorusso A; Malagigi V; Mangone I; Marcacci M; Puglia I; Rinaldi A; Savini G; Scialabba S; Tacconi P |
| EPI_ISL_2308718 | Laboratorio Analisi Osp. Città di Castello - Azienda USL Umbria9 | Istituto Zooprofilattico Sperimentale dell'Abruzzo e Molise "G. Caporale" | Ancora M; Calistri P; Cammà C; Curini V; Delli Compagni E; Di Domenico M; Di Pasquale A; Lorusso A; Malagigi V; Mangone I; Marcacci M; Puglia I; Rinaldi A; Savini G; Scialabba S; Tacconi P |
| EPI_ISL_5199353 | Laboratorio Angel ( Synlab) | Instituto Nacional de Salud- Dirección de Investigación en Salud Pública | Beatriz de Arco; Carlos Franco-Muñoz; Carmen Osorio; Diana Malo; Diego A. Álvarez-Díaz; Diego Andrés Prada; Gerardo Santamaría; Hector Alejandro Ruiz-Moreno; Jhonnatan Reales-González; Jorge Rivera; Juan Camilo Martinez; Julian Naizaque; Katherine Laiton-Donato; Lisseth Pardo; Magdalena Wiesner; Marcela Mercado-Reyes; Maria T. Herrera-Sepúlveda; Marta Lopez Blanco; Martha Lucia Ospina Martinez; Paola Rojas; Sergio Gomez; Sheryll Corchuelo; Tatiana Cobos; Ángela Alarcon Cruz |
| EPI_ISL_2340093, EPI_ISL_2340119, EPI_ISL_2340125, EPI_ISL_2340130, EPI_ISL_2340131, EPI_ISL_2340132, EPI_ISL_2340133, EPI_ISL_2340134 | | | |
| see above | Laboratorio Aziendale di Microbiologia e Virologia, Azienda Sanitaria dell'Alto Adige | Laboratorio Aziendale di Microbiologia e Virologia, Azienda Sanitaria dell'Alto Adige | Anne Picard; Chiara Cantaloni; Claudia Volpato; Elisa Masi; Elisabetta Giacobazzi; Elisabetta Pagani; Irene Bianconi; Stefanie Wieser |
| EPI_ISL_2534862, EPI_ISL_2534863, EPI_ISL_2534864, EPI_ISL_2534865, EPI_ISL_2534866, EPI_ISL_2534867, EPI_ISL_2534868, EPI_ISL_2534869, EPI_ISL_2534870, EPI_ISL_2534871, EPI_ISL_2534872, EPI_ISL_2534873, EPI_ISL_2534874, EPI_ISL_2534875, EPI_ISL_2534876, EPI_ISL_2534877, EPI_ISL_2534878, EPI_ISL_2534879, EPI_ISL_2534880, EPI_ISL_2534882, EPI_ISL_2534883, EPI_ISL_2534884, EPI_ISL_2534885, EPI_ISL_2534886, EPI_ISL_2534887, EPI_ISL_2534888, EPI_ISL_2534889, EPI_ISL_2534890, EPI_ISL_2534891, EPI_ISL_2534892, EPI_ISL_2534893, EPI_ISL_2534894, EPI_ISL_2534895, EPI_ISL_2534896, EPI_ISL_2534897, EPI_ISL_2534898, EPI_ISL_2534899, EPI_ISL_2534900, EPI_ISL_2534901, EPI_ISL_2534902, EPI_ISL_2534903, EPI_ISL_2534904, EPI_ISL_2534905, EPI_ISL_2534906, EPI_ISL_2534907, EPI_ISL_2534908, EPI_ISL_2534909, EPI_ISL_2534910, EPI_ISL_2534911, EPI_ISL_2534912, EPI_ISL_2534913, EPI_ISL_2534914, EPI_ISL_2534915, EPI_ISL_2534916, EPI_ISL_2534917, EPI_ISL_2534918, EPI_ISL_2534919, EPI_ISL_2534920, EPI_ISL_2534921, EPI_ISL_2534922, EPI_ISL_2534923, EPI_ISL_2534925, EPI_ISL_2534926, EPI_ISL_2534927, EPI_ISL_2534928, EPI_ISL_2534929, EPI_ISL_2534930, EPI_ISL_2534931, EPI_ISL_2534932, EPI_ISL_2534933, EPI_ISL_2534934, EPI_ISL_2534935, EPI_ISL_2534936, EPI_ISL_2534937, EPI_ISL_2534938, EPI_ISL_2534939, EPI_ISL_2534940, EPI_ISL_2534941, EPI_ISL_2534942, EPI_ISL_2534944, EPI_ISL_2534945, EPI_ISL_2534946, EPI_ISL_2534947, EPI_ISL_2534948, EPI_ISL_2534949, EPI_ISL_2534950, EPI_ISL_2534951, EPI_ISL_2534952, EPI_ISL_2534953, EPI_ISL_2534954, EPI_ISL_2534955, EPI_ISL_2534956, EPI_ISL_2534957, EPI_ISL_2534958, EPI_ISL_2534959, EPI_ISL_2534960, EPI_ISL_2534961, EPI_ISL_2534962, EPI_ISL_2534963, EPI_ISL_2534964, EPI_ISL_2534965, EPI_ISL_2534966, EPI_ISL_2534967, EPI_ISL_2534968, EPI_ISL_2534969, EPI_ISL_2534970, EPI_ISL_2534971, EPI_ISL_2534972, EPI_ISL_2534973, EPI_ISL_2534974, EPI_ISL_2534975, EPI_ISL_2534976, EPI_ISL_2534977, EPI_ISL_2534978, EPI_ISL_2534979, EPI_ISL_2534980, EPI_ISL_2534981, EPI_ISL_2534982, EPI_ISL_2534983, EPI_ISL_2534984, EPI_ISL_2534985, EPI_ISL_2534986, EPI_ISL_2534987, EPI_ISL_2534988, EPI_ISL_2534989, EPI_ISL_2534990, EPI_ISL_2534991, EPI_ISL_2534992, EPI_ISL_2534993, EPI_ISL_2534994, EPI_ISL_2534995, EPI_ISL_2534996, EPI_ISL_2534997, EPI_ISL_2534998, EPI_ISL_2534999, EPI_ISL_2535000, EPI_ISL_2535001, EPI_ISL_2535002, EPI_ISL_2535003, EPI_ISL_2535004, EPI_ISL_2535005, EPI_ISL_2535006, EPI_ISL_2535007, EPI_ISL_2535008, EPI_ISL_2535009, EPI_ISL_2535010, EPI_ISL_2535011, EPI_ISL_2535012, EPI_ISL_2535013, EPI_ISL_2535014, EPI_ISL_2535015, EPI_ISL_2535016, EPI_ISL_2535017, EPI_ISL_2535019, EPI_ISL_2535021, EPI_ISL_2535022, EPI_ISL_2535023, EPI_ISL_2535024, EPI_ISL_2535025, EPI_ISL_2535048, EPI_ISL_2535049, EPI_ISL_2535053, EPI_ISL_2535054, EPI_ISL_2535055, EPI_ISL_2535058, EPI_ISL_2535059, EPI_ISL_2535060, EPI_ISL_2535061, EPI_ISL_2535062, EPI_ISL_2535063, EPI_ISL_2535064, EPI_ISL_2535065, EPI_ISL_2535066, EPI_ISL_2535067, EPI_ISL_2535068, EPI_ISL_2535069, EPI_ISL_2535070 | | | |
| see above | Laboratorio Central Noel Nutels | Bioinformatics Laboratory / LNCC | Alessandra P Lamarca; Alexandra L Gerber; Amilcar Tanuri; Ana Paula de C Guimaraes; Ana Tereza R Vasconcelos; Andrea Cony Cavalcanti; Caio Luiz Pereira Ribeiro; Cassia Alves; Cintia Policarpo; Claudia Maria Braga de Mello; Cristiane Gomes da Silva; Diana Mariani; Douglas Terra Machado; Flavio Dias da Silva; Gleidson da Silva de Oliveira; Leandro Magalhaes de Souza; Liliane Cavalcante; Luiz G P de Almeida; Marcio Henrique de Oliveira Garcia; Mario Sergio Ribeiro; Ronaldo da Silva F Jr; Silvia Carvalho |
| EPI_ISL_2671728, EPI_ISL_2671729 | Laboratorio Central de Epidemiologia (LCE) | Unidad de Genomica Avanzada | ; Alejandra Garcia-Gasca; Alejandra Hernandez-Teran; Alejandro Sanchez-Flores; Alfredo Herrera-Estrella; Alicia Ocaña-Mondragon; Andreu Comas-Garcia; Angel Gustavo Salas-Lais; Antonio Loza Roman; Bernardo Martinez-Miguel; Blanca Taboada; Brenda Irasema Maldonado-Meza; Bruno Gomez-Gil; Carla Ivon Herrera-Najera; Carlos F. Arias; Celia Boukadida; Celida Duque Molina; Celida Martinez- Rodriguez; Clara Esperanza Santacruz-Tinoco; Concepcion Grajales-Muñiz; Consorcio Mexicano de Vigilancia Genomica (CoViGen-Mex). Authors (in alphabetical order): Julio Elias Alvarado-Yaah; Cristobal Chaidez-Quiroz; Daniel Fregoso-Rueda; Daniel Lira Morales; Eduardo Becerril-Vargas; Fernando Fontove-Herrera; Fidencio Mejia-Nepomuceno; Francisco Pulido; Gloria Elena Espinosa-Ayala; Gloria Maria Molina-Salinas; Gloria Vazquez; Hector Esteban Paz-Juarez; Hector Montoya-Fuentes; Helen Haydee Fernanda Ramirez-Plascencia; Irvin Gonzalez-Lopez; Jean Pierre Gonzalez; Jesus Hernandez; Joel Armando Vazquez-Perez.; Jorge Salas-Hernandez; Jose Antonio Enciso-Moreno; Jose Arturo Martinez-Orozco; Jose Esteban Muñoz-Medina; Jose de Jesus Nuñez-Contreras; Juan Bautista Chale-Dzul; Julissa Enciso-Ibarra; Luis Alberto Ochoa-Carrera; Margarita Matias-Florentino; Maria Guadalupe Santiago-Mauricio; Maria Guadalupe de Jesus Mireles-Rivera; Mario Mujica-Sanchez; Marissa Perez-Garcia; Nelly Selem-Mojica; Pavel Isa; Ricardo Ciria Merce; Ricardo Grande; Rosa Maria Gutierrez Rios; Santiago avila-Rios; Selene Zarate; Susana Lopez; Veronica Mata-Haro; Victor Eduardo Garcia-Arias; Victor Hugo Borja-Aburto |
| EPI_ISL_2490388, EPI_ISL_2490390, EPI_ISL_2490391, EPI_ISL_2490392 | Laboratorio Central de Epidemiología (LCE) | Centro de Investigación en Enfermedades Infecciosas (CIENI), Instituto Nacional de Enfermedades Respiratorias (INER) | ; Alejandra Garcia-Gasca; Alejandra Hernández-Terán; Alejandro Sanchez-Flores; Alfredo Herrera-Estrella; Alicia Ocaña-Mondragón; Andreu Comas-García; Angel Gustavo Salas-Lais; Antonio Loza Román; Bernardo Martínez-Miguel; Blanca Taboada; Brenda Irasema Maldonado-Meza; Bruno Gomez-Gil; Carla Ivón Herrera-Najera; Carlos F. Arias; Celia Boukadida; Clara Esperanza Santacruz-Tinoco; Concepción Grajales-Muñiz; Consorcio Mexicano de Vigilancia Genómica (CoViGen-Mex). Authors (in alphabetical order): Julio Elias Alvarado-Yaah; Cristóbal Cháidez-Quiróz; Célida Duque Molina; Célida Martínez- Rodríguez; Daniel Fregoso-Rueda; Daniel Lira Morales; Eduardo Becerril-Vargas; Fernando Fontove-Herrera; Fidencio Mejía-Nepomuceno; Francisco Pulido; Gloria Elena Espinosa-Ayala; Gloria María Molina-Salinas; Gloria Vazquez; Hector Esteban Paz-Juárez; Hector Montoya-Fuentes; Helen Haydee Fernanda Ramírez-Plascencia; Irvin González-López; Jean Pierre González; Joel Armando Vázquez-Pérez.; Jorge Salas-Hernández; José Antonio Enciso-Moreno; José Arturo Martínez-Orozco; José Esteban Muñoz-Medina; José de Jesús Nuñez-Contreras; Juan Bautista Chale-Dzul; Julissa Enciso-Ibarra; Luis Alberto Ochoa-Carrera; Margarita Matías-Florentino; Mario Mújica-Sánchez; Marissa Perez-Garcia; María Guadalupe Santiago-Mauricio; María Guadalupe de Jesús Mireles-Rivera; Nelly Sélem-Mojica; Pavel Isa; Ricardo Ciria Merce; Ricardo Grande; Rosa María Gutierrez Ríos; Santiago Ávila-Ríos; Selene Zárate; Susana Lopez; Victor Eduardo Garcia-Arias; Víctor Hugo Borja-Aburto |
| EPI_ISL_6944501, EPI_ISL_6944508, EPI_ISL_6944516, EPI_ISL_6944517, EPI_ISL_6944530, EPI_ISL_6944531, EPI_ISL_6944533, EPI_ISL_6944541, EPI_ISL_6944542 | | | |
| see above | Laboratorio Central de Salud Publica | Viral Special Pathogens Branch, Centers for Disease Control and Prevention | Andrea Gómez de la Fuente; Cynthia Vázquez; Emir Talundzic; Joel Montgomery; John Klena; Juan Torales; Justin Lee; María José Ortega; María Liz Gamarra; Shannon Whitmer; Shirley Villalba |
| EPI_ISL_2274072, EPI_ISL_2274075 | Laboratorio Central de Saude Publica do Estado da Bahia (LACEN/BA) | Laboratory of Respiratory Viruses and Measles, Oswaldo Cruz Institute, FIOCRUZ | Alice Sampaio Rocha; Ana Carolina Mendonca; Anna Carolina Paixao; Elisa Cavalcante Pereira; Felicidade Pereira; Fernando Motta; Luciana Appolinario; Marilda Siqueira on behalf of the Fiocruz COVID-19 Genomic Surveillance Network; Paola Resende; Renata Serrano Lopes; Taina Venas |
| EPI_ISL_2645384, EPI_ISL_2645389, EPI_ISL_2645390, EPI_ISL_2645391, EPI_ISL_2645407 | Laboratorio Central de Saude Publica do Estado de Alagoas (LACEN/AL) | Laboratory of Respiratory Viruses and Measles, Oswaldo Cruz Institute, FIOCRUZ | Alice Sampaio Rocha; Ana Carolina Mendonca; Anderson Brandao Leite; Anna Carolina Paixao; Elisa Cavalcante Pereira; Fernando Motta; Luciana Appolinario; Marilda Siqueira on behalf of the Fiocruz COVID-19 Genomic Surveillance Network; Paola Resende; Renata Serrano Lopes; Taina Venas |
| EPI_ISL_3832439, EPI_ISL_3832440, EPI_ISL_3832441, EPI_ISL_3832442, EPI_ISL_3832468 | Laboratorio Central de Saude Publica do Estado de Santa Catarina (LACEN/SC) | Laboratory of Respiratory Viruses and Measles, Oswaldo Cruz Institute, FIOCRUZ | Alice Sampaio Rocha; Ana Carolina Mendonca; Anna Carolina Paixao; Darcita Buerger Rovaris; Elisa Cavalcante Pereira; Fernando Motta; Luciana Appolinario; Marilda Siqueira on behalf of the Fiocruz COVID-19 Genomic Surveillance Network; Paola Resende; Renata Serrano Lopes; Sandra Bianchini Fernandes; Taina Venas |
| EPI_ISL_2863618 | Laboratorio Central de Saude Publica do Estado de Sergipe (LACEN/SE) | Laboratory of Respiratory Viruses and Measles, Oswaldo Cruz Institute, FIOCRUZ | Alice Sampaio Rocha; Ana Carolina Mendonca; Anna Carolina Paixao; Cliomar Alves dos Santos; Elisa Cavalcante Pereira; Fernando Motta; Luciana Appolinario; Marilda Siqueira on behalf of the Fiocruz COVID-19 Genomic Surveillance Network; Paola Resende; Renata Serrano Lopes; Tainá Moreira Martins Venas |
| EPI_ISL_2983240, EPI_ISL_2983241, EPI_ISL_2983300, EPI_ISL_2983326, EPI_ISL_2983327 | Laboratorio Central de Saude Publica do Estado do Maranhao (LACEN-MA) | Laboratory of Respiratory Viruses and Measles, Oswaldo Cruz Institute, FIOCRUZ | Agatha Cristinne Prudencio; Alice Sampaio Rocha; Ana Carolina Mendonca; Anna Carolina Paixao; Elisa Cavalcante Pereira; Fernando Motta; Ighor Leonardo Arantes Gomes; Lidio Gonçalves Lima Neto; Luciana Appolinario; Marilda Siqueira on behalf of the Fiocruz COVID-19 Genomic Surveillance Network; Paola Resende; Renata Serrano Lopes; Taina Moreira Venas |

| | | | |
|---|---|---|---|
| EPI_ISL_2645458, EPI_ISL_2645461, EPI_ISL_2645463, EPI_ISL_2645465, EPI_ISL_2645470, EPI_ISL_2645472, EPI_ISL_2645491, EPI_ISL_2645492, EPI_ISL_2645498, EPI_ISL_2983447, EPI_ISL_2983450, EPI_ISL_4061298, EPI_ISL_4061299, EPI_ISL_4061301, EPI_ISL_4061308, EPI_ISL_4061312, EPI_ISL_4061314, EPI_ISL_4061321, EPI_ISL_4061322 | | | |
| see above | Laboratorio Central de Saude Publica do Estado do Parana (LACEN/PR) | Laboratory of Respiratory Viruses and Measles, Oswaldo Cruz Institute, FIOCRUZ | Agatha Soares; Alice Sampaio Rocha; Ana Carolina Mendonca; Anna Carolina Paixao; Elisa Cavalcante Pereira; Fernando Motta; Ighor Leonardo Arantes; Irina Riediger; Luciana Appolinario; Marilda Siqueira on behalf of the Fiocruz COVID-19 Genomic Surveillance Network; Paola Resende; Renata Serrano Lopes; Taina Venas |
| EPI_ISL_2603442, EPI_ISL_2603443, EPI_ISL_2603447, EPI_ISL_2603536, EPI_ISL_2603537, EPI_ISL_2603538, EPI_ISL_2603539, EPI_ISL_2603540, EPI_ISL_2603541, EPI_ISL_2603542, EPI_ISL_2603547, EPI_ISL_2603548, EPI_ISL_2603549, EPI_ISL_2603550, EPI_ISL_2603551, EPI_ISL_2603552, EPI_ISL_2603553, EPI_ISL_2603554, EPI_ISL_2603557, EPI_ISL_2603558, EPI_ISL_2603559, EPI_ISL_2603560, EPI_ISL_2603561, EPI_ISL_2603562, EPI_ISL_2603563, EPI_ISL_2982770, EPI_ISL_2982771, EPI_ISL_2982773 | | | |
| see above | Laboratorio Central de Saude Publica do Estado do Rio Grande do Sul (LACEN-RS) | Laboratory of Respiratory Viruses and Measles, Oswaldo Cruz Institute, FIOCRUZ | Alice Sampaio Rocha; Ana Carolina Mendonca; Anderson Brandao Leite; Anna Carolina Paixao; Elisa Cavalcante Pereira; Fernando Motta; Luciana Appolinario; Marilda Siqueira on behalf of the Fiocruz COVID-19 Genomic Surveillance Network; Paola Resende; Renata Serrano Lopes; Taina Venas |
| EPI_ISL_3033060, EPI_ISL_3033062, EPI_ISL_3033063 | Laboratorio Central de la Ciudad de Santa Fe | Grupo de Genómica y Bioinformática del Instituto de Investigación de la Cadena Láctea CONICET-INTA on behalf of 'Proyecto Argentino Interinstitucional de genomica de SARS-CoV-2' (PAIS Consortium) | AF; Amadio; C; Eberhardt; G; Irazoqui; JM; MF; Mugna; Ojeda; Pastor; Rompato; V |
| EPI_ISL_2271687, EPI_ISL_2271694, EPI_ISL_2271697, EPI_ISL_2271699, EPI_ISL_2271704, EPI_ISL_2271705, EPI_ISL_2271706, EPI_ISL_2271708 | | | |
| see above | Laboratorio Central, Ministerio de Salud Cordoba | Instituto de Patologia Vegetal (CIAP-INTA) on behalf of 'Proyecto Argentino Interinstitucional de genomica de SARS-CoV-2' (PAIS Consortium) | Barbas, G.; Castro, G.; Debat, HJ.; FD; Fernandez; M; M.B.; Marquez, N.; Pisano; Re, V. |
| EPI_ISL_4878289 | Laboratorio Direccion Sanidad Policia Nacional | Instituto Nacional de Salud-Dirección de Investigación en Salud Pública | Carlos Franco-Muñoz; Carmen Osorio; Diana Malo; Diego A. Álvarez-Díaz; Diego Andrés Prada; Gerardo Santamaría; Hector Alejandro Ruiz-Moreno; Jhonnatan Reales-González; Jorge Rivera; Juan Camilo Martinez; Julian Naizaque; Katherine Laiton-Donato; Lisseth Pardo; Magdalena Wiesner; Marcela Mercado-Reyes; Maria T. Herrera-Sepúlveda; Marta Lopez Blanco; Martha Lucia Ospina Martinez; Paola Rojas; Sergio Gomez; Sheryll Corchuelo; Ángela Alarcon Cruz |
| EPI_ISL_4196716 | Laboratorio MICROBIOM | Laboratorio Central Mg. Luis Alfredo Pianciola on behalf of 'Proyecto Argentino Interinstitucional de genomica de SARS-CoV-2' (PAIS Consortium) | C Pintos; C Rastellini; C Ziehm; J Ousset; L Pianciola; M Fernandez; M Mazzeo; S Cecchi. |
| EPI_ISL_3275219 | Laboratorio Nacional de Salud, Ministerio de Salud Publica y Asistencia Social | Genomics and Proteomics Department, Gorgas Memorial Institute For Health Studies | Alexander A Martinez; Ambar Moreno; Claudia Estrada; Claudia Gonzalez V; César Roberto Conde Pereira; Jessica Gondola; Leyda Abrego; Marlene Castillo; Oris Chavarria |
| EPI_ISL_5199345 | Laboratorio SYNLAB Colombia | Instituto Nacional de Salud-Dirección de Investigación en Salud Pública | Beatriz de Arco; Carlos Franco-Muñoz; Carmen Osorio; Diana Malo; Diego A. Álvarez-Díaz; Diego Andrés Prada; Gerardo Santamaría; Hector Alejandro Ruiz-Moreno; Jhonnatan Reales-González; Jorge Rivera; Juan Camilo Martinez; Julian Naizaque; Katherine Laiton-Donato; Lisseth Pardo; Magdalena Wiesner; Marcela Mercado-Reyes; Maria T. Herrera-Sepúlveda; Marta Lopez Blanco; Martha Lucia Ospina Martinez; Paola Rojas; Sergio Gomez; Sheryll Corchuelo; Tatiana Cobos; Ángela Alarcon Cruz |
| EPI_ISL_5771879 | Laboratorio Salud Publica Mendoza | Nodo de Secuenciación Tierra del Fuego - Hospital Regional Ushuaia - Centro Austral De Investigaciones Científicas - Universidad Nacional De Tierra Del Fuego | AE; CF; Ceballos; F; Gallego; Gramundi; ID; Nardi; Rojas; SG |
| EPI_ISL_2240748 | Laboratorio analisi cliniche, ASST Valtellina e Alto Lario | Laboratory of Clinical Microbiology, Virology and Bioemergencies, ASST Fatebenefratelli Sacco - Sacco Hospital | Alberto Rizzo; Alessandro Mancon; Fiorenza Bracchitta; Luca Rizzuto; Maria Rita Gismondo; Valeria Micheli |
| EPI_ISL_2501854 | Laboratorio analisi, ASST Valtellina e Alto Lario | Laboratory of Clinical Microbiology, Virology and Bioemergencies, ASST Fatebenefratelli Sacco - Sacco Hospital | Alberto Rizzo; Alessandro Mancon; Fiorenza Bracchitta; Maria Rita Gismondo; Valeria Micheli |
| EPI_ISL_2462063 | Laboratorio de Biologia Molecular, Hospital San Pedro Claver | Microbiologia Molecular, Instituto SELADIS, Universidad Mayor de San Andrés | Aneth Vasquez Michel; Carmen Delgado Barrera; Oscar M. Rollano-Peñaloza; Sandra Miranda Sardon |
| EPI_ISL_2777435, EPI_ISL_2777436, EPI_ISL_2777437, EPI_ISL_2777438, EPI_ISL_4030325, EPI_ISL_4030352, EPI_ISL_4030355, EPI_ISL_4030358, EPI_ISL_4030363 | | | |
| see above | Laboratorio de Ecologia de Doencas Transmissiveis na Amazonia, Instituto Leonidas e Maria Deane - Fiocruz Amazonia | Laboratorio de Ecologia de Doencas Transmissiveis na Amazonia, Instituto Leonidas e Maria Deane - Fiocruz Amazonia | Alex Martins; André Corado; Debora Duarte; Fabiola Mendonça da Silva Chui; Felipe Naveca; Fernanda Nascimento; Fernando Fonseca de Almeida Val; George Silva; Gisely Cardoso de Melo; Karina Pessoa; Luciana Gonçalves; Marcus Vinicius Guimarães de Lacerda; Maria Júlia Brandão; Maria Paula Gomes Mourão; Mariana Xavier Simão; Matilde Mejía; Michele Jesus; Valdinete Nascimento; Vanderson de Souza Sampaio; Victor Souza; Ágatha Costa |
| EPI_ISL_2728561, EPI_ISL_3321109, EPI_ISL_3321111, EPI_ISL_3321270, EPI_ISL_3370427 | Laboratorio de Infectologia y Virologia Molecular | Laboratory of Molecular Virology, School of Medicine, Pontificia Universidad Catolica de Chile | Alejandro Bhrun; Ana Maria Contreras; Andres E. Munoz-Marcos; Carlos Palma; Catalina Pardo-Roa; Constanza Maldonado; Constanza Martinez-Valdevenito; Eileen Serrano; Erick Salinas; Estefany Poblete; Francisco Melo; Jennifer Angulo; Jorge Levican; Leonardo I. Almonacid; M. Belen Leyton; Magdalena Vera; Marcela Ferres; Maria Jose Avendano; Rafael A. Medina; Tamara Garcia-Salum |
| EPI_ISL_2674329, EPI_ISL_2674332, EPI_ISL_2674333 | Laboratorio de Inmunología y Biología Molecular | Instituto Nacional de Salud-Dirección de Investigación en Salud Pública | Carlos Franco-Muñoz; Carmen Osorio; Diana Malo; Diego A. Álvarez-Díaz; Diego Andrés Prada; Gerardo Santamaría; Hector Alejandro Ruiz-Moreno; Jhonnatan Reales-González; Jorge Rivera; Juan Camilo Martinez; Julian Naizaque; Katherine Laiton-Donato; Lisseth Pardo; Magdalena Wiesner; Marcela Mercado-Reyes; Maria T. Herrera-Sepúlveda; Marta Lopez Blanco; Martha Lucia Ospina Martinez; Paola Rojas; Sergio Gomez; Sheryll Corchuelo; Ángela Alarcon Cruz |
| EPI_ISL_3134277 | Laboratorio de Medicina Molecular, Universidad de Magallanes | Centro Asistencial Docente y de Investigacion, Universidad de Magallanes | Carolina Pérez; Constanza Ceroni; Diego Alvarez; Inés Cid; Jacqueline Aldridge; Jorge González; Juan Ríos; Marcelo Navarrete |
| EPI_ISL_3048968, EPI_ISL_3048969, EPI_ISL_3048970, EPI_ISL_3048971, EPI_ISL_3048972, EPI_ISL_3761576, EPI_ISL_3761613, EPI_ISL_3761616, EPI_ISL_3761646 | | | |
| see above | Laboratorio de Pesquisa em Virologia, FAMERP, SJRP | Laboratorio de Pesquisa em Virologia, FAMERP, SJRP | Beatriz de Carvalho Marques; Cecília Artico Banho; Cíntia Bittar; Fábio Sossai Possebon; Guilherme Campos; Helena Lage Ferreira; Jorge A. Petroli Marchesi; João Pessoa Araújo Jr.; Leila Sabrina Ullmann; Lívia Sacchetto; Maisa C. Pereira Parra; Marília Moraes; Maurício L. Nogueira.; Paula Rahal; Paulo Inacio da Costa |
| EPI_ISL_3672406, EPI_ISL_3672414, EPI_ISL_3672415, EPI_ISL_3672418, EPI_ISL_3826395, EPI_ISL_3826401, EPI_ISL_3826402, EPI_ISL_3826441, EPI_ISL_3991429, EPI_ISL_4052728, EPI_ISL_4052729, EPI_ISL_4052742, EPI_ISL_4052744, EPI_ISL_4052747, EPI_ISL_4052748, EPI_ISL_4052751, EPI_ISL_4052760, EPI_ISL_4052766, EPI_ISL_5934957 | | | |
| see above | Laboratorio de Referencia Nacional de Virus Respiratorios. Centro Nacional de Salud Publica. Instituto Nacional de Salud Peru. | Laboratorio de Referencia Nacional de Virus Respiratorios. Centro Nacional de Salud Publica. Instituto Nacional de Salud Peru. | Carlos Padilla Rojas; Henri Bailon Calderon; Iris Silva Molina; Joseph Huayra Niquen; Lely Solari Zerpa; Luis Barcena Flores; Marco Galarza Perez; Nancy Rojas Serrano; Nieves Sevilla Castañeda; Omar Caceres Rey; Orson Mestanza Millones; Princesa Medrano Alhuay; Priscila Lope Pari; Sandra Morales Ruiz; Sara Gordillo Vilchez; Steve Acedo Lazo; Veronica Hurtado Vela; Victor Jimenez Vasquez; Wendy Lizarraga Olivares |
| EPI_ISL_4405436 | Laboratorio de salud pública, Facultad de Ciencias Exactas, Universidad Nacional de La Plata | Área de Secuenciación del Laboratorio de Virología del Hospital de Niños Dr. Ricardo Gutierrez on behalf of 'Proyecto Argentino Interinstitucional de genomica de SARS-CoV-2' (PAIS Consortium) | A; Acuña; Angeletti; Cordero; D; Goya; LE; Lusso; MI; MS; Nabaes Jodar; Nadalich; Natale; R; S; Toro; V; Valinotto; Viegas, M. |
| EPI_ISL_7620375, EPI_ISL_7620376 | Laboratorio del Hospital Regional Ushuaia Gdor. Ernesto Campos | Nodo de Secuenciación Tierra del Fuego - Hospital Regional Ushuaia - Centro Austral De Investigaciones Científicas - Universidad Nacional De Tierra Del Fuego on behalf PAIS project and ANLIS-Malbran | AE; Boutureira, MF.; CA; CB; CF; Castro; Ceballos; Cáceres; De Roccis; F; G; Gallego; Gramundi; ID; Nardi; Rojas; SB; SG; Yulan |
| EPI_ISL_2180045 | Laboratorio di Microbiologia | Laboratorio di Microbiologia | Martinetti Lucchini Gladys; Valeria Spina |
| EPI_ISL_6368767, EPI_ISL_6368816, EPI_ISL_6368820, EPI_ISL_6368826 | Laboratorio nacional de salud publica | Viral Special Pathogens Branch, Centers for Disease Control and Prevention | Emir Talundzic; Isaac Miguel; Joel Montgomery; John Klena; Justin Lee; Melissa Mobley; Ronald Skewes-Ramm; Shannon Whitmer |
| EPI_ISL_2183402, EPI_ISL_2183411, EPI_ISL_2183432, EPI_ISL_2183464, EPI_ISL_2185473, EPI_ISL_2185474, EPI_ISL_2185479, EPI_ISL_2185508, EPI_ISL_2304829, EPI_ISL_2304837, EPI_ISL_2304850, EPI_ISL_2304856, EPI_ISL_2304882, EPI_ISL_2304889, EPI_ISL_2304913, EPI_ISL_2304918, EPI_ISL_2304920, EPI_ISL_2304921, EPI_ISL_2304923, EPI_ISL_2304925, EPI_ISL_2304926, EPI_ISL_2304939, EPI_ISL_2304944, EPI_ISL_2304945, EPI_ISL_2304959, EPI_ISL_2304961, EPI_ISL_2304963, EPI_ISL_2304964, EPI_ISL_2304968, EPI_ISL_2305035, EPI_ISL_2305043, EPI_ISL_2305051, EPI_ISL_2305053, EPI_ISL_2305056, EPI_ISL_2305057, EPI_ISL_2305059, EPI_ISL_2305060, EPI_ISL_2305063, EPI_ISL_2305074, EPI_ISL_2305075, EPI_ISL_2305080, EPI_ISL_2305095, EPI_ISL_2305179, EPI_ISL_2305190, EPI_ISL_2305191, EPI_ISL_2305197, EPI_ISL_2305202, EPI_ISL_2305243, EPI_ISL_2305257, EPI_ISL_2305286, EPI_ISL_2305293, EPI_ISL_2305336, EPI_ISL_2305339, EPI_ISL_2305373, EPI_ISL_2305385, EPI_ISL_2305766, EPI_ISL_2305771, EPI_ISL_2305789, EPI_ISL_2305797, EPI_ISL_2305802, EPI_ISL_2305805, EPI_ISL_2305810, EPI_ISL_2305812, EPI_ISL_2305818, EPI_ISL_2305827, EPI_ISL_2305833, EPI_ISL_2305834, EPI_ISL_2305835, EPI_ISL_2305850, EPI_ISL_2305856, EPI_ISL_2305860, EPI_ISL_2305866, EPI_ISL_2305874, EPI_ISL_2305904, EPI_ISL_2305905, EPI_ISL_2305906, EPI_ISL_2305974, EPI_ISL_2305977, EPI_ISL_2306009, EPI_ISL_2306026, EPI_ISL_2306028, EPI_ISL_2306030, EPI_ISL_2306032, EPI_ISL_2306038, EPI_ISL_2306051, EPI_ISL_2306054, EPI_ISL_2306057, EPI_ISL_2306059, EPI_ISL_2306071, EPI_ISL_2306072, EPI_ISL_2306082, EPI_ISL_2306120, EPI_ISL_2306131, EPI_ISL_2306173, EPI_ISL_2306186, EPI_ISL_2306193, EPI_ISL_2306218, EPI_ISL_2306255, EPI_ISL_2306275, EPI_ISL_2306276, EPI_ISL_2306286, EPI_ISL_2306301, EPI_ISL_2306315, EPI_ISL_2306329, EPI_ISL_2306335, EPI_ISL_2306336, EPI_ISL_2306414, EPI_ISL_2306417, EPI_ISL_2306422, EPI_ISL_2306426, | | | |

| | | | |
|---|---|---|---|
| EPI_ISL_2306429, EPI_ISL_2306432, EPI_ISL_2306433, EPI_ISL_2306435, EPI_ISL_2306439, EPI_ISL_2306442, EPI_ISL_2306455, EPI_ISL_2306459, EPI_ISL_2306461, EPI_ISL_2306468, EPI_ISL_2306471, EPI_ISL_2306475, EPI_ISL_2306478, EPI_ISL_2306481, EPI_ISL_2306489, EPI_ISL_2306504, EPI_ISL_2306540, EPI_ISL_2306550, EPI_ISL_2306552, EPI_ISL_2306555, EPI_ISL_2306557, EPI_ISL_2306564, EPI_ISL_2306565, EPI_ISL_2306566, EPI_ISL_2306574, EPI_ISL_2306599, EPI_ISL_2306603, EPI_ISL_2306616, EPI_ISL_2306621, EPI_ISL_2306623, EPI_ISL_2306634, EPI_ISL_2306640, EPI_ISL_2306671, EPI_ISL_2306681, EPI_ISL_2306700, EPI_ISL_2306705, EPI_ISL_2306718, EPI_ISL_2306727, EPI_ISL_2306729, EPI_ISL_2306737, EPI_ISL_2306743, EPI_ISL_2306748, EPI_ISL_2306759, EPI_ISL_2306761, EPI_ISL_2306762, EPI_ISL_2306777, EPI_ISL_2306804, EPI_ISL_2306811, EPI_ISL_2306849, EPI_ISL_2306857, EPI_ISL_2306858, EPI_ISL_2306885, EPI_ISL_2306917, EPI_ISL_2306919, EPI_ISL_2306930, EPI_ISL_2306942, EPI_ISL_2306976, EPI_ISL_2306979, EPI_ISL_2306984, EPI_ISL_2306988, EPI_ISL_2306993, EPI_ISL_2306997, EPI_ISL_2324462, EPI_ISL_2324465, EPI_ISL_2324468, EPI_ISL_2324469, EPI_ISL_2324474, EPI_ISL_2324481, EPI_ISL_2324485, EPI_ISL_2324487, EPI_ISL_2324493, EPI_ISL_2324495, EPI_ISL_2324519, EPI_ISL_2324530, EPI_ISL_2324536, EPI_ISL_2324538, EPI_ISL_2324544, EPI_ISL_2324565, EPI_ISL_2324567, EPI_ISL_2324570, EPI_ISL_2324577, EPI_ISL_2324580, EPI_ISL_2324588, EPI_ISL_2398076, EPI_ISL_2398079, EPI_ISL_2398083, EPI_ISL_2398097, EPI_ISL_2398100, EPI_ISL_2398111, EPI_ISL_2398114, EPI_ISL_2398115, EPI_ISL_2398153, EPI_ISL_2398158, EPI_ISL_2398177, EPI_ISL_2398220, EPI_ISL_2398618, EPI_ISL_2398624, EPI_ISL_2398632, EPI_ISL_2398647, EPI_ISL_2398703, EPI_ISL_2398725, EPI_ISL_2611254, EPI_ISL_2611255, EPI_ISL_2611256, EPI_ISL_2611264, EPI_ISL_2611267, EPI_ISL_2611268, EPI_ISL_2611271, EPI_ISL_4374194, EPI_ISL_4376799, EPI_ISL_4377573 | | | |
| see above | Laboratory Corporation of America | Centers for Disease Control and Prevention Division of Viral Diseases, Pathogen Discovery | Adrian Paskey; Amanda Douglas; Amanda Suchanek; Andrea Throop; Ayla Burns; Benjamin Rambo-Martin; Bobbi Croy; Brian Krueger; Brian Norvell; Christopher Gulvick; Christos Petropoulos; Clinton Paden; Clinton R. Paden; Craig Lukasik; Dakota Howard; Darlene Wagner; Debbie Boles; Dhwani Batra; Duncan MacCannell; Eyad Almasri; Goran Stevovic; Howard Engler; Hrushikesh Deshmukh; Jake Humphrey; Jana Schroth; Jason Caravas; Joe Voshell; John Pruitt; Jonathan Meltzer; Jonathan Williams; Kara Moser; Kimberly Wagner; Kristine Lacek; Lax Iyer; Lisa Pfefferle; Lyndon Tilson; Manoj Jain; Marcia Eisenberg; Mary Ann Cristobal; Mary Cristobal; Mary Williamson; Matthew Robinson; Matthew Schmerer; Michael Levandoski; Mike Sapeta; Mindy Nye; Minoo Agarwal; Mohan Kolli; Nuthawin Charoensri; Oren Cohen; Peter Cook; Peter W. Cook; Prashant Gupta; Qian Zeng; Rama Ghatti; Scott Parker; Scott Ryan; Scott Sammons; Shatavia Morrison; Stanley Letovsky; Steven Ragan; Suresh Babu Selvaraju; Suresh Selvaraju; Susan Countryman; Susan Hicks; Suzanne Dale; Thomas Urban; Tim Kuphal; Tricia Zwiefelhofer; Tymeckia Kendall; Victoria Caban Figueroa; Vincent Drouillon; Yvette Unoarumhi |
| EPI_ISL_3931704 | Laboratory of Microbiology and Virology, Ospedale Amedeo di Savoia, ASL "Città di Torino" | Laboratory of Microbiology and Virology, Ospedale Amedeo di Savoia, ASL "Città di Torino" | Debora Fiorito; Elisa Burdino; Francesco Cerutti; Gabriella Gregori; Maria Grazia Milia; Tiziano Allice; Valeria Ghisetti |
| EPI_ISL_2443634, EPI_ISL_2443659, EPI_ISL_2443660, EPI_ISL_2443661, EPI_ISL_3045471, EPI_ISL_4237260, EPI_ISL_4237344, EPI_ISL_4237345, EPI_ISL_4237346, EPI_ISL_4237347, EPI_ISL_4237417, EPI_ISL_4237428, EPI_ISL_6172747, EPI_ISL_6172748, EPI_ISL_6172749, EPI_ISL_6172750, EPI_ISL_6970988, EPI_ISL_6970991 | | | |
| see above | Laboratory of Respiratory Viruses and Measles, Oswaldo Cruz Institute, FIOCRUZ | Laboratory of Respiratory Viruses and Measles, Oswaldo Cruz Institute, FIOCRUZ | Agatha Cristinne Prudencio; Alice Sampaio Rocha; Ana Carolina Mendonca; Anna Carolina Paixao; Bruna Mendonça da Silva; Elisa Cavalcante Pereira; Fernando Motta; Ighor Arantes; Ighor Leonardo Arantes Gomes; Jéssica Graça Macedo de Carvalho; Larissa Macedo Pinto; Luciana Appolinario; Marilda Siqueira on behalf of the Fiocruz COVID-19 Genomic Surveillance Network; Paola Resende; Renata Serrano Lopes; Taina Venas; Victor Guimaraes; Vitor Guimaraes Ribeiro |
| EPI_ISL_2621576 | Laboratory, Gravity Diagnostics | Laboratory, Gravity Diagnostics | Diagnostics, G. |
| EPI_ISL_4600543, EPI_ISL_4600544, EPI_ISL_4600548, EPI_ISL_4600550, EPI_ISL_4600553, EPI_ISL_5017907 | Laboratório Central de Saúde Pública Dr. Costa Alvarenga | Coordenação Geral de Laboratórios de Saúde Pública (CGLAB/DAEVS/SVS/MS) | Vagner Fonseca; et al. |
| EPI_ISL_4600534 | Laboratório Central de Saúde Pública de Pernambuco | Coordenação Geral de Laboratórios de Saúde Pública (CGLAB/DAEVS/SVS/MS) | Vagner Fonseca; et al. |
| EPI_ISL_2777285, EPI_ISL_2777349, EPI_ISL_2777350, EPI_ISL_2777351, EPI_ISL_2777359, EPI_ISL_2777546, EPI_ISL_2777555, EPI_ISL_2777556, EPI_ISL_2777561, EPI_ISL_2777570, EPI_ISL_2777624, EPI_ISL_2777625, EPI_ISL_2777626, EPI_ISL_2777627, EPI_ISL_2777637, EPI_ISL_2777639, EPI_ISL_2777641, EPI_ISL_2777642, EPI_ISL_2777648, EPI_ISL_2777650, EPI_ISL_2777652, EPI_ISL_2777653, EPI_ISL_2777654, EPI_ISL_2777659, EPI_ISL_2777663, EPI_ISL_2777664, EPI_ISL_2777777, EPI_ISL_2777778, EPI_ISL_2777852, EPI_ISL_2777853, EPI_ISL_2777856, EPI_ISL_2777890, EPI_ISL_2777891, EPI_ISL_2777892, EPI_ISL_2777893, EPI_ISL_2777916, EPI_ISL_2777944, EPI_ISL_2777948, EPI_ISL_2777951, EPI_ISL_2777955 | | | |
| see above | Laboratório Central de Saúde Pública do Amazonas - LACEN-AM | Laboratorio de Ecologia de Doencas Transmissiveis na Amazonia, Instituto Leonidas e Maria Deane - Fiocruz Amazonia | André Corado; Debora Duarte; Felipe Naveca; Fernanda Nascimento; George Silva; Karina Pessoa; Luciana Gonçalves; Maria Júlia Brandão; Matilde Mejía; Michele Jesus; Valdinete Nascimento; Victor Souza; Ágatha Costa |
| EPI_ISL_4433691, EPI_ISL_4434012, EPI_ISL_4435003 | Laboratório de Baculovirus, Universidade de Brasília (UnB), Instituto de Ciências Biológicas (IB) | Laboratório de Virologia, Faculdade de Medicina, Universidade Federal de Mato Grosso (UFMT) | Bergman Morais Ribeiro; Fernando Lucas Melo; Francisco Scoffoni Kennedy de Azevedo; Gessica Fernanda Colnago de Lima; Renata Dezengrini Slhessarenko; Thais Campos Cruz |
| EPI_ISL_2731508, EPI_ISL_2731509, EPI_ISL_4543308 | Laboratório de Biotecnologia Aplicada (LBA) - Laboratório de Biologia Molecular - Hospital das Clínicas, Faculdade de Medicina de Botucatu, Departamento de Bioprocessos e Biotecnologia - Faculdade de Ciências Agronômicas, UNESP – Botucatu/SP | Laboratory of Respiratory Viruses and Measles, Oswaldo Cruz Institute, FIOCRUZ | Alice Sampaio Rocha; Ana Carolina Mendonca; Anna Carolina Paixao; Elisa Cavalcante Pereira; Felipe Allan da Silva Costa; Fernando Motta; Jayme Augusto de Souza Neto; Leonardo Nazario de Moraes; Luciana Appolinario; Marilda Siqueira on behalf of the Fiocruz COVID-19 Genomic Surveillance Network; Paola Resende; Patricia Akemi Assato; Rejane Maria Tommasini; Renata Serrano Lopes; Taina Venas |
| EPI_ISL_3873658, EPI_ISL_3873660, EPI_ISL_4051969 | Laboratório de Microbiologia Molecular - Universidade FEEVALE | Molecular Microbiology Laboratory | Alana Witt Hansen; Fernando Rosado Spilki; Fágner Henrique Heldt; Juliana Schons Gularte; Juliane Deise Fleck; Mariana Soares da Silva; Matheus Nunes Weber; Meriane Demoliner; Micheli Filippi; Paula Rodrigues de Almeida; Vyctoria Malayhka de Abreu Góes Pereira. |
| EPI_ISL_2536227, EPI_ISL_2536229, EPI_ISL_2536232, EPI_ISL_2536235, EPI_ISL_2536236, EPI_ISL_2536238, EPI_ISL_2536240, EPI_ISL_2536245, EPI_ISL_2536249, EPI_ISL_2536259, EPI_ISL_2557414 | | | |
| see above | Labortorio Central de Saude Publica do Estado de Santa Catarina (LACEN/SC) | Laboratory of Respiratory Viruses and Measles, Oswaldo Cruz Institute, FIOCRUZ | Alice Sampaio Rocha; Ana Carolina Mendonca; Anna Carolina Paixao; Darcita Buerger Rovaris; Elisa Cavalcante Pereira; Fernando Motta; Luciana Appolinario; Marilda Siqueira on behalf of the Fiocruz COVID-19 Genomic Surveillance Network; Paola Resende; Renata Serrano Lopes; Sandra Bianchini Fernandes; Taina Venas |
| EPI_ISL_2536287, EPI_ISL_2536288, EPI_ISL_2536289, EPI_ISL_2536290, EPI_ISL_2536291, EPI_ISL_2536292, EPI_ISL_2536293, EPI_ISL_2536294, EPI_ISL_2536295, EPI_ISL_2536296, EPI_ISL_2536297, EPI_ISL_2536298 | | | |
| see above | Labortorio Central de Saude Publica do Estado do Rio de Janeiro (LACEN/RJ) | Laboratory of Respiratory Viruses and Measles, Oswaldo Cruz Institute, FIOCRUZ | Alice Sampaio Rocha; Ana Carolina Mendonca; Andrea Cony Cavalcanti; Anna Carolina Paixao; Elisa Cavalcante Pereira; Fernando Motta; Luciana Appolinario; Marilda Siqueira on behalf of the Fiocruz COVID-19 Genomic Surveillance Network; Paola Resende; Renata Serrano Lopes; Taina Venas |
| EPI_ISL_2314838 | Landesgesundheitsamt Baden-Württemberg | Robert Koch Institute | |
| EPI_ISL_2557363, EPI_ISL_2557364, EPI_ISL_2557365, EPI_ISL_2557371, EPI_ISL_2557373, EPI_ISL_2557375, EPI_ISL_2863684, EPI_ISL_2863685, EPI_ISL_2863687, EPI_ISL_2863889 | | | |
| see above | Lboratorio Central de Saude Publica do Estado do Parana (LACEN/PR) | Laboratory of Respiratory Viruses and Measles, Oswaldo Cruz Institute, FIOCRUZ | Agatha Cristinne Prudencio; Agatha Cristinne Prudencio Soares; Alice Sampaio Rocha; Ana Carolina Mendonca; Anna Carolina Paixao; Elisa Cavalcante Pereira; Fernando Motta; Ighor Leonardo Arantes Gomes; Irina Riediger; Luciana Appolinario; Marilda Siqueira on behalf of the Fiocruz COVID-19 Genomic Surveillance Network; Paola Resende; Renata Serrano Lopes; Taina Venas |
| EPI_ISL_2199058, EPI_ISL_2199752, EPI_ISL_2237842 | Lighthouse Lab in Glasgow | Wellcome Sanger Institute for the COVID-19 Genomics UK (COG-UK) Consortium | Anna Dominiczak and Alex Alderton; Carol Clugston; Cordelia Langford; David Gray; David K. Jackson; Dominic Kwiatkowski; Ewan Harrison; Harper VanSteenhouse; Ian Johnston; Jeffrey Barrett; John Sillitoe on behalf of the Wellcome Sanger Institute COVID-19 Surveillance Team; Roberto Amato; Sonia Goncalves; Yumi Kasai |
| EPI_ISL_2118155, EPI_ISL_2236862, EPI_ISL_2236956, EPI_ISL_2253909, EPI_ISL_2253920 | Lighthouse Lab in Milton Keynes | Wellcome Sanger Institute for the COVID-19 Genomics UK (COG-UK) Consortium | Cordelia Langford; David K. Jackson; Dominic Kwiatkowski; Ewan Harrison; Ian Johnston; Jeffrey Barrett; John Sillitoe on behalf of the Wellcome Sanger Institute COVID-19 Surveillance Team; Roberto Amato; Sonia Goncalves; The Lighthouse Lab in Milton Keynes and Alex Alderton |
| EPI_ISL_2266545 | Limbach - MVZ Humangenetik Ulm | Robert Koch Institute | |
| EPI_ISL_2373100, EPI_ISL_2373103 | Lurie Children's Hospital of Chicago | Northwestern University - Ozer Lab | Egon A. Ozer; Judd F. Hultquist; Lacy M. Simons; Larry K. Kociolek; Michael G. Ison; Ramon Lorenzo-Redondo; Taylor J. Dean; William J. Muller; Xiaotian; Zheng |
| EPI_ISL_2376581, EPI_ISL_2425395 | M Health Fairview | Minnesota Department of Health, Public Health Laboratory | Alexandra Lorentz; Jacob Garfin; Matt Plumb; and Xiong Wang |
| EPI_ISL_2495422, EPI_ISL_2495424, EPI_ISL_2495608 | MB-Cadham Provincial laboratory | National Microbiology Laboratory (NML) | Anna Majer; Anneliese Landgraff; CanCOGeN's metadata curation team; Darian Hole; David Alexander; Elsie Grudeski; Gary Van Domselaar; Grace Seo; Jared Bullard; Jennifer Tanner; Kerry Dust; Kirsten Biggar; Madison Chapel; Morag Graham; Natalie Knox; Nathalie Bastien; Paul Van Caeseele; Philip Mabon; Public Health Agency of Canada CanCOGeN team; Rhiannon Huzarewich; Russell Mandes; Shari Tyson; Timothy Booth; Yan Li |
| EPI_ISL_2262825, EPI_ISL_2263075 | MDI Limbach Berlin GmbH; MVZ Labor Berlin | Robert Koch Institute | |
| EPI_ISL_2383931 | ME Health and Environmental Testing Laboratory | Centers for Disease Control and Prevention Division of Viral Diseases, Pathogen Discovery | Alison Laufer Halpin; Ben L. Rambo-Martin; Clinton R. Paden; Dakota Howard; Darlene Wagner; Dave Wentworth; Dhwani Batra; Jasmine Padilla; Justin Lee; Katie Dillon; Krista Queen; Kristen Knipe; Kristine Lacek; Mark Burroughs; Matthew Schmerer; Mili Sheth; Peter Cook; Sam Shepard; Sarah Nobles; Shoshona Le; Suxiang Tong; Vivien Dugan; Yvette Unoarumhi |
| EPI_ISL_2453673 | MEPHI, Aix Marseille University | MEPHI, Aix Marseille University | Anthony LEVASSEUR |
| EPI_ISL_2451618 | MI - Michigan Department of Health and Human Services - Bureau of Laboratories | Centers for Disease Control and Prevention Division of Viral Diseases, Pathogen Discovery | Alison Laufer Halpin; Ben L. Rambo-Martin; Clinton R. Paden; Dakota Howard; Darlene Wagner; Dave Wentworth; Dhwani Batra; Jasmine Padilla; Justin Lee; Katie Dillon; Krista Queen; Kristen Knipe; Kristine Lacek; Mark Burroughs; Matthew Schmerer; Mili Sheth; Peter Cook; Sam Shepard; Sarah Nobles; Shoshona Le; Suxiang Tong; Vivien Dugan; Yvette Unoarumhi |
| EPI_ISL_6492703, EPI_ISL_6492722, EPI_ISL_6492723, EPI_ISL_6492725, EPI_ISL_6492728 | MSHS Clinical Microbiology Laboratories | MSHS Pathogen Surveillance Program | Adolfo García-Sastre; Adriana van de Guchte; Ajay Obla; Alberto Paniz-Mondolfi; Ana S. Gonzalez-Reiche; Angela Amoako; Ashley Salimbangon; Betsaida Salom Melo; Bremy Alburquerque; Brianne Ciferri; Charles Gleason; Daniel Floda; Deena R. Altman; Denise Jurczyszak; Emilia Mia Sordillo; Gintaras Deikus; Giulio Kleiner; Gopi Patel; Hala Alshammary; Harm van Bakel; Irina Oussenko; Jayeeta Dutta; Juan Soto; Julia Matthews; Katherine Beach; Kathryn Twyman; Kayla Russo; Komal Srivastava; Levy Sominsky; Mahmoud Awawda; Marta Luksza; Matthew M. Hernandez; Melissa Gitman; Michael D. Nowak; Mitchell J. Sullivan; Nancy Francoeur; Robert Sebra; Sarah Schaefer; Shelcie Fabre; Shwetha Hara Sridhar; Viviana Simon; Ying-Chih Wang; Zenab Khan |
| EPI_ISL_2313131, EPI_ISL_2313155 | MVZ Labor Dr. Limbach & Kollegen GbR | Robert Koch Institute | |
| EPI_ISL_2245568, EPI_ISL_2245596, EPI_ISL_2802612, EPI_ISL_3298815, EPI_ISL_3298842, EPI_ISL_3298847, EPI_ISL_3298856, EPI_ISL_3298857, EPI_ISL_3298874, EPI_ISL_3298878, EPI_ISL_3298879, EPI_ISL_3298890, EPI_ISL_4083833, EPI_ISL_6532718 | | | |
| see above | Maine Health and Environmental Testing Laboratory | Tewhey Lab, The Jackson Laboratory | Barter, M.; Dewey, H.; H. and Tewhey, R.; Iosue, F.; Lynch, R.; Matluk, N.; Munger |
| EPI_ISL_2440926, EPI_ISL_2440979, EPI_ISL_2440993, EPI_ISL_2441051, EPI_ISL_2441063, EPI_ISL_2441067, EPI_ISL_2450946, EPI_ISL_2451027, EPI_ISL_2451029, EPI_ISL_2451123, EPI_ISL_2525127, EPI_ISL_2525324, EPI_ISL_2525328, EPI_ISL_2525355, EPI_ISL_2646541, EPI_ISL_2646544, EPI_ISL_2646564, EPI_ISL_2646573, EPI_ISL_2646578, EPI_ISL_2646614, EPI_ISL_2646619, EPI_ISL_2646658, | | | |

| | | | |
|---|---|---|---|
| EPI_ISL_2646685, EPI_ISL_2646692, EPI_ISL_2646724, EPI_ISL_2646732, EPI_ISL_2646801, EPI_ISL_2646872, EPI_ISL_2646888, EPI_ISL_4348256 | | | |
| see above | Mako Medical | Centers for Disease Control and Prevention Division of Viral Diseases, Pathogen Discovery | Adrian Paskey; Benjamin Rambo-Martin; Christopher Gulvick; Clinton Paden; Clinton R. Paden; Dakota Howard; Darlene Wagner; Dhwani Batra; Duncan MacCannell; Erisa Sula; Jason Caravas; Kara Moser; Kristine Lacek; Lauren Moon; Matthew Schmerer; Matthew Tugwell; Peter Cook; Peter W. Cook; Scott Sammons; Shatavia Morrison; Tymeckia Kendall; Victoria Caban Figueroa; Yvette Unoarumhi |
| EPI_ISL_2501754, EPI_ISL_2501831, EPI_ISL_2501832 | Maryland Genomics, Institute for Genome Sciences, University of Maryland School of Medicine | Maryland Genomics, Institute for Genome Sciences, University of Maryland School of Medicine | Claire M; Fraser; Hazen; Holly; Humphrys; Ivette; Jacques; Jonathan; Kranthi; Lim; Lisa D; Luke J; Mike; Ott; Ravel; Roussey; Sadzewicz; Sandra; Santana-Cruz; Tallon; Tracy; Vavikolanu |
| EPI_ISL_6680572, EPI_ISL_6680575, EPI_ISL_6680595, EPI_ISL_6680600, EPI_ISL_6680612, EPI_ISL_6680614, EPI_ISL_6680622, EPI_ISL_6680632, EPI_ISL_6680654, EPI_ISL_6680655, EPI_ISL_6680698, EPI_ISL_6680734, EPI_ISL_6680767, EPI_ISL_6680797, EPI_ISL_6680831, EPI_ISL_6680840, EPI_ISL_6680847, EPI_ISL_6680862, EPI_ISL_6680872, EPI_ISL_6680878, EPI_ISL_6680887, EPI_ISL_6680896, EPI_ISL_6680905, EPI_ISL_6680914, EPI_ISL_6680922, EPI_ISL_6680944, EPI_ISL_6680945, EPI_ISL_6680946, EPI_ISL_6680947, EPI_ISL_6680956, EPI_ISL_6680963, EPI_ISL_6681006, EPI_ISL_6681014, EPI_ISL_6681040 | | | |
| see above | Mass General Brigham | Mass General Brigham | A.E.; Adams, G.; Anahtar, M.; B.L.; B.W.; Bauer, M.; Birren; Branda, J.; Carter, A.; Cerrato, F.; Chaluvadi, S.; Chapman; Cusick, C.; D.J.; DeRuff, K.; E. and Sabeti; Flowers, K.; Gallagher, G.; Gladden-Young, A.; Gnirke, A.; Harris, J.; J.E.; K.J.; LaRocque, R.; Lagerborg, K.; Lemieux; Lin; Loreth, C.; MacInnis; Neumann, A.; Normandin, E.; P.C.; Park; Pierce, V.; Reilly, S.; Rosenberg; Rudy, M.; Ryan, E.; S.B.; Shaw, B.; Siddle; Slater, D.; Smole, S.; Tomkins-Tinch, C.; Turbett, S. |
| EPI_ISL_2229808, EPI_ISL_2483777, EPI_ISL_2483786, EPI_ISL_2483790, EPI_ISL_2483801, EPI_ISL_2483809, EPI_ISL_2484935, EPI_ISL_2484938, EPI_ISL_2484956 | | | |
| see above | Massachusetts State Public Health Laboratory | Massachusetts State Public Health Laboratory | Andrew Lang; Glen Gallagher; Sandra Smole; Timelia Fink |
| EPI_ISL_2389368 | Med. Labor Prof. Schenk Dr. Ansorge & Kollegen | Robert Koch Institute | |
| EPI_ISL_2372713 | Microbiologia CATLAB | Can Ruti SARS-CoV-2 Sequencing Hub (HUGTiP/IrsiCaixa/IGTP) | Anna Not; Antoni E Bordoy; Bonaventura Clotet; Cristina Casañ; Cristina Esteban; Francesc Catala-Moll; Gemma Clara; Ignacio Blanco; Marc Noguera-Julian; Maria Casadellà; Mercedes Guerrero; Montserrat Giménez; Pere-Joan Cardona; Pilar Armengol; Roger Pared |
| EPI_ISL_2285124, EPI_ISL_2285140, EPI_ISL_2285141, EPI_ISL_2285142, EPI_ISL_2285143, EPI_ISL_2365892, EPI_ISL_2967803 | | | |
| see above | Microbiology Department. Complexo Hospitalario Universitario de Vigo | Microbiology Department. Complexo Hospitalario Universitario de Vigo | Alfaya N; Alonso I; Alvarez M; Cabrera JJ; Carballo R; Cores O; Cortizo S; Davina C; Martinez L; Mediero G; Perez S; Potel C; Regueiro B; Rey S; Vasallo FJ; del-Campo V |
| EPI_ISL_2508406, EPI_ISL_2508453 | Microbiology Division, SC DHEC | Microbiology Division, SC DHEC | Flores, H.; Freeman, J. |
| EPI_ISL_2259140 | Microvida | Microvida | Jaco Verweij; Joep Stohr; Suzan D. Pas |
| EPI_ISL_2406491 | Middlemore Hospital | Institute of Environmental Science and Research (ESR) | Anja Werno; Antje van der Linden; Arlo Upton; Chris Mansell; David Hammer; Dragana Drinkovic; Erasmus Smit; Gary McAuliffe; Hana Sofia Andersson; Hermes Perez; James Ussher; Jill Sherwood; Jing Wang; Joep de Ligt; Josh Freeman; Julia Howard; Juliet Elvy; Lauren Jelly; Mary DeAlmeida; Matt Blakiston; Matt Storey; Matthew Rogers; Max Bloomfield; Michael Addidle; Michelle Balm; Muhammad Faisal; Nikki Freed; Olin Silander; Olivia Stroeven; Rachel Boyle; Sally Roberts; SallyAnn Harbison; Sarah Jefferies; Sharmini Muttaiyah; Susan Morpeth; Susan Taylor; Timothy Blackmore; Vani Sathyendran; Veronica Playle; Virginia Hope; Xiaoyun Ren |
| EPI_ISL_2178659, EPI_ISL_2178660, EPI_ISL_2178661, EPI_ISL_2232584, EPI_ISL_2244913, EPI_ISL_2249537, EPI_ISL_2249545, EPI_ISL_2249547, EPI_ISL_2249548, EPI_ISL_2325069, EPI_ISL_2403691 | | | |
| see above | Minnesota Department of Health, Public Health Laboratory | Minnesota Department of Health, Public Health Laboratory | Alexandra Lorentz; Jacob Garfin; Matt Plumb; and Xiong Wang |
| EPI_ISL_5599314, EPI_ISL_5599317, EPI_ISL_5599325 | Molecular Biology Laboratory, Vidant Medical Center | Brody Integrative Genomics Core, East Carolina University | Changhong Yin; Heather Duncan; John T. Fallon; Kimberly P. Briley; Weihua Huang |
| EPI_ISL_2567617 | Molecular Hepatology Laboratory AOU Messina | Molecular Hepatology Laboratory AOU Messina | Cristina Musolino; Daniele Lombardo; Giuseppina Raffa; Teresa Pollicino.; Valeria Chines |
| EPI_ISL_4025770, EPI_ISL_4025772, EPI_ISL_4025773, EPI_ISL_4025778, EPI_ISL_4025780, EPI_ISL_4025802, EPI_ISL_4025807 | | | |
| see above | NB-Hôpital Georges L. Dumont | New Brunswick - Vitalité Health Network | Allain E.; Chacko S.; Crapoulet N.; Desnoyers G.; Garceau R.; Lacroix J.; Shaw W. |
| EPI_ISL_3355029, EPI_ISL_3355032 | ND Dept. of Health Laboratory Services-Microbiology | Centers for Disease Control and Prevention Division of Viral Diseases, Pathogen Discovery | Alex Burgin; Ben Rambo-Martin; Clinton Paden; Dakota Howard; Dave Wentworth; Dhwani Batra; Jasmine Padilla; Justin Lee; Krista Queen; Kristen Knipe; Kristine Lacek; Mark Burroughs; Matthew Schmerer; Meghan Bentz; Mili Sheth; Peter Cook; Sam Shepard; Sarah Nobles; Suxiang Tong; Vivien Dugan; Yvette Unoarumhi |
| EPI_ISL_2384002 | NH Dept. of Health and Human Services Public Health Labs | Centers for Disease Control and Prevention Division of Viral Diseases, Pathogen Discovery | Alison Laufer Halpin; Ben L. Rambo-Martin; Clinton R. Paden; Dakota Howard; Darlene Wagner; Dave Wentworth; Dhwani Batra; Jasmine Padilla; Justin Lee; Katie Dillon; Krista Queen; Kristen Knipe; Kristine Lacek; Mark Burroughs; Matthew Schmerer; Mili Sheth; Peter Cook; Sam Shepard; Sarah Nobles; Shoshona Le; Suxiang Tong; Vivien Dugan; Yvette Unoarumhi |
| EPI_ISL_2557099 | NJDOH, Public Health and Environmental Laboratories | NJ_PHEL | Allison Roder; Byeong Jeong; Chelsea San Filippo; Dana Woell; Lindsey Bodnar; Ryan Pachucki; Shiv K. Verma |
| EPI_ISL_3025464 | NJDOH, Public Health and Environmental Laboratories | New Jersey Public Health and Environmental Laboratories (NJ PHEL) | Allison Roder; Byeong Jeong; Chelsea San Filippo; Dana Woell; Jacquelyn Deverell; Lindsey Bodnar; Ryan Pachucki; Shiv K. Verma |
| EPI_ISL_2451588, EPI_ISL_2451601 | NV-Southern Nevada Public Health Laboratory | Centers for Disease Control and Prevention Division of Viral Diseases, Pathogen Discovery | Alison Laufer Halpin; Ben L. Rambo-Martin; Clinton R. Paden; Dakota Howard; Darlene Wagner; Dave Wentworth; Dhwani Batra; Jasmine Padilla; Justin Lee; Katie Dillon; Krista Queen; Kristen Knipe; Kristine Lacek; Mark Burroughs; Matthew Schmerer; Mili Sheth; Peter Cook; Sam Shepard; Sarah Nobles; Shoshona Le; Suxiang Tong; Vivien Dugan; Yvette Unoarumhi |
| EPI_ISL_2427404, EPI_ISL_2427410, EPI_ISL_2427416, EPI_ISL_2427463 | NYU Langone Health | Departments of Pathology and Medicine, New York University School of Medicine | Adriana Heguy; Christian Marier; Dacia Dimartino; Emily Guzman; Gael Westby; Guiqing Wang; Paolo Cotzia; Paul Zappile; Peter Meyn; Sitharam Ramaswami; Yutong Zhang |
| EPI_ISL_2463605 | National Laboratory for Health, Environment and Food, OMM, Koper | NLZOH (National Laboratory for Health, Environment and Food) / CISLD (Clinical Institute of Special Laboratory Diagnostics), University Children's Hospital, University Medical Center Ljubljana | Aleksander Kocuvan; Aleksander Mahnic; Alenka Štorman; Ana Grom; Barbara Jenko Bizjan; Gašper Strugar; Kaja Tominc; Katarina Kozmos; Maja Rupnik; Marko Pokorn; Maruša Debeljak; Maša Jarčič; Mitja Rak / Jernej Kovač; Nika Gobec; Robert Šket; Sandra Janezic; Tadej Battelino; Tina Cvetković; Tine Tesovnik; Tjasa Žohar Čretnik |
| EPI_ISL_2179653, EPI_ISL_2179654, EPI_ISL_2179655, EPI_ISL_2179656, EPI_ISL_2179662, EPI_ISL_2179663, EPI_ISL_2228010, EPI_ISL_2228012, EPI_ISL_2228013, EPI_ISL_2228014, EPI_ISL_2271914, EPI_ISL_2271916, EPI_ISL_2271918, EPI_ISL_2271920, EPI_ISL_2271930, EPI_ISL_2271931, EPI_ISL_2271932, EPI_ISL_2271938, EPI_ISL_2271941, EPI_ISL_2271943, EPI_ISL_2271945, EPI_ISL_2271948, EPI_ISL_2687949, EPI_ISL_2687956 | | | |
| see above | National Platform bis UMONS/Jolimont | National Platform bis UMONS/Jolimont | Eric Tarantino; Florian Juszczak; François Dufrasne; Gautier Detry; Guillaume Bayon-Vicente; Ruddy Wattiez |
| EPI_ISL_2754074 | Nebraska Public Health Laboratory | NPHL COVID-19 Response Team | NPHL COVID-19 Response Team |
| EPI_ISL_2322848, EPI_ISL_2381943 | New Mexico Department of Health Scientific Laboratory | New Mexico Department of Health Scientific Laboratory | Anastacia Griego-Fisher; D'eldra Malone; Ellie Johnson; Jennifer Benoit; Keila Gutierrez; Linda Salazar; Ratheesh Rajan |
| EPI_ISL_2340043, EPI_ISL_2384275, EPI_ISL_2489955, EPI_ISL_2661612, EPI_ISL_2661624, EPI_ISL_2661626, EPI_ISL_2661632, EPI_ISL_2661633, EPI_ISL_2661637, EPI_ISL_2714611, EPI_ISL_2858825, EPI_ISL_2858828, EPI_ISL_2858829 | | | |
| see above | North Dakota Department of Health, Public Health Laboratory | North Dakota Department of Health, Public Health Laboratory | Lisa Wingerter |
| EPI_ISL_2536542, EPI_ISL_2536543, EPI_ISL_2536545, EPI_ISL_2536546, EPI_ISL_2536547, EPI_ISL_2536567, EPI_ISL_2536569, EPI_ISL_2536572, EPI_ISL_2536575, EPI_ISL_2536581, EPI_ISL_2536588, EPI_ISL_2536592, EPI_ISL_2536599, EPI_ISL_2536601, EPI_ISL_2536604, EPI_ISL_2536608, EPI_ISL_2536611, EPI_ISL_2536613, EPI_ISL_2536615, EPI_ISL_2536617, EPI_ISL_2536620, EPI_ISL_2536626, EPI_ISL_2692335, EPI_ISL_2692341, EPI_ISL_2692347, EPI_ISL_2692348, EPI_ISL_2692349, EPI_ISL_2692350, EPI_ISL_2692351, EPI_ISL_2692352, EPI_ISL_2692354, EPI_ISL_2692356, EPI_ISL_2692359, EPI_ISL_2692360, EPI_ISL_2692363, EPI_ISL_2692366, EPI_ISL_2692368, EPI_ISL_2692372, EPI_ISL_2692377, EPI_ISL_2692383, EPI_ISL_2692386, EPI_ISL_2692391, EPI_ISL_2692396, EPI_ISL_2692397, EPI_ISL_2692402, EPI_ISL_2692404, EPI_ISL_2692406, EPI_ISL_2692407, EPI_ISL_2692408, EPI_ISL_2692411 | | | |
| see above | Northwest Laboratory | Seattle Flu Study | Amanda Adler; Barry R. Lutz; Benjamin Pelle; Brian Hiatt; Caitlin R. Wolf; Chris D. Frazar; Deborah A. Nickerson; Elisabeth Brandstetter; Erica Ryke; Geoff Melly; Helen Y. Chu; Janet A. Englund; Jay Shendure; Jover Lee; Kairsten Fay; Kirsten Lacombe; Lea M. Starita; Mark J. Rieder; Matthew Richardson; Matthew Thompson; Melissa Truong; Michael Boeckh; Michael Famulare; Misja Ilcisin; Peter D. Han; Philip Dykema; Romesh Gautom; Scott Lindquist; Thomas R. Sibley; Trevor Bedford |
| EPI_ISL_2373002, EPI_ISL_2373003, EPI_ISL_2373008, EPI_ISL_2373009, EPI_ISL_2373011, EPI_ISL_2373019 | Northwestern Memorial Hospital | Northwestern University - Ozer Lab | Chad J. Achenbach; Chao Qi; Egon A. Ozer; Judd F. Hultquist; Lacy M. Simons; Lawrence J. Jennings; Michael G. Ison; Ramon Lorenzo-Redondo; Taylor J. Dean |
| EPI_ISL_2462253, EPI_ISL_2663619, EPI_ISL_2663736, EPI_ISL_2663738, EPI_ISL_2663740, EPI_ISL_2663741, EPI_ISL_2663752, EPI_ISL_2663755, EPI_ISL_2663764, EPI_ISL_2663765, EPI_ISL_2663769, EPI_ISL_2838825, EPI_ISL_2838826, EPI_ISL_2838827, EPI_ISL_2838828, EPI_ISL_2838829, EPI_ISL_2838830, EPI_ISL_2838831, EPI_ISL_2838832, EPI_ISL_2838833, EPI_ISL_2838834, EPI_ISL_2838835, EPI_ISL_2838836, EPI_ISL_2838837, EPI_ISL_2838838, EPI_ISL_2838839, EPI_ISL_2838840, EPI_ISL_2838841, EPI_ISL_2838842, EPI_ISL_2838843 | | | |
| see above | OHSU Lab Services Molecular Microbiology Lab | Oregon SARS-CoV-2 Genome Sequencing Center | '[UsersoroakBox SyncPHI_SARS-COV-2_Sample_Sheet_infoGISAID_Submission_Infometa_master_2021.xlsx]release20_final'!D:AC; 26; Alec J. Hirsch; Andrew C. Adey; Benjamin N. Bimber; Brendan L. O'Connell; Brian J. O'Roak; Cierra LaBlanc; Daniel N. Streblow; Donna Hansel; FALSE); Guang Fan; Ruth V. Nichols; Sonia Acharya; VLOOKUP(C378; VLOOKUP(C379; VLOOKUP(C389; VLOOKUP(C390; VLOOKUP(C391; VLOOKUP(C395; VLOOKUP(C396; VLOOKUP(C397; VLOOKUP(C398; VLOOKUP(C410; VLOOKUP(C412; VLOOKUP(C413; VLOOKUP(C416; VLOOKUP(C418; VLOOKUP(C419; VLOOKUP(C421; VLOOKUP(C422; VLOOKUP(C423; VLOOKUP(C424; William B. Messer; Xuan Qin |
| EPI_ISL_3065773, EPI_ISL_3065774, EPI_ISL_3065776, EPI_ISL_3065783, EPI_ISL_3065786, EPI_ISL_3065788, EPI_ISL_3065790, EPI_ISL_3065791, EPI_ISL_3065793, EPI_ISL_3065797, EPI_ISL_3065818, EPI_ISL_3065830, EPI_ISL_3065834, EPI_ISL_3065836, EPI_ISL_3065841, EPI_ISL_3065843, EPI_ISL_3065845, EPI_ISL_3065846, EPI_ISL_3065850, EPI_ISL_3065851 | | | |
| see above | OHSU-MM Lab | OHSU-MM Lab | Rebecca O'Gara |
| EPI_ISL_2228065, EPI_ISL_2228071, | OLVZ Aalst | OLVZ Aalst | Anne Vankeerberghen |

| | | | |
|---|---|---|---|
| EPI_ISL_2228072 | | | |
| EPI_ISL_2384211, EPI_ISL_2384217, EPI_ISL_2384220, EPI_ISL_2451994 | OR State PHL-Virology/Immunology Section | Centers for Disease Control and Prevention Division of Viral Diseases, Pathogen Discovery | Alison Laufer Halpin; Ben L. Rambo-Martin; Clinton R. Paden; Dakota Howard; Darlene Wagner; Dave Wentworth; Dhwani Batra; Jasmine Padilla; Justin Lee; Katie Dillon; Krista Queen; Kristen Knipe; Kristine Lacek; Mark Burroughs; Matthew Schmerer; Mili Sheth; Peter Cook; Sam Shepard; Sarah Nobles; Shoshona Le; Suxiang Tong; Vivien Dugan; Yvette Unoarumhi |
| EPI_ISL_3943627 | Olive View Medical Center | Los Angeles County Public Health Laboratories | P. Hemarajata et al. |
| EPI_ISL_2339321, EPI_ISL_2339322, EPI_ISL_2339323, EPI_ISL_2339324, EPI_ISL_2339325, EPI_ISL_2339327, EPI_ISL_2339335, EPI_ISL_2346112, EPI_ISL_2346113, EPI_ISL_2346120, EPI_ISL_2348931, EPI_ISL_2348933, EPI_ISL_2348949 | | | |
| see above | Oregon State Public Health Laboratory | Oregon State Public Health Laboratory | Eugene Yeboah; John Fontana and Shane Sevey; Laura Tsaknaridis; Rafia Razzaque; Vanda Makris |
| EPI_ISL_2483726 | Ospedale S.M. Goretti di Latina - ASL Latina presidio ospedaliero Nord | INMI Lazzaro Spallanzani IRCCS | A Di Caro; A Lucci; B Bartolini; CEM Gruber; E Giombini; F Messina; F Santini; G Bonfiglio; L Corso; M Rueca; MR Capobianchi; O Butera; S Pignalosa |
| EPI_ISL_2665404 | Ospedale Sandro Pertini ASL Roma 2 | National Institute for Infectious Diseases (INMI) L. Spallanzani I.R.C.C.S | A Di Caro; B Bartolini; CEM Gruber; E Giombini; F Messina; F Santini; G Bonfiglio; M Rueca; MR Capobianchi; O Butera |
| EPI_ISL_2536634, EPI_ISL_2536639, EPI_ISL_2536642, EPI_ISL_2536643 | Overlake Hospital | Seattle Flu Study | Amanda Adler; Barry R. Lutz; Benjamin Pelle; Brian Hiatt; Caitlin R. Wolf; Chris D. Frazar; Deborah A. Nickerson; Elisabeth Brandstetter; Erica Ryke; Geoff Melly; Helen Y. Chu; Janet A. Englund; Jay Shendure; Jover Lee; Kairsten Fay; Kirsten Lacombe; Lea M. Starita; Mark J. Rieder; Matthew Richardson; Matthew Thompson; Melissa Truong; Michael Boeckh; Michael Famulare; Misja Ilcisin; Peter D. Han; Philip Dykema; Romesh Gautom; Scott Lindquist; Thomas R. Sibley; Trevor Bedford |
| EPI_ISL_2445407, EPI_ISL_2445439 | PA CAPELA | Instituto Butantan | Antonio Jorge Martins; Claudia Renata dos Santos Barros; David Schlesinger; Debora Botequio Moretti; Dimas Tadeu Covas; Elaine Cristina Marqueze; Elaine Vieira Santos; Evandra Strazza Rodrigues; Heidge Fukumasu; Jayme Augusto de Souza-Neto; José Salvatore Leister Patané; Luiz Alcantara; Luiz Lehmann Coutinho; Maria Carolina Elias; Maurício Lacerda Nogueira; Rafael dos Santos Bezerra; Raul Machado Neto; Rejane Maria Tommasini Grotto; Ricardo Haddad; Sandra Coccuzzo Sampaio Vessoni; Simone Kashima; Svetoslav Nanev Slavov; Vincent Louis Viala |
| EPI_ISL_2445596, EPI_ISL_2445601 | POLICLINICA COVID 19 ITAPETININGA | Instituto Butantan | Antonio Jorge Martins; Claudia Renata dos Santos Barros; David Schlesinger; Debora Botequio Moretti; Dimas Tadeu Covas; Elaine Cristina Marqueze; Elaine Vieira Santos; Evandra Strazza Rodrigues; Heidge Fukumasu; Jayme Augusto de Souza-Neto; José Salvatore Leister Patané; Luiz Alcantara; Luiz Lehmann Coutinho; Maria Carolina Elias; Maurício Lacerda Nogueira; Rafael dos Santos Bezerra; Raul Machado Neto; Rejane Maria Tommasini Grotto; Ricardo Haddad; Sandra Coccuzzo Sampaio Vessoni; Simone Kashima; Svetoslav Nanev Slavov; Vincent Louis Viala |
| EPI_ISL_2445574, EPI_ISL_2445576 | POSTO DE SAUDE BENVIRA E S F 4 | Instituto Butantan | Antonio Jorge Martins; Claudia Renata dos Santos Barros; David Schlesinger; Debora Botequio Moretti; Dimas Tadeu Covas; Elaine Cristina Marqueze; Elaine Vieira Santos; Evandra Strazza Rodrigues; Heidge Fukumasu; Jayme Augusto de Souza-Neto; José Salvatore Leister Patané; Luiz Alcantara; Luiz Lehmann Coutinho; Maria Carolina Elias; Maurício Lacerda Nogueira; Rafael dos Santos Bezerra; Raul Machado Neto; Rejane Maria Tommasini Grotto; Ricardo Haddad; Sandra Coccuzzo Sampaio Vessoni; Simone Kashima; Svetoslav Nanev Slavov; Vincent Louis Viala |
| EPI_ISL_2445402 | POSTO DE SAUDE DO TANQUE ATIBAIA | Instituto Butantan | Antonio Jorge Martins; Claudia Renata dos Santos Barros; David Schlesinger; Debora Botequio Moretti; Dimas Tadeu Covas; Elaine Cristina Marqueze; Elaine Vieira Santos; Evandra Strazza Rodrigues; Heidge Fukumasu; Jayme Augusto de Souza-Neto; José Salvatore Leister Patané; Luiz Alcantara; Luiz Lehmann Coutinho; Maria Carolina Elias; Maurício Lacerda Nogueira; Rafael dos Santos Bezerra; Raul Machado Neto; Rejane Maria Tommasini Grotto; Ricardo Haddad; Sandra Coccuzzo Sampaio Vessoni; Simone Kashima; Svetoslav Nanev Slavov; Vincent Louis Viala |
| EPI_ISL_3102280 | POSTO DE SAUDE MARIA DE LOURDES MAGALHAES MAIA | Analytical Competence Molecular Epidemiology Lab/ACME, Oswaldo Cruz Foundation, Ceara (FIOCRUZ CE) | Cleber Furtado Aksenen; Fabio Miyajima; Fernando Braga Stehling; Francisco Eder de Moura Lopes; Jamille Maria Mendes Bezerra; Joaquim César do Nascimento Sousa Junior; Pedro Miguel Carneiro Jeronimo; Suzana Porto Almeida e Lucas Delerino; Thais Ferreira de Oliveira; Thais de Oliveira Costa; Ticiane Cavalcante de Souza; Veridiana Pessoa Miyajima |
| EPI_ISL_2621198 | PROANALAB | Universidad Nacional de Colombia - Laboratorio Genómico One Health | Andres F. Cardona-Rios; Carlos Franco-Muñoz; Carolina Muñoz-Arango; Celeny Ortiz; Daniel O. Maldonado-Perez; Diego A. Álvarez-Díaz; Hector Alejandro Ruiz-Moreno; Idabely Betancur Ortiz; Jorge E. Osorio; Juan P. Hernandez-Ortiz; Karl A Ciuoderis; Katherine Laiton-Donato; Laura Silvana Perez; Lina M. Hurtado; Marcela Mercado-Reyes; Maria Angélica Maya; Maria Stella López; Rita Almanza Payares; Sandra Ines Cano; Simón Villegas Velásquez |
| EPI_ISL_2445241, EPI_ISL_2445244, EPI_ISL_2445252, EPI_ISL_2445369 | PRONTO ATENDIMENTO DE VARGEM GRANDE PAULISTA | Instituto Butantan | Antonio Jorge Martins; Claudia Renata dos Santos Barros; David Schlesinger; Debora Botequio Moretti; Dimas Tadeu Covas; Elaine Cristina Marqueze; Elaine Vieira Santos; Evandra Strazza Rodrigues; Heidge Fukumasu; Jayme Augusto de Souza-Neto; José Salvatore Leister Patané; Luiz Alcantara; Luiz Lehmann Coutinho; Maria Carolina Elias; Maurício Lacerda Nogueira; Rafael dos Santos Bezerra; Raul Machado Neto; Rejane Maria Tommasini Grotto; Ricardo Haddad; Sandra Coccuzzo Sampaio Vessoni; Simone Kashima; Svetoslav Nanev Slavov; Vincent Louis Viala |
| EPI_ISL_2473695 | PRONTO SOCORRO MUNICIPAL DE SEVERINIA | Instituto Butantan | Antonio Jorge Martins; Claudia Renata dos Santos Barros; David Schlesinger; Debora Botequio Moretti; Dimas Tadeu Covas; Elaine Cristina Marqueze; Elaine Vieira Santos; Evandra Strazza Rodrigues; Heidge Fukumasu; Jayme Augusto de Souza-Neto; José Salvatore Leister Patané; Luiz Alcantara; Luiz Lehmann Coutinho; Maria Carolina Elias; Maurício Lacerda Nogueira; Rafael dos Santos Bezerra; Raul Machado Neto; Rejane Maria Tommasini Grotto; Ricardo Haddad; Sandra Coccuzzo Sampaio Vessoni; Simone Kashima; Svetoslav Nanev Slavov; Vincent Louis Viala |
| EPI_ISL_2106514, EPI_ISL_2106568, EPI_ISL_2106588, EPI_ISL_2234910, EPI_ISL_2234922, EPI_ISL_2234994, EPI_ISL_2235067, EPI_ISL_2235090, EPI_ISL_2235138, EPI_ISL_2235191, EPI_ISL_2235205, EPI_ISL_3933405, EPI_ISL_4096611, EPI_ISL_4096622, EPI_ISL_4096629 | | | |
| see above | Pandemic Response Lab - NYC | Pandemic Response Lab, R&D | Cybill del Castillo; Dylan Law; Haiping Hao; Henry Lee; Isabel Fernandez Escapa; Jon Laurent; Katharine Nelson; Melissa Hopkins; Michael Hammerling; Pradeep Bugga; Shinyoung Clair Kang; Sol Rey; William Ward |
| EPI_ISL_3192152 | Pathogen Genomics Center, National Institute of Infectious Diseases | Pathogen Genomics Center, National Institute of Infectious Diseases | Kentaro Itokawa; Makoto Kuroda; Masanori Hashino; Rina Tanaka; Tsuyoshi Sekizuka |
| EPI_ISL_2993757 | Pathology, UAB | Pathology, UAB | E.J.; Hendrickson; Hernandez, J.; Leal; Lefkowitz; Moates, D.; R.C.; S.M. |
| EPI_ISL_2663311, EPI_ISL_6907925, EPI_ISL_6907931, EPI_ISL_6907937, EPI_ISL_6907938, EPI_ISL_6907945, EPI_ISL_6907953, EPI_ISL_6907954, EPI_ISL_6907961, EPI_ISL_6908468, EPI_ISL_6908475, EPI_ISL_6908480, EPI_ISL_6908886, EPI_ISL_6908887, EPI_ISL_6908888, EPI_ISL_6908889, EPI_ISL_6908890, EPI_ISL_6908891, EPI_ISL_6908892, EPI_ISL_6908893, EPI_ISL_6908898, EPI_ISL_6908903, EPI_ISL_6908906, EPI_ISL_6908907, EPI_ISL_6908913, EPI_ISL_6908919, EPI_ISL_6908926, EPI_ISL_6908927 | | | |
| see above | Plataforma de Vigilancia Molecular (PVM) - FIOCRUZ/BA | Plataforma de Vigilancia Molecular (PVM) - FIOCRUZ/BA | Bruno Bezerril Andrade; Camila I. de Oliveira; Camila I. de Oliveira on behalf of the Fiocruz COVID-19 Genomic Surveillance Network.; Clarissa Araújo Gurgel; Eduardo Oyama; Francys A Rangel; Laila A da Silva; Laise de Moraes; Leonardo Paiva Farias; Marina Cucco; Pedro Silveira; Ricardo Khouri; Ricardo Khouri on behalf of the Fiocruz COVID-19 Genomic Surveillance Network.; Tiago Graf |
| EPI_ISL_2226166, EPI_ISL_2226168, EPI_ISL_2226170, EPI_ISL_2226174, EPI_ISL_2226176, EPI_ISL_2226181, EPI_ISL_2226243, EPI_ISL_2325032 | | | |
| see above | Plateforme de testing Namuroise | Plateforme de testing Namuroise | Degosserie Jonathan; Denis Olivier; Lesly Nyinkeu Kemamen; Mullier François; Otto Gaetan |
| EPI_ISL_2131311, EPI_ISL_2131313, EPI_ISL_2131316, EPI_ISL_2272691, EPI_ISL_2272709, EPI_ISL_2272771 | Platform BIS UZA/UAntwerpen | Labo Klinische Biologie, UZA | Basil Britto Xavier; Christine Lammens; Herman Goossens; Jasmine Coppens; Marie Le Mercier; Veerle Matheeussen |
| EPI_ISL_2405206 | Providence Saint Joseph Medical Center | Los Angeles County Public Health Laboratories | P. Hemarajata et al. |
| EPI_ISL_2699286, EPI_ISL_2699292, EPI_ISL_2699300, EPI_ISL_2699351, EPI_ISL_2699354, EPI_ISL_2699422 | Public Health Laboratory, Minnesota Department of Health | University of Minnesota Genomics Center | Corbin Dirkx; Daryl M. Gohl; Jaquelyn Kuriger-Laber; John Garbe; and Sean Wang |
| EPI_ISL_2280093, EPI_ISL_2280094, EPI_ISL_2280097, EPI_ISL_2280098, EPI_ISL_2280099, EPI_ISL_2280101, EPI_ISL_2307140, EPI_ISL_2307142, EPI_ISL_2307145, EPI_ISL_2307146, EPI_ISL_2307147, EPI_ISL_2307150, EPI_ISL_2307151, EPI_ISL_2307152, EPI_ISL_2307153, EPI_ISL_2371691, EPI_ISL_2371693, EPI_ISL_2371694, EPI_ISL_2371695, EPI_ISL_2371699, EPI_ISL_2371700, EPI_ISL_2371701, EPI_ISL_2371702, EPI_ISL_2371703, EPI_ISL_2371704, EPI_ISL_2371706, EPI_ISL_2371707, EPI_ISL_2371712, EPI_ISL_2371713, EPI_ISL_2371714, EPI_ISL_2371715, EPI_ISL_2371716, EPI_ISL_2371718, EPI_ISL_2371720, EPI_ISL_2371721, EPI_ISL_2371722, EPI_ISL_2371724, EPI_ISL_2371725, EPI_ISL_2371727, EPI_ISL_2371728, EPI_ISL_2371729, EPI_ISL_2371730, EPI_ISL_2371731, EPI_ISL_2371732, EPI_ISL_2371733, EPI_ISL_2371736, EPI_ISL_2372899, EPI_ISL_2372900, EPI_ISL_2372901, EPI_ISL_2372902, EPI_ISL_2372903, EPI_ISL_2372904, EPI_ISL_2372905, EPI_ISL_2372906, EPI_ISL_2372907, EPI_ISL_2372908, EPI_ISL_2372909, EPI_ISL_2372910, EPI_ISL_2372911, EPI_ISL_2372912, EPI_ISL_2372913, EPI_ISL_2372914, EPI_ISL_2372915, EPI_ISL_2372916, EPI_ISL_2372917, EPI_ISL_2372918, EPI_ISL_2372919, EPI_ISL_2372920, EPI_ISL_2372921, EPI_ISL_2372922, EPI_ISL_2372923, EPI_ISL_2372924, EPI_ISL_2372925, EPI_ISL_2372926, EPI_ISL_2372927, EPI_ISL_2372928, EPI_ISL_2372929, EPI_ISL_2372930, EPI_ISL_2372931, EPI_ISL_2372932, EPI_ISL_2372933, EPI_ISL_2372934, EPI_ISL_2372935, EPI_ISL_2372936, EPI_ISL_2372937, EPI_ISL_2372938, EPI_ISL_2372939, EPI_ISL_2372940, EPI_ISL_2372941, EPI_ISL_2372943, EPI_ISL_2372944, EPI_ISL_2372945, EPI_ISL_2372946, EPI_ISL_2372947, EPI_ISL_2372948, EPI_ISL_2372949, EPI_ISL_2372951, EPI_ISL_2372952, EPI_ISL_2399471, EPI_ISL_2399474, EPI_ISL_2399475, EPI_ISL_2399477, EPI_ISL_2399480, EPI_ISL_2399481, EPI_ISL_2399482, EPI_ISL_2399483, EPI_ISL_2399484, EPI_ISL_2399485, EPI_ISL_2399488, EPI_ISL_2399489, EPI_ISL_2399490, EPI_ISL_2399491, EPI_ISL_2399492, EPI_ISL_2399494, EPI_ISL_2399495, EPI_ISL_2399496, EPI_ISL_2399497, EPI_ISL_2399498, EPI_ISL_2399499, EPI_ISL_2399500, EPI_ISL_2399501, EPI_ISL_2399503, EPI_ISL_2399507, EPI_ISL_2399510, EPI_ISL_2451458, EPI_ISL_2451459, EPI_ISL_2451461, EPI_ISL_2451465, EPI_ISL_4366220, EPI_ISL_4366538, EPI_ISL_4370004, EPI_ISL_4370627, EPI_ISL_4372599, EPI_ISL_4373310 | | | |
| see above | Quest Diagnostics Incorporated | Centers for Disease Control and Prevention Division of Viral Diseases, Pathogen Discovery | A. Gerasimova; A. Perez; Adrian Paskey; B. Anderson; Benjamin Rambo-Martin; Christopher Gulvick; Clinton Paden; Clinton R. Paden; Dakota Howard; Darlene Wagner; Dhwani Batra; Duncan MacCannell; Erisa Sula; F. Lacbawan; I. A. Shlyakhter; I. Shlyakhter; Jason Caravas; K. Livingston; K.E. Livingston; Kara Moser; Kristine Lacek; L. Bernstein; L.E. Bernstein; M. Hua; Matthew Schmerer; P. Tanpaiboon; Peter Cook; Peter W. Cook; R. Kagan; R. M. Kagan; R. Owen; R. Rolando; R. V. Rolando; S. H. Rosenthal; S. Rosenthal; Scott Sammons; Shatavia Morrison; Tymeckia Kendall; Victoria Caban Figueroa; Y. Liu; Yvette Unoarumhi |
| EPI_ISL_6229875 | RAIMUNDO FERREIRA FILHO NOVO TEMPO | ACME Lab, Oswaldo Cruz Foundation, FIOCRUZ/CE | Carlos Leonardo de Aragao Araujo; Cecilia Leite Costa & Eduardo Ruback dos Santos on behalf of COVID-19 FIOCRUZ Genomic Network; Cleber Furtado Aksenen; Fabio Miyajima; Fernando Braga Stehling; Francisco Eder de Moura Lopes; Igor Oliveira Duarte; Jamille Maria Mendes Bezerra; Joaquim Cesar do Nascimento Sousa Junior; Pedro Miguel Carneiro Jeronimo; Suzana Porto Almeida; Thais Ferreira de Oliveira; Thais de Oliveira Costa; Ticiane Cavalcante de Souza; Veridiana Pessoa Miyajima |
| EPI_ISL_2452233, EPI_ISL_2452242, EPI_ISL_2452256, EPI_ISL_2452268, EPI_ISL_2463005 | RI State Health Laboratories | Centers for Disease Control and Prevention Division of Viral Diseases, Pathogen Discovery | Alison Laufer Halpin; Ben L. Rambo-Martin; Clinton R. Paden; Dakota Howard; Darlene Wagner; Dave Wentworth; Dhwani Batra; Jasmine Padilla; Justin Lee; Katie Dillon; Krista Queen; Kristen Knipe; Kristine Lacek; Mark Burroughs; Matthew Schmerer; Mili Sheth; Peter Cook; Sam Shepard; Sarah Nobles; Shoshona Le; Suxiang Tong; Vivien Dugan; Yvette Unoarumhi |
| EPI_ISL_2473877, EPI_ISL_2473892, EPI_ISL_2473923, EPI_ISL_2473934, EPI_ISL_2474050, EPI_ISL_2474065, EPI_ISL_2474066 | | | |
| see above | Reditus Laboratories | Reditus Laboratories | Alexa Eichelberger; Cassy Philips; Joshua J. Geltz; M.S.; Ph.D.; Rex Dyer; Robert M. Sgambelluri |
| EPI_ISL_2362382, EPI_ISL_2362390, EPI_ISL_2362397, EPI_ISL_2362403, EPI_ISL_2362406, EPI_ISL_2362409, EPI_ISL_2362416, EPI_ISL_2362429, EPI_ISL_2362440, EPI_ISL_2362452, EPI_ISL_2362458, EPI_ISL_2362461, EPI_ISL_2362472, EPI_ISL_2362473, EPI_ISL_2362474, EPI_ISL_2362475, EPI_ISL_2362482 | | | |
| see above | Rhode Island Department of Health | Infectious Disease Program, Broad Institute of Harvard and MIT | Adams, G.; Azevedo, K.; B.L.; B.W.; Bauer, M.; Birren; Carter, A.; Chaluvadi, S.; D.J.; DeRuff, K.; Gladden-Young, A.; Huard, R.; J.E.; K.J.; King, E.; Lagerborg, K.; Lemieux; Loreth, C.; Miller, A.; Normandin, E.; P.C.; Park; Pearlman, L.; Reilly, S.; Rudy, M.; Sabeti; Siddle; Tomkins-Tinch, C.; and MacInnis |
| EPI_ISL_2426818, EPI_ISL_2426820, EPI_ISL_2426821, EPI_ISL_2426847, EPI_ISL_2426848, EPI_ISL_2426849, EPI_ISL_2603331 | | | |
| see above | Rhode Island State Health Laboratory | Rhode Island State Health Laboratory | Ewa King; Kristin Carpenter-Azevedo; Richard C. Huard |
| EPI_ISL_2691916 | Rush University Medical Center | RIPHL at Rush University Medical Center | Diane Springer; Felix Araujo Perez; Joyce Houlihan; Kevin Kunstman; Laura Furtado; Marieta Hyde; Mary Hayden; Max Kolton; Stefan Green |
| EPI_ISL_2840617 | S.C. Laboratorio Analisi, ASL 3 Liguria | U.O. Igiene, Ospedale Policlinico San Martino | Bruzzone Bianca; Caligiuri Patrizia; De Pace Vanessa; Domnich Alexander; Icardi Giancarlo on behalf of SARS-CoV-2 ITALIAN RESEARCH ENTERPRISE-(SCIRE) Collaborative Group; Orsi Andrea; Ricucci Valentina; Spitaleri Antonino |

| | | | |
|---|---|---|---|
| EPI_ISL_2445409, EPI_ISL_2445411, EPI_ISL_2445413 | SAE CTA SERV ATEND ESPEC TESTAGEM E ACONSELHAMENTO | Instituto Butantan | Antonio Jorge Martins; Claudia Renata dos Santos Barros; David Schlesinger; Debora Botequio Moretti; Dimas Tadeu Covas; Elaine Cristina Marqueze; Elaine Vieira Santos; Evandra Strazza Rodrigues; Heidge Fukumasu; Jayme Augusto de Souza-Neto; José Salvatore Leister Patané; Luiz Alcantara; Luiz Lehmann Coutinho; Maria Carolina Elias; Maurício Lacerda Nogueira; Rafael dos Santos Bezerra; Raul Machado Neto; Rejane Maria Tommasini Grotto; Ricardo Haddad; Sandra Coccuzzo Sampaio Vessoni; Simone Kashima; Svetoslav Nanev Slavov; Vincent Louis Viala |
| EPI_ISL_2445604 | SANTA CASA DE MISERICORDIA DE TIETE | Instituto Butantan | Antonio Jorge Martins; Claudia Renata dos Santos Barros; David Schlesinger; Debora Botequio Moretti; Dimas Tadeu Covas; Elaine Cristina Marqueze; Elaine Vieira Santos; Evandra Strazza Rodrigues; Heidge Fukumasu; Jayme Augusto de Souza-Neto; José Salvatore Leister Patané; Luiz Alcantara; Luiz Lehmann Coutinho; Maria Carolina Elias; Maurício Lacerda Nogueira; Rafael dos Santos Bezerra; Raul Machado Neto; Rejane Maria Tommasini Grotto; Ricardo Haddad; Sandra Coccuzzo Sampaio Vessoni; Simone Kashima; Svetoslav Nanev Slavov; Vincent Louis Viala |
| EPI_ISL_2791931, EPI_ISL_2791934, EPI_ISL_2791935, EPI_ISL_2791938, EPI_ISL_2791942, EPI_ISL_2792257, EPI_ISL_2792258, EPI_ISL_2792271, EPI_ISL_2792292, EPI_ISL_2792301, EPI_ISL_2792491, EPI_ISL_2792495, EPI_ISL_2792501 | | | |
| see above | SARS-CoV-2 Sequencing Castilla y Leon-Spain Consortium | SARS-CoV-2 Sequencing Castilla y Leon-Spain Consortium | Antonio Orduña-Domingo; Carlos Fuster Foz; Carmen Aldea-Mansilla; Carmen Gimeno Crespo; David Abad; Gregoria Megías Lobón; Jose María Eiros Bouza; Laura Sánchez de Prada; M. Isabel Fernandez-Natal; Marta Dominguez-Gil; Marta Hernandez; María Antonia García Castro; Mª Fe Brezmes-Valdivieso; Noelia Arenal Andrés; Silvia Rojo; Sonsoles Garcinuño Pérez |
| EPI_ISL_2769807 | SARS-CoV-2 testing team, National Institute of Infectious Diseases | Pathogen Genomics Center, National Institute of Infectious Diseases | Hazuka Y Furihata; Hiromizu Takahashi; Kentaro Itokawa; Makoto Kuroda; Masanori Hashino; Masumichi Saito; Naomi Nojiri; Nozomu Hanaoka; Rina Tanaka; Tsuguto Fujimoto; Tsuyoshi Sekizuka |
| EPI_ISL_2445464, EPI_ISL_2445468, EPI_ISL_2445469, EPI_ISL_2445470, EPI_ISL_2445472, EPI_ISL_2445474, EPI_ISL_2445475, EPI_ISL_2445476, EPI_ISL_2445477 | | | |
| see above | SECRETARIA DE SAUDE DE LOUVEIRA | Instituto Butantan | Antonio Jorge Martins; Claudia Renata dos Santos Barros; David Schlesinger; Debora Botequio Moretti; Dimas Tadeu Covas; Elaine Cristina Marqueze; Elaine Vieira Santos; Evandra Strazza Rodrigues; Heidge Fukumasu; Jayme Augusto de Souza-Neto; José Salvatore Leister Patané; Luiz Alcantara; Luiz Lehmann Coutinho; Maria Carolina Elias; Maurício Lacerda Nogueira; Rafael dos Santos Bezerra; Raul Machado Neto; Rejane Maria Tommasini Grotto; Ricardo Haddad; Sandra Coccuzzo Sampaio Vessoni; Simone Kashima; Svetoslav Nanev Slavov; Vincent Louis Viala |
| EPI_ISL_2493447 | SECRETARIA DE SAUDE DE SANTA GERTRUDES | Instituto Butantan | Antonio Jorge Martins; Claudia Renata dos Santos Barros; David Schlesinger; Debora Botequio Moretti; Dimas Tadeu Covas; Elaine Cristina Marqueze; Elaine Vieira Santos; Evandra Strazza Rodrigues; Heidge Fukumasu; Jayme Augusto de Souza-Neto; José Salvatore Leister Patané; Luiz Alcantara; Luiz Lehmann Coutinho; Maria Carolina Elias; Maurício Lacerda Nogueira; Rafael dos Santos Bezerra; Raul Machado Neto; Rejane Maria Tommasini Grotto; Ricardo Haddad; Sandra Coccuzzo Sampaio Vessoni; Simone Kashima; Svetoslav Nanev Slavov; Vincent Louis Viala |
| EPI_ISL_2493444, EPI_ISL_2494280, EPI_ISL_2494286, EPI_ISL_2494287 | SECRETARIA DE SAUDE DE SAO PEDRO | Instituto Butantan | Antonio Jorge Martins; Claudia Renata dos Santos Barros; David Schlesinger; Debora Botequio Moretti; Dimas Tadeu Covas; Elaine Cristina Marqueze; Elaine Vieira Santos; Evandra Strazza Rodrigues; Heidge Fukumasu; Jayme Augusto de Souza-Neto; José Salvatore Leister Patané; Luiz Alcantara; Luiz Lehmann Coutinho; Maria Carolina Elias; Maurício Lacerda Nogueira; Rafael dos Santos Bezerra; Raul Machado Neto; Rejane Maria Tommasini Grotto; Ricardo Haddad; Sandra Coccuzzo Sampaio Vessoni; Simone Kashima; Svetoslav Nanev Slavov; Vincent Louis Viala |
| EPI_ISL_2445598, EPI_ISL_2445599 | SECRETARIA MUNICIPAL DA SAUDE DE ITARARE | Instituto Butantan | Antonio Jorge Martins; Claudia Renata dos Santos Barros; David Schlesinger; Debora Botequio Moretti; Dimas Tadeu Covas; Elaine Cristina Marqueze; Elaine Vieira Santos; Evandra Strazza Rodrigues; Heidge Fukumasu; Jayme Augusto de Souza-Neto; José Salvatore Leister Patané; Luiz Alcantara; Luiz Lehmann Coutinho; Maria Carolina Elias; Maurício Lacerda Nogueira; Rafael dos Santos Bezerra; Raul Machado Neto; Rejane Maria Tommasini Grotto; Ricardo Haddad; Sandra Coccuzzo Sampaio Vessoni; Simone Kashima; Svetoslav Nanev Slavov; Vincent Louis Viala |
| EPI_ISL_2493367, EPI_ISL_2493452 | SECRETARIA MUNICIPAL DE SAUDE DE CORDEIROPOLIS | Instituto Butantan | Antonio Jorge Martins; Claudia Renata dos Santos Barros; David Schlesinger; Debora Botequio Moretti; Dimas Tadeu Covas; Elaine Cristina Marqueze; Elaine Vieira Santos; Evandra Strazza Rodrigues; Heidge Fukumasu; Jayme Augusto de Souza-Neto; José Salvatore Leister Patané; Luiz Alcantara; Luiz Lehmann Coutinho; Maria Carolina Elias; Maurício Lacerda Nogueira; Rafael dos Santos Bezerra; Raul Machado Neto; Rejane Maria Tommasini Grotto; Ricardo Haddad; Sandra Coccuzzo Sampaio Vessoni; Simone Kashima; Svetoslav Nanev Slavov; Vincent Louis Viala |
| EPI_ISL_5825566 | SECRETARIA MUNICIPAL DE SAUDE DE ITAPIPOCA | Analytical Competence Molecular Epidemiology Lab/ACME, Oswaldo Cruz Foundation, Ceara (FIOCRUZ CE) | Carlos Leonardo de Aragao Araujo; Cecilia Leite Costa & Eduardo Ruback dos Santos on behalf of COVID-19 FIOCRUZ Genomic Network; Cleber Furtado Aksenen; Fabio Miyajima; Fernando Braga Stehling; Francisco Eder de Moura Lopes; Igor Oliveira Duarte; Jamille Maria Mendes Bezerra; Joaquim Cesar do Nascimento Sousa Junior; Pedro Miguel Carneiro Jeronimo; Suzana Porto Almeida; Thais Ferreira de Oliveira; Thais de Oliveira Costa; Ticiane Cavalcante de Souza; Veridiana Pessoa Miyajima |
| EPI_ISL_6229776, EPI_ISL_6229777 | SECRETARIA MUNICIPAL DE SAUDE DE JAGUARIBE | ACME Lab, Oswaldo Cruz Foundation, FIOCRUZ/CE | Carlos Leonardo de Aragao Araujo; Cecilia Leite Costa & Eduardo Ruback dos Santos on behalf of COVID-19 FIOCRUZ Genomic Network; Cleber Furtado Aksenen; Fabio Miyajima; Fernando Braga Stehling; Francisco Eder de Moura Lopes; Igor Oliveira Duarte; Jamille Maria Mendes Bezerra; Joaquim Cesar do Nascimento Sousa Junior; Pedro Miguel Carneiro Jeronimo; Suzana Porto Almeida; Thais Ferreira de Oliveira; Thais de Oliveira Costa; Ticiane Cavalcante de Souza; Veridiana Pessoa Miyajima |
| EPI_ISL_2801353 | SECRETARIA MUNICIPAL DE SAUDE DE TIANGUA | Analytical Competence Molecular Epidemiology Lab/ACME, Oswaldo Cruz Foundation, Ceara (FIOCRUZ CE) | Cleber Furtado Aksenen e Suzana Porto Almeida; Fabio Miyajima; Fernando Braga Stehling; Francisco Eder de Moura Lopes; Jamille Maria Mendes Bezerra; Joaquim César do Nascimento Sousa Junior; Pedro Miguel Carneiro Jeronimo; Thais Ferreira de Oliveira; Thais de Oliveira Costa; Ticiane Cavalcante de Souza; Veridiana Pessoa Miyajima |
| EPI_ISL_2445386 | SECRETARIA MUNICIPAL DE SAUDE DE VALPARAISO SP | Instituto Butantan | Antonio Jorge Martins; Claudia Renata dos Santos Barros; David Schlesinger; Debora Botequio Moretti; Dimas Tadeu Covas; Elaine Cristina Marqueze; Elaine Vieira Santos; Evandra Strazza Rodrigues; Heidge Fukumasu; Jayme Augusto de Souza-Neto; José Salvatore Leister Patané; Luiz Alcantara; Luiz Lehmann Coutinho; Maria Carolina Elias; Maurício Lacerda Nogueira; Rafael dos Santos Bezerra; Raul Machado Neto; Rejane Maria Tommasini Grotto; Ricardo Haddad; Sandra Coccuzzo Sampaio Vessoni; Simone Kashima; Svetoslav Nanev Slavov; Vincent Louis Viala |
| EPI_ISL_3102520 | SECRETARIA MUNICIPAL DE SAUDE DE VARZEA ALEGRE | Analytical Competence Molecular Epidemiology Lab/ACME, Oswaldo Cruz Foundation, Ceara (FIOCRUZ CE) | Cleber Furtado Aksenen; Fabio Miyajima; Fernando Braga Stehling; Francisco Eder de Moura Lopes; Jamille Maria Mendes Bezerra; Joaquim César do Nascimento Sousa Junior; Pedro Miguel Carneiro Jeronimo; Suzana Porto Almeida e Lucas Delerino; Thais Ferreira de Oliveira; Thais de Oliveira Costa; Ticiane Cavalcante de Souza; Veridiana Pessoa Miyajima |
| EPI_ISL_2445602, EPI_ISL_2473701 | SECRETARIA MUNICIPAL DE SAUDE SOROCABA | Instituto Butantan | Antonio Jorge Martins; Claudia Renata dos Santos Barros; David Schlesinger; Debora Botequio Moretti; Dimas Tadeu Covas; Elaine Cristina Marqueze; Elaine Vieira Santos; Evandra Strazza Rodrigues; Heidge Fukumasu; Jayme Augusto de Souza-Neto; José Salvatore Leister Patané; Luiz Alcantara; Luiz Lehmann Coutinho; Maria Carolina Elias; Maurício Lacerda Nogueira; Rafael dos Santos Bezerra; Raul Machado Neto; Rejane Maria Tommasini Grotto; Ricardo Haddad; Sandra Coccuzzo Sampaio Vessoni; Simone Kashima; Svetoslav Nanev Slavov; Vincent Louis Viala |
| EPI_ISL_2270897, EPI_ISL_2270917, EPI_ISL_2270926, EPI_ISL_2483578, EPI_ISL_2483579, EPI_ISL_2483580, EPI_ISL_2483581, EPI_ISL_2483582 | | | |
| see above | SIESP DIPARTIMENTO DI PREVENZIONE TERAMO | Istituto Zooprofilattico Sperimentale dell'Abruzzo e Molise "G. Caporale" | Ancora M; Calistri P; Cammà C; Caporale M; Curini V; Delli Compagni E; Di Domenico M; Di Lollo Valeria; Di Pasquale A; Lorusso A; Mangone I; Marcacci M; Puglia I; Rinaldi A; Savini G; Scialabba S |
| EPI_ISL_2587468, EPI_ISL_2587563, EPI_ISL_2587566, EPI_ISL_2587568, EPI_ISL_2587570, EPI_ISL_2587829, EPI_ISL_2587839, EPI_ISL_2587841, EPI_ISL_2587862, EPI_ISL_2587868, EPI_ISL_2587871, EPI_ISL_2587873, EPI_ISL_2587875, EPI_ISL_2587877, EPI_ISL_2587879 | | | |
| see above | SK-Roy Romanow Provincial Laboratory | National Microbiology Laboratory (NML) | Amanda Lang; Anna Majer; Anneliese Landgraff; CanCOGeN's metadata curation team; Darian Hole; Elsie Grudeski; Gary Van Domselaar; Grace Seo; Jennifer Tanner; Jessica Minion; Kirsten Biggar; Madison Chapel; Morag Graham; Natalie Knox; Nathalie Bastien; Philip Mabon; Public Health Agency of Canada CanCOGeN team; Rachel DePaulo; Rhiannon Huzarewich; Russell Mandes; Ryan McDonald; Shari Tyson; Timothy Booth; Yan Li |
| EPI_ISL_5844430, EPI_ISL_5844448, EPI_ISL_5844449, EPI_ISL_5844456, EPI_ISL_5844458, EPI_ISL_5844471, EPI_ISL_5844472, EPI_ISL_5844488, EPI_ISL_5844492, EPI_ISL_6124967, EPI_ISL_6124973, EPI_ISL_6124985, EPI_ISL_6125051 | | | |
| see above | SK-Roy Romanow Provincial Laboratory | Saskatchewan - Roy Romanow Provincial Laboratory (RRPL) | Alanna Senecal; Amanda Lang; Jessica Minion; Kara Loos; Keith MacKenzie; Meredith Faires; Rachel DePaulo; Roy Romanow Provincial Laboratory - Molecular Diagnostics; Ryan McDonald |
| EPI_ISL_2445541, EPI_ISL_2445580, EPI_ISL_2445600 | SMS SECRETARIA MUNICIPAL DE SAUDE DE BOITUVA | Instituto Butantan | Antonio Jorge Martins; Claudia Renata dos Santos Barros; David Schlesinger; Debora Botequio Moretti; Dimas Tadeu Covas; Elaine Cristina Marqueze; Elaine Vieira Santos; Evandra Strazza Rodrigues; Heidge Fukumasu; Jayme Augusto de Souza-Neto; José Salvatore Leister Patané; Luiz Alcantara; Luiz Lehmann Coutinho; Maria Carolina Elias; Maurício Lacerda Nogueira; Rafael dos Santos Bezerra; Raul Machado Neto; Rejane Maria Tommasini Grotto; Ricardo Haddad; Sandra Coccuzzo Sampaio Vessoni; Simone Kashima; Svetoslav Nanev Slavov; Vincent Louis Viala |
| EPI_ISL_2422317 | STONY BROOK UNIVERSITY HOSPITAL | Wadsworth Center, New York State Department of Health | Alexis Russell; Catharine Prussing; Daryl M. Lamson; Erasmus Schneider; Erica Lasek-Nesselquist; John Kelly; Jonathan Plitnick; Kirsten St. George; Matthew Shudt; Melissa A Leisner; Navjot Singh |
| EPI_ISL_5155736 | SUNY Upstate Medical University | 505 Irving Avenue, Syracuse, NY 13210 - Frank Middleton Lab | Brian Pavlovitz; Frank Middleton |
| EPI_ISL_2245236, EPI_ISL_2245265, EPI_ISL_2245424, EPI_ISL_2245430, EPI_ISL_2289019, EPI_ISL_2289033, EPI_ISL_2289037, EPI_ISL_2289065, EPI_ISL_2321660, EPI_ISL_2321661, EPI_ISL_2321662, EPI_ISL_2321664, EPI_ISL_2321665, EPI_ISL_2321666, EPI_ISL_2321667, EPI_ISL_2321717, EPI_ISL_2321718, EPI_ISL_2321720, EPI_ISL_2321721 | | | |
| see above | SYNLAB | GIGA Medical Genomics | Bouchra Boujemla; Cécile Meex; Keith Durkin; Maria Artesi; Marie-Pierre Hayette; Nathalie Renotte; Pierrette Melin; Raphaël Boreux; Sébastien Bontems; Vincent Bours |
| EPI_ISL_2445961, EPI_ISL_2536121 | SYNLAB | Instituto Nacional de Saude (INSA) | Borges et al |
| EPI_ISL_3276548 | SYNLAB | Universidad Nacional de Colombia - Laboratorio Genómico One Health | Andres F. Cardona-Rios; Carlos Franco-Muñoz; Carolina Muñoz-Arango; Celeny Ortiz; Daniel O. Maldonado-Perez; Diego A. Álvarez-Díaz; Hector Alejandro Ruiz-Moreno; Idabely Betancur Ortiz; Jorge E. Osorio; Juan P. Hernandez-Ortiz; Karl A Ciuoderis; Katherine Laiton-Donato; Laura Silvana Perez; Lina M. Hurtado; Marcela Mercado-Reyes; Maria Angélica Maya; Maria Stella López; Rita Almanza Payares; Sandra Ines Cano; Simón Villegas Velásquez |
| EPI_ISL_2265162, EPI_ISL_2316232 | SYNLAB MVZ Leinfelden-Echterdingen | Robert Koch Institute | |
| EPI_ISL_2319308, EPI_ISL_2349882, EPI_ISL_2349884, EPI_ISL_2349889, EPI_ISL_2349892, EPI_ISL_2349902, EPI_ISL_2349931, EPI_ISL_2349932, EPI_ISL_2349949, EPI_ISL_2349951, EPI_ISL_2349978, EPI_ISL_2349980, EPI_ISL_2349991, EPI_ISL_2349995, EPI_ISL_2349997, EPI_ISL_2349998, EPI_ISL_2350003, EPI_ISL_2350015, EPI_ISL_2350024, EPI_ISL_2350046, EPI_ISL_2350050, EPI_ISL_2350051, EPI_ISL_2350070, EPI_ISL_2350085, EPI_ISL_2350086, EPI_ISL_2350093, EPI_ISL_2881822, EPI_ISL_2881827, EPI_ISL_2881829 | | | |
| see above | Salud Digna | Instituto Nacional de Medicina Genomica | Abraham Campos-Romero; Cedro-Tanda A; Escobar-Arrazola; Gonzalez-Barrera D; Herrera-Montalvo LA.; Hidalgo-Miranda A; Luna-Ruiz Marco; M; Mendoza-Vargas A; Moreno-Camacho José Luis; Munguia-Garza P; Ramirez-Vega O; Rangel-DeLeon D; Reyes-Grajeda JP; Rodriguez-Gallegos Jorge |
| EPI_ISL_2628346, EPI_ISL_2628598, EPI_ISL_2628844 | San Diego County Public Health Laboratory | Andersen lab at Scripps Research | Brett Austin; Jovan Shephard; SEARCH Alliance San Diego with Ashleigh Murphy |
| EPI_ISL_2840867, EPI_ISL_2840895 | San Diego County Public Health Laboratory | San Diego County Public Health Laboratory | Ashleigh Murphy Schafer; Brett Austin |
| EPI_ISL_2631212, EPI_ISL_2650546, EPI_ISL_2650549, EPI_ISL_2650550, EPI_ISL_2650552, EPI_ISL_2650565 | Santa Clara Valley Medical Center | Santa Clara County Public Health Laboratory | Santa Clara County Public Health Department |
| EPI_ISL_2692314, EPI_ISL_2692317, EPI_ISL_2692318 | Seattle Flu Study | Seattle Flu Study | Amanda Adler; Barry R. Lutz; Benjamin Pelle; Caitlin R. Wolf; Chris D. Frazar; Deborah A. Nickerson; Elisabeth Brandstetter; Erica Ryke; Helen Y. Chu; Janet A. Englund; Jay Shendure; Jeff Duchin; Jover Lee; Kairsten Fay; Karen Cowgill; Kirsten Lacombe; Lea M. Starita; Mark J. Rieder; Matthew Richardson; Matthew Thompson; Melissa Truong; Michael Boeckh; Michael Famulare; Misja Ilcisin; Peter D. Han; Stephanie Schrag; Thomas R. Sibley; Trevor Bedford |
| EPI_ISL_2179796, EPI_ISL_2179797 | Servicio Microbiología Hospital La Paz | Servicio Microbiología Hospital La Paz | Elie Dahdouh; Fernando Lázaro; Jesús Mingorance Cruz; Rubén Cáceres |
| EPI_ISL_2158843, EPI_ISL_2158845, EPI_ISL_4082187, EPI_ISL_4082190, EPI_ISL_4082205, EPI_ISL_4082206, EPI_ISL_4082207, EPI_ISL_4082209, EPI_ISL_4082220, EPI_ISL_4082221, EPI_ISL_4082223, EPI_ISL_4082225, EPI_ISL_4082253, EPI_ISL_4082254, EPI_ISL_4082257, EPI_ISL_4082259, EPI_ISL_4082260, EPI_ISL_4082261, EPI_ISL_4082262, EPI_ISL_4082264, EPI_ISL_4082265, EPI_ISL_4082266, EPI_ISL_4082268, EPI_ISL_4082269, EPI_ISL_4082273, EPI_ISL_4082274, EPI_ISL_4082276, EPI_ISL_4082279, EPI_ISL_4082281, EPI_ISL_4082283, EPI_ISL_4082285, EPI_ISL_4082286, EPI_ISL_4082289, EPI_ISL_4082291, EPI_ISL_4082294, EPI_ISL_4082296, EPI_ISL_4082297, EPI_ISL_4082300, EPI_ISL_4082301, EPI_ISL_4082304, EPI_ISL_4082305, EPI_ISL_4082306, EPI_ISL_4082307, EPI_ISL_4082317, EPI_ISL_4082327, EPI_ISL_4082328, EPI_ISL_4082330, EPI_ISL_4082336, EPI_ISL_4082337, EPI_ISL_4082338, EPI_ISL_4082339, EPI_ISL_4082340, EPI_ISL_4082345, EPI_ISL_4082363, EPI_ISL_4082370, EPI_ISL_4082457, EPI_ISL_4082493, EPI_ISL_4082525, EPI_ISL_4082526, EPI_ISL_4082527, EPI_ISL_4082534, EPI_ISL_4082535, EPI_ISL_4082539, EPI_ISL_4082546, EPI_ISL_4082648, EPI_ISL_4082721, EPI_ISL_4082766, EPI_ISL_4082827, EPI_ISL_4082965, EPI_ISL_6694364, EPI_ISL_6694431 | | | |
| see above | Servicio Virosis Respiratorias-Departamento Virología-INEI | Instituto Nacional Enfermedades Infecciosas C.G.Malbran | Avaro M.; Baumeister E.; Benedetti E.; Campos J.; Cisterna D.; Dattero ME; De Belder D.; Haim MS.; Lorenzo F.; Molina V.; Perandones C.; Poklepovich T.; Pontoriero A.; Russo M.; Sanchez Loria J.; Tuduri E. |

| | | | |
|---|---|---|---|
| EPI_ISL_2426627, EPI_ISL_2426630, EPI_ISL_2426641, EPI_ISL_2558027, EPI_ISL_2558046, EPI_ISL_2861034 | Servicio de Microbiologia Hospital Ramon y Cajal | Servicio de Microbiologia Hospital Ramon y Cajal | Galan JC; Martinez L. Abreu M; Ponce M; y Gonzalez-Alba JM |
| EPI_ISL_2510576 | Servicio de Microbiología Clínica (Complejo Hospitalario de Navarra, Pamplona) | Centro de Secuenciación NASERTIC | Ana Miqueleiz; Ana Navascués; Carmen Ezpeleta Baquedano |
| EPI_ISL_2362319 | Servicio de Microbiología. Consorcio Hospital General Universitario de Valencia | SeqCOVID-SPAIN consortium/IBV(CSIC) | Begoña Fuster Escrivá; Carme Salvador García; Concepción Gimeno Cardona and SeqCOVID-SPAIN consortium; María Dolores Ocete; Rafael Medina González |
| EPI_ISL_2439985, EPI_ISL_2439986, EPI_ISL_2439987 | Servicio de Microbiología. Hospital Clínico Universitario de Valencia | SeqCOVID-SPAIN consortium/IBV(CSIC) | David Navarro Ortega; Eliseo Albert Vicent; Ignacio Torres and SeqCOVID-SPAIN consortium |
| EPI_ISL_5315786, EPI_ISL_5315789, EPI_ISL_5315792, EPI_ISL_5315793, EPI_ISL_5315794, EPI_ISL_5315795, EPI_ISL_5315796, EPI_ISL_5315797, EPI_ISL_5315799, EPI_ISL_5315800, EPI_ISL_5315801, EPI_ISL_5315802, EPI_ISL_5315803, EPI_ISL_5315804, EPI_ISL_5315805, EPI_ISL_5315807, EPI_ISL_5315810, EPI_ISL_5315814, EPI_ISL_5315815, EPI_ISL_5315818, EPI_ISL_5315820, EPI_ISL_5315822, EPI_ISL_5315826, EPI_ISL_5315827, EPI_ISL_5315830, EPI_ISL_5315839, EPI_ISL_5315857, EPI_ISL_5315859, EPI_ISL_6686903, EPI_ISL_6686904, EPI_ISL_6686905, EPI_ISL_6686906, EPI_ISL_6686907, EPI_ISL_6686910, EPI_ISL_6686911, EPI_ISL_6686912, EPI_ISL_6686913, EPI_ISL_6686915, EPI_ISL_6686917, EPI_ISL_6686918, EPI_ISL_6686920, EPI_ISL_6686923, EPI_ISL_6686924, EPI_ISL_6687389, EPI_ISL_6687400, EPI_ISL_6687403, EPI_ISL_6687444, EPI_ISL_6687447, EPI_ISL_6687659, EPI_ISL_6687673, EPI_ISL_6687737, EPI_ISL_6688132, EPI_ISL_6688244, EPI_ISL_6688305 | | | |
| see above | Shared Hospital Laboratory | Shared Hospital Laboratory | Christie Vermeiren; Finlay Maguire; Kevin Katz; Patryk Aftanas; Robert Kozak; Samira Mubareka |
| EPI_ISL_2422816 | Sharp HealthCare Laboratory | Andersen lab at Scripps Research | Art Mendoza; Cathy Woerle; Jacquelyn Berumen; Liam McGinnis; Omid Bakhtar; SEARCH Alliance San Diego with Aaron Harding |
| EPI_ISL_2801143, EPI_ISL_2801144, EPI_ISL_2801145 | Simple Laboratories | Gagnon Lab, Southern Illinois University | Keith Gagnon |
| EPI_ISL_2621881, EPI_ISL_2621883, EPI_ISL_2651208 | Somer | Universidad Nacional de Colombia - Laboratorio Genómico One Health | Andres F. Cardona-Rios; Carlos Franco-Muñoz; Carolina Muñoz-Arango; Celeny Ortiz; Daniel O. Maldonado-Perez; Diego A. Álvarez-Díaz; Hector Alejandro Ruiz-Moreno; Idabely Betancur Ortiz; Jorge E. Osorio; Juan P. Hernandez-Ortiz; Karl A Ciuoderis; Katherine Laiton-Donato; Laura Silvana Perez; Lina M. Hurtado; Marcela Mercado-Reyes; Maria Angélica Maya; Maria Stella López; Rita Almanza Payares; Sandra Ines Cano; Simón Villegas Velásquez |
| EPI_ISL_2449882 | Spital Limmattal | Institute of Medical Virology | Alexandra Trkola; Annette AudigŽ; Catharine Aquino; Cyril Shah; Daniel Ehrsam; Gabriela Ziltener; Guido Bloemberg; Hubert Rehrauer; Isabel StŸrmer; Joel Wirz; Jon Huder; JŸrg Bšni; Kevin Steiner; Maria GrŸnberg; Maryam Zaheri; Michael Huber; Riccarda Capaul; Stefan Schmutz; Verena Kufner; Weihong Qi |
| EPI_ISL_2449879 | Spital MŠnnedorf AG | Institute of Medical Virology | Alexandra Trkola; Annette AudigŽ; Catharine Aquino; Cyril Shah; Daniel Ehrsam; Gabriela Ziltener; Guido Bloemberg; Hubert Rehrauer; Isabel StŸrmer; Joel Wirz; Jon Huder; JŸrg Bšni; Kevin Steiner; Maria GrŸnberg; Maryam Zaheri; Michael Huber; Riccarda Capaul; Stefan Schmutz; Verena Kufner; Weihong Qi |
| EPI_ISL_4496866 | Stanford Health Care | Stanford University School of Medicine, Clinical Virology Laboratory | Becky Jiang; James Zehnder; Malaya K. Sahoo; Selamawit Bihon; and Benjamin A. Pinsky |
| EPI_ISL_2724635, EPI_ISL_2724639, EPI_ISL_2724643, EPI_ISL_2724646, EPI_ISL_2724651 | State Laboratories Division, Hawaii State Department of Health | State Laboratories Division, Hawaii State Department of Health | Ayana Garnet; Drew Kuwazaki; Edward Desmond; Pamela O'Brien; Razvan Sultana |
| EPI_ISL_2473299, EPI_ISL_2473310, EPI_ISL_2473321, EPI_ISL_2473335, EPI_ISL_2473336, EPI_ISL_2473351, EPI_ISL_2473363, EPI_ISL_2473366, EPI_ISL_2473368, EPI_ISL_2473382, EPI_ISL_2473390, EPI_ISL_2473401, EPI_ISL_2473409, EPI_ISL_2473413, EPI_ISL_2473423, EPI_ISL_2473426, EPI_ISL_2473448, EPI_ISL_2473468 | | | |
| see above | State Testing Facility | Altius Institute for Biomedical Research | Alex Isner; Alex Nguyen; Amanda Gale; Audra Johnson; Clem Green; Daniel Bates; Eric Thorland; Jacob Rodriguez; Jean Robinston; Jemma Nelson; Jessica Kunder; John Stamatoyannopoulos; Joshua Richards; Julia Wald; Kneshay Harper; Lauren Mitchell; Mark Frerker; Matt Hartman; Michael Buckley; Muhammad Halimun; Rebecca Bruders; Sadie Patraw; Sofia Olsson; Tobias Ragoczy |
| EPI_ISL_6794353 | State of New Hampshire Public Health Laboratories | State of New Hampshire Public Health Laboratories | Caitlin Mercier; Chris Benton; Jinfeng Li; Juan Bolanos; Xinglu Zhang |
| EPI_ISL_2278969, EPI_ISL_2414029, EPI_ISL_2414091, EPI_ISL_2581053 | Swedish national genomic surveillance program of SARS-CoV-2 | The Public Health Agency of Sweden | Alma Brolund; Maria Lind Karlberg; Maximilian Riess; Swedish national genomic surveillance program of SARS-CoV-2 |
| EPI_ISL_2438050, EPI_ISL_2621150, EPI_ISL_2621152, EPI_ISL_2621154, EPI_ISL_2621155, EPI_ISL_2621156 | Synlab | Universidad Nacional de Colombia - Laboratorio Genómico One Health | Andres F. Cardona-Rios; Carlos Franco-Muñoz; Carolina Muñoz-Arango; Celeny Ortiz; Daniel O. Maldonado-Perez; Diego A. Álvarez-Díaz; Hector Alejandro Ruiz-Moreno; Idabely Betancur Ortiz; Jorge E. Osorio; Juan P. Hernandez-Ortiz; Karl A Ciuoderis; Katherine Laiton-Donato; Laura Silvana Perez; Lina M. Hurtado; Marcela Mercado-Reyes; Maria Angélica Maya; Maria Stella López; Rita Almanza Payares; Sandra Ines Cano; Simón Villegas Velásquez |
| EPI_ISL_2263832, EPI_ISL_2266877 | Synlab MVZ Augsburg | Robert Koch Institute | |
| EPI_ISL_2423669, EPI_ISL_2491235, EPI_ISL_2491275, EPI_ISL_2600381, EPI_ISL_2600412, EPI_ISL_2671940, EPI_ISL_2671972, EPI_ISL_2672010, EPI_ISL_2672060, EPI_ISL_2672061, EPI_ISL_2672072, EPI_ISL_2672151, EPI_ISL_2672201, EPI_ISL_6253906, EPI_ISL_6253926 | | | |
| see above | TGen North | TGen North | "Jolene Bowers; Ashlyn Pfeiffer; Brett Van Tassel; Chris French; Darrin Lemmer; Dave Engelthaler; Dave Engelthaler"; Hayley Yaglom; Heather Centner; Jolene Bowers; The Arizona COVID Genomics Union (ACGU); The Arizona COVID Genomics Union (ACGU)" |
| EPI_ISL_2450366 | TN Division of Laboratory Services | Genomics and Discovery, Respiratory Viruses Branch, Division of Viral Diseases, Centers for Disease Control and Prevention | Adam Retchless; Anna Kelleher; Anna Uehara; Brian Lynch; Clinton R. Paden; Dhwani Batra; Haibin Wang; Han Jia Justin Ng; Jasmine Padilla; Jing Zhang; Justin Lee; Krista Queen; Mark Burroughs; Mili Sheth; Morgan Davis; Peter Cook; Rachel Marine; Sarah Nobles; Suxiang Tong; Tara Coalter; Yan Li; Ying Tao |
| EPI_ISL_2443313, EPI_ISL_2501739, EPI_ISL_2501740 | TXDSHS | TXDSHS | Anita Pokharel; Bonnie Oh; Chun Wang; Grace Kubin; Jenny Zhang; Karen Bobier; Lorraine Rodriguez; Maliha Rahman; Mayela Pedrueza; Myong Koag; Rachel Lee; Rashmi Tuladhar |
| EPI_ISL_2966815, EPI_ISL_2988615 | Tampa General Hospital Esoteric Lab | Tampa General Hospital Esoteric Research & Development Lab | Amorce Lima; Deanna Becker; Dominic Uy; Elaine Vendrone; Jon Faughn; Jorge Mecias-Frias; Suzane Silbert; Vicki Healer |
| EPI_ISL_2893503 | Texas Children's Hospital | Texas Children's Microbiome Center | Ila Singh; James Dunn; James Versalovic; Jennifer K. Spinler; Ruth Ann Luna |
| EPI_ISL_2678146, EPI_ISL_2678148, EPI_ISL_2678152, EPI_ISL_2678154, EPI_ISL_2678155, EPI_ISL_2678156, EPI_ISL_2678158, EPI_ISL_2716584, EPI_ISL_2716590, EPI_ISL_2716591, EPI_ISL_2716595, EPI_ISL_2716599, EPI_ISL_2716600, EPI_ISL_2756560, EPI_ISL_2756561, EPI_ISL_2756562, EPI_ISL_2756563, EPI_ISL_2756564, EPI_ISL_2756565, EPI_ISL_2756566, EPI_ISL_2756567, EPI_ISL_3642757 | | | |
| see above | The Caribbean Public Health Agency | Carrington Lab, Department of Preclinical Sciences, Faculty of Medical Sciences, The University of the West Indies, St Augustine Campus | Anushka Ramjag; Arianne Brown-Jordan; Avery Hinds; Christine V. F. Carrington; Christopher Oura; Gabriel Escobar; Nikita S. D. Sahadeo; Nuno Faria; Oliver Pybus; Risha Singh; Roshan Parasram; Sarah Hill; SueMin Nathaniel; Vernie Ramkissoon |
| EPI_ISL_2230024, EPI_ISL_2230026 | The Jackson Laboratory | The Jackson Laboratory | Adams M; Bergeron D; Kelly K; Li L; Long J; Omerza G; Renzette N |
| EPI_ISL_2534309, EPI_ISL_2758841, EPI_ISL_2834212 | The Ohio State University Applied Microbiology Services Laboratory | The Ohio State University Applied Microbiology Services Laboratory | Seth A. Faith PhD |
| EPI_ISL_3274693, EPI_ISL_3274694 | TriCore Reference Laboratories | Center for Global Health, University of New Mexico Health Sciences Center | Cecilia Thompson; Darrell Dinwiddie; Daryl Domman; Karissa Culbreath; Kendra Pesko; Kurt Schwalm; Valerie Morley |
| EPI_ISL_3185330 | U.O. Microbiologia Laboratorio Unico Centro Servizi - AUSL della Romagna | U.O. Microbiologia, Laboratorio Unico Centro Servizi - AUSL della Romagna | Giorgio Dirani |
| EPI_ISL_6229818, EPI_ISL_6229851, EPI_ISL_6229857 | UAPS CASEMIRO FILHO | ACME Lab, Oswaldo Cruz Foundation, FIOCRUZ/CE | Carlos Leonardo de Aragao Araujo; Cecilia Leite Costa & Eduardo Ruback dos Santos on behalf of COVID-19 FIOCRUZ Genomic Network; Cleber Furtado Aksenen; Fabio Miyajima; Fernando Braga Stehling; Francisco Eder de Moura Lopes; Igor Oliveira Duarte; Jamille Maria Mendes Bezerra; Joaquim Cesar do Nascimento Sousa Junior; Pedro Miguel Carneiro Jeronimo; Suzana Porto Almeida; Thais Ferreira de Oliveira; Thais de Oliveira Costa; Ticiane Cavalcante de Souza; Veridiana Pessoa Miyajima |
| EPI_ISL_2473693 | UBS ALCIMINIO DE ASSIS LOURENCO BADY BASSITT | Instituto Butantan | Antonio Jorge Martins; Claudia Renata dos Santos Barros; David Schlesinger; Debora Botequio Moretti; Dimas Tadeu Covas; Elaine Cristina Marqueze; Elaine Vieira Santos; Evandra Strazza Rodrigues; Heidge Fukumasu; Jayme Augusto de Souza-Neto; José Salvatore Leister Patané; Luiz Alcantara; Luiz Lehmann Coutinho; Maria Carolina Elias; Maurício Lacerda Nogueira; Rafael dos Santos Bezerra; Raul Machado Neto; Rejane Maria Tommasini Grotto; Ricardo Haddad; Sandra Coccuzzo Sampaio Vessoni; Simone Kashima; Svetoslav Nanev Slavov; Vincent Louis Viala |
| EPI_ISL_6229784 | UBS CENTRO | ACME Lab, Oswaldo Cruz Foundation, FIOCRUZ/CE | Carlos Leonardo de Aragao Araujo; Cecilia Leite Costa & Eduardo Ruback dos Santos on behalf of COVID-19 FIOCRUZ Genomic Network; Cleber Furtado Aksenen; Fabio Miyajima; Fernando Braga Stehling; Francisco Eder de Moura Lopes; Igor Oliveira Duarte; Jamille Maria Mendes Bezerra; Joaquim Cesar do Nascimento Sousa Junior; Pedro Miguel Carneiro Jeronimo; Suzana Porto Almeida; Thais Ferreira de Oliveira; Thais de Oliveira Costa; Ticiane Cavalcante de Souza; Veridiana Pessoa Miyajima |
| EPI_ISL_2445528 | UBS DE PEDRANOPOLIS | Instituto Butantan | Antonio Jorge Martins; Claudia Renata dos Santos Barros; David Schlesinger; Debora Botequio Moretti; Dimas Tadeu Covas; Elaine Cristina Marqueze; Elaine Vieira Santos; Evandra Strazza Rodrigues; Heidge Fukumasu; Jayme Augusto de Souza-Neto; José Salvatore Leister Patané; Luiz Alcantara; Luiz Lehmann Coutinho; Maria Carolina Elias; Maurício Lacerda Nogueira; Rafael dos Santos Bezerra; Raul Machado Neto; Rejane Maria Tommasini Grotto; Ricardo Haddad; Sandra Coccuzzo Sampaio Vessoni; Simone Kashima; Svetoslav Nanev Slavov; Vincent Louis Viala |
| EPI_ISL_2445536 | UBS DE UBARANA | Instituto Butantan | Antonio Jorge Martins; Claudia Renata dos Santos Barros; David Schlesinger; Debora Botequio Moretti; Dimas Tadeu Covas; Elaine Cristina Marqueze; Elaine Vieira Santos; Evandra Strazza Rodrigues; Heidge Fukumasu; Jayme Augusto de Souza-Neto; José Salvatore Leister Patané; Luiz Alcantara; Luiz Lehmann Coutinho; Maria Carolina Elias; Maurício Lacerda Nogueira; Rafael dos Santos Bezerra; Raul Machado Neto; Rejane Maria Tommasini Grotto; Ricardo Haddad; Sandra Coccuzzo Sampaio Vessoni; Simone Kashima; Svetoslav Nanev Slavov; Vincent Louis Viala |
| EPI_ISL_2445482, | UBS II DE REGENTE FEIJO | Instituto Butantan | Antonio Jorge Martins; Claudia Renata dos Santos Barros; David Schlesinger; Debora Botequio Moretti; Dimas Tadeu Covas; Elaine Cristina Marqueze; Elaine Vieira Santos; Evandra Strazza Rodrigues; Heidge Fukumasu; Jayme Augusto de Souza-Neto; José Salvatore Leister Patané; Luiz Alcantara; |

| | | | |
|---|---|---|---|
| EPI_ISL_2445484 | | | Luiz Lehmann Coutinho; Maria Carolina Elias; Maurício Lacerda Nogueira; Rafael dos Santos Bezerra; Raul Machado Neto; Rejane Maria Tommasini Grotto; Ricardo Haddad; Sandra Coccuzzo Sampaio Vessoni; Simone Kashima; Svetoslav Nanev Slavov; Vincent Louis Viala |
| EPI_ISL_2445516, EPI_ISL_2445518, EPI_ISL_2445519, EPI_ISL_2445520, EPI_ISL_2445534, EPI_ISL_2473696 | UBS II DE TANABI MILTON MARTINS PERCHES | Instituto Butantan | Antonio Jorge Martins; Claudia Renata dos Santos Barros; David Schlesinger; Debora Botequio Moretti; Dimas Tadeu Covas; Elaine Cristina Marqueze; Elaine Vieira Santos; Evandra Strazza Rodrigues; Heidge Fukumasu; Jayme Augusto de Souza-Neto; José Salvatore Leister Patané; Luiz Alcantara; Luiz Lehmann Coutinho; Maria Carolina Elias; Maurício Lacerda Nogueira; Rafael dos Santos Bezerra; Raul Machado Neto; Rejane Maria Tommasini Grotto; Ricardo Haddad; Sandra Coccuzzo Sampaio Vessoni; Simone Kashima; Svetoslav Nanev Slavov; Vincent Louis Viala |
| EPI_ISL_2445552, EPI_ISL_2445555, EPI_ISL_2473796 | UBS II DR JOSE CARLOS MACUCO JANINI | Instituto Butantan | Antonio Jorge Martins; Claudia Renata dos Santos Barros; David Schlesinger; Debora Botequio Moretti; Dimas Tadeu Covas; Elaine Cristina Marqueze; Elaine Vieira Santos; Evandra Strazza Rodrigues; Heidge Fukumasu; Jayme Augusto de Souza-Neto; José Salvatore Leister Patané; Luiz Alcantara; Luiz Lehmann Coutinho; Maria Carolina Elias; Maurício Lacerda Nogueira; Rafael dos Santos Bezerra; Raul Machado Neto; Rejane Maria Tommasini Grotto; Ricardo Haddad; Sandra Coccuzzo Sampaio Vessoni; Simone Kashima; Svetoslav Nanev Slavov; Vincent Louis Viala |
| EPI_ISL_2445543 | UBS III DR BENITO MALZONE | Instituto Butantan | Antonio Jorge Martins; Claudia Renata dos Santos Barros; David Schlesinger; Debora Botequio Moretti; Dimas Tadeu Covas; Elaine Cristina Marqueze; Elaine Vieira Santos; Evandra Strazza Rodrigues; Heidge Fukumasu; Jayme Augusto de Souza-Neto; José Salvatore Leister Patané; Luiz Alcantara; Luiz Lehmann Coutinho; Maria Carolina Elias; Maurício Lacerda Nogueira; Rafael dos Santos Bezerra; Raul Machado Neto; Rejane Maria Tommasini Grotto; Ricardo Haddad; Sandra Coccuzzo Sampaio Vessoni; Simone Kashima; Svetoslav Nanev Slavov; Vincent Louis Viala |
| EPI_ISL_5603211 | UBS NUBIA FERREIRA RAMOS | Analytical Competence Molecular Epidemiology Lab/ACME, Oswaldo Cruz Foundation, Ceara (FIOCRUZ CE) | Carlos Leonardo de Aragao Araujo; Cecilia Leite Costa & Eduardo Ruback dos Santos on behalf of COVID-19 FIOCRUZ Genomic Network; Cleber Furtado Aksenen; Fabio Miyajima; Fernando Braga Stehling; Francisco Eder de Moura Lopes; Igor Oliveira Duarte; Jamille Maria Mendes Bezerra; Joaquim Cesar do Nascimento Sousa Junior; Pedro Miguel Carneiro Jeronimo; Suzana Porto Almeida; Thais Ferreira de Oliveira; Thais de Oliveira Costa; Ticiane Cavalcante de Souza; Veridiana Pessoa Miyajima |
| EPI_ISL_2445522, EPI_ISL_2445524 | UBS PASTOR MARCELINO DEUNGARO RIO GRANDE | Instituto Butantan | Antonio Jorge Martins; Claudia Renata dos Santos Barros; David Schlesinger; Debora Botequio Moretti; Dimas Tadeu Covas; Elaine Cristina Marqueze; Elaine Vieira Santos; Evandra Strazza Rodrigues; Heidge Fukumasu; Jayme Augusto de Souza-Neto; José Salvatore Leister Patané; Luiz Alcantara; Luiz Lehmann Coutinho; Maria Carolina Elias; Maurício Lacerda Nogueira; Rafael dos Santos Bezerra; Raul Machado Neto; Rejane Maria Tommasini Grotto; Ricardo Haddad; Sandra Coccuzzo Sampaio Vessoni; Simone Kashima; Svetoslav Nanev Slavov; Vincent Louis Viala |
| EPI_ISL_2445254, EPI_ISL_2445276 | UBS SAO LOURENCO DA SERRA | Instituto Butantan | Antonio Jorge Martins; Claudia Renata dos Santos Barros; David Schlesinger; Debora Botequio Moretti; Dimas Tadeu Covas; Elaine Cristina Marqueze; Elaine Vieira Santos; Evandra Strazza Rodrigues; Heidge Fukumasu; Jayme Augusto de Souza-Neto; José Salvatore Leister Patané; Luiz Alcantara; Luiz Lehmann Coutinho; Maria Carolina Elias; Maurício Lacerda Nogueira; Rafael dos Santos Bezerra; Raul Machado Neto; Rejane Maria Tommasini Grotto; Ricardo Haddad; Sandra Coccuzzo Sampaio Vessoni; Simone Kashima; Svetoslav Nanev Slavov; Vincent Louis Viala |
| EPI_ISL_3104668, EPI_ISL_3104678, EPI_ISL_3104680, EPI_ISL_3104697, EPI_ISL_3104698, EPI_ISL_3104699, EPI_ISL_3104700 | | | |
| see above | UCC (Universidad Central del Caribe) | Grubaugh Lab - Yale School of Public Health | Anderson Brito; Andrea Firpo; Annie Watkins; Chaney Kalinich; Chantal Vogels; David Rivera Aponte; Ernesto Vazquez; Fabiola Cruz-Lopez; Fabiola Fontanet; Isabel Ott; Jessica Rothman; Joseph Fauver; Karolane Gonzalez; Kendall Billig; Mallery Breban; Mary Petrone; Natalie Machargo; Nathan Grubaugh; Patricia Serrano; Tania Mitwalli; Tara Alpert; Tobias Koch; Vianca Aponte |
| EPI_ISL_2617950 | UCLA Clinical Micro Lab | Los Angeles County Public Health Laboratories | P. Hemarajata et al. |
| EPI_ISL_2295457 | UCSC Genomics Institute | UCSC Genomics Institute | A. Marm Kilpatrick; Angie Hinrichs; Beth Shapiro; Bryan Thornlow; Ciara Wanket; David Haussler; Eric Beraut; Hugh Olsen; Ikenna Anigbogu; Isabel Bjork; Jakob McBroome; Jeremy Sanford; Joshua Kapp; Mark Akeson; Maximilian Haeussler; Michael Stone; Miten Jain; Molly Cassatt-Johnstone; Namrita Dhillon; Russell Corbett-Detig; Terren Chang; Yatish Turakhia |
| EPI_ISL_2438059 | UDEA | Universidad Nacional de Colombia - Laboratorio Genómico One Health | Andres F. Cardona-Rios; Carlos Franco-Muñoz; Carolina Muñoz-Arango; Celeny Ortiz; Daniel O. Maldonado-Perez; Diego A. Álvarez-Díaz; Hector Alejandro Ruiz-Moreno; Idabely Betancur Ortiz; Jorge E. Osorio; Juan P. Hernandez-Ortiz; Karl A Ciuoderis; Katherine Laiton-Donato; Laura Silvana Perez; Lina M. Hurtado; Marcela Mercado-Reyes; Maria Angélica Maya; Maria Stella López; Rita Almanza Payares; Sandra Ines Cano; Simón Villegas Velásquez |
| EPI_ISL_3409086, EPI_ISL_3409289, EPI_ISL_3409808, EPI_ISL_3409847 | UMass Memorial Medical Center | Center for Microbiome Research | Doyle V. Ward; Karl J. Simon; Richard T. Ellison III |
| EPI_ISL_6229799 | UNIDADE BASICA DE SAUDE DE LAGOA FUNDA | ACME Lab, Oswaldo Cruz Foundation, FIOCRUZ/CE | Carlos Leonardo de Aragao Araujo; Cecilia Leite Costa & Eduardo Ruback dos Santos on behalf of COVID-19 FIOCRUZ Genomic Network; Cleber Furtado Aksenen; Fabio Miyajima; Fernando Braga Stehling; Francisco Eder de Moura Lopes; Igor Oliveira Duarte; Jamille Maria Mendes Bezerra; Joaquim Cesar do Nascimento Sousa Junior; Pedro Miguel Carneiro Jeronimo; Suzana Porto Almeida; Thais Ferreira de Oliveira; Thais de Oliveira Costa; Ticiane Cavalcante de Souza; Veridiana Pessoa Miyajima |
| EPI_ISL_6229800 | UNIDADE BASICA DE SAUDE DE ROLDAO | ACME Lab, Oswaldo Cruz Foundation, FIOCRUZ/CE | Carlos Leonardo de Aragao Araujo; Cecilia Leite Costa & Eduardo Ruback dos Santos on behalf of COVID-19 FIOCRUZ Genomic Network; Cleber Furtado Aksenen; Fabio Miyajima; Fernando Braga Stehling; Francisco Eder de Moura Lopes; Igor Oliveira Duarte; Jamille Maria Mendes Bezerra; Joaquim Cesar do Nascimento Sousa Junior; Pedro Miguel Carneiro Jeronimo; Suzana Porto Almeida; Thais Ferreira de Oliveira; Thais de Oliveira Costa; Ticiane Cavalcante de Souza; Veridiana Pessoa Miyajima |
| EPI_ISL_6229801 | UNIDADE BASICA DE SAUDE DO CAMPO DE AVIACAO | ACME Lab, Oswaldo Cruz Foundation, FIOCRUZ/CE | Carlos Leonardo de Aragao Araujo; Cecilia Leite Costa & Eduardo Ruback dos Santos on behalf of COVID-19 FIOCRUZ Genomic Network; Cleber Furtado Aksenen; Fabio Miyajima; Fernando Braga Stehling; Francisco Eder de Moura Lopes; Igor Oliveira Duarte; Jamille Maria Mendes Bezerra; Joaquim Cesar do Nascimento Sousa Junior; Pedro Miguel Carneiro Jeronimo; Suzana Porto Almeida; Thais Ferreira de Oliveira; Thais de Oliveira Costa; Ticiane Cavalcante de Souza; Veridiana Pessoa Miyajima |
| EPI_ISL_2473794 | UNIDADE BASICA DE SAUDE II ESTRELA DO NORTE | Instituto Butantan | Antonio Jorge Martins; Claudia Renata dos Santos Barros; David Schlesinger; Debora Botequio Moretti; Dimas Tadeu Covas; Elaine Cristina Marqueze; Elaine Vieira Santos; Evandra Strazza Rodrigues; Heidge Fukumasu; Jayme Augusto de Souza-Neto; José Salvatore Leister Patané; Luiz Alcantara; Luiz Lehmann Coutinho; Maria Carolina Elias; Maurício Lacerda Nogueira; Rafael dos Santos Bezerra; Raul Machado Neto; Rejane Maria Tommasini Grotto; Ricardo Haddad; Sandra Coccuzzo Sampaio Vessoni; Simone Kashima; Svetoslav Nanev Slavov; Vincent Louis Viala |
| EPI_ISL_2445463 | UNIDADE BASICA SAUDE NATAL ABADIO LACERDA | Instituto Butantan | Antonio Jorge Martins; Claudia Renata dos Santos Barros; David Schlesinger; Debora Botequio Moretti; Dimas Tadeu Covas; Elaine Cristina Marqueze; Elaine Vieira Santos; Evandra Strazza Rodrigues; Heidge Fukumasu; Jayme Augusto de Souza-Neto; José Salvatore Leister Patané; Luiz Alcantara; Luiz Lehmann Coutinho; Maria Carolina Elias; Maurício Lacerda Nogueira; Rafael dos Santos Bezerra; Raul Machado Neto; Rejane Maria Tommasini Grotto; Ricardo Haddad; Sandra Coccuzzo Sampaio Vessoni; Simone Kashima; Svetoslav Nanev Slavov; Vincent Louis Viala |
| EPI_ISL_6229806, EPI_ISL_6229819 | UNIDADE DE PRONTO ATENDIMENTO DE IGUATU UPA | ACME Lab, Oswaldo Cruz Foundation, FIOCRUZ/CE | Carlos Leonardo de Aragao Araujo; Cecilia Leite Costa & Eduardo Ruback dos Santos on behalf of COVID-19 FIOCRUZ Genomic Network; Cleber Furtado Aksenen; Fabio Miyajima; Fernando Braga Stehling; Francisco Eder de Moura Lopes; Igor Oliveira Duarte; Jamille Maria Mendes Bezerra; Joaquim Cesar do Nascimento Sousa Junior; Pedro Miguel Carneiro Jeronimo; Suzana Porto Almeida; Thais Ferreira de Oliveira; Thais de Oliveira Costa; Ticiane Cavalcante de Souza; Veridiana Pessoa Miyajima |
| EPI_ISL_2445487, EPI_ISL_2445488, EPI_ISL_2445491 | UNIDADE DE SAUDE DR PHEBO DE OLIVEIRA ROGE FERREIRA | Instituto Butantan | Antonio Jorge Martins; Claudia Renata dos Santos Barros; David Schlesinger; Debora Botequio Moretti; Dimas Tadeu Covas; Elaine Cristina Marqueze; Elaine Vieira Santos; Evandra Strazza Rodrigues; Heidge Fukumasu; Jayme Augusto de Souza-Neto; José Salvatore Leister Patané; Luiz Alcantara; Luiz Lehmann Coutinho; Maria Carolina Elias; Maurício Lacerda Nogueira; Rafael dos Santos Bezerra; Raul Machado Neto; Rejane Maria Tommasini Grotto; Ricardo Haddad; Sandra Coccuzzo Sampaio Vessoni; Simone Kashima; Svetoslav Nanev Slavov; Vincent Louis Viala |
| EPI_ISL_2445480 | UNIDADE DE VIGILANCIA EM SAUDE | Instituto Butantan | Antonio Jorge Martins; Claudia Renata dos Santos Barros; David Schlesinger; Debora Botequio Moretti; Dimas Tadeu Covas; Elaine Cristina Marqueze; Elaine Vieira Santos; Evandra Strazza Rodrigues; Heidge Fukumasu; Jayme Augusto de Souza-Neto; José Salvatore Leister Patané; Luiz Alcantara; Luiz Lehmann Coutinho; Maria Carolina Elias; Maurício Lacerda Nogueira; Rafael dos Santos Bezerra; Raul Machado Neto; Rejane Maria Tommasini Grotto; Ricardo Haddad; Sandra Coccuzzo Sampaio Vessoni; Simone Kashima; Svetoslav Nanev Slavov; Vincent Louis Viala |
| EPI_ISL_2445234, EPI_ISL_2445236, EPI_ISL_2445237 | UNIDADE MISTA DE IGUAPE | Instituto Butantan | Antonio Jorge Martins; Claudia Renata dos Santos Barros; David Schlesinger; Debora Botequio Moretti; Dimas Tadeu Covas; Elaine Cristina Marqueze; Elaine Vieira Santos; Evandra Strazza Rodrigues; Heidge Fukumasu; Jayme Augusto de Souza-Neto; José Salvatore Leister Patané; Luiz Alcantara; Luiz Lehmann Coutinho; Maria Carolina Elias; Maurício Lacerda Nogueira; Rafael dos Santos Bezerra; Raul Machado Neto; Rejane Maria Tommasini Grotto; Ricardo Haddad; Sandra Coccuzzo Sampaio Vessoni; Simone Kashima; Svetoslav Nanev Slavov; Vincent Louis Viala |
| EPI_ISL_2493442, EPI_ISL_2493443 | UNIDADE MISTA DE SAUDE MOMBUCA | Instituto Butantan | Antonio Jorge Martins; Claudia Renata dos Santos Barros; David Schlesinger; Debora Botequio Moretti; Dimas Tadeu Covas; Elaine Cristina Marqueze; Elaine Vieira Santos; Evandra Strazza Rodrigues; Heidge Fukumasu; Jayme Augusto de Souza-Neto; José Salvatore Leister Patané; Luiz Alcantara; Luiz Lehmann Coutinho; Maria Carolina Elias; Maurício Lacerda Nogueira; Rafael dos Santos Bezerra; Raul Machado Neto; Rejane Maria Tommasini Grotto; Ricardo Haddad; Sandra Coccuzzo Sampaio Vessoni; Simone Kashima; Svetoslav Nanev Slavov; Vincent Louis Viala |
| EPI_ISL_2445585 | UNIDADE MISTA DE SAUDE ROSEIRA DR PAULO GUIMARAES CASTRO | Instituto Butantan | Antonio Jorge Martins; Claudia Renata dos Santos Barros; David Schlesinger; Debora Botequio Moretti; Dimas Tadeu Covas; Elaine Cristina Marqueze; Elaine Vieira Santos; Evandra Strazza Rodrigues; Heidge Fukumasu; Jayme Augusto de Souza-Neto; José Salvatore Leister Patané; Luiz Alcantara; Luiz Lehmann Coutinho; Maria Carolina Elias; Maurício Lacerda Nogueira; Rafael dos Santos Bezerra; Raul Machado Neto; Rejane Maria Tommasini Grotto; Ricardo Haddad; Sandra Coccuzzo Sampaio Vessoni; Simone Kashima; Svetoslav Nanev Slavov; Vincent Louis Viala |
| EPI_ISL_6229695 | UNIDADE PRONTO ATENDIMENTO CRISTO REDENTOR | ACME Lab, Oswaldo Cruz Foundation, FIOCRUZ/CE | Carlos Leonardo de Aragao Araujo; Cecilia Leite Costa & Eduardo Ruback dos Santos on behalf of COVID-19 FIOCRUZ Genomic Network; Cleber Furtado Aksenen; Fabio Miyajima; Fernando Braga Stehling; Francisco Eder de Moura Lopes; Igor Oliveira Duarte; Jamille Maria Mendes Bezerra; Joaquim Cesar do Nascimento Sousa Junior; Pedro Miguel Carneiro Jeronimo; Suzana Porto Almeida; Thais Ferreira de Oliveira; Thais de Oliveira Costa; Ticiane Cavalcante de Souza; Veridiana Pessoa Miyajima |
| EPI_ISL_6229811 | UNIDADE REFERENCIA COVID 19 | ACME Lab, Oswaldo Cruz Foundation, FIOCRUZ/CE | Carlos Leonardo de Aragao Araujo; Cecilia Leite Costa & Eduardo Ruback dos Santos on behalf of COVID-19 FIOCRUZ Genomic Network; Cleber Furtado Aksenen; Fabio Miyajima; Fernando Braga Stehling; Francisco Eder de Moura Lopes; Igor Oliveira Duarte; Jamille Maria Mendes Bezerra; Joaquim Cesar do Nascimento Sousa Junior; Pedro Miguel Carneiro Jeronimo; Suzana Porto Almeida; Thais Ferreira de Oliveira; Thais de Oliveira Costa; Ticiane Cavalcante de Souza; Veridiana Pessoa Miyajima |
| EPI_ISL_2434990 | UOC Microbiologia e Virologia, Azienda Ospedaliera Universitaria Senese, Siena, Italy | Dipartimento di Biotecnologie Mediche, University of Siena | Anichini G.; Coppi M.; Cusi M.G.; Gandolfo C.; Gori Savellini G.; Pinzauti D.; Rossolini G.M.; Santoro F. |
| EPI_ISL_2445243, EPI_ISL_2445245, EPI_ISL_2445248, EPI_ISL_2445249, EPI_ISL_2445250, EPI_ISL_2445251, EPI_ISL_2445253, EPI_ISL_2445256, EPI_ISL_2445260, EPI_ISL_2445261, EPI_ISL_2445264, EPI_ISL_2445269, EPI_ISL_2445270 | | | |
| see above | UPA DR AKIRA TADA | Instituto Butantan | Antonio Jorge Martins; Claudia Renata dos Santos Barros; David Schlesinger; Debora Botequio Moretti; Dimas Tadeu Covas; Elaine Cristina Marqueze; Elaine Vieira Santos; Evandra Strazza Rodrigues; Heidge Fukumasu; Jayme Augusto de Souza-Neto; José Salvatore Leister Patané; Luiz Alcantara; Luiz Lehmann Coutinho; Maria Carolina Elias; Maurício Lacerda Nogueira; Rafael dos Santos Bezerra; Raul Machado Neto; Rejane Maria Tommasini Grotto; Ricardo Haddad; Sandra Coccuzzo Sampaio Vessoni; Simone Kashima; Svetoslav Nanev Slavov; Vincent Louis Viala |
| EPI_ISL_2445272 | UPA DR FRANCO DA ROCHA | Instituto Butantan | Antonio Jorge Martins; Claudia Renata dos Santos Barros; David Schlesinger; Debora Botequio Moretti; Dimas Tadeu Covas; Elaine Cristina Marqueze; Elaine Vieira Santos; Evandra Strazza Rodrigues; Heidge Fukumasu; Jayme Augusto de Souza-Neto; José Salvatore Leister Patané; Luiz Alcantara; Luiz Lehmann Coutinho; Maria Carolina Elias; Maurício Lacerda Nogueira; Rafael dos Santos Bezerra; Raul Machado Neto; Rejane Maria Tommasini Grotto; Ricardo Haddad; Sandra Coccuzzo Sampaio Vessoni; Simone Kashima; Svetoslav Nanev Slavov; Vincent Louis Viala |
| EPI_ISL_2445336 | UPA GEISEL | Instituto Butantan | Antonio Jorge Martins; Claudia Renata dos Santos Barros; David Schlesinger; Debora Botequio Moretti; Dimas Tadeu Covas; Elaine Cristina Marqueze; Elaine Vieira Santos; Evandra Strazza Rodrigues; Heidge Fukumasu; Jayme Augusto de Souza-Neto; José Salvatore Leister Patané; Luiz Alcantara; Luiz Lehmann Coutinho; Maria Carolina Elias; Maurício Lacerda Nogueira; Rafael dos Santos Bezerra; Raul Machado Neto; Rejane Maria Tommasini Grotto; Ricardo Haddad; Sandra Coccuzzo Sampaio Vessoni; Simone Kashima; Svetoslav Nanev Slavov; Vincent Louis Viala |
| EPI_ISL_2445333, EPI_ISL_2445364 | UPA MARY DOTA | Instituto Butantan | Antonio Jorge Martins; Claudia Renata dos Santos Barros; David Schlesinger; Debora Botequio Moretti; Dimas Tadeu Covas; Elaine Cristina Marqueze; Elaine Vieira Santos; Evandra Strazza Rodrigues; Heidge Fukumasu; Jayme Augusto de Souza-Neto; José Salvatore Leister Patané; Luiz Alcantara; Luiz Lehmann Coutinho; Maria Carolina Elias; Maurício Lacerda Nogueira; Rafael dos Santos Bezerra; Raul Machado Neto; Rejane Maria Tommasini Grotto; Ricardo Haddad; Sandra Coccuzzo Sampaio Vessoni; Simone Kashima; Svetoslav Nanev Slavov; Vincent Louis Viala |
| EPI_ISL_5825567, EPI_ISL_5825568 | UPA PORTE II | Analytical Competence Molecular Epidemiology Lab/ACME, Oswaldo Cruz Foundation, Ceara (FIOCRUZ CE) | Carlos Leonardo de Aragao Araujo; Cecilia Leite Costa & Eduardo Ruback dos Santos on behalf of COVID-19 FIOCRUZ Genomic Network; Cleber Furtado Aksenen; Fabio Miyajima; Fernando Braga Stehling; Francisco Eder de Moura Lopes; Igor Oliveira Duarte; Jamille Maria Mendes Bezerra; Joaquim Cesar do Nascimento Sousa Junior; Pedro Miguel Carneiro Jeronimo; Suzana Porto Almeida; Thais Ferreira de Oliveira; Thais de Oliveira Costa; Ticiane Cavalcante de Souza; Veridiana Pessoa Miyajima |
| EPI_ISL_2445275, EPI_ISL_2445277, EPI_ISL_2445300, EPI_ISL_2445302, EPI_ISL_2445310 | UPA VILA SANTA CATARINA | Instituto Butantan | Antonio Jorge Martins; Claudia Renata dos Santos Barros; David Schlesinger; Debora Botequio Moretti; Dimas Tadeu Covas; Elaine Cristina Marqueze; Elaine Vieira Santos; Evandra Strazza Rodrigues; Heidge Fukumasu; Jayme Augusto de Souza-Neto; José Salvatore Leister Patané; Luiz Alcantara; Luiz Lehmann Coutinho; Maria Carolina Elias; Maurício Lacerda Nogueira; Rafael dos Santos Bezerra; Raul Machado Neto; Rejane Maria Tommasini Grotto; Ricardo Haddad; Sandra Coccuzzo Sampaio Vessoni; Simone Kashima; Svetoslav Nanev Slavov; Vincent Louis Viala |
| EPI_ISL_2450385, EPI_ISL_2450396 | US Air Force School of Aerospace Medicine | US Air Force School of Aerospace Medicine | Amanda Javorina; Anthony Fries; Carol Garrett; Clarise Starr; Elizabeth Macias; Jennifer Meyer; Sarah Purves; William Gruner |
| EPI_ISL_2445456 | USF II DR JOSE RODOLFO ZIMMERMANN DIAS | Instituto Butantan | Antonio Jorge Martins; Claudia Renata dos Santos Barros; David Schlesinger; Debora Botequio Moretti; Dimas Tadeu Covas; Elaine Cristina Marqueze; Elaine Vieira Santos; Evandra Strazza Rodrigues; Heidge Fukumasu; Jayme Augusto de Souza-Neto; José Salvatore Leister Patané; Luiz Alcantara; Luiz Lehmann Coutinho; Maria Carolina Elias; Maurício Lacerda Nogueira; Rafael dos Santos Bezerra; Raul Machado Neto; Rejane Maria Tommasini Grotto; Ricardo Haddad; Sandra Coccuzzo Sampaio Vessoni; Simone Kashima; Svetoslav Nanev Slavov; Vincent Louis Viala |
| EPI_ISL_2383988 | UT-Unified State Labs: Public Health Utah DOH | Centers for Disease Control and Prevention Division of Viral Diseases, Pathogen Discovery | Alison Laufer Halpin; Ben L. Rambo-Martin; Clinton R. Paden; Dakota Howard; Darlene Wagner; Dave Wentworth; Dhwani Batra; Jasmine Padilla; Justin Lee; Katie Dillon; Krista Queen; Kristen Knipe; Kristine Lacek; Mark Burroughs; Matthew Schmerer; Mili Sheth; Peter Cook; Sam Shepard; Sarah Nobles; Shoshona Le; Suxiang Tong; Vivien Dugan; Yvette Unoarumhi |
| EPI_ISL_2272609, EPI_ISL_2272617, EPI_ISL_2272656, EPI_ISL_2272660, EPI_ISL_2273727, EPI_ISL_2273742, EPI_ISL_2273758, EPI_ISL_2273764, EPI_ISL_2273771, EPI_ISL_2273779, EPI_ISL_2273781, EPI_ISL_2273783, EPI_ISL_2273790, EPI_ISL_2273794, EPI_ISL_2273801, EPI_ISL_2273807, EPI_ISL_2273809, EPI_ISL_2273810, EPI_ISL_2273831, EPI_ISL_2273832, EPI_ISL_2273833, EPI_ISL_2332846, EPI_ISL_2332880, EPI_ISL_2332883, EPI_ISL_2332901, EPI_ISL_2332909, EPI_ISL_2332913, EPI_ISL_2332921, EPI_ISL_2332923, EPI_ISL_2332940, EPI_ISL_2332946, EPI_ISL_2332960, EPI_ISL_2332965, EPI_ISL_2332970, EPI_ISL_2332973, EPI_ISL_2332974, EPI_ISL_2332988, EPI_ISL_2332989, EPI_ISL_2332992, EPI_ISL_2333008, EPI_ISL_2333022, EPI_ISL_2333034, EPI_ISL_2333036, EPI_ISL_2333038, EPI_ISL_2333061, EPI_ISL_2340694, EPI_ISL_2340695, EPI_ISL_2340705, EPI_ISL_2340711, EPI_ISL_2340712, EPI_ISL_2340721, EPI_ISL_2340743, EPI_ISL_2340790, EPI_ISL_2340804, EPI_ISL_2426399, EPI_ISL_2426408, EPI_ISL_2426424, EPI_ISL_2426430, EPI_ISL_2426433, EPI_ISL_2426434, EPI_ISL_2426436, EPI_ISL_2426461, EPI_ISL_2426484, EPI_ISL_2426500, EPI_ISL_2426501, EPI_ISL_2426503, EPI_ISL_2426507, EPI_ISL_2426515, EPI_ISL_2426518, EPI_ISL_2426529, EPI_ISL_2426530, EPI_ISL_2426539, EPI_ISL_2426549, EPI_ISL_2426563, EPI_ISL_2426570, EPI_ISL_2426572, EPI_ISL_2618668, EPI_ISL_2618669, EPI_ISL_2618671, EPI_ISL_2618702, EPI_ISL_2618709, EPI_ISL_2618728, EPI_ISL_2618730 | | | |

| | | | |
|---|---|---|---|
| see above | UW Virology Lab | UW Virology Lab | Alexander Greninger; Hong Xie; Keith R Jerome; Lasata Shrestha; Meei-Li Huang; Michelle Lin; Nathan Breit; Noah R. Baker; Patrick Mathias; Pavitra Roychoudhury; Ricardo Perez; Robert J. Livingston; Sean Ellis; Shah Mohamed Bakhash; Tien V. Nguyen |
| EPI_ISL_2671653 | Unidad de Investigacion Medica de Yucatan (UIMY) | Unidad de Genomica Avanzada | ; Alejandra Garcia-Gasca; Alejandra Hernandez-Teran; Alejandro Sanchez-Flores; Alfredo Herrera-Estrella; Alicia Ocaña-Mondragon; Andreu Comas-Garcia; Angel Gustavo Salas-Lais; Antonio Loza Roman; Bernardo Martinez-Miguel; Blanca Taboada; Brenda Irasema Maldonado-Meza; Bruno Gomez-Gil; Carla Ivon Herrera-Najera; Carlos F. Arias; Celia Boukadida; Celida Duque Molina; Celida Martinez- Rodriguez; Clara Esperanza Santacruz-Tinoco; Concepcion Grajales-Muñiz; Consorcio Mexicano de Vigilancia Genomica (CoViGen-Mex). Authors (in alphabetical order): Julio Elias Alvarado-Yaah; Cristobal Chaidez-Quiroz; Daniel Fregoso-Rueda; Daniel Lira Morales; Eduardo Becerril-Vargas; Fernando Fontove-Herrera; Fidencio Mejia-Nepomuceno; Francisco Pulido; Gloria Elena Espinosa-Ayala; Gloria Maria Molina-Salinas; Gloria Vazquez; Hector Esteban Paz-Juarez; Hector Montoya-Fuentes; Helen Haydee Fernanda Ramirez-Plascencia; Irvin Gonzalez-Lopez; Jean Pierre Gonzalez; Jesus Hernandez; Joel Armando Vazquez-Perez.; Jorge Salas-Hernandez; Jose Antonio Enciso-Moreno; Jose Arturo Martinez-Orozco; Jose Esteban Muñoz-Medina; Jose de Jesus Nuñez-Contreras; Juan Bautista Chale-Dzul; Julissa Enciso-Ibarra; Luis Alberto Ochoa-Carrera; Margarita Matias-Florentino; Maria Guadalupe Santiago-Mauricio; Maria Guadalupe de Jesus Mireles-Rivera; Mario Mujica-Sanchez; Marissa Perez-Garcia; Nelly Selem-Mojica; Pavel Isa; Ricardo Ciria Merce; Ricardo Grande; Rosa Maria Gutierrez Rios; Santiago avila-Rios; Selene Zarate; Susana Lopez; Veronica Mata-Haro; Victor Eduardo Garcia-Arias; Victor Hugo Borja-Aburto |
| EPI_ISL_2490386 | Unidad de Investigación Biomédica de Zacatecas (UIBZ) | Centro de Investigación en Enfermedades Infecciosas (CIENI), Instituto Nacional de Enfermedades Respiratorias (INER) | ; Alejandra Garcia-Gasca; Alejandra Hernández-Terán; Alejandro Sanchez-Flores; Alfredo Herrera-Estrella; Alicia Ocaña-Mondragón; Andreu Comas-García; Angel Gustavo Salas-Lais; Antonio Loza Román; Bernardo Martínez-Miguel; Blanca Taboada; Brenda Irasema Maldonado-Meza; Bruno Gomez-Gil; Carla Ivón Herrera-Najera; Carlos F. Arias; Celia Boukadida; Clara Esperanza Santacruz-Tinoco; Concepción Grajales-Muñiz; Consorcio Mexicano de Vigilancia Genómica (CoViGen-Mex). Authors (in alphabetical order): Julio Elias Alvarado-Yaah; Cristóbal Cháidez-Quiróz; Célida Duque Molina; Célida Martínez- Rodríguez; Daniel Fregoso-Rueda; Daniel Lira Morales; Eduardo Becerril-Vargas; Fernando Fontove-Herrera; Fidencio Mejía-Nepomuceno; Francisco Pulido; Gloria Elena Espinosa-Ayala; Gloria María Molina-Salinas; Gloria Vazquez; Hector Esteban Paz-Juárez; Hector Montoya-Fuentes; Helen Haydee Fernanda Ramírez-Plascencia; Irvin González-López; Jean Pierre González; Joel Armando Vázquez-Pérez.; Jorge Salas-Hernández; José Antonio Enciso-Moreno; José Arturo Martínez-Orozco; José Esteban Muñoz-Medina; José de Jesús Nuñez-Contreras; Juan Bautista Chale-Dzul; Julissa Enciso-Ibarra; Luis Alberto Ochoa-Carrera; Margarita Matías-Florentino; Mario Mújica-Sánchez; Marissa Pérez-García; María Guadalupe Santiago-Mauricio; María Guadalupe de Jesús Mireles-Rivera; Nelly Sélem-Mojica; Pavel Isa; Ricardo Ciria Merce; Ricardo Grande; Rosa María Gutierrez Ríos; Santiago Ávila-Ríos; Selene Zárate; Susana Lopez; Víctor Eduardo García-Arias; Víctor Hugo Borja-Aburto |
| EPI_ISL_2490378, EPI_ISL_2490379, EPI_ISL_2490380, EPI_ISL_2490381 | Unidad de Investigación Médica de Yucatán (UIMY) | Centro de Investigación en Enfermedades Infecciosas (CIENI), Instituto Nacional de Enfermedades Respiratorias (INER) | ; Alejandra Garcia-Gasca; Alejandra Hernández-Terán; Alejandro Sanchez-Flores; Alfredo Herrera-Estrella; Alicia Ocaña-Mondragón; Andreu Comas-García; Angel Gustavo Salas-Lais; Antonio Loza Román; Bernardo Martínez-Miguel; Blanca Taboada; Brenda Irasema Maldonado-Meza; Bruno Gomez-Gil; Carla Ivón Herrera-Najera; Carlos F. Arias; Celia Boukadida; Clara Esperanza Santacruz-Tinoco; Concepción Grajales-Muñiz; Consorcio Mexicano de Vigilancia Genómica (CoViGen-Mex). Authors (in alphabetical order): Julio Elias Alvarado-Yaah; Cristóbal Cháidez-Quiróz; Célida Duque Molina; Célida Martínez- Rodríguez; Daniel Fregoso-Rueda; Daniel Lira Morales; Eduardo Becerril-Vargas; Fernando Fontove-Herrera; Fidencio Mejía-Nepomuceno; Francisco Pulido; Gloria Elena Espinosa-Ayala; Gloria María Molina-Salinas; Gloria Vazquez; Hector Esteban Paz-Juárez; Hector Montoya-Fuentes; Helen Haydee Fernanda Ramírez-Plascencia; Irvin González-López; Jean Pierre González; Joel Armando Vázquez-Pérez.; Jorge Salas-Hernández; José Antonio Enciso-Moreno; José Arturo Martínez-Orozco; José Esteban Muñoz-Medina; José de Jesús Nuñez-Contreras; Juan Bautista Chale-Dzul; Julissa Enciso-Ibarra; Luis Alberto Ochoa-Carrera; Margarita Matías-Florentino; Mario Mújica-Sánchez; Marissa Pérez-García; María Guadalupe Santiago-Mauricio; María Guadalupe de Jesús Mireles-Rivera; Nelly Sélem-Mojica; Pavel Isa; Ricardo Ciria Merce; Ricardo Grande; Rosa María Gutierrez Ríos; Santiago Ávila-Ríos; Selene Zárate; Susana Lopez; Víctor Eduardo García-Arias; Víctor Hugo Borja-Aburto |
| EPI_ISL_2535174, EPI_ISL_2535181, EPI_ISL_2535197, EPI_ISL_2535198, EPI_ISL_2535199, EPI_ISL_2535200, EPI_ISL_2535201, EPI_ISL_2535202, EPI_ISL_2535203, EPI_ISL_2535204, EPI_ISL_2535205, EPI_ISL_2535206, EPI_ISL_2535212, EPI_ISL_2535215, EPI_ISL_2535216, EPI_ISL_2535217, EPI_ISL_2535218, EPI_ISL_2535219, EPI_ISL_2535228, EPI_ISL_2535229, EPI_ISL_2535231, EPI_ISL_2535232, EPI_ISL_2535233, EPI_ISL_2691626 | | | |
| see above | Unidade de apoio ao diagnostico da COVID - UNADIG | Bioinformatics Laboratory / LNCC | Alessandra P Lamarca; Alexandra L Gerber; Amilcar Tanuri; Ana Paula de C Guimaraes; Ana Tereza R Vasconcelos; Andrea Cony Cavalcanti; Caio Luiz Pereira Ribeiro; Cassia Alves; Cintia Policarpo; Claudia Maria Braga de Mello; Cristiane Gomes da Silva; Diana Mariani; Douglas Terra Machado; Flavio Dias da Silva; Gleidson da Silva de Oliveira; Leandro Magalhaes de Souza; Liliane Cavalcante; Luiz G P de Almeida; Marcio Henrique de Oliveira Garcia; Mario Sergio Ribeiro; Ronaldo da Silva F Jr; Silvia Carvalho |
| EPI_ISL_2204905 | United Hospital Center | WVU and Marshall University Combined Genomics Core Facilities | James Denvir; Peter Perrotta; Peter Stoilov; Ryan Percifield; Wesley Kimble |
| EPI_ISL_3216907, EPI_ISL_3477562 | Universidad del Valle, LDAB-Laboratorio de Diagnostico de Agentes Biologicos | Universidad del Valle, TAO-Lab, VIREM, NEAS Network | Andres Castillo; Beatriz Parra & COVID-19 Team Univalle; Diana López-Alvarez; Nelson Rivera Franco |
| EPI_ISL_2193384, EPI_ISL_2193388, EPI_ISL_2193389, EPI_ISL_2193390, EPI_ISL_2193391, EPI_ISL_2193393, EPI_ISL_2193396, EPI_ISL_2193400, EPI_ISL_2193401, EPI_ISL_2193402, EPI_ISL_2193403, EPI_ISL_2193404, EPI_ISL_2193405, EPI_ISL_2193406, EPI_ISL_2193407, EPI_ISL_2193411, EPI_ISL_2193414, EPI_ISL_2193417 | | | |
| see above | University Hospitals Translational Laboratory (UHTL), University Hospitals | University Hospitals Translational Laboratory (UHTL), University Hospitals | Alouani, D.; Sadri, N.; Song, X. |
| EPI_ISL_2931108, EPI_ISL_3388709 | University of Chicago Medicine | RIPHL at Rush University Medical Center | Edith Perez; Felix Araujo Perez; Kevin Kunstman; Laura Furtado; Lauren Megger; Marieta Hyde; Max Kolton; Stefan Green |
| EPI_ISL_2692096, EPI_ISL_2692100, EPI_ISL_2692101, EPI_ISL_2692122, EPI_ISL_2692125, EPI_ISL_2692131, EPI_ISL_2692132 | | | |
| see above | University of Illinois at Chicago | RIPHL at Rush University Medical Center | Felix Araujo Perez; Julie Bentea; Kevin Kunstman; Laura Furtado; Marieta Hyde; Max Kolton; Omar Perez; Stefan Green |
| EPI_ISL_2245221, EPI_ISL_2245251, EPI_ISL_2245260, EPI_ISL_2245261, EPI_ISL_2245405, EPI_ISL_2245425, EPI_ISL_2245426, EPI_ISL_2245427 | | | |
| see above | University of Liège COVID-19 testing center | GIGA Medical Genomics | Bouchra Boujemla; Cécile Meex; Fabrice Bureau; Keith Durkin; Laurent Gillet; Maria Artesi; Marie-Pierre Hayette; Nathalie Renotte; Sébastien Bontems; Vincent Bours; Wouter Coppieters |
| EPI_ISL_2376042, EPI_ISL_2400358 | University of Michigan Clinical Microbiology Laboratory | Lauring Lab, University of Michigan, Department of Microbiology and Immunology | Gilbert |
| EPI_ISL_2404840 | University of Oregon COVID-19 MAP Laboratory | University of Oregon Genomics and Cell Charactericzation Core Facility (GC3F) | Ariana White; Douglas Turnbull; Jason Carriere; Jason Sydes; Jeff Bishop; Peter Batzel |
| EPI_ISL_2340668, EPI_ISL_2340833, EPI_ISL_2340836 | University of Wisconsin-Madison AIDS Vaccine Research Laboratories | University of Wisconsin-Madison AIDS Vaccine Research Laboratories | Gage Moreno; Katarina Braun; et al. AIDS Vaccine Research Laboratories |
| EPI_ISL_2235442, EPI_ISL_2235443, EPI_ISL_2235445, EPI_ISL_2235451, EPI_ISL_2235452, EPI_ISL_2235454, EPI_ISL_2235455, EPI_ISL_2235456, EPI_ISL_2235457, EPI_ISL_2235462, EPI_ISL_2245014, EPI_ISL_2308705 | | | |
| see above | Università degli Studi di Perugia | Istituto Zooprofilattico Sperimentale dell'Abruzzo e Molise "G. Caporale" | Ancora M; Biagetti M; Calistri P; Camilloni B; Cammà C; Caporale M; Curini V; Delli Compagni E; Di Domenico M; Di Lollo Valeria; Di Pasquale A; GiammarioliM; Lorusso A; Mangone I; Marcacci M; Mencacci A; Puglia I; Rinaldi A; Savini G; Scialabba S |
| EPI_ISL_2449934, EPI_ISL_2449942 | UniversitŠtsSpital ZŸrich | Institute of Medical Virology | Alexandra Trkola; Annette AudigŽ; Catharine Aquino; Cyril Shah; Daniel Ehrsam; Gabriela Ziltener; Guido Bloemberg; Hubert Rehrauer; Isabel StŸrmer; Joel Wirz; Jon Huder; JŸrg Bšni; Kevin Steiner; Maria GrŸnberg; Maryam Zaheri; Michael Huber; Riccarda Capaul; Stefan Schmutz; Verena Kufner; Weihong Qi |
| EPI_ISL_2292200, EPI_ISL_2456321, EPI_ISL_2456598, EPI_ISL_2456676, EPI_ISL_2458490, EPI_ISL_2679351, EPI_ISL_2679353, EPI_ISL_2856068, EPI_ISL_2856095, EPI_ISL_2857979, EPI_ISL_2857990, EPI_ISL_2858009, EPI_ISL_2858015, EPI_ISL_2858020, EPI_ISL_2858028 | | | |
| see above | Utah Public Health Laboratory | Utah Public Health Laboratory | Erin L. Young; Kelly F. Oakeson; Olinto Linares-Perdomo; Tara Gallagher |
| EPI_ISL_2493366, EPI_ISL_2493450 | VIGILANCIA EPIDEMIOLOGICA DE LEME | Instituto Butantan | Antonio Jorge Martins; Claudia Renata dos Santos Barros; David Schlesinger; Debora Botequio Moretti; Dimas Tadeu Covas; Elaine Cristina Marqueze; Elaine Vieira Santos; Evandra Strazza Rodrigues; Heidge Fukumasu; Jayme Augusto de Souza-Neto; José Salvatore Leister Patané; Luiz Alcantara; Luiz Lehmann Coutinho; Maria Carolina Elias; Maurício Lacerda Nogueira; Rafael dos Santos Bezerra; Raul Machado Neto; Rejane Maria Tommasini Grotto; Ricardo Haddad; Sandra Coccuzzo Sampaio Vessoni; Simone Kashima; Svetoslav Nanev Slavov; Vincent Louis Viala |
| EPI_ISL_2493365, EPI_ISL_2493448, EPI_ISL_2500982 | VIGILANCIA EPIDEMIOLOGICA E CONTROLE DE VETORES PIRASSUNUN | Instituto Butantan | Antonio Jorge Martins; Claudia Renata dos Santos Barros; David Schlesinger; Debora Botequio Moretti; Dimas Tadeu Covas; Elaine Cristina Marqueze; Elaine Vieira Santos; Evandra Strazza Rodrigues; Heidge Fukumasu; Jayme Augusto de Souza-Neto; José Salvatore Leister Patané; Luiz Alcantara; Luiz Lehmann Coutinho; Maria Carolina Elias; Maurício Lacerda Nogueira; Rafael dos Santos Bezerra; Raul Machado Neto; Rejane Maria Tommasini Grotto; Ricardo Haddad; Sandra Coccuzzo Sampaio Vessoni; Simone Kashima; Svetoslav Nanev Slavov; Vincent Louis Viala |
| EPI_ISL_2445607 | VIGILANCIA EPIDIMIOLOGICA DE ARACARIGUAMA | Instituto Butantan | Antonio Jorge Martins; Claudia Renata dos Santos Barros; David Schlesinger; Debora Botequio Moretti; Dimas Tadeu Covas; Elaine Cristina Marqueze; Elaine Vieira Santos; Evandra Strazza Rodrigues; Heidge Fukumasu; Jayme Augusto de Souza-Neto; José Salvatore Leister Patané; Luiz Alcantara; Luiz Lehmann Coutinho; Maria Carolina Elias; Maurício Lacerda Nogueira; Rafael dos Santos Bezerra; Raul Machado Neto; Rejane Maria Tommasini Grotto; Ricardo Haddad; Sandra Coccuzzo Sampaio Vessoni; Simone Kashima; Svetoslav Nanev Slavov; Vincent Louis Viala |
| EPI_ISL_2445282, EPI_ISL_2445290, EPI_ISL_2445291, EPI_ISL_2445293, EPI_ISL_2445294, EPI_ISL_2445296, EPI_ISL_2445303, EPI_ISL_2473725, EPI_ISL_2473822 | | | |
| see above | VIGILANCIA SANITARIA E VIG EPIDEMIOLOGICA DE ITAPEVI | Instituto Butantan | Antonio Jorge Martins; Claudia Renata dos Santos Barros; David Schlesinger; Debora Botequio Moretti; Dimas Tadeu Covas; Elaine Cristina Marqueze; Elaine Vieira Santos; Evandra Strazza Rodrigues; Heidge Fukumasu; Jayme Augusto de Souza-Neto; José Salvatore Leister Patané; Luiz Alcantara; Luiz Lehmann Coutinho; Maria Carolina Elias; Maurício Lacerda Nogueira; Rafael dos Santos Bezerra; Raul Machado Neto; Rejane Maria Tommasini Grotto; Ricardo Haddad; Sandra Coccuzzo Sampaio Vessoni; Simone Kashima; Svetoslav Nanev Slavov; Vincent Louis Viala |
| EPI_ISL_5780268, EPI_ISL_5780272 | VUMC Molecular Infectious Diseases Laboratory (MIDL) | Dr. Suman Das Lab - Vanderbilt University Medical Center (VUMC) | Bookyung Park; Grant Vestal; Helen Boone; Hunter Brown; Jonathan E. Schmitz; Meghan Shilts; Seesandra Rajagopala; Suman B. Pakala; Suman Das |
| EPI_ISL_4089999 | Vermont Integrative Genomics Resource | Vermont Integrative Genomics Resource | Korin Eckstrom |
| EPI_ISL_2307457, EPI_ISL_2307461, EPI_ISL_2307612, EPI_ISL_2307706, EPI_ISL_2307750, EPI_ISL_2307771, EPI_ISL_2307844, EPI_ISL_2307878, EPI_ISL_2308026, EPI_ISL_2308074 | | | |
| see above | Viollier AG | Department of Biosystems Science and Engineering, ETH Zürich | Andrea Patrignani; Andreia Cabral de Gouvea; Catharine Aquino; Chaoran Chen; Christiane Beckmann; Christoph Noppen; Daniel Ehrsam; David Dreifuss; Doris Popovic; Griffin White; Isabel Stürmer; Ivan Topolsky; Jay Tracy; Katharina Jahn; Lara Fuhrmann; Laura Neff; Lennart Opitz; Maria Domenica Moccia; Maurice Redondo; Niko Beerenwinkel; Noemie Santamaria de Souza; Olivier Kobel; Philipp Jablonski; Ralph Schlapbach; Sarah Nadeau; Simon Grüter; Sophie Seidel; Tanja Stadler; Timothy Sykes |
| EPI_ISL_2226439 | Viollier AG | Viollier AG | Andrea Patrizia Salzmann; Christiane Beckmann; Christoph Noppen; Henriette Kurth; Maurice Redondo; Olivier Kobel |
| EPI_ISL_2884326 | Virology Laboratory, Scientific Department, Army Medical Center | Virology Laboratory, Scientific Department, Army Medical Center | Anella Monte; Anna Anselmo; Antonella Fortunato; Filippo Molinari; Florigio Lista; Francesco Giordani; Giancarlo Petralito; Giandomenico Cerreto; Giulia Campoli; Lucia Nicosia; Marzia Cavalli; Riccardo De Sanctis; Rossella Brandi; Silvia Fillo; Vanessa Vera Fain |
| EPI_ISL_2451952 | WA State Department of Health | Centers for Disease Control and Prevention Division of Viral Diseases, Pathogen Discovery | Alison Laufer Halpin; Ben L. Rambo-Martin; Clinton R. Paden; Dakota Howard; Darlene Wagner; Dave Wentworth; Dhwani Batra; Jasmine Padilla; Justin Lee; Katie Dillon; Krista Queen; Kristen Knipe; Kristine Lacek; Mark Burroughs; Matthew Schmerer; Mili Sheth; Peter Cook; Sam Shepard; Sarah Nobles; Shoshona Le; Suxiang Tong; Vivien Dugan; Yvette Unoarumhi |
| EPI_ISL_2336196, EPI_ISL_2336205, EPI_ISL_2336207, EPI_ISL_2336212, EPI_ISL_2336244, EPI_ISL_2378842, EPI_ISL_2378843, EPI_ISL_2378844, EPI_ISL_2378845, EPI_ISL_2378846, EPI_ISL_2378847, EPI_ISL_2378848, EPI_ISL_2378849, EPI_ISL_2378862 | | | |
| see above | Washington State Department of Health Public Health Laboratories | Washington State Department of Health Public Health Laboratories | Avi Singh; Darren Lucas; Denny Russell; Drew MacKellar; Geoff Melly; Hannah Gray; Joenice Gonzalez; JohnAric Peterson; Philip Dykema; Rebecca Cao; Vanessa De Los Santos |
| EPI_ISL_4212988 | Weill Cornell Medicine | New York Genome Center | Amy Baldwin; Andre Corvelo; Arryn Craney; Chris Mason; Dayna M. Oschwald; Hanna Rennert; Lars F Westblade; Margaret Elizabeth Ross; Melissa Cushing; Michael Zody; Olivier Elemento; Priya Velu; Samantha Fennessey; Tom Maniatis |
| EPI_ISL_2092835, EPI_ISL_2092836, EPI_ISL_2092839, EPI_ISL_2101360, EPI_ISL_2156023, EPI_ISL_2156025, EPI_ISL_2156026, EPI_ISL_2156028, EPI_ISL_2156033, EPI_ISL_2156034, EPI_ISL_2156044, EPI_ISL_2227378, EPI_ISL_2227379, EPI_ISL_2227380, EPI_ISL_2227384, EPI_ISL_2227391, EPI_ISL_2227393, EPI_ISL_2227396 | | | |
| see above | Wichita State University - Molecular Diagnostics Lab | Kansas Health and Environmental Lab | Ben Olsen; Jonathan Barnell; Katherine Wiggins; Mike Grose; and Phil Adam |
| EPI_ISL_2343266, EPI_ISL_2343269 | Willis-Knighton Medical Center Hospital Laboratory | LSUHS Emerging Viral Threat Laboratory | Alexander Mijalis; Andrew D. Yurochko; April N. Johnson; Christopher G. Kevil; Gregory L. Ware; Jennifer L. Carroll; Jeremy P. Kamil; John A. Vanchiere; Joseph A. Bocchini; Lorie M. Atkins; Maarten Van Diest; Rona S. Scott |
| EPI_ISL_2558085, EPI_ISL_2558171, EPI_ISL_2558269, EPI_ISL_2558332, EPI_ISL_2558340, EPI_ISL_2558361, EPI_ISL_2558384, EPI_ISL_2558436, EPI_ISL_2558447, EPI_ISL_2558481, EPI_ISL_2558484 | | | |
| see above | Wisconsin State Laboratory of Hygiene Communicable Disease Division | Wisconsin State Laboratory of Hygiene Communicable Disease Division | Abigail C. Shockey; Alicia J. Mooney; Erika M. Hanson; Kelsey R. Florek; Richard Griesser; Sara Wagner; Tonya Danz |

EPI_ISL_2193968, EPI_ISL_2226823, EPI_ISL_2444773, EPI_ISL_2728124, EPI_ISL_2842504, EPI_ISL_2842510, EPI_ISL_3758883

| | | | |
|---|---|---|---|
| see above | Wyoming Public Health Laboratory | Wyoming Public Health Laboratory | Ashley Norberg; Brian Dominguez; Brittany Oher; Cari Sloma; Channing Weber; Chayse Rowley; Elliot Thomasson; Jim Mildenberger; Lynette Gumbleton; Marley Goetz; Noah Hull; Robert Petit; Sam Britz; Taylor Fearing; Wanda Manley; and Rob Christensen |
| EPI_ISL_2426129, EPI_ISL_2501703 | Yale Clinical Virology Lab | Grubaugh Lab - Yale School of Public Health | Anderson Brito; Annie Watkins; Chaney Kalinich; Chantal Vogels; Isabel Ott; Jessica Rothman; Joseph Fauver; Mallery Breban; Marie L. Landry; Mary Petrone; Nathan Grubaugh; Tara Alpert |
| EPI_ISL_2381558 | ZOTZ KLIMAS MVZ Düsseldorf-Centrum GbR ÜBAG für Labormedizin, Genetik, Zytologie, Pathologie | Center of Medical Microbiology, Virology, and Hospital Hygiene, University of Duesseldorf | Alexander Dilthey; Andreas Walker; Daniel Strelow; Jessica Nicolai; Jörg Timm; Katrin Hoffmann; Klaus Pfeffer; Lisanna Hülse; Malte Kohns Vasconcelos; Maximilian Damagnez; Nadine Lübke; Patrick Finzer; Rainer Zotz; Tobias Wienemann; Torsten Houwaart |
| EPI_ISL_2445858 | unknown | Instituto Nacional de Saude (INSA) | Borges et al |
