## Supplement 3 for "Rapid and Accurate Identification of SARS-CoV-2 Omicron Variants Using Droplet Digital PCR (RT-ddPCR)": All_0517-0523.pdf

We gratefully acknowledge the following Authors from the Originating laboratories responsible for obtaining the specimens, as well as the Submitting laboratories where the genome data were generated and shared via GISAID, on which this research is based.

All Submitters of data may be contacted directly via www.gisaid.org

Authors are sorted alphabetically.

| Accession ID | Originating Laboratory | Submitting Laboratory | Authors |
|---|---|---|---|
| EPI_ISL_2689871, EPI_ISL_2689874 | "AK State Public Health Lab, State Health Department" | Centers for Disease Control and Prevention Division of Viral Diseases, Pathogen Discovery | Alison Laufer Halpin; Ben L. Rambo-Martin; Clinton R. Paden; Dakota Howard; Darlene Wagner; Dave Wentworth; Dhwani Batra; Jasmine Padilla; Justin Lee; Katie Dillon; Krista Queen; Kristen Knipe; Kristine Lacek; Mark Burroughs; Matthew Schmerer; Mili Sheth; Peter Cook; Sam Shepard; Sarah Nobles; Shoshona Le; Suxiang Tong; Vivien Dugan; Yvette Unoarumhi |
| EPI_ISL_2787716, EPI_ISL_2787732 | "HI Dept. of Health, State Laboratories Division" | Centers for Disease Control and Prevention Division of Viral Diseases, Pathogen Discovery | Alison Laufer Halpin; Ben L. Rambo-Martin; Clinton R. Paden; Dakota Howard; Darlene Wagner; Dave Wentworth; Dhwani Batra; Jasmine Padilla; Justin Lee; Katie Dillon; Krista Queen; Kristen Knipe; Kristine Lacek; Mark Burroughs; Matthew Schmerer; Mili Sheth; Peter Cook; Sam Shepard; Sarah Nobles; Shoshona Le; Suxiang Tong; Vivien Dugan; Yvette Unoarumhi |
| EPI_ISL_2651014 | A.S.L. BIELLA - OSPEDALE DEGLI INFERMI - SC LABORATORIO ANALISI - | Fondazione del Piemonte per l'Oncologia IRCCS | Antonino Sottile; Giorgio Giardina; Paola Marino; Silvia Brossa |
| EPI_ISL_2650995 | A.S.L. CITTA' DI TORINO - OSPEDALE AMEDEO DI SAVOIA - SC MICROBIOLOGIA E VIROLOGIA - | Fondazione del Piemonte per l'Oncologia IRCCS | Antonino Sottile; Giorgio Giardina; Paola Marino; Silvia Brossa |
| EPI_ISL_2651024, EPI_ISL_2651029 | A.S.L. NOVARA - ISTITUTO S.S. TRINITA' - BORGOMANERO - S.S. LABORATORIO ANALISI CHIMICO CLINICHE - | Fondazione del Piemonte per l'Oncologia IRCCS | Antonino Sottile; Giorgio Giardina; Paola Marino; Silvia Brossa |
| EPI_ISL_2621212, EPI_ISL_2621215 | ADILAB | Universidad Nacional de Colombia - Laboratorio Genómico One Health | Andres F. Cardona-Rios; Carlos Franco-Muñoz; Carolina Muñoz-Arango; Celeny Ortiz; Daniel O. Maldonado-Perez; Diego A. Álvarez-Díaz; Hector Alejandro Ruiz-Moreno; Idabely Betancur Ortiz; Jorge E. Osorio; Juan P. Hernandez-Ortiz; Karl A Ciuoderis; Katherine Laiton-Donato; Laura Silvana Perez; Lina M. Hurtado; Marcela Mercado-Reyes; Maria Angélica Maya; Maria Stella López; Rita Almanza Payares; Sandra Ines Cano; Simón Villegas Velásquez |
| EPI_ISL_2493256, EPI_ISL_2493285, EPI_ISL_2493739, EPI_ISL_2493740, EPI_ISL_2493741, EPI_ISL_2493742, EPI_ISL_2493743, EPI_ISL_2493744, EPI_ISL_2493745, EPI_ISL_2493746, EPI_ISL_2493747, EPI_ISL_2493748, EPI_ISL_2493749, EPI_ISL_2493750, EPI_ISL_2493751, EPI_ISL_2493752, EPI_ISL_2493753, EPI_ISL_2493754, EPI_ISL_2493755, EPI_ISL_2493756, EPI_ISL_2493757, EPI_ISL_2493758, EPI_ISL_2493759, EPI_ISL_2493760, EPI_ISL_2493761, EPI_ISL_2493762, EPI_ISL_2494293 | | | |
| see above | AFIP | Instituto Butantan | Antonio Jorge Martins; Claudia Renata dos Santos Barros; David Schlesinger; Debora Botequio Moretti; Dimas Tadeu Covas; Elaine Cristina Marqueze; Elaine Vieira Santos; Evandra Strazza Rodrigues; Heidge Fukumasu; Jayme Augusto de Souza-Neto; José Salvatore Leister Patané; Luiz Alcantara; Luiz Lehmann Coutinho; Maria Carolina Elias; Maurício Lacerda Nogueira; Rafael dos Santos Bezerra; Raul Machado Neto; Rejane Maria Tommasini Grotto; Ricardo Haddad; Sandra Coccuzzo Sampaio Vessoni; Simone Kashima; Svetoslav Nanev Slavov; Vincent Louis Viala |
| EPI_ISL_2627157 | ALBANY MEDICAL CENTER | Wadsworth Center, New York State Department of Health | Alexis Russell; Catharine Prussing; Daryl M. Lamson; Erasmus Schneider; Erica Lasek-Nesselquist; John Kelly; Jonathan Plitnick; Kirsten St. George; Matthew Shudt; Melissa A Leisner; Navjot Singh |
| EPI_ISL_5915162, EPI_ISL_5915188, EPI_ISL_5915195, EPI_ISL_5915211, EPI_ISL_5915212, EPI_ISL_5915226, EPI_ISL_5915260, EPI_ISL_5915261, EPI_ISL_5915293, EPI_ISL_5915294, EPI_ISL_5915311, EPI_ISL_5915312, EPI_ISL_5915313, EPI_ISL_5915319, EPI_ISL_6573790, EPI_ISL_6573874 | | | |
| see above | AMALAB/FACISA/UFRN | WallauLab on behalf of Fiocruz COVID-19 Genomic Surveillance Network | Alexandre Freitas da Silva; Allan Roberto Dias Nunes; Antonio Marinho da Silva Neto; Cassia Docena; Constância Flávia Junqueira Ayres; Filipe Zimmer Dezordi; Gabriel Luz Wallau; Gustavo Barbosa de Lima; Joana Cristina Medeiros Tavares Marques; Katya Anaya Jacinto; Lais Ceschini Machado; Lilian Caroliny Amorim Silva; Marcelo Henrique dos Santos Paiva; Mariane dos Santos Duarte; Matheus Filgueira Bezerra; Sinval Pinto Brandão Filho |
| EPI_ISL_2378739, EPI_ISL_2493553 | AMBULATORIO AVIACAO | Instituto Butantan | Antonio Jorge Martins; Claudia Renata dos Santos Barros; David Schlesinger; Debora Botequio Moretti; Dimas Tadeu Covas; Elaine Cristina Marqueze; Elaine Vieira Santos; Evandra Strazza Rodrigues; Heidge Fukumasu; Jayme Augusto de Souza-Neto; José Salvatore Leister Patané; Luiz Alcantara; Luiz Lehmann Coutinho; Maria Carolina Elias; Maurício Lacerda Nogueira; Rafael dos Santos Bezerra; Raul Machado Neto; Rejane Maria Tommasini Grotto; Ricardo Haddad; Sandra Coccuzzo Sampaio Vessoni; Simone Kashima; Svetoslav Nanev Slavov; Vincent Louis Viala |
| EPI_ISL_2493549, EPI_ISL_2493554 | AMBULATORIO BOQUEIRAO | Instituto Butantan | Antonio Jorge Martins; Claudia Renata dos Santos Barros; David Schlesinger; Debora Botequio Moretti; Dimas Tadeu Covas; Elaine Cristina Marqueze; Elaine Vieira Santos; Evandra Strazza Rodrigues; Heidge Fukumasu; Jayme Augusto de Souza-Neto; José Salvatore Leister Patané; Luiz Alcantara; Luiz Lehmann Coutinho; Maria Carolina Elias; Maurício Lacerda Nogueira; Rafael dos Santos Bezerra; Raul Machado Neto; Rejane Maria Tommasini Grotto; Ricardo Haddad; Sandra Coccuzzo Sampaio Vessoni; Simone Kashima; Svetoslav Nanev Slavov; Vincent Louis Viala |
| EPI_ISL_2493608, EPI_ISL_2493615 | AMBULATORIO COVID 19 | Instituto Butantan | Antonio Jorge Martins; Claudia Renata dos Santos Barros; David Schlesinger; Debora Botequio Moretti; Dimas Tadeu Covas; Elaine Cristina Marqueze; Elaine Vieira Santos; Evandra Strazza Rodrigues; Heidge Fukumasu; Jayme Augusto de Souza-Neto; José Salvatore Leister Patané; Luiz Alcantara; Luiz Lehmann Coutinho; Maria Carolina Elias; Maurício Lacerda Nogueira; Rafael dos Santos Bezerra; Raul Machado Neto; Rejane Maria Tommasini Grotto; Ricardo Haddad; Sandra Coccuzzo Sampaio Vessoni; Simone Kashima; Svetoslav Nanev Slavov; Vincent Louis Viala |
| EPI_ISL_2493701 | AMBULATORIO MEDICO E ODONTOLOGICO MUNICIPAL DE PIEDADE | Instituto Butantan | Antonio Jorge Martins; Claudia Renata dos Santos Barros; David Schlesinger; Debora Botequio Moretti; Dimas Tadeu Covas; Elaine Cristina Marqueze; Elaine Vieira Santos; Evandra Strazza Rodrigues; Heidge Fukumasu; Jayme Augusto de Souza-Neto; José Salvatore Leister Patané; Luiz Alcantara; Luiz Lehmann Coutinho; Maria Carolina Elias; Maurício Lacerda Nogueira; Rafael dos Santos Bezerra; Raul Machado Neto; Rejane Maria Tommasini Grotto; Ricardo Haddad; Sandra Coccuzzo Sampaio Vessoni; Simone Kashima; Svetoslav Nanev Slavov; Vincent Louis Viala |
| EPI_ISL_2473763, EPI_ISL_2473770, EPI_ISL_2473772, EPI_ISL_2473774, EPI_ISL_2493364 | AMBULATÓRIO INSTITUTO BUTANTAN | Instituto Butantan | Antonio Jorge Martins; Claudia Renata dos Santos Barros; David Schlesinger; Debora Botequio Moretti; Dimas Tadeu Covas; Elaine Cristina Marqueze; Elaine Vieira Santos; Evandra Strazza Rodrigues; Heidge Fukumasu; Jayme Augusto de Souza-Neto; José Salvatore Leister Patané; Luiz Alcantara; Luiz Lehmann Coutinho; Maria Carolina Elias; Maurício Lacerda Nogueira; Rafael dos Santos Bezerra; Raul Machado Neto; Rejane Maria Tommasini Grotto; Ricardo Haddad; Sandra Coccuzzo Sampaio Vessoni; Simone Kashima; Svetoslav Nanev Slavov; Vincent Louis Viala |
| EPI_ISL_2963428 | AOPD | Istituto Zooprofilattico Sperimentale delle Venezie | Adelaide Milani; Alessia Schivo; Alice Fusaro; Ambra Pastori; Annalisa Salviato; Antonia Ricci; Calogero Terregino; Edoardo Giussani; Elisa Palumbo; Erika Giorgia Quaranta; Isabella Monne; Luca Tassoni |
| EPI_ISL_2451779 | AR Dept. of Health-PHL, Molecular Diagnostics | Centers for Disease Control and Prevention Division of Viral Diseases, Pathogen Discovery | Alison Laufer Halpin; Ben L. Rambo-Martin; Clinton R. Paden; Dakota Howard; Darlene Wagner; Dave Wentworth; Dhwani Batra; Jasmine Padilla; Justin Lee; Katie Dillon; Krista Queen; Kristen Knipe; Kristine Lacek; Mark Burroughs; Matthew Schmerer; Mili Sheth; Peter Cook; Sam Shepard; Sarah Nobles; Shoshona Le; Suxiang Tong; Vivien Dugan; Yvette Unoarumhi |
| EPI_ISL_2467882, EPI_ISL_2534318, EPI_ISL_2658946 | AR Dept. of Health-PHL, Molecular Diagnostics | Genomics and Discovery, Respiratory Viruses Branch, Division of Viral Diseases, Centers for Disease Control and Prevention | Adam Retchless; Anna Kelleher; Anna Uehara; Brian Lynch; Clinton R. Paden; Dhwani Batra; Haibin Wang; Han Jia Justin Ng; Jasmine Padilla; Jing Zhang; Justin Lee; Krista Queen; Mark Burroughs; Mili Sheth; Morgan Davis; Peter Cook; Rachel Marine; Sarah Nobles; Suxiang Tong; Tara Coalter; Yan Li; Ying Tao |
| EPI_ISL_3122759, EPI_ISL_3122765, EPI_ISL_3122766 | ASST GOM NIGUARDA | ASST Grande ospedale Metropolitano Niguarda | Alice Nava |
| EPI_ISL_2963485 | AULSS 2 Marca Trevigiana | Istituto Zooprofilattico Sperimentale delle Venezie | Adelaide Milani; Alessia Schivo; Alice Fusaro; Ambra Pastori; Annalisa Salviato; Antonia Ricci; Calogero Terregino; Edoardo Giussani; Elisa Palumbo; Erika Giorgia Quaranta; Isabella Monne; Luca Tassoni |
| EPI_ISL_2322992 | AZ Jan Portaels | Imeda Hospital | Dagmar Obbels; Hanne Valgaeren; Johan Frans |
| EPI_ISL_2434165 | AZ Klina | AZ Klina | Carl Vael - Lynsey Berckmans |
| EPI_ISL_2323002, EPI_ISL_2323003 | AZ Sint Maarten | Imeda Hospital | Dagmar Obbels; Hanne Valgaeren; Johan Frans |
| EPI_ISL_2466351 | AZ Zeno Knokke-Blankenberge | AZ Sint Jan | Jorn Hellemans; Laurien Hoornaert; Marie Madeleine Chabert-Consen; Marijke Reynders; Patrick Descheemaeker; Thomas Van Landschoot |
| EPI_ISL_2280442, EPI_ISL_2280479, EPI_ISL_2280504, EPI_ISL_2280505, EPI_ISL_2280521, EPI_ISL_2280525, EPI_ISL_2280530, EPI_ISL_2281012, EPI_ISL_2281015, EPI_ISL_2281016, EPI_ISL_2281044, EPI_ISL_2281063, EPI_ISL_2281100, EPI_ISL_2281105, EPI_ISL_2281112, EPI_ISL_2281119, EPI_ISL_2281148, EPI_ISL_2281183, EPI_ISL_2281197, EPI_ISL_2281208, EPI_ISL_2281209, EPI_ISL_2281225, EPI_ISL_2283818, EPI_ISL_2283819, EPI_ISL_2283822, EPI_ISL_2283863, EPI_ISL_2283867, EPI_ISL_2283869, EPI_ISL_2283876, EPI_ISL_2368207, EPI_ISL_2368210, EPI_ISL_2368238, EPI_ISL_2368244, EPI_ISL_2368246, EPI_ISL_2368259, EPI_ISL_2368280, EPI_ISL_2368285, EPI_ISL_2368286, EPI_ISL_2368289, EPI_ISL_2368290, EPI_ISL_2368299, EPI_ISL_2368320, EPI_ISL_2368334, EPI_ISL_2368336, EPI_ISL_2368337, EPI_ISL_2368338, EPI_ISL_2368340, EPI_ISL_2368368, EPI_ISL_2368371, EPI_ISL_2368381, EPI_ISL_2368385, EPI_ISL_2368386, EPI_ISL_2368388, EPI_ISL_2368429, EPI_ISL_2368458, EPI_ISL_2368471, EPI_ISL_2368474, EPI_ISL_2368478, EPI_ISL_2368484, EPI_ISL_2368489, EPI_ISL_2368504, EPI_ISL_2368527, EPI_ISL_2368542, EPI_ISL_2368559, EPI_ISL_2368579, EPI_ISL_2368582, EPI_ISL_2368622, EPI_ISL_2368657, EPI_ISL_2368675, EPI_ISL_2368681, EPI_ISL_2368698, EPI_ISL_2368750, EPI_ISL_2368781, EPI_ISL_2368786, EPI_ISL_2368800, EPI_ISL_2368812, EPI_ISL_2368838, EPI_ISL_2368865, EPI_ISL_2368895, EPI_ISL_2368918, EPI_ISL_2368927, EPI_ISL_2368960, EPI_ISL_2368969, EPI_ISL_2368981, EPI_ISL_2369960, EPI_ISL_2369964, EPI_ISL_2369972, EPI_ISL_2369998, EPI_ISL_2370016, EPI_ISL_2370020, EPI_ISL_2370031, EPI_ISL_2370034, EPI_ISL_2370036, EPI_ISL_2370052, EPI_ISL_2370088, EPI_ISL_2370099, EPI_ISL_2370102, EPI_ISL_2370164, EPI_ISL_2370172, EPI_ISL_2370178, EPI_ISL_2370188, EPI_ISL_2370488, EPI_ISL_2370528, EPI_ISL_2370538, EPI_ISL_2370541, EPI_ISL_2370553, EPI_ISL_2370555, EPI_ISL_2370616, EPI_ISL_2370622, EPI_ISL_2370704, EPI_ISL_2372310, EPI_ISL_2372312, EPI_ISL_2372314, EPI_ISL_2372322, EPI_ISL_2372324, EPI_ISL_2372347, EPI_ISL_2372372, EPI_ISL_2372384, EPI_ISL_2372386, EPI_ISL_2372388, EPI_ISL_2372397, EPI_ISL_2372432, EPI_ISL_2372433, EPI_ISL_2372435, EPI_ISL_2372437, EPI_ISL_2372458, EPI_ISL_2372459, EPI_ISL_2421986, EPI_ISL_2421990, EPI_ISL_2421998, EPI_ISL_2422002, EPI_ISL_2422010, EPI_ISL_2422020, EPI_ISL_2422038, EPI_ISL_2422055, EPI_ISL_2422093, EPI_ISL_2422096, EPI_ISL_2422111, EPI_ISL_2489374, EPI_ISL_4360221, EPI_ISL_4362680, EPI_ISL_4364369 | | | |
| see above | Aegis Sciences Corporation | Centers for Disease Control and Prevention Division of Viral Diseases, Pathogen Discovery | Adrian Paskey; Alec Vest; Benjamin Rambo-Martin; Christopher Gulvick; Clinton Paden; Clinton R. Paden; Cyndi Clark; Dakota Howard; Darlene Wagner; Dhwani Batra; Dillon Nall; Duncan MacCannell; Erisa Sula; Ethan Sanders; Holly Houdeshell; Jason Caravas; Kara Moser; Kristine Lacek; Matthew Hardison; Matthew Schmerer; Ola Kvalvaag; Patrick Campbell; Peter Cook; Peter W. Cook; Rob Case; Scott Sammons; Shatavia Morrison; Shaun Westlund; Tymeckia Kendall; Victoria Caban Figueroa; Vikramsinha Ghorpade; Yvette Unoarumhi |
| EPI_ISL_2376008, EPI_ISL_2421717, EPI_ISL_2422100, EPI_ISL_2422101, EPI_ISL_2455425, EPI_ISL_2499921, EPI_ISL_2791461 | | | |
| see above | Alaska State Virology Laboratory | Alaska State Virology Laboratory | Elva House; Jack Chen; Jacob Zidek; Lisa Smith; Ph.D.; Stephanie DeRonde |
| EPI_ISL_2361724, EPI_ISL_2361726, EPI_ISL_2361729, EPI_ISL_2422125, EPI_ISL_2422127 | Algemeen Medisch Labo | Labo Klinische Biologie, UZA | Basil Britto Xavier; Christine Lammens; Herman Goossens; Jasmine Coppens; Marie Le Mercier; Veerle Matheeussen |
| EPI_ISL_3243026, EPI_ISL_3243032, EPI_ISL_3243033 | Applied DNA Sciences Inc | Applied DNA Sciences Inc | Huan Dong; Lucas Liu; Spencer William; Yuhua Sun |
| EPI_ISL_2331810, EPI_ISL_2331819 | Arcispedale Santa Maria Nuova, Autoimmunità, Allergologia e Biotecnologie Innovative | Istituto Zooprofilattico Sperimentale della Lombardia e dell'Emilia Romagna (IZSLER), Risk Analysis and Genomic Epidemiology Unit | Alessandro Zerbini; Erika Scaltriti; Ilaria Menozzi; Lucia Belloni; Marina Morganti; Stefania Croci; Stefano Pongolini |
| EPI_ISL_2339275, | Arizona State University | Arizona State University | Efrem S. Lim; Joshua LaBaer; Joy M. Blain; LaRinda A. Holland; Matthew F. Smith; Nicholas J. Mellor; Peter T. Skidmore; Rabia Maqsood; Valerie Harris; Vel Murugan |

| | | | |
|---|---|---|---|
| EPI_ISL_2339276, EPI_ISL_2339301, EPI_ISL_2484061, EPI_ISL_2484075, EPI_ISL_2484083 | | | |
| EPI_ISL_2931024 | Arkansas Department of Health | Center for Global Health, University of New Mexico Health Sciences Center | Atul Kothari; Darrell Dinwiddie; Daryl Domman; Joshua L. Kennedy; Kurt Schwalm; Valerie Morley |
| EPI_ISL_2692482, EPI_ISL_2692483, EPI_ISL_2692491, EPI_ISL_2692494, EPI_ISL_2692498, EPI_ISL_2715011, EPI_ISL_2715013, EPI_ISL_2715022, EPI_ISL_2715033, EPI_ISL_2715034 | | | |
| see above | Atlas Genomics | Seattle Flu Study | Amanda Adler; Barry R. Lutz; Benjamin Pelle; Brian Hiatt; Caitlin R. Wolf; Chris D. Frazar; Deborah A. Nickerson; Elisabeth Brandstetter; Erica Ryke; Geoff Melly; Helen Y. Chu; Janet A. Englund; Jay Shendure; Jover Lee; Kairsten Fay; Kirsten Lacombe; Lea M. Starita; Mark J. Rieder; Matthew Richardson; Matthew Thompson; Melissa Truong; Michael Boeckh; Michael Famulare; Misja Ilcisin; Peter D. Han; Philip Dykema; Romesh Gautom; Scott Lindquist; Thomas R. Sibley; Trevor Bedford |
| EPI_ISL_2427228, EPI_ISL_2485155, EPI_ISL_2485158, EPI_ISL_2616212, EPI_ISL_2616362 | Austrian Agency for Health and Food Safety (AGES) | Bergthaler laboratory, CeMM Research Center for Molecular Medicine of the Austrian Academy of Sciences | Andreas Bergthaler; Anna Schedl; Bekir Erguner; Benedikt Agerer; Christoph Bock; Fabian Amman; Jan Laine; Lukas Endler; Maelle Le Moing; Martin Senekowitsch; Matthew Thornton; Michael Schuster; Petr Triska; Thomas Penz |
| EPI_ISL_3231347 | Azarian Lab at Burnett School of Biomedical Sciences | Azarian Lab at Burnett School of Biomedical Sciences | Catherine Johnston; Eleonora Cella; Taj Azarian |
| EPI_ISL_2396878, EPI_ISL_2396882, EPI_ISL_2396883 | Azienda Ospedaliera Pugliese Ciaccio di Catanzaro SOC Microbiologia e Virologia | Azienda Ospedaliera Pugliese Ciaccio di Catanzaro SOC Microbiologia e Virologia | Rossana Tallerico Federica Pasceri Marco De Fazio Ilenia Talotta Giuseppina Panduri Pasquale Minchella |
| EPI_ISL_2600550, EPI_ISL_2600551, EPI_ISL_2600560 | Azienda Ospedaliera Terni | Istituto Zooprofilattico Sperimentale dell'Abruzzo e Molise "G. Caporale" | Ancora M; Calistri P; Cammà C; Curini V; Delli Compagni E; Di Domenico M; Di Pasquale A; Lorusso A; Mangone I; Marcacci M; Palumbo M; Puglia I; Rinaldi A; Savini G; Scaccetti A; Scialabba S |
| EPI_ISL_2331412, EPI_ISL_2331414, EPI_ISL_2331418, EPI_ISL_2331420, EPI_ISL_2331422, EPI_ISL_2331423, EPI_ISL_2331426, EPI_ISL_2331432, EPI_ISL_2331437 | | | |
| see above | Azienda Ospedaliero - Universitaria di Modena Policlinico - Virologia e Microbiologia Molecolare | Istituto Zooprofilattico Sperimentale della Lombardia e dell'Emilia Romagna (IZSLER), Risk Analysis and Genomic Epidemiology Unit | Erika Scaltriti; Giulia Fregni Serpini; Ilaria Menozzi; Marina Morganti; Monica Pecorari; Stefano Pongolini; William Gennari |
| EPI_ISL_2509112 | Azienda Sanitaria dell'Alto Adige - Laboratorio Aziendale di Microbiologia e Virologia | Azienda Sanitaria dell'Alto Adige | Irene Bianconi |
| EPI_ISL_2372816, EPI_ISL_2372820, EPI_ISL_2372822, EPI_ISL_2372826, EPI_ISL_2372827, EPI_ISL_2372828, EPI_ISL_2372835, EPI_ISL_2372838, EPI_ISL_2372839, EPI_ISL_2372840, EPI_ISL_2372845, EPI_ISL_2372846, EPI_ISL_2372850, EPI_ISL_2372854, EPI_ISL_2372855, EPI_ISL_2372860, EPI_ISL_2372867, EPI_ISL_2372871, EPI_ISL_2372873, EPI_ISL_2372875, EPI_ISL_2372876, EPI_ISL_2372881, EPI_ISL_2372890, EPI_ISL_2372891, EPI_ISL_2372893, EPI_ISL_2372897 | | | |
| see above | Azienda Sanitaria dell'Alto Adige Laboratorio Aziendale di Microbiologia e Virologia | Istituto di Genomica Applicata | Davide Scaglione; Eleonora Paparelli; Elisa Masi; Elisabetta Giacobazzi; Elisabetta Pagani; Gabriele Magris; Irena Jurman; Irene Bianconi; Michele Morgante; Stefanie Wieser; Vera Vendramin |
| EPI_ISL_2325020, EPI_ISL_2325021, EPI_ISL_2325024, EPI_ISL_2325029, EPI_ISL_2325031 | Azienda Sanitaria dell´Alto Adige - Laboratorio Aziendale di Microbiologia e Virologia | Azienda Sanitaria dell'Alto Adige | Irene Bianconi |
| EPI_ISL_2493187, EPI_ISL_2493199, EPI_ISL_2493763, EPI_ISL_2493764, EPI_ISL_2493765, EPI_ISL_2493766, EPI_ISL_2493767, EPI_ISL_2493768, EPI_ISL_2493769, EPI_ISL_2493770, EPI_ISL_2493771, EPI_ISL_2493772, EPI_ISL_2493773, EPI_ISL_2493774, EPI_ISL_2493775, EPI_ISL_2493776, EPI_ISL_2493777, EPI_ISL_2493778, EPI_ISL_2493780, EPI_ISL_2493781, EPI_ISL_2493782, EPI_ISL_2493783, EPI_ISL_2493784, EPI_ISL_2493785, EPI_ISL_2493786, EPI_ISL_2493787, EPI_ISL_2493788, EPI_ISL_2494314, EPI_ISL_2494315, EPI_ISL_2494319, EPI_ISL_2500974 | | | |
| see above | BIOFAST | Instituto Butantan | Antonio Jorge Martins; Claudia Renata dos Santos Barros; David Schlesinger; Debora Botequio Moretti; Dimas Tadeu Covas; Elaine Cristina Marqueze; Elaine Vieira Santos; Evandra Strazza Rodrigues; Heidge Fukumasu; Jayme Augusto de Souza-Neto; José Salvatore Leister Patané; Luiz Alcantara; Luiz Lehmann Coutinho; Maria Carolina Elias; Maurício Lacerda Nogueira; Rafael dos Santos Bezerra; Raul Machado Neto; Rejane Maria Tommasini Grotto; Ricardo Haddad; Sandra Coccuzzo Sampaio Vessoni; Simone Kashima; Svetoslav Nanev Slavov; Vincent Louis Viala |
| EPI_ISL_7672154, EPI_ISL_7672155, EPI_ISL_7672157, EPI_ISL_7672185 | Bambino Gesù Pediatric Hospital | Microbiology and Immunology Diagnosis Bambino Gesù Pediatric Hospital | Carlo Federico Perno; Claudia Alteri; Luna Colagrossi; Rossana Scutari; Valentino Costabile |
| EPI_ISL_2931172, EPI_ISL_2931176 | Baptist Health Medical Center | Center for Global Health, University of New Mexico Health Sciences Center | Amanda Novack; Darrell Dinwiddie; Daryl Domman; Dirk Haselow; Joshua L. Kennedy; Kurt Schwalm; Valerie Morley |
| EPI_ISL_2634980, EPI_ISL_2635010 | Bayerisches Landesamt für Gesundheit und Lebensmittelsicherheit (LGL) | Robert Koch Institute | |
| EPI_ISL_2401669, EPI_ISL_2401686, EPI_ISL_2401730, EPI_ISL_2401731 | BioneXt Lab | Laboratoire national de sante, Microbiology, Microbial Genomics Platform | Anke Wienecke-Baldacchino; Catherine Ragimbeau; Fatu Djabi; Jessica Tapp; Lise Pignon; Raoul Salmon; Tamir Abdelrahman; Thibault Ferrandon |
| EPI_ISL_3479077, EPI_ISL_3479078, EPI_ISL_3479079, EPI_ISL_3479084, EPI_ISL_3479088, EPI_ISL_3479092, EPI_ISL_3479093, EPI_ISL_3479094, EPI_ISL_3479101, EPI_ISL_3479105, EPI_ISL_3479106, EPI_ISL_3479107, EPI_ISL_3479109, EPI_ISL_3479111, EPI_ISL_3479113, EPI_ISL_3479114, EPI_ISL_3485747, EPI_ISL_3485761, EPI_ISL_3485768, EPI_ISL_3485769, EPI_ISL_3485771, EPI_ISL_3485781, EPI_ISL_3485783, EPI_ISL_3485880, EPI_ISL_3485888, EPI_ISL_3485915, EPI_ISL_3485918, EPI_ISL_3485920, EPI_ISL_3485955, EPI_ISL_3485972, EPI_ISL_3485991, EPI_ISL_3486000, EPI_ISL_3486001, EPI_ISL_3486003, EPI_ISL_3486004, EPI_ISL_3486007, EPI_ISL_3486008, EPI_ISL_3486009, EPI_ISL_3486013, EPI_ISL_3486015, EPI_ISL_3486019, EPI_ISL_3486023, EPI_ISL_3486026, EPI_ISL_3486027, EPI_ISL_3486034, EPI_ISL_3486035, EPI_ISL_3486036, EPI_ISL_3486040, EPI_ISL_3486042, EPI_ISL_3486044, EPI_ISL_3486052, EPI_ISL_3486059, EPI_ISL_3486063, EPI_ISL_3486065, EPI_ISL_3486070, EPI_ISL_3486074, EPI_ISL_3486079, EPI_ISL_3486082, EPI_ISL_3486083, EPI_ISL_3486084, EPI_ISL_3486086, EPI_ISL_3486088, EPI_ISL_3486089, EPI_ISL_3486092, EPI_ISL_3486095, EPI_ISL_3486096, EPI_ISL_3486097, EPI_ISL_3486100, EPI_ISL_3486102, EPI_ISL_3486105, EPI_ISL_3486149, EPI_ISL_3486150, EPI_ISL_3486152, EPI_ISL_3486153, EPI_ISL_3486156, EPI_ISL_3486184, EPI_ISL_3486185, EPI_ISL_3486187, EPI_ISL_3486193, EPI_ISL_3486194, EPI_ISL_3486195, EPI_ISL_3486196, EPI_ISL_3486197, EPI_ISL_3486200, EPI_ISL_3486204, EPI_ISL_3486205, EPI_ISL_3486208, EPI_ISL_3486209, EPI_ISL_3486210, EPI_ISL_3486211, EPI_ISL_3486215, EPI_ISL_3486216, EPI_ISL_3486217, EPI_ISL_3486221, EPI_ISL_3486222, EPI_ISL_3486230, EPI_ISL_3486233, EPI_ISL_3486244, EPI_ISL_3486246, EPI_ISL_3486250, EPI_ISL_3486262, EPI_ISL_3486327, EPI_ISL_3486331, EPI_ISL_3486335, EPI_ISL_3486338, EPI_ISL_3486365, EPI_ISL_3486366, EPI_ISL_3486367, EPI_ISL_3486379, EPI_ISL_3486381, EPI_ISL_3486382, EPI_ISL_3486383, EPI_ISL_3486406, EPI_ISL_3486407, EPI_ISL_3486408, EPI_ISL_3486417, EPI_ISL_3486418, EPI_ISL_3486419, EPI_ISL_3486425, EPI_ISL_3486426, EPI_ISL_3486429, EPI_ISL_3486431, EPI_ISL_3486432, EPI_ISL_3486433, EPI_ISL_3486437, EPI_ISL_3486440, EPI_ISL_3486441, EPI_ISL_3486447, EPI_ISL_3486448, EPI_ISL_3486450, EPI_ISL_3486460, EPI_ISL_3486473, EPI_ISL_3486481, EPI_ISL_3486482, EPI_ISL_3486484, EPI_ISL_3486487, EPI_ISL_3486488, EPI_ISL_3486489, EPI_ISL_3486490, EPI_ISL_3486491, EPI_ISL_3486492, EPI_ISL_3486493, EPI_ISL_3486498, EPI_ISL_3486499, EPI_ISL_3486500, EPI_ISL_3486502, EPI_ISL_3486503, EPI_ISL_3486505, EPI_ISL_3486507, EPI_ISL_3486508, EPI_ISL_3486509, EPI_ISL_3486512, EPI_ISL_3486513, EPI_ISL_3486515, EPI_ISL_3486516, EPI_ISL_3486518, EPI_ISL_3486522, EPI_ISL_3486523, EPI_ISL_3486525, EPI_ISL_3486527, EPI_ISL_3486530, EPI_ISL_3486531, EPI_ISL_3486534, EPI_ISL_3486535, EPI_ISL_3486536, EPI_ISL_3486538, EPI_ISL_3486539, EPI_ISL_3486542, EPI_ISL_3486543, EPI_ISL_3486545, EPI_ISL_3486553, EPI_ISL_3486555, EPI_ISL_3486556, EPI_ISL_3486559, EPI_ISL_3486560, EPI_ISL_3486563, EPI_ISL_3486566, EPI_ISL_3486568, EPI_ISL_3486570, EPI_ISL_3486571, EPI_ISL_3486573, EPI_ISL_3486576, EPI_ISL_3486577, EPI_ISL_3486586, EPI_ISL_3486590, EPI_ISL_3486591, EPI_ISL_3486592, EPI_ISL_3486594, EPI_ISL_3486595, EPI_ISL_3486597, EPI_ISL_3486598, EPI_ISL_3486600, EPI_ISL_3486601, EPI_ISL_3486602, EPI_ISL_3486606, EPI_ISL_3486608, EPI_ISL_3486613, EPI_ISL_3486614, EPI_ISL_3486615, EPI_ISL_3486616, EPI_ISL_3486621, EPI_ISL_3486628, EPI_ISL_3486629, EPI_ISL_3486630, EPI_ISL_3486631, EPI_ISL_3486633, EPI_ISL_3486636, EPI_ISL_3486637, EPI_ISL_3486638, EPI_ISL_3486639, EPI_ISL_3486640, EPI_ISL_3486642, EPI_ISL_3486643, EPI_ISL_3486644, EPI_ISL_3486645, EPI_ISL_3486656, EPI_ISL_3486661, EPI_ISL_3486662, EPI_ISL_3486663, EPI_ISL_3486664, EPI_ISL_3486667, EPI_ISL_3486671, EPI_ISL_3486677, EPI_ISL_3486680, EPI_ISL_3486684, EPI_ISL_3486685, EPI_ISL_3486688, EPI_ISL_3486690, EPI_ISL_3486692, EPI_ISL_3486696, EPI_ISL_3486698, EPI_ISL_3486699, EPI_ISL_3486702, EPI_ISL_3486705, EPI_ISL_3486707, EPI_ISL_3486708, EPI_ISL_3486712, EPI_ISL_3486713, EPI_ISL_3486715, EPI_ISL_3486716, EPI_ISL_3486720, EPI_ISL_3486721, EPI_ISL_3486731, EPI_ISL_3486737, EPI_ISL_3486738, EPI_ISL_3486743, EPI_ISL_3486745, EPI_ISL_3486746, EPI_ISL_3486747, EPI_ISL_3486748, EPI_ISL_3486750, EPI_ISL_3486754, EPI_ISL_3486757, EPI_ISL_3486758, EPI_ISL_3486760, EPI_ISL_3486762, EPI_ISL_3486767, EPI_ISL_3486769, EPI_ISL_3486771, EPI_ISL_3486778, EPI_ISL_3486779, EPI_ISL_3486780, EPI_ISL_3486781, EPI_ISL_3486783, EPI_ISL_3486786, EPI_ISL_3486787, EPI_ISL_3486788, EPI_ISL_3486789, EPI_ISL_3486799, EPI_ISL_3486800, EPI_ISL_3486807, EPI_ISL_3486814, EPI_ISL_3486816, EPI_ISL_3486820, EPI_ISL_3486824, EPI_ISL_3486826, EPI_ISL_3486828, EPI_ISL_3486831, EPI_ISL_3486832, EPI_ISL_3486833, EPI_ISL_3486835, EPI_ISL_3486838, EPI_ISL_3486839, EPI_ISL_3486841, EPI_ISL_3486846, EPI_ISL_3486848, EPI_ISL_3486849, EPI_ISL_3486851, EPI_ISL_3486853, EPI_ISL_3486855, EPI_ISL_3486856, EPI_ISL_3486857, EPI_ISL_3486859, EPI_ISL_3486864, EPI_ISL_3486865, EPI_ISL_3486869, EPI_ISL_3486879, EPI_ISL_3486885, EPI_ISL_3486886, EPI_ISL_3486891, EPI_ISL_3486901, EPI_ISL_3486923, EPI_ISL_3486925, EPI_ISL_3486926, EPI_ISL_3486928, EPI_ISL_3486934, EPI_ISL_3486935, EPI_ISL_3486936, EPI_ISL_3486937, EPI_ISL_3486938, EPI_ISL_3486946, EPI_ISL_3486948, EPI_ISL_3486950, EPI_ISL_3486951, EPI_ISL_3486956, EPI_ISL_3486959, EPI_ISL_3486962, EPI_ISL_3486970, EPI_ISL_3486972, EPI_ISL_3486975, EPI_ISL_3486979, EPI_ISL_3486981, EPI_ISL_3486985, EPI_ISL_3486986, EPI_ISL_3486996, EPI_ISL_3486999, EPI_ISL_3487008, EPI_ISL_3487010, EPI_ISL_3487014, EPI_ISL_3487018, EPI_ISL_3487023, EPI_ISL_3487025, EPI_ISL_3487033, EPI_ISL_3487035, EPI_ISL_3487044, EPI_ISL_3487052, EPI_ISL_3487057, EPI_ISL_3487065, EPI_ISL_3487066, EPI_ISL_3487067, EPI_ISL_3487071, EPI_ISL_3487074, EPI_ISL_3487075, EPI_ISL_3487085, EPI_ISL_3487089, EPI_ISL_3487090, EPI_ISL_3487093, EPI_ISL_3487096, EPI_ISL_3487098, EPI_ISL_3487108, EPI_ISL_3487117, EPI_ISL_3487121, EPI_ISL_3487128, EPI_ISL_3487130, EPI_ISL_3487131, EPI_ISL_3487136, EPI_ISL_3487140, EPI_ISL_3487141, EPI_ISL_3487143, EPI_ISL_3487144, EPI_ISL_3487146, EPI_ISL_3487149, EPI_ISL_3487150, EPI_ISL_3487151, EPI_ISL_3487152, EPI_ISL_3487158, EPI_ISL_3487160, EPI_ISL_3487167, EPI_ISL_3487168, EPI_ISL_3487169, EPI_ISL_3487173, EPI_ISL_3487174, EPI_ISL_3487177, EPI_ISL_3487178, EPI_ISL_3487179, EPI_ISL_3487182, EPI_ISL_3487183, EPI_ISL_3487184, EPI_ISL_3487185, EPI_ISL_3487186, EPI_ISL_3487187, EPI_ISL_3487191, EPI_ISL_3487193, EPI_ISL_3487194, EPI_ISL_3487196, EPI_ISL_3487197, EPI_ISL_3487198, EPI_ISL_3487205, EPI_ISL_3487206, EPI_ISL_3487207, EPI_ISL_3487210, EPI_ISL_3487211, EPI_ISL_3487212, EPI_ISL_3487213, EPI_ISL_3487217, EPI_ISL_3487218, EPI_ISL_3487220, EPI_ISL_3487222, EPI_ISL_3487225, EPI_ISL_3487232, EPI_ISL_3487233, EPI_ISL_3487234, EPI_ISL_3487235, EPI_ISL_3487236, EPI_ISL_3487242, EPI_ISL_3487244, EPI_ISL_3487248, EPI_ISL_3487249, EPI_ISL_3487250, EPI_ISL_3487251, EPI_ISL_3487252, EPI_ISL_3487253, EPI_ISL_3487256, EPI_ISL_3487257, EPI_ISL_3487263, EPI_ISL_3487264, EPI_ISL_3487268, EPI_ISL_3487269, EPI_ISL_3487270, EPI_ISL_3487275, EPI_ISL_3487278, EPI_ISL_3487279, EPI_ISL_3487281, EPI_ISL_3487284, EPI_ISL_3487290, EPI_ISL_3487291, EPI_ISL_3487293, EPI_ISL_3487294, EPI_ISL_3487297, EPI_ISL_3487298, EPI_ISL_3487299, EPI_ISL_3487300, EPI_ISL_3487302, EPI_ISL_3487310, EPI_ISL_3487314, EPI_ISL_3487315, EPI_ISL_3487318, EPI_ISL_3487325, EPI_ISL_3487326, EPI_ISL_3487328, EPI_ISL_3487333, EPI_ISL_3487334, EPI_ISL_3487345, EPI_ISL_3487348, EPI_ISL_3487349, EPI_ISL_3487352, EPI_ISL_3487353, EPI_ISL_3487354, EPI_ISL_3487355, EPI_ISL_3487356, EPI_ISL_3487357, EPI_ISL_3487358, EPI_ISL_3487359, EPI_ISL_3487360, EPI_ISL_3487361, EPI_ISL_3487363, EPI_ISL_3487369, EPI_ISL_3487371, EPI_ISL_3487375, EPI_ISL_3487377, EPI_ISL_3487379, EPI_ISL_3487383, EPI_ISL_3487384, EPI_ISL_3487391, EPI_ISL_3487394, EPI_ISL_3487395, EPI_ISL_3487396, EPI_ISL_3487397, EPI_ISL_3487398, EPI_ISL_3487399, EPI_ISL_3487403, EPI_ISL_3487404, EPI_ISL_3487411, EPI_ISL_3487412, EPI_ISL_3487413, EPI_ISL_3487415, EPI_ISL_3487416, EPI_ISL_3487418, EPI_ISL_3487422, EPI_ISL_3487423, EPI_ISL_3487424, EPI_ISL_3487427, EPI_ISL_3487438, EPI_ISL_3487439, EPI_ISL_3487451, EPI_ISL_3487452, EPI_ISL_3487454, EPI_ISL_3487457, EPI_ISL_3487459, EPI_ISL_3487463, EPI_ISL_3487465, EPI_ISL_3487466, EPI_ISL_3487467, EPI_ISL_3487470, EPI_ISL_3487472, EPI_ISL_3487473, EPI_ISL_3487474, EPI_ISL_3487478, EPI_ISL_3487479, EPI_ISL_3487483, EPI_ISL_3487485, EPI_ISL_3487487, EPI_ISL_3487489, EPI_ISL_3487490, EPI_ISL_3487491, EPI_ISL_3487497, EPI_ISL_3487498, EPI_ISL_3487499, EPI_ISL_3487503, EPI_ISL_3487504, EPI_ISL_3487506, EPI_ISL_3487507, EPI_ISL_3487508, EPI_ISL_3487509, EPI_ISL_3487510, EPI_ISL_3487520, EPI_ISL_3487521, EPI_ISL_3487523, EPI_ISL_3487524, EPI_ISL_3487526, EPI_ISL_3487527, EPI_ISL_3487535, EPI_ISL_3487536, EPI_ISL_3487542, EPI_ISL_3487545, EPI_ISL_3487548, EPI_ISL_3487549, EPI_ISL_3487550, EPI_ISL_3487551, EPI_ISL_3487555, EPI_ISL_3487576, EPI_ISL_3487577, EPI_ISL_3487578, EPI_ISL_3487579, EPI_ISL_3487596, EPI_ISL_3487599, EPI_ISL_3487605, EPI_ISL_3487635, EPI_ISL_3487776, EPI_ISL_3487780, EPI_ISL_3487781, EPI_ISL_3487782, EPI_ISL_3487783, EPI_ISL_3487784, EPI_ISL_3487785, EPI_ISL_3487786, EPI_ISL_3487787, EPI_ISL_3487788, EPI_ISL_3487789, EPI_ISL_3487790, EPI_ISL_3487791, EPI_ISL_3487792, EPI_ISL_3487795, EPI_ISL_3487797, EPI_ISL_3487798, EPI_ISL_3487957, EPI_ISL_3487958, EPI_ISL_3487959, EPI_ISL_3487960, EPI_ISL_3487963, EPI_ISL_3487968, EPI_ISL_3487970, EPI_ISL_3487971, EPI_ISL_3487978, EPI_ISL_3488131, EPI_ISL_3488135, EPI_ISL_3488136, EPI_ISL_3488149, EPI_ISL_3488151, EPI_ISL_3488153, EPI_ISL_3488161, EPI_ISL_3488175, EPI_ISL_3488177, EPI_ISL_3488182, EPI_ISL_3488183, EPI_ISL_3488184, EPI_ISL_3488190, EPI_ISL_3488199, EPI_ISL_3488200, EPI_ISL_3488201, EPI_ISL_3488202, EPI_ISL_3488205, EPI_ISL_3488216, EPI_ISL_3488217, EPI_ISL_3488225, EPI_ISL_3488228, EPI_ISL_3488233, EPI_ISL_3488239, EPI_ISL_3488243, EPI_ISL_3488247, EPI_ISL_3488256, EPI_ISL_3488274, EPI_ISL_3488277, EPI_ISL_3488279, EPI_ISL_3488286, EPI_ISL_3488290, EPI_ISL_3596532, EPI_ISL_3596549, EPI_ISL_3596552, EPI_ISL_3601447, EPI_ISL_3601448, EPI_ISL_3601452, EPI_ISL_3601453, EPI_ISL_3601472, EPI_ISL_3601473, EPI_ISL_3601474, EPI_ISL_3601475, EPI_ISL_3601476, EPI_ISL_3601478, EPI_ISL_3601479, EPI_ISL_3601490, EPI_ISL_3601492, EPI_ISL_3601499, EPI_ISL_3601500, EPI_ISL_3601501, EPI_ISL_3601505, EPI_ISL_3601506, EPI_ISL_3601509, EPI_ISL_3601511, EPI_ISL_3601513, EPI_ISL_3601525, EPI_ISL_3601527, EPI_ISL_3601529, EPI_ISL_3601530, EPI_ISL_3601531, EPI_ISL_3601534, EPI_ISL_3601537, EPI_ISL_3601545, EPI_ISL_3601546, EPI_ISL_3601547, EPI_ISL_3601552, EPI_ISL_3601553, EPI_ISL_3601555, EPI_ISL_3601562, EPI_ISL_3601574, EPI_ISL_3601593, EPI_ISL_3601596, EPI_ISL_3601601, EPI_ISL_3601602, EPI_ISL_3601603, EPI_ISL_3601606, EPI_ISL_3601872, EPI_ISL_3601876, EPI_ISL_3601880, EPI_ISL_3601881, EPI_ISL_3601886, EPI_ISL_3601887, EPI_ISL_3601890, EPI_ISL_3601891, EPI_ISL_3601894, EPI_ISL_3601895, EPI_ISL_3601896, EPI_ISL_3601897, EPI_ISL_3601898, EPI_ISL_3601902, EPI_ISL_3601903, EPI_ISL_3601907, EPI_ISL_3601908, EPI_ISL_3601909, EPI_ISL_3601910, EPI_ISL_3601912, EPI_ISL_3601913, EPI_ISL_3601914, EPI_ISL_3601915, EPI_ISL_3601918, EPI_ISL_3601919, EPI_ISL_3601920, EPI_ISL_3601921, EPI_ISL_3601927, EPI_ISL_3601930, EPI_ISL_3601931, EPI_ISL_3601936, EPI_ISL_3601937, EPI_ISL_3601938, EPI_ISL_3601939, EPI_ISL_3601943, EPI_ISL_3601944, EPI_ISL_3601947, EPI_ISL_3601948, EPI_ISL_3601950, EPI_ISL_3601953, EPI_ISL_3601955, EPI_ISL_3601957, EPI_ISL_3601958, EPI_ISL_3601959, EPI_ISL_3601960, EPI_ISL_3601961, EPI_ISL_3601963, EPI_ISL_3601969, EPI_ISL_3601971, EPI_ISL_3601978, EPI_ISL_3601979, EPI_ISL_3602006, EPI_ISL_3602016, EPI_ISL_3602398 | | | |
| see above | British Columbia Centre For Disease Control | BCCDC Public Health Laboratory | Ana Pacagnella; Corrinne Ng; Dan Fornika; John Tyson; Kim Macdonald; Kimia Kamelian; Linda Hoang; Loretta Janz; Mel Krajden; Prystajecky Natalie; Robert Azana; Shannon Russell |
| EPI_ISL_2991520, EPI_ISL_2991550, EPI_ISL_2991652, EPI_ISL_2991826, EPI_ISL_2991868, EPI_ISL_2991906 | Broad Institute Clinical Research Sequencing Platform | Infectious Disease Program, Broad Institute of Harvard and MIT | A.E.; Adams, G.; Anahtar, M.; B.L.; B.W.; Bauer, M.; Birren; Branda, J.; Carter, A.; Cerrato, F.; Chaluvadi, S.; Chapman; Cusick, C.; D.J.; DeRuff, K.; Flowers, K.; Gallagher, G.; Gladden-Young, A.; Gnirke, A.; Harris, J.; J.E.; K.J.; LaRocque, R.; Lagerborg, K.; Lemieux; Lin; Loreth, C.; MacInnis; Neumann, A.; Normandin, E.; P.C.; Park; Pierce, V.; Reilly, S.; Rosenberg, E.; Rudy, M.; Ryan, E.; S.B.; Sabeti; Shaw, B.; Siddle; Slater, D.; Smole, S.; Tomkins-Tinch, C.; Turbett, S. |

| | | | |
|---|---|---|---|
| EPI_ISL_2311868, EPI_ISL_2311872, EPI_ISL_2311877, EPI_ISL_2311887, EPI_ISL_2311901, EPI_ISL_2311915, EPI_ISL_2311917, EPI_ISL_2311918, EPI_ISL_2311924, EPI_ISL_2311927, EPI_ISL_2311929, EPI_ISL_2311932, EPI_ISL_2311937, EPI_ISL_2311942, EPI_ISL_2311946, EPI_ISL_2311947, EPI_ISL_2311956, EPI_ISL_2311960, EPI_ISL_2311963, EPI_ISL_2311986, EPI_ISL_2312004, EPI_ISL_2312015, EPI_ISL_2312027, EPI_ISL_2312040, EPI_ISL_2312041, EPI_ISL_2312068, EPI_ISL_2312069, EPI_ISL_2312073, EPI_ISL_2312087, EPI_ISL_2312094, EPI_ISL_2312108, EPI_ISL_2312110, EPI_ISL_2312119, EPI_ISL_2312121, EPI_ISL_2312122, EPI_ISL_2312124, EPI_ISL_2312130, EPI_ISL_2312135, EPI_ISL_2312144, EPI_ISL_2312148, EPI_ISL_2312149, EPI_ISL_2312154, EPI_ISL_2312155, EPI_ISL_2312156, EPI_ISL_2312158, EPI_ISL_2312174, EPI_ISL_2312187, EPI_ISL_2312195, EPI_ISL_2312204, EPI_ISL_2312208, EPI_ISL_2312227, EPI_ISL_2312235, EPI_ISL_2312237, EPI_ISL_2312247, EPI_ISL_2312254, EPI_ISL_2312263, EPI_ISL_2312271, EPI_ISL_2312279, EPI_ISL_2312280, EPI_ISL_2312285, EPI_ISL_2312288, EPI_ISL_2312293, EPI_ISL_2312294, EPI_ISL_2312296, EPI_ISL_2312298, EPI_ISL_2312326, EPI_ISL_2312330, EPI_ISL_2312331, EPI_ISL_2312333, EPI_ISL_2312334, EPI_ISL_2312343, EPI_ISL_2312350, EPI_ISL_2312351, EPI_ISL_2312352, EPI_ISL_2312357, EPI_ISL_2312359, EPI_ISL_2312360, EPI_ISL_2312370, EPI_ISL_2312379, EPI_ISL_2312388, EPI_ISL_2312393, EPI_ISL_2312395, EPI_ISL_2312409, EPI_ISL_2312416, EPI_ISL_2312422, EPI_ISL_2312424, EPI_ISL_2312426, EPI_ISL_2312442 | | | |
| see above | Broad Institute Clinical Research Sequencing Platform | Infectious Disease Program, Broad Institute of Harvard and MIT | Adams, G.; B.L.; B.W.; Bauer, M.; Birren; Blumenstiel, B.; Brown, C.; Carter, A.; Chaluvadi, S.; D.J.; DeFelice, M.; DeRuff, K.; Dodge, S.; Gabriel, S.; Gallagher, G.; Gladden-Young, A.; Granger, B.; J.E.; K.J.; Lagerborg, K.; Larkin, K.; Lee, M.; Lemieux; Lennon, N.; Loreth, C.; Madoff, L.; McGovern, S.; Meldrim, J.; Normandin, E.; P.C.; Park; Pearlman, L.; Reilly, S.; Rudy, M.; Sabeti; Siddle; Smole, S.; Tomkins-Tinch, C.; Vicente, G.; and MacInnis |
| EPI_ISL_2473723, EPI_ISL_2473818 | CADIP VILA REGINA FERNANDOPOLIS | Instituto Butantan | Antonio Jorge Martins; Claudia Renata dos Santos Barros; David Schlesinger; Debora Botequio Moretti; Dimas Tadeu Covas; Elaine Cristina Marqueze; Elaine Vieira Santos; Evandra Strazza Rodrigues; Heidge Fukumasu; Jayme Augusto de Souza-Neto; José Salvatore Leister Patané; Luiz Alcantara; Luiz Lehmann Coutinho; Maria Carolina Elias; Maurício Lacerda Nogueira; Rafael dos Santos Bezerra; Raul Machado Neto; Rejane Maria Tommasini Grotto; Ricardo Haddad; Sandra Coccuzzo Sampaio Vessoni; Simone Kashima; Svetoslav Nanev Slavov; Vincent Louis Viala |
| EPI_ISL_2890943, EPI_ISL_2891832, EPI_ISL_3244038, EPI_ISL_3244079, EPI_ISL_3244097, EPI_ISL_3244106, EPI_ISL_3244108, EPI_ISL_3244113, EPI_ISL_3244126, EPI_ISL_3244145, EPI_ISL_3244147, EPI_ISL_3244149, EPI_ISL_3244153, EPI_ISL_3244160, EPI_ISL_3244171, EPI_ISL_3244173, EPI_ISL_3244178, EPI_ISL_3244180, EPI_ISL_3244183, EPI_ISL_3244193, EPI_ISL_3244199, EPI_ISL_3244201, EPI_ISL_3244222, EPI_ISL_3244231, EPI_ISL_3244237, EPI_ISL_3244240, EPI_ISL_3244245, EPI_ISL_3244246, EPI_ISL_3244247, EPI_ISL_3244266, EPI_ISL_3244277, EPI_ISL_3244284, EPI_ISL_3244288, EPI_ISL_3244296, EPI_ISL_3244300, EPI_ISL_3244311, EPI_ISL_3244315, EPI_ISL_3244320, EPI_ISL_3244326, EPI_ISL_3244335, EPI_ISL_3244360, EPI_ISL_3244363, EPI_ISL_3244374, EPI_ISL_3244375, EPI_ISL_3244400, EPI_ISL_3244408, EPI_ISL_3244433, EPI_ISL_3244439, EPI_ISL_3244447, EPI_ISL_3244448, EPI_ISL_5696590 | | | |
| see above | CDPH VBL | California Department of Public Health | CDPH-COVIDNet; Emily Smith on behalf of CDPH-COVIDNet and Priva Path Labs Inc.; Emily Smith on behalf of CDPH-COVIDNet and UCSD EXCITE lab |
| EPI_ISL_2268284, EPI_ISL_2387174 | CENTOGENE Frankfurt Laboratory: Niederlassung Industriepark Höchst | Robert Koch Institute | |
| EPI_ISL_2801337, EPI_ISL_2801341, EPI_ISL_2801343, EPI_ISL_2801344, EPI_ISL_2801346, EPI_ISL_2801363, EPI_ISL_3102277, EPI_ISL_3102519 | | | |
| see above | CENTRO DE ATENDIMENTO PARA ENFRENTAMENTO AO COVID 19 | Analytical Competence Molecular Epidemiology Lab/ACME, Oswaldo Cruz Foundation, Ceara (FIOCRUZ CE) | Cleber Furtado Aksenen; Cleber Furtado Aksenen e Suzana Porto Almeida; Fabio Miyajima; Fernando Braga Stehling; Francisco Eder de Moura Lopes; Jamille Maria Mendes Bezerra; Joaquim César do Nascimento Sousa Junior; Pedro Miguel Carneiro Jeronimo; Suzana Porto Almeida e Lucas Delerino; Thais Ferreira de Oliveira; Thais de Oliveira Costa; Ticiane Cavalcante de Souza; Veridiana Pessoa Miyajima |
| EPI_ISL_2493375 | CENTRO DE ESPECIALIDDS MEDICAS IRMA LEOPOLDINA PIRASSUNUNGA | Instituto Butantan | Antonio Jorge Martins; Claudia Renata dos Santos Barros; David Schlesinger; Debora Botequio Moretti; Dimas Tadeu Covas; Elaine Cristina Marqueze; Elaine Vieira Santos; Evandra Strazza Rodrigues; Heidge Fukumasu; Jayme Augusto de Souza-Neto; José Salvatore Leister Patané; Luiz Alcantara; Luiz Lehmann Coutinho; Maria Carolina Elias; Maurício Lacerda Nogueira; Rafael dos Santos Bezerra; Raul Machado Neto; Rejane Maria Tommasini Grotto; Ricardo Haddad; Sandra Coccuzzo Sampaio Vessoni; Simone Kashima; Svetoslav Nanev Slavov; Vincent Louis Viala |
| EPI_ISL_2493155, EPI_ISL_2493435, EPI_ISL_2493436, EPI_ISL_2493437, EPI_ISL_2493438 | CENTRO DE GENOMAS SUL | Instituto Butantan | Antonio Jorge Martins; Claudia Renata dos Santos Barros; David Schlesinger; Debora Botequio Moretti; Dimas Tadeu Covas; Elaine Cristina Marqueze; Elaine Vieira Santos; Evandra Strazza Rodrigues; Heidge Fukumasu; Jayme Augusto de Souza-Neto; José Salvatore Leister Patané; Luiz Alcantara; Luiz Lehmann Coutinho; Maria Carolina Elias; Maurício Lacerda Nogueira; Rafael dos Santos Bezerra; Raul Machado Neto; Rejane Maria Tommasini Grotto; Ricardo Haddad; Sandra Coccuzzo Sampaio Vessoni; Simone Kashima; Svetoslav Nanev Slavov; Vincent Louis Viala |
| EPI_ISL_2493394 | CENTRO DE SAUDE | Instituto Butantan | Antonio Jorge Martins; Claudia Renata dos Santos Barros; David Schlesinger; Debora Botequio Moretti; Dimas Tadeu Covas; Elaine Cristina Marqueze; Elaine Vieira Santos; Evandra Strazza Rodrigues; Heidge Fukumasu; Jayme Augusto de Souza-Neto; José Salvatore Leister Patané; Luiz Alcantara; Luiz Lehmann Coutinho; Maria Carolina Elias; Maurício Lacerda Nogueira; Rafael dos Santos Bezerra; Raul Machado Neto; Rejane Maria Tommasini Grotto; Ricardo Haddad; Sandra Coccuzzo Sampaio Vessoni; Simone Kashima; Svetoslav Nanev Slavov; Vincent Louis Viala |
| EPI_ISL_2493282 | CENTRO DE SAUDE DE ANALANDIA | Instituto Butantan | Antonio Jorge Martins; Claudia Renata dos Santos Barros; David Schlesinger; Debora Botequio Moretti; Dimas Tadeu Covas; Elaine Cristina Marqueze; Elaine Vieira Santos; Evandra Strazza Rodrigues; Heidge Fukumasu; Jayme Augusto de Souza-Neto; José Salvatore Leister Patané; Luiz Alcantara; Luiz Lehmann Coutinho; Maria Carolina Elias; Maurício Lacerda Nogueira; Rafael dos Santos Bezerra; Raul Machado Neto; Rejane Maria Tommasini Grotto; Ricardo Haddad; Sandra Coccuzzo Sampaio Vessoni; Simone Kashima; Svetoslav Nanev Slavov; Vincent Louis Viala |
| EPI_ISL_2445165, EPI_ISL_2473820 | CENTRO DE SAUDE DE CANITAR | Instituto Butantan | Antonio Jorge Martins; Claudia Renata dos Santos Barros; David Schlesinger; Debora Botequio Moretti; Dimas Tadeu Covas; Elaine Cristina Marqueze; Elaine Vieira Santos; Evandra Strazza Rodrigues; Heidge Fukumasu; Jayme Augusto de Souza-Neto; José Salvatore Leister Patané; Luiz Alcantara; Luiz Lehmann Coutinho; Maria Carolina Elias; Maurício Lacerda Nogueira; Rafael dos Santos Bezerra; Raul Machado Neto; Rejane Maria Tommasini Grotto; Ricardo Haddad; Sandra Coccuzzo Sampaio Vessoni; Simone Kashima; Svetoslav Nanev Slavov; Vincent Louis Viala |
| EPI_ISL_2445197 | CENTRO DE SAUDE DE CRUZALIA | Instituto Butantan | Antonio Jorge Martins; Claudia Renata dos Santos Barros; David Schlesinger; Debora Botequio Moretti; Dimas Tadeu Covas; Elaine Cristina Marqueze; Elaine Vieira Santos; Evandra Strazza Rodrigues; Heidge Fukumasu; Jayme Augusto de Souza-Neto; José Salvatore Leister Patané; Luiz Alcantara; Luiz Lehmann Coutinho; Maria Carolina Elias; Maurício Lacerda Nogueira; Rafael dos Santos Bezerra; Raul Machado Neto; Rejane Maria Tommasini Grotto; Ricardo Haddad; Sandra Coccuzzo Sampaio Vessoni; Simone Kashima; Svetoslav Nanev Slavov; Vincent Louis Viala |
| EPI_ISL_2445153 | CENTRO DE SAUDE DE FLORIDA PAULISTA | Instituto Butantan | Antonio Jorge Martins; Claudia Renata dos Santos Barros; David Schlesinger; Debora Botequio Moretti; Dimas Tadeu Covas; Elaine Cristina Marqueze; Elaine Vieira Santos; Evandra Strazza Rodrigues; Heidge Fukumasu; Jayme Augusto de Souza-Neto; José Salvatore Leister Patané; Luiz Alcantara; Luiz Lehmann Coutinho; Maria Carolina Elias; Maurício Lacerda Nogueira; Rafael dos Santos Bezerra; Raul Machado Neto; Rejane Maria Tommasini Grotto; Ricardo Haddad; Sandra Coccuzzo Sampaio Vessoni; Simone Kashima; Svetoslav Nanev Slavov; Vincent Louis Viala |
| EPI_ISL_2473713 | CENTRO DE SAUDE DE IBIRAREMA | Instituto Butantan | Antonio Jorge Martins; Claudia Renata dos Santos Barros; David Schlesinger; Debora Botequio Moretti; Dimas Tadeu Covas; Elaine Cristina Marqueze; Elaine Vieira Santos; Evandra Strazza Rodrigues; Heidge Fukumasu; Jayme Augusto de Souza-Neto; José Salvatore Leister Patané; Luiz Alcantara; Luiz Lehmann Coutinho; Maria Carolina Elias; Maurício Lacerda Nogueira; Rafael dos Santos Bezerra; Raul Machado Neto; Rejane Maria Tommasini Grotto; Ricardo Haddad; Sandra Coccuzzo Sampaio Vessoni; Simone Kashima; Svetoslav Nanev Slavov; Vincent Louis Viala |
| EPI_ISL_2473744, EPI_ISL_2473745 | CENTRO DE SAUDE DE PIRAJUI | Instituto Butantan | Antonio Jorge Martins; Claudia Renata dos Santos Barros; David Schlesinger; Debora Botequio Moretti; Dimas Tadeu Covas; Elaine Cristina Marqueze; Elaine Vieira Santos; Evandra Strazza Rodrigues; Heidge Fukumasu; Jayme Augusto de Souza-Neto; José Salvatore Leister Patané; Luiz Alcantara; Luiz Lehmann Coutinho; Maria Carolina Elias; Maurício Lacerda Nogueira; Rafael dos Santos Bezerra; Raul Machado Neto; Rejane Maria Tommasini Grotto; Ricardo Haddad; Sandra Coccuzzo Sampaio Vessoni; Simone Kashima; Svetoslav Nanev Slavov; Vincent Louis Viala |
| EPI_ISL_2445194 | CENTRO DE SAUDE DE RIBEIRAO DO SUL | Instituto Butantan | Antonio Jorge Martins; Claudia Renata dos Santos Barros; David Schlesinger; Debora Botequio Moretti; Dimas Tadeu Covas; Elaine Cristina Marqueze; Elaine Vieira Santos; Evandra Strazza Rodrigues; Heidge Fukumasu; Jayme Augusto de Souza-Neto; José Salvatore Leister Patané; Luiz Alcantara; Luiz Lehmann Coutinho; Maria Carolina Elias; Maurício Lacerda Nogueira; Rafael dos Santos Bezerra; Raul Machado Neto; Rejane Maria Tommasini Grotto; Ricardo Haddad; Sandra Coccuzzo Sampaio Vessoni; Simone Kashima; Svetoslav Nanev Slavov; Vincent Louis Viala |
| EPI_ISL_2493354 | CENTRO DE SAUDE DR JOSE AGOSTINHO NOGUEIRA PAULICEIA | Instituto Butantan | Antonio Jorge Martins; Claudia Renata dos Santos Barros; David Schlesinger; Debora Botequio Moretti; Dimas Tadeu Covas; Elaine Cristina Marqueze; Elaine Vieira Santos; Evandra Strazza Rodrigues; Heidge Fukumasu; Jayme Augusto de Souza-Neto; José Salvatore Leister Patané; Luiz Alcantara; Luiz Lehmann Coutinho; Maria Carolina Elias; Maurício Lacerda Nogueira; Rafael dos Santos Bezerra; Raul Machado Neto; Rejane Maria Tommasini Grotto; Ricardo Haddad; Sandra Coccuzzo Sampaio Vessoni; Simone Kashima; Svetoslav Nanev Slavov; Vincent Louis Viala |
| EPI_ISL_2493377, EPI_ISL_2493378, EPI_ISL_2494150 | CENTRO DE SAUDE DR MARIO DIAS DE AGUIAR CAPIVARI | Instituto Butantan | Antonio Jorge Martins; Claudia Renata dos Santos Barros; David Schlesinger; Debora Botequio Moretti; Dimas Tadeu Covas; Elaine Cristina Marqueze; Elaine Vieira Santos; Evandra Strazza Rodrigues; Heidge Fukumasu; Jayme Augusto de Souza-Neto; José Salvatore Leister Patané; Luiz Alcantara; Luiz Lehmann Coutinho; Maria Carolina Elias; Maurício Lacerda Nogueira; Rafael dos Santos Bezerra; Raul Machado Neto; Rejane Maria Tommasini Grotto; Ricardo Haddad; Sandra Coccuzzo Sampaio Vessoni; Simone Kashima; Svetoslav Nanev Slavov; Vincent Louis Viala |
| EPI_ISL_5646027, EPI_ISL_5646030 | CENTRO DE SAUDE II BARRA BONITA | Instituto Butantan | Antonio Jorge Martins; Claudia Renata dos Santos Barros; David Schlesinger; Debora Botequio Moretti; Dimas Tadeu Covas; Elaine Cristina Marqueze; Elaine Vieira Santos; Evandra Strazza Rodrigues; Heidge Fukumasu; Jayme Augusto de Souza-Neto; José Salvatore Leister Patané; Luiz Alcantara; Luiz Lehmann Coutinho; Maria Carolina Elias; Maurício Lacerda Nogueira; Rafael dos Santos Bezerra; Raul Machado Neto; Rejane Maria Tommasini Grotto; Ricardo Haddad; Sandra Coccuzzo Sampaio Vessoni; Simone Kashima; Svetoslav Nanev Slavov; Vincent Louis Viala |
| EPI_ISL_5646049, EPI_ISL_5646073, EPI_ISL_5646078 | CENTRO DE SAUDE II DE CAFELANDIA | Instituto Butantan | Antonio Jorge Martins; Claudia Renata dos Santos Barros; David Schlesinger; Debora Botequio Moretti; Dimas Tadeu Covas; Elaine Cristina Marqueze; Elaine Vieira Santos; Evandra Strazza Rodrigues; Heidge Fukumasu; Jayme Augusto de Souza-Neto; José Salvatore Leister Patané; Luiz Alcantara; Luiz Lehmann Coutinho; Maria Carolina Elias; Maurício Lacerda Nogueira; Rafael dos Santos Bezerra; Raul Machado Neto; Rejane Maria Tommasini Grotto; Ricardo Haddad; Sandra Coccuzzo Sampaio Vessoni; Simone Kashima; Svetoslav Nanev Slavov; Vincent Louis Viala |
| EPI_ISL_2493643 | CENTRO DE SAUDE II DR GABRIEL MESQUITA VARGEM GDE DO SUL | Instituto Butantan | Antonio Jorge Martins; Claudia Renata dos Santos Barros; David Schlesinger; Debora Botequio Moretti; Dimas Tadeu Covas; Elaine Cristina Marqueze; Elaine Vieira Santos; Evandra Strazza Rodrigues; Heidge Fukumasu; Jayme Augusto de Souza-Neto; José Salvatore Leister Patané; Luiz Alcantara; Luiz Lehmann Coutinho; Maria Carolina Elias; Maurício Lacerda Nogueira; Rafael dos Santos Bezerra; Raul Machado Neto; Rejane Maria Tommasini Grotto; Ricardo Haddad; Sandra Coccuzzo Sampaio Vessoni; Simone Kashima; Svetoslav Nanev Slavov; Vincent Louis Viala |
| EPI_ISL_2494160 | CENTRO DE SAUDE II DR JOSE DE FELIPE E S PINHAL SP | Instituto Butantan | Antonio Jorge Martins; Claudia Renata dos Santos Barros; David Schlesinger; Debora Botequio Moretti; Dimas Tadeu Covas; Elaine Cristina Marqueze; Elaine Vieira Santos; Evandra Strazza Rodrigues; Heidge Fukumasu; Jayme Augusto de Souza-Neto; José Salvatore Leister Patané; Luiz Alcantara; Luiz Lehmann Coutinho; Maria Carolina Elias; Maurício Lacerda Nogueira; Rafael dos Santos Bezerra; Raul Machado Neto; Rejane Maria Tommasini Grotto; Ricardo Haddad; Sandra Coccuzzo Sampaio Vessoni; Simone Kashima; Svetoslav Nanev Slavov; Vincent Louis Viala |
| EPI_ISL_2493542, EPI_ISL_2493571, EPI_ISL_2493572, EPI_ISL_2493574, EPI_ISL_2493575, EPI_ISL_2493576, EPI_ISL_2493578, EPI_ISL_2493580, EPI_ISL_2493582, EPI_ISL_2493583, EPI_ISL_2493675, EPI_ISL_2493676, EPI_ISL_2493680, EPI_ISL_2493682, EPI_ISL_2493683 | | | |
| see above | CENTRO DE SAUDE II IBITINGA | Instituto Butantan | Antonio Jorge Martins; Claudia Renata dos Santos Barros; David Schlesinger; Debora Botequio Moretti; Dimas Tadeu Covas; Elaine Cristina Marqueze; Elaine Vieira Santos; Evandra Strazza Rodrigues; Heidge Fukumasu; Jayme Augusto de Souza-Neto; José Salvatore Leister Patané; Luiz Alcantara; Luiz Lehmann Coutinho; Maria Carolina Elias; Maurício Lacerda Nogueira; Rafael dos Santos Bezerra; Raul Machado Neto; Rejane Maria Tommasini Grotto; Ricardo Haddad; Sandra Coccuzzo Sampaio Vessoni; Simone Kashima; Svetoslav Nanev Slavov; Vincent Louis Viala |
| EPI_ISL_2493336, EPI_ISL_2493337, EPI_ISL_2493338, EPI_ISL_2493339, EPI_ISL_2493344, EPI_ISL_2493347 | CENTRO DE SAUDE II JUNQUEIROPOLIS | Instituto Butantan | Antonio Jorge Martins; Claudia Renata dos Santos Barros; David Schlesinger; Debora Botequio Moretti; Dimas Tadeu Covas; Elaine Cristina Marqueze; Elaine Vieira Santos; Evandra Strazza Rodrigues; Heidge Fukumasu; Jayme Augusto de Souza-Neto; José Salvatore Leister Patané; Luiz Alcantara; Luiz Lehmann Coutinho; Maria Carolina Elias; Maurício Lacerda Nogueira; Rafael dos Santos Bezerra; Raul Machado Neto; Rejane Maria Tommasini Grotto; Ricardo Haddad; Sandra Coccuzzo Sampaio Vessoni; Simone Kashima; Svetoslav Nanev Slavov; Vincent Louis Viala |
| EPI_ISL_2493263 | CENTRO DE SAUDE III AFFONSO LUZZI SANTA CRUZ DAS PALMEIRAS | Instituto Butantan | Antonio Jorge Martins; Claudia Renata dos Santos Barros; David Schlesinger; Debora Botequio Moretti; Dimas Tadeu Covas; Elaine Cristina Marqueze; Elaine Vieira Santos; Evandra Strazza Rodrigues; Heidge Fukumasu; Jayme Augusto de Souza-Neto; José Salvatore Leister Patané; Luiz Alcantara; Luiz Lehmann Coutinho; Maria Carolina Elias; Maurício Lacerda Nogueira; Rafael dos Santos Bezerra; Raul Machado Neto; Rejane Maria Tommasini Grotto; Ricardo Haddad; Sandra Coccuzzo Sampaio Vessoni; Simone Kashima; Svetoslav Nanev Slavov; Vincent Louis Viala |
| EPI_ISL_2493539, EPI_ISL_2493544, EPI_ISL_2493547, EPI_ISL_2493570 | CENTRO DE SAUDE III BORBOREMA | Instituto Butantan | Antonio Jorge Martins; Claudia Renata dos Santos Barros; David Schlesinger; Debora Botequio Moretti; Dimas Tadeu Covas; Elaine Cristina Marqueze; Elaine Vieira Santos; Evandra Strazza Rodrigues; Heidge Fukumasu; Jayme Augusto de Souza-Neto; José Salvatore Leister Patané; Luiz Alcantara; Luiz Lehmann Coutinho; Maria Carolina Elias; Maurício Lacerda Nogueira; Rafael dos Santos Bezerra; Raul Machado Neto; Rejane Maria Tommasini Grotto; Ricardo Haddad; Sandra Coccuzzo Sampaio Vessoni; Simone Kashima; Svetoslav Nanev Slavov; Vincent Louis Viala |
| EPI_ISL_2494317, EPI_ISL_2494332 | CENTRO DE SAUDE III DE AGUAS DE SAO PEDRO | Instituto Butantan | Antonio Jorge Martins; Claudia Renata dos Santos Barros; David Schlesinger; Debora Botequio Moretti; Dimas Tadeu Covas; Elaine Cristina Marqueze; Elaine Vieira Santos; Evandra Strazza Rodrigues; Heidge Fukumasu; Jayme Augusto de Souza-Neto; José Salvatore Leister Patané; Luiz Alcantara; Luiz Lehmann Coutinho; Maria Carolina Elias; Maurício Lacerda Nogueira; Rafael dos Santos Bezerra; Raul Machado Neto; Rejane Maria Tommasini Grotto; Ricardo Haddad; Sandra Coccuzzo Sampaio Vessoni; Simone Kashima; Svetoslav Nanev Slavov; Vincent Louis Viala |
| EPI_ISL_2493669 | CENTRO DE SAUDE III DOBRADA | Instituto Butantan | Antonio Jorge Martins; Claudia Renata dos Santos Barros; David Schlesinger; Debora Botequio Moretti; Dimas Tadeu Covas; Elaine Cristina Marqueze; Elaine Vieira Santos; Evandra Strazza Rodrigues; Heidge Fukumasu; Jayme Augusto de Souza-Neto; José Salvatore Leister Patané; Luiz Alcantara; Luiz Lehmann Coutinho; Maria Carolina Elias; Maurício Lacerda Nogueira; Rafael dos Santos Bezerra; Raul Machado Neto; Rejane Maria Tommasini Grotto; Ricardo Haddad; Sandra Coccuzzo Sampaio Vessoni; Simone Kashima; Svetoslav Nanev Slavov; Vincent Louis Viala |
| EPI_ISL_2493677, EPI_ISL_2493678 | CENTRO DE SAUDE III TABATINGA | Instituto Butantan | Antonio Jorge Martins; Claudia Renata dos Santos Barros; David Schlesinger; Debora Botequio Moretti; Dimas Tadeu Covas; Elaine Cristina Marqueze; Elaine Vieira Santos; Evandra Strazza Rodrigues; Heidge Fukumasu; Jayme Augusto de Souza-Neto; José Salvatore Leister Patané; Luiz Alcantara; Luiz Lehmann Coutinho; Maria Carolina Elias; Maurício Lacerda Nogueira; Rafael dos Santos Bezerra; Raul Machado Neto; Rejane Maria Tommasini Grotto; Ricardo Haddad; Sandra Coccuzzo Sampaio Vessoni; Simone Kashima; Svetoslav Nanev Slavov; Vincent Louis Viala |
| EPI_ISL_2494276, EPI_ISL_2494291 | CENTRO MEDICO DR NELSON SALOME DE CONCHAL | Instituto Butantan | Antonio Jorge Martins; Claudia Renata dos Santos Barros; David Schlesinger; Debora Botequio Moretti; Dimas Tadeu Covas; Elaine Cristina Marqueze; Elaine Vieira Santos; Evandra Strazza Rodrigues; Heidge Fukumasu; Jayme Augusto de Souza-Neto; José Salvatore Leister Patané; Luiz Alcantara; Luiz Lehmann Coutinho; Maria Carolina Elias; Maurício Lacerda Nogueira; Rafael dos Santos Bezerra; Raul Machado Neto; Rejane Maria Tommasini Grotto; Ricardo Haddad; Sandra Coccuzzo Sampaio Vessoni; Simone Kashima; Svetoslav Nanev Slavov; Vincent Louis Viala |
| EPI_ISL_2493182, EPI_ISL_2493292, EPI_ISL_2493300 | CENTRO MEDICO PMESP | Instituto Butantan | Antonio Jorge Martins; Claudia Renata dos Santos Barros; David Schlesinger; Debora Botequio Moretti; Dimas Tadeu Covas; Elaine Cristina Marqueze; Elaine Vieira Santos; Evandra Strazza Rodrigues; Heidge Fukumasu; Jayme Augusto de Souza-Neto; José Salvatore Leister Patané; Luiz Alcantara; Luiz Lehmann Coutinho; Maria Carolina Elias; Maurício Lacerda Nogueira; Rafael dos Santos Bezerra; Raul Machado Neto; Rejane Maria Tommasini Grotto; Ricardo Haddad; Sandra Coccuzzo Sampaio Vessoni; Simone Kashima; Svetoslav Nanev Slavov; Vincent Louis Viala |
| EPI_ISL_2493591, EPI_ISL_2493596, EPI_ISL_2493597, EPI_ISL_2493600, EPI_ISL_2493606, EPI_ISL_2493625 | CENTRO MEDICO SOCIAL COMUNITARIO JANUARIO TEODORO DE SOUZA | Instituto Butantan | Antonio Jorge Martins; Claudia Renata dos Santos Barros; David Schlesinger; Debora Botequio Moretti; Dimas Tadeu Covas; Elaine Cristina Marqueze; Elaine Vieira Santos; Evandra Strazza Rodrigues; Heidge Fukumasu; Jayme Augusto de Souza-Neto; José Salvatore Leister Patané; Luiz Alcantara; Luiz Lehmann Coutinho; Maria Carolina Elias; Maurício Lacerda Nogueira; Rafael dos Santos Bezerra; Raul Machado Neto; Rejane Maria Tommasini Grotto; Ricardo Haddad; Sandra Coccuzzo Sampaio Vessoni; Simone Kashima; Svetoslav Nanev Slavov; Vincent Louis Viala |
| EPI_ISL_2493433 | CENTRO SAUDE III DR JOSE TADEU MOURAO | Instituto Butantan | Antonio Jorge Martins; Claudia Renata dos Santos Barros; David Schlesinger; Debora Botequio Moretti; Dimas Tadeu Covas; Elaine Cristina Marqueze; Elaine Vieira Santos; Evandra Strazza Rodrigues; Heidge Fukumasu; Jayme Augusto de Souza-Neto; José Salvatore Leister Patané; Luiz Alcantara; Luiz Lehmann Coutinho; Maria Carolina Elias; Maurício Lacerda Nogueira; Rafael dos Santos Bezerra; Raul Machado Neto; Rejane Maria Tommasini Grotto; Ricardo Haddad; Sandra Coccuzzo Sampaio Vessoni; Simone Kashima; Svetoslav Nanev Slavov; Vincent Louis Viala |
| EPI_ISL_2621208 | CENTROLAB | Universidad Nacional de Colombia - Laboratorio Genómico One Health | Andres F. Cardona-Rios; Carlos Franco-Muñoz; Carolina Muñoz-Arango; Celeny Ortiz; Daniel O. Maldonado-Perez; Diego A. Álvarez-Díaz; Hector Alejandro Ruiz-Moreno; Idabely Betancur Ortiz; Jorge E. Osorio; Juan P. Hernandez-Ortiz; Karl A Ciuoderis; Katherine Laiton-Donato; Laura Silvana Perez; Lina M. Hurtado; Marcela Mercado-Reyes; Maria Angélica Maya; Maria Stella López; Rita Almanza Payares; Sandra Ines Cano; Simón Villegas Velásquez |

| | | | |
|---|---|---|---|
| EPI_ISL_2614613 | CETESB - Companhia Ambiental do Estado de Sao Paulo | Instituto Adolfo Lutz, Interdiciplinary Procedures Center, Strategic Laboratory | Caio Vinicius Dias Lopes; Claudia Regina Gonçalves; Claudio Tavares Sacchi; Erica Valessa Ramos Gomes; Karoline Rodrigues Campos; Leonardo Jose Tadeu de Araujo; Luisa Zanolli Moreno; Maria Ines Zanoli Sato; Mikaela R. F. Barbosa |
| EPI_ISL_2362539, EPI_ISL_2362540, EPI_ISL_2362557, EPI_ISL_2627846, EPI_ISL_2627847 | CHC Andrée Rosemon | Institut Pasteur de la Guyane | Anne Lavergne; Antoine Enfissi; Arielle Salmier; Dominique Rousset |
| EPI_ISL_2362105, EPI_ISL_2362114 | CHLC | Instituto Nacional de Saude (INSA) | Borges et al |
| EPI_ISL_2444361 | CHU LILLE | CHU Lille - Laboratoire de Virologie | AIT YAHYA Emilie; ALIDJINOU Enagnon Kazali; BOCKET Laurence; CREPIN Michel; DEMAY Christophe; ENGELMANN Ilka; GEFFROY Sandrine; GUIGON Aurélie; LAMBERT Valérie; LAZREK Mouna; NOBILLIAUX Florian; PREVOST Brigitte; THUILLIER Caroline; TINEZ Claire |
| EPI_ISL_2308117 | CHWAPI - SITE NOTRE DAME | Institut de Pathologie et Genetique (IPG) | Jérémie Gras; Pascale Hilbert |
| EPI_ISL_5645970 | CMSC VIL A MELHADO DR GIUSEPPE A SOBRINHO | Instituto Butantan | Antonio Jorge Martins; Claudia Renata dos Santos Barros; David Schlesinger; Debora Botequio Moretti; Dimas Tadeu Covas; Elaine Cristina Marqueze; Elaine Vieira Santos; Evandra Strazza Rodrigues; Heidge Fukumasu; Jayme Augusto de Souza-Neto; José Salvatore Leister Patané; Luiz Alcantara; Luiz Lehmann Coutinho; Maria Carolina Elias; Maurício Lacerda Nogueira; Rafael dos Santos Bezerra; Raul Machado Neto; Rejane Maria Tommasini Grotto; Ricardo Haddad; Sandra Coccuzzo Sampaio Vessoni; Simone Kashima; Svetoslav Nanev Slavov; Vincent Louis Viala |
| EPI_ISL_2362531, EPI_ISL_2362541, EPI_ISL_2362542, EPI_ISL_2362543, EPI_ISL_2362547, EPI_ISL_2362548, EPI_ISL_2362549, EPI_ISL_2362550, EPI_ISL_2362551, EPI_ISL_2362552 | | | |
| see above | CNR Institut Pasteur de la Guyane | Institut Pasteur de la Guyane | Anne Lavergne; Antoine Enfissi; Arielle Salmier; Dominique Rousset |
| EPI_ISL_2373756 | CNR Virus des Infections Respiratoires - France SUD | CNR Virus des Infections Respiratoires - France SUD | Antonin Bal; Bruno Lina; Bruno Simon; Gregory Destras; Gwendolyne Burfin; Hadrien Regue; Laurence Josset; Martine Valette; Quentin Semanas |
| EPI_ISL_2493384, EPI_ISL_2493385, EPI_ISL_2494153 | COORDENADORIA MUNICIPAL DE SAUDE DE IRACEMAPOLIS | Instituto Butantan | Antonio Jorge Martins; Claudia Renata dos Santos Barros; David Schlesinger; Debora Botequio Moretti; Dimas Tadeu Covas; Elaine Cristina Marqueze; Elaine Vieira Santos; Evandra Strazza Rodrigues; Heidge Fukumasu; Jayme Augusto de Souza-Neto; José Salvatore Leister Patané; Luiz Alcantara; Luiz Lehmann Coutinho; Maria Carolina Elias; Maurício Lacerda Nogueira; Rafael dos Santos Bezerra; Raul Machado Neto; Rejane Maria Tommasini Grotto; Ricardo Haddad; Sandra Coccuzzo Sampaio Vessoni; Simone Kashima; Svetoslav Nanev Slavov; Vincent Louis Viala |
| EPI_ISL_5646028 | CS DE ADOLFO | Instituto Butantan | Antonio Jorge Martins; Claudia Renata dos Santos Barros; David Schlesinger; Debora Botequio Moretti; Dimas Tadeu Covas; Elaine Cristina Marqueze; Elaine Vieira Santos; Evandra Strazza Rodrigues; Heidge Fukumasu; Jayme Augusto de Souza-Neto; José Salvatore Leister Patané; Luiz Alcantara; Luiz Lehmann Coutinho; Maria Carolina Elias; Maurício Lacerda Nogueira; Rafael dos Santos Bezerra; Raul Machado Neto; Rejane Maria Tommasini Grotto; Ricardo Haddad; Sandra Coccuzzo Sampaio Vessoni; Simone Kashima; Svetoslav Nanev Slavov; Vincent Louis Viala |
| EPI_ISL_2378744, EPI_ISL_2378753, EPI_ISL_2378754 | CS DE APARECIDA DOESTE | Instituto Butantan | Antonio Jorge Martins; Claudia Renata dos Santos Barros; David Schlesinger; Debora Botequio Moretti; Dimas Tadeu Covas; Elaine Cristina Marqueze; Elaine Vieira Santos; Evandra Strazza Rodrigues; Heidge Fukumasu; Jayme Augusto de Souza-Neto; José Salvatore Leister Patané; Luiz Alcantara; Luiz Lehmann Coutinho; Maria Carolina Elias; Maurício Lacerda Nogueira; Rafael dos Santos Bezerra; Raul Machado Neto; Rejane Maria Tommasini Grotto; Ricardo Haddad; Sandra Coccuzzo Sampaio Vessoni; Simone Kashima; Svetoslav Nanev Slavov; Vincent Louis Viala |
| EPI_ISL_2445209, EPI_ISL_2445210, EPI_ISL_2473720 | CS DE BALSAMO | Instituto Butantan | Antonio Jorge Martins; Claudia Renata dos Santos Barros; David Schlesinger; Debora Botequio Moretti; Dimas Tadeu Covas; Elaine Cristina Marqueze; Elaine Vieira Santos; Evandra Strazza Rodrigues; Heidge Fukumasu; Jayme Augusto de Souza-Neto; José Salvatore Leister Patané; Luiz Alcantara; Luiz Lehmann Coutinho; Maria Carolina Elias; Maurício Lacerda Nogueira; Rafael dos Santos Bezerra; Raul Machado Neto; Rejane Maria Tommasini Grotto; Ricardo Haddad; Sandra Coccuzzo Sampaio Vessoni; Simone Kashima; Svetoslav Nanev Slavov; Vincent Louis Viala |
| EPI_ISL_2493161 | CS DE CATIGUA | Instituto Butantan | Antonio Jorge Martins; Claudia Renata dos Santos Barros; David Schlesinger; Debora Botequio Moretti; Dimas Tadeu Covas; Elaine Cristina Marqueze; Elaine Vieira Santos; Evandra Strazza Rodrigues; Heidge Fukumasu; Jayme Augusto de Souza-Neto; José Salvatore Leister Patané; Luiz Alcantara; Luiz Lehmann Coutinho; Maria Carolina Elias; Maurício Lacerda Nogueira; Rafael dos Santos Bezerra; Raul Machado Neto; Rejane Maria Tommasini Grotto; Ricardo Haddad; Sandra Coccuzzo Sampaio Vessoni; Simone Kashima; Svetoslav Nanev Slavov; Vincent Louis Viala |
| EPI_ISL_2445212, EPI_ISL_2494297 | CS DE COSMORAMA | Instituto Butantan | Antonio Jorge Martins; Claudia Renata dos Santos Barros; David Schlesinger; Debora Botequio Moretti; Dimas Tadeu Covas; Elaine Cristina Marqueze; Elaine Vieira Santos; Evandra Strazza Rodrigues; Heidge Fukumasu; Jayme Augusto de Souza-Neto; José Salvatore Leister Patané; Luiz Alcantara; Luiz Lehmann Coutinho; Maria Carolina Elias; Maurício Lacerda Nogueira; Rafael dos Santos Bezerra; Raul Machado Neto; Rejane Maria Tommasini Grotto; Ricardo Haddad; Sandra Coccuzzo Sampaio Vessoni; Simone Kashima; Svetoslav Nanev Slavov; Vincent Louis Viala |
| EPI_ISL_2493424, EPI_ISL_5645969, EPI_ISL_5645974, EPI_ISL_5645976, EPI_ISL_5645981, EPI_ISL_5645982, EPI_ISL_5645986, EPI_ISL_5645987, EPI_ISL_5645988, EPI_ISL_5645989, EPI_ISL_5645993, EPI_ISL_5646005, EPI_ISL_5646007, EPI_ISL_5646011 | | | |
| see above | CS DE MONTE APRAZIVEL | Instituto Butantan | Antonio Jorge Martins; Claudia Renata dos Santos Barros; David Schlesinger; Debora Botequio Moretti; Dimas Tadeu Covas; Elaine Cristina Marqueze; Elaine Vieira Santos; Evandra Strazza Rodrigues; Heidge Fukumasu; Jayme Augusto de Souza-Neto; José Salvatore Leister Patané; Luiz Alcantara; Luiz Lehmann Coutinho; Maria Carolina Elias; Maurício Lacerda Nogueira; Rafael dos Santos Bezerra; Raul Machado Neto; Rejane Maria Tommasini Grotto; Ricardo Haddad; Sandra Coccuzzo Sampaio Vessoni; Simone Kashima; Svetoslav Nanev Slavov; Vincent Louis Viala |
| EPI_ISL_2493180, EPI_ISL_2493405, EPI_ISL_2493406, EPI_ISL_2493407, EPI_ISL_2493412, EPI_ISL_2493414, EPI_ISL_2493416 | | | |
| see above | CS DE NEVES PAULISTA | Instituto Butantan | Antonio Jorge Martins; Claudia Renata dos Santos Barros; David Schlesinger; Debora Botequio Moretti; Dimas Tadeu Covas; Elaine Cristina Marqueze; Elaine Vieira Santos; Evandra Strazza Rodrigues; Heidge Fukumasu; Jayme Augusto de Souza-Neto; José Salvatore Leister Patané; Luiz Alcantara; Luiz Lehmann Coutinho; Maria Carolina Elias; Maurício Lacerda Nogueira; Rafael dos Santos Bezerra; Raul Machado Neto; Rejane Maria Tommasini Grotto; Ricardo Haddad; Sandra Coccuzzo Sampaio Vessoni; Simone Kashima; Svetoslav Nanev Slavov; Vincent Louis Viala |
| EPI_ISL_2445213, EPI_ISL_2445214, EPI_ISL_2445215, EPI_ISL_2445217, EPI_ISL_2445218, EPI_ISL_2473721, EPI_ISL_2473722, EPI_ISL_2493661, EPI_ISL_2493662 | | | |
| see above | CS DE NOVO HORIZONTE | Instituto Butantan | Antonio Jorge Martins; Claudia Renata dos Santos Barros; David Schlesinger; Debora Botequio Moretti; Dimas Tadeu Covas; Elaine Cristina Marqueze; Elaine Vieira Santos; Evandra Strazza Rodrigues; Heidge Fukumasu; Jayme Augusto de Souza-Neto; José Salvatore Leister Patané; Luiz Alcantara; Luiz Lehmann Coutinho; Maria Carolina Elias; Maurício Lacerda Nogueira; Rafael dos Santos Bezerra; Raul Machado Neto; Rejane Maria Tommasini Grotto; Ricardo Haddad; Sandra Coccuzzo Sampaio Vessoni; Simone Kashima; Svetoslav Nanev Slavov; Vincent Louis Viala |
| EPI_ISL_5646025 | CS DE ONDA VERDE | Instituto Butantan | Antonio Jorge Martins; Claudia Renata dos Santos Barros; David Schlesinger; Debora Botequio Moretti; Dimas Tadeu Covas; Elaine Cristina Marqueze; Elaine Vieira Santos; Evandra Strazza Rodrigues; Heidge Fukumasu; Jayme Augusto de Souza-Neto; José Salvatore Leister Patané; Luiz Alcantara; Luiz Lehmann Coutinho; Maria Carolina Elias; Maurício Lacerda Nogueira; Rafael dos Santos Bezerra; Raul Machado Neto; Rejane Maria Tommasini Grotto; Ricardo Haddad; Sandra Coccuzzo Sampaio Vessoni; Simone Kashima; Svetoslav Nanev Slavov; Vincent Louis Viala |
| EPI_ISL_2493411, EPI_ISL_2493419, EPI_ISL_2493429 | CS DE ORINDIUVA | Instituto Butantan | Antonio Jorge Martins; Claudia Renata dos Santos Barros; David Schlesinger; Debora Botequio Moretti; Dimas Tadeu Covas; Elaine Cristina Marqueze; Elaine Vieira Santos; Evandra Strazza Rodrigues; Heidge Fukumasu; Jayme Augusto de Souza-Neto; José Salvatore Leister Patané; Luiz Alcantara; Luiz Lehmann Coutinho; Maria Carolina Elias; Maurício Lacerda Nogueira; Rafael dos Santos Bezerra; Raul Machado Neto; Rejane Maria Tommasini Grotto; Ricardo Haddad; Sandra Coccuzzo Sampaio Vessoni; Simone Kashima; Svetoslav Nanev Slavov; Vincent Louis Viala |
| EPI_ISL_2493410, EPI_ISL_2493413, EPI_ISL_2493415, EPI_ISL_2493417, EPI_ISL_2493420 | CS DE PALMARES PAULISTA | Instituto Butantan | Antonio Jorge Martins; Claudia Renata dos Santos Barros; David Schlesinger; Debora Botequio Moretti; Dimas Tadeu Covas; Elaine Cristina Marqueze; Elaine Vieira Santos; Evandra Strazza Rodrigues; Heidge Fukumasu; Jayme Augusto de Souza-Neto; José Salvatore Leister Patané; Luiz Alcantara; Luiz Lehmann Coutinho; Maria Carolina Elias; Maurício Lacerda Nogueira; Rafael dos Santos Bezerra; Raul Machado Neto; Rejane Maria Tommasini Grotto; Ricardo Haddad; Sandra Coccuzzo Sampaio Vessoni; Simone Kashima; Svetoslav Nanev Slavov; Vincent Louis Viala |
| EPI_ISL_2445211 | CS DE PLANALTO | Instituto Butantan | Antonio Jorge Martins; Claudia Renata dos Santos Barros; David Schlesinger; Debora Botequio Moretti; Dimas Tadeu Covas; Elaine Cristina Marqueze; Elaine Vieira Santos; Evandra Strazza Rodrigues; Heidge Fukumasu; Jayme Augusto de Souza-Neto; José Salvatore Leister Patané; Luiz Alcantara; Luiz Lehmann Coutinho; Maria Carolina Elias; Maurício Lacerda Nogueira; Rafael dos Santos Bezerra; Raul Machado Neto; Rejane Maria Tommasini Grotto; Ricardo Haddad; Sandra Coccuzzo Sampaio Vessoni; Simone Kashima; Svetoslav Nanev Slavov; Vincent Louis Viala |
| EPI_ISL_2493587 | CS DE QUINTANA | Instituto Butantan | Antonio Jorge Martins; Claudia Renata dos Santos Barros; David Schlesinger; Debora Botequio Moretti; Dimas Tadeu Covas; Elaine Cristina Marqueze; Elaine Vieira Santos; Evandra Strazza Rodrigues; Heidge Fukumasu; Jayme Augusto de Souza-Neto; José Salvatore Leister Patané; Luiz Alcantara; Luiz Lehmann Coutinho; Maria Carolina Elias; Maurício Lacerda Nogueira; Rafael dos Santos Bezerra; Raul Machado Neto; Rejane Maria Tommasini Grotto; Ricardo Haddad; Sandra Coccuzzo Sampaio Vessoni; Simone Kashima; Svetoslav Nanev Slavov; Vincent Louis Viala |
| EPI_ISL_2493362 | CS DE ROSANA | Instituto Butantan | Antonio Jorge Martins; Claudia Renata dos Santos Barros; David Schlesinger; Debora Botequio Moretti; Dimas Tadeu Covas; Elaine Cristina Marqueze; Elaine Vieira Santos; Evandra Strazza Rodrigues; Heidge Fukumasu; Jayme Augusto de Souza-Neto; José Salvatore Leister Patané; Luiz Alcantara; Luiz Lehmann Coutinho; Maria Carolina Elias; Maurício Lacerda Nogueira; Rafael dos Santos Bezerra; Raul Machado Neto; Rejane Maria Tommasini Grotto; Ricardo Haddad; Sandra Coccuzzo Sampaio Vessoni; Simone Kashima; Svetoslav Nanev Slavov; Vincent Louis Viala |
| EPI_ISL_2493400 | CS DE SANTA ALBERTINA | Instituto Butantan | Antonio Jorge Martins; Claudia Renata dos Santos Barros; David Schlesinger; Debora Botequio Moretti; Dimas Tadeu Covas; Elaine Cristina Marqueze; Elaine Vieira Santos; Evandra Strazza Rodrigues; Heidge Fukumasu; Jayme Augusto de Souza-Neto; José Salvatore Leister Patané; Luiz Alcantara; Luiz Lehmann Coutinho; Maria Carolina Elias; Maurício Lacerda Nogueira; Rafael dos Santos Bezerra; Raul Machado Neto; Rejane Maria Tommasini Grotto; Ricardo Haddad; Sandra Coccuzzo Sampaio Vessoni; Simone Kashima; Svetoslav Nanev Slavov; Vincent Louis Viala |
| EPI_ISL_2493421, EPI_ISL_2493422 | CS DE SANTA RITA DOESTE | Instituto Butantan | Antonio Jorge Martins; Claudia Renata dos Santos Barros; David Schlesinger; Debora Botequio Moretti; Dimas Tadeu Covas; Elaine Cristina Marqueze; Elaine Vieira Santos; Evandra Strazza Rodrigues; Heidge Fukumasu; Jayme Augusto de Souza-Neto; José Salvatore Leister Patané; Luiz Alcantara; Luiz Lehmann Coutinho; Maria Carolina Elias; Maurício Lacerda Nogueira; Rafael dos Santos Bezerra; Raul Machado Neto; Rejane Maria Tommasini Grotto; Ricardo Haddad; Sandra Coccuzzo Sampaio Vessoni; Simone Kashima; Svetoslav Nanev Slavov; Vincent Louis Viala |
| EPI_ISL_5645975, EPI_ISL_5645994, EPI_ISL_5646000, EPI_ISL_5646012, EPI_ISL_5646015, EPI_ISL_5646023 | CS DE URUPES | Instituto Butantan | Antonio Jorge Martins; Claudia Renata dos Santos Barros; David Schlesinger; Debora Botequio Moretti; Dimas Tadeu Covas; Elaine Cristina Marqueze; Elaine Vieira Santos; Evandra Strazza Rodrigues; Heidge Fukumasu; Jayme Augusto de Souza-Neto; José Salvatore Leister Patané; Luiz Alcantara; Luiz Lehmann Coutinho; Maria Carolina Elias; Maurício Lacerda Nogueira; Rafael dos Santos Bezerra; Raul Machado Neto; Rejane Maria Tommasini Grotto; Ricardo Haddad; Sandra Coccuzzo Sampaio Vessoni; Simone Kashima; Svetoslav Nanev Slavov; Vincent Louis Viala |
| EPI_ISL_2493430, EPI_ISL_2493431 | CS DE VALENTIM GENTIL | Instituto Butantan | Antonio Jorge Martins; Claudia Renata dos Santos Barros; David Schlesinger; Debora Botequio Moretti; Dimas Tadeu Covas; Elaine Cristina Marqueze; Elaine Vieira Santos; Evandra Strazza Rodrigues; Heidge Fukumasu; Jayme Augusto de Souza-Neto; José Salvatore Leister Patané; Luiz Alcantara; Luiz Lehmann Coutinho; Maria Carolina Elias; Maurício Lacerda Nogueira; Rafael dos Santos Bezerra; Raul Machado Neto; Rejane Maria Tommasini Grotto; Ricardo Haddad; Sandra Coccuzzo Sampaio Vessoni; Simone Kashima; Svetoslav Nanev Slavov; Vincent Louis Viala |
| EPI_ISL_2493299 | CS II DE IGARAPAVA | Instituto Butantan | Antonio Jorge Martins; Claudia Renata dos Santos Barros; David Schlesinger; Debora Botequio Moretti; Dimas Tadeu Covas; Elaine Cristina Marqueze; Elaine Vieira Santos; Evandra Strazza Rodrigues; Heidge Fukumasu; Jayme Augusto de Souza-Neto; José Salvatore Leister Patané; Luiz Alcantara; Luiz Lehmann Coutinho; Maria Carolina Elias; Maurício Lacerda Nogueira; Rafael dos Santos Bezerra; Raul Machado Neto; Rejane Maria Tommasini Grotto; Ricardo Haddad; Sandra Coccuzzo Sampaio Vessoni; Simone Kashima; Svetoslav Nanev Slavov; Vincent Louis Viala |
| EPI_ISL_2493543 | CS II DR SEBASTIAO RIBEIRO DO VALLE | Instituto Butantan | Antonio Jorge Martins; Claudia Renata dos Santos Barros; David Schlesinger; Debora Botequio Moretti; Dimas Tadeu Covas; Elaine Cristina Marqueze; Elaine Vieira Santos; Evandra Strazza Rodrigues; Heidge Fukumasu; Jayme Augusto de Souza-Neto; José Salvatore Leister Patané; Luiz Alcantara; Luiz Lehmann Coutinho; Maria Carolina Elias; Maurício Lacerda Nogueira; Rafael dos Santos Bezerra; Raul Machado Neto; Rejane Maria Tommasini Grotto; Ricardo Haddad; Sandra Coccuzzo Sampaio Vessoni; Simone Kashima; Svetoslav Nanev Slavov; Vincent Louis Viala |
| EPI_ISL_2493573 | CS III DR ANTONIO ANADAO SSGRAMA | Instituto Butantan | Antonio Jorge Martins; Claudia Renata dos Santos Barros; David Schlesinger; Debora Botequio Moretti; Dimas Tadeu Covas; Elaine Cristina Marqueze; Elaine Vieira Santos; Evandra Strazza Rodrigues; Heidge Fukumasu; Jayme Augusto de Souza-Neto; José Salvatore Leister Patané; Luiz Alcantara; Luiz Lehmann Coutinho; Maria Carolina Elias; Maurício Lacerda Nogueira; Rafael dos Santos Bezerra; Raul Machado Neto; Rejane Maria Tommasini Grotto; Ricardo Haddad; Sandra Coccuzzo Sampaio Vessoni; Simone Kashima; Svetoslav Nanev Slavov; Vincent Louis Viala |
| EPI_ISL_2493288 | CS LUIZ FIOD DE BURITIZAL | Instituto Butantan | Antonio Jorge Martins; Claudia Renata dos Santos Barros; David Schlesinger; Debora Botequio Moretti; Dimas Tadeu Covas; Elaine Cristina Marqueze; Elaine Vieira Santos; Evandra Strazza Rodrigues; Heidge Fukumasu; Jayme Augusto de Souza-Neto; José Salvatore Leister Patané; Luiz Alcantara; Luiz Lehmann Coutinho; Maria Carolina Elias; Maurício Lacerda Nogueira; Rafael dos Santos Bezerra; Raul Machado Neto; Rejane Maria Tommasini Grotto; Ricardo Haddad; Sandra Coccuzzo Sampaio Vessoni; Simone Kashima; Svetoslav Nanev Slavov; Vincent Louis Viala |
| EPI_ISL_2493301 | CSII DR JOSE FERREIRA TELLES | Instituto Butantan | Antonio Jorge Martins; Claudia Renata dos Santos Barros; David Schlesinger; Debora Botequio Moretti; Dimas Tadeu Covas; Elaine Cristina Marqueze; Elaine Vieira Santos; Evandra Strazza Rodrigues; Heidge Fukumasu; Jayme Augusto de Souza-Neto; José Salvatore Leister Patané; Luiz Alcantara; Luiz Lehmann Coutinho; Maria Carolina Elias; Maurício Lacerda Nogueira; Rafael dos Santos Bezerra; Raul Machado Neto; Rejane Maria Tommasini Grotto; Ricardo Haddad; Sandra Coccuzzo Sampaio Vessoni; Simone Kashima; Svetoslav Nanev Slavov; Vincent Louis Viala |
| EPI_ISL_2378743, EPI_ISL_2493622 | CSII DR WASHINGTON LUIS M RODRIGUES DA SILVA PITANGUEIRAS | Instituto Butantan | Antonio Jorge Martins; Claudia Renata dos Santos Barros; David Schlesinger; Debora Botequio Moretti; Dimas Tadeu Covas; Elaine Cristina Marqueze; Elaine Vieira Santos; Evandra Strazza Rodrigues; Heidge Fukumasu; Jayme Augusto de Souza-Neto; José Salvatore Leister Patané; Luiz Alcantara; Luiz Lehmann Coutinho; Maria Carolina Elias; Maurício Lacerda Nogueira; Rafael dos Santos Bezerra; Raul Machado Neto; Rejane Maria Tommasini Grotto; Ricardo Haddad; Sandra Coccuzzo Sampaio Vessoni; Simone Kashima; Svetoslav Nanev Slavov; Vincent Louis Viala |
| EPI_ISL_2501534, EPI_ISL_2501535, EPI_ISL_2501538 | CT Department of Public Health | CT Department of Public Health | Claire_Pearson; Tu_N_Nguyen |
| EPI_ISL_2787514 | CT-Dr. Katherine A. Kelley State Public Health Lab | Centers for Disease Control and Prevention Division of Viral Diseases, Pathogen Discovery | Alison Laufer Halpin; Ben L. Rambo-Martin; Clinton R. Paden; Dakota Howard; Darlene Wagner; Dave Wentworth; Dhwani Batra; Jasmine Padilla; Justin Lee; Katie Dillon; Krista Queen; Kristen Knipe; Kristine Lacek; Mark Burroughs; Matthew Schmerer; Mili Sheth; Peter Cook; Sam Shepard; Sarah Nobles; Shoshona Le; Suxiang Tong; Vivien Dugan; Yvette Unoarumhi |
| EPI_ISL_2892480, EPI_ISL_3536689, EPI_ISL_3536728, EPI_ISL_3536734, EPI_ISL_3536897, EPI_ISL_3536931, EPI_ISL_3536932, EPI_ISL_3536933, EPI_ISL_4005549, EPI_ISL_4005551 | | | |
| see above | California Department of Public Health | California Department of Public Health | CDPH IDLB COVIDNet et al |

| | | | |
|---|---|---|---|
| EPI_ISL_3134058, EPI_ISL_3134071, EPI_ISL_3134075, EPI_ISL_3134076, EPI_ISL_3134079, EPI_ISL_3134087 | California Department of Public Health Valencia Branch Laboratory | California Department of Public Health | CDPH-COVIDNet; UCSF Center for Advanced Technology |
| EPI_ISL_7507379, EPI_ISL_7507385 | Cedars-Sinai Medical Center, Molecular Pathology Laboratory of Department of Pathology & Laboratory Medicine and Genomic Core | Cedars-Sinai Medical Center, Molecular Pathology Laboratory of Department of Pathology & Laboratory Medicine and Genomic Core | Brian Davis; Eric Vail; Jasmine T Plummer; Jorge Mario Sincuir Martinez; Stephanie Chen; Wenjuan Zhang |
| EPI_ISL_2472901 | Cell-T | 1. Tricity SARS-CoV-2 sequencing consortium: University of Gdansk, Medical University of Gdansk, Vaxican Ltd., Invicta Ltd. 2. National Institute of Public Health - National Institute of Hygiene, Warsaw, Poland | Celina Cybulska; Karolina Gackowska; Katarzyna Groth; Katarzyna Zacharczuk; Krystyna Bienkowska Szewczyk; Lukasz Rabalski; Maciej Grzybek; Maciej Kosinski; Magdalena Nowakowska; Marcin Lubocki; Małgorzata Sadkowska-Todys; Tomasz Wołkowicz |
| EPI_ISL_3144947 | Center for Virology | Center for Virology | Jeremy V. Camp; Monika Redlberger-Fritz; Stephan W. Aberle |
| EPI_ISL_2502381, EPI_ISL_2502416, EPI_ISL_2502417, EPI_ISL_2502425, EPI_ISL_2502427, EPI_ISL_2502437, EPI_ISL_2502502, EPI_ISL_2502503, EPI_ISL_2502504, EPI_ISL_2502505, EPI_ISL_2502506, EPI_ISL_3462567, EPI_ISL_3462589, EPI_ISL_3462623, EPI_ISL_3462624, EPI_ISL_3462625, EPI_ISL_3462626 | | | |
| see above | Central Laboratory, Bureau of Public Health (BOG) and Academic Hospital Paramaribo | Erasmus Medical Center | Bas B Oude Munnink; Cherise Beek; Consuella Partowidjojo; Dion Gajadin; Ed PF IJzerman; Emmanuelle Munger; Gary Gummels; Ingrid SK Krishnadath; Lycke Woittiez; Marion PG Koopmans; Mireille Van de Veer; Phyllis Pinas; Princes Wongsowidjojo; Radjesh Ori; Ranisha Doerbalie; Rohma Banwari; Soeradj Harkisoen; Stephen Vreden; Tilotmadebie Ramlal; Verne Nanhoe |
| EPI_ISL_3255060, EPI_ISL_3255085, EPI_ISL_3255086, EPI_ISL_3255087, EPI_ISL_3255088, EPI_ISL_3255090, EPI_ISL_3255091, EPI_ISL_3255092, EPI_ISL_3255093, EPI_ISL_3255094, EPI_ISL_3255095, EPI_ISL_3255096, EPI_ISL_3255097, EPI_ISL_3255099, EPI_ISL_3255100, EPI_ISL_3255102, EPI_ISL_3255103, EPI_ISL_3255104, EPI_ISL_3255105, EPI_ISL_3255106, EPI_ISL_3255107, EPI_ISL_3255108, EPI_ISL_3255109, EPI_ISL_3255110, EPI_ISL_3255111, EPI_ISL_3255112, EPI_ISL_3255113, EPI_ISL_3255114, EPI_ISL_3255115, EPI_ISL_3255116, EPI_ISL_3255118, EPI_ISL_3255119, EPI_ISL_3255120, EPI_ISL_3255121, EPI_ISL_3255124, EPI_ISL_3255125, EPI_ISL_3255126, EPI_ISL_3255127, EPI_ISL_3255128, EPI_ISL_3255129, EPI_ISL_3255130, EPI_ISL_3255131, EPI_ISL_3255153, EPI_ISL_3255161, EPI_ISL_3255173, EPI_ISL_3255177, EPI_ISL_3255178, EPI_ISL_3255191, EPI_ISL_3255201, EPI_ISL_3255219, EPI_ISL_3255233 | | | |
| see above | Central Public Health Laboratory - LACEN - Bahia, Salvador, Brazil | Central Public Health Laboratory - LACEN -Bahia, Salvador, Brazil | Arabela Leal; Breno Dominguez; Felicidade Pereira; Jaqueline Gomes; Luciana Oliveira; Luiz Alcantara; Marcela Gómez; Marta Giovanetti; Patrícia Cajado; Stephane Tosta; Vagner Fonseca; Vanessa Nardy |
| EPI_ISL_5062577 | Centre Hospitalier Universitaire de Rouen Laboratoire de Virologie | Centre Hospitalier Universitaire de Rouen Laboratoire de Virologie | Alice Moisan; Fabienne De Oliveira; Marie Leoz |
| EPI_ISL_2842780, EPI_ISL_2842781 | Centro Medico ABC | Instituto Nacional de Medicina Genomica | Cedro-Tanda A; Cesar Lara; Daniel Aguirre; Escobar-Arrazola; Gonzalez-Barrera D; Herrera-Montalvo LA.; Hidalgo-Miranda A; Jacomine Reyes-Carrasco; M; Mendoza-Vargas A; Munguia-Garza P; Ramirez-Vega O; Rangel-DeLeon D; Reyes-Grajeda JP; Roxana Trejo-González |
| EPI_ISL_5926815, EPI_ISL_5926818, EPI_ISL_5926819, EPI_ISL_5926821, EPI_ISL_5926823, EPI_ISL_5926874, EPI_ISL_5926875, EPI_ISL_5926876, EPI_ISL_5926877, EPI_ISL_5926878, EPI_ISL_5926880, EPI_ISL_5926882, EPI_ISL_5926895, EPI_ISL_5926896 | | | |
| see above | Centro de Infectologia Charles Mérieux/ Laboratório Rodolphe Mérieux, FUNDHACRE | Bioinformatics Laboratory / LNCC | Alessandra P Lamarca; Alexandra L Gerber; Aline de Freitas Souza; Ana Paula de C Guimarães; Ana Tereza R Vasconcelos; Andreas Stocker; Cirley Maria de Oliveira Lobato; Douglas Terra Machado; Janaína Mazaro; Janete Taynã Nascimento Rodrigues; Luiz Fellype Alves de Souza; Luiz G P de Almeida; Ronaldo da Silva F Jr; Rutilene B. Souza |
| EPI_ISL_2671553, EPI_ISL_2671570 | Centro de Investigacion Biomedica de Occidente (CIBO) | Unidad de Genomica Avanzada | ; Alejandra Garcia-Gasca; Alejandra Hernandez-Teran; Alejandro Sanchez-Flores; Alfredo Herrera-Estrella; Alicia Ocaña-Mondragon; Andreu Comas-Garcia; Angel Gustavo Salas-Lais; Antonio Loza Roman; Bernardo Martinez-Miguel; Blanca Taboada; Brenda Irasema Maldonado-Meza; Bruno Gomez-Gil; Carla Ivon Herrera-Najera; Carlos F. Arias; Celia Boukadida; Celida Duque Molina; Celida Martinez- Rodriguez; Clara Esperanza Santacruz-Tinoco; Concepcion Grajales-Muñiz; Consorcio Mexicano de Vigilancia Genomica (CoViGen-Mex). Authors (in alphabetical order): Julio Elias Alvarado-Yaah; Cristobal Chaidez-Quiroz; Daniel Fregoso-Rueda; Daniel Lira Morales; Eduardo Becerril-Vargas; Fernando Fontove-Herrera; Fidencio Mejia-Nepomuceno; Francisco Pulido; Gloria Elena Espinosa-Ayala; Gloria Maria Molina-Salinas; Gloria Vazquez; Hector Esteban Paz-Juarez; Hector Montoya-Fuentes; Helen Haydee Fernanda Ramirez-Plascencia; Irvin Gonzalez-Lopez; Jean Pierre Gonzalez; Jesus Hernandez; Joel Armando Vazquez-Perez.; Jorge Salas-Hernandez; Jose Antonio Enciso-Moreno; Jose Arturo Martinez-Orozco; Jose Esteban Muñoz-Medina; Jose de Jesus Nuñez-Contreras; Juan Bautista Chale-Dzul; Julissa Enciso-Ibarra; Luis Alberto Ochoa-Carrera; Margarita Matias-Florentino; Maria Guadalupe Santiago-Mauricio; Maria Guadalupe de Jesus Mireles-Rivera; Mario Mujica-Sanchez; Marissa Perez-Garcia; Nelly Selem-Mojica; Pavel Isa; Ricardo Ciria Merce; Ricardo Grande; Rosa Maria Gutierrez Rios; Santiago avila-Rios; Selene Zarate; Susana Lopez; Veronica Mata-Haro; Victor Eduardo Garcia-Arias; Victor Hugo Borja-Aburto |
| EPI_ISL_2671543, EPI_ISL_2671544, EPI_ISL_2671619, EPI_ISL_2671624, EPI_ISL_2671716, EPI_ISL_2671719, EPI_ISL_2970005, EPI_ISL_2970007, EPI_ISL_2970050 | | | |
| see above | Centro de Investigacion Biomedica del Noreste (CIBIN) | Unidad de Genomica Avanzada | ; Alejandra Garcia-Gasca; Alejandra Hernandez-Teran; Alejandro Sanchez-Flores; Alfredo Herrera-Estrella; Alicia Ocaña-Mondragon; Andreu Comas-Garcia; Angel Gustavo Salas-Lais; Antonio Loza Roman; Bernardo Martinez-Miguel; Blanca Taboada; Brenda Irasema Maldonado-Meza; Bruno Gomez-Gil; Carla Ivon Herrera-Najera; Carlos F. Arias; Celia Boukadida; Celida Duque Molina; Celida Martinez- Rodriguez; Clara Esperanza Santacruz-Tinoco; Concepcion Grajales-Muñiz; Consorcio Mexicano de Vigilancia Genomica (CoViGen-Mex). Authors (in alphabetical order): Julio Elias Alvarado-Yaah; Cristobal Chaidez-Quiroz; Daniel Fregoso-Rueda; Daniel Lira Morales; Eduardo Becerril-Vargas; Fernando Fontove-Herrera; Fidencio Mejia-Nepomuceno; Francisco Pulido; Gloria Elena Espinosa-Ayala; Gloria Maria Molina-Salinas; Gloria Vazquez; Hector Esteban Paz-Juarez; Hector Montoya-Fuentes; Helen Haydee Fernanda Ramirez-Plascencia; Irvin Gonzalez-Lopez; Jean Pierre Gonzalez; Jesus Hernandez; Joel Armando Vazquez-Perez.; Jorge Salas-Hernandez; Jose Antonio Enciso-Moreno; Jose Arturo Martinez-Orozco; Jose Esteban Muñoz-Medina; Jose de Jesus Nuñez-Contreras; Juan Bautista Chale-Dzul; Julissa Enciso-Ibarra; Luis Alberto Ochoa-Carrera; Margarita Matias-Florentino; Maria Guadalupe Santiago-Mauricio; Maria Guadalupe de Jesus Mireles-Rivera; Mario Mujica-Sanchez; Marissa Perez-Garcia; Nelly Selem-Mojica; Pavel Isa; Ricardo Ciria Merce; Ricardo Grande; Rosa Maria Gutierrez Rios; Santiago avila-Rios; Selene Zarate; Susana Lopez; Veronica Mata-Haro; Victor Eduardo Garcia-Arias; Victor Hugo Borja-Aburto |
| EPI_ISL_4261896, EPI_ISL_4262029 | Chiu Laboratory, University of California, San Francisco | Chiu Laboratory, University of California, San Francisco | Alicia Sotomayor-Gonzalez; Baolin Wang; Candace Wang; Charles Chiu; Daniel Wong; Dustin Glasner; Venice Servellita; Xianding (Wayne) Deng; Yueyuan Zhang |
| EPI_ISL_2841838 | City of Milwaukee Health Department Laboratory | City of Milwaukee Health Department Laboratory | Amy Bauer; Jennifer Lentz; Manjeet Khubbar; Samantha Scott; Sanjib Bhattacharyya |
| EPI_ISL_2308246, EPI_ISL_2308247, EPI_ISL_2308248 | Clinical Emergency County Hospital Constanta | National Institute of Infectious Diseases-Prof. Dr. Matei Bals Molecular Diagnostics Laboratory | Andreea Tudor; Corina Casangiu; Dan Otelea; Leontina Banica; Marius Surleac; Ovidiu Vlaicu; Simona Paraschiv |
| EPI_ISL_4635665, EPI_ISL_4635666, EPI_ISL_4635667 | Clinical Microbiology, University of Texas Medical Branch (UTMB), Galveston | ClickSeq Lab, University of Texas Medical Branch, UTMB, Galveston | Dong J.; Jaworski E.; Judy B.; Ksiazek T.; Langsjoen R.M.; Machado R.R; McConnell A.J.; Menachery V.D.; Miller A.L.; Mitchell B.M.; Morris V.G.; Newman P.; Plante J.A.; Plante K.S.; Pyles R.B.; Ren P.; Routh A.; Saada N.I.; Sotcheff S.L.; Swetnam D.; Thompson J.K.; Weaver S.C; Widen S.; Zhou Y. |
| EPI_ISL_2380789, EPI_ISL_2380806 | Clinical Molecular Microbiology Laboratory, UNC Hospitals | Jeremy Wang | Alexander Rubinsteyn; Colleen Rice; Corbin Jones; Jason Smedberg; Jeremy Wang; Melissa Miller; Robert Hagan; Shawn Hawken |
| EPI_ISL_2408427, EPI_ISL_2408442, EPI_ISL_2466571 | Clinical Virology | Clinical Bacteriology | Adrian Egli; Alfredo Mari; Fanny Wegner; Hans Hirsch; Helena MB Seth-Smith; Julia Bielicki; Karoline Leuzinger; Manuel Battegay; Tim Roloff |
| EPI_ISL_2427393, EPI_ISL_2427395, EPI_ISL_2427401 | Clinique Saint-Pierre Ottignies | UCLouvain/IREC/MBLG | Benoit Kabamba Mukadi; Bertrand Bearzatto; Jean Ruelle; Ophélie Simon |
| EPI_ISL_2348842, EPI_ISL_2348849, EPI_ISL_2348863, EPI_ISL_2348868, EPI_ISL_2348871, EPI_ISL_2348874, EPI_ISL_2348878, EPI_ISL_2348884, EPI_ISL_2348889, EPI_ISL_2348897, EPI_ISL_2348900, EPI_ISL_2348915, EPI_ISL_2427333, EPI_ISL_2427339, EPI_ISL_2778039, EPI_ISL_2778084 | | | |
| see above | Cliniques universitaires Saint-Luc | UCLouvain/IREC/MBLG | Benoit Kabamba Mukadi; Bertrand Bearzatto; Jean Ruelle; Nicolas Pinte; Ophélie Simon |
| EPI_ISL_5260659, EPI_ISL_5323524, EPI_ISL_5323526 | Color Genomics | Chiu Laboratory, University of California, San Francisco | Alicia Sotomayor-Gonzalez; Alicia Zhou; Baolin Wang; Charles Chiu; Daniel Wong; Erika Torres; Noah Brazer; Scott Topper; Venice Servellita |
| EPI_ISL_2550016, EPI_ISL_2550017, EPI_ISL_2550023, EPI_ISL_2550261, EPI_ISL_2550275, EPI_ISL_2550311, EPI_ISL_2550457, EPI_ISL_2987416, EPI_ISL_2987417, EPI_ISL_2987418, EPI_ISL_2987419, EPI_ISL_3159538, EPI_ISL_3159664, EPI_ISL_3160137, EPI_ISL_3404355, EPI_ISL_6211477 | | | |
| see above | Colorado Department of Public Health and Environment | Colorado Department of Public Health and Environment | Alexandria Rossheim; Diana Ir; Emily A. Travanty; Laura Bankers; Mandy Waters; Michael Martin; Molly C. Hetherington-Rauth; Sarah Elizabeth Totten; Shannon R. Matzinger |
| EPI_ISL_3368476 | Compensar Calle 63 | Centro de Investigaciones en Microbiología y Biotecnología-UR (CIMBIUR), Facultad de Ciencias Naturales, Universidad del Rosario, Bogotá, Colombia | Angie Ramírez; Juan David Ramírez; Luz H. Patiño; Marcela Mercado-Reyes; Marina Muñoz; Nathalia Ballesteros; Nicolas Niño; Sergio Castañeda |
| EPI_ISL_2562711 | Creighton University Medical Center | Creighton COVID Consortium | Cynthia Watson; Holly Stessman; Jake Siedlik; Michael Belshan; Richard Goering |
| EPI_ISL_2755328, EPI_ISL_2755332, EPI_ISL_2755363, EPI_ISL_2755366, EPI_ISL_2755396, EPI_ISL_2755702, EPI_ISL_2755704, EPI_ISL_2755945, EPI_ISL_2755965, EPI_ISL_2755982, EPI_ISL_2755983, EPI_ISL_2755986, EPI_ISL_2783542, EPI_ISL_2783553, EPI_ISL_2783565, EPI_ISL_2783577, EPI_ISL_2783583, EPI_ISL_2783587, EPI_ISL_2783602, EPI_ISL_2783603, EPI_ISL_2783605, EPI_ISL_2783611, EPI_ISL_2783614, EPI_ISL_2783619, EPI_ISL_2783623, EPI_ISL_2783628, EPI_ISL_2783630, EPI_ISL_2783631, EPI_ISL_2783637, EPI_ISL_2783639, EPI_ISL_2783641, EPI_ISL_2783642, EPI_ISL_2783646, EPI_ISL_2783652, EPI_ISL_2783697, EPI_ISL_2783704, EPI_ISL_2783705, EPI_ISL_2783707, EPI_ISL_2783711, EPI_ISL_2783713, EPI_ISL_2783715, EPI_ISL_2783726, EPI_ISL_2783727, EPI_ISL_2783753, EPI_ISL_2783754, EPI_ISL_2783763, EPI_ISL_2783776, EPI_ISL_2783839, EPI_ISL_2783841, EPI_ISL_2783842, EPI_ISL_2783851, EPI_ISL_2783852, EPI_ISL_2783853, EPI_ISL_2783863, EPI_ISL_2783880, EPI_ISL_2783917, EPI_ISL_2783922, EPI_ISL_2783923, EPI_ISL_2783924, EPI_ISL_2783928, EPI_ISL_2783929, EPI_ISL_2783933, EPI_ISL_2783936, EPI_ISL_2783946, EPI_ISL_2783947, EPI_ISL_2783948, EPI_ISL_2783961, EPI_ISL_2783963, EPI_ISL_2783967, EPI_ISL_2783968, EPI_ISL_2783972, EPI_ISL_2784012, EPI_ISL_2784013, EPI_ISL_2784015, EPI_ISL_2784019, EPI_ISL_2784021, EPI_ISL_2784028, EPI_ISL_2784033, EPI_ISL_2784034, EPI_ISL_2784038, EPI_ISL_2784040, EPI_ISL_2784071, EPI_ISL_2784074, EPI_ISL_2784080, EPI_ISL_2839905, EPI_ISL_2941943, EPI_ISL_2942115, EPI_ISL_2942122, EPI_ISL_2942124, EPI_ISL_2942126, EPI_ISL_3064391, EPI_ISL_3270167, EPI_ISL_3270193, EPI_ISL_3270317, EPI_ISL_3270350, EPI_ISL_3270362, EPI_ISL_3270389 | | | |
| see above | Curative Labs | Curative Labs | Elias L. Salfati; Eugenia Khorosheva; George Way; J.Cesar Ignacio-Espinoza; Janet Chen; Mikhail Hanewich-Hollatz; Nabjot Sandhu; Sophia Quasem; Vladimir Slepnev; Zhiyi Xie |
| EPI_ISL_4422790, EPI_ISL_4422792, EPI_ISL_4422995, EPI_ISL_4422997, EPI_ISL_4423002, EPI_ISL_4427878, EPI_ISL_4427880, EPI_ISL_4427882 | | | |
| see above | DASA | DASA | Adriano Bonaldi; Angelica Hristov; Annelise Lopes; Bianca Cota; Cristina Oliveira; Jose Levi; Lidia Yamamoto; Paulo Pierry; Rodrigo Guarischi; Rodrigo Salazar |
| EPI_ISL_3899242 | DC Public Health Lab/ Dept. of Forensic Sciences | DC Public Health Lab/ Dept. of Forensic Sciences | Brittany Hamilton; Connie Maza; Elizabeth Zelaya; Janis Doss; Jocelyn Hauser; Monica Mann; Nathan Bruns; Sarah Scott; Scott Nguyen |
| EPI_ISL_2840444 | DE Public Health Laboratory | Centers for Disease Control and Prevention Division of Viral | Alison Laufer Halpin; Ben L. Rambo-Martin; Clinton R. Paden; Dakota Howard; Darlene Wagner; Dave Wentworth; Dhwani Batra; Jasmine Padilla; Justin Lee; Katie Dillon; Krista Queen; Kristen Knipe; Kristine Lacek; Mark Burroughs; Matthew Schmerer; Mili Sheth; Peter Cook; Sam Shepard; Sarah Nobles; Shoshona Le; Suxiang Tong; Vivien Dugan; Yvette Unoarumhi |

| | | | |
|---|---|---|---|
| | | Diseases, Pathogen Discovery | |
| EPI_ISL_2493631, EPI_ISL_2493632, EPI_ISL_2493634, EPI_ISL_2493636, EPI_ISL_2493638, EPI_ISL_2493639, EPI_ISL_2493641, EPI_ISL_2493642 | | | |
| see above | DEPARTAMENTO DE SAUDE MUNICIPAL SOCORRO SP | Instituto Butantan | Antonio Jorge Martins; Claudia Renata dos Santos Barros; David Schlesinger; Debora Botequio Moretti; Dimas Tadeu Covas; Elaine Cristina Marqueze; Elaine Vieira Santos; Evandra Strazza Rodrigues; Heidge Fukumasu; Jayme Augusto de Souza-Neto; José Salvatore Leister Patané; Luiz Lehmann Coutinho; Maria Carolina Elias; Maurício Lacerda Nogueira; Rafael dos Santos Bezerra; Raul Machado Neto; Rejane Maria Tommasini Grotto; Ricardo Haddad; Sandra Coccuzzo Sampaio Vessoni; Simone Kashima; Svetoslav Nanev Slavov; Vincent Louis Viala |
| EPI_ISL_2445177, EPI_ISL_2445178, EPI_ISL_2445180, EPI_ISL_2445182, EPI_ISL_2445183, EPI_ISL_2445184, EPI_ISL_2445186 | | | |
| see above | DIRETORIA MUNICIPAL DE SAUDE | Instituto Butantan | Antonio Jorge Martins; Claudia Renata dos Santos Barros; David Schlesinger; Debora Botequio Moretti; Dimas Tadeu Covas; Elaine Cristina Marqueze; Elaine Vieira Santos; Evandra Strazza Rodrigues; Heidge Fukumasu; Jayme Augusto de Souza-Neto; José Salvatore Leister Patané; Luiz Alcantara; Luiz Lehmann Coutinho; Maria Carolina Elias; Maurício Lacerda Nogueira; Rafael dos Santos Bezerra; Raul Machado Neto; Rejane Maria Tommasini Grotto; Ricardo Haddad; Sandra Coccuzzo Sampaio Vessoni; Simone Kashima; Svetoslav Nanev Slavov; Vincent Louis Viala |
| EPI_ISL_2466682, EPI_ISL_2466683 | DOHMH Corona | New York City Public Health Laboratory | Jade Wang; et al. |
| EPI_ISL_2383834, EPI_ISL_2383835, EPI_ISL_2383849, EPI_ISL_2383852, EPI_ISL_2383860, EPI_ISL_2383864, EPI_ISL_2662782, EPI_ISL_3692674, EPI_ISL_3692687, EPI_ISL_4556697, EPI_ISL_4556702 | | | |
| see above | DPHL | Delaware Public Health Lab | Rebecca Savage |
| EPI_ISL_4405178, EPI_ISL_4405181 | Departamento de Biología y genética molecular, IACA Laboratorios. | Área de Secuenciación del Laboratorio de Virología del Hospital de Niños Dr. Ricardo Gutierrez on behalf of 'Proyecto Argentino Interinstitucional de genomica de SARS-CoV-2' (PAIS Consortium) | ; A; Acuña; D; Goya; LE; Lusso; MI; MS; Masciovecchio MV; Nabaes Jodar; Natale; S; Streitenberger ER; Suárez; Valinotto; Viegas, M. |
| EPI_ISL_2429555, EPI_ISL_2430305, EPI_ISL_2430615, EPI_ISL_2431115, EPI_ISL_2431254, EPI_ISL_2431486, EPI_ISL_2431537 | | | |
| see above | Department of Bacteria, Parasites and Fungi, Statens Serum Institut, Copenhagen, Denmark | Statens Serum Institut Bioinformatics and Microbial Genomics | Danish Covid-19 Genome Consortium |
| EPI_ISL_2245263, EPI_ISL_2245264, EPI_ISL_2289048, EPI_ISL_2348563, EPI_ISL_2348564 | Department of Clinical Microbiology | GIGA Medical Genomics | Bouchra Boujemla; Cécile Meex; Keith Durkin; Maria Artesi; Marie-Pierre Hayette; Nathalie Renotte; Pierrette Melin; Raphaël Boreux; Sébastien Bontems; Vincent Bours |
| EPI_ISL_2894980 | Department of Virology | Department of Virology | Aamer Ikram; Massab Umair; Muhammad Ammar; Muhammad Salman; Nazish Badar; Syed Adnan Haider; Zaira Rehman |
| EPI_ISL_2246280, EPI_ISL_2246283, EPI_ISL_2420647, EPI_ISL_2420648, EPI_ISL_2544515, EPI_ISL_2544518, EPI_ISL_2544519, EPI_ISL_2544523 | | | |
| see above | Department of Virology, Istituto Zooprofilattico Sperimentale del Lazio e della Toscana (IZSLT) | Department of General Diagnostics; Department of Virology; Istituto Zooprofilattico Sperimentale del Lazio e della Toscana (IZSLT) | Alessia Franco; Antonella Cersini; Antonio Battisti.; Elena L. Diaconu; Fabiola Feltrin; Giuseppe Manna; Patricia Alba; Raffaela Conti; Teresa Scicluna; Virginia Carfora |
| EPI_ISL_2423034 | Diagnostyka. Laboratoria Medyczne. | 1. ViroGenetics - BSL3 Laboratory of Virology, Małopolska Centre of Biotechnology, Jagiellonian University; 2. Diagtron Laboratoria Lukasz Rabalski | Foremny J; Gromowski, T.; Kowalski, M.; Labaj; Maciej Kosinski; Mazur-Panasiuk, N.; Natalia Derewonko; P.P.; Pyrc, K.; Rabalski L.; Rogalska-Kupiec M.; Swadzba J.; Sylwia Januszczak; Szulc, P.; Wydmanski, W.; Zielinska M |
| EPI_ISL_2403852, EPI_ISL_2403858, EPI_ISL_2403859 | Dianalabs SA | Genesupport | Tanguy ARAUD |
| EPI_ISL_3578151, EPI_ISL_3578162, EPI_ISL_3578168, EPI_ISL_3578189, EPI_ISL_3578236, EPI_ISL_3578240, EPI_ISL_3635733, EPI_ISL_3635740 | | | |
| see above | Dr. Risch Ostschweiz AG | Microbiology, Dr. Risch | Dominique Fabien Hilti; Faina Wehrli; Lorenz Risch; Martin Risch; Nadia Wohlwend; Sinem Kas; Thomas Bodmer |
| EPI_ISL_2405658, EPI_ISL_2405834, EPI_ISL_2405886, EPI_ISL_2406008, EPI_ISL_2406028, EPI_ISL_2406085, EPI_ISL_2406108, EPI_ISL_2406122, EPI_ISL_2406198, EPI_ISL_2406199, EPI_ISL_2406225, EPI_ISL_2406236, EPI_ISL_2406247, EPI_ISL_2406269, EPI_ISL_2406320, EPI_ISL_2406344, EPI_ISL_2406366, EPI_ISL_2406374, EPI_ISL_2406413, EPI_ISL_2406434, EPI_ISL_2406445, EPI_ISL_2406451, EPI_ISL_2406457, EPI_ISL_2475676, EPI_ISL_2475681, EPI_ISL_2475682, EPI_ISL_2475683, EPI_ISL_2475684, EPI_ISL_2475685, EPI_ISL_2475686, EPI_ISL_2475688, EPI_ISL_2475689, EPI_ISL_2475690, EPI_ISL_2475696, EPI_ISL_2475697, EPI_ISL_2475698, EPI_ISL_2475699, EPI_ISL_2475700, EPI_ISL_2475705, EPI_ISL_2475706, EPI_ISL_2475708, EPI_ISL_2609957, EPI_ISL_2610746, EPI_ISL_2672800 | | | |
| see above | Dutch COVID-19 response team | National Institute for Public Health and the Environment (RIVM) | Adam Meijer; AnneMarie van den Brandt; Annelies Kroneman; Bas van der Veer; Chantal Reusken; Dennis Schmitz; Dirk Eggink; Euníce Then; Florian Zwagemaker; Harry Vennema; James Groot; Jeroen Cremer; Karim Hajji; Kim Freriks; Linda van de Nes; Lisa Wijsman; Lynn Aarts; Melissa van Tuil; Ryanne Jaarsma; Sanne Bos; Sharon van den Brink; Sjoerd Kuiling; on behalf of the national COVID-19 response team |
| EPI_ISL_2493401, EPI_ISL_2493408, EPI_ISL_5646029, EPI_ISL_5646031 | EMERGENCIA RESPIRATORIA DE NOVA GRANADA | Instituto Butantan | Antonio Jorge Martins; Claudia Renata dos Santos Barros; David Schlesinger; Debora Botequio Moretti; Dimas Tadeu Covas; Elaine Cristina Marqueze; Elaine Vieira Santos; Evandra Strazza Rodrigues; Heidge Fukumasu; Jayme Augusto de Souza-Neto; José Salvatore Leister Patané; Luiz Alcantara; Luiz Lehmann Coutinho; Maria Carolina Elias; Maurício Lacerda Nogueira; Rafael dos Santos Bezerra; Raul Machado Neto; Rejane Maria Tommasini Grotto; Ricardo Haddad; Sandra Coccuzzo Sampaio Vessoni; Simone Kashima; Svetoslav Nanev Slavov; Vincent Louis Viala |
| EPI_ISL_5645998 | ESF NOVA TANABI II | Instituto Butantan | Antonio Jorge Martins; Claudia Renata dos Santos Barros; David Schlesinger; Debora Botequio Moretti; Dimas Tadeu Covas; Elaine Cristina Marqueze; Elaine Vieira Santos; Evandra Strazza Rodrigues; Heidge Fukumasu; Jayme Augusto de Souza-Neto; José Salvatore Leister Patané; Luiz Alcantara; Luiz Lehmann Coutinho; Maria Carolina Elias; Maurício Lacerda Nogueira; Rafael dos Santos Bezerra; Raul Machado Neto; Rejane Maria Tommasini Grotto; Ricardo Haddad; Sandra Coccuzzo Sampaio Vessoni; Simone Kashima; Svetoslav Nanev Slavov; Vincent Louis Viala |
| EPI_ISL_2445157, EPI_ISL_2473706 | ESF PARQUE COLINAS | Instituto Butantan | Antonio Jorge Martins; Claudia Renata dos Santos Barros; David Schlesinger; Debora Botequio Moretti; Dimas Tadeu Covas; Elaine Cristina Marqueze; Elaine Vieira Santos; Evandra Strazza Rodrigues; Heidge Fukumasu; Jayme Augusto de Souza-Neto; José Salvatore Leister Patané; Luiz Alcantara; Luiz Lehmann Coutinho; Maria Carolina Elias; Maurício Lacerda Nogueira; Rafael dos Santos Bezerra; Raul Machado Neto; Rejane Maria Tommasini Grotto; Ricardo Haddad; Sandra Coccuzzo Sampaio Vessoni; Simone Kashima; Svetoslav Nanev Slavov; Vincent Louis Viala |
| EPI_ISL_2445191, EPI_ISL_2445192, EPI_ISL_2473712 | ESF PQ UNIVERSITARIO DE ASSIS | Instituto Butantan | Antonio Jorge Martins; Claudia Renata dos Santos Barros; David Schlesinger; Debora Botequio Moretti; Dimas Tadeu Covas; Elaine Cristina Marqueze; Elaine Vieira Santos; Evandra Strazza Rodrigues; Heidge Fukumasu; Jayme Augusto de Souza-Neto; José Salvatore Leister Patané; Luiz Alcantara; Luiz Lehmann Coutinho; Maria Carolina Elias; Maurício Lacerda Nogueira; Rafael dos Santos Bezerra; Raul Machado Neto; Rejane Maria Tommasini Grotto; Ricardo Haddad; Sandra Coccuzzo Sampaio Vessoni; Simone Kashima; Svetoslav Nanev Slavov; Vincent Louis Viala |
| EPI_ISL_2445169, EPI_ISL_2445176 | ESF VILA GLORIA I | Instituto Butantan | Antonio Jorge Martins; Claudia Renata dos Santos Barros; David Schlesinger; Debora Botequio Moretti; Dimas Tadeu Covas; Elaine Cristina Marqueze; Elaine Vieira Santos; Evandra Strazza Rodrigues; Heidge Fukumasu; Jayme Augusto de Souza-Neto; José Salvatore Leister Patané; Luiz Alcantara; Luiz Lehmann Coutinho; Maria Carolina Elias; Maurício Lacerda Nogueira; Rafael dos Santos Bezerra; Raul Machado Neto; Rejane Maria Tommasini Grotto; Ricardo Haddad; Sandra Coccuzzo Sampaio Vessoni; Simone Kashima; Svetoslav Nanev Slavov; Vincent Louis Viala |
| EPI_ISL_2445171, EPI_ISL_2473816 | ESF VILA PROGRESSO | Instituto Butantan | Antonio Jorge Martins; Claudia Renata dos Santos Barros; David Schlesinger; Debora Botequio Moretti; Dimas Tadeu Covas; Elaine Cristina Marqueze; Elaine Vieira Santos; Evandra Strazza Rodrigues; Heidge Fukumasu; Jayme Augusto de Souza-Neto; José Salvatore Leister Patané; Luiz Alcantara; Luiz Lehmann Coutinho; Maria Carolina Elias; Maurício Lacerda Nogueira; Rafael dos Santos Bezerra; Raul Machado Neto; Rejane Maria Tommasini Grotto; Ricardo Haddad; Sandra Coccuzzo Sampaio Vessoni; Simone Kashima; Svetoslav Nanev Slavov; Vincent Louis Viala |
| EPI_ISL_2445158, EPI_ISL_2445160 | ESF VITORIA | Instituto Butantan | Antonio Jorge Martins; Claudia Renata dos Santos Barros; David Schlesinger; Debora Botequio Moretti; Dimas Tadeu Covas; Elaine Cristina Marqueze; Elaine Vieira Santos; Evandra Strazza Rodrigues; Heidge Fukumasu; Jayme Augusto de Souza-Neto; José Salvatore Leister Patané; Luiz Alcantara; Luiz Lehmann Coutinho; Maria Carolina Elias; Maurício Lacerda Nogueira; Rafael dos Santos Bezerra; Raul Machado Neto; Rejane Maria Tommasini Grotto; Ricardo Haddad; Sandra Coccuzzo Sampaio Vessoni; Simone Kashima; Svetoslav Nanev Slavov; Vincent Louis Viala |
| EPI_ISL_3215305 | EXCITE Lab | Andersen lab at Scripps Research | Abigail Schnapper; Angela Scioscia; Chip Schooley; David Pride; Helena Tubb; Natasha Martin Cheryl Anderson; Sawyer Farmer; Sharon Reed; Tommy Valles + SEARCH |
| EPI_ISL_2940608, EPI_ISL_2940632, EPI_ISL_2940633, EPI_ISL_2940801, EPI_ISL_2940803, EPI_ISL_2940805, EPI_ISL_2940807, EPI_ISL_2940811, EPI_ISL_2940812, EPI_ISL_2940813, EPI_ISL_2940814, EPI_ISL_2940815, EPI_ISL_2940816, EPI_ISL_2940818, EPI_ISL_2940819, EPI_ISL_2940820, EPI_ISL_2940821, EPI_ISL_2940832, EPI_ISL_2940833, EPI_ISL_2940849, EPI_ISL_6661152 | | | |
| see above | Edmonton Provincial Lab | Public Health Agency of Canada (PHAC) National Microbiology Laboratory | Buss; Croxen M; Deo A; Dieu P; E; Ferrato C; Gill K; Khan F; Koleva P; Li V; Lloyd C; Lynch T; Ma R; Murphy S; Pabbaraju K; Shokoples S; Thayer J; Tipples G; Whitehouse M; Wong A; Yu C; Zelyas N |
| EPI_ISL_3758746 | Emory Molecular Diagnostics Laboratory, Emory Healthcare | Piantadosi Lab, Emory Department of Pathology | Ahmed Babiker; Anne Piantadosi; Dara Khosravi |
| EPI_ISL_2362558 | Eurofins Labazur Guyane SELAS | Institut Pasteur de la Guyane | Anne Lavergne; Antoine Enfissi; Arielle Salmier; Dominique Rousset |
| EPI_ISL_2631904, EPI_ISL_2631911 | Eurofins LifeCodexx GmbH | Robert Koch Institute | |
| EPI_ISL_2445114, EPI_ISL_2445115, EPI_ISL_2445116, EPI_ISL_2493708, EPI_ISL_2493709, EPI_ISL_2493710, EPI_ISL_2493711, EPI_ISL_2493712, EPI_ISL_2494270, EPI_ISL_2494296 | | | |
| see above | FACULDADE DE MEDICINA ABC | Instituto Butantan | Antonio Jorge Martins; Claudia Renata dos Santos Barros; David Schlesinger; Debora Botequio Moretti; Dimas Tadeu Covas; Elaine Cristina Marqueze; Elaine Vieira Santos; Evandra Strazza Rodrigues; Heidge Fukumasu; Jayme Augusto de Souza-Neto; José Salvatore Leister Patané; Luiz Alcantara; Luiz Lehmann Coutinho; Maria Carolina Elias; Maurício Lacerda Nogueira; Rafael dos Santos Bezerra; Raul Machado Neto; Rejane Maria Tommasini Grotto; Ricardo Haddad; Sandra Coccuzzo Sampaio Vessoni; Simone Kashima; Svetoslav Nanev Slavov; Vincent Louis Viala |
| EPI_ISL_2443017, EPI_ISL_2443019, EPI_ISL_2443032, EPI_ISL_2443033, EPI_ISL_2443034, EPI_ISL_2443040, EPI_ISL_2443041, EPI_ISL_2567201, EPI_ISL_2567202, EPI_ISL_2567214, EPI_ISL_2567215, EPI_ISL_2567219, EPI_ISL_2567238, EPI_ISL_2836193, EPI_ISL_2836214, EPI_ISL_2836219, EPI_ISL_2836220, EPI_ISL_2836236, EPI_ISL_2836242, EPI_ISL_2836243, EPI_ISL_2836319, EPI_ISL_2836330, EPI_ISL_2836499, EPI_ISL_2836820, EPI_ISL_5866263, EPI_ISL_6050033, EPI_ISL_6050034, EPI_ISL_6050035, EPI_ISL_6050036, EPI_ISL_6050037, EPI_ISL_6050038, EPI_ISL_6050042, EPI_ISL_6050044, EPI_ISL_6050045, EPI_ISL_6050098, EPI_ISL_6050665, EPI_ISL_6050726, EPI_ISL_6050746 | | | |
| see above | Florida Bureau of Public Health Laboratories | Florida Bureau of Public Health Laboratories | Jason Blanton; Namratha Tarigopula; Sarah Schmedes; Tiffany Splatt |
| EPI_ISL_2269311, EPI_ISL_2269314, EPI_ISL_2269318, EPI_ISL_2269319, EPI_ISL_2269323, EPI_ISL_2269335, EPI_ISL_2269352, EPI_ISL_2269358, EPI_ISL_2269360, EPI_ISL_2269368, EPI_ISL_2269371, EPI_ISL_2368135, EPI_ISL_2368140, EPI_ISL_2368143, EPI_ISL_2368154, EPI_ISL_2368177, EPI_ISL_2371518, EPI_ISL_2371573, EPI_ISL_2371575, EPI_ISL_2371576, EPI_ISL_2371577, EPI_ISL_2371578, EPI_ISL_2371579, EPI_ISL_2371580, EPI_ISL_2371584, EPI_ISL_2371585, EPI_ISL_2371586, EPI_ISL_2371587, EPI_ISL_2371617, EPI_ISL_2371626, EPI_ISL_2371639, EPI_ISL_2371650, EPI_ISL_2371654, EPI_ISL_2371657, EPI_ISL_2371664, EPI_ISL_2371672, EPI_ISL_2371673, EPI_ISL_2371674, EPI_ISL_2480073, EPI_ISL_2480079, EPI_ISL_2525446, EPI_ISL_2525459, EPI_ISL_2525464, EPI_ISL_2525468, EPI_ISL_2525472, EPI_ISL_2525480, EPI_ISL_2525482, EPI_ISL_2525497, EPI_ISL_2525503, EPI_ISL_2525517, EPI_ISL_2525521, EPI_ISL_2525525, EPI_ISL_2525544, EPI_ISL_2525551, EPI_ISL_2525557, EPI_ISL_2525633, EPI_ISL_2525785, EPI_ISL_2525787, EPI_ISL_2525831, EPI_ISL_2525836, EPI_ISL_2525885, EPI_ISL_2525910, EPI_ISL_2525921, EPI_ISL_2525951, EPI_ISL_2525962, EPI_ISL_2525993, EPI_ISL_2526024, EPI_ISL_2526057, EPI_ISL_4368887, EPI_ISL_4371203, EPI_ISL_4371871, EPI_ISL_4372271 | | | |
| see above | Fulgent Genetics | Centers for Disease Control and Prevention Division of Viral Diseases, Pathogen Discovery | Adrian Paskey; Becky Tsai; Benafsh Sapra; Benjamin Rambo-Martin; Christopher Gulvick; Clinton Paden; Clinton R. Paden; Dakota Howard; Darlene Wagner; Dhwani Batra; Doreen Ng; Duncan MacCannell; Erisa Sula; Harry Gao; James Xie; Jason Caravas; John Gao; Joseph Fierro; Kara Moser; Kristine Lacek; Matthew Schmerer; Mickey Li; Peter Cook; Peter W. Cook; Scott Sammons; Shatavia Morrison; Tymeckia Kendall; Victoria Caban Figueroa; Yan Meng; Yvette Unoarumhi |
| EPI_ISL_2550708 | GA Department of Public Health | GA Department of Public Health | Aliyah Fields; Cynthia Dixey; Jonathan Edwards; Stacy Reeves; Taylor Smith; Tonia Parrott |
| EPI_ISL_3031056 | GLENS FALLS HOSPITAL LABORATORY | Wadsworth Center, New York State Department of Health | Alexis Russell; Catharine Prussing; Daryl M. Lamson; Erasmus Schneider; Erica Lasek-Nesselquist; John Kelly; Jonathan Plitnick; Kirsten St. George; Matthew Shudt; Melissa A Leisner; Navjot Singh |

| | | | |
|---|---|---|---|
| EPI_ISL_3491813, EPI_ISL_3491816, EPI_ISL_3491817, EPI_ISL_3491834, EPI_ISL_3491835 | Gencore - Universidad de los Andes | Gencore - Universidad de los Andes | Catherine Jaller; Cristian Barrera; David González; Luisa Sacristan; Marcela Guevara; Silvia Restrepo |
| EPI_ISL_2509163, EPI_ISL_2509166, EPI_ISL_2509168, EPI_ISL_2509185, EPI_ISL_2509282, EPI_ISL_2509292, EPI_ISL_2509294, EPI_ISL_2509296, EPI_ISL_2509297, EPI_ISL_2509300, EPI_ISL_2509302, EPI_ISL_2509303, EPI_ISL_2509306, EPI_ISL_2509308, EPI_ISL_2509309, EPI_ISL_2509310, EPI_ISL_2509316, EPI_ISL_2509318, EPI_ISL_2509319, EPI_ISL_2509320, EPI_ISL_2509321, EPI_ISL_2509322, EPI_ISL_2509323, EPI_ISL_2543436, EPI_ISL_2543438, EPI_ISL_2543447, EPI_ISL_2543449, EPI_ISL_2543450, EPI_ISL_2543451, EPI_ISL_2543452, EPI_ISL_2543453, EPI_ISL_2543454, EPI_ISL_2543455, EPI_ISL_2543456, EPI_ISL_2543457, EPI_ISL_2543458, EPI_ISL_2543461, EPI_ISL_2543464, EPI_ISL_2543465, EPI_ISL_2543469, EPI_ISL_2543470, EPI_ISL_2543471, EPI_ISL_2543472, EPI_ISL_2543473, EPI_ISL_2543475, EPI_ISL_2543476, EPI_ISL_2543477, EPI_ISL_2543478, EPI_ISL_2543482, EPI_ISL_2543483, EPI_ISL_2543484, EPI_ISL_2543488, EPI_ISL_2543489, EPI_ISL_2543490, EPI_ISL_2543491, EPI_ISL_2543492, EPI_ISL_2543493, EPI_ISL_2543494, EPI_ISL_2543495, EPI_ISL_2543496, EPI_ISL_2543497, EPI_ISL_2543499, EPI_ISL_2543500, EPI_ISL_2543501, EPI_ISL_2543502, EPI_ISL_2543504, EPI_ISL_2543505, EPI_ISL_2543509, EPI_ISL_2543510, EPI_ISL_2543511, EPI_ISL_2543512, EPI_ISL_2543513, EPI_ISL_2543515, EPI_ISL_2543526, EPI_ISL_2543527, EPI_ISL_2543542, EPI_ISL_2543543, EPI_ISL_2543544, EPI_ISL_2543545, EPI_ISL_2543547, EPI_ISL_2543548, EPI_ISL_2543549, EPI_ISL_2543551, EPI_ISL_2543553, EPI_ISL_2543554, EPI_ISL_2543557, EPI_ISL_2543558, EPI_ISL_2543571, EPI_ISL_2543573, EPI_ISL_2543577, EPI_ISL_2543583, EPI_ISL_2543584, EPI_ISL_2543585, EPI_ISL_2543586, EPI_ISL_2543617, EPI_ISL_2543619, EPI_ISL_2543620, EPI_ISL_2557264, EPI_ISL_2557268, EPI_ISL_2892595, EPI_ISL_2892608 | | | |
| see above | Genetica Molecular and Subdepartamento de Virologia ISP Chile | Instituto de Salud Publica de Chile | Andres Castillo; Barbara Parra; Constanza Campano; Gisselle Barra; Javier Tognarelli; Jorge Fernandez; Karen Orostica; Loredana Arata; Patricia Bustos; Rodrigo Fasce; Soledad Ulloa |
| EPI_ISL_2535904, EPI_ISL_2535947, EPI_ISL_2535961, EPI_ISL_2535965, EPI_ISL_2535968, EPI_ISL_2535977, EPI_ISL_2535981, EPI_ISL_2535988, EPI_ISL_2536002, EPI_ISL_2536023, EPI_ISL_2536025, EPI_ISL_2536030, EPI_ISL_2536036, EPI_ISL_2537091, EPI_ISL_2537097, EPI_ISL_2537098, EPI_ISL_2537099, EPI_ISL_2537108, EPI_ISL_2537109, EPI_ISL_2537126, EPI_ISL_2537129, EPI_ISL_2613921, EPI_ISL_2614004, EPI_ISL_2614052, EPI_ISL_2614055, EPI_ISL_2614056 | | | |
| see above | Gravity Diagnostics, LLC | Gravity Diagnostics, LLC | Gravity Diagnostics |
| EPI_ISL_4511673 | Gravity Diagnostics, LLC | University of Louisville Sequencing Technology Center | Elizabeth Hudson; Eric C. Rouchka; James P. Canner; Julia H. Chariker; Melissa L. Smith; Ryan Walker; William Lauer |
| EPI_ISL_2376243, EPI_ISL_2376247, EPI_ISL_2376249, EPI_ISL_2500991 | Gundersen Clinical Microbiology Laboratory | Kabara Cancer Research Institute | Craig S. Richmond; Paraic A. Kenny |
| EPI_ISL_6229715 | HGF HOSPITAL GERAL DE FORTALEZA | ACME Lab, Oswaldo Cruz Foundation, FIOCRUZ/CE | Carlos Leonardo de Aragao Araujo; Cecilia Leite Costa & Eduardo Ruback dos Santos on behalf of COVID-19 FIOCRUZ Genomic Network; Cleber Furtado Aksenen; Fabio Miyajima; Fernando Braga Stehling; Francisco Eder de Moura Lopes; Igor Oliveira Duarte; Jamille Maria Mendes Bezerra; Joaquim Cesar do Nascimento Sousa Junior; Pedro Miguel Carneiro Jeronimo; Suzana Porto Almeida; Thais Ferreira de Oliveira; Thais de Oliveira Costa; Ticiane Cavalcante de Souza; Veridiana Pessoa Miyajima |
| EPI_ISL_2452142 | HI Dept. of Health, State Laboratories Division | Centers for Disease Control and Prevention Division of Viral Diseases, Pathogen Discovery | Alison Laufer Halpin; Ben L. Rambo-Martin; Clinton R. Paden; Dakota Howard; Darlene Wagner; Dave Wentworth; Dhwani Batra; Jasmine Padilla; Justin Lee; Katie Dillon; Krista Queen; Kristen Knipe; Kristine Lacek; Mark Burroughs; Matthew Schmerer; Mili Sheth; Peter Cook; Sam Shepard; Sarah Nobles; Shoshona Le; Suxiang Tong; Vivien Dugan; Yvette Unoarumhi |
| EPI_ISL_2466447, EPI_ISL_2466448, EPI_ISL_2466449, EPI_ISL_2466450, EPI_ISL_2466451, EPI_ISL_2466452, EPI_ISL_2466453, EPI_ISL_2466454, EPI_ISL_2466455, EPI_ISL_2466456, EPI_ISL_2466457, EPI_ISL_2466458, EPI_ISL_2466459, EPI_ISL_2466460, EPI_ISL_2466461, EPI_ISL_2466462, EPI_ISL_2466463, EPI_ISL_2466466, EPI_ISL_2497441, EPI_ISL_2497442, EPI_ISL_2497465, EPI_ISL_2617586, EPI_ISL_2617607, EPI_ISL_3254316, EPI_ISL_3254321, EPI_ISL_3386138, EPI_ISL_5438934 | | | |
| see above | HLAGYN - Laboratorio de Imunologia de Transplantes de Goias | HLAGYN - Laboratorio de Imunologia de Transplantes de Goias | Alessandro Leonardo Alvares Magalhaes; Daniel Ferreira de Sousa; Erika Lopes Rocha Batista; Fernando Antonio Vinhal dos Santos; Frederico Rodrigues Vinhal; Kamila Oliveira Reis De Freitas.; Lucas Carlos Gomes Pereira; Sabrina Sara Moreira Duarte |
| EPI_ISL_2473702, EPI_ISL_2473749, EPI_ISL_2473758, EPI_ISL_2473759 | HOSP GERAL DE SAO PAULO | Instituto Butantan | Antonio Jorge Martins; Claudia Renata dos Santos Barros; David Schlesinger; Debora Botequio Moretti; Dimas Tadeu Covas; Elaine Cristina Marqueze; Elaine Vieira Santos; Evandra Strazza Rodrigues; Heidge Fukumasu; Jayme Augusto de Souza-Neto; José Salvatore Leister Patané; Luiz Alcantara; Luiz Lehmann Coutinho; Maria Carolina Elias; Maurício Lacerda Nogueira; Rafael dos Santos Bezerra; Raul Machado Neto; Rejane Maria Tommasini Grotto; Ricardo Haddad; Sandra Coccuzzo Sampaio Vessoni; Simone Kashima; Svetoslav Nanev Slavov; Vincent Louis Viala |
| EPI_ISL_3102255 | HOSPITAL CURA DARS | Analytical Competence Molecular Epidemiology Lab/ACME, Oswaldo Cruz Foundation, Ceara (FIOCRUZ CE) | Cleber Furtado Aksenen; Fabio Miyajima; Fernando Braga Stehling; Francisco Eder de Moura Lopes; Jamille Maria Mendes Bezerra; Joaquim César do Nascimento Sousa Junior; Pedro Miguel Carneiro Jeronimo; Suzana Porto Almeida e Lucas Delerino; Thais Ferreira de Oliveira; Thais de Oliveira Costa; Ticiane Cavalcante de Souza; Veridiana Pessoa Miyajima |
| EPI_ISL_2378745, EPI_ISL_2493192, EPI_ISL_2493303, EPI_ISL_2493304, EPI_ISL_2493305, EPI_ISL_2493306, EPI_ISL_2493307, EPI_ISL_2493308, EPI_ISL_2493309, EPI_ISL_2493310, EPI_ISL_2493312, EPI_ISL_2493316, EPI_ISL_2493318, EPI_ISL_2493320, EPI_ISL_2493321, EPI_ISL_2493322, EPI_ISL_2493323, EPI_ISL_2493324, EPI_ISL_2493325, EPI_ISL_2493330, EPI_ISL_2493331, EPI_ISL_2493332, EPI_ISL_2493334 | | | |
| see above | HOSPITAL DO RIM E HIPERTENSAO | Instituto Butantan | Antonio Jorge Martins; Claudia Renata dos Santos Barros; David Schlesinger; Debora Botequio Moretti; Dimas Tadeu Covas; Elaine Cristina Marqueze; Elaine Vieira Santos; Evandra Strazza Rodrigues; Heidge Fukumasu; Jayme Augusto de Souza-Neto; José Salvatore Leister Patané; Luiz Alcantara; Luiz Lehmann Coutinho; Maria Carolina Elias; Maurício Lacerda Nogueira; Rafael dos Santos Bezerra; Raul Machado Neto; Rejane Maria Tommasini Grotto; Ricardo Haddad; Sandra Coccuzzo Sampaio Vessoni; Simone Kashima; Svetoslav Nanev Slavov; Vincent Louis Viala |
| EPI_ISL_3102290, EPI_ISL_3102292, EPI_ISL_3102317, EPI_ISL_3102522 | HOSPITAL ESTADUAL LEONARDO DA VINCI | Analytical Competence Molecular Epidemiology Lab/ACME, Oswaldo Cruz Foundation, Ceara (FIOCRUZ CE) | Cleber Furtado Aksenen; Fabio Miyajima; Fernando Braga Stehling; Francisco Eder de Moura Lopes; Jamille Maria Mendes Bezerra; Joaquim César do Nascimento Sousa Junior; Pedro Miguel Carneiro Jeronimo; Suzana Porto Almeida e Lucas Delerino; Thais Ferreira de Oliveira; Thais de Oliveira Costa; Ticiane Cavalcante de Souza; Veridiana Pessoa Miyajima |
| EPI_ISL_3260869 | HOSPITAL GENERAL UNIVERSITARIO DE ALBACETE | Instituto de Salud Carlos III | A. Monzón; CARIDAD; F. Casas; I. Jiménez; I. SAINZ DE BARANDA CAMINO; M. Camarero; P. Zaballos; S. Cuesta; S. Iglesias-Caballero; S. Pozo; S. Varona; Sandonís; V. Vázquez-Morón |
| EPI_ISL_3260876 | HOSPITAL GENERAL UNIVERSITARIO DE GUADALAJARA | Instituto de Salud Carlos III | A. Monzón; ALEJANDRO; F. Casas; I. Jiménez; I.GONZALEZ PRAETORIUS; Iglesias-Caballero; M. Sandonís; P. Zaballos; S. Camarero; S. Cuesta; S. Pozo; S. Varona; V. Vázquez-Morón |
| EPI_ISL_3260838, EPI_ISL_3260840 | HOSPITAL GENERAL UNIVERSITARIO SANTA LUCIA | Instituto de Salud Carlos III | A. Monzón; ANA; F. Casas; I. Jiménez; I.BLAZQUEZ ABELLAN; M. Sandonís; P. Zaballos; S. Cuesta; S. Iglesias-Caballero; S. Pozo; S. Varona; V. Camarero; Vázquez-Morón |
| EPI_ISL_5645965, EPI_ISL_5646020 | HOSPITAL MATERNIDADE SAO JOSE ITAPUI | Instituto Butantan | Antonio Jorge Martins; Claudia Renata dos Santos Barros; David Schlesinger; Debora Botequio Moretti; Dimas Tadeu Covas; Elaine Cristina Marqueze; Elaine Vieira Santos; Evandra Strazza Rodrigues; Heidge Fukumasu; Jayme Augusto de Souza-Neto; José Salvatore Leister Patané; Luiz Alcantara; Luiz Lehmann Coutinho; Maria Carolina Elias; Maurício Lacerda Nogueira; Rafael dos Santos Bezerra; Raul Machado Neto; Rejane Maria Tommasini Grotto; Ricardo Haddad; Sandra Coccuzzo Sampaio Vessoni; Simone Kashima; Svetoslav Nanev Slavov; Vincent Louis Viala |
| EPI_ISL_2493685, EPI_ISL_2493687, EPI_ISL_2493689 | HOSPITAL MUNICIPAL DE IBIUNA IBIUNA SP | Instituto Butantan | Antonio Jorge Martins; Claudia Renata dos Santos Barros; David Schlesinger; Debora Botequio Moretti; Dimas Tadeu Covas; Elaine Cristina Marqueze; Elaine Vieira Santos; Evandra Strazza Rodrigues; Heidge Fukumasu; Jayme Augusto de Souza-Neto; José Salvatore Leister Patané; Luiz Alcantara; Luiz Lehmann Coutinho; Maria Carolina Elias; Maurício Lacerda Nogueira; Rafael dos Santos Bezerra; Raul Machado Neto; Rejane Maria Tommasini Grotto; Ricardo Haddad; Sandra Coccuzzo Sampaio Vessoni; Simone Kashima; Svetoslav Nanev Slavov; Vincent Louis Viala |
| EPI_ISL_2493592, EPI_ISL_2493593, EPI_ISL_2493609, EPI_ISL_2493650, EPI_ISL_2493651, EPI_ISL_2493652 | HOSPITAL MUNICIPAL DE ILHABELA GOV MARIO COVAS JR | Instituto Butantan | Antonio Jorge Martins; Claudia Renata dos Santos Barros; David Schlesinger; Debora Botequio Moretti; Dimas Tadeu Covas; Elaine Cristina Marqueze; Elaine Vieira Santos; Evandra Strazza Rodrigues; Heidge Fukumasu; Jayme Augusto de Souza-Neto; José Salvatore Leister Patané; Luiz Alcantara; Luiz Lehmann Coutinho; Maria Carolina Elias; Maurício Lacerda Nogueira; Rafael dos Santos Bezerra; Raul Machado Neto; Rejane Maria Tommasini Grotto; Ricardo Haddad; Sandra Coccuzzo Sampaio Vessoni; Simone Kashima; Svetoslav Nanev Slavov; Vincent Louis Viala |
| EPI_ISL_2493628 | HOSPITAL MUNICIPAL DE ITAPIRA | Instituto Butantan | Antonio Jorge Martins; Claudia Renata dos Santos Barros; David Schlesinger; Debora Botequio Moretti; Dimas Tadeu Covas; Elaine Cristina Marqueze; Elaine Vieira Santos; Evandra Strazza Rodrigues; Heidge Fukumasu; Jayme Augusto de Souza-Neto; José Salvatore Leister Patané; Luiz Alcantara; Luiz Lehmann Coutinho; Maria Carolina Elias; Maurício Lacerda Nogueira; Rafael dos Santos Bezerra; Raul Machado Neto; Rejane Maria Tommasini Grotto; Ricardo Haddad; Sandra Coccuzzo Sampaio Vessoni; Simone Kashima; Svetoslav Nanev Slavov; Vincent Louis Viala |
| EPI_ISL_2493392, EPI_ISL_2500976 | HOSPITAL MUNICIPAL DR TABAJARA RAMOS | Instituto Butantan | Antonio Jorge Martins; Claudia Renata dos Santos Barros; David Schlesinger; Debora Botequio Moretti; Dimas Tadeu Covas; Elaine Cristina Marqueze; Elaine Vieira Santos; Evandra Strazza Rodrigues; Heidge Fukumasu; Jayme Augusto de Souza-Neto; José Salvatore Leister Patané; Luiz Alcantara; Luiz Lehmann Coutinho; Maria Carolina Elias; Maurício Lacerda Nogueira; Rafael dos Santos Bezerra; Raul Machado Neto; Rejane Maria Tommasini Grotto; Ricardo Haddad; Sandra Coccuzzo Sampaio Vessoni; Simone Kashima; Svetoslav Nanev Slavov; Vincent Louis Viala |
| EPI_ISL_6229727, EPI_ISL_6229728 | HOSPITAL MUNICIPAL HUMBERTO DE QUEIROZ | ACME Lab, Oswaldo Cruz Foundation, FIOCRUZ/CE | Carlos Leonardo de Aragao Araujo; Cecilia Leite Costa & Eduardo Ruback dos Santos on behalf of COVID-19 FIOCRUZ Genomic Network; Cleber Furtado Aksenen; Fabio Miyajima; Fernando Braga Stehling; Francisco Eder de Moura Lopes; Igor Oliveira Duarte; Jamille Maria Mendes Bezerra; Joaquim Cesar do Nascimento Sousa Junior; Pedro Miguel Carneiro Jeronimo; Suzana Porto Almeida; Thais Ferreira de Oliveira; Thais de Oliveira Costa; Ticiane Cavalcante de Souza; Veridiana Pessoa Miyajima |
| EPI_ISL_2493688 | HOSPITAL MUNICIPAL LEONARDUS VAN MELLIS | Instituto Butantan | Antonio Jorge Martins; Claudia Renata dos Santos Barros; David Schlesinger; Debora Botequio Moretti; Dimas Tadeu Covas; Elaine Cristina Marqueze; Elaine Vieira Santos; Evandra Strazza Rodrigues; Heidge Fukumasu; Jayme Augusto de Souza-Neto; José Salvatore Leister Patané; Luiz Alcantara; Luiz Lehmann Coutinho; Maria Carolina Elias; Maurício Lacerda Nogueira; Rafael dos Santos Bezerra; Raul Machado Neto; Rejane Maria Tommasini Grotto; Ricardo Haddad; Sandra Coccuzzo Sampaio Vessoni; Simone Kashima; Svetoslav Nanev Slavov; Vincent Louis Viala |
| EPI_ISL_2801335, EPI_ISL_2801336, EPI_ISL_2801345 | HOSPITAL MUNICIPAL NOSSA SENHORA DOS MILAGRES | Analytical Competence Molecular Epidemiology Lab/ACME, Oswaldo Cruz Foundation, Ceara (FIOCRUZ CE) | Cleber Furtado Aksenen e Suzana Porto Almeida; Fabio Miyajima; Fernando Braga Stehling; Francisco Eder de Moura Lopes; Jamille Maria Mendes Bezerra; Joaquim César do Nascimento Sousa Junior; Pedro Miguel Carneiro Jeronimo; Thais Ferreira de Oliveira; Thais de Oliveira Costa; Ticiane Cavalcante de Souza; Veridiana Pessoa Miyajima |
| EPI_ISL_2493296 | HOSPITAL MUNICIPAL SANTO ANTONIO | Instituto Butantan | Antonio Jorge Martins; Claudia Renata dos Santos Barros; David Schlesinger; Debora Botequio Moretti; Dimas Tadeu Covas; Elaine Cristina Marqueze; Elaine Vieira Santos; Evandra Strazza Rodrigues; Heidge Fukumasu; Jayme Augusto de Souza-Neto; José Salvatore Leister Patané; Luiz Alcantara; Luiz Lehmann Coutinho; Maria Carolina Elias; Maurício Lacerda Nogueira; Rafael dos Santos Bezerra; Raul Machado Neto; Rejane Maria Tommasini Grotto; Ricardo Haddad; Sandra Coccuzzo Sampaio Vessoni; Simone Kashima; Svetoslav Nanev Slavov; Vincent Louis Viala |
| EPI_ISL_6229741, EPI_ISL_6229742, EPI_ISL_6229743, EPI_ISL_6229744, EPI_ISL_6229745, EPI_ISL_6229746, EPI_ISL_6229821 | | | |
| see above | HOSPITAL REGIONAL DO CARIRI | ACME Lab, Oswaldo Cruz Foundation, FIOCRUZ/CE | Carlos Leonardo de Aragao Araujo; Cecilia Leite Costa & Eduardo Ruback dos Santos on behalf of COVID-19 FIOCRUZ Genomic Network; Cleber Furtado Aksenen; Fabio Miyajima; Fernando Braga Stehling; Francisco Eder de Moura Lopes; Igor Oliveira Duarte; Jamille Maria Mendes Bezerra; Joaquim Cesar do Nascimento Sousa Junior; Pedro Miguel Carneiro Jeronimo; Suzana Porto Almeida; Thais Ferreira de Oliveira; Thais de Oliveira Costa; Ticiane Cavalcante de Souza; Veridiana Pessoa Miyajima |
| EPI_ISL_6229749 | HOSPITAL REGIONAL NORTE | ACME Lab, Oswaldo Cruz Foundation, FIOCRUZ/CE | Carlos Leonardo de Aragao Araujo; Cecilia Leite Costa & Eduardo Ruback dos Santos on behalf of COVID-19 FIOCRUZ Genomic Network; Cleber Furtado Aksenen; Fabio Miyajima; Fernando Braga Stehling; Francisco Eder de Moura Lopes; Igor Oliveira Duarte; Jamille Maria Mendes Bezerra; Joaquim Cesar do Nascimento Sousa Junior; Pedro Miguel Carneiro Jeronimo; Suzana Porto Almeida; Thais Ferreira de Oliveira; Thais de Oliveira Costa; Ticiane Cavalcante de Souza; Veridiana Pessoa Miyajima |
| EPI_ISL_2502760, EPI_ISL_2502762 | HOSPITAL SAN JUAN DE DIOS | Inciensa, Instituto Costarricense de InvestigaciOn y Enseñanza en NutriciOn y Salud | Adriana Godínez; Claudio Soto-Garita; Estela Cordero; Francisco Duarte; Hebleen Porras; Jose Luis Vargas; Joselyn Prado & Marco Cháves-Otárola; Mariela Gutierrez; Melany CalderOn |
| EPI_ISL_3502489, EPI_ISL_3502591, EPI_ISL_3502593 | HOSPITAL UNIVERSITARIO 12 DE OCTUBRE | HOSPITAL UNIVERSITARIO 12 DE OCTUBRE | Carmen Martín-Higuera; Esther Viedma; Irene Múñoz-Gallego; M.ª Dolores Folgueira; Mar Aguilera; Noelia Moral; Rafael Delgado; Sagrario Zurita |
| EPI_ISL_3235348, EPI_ISL_3235352, EPI_ISL_3235354 | Hackensack Medical Center | New York Genome Center | Amy Baldwin; Andre Corvelo; Barry Kreiswirth; David Perlin; Dayna M. Oschwald; Jose Mediavilla; Kaelea Composto; Kar Chow; Liang Chen; Marcus Cunningham; Michael Zody; Samantha Fennessey; Tom Maniatis |
| EPI_ISL_2235694 | Health Services Laboratories | Wellcome Sanger Institute for the COVID-19 Genomics UK (COG-UK) Consortium | Cordelia Langford; David K. Jackson; Dominic Kwiatkowski; Ewan Harrison; Health Services Laboratories and Alex Alderton; Ian Johnston; Jeffrey Barrett; John Sillitoe on behalf of the Wellcome Sanger Institute COVID-19 Surveillance Team; Roberto Amato; Sonia Goncalves |

| | | | |
|---|---|---|---|
| EPI_ISL_5771881 | Hospital Lagomaggiore Mendoza | Nodo de Secuenciación Tierra del Fuego - Hospital Regional Ushuaia - Centro Austral De Investigaciones Científicas - Universidad Nacional De Tierra Del Fuego | AE; CF; Ceballos; F; Gallego; Gramundi; ID; Nardi; Rojas; SG |
| EPI_ISL_2464080 | Hospital | National Reference Center for Viruses of Respiratory Infections, Institut Pasteur, Paris | Angela Brisebarre; Camille Capel; Christophe Malabat; Corinne Maufrais; Emmanuelle Permal; Etienne Simon-Lorière; Frédéric Lemoine; Louise Lefrançois; Marion Barbet; Maud Vanpeene; Méline Bizard; Patricia Stoessel-Thouvenin; Sylvie Behillil; Sylvie Van der Werf; Vincent Enouf |
| EPI_ISL_2812290 | Hospital Cangas y Severo Ochoa | Laboratorio de Virología HUCA | Abreu F; Alvarez-Arguelles ME; Boga JA; Castelló C; Costales I; Coto E; Gómez de Oña J; Martín-Rodríguez G; Melón S; Perez-Martínez Z; Rojo S; Sandoval M |
| EPI_ISL_2812439 | Hospital Carmen y Severo Ochoa | Laboratorio de Virología HUCA | Abreu F; Alvarez-Arguelles ME; Boga JA; Castelló C; Costales I; Coto E; Gómez de Oña J; Martín-Rodríguez G; Melón S; Perez-Martínez Z; Rojo S; Sandoval M |
| EPI_ISL_5771883 | Hospital Central Mendoza | Nodo de Secuenciación Tierra del Fuego - Hospital Regional Ushuaia - Centro Austral De Investigaciones Científicas - Universidad Nacional De Tierra Del Fuego | AE; CF; Ceballos; F; Gallego; Gramundi; ID; Nardi; Rojas; SG |
| EPI_ISL_2304183, EPI_ISL_2304184, EPI_ISL_2304185, EPI_ISL_2304186, EPI_ISL_3505960, EPI_ISL_3505961, EPI_ISL_3505962, EPI_ISL_3505963, EPI_ISL_3505967, EPI_ISL_3505969, EPI_ISL_3505975, EPI_ISL_3505976, EPI_ISL_3505978 | | | |
| see above | Hospital General Universitario Gregorio Marañón | Hospital General Universitario Gregorio Marañón | Cristina Rodriguez-Grande; Darío García de Viedma; Julia Suárez; Laura Pérez-Lago; Marta Herranz Martin; Patricia Muñoz; Pedro Sola Campoy; Pilar Catalán; Sergio Buenestado Serrano; Victor Manuel de la Cueva |
| EPI_ISL_2598381, EPI_ISL_2598385 | Hospital General Universitario de Alicante - Instituto de Investigación Sanitaria y Biomédica de Alicante | SeqCOVID-SPAIN consortium/IBV(CSIC) | Carmen Molina Pardines and SeqCOVID-SPAIN consortium; Maripaz Ventero Martín |
| EPI_ISL_3033046, EPI_ISL_3033047, EPI_ISL_3033048, EPI_ISL_3260091 | Hospital Jaime Ferre - SAMCO Rafaela | Grupo de Genómica y Bioinformática del Instituto de Investigación de la Cadena Láctea CONICET-INTA on behalf of 'Proyecto Argentino Interinstitucional de genomica de SARS-CoV-2' (PAIS Consortium) | AF; Aliprandi D; Amadio; C; Eberhardt; Irazoqui; Isaia; JF; JM; MF; Pandolfi; Quaranta; Soratti R; V |
| EPI_ISL_2284974, EPI_ISL_2284975, EPI_ISL_2284976, EPI_ISL_2391949, EPI_ISL_2391950, EPI_ISL_2391951 | Hospital Universitari Arnau de Vilanova | Hospital Universitari Vall d'Hebron - Vall d'Hebron Institut de Recerca | Alejandra González-Sánchez; Andrés Antón; Ariadna Rando; Carla Castillo; Cristina Andrés; Damir Garcia-Cehic; Josep Quer; Juliana Esperalba; Maria Carmen Martin; Maria Gema Codina; Maria Piñana; Tomàs Pumarola |
| EPI_ISL_2516572, EPI_ISL_2516580, EPI_ISL_2516581, EPI_ISL_2516582, EPI_ISL_2637297 | Hospital Universitari Bellvitge | Microbiology Department | Aida Gonzalez-Diaz; Carmen Ardanuy; Jordi Camara; Jordi Niubó; Laura Calatayud; M Angeles Domínguez; Miguel Fernandez-Huerta; Sara Marti |
| EPI_ISL_2652049, EPI_ISL_2652054 | Hospital Universitari Joan XXIII | Hospital Universitari Joan XXIII | Carla Martín; Clara Benavent; Cristina Gutiérrez; Ester Picó; Gemma Recio; Margarida Terrón; Natalia Bastón |
| EPI_ISL_2284977, EPI_ISL_2391939 | Hospital Universitari Joan XXIII de Tarragona | Hospital Universitari Vall d'Hebron - Vall d'Hebron Institut de Recerca | Alejandra González-Sánchez; Andrés Antón; Ariadna Rando; Carla Castillo; Cristina Andrés; Damir Garcia-Cehic; Josep Quer; Juliana Esperalba; Maria Carmen Martin; Maria Gema Codina; Maria Piñana; Tomàs Pumarola |
| EPI_ISL_7339451, EPI_ISL_7339475, EPI_ISL_7339477 | Hospital Universitario San Ignacio | Centro de Investigaciones en Microbiología y Biotecnología-UR (CIMBIUR), Facultad de Ciencias Naturales, Universidad del Rosario, Bogotá, Colombia | Alberto Paniz-Mondolfi; Angie Ramírez; Beatriz Ariza; Camilo A. Correa-Cárdenas; Carlos Gómez-Restrepo; Claudia Cardozo-Romero; Claudia Méndez; David-Santiago Quevedo; Guido España; Hernando Díaz; Juan David Ramírez; Juliana Cuervo-Rojas; Julie Pérez; Luz H. Patiño; Manuel-Antonio Franco; Maria-Clara Duque; Marina Muñoz; Nathalia Ballesteros; Nicolas Luna; Sergio Castañeda; Zulma M. Cucunubá |
| EPI_ISL_4196733 | Hospital zonal Francisco Lopez Lima General Roca | Laboratorio Central Mg. Luis Alfredo Pianciola on behalf of 'Proyecto Argentino Interinstitucional de genomica de SARS-CoV-2' (PAIS Consortium) | A Montecinos.; C Pintos; C Rastellini; C Ziehm; D Durany; J Laher; J Ousset; L Pianciola; M Fernandez; M Mazzeo; T Paniceres |
| EPI_ISL_5070650, EPI_ISL_5070653, EPI_ISL_5070673, EPI_ISL_5070684, EPI_ISL_5070765, EPI_ISL_5070799, EPI_ISL_5070847 | | | |
| see above | Houston Methodist Hospital | Houston Methodist Hospital | Ilya J. Finkelstein; James J. Davis; Jessica Cambric; Jimmy Gollihar; Kristina Reppond; Layne Pruitt; Madison N. Shyer; Marcus Nguyen; Matthew Ojeda Saavedra; Paul A. Christensen; Prasanti Yerramilli; Randall J. Olsen; Robert Olson; Ryan Gadd; S. Wesley Long; Sishir Subedi; and James M. Musser |
| EPI_ISL_2497653, EPI_ISL_2497660, EPI_ISL_2497663, EPI_ISL_2497664 | Humboldt County Public Health Laboratory | Chan-Zuckerberg Biohub | CZB Cliahub Consortium |
| EPI_ISL_2680474, EPI_ISL_2680483 | Humboldt County Public Health Laboratory | Humboldt County Public Health Laboratory | Jeremy Corrigan |
| EPI_ISL_3276632 | ICMT-Apartado | Universidad Nacional de Colombia - Laboratorio Genómico One Health | Andres F. Cardona-Rios; Carlos Franco-Muñoz; Carolina Muñoz-Arango; Celeny Ortiz; Daniel O. Maldonado-Perez; Diego A. Álvarez-Díaz; Hector Alejandro Ruiz-Moreno; Idabely Betancur Ortiz; Jorge E. Osorio; Juan P. Hernandez-Ortiz; Karl A Ciuoderis; Katherine Laiton-Donato; Laura Silvana Perez; Lina M. Hurtado; Marcela Mercado-Reyes; Maria Angélica Maya; Maria Stella López; Rita Almanza Payares; Sandra Ines Cano; Simón Villegas Velásquez |
| EPI_ISL_2690066, EPI_ISL_2690070, EPI_ISL_2690072, EPI_ISL_2690082, EPI_ISL_2690083 | ID Bureau of Laboratories | Centers for Disease Control and Prevention Division of Viral Diseases, Pathogen Discovery | Alison Laufer Halpin; Ben L. Rambo-Martin; Clinton R. Paden; Dakota Howard; Darlene Wagner; Dave Wentworth; Dhwani Batra; Jasmine Padilla; Justin Lee; Katie Dillon; Krista Queen; Kristen Knipe; Kristine Lacek; Mark Burroughs; Matthew Schmerer; Mili Sheth; Peter Cook; Sam Shepard; Sarah Nobles; Shoshona Le; Suxiang Tong; Vivien Dugan; Yvette Unoarumhi |
| EPI_ISL_2451787, EPI_ISL_2451789, EPI_ISL_2451792, EPI_ISL_2451798, EPI_ISL_2451804, EPI_ISL_2451808, EPI_ISL_2451811 | | | |
| see above | IL Dept. of Public Health Springfield Laboratory | Centers for Disease Control and Prevention Division of Viral Diseases, Pathogen Discovery | Alison Laufer Halpin; Ben L. Rambo-Martin; Clinton R. Paden; Dakota Howard; Darlene Wagner; Dave Wentworth; Dhwani Batra; Jasmine Padilla; Justin Lee; Katie Dillon; Krista Queen; Kristen Knipe; Kristine Lacek; Mark Burroughs; Matthew Schmerer; Mili Sheth; Peter Cook; Sam Shepard; Sarah Nobles; Shoshona Le; Suxiang Tong; Vivien Dugan; Yvette Unoarumhi |
| EPI_ISL_2389995 | IMD - Institut für Medizinische Diagnostik Berlin-Potsdam | Robert Koch Institute | |
| EPI_ISL_2313977 | IMD Labor Frankfurt | Robert Koch Institute | |
| EPI_ISL_2689991 | IN State Department of Health Laboratory Services | Centers for Disease Control and Prevention Division of Viral Diseases, Pathogen Discovery | Alison Laufer Halpin; Ben L. Rambo-Martin; Clinton R. Paden; Dakota Howard; Darlene Wagner; Dave Wentworth; Dhwani Batra; Jasmine Padilla; Justin Lee; Katie Dillon; Krista Queen; Kristen Knipe; Kristine Lacek; Mark Burroughs; Matthew Schmerer; Mili Sheth; Peter Cook; Sam Shepard; Sarah Nobles; Shoshona Le; Suxiang Tong; Vivien Dugan; Yvette Unoarumhi |
| EPI_ISL_2439152, EPI_ISL_2439163, EPI_ISL_2439164, EPI_ISL_2439166, EPI_ISL_2439175, EPI_ISL_2439177, EPI_ISL_2439178, EPI_ISL_2439179, EPI_ISL_2439181, EPI_ISL_2544554, EPI_ISL_2544556, EPI_ISL_2544607, EPI_ISL_2544609, EPI_ISL_2544622, EPI_ISL_2544638, EPI_ISL_2768054 | | | |
| see above | IN State Department of Health Laboratory Services | IN State Department of Health Laboratory Services | Brian Pope; Cassandra Campion; Jamie Yeadon; Kyle Brownlee; Lixia Liu; Mark Glazier; Melissa Hindenlang |
| EPI_ISL_2602399, EPI_ISL_2602893 | INMI Lazzaro Spallanzani IRCCS | INMI Lazzaro Spallanzani IRCCS | A Massacci; B Bartolini; CEM Gruber; E Giombini; F Messina; F Santini; G Bonfiglio; G Orlandi; M Pallocca; M Rueca; O Butera; S Donzelli |
| EPI_ISL_2494022 | INSIDE CRSN CACHOEIRINHA/CASA VERDE | Instituto Butantan | Antonio Jorge Martins; Claudia Renata dos Santos Barros; David Schlesinger; Debora Botequio Moretti; Dimas Tadeu Covas; Elaine Cristina Marqueze; Elaine Vieira Santos; Evandra Strazza Rodrigues; Heidge Fukumasu; Jayme Augusto de Souza-Neto; José Salvatore Leister Patané; Luiz Alcantara; Luiz Lehmann Coutinho; Maria Carolina Elias; Maurício Lacerda Nogueira; Rafael dos Santos Bezerra; Raul Machado Neto; Rejane Maria Tommasini Grotto; Ricardo Haddad; Sandra Coccuzzo Sampaio Vessoni; Simone Kashima; Svetoslav Nanev Slavov; Vincent Louis Viala |
| EPI_ISL_2493969, EPI_ISL_2494037 | INSIDE CRSN FREGUESIA/BRASILÂNDIA | Instituto Butantan | Antonio Jorge Martins; Claudia Renata dos Santos Barros; David Schlesinger; Debora Botequio Moretti; Dimas Tadeu Covas; Elaine Cristina Marqueze; Elaine Vieira Santos; Evandra Strazza Rodrigues; Heidge Fukumasu; Jayme Augusto de Souza-Neto; José Salvatore Leister Patané; Luiz Alcantara; Luiz Lehmann Coutinho; Maria Carolina Elias; Maurício Lacerda Nogueira; Rafael dos Santos Bezerra; Raul Machado Neto; Rejane Maria Tommasini Grotto; Ricardo Haddad; Sandra Coccuzzo Sampaio Vessoni; Simone Kashima; Svetoslav Nanev Slavov; Vincent Louis Viala |
| EPI_ISL_2493974, EPI_ISL_2494274 | INSIDE CRSN PERUS | Instituto Butantan | Antonio Jorge Martins; Claudia Renata dos Santos Barros; David Schlesinger; Debora Botequio Moretti; Dimas Tadeu Covas; Elaine Cristina Marqueze; Elaine Vieira Santos; Evandra Strazza Rodrigues; Heidge Fukumasu; Jayme Augusto de Souza-Neto; José Salvatore Leister Patané; Luiz Alcantara; Luiz Lehmann Coutinho; Maria Carolina Elias; Maurício Lacerda Nogueira; Rafael dos Santos Bezerra; Raul Machado Neto; Rejane Maria Tommasini Grotto; Ricardo Haddad; Sandra Coccuzzo Sampaio Vessoni; Simone Kashima; Svetoslav Nanev Slavov; Vincent Louis Viala |
| EPI_ISL_2493966 | INSIDE CRSN VILA MARIA/VILA GUILHERME | Instituto Butantan | Antonio Jorge Martins; Claudia Renata dos Santos Barros; David Schlesinger; Debora Botequio Moretti; Dimas Tadeu Covas; Elaine Cristina Marqueze; Elaine Vieira Santos; Evandra Strazza Rodrigues; Heidge Fukumasu; Jayme Augusto de Souza-Neto; José Salvatore Leister Patané; Luiz Alcantara; Luiz Lehmann Coutinho; Maria Carolina Elias; Maurício Lacerda Nogueira; Rafael dos Santos Bezerra; Raul Machado Neto; Rejane Maria Tommasini Grotto; Ricardo Haddad; Sandra Coccuzzo Sampaio Vessoni; Simone Kashima; Svetoslav Nanev Slavov; Vincent Louis Viala |
| EPI_ISL_2493981, EPI_ISL_2494014 | INSIDE CRSO LAPA/PINHEIROS | Instituto Butantan | Antonio Jorge Martins; Claudia Renata dos Santos Barros; David Schlesinger; Debora Botequio Moretti; Dimas Tadeu Covas; Elaine Cristina Marqueze; Elaine Vieira Santos; Evandra Strazza Rodrigues; Heidge Fukumasu; Jayme Augusto de Souza-Neto; José Salvatore Leister Patané; Luiz Alcantara; Luiz Lehmann Coutinho; Maria Carolina Elias; Maurício Lacerda Nogueira; Rafael dos Santos Bezerra; Raul Machado Neto; Rejane Maria Tommasini Grotto; Ricardo Haddad; Sandra Coccuzzo Sampaio Vessoni; Simone Kashima; Svetoslav Nanev Slavov; Vincent Louis Viala |
| EPI_ISL_2493991 | INSIDE CRSSU MOOCA/ARICANDUVA | Instituto Butantan | Antonio Jorge Martins; Claudia Renata dos Santos Barros; David Schlesinger; Debora Botequio Moretti; Dimas Tadeu Covas; Elaine Cristina Marqueze; Elaine Vieira Santos; Evandra Strazza Rodrigues; Heidge Fukumasu; Jayme Augusto de Souza-Neto; José Salvatore Leister Patané; Luiz Alcantara; Luiz Lehmann Coutinho; Maria Carolina Elias; Maurício Lacerda Nogueira; Rafael dos Santos Bezerra; Raul Machado Neto; Rejane Maria Tommasini Grotto; Ricardo Haddad; Sandra Coccuzzo Sampaio Vessoni; Simone Kashima; Svetoslav Nanev Slavov; Vincent Louis Viala |

| | | | |
|---|---|---|---|
| EPI_ISL_2494013, EPI_ISL_2494021 | INSIDE CRSSU SAPOPEMBA/VILA PRUDENTE | Instituto Butantan | Antonio Jorge Martins; Claudia Renata dos Santos Barros; David Schlesinger; Debora Botequio Moretti; Dimas Tadeu Covas; Elaine Cristina Marqueze; Elaine Vieira Santos; Evandra Strazza Rodrigues; Heidge Fukumasu; Jayme Augusto de Souza-Neto; José Salvatore Leister Patané; Luiz Alcantara; Luiz Lehmann Coutinho; Maria Carolina Elias; Maurício Lacerda Nogueira; Rafael dos Santos Bezerra; Raul Machado Neto; Rejane Maria Tommasini Grotto; Ricardo Haddad; Sandra Coccuzzo Sampaio Vessoni; Simone Kashima; Svetoslav Nanev Slavov; Vincent Louis Viala |
| EPI_ISL_2493178, EPI_ISL_2493713, EPI_ISL_2493714, EPI_ISL_2493716, EPI_ISL_2493717, EPI_ISL_2493718, EPI_ISL_2493719, EPI_ISL_2493720, EPI_ISL_2493722, EPI_ISL_2493724, EPI_ISL_2493725, EPI_ISL_2493727, EPI_ISL_2493728, EPI_ISL_2493729, EPI_ISL_2493730, EPI_ISL_2493731, EPI_ISL_2493732, EPI_ISL_2493733, EPI_ISL_2493734, EPI_ISL_2493735, EPI_ISL_2493736, EPI_ISL_2493737, EPI_ISL_2493738, EPI_ISL_2494318, EPI_ISL_2500972, EPI_ISL_2500973 | | | |
| see above | INSIDE DIAGNÓSTICOS | Instituto Butantan | Antonio Jorge Martins; Claudia Renata dos Santos Barros; David Schlesinger; Debora Botequio Moretti; Dimas Tadeu Covas; Elaine Cristina Marqueze; Elaine Vieira Santos; Evandra Strazza Rodrigues; Heidge Fukumasu; Jayme Augusto de Souza-Neto; José Salvatore Leister Patané; Luiz Alcantara; Luiz Lehmann Coutinho; Maria Carolina Elias; Maurício Lacerda Nogueira; Rafael dos Santos Bezerra; Raul Machado Neto; Rejane Maria Tommasini Grotto; Ricardo Haddad; Sandra Coccuzzo Sampaio Vessoni; Simone Kashima; Svetoslav Nanev Slavov; Vincent Louis Viala |
| EPI_ISL_2493289, EPI_ISL_2493704, EPI_ISL_2493705, EPI_ISL_2493707 | INSIDE DIAGNÓSTICOS NORTE BRASILÂNDIA/FREGUESIA | Instituto Butantan | Antonio Jorge Martins; Claudia Renata dos Santos Barros; David Schlesinger; Debora Botequio Moretti; Dimas Tadeu Covas; Elaine Cristina Marqueze; Elaine Vieira Santos; Evandra Strazza Rodrigues; Heidge Fukumasu; Jayme Augusto de Souza-Neto; José Salvatore Leister Patané; Luiz Alcantara; Luiz Lehmann Coutinho; Maria Carolina Elias; Maurício Lacerda Nogueira; Rafael dos Santos Bezerra; Raul Machado Neto; Rejane Maria Tommasini Grotto; Ricardo Haddad; Sandra Coccuzzo Sampaio Vessoni; Simone Kashima; Svetoslav Nanev Slavov; Vincent Louis Viala |
| EPI_ISL_2493706 | INSIDE DIAGNÓSTICOS NORTE CACHOEIRINHA/CASA VERDE | Instituto Butantan | Antonio Jorge Martins; Claudia Renata dos Santos Barros; David Schlesinger; Debora Botequio Moretti; Dimas Tadeu Covas; Elaine Cristina Marqueze; Elaine Vieira Santos; Evandra Strazza Rodrigues; Heidge Fukumasu; Jayme Augusto de Souza-Neto; José Salvatore Leister Patané; Luiz Alcantara; Luiz Lehmann Coutinho; Maria Carolina Elias; Maurício Lacerda Nogueira; Rafael dos Santos Bezerra; Raul Machado Neto; Rejane Maria Tommasini Grotto; Ricardo Haddad; Sandra Coccuzzo Sampaio Vessoni; Simone Kashima; Svetoslav Nanev Slavov; Vincent Louis Viala |
| EPI_ISL_2488728, EPI_ISL_2757619, EPI_ISL_2757630 | INSPI-CRN DE INFLUENZA Y OTROS VIRUS RESPIRATORIOS | NIC-INSPI | Alfredo Bruno; Domenica de Mora.; Jimmy Garcés; Johanna Laines; Lizbeth Patiño; Manuel Gonzalez; Maritza Olmedo; Michelle Páez |
| EPI_ISL_2547524, EPI_ISL_2547525 | INT Fondazione Pascale | INT Fondazione Pascale | INT Fondazione Pascale |
| EPI_ISL_2716832, EPI_ISL_2716833, EPI_ISL_2774479, EPI_ISL_2774480, EPI_ISL_2774486, EPI_ISL_2774487, EPI_ISL_2774488, EPI_ISL_2774491, EPI_ISL_2774492 | | | |
| see above | IRCCS San Gallicano Dermatological Institute | IRCCS Regina Elena National Cancer Institute | Aldo Morrone; Alice Massacci; Antonio Federico; Carla Mottini; Cesare Ernesto Maria Gruber; Eleonora Sperandio; Emanuela Giombini; Fabrizio Ensoli; Francesca De Nicola; Francesca Maione; Francisco Obregon; Frauke Goeman; Fulvia Fraticelli; Fulvia Pimpinelli; Gennaro Ciliberto; Giovanni Blandino; Giulia Orlandi; Maria Rosaria Capobianchi; Martina Diano; Martina Pontone; Matteo Pallocca; Maurizio Fanciulli; Sabrina Strano; Sara Donzelli |
| EPI_ISL_2693359, EPI_ISL_2693365, EPI_ISL_2693367 | ISDH | Quantigen Biosciences | Paul J. Childress |
| EPI_ISL_2789974, EPI_ISL_2789975, EPI_ISL_2789977, EPI_ISL_2801467 | IVY3 Central Lab, Vanderbilt University Medical Center | Lauring Lab, University of Michigan, Department of Microbiology and Immunology | Valesano |
| EPI_ISL_3064189 | Idaho Bureau of Laboratories | Idaho Bureau of Laboratories | "R. Beukelman; Aimee Ceniseros; Christian Loera; Christopher Ball"; Matthew Charles Burns; Robert L. Voermans |
| EPI_ISL_2801137, EPI_ISL_2801142 | Illinois Department of Public Health | Gagnon Lab, Southern Illinois University | Keith Gagnon |
| EPI_ISL_2300549, EPI_ISL_2467386, EPI_ISL_2467450, EPI_ISL_2467451, EPI_ISL_2467479, EPI_ISL_2467480, EPI_ISL_2467483, EPI_ISL_2467486, EPI_ISL_2467488, EPI_ISL_2467490, EPI_ISL_2467491, EPI_ISL_2467501, EPI_ISL_2467502, EPI_ISL_2467505, EPI_ISL_2467506, EPI_ISL_2467510, EPI_ISL_2800747, EPI_ISL_2800754, EPI_ISL_2800760, EPI_ISL_2800822, EPI_ISL_2800839, EPI_ISL_2800858, EPI_ISL_2800866 | | | |
| see above | Illinois Department of Public Health | Illinois Department of Public Health - Chicago Lab | Ira Heimler; Joel Price; Vineet K. Dhiman |
| EPI_ISL_2339346, EPI_ISL_2339402, EPI_ISL_2339405, EPI_ISL_2339410, EPI_ISL_2339411, EPI_ISL_2339412, EPI_ISL_2339413, EPI_ISL_2339416, EPI_ISL_2339419, EPI_ISL_2339424, EPI_ISL_2339425, EPI_ISL_2339426, EPI_ISL_2339432, EPI_ISL_2339433, EPI_ISL_2339434, EPI_ISL_2339435, EPI_ISL_2403013, EPI_ISL_2403027, EPI_ISL_2403057, EPI_ISL_2403058, EPI_ISL_2403059, EPI_ISL_2403060, EPI_ISL_2403065, EPI_ISL_2403072, EPI_ISL_2403073, EPI_ISL_2403090, EPI_ISL_2403091, EPI_ISL_2403092, EPI_ISL_2403097, EPI_ISL_2403098, EPI_ISL_2403099, EPI_ISL_2403101, EPI_ISL_2403103, EPI_ISL_2403110, EPI_ISL_2403117, EPI_ISL_2403127, EPI_ISL_2403128, EPI_ISL_2403131, EPI_ISL_2403137, EPI_ISL_2403145, EPI_ISL_2403185, EPI_ISL_2598182 | | | |
| see above | Illinois Department of Public Health - Springfield Lab | Illinois Department of Public Health - Springfield Lab | Bryan Sim; Gordon McCall |
| EPI_ISL_2533928 | Imágenes Diagnósticas de Mazatlán | Microbial Genomics Laboratory | ; Alejandra García-Gasca; Alejandra Hernández-Terán; Alejandro Sánchez-Flores; Alfredo Herrera-Estrella; Alicia Ocaña-Mondragón; Andreu Comas-García; Angel Gustavo Salas-Lais; Antonio Loza Román; Bernardo Martínez-Miguel; Blanca Taboada; Brenda Irasema Maldonado-Meza; Bruno Gómez-Gil; Carla Ivón Herrera-Najera; Carlos F. Arias; Celia Boukadida; Clara Esperanza Santacruz-Tinoco; Concepción Grajales-Muñiz; Consorcio Mexicano de Vigilancia Genómica (CoViGen-Mex). Authors (in alphabetical order): Julio Elias Alvarado-Yaah; Cristóbal Cháidez-Quiróz; Célida Duque Molina; Célida Martínez- Rodríguez; Daniel Fregoso-Rueda; Daniel Lira Morales; Eduardo Becerril-Vargas; Fernando Fontove-Herrera; Fidencio Mejía-Nepomuceno; Francisco Pulido; Gloria Elena Espinosa-Ayala; Gloria María Molina-Salinas; Gloria Vazquez; Hector Esteban Paz-Juárez; Hector Montoya-Fuentes; Helen Haydee Fernanda Ramírez-Plascencia; Irvin González-López; Jean Pierre González; Jesús Hernández; Joel Armando Vázquez-Pérez.; Jorge Salas-Hernández; José Antonio Enciso-Moreno; José Arturo Martínez-Orozco; José Esteban Muñoz-Medina; José de Jesús Nuñez-Contreras; Juan Bautista Chale-Dzul; Julissa Enciso-Ibarra; Luis Alberto Ochoa-Carrera; Margarita Matías-Florentino; Mario Mújica-Sánchez; Marissa Perez-Garcia; María Guadalupe Santiago-Mauricio; María Guadalupe de Jesús Mireles-Rivera; Nelly Sélem-Mojica; Pavel Isa; Ricardo Ciria Merce; Ricardo Grande; Rosa María Gutiérrez Rios; Santiago Ávila-Ríos; Selene Zárate; Susana Lopez; Verónica Mata-Haro; Victor Eduardo García-Arias; Víctor Hugo Borja-Aburto |
| EPI_ISL_2715085, EPI_ISL_2715093 | Incyte Diagnostics Spokane | Seattle Flu Study | Amanda Adler; Barry R. Lutz; Benjamin Pelle; Brian Hiatt; Caitlin R. Wolf; Chris D. Frazar; Deborah A. Nickerson; Elisabeth Brandstetter; Erica Ryke; Geoff Melly; Helen Y. Chu; Janet A. Englund; Jay Shendure; Jover Lee; Kairsten Fay; Kirsten Lacombe; Lea M. Starita; Mark J. Rieder; Matthew Richardson; Matthew Thompson; Melissa Truong; Michael Boeckh; Michael Famulare; Misja Ilcisin; Peter D. Han; Philip Dykema; Romesh Gautom; Scott Lindquist; Thomas R. Sibley; Trevor Bedford |
| EPI_ISL_2660337, EPI_ISL_2660339, EPI_ISL_2660351, EPI_ISL_2660355, EPI_ISL_2660360, EPI_ISL_2660363, EPI_ISL_2660370, EPI_ISL_2660374 | | | |
| see above | Infectious Disease Program, Broad Institute of Harvard and MIT | Infectious Disease Program, Broad Institute of Harvard and MIT | A.E.; Adams, G.; Anahtar, M.; B.L.; B.W.; Bauer, M.; Birren; Branda, J.; Carter, A.; Cerrato, F.; Chaluvadi, S.; Chapman; Cusick, C.; D.J.; DeRuff, K.; Flowers, K.; Gallagher, G.; Gladden-Young, A.; Gnirke, A.; Harris, J.; J.E.; K.J.; LaRocque, R.; Lagerborg, K.; Lemieux; Lin; Loreth, C.; MacInnis; Neumann, A.; Normandin, E.; P.C.; Park; Pierce, V.; Reilly, S.; Rosenberg, E.; Rudy, M.; Ryan, E.; S.B.; Sabeti; Shaw, B.; Siddle; Slater, D.; Smole, S.; Tomkins-Tinch, C.; Turbett, S. |
| EPI_ISL_2323498, EPI_ISL_2375555, EPI_ISL_2375615, EPI_ISL_2375624, EPI_ISL_2375679, EPI_ISL_2375707, EPI_ISL_2375785, EPI_ISL_2652549, EPI_ISL_2652627, EPI_ISL_2652711, EPI_ISL_2652728, EPI_ISL_2652778, EPI_ISL_2652797, EPI_ISL_2652818, EPI_ISL_2652854, EPI_ISL_2652870, EPI_ISL_2652884, EPI_ISL_2652907, EPI_ISL_2652924, EPI_ISL_2652938, EPI_ISL_2652968 | | | |
| see above | Infinity Biologix | Centers for Disease Control and Prevention Division of Viral Diseases, Pathogen Discovery | Adrian Paskey; Benjamin Rambo-Martin; Chirayu Goswami; Christian Bixby; Christopher Gulvick; Clinton R. Paden; Dakota Howard; Darlene Wagner; Dhwani Batra; Duncan MacCannell; Jason Caravas; Jonathan Schultz; Kara Moser; Matthew Schmerer; Peter W. Cook; Robin Grimwood; Russ Hager; Scott Sammons; Shatavia Morrison; Yihe Wang; Yvette Unoarumhi |
| EPI_ISL_2828663, EPI_ISL_2828664, EPI_ISL_2828665 | Instituo Adolfo Lutz - Reginal de Ribeirao Preto | Instituto Adolfo Lutz, Interdiciplinary Procedures Center, Strategic Laboratory | Caio Vinicius Dias Lopes; Claudia Regina Gonçalves; Claudio Tavares Sacchi; Erica Valessa Ramos Gomes; Karoline Rodrigues Campos |
| EPI_ISL_2691151, EPI_ISL_2691152, EPI_ISL_2691153, EPI_ISL_2691154, EPI_ISL_2691156, EPI_ISL_2691159, EPI_ISL_2691160, EPI_ISL_2691161, EPI_ISL_2691162, EPI_ISL_2691163, EPI_ISL_2691164, EPI_ISL_2691165, EPI_ISL_2691166, EPI_ISL_2691167, EPI_ISL_2691168, EPI_ISL_2691169, EPI_ISL_2691170, EPI_ISL_2828658, EPI_ISL_2828660 | | | |
| see above | Instituo Adolfo Lutz Central | Instituto Adolfo Lutz, Interdiciplinary Procedures Center, Strategic Laboratory | Caio Vinicius Dias Lopes; Claudia Regina Gonçalves; Claudio Tavares Sacchi; Erica Valessa Ramos Gomes; Karoline Rodrigues Campos; Leonardo Jose Tadeu de Araujo |
| EPI_ISL_2691173, EPI_ISL_2691174, EPI_ISL_2691175, EPI_ISL_2691176, EPI_ISL_2691177, EPI_ISL_2691178, EPI_ISL_2691180, EPI_ISL_2691181, EPI_ISL_2691182, EPI_ISL_2691183, EPI_ISL_2691184, EPI_ISL_2691185, EPI_ISL_2691186, EPI_ISL_2691187, EPI_ISL_2691188, EPI_ISL_2691189, EPI_ISL_2691190, EPI_ISL_2691191, EPI_ISL_2691192, EPI_ISL_2691193, EPI_ISL_2691194, EPI_ISL_2691195, EPI_ISL_2691196, EPI_ISL_2691197, EPI_ISL_2691198, EPI_ISL_2691199, EPI_ISL_2691200, EPI_ISL_2691201, EPI_ISL_2691202, EPI_ISL_2691203, EPI_ISL_2828647, EPI_ISL_2828648, EPI_ISL_2828649, EPI_ISL_2828650, EPI_ISL_2828651, EPI_ISL_2828652, EPI_ISL_2828653 | | | |
| see above | Instituto Adolfo Lutz - Reginal de Sao José do Rio Preto | Instituto Adolfo Lutz, Interdiciplinary Procedures Center, Strategic Laboratory | Caio Vinicius Dias Lopes; Claudia Regina Gonçalves; Claudio Tavares Sacchi; Erica Valessa Ramos Gomes; Karoline Rodrigues Campos; Leonardo Jose Tadeu de Araujo |
| EPI_ISL_2919243, EPI_ISL_2919244, EPI_ISL_2919245, EPI_ISL_2919246, EPI_ISL_2919254, EPI_ISL_3316169, EPI_ISL_5863741 | | | |
| see above | Instituto Adolfo Lutz - Regional de Bauru | Instituto Adolfo Lutz, Interdiciplinary Procedures Center, Strategic Laboratory | Ariadne Ferreira Amarante; Caio Vinicius Dias Lopes; Claudia Regina Gonçalves; Claudio Tavares Sacchi; Erica Valessa Ramos Gomes; Karoline Rodrigues Campos; Leonardo Tadeu de Araujo; Marlon Benedito Nascimento Santos; Marlon Benedito Nascomento Santos |
| EPI_ISL_3316221 | Instituto Adolfo Lutz - Regional de Campinas | Instituto Adolfo Lutz, Interdiciplinary Procedures Center, Strategic Laboratory | Caio Vinicius Dias Lopes; Claudia Regina Gonçalves; Claudio Tavares Sacchi; Karoline Rodrigues Campos; Leonardo Tadeu de Araujo; Marlon Benedito Nascomento Santos |
| EPI_ISL_3316174, EPI_ISL_3316228 | Instituto Adolfo Lutz - Regional de Presidente Prudente | Instituto Adolfo Lutz, Interdiciplinary Procedures Center, Strategic Laboratory | Caio Vinicius Dias Lopes; Claudia Regina Gonçalves; Claudio Tavares Sacchi; Karoline Rodrigues Campos; Leonardo Tadeu de Araujo; Marlon Benedito Nascomento Santos |
| EPI_ISL_2756431, EPI_ISL_2958848, EPI_ISL_2958849, EPI_ISL_2958850, EPI_ISL_2958851, EPI_ISL_2958852, EPI_ISL_2958853 | | | |
| see above | Instituto Adolfo Lutz Central | Instituto Adolfo Lutz, Interdiciplinary Procedures Center, Strategic Laboratory | Caio Vinicius Dias Lopes; Claudia Regina Gonçalves; Claudio Tavares Sacchi; Erica Valessa Ramos Gomes; Karoline Rodrigues Campos; Leonardo Jose Tadeu de Araujo |
| EPI_ISL_2646123, EPI_ISL_2646131 | Instituto Nacional de Medicina Genomica | Centro de Investigación en Enfermedades Infecciosas (CIENI), Instituto Nacional de Enfermedades Respiratorias (INER) | Avila-Rios S; Boukadida C; Cedro-Tanda A; Herrera-Montalvo LA; Hidalgo-Miranda A; Matias-Florentino M; Mendoza-Vargas A; Perez-Garcia M; Reyes-Grajeda JP; Reyes-Teran G |
| EPI_ISL_2350106, EPI_ISL_2350111, EPI_ISL_2350113 | Instituto Nacional de Medicina Genomica | Instituto Nacional de Medicina Genomica | Cedro-Tanda A; Cisneros- Villanueva M; Escobar-Arrazola; Gonzalez-Barrera D; Herrera-Montalvo LA.; Hidalgo-Miranda A; M; Mendoza-Vargas A; Munguia-Garza P; Ramirez-Vega O; Rangel-DeLeon D; Reyes-Grajeda JP |
| EPI_ISL_2500971 | Instituto Nacional de Salud | Instituto Nacional de Salud-Dirección de Investigación en Salud Pública | Carlos Franco-Muñoz; Carmen Osorio; Diana Malo; Diego A. Álvarez-Díaz; Diego Andrés Prada; Gerardo Santamaría; Hector Alejandro Ruiz-Moreno; Jhonnatan Reales-González; Jorge Rivera; Juan Camilo Martinez; Julian Naizaque; Katherine Laiton-Donato; Lisseth Pardo; Magdalena Wiesner; Marcela Mercado-Reyes; Maria T. Herrera-Sepúlveda; Marta Lopez Blanco; Martha Lucia Ospina Martinez; Paola Rojas; Sergio Gomez; Sheryll Corchuelo; Ángela Alarcon Cruz |
| EPI_ISL_2378698, EPI_ISL_3118816, EPI_ISL_3118817, EPI_ISL_3118819, EPI_ISL_3118820, EPI_ISL_3118828, EPI_ISL_3118830, EPI_ISL_3118833, EPI_ISL_3118834, EPI_ISL_3118835, EPI_ISL_3118836, EPI_ISL_3118840, EPI_ISL_3118842, EPI_ISL_3118844, EPI_ISL_3118846 | | | |
| see above | Instituto de Biotecnologia - UNESP-Botucatu-SP | Instituto de Biotecnologia - UNESP-Botucatu-SP | Cecília Artico Banho; Cíntia Bittar; Fábio Sossai Possebon; Guilherme Campos; Helena Lage Ferreira; Jorge A. Petroli Marchesi; João Pessoa Araújo Jr.; Leila Sabrina Ullmann; Lívia Sacchetto; Maisa C. Pereira Parra; Marília Moraes; Maurício L. Nogueira; Paula Rahal; Paulo Inacio da Costa |

| | | | |
|---|---|---|---|
| EPI_ISL_3104702, EPI_ISL_3236435, EPI_ISL_3236436 | Instituto de Medicina Tropical & Salud Global Universidad Iberoamericana | Grubaugh Lab - Yale School of Public Health | Alejandaro Vallejo Degaudenzi; Anderson Brito; Andrea Firpo; Annie Watkins; Chaney Kalinich; Chantal Vogels; David Rivera Aponte; Elisa Contreras; Ernesto Vazquez; Esperanza Mendoza; Fabiola Cruz-Lopez; Fabiola Fontanet; Isabel Ott; Jessica Rothman; Joseph Fauver; Karolane Gonzalez; Kendall Billig; Mallery Breban; Mary Petrone; Natalie Machargo; Nathan Grubaugh; Patricia Serrano; Robert Paulino-Ramirez; Tania Mitwalli; Tara Alpert; Tobias Koch; Vianca Aponte; Victor Virgilio Calderon |
| EPI_ISL_5147025 | Instituto de Medicina Tropical Alexander Von Humboldt, Sede Cusco | Laboratorio de Genómica Microbiana, Universidad Peruana Cayetano Heredia | Alejandra Dávila-Barclay; Diego Cuicapuza; Fátima Concha; Guillermo Salvatierra; Janet Huancachoque; Luis González; Maria Luisa Morales; Martha Fernández; Miguel Cabada; Pablo Tsukayama; Pedro E. Romero; Pool Marcos |
| EPI_ISL_2636258 | Israel Central Virology laboratory | Israel National Consortium for SARS-CoV-2 sequencing | Dana Bar-Ilan; Efrat Dahan Bucris; Efrat Glick-Saar; Ella Mendelson; Gideon Rechavi; Michal Mandelboim; Miranda Geva; Neta Zuckerman; Omri Nayshool; Oran Erster; Orna Mor |
| EPI_ISL_2275794, EPI_ISL_2275799, EPI_ISL_2275802, EPI_ISL_2275803, EPI_ISL_2275892, EPI_ISL_2275898, EPI_ISL_2275900, EPI_ISL_2275934, EPI_ISL_2276034, EPI_ISL_2276046, EPI_ISL_2276048, EPI_ISL_2276051, EPI_ISL_2276056, EPI_ISL_2282142, EPI_ISL_2282143, EPI_ISL_2282148, EPI_ISL_2282209, EPI_ISL_2282210, EPI_ISL_2282211, EPI_ISL_2282223, EPI_ISL_2282224, EPI_ISL_2282227, EPI_ISL_2357159, EPI_ISL_2357181, EPI_ISL_2357186, EPI_ISL_2357189, EPI_ISL_2357199, EPI_ISL_2357211, EPI_ISL_2357214, EPI_ISL_2357217, EPI_ISL_2357220, EPI_ISL_2455740, EPI_ISL_2455745, EPI_ISL_2455757, EPI_ISL_2455821, EPI_ISL_2455854, EPI_ISL_2455855 | | | |
| see above | Istituto Zooprofilattico Sperimentale del Mezzogiorno | TIGEM | Antonio Grimaldi Patrizia Annunziata Francesco Panariello Biancamaria Pierri Claudia Tiberio Teresa Giuliano Valentina Bouche Chiara Colantuono Maria Concetta Cuomo Denise Di Concilio Lucio Di Filippo Anna Manfredi Marcello Salvi Antonio Limone Luigi Atripaldi Pellegrino Cerino Andrea Ballabio Davide Cacchiarelli |
| EPI_ISL_2275950, EPI_ISL_2275951, EPI_ISL_2275952 | Istituto Zooprofilattico Sperimentale del Mezzogiorno - Azienda Ospedaliera "Pugliese Ciaccio" di Catanzaro | TIGEM | Antonio Grimaldi Patrizia Annunziata Francesco Panariello Biancamaria Pierri Claudia Tiberio Teresa Giuliano Valentina Bouche Chiara Colantuono Maria Concetta Cuomo Denise Di Concilio Lucio Di Filippo Anna Manfredi Pasquale Minchella Marcello Salvi Antonio Limone Luigi Atripaldi Pellegrino Cerino Andrea Ballabio Davide Cacchiarelli |
| EPI_ISL_2322376, EPI_ISL_2322377 | Jessa | Jessa | Berden et al. on behalf of the Jessa_cmdLab |
| EPI_ISL_2692070, EPI_ISL_2692072, EPI_ISL_2692074, EPI_ISL_2692076 | John H Stroger Jr Hospital of Cook County | RIPHL at Rush University Medical Center | Ah-Young Sung; Felix Araujo Perez; Kevin Kunstman; Laura Furtado; Marieta Hyde; Max Kolton; Sharon Welbel; Stefan Green |
| EPI_ISL_2451581, EPI_ISL_2645292, EPI_ISL_2645297 | KS Health and Environmental Laboratories | Centers for Disease Control and Prevention Division of Viral Diseases, Pathogen Discovery | Alison Laufer Halpin; Ben L. Rambo-Martin; Clinton R. Paden; Dakota Howard; Darlene Wagner; Dave Wentworth; Dhwani Batra; Jasmine Padilla; Justin Lee; Katie Dillon; Krista Queen; Kristen Knipe; Kristine Lacek; Mark Burroughs; Matthew Schmerer; Mili Sheth; Peter Cook; Sam Shepard; Sarah Nobles; Shoshona Le; Suxiang Tong; Vivien Dugan; Yvette Unoarumhi |
| EPI_ISL_2725618, EPI_ISL_2725651, EPI_ISL_2726466, EPI_ISL_2726474, EPI_ISL_2834757, EPI_ISL_2834791, EPI_ISL_2834831, EPI_ISL_2834840, EPI_ISL_2839469, EPI_ISL_2839473, EPI_ISL_2839488, EPI_ISL_2839490, EPI_ISL_2840450, EPI_ISL_2840452, EPI_ISL_2864231, EPI_ISL_2864243, EPI_ISL_2864244 | | | |
| see above | KU Leuven, Rega Institute, Clinical and Epidemiological Virology | KU Leuven, Rega Institute, Clinical and Epidemiological Virology | Bert Vanmechelen; Joan Marti-Carerras; Piet Maes; Tony Wawina-Bokalanga |
| EPI_ISL_3547385, EPI_ISL_3561008, EPI_ISL_3561166, EPI_ISL_3561299, EPI_ISL_3561350, EPI_ISL_3561383, EPI_ISL_3561386, EPI_ISL_3561387, EPI_ISL_3561389, EPI_ISL_3561390, EPI_ISL_3561400, EPI_ISL_3561403, EPI_ISL_3561413, EPI_ISL_3561527, EPI_ISL_3561531, EPI_ISL_3561549, EPI_ISL_3561558, EPI_ISL_3561576, EPI_ISL_3561614, EPI_ISL_3561809, EPI_ISL_3561836, EPI_ISL_3561840, EPI_ISL_3561955, EPI_ISL_3562007, EPI_ISL_3562049, EPI_ISL_3562077, EPI_ISL_3562079, EPI_ISL_3562085, EPI_ISL_3562090, EPI_ISL_4005454 | | | |
| see above | Kaiser Permanente NW Reginal Lab | OHSU MM Lab | Yun Wu |
| EPI_ISL_3504861, EPI_ISL_3504868, EPI_ISL_3504870, EPI_ISL_3504873, EPI_ISL_3504886, EPI_ISL_3504887, EPI_ISL_3504889, EPI_ISL_3504905, EPI_ISL_3504907, EPI_ISL_3504914, EPI_ISL_3504923, EPI_ISL_3504930, EPI_ISL_3504947, EPI_ISL_3504962, EPI_ISL_3504965, EPI_ISL_3504969, EPI_ISL_3504980, EPI_ISL_3504983, EPI_ISL_3505060, EPI_ISL_3505063, EPI_ISL_3505065, EPI_ISL_3505071, EPI_ISL_3505080, EPI_ISL_3505086, EPI_ISL_3505087, EPI_ISL_3505091, EPI_ISL_3505109, EPI_ISL_3505112, EPI_ISL_3505117, EPI_ISL_3505123, EPI_ISL_3505124, EPI_ISL_3505125, EPI_ISL_3505127, EPI_ISL_3505128, EPI_ISL_3505130 | | | |
| see above | Kaiser Permanente NW Reginal Lab | OHSU-MM Lab | Yun Wu |
| EPI_ISL_2933983, EPI_ISL_2934066 | Kaiser Regional | Santa Clara County Public Health Laboratory | Santa Clara County Public Health Department |
| EPI_ISL_2650540 | Kaiser San Jose | Santa Clara County Public Health Laboratory | Santa Clara County Public Health Department |
| EPI_ISL_2230470, EPI_ISL_2230472, EPI_ISL_2248751, EPI_ISL_2276095, EPI_ISL_2289071, EPI_ISL_2289072, EPI_ISL_2289073, EPI_ISL_2289074 | | | |
| see above | Kansas Health and Environmental Lab | Kansas Health and Environmental Lab | Ben Olsen; Jonathan Barnell; Katherine Wiggins; Mike Grose; and Phil Adam |
| EPI_ISL_2227442, EPI_ISL_2227446, EPI_ISL_2227451, EPI_ISL_2227454 | Kansas Health and Environmental Lab | Wichita State University - Molecular Diagnostics Lab | Ben Olsen; Jonathan Barnell; Katherine Wiggins; Mike Grose; and Phil Adam |
| EPI_ISL_2898360, EPI_ISL_2898414 | Karolinska University Hospital Solna | Karolinska University Hospital | Annelie Bjerkner; Isak Sylvin; Jan Albert; Karolina Ininbergs; Lina Guerra Blomqvist; Lynda Eneh; Martin Ekman; Martina Wahlund; Robert Dyrdak; Sandra Broddesson; Tanja Normark; Tobias Allander; Valtteri Wirta; Zhibing Yun |
| EPI_ISL_4080577, EPI_ISL_4080594, EPI_ISL_4195028, EPI_ISL_4348307 | Kentucky State Public Health Lab | Kentucky State Public Health Lab | Anjayya Tummala; Chitralipi Kattunga; Joshua Tobias; Karim George; Matthew Johnson; Rachel Zinner; Rhonda Lucas; Vaneet Arora; William Grooms |
| EPI_ISL_2482670, EPI_ISL_2482673 | LABORATOIRE CREAVALLEE | CNR Virus des Infections Respiratoires - France SUD | Antonin Bal; Bruno Lina; Bruno Simon; Gregory Destras; Gwendolyne Burfin; Hadrien Regue; Laurence Josset; Martine Valette; Quentin Semanas |
| EPI_ISL_2482522, EPI_ISL_2482523 | LABORATOIRE DE BIOLOGIE MEDICALE | CNR Virus des Infections Respiratoires - France SUD | Antonin Bal; Bruno Lina; Bruno Simon; Gregory Destras; Gwendolyne Burfin; Hadrien Regue; Laurence Josset; Martine Valette; Quentin Semanas |
| EPI_ISL_2493646, EPI_ISL_2493647, EPI_ISL_2493648 | LABORATORIO DE FRANCA | Instituto Butantan | Antonio Jorge Martins; Claudia Renata dos Santos Barros; David Schlesinger; Debora Botequio Moretti; Dimas Tadeu Covas; Elaine Cristina Marqueze; Elaine Vieira Santos; Evandra Strazza Rodrigues; Heidge Fukumasu; Jayme Augusto de Souza-Neto; José Salvatore Leister Patané; Luiz Alcantara; Luiz Lehmann Coutinho; Maria Carolina Elias; Maurício Lacerda Nogueira; Rafael dos Santos Bezerra; Raul Machado Neto; Rejane Maria Tommasini Grotto; Ricardo Haddad; Sandra Coccuzzo Sampaio Vessoni; Simone Kashima; Svetoslav Nanev Slavov; Vincent Louis Viala |
| EPI_ISL_2651229 | LABORATORIO HEMATOLOGICO | Universidad Nacional de Colombia - Laboratorio Genómico One Health | Andres F. Cardona-Rios; Carlos Franco-Muñoz; Carolina Muñoz-Arango; Celeny Ortiz; Daniel O. Maldonado-Perez; Diego A. Álvarez-Díaz; Hector Alejandro Ruiz-Moreno; Idabely Betancur Ortiz; Jorge E. Osorio; Juan P. Hernandez-Ortiz; Karl A Ciuoderis; Katherine Laiton-Donato; Laura Silvana Perez; Lina M. Hurtado; Marcela Mercado-Reyes; Maria Angélica Maya; Maria Stella López; Rita Almanza Payares; Sandra Ines Cano; Simón Villegas Velásquez |
| EPI_ISL_2493684, EPI_ISL_2493702, EPI_ISL_2493703 | LABORATORIO MUNICIPAL DE ANALISES CLINICAS DE ITANHAEM | Instituto Butantan | Antonio Jorge Martins; Claudia Renata dos Santos Barros; David Schlesinger; Debora Botequio Moretti; Dimas Tadeu Covas; Elaine Cristina Marqueze; Elaine Vieira Santos; Evandra Strazza Rodrigues; Heidge Fukumasu; Jayme Augusto de Souza-Neto; José Salvatore Leister Patané; Luiz Alcantara; Luiz Lehmann Coutinho; Maria Carolina Elias; Maurício Lacerda Nogueira; Rafael dos Santos Bezerra; Raul Machado Neto; Rejane Maria Tommasini Grotto; Ricardo Haddad; Sandra Coccuzzo Sampaio Vessoni; Simone Kashima; Svetoslav Nanev Slavov; Vincent Louis Viala |
| EPI_ISL_2493379, EPI_ISL_2493380, EPI_ISL_2493381, EPI_ISL_2493397, EPI_ISL_2493398, EPI_ISL_2493403, EPI_ISL_2494151, EPI_ISL_2494299 | | | |
| see above | LABORATORIO MUNICIPAL DE ANALISES CLINICAS DE RIO CLARO | Instituto Butantan | Antonio Jorge Martins; Claudia Renata dos Santos Barros; David Schlesinger; Debora Botequio Moretti; Dimas Tadeu Covas; Elaine Cristina Marqueze; Elaine Vieira Santos; Evandra Strazza Rodrigues; Heidge Fukumasu; Jayme Augusto de Souza-Neto; José Salvatore Leister Patané; Luiz Alcantara; Luiz Lehmann Coutinho; Maria Carolina Elias; Maurício Lacerda Nogueira; Rafael dos Santos Bezerra; Raul Machado Neto; Rejane Maria Tommasini Grotto; Ricardo Haddad; Sandra Coccuzzo Sampaio Vessoni; Simone Kashima; Svetoslav Nanev Slavov; Vincent Louis Viala |
| EPI_ISL_2493150, EPI_ISL_2493186, EPI_ISL_2493190, EPI_ISL_2493649, EPI_ISL_2493653, EPI_ISL_2493696, EPI_ISL_2494154, EPI_ISL_2494157 | | | |
| see above | LABORATORIO MUNICIPAL DE PIRACICABA | Instituto Butantan | Antonio Jorge Martins; Claudia Renata dos Santos Barros; David Schlesinger; Debora Botequio Moretti; Dimas Tadeu Covas; Elaine Cristina Marqueze; Elaine Vieira Santos; Evandra Strazza Rodrigues; Heidge Fukumasu; Jayme Augusto de Souza-Neto; José Salvatore Leister Patané; Luiz Alcantara; Luiz Lehmann Coutinho; Maria Carolina Elias; Maurício Lacerda Nogueira; Rafael dos Santos Bezerra; Raul Machado Neto; Rejane Maria Tommasini Grotto; Ricardo Haddad; Sandra Coccuzzo Sampaio Vessoni; Simone Kashima; Svetoslav Nanev Slavov; Vincent Louis Viala |
| EPI_ISL_3404821, EPI_ISL_3404822, EPI_ISL_3404823, EPI_ISL_3404824, EPI_ISL_3404826, EPI_ISL_3404827, EPI_ISL_3404828, EPI_ISL_3404829, EPI_ISL_3404835, EPI_ISL_3404836, EPI_ISL_3404837, EPI_ISL_4395331 | | | |
| see above | LACEN | LGBio (Laboratorio de Genetica & Biodiversidade) | Alex Honda Bernardes; Amanda Alves de Melo; Aparecido Divino da Cruz; Cintia Pelegrineti Targueta de Azevedo Brito; Daniela de Melo e Silva; Elisangela de Paula Silveira Lacerda; Juliana Santana de Curcio; Luiz Augusto Pereira; Marc Alexandre Duarte Gigonzac; Mariana Pires de Campos Telles; Ramilla dos Santos Braga; Renata de Oliveira Dias; Rhewter Nunes; Thais Cidália Vieira Gigonzac; Thais Guimarães Castro; Thays Millena Alves Pedroso |
| EPI_ISL_2488814, EPI_ISL_2488815, EPI_ISL_2488816, EPI_ISL_2488817, EPI_ISL_2488818, EPI_ISL_2488819 | LACEN - Laboratório Central de Saúde Pública do Para | Evandro Chagas Institute | A.M.; Barbagelata; E.C.; E.M.A.; Ferreira; J.A.; Junior; K.C.; L.C.; L.S.; M.C.; P.S.; Pinheiro; Santos; Silva; Sousa; Sousa Junior; W.D.C.; da Silva |
| EPI_ISL_3316308, EPI_ISL_3316309, EPI_ISL_3316310, EPI_ISL_3671906 | LACEN do Distrito Federal | Instituto Adolfo Lutz, Interdiciplinary Procedures Center, Strategic Laboratory | Caio Vinicius Dias Lopes; Claudia Regina Gonçalves; Claudio Tavares Sacchi; Karoline Rodrigues Campos; Leonardo Tadeu de Araujo; Marlon Benedito Nascomento Santos |
| EPI_ISL_3316160, EPI_ISL_3316176, EPI_ISL_3316177, EPI_ISL_3316180, EPI_ISL_3316188, EPI_ISL_3316217, EPI_ISL_3316219, EPI_ISL_3316230 | | | |
| see above | LACEN do Estado de Mato Grosso | Instituto Adolfo Lutz, Interdiciplinary Procedures Center, Strategic Laboratory | Caio Vinicius Dias Lopes; Claudia Regina Gonçalves; Claudio Tavares Sacchi; Karoline Rodrigues Campos; Leonardo Tadeu de Araujo; Marlon Benedito Nascomento Santos |
| EPI_ISL_2919205, EPI_ISL_2919208, EPI_ISL_2919209, EPI_ISL_3316213 | LACEN do Estado de Tocantins | Instituto Adolfo Lutz, Interdiciplinary Procedures Center, Strategic Laboratory | Caio Vinicius Dias Lopes; Claudia Regina Gonçalves; Claudio Tavares Sacchi; Erica Valessa Ramos Gomes; Karoline Rodrigues Campos; Leonardo Tadeu de Araujo; Marlon Benedito Nascomento Santos |
| EPI_ISL_3545806, EPI_ISL_3545807, EPI_ISL_3545808, EPI_ISL_3691363, EPI_ISL_3691364, EPI_ISL_3691365, EPI_ISL_3691366, EPI_ISL_3691367 | | | |
| see above | LACEN do Estado do Mato Grosso do Sul | Instituto Adolfo Lutz, Interdiciplinary Procedures Center, Strategic Laboratory | Caio Vinicius Dias Lopes; Claudia Regina Gonçalves; Claudio Tavares Sacchi; Karoline Rodrigues Campos; Leonardo Tadeu de Araujo; Marlon Benedito Nascomento Santos |

| | | | |
|---|---|---|---|
| EPI_ISL_3835228, EPI_ISL_3835236 | LACEN/PB | WallauLab on behalf of Fiocruz COVID-19 Genomic Surveillance Network | Alexandre Freitas da Silva; Cassia Docena; Constância Flávia Junqueira Ayres; Filipe Zimmer Dezordi; Gabriel Luz Wallau; Gustavo Barbosa de Lima; Lais Ceschini Machado; Lilian Caroliny Amorim Silva; Marcelo Henrique dos Santos Paiva; Matheus Filgueira Bezerra; Sinval Pinto Brandão Filho |
| EPI_ISL_3134859, EPI_ISL_3447590 | LACEN/PE | WallauLab on behalf of Fiocruz COVID-19 Genomic Surveillance Network | Alexandre Freitas da Silva; Cassia Docena; Constância Flávia Junqueira Ayres; Filipe Zimmer Dezordi; Gabriel Luz Wallau; Gustavo Barbosa de Lima; Lais Ceschini Machado; Lilian Caroliny Amorim Silva; Marcelo Henrique dos Santos Paiva; Matheus Filgueira Bezerra; Sinval Pinto Brandão Filho |
| EPI_ISL_4487828, EPI_ISL_4487840, EPI_ISL_4487854, EPI_ISL_4487864, EPI_ISL_4487875, EPI_ISL_4487880, EPI_ISL_4487885 | | | |
| see above | LACEN/PI | WallauLab on behalf of Fiocruz COVID-19 Genomic Surveillance Network | Adelino Soares Lima Neto; Alexandre Freitas da Silva; Antônio Marinho; Cassia Docena; Constância Flávia Junqueira Ayres; Filipe Zimmer Dezordi; Gabriel Luz Wallau; Gustavo Barbosa de Lima; Hellen de Oliveira Amaral; Jacenir Reis dos Santos Mallet; Joana Carolina Viana Lima; Lais Ceschini Machado; Leandro de Mattos; Lilian Caroliny Amorim Silva; Marcela de Lacerda Valença Queiroz; Marcelo Adriano da Cunha e Silva Vieira; Marcelo Henrique dos Santos Paiva; Matheus Filgueira Bezerra; Sinval Pinto Brandão Filho; Túlio de Lima Campos; Vladimir Costa Silva; Walterlene de Carvalho Goçalves |
| EPI_ISL_5193555, EPI_ISL_5193556, EPI_ISL_5193557, EPI_ISL_5193558, EPI_ISL_5193559, EPI_ISL_5193560, EPI_ISL_5193561, EPI_ISL_5193562, EPI_ISL_5193563, EPI_ISL_5193595, EPI_ISL_5193600, EPI_ISL_5193617, EPI_ISL_5193618, EPI_ISL_5193619, EPI_ISL_5193620 | | | |
| see above | LACEN/RR | Laboratorio de Ecologia de Doencas Transmissiveis na Amazonia, Instituto Leonidas e Maria Deane - Fiocruz Amazonia | André Corado; Cátia Alexandra Ribeiro Meneses; Felipe Naveca; Fernanda Nascimento; George Silva; Ilma de Aguiar Antony; Janis Cassimiro Lopes; Luciana Gonçalves; Maria Júlia Brandão; Matilde Mejía; Pricila Carlos Veloso; Rodrigo Melo Maito; Valdinete Nascimento; Victor Souza |
| EPI_ISL_3010762, EPI_ISL_3010770, EPI_ISL_3010773, EPI_ISL_3010774, EPI_ISL_3010778, EPI_ISL_3010785, EPI_ISL_3010786, EPI_ISL_3010792, EPI_ISL_3010795, EPI_ISL_3010796, EPI_ISL_3010814, EPI_ISL_3010815, EPI_ISL_3010820, EPI_ISL_3010821, EPI_ISL_3010829, EPI_ISL_3010834, EPI_ISL_3010838, EPI_ISL_3010844, EPI_ISL_3010850, EPI_ISL_3010851, EPI_ISL_3010857, EPI_ISL_3010865, EPI_ISL_3010867, EPI_ISL_3010869, EPI_ISL_3010872, EPI_ISL_3010878, EPI_ISL_3010880, EPI_ISL_3010883, EPI_ISL_3010884, EPI_ISL_3010887, EPI_ISL_3010908, EPI_ISL_3010916, EPI_ISL_3010919, EPI_ISL_3010927, EPI_ISL_3010929, EPI_ISL_3261103 | | | |
| see above | LATE - Laboratório de Técnicas Especiais - Hospital Israelita Albert Einstein | LATE - Laboratório de Técnicas Especiais - Hospital Israelita Albert Einstein | Alexandre Hideaki Takara; Ana Paula Moreira Salles; Anelisie da Silva Santos; Deyvid Amgarten; Erick Gustavo Dorlass; Fernanda de Mello Malta; João Renato Rebello Pinho; Marcio Anunciacao Menezes; Pedro Henrique Sebe Rodrigues; Raquel Riyuzo |
| EPI_ISL_2476439 | LESP Baja California | Instituto de Diagnostico y Referencia Epidemiologicos (INDRE) | Abril Rodriguez-Maldonado; Ariadna Medina-Benitez; Claudia Wong-Arambula; Ernesto Ramirez-Gonzalez.; Gisela Barrera-Badillo; Irma Lopez-Martinez; Joaquin Quiroz-Mercado; Lucia Hernandez-Rivas; Natividad Cruz-Ortiz; Sergio Rangel-Guerrero; Tatiana Nunez-Garcia; Vanessa Rivero-Arredondo |
| EPI_ISL_2545729, EPI_ISL_2545730 | LESP Baja California Sur | Instituto de Diagnostico y Referencia Epidemiologicos (INDRE) | Abril Rodriguez-Maldonado; Ariadna Medina-Benitez; Claudia Wong-Arambula; Ernesto Ramirez-Gonzalez.; Gisela Barrera-Badillo; Irma Lopez-Martinez; Joaquin Quiroz-Mercado; Lucia Hernandez-Rivas; Natividad Cruz-Ortiz; Sergio Rangel-Guerrero; Tatiana Nunez-Garcia; Vanessa Rivero-Arredondo |
| EPI_ISL_2545726, EPI_ISL_2545727 | LESP Campeche | Instituto de Diagnostico y Referencia Epidemiologicos (INDRE) | Abril Rodriguez-Maldonado; Ariadna Medina-Benitez; Claudia Wong-Arambula; Ernesto Ramirez-Gonzalez.; Gisela Barrera-Badillo; Irma Lopez-Martinez; Joaquin Quiroz-Mercado; Lucia Hernandez-Rivas; Natividad Cruz-Ortiz; Sergio Rangel-Guerrero; Tatiana Nunez-Garcia; Vanessa Rivero-Arredondo |
| EPI_ISL_2476409, EPI_ISL_2476410, EPI_ISL_2476411, EPI_ISL_2476427, EPI_ISL_2479882, EPI_ISL_2545695, EPI_ISL_2545696, EPI_ISL_2545697, EPI_ISL_2545722, EPI_ISL_2674632 | | | |
| see above | LESP Ciudad de Mexico | Instituto de Diagnostico y Referencia Epidemiologicos (INDRE) | Abril Rodriguez-Maldonado; Ariadna Medina-Benitez; Claudia Wong-Arambula; Ernesto Ramirez-Gonzalez.; Gisela Barrera-Badillo; Irma Lopez-Martinez; Joaquin Quiroz-Mercado; Lucia Hernandez-Rivas; Natividad Cruz-Ortiz; Sergio Rangel-Guerrero; Tatiana Nunez-Garcia; Vanessa Rivero-Arredondo |
| EPI_ISL_2545728 | LESP Coahuila | Instituto de Diagnostico y Referencia Epidemiologicos (INDRE) | Abril Rodriguez-Maldonado; Ariadna Medina-Benitez; Claudia Wong-Arambula; Ernesto Ramirez-Gonzalez.; Gisela Barrera-Badillo; Irma Lopez-Martinez; Joaquin Quiroz-Mercado; Lucia Hernandez-Rivas; Natividad Cruz-Ortiz; Sergio Rangel-Guerrero; Tatiana Nunez-Garcia; Vanessa Rivero-Arredondo |
| EPI_ISL_2545698 | LESP Estado de Mexico | Instituto de Diagnostico y Referencia Epidemiologicos (INDRE) | Abril Rodriguez-Maldonado; Ariadna Medina-Benitez; Claudia Wong-Arambula; Ernesto Ramirez-Gonzalez.; Gisela Barrera-Badillo; Irma Lopez-Martinez; Joaquin Quiroz-Mercado; Lucia Hernandez-Rivas; Natividad Cruz-Ortiz; Sergio Rangel-Guerrero; Tatiana Nunez-Garcia; Vanessa Rivero-Arredondo |
| EPI_ISL_2476419, EPI_ISL_2476420, EPI_ISL_2476421, EPI_ISL_2476422 | LESP Guanajuato | Instituto de Diagnostico y Referencia Epidemiologicos (INDRE) | Abril Rodriguez-Maldonado; Ariadna Medina-Benitez; Claudia Wong-Arambula; Ernesto Ramirez-Gonzalez.; Gisela Barrera-Badillo; Irma Lopez-Martinez; Joaquin Quiroz-Mercado; Lucia Hernandez-Rivas; Natividad Cruz-Ortiz; Sergio Rangel-Guerrero; Tatiana Nunez-Garcia; Vanessa Rivero-Arredondo |
| EPI_ISL_2479895 | LESP Nuevo Leon | Instituto de Diagnostico y Referencia Epidemiologicos (INDRE) | Abril Rodriguez-Maldonado; Ariadna Medina-Benitez; Claudia Wong-Arambula; Ernesto Ramirez-Gonzalez.; Gisela Barrera-Badillo; Irma Lopez-Martinez; Joaquin Quiroz-Mercado; Lucia Hernandez-Rivas; Natividad Cruz-Ortiz; Sergio Rangel-Guerrero; Tatiana Nunez-Garcia; Vanessa Rivero-Arredondo |
| EPI_ISL_2476430 | LESP Oaxaca | Instituto de Diagnostico y Referencia Epidemiologicos (INDRE) | Abril Rodriguez-Maldonado; Ariadna Medina-Benitez; Claudia Wong-Arambula; Ernesto Ramirez-Gonzalez.; Gisela Barrera-Badillo; Irma Lopez-Martinez; Joaquin Quiroz-Mercado; Lucia Hernandez-Rivas; Natividad Cruz-Ortiz; Sergio Rangel-Guerrero; Tatiana Nunez-Garcia; Vanessa Rivero-Arredondo |
| EPI_ISL_2479914, EPI_ISL_2859052 | LESP Quintana Roo | Instituto de Diagnostico y Referencia Epidemiologicos (INDRE) | Abril Rodriguez-Maldonado; Ariadna Medina-Benitez; Claudia Wong-Arambula; Ernesto Ramirez-Gonzalez.; Gisela Barrera-Badillo; Irma Lopez-Martinez; Joaquin Quiroz-Mercado; Lucia Hernandez-Rivas; Maribel Gonzalez-Villa; Natividad Cruz-Ortiz; Sergio Rangel-Guerrero; Tatiana Nunez-Garcia; Vanessa Rivero-Arredondo |
| EPI_ISL_2476437 | LESP Sinaloa | Instituto de Diagnostico y Referencia Epidemiologicos (INDRE) | Abril Rodriguez-Maldonado; Ariadna Medina-Benitez; Claudia Wong-Arambula; Ernesto Ramirez-Gonzalez.; Gisela Barrera-Badillo; Irma Lopez-Martinez; Joaquin Quiroz-Mercado; Lucia Hernandez-Rivas; Natividad Cruz-Ortiz; Sergio Rangel-Guerrero; Tatiana Nunez-Garcia; Vanessa Rivero-Arredondo |
| EPI_ISL_2545689, EPI_ISL_2545690, EPI_ISL_2545691 | LESP Tamaulipas | Instituto de Diagnostico y Referencia Epidemiologicos (INDRE) | Abril Rodriguez-Maldonado; Ariadna Medina-Benitez; Claudia Wong-Arambula; Ernesto Ramirez-Gonzalez.; Gisela Barrera-Badillo; Irma Lopez-Martinez; Joaquin Quiroz-Mercado; Lucia Hernandez-Rivas; Natividad Cruz-Ortiz; Sergio Rangel-Guerrero; Tatiana Nunez-Garcia; Vanessa Rivero-Arredondo |
| EPI_ISL_2479887, EPI_ISL_2545688, EPI_ISL_2545735, EPI_ISL_2545736, EPI_ISL_2545737, EPI_ISL_2545738, EPI_ISL_2545739, EPI_ISL_2545740, EPI_ISL_2545741, EPI_ISL_2545742, EPI_ISL_2545743, EPI_ISL_2545744, EPI_ISL_2545745, EPI_ISL_2545746, EPI_ISL_2545747, EPI_ISL_2545748, EPI_ISL_2545750, EPI_ISL_2545751, EPI_ISL_2545752, EPI_ISL_2545753, EPI_ISL_2545754, EPI_ISL_2545755, EPI_ISL_2545756, EPI_ISL_2545757, EPI_ISL_2545758, EPI_ISL_2545759, EPI_ISL_2545760, EPI_ISL_2545762, EPI_ISL_2545763, EPI_ISL_2545764, EPI_ISL_2545765, EPI_ISL_2545766, EPI_ISL_2545767, EPI_ISL_2545768, EPI_ISL_2545769, EPI_ISL_2545771, EPI_ISL_2545772, EPI_ISL_2545773, EPI_ISL_2545774, EPI_ISL_2545775, EPI_ISL_2545776, EPI_ISL_2545777 | | | |
| see above | LESP Yucatan | Instituto de Diagnostico y Referencia Epidemiologicos (INDRE) | Abril Rodriguez-Maldonado; Ariadna Medina-Benitez; Claudia Wong-Arambula; Ernesto Ramirez-Gonzalez.; Gisela Barrera-Badillo; Irma Lopez-Martinez; Joaquin Quiroz-Mercado; Lucia Hernandez-Rivas; Natividad Cruz-Ortiz; Sergio Rangel-Guerrero; Tatiana Nunez-Garcia; Vanessa Rivero-Arredondo |
| EPI_ISL_2331863, EPI_ISL_2362646, EPI_ISL_2362649, EPI_ISL_2362650, EPI_ISL_2362657, EPI_ISL_2362658, EPI_ISL_2362661, EPI_ISL_2362665 | | | |
| see above | LHUB-ULB | Labo Klinische Biologie, UZA | Basil Britto Xavier; Christine Lammens; Herman Goossens; Jasmine Coppens; Marie Le Mercier; Veerle Matheeussen |
| EPI_ISL_2362942, EPI_ISL_2362949, EPI_ISL_2362979, EPI_ISL_2362980, EPI_ISL_2604980, EPI_ISL_2605026, EPI_ISL_2605036, EPI_ISL_2605038, EPI_ISL_2605145 | | | |
| see above | Lab voor klinische biologie | Lab voor klinische biologie | Bruno Verhasselt; Hannelore Hamerlinck; Marija Janevska |
| EPI_ISL_2275825, EPI_ISL_2275826, EPI_ISL_2275827, EPI_ISL_2275838, EPI_ISL_2275841, EPI_ISL_2275843, EPI_ISL_2275846, EPI_ISL_2275860, EPI_ISL_2275870 | | | |
| see above | Lab. Microbiologia e Virologia Cotugno A.O. dei Colli - Istituto Zooprofilattico Sperimentale del Mezzogiorno | TIGEM | Antonio Grimaldi Patrizia Annunziata Francesco Panariello Biancamaria Pierri Claudia Tiberio Teresa Giuliano Valentina Bouche Chiara Colantuono Maria Concetta Cuomo Denise Di Concilio Lucio Di Filippo Anna Manfredi Marcello Salvi Antonio Limone Luigi Atripaldi Pellegrino Cerino Andrea Ballabio Davide Cacchiarelli |
| EPI_ISL_2635918, EPI_ISL_2845704 | Labor Becker & Kollegen (Standort München) | Robert Koch Institute | |
| EPI_ISL_2339475 | Labor Berlin Charité Vivantes GmbH / Institut für Virologie | Charité Universitätsmedizin Berlin, Institut für Virologie/Labor Berlin | Barbara Mühlemann; Christian Drosten; Christine Stephan; Peter Menzel; Rolf Schwarzer; Terry Jones; Victor M Corman |
| EPI_ISL_2389341 | Labor Dr. Heidrich & Kollegen MVZ GmbH Hamburg | Robert Koch Institute | |
| EPI_ISL_2386549, EPI_ISL_2469752 | Labor Dr. Wisplinghoff - Köln | Robert Koch Institute | |
| EPI_ISL_2387434, EPI_ISL_2387446 | Labor Lübeck bzw. Laborärztliche Gemeinschaftspraxis Lübeck | Robert Koch Institute | |
| EPI_ISL_2314742 | Labor Prof. Dr. G. Enders MVZ GbR | Robert Koch Institute | |
| EPI_ISL_2316147 | Labor für medizinische Mikrobiologie Bakteriologie Mykologie Virologie & Infektionsserologie - Labor Mölbis | Robert Koch Institute | |
| EPI_ISL_2362544, EPI_ISL_2362545, EPI_ISL_2362546, EPI_ISL_2362553, EPI_ISL_2362554, EPI_ISL_2362555, EPI_ISL_2426975 | | | |
| see above | Laboratoire Carage | Institut Pasteur de la Guyane | Anne Lavergne; Antoine Enfissi; Arielle Salmier; Dominique Rousset |
| EPI_ISL_2492506, EPI_ISL_2492507, EPI_ISL_2492508, EPI_ISL_2492509, EPI_ISL_2492510, EPI_ISL_2492511, EPI_ISL_2492513, EPI_ISL_2492514, EPI_ISL_2492515, EPI_ISL_2492516, EPI_ISL_2492517, EPI_ISL_2492518, EPI_ISL_2492519, EPI_ISL_2492520, EPI_ISL_2492521, EPI_ISL_2492522, EPI_ISL_2492523, EPI_ISL_2492524, EPI_ISL_2492525, EPI_ISL_2492526, EPI_ISL_2492527, EPI_ISL_2492528, EPI_ISL_2492529, EPI_ISL_2492530, EPI_ISL_2492531, EPI_ISL_2492533, EPI_ISL_2492534, EPI_ISL_2492535, EPI_ISL_2492536, EPI_ISL_2492537, EPI_ISL_2492538, EPI_ISL_2492539, EPI_ISL_2492540, EPI_ISL_2492541, EPI_ISL_2492542, EPI_ISL_2492543, EPI_ISL_2492544, EPI_ISL_2492545 | | | |
| see above | Laboratoire National de Santé Publique – LNSP (HAITI - LNSP) | Laboratory of Respiratory Viruses and Measles, Oswaldo Cruz Institute, FIOCRUZ | Alice Sampaio Rocha; Ana Carolina Mendonca; Anna Carolina Paixao; Elisa Cavalcante Pereira; Fernando Motta; Ito Journel; Jaques Boncy; Luciana Appolinario; Marilda Siqueira on behalf of the Fiocruz COVID-19 Genomic Surveillance Network; Paola Resende; Patrick Delly; Renata Serrano Lopes; Taina Venas |

| | | | |
|---|---|---|---|
| EPI_ISL_3458669, EPI_ISL_3458670, EPI_ISL_3458702, EPI_ISL_5872954, EPI_ISL_5873012, EPI_ISL_7652043 | Laboratoire de santé publique du Québec | Laboratoire de santé publique du Québec | Guillaume Bourque; Ioannis Ragoussis; Jesse Shapiro; Mark Lathrop and Judith Fafard on behalf of the CoVSeQ research group; Mark Lathrop and Michel Roger on behalf of the CoVSeQ research group; Sandrine Moreira |
| EPI_ISL_2401180, EPI_ISL_2401181, EPI_ISL_2401183 | Laboratoire national de sante, Microbiology, Virology | Laboratoire national de sante, Microbiology, Microbial Genomics Platform | Anke Wienecke-Baldacchino; Catherine Ragimbeau; Fatu Djabi; Jessica Tapp; Lise Pignon; Raoul Salmon; Tamir Abdelrahman; Trung Nguyen Nguyen |
| EPI_ISL_2983179, EPI_ISL_3190350 | Laboratorio Central de Saude Publica do Estado de Goias (LACEN/GO) | Laboratory of Respiratory Viruses and Measles, Oswaldo Cruz Institute, FIOCRUZ | Agatha Cristinne Prudencio; Alice Sampaio Rocha; Ana Carolina Mendonca; Ana Flavia Mendonça; Anna Carolina Paixao; Carmen Helena Ramos; Cassiane Casanova; Elisa Cavalcante Pereira; Fernando Motta; Flavia Pereira Amorim da Silva; Ighor Leonardo Arantes Gomes; Luciana Appolinario; Luiz Augusto Pereira; Marilda Siqueira on behalf of the Fiocruz COVID-19 Genomic Surveillance Network; Paola Resende; Rafael Souza Guedes; Renata Serrano Lopes; Taina Venas; Vinicius Lemes da Silva |
| EPI_ISL_2645430 | Laboratorio Central de Saude Publica do Estado do Para (LACEN/PA) | Laboratory of Respiratory Viruses and Measles, Oswaldo Cruz Institute, FIOCRUZ | Alice Sampaio Rocha; Ana Carolina Mendonca; Anna Carolina Paixao; Elisa Cavalcante Pereira; Fernando Motta; Luciana Appolinario; Marilda Siqueira on behalf of the Fiocruz COVID-19 Genomic Surveillance Network; Paola Resende; Renata Serrano Lopes; Taina Venas; Valnete Andrade |
| EPI_ISL_2308637 | Laboratorio Analisi Osp. Città di Castello - Azienda USL Umbria2 | Istituto Zooprofilattico Sperimentale dell'Abruzzo e Molise "G. Caporale" | Ancora M; Calistri P; Cammà C; Curini V; Delli Compagni E; Di Domenico M; Di Pasquale A; Lorusso A; Malagigi V; Mangone I; Marcacci M; Puglia I; Rinaldi A; Savini G; Scialabba S; Taccani P |
| EPI_ISL_4196767 | Laboratorio Central Mg. Luis Alfredo Pianciola | Laboratorio Central Mg. Luis Alfredo Pianciola on behalf of 'Proyecto Argentino Interinstitucional de genomica de SARS-CoV-2' (PAIS Consortium) | C Pintos; C Rastellini.; C Ziehm; J Ousset; L Pianciola; M Fernandez; M Mazzeo |
| EPI_ISL_2535020, EPI_ISL_2535026, EPI_ISL_2535027, EPI_ISL_2535028, EPI_ISL_2535029, EPI_ISL_2535030, EPI_ISL_2535031, EPI_ISL_2535032, EPI_ISL_2535033, EPI_ISL_2535034, EPI_ISL_2535035, EPI_ISL_2535036, EPI_ISL_2535037, EPI_ISL_2535038, EPI_ISL_2535039, EPI_ISL_2535040, EPI_ISL_2535041, EPI_ISL_2535042, EPI_ISL_2535043, EPI_ISL_2535044, EPI_ISL_2535045, EPI_ISL_2535046, EPI_ISL_2535047, EPI_ISL_2535050, EPI_ISL_2535051, EPI_ISL_2535052, EPI_ISL_2535057, EPI_ISL_2535073, EPI_ISL_2535074, EPI_ISL_2535075, EPI_ISL_2535076, EPI_ISL_2535077, EPI_ISL_2535079, EPI_ISL_2535080, EPI_ISL_2535081, EPI_ISL_2535082, EPI_ISL_2535083, EPI_ISL_2535084, EPI_ISL_2535085, EPI_ISL_2535086, EPI_ISL_2535089, EPI_ISL_2535090, EPI_ISL_2535091, EPI_ISL_2535092, EPI_ISL_2535093, EPI_ISL_2535094, EPI_ISL_2535095, EPI_ISL_2535096, EPI_ISL_2535097, EPI_ISL_2535098, EPI_ISL_2535099, EPI_ISL_2535100, EPI_ISL_2535102, EPI_ISL_2535103, EPI_ISL_2535104, EPI_ISL_2535105, EPI_ISL_2535106, EPI_ISL_2691353, EPI_ISL_2691354, EPI_ISL_2691355, EPI_ISL_2691356 | | | |
| see above | Laboratorio Central Noel Nutels | Bioinformatics Laboratory / LNCC | Alessandra P Lamarca; Alexandra L Gerber; Amilcar Tanuri; Ana Paula de C Guimaraes; Ana Tereza R Vasconcelos; Andrea Cony Cavalcanti; Caio Luiz Pereira Ribeiro; Cassia Alves; Cintia Policarpo; Claudia Maria Braga de Mello; Cristiane Gomes da Silva; Diana Mariani; Douglas Terra Machado; Flavio Dias da Silva; Gleidson da Silva de Oliveira; Leandro Magalhaes de Souza; Liliane Cavalcante; Luiz G P de Almeida; Marcio Henrique de Oliveira Garcia; Mario Sergio Ribeiro; Ronaldo da Silva F Jr; Silvia Carvalho |
| EPI_ISL_2969862, EPI_ISL_2969900, EPI_ISL_2969935 | Laboratorio Central de Epidemiologia (LCE) | Unidad de Genomica Avanzada | ; Alejandra Garcia-Gasca; Alejandra Hernandez-Teran; Alejandro Sanchez-Flores; Alfredo Herrera-Estrella; Alicia Ocaña-Mondragon; Andreu Comas-Garcia; Angel Gustavo Salas-Lais; Antonio Loza Roman; Bernardo Martinez-Miguel; Blanca Taboada; Brenda Irasema Maldonado-Meza; Bruno Gomez-Gil; Carla Ivon Herrera-Najera; Carlos F. Arias; Celia Boukadida; Celida Duque Molina; Celida Martinez- Rodriguez; Clara Esperanza Santacruz-Tinoco; Concepcion Grajales-Muñiz; Consorcio Mexicano de Vigilancia Genomica (CoViGen-Mex). Authors (in alphabetical order): Julio Elias Alvarado-Yaah; Cristobal Chaidez-Quiroz; Daniel Fregoso-Rueda; Daniel Lira Morales; Eduardo Becerril-Vargas; Fernando Fontove-Herrera; Fidencio Mejia-Nepomuceno; Francisco Pulido; Gloria Elena Espinosa-Ayala; Gloria Maria Molina-Salinas; Gloria Vazquez; Hector Esteban Paz-Juarez; Hector Montoya-Fuentes; Helen Haydee Fernanda Ramirez-Plascencia; Irvin Gonzalez-Lopez; Jean Pierre Gonzalez; Jesus Hernandez; Joel Armando Vazquez-Perez.; Jorge Salas-Hernandez; Jose Antonio Enciso-Moreno; Jose Arturo Martinez-Orozco; Jose Esteban Muñoz-Medina; Jose de Jesus Nuñez-Contreras; Juan Bautista Chale-Dzul; Julissa Enciso-Ibarra; Luis Alberto Ochoa-Carrera; Margarita Matias-Florentino; Maria Guadalupe Santiago-Mauricio; Maria Guadalupe de Jesus Mireles-Rivera; Mario Mujica-Sanchez; Marissa Perez-Garcia; Nelly Selem-Mojica; Pavel Isa; Ricardo Ciria Merce; Ricardo Grande; Rosa Maria Gutierrez Rios; Santiago avila-Rios; Selene Zarate; Susana Lopez; Veronica Mata-Haro; Victor Eduardo Garcia-Arias; Victor Hugo Borja-Aburto |
| EPI_ISL_2671487, EPI_ISL_2671489, EPI_ISL_2671491, EPI_ISL_2671492, EPI_ISL_2671529, EPI_ISL_2671533, EPI_ISL_2671535, EPI_ISL_2671540, EPI_ISL_2671569, EPI_ISL_2671645, EPI_ISL_2671736, EPI_ISL_2671741 | | | |
| see above | Laboratorio Central de Epidemiologia (LCE) | Unidad de Genomica Avanzada | ; Alejandra Garcia-Gasca; Alejandra Hernandez-Teran; Alejandro Sanchez-Flores; Alfredo Herrera-Estrella; Alicia Ocaña-Mondragon; Andreu Comas-Garcia; Angel Gustavo Salas-Lais; Antonio Loza Roman; Bernardo Martinez-Miguel; Blanca Taboada; Brenda Irasema Maldonado-Meza; Bruno Gomez-Gil; Carla Ivon Herrera-Najera; Carlos F. Arias; Celia Boukadida; Celida Duque Molina; Celida Martinez- Rodriguez; Clara Esperanza Santacruz-Tinoco; Concepcion Grajales-Muñiz; Consorcio Mexicano de Vigilancia Genomica (CoViGen-Mex). Authors (in alphabetical order): Julio Elias Alvarado-Yaah; Cristobal Chaidez-Quiroz; Daniel Fregoso-Rueda; Daniel Lira Morales; Eduardo Becerril-Vargas; Fernando Fontove-Herrera; Fidencio Mejia-Nepomuceno; Francisco Pulido; Gloria Elena Espinosa-Ayala; Gloria Maria Molina-Salinas; Gloria Vazquez; Hector Esteban Paz-Juarez; Hector Montoya-Fuentes; Helen Haydee Fernanda Ramirez-Plascencia; Irvin Gonzalez-Lopez; Jean Pierre Gonzalez; Jesus Hernandez; Joel Armando Vazquez-Perez.; Jorge Salas-Hernandez; Jose Antonio Enciso-Moreno; Jose Arturo Martinez-Orozco; Jose Esteban Muñoz-Medina; Jose de Jesus Nuñez-Contreras; Juan Bautista Chale-Dzul; Julissa Enciso-Ibarra; Luis Alberto Ochoa-Carrera; Margarita Matias-Florentino; Maria Guadalupe Santiago-Mauricio; Maria Guadalupe de Jesus Mireles-Rivera; Mario Mujica-Sanchez; Marissa Perez-Garcia; Nelly Selem-Mojica; Pavel Isa; Ricardo Ciria Merce; Ricardo Grande; Rosa Maria Gutierrez Rios; Santiago avila-Rios; Selene Zarate; Susana Lopez; Veronica Mata-Haro; Victor Eduardo Garcia-Arias; Victor Hugo Borja-Aburto |
| EPI_ISL_2681081, EPI_ISL_2681131, EPI_ISL_2681152, EPI_ISL_2681237, EPI_ISL_2681332, EPI_ISL_2681386 | Laboratorio Central de Epidemiología (LCE) | Instituto de Biotecnología de la UNAM | ; Alejandra García-Gasca; Alejandra Hernández-Terán; Alejandro Sánchez-Flores; Alfredo Herrera-Estrella; Alicia Ocaña-Mondragón; Andreu Comas-García; Angel Gustavo Salas-Lais; Antonio Loza Román; Bernardo Martínez-Miguel; Blanca Taboada; Brenda Irasema Maldonado-Meza; Bruno Gómez-Gil; Carla Ivón Herrera-Najera; Carlos F. Arias; Celia Boukadida; Clara Esperanza Santacruz-Tinoco; Concepción Grajales-Muñiz; Consorcio Mexicano de Vigilancia Genómica (CoViGen-Mex). Authors (in alphabetical order): Julio Elias Alvarado-Yaah; Cristóbal Cháidez-Quiróz; Célida Duque Molina; Célida Martínez- Rodríguez; Daniel Fregoso-Rueda; Daniel Lira Morales; Eduardo Becerril-Vargas; Fernando Fontove-Herrera; Fidencio Mejía-Nepomuceno; Francisco Pulido; Gloria Elena Espinosa-Ayala; Gloria María Molina-Salinas; Gloria Vazquez; Hector Esteban Paz-Juárez; Hector Montoya-Fuentes; Helen Haydee Fernanda Ramírez-Plascencia; Irvin González-López; Jean Pierre González; Jesús Hernández; Joel Armando Vázquez-Pérez.; Jorge Salas-Hernández; José Antonio Enciso-Moreno; José Arturo Martínez-Orozco; José Esteban Muñoz-Medina; José de Jesús Nuñez-Contreras; Juan Bautista Chale-Dzul; Julissa Enciso-Ibarra; Luis Alberto Ochoa-Carrera; Margarita Matías-Florentino; Mario Mújica-Sánchez; Marissa Perez-García; María Guadalupe Santiago-Mauricio; María Guadalupe de Jesús Mireles-Rivera; Nelly Sélem-Mojica; Pavel Isa; Ricardo Ciria Mercé; Ricardo Grande; Rosa María Gutiérrez Ríos; Santiago Ávila-Ríos; Selene Zárate; Susana Lopez; Verónica Mata-Haro; Victor Eduardo García-Arias; Víctor Hugo Borja-Aburto |
| EPI_ISL_3388827 | Laboratorio Central de Salud Publica | Laboratorio de Biología Molecular, Instituto de Medicina Regional on behalf of 'Proyecto Argentino Interinstitucional de genomica de SARS-CoV-2' (PAIS Consortium) | Andrea Ayala; Bettina Brusés; Erica Struss; Esteban Paredes; Griselda Oria; Horacio Lucero.; Javier Mussin; Laura Formichelli; Melina Lorenzini Campos; Raúl Maximiliano Acevedo; Verónica Gómez; Victoria Femenías |
| EPI_ISL_6944507, EPI_ISL_6944515, EPI_ISL_6944519, EPI_ISL_6944521, EPI_ISL_6944524, EPI_ISL_6944535 | Laboratorio Central de Salud Publica | Viral Special Pathogens Branch, Centers for Disease Control and Prevention | Andrea Gómez de la Fuente; Cynthia Vázquez; Emir Talundzic; Joel Montgomery; John Klena; Juan Torales; Justin Lee; María José Ortega; María Liz Gamarra; Shannon Whitmer; Shirley Villalba |
| EPI_ISL_4518739, EPI_ISL_4518751 | Laboratorio Central de Salud Publica de Paraguay | Fundação Ezequiel Dias | Andre Leal; Andrea Gómez de la Fuente; Cynthia Vazquez; Elaine Cristina; Felipe Iani; Flavia Aburjaile; Gislene Garcia de Castro Lichs; Glauco Carvalho; Hegger Fritsch; Joilson Xavier; Juan Torales; Luiz Alcantara.; Luiz Henrique Ferraz Demarchi; Luiz Takao Watanabe; Marina Castilhos Souza Umaki Zardin; Marta Giovanetti; María José Ortega; María Liz Gamarra; Natalia Guimaraes; Raquel da Silva Ferreira; Shirley Villalba; Talita Adelino; Vagner Fonseca; de Oliveira |
| EPI_ISL_2466262, EPI_ISL_2466263, EPI_ISL_2466264, EPI_ISL_2557337, EPI_ISL_2557338, EPI_ISL_2557339 | Laboratorio Central de Saude Publica do Estado da Bahia (LACEN/BA) | Laboratory of Respiratory Viruses and Measles, Oswaldo Cruz Institute, FIOCRUZ | Alice Sampaio Rocha; Ana Carolina Mendonca; Anna Carolina Paixao; Elisa Cavalcante Pereira; Felicidade Pereira; Fernando Motta; Luciana Appolinario; Marilda Siqueira on behalf of the Fiocruz COVID-19 Genomic Surveillance Network; Paola Resende; Renata Serrano Lopes; Taina Venas |
| EPI_ISL_2645392, EPI_ISL_2645394, EPI_ISL_2645395, EPI_ISL_2645396, EPI_ISL_2645402, EPI_ISL_2645406 | Laboratorio Central de Saude Publica do Estado de Alagoas (LACEN/AL) | Laboratory of Respiratory Viruses and Measles, Oswaldo Cruz Institute, FIOCRUZ | Alice Sampaio Rocha; Ana Carolina Mendonca; Anderson Brandao Leite; Anna Carolina Paixao; Elisa Cavalcante Pereira; Fernando Motta; Luciana Appolinario; Marilda Siqueira on behalf of the Fiocruz COVID-19 Genomic Surveillance Network; Paola Resende; Renata Serrano Lopes; Taina Venas |
| EPI_ISL_3045462, EPI_ISL_3190239, EPI_ISL_3190241 | Laboratorio Central de Saude Publica do Estado do Espirito Santo (LACEN/ES) | Laboratory of Respiratory Viruses and Measles, Oswaldo Cruz Institute, FIOCRUZ | Alice Sampaio Rocha; Ana Carolina Mendonca; Anna Carolina Paixao; Elisa Cavalcante Pereira; Fernando Motta; Luciana Appolinario; Marilda Siqueira on behalf of the Fiocruz COVID-19 Genomic Surveillance Network; Paola Resende; Renata Serrano Lopes; Rodrigo Ribeiro Rodrigues; Taina Venas |
| EPI_ISL_2983188, EPI_ISL_2983189, EPI_ISL_2983190, EPI_ISL_2983191, EPI_ISL_2983192, EPI_ISL_2983193, EPI_ISL_2983194, EPI_ISL_2983195, EPI_ISL_2983301, EPI_ISL_2983302, EPI_ISL_2983303, EPI_ISL_2983328, EPI_ISL_3190254, EPI_ISL_3190255, EPI_ISL_3190256, EPI_ISL_3190257, EPI_ISL_3190258, EPI_ISL_3190259, EPI_ISL_3190260, EPI_ISL_3539935 | | | |
| see above | Laboratorio Central de Saude Publica do Estado do Maranhao (LACEN-MA) | Laboratory of Respiratory Viruses and Measles, Oswaldo Cruz Institute, FIOCRUZ | Agatha Cristinne Prudencio; Agatha Soares; Alice Sampaio Rocha; Ana Carolina Mendonca; Anna Carolina Paixao; Elisa Cavalcante Pereira; Fernando Motta; Ighor Arantes; Ighor Leonardo Arantes Gomes; Lidio Gonçalves Lima Neto; Luciana Appolinario; Marilda Siqueira on behalf of the Fiocruz COVID-19 Genomic Surveillance Network; Paola Resende; Renata Serrano Lopes; Taina Moreira Venas; Tainá Venas |
| EPI_ISL_2645466, EPI_ISL_2645471, EPI_ISL_2645474, EPI_ISL_2645482, EPI_ISL_2645484, EPI_ISL_2645485, EPI_ISL_2645487, EPI_ISL_2645494, EPI_ISL_2645495, EPI_ISL_2645497, EPI_ISL_2645499, EPI_ISL_2645500, EPI_ISL_2645501, EPI_ISL_2645503, EPI_ISL_2645504, EPI_ISL_2983448, EPI_ISL_4633885 | | | |
| see above | Laboratorio Central de Saude Publica do Estado do Parana (LACEN/PR) | Laboratory of Respiratory Viruses and Measles, Oswaldo Cruz Institute, FIOCRUZ | Agatha Soares; Alice Sampaio Rocha; Ana Carolina Mendonca; Anna Carolina Paixao; Elisa Cavalcante Pereira; Fernando Motta; Ighor Leonardo Arantes; Irina Riediger; Luciana Appolinario; Marilda Siqueira on behalf of the Fiocruz COVID-19 Genomic Surveillance Network; Paola Resende; Renata Serrano Lopes; Taina Venas |
| EPI_ISL_2603432, EPI_ISL_2982758, EPI_ISL_2982759, EPI_ISL_2982760, EPI_ISL_2982761, EPI_ISL_2982774, EPI_ISL_2982778, EPI_ISL_2982779, EPI_ISL_2982782, EPI_ISL_2982783, EPI_ISL_2982785, EPI_ISL_2982787, EPI_ISL_2982789, EPI_ISL_2982790, EPI_ISL_2982791, EPI_ISL_2982793, EPI_ISL_2982794 | | | |
| see above | Laboratorio Central de Saude Publica do Estado do Rio Grande do Sul (LACEN-RS) | Laboratory of Respiratory Viruses and Measles, Oswaldo Cruz Institute, FIOCRUZ | Alice Sampaio Rocha; Ana Carolina Mendonca; Anderson Brandao Leite; Anna Carolina Paixao; Elisa Cavalcante Pereira; Fernando Motta; Luciana Appolinario; Marilda Siqueira on behalf of the Fiocruz COVID-19 Genomic Surveillance Network; Paola Resende; Renata Serrano Lopes; Taina Venas |
| EPI_ISL_4196715 | Laboratorio MICROBIOM | Laboratorio Central Mg. Luis Alfredo Pianciola on behalf of 'Proyecto Argentino Interinstitucional de genomica de SARS-CoV-2' (PAIS Consortium) | C Pintos; C Rastellini; C Ziehm; J Ousset; L Pianciola; M Fernandez; M Mazzeo; S Cecchi. |
| EPI_ISL_2193289, EPI_ISL_2193290, EPI_ISL_2193292, EPI_ISL_2193294, EPI_ISL_2193295, EPI_ISL_2193296, EPI_ISL_2193297, EPI_ISL_2193298, EPI_ISL_2193299, EPI_ISL_2193300, EPI_ISL_2193301 | | | |
| see above | Laboratorio Microbiologia PO Cardarelli | Laboratorio Microbiologia PO Cardarelli | Felice V.; Niro G. Scutella' M. |
| EPI_ISL_3045388, EPI_ISL_3045389, EPI_ISL_3045404, EPI_ISL_3045409, EPI_ISL_3045413, EPI_ISL_3045415, EPI_ISL_3045418, EPI_ISL_3045419, EPI_ISL_3045420 | | | |
| see above | Laboratorio Nacional de Salud Pública Dr. Defilló - LNSPDD | Laboratory of Respiratory Viruses and Measles, Oswaldo Cruz | Alice Sampaio Rocha; Ana Carolina Mendonca; Anna Carolina Paixao; Elisa Cavalcante Pereira; Fernando Motta; Grey Benoit Vasquez; Isaac Miguel Sanchez; Ivonne Imbert; Lucia de la Cruz; Luciana Appolinario; Marilda Siqueira on behalf of the Fiocruz COVID-19 Genomic Surveillance Network; Nurys de Castro; Paola Resende; Renata Serrano Lopes; Ronald Skewes; Taina Venas |

| | | | |
|---|---|---|---|
| | | Institute, FIOCRUZ | |
| EPI_ISL_4029413, EPI_ISL_4029414 | Laboratorio Nacional de Vigilancia de la Salud – Sección de Virologia | Genomics and Proteomics Departament, Gorgas Memorial Institute For Health Studies | Alexander Martinez; Ambar Moreno; Claudia Díaz; Claudia Gonzalez; Jessica Gondola; Leyda Abrego; Marlene Castillo; Mitzi Castro; Nelly Amador; Oris Chavarria; Sandra Paola Paz; Sofia Carolina Alvarado |
| EPI_ISL_4405298 | Laboratorio de Biología Molecular Bolívar | Área de Secuenciación del Laboratorio de Virología del Hospital de Niños Dr. Ricardo Gutierrez on behalf of 'Proyecto Argentino Interinstitucional de genomica de SARS-CoV-2' (PAIS Consortium) | Acuña; Cabrerizo; Caviglia; D; Di Claudio; D'Andrea; F; Goya; Jofré; LE; Larreche; Lescano; Lusso; M; ME; MI; ML; MR; MS; Martínez; Nabaes Jodar; Natale; R; S; Ticeira; Valinotto; Viegas, M. |
| EPI_ISL_2777439, EPI_ISL_2777440, EPI_ISL_2777441, EPI_ISL_2777442, EPI_ISL_2777443, EPI_ISL_2777444, EPI_ISL_2777445, EPI_ISL_4030383 | | | |
| see above | Laboratorio de Ecologia de Doencas Transmissiveis na Amazonia, Instituto Leonidas e Maria Deane - Fiocruz Amazonia | Laboratorio de Ecologia de Doencas Transmissiveis na Amazonia, Instituto Leonidas e Maria Deane - Fiocruz Amazonia | André Corado; Debora Duarte; Felipe Naveca; Fernanda Nascimento; George Silva; Karina Pessoa; Luciana Gonçalves; Maria Júlia Brandão; Matilde Mejía; Michele Jesus; Valdinete Nascimento; Victor Souza; Ágatha Costa |
| EPI_ISL_3535758 | Laboratorio de Infectologia y Virologia Molecular | Laboratory of Molecular Virology, School of Medicine, Pontificia Universidad Catolica de Chile | Ana Maira Guzman; Andres E. Munoz-Marcos; Catalina Pardo-Roa; Eileen Serrano; Erick Salinas; Estefany Poblete; Francisco Melo; Jorge Levican; Leonardo I. Almonacid; Maria Jose Avendano; Maria Patricia Vega; Rafael A. Medina; Ricardo Enrique de la Barra; Tamara Garcia-Salum |
| EPI_ISL_2907517, EPI_ISL_3134274, EPI_ISL_3134276 | Laboratorio de Medicina Molecular, Universidad de Magallanes | Centro Asistencial Docente y de Investigacion, Universidad de Magallanes | Carolina Pérez; Constanza Ceroni; Diego Alvarez; Inés Cid; Jacqueline Aldridge; Jorge González; Juan Ríos; Marcelo Navarrete; Roberto Uribe-Paredes |
| EPI_ISL_3048973, EPI_ISL_3048974, EPI_ISL_3048975, EPI_ISL_3048976, EPI_ISL_3048977, EPI_ISL_3048978, EPI_ISL_3761570, EPI_ISL_3761645 | | | |
| see above | Laboratorio de Pesquisa em Virologia, FAMERP, SJRP | Laboratorio de Pesquisa em Virologia, FAMERP, SJRP | Beatriz de Carvalho Marques; Cecília Artico Banho; Cíntia Bittar; Fábio Sossai Possebon; Guilherme Campos; Helena Lage Ferreira; Jorge A. Petroli Marchesi; João Pessoa Araújo Jr.; Leila Sabrina Ullmann; Lívia Sacchetto; Maisa C. Pereira Parra; Marília Moraes; Maurício L. Nogueira.; Paula Rahal; Paulo Inacio da Costa |
| EPI_ISL_3401433, EPI_ISL_3672410, EPI_ISL_3826396, EPI_ISL_3826403, EPI_ISL_3826404, EPI_ISL_3826405, EPI_ISL_3826406, EPI_ISL_3826407, EPI_ISL_3826408, EPI_ISL_3826409, EPI_ISL_3826410, EPI_ISL_3826411, EPI_ISL_3991433, EPI_ISL_3991438, EPI_ISL_3991442, EPI_ISL_3991445, EPI_ISL_3991449, EPI_ISL_3991655, EPI_ISL_3991661, EPI_ISL_3991672, EPI_ISL_3991685, EPI_ISL_3991688, EPI_ISL_3991692, EPI_ISL_3991694, EPI_ISL_4052702, EPI_ISL_4052712, EPI_ISL_4052713, EPI_ISL_4052714, EPI_ISL_4052719, EPI_ISL_4052720, EPI_ISL_4052721, EPI_ISL_4052722, EPI_ISL_4052723, EPI_ISL_4052724, EPI_ISL_4052725, EPI_ISL_4052733, EPI_ISL_4052734, EPI_ISL_4052736, EPI_ISL_4052746, EPI_ISL_4052754, EPI_ISL_4052757, EPI_ISL_4052759, EPI_ISL_4052763, EPI_ISL_4052765 | | | |
| see above | Laboratorio de Referencia Nacional de Virus Respiratorios. Centro Nacional de Salud Publica. Instituto Nacional de Salud Peru. | Laboratorio de Referencia Nacional de Virus Respiratorios. Centro Nacional de Salud Publica. Instituto Nacional de Salud Peru. | Carlos Padilla Rojas; Henri Bailon Calderon; Iris Silva Molina; Joseph Huayra Niquen; Lely Solari Zerpa; Luis Barcena Flores; Marco Galarza Perez; Nancy Rojas Serrano; Nieves Sevilla Castañeda; Omar Caceres Rey; Orson Mestanza Millones; Princesa Medrano Alhuay; Priscila Lope Pari; Sandra Morales Ruiz; Sara Gordillo Vilchez; Steve Acedo Lazo; Veronica Hurtado Vela; Victor Jimenez Vasquez; Wendy Lizarraga Olivares |
| EPI_ISL_2657860, EPI_ISL_2657861 | Laboratorio de Salud Pública Fronterizo de Arauca | Instituto Nacional de Salud- Dirección de Investigación en Salud Pública | Carlos Franco-Muñoz; Carmen Osorio; Diana Malo; Diego A. Álvarez-Díaz; Diego Andrés Prada; Gerardo Santamaría; Hector Alejandro Ruiz-Moreno; Jhonnatan Reales-González; Jorge Rivera; Juan Camilo Martinez; Julian Naizaque; Katherine Laiton-Donato; Lisseth Pardo; Magdalena Wiesner; Marcela Mercado-Reyes; Maria T. Herrera-Sepúlveda; Marta Lopez Blanco; Martha Lucia Ospina Martinez; Paola Rojas; Sergio Gomez; Sheryll Corchuelo; Ángela Alarcon Cruz |
| EPI_ISL_4405560 | Laboratorio del Hospital Interzonal General de Agudos " Evita " | Área de Secuenciación del Laboratorio de Virología del Hospital de Niños Dr. Ricardo Gutierrez on behalf of 'Proyecto Argentino Interinstitucional de genomica de SARS-CoV-2' (PAIS Consortium) | A; Acuña; D; Desimone; E; Goya; Grossi; I; L; LE; Luczac; Lusso; MI; MS; Musto; Nabaes Jodar; Natale; O; S; Serrano; Valinotto; Viegas, M. |
| EPI_ISL_4405225, EPI_ISL_4405228 | Laboratorio del Hospital Interzonal General de Agudos "Eva Perón" | Área de Secuenciación del Laboratorio de Virología del Hospital de Niños Dr. Ricardo Gutierrez on behalf of 'Proyecto Argentino Interinstitucional de genomica de SARS-CoV-2' (PAIS Consortium) | Acuña; C; Carulla; D; Goya; Kairiyama; LE; Lusso; M; MI; MS; Nabaes Jodar; Natale; Pengue; Piñeyro C; S; Valinotto; Viegas, M. |
| EPI_ISL_7620350, EPI_ISL_7620351, EPI_ISL_7620378, EPI_ISL_7620379, EPI_ISL_7620380, EPI_ISL_7620381, EPI_ISL_7620382 | | | |
| see above | Laboratorio del Hospital Regional Ushuaia Gdor. Ernesto Campos | Nodo de Secuenciación Tierra del Fuego - Hospital Regional Ushuaia - Centro Austral De Investigaciones Científicas - Universidad Nacional De Tierra Del Fuego on behalf PAIS project and ANLIS-Malbran | AE; Boutureira, MF.; CA; CB; CF; Castro; Ceballos; Cáceres; De Roccis; F; G; Gallego; Gramundi; ID; Nardi; Rojas; SB; SG; Yulan |
| EPI_ISL_2360296, EPI_ISL_2360308, EPI_ISL_2360333, EPI_ISL_2361739 | Laboratorio di Microbiologia | Laboratorio di Microbiologia | Martinetti Lucchini Gladys; Valeria Spina |
| EPI_ISL_3356287, EPI_ISL_3356288 | Laboratorio di Patologia Clinica, Ospedale San Paolo in Valloria, ASL 2 Liguria | U.O. Igiene, Ospedale Policlinico San Martino | Bruzzone Bianca; Caligiuri Patrizia; De Pace Vanessa; Domnich Alexander; Icardi Giancarlo on behalf of SARS-CoV-2 ITALIAN RESEARCH ENTERPRISE-(SCIRE) Collaborative Group; Lillo Flavia; Orsi Andrea; Ricucci Valentina |
| EPI_ISL_6368812, EPI_ISL_6368813, EPI_ISL_6368814, EPI_ISL_6368815, EPI_ISL_6368817, EPI_ISL_6368818, EPI_ISL_6368819, EPI_ISL_6368821, EPI_ISL_6368822, EPI_ISL_6368823, EPI_ISL_6368824, EPI_ISL_6368825, EPI_ISL_6368827, EPI_ISL_6368828 | | | |
| see above | Laboratorio nacional de salud publica | Viral Special Pathogens Branch, Centers for Disease Control and Prevention | Emir Talundzic; Isaac Miguel; Joel Montgomery; John Klena; Justin Lee; Melissa Mobley; Ronald Skewes-Ramm; Shannon Whitmer |
| EPI_ISL_2305583, EPI_ISL_2305585, EPI_ISL_2305589, EPI_ISL_2305622, EPI_ISL_2307041, EPI_ISL_2307043, EPI_ISL_2307045, EPI_ISL_2307053, EPI_ISL_2397838, EPI_ISL_2397866, EPI_ISL_2397878, EPI_ISL_2397903, EPI_ISL_2397913, EPI_ISL_2397955, EPI_ISL_2397956, EPI_ISL_2397957, EPI_ISL_2397960, EPI_ISL_2397962, EPI_ISL_2397965, EPI_ISL_2397967, EPI_ISL_2397982, EPI_ISL_2397998, EPI_ISL_2398009, EPI_ISL_2398020, EPI_ISL_2398055, EPI_ISL_2398208, EPI_ISL_2398222, EPI_ISL_2398225, EPI_ISL_2398235, EPI_ISL_2398244, EPI_ISL_2398245, EPI_ISL_2398252, EPI_ISL_2398255, EPI_ISL_2398256, EPI_ISL_2398276, EPI_ISL_2398281, EPI_ISL_2398316, EPI_ISL_2398318, EPI_ISL_2398319, EPI_ISL_2398324, EPI_ISL_2398325, EPI_ISL_2398334, EPI_ISL_2398338, EPI_ISL_2398341, EPI_ISL_2398343, EPI_ISL_2398351, EPI_ISL_2398352, EPI_ISL_2398355, EPI_ISL_2398364, EPI_ISL_2398370, EPI_ISL_2398372, EPI_ISL_2398373, EPI_ISL_2398376, EPI_ISL_2398381, EPI_ISL_2398386, EPI_ISL_2398390, EPI_ISL_2398401, EPI_ISL_2398414, EPI_ISL_2398418, EPI_ISL_2398428, EPI_ISL_2398444, EPI_ISL_2398451, EPI_ISL_2398454, EPI_ISL_2398458, EPI_ISL_2398459, EPI_ISL_2398476, EPI_ISL_2398479, EPI_ISL_2398481, EPI_ISL_2398482, EPI_ISL_2398483, EPI_ISL_2398491, EPI_ISL_2398492, EPI_ISL_2398502, EPI_ISL_2398503, EPI_ISL_2398508, EPI_ISL_2398513, EPI_ISL_2398520, EPI_ISL_2398523, EPI_ISL_2398526, EPI_ISL_2398530, EPI_ISL_2398531, EPI_ISL_2398567, EPI_ISL_2398769, EPI_ISL_2398803, EPI_ISL_2398804, EPI_ISL_2398822, EPI_ISL_2398841, EPI_ISL_2398842, EPI_ISL_2398845, EPI_ISL_2398854, EPI_ISL_2398859, EPI_ISL_2398876, EPI_ISL_2398884, EPI_ISL_2398902, EPI_ISL_2398910, EPI_ISL_2398917, EPI_ISL_2398922, EPI_ISL_2398928, EPI_ISL_2398929, EPI_ISL_2398935, EPI_ISL_2398936, EPI_ISL_2398941, EPI_ISL_2398948, EPI_ISL_2398950, EPI_ISL_2398957, EPI_ISL_2398969, EPI_ISL_2398979, EPI_ISL_2398981, EPI_ISL_2398986, EPI_ISL_2399041, EPI_ISL_2399050, EPI_ISL_2399052, EPI_ISL_2399061, EPI_ISL_2399065, EPI_ISL_2399073, EPI_ISL_2399081, EPI_ISL_2399082, EPI_ISL_2399092, EPI_ISL_2399148, EPI_ISL_2399149, EPI_ISL_2399155, EPI_ISL_2399166, EPI_ISL_2399193, EPI_ISL_2399204, EPI_ISL_2399221, EPI_ISL_2399228, EPI_ISL_2399233, EPI_ISL_2399234, EPI_ISL_2399249, EPI_ISL_2399255, EPI_ISL_2399258, EPI_ISL_2399265, EPI_ISL_2399267, EPI_ISL_2399268, EPI_ISL_2399270, EPI_ISL_2399272, EPI_ISL_2399290, EPI_ISL_2399296, EPI_ISL_2399302, EPI_ISL_2399336, EPI_ISL_2399338, EPI_ISL_2399351, EPI_ISL_2399355, EPI_ISL_2399367, EPI_ISL_2480199, EPI_ISL_2480214, EPI_ISL_2480638, EPI_ISL_2480639, EPI_ISL_2480646, EPI_ISL_2480666, EPI_ISL_2480702, EPI_ISL_2480713, EPI_ISL_2480721, EPI_ISL_2480758, EPI_ISL_2480759, EPI_ISL_2480762, EPI_ISL_2480789, EPI_ISL_2480790, EPI_ISL_2480791, EPI_ISL_2480793, EPI_ISL_2480794, EPI_ISL_2481019, EPI_ISL_2481039, EPI_ISL_2481044, EPI_ISL_2481058, EPI_ISL_2481062, EPI_ISL_2481084, EPI_ISL_2481110, EPI_ISL_2481130, EPI_ISL_2481143, EPI_ISL_2481144, EPI_ISL_2481500, EPI_ISL_2481514, EPI_ISL_2481519, EPI_ISL_2481521, EPI_ISL_2481578, EPI_ISL_2481597, EPI_ISL_2481875, EPI_ISL_4373760, EPI_ISL_4374073, EPI_ISL_4374082, EPI_ISL_4375799, EPI_ISL_4379207, EPI_ISL_4381500 | | | |
| see above | Laboratory Corporation of America | Centers for Disease Control and Prevention Division of Viral Diseases, Pathogen Discovery | Adrian Paskey; Amanda Douglas; Amanda Suchanek; Andrea Throop; Ayla Burns; Benjamin Rambo-Martin; Bobbi Croy; Brian Krueger; Brian Norvell; Christopher Gulvick; Christos Petropoulos; Clinton Paden; Clinton R. Paden; Craig Lukasik; Dakota Howard; Darlene Wagner; Debbie Boles; Dhwani Batra; Duncan MacCannell; Eyad Almasri; Goran Stevovic; Howard Engler; Hrushikesh Deshmukh; Jake Humphrey; Jana Schroth; Jason Caravas; Joe Voshell; John Pruitt; Jonathan Meltzer; Jonathan Williams; Kara Moser; Kimberly Wagner; Kristine Lacek; Lax Iyer; Lisa Pfefferle; Lyndon Tilson; Manoj Jain; Marcia Eisenberg; Mary Ann Cristobal; Mary Cristobal; Mary Williamson; Matthew Robinson; Matthew Schmerer; Michael Levandoski; Mike Sapeta; Mindy Nye; Minoo Agarwal; Mohan Kolli; Nuthawin Charoensri; Oren Cohen; Peter Cook; Peter W. Cook; Prashant Gupta; Qian Zeng; Rama Ghatti; Scott Parker; Scott Ryan; Scott Sammons; Shatavia Morrison; Stanley Letovsky; Steven Ragan; Suresh Babu Selvaraju; Suresh Selvaraju; Susan Countryman; Susan Hicks; Suzanne Dale; Thomas Urban; Tim Kuphal; Tricia Zwiefelhofer; Tymeckia Kendall; Victoria Caban Figueroa; Vincent Drouillon; Yvette Unoarumhi |
| EPI_ISL_2443662, EPI_ISL_3539937, EPI_ISL_4237340, EPI_ISL_4237341, EPI_ISL_4237342, EPI_ISL_4237343, EPI_ISL_4237439, EPI_ISL_6172751, EPI_ISL_6172752, EPI_ISL_6172753, EPI_ISL_6172754, EPI_ISL_6172755, EPI_ISL_6172756, EPI_ISL_6172757, EPI_ISL_7105923, EPI_ISL_7105959, EPI_ISL_7105988 | | | |
| see above | Laboratory of Respiratory Viruses and Measles, Oswaldo Cruz Institute, FIOCRUZ | Laboratory of Respiratory Viruses and Measles, Oswaldo Cruz Institute, FIOCRUZ | Agatha Cristinne Prudencio; Alice Sampaio Rocha; Ana Carolina Mendonca; Anna Carolina Paixao; Bruna Mendonça da Silva; Elisa Cavalcante Pereira; Fernando Motta; Ighor Arantes; Ighor Leonardo Arantes Gomes; Jéssica Graça Macedo de Carvalho; Larissa Macedo Pinto; Luciana Appolinario; Marilda Siqueira on behalf of the Fiocruz COVID-19 Genomic Surveillance Network; Paola Resende; Renata Serrano Lopes; Taina Venas; Victor Guimaraes; Vitor Guimaraes Ribeiro |
| EPI_ISL_5017906 | Laboratório Central de Saúde Pública de Alagoas | Coordenação Geral de Laboratórios de Saúde Pública (CGLAB/DAEVS/SVS/MS) | Vagner Fonseca; et al. |
| EPI_ISL_4600508, EPI_ISL_4600514 | Laboratório Central de Saúde Pública de Pernambuco | Coordenação Geral de Laboratórios de Saúde Pública (CGLAB/DAEVS/SVS/MS) | Vagner Fonseca; et al. |
| EPI_ISL_2777244, EPI_ISL_2777245, EPI_ISL_2777284, EPI_ISL_2777297, EPI_ISL_2777299, EPI_ISL_2777316, EPI_ISL_2777338, EPI_ISL_2777339, EPI_ISL_2777344, EPI_ISL_2777353, EPI_ISL_2777354, EPI_ISL_2777511, EPI_ISL_2777562, EPI_ISL_2777584, EPI_ISL_2777640, EPI_ISL_2777645, EPI_ISL_2777671, EPI_ISL_2777672, EPI_ISL_2777674, EPI_ISL_2777675, EPI_ISL_2777678, EPI_ISL_2777679, EPI_ISL_2777682, EPI_ISL_2777711, EPI_ISL_2777826, EPI_ISL_2777901, EPI_ISL_2777902, EPI_ISL_2777903, EPI_ISL_2777904, EPI_ISL_2777905, EPI_ISL_2777906, EPI_ISL_2777908, EPI_ISL_2777909, EPI_ISL_2777911, EPI_ISL_2777912, EPI_ISL_2777921, EPI_ISL_2777924, EPI_ISL_2777925, EPI_ISL_2777926, EPI_ISL_2777929, EPI_ISL_2777930, EPI_ISL_2777933, EPI_ISL_2777934, EPI_ISL_2777935, EPI_ISL_2777938, EPI_ISL_2777939, EPI_ISL_2777940, EPI_ISL_2777941, EPI_ISL_2777946, EPI_ISL_2777947, EPI_ISL_2777949, EPI_ISL_2777952, EPI_ISL_2777956, EPI_ISL_2777957, EPI_ISL_2777958, EPI_ISL_2777959, EPI_ISL_2777960, EPI_ISL_2777961, EPI_ISL_2777965, EPI_ISL_2777966, EPI_ISL_2777985, EPI_ISL_2777991, EPI_ISL_2778006, EPI_ISL_2778007, EPI_ISL_2778009 | | | |
| see above | Laboratório Central de Saúde Pública do Amazonas - LACEN-AM | Laboratorio de Ecologia de Doencas Transmissiveis na Amazonia, Instituto Leonidas e Maria Deane - Fiocruz Amazonia | André Corado; Debora Duarte; Felipe Naveca; Fernanda Nascimento; George Silva; Karina Pessoa; Luciana Gonçalves; Maria Júlia Brandão; Matilde Mejía; Michele Jesus; Valdinete Nascimento; Victor Souza; Ágatha Costa |
| EPI_ISL_7261615, EPI_ISL_7261616 | Laboratório Central de Saúde Pública do Estado do Tocantins (LACEN/TO) | Bioinformatics and Biotechnology Laboratory (Labinftec/Federal | Fabrício Souza Campos; Jucimária Dantas Galvão; Raíssa Nunes dos Santos; Ueric José Borges de Souza |

| | | | |
|---|---|---|---|
| | | University of Tocantins, Campus of Gurupi) | |
| EPI_ISL_2731510, EPI_ISL_2731511, EPI_ISL_2731512, EPI_ISL_2731513, EPI_ISL_2731514, EPI_ISL_2731515, EPI_ISL_2731516, EPI_ISL_2731517, EPI_ISL_2731518, EPI_ISL_2731519, EPI_ISL_2731520, EPI_ISL_2731521, EPI_ISL_2731522, EPI_ISL_2731523, EPI_ISL_2731524, EPI_ISL_2731525, EPI_ISL_2731526, EPI_ISL_2731527, EPI_ISL_2731528, EPI_ISL_2731529, EPI_ISL_2731530, EPI_ISL_2731531, EPI_ISL_2731532, EPI_ISL_2731533, EPI_ISL_2731534, EPI_ISL_2731535, EPI_ISL_2731536, EPI_ISL_2731537, EPI_ISL_2731538, EPI_ISL_2731539, EPI_ISL_2731540, EPI_ISL_2731541, EPI_ISL_2731542, EPI_ISL_2731543, EPI_ISL_2731544, EPI_ISL_2731545, EPI_ISL_2731546, EPI_ISL_2731547, EPI_ISL_2731548, EPI_ISL_2731549, EPI_ISL_2731550, EPI_ISL_2731551, EPI_ISL_2731552, EPI_ISL_2731553, EPI_ISL_2731554, EPI_ISL_2731555, EPI_ISL_2731556, EPI_ISL_2731557, EPI_ISL_2731558, EPI_ISL_2731559, EPI_ISL_2731560, EPI_ISL_2731561, EPI_ISL_2731562, EPI_ISL_2731563, EPI_ISL_2731564, EPI_ISL_2731565, EPI_ISL_2731566, EPI_ISL_2731567, EPI_ISL_2731568, EPI_ISL_2731569, EPI_ISL_2731570, EPI_ISL_2731571, EPI_ISL_2731572, EPI_ISL_2731573, EPI_ISL_2731574, EPI_ISL_2731575, EPI_ISL_2731576, EPI_ISL_2731577, EPI_ISL_2731578, EPI_ISL_2731579, EPI_ISL_2731580, EPI_ISL_2731581, EPI_ISL_2731582, EPI_ISL_2731583, EPI_ISL_2731584, EPI_ISL_2731585, EPI_ISL_2731586, EPI_ISL_2731587, EPI_ISL_2731588, EPI_ISL_2731589, EPI_ISL_2731590, EPI_ISL_2731591, EPI_ISL_2731592, EPI_ISL_2731593, EPI_ISL_2731594, EPI_ISL_2731595, EPI_ISL_2731596, EPI_ISL_2731597, EPI_ISL_2731598, EPI_ISL_2731599, EPI_ISL_2731600, EPI_ISL_2731601, EPI_ISL_2731602, EPI_ISL_2731603, EPI_ISL_2731604, EPI_ISL_2731605, EPI_ISL_2731606, EPI_ISL_2731607, EPI_ISL_2731608, EPI_ISL_2731609, EPI_ISL_2731610, EPI_ISL_2731611, EPI_ISL_2731612, EPI_ISL_2731613, EPI_ISL_2731614, EPI_ISL_2731615, EPI_ISL_2731616, EPI_ISL_2731617, EPI_ISL_2731618, EPI_ISL_2731619, EPI_ISL_2731620, EPI_ISL_2731621, EPI_ISL_2731622, EPI_ISL_2731623, EPI_ISL_2731625, EPI_ISL_2731626, EPI_ISL_2731627, EPI_ISL_2731628, EPI_ISL_2731629, EPI_ISL_2731630, EPI_ISL_2731631, EPI_ISL_2731632, EPI_ISL_2731633, EPI_ISL_2731634, EPI_ISL_2731637, EPI_ISL_2731638, EPI_ISL_2731639, EPI_ISL_2731640, EPI_ISL_2731641, EPI_ISL_2731642, EPI_ISL_2731643, EPI_ISL_2731644, EPI_ISL_2731645, EPI_ISL_2731646, EPI_ISL_2731647, EPI_ISL_2731648, EPI_ISL_2731649, EPI_ISL_2731650, EPI_ISL_2731651, EPI_ISL_2731652, EPI_ISL_2731653, EPI_ISL_2731654, EPI_ISL_2731655, EPI_ISL_2731656, EPI_ISL_2731657, EPI_ISL_2731658, EPI_ISL_2731659, EPI_ISL_2731660, EPI_ISL_2731661, EPI_ISL_2731662, EPI_ISL_2731663, EPI_ISL_2731664, EPI_ISL_2731665, EPI_ISL_2731667, EPI_ISL_2731668, EPI_ISL_2731669, EPI_ISL_2731671, EPI_ISL_2731672, EPI_ISL_2731673, EPI_ISL_2731674, EPI_ISL_2731675, EPI_ISL_2731676, EPI_ISL_2731677, EPI_ISL_2790137, EPI_ISL_2790138, EPI_ISL_2790139, EPI_ISL_2790140, EPI_ISL_2790142, EPI_ISL_2790143, EPI_ISL_2790144, EPI_ISL_2790145, EPI_ISL_2790146, EPI_ISL_2790147, EPI_ISL_2790148, EPI_ISL_2790149, EPI_ISL_2790150, EPI_ISL_2790151, EPI_ISL_2790152, EPI_ISL_2790153, EPI_ISL_2790154, EPI_ISL_2790155, EPI_ISL_2790156, EPI_ISL_2790157, EPI_ISL_2790158, EPI_ISL_2790159, EPI_ISL_2790160, EPI_ISL_2790161, EPI_ISL_2790162, EPI_ISL_2790163, EPI_ISL_2790164, EPI_ISL_2790165, EPI_ISL_2790166, EPI_ISL_2790167, EPI_ISL_2790168, EPI_ISL_2790169, EPI_ISL_2790170, EPI_ISL_2790171, EPI_ISL_2790172, EPI_ISL_2790173, EPI_ISL_2790174, EPI_ISL_2835107, EPI_ISL_2835108, EPI_ISL_2835109, EPI_ISL_2835110, EPI_ISL_2835111, EPI_ISL_2835112, EPI_ISL_2835113, EPI_ISL_2835114, EPI_ISL_2835116, EPI_ISL_2835118, EPI_ISL_2835119, EPI_ISL_2835120, EPI_ISL_2835121, EPI_ISL_2835122, EPI_ISL_2835123, EPI_ISL_2835124, EPI_ISL_2835125, EPI_ISL_2835126, EPI_ISL_2835127, EPI_ISL_2835128, EPI_ISL_2835129, EPI_ISL_2835130, EPI_ISL_2835131, EPI_ISL_2835132, EPI_ISL_2835133, EPI_ISL_2835134, EPI_ISL_2835135, EPI_ISL_2835136, EPI_ISL_2835137, EPI_ISL_2835138, EPI_ISL_2835139, EPI_ISL_2835140, EPI_ISL_2835141, EPI_ISL_2835142, EPI_ISL_2835143, EPI_ISL_2835144, EPI_ISL_2835145, EPI_ISL_2835146, EPI_ISL_2835147, EPI_ISL_2835148, EPI_ISL_2835149, EPI_ISL_2835150, EPI_ISL_2835151, EPI_ISL_2835152, EPI_ISL_2835153, EPI_ISL_2835154, EPI_ISL_2835155, EPI_ISL_2835156, EPI_ISL_2835157, EPI_ISL_2835158, EPI_ISL_2835159, EPI_ISL_2835160, EPI_ISL_2835161, EPI_ISL_2835162, EPI_ISL_2835163, EPI_ISL_2835164, EPI_ISL_2835165, EPI_ISL_2835167, EPI_ISL_2835169, EPI_ISL_2835170, EPI_ISL_2835171, EPI_ISL_2835172, EPI_ISL_2835173, EPI_ISL_2835174, EPI_ISL_2835175, EPI_ISL_2835176, EPI_ISL_2835177, EPI_ISL_2835178, EPI_ISL_2835179, EPI_ISL_2835180, EPI_ISL_2835181, EPI_ISL_2835182, EPI_ISL_2835183, EPI_ISL_2835184, EPI_ISL_2835185, EPI_ISL_2835186, EPI_ISL_2835188, EPI_ISL_2835189, EPI_ISL_2835191, EPI_ISL_2835192, EPI_ISL_2835193, EPI_ISL_2835194, EPI_ISL_2835195, EPI_ISL_2835196, EPI_ISL_2835197, EPI_ISL_2835198, EPI_ISL_2835199, EPI_ISL_2835200, EPI_ISL_2835201, EPI_ISL_2835202, EPI_ISL_2835203, EPI_ISL_2835204, EPI_ISL_2835205, EPI_ISL_2835206, EPI_ISL_2835207, EPI_ISL_2835208, EPI_ISL_2835209, EPI_ISL_2835210, EPI_ISL_2835211, EPI_ISL_2835212, EPI_ISL_2835213, EPI_ISL_2835214, EPI_ISL_2835215, EPI_ISL_2983471, EPI_ISL_4543263, EPI_ISL_4543264, EPI_ISL_4543287, EPI_ISL_4543288, EPI_ISL_4543289, EPI_ISL_4543290, EPI_ISL_4543291, EPI_ISL_4543292, EPI_ISL_4543293, EPI_ISL_4543294, EPI_ISL_4543295, EPI_ISL_4543296, EPI_ISL_4543297, EPI_ISL_4543298, EPI_ISL_4543299, EPI_ISL_4543300, EPI_ISL_4543301, EPI_ISL_4543302, EPI_ISL_4543303, EPI_ISL_4543304, EPI_ISL_4543311, EPI_ISL_4543316, EPI_ISL_4543344, EPI_ISL_4543357, EPI_ISL_4543364, EPI_ISL_4543366, EPI_ISL_4543369 | | | |
| see above | Laboratório de Biotecnologia Aplicada (LBA) - Laboratório de Biologia Molecular - Hospital das Clínicas, Faculdade de Medicina de Botucatu, Departamento de Bioprocessos e Biotecnologia - Faculdade de Ciências Agronômicas, UNESP – Botucatu/SP | Laboratory of Respiratory Viruses and Measles, Oswaldo Cruz Institute, FIOCRUZ | Alice Sampaio Rocha; Ana Carolina Mendonca; Anna Carolina Paixao; Elisa Cavalcante Pereira; Felipe Allan da Silva Costa; Fernando Motta; Jayme Augusto de Souza Neto; Leonardo Nazario de Moraes; Luciana Appolinario; Marilda Siqueira on behalf of the Fiocruz COVID-19 Genomic Surveillance Network; Paola Resende; Patricia Akemi Assato; Rejane Maria Tommasini; Renata Serrano Lopes; Taina Venas |
| EPI_ISL_4051962 | Laboratório de Microbiologia Molecular - Universidade FEEVALE | Molecular Microbiology Laboratory | Alana Witt Hansen; Fernando Rosado Spilki; Fágner Henrique Heldt; Juliana Schons Gularte; Juliane Deise Fleck; Mariana Soares da Silva; Matheus Nunes Weber; Meriane Demoliner; Micheli Filippi; Paula Rodrigues de Almeida; Vyctoria Malayhka de Abreu Góes Pereira. |
| EPI_ISL_4520593 | Laboratório de Virologia e Cultivo Celular - LABVICC | Laboratório Baculovírus | Aline Belmok; Bergmann Morais Ribeiro; Clayton Luiz Borges; Déborah Anjos; Elen Regozino Muniz; Fabiola Souza Fiaccadori; Fernando L Melo; Gabriela Silvério Bazilio; Juliana Alves Parente Rocha; Keila Correia de Alcântara; Menira Souza; Nathânia Dábilla; Valéria Christina de Rezende Feres |
| EPI_ISL_2557416, EPI_ISL_2557418, EPI_ISL_2557419, EPI_ISL_2557421 | Labortorio Central de Saude Publica do Estado de Santa Catarina (LACEN/SC) | Laboratory of Respiratory Viruses and Measles, Oswaldo Cruz Institute, FIOCRUZ | Alice Sampaio Rocha; Ana Carolina Mendonca; Anna Carolina Paixao; Darcita Buerger Rovaris; Elisa Cavalcante Pereira; Fernando Motta; Luciana Appolinario; Marilda Siqueira on behalf of the Fiocruz COVID-19 Genomic Surveillance Network; Paola Resende; Renata Serrano Lopes; Sandra Bianchini Fernandes; Taina Venas |
| EPI_ISL_2536277, EPI_ISL_2536278, EPI_ISL_2536279, EPI_ISL_2536280, EPI_ISL_2536281, EPI_ISL_2536282, EPI_ISL_2536283, EPI_ISL_2536284, EPI_ISL_2536285, EPI_ISL_2536286, EPI_ISL_2557380, EPI_ISL_2557381, EPI_ISL_2557382, EPI_ISL_2557383, EPI_ISL_2557406 | | | |
| see above | Labortorio Central de Saude Publica do Estado do Rio de Janeiro (LACEN/RJ) | Laboratory of Respiratory Viruses and Measles, Oswaldo Cruz Institute, FIOCRUZ | Alice Sampaio Rocha; Ana Carolina Mendonca; Andrea Cony Cavalcanti; Anna Carolina Paixao; Elisa Cavalcante Pereira; Fernando Motta; Luciana Appolinario; Marilda Siqueira on behalf of the Fiocruz COVID-19 Genomic Surveillance Network; Paola Resende; Renata Serrano Lopes; Taina Venas |
| EPI_ISL_2645509, EPI_ISL_2645510 | Labortorio Central de Saude Publica do Estado do Tocantins (LACEN/TO) | Laboratory of Respiratory Viruses and Measles, Oswaldo Cruz Institute, FIOCRUZ | Alice Sampaio Rocha; Ana Carolina Mendonca; Anna Carolina Paixao; Elisa Cavalcante Pereira; Fernando Motta; Jucimaria Dantas Galvao; Luciana Appolinario; Marilda Siqueira on behalf of the Fiocruz COVID-19 Genomic Surveillance Network; Paola Resende; Renata Serrano Lopes; Taina Venas |
| EPI_ISL_2557362 | Lboratorio Central de Saude Publica do Estado do Parana (LACEN/PR) | Laboratory of Respiratory Viruses and Measles, Oswaldo Cruz Institute, FIOCRUZ | Alice Sampaio Rocha; Ana Carolina Mendonca; Anna Carolina Paixao; Elisa Cavalcante Pereira; Fernando Motta; Irina Riediger; Luciana Appolinario; Marilda Siqueira on behalf of the Fiocruz COVID-19 Genomic Surveillance Network; Paola Resende; Renata Serrano Lopes; Taina Venas |
| EPI_ISL_2235591, EPI_ISL_2316530, EPI_ISL_2317615, EPI_ISL_2348483 | Lighthouse Lab in Milton Keynes | Wellcome Sanger Institute for the COVID-19 Genomics UK (COG-UK) Consortium | Cordelia Langford; David K. Jackson; Dominic Kwiatkowski; Ewan Harrison; Ian Johnston; Jeffrey Barrett; John Sillitoe on behalf of the Wellcome Sanger Institute COVID-19 Surveillance Team; Roberto Amato; Sonia Goncalves; The Lighthouse Lab in Milton Keynes and Alex Alderton |
| EPI_ISL_2266523, EPI_ISL_2389148 | Limbach - MVZ Humangenetik Ulm | Robert Koch Institute | |
| EPI_ISL_2373107, EPI_ISL_2842585, EPI_ISL_2842589 | Lurie Children's Hospital of Chicago | Northwestern University - Ozer Lab | Egon A. Ozer; Judd F. Hultquist; Lacy M. Simons; Larry K. Kociolek; Michael G. Ison; Ramon Lorenzo-Redondo; Taylor J. Dean; William J. Muller; Xiaotian; Zheng |
| EPI_ISL_2801333 | MATERNIDADE ESCOLA ASSIS CHATEAUBRIAND | Analytical Competence Molecular Epidemiology Lab/ACME, Oswaldo Cruz Foundation, Ceara (FIOCRUZ CE) | Cleber Furtado Aksenen e Suzana Porto Almeida; Fabio Miyajima; Fernando Braga Stehling; Francisco Eder de Moura Lopes; Jamille Maria Mendes Bezerra; Joaquim César do Nascimento Sousa Junior; Pedro Miguel Carneiro Jeronimo; Thais Ferreira de Oliveira; Thais de Oliveira Costa; Ticiane Cavalcante de Souza; Veridiana Pessoa Miyajima |
| EPI_ISL_2495429, EPI_ISL_2495430, EPI_ISL_2495431, EPI_ISL_2495432, EPI_ISL_2495433, EPI_ISL_2495617 | MB-Cadham Provincial laboratory | National Microbiology Laboratory (NML) | Anna Majer; Anneliese Landgraff; CanCOGeN's metadata curation team; Darian Hole; David Alexander; Elsie Grudeski; Gary Van Domselaar; Grace Seo; Jared Bullard; Jennifer Tanner; Kerry Dust; Kirsten Biggar; Madison Chapel; Morag Graham; Natalie Knox; Nathalie Bastien; Paul Van Caeseele; Philip Mabon; Public Health Agency of Canada CanCOGeN team; Rhiannon Huzarewich; Russell Mandes; Shari Tyson; Timothy Booth; Yan Li |
| EPI_ISL_2453622, EPI_ISL_2453794, EPI_ISL_2453796, EPI_ISL_2453806, EPI_ISL_2453807, EPI_ISL_2453810, EPI_ISL_2453816, EPI_ISL_2453910, EPI_ISL_2453951, EPI_ISL_2453960, EPI_ISL_2453961, EPI_ISL_2453970, EPI_ISL_2843644 | | | |
| see above | MEPHI, Aix Marseille University | MEPHI, Aix Marseille University | Anthony LEVASSEUR |
| EPI_ISL_2787676 | MI - Michigan Department of Health and Human Services - Bureau of Laboratories | Centers for Disease Control and Prevention Division of Viral Diseases, Pathogen Discovery | Alison Laufer Halpin; Ben L. Rambo-Martin; Clinton R. Paden; Dakota Howard; Darlene Wagner; Dave Wentworth; Dhwani Batra; Jasmine Padilla; Justin Lee; Katie Dillon; Krista Queen; Kristen Knipe; Kristine Lacek; Mark Burroughs; Matthew Schmerer; Mili Sheth; Peter Cook; Sam Shepard; Sarah Nobles; Shoshona Le; Suxiang Tong; Vivien Dugan; Yvette Unoarumhi |
| EPI_ISL_2567346 | MIRIALIS CLUSES BECHET | CNR Virus des Infections Respiratoires - France SUD | Antonin Bal; Bruno Lina; Bruno Simon; Gregory Destras; Gwendolyne Burfin; Hadrien Regue; Laurence Josset; Martine Valette; Quentin Semanas |
| EPI_ISL_6492740, EPI_ISL_6492741, EPI_ISL_6492742, EPI_ISL_6492743, EPI_ISL_6492747, EPI_ISL_6492755, EPI_ISL_6492759, EPI_ISL_6492760, EPI_ISL_6492761, EPI_ISL_6492762, EPI_ISL_6492774 | | | |
| see above | MSHS Clinical Microbiology Laboratories | MSHS Pathogen Surveillance Program | Adolfo García-Sastre; Adriana van de Guchte; Ajay Obla; Alberto Paniz-Mondolfi; Ana S. Gonzalez-Reiche; Angela Amoako; Ashley Salimbangon; Betsaida Salom Melo; Bremy Alburquerque; Brianne Ciferri; Charles Gleason; Daniel Floda; Deena R. Altman; Denise Jurczyszak; Emilia Mia Sordillo; Gintaras Deikus; Giulio Kleiner; Gopi Patel; Hala Alshammary; Harm van Bakel; Irina Oussenko; Jayeeta Dutta; Juan Soto; Julia Matthews; Katherine Beach; Kathryn Twyman; Kayla Russo; Komal Srivastava; Levy Sominsky; Mahmoud Awawda; Marta Luksza; Matthew M. Hernandez; Melissa Gitman; Michael D. Nowak; Mitchell J. Sullivan; Nancy Francoeur; Robert Sebra; Sarah Schaefer; Shelcie Fabre; Shwetha Hara Sridhar; Viviana Simon; Ying-Chih Wang; Zenab Khan |
| EPI_ISL_2451640, EPI_ISL_2463001 | MT Public Health Laboratory | Centers for Disease Control and Prevention Division of Viral Diseases, Pathogen Discovery | Alison Laufer Halpin; Ben L. Rambo-Martin; Clinton R. Paden; Dakota Howard; Darlene Wagner; Dave Wentworth; Dhwani Batra; Jasmine Padilla; Justin Lee; Katie Dillon; Krista Queen; Kristen Knipe; Kristine Lacek; Mark Burroughs; Matthew Schmerer; Mili Sheth; Peter Cook; Sam Shepard; Sarah Nobles; Shoshona Le; Suxiang Tong; Vivien Dugan; Yvette Unoarumhi |
| EPI_ISL_2388186 | MVZ Dr. Eberhard & Partner Dortmund | Robert Koch Institute | |
| EPI_ISL_2386240 | MVZ Labor Dr. Reising-Ackermann und Kollegen | Robert Koch Institute | |
| EPI_ISL_2314745, EPI_ISL_2314769, EPI_ISL_2386276 | MVZ für Laboratoriumsmedizin und Mikrobiologie Koblenz-Mittelrhein (Labor Koblenz) | Robert Koch Institute | |
| EPI_ISL_2501161, EPI_ISL_2501545 | Maine HETL | Health and Environmental Testing Laboratory | Barter, M.; Dewey, H.; Grieser, H.; H. and Tewhey, R.; Lynch, R.; Martha, J.; Matluk, N.; Meak, S.; Munger |
| EPI_ISL_2802687, EPI_ISL_2802691, EPI_ISL_3298895, EPI_ISL_4083849 | Maine Health and Environmental Testing Laboratory | Tewhey Lab, The Jackson Laboratory | Barter, M.; Dewey, H.; H. and Tewhey, R.; Iosue, F.; Lynch, R.; Matluk, N.; Munger |
| EPI_ISL_2451212, EPI_ISL_2646930, EPI_ISL_2646959, EPI_ISL_2646981, EPI_ISL_2647041, EPI_ISL_2647100, EPI_ISL_2647267, EPI_ISL_2647276, EPI_ISL_2647279, EPI_ISL_2647285, EPI_ISL_2647303, EPI_ISL_2647314, EPI_ISL_2647321, EPI_ISL_2647323, EPI_ISL_2647325, EPI_ISL_2647376, EPI_ISL_2647434, EPI_ISL_2647439, EPI_ISL_2647450, EPI_ISL_2647451, EPI_ISL_2647463, EPI_ISL_2647917, EPI_ISL_2647965, EPI_ISL_2786548 | | | |
| see above | Mako Medical | Centers for Disease Control and Prevention Division of Viral Diseases, Pathogen Discovery | Adrian Paskey; Benjamin Rambo-Martin; Christopher Gulvick; Clinton R. Paden; Dakota Howard; Darlene Wagner; Dhwani Batra; Duncan MacCannell; Jason Caravas; Kara Moser; Lauren Moon; Matthew Schmerer; Matthew Tugwell; Peter W. Cook; Scott Sammons; Shatavia Morrison; Yvette Unoarumhi |
| EPI_ISL_2598490, EPI_ISL_3012133, EPI_ISL_3012137, EPI_ISL_3012148 | Maryland Genomics, Institute for Genome Sciences, University of Maryland School of Medicine | Maryland Genomics, Institute for Genome Sciences, University of Maryland School of Medicine | Claire M; Fraser; Hazen; Holly; Humphrys; Ivette; Jacques; Jonathan; Kranthi; Lim; Lisa D; Luke J; Mike; Ott; Ravel; Roussey; Sadzewicz; Sandra; Santana-Cruz; Tallon; Tracy; Vavikolanu |

| | | | |
|---|---|---|---|
| EPI_ISL_2550760, EPI_ISL_2550761, EPI_ISL_2550782 | Maryland Public Health Laboratory (MD PHL) | Maryland Public Health Laboratory (MD PHL) | Maryland Department of Health Laboratories Administration |
| EPI_ISL_6680623, EPI_ISL_6680691, EPI_ISL_6680773, EPI_ISL_6680776, EPI_ISL_6680787, EPI_ISL_6680909, EPI_ISL_6680971, EPI_ISL_6680994, EPI_ISL_6680995 | | | |
| see above | Mass General Brigham | Mass General Brigham | A.E.; Adams, G.; Anahtar, M.; B.L.; B.W.; Bauer, M.; Birren; Branda, J.; Carter, A.; Cerrato, F.; Chaluvadi, S.; Chapman; Cusick, C.; D.J.; DeRuff, K.; E. and Sabeti; Flowers, K.; Gallagher, G.; Gladden-Young, A.; Gnirke, A.; Harris, J.; J.E.; K.J.; LaRocque, R.; Lagerborg, K.; Lemieux; Lin; Loreth, C.; MacInnis; Neumann, A.; Normandin, E.; P.C.; Park; Pierce, V.; Reilly, S.; Rosenberg; Rudy, M.; Ryan, E.; S.B.; Shaw, B.; Siddle; Slater, D.; Smole, S.; Tomkins-Tinch, C.; Turbett, S. |
| EPI_ISL_2483791, EPI_ISL_2483803, EPI_ISL_2484937, EPI_ISL_2484959 | Massachusetts State Public Health Laboratory | Massachusetts State Public Health Laboratory | Andrew Lang; Glen Gallagher; Sandra Smole; Timelia Fink |
| EPI_ISL_2450322 | Max von Pettenkofer Institute, Virology, National Reference Center for Retroviruses, LMU Munich | Laboratory for Functional Genome Analysis; Dept. Genomics; Gene Center of the LMU Munich | Alexander Graf; Helmut Blum; Max Muenchhoff; Oliver Keppler; Stefan Krebs |
| EPI_ISL_2501145, EPI_ISL_2663230 | Mayo Clinic & Mayo Clinic Laboratories | Minnesota Department of Health, Public Health Laboratory | Alexandra Lorentz; Jacob Garfin; Matt Plumb; and Xiong Wang |
| EPI_ISL_2387348 | Medizinisches Labor Wahl Lüdenscheid | Robert Koch Institute | |
| EPI_ISL_2533757 | Michael E. Debakey VA Medical Center | Genomics and Discovery, Respiratory Viruses Branch, Division of Viral Diseases, Centers for Disease Control and Prevention | Adam Retchless; Anna Kelleher; Anna Uehara; Brian Lynch; Clinton R. Paden; Dhwani Batra; Haibin Wang; Han Jia Justin Ng; Jasmine Padilla; Jing Zhang; Justin Lee; Krista Queen; Mark Burroughs; Mili Sheth; Morgan Davis; Peter Cook; Rachel Marine; Sarah Nobles; Suxiang Tong; Tara Coalter; Yan Li; Ying Tao |
| EPI_ISL_2433981, EPI_ISL_2434027, EPI_ISL_2434039 | Microbiologia CATLAB | Can Ruti SARS-CoV-2 Sequencing Hub (HUGTiP/IrsiCaixa/IGTP) | Alba Sánchez; Anna Not; Antoni E Bordoy; Bonaventura Clotet; Cristina Casañ; Cristina Esteban; Francesc Catala-Moll; Gemma Clara; Ignacio Blanco; Marc Noguera-Julian; Maria Casadellà; Mariona Parera; Mercedes Guerrero; Montserrat Giménez; Pere-Joan Cardona; Pilar Armengol; Roger Paredes; Verónica Saludes; and Elisa Martró on behalf of the Can Ruti SARS-CoV-2 Sequencing Hub. |
| EPI_ISL_2448032 | Microbiology Department, Laboratori Clínic Metropolitana Nord. Hospital Universitari Germans Trias i Pujol | Can Ruti SARS-CoV-2 Sequencing Hub (HUGTiP/IrsiCaixa/IGTP) | Alba Snchez; Anna Not; Antoni E Bordoy; Bonaventura Clotet; Cristina Casa; Cristina Esteban; Francesc Catala-Moll; Gemma Clara; Ignacio Blanco; Marc Noguera-Julian; Maria Casadell; Mariona Parera; Mercedes Guerrero; Montserrat Gimnez; Pere-Joan Cardona; Pilar Armengol; Roger Paredes; Vernica Saludes; and Elisa Martr on behalf of the Can Ruti SARS-CoV-2 Sequencing Hub. |
| EPI_ISL_2508520, EPI_ISL_2508524, EPI_ISL_2508525, EPI_ISL_2508526, EPI_ISL_2508527 | Microbiology Division, SC DHEC | Microbiology Division, SC DHEC | Flores, H.; Freeman, J. |
| EPI_ISL_2325066, EPI_ISL_2325068, EPI_ISL_2325096, EPI_ISL_2325100, EPI_ISL_2325105, EPI_ISL_2376558, EPI_ISL_2376567, EPI_ISL_2403724 | | | |
| see above | Minnesota Department of Health, Public Health Laboratory | Minnesota Department of Health, Public Health Laboratory | Alexandra Lorentz; Jacob Garfin; Matt Plumb; and Xiong Wang |
| EPI_ISL_2501922, EPI_ISL_2501926, EPI_ISL_2501927 | Missouri State Public Health Laboratory | Missouri State Public Health Laboratory | Ashley New; Joshua Barry; Matthew Sinn |
| EPI_ISL_2399518 | Molecular Diagnostics Pathology Department Mater Dei Hospital Malta | Molecular Diagnostics Pathology Department Mater Dei Hospital Malta | C Cilia; G Zahra; L Grech; M Briffa; R Borg |
| EPI_ISL_2612458 | Montana Public Health Laboratory | Montana Public Health Laboratory | Carrie Biskupiak; Deborah Gibson; Joy Ritter; Michael Dills; Michelle Mozer |
| EPI_ISL_4025823 | NB-Hôpital Georges L. Dumont | New Brunswick - Vitalité Health Network | Allain E.; Chacko S.; Crapoulet N.; Desnoyers G.; Garceau R.; Lacroix J.; Shaw W. |
| EPI_ISL_2452194, EPI_ISL_2452197 | NH Dept. of Health and Human Services Public Health Labs | Centers for Disease Control and Prevention Division of Viral Diseases, Pathogen Discovery | Alison Laufer Halpin; Ben L. Rambo-Martin; Clinton R. Paden; Dakota Howard; Darlene Wagner; Dave Wentworth; Dhwani Batra; Jasmine Padilla; Justin Lee; Katie Dillon; Krista Queen; Kristen Knipe; Kristine Lacek; Mark Burroughs; Matthew Schmerer; Mili Sheth; Peter Cook; Sam Shepard; Sarah Nobles; Shoshona Le; Suxiang Tong; Vivien Dugan; Yvette Unoarumhi |
| EPI_ISL_2376125, EPI_ISL_2557119 | NJDOH, Public Health and Environmental Laboratories | NJ_PHEL | Allison Roder; Byeong Jeong; Chelsea San Filippo; Dana Woell; Jacquelyn Deverell; Lindsey Bodnar; Ryan Pachucki; Shiv K. Verma |
| EPI_ISL_2484592, EPI_ISL_2484594 | NL-Dr. Leonard A. Miller Centre for Health Services | National Microbiology Laboratory (NML) | Adel Malek; Anna Majer; Anneliese Landgraff; CanCOGeN's metadata curation team; Darian Hole; Elsie Grudeski; Gary Van Domselaar; George Zahariadis; Grace Seo; Jennifer Tanner; Kerri Smith; Kirsten Biggar; Laura Gilbert; Madison Chapel; Morag Graham; Natalie Knox; Nathalie Bastien; Philip Mabon; Public Health Agency of Canada CanCOGeN team; Rhiannon Huzarewich; Robert Needle; Russell Mandes; Shari Tyson; Timothy Booth; Yan Li; Yang Yu |
| EPI_ISL_2627153, EPI_ISL_2627154 | NORTHWELL HEALTH LABORATORIES | Wadsworth Center, New York State Department of Health | Alexis Russell; Catharine Prussing; Daryl M. Lamson; Erasmus Schneider; Erica Lasek-Nesselquist; John Kelly; Jonathan Plitnick; Kirsten St. George; Matthew Shudt; Melissa A Leisner; Navjot Singh |
| EPI_ISL_2445131, EPI_ISL_2445138, EPI_ISL_2473738, EPI_ISL_2473743, EPI_ISL_2473750, EPI_ISL_2473751, EPI_ISL_5645996, EPI_ISL_5645999, EPI_ISL_5646026 | | | |
| see above | NUCLEO DE SAUDE VILA FALCAO DE BAURU | Instituto Butantan | Antonio Jorge Martins; Claudia Renata dos Santos Barros; David Schlesinger; Debora Botequio Moretti; Dimas Tadeu Covas; Elaine Cristina Marqueze; Elaine Vieira Santos; Evandra Strazza Rodrigues; Heidge Fukumasu; Jayme Augusto de Souza-Neto; José Salvatore Leister Patané; Luiz Alcantara; Luiz Lehmann Coutinho; Maria Carolina Elias; Maurício Lacerda Nogueira; Rafael dos Santos Bezerra; Raul Machado Neto; Rejane Maria Tommasini Grotto; Ricardo Haddad; Sandra Coccuzzo Sampaio Vessoni; Simone Kashima; Svetoslav Nanev Slavov; Vincent Louis Viala |
| EPI_ISL_5645963, EPI_ISL_5646022 | NUCLEO SAUDE III N HABITACIONAL PRESIDENTE GEISEL BAURU | Instituto Butantan | Antonio Jorge Martins; Claudia Renata dos Santos Barros; David Schlesinger; Debora Botequio Moretti; Dimas Tadeu Covas; Elaine Cristina Marqueze; Elaine Vieira Santos; Evandra Strazza Rodrigues; Heidge Fukumasu; Jayme Augusto de Souza-Neto; José Salvatore Leister Patané; Luiz Alcantara; Luiz Lehmann Coutinho; Maria Carolina Elias; Maurício Lacerda Nogueira; Rafael dos Santos Bezerra; Raul Machado Neto; Rejane Maria Tommasini Grotto; Ricardo Haddad; Sandra Coccuzzo Sampaio Vessoni; Simone Kashima; Svetoslav Nanev Slavov; Vincent Louis Viala |
| EPI_ISL_2427468, EPI_ISL_2427477, EPI_ISL_2458161 | NYU Langone Health | Departments of Pathology and Medicine, New York University School of Medicine | Adriana Heguy; Christian Marier; Dacia Dimartino; Emily Guzman; Gael Westby; Guiqing Wang; Paolo Cotzia; Paul Zappile; Peter Meyn; Sitharam Ramaswami; Yutong Zhang |
| EPI_ISL_2726981, EPI_ISL_2726982 | National Institute of Public Health | National Institute of Public Health | Alexander Nagy; Dusan Trnka; Helena Jirincova; Jaromira Vecerova; Timotej Suri |
| EPI_ISL_2463454 | National Laboratory for Health, Environment and Food, OMM, Maribor | NLZOH (National Laboratory for Health, Environment and Food) / CISLD (Clinical Institute of Special Laboratory Diagnostics), University Children's Hospital, University Medical Center Ljubljana | Aleksander Kocuvan; Aleksander Mahnic; Alenka Štorman; Ana Grom; Andrej Golle / Jernej Kovač; Barbara Jenko Bizjan; Kaja Tominc; Katarina Kozmos; Maja Rupnik; Marko Pokorn; Maruša Debeljak; Maša Jarčič; Mojca Cimerman; Nika Gobec; Nika Volmajer; Robert Šket; Sandra Janezic; Tadej Battelino; Tine Tesovnik; Tjasa Žohar Čretnik |
| EPI_ISL_2271946, EPI_ISL_2379264, EPI_ISL_2466329, EPI_ISL_2466332, EPI_ISL_2466333, EPI_ISL_2466337, EPI_ISL_2483848, EPI_ISL_2483849, EPI_ISL_2483851, EPI_ISL_2483852, EPI_ISL_2483853, EPI_ISL_2483854, EPI_ISL_2545463, EPI_ISL_2545464, EPI_ISL_2545471, EPI_ISL_2549145, EPI_ISL_2687938, EPI_ISL_2687941, EPI_ISL_2687942, EPI_ISL_2687943, EPI_ISL_2687947, EPI_ISL_2687948, EPI_ISL_2687953 | | | |
| see above | National Platform bis UMONS/Jolimont | National Platform bis UMONS/Jolimont | Florian Juszczak; François Dufrasne; Gautier Detry; Guillaume Bayon-Vicente; Ruddy Wattiez |
| EPI_ISL_2233081 | Nebraska Public Health Laboratory | NPHL COVID-19 Response Team | NPHL COVID-19 Response Team |
| EPI_ISL_2277638 | Nevada State Public Health Laboratory | Nevada State Public Health Laboratory | Andrew Gorzalski; Mark Pandori |
| EPI_ISL_2652062 | New Mexico Department of Health Scientific Laboratory | New Mexico Department of Health Scientific Laboratory | Anastacia Griego-Fisher; D'eldra Malone; Ellie Johnson; Jennifer Benoit; Keila Gutierrez; Linda Salazar; Ratheesh Rajan |
| EPI_ISL_2489949, EPI_ISL_2489951, EPI_ISL_2661645, EPI_ISL_2661649, EPI_ISL_2661650, EPI_ISL_2661652, EPI_ISL_2661653, EPI_ISL_2661655, EPI_ISL_2661662, EPI_ISL_2714563 | | | |
| see above | North Dakota Department of Health, Public Health Laboratory | North Dakota Department of Health, Public Health Laboratory | Lisa Wingerter |
| EPI_ISL_2692388, EPI_ISL_2715102, EPI_ISL_2715106, EPI_ISL_2715108, EPI_ISL_2715111, EPI_ISL_2715112, EPI_ISL_2715117, EPI_ISL_2715120, EPI_ISL_2715124, EPI_ISL_2715126, EPI_ISL_2715127, EPI_ISL_2715149, EPI_ISL_2715151, EPI_ISL_2715152, EPI_ISL_2715156, EPI_ISL_2715162, EPI_ISL_2715164, EPI_ISL_2715169, EPI_ISL_2715171, EPI_ISL_2715175 | | | |
| see above | Northwest Laboratory | Seattle Flu Study | Amanda Adler; Barry R. Lutz; Benjamin Pelle; Brian Hiatt; Caitlin R. Wolf; Chris D. Frazar; Deborah A. Nickerson; Elisabeth Brandstetter; Erica Ryke; Geoff Melly; Helen Y. Chu; Janet A. Englund; Jay Shendure; Jover Lee; Kairsten Fay; Kirsten Lacombe; Lea M. Starita; Mark J. Rieder; Matthew Richardson; Matthew Thompson; Melissa Truong; Michael Boeckh; Michael Famulare; Misja Ilcisin; Peter D. Han; Philip Dykema; Romesh Gautom; Scott Lindquist; Thomas R. Sibley; Trevor Bedford |
| EPI_ISL_2373022, EPI_ISL_2373023, EPI_ISL_2373024, EPI_ISL_2373031, EPI_ISL_2373032, EPI_ISL_2373040, EPI_ISL_2373041, EPI_ISL_2373042 | | | |
| see above | Northwestern Memorial Hospital | Northwestern University - Ozer Lab | Chad J. Achenbach; Chao Qi; Egon A. Ozer; Judd F. Hultquist; Lacy M. Simons; Lawrence J. Jennings; Michael G. Ison; Ramon Lorenzo-Redondo; Taylor J. Dean |
| EPI_ISL_2692046, EPI_ISL_2692051 | Northwestern Memorial Hospital | RIPHL at Rush University Medical Center | Amber Kimble; Chao Qi; Felix Araujo Perez; Kevin Kunstman; Laura Furtado; Marieta Hyde; Max Kolton; Stefan Green |
| EPI_ISL_2950658, EPI_ISL_2950961 | Novogenia GmbH | Elling and Cochella laboratories, IMBA/IMP | Alexander Stark; Ezgi Oezkan; Luisa Cochella and Ulrich Elling; Marcus Strobl; Olga Frank; Ramesh Yelagandula; Tanino Albanese |
| EPI_ISL_2838615, EPI_ISL_2838844, EPI_ISL_2838845, EPI_ISL_2838846, EPI_ISL_2838847, EPI_ISL_2838848, EPI_ISL_2838849, EPI_ISL_2838850, EPI_ISL_2838851, EPI_ISL_2838852, EPI_ISL_2838853, EPI_ISL_2838854, EPI_ISL_2838855 | | | |
| see above | OHSU Lab Services Molecular Microbiology Lab | Oregon SARS-CoV-2 Genome Sequencing Center | '[UsersoroakBox SyncPHI_SARS-COV-2_Sample_Sheet_infoGISAID_Submission_Infometa_master_2021.xlsx]release20_final'!D:AC; 26; FALSE); VLOOKUP(C427; VLOOKUP(C428; VLOOKUP(C429; VLOOKUP(C430; VLOOKUP(C433; VLOOKUP(C434; VLOOKUP(C438; VLOOKUP(C440; VLOOKUP(C441; VLOOKUP(C454; VLOOKUP(C455; VLOOKUP(C459; VLOOKUP(C463 |

| | | | |
|---|---|---|---|
| EPI_ISL_2467884 | OK Public Health Laboratory, Oklahoma State DOH | Genomics and Discovery, Respiratory Viruses Branch, Division of Viral Diseases, Centers for Disease Control and Prevention | Adam Retchless; Anna Kelleher; Anna Uehara; Brian Lynch; Clinton R. Paden; Dhwani Batra; Haibin Wang; Han Jia Justin Ng; Jasmine Padilla; Jing Zhang; Justin Lee; Krista Queen; Mark Burroughs; Mili Sheth; Morgan Davis; Peter Cook; Rachel Marine; Sarah Nobles; Suxiang Tong; Tara Coalter; Yan Li; Ying Tao |
| EPI_ISL_2332806, EPI_ISL_2332807, EPI_ISL_2332808, EPI_ISL_5331894, EPI_ISL_5331895, EPI_ISL_5331896 | OLVZ Aalst | OLVZ Aalst | Anne Vankeerberghen; Astrid Holderbeke |
| EPI_ISL_2451980, EPI_ISL_2451989, EPI_ISL_2689978, EPI_ISL_2689988 | OR State PHL-Virology/Immunology Section | Centers for Disease Control and Prevention Division of Viral Diseases, Pathogen Discovery | Alison Laufer Halpin; Ben L. Rambo-Martin; Clinton R. Paden; Dakota Howard; Darlene Wagner; Dave Wentworth; Dhwani Batra; Jasmine Padilla; Justin Lee; Katie Dillon; Krista Queen; Kristen Knipe; Kristine Lacek; Mark Burroughs; Matthew Schmerer; Mili Sheth; Peter Cook; Sam Shepard; Sarah Nobles; Shoshona Le; Suxiang Tong; Vivien Dugan; Yvette Unoarumhi |
| EPI_ISL_2566359 | Operation Infectious Diseases, Naval Health Research Center | Operation Infectious Diseases, Naval Health Research Center | A.E.; Balansay-Ames; C.A.; Fierro, M.; Iniguez-Stevens, E.; Kriner, P.; M.S.; Moser, K.; Myers; Pan; Phippard; R.S.; R.W.; Underwood |
| EPI_ISL_2348953, EPI_ISL_2382530, EPI_ISL_2423509, EPI_ISL_2424248, EPI_ISL_2424255, EPI_ISL_2473661, EPI_ISL_2473662, EPI_ISL_2473663, EPI_ISL_2473672, EPI_ISL_2727477 | | | |
| see above | Oregon State Public Health Laboratory | Oregon State Public Health Laboratory | Eugene Yeboah; John Fontana and Shane Sevey; Laura Tsaknaridis; Rafia Razzaque; Vanda Makris |
| EPI_ISL_2483767 | Ospedale "F. Spaziani" Frosinone | INMI Lazzaro Spallanzani IRCCS | B Bartolini; C Gargiulo; CEM Gruber; F Messina; GF Inger; M Pallocca; M Rueca; O Butera; R Pulselli |
| EPI_ISL_2602867, EPI_ISL_2602868 | Ospedale S.M. Goretti di Latina - ASL Latina presidio ospedaliero Nord | INMI Lazzaro Spallanzani IRCCS | A Lucci; A Massacci; B Bartolini; CEM Gruber; E Giombini; F De Nicola; F Messina; F Santini; G Bonfiglio; G Orlandi; L Corso; L Di Biase; M Pallocca; M Rueca; O Butera |
| EPI_ISL_2602889 | Ospedale San Camillo De Lellis | INMI Lazzaro Spallanzani IRCCS | A De Luca; CEM Gruber; E Giombini; F Santini; GF Inger; M Pallocca; M Rueca; O Butera; S Venarubea |
| EPI_ISL_2602406, EPI_ISL_2602409 | Ospedale San Filippo Neri | INMI Lazzaro Spallanzani IRCCS | A Tamburro; B Bartolini; CEM Gruber; E Sperandio; F De Nicola; F Santini; G Bonfiglio; G Orlandi; M Meledandri; O Butera |
| EPI_ISL_2602421, EPI_ISL_2602427, EPI_ISL_2602430, EPI_ISL_2602431 | Ospedale San Giovanni Evangelista | INMI Lazzaro Spallanzani IRCCS | A Massacci; B Bartolini; CEM Gruber; D Cerini; D Di Fusco; E Giombini; F Messina; F Santini; G Bonfiglio; G Orlandi; GF Inger; M Pallocca; M Rueca; O Butera; S Donzelli |
| EPI_ISL_2483733, EPI_ISL_2483736, EPI_ISL_2483737, EPI_ISL_2483741, EPI_ISL_2483742, EPI_ISL_2483743, EPI_ISL_2483747, EPI_ISL_2483749, EPI_ISL_2483750, EPI_ISL_2483753, EPI_ISL_2483756, EPI_ISL_2483759, EPI_ISL_2483760, EPI_ISL_2602439, EPI_ISL_2602885, EPI_ISL_2602886, EPI_ISL_2602888 | | | |
| see above | Ospedale Sandro Pertini ASL Roma 2 | INMI Lazzaro Spallanzani IRCCS | A Massacci; B Bartolini; CEM Gruber; E Giombini; E Sperandio; F De Nicola; F Messina; F Santini; G Bonfiglio; G Cappiello; G Orlandi; GF Inger; L Ciuffreda; M Pallocca; M Rueca; MC Cava; O Butera; R Longo; S Donzelli; S Romano; V Michela |
| EPI_ISL_2602845, EPI_ISL_2602850, EPI_ISL_2602853, EPI_ISL_2602857 | Ospedale di Genzano - ASL RM 6 | INMI Lazzaro Spallanzani IRCCS | A Massacci; CEM Gruber; E Conti; E Giombini; F De Nicola; F Messina; F Santini; G Bonfiglio; G Orlandi; G Tramini; GF Inger; M Pallocca; M Rueca; O Butera; S Donzelli |
| EPI_ISL_2692514 | Overlake Hospital | Seattle Flu Study | Amanda Adler; Barry R. Lutz; Benjamin Pelle; Brian Hiatt; Caitlin R. Wolf; Chris D. Frazar; Deborah A. Nickerson; Elisabeth Brandstetter; Erica Ryke; Geoff Melly; Helen Y. Chu; Janet A. Englund; Jay Shendure; Jover Lee; Kairsten Fay; Kirsten Lacombe; Lea M. Starita; Mark J. Rieder; Matthew Richardson; Matthew Thompson; Melissa Truong; Michael Boeckh; Michael Famulare; Misja Ilcisin; Peter D. Han; Philip Dykema; Romesh Gautom; Scott Lindquist; Thomas R. Sibley; Trevor Bedford |
| EPI_ISL_2378748, EPI_ISL_2378749, EPI_ISL_2493356, EPI_ISL_2493357, EPI_ISL_2493358, EPI_ISL_2493359 | PAS JOAO ANTONIO DO NASCIMENTO | Instituto Butantan | Antonio Jorge Martins; Claudia Renata dos Santos Barros; David Schlesinger; Debora Botequio Moretti; Dimas Tadeu Covas; Elaine Cristina Marqueze; Elaine Vieira Santos; Evandra Strazza Rodrigues; Heidge Fukumasu; Jayme Augusto de Souza-Neto; José Salvatore Leister Patané; Luiz Alcantara; Luiz Lehmann Coutinho; Maria Carolina Elias; Maurício Lacerda Nogueira; Rafael dos Santos Bezerra; Raul Machado Neto; Rejane Maria Tommasini Grotto; Ricardo Haddad; Sandra Coccuzzo Sampaio Vessoni; Simone Kashima; Svetoslav Nanev Slavov; Vincent Louis Viala |
| EPI_ISL_2500981 | PAS JULIANIA DE HERCULANDIA | Instituto Butantan | Antonio Jorge Martins; Claudia Renata dos Santos Barros; David Schlesinger; Debora Botequio Moretti; Dimas Tadeu Covas; Elaine Cristina Marqueze; Elaine Vieira Santos; Evandra Strazza Rodrigues; Heidge Fukumasu; Jayme Augusto de Souza-Neto; José Salvatore Leister Patané; Luiz Alcantara; Luiz Lehmann Coutinho; Maria Carolina Elias; Maurício Lacerda Nogueira; Rafael dos Santos Bezerra; Raul Machado Neto; Rejane Maria Tommasini Grotto; Ricardo Haddad; Sandra Coccuzzo Sampaio Vessoni; Simone Kashima; Svetoslav Nanev Slavov; Vincent Louis Viala |
| EPI_ISL_2493667, EPI_ISL_2493673 | POLICLINICA COVID 19 ITAPETININGA | Instituto Butantan | Antonio Jorge Martins; Claudia Renata dos Santos Barros; David Schlesinger; Debora Botequio Moretti; Dimas Tadeu Covas; Elaine Cristina Marqueze; Elaine Vieira Santos; Evandra Strazza Rodrigues; Heidge Fukumasu; Jayme Augusto de Souza-Neto; José Salvatore Leister Patané; Luiz Alcantara; Luiz Lehmann Coutinho; Maria Carolina Elias; Maurício Lacerda Nogueira; Rafael dos Santos Bezerra; Raul Machado Neto; Rejane Maria Tommasini Grotto; Ricardo Haddad; Sandra Coccuzzo Sampaio Vessoni; Simone Kashima; Svetoslav Nanev Slavov; Vincent Louis Viala |
| EPI_ISL_6229763 | POSTO DE SAUDE DE CRIOULAS | ACME Lab, Oswaldo Cruz Foundation, FIOCRUZ/CE | Carlos Leonardo de Aragao Araujo; Cecilia Leite Costa & Eduardo Ruback dos Santos on behalf of COVID-19 FIOCRUZ Genomic Network; Cleber Furtado Aksenen; Fabio Miyajima; Fernando Braga Stehling; Francisco Eder de Moura Lopes; Igor Oliveira Duarte; Jamille Maria Mendes Bezerra; Joaquim Cesar do Nascimento Sousa Junior; Pedro Miguel Carneiro Jeronimo; Suzana Porto Almeida; Thais Ferreira de Oliveira; Thais de Oliveira Costa; Ticiane Cavalcante de Souza; Veridiana Pessoa Miyajima |
| EPI_ISL_6229764 | POSTO DE SAUDE DONA LILI | ACME Lab, Oswaldo Cruz Foundation, FIOCRUZ/CE | Carlos Leonardo de Aragao Araujo; Cecilia Leite Costa & Eduardo Ruback dos Santos on behalf of COVID-19 FIOCRUZ Genomic Network; Cleber Furtado Aksenen; Fabio Miyajima; Fernando Braga Stehling; Francisco Eder de Moura Lopes; Igor Oliveira Duarte; Jamille Maria Mendes Bezerra; Joaquim Cesar do Nascimento Sousa Junior; Pedro Miguel Carneiro Jeronimo; Suzana Porto Almeida; Thais Ferreira de Oliveira; Thais de Oliveira Costa; Ticiane Cavalcante de Souza; Veridiana Pessoa Miyajima |
| EPI_ISL_6229696 | POSTO DE SAUDE MAE OTAVIA | ACME Lab, Oswaldo Cruz Foundation, FIOCRUZ/CE | Carlos Leonardo de Aragao Araujo; Cecilia Leite Costa & Eduardo Ruback dos Santos on behalf of COVID-19 FIOCRUZ Genomic Network; Cleber Furtado Aksenen; Fabio Miyajima; Fernando Braga Stehling; Francisco Eder de Moura Lopes; Igor Oliveira Duarte; Jamille Maria Mendes Bezerra; Joaquim Cesar do Nascimento Sousa Junior; Pedro Miguel Carneiro Jeronimo; Suzana Porto Almeida; Thais Ferreira de Oliveira; Thais de Oliveira Costa; Ticiane Cavalcante de Souza; Veridiana Pessoa Miyajima |
| EPI_ISL_6229850 | POSTO DE SAUDE MONSENHOR DIOMEDES | ACME Lab, Oswaldo Cruz Foundation, FIOCRUZ/CE | Carlos Leonardo de Aragao Araujo; Cecilia Leite Costa & Eduardo Ruback dos Santos on behalf of COVID-19 FIOCRUZ Genomic Network; Cleber Furtado Aksenen; Fabio Miyajima; Fernando Braga Stehling; Francisco Eder de Moura Lopes; Igor Oliveira Duarte; Jamille Maria Mendes Bezerra; Joaquim Cesar do Nascimento Sousa Junior; Pedro Miguel Carneiro Jeronimo; Suzana Porto Almeida; Thais Ferreira de Oliveira; Thais de Oliveira Costa; Ticiane Cavalcante de Souza; Veridiana Pessoa Miyajima |
| EPI_ISL_2493393 | POSTO MEDICO CIDADE NOVA | Instituto Butantan | Antonio Jorge Martins; Claudia Renata dos Santos Barros; David Schlesinger; Debora Botequio Moretti; Dimas Tadeu Covas; Elaine Cristina Marqueze; Elaine Vieira Santos; Evandra Strazza Rodrigues; Heidge Fukumasu; Jayme Augusto de Souza-Neto; José Salvatore Leister Patané; Luiz Alcantara; Luiz Lehmann Coutinho; Maria Carolina Elias; Maurício Lacerda Nogueira; Rafael dos Santos Bezerra; Raul Machado Neto; Rejane Maria Tommasini Grotto; Ricardo Haddad; Sandra Coccuzzo Sampaio Vessoni; Simone Kashima; Svetoslav Nanev Slavov; Vincent Louis Viala |
| EPI_ISL_5645968 | POSTO MEDICO EUROPA | Instituto Butantan | Antonio Jorge Martins; Claudia Renata dos Santos Barros; David Schlesinger; Debora Botequio Moretti; Dimas Tadeu Covas; Elaine Cristina Marqueze; Elaine Vieira Santos; Evandra Strazza Rodrigues; Heidge Fukumasu; Jayme Augusto de Souza-Neto; José Salvatore Leister Patané; Luiz Alcantara; Luiz Lehmann Coutinho; Maria Carolina Elias; Maurício Lacerda Nogueira; Rafael dos Santos Bezerra; Raul Machado Neto; Rejane Maria Tommasini Grotto; Ricardo Haddad; Sandra Coccuzzo Sampaio Vessoni; Simone Kashima; Svetoslav Nanev Slavov; Vincent Louis Viala |
| EPI_ISL_2493589, EPI_ISL_2493590, EPI_ISL_2493629 | POSTO MEDICO NATAL DIEGUES DE ESTIVA GERBI | Instituto Butantan | Antonio Jorge Martins; Claudia Renata dos Santos Barros; David Schlesinger; Debora Botequio Moretti; Dimas Tadeu Covas; Elaine Cristina Marqueze; Elaine Vieira Santos; Evandra Strazza Rodrigues; Heidge Fukumasu; Jayme Augusto de Souza-Neto; José Salvatore Leister Patané; Luiz Alcantara; Luiz Lehmann Coutinho; Maria Carolina Elias; Maurício Lacerda Nogueira; Rafael dos Santos Bezerra; Raul Machado Neto; Rejane Maria Tommasini Grotto; Ricardo Haddad; Sandra Coccuzzo Sampaio Vessoni; Simone Kashima; Svetoslav Nanev Slavov; Vincent Louis Viala |
| EPI_ISL_2621181 | PRIME DIAGNOSTICS | Universidad Nacional de Colombia - Laboratorio Genómico One Health | Andres F. Cardona-Rios; Carlos Franco-Muñoz; Carolina Muñoz-Arango; Celeny Ortiz; Daniel O. Maldonado-Perez; Diego A. Álvarez-Díaz; Hector Alejandro Ruiz-Moreno; Idabely Betancur Ortiz; Jorge E. Osorio; Juan P. Hernandez-Ortiz; Karl A Ciuoderis; Katherine Laiton-Donato; Laura Silvana Perez; Lina M. Hurtado; Marcela Mercado-Reyes; Maria Angélica Maya; Maria Stella López; Rita Almanza Payares; Sandra Ines Cano; Simón Villegas Velásquez |
| EPI_ISL_2378747, EPI_ISL_2493277, EPI_ISL_2493355, EPI_ISL_2493694, EPI_ISL_2493695 | PROGRAMA SAUDE DA FAMILIA I IEPE | Instituto Butantan | Antonio Jorge Martins; Claudia Renata dos Santos Barros; David Schlesinger; Debora Botequio Moretti; Dimas Tadeu Covas; Elaine Cristina Marqueze; Elaine Vieira Santos; Evandra Strazza Rodrigues; Heidge Fukumasu; Jayme Augusto de Souza-Neto; José Salvatore Leister Patané; Luiz Alcantara; Luiz Lehmann Coutinho; Maria Carolina Elias; Maurício Lacerda Nogueira; Rafael dos Santos Bezerra; Raul Machado Neto; Rejane Maria Tommasini Grotto; Ricardo Haddad; Sandra Coccuzzo Sampaio Vessoni; Simone Kashima; Svetoslav Nanev Slavov; Vincent Louis Viala |
| EPI_ISL_2493552, EPI_ISL_2493556, EPI_ISL_2493564 | PRONTO SOCORRO DE AGENOR DE CAMPOS MONGAGUA | Instituto Butantan | Antonio Jorge Martins; Claudia Renata dos Santos Barros; David Schlesinger; Debora Botequio Moretti; Dimas Tadeu Covas; Elaine Cristina Marqueze; Elaine Vieira Santos; Evandra Strazza Rodrigues; Heidge Fukumasu; Jayme Augusto de Souza-Neto; José Salvatore Leister Patané; Luiz Alcantara; Luiz Lehmann Coutinho; Maria Carolina Elias; Maurício Lacerda Nogueira; Rafael dos Santos Bezerra; Raul Machado Neto; Rejane Maria Tommasini Grotto; Ricardo Haddad; Sandra Coccuzzo Sampaio Vessoni; Simone Kashima; Svetoslav Nanev Slavov; Vincent Louis Viala |
| EPI_ISL_2493594, EPI_ISL_2493595, EPI_ISL_2493613, EPI_ISL_2493618, EPI_ISL_2493624, EPI_ISL_2493626 | PRONTO SOCORRO MUNICIPAL DE CRAVINHOS | Instituto Butantan | Antonio Jorge Martins; Claudia Renata dos Santos Barros; David Schlesinger; Debora Botequio Moretti; Dimas Tadeu Covas; Elaine Cristina Marqueze; Elaine Vieira Santos; Evandra Strazza Rodrigues; Heidge Fukumasu; Jayme Augusto de Souza-Neto; José Salvatore Leister Patané; Luiz Alcantara; Luiz Lehmann Coutinho; Maria Carolina Elias; Maurício Lacerda Nogueira; Rafael dos Santos Bezerra; Raul Machado Neto; Rejane Maria Tommasini Grotto; Ricardo Haddad; Sandra Coccuzzo Sampaio Vessoni; Simone Kashima; Svetoslav Nanev Slavov; Vincent Louis Viala |
| EPI_ISL_2493179, EPI_ISL_2493189, EPI_ISL_2493640, EPI_ISL_2493644, EPI_ISL_2493645 | PSF VIRGINIO BASSO | Instituto Butantan | Antonio Jorge Martins; Claudia Renata dos Santos Barros; David Schlesinger; Debora Botequio Moretti; Dimas Tadeu Covas; Elaine Cristina Marqueze; Elaine Vieira Santos; Evandra Strazza Rodrigues; Heidge Fukumasu; Jayme Augusto de Souza-Neto; José Salvatore Leister Patané; Luiz Alcantara; Luiz Lehmann Coutinho; Maria Carolina Elias; Maurício Lacerda Nogueira; Rafael dos Santos Bezerra; Raul Machado Neto; Rejane Maria Tommasini Grotto; Ricardo Haddad; Sandra Coccuzzo Sampaio Vessoni; Simone Kashima; Svetoslav Nanev Slavov; Vincent Louis Viala |
| EPI_ISL_2234960, EPI_ISL_2235006, EPI_ISL_2235014, EPI_ISL_2235050, EPI_ISL_2235081, EPI_ISL_2235158, EPI_ISL_2235161, EPI_ISL_2235194, EPI_ISL_2349620, EPI_ISL_2349634, EPI_ISL_2349646, EPI_ISL_2349647, EPI_ISL_2349660, EPI_ISL_2349661, EPI_ISL_4096645, EPI_ISL_5438558, EPI_ISL_5438560, EPI_ISL_5438561, EPI_ISL_5438562, EPI_ISL_5438568, EPI_ISL_5438608 | | | |
| see above | Pandemic Response Lab - NYC | Pandemic Response Lab, R&D | Alex Carpio; Cybill del Castillo; Dylan Law; Haiping Hao; Henry Lee; Isabel Fernandez Escapa; Jon Laurent; Katharine Nelson; Melissa Hopkins; Michael Hammerling; Pradeep Bugga; Shinyoung Clair Kang; Sol Rey; William Ward |
| EPI_ISL_2451606 | Philadelphia Department of Public Health | Centers for Disease Control and Prevention Division of Viral Diseases, Pathogen Discovery | Alison Laufer Halpin; Ben L. Rambo-Martin; Clinton R. Paden; Dakota Howard; Darlene Wagner; Dave Wentworth; Dhwani Batra; Jasmine Padilla; Justin Lee; Katie Dillon; Krista Queen; Kristen Knipe; Kristine Lacek; Mark Burroughs; Matthew Schmerer; Mili Sheth; Peter Cook; Sam Shepard; Sarah Nobles; Shoshona Le; Suxiang Tong; Vivien Dugan; Yvette Unoarumhi |
| EPI_ISL_6907363, EPI_ISL_6907368, EPI_ISL_6907377, EPI_ISL_6907378, EPI_ISL_6907385, EPI_ISL_6907390, EPI_ISL_6907392, EPI_ISL_6907967, EPI_ISL_6907973, EPI_ISL_6907980, EPI_ISL_6907986 | | | |
| see above | Plataforma de Vigilancia Molecular (PVM) - FIOCRUZ/BA | Plataforma de Vigilancia Molecular (PVM) - FIOCRUZ/BA | Bruno Bezerril Andrade; Camila I. de Oliveira; Clarissa Araújo Gurgel; Eduardo Oyama; Francys A Rangel; Laila A da Silva; Laise de Moraes; Leonardo Paiva Farias; Marina Cucco; Pedro Silveira; Ricardo Khouri on behalf of the Fiocruz COVID-19 Genomic Surveillance Network.; Tiago Graf |
| EPI_ISL_2264252, EPI_ISL_2285226, EPI_ISL_2285227, EPI_ISL_2285822, EPI_ISL_2285825, EPI_ISL_2325040, EPI_ISL_2325041, EPI_ISL_2325046, EPI_ISL_2325048 | | | |
| see above | Plateforme de testing Namuroise | Plateforme de testing Namuroise | Degosserie Jonathan; Denis Olivier; Lesly Nyinkeu Kemamen; Mullier François; Otto Gaetan |

| | | | |
|---|---|---|---|
| EPI_ISL_2246082, EPI_ISL_2246113, EPI_ISL_2272758, EPI_ISL_2331844, EPI_ISL_2362593, EPI_ISL_2362594, EPI_ISL_2362610, EPI_ISL_2362613 | | | |
| see above | Platform BIS UZA/UAntwerpen | Labo Klinische Biologie, UZA | Basil Britto Xavier; Christine Lammens; Herman Goossens; Jasmine Coppens; Marie Le Mercier; Veerle Matheeussen |
| EPI_ISL_2759149 | Population Medicine and Diagnostic Sciences, Cornell University | Population Medicine and Diagnostic Sciences, Cornell University | Anderson; B.D.; Caserta; Cronk; D.G.; Diel; L.C.; Laverack, M.; Mitchell; P.K.; Plocharczyk, E.; R.R.; Venugopalan, R. |
| EPI_ISL_7406391, EPI_ISL_7406400, EPI_ISL_7406407 | Programa Laboratorio de Salud Pública "Dr Dalmiro Pérez Laborda" | Área de Secuenciación del Laboratorio de Virología del Hospital de Niños Dr. Ricardo Gutierrez on behalf of 'Proyecto Argentino Interinstitucional de genomica de SARS-CoV-2' (PAIS Consortium) | A; AM; Acuña; Aguilera; BH; Bohn; Cabral Bombardieri; Campos; Carrizo; D; E; Esteves; F; G; Goya; J; JM; JP; Jofré; L; LE; Lacaze; Lusso; M; MA; MI; MS; Mastrodonato; Molina; Nabaes Jodar; Natale; Olivera; Perez Diaz; Peñalva; Quijano; Rivero; Rosales; S; Talia; Valinotto; Viegas, M. |
| EPI_ISL_2699437, EPI_ISL_2709021, EPI_ISL_2709031, EPI_ISL_2709034, EPI_ISL_2709048, EPI_ISL_2709092, EPI_ISL_2709099, EPI_ISL_2709110, EPI_ISL_2709119, EPI_ISL_2709123, EPI_ISL_2709124, EPI_ISL_2709126, EPI_ISL_2709130, EPI_ISL_2709145, EPI_ISL_2709149, EPI_ISL_2709151 | | | |
| see above | Public Health Laboratory, Minnesota Department of Health | University of Minnesota Genomics Center | Corbin Dirkx; Daryl M. Gohl; Jaquelyn Kuriger-Laber; John Garbe; and Sean Wang |
| EPI_ISL_2399465, EPI_ISL_2399466, EPI_ISL_2399467, EPI_ISL_2399468, EPI_ISL_2399469, EPI_ISL_2399472, EPI_ISL_2399473, EPI_ISL_2399486, EPI_ISL_2399487, EPI_ISL_2399502, EPI_ISL_2399504, EPI_ISL_2399505, EPI_ISL_2399508, EPI_ISL_2399511, EPI_ISL_2399512, EPI_ISL_2399514, EPI_ISL_2399515, EPI_ISL_2439774, EPI_ISL_2451402, EPI_ISL_2451403, EPI_ISL_2451404, EPI_ISL_2451405, EPI_ISL_2451406, EPI_ISL_2451407, EPI_ISL_2451408, EPI_ISL_2451409, EPI_ISL_2451410, EPI_ISL_2451411, EPI_ISL_2451412, EPI_ISL_2451413, EPI_ISL_2451414, EPI_ISL_2451415, EPI_ISL_2451416, EPI_ISL_2451417, EPI_ISL_2451418, EPI_ISL_2451419, EPI_ISL_2451420, EPI_ISL_2451421, EPI_ISL_2451422, EPI_ISL_2451423, EPI_ISL_2451424, EPI_ISL_2451425, EPI_ISL_2451426, EPI_ISL_2451427, EPI_ISL_2451429, EPI_ISL_2451430, EPI_ISL_2451431, EPI_ISL_2451432, EPI_ISL_2451433, EPI_ISL_2451434, EPI_ISL_2451435, EPI_ISL_2451436, EPI_ISL_2451437, EPI_ISL_2451438, EPI_ISL_2451439, EPI_ISL_2451440, EPI_ISL_2451441, EPI_ISL_2451442, EPI_ISL_2451443, EPI_ISL_2451444, EPI_ISL_2451445, EPI_ISL_2451446, EPI_ISL_2451447, EPI_ISL_2451448, EPI_ISL_2451449, EPI_ISL_2451450, EPI_ISL_2451451, EPI_ISL_2451453, EPI_ISL_2451454, EPI_ISL_2451455, EPI_ISL_2451456, EPI_ISL_2451457, EPI_ISL_2451460, EPI_ISL_2451464, EPI_ISL_2451466, EPI_ISL_2451467, EPI_ISL_2451468, EPI_ISL_2451469, EPI_ISL_2451471, EPI_ISL_2451472, EPI_ISL_2451474, EPI_ISL_2451476, EPI_ISL_2451477, EPI_ISL_2451478, EPI_ISL_2451479, EPI_ISL_2451480, EPI_ISL_2451481, EPI_ISL_2451482, EPI_ISL_2451484, EPI_ISL_2451485, EPI_ISL_2451486, EPI_ISL_2451487, EPI_ISL_2451488, EPI_ISL_2451489, EPI_ISL_2451490, EPI_ISL_2451507, EPI_ISL_2451508, EPI_ISL_2451510, EPI_ISL_2451511, EPI_ISL_2451512, EPI_ISL_2451513, EPI_ISL_2451514, EPI_ISL_2451515, EPI_ISL_2451516, EPI_ISL_2451517, EPI_ISL_2451519, EPI_ISL_2451520, EPI_ISL_2451521, EPI_ISL_2451522, EPI_ISL_2451523, EPI_ISL_2451525, EPI_ISL_2451526, EPI_ISL_2451528, EPI_ISL_2451531, EPI_ISL_2451532, EPI_ISL_2451534, EPI_ISL_2451536, EPI_ISL_2451537, EPI_ISL_2451539, EPI_ISL_2451540, EPI_ISL_2451542, EPI_ISL_2451544, EPI_ISL_2451545, EPI_ISL_2451547, EPI_ISL_2451548, EPI_ISL_2451553, EPI_ISL_2451554, EPI_ISL_2451556, EPI_ISL_2451560, EPI_ISL_2451561, EPI_ISL_2451562, EPI_ISL_4368888, EPI_ISL_4372333 | | | |
| see above | Quest Diagnostics Incorporated | Centers for Disease Control and Prevention Division of Viral Diseases, Pathogen Discovery | A. Gerasimova; A. Perez; Adrian Paskey; B. Anderson; Benjamin Rambo-Martin; Christopher Gulvick; Clinton Paden; Clinton R. Paden; Dakota Howard; Darlene Wagner; Dhwani Batra; Duncan MacCannell; Erisa Sula; F. Lacbawan; I. A. Shlyakhter; I. Shlyakhter; Jason Caravas; K. Livingston; K.E. Livingston; Kara Moser; Kristine Lacek; L. Bernstein; L.E. Bernstein; M. Hua; Matthew Schmerer; P. Tanpaiboon; Peter Cook; Peter W. Cook; R. Kagan; R. M. Kagan; R. Owen; R. Rolando; R. V. Rolando; S. H. Rosenthal; S. Rosenthal; Scott Sammons; Shatavia Morrison; Tymeckia Kendall; Victoria Caban Figueroa; Y. Liu; Yvette Unoarumhi |
| EPI_ISL_2603312, EPI_ISL_2725578, EPI_ISL_2725579 | Rhode Island State Health Laboratory | Rhode Island State Health Laboratory | Ewa King; Kristin Carpenter-Azevedo; Richard C. Huard |
| EPI_ISL_2313342, EPI_ISL_2313344, EPI_ISL_2313348 | Robert Koch-Institut ZBS1 | Robert Koch Institute | Annika Brinkmann |
| EPI_ISL_2691936, EPI_ISL_2691937, EPI_ISL_2691939, EPI_ISL_2691940, EPI_ISL_2691944, EPI_ISL_2691945, EPI_ISL_2691948, EPI_ISL_2691952, EPI_ISL_2691957 | | | |
| see above | Rush University Medical Center | RIPHL at Rush University Medical Center | Diane Springer; Felix Araujo Perez; Joyce Houlihan; Kevin Kunstman; Laura Furtado; Marieta Hyde; Mary Hayden; Max Kolton; Stefan Green |
| EPI_ISL_3356302 | S.C. Laboratorio Analisi, ASL 3 Liguria | U.O. Igiene, Ospedale Policlinico San Martino | Bruzzone Bianca; Caligiuri Patrizia; De Pace Vanessa; Domnich Alexander; Icardi Giancarlo on behalf of SARS-CoV-2 ITALIAN RESEARCH ENTERPRISE-(SCIRE) Collaborative Group; Orsi Andrea; Ricucci Valentina; Spitaleri Antonino |
| EPI_ISL_2445155, EPI_ISL_2445156, EPI_ISL_2445159, EPI_ISL_2445161, EPI_ISL_2445162, EPI_ISL_2445163, EPI_ISL_2445166, EPI_ISL_2445167, EPI_ISL_2445170, EPI_ISL_2445172, EPI_ISL_2445174, EPI_ISL_2473708, EPI_ISL_2473709 | | | |
| see above | SAE CTA SERV ATEND ESPEC TESTAGEM E ACONSELHAMENTO | Instituto Butantan | Antonio Jorge Martins; Claudia Renata dos Santos Barros; David Schlesinger; Debora Botequio Moretti; Dimas Tadeu Covas; Elaine Cristina Marqueze; Elaine Vieira Santos; Evandra Strazza Rodrigues; Heidge Fukumasu; Jayme Augusto de Souza-Neto; José Salvatore Leister Patané; Luiz Lehmann Coutinho; Maria Carolina Elias; Maurício Lacerda Nogueira; Rafael dos Santos Bezerra; Raul Machado Neto; Rejane Maria Tommasini Grotto; Ricardo Haddad; Sandra Coccuzzo Sampaio Vessoni; Simone Kashima; Svetoslav Nanev Slavov; Vincent Louis Viala |
| EPI_ISL_2621209 | SAN VICENTE FUNDACION | Universidad Nacional de Colombia - Laboratorio Genómico One Health | Andres F. Cardona-Rios; Carlos Franco-Muñoz; Carolina Muñoz-Arango; Celeny Ortiz; Daniel O. Maldonado-Perez; Diego A. Álvarez-Díaz; Hector Alejandro Ruiz-Moreno; Idabely Betancur Ortiz; Jorge E. Osorio; Juan P. Hernandez-Ortiz; Karl A Ciuoderis; Katherine Laiton-Donato; Laura Silvana Perez; Lina M. Hurtado; Marcela Mercado-Reyes; Maria Angélica Maya; Maria Stella López; Rita Almanza Payares; Sandra Ines Cano; Simón Villegas Velásquez |
| EPI_ISL_2445139, EPI_ISL_2445140, EPI_ISL_2445141, EPI_ISL_2473755 | SANTA CASA DE DOIS CORREGOS | Instituto Butantan | Antonio Jorge Martins; Claudia Renata dos Santos Barros; David Schlesinger; Debora Botequio Moretti; Dimas Tadeu Covas; Elaine Cristina Marqueze; Elaine Vieira Santos; Evandra Strazza Rodrigues; Heidge Fukumasu; Jayme Augusto de Souza-Neto; José Salvatore Leister Patané; Luiz Lehmann Coutinho; Maria Carolina Elias; Maurício Lacerda Nogueira; Rafael dos Santos Bezerra; Raul Machado Neto; Rejane Maria Tommasini Grotto; Ricardo Haddad; Sandra Coccuzzo Sampaio Vessoni; Simone Kashima; Svetoslav Nanev Slavov; Vincent Louis Viala |
| EPI_ISL_2493681 | SANTA CASA DE MISERICORDIA DE TIETE | Instituto Butantan | Antonio Jorge Martins; Claudia Renata dos Santos Barros; David Schlesinger; Debora Botequio Moretti; Dimas Tadeu Covas; Elaine Cristina Marqueze; Elaine Vieira Santos; Evandra Strazza Rodrigues; Heidge Fukumasu; Jayme Augusto de Souza-Neto; José Salvatore Leister Patané; Luiz Lehmann Coutinho; Maria Carolina Elias; Maurício Lacerda Nogueira; Rafael dos Santos Bezerra; Raul Machado Neto; Rejane Maria Tommasini Grotto; Ricardo Haddad; Sandra Coccuzzo Sampaio Vessoni; Simone Kashima; Svetoslav Nanev Slavov; Vincent Louis Viala |
| EPI_ISL_2986088 | SARS-CoV-2 Sequencing Castilla y Leon-Spain Consortium | SARS-CoV-2 Sequencing Castilla y Leon-Spain Consortium | Antonio Orduña-Domingo; Carlos Fuster Foz; Carmen Aldea-Mansilla; Carmen Gimeno Crespo; David Abad; Gabriel March Rossello; Gregoria Megías Lobón; Jose María Eiros Bouza; M. Isabel Fernandez-Natal; Marta Dominguez-Gil; Marta Hernandez; María Antonia García Castro; Mª Fe Brezmes-Valdivieso; Noelia Arenal Andrés; Silvia Rojo; Sonsoles Garcinuño Pérez |
| EPI_ISL_2447611, EPI_ISL_2447612, EPI_ISL_2764736 | SARS-CoV-2 testing team, National Institute of Infectious Diseases | Pathogen Genomics Center, National Institute of Infectious Diseases | Hazuka Y Furihata; Hiromizu Takahashi; Kentaro Itokawa; Makoto Kuroda; Masanori Hashino; Masumichi Saito; Naomi Nojiri; Nozomu Hanaoka; Rina Tanaka; Sana Uchikoba; Tsuguto Fujimoto; Tsuyoshi Sekizuka |
| EPI_ISL_2451972, EPI_ISL_2645318, EPI_ISL_2645320 | SC Dept of Health and Env. Control-Bureau of Laboratories | Centers for Disease Control and Prevention Division of Viral Diseases, Pathogen Discovery | Alison Laufer Halpin; Ben L. Rambo-Martin; Clinton R. Paden; Dakota Howard; Darlene Wagner; Dave Wentworth; Dhwani Batra; Jasmine Padilla; Justin Lee; Katie Dillon; Krista Queen; Kristen Knipe; Kristine Lacek; Mark Burroughs; Matthew Schmerer; Mili Sheth; Peter Cook; Sam Shepard; Sarah Nobles; Shoshona Le; Suxiang Tong; Vivien Dugan; Yvette Unoarumhi |
| EPI_ISL_3355017 | SD Public Health Laboratory | Centers for Disease Control and Prevention Division of Viral Diseases, Pathogen Discovery | Alex Burgin; Ben Rambo-Martin; Clinton Paden; Dakota Howard; Dave Wentworth; Dhwani Batra; Jasmine Padilla; Justin Lee; Krista Queen; Kristen Knipe; Kristine Lacek; Mark Burroughs; Matthew Schmerer; Meghan Bentz; Mili Sheth; Peter Cook; Sam Shepard; Sarah Nobles; Suxiang Tong; Vivien Dugan; Yvette Unoarumhi |
| EPI_ISL_2493619, EPI_ISL_2493621, EPI_ISL_2493627 | SECRETARIA DE SAUDE | Instituto Butantan | Antonio Jorge Martins; Claudia Renata dos Santos Barros; David Schlesinger; Debora Botequio Moretti; Dimas Tadeu Covas; Elaine Cristina Marqueze; Elaine Vieira Santos; Evandra Strazza Rodrigues; Heidge Fukumasu; Jayme Augusto de Souza-Neto; José Salvatore Leister Patané; Luiz Lehmann Coutinho; Maria Carolina Elias; Maurício Lacerda Nogueira; Rafael dos Santos Bezerra; Raul Machado Neto; Rejane Maria Tommasini Grotto; Ricardo Haddad; Sandra Coccuzzo Sampaio Vessoni; Simone Kashima; Svetoslav Nanev Slavov; Vincent Louis Viala |
| EPI_ISL_2494147, EPI_ISL_2494148 | SECRETARIA DE SAUDE DE RAFARD | Instituto Butantan | Antonio Jorge Martins; Claudia Renata dos Santos Barros; David Schlesinger; Debora Botequio Moretti; Dimas Tadeu Covas; Elaine Cristina Marqueze; Elaine Vieira Santos; Evandra Strazza Rodrigues; Heidge Fukumasu; Jayme Augusto de Souza-Neto; José Salvatore Leister Patané; Luiz Lehmann Coutinho; Maria Carolina Elias; Maurício Lacerda Nogueira; Rafael dos Santos Bezerra; Raul Machado Neto; Rejane Maria Tommasini Grotto; Ricardo Haddad; Sandra Coccuzzo Sampaio Vessoni; Simone Kashima; Svetoslav Nanev Slavov; Vincent Louis Viala |
| EPI_ISL_2493373, EPI_ISL_2493389, EPI_ISL_2493390, EPI_ISL_2494149, EPI_ISL_2494156 | SECRETARIA DE SAUDE DE SANTA GERTRUDES | Instituto Butantan | Antonio Jorge Martins; Claudia Renata dos Santos Barros; David Schlesinger; Debora Botequio Moretti; Dimas Tadeu Covas; Elaine Cristina Marqueze; Elaine Vieira Santos; Evandra Strazza Rodrigues; Heidge Fukumasu; Jayme Augusto de Souza-Neto; José Salvatore Leister Patané; Luiz Lehmann Coutinho; Maria Carolina Elias; Maurício Lacerda Nogueira; Rafael dos Santos Bezerra; Raul Machado Neto; Rejane Maria Tommasini Grotto; Ricardo Haddad; Sandra Coccuzzo Sampaio Vessoni; Simone Kashima; Svetoslav Nanev Slavov; Vincent Louis Viala |
| EPI_ISL_2493281, EPI_ISL_2493371, EPI_ISL_2493388, EPI_ISL_2494155 | SECRETARIA DE SAUDE DE SAO PEDRO | Instituto Butantan | Antonio Jorge Martins; Claudia Renata dos Santos Barros; David Schlesinger; Debora Botequio Moretti; Dimas Tadeu Covas; Elaine Cristina Marqueze; Elaine Vieira Santos; Evandra Strazza Rodrigues; Heidge Fukumasu; Jayme Augusto de Souza-Neto; José Salvatore Leister Patané; Luiz Lehmann Coutinho; Maria Carolina Elias; Maurício Lacerda Nogueira; Rafael dos Santos Bezerra; Raul Machado Neto; Rejane Maria Tommasini Grotto; Ricardo Haddad; Sandra Coccuzzo Sampaio Vessoni; Simone Kashima; Svetoslav Nanev Slavov; Vincent Louis Viala |
| EPI_ISL_2493598, EPI_ISL_2493601, EPI_ISL_2493603, EPI_ISL_2493612 | SECRETARIA MUNICIPAL DA SAUDE DE GUARIBA | Instituto Butantan | Antonio Jorge Martins; Claudia Renata dos Santos Barros; David Schlesinger; Debora Botequio Moretti; Dimas Tadeu Covas; Elaine Cristina Marqueze; Elaine Vieira Santos; Evandra Strazza Rodrigues; Heidge Fukumasu; Jayme Augusto de Souza-Neto; José Salvatore Leister Patané; Luiz Lehmann Coutinho; Maria Carolina Elias; Maurício Lacerda Nogueira; Rafael dos Santos Bezerra; Raul Machado Neto; Rejane Maria Tommasini Grotto; Ricardo Haddad; Sandra Coccuzzo Sampaio Vessoni; Simone Kashima; Svetoslav Nanev Slavov; Vincent Louis Viala |
| EPI_ISL_2493577, EPI_ISL_2493579, EPI_ISL_2493581, EPI_ISL_2493630, EPI_ISL_2493633, EPI_ISL_2493635, EPI_ISL_2493637, EPI_ISL_2494277, EPI_ISL_2494278 | | | |
| see above | SECRETARIA MUNICIPAL DE SAUDE | Instituto Butantan | Antonio Jorge Martins; Claudia Renata dos Santos Barros; David Schlesinger; Debora Botequio Moretti; Dimas Tadeu Covas; Elaine Cristina Marqueze; Elaine Vieira Santos; Evandra Strazza Rodrigues; Heidge Fukumasu; Jayme Augusto de Souza-Neto; José Salvatore Leister Patané; Luiz Lehmann Coutinho; Maria Carolina Elias; Maurício Lacerda Nogueira; Rafael dos Santos Bezerra; Raul Machado Neto; Rejane Maria Tommasini Grotto; Ricardo Haddad; Sandra Coccuzzo Sampaio Vessoni; Simone Kashima; Svetoslav Nanev Slavov; Vincent Louis Viala |
| EPI_ISL_2493184, EPI_ISL_2493283, EPI_ISL_2493382, EPI_ISL_2493396, EPI_ISL_2493427, EPI_ISL_2493428, EPI_ISL_2494158 | | | |
| see above | SECRETARIA MUNICIPAL DE SAUDE DE CORDEIROPOLIS | Instituto Butantan | Antonio Jorge Martins; Claudia Renata dos Santos Barros; David Schlesinger; Debora Botequio Moretti; Dimas Tadeu Covas; Elaine Cristina Marqueze; Elaine Vieira Santos; Evandra Strazza Rodrigues; Heidge Fukumasu; Jayme Augusto de Souza-Neto; José Salvatore Leister Patané; Luiz Lehmann Coutinho; Maria Carolina Elias; Maurício Lacerda Nogueira; Rafael dos Santos Bezerra; Raul Machado Neto; Rejane Maria Tommasini Grotto; Ricardo Haddad; Sandra Coccuzzo Sampaio Vessoni; Simone Kashima; Svetoslav Nanev Slavov; Vincent Louis Viala |
| EPI_ISL_5645964, EPI_ISL_5645971, EPI_ISL_5645980, EPI_ISL_5646001, EPI_ISL_5646002, EPI_ISL_5646008, EPI_ISL_5646009, EPI_ISL_5646017 | | | |
| see above | SECRETARIA MUNICIPAL DE SAUDE DE PEDERNEIRAS | Instituto Butantan | Antonio Jorge Martins; Claudia Renata dos Santos Barros; David Schlesinger; Debora Botequio Moretti; Dimas Tadeu Covas; Elaine Cristina Marqueze; Elaine Vieira Santos; Evandra Strazza Rodrigues; Heidge Fukumasu; Jayme Augusto de Souza-Neto; José Salvatore Leister Patané; Luiz Lehmann Coutinho; Maria Carolina Elias; Maurício Lacerda Nogueira; Rafael dos Santos Bezerra; Raul Machado Neto; Rejane Maria Tommasini Grotto; Ricardo Haddad; Sandra Coccuzzo Sampaio Vessoni; Simone Kashima; Svetoslav Nanev Slavov; Vincent Louis Viala |
| EPI_ISL_2801338, EPI_ISL_2801339, EPI_ISL_2801340, EPI_ISL_2801342, EPI_ISL_2801351, EPI_ISL_2801354 | SECRETARIA MUNICIPAL DE SAUDE DE TIANGUA | Analytical Competence Molecular Epidemiology Lab/ACME, Oswaldo Cruz Foundation, Ceara (FIOCRUZ CE) | Cleber Furtado Aksenen e Suzana Porto Almeida; Fabio Miyajima; Fernando Braga Stehling; Francisco Eder de Moura Lopes; Jamille Maria Mendes Bezerra; Joaquim César do Nascimento Sousa Junior; Pedro Miguel Carneiro Jeronimo; Thais Ferreira de Oliveira; Thais de Oliveira Costa; Ticiane Cavalcante de Souza; Veridiana Pessoa Miyajima |
| EPI_ISL_2493679 | SECRETARIA MUNICIPAL DE SAUDE PORTO FERREIRA | Instituto Butantan | Antonio Jorge Martins; Claudia Renata dos Santos Barros; David Schlesinger; Debora Botequio Moretti; Dimas Tadeu Covas; Elaine Cristina Marqueze; Elaine Vieira Santos; Evandra Strazza Rodrigues; Heidge Fukumasu; Jayme Augusto de Souza-Neto; José Salvatore Leister Patané; Luiz Lehmann Coutinho; Maria Carolina Elias; Maurício Lacerda Nogueira; Rafael dos Santos Bezerra; Raul Machado Neto; Rejane Maria Tommasini Grotto; Ricardo Haddad; Sandra Coccuzzo Sampaio Vessoni; Simone Kashima; Svetoslav Nanev Slavov; Vincent Louis Viala |
| EPI_ISL_2493654, EPI_ISL_2493657, EPI_ISL_2493660, EPI_ISL_2493668, EPI_ISL_2493674, EPI_ISL_2493690, EPI_ISL_2493691, EPI_ISL_2493692, EPI_ISL_2493693, EPI_ISL_2493697, EPI_ISL_2493698, EPI_ISL_2493700, EPI_ISL_2500979, EPI_ISL_2500983 | | | |
| see above | SECRETARIA MUNICIPAL DE SAUDE | Instituto Butantan | Antonio Jorge Martins; Claudia Renata dos Santos Barros; David Schlesinger; Debora Botequio Moretti; Dimas Tadeu Covas; Elaine Cristina Marqueze; Elaine Vieira Santos; Evandra Strazza Rodrigues; Heidge Fukumasu; Jayme Augusto de Souza-Neto; José Salvatore Leister Patané; Luiz Alcantara; Luiz |

| | | | |
|---|---|---|---|
| | SOROCABA | | Lehmann Coutinho; Maria Carolina Elias; Maurício Lacerda Nogueira; Rafael dos Santos Bezerra; Raul Machado Neto; Rejane Maria Tommasini Grotto; Ricardo Haddad; Sandra Coccuzzo Sampaio Vessoni; Simone Kashima; Svetoslav Nanev Slavov; Vincent Louis Viala |
| EPI_ISL_2493568 | SERV DE VIG SANITARIA EPIDEMIO E CTRL DE ZOONOZES GUARUJA | Instituto Butantan | Antonio Jorge Martins; Claudia Renata dos Santos Barros; David Schlesinger; Debora Botequio Moretti; Dimas Tadeu Covas; Elaine Cristina Marqueze; Elaine Vieira Santos; Evandra Strazza Rodrigues; Heidge Fukumasu; Jayme Augusto de Souza-Neto; José Salvatore Leister Patané; Luiz Alcantara; Luiz Lehmann Coutinho; Maria Carolina Elias; Maurício Lacerda Nogueira; Rafael dos Santos Bezerra; Raul Machado Neto; Rejane Maria Tommasini Grotto; Ricardo Haddad; Sandra Coccuzzo Sampaio Vessoni; Simone Kashima; Svetoslav Nanev Slavov; Vincent Louis Viala |
| EPI_ISL_2483583, EPI_ISL_2483588 | SIESP CHIETI - DRIVE IN ORTONA | Istituto Zooprofilattico Sperimentale dell'Abruzzo e Molise "G. Caporale" | Ancora M; Calistri P; Cammà C; Caporale M; Curini V; Delli Compagni E; Di Domenico M; Di Lollo Valeria; Di Pasquale A; Lorusso A; Mangone I; Marcacci M; Puglia I; Rinaldi A; Savini G; Scialabba S |
| EPI_ISL_2483590 | SIESP DIPARTIMENTO DI PREVENZIONE CHIETI | Istituto Zooprofilattico Sperimentale dell'Abruzzo e Molise "G. Caporale" | Ancora M; Calistri P; Cammà C; Caporale M; Curini V; Delli Compagni E; Di Domenico M; Di Lollo Valeria; Di Pasquale A; Lorusso A; Mangone I; Marcacci M; Puglia I; Rinaldi A; Savini G; Scialabba S |
| EPI_ISL_2308558, EPI_ISL_2308560, EPI_ISL_2308564, EPI_ISL_2308587, EPI_ISL_2308599, EPI_ISL_2483584, EPI_ISL_2483585, EPI_ISL_2483586, EPI_ISL_2483587, EPI_ISL_2483589 | | | |
| see above | SIESP DIPARTIMENTO DI PREVENZIONE TERAMO | Istituto Zooprofilattico Sperimentale dell'Abruzzo e Molise "G. Caporale" | Ancora M; Calistri P; Cammà C; Caporale M; Curini V; Delli Compagni E; Di Domenico M; Di Lollo Valeria; Di Pasquale A; Lorusso A; Mangone I; Marcacci M; Puglia I; Rinaldi A; Savini G; Scialabba S |
| EPI_ISL_5844439, EPI_ISL_5844496, EPI_ISL_5844513, EPI_ISL_5844518, EPI_ISL_5844519, EPI_ISL_5844560, EPI_ISL_5844567, EPI_ISL_5844568, EPI_ISL_5844573, EPI_ISL_5844581, EPI_ISL_5844588, EPI_ISL_5844593, EPI_ISL_5844596, EPI_ISL_5844597, EPI_ISL_5844604, EPI_ISL_5844608, EPI_ISL_5844621, EPI_ISL_5844623, EPI_ISL_5844625, EPI_ISL_5844627, EPI_ISL_5844633, EPI_ISL_5844642, EPI_ISL_5844665, EPI_ISL_5844666, EPI_ISL_5844676 | | | |
| see above | SK-Roy Romanow Provincial Laboratory | Saskatchewan - Roy Romanow Provincial Laboratory (RRPL) | Alanna Senecal; Amanda Lang; Jessica Minion; Kara Loos; Keith MacKenzie; Meredith Faires; Rachel DePaulo; Roy Romanow Provincial Laboratory - Molecular Diagnostics; Ryan McDonald |
| EPI_ISL_2493671 | SMS SECRETARIA MUNICIPAL DE SAUDE DE BOITUVA | Instituto Butantan | Antonio Jorge Martins; Claudia Renata dos Santos Barros; David Schlesinger; Debora Botequio Moretti; Dimas Tadeu Covas; Elaine Cristina Marqueze; Elaine Vieira Santos; Evandra Strazza Rodrigues; Heidge Fukumasu; Jayme Augusto de Souza-Neto; José Salvatore Leister Patané; Luiz Alcantara; Luiz Lehmann Coutinho; Maria Carolina Elias; Maurício Lacerda Nogueira; Rafael dos Santos Bezerra; Raul Machado Neto; Rejane Maria Tommasini Grotto; Ricardo Haddad; Sandra Coccuzzo Sampaio Vessoni; Simone Kashima; Svetoslav Nanev Slavov; Vincent Louis Viala |
| EPI_ISL_2621829, EPI_ISL_2621832, EPI_ISL_2621834, EPI_ISL_2621835, EPI_ISL_2621836, EPI_ISL_2621838 | SURA | Universidad Nacional de Colombia - Laboratorio Genómico One Health | Andres F. Cardona-Rios; Carlos Franco-Muñoz; Carolina Muñoz-Arango; Celeny Ortiz; Daniel O. Maldonado-Perez; Diego A. Álvarez-Díaz; Hector Alejandro Ruiz-Moreno; Idabely Betancur Ortiz; Jorge E. Osorio; Juan P. Hernandez-Ortiz; Karl A Ciuoderis; Katherine Laiton-Donato; Laura Silvana Perez; Lina M. Hurtado; Marcela Mercado-Reyes; Maria Angélica Maya; Maria Stella López; Rita Almanza Payares; Sandra Ines Cano; Simón Villegas Velásquez |
| EPI_ISL_2289057, EPI_ISL_2321659, EPI_ISL_2321716, EPI_ISL_2321719, EPI_ISL_2321722, EPI_ISL_2348517, EPI_ISL_2348518, EPI_ISL_2348520, EPI_ISL_2348527, EPI_ISL_2348532, EPI_ISL_2348534, EPI_ISL_2348536, EPI_ISL_2348537, EPI_ISL_2348540, EPI_ISL_2348544, EPI_ISL_2348546, EPI_ISL_2348570, EPI_ISL_2382807, EPI_ISL_2382814, EPI_ISL_2382815, EPI_ISL_2382816, EPI_ISL_2382819, EPI_ISL_2402385, EPI_ISL_2402393, EPI_ISL_2402394, EPI_ISL_2402396, EPI_ISL_2402404, EPI_ISL_2402406, EPI_ISL_2402409, EPI_ISL_2402411, EPI_ISL_2402412 | | | |
| see above | SYNLAB | GIGA Medical Genomics | Bouchra Boujemla; Cécile Meex; Keith Durkin; Maria Artesi; Marie-Pierre Hayette; Nathalie Renotte; Pierrette Melin; Raphaël Boreux; Sébastien Bontems; Vincent Bours |
| EPI_ISL_3276547 | SYNLAB | Universidad Nacional de Colombia - Laboratorio Genómico One Health | Andres F. Cardona-Rios; Carlos Franco-Muñoz; Carolina Muñoz-Arango; Celeny Ortiz; Daniel O. Maldonado-Perez; Diego A. Álvarez-Díaz; Hector Alejandro Ruiz-Moreno; Idabely Betancur Ortiz; Jorge E. Osorio; Juan P. Hernandez-Ortiz; Karl A Ciuoderis; Katherine Laiton-Donato; Laura Silvana Perez; Lina M. Hurtado; Marcela Mercado-Reyes; Maria Angélica Maya; Maria Stella López; Rita Almanza Payares; Sandra Ines Cano; Simón Villegas Velásquez |
| EPI_ISL_2316219 | SYNLAB MVZ Leinfelden-Echterdingen | Robert Koch Institute | |
| EPI_ISL_2267851, EPI_ISL_2315311, EPI_ISL_2386398, EPI_ISL_2386432 | SYNLAB MVZ Leverkusen | Robert Koch Institute | |
| EPI_ISL_2386315 | SYNLAB MVZ Weiden | Robert Koch Institute | |
| EPI_ISL_2350033, EPI_ISL_2350037, EPI_ISL_2350055, EPI_ISL_2350132, EPI_ISL_2350133, EPI_ISL_2350135, EPI_ISL_2444400, EPI_ISL_2444402, EPI_ISL_2444408, EPI_ISL_2444422, EPI_ISL_2444427, EPI_ISL_2444433, EPI_ISL_2444451, EPI_ISL_2444458, EPI_ISL_2444463, EPI_ISL_2444467, EPI_ISL_2444474, EPI_ISL_2444477, EPI_ISL_2444479, EPI_ISL_2444481, EPI_ISL_2444482, EPI_ISL_2444485, EPI_ISL_2444486, EPI_ISL_2444488, EPI_ISL_2444491, EPI_ISL_2444506, EPI_ISL_2444507, EPI_ISL_2444516, EPI_ISL_2444518, EPI_ISL_2444519, EPI_ISL_2444524, EPI_ISL_2444527, EPI_ISL_2444531, EPI_ISL_2444533, EPI_ISL_2444540, EPI_ISL_2444545, EPI_ISL_2444548, EPI_ISL_2444552, EPI_ISL_2550734, EPI_ISL_2550735, EPI_ISL_2550736, EPI_ISL_2550737, EPI_ISL_2550738, EPI_ISL_2550739, EPI_ISL_2550740, EPI_ISL_2550742, EPI_ISL_2550743, EPI_ISL_2550744, EPI_ISL_2550749, EPI_ISL_2603663, EPI_ISL_2603669, EPI_ISL_2881799, EPI_ISL_2881802, EPI_ISL_2881806, EPI_ISL_2881808 | | | |
| see above | Salud Digna | Instituto Nacional de Medicina Genomica | Abraham Campos-Romero; Cedro-Tanda A; Escobar-Arrazola; Gonzalez-Barrera D; Herrera-Montalvo LA.; Hidalgo-Miranda A; Luna-Ruiz Marco; M; Mendoza-Vargas A; Moreno-Camacho José Luis; Munguia-Garza P; Ramirez-Vega O; Rangel-DeLeon D; Reyes-Grajeda JP; Rodriguez-Gallegos Jorge |
| EPI_ISL_2628904 | San Diego County Public Health Laboratory | Andersen lab at Scripps Research | Brett Austin; Jovan Shephard; SEARCH Alliance San Diego with Ashleigh Murphy |
| EPI_ISL_2735874, EPI_ISL_2735882, EPI_ISL_2735893, EPI_ISL_2735895, EPI_ISL_2735899, EPI_ISL_2735916 | Seattle Flu Study | Seattle Flu Study | Amanda Adler; Barry R. Lutz; Benjamin Pelle; Caitlin R. Wolf; Chris D. Frazar; Deborah A. Nickerson; Elisabeth Brandstetter; Erica Ryke; Helen Y. Chu; Janet A. Englund; Jay Shendure; Jeff Duchin; Jover Lee; Kairsten Fay; Karen Cowgill; Kirsten Lacombe; Lea M. Starita; Mark J. Rieder; Matthew Richardson; Matthew Thompson; Melissa Truong; Michael Boeckh; Michael Famulare; Misja Ilcisin; Peter D. Han; Stephanie Schrag; Thomas R. Sibley; Trevor Bedford |
| EPI_ISL_4082247, EPI_ISL_4082282, EPI_ISL_4082287, EPI_ISL_4082298, EPI_ISL_4082299, EPI_ISL_4082310, EPI_ISL_4082319, EPI_ISL_4082320, EPI_ISL_4082321, EPI_ISL_4082335, EPI_ISL_4082341, EPI_ISL_4082342, EPI_ISL_4082344, EPI_ISL_4082346, EPI_ISL_4082351, EPI_ISL_4082353, EPI_ISL_4082354, EPI_ISL_4082358, EPI_ISL_4082361, EPI_ISL_4082364, EPI_ISL_4082372, EPI_ISL_4082382, EPI_ISL_4082383, EPI_ISL_4082385, EPI_ISL_4082388, EPI_ISL_4082390, EPI_ISL_4082392, EPI_ISL_4082393, EPI_ISL_4082394, EPI_ISL_4082398, EPI_ISL_4082405, EPI_ISL_4082418, EPI_ISL_4082422, EPI_ISL_4082423, EPI_ISL_4082426, EPI_ISL_4082438, EPI_ISL_4082446, EPI_ISL_4082452, EPI_ISL_4082484, EPI_ISL_4082489, EPI_ISL_4082538, EPI_ISL_4082562, EPI_ISL_4082607, EPI_ISL_4082623, EPI_ISL_4082624, EPI_ISL_4082653, EPI_ISL_4082655, EPI_ISL_4082659, EPI_ISL_4082661, EPI_ISL_4082669, EPI_ISL_4082701, EPI_ISL_4082788, EPI_ISL_4082884, EPI_ISL_4082992, EPI_ISL_4083068, EPI_ISL_4083069, EPI_ISL_4083151, EPI_ISL_4083364 | | | |
| see above | Servicio Virosis Respiratorias-Departamento Virología-INEI | Instituto Nacional Enfermedades Infecciosas C.G.Malbran | Avaro M.; Baumeister E.; Benedetti E.; Campos J.; Cisterna D.; Dattero ME; De Belder D.; Haim MS.; Lorenzo F.; Molina V.; Perandones C.; Poklepovich T.; Pontoriero A.; Russo M.; Sanchez Loria J.; Tuduri E. |
| EPI_ISL_2426635, EPI_ISL_2426651, EPI_ISL_2861036 | Servicio de Microbiologia Hospital Ramon y Cajal | Servicio de Microbiologia Hospital Ramon y Cajal | Galan JC; Martinez L. Abreu M; Ponce M; y Gonzalez-Alba JM |
| EPI_ISL_2510590, EPI_ISL_2510591, EPI_ISL_2510593, EPI_ISL_2510594, EPI_ISL_2510663 | Servicio de Microbiología Clínica (Complejo Hospitalario de Navarra, Pamplona) | Centro de Secuenciación NASERTIC | Ana Miqueleiz; Ana Navascués; Carmen Ezpeleta Baquedano |
| EPI_ISL_2658196, EPI_ISL_2658198, EPI_ISL_2658199 | Servicio de Microbiología. Consorcio Hospital General Universitario de Valencia | SeqCOVID-SPAIN consortium/IBV(CSIC) | Begoña Fuster Escrivá; Carme Salvador García; Concepción Gimeno Cardona and SeqCOVID-SPAIN consortium; María Dolores Ocete; Rafael Medina González |
| EPI_ISL_2439988, EPI_ISL_2439989 | Servicio de Microbiología. Hospital Clínico Universitario de Valencia | SeqCOVID-SPAIN consortium/IBV(CSIC) | David Navarro Ortega; Eliseo Albert Vicent; Ignacio Torres and SeqCOVID-SPAIN consortium |
| EPI_ISL_2627507 | Servicio de Microbiología. Hospital Universitario Doctor Peset | SeqCOVID-SPAIN consortium/IBV(CSIC) | José Miguel Nogueira Coito and SeqCOVID-SPAIN consortium; Juan Alberola Enguídanos; Juan José Camarena Miñana; Rosa González Pellicer |
| EPI_ISL_5315610, EPI_ISL_5315619, EPI_ISL_5315625, EPI_ISL_5315636, EPI_ISL_5315637, EPI_ISL_5315638, EPI_ISL_5315654, EPI_ISL_5315660, EPI_ISL_5315662, EPI_ISL_5315664, EPI_ISL_5315665, EPI_ISL_5315685, EPI_ISL_5315686, EPI_ISL_5315687, EPI_ISL_5315689, EPI_ISL_5315692, EPI_ISL_5315693, EPI_ISL_5315696, EPI_ISL_5315715, EPI_ISL_5315717, EPI_ISL_5315718, EPI_ISL_5315719, EPI_ISL_5315721, EPI_ISL_5315724, EPI_ISL_5315725, EPI_ISL_5315726, EPI_ISL_5315727, EPI_ISL_5315728, EPI_ISL_5315729, EPI_ISL_5315731, EPI_ISL_5315733, EPI_ISL_5315734, EPI_ISL_5315736, EPI_ISL_5315748, EPI_ISL_5315749, EPI_ISL_5315750, EPI_ISL_5315751, EPI_ISL_5315755, EPI_ISL_5315757, EPI_ISL_5315758, EPI_ISL_5315759, EPI_ISL_5315760, EPI_ISL_5315761, EPI_ISL_5315762, EPI_ISL_5315763, EPI_ISL_5315764, EPI_ISL_5315765, EPI_ISL_5315766, EPI_ISL_5315767, EPI_ISL_5315768, EPI_ISL_6686926, EPI_ISL_6686930, EPI_ISL_6686938, EPI_ISL_6687404 | | | |
| see above | Shared Hospital Laboratory | Shared Hospital Laboratory | Christie Vermeiren; Finlay Maguire; Kevin Katz; Patryk Aftanas; Robert Kozak; Samira Mubareka |
| EPI_ISL_2628392, EPI_ISL_2628394 | Sharp HealthCare Laboratory | Andersen lab at Scripps Research | Art Mendoza; Cathy Woerle; Jacquelyn Berumen; Liam McGinnis; Omid Bakhtar; SEARCH Alliance San Diego with Aaron Harding |
| EPI_ISL_2801031, EPI_ISL_2801036, EPI_ISL_2801146, EPI_ISL_2801147, EPI_ISL_2801148, EPI_ISL_2801149, EPI_ISL_2801150, EPI_ISL_2801151 | | | |
| see above | Simple Laboratories | Gagnon Lab, Southern Illinois University | Keith Gagnon |
| EPI_ISL_2621184, EPI_ISL_2621188, EPI_ISL_2621193, EPI_ISL_2621195, EPI_ISL_2621196 | Somer | Universidad Nacional de Colombia - Laboratorio Genómico One Health | Andres F. Cardona-Rios; Carlos Franco-Muñoz; Carolina Muñoz-Arango; Celeny Ortiz; Daniel O. Maldonado-Perez; Diego A. Álvarez-Díaz; Hector Alejandro Ruiz-Moreno; Idabely Betancur Ortiz; Jorge E. Osorio; Juan P. Hernandez-Ortiz; Karl A Ciuoderis; Katherine Laiton-Donato; Laura Silvana Perez; Lina M. Hurtado; Marcela Mercado-Reyes; Maria Angélica Maya; Maria Stella López; Rita Almanza Payares; Sandra Ines Cano; Simón Villegas Velásquez |
| EPI_ISL_2688001, EPI_ISL_2688002, EPI_ISL_2688003, EPI_ISL_3394091, EPI_ISL_3394111 | Sonic Reference Laboratory | Sonic Reference Laboratory | "Robert Moller; Bijaya Dhakal; Cora Lahey; Ellen Pizarek"; John Woody; Krishna Maddula |
| EPI_ISL_2484885, EPI_ISL_2484886, EPI_ISL_2484887 | Southern Nevada Public Health Laboratory | Southern Nevada Public Health Laboratory | Michael Picker |
| EPI_ISL_2420374, EPI_ISL_2510856, EPI_ISL_2510888, EPI_ISL_2510973, | Spital Limmattal | Institute of Medical Virology | Alexandra Trkola; Annette Audigé; Annette AudigŽ; Catharine Aquino; Cyril Shah; Daniel Ehrsam; Gabriela Ziltener; Guido Bloemberg; Hubert Rehrauer; Isabel StŸrmer; Joel Wirz; Jon Huder; Jürg Böni; JŸrg Bšni; Kevin Steiner; Maria Grünberg; Maria GrŸnberg; Maryam Zaheri; Michael Huber; Riccarda Capaul; Stefan Schmutz; Verena Kufner; Weihong Qi |

| | | | |
|---|---|---|---|
| EPI_ISL_2510976 | | | |
| EPI_ISL_4496871, EPI_ISL_4496875, EPI_ISL_4496876, EPI_ISL_4496877, EPI_ISL_4496883 | Stanford Health Care | Stanford University School of Medicine, Clinical Virology Laboratory | Becky Jiang; Bernadette Troung; James Zehnder; Malaya K. Sahoo; Selamawit Bihon; and Benjamin A. Pinsky |
| EPI_ISL_2724657, EPI_ISL_2724669, EPI_ISL_2724719, EPI_ISL_2724720, EPI_ISL_2724721, EPI_ISL_2724725, EPI_ISL_2724730, EPI_ISL_2724807 | | | |
| see above | State Laboratories Division, Hawaii State Department of Health | State Laboratories Division, Hawaii State Department of Health | Ayana Garnet; Drew Kuwazaki; Edward Desmond; Pamela O'Brien; Razvan Sultana |
| EPI_ISL_2473474, EPI_ISL_2473478, EPI_ISL_2473481, EPI_ISL_2473489, EPI_ISL_2473495, EPI_ISL_2473518, EPI_ISL_2473523, EPI_ISL_2473526, EPI_ISL_2473537, EPI_ISL_2473541, EPI_ISL_2473557, EPI_ISL_2473560, EPI_ISL_2473561, EPI_ISL_2800583, EPI_ISL_2800584 | | | |
| see above | State Testing Facility | Altius Institute for Biomedical Research | Alex Isner; Alex Nguyen; Amanda Gale; Audra Johnson; Clem Green; Daniel Bates; Eric Thorland; Jacob Rodriguez; Jean Robinston; Jemma Nelson; Jessica Kunder; John Stamatoyannopoulos; Joshua Richards; Julia Wald; Kneshay Harper; Lauren Mitchell; Mark Frerker; Matt Hartman; Michael Buckley; Muhammad Halimun; Rebecca Bruders; Sadie Patraw; Sofia Olsson; Tobias Ragoczy |
| EPI_ISL_2581084, EPI_ISL_2581124, EPI_ISL_2581130, EPI_ISL_2581444, EPI_ISL_2980127, EPI_ISL_3157893 | Swedish national genomic surveillance program of SARS-CoV-2 | The Public Health Agency of Sweden | Alma Brolund; Maria Lind Karlberg; Maximilian Riess; Swedish national genomic surveillance program of SARS-CoV-2 |
| EPI_ISL_2421182 | TEMPUS LABS INC | Wadsworth Center, New York State Department of Health | Alexis Russell; Catharine Prussing; Daryl M. Lamson; Erasmus Schneider; Erica Lasek-Nesselquist; John Kelly; Jonathan Plitnick; Kirsten St. George; Matthew Shudt; Melissa A Leisner; Navjot Singh |
| EPI_ISL_2491118, EPI_ISL_2491144, EPI_ISL_2491182, EPI_ISL_2491218, EPI_ISL_2491282, EPI_ISL_2491305, EPI_ISL_2600429, EPI_ISL_2600436, EPI_ISL_2600527, EPI_ISL_2671915, EPI_ISL_2671917, EPI_ISL_2672016, EPI_ISL_2672027 | | | |
| see above | TGen North | TGen North | "Jolene Bowers; Ashlyn Pfeiffer; Chris French; Darrin Lemmer; Dave Engelthaler; Hayley Yaglom; Heather Centner; Jolene Bowers; The Arizona COVID Genomics Union (ACGU); The Arizona COVID Genomics Union (ACGU)" |
| EPI_ISL_2451912 | TX DSHS, Lab Services Section MC 1947 | Centers for Disease Control and Prevention Division of Viral Diseases, Pathogen Discovery | Alison Laufer Halpin; Ben L. Rambo-Martin; Clinton R. Paden; Dakota Howard; Darlene Wagner; Dave Wentworth; Dhwani Batra; Jasmine Padilla; Justin Lee; Katie Dillon; Krista Queen; Kristen Knipe; Kristine Lacek; Mark Burroughs; Matthew Schmerer; Mili Sheth; Peter Cook; Sam Shepard; Sarah Nobles; Shoshona Le; Suxiang Tong; Vivien Dugan; Yvette Unoarumhi |
| EPI_ISL_2443299, EPI_ISL_2565761, EPI_ISL_2565785 | TXDSHS | TXDSHS | Anita Pokharel; Bonnie Oh; Chun Wang; Grace Kubin; Jenny Zhang; Karen Bobier; Lorraine Rodriguez; Maliha Rahman; Mayela Pedrueza; Myong Koag; Rachel Lee; Rashmi Tuladhar |
| EPI_ISL_2790532, EPI_ISL_2966835 | Tampa General Hospital Esoteric Lab | Tampa General Hospital Esoteric Research & Development Lab | Amorce Lima; Deanna Becker; Dominic Uy; Elaine Vendrone; Jon Faughn; Jorge Mecias-Frias; Suzane Silbert; Vicki Healer |
| EPI_ISL_2716586 | The Caribbean Public Health Agency | Carrington Lab, Department of Preclinical Sciences, Faculty of Medical Sciences, The University of the West Indies, St Augustine Campus | Anushka Ramjag; Arianne Brown-Jordan; Avery Hinds; Christine V. F. Carrington; Gabriel Escobar; Nikita S. D. Sahadeo; Nuno Faria; Oliver Pybus; Risha Singh; Roshan Parasram; Sarah Hill; SueMin Nathaniel; Vernie Ramkissoon |
| EPI_ISL_2346333, EPI_ISL_2346334, EPI_ISL_2346335, EPI_ISL_2346338 | The Jackson Laboratory | The Jackson Laboratory | Adams M; Bergeron D; Kelly K; Li L; Long J; Omerza G; Renzette N |
| EPI_ISL_2534312, EPI_ISL_2758850, EPI_ISL_2758859, EPI_ISL_2834236 | The Ohio State University Applied Microbiology Services Laboratory | The Ohio State University Applied Microbiology Services Laboratory | Seth A. Faith PhD |
| EPI_ISL_3152443 | TriCore Reference Laboratories | Center for Global Health, University of New Mexico Health Sciences Center | Cecilia Thompson; Darrell Dinwiddie; Daryl Domman; Karissa Culbreath; Kendra Pesko; Kurt Schwalm; Valerie Morley |
| EPI_ISL_3185340, EPI_ISL_3185341, EPI_ISL_3185345, EPI_ISL_3185347, EPI_ISL_3185356, EPI_ISL_3215016 | U.O. Microbiologia Laboratorio Unico Centro Servizi - AUSL della Romagna | U.O. Microbiologia, Laboratorio Unico Centro Servizi - AUSL della Romagna | Giorgio Dirani |
| EPI_ISL_2978354, EPI_ISL_2978358 | U.O. Microbiologia Laboratorio Unico Centro Servizi AUSL della Romagna | U.O. Microbiologia Laboratorio Unico Centro Servizi AUSL della Romagna | Francesca Taddei; Giorgio Dirani; Giulia Gatti; Ilaria Poggianti; Silvia Zannoli; Vittorio Sambri |
| EPI_ISL_6229781 | UAPS HUMBERTO BEZERRA | ACME Lab, Oswaldo Cruz Foundation, FIOCRUZ/CE | Carlos Leonardo de Aragao Araujo; Cecilia Leite Costa & Eduardo Ruback dos Santos on behalf of COVID-19 FIOCRUZ Genomic Network; Cleber Furtado Aksenen; Fabio Miyajima; Fernando Braga Stehling; Francisco Eder de Moura Lopes; Igor Oliveira Duarte; Jamille Maria Mendes Bezerra; Joaquim Cesar do Nascimento Sousa Junior; Pedro Miguel Carneiro Jeronimo; Suzana Porto Almeida; Thais Ferreira de Oliveira; Thais de Oliveira Costa; Ticiane Cavalcante de Souza; Veridiana Pessoa Miyajima |
| EPI_ISL_6229782 | UAPS MELO JABORANDI | ACME Lab, Oswaldo Cruz Foundation, FIOCRUZ/CE | Carlos Leonardo de Aragao Araujo; Cecilia Leite Costa & Eduardo Ruback dos Santos on behalf of COVID-19 FIOCRUZ Genomic Network; Cleber Furtado Aksenen; Fabio Miyajima; Fernando Braga Stehling; Francisco Eder de Moura Lopes; Igor Oliveira Duarte; Jamille Maria Mendes Bezerra; Joaquim Cesar do Nascimento Sousa Junior; Pedro Miguel Carneiro Jeronimo; Suzana Porto Almeida; Thais Ferreira de Oliveira; Thais de Oliveira Costa; Ticiane Cavalcante de Souza; Veridiana Pessoa Miyajima |
| EPI_ISL_6229783 | UAPS RECAMONDE CAPELO | ACME Lab, Oswaldo Cruz Foundation, FIOCRUZ/CE | Carlos Leonardo de Aragao Araujo; Cecilia Leite Costa & Eduardo Ruback dos Santos on behalf of COVID-19 FIOCRUZ Genomic Network; Cleber Furtado Aksenen; Fabio Miyajima; Fernando Braga Stehling; Francisco Eder de Moura Lopes; Igor Oliveira Duarte; Jamille Maria Mendes Bezerra; Joaquim Cesar do Nascimento Sousa Junior; Pedro Miguel Carneiro Jeronimo; Suzana Porto Almeida; Thais Ferreira de Oliveira; Thais de Oliveira Costa; Ticiane Cavalcante de Souza; Veridiana Pessoa Miyajima |
| EPI_ISL_2445173 | UBS CAIC OURINHOS | Instituto Butantan | Antonio Jorge Martins; Claudia Renata dos Santos Barros; David Schlesinger; Debora Botequio Moretti; Dimas Tadeu Covas; Elaine Cristina Marqueze; Elaine Vieira Santos; Evandra Strazza Rodrigues; Heidge Fukumasu; Jayme Augusto de Souza-Neto; José Salvatore Leister Patané; Luiz Alcantara; Luiz Lehmann Coutinho; Maria Carolina Elias; Maurício Lacerda Nogueira; Rafael dos Santos Bezerra; Raul Machado Neto; Rejane Maria Tommasini Grotto; Ricardo Haddad; Sandra Coccuzzo Sampaio Vessoni; Simone Kashima; Svetoslav Nanev Slavov; Vincent Louis Viala |
| EPI_ISL_2445207, EPI_ISL_2473718, EPI_ISL_5645966, EPI_ISL_5646010, EPI_ISL_5646032 | UBS CELESTINO CARNIELO | Instituto Butantan | Antonio Jorge Martins; Claudia Renata dos Santos Barros; David Schlesinger; Debora Botequio Moretti; Dimas Tadeu Covas; Elaine Cristina Marqueze; Elaine Vieira Santos; Evandra Strazza Rodrigues; Heidge Fukumasu; Jayme Augusto de Souza-Neto; José Salvatore Leister Patané; Luiz Alcantara; Luiz Lehmann Coutinho; Maria Carolina Elias; Maurício Lacerda Nogueira; Rafael dos Santos Bezerra; Raul Machado Neto; Rejane Maria Tommasini Grotto; Ricardo Haddad; Sandra Coccuzzo Sampaio Vessoni; Simone Kashima; Svetoslav Nanev Slavov; Vincent Louis Viala |
| EPI_ISL_2445208, EPI_ISL_2445216, EPI_ISL_2473719, EPI_ISL_2493655, EPI_ISL_2493656, EPI_ISL_2493658 | UBS DE PARAISO | Instituto Butantan | Antonio Jorge Martins; Claudia Renata dos Santos Barros; David Schlesinger; Debora Botequio Moretti; Dimas Tadeu Covas; Elaine Cristina Marqueze; Elaine Vieira Santos; Evandra Strazza Rodrigues; Heidge Fukumasu; Jayme Augusto de Souza-Neto; José Salvatore Leister Patané; Luiz Alcantara; Luiz Lehmann Coutinho; Maria Carolina Elias; Maurício Lacerda Nogueira; Rafael dos Santos Bezerra; Raul Machado Neto; Rejane Maria Tommasini Grotto; Ricardo Haddad; Sandra Coccuzzo Sampaio Vessoni; Simone Kashima; Svetoslav Nanev Slavov; Vincent Louis Viala |
| EPI_ISL_2493418 | UBS DE PEDRANOPOLIS | Instituto Butantan | Antonio Jorge Martins; Claudia Renata dos Santos Barros; David Schlesinger; Debora Botequio Moretti; Dimas Tadeu Covas; Elaine Cristina Marqueze; Elaine Vieira Santos; Evandra Strazza Rodrigues; Heidge Fukumasu; Jayme Augusto de Souza-Neto; José Salvatore Leister Patané; Luiz Alcantara; Luiz Lehmann Coutinho; Maria Carolina Elias; Maurício Lacerda Nogueira; Rafael dos Santos Bezerra; Raul Machado Neto; Rejane Maria Tommasini Grotto; Ricardo Haddad; Sandra Coccuzzo Sampaio Vessoni; Simone Kashima; Svetoslav Nanev Slavov; Vincent Louis Viala |
| EPI_ISL_5645997 | UBS DE SANTA SALETE | Instituto Butantan | Antonio Jorge Martins; Claudia Renata dos Santos Barros; David Schlesinger; Debora Botequio Moretti; Dimas Tadeu Covas; Elaine Cristina Marqueze; Elaine Vieira Santos; Evandra Strazza Rodrigues; Heidge Fukumasu; Jayme Augusto de Souza-Neto; José Salvatore Leister Patané; Luiz Alcantara; Luiz Lehmann Coutinho; Maria Carolina Elias; Maurício Lacerda Nogueira; Rafael dos Santos Bezerra; Raul Machado Neto; Rejane Maria Tommasini Grotto; Ricardo Haddad; Sandra Coccuzzo Sampaio Vessoni; Simone Kashima; Svetoslav Nanev Slavov; Vincent Louis Viala |
| EPI_ISL_2445195 | UBS DE SUD MENNUCCI | Instituto Butantan | Antonio Jorge Martins; Claudia Renata dos Santos Barros; David Schlesinger; Debora Botequio Moretti; Dimas Tadeu Covas; Elaine Cristina Marqueze; Elaine Vieira Santos; Evandra Strazza Rodrigues; Heidge Fukumasu; Jayme Augusto de Souza-Neto; José Salvatore Leister Patané; Luiz Alcantara; Luiz Lehmann Coutinho; Maria Carolina Elias; Maurício Lacerda Nogueira; Rafael dos Santos Bezerra; Raul Machado Neto; Rejane Maria Tommasini Grotto; Ricardo Haddad; Sandra Coccuzzo Sampaio Vessoni; Simone Kashima; Svetoslav Nanev Slavov; Vincent Louis Viala |
| EPI_ISL_2445206, EPI_ISL_2493177, EPI_ISL_2493402, EPI_ISL_2493404, EPI_ISL_2493409, EPI_ISL_2493423, EPI_ISL_2493425, EPI_ISL_2493426, EPI_ISL_2494337, EPI_ISL_2500975 | | | |
| see above | UBS DE TABAPUA | Instituto Butantan | Antonio Jorge Martins; Claudia Renata dos Santos Barros; David Schlesinger; Debora Botequio Moretti; Dimas Tadeu Covas; Elaine Cristina Marqueze; Elaine Vieira Santos; Evandra Strazza Rodrigues; Heidge Fukumasu; Jayme Augusto de Souza-Neto; José Salvatore Leister Patané; Luiz Alcantara; Luiz Lehmann Coutinho; Maria Carolina Elias; Maurício Lacerda Nogueira; Rafael dos Santos Bezerra; Raul Machado Neto; Rejane Maria Tommasini Grotto; Ricardo Haddad; Sandra Coccuzzo Sampaio Vessoni; Simone Kashima; Svetoslav Nanev Slavov; Vincent Louis Viala |
| EPI_ISL_2493659, EPI_ISL_2493665, EPI_ISL_2493666 | UBS DE UBARANA | Instituto Butantan | Antonio Jorge Martins; Claudia Renata dos Santos Barros; David Schlesinger; Debora Botequio Moretti; Dimas Tadeu Covas; Elaine Cristina Marqueze; Elaine Vieira Santos; Evandra Strazza Rodrigues; Heidge Fukumasu; Jayme Augusto de Souza-Neto; José Salvatore Leister Patané; Luiz Alcantara; Luiz Lehmann Coutinho; Maria Carolina Elias; Maurício Lacerda Nogueira; Rafael dos Santos Bezerra; Raul Machado Neto; Rejane Maria Tommasini Grotto; Ricardo Haddad; Sandra Coccuzzo Sampaio Vessoni; Simone Kashima; Svetoslav Nanev Slavov; Vincent Louis Viala |
| EPI_ISL_6229785, EPI_ISL_6229786, EPI_ISL_6229787, EPI_ISL_6229788 | UBS DO BAIRRO SAO JOSE | ACME Lab, Oswaldo Cruz Foundation, FIOCRUZ/CE | Carlos Leonardo de Aragao Araujo; Cecilia Leite Costa & Eduardo Ruback dos Santos on behalf of COVID-19 FIOCRUZ Genomic Network; Cleber Furtado Aksenen; Fabio Miyajima; Fernando Braga Stehling; Francisco Eder de Moura Lopes; Igor Oliveira Duarte; Jamille Maria Mendes Bezerra; Joaquim Cesar do Nascimento Sousa Junior; Pedro Miguel Carneiro Jeronimo; Suzana Porto Almeida; Thais Ferreira de Oliveira; Thais de Oliveira Costa; Ticiane Cavalcante de Souza; Veridiana Pessoa Miyajima |
| EPI_ISL_2378746, EPI_ISL_2493168, EPI_ISL_2493278, EPI_ISL_2493326, EPI_ISL_2493327, EPI_ISL_2493328, EPI_ISL_2493340, EPI_ISL_2493341, EPI_ISL_2493343, EPI_ISL_2493346, EPI_ISL_2493348, EPI_ISL_2493349, EPI_ISL_2493350, EPI_ISL_2493351, EPI_ISL_2493352, EPI_ISL_2493353, EPI_ISL_2500984 | | | |
| see above | UBS II DE PIRAPOZINHO C SERVICO DE EACS E ESF | Instituto Butantan | Antonio Jorge Martins; Claudia Renata dos Santos Barros; David Schlesinger; Debora Botequio Moretti; Dimas Tadeu Covas; Elaine Cristina Marqueze; Elaine Vieira Santos; Evandra Strazza Rodrigues; Heidge Fukumasu; Jayme Augusto de Souza-Neto; José Salvatore Leister Patané; Luiz Alcantara; Luiz Lehmann Coutinho; Maria Carolina Elias; Maurício Lacerda Nogueira; Rafael dos Santos Bezerra; Raul Machado Neto; Rejane Maria Tommasini Grotto; Ricardo Haddad; Sandra Coccuzzo Sampaio Vessoni; Simone Kashima; Svetoslav Nanev Slavov; Vincent Louis Viala |
| EPI_ISL_2445202, EPI_ISL_5645977, EPI_ISL_5645979, EPI_ISL_5645984, EPI_ISL_5645995, EPI_ISL_5646004, EPI_ISL_5646013, EPI_ISL_5646014 | | | |
| see above | UBS II DE TANABI MILTON MARTINS | Instituto Butantan | Antonio Jorge Martins; Claudia Renata dos Santos Barros; David Schlesinger; Debora Botequio Moretti; Dimas Tadeu Covas; Elaine Cristina Marqueze; Elaine Vieira Santos; Evandra Strazza Rodrigues; Heidge Fukumasu; Jayme Augusto de Souza-Neto; José Salvatore Leister Patané; Luiz Alcantara; Luiz |

| | | | |
|---|---|---|---|
| | PERCHES | | Lehmann Coutinho; Maria Carolina Elias; Maurício Lacerda Nogueira; Rafael dos Santos Bezerra; Raul Machado Neto; Rejane Maria Tommasini Grotto; Ricardo Haddad; Sandra Coccuzzo Sampaio Vessoni; Simone Kashima; Svetoslav Nanev Slavov; Vincent Louis Viala |
| EPI_ISL_2493360 | UBS II DR JOSE CARLOS MACUCO JANINI | Instituto Butantan | Antonio Jorge Martins; Claudia Renata dos Santos Barros; David Schlesinger; Debora Botequio Moretti; Dimas Tadeu Covas; Elaine Cristina Marqueze; Elaine Vieira Santos; Evandra Strazza Rodrigues; Heidge Fukumasu; Jayme Augusto de Souza-Neto; José Salvatore Leister Patané; Luiz Alcantara; Luiz Lehmann Coutinho; Maria Carolina Elias; Maurício Lacerda Nogueira; Rafael dos Santos Bezerra; Raul Machado Neto; Rejane Maria Tommasini Grotto; Ricardo Haddad; Sandra Coccuzzo Sampaio Vessoni; Simone Kashima; Svetoslav Nanev Slavov; Vincent Louis Viala |
| EPI_ISL_2493531, EPI_ISL_2493545, EPI_ISL_2493550 | UBS III DE PARIQUERA ACU PARIQUERA ACU | Instituto Butantan | Antonio Jorge Martins; Claudia Renata dos Santos Barros; David Schlesinger; Debora Botequio Moretti; Dimas Tadeu Covas; Elaine Cristina Marqueze; Elaine Vieira Santos; Evandra Strazza Rodrigues; Heidge Fukumasu; Jayme Augusto de Souza-Neto; José Salvatore Leister Patané; Luiz Alcantara; Luiz Lehmann Coutinho; Maria Carolina Elias; Maurício Lacerda Nogueira; Rafael dos Santos Bezerra; Raul Machado Neto; Rejane Maria Tommasini Grotto; Ricardo Haddad; Sandra Coccuzzo Sampaio Vessoni; Simone Kashima; Svetoslav Nanev Slavov; Vincent Louis Viala |
| EPI_ISL_2493686, EPI_ISL_2493699 | UBS III DE RANCHARIA | Instituto Butantan | Antonio Jorge Martins; Claudia Renata dos Santos Barros; David Schlesinger; Debora Botequio Moretti; Dimas Tadeu Covas; Elaine Cristina Marqueze; Elaine Vieira Santos; Evandra Strazza Rodrigues; Heidge Fukumasu; Jayme Augusto de Souza-Neto; José Salvatore Leister Patané; Luiz Alcantara; Luiz Lehmann Coutinho; Maria Carolina Elias; Maurício Lacerda Nogueira; Rafael dos Santos Bezerra; Raul Machado Neto; Rejane Maria Tommasini Grotto; Ricardo Haddad; Sandra Coccuzzo Sampaio Vessoni; Simone Kashima; Svetoslav Nanev Slavov; Vincent Louis Viala |
| EPI_ISL_2493663, EPI_ISL_2493664 | UBS LUIS FACHIN IPIGUA | Instituto Butantan | Antonio Jorge Martins; Claudia Renata dos Santos Barros; David Schlesinger; Debora Botequio Moretti; Dimas Tadeu Covas; Elaine Cristina Marqueze; Elaine Vieira Santos; Evandra Strazza Rodrigues; Heidge Fukumasu; Jayme Augusto de Souza-Neto; José Salvatore Leister Patané; Luiz Alcantara; Luiz Lehmann Coutinho; Maria Carolina Elias; Maurício Lacerda Nogueira; Rafael dos Santos Bezerra; Raul Machado Neto; Rejane Maria Tommasini Grotto; Ricardo Haddad; Sandra Coccuzzo Sampaio Vessoni; Simone Kashima; Svetoslav Nanev Slavov; Vincent Louis Viala |
| EPI_ISL_6229790, EPI_ISL_6229791, EPI_ISL_6229792, EPI_ISL_6229794, EPI_ISL_6229831 | UBS MARATOAN | ACME Lab, Oswaldo Cruz Foundation, FIOCRUZ/CE | Carlos Leonardo de Aragao Araujo; Cecilia Leite Costa & Eduardo Ruback dos Santos on behalf of COVID-19 FIOCRUZ Genomic Network; Cleber Furtado Aksenen; Fabio Miyajima; Fernando Braga Stehling; Francisco Eder de Moura Lopes; Igor Oliveira Duarte; Jamille Maria Mendes Bezerra; Joaquim Cesar do Nascimento Sousa Junior; Pedro Miguel Carneiro Jeronimo; Suzana Porto Almeida; Thais Ferreira de Oliveira; Thais de Oliveira Costa; Ticiane Cavalcante de Souza; Veridiana Pessoa Miyajima |
| EPI_ISL_2493584 | UBS MARIA IZABEL | Instituto Butantan | Antonio Jorge Martins; Claudia Renata dos Santos Barros; David Schlesinger; Debora Botequio Moretti; Dimas Tadeu Covas; Elaine Cristina Marqueze; Elaine Vieira Santos; Evandra Strazza Rodrigues; Heidge Fukumasu; Jayme Augusto de Souza-Neto; José Salvatore Leister Patané; Luiz Alcantara; Luiz Lehmann Coutinho; Maria Carolina Elias; Maurício Lacerda Nogueira; Rafael dos Santos Bezerra; Raul Machado Neto; Rejane Maria Tommasini Grotto; Ricardo Haddad; Sandra Coccuzzo Sampaio Vessoni; Simone Kashima; Svetoslav Nanev Slavov; Vincent Louis Viala |
| EPI_ISL_2445198, EPI_ISL_2445199, EPI_ISL_2445200, EPI_ISL_2445201, EPI_ISL_2473714, EPI_ISL_2473715 | UBS PASTOR MARCELINO DEUNGARO RIO GRANDE | Instituto Butantan | Antonio Jorge Martins; Claudia Renata dos Santos Barros; David Schlesinger; Debora Botequio Moretti; Dimas Tadeu Covas; Elaine Cristina Marqueze; Elaine Vieira Santos; Evandra Strazza Rodrigues; Heidge Fukumasu; Jayme Augusto de Souza-Neto; José Salvatore Leister Patané; Luiz Alcantara; Luiz Lehmann Coutinho; Maria Carolina Elias; Maurício Lacerda Nogueira; Rafael dos Santos Bezerra; Raul Machado Neto; Rejane Maria Tommasini Grotto; Ricardo Haddad; Sandra Coccuzzo Sampaio Vessoni; Simone Kashima; Svetoslav Nanev Slavov; Vincent Louis Viala |
| EPI_ISL_2493293 | UBS PAULO HUMBERTO DE FIGUEIREDO | Instituto Butantan | Antonio Jorge Martins; Claudia Renata dos Santos Barros; David Schlesinger; Debora Botequio Moretti; Dimas Tadeu Covas; Elaine Cristina Marqueze; Elaine Vieira Santos; Evandra Strazza Rodrigues; Heidge Fukumasu; Jayme Augusto de Souza-Neto; José Salvatore Leister Patané; Luiz Alcantara; Luiz Lehmann Coutinho; Maria Carolina Elias; Maurício Lacerda Nogueira; Rafael dos Santos Bezerra; Raul Machado Neto; Rejane Maria Tommasini Grotto; Ricardo Haddad; Sandra Coccuzzo Sampaio Vessoni; Simone Kashima; Svetoslav Nanev Slavov; Vincent Louis Viala |
| EPI_ISL_2445188, EPI_ISL_2473710 | UBS VILA RIBEIRO | Instituto Butantan | Antonio Jorge Martins; Claudia Renata dos Santos Barros; David Schlesinger; Debora Botequio Moretti; Dimas Tadeu Covas; Elaine Cristina Marqueze; Elaine Vieira Santos; Evandra Strazza Rodrigues; Heidge Fukumasu; Jayme Augusto de Souza-Neto; José Salvatore Leister Patané; Luiz Alcantara; Luiz Lehmann Coutinho; Maria Carolina Elias; Maurício Lacerda Nogueira; Rafael dos Santos Bezerra; Raul Machado Neto; Rejane Maria Tommasini Grotto; Ricardo Haddad; Sandra Coccuzzo Sampaio Vessoni; Simone Kashima; Svetoslav Nanev Slavov; Vincent Louis Viala |
| EPI_ISL_3321306, EPI_ISL_3370429 | UC-Christus Clinical Hospital Laboratory | Laboratory of Molecular Virology, School of Medicine, Pontificia Universidad Catolica de Chile | Ana Maira Guzman; Andres E. Munoz-Marcos; Catalina Pardo-Roa; Eileen Serrano; Erick Salinas; Estefany Poblete; Francisco Melo; Jorge Levican; Leonardo I. Almonacid; Maria Jose Avendano; Maria Patricia Vega; Rafael A. Medina; Ricardo Enrique de la Barra; Tamara Garcia-Salum |
| EPI_ISL_2547290, EPI_ISL_2547291, EPI_ISL_2547292, EPI_ISL_2547299, EPI_ISL_2547312, EPI_ISL_2547314, EPI_ISL_2547315, EPI_ISL_2547316 | | | |
| see above | UHTL, University Hospitals | UHTL, University Hospitals | Alouani, D.; Sadri, N.; Song, X. |
| EPI_ISL_2448598 | ULSS 2 Marca Trevigiana | Istituto Zooprofilattico Sperimentale delle Venezie | Adelaide Milani; Alessia Schivo; Alice Fusaro; Ambra Pastori; Annalisa Salviato; Antonia Ricci; Calogero Terregino; Edoardo Giussani; Elisa Palumbo; Erika Giorgia Quaranta; Isabella Monne; Luca Tassoni |
| EPI_ISL_3409199, EPI_ISL_3410104 | UMass Memorial Medical Center | Center for Microbiome Research | Doyle V. Ward; Karl J. Simon; Richard T. Ellison III |
| EPI_ISL_2493585, EPI_ISL_2493588 | UNIDADE BASICA DE SAUDE DE TARUMA | Instituto Butantan | Antonio Jorge Martins; Claudia Renata dos Santos Barros; David Schlesinger; Debora Botequio Moretti; Dimas Tadeu Covas; Elaine Cristina Marqueze; Elaine Vieira Santos; Evandra Strazza Rodrigues; Heidge Fukumasu; Jayme Augusto de Souza-Neto; José Salvatore Leister Patané; Luiz Alcantara; Luiz Lehmann Coutinho; Maria Carolina Elias; Maurício Lacerda Nogueira; Rafael dos Santos Bezerra; Raul Machado Neto; Rejane Maria Tommasini Grotto; Ricardo Haddad; Sandra Coccuzzo Sampaio Vessoni; Simone Kashima; Svetoslav Nanev Slavov; Vincent Louis Viala |
| EPI_ISL_2383035, EPI_ISL_2493383, EPI_ISL_2494152 | UNIDADE BASICA DE SAUDE DR MATHEUS GABRIEL BONASSA | Instituto Butantan | Antonio Jorge Martins; Claudia Renata dos Santos Barros; David Schlesinger; Debora Botequio Moretti; Dimas Tadeu Covas; Elaine Cristina Marqueze; Elaine Vieira Santos; Evandra Strazza Rodrigues; Heidge Fukumasu; Jayme Augusto de Souza-Neto; José Salvatore Leister Patané; Luiz Alcantara; Luiz Lehmann Coutinho; Maria Carolina Elias; Maurício Lacerda Nogueira; Rafael dos Santos Bezerra; Raul Machado Neto; Rejane Maria Tommasini Grotto; Ricardo Haddad; Sandra Coccuzzo Sampaio Vessoni; Simone Kashima; Svetoslav Nanev Slavov; Vincent Louis Viala |
| EPI_ISL_2493311, EPI_ISL_2493313, EPI_ISL_2493314, EPI_ISL_2493315, EPI_ISL_2493317, EPI_ISL_2493319 | UNIDADE BASICA DE SAUDE HORTOLANDIA PACS | Instituto Butantan | Antonio Jorge Martins; Claudia Renata dos Santos Barros; David Schlesinger; Debora Botequio Moretti; Dimas Tadeu Covas; Elaine Cristina Marqueze; Elaine Vieira Santos; Evandra Strazza Rodrigues; Heidge Fukumasu; Jayme Augusto de Souza-Neto; José Salvatore Leister Patané; Luiz Alcantara; Luiz Lehmann Coutinho; Maria Carolina Elias; Maurício Lacerda Nogueira; Rafael dos Santos Bezerra; Raul Machado Neto; Rejane Maria Tommasini Grotto; Ricardo Haddad; Sandra Coccuzzo Sampaio Vessoni; Simone Kashima; Svetoslav Nanev Slavov; Vincent Louis Viala |
| EPI_ISL_2493294, EPI_ISL_2493295 | UNIDADE BASICA SAUDE NATAL ABADIO LACERDA | Instituto Butantan | Antonio Jorge Martins; Claudia Renata dos Santos Barros; David Schlesinger; Debora Botequio Moretti; Dimas Tadeu Covas; Elaine Cristina Marqueze; Elaine Vieira Santos; Evandra Strazza Rodrigues; Heidge Fukumasu; Jayme Augusto de Souza-Neto; José Salvatore Leister Patané; Luiz Alcantara; Luiz Lehmann Coutinho; Maria Carolina Elias; Maurício Lacerda Nogueira; Rafael dos Santos Bezerra; Raul Machado Neto; Rejane Maria Tommasini Grotto; Ricardo Haddad; Sandra Coccuzzo Sampaio Vessoni; Simone Kashima; Svetoslav Nanev Slavov; Vincent Louis Viala |
| EPI_ISL_2378750 | UNIDADE DE SAUDE DA FAMILIA DE FLORIDA PAULISTA II | Instituto Butantan | Antonio Jorge Martins; Claudia Renata dos Santos Barros; David Schlesinger; Debora Botequio Moretti; Dimas Tadeu Covas; Elaine Cristina Marqueze; Elaine Vieira Santos; Evandra Strazza Rodrigues; Heidge Fukumasu; Jayme Augusto de Souza-Neto; José Salvatore Leister Patané; Luiz Alcantara; Luiz Lehmann Coutinho; Maria Carolina Elias; Maurício Lacerda Nogueira; Rafael dos Santos Bezerra; Raul Machado Neto; Rejane Maria Tommasini Grotto; Ricardo Haddad; Sandra Coccuzzo Sampaio Vessoni; Simone Kashima; Svetoslav Nanev Slavov; Vincent Louis Viala |
| EPI_ISL_6229809 | UNIDADE DE SAUDE DA FAMILIA MONSENHOR SABINO | ACME Lab, Oswaldo Cruz Foundation, FIOCRUZ/CE | Carlos Leonardo de Aragao Araujo; Cecilia Leite Costa & Eduardo Ruback dos Santos on behalf of COVID-19 FIOCRUZ Genomic Network; Cleber Furtado Aksenen; Fabio Miyajima; Fernando Braga Stehling; Francisco Eder de Moura Lopes; Igor Oliveira Duarte; Jamille Maria Mendes Bezerra; Joaquim Cesar do Nascimento Sousa Junior; Pedro Miguel Carneiro Jeronimo; Suzana Porto Almeida; Thais Ferreira de Oliveira; Thais de Oliveira Costa; Ticiane Cavalcante de Souza; Veridiana Pessoa Miyajima |
| EPI_ISL_2494305 | UNIDADE DE SAUDE DA FAMILIA NOVO HORIZONTE | Instituto Butantan | Antonio Jorge Martins; Claudia Renata dos Santos Barros; David Schlesinger; Debora Botequio Moretti; Dimas Tadeu Covas; Elaine Cristina Marqueze; Elaine Vieira Santos; Evandra Strazza Rodrigues; Heidge Fukumasu; Jayme Augusto de Souza-Neto; José Salvatore Leister Patané; Luiz Alcantara; Luiz Lehmann Coutinho; Maria Carolina Elias; Maurício Lacerda Nogueira; Rafael dos Santos Bezerra; Raul Machado Neto; Rejane Maria Tommasini Grotto; Ricardo Haddad; Sandra Coccuzzo Sampaio Vessoni; Simone Kashima; Svetoslav Nanev Slavov; Vincent Louis Viala |
| EPI_ISL_2493526 | UNIDADE DE SAUDE DE ITOBI ALCIBIADES PIRES | Instituto Butantan | Antonio Jorge Martins; Claudia Renata dos Santos Barros; David Schlesinger; Debora Botequio Moretti; Dimas Tadeu Covas; Elaine Cristina Marqueze; Elaine Vieira Santos; Evandra Strazza Rodrigues; Heidge Fukumasu; Jayme Augusto de Souza-Neto; José Salvatore Leister Patané; Luiz Alcantara; Luiz Lehmann Coutinho; Maria Carolina Elias; Maurício Lacerda Nogueira; Rafael dos Santos Bezerra; Raul Machado Neto; Rejane Maria Tommasini Grotto; Ricardo Haddad; Sandra Coccuzzo Sampaio Vessoni; Simone Kashima; Svetoslav Nanev Slavov; Vincent Louis Viala |
| EPI_ISL_2378742, EPI_ISL_2493297, EPI_ISL_2493298 | UNIDADE DE SAUDE DR PHEBO DE OLIVEIRA ROGE FERREIRA | Instituto Butantan | Antonio Jorge Martins; Claudia Renata dos Santos Barros; David Schlesinger; Debora Botequio Moretti; Dimas Tadeu Covas; Elaine Cristina Marqueze; Elaine Vieira Santos; Evandra Strazza Rodrigues; Heidge Fukumasu; Jayme Augusto de Souza-Neto; José Salvatore Leister Patané; Luiz Alcantara; Luiz Lehmann Coutinho; Maria Carolina Elias; Maurício Lacerda Nogueira; Rafael dos Santos Bezerra; Raul Machado Neto; Rejane Maria Tommasini Grotto; Ricardo Haddad; Sandra Coccuzzo Sampaio Vessoni; Simone Kashima; Svetoslav Nanev Slavov; Vincent Louis Viala |
| EPI_ISL_2493167, EPI_ISL_2493376 | UNIDADE DE VIGILANCIA EPIDEMIOLOGICA | Instituto Butantan | Antonio Jorge Martins; Claudia Renata dos Santos Barros; David Schlesinger; Debora Botequio Moretti; Dimas Tadeu Covas; Elaine Cristina Marqueze; Elaine Vieira Santos; Evandra Strazza Rodrigues; Heidge Fukumasu; Jayme Augusto de Souza-Neto; José Salvatore Leister Patané; Luiz Alcantara; Luiz Lehmann Coutinho; Maria Carolina Elias; Maurício Lacerda Nogueira; Rafael dos Santos Bezerra; Raul Machado Neto; Rejane Maria Tommasini Grotto; Ricardo Haddad; Sandra Coccuzzo Sampaio Vessoni; Simone Kashima; Svetoslav Nanev Slavov; Vincent Louis Viala |
| EPI_ISL_2493181, EPI_ISL_2493602, EPI_ISL_2493604, EPI_ISL_2493605, EPI_ISL_2493607, EPI_ISL_2493611, EPI_ISL_2493614, EPI_ISL_2493616, EPI_ISL_2493617, EPI_ISL_2493620, EPI_ISL_2493623 | | | |
| see above | UNIDADE MISTA DE LUIZ ANTONIO | Instituto Butantan | Antonio Jorge Martins; Claudia Renata dos Santos Barros; David Schlesinger; Debora Botequio Moretti; Dimas Tadeu Covas; Elaine Cristina Marqueze; Elaine Vieira Santos; Evandra Strazza Rodrigues; Heidge Fukumasu; Jayme Augusto de Souza-Neto; José Salvatore Leister Patané; Luiz Alcantara; Luiz Lehmann Coutinho; Maria Carolina Elias; Maurício Lacerda Nogueira; Rafael dos Santos Bezerra; Raul Machado Neto; Rejane Maria Tommasini Grotto; Ricardo Haddad; Sandra Coccuzzo Sampaio Vessoni; Simone Kashima; Svetoslav Nanev Slavov; Vincent Louis Viala |
| EPI_ISL_2445196 | UNIDADE MISTA DE SAUDE DE ORIENTE | Instituto Butantan | Antonio Jorge Martins; Claudia Renata dos Santos Barros; David Schlesinger; Debora Botequio Moretti; Dimas Tadeu Covas; Elaine Cristina Marqueze; Elaine Vieira Santos; Evandra Strazza Rodrigues; Heidge Fukumasu; Jayme Augusto de Souza-Neto; José Salvatore Leister Patané; Luiz Alcantara; Luiz Lehmann Coutinho; Maria Carolina Elias; Maurício Lacerda Nogueira; Rafael dos Santos Bezerra; Raul Machado Neto; Rejane Maria Tommasini Grotto; Ricardo Haddad; Sandra Coccuzzo Sampaio Vessoni; Simone Kashima; Svetoslav Nanev Slavov; Vincent Louis Viala |
| EPI_ISL_2493386, EPI_ISL_2493387 | UNIDADE MISTA DE SAUDE MOMBUCA | Instituto Butantan | Antonio Jorge Martins; Claudia Renata dos Santos Barros; David Schlesinger; Debora Botequio Moretti; Dimas Tadeu Covas; Elaine Cristina Marqueze; Elaine Vieira Santos; Evandra Strazza Rodrigues; Heidge Fukumasu; Jayme Augusto de Souza-Neto; José Salvatore Leister Patané; Luiz Alcantara; Luiz Lehmann Coutinho; Maria Carolina Elias; Maurício Lacerda Nogueira; Rafael dos Santos Bezerra; Raul Machado Neto; Rejane Maria Tommasini Grotto; Ricardo Haddad; Sandra Coccuzzo Sampaio Vessoni; Simone Kashima; Svetoslav Nanev Slavov; Vincent Louis Viala |
| EPI_ISL_2493372 | UNIDADE MISTA OSVALDO P CAMARGO ELIAS FAUSTO | Instituto Butantan | Antonio Jorge Martins; Claudia Renata dos Santos Barros; David Schlesinger; Debora Botequio Moretti; Dimas Tadeu Covas; Elaine Cristina Marqueze; Elaine Vieira Santos; Evandra Strazza Rodrigues; Heidge Fukumasu; Jayme Augusto de Souza-Neto; José Salvatore Leister Patané; Luiz Alcantara; Luiz Lehmann Coutinho; Maria Carolina Elias; Maurício Lacerda Nogueira; Rafael dos Santos Bezerra; Raul Machado Neto; Rejane Maria Tommasini Grotto; Ricardo Haddad; Sandra Coccuzzo Sampaio Vessoni; Simone Kashima; Svetoslav Nanev Slavov; Vincent Louis Viala |
| EPI_ISL_2378752, EPI_ISL_2445154, EPI_ISL_2445164, EPI_ISL_2445168, EPI_ISL_2445175, EPI_ISL_2445181, EPI_ISL_2445185, EPI_ISL_2473707, EPI_ISL_2473724, EPI_ISL_2473812 | | | |
| see above | UNIDADE REFERENCIAL SUDOESTE | Instituto Butantan | Antonio Jorge Martins; Claudia Renata dos Santos Barros; David Schlesinger; Debora Botequio Moretti; Dimas Tadeu Covas; Elaine Cristina Marqueze; Elaine Vieira Santos; Evandra Strazza Rodrigues; Heidge Fukumasu; Jayme Augusto de Souza-Neto; José Salvatore Leister Patané; Luiz Alcantara; Luiz Lehmann Coutinho; Maria Carolina Elias; Maurício Lacerda Nogueira; Rafael dos Santos Bezerra; Raul Machado Neto; Rejane Maria Tommasini Grotto; Ricardo Haddad; Sandra Coccuzzo Sampaio Vessoni; Simone Kashima; Svetoslav Nanev Slavov; Vincent Louis Viala |
| EPI_ISL_2820956 | UOC Microbiologia e Virologia, Azienda Ospedaliera Universitaria Senese, Siena, Italy | Dipartimento di Biotecnologie Mediche | Claudia Gandolfo; David Pinzauti; Francesco Santoro; Gabriele Anichini; Gianni Gori Savellini; Gianni Pozzi; Maria Grazia Cusi |
| EPI_ISL_6229813, EPI_ISL_6229814, EPI_ISL_6229815 | UPA 24H DE QUIXERAMOBIM | ACME Lab, Oswaldo Cruz Foundation, FIOCRUZ/CE | Carlos Leonardo de Aragao Araujo; Cecilia Leite Costa & Eduardo Ruback dos Santos on behalf of COVID-19 FIOCRUZ Genomic Network; Cleber Furtado Aksenen; Fabio Miyajima; Fernando Braga Stehling; Francisco Eder de Moura Lopes; Igor Oliveira Duarte; Jamille Maria Mendes Bezerra; Joaquim Cesar do Nascimento Sousa Junior; Pedro Miguel Carneiro Jeronimo; Suzana Porto Almeida; Thais Ferreira de Oliveira; Thais de Oliveira Costa; Ticiane Cavalcante de Souza; Veridiana Pessoa Miyajima |
| EPI_ISL_5645973, EPI_ISL_5645978, EPI_ISL_5645990 | UPA BELA VISTA | Instituto Butantan | Antonio Jorge Martins; Claudia Renata dos Santos Barros; David Schlesinger; Debora Botequio Moretti; Dimas Tadeu Covas; Elaine Cristina Marqueze; Elaine Vieira Santos; Evandra Strazza Rodrigues; Heidge Fukumasu; Jayme Augusto de Souza-Neto; José Salvatore Leister Patané; Luiz Alcantara; Luiz Lehmann Coutinho; Maria Carolina Elias; Maurício Lacerda Nogueira; Rafael dos Santos Bezerra; Raul Machado Neto; Rejane Maria Tommasini Grotto; Ricardo Haddad; Sandra Coccuzzo Sampaio Vessoni; Simone Kashima; Svetoslav Nanev Slavov; Vincent Louis Viala |
| EPI_ISL_2378740, EPI_ISL_2378741, EPI_ISL_2493566, EPI_ISL_2494298 | UPA DE BEBEDOURO | Instituto Butantan | Antonio Jorge Martins; Claudia Renata dos Santos Barros; David Schlesinger; Debora Botequio Moretti; Dimas Tadeu Covas; Elaine Cristina Marqueze; Elaine Vieira Santos; Evandra Strazza Rodrigues; Heidge Fukumasu; Jayme Augusto de Souza-Neto; José Salvatore Leister Patané; Luiz Alcantara; Luiz Lehmann Coutinho; Maria Carolina Elias; Maurício Lacerda Nogueira; Rafael dos Santos Bezerra; Raul Machado Neto; Rejane Maria Tommasini Grotto; Ricardo Haddad; Sandra Coccuzzo Sampaio Vessoni; Simone Kashima; Svetoslav Nanev Slavov; Vincent Louis Viala |
| EPI_ISL_5645985, EPI_ISL_5646021 | UPA GEISEL | Instituto Butantan | Antonio Jorge Martins; Claudia Renata dos Santos Barros; David Schlesinger; Debora Botequio Moretti; Dimas Tadeu Covas; Elaine Cristina Marqueze; Elaine Vieira Santos; Evandra Strazza Rodrigues; Heidge Fukumasu; Jayme Augusto de Souza-Neto; José Salvatore Leister Patané; Luiz Alcantara; Luiz Lehmann Coutinho; Maria Carolina Elias; Maurício Lacerda Nogueira; Rafael dos Santos Bezerra; Raul Machado Neto; Rejane Maria Tommasini Grotto; Ricardo Haddad; Sandra Coccuzzo Sampaio Vessoni; Simone Kashima; Svetoslav Nanev Slavov; Vincent Louis Viala |
| EPI_ISL_5645972, EPI_ISL_5645983 | UPA IPIRANGA | Instituto Butantan | Antonio Jorge Martins; Claudia Renata dos Santos Barros; David Schlesinger; Debora Botequio Moretti; Dimas Tadeu Covas; Elaine Cristina Marqueze; Elaine Vieira Santos; Evandra Strazza Rodrigues; Heidge Fukumasu; Jayme Augusto de Souza-Neto; José Salvatore Leister Patané; Luiz Alcantara; Luiz Lehmann Coutinho; Maria Carolina Elias; Maurício Lacerda Nogueira; Rafael dos Santos Bezerra; Raul Machado Neto; Rejane Maria Tommasini Grotto; Ricardo Haddad; Sandra Coccuzzo Sampaio Vessoni; Simone Kashima; Svetoslav Nanev Slavov; Vincent Louis Viala |
| EPI_ISL_5646019, EPI_ISL_5646034 | UPA MARY DOTA | Instituto Butantan | Antonio Jorge Martins; Claudia Renata dos Santos Barros; David Schlesinger; Debora Botequio Moretti; Dimas Tadeu Covas; Elaine Cristina Marqueze; Elaine Vieira Santos; Evandra Strazza Rodrigues; Heidge Fukumasu; Jayme Augusto de Souza-Neto; José Salvatore Leister Patané; Luiz Alcantara; Luiz Lehmann Coutinho; Maria Carolina Elias; Maurício Lacerda Nogueira; Rafael dos Santos Bezerra; Raul Machado Neto; Rejane Maria Tommasini Grotto; Ricardo Haddad; Sandra Coccuzzo Sampaio Vessoni; Simone Kashima; Svetoslav Nanev Slavov; Vincent Louis Viala |

| | | | |
|---|---|---|---|
| EPI_ISL_2661479, EPI_ISL_2661486 | US Air Force School of Aerospace Medicine | US Air Force School of Aerospace Medicine | Amanda Javorina; Anthony Fries; Carol Garrett; Clarise Starr; Elizabeth Macias; Jennifer Meyer; Sarah Purves; William Gruner |
| EPI_ISL_2378738, EPI_ISL_2493546 | USAFA FORTE | Instituto Butantan | Antonio Jorge Martins; Claudia Renata dos Santos Barros; David Schlesinger; Debora Botequio Moretti; Dimas Tadeu Covas; Elaine Cristina Marqueze; Elaine Vieira Santos; Evandra Strazza Rodrigues; Heidge Fukumasu; Jayme Augusto de Souza-Neto; José Salvatore Leister Patané; Luiz Alcantara; Luiz Lehmann Coutinho; Maria Carolina Elias; Maurício Lacerda Nogueira; Rafael dos Santos Bezerra; Raul Machado Neto; Rejane Maria Tommasini Grotto; Ricardo Haddad; Sandra Coccuzzo Sampaio Vessoni; Simone Kashima; Svetoslav Nanev Slavov; Vincent Louis Viala |
| EPI_ISL_2493536, EPI_ISL_2494304 | USAFA GUILHERMINA | Instituto Butantan | Antonio Jorge Martins; Claudia Renata dos Santos Barros; David Schlesinger; Debora Botequio Moretti; Dimas Tadeu Covas; Elaine Cristina Marqueze; Elaine Vieira Santos; Evandra Strazza Rodrigues; Heidge Fukumasu; Jayme Augusto de Souza-Neto; José Salvatore Leister Patané; Luiz Alcantara; Luiz Lehmann Coutinho; Maria Carolina Elias; Maurício Lacerda Nogueira; Rafael dos Santos Bezerra; Raul Machado Neto; Rejane Maria Tommasini Grotto; Ricardo Haddad; Sandra Coccuzzo Sampaio Vessoni; Simone Kashima; Svetoslav Nanev Slavov; Vincent Louis Viala |
| EPI_ISL_2493555 | USAFA MELVI | Instituto Butantan | Antonio Jorge Martins; Claudia Renata dos Santos Barros; David Schlesinger; Debora Botequio Moretti; Dimas Tadeu Covas; Elaine Cristina Marqueze; Elaine Vieira Santos; Evandra Strazza Rodrigues; Heidge Fukumasu; Jayme Augusto de Souza-Neto; José Salvatore Leister Patané; Luiz Alcantara; Luiz Lehmann Coutinho; Maria Carolina Elias; Maurício Lacerda Nogueira; Rafael dos Santos Bezerra; Raul Machado Neto; Rejane Maria Tommasini Grotto; Ricardo Haddad; Sandra Coccuzzo Sampaio Vessoni; Simone Kashima; Svetoslav Nanev Slavov; Vincent Louis Viala |
| EPI_ISL_2493558 | USAFA MIRIM | Instituto Butantan | Antonio Jorge Martins; Claudia Renata dos Santos Barros; David Schlesinger; Debora Botequio Moretti; Dimas Tadeu Covas; Elaine Cristina Marqueze; Elaine Vieira Santos; Evandra Strazza Rodrigues; Heidge Fukumasu; Jayme Augusto de Souza-Neto; José Salvatore Leister Patané; Luiz Alcantara; Luiz Lehmann Coutinho; Maria Carolina Elias; Maurício Lacerda Nogueira; Rafael dos Santos Bezerra; Raul Machado Neto; Rejane Maria Tommasini Grotto; Ricardo Haddad; Sandra Coccuzzo Sampaio Vessoni; Simone Kashima; Svetoslav Nanev Slavov; Vincent Louis Viala |
| EPI_ISL_2493557 | USAFA OCIAN | Instituto Butantan | Antonio Jorge Martins; Claudia Renata dos Santos Barros; David Schlesinger; Debora Botequio Moretti; Dimas Tadeu Covas; Elaine Cristina Marqueze; Elaine Vieira Santos; Evandra Strazza Rodrigues; Heidge Fukumasu; Jayme Augusto de Souza-Neto; José Salvatore Leister Patané; Luiz Alcantara; Luiz Lehmann Coutinho; Maria Carolina Elias; Maurício Lacerda Nogueira; Rafael dos Santos Bezerra; Raul Machado Neto; Rejane Maria Tommasini Grotto; Ricardo Haddad; Sandra Coccuzzo Sampaio Vessoni; Simone Kashima; Svetoslav Nanev Slavov; Vincent Louis Viala |
| EPI_ISL_2493559, EPI_ISL_2493561 | USAFA PRINCESA | Instituto Butantan | Antonio Jorge Martins; Claudia Renata dos Santos Barros; David Schlesinger; Debora Botequio Moretti; Dimas Tadeu Covas; Elaine Cristina Marqueze; Elaine Vieira Santos; Evandra Strazza Rodrigues; Heidge Fukumasu; Jayme Augusto de Souza-Neto; José Salvatore Leister Patané; Luiz Alcantara; Luiz Lehmann Coutinho; Maria Carolina Elias; Maurício Lacerda Nogueira; Rafael dos Santos Bezerra; Raul Machado Neto; Rejane Maria Tommasini Grotto; Ricardo Haddad; Sandra Coccuzzo Sampaio Vessoni; Simone Kashima; Svetoslav Nanev Slavov; Vincent Louis Viala |
| EPI_ISL_2493535, EPI_ISL_2493541, EPI_ISL_2494339 | USAFA SAMAMBAIA | Instituto Butantan | Antonio Jorge Martins; Claudia Renata dos Santos Barros; David Schlesinger; Debora Botequio Moretti; Dimas Tadeu Covas; Elaine Cristina Marqueze; Elaine Vieira Santos; Evandra Strazza Rodrigues; Heidge Fukumasu; Jayme Augusto de Souza-Neto; José Salvatore Leister Patané; Luiz Alcantara; Luiz Lehmann Coutinho; Maria Carolina Elias; Maurício Lacerda Nogueira; Rafael dos Santos Bezerra; Raul Machado Neto; Rejane Maria Tommasini Grotto; Ricardo Haddad; Sandra Coccuzzo Sampaio Vessoni; Simone Kashima; Svetoslav Nanev Slavov; Vincent Louis Viala |
| EPI_ISL_2493560, EPI_ISL_2493563 | USAFA SAO JORGE | Instituto Butantan | Antonio Jorge Martins; Claudia Renata dos Santos Barros; David Schlesinger; Debora Botequio Moretti; Dimas Tadeu Covas; Elaine Cristina Marqueze; Elaine Vieira Santos; Evandra Strazza Rodrigues; Heidge Fukumasu; Jayme Augusto de Souza-Neto; José Salvatore Leister Patané; Luiz Alcantara; Luiz Lehmann Coutinho; Maria Carolina Elias; Maurício Lacerda Nogueira; Rafael dos Santos Bezerra; Raul Machado Neto; Rejane Maria Tommasini Grotto; Ricardo Haddad; Sandra Coccuzzo Sampaio Vessoni; Simone Kashima; Svetoslav Nanev Slavov; Vincent Louis Viala |
| EPI_ISL_2493551 | USAFA TUDE BASTOS | Instituto Butantan | Antonio Jorge Martins; Claudia Renata dos Santos Barros; David Schlesinger; Debora Botequio Moretti; Dimas Tadeu Covas; Elaine Cristina Marqueze; Elaine Vieira Santos; Evandra Strazza Rodrigues; Heidge Fukumasu; Jayme Augusto de Souza-Neto; José Salvatore Leister Patané; Luiz Alcantara; Luiz Lehmann Coutinho; Maria Carolina Elias; Maurício Lacerda Nogueira; Rafael dos Santos Bezerra; Raul Machado Neto; Rejane Maria Tommasini Grotto; Ricardo Haddad; Sandra Coccuzzo Sampaio Vessoni; Simone Kashima; Svetoslav Nanev Slavov; Vincent Louis Viala |
| EPI_ISL_2473711 | USF ITAJUBY | Instituto Butantan | Antonio Jorge Martins; Claudia Renata dos Santos Barros; David Schlesinger; Debora Botequio Moretti; Dimas Tadeu Covas; Elaine Cristina Marqueze; Elaine Vieira Santos; Evandra Strazza Rodrigues; Heidge Fukumasu; Jayme Augusto de Souza-Neto; José Salvatore Leister Patané; Luiz Alcantara; Luiz Lehmann Coutinho; Maria Carolina Elias; Maurício Lacerda Nogueira; Rafael dos Santos Bezerra; Raul Machado Neto; Rejane Maria Tommasini Grotto; Ricardo Haddad; Sandra Coccuzzo Sampaio Vessoni; Simone Kashima; Svetoslav Nanev Slavov; Vincent Louis Viala |
| EPI_ISL_2378751, EPI_ISL_2445203, EPI_ISL_2445204, EPI_ISL_2445205, EPI_ISL_2473716, EPI_ISL_2473717, EPI_ISL_2473817 | | | |
| see above | USF PAULISTA FERNANDOPOLIS ANTONIO PIVATO | Instituto Butantan | Antonio Jorge Martins; Claudia Renata dos Santos Barros; David Schlesinger; Debora Botequio Moretti; Dimas Tadeu Covas; Elaine Cristina Marqueze; Elaine Vieira Santos; Evandra Strazza Rodrigues; Heidge Fukumasu; Jayme Augusto de Souza-Neto; José Salvatore Leister Patané; Luiz Alcantara; Luiz Lehmann Coutinho; Maria Carolina Elias; Maurício Lacerda Nogueira; Rafael dos Santos Bezerra; Raul Machado Neto; Rejane Maria Tommasini Grotto; Ricardo Haddad; Sandra Coccuzzo Sampaio Vessoni; Simone Kashima; Svetoslav Nanev Slavov; Vincent Louis Viala |
| EPI_ISL_2332832, EPI_ISL_2332835, EPI_ISL_2426260, EPI_ISL_2426281, EPI_ISL_2426285, EPI_ISL_2426288, EPI_ISL_2426293, EPI_ISL_2426327, EPI_ISL_2426347, EPI_ISL_2426351, EPI_ISL_2426357, EPI_ISL_2426374, EPI_ISL_2426376, EPI_ISL_2426379, EPI_ISL_2426390, EPI_ISL_2432638, EPI_ISL_2432639, EPI_ISL_2432647, EPI_ISL_2432656, EPI_ISL_2432665, EPI_ISL_2432666, EPI_ISL_2432675, EPI_ISL_2432676, EPI_ISL_2432688, EPI_ISL_2432707, EPI_ISL_2432708, EPI_ISL_2432709, EPI_ISL_2432711, EPI_ISL_2432712, EPI_ISL_2432716, EPI_ISL_2432718, EPI_ISL_2432721, EPI_ISL_2432723, EPI_ISL_2432724, EPI_ISL_2432726, EPI_ISL_2432732, EPI_ISL_2432736, EPI_ISL_2432738, EPI_ISL_2432739, EPI_ISL_2432741, EPI_ISL_2432743, EPI_ISL_2432744, EPI_ISL_2432766, EPI_ISL_2432776, EPI_ISL_2432783, EPI_ISL_2432785, EPI_ISL_2432792, EPI_ISL_2432799, EPI_ISL_2432800, EPI_ISL_2432801, EPI_ISL_2432827, EPI_ISL_2432844, EPI_ISL_2432847, EPI_ISL_2432849, EPI_ISL_2432853, EPI_ISL_2432858, EPI_ISL_2432859, EPI_ISL_2432868, EPI_ISL_2432874, EPI_ISL_2432876, EPI_ISL_2432877, EPI_ISL_2432889, EPI_ISL_2432899, EPI_ISL_2432902, EPI_ISL_2432904, EPI_ISL_2432907, EPI_ISL_2432911, EPI_ISL_2544921, EPI_ISL_2544938, EPI_ISL_2544942, EPI_ISL_2544949, EPI_ISL_2544952, EPI_ISL_2544954, EPI_ISL_2544958, EPI_ISL_2544968 | | | |
| see above | UW Virology Lab | UW Virology Lab | Alexander Greninger; Hong Xie; Keith R Jerome; Lasata Shrestha; Meei-Li Huang; Michelle Lin; Nathan Breit; Noah R. Baker; Patrick Mathias; Pavitra Roychoudhury; Ricardo Perez; Robert J. Livingston; Sean Ellis; Shah Mohamed Bakhash; Tien V. Nguyen |
| EPI_ISL_2671483, EPI_ISL_2671769, EPI_ISL_2671777, EPI_ISL_2671780 | Unidad de Investigacion Biomedica de Zacatecas (UIBZ) | Unidad de Genomica Avanzada | ; Alejandra Garcia-Gasca; Alejandra Hernandez-Teran; Alejandro Sanchez-Flores; Alfredo Herrera-Estrella; Alicia Ocaña-Mondragon; Andreu Comas-Garcia; Angel Gustavo Salas-Lais; Antonio Loza Roman; Bernardo Martinez-Miguel; Blanca Taboada; Brenda Irasema Maldonado-Meza; Bruno Gomez-Gil; Carla Ivon Herrera-Najera; Carlos F. Arias; Celia Boukadida; Celida Duque Molina; Celida Martinez- Rodriguez; Clara Esperanza Santacruz-Tinoco; Concepcion Grajales-Muñiz; Consorcio Mexicano de Vigilancia Genomica (CoViGen-Mex). Authors (in alphabetical order): Julio Elias Alvarado-Yaah; Cristobal Chaidez-Quiroz; Daniel Fregoso-Rueda; Daniel Lira Morales; Eduardo Becerril-Vargas; Fernando Fontove-Herrera; Fidencio Mejia-Nepomuceno; Francisco Pulido; Gloria Elena Espinosa-Ayala; Gloria Maria Molina-Salinas; Gloria Vazquez; Hector Esteban Paz-Juarez; Hector Montoya-Fuentes; Helen Haydee Fernanda Ramirez-Plascencia; Irvin Gonzalez-Lopez; Jean Pierre Gonzalez; Jesus Hernandez; Joel Armando Vazquez-Perez.; Jorge Salas-Hernandez; Jose Antonio Enciso-Moreno; Jose Arturo Martinez-Orozco; Jose Esteban Muñoz-Medina; Jose de Jesus Nuñez-Contreras; Juan Bautista Chale-Dzul; Julissa Enciso-Ibarra; Luis Alberto Ochoa-Carrera; Margarita Matias-Florentino; Maria Guadalupe Santiago-Mauricio; Maria Guadalupe de Jesus Mireles-Rivera; Mario Mujica-Sanchez; Marissa Perez-Garcia; Nelly Selem-Mojica; Pavel Isa; Ricardo Ciria Merce; Ricardo Grande; Rosa Maria Gutierrez Rios; Santiago avila-Rios; Selene Zarate; Susana Lopez; Veronica Mata-Haro; Victor Eduardo Garcia-Arias; Victor Hugo Borja-Aburto |
| EPI_ISL_2671494, EPI_ISL_2671495, EPI_ISL_2671498, EPI_ISL_2671499, EPI_ISL_2671654, EPI_ISL_2671656, EPI_ISL_2671657, EPI_ISL_2671658, EPI_ISL_2671661, EPI_ISL_2671662, EPI_ISL_2671663, EPI_ISL_2671664, EPI_ISL_2671665, EPI_ISL_2671666, EPI_ISL_2671667, EPI_ISL_2671668, EPI_ISL_2671669, EPI_ISL_2671670, EPI_ISL_2671672, EPI_ISL_2671674, EPI_ISL_2671675, EPI_ISL_2671676, EPI_ISL_2671677, EPI_ISL_2671678, EPI_ISL_2671681, EPI_ISL_2671683, EPI_ISL_2671684, EPI_ISL_2671693, EPI_ISL_2671710, EPI_ISL_2671715, EPI_ISL_2671746, EPI_ISL_2671748, EPI_ISL_2671749, EPI_ISL_2671781, EPI_ISL_2671782, EPI_ISL_2671783, EPI_ISL_2970028, EPI_ISL_5332882, EPI_ISL_5332891 | | | |
| see above | Unidad de Investigacion Medica de Yucatan (UIMY) | Unidad de Genomica Avanzada | ; Alejandra Garcia-Gasca; Alejandra Hernandez-Teran; Alejandro Sanchez-Flores; Alfredo Herrera-Estrella; Alicia OcaÃ±a-Mondragon; Alicia Ocaña-Mondragon; Andreu Comas-Garcia; Angel Gustavo Salas-Lais; Antonio Loza Roman; Bernardo Martinez-Miguel; Blanca Taboada; Brenda Irasema Maldonado-Meza; Bruno Gomez-Gil; Carla Ivon Herrera-Najera; Carlos F. Arias; Celia Boukadida; Celida Duque Molina; Celida Martinez- Rodriguez; Clara Esperanza Santacruz-Tinoco; Concepcion Grajales-MuÃ±iz; Concepcion Grajales-Muñiz; Consorcio Mexicano de Vigilancia Genomica (CoViGen-Mex). Authors (in alphabetical order): Julio Elias Alvarado-Yaah; Cristobal Chaidez-Quiroz; Daniel Fregoso-Rueda; Daniel Lira Morales; Eduardo Becerril-Vargas; Fernando Fontove-Herrera; Fidencio Mejia-Nepomuceno; Francisco Pulido; Gloria Elena Espinosa-Ayala; Gloria Maria Molina-Salinas; Gloria Vazquez; Hector Esteban Paz-Juarez; Hector Montoya-Fuentes; Helen Haydee Fernanda Ramirez-Plascencia; Irvin Gonzalez-Lopez; Jean Pierre Gonzalez; Jesus Hernandez; Joel Armando Vazquez-Perez.; Jorge Salas-Hernandez; Jose Antonio Enciso-Moreno; Jose Arturo Martinez-Orozco; Jose Esteban MuÃ±oz-Medina; Jose Esteban Muñoz-Medina; Jose de Jesus NuÃ±ez-Contreras; Jose de Jesus Nuñez-Contreras; Juan Bautista Chale-Dzul; Julissa Enciso-Ibarra; Luis Alberto Ochoa-Carrera; Margarita Matias-Florentino; Maria Guadalupe Santiago-Mauricio; Maria Guadalupe de Jesus Mireles-Rivera; Mario Mujica-Sanchez; Marissa Perez-Garcia; Nelly Selem-Mojica; Pavel Isa; Ricardo Ciria Merce; Ricardo Grande; Rosa Maria Gutierrez Rios; Santiago avila-Rios; Selene Zarate; Susana Lopez; Veronica Mata-Haro; Victor Eduardo Garcia-Arias; Victor Hugo Borja-Aburto |
| EPI_ISL_2681367 | Unidad de Investigación Biomédica de Zacatecas (UIBZ) | Instituto de Biotecnología de la UNAM | ; Alejandra García-Gasca; Alejandra Hernández-Terán; Alejandro Sánchez-Flores; Alfredo Herrera-Estrella; Alicia Ocaña-Mondragón; Andreu Comas-García; Angel Gustavo Salas-Lais; Antonio Loza Román; Bernardo Martínez-Miguel; Blanca Taboada; Brenda Irasema Maldonado-Meza; Bruno Gómez-Gil; Carla Ivón Herrera-Najera; Carlos F. Arias; Celia Boukadida; Clara Esperanza Santacruz-Tinoco; Concepción Grajales-Muñiz; Consorcio Mexicano de Vigilancia Genómica (CoViGen-Mex). Authors (in alphabetical order): Julio Elias Alvarado-Yaah; Cristóbal Cháidez-Quiróz; Célida Duque Molina; Célida Martínez- Rodríguez; Daniel Fregoso-Rueda; Daniel Lira Morales; Eduardo Becerril-Vargas; Fernando Fontove-Herrera; Fidencio Mejía-Nepomuceno; Francisco Pulido; Gloria Elena Espinosa-Ayala; Gloria María Molina-Salinas; Gloria Vazquez; Hector Esteban Paz-Juárez; Hector Montoya-Fuentes; Helen Haydee Fernanda Ramírez-Plascencia; Irvin González-López; Jean Pierre González; Jesús Hernández; Joel Armando Vázquez-Pérez.; Jorge Salas-Hernández; José Antonio Enciso-Moreno; José Arturo Martínez-Orozco; José Esteban Muñoz-Medina; José de Jesús Nuñez-Contreras; Juan Bautista Chale-Dzul; Julissa Enciso-Ibarra; Luis Alberto Ochoa-Carrera; Margarita Matías-Florentino; Mario Mújica-Sánchez; Marissa Perez-Garcia; María Guadalupe Santiago-Mauricio; María Guadalupe de Jesús Mireles-Rivera; Nelly Sélem-Mojica; Pavel Isa; Ricardo Ciria Merce; Ricardo Grande; Rosa María Gutiérrez Rios; Santiago Ávila-Ríos; Selene Zárate; Susana Lopez; Verónica Mata-Haro; Victor Eduardo García-Arias; Víctor Hugo Borja-Aburto |
| EPI_ISL_2535111, EPI_ISL_2535114, EPI_ISL_2535115, EPI_ISL_2535116, EPI_ISL_2535117, EPI_ISL_2535118, EPI_ISL_2535119, EPI_ISL_2535120, EPI_ISL_2535121, EPI_ISL_2535122, EPI_ISL_2535123, EPI_ISL_2535124, EPI_ISL_2535125, EPI_ISL_2535126, EPI_ISL_2535127, EPI_ISL_2535128, EPI_ISL_2535129, EPI_ISL_2535130, EPI_ISL_2535131, EPI_ISL_2535132, EPI_ISL_2535133, EPI_ISL_2535134, EPI_ISL_2535135, EPI_ISL_2535136, EPI_ISL_2535139, EPI_ISL_2535140, EPI_ISL_2535141, EPI_ISL_2535142, EPI_ISL_2535143, EPI_ISL_2535144, EPI_ISL_2535145, EPI_ISL_2535146, EPI_ISL_2535147, EPI_ISL_2535148, EPI_ISL_2535149, EPI_ISL_2535150, EPI_ISL_2535151, EPI_ISL_2535152, EPI_ISL_2535153, EPI_ISL_2535154, EPI_ISL_2535155, EPI_ISL_2535156, EPI_ISL_2535157, EPI_ISL_2535158, EPI_ISL_2535159, EPI_ISL_2535160, EPI_ISL_2535161, EPI_ISL_2535162, EPI_ISL_2535163, EPI_ISL_2535164, EPI_ISL_2535165, EPI_ISL_2535166, EPI_ISL_2535167, EPI_ISL_2535168, EPI_ISL_2535169, EPI_ISL_2535170, EPI_ISL_2535171, EPI_ISL_2535172, EPI_ISL_2535173, EPI_ISL_2535176, EPI_ISL_2535177, EPI_ISL_2535178, EPI_ISL_2535179, EPI_ISL_2535180, EPI_ISL_2535182, EPI_ISL_2535183, EPI_ISL_2535184, EPI_ISL_2535185, EPI_ISL_2535186, EPI_ISL_2535187, EPI_ISL_2535188, EPI_ISL_2535190, EPI_ISL_2535191, EPI_ISL_2535192, EPI_ISL_2535193, EPI_ISL_2535194, EPI_ISL_2535195, EPI_ISL_2535196, EPI_ISL_2535207, EPI_ISL_2535208, EPI_ISL_2535209, EPI_ISL_2535210, EPI_ISL_2535211, EPI_ISL_2535213, EPI_ISL_2535221, EPI_ISL_2535222, EPI_ISL_2535223, EPI_ISL_2535224, EPI_ISL_2535225, EPI_ISL_2535226, EPI_ISL_2535227, EPI_ISL_2535234, EPI_ISL_2535235, EPI_ISL_2691623, EPI_ISL_2691624, EPI_ISL_2691625, EPI_ISL_2691645, EPI_ISL_2691646 | | | |
| see above | Unidade de apoio ao diagnostico da COVID - UNADIG | Bioinformatics Laboratory / LNCC | Alessandra P Lamarca; Alexandra L Gerber; Amilcar Tanuri; Ana Paula de C Guimaraes; Ana Tereza R Vasconcelos; Andrea Cony Cavalcanti; Caio Luiz Pereira Ribeiro; Cassia Alves; Cintia Policarpo; Claudia Maria Braga de Mello; Cristiane Gomes da Silva; Diana Mariani; Douglas Terra Machado; Flavio Dias da Silva; Gleidson da Silva de Oliveira; Leandro Magalhaes de Souza; Liliane Cavalcante; Luiz G P de Almeida; Marcio Henrique de Oliveira Garcia; Mario Sergio Ribeiro; Ronaldo da Silva F Jr; Silvia Carvalho |
| EPI_ISL_2501855 | Unità Operativa Complessa Analisi Chimico Cliniche - ASST Brianza | Laboratory of Clinical Microbiology, Virology and Bioemergencies, ASST Fatebenefratelli Sacco - Sacco Hospital | Alberto Rizzo; Alessandro Mancon; Fiorenza Bracchitta; Maria Rita Gismondo; Valeria Micheli |
| EPI_ISL_3216908, EPI_ISL_3477566 | Universidad del Valle, LDAB-Laboratorio de Diagnostico de Agentes Biologicos | Universidad del Valle, TAO-Lab, VIREM, NEAS Network | Andres Castillo; Beatriz Parra & COVID-19 Team Univalle; Diana López-Alvarez; Nelson Rivera Franco |
| EPI_ISL_2691988, EPI_ISL_2691991, EPI_ISL_2691994 | University of Chicago Medicine | RIPHL at Rush University Medical Center | Felix Araujo Perez; Kevin Kunstman; Laura Furtado; Lauren Megger; Marieta Hyde; Max Kolton; Stefan Green |
| EPI_ISL_2692103, EPI_ISL_2692104, EPI_ISL_2692107, EPI_ISL_2692108, EPI_ISL_2692113, EPI_ISL_2692114, EPI_ISL_2692129 | | | |
| see above | University of Illinois at Chicago | RIPHL at Rush University Medical Center | Felix Araujo Perez; Julie Bentea; Kevin Kunstman; Laura Furtado; Marieta Hyde; Max Kolton; Omar Perez; Stefan Green |
| EPI_ISL_2288991, EPI_ISL_2483947 | University of Liège COVID-19 testing center | GIGA Medical Genomics | Bouchra Boujemla; Cécile Meex; Fabrice Bureau; Keith Durkin; Laurent Gillet; Maria Artesi; Marie-Pierre Hayette; Nathalie Renotte; Sébastien Bontems; Vincent Bours; Wouter Coppieters |
| EPI_ISL_2400362, EPI_ISL_2400365, EPI_ISL_2400366, EPI_ISL_2400372 | University of Michigan Clinical Microbiology Laboratory | Lauring Lab, University of Michigan, Department of Microbiology and Immunology | Gilbert |
| EPI_ISL_2426793 | University of Mississippi Medical Center, Department of Pathology | University of Mississippi Medical Center, Molecular and Genomics Core Facility | Ashley C. Johnson; D. Ashley Robinson; Ithiel J. Frame; Krishna K. Ayyalasomayajula; Michael R. Garrett |
| EPI_ISL_2404846 | University of Oregon COVID-19 MAP Laboratory | University of Oregon Genomics and Cell Characterizcation Core Facility (GC3F) | Ariana White; Douglas Turnbull; Jason Carriere; Jason Sydes; Jeff Bishop; Peter Batzel |
| EPI_ISL_7635965, EPI_ISL_7635966, EPI_ISL_7635967, EPI_ISL_7635968, EPI_ISL_7635974, EPI_ISL_7635976, EPI_ISL_7635983 | | | |
| see above | University of Rome Tor Vergata: Departm Experim Medicine Chair of Virology | University of Rome Tor Vergata: Departm Experim Medicine Chair of Virology | Francesca Ceccherini-Silberstein; Loredana Sarmati; Lorenzo Piermatteo; Luca Carioti; Marco Iannetta; Maria Concetta Bellocchi; Massimo Andreoni |
| EPI_ISL_2340675, EPI_ISL_2340682, EPI_ISL_2340828, EPI_ISL_2340839, EPI_ISL_2340840, EPI_ISL_2463288, EPI_ISL_2467795, EPI_ISL_2467796 | | | |
| see above | University of Wisconsin-Madison AIDS Vaccine Research Laboratories | University of Wisconsin-Madison AIDS Vaccine Research Laboratories | Gage Moreno; Katarina Braun; et al. AIDS Vaccine Research Laboratories |
| EPI_ISL_3014192, EPI_ISL_3014299 | Università Federico II - Dipartimento di scienze mediche traslazionali - Napoli | TIGEM | Antonio Grimaldi Patrizia Annunziata Francesco Panariello Teresa Giuliano Michele Cennamo Valentina Bouche Chiara Colantuono Lucio Di Filippo Mariano Fiorenza Anna Manfredi Marcello Salvi Giuseppe Portella Andrea Ballabio Davide Cacchiarelli |
| EPI_ISL_2308622, EPI_ISL_2308623, | Università degli Studi di Perugia | Istituto Zooprofilattico Sperimentale dell'Abruzzo e Molise | Ancora M; Biagetti M; Calistri P; Camilloni B; Cammà C; Curini V; Delli Compagni E; Di Domenico M; Di Pasquale A; Giammarioli M; Lorusso A; Mangone I; Marcacci M; Mencacci A; Puglia I; Rinaldi A; Savini G; Scialabba S |

| | | | |
|---|---|---|---|
| EPI_ISL_2308626 | | “G. Caporale” | |
| EPI_ISL_5397430 | Universitätsklinikum Köln; Institut für Virologie | Robert Koch Institute | |
| EPI_ISL_2510857, EPI_ISL_2510880, EPI_ISL_2510894, EPI_ISL_2510900, EPI_ISL_2510943, EPI_ISL_2510999, EPI_ISL_2511002, EPI_ISL_2511008 | | | |
| see above | UniversitŠtsSpital ZŸrich | Institute of Medical Virology | Alexandra Trkola; Annette AudigŽ; Catharine Aquino; Cyril Shah; Daniel Ehrsam; Gabriela Ziltener; Guido Bloemberg; Hubert Rehrauer; Isabel StŸrmer; Joel Wirz; Jon Huder; JŸrg Bšni; Kevin Steiner; Maria GrŸnberg; Maryam Zaheri; Michael Huber; Riccarda Capaul; Stefan Schmutz; Verena Kufner; Weihong Qi |
| EPI_ISL_2483459 | Usansolo-Galdakao University Hospital | Cruces University Hospital | Ana Belén de la Hoz; Ana Gual-de-Torrella; Izaskun Alejo-Cancho; Mikel Gallego |
| EPI_ISL_2456657, EPI_ISL_2456732, EPI_ISL_2458487, EPI_ISL_2679601, EPI_ISL_2679632, EPI_ISL_2855894, EPI_ISL_2856073, EPI_ISL_2856080, EPI_ISL_2856141, EPI_ISL_2856152, EPI_ISL_2856452, EPI_ISL_2856473, EPI_ISL_2856496, EPI_ISL_2857920, EPI_ISL_2857948, EPI_ISL_2857955 | | | |
| see above | Utah Public Health Laboratory | Utah Public Health Laboratory | Erin L. Young; Kelly F. Oakeson; Olinto Linares-Perdomo; Tara Gallagher |
| EPI_ISL_2493670, EPI_ISL_2493672 | VIGILANCIA EPIDEMIOLOGICA DE IBATE | Instituto Butantan | Antonio Jorge Martins; Claudia Renata dos Santos Barros; David Schlesinger; Debora Botequio Moretti; Dimas Tadeu Covas; Elaine Cristina Marqueze; Elaine Vieira Santos; Evandra Strazza Rodrigues; Heidge Fukumasu; Jayme Augusto de Souza-Neto; José Salvatore Leister Patané; Luiz Alcantara; Luiz Lehmann Coutinho; Maria Carolina Elias; Maurício Lacerda Nogueira; Rafael dos Santos Bezerra; Raul Machado Neto; Rejane Maria Tommasini Grotto; Ricardo Haddad; Sandra Coccuzzo Sampaio Vessoni; Simone Kashima; Svetoslav Nanev Slavov; Vincent Louis Viala |
| EPI_ISL_2493395, EPI_ISL_2493399, EPI_ISL_2494335 | VIGILANCIA EPIDEMIOLOGICA DE LEME | Instituto Butantan | Antonio Jorge Martins; Claudia Renata dos Santos Barros; David Schlesinger; Debora Botequio Moretti; Dimas Tadeu Covas; Elaine Cristina Marqueze; Elaine Vieira Santos; Evandra Strazza Rodrigues; Heidge Fukumasu; Jayme Augusto de Souza-Neto; José Salvatore Leister Patané; Luiz Alcantara; Luiz Lehmann Coutinho; Maria Carolina Elias; Maurício Lacerda Nogueira; Rafael dos Santos Bezerra; Raul Machado Neto; Rejane Maria Tommasini Grotto; Ricardo Haddad; Sandra Coccuzzo Sampaio Vessoni; Simone Kashima; Svetoslav Nanev Slavov; Vincent Louis Viala |
| EPI_ISL_2383034, EPI_ISL_2493391, EPI_ISL_2493432, EPI_ISL_2493434, EPI_ISL_2494283 | VIGILANCIA EPIDEMIOLOGICA E CONTROLE DE VETORES PIRASSUNUN | Instituto Butantan | Antonio Jorge Martins; Claudia Renata dos Santos Barros; David Schlesinger; Debora Botequio Moretti; Dimas Tadeu Covas; Elaine Cristina Marqueze; Elaine Vieira Santos; Evandra Strazza Rodrigues; Heidge Fukumasu; Jayme Augusto de Souza-Neto; José Salvatore Leister Patané; Luiz Alcantara; Luiz Lehmann Coutinho; Maria Carolina Elias; Maurício Lacerda Nogueira; Rafael dos Santos Bezerra; Raul Machado Neto; Rejane Maria Tommasini Grotto; Ricardo Haddad; Sandra Coccuzzo Sampaio Vessoni; Simone Kashima; Svetoslav Nanev Slavov; Vincent Louis Viala |
| EPI_ISL_2493599, EPI_ISL_2493610 | VIGILANCIA EPIDEMIOLOGICA JARDINOPOLIS SP | Instituto Butantan | Antonio Jorge Martins; Claudia Renata dos Santos Barros; David Schlesinger; Debora Botequio Moretti; Dimas Tadeu Covas; Elaine Cristina Marqueze; Elaine Vieira Santos; Evandra Strazza Rodrigues; Heidge Fukumasu; Jayme Augusto de Souza-Neto; José Salvatore Leister Patané; Luiz Alcantara; Luiz Lehmann Coutinho; Maria Carolina Elias; Maurício Lacerda Nogueira; Rafael dos Santos Bezerra; Raul Machado Neto; Rejane Maria Tommasini Grotto; Ricardo Haddad; Sandra Coccuzzo Sampaio Vessoni; Simone Kashima; Svetoslav Nanev Slavov; Vincent Louis Viala |
| EPI_ISL_2375422, EPI_ISL_2462653 | Viollier AG | Department of Biosystems Science and Engineering, ETH Zürich | Andrea Patrignani; Andreia Cabral de Gouvea; Catharine Aquino; Chaoran Chen; Christiane Beckmann; Christoph Noppen; Daniel Ehrsam; David Dreifuss; Doris Popovic; Griffin White; Isabel Stürmer; Ivan Topolsky; Jay Tracy; Katharina Jahn; Kim Philipp Jablonski; Lara Fuhrmann; Laura Neff; Lennart Opitz; Maria Domenica Moccia; Maurice Redondo; Niko Beerenwinkel; Noemie Santamaria de Souza; Olivier Kobel; Philipp Jablonski; Ralph Schlapbach; Sarah Nadeau; Simon Grüter; Sophie Seidel; Tanja Stadler; Timothy Sykes |
| EPI_ISL_2344538, EPI_ISL_2603207, EPI_ISL_2603208, EPI_ISL_2603217 | Virginia Division of Consolidated Laboratory Services | Virginia Division of Consolidated Laboratory Services | Virginia DCLS |
| EPI_ISL_2884069, EPI_ISL_2884409, EPI_ISL_2884410, EPI_ISL_2884412, EPI_ISL_2884414 | Virology Laboratory, Scientific Department, Army Medical Center | Virology Laboratory, Scientific Department, Army Medical Center | Anella Monte; Anna Anselmo; Antonella Fortunato; Filippo Molinari; Florigio Lista; Francesco Giordani; Giancarlo Petralito; Giandomenico Cerreto; Giulia Campoli; Lucia Nicosia; Marzia Cavalli; Riccardo De Sanctis; Rossella Brandi; Silvia Fillo; Vanessa Vera Fain |
| EPI_ISL_2274977 | Virology Unit, AOUP | Virology Unit, AOUP | Marialinda Vatteroni; Mauro Pistello; Susi Frateschi |
| EPI_ISL_2451955 | WA State Department of Health | Centers for Disease Control and Prevention Division of Viral Diseases, Pathogen Discovery | Alison Laufer Halpin; Ben L. Rambo-Martin; Clinton R. Paden; Dakota Howard; Darlene Wagner; Dave Wentworth; Dhwani Batra; Jasmine Padilla; Justin Lee; Katie Dillon; Krista Queen; Kristen Knipe; Kristine Lacek; Mark Burroughs; Matthew Schmerer; Mili Sheth; Peter Cook; Sam Shepard; Sarah Nobles; Shoshona Le; Suxiang Tong; Vivien Dugan; Yvette Unoarumhi |
| EPI_ISL_2627011 | WESTCHESTER MEDICAL CENTER | Wadsworth Center, New York State Department of Health | Alexis Russell; Catharine Prussing; Daryl M. Lamson; Erasmus Schneider; Erica Lasek-Nesselquist; John Kelly; Jonathan Plitnick; Kirsten St. George; Matthew Shudt; Melissa A Leisner; Navjot Singh |
| EPI_ISL_2452015 | WI State Laboratory of Hygiene | Centers for Disease Control and Prevention Division of Viral Diseases, Pathogen Discovery | Alison Laufer Halpin; Ben L. Rambo-Martin; Clinton R. Paden; Dakota Howard; Darlene Wagner; Dave Wentworth; Dhwani Batra; Jasmine Padilla; Justin Lee; Katie Dillon; Krista Queen; Kristen Knipe; Kristine Lacek; Mark Burroughs; Matthew Schmerer; Mili Sheth; Peter Cook; Sam Shepard; Sarah Nobles; Shoshona Le; Suxiang Tong; Vivien Dugan; Yvette Unoarumhi |
| EPI_ISL_2452097 | WY Public Health Laboratory | Centers for Disease Control and Prevention Division of Viral Diseases, Pathogen Discovery | Alison Laufer Halpin; Ben L. Rambo-Martin; Clinton R. Paden; Dakota Howard; Darlene Wagner; Dave Wentworth; Dhwani Batra; Jasmine Padilla; Justin Lee; Katie Dillon; Krista Queen; Kristen Knipe; Kristine Lacek; Mark Burroughs; Matthew Schmerer; Mili Sheth; Peter Cook; Sam Shepard; Sarah Nobles; Shoshona Le; Suxiang Tong; Vivien Dugan; Yvette Unoarumhi |
| EPI_ISL_2336247, EPI_ISL_2378831, EPI_ISL_2378832, EPI_ISL_2378881, EPI_ISL_2378883, EPI_ISL_2378884, EPI_ISL_2378885, EPI_ISL_2378886, EPI_ISL_2443465, EPI_ISL_2557050, EPI_ISL_2651041 | | | |
| see above | Washington State Department of Health Public Health Laboratories | Washington State Department of Health Public Health Laboratories | Avi Singh; Darren Lucas; Denny Russell; Drew MacKellar; Geoff Melly; Hannah Gray; Joenice Gonzalez; JohnAric Peterson; Philip Dykema; Rebecca Cao; Vanessa De Los Santos |
| EPI_ISL_2402825, EPI_ISL_2402894 | Weill Cornell Medicine | New York Genome Center | Andre Corvelo; Arryn Craney; Chris Mason; Dayna M. Oschwald; Hanna Rennert; Lars F Westblade; Margaret Elizabeth Ross; Melissa Cushing; Michael Zody; Olivier Elemento; Priya Velu; Samantha Fennessey; Tom Maniatis |
| EPI_ISL_2657692, EPI_ISL_2657693, EPI_ISL_2657695 | Wexner Medical Center | The Ohio State University College of Medicine | Koenig, S.; Seminetta, J. |
| EPI_ISL_2248756, EPI_ISL_2261797, EPI_ISL_2261811, EPI_ISL_2261828, EPI_ISL_2261829, EPI_ISL_2276083, EPI_ISL_2276085, EPI_ISL_2284855, EPI_ISL_2284856, EPI_ISL_2284858, EPI_ISL_2284860, EPI_ISL_2284864, EPI_ISL_2284866, EPI_ISL_2284867, EPI_ISL_2289075, EPI_ISL_2289077 | | | |
| see above | Wichita State University - Molecular Diagnostics Lab | Kansas Health and Environmental Lab | Ben Olsen; Jonathan Barnell; Katherine Wiggins; Mike Grose; and Phil Adam |
| EPI_ISL_2382545, EPI_ISL_2382547, EPI_ISL_2382550 | Wichita State University- Molecular Diagonstic lab | Kansas Health and Environmental Lab | Ben Olsen; Jonathan Barnell; Katherine Wiggins; Mike Grose; and Phil Adam |
| EPI_ISL_2383172 | Willis-Knighton Medical Center Hospital Laboratory | LSUHS Emerging Viral Threat Laboratory | Alexander Mijalis; Andrew D. Yurochko; April N. Johnson; Christopher G. Kevil; Gregory L. Ware; Jennifer L. Carroll; Jeremy P. Kamil; John A. Vanchiere; Joseph A. Bocchini; Lorie M. Atkins; Maarten Van Diest; Rona S. Scott |
| EPI_ISL_2558268, EPI_ISL_2558326, EPI_ISL_2558336, EPI_ISL_2558375, EPI_ISL_2558377, EPI_ISL_2558378, EPI_ISL_2558379, EPI_ISL_2558562, EPI_ISL_2558569, EPI_ISL_2558572, EPI_ISL_2558605, EPI_ISL_2558627 | | | |
| see above | Wisconsin State Laboratory of Hygiene Communicable Disease Division | Wisconsin State Laboratory of Hygiene Communicable Disease Division | Abigail C. Shockey; Alicia J. Mooney; Erika M. Hanson; Kelsey R. Florek; Richard Griesser; Sara Wagner; Tonya Danz |
| EPI_ISL_2423051, EPI_ISL_2616539, EPI_ISL_3010473, EPI_ISL_3010476 | Wyoming Public Health Laboratory | Wyoming Public Health Laboratory | Ashley Norberg; Brian Dominguez; Brittany Oher; Cari Sloma; Channing Weber; Chayse Rowley; Elliot Thomasson; Jim Mildenberger; Marley Goetz; Robert Petit; Sam Britz; Taylor Fearing; and Rob Christensen |
| EPI_ISL_2501685, EPI_ISL_2501704, EPI_ISL_2501705 | Yale Clinical Virology Lab | Grubaugh Lab - Yale School of Public Health | Anderson Brito; Annie Watkins; Chaney Kalinich; Chantal Vogels; Isabel Ott; Jessica Rothman; Joseph Fauver; Mallery Breban; Marie L. Landry; Mary Petrone; Nathan Grubaugh; Tara Alpert |
| EPI_ISL_2360232 | Zentrallabor Zürich | Institute of Medical Virology | Alexandra Trkola; Annette Audigé; Cyril Shah; Gabriela Ziltener; Guido Bloemberg; Jon Huder; Jürg Böni; Kevin Steiner; Maria Grünberg; Maryam Zaheri; Michael Huber; Riccarda Capaul; Stefan Schmutz; Verena Kufner |
| EPI_ISL_2387321 | amedes MVZ für Laboratoriumsdiagnostik Raubling GmbH | Robert Koch Institute | |
| EPI_ISL_2488813 | Evandro Chagas Institute | Evandro Chagas Institute | A.M.; Barbagelata; E.C.; E.M.A.; Ferreira; J.A.; Junior; K.C.; L.C.; L.S.; M.C.; P.S.; Pinheiro; Santos; Silva; Sousa; Sousa Junior; W.D.C.; da Silva |
