## Supplement 3 for "Rapid and Accurate Identification of SARS-CoV-2 Omicron Variants Using Droplet Digital PCR (RT-ddPCR)": All_0524-0530.pdf

We gratefully acknowledge the following Authors from the Originating laboratories responsible for obtaining the specimens, as well as the Submitting laboratories where the genome data were generated and shared via GISAID, on which this research is based.

All Submitters of data may be contacted directly via www.gisaid.org

Authors are sorted alphabetically.

| Accession ID | Originating Laboratory | Submitting Laboratory | Authors |
|---|---|---|---|
| EPI_ISL_2787711 | "HI Dept. of Health, State Laboratories Division" | Centers for Disease Control and Prevention Division of Viral Diseases, Pathogen Discovery | Alison Laufer Halpin; Ben L. Rambo-Martin; Clinton R. Paden; Dakota Howard; Darlene Wagner; Dave Wentworth; Dhwani Batra; Jasmine Padilla; Justin Lee; Katie Dillon; Krista Queen; Kristen Knipe; Kristine Lacek; Mark Burroughs; Matthew Schmerer; Mili Sheth; Peter Cook; Sam Shepard; Sarah Nobles; Shoshona Le; Suxiang Tong; Vivien Dugan; Yvette Unoarumhi |
| EPI_ISL_2690045 | "NM Dept. Health, Scientific Laboratory Division " | Centers for Disease Control and Prevention Division of Viral Diseases, Pathogen Discovery | Alison Laufer Halpin; Ben L. Rambo-Martin; Clinton R. Paden; Dakota Howard; Darlene Wagner; Dave Wentworth; Dhwani Batra; Jasmine Padilla; Justin Lee; Katie Dillon; Krista Queen; Kristen Knipe; Kristine Lacek; Mark Burroughs; Matthew Schmerer; Mili Sheth; Peter Cook; Sam Shepard; Sarah Nobles; Shoshona Le; Suxiang Tong; Vivien Dugan; Yvette Unoarumhi |
| EPI_ISL_2721200, EPI_ISL_2721202, EPI_ISL_2721203, EPI_ISL_2721205 | A.S.L. ALESSANDRIA | Fondazione del Piemonte per l'Oncologia IRCCS | Antonino Sottile; Giorgio Giardina; Paola Marino; Silvia Brossa |
| EPI_ISL_2621214, EPI_ISL_2621217, EPI_ISL_2651234, EPI_ISL_3276541, EPI_ISL_3276542 | ADILAB | Universidad Nacional de Colombia - Laboratorio Genómico One Health | Andres F. Cardona-Rios; Carlos Franco-Muñoz; Carolina Muñoz-Arango; Celeny Ortiz; Daniel O. Maldonado-Perez; Diego A. Álvarez-Díaz; Hector Alejandro Ruiz-Moreno; Idabely Betancur Ortiz; Jorge E. Osorio; Juan P. Hernandez-Ortiz; Karl A Ciuoderis; Katherine Laiton-Donato; Laura Silvana Perez; Lina M. Hurtado; Marcela Mercado-Reyes; Maria Angélica Maya; Maria Stella López; Rita Almanza Payares; Sandra Ines Cano; Simón Villegas Velásquez |
| EPI_ISL_2494052, EPI_ISL_2494053, EPI_ISL_2494054, EPI_ISL_2494055, EPI_ISL_2494056, EPI_ISL_2494057 | AFIP LESTE | Instituto Butantan | Antonio Jorge Martins; Claudia Renata dos Santos Barros; David Schlesinger; Debora Botequio Moretti; Dimas Tadeu Covas; Elaine Cristina Marqueze; Elaine Vieira Santos; Evandra Strazza Rodrigues; Heidge Fukumasu; Jayme Augusto de Souza-Neto; José Salvatore Leister Patané; Luiz Alcantara; Luiz Lehmann Coutinho; Maria Carolina Elias; Maurício Lacerda Nogueira; Rafael dos Santos Bezerra; Raul Machado Neto; Rejane Maria Tommasini Grotto; Ricardo Haddad; Sandra Coccuzzo Sampaio Vessoni; Simone Kashima; Svetoslav Nanev Slavov; Vincent Louis Viala |
| EPI_ISL_2493183, EPI_ISL_2493266, EPI_ISL_2493269, EPI_ISL_2494058, EPI_ISL_2494059, EPI_ISL_2494060, EPI_ISL_2494061, EPI_ISL_2494062, EPI_ISL_2494063, EPI_ISL_2494064, EPI_ISL_2494065, EPI_ISL_2494066 | | | |
| see above | AFIP SUDESTE | Instituto Butantan | Antonio Jorge Martins; Claudia Renata dos Santos Barros; David Schlesinger; Debora Botequio Moretti; Dimas Tadeu Covas; Elaine Cristina Marqueze; Elaine Vieira Santos; Evandra Strazza Rodrigues; Heidge Fukumasu; Jayme Augusto de Souza-Neto; José Salvatore Leister Patané; Luiz Alcantara; Luiz Lehmann Coutinho; Maria Carolina Elias; Maurício Lacerda Nogueira; Rafael dos Santos Bezerra; Raul Machado Neto; Rejane Maria Tommasini Grotto; Ricardo Haddad; Sandra Coccuzzo Sampaio Vessoni; Simone Kashima; Svetoslav Nanev Slavov; Vincent Louis Viala |
| EPI_ISL_2493169, EPI_ISL_2493279, EPI_ISL_2493287, EPI_ISL_2494067, EPI_ISL_2494068, EPI_ISL_2494069, EPI_ISL_2494070, EPI_ISL_2494071, EPI_ISL_2494072, EPI_ISL_2494073, EPI_ISL_2494074, EPI_ISL_2494075, EPI_ISL_2494076, EPI_ISL_2494077, EPI_ISL_2494078, EPI_ISL_2494079, EPI_ISL_2494080, EPI_ISL_2494081, EPI_ISL_2494082, EPI_ISL_2494083, EPI_ISL_2494084, EPI_ISL_2494085, EPI_ISL_2494086, EPI_ISL_2494087, EPI_ISL_2494088, EPI_ISL_2494089, EPI_ISL_2494090, EPI_ISL_2494091, EPI_ISL_2494092, EPI_ISL_2494093, EPI_ISL_2494094, EPI_ISL_2494095, EPI_ISL_2494096, EPI_ISL_2494097, EPI_ISL_2494098, EPI_ISL_2494311, EPI_ISL_2494312, EPI_ISL_2494330 | | | |
| see above | AFIP SUL | Instituto Butantan | Antonio Jorge Martins; Claudia Renata dos Santos Barros; David Schlesinger; Debora Botequio Moretti; Dimas Tadeu Covas; Elaine Cristina Marqueze; Elaine Vieira Santos; Evandra Strazza Rodrigues; Heidge Fukumasu; Jayme Augusto de Souza-Neto; José Salvatore Leister Patané; Luiz Alcantara; Luiz Lehmann Coutinho; Maria Carolina Elias; Maurício Lacerda Nogueira; Rafael dos Santos Bezerra; Raul Machado Neto; Rejane Maria Tommasini Grotto; Ricardo Haddad; Sandra Coccuzzo Sampaio Vessoni; Simone Kashima; Svetoslav Nanev Slavov; Vincent Louis Viala |
| EPI_ISL_2627161 | ALBANY MEDICAL CENTER | Wadsworth Center, New York State Department of Health | Alexis Russell; Catharine Prussing; Daryl M. Lamson; Erasmus Schneider; Erica Lasek-Nesselquist; John Kelly; Jonathan Plitnick; Kirsten St. George; Matthew Shudt; Melissa A Leisner; Navjot Singh |
| EPI_ISL_5915181, EPI_ISL_5915191, EPI_ISL_5915196, EPI_ISL_5915203, EPI_ISL_5915262, EPI_ISL_5915263, EPI_ISL_5915295, EPI_ISL_5915300, EPI_ISL_5915303, EPI_ISL_5915305, EPI_ISL_5915307, EPI_ISL_6573945 | | | |
| see above | AMALAB/FACISA/UFRN | WallauLab on behalf of Fiocruz COVID-19 Genomic Surveillance Network | Alexandre Freitas da Silva; Allan Roberto Dias Nunes; Antonio Marinho da Silva Neto; Cassia Docena; Constância Flávia Junqueira Ayres; Filipe Zimmer Dezordi; Gabriel Luz Wallau; Gustavo Barbosa de Lima; Joana Cristina Medeiros Tavares Marques; Katya Anaya Jacinto; Lais Ceschini Machado; Lilian Caroliny Amorim Silva; Marcelo Henrique dos Santos Paiva; Mariane dos Santos Duarte; Matheus Filgueira Bezerra; Sinval Pinto Brandão Filho |
| EPI_ISL_2462327, EPI_ISL_2493170, EPI_ISL_2494133, EPI_ISL_2494134, EPI_ISL_2494137, EPI_ISL_2494138, EPI_ISL_2494331 | | | |
| see above | AMBULATORIO DE ESPECIALIDADE V E MOGI MIRIM | Instituto Butantan | Antonio Jorge Martins; Claudia Renata dos Santos Barros; David Schlesinger; Debora Botequio Moretti; Dimas Tadeu Covas; Elaine Cristina Marqueze; Elaine Vieira Santos; Evandra Strazza Rodrigues; Heidge Fukumasu; Jayme Augusto de Souza-Neto; José Salvatore Leister Patané; Luiz Alcantara; Luiz Lehmann Coutinho; Maria Carolina Elias; Maurício Lacerda Nogueira; Rafael dos Santos Bezerra; Raul Machado Neto; Rejane Maria Tommasini Grotto; Ricardo Haddad; Sandra Coccuzzo Sampaio Vessoni; Simone Kashima; Svetoslav Nanev Slavov; Vincent Louis Viala |
| EPI_ISL_5649587, EPI_ISL_5649636, EPI_ISL_5649667, EPI_ISL_5650177 | AMBULATORIO DR JOAO LANIA CENTRO JOSE BONIFACIO | Instituto Butantan | Antonio Jorge Martins; Claudia Renata dos Santos Barros; David Schlesinger; Debora Botequio Moretti; Dimas Tadeu Covas; Elaine Cristina Marqueze; Elaine Vieira Santos; Evandra Strazza Rodrigues; Heidge Fukumasu; Jayme Augusto de Souza-Neto; José Salvatore Leister Patané; Luiz Alcantara; Luiz Lehmann Coutinho; Maria Carolina Elias; Maurício Lacerda Nogueira; Rafael dos Santos Bezerra; Raul Machado Neto; Rejane Maria Tommasini Grotto; Ricardo Haddad; Sandra Coccuzzo Sampaio Vessoni; Simone Kashima; Svetoslav Nanev Slavov; Vincent Louis Viala |
| EPI_ISL_5646085, EPI_ISL_5646096, EPI_ISL_5646108 | AMBULATORIO MEDICO MUNICIPAL DE AGUDOS | Instituto Butantan | Antonio Jorge Martins; Claudia Renata dos Santos Barros; David Schlesinger; Debora Botequio Moretti; Dimas Tadeu Covas; Elaine Cristina Marqueze; Elaine Vieira Santos; Evandra Strazza Rodrigues; Heidge Fukumasu; Jayme Augusto de Souza-Neto; José Salvatore Leister Patané; Luiz Alcantara; Luiz Lehmann Coutinho; Maria Carolina Elias; Maurício Lacerda Nogueira; Rafael dos Santos Bezerra; Raul Machado Neto; Rejane Maria Tommasini Grotto; Ricardo Haddad; Sandra Coccuzzo Sampaio Vessoni; Simone Kashima; Svetoslav Nanev Slavov; Vincent Louis Viala |
| EPI_ISL_2827985 | AREA DE SALUD ESPARZA | Inciensa, Instituto Costarricense de Investigación y Enseñanza en Nutrición y Salud | Adriana Godínez; Claudio Soto-Garita; Estela Cordero; Francisco Duarte; Hebleen Porras; Joselyn Prado & María José Gómez; José Luis Vargas; Mariela Gutiérrez; Melany Calderón |
| EPI_ISL_3061203 | ASST FBF Sacco | Laboratory of Clinical Microbiology, Virology and Bioemergencies, ASST Fatebenefratelli Sacco - Sacco Hospital | Valeria Micheli |
| EPI_ISL_3495339, EPI_ISL_3495353 | ASST GOM NIGUARDA | ASST Grande ospedale Metropolitano Niguarda | Alice Nava |
| EPI_ISL_2638107, EPI_ISL_2638115, EPI_ISL_2638120, EPI_ISL_2638124, EPI_ISL_2638125, EPI_ISL_2638139 | AULSS 2 Marca Trevigiana | Istituto Zooprofilattico Sperimentale delle Venezie | Adelaide Milani; Alessia Schivo; Alice Fusaro; Ambra Pastori; Annalisa Salviato; Antonia Ricci; Calogero Terregino; Edoardo Giussani; Elisa Palumbo; Erika Giorgia Quaranta; Isabella Monne; Luca Tassoni |
| EPI_ISL_2651227 | AYUDAS DIAGNOSTICAS SURA | Universidad Nacional de Colombia - Laboratorio Genómico One Health | Andres F. Cardona-Rios; Carlos Franco-Muñoz; Carolina Muñoz-Arango; Celeny Ortiz; Daniel O. Maldonado-Perez; Diego A. Álvarez-Díaz; Hector Alejandro Ruiz-Moreno; Idabely Betancur Ortiz; Jorge E. Osorio; Juan P. Hernandez-Ortiz; Karl A Ciuoderis; Katherine Laiton-Donato; Laura Silvana Perez; Lina M. Hurtado; Marcela Mercado-Reyes; Maria Angélica Maya; Maria Stella López; Rita Almanza Payares; Sandra Ines Cano; Simón Villegas Velásquez |
| EPI_ISL_2566921 | AZ Klina | AZ Klina | Carl Vael - Lynsey Berckmans |
| EPI_ISL_2377054 | AZDelta | AZ Delta Medical Laboratories in Roeselare, Belgium | Dieter De Smet; Geert Martens; Merijn Vanhee; on behalf of AZ Delta COVID-19 Genomics core (member of Genomic surveillance of SARS-CoV-2 in Belgium network) |
| EPI_ISL_2370340, EPI_ISL_2370392, EPI_ISL_2370415, EPI_ISL_2370609, EPI_ISL_2370627, EPI_ISL_2370713, EPI_ISL_2370718, EPI_ISL_2372466, EPI_ISL_2489069, EPI_ISL_2489079, EPI_ISL_2489080, EPI_ISL_2489097, EPI_ISL_2489100, EPI_ISL_2489101, EPI_ISL_2489107, EPI_ISL_2489123, EPI_ISL_2489126, EPI_ISL_2489130, EPI_ISL_2489138, EPI_ISL_2489139, EPI_ISL_2489161, EPI_ISL_2489172, EPI_ISL_2489175, EPI_ISL_2489274, EPI_ISL_2489280, EPI_ISL_2489281, EPI_ISL_2489282, EPI_ISL_2489285, EPI_ISL_2489301, EPI_ISL_2489306, EPI_ISL_2489307, EPI_ISL_2489308, EPI_ISL_2489309, EPI_ISL_2489320, EPI_ISL_2489332, EPI_ISL_2489342, EPI_ISL_2489345, EPI_ISL_2489367, EPI_ISL_2489369, EPI_ISL_2489370, EPI_ISL_2489373, EPI_ISL_2489382, EPI_ISL_2489403, EPI_ISL_2489405, EPI_ISL_2489407, EPI_ISL_2489438, EPI_ISL_2489453, EPI_ISL_2489457, EPI_ISL_2489459, EPI_ISL_2489471, EPI_ISL_2489479, EPI_ISL_2489530, EPI_ISL_2489533, EPI_ISL_2489535, EPI_ISL_2489541, EPI_ISL_2489587, EPI_ISL_2489589, EPI_ISL_2489605, EPI_ISL_2489615, EPI_ISL_2489631, EPI_ISL_2489634, EPI_ISL_2489651, EPI_ISL_2489672, EPI_ISL_2489694, EPI_ISL_2489695, EPI_ISL_2489696, EPI_ISL_2489702, EPI_ISL_2489719, EPI_ISL_2489728, EPI_ISL_2489732, EPI_ISL_2489754, EPI_ISL_2489758, EPI_ISL_2489759, EPI_ISL_2489762, EPI_ISL_2489767, EPI_ISL_2489802, EPI_ISL_2489805, EPI_ISL_2528537, EPI_ISL_2528737, EPI_ISL_2530932, EPI_ISL_2530936, EPI_ISL_2530940, EPI_ISL_2530948, EPI_ISL_2530957, EPI_ISL_2530959, EPI_ISL_2530970, EPI_ISL_2531006, EPI_ISL_2531014, EPI_ISL_2531027, EPI_ISL_2531040, EPI_ISL_2531047, EPI_ISL_2531055, EPI_ISL_2531065, EPI_ISL_2531070, EPI_ISL_4360419, EPI_ISL_4362719, EPI_ISL_4363019, EPI_ISL_4366474 | | | |
| see above | Aegis Sciences Corporation | Centers for Disease Control and Prevention Division of Viral Diseases, Pathogen Discovery | Adrian Paskey; Alec Vest; Benjamin Rambo-Martin; Christopher Gulvick; Clinton Paden; Clinton R. Paden; Cyndi Clark; Dakota Howard; Darlene Wagner; Dhwani Batra; Dillon Nall; Duncan MacCannell; Erisa Sula; Ethan Sanders; Holly Houdeshell; Jason Caravas; Kara Moser; Kristine Lacek; Matthew Hardison; Matthew Schmerer; Ola Kvalvaag; Patrick Campbell; Peter Cook; Peter W. Cook; Rob Case; Scott Sammons; Shatavia Morrison; Shaun Westlund; Tymeckia Kendall; Victoria Caban Figueroa; Vikramsinha Ghorpade; Yvette Unoarumhi |
| EPI_ISL_2445953 | Affidea | Instituto Nacional de Saude (INSA) | Borges et al |
| EPI_ISL_2423413, EPI_ISL_2455421, EPI_ISL_2455428, EPI_ISL_2455429 | Alaska State Virology Laboratory | Alaska State Virology Laboratory | Elva House; Jack Chen; Jacob Zidek; Lisa Smith; Ph.D.; Stephanie DeRonde |
| EPI_ISL_2493193, EPI_ISL_2493286, EPI_ISL_2494039, EPI_ISL_2494040, EPI_ISL_2494041, EPI_ISL_2494042, EPI_ISL_2494043, EPI_ISL_2494044, EPI_ISL_2494045, EPI_ISL_2494046, EPI_ISL_2494047, EPI_ISL_2494048, EPI_ISL_2494049, EPI_ISL_2494050, EPI_ISL_2494051, EPI_ISL_2494329, EPI_ISL_2500985 | | | |
| see above | Ambulatório IB | Instituto Butantan | Antonio Jorge Martins; Claudia Renata dos Santos Barros; David Schlesinger; Debora Botequio Moretti; Dimas Tadeu Covas; Elaine Cristina Marqueze; Elaine Vieira Santos; Evandra Strazza Rodrigues; Heidge Fukumasu; Jayme Augusto de Souza-Neto; José Salvatore Leister Patané; Luiz Alcantara; Luiz Lehmann Coutinho; Maria Carolina Elias; Maurício Lacerda Nogueira; Rafael dos Santos Bezerra; Raul Machado Neto; Rejane Maria Tommasini Grotto; Ricardo Haddad; Sandra Coccuzzo Sampaio Vessoni; Simone Kashima; Svetoslav Nanev Slavov; Vincent Louis Viala |
| EPI_ISL_2484085, EPI_ISL_2484086, EPI_ISL_2484102, EPI_ISL_2484111, EPI_ISL_2484113, EPI_ISL_2484124 | Arizona State University | Arizona State University | Efrem S. Lim; Joshua LaBaer; Joy M. Blain; LaRinda A. Holland; Matthew F. Smith; Nicholas J. Mellor; Peter T. Skidmore; Rabia Maqsood; Valerie Harris; Vel Murugan |

| | | | |
|---|---|---|---|
| EPI_ISL_3030303 | Ascoli Piceno | Microbiology University Politecnica delle Marche | Anna Valenza; Carla Acciarri; Katia Marinelli; Monica Lucia Ferreri; Patrizia Bagnarelli; Roberta Longo; Sara Caucci; Stefano Menzo |
| EPI_ISL_2715055, EPI_ISL_2715059, EPI_ISL_2715069, EPI_ISL_2715073, EPI_ISL_2715182, EPI_ISL_2715183, EPI_ISL_2715184, EPI_ISL_2715186, EPI_ISL_2715194, EPI_ISL_2715195, EPI_ISL_2715196, EPI_ISL_2715197, EPI_ISL_2715198, EPI_ISL_2715199 | | | |
| see above | Atlas Genomics | Seattle Flu Study | Amanda Adler; Barry R. Lutz; Benjamin Pelle; Brian Hiatt; Caitlin R. Wolf; Chris D. Frazar; Deborah A. Nickerson; Elisabeth Brandstetter; Erica Ryke; Geoff Melly; Helen Y. Chu; Janet A. Englund; Jay Shendure; Jover Lee; Kairsten Fay; Kirsten Lacombe; Lea M. Starita; Mark J. Rieder; Matthew Richardson; Matthew Thompson; Melissa Truong; Michael Boeckh; Michael Famulare; Misja Ilcisin; Peter D. Han; Philip Dykema; Romesh Gautom; Scott Lindquist; Thomas R. Sibley; Trevor Bedford |
| EPI_ISL_2657455, EPI_ISL_2657456, EPI_ISL_2657472 | Austrian Agency for Health and Food Safety (AGES) | Bergthaler laboratory, CeMM Research Center for Molecular Medicine of the Austrian Academy of Sciences | Andreas Bergthaler; Anna Schedl; Bekir Erguner; Benedikt Agerer; Christoph Bock; Fabian Amman; Jan Laine; Lukas Endler; Maelle Le Moing; Martin Senekowitsch; Matthew Thornton; Michael Schuster; Petr Triska; Thomas Penz |
| EPI_ISL_2522106 | Azienda Ospedaliera San Camillo Forlanini | INMI Lazzaro Spallanzani IRCCS | A D'Agostino; A Di Caro; B Bartolini; CEM Gruber; E Giombini; F Messina; F Santini; G Bonfiglio; G Parisi; M Rueca; ML Guarino; MR Capobianchi; O Butera |
| EPI_ISL_2600785, EPI_ISL_2600786 | Azienda Ospedaliera Terni | Istituto Zooprofilattico Sperimentale dell'Abruzzo e Molise "G. Caporale" | Ancora M; Calistri P; Cammà C; Curini V; Delli Compagni E; Di Domenico M; Di Pasquale A; Lorusso A; Mangone I; Marcacci M; Palumbo M; Puglia I; Rinaldi A; Savini G; Scaccetti A; Scialabba S |
| EPI_ISL_2517092, EPI_ISL_2517093, EPI_ISL_2517105, EPI_ISL_2517106, EPI_ISL_2517108, EPI_ISL_2517118, EPI_ISL_2517120, EPI_ISL_2517121, EPI_ISL_2517123, EPI_ISL_2517128, EPI_ISL_2517132, EPI_ISL_2517153 | | | |
| see above | Azienda Sanitaria dell'Alto Adige Laboratorio Aziendale di Microbiologia e Virologia | Istituto di Genomica Applicata | Davide Scaglione; Eleonora Paparelli; Elisa Masi; Elisabetta Giacobazzi; Elisabetta Pagani; Gabriele Magris; Irena Jurman; Irene Bianconi; Michele Morgante; Stefanie Wieser; Vera Vendramin |
| EPI_ISL_2600796, EPI_ISL_2600797, EPI_ISL_2600799 | Azienda USL Umbria 2 | Istituto Zooprofilattico Sperimentale dell'Abruzzo e Molise "G. Caporale" | Ancora M; Calistri P; Cammà C; Curini V; Delli Compagni E; Di Domenico M; Di Pasquale A; Lorusso A; Mangone I; Marcacci M; Pistoni E; Proietti A; Puglia I; Rinaldi A; Savini G; Scialabba S |
| EPI_ISL_2493261, EPI_ISL_2493262, EPI_ISL_2493268, EPI_ISL_2494220, EPI_ISL_2494221, EPI_ISL_2494222, EPI_ISL_2494223, EPI_ISL_2494224 | | | |
| see above | BIOFAST CENTRO | Instituto Butantan | Antonio Jorge Martins; Claudia Renata dos Santos Barros; David Schlesinger; Debora Botequio Moretti; Dimas Tadeu Covas; Elaine Cristina Marqueze; Elaine Vieira Santos; Evandra Strazza Rodrigues; Heidge Fukumasu; Jayme Augusto de Souza-Neto; José Salvatore Leister Patané; Luiz Alcantara; Luiz Lehmann Coutinho; Maria Carolina Elias; Maurício Lacerda Nogueira; Rafael dos Santos Bezerra; Raul Machado Neto; Rejane Maria Tommasini Grotto; Ricardo Haddad; Sandra Coccuzzo Sampaio Vessoni; Simone Kashima; Svetoslav Nanev Slavov; Vincent Louis Viala |
| EPI_ISL_2493191, EPI_ISL_2493248, EPI_ISL_2493291, EPI_ISL_2494225, EPI_ISL_2494226, EPI_ISL_2494227, EPI_ISL_2494228, EPI_ISL_2494229, EPI_ISL_2494230, EPI_ISL_2494231, EPI_ISL_2494232, EPI_ISL_2494233, EPI_ISL_2494234, EPI_ISL_2494235, EPI_ISL_2494236, EPI_ISL_2494237, EPI_ISL_2494238, EPI_ISL_2494239, EPI_ISL_2494240, EPI_ISL_2494241, EPI_ISL_2494242, EPI_ISL_2494243, EPI_ISL_2494244, EPI_ISL_2494245, EPI_ISL_2494246, EPI_ISL_2494247, EPI_ISL_2494248, EPI_ISL_2494249, EPI_ISL_2494250, EPI_ISL_2494251, EPI_ISL_2494252, EPI_ISL_2494253, EPI_ISL_2494254, EPI_ISL_2494255, EPI_ISL_2494256, EPI_ISL_2494257, EPI_ISL_2494258, EPI_ISL_2494259, EPI_ISL_2494260, EPI_ISL_2494261, EPI_ISL_2494262, EPI_ISL_2494263, EPI_ISL_2494264, EPI_ISL_2494265, EPI_ISL_2494266, EPI_ISL_2494301 | | | |
| see above | BIOFAST LESTE | Instituto Butantan | Antonio Jorge Martins; Claudia Renata dos Santos Barros; David Schlesinger; Debora Botequio Moretti; Dimas Tadeu Covas; Elaine Cristina Marqueze; Elaine Vieira Santos; Evandra Strazza Rodrigues; Heidge Fukumasu; Jayme Augusto de Souza-Neto; José Salvatore Leister Patané; Luiz Alcantara; Luiz Lehmann Coutinho; Maria Carolina Elias; Maurício Lacerda Nogueira; Rafael dos Santos Bezerra; Raul Machado Neto; Rejane Maria Tommasini Grotto; Ricardo Haddad; Sandra Coccuzzo Sampaio Vessoni; Simone Kashima; Svetoslav Nanev Slavov; Vincent Louis Viala |
| EPI_ISL_2493188, EPI_ISL_2493270, EPI_ISL_2494213, EPI_ISL_2494214, EPI_ISL_2494215, EPI_ISL_2494216, EPI_ISL_2494217, EPI_ISL_2494218, EPI_ISL_2494219, EPI_ISL_2494333 | | | |
| see above | BIOFAST NORTE | Instituto Butantan | Antonio Jorge Martins; Claudia Renata dos Santos Barros; David Schlesinger; Debora Botequio Moretti; Dimas Tadeu Covas; Elaine Cristina Marqueze; Elaine Vieira Santos; Evandra Strazza Rodrigues; Heidge Fukumasu; Jayme Augusto de Souza-Neto; José Salvatore Leister Patané; Luiz Alcantara; Luiz Lehmann Coutinho; Maria Carolina Elias; Maurício Lacerda Nogueira; Rafael dos Santos Bezerra; Raul Machado Neto; Rejane Maria Tommasini Grotto; Ricardo Haddad; Sandra Coccuzzo Sampaio Vessoni; Simone Kashima; Svetoslav Nanev Slavov; Vincent Louis Viala |
| EPI_ISL_2493260, EPI_ISL_2493267, EPI_ISL_2494202, EPI_ISL_2494203, EPI_ISL_2494204, EPI_ISL_2494205, EPI_ISL_2494206, EPI_ISL_2494207, EPI_ISL_2494208, EPI_ISL_2494209, EPI_ISL_2494210, EPI_ISL_2494211, EPI_ISL_2494212, EPI_ISL_2494275, EPI_ISL_2494313 | | | |
| see above | BIOFAST SUL | Instituto Butantan | Antonio Jorge Martins; Claudia Renata dos Santos Barros; David Schlesinger; Debora Botequio Moretti; Dimas Tadeu Covas; Elaine Cristina Marqueze; Elaine Vieira Santos; Evandra Strazza Rodrigues; Heidge Fukumasu; Jayme Augusto de Souza-Neto; José Salvatore Leister Patané; Luiz Alcantara; Luiz Lehmann Coutinho; Maria Carolina Elias; Maurício Lacerda Nogueira; Rafael dos Santos Bezerra; Raul Machado Neto; Rejane Maria Tommasini Grotto; Ricardo Haddad; Sandra Coccuzzo Sampaio Vessoni; Simone Kashima; Svetoslav Nanev Slavov; Vincent Louis Viala |
| EPI_ISL_7672200, EPI_ISL_7672504 | Bambino Gesù Pediatric Hospital | Microbiology and Immunology Diagnosis Bambino Gesù Pediatric Hospital | Carlo Federico Perno; Claudia Alteri; Luna Colagrossi; Rossana Scutari; Valentino Costabile |
| EPI_ISL_2464480 | Biogroup Bio Lam-LCD Saint-Denis | Department of Virology, Henri Mondor University Hospital, Assistance Publique Hôpitaux de Paris, Université Paris-Est Créteil, INSERM U955 | Alexandre Soulier; Christophe Rodriguez; Elisabeth Trawinski; Guillaume Gricourt; Jean-Michel Pawlotsky; Melissa N'Debi; Slim Fourati; Vanessa Demontant |
| EPI_ISL_3145457, EPI_ISL_3145475, EPI_ISL_3145476, EPI_ISL_3145477 | BioneXt Lab | Laboratoire national de sante, Microbiology, Microbial Genomics Platform | Anke Wienecke-Baldacchino; Catherine Ragimbeau; Elodie Solarino; Fatu Djabi; Jessica Tapp; Lise Pignon; Raoul Salmon; Tamir Abdelrahman; Thibault Ferrandon; Virginie Jover |
| EPI_ISL_3479117, EPI_ISL_3479120, EPI_ISL_3479123, EPI_ISL_3479125, EPI_ISL_3479126, EPI_ISL_3479128, EPI_ISL_3479129, EPI_ISL_3479132, EPI_ISL_3479133, EPI_ISL_3479134, EPI_ISL_3479135, EPI_ISL_3487709, EPI_ISL_3487711, EPI_ISL_3487713, EPI_ISL_3487722, EPI_ISL_3487730, EPI_ISL_3487731, EPI_ISL_3487732, EPI_ISL_3487733, EPI_ISL_3487736, EPI_ISL_3487740, EPI_ISL_3487741, EPI_ISL_3487742, EPI_ISL_3487746, EPI_ISL_3487748, EPI_ISL_3487752, EPI_ISL_3487755, EPI_ISL_3487756, EPI_ISL_3487764, EPI_ISL_3487765, EPI_ISL_3487767, EPI_ISL_3487769, EPI_ISL_3487799, EPI_ISL_3487801, EPI_ISL_3487802, EPI_ISL_3487803, EPI_ISL_3487804, EPI_ISL_3487805, EPI_ISL_3487807, EPI_ISL_3487808, EPI_ISL_3487809, EPI_ISL_3487811, EPI_ISL_3487816, EPI_ISL_3487819, EPI_ISL_3487820, EPI_ISL_3487824, EPI_ISL_3487827, EPI_ISL_3487829, EPI_ISL_3487831, EPI_ISL_3487832, EPI_ISL_3487835, EPI_ISL_3487836, EPI_ISL_3487840, EPI_ISL_3487844, EPI_ISL_3487851, EPI_ISL_3487853, EPI_ISL_3487863, EPI_ISL_3487866, EPI_ISL_3487875, EPI_ISL_3487876, EPI_ISL_3487877, EPI_ISL_3487878, EPI_ISL_3487879, EPI_ISL_3487880, EPI_ISL_3487882, EPI_ISL_3487884, EPI_ISL_3487886, EPI_ISL_3487887, EPI_ISL_3487888, EPI_ISL_3487889, EPI_ISL_3487892, EPI_ISL_3487900, EPI_ISL_3487904, EPI_ISL_3487908, EPI_ISL_3487913, EPI_ISL_3487914, EPI_ISL_3487917, EPI_ISL_3487933, EPI_ISL_3487946, EPI_ISL_3487947, EPI_ISL_3487948, EPI_ISL_3487952, EPI_ISL_3487955, EPI_ISL_3487973, EPI_ISL_3487974, EPI_ISL_3487975, EPI_ISL_3487976, EPI_ISL_3487977, EPI_ISL_3487980, EPI_ISL_3487988, EPI_ISL_3487990, EPI_ISL_3487996, EPI_ISL_3487997, EPI_ISL_3487999, EPI_ISL_3488000, EPI_ISL_3488006, EPI_ISL_3488020, EPI_ISL_3488021, EPI_ISL_3488030, EPI_ISL_3488031, EPI_ISL_3488032, EPI_ISL_3488033, EPI_ISL_3488034, EPI_ISL_3488046, EPI_ISL_3488049, EPI_ISL_3488053, EPI_ISL_3488054, EPI_ISL_3488055, EPI_ISL_3488056, EPI_ISL_3488058, EPI_ISL_3488059, EPI_ISL_3488060, EPI_ISL_3488061, EPI_ISL_3488064, EPI_ISL_3488066, EPI_ISL_3488068, EPI_ISL_3488070, EPI_ISL_3488074, EPI_ISL_3488075, EPI_ISL_3488076, EPI_ISL_3488077, EPI_ISL_3488079, EPI_ISL_3488080, EPI_ISL_3488083, EPI_ISL_3488085, EPI_ISL_3488086, EPI_ISL_3488087, EPI_ISL_3488088, EPI_ISL_3488090, EPI_ISL_3488092, EPI_ISL_3488093, EPI_ISL_3488097, EPI_ISL_3488098, EPI_ISL_3488100, EPI_ISL_3488101, EPI_ISL_3488102, EPI_ISL_3488106, EPI_ISL_3488110, EPI_ISL_3488111, EPI_ISL_3488115, EPI_ISL_3488116, EPI_ISL_3488120, EPI_ISL_3488122, EPI_ISL_3488126, EPI_ISL_3488207, EPI_ISL_3488208, EPI_ISL_3488261, EPI_ISL_3488287, EPI_ISL_3488294, EPI_ISL_3488312, EPI_ISL_3488313, EPI_ISL_3488316, EPI_ISL_3488318, EPI_ISL_3488319, EPI_ISL_3488323, EPI_ISL_3488324, EPI_ISL_3488326, EPI_ISL_3488327, EPI_ISL_3488331, EPI_ISL_3488337, EPI_ISL_3488340, EPI_ISL_3488349, EPI_ISL_3488350, EPI_ISL_3488357, EPI_ISL_3488359, EPI_ISL_3488360, EPI_ISL_3488398, EPI_ISL_3488404, EPI_ISL_3488406, EPI_ISL_3488416, EPI_ISL_3488419, EPI_ISL_3488421, EPI_ISL_3488423, EPI_ISL_3488424, EPI_ISL_3488425, EPI_ISL_3488429, EPI_ISL_3488430, EPI_ISL_3488431, EPI_ISL_3488432, EPI_ISL_3488434, EPI_ISL_3488435, EPI_ISL_3488436, EPI_ISL_3488437, EPI_ISL_3488439, EPI_ISL_3488440, EPI_ISL_3488451, EPI_ISL_3488452, EPI_ISL_3488460, EPI_ISL_3488464, EPI_ISL_3488466, EPI_ISL_3488470, EPI_ISL_3488471, EPI_ISL_3488472, EPI_ISL_3488473, EPI_ISL_3488474, EPI_ISL_3488479, EPI_ISL_3488485, EPI_ISL_3488486, EPI_ISL_3488487, EPI_ISL_3488488, EPI_ISL_3488489, EPI_ISL_3488490, EPI_ISL_3488495, EPI_ISL_3488496, EPI_ISL_3488498, EPI_ISL_3488499, EPI_ISL_3488501, EPI_ISL_3488511, EPI_ISL_3488514, EPI_ISL_3488515, EPI_ISL_3488516, EPI_ISL_3488517, EPI_ISL_3488519, EPI_ISL_3488522, EPI_ISL_3488525, EPI_ISL_3488526, EPI_ISL_3488529, EPI_ISL_3488531, EPI_ISL_3488532, EPI_ISL_3488543, EPI_ISL_3488544, EPI_ISL_3488592, EPI_ISL_3488593, EPI_ISL_3488594, EPI_ISL_3488597, EPI_ISL_3488598, EPI_ISL_3488599, EPI_ISL_3488600, EPI_ISL_3488604, EPI_ISL_3488605, EPI_ISL_3488612, EPI_ISL_3488613, EPI_ISL_3488616, EPI_ISL_3488622, EPI_ISL_3488624, EPI_ISL_3488628, EPI_ISL_3488629, EPI_ISL_3488633, EPI_ISL_3488634, EPI_ISL_3488636, EPI_ISL_3488637, EPI_ISL_3488638, EPI_ISL_3488639, EPI_ISL_3488641, EPI_ISL_3488644, EPI_ISL_3488647, EPI_ISL_3488648, EPI_ISL_3488649, EPI_ISL_3488652, EPI_ISL_3488655, EPI_ISL_3488658, EPI_ISL_3488660, EPI_ISL_3488661, EPI_ISL_3488663, EPI_ISL_3488666, EPI_ISL_3488667, EPI_ISL_3488671, EPI_ISL_3488674, EPI_ISL_3488681, EPI_ISL_3488683, EPI_ISL_3488696, EPI_ISL_3488697, EPI_ISL_3488700, EPI_ISL_3488701, EPI_ISL_3488703, EPI_ISL_3488707, EPI_ISL_3488708, EPI_ISL_3488716, EPI_ISL_3488717, EPI_ISL_3488718, EPI_ISL_3488724, EPI_ISL_3488725, EPI_ISL_3488726, EPI_ISL_3488727, EPI_ISL_3488729, EPI_ISL_3488730, EPI_ISL_3488738, EPI_ISL_3488746, EPI_ISL_3488750, EPI_ISL_3488754, EPI_ISL_3488755, EPI_ISL_3488761, EPI_ISL_3488788, EPI_ISL_3488790, EPI_ISL_3488791, EPI_ISL_3488794, EPI_ISL_3488800, EPI_ISL_3488802, EPI_ISL_3488803, EPI_ISL_3488804, EPI_ISL_3488806, EPI_ISL_3488808, EPI_ISL_3488809, EPI_ISL_3488814, EPI_ISL_3488815, EPI_ISL_3488816, EPI_ISL_3488817, EPI_ISL_3488819, EPI_ISL_3488820, EPI_ISL_3488821, EPI_ISL_3488822, EPI_ISL_3488825, EPI_ISL_3488826, EPI_ISL_3488827, EPI_ISL_3488828, EPI_ISL_3488830, EPI_ISL_3488831, EPI_ISL_3488832, EPI_ISL_3488834, EPI_ISL_3488838, EPI_ISL_3488840, EPI_ISL_3488847, EPI_ISL_3488848, EPI_ISL_3488853, EPI_ISL_3488854, EPI_ISL_3488856, EPI_ISL_3488857, EPI_ISL_3488858, EPI_ISL_3488859, EPI_ISL_3488860, EPI_ISL_3488862, EPI_ISL_3488864, EPI_ISL_3488867, EPI_ISL_3488870, EPI_ISL_3488871, EPI_ISL_3488872, EPI_ISL_3488873, EPI_ISL_3488880, EPI_ISL_3488881, EPI_ISL_3488886, EPI_ISL_3488890, EPI_ISL_3488891, EPI_ISL_3488894, EPI_ISL_3488899, EPI_ISL_3488900, EPI_ISL_3488901, EPI_ISL_3488902, EPI_ISL_3488909, EPI_ISL_3488913, EPI_ISL_3488916, EPI_ISL_3488917, EPI_ISL_3488918, EPI_ISL_3488920, EPI_ISL_3488921, EPI_ISL_3488923, EPI_ISL_3488924, EPI_ISL_3488927, EPI_ISL_3488930, EPI_ISL_3488931, EPI_ISL_3488932, EPI_ISL_3488933, EPI_ISL_3488940, EPI_ISL_3488941, EPI_ISL_3488942, EPI_ISL_3488945, EPI_ISL_3488946, EPI_ISL_3488947, EPI_ISL_3488950, EPI_ISL_3488954, EPI_ISL_3488955, EPI_ISL_3488960, EPI_ISL_3488965, EPI_ISL_3488968, EPI_ISL_3488969, EPI_ISL_3488970, EPI_ISL_3488972, EPI_ISL_3488973, EPI_ISL_3488975, EPI_ISL_3488979, EPI_ISL_3488984, EPI_ISL_3488989, EPI_ISL_3488991, EPI_ISL_3488996, EPI_ISL_3488998, EPI_ISL_3488999, EPI_ISL_3489009, EPI_ISL_3489013, EPI_ISL_3489014, EPI_ISL_3489015, EPI_ISL_3489016, EPI_ISL_3489019, EPI_ISL_3489023, EPI_ISL_3489025, EPI_ISL_3489026, EPI_ISL_3489036, EPI_ISL_3489037, EPI_ISL_3489040, EPI_ISL_3489042, EPI_ISL_3489043, EPI_ISL_3489044, EPI_ISL_3489046, EPI_ISL_3489047, EPI_ISL_3489050, EPI_ISL_3489064, EPI_ISL_3489066, EPI_ISL_3489067, EPI_ISL_3489069, EPI_ISL_3489072, EPI_ISL_3489077, EPI_ISL_3489078, EPI_ISL_3489080, EPI_ISL_3489082, EPI_ISL_3489088, EPI_ISL_3489089, EPI_ISL_3489091, EPI_ISL_3489092, EPI_ISL_3489145, EPI_ISL_3489189, EPI_ISL_3489199, EPI_ISL_3489375, EPI_ISL_3489385, EPI_ISL_3489386, EPI_ISL_3489389, EPI_ISL_3489391, EPI_ISL_3489392, EPI_ISL_3489393, EPI_ISL_3489394, EPI_ISL_3489395, EPI_ISL_3489397, EPI_ISL_3489403, EPI_ISL_3489406, EPI_ISL_3489407, EPI_ISL_3489408, EPI_ISL_3489413, EPI_ISL_3489415, EPI_ISL_3489418, EPI_ISL_3489419, EPI_ISL_3489421, EPI_ISL_3489422, EPI_ISL_3489423, EPI_ISL_3489424, EPI_ISL_3489426, EPI_ISL_3489428, EPI_ISL_3489430, EPI_ISL_3489431, EPI_ISL_3489432, EPI_ISL_3489433, EPI_ISL_3489435, EPI_ISL_3489441, EPI_ISL_3489443, EPI_ISL_3489446, EPI_ISL_3489447, EPI_ISL_3489448, EPI_ISL_3489449, EPI_ISL_3489464, EPI_ISL_3489468, EPI_ISL_3489471, EPI_ISL_3489472, EPI_ISL_3489478, EPI_ISL_3489579, EPI_ISL_3489587, EPI_ISL_3489590, EPI_ISL_3489600, EPI_ISL_3489603, EPI_ISL_3489606, EPI_ISL_3489610, EPI_ISL_3489611, EPI_ISL_3489614, EPI_ISL_3489619, EPI_ISL_3489627, EPI_ISL_3489628, EPI_ISL_3489630, EPI_ISL_3489632, EPI_ISL_3489633, EPI_ISL_3489634, EPI_ISL_3489635, EPI_ISL_3489636, EPI_ISL_3489637, EPI_ISL_3489640, EPI_ISL_3489668, EPI_ISL_3489688, EPI_ISL_3489778 | | | |
| see above | British Columbia Centre For Disease Control | BCCDC Public Health Laboratory | Ana Pacagnella; Corrinne Ng; Dan Fornika; John Tyson; Kim Macdonald; Kimia Kamelian; Linda Hoang; Loretta Janz; Mel Krajden; Prystajecky Natalie; Robert Azana; Shannon Russell |
| EPI_ISL_2991511, EPI_ISL_2991521, EPI_ISL_2991527, EPI_ISL_2991530, EPI_ISL_2991535, EPI_ISL_2991544, EPI_ISL_2991549, EPI_ISL_2991555, EPI_ISL_2991565, EPI_ISL_2991573, EPI_ISL_2991612, EPI_ISL_2991630, EPI_ISL_2991634, EPI_ISL_2991643, EPI_ISL_2991647, EPI_ISL_2991659, EPI_ISL_2991676, EPI_ISL_2991687, EPI_ISL_2991720, EPI_ISL_2991727, EPI_ISL_2991728, EPI_ISL_2991738, EPI_ISL_2991750, EPI_ISL_2991755, EPI_ISL_2991767, EPI_ISL_2991781, EPI_ISL_2991790, EPI_ISL_2991794, EPI_ISL_2991829, EPI_ISL_2991841, EPI_ISL_2991846, EPI_ISL_2991854, EPI_ISL_2991857, EPI_ISL_2991879, EPI_ISL_2991887, EPI_ISL_2991893, EPI_ISL_2991896, EPI_ISL_2991897, EPI_ISL_2991914, EPI_ISL_2991940, EPI_ISL_2991941, EPI_ISL_2991942, EPI_ISL_2991958 | | | |
| see above | Broad Institute Clinical Research Sequencing Platform | Infectious Disease Program, Broad Institute of Harvard and MIT | A.E.; Adams, G.; Anahtar, M.; B.L.; B.W.; Bauer, M.; Birren; Branda, J.; Carter, A.; Cerrato, F.; Chaluvadi, S.; Chapman; Cusick, C.; D.J.; DeRuff, K.; Flowers, K.; Gallagher, G.; Gladden-Young, A.; Gnirke, A.; Harris, J.; J.E.; K.J.; LaRocque, R.; Lagerborg, K.; Lemieux; Lin; Loreth, C.; MacInnis; Neumann, A.; Normandin, E.; P.C.; Park; Pierce, V.; Reilly, S.; Rosenberg, E.; Rudy, M.; Ryan, E.; S.B.; Sabeti; Shaw, B.; Siddle; Slater, D.; Smole, S.; Tomkins-Tinch, C.; Turbett, S. |
| EPI_ISL_2483916, EPI_ISL_2483917 | C.H.Bois de l'Abbaye | GIGA Medical Genomics | Bouchra Boujemla; Cécile Meex; Keith Durkin; Maria Artesi; Marie-Pierre Hayette; Nathalie Renotte; Pierrette Melin; Raphaël Boreux; Sébastien Bontems; Vincent Bours |
| EPI_ISL_2709557, EPI_ISL_2787323 | CA-Los Angeles County Public Health Laboratory | Centers for Disease Control and Prevention Division of Viral Diseases, Pathogen Discovery | Alison Laufer Halpin; Ben L. Rambo-Martin; Clinton R. Paden; Dakota Howard; Darlene Wagner; Dave Wentworth; Dhwani Batra; Jasmine Padilla; Justin Lee; Katie Dillon; Krista Queen; Kristen Knipe; Kristine Lacek; Mark Burroughs; Matthew Schmerer; Mili Sheth; Peter Cook; Sam Shepard; Sarah Nobles; Shoshona Le; Suxiang Tong; Vivien Dugan; Yvette Unoarumhi |
| EPI_ISL_5650199 | CADIP VILA REGINA FERNANDOPOLIS | Instituto Butantan | Antonio Jorge Martins; Claudia Renata dos Santos Barros; David Schlesinger; Debora Botequio Moretti; Dimas Tadeu Covas; Elaine Cristina Marqueze; Elaine Vieira Santos; Evandra Strazza Rodrigues; Heidge Fukumasu; Jayme Augusto de Souza-Neto; José Salvatore Leister Patané; Luiz Alcantara; Luiz Lehmann Coutinho; Maria Carolina Elias; Maurício Lacerda Nogueira; Rafael dos Santos Bezerra; Raul Machado Neto; Rejane Maria Tommasini Grotto; Ricardo Haddad; Sandra Coccuzzo Sampaio Vessoni; Simone Kashima; Svetoslav Nanev Slavov; Vincent Louis Viala |
| EPI_ISL_2891968, EPI_ISL_2891977, EPI_ISL_2892078, EPI_ISL_2892202, EPI_ISL_2892208, EPI_ISL_2892267, EPI_ISL_2892277, EPI_ISL_2892318, EPI_ISL_2892353, EPI_ISL_2892354, EPI_ISL_2892355, EPI_ISL_2892356, EPI_ISL_2922119, EPI_ISL_3244455, EPI_ISL_3244457, EPI_ISL_3244465, EPI_ISL_3244470, EPI_ISL_3244476, EPI_ISL_3244477, EPI_ISL_3244482, EPI_ISL_3244489, EPI_ISL_3671255, EPI_ISL_3671256, EPI_ISL_3671257, EPI_ISL_3671258, EPI_ISL_3671259, EPI_ISL_3671260, EPI_ISL_3671261, EPI_ISL_3671262, EPI_ISL_3671263, EPI_ISL_3671265 | | | |
| see above | CDPH VBL | California Department of Public Health | CDPH-COVIDNet; Emily Smith on behalf of CDPH-COVIDNet and UCSD EXCITE lab |
| EPI_ISL_2471227, EPI_ISL_2471347 | CENTOGENE Frankfurt Laboratory: Niederlassung Industriepark Höchst | Robert Koch Institute | |
| EPI_ISL_2801364, EPI_ISL_3102495, EPI_ISL_3102496, EPI_ISL_3102497, EPI_ISL_3102498, EPI_ISL_3102499, EPI_ISL_3102501, EPI_ISL_3102502, EPI_ISL_3102503, EPI_ISL_3102504, EPI_ISL_3102523, EPI_ISL_3102524, EPI_ISL_3102525 | | | |
| see above | CENTRO DE ATENDIMENTO PARA ENFRENTAMENTO AO COVID 19 | Analytical Competence Molecular Epidemiology Lab/ACME, Oswaldo Cruz Foundation, Ceara (FIOCRUZ CE) | Cleber Furtado Aksenen; Cleber Furtado Aksenen e Suzana Porto Almeida; Fabio Miyajima; Fernando Braga Stehling; Francisco Eder de Moura Lopes; Jamille Maria Mendes Bezerra; Joaquim César do Nascimento Sousa Junior; Pedro Miguel Carneiro Jeronimo; Suzana Porto Almeida e Lucas Delerino; Thais Ferreira de Oliveira; Thais de Oliveira Costa; Ticiane Cavalcante de Souza; Veridiana Pessoa Miyajima |
| EPI_ISL_5649678, EPI_ISL_5649679, EPI_ISL_5650107, EPI_ISL_5650174 | CENTRO DE ESPECIALIDADES MARIAS DA GLORIA | Instituto Butantan | Antonio Jorge Martins; Claudia Renata dos Santos Barros; David Schlesinger; Debora Botequio Moretti; Dimas Tadeu Covas; Elaine Cristina Marqueze; Elaine Vieira Santos; Evandra Strazza Rodrigues; Heidge Fukumasu; Jayme Augusto de Souza-Neto; José Salvatore Leister Patané; Luiz Alcantara; Luiz Lehmann Coutinho; Maria Carolina Elias; Maurício Lacerda Nogueira; Rafael dos Santos Bezerra; Raul Machado Neto; Rejane Maria Tommasini Grotto; Ricardo Haddad; Sandra Coccuzzo Sampaio Vessoni; Simone Kashima; Svetoslav Nanev Slavov; Vincent Louis Viala |
| EPI_ISL_2493280, EPI_ISL_2493789, EPI_ISL_2493790, | CENTRO DE GENOMAS | Instituto Butantan | Antonio Jorge Martins; Claudia Renata dos Santos Barros; David Schlesinger; Debora Botequio Moretti; Dimas Tadeu Covas; Elaine Cristina Marqueze; Elaine Vieira Santos; Evandra Strazza Rodrigues; Heidge Fukumasu; Jayme Augusto de Souza-Neto; José Salvatore Leister Patané; Luiz Alcantara; Luiz Lehmann Coutinho; Maria Carolina Elias; Maurício Lacerda Nogueira; Rafael dos Santos Bezerra; Raul Machado Neto; Rejane Maria Tommasini Grotto; Ricardo Haddad; Sandra Coccuzzo Sampaio Vessoni; Simone Kashima; Svetoslav Nanev Slavov; Vincent Louis Viala |

| | | | |
|---|---|---|---|
| EPI_ISL_2493791 | | | |
| EPI_ISL_2493153, EPI_ISL_2493824, EPI_ISL_2493826, EPI_ISL_2494127, EPI_ISL_2494128 | CENTRO DE REFERENCIA DO IDOSO DR HUMBERTO MENDES DE CARVALHO | Instituto Butantan | Antonio Jorge Martins; Claudia Renata dos Santos Barros; David Schlesinger; Debora Botequio Moretti; Dimas Tadeu Covas; Elaine Cristina Marqueze; Elaine Vieira Santos; Evandra Strazza Rodrigues; Heidge Fukumasu; Jayme Augusto de Souza-Neto; José Salvatore Leister Patané; Luiz Alcantara; Luiz Lehmann Coutinho; Maria Carolina Elias; Maurício Lacerda Nogueira; Rafael dos Santos Bezerra; Raul Machado Neto; Rejane Maria Tommasini Grotto; Ricardo Haddad; Sandra Coccuzzo Sampaio Vessoni; Simone Kashima; Svetoslav Nanev Slavov; Vincent Louis Viala |
| EPI_ISL_2493917, EPI_ISL_2494309 | CENTRO DE REFERENCIA E TREINAMENTO DSTAIDS SAO PAULO | Instituto Butantan | Antonio Jorge Martins; Claudia Renata dos Santos Barros; David Schlesinger; Debora Botequio Moretti; Dimas Tadeu Covas; Elaine Cristina Marqueze; Elaine Vieira Santos; Evandra Strazza Rodrigues; Heidge Fukumasu; Jayme Augusto de Souza-Neto; José Salvatore Leister Patané; Luiz Alcantara; Luiz Lehmann Coutinho; Maria Carolina Elias; Maurício Lacerda Nogueira; Rafael dos Santos Bezerra; Raul Machado Neto; Rejane Maria Tommasini Grotto; Ricardo Haddad; Sandra Coccuzzo Sampaio Vessoni; Simone Kashima; Svetoslav Nanev Slavov; Vincent Louis Viala |
| EPI_ISL_2494179 | CENTRO DE SAUDE DE ANALANDIA | Instituto Butantan | Antonio Jorge Martins; Claudia Renata dos Santos Barros; David Schlesinger; Debora Botequio Moretti; Dimas Tadeu Covas; Elaine Cristina Marqueze; Elaine Vieira Santos; Evandra Strazza Rodrigues; Heidge Fukumasu; Jayme Augusto de Souza-Neto; José Salvatore Leister Patané; Luiz Alcantara; Luiz Lehmann Coutinho; Maria Carolina Elias; Maurício Lacerda Nogueira; Rafael dos Santos Bezerra; Raul Machado Neto; Rejane Maria Tommasini Grotto; Ricardo Haddad; Sandra Coccuzzo Sampaio Vessoni; Simone Kashima; Svetoslav Nanev Slavov; Vincent Louis Viala |
| EPI_ISL_5649719, EPI_ISL_5649803, EPI_ISL_5649947 | CENTRO DE SAUDE DE LUCELIA | Instituto Butantan | Antonio Jorge Martins; Claudia Renata dos Santos Barros; David Schlesinger; Debora Botequio Moretti; Dimas Tadeu Covas; Elaine Cristina Marqueze; Elaine Vieira Santos; Evandra Strazza Rodrigues; Heidge Fukumasu; Jayme Augusto de Souza-Neto; José Salvatore Leister Patané; Luiz Alcantara; Luiz Lehmann Coutinho; Maria Carolina Elias; Maurício Lacerda Nogueira; Rafael dos Santos Bezerra; Raul Machado Neto; Rejane Maria Tommasini Grotto; Ricardo Haddad; Sandra Coccuzzo Sampaio Vessoni; Simone Kashima; Svetoslav Nanev Slavov; Vincent Louis Viala |
| EPI_ISL_5646046 | CENTRO DE SAUDE DE NANTES | Instituto Butantan | Antonio Jorge Martins; Claudia Renata dos Santos Barros; David Schlesinger; Debora Botequio Moretti; Dimas Tadeu Covas; Elaine Cristina Marqueze; Elaine Vieira Santos; Evandra Strazza Rodrigues; Heidge Fukumasu; Jayme Augusto de Souza-Neto; José Salvatore Leister Patané; Luiz Alcantara; Luiz Lehmann Coutinho; Maria Carolina Elias; Maurício Lacerda Nogueira; Rafael dos Santos Bezerra; Raul Machado Neto; Rejane Maria Tommasini Grotto; Ricardo Haddad; Sandra Coccuzzo Sampaio Vessoni; Simone Kashima; Svetoslav Nanev Slavov; Vincent Louis Viala |
| EPI_ISL_6229703, EPI_ISL_6229854 | CENTRO DE SAUDE DE PENTECOSTE | ACME Lab, Oswaldo Cruz Foundation, FIOCRUZ/CE | Carlos Leonardo de Aragao Araujo; Cecilia Leite Costa & Eduardo Ruback dos Santos on behalf of COVID-19 FIOCRUZ Genomic Network; Cleber Furtado Aksenen; Fabio Miyajima; Fernando Braga Stehling; Francisco Eder de Moura Lopes; Igor Oliveira Duarte; Jamille Maria Mendes Bezerra; Joaquim Cesar do Nascimento Sousa Junior; Pedro Miguel Carneiro Jeronimo; Suzana Porto Almeida; Thais Ferreira de Oliveira; Thais de Oliveira Costa; Ticiane Cavalcante de Souza; Veridiana Pessoa Miyajima |
| EPI_ISL_2462314, EPI_ISL_2493792, EPI_ISL_2493793, EPI_ISL_2493794, EPI_ISL_2493795, EPI_ISL_2493796, EPI_ISL_2493797 | | | |
| see above | CENTRO DE SAUDE DE PIRAJUI | Instituto Butantan | Antonio Jorge Martins; Claudia Renata dos Santos Barros; David Schlesinger; Debora Botequio Moretti; Dimas Tadeu Covas; Elaine Cristina Marqueze; Elaine Vieira Santos; Evandra Strazza Rodrigues; Heidge Fukumasu; Jayme Augusto de Souza-Neto; José Salvatore Leister Patané; Luiz Alcantara; Luiz Lehmann Coutinho; Maria Carolina Elias; Maurício Lacerda Nogueira; Rafael dos Santos Bezerra; Raul Machado Neto; Rejane Maria Tommasini Grotto; Ricardo Haddad; Sandra Coccuzzo Sampaio Vessoni; Simone Kashima; Svetoslav Nanev Slavov; Vincent Louis Viala |
| EPI_ISL_5649630, EPI_ISL_5649643 | CENTRO DE SAUDE DE SPEDRO DO TURVO | Instituto Butantan | Antonio Jorge Martins; Claudia Renata dos Santos Barros; David Schlesinger; Debora Botequio Moretti; Dimas Tadeu Covas; Elaine Cristina Marqueze; Elaine Vieira Santos; Evandra Strazza Rodrigues; Heidge Fukumasu; Jayme Augusto de Souza-Neto; José Salvatore Leister Patané; Luiz Alcantara; Luiz Lehmann Coutinho; Maria Carolina Elias; Maurício Lacerda Nogueira; Rafael dos Santos Bezerra; Raul Machado Neto; Rejane Maria Tommasini Grotto; Ricardo Haddad; Sandra Coccuzzo Sampaio Vessoni; Simone Kashima; Svetoslav Nanev Slavov; Vincent Louis Viala |
| EPI_ISL_5646054, EPI_ISL_5646057, EPI_ISL_5646059, EPI_ISL_5646062, EPI_ISL_5646064, EPI_ISL_5646066, EPI_ISL_5646068, EPI_ISL_5646072, EPI_ISL_5646074, EPI_ISL_5646079, EPI_ISL_5646080, EPI_ISL_5646081, EPI_ISL_5646084, EPI_ISL_5646086, EPI_ISL_5646093, EPI_ISL_5646098, EPI_ISL_5646106 | | | |
| see above | CENTRO DE SAUDE II BARRA BONITA | Instituto Butantan | Antonio Jorge Martins; Claudia Renata dos Santos Barros; David Schlesinger; Debora Botequio Moretti; Dimas Tadeu Covas; Elaine Cristina Marqueze; Elaine Vieira Santos; Evandra Strazza Rodrigues; Heidge Fukumasu; Jayme Augusto de Souza-Neto; José Salvatore Leister Patané; Luiz Alcantara; Luiz Lehmann Coutinho; Maria Carolina Elias; Maurício Lacerda Nogueira; Rafael dos Santos Bezerra; Raul Machado Neto; Rejane Maria Tommasini Grotto; Ricardo Haddad; Sandra Coccuzzo Sampaio Vessoni; Simone Kashima; Svetoslav Nanev Slavov; Vincent Louis Viala |
| EPI_ISL_2494195 | CENTRO DE SAUDE II DR JOSE DE FELIPE E S PINHAL SP | Instituto Butantan | Antonio Jorge Martins; Claudia Renata dos Santos Barros; David Schlesinger; Debora Botequio Moretti; Dimas Tadeu Covas; Elaine Cristina Marqueze; Elaine Vieira Santos; Evandra Strazza Rodrigues; Heidge Fukumasu; Jayme Augusto de Souza-Neto; José Salvatore Leister Patané; Luiz Alcantara; Luiz Lehmann Coutinho; Maria Carolina Elias; Maurício Lacerda Nogueira; Rafael dos Santos Bezerra; Raul Machado Neto; Rejane Maria Tommasini Grotto; Ricardo Haddad; Sandra Coccuzzo Sampaio Vessoni; Simone Kashima; Svetoslav Nanev Slavov; Vincent Louis Viala |
| EPI_ISL_2494192 | CENTRO DE SAUDE II DR JOSE PAIONE MOCOCA | Instituto Butantan | Antonio Jorge Martins; Claudia Renata dos Santos Barros; David Schlesinger; Debora Botequio Moretti; Dimas Tadeu Covas; Elaine Cristina Marqueze; Elaine Vieira Santos; Evandra Strazza Rodrigues; Heidge Fukumasu; Jayme Augusto de Souza-Neto; José Salvatore Leister Patané; Luiz Alcantara; Luiz Lehmann Coutinho; Maria Carolina Elias; Maurício Lacerda Nogueira; Rafael dos Santos Bezerra; Raul Machado Neto; Rejane Maria Tommasini Grotto; Ricardo Haddad; Sandra Coccuzzo Sampaio Vessoni; Simone Kashima; Svetoslav Nanev Slavov; Vincent Louis Viala |
| EPI_ISL_2494165, EPI_ISL_2494186 | CENTRO DE SAUDE III DE DIVINOLANDIA | Instituto Butantan | Antonio Jorge Martins; Claudia Renata dos Santos Barros; David Schlesinger; Debora Botequio Moretti; Dimas Tadeu Covas; Elaine Cristina Marqueze; Elaine Vieira Santos; Evandra Strazza Rodrigues; Heidge Fukumasu; Jayme Augusto de Souza-Neto; José Salvatore Leister Patané; Luiz Alcantara; Luiz Lehmann Coutinho; Maria Carolina Elias; Maurício Lacerda Nogueira; Rafael dos Santos Bezerra; Raul Machado Neto; Rejane Maria Tommasini Grotto; Ricardo Haddad; Sandra Coccuzzo Sampaio Vessoni; Simone Kashima; Svetoslav Nanev Slavov; Vincent Louis Viala |
| EPI_ISL_2494108 | CENTRO DE SAUDE III DOBRADA | Instituto Butantan | Antonio Jorge Martins; Claudia Renata dos Santos Barros; David Schlesinger; Debora Botequio Moretti; Dimas Tadeu Covas; Elaine Cristina Marqueze; Elaine Vieira Santos; Evandra Strazza Rodrigues; Heidge Fukumasu; Jayme Augusto de Souza-Neto; José Salvatore Leister Patané; Luiz Alcantara; Luiz Lehmann Coutinho; Maria Carolina Elias; Maurício Lacerda Nogueira; Rafael dos Santos Bezerra; Raul Machado Neto; Rejane Maria Tommasini Grotto; Ricardo Haddad; Sandra Coccuzzo Sampaio Vessoni; Simone Kashima; Svetoslav Nanev Slavov; Vincent Louis Viala |
| EPI_ISL_5646038, EPI_ISL_5646050, EPI_ISL_5646053, EPI_ISL_5646056, EPI_ISL_5646069, EPI_ISL_5646082 | CENTRO DE SAUDE III FLORENCIO CAMARGO GUIMARAES ITAJU | Instituto Butantan | Antonio Jorge Martins; Claudia Renata dos Santos Barros; David Schlesinger; Debora Botequio Moretti; Dimas Tadeu Covas; Elaine Cristina Marqueze; Elaine Vieira Santos; Evandra Strazza Rodrigues; Heidge Fukumasu; Jayme Augusto de Souza-Neto; José Salvatore Leister Patané; Luiz Alcantara; Luiz Lehmann Coutinho; Maria Carolina Elias; Maurício Lacerda Nogueira; Rafael dos Santos Bezerra; Raul Machado Neto; Rejane Maria Tommasini Grotto; Ricardo Haddad; Sandra Coccuzzo Sampaio Vessoni; Simone Kashima; Svetoslav Nanev Slavov; Vincent Louis Viala |
| EPI_ISL_2493822 | CENTRO DE SAUDE III SANTA LUCIA | Instituto Butantan | Antonio Jorge Martins; Claudia Renata dos Santos Barros; David Schlesinger; Debora Botequio Moretti; Dimas Tadeu Covas; Elaine Cristina Marqueze; Elaine Vieira Santos; Evandra Strazza Rodrigues; Heidge Fukumasu; Jayme Augusto de Souza-Neto; José Salvatore Leister Patané; Luiz Alcantara; Luiz Lehmann Coutinho; Maria Carolina Elias; Maurício Lacerda Nogueira; Rafael dos Santos Bezerra; Raul Machado Neto; Rejane Maria Tommasini Grotto; Ricardo Haddad; Sandra Coccuzzo Sampaio Vessoni; Simone Kashima; Svetoslav Nanev Slavov; Vincent Louis Viala |
| EPI_ISL_2493154, EPI_ISL_2494109, EPI_ISL_2494115, EPI_ISL_2494123 | CENTRO DE SAUDE III TABATINGA | Instituto Butantan | Antonio Jorge Martins; Claudia Renata dos Santos Barros; David Schlesinger; Debora Botequio Moretti; Dimas Tadeu Covas; Elaine Cristina Marqueze; Elaine Vieira Santos; Evandra Strazza Rodrigues; Heidge Fukumasu; Jayme Augusto de Souza-Neto; José Salvatore Leister Patané; Luiz Alcantara; Luiz Lehmann Coutinho; Maria Carolina Elias; Maurício Lacerda Nogueira; Rafael dos Santos Bezerra; Raul Machado Neto; Rejane Maria Tommasini Grotto; Ricardo Haddad; Sandra Coccuzzo Sampaio Vessoni; Simone Kashima; Svetoslav Nanev Slavov; Vincent Louis Viala |
| EPI_ISL_2493950, EPI_ISL_2493951 | CENTRO DE SAUDE SAO ROQUE DR JOSE CARVALHO BRITO | Instituto Butantan | Antonio Jorge Martins; Claudia Renata dos Santos Barros; David Schlesinger; Debora Botequio Moretti; Dimas Tadeu Covas; Elaine Cristina Marqueze; Elaine Vieira Santos; Evandra Strazza Rodrigues; Heidge Fukumasu; Jayme Augusto de Souza-Neto; José Salvatore Leister Patané; Luiz Alcantara; Luiz Lehmann Coutinho; Maria Carolina Elias; Maurício Lacerda Nogueira; Rafael dos Santos Bezerra; Raul Machado Neto; Rejane Maria Tommasini Grotto; Ricardo Haddad; Sandra Coccuzzo Sampaio Vessoni; Simone Kashima; Svetoslav Nanev Slavov; Vincent Louis Viala |
| EPI_ISL_2462325, EPI_ISL_2494130 | CENTRO MUNICIPAL DE SAUDE RUY DE CAMARGO | Instituto Butantan | Antonio Jorge Martins; Claudia Renata dos Santos Barros; David Schlesinger; Debora Botequio Moretti; Dimas Tadeu Covas; Elaine Cristina Marqueze; Elaine Vieira Santos; Evandra Strazza Rodrigues; Heidge Fukumasu; Jayme Augusto de Souza-Neto; José Salvatore Leister Patané; Luiz Alcantara; Luiz Lehmann Coutinho; Maria Carolina Elias; Maurício Lacerda Nogueira; Rafael dos Santos Bezerra; Raul Machado Neto; Rejane Maria Tommasini Grotto; Ricardo Haddad; Sandra Coccuzzo Sampaio Vessoni; Simone Kashima; Svetoslav Nanev Slavov; Vincent Louis Viala |
| EPI_ISL_2494159, EPI_ISL_2494188 | CENTRO SAUDE III DR JOSE TADEU MOURAO | Instituto Butantan | Antonio Jorge Martins; Claudia Renata dos Santos Barros; David Schlesinger; Debora Botequio Moretti; Dimas Tadeu Covas; Elaine Cristina Marqueze; Elaine Vieira Santos; Evandra Strazza Rodrigues; Heidge Fukumasu; Jayme Augusto de Souza-Neto; José Salvatore Leister Patané; Luiz Alcantara; Luiz Lehmann Coutinho; Maria Carolina Elias; Maurício Lacerda Nogueira; Rafael dos Santos Bezerra; Raul Machado Neto; Rejane Maria Tommasini Grotto; Ricardo Haddad; Sandra Coccuzzo Sampaio Vessoni; Simone Kashima; Svetoslav Nanev Slavov; Vincent Louis Viala |
| EPI_ISL_2444390 | CH SAINT-OMER | CHU Lille - Laboratoire de Virologie | AIT YAHYA Emilie; ALIDJINOU Enagnon Kazali; BOCKET Laurence; CREPIN Michel; DEMAY Christophe; ENGELMANN Ilka; GEFFROY Sandrine; GUIGON Aurélie; LAMBERT Valérie; LAZREK Mouna; NOBILLIAUX Florian; PREVOST Brigitte; THUILLIER Caroline; TINEZ Claire |
| EPI_ISL_2637474 | CH.TROYES | Department of Virology, Henri Mondor University Hospital, Assistance Publique Hôpitaux de Paris, Université Paris-Est Créteil, INSERM U955 | Alexandre Soulier; Christophe Rodriguez; Elisabeth Trawinski; Guillaume Gricourt; Jean-Michel Pawlotsky; Melissa N'Debi; Slim Fourati; Vanessa Demontant |
| EPI_ISL_2426967, EPI_ISL_2426971, EPI_ISL_2426982, EPI_ISL_2627848 | CHC Andrée Rosemon | Institut Pasteur de la Guyane | Anne Lavergne; Antoine Enfissi; Arielle Salmier; Dominique Rousset |
| EPI_ISL_2536084, EPI_ISL_2536085 | CHMT | Instituto Nacional de Saude (INSA) | Borges et al |
| EPI_ISL_2493898 | CLINICA SALTO SAUDE BELA VISTA | Instituto Butantan | Antonio Jorge Martins; Claudia Renata dos Santos Barros; David Schlesinger; Debora Botequio Moretti; Dimas Tadeu Covas; Elaine Cristina Marqueze; Elaine Vieira Santos; Evandra Strazza Rodrigues; Heidge Fukumasu; Jayme Augusto de Souza-Neto; José Salvatore Leister Patané; Luiz Alcantara; Luiz Lehmann Coutinho; Maria Carolina Elias; Maurício Lacerda Nogueira; Rafael dos Santos Bezerra; Raul Machado Neto; Rejane Maria Tommasini Grotto; Ricardo Haddad; Sandra Coccuzzo Sampaio Vessoni; Simone Kashima; Svetoslav Nanev Slavov; Vincent Louis Viala |
| EPI_ISL_2426968, EPI_ISL_2426969, EPI_ISL_2426972, EPI_ISL_2426973, EPI_ISL_2426974, EPI_ISL_2426983, EPI_ISL_2426984 | | | |
| see above | CNR Institut Pasteur de la Guyane | Institut Pasteur de la Guyane | Anne Lavergne; Antoine Enfissi; Arielle Salmier; Dominique Rousset |
| EPI_ISL_5649619, EPI_ISL_5650171 | CS DE BALSAMO | Instituto Butantan | Antonio Jorge Martins; Claudia Renata dos Santos Barros; David Schlesinger; Debora Botequio Moretti; Dimas Tadeu Covas; Elaine Cristina Marqueze; Elaine Vieira Santos; Evandra Strazza Rodrigues; Heidge Fukumasu; Jayme Augusto de Souza-Neto; José Salvatore Leister Patané; Luiz Alcantara; Luiz Lehmann Coutinho; Maria Carolina Elias; Maurício Lacerda Nogueira; Rafael dos Santos Bezerra; Raul Machado Neto; Rejane Maria Tommasini Grotto; Ricardo Haddad; Sandra Coccuzzo Sampaio Vessoni; Simone Kashima; Svetoslav Nanev Slavov; Vincent Louis Viala |
| EPI_ISL_5649594, EPI_ISL_5649607, EPI_ISL_5649654, EPI_ISL_5650076, EPI_ISL_5650081, EPI_ISL_5650170 | CS DE CATIGUA | Instituto Butantan | Antonio Jorge Martins; Claudia Renata dos Santos Barros; David Schlesinger; Debora Botequio Moretti; Dimas Tadeu Covas; Elaine Cristina Marqueze; Elaine Vieira Santos; Evandra Strazza Rodrigues; Heidge Fukumasu; Jayme Augusto de Souza-Neto; José Salvatore Leister Patané; Luiz Alcantara; Luiz Lehmann Coutinho; Maria Carolina Elias; Maurício Lacerda Nogueira; Rafael dos Santos Bezerra; Raul Machado Neto; Rejane Maria Tommasini Grotto; Ricardo Haddad; Sandra Coccuzzo Sampaio Vessoni; Simone Kashima; Svetoslav Nanev Slavov; Vincent Louis Viala |
| EPI_ISL_5649576, EPI_ISL_5649585, EPI_ISL_5649601, EPI_ISL_5649626, EPI_ISL_5649653, EPI_ISL_5650067, EPI_ISL_5650112, EPI_ISL_5650141, EPI_ISL_5650175, EPI_ISL_5650180 | | | |
| see above | CS DE MONTE APRAZIVEL | Instituto Butantan | Antonio Jorge Martins; Claudia Renata dos Santos Barros; David Schlesinger; Debora Botequio Moretti; Dimas Tadeu Covas; Elaine Cristina Marqueze; Elaine Vieira Santos; Evandra Strazza Rodrigues; Heidge Fukumasu; Jayme Augusto de Souza-Neto; José Salvatore Leister Patané; Luiz Alcantara; Luiz Lehmann Coutinho; Maria Carolina Elias; Maurício Lacerda Nogueira; Rafael dos Santos Bezerra; Raul Machado Neto; Rejane Maria Tommasini Grotto; Ricardo Haddad; Sandra Coccuzzo Sampaio Vessoni; Simone Kashima; Svetoslav Nanev Slavov; Vincent Louis Viala |
| EPI_ISL_5649579, EPI_ISL_5649586, EPI_ISL_5650200 | CS DE NEVES PAULISTA | Instituto Butantan | Antonio Jorge Martins; Claudia Renata dos Santos Barros; David Schlesinger; Debora Botequio Moretti; Dimas Tadeu Covas; Elaine Cristina Marqueze; Elaine Vieira Santos; Evandra Strazza Rodrigues; Heidge Fukumasu; Jayme Augusto de Souza-Neto; José Salvatore Leister Patané; Luiz Alcantara; Luiz Lehmann Coutinho; Maria Carolina Elias; Maurício Lacerda Nogueira; Rafael dos Santos Bezerra; Raul Machado Neto; Rejane Maria Tommasini Grotto; Ricardo Haddad; Sandra Coccuzzo Sampaio Vessoni; Simone Kashima; Svetoslav Nanev Slavov; Vincent Louis Viala |
| EPI_ISL_5649584, EPI_ISL_5649677, EPI_ISL_5650169, EPI_ISL_5650410 | CS DE NOVAIS | Instituto Butantan | Antonio Jorge Martins; Claudia Renata dos Santos Barros; David Schlesinger; Debora Botequio Moretti; Dimas Tadeu Covas; Elaine Cristina Marqueze; Elaine Vieira Santos; Evandra Strazza Rodrigues; Heidge Fukumasu; Jayme Augusto de Souza-Neto; José Salvatore Leister Patané; Luiz Alcantara; Luiz Lehmann Coutinho; Maria Carolina Elias; Maurício Lacerda Nogueira; Rafael dos Santos Bezerra; Raul Machado Neto; Rejane Maria Tommasini Grotto; Ricardo Haddad; Sandra Coccuzzo Sampaio Vessoni; Simone Kashima; Svetoslav Nanev Slavov; Vincent Louis Viala |
| EPI_ISL_5650166 | CS DE ORINDIUVA | Instituto Butantan | Antonio Jorge Martins; Claudia Renata dos Santos Barros; David Schlesinger; Debora Botequio Moretti; Dimas Tadeu Covas; Elaine Cristina Marqueze; Elaine Vieira Santos; Evandra Strazza Rodrigues; Heidge Fukumasu; Jayme Augusto de Souza-Neto; José Salvatore Leister Patané; Luiz Alcantara; Luiz Lehmann Coutinho; Maria Carolina Elias; Maurício Lacerda Nogueira; Rafael dos Santos Bezerra; Raul Machado Neto; Rejane Maria Tommasini Grotto; Ricardo Haddad; Sandra Coccuzzo Sampaio Vessoni; Simone Kashima; Svetoslav Nanev Slavov; Vincent Louis Viala |
| EPI_ISL_5650085, EPI_ISL_5650182 | CS DE SANTA ALBERTINA | Instituto Butantan | Antonio Jorge Martins; Claudia Renata dos Santos Barros; David Schlesinger; Debora Botequio Moretti; Dimas Tadeu Covas; Elaine Cristina Marqueze; Elaine Vieira Santos; Evandra Strazza Rodrigues; Heidge Fukumasu; Jayme Augusto de Souza-Neto; José Salvatore Leister Patané; Luiz Alcantara; Luiz Lehmann Coutinho; Maria Carolina Elias; Maurício Lacerda Nogueira; Rafael dos Santos Bezerra; Raul Machado Neto; Rejane Maria Tommasini Grotto; Ricardo Haddad; Sandra Coccuzzo Sampaio Vessoni; Simone Kashima; Svetoslav Nanev Slavov; Vincent Louis Viala |
| EPI_ISL_5649609, EPI_ISL_5650074, EPI_ISL_5650120 | CS DE VALENTIM GENTIL | Instituto Butantan | Antonio Jorge Martins; Claudia Renata dos Santos Barros; David Schlesinger; Debora Botequio Moretti; Dimas Tadeu Covas; Elaine Cristina Marqueze; Elaine Vieira Santos; Evandra Strazza Rodrigues; Heidge Fukumasu; Jayme Augusto de Souza-Neto; José Salvatore Leister Patané; Luiz Alcantara; Luiz Lehmann Coutinho; Maria Carolina Elias; Maurício Lacerda Nogueira; Rafael dos Santos Bezerra; Raul Machado Neto; Rejane Maria Tommasini Grotto; Ricardo Haddad; Sandra Coccuzzo Sampaio Vessoni; Simone Kashima; Svetoslav Nanev Slavov; Vincent Louis Viala |
| EPI_ISL_5649716, | CS II DR ANTONIO VICOSO MOREIRA DE | Instituto Butantan | Antonio Jorge Martins; Claudia Renata dos Santos Barros; David Schlesinger; Debora Botequio Moretti; Dimas Tadeu Covas; Elaine Cristina Marqueze; Elaine Vieira Santos; Evandra Strazza Rodrigues; Heidge Fukumasu; Jayme Augusto de Souza-Neto; José Salvatore Leister Patané; Luiz Alcantara; |

| | | | |
|---|---|---|---|
| EPI_ISL_5649918 | REZENDE SUMARE | | Luiz Lehmann Coutinho; Maria Carolina Elias; Maurício Lacerda Nogueira; Rafael dos Santos Bezerra; Raul Machado Neto; Rejane Maria Tommasini Grotto; Ricardo Haddad; Sandra Coccuzzo Sampaio Vessoni; Simone Kashima; Svetoslav Nanev Slavov; Vincent Louis Viala |
| EPI_ISL_5649867, EPI_ISL_5649996 | CS II DR JOSE CARQUEIJO SCRPARDO | Instituto Butantan | Antonio Jorge Martins; Claudia Renata dos Santos Barros; David Schlesinger; Debora Botequio Moretti; Dimas Tadeu Covas; Elaine Cristina Marqueze; Elaine Vieira Santos; Evandra Strazza Rodrigues; Heidge Fukumasu; Jayme Augusto de Souza-Neto; José Salvatore Leister Patané; Luiz Alcantara; Luiz Lehmann Coutinho; Maria Carolina Elias; Maurício Lacerda Nogueira; Rafael dos Santos Bezerra; Raul Machado Neto; Rejane Maria Tommasini Grotto; Ricardo Haddad; Sandra Coccuzzo Sampaio Vessoni; Simone Kashima; Svetoslav Nanev Slavov; Vincent Louis Viala |
| EPI_ISL_2494183 | CS III DR ANTONIO ANADAO SSGRAMA | Instituto Butantan | Antonio Jorge Martins; Claudia Renata dos Santos Barros; David Schlesinger; Debora Botequio Moretti; Dimas Tadeu Covas; Elaine Cristina Marqueze; Elaine Vieira Santos; Evandra Strazza Rodrigues; Heidge Fukumasu; Jayme Augusto de Souza-Neto; José Salvatore Leister Patané; Luiz Alcantara; Luiz Lehmann Coutinho; Maria Carolina Elias; Maurício Lacerda Nogueira; Rafael dos Santos Bezerra; Raul Machado Neto; Rejane Maria Tommasini Grotto; Ricardo Haddad; Sandra Coccuzzo Sampaio Vessoni; Simone Kashima; Svetoslav Nanev Slavov; Vincent Louis Viala |
| EPI_ISL_2493829, EPI_ISL_2493830, EPI_ISL_2494321 | CSII DR WASHINGTON LUIS M RODRIGUES DA SILVA PITANGUEIRAS | Instituto Butantan | Antonio Jorge Martins; Claudia Renata dos Santos Barros; David Schlesinger; Debora Botequio Moretti; Dimas Tadeu Covas; Elaine Cristina Marqueze; Elaine Vieira Santos; Evandra Strazza Rodrigues; Heidge Fukumasu; Jayme Augusto de Souza-Neto; José Salvatore Leister Patané; Luiz Alcantara; Luiz Lehmann Coutinho; Maria Carolina Elias; Maurício Lacerda Nogueira; Rafael dos Santos Bezerra; Raul Machado Neto; Rejane Maria Tommasini Grotto; Ricardo Haddad; Sandra Coccuzzo Sampaio Vessoni; Simone Kashima; Svetoslav Nanev Slavov; Vincent Louis Viala |
| EPI_ISL_2626592, EPI_ISL_3023773 | CT Department of Public Health | CT Department of Public Health | Claire_Pearson; Tu_N_Nguyen |
| EPI_ISL_2892470, EPI_ISL_2892516, EPI_ISL_2892517, EPI_ISL_2892533, EPI_ISL_3536905 | California Department of Public Health | California Department of Public Health | CDPH IDLB COVIDNet et al |
| EPI_ISL_3134059, EPI_ISL_3134061, EPI_ISL_3134074, EPI_ISL_3134084, EPI_ISL_3134090, EPI_ISL_3134097, EPI_ISL_3134102, EPI_ISL_3134105, EPI_ISL_3134106, EPI_ISL_3134107, EPI_ISL_3134109, EPI_ISL_3134116, EPI_ISL_3134118, EPI_ISL_3134119, EPI_ISL_3134121, EPI_ISL_3134123, EPI_ISL_3134124, EPI_ISL_3134125, EPI_ISL_3134128, EPI_ISL_3134129, EPI_ISL_3134131, EPI_ISL_3134136, EPI_ISL_3134140, EPI_ISL_3134145, EPI_ISL_3134147, EPI_ISL_3134151, EPI_ISL_3134154, EPI_ISL_3134155, EPI_ISL_3134158, EPI_ISL_3134160, EPI_ISL_3134162 | | | |
| see above | California Department of Public Health Valencia Branch Laboratory | California Department of Public Health | CDPH-COVIDNet; UCSF Center for Advanced Technology |
| EPI_ISL_2774013 | Cantacuzino National Military-Medical Institute, Viral Respiratory Infections Laboratory | Cantacuzino Institute Virology | Luiza Ustea; Mihaela Lazar; Mihaela Oprea; Nicoleta Paraschiv; Sorin Dinu |
| EPI_ISL_7507361 | Cedars-Sinai Medical Center, Molecular Pathology Laboratory of Department of Pathology & Laboratory Medicine and Genomic Core | Cedars-Sinai Medical Center, Molecular Pathology Laboratory of Department of Pathology & Laboratory Medicine and Genomic Core | Brian Davis; Eric Vail; Jasmine T Plummer; Jorge Mario Sincuir Martinez; Stephanie Chen; Wenjuan Zhang |
| EPI_ISL_2450018 | Center for Laboratory Medicine | Center for Laboratory Medicine | Yannick Gerth |
| EPI_ISL_2502507, EPI_ISL_3462530, EPI_ISL_3462541, EPI_ISL_3462556, EPI_ISL_3462557, EPI_ISL_3462574, EPI_ISL_3462590, EPI_ISL_3462606, EPI_ISL_3462630, EPI_ISL_3462631, EPI_ISL_3462632, EPI_ISL_3462633, EPI_ISL_3462634, EPI_ISL_3462635, EPI_ISL_3462636, EPI_ISL_3462637, EPI_ISL_3462638, EPI_ISL_3462639, EPI_ISL_3462640, EPI_ISL_3462641, EPI_ISL_3462642, EPI_ISL_3462643, EPI_ISL_3462644, EPI_ISL_3462645, EPI_ISL_3462727, EPI_ISL_3462728, EPI_ISL_3464736 | | | |
| see above | Central Laboratory, Bureau of Public Health (BOG) and Academic Hospital Paramaribo | Erasmus Medical Center | Bas B Oude Munnink; Cherise Beek; Consuella Partowidjojo; Dion Gajadin; Ed PF IJzerman; Emmanuelle Munger; Gary Gummels; Ingrid SK Krishnadath; Lycke Woittiez; Marion PG Koopmans; Mireille Van de Veer; Phyllis Pinas; Princes Wongsowidjojo; Radjesh Ori; Ranisha Doerbalie; Rohma Banwari; Soeradj Harkisoen; Stephen Vreden; Tilotmadebie Ramlal; Verne Nanhoe |
| EPI_ISL_3255137, EPI_ISL_3255155, EPI_ISL_3255163, EPI_ISL_3255171, EPI_ISL_3255174, EPI_ISL_3255176, EPI_ISL_3255182, EPI_ISL_3255190, EPI_ISL_3255196, EPI_ISL_3255213, EPI_ISL_3255281 | | | |
| see above | Central Public Health Laboratory - LACEN - Bahia, Salvador, Brazil | Central Public Health Laboratory - LACEN -Bahia, Salvador, Brazil | Arabela Leal; Breno Dominguez; Felicidade Pereira; Jaqueline Gomes; Luciana Oliveira; Luiz Alcantara; Marcela Gómez; Marta Giovanetti; Patrícia Cajado; Stephane Tosta; Vagner Fonseca; Vanessa Nardy |
| EPI_ISL_5926866, EPI_ISL_5926883, EPI_ISL_5926884, EPI_ISL_5926898 | Centro de Infectologia Charles Mérieux/ Laboratório Rodolphe Mérieux, FUNDHACRE | Bioinformatics Laboratory / LNCC | Alessandra P Lamarca; Alexandra L Gerber; Aline de Freitas Souza; Ana Paula de C Guimarães; Ana Tereza R Vasconcelos; Andreas Stocker; Cirley Maria de Oliveira Lobato; Douglas Terra Machado; Janaína Mazaro; Janete Taynã Nascimento Rodrigues; Luiz Fellype Alves de Souza; Luiz G P de Almeida; Ronaldo da Silva F Jr; Rutilene B. Souza |
| EPI_ISL_2671568, EPI_ISL_2671575, EPI_ISL_2671576, EPI_ISL_2671582, EPI_ISL_2671585, EPI_ISL_2671586, EPI_ISL_2671703 | | | |
| see above | Centro de Investigacion Biomedica de Occidente (CIBO) | Unidad de Genomica Avanzada | ; Alejandra Garcia-Gasca; Alejandra Hernandez-Teran; Alejandro Sanchez-Flores; Alfredo Herrera-Estrella; Alicia Ocaña-Mondragon; Andreu Comas-Garcia; Angel Gustavo Salas-Lais; Antonio Loza Roman; Bernardo Martinez-Miguel; Blanca Taboada; Brenda Irasema Maldonado-Meza; Bruno Gomez-Gil; Carla Ivon Herrera-Najera; Carlos F. Arias; Celia Boukadida; Celida Duque Molina; Celida Martinez- Rodriguez; Clara Esperanza Santacruz-Tinoco; Concepcion Grajales-Muñiz; Consorcio Mexicano de Vigilancia Genomica (CoViGen-Mex). Authors (in alphabetical order): Julio Elias Alvarado-Yaah; Cristobal Chaidez-Quiroz; Daniel Fregoso-Rueda; Daniel Lira Morales; Eduardo Becerril-Vargas; Fernando Fontove-Herrera; Fidencio Mejia-Nepomuceno; Francisco Pulido; Gloria Elena Espinosa-Ayala; Gloria Maria Molina-Salinas; Gloria Vazquez; Hector Esteban Paz-Juarez; Hector Montoya-Fuentes; Helen Haydee Fernanda Ramirez-Plascencia; Irvin Gonzalez-Lopez; Jean Pierre Gonzalez; Jesus Hernandez; Joel Armando Vazquez-Perez.; Jorge Salas-Hernandez; Jose Antonio Enciso-Moreno; Jose Arturo Martinez-Orozco; Jose Esteban Muñoz-Medina; Jose de Jesus Nuñez-Contreras; Juan Bautista Chale-Dzul; Julissa Enciso-Ibarra; Luis Alberto Ochoa-Carrera; Margarita Matias-Florentino; Maria Guadalupe Santiago-Mauricio; Maria Guadalupe de Jesus Mireles-Rivera; Mario Mujica-Sanchez; Marissa Perez-Garcia; Nelly Selem-Mojica; Pavel Isa; Ricardo Ciria Merce; Ricardo Grande; Rosa Maria Gutierrez Rios; Santiago avila-Rios; Selene Zarate; Susana Lopez; Veronica Mata-Haro; Victor Eduardo Garcia-Arias; Victor Hugo Borja-Aburto |
| EPI_ISL_2671560 | Centro de Investigacion Biomedica del Noreste (CIBIN) | Unidad de Genomica Avanzada | ; Alejandra Garcia-Gasca; Alejandra Hernandez-Teran; Alejandro Sanchez-Flores; Alfredo Herrera-Estrella; Alicia Ocaña-Mondragon; Andreu Comas-Garcia; Angel Gustavo Salas-Lais; Antonio Loza Roman; Bernardo Martinez-Miguel; Blanca Taboada; Brenda Irasema Maldonado-Meza; Bruno Gomez-Gil; Carla Ivon Herrera-Najera; Carlos F. Arias; Celia Boukadida; Celida Duque Molina; Celida Martinez- Rodriguez; Clara Esperanza Santacruz-Tinoco; Concepcion Grajales-Muñiz; Consorcio Mexicano de Vigilancia Genomica (CoViGen-Mex). Authors (in alphabetical order): Julio Elias Alvarado-Yaah; Cristobal Chaidez-Quiroz; Daniel Fregoso-Rueda; Daniel Lira Morales; Eduardo Becerril-Vargas; Fernando Fontove-Herrera; Fidencio Mejia-Nepomuceno; Francisco Pulido; Gloria Elena Espinosa-Ayala; Gloria Maria Molina-Salinas; Gloria Vazquez; Hector Esteban Paz-Juarez; Hector Montoya-Fuentes; Helen Haydee Fernanda Ramirez-Plascencia; Irvin Gonzalez-Lopez; Jean Pierre Gonzalez; Jesus Hernandez; Joel Armando Vazquez-Perez.; Jorge Salas-Hernandez; Jose Antonio Enciso-Moreno; Jose Arturo Martinez-Orozco; Jose Esteban Muñoz-Medina; Jose de Jesus Nuñez-Contreras; Juan Bautista Chale-Dzul; Julissa Enciso-Ibarra; Luis Alberto Ochoa-Carrera; Margarita Matias-Florentino; Maria Guadalupe Santiago-Mauricio; Maria Guadalupe de Jesus Mireles-Rivera; Mario Mujica-Sanchez; Marissa Perez-Garcia; Nelly Selem-Mojica; Pavel Isa; Ricardo Ciria Merce; Ricardo Grande; Rosa Maria Gutierrez Rios; Santiago avila-Rios; Selene Zarate; Susana Lopez; Veronica Mata-Haro; Victor Eduardo Garcia-Arias; Victor Hugo Borja-Aburto |
| EPI_ISL_4405201, EPI_ISL_4405419, EPI_ISL_4405462 | Centro de Investigaciones Básicas y Aplicadas, UNNOBA | Área de Secuenciación del Laboratorio de Virología del Hospital de Niños Dr. Ricardo Gutierrez on behalf of 'Proyecto Argentino Interinstitucional de genomica de SARS-CoV-2' (PAIS Consortium) | A; Acuña; Alaniz; Bagnis; Barbero; Bonadeo; Brandone; C; Cassarini; Castro; Chimento; Cristina; D; Demarchi; Español; F; Fernández; G; Goya; Gracia Balbi; Hernández del Pin; I; Icardi; L; LE; Lusso; M; MI; MS; Menite; Moroni; Morro; N; Nabaes Jodar; Natale; P; Palumbo; Pasquinelli; Perrone; R; Romano; Rosales; S; Sevic; Spinelli; V; Valinotto; Valla; Viegas, M.; Villafañe; Vitale |
| EPI_ISL_2801716 | Centro de Investigación Biomédica de Occidente (CIBO) | Instituto de Biotecnología de la UNAM | ; Alejandra García-Gasca; Alejandra Hernández-Terán; Alejandro Sánchez-Flores; Alfredo Herrera-Estrella; Alicia Ocaña-Mondragón; Andreu Comas-García; Angel Gustavo Salas-Lais; Antonio Loza Román; Bernardo Martínez-Miguel; Blanca Taboada; Brenda Irasema Maldonado-Meza; Bruno Gómez-Gil; Carla Ivón Herrera-Najera; Carlos F. Arias; Celia Boukadida; Clara Esperanza Santacruz-Tinoco; Concepción Grajales-Muñiz; Consorcio Mexicano de Vigilancia Genómica (CoViGen-Mex). Authors (in alphabetical order): Julio Elias Alvarado-Yaah; Cristóbal Cháidez-Quiróz; Célida Duque Molina; Célida Martínez- Rodríguez; Daniel Fregoso-Rueda; Daniel Lira Morales; Eduardo Becerril-Vargas; Fernando Fontove-Herrera; Fidencio Mejía-Nepomuceno; Francisco Pulido; Gloria Elena Espinosa-Ayala; Gloria María Molina-Salinas; Gloria Vazquez; Hector Esteban Paz-Juárez; Hector Montoya-Fuentes; Helen Haydee Fernanda Ramírez-Plascencia; Irvin González-López; Jean Pierre González; Jesús Hernández; Joel Armando Vázquez-Pérez.; Jorge Salas-Hernández; José Antonio Enciso-Moreno; José Arturo Martínez-Orozco; José Esteban Muñoz-Medina; José de Jesús Nuñez-Contreras; Juan Bautista Chale-Dzul; Julissa Enciso-Ibarra; Luis Alberto Ochoa-Carrera; Margarita Matías-Florentino; Mario Mújica-Sánchez; Marissa Perez-Garcia; María Guadalupe Santiago-Mauricio; María Guadalupe de Jesús Mireles-Rivera; Nelly Sélem-Mojica; Pavel Isa; Ricardo Ciria Merce; Ricardo Grande; Rosa María Gutiérrez Ríos; Santiago Ávila-Ríos; Selene Zárate; Susana Lopez; Verónica Mata-Haro; Víctor Eduardo García-Arias; Víctor Hugo Borja-Aburto |
| EPI_ISL_4261909, EPI_ISL_4261912, EPI_ISL_4261913, EPI_ISL_5055013 | Chiu Laboratory, University of California, San Francisco | Chiu Laboratory, University of California, San Francisco | Alicia Sotomayor-Gonzalez; Baolin Wang; Candace Wang; Charles Chiu; Daniel Wong; Dustin Glasner; Venice Servellita; Xianding (Wayne) Deng; Yueyuan Zhang |
| EPI_ISL_4635670, EPI_ISL_4635671 | Clinical Microbiology, University of Texas Medical Branch (UTMB), Galveston | ClickSeq Lab, University of Texas Medical Branch, UTMB, Galveston | Dong J.; Jaworski E.; Judy B.; Ksiazek T.; Langsjoen R.M.; Machado R.R; McConnell A.J.; Menachery V.D.; Miller A.L.; Mitchell B.M.; Morris V.G.; Newman P.; Plante J.A.; Plante K.S.; Pyles R.B.; Ren P.; Routh A.; Saada N.I.; Sotcheff S.L.; Swetnam D.; Thompson J.K.; Weaver S.C; Widen S.; Zhou Y. |
| EPI_ISL_3061525 | Clinical Molecular Microbiology Laboratory, UNC Hospitals | Jeremy Wang | Corbin Jones; Jeremy Wang; Melissa Miller; Shawn Hawken |
| EPI_ISL_2610956, EPI_ISL_2610962 | Clinical Virology | Clinical Bacteriology | Adrian Egli; Alfredo Mari; Fanny Wegner; Hans Hirsch; Helena MB Seth-Smith; Julia Bielicki; Karoline Leuzinger; Manuel Battegay; Tim Roloff |
| EPI_ISL_2402436 | Clinique André Renard Herstal | GIGA Medical Genomics | Bouchra Boujemla; Cécile Meex; Keith Durkin; Maria Artesi; Marie-Pierre Hayette; Nathalie Renotte; Pierrette Melin; Raphaël Boreux; Sébastien Bontems; Vincent Bours |
| EPI_ISL_2492365 | Clinique Saint-Pierre Ottignies | UCLouvain/IREC/MBLG | Benoit Kabamba Mukadi; Jean Ruelle |
| EPI_ISL_2427324, EPI_ISL_2427364, EPI_ISL_2492421, EPI_ISL_2492426, EPI_ISL_2778062, EPI_ISL_2778087 | Cliniques universitaires Saint-Luc | UCLouvain/IREC/MBLG | Benoit Kabamba Mukadi; Bertrand Bearzatto; Jean Ruelle; Ophélie Simon |
| EPI_ISL_4300384, EPI_ISL_4300387, EPI_ISL_4300388, EPI_ISL_4300390, EPI_ISL_4300391, EPI_ISL_4300392, EPI_ISL_4300403 | | | |
| see above | Color Genomics | Chiu Laboratory, University of California, San Francisco | Alicia Sotomayor-Gonzalez; Alicia Zhou; Baolin Wang; Charles Chiu; Daniel Wong; Erika Torres; Madyson Wanner; Noah Brazer; Scott Topper; Venice Servellita; Yueyuan Zhang |
| EPI_ISL_2550357, EPI_ISL_2550387, EPI_ISL_2550395, EPI_ISL_2550406, EPI_ISL_2550478, EPI_ISL_2550500, EPI_ISL_2550506, EPI_ISL_2790625, EPI_ISL_2790667, EPI_ISL_2941594, EPI_ISL_2941636, EPI_ISL_3159610, EPI_ISL_3159620, EPI_ISL_3159662, EPI_ISL_3404121, EPI_ISL_3988717 | | | |
| see above | Colorado Department of Public Health and Environment | Colorado Department of Public Health and Environment | Alexandria Rossheim; Diana Ir; Emily A. Travanty; Laura Bankers; Mandy Waters; Michael A. Martin; Michael Martin; Molly C. Hetherington-Rauth; Sarah Elizabeth Totten; Shannon R. Matzinger |
| EPI_ISL_2659116 | County of San Luis Obispo Public Health Laboratory | County of San Luis Obispo Public Health Laboratory | Frances Sidhu; Katrina Erwin |
| EPI_ISL_2507361, EPI_ISL_2755414, EPI_ISL_2755445, EPI_ISL_2755712, EPI_ISL_2755723, EPI_ISL_2755724, EPI_ISL_2755725, EPI_ISL_2755727, EPI_ISL_2755728, EPI_ISL_2756016, EPI_ISL_2756056, EPI_ISL_2784235, EPI_ISL_2941964, EPI_ISL_2942145, EPI_ISL_2942152, EPI_ISL_3270253, EPI_ISL_3270273 | | | |
| see above | Curative Labs | Curative Labs | Elias L. Salfati; Eugenia Khorosheva; George Way; J.Cesar Ignacio-Espinoza; Janet Chen; Mikhail Hanewich-Hollatz; Nabjot Sandhu; Sophia Quasem; Vladimir Slepnev; Zhiyi Xie |
| EPI_ISL_3664275, EPI_ISL_4415776, EPI_ISL_4415777, EPI_ISL_4415843, EPI_ISL_4422787, EPI_ISL_4422793, EPI_ISL_4422796, EPI_ISL_4423004, EPI_ISL_4423007, EPI_ISL_4423009, EPI_ISL_4426837, EPI_ISL_4427864, EPI_ISL_4427866, EPI_ISL_4427868, EPI_ISL_4427869, EPI_ISL_4427871, EPI_ISL_4427873, EPI_ISL_4427875, EPI_ISL_4427876, EPI_ISL_4428164, EPI_ISL_6851409 | | | |
| see above | DASA | DASA | Adriano Bonaldi; Angelica Hristov; Annelise Lopes; Bianca Cota; Camila Romano; Cristina Oliveira; Jose Levi; Lidia Yamamoto; Luciane Sussuchi; Paulo Pierry; Rodrigo Guarischi; Rodrigo Salazar |

| | | | |
|---|---|---|---|
| EPI_ISL_2662745, EPI_ISL_2662750, EPI_ISL_2662763, EPI_ISL_2662807, EPI_ISL_3692608 | DPHL | Delaware Public Health Lab | Rebecca Savage |
| EPI_ISL_2759813 | Department für Labormedizin Abteilung III Bereich Molekulare Diagnostik Universitätsklinikum Halle(Saale) | Robert Koch Institute | |
| EPI_ISL_2429493, EPI_ISL_2431024, EPI_ISL_2432301 | Department of Bacteria, Parasites and Fungi, Statens Serum Institut, Copenhagen, Denmark | Statens Serum Institut Bioinformatics and Microbial Genomics | Danish Covid-19 Genome Consortium |
| EPI_ISL_2636598, EPI_ISL_2636627, EPI_ISL_2636632, EPI_ISL_2636642 | Diagnosticum - Labor Neukirchen | Robert Koch Institute | |
| EPI_ISL_2503963 | Diagnostyka | 1. Academic Center for Pathomorphological and Genetic-Molecular Diagnostics ltd, Bialystok, Poland 2. National Institute of Public Health - National Institute of Hygiene, Warsaw, Poland | Anetta Sulewska; Jacek Nikliński; Janusz Dzięcioł; Joanna Kiśluk; Katarzyna Zacharczuk; Konrad Raczkowski; Magdalena Nowakowska; Małgorzata Sadkowska-Todys; Piotr Karabowicz; Piotr Majewski; Przemysław Biecek. Joanna Reszeć; Radosław Charkiewicz; Tomasz Wołkowicz |
| EPI_ISL_2597808, EPI_ISL_2597811 | Diagnostyka | 1. Tricity SARS-CoV-2 sequencing consortium: University of Gdansk, Medical University of Gdansk, Vaxican Ltd., Invicta Ltd. 2. National Institute of Public Health - National Institute of Hygiene, Warsaw, Poland | Celina Cybulska; Karolina Gackowska; Katarzyna Groth; Katarzyna Zacharczuk; Krystyna Bienkowska Szewczyk; Lukasz Rabalski; Maciej Grzybek; Maciej Kosinski; Magdalena Nowakowska; Marcin Lubocki; Małgorzata Sadkowska-Todys; Tomasz Wołkowicz |
| EPI_ISL_2504119 | Diagnostyka Sp. z o.o. | 1. Academic Center for Pathomorphological and Genetic-Molecular Diagnostics ltd, Bialystok, Poland 2. National Institute of Public Health - National Institute of Hygiene, Warsaw, Poland | Anetta Sulewska; Jacek Nikliński; Janusz Dzięcioł; Joanna Kiśluk; Katarzyna Zacharczuk; Konrad Raczkowski; Magdalena Nowakowska; Małgorzata Sadkowska-Todys; Piotr Karabowicz; Piotr Majewski; Przemysław Biecek. Joanna Reszeć; Radosław Charkiewicz; Tomasz Wołkowicz |
| EPI_ISL_3050651 | Dirección regional de salud del Callao (DIRESA-CALLAO) | Centro de Investigaciones Tecnológicas, Biomédicas y Medioambientales (CITBM) | B; Huaman; J. Alarcon; M. Cuellar; M. Ramirez; M. Sovero |
| EPI_ISL_3474267 | Dutch COVID-19 response team | Medical Microbiology, Maastricht University Medical Centre | Brian van der Veer*; Carmen Reumkens; Christian Hoebe; Erik Beuken; Jozef Dingemans*; Lieke van Alphen; Paul Savelkoul |
| EPI_ISL_2405546, EPI_ISL_2405560, EPI_ISL_2405571, EPI_ISL_2405572, EPI_ISL_2405606, EPI_ISL_2474206, EPI_ISL_2474245, EPI_ISL_2475678, EPI_ISL_2475679, EPI_ISL_2475680, EPI_ISL_2475687, EPI_ISL_2475691, EPI_ISL_2475692, EPI_ISL_2475693, EPI_ISL_2475694, EPI_ISL_2475695, EPI_ISL_2475701, EPI_ISL_2475702, EPI_ISL_2475703, EPI_ISL_2475707, EPI_ISL_2475709, EPI_ISL_2475711, EPI_ISL_2475712, EPI_ISL_2475713, EPI_ISL_2475714, EPI_ISL_2475715, EPI_ISL_2475716, EPI_ISL_2475717, EPI_ISL_2475718, EPI_ISL_2475719, EPI_ISL_2475720, EPI_ISL_2475721, EPI_ISL_2475722, EPI_ISL_2475723, EPI_ISL_2475724, EPI_ISL_2475725, EPI_ISL_2475726, EPI_ISL_2475728, EPI_ISL_2609822, EPI_ISL_2610051, EPI_ISL_2610092, EPI_ISL_2610234, EPI_ISL_2610296, EPI_ISL_2610814, EPI_ISL_2610823, EPI_ISL_2610854, EPI_ISL_2672546, EPI_ISL_2673399 | | | |
| see above | Dutch COVID-19 response team | National Institute for Public Health and the Environment (RIVM) | Adam Meijer; AnneMarie van den Brandt; Annelies Kroneman; Bas van der Veer; Chantal Reusken; Dennis Schmitz; Dirk Eggink; Euníce Then; Florian Zwagemaker; Harry Vennema; James Groot; Jeroen Cremer; Karim Hajji; Kim Freriks; Linda van de Nes; Lisa Wijsman; Lynn Aarts; Melissa van Tuil; Ryanne Jaarsma; Sanne Bos; Sharon van den Brink; Sjoerd Kuiling; on behalf of the national COVID-19 response team |
| EPI_ISL_5646094 | ESTRATEGIA DE SAUDE DA FAMILIA II | Instituto Butantan | Antonio Jorge Martins; Claudia Renata dos Santos Barros; David Schlesinger; Debora Botequio Moretti; Dimas Tadeu Covas; Elaine Cristina Marqueze; Elaine Vieira Santos; Evandra Strazza Rodrigues; Heidge Fukumasu; Jayme Augusto de Souza-Neto; José Salvatore Leister Patané; Luiz Alcantara; Luiz Lehmann Coutinho; Maria Carolina Elias; Maurício Lacerda Nogueira; Rafael dos Santos Bezerra; Raul Machado Neto; Rejane Maria Tommasini Grotto; Ricardo Haddad; Sandra Coccuzzo Sampaio Vessoni; Simone Kashima; Svetoslav Nanev Slavov; Vincent Louis Viala |
| EPI_ISL_2940607, EPI_ISL_2940724, EPI_ISL_2940736, EPI_ISL_3117259, EPI_ISL_3117270, EPI_ISL_3117271, EPI_ISL_3117403, EPI_ISL_3117415 | | | |
| see above | Edmonton Provincial Lab | Public Health Agency of Canada (PHAC) National Microbiology Laboratory | Buss; Croxen M; Deo A; Dieu P; E; Ferrato C; Gill K; Khan F; Koleva P; Li V; Lloyd C; Lynch T; Ma R; Murphy S; Pabbaraju K; Shokoples S; Thayer J; Tipples G; Whitehouse M; Wong A; Yu C; Zelyas N |
| EPI_ISL_2842776 | Emory Molecular Diagnostics Laboratory, Emory Healthcare | Piantadosi Lab, Emory Department of Pathology | Ahmed Babiker; Anne Piantadosi; Ludy Registre Carmola |
| EPI_ISL_2634510, EPI_ISL_2634515, EPI_ISL_2634522, EPI_ISL_2634524, EPI_ISL_2634526, EPI_ISL_2634534, EPI_ISL_2634536, EPI_ISL_2634551, EPI_ISL_2634563, EPI_ISL_2634565, EPI_ISL_2634567, EPI_ISL_2634581, EPI_ISL_2634583, EPI_ISL_2634585, EPI_ISL_2634605, EPI_ISL_2634608, EPI_ISL_2634610, EPI_ISL_2634612, EPI_ISL_2634619, EPI_ISL_2634621, EPI_ISL_2634628, EPI_ISL_2634637, EPI_ISL_2634652, EPI_ISL_2634654, EPI_ISL_2634667, EPI_ISL_2634669, EPI_ISL_2634671, EPI_ISL_2634687 | | | |
| see above | Eurofins LifeCodexx GmbH | Robert Koch Institute | |
| EPI_ISL_2494267, EPI_ISL_2494268, EPI_ISL_2494269, EPI_ISL_2494271, EPI_ISL_2494272, EPI_ISL_2494273, EPI_ISL_2494302, EPI_ISL_5646241 | | | |
| see above | FACULDADE DE MEDICINA ABC | Instituto Butantan | Antonio Jorge Martins; Claudia Renata dos Santos Barros; David Schlesinger; Debora Botequio Moretti; Dimas Tadeu Covas; Elaine Cristina Marqueze; Elaine Vieira Santos; Evandra Strazza Rodrigues; Heidge Fukumasu; Jayme Augusto de Souza-Neto; José Salvatore Leister Patané; Luiz Alcantara; Luiz Lehmann Coutinho; Maria Carolina Elias; Maurício Lacerda Nogueira; Rafael dos Santos Bezerra; Raul Machado Neto; Rejane Maria Tommasini Grotto; Ricardo Haddad; Sandra Coccuzzo Sampaio Vessoni; Simone Kashima; Svetoslav Nanev Slavov; Vincent Louis Viala |
| EPI_ISL_2787209, EPI_ISL_2787215, EPI_ISL_2787217, EPI_ISL_2787223 | FL Bureau of Public Health Laboratories-Tampa | Centers for Disease Control and Prevention Division of Viral Diseases, Pathogen Discovery | Alison Laufer Halpin; Ben L. Rambo-Martin; Clinton R. Paden; Dakota Howard; Darlene Wagner; Dave Wentworth; Dhwani Batra; Jasmine Padilla; Justin Lee; Katie Dillon; Krista Queen; Kristen Knipe; Kristine Lacek; Mark Burroughs; Matthew Schmerer; Mili Sheth; Peter Cook; Sam Shepard; Sarah Nobles; Shoshona Le; Suxiang Tong; Vivien Dugan; Yvette Unoarumhi |
| EPI_ISL_2836332, EPI_ISL_2836414, EPI_ISL_2836422, EPI_ISL_2836488, EPI_ISL_2836489, EPI_ISL_2836505, EPI_ISL_2836511, EPI_ISL_2836512, EPI_ISL_2836515, EPI_ISL_2836579, EPI_ISL_2836583, EPI_ISL_3100709, EPI_ISL_6049965, EPI_ISL_6049967, EPI_ISL_6050043, EPI_ISL_6050057, EPI_ISL_6050058, EPI_ISL_6050059, EPI_ISL_6050060, EPI_ISL_6050061, EPI_ISL_6050062, EPI_ISL_6050067, EPI_ISL_6050695 | | | |
| see above | Florida Bureau of Public Health Laboratories | Florida Bureau of Public Health Laboratories | Jason Blanton; Namratha Tarigopula; Sarah Schmedes; Tiffany Splatt |
| EPI_ISL_2371530, EPI_ISL_2371531, EPI_ISL_2371536, EPI_ISL_2371548, EPI_ISL_2371558, EPI_ISL_2441851, EPI_ISL_2441852, EPI_ISL_2441875, EPI_ISL_2441877, EPI_ISL_2441886, EPI_ISL_2441887, EPI_ISL_2441904, EPI_ISL_2441907, EPI_ISL_2441917, EPI_ISL_2441919, EPI_ISL_2441935, EPI_ISL_2441950, EPI_ISL_2479997, EPI_ISL_2479998, EPI_ISL_2479999, EPI_ISL_2480000, EPI_ISL_2480002, EPI_ISL_2480036, EPI_ISL_2480052, EPI_ISL_2480152, EPI_ISL_2480153, EPI_ISL_2480157, EPI_ISL_2480169, EPI_ISL_2480171, EPI_ISL_2480173, EPI_ISL_2480175, EPI_ISL_2480176, EPI_ISL_2480178, EPI_ISL_2480181, EPI_ISL_2480182, EPI_ISL_2480189, EPI_ISL_2480193, EPI_ISL_2480194, EPI_ISL_2480195, EPI_ISL_2525648, EPI_ISL_2525652, EPI_ISL_2525660, EPI_ISL_2525669, EPI_ISL_2525671, EPI_ISL_2525687, EPI_ISL_2525702, EPI_ISL_2525704, EPI_ISL_2525706, EPI_ISL_2525708, EPI_ISL_2525725, EPI_ISL_2525727, EPI_ISL_2525729, EPI_ISL_2525879, EPI_ISL_2525883, EPI_ISL_2525895, EPI_ISL_2525905, EPI_ISL_2525907, EPI_ISL_2525916, EPI_ISL_2525927, EPI_ISL_2525931, EPI_ISL_2686477, EPI_ISL_2686486, EPI_ISL_4371447 | | | |
| see above | Fulgent Genetics | Centers for Disease Control and Prevention Division of Viral Diseases, Pathogen Discovery | Adrian Paskey; Becky Tsai; Benafsh Sapra; Benjamin Rambo-Martin; Christopher Gulvick; Clinton Paden; Clinton R. Paden; Dakota Howard; Darlene Wagner; Dhwani Batra; Doreen Ng; Duncan MacCannell; Erisa Sula; Harry Gao; James Xie; Jason Caravas; John Gao; Joseph Fierro; Kara Moser; Kristine Lacek; Matthew Schmerer; Mickey Li; Peter Cook; Peter W. Cook; Scott Sammons; Shatavia Morrison; Tymeckia Kendall; Victoria Caban Figueroa; Yan Meng; Yvette Unoarumhi |
| EPI_ISL_2645769 | GA Department of Public Health | GA Department of Public Health | Aliyah Fields; Cynthia Dixey; Jonathan Edwards; Stacy Reeves; Taylor Smith; Tonia Parrott |
| EPI_ISL_2690029, EPI_ISL_2840409 | GA Department of Public Health Laboratory | Centers for Disease Control and Prevention Division of Viral Diseases, Pathogen Discovery | Alison Laufer Halpin; Ben L. Rambo-Martin; Clinton R. Paden; Dakota Howard; Darlene Wagner; Dave Wentworth; Dhwani Batra; Jasmine Padilla; Justin Lee; Katie Dillon; Krista Queen; Kristen Knipe; Kristine Lacek; Mark Burroughs; Matthew Schmerer; Mili Sheth; Peter Cook; Sam Shepard; Sarah Nobles; Shoshona Le; Suxiang Tong; Vivien Dugan; Yvette Unoarumhi |
| EPI_ISL_2450171 | GHDC - SITE ST JOSEPH | Institut de Pathologie et Genetique (IPG) | Jérémie Gras; Pascale Hilbert |
| EPI_ISL_2493943 | GRUPO TECNICO DE VIGILANCIA SANITARIA E EPIDEMIOLOGICA | Instituto Butantan | Antonio Jorge Martins; Claudia Renata dos Santos Barros; David Schlesinger; Debora Botequio Moretti; Dimas Tadeu Covas; Elaine Cristina Marqueze; Elaine Vieira Santos; Evandra Strazza Rodrigues; Heidge Fukumasu; Jayme Augusto de Souza-Neto; José Salvatore Leister Patané; Luiz Alcantara; Luiz Lehmann Coutinho; Maria Carolina Elias; Maurício Lacerda Nogueira; Rafael dos Santos Bezerra; Raul Machado Neto; Rejane Maria Tommasini Grotto; Ricardo Haddad; Sandra Coccuzzo Sampaio Vessoni; Simone Kashima; Svetoslav Nanev Slavov; Vincent Louis Viala |
| EPI_ISL_2543522, EPI_ISL_2543523, EPI_ISL_2543524, EPI_ISL_2543529, EPI_ISL_2543552, EPI_ISL_2543580, EPI_ISL_2543581, EPI_ISL_2543582, EPI_ISL_2543616, EPI_ISL_2543623, EPI_ISL_2543624, EPI_ISL_2543625, EPI_ISL_2543626, EPI_ISL_2543627, EPI_ISL_2543628, EPI_ISL_2543630, EPI_ISL_2543631, EPI_ISL_2543632, EPI_ISL_2543633, EPI_ISL_2543634, EPI_ISL_2543635, EPI_ISL_2543638, EPI_ISL_2543639, EPI_ISL_2543640, EPI_ISL_2543641, EPI_ISL_2543642, EPI_ISL_2543644, EPI_ISL_2543645, EPI_ISL_2543646, EPI_ISL_2543647, EPI_ISL_2543648, EPI_ISL_2543649, EPI_ISL_2543651, EPI_ISL_2543653, EPI_ISL_2543655, EPI_ISL_2543657, EPI_ISL_2543661, EPI_ISL_2543662, EPI_ISL_2543664, EPI_ISL_2543665, EPI_ISL_2557252, EPI_ISL_2557254, EPI_ISL_2557255, EPI_ISL_2557256, EPI_ISL_2557257, EPI_ISL_2557258, EPI_ISL_2557259, EPI_ISL_2557260, EPI_ISL_2557261, EPI_ISL_2557262, EPI_ISL_2557269, EPI_ISL_2557274, EPI_ISL_2557276, EPI_ISL_2557279, EPI_ISL_2557280, EPI_ISL_2557282, EPI_ISL_2557283, EPI_ISL_2557284, EPI_ISL_2557285, EPI_ISL_2557286, EPI_ISL_2557287, EPI_ISL_2557288, EPI_ISL_2557289, EPI_ISL_2557290, EPI_ISL_2557291, EPI_ISL_2557294, EPI_ISL_2557296, EPI_ISL_2557297, EPI_ISL_2557298, EPI_ISL_2557299, EPI_ISL_2557300, EPI_ISL_2557301, EPI_ISL_2659117, EPI_ISL_2659119, EPI_ISL_2659132, EPI_ISL_2659170, EPI_ISL_2659171, EPI_ISL_2663410, EPI_ISL_2663411, EPI_ISL_2663416, EPI_ISL_2663420, EPI_ISL_2663422, EPI_ISL_2663456, EPI_ISL_2663457, EPI_ISL_2663458, EPI_ISL_2663459, EPI_ISL_2663460, EPI_ISL_2663461, EPI_ISL_2663554, EPI_ISL_2663557, EPI_ISL_2663559, EPI_ISL_2663560, EPI_ISL_2663561, EPI_ISL_2663562, EPI_ISL_2663563, EPI_ISL_2663564, EPI_ISL_2663565, EPI_ISL_2663568, EPI_ISL_2663569, EPI_ISL_2663570, EPI_ISL_2663571, EPI_ISL_2663572, EPI_ISL_2663573, EPI_ISL_2663574, EPI_ISL_2663575, EPI_ISL_2663576 | | | |
| see above | Genetica Molecular and Subdepartamento de Virologia ISP Chile | Instituto de Salud Publica de Chile | Andres Castillo; Barbara Parra; Constanza Campano; Gisselle Barra; Javier Tognarelli; Jorge Fernandez; Karen Orostica; Loredana Arata; Patricia Bustos; Rodrigo Fasce; Soledad Ulloa |
| EPI_ISL_4054613, EPI_ISL_4054615 | Grady Memorial Hospital, Clinical Microbiology Laboratory | Piantadosi Lab, Emory Department of Pathology | Anne Piantadosi; Ludy Registre Carmola; Yun F (Wayne) Wang |
| EPI_ISL_2537140, EPI_ISL_2537141, EPI_ISL_2537142, EPI_ISL_2537146, EPI_ISL_2537191, EPI_ISL_2537202, EPI_ISL_2537203, EPI_ISL_2537207, EPI_ISL_2537217, EPI_ISL_2537219, EPI_ISL_2537221, EPI_ISL_2537223, EPI_ISL_2537228, EPI_ISL_2537233 | | | |
| see above | Gravity Diagnostics, LLC | Gravity Diagnostics, LLC | Gravity Diagnostics |
| EPI_ISL_2376254, EPI_ISL_2376255, EPI_ISL_2501001 | Gundersen Clinical Microbiology Laboratory | Kabara Cancer Research Institute | Craig S. Richmond; Paraic A. Kenny |
| EPI_ISL_2536097 | H Braga | Instituto Nacional de Saude (INSA) | Borges et al |
| EPI_ISL_5646131, | HC-UNESP | Instituto Butantan | Antonio Jorge Martins; Claudia Renata dos Santos Barros; David Schlesinger; Debora Botequio Moretti; Dimas Tadeu Covas; Elaine Cristina Marqueze; Elaine Vieira Santos; Evandra Strazza Rodrigues; Heidge Fukumasu; Jayme Augusto de Souza-Neto; José Salvatore Leister Patané; Luiz Alcantara; |

| | | | |
|---|---|---|---|
| EPI_ISL_5646162, EPI_ISL_5646190 | | | Luiz Lehmann Coutinho; Maria Carolina Elias; Maurício Lacerda Nogueira; Rafael dos Santos Bezerra; Raul Machado Neto; Rejane Maria Tommasini Grotto; Ricardo Haddad; Sandra Coccuzzo Sampaio Vessoni; Simone Kashima; Svetoslav Nanev Slavov; Vincent Louis Viala |
| EPI_ISL_6229716, EPI_ISL_6229717, EPI_ISL_6229824 | HGF HOSPITAL GERAL DE FORTALEZA | ACME Lab, Oswaldo Cruz Foundation, FIOCRUZ/CE | Carlos Leonardo de Aragao Araujo; Cecilia Leite Costa & Eduardo Ruback dos Santos on behalf of COVID-19 FIOCRUZ Genomic Network; Cleber Furtado Aksenen; Fabio Miyajima; Fernando Braga Stehling; Francisco Eder de Moura Lopes; Igor Oliveira Duarte; Jamille Maria Mendes Bezerra; Joaquim Cesar do Nascimento Sousa Junior; Pedro Miguel Carneiro Jeronimo; Suzana Porto Almeida; Thais Ferreira de Oliveira; Thais de Oliveira Costa; Ticiane Cavalcante de Souza; Veridiana Pessoa Miyajima |
| EPI_ISL_2497443, EPI_ISL_2497444, EPI_ISL_2497445, EPI_ISL_2497446, EPI_ISL_2497447, EPI_ISL_2497448, EPI_ISL_2497449, EPI_ISL_2497450, EPI_ISL_2497451, EPI_ISL_2497452, EPI_ISL_2497453, EPI_ISL_2497454, EPI_ISL_2497455, EPI_ISL_2497456, EPI_ISL_2497457, EPI_ISL_2497458, EPI_ISL_2497459, EPI_ISL_2497460, EPI_ISL_2497461, EPI_ISL_2497462, EPI_ISL_2497463, EPI_ISL_2497464, EPI_ISL_2497466, EPI_ISL_2497467, EPI_ISL_2497468, EPI_ISL_2497469, EPI_ISL_2497470, EPI_ISL_2497471, EPI_ISL_2497472, EPI_ISL_2497473, EPI_ISL_2497474, EPI_ISL_2497475, EPI_ISL_2497476, EPI_ISL_2617587, EPI_ISL_2617588, EPI_ISL_2617608, EPI_ISL_2617609, EPI_ISL_2617627, EPI_ISL_2921533, EPI_ISL_3254322, EPI_ISL_3254323, EPI_ISL_3254324, EPI_ISL_3254325, EPI_ISL_3254326, EPI_ISL_3254327, EPI_ISL_3254348, EPI_ISL_4061647, EPI_ISL_4890088, EPI_ISL_4890102 | | | |
| see above | HLAGYN - Laboratorio de Imunologia de Transplantes de Goias | HLAGYN - Laboratorio de Imunologia de Transplantes de Goias | Alessandro Leonardo Alvares Magalhaes; Daniel Ferreira de Sousa; Erika Lopes Rocha Batista; Fernando Antonio Vinhal dos Santos; Frederico Rodrigues Vinhal; Kamila Oliveira Reis De Freitas.; Lucas Carlos Gomes Pereira; Sabrina Sara Moreira Duarte |
| EPI_ISL_2482830, EPI_ISL_2482832 | HOPITAL ALBI | CNR Virus des Infections Respiratoires - France SUD | Antonin Bal; Bruno Lina; Bruno Simon; Gregory Destras; Gwendolyne Burfin; Hadrien Regue; Laurence Josset; Martine Valette; Quentin Semanas |
| EPI_ISL_6229843 | HOSP DR THADEU DE PAULO BRITO | ACME Lab, Oswaldo Cruz Foundation, FIOCRUZ/CE | Carlos Leonardo de Aragao Araujo; Cecilia Leite Costa & Eduardo Ruback dos Santos on behalf of COVID-19 FIOCRUZ Genomic Network; Cleber Furtado Aksenen; Fabio Miyajima; Fernando Braga Stehling; Francisco Eder de Moura Lopes; Igor Oliveira Duarte; Jamille Maria Mendes Bezerra; Joaquim Cesar do Nascimento Sousa Junior; Pedro Miguel Carneiro Jeronimo; Suzana Porto Almeida; Thais Ferreira de Oliveira; Thais de Oliveira Costa; Ticiane Cavalcante de Souza; Veridiana Pessoa Miyajima |
| EPI_ISL_3912084 | HOSP MATERN LIA LOIOLA DE ALENCAR | Analytical Competence Molecular Epidemiology Lab/ACME, Oswaldo Cruz Foundation, Ceara (FIOCRUZ CE) | Cleber Furtado Aksenen; Fabio Miyajima; Fernando Braga Stehling; Francisco Eder de Moura Lopes; Jamille Maria Mendes Bezerra; Joaquim Cesar do Nascimento Sousa Junior; Pedro Miguel Carneiro Jeronimo; Suzana Porto Almeida & Lucas Delerino on behalf of COVID-19 FIOCRUZ Genomic Network; Thais Ferreira de Oliveira; Thais de Oliveira Costa; Ticiane Cavalcante de Souza; Veridiana Pessoa Miyajima |
| EPI_ISL_2493845, EPI_ISL_2493850, EPI_ISL_2493851, EPI_ISL_2493853, EPI_ISL_2493854, EPI_ISL_2493855, EPI_ISL_2493856, EPI_ISL_2493857, EPI_ISL_2493859, EPI_ISL_2493860, EPI_ISL_2493905, EPI_ISL_2493918, EPI_ISL_2493919, EPI_ISL_2493920, EPI_ISL_2493921, EPI_ISL_2493922, EPI_ISL_2493923, EPI_ISL_2493924, EPI_ISL_2493925, EPI_ISL_2493926, EPI_ISL_2493927, EPI_ISL_2493928, EPI_ISL_2493929, EPI_ISL_2493930, EPI_ISL_2493937, EPI_ISL_2493938, EPI_ISL_2493939, EPI_ISL_2493954, EPI_ISL_2494300 | | | |
| see above | HOSP MUN DE MOGI DAS CRUZES PREF WALDEMAR COSTA FILHO | Instituto Butantan | Antonio Jorge Martins; Claudia Renata dos Santos Barros; David Schlesinger; Debora Botequio Moretti; Dimas Tadeu Covas; Elaine Cristina Marqueze; Elaine Vieira Santos; Evandra Strazza Rodrigues; Heidge Fukumasu; Jayme Augusto de Souza-Neto; José Salvatore Leister Patané; Luiz Alcantara; Luiz Lehmann Coutinho; Maria Carolina Elias; Maurício Lacerda Nogueira; Rafael dos Santos Bezerra; Raul Machado Neto; Rejane Maria Tommasini Grotto; Ricardo Haddad; Sandra Coccuzzo Sampaio Vessoni; Simone Kashima; Svetoslav Nanev Slavov; Vincent Louis Viala |
| EPI_ISL_2493941 | HOSP MUN JOSANIAS CASTANHA BRAGA | Instituto Butantan | Antonio Jorge Martins; Claudia Renata dos Santos Barros; David Schlesinger; Debora Botequio Moretti; Dimas Tadeu Covas; Elaine Cristina Marqueze; Elaine Vieira Santos; Evandra Strazza Rodrigues; Heidge Fukumasu; Jayme Augusto de Souza-Neto; José Salvatore Leister Patané; Luiz Alcantara; Luiz Lehmann Coutinho; Maria Carolina Elias; Maurício Lacerda Nogueira; Rafael dos Santos Bezerra; Raul Machado Neto; Rejane Maria Tommasini Grotto; Ricardo Haddad; Sandra Coccuzzo Sampaio Vessoni; Simone Kashima; Svetoslav Nanev Slavov; Vincent Louis Viala |
| EPI_ISL_2493956, EPI_ISL_2493959 | HOSPITAL DE CAMPANHA COVID 19 DE MAIRIPORA | Instituto Butantan | Antonio Jorge Martins; Claudia Renata dos Santos Barros; David Schlesinger; Debora Botequio Moretti; Dimas Tadeu Covas; Elaine Cristina Marqueze; Elaine Vieira Santos; Evandra Strazza Rodrigues; Heidge Fukumasu; Jayme Augusto de Souza-Neto; José Salvatore Leister Patané; Luiz Alcantara; Luiz Lehmann Coutinho; Maria Carolina Elias; Maurício Lacerda Nogueira; Rafael dos Santos Bezerra; Raul Machado Neto; Rejane Maria Tommasini Grotto; Ricardo Haddad; Sandra Coccuzzo Sampaio Vessoni; Simone Kashima; Svetoslav Nanev Slavov; Vincent Louis Viala |
| EPI_ISL_2493873, EPI_ISL_2493876, EPI_ISL_2493877 | HOSPITAL DE CAMPANHA COVID 19 MUNICIPIO DE TAUBATE | Instituto Butantan | Antonio Jorge Martins; Claudia Renata dos Santos Barros; David Schlesinger; Debora Botequio Moretti; Dimas Tadeu Covas; Elaine Cristina Marqueze; Elaine Vieira Santos; Evandra Strazza Rodrigues; Heidge Fukumasu; Jayme Augusto de Souza-Neto; José Salvatore Leister Patané; Luiz Alcantara; Luiz Lehmann Coutinho; Maria Carolina Elias; Maurício Lacerda Nogueira; Rafael dos Santos Bezerra; Raul Machado Neto; Rejane Maria Tommasini Grotto; Ricardo Haddad; Sandra Coccuzzo Sampaio Vessoni; Simone Kashima; Svetoslav Nanev Slavov; Vincent Louis Viala |
| EPI_ISL_2827977 | HOSPITAL DR. ENRIQUE BALTODANO BRICEÑO | Inciensa, Instituto Costarricense de Investigación y Enseñanza en Nutrición y Salud | Adriana Godínez; Claudio Soto-Garita; Estela Cordero; Francisco Duarte; Hebleen Porras; Joselyn Prado & Adriana Bermúdez; José Luis Vargas; Mariela Gutiérrez; Melany Calderón |
| EPI_ISL_2493257, EPI_ISL_2493258, EPI_ISL_2493863, EPI_ISL_2493864, EPI_ISL_2493865, EPI_ISL_2493870, EPI_ISL_2493871, EPI_ISL_2500986 | | | |
| see above | HOSPITAL E MATERNIDADE NOSSA SENHORA DA AJUDA | Instituto Butantan | Antonio Jorge Martins; Claudia Renata dos Santos Barros; David Schlesinger; Debora Botequio Moretti; Dimas Tadeu Covas; Elaine Cristina Marqueze; Elaine Vieira Santos; Evandra Strazza Rodrigues; Heidge Fukumasu; Jayme Augusto de Souza-Neto; José Salvatore Leister Patané; Luiz Alcantara; Luiz Lehmann Coutinho; Maria Carolina Elias; Maurício Lacerda Nogueira; Rafael dos Santos Bezerra; Raul Machado Neto; Rejane Maria Tommasini Grotto; Ricardo Haddad; Sandra Coccuzzo Sampaio Vessoni; Simone Kashima; Svetoslav Nanev Slavov; Vincent Louis Viala |
| EPI_ISL_3102506 | HOSPITAL ESTADUAL LEONARDO DA VINCI | Analytical Competence Molecular Epidemiology Lab/ACME, Oswaldo Cruz Foundation, Ceara (FIOCRUZ CE) | Cleber Furtado Aksenen; Fabio Miyajima; Fernando Braga Stehling; Francisco Eder de Moura Lopes; Jamille Maria Mendes Bezerra; Joaquim César do Nascimento Sousa Junior; Pedro Miguel Carneiro Jeronimo; Suzana Porto Almeida e Lucas Delerino; Thais Ferreira de Oliveira; Thais de Oliveira Costa; Ticiane Cavalcante de Souza; Veridiana Pessoa Miyajima |
| EPI_ISL_4425142 | HOSPITAL GENERAL UNIVERSITARIO DE ALBACETE | Instituto de Salud Carlos III | A. Monzón; CARIDAD; F. Casas; I. Jiménez; I.SAINZ DE BARANDA CAMINO; M. Sandonís; P. Zaballos; S. Cuesta; S. Iglesias-Caballero; S. Pozo; S. Varona; V. Camarero; Vázquez-Morón |
| EPI_ISL_2493940 | HOSPITAL GERAL JESUS TEIXEIRA DA COSTA GUAIANASES SAO PAULO | Instituto Butantan | Antonio Jorge Martins; Claudia Renata dos Santos Barros; David Schlesinger; Debora Botequio Moretti; Dimas Tadeu Covas; Elaine Cristina Marqueze; Elaine Vieira Santos; Evandra Strazza Rodrigues; Heidge Fukumasu; Jayme Augusto de Souza-Neto; José Salvatore Leister Patané; Luiz Alcantara; Luiz Lehmann Coutinho; Maria Carolina Elias; Maurício Lacerda Nogueira; Rafael dos Santos Bezerra; Raul Machado Neto; Rejane Maria Tommasini Grotto; Ricardo Haddad; Sandra Coccuzzo Sampaio Vessoni; Simone Kashima; Svetoslav Nanev Slavov; Vincent Louis Viala |
| EPI_ISL_2493861, EPI_ISL_2493957 | HOSPITAL HELIOPOLIS UNIDADE DE GESTAO ASSISTENCIAL I SP | Instituto Butantan | Antonio Jorge Martins; Claudia Renata dos Santos Barros; David Schlesinger; Debora Botequio Moretti; Dimas Tadeu Covas; Elaine Cristina Marqueze; Elaine Vieira Santos; Evandra Strazza Rodrigues; Heidge Fukumasu; Jayme Augusto de Souza-Neto; José Salvatore Leister Patané; Luiz Alcantara; Luiz Lehmann Coutinho; Maria Carolina Elias; Maurício Lacerda Nogueira; Rafael dos Santos Bezerra; Raul Machado Neto; Rejane Maria Tommasini Grotto; Ricardo Haddad; Sandra Coccuzzo Sampaio Vessoni; Simone Kashima; Svetoslav Nanev Slavov; Vincent Louis Viala |
| EPI_ISL_3102330 | HOSPITAL MATERNIDADE AGENOR ARAUJO | Analytical Competence Molecular Epidemiology Lab/ACME, Oswaldo Cruz Foundation, Ceara (FIOCRUZ CE) | Cleber Furtado Aksenen; Fabio Miyajima; Fernando Braga Stehling; Francisco Eder de Moura Lopes; Jamille Maria Mendes Bezerra; Joaquim César do Nascimento Sousa Junior; Pedro Miguel Carneiro Jeronimo; Suzana Porto Almeida e Lucas Delerino; Thais Ferreira de Oliveira; Thais de Oliveira Costa; Ticiane Cavalcante de Souza; Veridiana Pessoa Miyajima |
| EPI_ISL_5646121, EPI_ISL_5646128 | HOSPITAL MATERNIDADE SAO JOSE ITAPUI | Instituto Butantan | Antonio Jorge Martins; Claudia Renata dos Santos Barros; David Schlesinger; Debora Botequio Moretti; Dimas Tadeu Covas; Elaine Cristina Marqueze; Elaine Vieira Santos; Evandra Strazza Rodrigues; Heidge Fukumasu; Jayme Augusto de Souza-Neto; José Salvatore Leister Patané; Luiz Alcantara; Luiz Lehmann Coutinho; Maria Carolina Elias; Maurício Lacerda Nogueira; Rafael dos Santos Bezerra; Raul Machado Neto; Rejane Maria Tommasini Grotto; Ricardo Haddad; Sandra Coccuzzo Sampaio Vessoni; Simone Kashima; Svetoslav Nanev Slavov; Vincent Louis Viala |
| EPI_ISL_2827981 | HOSPITAL MONSEÑOR SANABRIA | Inciensa, Instituto Costarricense de Investigación y Enseñanza en Nutrición y Salud | Adriana Godínez; Claudio Soto-Garita; Estela Cordero; Francisco Duarte; Hebleen Porras; Joselyn Prado & María José Gómez; José Luis Vargas; Mariela Gutiérrez; Melany Calderón |
| EPI_ISL_2493882 | HOSPITAL MUN DR GUIDO GUIDA | Instituto Butantan | Antonio Jorge Martins; Claudia Renata dos Santos Barros; David Schlesinger; Debora Botequio Moretti; Dimas Tadeu Covas; Elaine Cristina Marqueze; Elaine Vieira Santos; Evandra Strazza Rodrigues; Heidge Fukumasu; Jayme Augusto de Souza-Neto; José Salvatore Leister Patané; Luiz Alcantara; Luiz Lehmann Coutinho; Maria Carolina Elias; Maurício Lacerda Nogueira; Rafael dos Santos Bezerra; Raul Machado Neto; Rejane Maria Tommasini Grotto; Ricardo Haddad; Sandra Coccuzzo Sampaio Vessoni; Simone Kashima; Svetoslav Nanev Slavov; Vincent Louis Viala |
| EPI_ISL_2493955 | HOSPITAL MUNICIPAL DE IBIUNA IBIUNA SP | Instituto Butantan | Antonio Jorge Martins; Claudia Renata dos Santos Barros; David Schlesinger; Debora Botequio Moretti; Dimas Tadeu Covas; Elaine Cristina Marqueze; Elaine Vieira Santos; Evandra Strazza Rodrigues; Heidge Fukumasu; Jayme Augusto de Souza-Neto; José Salvatore Leister Patané; Luiz Alcantara; Luiz Lehmann Coutinho; Maria Carolina Elias; Maurício Lacerda Nogueira; Rafael dos Santos Bezerra; Raul Machado Neto; Rejane Maria Tommasini Grotto; Ricardo Haddad; Sandra Coccuzzo Sampaio Vessoni; Simone Kashima; Svetoslav Nanev Slavov; Vincent Louis Viala |
| EPI_ISL_2462326, EPI_ISL_2493149, EPI_ISL_2493158, EPI_ISL_2493172, EPI_ISL_2493174, EPI_ISL_2494132, EPI_ISL_2494135, EPI_ISL_2494136, EPI_ISL_2494139, EPI_ISL_2494140 | | | |
| see above | HOSPITAL MUNICIPAL DE ITAPIRA | Instituto Butantan | Antonio Jorge Martins; Claudia Renata dos Santos Barros; David Schlesinger; Debora Botequio Moretti; Dimas Tadeu Covas; Elaine Cristina Marqueze; Elaine Vieira Santos; Evandra Strazza Rodrigues; Heidge Fukumasu; Jayme Augusto de Souza-Neto; José Salvatore Leister Patané; Luiz Alcantara; Luiz Lehmann Coutinho; Maria Carolina Elias; Maurício Lacerda Nogueira; Rafael dos Santos Bezerra; Raul Machado Neto; Rejane Maria Tommasini Grotto; Ricardo Haddad; Sandra Coccuzzo Sampaio Vessoni; Simone Kashima; Svetoslav Nanev Slavov; Vincent Louis Viala |
| EPI_ISL_2801347 | HOSPITAL MUNICIPAL DR EDUARDO DIAS | Analytical Competence Molecular Epidemiology Lab/ACME, Oswaldo Cruz Foundation, Ceara (FIOCRUZ CE) | Cleber Furtado Aksenen e Suzana Porto Almeida; Fabio Miyajima; Fernando Braga Stehling; Francisco Eder de Moura Lopes; Jamille Maria Mendes Bezerra; Joaquim César do Nascimento Sousa Junior; Pedro Miguel Carneiro Jeronimo; Thais Ferreira de Oliveira; Thais de Oliveira Costa; Ticiane Cavalcante de Souza; Veridiana Pessoa Miyajima |
| EPI_ISL_2462315, EPI_ISL_2493821, EPI_ISL_2493827, EPI_ISL_5649648, EPI_ISL_5649754, EPI_ISL_5649826, EPI_ISL_5649885, EPI_ISL_5649894 | | | |
| see above | HOSPITAL MUNICIPAL DR MARIO GATTI CAMPINAS | Instituto Butantan | Antonio Jorge Martins; Claudia Renata dos Santos Barros; David Schlesinger; Debora Botequio Moretti; Dimas Tadeu Covas; Elaine Cristina Marqueze; Elaine Vieira Santos; Evandra Strazza Rodrigues; Heidge Fukumasu; Jayme Augusto de Souza-Neto; José Salvatore Leister Patané; Luiz Alcantara; Luiz Lehmann Coutinho; Maria Carolina Elias; Maurício Lacerda Nogueira; Rafael dos Santos Bezerra; Raul Machado Neto; Rejane Maria Tommasini Grotto; Ricardo Haddad; Sandra Coccuzzo Sampaio Vessoni; Simone Kashima; Svetoslav Nanev Slavov; Vincent Louis Viala |
| EPI_ISL_2494164 | HOSPITAL MUNICIPAL JORGE CURY DE SANTA MARIA DA SERRA | Instituto Butantan | Antonio Jorge Martins; Claudia Renata dos Santos Barros; David Schlesinger; Debora Botequio Moretti; Dimas Tadeu Covas; Elaine Cristina Marqueze; Elaine Vieira Santos; Evandra Strazza Rodrigues; Heidge Fukumasu; Jayme Augusto de Souza-Neto; José Salvatore Leister Patané; Luiz Alcantara; Luiz Lehmann Coutinho; Maria Carolina Elias; Maurício Lacerda Nogueira; Rafael dos Santos Bezerra; Raul Machado Neto; Rejane Maria Tommasini Grotto; Ricardo Haddad; Sandra Coccuzzo Sampaio Vessoni; Simone Kashima; Svetoslav Nanev Slavov; Vincent Louis Viala |
| EPI_ISL_2801348, EPI_ISL_2801349, EPI_ISL_2801355, EPI_ISL_2801370 | HOSPITAL MUNICIPAL NOSSA SENHORA DOS MILAGRES | Analytical Competence Molecular Epidemiology Lab/ACME, Oswaldo Cruz Foundation, Ceara (FIOCRUZ CE) | Cleber Furtado Aksenen e Suzana Porto Almeida; Fabio Miyajima; Fernando Braga Stehling; Francisco Eder de Moura Lopes; Jamille Maria Mendes Bezerra; Joaquim César do Nascimento Sousa Junior; Pedro Miguel Carneiro Jeronimo; Thais Ferreira de Oliveira; Thais de Oliveira Costa; Ticiane Cavalcante de Souza; Veridiana Pessoa Miyajima |
| EPI_ISL_6229738, EPI_ISL_6229739, EPI_ISL_6229740 | HOSPITAL REGIONAL DE IGUATU | ACME Lab, Oswaldo Cruz Foundation, FIOCRUZ/CE | Carlos Leonardo de Aragao Araujo; Cecilia Leite Costa & Eduardo Ruback dos Santos on behalf of COVID-19 FIOCRUZ Genomic Network; Cleber Furtado Aksenen; Fabio Miyajima; Fernando Braga Stehling; Francisco Eder de Moura Lopes; Igor Oliveira Duarte; Jamille Maria Mendes Bezerra; Joaquim Cesar do Nascimento Sousa Junior; Pedro Miguel Carneiro Jeronimo; Suzana Porto Almeida; Thais Ferreira de Oliveira; Thais de Oliveira Costa; Ticiane Cavalcante de Souza; Veridiana Pessoa Miyajima |
| EPI_ISL_3102505 | HOSPITAL REGIONAL DE IGUATU | Analytical Competence Molecular Epidemiology Lab/ACME, Oswaldo Cruz Foundation, Ceara (FIOCRUZ CE) | Cleber Furtado Aksenen; Fabio Miyajima; Fernando Braga Stehling; Francisco Eder de Moura Lopes; Jamille Maria Mendes Bezerra; Joaquim César do Nascimento Sousa Junior; Pedro Miguel Carneiro Jeronimo; Suzana Porto Almeida e Lucas Delerino; Thais Ferreira de Oliveira; Thais de Oliveira Costa; Ticiane Cavalcante de Souza; Veridiana Pessoa Miyajima |
| EPI_ISL_3102304 | HOSPITAL REGIONAL DO SERTAO CENTRAL | Analytical Competence Molecular Epidemiology Lab/ACME, Oswaldo Cruz Foundation, Ceara (FIOCRUZ CE) | Cleber Furtado Aksenen; Fabio Miyajima; Fernando Braga Stehling; Francisco Eder de Moura Lopes; Jamille Maria Mendes Bezerra; Joaquim César do Nascimento Sousa Junior; Pedro Miguel Carneiro Jeronimo; Suzana Porto Almeida e Lucas Delerino; Thais Ferreira de Oliveira; Thais de Oliveira Costa; Ticiane Cavalcante de Souza; Veridiana Pessoa Miyajima |
| EPI_ISL_2827973 | HOSPITAL SAN JUAN DE DIOS | Inciensa, Instituto Costarricense de Investigación y Enseñanza en Nutrición y Salud | Adriana Godínez; Claudio Soto-Garita; Estela Cordero; Francisco Duarte; Hebleen Porras; Joselyn Prado & Paola Sánchez; José Luis Vargas; Mariela Gutiérrez; Melany Calderón |
| EPI_ISL_2827986 | HOSPITAL SAN VICENTE DE PAUL | Inciensa, Instituto Costarricense de Investigación y Enseñanza en Nutrición y Salud | Adriana Godínez; Claudio Soto-Garita; Estela Cordero; Francisco Duarte; Hebleen Porras; Joselyn Prado & Josiany Arce; José Luis Vargas; Mariela Gutiérrez; Melany Calderón |
| EPI_ISL_2494168, EPI_ISL_2494172 | HOSPITAL SAO JOSE ITIRAPINA | Instituto Butantan | Antonio Jorge Martins; Claudia Renata dos Santos Barros; David Schlesinger; Debora Botequio Moretti; Dimas Tadeu Covas; Elaine Cristina Marqueze; Elaine Vieira Santos; Evandra Strazza Rodrigues; Heidge Fukumasu; Jayme Augusto de Souza-Neto; José Salvatore Leister Patané; Luiz Alcantara; Luiz Lehmann Coutinho; Maria Carolina Elias; Maurício Lacerda Nogueira; Rafael dos Santos Bezerra; Raul Machado Neto; Rejane Maria Tommasini Grotto; Ricardo Haddad; Sandra Coccuzzo Sampaio Vessoni; Simone Kashima; Svetoslav Nanev Slavov; Vincent Louis Viala |
| EPI_ISL_3532488 | HOSPITAL UNIVERSITARIO 12 DE OCTUBRE | HOSPITAL UNIVERSITARIO 12 DE OCTUBRE | Carmen Martín-Higuera; Esther Viedma; Irene Múñoz-Gallego; M.ª Dolores Folgueira; Mar Aguilera; Noelia Moral; Rafael Delgado; Sagrario Zurita |
| EPI_ISL_4425079 | HOSPITAL UNIVERSITARIO VIRGEN DE LA ARRIXACA | Instituto de Salud Carlos III | A. Monzón; F. Casas; I. Jiménez; I.MORENO PARRADO; LAURA; M. Sandonís; P. Zaballos; S. Cuesta; S. Iglesias-Caballero; S. Pozo; S. Varona; V. Camarero; Vázquez-Morón |
| EPI_ISL_3235332, EPI_ISL_3235349, | Hackensack Medical Center | New York Genome Center | Amy Baldwin; Andre Corvelo; Barry Kreiswirth; David Perlin; Dayna M. Oschwald; Jose Mediavilla; Kaelea Composto; Kar Chow; Liang Chen; Marcus Cunningham; Michael Zody; Samantha Fennessey; Tom Maniatis |

| | | | |
|---|---|---|---|
| EPI_ISL_3235350, EPI_ISL_3235357 | | | |
| EPI_ISL_3388832 | Hosp. Pediatrico Dr. Avelino Castelán | Laboratorio de Biología Molecular, Instituto de Medicina Regional on behalf of 'Proyecto Argentino Interinstitucional de genomica de SARS-CoV-2' (PAIS Consortium) | Bettina Brusés; Gioia Lucia Marino; Griselda Oria; Horacio Lucero.; Javier Mussin; Julieta Ronchi; Laura Formichelli; María Andrea Gili; María Cecilia López; Melina Lorenzini Campos; Raúl Maximiliano Acevedo |
| EPI_ISL_2812647 | Hospital Carmen y Severo Ochoa | Laboratorio de Virología HUCA | Abreu F; Alvarez-Arguelles ME; Boga JA; Castelló C; Costales I; Coto E; Gómez de Oña J; Martín-Rodríguez G; Melón S; Perez-Martínez Z; Rojo S; Sandoval M |
| EPI_ISL_3147312, EPI_ISL_3147475 | Hospital Center Emile Mayrisch | Laboratoire national de sante, Microbiology, Microbial Genomics Platform | Anke Wienecke-Baldacchino; Catherine Ragimbeau; Cynthia Oxacelay; Elodie Solarino; Fatu Djabi; Jessica Tapp; Lise Pignon; Raoul Salmon; Tamir Abdelrahman; Virginie Jover |
| EPI_ISL_2402499, EPI_ISL_3505984, EPI_ISL_3505988 | Hospital General Universitario Gregorio Marañón | Hospital General Universitario Gregorio Marañón | Cristina Rodriguez-Grande; Darío García de Viedma; Julia Suárez; Laura Pérez-Lago; Marta Herranz Martin; Patricia Muñoz; Pedro Sola Campoy; Pilar Catalán; Sergio Buenestado Serrano; Victor Manuel de la Cueva |
| EPI_ISL_2598370, EPI_ISL_2598378, EPI_ISL_2598379, EPI_ISL_2598380, EPI_ISL_3216189, EPI_ISL_3216385, EPI_ISL_3216392 | | | |
| see above | Hospital General Universitario de Alicante - Instituto de Investigación Sanitaria y Biomédica de Alicante | SeqCOVID-SPAIN consortium/IBV(CSIC) | Carmen Molina Pardines and SeqCOVID-SPAIN consortium; Maripaz Ventero Martín |
| EPI_ISL_4405119 | Hospital General de Agudos Dr. Cosme Argerich | Área de Secuenciación del Laboratorio de Virología del Hospital de Niños Dr. Ricardo Gutierrez on behalf of 'Proyecto Argentino Interinstitucional de genomica de SARS-CoV-2' (PAIS Consortium) | Acuña; Andrea Fernández; D; Florencia Funez; Florencia Rodríguez; Goya; Jésica Galeano; Karina Polanski; LE; Lusso; MI; MS; Marcia Pozzati; Nabaes Jodar; Natale; S; Valinotto; Viegas, M. |
| EPI_ISL_3260092, EPI_ISL_3260093 | Hospital Jaime Ferre - SAMCO Rafaela | Grupo de Genómica y Bioinformática del Instituto de Investigación de la Cadena Láctea CONICET-INTA on behalf of 'Proyecto Argentino Interinstitucional de genomica de SARS-CoV-2' (PAIS Consortium) | AF; Aliprandi D; Amadio; C; Eberhardt; Irazoqui; Isaia; JF; JM; MF; Pandolfi; Quaranta; Soratti R; V |
| EPI_ISL_7711735 | Hospital Psiquiatrico Dr. Manuel Chapui | Universidad de Costa Rica e Instituto Costarricense de Investigacion y Ensenanza en Nutricion y Salud | Brenes Hebleen; Camacho Erwin; Campos-Sanchez Rebeca; Ceballos Ana; Cordero Estela; Cristancho Marco; Duarte Francisco; Fernandez-Do Porto Dario; Herrera-Estrella Alfredo; Jimenez-Moraila Beatriz; Kreuze Jan; Mireles-Rivera Guadalupe; Molina-Mora Jose; Munoz-Medina Jose Esteban; Negri Tatiana; Nunes Gisele; Oliveira Guilherme; Oliveira Renato; Reales-Gonzalez Jhonnatan; Remes-Lenicov Federico; Reyes Alejandro; Sosa Ezequiel; Soto Claudio; Tsukayama Pablo; Turjanski Adrian; and Zayat Jonathan |
| EPI_ISL_7711719 | Hospital San Juan De Dios [San Jose/San Jose] | Universidad de Costa Rica e Instituto Costarricense de Investigacion y Ensenanza en Nutricion y Salud | Brenes Hebleen; Camacho Erwin; Campos-Sanchez Rebeca; Ceballos Ana; Cordero Estela; Cristancho Marco; Duarte Francisco; Fernandez-Do Porto Dario; Herrera-Estrella Alfredo; Jimenez-Moraila Beatriz; Kreuze Jan; Mireles-Rivera Guadalupe; Molina-Mora Jose; Munoz-Medina Jose Esteban; Negri Tatiana; Nunes Gisele; Oliveira Guilherme; Oliveira Renato; Reales-Gonzalez Jhonnatan; Remes-Lenicov Federico; Reyes Alejandro; Sosa Ezequiel; Soto Claudio; Tsukayama Pablo; Turjanski Adrian; and Zayat Jonathan |
| EPI_ISL_2516536, EPI_ISL_2637300 | Hospital Universitari Bellvitge | Microbiology Department | Aida Gonzalez-Diaz; Carmen Ardanuy; Jordi Camara; Jordi Niubó; Laura Calatayud; M Angeles Domínguez; Miguel Fernandez-Huerta; Sara Marti |
| EPI_ISL_2566592, EPI_ISL_2566593, EPI_ISL_2566594, EPI_ISL_2566596, EPI_ISL_2652058, EPI_ISL_2652059, EPI_ISL_2652064 | | | |
| see above | Hospital Universitari Joan XXIII | Hospital Universitari Joan XXIII | Carla Martín; Clara Benavent; Cristina Gutiérrez; Ester Picó; Gemma Recio; Margarida Terrón; Natalia Bastón |
| EPI_ISL_2600854, EPI_ISL_2600865, EPI_ISL_2820650, EPI_ISL_2820661 | Hospital Universitari i Politècnic La Fe de València | SeqCOVID-SPAIN consortium/IBV(CSIC) | Ana Gil Brusola; Eva González Barberá; José Luis López Hontangas and SeqCOVID-SPAIN consortium; María Dolores Gómez Ruiz; Salvador Giner Almaraz |
| EPI_ISL_7339460, EPI_ISL_7339472, EPI_ISL_7339473, EPI_ISL_7339474 | Hospital Universitario San Ignacio | Centro de Investigaciones en Microbiología y Biotecnología-UR (CIMBIUR), Facultad de Ciencias Naturales, Universidad del Rosario, Bogotá, Colombia | Alberto Paniz-Mondolfi; Angie Ramírez; Beatriz Ariza; Camilo A. Correa-Cárdenas; Carlos Gómez-Restrepo; Claudia Cardozo-Romero; Claudia Méndez; David-Santiago Quevedo; Guido España; Hernando Díaz; Juan David Ramírez; Juliana Cuervo-Rojas; Julie Pérez; Luz H. Patiño; Manuel-Antonio Franco; Maria-Clara Duque; Marina Muñoz; Nathalia Ballesteros; Nicolas Luna; Sergio Castañeda; Zulma M. Cucunubá |
| EPI_ISL_2545330 | Hospital Universitario Virgen de las Nieves de Granada-SAS | SeqCOVID-SPAIN consortium/IBV(CSIC) | Irene Pedrosa Corral; José M. Navarro-Marí and SeqCOVID-SPAIN consortium; Sara Sanbonmatsu Gámez |
| EPI_ISL_4196727 | Hospital Zonal Dr. Ramón Carrillo | Laboratorio Central Mg. Luis Alfredo Pianciola on behalf of 'Proyecto Argentino Interinstitucional de genomica de SARS-CoV-2' (PAIS Consortium) | C Pintos; C Rastellini; C Ziehm; J Ousset; L Pianciola; M Fernandez; M L Álvarez; M Mazzeo; P V Blanco.; Y Espasandin |
| EPI_ISL_5195523, EPI_ISL_5195528 | Hospital da Baleia | Instituto René Rachou / Fiocruz Minas | Alana Oliveira; Anna Salim; Camila Corsini; Daniel Miranda; Gabriel Fernandes; Mozar de Castro; Nathalie Almeida; Pedro Alves; Priscilla Filgueiras; Rafaella Fortini; Raphael Silva; Raquel Vilela; Rubens do Monte Neto; Sarah Gomes; Thaís Silva; Wander Jeremias |
| EPI_ISL_3412036 | Hospital of the University of Pennsylvania Molecular Pathology Lab | Bushman | Andrew D. Marques; Aoife M. Roche; Arupa Ganguly; Ayannah S. Fitzgerald; Brendan Kelly; Jevon Graham-Wooten; John K. Everett; Kyle Rodino; Layla A. Khatib; Mike Feldman; Ronald G. Collman and Frederic Bushan; Samantha A. Whiteside; Scott Sherrill-Mix; Shantan Reddy; Young Hwang |
| EPI_ISL_2597697, EPI_ISL_2653039, EPI_ISL_2653041, EPI_ISL_2653042, EPI_ISL_3023276, EPI_ISL_3023284, EPI_ISL_3023316 | | | |
| see above | Hospital of the University of Pennsylvania Molecular Pathology Lab | Bushman Lab - University of Pennsylvania | Aoife M. Roche; Arupa Ganguly; Ayannah S. Fitzgerald; Brendan Kelly; Jevon Graham-Wooten; John K. Everett; Kyle Rodino; Layla A. Khatib; Mike Feldman; Ronald G. Collman and Frederic Bushan; Samantha A. Whiteside; Scott Sherrill-Mix; Shantan Reddy; Young Hwang |
| EPI_ISL_4196734 | Hospital zonal Francisco Lopez Lima General Roca | Laboratorio Central Mg. Luis Alfredo Pianciola on behalf of 'Proyecto Argentino Interinstitucional de genomica de SARS-CoV-2' (PAIS Consortium) | A Montecinos.; C Pintos; C Rastellini; C Ziehm; D Durany; J Laher; J Ousset; L Pianciola; M Fernandez; M Mazzeo; T Paniceres |
| EPI_ISL_4196705, EPI_ISL_4196706 | Hospital Programa Area Cipolletti, Dr Pedro Moguillansky | Laboratorio Central Mg. Luis Alfredo Pianciola on behalf of 'Proyecto Argentino Interinstitucional de genomica de SARS-CoV-2' (PAIS Consortium) | C Pintos; C Rastellini; C Utreras.; C Ziehm; J Ousset; L Pianciola; L Pinoche; M Fernandez; M Mazzeo; M Roncallo; R Vilugron; S Marinelli; Y Guerrero |
| EPI_ISL_2616770 | Houston Health Dept. | Houston Health Dept. | Adolpho Lara; Pamela Brown; Ryker Penn; Yanlai Lai |
| EPI_ISL_5070875, EPI_ISL_5070876, EPI_ISL_5070885, EPI_ISL_5070889, EPI_ISL_5070909, EPI_ISL_5070936, EPI_ISL_5070937, EPI_ISL_5070950, EPI_ISL_5070964, EPI_ISL_5071071 | | | |
| see above | Houston Methodist Hospital | Houston Methodist Hospital | Ilya J. Finkelstein; James J. Davis; Jessica Cambric; Jimmy Gollihar; Kristina Reppond; Layne Pruitt; Madison N. Shyer; Marcus Nguyen; Matthew Ojeda Saavedra; Paul A. Christensen; Prasanti Yerramilli; Randall J. Olsen; Robert Olson; Ryan Gadd; S. Wesley Long; Sishir Subedi; and James M. Musser |
| EPI_ISL_2680482 | Humboldt County Public Health Laboratory | Humboldt County Public Health Laboratory | Jeremy Corrigan |
| EPI_ISL_2464497 | Hôpital Avicenne | Department of Virology, Henri Mondor University Hospital, Assistance Publique Hôpitaux de Paris, Université Paris-Est Créteil, INSERM U955 | Alexandre Soulier; Christophe Rodriguez; Elisabeth Trawinski; Guillaume Gricourt; Jean-Michel Pawlotsky; Melissa N'Debi; Slim Fourati; Vanessa Demontant |
| EPI_ISL_2689890 | IA State Hygienic Laboratory | Centers for Disease Control and Prevention Division of Viral Diseases, Pathogen Discovery | Alison Laufer Halpin; Ben L. Rambo-Martin; Clinton R. Paden; Dakota Howard; Darlene Wagner; Dave Wentworth; Dhwani Batra; Jasmine Padilla; Justin Lee; Katie Dillon; Krista Queen; Kristen Knipe; Kristine Lacek; Mark Burroughs; Matthew Schmerer; Mili Sheth; Peter Cook; Sam Shepard; Sarah Nobles; Shoshona Le; Suxiang Tong; Vivien Dugan; Yvette Unoarumhi |
| EPI_ISL_2690063, EPI_ISL_2690064 | ID Bureau of Laboratories | Centers for Disease Control and Prevention Division of Viral Diseases, Pathogen Discovery | Alison Laufer Halpin; Ben L. Rambo-Martin; Clinton R. Paden; Dakota Howard; Darlene Wagner; Dave Wentworth; Dhwani Batra; Jasmine Padilla; Justin Lee; Katie Dillon; Krista Queen; Kristen Knipe; Kristine Lacek; Mark Burroughs; Matthew Schmerer; Mili Sheth; Peter Cook; Sam Shepard; Sarah Nobles; Shoshona Le; Suxiang Tong; Vivien Dugan; Yvette Unoarumhi |
| EPI_ISL_2632151 | IMD - MVZ Labor Martinsried | Robert Koch Institute | |
| EPI_ISL_2544532, EPI_ISL_2544533, EPI_ISL_2544573, EPI_ISL_2544582, EPI_ISL_2544584, EPI_ISL_2544592, EPI_ISL_2544640, EPI_ISL_2544660 | | | |
| see above | IN State Department of Health Laboratory Services | IN State Department of Health Laboratory Services | Brian Pope; Cassandra Campion; Jamie Yeadon; Kyle Brownlee; Lixia Liu; Mark Glazier; Melissa Hindenlang |
| EPI_ISL_6115482 | INSA | Instituto Nacional de Saude (INSA) | Borges et al |

| | | | |
|---|---|---|---|
| EPI_ISL_2493987, EPI_ISL_2493994, EPI_ISL_2494001, EPI_ISL_2494327 | INSIDE CRSN CACHOEIRINHA/CASA VERDE | Instituto Butantan | Antonio Jorge Martins; Claudia Renata dos Santos Barros; David Schlesinger; Debora Botequio Moretti; Dimas Tadeu Covas; Elaine Cristina Marqueze; Elaine Vieira Santos; Evandra Strazza Rodrigues; Heidge Fukumasu; Jayme Augusto de Souza-Neto; José Salvatore Leister Patané; Luiz Alcantara; Luiz Lehmann Coutinho; Maria Carolina Elias; Maurício Lacerda Nogueira; Rafael dos Santos Bezerra; Raul Machado Neto; Rejane Maria Tommasini Grotto; Ricardo Haddad; Sandra Coccuzzo Sampaio Vessoni; Simone Kashima; Svetoslav Nanev Slavov; Vincent Louis Viala |
| EPI_ISL_2493973, EPI_ISL_2493982, EPI_ISL_2493988, EPI_ISL_2493992, EPI_ISL_2493998, EPI_ISL_2494024, EPI_ISL_2494035 | | | |
| see above | INSIDE CRSN FREGUESIA/BRASILÂNDIA | Instituto Butantan | Antonio Jorge Martins; Claudia Renata dos Santos Barros; David Schlesinger; Debora Botequio Moretti; Dimas Tadeu Covas; Elaine Cristina Marqueze; Elaine Vieira Santos; Evandra Strazza Rodrigues; Heidge Fukumasu; Jayme Augusto de Souza-Neto; José Salvatore Leister Patané; Luiz Alcantara; Luiz Lehmann Coutinho; Maria Carolina Elias; Maurício Lacerda Nogueira; Rafael dos Santos Bezerra; Raul Machado Neto; Rejane Maria Tommasini Grotto; Ricardo Haddad; Sandra Coccuzzo Sampaio Vessoni; Simone Kashima; Svetoslav Nanev Slavov; Vincent Louis Viala |
| EPI_ISL_2493979, EPI_ISL_2493986, EPI_ISL_2494026 | INSIDE CRSN PERUS | Instituto Butantan | Antonio Jorge Martins; Claudia Renata dos Santos Barros; David Schlesinger; Debora Botequio Moretti; Dimas Tadeu Covas; Elaine Cristina Marqueze; Elaine Vieira Santos; Evandra Strazza Rodrigues; Heidge Fukumasu; Jayme Augusto de Souza-Neto; José Salvatore Leister Patané; Luiz Alcantara; Luiz Lehmann Coutinho; Maria Carolina Elias; Maurício Lacerda Nogueira; Rafael dos Santos Bezerra; Raul Machado Neto; Rejane Maria Tommasini Grotto; Ricardo Haddad; Sandra Coccuzzo Sampaio Vessoni; Simone Kashima; Svetoslav Nanev Slavov; Vincent Louis Viala |
| EPI_ISL_2493201, EPI_ISL_2493967, EPI_ISL_2493971, EPI_ISL_2493993, EPI_ISL_2494003, EPI_ISL_2494008, EPI_ISL_2494011, EPI_ISL_2494027 | | | |
| see above | INSIDE CRSN PIRITUBA | Instituto Butantan | Antonio Jorge Martins; Claudia Renata dos Santos Barros; David Schlesinger; Debora Botequio Moretti; Dimas Tadeu Covas; Elaine Cristina Marqueze; Elaine Vieira Santos; Evandra Strazza Rodrigues; Heidge Fukumasu; Jayme Augusto de Souza-Neto; José Salvatore Leister Patané; Luiz Alcantara; Luiz Lehmann Coutinho; Maria Carolina Elias; Maurício Lacerda Nogueira; Rafael dos Santos Bezerra; Raul Machado Neto; Rejane Maria Tommasini Grotto; Ricardo Haddad; Sandra Coccuzzo Sampaio Vessoni; Simone Kashima; Svetoslav Nanev Slavov; Vincent Louis Viala |
| EPI_ISL_2493259, EPI_ISL_2493984, EPI_ISL_2494006, EPI_ISL_2494007 | INSIDE CRSN VILA MARIA/VILA GUILHERME | Instituto Butantan | Antonio Jorge Martins; Claudia Renata dos Santos Barros; David Schlesinger; Debora Botequio Moretti; Dimas Tadeu Covas; Elaine Cristina Marqueze; Elaine Vieira Santos; Evandra Strazza Rodrigues; Heidge Fukumasu; Jayme Augusto de Souza-Neto; José Salvatore Leister Patané; Luiz Alcantara; Luiz Lehmann Coutinho; Maria Carolina Elias; Maurício Lacerda Nogueira; Rafael dos Santos Bezerra; Raul Machado Neto; Rejane Maria Tommasini Grotto; Ricardo Haddad; Sandra Coccuzzo Sampaio Vessoni; Simone Kashima; Svetoslav Nanev Slavov; Vincent Louis Viala |
| EPI_ISL_2493265, EPI_ISL_2493968, EPI_ISL_2494000, EPI_ISL_2494002, EPI_ISL_2494010, EPI_ISL_2494023, EPI_ISL_2494030, EPI_ISL_2494033 | | | |
| see above | INSIDE CRSO BUTANTÃ | Instituto Butantan | Antonio Jorge Martins; Claudia Renata dos Santos Barros; David Schlesinger; Debora Botequio Moretti; Dimas Tadeu Covas; Elaine Cristina Marqueze; Elaine Vieira Santos; Evandra Strazza Rodrigues; Heidge Fukumasu; Jayme Augusto de Souza-Neto; José Salvatore Leister Patané; Luiz Alcantara; Luiz Lehmann Coutinho; Maria Carolina Elias; Maurício Lacerda Nogueira; Rafael dos Santos Bezerra; Raul Machado Neto; Rejane Maria Tommasini Grotto; Ricardo Haddad; Sandra Coccuzzo Sampaio Vessoni; Simone Kashima; Svetoslav Nanev Slavov; Vincent Louis Viala |
| EPI_ISL_2493200, EPI_ISL_2494015 | INSIDE CRSO LAPA/PINHEIROS | Instituto Butantan | Antonio Jorge Martins; Claudia Renata dos Santos Barros; David Schlesinger; Debora Botequio Moretti; Dimas Tadeu Covas; Elaine Cristina Marqueze; Elaine Vieira Santos; Evandra Strazza Rodrigues; Heidge Fukumasu; Jayme Augusto de Souza-Neto; José Salvatore Leister Patané; Luiz Alcantara; Luiz Lehmann Coutinho; Maria Carolina Elias; Maurício Lacerda Nogueira; Rafael dos Santos Bezerra; Raul Machado Neto; Rejane Maria Tommasini Grotto; Ricardo Haddad; Sandra Coccuzzo Sampaio Vessoni; Simone Kashima; Svetoslav Nanev Slavov; Vincent Louis Viala |
| EPI_ISL_2493964, EPI_ISL_2493972, EPI_ISL_2493976, EPI_ISL_2493978, EPI_ISL_2493980, EPI_ISL_2493995, EPI_ISL_2494012, EPI_ISL_2494016, EPI_ISL_2494017, EPI_ISL_2494018, EPI_ISL_2494029, EPI_ISL_2494034, EPI_ISL_2494038 | | | |
| see above | INSIDE CRSS CAPELA DO SOCORRO | Instituto Butantan | Antonio Jorge Martins; Claudia Renata dos Santos Barros; David Schlesinger; Debora Botequio Moretti; Dimas Tadeu Covas; Elaine Cristina Marqueze; Elaine Vieira Santos; Evandra Strazza Rodrigues; Heidge Fukumasu; Jayme Augusto de Souza-Neto; José Salvatore Leister Patané; Luiz Alcantara; Luiz Lehmann Coutinho; Maria Carolina Elias; Maurício Lacerda Nogueira; Rafael dos Santos Bezerra; Raul Machado Neto; Rejane Maria Tommasini Grotto; Ricardo Haddad; Sandra Coccuzzo Sampaio Vessoni; Simone Kashima; Svetoslav Nanev Slavov; Vincent Louis Viala |
| EPI_ISL_2462331, EPI_ISL_2493960, EPI_ISL_2493961, EPI_ISL_2493977, EPI_ISL_2493989, EPI_ISL_2493990, EPI_ISL_2493997, EPI_ISL_2494005, EPI_ISL_2494036 | | | |
| see above | INSIDE CRSS PARELHEIROS | Instituto Butantan | Antonio Jorge Martins; Claudia Renata dos Santos Barros; David Schlesinger; Debora Botequio Moretti; Dimas Tadeu Covas; Elaine Cristina Marqueze; Elaine Vieira Santos; Evandra Strazza Rodrigues; Heidge Fukumasu; Jayme Augusto de Souza-Neto; José Salvatore Leister Patané; Luiz Alcantara; Luiz Lehmann Coutinho; Maria Carolina Elias; Maurício Lacerda Nogueira; Rafael dos Santos Bezerra; Raul Machado Neto; Rejane Maria Tommasini Grotto; Ricardo Haddad; Sandra Coccuzzo Sampaio Vessoni; Simone Kashima; Svetoslav Nanev Slavov; Vincent Louis Viala |
| EPI_ISL_2493274, EPI_ISL_2493975, EPI_ISL_2493996, EPI_ISL_2494025, EPI_ISL_2494279, EPI_ISL_2494316 | INSIDE CRSSU IPIRANGA | Instituto Butantan | Antonio Jorge Martins; Claudia Renata dos Santos Barros; David Schlesinger; Debora Botequio Moretti; Dimas Tadeu Covas; Elaine Cristina Marqueze; Elaine Vieira Santos; Evandra Strazza Rodrigues; Heidge Fukumasu; Jayme Augusto de Souza-Neto; José Salvatore Leister Patané; Luiz Alcantara; Luiz Lehmann Coutinho; Maria Carolina Elias; Maurício Lacerda Nogueira; Rafael dos Santos Bezerra; Raul Machado Neto; Rejane Maria Tommasini Grotto; Ricardo Haddad; Sandra Coccuzzo Sampaio Vessoni; Simone Kashima; Svetoslav Nanev Slavov; Vincent Louis Viala |
| EPI_ISL_2493963, EPI_ISL_2493970, EPI_ISL_2494328 | INSIDE CRSSU JABAQUARA/VILA MARIANA | Instituto Butantan | Antonio Jorge Martins; Claudia Renata dos Santos Barros; David Schlesinger; Debora Botequio Moretti; Dimas Tadeu Covas; Elaine Cristina Marqueze; Elaine Vieira Santos; Evandra Strazza Rodrigues; Heidge Fukumasu; Jayme Augusto de Souza-Neto; José Salvatore Leister Patané; Luiz Alcantara; Luiz Lehmann Coutinho; Maria Carolina Elias; Maurício Lacerda Nogueira; Rafael dos Santos Bezerra; Raul Machado Neto; Rejane Maria Tommasini Grotto; Ricardo Haddad; Sandra Coccuzzo Sampaio Vessoni; Simone Kashima; Svetoslav Nanev Slavov; Vincent Louis Viala |
| EPI_ISL_2494004, EPI_ISL_2494028, EPI_ISL_2494031, EPI_ISL_2494032, EPI_ISL_2494310 | INSIDE CRSSU MOOCA/ARICANDUVA | Instituto Butantan | Antonio Jorge Martins; Claudia Renata dos Santos Barros; David Schlesinger; Debora Botequio Moretti; Dimas Tadeu Covas; Elaine Cristina Marqueze; Elaine Vieira Santos; Evandra Strazza Rodrigues; Heidge Fukumasu; Jayme Augusto de Souza-Neto; José Salvatore Leister Patané; Luiz Alcantara; Luiz Lehmann Coutinho; Maria Carolina Elias; Maurício Lacerda Nogueira; Rafael dos Santos Bezerra; Raul Machado Neto; Rejane Maria Tommasini Grotto; Ricardo Haddad; Sandra Coccuzzo Sampaio Vessoni; Simone Kashima; Svetoslav Nanev Slavov; Vincent Louis Viala |
| EPI_ISL_2493962, EPI_ISL_2493965, EPI_ISL_2493983, EPI_ISL_2493999, EPI_ISL_2494009, EPI_ISL_2494019, EPI_ISL_2494020 | | | |
| see above | INSIDE CRSSU SAPOPEMBA/VILA PRUDENTE | Instituto Butantan | Antonio Jorge Martins; Claudia Renata dos Santos Barros; David Schlesinger; Debora Botequio Moretti; Dimas Tadeu Covas; Elaine Cristina Marqueze; Elaine Vieira Santos; Evandra Strazza Rodrigues; Heidge Fukumasu; Jayme Augusto de Souza-Neto; José Salvatore Leister Patané; Luiz Alcantara; Luiz Lehmann Coutinho; Maria Carolina Elias; Maurício Lacerda Nogueira; Rafael dos Santos Bezerra; Raul Machado Neto; Rejane Maria Tommasini Grotto; Ricardo Haddad; Sandra Coccuzzo Sampaio Vessoni; Simone Kashima; Svetoslav Nanev Slavov; Vincent Louis Viala |
| EPI_ISL_5645609, EPI_ISL_5645618, EPI_ISL_5645628, EPI_ISL_5645630, EPI_ISL_5645632, EPI_ISL_5645653, EPI_ISL_5645671, EPI_ISL_5645678, EPI_ISL_5645685, EPI_ISL_5645711, EPI_ISL_5645716, EPI_ISL_5645731, EPI_ISL_5645735, EPI_ISL_5645752, EPI_ISL_5645768, EPI_ISL_5645773, EPI_ISL_5645803, EPI_ISL_5645807, EPI_ISL_5645822, EPI_ISL_5645823, EPI_ISL_5645842, EPI_ISL_5645844, EPI_ISL_5645849, EPI_ISL_5645866, EPI_ISL_5645868, EPI_ISL_5645869, EPI_ISL_5645889, EPI_ISL_5645890, EPI_ISL_5645906, EPI_ISL_5645934, EPI_ISL_5645940, EPI_ISL_5645942, EPI_ISL_5645959 | | | |
| see above | INSIDE DIAGNOSTICOS | Instituto Butantan | Antonio Jorge Martins; Claudia Renata dos Santos Barros; David Schlesinger; Debora Botequio Moretti; Dimas Tadeu Covas; Elaine Cristina Marqueze; Elaine Vieira Santos; Evandra Strazza Rodrigues; Heidge Fukumasu; Jayme Augusto de Souza-Neto; José Salvatore Leister Patané; Luiz Alcantara; Luiz Lehmann Coutinho; Maria Carolina Elias; Maurício Lacerda Nogueira; Rafael dos Santos Bezerra; Raul Machado Neto; Rejane Maria Tommasini Grotto; Ricardo Haddad; Sandra Coccuzzo Sampaio Vessoni; Simone Kashima; Svetoslav Nanev Slavov; Vincent Louis Viala |
| EPI_ISL_2774144 | IRCCS San Gallicano Dermatological Institute | IRCCS Regina Elena National Cancer Institute | Aldo Morrone; Fabrizio Ensoli; Fulvia Pimpinelli; Gennaro Ciliberto; Giovanni Blandino; Grazia Prignano; Matteo Pallocca; Maurizio Fanciulli; Sabrina Strano; Sara Donzelli |
| EPI_ISL_2757934, EPI_ISL_2757935, EPI_ISL_2757937 | ISDH | Quantigen Biosciences | Paul J. Childress |
| EPI_ISL_2628209, EPI_ISL_2628300, EPI_ISL_2628305 | IVIC | Laboratorio de Virología Molecular | Carmen L Loureiro; CoViVen Group; Domingo J Garzaro; Esmeralda Vizzi; Flor H Pujol; Héctor R Rangel; José Luis Zambrano; Lieska Rodríguez; Mariana Hidalgo; Pierina D´Angelo; Rossana C Jaspe; Víctor Alarcón; Yoneira Sulbaran; Zoila Moros |
| EPI_ISL_2789980, EPI_ISL_2801468 | IVY3 Central Lab, Vanderbilt University Medical Center | Lauring Lab, University of Michigan, Department of Microbiology and Immunology | Valesano |
| EPI_ISL_2934950, EPI_ISL_2934954, EPI_ISL_2934961, EPI_ISL_2934962, EPI_ISL_2934964, EPI_ISL_2934985, EPI_ISL_2934987 | | | |
| see above | Idaho Bureau of Laboratories | Center for Global Health, University of New Mexico Health Sciences Center | Christopher Ball; Darrell Dinwiddie; Daryl Domman; Kurt Schwalm; Matthew Burns; Robert Voermans; Valerie Morley |
| EPI_ISL_3191039 | Idaho Bureau of Laboratories | Idaho Bureau of Laboratories | "R. Beukelman; Aimee Ceniseros; Christian Loera; Christopher Ball"; Matthew Charles Burns; Robert L. Voermans |
| EPI_ISL_2801141 | Illinois Department of Public Health | Gagnon Lab, Southern Illinois University | Keith Gagnon |
| EPI_ISL_2800710, EPI_ISL_2800718, EPI_ISL_2800720, EPI_ISL_2800745, EPI_ISL_2800852, EPI_ISL_2800856, EPI_ISL_2800859, EPI_ISL_2800863, EPI_ISL_3231891, EPI_ISL_3231929 | | | |
| see above | Illinois Department of Public Health | Illinois Department of Public Health - Chicago Lab | Ira Heimler; Joel Price; Vineet K. Dhiman |
| EPI_ISL_2403009, EPI_ISL_2403012, EPI_ISL_2403053, EPI_ISL_2403055, EPI_ISL_2403074, EPI_ISL_2403083, EPI_ISL_2403148, EPI_ISL_2403158, EPI_ISL_2403165, EPI_ISL_2403168, EPI_ISL_2403173, EPI_ISL_2403174, EPI_ISL_2403175, EPI_ISL_2403176, EPI_ISL_2403177, EPI_ISL_2403178, EPI_ISL_2403179, EPI_ISL_2403183, EPI_ISL_2598169, EPI_ISL_2598171, EPI_ISL_2598176, EPI_ISL_2598178, EPI_ISL_2598181, EPI_ISL_2598192, EPI_ISL_2598198, EPI_ISL_2598200, EPI_ISL_2598201, EPI_ISL_2598202, EPI_ISL_2598203, EPI_ISL_2598204, EPI_ISL_2598207, EPI_ISL_2598219, EPI_ISL_2598226, EPI_ISL_2885459 | | | |
| see above | Illinois Department of Public Health - Springfield Lab | Illinois Department of Public Health - Springfield Lab | Bryan Sim; Gordon McCall |
| EPI_ISL_2533929 | Imágenes Diagnósticas de Mazatlán | Microbial Genomics Laboratory | ; Alejandra García-Gasca; Alejandra Hernández-Terán; Alejandro Sánchez-Flores; Alfredo Herrera-Estrella; Alicia Ocaña-Mondragón; Andreu Comas-García; Angel Gustavo Salas-Lais; Antonio Loza Román; Bernardo Martínez-Miguel; Blanca Taboada; Brenda Irasema Maldonado-Meza; Bruno Gómez-Gil; Carla Ivón Herrera-Najera; Carlos F. Arias; Celia Boukadida; Clara Esperanza Santacruz-Tinoco; Concepción Grajales-Muñiz; Consorcio Mexicano de Vigilancia Genómica (CoViGen-Mex). Authors (in alphabetical order): Julio Elias Alvarado-Yaah; Cristóbal Cháidez-Quiróz; Célida Duque Molina; Célida Martínez- Rodríguez; Daniel Fregoso-Rueda; Daniel Lira Morales; Eduardo Becerril-Vargas; Fernando Fontove-Herrera; Fidencio Mejía-Nepomuceno; Francisco Pulido; Gloria Elena Espinosa-Ayala; Gloria María Molina-Salinas; Gloria Vazquez; Hector Esteban Paz-Juárez; Hector Montoya-Fuentes; Helen Haydee Fernanda Ramírez-Plascencia; Irvin González-López; Jean Pierre González; Jesús Hernández; Joel Armando Vázquez-Pérez.; Jorge Salas-Hernández; José Antonio Enciso-Moreno; José Arturo Martínez-Orozco; José Esteban Muñoz-Medina; José de Jesús Nuñez-Contreras; Juan Bautista Chale-Dzul; Julissa Enciso-Ibarra; Luis Alberto Ochoa-Carrera; Margarita Matías-Florentino; Mario Mújica-Sánchez; Marissa Perez-Garcia; María Guadalupe Santiago-Mauricio; María Guadalupe de Jesús Mireles-Rivera; Nelly Sélem-Mojica; Pavel Isa; Ricardo Ciria Merce; Ricardo Grande; Rosa María Gutiérrez Rios; Santiago Ávila-Ríos; Selene Zárate; Susana Lopez; Verónica Mata-Haro; Victor Eduardo García-Arias; Víctor Hugo Borja-Aburto |
| EPI_ISL_2603609, EPI_ISL_2603610, EPI_ISL_2603618, EPI_ISL_2603620, EPI_ISL_2603622, EPI_ISL_2603631, EPI_ISL_2603633, EPI_ISL_2603638 | | | |
| see above | InDRE | Instituto Nacional de Medicina Genomica | Cedro-Tanda A; Garcia-Cardenas FJ; Gisela Barrera-Badillo; Gonzalez-Barrera D; Gonzalez-Woge MA; Herrera-Montalvo LA.; Hidalgo-Miranda A; Irma Lopez-Martinez; Jose Ernesto Ramirez González; Mendoza-Vargas A; Munguia-Garza P; Rangel-DeLeon D; Reyes-Grajeda JP |
| EPI_ISL_2735955, EPI_ISL_2735971, EPI_ISL_2735978, EPI_ISL_2735980, EPI_ISL_2736006 | Incyte Diagnostics Spokane | Seattle Flu Study | Amanda Adler; Barry R. Lutz; Benjamin Pelle; Brian Hiatt; Caitlin R. Wolf; Chris D. Frazar; Deborah A. Nickerson; Elisabeth Brandstetter; Erica Ryke; Geoff Melly; Helen Y. Chu; Janet A. Englund; Jay Shendure; Jover Lee; Kairsten Fay; Kirsten Lacombe; Lea M. Starita; Mark J. Rieder; Matthew Richardson; Matthew Thompson; Melissa Truong; Michael Boeckh; Michael Famulare; Misja Ilcisin; Peter D. Han; Philip Dykema; Romesh Gautom; Scott Lindquist; Thomas R. Sibley; Trevor Bedford |
| EPI_ISL_2660399, EPI_ISL_2660401, EPI_ISL_2660405, EPI_ISL_2660431, EPI_ISL_2660445, | Infectious Disease Program, Broad Institute of Harvard and MIT | Infectious Disease Program, Broad Institute of Harvard and MIT | A.E.; Adams, G.; Anahtar, M.; B.L.; B.W.; Bauer, M.; Birren; Branda, J.; Carter, A.; Cerrato, F.; Chaluvadi, S.; Chapman; Cusick, C.; D.J.; DeRuff, K.; Flowers, K.; Gallagher, G.; Gladden-Young, A.; Gnirke, A.; Harris, J.; J.E.; K.J.; LaRocque, R.; Lagerborg, K.; Lemieux; Lin; Loreth, C.; MacInnis; Neumann, A.; Normandin, E.; P.C.; Park; Pierce, V.; Reilly, S.; Rosenberg, E.; Rudy, M.; Ryan, E.; S.B.; Sabeti; Shaw, B.; Siddle; Slater, D.; Smole, S.; Tomkins-Tinch, C.; Turbett, S. |

| | | | |
|---|---|---|---|
| EPI_ISL_2660449 | | | |
| EPI_ISL_2557441, EPI_ISL_2557445, EPI_ISL_2557448, EPI_ISL_2557465, EPI_ISL_2557478, EPI_ISL_2557518, EPI_ISL_2557519, EPI_ISL_2557527, EPI_ISL_2557538, EPI_ISL_2557541, EPI_ISL_2557560, EPI_ISL_2557574, EPI_ISL_2557588, EPI_ISL_2557593, EPI_ISL_2557598, EPI_ISL_2557601, EPI_ISL_2557626, EPI_ISL_2652607, EPI_ISL_2652780, EPI_ISL_2652967 | | | |
| see above | Infinity Biologix | Centers for Disease Control and Prevention Division of Viral Diseases, Pathogen Discovery | Adrian Paskey; Benjamin Rambo-Martin; Chirayu Goswami; Christian Bixby; Christopher Gulvick; Clinton R. Paden; Dakota Howard; Darlene Wagner; Dhwani Batra; Duncan MacCannell; Jason Caravas; Jonathan Schultz; Kara Moser; Matthew Schmerer; Peter W. Cook; Robin Grimwood; Russ Hager; Scott Sammons; Shatavia Morrison; Yihe Wang; Yvette Unoarumhi |
| EPI_ISL_2492351 | Institute of Microbiology, Universidad San Francisco de Quito | Institute of Microbiology, Universidad San Francisco de Quito | Belén Prado-Vivar; Bernardo Gutiérrez; Fernanda Zurita; Gabriel Trueba; Juan José Guadalupe; Lofrein Vera; Michelle Grunauer; Monica Becerra-Wong; Patricio Rojas-Silva; Paúl Cárdenas; Sully Márquez; Verónica Barragán |
| EPI_ISL_2919255, EPI_ISL_2919257 | Instituto Adolfo Lutz - Regional de Bauru | Instituto Adolfo Lutz, Interdiciplinary Procedures Center, Strategic Laboratory | Caio Vinicius Dias Lopes; Claudia Regina Gonçalves; Claudio Tavares Sacchi; Erica Valessa Ramos Gomes; Karoline Rodrigues Campos |
| EPI_ISL_2828684, EPI_ISL_2828685 | Instituto Adolfo Lutz - Regional de Campinas | Instituto Adolfo Lutz, Interdiciplinary Procedures Center, Strategic Laboratory | Caio Vinicius Dias Lopes; Claudia Regina Gonçalves; Claudio Tavares Sacchi; Erica Valessa Ramos Gomes; Karoline Rodrigues Campos |
| EPI_ISL_2828669, EPI_ISL_2828670, EPI_ISL_2828671, EPI_ISL_2828672, EPI_ISL_2828673, EPI_ISL_2828674, EPI_ISL_2828675, EPI_ISL_2828676, EPI_ISL_2828677, EPI_ISL_2828678, EPI_ISL_2828679, EPI_ISL_2828680, EPI_ISL_2828681, EPI_ISL_2828682, EPI_ISL_2828683 | | | |
| see above | Instituto Adolfo Lutz - Regional de Santo Andre | Instituto Adolfo Lutz, Interdiciplinary Procedures Center, Strategic Laboratory | Caio Vinicius Dias Lopes; Claudia Regina Gonçalves; Claudio Tavares Sacchi; Erica Valessa Ramos Gomes; Karoline Rodrigues Campos |
| EPI_ISL_2919267 | Instituto Adolfo Lutz - Regional de Sao Jose do Rio Preto | Instituto Adolfo Lutz, Interdiciplinary Procedures Center, Strategic Laboratory | Caio Vinicius Dias Lopes; Claudia Regina Gonçalves; Claudio Tavares Sacchi; Erica Valessa Ramos Gomes; Karoline Rodrigues Campos |
| EPI_ISL_2646135, EPI_ISL_2646136 | Instituto Nacional de Medicina Genomica | Centro de Investigación en Enfermedades Infecciosas (CIENI), Instituto Nacional de Enfermedades Respiratorias (INER) | Avila-Rios S; Boukadida C; Cedro-Tanda A; Herrera-Montalvo LA; Hidalgo-Miranda A; Matias-Florentino M; Mendoza-Vargas A; Perez-Garcia M; Reyes-Grajeda JP; Reyes-Teran G |
| EPI_ISL_2843903, EPI_ISL_2843904, EPI_ISL_2843905, EPI_ISL_2843907, EPI_ISL_2843908, EPI_ISL_2843910, EPI_ISL_2843911, EPI_ISL_2843912, EPI_ISL_2843913, EPI_ISL_2843914, EPI_ISL_2843915, EPI_ISL_2843916, EPI_ISL_2843917, EPI_ISL_2843918, EPI_ISL_2843919, EPI_ISL_2843920, EPI_ISL_2843921, EPI_ISL_2843922, EPI_ISL_2843923, EPI_ISL_2843924, EPI_ISL_3118847, EPI_ISL_3118848, EPI_ISL_3118849, EPI_ISL_3118851, EPI_ISL_3118852, EPI_ISL_3118853, EPI_ISL_3118855, EPI_ISL_3118857, EPI_ISL_3118858, EPI_ISL_3118859, EPI_ISL_3118860, EPI_ISL_3118861, EPI_ISL_3118862, EPI_ISL_3244532, EPI_ISL_3244533, EPI_ISL_3244535, EPI_ISL_3244538, EPI_ISL_3244539, EPI_ISL_3244540 | | | |
| see above | Instituto de Biotecnologia - UNESP-Botucatu-SP | Instituto de Biotecnologia - UNESP-Botucatu-SP | Cecília Artico Banho; Cíntia Bittar; Fábio Sossai Possebon; Guilherme Campos; Helena Lage Ferreira; Jorge A. Petroli Marchesi; João Pessoa Araújo Jr.; Leila Sabrina Ullmann; Lívia Sacchetto; Maisa C. Pereira Parra; Marília Moraes; Maurício L. Nogueira; Paula Rahal; Paulo Inacio da Costa |
| EPI_ISL_3020244 | Instituto de Medicina Tropical Alexander Von Humboldt, Sede Cusco | Laboratorio de Genómica Microbiana, Universidad Peruana Cayetano Heredia | Alejandra Dávila-Barclay; Diego Cuicapuza; Guillermo Salvatierra; Janet Huancachoque; Luis González; Pablo Tsukayama; Pedro E. Romero; Pool Marcos |
| EPI_ISL_2455808, EPI_ISL_2455811, EPI_ISL_2455824, EPI_ISL_2455825, EPI_ISL_2455828, EPI_ISL_2455829, EPI_ISL_2455832, EPI_ISL_2455840 | | | |
| see above | Istituto Zooprofilattico Sperimentale del Mezzogiorno | TIGEM | Antonio Grimaldi Patrizia Annunziata Francesco Panariello Biancamaria Pierri Claudia Tiberio Teresa Giuliano Valentina Bouche Chiara Colantuono Maria Concetta Cuomo Denise Di Concilio Lucio Di Filippo Anna Manfredi Marcello Salvi Antonio Limone Luigi Atripaldi Pellegrino Cerino Andrea Ballabio Davide Cacchiarelli |
| EPI_ISL_2465123, EPI_ISL_2465124, EPI_ISL_2465148, EPI_ISL_2465152, EPI_ISL_2465153, EPI_ISL_2465184, EPI_ISL_2465195 | | | |
| see above | Istituto Zooprofilattico Sperimentale del Mezzogiorno | Telethon Institute of Genetics and Medicine (TIGEM) | Antonio Grimaldi Patrizia Annunziata Francesco Panariello Biancamaria Pierri Claudia Tiberio Teresa Giuliano Valentina Bouche Chiara Colantuono Maria Concetta Cuomo Denise Di Concilio Lucio Di Filippo Anna Manfredi Marcello Salvi Antonio Limone Luigi Atripaldi Pellegrino Cerino Andrea Ballabio Davide Cacchiarelli |
| EPI_ISL_2382901, EPI_ISL_2382907 | Jessa | Jessa | Raymaekers et al. on behalf of Jessa_cmdLab |
| EPI_ISL_2690053 | KS Health and Environmental Laboratories | Centers for Disease Control and Prevention Division of Viral Diseases, Pathogen Discovery | Alison Laufer Halpin; Ben L. Rambo-Martin; Clinton R. Paden; Dakota Howard; Darlene Wagner; Dave Wentworth; Dhwani Batra; Jasmine Padilla; Justin Lee; Katie Dillon; Krista Queen; Kristen Knipe; Kristine Lacek; Mark Burroughs; Matthew Schmerer; Mili Sheth; Peter Cook; Sam Shepard; Sarah Nobles; Shoshona Le; Suxiang Tong; Vivien Dugan; Yvette Unoarumhi |
| EPI_ISL_2834766, EPI_ISL_2834767, EPI_ISL_2834897, EPI_ISL_2840470, EPI_ISL_2840537, EPI_ISL_2864219, EPI_ISL_2864293, EPI_ISL_2864294, EPI_ISL_2864314, EPI_ISL_2864461, EPI_ISL_2864466, EPI_ISL_2864476, EPI_ISL_2864486, EPI_ISL_2886238 | | | |
| see above | KU Leuven, Rega Institute, Clinical and Epidemiological Virology | KU Leuven, Rega Institute, Clinical and Epidemiological Virology | Bert Vanmechelen; Joan Marti-Carerras; Piet Maes; Tony Wawina-Bokalanga |
| EPI_ISL_3561106, EPI_ISL_3561409, EPI_ISL_3561424, EPI_ISL_3561441, EPI_ISL_3561780, EPI_ISL_3561827, EPI_ISL_3561946, EPI_ISL_3562021, EPI_ISL_3825501, EPI_ISL_3825502, EPI_ISL_3825515, EPI_ISL_3825535, EPI_ISL_3825545, EPI_ISL_3825546, EPI_ISL_3825550, EPI_ISL_3825551, EPI_ISL_3825557, EPI_ISL_3825568, EPI_ISL_3825573, EPI_ISL_3825589, EPI_ISL_3825595, EPI_ISL_3825600, EPI_ISL_3825603, EPI_ISL_3825606, EPI_ISL_3825624, EPI_ISL_3825627, EPI_ISL_3825645, EPI_ISL_3825671, EPI_ISL_3825679, EPI_ISL_3825680, EPI_ISL_3825703, EPI_ISL_3825705, EPI_ISL_3825706, EPI_ISL_3825732, EPI_ISL_3825766, EPI_ISL_3825771, EPI_ISL_3825774, EPI_ISL_3825789, EPI_ISL_3825794, EPI_ISL_3825803, EPI_ISL_3826986, EPI_ISL_3826987, EPI_ISL_3826990, EPI_ISL_3827008, EPI_ISL_3940992, EPI_ISL_3941050 | | | |
| see above | Kaiser Permanente NW Reginal Lab | OHSU MM Lab | Xuan Qin; Yun Wu |
| EPI_ISL_2934071 | Kaiser Regional | Santa Clara County Public Health Laboratory | Santa Clara County Public Health Department |
| EPI_ISL_2400295, EPI_ISL_2400296, EPI_ISL_2443142 | Kansas Health and Environmental Lab | Kansas Health and Environmental Lab | Ben Olsen; Jonathan Barnell; Katherine Wiggins; Mike Grose; and Phil Adam |
| EPI_ISL_2898377, EPI_ISL_2898394 | Karolinska University Hospital Solna | Karolinska University Hospital | Annelie Bjerkner; Isak Sylvin; Jan Albert; Karolina Ininbergs; Lina Guerra Blomqvist; Lynda Eneh; Martin Ekman; Martina Wahlund; Robert Dyrdak; Sandra Broddesson; Tanja Normark; Tobias Allander; Valtteri Wirta; Zhibing Yun |
| EPI_ISL_4080592, EPI_ISL_4080595, EPI_ISL_4080598, EPI_ISL_4172118, EPI_ISL_4195033 | Kentucky State Public Health Lab | Kentucky State Public Health Lab | Anjayya Tummala; Chitralipi Kattunga; Joshua Tobias; Karim George; Matthew Johnson; Rachel Zinner; Rhonda Lucas; Vaneet Arora; William Grooms |
| EPI_ISL_2483830, EPI_ISL_2483832 | Klinisch Laboratorium ZNA | Klinisch Laboratorium ZNA | Verstrepen et al. |
| EPI_ISL_2482912 | LABORATOIRE BIOESTEREL | CNR Virus des Infections Respiratoires - France SUD | Antonin Bal; Bruno Lina; Bruno Simon; Gregory Destras; Gwendolyne Burfin; Hadrien Regue; Laurence Josset; Martine Valette; Quentin Semanas |
| EPI_ISL_2462323, EPI_ISL_2493151, EPI_ISL_2493165, EPI_ISL_2493185, EPI_ISL_2494141, EPI_ISL_2494142, EPI_ISL_2494143 | | | |
| see above | LABORATORIO DE FRANCA | Instituto Butantan | Antonio Jorge Martins; Claudia Renata dos Santos Barros; David Schlesinger; Debora Botequio Moretti; Dimas Tadeu Covas; Elaine Cristina Marqueze; Elaine Vieira Santos; Evandra Strazza Rodrigues; Heidge Fukumasu; Jayme Augusto de Souza-Neto; José Salvatore Leister Patané; Luiz Alcantara; Luiz Lehmann Coutinho; Maria Carolina Elias; Maurício Lacerda Nogueira; Rafael dos Santos Bezerra; Raul Machado Neto; Rejane Maria Tommasini Grotto; Ricardo Haddad; Sandra Coccuzzo Sampaio Vessoni; Simone Kashima; Svetoslav Nanev Slavov; Vincent Louis Viala |
| EPI_ISL_2651231 | LABORATORIO HEMATOLOGICO | Universidad Nacional de Colombia - Laboratorio Genómico One Health | Andres F. Cardona-Rios; Carlos Franco-Muñoz; Carolina Muñoz-Arango; Celeny Ortiz; Daniel O. Maldonado-Perez; Diego A. Álvarez-Díaz; Hector Alejandro Ruiz-Moreno; Idabely Betancur Ortiz; Jorge E. Osorio; Juan P. Hernandez-Ortiz; Karl A Ciuoderis; Katherine Laiton-Donato; Laura Silvana Perez; Lina M. Hurtado; Marcela Mercado-Reyes; Maria Angélica Maya; Maria Stella López; Rita Almanza Payares; Sandra Ines Cano; Simón Villegas Velásquez |
| EPI_ISL_2494166, EPI_ISL_2494176, EPI_ISL_2494184 | LABORATORIO MUNICIPAL DE ANALISES CLINICAS DE RIO CLARO | Instituto Butantan | Antonio Jorge Martins; Claudia Renata dos Santos Barros; David Schlesinger; Debora Botequio Moretti; Dimas Tadeu Covas; Elaine Cristina Marqueze; Elaine Vieira Santos; Evandra Strazza Rodrigues; Heidge Fukumasu; Jayme Augusto de Souza-Neto; José Salvatore Leister Patané; Luiz Alcantara; Luiz Lehmann Coutinho; Maria Carolina Elias; Maurício Lacerda Nogueira; Rafael dos Santos Bezerra; Raul Machado Neto; Rejane Maria Tommasini Grotto; Ricardo Haddad; Sandra Coccuzzo Sampaio Vessoni; Simone Kashima; Svetoslav Nanev Slavov; Vincent Louis Viala |
| EPI_ISL_2494163, EPI_ISL_2494180, EPI_ISL_2494182, EPI_ISL_2494193, EPI_ISL_2494200, EPI_ISL_2494338, EPI_ISL_5645591, EPI_ISL_5645733, EPI_ISL_5645836 | | | |
| see above | LABORATORIO MUNICIPAL DE PIRACICABA | Instituto Butantan | Antonio Jorge Martins; Claudia Renata dos Santos Barros; David Schlesinger; Debora Botequio Moretti; Dimas Tadeu Covas; Elaine Cristina Marqueze; Elaine Vieira Santos; Evandra Strazza Rodrigues; Heidge Fukumasu; Jayme Augusto de Souza-Neto; José Salvatore Leister Patané; Luiz Alcantara; Luiz Lehmann Coutinho; Maria Carolina Elias; Maurício Lacerda Nogueira; Rafael dos Santos Bezerra; Raul Machado Neto; Rejane Maria Tommasini Grotto; Ricardo Haddad; Sandra Coccuzzo Sampaio Vessoni; Simone Kashima; Svetoslav Nanev Slavov; Vincent Louis Viala |
| EPI_ISL_2493881, EPI_ISL_2493884, EPI_ISL_2493885, EPI_ISL_2493886, EPI_ISL_2493887, EPI_ISL_2493888, EPI_ISL_2493906, EPI_ISL_2493908, EPI_ISL_2493948, EPI_ISL_2494323, EPI_ISL_2500987 | | | |
| see above | LABORATORIO MUNICIPAL DE SUZANO | Instituto Butantan | Antonio Jorge Martins; Claudia Renata dos Santos Barros; David Schlesinger; Debora Botequio Moretti; Dimas Tadeu Covas; Elaine Cristina Marqueze; Elaine Vieira Santos; Evandra Strazza Rodrigues; Heidge Fukumasu; Jayme Augusto de Souza-Neto; José Salvatore Leister Patané; Luiz Alcantara; Luiz Lehmann Coutinho; Maria Carolina Elias; Maurício Lacerda Nogueira; Rafael dos Santos Bezerra; Raul Machado Neto; Rejane Maria Tommasini Grotto; Ricardo Haddad; Sandra Coccuzzo Sampaio Vessoni; Simone Kashima; Svetoslav Nanev Slavov; Vincent Louis Viala |
| EPI_ISL_3404830, EPI_ISL_3404831, EPI_ISL_3404832, EPI_ISL_3404833, EPI_ISL_3404834, EPI_ISL_3404838, EPI_ISL_3404839, EPI_ISL_3404840, EPI_ISL_3404841, EPI_ISL_3404842, EPI_ISL_3404843, EPI_ISL_3404846, EPI_ISL_3404847, EPI_ISL_3404879, EPI_ISL_3404880, EPI_ISL_3404881 | | | |
| see above | LACEN | LGBio (Laboratorio de Genetica & Biodiversidade) | Alex Honda Bernardes; Amanda Alves de Melo; Aparecido Divino da Cruz; Cintia Pelegrineti Targueta de Azevedo Brito; Daniela de Melo e Silva; Elisangela de Paula Silveira Lacerda; Juliana Santana de Curcio; Luiz Augusto Pereira; Marc Alexandre Duarte Gigonzac; Mariana Pires de Campos Telles; Ramilla dos Santos Braga; Renata de Oliveira Dias; Rhewter Nunes; Thais Cidália Vieira Gigonzac; Thais Guimarães Castro; Thays Millena Alves Pedroso |
| EPI_ISL_6721834 | LACEN - Laboratório Central de Saúde Pública do Ceara | Evandro Chagas Institute | Delana Andreza Melo Bezerra; Dielle Monteiro Teixeira; Jedson Ferreira Cardoso; Jessylene de Almeida Ferreira; Kenny da Costa Pinheiro; Luana Soares Barbagelata; Luana da Silva Soares; Mirleide Cordeiro dos Santos; Patrícia dos Santos Lobo; Rayssa Layna da Silva Bedran; Sandro Patroca da Silva |
| EPI_ISL_3946592, EPI_ISL_3946593, EPI_ISL_3946594 | LACEN do Distrito Federal | Instituto Adolfo Lutz, Interdiciplinary Procedures Center, Strategic Laboratory | Claudio Tavares Sacchi; Karoline Rodrigues Campos |
| EPI_ISL_3545805, EPI_ISL_3691360, EPI_ISL_3691361 | LACEN do Estado do Mato Grosso do Sul | Instituto Adolfo Lutz, Interdiciplinary Procedures Center, Strategic Laboratory | Caio Vinicius Dias Lopes; Claudia Regina Gonçalves; Claudio Tavares Sacchi; Karoline Rodrigues Campos; Leonardo Tadeu de Araujo; Marlon Benedito Nascomento Santos |
| EPI_ISL_3835195, EPI_ISL_3835196, EPI_ISL_3835197, EPI_ISL_3835198, EPI_ISL_3835199, EPI_ISL_3835200, EPI_ISL_3835201, EPI_ISL_3835202, EPI_ISL_3835203, EPI_ISL_3835204, EPI_ISL_3835205, EPI_ISL_3835206, EPI_ISL_3835207, EPI_ISL_3835208, EPI_ISL_3835209, EPI_ISL_3835210, EPI_ISL_3835211, EPI_ISL_3835212, EPI_ISL_3835213, EPI_ISL_3835214, EPI_ISL_3835215, EPI_ISL_3835216, | | | |

| | | | |
|---|---|---|---|
| EPI_ISL_3835217, EPI_ISL_3835218, EPI_ISL_3835219, EPI_ISL_3835220, EPI_ISL_3835221, EPI_ISL_3835222, EPI_ISL_3835223, EPI_ISL_3835224, EPI_ISL_3835225, EPI_ISL_3835226, EPI_ISL_3835227, EPI_ISL_3835229, EPI_ISL_3835230, EPI_ISL_3835231, EPI_ISL_3835232, EPI_ISL_3835233, EPI_ISL_3835234, EPI_ISL_3835235, EPI_ISL_3835237, EPI_ISL_3835238, EPI_ISL_3835239 | | | |
| see above | LACEN/PB | WallauLab on behalf of Fiocruz COVID-19 Genomic Surveillance Network | Alexandre Freitas da Silva; Cassia Docena; Constância Flávia Junqueira Ayres; Filipe Zimmer Dezordi; Gabriel Luz Wallau; Gustavo Barbosa de Lima; Lais Ceschini Machado; Lilian Caroliny Amorim Silva; Maira Galdino da Rocha Pitta; Marcelo Henrique dos Santos Paiva; Matheus Filgueira Bezerra; Michelly Cristiny Pereira; Rômulo Pessoa e Silva; Sinval Pinto Brandão Filho |
| EPI_ISL_4487764, EPI_ISL_4487774, EPI_ISL_4487785, EPI_ISL_4487796, EPI_ISL_4487806 | LACEN/PI | WallauLab on behalf of Fiocruz COVID-19 Genomic Surveillance Network | Adelino Soares Lima Neto; Alexandre Freitas da Silva; Antônio Marinho; Cassia Docena; Constância Flávia Junqueira Ayres; Filipe Zimmer Dezordi; Gabriel Luz Wallau; Gustavo Barbosa de Lima; Hellen de Oliveira Amaral; Jacenir Reis dos Santos Mallet; Joana Carolina Viana Lima; Lais Ceschini Machado; Leandro de Mattos; Lilian Caroliny Amorim Silva; Marcela de Lacerda Valença Queiroz; Marcelo Adriano da Cunha e Silva Vieira; Marcelo Henrique dos Santos Paiva; Matheus Filgueira Bezerra; Sinval Pinto Brandão Filho; Túlio de Lima Campos; Vladimir Costa Silva; Walterlene de Carvalho Goçalves |
| EPI_ISL_5193462, EPI_ISL_5193547, EPI_ISL_5193548, EPI_ISL_5193549, EPI_ISL_5193550, EPI_ISL_5193551, EPI_ISL_5193552, EPI_ISL_5193553, EPI_ISL_5193554, EPI_ISL_5193594, EPI_ISL_5193646 | | | |
| see above | LACEN/RR | Laboratorio de Ecologia de Doencas Transmissiveis na Amazonia, Instituto Leonidas e Maria Deane - Fiocruz Amazonia | André Corado; Cátia Alexandra Ribeiro Meneses; Felipe Naveca; Fernanda Nascimento; George Silva; Ilma de Aguiar Antony; Janis Cassimiro Lopes; Luciana Gonçalves; Maria Júlia Brandão; Matilde Mejía; Pricila Carlos Veloso; Rodrigo Melo Maito; Valdinete Nascimento; Victor Souza |
| EPI_ISL_3010759, EPI_ISL_3010769, EPI_ISL_3010776, EPI_ISL_3010783, EPI_ISL_3010794, EPI_ISL_3010803, EPI_ISL_3010808, EPI_ISL_3010809, EPI_ISL_3010810, EPI_ISL_3010818, EPI_ISL_3010819, EPI_ISL_3010827, EPI_ISL_3010828, EPI_ISL_3010830, EPI_ISL_3010832, EPI_ISL_3010833, EPI_ISL_3010836, EPI_ISL_3010837, EPI_ISL_3010843, EPI_ISL_3010847, EPI_ISL_3010848, EPI_ISL_3010856, EPI_ISL_3010860, EPI_ISL_3010861, EPI_ISL_3010863, EPI_ISL_3010879, EPI_ISL_3010886, EPI_ISL_3010891, EPI_ISL_3010892, EPI_ISL_3010896, EPI_ISL_3010897, EPI_ISL_3010899, EPI_ISL_3010904, EPI_ISL_3010906, EPI_ISL_3010910, EPI_ISL_3010911, EPI_ISL_3010915, EPI_ISL_3010918, EPI_ISL_3010923, EPI_ISL_3010924, EPI_ISL_3010925, EPI_ISL_3010926, EPI_ISL_3010933, EPI_ISL_3010934, EPI_ISL_3261078, EPI_ISL_3261115 | | | |
| see above | LATE - Laboratório de Técnicas Especiais - Hospital Israelita Albert Einstein | LATE - Laboratório de Técnicas Especiais - Hospital Israelita Albert Einstein | Alexandre Hideaki Takara; Ana Paula Moreira Salles; Anelisie da Silva Santos; Deyvid Amgarten; Erick Gustavo Dorlass; Fernanda de Mello Malta; João Renato Rebello Pinho; Marcio Anunciacao Menezes; Pedro Henrique Sebe Rodrigues; Raquel Riyuzo |
| EPI_ISL_2545806 | LESP Campeche | Instituto de Diagnostico y Referencia Epidemiologicos (INDRE) | Abril Rodriguez-Maldonado; Ariadna Medina-Benitez; Claudia Wong-Arambula; Ernesto Ramirez-Gonzalez.; Gisela Barrera-Badillo; Irma Lopez-Martinez; Joaquin Quiroz-Mercado; Lucia Hernandez-Rivas; Natividad Cruz-Ortiz; Sergio Rangel-Guerrero; Tatiana Nunez-Garcia; Vanessa Rivero-Arredondo |
| EPI_ISL_2674664 | LESP Estado de Mexico | Instituto de Diagnostico y Referencia Epidemiologicos (INDRE) | Abril Rodriguez-Maldonado; Ariadna Medina-Benitez; Claudia Wong-Arambula; Ernesto Ramirez-Gonzalez.; Gisela Barrera-Badillo; Irma Lopez-Martinez; Joaquin Quiroz-Mercado; Lucia Hernandez-Rivas; Natividad Cruz-Ortiz; Sergio Rangel-Guerrero; Tatiana Nunez-Garcia; Vanessa Rivero-Arredondo |
| EPI_ISL_2779057, EPI_ISL_2779058, EPI_ISL_2779120, EPI_ISL_2800917 | LESP Guanajuato | Instituto de Diagnostico y Referencia Epidemiologicos (INDRE) | Abril Rodriguez-Maldonado; Ariadna Medina-Benitez; Claudia Wong-Arambula; Ernesto Ramirez-Gonzalez.; Gisela Barrera-Badillo; Irma Lopez-Martinez; Joaquin Quiroz-Mercado; Lucia Hernandez-Rivas; Maribel Gonzalez-Villa; Natividad Cruz-Ortiz; Sergio Rangel-Guerrero; Tatiana Nunez-Garcia; Vanessa Rivero-Arredondo |
| EPI_ISL_2674665, EPI_ISL_2674667 | LESP Hidalgo | Instituto de Diagnostico y Referencia Epidemiologicos (INDRE) | Abril Rodriguez-Maldonado; Ariadna Medina-Benitez; Claudia Wong-Arambula; Ernesto Ramirez-Gonzalez.; Gisela Barrera-Badillo; Irma Lopez-Martinez; Joaquin Quiroz-Mercado; Lucia Hernandez-Rivas; Natividad Cruz-Ortiz; Sergio Rangel-Guerrero; Tatiana Nunez-Garcia; Vanessa Rivero-Arredondo |
| EPI_ISL_2479916, EPI_ISL_2545683 | LESP Jalisco | Instituto de Diagnostico y Referencia Epidemiologicos (INDRE) | Abril Rodriguez-Maldonado; Ariadna Medina-Benitez; Claudia Wong-Arambula; Ernesto Ramirez-Gonzalez.; Gisela Barrera-Badillo; Irma Lopez-Martinez; Joaquin Quiroz-Mercado; Lucia Hernandez-Rivas; Natividad Cruz-Ortiz; Sergio Rangel-Guerrero; Tatiana Nunez-Garcia; Vanessa Rivero-Arredondo |
| EPI_ISL_2674629, EPI_ISL_2674630, EPI_ISL_2674631 | LESP Nuevo Leon | Instituto de Diagnostico y Referencia Epidemiologicos (INDRE) | Abril Rodriguez-Maldonado; Ariadna Medina-Benitez; Claudia Wong-Arambula; Ernesto Ramirez-Gonzalez.; Gisela Barrera-Badillo; Irma Lopez-Martinez; Joaquin Quiroz-Mercado; Lucia Hernandez-Rivas; Natividad Cruz-Ortiz; Sergio Rangel-Guerrero; Tatiana Nunez-Garcia; Vanessa Rivero-Arredondo |
| EPI_ISL_2545732 | LESP Queretaro | Instituto de Diagnostico y Referencia Epidemiologicos (INDRE) | Abril Rodriguez-Maldonado; Ariadna Medina-Benitez; Claudia Wong-Arambula; Ernesto Ramirez-Gonzalez.; Gisela Barrera-Badillo; Irma Lopez-Martinez; Joaquin Quiroz-Mercado; Lucia Hernandez-Rivas; Natividad Cruz-Ortiz; Sergio Rangel-Guerrero; Tatiana Nunez-Garcia; Vanessa Rivero-Arredondo |
| EPI_ISL_2545693, EPI_ISL_2545694, EPI_ISL_2545808, EPI_ISL_2545809, EPI_ISL_2545810, EPI_ISL_2545811 | LESP Quintana Roo | Instituto de Diagnostico y Referencia Epidemiologicos (INDRE) | Abril Rodriguez-Maldonado; Ariadna Medina-Benitez; Claudia Wong-Arambula; Ernesto Ramirez-Gonzalez.; Gisela Barrera-Badillo; Irma Lopez-Martinez; Joaquin Quiroz-Mercado; Lucia Hernandez-Rivas; Natividad Cruz-Ortiz; Sergio Rangel-Guerrero; Tatiana Nunez-Garcia; Vanessa Rivero-Arredondo |
| EPI_ISL_2545692, EPI_ISL_2674634 | LESP Sinaloa | Instituto de Diagnostico y Referencia Epidemiologicos (INDRE) | Abril Rodriguez-Maldonado; Ariadna Medina-Benitez; Claudia Wong-Arambula; Ernesto Ramirez-Gonzalez.; Gisela Barrera-Badillo; Irma Lopez-Martinez; Joaquin Quiroz-Mercado; Lucia Hernandez-Rivas; Natividad Cruz-Ortiz; Sergio Rangel-Guerrero; Tatiana Nunez-Garcia; Vanessa Rivero-Arredondo |
| EPI_ISL_2545782, EPI_ISL_2545783, EPI_ISL_2545784, EPI_ISL_2545785, EPI_ISL_2545786, EPI_ISL_2545787, EPI_ISL_2545788, EPI_ISL_2545790, EPI_ISL_2545791, EPI_ISL_2545792, EPI_ISL_2545794, EPI_ISL_2545795, EPI_ISL_2545796, EPI_ISL_2545797 | | | |
| see above | LESP Yucatan | Instituto de Diagnostico y Referencia Epidemiologicos (INDRE) | Abril Rodriguez-Maldonado; Ariadna Medina-Benitez; Claudia Wong-Arambula; Ernesto Ramirez-Gonzalez.; Gisela Barrera-Badillo; Irma Lopez-Martinez; Joaquin Quiroz-Mercado; Lucia Hernandez-Rivas; Natividad Cruz-Ortiz; Sergio Rangel-Guerrero; Tatiana Nunez-Garcia; Vanessa Rivero-Arredondo |
| EPI_ISL_2443442 | LHUB-ULB | Labo Klinische Biologie, UZA | Basil Britto Xavier; Christine Lammens; Herman Goossens; Jasmine Coppens; Marie Le Mercier; Veerle Matheeussen |
| EPI_ISL_2605070, EPI_ISL_2605071, EPI_ISL_2605072, EPI_ISL_2605102 | Lab voor klinische biologie | Lab voor klinische biologie | Bruno Verhasselt; Hannelore Hamerlinck; Marija Janevska |
| EPI_ISL_2551033, EPI_ISL_2551034, EPI_ISL_2551041, EPI_ISL_2551050, EPI_ISL_2551052, EPI_ISL_2551055, EPI_ISL_2551087 | | | |
| see above | Lab. Microbiologia e Virologia Cotugno A.O. dei Colli - Istituto Zooprofilattico Sperimentale del Mezzogiorno | TIGEM | Antonio Grimaldi Patrizia Annunziata Francesco Panariello Biancamaria Pierri Claudia Tiberio Teresa Giuliano Valentina Bouche Chiara Colantuono Maria Concetta Cuomo Denise Di Concilio Lucio Di Filippo Anna Manfredi Marcello Salvi Antonio Limone Luigi Atripaldi Pellegrino Cerino Andrea Ballabio Davide Cacchiarelli |
| EPI_ISL_2464257, EPI_ISL_2464260, EPI_ISL_2532367, EPI_ISL_2535296, EPI_ISL_2535450, EPI_ISL_2535453, EPI_ISL_2535459 | | | |
| see above | Labo Analyses Med | National Reference Center for Viruses of Respiratory Infections, Institut Pasteur, Paris | Amaury Vaysse; Angela Brisebarre; Camille Capel; Christophe Malabat; Claire Felloni; Corinne Maufrais; Domitille Leman; Etienne Simon-Lorière; Frédéric Lemoine; Hervé Dupont; Louise Lefrançois; Marion Barbet; Maud Vanpeene; Méline Bizard; OphéLie Said-Delattre; Sylvie Behillil; Sylvie Van der Werf; Victoire Baillet; Vincent Enouf |
| EPI_ISL_2845699, EPI_ISL_2845700, EPI_ISL_2845710, EPI_ISL_2845717 | Labor Becker & Kollegen (Standort München) | Robert Koch Institute | |
| EPI_ISL_2689539 | Labor Dr. Fenner und Kollegen | Heinrich Pette Institute, Leibniz Institute for Experimental Virology | Adam Grundhoff; Alexis Robitaille; Johannes Knobloch; Martin Aepfelbacher; Nicole Fischer; Thomas Günther |
| EPI_ISL_2426981, EPI_ISL_2627850, EPI_ISL_2627851, EPI_ISL_2629225, EPI_ISL_2629227 | Laboratoire Carage | Institut Pasteur de la Guyane | Anne Lavergne; Antoine Enfissi; Arielle Salmier; Dominique Rousset |
| EPI_ISL_3458779, EPI_ISL_3458780, EPI_ISL_3458796, EPI_ISL_3458798, EPI_ISL_5867806, EPI_ISL_5867807, EPI_ISL_5867808 | | | |
| see above | Laboratoire de santé publique du Québec | Laboratoire de santé publique du Québec | Guillaume Bourque; Ioannis Ragoussis; Jesse Shapiro; Mark Lathrop and Michel Roger on behalf of the CoVSeQ research group; Sandrine Moreira |
| EPI_ISL_3147251 | Laboratoires Reunis | Laboratoire national de sante, Microbiology, Microbial Genomics Platform | Anke Wienecke-Baldacchino; Bernard Weber; Catherine Ragimbeau; Elodie Solarino; Fatu Djabi; Jessica Tapp; Lise Pignon; Raoul Salmon; Tamir Abdelrahman; Virginie Jover |
| EPI_ISL_3147801, EPI_ISL_3147804, EPI_ISL_3147820, EPI_ISL_3148065, EPI_ISL_3148150 | Laboratoires d'analyses medicales - Ketterthill | Laboratoire national de sante, Microbiology, Microbial Genomics Platform | Anke Wienecke-Baldacchino; Caroline Scheiber; Catherine Ragimbeau; Elodie Solarino; Fatu Djabi; Jessica Tapp; Lise Pignon; Raoul Salmon; Serge Vedy; Tamir Abdelrahman; Virginie Jover |
| EPI_ISL_2983181, EPI_ISL_3190357 | Laboratorio Central de Saude Publica do Estado de Goias (LACEN/GO) | Laboratory of Respiratory Viruses and Measles, Oswaldo Cruz Institute, FIOCRUZ | Agatha Cristinne Prudencio; Alice Sampaio Rocha; Ana Carolina Mendonca; Ana Flavia Mendonça; Anna Carolina Paixao; Carmen Helena Ramos; Cassiane Casanova; Elisa Cavalcante Pereira; Fernando Motta; Flavia Pereira Amorim da Silva; Ighor Leonardo Arantes Gomes; Luciana Appolinario; Luiz Augusto Pereira; Marilda Siqueira on behalf of the Fiocruz COVID-19 Genomic Surveillance Network; Paola Resende; Rafael Souza Guedes; Renata Serrano Lopes; Taina Venas; Vinicius Lemes da Silva |
| EPI_ISL_2645434, EPI_ISL_2645435 | Laboratorio Central de Saude Publica do Estado do Para (LACEN/PA) | Laboratory of Respiratory Viruses and Measles, Oswaldo Cruz Institute, FIOCRUZ | Alice Sampaio Rocha; Ana Carolina Mendonca; Anna Carolina Paixao; Elisa Cavalcante Pereira; Fernando Motta; Luciana Appolinario; Marilda Siqueira on behalf of the Fiocruz COVID-19 Genomic Surveillance Network; Paola Resende; Renata Serrano Lopes; Taina Venas; Valnete Andrade |
| EPI_ISL_4946234 | Laboratorio Angel ( Synlab) | Instituto Nacional de Salud- Dirección de Investigación en Salud Pública | Carlos Franco-Muñoz; Carmen Osorio; Diana Malo; Diego A. Álvarez-Díaz; Diego Andrés Prada; Gerardo Santamaría; Hector Alejandro Ruiz-Moreno; Jhonnatan Reales-González; Jorge Rivera; Juan Camilo Martinez; Julian Naizaque; Katherine Laiton-Donato; Lisseth Pardo; Magdalena Wiesner; Marcela Mercado-Reyes; Maria T. Herrera-Sepúlveda; Marta Lopez Blanco; Martha Lucia Ospina Martinez; Paola Rojas; Sergio Gomez; Sheryll Corchuelo; Ángela Alarcon Cruz |
| EPI_ISL_2535087, EPI_ISL_2535088, EPI_ISL_2535108, EPI_ISL_2535109, EPI_ISL_2535110, EPI_ISL_2691315, EPI_ISL_2691316, EPI_ISL_2691317, EPI_ISL_2691318, EPI_ISL_2691319, EPI_ISL_2691320, EPI_ISL_2691321, EPI_ISL_2691322, EPI_ISL_2691323, EPI_ISL_2691324, EPI_ISL_2691325, EPI_ISL_2691326, EPI_ISL_2691327, EPI_ISL_2691328, EPI_ISL_2691331, EPI_ISL_2691332, EPI_ISL_2691333, EPI_ISL_2691334, EPI_ISL_2691335, EPI_ISL_2691336, EPI_ISL_2691337, EPI_ISL_2691338, EPI_ISL_2691339, EPI_ISL_2691340, EPI_ISL_2691341, EPI_ISL_2691342, EPI_ISL_2691343, EPI_ISL_2691344, EPI_ISL_2691345, EPI_ISL_2691346, EPI_ISL_2691347, EPI_ISL_2691348, EPI_ISL_2691349, EPI_ISL_2691350, EPI_ISL_2691351, EPI_ISL_2691352, EPI_ISL_2691357, EPI_ISL_2691358, EPI_ISL_2691359, EPI_ISL_2691360, EPI_ISL_2691361, EPI_ISL_2691362, EPI_ISL_2691363, EPI_ISL_2691364, EPI_ISL_2691365, EPI_ISL_2691366, EPI_ISL_2691367, EPI_ISL_2691368, EPI_ISL_2691369, EPI_ISL_2691370, EPI_ISL_2691371, EPI_ISL_2691372, EPI_ISL_2691374, EPI_ISL_2691375, EPI_ISL_2691376, EPI_ISL_2691377, EPI_ISL_2691378, EPI_ISL_2691380, EPI_ISL_2691381, EPI_ISL_2691382, EPI_ISL_2691383, EPI_ISL_2691384, EPI_ISL_2691385, EPI_ISL_2691386, EPI_ISL_2691387, EPI_ISL_2691388, EPI_ISL_2691389, EPI_ISL_2691390, EPI_ISL_2691391, EPI_ISL_2691392, EPI_ISL_2691393, EPI_ISL_2691394, EPI_ISL_2691395, EPI_ISL_2691396, EPI_ISL_2691397, EPI_ISL_2691398, EPI_ISL_2691399, EPI_ISL_2691400, EPI_ISL_2691401, EPI_ISL_2691402, EPI_ISL_2691403, EPI_ISL_2691404, EPI_ISL_2691405, | | | |

| | | | |
|---|---|---|---|
| EPI_ISL_2691406, EPI_ISL_2691407, EPI_ISL_2691408, EPI_ISL_2691409, EPI_ISL_2691410, EPI_ISL_2691411, EPI_ISL_2691412, EPI_ISL_2691413, EPI_ISL_2691414, EPI_ISL_2691415, EPI_ISL_2691416, EPI_ISL_2691417, EPI_ISL_2691418, EPI_ISL_2691419, EPI_ISL_2691420, EPI_ISL_2691421, EPI_ISL_2691422, EPI_ISL_2691423, EPI_ISL_2691424, EPI_ISL_2691425, EPI_ISL_2691426, EPI_ISL_2691427, EPI_ISL_2691428, EPI_ISL_2691429, EPI_ISL_2691430, EPI_ISL_2691431, EPI_ISL_2691432, EPI_ISL_2691433, EPI_ISL_2691434, EPI_ISL_2691435, EPI_ISL_2691436, EPI_ISL_2691437, EPI_ISL_2691438, EPI_ISL_2691439, EPI_ISL_2691441, EPI_ISL_2691442, EPI_ISL_2691443, EPI_ISL_2691444, EPI_ISL_2691445, EPI_ISL_2691446, EPI_ISL_2691447, EPI_ISL_2691448, EPI_ISL_2691449, EPI_ISL_2691450, EPI_ISL_2691451, EPI_ISL_2691452, EPI_ISL_2691453, EPI_ISL_2691454, EPI_ISL_2691455, EPI_ISL_2691456, EPI_ISL_2691457, EPI_ISL_2691458, EPI_ISL_2691459, EPI_ISL_2691460, EPI_ISL_2691461, EPI_ISL_2691462, EPI_ISL_2691463, EPI_ISL_2691464, EPI_ISL_2691465, EPI_ISL_2691467, EPI_ISL_2691468, EPI_ISL_2691469, EPI_ISL_2691470, EPI_ISL_2691471, EPI_ISL_2691472, EPI_ISL_2691473, EPI_ISL_2691474, EPI_ISL_2691475, EPI_ISL_2691476, EPI_ISL_2691477, EPI_ISL_2691478, EPI_ISL_2691479, EPI_ISL_2691480, EPI_ISL_2691481, EPI_ISL_2691482, EPI_ISL_2691483, EPI_ISL_2691484, EPI_ISL_2691487, EPI_ISL_2691488, EPI_ISL_2691489, EPI_ISL_2691490, EPI_ISL_2691491, EPI_ISL_2691492, EPI_ISL_2691493, EPI_ISL_2691494, EPI_ISL_2691495, EPI_ISL_2691496, EPI_ISL_2691545, EPI_ISL_2691564, EPI_ISL_2691565, EPI_ISL_2691566, EPI_ISL_2691567 | | | |
| see above | Laboratorio Central Noel Nutels | Bioinformatics Laboratory / LNCC | Alessandra P Lamarca; Alexandra L Gerber; Amilcar Tanuri; Ana Paula de C Guimaraes; Ana Tereza R Vasconcelos; Andrea Cony Cavalcanti; Caio Luiz Pereira Ribeiro; Cassia Alves; Cintia Policarpo; Claudia Maria Braga de Mello; Cristiane Gomes da Silva; Diana Mariani; Douglas Terra Machado; Flavio Dias da Silva; Gleidson da Silva de Oliveira; Leandro Magalhaes de Souza; Liliane Cavalcante; Luiz G P de Almeida; Marcio Henrique de Oliveira Garcia; Mario Sergio Ribeiro; Ronaldo da Silva F Jr; Silvia Carvalho |
| EPI_ISL_2681076, EPI_ISL_2681093, EPI_ISL_2681112, EPI_ISL_2681125, EPI_ISL_2681129, EPI_ISL_2681139, EPI_ISL_2681144, EPI_ISL_2681146, EPI_ISL_2681154, EPI_ISL_2681157, EPI_ISL_2681158, EPI_ISL_2681163, EPI_ISL_2681166, EPI_ISL_2681172, EPI_ISL_2681187, EPI_ISL_2681192, EPI_ISL_2681204, EPI_ISL_2681210, EPI_ISL_2681230, EPI_ISL_2681240, EPI_ISL_2681245, EPI_ISL_2681254, EPI_ISL_2681258, EPI_ISL_2681269, EPI_ISL_2681284, EPI_ISL_2681297, EPI_ISL_2681314, EPI_ISL_2681337, EPI_ISL_2801762, EPI_ISL_2801763, EPI_ISL_2801764, EPI_ISL_2801768, EPI_ISL_2801769 | | | |
| see above | Laboratorio Central de Epidemiología (LCE) | Instituto de Biotecnología de la UNAM | ; Alejandra García-Gasca; Alejandra Hernández-Terán; Alejandro Sánchez-Flores; Alfredo Herrera-Estrella; Alicia Ocaña-Mondragón; Andreu Comas-García; Angel Gustavo Salas-Lais; Antonio Loza Román; Bernardo Martínez-Miguel; Blanca Taboada; Brenda Irasema Maldonado-Meza; Bruno Gómez-Gil; Carla Ivón Herrera-Najera; Carlos F. Arias; Celia Boukadida; Clara Esperanza Santacruz-Tinoco; Concepción Grajales-Muñiz; Consorcio Mexicano de Vigilancia Genómica (CoViGen-Mex). Authors (in alphabetical order): Julio Elias Alvarado-Yaah; Cristóbal Cháidez-Quiróz; Célida Duque Molina; Célida Martínez- Rodríguez; Daniel Fregoso-Rueda; Daniel Lira Morales; Eduardo Becerril-Vargas; Fernando Fontove-Herrera; Fidencio Mejía-Nepomuceno; Francisco Pulido; Gloria Elena Espinosa-Ayala; Gloria María Molina-Salinas; Gloria Vazquez; Hector Esteban Paz-Juárez; Hector Montoya-Fuentes; Helen Haydee Fernanda Ramírez-Plascencia; Irvin González-López; Jean Pierre González; Jesús Hernández; Joel Armando Vázquez-Pérez.; Jorge Salas-Hernández; José Antonio Enciso-Moreno; José Arturo Martínez-Orozco; José Esteban Muñoz-Medina; José de Jesús Nuñez-Contreras; Juan Bautista Chale-Dzul; Julissa Enciso-Ibarra; Luis Alberto Ochoa-Carrera; Margarita Matías-Florentino; Mario Mújica-Sánchez; Marissa Perez-Garcia; María Guadalupe Santiago-Mauricio; María Guadalupe de Jesús Mireles-Rivera; Nelly Sélem-Mojica; Pavel Isa; Ricardo Ciria Merce; Ricardo Grande; Rosa María Gutiérrez Rios; Santiago Ávila-Ríos; Selene Zárate; Susana Lopez; Verónica Mata-Haro; Victor Eduardo García-Arias; Víctor Hugo Borja-Aburto |
| EPI_ISL_6944497, EPI_ISL_6944498, EPI_ISL_6944509, EPI_ISL_6944525, EPI_ISL_6944545 | Laboratorio Central de Salud Publica | Viral Special Pathogens Branch, Centers for Disease Control and Prevention | Andrea Gómez de la Fuente; Cynthia Vázquez; Emir Talundzic; Joel Montgomery; John Klena; Juan Torales; Justin Lee; María José Ortega; María Liz Gamarra; Shannon Whitmer; Shirley Villalba |
| EPI_ISL_4030305, EPI_ISL_4030311, EPI_ISL_4030320 | Laboratorio Central de Saude Publica do Amazonas - LACEN-AM | Laboratorio de Ecologia de Doencas Transmissiveis na Amazonia, Instituto Leonidas e Maria Deane - Fiocruz Amazonia | André Corado; Debora Duarte; Felipe Naveca; Fernanda Nascimento; George Silva; Karina Pessoa; Luciana Gonçalves; Maria Júlia Brandão; Matilde Mejía; Michele Jesus; Valdinete Nascimento; Victor Souza; Ágatha Costa |
| EPI_ISL_3434726, EPI_ISL_3434762, EPI_ISL_3434763, EPI_ISL_3434764, EPI_ISL_3434765, EPI_ISL_3434767, EPI_ISL_3434768, EPI_ISL_3434775, EPI_ISL_3434777, EPI_ISL_3539936 | | | |
| see above | Laboratorio Central de Saude Publica do Estado da Paraiba (LACEN/PB) | Laboratory of Respiratory Viruses and Measles, Oswaldo Cruz Institute, FIOCRUZ | Alice Sampaio Rocha; Ana Carolina Mendonca; Anna Carolina Paixao; Dalane Loudal Florentino Teixeira; Elisa Cavalcante Pereira; Fernando Motta; Joao Felipe Bezerra; Luciana Appolinario; Marilda Siqueira on behalf of the Fiocruz COVID-19 Genomic Surveillance Network; Paola Resende; Renata Serrano Lopes; Taina Venas |
| EPI_ISL_2645404, EPI_ISL_2983064, EPI_ISL_2983065, EPI_ISL_2983070, EPI_ISL_2983338, EPI_ISL_2983453, EPI_ISL_2983454, EPI_ISL_2983478 | | | |
| see above | Laboratorio Central de Saude Publica do Estado de Alagoas (LACEN/AL) | Laboratory of Respiratory Viruses and Measles, Oswaldo Cruz Institute, FIOCRUZ | Agatha Cristinne Prudencio; Alice Sampaio Rocha; Ana Carolina Mendonca; Anderson Brandao Leite; Anna Carolina Paixao; Elisa Cavalcante Pereira; Fernando Motta; Ighor Leonardo Arantes Gomes; Luciana Appolinario; Marilda Siqueira on behalf of the Fiocruz COVID-19 Genomic Surveillance Network; Paola Resende; Renata Serrano Lopes; Taina Venas |
| EPI_ISL_2863647, EPI_ISL_2863648, EPI_ISL_2863651, EPI_ISL_2863653, EPI_ISL_2863655, EPI_ISL_2863658, EPI_ISL_2863660, EPI_ISL_2863662, EPI_ISL_2863664, EPI_ISL_2863666, EPI_ISL_2863712 | | | |
| see above | Laboratorio Central de Saude Publica do Estado de Santa Catarina (LACEN-SC) | Laboratory of Respiratory Viruses and Measles, Oswaldo Cruz Institute, FIOCRUZ | Alice Sampaio Rocha; Ana Carolina Mendonca; Anna Carolina Paixao; Darcita Buerger Rovaris; Elisa Cavalcante Pereira; Fernando Motta; Luciana Appolinario; Marilda Siqueira on behalf of the Fiocruz COVID-19 Genomic Surveillance Network; Paola Resende; Renata Serrano Lopes; Sandra Bianchini Fernandes; Taina Venas |
| EPI_ISL_3190167 | Laboratorio Central de Saude Publica do Estado de Santa Catarina (LACEN/SC) | Laboratory of Respiratory Viruses and Measles, Oswaldo Cruz Institute, FIOCRUZ | Alice Sampaio Rocha; Ana Carolina Mendonca; Anna Carolina Paixao; Darcita Buerger Rovaris; Elisa Cavalcante Pereira; Fernando Motta; Luciana Appolinario; Marilda Siqueira on behalf of the Fiocruz COVID-19 Genomic Surveillance Network; Paola Resende; Renata Serrano Lopes; Sandra Bianchini Fernandes; Taina Venas |
| EPI_ISL_2863573, EPI_ISL_2863575, EPI_ISL_2863621, EPI_ISL_2863623, EPI_ISL_2863626, EPI_ISL_2863630, EPI_ISL_2863634, EPI_ISL_2863637, EPI_ISL_2863643 | | | |
| see above | Laboratorio Central de Saude Publica do Estado de Sergipe (LACEN/SE) | Laboratory of Respiratory Viruses and Measles, Oswaldo Cruz Institute, FIOCRUZ | Alice Sampaio Rocha; Ana Carolina Mendonca; Anna Carolina Paixao; Cliomar Alves dos Santos; Elisa Cavalcante Pereira; Fernando Motta; Luciana Appolinario; Marilda Siqueira on behalf of the Fiocruz COVID-19 Genomic Surveillance Network; Paola Resende; Renata Serrano Lopes; Tainá Moreira Martins Venas |
| EPI_ISL_2375476, EPI_ISL_2375477, EPI_ISL_3045370 | Laboratorio Central de Saude Publica do Estado do Espirito Santo (LACEN/ES) | Laboratory of Respiratory Viruses and Measles, Oswaldo Cruz Institute, FIOCRUZ | Alice Sampaio Rocha; Ana Carolina Mendonca; Anna Carolina Paixao; Eilisa Cavalcante Pereira; Elisa Cavalcante Pereira; Fernando Motta; Luciana Appolinario; Marilda Siqueira on behalf of the Fiocruz COVID-19 Genomic Surveillance Network; Paola Resende; Renata Serrano Lopes; Rodrigo Ribeiro Rodrigues; Taina Venas |
| EPI_ISL_2645445, EPI_ISL_2645447, EPI_ISL_2645449, EPI_ISL_2645450, EPI_ISL_2645451, EPI_ISL_2645454, EPI_ISL_2645457, EPI_ISL_2645460, EPI_ISL_2645462, EPI_ISL_2645464, EPI_ISL_2645467, EPI_ISL_2645468, EPI_ISL_2645469, EPI_ISL_2645473, EPI_ISL_2645475, EPI_ISL_2645477, EPI_ISL_2645479, EPI_ISL_2645480, EPI_ISL_2645481, EPI_ISL_2645493, EPI_ISL_2645502, EPI_ISL_2983428 | | | |
| see above | Laboratorio Central de Saude Publica do Estado do Parana (LACEN/PR) | Laboratory of Respiratory Viruses and Measles, Oswaldo Cruz Institute, FIOCRUZ | Alice Sampaio Rocha; Ana Carolina Mendonca; Anna Carolina Paixao; Elisa Cavalcante Pereira; Fernando Motta; Irina Riediger; Luciana Appolinario; Marilda Siqueira on behalf of the Fiocruz COVID-19 Genomic Surveillance Network; Paola Resende; Renata Serrano Lopes; Taina Venas |
| EPI_ISL_2603441, EPI_ISL_2603529, EPI_ISL_2603530, EPI_ISL_2603531, EPI_ISL_2603532, EPI_ISL_2603533, EPI_ISL_2603534, EPI_ISL_2982762, EPI_ISL_2982776, EPI_ISL_2982795, EPI_ISL_2982798, EPI_ISL_2982799, EPI_ISL_2982801, EPI_ISL_2982803, EPI_ISL_2982805, EPI_ISL_2982807, EPI_ISL_2982809, EPI_ISL_2982810, EPI_ISL_2982811, EPI_ISL_2982812, EPI_ISL_2982813 | | | |
| see above | Laboratorio Central de Saude Publica do Estado do Rio Grande do Sul (LACEN-RS) | Laboratory of Respiratory Viruses and Measles, Oswaldo Cruz Institute, FIOCRUZ | Alice Sampaio Rocha; Ana Carolina Mendonca; Anderson Brandao Leite; Anna Carolina Paixao; Elisa Cavalcante Pereira; Fernando Motta; Luciana Appolinario; Marilda Siqueira on behalf of the Fiocruz COVID-19 Genomic Surveillance Network; Paola Resende; Renata Serrano Lopes; Taina Venas |
| EPI_ISL_2863584, EPI_ISL_2863585, EPI_ISL_2863586, EPI_ISL_2863587, EPI_ISL_2863588 | Laboratorio Central de Saude Publica do Estado do Rio de Janeiro (LACEN-RJ) | Laboratory of Respiratory Viruses and Measles, Oswaldo Cruz Institute, FIOCRUZ | Agatha Cristinne Prudencio; Alice Sampaio Rocha; Ana Carolina Mendonca; Andrea Cony Cavalcanti; Anna Carolina Paixao; Elisa Cavalcante Pereira; Fernando Motta; Ighor Leonardo Arantes Gomes; Luciana Appolinario; Marilda Siqueira on behalf of the Fiocruz COVID-19 Genomic Surveillance Network; Paola Resende; Renata Serrano Lopes; Taina Venas |
| EPI_ISL_3033064, EPI_ISL_3033065, EPI_ISL_3033067, EPI_ISL_3033068, EPI_ISL_3033071, EPI_ISL_3033072 | Laboratorio Central de la Ciudad de Santa Fe | Grupo de Genómica y Bioinformática del Instituto de Investigación de la Cadena Láctea CONICET-INTA on behalf of 'Proyecto Argentino Interinstitucional de genomica de SARS-CoV-2' (PAIS Consortium) | AF; Amadio; C; Eberhardt; G; Irazoqui; JM; MF; Mugna; Ojeda; Pastor; Rompato; V |
| EPI_ISL_4878296 | Laboratorio Direccion Sanidad Policia Nacional | Instituto Nacional de Salud- Dirección de Investigación en Salud Pública | Carlos Franco-Muñoz; Carmen Osorio; Diana Malo; Diego A. Álvarez-Díaz; Diego Andrés Prada; Gerardo Santamaría; Hector Alejandro Ruiz-Moreno; Jhonnatan Reales-González; Jorge Rivera; Juan Camilo Martinez; Julian Naizaque; Katherine Laiton-Donato; Lisseth Pardo; Magdalena Wiesner; Marcela Mercado-Reyes; Maria T. Herrera-Sepúlveda; Marta Lopez Blanco; Martha Lucia Ospina Martinez; Paola Rojas; Sergio Gomez; Sheryll Corchuelo; Ángela Alarcon Cruz |
| EPI_ISL_5199342 | Laboratorio SYNLAB Colombia | Instituto Nacional de Salud- Dirección de Investigación en Salud Pública | Beatriz de Arco; Carlos Franco-Muñoz; Carmen Osorio; Diana Malo; Diego A. Álvarez-Díaz; Diego Andrés Prada; Gerardo Santamaría; Hector Alejandro Ruiz-Moreno; Jhonnatan Reales-González; Jorge Rivera; Juan Camilo Martinez; Julian Naizaque; Katherine Laiton-Donato; Lisseth Pardo; Magdalena Wiesner; Marcela Mercado-Reyes; Maria T. Herrera-Sepúlveda; Marta Lopez Blanco; Martha Lucia Ospina Martinez; Paola Rojas; Sergio Gomez; Sheryll Corchuelo; Tatiana Cobos; Ángela Alarcon Cruz |
| EPI_ISL_2777446, EPI_ISL_2777447, EPI_ISL_2777448, EPI_ISL_2777449, EPI_ISL_2777450, EPI_ISL_2777451, EPI_ISL_2777452, EPI_ISL_2777453, EPI_ISL_4030329, EPI_ISL_4030332, EPI_ISL_4030350, EPI_ISL_4030353, EPI_ISL_4030356, EPI_ISL_4030367 | | | |
| see above | Laboratorio de Ecologia de Doencas Transmissiveis na Amazonia, Instituto Leonidas e Maria Deane - Fiocruz Amazonia | Laboratorio de Ecologia de Doencas Transmissiveis na Amazonia, Instituto Leonidas e Maria Deane - Fiocruz Amazonia | Alex Martins; André Corado; Debora Duarte; Fabiola Mendonça da Silva Chui; Felipe Naveca; Fernanda Nascimento; Fernando Fonseca de Almeida Val; George Silva; Gisely Cardoso de Melo; Karina Pessoa; Luciana Gonçalves; Marcus Vinicius Guimarães de Lacerda; Maria Júlia Brandão; Maria Paula Gomes Mourão; Mariana Xavier Simão; Matilde Mejía; Michele Jesus; Valdinete Nascimento; Vanderson de Souza Sampaio; Victor Souza; Ágatha Costa |
| EPI_ISL_2728574, EPI_ISL_2728575, EPI_ISL_2728576, EPI_ISL_2728578, EPI_ISL_2728579, EPI_ISL_3321117, EPI_ISL_3321276 | | | |
| see above | Laboratorio de Infectologia y Virologia Molecular | Laboratory of Molecular Virology, School of Medicine, Pontificia Universidad Catolica de Chile | Alejandro Bhrun; Ana Maria Contreras; Andres E. Munoz-Marcos; Carlos Palma; Catalina Pardo-Roa; Constanza Maldonado; Constanza Martinez-Valdevenito; Eileen Serrano; Erick Salinas; Estefany Poblete; Francisco Melo; Jennifer Angulo; Jorge Levican; Leonardo I. Almonacid; M. Belen Leyton; Magdalena Vera; Marcela Ferres; Maria Jose Avendano; Rafael A. Medina; Tamara Garcia-Salum |
| EPI_ISL_3134273, EPI_ISL_3134275 | Laboratorio de Medicina Molecular, Universidad de Magallanes | Centro Asistencial Docente y de Investigacion, Universidad de Magallanes | Carolina Pérez; Constanza Ceroni; Diego Alvarez; Inés Cid; Jacqueline Aldridge; Jorge González; Juan Ríos; Marcelo Navarrete |
| EPI_ISL_3048979, EPI_ISL_3048980, EPI_ISL_3761520, EPI_ISL_3761617, EPI_ISL_3761637, EPI_ISL_3761638, EPI_ISL_3761639, EPI_ISL_3761640, EPI_ISL_3761641, EPI_ISL_3761642, EPI_ISL_3761643, EPI_ISL_3761644 | | | |
| see above | Laboratorio de Pesquisa em Virologia, FAMERP, SJRP | Laboratorio de Pesquisa em Virologia, FAMERP, SJRP | Beatriz de Carvalho Marques; Cecília Artico Banho; Cíntia Bittar; Fábio Sossai Possebon; Guilherme Campos; Helena Lage Ferreira; Jorge A. Petroli Marchesi; João Pessoa Araújo Jr.; Leila Sabrina Ullmann; Lívia Sacchetto; Maisa C. Pereira Parra; Marília Moraes; Maurício L. Nogueira.; Paula Rahal; Paulo Inacio da Costa |
| EPI_ISL_3672436, EPI_ISL_3826440, EPI_ISL_3991452, EPI_ISL_4052704, EPI_ISL_4052707, EPI_ISL_4052727, EPI_ISL_4052755, EPI_ISL_4080451, EPI_ISL_4080452 | | | |
| see above | Laboratorio de Referencia Nacional de Virus Respiratorios. Centro Nacional de Salud Publica. Instituto Nacional de Salud Peru. | Laboratorio de Referencia Nacional de Virus Respiratorios. Centro Nacional de Salud Publica. Instituto Nacional de Salud Peru. | Carlos Padilla Rojas; Henri Bailon Calderon; Iris Silva Molina; Joseph Huayra Niquen; Lely Solari Zerpa; Luis Barcena Flores; Marco Galarza Perez; Nancy Rojas Serrano; Nieves Sevilla Castañeda; Omar Caceres Rey; Orson Mestanza Millones; Princesa Medrano Alhuay; Priscila Lope Pari; Sandra Morales Ruiz; Sara Gordillo Vilchez; Steve Acedo Lazo; Veronica Hurtado Vela; Victor Jimenez Vasquez; Wendy Lizarraga Olivares |
| EPI_ISL_2608291, EPI_ISL_2812650 | Laboratorio de Virología HUCA | Laboratorio de Virología HUCA | Abreu F; Alvarez-Arguelles ME; Boga JA; Castelló C; Costales I; Coto E; Gómez de Oña J; Martín-Rodríguez G; Melón S; Perez-Martínez Z; Rojo S; Sandoval M |
| EPI_ISL_4405112, EPI_ISL_4405116, EPI_ISL_4405132 | Laboratorio de Virología del Hospital de Niños Dr. Ricardo Gutierrez | Área de Secuenciación del Laboratorio de Virología del Hospital de Niños Dr. Ricardo Gutierrez on behalf of 'Proyecto Argentino | AS; Acevedo; Acuña; Alexay; Alvarez Lopez; C; D; E; Goya; Gravis; Jacques; LE; Lusso; ME; MI; MS; Mistchenko; Nabaes Jodar; Natale; O; S; Valinotto; Viegas, M. |

| | | | |
|---|---|---|---|
| | | Interinstitucional de genomica de SARS-CoV-2' (PAIS Consortium) | |
| EPI_ISL_4405563 | Laboratorio del Hospital Interzonal General de Agudos " Evita " | Área de Secuenciación del Laboratorio de Virología del Hospital de Niños Dr. Ricardo Gutierrez on behalf of 'Proyecto Argentino Interinstitucional de genomica de SARS-CoV-2' (PAIS Consortium) | A; Acuña; D; Desimone; E; Goya; Grossi; I; L; LE; Luczac; Lusso; MI; MS; Musto; Nabaes Jodar; Natale; O; S; Serrano; Valinotto; Viegas, M. |
| EPI_ISL_5771884 | Laboratorio del Hospital Regional Ushuaia Gdor. Ernesto Campos | Nodo de Secuenciación Tierra del Fuego - Hospital Regional Ushuaia - Centro Austral De Investigaciones Científicas - Universidad Nacional De Tierra Del Fuego | AE; Boutureira, MF.; CA; CB; CF; Castro; Ceballos; Cáceres; De Roccis; F; G; Gallego; Gramundi; ID; Nardi; Rojas; SB; SG; Yulan |
| EPI_ISL_2399376, EPI_ISL_2399377, EPI_ISL_2399380, EPI_ISL_2399399, EPI_ISL_2399403, EPI_ISL_2480218, EPI_ISL_2480220, EPI_ISL_2480232, EPI_ISL_2480233, EPI_ISL_2480238, EPI_ISL_2480251, EPI_ISL_2480262, EPI_ISL_2480272, EPI_ISL_2480274, EPI_ISL_2480277, EPI_ISL_2480312, EPI_ISL_2480363, EPI_ISL_2480366, EPI_ISL_2480368, EPI_ISL_2480378, EPI_ISL_2480380, EPI_ISL_2480382, EPI_ISL_2480402, EPI_ISL_2480404, EPI_ISL_2480405, EPI_ISL_2480418, EPI_ISL_2480509, EPI_ISL_2480531, EPI_ISL_2480858, EPI_ISL_2480859, EPI_ISL_2480860, EPI_ISL_2480861, EPI_ISL_2480922, EPI_ISL_2480923, EPI_ISL_2480924, EPI_ISL_2480935, EPI_ISL_2480936, EPI_ISL_2480944, EPI_ISL_2480970, EPI_ISL_2480976, EPI_ISL_2481189, EPI_ISL_2481190, EPI_ISL_2481191, EPI_ISL_2481203, EPI_ISL_2481220, EPI_ISL_2481221, EPI_ISL_2481224, EPI_ISL_2481225, EPI_ISL_2481252, EPI_ISL_2481293, EPI_ISL_2481301, EPI_ISL_2481445, EPI_ISL_2481448, EPI_ISL_2481449, EPI_ISL_2481454, EPI_ISL_2481460, EPI_ISL_2481461, EPI_ISL_2481472, EPI_ISL_2481612, EPI_ISL_2481647, EPI_ISL_2481651, EPI_ISL_2481652, EPI_ISL_2481658, EPI_ISL_2481698, EPI_ISL_2481706, EPI_ISL_2481710, EPI_ISL_2481726, EPI_ISL_2481727, EPI_ISL_2481736, EPI_ISL_2481748, EPI_ISL_2481761, EPI_ISL_2481764, EPI_ISL_2481785, EPI_ISL_2481797, EPI_ISL_2481801, EPI_ISL_2481805, EPI_ISL_2481808, EPI_ISL_2481838, EPI_ISL_2481846, EPI_ISL_2481847, EPI_ISL_2481856, EPI_ISL_2481857, EPI_ISL_2481879, EPI_ISL_2481892, EPI_ISL_2481917, EPI_ISL_2481919, EPI_ISL_2481939, EPI_ISL_2481961, EPI_ISL_2481962, EPI_ISL_2481964, EPI_ISL_2481965, EPI_ISL_2481967, EPI_ISL_2481968, EPI_ISL_2481969, EPI_ISL_2481970, EPI_ISL_2481981, EPI_ISL_2481982, EPI_ISL_2481998, EPI_ISL_2482020, EPI_ISL_2482027, EPI_ISL_2482037, EPI_ISL_2482038, EPI_ISL_2482039, EPI_ISL_2482040, EPI_ISL_2482043, EPI_ISL_2482045, EPI_ISL_2482048, EPI_ISL_2482050, EPI_ISL_2482055, EPI_ISL_2482057, EPI_ISL_2482067, EPI_ISL_2482075, EPI_ISL_2482078, EPI_ISL_2482094, EPI_ISL_2482117, EPI_ISL_2482118, EPI_ISL_2482127, EPI_ISL_2482128, EPI_ISL_2482130, EPI_ISL_2482138, EPI_ISL_2482142, EPI_ISL_2482146, EPI_ISL_2482150, EPI_ISL_2482151, EPI_ISL_2482161, EPI_ISL_2482164, EPI_ISL_2482175, EPI_ISL_2482181, EPI_ISL_2482183, EPI_ISL_2482185, EPI_ISL_2482192, EPI_ISL_2482209, EPI_ISL_2482210, EPI_ISL_2482215, EPI_ISL_2482216, EPI_ISL_2482222, EPI_ISL_2482239, EPI_ISL_2611281, EPI_ISL_2611286, EPI_ISL_2611290, EPI_ISL_2611447, EPI_ISL_2611450, EPI_ISL_2611451, EPI_ISL_2611459, EPI_ISL_4374802, EPI_ISL_4375364, EPI_ISL_4375433, EPI_ISL_4375951, EPI_ISL_4376423, EPI_ISL_4377396, EPI_ISL_4379320, EPI_ISL_4379413, EPI_ISL_4380313, EPI_ISL_4381190 | | | |
| see above | Laboratory Corporation of America | Centers for Disease Control and Prevention Division of Viral Diseases, Pathogen Discovery | ; Adrian Paskey; Amanda Douglas; Amanda Suchanek; Andrea Throop; Ayla Burns; Benjamin Rambo-Martin; Bobbi Croy; Brian Krueger; Brian Norvell; Christopher Gulvick; Christos Petropoulos; Clinton Paden; Clinton R. Paden; Craig Lukasik; Dakota Howard; Darlene Wagner; Debbie Boles; Dhwani Batra; Duncan MacCannell; Eyad Almasri; Goran Stevovic; Howard Engler; Hrushikesh Deshmukh; Jake Humphrey; Jana Schroth; Jason Caravas; Joe Voshell; John Pruitt; Jonathan Meltzer; Jonathan Williams; Kara Moser; Kimberly Wagner; Kristine Lacek; Lax Iyer; Lisa Pfefferle; Lyndon Tilson; Manoj Jain; Marcia Eisenberg; Mary Ann Cristobal; Mary Cristobal; Mary Williamson; Matthew Robinson; Matthew Schmerer; Michael Levandoski; Mike Sapeta; Mindy Nye; Minoo Agarwal; Mohan Kolli; Nuthawin Charoensri; Oren Cohen; Peter Cook; Peter W. Cook; Prashant Gupta; Qian Zeng; Rama Ghatti; Scott Parker; Scott Ryan; Scott Sammons; Shatavia Morrison; Stanley Letovsky; Steven Ragan; Suresh Babu Selvaraju; Suresh Selvaraju; Susan Countryman; Susan Hicks; Suzanne Dale; Thomas Urban; Tim Kuphal; Tricia Zwiefelhofer; Tymeckia Kendall; Victoria Caban Figueroa; Vincent Drouillon; Yvette Unoarumhi |
| EPI_ISL_2501861 | Laboratory of Clinical Microbiology, Virology and Bioemergencies, ASST Fatebenefratelli Sacco - Sacco Hospital | Laboratory of Clinical Microbiology, Virology and Bioemergencies, ASST Fatebenefratelli Sacco - Sacco Hospital | Alberto Rizzo; Alessandro Mancon; Fiorenza Bracchitta; Maria Rita Gismondo; Valeria Micheli |
| EPI_ISL_2557405, EPI_ISL_2863569, EPI_ISL_2863605, EPI_ISL_2863607, EPI_ISL_2863609, EPI_ISL_2863611, EPI_ISL_2863613, EPI_ISL_2863614, EPI_ISL_2863616, EPI_ISL_4212881, EPI_ISL_5158278, EPI_ISL_5158280, EPI_ISL_5158284, EPI_ISL_6172758, EPI_ISL_6172759, EPI_ISL_7106013, EPI_ISL_7106044, EPI_ISL_7111361, EPI_ISL_7111372 | | | |
| see above | Laboratory of Respiratory Viruses and Measles, Oswaldo Cruz Institute, FIOCRUZ | Laboratory of Respiratory Viruses and Measles, Oswaldo Cruz Institute, FIOCRUZ | Agatha Cristinne Prudencio; Alice Sampaio Rocha; Ana Carolina Mendonca; Anna Carolina Paixao; Bruna Mendonça da Silva; Elisa Cavalcante Pereira; Fernando Motta; Ighor Arantes; Ighor Leonardo Arantes Gomes; Jéssica Graça Macedo de Carvalho; Larissa Macedo Pinto; Luciana Appolinario; Marilda Siqueira on behalf of the Fiocruz COVID-19 Genomic Surveillance Network; Paola Resende; Patricia Brasil; Renata Serrano Lopes; Taina Venas; Thais Aguiar; Victor Guimaraes; Vitor Guimaraes Ribeiro |
| EPI_ISL_2777293, EPI_ISL_2777341, EPI_ISL_2777342, EPI_ISL_2777343, EPI_ISL_2777355, EPI_ISL_2777512, EPI_ISL_2777564, EPI_ISL_2777590, EPI_ISL_2777592, EPI_ISL_2777598, EPI_ISL_2777602, EPI_ISL_2777607, EPI_ISL_2777608, EPI_ISL_2777611, EPI_ISL_2777613, EPI_ISL_2777616, EPI_ISL_2777644, EPI_ISL_2777656, EPI_ISL_2777658, EPI_ISL_2777661, EPI_ISL_2777665, EPI_ISL_2777669, EPI_ISL_2777673, EPI_ISL_2777677, EPI_ISL_2777680, EPI_ISL_2777681, EPI_ISL_2777684, EPI_ISL_2777685, EPI_ISL_2777688, EPI_ISL_2777690, EPI_ISL_2777692, EPI_ISL_2777694, EPI_ISL_2777695, EPI_ISL_2777712, EPI_ISL_2777806, EPI_ISL_2777810, EPI_ISL_2777814, EPI_ISL_2777831, EPI_ISL_2777863, EPI_ISL_2777865, EPI_ISL_2777866, EPI_ISL_2777895, EPI_ISL_2777896, EPI_ISL_2777897, EPI_ISL_2777910, EPI_ISL_2777942, EPI_ISL_2777953, EPI_ISL_2777967, EPI_ISL_3050295, EPI_ISL_3050296, EPI_ISL_3050297, EPI_ISL_3050298 | | | |
| see above | Laboratório Central de Saúde Pública do Amazonas - LACEN-AM | Laboratorio de Ecologia de Doencas Transmissiveis na Amazonia, Instituto Leonidas e Maria Deane - Fiocruz Amazonia | André Corado; Debora Duarte; Felipe Naveca; Fernanda Nascimento; George Silva; Karina Pessoa; Luciana Gonçalves; Maria Júlia Brandão; Matilde Mejía; Michele Jesus; Valdinete Nascimento; Victor Souza; Ágatha Costa |
| EPI_ISL_3584949, EPI_ISL_3585297, EPI_ISL_3585952, EPI_ISL_3586500, EPI_ISL_3587057, EPI_ISL_3587490, EPI_ISL_3598262, EPI_ISL_3602956, EPI_ISL_3602957, EPI_ISL_3602958, EPI_ISL_3602959, EPI_ISL_3602960, EPI_ISL_3602961, EPI_ISL_3602964, EPI_ISL_3610172, EPI_ISL_3610352, EPI_ISL_3610353 | | | |
| see above | Laboratório Central de Saúde Pública do Estado do Tocantins (LACEN/TO) | Bioinformatics and Biotechnology Laboratory (Labinftec/Federal University of Tocantins, Campus of Gurupi) | Bergmann Morais Ribeiro; Fabrício Souza Campos; Fernando Lucas Melo; Jucimária Dantas Galvão; Raíssa Nunes dos Santos; Ueric José Borges de Souza |
| EPI_ISL_2835216, EPI_ISL_2835217, EPI_ISL_2835218, EPI_ISL_2835219, EPI_ISL_2835220, EPI_ISL_2835221, EPI_ISL_2835222, EPI_ISL_2835223, EPI_ISL_2983455, EPI_ISL_2983456, EPI_ISL_2983457, EPI_ISL_2983458, EPI_ISL_2983459, EPI_ISL_2983460, EPI_ISL_2983461, EPI_ISL_2983462, EPI_ISL_2983463, EPI_ISL_2983464, EPI_ISL_2983465, EPI_ISL_2983466, EPI_ISL_2983467, EPI_ISL_2983468, EPI_ISL_2983469, EPI_ISL_4543261, EPI_ISL_4543262, EPI_ISL_4543286, EPI_ISL_4543338, EPI_ISL_4543342, EPI_ISL_4543348 | | | |
| see above | Laboratório de Biotecnologia Aplicada (LBA) - Laboratório de Biologia Molecular - Hospital das Clínicas, Faculdade de Medicina de Botucatu, Departamento de Bioprocessos e Biotecnologia - Faculdade de Ciências Agronômicas, UNESP – Botucatu/SP | Laboratory of Respiratory Viruses and Measles, Oswaldo Cruz Institute, FIOCRUZ | Alice Sampaio Rocha; Ana Carolina Mendonca; Anna Carolina Paixao; Elisa Cavalcante Pereira; Felipe Allan da Silva Costa; Fernando Motta; Jayme Augusto de Souza Neto; Leonardo Nazario de Moraes; Luciana Appolinario; Marilda Siqueira on behalf of the Fiocruz COVID-19 Genomic Surveillance Network; Paola Resende; Patricia Akemi Assato; Rejane Maria Tommasini; Renata Serrano Lopes; Taina Venas |
| EPI_ISL_4520574, EPI_ISL_4520575, EPI_ISL_4520594, EPI_ISL_4520595 | Laboratório de Virologia e Cultivo Celular - LABVICC | Laboratório Baculovírus | Aline Belmok; Bergmann Morais Ribeiro; Clayton Luiz Borges; Déborah Anjos; Elen Regozino Muniz; Fabiola Souza Fiaccadori; Fernando L Melo; Gabriela Silvério Bazilio; Juliana Alves Parente Rocha; Keila Correia de Alcântara; Menira Souza; Nathânia Dábilla; Valéria Christina de Rezende Feres |
| EPI_ISL_2557411, EPI_ISL_2557412, EPI_ISL_2557413 | Labortorio Central de Saude Publica do Estado de Santa Catarina (LACEN/SC) | Laboratory of Respiratory Viruses and Measles, Oswaldo Cruz Institute, FIOCRUZ | Alice Sampaio Rocha; Ana Carolina Mendonca; Anna Carolina Paixao; Darcita Buerger Rovaris; Elisa Cavalcante Pereira; Fernando Motta; Luciana Appolinario; Marilda Siqueira on behalf of the Fiocruz COVID-19 Genomic Surveillance Network; Paola Resende; Renata Serrano Lopes; Sandra Bianchini Fernandes; Taina Venas |
| EPI_ISL_2645505 | Labortorio Central de Saude Publica do Estado do Rio de Janeiro (LACEN/RJ) | Laboratory of Respiratory Viruses and Measles, Oswaldo Cruz Institute, FIOCRUZ | Alice Sampaio Rocha; Ana Carolina Mendonca; Andrea Cony Cavalcanti; Anna Carolina Paixao; Elisa Cavalcante Pereira; Fernando Motta; Luciana Appolinario; Marilda Siqueira on behalf of the Fiocruz COVID-19 Genomic Surveillance Network; Paola Resende; Renata Serrano Lopes; Taina Venas |
| EPI_ISL_2645508, EPI_ISL_2645511, EPI_ISL_2645512, EPI_ISL_2645513 | Labortorio Central de Saude Publica do Estado do Tocantins (LACEN/TO) | Laboratory of Respiratory Viruses and Measles, Oswaldo Cruz Institute, FIOCRUZ | Alice Sampaio Rocha; Ana Carolina Mendonca; Anna Carolina Paixao; Elisa Cavalcante Pereira; Fernando Motta; Jucimaria Dantas Galvao; Luciana Appolinario; Marilda Siqueira on behalf of the Fiocruz COVID-19 Genomic Surveillance Network; Paola Resende; Renata Serrano Lopes; Taina Venas |
| EPI_ISL_2613809, EPI_ISL_2613810, EPI_ISL_2613811, EPI_ISL_2613812, EPI_ISL_2613813, EPI_ISL_2613814, EPI_ISL_2613815 | | | |
| see above | Lboratorio Central de Saude Publica do Estado do Parana (LACEN/PR) | Laboratory of Respiratory Viruses and Measles, Oswaldo Cruz Institute, FIOCRUZ | Alice Sampaio Rocha; Ana Carolina Mendonca; Anna Carolina Paixao; Elisa Cavalcante Pereira; Fernando Motta; Irina Riediger; Luciana Appolinario; Marilda Siqueira on behalf of the Fiocruz COVID-19 Genomic Surveillance Network; Paola Resende; Renata Serrano Lopes; Taina Venas |
| EPI_ISL_2532777 | Lifebrain Covid Labor GmbH | Dept. of Laboratory Medicine | Anna Gschaider; Claudia Weber; Fabian Konig; Harald Esterbauer; Oswald Wagner; Petra Jurkowitsch; Robert Strassl; Sabina Plumer |
| EPI_ISL_2446852, EPI_ISL_2446920, EPI_ISL_2515298 | Lighthouse Lab in Alderley Park | Wellcome Sanger Institute for the COVID-19 Genomics UK (COG-UK) Consortium | Cordelia Langford; David K. Jackson; Dominic Kwiatkowski; Ewan Harrison; Ian Johnston; Jacquelyn Wynn; Jeffrey Barrett; John Sillitoe on behalf of the Wellcome Sanger Institute COVID-19 Surveillance Team; Mairead Hyland; Roberto Amato; Sonia Goncalves; The Lighthouse Lab in Alderley Park and Alex Alderton |
| EPI_ISL_2352978, EPI_ISL_2392333, EPI_ISL_2395953, EPI_ISL_2437005, EPI_ISL_2446442 | Lighthouse Lab in Milton Keynes | Wellcome Sanger Institute for the COVID-19 Genomics UK (COG-UK) Consortium | Cordelia Langford; David K. Jackson; Dominic Kwiatkowski; Ewan Harrison; Ian Johnston; Jeffrey Barrett; John Sillitoe on behalf of the Wellcome Sanger Institute COVID-19 Surveillance Team; Roberto Amato; Sonia Goncalves; The Lighthouse Lab in Milton Keynes and Alex Alderton |
| EPI_ISL_2390202 | Limbach - MVZ Labor Ravensburg Labor Dr. Gärtner | Robert Koch Institute | |
| EPI_ISL_3372610 | Lincoln Diagnostics | New York City Public Health Laboratory | Jade Wang; et al. |
| EPI_ISL_5778156 | Louisville Metro Public Health and Wellness Department/Louisville Metro Department of Corrections | University of Louisville Sequencing Technology Center | Elizabeth Hudson; Eric C. Rouchka; Julia H. Chariker; Leslie A. Wolf; Melissa L. Smith; William Lauer |
| EPI_ISL_2350819 | M.Sc Medical bioinformatics, Shri Ramachandra University | M.Sc Medical bioinformatics, Shri Ramachandra University | Thara, D. |
| EPI_ISL_6229817, EPI_ISL_6229848, EPI_ISL_6229867 | MATERNIDADE E HOSPITAL SANTA ISABEL | ACME Lab, Oswaldo Cruz Foundation, FIOCRUZ/CE | Carlos Leonardo de Aragao Araujo; Cecilia Leite Costa & Eduardo Ruback dos Santos on behalf of COVID-19 FIOCRUZ Genomic Network; Cleber Furtado Aksenen; Fabio Miyajima; Fernando Braga Stehling; Francisco Eder de Moura Lopes; Igor Oliveira Duarte; Jamille Maria Mendes Bezerra; Joaquim Cesar do Nascimento Sousa Junior; Pedro Miguel Carneiro Jeronimo; Suzana Porto Almeida; Thais Ferreira de Oliveira; Thais de Oliveira Costa; Ticiane Cavalcante de Souza; Veridiana Pessoa Miyajima |
| EPI_ISL_3356915, EPI_ISL_3356916, EPI_ISL_3356917 | MB-Cadham Provincial laboratory | Manitoba Cadham Provincial Laboratory | Anna Majer; Anneliese Landgraff; CanCOGeN's metadata curation team; Darian Hole; David Alexander; Elsie Grudeski; Gary Van Domselaar; Grace Seo; Jared Bullard; Jennifer Tanner; Kerry Dust; Kirsten Biggar; Madison Chapel; Morag Graham; Natalie Knox; Nathalie Bastien; Paul Van Caeseele; Philip Mabon; Public Health Agency of Canada CanCOGeN team; Rhiannon Huzarewich; Russell Mandes; Shari Tyson; Timothy Booth; Yan Li |
| EPI_ISL_4494278, EPI_ISL_4494286, | MB-Cadham Provincial laboratory | National Microbiology Laboratory (NML) | Anna Majer; Anneliese Landgraff; David Alexander; Elsie Grudeski; Gary Van Domselaar; Grace Seo; Jared Bullard; Jennifer Tanner; Kerry Dust; Morag Graham; Nat; Natalie Knox; Paul Van Caeseele; Philip Mabon; Rhiannon Huzarewich; Russell Mandes; Shari Tyson |

| | | | |
|---|---|---|---|
| EPI_ISL_4494292 | | | |
| EPI_ISL_2595748 | MEPHI, Aix Marseille University | MEPHI, Aix Marseille University | Anthony LEVASSEUR |
| EPI_ISL_6492777, EPI_ISL_6492778, EPI_ISL_6492779, EPI_ISL_6492782, EPI_ISL_6492783, EPI_ISL_6492786, EPI_ISL_6492790, EPI_ISL_6492791, EPI_ISL_6492792, EPI_ISL_6492793, EPI_ISL_6492796 | | | |
| see above | MSHS Clinical Microbiology Laboratories | MSHS Pathogen Surveillance Program | Adolfo García-Sastre; Adriana van de Guchte; Ajay Obla; Alberto Paniz-Mondolfi; Ana S. Gonzalez-Reiche; Angela Amoako; Ashley Salimbangon; Betsaida Salom Melo; Bremy Alburquerque; Brianne Ciferri; Charles Gleason; Daniel Floda; Deena R. Altman; Denise Jurczyszak; Emilia Mia Sordillo; Gintaras Deikus; Giulio Kleiner; Gopi Patel; Hala Alshammary; Harm van Bakel; Irina Oussenko; Jayeeta Dutta; Juan Soto; Julia Matthews; Katherine Beach; Kathryn Twyman; Kayla Russo; Komal Srivastava; Levy Sominsky; Mahmoud Awawda; Marta Luksza; Matthew M. Hernandez; Melissa Gitman; Michael D. Nowak; Mitchell J. Sullivan; Nancy Francoeur; Robert Sebra; Sarah Schaefer; Shelcie Fabre; Shwetha Hara Sridhar; Viviana Simon; Ying-Chih Wang; Zenab Khan |
| EPI_ISL_2690128 | MT Public Health Laboratory | Centers for Disease Control and Prevention Division of Viral Diseases, Pathogen Discovery | Alison Laufer Halpin; Ben L. Rambo-Martin; Clinton R. Paden; Dakota Howard; Darlene Wagner; Dave Wentworth; Dhwani Batra; Jasmine Padilla; Justin Lee; Katie Dillon; Krista Queen; Kristen Knipe; Kristine Lacek; Mark Burroughs; Matthew Schmerer; Mili Sheth; Peter Cook; Sam Shepard; Sarah Nobles; Shoshona Le; Suxiang Tong; Vivien Dugan; Yvette Unoarumhi |
| EPI_ISL_2471539 | MVZ Labor Dr. Fenner und Kollegen (Standort Hamburg) | Robert Koch Institute | |
| EPI_ISL_2472415, EPI_ISL_2472416, EPI_ISL_2472447, EPI_ISL_2472482 | MVZ Labor Dr. Limbach & Kollegen GbR | Robert Koch Institute | |
| EPI_ISL_2390096, EPI_ISL_2471465, EPI_ISL_2472437 | MVZ Labor Dr. Reising-Ackermann und Kollegen | Robert Koch Institute | |
| EPI_ISL_2802713, EPI_ISL_3085932 | Maine Health and Environmental Testing Laboratory | Tewhey Lab, The Jackson Laboratory | Barter, M.; Dewey, H.; H. and Tewhey, R.; Iosue, F.; Lynch, R.; Matluk, N.; Munger |
| EPI_ISL_2647466, EPI_ISL_2647483, EPI_ISL_2647487, EPI_ISL_2647528, EPI_ISL_2647529, EPI_ISL_2647588, EPI_ISL_2647651, EPI_ISL_2647655, EPI_ISL_2647666, EPI_ISL_2647694, EPI_ISL_2647700, EPI_ISL_2647702, EPI_ISL_2647709, EPI_ISL_2647714, EPI_ISL_2647716, EPI_ISL_2647737, EPI_ISL_2647823, EPI_ISL_2647845, EPI_ISL_2647854, EPI_ISL_2647866, EPI_ISL_2647869, EPI_ISL_2647889, EPI_ISL_2647890, EPI_ISL_2647936, EPI_ISL_2647956, EPI_ISL_2647960, EPI_ISL_2647968, EPI_ISL_2647981 | | | |
| see above | Mako Medical | Centers for Disease Control and Prevention Division of Viral Diseases, Pathogen Discovery | Adrian Paskey; Benjamin Rambo-Martin; Christopher Gulvick; Clinton R. Paden; Dakota Howard; Darlene Wagner; Dhwani Batra; Duncan MacCannell; Jason Caravas; Kara Moser; Lauren Moon; Matthew Schmerer; Matthew Tugwell; Peter W. Cook; Scott Sammons; Shatavia Morrison; Yvette Unoarumhi |
| EPI_ISL_2598425, EPI_ISL_3009153, EPI_ISL_3012165, EPI_ISL_3012168, EPI_ISL_3012169, EPI_ISL_3012170, EPI_ISL_3012175, EPI_ISL_3435356 | | | |
| see above | Maryland Genomics, Institute for Genome Sciences, University of Maryland School of Medicine | Maryland Genomics, Institute for Genome Sciences, University of Maryland School of Medicine | Claire M; Fraser; Hazen; Holly; Humphrys; Ivette; Jacques; Jonathan; Kranthi; Lim; Lisa D; Luke J; Mike; Ott; Ravel; Roussey; Sadzewicz; Sandra; Santana-Cruz; Tallon; Tracy; Vavikolanu |
| EPI_ISL_2550770 | Maryland Public Health Laboratory (MD PHL) | Maryland Public Health Laboratory (MD PHL) | Maryland Department of Health Laboratories Administration |
| EPI_ISL_2688815 | Massachusetts State Public Health Laboratory | Massachusetts State Public Health Laboratory | Andrew Lang; Glen Gallagher; Sandra Smole; Timelia Fink |
| EPI_ISL_2501146 | Mayo Clinic & Mayo Clinic Laboratories | Minnesota Department of Health, Public Health Laboratory | Alexandra Lorentz; Jacob Garfin; Matt Plumb; and Xiong Wang |
| EPI_ISL_2389363 | Med. Labor Prof. Schenk Dr. Ansorge & Kollegen | Robert Koch Institute | |
| EPI_ISL_2952250, EPI_ISL_2952265, EPI_ISL_2952266, EPI_ISL_2952267, EPI_ISL_2952414, EPI_ISL_2952432, EPI_ISL_2952437, EPI_ISL_2952441, EPI_ISL_2952463, EPI_ISL_2952509, EPI_ISL_2952523, EPI_ISL_2952527 | | | |
| see above | Medizinische Universitaet Innsbruck Institut fuer Virologie | Elling and Cochella laboratories, IMBA/IMP | Alexander Stark; Ezgi Oezkan; Luisa Cochella and Ulrich Elling; Marcus Strobl; Olga Frank; Ramesh Yelagandula; Tanino Albanese |
| EPI_ISL_2388225 | Medizinisches Labor Wahl Lüdenscheid | Robert Koch Institute | |
| EPI_ISL_2433970, EPI_ISL_2433982, EPI_ISL_2433993 | Microbiologia CATLAB | Can Ruti SARS-CoV-2 Sequencing Hub (HUGTiP/IrsiCaixa/IGTP) | Alba Sánchez; Anna Not; Antoni E Bordoy; Bonaventura Clotet; Cristina Casañ; Cristina Esteban; Francesc Catala-Moll; Gemma Clara; Ignacio Blanco; Marc Noguera-Julian; Maria Casadellà; Mariona Parera; Mercedes Guerrero; Montserrat Giménez; Pere-Joan Cardona; Pilar Armengol; Roger Paredes; Verónica Saludes; and Elisa Martró on behalf of the Can Ruti SARS-CoV-2 Sequencing Hub. |
| EPI_ISL_2448064, EPI_ISL_2650271, EPI_ISL_2650283, EPI_ISL_2650293 | Microbiologia CATLAB | Can Ruti SARS-CoV-2 Sequencing Hub (HUGTiP/IrsiCaixa/IGTP) | Alba Snchez; Alba Sánchez; Anna Not; Antoni E Bordoy; Bonaventura Clotet; Cristina Casa; Cristina Casañ; Cristina Esteban; Francesc Catala-Moll; Gemma Clara; Ignacio Blanco; Marc Noguera-Julian; Maria Casadell; Maria Casadellà; Mariona Parera; Mercedes Guerrero; Montserrat Gimnez; Montserrat Giménez; Pere-Joan Cardona; Pilar Armengol; Roger Paredes; Vernica Saludes; Verónica Saludes; and Elisa Martr on behalf of the Can Ruti SARS-CoV-2 Sequencing Hub.; and Elisa Martró on behalf of the Can Ruti SARS-CoV-2 Sequencing Hub |
| EPI_ISL_2955976 | Microbiology Department & Molecular Biology CORE \| CDB \| Hospital Clinic \| Barcelona | Microbiology Department & Molecular Biology CORE \| CDB \| Hospital Clinic \| Barcelona | Andrea Vergara Gomez; Jose Luis Villanueva Cañas; Miguel Julian Martínez Yoldi; Mª Angeles Marcos Maeso; Mª del Mar Mosquera Gutierrez |
| EPI_ISL_2448054 | Microbiology Department, Laboratori Clínic Metropolitana Nord. Hospital Universitari Germans Trias i Pujol | Can Ruti SARS-CoV-2 Sequencing Hub (HUGTiP/IrsiCaixa/IGTP) | Alba Snchez; Anna Not; Antoni E Bordoy; Bonaventura Clotet; Cristina Casa; Cristina Esteban; Francesc Catala-Moll; Gemma Clara; Ignacio Blanco; Marc Noguera-Julian; Maria Casadell; Mariona Parera; Mercedes Guerrero; Montserrat Gimnez; Pere-Joan Cardona; Pilar Armengol; Roger Paredes; Vernica Saludes; and Elisa Martr on behalf of the Can Ruti SARS-CoV-2 Sequencing Hub. |
| EPI_ISL_2756396 | Microbiology Department, University Hospital Donostia | Microbiology Department, University Hospital Donostia | Cilla G.; Gomez M; Marimon JM; Martin-Peñaranda T; Montes M; Piñeiro L; Sorarrain A |
| EPI_ISL_2501366, EPI_ISL_2501367, EPI_ISL_2501368 | Microbiology Department. Complexo Hospitalario Universitario de Vigo | Microbiology Department. Complexo Hospitalario Universitario de Vigo | Alfaya N; Alonso I; Alvarez M; Cabrera JJ; Carballo R; Cores O; Cortizo S; Martinez L; Mediero G; Perez S; Potel C; Regueiro B; Rey S; Vasallo FJ; del-Campo V |
| EPI_ISL_2508530 | Microbiology Division, SC DHEC | Microbiology Division, SC DHEC | Flores, H.; Freeman, J. |
| EPI_ISL_2629189 | Microbiology Division, South Carolina Department of Health and Environmental Control | Microbiology Division, South Carolina Department of Health and Environmental Control | Flores; Freeman; H.V.; J.S. |
| EPI_ISL_2936682 | Ministry of Health Turkey | Ministry of Health Turkey | Fatma Bayrakdar; Gulay Korukluoglu; Suleyman Yalcin; Yasemin Cosgun |
| EPI_ISL_2443839, EPI_ISL_2455343, EPI_ISL_2565864, EPI_ISL_2565881, EPI_ISL_2598153, EPI_ISL_2658515, EPI_ISL_2663196 | | | |
| see above | Minnesota Department of Health, Public Health Laboratory | Minnesota Department of Health, Public Health Laboratory | Alexandra Lorentz; Jacob Garfin; Matt Plumb; and Xiong Wang |
| EPI_ISL_2635085 | Missouri State Public Health Laboratory | Missouri State Public Health Laboratory | Ashley New; Joshua Barry; Matthew Sinn |
| EPI_ISL_3142935 | Molecular & Genomic Pathology Laboratory, Thomas Jefferson University Hospital | Molecular & Genomic Pathology Laboratory, Thomas Jefferson University Hospital | Nitika Badjatia; Run Jin; Shannon Mahoney; Zi-xuan Wang |
| EPI_ISL_5598703 | Molecular Biology Laboratory, Vidant Medical Center | Brody Integrative Genomics Core, East Carolina University | Changhong Yin; Heather Duncan; John T. Fallon; Kimberly P. Briley; Weihua Huang |
| EPI_ISL_2689917, EPI_ISL_2689927, EPI_ISL_2689930 | NE Public Health Laboratory | Centers for Disease Control and Prevention Division of Viral Diseases, Pathogen Discovery | Alison Laufer Halpin; Ben L. Rambo-Martin; Clinton R. Paden; Dakota Howard; Darlene Wagner; Dave Wentworth; Dhwani Batra; Jasmine Padilla; Justin Lee; Katie Dillon; Krista Queen; Kristen Knipe; Kristine Lacek; Mark Burroughs; Matthew Schmerer; Mili Sheth; Peter Cook; Sam Shepard; Sarah Nobles; Shoshona Le; Suxiang Tong; Vivien Dugan; Yvette Unoarumhi |
| EPI_ISL_2689938 | NJ Public Health and Environmental Laboratories | Centers for Disease Control and Prevention Division of Viral Diseases, Pathogen Discovery | Alison Laufer Halpin; Ben L. Rambo-Martin; Clinton R. Paden; Dakota Howard; Darlene Wagner; Dave Wentworth; Dhwani Batra; Jasmine Padilla; Justin Lee; Katie Dillon; Krista Queen; Kristen Knipe; Kristine Lacek; Mark Burroughs; Matthew Schmerer; Mili Sheth; Peter Cook; Sam Shepard; Sarah Nobles; Shoshona Le; Suxiang Tong; Vivien Dugan; Yvette Unoarumhi |
| EPI_ISL_2443124, EPI_ISL_2443127, EPI_ISL_2557131 | NJDOH, Public Health and Environmental Laboratories | NJ_PHEL | Allison Roder; Byeong Jeong; Chelsea San Filippo; Dana Woell; Lindsey Bodnar; Ryan Pachucki; Shiv K. Verma |
| EPI_ISL_2627177, EPI_ISL_2627179 | NORTHWELL HEALTH LABORATORIES | Wadsworth Center, New York State Department of Health | Alexis Russell; Catharine Prussing; Daryl M. Lamson; Erasmus Schneider; Erica Lasek-Nesselquist; John Kelly; Jonathan Plitnick; Kirsten St. George; Matthew Shudt; Melissa A Leisner; Navjot Singh |
| EPI_ISL_5646087, EPI_ISL_5646089 | NUCLEO DE SAUDE VILA FALCAO DE BAURU | Instituto Butantan | Antonio Jorge Martins; Claudia Renata dos Santos Barros; David Schlesinger; Debora Botequio Moretti; Dimas Tadeu Covas; Elaine Cristina Marqueze; Elaine Vieira Santos; Evandra Strazza Rodrigues; Heidge Fukumasu; Jayme Augusto de Souza-Neto; José Salvatore Leister Patané; Luiz Alcantara; Luiz Lehmann Coutinho; Maria Carolina Elias; Maurício Lacerda Nogueira; Rafael dos Santos Bezerra; Raul Machado Neto; Rejane Maria Tommasini Grotto; Ricardo Haddad; Sandra Coccuzzo Sampaio Vessoni; Simone Kashima; Svetoslav Nanev Slavov; Vincent Louis Viala |
| EPI_ISL_3134752, EPI_ISL_3134753, EPI_ISL_3134754, EPI_ISL_3134755, EPI_ISL_3134756, EPI_ISL_3134758, EPI_ISL_3134759, EPI_ISL_3134760, EPI_ISL_3134761, EPI_ISL_3134762, EPI_ISL_3134763, EPI_ISL_3134764, EPI_ISL_3134765, EPI_ISL_3134766, EPI_ISL_3134767, EPI_ISL_3134768, EPI_ISL_3134769, EPI_ISL_3134770, EPI_ISL_3134771, EPI_ISL_3134772, EPI_ISL_3134773, EPI_ISL_3134774, EPI_ISL_3134775, EPI_ISL_3134776 | | | |
| see above | NUPIT/UFPE | WallauLab on behalf of Fiocruz COVID-19 Genomic Surveillance Network | Alexandre Freitas da Silva; Cassia Docena; Constância Flávia Junqueira Ayres; Filipe Zimmer Dezordi; Gabriel Luz Wallau; Gustavo Barbosa de Lima; Lais Ceschini Machado; Lilian Caroliny Amorim Silva; Maira Galdino da Rocha Pitta; Marcelo Henrique dos Santos Paiva; Matheus Filgueira Bezerra; Michelly Cristiny Pereira; Rômulo Pessoa e Silva; Sinval Pinto Brandão Filho |

| | | | |
|---|---|---|---|
| EPI_ISL_2663515, EPI_ISL_2663535 | NYU Langone Health | Departments of Pathology and Medicine, New York University School of Medicine | Adriana Heguy; Christian Marier; Dacia Dimartino; Emily Guzman; Gael Westby; Guiqing Wang; Paolo Cotzia; Paul Zappile; Peter Meyn; Sitharam Ramaswami; Yutong Zhang |
| EPI_ISL_2726984, EPI_ISL_2726985 | National Institute of Public Health | National Institute of Public Health | Alexander Nagy; Dusan Trnka; Helena Jirincova; Jaromira Vecerova; Timotej Suri |
| EPI_ISL_2382345, EPI_ISL_2382348, EPI_ISL_2483709, EPI_ISL_2483714, EPI_ISL_2483715, EPI_ISL_2545437, EPI_ISL_2545439, EPI_ISL_2545455, EPI_ISL_2545457, EPI_ISL_2545460, EPI_ISL_2687955, EPI_ISL_3133025 | | | |
| see above | National Platform bis UMONS/Jolimont | National Platform bis UMONS/Jolimont | Eric Tarantino; Florian Juszczak; François Dufrasne; Gautier Detry; Guillaume Bayon-Vicente; Laetitia Gheysen; Ruddy Wattiez |
| EPI_ISL_2450054 | National Virus Reference Laboratory | National Virus Reference Laboratory | Charlene Bennett; Cillian F De Gascun; Gabriel Gonzalez; Jonathan Dean; Michael Carr; Zoe Yandle |
| EPI_ISL_2308536, EPI_ISL_2382424, EPI_ISL_2382430 | Nebraska Public Health Laboratory | NPHL COVID-19 Response Team | NPHL COVID-19 Response Team |
| EPI_ISL_2661676, EPI_ISL_2776371, EPI_ISL_2858718 | North Dakota Department of Health, Public Health Laboratory | North Dakota Department of Health, Public Health Laboratory | Lisa Wingerter |
| EPI_ISL_2715099, EPI_ISL_2715129, EPI_ISL_2715133, EPI_ISL_2715138, EPI_ISL_2715139, EPI_ISL_2715140, EPI_ISL_2736013, EPI_ISL_2736015, EPI_ISL_2736019, EPI_ISL_2736024, EPI_ISL_2736062, EPI_ISL_2736065, EPI_ISL_2736186, EPI_ISL_2736189, EPI_ISL_2736201, EPI_ISL_2736205, EPI_ISL_2736208, EPI_ISL_2736210, EPI_ISL_2736212, EPI_ISL_3015662, EPI_ISL_3015699 | | | |
| see above | Northwest Laboratory | Seattle Flu Study | Amanda Adler; Barry R. Lutz; Benjamin Pelle; Brian Hiatt; Caitlin R. Wolf; Chris D. Frazar; Deborah A. Nickerson; Elisabeth Brandstetter; Erica Ryke; Geoff Melly; Helen Y. Chu; Janet A. Englund; Jay Shendure; Jover Lee; Kairsten Fay; Kirsten Lacombe; Lea M. Starita; Mark J. Rieder; Matthew Richardson; Matthew Thompson; Melissa Truong; Michael Boeckh; Michael Famulare; Misja Ilcisin; Peter D. Han; Philip Dykema; Romesh Gautom; Scott Lindquist; Thomas R. Sibley; Trevor Bedford |
| EPI_ISL_2373049, EPI_ISL_2373050, EPI_ISL_2842596, EPI_ISL_2842597 | Northwestern Memorial Hospital | Northwestern University - Ozer Lab | Chad J. Achenbach; Chao Qi; Egon A. Ozer; Judd F. Hultquist; Lacy M. Simons; Lawrence J. Jennings; Michael G. Ison; Ramon Lorenzo-Redondo; Taylor J. Dean |
| EPI_ISL_3388710 | Northwestern Memorial Hospital | RIPHL at Rush University Medical Center | Amber Kimble; Chao Qi; Edith Perez; Felix Araujo Perez; Kevin Kunstman; Laura Furtado; Marieta Hyde; Max Kolton; Stefan Green |
| EPI_ISL_2838571, EPI_ISL_2838591, EPI_ISL_2838606, EPI_ISL_2838619, EPI_ISL_2838793, EPI_ISL_2838809, EPI_ISL_2838856, EPI_ISL_2838857, EPI_ISL_2838858, EPI_ISL_2838859, EPI_ISL_2838860, EPI_ISL_2838861, EPI_ISL_2838862, EPI_ISL_2838863, EPI_ISL_2838864, EPI_ISL_2838865, EPI_ISL_2838866, EPI_ISL_2838867, EPI_ISL_2838868, EPI_ISL_2838869, EPI_ISL_2838870, EPI_ISL_2838871, EPI_ISL_2838872, EPI_ISL_2838873, EPI_ISL_2838874, EPI_ISL_2838875, EPI_ISL_2838876 | | | |
| see above | OHSU Lab Services Molecular Microbiology Lab | Oregon SARS-CoV-2 Genome Sequencing Center | '[UsersoroakBox SyncPHI_SARS-COV-2_Sample_Sheet_infoGISAID_Submission_Infometa_master_2021.xlsx]release20_final'!D:AC; 26; FALSE); VLOOKUP(C466; VLOOKUP(C468; VLOOKUP(C474; VLOOKUP(C477; VLOOKUP(C479; VLOOKUP(C483; VLOOKUP(C488; VLOOKUP(C489; VLOOKUP(C492; VLOOKUP(C502; VLOOKUP(C503; VLOOKUP(C504; VLOOKUP(C505; VLOOKUP(C506; VLOOKUP(C508; VLOOKUP(C509; VLOOKUP(C510; VLOOKUP(C513; VLOOKUP(C516; VLOOKUP(C517; VLOOKUP(C518; VLOOKUP(C519; VLOOKUP(C520; VLOOKUP(C521; VLOOKUP(C522; VLOOKUP(C525; VLOOKUP(C526 |
| EPI_ISL_2402536, EPI_ISL_2402542, EPI_ISL_2402543, EPI_ISL_5331897 | OLVZ Aalst | OLVZ Aalst | Astrid Holderbeke |
| EPI_ISL_2689979, EPI_ISL_2689981, EPI_ISL_2689984, EPI_ISL_2787421 | OR State PHL-Virology/Immunology Section | Centers for Disease Control and Prevention Division of Viral Diseases, Pathogen Discovery | Alison Laufer Halpin; Ben L. Rambo-Martin; Clinton R. Paden; Dakota Howard; Darlene Wagner; Dave Wentworth; Dhwani Batra; Jasmine Padilla; Justin Lee; Katie Dillon; Krista Queen; Kristen Knipe; Kristine Lacek; Mark Burroughs; Matthew Schmerer; Mili Sheth; Peter Cook; Sam Shepard; Sarah Nobles; Shoshona Le; Suxiang Tong; Vivien Dugan; Yvette Unoarumhi |
| EPI_ISL_2621529 | Omega Diagnostics at Mounes | Omega Diagnostics at Mounes | Ashleigh McGrail; Cynthia Corley; Latira Haynes-Jacob; MD; Vivek Khare |
| EPI_ISL_2423535 | Omics Sciences Laboratory | Omics Sciences Laboratory | Darlyn Amaya; Derly Andrade Molina; Gabriel Morey León; Juan Carlos Fernández Cadena; Rubén Armas González |
| EPI_ISL_2424249, EPI_ISL_2424250, EPI_ISL_2473666, EPI_ISL_2557923, EPI_ISL_2557926, EPI_ISL_2604205 | Oregon State Public Health Laboratory | Oregon State Public Health Laboratory | Eugene Yeboah; John Fontana and Shane Sevey; Laura Tsaknaridis; Rafia Razzaque; Vanda Makris |
| EPI_ISL_2665406 | Ospedale S.M. Goretti di Latina - ASL Latina presidio ospedaliero Nord | National Institute for Infectious Diseases (INMI) L. Spallanzani I.R.C.C.S | A Di Caro; B Bartolini; CEM Gruber; E Giombini; F Messina; F Santini; G Bonfiglio; M Rueca; MR Capobianchi; O Butera |
| EPI_ISL_2665409 | Ospedale San Filippo Neri | National Institute for Infectious Diseases (INMI) L. Spallanzani I.R.C.C.S | A Di Caro; B Bartolini; CEM Gruber; E Giombini; F Messina; F Santini; G Bonfiglio; M Rueca; MR Capobianchi; O Butera |
| EPI_ISL_3104899, EPI_ISL_3105045 | Overlake Hospital | Seattle Flu Study | Amanda Adler; Barry R. Lutz; Benjamin Pelle; Brian Hiatt; Caitlin R. Wolf; Chris D. Frazar; Deborah A. Nickerson; Elisabeth Brandstetter; Erica Ryke; Geoff Melly; Helen Y. Chu; Janet A. Englund; Jay Shendure; Jover Lee; Kairsten Fay; Kirsten Lacombe; Lea M. Starita; Mark J. Rieder; Matthew Richardson; Matthew Thompson; Melissa Truong; Michael Boeckh; Michael Famulare; Misja Ilcisin; Peter D. Han; Philip Dykema; Romesh Gautom; Scott Lindquist; Thomas R. Sibley; Trevor Bedford |
| EPI_ISL_2493828 | PA DE IBITIUVA DR OTAVIO BENETTI PITANGUEIRAS | Instituto Butantan | Antonio Jorge Martins; Claudia Renata dos Santos Barros; David Schlesinger; Debora Botequio Moretti; Dimas Tadeu Covas; Elaine Cristina Marqueze; Elaine Vieira Santos; Evandra Strazza Rodrigues; Heidge Fukumasu; Jayme Augusto de Souza-Neto; José Salvatore Leister Patané; Luiz Alcantara; Luiz Lehmann Coutinho; Maria Carolina Elias; Maurício Lacerda Nogueira; Rafael dos Santos Bezerra; Raul Machado Neto; Rejane Maria Tommasini Grotto; Ricardo Haddad; Sandra Coccuzzo Sampaio Vessoni; Simone Kashima; Svetoslav Nanev Slavov; Vincent Louis Viala |
| EPI_ISL_5649639 | PACS JOAO FRANCISCO MARQUES SANTA LUZIA JOSE BONIFACIO | Instituto Butantan | Antonio Jorge Martins; Claudia Renata dos Santos Barros; David Schlesinger; Debora Botequio Moretti; Dimas Tadeu Covas; Elaine Cristina Marqueze; Elaine Vieira Santos; Evandra Strazza Rodrigues; Heidge Fukumasu; Jayme Augusto de Souza-Neto; José Salvatore Leister Patané; Luiz Alcantara; Luiz Lehmann Coutinho; Maria Carolina Elias; Maurício Lacerda Nogueira; Rafael dos Santos Bezerra; Raul Machado Neto; Rejane Maria Tommasini Grotto; Ricardo Haddad; Sandra Coccuzzo Sampaio Vessoni; Simone Kashima; Svetoslav Nanev Slavov; Vincent Louis Viala |
| EPI_ISL_2493913 | POLICLINICA COVID 19 ITAPETININGA | Instituto Butantan | Antonio Jorge Martins; Claudia Renata dos Santos Barros; David Schlesinger; Debora Botequio Moretti; Dimas Tadeu Covas; Elaine Cristina Marqueze; Elaine Vieira Santos; Evandra Strazza Rodrigues; Heidge Fukumasu; Jayme Augusto de Souza-Neto; José Salvatore Leister Patané; Luiz Alcantara; Luiz Lehmann Coutinho; Maria Carolina Elias; Maurício Lacerda Nogueira; Rafael dos Santos Bezerra; Raul Machado Neto; Rejane Maria Tommasini Grotto; Ricardo Haddad; Sandra Coccuzzo Sampaio Vessoni; Simone Kashima; Svetoslav Nanev Slavov; Vincent Louis Viala |
| EPI_ISL_5646041, EPI_ISL_5646045, EPI_ISL_5646052, EPI_ISL_5646071, EPI_ISL_5646088, EPI_ISL_5646090, EPI_ISL_5646091, EPI_ISL_5646092, EPI_ISL_5646100, EPI_ISL_5646104, EPI_ISL_5646105, EPI_ISL_5646109, EPI_ISL_5646110, EPI_ISL_5646111, EPI_ISL_5646112, EPI_ISL_5646113 | | | |
| see above | PROGRAMA SAUDE DA FAMILIA I IEPE | Instituto Butantan | Antonio Jorge Martins; Claudia Renata dos Santos Barros; David Schlesinger; Debora Botequio Moretti; Dimas Tadeu Covas; Elaine Cristina Marqueze; Elaine Vieira Santos; Evandra Strazza Rodrigues; Heidge Fukumasu; Jayme Augusto de Souza-Neto; José Salvatore Leister Patané; Luiz Alcantara; Luiz Lehmann Coutinho; Maria Carolina Elias; Maurício Lacerda Nogueira; Rafael dos Santos Bezerra; Raul Machado Neto; Rejane Maria Tommasini Grotto; Ricardo Haddad; Sandra Coccuzzo Sampaio Vessoni; Simone Kashima; Svetoslav Nanev Slavov; Vincent Louis Viala |
| EPI_ISL_2493875, EPI_ISL_2493878, EPI_ISL_2493883, EPI_ISL_2493891, EPI_ISL_2493896, EPI_ISL_2493899, EPI_ISL_2493901, EPI_ISL_2493902, EPI_ISL_2493904, EPI_ISL_2493910, EPI_ISL_2493911, EPI_ISL_2493915, EPI_ISL_2493931, EPI_ISL_2493932, EPI_ISL_2493933, EPI_ISL_2493935, EPI_ISL_2493936, EPI_ISL_2494295, EPI_ISL_2494325, EPI_ISL_2494326 | | | |
| see above | PRONTO ATENDIMENTO DE VARGEM GRANDE PAULISTA | Instituto Butantan | Antonio Jorge Martins; Claudia Renata dos Santos Barros; David Schlesinger; Debora Botequio Moretti; Dimas Tadeu Covas; Elaine Cristina Marqueze; Elaine Vieira Santos; Evandra Strazza Rodrigues; Heidge Fukumasu; Jayme Augusto de Souza-Neto; José Salvatore Leister Patané; Luiz Alcantara; Luiz Lehmann Coutinho; Maria Carolina Elias; Maurício Lacerda Nogueira; Rafael dos Santos Bezerra; Raul Machado Neto; Rejane Maria Tommasini Grotto; Ricardo Haddad; Sandra Coccuzzo Sampaio Vessoni; Simone Kashima; Svetoslav Nanev Slavov; Vincent Louis Viala |
| EPI_ISL_2493846, EPI_ISL_2493847 | PS E AMBULATORIO ENGENHEIRO CARDOSO | Instituto Butantan | Antonio Jorge Martins; Claudia Renata dos Santos Barros; David Schlesinger; Debora Botequio Moretti; Dimas Tadeu Covas; Elaine Cristina Marqueze; Elaine Vieira Santos; Evandra Strazza Rodrigues; Heidge Fukumasu; Jayme Augusto de Souza-Neto; José Salvatore Leister Patané; Luiz Alcantara; Luiz Lehmann Coutinho; Maria Carolina Elias; Maurício Lacerda Nogueira; Rafael dos Santos Bezerra; Raul Machado Neto; Rejane Maria Tommasini Grotto; Ricardo Haddad; Sandra Coccuzzo Sampaio Vessoni; Simone Kashima; Svetoslav Nanev Slavov; Vincent Louis Viala |
| EPI_ISL_2493900 | PSF DR ANTONIO PIRES DE ALMEIDA PORTO FELIZ | Instituto Butantan | Antonio Jorge Martins; Claudia Renata dos Santos Barros; David Schlesinger; Debora Botequio Moretti; Dimas Tadeu Covas; Elaine Cristina Marqueze; Elaine Vieira Santos; Evandra Strazza Rodrigues; Heidge Fukumasu; Jayme Augusto de Souza-Neto; José Salvatore Leister Patané; Luiz Alcantara; Luiz Lehmann Coutinho; Maria Carolina Elias; Maurício Lacerda Nogueira; Rafael dos Santos Bezerra; Raul Machado Neto; Rejane Maria Tommasini Grotto; Ricardo Haddad; Sandra Coccuzzo Sampaio Vessoni; Simone Kashima; Svetoslav Nanev Slavov; Vincent Louis Viala |
| EPI_ISL_5649635 | PSF PARQUE EMILIA | Instituto Butantan | Antonio Jorge Martins; Claudia Renata dos Santos Barros; David Schlesinger; Debora Botequio Moretti; Dimas Tadeu Covas; Elaine Cristina Marqueze; Elaine Vieira Santos; Evandra Strazza Rodrigues; Heidge Fukumasu; Jayme Augusto de Souza-Neto; José Salvatore Leister Patané; Luiz Alcantara; Luiz Lehmann Coutinho; Maria Carolina Elias; Maurício Lacerda Nogueira; Rafael dos Santos Bezerra; Raul Machado Neto; Rejane Maria Tommasini Grotto; Ricardo Haddad; Sandra Coccuzzo Sampaio Vessoni; Simone Kashima; Svetoslav Nanev Slavov; Vincent Louis Viala |
| EPI_ISL_2349625, EPI_ISL_2349630, EPI_ISL_2349631, EPI_ISL_2349675, EPI_ISL_2446041, EPI_ISL_2446045, EPI_ISL_2446051, EPI_ISL_2446057, EPI_ISL_2446076 | | | |
| see above | Pandemic Response Lab - NYC | Pandemic Response Lab, R&D | Cybill del Castillo; Dylan Law; Haiping Hao; Henry Lee; Jon Laurent; Katharine Nelson; Melissa Hopkins; Michael Hammerling; Pradeep Bugga; Shinyoung Clair Kang; Sol Rey; William Ward |
| EPI_ISL_3050138 | Pathology, UAB | Pathology, UAB | E.J.; Hendrickson; Hernandez, J.; Leal; Lefkowitz; Moates, D.; R.C.; S.M. |
| EPI_ISL_3030327 | Pesaro | Microbiology University Politecnica delle Marche | Anna Valenza; Carla Acciarri; Katia Marinelli; Monica Lucia Ferreri; Patrizia Bagnarelli; Roberta Longo; Sara Caucci; Stefano Menzo |
| EPI_ISL_2642050 | Pharmgenetix GmbH | Pharmgenetix GmbH | Arnoldas Matulevicius; Besard Avdiu; Simone Vanoni |
| EPI_ISL_6907397, EPI_ISL_6907400, EPI_ISL_6907401, EPI_ISL_6907404, EPI_ISL_6907407, EPI_ISL_6907414, EPI_ISL_6907415, EPI_ISL_6907423, EPI_ISL_6907432, EPI_ISL_6907990, EPI_ISL_6907992 | | | |
| see above | Plataforma de Vigilancia Molecular (PVM) - FIOCRUZ/BA | Plataforma de Vigilancia Molecular (PVM) - FIOCRUZ/BA | Bruno Bezerril Andrade; Camila I. de Oliveira; Clarissa Araújo Gurgel; Eduardo Oyama; Francys A Rangel; Laila A da Silva; Laise de Moraes; Leonardo Paiva Farias; Marina Cucco; Pedro Silveira; Ricardo Khouri on behalf of the Fiocruz COVID-19 Genomic Surveillance Network.; Tiago Graf |
| EPI_ISL_2346313, EPI_ISL_2346314, EPI_ISL_2346316, EPI_ISL_2346319, EPI_ISL_2379747, EPI_ISL_2382270, EPI_ISL_2382281, EPI_ISL_2382286, EPI_ISL_2382291, EPI_ISL_2382292, EPI_ISL_2391766, EPI_ISL_2397215 | | | |
| see above | Plateforme de testing Namuroise | Plateforme de testing Namuroise | Degosserie Jonathan; Denis Olivier; Lesly Nyinkeu Kemamen; Mullier François; Nicolas Gilliard; Otto Gaetan |
| EPI_ISL_2362633, EPI_ISL_2422169 | Platform BIS UZA/UAntwerpen | Labo Klinische Biologie, UZA | Basil Britto Xavier; Christine Lammens; Herman Goossens; Jasmine Coppens; Marie Le Mercier; Veerle Matheeussen |
| EPI_ISL_2665408 | Policlinico Universitario Campus Bio-Medico | National Institute for Infectious Diseases (INMI) L. Spallanzani I.R.C.C.S | A Di Caro; B Bartolini; CEM Gruber; E Giombini; F Messina; F Santini; G Bonfiglio; M Rueca; MR Capobianchi; O Butera |
| EPI_ISL_7406401, EPI_ISL_7406405 | Programa Laboratorio de Salud Pública "Dr Dalmiro Pérez Laborda" | Área de Secuenciación del Laboratorio de Virología del Hospital de Niños Dr. Ricardo Gutierrez on behalf of 'Proyecto Argentino Interinstitucional de genomica de SARS-CoV-2' (PAIS Consortium) | A; AM; Acuña; Aguilera; BH; Bohn; Cabral Bombardieri; Campos; Carrizo; D; E; Esteves; F; G; Goya; J; JM; JP; Jofré; L; LE; Lacaze; Lusso; M; MA; MI; MS; Mastrodonato; Molina; Nabaes Jodar; Natale; Olivera; Perez Diaz; Peñalva; Quijano; Rivero; Rosales; S; Talia; Valinotto; Viegas, M. |

| | | | |
|---|---|---|---|
| EPI_ISL_2699447, EPI_ISL_2699454, EPI_ISL_2699462, EPI_ISL_2699468, EPI_ISL_2699470, EPI_ISL_2699479, EPI_ISL_2699485, EPI_ISL_2699490, EPI_ISL_2699491, EPI_ISL_2699496, EPI_ISL_2699498 | | | |
| see above | Public Health Laboratory, Minnesota Department of Health | University of Minnesota Genomics Center | Corbin Dirkx; Daryl M. Gohl; Jaquelyn Kuriger-Laber; John Garbe; and Sean Wang |
| EPI_ISL_2615920 | QLabs | WVU and Marshall University Combined Genomics Core Facilities | James Denvir; Peter Perrotta; Peter Stoilov; Ryan Percifield; Wesley Kimble |
| EPI_ISL_2624021, EPI_ISL_2624022, EPI_ISL_2624052, EPI_ISL_2706658, EPI_ISL_2706882, EPI_ISL_2706917 | Quadram Institute Bioscience | COVID-19 Genomics UK (COG-UK) Consortium | Alexander J Trotter; Alison E. Mather; Alp Aydin; Ana P. Tedim; Anastasia Kolyva; Andrew Bell; Andrew J. Page; Claire Stuart; Dave J. Baker; Gemma L. Kay; John Wain; Justin O'Grady; Leonardo de Oliveira Martins; Lizzie Meadows; Maria Diaz; Mark Webber; Muhammed Yasir; Nabil-Fareed Alikhan; Ngozi Elumogo; Nicholas M. Thomson; Rachael Stanley; Rachel Gilroy; Reenesh Prakash; Samir Dervisevic; Samuel Bloomfield; Steven Rudder; Thanh Le-Viet |
| EPI_ISL_2451316, EPI_ISL_2451509, EPI_ISL_2451518, EPI_ISL_2451527, EPI_ISL_2451533, EPI_ISL_2451535, EPI_ISL_2451543, EPI_ISL_2451546, EPI_ISL_2451555, EPI_ISL_2451558, EPI_ISL_2451564, EPI_ISL_2451565, EPI_ISL_2463007, EPI_ISL_2463008, EPI_ISL_2463009, EPI_ISL_2463010, EPI_ISL_2463011, EPI_ISL_2463012, EPI_ISL_2463013, EPI_ISL_2463014, EPI_ISL_2463015, EPI_ISL_2463016, EPI_ISL_2463017, EPI_ISL_2463018, EPI_ISL_2463019, EPI_ISL_2463138, EPI_ISL_2488820, EPI_ISL_2488821, EPI_ISL_2488822, EPI_ISL_2488823, EPI_ISL_2488824, EPI_ISL_2488825, EPI_ISL_2488826, EPI_ISL_2488827, EPI_ISL_2488828, EPI_ISL_2488829, EPI_ISL_2488830, EPI_ISL_2488831, EPI_ISL_2488832, EPI_ISL_2488833, EPI_ISL_2488834, EPI_ISL_2488835, EPI_ISL_2488836, EPI_ISL_2488837, EPI_ISL_2488838, EPI_ISL_2488839, EPI_ISL_2488840, EPI_ISL_2488841, EPI_ISL_2488842, EPI_ISL_2488843, EPI_ISL_2488844, EPI_ISL_2488845, EPI_ISL_2488846, EPI_ISL_2488847, EPI_ISL_2488848, EPI_ISL_2488849, EPI_ISL_2488850, EPI_ISL_2488853, EPI_ISL_2488854, EPI_ISL_2488855, EPI_ISL_2488856, EPI_ISL_2488858, EPI_ISL_2488859, EPI_ISL_2488860, EPI_ISL_2488861, EPI_ISL_2534520, EPI_ISL_2534521, EPI_ISL_2534522, EPI_ISL_2534525, EPI_ISL_2534526, EPI_ISL_2534527, EPI_ISL_2534528, EPI_ISL_2534529, EPI_ISL_2534531, EPI_ISL_2534532, EPI_ISL_2649949, EPI_ISL_2649950, EPI_ISL_2649956, EPI_ISL_2649961, EPI_ISL_2649962, EPI_ISL_2649985, EPI_ISL_4367580, EPI_ISL_4367678, EPI_ISL_4372717 | | | |
| see above | Quest Diagnostics Incorporated | Centers for Disease Control and Prevention Division of Viral Diseases, Pathogen Discovery | A. Gerasimova; A. Perez; Adrian Paskey; B. Anderson; Benjamin Rambo-Martin; Christopher Gulvick; Clinton Paden; Clinton R. Paden; Dakota Howard; Darlene Wagner; Dhwani Batra; Duncan MacCannell; Erisa Sula; F. Lacbawan; I. A. Shlyakhter; I. Shlyakhter; Jason Caravas; K. Livingston; K.E. Livingston; Kara Moser; Kristine Lacek; L. Bernstein; L.E. Bernstein; M. Hua; Matthew Schmerer; P. Tanpaiboon; Peter Cook; Peter W. Cook; R. Kagan; R. M. Kagan; R. Owen; R. Rolando; R. V. Rolando; S. H. Rosenthal; S. Rosenthal; Scott Sammons; Shatavia Morrison; Tymeckia Kendall; Victoria Caban Figueroa; Y. Liu; Yvette Unoarumhi |
| EPI_ISL_2691942, EPI_ISL_2931090, EPI_ISL_2931091, EPI_ISL_2931092, EPI_ISL_2931093 | Rush University Medical Center | RIPHL at Rush University Medical Center | Diane Springer; Felix Araujo Perez; Joyce Houlihan; Kevin Kunstman; Laura Furtado; Marieta Hyde; Mary Hayden; Max Kolton; Stefan Green |
| EPI_ISL_2840622 | S.C. Laboratorio Analisi, ASL 3 Liguria | U.O. Igiene, Ospedale Policlinico San Martino | Bruzzone Bianca; Caligiuri Patrizia; De Pace Vanessa; Domnich Alexander; Icardi Giancarlo on behalf of SARS-CoV-2 ITALIAN RESEARCH ENTERPRISE-(SCIRE) Collaborative Group; Orsi Andrea; Ricucci Valentina; Spitaleri Antonino |
| EPI_ISL_5650058, EPI_ISL_5650126 | SANTA CASA DE JOSE BONIFACIO | Instituto Butantan | Antonio Jorge Martins; Claudia Renata dos Santos Barros; David Schlesinger; Debora Botequio Moretti; Dimas Tadeu Covas; Elaine Cristina Marqueze; Elaine Vieira Santos; Evandra Strazza Rodrigues; Heidge Fukumasu; Jayme Augusto de Souza-Neto; José Salvatore Leister Patané; Luiz Alcantara; Luiz Lehmann Coutinho; Maria Carolina Elias; Maurício Lacerda Nogueira; Rafael dos Santos Bezerra; Raul Machado Neto; Rejane Maria Tommasini Grotto; Ricardo Haddad; Sandra Coccuzzo Sampaio Vessoni; Simone Kashima; Svetoslav Nanev Slavov; Vincent Louis Viala |
| EPI_ISL_2493867 | SANTA CASA DE MISERICORDIA DE TIETE | Instituto Butantan | Antonio Jorge Martins; Claudia Renata dos Santos Barros; David Schlesinger; Debora Botequio Moretti; Dimas Tadeu Covas; Elaine Cristina Marqueze; Elaine Vieira Santos; Evandra Strazza Rodrigues; Heidge Fukumasu; Jayme Augusto de Souza-Neto; José Salvatore Leister Patané; Luiz Alcantara; Luiz Lehmann Coutinho; Maria Carolina Elias; Maurício Lacerda Nogueira; Rafael dos Santos Bezerra; Raul Machado Neto; Rejane Maria Tommasini Grotto; Ricardo Haddad; Sandra Coccuzzo Sampaio Vessoni; Simone Kashima; Svetoslav Nanev Slavov; Vincent Louis Viala |
| EPI_ISL_5646076 | SANTA CASA DE PIRATININGA | Instituto Butantan | Antonio Jorge Martins; Claudia Renata dos Santos Barros; David Schlesinger; Debora Botequio Moretti; Dimas Tadeu Covas; Elaine Cristina Marqueze; Elaine Vieira Santos; Evandra Strazza Rodrigues; Heidge Fukumasu; Jayme Augusto de Souza-Neto; José Salvatore Leister Patané; Luiz Alcantara; Luiz Lehmann Coutinho; Maria Carolina Elias; Maurício Lacerda Nogueira; Rafael dos Santos Bezerra; Raul Machado Neto; Rejane Maria Tommasini Grotto; Ricardo Haddad; Sandra Coccuzzo Sampaio Vessoni; Simone Kashima; Svetoslav Nanev Slavov; Vincent Louis Viala |
| EPI_ISL_2493880 | SANTA CASA DE PORTO FELIZ | Instituto Butantan | Antonio Jorge Martins; Claudia Renata dos Santos Barros; David Schlesinger; Debora Botequio Moretti; Dimas Tadeu Covas; Elaine Cristina Marqueze; Elaine Vieira Santos; Evandra Strazza Rodrigues; Heidge Fukumasu; Jayme Augusto de Souza-Neto; José Salvatore Leister Patané; Luiz Alcantara; Luiz Lehmann Coutinho; Maria Carolina Elias; Maurício Lacerda Nogueira; Rafael dos Santos Bezerra; Raul Machado Neto; Rejane Maria Tommasini Grotto; Ricardo Haddad; Sandra Coccuzzo Sampaio Vessoni; Simone Kashima; Svetoslav Nanev Slavov; Vincent Louis Viala |
| EPI_ISL_2493909 | SANTA CASA SALTO DE PIRAPORA | Instituto Butantan | Antonio Jorge Martins; Claudia Renata dos Santos Barros; David Schlesinger; Debora Botequio Moretti; Dimas Tadeu Covas; Elaine Cristina Marqueze; Elaine Vieira Santos; Evandra Strazza Rodrigues; Heidge Fukumasu; Jayme Augusto de Souza-Neto; José Salvatore Leister Patané; Luiz Alcantara; Luiz Lehmann Coutinho; Maria Carolina Elias; Maurício Lacerda Nogueira; Rafael dos Santos Bezerra; Raul Machado Neto; Rejane Maria Tommasini Grotto; Ricardo Haddad; Sandra Coccuzzo Sampaio Vessoni; Simone Kashima; Svetoslav Nanev Slavov; Vincent Louis Viala |
| EPI_ISL_2494169, EPI_ISL_2494177, EPI_ISL_2494189, EPI_ISL_2494196 | SANTA CASA SANTA RITA DO PASSA QUATRO | Instituto Butantan | Antonio Jorge Martins; Claudia Renata dos Santos Barros; David Schlesinger; Debora Botequio Moretti; Dimas Tadeu Covas; Elaine Cristina Marqueze; Elaine Vieira Santos; Evandra Strazza Rodrigues; Heidge Fukumasu; Jayme Augusto de Souza-Neto; José Salvatore Leister Patané; Luiz Alcantara; Luiz Lehmann Coutinho; Maria Carolina Elias; Maurício Lacerda Nogueira; Rafael dos Santos Bezerra; Raul Machado Neto; Rejane Maria Tommasini Grotto; Ricardo Haddad; Sandra Coccuzzo Sampaio Vessoni; Simone Kashima; Svetoslav Nanev Slavov; Vincent Louis Viala |
| EPI_ISL_2645312 | SC Dept of Health and Env. Control-Bureau of Laboratories | Centers for Disease Control and Prevention Division of Viral Diseases, Pathogen Discovery | Alison Laufer Halpin; Ben L. Rambo-Martin; Clinton R. Paden; Dakota Howard; Darlene Wagner; Dave Wentworth; Dhwani Batra; Jasmine Padilla; Justin Lee; Katie Dillon; Krista Queen; Kristen Knipe; Kristine Lacek; Mark Burroughs; Matthew Schmerer; Mili Sheth; Peter Cook; Sam Shepard; Sarah Nobles; Shoshona Le; Suxiang Tong; Vivien Dugan; Yvette Unoarumhi |
| EPI_ISL_2494161, EPI_ISL_2494170, EPI_ISL_2494178 | SECRETARIA DE SAUDE DE SANTA GERTRUDES | Instituto Butantan | Antonio Jorge Martins; Claudia Renata dos Santos Barros; David Schlesinger; Debora Botequio Moretti; Dimas Tadeu Covas; Elaine Cristina Marqueze; Elaine Vieira Santos; Evandra Strazza Rodrigues; Heidge Fukumasu; Jayme Augusto de Souza-Neto; José Salvatore Leister Patané; Luiz Alcantara; Luiz Lehmann Coutinho; Maria Carolina Elias; Maurício Lacerda Nogueira; Rafael dos Santos Bezerra; Raul Machado Neto; Rejane Maria Tommasini Grotto; Ricardo Haddad; Sandra Coccuzzo Sampaio Vessoni; Simone Kashima; Svetoslav Nanev Slavov; Vincent Louis Viala |
| EPI_ISL_2494167, EPI_ISL_2494190 | SECRETARIA DE SAUDE DE SAO PEDRO | Instituto Butantan | Antonio Jorge Martins; Claudia Renata dos Santos Barros; David Schlesinger; Debora Botequio Moretti; Dimas Tadeu Covas; Elaine Cristina Marqueze; Elaine Vieira Santos; Evandra Strazza Rodrigues; Heidge Fukumasu; Jayme Augusto de Souza-Neto; José Salvatore Leister Patané; Luiz Alcantara; Luiz Lehmann Coutinho; Maria Carolina Elias; Maurício Lacerda Nogueira; Rafael dos Santos Bezerra; Raul Machado Neto; Rejane Maria Tommasini Grotto; Ricardo Haddad; Sandra Coccuzzo Sampaio Vessoni; Simone Kashima; Svetoslav Nanev Slavov; Vincent Louis Viala |
| EPI_ISL_2493290 | SECRETARIA MUNICIPAL DA SAUDE DE ITAPEVA SP | Instituto Butantan | Antonio Jorge Martins; Claudia Renata dos Santos Barros; David Schlesinger; Debora Botequio Moretti; Dimas Tadeu Covas; Elaine Cristina Marqueze; Elaine Vieira Santos; Evandra Strazza Rodrigues; Heidge Fukumasu; Jayme Augusto de Souza-Neto; José Salvatore Leister Patané; Luiz Alcantara; Luiz Lehmann Coutinho; Maria Carolina Elias; Maurício Lacerda Nogueira; Rafael dos Santos Bezerra; Raul Machado Neto; Rejane Maria Tommasini Grotto; Ricardo Haddad; Sandra Coccuzzo Sampaio Vessoni; Simone Kashima; Svetoslav Nanev Slavov; Vincent Louis Viala |
| EPI_ISL_2493903 | SECRETARIA MUNICIPAL DA SAUDE DE ITARARE | Instituto Butantan | Antonio Jorge Martins; Claudia Renata dos Santos Barros; David Schlesinger; Debora Botequio Moretti; Dimas Tadeu Covas; Elaine Cristina Marqueze; Elaine Vieira Santos; Evandra Strazza Rodrigues; Heidge Fukumasu; Jayme Augusto de Souza-Neto; José Salvatore Leister Patané; Luiz Alcantara; Luiz Lehmann Coutinho; Maria Carolina Elias; Maurício Lacerda Nogueira; Rafael dos Santos Bezerra; Raul Machado Neto; Rejane Maria Tommasini Grotto; Ricardo Haddad; Sandra Coccuzzo Sampaio Vessoni; Simone Kashima; Svetoslav Nanev Slavov; Vincent Louis Viala |
| EPI_ISL_2493176, EPI_ISL_2493275, EPI_ISL_2493276, EPI_ISL_2494110 | SECRETARIA MUNICIPAL DE SAUDE | Instituto Butantan | Antonio Jorge Martins; Claudia Renata dos Santos Barros; David Schlesinger; Debora Botequio Moretti; Dimas Tadeu Covas; Elaine Cristina Marqueze; Elaine Vieira Santos; Evandra Strazza Rodrigues; Heidge Fukumasu; Jayme Augusto de Souza-Neto; José Salvatore Leister Patané; Luiz Alcantara; Luiz Lehmann Coutinho; Maria Carolina Elias; Maurício Lacerda Nogueira; Rafael dos Santos Bezerra; Raul Machado Neto; Rejane Maria Tommasini Grotto; Ricardo Haddad; Sandra Coccuzzo Sampaio Vessoni; Simone Kashima; Svetoslav Nanev Slavov; Vincent Louis Viala |
| EPI_ISL_2493156, EPI_ISL_2494185, EPI_ISL_2494194 | SECRETARIA MUNICIPAL DE SAUDE DE CORDEIROPOLIS | Instituto Butantan | Antonio Jorge Martins; Claudia Renata dos Santos Barros; David Schlesinger; Debora Botequio Moretti; Dimas Tadeu Covas; Elaine Cristina Marqueze; Elaine Vieira Santos; Evandra Strazza Rodrigues; Heidge Fukumasu; Jayme Augusto de Souza-Neto; José Salvatore Leister Patané; Luiz Alcantara; Luiz Lehmann Coutinho; Maria Carolina Elias; Maurício Lacerda Nogueira; Rafael dos Santos Bezerra; Raul Machado Neto; Rejane Maria Tommasini Grotto; Ricardo Haddad; Sandra Coccuzzo Sampaio Vessoni; Simone Kashima; Svetoslav Nanev Slavov; Vincent Louis Viala |
| EPI_ISL_2493825, EPI_ISL_5649737 | SECRETARIA MUNICIPAL DE SAUDE DE MORUNGABA | Instituto Butantan | Antonio Jorge Martins; Claudia Renata dos Santos Barros; David Schlesinger; Debora Botequio Moretti; Dimas Tadeu Covas; Elaine Cristina Marqueze; Elaine Vieira Santos; Evandra Strazza Rodrigues; Heidge Fukumasu; Jayme Augusto de Souza-Neto; José Salvatore Leister Patané; Luiz Alcantara; Luiz Lehmann Coutinho; Maria Carolina Elias; Maurício Lacerda Nogueira; Rafael dos Santos Bezerra; Raul Machado Neto; Rejane Maria Tommasini Grotto; Ricardo Haddad; Sandra Coccuzzo Sampaio Vessoni; Simone Kashima; Svetoslav Nanev Slavov; Vincent Louis Viala |
| EPI_ISL_2493802, EPI_ISL_2493823, EPI_ISL_2494306, EPI_ISL_2494307, EPI_ISL_2494308 | SECRETARIA MUNICIPAL DE SAUDE DE PEDERNEIRAS | Instituto Butantan | Antonio Jorge Martins; Claudia Renata dos Santos Barros; David Schlesinger; Debora Botequio Moretti; Dimas Tadeu Covas; Elaine Cristina Marqueze; Elaine Vieira Santos; Evandra Strazza Rodrigues; Heidge Fukumasu; Jayme Augusto de Souza-Neto; José Salvatore Leister Patané; Luiz Alcantara; Luiz Lehmann Coutinho; Maria Carolina Elias; Maurício Lacerda Nogueira; Rafael dos Santos Bezerra; Raul Machado Neto; Rejane Maria Tommasini Grotto; Ricardo Haddad; Sandra Coccuzzo Sampaio Vessoni; Simone Kashima; Svetoslav Nanev Slavov; Vincent Louis Viala |
| EPI_ISL_2462329, EPI_ISL_2494146, EPI_ISL_5646040, EPI_ISL_5646042, EPI_ISL_5646043, EPI_ISL_5646044, EPI_ISL_5646047, EPI_ISL_5646048, EPI_ISL_5646051, EPI_ISL_5646055, EPI_ISL_5646058, EPI_ISL_5646060, EPI_ISL_5646061, EPI_ISL_5646063, EPI_ISL_5646070, EPI_ISL_5646075, EPI_ISL_5646095, EPI_ISL_5646097, EPI_ISL_5646099, EPI_ISL_5646101, EPI_ISL_5646107 | | | |
| see above | SECRETARIA MUNICIPAL DE SAUDE DE PRES BERNARDES SP | Instituto Butantan | Antonio Jorge Martins; Claudia Renata dos Santos Barros; David Schlesinger; Debora Botequio Moretti; Dimas Tadeu Covas; Elaine Cristina Marqueze; Elaine Vieira Santos; Evandra Strazza Rodrigues; Heidge Fukumasu; Jayme Augusto de Souza-Neto; José Salvatore Leister Patané; Luiz Alcantara; Luiz Lehmann Coutinho; Maria Carolina Elias; Maurício Lacerda Nogueira; Rafael dos Santos Bezerra; Raul Machado Neto; Rejane Maria Tommasini Grotto; Ricardo Haddad; Sandra Coccuzzo Sampaio Vessoni; Simone Kashima; Svetoslav Nanev Slavov; Vincent Louis Viala |
| EPI_ISL_2801350, EPI_ISL_2801359, EPI_ISL_2801366 | SECRETARIA MUNICIPAL DE SAUDE DE TIANGUA | Analytical Competence Molecular Epidemiology Lab/ACME, Oswaldo Cruz Foundation, Ceara (FIOCRUZ CE) | Cleber Furtado Aksenen e Suzana Porto Almeida; Fabio Miyajima; Fernando Braga Stehling; Francisco Eder de Moura Lopes; Jamille Maria Mendes Bezerra; Joaquim César do Nascimento Sousa Junior; Pedro Miguel Carneiro Jeronimo; Thais Ferreira de Oliveira; Thais de Oliveira Costa; Ticiane Cavalcante de Souza; Veridiana Pessoa Miyajima |
| EPI_ISL_2462330, EPI_ISL_2493832, EPI_ISL_2493833, EPI_ISL_2493834, EPI_ISL_2493837, EPI_ISL_2493838, EPI_ISL_2493839, EPI_ISL_2493840, EPI_ISL_2493842, EPI_ISL_2493889, EPI_ISL_2493890, EPI_ISL_2493892, EPI_ISL_2493893, EPI_ISL_2493894, EPI_ISL_2494294, EPI_ISL_2494322 | | | |
| see above | SERV DE VIG SANITARIA EPIDEMIO E CTRL DE ZOONOZES GUARUJA | Instituto Butantan | Antonio Jorge Martins; Claudia Renata dos Santos Barros; David Schlesinger; Debora Botequio Moretti; Dimas Tadeu Covas; Elaine Cristina Marqueze; Elaine Vieira Santos; Evandra Strazza Rodrigues; Heidge Fukumasu; Jayme Augusto de Souza-Neto; José Salvatore Leister Patané; Luiz Alcantara; Luiz Lehmann Coutinho; Maria Carolina Elias; Maurício Lacerda Nogueira; Rafael dos Santos Bezerra; Raul Machado Neto; Rejane Maria Tommasini Grotto; Ricardo Haddad; Sandra Coccuzzo Sampaio Vessoni; Simone Kashima; Svetoslav Nanev Slavov; Vincent Louis Viala |
| EPI_ISL_2483592 | SIESP DIPARTIMENTO DI PREVENZIONE CHIETI | Istituto Zooprofilattico Sperimentale dell'Abruzzo e Molise "G. Caporale" | Ancora M; Calistri P; Cammà C; Caporale M; Curini V; Delli Compagni E; Di Domenico M; Di Lollo Valeria; Di Pasquale A; Lorusso A; Mangone I; Marcacci M; Puglia I; Rinaldi A; Savini G; Scialabba S |
| EPI_ISL_2483591, EPI_ISL_2483593 | SIESP DIPARTIMENTO DI PREVENZIONE TERAMO | Istituto Zooprofilattico Sperimentale dell'Abruzzo e Molise "G. Caporale" | Ancora M; Calistri P; Cammà C; Caporale M; Curini V; Delli Compagni E; Di Domenico M; Di Lollo Valeria; Di Pasquale A; Lorusso A; Mangone I; Marcacci M; Puglia I; Rinaldi A; Savini G; Scialabba S |
| EPI_ISL_2889273, EPI_ISL_2889315, EPI_ISL_2889331, EPI_ISL_2889332, EPI_ISL_2889333, EPI_ISL_2889395, EPI_ISL_2889404, EPI_ISL_2889423, EPI_ISL_2889432, EPI_ISL_2889450, EPI_ISL_2889451, EPI_ISL_2889452, EPI_ISL_2889525, EPI_ISL_2889526, EPI_ISL_2889554, EPI_ISL_2889577, EPI_ISL_2889584, EPI_ISL_2889618, EPI_ISL_2889632, EPI_ISL_2889633, EPI_ISL_2889634 | | | |
| see above | SK-Roy Romanow Provincial Laboratory | National Microbiology Laboratory (NML) | Alanna Senecal; Amanda Lang; Anna Majer; Anneliese Landgraff; CanCOGeN's metadata curation team; Darian Hole; Elsie Grudeski; Gary Van Domselaar; Grace Seo; Jennifer Tanner; Jessica Minion; Kara Loos; Keith MacKenzie; Kirsten Biggar; Madison Chapel; Meredith Faires; Morag Graham; Natalie Knox; Nathalie Bastien; Philip Mabon; Public Health Agency of Canada CanCOGeN team; Rachel DePaulo; Rhiannon Huzarewich; Russell Mandes; Ryan McDonald; Shari Tyson; Timothy Booth; Yan Li |
| EPI_ISL_5844705, EPI_ISL_5844718 | SK-Roy Romanow Provincial Laboratory | Saskatchewan - Roy Romanow Provincial Laboratory (RRPL) | Alanna Senecal; Amanda Lang; Jessica Minion; Kara Loos; Keith MacKenzie; Meredith Faires; Rachel DePaulo; Roy Romanow Provincial Laboratory - Molecular Diagnostics; Ryan McDonald |
| EPI_ISL_2462317, EPI_ISL_2493862, EPI_ISL_2493872, EPI_ISL_2493874, EPI_ISL_2493942 | SMS ARUJA | Instituto Butantan | Antonio Jorge Martins; Claudia Renata dos Santos Barros; David Schlesinger; Debora Botequio Moretti; Dimas Tadeu Covas; Elaine Cristina Marqueze; Elaine Vieira Santos; Evandra Strazza Rodrigues; Heidge Fukumasu; Jayme Augusto de Souza-Neto; José Salvatore Leister Patané; Luiz Alcantara; Luiz Lehmann Coutinho; Maria Carolina Elias; Maurício Lacerda Nogueira; Rafael dos Santos Bezerra; Raul Machado Neto; Rejane Maria Tommasini Grotto; Ricardo Haddad; Sandra Coccuzzo Sampaio Vessoni; Simone Kashima; Svetoslav Nanev Slavov; Vincent Louis Viala |
| EPI_ISL_2402376, EPI_ISL_2402418, EPI_ISL_2402419, EPI_ISL_2402420, EPI_ISL_2402423, EPI_ISL_2402425, EPI_ISL_2402434, EPI_ISL_2402441, EPI_ISL_2402442, EPI_ISL_2402450, EPI_ISL_2402451, EPI_ISL_2402453, EPI_ISL_2402454, EPI_ISL_2402455, EPI_ISL_2402456, EPI_ISL_2402458, EPI_ISL_2426581, EPI_ISL_2426582, EPI_ISL_2426584, EPI_ISL_2426594, EPI_ISL_2426597, EPI_ISL_2426600, EPI_ISL_2483895 | | | |
| see above | SYNLAB | GIGA Medical Genomics | Bouchra Boujemla; Cécile Meex; Keith Durkin; Maria Artesi; Marie-Pierre Hayette; Nathalie Renotte; Pierrette Melin; Raphaël Boreux; Sébastien Bontems; Vincent Bours |
| EPI_ISL_2472365 | SYNLAB MVZ Berlin | Robert Koch Institute | |
| EPI_ISL_2389554, EPI_ISL_2389772, | SYNLAB MVZ Heidelberg | Robert Koch Institute | |

| | | | |
|---|---|---|---|
| EPI_ISL_2389779, EPI_ISL_2389794, EPI_ISL_2389799, EPI_ISL_2389805 | | | |
| EPI_ISL_2387454, EPI_ISL_2387564, EPI_ISL_2387700, EPI_ISL_2387944, EPI_ISL_2389534, EPI_ISL_2389539, EPI_ISL_2389667, EPI_ISL_2389701, EPI_ISL_2389816 | | | |
| see above | SYNLAB MVZ Leinfelden-Echterdingen | Robert Koch Institute | |
| EPI_ISL_2387475, EPI_ISL_2387596, EPI_ISL_2389617 | SYNLAB MVZ Leverkusen | Robert Koch Institute | |
| EPI_ISL_2469537 | SYNLAB MVZ Trier | Robert Koch Institute | |
| EPI_ISL_2469483 | SYNLAB MVZ Weiden | Robert Koch Institute | |
| EPI_ISL_2603673, EPI_ISL_2603674, EPI_ISL_2603678, EPI_ISL_2603683, EPI_ISL_2603684, EPI_ISL_2603692, EPI_ISL_2603710, EPI_ISL_2603722, EPI_ISL_2603727, EPI_ISL_2603733, EPI_ISL_2603734, EPI_ISL_2603744, EPI_ISL_2603758, EPI_ISL_2603766, EPI_ISL_2603770, EPI_ISL_2603771, EPI_ISL_2603782, EPI_ISL_2603783, EPI_ISL_2603784, EPI_ISL_2603795, EPI_ISL_2603797, EPI_ISL_2603798, EPI_ISL_2603807, EPI_ISL_2603809, EPI_ISL_2692785, EPI_ISL_2692790, EPI_ISL_2692792, EPI_ISL_2692793, EPI_ISL_2692797, EPI_ISL_2692799, EPI_ISL_2692800, EPI_ISL_2692802, EPI_ISL_2692803 | | | |
| see above | Salud Digna | Instituto Nacional de Medicina Genomica | Abraham Campos-Romero; Cedro-Tanda A; Escobar-Arrazola; Gonzalez-Barrera D; Herrera-Montalvo LA.; Hidalgo-Miranda A; Luna-Ruiz Marco; M; Mendoza-Vargas A; Moreno-Camacho José Luis; Munguia-Garza P; Ramirez-Vega O; Rangel-DeLeon D; Reyes-Grajeda JP; Rodriguez-Gallegos Jorge |
| EPI_ISL_2840638, EPI_ISL_2842703, EPI_ISL_2842704, EPI_ISL_2842705 | San Diego County Public Health Laboratory | San Diego County Public Health Laboratory | Ashleigh Murphy Schafer; Brett Austin |
| EPI_ISL_2650572, EPI_ISL_2650573, EPI_ISL_2650574, EPI_ISL_2691314 | Santa Clara Valley Medical Center | Santa Clara County Public Health Laboratory | Santa Clara County Public Health Department |
| EPI_ISL_2627004 | Saratoga Hospital Laboratory | Wadsworth Center, New York State Department of Health | Alexis Russell; Catharine Prussing; Daryl M. Lamson; Erasmus Schneider; Erica Lasek-Nesselquist; John Kelly; Jonathan Plitnick; Kirsten St. George; Matthew Shudt; Melissa A Leisner; Navjot Singh |
| EPI_ISL_2736549, EPI_ISL_2736569 | Seattle Flu Study | Seattle Flu Study | Amanda Adler; Barry R. Lutz; Benjamin Pelle; Caitlin R. Wolf; Chris D. Frazar; Deborah A. Nickerson; Elisabeth Brandstetter; Erica Ryke; Helen Y. Chu; Janet A. Englund; Jay Shendure; Jover Lee; Kairsten Fay; Kirsten Lacombe; Lea M. Starita; Mark J. Rieder; Matthew Richardson; Matthew Thompson; Melissa Truong; Michael Boeckh; Michael Famulare; Misja Ilcisin; Peter D. Han; Thomas R. Sibley; Trevor Bedford |
| EPI_ISL_2680930 | Servicio Diagnóstico Clínico | Laboratorio nacional-LANIIA-CIAD | ; Alejandra García-Gasca; Alejandra Hernández-Terán; Alejandro Sánchez-Flores; Alfredo Herrera-Estrella; Alicia Ocaña-Mondragón; Andreu Comas-García; Angel Gustavo Salas-Lais; Antonio Loza Román; Bernardo Martínez-Miguel; Blanca Taboada; Brenda Irasema Maldonado-Meza; Bruno Gómez-Gil; Carla Ivón Herrera-Najera; Carlos F. Arias; Celia Boukadida; Clara Esperanza Santacruz-Tinoco; Concepción Grajales-Muñiz; Consorcio Mexicano de Vigilancia Genómica (CoViGen-Mex). Authors (in alphabetical order): Julio Elias Alvarado-Yaah; Cristóbal Cháidez-Quiróz; Célida Duque Molina; Célida Martínez- Rodríguez; Daniel Fregoso-Rueda; Daniel Lira Morales; Eduardo Becerril-Vargas; Fernando Fontove-Herrera; Fidencio Mejía-Nepomuceno; Francisco Pulido; Gloria Elena Espinosa-Ayala; Gloria María Molina-Salinas; Gloria Vazquez; Hector Esteban Paz-Juárez; Hector Montoya-Fuentes; Helen Haydee Fernanda Ramírez-Plascencia; Irvin González-López; Jean Pierre González; Jesús Hernández; Joel Armando Vázquez-Pérez.; Jorge Salas-Hernández; José Antonio Enciso-Moreno; José Arturo Martínez-Orozco; José Esteban Muñoz-Medina; José de Jesús Nuñez-Contreras; Juan Bautista Chale-Dzul; Julissa Enciso-Ibarra; Luis Alberto Ochoa-Carrera; Margarita Matías-Florentino; Mario Mújica-Sánchez; Marissa Perez-Garcia; María Guadalupe Santiago-Mauricio; María Guadalupe de Jesús Mireles-Rivera; Nelly Sélem-Mojica; Pavel Isa; Ricardo Ciria Merce; Ricardo Grande; Rosa María Gutiérrez Rios; Santiago Ávila-Ríos; Selene Zárate; Susana Lopez; Verónica Mata-Haro; Victor Eduardo García-Arias; Víctor Hugo Borja-Aburto |
| EPI_ISL_4082324, EPI_ISL_4082325, EPI_ISL_4082359, EPI_ISL_4082362, EPI_ISL_4082365, EPI_ISL_4082368, EPI_ISL_4082377, EPI_ISL_4082378, EPI_ISL_4082384, EPI_ISL_4082401, EPI_ISL_4082410, EPI_ISL_4082412, EPI_ISL_4082413, EPI_ISL_4082414, EPI_ISL_4082415, EPI_ISL_4082416, EPI_ISL_4082425, EPI_ISL_4082427, EPI_ISL_4082429, EPI_ISL_4082432, EPI_ISL_4082433, EPI_ISL_4082434, EPI_ISL_4082435, EPI_ISL_4082437, EPI_ISL_4082448, EPI_ISL_4082450, EPI_ISL_4082464, EPI_ISL_4082470, EPI_ISL_4082475, EPI_ISL_4082476, EPI_ISL_4082478, EPI_ISL_4082481, EPI_ISL_4082530, EPI_ISL_4082536, EPI_ISL_4082553, EPI_ISL_4082554, EPI_ISL_4082564, EPI_ISL_4082565, EPI_ISL_4082566, EPI_ISL_4082583, EPI_ISL_4082615, EPI_ISL_4082616, EPI_ISL_4082639, EPI_ISL_4082650, EPI_ISL_4082652, EPI_ISL_4082670, EPI_ISL_4082765, EPI_ISL_4083023, EPI_ISL_4083024, EPI_ISL_5158410, EPI_ISL_6694365 | | | |
| see above | Servicio Virosis Respiratorias-Departamento Virología-INEI | Instituto Nacional Enfermedades Infecciosas C.G.Malbran | Avaro M.; Baumeister E.; Benedetti E.; Campos J.; Cisterna D.; Dattero ME; De Belder D.; Haim MS.; Lorenzo F.; Molina V.; Perandones C.; Poklepovich T.; Pontoriero A.; Russo M.; Sanchez Loria J.; Tuduri E. |
| EPI_ISL_2558022, EPI_ISL_2558028, EPI_ISL_2558039, EPI_ISL_2558044 | Servicio de Microbiologia Hospital Ramon y Cajal | Servicio de Microbiologia Hospital Ramon y Cajal | Galan JC; Martinez L. Abreu M; Ponce M; y Gonzalez-Alba JM |
| EPI_ISL_2516674, EPI_ISL_2516683, EPI_ISL_2516693, EPI_ISL_2516694, EPI_ISL_2516695 | Servicio de Microbiología Clínica (Complejo Hospitalario de Navarra, Pamplona) | Centro de Secuenciación NASERTIC | Ana Miqueleiz; Ana Navascués; Carmen Ezpeleta Baquedano |
| EPI_ISL_2658202, EPI_ISL_2658207, EPI_ISL_2658212, EPI_ISL_2658215, EPI_ISL_2658227 | Servicio de Microbiología. Consorcio Hospital General Universitario de Valencia | SeqCOVID-SPAIN consortium/IBV(CSIC) | Begoña Fuster Escrivá; Carme Salvador García; Concepción Gimeno Cardona and SeqCOVID-SPAIN consortium; María Dolores Ocete; Rafael Medina González |
| EPI_ISL_2790433 | Servizo de Microbioloxía. Complexo Hospitalario de Santiago de Compostela | Servizo de Microbioloxía. Complexo Hospitalario de Santiago de Compostela | Amparo Coira; Antonio Aguilera; Carlos García_Riestra; Daniel Navarro; Gema Barbeito; Iria Roca; José Llovo; Laura Millán; Manuela Hernández; María Luisa Pérez_del_Molino; Mercedes Treviño; Rocío trastoy; Teresa Lopez_Valiño; Xosé Costa |
| EPI_ISL_5315550, EPI_ISL_5315553, EPI_ISL_5315561, EPI_ISL_5315575, EPI_ISL_5315593 | Shared Hospital Laboratory | Shared Hospital Laboratory | Christie Vermeiren; Finlay Maguire; Kevin Katz; Patryk Aftanas; Robert Kozak; Samira Mubareka |
| EPI_ISL_2688005, EPI_ISL_3394032, EPI_ISL_3394055 | Sonic Reference Laboratory | Sonic Reference Laboratory | "Robert Moller; Bijaya Dhakal; Cora Lahey; Ellen Pizarek"; John Woody; Krishna Maddula |
| EPI_ISL_2484888, EPI_ISL_2484889, EPI_ISL_2484890, EPI_ISL_2484891, EPI_ISL_2484892, EPI_ISL_2716898 | Southern Nevada Public Health Laboratory | Southern Nevada Public Health Laboratory | Michael Picker |
| EPI_ISL_2918821 | Spital Limmattal | Institute of Medical Virology | Alexandra Trkola; Annette Audigé; Catharine Aquino; Cyril Shah; Daniel Ehrsam; Gabriela Ziltener; Guido Bloemberg; Hubert Rehrauer; Isabel Stürmer; Joel Wirz; Jon Huder; Jürg Böni; Kevin Steiner; Maria Grünberg; Maryam Zaheri; Michael Huber; Riccarda Capaul; Stefan Schmutz; Verena Kufner; Weihong Qi |
| EPI_ISL_4496901 | Stanford Health Care | Stanford University School of Medicine, Clinical Virology Laboratory | Becky Jiang; Bernadette Troung; James Zehnder; Malaya K. Sahoo; Selamawit Bihon; and Benjamin A. Pinsky |
| EPI_ISL_2724699, EPI_ISL_2724711, EPI_ISL_2724748, EPI_ISL_2724749, EPI_ISL_2724766, EPI_ISL_2724777, EPI_ISL_2724783, EPI_ISL_2724788, EPI_ISL_2724801, EPI_ISL_2724825, EPI_ISL_2724826, EPI_ISL_2724828, EPI_ISL_2724830, EPI_ISL_2724833, EPI_ISL_2724850, EPI_ISL_2724884 | | | |
| see above | State Laboratories Division, Hawaii State Department of Health | State Laboratories Division, Hawaii State Department of Health | Ayana Garnet; Drew Kuwazaki; Edward Desmond; Pamela O'Brien; Razvan Sultana |
| EPI_ISL_2800574, EPI_ISL_2800575, EPI_ISL_2800576, EPI_ISL_2800580, EPI_ISL_2800585, EPI_ISL_2800586, EPI_ISL_2800587, EPI_ISL_2800588, EPI_ISL_2800589, EPI_ISL_2800590, EPI_ISL_2800591, EPI_ISL_2800592, EPI_ISL_2835236, EPI_ISL_2835237 | | | |
| see above | State Testing Facility | Altius Institute for Biomedical Research | Alex Isner; Alex Nguyen; Amanda Gale; Audra Johnson; Clem Green; Daniel Bates; Eric Thorland; Jacob Rodriguez; Jean Robinston; Jemma Nelson; Jessica Kunder; John Stamatoyannopoulos; Joshua Richards; Julia Wald; Kneshay Harper; Lauren Mitchell; Mark Frerker; Matt Hartman; Michael Buckley; Muhammad Halimun; Rebecca Bruders; Sadie Patraw; Sofia Olsson; Tobias Ragoczy |
| EPI_ISL_2389576, EPI_ISL_2389730 | Synlab MVZ Augsburg | Robert Koch Institute | |
| EPI_ISL_2626963, EPI_ISL_2626968 | TEMPUS LABS INC | Wadsworth Center, New York State Department of Health | Alexis Russell; Catharine Prussing; Daryl M. Lamson; Erasmus Schneider; Erica Lasek-Nesselquist; John Kelly; Jonathan Plitnick; Kirsten St. George; Matthew Shudt; Melissa A Leisner; Navjot Singh |
| EPI_ISL_2490995, EPI_ISL_2600393, EPI_ISL_2600454, EPI_ISL_2769277, EPI_ISL_2769300, EPI_ISL_2769303, EPI_ISL_2769339, EPI_ISL_3325992, EPI_ISL_3326594 | | | |
| see above | TGen North | TGen North | "Jolene Bowers; Ashlyn Pfeiffer; Brett Van Tassel; Chris French; Darrin Lemmer; Dave Engelthaler; Dave Engelthaler"; Hayley Yaglom; Heather Centner; Jolene Bowers; The Arizona COVID Genomics Union (ACGU)" |
| EPI_ISL_2565760, EPI_ISL_2565762, EPI_ISL_2658961 | TXDSHS | TXDSHS | Anita Pokharel; Bonnie Oh; Chun Wang; Grace Kubin; Jenny Zhang; Karen Bobier; Lorraine Rodriguez; Maliha Rahman; Mayela Pedrueza; Myong Koag; Rachel Lee; Rashmi Tuladhar |
| EPI_ISL_2790535, EPI_ISL_2790536 | Tampa General Hospital Esoteric Lab | Tampa General Hospital Esoteric Research & Development Lab | Amorce Lima; Deanna Becker; Dominic Uy; Elaine Vendrone; Jon Faughn; Jorge Mecias-Frias; Suzane Silbert; Vicki Healer |
| EPI_ISL_2716588, EPI_ISL_2716589, EPI_ISL_2756569, EPI_ISL_2756570, EPI_ISL_2756571, EPI_ISL_2756572, EPI_ISL_2756573, EPI_ISL_2756574, EPI_ISL_2756585 | | | |
| see above | The Caribbean Public Health Agency | Carrington Lab, Department of Preclinical Sciences, Faculty of Medical Sciences, The University of the West Indies, St Augustine | Anushka Ramjag; Arianne Brown-Jordan; Avery Hinds; Christine V. F. Carrington; Gabriel Escobar; Nikita S. D. Sahadeo; Nuno Faria; Oliver Pybus; Risha Singh; Roshan Parasram; Sarah Hill; SueMin Nathaniel; Vernie Ramkissoon |

| | | | |
|---|---|---|---|
| | | Campus | |
| EPI_ISL_2466707 | The Jackson Laboratory | The Jackson Laboratory | Adams M; Bergeron D; Kelly K; Li L; Long J; Omerza G; Renzette N |
| EPI_ISL_3930809 | The National University Hospital of Iceland | deCODE genetics | Agnar Helgason; Alma Moller; Arna B Agustsdottir; Arnaldur Gylfason; Asgeir Sigurdsson; Aslaug Jonasdottir; Berglind Eiriksdottir; Bjarni Thorbjornsson; Brynjar O Jensson; Daniel F Gudbjartsson; Droplaug N Magnusdottir; Elisabet E Gardarsdottir; Emil A Thorarensen; Gardar Sveinbjornsson; Gisli Masson; Gudmundur Georgsson; Gudmundur L Norddahl; Gudrun Sigmundsdottir; Hakon Jonsson; Hannes Eggertsson; Hilma Holm; Ingileif Jonsdottir; Jona Saemundsdottir; Kamilla S Josefsdottir; Kari Stefansson; Karl G Kristinsson; Kjartan R Gudmundsson; Kristin E Sveinsdottir; Kristjan E Hjorleifsson; Louise le Roux; Maney Sveinsdottir; Olafia S Gretarsdottir; Olafur T Magnusson; Pall Melsted; Patrick Sulem; Run Fridriksdottir; Solvi Rognvaldsson; Thora R Gunnarsdottir; Thordur Kristjansson; Thorolfur Gudnason; Unnur Thorsteinsdottir |
| EPI_ISL_2758876 | The Ohio State University Applied Microbiology Services Laboratory | The Ohio State University Applied Microbiology Services Laboratory | Seth A. Faith PhD |
| EPI_ISL_3152415, EPI_ISL_3152417 | TriCore Reference Laboratories | Center for Global Health, University of New Mexico Health Sciences Center | Cecilia Thompson; Darrell Dinwiddie; Daryl Domman; Karissa Culbreath; Kendra Pesko; Kurt Schwalm; Valerie Morley |
| EPI_ISL_3214690 | U.O. Microbiologia Laboratorio Unico Centro Servizi - AUSL della Romagna | U.O. Microbiologia, Laboratorio Unico Centro Servizi - AUSL della Romagna | Giorgio Dirani |
| EPI_ISL_6229837 | UBASF DR ERNANI BARREIRA FLORES PARANA E BRASILIA | ACME Lab, Oswaldo Cruz Foundation, FIOCRUZ/CE | Carlos Leonardo de Aragao Araujo; Cecilia Leite Costa & Eduardo Ruback dos Santos on behalf of COVID-19 FIOCRUZ Genomic Network; Cleber Furtado Aksenen; Fabio Miyajima; Fernando Braga Stehling; Francisco Eder de Moura Lopes; Igor Oliveira Duarte; Jamille Maria Mendes Bezerra; Joaquim Cesar do Nascimento Sousa Junior; Pedro Miguel Carneiro Jeronimo; Suzana Porto Almeida; Thais Ferreira de Oliveira; Thais de Oliveira Costa; Ticiane Cavalcante de Souza; Veridiana Pessoa Miyajima |
| EPI_ISL_6229839 | UBS CAIC II | ACME Lab, Oswaldo Cruz Foundation, FIOCRUZ/CE | Carlos Leonardo de Aragao Araujo; Cecilia Leite Costa & Eduardo Ruback dos Santos on behalf of COVID-19 FIOCRUZ Genomic Network; Cleber Furtado Aksenen; Fabio Miyajima; Fernando Braga Stehling; Francisco Eder de Moura Lopes; Igor Oliveira Duarte; Jamille Maria Mendes Bezerra; Joaquim Cesar do Nascimento Sousa Junior; Pedro Miguel Carneiro Jeronimo; Suzana Porto Almeida; Thais Ferreira de Oliveira; Thais de Oliveira Costa; Ticiane Cavalcante de Souza; Veridiana Pessoa Miyajima |
| EPI_ISL_3912271, EPI_ISL_3912272, EPI_ISL_3912273, EPI_ISL_3912274, EPI_ISL_3912275, EPI_ISL_3912276, EPI_ISL_3912277 | | | |
| see above | UBS CAIC II | Analytical Competence Molecular Epidemiology Lab/ACME, Oswaldo Cruz Foundation, Ceara (FIOCRUZ CE) | Cleber Furtado Aksenen; Fabio Miyajima; Fernando Braga Stehling; Francisco Eder de Moura Lopes; Jamille Maria Mendes Bezerra; Joaquim Cesar do Nascimento Sousa Junior; Pedro Miguel Carneiro Jeronimo; Suzana Porto Almeida & Lucas Delerino on behalf of COVID-19 FIOCRUZ Genomic Network; Thais Ferreira de Oliveira; Thais de Oliveira Costa; Ticiane Cavalcante de Souza; Veridiana Pessoa Miyajima |
| EPI_ISL_2462328, EPI_ISL_2494144, EPI_ISL_5649624 | UBS CELESTINO CARNIELO | Instituto Butantan | Antonio Jorge Martins; Claudia Renata dos Santos Barros; David Schlesinger; Debora Botequio Moretti; Dimas Tadeu Covas; Elaine Cristina Marqueze; Elaine Vieira Santos; Evandra Strazza Rodrigues; Heidge Fukumasu; Jayme Augusto de Souza-Neto; José Salvatore Leister Patané; Luiz Alcantara; Luiz Lehmann Coutinho; Maria Carolina Elias; Maurício Lacerda Nogueira; Rafael dos Santos Bezerra; Raul Machado Neto; Rejane Maria Tommasini Grotto; Ricardo Haddad; Sandra Coccuzzo Sampaio Vessoni; Simone Kashima; Svetoslav Nanev Slavov; Vincent Louis Viala |
| EPI_ISL_2493866, EPI_ISL_2493868, EPI_ISL_2493869 | UBS DE JAMBEIRO JOCELINA MEIRELES | Instituto Butantan | Antonio Jorge Martins; Claudia Renata dos Santos Barros; David Schlesinger; Debora Botequio Moretti; Dimas Tadeu Covas; Elaine Cristina Marqueze; Elaine Vieira Santos; Evandra Strazza Rodrigues; Heidge Fukumasu; Jayme Augusto de Souza-Neto; José Salvatore Leister Patané; Luiz Alcantara; Luiz Lehmann Coutinho; Maria Carolina Elias; Maurício Lacerda Nogueira; Rafael dos Santos Bezerra; Raul Machado Neto; Rejane Maria Tommasini Grotto; Ricardo Haddad; Sandra Coccuzzo Sampaio Vessoni; Simone Kashima; Svetoslav Nanev Slavov; Vincent Louis Viala |
| EPI_ISL_5650084, EPI_ISL_5650179 | UBS DE TABAPUA | Instituto Butantan | Antonio Jorge Martins; Claudia Renata dos Santos Barros; David Schlesinger; Debora Botequio Moretti; Dimas Tadeu Covas; Elaine Cristina Marqueze; Elaine Vieira Santos; Evandra Strazza Rodrigues; Heidge Fukumasu; Jayme Augusto de Souza-Neto; José Salvatore Leister Patané; Luiz Alcantara; Luiz Lehmann Coutinho; Maria Carolina Elias; Maurício Lacerda Nogueira; Rafael dos Santos Bezerra; Raul Machado Neto; Rejane Maria Tommasini Grotto; Ricardo Haddad; Sandra Coccuzzo Sampaio Vessoni; Simone Kashima; Svetoslav Nanev Slavov; Vincent Louis Viala |
| EPI_ISL_5649566, EPI_ISL_5649567 | UBS DE UBARANA | Instituto Butantan | Antonio Jorge Martins; Claudia Renata dos Santos Barros; David Schlesinger; Debora Botequio Moretti; Dimas Tadeu Covas; Elaine Cristina Marqueze; Elaine Vieira Santos; Evandra Strazza Rodrigues; Heidge Fukumasu; Jayme Augusto de Souza-Neto; José Salvatore Leister Patané; Luiz Alcantara; Luiz Lehmann Coutinho; Maria Carolina Elias; Maurício Lacerda Nogueira; Rafael dos Santos Bezerra; Raul Machado Neto; Rejane Maria Tommasini Grotto; Ricardo Haddad; Sandra Coccuzzo Sampaio Vessoni; Simone Kashima; Svetoslav Nanev Slavov; Vincent Louis Viala |
| EPI_ISL_6229789, EPI_ISL_6229816, EPI_ISL_6229874 | UBS DO BAIRRO SAO JOSE | ACME Lab, Oswaldo Cruz Foundation, FIOCRUZ/CE | Carlos Leonardo de Aragao Araujo; Cecilia Leite Costa & Eduardo Ruback dos Santos on behalf of COVID-19 FIOCRUZ Genomic Network; Cleber Furtado Aksenen; Fabio Miyajima; Fernando Braga Stehling; Francisco Eder de Moura Lopes; Igor Oliveira Duarte; Jamille Maria Mendes Bezerra; Joaquim Cesar do Nascimento Sousa Junior; Pedro Miguel Carneiro Jeronimo; Suzana Porto Almeida; Thais Ferreira de Oliveira; Thais de Oliveira Costa; Ticiane Cavalcante de Souza; Veridiana Pessoa Miyajima |
| EPI_ISL_3912284, EPI_ISL_3912285, EPI_ISL_3912286, EPI_ISL_3912287 | UBS DO BAIRRO SAO JOSE | Analytical Competence Molecular Epidemiology Lab/ACME, Oswaldo Cruz Foundation, Ceara (FIOCRUZ CE) | Cleber Furtado Aksenen; Fabio Miyajima; Fernando Braga Stehling; Francisco Eder de Moura Lopes; Jamille Maria Mendes Bezerra; Joaquim Cesar do Nascimento Sousa Junior; Pedro Miguel Carneiro Jeronimo; Suzana Porto Almeida & Lucas Delerino on behalf of COVID-19 FIOCRUZ Genomic Network; Thais Ferreira de Oliveira; Thais de Oliveira Costa; Ticiane Cavalcante de Souza; Veridiana Pessoa Miyajima |
| EPI_ISL_2462320, EPI_ISL_2462321, EPI_ISL_2493163, EPI_ISL_2494114, EPI_ISL_2494118, EPI_ISL_2494119, EPI_ISL_2494120, EPI_ISL_2494121, EPI_ISL_2494122, EPI_ISL_2494126, EPI_ISL_2494129 | | | |
| see above | UBS DR ALFREDO DANTAS DE SOUZA UMUARAMA | Instituto Butantan | Antonio Jorge Martins; Claudia Renata dos Santos Barros; David Schlesinger; Debora Botequio Moretti; Dimas Tadeu Covas; Elaine Cristina Marqueze; Elaine Vieira Santos; Evandra Strazza Rodrigues; Heidge Fukumasu; Jayme Augusto de Souza-Neto; José Salvatore Leister Patané; Luiz Alcantara; Luiz Lehmann Coutinho; Maria Carolina Elias; Maurício Lacerda Nogueira; Rafael dos Santos Bezerra; Raul Machado Neto; Rejane Maria Tommasini Grotto; Ricardo Haddad; Sandra Coccuzzo Sampaio Vessoni; Simone Kashima; Svetoslav Nanev Slavov; Vincent Louis Viala |
| EPI_ISL_5649649, EPI_ISL_5649758, EPI_ISL_5649774, EPI_ISL_5649782, EPI_ISL_5649958 | UBS DR WALDOMIRO FERREIRA NEVES SCRPARDO | Instituto Butantan | Antonio Jorge Martins; Claudia Renata dos Santos Barros; David Schlesinger; Debora Botequio Moretti; Dimas Tadeu Covas; Elaine Cristina Marqueze; Elaine Vieira Santos; Evandra Strazza Rodrigues; Heidge Fukumasu; Jayme Augusto de Souza-Neto; José Salvatore Leister Patané; Luiz Alcantara; Luiz Lehmann Coutinho; Maria Carolina Elias; Maurício Lacerda Nogueira; Rafael dos Santos Bezerra; Raul Machado Neto; Rejane Maria Tommasini Grotto; Ricardo Haddad; Sandra Coccuzzo Sampaio Vessoni; Simone Kashima; Svetoslav Nanev Slavov; Vincent Louis Viala |
| EPI_ISL_5649637 | UBS ESPIRITO SANTO DO TURVO | Instituto Butantan | Antonio Jorge Martins; Claudia Renata dos Santos Barros; David Schlesinger; Debora Botequio Moretti; Dimas Tadeu Covas; Elaine Cristina Marqueze; Elaine Vieira Santos; Evandra Strazza Rodrigues; Heidge Fukumasu; Jayme Augusto de Souza-Neto; José Salvatore Leister Patané; Luiz Alcantara; Luiz Lehmann Coutinho; Maria Carolina Elias; Maurício Lacerda Nogueira; Rafael dos Santos Bezerra; Raul Machado Neto; Rejane Maria Tommasini Grotto; Ricardo Haddad; Sandra Coccuzzo Sampaio Vessoni; Simone Kashima; Svetoslav Nanev Slavov; Vincent Louis Viala |
| EPI_ISL_5649695, EPI_ISL_5649697, EPI_ISL_5649808, EPI_ISL_5649942, EPI_ISL_5650017 | UBS GLENDA RODELLA DUMAS | Instituto Butantan | Antonio Jorge Martins; Claudia Renata dos Santos Barros; David Schlesinger; Debora Botequio Moretti; Dimas Tadeu Covas; Elaine Cristina Marqueze; Elaine Vieira Santos; Evandra Strazza Rodrigues; Heidge Fukumasu; Jayme Augusto de Souza-Neto; José Salvatore Leister Patané; Luiz Alcantara; Luiz Lehmann Coutinho; Maria Carolina Elias; Maurício Lacerda Nogueira; Rafael dos Santos Bezerra; Raul Machado Neto; Rejane Maria Tommasini Grotto; Ricardo Haddad; Sandra Coccuzzo Sampaio Vessoni; Simone Kashima; Svetoslav Nanev Slavov; Vincent Louis Viala |
| EPI_ISL_2462332 | UBS III DE ELDORADO ELDORADO S P | Instituto Butantan | Antonio Jorge Martins; Claudia Renata dos Santos Barros; David Schlesinger; Debora Botequio Moretti; Dimas Tadeu Covas; Elaine Cristina Marqueze; Elaine Vieira Santos; Evandra Strazza Rodrigues; Heidge Fukumasu; Jayme Augusto de Souza-Neto; José Salvatore Leister Patané; Luiz Alcantara; Luiz Lehmann Coutinho; Maria Carolina Elias; Maurício Lacerda Nogueira; Rafael dos Santos Bezerra; Raul Machado Neto; Rejane Maria Tommasini Grotto; Ricardo Haddad; Sandra Coccuzzo Sampaio Vessoni; Simone Kashima; Svetoslav Nanev Slavov; Vincent Louis Viala |
| EPI_ISL_2466733 | UBS III DE RANCHARIA | Instituto Butantan | Antonio Jorge Martins; Claudia Renata dos Santos Barros; David Schlesinger; Debora Botequio Moretti; Dimas Tadeu Covas; Elaine Cristina Marqueze; Elaine Vieira Santos; Evandra Strazza Rodrigues; Heidge Fukumasu; Jayme Augusto de Souza-Neto; José Salvatore Leister Patané; Luiz Alcantara; Luiz Lehmann Coutinho; Maria Carolina Elias; Maurício Lacerda Nogueira; Rafael dos Santos Bezerra; Raul Machado Neto; Rejane Maria Tommasini Grotto; Ricardo Haddad; Sandra Coccuzzo Sampaio Vessoni; Simone Kashima; Svetoslav Nanev Slavov; Vincent Louis Viala |
| EPI_ISL_2493820, EPI_ISL_2494320 | UBS JARDIM CAIUBY | Instituto Butantan | Antonio Jorge Martins; Claudia Renata dos Santos Barros; David Schlesinger; Debora Botequio Moretti; Dimas Tadeu Covas; Elaine Cristina Marqueze; Elaine Vieira Santos; Evandra Strazza Rodrigues; Heidge Fukumasu; Jayme Augusto de Souza-Neto; José Salvatore Leister Patané; Luiz Alcantara; Luiz Lehmann Coutinho; Maria Carolina Elias; Maurício Lacerda Nogueira; Rafael dos Santos Bezerra; Raul Machado Neto; Rejane Maria Tommasini Grotto; Ricardo Haddad; Sandra Coccuzzo Sampaio Vessoni; Simone Kashima; Svetoslav Nanev Slavov; Vincent Louis Viala |
| EPI_ISL_2493849, EPI_ISL_2494324 | UBS JARDIM ODETE | Instituto Butantan | Antonio Jorge Martins; Claudia Renata dos Santos Barros; David Schlesinger; Debora Botequio Moretti; Dimas Tadeu Covas; Elaine Cristina Marqueze; Elaine Vieira Santos; Evandra Strazza Rodrigues; Heidge Fukumasu; Jayme Augusto de Souza-Neto; José Salvatore Leister Patané; Luiz Alcantara; Luiz Lehmann Coutinho; Maria Carolina Elias; Maurício Lacerda Nogueira; Rafael dos Santos Bezerra; Raul Machado Neto; Rejane Maria Tommasini Grotto; Ricardo Haddad; Sandra Coccuzzo Sampaio Vessoni; Simone Kashima; Svetoslav Nanev Slavov; Vincent Louis Viala |
| EPI_ISL_6229795, EPI_ISL_6229796, EPI_ISL_6229797, EPI_ISL_6229798 | UBS MARATOAN | ACME Lab, Oswaldo Cruz Foundation, FIOCRUZ/CE | Carlos Leonardo de Aragao Araujo; Cecilia Leite Costa & Eduardo Ruback dos Santos on behalf of COVID-19 FIOCRUZ Genomic Network; Cleber Furtado Aksenen; Fabio Miyajima; Fernando Braga Stehling; Francisco Eder de Moura Lopes; Igor Oliveira Duarte; Jamille Maria Mendes Bezerra; Joaquim Cesar do Nascimento Sousa Junior; Pedro Miguel Carneiro Jeronimo; Suzana Porto Almeida; Thais Ferreira de Oliveira; Thais de Oliveira Costa; Ticiane Cavalcante de Souza; Veridiana Pessoa Miyajima |
| EPI_ISL_3912289 | UBS MARATOAN | Analytical Competence Molecular Epidemiology Lab/ACME, Oswaldo Cruz Foundation, Ceara (FIOCRUZ CE) | Cleber Furtado Aksenen; Fabio Miyajima; Fernando Braga Stehling; Francisco Eder de Moura Lopes; Jamille Maria Mendes Bezerra; Joaquim Cesar do Nascimento Sousa Junior; Pedro Miguel Carneiro Jeronimo; Suzana Porto Almeida & Lucas Delerino on behalf of COVID-19 FIOCRUZ Genomic Network; Thais Ferreira de Oliveira; Thais de Oliveira Costa; Ticiane Cavalcante de Souza; Veridiana Pessoa Miyajima |
| EPI_ISL_5650052, EPI_ISL_5650193 | UBS PASTOR MARCELINO DEUNGARO RIO GRANDE | Instituto Butantan | Antonio Jorge Martins; Claudia Renata dos Santos Barros; David Schlesinger; Debora Botequio Moretti; Dimas Tadeu Covas; Elaine Cristina Marqueze; Elaine Vieira Santos; Evandra Strazza Rodrigues; Heidge Fukumasu; Jayme Augusto de Souza-Neto; José Salvatore Leister Patané; Luiz Alcantara; Luiz Lehmann Coutinho; Maria Carolina Elias; Maurício Lacerda Nogueira; Rafael dos Santos Bezerra; Raul Machado Neto; Rejane Maria Tommasini Grotto; Ricardo Haddad; Sandra Coccuzzo Sampaio Vessoni; Simone Kashima; Svetoslav Nanev Slavov; Vincent Louis Viala |
| EPI_ISL_3321308, EPI_ISL_3370430, EPI_ISL_3370431, EPI_ISL_3370432, EPI_ISL_3370433 | UC-Christus Clinical Hospital Laboratory | Laboratory of Molecular Virology, School of Medicine, Pontificia Universidad Catolica de Chile | Ana Maira Guzman; Andres E. Munoz-Marcos; Catalina Pardo-Roa; Eileen Serrano; Erick Salinas; Estefany Poblete; Francisco Melo; Jorge Levican; Leonardo I. Almonacid; Maria Jose Avendano; Maria Patricia Vega; Rafael A. Medina; Ricardo Enrique de la Barra; Tamara Garcia-Salum |
| EPI_ISL_2960091 | UCC (Universidad Central del Caribe) | Grubaugh Lab - Yale School of Public Health | Anderson Brito; Andrea Firpo; Annie Watkins; Chaney Kalinich; Chantal Vogels; David Rivera Aponte; Ernesto Vazquez; Fabiola Cruz-Lopez; Fabiola Fontanet; Isabel Ott; Jessica Rothman; Joseph Fauver; Karolane Gonzalez; Kendall Billig; Mallery Breban; Mary Petrone; Natalie Machargo; Nathan Grubaugh; Patricia Serrano; Tania Mitwalli; Tara Alpert; Tobias Koch; Vianca Aponte |
| EPI_ISL_2617953 | UCLA Clinical Micro Lab | Los Angeles County Public Health Laboratories | P. Hemarajata et al. |
| EPI_ISL_2547293, EPI_ISL_2547300, EPI_ISL_2547301, EPI_ISL_2547302, EPI_ISL_2547303, EPI_ISL_2547313, EPI_ISL_2547319, EPI_ISL_2547320, EPI_ISL_2547321 | | | |
| see above | UHTL, University Hospitals | UHTL, University Hospitals | Alouani, D.; Sadri, N.; Song, X. |
| EPI_ISL_3385787 | UNIDAD DE DIAGNOSTICO HEMATO ONCOLOGICA | Instituto Nacional de Salud- Dirección de Investigación en Salud Pública | Carlos Franco-Muñoz; Carmen Osorio; Diana Malo; Diego A. Álvarez-Díaz; Diego Andrés Prada; Gerardo Santamaría; Hector Alejandro Ruiz-Moreno; Jhonnatan Reales-González; Jorge Rivera; Juan Camilo Martinez; Julian Naizaque; Katherine Laiton-Donato; Lisseth Pardo; Magdalena Wiesner; Marcela Mercado-Reyes; Maria T. Herrera-Sepúlveda; Marta Lopez Blanco; Martha Lucia Ospina Martinez; Paola Rojas; Sergio Gomez; Sheryll Corchuelo; Ángela Alarcon Cruz |
| EPI_ISL_5649650, EPI_ISL_5649671, EPI_ISL_5649816 | UNIDADE BASICA DE SAUDE ALBERTO CARLOS BOINTONI | Instituto Butantan | Antonio Jorge Martins; Claudia Renata dos Santos Barros; David Schlesinger; Debora Botequio Moretti; Dimas Tadeu Covas; Elaine Cristina Marqueze; Elaine Vieira Santos; Evandra Strazza Rodrigues; Heidge Fukumasu; Jayme Augusto de Souza-Neto; José Salvatore Leister Patané; Luiz Alcantara; Luiz Lehmann Coutinho; Maria Carolina Elias; Maurício Lacerda Nogueira; Rafael dos Santos Bezerra; Raul Machado Neto; Rejane Maria Tommasini Grotto; Ricardo Haddad; Sandra Coccuzzo Sampaio Vessoni; Simone Kashima; Svetoslav Nanev Slavov; Vincent Louis Viala |
| EPI_ISL_3912298 | UNIDADE BASICA DE SAUDE DIVINO ESPIRITO SANTO | Analytical Competence Molecular Epidemiology Lab/ACME, Oswaldo Cruz Foundation, Ceara (FIOCRUZ CE) | Cleber Furtado Aksenen; Fabio Miyajima; Fernando Braga Stehling; Francisco Eder de Moura Lopes; Jamille Maria Mendes Bezerra; Joaquim Cesar do Nascimento Sousa Junior; Pedro Miguel Carneiro Jeronimo; Suzana Porto Almeida & Lucas Delerino on behalf of COVID-19 FIOCRUZ Genomic Network; Thais Ferreira de Oliveira; Thais de Oliveira Costa; Ticiane Cavalcante de Souza; Veridiana Pessoa Miyajima |
| EPI_ISL_3912300 | UNIDADE BASICA DE SAUDE DO CAMPO DE AVIACAO | Analytical Competence Molecular Epidemiology Lab/ACME, Oswaldo Cruz Foundation, Ceara (FIOCRUZ CE) | Cleber Furtado Aksenen; Fabio Miyajima; Fernando Braga Stehling; Francisco Eder de Moura Lopes; Jamille Maria Mendes Bezerra; Joaquim Cesar do Nascimento Sousa Junior; Pedro Miguel Carneiro Jeronimo; Suzana Porto Almeida & Lucas Delerino on behalf of COVID-19 FIOCRUZ Genomic Network; Thais Ferreira de Oliveira; Thais de Oliveira Costa; Ticiane Cavalcante de Souza; Veridiana Pessoa Miyajima |
| EPI_ISL_3912301 | UNIDADE BASICA DE SAUDE DO SAO FRANCISCO | Analytical Competence Molecular Epidemiology Lab/ACME, Oswaldo Cruz Foundation, Ceara (FIOCRUZ CE) | Cleber Furtado Aksenen; Fabio Miyajima; Fernando Braga Stehling; Francisco Eder de Moura Lopes; Jamille Maria Mendes Bezerra; Joaquim Cesar do Nascimento Sousa Junior; Pedro Miguel Carneiro Jeronimo; Suzana Porto Almeida & Lucas Delerino on behalf of COVID-19 FIOCRUZ Genomic Network; Thais Ferreira de Oliveira; Thais de Oliveira Costa; Ticiane Cavalcante de Souza; Veridiana Pessoa Miyajima |

| | | | |
|---|---|---|---|
| EPI_ISL_2494175, EPI_ISL_2494187 | UNIDADE BASICA DE SAUDE DR MATHEUS GABRIEL BONASSA | Instituto Butantan | Antonio Jorge Martins; Claudia Renata dos Santos Barros; David Schlesinger; Debora Botequio Moretti; Dimas Tadeu Covas; Elaine Cristina Marqueze; Elaine Vieira Santos; Evandra Strazza Rodrigues; Heidge Fukumasu; Jayme Augusto de Souza-Neto; José Salvatore Leister Patané; Luiz Alcantara; Luiz Lehmann Coutinho; Maria Carolina Elias; Maurício Lacerda Nogueira; Rafael dos Santos Bezerra; Raul Machado Neto; Rejane Maria Tommasini Grotto; Ricardo Haddad; Sandra Coccuzzo Sampaio Vessoni; Simone Kashima; Svetoslav Nanev Slavov; Vincent Louis Viala |
| EPI_ISL_5646065, EPI_ISL_5646067 | UNIDADE BASICA DE SAUDE GUAICARA | Instituto Butantan | Antonio Jorge Martins; Claudia Renata dos Santos Barros; David Schlesinger; Debora Botequio Moretti; Dimas Tadeu Covas; Elaine Cristina Marqueze; Elaine Vieira Santos; Evandra Strazza Rodrigues; Heidge Fukumasu; Jayme Augusto de Souza-Neto; José Salvatore Leister Patané; Luiz Alcantara; Luiz Lehmann Coutinho; Maria Carolina Elias; Maurício Lacerda Nogueira; Rafael dos Santos Bezerra; Raul Machado Neto; Rejane Maria Tommasini Grotto; Ricardo Haddad; Sandra Coccuzzo Sampaio Vessoni; Simone Kashima; Svetoslav Nanev Slavov; Vincent Louis Viala |
| EPI_ISL_5646083 | UNIDADE BASICA DE SAUDE II DE TARABAI | Instituto Butantan | Antonio Jorge Martins; Claudia Renata dos Santos Barros; David Schlesinger; Debora Botequio Moretti; Dimas Tadeu Covas; Elaine Cristina Marqueze; Elaine Vieira Santos; Evandra Strazza Rodrigues; Heidge Fukumasu; Jayme Augusto de Souza-Neto; José Salvatore Leister Patané; Luiz Alcantara; Luiz Lehmann Coutinho; Maria Carolina Elias; Maurício Lacerda Nogueira; Rafael dos Santos Bezerra; Raul Machado Neto; Rejane Maria Tommasini Grotto; Ricardo Haddad; Sandra Coccuzzo Sampaio Vessoni; Simone Kashima; Svetoslav Nanev Slavov; Vincent Louis Viala |
| EPI_ISL_3912309 | UNIDADE DE PRONTO ATENDIMENTO ARACATI UPA ARACATI | Analytical Competence Molecular Epidemiology Lab/ACME, Oswaldo Cruz Foundation, Ceara (FIOCRUZ CE) | Cleber Furtado Aksenen; Fabio Miyajima; Fernando Braga Stehling; Francisco Eder de Moura Lopes; Jamille Maria Mendes Bezerra; Joaquim Cesar do Nascimento Sousa Junior; Pedro Miguel Carneiro Jeronimo; Suzana Porto Almeida & Lucas Delerino on behalf of COVID-19 FIOCRUZ Genomic Network; Thais Ferreira de Oliveira; Thais de Oliveira Costa; Ticiane Cavalcante de Souza; Veridiana Pessoa Miyajima |
| EPI_ISL_6229807 | UNIDADE DE PRONTO ATENDIMENTO DE IGUATU UPA | ACME Lab, Oswaldo Cruz Foundation, FIOCRUZ/CE | Carlos Leonardo de Aragao Araujo; Cecilia Leite Costa & Eduardo Ruback dos Santos on behalf of COVID-19 FIOCRUZ Genomic Network; Cleber Furtado Aksenen; Fabio Miyajima; Fernando Braga Stehling; Francisco Eder de Moura Lopes; Igor Oliveira Duarte; Jamille Maria Mendes Bezerra; Joaquim Cesar do Nascimento Sousa Junior; Pedro Miguel Carneiro Jeronimo; Suzana Porto Almeida; Thais Ferreira de Oliveira; Thais de Oliveira Costa; Ticiane Cavalcante de Souza; Veridiana Pessoa Miyajima |
| EPI_ISL_2493835 | UNIDADE DE PRONTO ATENDIMENTO DE PITANGUEIRAS | Instituto Butantan | Antonio Jorge Martins; Claudia Renata dos Santos Barros; David Schlesinger; Debora Botequio Moretti; Dimas Tadeu Covas; Elaine Cristina Marqueze; Elaine Vieira Santos; Evandra Strazza Rodrigues; Heidge Fukumasu; Jayme Augusto de Souza-Neto; José Salvatore Leister Patané; Luiz Alcantara; Luiz Lehmann Coutinho; Maria Carolina Elias; Maurício Lacerda Nogueira; Rafael dos Santos Bezerra; Raul Machado Neto; Rejane Maria Tommasini Grotto; Ricardo Haddad; Sandra Coccuzzo Sampaio Vessoni; Simone Kashima; Svetoslav Nanev Slavov; Vincent Louis Viala |
| EPI_ISL_6229808 | UNIDADE DE PSF VILA CENTENARIO | ACME Lab, Oswaldo Cruz Foundation, FIOCRUZ/CE | Carlos Leonardo de Aragao Araujo; Cecilia Leite Costa & Eduardo Ruback dos Santos on behalf of COVID-19 FIOCRUZ Genomic Network; Cleber Furtado Aksenen; Fabio Miyajima; Fernando Braga Stehling; Francisco Eder de Moura Lopes; Igor Oliveira Duarte; Jamille Maria Mendes Bezerra; Joaquim Cesar do Nascimento Sousa Junior; Pedro Miguel Carneiro Jeronimo; Suzana Porto Almeida; Thais Ferreira de Oliveira; Thais de Oliveira Costa; Ticiane Cavalcante de Souza; Veridiana Pessoa Miyajima |
| EPI_ISL_2493803, EPI_ISL_2493804, EPI_ISL_2493805, EPI_ISL_2493806, EPI_ISL_5646039, EPI_ISL_5646077 | UNIDADE DE SAUDE DA FAMILIA DR AMELIO TANGANELLI | Instituto Butantan | Antonio Jorge Martins; Claudia Renata dos Santos Barros; David Schlesinger; Debora Botequio Moretti; Dimas Tadeu Covas; Elaine Cristina Marqueze; Elaine Vieira Santos; Evandra Strazza Rodrigues; Heidge Fukumasu; Jayme Augusto de Souza-Neto; José Salvatore Leister Patané; Luiz Alcantara; Luiz Lehmann Coutinho; Maria Carolina Elias; Maurício Lacerda Nogueira; Rafael dos Santos Bezerra; Raul Machado Neto; Rejane Maria Tommasini Grotto; Ricardo Haddad; Sandra Coccuzzo Sampaio Vessoni; Simone Kashima; Svetoslav Nanev Slavov; Vincent Louis Viala |
| EPI_ISL_2493844 | UNIDADE DE SAUDE DA FAMILIA JARDIM AMERICA ITAQUAQUECETUBA | Instituto Butantan | Antonio Jorge Martins; Claudia Renata dos Santos Barros; David Schlesinger; Debora Botequio Moretti; Dimas Tadeu Covas; Elaine Cristina Marqueze; Elaine Vieira Santos; Evandra Strazza Rodrigues; Heidge Fukumasu; Jayme Augusto de Souza-Neto; José Salvatore Leister Patané; Luiz Alcantara; Luiz Lehmann Coutinho; Maria Carolina Elias; Maurício Lacerda Nogueira; Rafael dos Santos Bezerra; Raul Machado Neto; Rejane Maria Tommasini Grotto; Ricardo Haddad; Sandra Coccuzzo Sampaio Vessoni; Simone Kashima; Svetoslav Nanev Slavov; Vincent Louis Viala |
| EPI_ISL_2494181 | UNIDADE DE VIGILANCIA EPIDEMIOLOGICA | Instituto Butantan | Antonio Jorge Martins; Claudia Renata dos Santos Barros; David Schlesinger; Debora Botequio Moretti; Dimas Tadeu Covas; Elaine Cristina Marqueze; Elaine Vieira Santos; Evandra Strazza Rodrigues; Heidge Fukumasu; Jayme Augusto de Souza-Neto; José Salvatore Leister Patané; Luiz Alcantara; Luiz Lehmann Coutinho; Maria Carolina Elias; Maurício Lacerda Nogueira; Rafael dos Santos Bezerra; Raul Machado Neto; Rejane Maria Tommasini Grotto; Ricardo Haddad; Sandra Coccuzzo Sampaio Vessoni; Simone Kashima; Svetoslav Nanev Slavov; Vincent Louis Viala |
| EPI_ISL_2493947 | UNIDADE MISTA ARACOIABA DA SERRA | Instituto Butantan | Antonio Jorge Martins; Claudia Renata dos Santos Barros; David Schlesinger; Debora Botequio Moretti; Dimas Tadeu Covas; Elaine Cristina Marqueze; Elaine Vieira Santos; Evandra Strazza Rodrigues; Heidge Fukumasu; Jayme Augusto de Souza-Neto; José Salvatore Leister Patané; Luiz Alcantara; Luiz Lehmann Coutinho; Maria Carolina Elias; Maurício Lacerda Nogueira; Rafael dos Santos Bezerra; Raul Machado Neto; Rejane Maria Tommasini Grotto; Ricardo Haddad; Sandra Coccuzzo Sampaio Vessoni; Simone Kashima; Svetoslav Nanev Slavov; Vincent Louis Viala |
| EPI_ISL_2494174 | UNIDADE MISTA DE SAUDE RAFARD | Instituto Butantan | Antonio Jorge Martins; Claudia Renata dos Santos Barros; David Schlesinger; Debora Botequio Moretti; Dimas Tadeu Covas; Elaine Cristina Marqueze; Elaine Vieira Santos; Evandra Strazza Rodrigues; Heidge Fukumasu; Jayme Augusto de Souza-Neto; José Salvatore Leister Patané; Luiz Alcantara; Luiz Lehmann Coutinho; Maria Carolina Elias; Maurício Lacerda Nogueira; Rafael dos Santos Bezerra; Raul Machado Neto; Rejane Maria Tommasini Grotto; Ricardo Haddad; Sandra Coccuzzo Sampaio Vessoni; Simone Kashima; Svetoslav Nanev Slavov; Vincent Louis Viala |
| EPI_ISL_2493807, EPI_ISL_2493808 | UNIDADE SAUDE FAMILIA DR PIERRE ANTONIO COPPIETERS AREALVA | Instituto Butantan | Antonio Jorge Martins; Claudia Renata dos Santos Barros; David Schlesinger; Debora Botequio Moretti; Dimas Tadeu Covas; Elaine Cristina Marqueze; Elaine Vieira Santos; Evandra Strazza Rodrigues; Heidge Fukumasu; Jayme Augusto de Souza-Neto; José Salvatore Leister Patané; Luiz Alcantara; Luiz Lehmann Coutinho; Maria Carolina Elias; Maurício Lacerda Nogueira; Rafael dos Santos Bezerra; Raul Machado Neto; Rejane Maria Tommasini Grotto; Ricardo Haddad; Sandra Coccuzzo Sampaio Vessoni; Simone Kashima; Svetoslav Nanev Slavov; Vincent Louis Viala |
| EPI_ISL_2493897, EPI_ISL_2493912 | UNIDADE SENTINELA COVID19 | Instituto Butantan | Antonio Jorge Martins; Claudia Renata dos Santos Barros; David Schlesinger; Debora Botequio Moretti; Dimas Tadeu Covas; Elaine Cristina Marqueze; Elaine Vieira Santos; Evandra Strazza Rodrigues; Heidge Fukumasu; Jayme Augusto de Souza-Neto; José Salvatore Leister Patané; Luiz Alcantara; Luiz Lehmann Coutinho; Maria Carolina Elias; Maurício Lacerda Nogueira; Rafael dos Santos Bezerra; Raul Machado Neto; Rejane Maria Tommasini Grotto; Ricardo Haddad; Sandra Coccuzzo Sampaio Vessoni; Simone Kashima; Svetoslav Nanev Slavov; Vincent Louis Viala |
| EPI_ISL_2493907 | UNIDADE SENTINELA VILA APARECIDA | Instituto Butantan | Antonio Jorge Martins; Claudia Renata dos Santos Barros; David Schlesinger; Debora Botequio Moretti; Dimas Tadeu Covas; Elaine Cristina Marqueze; Elaine Vieira Santos; Evandra Strazza Rodrigues; Heidge Fukumasu; Jayme Augusto de Souza-Neto; José Salvatore Leister Patané; Luiz Alcantara; Luiz Lehmann Coutinho; Maria Carolina Elias; Maurício Lacerda Nogueira; Rafael dos Santos Bezerra; Raul Machado Neto; Rejane Maria Tommasini Grotto; Ricardo Haddad; Sandra Coccuzzo Sampaio Vessoni; Simone Kashima; Svetoslav Nanev Slavov; Vincent Louis Viala |
| EPI_ISL_5649696 | UPA 24 HS UNIDADE DE PRONTO ATENDIMENTO DR MARIO NUNES MIRAN | Instituto Butantan | Antonio Jorge Martins; Claudia Renata dos Santos Barros; David Schlesinger; Debora Botequio Moretti; Dimas Tadeu Covas; Elaine Cristina Marqueze; Elaine Vieira Santos; Evandra Strazza Rodrigues; Heidge Fukumasu; Jayme Augusto de Souza-Neto; José Salvatore Leister Patané; Luiz Alcantara; Luiz Lehmann Coutinho; Maria Carolina Elias; Maurício Lacerda Nogueira; Rafael dos Santos Bezerra; Raul Machado Neto; Rejane Maria Tommasini Grotto; Ricardo Haddad; Sandra Coccuzzo Sampaio Vessoni; Simone Kashima; Svetoslav Nanev Slavov; Vincent Louis Viala |
| EPI_ISL_2462322, EPI_ISL_2494116, EPI_ISL_2494124, EPI_ISL_2494125 | UPA DE BEBEDOURO | Instituto Butantan | Antonio Jorge Martins; Claudia Renata dos Santos Barros; David Schlesinger; Debora Botequio Moretti; Dimas Tadeu Covas; Elaine Cristina Marqueze; Elaine Vieira Santos; Evandra Strazza Rodrigues; Heidge Fukumasu; Jayme Augusto de Souza-Neto; José Salvatore Leister Patané; Luiz Alcantara; Luiz Lehmann Coutinho; Maria Carolina Elias; Maurício Lacerda Nogueira; Rafael dos Santos Bezerra; Raul Machado Neto; Rejane Maria Tommasini Grotto; Ricardo Haddad; Sandra Coccuzzo Sampaio Vessoni; Simone Kashima; Svetoslav Nanev Slavov; Vincent Louis Viala |
| EPI_ISL_5646370 | UPA IPIRANGA | Instituto Butantan | Antonio Jorge Martins; Claudia Renata dos Santos Barros; David Schlesinger; Debora Botequio Moretti; Dimas Tadeu Covas; Elaine Cristina Marqueze; Elaine Vieira Santos; Evandra Strazza Rodrigues; Heidge Fukumasu; Jayme Augusto de Souza-Neto; José Salvatore Leister Patané; Luiz Alcantara; Luiz Lehmann Coutinho; Maria Carolina Elias; Maurício Lacerda Nogueira; Rafael dos Santos Bezerra; Raul Machado Neto; Rejane Maria Tommasini Grotto; Ricardo Haddad; Sandra Coccuzzo Sampaio Vessoni; Simone Kashima; Svetoslav Nanev Slavov; Vincent Louis Viala |
| EPI_ISL_5646313 | UPA MARY DOTA | Instituto Butantan | Antonio Jorge Martins; Claudia Renata dos Santos Barros; David Schlesinger; Debora Botequio Moretti; Dimas Tadeu Covas; Elaine Cristina Marqueze; Elaine Vieira Santos; Evandra Strazza Rodrigues; Heidge Fukumasu; Jayme Augusto de Souza-Neto; José Salvatore Leister Patané; Luiz Alcantara; Luiz Lehmann Coutinho; Maria Carolina Elias; Maurício Lacerda Nogueira; Rafael dos Santos Bezerra; Raul Machado Neto; Rejane Maria Tommasini Grotto; Ricardo Haddad; Sandra Coccuzzo Sampaio Vessoni; Simone Kashima; Svetoslav Nanev Slavov; Vincent Louis Viala |
| EPI_ISL_2493816, EPI_ISL_2493848, EPI_ISL_2493852, EPI_ISL_2493858 | UPA SADAKO SEDOGUTI | Instituto Butantan | Antonio Jorge Martins; Claudia Renata dos Santos Barros; David Schlesinger; Debora Botequio Moretti; Dimas Tadeu Covas; Elaine Cristina Marqueze; Elaine Vieira Santos; Evandra Strazza Rodrigues; Heidge Fukumasu; Jayme Augusto de Souza-Neto; José Salvatore Leister Patané; Luiz Alcantara; Luiz Lehmann Coutinho; Maria Carolina Elias; Maurício Lacerda Nogueira; Rafael dos Santos Bezerra; Raul Machado Neto; Rejane Maria Tommasini Grotto; Ricardo Haddad; Sandra Coccuzzo Sampaio Vessoni; Simone Kashima; Svetoslav Nanev Slavov; Vincent Louis Viala |
| EPI_ISL_2868467, EPI_ISL_2868527 | URMC LABS | Wadsworth Center, New York State Department of Health | Alexis Russell; Catharine Prussing; Daryl M. Lamson; Erasmus Schneider; Erica Lasek-Nesselquist; John Kelly; Jonathan Plitnick; Kirsten St. George; Matthew Shudt; Melissa A Leisner; Navjot Singh |
| EPI_ISL_2715302, EPI_ISL_2715306, EPI_ISL_2715316 | US Air Force School of Aerospace Medicine | US Air Force School of Aerospace Medicine | Amanda Javorina; Anthony Fries; Carol Garrett; Clarise Starr; Elizabeth Macias; Jennifer Meyer; Sarah Purves; William Gruner |
| EPI_ISL_2493879 | USAFA MARACANA | Instituto Butantan | Antonio Jorge Martins; Claudia Renata dos Santos Barros; David Schlesinger; Debora Botequio Moretti; Dimas Tadeu Covas; Elaine Cristina Marqueze; Elaine Vieira Santos; Evandra Strazza Rodrigues; Heidge Fukumasu; Jayme Augusto de Souza-Neto; José Salvatore Leister Patané; Luiz Alcantara; Luiz Lehmann Coutinho; Maria Carolina Elias; Maurício Lacerda Nogueira; Rafael dos Santos Bezerra; Raul Machado Neto; Rejane Maria Tommasini Grotto; Ricardo Haddad; Sandra Coccuzzo Sampaio Vessoni; Simone Kashima; Svetoslav Nanev Slavov; Vincent Louis Viala |
| EPI_ISL_2493895 | USAFA MIRIM | Instituto Butantan | Antonio Jorge Martins; Claudia Renata dos Santos Barros; David Schlesinger; Debora Botequio Moretti; Dimas Tadeu Covas; Elaine Cristina Marqueze; Elaine Vieira Santos; Evandra Strazza Rodrigues; Heidge Fukumasu; Jayme Augusto de Souza-Neto; José Salvatore Leister Patané; Luiz Alcantara; Luiz Lehmann Coutinho; Maria Carolina Elias; Maurício Lacerda Nogueira; Rafael dos Santos Bezerra; Raul Machado Neto; Rejane Maria Tommasini Grotto; Ricardo Haddad; Sandra Coccuzzo Sampaio Vessoni; Simone Kashima; Svetoslav Nanev Slavov; Vincent Louis Viala |
| EPI_ISL_2493831 | USAFA SAO JORGE | Instituto Butantan | Antonio Jorge Martins; Claudia Renata dos Santos Barros; David Schlesinger; Debora Botequio Moretti; Dimas Tadeu Covas; Elaine Cristina Marqueze; Elaine Vieira Santos; Evandra Strazza Rodrigues; Heidge Fukumasu; Jayme Augusto de Souza-Neto; José Salvatore Leister Patané; Luiz Alcantara; Luiz Lehmann Coutinho; Maria Carolina Elias; Maurício Lacerda Nogueira; Rafael dos Santos Bezerra; Raul Machado Neto; Rejane Maria Tommasini Grotto; Ricardo Haddad; Sandra Coccuzzo Sampaio Vessoni; Simone Kashima; Svetoslav Nanev Slavov; Vincent Louis Viala |
| EPI_ISL_5649628, EPI_ISL_5649631 | USF DR SAMUEL MARTINS FIGUEIRA SCRPARDO | Instituto Butantan | Antonio Jorge Martins; Claudia Renata dos Santos Barros; David Schlesinger; Debora Botequio Moretti; Dimas Tadeu Covas; Elaine Cristina Marqueze; Elaine Vieira Santos; Evandra Strazza Rodrigues; Heidge Fukumasu; Jayme Augusto de Souza-Neto; José Salvatore Leister Patané; Luiz Alcantara; Luiz Lehmann Coutinho; Maria Carolina Elias; Maurício Lacerda Nogueira; Rafael dos Santos Bezerra; Raul Machado Neto; Rejane Maria Tommasini Grotto; Ricardo Haddad; Sandra Coccuzzo Sampaio Vessoni; Simone Kashima; Svetoslav Nanev Slavov; Vincent Louis Viala |
| EPI_ISL_2493801 | USF JARDIM AMERICA FIGUEIRA | Instituto Butantan | Antonio Jorge Martins; Claudia Renata dos Santos Barros; David Schlesinger; Debora Botequio Moretti; Dimas Tadeu Covas; Elaine Cristina Marqueze; Elaine Vieira Santos; Evandra Strazza Rodrigues; Heidge Fukumasu; Jayme Augusto de Souza-Neto; José Salvatore Leister Patané; Luiz Alcantara; Luiz Lehmann Coutinho; Maria Carolina Elias; Maurício Lacerda Nogueira; Rafael dos Santos Bezerra; Raul Machado Neto; Rejane Maria Tommasini Grotto; Ricardo Haddad; Sandra Coccuzzo Sampaio Vessoni; Simone Kashima; Svetoslav Nanev Slavov; Vincent Louis Viala |
| EPI_ISL_2493811, EPI_ISL_2493814, EPI_ISL_2493817 | USF JARDIM CAMANDUCAIA | Instituto Butantan | Antonio Jorge Martins; Claudia Renata dos Santos Barros; David Schlesinger; Debora Botequio Moretti; Dimas Tadeu Covas; Elaine Cristina Marqueze; Elaine Vieira Santos; Evandra Strazza Rodrigues; Heidge Fukumasu; Jayme Augusto de Souza-Neto; José Salvatore Leister Patané; Luiz Alcantara; Luiz Lehmann Coutinho; Maria Carolina Elias; Maurício Lacerda Nogueira; Rafael dos Santos Bezerra; Raul Machado Neto; Rejane Maria Tommasini Grotto; Ricardo Haddad; Sandra Coccuzzo Sampaio Vessoni; Simone Kashima; Svetoslav Nanev Slavov; Vincent Louis Viala |
| EPI_ISL_2493799, EPI_ISL_2493810, EPI_ISL_2493812, EPI_ISL_2493813 | USF JARDIM DAS AVES MOREIRINHA | Instituto Butantan | Antonio Jorge Martins; Claudia Renata dos Santos Barros; David Schlesinger; Debora Botequio Moretti; Dimas Tadeu Covas; Elaine Cristina Marqueze; Elaine Vieira Santos; Evandra Strazza Rodrigues; Heidge Fukumasu; Jayme Augusto de Souza-Neto; José Salvatore Leister Patané; Luiz Alcantara; Luiz Lehmann Coutinho; Maria Carolina Elias; Maurício Lacerda Nogueira; Rafael dos Santos Bezerra; Raul Machado Neto; Rejane Maria Tommasini Grotto; Ricardo Haddad; Sandra Coccuzzo Sampaio Vessoni; Simone Kashima; Svetoslav Nanev Slavov; Vincent Louis Viala |
| EPI_ISL_2493819 | USF JARDIM JOSELY | Instituto Butantan | Antonio Jorge Martins; Claudia Renata dos Santos Barros; David Schlesinger; Debora Botequio Moretti; Dimas Tadeu Covas; Elaine Cristina Marqueze; Elaine Vieira Santos; Evandra Strazza Rodrigues; Heidge Fukumasu; Jayme Augusto de Souza-Neto; José Salvatore Leister Patané; Luiz Alcantara; Luiz Lehmann Coutinho; Maria Carolina Elias; Maurício Lacerda Nogueira; Rafael dos Santos Bezerra; Raul Machado Neto; Rejane Maria Tommasini Grotto; Ricardo Haddad; Sandra Coccuzzo Sampaio Vessoni; Simone Kashima; Svetoslav Nanev Slavov; Vincent Louis Viala |
| EPI_ISL_2493798, EPI_ISL_2493809 | USF JARDIM SAO DIMAS | Instituto Butantan | Antonio Jorge Martins; Claudia Renata dos Santos Barros; David Schlesinger; Debora Botequio Moretti; Dimas Tadeu Covas; Elaine Cristina Marqueze; Elaine Vieira Santos; Evandra Strazza Rodrigues; Heidge Fukumasu; Jayme Augusto de Souza-Neto; José Salvatore Leister Patané; Luiz Alcantara; Luiz Lehmann Coutinho; Maria Carolina Elias; Maurício Lacerda Nogueira; Rafael dos Santos Bezerra; Raul Machado Neto; Rejane Maria Tommasini Grotto; Ricardo Haddad; Sandra Coccuzzo Sampaio Vessoni; Simone Kashima; Svetoslav Nanev Slavov; Vincent Louis Viala |
| EPI_ISL_2493815 | USF JARDIM SILVESTRE II | Instituto Butantan | Antonio Jorge Martins; Claudia Renata dos Santos Barros; David Schlesinger; Debora Botequio Moretti; Dimas Tadeu Covas; Elaine Cristina Marqueze; Elaine Vieira Santos; Evandra Strazza Rodrigues; Heidge Fukumasu; Jayme Augusto de Souza-Neto; José Salvatore Leister Patané; Luiz Alcantara; Luiz Lehmann Coutinho; Maria Carolina Elias; Maurício Lacerda Nogueira; Rafael dos Santos Bezerra; Raul Machado Neto; Rejane Maria Tommasini Grotto; Ricardo Haddad; Sandra Coccuzzo Sampaio Vessoni; Simone Kashima; Svetoslav Nanev Slavov; Vincent Louis Viala |
| EPI_ISL_5649686 | USF PAULISTA FERNANDOPOLIS ANTONIO PIVATO | Instituto Butantan | Antonio Jorge Martins; Claudia Renata dos Santos Barros; David Schlesinger; Debora Botequio Moretti; Dimas Tadeu Covas; Elaine Cristina Marqueze; Elaine Vieira Santos; Evandra Strazza Rodrigues; Heidge Fukumasu; Jayme Augusto de Souza-Neto; José Salvatore Leister Patané; Luiz Alcantara; Luiz Lehmann Coutinho; Maria Carolina Elias; Maurício Lacerda Nogueira; Rafael dos Santos Bezerra; Raul Machado Neto; Rejane Maria Tommasini Grotto; Ricardo Haddad; Sandra Coccuzzo Sampaio Vessoni; Simone Kashima; Svetoslav Nanev Slavov; Vincent Louis Viala |
| EPI_ISL_5649588, EPI_ISL_5649606, EPI_ISL_5649620, EPI_ISL_5649710, EPI_ISL_5649744, EPI_ISL_5649749, EPI_ISL_5649790, EPI_ISL_5649877 | | | |
| see above | USF SALERNO | Instituto Butantan | Antonio Jorge Martins; Claudia Renata dos Santos Barros; David Schlesinger; Debora Botequio Moretti; Dimas Tadeu Covas; Elaine Cristina Marqueze; Elaine Vieira Santos; Evandra Strazza Rodrigues; Heidge Fukumasu; Jayme Augusto de Souza-Neto; José Salvatore Leister Patané; Luiz Alcantara; Luiz Lehmann Coutinho; Maria Carolina Elias; Maurício Lacerda Nogueira; Rafael dos Santos Bezerra; Raul Machado Neto; Rejane Maria Tommasini Grotto; Ricardo Haddad; Sandra Coccuzzo Sampaio Vessoni; Simone Kashima; Svetoslav Nanev Slavov; Vincent Louis Viala |
| EPI_ISL_2493818 | USF SANTA MARIA DO AMPARO | Instituto Butantan | Antonio Jorge Martins; Claudia Renata dos Santos Barros; David Schlesinger; Debora Botequio Moretti; Dimas Tadeu Covas; Elaine Cristina Marqueze; Elaine Vieira Santos; Evandra Strazza Rodrigues; Heidge Fukumasu; Jayme Augusto de Souza-Neto; José Salvatore Leister Patané; Luiz Alcantara; Luiz Lehmann Coutinho; Maria Carolina Elias; Maurício Lacerda Nogueira; Rafael dos Santos Bezerra; Raul Machado Neto; Rejane Maria Tommasini Grotto; Ricardo Haddad; Sandra Coccuzzo Sampaio Vessoni; Simone Kashima; Svetoslav Nanev Slavov; Vincent Louis Viala |
| EPI_ISL_2494145 | USF SERROTE REGISTRO | Instituto Butantan | Antonio Jorge Martins; Claudia Renata dos Santos Barros; David Schlesinger; Debora Botequio Moretti; Dimas Tadeu Covas; Elaine Cristina Marqueze; Elaine Vieira Santos; Evandra Strazza Rodrigues; Heidge Fukumasu; Jayme Augusto de Souza-Neto; José Salvatore Leister Patané; Luiz Alcantara; Luiz Lehmann Coutinho; Maria Carolina Elias; Maurício Lacerda Nogueira; Rafael dos Santos Bezerra; Raul Machado Neto; Rejane Maria Tommasini Grotto; Ricardo Haddad; Sandra Coccuzzo Sampaio Vessoni; Simone Kashima; Svetoslav Nanev Slavov; Vincent Louis Viala |
| EPI_ISL_2493800 | USF TRES PONTES | Instituto Butantan | Antonio Jorge Martins; Claudia Renata dos Santos Barros; David Schlesinger; Debora Botequio Moretti; Dimas Tadeu Covas; Elaine Cristina Marqueze; Elaine Vieira Santos; Evandra Strazza Rodrigues; Heidge Fukumasu; Jayme Augusto de Souza-Neto; José Salvatore Leister Patané; Luiz Alcantara; Luiz Lehmann Coutinho; Maria Carolina Elias; Maurício Lacerda Nogueira; Rafael dos Santos Bezerra; Raul Machado Neto; Rejane Maria Tommasini Grotto; Ricardo Haddad; Sandra Coccuzzo Sampaio Vessoni; Simone Kashima; Svetoslav Nanev Slavov; Vincent Louis Viala |
| EPI_ISL_2493914 | USF YEN WANN SHENG | Instituto Butantan | Antonio Jorge Martins; Claudia Renata dos Santos Barros; David Schlesinger; Debora Botequio Moretti; Dimas Tadeu Covas; Elaine Cristina Marqueze; Elaine Vieira Santos; Evandra Strazza Rodrigues; Heidge Fukumasu; Jayme Augusto de Souza-Neto; José Salvatore Leister Patané; Luiz Alcantara; Luiz Lehmann Coutinho; Maria Carolina Elias; Maurício Lacerda Nogueira; Rafael dos Santos Bezerra; Raul Machado Neto; Rejane Maria Tommasini Grotto; Ricardo Haddad; Sandra Coccuzzo Sampaio Vessoni; Simone Kashima; Svetoslav Nanev Slavov; Vincent Louis Viala |
| EPI_ISL_2432601, EPI_ISL_2432602, EPI_ISL_2432619, EPI_ISL_2497190, EPI_ISL_2497191, EPI_ISL_2497193, EPI_ISL_2497196, EPI_ISL_2497200, EPI_ISL_2497208, EPI_ISL_2497213, EPI_ISL_2497214, EPI_ISL_2497215, EPI_ISL_2497216, EPI_ISL_2497220, EPI_ISL_2497221, EPI_ISL_2497222, EPI_ISL_2497230, EPI_ISL_2497231, EPI_ISL_2497232, EPI_ISL_2497237, EPI_ISL_2497245, EPI_ISL_2497246, EPI_ISL_2497247, EPI_ISL_2497250, EPI_ISL_2497261, EPI_ISL_2497263, EPI_ISL_2497268, EPI_ISL_2497269, EPI_ISL_2497275, EPI_ISL_2497280, EPI_ISL_2497287, EPI_ISL_2497289, EPI_ISL_2497292, EPI_ISL_2497295, EPI_ISL_2497297, EPI_ISL_2497299, EPI_ISL_2497303, EPI_ISL_2497308, EPI_ISL_2497309, EPI_ISL_2497310, EPI_ISL_2497316, EPI_ISL_2497321, EPI_ISL_2497325, EPI_ISL_2497327, EPI_ISL_2497338, EPI_ISL_2497344, EPI_ISL_2497350, EPI_ISL_2497356, EPI_ISL_2497365, EPI_ISL_2497372, EPI_ISL_2497383, EPI_ISL_2497384, EPI_ISL_2497392, EPI_ISL_2497396, EPI_ISL_2497397, EPI_ISL_2497398, EPI_ISL_2497414, EPI_ISL_2497419, EPI_ISL_2497932, EPI_ISL_2497933, EPI_ISL_2497934, EPI_ISL_2497935, EPI_ISL_2497936, EPI_ISL_2497937, EPI_ISL_2497938, EPI_ISL_2497939, | | | |

| | | | |
|---|---|---|---|
| EPI_ISL_2497940, EPI_ISL_2545020, EPI_ISL_2545035, EPI_ISL_2545036, EPI_ISL_2545045, EPI_ISL_2545058, EPI_ISL_2545061, EPI_ISL_2545068, EPI_ISL_2545073, EPI_ISL_2545075, EPI_ISL_2545082, EPI_ISL_2545087, EPI_ISL_2545089, EPI_ISL_2545090, EPI_ISL_2545093, EPI_ISL_2545099, EPI_ISL_2545104, EPI_ISL_2545107, EPI_ISL_2545119, EPI_ISL_2545123, EPI_ISL_2545127, EPI_ISL_2545139, EPI_ISL_2545143, EPI_ISL_2545150, EPI_ISL_2545153, EPI_ISL_2545154, EPI_ISL_2545155, EPI_ISL_2545156, EPI_ISL_2545157, EPI_ISL_2545158, EPI_ISL_2545159, EPI_ISL_2545160, EPI_ISL_2545161, EPI_ISL_2545162, EPI_ISL_2545163, EPI_ISL_2545164, EPI_ISL_2545165, EPI_ISL_2545166, EPI_ISL_2545167, EPI_ISL_2545168, EPI_ISL_2545169, EPI_ISL_2545170, EPI_ISL_2545171, EPI_ISL_2545172, EPI_ISL_2545173, EPI_ISL_2545174, EPI_ISL_2545175, EPI_ISL_2545176, EPI_ISL_2545177, EPI_ISL_3411451 | | | |
| see above | UW Virology Lab | UW Virology Lab | Alexander Greninger; Hong Xie; Keith R Jerome; Lasata Shrestha; Maria Lukes; Meei-Li Huang; Michelle Lin; Nathan Breit; Noah R. Baker; Patrick Mathias; Pavitra Roychoudhury; Ricardo Perez; Robert J. Livingston; Sean Ellis; Shah Mohamed Bakhash; Tien V. Nguyen |
| EPI_ISL_2671502, EPI_ISL_2671503, EPI_ISL_2671504, EPI_ISL_2671685, EPI_ISL_2671687, EPI_ISL_2671688, EPI_ISL_2671689, EPI_ISL_2671692, EPI_ISL_2671694, EPI_ISL_2671695, EPI_ISL_2671697, EPI_ISL_2671751, EPI_ISL_2671753, EPI_ISL_2671755, EPI_ISL_2671756, EPI_ISL_2671757, EPI_ISL_2671784, EPI_ISL_2671785, EPI_ISL_2970029 | | | |
| see above | Unidad de Investigacion Medica de Yucatan (UIMY) | Unidad de Genomica Avanzada | ; Alejandra Garcia-Gasca; Alejandra Hernandez-Teran; Alejandro Sanchez-Flores; Alfredo Herrera-Estrella; Alicia Ocaña-Mondragon; Andreu Comas-Garcia; Angel Gustavo Salas-Lais; Antonio Loza Roman; Bernardo Martinez-Miguel; Blanca Taboada; Brenda Irasema Maldonado-Meza; Bruno Gomez-Gil; Carla Ivon Herrera-Najera; Carlos F. Arias; Celia Boukadida; Celida Duque Molina; Celida Martinez- Rodriguez; Clara Esperanza Santacruz-Tinoco; Concepcion Grajales-Muñiz; Consorcio Mexicano de Vigilancia Genomica (CoViGen-Mex). Authors (in alphabetical order): Julio Elias Alvarado-Yaah; Cristobal Chaidez-Quiroz; Daniel Fregoso-Rueda; Daniel Lira Morales; Eduardo Becerril-Vargas; Fernando Fontove-Herrera; Fidencio Mejia-Nepomuceno; Francisco Pulido; Gloria Elena Espinosa-Ayala; Gloria Maria Molina-Salinas; Gloria Vazquez; Hector Esteban Paz-Juarez; Hector Montoya-Fuentes; Helen Haydee Fernanda Ramirez-Plascencia; Irvin Gonzalez-Lopez; Jean Pierre Gonzalez; Jesus Hernandez; Joel Armando Vazquez-Perez.; Jorge Salas-Hernandez; Jose Antonio Enciso-Moreno; Jose Arturo Martinez-Orozco; Jose Esteban Muñoz-Medina; Jose de Jesus Nuñez-Contreras; Juan Bautista Chale-Dzul; Julissa Enciso-Ibarra; Luis Alberto Ochoa-Carrera; Margarita Matias-Florentino; Maria Guadalupe Santiago-Mauricio; Maria Guadalupe de Jesus Mireles-Rivera; Mario Mujica-Sanchez; Marissa Perez-Garcia; Nelly Selem-Mojica; Pavel Isa; Ricardo Ciria Merce; Ricardo Grande; Rosa Maria Gutierrez Rios; Santiago avila-Rios; Selene Zarate; Susana Lopez; Veronica Mata-Haro; Victor Eduardo Garcia-Arias; Victor Hugo Borja-Aburto |
| EPI_ISL_2681346 | Unidad de Investigación Biomédica de Zacatecas (UIBZ) | Instituto de Biotecnología de la UNAM | ; Alejandra García-Gasca; Alejandra Hernández-Terán; Alejandro Sánchez-Flores; Alfredo Herrera-Estrella; Alicia Ocaña-Mondragón; Andreu Comas-García; Angel Gustavo Salas-Lais; Antonio Loza Román; Bernardo Martínez-Miguel; Blanca Taboada; Brenda Irasema Maldonado-Meza; Bruno Gómez-Gil; Carla Ivón Herrera-Najera; Carlos F. Arias; Celia Boukadida; Clara Esperanza Santacruz-Tinoco; Concepción Grajales-Muñiz; Consorcio Mexicano de Vigilancia Genómica (CoViGen-Mex). Authors (in alphabetical order): Julio Elias Alvarado-Yaah; Cristóbal Cháidez-Quiróz; Célida Duque Molina; Célida Martínez- Rodríguez; Daniel Fregoso-Rueda; Daniel Lira Morales; Eduardo Becerril-Vargas; Fernando Fontove-Herrera; Fidencio Mejía-Nepomuceno; Francisco Pulido; Gloria Elena Espinosa-Ayala; Gloria María Molina-Salinas; Gloria Vazquez; Hector Esteban Paz-Juárez; Hector Montoya-Fuentes; Helen Haydee Fernanda Ramírez-Plascencia; Irvin González-López; Jean Pierre González; Jesús Hernández; Joel Armando Vázquez-Pérez.; Jorge Salas-Hernández; José Antonio Enciso-Moreno; José Arturo Martínez-Orozco; José Esteban Muñoz-Medina; José de Jesús Nuñez-Contreras; Juan Bautista Chale-Dzul; Julissa Enciso-Ibarra; Luis Alberto Ochoa-Carrera; Margarita Matías-Florentino; Mario Mújica-Sánchez; Marissa Perez-Garcia; María Guadalupe Santiago-Mauricio; María Guadalupe de Jesús Mireles-Rivera; Nelly Sélem-Mojica; Pavel Isa; Ricardo Ciria Merce; Ricardo Grande; Rosa María Gutiérrez Rios; Santiago Ávila-Ríos; Selene Zárate; Susana Lopez; Verónica Mata-Haro; Victor Eduardo García-Arias; Víctor Hugo Borja-Aburto |
| EPI_ISL_2681055, EPI_ISL_2681077, EPI_ISL_2681078, EPI_ISL_2681084, EPI_ISL_2681090, EPI_ISL_2681292, EPI_ISL_2681304, EPI_ISL_2681338, EPI_ISL_2681353 | | | |
| see above | Unidad de Investigación Médica de Yucatán (UIMY) | Instituto de Biotecnología de la UNAM | ; Alejandra García-Gasca; Alejandra Hernández-Terán; Alejandro Sánchez-Flores; Alfredo Herrera-Estrella; Alicia Ocaña-Mondragón; Andreu Comas-García; Angel Gustavo Salas-Lais; Antonio Loza Román; Bernardo Martínez-Miguel; Blanca Taboada; Brenda Irasema Maldonado-Meza; Bruno Gómez-Gil; Carla Ivón Herrera-Najera; Carlos F. Arias; Celia Boukadida; Clara Esperanza Santacruz-Tinoco; Concepción Grajales-Muñiz; Consorcio Mexicano de Vigilancia Genómica (CoViGen-Mex). Authors (in alphabetical order): Julio Elias Alvarado-Yaah; Cristóbal Cháidez-Quiróz; Célida Duque Molina; Célida Martínez- Rodríguez; Daniel Fregoso-Rueda; Daniel Lira Morales; Eduardo Becerril-Vargas; Fernando Fontove-Herrera; Fidencio Mejía-Nepomuceno; Francisco Pulido; Gloria Elena Espinosa-Ayala; Gloria María Molina-Salinas; Gloria Vazquez; Hector Esteban Paz-Juárez; Hector Montoya-Fuentes; Helen Haydee Fernanda Ramírez-Plascencia; Irvin González-López; Jean Pierre González; Jesús Hernández; Joel Armando Vázquez-Pérez.; Jorge Salas-Hernández; José Antonio Enciso-Moreno; José Arturo Martínez-Orozco; José Esteban Muñoz-Medina; José de Jesús Nuñez-Contreras; Juan Bautista Chale-Dzul; Julissa Enciso-Ibarra; Luis Alberto Ochoa-Carrera; Margarita Matías-Florentino; Mario Mújica-Sánchez; Marissa Perez-Garcia; María Guadalupe Santiago-Mauricio; María Guadalupe de Jesús Mireles-Rivera; Nelly Sélem-Mojica; Pavel Isa; Ricardo Ciria Merce; Ricardo Grande; Rosa María Gutiérrez Rios; Santiago Ávila-Ríos; Selene Zárate; Susana Lopez; Verónica Mata-Haro; Victor Eduardo García-Arias; Víctor Hugo Borja-Aburto |
| EPI_ISL_2691569, EPI_ISL_2691574, EPI_ISL_2691575, EPI_ISL_2691576, EPI_ISL_2691577, EPI_ISL_2691578, EPI_ISL_2691579, EPI_ISL_2691580, EPI_ISL_2691581, EPI_ISL_2691582, EPI_ISL_2691583, EPI_ISL_2691584, EPI_ISL_2691585, EPI_ISL_2691586, EPI_ISL_2691587, EPI_ISL_2691588, EPI_ISL_2691589, EPI_ISL_2691590, EPI_ISL_2691591, EPI_ISL_2691592, EPI_ISL_2691593, EPI_ISL_2691594, EPI_ISL_2691595, EPI_ISL_2691596, EPI_ISL_2691597, EPI_ISL_2691598, EPI_ISL_2691599, EPI_ISL_2691600, EPI_ISL_2691601, EPI_ISL_2691602, EPI_ISL_2691603, EPI_ISL_2691604, EPI_ISL_2691605, EPI_ISL_2691606, EPI_ISL_2691607, EPI_ISL_2691608, EPI_ISL_2691609, EPI_ISL_2691610, EPI_ISL_2691611, EPI_ISL_2691612, EPI_ISL_2691613, EPI_ISL_2691614, EPI_ISL_2691615, EPI_ISL_2691616, EPI_ISL_2691617, EPI_ISL_2691618, EPI_ISL_2691619, EPI_ISL_2691620, EPI_ISL_2691621, EPI_ISL_2691622, EPI_ISL_2691627, EPI_ISL_2691628, EPI_ISL_2691629, EPI_ISL_2691630, EPI_ISL_2691631, EPI_ISL_2691632, EPI_ISL_2691633, EPI_ISL_2691634, EPI_ISL_2691635, EPI_ISL_2691636, EPI_ISL_2691637, EPI_ISL_2691638, EPI_ISL_2691639, EPI_ISL_2691640, EPI_ISL_2691643, EPI_ISL_2691644, EPI_ISL_2691647, EPI_ISL_2691648, EPI_ISL_2691649, EPI_ISL_2691650, EPI_ISL_2691651, EPI_ISL_2691652, EPI_ISL_2691653, EPI_ISL_2691654, EPI_ISL_2691655, EPI_ISL_2691656, EPI_ISL_2691657, EPI_ISL_2691658, EPI_ISL_2691659, EPI_ISL_2691660, EPI_ISL_2691661, EPI_ISL_2691662, EPI_ISL_2691663, EPI_ISL_2691664, EPI_ISL_2691665, EPI_ISL_2691666, EPI_ISL_2691667, EPI_ISL_2691668, EPI_ISL_2691669, EPI_ISL_2691670, EPI_ISL_2691671, EPI_ISL_2691672, EPI_ISL_2691673, EPI_ISL_2691674, EPI_ISL_2691675, EPI_ISL_2691676, EPI_ISL_2691677, EPI_ISL_2691678, EPI_ISL_2691679, EPI_ISL_2691680, EPI_ISL_2691681, EPI_ISL_2691682, EPI_ISL_2691683, EPI_ISL_2691684 | | | |
| see above | Unidade de apoio ao diagnostico da COVID - UNADIG | Bioinformatics Laboratory / LNCC | Alessandra P Lamarca; Alexandra L Gerber; Amilcar Tanuri; Ana Paula de C Guimaraes; Ana Tereza R Vasconcelos; Andrea Cony Cavalcanti; Caio Luiz Pereira Ribeiro; Cassia Alves; Cintia Policarpo; Claudia Maria Braga de Mello; Cristiane Gomes da Silva; Diana Mariani; Douglas Terra Machado; Flavio Dias da Silva; Gleidson da Silva de Oliveira; Leandro Magalhaes de Souza; Liliane Cavalcante; Luiz G P de Almeida; Marcio Henrique de Oliveira Garcia; Mario Sergio Ribeiro; Ronaldo da Silva F Jr; Silvia Carvalho |
| EPI_ISL_2544253 | Unilabs | Institute of Medical Virology | Alexandra Trkola; Annette Audigé; Cyril Shah; Gabriela Ziltener; Guido Bloemberg; Jon Huder; Jürg Böni; Kevin Steiner; Maria Grünberg; Maryam Zaheri; Michael Huber; Riccarda Capaul; Stefan Schmutz; Verena Kufner |
| EPI_ISL_2898512 | Unilabs | Karolinska University Hospital | Annelie Bjerkner; Isak Sylvin; Jan Albert; Karolina Ininbergs; Lina Guerra Blomqvist; Lynda Eneh; Martin Ekman; Martina Wahlund; Robert Dyrdak; Sandra Broddesson; Tanja Normark; Tobias Allander; Valtteri Wirta; Zhibing Yun |
| EPI_ISL_3477567 | Universidad del Valle, LDAB-Laboratorio de Diagnostico de Agentes Biologicos | Universidad del Valle, TAO-Lab, VIREM, NEAS Network | Andres Castillo; Beatriz Parra & COVID-19 Team Univalle; Diana López-Alvarez; Nelson Rivera Franco |
| EPI_ISL_2405336 | University Hospitals of Geneva, Laboratory of Virology | HUG, Laboratory of Virology and the Health2030 Genome Center | Ana Rita Goncalves; Deborah Penet; Emmanouil Dermitzakis; Henri Pegeot; Ioannis Xenarios; Keith Harshman; Laurent Kaiser; Lorenzo Cerutti; Melyssa Elies; Samuel Cordey |
| EPI_ISL_2692019, EPI_ISL_2692022, EPI_ISL_2692023, EPI_ISL_2931105 | University of Chicago Medicine | RIPHL at Rush University Medical Center | Felix Araujo Perez; Kevin Kunstman; Laura Furtado; Lauren Megger; Marieta Hyde; Max Kolton; Stefan Green |
| EPI_ISL_2692112, EPI_ISL_2692121 | University of Illinois at Chicago | RIPHL at Rush University Medical Center | Felix Araujo Perez; Julie Bentea; Kevin Kunstman; Laura Furtado; Marieta Hyde; Max Kolton; Omar Perez; Stefan Green |
| EPI_ISL_2382835, EPI_ISL_2382847, EPI_ISL_2382857, EPI_ISL_2382858, EPI_ISL_2382862, EPI_ISL_2483874, EPI_ISL_2483875, EPI_ISL_2483882 | | | |
| see above | University of Liège COVID-19 testing center | GIGA Medical Genomics | Bouchra Boujemla; Cécile Meex; Fabrice Bureau; Keith Durkin; Laurent Gillet; Maria Artesi; Marie-Pierre Hayette; Nathalie Renotte; Sébastien Bontems; Vincent Bours; Wouter Coppieters |
| EPI_ISL_2497895 | University of Mississippi Medical Center, Department of Pathology | University of Mississippi Medical Center, Molecular and Genomics Core Facility | Ashley C. Johnson; D. Ashley Robinson; Ithiel J. Frame; Krishna K. Ayyalasomayajula; Michael R. Garrett |
| EPI_ISL_2650508, EPI_ISL_2650510, EPI_ISL_2650512, EPI_ISL_2650515 | University of Oregon COVID-19 MAP Laboratory | University of Oregon Genomics and Cell Charactericzation Core Facility (GC3F) | Ariana White; Douglas Turnbull; Jason Carriere; Jason Sydes; Jeff Bishop; Peter Batzel |
| EPI_ISL_7635970, EPI_ISL_7635971, EPI_ISL_7635972, EPI_ISL_7635975, EPI_ISL_7635982 | University of Rome Tor Vergata: Departm Experim Medicine Chair of Virology | University of Rome Tor Vergata: Departm Experim Medicine Chair of Virology | Francesca Ceccherini-Silberstein; Loredana Sarmati; Lorenzo Piermatteo; Luca Carioti; Marco Iannetta; Maria Concetta Bellocchi; Massimo Andreoni |
| EPI_ISL_2467792, EPI_ISL_2467793 | University of Wisconsin-Madison AIDS Vaccine Research Laboratories | University of Wisconsin-Madison AIDS Vaccine Research Laboratories | Gage Moreno; Katarina Braun; et al. AIDS Vaccine Research Laboratories |
| EPI_ISL_2600748, EPI_ISL_2600749, EPI_ISL_2600750, EPI_ISL_2600751, EPI_ISL_2600752, EPI_ISL_2600753, EPI_ISL_2600754, EPI_ISL_2600755, EPI_ISL_2600756, EPI_ISL_2600760, EPI_ISL_2600766, EPI_ISL_2600768, EPI_ISL_2600769, EPI_ISL_2600770, EPI_ISL_2600771, EPI_ISL_2600772, EPI_ISL_2600773, EPI_ISL_2600774, EPI_ISL_2600775, EPI_ISL_2612314 | | | |
| see above | Università degli Studi di Perugia | Istituto Zooprofilattico Sperimentale dell'Abruzzo e Molise "G. Caporale" | Ancora M; Biagetti M; Calistri P; Camilloni B; Cammà C; Curini V; Delli Compagni E; Di Domenico M; Di Pasquale A; Giammarioli M; Lorusso A; Mangone I; Marcacci M; Mencacci A; Puglia I; Rinaldi A; Savini G; Scialabba S |
| EPI_ISL_2484682, EPI_ISL_2918786 | UniversitätsSpital Zürich | Institute of Medical Virology | Alexandra Trkola; Annette Audigé; Catharine Aquino; Cyril Shah; Daniel Ehrsam; Gabriela Ziltener; Guido Bloemberg; Hubert Rehrauer; Isabel Stürmer; Joel Wirz; Jon Huder; Jürg Böni; Kevin Steiner; Maria Grünberg; Maryam Zaheri; Michael Huber; Riccarda Capaul; Stefan Schmutz; Verena Kufner; Weihong Qi |
| EPI_ISL_2845768 | Universitätsklinikum Heidelberg | Robert Koch Institute | |
| EPI_ISL_5904936 | Universitätsklinikum Ulm - Institut für Virologie | Robert Koch Institute | |
| EPI_ISL_3030354 | Urbino | Microbiology University Politecnica delle Marche | Anna Valenza; Carla Acciarri; Katia Marinelli; Monica Lucia Ferreri; Patrizia Bagnarelli; Roberta Longo; Sara Caucci; Stefano Menzo |
| EPI_ISL_2678933, EPI_ISL_2678949, EPI_ISL_2678952, EPI_ISL_2678954, EPI_ISL_2678965, EPI_ISL_2679234, EPI_ISL_2679289, EPI_ISL_2679299, EPI_ISL_2679344, EPI_ISL_2856371, EPI_ISL_2856376, EPI_ISL_2856380, EPI_ISL_2856394, EPI_ISL_2857336, EPI_ISL_2857346, EPI_ISL_2857347, EPI_ISL_2857348 | | | |
| see above | Utah Public Health Laboratory | Utah Public Health Laboratory | Erin L. Young; Kelly F. Oakeson; Olinto Linares-Perdomo; Tara Gallagher |
| EPI_ISL_3355159, EPI_ISL_3355191 | VA-Division of Consolidated Laboratory Services | Centers for Disease Control and Prevention Division of Viral Diseases, Pathogen Discovery | Alex Burgin; Ben Rambo-Martin; Clinton Paden; Dakota Howard; Dave Wentworth; Dhwani Batra; Jasmine Padilla; Justin Lee; Krista Queen; Kristen Knipe; Kristine Lacek; Mark Burroughs; Matthew Schmerer; Meghan Bentz; Mili Sheth; Peter Cook; Sam Shepard; Sarah Nobles; Suxiang Tong; Vivien Dugan; Yvette Unoarumhi |
| EPI_ISL_2462333 | VIGILANCIA EM SAUDE | Instituto Butantan | Antonio Jorge Martins; Claudia Renata dos Santos Barros; David Schlesinger; Debora Botequio Moretti; Dimas Tadeu Covas; Elaine Cristina Marqueze; Elaine Vieira Santos; Evandra Strazza Rodrigues; Heidge Fukumasu; Jayme Augusto de Souza-Neto; José Salvatore Leister Patané; Luiz Alcantara; Luiz Lehmann Coutinho; Maria Carolina Elias; Maurício Lacerda Nogueira; Rafael dos Santos Bezerra; Raul Machado Neto; Rejane Maria Tommasini Grotto; Ricardo Haddad; Sandra Coccuzzo Sampaio Vessoni; Simone Kashima; Svetoslav Nanev Slavov; Vincent Louis Viala |
| EPI_ISL_2462316, EPI_ISL_2493841, EPI_ISL_2493843 | VIGILANCIA EM SAUDE DE FCO MORATO | Instituto Butantan | Antonio Jorge Martins; Claudia Renata dos Santos Barros; David Schlesinger; Debora Botequio Moretti; Dimas Tadeu Covas; Elaine Cristina Marqueze; Elaine Vieira Santos; Evandra Strazza Rodrigues; Heidge Fukumasu; Jayme Augusto de Souza-Neto; José Salvatore Leister Patané; Luiz Alcantara; Luiz Lehmann Coutinho; Maria Carolina Elias; Maurício Lacerda Nogueira; Rafael dos Santos Bezerra; Raul Machado Neto; Rejane Maria Tommasini Grotto; Ricardo Haddad; Sandra Coccuzzo Sampaio Vessoni; Simone Kashima; Svetoslav Nanev Slavov; Vincent Louis Viala |
| EPI_ISL_2493159, EPI_ISL_2493166, EPI_ISL_2494111, EPI_ISL_2494112, EPI_ISL_2494113 | VIGILANCIA EM SAUDE SAE VIGILANCIA EPIDEMIOLOGICA | Instituto Butantan | Antonio Jorge Martins; Claudia Renata dos Santos Barros; David Schlesinger; Debora Botequio Moretti; Dimas Tadeu Covas; Elaine Cristina Marqueze; Elaine Vieira Santos; Evandra Strazza Rodrigues; Heidge Fukumasu; Jayme Augusto de Souza-Neto; José Salvatore Leister Patané; Luiz Alcantara; Luiz Lehmann Coutinho; Maria Carolina Elias; Maurício Lacerda Nogueira; Rafael dos Santos Bezerra; Raul Machado Neto; Rejane Maria Tommasini Grotto; Ricardo Haddad; Sandra Coccuzzo Sampaio Vessoni; Simone Kashima; Svetoslav Nanev Slavov; Vincent Louis Viala |
| EPI_ISL_2493916 | VIGILANCIA EPIDEMIOLOGICA DE FERRAZ DE VASCONCELOS | Instituto Butantan | Antonio Jorge Martins; Claudia Renata dos Santos Barros; David Schlesinger; Debora Botequio Moretti; Dimas Tadeu Covas; Elaine Cristina Marqueze; Elaine Vieira Santos; Evandra Strazza Rodrigues; Heidge Fukumasu; Jayme Augusto de Souza-Neto; José Salvatore Leister Patané; Luiz Alcantara; Luiz Lehmann Coutinho; Maria Carolina Elias; Maurício Lacerda Nogueira; Rafael dos Santos Bezerra; Raul Machado Neto; Rejane Maria Tommasini Grotto; Ricardo Haddad; Sandra Coccuzzo Sampaio Vessoni; Simone Kashima; Svetoslav Nanev Slavov; Vincent Louis Viala |
| EPI_ISL_2494162, EPI_ISL_2494173, EPI_ISL_2494336 | VIGILANCIA EPIDEMIOLOGICA DE LEME | Instituto Butantan | Antonio Jorge Martins; Claudia Renata dos Santos Barros; David Schlesinger; Debora Botequio Moretti; Dimas Tadeu Covas; Elaine Cristina Marqueze; Elaine Vieira Santos; Evandra Strazza Rodrigues; Heidge Fukumasu; Jayme Augusto de Souza-Neto; José Salvatore Leister Patané; Luiz Alcantara; Luiz Lehmann Coutinho; Maria Carolina Elias; Maurício Lacerda Nogueira; Rafael dos Santos Bezerra; Raul Machado Neto; Rejane Maria Tommasini Grotto; Ricardo Haddad; Sandra Coccuzzo Sampaio Vessoni; Simone Kashima; Svetoslav Nanev Slavov; Vincent Louis Viala |
| EPI_ISL_2493284, EPI_ISL_2494171, EPI_ISL_2494191, EPI_ISL_2494197, EPI_ISL_2494198, | VIGILANCIA EPIDEMIOLOGICA E CONTROLE DE VETORES PIRASSUNUN | Instituto Butantan | Antonio Jorge Martins; Claudia Renata dos Santos Barros; David Schlesinger; Debora Botequio Moretti; Dimas Tadeu Covas; Elaine Cristina Marqueze; Elaine Vieira Santos; Evandra Strazza Rodrigues; Heidge Fukumasu; Jayme Augusto de Souza-Neto; José Salvatore Leister Patané; Luiz Alcantara; Luiz Lehmann Coutinho; Maria Carolina Elias; Maurício Lacerda Nogueira; Rafael dos Santos Bezerra; Raul Machado Neto; Rejane Maria Tommasini Grotto; Ricardo Haddad; Sandra Coccuzzo Sampaio Vessoni; Simone Kashima; Svetoslav Nanev Slavov; Vincent Louis Viala |

| | | | |
|---|---|---|---|
| EPI_ISL_2494199 | | | |
| EPI_ISL_2493944, EPI_ISL_2493945, EPI_ISL_2493946, EPI_ISL_2493949, EPI_ISL_2493952, EPI_ISL_2493953, EPI_ISL_2493958 | | | |
| see above | VIGILANCIA SANITARIA E VIG EPIDEMIOLOGICA DE ITAPEVI | Instituto Butantan | Antonio Jorge Martins; Claudia Renata dos Santos Barros; David Schlesinger; Debora Botequio Moretti; Dimas Tadeu Covas; Elaine Cristina Marqueze; Elaine Vieira Santos; Evandra Strazza Rodrigues; Heidge Fukumasu; Jayme Augusto de Souza-Neto; José Salvatore Leister Patané; Luiz Alcantara; Luiz Lehmann Coutinho; Maria Carolina Elias; Maurício Lacerda Nogueira; Rafael dos Santos Bezerra; Raul Machado Neto; Rejane Maria Tommasini Grotto; Ricardo Haddad; Sandra Coccuzzo Sampaio Vessoni; Simone Kashima; Svetoslav Nanev Slavov; Vincent Louis Viala |
| EPI_ISL_4090004 | Vermont Integrative Genomics Resource | Vermont Integrative Genomics Resource | Korin Eckstrom |
| EPI_ISL_2462557, EPI_ISL_2462716, EPI_ISL_2544406, EPI_ISL_2544492 | Viollier AG | Department of Biosystems Science and Engineering, ETH Zürich | Andrea Patrignani; Andreia Cabral de Gouvea; Catharine Aquino; Chaoran Chen; Christian Beisel; Christiane Beckmann; Christoph Noppen; Daniel Ehrsam; Doris Popovic; Elodie Burcklen; Griffin White; Ina Nissen; Isabel Stürmer; Ivan Topolsky; Jay Tracy; Kim Philipp Jablonski; Lara Fuhrmann; Laura Neff; Lennart Opitz; Maria Domenica Moccia; Maurice Redondo; Mirjam Feldkamp; Natascha Santacroce; Niko Beerenwinkel; Olivier Kobel; Ralph Schlapbach; Rebecca Denes; Sarah Nadeau; Simon Grüter; Tanja Stadler; Timothy Sykes |
| EPI_ISL_2420652 | Viollier AG | Viollier AG | Andrea Patrizia Salzmann; Christiane Beckmann; Christoph Noppen; Henriette Kurth; Maurice Redondo; Olivier Kobel |
| EPI_ISL_2603344, EPI_ISL_2603346, EPI_ISL_2603359, EPI_ISL_2689849, EPI_ISL_2689853, EPI_ISL_2689854, EPI_ISL_2689855 | | | |
| see above | Virginia Division of Consolidated Laboratory Services | Virginia Division of Consolidated Laboratory Services | Virginia DCLS; Virginia Division of Consolidated Laboratory Services |
| EPI_ISL_2884416, EPI_ISL_2884595 | Virology Laboratory, Scientific Department, Army Medical Center | Virology Laboratory, Scientific Department, Army Medical Center | Anella Monte; Anna Anselmo; Antonella Fortunato; Filippo Molinari; Florigio Lista; Francesco Giordani; Giancarlo Petralito; Giandomenico Cerreto; Giulia Campoli; Lucia Nicosia; Marzia Cavalli; Riccardo De Sanctis; Rossella Brandi; Silvia Fillo; Vanessa Vera Fain |
| EPI_ISL_2690105, EPI_ISL_2690106, EPI_ISL_2690110 | WA State Department of Health | Centers for Disease Control and Prevention Division of Viral Diseases, Pathogen Discovery | Alison Laufer Halpin; Ben L. Rambo-Martin; Clinton R. Paden; Dakota Howard; Darlene Wagner; Dave Wentworth; Dhwani Batra; Jasmine Padilla; Justin Lee; Katie Dillon; Krista Queen; Kristen Knipe; Kristine Lacek; Mark Burroughs; Matthew Schmerer; Mili Sheth; Peter Cook; Sam Shepard; Sarah Nobles; Shoshona Le; Suxiang Tong; Vivien Dugan; Yvette Unoarumhi |
| EPI_ISL_2615464 | WV Labs | WVU and Marshall University Combined Genomics Core Facilities | James Denvir; Peter Perrotta; Peter Stoilov; Ryan Percifield; Wesley Kimble |
| EPI_ISL_2378872, EPI_ISL_2557055, EPI_ISL_2557056, EPI_ISL_2557057, EPI_ISL_2557066, EPI_ISL_2557067, EPI_ISL_2557075, EPI_ISL_2557079 | | | |
| see above | Washington State Department of Health Public Health Laboratories | Washington State Department of Health Public Health Laboratories | Avi Singh; Darren Lucas; Denny Russell; Drew MacKellar; Geoff Melly; Hannah Gray; Joenice Gonzalez; JohnAric Peterson; Philip Dykema; Rebecca Cao; Vanessa De Los Santos |
| EPI_ISL_2774524, EPI_ISL_6678510 | Weill Cornell Medicine | New York Genome Center | Amy Baldwin; Andre Corvelo; Arryn Craney; Chris Mason; Dayna M. Oschwald; Hanna Rennert; Lars F Westblade; Margaret Elizabeth Ross; Melissa Cushing; Michael Zody; Olivier Elemento; Priya Velu; Samantha Fennessey; Tom Maniatis |
| EPI_ISL_2400234 | Wichita State University - Molecular Diagnostics Lab | Kansas Health and Environmental Lab | Ben Olsen; Jonathan Barnell; Katherine Wiggins; Mike Grose; and Phil Adam |
| EPI_ISL_2382560, EPI_ISL_2382562, EPI_ISL_2382563, EPI_ISL_2383016, EPI_ISL_2383017, EPI_ISL_2383018 | Wichita State University- Molecular Diagonstic lab | Kansas Health and Environmental Lab | Ben Olsen; Jonathan Barnell; Katherine Wiggins; Mike Grose; and Phil Adam |
| EPI_ISL_2383180, EPI_ISL_2561917, EPI_ISL_2561924, EPI_ISL_2561926 | Willis-Knighton Medical Center Hospital Laboratory | LSUHS Emerging Viral Threat Laboratory | Alexander Mijalis; Andrew D. Yurochko; April N. Johnson; Christopher G. Kevil; Gregory L. Ware; Jennifer L. Carroll; Jeremy P. Kamil; John A. Vanchiere; Joseph A. Bocchini; Lorie M. Atkins; Maarten Van Diest; Rona S. Scott |
| EPI_ISL_2558601, EPI_ISL_2558668, EPI_ISL_2727681, EPI_ISL_2727705, EPI_ISL_2727708, EPI_ISL_2727712 | Wisconsin State Laboratory of Hygiene Communicable Disease Division | Wisconsin State Laboratory of Hygiene Communicable Disease Division | Abigail C. Shockey; Alicia J. Mooney; Erika M. Hanson; Kelsey R. Florek; Richard Griesser; Sara Wagner; Tonya Danz |
| EPI_ISL_2383131, EPI_ISL_2383147, EPI_ISL_2455253, EPI_ISL_2603278, EPI_ISL_2603285, EPI_ISL_2603291, EPI_ISL_2728125 | | | |
| see above | Wyoming Public Health Laboratory | Wyoming Public Health Laboratory | Ashley Norberg; Brian Dominguez; Brittany Oher; Cari Sloma; Channing Weber; Chayse Rowley; Elliot Thomasson; Jim Mildenberger; Lynette Gumbleton; Marley Goetz; Noah Hull; Robert Petit; Sam Britz; Taylor Fearing; Wanda Manley; and Rob Christensen |
| EPI_ISL_2501663, EPI_ISL_2716232 | Yale Clinical Virology Lab | Grubaugh Lab - Yale School of Public Health | Anderson Brito; Annie Watkins; Chaney Kalinich; Chantal Vogels; Isabel Ott; Jessica Rothman; Joseph Fauver; Mallery Breban; Marie L. Landry; Mary Petrone; Nathan Grubaugh; Tara Alpert |
| EPI_ISL_2382881, EPI_ISL_2382888 | ZOL | Jessa | Raymaekers et al. on behalf of Jessa_cmdLab |
| EPI_ISL_2566794 | Zakład Mikrobiologii USK Opole | 1. National Institute of Public Health - National Institute of Hygiene; 2. Eurofins Genomics Europe Sequencing GmbH | ECDC COVID-19 WGS support team; Eurofins Genomics Europe Sequencing Team; Gierczyńki Rafał; Sadkowska-Todys Małgorzata; Wołkowicz Tomasz; Zacharczuk Katarzyna |
| EPI_ISL_2470909 | amedes MVZ für Laboratoriumsmedizin Fürstenfeldbruck | Robert Koch Institute | |
