## Supplement 3 for "Rapid and Accurate Identification of SARS-CoV-2 Omicron Variants Using Droplet Digital PCR (RT-ddPCR)": All_0531-0606.pdf

We gratefully acknowledge the following Authors from the Originating laboratories responsible for obtaining the specimens, as well as the Submitting laboratories where the genome data were generated and shared via GISAID, on which this research is based.

All Submitters of data may be contacted directly via www.gisaid.org

Authors are sorted alphabetically.

| Accession ID | Originating Laboratory | Submitting Laboratory | Authors |
|---|---|---|---|
| EPI_ISL_2987104, EPI_ISL_2987105, EPI_ISL_2987111 | "CDPH, Viral and Rickettsial Disease Laboratory" | Centers for Disease Control and Prevention Division of Viral Diseases, Pathogen Discovery | Alison Laufer Halpin; Ben L. Rambo-Martin; Clinton R. Paden; Dakota Howard; Darlene Wagner; Dave Wentworth; Dhwani Batra; Jasmine Padilla; Justin Lee; Katie Dillon; Krista Queen; Kristen Knipe; Kristine Lacek; Mark Burroughs; Matthew Schmerer; Mili Sheth; Peter Cook; Sam Shepard; Sarah Nobles; Shoshona Le; Suxiang Tong; Vivien Dugan; Yvette Unoarumhi |
| EPI_ISL_5646317 | ADECIO POMPEU CARVALHO | Instituto Butantan | Antonio Jorge Martins; Claudia Renata dos Santos Barros; David Schlesinger; Debora Botequio Moretti; Dimas Tadeu Covas; Elaine Cristina Marqueze; Elaine Vieira Santos; Evandra Strazza Rodrigues; Heidge Fukumasu; Jayme Augusto de Souza-Neto; José Salvatore Leister Patané; Luiz Alcantara; Luiz Lehmann Coutinho; Maria Carolina Elias; Maurício Lacerda Nogueira; Rafael dos Santos Bezerra; Raul Machado Neto; Rejane Maria Tommasini Grotto; Ricardo Haddad; Sandra Coccuzzo Sampaio Vessoni; Simone Kashima; Svetoslav Nanev Slavov; Vincent Louis Viala |
| EPI_ISL_5645634, EPI_ISL_5645644, EPI_ISL_5645645, EPI_ISL_5645657, EPI_ISL_5645662, EPI_ISL_5645667, EPI_ISL_5645676, EPI_ISL_5645677, EPI_ISL_5645696, EPI_ISL_5645697, EPI_ISL_5645698, EPI_ISL_5645699, EPI_ISL_5645702, EPI_ISL_5645706, EPI_ISL_5645740, EPI_ISL_5645750, EPI_ISL_5645751, EPI_ISL_5645756, EPI_ISL_5645764, EPI_ISL_5645772, EPI_ISL_5645789, EPI_ISL_5645790, EPI_ISL_5645797, EPI_ISL_5645798, EPI_ISL_5645806, EPI_ISL_5645839, EPI_ISL_5645853, EPI_ISL_5645863, EPI_ISL_5645864, EPI_ISL_5645873, EPI_ISL_5645880, EPI_ISL_5645881, EPI_ISL_5645888, EPI_ISL_5645895, EPI_ISL_5645896, EPI_ISL_5645904, EPI_ISL_5645905, EPI_ISL_5645911, EPI_ISL_5645912, EPI_ISL_5645920, EPI_ISL_5645929, EPI_ISL_5645933, EPI_ISL_5645937, EPI_ISL_5645948, EPI_ISL_5645949, EPI_ISL_5645950, EPI_ISL_5645951 | | | |
| see above | AFIP | Instituto Butantan | Antonio Jorge Martins; Claudia Renata dos Santos Barros; David Schlesinger; Debora Botequio Moretti; Dimas Tadeu Covas; Elaine Cristina Marqueze; Elaine Vieira Santos; Evandra Strazza Rodrigues; Heidge Fukumasu; Jayme Augusto de Souza-Neto; José Salvatore Leister Patané; Luiz Alcantara; Luiz Lehmann Coutinho; Maria Carolina Elias; Maurício Lacerda Nogueira; Rafael dos Santos Bezerra; Raul Machado Neto; Rejane Maria Tommasini Grotto; Ricardo Haddad; Sandra Coccuzzo Sampaio Vessoni; Simone Kashima; Svetoslav Nanev Slavov; Vincent Louis Viala |
| EPI_ISL_5915146, EPI_ISL_5915197, EPI_ISL_5915214, EPI_ISL_5915265, EPI_ISL_5915266, EPI_ISL_5915314, EPI_ISL_6573787, EPI_ISL_6573788, EPI_ISL_6573854, EPI_ISL_6573924, EPI_ISL_6573938 | | | |
| see above | AMALAB/FACISA/UFRN | WallauLab on behalf of Fiocruz COVID-19 Genomic Surveillance Network | Alexandre Freitas da Silva; Allan Roberto Dias Nunes; Antonio Marinho da Silva Neto; Cassia Docena; Constância Flávia Junqueira Ayres; Filipe Zimmer Dezordi; Gabriel Luz Wallau; Gustavo Barbosa de Lima; Joana Cristina Medeiros Tavares Marques; Katya Anaya Jacinto; Lais Ceschini Machado; Lilian Caroliny Amorim Silva; Marcelo Henrique dos Santos Paiva; Mariane dos Santos Duarte; Matheus Filgueira Bezerra; Sinval Pinto Brandão Filho |
| EPI_ISL_5645598, EPI_ISL_5645602, EPI_ISL_5645610, EPI_ISL_5645614, EPI_ISL_5645627, EPI_ISL_5645631, EPI_ISL_5645646, EPI_ISL_5645656, EPI_ISL_5645700, EPI_ISL_5645707, EPI_ISL_5645748 | | | |
| see above | AMBULATORIO IB | Instituto Butantan | Antonio Jorge Martins; Claudia Renata dos Santos Barros; David Schlesinger; Debora Botequio Moretti; Dimas Tadeu Covas; Elaine Cristina Marqueze; Elaine Vieira Santos; Evandra Strazza Rodrigues; Heidge Fukumasu; Jayme Augusto de Souza-Neto; José Salvatore Leister Patané; Luiz Alcantara; Luiz Lehmann Coutinho; Maria Carolina Elias; Maurício Lacerda Nogueira; Rafael dos Santos Bezerra; Raul Machado Neto; Rejane Maria Tommasini Grotto; Ricardo Haddad; Sandra Coccuzzo Sampaio Vessoni; Simone Kashima; Svetoslav Nanev Slavov; Vincent Louis Viala |
| EPI_ISL_5645655, EPI_ISL_5645809, EPI_ISL_5645837 | AMBULATORIO MEDICO E ODONTOLOGICO MUNICIPAL DE PIEDADE | Instituto Butantan | Antonio Jorge Martins; Claudia Renata dos Santos Barros; David Schlesinger; Debora Botequio Moretti; Dimas Tadeu Covas; Elaine Cristina Marqueze; Elaine Vieira Santos; Evandra Strazza Rodrigues; Heidge Fukumasu; Jayme Augusto de Souza-Neto; José Salvatore Leister Patané; Luiz Alcantara; Luiz Lehmann Coutinho; Maria Carolina Elias; Maurício Lacerda Nogueira; Rafael dos Santos Bezerra; Raul Machado Neto; Rejane Maria Tommasini Grotto; Ricardo Haddad; Sandra Coccuzzo Sampaio Vessoni; Simone Kashima; Svetoslav Nanev Slavov; Vincent Louis Viala |
| EPI_ISL_5646143, EPI_ISL_5646155, EPI_ISL_5646164, EPI_ISL_5646171, EPI_ISL_5646172, EPI_ISL_5646175, EPI_ISL_5646183, EPI_ISL_5646191, EPI_ISL_5646199, EPI_ISL_5646201, EPI_ISL_5646342, EPI_ISL_5646343, EPI_ISL_5646346, EPI_ISL_5646349, EPI_ISL_5646354, EPI_ISL_5646367 | | | |
| see above | AMBULATORIO MEDICO MUNICIPAL DE AGUDOS | Instituto Butantan | Antonio Jorge Martins; Claudia Renata dos Santos Barros; David Schlesinger; Debora Botequio Moretti; Dimas Tadeu Covas; Elaine Cristina Marqueze; Elaine Vieira Santos; Evandra Strazza Rodrigues; Heidge Fukumasu; Jayme Augusto de Souza-Neto; José Salvatore Leister Patané; Luiz Alcantara; Luiz Lehmann Coutinho; Maria Carolina Elias; Maurício Lacerda Nogueira; Rafael dos Santos Bezerra; Raul Machado Neto; Rejane Maria Tommasini Grotto; Ricardo Haddad; Sandra Coccuzzo Sampaio Vessoni; Simone Kashima; Svetoslav Nanev Slavov; Vincent Louis Viala |
| EPI_ISL_2827997, EPI_ISL_2827998 | AREA DE SALUD BAGACES | Inciensa, Instituto Costarricense de Investigación y Enseñanza en Nutrición y Salud | Adriana Godínez; Claudio Soto-Garita; Estela Cordero; Francisco Duarte; Hebleen Porras; Joselyn Prado & Karol Matamoros; José Luis Vargas; Mariela Gutiérrez; Melany Calderón |
| EPI_ISL_2827996 | AREA DE SALUD CARTAGO | Inciensa, Instituto Costarricense de Investigación y Enseñanza en Nutrición y Salud | Adriana Godínez; Claudio Soto-Garita; Estela Cordero; Francisco Duarte; Hebleen Porras; Joselyn Prado & Monserrat Segura; José Luis Vargas; Mariela Gutiérrez; Melany Calderón |
| EPI_ISL_2827988, EPI_ISL_2827989, EPI_ISL_2827990 | AREA DE SALUD CIUDAD QUESADA | Inciensa, Instituto Costarricense de Investigación y Enseñanza en Nutrición y Salud | Adriana Godínez; Claudio Soto-Garita; Estela Cordero; Francisco Duarte; Hebleen Porras; Joselyn Prado & Juan Carlos Villalobos; José Luis Vargas; Mariela Gutiérrez; Melany Calderón |
| EPI_ISL_2827999 | AREA DE SALUD DESAMPARADOS 2 (COOPESALUD) | Inciensa, Instituto Costarricense de Investigación y Enseñanza en Nutrición y Salud | Adriana Godínez; Claudio Soto-Garita; Estela Cordero; Francisco Duarte; Hebleen Porras; Joselyn Prado & Alberto Quirós; José Luis Vargas; Mariela Gutiérrez; Melany Calderón |
| EPI_ISL_2827970 | AREA DE SALUD GUATUSO | Inciensa, Instituto Costarricense de Investigación y Enseñanza en Nutrición y Salud | Adriana Godínez; Claudio Soto-Garita; Estela Cordero; Francisco Duarte; Hebleen Porras; Joselyn Prado & Francisco Chacón; José Luis Vargas; Mariela Gutiérrez; Melany Calderón |
| EPI_ISL_2695447 | ARS Algarve - Laboratorio Laura Ayres | Instituto Nacional de Saude (INSA) | Borges et al |
| EPI_ISL_2815366 | ASL BARI | University of Bari Biomedical Sciences and Human Oncology | Maria Chironna |
| EPI_ISL_5647175 | ASSOCIACAO HOSPITAL BENEFICENTE SAGRADO CORACAO DE JESUS | Instituto Butantan | Antonio Jorge Martins; Claudia Renata dos Santos Barros; David Schlesinger; Debora Botequio Moretti; Dimas Tadeu Covas; Elaine Cristina Marqueze; Elaine Vieira Santos; Evandra Strazza Rodrigues; Heidge Fukumasu; Jayme Augusto de Souza-Neto; José Salvatore Leister Patané; Luiz Alcantara; Luiz Lehmann Coutinho; Maria Carolina Elias; Maurício Lacerda Nogueira; Rafael dos Santos Bezerra; Raul Machado Neto; Rejane Maria Tommasini Grotto; Ricardo Haddad; Sandra Coccuzzo Sampaio Vessoni; Simone Kashima; Svetoslav Nanev Slavov; Vincent Louis Viala |
| EPI_ISL_2638097, EPI_ISL_2638103, EPI_ISL_2638105, EPI_ISL_2638106, EPI_ISL_2925857 | AULSS 2 Marca Trevigiana | Istituto Zooprofilattico Sperimentale delle Venezie | Adelaide Milani; Alessia Schivo; Alice Fusaro; Ambra Pastori; Annalisa Salviato; Antonia Ricci; Calogero Terregino; Edoardo Giussani; Elisa Palumbo; Erika Giorgia Quaranta; Isabella Monne; Luca Tassoni |
| EPI_ISL_3006782 | AULSS 9 Scaligera | Istituto Zooprofilattico Sperimentale delle Venezie | Adelaide Milani; Alessia Schivo; Alice Fusaro; Ambra Pastori; Annalisa Salviato; Antonia Ricci; Calogero Terregino; Edoardo Giussani; Elisa Palumbo; Erika Giorgia Quaranta; Isabella Monne; Luca Tassoni |
| EPI_ISL_2489225, EPI_ISL_2527882, EPI_ISL_2527899, EPI_ISL_2527906, EPI_ISL_2527909, EPI_ISL_2527911, EPI_ISL_2527937, EPI_ISL_2527954, EPI_ISL_2527961, EPI_ISL_2527972, EPI_ISL_2527974, EPI_ISL_2527995, EPI_ISL_2528002, EPI_ISL_2528006, EPI_ISL_2528019, EPI_ISL_2528037, EPI_ISL_2528052, EPI_ISL_2528062, EPI_ISL_2528065, EPI_ISL_2528074, EPI_ISL_2528076, EPI_ISL_2528087, EPI_ISL_2528104, EPI_ISL_2528106, EPI_ISL_2528111, EPI_ISL_2528132, EPI_ISL_2528139, EPI_ISL_2528155, EPI_ISL_2528163, EPI_ISL_2528165, EPI_ISL_2528216, EPI_ISL_2528218, EPI_ISL_2528226, EPI_ISL_2528230, EPI_ISL_2528233, EPI_ISL_2528235, EPI_ISL_2528237, EPI_ISL_2528241, EPI_ISL_2528283, EPI_ISL_2528299, EPI_ISL_2528303, EPI_ISL_2528310, EPI_ISL_2528318, EPI_ISL_2528372, EPI_ISL_2528376, EPI_ISL_2528380, EPI_ISL_2528383, EPI_ISL_2528387, EPI_ISL_2528394, EPI_ISL_2528404, EPI_ISL_2528419, EPI_ISL_2528425, EPI_ISL_2528428, EPI_ISL_2528451, EPI_ISL_2528453, EPI_ISL_2528455, EPI_ISL_2528458, EPI_ISL_2528498, EPI_ISL_2528504, EPI_ISL_2528506, EPI_ISL_2528519, EPI_ISL_2528541, EPI_ISL_2528546, EPI_ISL_2528550, EPI_ISL_2528556, EPI_ISL_2528582, EPI_ISL_2528624, EPI_ISL_2528642, EPI_ISL_2528654, EPI_ISL_2528666, EPI_ISL_2528734, EPI_ISL_2528739, EPI_ISL_2528747, EPI_ISL_2528753, EPI_ISL_2528760, EPI_ISL_2528768, EPI_ISL_2528779, EPI_ISL_2528783, EPI_ISL_2528795, EPI_ISL_2528836, EPI_ISL_2528845, EPI_ISL_2528862, EPI_ISL_2528866, EPI_ISL_2528871, EPI_ISL_2528875, EPI_ISL_2528877, EPI_ISL_2528971, EPI_ISL_2528976, EPI_ISL_2528978, EPI_ISL_2528980, EPI_ISL_2528987, EPI_ISL_2528990, EPI_ISL_2529005, EPI_ISL_2529013, EPI_ISL_2534534, EPI_ISL_2534541, EPI_ISL_2534542, EPI_ISL_2534550, EPI_ISL_2534552, EPI_ISL_2686906, EPI_ISL_2686908, EPI_ISL_2686911, EPI_ISL_2686917, EPI_ISL_2686919, EPI_ISL_2686920, EPI_ISL_2686921, EPI_ISL_2686950, EPI_ISL_2687006, EPI_ISL_2687021, EPI_ISL_2687034, EPI_ISL_2687035, EPI_ISL_2687047, EPI_ISL_2687061, EPI_ISL_2687093, EPI_ISL_2687094, EPI_ISL_2687202, EPI_ISL_2687213, EPI_ISL_2709498, EPI_ISL_4360971, EPI_ISL_4362395, EPI_ISL_4363898, EPI_ISL_4364732 | | | |
| see above | Aegis Sciences Corporation | Centers for Disease Control and Prevention Division of Viral Diseases, Pathogen Discovery | Adrian Paskey; Alec Vest; Benjamin Rambo-Martin; Christopher Gulvick; Clinton Paden; Clinton R. Paden; Cyndi Clark; Dakota Howard; Darlene Wagner; Dhwani Batra; Dillon Nall; Duncan MacCannell; Erisa Sula; Ethan Sanders; Holly Houdeshell; Jason Caravas; Kara Moser; Kristine Lacek; Matthew Hardison; Matthew Schmerer; Ola Kvalvaag; Patrick Campbell; Peter Cook; Peter W. Cook; Rob Case; Scott Sammons; Shatavia Morrison; Shaun Westlund; Tymeckia Kendall; Victoria Caban Figueroa; Vikramsinha Ghorpade; Yvette Unoarumhi |
| EPI_ISL_2503981, EPI_ISL_2503982, EPI_ISL_2504003, EPI_ISL_2504004 | Alaska State Virology Laboratory | Alaska State Virology Laboratory | Elva House; Jack Chen; Jacob Zidek; Lisa Smith; Ph.D.; Stephanie DeRonde |
| EPI_ISL_3030252, EPI_ISL_3030253, EPI_ISL_3030254, EPI_ISL_3030267, EPI_ISL_3030268, EPI_ISL_3030270, EPI_ISL_3030329 | | | |
| see above | Ancona | Microbiology University Politecnica delle Marche | Anna Valenza; Carla Acciarri; Katia Marinelli; Monica Lucia Ferreri; Patrizia Bagnarelli; Roberta Longo; Sara Caucci; Stefano Menzo |
| EPI_ISL_2650135, EPI_ISL_2650136, EPI_ISL_2650137, EPI_ISL_2650141, EPI_ISL_2650142, EPI_ISL_2650151 | Arizona State Public Health Laboratory | Arizona State Public Health Laboratory | Jessica Escobar; Katherine Fullerton; Linda Getsinger; Matthew Contursi; Nobuko Fukushima; Stacy White; Trung Huynh; Victor Waddell |
| EPI_ISL_2484109, EPI_ISL_2484131, EPI_ISL_2604137, EPI_ISL_2604154 | Arizona State University | Arizona State University | Efrem S. Lim; Joshua LaBaer; Joy M. Blain; LaRinda A. Holland; Matthew F. Smith; Nicholas J. Mellor; Peter T. Skidmore; Rabia Maqsood; Valerie Harris; Vel Murugan |
| EPI_ISL_2810107 | Arión Genética | Instituto Nacional de Medicina Genomica | Cedro-Tanda A; Díaz-Quiñonez José Alberto; Escobar-Arrazola M; Gonzalez-Barrera D; Herrera-Montalvo LA.; Hidalgo-Miranda A; Mendoza-Vargas A; Munguia-Garza P; Ramirez-Vega O; Rangel-DeLeon D; Reyes-Grajeda JP; Roldan-Castillo Magaly; Uribe-Figueroa Laura; Verea Jazmin |
| EPI_ISL_2715207, EPI_ISL_2736221, EPI_ISL_2736256, EPI_ISL_2736276 | Atlas Genomics | Seattle Flu Study | Amanda Adler; Barry R. Lutz; Benjamin Pelle; Brian Hiatt; Caitlin R. Wolf; Chris D. Frazar; Deborah A. Nickerson; Elisabeth Brandstetter; Erica Ryke; Geoff Melly; Helen Y. Chu; Janet A. Englund; Jay Shendure; Jover Lee; Kairsten Fay; Kirsten Lacombe; Lea M. Starita; Mark J. Rieder; Matthew Richardson; Matthew Thompson; Melissa Truong; Michael Boeckh; Michael Famulare; Misja Ilcisin; Peter D. Han; Philip Dykema; Romesh Gautom; Scott Lindquist; Thomas R. Sibley; Trevor Bedford |
| EPI_ISL_2616206, EPI_ISL_2657459, | Austrian Agency for Health and Food Safety (AGES) | Bergthaler laboratory, CeMM Research Center for Molecular | Andreas Bergthaler; Anna Schedl; Bekir Erguner; Benedikt Agerer; Christoph Bock; Fabian Amman; Jan Laine; Lukas Endler; Maelle Le Moing; Martin Senekowitsch; Matthew Thornton; Michael Schuster; Petr Triska; Thomas Penz |

| | | | |
|---|---|---|---|
| EPI_ISL_2657460 | | Medicine of the Austrian Academy of Sciences | |
| EPI_ISL_2622160 | Azienda Ospedaliera Pugliese Ciaccio di Catanzaro SOC Microbiologia e Virologia | Azienda Ospedaliera Pugliese Ciaccio di Catanzaro SOC Microbiologia e Virologia | Rossana Tallerico Federica Pasceri Marco De Fazio Ilenia Talotta Giuseppina Panduri Pasquale Minchella |
| EPI_ISL_2686081, EPI_ISL_2686084 | Azienda Ospedaliera Terni | Istituto Zooprofilattico Sperimentale dell'Abruzzo e Molise "G. Caporale" | Ancora M; Calistri P; Cammà C; Curini V; Delli Compagni E; Di Domenico M; Di Pasquale A; Lorusso A; Mangone I; Marcacci M; Palumbo M; Puglia I; Rinaldi A; Savini G; Scaccetti A; Scialabba S |
| EPI_ISL_2550615, EPI_ISL_2550617, EPI_ISL_2550623, EPI_ISL_2550630, EPI_ISL_2550633, EPI_ISL_2550634, EPI_ISL_2550635, EPI_ISL_2550642, EPI_ISL_2550661, EPI_ISL_2550665, EPI_ISL_2550670, EPI_ISL_2550672, EPI_ISL_2550673, EPI_ISL_2550675, EPI_ISL_2550686 | | | |
| see above | Azienda Sanitaria dell'Alto Adige Laboratorio Aziendale di Microbiologia e Virologia | Istituto di Genomica Applicata | Davide Scaglione; Eleonora Paparelli; Elisa Masi; Elisabetta Giacobazzi; Elisabetta Pagani; Gabriele Magris; Irena Jurman; Irene Bianconi; Michele Morgante; Stefanie Wieser; Vera Vendramin |
| EPI_ISL_2686076, EPI_ISL_2686079, EPI_ISL_2686080 | Azienda USL Umbria 2 | Istituto Zooprofilattico Sperimentale dell'Abruzzo e Molise "G. Caporale" | Ancora M; Calistri P; Cammà C; Curini V; Delli Compagni E; Di Domenico M; Di Pasquale A; Lorusso A; Mangone I; Marcacci M; Pistoni E; Proietti A; Puglia I; Rinaldi A; Savini G; Scialabba S |
| EPI_ISL_5646208, EPI_ISL_5646209, EPI_ISL_5646211, EPI_ISL_5646212, EPI_ISL_5646213, EPI_ISL_5646215, EPI_ISL_5646216, EPI_ISL_5646217, EPI_ISL_5646218, EPI_ISL_5646221, EPI_ISL_5646222, EPI_ISL_5646223, EPI_ISL_5646224, EPI_ISL_5646225, EPI_ISL_5646226, EPI_ISL_5646227, EPI_ISL_5646228, EPI_ISL_5646229, EPI_ISL_5646231, EPI_ISL_5646233, EPI_ISL_5646236, EPI_ISL_5646237, EPI_ISL_5646238, EPI_ISL_5646239, EPI_ISL_5646240, EPI_ISL_5646243, EPI_ISL_5646245, EPI_ISL_5646246, EPI_ISL_5646247, EPI_ISL_5646249, EPI_ISL_5646250, EPI_ISL_5646251, EPI_ISL_5646252, EPI_ISL_5646253, EPI_ISL_5646254, EPI_ISL_5646255, EPI_ISL_5646256, EPI_ISL_5646257, EPI_ISL_5646258, EPI_ISL_5646259, EPI_ISL_5646260, EPI_ISL_5646261, EPI_ISL_5646264, EPI_ISL_5646265, EPI_ISL_5646266, EPI_ISL_5646268, EPI_ISL_5646269, EPI_ISL_5646270, EPI_ISL_5646271, EPI_ISL_5646272, EPI_ISL_5646273, EPI_ISL_5646274, EPI_ISL_5646275, EPI_ISL_5646276, EPI_ISL_5646277, EPI_ISL_5646278, EPI_ISL_5646280, EPI_ISL_5646281, EPI_ISL_5646282, EPI_ISL_5646284, EPI_ISL_5646285, EPI_ISL_5646286, EPI_ISL_5646287, EPI_ISL_5646288, EPI_ISL_5647956, EPI_ISL_5647961 | | | |
| see above | BIOFAST | Instituto Butantan | Antonio Jorge Martins; Claudia Renata dos Santos Barros; David Schlesinger; Debora Botequio Moretti; Dimas Tadeu Covas; Elaine Cristina Marqueze; Elaine Vieira Santos; Evandra Strazza Rodrigues; Heidge Fukumasu; Jayme Augusto de Souza-Neto; José Salvatore Leister Patané; Luiz Alcantara; Luiz Lehmann Coutinho; Maria Carolina Elias; Maurício Lacerda Nogueira; Rafael dos Santos Bezerra; Raul Machado Neto; Rejane Maria Tommasini Grotto; Ricardo Haddad; Sandra Coccuzzo Sampaio Vessoni; Simone Kashima; Svetoslav Nanev Slavov; Vincent Louis Viala |
| EPI_ISL_7672153, EPI_ISL_7672174, EPI_ISL_7672481 | Bambino Gesù Pediatric Hospital | Microbiology and Immunology Diagnosis Bambino Gesù Pediatric Hospital | Carlo Federico Perno; Claudia Alteri; Luna Colagrossi; Rossana Scutari; Valentino Costabile |
| EPI_ISL_3504551, EPI_ISL_3504577 | Baptist Health Medical Center | Center for Global Health, University of New Mexico Health Sciences Center | Amanda Novack; Darrell Dinwiddie; Daryl Domman; Dirk Haselow; Joshua L. Kennedy; Kurt Schwalm; Valerie Morley |
| EPI_ISL_2760052 | Bayerisches Landesamt für Gesundheit und Lebensmittelsicherheit (LGL) | Robert Koch Institute | |
| EPI_ISL_2626816 | Baylor Scott & White-Temple | Baylor Scott & White-Temple | Ari Rao; Kimberly Walker; Linden Morales; Marcus Volz; Shelby Johnson |
| EPI_ISL_3144989, EPI_ISL_3145478, EPI_ISL_3145479, EPI_ISL_3145480, EPI_ISL_3145481 | BioneXt Lab | Laboratoire national de sante, Microbiology, Microbial Genomics Platform | Anke Wienecke-Baldacchino; Catherine Ragimbeau; Elodie Solarino; Fatu Djabi; Jessica Tapp; Lise Pignon; Raoul Salmon; Tamir Abdelrahman; Thibault Ferrandon; Virginie Jover |
| EPI_ISL_2633914 | Bioscientia MVZ Labor Karlsruhe GmbH | Robert Koch Institute | |
| EPI_ISL_3479146, EPI_ISL_3479147, EPI_ISL_3479148, EPI_ISL_3479149, EPI_ISL_3479151, EPI_ISL_3479155, EPI_ISL_3479156, EPI_ISL_3479159, EPI_ISL_3479161, EPI_ISL_3489362, EPI_ISL_3489363, EPI_ISL_3489364, EPI_ISL_3489366, EPI_ISL_3489390, EPI_ISL_3489454, EPI_ISL_3489461, EPI_ISL_3489462, EPI_ISL_3489476, EPI_ISL_3489477, EPI_ISL_3489491, EPI_ISL_3489498, EPI_ISL_3489504, EPI_ISL_3489505, EPI_ISL_3489507, EPI_ISL_3489509, EPI_ISL_3489510, EPI_ISL_3489511, EPI_ISL_3489513, EPI_ISL_3489515, EPI_ISL_3489518, EPI_ISL_3489519, EPI_ISL_3489521, EPI_ISL_3489525, EPI_ISL_3489526, EPI_ISL_3489527, EPI_ISL_3489528, EPI_ISL_3489534, EPI_ISL_3489535, EPI_ISL_3489536, EPI_ISL_3489537, EPI_ISL_3489538, EPI_ISL_3489539, EPI_ISL_3489543, EPI_ISL_3489545, EPI_ISL_3489546, EPI_ISL_3489550, EPI_ISL_3489554, EPI_ISL_3489558, EPI_ISL_3489559, EPI_ISL_3489560, EPI_ISL_3489561, EPI_ISL_3489564, EPI_ISL_3489568, EPI_ISL_3489569, EPI_ISL_3489570, EPI_ISL_3489643, EPI_ISL_3489644, EPI_ISL_3489645, EPI_ISL_3489649, EPI_ISL_3489654, EPI_ISL_3489655, EPI_ISL_3489657, EPI_ISL_3489658, EPI_ISL_3489694, EPI_ISL_3489695, EPI_ISL_3489696, EPI_ISL_3489697, EPI_ISL_3489699, EPI_ISL_3489700, EPI_ISL_3489701, EPI_ISL_3489706, EPI_ISL_3489707, EPI_ISL_3489712, EPI_ISL_3489716, EPI_ISL_3489718, EPI_ISL_3489719, EPI_ISL_3489720, EPI_ISL_3489721, EPI_ISL_3489722, EPI_ISL_3489724, EPI_ISL_3489725, EPI_ISL_3489726, EPI_ISL_3489727, EPI_ISL_3489730, EPI_ISL_3489733, EPI_ISL_3489734, EPI_ISL_3489741, EPI_ISL_3489743, EPI_ISL_3489744, EPI_ISL_3489747, EPI_ISL_3489748, EPI_ISL_3489751, EPI_ISL_3489755, EPI_ISL_3489784, EPI_ISL_3489791, EPI_ISL_3489796, EPI_ISL_3489799, EPI_ISL_3489802, EPI_ISL_3489803, EPI_ISL_3489804, EPI_ISL_3489808, EPI_ISL_3489809, EPI_ISL_3489810, EPI_ISL_3489811, EPI_ISL_3489812, EPI_ISL_3489813, EPI_ISL_3489816, EPI_ISL_3489817, EPI_ISL_3489825, EPI_ISL_3489826, EPI_ISL_3489827, EPI_ISL_3489829, EPI_ISL_3489830, EPI_ISL_3489831, EPI_ISL_3489832, EPI_ISL_3489837, EPI_ISL_3489838, EPI_ISL_3489839, EPI_ISL_3489840, EPI_ISL_3489842, EPI_ISL_3489851, EPI_ISL_3489852, EPI_ISL_3489856, EPI_ISL_3489857, EPI_ISL_3489871, EPI_ISL_3489872, EPI_ISL_3489873, EPI_ISL_3489875, EPI_ISL_3489876, EPI_ISL_3489879, EPI_ISL_3489882, EPI_ISL_3489885, EPI_ISL_3489886, EPI_ISL_3489887, EPI_ISL_3489888, EPI_ISL_3489889, EPI_ISL_3489890, EPI_ISL_3489892, EPI_ISL_3489893, EPI_ISL_3489894, EPI_ISL_3489897, EPI_ISL_3489899, EPI_ISL_3489902, EPI_ISL_3489903, EPI_ISL_3489905, EPI_ISL_3489906, EPI_ISL_3489913, EPI_ISL_3489914, EPI_ISL_3489915, EPI_ISL_3489917, EPI_ISL_3489918, EPI_ISL_3489919, EPI_ISL_3489920, EPI_ISL_3489921, EPI_ISL_3489923, EPI_ISL_3489928, EPI_ISL_3489929, EPI_ISL_3489930, EPI_ISL_3489931, EPI_ISL_3489932, EPI_ISL_3489936, EPI_ISL_3489937, EPI_ISL_3489940, EPI_ISL_3489946, EPI_ISL_3489948, EPI_ISL_3489951, EPI_ISL_3489952, EPI_ISL_3489957, EPI_ISL_3489959, EPI_ISL_3489960, EPI_ISL_3489963, EPI_ISL_3489966, EPI_ISL_3489969, EPI_ISL_3489970, EPI_ISL_3489971, EPI_ISL_3489974, EPI_ISL_3489976, EPI_ISL_3489977, EPI_ISL_3489980, EPI_ISL_3489981, EPI_ISL_3489982, EPI_ISL_3489983, EPI_ISL_3489984, EPI_ISL_3489985, EPI_ISL_3489994, EPI_ISL_3489996, EPI_ISL_3489999, EPI_ISL_3490000, EPI_ISL_3490001, EPI_ISL_3490015, EPI_ISL_3490016, EPI_ISL_3490017, EPI_ISL_3490019, EPI_ISL_3490020, EPI_ISL_3490022, EPI_ISL_3490023, EPI_ISL_3490025, EPI_ISL_3490026, EPI_ISL_3490027, EPI_ISL_3490031, EPI_ISL_3490032, EPI_ISL_3490036, EPI_ISL_3490037, EPI_ISL_3490038, EPI_ISL_3490039, EPI_ISL_3490040, EPI_ISL_3490041, EPI_ISL_3490043, EPI_ISL_3490045, EPI_ISL_3490047, EPI_ISL_3490052, EPI_ISL_3490054, EPI_ISL_3490055, EPI_ISL_3490060, EPI_ISL_3490065, EPI_ISL_3490067, EPI_ISL_3490068, EPI_ISL_3490071, EPI_ISL_3490072, EPI_ISL_3490073, EPI_ISL_3490074, EPI_ISL_3490075, EPI_ISL_3490076, EPI_ISL_3490078, EPI_ISL_3490079, EPI_ISL_3490085, EPI_ISL_3490086, EPI_ISL_3490088, EPI_ISL_3490091, EPI_ISL_3490096, EPI_ISL_3490097, EPI_ISL_3490099, EPI_ISL_3490107, EPI_ISL_3490108, EPI_ISL_3490112, EPI_ISL_3490113, EPI_ISL_3490114, EPI_ISL_3490116, EPI_ISL_3490117, EPI_ISL_3490120, EPI_ISL_3490121, EPI_ISL_3490122, EPI_ISL_3490123, EPI_ISL_3490124, EPI_ISL_3490126, EPI_ISL_3490127, EPI_ISL_3490128, EPI_ISL_3490130, EPI_ISL_3490131, EPI_ISL_3490132, EPI_ISL_3490133, EPI_ISL_3490134, EPI_ISL_3490135, EPI_ISL_3490136, EPI_ISL_3490137, EPI_ISL_3490139, EPI_ISL_3490140, EPI_ISL_3490141, EPI_ISL_3490144, EPI_ISL_3490145, EPI_ISL_3490150, EPI_ISL_3490151, EPI_ISL_3490152, EPI_ISL_3490153, EPI_ISL_3490156, EPI_ISL_3490157, EPI_ISL_3490163, EPI_ISL_3490164, EPI_ISL_3490165, EPI_ISL_3490166, EPI_ISL_3490167, EPI_ISL_3490169, EPI_ISL_3490170, EPI_ISL_3490174, EPI_ISL_3490175, EPI_ISL_3490176, EPI_ISL_3490181, EPI_ISL_3490184, EPI_ISL_3490185, EPI_ISL_3490189, EPI_ISL_3490190, EPI_ISL_3490191, EPI_ISL_3490197, EPI_ISL_3490198, EPI_ISL_3490207, EPI_ISL_3490214, EPI_ISL_3490215, EPI_ISL_3490216, EPI_ISL_3490217, EPI_ISL_3490220, EPI_ISL_3490224, EPI_ISL_3490228, EPI_ISL_3490229, EPI_ISL_3490230, EPI_ISL_3490231, EPI_ISL_3490232, EPI_ISL_3490234, EPI_ISL_3490236, EPI_ISL_3490238, EPI_ISL_3490239, EPI_ISL_3490240, EPI_ISL_3490244, EPI_ISL_3490245, EPI_ISL_3490250, EPI_ISL_3490255, EPI_ISL_3490256, EPI_ISL_3490257, EPI_ISL_3490258, EPI_ISL_3490259, EPI_ISL_3490261, EPI_ISL_3490262, EPI_ISL_3490264, EPI_ISL_3490273, EPI_ISL_3490275, EPI_ISL_3490276, EPI_ISL_3490277, EPI_ISL_3490278, EPI_ISL_3490281, EPI_ISL_3490282, EPI_ISL_3490283, EPI_ISL_3490284, EPI_ISL_3490289, EPI_ISL_3490292, EPI_ISL_3490293, EPI_ISL_3490294, EPI_ISL_3490295, EPI_ISL_3490297, EPI_ISL_3490298, EPI_ISL_3490303, EPI_ISL_3490306, EPI_ISL_3490310, EPI_ISL_3490313, EPI_ISL_3490314, EPI_ISL_3490315, EPI_ISL_3490316, EPI_ISL_3490317, EPI_ISL_3490318, EPI_ISL_3490320, EPI_ISL_3490321, EPI_ISL_3490322, EPI_ISL_3490329, EPI_ISL_3490331, EPI_ISL_3490332, EPI_ISL_3490333, EPI_ISL_3490336, EPI_ISL_3490339, EPI_ISL_3490346, EPI_ISL_3490348, EPI_ISL_3490355, EPI_ISL_3490371, EPI_ISL_3490373, EPI_ISL_3490374, EPI_ISL_3490383, EPI_ISL_3490437, EPI_ISL_3490438, EPI_ISL_3490444, EPI_ISL_3490445, EPI_ISL_3490446, EPI_ISL_3490447, EPI_ISL_3490448, EPI_ISL_3490450, EPI_ISL_3490451, EPI_ISL_3490456, EPI_ISL_3490458, EPI_ISL_3490461, EPI_ISL_3490462, EPI_ISL_3490465, EPI_ISL_3490467, EPI_ISL_3490469, EPI_ISL_3490470, EPI_ISL_3490509, EPI_ISL_3490510, EPI_ISL_3490513, EPI_ISL_3490516, EPI_ISL_3490517, EPI_ISL_3490522, EPI_ISL_3490524, EPI_ISL_3490526, EPI_ISL_3490535, EPI_ISL_3490548, EPI_ISL_3490551, EPI_ISL_3490556, EPI_ISL_3490557, EPI_ISL_3490566, EPI_ISL_3490574, EPI_ISL_3490575, EPI_ISL_3490578, EPI_ISL_3490582, EPI_ISL_3490585, EPI_ISL_3490586, EPI_ISL_3490588 | | | |
| see above | British Columbia Centre For Disease Control | BCCDC Public Health Laboratory | Ana Pacagnella; Corrinne Ng; Dan Fornika; John Tyson; Kim Macdonald; Kimia Kamelian; Linda Hoang; Loretta Janz; Mel Krajden; Prystajecky Natalie; Robert Azana; Shannon Russell |
| EPI_ISL_2991529, EPI_ISL_2991540, EPI_ISL_2991577, EPI_ISL_2991593, EPI_ISL_2991613, EPI_ISL_2991625, EPI_ISL_2991631, EPI_ISL_2991656, EPI_ISL_2991721, EPI_ISL_2991732, EPI_ISL_2991769, EPI_ISL_2991780, EPI_ISL_2991795, EPI_ISL_2991815, EPI_ISL_2991836, EPI_ISL_2991926, EPI_ISL_2991930, EPI_ISL_2991985 | | | |
| see above | Broad Institute Clinical Research Sequencing Platform | Infectious Disease Program, Broad Institute of Harvard and MIT | A.E.; Adams, G.; Anahtar, M.; B.L.; B.W.; Bauer, M.; Birren; Branda, J.; Carter, A.; Cerrato, F.; Chaluvadi, S.; Chapman; Cusick, C.; D.J.; DeRuff, K.; Flowers, K.; Gallagher, G.; Gladden-Young, A.; Gnirke, A.; Harris, J.; J.E.; K.J.; LaRocque, R.; Lagerborg, K.; Lemieux; Lin; Loreth, C.; MacInnis; Neumann, A.; Normandin, E.; P.C.; Park; Pierce, V.; Reilly, S.; Rosenberg, E.; Rudy, M.; Ryan, E.; S.B.; Sabeti; Shaw, B.; Siddle; Slater, D.; Smole, S.; Tomkins-Tinch, C.; Turbett, S. |
| EPI_ISL_2685871 | C.H.Bois de l'Abbaye | GIGA Medical Genomics | Bouchra Boujemla; Cécile Meex; Keith Durkin; Maria Artesi; Marie-Pierre Hayette; Nathalie Renotte; Pierrette Melin; Raphaël Boreux; Sébastien Bontems; Vincent Bours |
| EPI_ISL_2787321 | CA-Los Angeles County Public Health Laboratory | Centers for Disease Control and Prevention Division of Viral Diseases, Pathogen Discovery | Alison Laufer Halpin; Ben L. Rambo-Martin; Clinton R. Paden; Dakota Howard; Darlene Wagner; Dave Wentworth; Dhwani Batra; Jasmine Padilla; Justin Lee; Katie Dillon; Krista Queen; Kristen Knipe; Kristine Lacek; Mark Burroughs; Matthew Schmerer; Mili Sheth; Peter Cook; Sam Shepard; Sarah Nobles; Shoshona Le; Suxiang Tong; Vivien Dugan; Yvette Unoarumhi |
| EPI_ISL_2923285, EPI_ISL_2923355, EPI_ISL_2923360, EPI_ISL_2923361, EPI_ISL_2923368, EPI_ISL_2923371, EPI_ISL_2923372, EPI_ISL_2923388, EPI_ISL_2923436, EPI_ISL_2923525, EPI_ISL_2923566, EPI_ISL_2923633, EPI_ISL_2923635, EPI_ISL_2923638, EPI_ISL_2923659, EPI_ISL_2923663, EPI_ISL_2923741, EPI_ISL_2923742, EPI_ISL_2923756, EPI_ISL_3244492, EPI_ISL_3244501, EPI_ISL_3244519, EPI_ISL_3671066, EPI_ISL_3671070, EPI_ISL_3671253, EPI_ISL_3671254, EPI_ISL_3671266, EPI_ISL_3671267, EPI_ISL_3671268, EPI_ISL_3671269, EPI_ISL_3671270, EPI_ISL_3671271, EPI_ISL_3671272, EPI_ISL_3671273, EPI_ISL_3671274, EPI_ISL_3671275, EPI_ISL_3671276, EPI_ISL_3671277, EPI_ISL_3671278, EPI_ISL_3671279, EPI_ISL_3671280, EPI_ISL_3671281, EPI_ISL_3672280, EPI_ISL_3672287 | | | |
| see above | CDPH VBL | California Department of Public Health | CDPH-COVIDNet; Emily Smith on behalf of CDPH-COVIDNet and UCSD EXCITE lab |
| EPI_ISL_3102257, EPI_ISL_3102261, EPI_ISL_3102268, EPI_ISL_3102274, EPI_ISL_3102278, EPI_ISL_3102288, EPI_ISL_3102295, EPI_ISL_3102305, EPI_ISL_3912144 | | | |
| see above | CENTRO DE ATENDIMENTO PARA ENFRENTAMENTO AO COVID 19 | Analytical Competence Molecular Epidemiology Lab/ACME, Oswaldo Cruz Foundation, Ceara (FIOCRUZ CE) | Cleber Furtado Aksenen; Fabio Miyajima; Fernando Braga Stehling; Francisco Eder de Moura Lopes; Jamille Maria Mendes Bezerra; Joaquim Cesar do Nascimento Sousa Junior; Joaquim César do Nascimento Sousa Junior; Pedro Miguel Carneiro Jeronimo; Suzana Porto Almeida & Lucas Delerino on behalf of COVID-19 FIOCRUZ Genomic Network; Suzana Porto Almeida e Lucas Delerino; Thais Ferreira de Oliveira; Thais de Oliveira Costa; Ticiane Cavalcante de Souza; Veridiana Pessoa Miyajima |
| EPI_ISL_5647110, EPI_ISL_5647115, EPI_ISL_5647147, EPI_ISL_5647158, EPI_ISL_5647197 | CENTRO DE ESPECIALIDADES MARIAS DA GLORIA | Instituto Butantan | Antonio Jorge Martins; Claudia Renata dos Santos Barros; David Schlesinger; Debora Botequio Moretti; Dimas Tadeu Covas; Elaine Cristina Marqueze; Elaine Vieira Santos; Evandra Strazza Rodrigues; Heidge Fukumasu; Jayme Augusto de Souza-Neto; José Salvatore Leister Patané; Luiz Alcantara; Luiz Lehmann Coutinho; Maria Carolina Elias; Maurício Lacerda Nogueira; Rafael dos Santos Bezerra; Raul Machado Neto; Rejane Maria Tommasini Grotto; Ricardo Haddad; Sandra Coccuzzo Sampaio Vessoni; Simone Kashima; Svetoslav Nanev Slavov; Vincent Louis Viala |
| EPI_ISL_5646379 | CENTRO DE SAUDE DE ADAMANTINA | Instituto Butantan | Antonio Jorge Martins; Claudia Renata dos Santos Barros; David Schlesinger; Debora Botequio Moretti; Dimas Tadeu Covas; Elaine Cristina Marqueze; Elaine Vieira Santos; Evandra Strazza Rodrigues; Heidge Fukumasu; Jayme Augusto de Souza-Neto; José Salvatore Leister Patané; Luiz Alcantara; Luiz Lehmann Coutinho; Maria Carolina Elias; Maurício Lacerda Nogueira; Rafael dos Santos Bezerra; Raul Machado Neto; Rejane Maria Tommasini Grotto; Ricardo Haddad; Sandra Coccuzzo Sampaio Vessoni; Simone Kashima; Svetoslav Nanev Slavov; Vincent Louis Viala |
| EPI_ISL_5646119, EPI_ISL_5646135, EPI_ISL_5646139, EPI_ISL_5646144, EPI_ISL_5646165, EPI_ISL_5646173, EPI_ISL_5646179, EPI_ISL_5646182, EPI_ISL_5646187, EPI_ISL_5646188, EPI_ISL_5646200, EPI_ISL_5646203 | | | |
| see above | CENTRO DE SAUDE DE AVAI | Instituto Butantan | Antonio Jorge Martins; Claudia Renata dos Santos Barros; David Schlesinger; Debora Botequio Moretti; Dimas Tadeu Covas; Elaine Cristina Marqueze; Elaine Vieira Santos; Evandra Strazza Rodrigues; Heidge Fukumasu; Jayme Augusto de Souza-Neto; José Salvatore Leister Patané; Luiz Alcantara; Luiz Lehmann Coutinho; Maria Carolina Elias; Maurício Lacerda Nogueira; Rafael dos Santos Bezerra; Raul Machado Neto; Rejane Maria Tommasini Grotto; Ricardo Haddad; Sandra Coccuzzo Sampaio Vessoni; Simone Kashima; Svetoslav Nanev Slavov; Vincent Louis Viala |
| EPI_ISL_5649627, EPI_ISL_5649689 | CENTRO DE SAUDE DE CAJOBI | Instituto Butantan | Antonio Jorge Martins; Claudia Renata dos Santos Barros; David Schlesinger; Debora Botequio Moretti; Dimas Tadeu Covas; Elaine Cristina Marqueze; Elaine Vieira Santos; Evandra Strazza Rodrigues; Heidge Fukumasu; Jayme Augusto de Souza-Neto; José Salvatore Leister Patané; Luiz Alcantara; Luiz Lehmann Coutinho; Maria Carolina Elias; Maurício Lacerda Nogueira; Rafael dos Santos Bezerra; Raul Machado Neto; Rejane Maria Tommasini Grotto; Ricardo Haddad; Sandra Coccuzzo Sampaio Vessoni; Simone Kashima; Svetoslav Nanev Slavov; Vincent Louis Viala |
| EPI_ISL_5646290 | CENTRO DE SAUDE DE CHAVANTES | Instituto Butantan | Antonio Jorge Martins; Claudia Renata dos Santos Barros; David Schlesinger; Debora Botequio Moretti; Dimas Tadeu Covas; Elaine Cristina Marqueze; Elaine Vieira Santos; Evandra Strazza Rodrigues; Heidge Fukumasu; Jayme Augusto de Souza-Neto; José Salvatore Leister Patané; Luiz Alcantara; Luiz Lehmann Coutinho; Maria Carolina Elias; Maurício Lacerda Nogueira; Rafael dos Santos Bezerra; Raul Machado Neto; Rejane Maria Tommasini Grotto; Ricardo Haddad; Sandra Coccuzzo Sampaio Vessoni; Simone Kashima; Svetoslav Nanev Slavov; Vincent Louis Viala |
| EPI_ISL_5646362 | CENTRO DE SAUDE DE FLORINEA | Instituto Butantan | Antonio Jorge Martins; Claudia Renata dos Santos Barros; David Schlesinger; Debora Botequio Moretti; Dimas Tadeu Covas; Elaine Cristina Marqueze; Elaine Vieira Santos; Evandra Strazza Rodrigues; Heidge Fukumasu; Jayme Augusto de Souza-Neto; José Salvatore Leister Patané; Luiz Alcantara; Luiz Lehmann Coutinho; Maria Carolina Elias; Maurício Lacerda Nogueira; Rafael dos Santos Bezerra; Raul Machado Neto; Rejane Maria Tommasini Grotto; Ricardo Haddad; Sandra Coccuzzo Sampaio Vessoni; Simone Kashima; Svetoslav Nanev Slavov; Vincent Louis Viala |
| EPI_ISL_5645581, EPI_ISL_5649618, EPI_ISL_5649663, EPI_ISL_5650219 | CENTRO DE SAUDE DE LUCELIA | Instituto Butantan | Antonio Jorge Martins; Claudia Renata dos Santos Barros; David Schlesinger; Debora Botequio Moretti; Dimas Tadeu Covas; Elaine Cristina Marqueze; Elaine Vieira Santos; Evandra Strazza Rodrigues; Heidge Fukumasu; Jayme Augusto de Souza-Neto; José Salvatore Leister Patané; Luiz Alcantara; Luiz Lehmann Coutinho; Maria Carolina Elias; Maurício Lacerda Nogueira; Rafael dos Santos Bezerra; Raul Machado Neto; Rejane Maria Tommasini Grotto; Ricardo Haddad; Sandra Coccuzzo Sampaio Vessoni; Simone Kashima; Svetoslav Nanev Slavov; Vincent Louis Viala |
| EPI_ISL_5646376 | CENTRO DE SAUDE DE PLATINA | Instituto Butantan | Antonio Jorge Martins; Claudia Renata dos Santos Barros; David Schlesinger; Debora Botequio Moretti; Dimas Tadeu Covas; Elaine Cristina Marqueze; Elaine Vieira Santos; Evandra Strazza Rodrigues; Heidge Fukumasu; Jayme Augusto de Souza-Neto; José Salvatore Leister Patané; Luiz Alcantara; Luiz Lehmann Coutinho; Maria Carolina Elias; Maurício Lacerda Nogueira; Rafael dos Santos Bezerra; Raul Machado Neto; Rejane Maria Tommasini Grotto; Ricardo Haddad; Sandra Coccuzzo Sampaio Vessoni; Simone Kashima; Svetoslav Nanev Slavov; Vincent Louis Viala |
| EPI_ISL_5647170 | CENTRO DE SAUDE DR MARIO DIAS DE AGUIAR CAPIVARI | Instituto Butantan | Antonio Jorge Martins; Claudia Renata dos Santos Barros; David Schlesinger; Debora Botequio Moretti; Dimas Tadeu Covas; Elaine Cristina Marqueze; Elaine Vieira Santos; Evandra Strazza Rodrigues; Heidge Fukumasu; Jayme Augusto de Souza-Neto; José Salvatore Leister Patané; Luiz Alcantara; Luiz Lehmann Coutinho; Maria Carolina Elias; Maurício Lacerda Nogueira; Rafael dos Santos Bezerra; Raul Machado Neto; Rejane Maria Tommasini Grotto; Ricardo Haddad; Sandra Coccuzzo Sampaio Vessoni; Simone Kashima; Svetoslav Nanev Slavov; Vincent Louis Viala |

| | | | |
|---|---|---|---|
| EPI_ISL_5646316, EPI_ISL_5646325, EPI_ISL_5646335 | CENTRO DE SAUDE DR NELSON DA CUNHA BASTOS | Instituto Butantan | Antonio Jorge Martins; Claudia Renata dos Santos Barros; David Schlesinger; Debora Botequio Moretti; Dimas Tadeu Covas; Elaine Cristina Marqueze; Elaine Vieira Santos; Evandra Strazza Rodrigues; Heidge Fukumasu; Jayme Augusto de Souza-Neto; José Salvatore Leister Patané; Luiz Alcantara; Luiz Lehmann Coutinho; Maria Carolina Elias; Maurício Lacerda Nogueira; Rafael dos Santos Bezerra; Raul Machado Neto; Rejane Maria Tommasini Grotto; Ricardo Haddad; Sandra Coccuzzo Sampaio Vessoni; Simone Kashima; Svetoslav Nanev Slavov; Vincent Louis Viala |
| EPI_ISL_5645727 | CENTRO DE SAUDE DR RENATO DE CARVALHO RIBEIRO ANGATUBA | Instituto Butantan | Antonio Jorge Martins; Claudia Renata dos Santos Barros; David Schlesinger; Debora Botequio Moretti; Dimas Tadeu Covas; Elaine Cristina Marqueze; Elaine Vieira Santos; Evandra Strazza Rodrigues; Heidge Fukumasu; Jayme Augusto de Souza-Neto; José Salvatore Leister Patané; Luiz Alcantara; Luiz Lehmann Coutinho; Maria Carolina Elias; Maurício Lacerda Nogueira; Rafael dos Santos Bezerra; Raul Machado Neto; Rejane Maria Tommasini Grotto; Ricardo Haddad; Sandra Coccuzzo Sampaio Vessoni; Simone Kashima; Svetoslav Nanev Slavov; Vincent Louis Viala |
| EPI_ISL_5645637, EPI_ISL_5645709, EPI_ISL_5645722, EPI_ISL_5645741 | CENTRO DE SAUDE ESF IV ZONA RURAL DOMINGOS DE S SJRIOPARDO | Instituto Butantan | Antonio Jorge Martins; Claudia Renata dos Santos Barros; David Schlesinger; Debora Botequio Moretti; Dimas Tadeu Covas; Elaine Cristina Marqueze; Elaine Vieira Santos; Evandra Strazza Rodrigues; Heidge Fukumasu; Jayme Augusto de Souza-Neto; José Salvatore Leister Patané; Luiz Alcantara; Luiz Lehmann Coutinho; Maria Carolina Elias; Maurício Lacerda Nogueira; Rafael dos Santos Bezerra; Raul Machado Neto; Rejane Maria Tommasini Grotto; Ricardo Haddad; Sandra Coccuzzo Sampaio Vessoni; Simone Kashima; Svetoslav Nanev Slavov; Vincent Louis Viala |
| EPI_ISL_5645796, EPI_ISL_5645946, EPI_ISL_5645956 | CENTRO DE SAUDE II | Instituto Butantan | Antonio Jorge Martins; Claudia Renata dos Santos Barros; David Schlesinger; Debora Botequio Moretti; Dimas Tadeu Covas; Elaine Cristina Marqueze; Elaine Vieira Santos; Evandra Strazza Rodrigues; Heidge Fukumasu; Jayme Augusto de Souza-Neto; José Salvatore Leister Patané; Luiz Alcantara; Luiz Lehmann Coutinho; Maria Carolina Elias; Maurício Lacerda Nogueira; Rafael dos Santos Bezerra; Raul Machado Neto; Rejane Maria Tommasini Grotto; Ricardo Haddad; Sandra Coccuzzo Sampaio Vessoni; Simone Kashima; Svetoslav Nanev Slavov; Vincent Louis Viala |
| EPI_ISL_5647155 | CENTRO DE SAUDE II DR JOSE DE FELIPE E S PINHAL SP | Instituto Butantan | Antonio Jorge Martins; Claudia Renata dos Santos Barros; David Schlesinger; Debora Botequio Moretti; Dimas Tadeu Covas; Elaine Cristina Marqueze; Elaine Vieira Santos; Evandra Strazza Rodrigues; Heidge Fukumasu; Jayme Augusto de Souza-Neto; José Salvatore Leister Patané; Luiz Alcantara; Luiz Lehmann Coutinho; Maria Carolina Elias; Maurício Lacerda Nogueira; Rafael dos Santos Bezerra; Raul Machado Neto; Rejane Maria Tommasini Grotto; Ricardo Haddad; Sandra Coccuzzo Sampaio Vessoni; Simone Kashima; Svetoslav Nanev Slavov; Vincent Louis Viala |
| EPI_ISL_5647108, EPI_ISL_5647181 | CENTRO DE SAUDE II DR JOSE PAIONE MOCOCA | Instituto Butantan | Antonio Jorge Martins; Claudia Renata dos Santos Barros; David Schlesinger; Debora Botequio Moretti; Dimas Tadeu Covas; Elaine Cristina Marqueze; Elaine Vieira Santos; Evandra Strazza Rodrigues; Heidge Fukumasu; Jayme Augusto de Souza-Neto; José Salvatore Leister Patané; Luiz Alcantara; Luiz Lehmann Coutinho; Maria Carolina Elias; Maurício Lacerda Nogueira; Rafael dos Santos Bezerra; Raul Machado Neto; Rejane Maria Tommasini Grotto; Ricardo Haddad; Sandra Coccuzzo Sampaio Vessoni; Simone Kashima; Svetoslav Nanev Slavov; Vincent Louis Viala |
| EPI_ISL_5649611, EPI_ISL_5649690 | CENTRO DE SAUDE II IBITINGA | Instituto Butantan | Antonio Jorge Martins; Claudia Renata dos Santos Barros; David Schlesinger; Debora Botequio Moretti; Dimas Tadeu Covas; Elaine Cristina Marqueze; Elaine Vieira Santos; Evandra Strazza Rodrigues; Heidge Fukumasu; Jayme Augusto de Souza-Neto; José Salvatore Leister Patané; Luiz Alcantara; Luiz Lehmann Coutinho; Maria Carolina Elias; Maurício Lacerda Nogueira; Rafael dos Santos Bezerra; Raul Machado Neto; Rejane Maria Tommasini Grotto; Ricardo Haddad; Sandra Coccuzzo Sampaio Vessoni; Simone Kashima; Svetoslav Nanev Slavov; Vincent Louis Viala |
| EPI_ISL_5645640 | CENTRO DE SAUDE II SAO MIGUEL ARCANJO | Instituto Butantan | Antonio Jorge Martins; Claudia Renata dos Santos Barros; David Schlesinger; Debora Botequio Moretti; Dimas Tadeu Covas; Elaine Cristina Marqueze; Elaine Vieira Santos; Evandra Strazza Rodrigues; Heidge Fukumasu; Jayme Augusto de Souza-Neto; José Salvatore Leister Patané; Luiz Alcantara; Luiz Lehmann Coutinho; Maria Carolina Elias; Maurício Lacerda Nogueira; Rafael dos Santos Bezerra; Raul Machado Neto; Rejane Maria Tommasini Grotto; Ricardo Haddad; Sandra Coccuzzo Sampaio Vessoni; Simone Kashima; Svetoslav Nanev Slavov; Vincent Louis Viala |
| EPI_ISL_5645682 | CENTRO DE SAUDE II TIETE DR RUI SILVEIRA MELO | Instituto Butantan | Antonio Jorge Martins; Claudia Renata dos Santos Barros; David Schlesinger; Debora Botequio Moretti; Dimas Tadeu Covas; Elaine Cristina Marqueze; Elaine Vieira Santos; Evandra Strazza Rodrigues; Heidge Fukumasu; Jayme Augusto de Souza-Neto; José Salvatore Leister Patané; Luiz Alcantara; Luiz Lehmann Coutinho; Maria Carolina Elias; Maurício Lacerda Nogueira; Rafael dos Santos Bezerra; Raul Machado Neto; Rejane Maria Tommasini Grotto; Ricardo Haddad; Sandra Coccuzzo Sampaio Vessoni; Simone Kashima; Svetoslav Nanev Slavov; Vincent Louis Viala |
| EPI_ISL_5650372 | CENTRO DE SAUDE III ALTO ALEGRE | Instituto Butantan | Antonio Jorge Martins; Claudia Renata dos Santos Barros; David Schlesinger; Debora Botequio Moretti; Dimas Tadeu Covas; Elaine Cristina Marqueze; Elaine Vieira Santos; Evandra Strazza Rodrigues; Heidge Fukumasu; Jayme Augusto de Souza-Neto; José Salvatore Leister Patané; Luiz Alcantara; Luiz Lehmann Coutinho; Maria Carolina Elias; Maurício Lacerda Nogueira; Rafael dos Santos Bezerra; Raul Machado Neto; Rejane Maria Tommasini Grotto; Ricardo Haddad; Sandra Coccuzzo Sampaio Vessoni; Simone Kashima; Svetoslav Nanev Slavov; Vincent Louis Viala |
| EPI_ISL_5646154 | CENTRO DE SAUDE III DE BORACEIA | Instituto Butantan | Antonio Jorge Martins; Claudia Renata dos Santos Barros; David Schlesinger; Debora Botequio Moretti; Dimas Tadeu Covas; Elaine Cristina Marqueze; Elaine Vieira Santos; Evandra Strazza Rodrigues; Heidge Fukumasu; Jayme Augusto de Souza-Neto; José Salvatore Leister Patané; Luiz Alcantara; Luiz Lehmann Coutinho; Maria Carolina Elias; Maurício Lacerda Nogueira; Rafael dos Santos Bezerra; Raul Machado Neto; Rejane Maria Tommasini Grotto; Ricardo Haddad; Sandra Coccuzzo Sampaio Vessoni; Simone Kashima; Svetoslav Nanev Slavov; Vincent Louis Viala |
| EPI_ISL_5646725, EPI_ISL_5646735 | CENTRO DE SAUDE III DE DIVINOLANDIA | Instituto Butantan | Antonio Jorge Martins; Claudia Renata dos Santos Barros; David Schlesinger; Debora Botequio Moretti; Dimas Tadeu Covas; Elaine Cristina Marqueze; Elaine Vieira Santos; Evandra Strazza Rodrigues; Heidge Fukumasu; Jayme Augusto de Souza-Neto; José Salvatore Leister Patané; Luiz Alcantara; Luiz Lehmann Coutinho; Maria Carolina Elias; Maurício Lacerda Nogueira; Rafael dos Santos Bezerra; Raul Machado Neto; Rejane Maria Tommasini Grotto; Ricardo Haddad; Sandra Coccuzzo Sampaio Vessoni; Simone Kashima; Svetoslav Nanev Slavov; Vincent Louis Viala |
| EPI_ISL_5650342, EPI_ISL_5650364 | CENTRO DE SAUDE III GLICERIO GIMAIEL ELIAS GIMAIEL | Instituto Butantan | Antonio Jorge Martins; Claudia Renata dos Santos Barros; David Schlesinger; Debora Botequio Moretti; Dimas Tadeu Covas; Elaine Cristina Marqueze; Elaine Vieira Santos; Evandra Strazza Rodrigues; Heidge Fukumasu; Jayme Augusto de Souza-Neto; José Salvatore Leister Patané; Luiz Alcantara; Luiz Lehmann Coutinho; Maria Carolina Elias; Maurício Lacerda Nogueira; Rafael dos Santos Bezerra; Raul Machado Neto; Rejane Maria Tommasini Grotto; Ricardo Haddad; Sandra Coccuzzo Sampaio Vessoni; Simone Kashima; Svetoslav Nanev Slavov; Vincent Louis Viala |
| EPI_ISL_5650348, EPI_ISL_5650376, EPI_ISL_5650380 | CENTRO DE SAUDE III SANTA ERNESTINA | Instituto Butantan | Antonio Jorge Martins; Claudia Renata dos Santos Barros; David Schlesinger; Debora Botequio Moretti; Dimas Tadeu Covas; Elaine Cristina Marqueze; Elaine Vieira Santos; Evandra Strazza Rodrigues; Heidge Fukumasu; Jayme Augusto de Souza-Neto; José Salvatore Leister Patané; Luiz Alcantara; Luiz Lehmann Coutinho; Maria Carolina Elias; Maurício Lacerda Nogueira; Rafael dos Santos Bezerra; Raul Machado Neto; Rejane Maria Tommasini Grotto; Ricardo Haddad; Sandra Coccuzzo Sampaio Vessoni; Simone Kashima; Svetoslav Nanev Slavov; Vincent Louis Viala |
| EPI_ISL_5649613, EPI_ISL_5649688, EPI_ISL_5650238, EPI_ISL_5650343, EPI_ISL_5650349, EPI_ISL_5650369, EPI_ISL_5650382, EPI_ISL_5650389 | | | |
| see above | CENTRO DE SAUDE III TABATINGA | Instituto Butantan | Antonio Jorge Martins; Claudia Renata dos Santos Barros; David Schlesinger; Debora Botequio Moretti; Dimas Tadeu Covas; Elaine Cristina Marqueze; Elaine Vieira Santos; Evandra Strazza Rodrigues; Heidge Fukumasu; Jayme Augusto de Souza-Neto; José Salvatore Leister Patané; Luiz Alcantara; Luiz Lehmann Coutinho; Maria Carolina Elias; Maurício Lacerda Nogueira; Rafael dos Santos Bezerra; Raul Machado Neto; Rejane Maria Tommasini Grotto; Ricardo Haddad; Sandra Coccuzzo Sampaio Vessoni; Simone Kashima; Svetoslav Nanev Slavov; Vincent Louis Viala |
| EPI_ISL_5645626, EPI_ISL_5645729 | CENTRO DE SAUDE TEREZINHA DE MORAES ARSILLA | Instituto Butantan | Antonio Jorge Martins; Claudia Renata dos Santos Barros; David Schlesinger; Debora Botequio Moretti; Dimas Tadeu Covas; Elaine Cristina Marqueze; Elaine Vieira Santos; Evandra Strazza Rodrigues; Heidge Fukumasu; Jayme Augusto de Souza-Neto; José Salvatore Leister Patané; Luiz Alcantara; Luiz Lehmann Coutinho; Maria Carolina Elias; Maurício Lacerda Nogueira; Rafael dos Santos Bezerra; Raul Machado Neto; Rejane Maria Tommasini Grotto; Ricardo Haddad; Sandra Coccuzzo Sampaio Vessoni; Simone Kashima; Svetoslav Nanev Slavov; Vincent Louis Viala |
| EPI_ISL_5645701, EPI_ISL_5645723, EPI_ISL_5645736, EPI_ISL_5645851 | CENTRO DE TRIAGEM COVID 19 | Instituto Butantan | Antonio Jorge Martins; Claudia Renata dos Santos Barros; David Schlesinger; Debora Botequio Moretti; Dimas Tadeu Covas; Elaine Cristina Marqueze; Elaine Vieira Santos; Evandra Strazza Rodrigues; Heidge Fukumasu; Jayme Augusto de Souza-Neto; José Salvatore Leister Patané; Luiz Alcantara; Luiz Lehmann Coutinho; Maria Carolina Elias; Maurício Lacerda Nogueira; Rafael dos Santos Bezerra; Raul Machado Neto; Rejane Maria Tommasini Grotto; Ricardo Haddad; Sandra Coccuzzo Sampaio Vessoni; Simone Kashima; Svetoslav Nanev Slavov; Vincent Louis Viala |
| EPI_ISL_5647188 | CENTRO MEDICO DR NELSON SALOME DE CONCHAL | Instituto Butantan | Antonio Jorge Martins; Claudia Renata dos Santos Barros; David Schlesinger; Debora Botequio Moretti; Dimas Tadeu Covas; Elaine Cristina Marqueze; Elaine Vieira Santos; Evandra Strazza Rodrigues; Heidge Fukumasu; Jayme Augusto de Souza-Neto; José Salvatore Leister Patané; Luiz Alcantara; Luiz Lehmann Coutinho; Maria Carolina Elias; Maurício Lacerda Nogueira; Rafael dos Santos Bezerra; Raul Machado Neto; Rejane Maria Tommasini Grotto; Ricardo Haddad; Sandra Coccuzzo Sampaio Vessoni; Simone Kashima; Svetoslav Nanev Slavov; Vincent Louis Viala |
| EPI_ISL_2637392 | CH.INTERCOMMUNAL DE CRETEIL | Department of Virology, Henri Mondor University Hospital, Assistance Publique Hôpitaux de Paris, Université Paris-Est Créteil, INSERM U955 | Alexandre Soulier; Christophe Rodriguez; Elisabeth Trawinski; Guillaume Gricourt; Jean-Michel Pawlotsky; Melissa N'Debi; Slim Fourati; Vanessa Demontant |
| EPI_ISL_2627852, EPI_ISL_2627853, EPI_ISL_2627854, EPI_ISL_2627855, EPI_ISL_2627856, EPI_ISL_2627857, EPI_ISL_2627858, EPI_ISL_2627859, EPI_ISL_2629223, EPI_ISL_2629224 | | | |
| see above | CHC Andrée Rosemon | Institut Pasteur de la Guyane | Anne Lavergne; Antoine Enfissi; Arielle Salmier; Dominique Rousset |
| EPI_ISL_2695339, EPI_ISL_2695345 | CHEDV | Instituto Nacional de Saude (INSA) | Borges et al |
| EPI_ISL_2695098 | CHULN - H Santa Maria | Instituto Nacional de Saude (INSA) | Borges et al |
| EPI_ISL_2637260 | CLILAB | Microbiology Department | Aida Gonzalez-Diaz; Carmen Ardanuy; Jordi Camara; Jordi Niubó; Laura Calatayud; M Angeles Domínguez; Miguel Fernandez-Huerta; Sara Marti |
| EPI_ISL_2627860, EPI_ISL_2627861, EPI_ISL_2627862, EPI_ISL_2627863, EPI_ISL_2627864, EPI_ISL_2627865, EPI_ISL_2627866 | | | |
| see above | CNR Institut Pasteur de la Guyane | Institut Pasteur de la Guyane | Anne Lavergne; Antoine Enfissi; Arielle Salmier; Dominique Rousset |
| EPI_ISL_5647162, EPI_ISL_5647179 | COORDENADORIA MUNICIPAL DE SAUDE DE IRACEMAPOLIS | Instituto Butantan | Antonio Jorge Martins; Claudia Renata dos Santos Barros; David Schlesinger; Debora Botequio Moretti; Dimas Tadeu Covas; Elaine Cristina Marqueze; Elaine Vieira Santos; Evandra Strazza Rodrigues; Heidge Fukumasu; Jayme Augusto de Souza-Neto; José Salvatore Leister Patané; Luiz Alcantara; Luiz Lehmann Coutinho; Maria Carolina Elias; Maurício Lacerda Nogueira; Rafael dos Santos Bezerra; Raul Machado Neto; Rejane Maria Tommasini Grotto; Ricardo Haddad; Sandra Coccuzzo Sampaio Vessoni; Simone Kashima; Svetoslav Nanev Slavov; Vincent Louis Viala |
| EPI_ISL_5647113, EPI_ISL_5647124, EPI_ISL_5647127 | CS DE ADOLFO | Instituto Butantan | Antonio Jorge Martins; Claudia Renata dos Santos Barros; David Schlesinger; Debora Botequio Moretti; Dimas Tadeu Covas; Elaine Cristina Marqueze; Elaine Vieira Santos; Evandra Strazza Rodrigues; Heidge Fukumasu; Jayme Augusto de Souza-Neto; José Salvatore Leister Patané; Luiz Alcantara; Luiz Lehmann Coutinho; Maria Carolina Elias; Maurício Lacerda Nogueira; Rafael dos Santos Bezerra; Raul Machado Neto; Rejane Maria Tommasini Grotto; Ricardo Haddad; Sandra Coccuzzo Sampaio Vessoni; Simone Kashima; Svetoslav Nanev Slavov; Vincent Louis Viala |
| EPI_ISL_5645578 | CS DE CAMPOS NOVOS PTA | Instituto Butantan | Antonio Jorge Martins; Claudia Renata dos Santos Barros; David Schlesinger; Debora Botequio Moretti; Dimas Tadeu Covas; Elaine Cristina Marqueze; Elaine Vieira Santos; Evandra Strazza Rodrigues; Heidge Fukumasu; Jayme Augusto de Souza-Neto; José Salvatore Leister Patané; Luiz Alcantara; Luiz Lehmann Coutinho; Maria Carolina Elias; Maurício Lacerda Nogueira; Rafael dos Santos Bezerra; Raul Machado Neto; Rejane Maria Tommasini Grotto; Ricardo Haddad; Sandra Coccuzzo Sampaio Vessoni; Simone Kashima; Svetoslav Nanev Slavov; Vincent Louis Viala |
| EPI_ISL_5650334, EPI_ISL_5650361, EPI_ISL_5650385, EPI_ISL_5650386 | CS DE MARAPOAMA | Instituto Butantan | Antonio Jorge Martins; Claudia Renata dos Santos Barros; David Schlesinger; Debora Botequio Moretti; Dimas Tadeu Covas; Elaine Cristina Marqueze; Elaine Vieira Santos; Evandra Strazza Rodrigues; Heidge Fukumasu; Jayme Augusto de Souza-Neto; José Salvatore Leister Patané; Luiz Alcantara; Luiz Lehmann Coutinho; Maria Carolina Elias; Maurício Lacerda Nogueira; Rafael dos Santos Bezerra; Raul Machado Neto; Rejane Maria Tommasini Grotto; Ricardo Haddad; Sandra Coccuzzo Sampaio Vessoni; Simone Kashima; Svetoslav Nanev Slavov; Vincent Louis Viala |
| EPI_ISL_5649569, EPI_ISL_5649571, EPI_ISL_5649581, EPI_ISL_5649583, EPI_ISL_5649603, EPI_ISL_5649629, EPI_ISL_5649634, EPI_ISL_5649642, EPI_ISL_5649683, EPI_ISL_5650203, EPI_ISL_5650208, EPI_ISL_5650216, EPI_ISL_5650222, EPI_ISL_5650224, EPI_ISL_5650226, EPI_ISL_5650230, EPI_ISL_5650415, EPI_ISL_5650416 | | | |
| see above | CS DE MONTE APRAZIVEL | Instituto Butantan | Antonio Jorge Martins; Claudia Renata dos Santos Barros; David Schlesinger; Debora Botequio Moretti; Dimas Tadeu Covas; Elaine Cristina Marqueze; Elaine Vieira Santos; Evandra Strazza Rodrigues; Heidge Fukumasu; Jayme Augusto de Souza-Neto; José Salvatore Leister Patané; Luiz Alcantara; Luiz Lehmann Coutinho; Maria Carolina Elias; Maurício Lacerda Nogueira; Rafael dos Santos Bezerra; Raul Machado Neto; Rejane Maria Tommasini Grotto; Ricardo Haddad; Sandra Coccuzzo Sampaio Vessoni; Simone Kashima; Svetoslav Nanev Slavov; Vincent Louis Viala |
| EPI_ISL_5646321, EPI_ISL_5646368, EPI_ISL_5646374 | CS DE NEVES PAULISTA | Instituto Butantan | Antonio Jorge Martins; Claudia Renata dos Santos Barros; David Schlesinger; Debora Botequio Moretti; Dimas Tadeu Covas; Elaine Cristina Marqueze; Elaine Vieira Santos; Evandra Strazza Rodrigues; Heidge Fukumasu; Jayme Augusto de Souza-Neto; José Salvatore Leister Patané; Luiz Alcantara; Luiz Lehmann Coutinho; Maria Carolina Elias; Maurício Lacerda Nogueira; Rafael dos Santos Bezerra; Raul Machado Neto; Rejane Maria Tommasini Grotto; Ricardo Haddad; Sandra Coccuzzo Sampaio Vessoni; Simone Kashima; Svetoslav Nanev Slavov; Vincent Louis Viala |
| EPI_ISL_5646355 | CS DE POPULINA | Instituto Butantan | Antonio Jorge Martins; Claudia Renata dos Santos Barros; David Schlesinger; Debora Botequio Moretti; Dimas Tadeu Covas; Elaine Cristina Marqueze; Elaine Vieira Santos; Evandra Strazza Rodrigues; Heidge Fukumasu; Jayme Augusto de Souza-Neto; José Salvatore Leister Patané; Luiz Alcantara; Luiz Lehmann Coutinho; Maria Carolina Elias; Maurício Lacerda Nogueira; Rafael dos Santos Bezerra; Raul Machado Neto; Rejane Maria Tommasini Grotto; Ricardo Haddad; Sandra Coccuzzo Sampaio Vessoni; Simone Kashima; Svetoslav Nanev Slavov; Vincent Louis Viala |
| EPI_ISL_5645652 | CS GILBERTO MIRANDA CABRAL DE PARAPUA | Instituto Butantan | Antonio Jorge Martins; Claudia Renata dos Santos Barros; David Schlesinger; Debora Botequio Moretti; Dimas Tadeu Covas; Elaine Cristina Marqueze; Elaine Vieira Santos; Evandra Strazza Rodrigues; Heidge Fukumasu; Jayme Augusto de Souza-Neto; José Salvatore Leister Patané; Luiz Alcantara; Luiz Lehmann Coutinho; Maria Carolina Elias; Maurício Lacerda Nogueira; Rafael dos Santos Bezerra; Raul Machado Neto; Rejane Maria Tommasini Grotto; Ricardo Haddad; Sandra Coccuzzo Sampaio Vessoni; Simone Kashima; Svetoslav Nanev Slavov; Vincent Louis Viala |
| EPI_ISL_5645734 | CSA COMPLEXO DE SAUDE ANHANGUERA TUCURUVI | Instituto Butantan | Antonio Jorge Martins; Claudia Renata dos Santos Barros; David Schlesinger; Debora Botequio Moretti; Dimas Tadeu Covas; Elaine Cristina Marqueze; Elaine Vieira Santos; Evandra Strazza Rodrigues; Heidge Fukumasu; Jayme Augusto de Souza-Neto; José Salvatore Leister Patané; Luiz Alcantara; Luiz Lehmann Coutinho; Maria Carolina Elias; Maurício Lacerda Nogueira; Rafael dos Santos Bezerra; Raul Machado Neto; Rejane Maria Tommasini Grotto; Ricardo Haddad; Sandra Coccuzzo Sampaio Vessoni; Simone Kashima; Svetoslav Nanev Slavov; Vincent Louis Viala |
| EPI_ISL_5645595 | CSA COMPLEXO DE SAUDE ANHANGUERA VILA JAGUARA | Instituto Butantan | Antonio Jorge Martins; Claudia Renata dos Santos Barros; David Schlesinger; Debora Botequio Moretti; Dimas Tadeu Covas; Elaine Cristina Marqueze; Elaine Vieira Santos; Evandra Strazza Rodrigues; Heidge Fukumasu; Jayme Augusto de Souza-Neto; José Salvatore Leister Patané; Luiz Alcantara; Luiz Lehmann Coutinho; Maria Carolina Elias; Maurício Lacerda Nogueira; Rafael dos Santos Bezerra; Raul Machado Neto; Rejane Maria Tommasini Grotto; Ricardo Haddad; Sandra Coccuzzo Sampaio Vessoni; Simone Kashima; Svetoslav Nanev Slavov; Vincent Louis Viala |
| EPI_ISL_2626591, EPI_ISL_2626593 | CT Department of Public Health | CT Department of Public Health | Claire_Pearson; Tu_N_Nguyen |
| EPI_ISL_2892525, EPI_ISL_3536715, EPI_ISL_3536907, EPI_ISL_3536934, EPI_ISL_4005556, EPI_ISL_4005559, EPI_ISL_4005560, EPI_ISL_4005561, EPI_ISL_4005562 | | | |
| see above | California Department of Public Health | California Department of Public Health | CDPH IDLB COVIDNet et al |
| EPI_ISL_3911439, | Cantacuzino National Military- | Cantacuzino Institute Virology | Carmen Cherciu; Luiza Ustea; Mihaela Lazar; Mihaela Oprea; Nicoleta Paraschiv; Sorin Dinu |

| | | | |
|---|---|---|---|
| EPI_ISL_3911440, EPI_ISL_3911441 | Medical Institute, Viral Respiratory Infections Laboratory | | |
| EPI_ISL_7507366, EPI_ISL_7507378 | Cedars-Sinai Medical Center, Molecular Pathology Laboratory of Department of Pathology & Laboratory Medicine and Genomic Core | Cedars-Sinai Medical Center, Molecular Pathology Laboratory of Department of Pathology & Laboratory Medicine and Genomic Core | Brian Davis; Eric Vail; Jasmine T Plummer; Jorge Mario Sincuir Martinez; Stephanie Chen; Wenjuan Zhang |
| EPI_ISL_3462553, EPI_ISL_3462558, EPI_ISL_3462562, EPI_ISL_3462571, EPI_ISL_3462585, EPI_ISL_3462592, EPI_ISL_3462597, EPI_ISL_3462603, EPI_ISL_3462607, EPI_ISL_3462647, EPI_ISL_3462648, EPI_ISL_3462649, EPI_ISL_3462650, EPI_ISL_3462651, EPI_ISL_3462652, EPI_ISL_3462653 | | | |
| see above | Central Laboratory, Bureau of Public Health (BOG) and Academic Hospital Paramaribo | Erasmus Medical Center | Bas B Oude Munnink; Cherise Beek; Consuella Partowidjojo; Dion Gajadin; Ed PF IJzerman; Emmanuelle Munger; Gary Gummels; Ingrid SK Krishnadath; Lycke Woittiez; Marion PG Koopmans; Mireille Van de Veer; Phyllis Pinas; Princes Wongsowidjojo; Radjesh Ori; Ranisha Doerbalie; Rohma Banwari; Soeradj Harkisoen; Stephen Vreden; Tilotmadebie Ramlal; Verne Nanhoe |
| EPI_ISL_3255135, EPI_ISL_3255138, EPI_ISL_3255140, EPI_ISL_3255144, EPI_ISL_3255145, EPI_ISL_3255146, EPI_ISL_3255148, EPI_ISL_3255149, EPI_ISL_3255151, EPI_ISL_3255154, EPI_ISL_3255156, EPI_ISL_3255157, EPI_ISL_3255159, EPI_ISL_3255160, EPI_ISL_3255164, EPI_ISL_3255166, EPI_ISL_3255168, EPI_ISL_3255180, EPI_ISL_3255181, EPI_ISL_3255183, EPI_ISL_3255184, EPI_ISL_3255188, EPI_ISL_3255189, EPI_ISL_3255220 | | | |
| see above | Central Public Health Laboratory - LACEN -Bahia, Salvador, Brazil | Central Public Health Laboratory - LACEN -Bahia, Salvador, Brazil | Arabela Leal; Breno Dominguez; Felicidade Pereira; Jaqueline Gomes; Luciana Oliveira; Luiz Alcantara; Marcela Gómez; Marta Giovanetti; Patrícia Cajado; Stephane Tosta; Vagner Fonseca; Vanessa Nardy |
| EPI_ISL_2491661 | Centre Hospitalier Universitaire Clermont-Ferrand | CHU Clermont-Ferrand, service de virologie | Bisseux Maxime; Combes Patricia; Henquell Cécile; Mirand Audrey |
| EPI_ISL_4405267 | Centro Provincial de Referencia VIH/SIDA y Hepatitis Virales – Instituto Biológico Dr Tomás Perón | Área de Secuenciación del Laboratorio de Virología del Hospital de Niños Dr. Ricardo Gutierrez on behalf of 'Proyecto Argentino Interinstitucional de genomica de SARS-CoV-2' (PAIS Consortium) | A; Acuña; C; Carlos; Corazza; D; Dehaut; Dolcini; F; Fresina; Gatti; Gimenez; González; Goya; Gutierrez; K; L; LE; Lusso; M; MI; MS; Nabaes Jodar; Nardone; Natale; Nogueira Laspuir; Pagliari; R; Real; Romanko; S; Sanchez; Suarez; Szymanowski; Talavera; Ten Huver; V; Valinotto; Valle; Viegas, M. |
| EPI_ISL_5926831, EPI_ISL_5926840, EPI_ISL_5926841, EPI_ISL_5926868, EPI_ISL_5926869, EPI_ISL_5926870, EPI_ISL_5926871, EPI_ISL_5926893, EPI_ISL_5926894 | | | |
| see above | Centro de Infectologia Charles Mérieux/ Laboratório Rodolphe Mérieux, FUNDHACRE | Bioinformatics Laboratory / LNCC | Alessandra P Lamarca; Alexandra L Gerber; Aline de Freitas Souza; Ana Paula de C Guimarães; Ana Tereza R Vasconcelos; Andreas Stocker; Cirley Maria de Oliveira Lobato; Douglas Terra Machado; Janaína Mazaro; Janete Taynã Nascimento Rodrigues; Luiz Fellype Alves de Souza; Luiz G P de Almeida; Ronaldo da Silva F Jr; Rutilene B. Souza |
| EPI_ISL_4405465 | Centro de Investigaciones Básicas y Aplicadas, UNNOBA | Área de Secuenciación del Laboratorio de Virología del Hospital de Niños Dr. Ricardo Gutierrez on behalf of 'Proyecto Argentino Interinstitucional de genomica de SARS-CoV-2' (PAIS Consortium) | A; Acuña; Alaniz; Bagnis; Barbero; Bonadeo; Brandone; C; Cassarini; Castro; Chimento; Cristina; D; Demarchi; Español; F; Fernández; G; Goya; Gracia Balbi; Hernández del Pin; I; Icardi; L; LE; Lusso; M; MI; MS; Menite; Moroni; Morro; N; Nabaes Jodar; Natale; P; Palumbo; Pasquinelli; Perrone; R; Romano; Rosales; S; Sevic; Spinelli; V; Valinotto; Valla; Viegas, M.; Villafañe; Vitale |
| EPI_ISL_2801712, EPI_ISL_2801721, EPI_ISL_2801726, EPI_ISL_2801729, EPI_ISL_2801731, EPI_ISL_2801733, EPI_ISL_2801734 | | | |
| see above | Centro de Investigación Biomédica de Occidente (CIBO) | Instituto de Biotecnología de la UNAM | ; Alejandra García-Gasca; Alejandra Hernández-Terán; Alejandro Sánchez-Flores; Alfredo Herrera-Estrella; Alicia Ocaña-Mondragón; Andreu Comas-García; Angel Gustavo Salas-Lais; Antonio Loza Román; Bernardo Martínez-Miguel; Blanca Taboada; Brenda Irasema Maldonado-Meza; Bruno Gómez-Gil; Carla Ivón Herrera-Najera; Carlos F. Arias; Celia Boukadida; Clara Esperanza Santacruz-Tinoco; Concepción Grajales-Muñiz; Consorcio Mexicano de Vigilancia Genómica (CoViGen-Mex). Authors (in alphabetical order): Julio Elias Alvarado-Yaah; Cristóbal Cháidez-Quiróz; Célida Duque Molina; Célida Martínez- Rodríguez; Daniel Fregoso-Rueda; Daniel Lira Morales; Eduardo Becerril-Vargas; Fernando Fontove-Herrera; Fidencio Mejía-Nepomuceno; Francisco Pulido; Gloria Elena Espinosa-Ayala; Gloria María Molina-Salinas; Gloria Vazquez; Hector Esteban Paz-Juárez; Hector Montoya-Fuentes; Helen Haydee Fernanda Ramírez-Plascencia; Irvin González-López; Jean Pierre González; Jesús Hernández; Joel Armando Vázquez-Pérez.; Jorge Salas-Hernández; José Antonio Enciso-Moreno; José Arturo Martínez-Orozco; José Esteban Muñoz-Medina; José de Jesús Nuñez-Contreras; Juan Bautista Chale-Dzul; Julissa Enciso-Ibarra; Luis Alberto Ochoa-Carrera; Margarita Matías-Florentino; Mario Mújica-Sánchez; Marissa Perez-Garcia; María Guadalupe Santiago-Mauricio; María Guadalupe de Jesús Mireles-Rivera; Nelly Sélem-Mojica; Pavel Isa; Ricardo Ciria Merce; Ricardo Grande; Rosa María Gutiérrez Rios; Santiago Ávila-Ríos; Selene Zárate; Susana Lopez; Verónica Mata-Haro; Victor Eduardo García-Arias; Víctor Hugo Borja-Aburto |
| EPI_ISL_2681227, EPI_ISL_2681260, EPI_ISL_2681296, EPI_ISL_2681348, EPI_ISL_2801658, EPI_ISL_2801659, EPI_ISL_2801660, EPI_ISL_2801664 | | | |
| see above | Centro de Investigación Biomédica del Noreste (CIBIN) | Instituto de Biotecnología de la UNAM | ; Alejandra García-Gasca; Alejandra Hernández-Terán; Alejandro Sánchez-Flores; Alfredo Herrera-Estrella; Alicia Ocaña-Mondragón; Andreu Comas-García; Angel Gustavo Salas-Lais; Antonio Loza Román; Bernardo Martínez-Miguel; Blanca Taboada; Brenda Irasema Maldonado-Meza; Bruno Gómez-Gil; Carla Ivón Herrera-Najera; Carlos F. Arias; Celia Boukadida; Clara Esperanza Santacruz-Tinoco; Concepción Grajales-Muñiz; Consorcio Mexicano de Vigilancia Genómica (CoViGen-Mex). Authors (in alphabetical order): Julio Elias Alvarado-Yaah; Cristóbal Cháidez-Quiróz; Célida Duque Molina; Célida Martínez- Rodríguez; Daniel Fregoso-Rueda; Daniel Lira Morales; Eduardo Becerril-Vargas; Fernando Fontove-Herrera; Fidencio Mejía-Nepomuceno; Francisco Pulido; Gloria Elena Espinosa-Ayala; Gloria María Molina-Salinas; Gloria Vazquez; Hector Esteban Paz-Juárez; Hector Montoya-Fuentes; Helen Haydee Fernanda Ramírez-Plascencia; Irvin González-López; Jean Pierre González; Jesús Hernández; Joel Armando Vázquez-Pérez.; Jorge Salas-Hernández; José Antonio Enciso-Moreno; José Arturo Martínez-Orozco; José Esteban Muñoz-Medina; José de Jesús Nuñez-Contreras; Juan Bautista Chale-Dzul; Julissa Enciso-Ibarra; Luis Alberto Ochoa-Carrera; Margarita Matías-Florentino; Mario Mújica-Sánchez; Marissa Perez-Garcia; María Guadalupe Santiago-Mauricio; María Guadalupe de Jesús Mireles-Rivera; Nelly Sélem-Mojica; Pavel Isa; Ricardo Ciria Merce; Ricardo Grande; Rosa María Gutiérrez Rios; Santiago Ávila-Ríos; Selene Zárate; Susana Lopez; Verónica Mata-Haro; Victor Eduardo García-Arias; Víctor Hugo Borja-Aburto |
| EPI_ISL_4261888, EPI_ISL_4261914, EPI_ISL_4261915, EPI_ISL_4261925 | Chiu Laboratory, University of California, San Francisco | Chiu Laboratory, University of California, San Francisco | Alicia Sotomayor-Gonzalez; Baolin Wang; Candace Wang; Charles Chiu; Daniel Wong; Dustin Glasner; Venice Servellita; Xianding (Wayne) Deng; Yueyuan Zhang |
| EPI_ISL_3242518, EPI_ISL_3242523, EPI_ISL_3242530, EPI_ISL_3242535, EPI_ISL_3242538, EPI_ISL_3242545, EPI_ISL_3242553, EPI_ISL_3242703, EPI_ISL_3242704, EPI_ISL_3242705, EPI_ISL_3242706, EPI_ISL_3242707, EPI_ISL_3242708 | | | |
| see above | Clinica INDISA | "Facultad de Ciencias de la Vida, UNAB" | "Claudio Meneses; Ariel Orellana"; Claudio Olmos; Daniel Leon; Dayan Sanhueza; Eduardo Castro; Gonzalo Campaña; Macarena Bastias; Paola Pidal; Ricardo Yusta; Sebastian Wolter; Susana Saez; Víctor Monreal; Waldo Diaz |
| EPI_ISL_3061549, EPI_ISL_3061581 | Clinical Molecular Microbiology Laboratory, UNC Hospitals | Jeremy Wang | Corbin Jones; Jeremy Wang; Melissa Miller; Shawn Hawken |
| EPI_ISL_2466606 | Clinical Virology | Clinical Bacteriology | Adrian Egli; Alfredo Mari; Fanny Wegner; Hans Hirsch; Helena MB Seth-Smith; Julia Bielicki; Karoline Leuzinger; Manuel Battegay; Tim Roloff |
| EPI_ISL_2626066, EPI_ISL_2626071, EPI_ISL_2626077 | Clinique Saint-Pierre Ottignies | UCLouvain/IREC/MBLG | Benoit Kabamba Mukadi; Bertrand Bearzatto; Jean Ruelle |
| EPI_ISL_2492422, EPI_ISL_2492423, EPI_ISL_2492425 | Cliniques universitaires Saint-Luc | UCLouvain/IREC/MBLG | Benoit Kabamba Mukadi; Jean Ruelle |
| EPI_ISL_4300423 | Color Genomics | Chiu Laboratory, University of California, San Francisco | Alicia Sotomayor-Gonzalez; Alicia Zhou; Baolin Wang; Charles Chiu; Daniel Wong; Erika Torres; Madyson Wanner; Noah Brazer; Scott Topper; Venice Servellita; Yueyuan Zhang |
| EPI_ISL_2790645, EPI_ISL_2941608, EPI_ISL_3159665, EPI_ISL_3160140, EPI_ISL_3404119, EPI_ISL_3404120 | Colorado Department of Public Health and Environment | Colorado Department of Public Health and Environment | Alexandria Rossheim; Diana Ir; Emily A. Travanty; Laura Bankers; Mandy Waters; Michael A. Martin; Michael Martin; Molly C. Hetherington-Rauth; Sarah Elizabeth Totten; Shannon R. Matzinger |
| EPI_ISL_2755463, EPI_ISL_2784119, EPI_ISL_2784127, EPI_ISL_2784128, EPI_ISL_2784129, EPI_ISL_2784145, EPI_ISL_2784146, EPI_ISL_2784148, EPI_ISL_2784155, EPI_ISL_2784162, EPI_ISL_2784164, EPI_ISL_2784165, EPI_ISL_2784170, EPI_ISL_2784179, EPI_ISL_2784184, EPI_ISL_2784189, EPI_ISL_2784240, EPI_ISL_2784257, EPI_ISL_2784273, EPI_ISL_2784287, EPI_ISL_2839767, EPI_ISL_2839897, EPI_ISL_2941972, EPI_ISL_2941981, EPI_ISL_2942000, EPI_ISL_2942002, EPI_ISL_2942156, EPI_ISL_2942171, EPI_ISL_2942175, EPI_ISL_3064653, EPI_ISL_3270295, EPI_ISL_3270300, EPI_ISL_3270372 | | | |
| see above | Curative Labs | Curative Labs | Elias L. Salfati; Eugenia Khorosheva; George Way; J.Cesar Ignacio-Espinoza; Janet Chen; Mikhail Hanewich-Hollatz; Nabjot Sandhu; Sophia Quasem; Vladimir Slepnev; Zhiyi Xie |
| EPI_ISL_2832474 | Curative Labs | New Mexico Department of Health Scientific Laboratory | Anastacia Griego-Fisher; D'eldra Malone; Ellie Johnson; Jennifer Benoit; Keila Gutierrez; Linda Salazar; Ratheesh Rajan |
| EPI_ISL_4415765, EPI_ISL_4415766, EPI_ISL_4415769, EPI_ISL_4415771, EPI_ISL_4415773, EPI_ISL_4415774, EPI_ISL_4415778, EPI_ISL_4415779, EPI_ISL_4415780, EPI_ISL_4415781, EPI_ISL_4415782, EPI_ISL_4415783, EPI_ISL_4415784, EPI_ISL_4415785, EPI_ISL_4415786, EPI_ISL_4415787, EPI_ISL_4415788, EPI_ISL_4415789, EPI_ISL_4415792, EPI_ISL_4415793, EPI_ISL_4415795, EPI_ISL_4415796, EPI_ISL_4415797, EPI_ISL_4415798, EPI_ISL_4415810, EPI_ISL_4415815, EPI_ISL_4415816, EPI_ISL_4415818, EPI_ISL_4415821, EPI_ISL_4415822, EPI_ISL_4415823, EPI_ISL_4415825, EPI_ISL_4415826, EPI_ISL_4415827, EPI_ISL_4415828, EPI_ISL_4415829, EPI_ISL_4415832, EPI_ISL_4415833, EPI_ISL_4415834, EPI_ISL_4415837, EPI_ISL_4415838, EPI_ISL_4415839, EPI_ISL_4415840, EPI_ISL_4415841, EPI_ISL_4415842, EPI_ISL_4415844, EPI_ISL_4415845, EPI_ISL_4415846, EPI_ISL_4415849, EPI_ISL_4415850, EPI_ISL_4415853, EPI_ISL_4415856, EPI_ISL_4415857, EPI_ISL_4415858, EPI_ISL_4415859, EPI_ISL_4415863, EPI_ISL_4415864, EPI_ISL_4415865, EPI_ISL_4415866, EPI_ISL_4415867, EPI_ISL_4415868, EPI_ISL_4415869, EPI_ISL_4415870, EPI_ISL_4415871, EPI_ISL_4415872, EPI_ISL_4415873, EPI_ISL_4415874, EPI_ISL_4415875, EPI_ISL_4415881, EPI_ISL_4415882, EPI_ISL_4415883, EPI_ISL_4415884, EPI_ISL_4415885, EPI_ISL_4415895, EPI_ISL_4415896, EPI_ISL_4415907, EPI_ISL_4415908, EPI_ISL_4415909, EPI_ISL_4415910, EPI_ISL_4415911, EPI_ISL_4415912, EPI_ISL_4415913, EPI_ISL_4415919, EPI_ISL_4415920, EPI_ISL_4415921, EPI_ISL_4415922, EPI_ISL_4415923, EPI_ISL_4415924, EPI_ISL_4415925, EPI_ISL_4415926, EPI_ISL_4415927, EPI_ISL_4415928, EPI_ISL_4415929, EPI_ISL_4415930, EPI_ISL_4415931, EPI_ISL_4415932, EPI_ISL_4415933, EPI_ISL_4415935, EPI_ISL_4415942, EPI_ISL_4415947, EPI_ISL_4415948, EPI_ISL_4415949, EPI_ISL_4415950, EPI_ISL_4415951, EPI_ISL_4415954, EPI_ISL_4415955, EPI_ISL_4415965, EPI_ISL_4415978, EPI_ISL_4415991, EPI_ISL_4416013, EPI_ISL_4416017, EPI_ISL_4416020, EPI_ISL_4416021, EPI_ISL_4416024, EPI_ISL_4416029, EPI_ISL_4416030, EPI_ISL_4416038, EPI_ISL_4416041, EPI_ISL_4416043, EPI_ISL_4416046, EPI_ISL_4416048, EPI_ISL_4416051, EPI_ISL_4416055, EPI_ISL_4416056, EPI_ISL_4416058, EPI_ISL_4416059, EPI_ISL_4416061, EPI_ISL_4416063, EPI_ISL_4416065, EPI_ISL_4416070, EPI_ISL_4416073, EPI_ISL_4416080, EPI_ISL_4416081, EPI_ISL_4416085, EPI_ISL_4416086, EPI_ISL_4416087, EPI_ISL_4416089, EPI_ISL_4416090, EPI_ISL_4416094, EPI_ISL_4416095, EPI_ISL_4422498, EPI_ISL_4422515, EPI_ISL_4422518, EPI_ISL_4422520, EPI_ISL_4422527, EPI_ISL_4422532, EPI_ISL_4422539, EPI_ISL_4422545, EPI_ISL_4422548, EPI_ISL_4422551, EPI_ISL_4422557, EPI_ISL_4422560, EPI_ISL_4422563, EPI_ISL_4422565, EPI_ISL_4422571, EPI_ISL_4422574, EPI_ISL_4422577, EPI_ISL_4422582, EPI_ISL_4422588, EPI_ISL_4422593, EPI_ISL_4422610, EPI_ISL_4422623, EPI_ISL_4422625, EPI_ISL_4422627, EPI_ISL_4422629, EPI_ISL_4422634, EPI_ISL_4422641, EPI_ISL_4422648, EPI_ISL_4422652, EPI_ISL_4422660, EPI_ISL_4422663, EPI_ISL_4422671, EPI_ISL_4422673, EPI_ISL_4422685, EPI_ISL_4422704, EPI_ISL_4422706, EPI_ISL_4422726, EPI_ISL_4422746, EPI_ISL_4422748, EPI_ISL_4422769, EPI_ISL_4422771, EPI_ISL_4422774, EPI_ISL_4422777, EPI_ISL_4422784, EPI_ISL_4422802, EPI_ISL_4422812, EPI_ISL_4422818, EPI_ISL_4422823, EPI_ISL_4422828, EPI_ISL_4422837, EPI_ISL_4422842, EPI_ISL_4422843, EPI_ISL_4422851, EPI_ISL_4422860, EPI_ISL_4422862, EPI_ISL_4422865, EPI_ISL_4422869, EPI_ISL_4422872, EPI_ISL_4422874, EPI_ISL_4422878, EPI_ISL_4422881, EPI_ISL_4422884, EPI_ISL_4422888, EPI_ISL_4422890, EPI_ISL_4422891, EPI_ISL_4422893, EPI_ISL_4422895, EPI_ISL_4422903, EPI_ISL_4422908, EPI_ISL_4422912, EPI_ISL_4422915, EPI_ISL_4422931, EPI_ISL_4422942, EPI_ISL_4422947, EPI_ISL_4423011, EPI_ISL_4423014, EPI_ISL_4423018, EPI_ISL_4423020, EPI_ISL_4423021, EPI_ISL_4423027, EPI_ISL_4423045, EPI_ISL_4423046, EPI_ISL_4423048, EPI_ISL_4423050, EPI_ISL_4423052, EPI_ISL_4423054, EPI_ISL_4423056, EPI_ISL_4423058, EPI_ISL_4423059, EPI_ISL_4423065, EPI_ISL_4423080, EPI_ISL_4423082, EPI_ISL_4423084, EPI_ISL_4423098, EPI_ISL_4423100, EPI_ISL_4423102, EPI_ISL_4423105, EPI_ISL_4423113, EPI_ISL_4423117, EPI_ISL_4423129, EPI_ISL_4423136, EPI_ISL_4423138, EPI_ISL_4423140, EPI_ISL_4423260, EPI_ISL_4423269, EPI_ISL_4423271, EPI_ISL_4423272, EPI_ISL_4423274, EPI_ISL_4423280, EPI_ISL_4423283, EPI_ISL_4423290, EPI_ISL_4423292, EPI_ISL_4423296, EPI_ISL_4423299, EPI_ISL_4423301, EPI_ISL_4423306, EPI_ISL_4423308, EPI_ISL_4423312, EPI_ISL_4423317, EPI_ISL_4423318, EPI_ISL_4423320, EPI_ISL_4423322, EPI_ISL_4423324, EPI_ISL_4423326, EPI_ISL_4423327, EPI_ISL_4423329, EPI_ISL_4423331, EPI_ISL_4423333, EPI_ISL_4423391, EPI_ISL_4423393, EPI_ISL_4423394, EPI_ISL_4423396, EPI_ISL_4423398, EPI_ISL_4423400, EPI_ISL_4423402, EPI_ISL_4423403, EPI_ISL_4423405, EPI_ISL_4423407, EPI_ISL_4423409, EPI_ISL_4423410, EPI_ISL_4423412, EPI_ISL_4423414, EPI_ISL_4423415, EPI_ISL_4423417, EPI_ISL_4423419, EPI_ISL_4423421, EPI_ISL_4423423, EPI_ISL_4423424, EPI_ISL_4423426, EPI_ISL_4423428, EPI_ISL_4423429, EPI_ISL_4423431, EPI_ISL_4423433, EPI_ISL_4423435, EPI_ISL_4423436, EPI_ISL_4423438, EPI_ISL_4423440, EPI_ISL_4423442, EPI_ISL_4423443, EPI_ISL_4423445, EPI_ISL_4423449, EPI_ISL_4423451, EPI_ISL_4423453, EPI_ISL_4423455, EPI_ISL_4423457, EPI_ISL_4423458, EPI_ISL_4423460, EPI_ISL_4423463, EPI_ISL_4423466, EPI_ISL_4423468, EPI_ISL_4423471, EPI_ISL_4423473, EPI_ISL_4423475, EPI_ISL_4423477, EPI_ISL_4423478, EPI_ISL_4423480, EPI_ISL_4423483, EPI_ISL_4423485, EPI_ISL_4423487, EPI_ISL_4423488, EPI_ISL_4423490, EPI_ISL_4423493, EPI_ISL_4423496, EPI_ISL_4423499, EPI_ISL_4423528, EPI_ISL_4423529, EPI_ISL_4423531, EPI_ISL_4423533, EPI_ISL_4423536, EPI_ISL_4423539, EPI_ISL_4423540, EPI_ISL_4423542, EPI_ISL_4423545, EPI_ISL_4423548, EPI_ISL_4423550, EPI_ISL_4423553, EPI_ISL_4423556, EPI_ISL_4423559, EPI_ISL_4423561, EPI_ISL_4423564, EPI_ISL_4423568, EPI_ISL_4423571, EPI_ISL_4423573, EPI_ISL_4423575, EPI_ISL_4423577, EPI_ISL_4423580, EPI_ISL_4426749, EPI_ISL_4426774, EPI_ISL_4426843, EPI_ISL_4427554, EPI_ISL_4427557, EPI_ISL_4427558, EPI_ISL_4427560, EPI_ISL_4427562, EPI_ISL_4427564, EPI_ISL_4427566, EPI_ISL_4427568, EPI_ISL_4427570, EPI_ISL_4427573, EPI_ISL_4427577, EPI_ISL_4427579, EPI_ISL_4427581, EPI_ISL_4427583, EPI_ISL_4427584, EPI_ISL_4427586, EPI_ISL_4427590, EPI_ISL_4427593, EPI_ISL_4427594, EPI_ISL_4427596, EPI_ISL_4427598, EPI_ISL_4427600, EPI_ISL_4427603, EPI_ISL_4427606, EPI_ISL_4427608, EPI_ISL_4427610, EPI_ISL_4427613, EPI_ISL_4427616, EPI_ISL_4427617, EPI_ISL_4427619, EPI_ISL_4427622, EPI_ISL_4427625, EPI_ISL_4427628, EPI_ISL_4427631, EPI_ISL_4427634, EPI_ISL_4427637, EPI_ISL_4427639, EPI_ISL_4427641, EPI_ISL_4427643, EPI_ISL_4427645, EPI_ISL_4427647, EPI_ISL_4427648, EPI_ISL_4427650, EPI_ISL_4427652, EPI_ISL_4427654, EPI_ISL_4427655, EPI_ISL_4427657, EPI_ISL_4427661, EPI_ISL_4427663, EPI_ISL_4427665, EPI_ISL_4427666, EPI_ISL_4427670, EPI_ISL_4427672, EPI_ISL_4427675, EPI_ISL_4427677, EPI_ISL_4427679, EPI_ISL_4427681, EPI_ISL_4427682, EPI_ISL_4427684, EPI_ISL_4427686, EPI_ISL_4427687, EPI_ISL_4427689, EPI_ISL_4427691, EPI_ISL_4427693, EPI_ISL_4427694, EPI_ISL_4427696, EPI_ISL_4427698, EPI_ISL_4427700, EPI_ISL_4427701, EPI_ISL_4427703, EPI_ISL_4427705, EPI_ISL_4427707, EPI_ISL_4427709, EPI_ISL_4427712, EPI_ISL_4427715, EPI_ISL_4427718, EPI_ISL_4427720, EPI_ISL_4427722, EPI_ISL_4427724, EPI_ISL_4427726, EPI_ISL_4427728, EPI_ISL_4427729, EPI_ISL_4427731, EPI_ISL_4427733, EPI_ISL_4427735, EPI_ISL_4427737, EPI_ISL_4427739, EPI_ISL_4427740, EPI_ISL_4427742, EPI_ISL_4427744, EPI_ISL_4427746, | | | |

| | | | |
|---|---|---|---|
| EPI_ISL_4427748, EPI_ISL_4427751, EPI_ISL_4427755, EPI_ISL_4427756, EPI_ISL_4427758, EPI_ISL_4427760, EPI_ISL_4427762, EPI_ISL_4427764, EPI_ISL_4427766, EPI_ISL_4427767, EPI_ISL_4427769, EPI_ISL_4427771, EPI_ISL_4427773, EPI_ISL_4427775, EPI_ISL_4427776, EPI_ISL_4427778, EPI_ISL_4427780, EPI_ISL_4427782, EPI_ISL_4427784, EPI_ISL_4427785, EPI_ISL_4427787, EPI_ISL_4427789, EPI_ISL_4427791, EPI_ISL_4427793, EPI_ISL_4427795, EPI_ISL_4427797, EPI_ISL_4427799, EPI_ISL_4427800, EPI_ISL_4427802, EPI_ISL_4427804, EPI_ISL_4427806, EPI_ISL_4427808, EPI_ISL_4427810, EPI_ISL_4427811, EPI_ISL_4427813, EPI_ISL_4427815, EPI_ISL_4427817, EPI_ISL_4427819, EPI_ISL_4427820, EPI_ISL_4427822, EPI_ISL_4427824, EPI_ISL_4427826, EPI_ISL_4427828, EPI_ISL_4427829, EPI_ISL_4427831, EPI_ISL_4427833, EPI_ISL_4427835, EPI_ISL_4427837, EPI_ISL_4427838, EPI_ISL_4427840, EPI_ISL_4427842, EPI_ISL_4427844, EPI_ISL_4427846, EPI_ISL_4427848, EPI_ISL_4427850, EPI_ISL_4427852, EPI_ISL_4427854, EPI_ISL_4427855, EPI_ISL_4427857, EPI_ISL_4427859, EPI_ISL_4428162, EPI_ISL_4428173, EPI_ISL_4428188, EPI_ISL_4428201, EPI_ISL_4428204, EPI_ISL_4428226, EPI_ISL_4428238, EPI_ISL_4428242, EPI_ISL_4428252, EPI_ISL_4428302, EPI_ISL_4428305, EPI_ISL_4428309, EPI_ISL_4428311, EPI_ISL_4428314, EPI_ISL_7743846, EPI_ISL_7743905, EPI_ISL_7743908, EPI_ISL_7743910, EPI_ISL_7743911, EPI_ISL_7743912, EPI_ISL_7743916 | | | |
| see above | DASA | DASA | Adriano Bonaldi; Angelica Hristov; Annelise Lopes; Bianca Cota; Cristina Oliveira; Jose Levi; Lidia Yamamoto; Paulo Pierry; Rodrigo Guarischi; Rodrigo Salazar |
| EPI_ISL_2689001, EPI_ISL_2689008, EPI_ISL_3547445 | DC Public Health Lab/ Dept. of Forensic Sciences | DC Public Health Lab/ Dept. of Forensic Sciences | Brittany Hamilton; Connie Maza; David Payne; Elizabeth Zelaya; Janis Doss; Jocelyn Hauser; Monica Mann; Sarah Scott; Scott Nguyen |
| EPI_ISL_5646344, EPI_ISL_5647116, EPI_ISL_5647122, EPI_ISL_5647131, EPI_ISL_5647140, EPI_ISL_5647184, EPI_ISL_5647185 | | | |
| see above | DEPARTAMENTO DE SAUDE MUNICIPAL SOCORRO SP | Instituto Butantan | Antonio Jorge Martins; Claudia Renata dos Santos Barros; David Schlesinger; Debora Botequio Moretti; Dimas Tadeu Covas; Elaine Cristina Marqueze; Elaine Vieira Santos; Evandra Strazza Rodrigues; Heidge Fukumasu; Jayme Augusto de Souza-Neto; José Salvatore Leister Patané; Luiz Alcantara; Luiz Lehmann Coutinho; Maria Carolina Elias; Maurício Lacerda Nogueira; Rafael dos Santos Bezerra; Raul Machado Neto; Rejane Maria Tommasini Grotto; Ricardo Haddad; Sandra Coccuzzo Sampaio Vessoni; Simone Kashima; Svetoslav Nanev Slavov; Vincent Louis Viala |
| EPI_ISL_5650354 | DEPARTAMENTO MUNICIPAL DE SAUDE DE BURITAMA | Instituto Butantan | Antonio Jorge Martins; Claudia Renata dos Santos Barros; David Schlesinger; Debora Botequio Moretti; Dimas Tadeu Covas; Elaine Cristina Marqueze; Elaine Vieira Santos; Evandra Strazza Rodrigues; Heidge Fukumasu; Jayme Augusto de Souza-Neto; José Salvatore Leister Patané; Luiz Alcantara; Luiz Lehmann Coutinho; Maria Carolina Elias; Maurício Lacerda Nogueira; Rafael dos Santos Bezerra; Raul Machado Neto; Rejane Maria Tommasini Grotto; Ricardo Haddad; Sandra Coccuzzo Sampaio Vessoni; Simone Kashima; Svetoslav Nanev Slavov; Vincent Louis Viala |
| EPI_ISL_5646315, EPI_ISL_5646341 | DIRETORIA MUNICIPAL DE SAUDE | Instituto Butantan | Antonio Jorge Martins; Claudia Renata dos Santos Barros; David Schlesinger; Debora Botequio Moretti; Dimas Tadeu Covas; Elaine Cristina Marqueze; Elaine Vieira Santos; Evandra Strazza Rodrigues; Heidge Fukumasu; Jayme Augusto de Souza-Neto; José Salvatore Leister Patané; Luiz Alcantara; Luiz Lehmann Coutinho; Maria Carolina Elias; Maurício Lacerda Nogueira; Rafael dos Santos Bezerra; Raul Machado Neto; Rejane Maria Tommasini Grotto; Ricardo Haddad; Sandra Coccuzzo Sampaio Vessoni; Simone Kashima; Svetoslav Nanev Slavov; Vincent Louis Viala |
| EPI_ISL_5646531, EPI_ISL_5646658, EPI_ISL_5646659, EPI_ISL_5646717, EPI_ISL_5647167 | DIRETORIA MUNICIPAL DE SAUDE DE ENGENHEIRO COELHO | Instituto Butantan | Antonio Jorge Martins; Claudia Renata dos Santos Barros; David Schlesinger; Debora Botequio Moretti; Dimas Tadeu Covas; Elaine Cristina Marqueze; Elaine Vieira Santos; Evandra Strazza Rodrigues; Heidge Fukumasu; Jayme Augusto de Souza-Neto; José Salvatore Leister Patané; Luiz Alcantara; Luiz Lehmann Coutinho; Maria Carolina Elias; Maurício Lacerda Nogueira; Rafael dos Santos Bezerra; Raul Machado Neto; Rejane Maria Tommasini Grotto; Ricardo Haddad; Sandra Coccuzzo Sampaio Vessoni; Simone Kashima; Svetoslav Nanev Slavov; Vincent Louis Viala |
| EPI_ISL_2834728 | DISAN POLICIA NACIONAL | Instituto Nacional de Salud-Dirección de Investigación en Salud Pública | Carlos Franco-Muñoz; Carmen Osorio; Diana Malo; Diego A. Álvarez-Díaz; Diego Andrés Prada; Gerardo Santamaría; Hector Alejandro Ruiz-Moreno; Jhonnatan Reales-González; Jorge Rivera; Juan Camilo Martinez; Julian Naizaque; Katherine Laiton-Donato; Lisseth Pardo; Magdalena Wiesner; Marcela Mercado-Reyes; Maria T. Herrera-Sepúlveda; Marta Lopez Blanco; Martha Lucia Ospina Martinez; Paola Rojas; Sergio Gomez; Sheryll Corchuelo; Ángela Alarcon Cruz |
| EPI_ISL_2662776, EPI_ISL_3692597 | DPHL | Delaware Public Health Lab | Rebecca Savage |
| EPI_ISL_2666116, EPI_ISL_2666225, EPI_ISL_2669007, EPI_ISL_2669330, EPI_ISL_2675490 | Department of Bacteria, Parasites and Fungi, Statens Serum Institut, Copenhagen, Denmark | Statens Serum Institut Bioinformatics and Microbial Genomics | Danish Covid-19 Genome Consortium |
| EPI_ISL_2685865 | Department of Clinical Microbiology | GIGA Medical Genomics | Bouchra Boujemla; Cécile Meex; Keith Durkin; Maria Artesi; Marie-Pierre Hayette; Nathalie Renotte; Pierrette Melin; Raphaël Boreux; Sébastien Bontems; Vincent Bours |
| EPI_ISL_2673507, EPI_ISL_2673508, EPI_ISL_2673511, EPI_ISL_2673513, EPI_ISL_2673514, EPI_ISL_2673515 | Department of Virology, Istituto Zooprofilattico Sperimentale del Lazio e della Toscana (IZSLT) | Department of General Diagnostics; Department of Virology; Istituto Zooprofilattico Sperimentale del Lazio e della Toscana (IZSLT) | Alessia Franco; Antonella Cersini; Antonio Battisti.; Elena L. Diaconu; Fabiola Feltrin; Giuseppe Manna; Patricia Alba; Raffaela Conti; Teresa Scicluna; Virginia Carfora |
| EPI_ISL_2636681, EPI_ISL_2636682, EPI_ISL_2636683, EPI_ISL_2636685, EPI_ISL_2636711, EPI_ISL_2762347 | Diagnosticum - Labor Neukirchen | Robert Koch Institute | |
| EPI_ISL_2657191 | Diagnostyka | 1. Tricity SARS-CoV-2 sequencing consortium: University of Gdansk, Medical University of Gdansk, Vaxican Ltd., Invicta Ltd. 2. National Institute of Public Health - National Institute of Hygiene, Warsaw, Poland | Celina Cybulska; Karolina Gackowska; Katarzyna Groth; Katarzyna Zacharczuk; Krystyna Bienkowska Szewczyk; Lukasz Rabalski; Maciej Grzybek; Maciej Kosinski; Magdalena Nowakowska; Marcin Lubocki; Małgorzata Sadkowska-Todys; Tomasz Wołkowicz |
| EPI_ISL_2966246 | Dipartimento di Medicina di Laboratorio, Azienda sanitaria universitaria Friuli Centrale (ASU FC) | Dipartimento di Medicina di Laboratorio, Azienda sanitaria universitaria Friuli Centrale (ASU FC) | Catia Mio; Chiara Dal Secco; Corrado Pipan; Francesco Curcio; Stefania Marzinotto |
| EPI_ISL_3050667, EPI_ISL_3050671, EPI_ISL_3050674, EPI_ISL_3050682 | Dirección regional de salud del Callao (DIRESA-CALLAO) | Centro de Investigaciones Tecnológicas, Biomédicas y Medioambientales (CITBM) | B; Huaman; J. Alarcon; M. Cuellar; M. Ramirez; M. Sovero |
| EPI_ISL_3578237, EPI_ISL_3578245, EPI_ISL_3578257, EPI_ISL_3578264 | Dr. Risch Ostschweiz AG | Microbiology, Dr. Risch | Dominique Fabien Hilti; Faina Wehrli; Lorenz Risch; Martin Risch; Nadia Wohlwend; Sinem Kas; Thomas Bodmer |
| EPI_ISL_3474305 | Dutch COVID-19 response team | Medical Microbiology, Maastricht University Medical Centre | Brian van der Veer*; Carmen Reumkens; Christian Hoebe; Erik Beuken; Jozef Dingemans*; Lieke van Alphen; Paul Savelkoul |
| EPI_ISL_2610115, EPI_ISL_2610117, EPI_ISL_2610138, EPI_ISL_2610149, EPI_ISL_2610150, EPI_ISL_2610169, EPI_ISL_2610177, EPI_ISL_2610295, EPI_ISL_2610322, EPI_ISL_2610331, EPI_ISL_2610341, EPI_ISL_2610351, EPI_ISL_2610352, EPI_ISL_2610359, EPI_ISL_2610556, EPI_ISL_2610598, EPI_ISL_2610609, EPI_ISL_2610641, EPI_ISL_2610644, EPI_ISL_2610647, EPI_ISL_2610655, EPI_ISL_2610656, EPI_ISL_2610657, EPI_ISL_2610663, EPI_ISL_2610665, EPI_ISL_2610667, EPI_ISL_2610671, EPI_ISL_2610672, EPI_ISL_2610673, EPI_ISL_2610679, EPI_ISL_2610691, EPI_ISL_2610694, EPI_ISL_2610697, EPI_ISL_2610698, EPI_ISL_2610699, EPI_ISL_2610700, EPI_ISL_2610733, EPI_ISL_2610743, EPI_ISL_2610753, EPI_ISL_2610754, EPI_ISL_2610755, EPI_ISL_2610765, EPI_ISL_2610766, EPI_ISL_2610776, EPI_ISL_2610834, EPI_ISL_2610844, EPI_ISL_2610853, EPI_ISL_2672303, EPI_ISL_2672423, EPI_ISL_2672626, EPI_ISL_2672638, EPI_ISL_2672650, EPI_ISL_2672728, EPI_ISL_2672902, EPI_ISL_2673181 | | | |
| see above | Dutch COVID-19 response team | National Institute for Public Health and the Environment (RIVM) | Adam Meijer; AnneMarie van den Brandt; Annelies Kroneman; Bas van der Veer; Chantal Reusken; Dennis Schmitz; Dirk Eggink; Euníce Then; Florian Zwagemaker; Harry Vennema; Jeroen Cremer; Karim Hajji; Kim Freriks; Linda van de Nes; Lisa Wijsman; Lynn Aarts; Melissa van Tuil; Ryanne Jaarsma; Sanne Bos; Sharon van den Brink; on behalf of the national COVID-19 response team |
| EPI_ISL_4494948, EPI_ISL_4494950, EPI_ISL_4494952, EPI_ISL_4494954, EPI_ISL_4494956, EPI_ISL_4494958, EPI_ISL_4494959, EPI_ISL_4494961, EPI_ISL_4494963, EPI_ISL_4494967, EPI_ISL_4494969, EPI_ISL_4494974, EPI_ISL_4494978, EPI_ISL_4494980, EPI_ISL_4494982 | | | |
| see above | Dynacare Manitoba | Dynacare Manitoba | David Alexander; Hilary Racher; Hongbin Li; Melissa Desaulnier; Tintu Abraham |
| EPI_ISL_5647114, EPI_ISL_5647117, EPI_ISL_5647119, EPI_ISL_5647121, EPI_ISL_5647129, EPI_ISL_5647132, EPI_ISL_5647137, EPI_ISL_5647150, EPI_ISL_5647156, EPI_ISL_5647157, EPI_ISL_5647171, EPI_ISL_5647177, EPI_ISL_5647187 | | | |
| see above | ESF NOVA TANABI II | Instituto Butantan | Antonio Jorge Martins; Claudia Renata dos Santos Barros; David Schlesinger; Debora Botequio Moretti; Dimas Tadeu Covas; Elaine Cristina Marqueze; Elaine Vieira Santos; Evandra Strazza Rodrigues; Heidge Fukumasu; Jayme Augusto de Souza-Neto; José Salvatore Leister Patané; Luiz Alcantara; Luiz Lehmann Coutinho; Maria Carolina Elias; Maurício Lacerda Nogueira; Rafael dos Santos Bezerra; Raul Machado Neto; Rejane Maria Tommasini Grotto; Ricardo Haddad; Sandra Coccuzzo Sampaio Vessoni; Simone Kashima; Svetoslav Nanev Slavov; Vincent Louis Viala |
| EPI_ISL_3215320, EPI_ISL_3215323, EPI_ISL_3215763 | EXCITE Lab | Andersen lab at Scripps Research | Abigail Schnapper; Angela Scioscia; Cheryl Anderson; Chip Schooley; David Pride; Greg Humphrey; Helena Tubb; Natasha Martin; Natasha Martin Cheryl Anderson; Sawyer Farmer; Sharon Reed; Smruthi Karthikeyan; Tommy Valles + SEARCH; Tommy Valles + SEARCH |
| EPI_ISL_3117260, EPI_ISL_3117261, EPI_ISL_3117262, EPI_ISL_3117405, EPI_ISL_3117406, EPI_ISL_3117408 | Edmonton Provincial Lab | Public Health Agency of Canada (PHAC) National Microbiology Laboratory | Buss; Croxen M; Deo A; Dieu P; E; Ferrato C; Gill K; Khan F; Koleva P; Li V; Lloyd C; Lynch T; Ma R; Murphy S; Pabbaraju K; Shokoples S; Thayer J; Tipples G; Whitehouse M; Wong A; Yu C; Zelyas N |
| EPI_ISL_5646214, EPI_ISL_5646220, EPI_ISL_5646242, EPI_ISL_5646244, EPI_ISL_5646263, EPI_ISL_5646283, EPI_ISL_5646289 | | | |
| see above | FACULDADE DE MEDICINA ABC | Instituto Butantan | Antonio Jorge Martins; Claudia Renata dos Santos Barros; David Schlesinger; Debora Botequio Moretti; Dimas Tadeu Covas; Elaine Cristina Marqueze; Elaine Vieira Santos; Evandra Strazza Rodrigues; Heidge Fukumasu; Jayme Augusto de Souza-Neto; José Salvatore Leister Patané; Luiz Alcantara; Luiz Lehmann Coutinho; Maria Carolina Elias; Maurício Lacerda Nogueira; Rafael dos Santos Bezerra; Raul Machado Neto; Rejane Maria Tommasini Grotto; Ricardo Haddad; Sandra Coccuzzo Sampaio Vessoni; Simone Kashima; Svetoslav Nanev Slavov; Vincent Louis Viala |
| EPI_ISL_3474175, EPI_ISL_3525029 | FYR Diagnostics | FYR Diagnostics | Blohm; Ghai; Tre; Vikas |
| EPI_ISL_2919147, EPI_ISL_2919148 | Fakultní nemocnice Hradec Králové | Fakultní nemocnice Hradec Králové | Helena Kovarikova; Marketa Gancarcikova |
| EPI_ISL_2567245, EPI_ISL_2567246, EPI_ISL_2567247, EPI_ISL_2567252, EPI_ISL_2567253, EPI_ISL_2567257, EPI_ISL_2567258, EPI_ISL_2567259, EPI_ISL_2567260, EPI_ISL_2567263, EPI_ISL_2836502, EPI_ISL_2836523, EPI_ISL_2836551, EPI_ISL_2836581, EPI_ISL_2836835, EPI_ISL_2920455, EPI_ISL_2920457, EPI_ISL_2920458, EPI_ISL_2920461, EPI_ISL_2920463, EPI_ISL_2920464, EPI_ISL_2920465, EPI_ISL_2920466, EPI_ISL_2920477, EPI_ISL_2920479, EPI_ISL_3100715, EPI_ISL_3100717, EPI_ISL_3100718, EPI_ISL_3100719, EPI_ISL_3100721, EPI_ISL_3100728, EPI_ISL_3100736, EPI_ISL_3100740, EPI_ISL_3100745, EPI_ISL_3100746, EPI_ISL_3100747, EPI_ISL_3100750, EPI_ISL_5866264, EPI_ISL_6050063, EPI_ISL_6050064, EPI_ISL_6050065, EPI_ISL_6050066, EPI_ISL_6050682 | | | |
| see above | Florida Bureau of Public Health Laboratories | Florida Bureau of Public Health Laboratories | Jason Blanton; Namratha Tarigopula; Sarah Schmedes; Tiffany Splatt |
| EPI_ISL_2480093, EPI_ISL_2480095, EPI_ISL_2480096, EPI_ISL_2480099, EPI_ISL_2480107, EPI_ISL_2480125, EPI_ISL_2480126, EPI_ISL_2480135, EPI_ISL_2480139, EPI_ISL_2480143, EPI_ISL_2527565, EPI_ISL_2527567, EPI_ISL_2527570, EPI_ISL_2527573, EPI_ISL_2527581, EPI_ISL_2527586, EPI_ISL_2527592, EPI_ISL_2527605, EPI_ISL_2527608, EPI_ISL_2527636, EPI_ISL_2527643, EPI_ISL_2527645, | | | |

| | | | |
|---|---|---|---|
| EPI_ISL_2527650, EPI_ISL_2527656, EPI_ISL_2527660, EPI_ISL_2527680, EPI_ISL_2527735, EPI_ISL_2527739, EPI_ISL_2527741, EPI_ISL_2527753, EPI_ISL_2527757, EPI_ISL_2527760, EPI_ISL_2527811, EPI_ISL_2598710, EPI_ISL_2598808, EPI_ISL_2598814, EPI_ISL_2598822, EPI_ISL_2598828, EPI_ISL_2599922, EPI_ISL_2599923, EPI_ISL_2686449, EPI_ISL_2686452, EPI_ISL_2686453, EPI_ISL_2686472, EPI_ISL_2686496, EPI_ISL_2686518 | | | |
| see above | Fulgent Genetics | Centers for Disease Control and Prevention Division of Viral Diseases, Pathogen Discovery | Adrian Paskey; Becky Tsai; Benafsh Sapra; Benjamin Rambo-Martin; Christopher Gulvick; Clinton R. Paden; Dakota Howard; Darlene Wagner; Dhwani Batra; Doreen Ng; Duncan MacCannell; Harry Gao; James Xie; Jason Caravas; John Gao; Joseph Fierro; Kara Moser; Matthew Schmerer; Mickey Li; Peter W. Cook; Scott Sammons; Shatavia Morrison; Yan Meng; Yvette Unoarumhi |
| EPI_ISL_2644693 | Furst Medical Laboratory | Norwegian Institute of Public Health, Department of Virology | Atiya R Ali; Debech Nadia; Engebretsen Serina Beate; Garcia Llorente Ignacio; Hilde Elshaug; Hilde Vollan; Jon Bråte; Kamilla Heddeland Instefjord; Karoline Bragstad; Kathrine Stene-Johansen; Line Victoria Moen; Marie Paulsen Madsen; Olav Hungnes; Pedersen Benedikte Nevjen; Rasmus Riis Kopperud |
| EPI_ISL_3800440 | GA Department of Public Health Laboratory | Centers for Disease Control and Prevention Division of Viral Diseases, Pathogen Discovery | Alex Burgin; Ben Rambo-Martin; Clinton Paden; Dakota Howard; Dave Wentworth; Dhwani Batra; Jasmine Padilla; Justin Lee; Krista Queen; Kristen Knipe; Kristine Lacek; Mark Burroughs; Matthew Schmerer; Meghan Bentz; Mili Sheth; Peter Cook; Sam Shepard; Sarah Nobles; Suxiang Tong; Vivien Dugan; Yvette Unoarumhi |
| EPI_ISL_2754920 | GA Department of Public Health Laboratory | Genomics and Discovery, Respiratory Viruses Branch, Division of Viral Diseases, Centers for Disease Control and Prevention | Adam Retchless; Anna Kelleher; Anna Uehara; Brian Lynch; Clinton R. Paden; Dhwani Batra; Haibin Wang; Han Jia Justin Ng; Jasmine Padilla; Jing Zhang; Justin Lee; Krista Queen; Mark Burroughs; Mili Sheth; Morgan Davis; Peter Cook; Rachel Marine; Sarah Nobles; Suxiang Tong; Tara Coalter; Yan Li; Ying Tao |
| EPI_ISL_4196870, EPI_ISL_4196872, EPI_ISL_4196874, EPI_ISL_4196875, EPI_ISL_4196876 | Gencore - Universidad de los Andes | Gencore - Universidad de los Andes | Cristian Barrera; David González; Gabriela Ariza; Luisa Sacristan; Marcela Guevara; Silvia Restrepo |
| EPI_ISL_2659120, EPI_ISL_2659122, EPI_ISL_2659125, EPI_ISL_2659127, EPI_ISL_2659128, EPI_ISL_2659129, EPI_ISL_2659130, EPI_ISL_2659131, EPI_ISL_2659136, EPI_ISL_2659137, EPI_ISL_2659138, EPI_ISL_2659139, EPI_ISL_2659140, EPI_ISL_2659141, EPI_ISL_2659142, EPI_ISL_2659143, EPI_ISL_2659144, EPI_ISL_2659145, EPI_ISL_2659147, EPI_ISL_2659148, EPI_ISL_2659149, EPI_ISL_2659150, EPI_ISL_2659155, EPI_ISL_2659156, EPI_ISL_2659157, EPI_ISL_2659158, EPI_ISL_2659159, EPI_ISL_2659160, EPI_ISL_2659161, EPI_ISL_2659167, EPI_ISL_2659172, EPI_ISL_2659173, EPI_ISL_2659174, EPI_ISL_2659175, EPI_ISL_2659178, EPI_ISL_2659179, EPI_ISL_2659180, EPI_ISL_2659181, EPI_ISL_2659182, EPI_ISL_2659183, EPI_ISL_2659184, EPI_ISL_2659185, EPI_ISL_2659186, EPI_ISL_2659187, EPI_ISL_2659189, EPI_ISL_2659191, EPI_ISL_2659192, EPI_ISL_2659193, EPI_ISL_2659194, EPI_ISL_2659195, EPI_ISL_2663447, EPI_ISL_2663448, EPI_ISL_2663449, EPI_ISL_2663450, EPI_ISL_2663451, EPI_ISL_2663452, EPI_ISL_2663583, EPI_ISL_2663584, EPI_ISL_2663586, EPI_ISL_2663587, EPI_ISL_2663588, EPI_ISL_2663601, EPI_ISL_2663605, EPI_ISL_2663607, EPI_ISL_2663611, EPI_ISL_2663612, EPI_ISL_2663614, EPI_ISL_2663615, EPI_ISL_2663616, EPI_ISL_2663617, EPI_ISL_2663618, EPI_ISL_2756057, EPI_ISL_2756058, EPI_ISL_2756060, EPI_ISL_2756061, EPI_ISL_2756062, EPI_ISL_2756063, EPI_ISL_2756064, EPI_ISL_2756066, EPI_ISL_2756067, EPI_ISL_2756068, EPI_ISL_2756069, EPI_ISL_2756070, EPI_ISL_2756114, EPI_ISL_2756220, EPI_ISL_2756246, EPI_ISL_2756247, EPI_ISL_2756276, EPI_ISL_2756277, EPI_ISL_2756278, EPI_ISL_2756279, EPI_ISL_2756280, EPI_ISL_2756281, EPI_ISL_2756282, EPI_ISL_2756283, EPI_ISL_2756284, EPI_ISL_2756286, EPI_ISL_2756287, EPI_ISL_2756288, EPI_ISL_2756289, EPI_ISL_2756291, EPI_ISL_2756293, EPI_ISL_2756306, EPI_ISL_2756313, EPI_ISL_2756315, EPI_ISL_2756323, EPI_ISL_2756325, EPI_ISL_2756327, EPI_ISL_2756328, EPI_ISL_2756331, EPI_ISL_2892609, EPI_ISL_2892610, EPI_ISL_2892706, EPI_ISL_3185455 | | | |
| see above | Genetica Molecular and Subdepartamento de Virologia ISP Chile | Instituto de Salud Publica de Chile | Andres Castillo; Barbara Parra; Constanza Campano; Gisselle Barra; Javier Tognarelli; Jorge Fernandez; Karen Orostica; Loredana Arata; Patricia Bustos; Rodrigo Fasce; Soledad Ulloa |
| EPI_ISL_2621554, EPI_ISL_2621555 | Genomics, Poplar Healthcare | Genomics, Poplar Healthcare | Truong, M. |
| EPI_ISL_2537241, EPI_ISL_2537257, EPI_ISL_2537259, EPI_ISL_2537272, EPI_ISL_2854941, EPI_ISL_2854951, EPI_ISL_2854955, EPI_ISL_2854961 | | | |
| see above | Gravity Diagnostics, LLC | Gravity Diagnostics, LLC | Gravity Diagnostics |
| EPI_ISL_7476556, EPI_ISL_7476635 | Grupo de Investigación en Enfermedades Tropicales del Ejército (GINETEJ), Laboratorio de Referencia e Investigación, Dirección de Sanidad Ejército, Bogotá, Colombia | Centro de Investigaciones en Microbiología y Biotecnología-UR (CIMBIUR), Facultad de Ciencias Naturales, Universidad del Rosario, Bogotá, Colombia | Alberto E. Paniz-Mondolfi; Angie L. Ramírez; Camilo Correa-Cardenas; Claudia Mendez; Enzo Guerrero-Araya; Juan David Ramírez; Julie Perez; Luz H. Patiño; Maria Clara Duque; Marina Muñoz; Nathalia Ballesteros; Nicolás Luna; Sergio Castañeda |
| EPI_ISL_2695275 | H Braga | Instituto Nacional de Saude (INSA) | Borges et al |
| EPI_ISL_3102422, EPI_ISL_3536363 | H J M A HOSPITAL JOSE MARTINIANO DE ALENCAR | Analytical Competence Molecular Epidemiology Lab/ACME, Oswaldo Cruz Foundation, Ceara (FIOCRUZ CE) | Cleber Furtado Aksenen; Fabio Miyajima; Fernando Braga Stehling; Francisco Eder de Moura Lopes; Jamille Maria Mendes Bezerra; Joaquim César do Nascimento Sousa Junior; Pedro Miguel Carneiro Jeronimo; Suzana Porto Almeida e Lucas Delerino; Thais Ferreira de Oliveira; Thais de Oliveira Costa; Ticiane Cavalcante de Souza; Veridiana Pessoa Miyajima |
| EPI_ISL_5646126 | HC-UNESP | Instituto Butantan | Antonio Jorge Martins; Claudia Renata dos Santos Barros; David Schlesinger; Debora Botequio Moretti; Dimas Tadeu Covas; Elaine Cristina Marqueze; Elaine Vieira Santos; Evandra Strazza Rodrigues; Heidge Fukumasu; Jayme Augusto de Souza-Neto; José Salvatore Leister Patané; Luiz Alcantara; Luiz Lehmann Coutinho; Maria Carolina Elias; Maurício Lacerda Nogueira; Rafael dos Santos Bezerra; Raul Machado Neto; Rejane Maria Tommasini Grotto; Ricardo Haddad; Sandra Coccuzzo Sampaio Vessoni; Simone Kashima; Svetoslav Nanev Slavov; Vincent Louis Viala |
| EPI_ISL_3102348, EPI_ISL_3102427, EPI_ISL_3536213, EPI_ISL_3536311 | HEMOCE CENTRO DE HEMATOLOGIA E HEMOTERAPIA DO CEARA | Analytical Competence Molecular Epidemiology Lab/ACME, Oswaldo Cruz Foundation, Ceara (FIOCRUZ CE) | Cleber Furtado Aksenen; Fabio Miyajima; Fernando Braga Stehling; Francisco Eder de Moura Lopes; Jamille Maria Mendes Bezerra; Joaquim César do Nascimento Sousa Junior; Pedro Miguel Carneiro Jeronimo; Suzana Porto Almeida e Lucas Delerino; Thais Ferreira de Oliveira; Thais de Oliveira Costa; Ticiane Cavalcante de Souza; Veridiana Pessoa Miyajima |
| EPI_ISL_3102345, EPI_ISL_3102381, EPI_ISL_3102392, EPI_ISL_3536166, EPI_ISL_3536214 | HGCC HOSPITAL GERAL DR CESAR CALS | Analytical Competence Molecular Epidemiology Lab/ACME, Oswaldo Cruz Foundation, Ceara (FIOCRUZ CE) | Cleber Furtado Aksenen; Fabio Miyajima; Fernando Braga Stehling; Francisco Eder de Moura Lopes; Jamille Maria Mendes Bezerra; Joaquim César do Nascimento Sousa Junior; Pedro Miguel Carneiro Jeronimo; Suzana Porto Almeida e Lucas Delerino; Thais Ferreira de Oliveira; Thais de Oliveira Costa; Ticiane Cavalcante de Souza; Veridiana Pessoa Miyajima |
| EPI_ISL_3102340, EPI_ISL_3536217, EPI_ISL_3536221 | HIAS HOSPITAL INFANTIL ALBERT SABIN | Analytical Competence Molecular Epidemiology Lab/ACME, Oswaldo Cruz Foundation, Ceara (FIOCRUZ CE) | Cleber Furtado Aksenen; Fabio Miyajima; Fernando Braga Stehling; Francisco Eder de Moura Lopes; Jamille Maria Mendes Bezerra; Joaquim César do Nascimento Sousa Junior; Pedro Miguel Carneiro Jeronimo; Suzana Porto Almeida e Lucas Delerino; Thais Ferreira de Oliveira; Thais de Oliveira Costa; Ticiane Cavalcante de Souza; Veridiana Pessoa Miyajima |
| EPI_ISL_2617589, EPI_ISL_2617590, EPI_ISL_2617592, EPI_ISL_2617593, EPI_ISL_2617594, EPI_ISL_2617595, EPI_ISL_2617596, EPI_ISL_2617597, EPI_ISL_2617598, EPI_ISL_2617599, EPI_ISL_2617600, EPI_ISL_2617601, EPI_ISL_2617602, EPI_ISL_2617603, EPI_ISL_2617610, EPI_ISL_2617611, EPI_ISL_2617612, EPI_ISL_2617613, EPI_ISL_2617614, EPI_ISL_2617615, EPI_ISL_2617616, EPI_ISL_2617617, EPI_ISL_2617618, EPI_ISL_2617619, EPI_ISL_2617620, EPI_ISL_2617621, EPI_ISL_2680895, EPI_ISL_2680896, EPI_ISL_2680897, EPI_ISL_2680898, EPI_ISL_2680899, EPI_ISL_2680900, EPI_ISL_2680901, EPI_ISL_2921534, EPI_ISL_2921535, EPI_ISL_2921536, EPI_ISL_2921537, EPI_ISL_2921602, EPI_ISL_2921606, EPI_ISL_2928511, EPI_ISL_3254317, EPI_ISL_3254328, EPI_ISL_3254349, EPI_ISL_3254350, EPI_ISL_3254351, EPI_ISL_3254352, EPI_ISL_3254353, EPI_ISL_3254354, EPI_ISL_3254355, EPI_ISL_3254356, EPI_ISL_3254357, EPI_ISL_3254358, EPI_ISL_3254359, EPI_ISL_3254360, EPI_ISL_3254361, EPI_ISL_3254362, EPI_ISL_3254363, EPI_ISL_3254364, EPI_ISL_3254365, EPI_ISL_3254366, EPI_ISL_3254368, EPI_ISL_3254369, EPI_ISL_3254370, EPI_ISL_3254371, EPI_ISL_3254372, EPI_ISL_3254373, EPI_ISL_3254374, EPI_ISL_3254375, EPI_ISL_3254376, EPI_ISL_3254377, EPI_ISL_3254378, EPI_ISL_3254379, EPI_ISL_3254380, EPI_ISL_3254381, EPI_ISL_3254382, EPI_ISL_3254383, EPI_ISL_3254384, EPI_ISL_3254385, EPI_ISL_3254386, EPI_ISL_3254387, EPI_ISL_3254388, EPI_ISL_3254389, EPI_ISL_3254390, EPI_ISL_3254391, EPI_ISL_3254392, EPI_ISL_3254393, EPI_ISL_3254394, EPI_ISL_3254395, EPI_ISL_3254396, EPI_ISL_3254397, EPI_ISL_3254398, EPI_ISL_3254399, EPI_ISL_3254400, EPI_ISL_3254401, EPI_ISL_3254402, EPI_ISL_3254403, EPI_ISL_3254404, EPI_ISL_3254405, EPI_ISL_3254406, EPI_ISL_3254407, EPI_ISL_3254408, EPI_ISL_3254409, EPI_ISL_3254410, EPI_ISL_3254411, EPI_ISL_3254412, EPI_ISL_3254413, EPI_ISL_3254414, EPI_ISL_3254415, EPI_ISL_3254416, EPI_ISL_3254417, EPI_ISL_4890090, EPI_ISL_4890108, EPI_ISL_5438927, EPI_ISL_5438949 | | | |
| see above | HLAGYN - Laboratorio de Imunologia de Transplantes de Goias | HLAGYN - Laboratorio de Imunologia de Transplantes de Goias | Alessandro Leonardo Alvares Magalhaes; Daniel Ferreira de Sousa; Danielle de Paiva Rezende; Erika Lopes Rocha Batista; Fernando Antonio Vinhal dos Santos; Frederico Rodrigues Vinhal; Kamila Oliveira Reis De Freitas.; Lucas Carlos Gomes Pereira; Sabrina Sara Moreira Duarte |
| EPI_ISL_3102424, EPI_ISL_3102436, EPI_ISL_3102448, EPI_ISL_3102469, EPI_ISL_3536227, EPI_ISL_3536228, EPI_ISL_3536239, EPI_ISL_3536247, EPI_ISL_3536249 | | | |
| see above | HM HOSPITAL DE MESSEJANA DR CARLOS ALBERTO STUDART GOMES | Analytical Competence Molecular Epidemiology Lab/ACME, Oswaldo Cruz Foundation, Ceara (FIOCRUZ CE) | Cleber Furtado Aksenen; Fabio Miyajima; Fernando Braga Stehling; Francisco Eder de Moura Lopes; Jamille Maria Mendes Bezerra; Joaquim César do Nascimento Sousa Junior; Pedro Miguel Carneiro Jeronimo; Suzana Porto Almeida e Lucas Delerino; Thais Ferreira de Oliveira; Thais de Oliveira Costa; Ticiane Cavalcante de Souza; Veridiana Pessoa Miyajima |
| EPI_ISL_5645861 | HOSPITAL DE CAMPANHA COVID 19 | Instituto Butantan | Antonio Jorge Martins; Claudia Renata dos Santos Barros; David Schlesinger; Debora Botequio Moretti; Dimas Tadeu Covas; Elaine Cristina Marqueze; Elaine Vieira Santos; Evandra Strazza Rodrigues; Heidge Fukumasu; Jayme Augusto de Souza-Neto; José Salvatore Leister Patané; Luiz Alcantara; Luiz Lehmann Coutinho; Maria Carolina Elias; Maurício Lacerda Nogueira; Rafael dos Santos Bezerra; Raul Machado Neto; Rejane Maria Tommasini Grotto; Ricardo Haddad; Sandra Coccuzzo Sampaio Vessoni; Simone Kashima; Svetoslav Nanev Slavov; Vincent Louis Viala |
| EPI_ISL_5645621, EPI_ISL_5645635, EPI_ISL_5645636, EPI_ISL_5645763, EPI_ISL_5645766, EPI_ISL_5645787, EPI_ISL_5645804, EPI_ISL_5645817, EPI_ISL_5645832, EPI_ISL_5645903, EPI_ISL_5645918, EPI_ISL_5645932, EPI_ISL_5646142, EPI_ISL_5646148, EPI_ISL_5646149, EPI_ISL_5646150, EPI_ISL_5646153, EPI_ISL_5646157, EPI_ISL_5646159, EPI_ISL_5646163, EPI_ISL_5646168, EPI_ISL_5646169, EPI_ISL_5646180, EPI_ISL_5646181, EPI_ISL_5646185, EPI_ISL_5646189, EPI_ISL_5646196, EPI_ISL_5646197 | | | |
| see above | HOSPITAL DE CAMPANHA COVID 19 MUNICIPIO DE TAUBATE | Instituto Butantan | Antonio Jorge Martins; Claudia Renata dos Santos Barros; David Schlesinger; Debora Botequio Moretti; Dimas Tadeu Covas; Elaine Cristina Marqueze; Elaine Vieira Santos; Evandra Strazza Rodrigues; Heidge Fukumasu; Jayme Augusto de Souza-Neto; José Salvatore Leister Patané; Luiz Alcantara; Luiz Lehmann Coutinho; Maria Carolina Elias; Maurício Lacerda Nogueira; Rafael dos Santos Bezerra; Raul Machado Neto; Rejane Maria Tommasini Grotto; Ricardo Haddad; Sandra Coccuzzo Sampaio Vessoni; Simone Kashima; Svetoslav Nanev Slavov; Vincent Louis Viala |
| EPI_ISL_2827976 | HOSPITAL DR. ENRIQUE BALTODANO BRICEÑO | Inciensa, Instituto Costarricense de Investigación y Enseñanza en Nutrición y Salud | Adriana Godínez; Claudio Soto-Garita; Estela Cordero; Francisco Duarte; Hebleen Porras; Joselyn Prado & Adriana Bermúdez; José Luis Vargas; Mariela Gutiérrez; Melany Calderón |
| EPI_ISL_2827967 | HOSPITAL DR. RAFAEL A. CALDERON GUARDIA | Inciensa, Instituto Costarricense de Investigación y Enseñanza en Nutrición y Salud | Adriana Godínez; Claudio Soto-Garita; Estela Cordero; Francisco Duarte; Hebleen Porras; Joselyn Prado & Fabian Salas; José Luis Vargas; Mariela Gutiérrez; Melany Calderón |
| EPI_ISL_2827963, EPI_ISL_2827964 | HOSPITAL DR. WILLIAM ALLEN | Inciensa, Instituto Costarricense de Investigación y Enseñanza en Nutrición y Salud | Adriana Godínez; Claudio Soto-Garita; Estela Cordero; Francisco Duarte; Hebleen Porras; José Luis Vargas; Mariela Gutiérrez & Joselyn Prado; Melany Calderón |
| EPI_ISL_3102412, EPI_ISL_3102442, EPI_ISL_3536304 | HOSPITAL E MATERNIDADE DRA ZILDA ARNS NEUMANN | Analytical Competence Molecular Epidemiology Lab/ACME, Oswaldo Cruz Foundation, Ceara (FIOCRUZ CE) | Cleber Furtado Aksenen; Fabio Miyajima; Fernando Braga Stehling; Francisco Eder de Moura Lopes; Jamille Maria Mendes Bezerra; Joaquim César do Nascimento Sousa Junior; Pedro Miguel Carneiro Jeronimo; Suzana Porto Almeida e Lucas Delerino; Thais Ferreira de Oliveira; Thais de Oliveira Costa; Ticiane Cavalcante de Souza; Veridiana Pessoa Miyajima |
| EPI_ISL_3102354, EPI_ISL_3102364, EPI_ISL_3102376, EPI_ISL_3102383, EPI_ISL_3102387, EPI_ISL_3102396, EPI_ISL_3102405, EPI_ISL_3536173, EPI_ISL_3536187, EPI_ISL_3536190, EPI_ISL_3536267, EPI_ISL_3536273, EPI_ISL_3536275, EPI_ISL_3536278, EPI_ISL_3536281, EPI_ISL_3536288, EPI_ISL_3536324, EPI_ISL_3536327, EPI_ISL_3536328, EPI_ISL_3536332, EPI_ISL_3536334, EPI_ISL_3536336, EPI_ISL_3536341, EPI_ISL_3536349, EPI_ISL_3536393 | | | |
| see above | HOSPITAL ESTADUAL LEONARDO DA VINCI | Analytical Competence Molecular Epidemiology Lab/ACME, Oswaldo | Cleber Furtado Aksenen; Fabio Miyajima; Fernando Braga Stehling; Francisco Eder de Moura Lopes; Jamille Maria Mendes Bezerra; Joaquim César do Nascimento Sousa Junior; Pedro Miguel Carneiro Jeronimo; Suzana Porto Almeida e Lucas Delerino; Thais Ferreira de Oliveira; Thais de Oliveira Costa; Ticiane Cavalcante de Souza; Veridiana Pessoa Miyajima |

| | | | |
|---|---|---|---|
| | | Cruz Foundation, Ceara (FIOCRUZ CE) | |
| EPI_ISL_4425156, EPI_ISL_4425158, EPI_ISL_4425162 | HOSPITAL GENERAL UNIVERSITARIO DE ALBACETE | Instituto de Salud Carlos III | A. Monzón; CARIDAD; F. Casas; I. Jiménez; I. SAINZ DE BARANDA CAMINO; I.SAINZ DE BARANDA CAMINO; M. Camarero; M. Sandonís; P. Zaballos; S. Cuesta; S. Iglesias-Caballero; S. Pozo; S. Varona; Sandonís; V. Camarero; V. Vázquez-Morón; Vázquez-Morón |
| EPI_ISL_4425194 | HOSPITAL GENERAL UNIVERSITARIO DE GUADALAJARA | Instituto de Salud Carlos III | A. Monzón; ALEJANDRO; F. Casas; I. GONZALEZ PRAETORIUS; I. Jiménez; M. Camarero; P. Zaballos; S. Cuesta; S. Iglesias-Caballero; S. Pozo; S. Varona; Sandonís; V. Vázquez-Morón |
| EPI_ISL_2827975 | HOSPITAL LA ANEXION | Inciensa, Instituto Costarricense de Investigación y Enseñanza en Nutrición y Salud | Adriana Godínez; Claudio Soto-Garita; Estela Cordero; Francisco Duarte; Hebleen Porras; Joselyn Prado & Adriana Bermúdez; José Luis Vargas; Mariela Gutiérrez; Melany Calderón |
| EPI_ISL_4601280 | HOSPITAL LOS ARCOS | Instituto de Salud Carlos III | A. Monzón; F. Casas; I. Jiménez; I.CAMARA SIMON; M. Sandonís; MARGARITA; P. Zaballos; S. Cuesta; S. Iglesias-Caballero; S. Pozo; S. Varona; V. Camarero; Vázquez-Morón |
| EPI_ISL_2534391, EPI_ISL_2534402 | HOSPITAL MANACOR | HOSPITAL UNIVERSITARIO SON ESPASES | Hospital Universitario Son Espases; Servicio de Microbiología |
| EPI_ISL_5646127, EPI_ISL_5646193, EPI_ISL_5646194 | HOSPITAL MATERNIDADE SAO JOSE ITAPUI | Instituto Butantan | Antonio Jorge Martins; Claudia Renata dos Santos Barros; David Schlesinger; Debora Botequio Moretti; Dimas Tadeu Covas; Elaine Cristina Marqueze; Elaine Vieira Santos; Evandra Strazza Rodrigues; Heidge Fukumasu; Jayme Augusto de Souza-Neto; José Salvatore Leister Patané; Luiz Alcantara; Luiz Lehmann Coutinho; Maria Carolina Elias; Maurício Lacerda Nogueira; Rafael dos Santos Bezerra; Raul Machado Neto; Rejane Maria Tommasini Grotto; Ricardo Haddad; Sandra Coccuzzo Sampaio Vessoni; Simone Kashima; Svetoslav Nanev Slavov; Vincent Louis Viala |
| EPI_ISL_5645761, EPI_ISL_5645927, EPI_ISL_5646145 | HOSPITAL MUNICIPAL DE IBIUNA IBIUNA SP | Instituto Butantan | Antonio Jorge Martins; Claudia Renata dos Santos Barros; David Schlesinger; Debora Botequio Moretti; Dimas Tadeu Covas; Elaine Cristina Marqueze; Elaine Vieira Santos; Evandra Strazza Rodrigues; Heidge Fukumasu; Jayme Augusto de Souza-Neto; José Salvatore Leister Patané; Luiz Alcantara; Luiz Lehmann Coutinho; Maria Carolina Elias; Maurício Lacerda Nogueira; Rafael dos Santos Bezerra; Raul Machado Neto; Rejane Maria Tommasini Grotto; Ricardo Haddad; Sandra Coccuzzo Sampaio Vessoni; Simone Kashima; Svetoslav Nanev Slavov; Vincent Louis Viala |
| EPI_ISL_5649580, EPI_ISL_5649623 | HOSPITAL MUNICIPAL DR MARIO GATTI CAMPINAS | Instituto Butantan | Antonio Jorge Martins; Claudia Renata dos Santos Barros; David Schlesinger; Debora Botequio Moretti; Dimas Tadeu Covas; Elaine Cristina Marqueze; Elaine Vieira Santos; Evandra Strazza Rodrigues; Heidge Fukumasu; Jayme Augusto de Souza-Neto; José Salvatore Leister Patané; Luiz Alcantara; Luiz Lehmann Coutinho; Maria Carolina Elias; Maurício Lacerda Nogueira; Rafael dos Santos Bezerra; Raul Machado Neto; Rejane Maria Tommasini Grotto; Ricardo Haddad; Sandra Coccuzzo Sampaio Vessoni; Simone Kashima; Svetoslav Nanev Slavov; Vincent Louis Viala |
| EPI_ISL_5645617, EPI_ISL_5645633, EPI_ISL_5645659, EPI_ISL_5645673, EPI_ISL_5645742, EPI_ISL_5645907, EPI_ISL_5645924 | | | |
| see above | HOSPITAL MUNICIPAL DR TABAJARA RAMOS | Instituto Butantan | Antonio Jorge Martins; Claudia Renata dos Santos Barros; David Schlesinger; Debora Botequio Moretti; Dimas Tadeu Covas; Elaine Cristina Marqueze; Elaine Vieira Santos; Evandra Strazza Rodrigues; Heidge Fukumasu; Jayme Augusto de Souza-Neto; José Salvatore Leister Patané; Luiz Alcantara; Luiz Lehmann Coutinho; Maria Carolina Elias; Maurício Lacerda Nogueira; Rafael dos Santos Bezerra; Raul Machado Neto; Rejane Maria Tommasini Grotto; Ricardo Haddad; Sandra Coccuzzo Sampaio Vessoni; Simone Kashima; Svetoslav Nanev Slavov; Vincent Louis Viala |
| EPI_ISL_5647183 | HOSPITAL MUNICIPAL JORGE CURY DE SANTA MARIA DA SERRA | Instituto Butantan | Antonio Jorge Martins; Claudia Renata dos Santos Barros; David Schlesinger; Debora Botequio Moretti; Dimas Tadeu Covas; Elaine Cristina Marqueze; Elaine Vieira Santos; Evandra Strazza Rodrigues; Heidge Fukumasu; Jayme Augusto de Souza-Neto; José Salvatore Leister Patané; Luiz Alcantara; Luiz Lehmann Coutinho; Maria Carolina Elias; Maurício Lacerda Nogueira; Rafael dos Santos Bezerra; Raul Machado Neto; Rejane Maria Tommasini Grotto; Ricardo Haddad; Sandra Coccuzzo Sampaio Vessoni; Simone Kashima; Svetoslav Nanev Slavov; Vincent Louis Viala |
| EPI_ISL_3102265, EPI_ISL_3102299, EPI_ISL_3536345 | HOSPITAL MUNICIPAL NOSSA SENHORA DOS MILAGRES | Analytical Competence Molecular Epidemiology Lab/ACME, Oswaldo Cruz Foundation, Ceara (FIOCRUZ CE) | Cleber Furtado Aksenen; Fabio Miyajima; Fernando Braga Stehling; Francisco Eder de Moura Lopes; Jamille Maria Mendes Bezerra; Joaquim César do Nascimento Sousa Junior; Pedro Miguel Carneiro Jeronimo; Suzana Porto Almeida e Lucas Delerino; Thais Ferreira de Oliveira; Thais de Oliveira Costa; Ticiane Cavalcante de Souza; Veridiana Pessoa Miyajima |
| EPI_ISL_2597993, EPI_ISL_5646177 | HOSPITAL REGIONAL DE ITAPETININGA | Instituto Butantan | Antonio Jorge Martins; Claudia Renata dos Santos Barros; David Schlesinger; Debora Botequio Moretti; Dimas Tadeu Covas; Elaine Cristina Marqueze; Elaine Vieira Santos; Evandra Strazza Rodrigues; Heidge Fukumasu; Jayme Augusto de Souza-Neto; José Salvatore Leister Patané; Luiz Alcantara; Luiz Lehmann Coutinho; Maria Carolina Elias; Maurício Lacerda Nogueira; Rafael dos Santos Bezerra; Raul Machado Neto; Rejane Maria Tommasini Grotto; Ricardo Haddad; Sandra Coccuzzo Sampaio Vessoni; Simone Kashima; Svetoslav Nanev Slavov; Vincent Louis Viala |
| EPI_ISL_3536397 | HOSPITAL REGIONAL UNIMED | Analytical Competence Molecular Epidemiology Lab/ACME, Oswaldo Cruz Foundation, Ceara (FIOCRUZ CE) | Cleber Furtado Aksenen; Fabio Miyajima; Fernando Braga Stehling; Francisco Eder de Moura Lopes; Jamille Maria Mendes Bezerra; Joaquim César do Nascimento Sousa Junior; Pedro Miguel Carneiro Jeronimo; Suzana Porto Almeida e Lucas Delerino; Thais Ferreira de Oliveira; Thais de Oliveira Costa; Ticiane Cavalcante de Souza; Veridiana Pessoa Miyajima |
| EPI_ISL_3536153, EPI_ISL_3536289, EPI_ISL_3536308 | HOSPITAL SAO JOSE DE DOENCAS INFECCIOSAS | Analytical Competence Molecular Epidemiology Lab/ACME, Oswaldo Cruz Foundation, Ceara (FIOCRUZ CE) | Cleber Furtado Aksenen; Fabio Miyajima; Fernando Braga Stehling; Francisco Eder de Moura Lopes; Jamille Maria Mendes Bezerra; Joaquim César do Nascimento Sousa Junior; Pedro Miguel Carneiro Jeronimo; Suzana Porto Almeida e Lucas Delerino; Thais Ferreira de Oliveira; Thais de Oliveira Costa; Ticiane Cavalcante de Souza; Veridiana Pessoa Miyajima |
| EPI_ISL_5647196 | HOSPITAL SAO JOSE ITIRAPINA | Instituto Butantan | Antonio Jorge Martins; Claudia Renata dos Santos Barros; David Schlesinger; Debora Botequio Moretti; Dimas Tadeu Covas; Elaine Cristina Marqueze; Elaine Vieira Santos; Evandra Strazza Rodrigues; Heidge Fukumasu; Jayme Augusto de Souza-Neto; José Salvatore Leister Patané; Luiz Alcantara; Luiz Lehmann Coutinho; Maria Carolina Elias; Maurício Lacerda Nogueira; Rafael dos Santos Bezerra; Raul Machado Neto; Rejane Maria Tommasini Grotto; Ricardo Haddad; Sandra Coccuzzo Sampaio Vessoni; Simone Kashima; Svetoslav Nanev Slavov; Vincent Louis Viala |
| EPI_ISL_3536335 | HOSPITAL SAO MATEUS | Analytical Competence Molecular Epidemiology Lab/ACME, Oswaldo Cruz Foundation, Ceara (FIOCRUZ CE) | Cleber Furtado Aksenen; Fabio Miyajima; Fernando Braga Stehling; Francisco Eder de Moura Lopes; Jamille Maria Mendes Bezerra; Joaquim César do Nascimento Sousa Junior; Pedro Miguel Carneiro Jeronimo; Suzana Porto Almeida e Lucas Delerino; Thais Ferreira de Oliveira; Thais de Oliveira Costa; Ticiane Cavalcante de Souza; Veridiana Pessoa Miyajima |
| EPI_ISL_5646137 | HOSPITAL SAO MIGUEL ARCANJO | Instituto Butantan | Antonio Jorge Martins; Claudia Renata dos Santos Barros; David Schlesinger; Debora Botequio Moretti; Dimas Tadeu Covas; Elaine Cristina Marqueze; Elaine Vieira Santos; Evandra Strazza Rodrigues; Heidge Fukumasu; Jayme Augusto de Souza-Neto; José Salvatore Leister Patané; Luiz Alcantara; Luiz Lehmann Coutinho; Maria Carolina Elias; Maurício Lacerda Nogueira; Rafael dos Santos Bezerra; Raul Machado Neto; Rejane Maria Tommasini Grotto; Ricardo Haddad; Sandra Coccuzzo Sampaio Vessoni; Simone Kashima; Svetoslav Nanev Slavov; Vincent Louis Viala |
| EPI_ISL_4425145 | HOSPITAL UNIVERSITARIO DE CEUTA | Instituto de Salud Carlos III | A. Monzón; F. Casas; I. Jiménez; I.HIJANO VILLEGAS; M. Sandonís; P. Zaballos; S. Cuesta; S. Iglesias-Caballero; S. Pozo; S. Varona; SALOME; V. Camarero; Vázquez-Morón |
| EPI_ISL_3102454, EPI_ISL_3536326 | HOSPITAL UNIVERSITARIO WALTER CANTIDIO | Analytical Competence Molecular Epidemiology Lab/ACME, Oswaldo Cruz Foundation, Ceara (FIOCRUZ CE) | Cleber Furtado Aksenen; Fabio Miyajima; Fernando Braga Stehling; Francisco Eder de Moura Lopes; Jamille Maria Mendes Bezerra; Joaquim César do Nascimento Sousa Junior; Pedro Miguel Carneiro Jeronimo; Suzana Porto Almeida e Lucas Delerino; Thais Ferreira de Oliveira; Thais de Oliveira Costa; Ticiane Cavalcante de Souza; Veridiana Pessoa Miyajima |
| EPI_ISL_4425307 | HOSPITAL VIRGEN DE LA SALUD DE TOLEDO | Instituto de Salud Carlos III | A. Monzón; EVA; F. Casas; I. Jiménez; I.HEREDERO GALVEZ; Iglesias-Caballero; M. Sandonís; P. Zaballos; S. Camarero; S. Cuesta; S. Pozo; S. Varona; V. Vázquez-Morón |
| EPI_ISL_4743413 | HPTU | Laboratorio Departamental de Salud Publica de Antioquia | Andres F. Cardona-Rios; Cristian Arbey Velarde Hoyos; Gloria Isabel Escobar; Idabely Betancur Ortiz; Juan P. Hernandez-Ortiz; Maria Stella López |
| EPI_ISL_3276546 | HPTU | Universidad Nacional de Colombia - Laboratorio Genómico One Health | Andres F. Cardona-Rios; Carlos Franco-Muñoz; Carolina Muñoz-Arango; Celeny Ortiz; Daniel O. Maldonado-Perez; Diego A. Álvarez-Díaz; Hector Alejandro Ruiz-Moreno; Idabely Betancur Ortiz; Jorge E. Osorio; Juan P. Hernandez-Ortiz; Karl A Ciuoderis; Katherine Laiton-Donato; Laura Silvana Perez; Lina M. Hurtado; Marcela Mercado-Reyes; Maria Angélica Maya; Maria Stella López; Rita Almanza Payares; Sandra Ines Cano; Simón Villegas Velásquez |
| EPI_ISL_3235338, EPI_ISL_3235351, EPI_ISL_3235353 | Hackensack Medical Center | New York Genome Center | Amy Baldwin; Andre Corvelo; Barry Kreiswirth; David Perlin; Dayna M. Oschwald; Jose Mediavilla; Kaelea Composto; Kar Chow; Liang Chen; Marcus Cunningham; Michael Zody; Samantha Fennessey; Tom Maniatis |
| EPI_ISL_2727869 | HealthPartners Central Lab | Minnesota Department of Health, Public Health Laboratory | Alexandra Lorentz; Jacob Garfin; Matt Plumb; and Xiong Wang |
| EPI_ISL_3149723, EPI_ISL_3149731 | Hosp. Pediatrico Dr. Avelino Castelán | Laboratorio de Biología Molecular, Instituto de Medicina Regional on behalf of 'Proyecto Argentino Interinstitucional de genomica de SARS-CoV-2' (PAIS Consortium) | Bettina Brusés; Gioia Lucia Marino; Griselda Oria; Horacio Lucero.; Javier Mussin; Julieta Ronchi; Laura Formichelli; María Andrea Gili; María Cecilia López; Melina Lorenzini Campos; Raúl Maximiliano Acevedo |
| EPI_ISL_2627942 | Hospital | National Reference Center for Viruses of Respiratory Infections, Institut Pasteur, Paris | Angela Brisebarre; Camille Capel; Christophe Malabat; Corinne Maufrais; Etienne Simon-Lorière; Frédéric Lemoine; Louise Lefrançois; Marion Barbet; Maud Vanpeene; Méline Bizard; Patricia Stoessel-Thouvenin; Pierre Lechat; Sylvie Behillil; Sylvie Van der Werf; Vincent Enouf |
| EPI_ISL_2484017, EPI_ISL_2484018 | Hospital General Universitario Gregorio Marañón | Hospital General Universitario Gregorio Marañón | Cristina Rodriguez-Grande; Darío García de Viedma; Julia Suárez; Laura Pérez-Lago; Marta Herranz Martin; Patricia Muñoz; Pedro Sola Campoy; Pilar Catalán; Sergio Buenestado Serrano; Victor Manuel de la Cueva |
| EPI_ISL_3033080, EPI_ISL_3033082, EPI_ISL_3033084, EPI_ISL_3260095 | Hospital Jaime Ferre - SAMCO Rafaela | Grupo de Genómica y Bioinformática del Instituto de Investigación de la Cadena Láctea CONICET-INTA on behalf of 'Proyecto Argentino Interinstitucional de genomica de SARS-CoV-2' (PAIS Consortium) | AF; Aliprandi D; Amadio; C; Eberhardt; Irazoqui; Isaia; JF; JM; MF; Pandolfi; Quaranta; Soratti R; V |
| EPI_ISL_2894555 | Hospital Margarita Maza de Juárez | Microbial Genomics Laboratory | ; Alejandra García-Gasca; Alejandra Hernández-Terán; Alejandro Sánchez-Flores; Alfredo Herrera-Estrella; Alicia Ocaña-Mondragón; Andreu Comas-García; Angel Gustavo Salas-Lais; Antonio Loza Román; Bernardo Martínez-Miguel; Blanca Taboada; Brenda Irasema Maldonado-Meza; Bruno Gómez-Gil; Carla Ivón Herrera-Najera; Carlos F. Arias; Celia Boukadida; Clara Esperanza Santacruz-Tinoco; Concepción Grajales-Muñiz; Consorcio Mexicano de Vigilancia Genómica (CoViGen-Mex). Authors (in alphabetical order): Julio Elias Alvarado-Yaah; Cristóbal Cháidez-Quiróz; Célida Duque Molina; Célida Martínez- Rodríguez; Daniel Fregoso-Rueda; Daniel Lira Morales; Eduardo Becerril-Vargas; Fernando Fontove-Herrera; Fidencio Mejía-Nepomuceno; Francisco Pulido; Gloria Elena Espinosa-Ayala; Gloria María Molina-Salinas; Gloria Vazquez; Hector Esteban Paz-Juárez; Hector Montoya-Fuentes; Helen Haydee Fernanda Ramírez-Plascencia; Irvin González-López; Jean Pierre González; Jesús Hernández; Joel Armando Vázquez-Pérez.; Jorge Salas-Hernández; José Antonio Enciso-Moreno; José Arturo Martínez-Orozco; José Esteban Muñoz-Medina; José de Jesús Nuñez-Contreras; Juan Bautista Chale-Dzul; Julissa Enciso-Ibarra; Luis Alberto Ochoa-Carrera; Margarita Matías-Florentino; Mario Mújica-Sánchez; Marissa Perez-Garcia; María Guadalupe Santiago-Mauricio; María Guadalupe de Jesús Mireles-Rivera; Nelly Sélem-Mojica; Pavel Isa; Ricardo Ciria Merce; Ricardo Grande; Rosa María Gutiérrez Rios; Santiago Ávila-Ríos; Selene Zárate; Susana Lopez; Verónica Mata-Haro; Victor Eduardo García-Arias; Víctor Hugo Borja-Aburto |
| EPI_ISL_2680935 | Hospital Sharp | Microbial Genomics Laboratory | ; Alejandra García-Gasca; Alejandra Hernández-Terán; Alejandro Sánchez-Flores; Alfredo Herrera-Estrella; Alicia Ocaña-Mondragón; Andreu Comas-García; Angel Gustavo Salas-Lais; Antonio Loza Román; Bernardo Martínez-Miguel; Blanca Taboada; Brenda Irasema Maldonado-Meza; Bruno Gómez-Gil; Carla Ivón Herrera-Najera; Carlos F. Arias; Celia Boukadida; Clara Esperanza Santacruz-Tinoco; Concepción Grajales-Muñiz; Consorcio Mexicano de Vigilancia Genómica (CoViGen-Mex). Authors (in alphabetical order): Julio Elias Alvarado-Yaah; Cristóbal Cháidez-Quiróz; Célida Duque Molina; Célida Martínez- Rodríguez; Daniel Fregoso-Rueda; Daniel Lira Morales; Eduardo Becerril-Vargas; Fernando Fontove-Herrera; Fidencio Mejía-Nepomuceno; Francisco Pulido; Gloria Elena Espinosa-Ayala; Gloria María Molina-Salinas; Gloria Vazquez; Hector Esteban Paz-Juárez; Hector Montoya-Fuentes; Helen Haydee Fernanda Ramírez-Plascencia; Irvin González-López; Jean Pierre González; Jesús Hernández; Joel Armando Vázquez-Pérez.; Jorge Salas-Hernández; José Antonio Enciso-Moreno; José Arturo Martínez-Orozco; José Esteban Muñoz-Medina; José de Jesús Nuñez-Contreras; Juan Bautista Chale-Dzul; Julissa Enciso-Ibarra; Luis Alberto Ochoa-Carrera; Margarita Matías-Florentino; Mario Mújica-Sánchez; Marissa Perez-Garcia; María Guadalupe Santiago-Mauricio; María Guadalupe de Jesús Mireles-Rivera; Nelly Sélem-Mojica; Pavel Isa; Ricardo Ciria Merce; Ricardo Grande; Rosa María Gutiérrez Rios; Santiago Ávila-Ríos; Selene Zárate; Susana Lopez; Verónica Mata-Haro; Victor Eduardo García-Arias; Víctor Hugo Borja-Aburto |
| EPI_ISL_2493148 | Hospital Universitari Arnau de | Hospital Universitari Vall d'Hebron - | Alejandra González-Sánchez; Andrés Antón; Ariadna Rando; Carla Castillo; Cristina Andrés; Damir Garcia-Cehic; Josep Quer; Juliana Esperalba; Maria Carmen Martin; Maria Gema Codina; Maria Piñana; Tomàs Pumarola |

| | | | |
|---|---|---|---|
| | Vilanova | Vall d'Hebron Institut de Recerca | |
| EPI_ISL_2566595 | Hospital Universitari Joan XXIII | Hospital Universitari Joan XXIII | Carla Martín; Clara Benavent; Cristina Gutiérrez; Ester Picó; Gemma Recio; Margarida Terrón; Natalia Bastón |
| EPI_ISL_2775934, EPI_ISL_2775946, EPI_ISL_2820672, EPI_ISL_2820673 | Hospital Universitari i Politècnic La Fe de València | SeqCOVID-SPAIN consortium/IBV(CSIC) | Ana Gil Brusola; Eva González Barberá; José Luis López Hontangas and SeqCOVID-SPAIN consortium; María Dolores Gómez Ruiz; Salvador Giner Almaraz |
| EPI_ISL_7339459, EPI_ISL_7339467 | Hospital Universitario San Ignacio | Centro de Investigaciones en Microbiología y Biotecnología-UR (CIMBIUR), Facultad de Ciencias Naturales, Universidad del Rosario, Bogotá, Colombia | Alberto Paniz-Mondolfi; Angie Ramírez; Beatriz Ariza; Camilo A. Correa-Cárdenas; Carlos Gómez-Restrepo; Claudia Cardozo-Romero; Claudia Méndez; David-Santiago Quevedo; Guido España; Hernando Díaz; Juan David Ramírez; Juliana Cuervo-Rojas; Julie Pérez; Luz H. Patiño; Manuel-Antonio Franco; Maria-Clara Duque; Marina Muñoz; Nathalia Ballesteros; Nicolas Luna; Sergio Castañeda; Zulma M. Cucunubá |
| EPI_ISL_2620962, EPI_ISL_2620963 | Hospital Universitario Virgen de las Nieves de Granada-SAS | SeqCOVID-SPAIN consortium/IBV(CSIC) | Irene Pedrosa Corral; José M. Navarro-Marí and SeqCOVID-SPAIN consortium; Sara Sanbonmatsu Gámez |
| EPI_ISL_2885999, EPI_ISL_2886010 | Hospital Universitario de Guadalajara | SeqCOVID-SPAIN consortium/IBV(CSIC) | Alejandro González Praetorius y Mariela Martínez Ramírez and SeqCOVID-SPAIN consortium |
| EPI_ISL_2925674, EPI_ISL_2925675, EPI_ISL_2925676, EPI_ISL_2925677 | Hospital de Cabueñes | Laboratorio de Virología HUCA | Abreu F; Alvarez-Arguelles ME; Boga JA; Castelló C; Costales I; Coto E; Gómez de Oña J; Martín-Rodríguez G; Melón; Melón S; Perez-Martínez Z; Rojo S; Sandoval M |
| EPI_ISL_3023343 | Hospital of the University of Pennsylvania Molecular Pathology Lab | Bushman Lab - University of Pennsylvania | Aoife M. Roche; Arupa Ganguly; Ayannah S. Fitzgerald; Brendan Kelly; Jevon Graham-Wooten; John K. Everett; Kyle Rodino; Layla A. Khatib; Mike Feldman; Ronald G. Collman and Frederic Bushan; Samantha A. Whiteside; Scott Sherrill-Mix; Shantan Reddy; Young Hwang |
| EPI_ISL_4196707, EPI_ISL_4196708, EPI_ISL_4196709, EPI_ISL_4196710, EPI_ISL_4196711, EPI_ISL_4196712 | Hospital Programa Area Cipolletti, Dr Pedro Moguillansky | Laboratorio Central Mg. Luis Alfredo Pianciola on behalf of 'Proyecto Argentino Interinstitucional de genomica de SARS-CoV-2' (PAIS Consortium) | C Pintos; C Rastellini; C Utreras.; C Ziehm; J Ousset; L Pianciola; L Pinoche; M Fernandez; M Mazzeo; M Roncallo; R Vilugron; S Marinelli; Y Guerrero |
| EPI_ISL_2968636, EPI_ISL_3398946 | Houston Health Dept. | Houston Health Dept. | Adolpho Lara; Pamela Brown; Ryker Penn; Yanlai Lai |
| EPI_ISL_5070940, EPI_ISL_5070968, EPI_ISL_5070983, EPI_ISL_5070999, EPI_ISL_5071006, EPI_ISL_5071075, EPI_ISL_5071077, EPI_ISL_5072624, EPI_ISL_5072648, EPI_ISL_5072650, EPI_ISL_5072654, EPI_ISL_5072705, EPI_ISL_5072753, EPI_ISL_5072770, EPI_ISL_5072792, EPI_ISL_5072960 | | | |
| see above | Houston Methodist Hospital | Houston Methodist Hospital | Ilya J. Finkelstein; James J. Davis; Jessica Cambric; Jimmy Gollihar; Kristina Reppond; Layne Pruitt; Madison N. Shyer; Marcus Nguyen; Matthew Ojeda Saavedra; Paul A. Christensen; Prasanti Yerramilli; Randall J. Olsen; Robert Olson; Ryan Gadd; S. Wesley Long; Sishir Subedi; and James M. Musser |
| EPI_ISL_2778970 | Humboldt County Public Health Laboratory | Humboldt County Public Health Laboratory | Jeremy Corrigan |
| EPI_ISL_2787361, EPI_ISL_2787367, EPI_ISL_2787369, EPI_ISL_2787380, EPI_ISL_2787389, EPI_ISL_2787392, EPI_ISL_3355076 | | | |
| see above | IA State Hygienic Laboratory | Centers for Disease Control and Prevention Division of Viral Diseases, Pathogen Discovery | Alex Burgin; Alison Laufer Halpin; Ben L. Rambo-Martin; Ben Rambo-Martin; Clinton Paden; Clinton R. Paden; Dakota Howard; Darlene Wagner; Dave Wentworth; Dhwani Batra; Jasmine Padilla; Justin Lee; Katie Dillon; Krista Queen; Kristen Knipe; Kristine Lacek; Mark Burroughs; Matthew Schmerer; Meghan Bentz; Mili Sheth; Peter Cook; Sam Shepard; Sarah Nobles; Shoshona Le; Suxiang Tong; Vivien Dugan; Yvette Unoarumhi |
| EPI_ISL_2788130, EPI_ISL_2788133, EPI_ISL_2788138, EPI_ISL_2788144, EPI_ISL_2788147 | IL Dept. of Public Health Springfield Laboratory | Centers for Disease Control and Prevention Division of Viral Diseases, Pathogen Discovery | Alison Laufer Halpin; Ben L. Rambo-Martin; Clinton R. Paden; Dakota Howard; Darlene Wagner; Dave Wentworth; Dhwani Batra; Jasmine Padilla; Justin Lee; Katie Dillon; Krista Queen; Kristen Knipe; Kristine Lacek; Mark Burroughs; Matthew Schmerer; Mili Sheth; Peter Cook; Sam Shepard; Sarah Nobles; Shoshona Le; Suxiang Tong; Vivien Dugan; Yvette Unoarumhi |
| EPI_ISL_2443175, EPI_ISL_2544602, EPI_ISL_2544652, EPI_ISL_2544664, EPI_ISL_2544674, EPI_ISL_2557221, EPI_ISL_2557242, EPI_ISL_2725042, EPI_ISL_2725070 | | | |
| see above | IN State Department of Health Laboratory Services | IN State Department of Health Laboratory Services | Brian Pope; Cassandra Campion; Jamie Yeadon; Kyle Brownlee; Lixia Liu; Mark Glazier; Melissa Hindenlang |
| EPI_ISL_6115483 | INSA | Instituto Nacional de Saude (INSA) | Borges et al |
| EPI_ISL_5645592, EPI_ISL_5645603, EPI_ISL_5645608, EPI_ISL_5645613, EPI_ISL_5645615, EPI_ISL_5645625, EPI_ISL_5645641, EPI_ISL_5645642, EPI_ISL_5645643, EPI_ISL_5645649, EPI_ISL_5645654, EPI_ISL_5645658, EPI_ISL_5645660, EPI_ISL_5645661, EPI_ISL_5645663, EPI_ISL_5645665, EPI_ISL_5645670, EPI_ISL_5645675, EPI_ISL_5645680, EPI_ISL_5645681, EPI_ISL_5645686, EPI_ISL_5645687, EPI_ISL_5645703, EPI_ISL_5645704, EPI_ISL_5645705, EPI_ISL_5645708, EPI_ISL_5645710, EPI_ISL_5645713, EPI_ISL_5645719, EPI_ISL_5645720, EPI_ISL_5645721, EPI_ISL_5645732, EPI_ISL_5645744, EPI_ISL_5645749, EPI_ISL_5645757, EPI_ISL_5645758, EPI_ISL_5645759, EPI_ISL_5645767, EPI_ISL_5645769, EPI_ISL_5645771, EPI_ISL_5645774, EPI_ISL_5645781, EPI_ISL_5645784, EPI_ISL_5645785, EPI_ISL_5645791, EPI_ISL_5645792, EPI_ISL_5645793, EPI_ISL_5645794, EPI_ISL_5645799, EPI_ISL_5645801, EPI_ISL_5645802, EPI_ISL_5645812, EPI_ISL_5645813, EPI_ISL_5645814, EPI_ISL_5645815, EPI_ISL_5645816, EPI_ISL_5645819, EPI_ISL_5645820, EPI_ISL_5645821, EPI_ISL_5645829, EPI_ISL_5645835, EPI_ISL_5645840, EPI_ISL_5645841, EPI_ISL_5645843, EPI_ISL_5645845, EPI_ISL_5645850, EPI_ISL_5645854, EPI_ISL_5645855, EPI_ISL_5645865, EPI_ISL_5645867, EPI_ISL_5645882, EPI_ISL_5645891, EPI_ISL_5645897, EPI_ISL_5645913, EPI_ISL_5645914, EPI_ISL_5645921, EPI_ISL_5645922, EPI_ISL_5645930, EPI_ISL_5645931, EPI_ISL_5645938, EPI_ISL_5645939, EPI_ISL_5645941, EPI_ISL_5645943, EPI_ISL_5645944, EPI_ISL_5645945, EPI_ISL_5645957, EPI_ISL_5645960, EPI_ISL_5646210, EPI_ISL_5646219, EPI_ISL_5646230, EPI_ISL_5646232, EPI_ISL_5646234, EPI_ISL_5646235, EPI_ISL_5646248, EPI_ISL_5646262, EPI_ISL_5646267, EPI_ISL_5646279, EPI_ISL_5646499, EPI_ISL_5646565, EPI_ISL_5646777, EPI_ISL_5646805, EPI_ISL_5646812, EPI_ISL_5646826, EPI_ISL_5646828, EPI_ISL_5646853, EPI_ISL_5647017, EPI_ISL_5647034 | | | |
| see above | INSIDE DIAGNOSTICOS | Instituto Butantan | Antonio Jorge Martins; Claudia Renata dos Santos Barros; David Schlesinger; Debora Botequio Moretti; Dimas Tadeu Covas; Elaine Cristina Marqueze; Elaine Vieira Santos; Evandra Strazza Rodrigues; Heidge Fukumasu; Jayme Augusto de Souza-Neto; José Salvatore Leister Patané; Luiz Alcantara; Luiz Lehmann Coutinho; Maria Carolina Elias; Maurício Lacerda Nogueira; Rafael dos Santos Bezerra; Raul Machado Neto; Rejane Maria Tommasini Grotto; Ricardo Haddad; Sandra Coccuzzo Sampaio Vessoni; Simone Kashima; Svetoslav Nanev Slavov; Vincent Louis Viala |
| EPI_ISL_2893559, EPI_ISL_2893603, EPI_ISL_2893605 | INSPI-CRN DE INFLUENZA Y OTROS VIRUS RESPIRATORIOS | NIC-INSPI | Alfredo Bruno; Domenica de Mora.; Jimmy Garcés; Johanna Laines; Lizbeth Patiño; Manuel Gonzalez; Maritza Olmedo; Michelle Páez |
| EPI_ISL_2953033, EPI_ISL_2953043, EPI_ISL_2953053 | IPF (Institut fr Pathologie/Landeskrankenhaus Feldkirch) | Elling and Cochella laboratories, IMBA/IMP | Alexander Stark; Ezgi Oezkan; Luisa Cochella and Ulrich Elling; Marcus Strobl; Olga Frank; Ramesh Yelagandula; Tanino Albanese |
| EPI_ISL_2774146, EPI_ISL_2774147, EPI_ISL_2774150, EPI_ISL_2774155, EPI_ISL_2774157, EPI_ISL_2774158, EPI_ISL_2774160 | | | |
| see above | IRCCS San Gallicano Dermatological Institute | IRCCS Regina Elena National Cancer Institute | Aldo Morrone; Alice Massacci; Arianna Mastrofrancesco; Eleonora Sperandio; Fabrizio Ensoli; Francesca De Nicola; Frauke Goeman; Fulvia Pimpinelli; Gennaro Ciliberto; Giovanna D'agosto; Giovanni Blandino; Giulia Orlandi; Ludovica Ciuffreda; Martina Diano; Matteo Pallocca; Maurizio Fanciulli; Sara Donzelli; Sara Petrolo; Serena Salvo; Silvia Paluzzi; di Domenico Enea Gino |
| EPI_ISL_2726937 | ISDH | Quantigen Biosciences | Paul J. Childress |
| EPI_ISL_3010167, EPI_ISL_3010169 | IVY3 Central Lab, Vanderbilt University Medical Center | Lauring Lab, University of Michigan, Department of Microbiology and Immunology | Valesano |
| EPI_ISL_2934974, EPI_ISL_3066923, EPI_ISL_3066925, EPI_ISL_3066974 | Idaho Bureau of Laboratories | Center for Global Health, University of New Mexico Health Sciences Center | Christopher Ball; Darrell Dinwiddie; Daryl Domman; Kurt Schwalm; Matthew Burns; Robert Voermans; Valerie Morley |
| EPI_ISL_3190989, EPI_ISL_3190993 | Idaho Bureau of Laboratories | Idaho Bureau of Laboratories | "R. Beukelman; Aimee Ceniseros; Christian Loera; Christopher Ball"; Matthew Charles Burns; Robert L. Voermans |
| EPI_ISL_2800711, EPI_ISL_2800716, EPI_ISL_2800721, EPI_ISL_2800723, EPI_ISL_2800748, EPI_ISL_2800826, EPI_ISL_2800830, EPI_ISL_2800849, EPI_ISL_2800850, EPI_ISL_2800857, EPI_ISL_2800861, EPI_ISL_2800862, EPI_ISL_2800865 | | | |
| see above | Illinois Department of Public Health | Illinois Department of Public Health - Chicago Lab | Ira Heimler; Joel Price; Vineet K. Dhiman |
| EPI_ISL_2403180, EPI_ISL_2598232, EPI_ISL_2598233, EPI_ISL_2598234, EPI_ISL_2598236, EPI_ISL_2598238, EPI_ISL_2598246, EPI_ISL_2774325, EPI_ISL_2774339, EPI_ISL_2774345, EPI_ISL_2774352, EPI_ISL_2774358, EPI_ISL_2774368, EPI_ISL_2774371 | | | |
| see above | Illinois Department of Public Health - Springfield Lab | Illinois Department of Public Health - Springfield Lab | Bryan Sim; Gordon McCall |
| EPI_ISL_2680934 | Imágenes Diagnósticas de Mazatlán | Microbial Genomics Laboratory | ; Alejandra García-Gasca; Alejandra Hernández-Terán; Alejandro Sánchez-Flores; Alfredo Herrera-Estrella; Alicia Ocaña-Mondragón; Andreu Comas-García; Angel Gustavo Salas-Lais; Antonio Loza Román; Bernardo Martínez-Miguel; Blanca Taboada; Brenda Irasema Maldonado-Meza; Bruno Gómez-Gil; Carla Ivón Herrera-Najera; Carlos F. Arias; Celia Boukadida; Clara Esperanza Santacruz-Tinoco; Concepción Grajales-Muñiz; Consorcio Mexicano de Vigilancia Genómica (CoViGen-Mex). Authors (in alphabetical order): Julio Elias Alvarado-Yaah; Cristóbal Cháidez-Quiróz; Célida Duque Molina; Célida Martínez- Rodríguez; Daniel Fregoso-Rueda; Daniel Lira Morales; Eduardo Becerril-Vargas; Fernando Fontove-Herrera; Fidencio Mejía-Nepomuceno; Francisco Pulido; Gloria Elena Espinosa-Ayala; Gloria María Molina-Salinas; Gloria Vazquez; Hector Esteban Paz-Juárez; Hector Montoya-Fuentes; Helen Haydee Fernanda Ramírez-Plascencia; Irvin González-López; Jean Pierre González; Jesús Hernández; Joel Armando Vázquez-Pérez.; Jorge Salas-Hernández; José Antonio Enciso-Moreno; José Arturo Martínez-Orozco; José Esteban Muñoz-Medina; José de Jesús Nuñez-Contreras; Juan Bautista Chale-Dzul; Julissa Enciso-Ibarra; Luis Alberto Ochoa-Carrera; Margarita Matías-Florentino; Mario Mújica-Sánchez; Marissa Perez-Garcia; María Guadalupe Santiago-Mauricio; María Guadalupe de Jesús Mireles-Rivera; Nelly Sélem-Mojica; Pavel Isa; Ricardo Ciria Merce; Ricardo Grande; Rosa María Gutiérrez Rios; Santiago Ávila-Ríos; Selene Zárate; Susana Lopez; Verónica Mata-Haro; Victor Eduardo García-Arias; Víctor Hugo Borja-Aburto |
| EPI_ISL_2715218, EPI_ISL_2715220, EPI_ISL_2715226 | Incyte Diagnostics Spokane | Seattle Flu Study | Amanda Adler; Barry R. Lutz; Benjamin Pelle; Brian Hiatt; Caitlin R. Wolf; Chris D. Frazar; Deborah A. Nickerson; Elisabeth Brandstetter; Erica Ryke; Geoff Melly; Helen Y. Chu; Janet A. Englund; Jay Shendure; Jover Lee; Kairsten Fay; Kirsten Lacombe; Lea M. Starita; Mark J. Rieder; Matthew Richardson; Matthew Thompson; Melissa Truong; Michael Boeckh; Michael Famulare; Misja Ilcisin; Peter D. Han; Philip Dykema; Romesh Gautom; Scott Lindquist; Thomas R. Sibley; Trevor Bedford |
| EPI_ISL_2713268, EPI_ISL_2713270, EPI_ISL_2713283, EPI_ISL_2713291, EPI_ISL_2992067 | Infectious Disease Program, Broad Institute of Harvard and MIT | Infectious Disease Program, Broad Institute of Harvard and MIT | A.E.; Adams, G.; Anahtar, M.; B.L.; B.W.; Bauer, M.; Birren; Branda, J.; Carter, A.; Cerrato, F.; Chaluvadi, S.; Chapman; Cusick, C.; D.J.; DeRuff, K.; Flowers, K.; Gallagher, G.; Gladden-Young, A.; Gnirke, A.; Harris, J.; J.E.; K.J.; LaRocque, R.; Lagerborg, K.; Lemieux; Lin; Loreth, C.; MacInnis; Neumann, A.; Normandin, E.; P.C.; Park; Pierce, V.; Reilly, S.; Rosenberg, E.; Rudy, M.; Ryan, E.; S.B.; Sabeti; Shaw, B.; Siddle; Slater, D.; Smole, S.; Tomkins-Tinch, C.; Turbett, S. |
| EPI_ISL_2557614, EPI_ISL_2653011, EPI_ISL_2653014, EPI_ISL_2653015, EPI_ISL_2653035, EPI_ISL_2653038, EPI_ISL_2653049, EPI_ISL_2653056, EPI_ISL_2653059, EPI_ISL_2686889, EPI_ISL_2686890, EPI_ISL_2686893 | | | |

| | | | |
|---|---|---|---|
| see above | Infinity Biologix | Centers for Disease Control and Prevention Division of Viral Diseases, Pathogen Discovery | Adrian Paskey; Benjamin Rambo-Martin; Chirayu Goswami; Christian Bixby; Christopher Gulvick; Clinton R. Paden; Dakota Howard; Darlene Wagner; Dhwani Batra; Duncan MacCannell; Jason Caravas; Jonathan Schultz; Kara Moser; Matthew Schmerer; Peter W. Cook; Robin Grimwood; Russ Hager; Scott Sammons; Shatavia Morrison; Yihe Wang; Yvette Unoarumhi |
| EPI_ISL_2549253 | Institut de Virologie du CHU de Strasbourg | Institut de Virologie du CHU de Strasbourg | Fafi-Kremer Samira; Gallais Floriane; Gantner Pierre; Laugel Elodie; Solis Morgane; Velay Aurélie; Wendling Marie-Josée |
| EPI_ISL_2522055 | Institute of Microbiology and Immunology, Faculty of Medicine, University of Ljubljana | Institute of Microbiology and Immunology, Faculty of Medicine, University of Ljubljana | Alen Suljič; Andraž Celar; Cene Skubic; Dominika Šturm; Doroteja Vlaj; Mario Poljak; Matic Brvar; Miša Korva; Patricija Pozvek; Samo Zakotnik; Tatjana Avšič – Županc; Tomaž Mark Zorec; Špela Pleh |
| EPI_ISL_3316157, EPI_ISL_3316158, EPI_ISL_3316193, EPI_ISL_3316197 | Instituto Adolfo Lutz - Regional de Bauru | Instituto Adolfo Lutz, Interdiciplinary Procedures Center, Strategic Laboratory | Caio Vinicius Dias Lopes; Claudia Regina Gonçalves; Claudio Tavares Sacchi; Karoline Rodrigues Campos; Leonardo Tadeu de Araujo; Marlon Benedito Nascomento Santos |
| EPI_ISL_3259696 | Instituto Adolfo Lutz - Regional de Campinas | Instituto Adolfo Lutz, Interdiciplinary Procedures Center, Strategic Laboratory | Caio Vinicius Dias Lopes; Claudia Regina Gonçalves; Claudio Tavares Sacchi; Erica Valessa Ramos Gomes; Karoline Rodrigues Campos; Leonardo Tadeu de Araujo; Marlon Benedito Nascomento Santos |
| EPI_ISL_3717850, EPI_ISL_3717852, EPI_ISL_3717887 | Instituto Adolfo Lutz - Regional de Marilia | Instituto Adolfo Lutz, Interdiciplinary Procedures Center, Strategic Laboratory | Caio Vinicius Dias Lopes; Claudia Regina Gonçalves; Claudio Tavares Sacchi; Karoline Rodrigues Campos; Leonardo Tadeu de Araujo; Marlon Benedito Nascimento Santos |
| EPI_ISL_5640217 | Instituto Adolfo Lutz - Regional de Rio Claro | Instituto Adolfo Lutz, Interdiciplinary Procedures Center, Strategic Laboratory | Caio Vinicius Dias Lopes; Claudia Regina Gonçalves; Claudio Tavares Sacchi; Karoline Rodrigues Campos; Leonardo Tadeu de Araujo; Marlon Benedito Nascimento Santos |
| EPI_ISL_4888042, EPI_ISL_4888043 | Instituto Adolfo Lutz Central | Instituto Adolfo Lutz, Interdiciplinary Procedures Center, Strategic Laboratory | Claudio Tavares Sacchi; Karoline Rodrigues Campos; Leonardo Tadeu de Araujo; Marlon Benedito Nascimento Santos |
| EPI_ISL_2648116, EPI_ISL_2648117, EPI_ISL_2777212, EPI_ISL_2777213, EPI_ISL_2777217 | Instituto Nacional de Enfermedades Respiratorias (INER) | Centro de Investigación en Enfermedades Infecciosas (CIENI), Instituto Nacional de Enfermedades Respiratorias (INER) | ; Alejandra Garcia-Gasca; Alejandra Hernández-Terán; Alejandro Sanchez-Flores; Alfredo Herrera-Estrella; Alicia Ocaña-Mondragón; Andreu Comas-García; Angel Gustavo Salas-Lais; Antonio Loza Román; Bernardo Martínez-Miguel; Blanca Taboada; Brenda Irasema Maldonado-Meza; Bruno Gomez-Gil; Carla Ivón Herrera-Najera; Carlos F. Arias; Celia Boukadida; Clara Esperanza Santacruz-Tinoco; Concepción Grajales-Muñiz; Consorcio Mexicano de Vigilancia Genómica (CoViGen-Mex). Authors (in alphabetical order): Julio Elias Alvarado-Yaah; Cristóbal Cháidez-Quiróz; Célida Duque Molina; Célida Martínez- Rodríguez; Daniel Fregoso-Rueda; Daniel Lira Morales; Eduardo Becerril-Vargas; Fernando Fontove-Herrera; Fidencio Mejía-Nepomuceno; Francisco Pulido; Gloria Elena Espinosa-Ayala; Gloria María Molina-Salinas; Gloria Vazquez; Hector Esteban Paz-Juárez; Hector Montoya-Fuentes; Helen Haydee Fernanda Ramírez-Plascencia; Irvin González-López; Jean Pierre González; Joel Armando Vázquez-Pérez.; Jorge Salas-Hernández; José Antonio Enciso-Moreno; José Arturo Martínez-Orozco; José Esteban Muñoz-Medina; José de Jesús Nuñez-Contreras; Juan Bautista Chale-Dzul; Julissa Enciso-Ibarra; Kathia Elizabeth Tapia-Diaz; Luis Alberto Ochoa-Carrera; Margarita Matías-Florentino; Mario Mújica-Sánchez; Marissa Pérez-García; María Guadalupe Santiago-Mauricio; María Guadalupe de Jesús Mireles-Rivera; Nelly Sélem-Mojica; Pavel Isa; Ricardo Ciria Merce; Ricardo Grande; Rosa María Gutierrez Rios; Santiago Ávila-Ríos; Selene Zárate; Susana Lopez; Victor Eduardo Garcia-Arias; Víctor Hugo Borja-Aburto |
| EPI_ISL_2843909, EPI_ISL_2843925, EPI_ISL_2843926, EPI_ISL_2843927, EPI_ISL_2843928, EPI_ISL_2843929, EPI_ISL_2843930, EPI_ISL_2843931, EPI_ISL_2843932, EPI_ISL_2843933, EPI_ISL_2843934, EPI_ISL_2843935, EPI_ISL_2843936, EPI_ISL_2843937, EPI_ISL_2843938, EPI_ISL_2843939, EPI_ISL_2843940, EPI_ISL_2843941, EPI_ISL_2843942, EPI_ISL_2843943, EPI_ISL_2843944, EPI_ISL_2843945, EPI_ISL_2843946, EPI_ISL_2843947, EPI_ISL_2843949, EPI_ISL_2843950, EPI_ISL_2843951, EPI_ISL_2843952, EPI_ISL_2843953, EPI_ISL_2843954, EPI_ISL_2843955, EPI_ISL_2843956, EPI_ISL_2843957, EPI_ISL_2843958, EPI_ISL_2843959, EPI_ISL_2843960, EPI_ISL_2843961, EPI_ISL_2843962, EPI_ISL_2843963, EPI_ISL_2843964, EPI_ISL_2843965, EPI_ISL_2843967, EPI_ISL_2843968, EPI_ISL_2843969, EPI_ISL_2843970, EPI_ISL_2843971, EPI_ISL_2843972, EPI_ISL_2843973, EPI_ISL_2843974, EPI_ISL_2843975, EPI_ISL_2843976, EPI_ISL_2843977, EPI_ISL_2843978, EPI_ISL_2843979, EPI_ISL_2843980, EPI_ISL_2843981, EPI_ISL_2843983, EPI_ISL_2843984, EPI_ISL_2843985, EPI_ISL_2843986, EPI_ISL_2843987, EPI_ISL_2843988, EPI_ISL_2843989, EPI_ISL_2843990, EPI_ISL_2843991, EPI_ISL_2843992, EPI_ISL_2843993, EPI_ISL_2843994, EPI_ISL_2843995, EPI_ISL_2843996, EPI_ISL_2843997, EPI_ISL_2843998, EPI_ISL_2843999, EPI_ISL_2844000, EPI_ISL_2844001, EPI_ISL_2844002, EPI_ISL_2844003, EPI_ISL_2844004, EPI_ISL_2844005, EPI_ISL_2844006, EPI_ISL_2844007, EPI_ISL_2844008, EPI_ISL_2844009, EPI_ISL_2844010, EPI_ISL_2844011, EPI_ISL_2844012, EPI_ISL_2844013, EPI_ISL_2844014, EPI_ISL_2844015, EPI_ISL_2844016, EPI_ISL_2844017, EPI_ISL_2844018, EPI_ISL_2844019, EPI_ISL_2844020, EPI_ISL_2844021, EPI_ISL_2844022, EPI_ISL_2844023, EPI_ISL_2844024, EPI_ISL_2844025, EPI_ISL_2844026, EPI_ISL_2844027, EPI_ISL_2844028, EPI_ISL_2844029, EPI_ISL_2844030, EPI_ISL_2844031, EPI_ISL_2844032, EPI_ISL_2844033, EPI_ISL_2844034, EPI_ISL_2844035, EPI_ISL_2844036, EPI_ISL_2844037, EPI_ISL_2844038, EPI_ISL_2844039, EPI_ISL_2844040, EPI_ISL_2844041, EPI_ISL_2844044, EPI_ISL_2844045, EPI_ISL_2844046, EPI_ISL_2844047, EPI_ISL_2844048, EPI_ISL_2844050, EPI_ISL_2844051, EPI_ISL_2844052, EPI_ISL_2844053, EPI_ISL_2844054, EPI_ISL_2844055, EPI_ISL_2844056, EPI_ISL_3118865, EPI_ISL_3118868, EPI_ISL_3118871, EPI_ISL_3118872, EPI_ISL_3118874, EPI_ISL_3118875, EPI_ISL_3118878, EPI_ISL_3118879, EPI_ISL_3118882, EPI_ISL_3118883, EPI_ISL_3118884, EPI_ISL_3118886, EPI_ISL_3244536, EPI_ISL_3244537, EPI_ISL_3244541, EPI_ISL_3244542, EPI_ISL_3244543, EPI_ISL_3244544, EPI_ISL_3244545, EPI_ISL_3244546, EPI_ISL_3244547, EPI_ISL_3244548, EPI_ISL_3244549, EPI_ISL_3244550, EPI_ISL_3244551, EPI_ISL_3244552, EPI_ISL_3244553, EPI_ISL_3244554, EPI_ISL_3244555, EPI_ISL_3244556, EPI_ISL_3244557, EPI_ISL_3244558, EPI_ISL_3244559, EPI_ISL_3244560, EPI_ISL_3244561, EPI_ISL_3244562, EPI_ISL_3244564, EPI_ISL_3244566, EPI_ISL_3244567 | | | |
| see above | Instituto de Biotecnologia - UNESP-Botucatu-SP | Instituto de Biotecnologia - UNESP-Botucatu-SP | Cecília Artico Banho; Cíntia Bittar; Fábio Sossai Possebon; Guilherme Campos; Helena Lage Ferreira; Jorge A. Petroli Marchesi; João Pessoa Araújo Jr.; Leila Sabrina Ullmann; Lívia Sacchetto; Maisa C. Pereira Parra; Marília Moraes; Maurício L. Nogueira; Paula Rahal; Paulo Inacio da Costa |
| EPI_ISL_3236442, EPI_ISL_3236443 | Instituto de Medicina Tropical & Salud Global Universidad Iberoamericana | Grubaugh Lab - Yale School of Public Health | Alejandaro Vallejo Degaudenzi; Anderson Brito; Annie Watkins; Chaney Kalinich; Chantal Vogels; Elisa Contreras; Esperanza Mendoza; Isabel Ott; Jessica Rothman; Joseph Fauver; Kendall Billig; Mallery Breban; Mary Petrone; Nathan Grubaugh; Robert Paulino-Ramirez; Tara Alpert; Tobias Koch; Victor Virgilio Calderon |
| EPI_ISL_2550983, EPI_ISL_2550985, EPI_ISL_2551109, EPI_ISL_2551114, EPI_ISL_2551117, EPI_ISL_2551127, EPI_ISL_2551139, EPI_ISL_2551142, EPI_ISL_2551145, EPI_ISL_2551148 | | | |
| see above | Istituto Zooprofilattico Sperimentale del Mezzogiorno | TIGEM | Antonio Grimaldi Patrizia Annunziata Francesco Panariello Biancamaria Pierri Claudia Tiberio Teresa Giuliano Valentina Bouche Chiara Colantuono Maria Concetta Cuomo Denise Di Concilio Lucio Di Filippo Anna Manfredi Marcello Salvi Antonio Limone Luigi Atripaldi Pellegrino Cerino Andrea Ballabio Davide Cacchiarelli |
| EPI_ISL_2641746, EPI_ISL_2641750, EPI_ISL_2641751, EPI_ISL_2641753, EPI_ISL_2641754, EPI_ISL_2641771, EPI_ISL_2641775, EPI_ISL_2641816, EPI_ISL_2641817, EPI_ISL_2641818, EPI_ISL_2641820 | | | |
| see above | Istituto Zooprofilattico Sperimentale del Mezzogiorno | Telethon Institute of Genetics and Medicine (TIGEM) | Antonio Grimaldi Patrizia Annunziata Francesco Panariello Biancamaria Pierri Claudia Tiberio Teresa Giuliano Valentina Bouche Chiara Colantuono Maria Concetta Cuomo Denise Di Concilio Lucio Di Filippo Anna Manfredi Marcello Salvi Antonio Limone Luigi Atripaldi Pellegrino Cerino Andrea Ballabio Davide Cacchiarelli |
| EPI_ISL_2611997 | Istituto Zooprofilattico Sperimentale della Puglia e della Basilicata | Istituto Zooprofilattico Sperimentale della Puglia e della Basilicata | Bianco A.; Capozzi L.; Del Sambro L.; Difato L.; Galante D.; Pace L.; Parisi A.; Pennuzzi G.; Simone D. |
| EPI_ISL_2462990, EPI_ISL_2462991 | Jessa | Jessa | Berden et al. on behalf of the Jessa_cmdLab |
| EPI_ISL_2692081, EPI_ISL_2692084 | John H Stroger Jr Hospital of Cook County | RIPHL at Rush University Medical Center | Ah-Young Sung; Felix Araujo Perez; Kevin Kunstman; Laura Furtado; Marieta Hyde; Max Kolton; Sharon Welbel; Stefan Green |
| EPI_ISL_2690047, EPI_ISL_2690054 | KS Health and Environmental Laboratories | Centers for Disease Control and Prevention Division of Viral Diseases, Pathogen Discovery | Alison Laufer Halpin; Ben L. Rambo-Martin; Clinton R. Paden; Dakota Howard; Darlene Wagner; Dave Wentworth; Dhwani Batra; Jasmine Padilla; Justin Lee; Katie Dillon; Krista Queen; Kristen Knipe; Kristine Lacek; Mark Burroughs; Matthew Schmerer; Mili Sheth; Peter Cook; Sam Shepard; Sarah Nobles; Shoshona Le; Suxiang Tong; Vivien Dugan; Yvette Unoarumhi |
| EPI_ISL_2864593, EPI_ISL_2864594, EPI_ISL_2864596, EPI_ISL_2864598, EPI_ISL_2864601, EPI_ISL_2864603, EPI_ISL_2864605, EPI_ISL_2864610, EPI_ISL_2864612, EPI_ISL_2864654, EPI_ISL_2864656, EPI_ISL_2864666, EPI_ISL_2864668, EPI_ISL_2864670, EPI_ISL_2864672, EPI_ISL_2864693, EPI_ISL_2864694, EPI_ISL_2864698, EPI_ISL_2864725, EPI_ISL_2864731, EPI_ISL_2864735, EPI_ISL_2877046, EPI_ISL_2877443, EPI_ISL_2877445, EPI_ISL_2877447, EPI_ISL_2877506, EPI_ISL_2877522, EPI_ISL_2877524, EPI_ISL_2877526, EPI_ISL_2964776, EPI_ISL_2964777, EPI_ISL_2964778, EPI_ISL_2964909 | | | |
| see above | KU Leuven, Rega Institute, Clinical and Epidemiological Virology | KU Leuven, Rega Institute, Clinical and Epidemiological Virology | Bert Vanmechelen; Joan Marti-Carerras; Piet Maes; Tony Wawina-Bokalanga |
| EPI_ISL_3561384, EPI_ISL_3561823, EPI_ISL_3561852, EPI_ISL_3561917, EPI_ISL_3562091, EPI_ISL_3939815, EPI_ISL_3939817, EPI_ISL_3939821, EPI_ISL_3939835, EPI_ISL_3940785, EPI_ISL_3940787, EPI_ISL_3940797, EPI_ISL_3940808, EPI_ISL_3940811, EPI_ISL_3940817, EPI_ISL_3940821, EPI_ISL_3940822, EPI_ISL_3940827, EPI_ISL_3940833, EPI_ISL_3940842, EPI_ISL_3940849, EPI_ISL_3940850, EPI_ISL_3940853, EPI_ISL_3940909, EPI_ISL_3940915, EPI_ISL_3940938, EPI_ISL_3941001, EPI_ISL_3941031, EPI_ISL_3941046, EPI_ISL_3941052, EPI_ISL_3941054, EPI_ISL_3941067, EPI_ISL_3941069, EPI_ISL_3941073, EPI_ISL_4102660, EPI_ISL_4102661 | | | |
| see above | Kaiser Permanente NW Reginal Lab | OHSU MM Lab | Amber Halse; Jeannine Lama; Xuan Qin; Yun Wu |
| EPI_ISL_2754010 | Kaiser Regional | Santa Clara County Public Health Laboratory | Santa Clara County Public Health Department |
| EPI_ISL_2443131, EPI_ISL_2455455 | Kansas Health and Environmental Lab | Kansas Health and Environmental Lab | Ben Olsen; Jonathan Barnell; Katherine Wiggins; Mike Grose; and Phil Adam |
| EPI_ISL_2898793 | Karolinska University Hospital Huddinge | Karolinska University Hospital | Annelie Bjerkner; Isak Sylvin; Jan Albert; Karolina Ininbergs; Lina Guerra Blomqvist; Lynda Eneh; Martin Ekman; Martina Wahlund; Robert Dyrdak; Sandra Broddesson; Tanja Normark; Tobias Allander; Valtteri Wirta; Zhibing Yun |
| EPI_ISL_4080603, EPI_ISL_4080608 | Kentucky State Public Health Lab | Kentucky State Public Health Lab | Anjayya Tummala; Chitralipi Kattunga; Joshua Tobias; Karim George; Matthew Johnson; Rachel Zinner; Rhonda Lucas; Vaneet Arora; William Grooms |
| EPI_ISL_2627495 | Klinisch Laboratorium ZNA | Klinisch Laboratorium ZNA | Verstrepen et al. |
| EPI_ISL_5646758, EPI_ISL_5647142, EPI_ISL_5647166, EPI_ISL_5647194 | LABORATORIO MUNICIPAL DE ANALISES CLINICAS DE RIO CLARO | Instituto Butantan | Antonio Jorge Martins; Claudia Renata dos Santos Barros; David Schlesinger; Debora Botequio Moretti; Dimas Tadeu Covas; Elaine Cristina Marqueze; Elaine Vieira Santos; Evandra Strazza Rodrigues; Heidge Fukumasu; Jayme Augusto de Souza-Neto; José Salvatore Leister Patané; Luiz Alcantara; Luiz Lehmann Coutinho; Maria Carolina Elias; Maurício Lacerda Nogueira; Rafael dos Santos Bezerra; Raul Machado Neto; Rejane Maria Tommasini Grotto; Ricardo Haddad; Sandra Coccuzzo Sampaio Vessoni; Simone Kashima; Svetoslav Nanev Slavov; Vincent Louis Viala |
| EPI_ISL_5645606, EPI_ISL_5645674, EPI_ISL_5645689, EPI_ISL_5645782, EPI_ISL_5646529 | LABORATORIO MUNICIPAL DE PIRACICABA | Instituto Butantan | Antonio Jorge Martins; Claudia Renata dos Santos Barros; David Schlesinger; Debora Botequio Moretti; Dimas Tadeu Covas; Elaine Cristina Marqueze; Elaine Vieira Santos; Evandra Strazza Rodrigues; Heidge Fukumasu; Jayme Augusto de Souza-Neto; José Salvatore Leister Patané; Luiz Alcantara; Luiz Lehmann Coutinho; Maria Carolina Elias; Maurício Lacerda Nogueira; Rafael dos Santos Bezerra; Raul Machado Neto; Rejane Maria Tommasini Grotto; Ricardo Haddad; Sandra Coccuzzo Sampaio Vessoni; Simone Kashima; Svetoslav Nanev Slavov; Vincent Louis Viala |
| EPI_ISL_3404845, EPI_ISL_3404851, EPI_ISL_3404852, EPI_ISL_3404853, EPI_ISL_3404854, EPI_ISL_3404855, EPI_ISL_3404860 | | | |
| see above | LACEN | LGBio (Laboratorio de Genetica & Biodiversidade) | Alex Honda Bernardes; Amanda Alves de Melo; Aparecido Divino da Cruz; Cintia Pelegrineti Targueta de Azevedo Brito; Daniela de Melo e Silva; Elisangela de Paula Silveira Lacerda; Juliana Santana de Curcio; Luiz Augusto Pereira; Marc Alexandre Duarte Gigonzac; Mariana Pires de Campos Telles; Ramilla dos Santos Braga; Renata de Oliveira Dias; Rhewter Nunes; Thais Cidália Vieira Gigonzac; Thais Guimarães Castro; Thays Millena Alves Pedroso |
| EPI_ISL_6721500, EPI_ISL_6721809 | LACEN - Laboratório Central de Saúde Pública de Pernambuco | Evandro Chagas Institute | Delana Andreza Melo Bezerra; Dielle Monteiro Teixeira; Jedson Ferreira Cardoso; Jessylene de Almeida Ferreira; Kenny da Costa Pinheiro; Luana Soares Barbagelata; Luana da Silva Soares; Mirleide Cordeiro dos Santos; Patrícia dos Santos Lobo; Rayssa Layna da Silva Bedran; Sandro Patroca da Silva |
| EPI_ISL_6721542 | LACEN - Laboratório Central de Saúde Pública do Amazonas | Evandro Chagas Institute | Delana Andreza Melo Bezerra; Dielle Monteiro Teixeira; Jedson Ferreira Cardoso; Jessylene de Almeida Ferreira; Kenny da Costa Pinheiro; Luana Soares Barbagelata; Luana da Silva Soares; Mirleide Cordeiro dos Santos; Patrícia dos Santos Lobo; Rayssa Layna da Silva Bedran; Sandro Patroca da Silva |

| | | | |
|---|---|---|---|
| EPI_ISL_6721776 | LACEN - Laboratório Central de Saúde Pública do Rio Grande do Norte | Evandro Chagas Institute | Delana Andreza Melo Bezerra; Dielle Monteiro Teixeira; Jedson Ferreira Cardoso; Jessylene de Almeida Ferreira; Kenny da Costa Pinheiro; Luana Soares Barbagelata; Luana da Silva Soares; Mirleide Cordeiro dos Santos; Patrícia dos Santos Lobo; Rayssa Layna da Silva Bedran; Sandro Patroca da Silva |
| EPI_ISL_6840903, EPI_ISL_6840923, EPI_ISL_6840953 | LACEN do Estado de Rondonia | Instituto Adolfo Lutz, Interdiciplinary Procedures Center, Strategic Laboratory | Ariadne Ferreira Amarante; Caio Vinicius Dias Lopes; Claudia Regina Gonçalves; Claudio Tavares Sacchi; Karoline Rodrigues Campos; Marlon Benedito Nascimento Santos |
| EPI_ISL_5193543, EPI_ISL_5193544, EPI_ISL_5193545, EPI_ISL_5193546, EPI_ISL_5193584, EPI_ISL_5193634, EPI_ISL_5193635 | | | |
| see above | LACEN/RR | Laboratorio de Ecologia de Doencas Transmissiveis na Amazonia, Instituto Leonidas e Maria Deane - Fiocruz Amazonia | André Corado; Cátia Alexandra Ribeiro Meneses; Felipe Naveca; Fernanda Nascimento; George Silva; Ilma de Aguiar Antony; Janis Cassimiro Lopes; Luciana Gonçalves; Maria Júlia Brandão; Matilde Mejía; Pricila Carlos Veloso; Rodrigo Melo Maito; Valdinete Nascimento; Victor Souza |
| EPI_ISL_3010761, EPI_ISL_3010767, EPI_ISL_3010768, EPI_ISL_3010800, EPI_ISL_3010801, EPI_ISL_3010805, EPI_ISL_3010806, EPI_ISL_3010817, EPI_ISL_3010823, EPI_ISL_3010824, EPI_ISL_3010825, EPI_ISL_3010842, EPI_ISL_3010859, EPI_ISL_3010866, EPI_ISL_3010870, EPI_ISL_3010871, EPI_ISL_3010881, EPI_ISL_3010889, EPI_ISL_3010890, EPI_ISL_3010900, EPI_ISL_3010905, EPI_ISL_3010907, EPI_ISL_3010909, EPI_ISL_3010912, EPI_ISL_3010917, EPI_ISL_3010920, EPI_ISL_3010928, EPI_ISL_3010931, EPI_ISL_3134946 | | | |
| see above | LATE - Laboratório de Técnicas Especiais - Hospital Israelita Albert Einstein | LATE - Laboratório de Técnicas Especiais - Hospital Israelita Albert Einstein | Alexandre Hideaki Takara; Ana Paula Moreira Salles; Anelisie da Silva Santos; Deyvid Amgarten; Erick Gustavo Dorlass; Fernanda de Mello Malta; João Renato Rebello Pinho; Marcio Anunciacao Menezes; Pedro Henrique Sebe Rodrigues; Raquel Riyuzo |
| EPI_ISL_4666426 | LDSP | Laboratorio Departamental de Salud Publica de Antioquia | Andres F. Cardona-Rios; Cristian Arbey Velarde Hoyos; Gloria Isabel Escobar; Idabely Betancur Ortiz; Juan P. Hernandez-Ortiz; Maria Stella López |
| EPI_ISL_2674699 | LESP Chihuahua | Instituto de Diagnostico y Referencia Epidemiologicos (INDRE) | Abril Rodriguez-Maldonado; Ariadna Medina-Benitez; Claudia Wong-Arambula; Ernesto Ramirez-Gonzalez.; Gisela Barrera-Badillo; Irma Lopez-Martinez; Joaquin Quiroz-Mercado; Lucia Hernandez-Rivas; Natividad Cruz-Ortiz; Sergio Rangel-Guerrero; Tatiana Nunez-Garcia; Vanessa Rivero-Arredondo |
| EPI_ISL_2937770 | LESP Ciudad de Mexico | Instituto de Diagnostico y Referencia Epidemiologicos (INDRE) | Abril Rodriguez-Maldonado; Ariadna Medina-Benitez; Claudia Wong-Arambula; Ernesto Ramirez-Gonzalez.; Gisela Barrera-Badillo; Irma Lopez-Martinez; Joaquin Quiroz-Mercado; Lucia Hernandez-Rivas; Maribel Gonzalez-Villa; Natividad Cruz-Ortiz; Sergio Rangel-Guerrero; Tatiana Nunez-Garcia; Vanessa Rivero-Arredondo |
| EPI_ISL_2674642 | LESP Durango | Instituto de Diagnostico y Referencia Epidemiologicos (INDRE) | Abril Rodriguez-Maldonado; Ariadna Medina-Benitez; Claudia Wong-Arambula; Ernesto Ramirez-Gonzalez.; Gisela Barrera-Badillo; Irma Lopez-Martinez; Joaquin Quiroz-Mercado; Lucia Hernandez-Rivas; Natividad Cruz-Ortiz; Sergio Rangel-Guerrero; Tatiana Nunez-Garcia; Vanessa Rivero-Arredondo |
| EPI_ISL_2674695 | LESP Guanajuato | Instituto de Diagnostico y Referencia Epidemiologicos (INDRE) | Abril Rodriguez-Maldonado; Ariadna Medina-Benitez; Claudia Wong-Arambula; Ernesto Ramirez-Gonzalez.; Gisela Barrera-Badillo; Irma Lopez-Martinez; Joaquin Quiroz-Mercado; Lucia Hernandez-Rivas; Natividad Cruz-Ortiz; Sergio Rangel-Guerrero; Tatiana Nunez-Garcia; Vanessa Rivero-Arredondo |
| EPI_ISL_2779050 | LESP Guerrero | Instituto de Diagnostico y Referencia Epidemiologicos (INDRE) | Abril Rodriguez-Maldonado; Ariadna Medina-Benitez; Claudia Wong-Arambula; Ernesto Ramirez-Gonzalez.; Gisela Barrera-Badillo; Irma Lopez-Martinez; Joaquin Quiroz-Mercado; Lucia Hernandez-Rivas; Maribel Gonzalez-Villa; Natividad Cruz-Ortiz; Sergio Rangel-Guerrero; Tatiana Nunez-Garcia; Vanessa Rivero-Arredondo |
| EPI_ISL_2674671 | LESP Hidalgo | Instituto de Diagnostico y Referencia Epidemiologicos (INDRE) | Abril Rodriguez-Maldonado; Ariadna Medina-Benitez; Claudia Wong-Arambula; Ernesto Ramirez-Gonzalez.; Gisela Barrera-Badillo; Irma Lopez-Martinez; Joaquin Quiroz-Mercado; Lucia Hernandez-Rivas; Natividad Cruz-Ortiz; Sergio Rangel-Guerrero; Tatiana Nunez-Garcia; Vanessa Rivero-Arredondo |
| EPI_ISL_3033575 | LESP Michoacan | Instituto de Diagnostico y Referencia Epidemiologicos (INDRE) | Abril Rodriguez-Maldonado; Ariadna Medina-Benitez; Claudia Wong-Arambula; Ernesto Ramirez-Gonzalez.; Gisela Barrera-Badillo; Irma Lopez-Martinez; Joaquin Quiroz-Mercado; Lucia Hernandez-Rivas; Maribel Gonzalez-Villa; Natividad Cruz-Ortiz; Sergio Rangel-Guerrero; Tatiana Nunez-Garcia; Vanessa Rivero-Arredondo |
| EPI_ISL_2674673, EPI_ISL_2674700 | LESP Puebla | Instituto de Diagnostico y Referencia Epidemiologicos (INDRE) | Abril Rodriguez-Maldonado; Ariadna Medina-Benitez; Claudia Wong-Arambula; Ernesto Ramirez-Gonzalez.; Gisela Barrera-Badillo; Irma Lopez-Martinez; Joaquin Quiroz-Mercado; Lucia Hernandez-Rivas; Natividad Cruz-Ortiz; Sergio Rangel-Guerrero; Tatiana Nunez-Garcia; Vanessa Rivero-Arredondo |
| EPI_ISL_2674655, EPI_ISL_2779051, EPI_ISL_2779052, EPI_ISL_2779176, EPI_ISL_2779177, EPI_ISL_2937765 | LESP Quintana Roo | Instituto de Diagnostico y Referencia Epidemiologicos (INDRE) | Abril Rodriguez-Maldonado; Ariadna Medina-Benitez; Claudia Wong-Arambula; Ernesto Ramirez-Gonzalez.; Gisela Barrera-Badillo; Irma Lopez-Martinez; Joaquin Quiroz-Mercado; Lucia Hernandez-Rivas; Maribel Gonzalez-Villa; Natividad Cruz-Ortiz; Sergio Rangel-Guerrero; Tatiana Nunez-Garcia; Vanessa Rivero-Arredondo |
| EPI_ISL_2674648, EPI_ISL_2674649 | LESP Sonora | Instituto de Diagnostico y Referencia Epidemiologicos (INDRE) | Abril Rodriguez-Maldonado; Ariadna Medina-Benitez; Claudia Wong-Arambula; Ernesto Ramirez-Gonzalez.; Gisela Barrera-Badillo; Irma Lopez-Martinez; Joaquin Quiroz-Mercado; Lucia Hernandez-Rivas; Natividad Cruz-Ortiz; Sergio Rangel-Guerrero; Tatiana Nunez-Garcia; Vanessa Rivero-Arredondo |
| EPI_ISL_2937814 | LESP Tabasco | Instituto de Diagnostico y Referencia Epidemiologicos (INDRE) | Abril Rodriguez-Maldonado; Ariadna Medina-Benitez; Claudia Wong-Arambula; Ernesto Ramirez-Gonzalez.; Gisela Barrera-Badillo; Irma Lopez-Martinez; Joaquin Quiroz-Mercado; Lucia Hernandez-Rivas; Maribel Gonzalez-Villa; Natividad Cruz-Ortiz; Sergio Rangel-Guerrero; Tatiana Nunez-Garcia; Vanessa Rivero-Arredondo |
| EPI_ISL_2779060, EPI_ISL_2779062, EPI_ISL_2779063, EPI_ISL_2779064, EPI_ISL_2779065, EPI_ISL_2858997, EPI_ISL_2858999 | | | |
| see above | LESP Tamaulipas | Instituto de Diagnostico y Referencia Epidemiologicos (INDRE) | Abril Rodriguez-Maldonado; Ariadna Medina-Benitez; Claudia Wong-Arambula; Ernesto Ramirez-Gonzalez.; Gisela Barrera-Badillo; Irma Lopez-Martinez; Joaquin Quiroz-Mercado; Lucia Hernandez-Rivas; Maribel Gonzalez-Villa; Natividad Cruz-Ortiz; Sergio Rangel-Guerrero; Tatiana Nunez-Garcia; Vanessa Rivero-Arredondo |
| EPI_ISL_2674686, EPI_ISL_2674704, EPI_ISL_2779066, EPI_ISL_2779067, EPI_ISL_2779068, EPI_ISL_2779069, EPI_ISL_2779070, EPI_ISL_2779071, EPI_ISL_2779072, EPI_ISL_2779073, EPI_ISL_2779074, EPI_ISL_2779075, EPI_ISL_2779076, EPI_ISL_2779077, EPI_ISL_2779078, EPI_ISL_2779079, EPI_ISL_2779080, EPI_ISL_2779081, EPI_ISL_2779082, EPI_ISL_2779083, EPI_ISL_2779084, EPI_ISL_2779085, EPI_ISL_2779086, EPI_ISL_2779087, EPI_ISL_2779088, EPI_ISL_2779089, EPI_ISL_2779090, EPI_ISL_2779091, EPI_ISL_2779092, EPI_ISL_2779093, EPI_ISL_2779094, EPI_ISL_2779095, EPI_ISL_2779096, EPI_ISL_2779097, EPI_ISL_2779098, EPI_ISL_2779099, EPI_ISL_2779100, EPI_ISL_2779101, EPI_ISL_2779102, EPI_ISL_2779103, EPI_ISL_2779104, EPI_ISL_2779105, EPI_ISL_2779106, EPI_ISL_2779107, EPI_ISL_2779108, EPI_ISL_2779109, EPI_ISL_2779110, EPI_ISL_2779111, EPI_ISL_2779112, EPI_ISL_2779113, EPI_ISL_2779158, EPI_ISL_3033478 | | | |
| see above | LESP Yucatan | Instituto de Diagnostico y Referencia Epidemiologicos (INDRE) | Abril Rodriguez-Maldonado; Ariadna Medina-Benitez; Claudia Wong-Arambula; Ernesto Ramirez-Gonzalez.; Gisela Barrera-Badillo; Irma Lopez-Martinez; Joaquin Quiroz-Mercado; Lucia Hernandez-Rivas; Maribel Gonzalez-Villa; Natividad Cruz-Ortiz; Sergio Rangel-Guerrero; Tatiana Nunez-Garcia; Vanessa Rivero-Arredondo |
| EPI_ISL_2443462, EPI_ISL_2443464, EPI_ISL_2545646, EPI_ISL_2545649, EPI_ISL_2545656, EPI_ISL_2545657, EPI_ISL_2545658 | | | |
| see above | LHUB-ULB | Labo Klinische Biologie, UZA | Basil Britto Xavier; Christine Lammens; Herman Goossens; Jasmine Coppens; Marie Le Mercier; Veerle Matheeussen |
| EPI_ISL_2422367 | LKO | Jessa | Berden et al. on behalf of the Jessa_cmdLab |
| EPI_ISL_2695394 | Lab La Salete Robles - VN Famalicao | Instituto Nacional de Saude (INSA) | Borges et al |
| EPI_ISL_2605277, EPI_ISL_2605287, EPI_ISL_2605402, EPI_ISL_2605459, EPI_ISL_2605480, EPI_ISL_2605602 | Lab voor klinische biologie | Lab voor klinische biologie | Bruno Verhasselt; Hannelore Hamerlinck; Marija Janevska |
| EPI_ISL_2551023 | Lab. Microbiologia e Virologia Cotugno A.O. dei Colli - Istituto Zooprofilattico Sperimentale del Mezzogiorno | TIGEM | Antonio Grimaldi Patrizia Annunziata Francesco Panariello Biancamaria Pierri Claudia Tiberio Teresa Giuliano Valentina Bouche Chiara Colantuono Maria Concetta Cuomo Denise Di Concilio Lucio Di Filippo Anna Manfredi Marcello Salvi Antonio Limone Luigi Atripaldi Pellegrino Cerino Andrea Ballabio Davide Cacchiarelli |
| EPI_ISL_2725250, EPI_ISL_2725252, EPI_ISL_2725257, EPI_ISL_2725263, EPI_ISL_2725265, EPI_ISL_2725266, EPI_ISL_2725268, EPI_ISL_2725271, EPI_ISL_2725275, EPI_ISL_2725276, EPI_ISL_2725281, EPI_ISL_2725282, EPI_ISL_2725294 | | | |
| see above | Lab. Microbiologia e Virologia Cotugno A.O. dei Colli - Istituto Zooprofilattico Sperimentale del Mezzogiorno | Telethon Institute of Genetics and Medicine (TIGEM) | Antonio Grimaldi Patrizia Annunziata Francesco Panariello Biancamaria Pierri Claudia Tiberio Teresa Giuliano Valentina Bouche Chiara Colantuono Maria Concetta Cuomo Denise Di Concilio Lucio Di Filippo Anna Manfredi Marcello Salvi Antonio Limone Luigi Atripaldi Pellegrino Cerino Andrea Ballabio Davide Cacchiarelli |
| EPI_ISL_2650016 | LabPLUS | Institute of Environmental Science and Research (ESR) | Anja Werno; Antje van der Linden; Arlo Upton; Chris Mansell; David Hammer; Dragana Drinkovic; Erasmus Smit; Gary McAuliffe; Hana Sofia Andersson; Hermes Perez; James Ussher; Jill Sherwood; Jing Wang; Joep de Ligt; Josh Freeman; Julia Howard; Juliet Elvy; Lauren Jelly; Mary DeAlmeida; Matt Blakiston; Matt Storey; Matthew Rogers; Max Bloomfield; Michael Addidle; Michelle Balm; Muhammad Faisal; Nikki Freed; Olin Silander; Olivia Stroeven; Rachel Boyle; Sally Roberts; SallyAnn Harbison; Sarah Jefferies; Sharmini Muttaiyah; Susan Morpeth; Susan Taylor; Timothy Blackmore; Vani Sathyendran; Veronica Playle; Virginia Hope; Xiaoyun Ren |
| EPI_ISL_2709274, EPI_ISL_2835853 | Labo Analyses Med | National Reference Center for Viruses of Respiratory Infections, Institut Pasteur, Paris | Angela Brisebarre; Anne-Laure Garand; Camille Capel; Christophe Malabat; Corinne Maufrais; Damien Mornico; Etienne Simon-Lorière; Frédéric Lemoine; Louise Lefrançois; Marion Barbet; Maud Vanpeene; Méline Bizard; Sabine Trombert; Sylvie Behillil; Sylvie Van der Werf; Vincent Enouf |
| EPI_ISL_2845792, EPI_ISL_2845794, EPI_ISL_2845803 | Labor Becker & Kollegen (Standort München) | Robert Koch Institute | |
| EPI_ISL_2504031, EPI_ISL_2504046 | Labor Berlin Charité Vivantes GmbH / Institut für Virologie | Charité Universitätsmedizin Berlin, Institut für Virologie/Labor Berlin | Barbara Mühlemann; Christian Drosten; Christine Stephan; Peter Menzel; Rolf Schwarzer; Terry Jones; Victor M Corman |
| EPI_ISL_2678554 | Labor Blackholm MVZ | Robert Koch Institute | |
| EPI_ISL_2631832 | Labor Dr. Wisplinghoff - Köln | Robert Koch Institute | |
| EPI_ISL_2629226, EPI_ISL_2629228, EPI_ISL_2629229, EPI_ISL_2629230, EPI_ISL_2629231, EPI_ISL_2835985, EPI_ISL_2835986, EPI_ISL_2835987, EPI_ISL_2835988 | | | |
| see above | Laboratoire Carage | Institut Pasteur de la Guyane | Anne Lavergne; Antoine Enfissi; Arielle Salmier; Dominique Rousset |
| EPI_ISL_5135803, EPI_ISL_5136250 | Laboratoire de Virologie – CHU François Mitterrand - Dijon – France | INSERM U1231 - Equipe GAD | Ali SI-MOHAMMED; Hélène GIRAUDON; Lionel PIROTH; Yannis DUFFOURD |
| EPI_ISL_3458806, EPI_ISL_3458809, EPI_ISL_3458826, EPI_ISL_3458827, EPI_ISL_3458904, EPI_ISL_3458905, EPI_ISL_3458907 | | | |
| see above | Laboratoire de santé publique | Laboratoire de santé publique du | Guillaume Bourque; Ioannis Ragoussis; Jesse Shapiro; Mark Lathrop and Michel Roger on behalf of the CoVSeQ research group; Sandrine Moreira |

| | | | |
|---|---|---|---|
| | du Québec | Québec | |
| EPI_ISL_3147555 | Laboratoire national de sante, Microbiology, Virology | Laboratoire national de sante, Microbiology, Microbial Genomics Platform | Anke Wienecke-Baldacchino; Catherine Ragimbeau; Elodie Solarino; Fatu Djabi; Jessica Tapp; Lise Pignon; Raoul Salmon; Tamir Abdelrahman; Trung Nguyen Nguyen; Virginie Jover |
| EPI_ISL_3148095, EPI_ISL_3148151, EPI_ISL_3148152 | Laboratoires d'analyses medicales - Ketterthill | Laboratoire national de sante, Microbiology, Microbial Genomics Platform | Anke Wienecke-Baldacchino; Caroline Scheiber; Catherine Ragimbeau; Elodie Solarino; Fatu Djabi; Jessica Tapp; Lise Pignon; Raoul Salmon; Serge Vedy; Tamir Abdelrahman; Virginie Jover |
| EPI_ISL_4946218 | Laboratorio Angel ( Synlab) | Instituto Nacional de Salud-Dirección de Investigación en Salud Pública | Carlos Franco-Muñoz; Carmen Osorio; Diana Malo; Diego A. Álvarez-Díaz; Diego Andrés Prada; Gerardo Santamaría; Hector Alejandro Ruiz-Moreno; Jhonnatan Reales-González; Jorge Rivera; Juan Camilo Martinez; Julian Naizaque; Katherine Laiton-Donato; Lisseth Pardo; Magdalena Wiesner; Marcela Mercado-Reyes; Maria T. Herrera-Sepúlveda; Marta Lopez Blanco; Martha Lucia Ospina Martinez; Paola Rojas; Sergio Gomez; Sheryll Corchuelo; Ángela Alarcon Cruz |
| EPI_ISL_2691373, EPI_ISL_2691466, EPI_ISL_2691497, EPI_ISL_2691498, EPI_ISL_2691499, EPI_ISL_2691500, EPI_ISL_2691501, EPI_ISL_2691502, EPI_ISL_2691503, EPI_ISL_2691504, EPI_ISL_2691505, EPI_ISL_2691506, EPI_ISL_2691507, EPI_ISL_2691508, EPI_ISL_2691509, EPI_ISL_2691510, EPI_ISL_2691511, EPI_ISL_2691512, EPI_ISL_2691513, EPI_ISL_2691514, EPI_ISL_2691515, EPI_ISL_2691516, EPI_ISL_2691517, EPI_ISL_2691518, EPI_ISL_2691519, EPI_ISL_2691520, EPI_ISL_2691521, EPI_ISL_2691522, EPI_ISL_2691523, EPI_ISL_2691524, EPI_ISL_2691525, EPI_ISL_2691526, EPI_ISL_2691527, EPI_ISL_2691528, EPI_ISL_2691529, EPI_ISL_2691530, EPI_ISL_2691531, EPI_ISL_2691533, EPI_ISL_2691534, EPI_ISL_2691535, EPI_ISL_2691536, EPI_ISL_2691537, EPI_ISL_2691538, EPI_ISL_2691539, EPI_ISL_2691540, EPI_ISL_2691541, EPI_ISL_2691542, EPI_ISL_2691543, EPI_ISL_2691544, EPI_ISL_2691546, EPI_ISL_2691547, EPI_ISL_2691548, EPI_ISL_2691549, EPI_ISL_2691550, EPI_ISL_2691551, EPI_ISL_2691553, EPI_ISL_2691554, EPI_ISL_2691555, EPI_ISL_2691556, EPI_ISL_2691557, EPI_ISL_2691558, EPI_ISL_2691560, EPI_ISL_2691561, EPI_ISL_2691562, EPI_ISL_2691563, EPI_ISL_2691568, EPI_ISL_2836973, EPI_ISL_2836974, EPI_ISL_2837043, EPI_ISL_2837053 | | | |
| see above | Laboratorio Central Noel Nutels | Bioinformatics Laboratory / LNCC | Alessandra P Lamarca; Alexandra L Gerber; Amilcar Tanuri; Ana Paula de C Guimaraes; Ana Tereza R Vasconcelos; Andrea Cony Cavalcanti; Caio Luiz Pereira Ribeiro; Cassia Alves; Cintia Policarpo; Claudia Maria Braga de Mello; Cristiane Gomes da Silva; Diana Mariani; Douglas Terra Machado; Flavio Dias da Silva; Gleidson da Silva de Oliveira; Leandro Magalhaes de Souza; Liliane Cavalcante; Luiz G P de Almeida; Marcio Henrique de Oliveira Garcia; Mario Sergio Ribeiro; Ronaldo da Silva F Jr; Silvia Carvalho |
| EPI_ISL_2942577 | Laboratorio Central de Epidemiologia (LCE) | Unidad de Genomica Avanzada | ; Alejandra Garcia-Gasca; Alejandra Hernandez-Teran; Alejandro Sanchez-Flores; Alfredo Herrera-Estrella; Alicia Ocaña-Mondragon; Andreu Comas-Garcia; Angel Gustavo Salas-Lais; Antonio Loza Roman; Bernardo Martinez-Miguel; Blanca Taboada; Brenda Irasema Maldonado-Meza; Bruno Gomez-Gil; Carla Ivon Herrera-Najera; Carlos F. Arias; Celia Boukadida; Celida Duque Molina; Celida Martinez- Rodriguez; Clara Esperanza Santacruz-Tinoco; Concepcion Grajales-Muñiz; Consorcio Mexicano de Vigilancia Genomica (CoViGen-Mex). Authors (in alphabetical order): Julio Elias Alvarado-Yaah; Cristobal Chaidez-Quiroz; Daniel Fregoso-Rueda; Daniel Lira Morales; Eduardo Becerril-Vargas; Fernando Fontove-Herrera; Fidencio Mejia-Nepomuceno; Francisco Pulido; Gloria Elena Espinosa-Ayala; Gloria Maria Molina-Salinas; Gloria Vazquez; Hector Esteban Paz-Juarez; Hector Montoya-Fuentes; Helen Haydee Fernanda Ramirez-Plascencia; Irvin Gonzalez-Lopez; Jean Pierre Gonzalez; Jesus Hernandez; Joel Armando Vazquez-Perez.; Jorge Salas-Hernandez; Jose Antonio Enciso-Moreno; Jose Arturo Martinez-Orozco; Jose Esteban Muñoz-Medina; Jose de Jesus Nuñez-Contreras; Juan Bautista Chale-Dzul; Julissa Enciso-Ibarra; Luis Alberto Ochoa-Carrera; Margarita Matias-Florentino; Maria Guadalupe Santiago-Mauricio; Maria Guadalupe de Jesus Mireles-Rivera; Mario Mujica-Sanchez; Marissa Perez-Garcia; Nelly Selem-Mojica; Pavel Isa; Ricardo Ciria Merce; Ricardo Grande; Rosa Maria Gutierrez Rios; Santiago avila-Rios; Selene Zarate; Susana Lopez; Veronica Mata-Haro; Victor Eduardo Garcia-Arias; Victor Hugo Borja-Aburto |
| EPI_ISL_2681179, EPI_ISL_2681182, EPI_ISL_2681217, EPI_ISL_2681264, EPI_ISL_2681280, EPI_ISL_2801774, EPI_ISL_2801775, EPI_ISL_2801786, EPI_ISL_2801787, EPI_ISL_2801796, EPI_ISL_2801801, EPI_ISL_2801802, EPI_ISL_2801803, EPI_ISL_2801805, EPI_ISL_2801817, EPI_ISL_2801824, EPI_ISL_2801825, EPI_ISL_2801827, EPI_ISL_2801831, EPI_ISL_2801832, EPI_ISL_2801834, EPI_ISL_2801842, EPI_ISL_2801846, EPI_ISL_2801872, EPI_ISL_3133777, EPI_ISL_3355557 | | | |
| see above | Laboratorio Central de Epidemiología (LCE) | Instituto de Biotecnología de la UNAM | ; Alejandra García-Gasca; Alejandra Hernández-Terán; Alejandro Sánchez-Flores; Alfredo Herrera-Estrella; Alicia Ocaña-Mondragón; Andreu Comas-García; Angel Gustavo Salas-Lais; Antonio Loza Román; Bernardo Martínez-Miguel; Blanca Taboada; Brenda Irasema Maldonado-Meza; Bruno Gómez-Gil; Carla Ivón Herrera-Najera; Carlos F. Arias; Celia Boukadida; Clara Esperanza Santacruz-Tinoco; Concepción Grajales-Muñiz; Consorcio Mexicano de Vigilancia Genómica (CoViGen-Mex). Authors (in alphabetical order): Julio Elias Alvarado-Yaah; Cristóbal Cháidez-Quiróz; Célida Duque Molina; Célida Martínez- Rodríguez; Daniel Fregoso-Rueda; Daniel Lira Morales; Eduardo Becerril-Vargas; Fernando Fontove-Herrera; Fidencio Mejía-Nepomuceno; Francisco Pulido; Gloria Elena Espinosa-Ayala; Gloria María Molina-Salinas; Gloria Vazquez; Hector Esteban Paz-Juárez; Hector Montoya-Fuentes; Helen Haydee Fernanda Ramírez-Plascencia; Irvin González-López; Jean Pierre González; Jesús Hernández; Joel Armando Vázquez-Pérez.; Jorge Salas-Hernández; José Antonio Enciso-Moreno; José Arturo Martínez-Orozco; José Esteban Muñoz-Medina; José de Jesús Nuñez-Contreras; Juan Bautista Chale-Dzul; Julissa Enciso-Ibarra; Luis Alberto Ochoa-Carrera; Margarita Matías-Florentino; Mario Mújica-Sánchez; Marissa Perez-Garcia; María Guadalupe Santiago-Mauricio; María Guadalupe de Jesús Mireles-Rivera; Nelly Sélem-Mojica; Pavel Isa; Ricardo Ciria Merce; Ricardo Grande; Rosa María Gutiérrez Rios; Santiago Ávila-Ríos; Selene Zárate; Susana Lopez; Verónica Mata-Haro; Victor Eduardo García-Arias; Víctor Hugo Borja-Aburto |
| EPI_ISL_6944494, EPI_ISL_6944504, EPI_ISL_6944518, EPI_ISL_6944520, EPI_ISL_6944543, EPI_ISL_6944552, EPI_ISL_6944568, EPI_ISL_6944569, EPI_ISL_6944585, EPI_ISL_6944603 | | | |
| see above | Laboratorio Central de Salud Publica | Viral Special Pathogens Branch, Centers for Disease Control and Prevention | Andrea Gómez de la Fuente; Cynthia Vázquez; Emir Talundzic; Joel Montgomery; John Klena; Juan Torales; Justin Lee; María José Ortega; María Liz Gamarra; Shannon Whitmer; Shirley Villalba |
| EPI_ISL_4303367, EPI_ISL_4303368, EPI_ISL_4303369, EPI_ISL_4303370, EPI_ISL_4303371, EPI_ISL_4303372, EPI_ISL_4303373, EPI_ISL_4303374, EPI_ISL_4303375, EPI_ISL_4303376, EPI_ISL_4303377, EPI_ISL_4303378, EPI_ISL_4303379, EPI_ISL_4303380 | | | |
| see above | Laboratorio Central de Salud Pública | Laboratorio Central de Salud Pública | Alexander Martinez; Andrea Gómez de la Fuente; Cesar Cantero; Cynthia Vazquez; Juan Torales; Julio Barrios; Maria Liz Gamarra; Sandra Gonzalez; Shirley Villalba; Tania Alfonzo |
| EPI_ISL_3434753, EPI_ISL_3434758 | Laboratorio Central de Saude Publica do Estado Maranhao (LACEN-MA) | Laboratory of Respiratory Viruses and Measles, Oswaldo Cruz Institute, FIOCRUZ | Alice Sampaio Rocha; Ana Carolina Mendonca; Anna Carolina Paixao; Elisa Cavalcante Pereira; Fernando Motta; Lidio Gonçalves Lima Neto; Luciana Appolinario; Marilda Siqueira on behalf of the Fiocruz COVID-19 Genomic Surveillance Network; Paola Resende; Renata Serrano Lopes; Taina Venas |
| EPI_ISL_2645408, EPI_ISL_2645409, EPI_ISL_2645410 | Laboratorio Central de Saude Publica do Estado da Bahia (LACEN/BA) | Laboratory of Respiratory Viruses and Measles, Oswaldo Cruz Institute, FIOCRUZ | Alice Sampaio Rocha; Ana Carolina Mendonca; Anna Carolina Paixao; Elisa Cavalcante Pereira; Felicidade Pereira; Fernando Motta; Luciana Appolinario; Marilda Siqueira on behalf of the Fiocruz COVID-19 Genomic Surveillance Network; Paola Resende; Renata Serrano Lopes; Taina Venas |
| EPI_ISL_3190176, EPI_ISL_3190178, EPI_ISL_3235238, EPI_ISL_3235239, EPI_ISL_3235240, EPI_ISL_3235241, EPI_ISL_3235242, EPI_ISL_3235243, EPI_ISL_3235245, EPI_ISL_3235246, EPI_ISL_3235247, EPI_ISL_3235248, EPI_ISL_3235249, EPI_ISL_3235250, EPI_ISL_3235251, EPI_ISL_3235252, EPI_ISL_3235253, EPI_ISL_3235254, EPI_ISL_3235255, EPI_ISL_3434783, EPI_ISL_3434784, EPI_ISL_3434785, EPI_ISL_3434786, EPI_ISL_3434787, EPI_ISL_3434788 | | | |
| see above | Laboratorio Central de Saude Publica do Estado da Paraiba (LACEN/PB) | Laboratory of Respiratory Viruses and Measles, Oswaldo Cruz Institute, FIOCRUZ | Alice Sampaio Rocha; Ana Carolina Mendonca; Anna Carolina Paixao; Dalane Loudal Florentino Teixeira; Elisa Cavalcante Pereira; Fernando Motta; Joao Felipe Bezerra; Luciana Appolinario; Marilda Siqueira on behalf of the Fiocruz COVID-19 Genomic Surveillance Network; Paola Resende; Renata Serrano Lopes; Taina Venas |
| EPI_ISL_2983071, EPI_ISL_2983072, EPI_ISL_2983073, EPI_ISL_2983074, EPI_ISL_2983075 | Laboratorio Central de Saude Publica do Estado de Alagoas (LACEN/AL) | Laboratory of Respiratory Viruses and Measles, Oswaldo Cruz Institute, FIOCRUZ | Agatha Cristinne Prudencio; Alice Sampaio Rocha; Ana Carolina Mendonca; Anderson Brandao Leite; Anna Carolina Paixao; Elisa Cavalcante Pereira; Fernando Motta; Ighor Leonardo Arantes Gomes; Luciana Appolinario; Marilda Siqueira on behalf of the Fiocruz COVID-19 Genomic Surveillance Network; Paola Resende; Renata Serrano Lopes; Taina Venas |
| EPI_ISL_2983218, EPI_ISL_2983354 | Laboratorio Central de Saude Publica do Estado de Santa Catarina (LACEN-SC) | Laboratory of Respiratory Viruses and Measles, Oswaldo Cruz Institute, FIOCRUZ | Agatha Cristinne Prudencio; Alice Sampaio Rocha; Ana Carolina Mendonca; Anna Carolina Paixao; Darcita Buerger Rovaris; Elisa Cavalcante Pereira; Fernando Motta; Ighor Leonardo Arantes Gomes; Luciana Appolinario; Marilda Siqueira on behalf of the Fiocruz COVID-19 Genomic Surveillance Network; Paola Resende; Renata Serrano Lopes; Sandra Bianchini Fernandes; Taina Venas |
| EPI_ISL_2983385, EPI_ISL_2983404, EPI_ISL_2983406, EPI_ISL_2983407, EPI_ISL_2983408, EPI_ISL_2983409, EPI_ISL_2983411, EPI_ISL_2983413, EPI_ISL_2983414, EPI_ISL_2983415, EPI_ISL_4563061 | | | |
| see above | Laboratorio Central de Saude Publica do Estado de Santa Catarina (LACEN/SC) | Laboratory of Respiratory Viruses and Measles, Oswaldo Cruz Institute, FIOCRUZ | Alice Sampaio Rocha; Ana Carolina Mendonca; Anna Carolina Paixao; Darcita Buerger Rovaris; Elisa Cavalcante Pereira; Fernando Motta; Luciana Appolinario; Marilda Siqueira on behalf of the Fiocruz COVID-19 Genomic Surveillance Network; Paola Resende; Renata Serrano Lopes; Sandra Bianchini Fernandes; Taina Venas |
| EPI_ISL_4061452, EPI_ISL_4061453 | Laboratorio Central de Saude Publica do Estado de Sergipe (LACEN/SE) | Laboratory of Respiratory Viruses and Measles, Oswaldo Cruz Institute, FIOCRUZ | Agatha Soares; Alice Sampaio Rocha; Ana Carolina Mendonca; Anna Carolina Paixao; Cliomar Alves dos Santos; Elisa Cavalcante Pereira; Fernando Motta; Ighor Arantes; Luciana Appolinario; Marilda Siqueira on behalf of the Fiocruz COVID-19 Genomic Surveillance Network; Paola Resende; Renata Serrano Lopes; Tainá Moreira Martins Venas |
| EPI_ISL_3190261, EPI_ISL_3190262, EPI_ISL_3190263 | Laboratorio Central de Saude Publica do Estado do Maranhao (LACEN-MA) | Laboratory of Respiratory Viruses and Measles, Oswaldo Cruz Institute, FIOCRUZ | Agatha Soares; Alice Sampaio Rocha; Ana Carolina Mendonca; Anna Carolina Paixao; Elisa Cavalcante Pereira; Fernando Motta; Ighor Arantes; Lidio Gonçalves Lima Neto; Luciana Appolinario; Marilda Siqueira on behalf of the Fiocruz COVID-19 Genomic Surveillance Network; Paola Resende; Renata Serrano Lopes; Tainá Venas |
| EPI_ISL_3190172, EPI_ISL_3190173 | Laboratorio Central de Saude Publica do Estado do Maranhao (LACEN/MA) | Laboratory of Respiratory Viruses and Measles, Oswaldo Cruz Institute, FIOCRUZ | Agatha Soares; Alice Sampaio Rocha; Ana Carolina Mendonca; Anna Carolina Paixao; Elisa Cavalcante Pereira; Fernando Motta; Ighor Arantes; Lidio Gonçalves Lima Neto; Luciana Appolinario; Marilda Siqueira on behalf of the Fiocruz COVID-19 Genomic Surveillance Network; Paola Resende; Renata Serrano Lopes; Tainá Venas |
| EPI_ISL_2645446, EPI_ISL_2645448, EPI_ISL_2645452, EPI_ISL_2645453, EPI_ISL_2645455, EPI_ISL_2645456, EPI_ISL_2645459, EPI_ISL_2645476, EPI_ISL_2645478, EPI_ISL_2645483, EPI_ISL_2645486, EPI_ISL_2983429, EPI_ISL_2983430, EPI_ISL_2983431, EPI_ISL_2983432, EPI_ISL_2983433, EPI_ISL_2983434, EPI_ISL_2983435, EPI_ISL_2983436, EPI_ISL_2983437, EPI_ISL_2983438, EPI_ISL_2983439, EPI_ISL_2983441, EPI_ISL_2983442, EPI_ISL_2983451, EPI_ISL_3190267 | | | |
| see above | Laboratorio Central de Saude Publica do Estado do Parana (LACEN/PR) | Laboratory of Respiratory Viruses and Measles, Oswaldo Cruz Institute, FIOCRUZ | Alice Sampaio Rocha; Ana Carolina Mendonca; Anna Carolina Paixao; Elisa Cavalcante Pereira; Fernando Motta; Irina Riediger; Luciana Appolinario; Marilda Siqueira on behalf of the Fiocruz COVID-19 Genomic Surveillance Network; Paola Resende; Renata Serrano Lopes; Taina Venas |
| EPI_ISL_2982763, EPI_ISL_2982764, EPI_ISL_2982767, EPI_ISL_2982769, EPI_ISL_2982784, EPI_ISL_2982814, EPI_ISL_2982815, EPI_ISL_2982816, EPI_ISL_2982817, EPI_ISL_2982818, EPI_ISL_2982819, EPI_ISL_2982820, EPI_ISL_2982821 | | | |
| see above | Laboratorio Central de Saude Publica do Estado do Rio Grande do Sul (LACEN-RS) | Laboratory of Respiratory Viruses and Measles, Oswaldo Cruz Institute, FIOCRUZ | Alice Sampaio Rocha; Ana Carolina Mendonca; Anderson Brandao Leite; Anna Carolina Paixao; Elisa Cavalcante Pereira; Fernando Motta; Luciana Appolinario; Marilda Siqueira on behalf of the Fiocruz COVID-19 Genomic Surveillance Network; Paola Resende; Renata Serrano Lopes; Taina Venas |
| EPI_ISL_2863667, EPI_ISL_2863669, EPI_ISL_2863671, EPI_ISL_2863673, EPI_ISL_2863675, EPI_ISL_2863727 | Laboratorio Central de Saude Publica do Estado do Rio de Janeiro (LACEN-RJ) | Laboratory of Respiratory Viruses and Measles, Oswaldo Cruz Institute, FIOCRUZ | Agatha Cristinne Prudencio; Alice Sampaio Rocha; Ana Carolina Mendonca; Andrea Cony Cavalcanti; Anna Carolina Paixao; Elisa Cavalcante Pereira; Fernando Motta; Ighor Leonardo Arantes Gomes; Luciana Appolinario; Marilda Siqueira on behalf of the Fiocruz COVID-19 Genomic Surveillance Network; Paola Resende; Renata Serrano Lopes; Taina Venas |
| EPI_ISL_3033074, EPI_ISL_3033075, EPI_ISL_3260096, EPI_ISL_3260097 | Laboratorio Central de la Ciudad de Santa Fe | Grupo de Genómica y Bioinformática del Instituto de Investigación de la Cadena Láctea CONICET-INTA on behalf of 'Proyecto Argentino Interinstitucional de genomica de SARS-CoV-2' (PAIS Consortium) | AF; Amadio; C; Eberhardt; G; Irazoqui; JM; MF; Mugna; Ojeda; Pastor; Rompato; V |
| EPI_ISL_2940269 | Laboratorio Central, Ministerio de Salud Córdoba | Instituto de Patologia Vegetal (CIAP-INTA) on behalf of 'Proyecto Argentino Interinstitucional de genomica de SARS-CoV-2' (PAIS Consortium) | Barbas, G.; Castro, G.; Debat, HJ.; FD; Fernández; M; M.B.; Marquez, N.; Pisano; Re, V. |
| EPI_ISL_7740603 | Laboratorio Nacional de Salud | Asociación de Salud Integral / Clínica Familiar "Luis Ángel García" | Ana S. Gonzalez-Reiche; Claudia Rangel; Danicela Mercado; Eduardo Arathon; Hilda Ruiz; Luis Aguirre; Luis Rivas; Narda Medina; Oscar Bonilla; Osmar Gamboa. |

| | | | |
|---|---|---|---|
| EPI_ISL_5332833, EPI_ISL_5332836, EPI_ISL_5332846, EPI_ISL_6121560, EPI_ISL_6121565 | Laboratorio de Alta Complejidad (LACMI), Instituto de Genética Humana (IgeHM) Parque de la Salud de la Provincia de Misiones, Universidad Nacional de Misiones | Grupo de Genómica y Bioinformática del Instituto de Investigación de la Cadena Láctea CONICET-INTA on behalf of 'Proyecto Argentino Interinstitucional de genomica de SARS-CoV-2' (PAIS Consortium) | AF; Aguirre; Amadio; Benitez; C; D; Eberhardt; Fontana; G; Gamarra; Gonzales; Irazoqui; JM; K; Ludojoski; M; MF; R; Salvatierra; Silva; TA; Urquiza; Vogel |
| EPI_ISL_3050307, EPI_ISL_4030344, EPI_ISL_4030365 | Laboratorio de Ecologia de Doencas Transmissiveis na Amazonia, Instituto Leonidas e Maria Deane - Fiocruz Amazonia | Laboratorio de Ecologia de Doencas Transmissiveis na Amazonia, Instituto Leonidas e Maria Deane - Fiocruz Amazonia | Alex Martins; André Corado; Debora Duarte; Fabiola Mendonça da Silva Chui; Felipe Naveca; Fernanda Nascimento; Fernando Fonseca de Almeida Val; George Silva; Gisely Cardoso de Melo; Karina Pessoa; Luciana Gonçalves; Marcus Vinicius Guimarães de Lacerda; Maria Júlia Brandão; Maria Paula Gomes Mourão; Mariana Xavier Simão; Matilde Mejía; Michele Jesus; Valdinete Nascimento; Vanderson de Souza Sampaio; Victor Souza; Ágatha Costa |
| EPI_ISL_4889734, EPI_ISL_4889775 | Laboratorio de Infectología, Servicio de Infectología, Hospital Universitario Dr. José Eleuterio González - Universidad Autónoma de Nuevo León | Centro de Investigación e Innovación en Virología Médica, Departamento de Bioquímica y Medicina Molecular,Facultad de Medicina - Universidad Autónoma de Nuevo León | Adrian Camacho-Ortiz; Ana M. Rivas-Estilla; Daniel Arellanos-Soto; Daniel Salas-Treviño; Eduardo Perez-Alba; Javier Ramos-Jiménez; Kame A. Galán-Huerta; Laura Nuzzolo-Shihadeh; Paola Bocanegra-Ibarias; Samantha M. Flores-Treviño; Sonia A. Lozano-Sepúlveda |
| EPI_ISL_2907519, EPI_ISL_3134269, EPI_ISL_3134270, EPI_ISL_3134271, EPI_ISL_3134272 | Laboratorio de Medicina Molecular, Universidad de Magallanes | Centro Asistencial Docente y de Investigacion, Universidad de Magallanes | Carolina Pérez; Constanza Ceroni; Diego Alvarez; Inés Cid; Jacqueline Aldridge; Jorge González; Juan Ríos; Marcelo Navarrete; Roberto Uribe-Paredes |
| EPI_ISL_3048981, EPI_ISL_3048982, EPI_ISL_3048983, EPI_ISL_3048984, EPI_ISL_3048985, EPI_ISL_3048986, EPI_ISL_3048987, EPI_ISL_3048988, EPI_ISL_3048989, EPI_ISL_3048990, EPI_ISL_3048991, EPI_ISL_3048992, EPI_ISL_3048993, EPI_ISL_3048994, EPI_ISL_3048995, EPI_ISL_3048996 | | | |
| see above | Laboratorio de Pesquisa em Virologia, FAMERP, SJRP | Laboratorio de Pesquisa em Virologia, FAMERP, SJRP | Cecília Artico Banho; Cíntia Bittar; Fábio Sossai Possebon; Guilherme Campos; Helena Lage Ferreira; Jorge A. Petroli Marchesi; João Pessoa Araújo Jr.; Leila Sabrina Ullmann; Lívia Sacchetto; Maisa C. Pereira Parra; Marília Moraes; Maurício L. Nogueira.; Paula Rahal; Paulo Inacio da Costa |
| EPI_ISL_3672425, EPI_ISL_3672428, EPI_ISL_3672429, EPI_ISL_3672430, EPI_ISL_3672435, EPI_ISL_3826385, EPI_ISL_3826386, EPI_ISL_3826387, EPI_ISL_3826388, EPI_ISL_3826419, EPI_ISL_3826429, EPI_ISL_3826434, EPI_ISL_3826435, EPI_ISL_3991455, EPI_ISL_3991458, EPI_ISL_3991462, EPI_ISL_3991465, EPI_ISL_3991468, EPI_ISL_4052706, EPI_ISL_4052708, EPI_ISL_4052709, EPI_ISL_4052710, EPI_ISL_4052731, EPI_ISL_4052732, EPI_ISL_4052735, EPI_ISL_4052738, EPI_ISL_4052739, EPI_ISL_4052749, EPI_ISL_4052756, EPI_ISL_4052764, EPI_ISL_4080443, EPI_ISL_4080444, EPI_ISL_4080446, EPI_ISL_4080457, EPI_ISL_4080458, EPI_ISL_4080459, EPI_ISL_4080460, EPI_ISL_4080472, EPI_ISL_4080503, EPI_ISL_4080515, EPI_ISL_4080518, EPI_ISL_4080519, EPI_ISL_4172679, EPI_ISL_4172681, EPI_ISL_4172683, EPI_ISL_4172684, EPI_ISL_4172686, EPI_ISL_4172688, EPI_ISL_4172690, EPI_ISL_4172707, EPI_ISL_4172717, EPI_ISL_4172732, EPI_ISL_4172734, EPI_ISL_4172735, EPI_ISL_4172737, EPI_ISL_4172739, EPI_ISL_4172894, EPI_ISL_4172905, EPI_ISL_5500122 | | | |
| see above | Laboratorio de Referencia Nacional de Virus Respiratorios. Centro Nacional de Salud Publica. Instituto Nacional de Salud Peru. | Laboratorio de Referencia Nacional de Virus Respiratorios. Centro Nacional de Salud Publica. Instituto Nacional de Salud Peru. | Carlos Padilla Rojas; Henri Bailon Calderon; Iris Silva Molina; Joseph Huayra Niquen; Lely Solari Zerpa; Luis Barcena Flores; Marco Galarza Perez; Nancy Rojas Serrano; Nieves Sevilla Castañeda; Omar Caceres Rey; Orson Mestanza Millones; Princesa Medrano Alhuay; Priscila Lope Pari; Sandra Morales Ruiz; Sara Gordillo Vilchez; Steve Acedo Lazo; Veronica Hurtado Vela; Victor Jimenez Vasquez; Wendy Lizarraga Olivares |
| EPI_ISL_2812709 | Laboratorio de Virología HUCA | Laboratorio de Virología HUCA | Abreu F; Alvarez-Arguelles ME; Boga JA; Castelló C; Costales I; Coto E; Gómez de Oña J; Martín-Rodríguez G; Melón S; Perez-Martínez Z; Rojo S; Sandoval M |
| EPI_ISL_4405480 | Laboratorio del Hospital Interzonal General de Agudos " Evita " | Área de Secuenciación del Laboratorio de Virología del Hospital de Niños Dr. Ricardo Gutierrez on behalf of 'Proyecto Argentino Interinstitucional de genomica de SARS-CoV-2' (PAIS Consortium) | A; Acuña; D; Desimone; E; Goya; Grossi; I; L; LE; Luczac; Lusso; MI; MS; Musto; Nabaes Jodar; Natale; O; S; Serrano; Valinotto; Viegas, M. |
| EPI_ISL_5771885 | Laboratorio del Hospital Regional Ushuaia Gdor. Ernesto Campos | Nodo de Secuenciación Tierra del Fuego - Hospital Regional Ushuaia - Centro Austral De Investigaciones Científicas - Universidad Nacional De Tierra Del Fuego | AE; Boutureira, MF.; CA; CB; CF; Castro; Ceballos; Cáceres; De Roccis; F; G; Gallego; Gramundi; ID; Nardi; Rojas; SB; SG; Yulan |
| EPI_ISL_7620383 | Laboratorio del Hospital Regional Ushuaia Gdor. Ernesto Campos | Nodo de Secuenciación Tierra del Fuego - Hospital Regional Ushuaia - Centro Austral De Investigaciones Científicas - Universidad Nacional De Tierra Del Fuego on behalf PAIS project and ANLIS-Malbran | AE; Boutureira, MF.; CA; CB; CF; Castro; Ceballos; Cáceres; De Roccis; F; G; Gallego; Gramundi; ID; Nardi; Rojas; SB; SG; Yulan |
| EPI_ISL_2680918, EPI_ISL_2680925, EPI_ISL_2894551, EPI_ISL_2987688, EPI_ISL_2987689 | Laboratorios Delia Barraza | Laboratorio nacional-LANIIA-CIAD | ; Alejandra García-Gasca; Alejandra Hernández-Terán; Alejandro Sánchez-Flores; Alfredo Herrera-Estrella; Alicia Ocaña-Mondragón; Andreu Comas-García; Angel Gustavo Salas-Lais; Antonio Loza Román; Bernardo Martínez-Miguel; Blanca Taboada; Brenda Irasema Maldonado-Meza; Bruno Gómez-Gil; Carla Ivón Herrera-Najera; Carlos F. Arias; Celia Boukadida; Clara Esperanza Santacruz-Tinoco; Claudia Soto Félix; Concepción Grajales-Muñiz; Consorcio Mexicano de Vigilancia Genómica (CoViGen-Mex). Authors (in alphabetical order): Julio Elias Alvarado-Yaah; Cristóbal Cháidez-Quiróz; Célida Duque Molina; Célida Martínez- Rodríguez; Daniel Fregoso-Rueda; Daniel Lira Morales; Eduardo Becerril-Vargas; Fernando Fontove-Herrera; Fidencio Mejía-Nepomuceno; Francisco Pulido; Gloria Elena Espinosa-Ayala; Gloria María Molina-Salinas; Gloria Vazquez; Hector Esteban Paz-Juárez; Hector Montoya-Fuentes; Helen Haydee Fernanda Ramírez-Plascencia; Irvin González-López; Jean Pierre González; Jesús Hernández; Joel Armando Vázquez-Pérez. Laboratorios Delia Barraza: María Delia Barraza Sámano; Jorge Salas-Hernández; José Antonio Enciso-Moreno; José Arturo Martínez-Orozco; José Esteban Muñoz-Medina; José de Jesús Nuñez-Contreras; Juan Bautista Chale-Dzul; Julissa Enciso-Ibarra; Luis Alberto Ochoa-Carrera; Margarita Matías-Florentino; Mario Mújica-Sánchez; Marissa Perez-Garcia; María Guadalupe Santiago-Mauricio; María Guadalupe de Jesús Mireles-Rivera; Melissa García Angulo.; Nelly Sélem-Mojica; Pavel Isa; Ricardo Ciria Merce; Ricardo Grande; Rosa María Gutiérrez Rios; Santiago Ávila-Ríos; Selene Zárate; Susana Lopez; Verónica Mata-Haro; Victor Eduardo García-Arias; Víctor Hugo Borja-Aburto |
| EPI_ISL_2480558, EPI_ISL_2480565, EPI_ISL_2481485, EPI_ISL_2611120, EPI_ISL_2611122, EPI_ISL_2611124, EPI_ISL_2611135, EPI_ISL_2611136, EPI_ISL_2611147, EPI_ISL_2611151, EPI_ISL_2611154, EPI_ISL_2611177, EPI_ISL_2611183, EPI_ISL_2611187, EPI_ISL_2611190, EPI_ISL_2611194, EPI_ISL_2611209, EPI_ISL_2611220, EPI_ISL_2611229, EPI_ISL_2611230, EPI_ISL_2611232, EPI_ISL_2611233, EPI_ISL_2611238, EPI_ISL_2611239, EPI_ISL_2611240, EPI_ISL_2611242, EPI_ISL_2611301, EPI_ISL_2611304, EPI_ISL_2611313, EPI_ISL_2611314, EPI_ISL_2611321, EPI_ISL_2611323, EPI_ISL_2611324, EPI_ISL_2611332, EPI_ISL_2611334, EPI_ISL_2611338, EPI_ISL_2611342, EPI_ISL_2611343, EPI_ISL_2611344, EPI_ISL_2611349, EPI_ISL_2611364, EPI_ISL_2611482, EPI_ISL_2611488, EPI_ISL_2611512, EPI_ISL_2611516, EPI_ISL_2611522, EPI_ISL_2611532, EPI_ISL_2611539, EPI_ISL_2611552, EPI_ISL_2611555, EPI_ISL_2611557, EPI_ISL_2611558, EPI_ISL_2611561, EPI_ISL_2611592, EPI_ISL_2611593, EPI_ISL_2611607, EPI_ISL_2611626, EPI_ISL_2611627, EPI_ISL_2611631, EPI_ISL_2611641, EPI_ISL_2611645, EPI_ISL_2611646, EPI_ISL_2611655, EPI_ISL_2611656, EPI_ISL_2611657, EPI_ISL_2611685, EPI_ISL_2611692, EPI_ISL_2611734, EPI_ISL_2611767, EPI_ISL_4372600, EPI_ISL_4372635, EPI_ISL_4372782, EPI_ISL_4372944, EPI_ISL_4372981, EPI_ISL_4373279, EPI_ISL_4373868, EPI_ISL_4373876, EPI_ISL_4374069, EPI_ISL_4374295, EPI_ISL_4374312, EPI_ISL_4374316, EPI_ISL_4374408, EPI_ISL_4374539, EPI_ISL_4374643, EPI_ISL_4374691, EPI_ISL_4374806, EPI_ISL_4374872, EPI_ISL_4374925, EPI_ISL_4374999, EPI_ISL_4375181, EPI_ISL_4375240, EPI_ISL_4375600, EPI_ISL_4375720, EPI_ISL_4375854, EPI_ISL_4376008, EPI_ISL_4376072, EPI_ISL_4376135, EPI_ISL_4376419, EPI_ISL_4376640, EPI_ISL_4376988, EPI_ISL_4377086, EPI_ISL_4377443, EPI_ISL_4377612, EPI_ISL_4377643, EPI_ISL_4377739, EPI_ISL_4377800, EPI_ISL_4377845, EPI_ISL_4378171, EPI_ISL_4378200, EPI_ISL_4378312, EPI_ISL_4378884, EPI_ISL_4378899, EPI_ISL_4379176, EPI_ISL_4379478, EPI_ISL_4379661, EPI_ISL_4379784, EPI_ISL_4379898, EPI_ISL_4379902, EPI_ISL_4380118, EPI_ISL_4380559, EPI_ISL_4380587, EPI_ISL_4380609, EPI_ISL_4380773, EPI_ISL_4380862, EPI_ISL_4381246, EPI_ISL_4381352, EPI_ISL_4381436, EPI_ISL_4381450, EPI_ISL_4381568, EPI_ISL_4381673, EPI_ISL_4381766, EPI_ISL_4382083, EPI_ISL_4382293 | | | |
| see above | Laboratory Corporation of America | Centers for Disease Control and Prevention Division of Viral Diseases, Pathogen Discovery | Adrian Paskey; Amanda Douglas; Amanda Suchanek; Andrea Throop; Ayla Burns; Benjamin Rambo-Martin; Bobbi Croy; Brian Krueger; Brian Norvell; Christopher Gulvick; Christos Petropoulos; Clinton Paden; Clinton R. Paden; Craig Lukasik; Dakota Howard; Darlene Wagner; Debbie Boles; Dhwani Batra; Duncan MacCannell; Eyad Almasri; Goran Stevovic; Howard Engler; Hrushikesh Deshmukh; Jake Humphrey; Jana Schroth; Jason Caravas; Joe Voshell; John Pruitt; Jonathan Meltzer; Jonathan Williams; Kara Moser; Kimberly Wagner; Kristine Lacek; Lax Iyer; Lisa Pfefferle; Lyndon Tilson; Manoj Jain; Marcia Eisenberg; Mary Ann Cristobal; Mary Cristobal; Mary Williamson; Matthew Robinson; Matthew Schmerer; Michael Levandoski; Mike Sapeta; Mindy Nye; Minoo Agarwal; Mohan Kolli; Nuthawin Charoensri; Oren Cohen; Peter Cook; Peter W. Cook; Prashant Gupta; Qian Zeng; Rama Ghatti; Scott Parker; Scott Ryan; Scott Sammons; Shatavia Morrison; Stanley Letovsky; Steven Ragan; Suresh Babu Selvaraju; Suresh Selvaraju; Susan Countryman; Susan Hicks; Suzanne Dale; Thomas Urban; Tim Kuphal; Tricia Zwiefelhofer; Tymeckia Kendall; Victoria Caban Figueroa; Vincent Drouillon; Yvette Unoarumhi |
| EPI_ISL_3931705 | Laboratory of Microbiology and Virology, Ospedale Amedeo di Savoia, ASL "Città di Torino" | Laboratory of Microbiology and Virology, Ospedale Amedeo di Savoia, ASL "Città di Torino" | Debora Fiorito; Elisa Burdino; Francesco Cerutti; Gabriella Gregori; Maria Grazia Milia; Tiziano Allice; Valeria Ghisetti |
| EPI_ISL_2557407, EPI_ISL_2982716, EPI_ISL_2982717, EPI_ISL_2982718, EPI_ISL_2982719, EPI_ISL_2982864, EPI_ISL_2982865, EPI_ISL_2982866, EPI_ISL_3045383, EPI_ISL_7106077 | | | |
| see above | Laboratory of Respiratory Viruses and Measles, Oswaldo Cruz Institute, FIOCRUZ | Laboratory of Respiratory Viruses and Measles, Oswaldo Cruz Institute, FIOCRUZ | Agatha Cristinne Prudencio; Alice Sampaio Rocha; Ana Carolina Mendonca; Anna Carolina Paixao; Bruna Mendonça da Silva; Elisa Cavalcante Pereira; Fernando Motta; Ighor Arantes; Ighor Leonardo Arantes Gomes; Jéssica Graça Macedo de Carvalho; Larissa Macedo Pinto; Luciana Appolinario; Marilda Siqueira on behalf of the Fiocruz COVID-19 Genomic Surveillance Network; Paola Resende; Renata Serrano Lopes; Taina Venas; Victor Guimaraes |
| EPI_ISL_3025606, EPI_ISL_3025615, EPI_ISL_3025620, EPI_ISL_3025626 | Laboratory, Gravity Diagnostics | Laboratory, Gravity Diagnostics | Diagnostics, G. |
| EPI_ISL_4600484 | Laboratório Central de Saúde Pública Profº Gonçalo Moniz | Coordenação Geral de Laboratórios de Saúde Pública (CGLAB/DAEVS/SVS/MS) | Vagner Fonseca; et al. |
| EPI_ISL_4600513, EPI_ISL_4600520, EPI_ISL_4600528 | Laboratório Central de Saúde Pública de Pernambuco | Coordenação Geral de Laboratórios de Saúde Pública (CGLAB/DAEVS/SVS/MS) | Vagner Fonseca; et al. |
| EPI_ISL_2777898, EPI_ISL_2777968, EPI_ISL_3050301, EPI_ISL_3050302, EPI_ISL_3050303, EPI_ISL_3050304, EPI_ISL_3050305, EPI_ISL_3050306, EPI_ISL_3050308, EPI_ISL_3050309, EPI_ISL_3050310, EPI_ISL_3050311, EPI_ISL_3050312, EPI_ISL_3050313, EPI_ISL_3050314, EPI_ISL_3050315, EPI_ISL_3050316, EPI_ISL_3050317, EPI_ISL_3050318, EPI_ISL_3050319, EPI_ISL_3050320, EPI_ISL_3050321, EPI_ISL_3050322, EPI_ISL_3050323, EPI_ISL_3050324, EPI_ISL_3050325, EPI_ISL_3050326, EPI_ISL_3050327, EPI_ISL_3050328, EPI_ISL_3050329, EPI_ISL_3050330, EPI_ISL_3050331, EPI_ISL_3050332, EPI_ISL_3050333 | | | |
| see above | Laboratório Central de Saúde Pública do Amazonas - LACEN-AM | Laboratorio de Ecologia de Doencas Transmissiveis na Amazonia, Instituto Leonidas e Maria Deane - Fiocruz Amazonia | André Corado; Debora Duarte; Felipe Naveca; Fernanda Nascimento; George Silva; Karina Pessoa; Luciana Gonçalves; Maria Júlia Brandão; Matilde Mejía; Michele Jesus; Valdinete Nascimento; Victor Souza; Ágatha Costa |
| EPI_ISL_3426438, EPI_ISL_3610354, EPI_ISL_3610355, EPI_ISL_3610356, EPI_ISL_3610357, EPI_ISL_3610358, EPI_ISL_3610359 | | | |
| see above | Laboratório Central de Saúde Pública do Estado do Tocantins (LACEN/TO) | Bioinformatics and Biotechnology Laboratory (Labinftec/Federal University of Tocantins, Campus of Gurupi) | Bergmann Morais Ribeiro; Chácaras 69/72; Fabrício Souza Campos; Fernando Lucas Melo; Gurupi/TO Ueric José Borges de Souza; Jucimária Dantas Galvão; Laboratório de Bioinformática e Biotecnologia; Lote 7; Raíssa Nunes dos Santos; Rua Badejós; Ueric José Borges de Souza; Zona Rural \| 77.402-970\| UFT Campus de Gurupi |
| EPI_ISL_3873632, EPI_ISL_3873654 | Laboratório de Microbiologia Molecular - Universidade | Molecular Microbiology Laboratory | Alana Witt Hansen; Fernando Rosado Spilki; Fágner Henrique Heldt; Juliana Schons Gularte; Juliane Deise Fleck; Mariana Soares da Silva; Matheus Nunes Weber; Meriane Demoliner; Micheli Filippi; Paula Rodrigues de Almeida; Vyctoria Malayhka de Abreu Góes Pereira. |

| | | | |
|---|---|---|---|
| | FEEVALE | | |
| EPI_ISL_4520596 | Laboratório de Virologia e Cultivo Celular - LABVICC | Laboratório Baculovírus | Aline Belmok; Bergmann Morais Ribeiro; Clayton Luiz Borges; Déborah Anjos; Elen Regozino Muniz; Fabiola Souza Fiaccadori; Fernando L Melo; Gabriela Silvério Bazilio; Juliana Alves Parente Rocha; Keila Correia de Alcântara; Menira Souza; Nathânia Dábilla; Valéria Christina de Rezende Feres |
| EPI_ISL_2863891, EPI_ISL_3045378 | Labortorio Central de Saude Publica do Estado do Rio de Janeiro (LACEN/RJ) | Laboratory of Respiratory Viruses and Measles, Oswaldo Cruz Institute, FIOCRUZ | Agatha Cristinne Prudencio Soares; Alice Sampaio Rocha; Ana Carolina Mendonca; Andrea Cony Cavalcanti; Anna Carolina Paixao; Elisa Cavalcante Pereira; Fernando Motta; Ighor Leonardo Arantes Gomes; Luciana Appolinario; Marilda Siqueira on behalf of the Fiocruz COVID-19 Genomic Surveillance Network; Paola Resende; Renata Serrano Lopes; Taina Venas |
| EPI_ISL_2645507, EPI_ISL_2863596 | Labortorio Central de Saude Publica do Estado do Tocantins (LACEN/TO) | Laboratory of Respiratory Viruses and Measles, Oswaldo Cruz Institute, FIOCRUZ | Alice Sampaio Rocha; Ana Carolina Mendonca; Anna Carolina Paixao; Elisa Cavalcante Pereira; Fernando Motta; Jucimaria Dantas Galvao; Luciana Appolinario; Marilda Siqueira on behalf of the Fiocruz COVID-19 Genomic Surveillance Network; Paola Resende; Renata Serrano Lopes; Taina Venas |
| EPI_ISL_2515605 | Lighthouse Lab in Alderley Park | Wellcome Sanger Institute for the COVID-19 Genomics UK (COG-UK) Consortium | Cordelia Langford; David K. Jackson; Dominic Kwiatkowski; Ewan Harrison; Ian Johnston; Jacquelyn Wynn; Jeffrey Barrett; John Sillitoe on behalf of the Wellcome Sanger Institute COVID-19 Surveillance Team; Mairead Hyland; Roberto Amato; Sonia Goncalves; The Lighthouse Lab in Alderley Park and Alex Alderton |
| EPI_ISL_2457435 | Lighthouse Lab in Glasgow | Wellcome Sanger Institute for the COVID-19 Genomics UK (COG-UK) Consortium | Anna Dominiczak and Alex Alderton; Carol Clugston; Cordelia Langford; David Gray; David K. Jackson; Dominic Kwiatkowski; Ewan Harrison; Harper VanSteenhouse; Ian Johnston; Jeffrey Barrett; John Sillitoe on behalf of the Wellcome Sanger Institute COVID-19 Surveillance Team; Roberto Amato; Sonia Goncalves; Yumi Kasai |
| EPI_ISL_2512526, EPI_ISL_2514791, EPI_ISL_2539223 | Lighthouse Lab in Milton Keynes | Wellcome Sanger Institute for the COVID-19 Genomics UK (COG-UK) Consortium | Cordelia Langford; David K. Jackson; Dominic Kwiatkowski; Ewan Harrison; Ian Johnston; Jeffrey Barrett; John Sillitoe on behalf of the Wellcome Sanger Institute COVID-19 Surveillance Team; Roberto Amato; Sonia Goncalves; The Lighthouse Lab in Milton Keynes and Alex Alderton |
| EPI_ISL_2632968 | Limbach - MVZ Labor Ludwigsburg | Robert Koch Institute | |
| EPI_ISL_2842581 | Lurie Children's Hospital of Chicago | Northwestern University - Ozer Lab | Egon A. Ozer; Judd F. Hultquist; Lacy M. Simons; Larry K. Kociolek; Michael G. Ison; Ramon Lorenzo-Redondo; Taylor J. Dean; William J. Muller; Xiaotian; Zheng |
| EPI_ISL_3356857 | MB-Cadham Provincial laboratory | National Microbiology Laboratory (NML) | Anna Majer; Anneliese Landgraff; CanCOGeN's metadata curation team; Darian Hole; David Alexander; Elsie Grudeski; Gary Van Domselaar; Grace Seo; Jared Bullard; Jennifer Tanner; Kerry Dust; Kirsten Biggar; Madison Chapel; Morag Graham; Natalie Knox; Nathalie Bastien; Paul Van Caeseele; Philip Mabon; Public Health Agency of Canada CanCOGeN team; Rhiannon Huzarewich; Russell Mandes; Shari Tyson; Timothy Booth; Yan Li |
| EPI_ISL_2598083 | MD PHL | MD PHL | Maryland Department of Health Laboratories Administration |
| EPI_ISL_2471925 | MDI Limbach Berlin GmbH; MVZ Labor Berlin | Robert Koch Institute | |
| EPI_ISL_2595749 | MEPHI, Aix Marseille University | MEPHI, Aix Marseille University | Anthony LEVASSEUR |
| EPI_ISL_2626996 | MONTEFIORE MEDICAL CENTER LABORATORIES | Wadsworth Center, New York State Department of Health | Alexis Russell; Catharine Prussing; Daryl M. Lamson; Erasmus Schneider; Erica Lasek-Nesselquist; John Kelly; Jonathan Plitnick; Kirsten St. George; Matthew Shudt; Melissa A Leisner; Navjot Singh |
| EPI_ISL_2787554 | MS Public Health Laboratory | Centers for Disease Control and Prevention Division of Viral Diseases, Pathogen Discovery | Alison Laufer Halpin; Ben L. Rambo-Martin; Clinton R. Paden; Dakota Howard; Darlene Wagner; Dave Wentworth; Dhwani Batra; Jasmine Padilla; Justin Lee; Katie Dillon; Krista Queen; Kristen Knipe; Kristine Lacek; Mark Burroughs; Matthew Schmerer; Mili Sheth; Peter Cook; Sam Shepard; Sarah Nobles; Shoshona Le; Suxiang Tong; Vivien Dugan; Yvette Unoarumhi |
| EPI_ISL_6492802, EPI_ISL_6492803, EPI_ISL_6492805, EPI_ISL_6492806, EPI_ISL_6492811, EPI_ISL_6492813, EPI_ISL_6492816 | | | |
| see above | MSHS Clinical Microbiology Laboratories | MSHS Pathogen Surveillance Program | Adolfo García-Sastre; Adriana van de Guchte; Ajay Obla; Alberto Paniz-Mondolfi; Ana S. Gonzalez-Reiche; Angela Amoako; Ashley Salimbangon; Betsaida Salom Melo; Bremy Alburquerque; Brianne Ciferri; Charles Gleason; Daniel Floda; Deena R. Altman; Denise Jurczyszak; Emilia Mia Sordillo; Gintaras Deikus; Giulio Kleiner; Gopi Patel; Hala Alshammary; Harm van Bakel; Irina Oussenko; Jayeeta Dutta; Juan Soto; Julia Matthews; Katherine Beach; Kathryn Twyman; Kayla Russo; Komal Srivastava; Levy Sominsky; Mahmoud Awawda; Marta Luksza; Matthew M. Hernandez; Melissa Gitman; Michael D. Nowak; Mitchell J. Sullivan; Nancy Francoeur; Robert Sebra; Sarah Schaefer; Shelcie Fabre; Shwetha Hara Sridhar; Viviana Simon; Ying-Chih Wang; Zain Khalil; Zenab Khan |
| EPI_ISL_2690127 | MT Public Health Laboratory | Centers for Disease Control and Prevention Division of Viral Diseases, Pathogen Discovery | Alison Laufer Halpin; Ben L. Rambo-Martin; Clinton R. Paden; Dakota Howard; Darlene Wagner; Dave Wentworth; Dhwani Batra; Jasmine Padilla; Justin Lee; Katie Dillon; Krista Queen; Kristen Knipe; Kristine Lacek; Mark Burroughs; Matthew Schmerer; Mili Sheth; Peter Cook; Sam Shepard; Sarah Nobles; Shoshona Le; Suxiang Tong; Vivien Dugan; Yvette Unoarumhi |
| EPI_ISL_2632797, EPI_ISL_2632803, EPI_ISL_2632810, EPI_ISL_2632812, EPI_ISL_2632831 | MVZ Labor Dr. Limbach & Kollegen GbR | Robert Koch Institute | |
| EPI_ISL_3085947 | Maine Health and Environmental Testing Laboratory | Tewhey Lab, The Jackson Laboratory | Barter, M.; Dewey, H.; H. and Tewhey, R.; Iosue, F.; Lynch, R.; Matluk, N.; Munger |
| EPI_ISL_2786002, EPI_ISL_2786004, EPI_ISL_2786064, EPI_ISL_2786086, EPI_ISL_2786093, EPI_ISL_2786115, EPI_ISL_2786129, EPI_ISL_2786143, EPI_ISL_2786191, EPI_ISL_2875242, EPI_ISL_2875351 | | | |
| see above | Mako Medical | Centers for Disease Control and Prevention Division of Viral Diseases, Pathogen Discovery | Adrian Paskey; Benjamin Rambo-Martin; Christopher Gulvick; Clinton R. Paden; Dakota Howard; Darlene Wagner; Dhwani Batra; Duncan MacCannell; Jason Caravas; Kara Moser; Lauren Moon; Matthew Schmerer; Matthew Tugwell; Peter W. Cook; Scott Sammons; Shatavia Morrison; Yvette Unoarumhi |
| EPI_ISL_6666013 | Mass General Brigham | Mass General Brigham | A.E.; Adams, G.; Anahtar, M.; B.L.; B.W.; Bauer, M.; Birren; Branda, J.; Carter, A.; Cerrato, F.; Chaluvadi, S.; Chapman; Cusick, C.; D.J.; DeRuff, K.; E. and Sabeti; Flowers, K.; Gallagher, G.; Gladden-Young, A.; Gnirke, A.; Harris, J.; J.E.; K.J.; LaRocque, R.; Lagerborg, K.; Lemieux; Lin; Loreth, C.; MacInnis; Neumann, A.; Normandin, E.; P.C.; Park; Pierce, V.; Reilly, S.; Rosenberg; Rudy, M.; Ryan, E.; S.B.; Shaw, B.; Siddle; Slater, D.; Smole, S.; Tomkins-Tinch, C.; Turbett, S. |
| EPI_ISL_2688808, EPI_ISL_2688819, EPI_ISL_2920253, EPI_ISL_2920262, EPI_ISL_2920263, EPI_ISL_2920266 | Massachusetts State Public Health Laboratory | Massachusetts State Public Health Laboratory | Andrew Lang; Glen Gallagher; Sandra Smole; Timelia Fink |
| EPI_ISL_2650316, EPI_ISL_2650328 | Microbiologia CATLAB | Can Ruti SARS-CoV-2 Sequencing Hub (HUGTiP/IrsiCaixa/IGTP) | Alba Sánchez; Anna Not; Antoni E Bordoy; Bonaventura Clotet; Cristina Casañ; Cristina Esteban; Francesc Catala-Moll; Gemma Clara; Ignacio Blanco; Marc Noguera-Julian; Maria Casadellà; Mariona Parera; Mercedes Guerrero; Montserrat Giménez; Pere-Joan Cardona; Pilar Armengol; Roger Paredes; Verónica Saludes; and Elisa Martró on behalf of the Can Ruti SARS-CoV-2 Sequencing Hub |
| EPI_ISL_2448086 | Microbiology Department, Laboratori Clínic Metropolitana Nord. Hospital Universitari Germans Trias i Pujol | Can Ruti SARS-CoV-2 Sequencing Hub (HUGTiP/IrsiCaixa/IGTP) | Alba Snchez; Anna Not; Antoni E Bordoy; Bonaventura Clotet; Cristina Casa; Cristina Esteban; Francesc Catala-Moll; Gemma Clara; Ignacio Blanco; Marc Noguera-Julian; Maria Casadell; Mariona Parera; Mercedes Guerrero; Montserrat Gimnez; Pere-Joan Cardona; Pilar Armengol; Roger Paredes; Vernica Saludes; and Elisa Martr on behalf of the Can Ruti SARS-CoV-2 Sequencing Hub. |
| EPI_ISL_2774104 | Microbiology Department, University Hospital Donostia | Microbiology Department, University Hospital Donostia | Cilla G.; Gomez M; Marimon JM; Martin-Peñaranda T; Montes M; Piñeiro L; Sorarrain A |
| EPI_ISL_2968531, EPI_ISL_3066700, EPI_ISL_3066701, EPI_ISL_3066703, EPI_ISL_3066705, EPI_ISL_3066706, EPI_ISL_3066707, EPI_ISL_3066709, EPI_ISL_3066710, EPI_ISL_3066715, EPI_ISL_3066726, EPI_ISL_3066727, EPI_ISL_3066729, EPI_ISL_3066730 | | | |
| see above | Ministry of Health Turkey | Ministry of Health Turkey | Fatma Bayrakdar; Gulay Korukluoglu; Suleyman Yalcin; Yasemin Cosgun |
| EPI_ISL_2443840, EPI_ISL_2501152, EPI_ISL_2652483 | Minnesota Department of Health, Public Health Laboratory | Minnesota Department of Health, Public Health Laboratory | Alexandra Lorentz; Jacob Garfin; Matt Plumb; and Xiong Wang |
| EPI_ISL_2938308 | Mobile County Health Department | Synergy Laboratories | Megan Cornwell |
| EPI_ISL_3142934 | Molecular & Genomic Pathology Laboratory, Thomas Jefferson University Hospital | Molecular & Genomic Pathology Laboratory, Thomas Jefferson University Hospital | Nitika Badjatia; Run Jin; Shannon Mahoney; Zi-xuan Wang |
| EPI_ISL_5598228 | Molecular Biology Laboratory, Vidant Medical Center | Brody Integrative Genomics Core, East Carolina University | Changhong Yin; Heather Duncan; John T. Fallon; Kimberly P. Briley; Weihua Huang |
| EPI_ISL_3060283 | Molecular Biology Laboratory, Westchester Medical Center | Genomics Core Laboratory, New York Medical College | Changhong Yin; Jian Zhuge; Utsav Pandey; Weihua Huang |
| EPI_ISL_5681776 | NAMRU-6 | NAMRU-6 | Cristhopher Cruz; Eugenio Abente; Gilda Troncos; Marita Silva; Paul Graf.; Sonia Ampuero; Victor Herrera; Victoria Espejo; Yeny Tinoco |
| EPI_ISL_2627767, EPI_ISL_2627768 | NCSLPH | NCSLPH | Chase K; Glover W; Greene S; Miller MC |
| EPI_ISL_2689925 | NE Public Health Laboratory | Centers for Disease Control and Prevention Division of Viral Diseases, Pathogen Discovery | Alison Laufer Halpin; Ben L. Rambo-Martin; Clinton R. Paden; Dakota Howard; Darlene Wagner; Dave Wentworth; Dhwani Batra; Jasmine Padilla; Justin Lee; Katie Dillon; Krista Queen; Kristen Knipe; Kristine Lacek; Mark Burroughs; Matthew Schmerer; Mili Sheth; Peter Cook; Sam Shepard; Sarah Nobles; Shoshona Le; Suxiang Tong; Vivien Dugan; Yvette Unoarumhi |
| EPI_ISL_2787928 | NJ Public Health and Environmental Laboratories | Centers for Disease Control and Prevention Division of Viral Diseases, Pathogen Discovery | Alison Laufer Halpin; Ben L. Rambo-Martin; Clinton R. Paden; Dakota Howard; Darlene Wagner; Dave Wentworth; Dhwani Batra; Jasmine Padilla; Justin Lee; Katie Dillon; Krista Queen; Kristen Knipe; Kristine Lacek; Mark Burroughs; Matthew Schmerer; Mili Sheth; Peter Cook; Sam Shepard; Sarah Nobles; Shoshona Le; Suxiang Tong; Vivien Dugan; Yvette Unoarumhi |
| EPI_ISL_2557117, EPI_ISL_2557134, | NJDOH, Public Health and Environmental Laboratories | NJ_PHEL | Allison Roder; Byeong Jeong; Chelsea San Filippo; Dana Woell; Jacquelyn Deverell; Lindsey Bodnar; Ryan Pachucki; Shiv K. Verma |

| | | | |
|---|---|---|---|
| EPI_ISL_2649830, EPI_ISL_2649831, EPI_ISL_2842676 | | | |
| EPI_ISL_5646318, EPI_ISL_5646330, EPI_ISL_5646348 | NUCLEO DE SAUDE MARY DOTA DE BAURU | Instituto Butantan | Antonio Jorge Martins; Claudia Renata dos Santos Barros; David Schlesinger; Debora Botequio Moretti; Dimas Tadeu Covas; Elaine Cristina Marqueze; Elaine Vieira Santos; Evandra Strazza Rodrigues; Heidge Fukumasu; Jayme Augusto de Souza-Neto; José Salvatore Leister Patané; Luiz Alcantara; Luiz Lehmann Coutinho; Maria Carolina Elias; Maurício Lacerda Nogueira; Rafael dos Santos Bezerra; Raul Machado Neto; Rejane Maria Tommasini Grotto; Ricardo Haddad; Sandra Coccuzzo Sampaio Vessoni; Simone Kashima; Svetoslav Nanev Slavov; Vincent Louis Viala |
| EPI_ISL_2821280, EPI_ISL_2821281, EPI_ISL_2821282, EPI_ISL_2821283, EPI_ISL_2821284, EPI_ISL_3046158, EPI_ISL_3046159 | | | |
| see above | NUPIT/UFPE | WallauLab on behalf of Fiocruz COVID-19 Genomic Surveillance Network | Alexandre Freitas da Silva; Antonio Mauro Rezende; Cassia Docena; Constância Flávia Junqueira Ayres; Cássia Docena; Duschinka Ribeiro Duarte Guedes; Elisama Helvecio; Filipe Zimmer Dezordi; Gabriel Luz Wallau; Gustavo Barbosa de Lima; Lais Ceschini Machado; Larissa Krokovsky; Laís Ceschini Machado; Lilian Caroliny Amorim Silva; Maira Galdino da Rocha Pitta; Marcelo Henrique dos Santos Paiva; Matheus Filgueira Bezerra; Michelly Cristiny Pereira; Rômulo Pessoa e Silva; Sinval Pinto Brandão Filho |
| EPI_ISL_2663528, EPI_ISL_3245467 | NYU Langone Health | Departments of Pathology and Medicine, New York University School of Medicine | Adriana Heguy; Christian Marier; Dacia Dimartino; Emily Guzman; Gael Westby; Guiqing Wang; Paolo Cotzia; Paul Zappile; Peter Meyn; Sitharam Ramaswami; Yutong Zhang |
| EPI_ISL_2466319, EPI_ISL_2466320, EPI_ISL_2483717, EPI_ISL_2483718, EPI_ISL_2483719, EPI_ISL_2483838, EPI_ISL_2545466, EPI_ISL_2545467, EPI_ISL_2545468, EPI_ISL_2549195, EPI_ISL_2687969, EPI_ISL_2687975 | | | |
| see above | National Platform bis UMONS/Jolimont | National Platform bis UMONS/Jolimont | Florian Juszczak; François Dufrasne; Gautier Detry; Guillaume Bayon-Vicente; Ruddy Wattiez |
| EPI_ISL_2400264, EPI_ISL_2442390, EPI_ISL_2442394, EPI_ISL_2466750, EPI_ISL_2754891 | Nebraska Public Health Laboratory | NPHL COVID-19 Response Team | NPHL COVID-19 Response Team |
| EPI_ISL_2652076 | New Mexico Department of Health Scientific Laboratory | New Mexico Department of Health Scientific Laboratory | Anastacia Griego-Fisher; D'eldra Malone; Ellie Johnson; Jennifer Benoit; Keila Gutierrez; Linda Salazar; Ratheesh Rajan |
| EPI_ISL_2714620 | North Dakota Department of Health, Public Health Laboratory | North Dakota Department of Health, Public Health Laboratory | Lisa Wingerter |
| EPI_ISL_2736086, EPI_ISL_2736088, EPI_ISL_2736090, EPI_ISL_2736091, EPI_ISL_2736095, EPI_ISL_2736101, EPI_ISL_2736106, EPI_ISL_2736115, EPI_ISL_2736125, EPI_ISL_2736128, EPI_ISL_2736137, EPI_ISL_2736147, EPI_ISL_2736150, EPI_ISL_2736154, EPI_ISL_2736162, EPI_ISL_2736163, EPI_ISL_2736172, EPI_ISL_3015638, EPI_ISL_3015641, EPI_ISL_3015644, EPI_ISL_3015645, EPI_ISL_3015651, EPI_ISL_3015654, EPI_ISL_3015663, EPI_ISL_3015694, EPI_ISL_3015696, EPI_ISL_3015697, EPI_ISL_3015702, EPI_ISL_3015704, EPI_ISL_3015705, EPI_ISL_3015706, EPI_ISL_3015709, EPI_ISL_3015710, EPI_ISL_3015714 | | | |
| see above | Northwest Laboratory | Seattle Flu Study | Amanda Adler; Barry R. Lutz; Benjamin Pelle; Brian Hiatt; Caitlin R. Wolf; Chris D. Frazar; Deborah A. Nickerson; Elisabeth Brandstetter; Erica Ryke; Geoff Melly; Helen Y. Chu; Janet A. Englund; Jay Shendure; Jover Lee; Kairsten Fay; Kirsten Lacombe; Lea M. Starita; Mark J. Rieder; Matthew Richardson; Matthew Thompson; Melissa Truong; Michael Boeckh; Michael Famulare; Misja Ilcisin; Peter D. Han; Philip Dykema; Romesh Gautom; Scott Lindquist; Thomas R. Sibley; Trevor Bedford |
| EPI_ISL_2842605, EPI_ISL_2842608 | Northwestern Memorial Hospital | Northwestern University - Ozer Lab | Chad J. Achenbach; Chao Qi; Egon A. Ozer; Judd F. Hultquist; Lacy M. Simons; Lawrence J. Jennings; Michael G. Ison; Ramon Lorenzo-Redondo; Taylor J. Dean |
| EPI_ISL_2838814, EPI_ISL_2838877, EPI_ISL_2838878, EPI_ISL_2838879, EPI_ISL_2838880, EPI_ISL_2838881, EPI_ISL_2838882 | | | |
| see above | OHSU Lab Services Molecular Microbiology Lab | Oregon SARS-CoV-2 Genome Sequencing Center | '[UsersoroakBox SyncPHI_SARS-COV-2_Sample_Sheet_infoGISAID_Submission_Infometa_master_2021.xlsx]release20_final'!D:AC; 26; FALSE); VLOOKUP(C528; VLOOKUP(C529; VLOOKUP(C530; VLOOKUP(C533; VLOOKUP(C534; VLOOKUP(C536; VLOOKUP(C539 |
| EPI_ISL_2402548, EPI_ISL_2402549, EPI_ISL_2544902 | OLVZ Aalst | OLVZ Aalst | Anne Vankeerberghen; Astrid Holderbeke |
| EPI_ISL_2787447, EPI_ISL_2787450, EPI_ISL_2787453, EPI_ISL_2787459 | OR State PHL-Virology/Immunology Section | Centers for Disease Control and Prevention Division of Viral Diseases, Pathogen Discovery | Alison Laufer Halpin; Ben L. Rambo-Martin; Clinton R. Paden; Dakota Howard; Darlene Wagner; Dave Wentworth; Dhwani Batra; Jasmine Padilla; Justin Lee; Katie Dillon; Krista Queen; Kristen Knipe; Kristine Lacek; Mark Burroughs; Matthew Schmerer; Mili Sheth; Peter Cook; Sam Shepard; Sarah Nobles; Shoshona Le; Suxiang Tong; Vivien Dugan; Yvette Unoarumhi |
| EPI_ISL_2600834 | OSPEDALE CIVILE TERAMO | Istituto Zooprofilattico Sperimentale dell'Abruzzo e Molise "G. Caporale" | Ancora M; Calistri P; Cammà C; Caporale M; Curini V; Delli Compagni E; Di Domenico M; Di Lollo Valeria; Di Pasquale A; Lorusso A; Mangone I; Marcacci M; Puglia I; Rinaldi A; Savini G; Scialabba S |
| EPI_ISL_2928154, EPI_ISL_2928156, EPI_ISL_2928158, EPI_ISL_2928161 | Orange County Public Health Laboratory | Orange County Public Health Laboratory | Angelica Torres; Julia Wolfe |
| EPI_ISL_2473664, EPI_ISL_2473665, EPI_ISL_2604198, EPI_ISL_2604204, EPI_ISL_2612541, EPI_ISL_2631151, EPI_ISL_2631187 | | | |
| see above | Oregon State Public Health Laboratory | Oregon State Public Health Laboratory | Eugene Yeboah; John Fontana and Shane Sevey; Laura Tsaknaridis; Rafia Razzaque; Vanda Makris |
| EPI_ISL_2665399 | Ospedale S.M. Goretti di Latina - ASL Latina presidio ospedaliero Nord | National Institute for Infectious Diseases (INMI) L. Spallanzani I.R.C.C.S | A Di Caro; B Bartolini; CEM Gruber; E Giombini; F Messina; F Santini; G Bonfiglio; M Rueca; MR Capobianchi; O Butera |
| EPI_ISL_2665398 | Ospedale San Filippo Neri | National Institute for Infectious Diseases (INMI) L. Spallanzani I.R.C.C.S | A Di Caro; B Bartolini; CEM Gruber; E Giombini; F Messina; F Santini; G Bonfiglio; M Rueca; MR Capobianchi; O Butera |
| EPI_ISL_3104860, EPI_ISL_3104875, EPI_ISL_3104972, EPI_ISL_3105032 | Overlake Hospital | Seattle Flu Study | Amanda Adler; Barry R. Lutz; Benjamin Pelle; Brian Hiatt; Caitlin R. Wolf; Chris D. Frazar; Deborah A. Nickerson; Elisabeth Brandstetter; Erica Ryke; Geoff Melly; Helen Y. Chu; Janet A. Englund; Jay Shendure; Jover Lee; Kairsten Fay; Kirsten Lacombe; Lea M. Starita; Mark J. Rieder; Matthew Richardson; Matthew Thompson; Melissa Truong; Michael Boeckh; Michael Famulare; Misja Ilcisin; Peter D. Han; Philip Dykema; Romesh Gautom; Scott Lindquist; Thomas R. Sibley; Trevor Bedford |
| EPI_ISL_5646312, EPI_ISL_5646331, EPI_ISL_5646334, EPI_ISL_5646337, EPI_ISL_5646339, EPI_ISL_5646373, EPI_ISL_5646377 | | | |
| see above | PA MATAO | Instituto Butantan | Antonio Jorge Martins; Claudia Renata dos Santos Barros; David Schlesinger; Debora Botequio Moretti; Dimas Tadeu Covas; Elaine Cristina Marqueze; Elaine Vieira Santos; Evandra Strazza Rodrigues; Heidge Fukumasu; Jayme Augusto de Souza-Neto; José Salvatore Leister Patané; Luiz Alcantara; Luiz Lehmann Coutinho; Maria Carolina Elias; Maurício Lacerda Nogueira; Rafael dos Santos Bezerra; Raul Machado Neto; Rejane Maria Tommasini Grotto; Ricardo Haddad; Sandra Coccuzzo Sampaio Vessoni; Simone Kashima; Svetoslav Nanev Slavov; Vincent Louis Viala |
| EPI_ISL_2691298 | PAMF Palo Alto | Santa Clara County Public Health Laboratory | Santa Clara County Public Health Department |
| EPI_ISL_5645786, EPI_ISL_5645872, EPI_ISL_5645916, EPI_ISL_5645926, EPI_ISL_5645958 | PAS JOAO ANTONIO DO NASCIMENTO | Instituto Butantan | Antonio Jorge Martins; Claudia Renata dos Santos Barros; David Schlesinger; Debora Botequio Moretti; Dimas Tadeu Covas; Elaine Cristina Marqueze; Elaine Vieira Santos; Evandra Strazza Rodrigues; Heidge Fukumasu; Jayme Augusto de Souza-Neto; José Salvatore Leister Patané; Luiz Alcantara; Luiz Lehmann Coutinho; Maria Carolina Elias; Maurício Lacerda Nogueira; Rafael dos Santos Bezerra; Raul Machado Neto; Rejane Maria Tommasini Grotto; Ricardo Haddad; Sandra Coccuzzo Sampaio Vessoni; Simone Kashima; Svetoslav Nanev Slavov; Vincent Louis Viala |
| EPI_ISL_2626972 | PECONIC BAY MEDICAL CENTER LABORATORY | Wadsworth Center, New York State Department of Health | Alexis Russell; Catharine Prussing; Daryl M. Lamson; Erasmus Schneider; Erica Lasek-Nesselquist; John Kelly; Jonathan Plitnick; Kirsten St. George; Matthew Shudt; Melissa A Leisner; Navjot Singh |
| EPI_ISL_5645639, EPI_ISL_5645651, EPI_ISL_5645664, EPI_ISL_5645695, EPI_ISL_5645765, EPI_ISL_5645876, EPI_ISL_5645878, EPI_ISL_5645902 | | | |
| see above | POLICLINICA ALCIA LINEIA VITTI REJANI | Instituto Butantan | Antonio Jorge Martins; Claudia Renata dos Santos Barros; David Schlesinger; Debora Botequio Moretti; Dimas Tadeu Covas; Elaine Cristina Marqueze; Elaine Vieira Santos; Evandra Strazza Rodrigues; Heidge Fukumasu; Jayme Augusto de Souza-Neto; José Salvatore Leister Patané; Luiz Alcantara; Luiz Lehmann Coutinho; Maria Carolina Elias; Maurício Lacerda Nogueira; Rafael dos Santos Bezerra; Raul Machado Neto; Rejane Maria Tommasini Grotto; Ricardo Haddad; Sandra Coccuzzo Sampaio Vessoni; Simone Kashima; Svetoslav Nanev Slavov; Vincent Louis Viala |
| EPI_ISL_5645622, EPI_ISL_5645712, EPI_ISL_5645730, EPI_ISL_5646152 | POLICLINICA COVID 19 ITAPETININGA | Instituto Butantan | Antonio Jorge Martins; Claudia Renata dos Santos Barros; David Schlesinger; Debora Botequio Moretti; Dimas Tadeu Covas; Elaine Cristina Marqueze; Elaine Vieira Santos; Evandra Strazza Rodrigues; Heidge Fukumasu; Jayme Augusto de Souza-Neto; José Salvatore Leister Patané; Luiz Alcantara; Luiz Lehmann Coutinho; Maria Carolina Elias; Maurício Lacerda Nogueira; Rafael dos Santos Bezerra; Raul Machado Neto; Rejane Maria Tommasini Grotto; Ricardo Haddad; Sandra Coccuzzo Sampaio Vessoni; Simone Kashima; Svetoslav Nanev Slavov; Vincent Louis Viala |
| EPI_ISL_5645597, EPI_ISL_5645601, EPI_ISL_5645856, EPI_ISL_5645870 | POSTO MEDICO NATAL DIEGUES DE ESTIVA GERBI | Instituto Butantan | Antonio Jorge Martins; Claudia Renata dos Santos Barros; David Schlesinger; Debora Botequio Moretti; Dimas Tadeu Covas; Elaine Cristina Marqueze; Elaine Vieira Santos; Evandra Strazza Rodrigues; Heidge Fukumasu; Jayme Augusto de Souza-Neto; José Salvatore Leister Patané; Luiz Alcantara; Luiz Lehmann Coutinho; Maria Carolina Elias; Maurício Lacerda Nogueira; Rafael dos Santos Bezerra; Raul Machado Neto; Rejane Maria Tommasini Grotto; Ricardo Haddad; Sandra Coccuzzo Sampaio Vessoni; Simone Kashima; Svetoslav Nanev Slavov; Vincent Louis Viala |
| EPI_ISL_5647125 | PRONTO ATENDIMENTO DR SERGIO AROUCA CAMPO GRANDE | Instituto Butantan | Antonio Jorge Martins; Claudia Renata dos Santos Barros; David Schlesinger; Debora Botequio Moretti; Dimas Tadeu Covas; Elaine Cristina Marqueze; Elaine Vieira Santos; Evandra Strazza Rodrigues; Heidge Fukumasu; Jayme Augusto de Souza-Neto; José Salvatore Leister Patané; Luiz Alcantara; Luiz Lehmann Coutinho; Maria Carolina Elias; Maurício Lacerda Nogueira; Rafael dos Santos Bezerra; Raul Machado Neto; Rejane Maria Tommasini Grotto; Ricardo Haddad; Sandra Coccuzzo Sampaio Vessoni; Simone Kashima; Svetoslav Nanev Slavov; Vincent Louis Viala |
| EPI_ISL_5649595, EPI_ISL_5649640, EPI_ISL_5649661, EPI_ISL_5649691 | PRONTO SOCORRO MUNICIPAL DE SEVERINIA | Instituto Butantan | Antonio Jorge Martins; Claudia Renata dos Santos Barros; David Schlesinger; Debora Botequio Moretti; Dimas Tadeu Covas; Elaine Cristina Marqueze; Elaine Vieira Santos; Evandra Strazza Rodrigues; Heidge Fukumasu; Jayme Augusto de Souza-Neto; José Salvatore Leister Patané; Luiz Alcantara; Luiz Lehmann Coutinho; Maria Carolina Elias; Maurício Lacerda Nogueira; Rafael dos Santos Bezerra; Raul Machado Neto; Rejane Maria Tommasini Grotto; Ricardo Haddad; Sandra Coccuzzo Sampaio Vessoni; Simone Kashima; Svetoslav Nanev Slavov; Vincent Louis Viala |
| EPI_ISL_5646133 | PSF DR ANTONIO PIRES DE ALMEIDA PORTO FELIZ | Instituto Butantan | Antonio Jorge Martins; Claudia Renata dos Santos Barros; David Schlesinger; Debora Botequio Moretti; Dimas Tadeu Covas; Elaine Cristina Marqueze; Elaine Vieira Santos; Evandra Strazza Rodrigues; Heidge Fukumasu; Jayme Augusto de Souza-Neto; José Salvatore Leister Patané; Luiz Alcantara; Luiz Lehmann Coutinho; Maria Carolina Elias; Maurício Lacerda Nogueira; Rafael dos Santos Bezerra; Raul Machado Neto; Rejane Maria Tommasini Grotto; Ricardo Haddad; Sandra Coccuzzo Sampaio Vessoni; Simone Kashima; Svetoslav Nanev Slavov; Vincent Louis Viala |
| EPI_ISL_5645737 | PSF MARIA JOSE SALTO DE | Instituto Butantan | Antonio Jorge Martins; Claudia Renata dos Santos Barros; David Schlesinger; Debora Botequio Moretti; Dimas Tadeu Covas; Elaine Cristina Marqueze; Elaine Vieira Santos; Evandra Strazza Rodrigues; Heidge Fukumasu; Jayme Augusto de Souza-Neto; José Salvatore Leister Patané; Luiz Alcantara; Luiz Lehmann |

| | | | |
|---|---|---|---|
| | PIRAPORA | | Coutinho; Maria Carolina Elias; Maurício Lacerda Nogueira; Rafael dos Santos Bezerra; Raul Machado Neto; Rejane Maria Tommasini Grotto; Ricardo Haddad; Sandra Coccuzzo Sampaio Vessoni; Simone Kashima; Svetoslav Nanev Slavov; Vincent Louis Viala |
| EPI_ISL_2446088, EPI_ISL_2446091, EPI_ISL_2446094, EPI_ISL_2631339, EPI_ISL_2631340, EPI_ISL_2631343, EPI_ISL_2631349, EPI_ISL_2631350, EPI_ISL_2631354, EPI_ISL_2631356, EPI_ISL_2631359, EPI_ISL_2631360, EPI_ISL_2631366, EPI_ISL_2631371, EPI_ISL_2631382, EPI_ISL_2631386, EPI_ISL_3933321 | | | |
| see above | Pandemic Response Lab - NYC | Pandemic Response Lab, R&D | Cybill del Castillo; Dylan Law; Haiping Hao; Henry Lee; Isabel Fernandez Escapa; Jon Laurent; Katharine Nelson; Melissa Hopkins; Michael Hammerling; Pradeep Bugga; Shinyoung Clair Kang; Sol Rey; William Ward |
| EPI_ISL_3030263, EPI_ISL_3030264, EPI_ISL_3030265 | Pesaro | Microbiology University Politecnica delle Marche | Anna Valenza; Carla Acciarri; Katia Marinelli; Monica Lucia Ferreri; Patrizia Bagnarelli; Roberta Longo; Sara Caucci; Stefano Menzo |
| EPI_ISL_6907433 | Plataforma de Vigilancia Molecular (PVM) - FIOCRUZ/BA | Plataforma de Vigilancia Molecular (PVM) - FIOCRUZ/BA | Bruno Bezerril Andrade; Camila I. de Oliveira; Clarissa Araújo Gurgel; Eduardo Oyama; Francys A Rangel; Laila A da Silva; Laise de Moraes; Leonardo Paiva Farias; Marina Cucco; Pedro Silveira; Ricardo Khouri on behalf of the Fiocruz COVID-19 Genomic Surveillance Network.; Tiago Graf |
| EPI_ISL_2521960, EPI_ISL_2521961, EPI_ISL_2521969, EPI_ISL_2521975, EPI_ISL_2532759, EPI_ISL_2544717, EPI_ISL_2544721 | | | |
| see above | Plateforme de testing Namuroise | Plateforme de testing Namuroise | Degosserie Jonathan; Demars Aurore; Denis Olivier; Maschietto Céline; Mullier François; Nicolas GILLIARD; Nicolas Gilliard; Nobis Chloé; Otto Gaetan |
| EPI_ISL_2422139, EPI_ISL_2443391, EPI_ISL_2483256, EPI_ISL_2483279, EPI_ISL_2483291, EPI_ISL_2483294 | Platform BIS UZA/UAntwerpen | Labo Klinische Biologie, UZA | Basil Britto Xavier; Christine Lammens; Herman Goossens; Jasmine Coppens; Marie Le Mercier; Veerle Matheeussen |
| EPI_ISL_5646118, EPI_ISL_5646120, EPI_ISL_5646124, EPI_ISL_5646125, EPI_ISL_5646129, EPI_ISL_5646130, EPI_ISL_5646132, EPI_ISL_5646134, EPI_ISL_5646136, EPI_ISL_5646138, EPI_ISL_5646141, EPI_ISL_5646160, EPI_ISL_5646166, EPI_ISL_5646176, EPI_ISL_5646178 | | | |
| see above | Prefeitura de Botucatu | Instituto Butantan | Antonio Jorge Martins; Claudia Renata dos Santos Barros; David Schlesinger; Debora Botequio Moretti; Dimas Tadeu Covas; Elaine Cristina Marqueze; Elaine Vieira Santos; Evandra Strazza Rodrigues; Heidge Fukumasu; Jayme Augusto de Souza-Neto; José Salvatore Leister Patané; Luiz Alcantara; Luiz Lehmann Coutinho; Maria Carolina Elias; Maurício Lacerda Nogueira; Rafael dos Santos Bezerra; Raul Machado Neto; Rejane Maria Tommasini Grotto; Ricardo Haddad; Sandra Coccuzzo Sampaio Vessoni; Simone Kashima; Svetoslav Nanev Slavov; Vincent Louis Viala |
| EPI_ISL_7406399 | Programa Laboratorio de Salud Pública "Dr Dalmiro Pérez Laborda" | Área de Secuenciación del Laboratorio de Virología del Hospital de Niños Dr. Ricardo Gutierrez on behalf of 'Proyecto Argentino Interinstitucional de genomica de SARS-CoV-2' (PAIS Consortium) | A; AM; Acuña; Aguilera; BH; Bohn; Cabral Bombardieri; Campos; Carrizo; D; E; Esteves; F; G; Goya; J; JM; JP; Jofré; L; LE; Lacaze; Lusso; M; MA; MI; MS; Mastrodonato; Molina; Nabaes Jodar; Natale; Olivera; Perez Diaz; Peñalva; Quijano; Rivero; Rosales; S; Talia; Valinotto; Viegas, M. |
| EPI_ISL_3132189 | Puerto Rico Department of Health | Centers for Disease Control and Prevention, Dengue Branch | Betzabel Flores; Gabriela Paz-Bailey; Gilberto A. Santiago; Glenda Gonzalez; Jorge L. Munoz-Jordan; Keyla Charriez |
| EPI_ISL_2758341 | QLabs | WVU and Marshall University Combined Genomics Core Facilities | James Denvir; Peter Perrotta; Peter Stoilov; Ryan Percifield; Wesley Kimble |
| EPI_ISL_2520234, EPI_ISL_2706726 | Quadram Institute Bioscience | COVID-19 Genomics UK (COG-UK) Consortium | Alexander J Trotter; Alison E. Mather; Alp Aydin; Ana P. Tedim; Anastasia Kolyva; Andrew Bell; Andrew J. Page; Claire Stuart; Dave J. Baker; Gemma L. Kay; John Wain; Justin O'Grady; Leonardo de Oliveira Martins; Lizzie Meadows; Maria Diaz; Mark Webber; Muhammed Yasir; Nabil-Fareed Alikhan; Ngozi Elumogo; Nicholas M. Thomson; Rachael Stanley; Rachel Gilroy; Reenesh Prakash; Samir Dervisevic; Samuel Bloomfield; Steven Rudder; Thanh Le-Viet |
| EPI_ISL_2529170, EPI_ISL_2534553, EPI_ISL_2534554, EPI_ISL_2534555, EPI_ISL_2534557, EPI_ISL_2534558, EPI_ISL_2534559, EPI_ISL_2534560, EPI_ISL_2534561, EPI_ISL_2534563, EPI_ISL_2534565, EPI_ISL_2649948, EPI_ISL_2649951, EPI_ISL_2649953, EPI_ISL_2649954, EPI_ISL_2649955, EPI_ISL_2649957, EPI_ISL_2649958, EPI_ISL_2649960, EPI_ISL_2649963, EPI_ISL_2649964, EPI_ISL_2649965, EPI_ISL_2649966, EPI_ISL_2649967, EPI_ISL_2649968, EPI_ISL_2649976, EPI_ISL_2649977, EPI_ISL_2649978, EPI_ISL_2649979, EPI_ISL_2649980, EPI_ISL_2649981, EPI_ISL_2649982, EPI_ISL_2649983, EPI_ISL_2649984, EPI_ISL_2649986, EPI_ISL_2649987, EPI_ISL_2649988, EPI_ISL_2649989, EPI_ISL_2649990, EPI_ISL_2686368, EPI_ISL_2686373, EPI_ISL_2686378, EPI_ISL_2876143, EPI_ISL_4370349 | | | |
| see above | Quest Diagnostics Incorporated | Centers for Disease Control and Prevention Division of Viral Diseases, Pathogen Discovery | A. Gerasimova; A. Perez; Adrian Paskey; B. Anderson; Benjamin Rambo-Martin; Christopher Gulvick; Clinton Paden; Clinton R. Paden; Dakota Howard; Darlene Wagner; Dhwani Batra; Duncan MacCannell; Erisa Sula; F. Lacbawan; I. A. Shlyakhter; I. Shlyakhter; Jason Caravas; K. Livingston; K.E. Livingston; Kara Moser; Kristine Lacek; L. Bernstein; L.E. Bernstein; M. Hua; Matthew Schmerer; P. Tanpaiboon; Peter Cook; Peter W. Cook; R. Kagan; R. M. Kagan; R. Owen; R. Rolando; R. V. Rolando; S. H. Rosenthal; S. Rosenthal; Scott Sammons; Shatavia Morrison; Tymeckia Kendall; Victoria Caban Figueroa; Y. Liu; Yvette Unoarumhi |
| EPI_ISL_2787984, EPI_ISL_2787987 | RI State Health Laboratories | Centers for Disease Control and Prevention Division of Viral Diseases, Pathogen Discovery | Alison Laufer Halpin; Ben L. Rambo-Martin; Clinton R. Paden; Dakota Howard; Darlene Wagner; Dave Wentworth; Dhwani Batra; Jasmine Padilla; Justin Lee; Katie Dillon; Krista Queen; Kristen Knipe; Kristine Lacek; Mark Burroughs; Matthew Schmerer; Mili Sheth; Peter Cook; Sam Shepard; Sarah Nobles; Shoshona Le; Suxiang Tong; Vivien Dugan; Yvette Unoarumhi |
| EPI_ISL_2931094 | Rush University Medical Center | RIPHL at Rush University Medical Center | Diane Springer; Felix Araujo Perez; Joyce Houlihan; Kevin Kunstman; Laura Furtado; Marieta Hyde; Mary Hayden; Max Kolton; Stefan Green |
| EPI_ISL_2925707 | S. Camillo | National Institute for Infectious Diseases (INMI) L. Spallanzani I.R.C.C.S | A Di Caro; B Bartolini; CEM Gruber; E Giombini; F Messina; F Santini; G Bonfiglio; M Rueca; MR Capobianchi; O Butera |
| EPI_ISL_5645600, EPI_ISL_5645611, EPI_ISL_5645620, EPI_ISL_5645669, EPI_ISL_5645691, EPI_ISL_5645725, EPI_ISL_5645780, EPI_ISL_5645783, EPI_ISL_5645788, EPI_ISL_5645818, EPI_ISL_5645834, EPI_ISL_5645852, EPI_ISL_5645862, EPI_ISL_5645894, EPI_ISL_5645952, EPI_ISL_5645953, EPI_ISL_5645955 | | | |
| see above | SANTA CASA DE GUARAREMA | Instituto Butantan | Antonio Jorge Martins; Claudia Renata dos Santos Barros; David Schlesinger; Debora Botequio Moretti; Dimas Tadeu Covas; Elaine Cristina Marqueze; Elaine Vieira Santos; Evandra Strazza Rodrigues; Heidge Fukumasu; Jayme Augusto de Souza-Neto; José Salvatore Leister Patané; Luiz Alcantara; Luiz Lehmann Coutinho; Maria Carolina Elias; Maurício Lacerda Nogueira; Rafael dos Santos Bezerra; Raul Machado Neto; Rejane Maria Tommasini Grotto; Ricardo Haddad; Sandra Coccuzzo Sampaio Vessoni; Simone Kashima; Svetoslav Nanev Slavov; Vincent Louis Viala |
| EPI_ISL_5645746, EPI_ISL_5645901, EPI_ISL_5646192 | SANTA CASA DE MISERICORDIA DE TIETE | Instituto Butantan | Antonio Jorge Martins; Claudia Renata dos Santos Barros; David Schlesinger; Debora Botequio Moretti; Dimas Tadeu Covas; Elaine Cristina Marqueze; Elaine Vieira Santos; Evandra Strazza Rodrigues; Heidge Fukumasu; Jayme Augusto de Souza-Neto; José Salvatore Leister Patané; Luiz Alcantara; Luiz Lehmann Coutinho; Maria Carolina Elias; Maurício Lacerda Nogueira; Rafael dos Santos Bezerra; Raul Machado Neto; Rejane Maria Tommasini Grotto; Ricardo Haddad; Sandra Coccuzzo Sampaio Vessoni; Simone Kashima; Svetoslav Nanev Slavov; Vincent Louis Viala |
| EPI_ISL_5645599, EPI_ISL_5645604, EPI_ISL_5645607, EPI_ISL_5645624, EPI_ISL_5645724, EPI_ISL_5645833, EPI_ISL_5645860, EPI_ISL_5645919, EPI_ISL_5646170, EPI_ISL_5646186 | | | |
| see above | SANTA CASA DE MISERICORDIA DE UBATUBA | Instituto Butantan | Antonio Jorge Martins; Claudia Renata dos Santos Barros; David Schlesinger; Debora Botequio Moretti; Dimas Tadeu Covas; Elaine Cristina Marqueze; Elaine Vieira Santos; Evandra Strazza Rodrigues; Heidge Fukumasu; Jayme Augusto de Souza-Neto; José Salvatore Leister Patané; Luiz Alcantara; Luiz Lehmann Coutinho; Maria Carolina Elias; Maurício Lacerda Nogueira; Rafael dos Santos Bezerra; Raul Machado Neto; Rejane Maria Tommasini Grotto; Ricardo Haddad; Sandra Coccuzzo Sampaio Vessoni; Simone Kashima; Svetoslav Nanev Slavov; Vincent Louis Viala |
| EPI_ISL_5647180 | SANTA CASA SANTA RITA DO PASSA QUATRO | Instituto Butantan | Antonio Jorge Martins; Claudia Renata dos Santos Barros; David Schlesinger; Debora Botequio Moretti; Dimas Tadeu Covas; Elaine Cristina Marqueze; Elaine Vieira Santos; Evandra Strazza Rodrigues; Heidge Fukumasu; Jayme Augusto de Souza-Neto; José Salvatore Leister Patané; Luiz Alcantara; Luiz Lehmann Coutinho; Maria Carolina Elias; Maurício Lacerda Nogueira; Rafael dos Santos Bezerra; Raul Machado Neto; Rejane Maria Tommasini Grotto; Ricardo Haddad; Sandra Coccuzzo Sampaio Vessoni; Simone Kashima; Svetoslav Nanev Slavov; Vincent Louis Viala |
| EPI_ISL_2957883, EPI_ISL_2958106, EPI_ISL_2986098 | SARS-CoV-2 Sequencing Castilla y Leon-Spain Consortium | SARS-CoV-2 Sequencing Castilla y Leon-Spain Consortium | Antonio Orduña-Domingo; Carlos Fuster Foz; Carmen Aldea-Mansilla; Carmen Gimeno Crespo; David Abad; Gabriel March Rossello; Gregoria Megías Lobón; Jose María Eiros Bouza; Laura Sánchez de Prada; M. Isabel Fernandez-Natal; Marta Dominguez-Gil; Marta Hernandez; María Antonia García Castro; Mª Fe Brezmes-Valdivieso; Noelia Arenal Andrés; Silvia Rojo; Sonsoles Garcinuño Pérez |
| EPI_ISL_2828387, EPI_ISL_2828388 | SC (UCO) Igiene e Sanità Pubblica, ASUGI, Trieste | ARGO Laboratorio Genomica ed Epigenomica | Barbone F; Breda C; Busetti M; D'Agaro P; Dal Monego S; Degasperi M; Fontana F; Licastro D; Marcello A; Piscianz E; Segat L |
| EPI_ISL_2828384 | SC (UCO) Igiene e Sanità Pubblica, ASUGI, Trieste and SC Microbiologia e Virologia, ASFO, Pordenone | ARGO Laboratorio Genomica ed Epigenomica | Barbone F; Breda C; D'Agaro P; Dal Monego S; De Rosa R; Degasperi M; Licastro D; Marcello A; Segat L |
| EPI_ISL_5647149, EPI_ISL_5647182 | SECRETARIA DE SAUDE DE SANTA GERTRUDES | Instituto Butantan | Antonio Jorge Martins; Claudia Renata dos Santos Barros; David Schlesinger; Debora Botequio Moretti; Dimas Tadeu Covas; Elaine Cristina Marqueze; Elaine Vieira Santos; Evandra Strazza Rodrigues; Heidge Fukumasu; Jayme Augusto de Souza-Neto; José Salvatore Leister Patané; Luiz Alcantara; Luiz Lehmann Coutinho; Maria Carolina Elias; Maurício Lacerda Nogueira; Rafael dos Santos Bezerra; Raul Machado Neto; Rejane Maria Tommasini Grotto; Ricardo Haddad; Sandra Coccuzzo Sampaio Vessoni; Simone Kashima; Svetoslav Nanev Slavov; Vincent Louis Viala |
| EPI_ISL_5647186 | SECRETARIA DE SAUDE DE SAO PEDRO | Instituto Butantan | Antonio Jorge Martins; Claudia Renata dos Santos Barros; David Schlesinger; Debora Botequio Moretti; Dimas Tadeu Covas; Elaine Cristina Marqueze; Elaine Vieira Santos; Evandra Strazza Rodrigues; Heidge Fukumasu; Jayme Augusto de Souza-Neto; José Salvatore Leister Patané; Luiz Alcantara; Luiz Lehmann Coutinho; Maria Carolina Elias; Maurício Lacerda Nogueira; Rafael dos Santos Bezerra; Raul Machado Neto; Rejane Maria Tommasini Grotto; Ricardo Haddad; Sandra Coccuzzo Sampaio Vessoni; Simone Kashima; Svetoslav Nanev Slavov; Vincent Louis Viala |
| EPI_ISL_5646314, EPI_ISL_5646385 | SECRETARIA MUNICIPAL DE SAUDE DE SANTA BARBARA D OESTE | Instituto Butantan | Antonio Jorge Martins; Claudia Renata dos Santos Barros; David Schlesinger; Debora Botequio Moretti; Dimas Tadeu Covas; Elaine Cristina Marqueze; Elaine Vieira Santos; Evandra Strazza Rodrigues; Heidge Fukumasu; Jayme Augusto de Souza-Neto; José Salvatore Leister Patané; Luiz Alcantara; Luiz Lehmann Coutinho; Maria Carolina Elias; Maurício Lacerda Nogueira; Rafael dos Santos Bezerra; Raul Machado Neto; Rejane Maria Tommasini Grotto; Ricardo Haddad; Sandra Coccuzzo Sampaio Vessoni; Simone Kashima; Svetoslav Nanev Slavov; Vincent Louis Viala |
| EPI_ISL_5646516, EPI_ISL_5646562, EPI_ISL_5646808, EPI_ISL_5647152, EPI_ISL_5647163 | SECRETARIA MUNICIPAL DE SAUDE DE CORDEIROPOLIS | Instituto Butantan | Antonio Jorge Martins; Claudia Renata dos Santos Barros; David Schlesinger; Debora Botequio Moretti; Dimas Tadeu Covas; Elaine Cristina Marqueze; Elaine Vieira Santos; Evandra Strazza Rodrigues; Heidge Fukumasu; Jayme Augusto de Souza-Neto; José Salvatore Leister Patané; Luiz Alcantara; Luiz Lehmann Coutinho; Maria Carolina Elias; Maurício Lacerda Nogueira; Rafael dos Santos Bezerra; Raul Machado Neto; Rejane Maria Tommasini Grotto; Ricardo Haddad; Sandra Coccuzzo Sampaio Vessoni; Simone Kashima; Svetoslav Nanev Slavov; Vincent Louis Viala |
| EPI_ISL_5646140, EPI_ISL_5646158 | SECRETARIA MUNICIPAL DE SAUDE DE MACATUBA | Instituto Butantan | Antonio Jorge Martins; Claudia Renata dos Santos Barros; David Schlesinger; Debora Botequio Moretti; Dimas Tadeu Covas; Elaine Cristina Marqueze; Elaine Vieira Santos; Evandra Strazza Rodrigues; Heidge Fukumasu; Jayme Augusto de Souza-Neto; José Salvatore Leister Patané; Luiz Alcantara; Luiz Lehmann Coutinho; Maria Carolina Elias; Maurício Lacerda Nogueira; Rafael dos Santos Bezerra; Raul Machado Neto; Rejane Maria Tommasini Grotto; Ricardo Haddad; Sandra Coccuzzo Sampaio Vessoni; Simone Kashima; Svetoslav Nanev Slavov; Vincent Louis Viala |
| EPI_ISL_3102318, EPI_ISL_3536383, EPI_ISL_3536386, EPI_ISL_3536387, EPI_ISL_3536389, EPI_ISL_3536391 | SECRETARIA MUNICIPAL DE SAUDE DE TIANGUA | Analytical Competence Molecular Epidemiology Lab/ACME, Oswaldo Cruz Foundation, Ceara (FIOCRUZ CE) | Cleber Furtado Aksenen; Fabio Miyajima; Fernando Braga Stehling; Francisco Eder de Moura Lopes; Jamille Maria Mendes Bezerra; Joaquim César do Nascimento Sousa Junior; Pedro Miguel Carneiro Jeronimo; Suzana Porto Almeida e Lucas Delerino; Thais Ferreira de Oliveira; Thais de Oliveira Costa; Ticiane Cavalcante de Souza; Veridiana Pessoa Miyajima |
| EPI_ISL_5647207, EPI_ISL_5647208, EPI_ISL_5647235 | SECRETARIA MUNICIPAL DE SAUDE DE TORRINHA | Instituto Butantan | Antonio Jorge Martins; Claudia Renata dos Santos Barros; David Schlesinger; Debora Botequio Moretti; Dimas Tadeu Covas; Elaine Cristina Marqueze; Elaine Vieira Santos; Evandra Strazza Rodrigues; Heidge Fukumasu; Jayme Augusto de Souza-Neto; José Salvatore Leister Patané; Luiz Alcantara; Luiz Lehmann Coutinho; Maria Carolina Elias; Maurício Lacerda Nogueira; Rafael dos Santos Bezerra; Raul Machado Neto; Rejane Maria Tommasini Grotto; Ricardo Haddad; Sandra Coccuzzo Sampaio Vessoni; Simone Kashima; Svetoslav Nanev Slavov; Vincent Louis Viala |
| EPI_ISL_5645594, EPI_ISL_5645596, EPI_ISL_5645605, EPI_ISL_5645616, EPI_ISL_5645629, EPI_ISL_5645638, EPI_ISL_5645648, EPI_ISL_5645650, EPI_ISL_5645666, EPI_ISL_5645668, EPI_ISL_5645679, EPI_ISL_5645683, EPI_ISL_5645684, EPI_ISL_5645688, EPI_ISL_5645692, EPI_ISL_5645694, EPI_ISL_5645714, EPI_ISL_5645717, EPI_ISL_5645728, EPI_ISL_5645738, EPI_ISL_5645760, EPI_ISL_5645775, EPI_ISL_5645778, EPI_ISL_5645830, EPI_ISL_5645831, EPI_ISL_5645846, EPI_ISL_5645857, EPI_ISL_5645858, EPI_ISL_5645871, EPI_ISL_5645875, EPI_ISL_5645885, EPI_ISL_5645900, EPI_ISL_5645915, EPI_ISL_5645925, EPI_ISL_5645935, EPI_ISL_5645954 | | | |
| see above | SECRETARIA MUNICIPAL DE SAUDE SOROCABA | Instituto Butantan | Antonio Jorge Martins; Claudia Renata dos Santos Barros; David Schlesinger; Debora Botequio Moretti; Dimas Tadeu Covas; Elaine Cristina Marqueze; Elaine Vieira Santos; Evandra Strazza Rodrigues; Heidge Fukumasu; Jayme Augusto de Souza-Neto; José Salvatore Leister Patané; Luiz Alcantara; Luiz Lehmann Coutinho; Maria Carolina Elias; Maurício Lacerda Nogueira; Rafael dos Santos Bezerra; Raul Machado Neto; Rejane Maria Tommasini Grotto; Ricardo Haddad; Sandra Coccuzzo Sampaio Vessoni; Simone Kashima; Svetoslav Nanev Slavov; Vincent Louis Viala |
| EPI_ISL_5646303, EPI_ISL_5646327, | SERV DE VIG SANITARIA EPIDEMIO E CTRL DE | Instituto Butantan | Antonio Jorge Martins; Claudia Renata dos Santos Barros; David Schlesinger; Debora Botequio Moretti; Dimas Tadeu Covas; Elaine Cristina Marqueze; Elaine Vieira Santos; Evandra Strazza Rodrigues; Heidge Fukumasu; Jayme Augusto de Souza-Neto; José Salvatore Leister Patané; Luiz Alcantara; Luiz Lehmann Coutinho; Maria Carolina Elias; Maurício Lacerda Nogueira; Rafael dos Santos Bezerra; Raul Machado Neto; Rejane Maria Tommasini Grotto; Ricardo Haddad; Sandra Coccuzzo Sampaio Vessoni; Simone Kashima; Svetoslav Nanev Slavov; Vincent Louis Viala |

| | | | |
|---|---|---|---|
| EPI_ISL_5646338, EPI_ISL_5646357, EPI_ISL_5646364 | ZOONOZES GUARUJA | | |
| EPI_ISL_2600802, EPI_ISL_2600822 | SIESP DIPARTIMENTO DI PREVENZIONE CHIETI | Istituto Zooprofilattico Sperimentale dell'Abruzzo e Molise "G. Caporale" | Ancora M; Calistri P; Cammà C; Caporale M; Curini V; Delli Compagni E; Di Domenico M; Di Lollo Valeria; Di Pasquale A; Lorusso A; Mangone I; Marcacci M; Puglia I; Rinaldi A; Savini G; Scialabba S |
| EPI_ISL_2889518, EPI_ISL_2889519, EPI_ISL_2889521, EPI_ISL_2889522, EPI_ISL_2889528, EPI_ISL_2889535, EPI_ISL_2889585, EPI_ISL_2889586, EPI_ISL_2889587, EPI_ISL_2889588, EPI_ISL_2889607, EPI_ISL_2889644, EPI_ISL_2889647, EPI_ISL_2889649, EPI_ISL_2889654, EPI_ISL_2889656, EPI_ISL_2889657, EPI_ISL_2889663, EPI_ISL_2889678, EPI_ISL_2889679, EPI_ISL_2889680, EPI_ISL_2889681, EPI_ISL_2889705, EPI_ISL_2889707, EPI_ISL_2889719, EPI_ISL_2889727, EPI_ISL_2889771, EPI_ISL_2889772, EPI_ISL_2889777, EPI_ISL_2889778, EPI_ISL_2889781, EPI_ISL_2889783, EPI_ISL_2889802, EPI_ISL_2889805, EPI_ISL_2889806, EPI_ISL_2889807, EPI_ISL_2889808, EPI_ISL_2889818, EPI_ISL_2889819, EPI_ISL_2889820, EPI_ISL_2889824, EPI_ISL_3255963, EPI_ISL_3255968 | | | |
| see above | SK-Roy Romanow Provincial Laboratory | National Microbiology Laboratory (NML) | Alanna Senecal; Amanda Lang; Anna Majer; Anneliese Landgraff; CanCOGeN's metadata curation team; Darian Hole; Elsie Grudeski; Gary Van Domselaar; Grace Seo; Jennifer Tanner; Jessica Minion; Kara Loos; Keith MacKenzie; Kirsten Biggar; Madison Chapel; Meredith Faires; Morag Graham; Natalie Knox; Nathalie Bastien; Philip Mabon; Public Health Agency of Canada CanCOGeN team; Rachel DePaulo; Rhiannon Huzarewich; Russell Mandes; Ryan McDonald; Shari Tyson; Timothy Booth; Yan Li |
| EPI_ISL_5844743, EPI_ISL_5844750, EPI_ISL_5844762, EPI_ISL_5844763, EPI_ISL_5844807 | SK-Roy Romanow Provincial Laboratory | Saskatchewan - Roy Romanow Provincial Laboratory (RRPL) | Alanna Senecal; Amanda Lang; Jessica Minion; Kara Loos; Keith MacKenzie; Meredith Faires; Rachel DePaulo; Roy Romanow Provincial Laboratory - Molecular Diagnostics; Ryan McDonald |
| EPI_ISL_3276557 | SOMER | Universidad Nacional de Colombia - Laboratorio Genómico One Health | Andres F. Cardona-Rios; Carlos Franco-Muñoz; Carolina Muñoz-Arango; Celeny Ortiz; Daniel O. Maldonado-Perez; Diego A. Álvarez-Díaz; Hector Alejandro Ruiz-Moreno; Idabely Betancur Ortiz; Jorge E. Osorio; Juan P. Hernandez-Ortiz; Karl A Ciuoderis; Katherine Laiton-Donato; Laura Silvana Perez; Lina M. Hurtado; Marcela Mercado-Reyes; Maria Angélica Maya; Maria Stella López; Rita Almanza Payares; Sandra Ines Cano; Simón Villegas Velásquez |
| EPI_ISL_2426606, EPI_ISL_2483922, EPI_ISL_2483924, EPI_ISL_2483925, EPI_ISL_2483926, EPI_ISL_2483929, EPI_ISL_2499973, EPI_ISL_2499975, EPI_ISL_2499976, EPI_ISL_2499977, EPI_ISL_2499978, EPI_ISL_2499979, EPI_ISL_2499981, EPI_ISL_2499983, EPI_ISL_2612779, EPI_ISL_2612780, EPI_ISL_2612801, EPI_ISL_2612831, EPI_ISL_2612832, EPI_ISL_2685874, EPI_ISL_2685876, EPI_ISL_2685878, EPI_ISL_2685879, EPI_ISL_2685880, EPI_ISL_2685883 | | | |
| see above | SYNLAB | GIGA Medical Genomics | Bouchra Boujemla; Cécile Meex; Keith Durkin; Maria Artesi; Marie-Pierre Hayette; Nathalie Renotte; Pierrette Melin; Raphaël Boreux; Sébastien Bontems; Vincent Bours |
| EPI_ISL_2633301 | SYNLAB MVZ Heidelberg | Robert Koch Institute | |
| EPI_ISL_2469512, EPI_ISL_2469528, EPI_ISL_2469561, EPI_ISL_2469734, EPI_ISL_2631572, EPI_ISL_2631604, EPI_ISL_2631695, EPI_ISL_3120754 | | | |
| see above | SYNLAB MVZ Leinfelden-Echterdingen | Robert Koch Institute | |
| EPI_ISL_2631725, EPI_ISL_3120799 | SYNLAB MVZ Leverkusen | Robert Koch Institute | |
| EPI_ISL_2472105, EPI_ISL_2633024 | SYNLAB MVZ Weiden | Robert Koch Institute | |
| EPI_ISL_6692745 | Saint Louise Hospital | Santa Clara County Public Health Laboratory | Santa Clara County Public Health Department |
| EPI_ISL_2603679, EPI_ISL_2603686, EPI_ISL_2603687, EPI_ISL_2603693, EPI_ISL_2603695, EPI_ISL_2603701, EPI_ISL_2603702, EPI_ISL_2603703, EPI_ISL_2603707, EPI_ISL_2603711, EPI_ISL_2603712, EPI_ISL_2603717, EPI_ISL_2603721, EPI_ISL_2603731, EPI_ISL_2603745, EPI_ISL_2616784, EPI_ISL_2616785, EPI_ISL_2616786, EPI_ISL_2616788, EPI_ISL_2616789, EPI_ISL_2616790, EPI_ISL_2616792, EPI_ISL_2616793, EPI_ISL_2616794, EPI_ISL_2616797, EPI_ISL_2616798, EPI_ISL_2616799, EPI_ISL_2616804, EPI_ISL_2616806, EPI_ISL_2616807, EPI_ISL_2616808, EPI_ISL_2616809, EPI_ISL_2616810, EPI_ISL_2616812, EPI_ISL_2616813, EPI_ISL_2616816, EPI_ISL_2616818, EPI_ISL_2616819, EPI_ISL_2616822, EPI_ISL_2616823, EPI_ISL_2616826, EPI_ISL_2616827, EPI_ISL_2616830, EPI_ISL_2616831, EPI_ISL_2616832, EPI_ISL_2616837, EPI_ISL_2616838, EPI_ISL_2616843, EPI_ISL_2616845, EPI_ISL_2616848, EPI_ISL_2616852, EPI_ISL_2616853, EPI_ISL_2616854, EPI_ISL_2616858, EPI_ISL_2616859, EPI_ISL_2616864, EPI_ISL_2616865, EPI_ISL_2616866, EPI_ISL_2616869, EPI_ISL_2616872, EPI_ISL_2616877, EPI_ISL_2616878, EPI_ISL_2616879, EPI_ISL_2616881, EPI_ISL_2616890, EPI_ISL_2616892, EPI_ISL_2616893, EPI_ISL_2616895, EPI_ISL_2616900, EPI_ISL_2616903, EPI_ISL_2616904, EPI_ISL_2616906, EPI_ISL_2616907, EPI_ISL_2616913, EPI_ISL_2616916, EPI_ISL_2616920, EPI_ISL_2616921, EPI_ISL_2616922, EPI_ISL_2616929, EPI_ISL_2616933, EPI_ISL_2616935, EPI_ISL_2616936, EPI_ISL_2616937, EPI_ISL_2616942, EPI_ISL_2616945, EPI_ISL_2616950, EPI_ISL_2616952, EPI_ISL_2616954, EPI_ISL_2616955, EPI_ISL_2616956, EPI_ISL_2616959, EPI_ISL_2616960, EPI_ISL_2616962, EPI_ISL_2616975, EPI_ISL_2616977, EPI_ISL_2616982, EPI_ISL_2616983, EPI_ISL_2616985, EPI_ISL_2616987, EPI_ISL_2616989, EPI_ISL_2616990, EPI_ISL_2616991, EPI_ISL_2616992, EPI_ISL_2616993, EPI_ISL_2616997, EPI_ISL_2616999, EPI_ISL_2617000, EPI_ISL_2617002, EPI_ISL_2617003, EPI_ISL_2617004, EPI_ISL_2617006, EPI_ISL_2617007, EPI_ISL_2617008, EPI_ISL_2617009, EPI_ISL_2617010, EPI_ISL_2617012, EPI_ISL_2617013, EPI_ISL_2617014, EPI_ISL_2617015, EPI_ISL_2617016, EPI_ISL_2617017, EPI_ISL_2692540, EPI_ISL_2692541, EPI_ISL_2692553, EPI_ISL_2692554, EPI_ISL_2692559, EPI_ISL_2692560, EPI_ISL_2692561, EPI_ISL_2692567, EPI_ISL_2692568, EPI_ISL_2692569, EPI_ISL_2692576, EPI_ISL_2692577, EPI_ISL_2692584, EPI_ISL_2692590, EPI_ISL_2692600, EPI_ISL_2692603, EPI_ISL_2692605, EPI_ISL_2692610, EPI_ISL_2692611, EPI_ISL_2692619, EPI_ISL_2692804, EPI_ISL_2692805, EPI_ISL_2692806, EPI_ISL_2692808, EPI_ISL_2692814, EPI_ISL_2692815, EPI_ISL_2692816, EPI_ISL_2692819, EPI_ISL_2692820, EPI_ISL_2692821, EPI_ISL_2692822, EPI_ISL_2692823, EPI_ISL_2692824, EPI_ISL_2692830, EPI_ISL_2810103, EPI_ISL_2810105, EPI_ISL_2810135, EPI_ISL_2810136, EPI_ISL_2810148, EPI_ISL_2881842, EPI_ISL_2881851, EPI_ISL_6437697 | | | |
| see above | Salud Digna | Instituto Nacional de Medicina Genomica | Abraham Campos-Romero; Cedro-Tanda A; Cruz-Islas Jazmin; Escobar-Arrazola; Gonzalez-Barrera D; Herrera-Montalvo LA.; Hidalgo-Miranda A; Luna-Ruiz Marco; M; Mendoza-Vargas A; Moreno-Camacho José Luis; Munguia-Garza P; Ramirez-Vega O; Rangel-DeLeon D; Reyes-Grajeda JP; Rodriguez-Gallegos Jorge |
| EPI_ISL_2840862, EPI_ISL_2840863, EPI_ISL_2840864 | San Diego County Public Health Laboratory | San Diego County Public Health Laboratory | Ashleigh Murphy Schafer; Brett Austin |
| EPI_ISL_3219154 | Santa Clara Valley Medical Center | Santa Clara County Public Health Laboratory | Santa Clara County Public Health Department |
| EPI_ISL_3015780, EPI_ISL_3015785 | Seattle Flu Study | Seattle Flu Study | Amanda Adler; Barry R. Lutz; Benjamin Pelle; Caitlin R. Wolf; Chris D. Frazar; Deborah A. Nickerson; Elisabeth Brandstetter; Erica Ryke; Helen Y. Chu; Janet A. Englund; Jay Shendure; Jover Lee; Kairsten Fay; Kirsten Lacombe; Lea M. Starita; Mark J. Rieder; Matthew Richardson; Matthew Thompson; Melissa Truong; Michael Boeckh; Michael Famulare; Misja Ilcisin; Peter D. Han; Thomas R. Sibley; Trevor Bedford |
| EPI_ISL_4082381, EPI_ISL_4082397, EPI_ISL_4082441, EPI_ISL_4082442, EPI_ISL_4082443, EPI_ISL_4082445, EPI_ISL_4082479, EPI_ISL_4082497, EPI_ISL_4082499, EPI_ISL_4082507, EPI_ISL_4082510, EPI_ISL_4082514, EPI_ISL_4082517, EPI_ISL_4082521, EPI_ISL_4082522, EPI_ISL_4082524, EPI_ISL_4082531, EPI_ISL_4082541, EPI_ISL_4082543, EPI_ISL_4082544, EPI_ISL_4082545, EPI_ISL_4082547, EPI_ISL_4082548, EPI_ISL_4082549, EPI_ISL_4082550, EPI_ISL_4082551, EPI_ISL_4082557, EPI_ISL_4082558, EPI_ISL_4082559, EPI_ISL_4082567, EPI_ISL_4082572, EPI_ISL_4082573, EPI_ISL_4082576, EPI_ISL_4082577, EPI_ISL_4082580, EPI_ISL_4082581, EPI_ISL_4082582, EPI_ISL_4082584, EPI_ISL_4082585, EPI_ISL_4082593, EPI_ISL_4082595, EPI_ISL_4082602, EPI_ISL_4082605, EPI_ISL_4082606, EPI_ISL_4082611, EPI_ISL_4082630, EPI_ISL_4082631, EPI_ISL_4082634, EPI_ISL_4082637, EPI_ISL_4082638, EPI_ISL_4082640, EPI_ISL_4082641, EPI_ISL_4082642, EPI_ISL_4082643, EPI_ISL_4082644, EPI_ISL_4082656, EPI_ISL_4082672, EPI_ISL_4082673, EPI_ISL_4082674, EPI_ISL_4082675, EPI_ISL_4082676, EPI_ISL_4082677, EPI_ISL_4082678, EPI_ISL_4082679, EPI_ISL_4082689, EPI_ISL_4082769, EPI_ISL_4082814, EPI_ISL_4082815, EPI_ISL_4082850, EPI_ISL_4082866, EPI_ISL_4082990, EPI_ISL_4083019, EPI_ISL_4083025, EPI_ISL_4083026, EPI_ISL_4083086, EPI_ISL_4083097, EPI_ISL_4083197, EPI_ISL_4083198, EPI_ISL_4083200, EPI_ISL_4083201, EPI_ISL_4083320, EPI_ISL_6694367 | | | |
| see above | Servicio Virosis Respiratorias-Departamento Virología-INEI | Instituto Nacional Enfermedades Infecciosas C.G.Malbran | Avaro M.; Baumeister E.; Benedetti E.; Campos J.; Cisterna D.; Dattero ME; De Belder D.; Haim MS.; Lorenzo F.; Molina V.; Perandones C.; Poklepovich T.; Pontoriero A.; Russo M.; Sanchez Loria J.; Tuduri E. |
| EPI_ISL_2689718 | Servicio de Microbiologia Hospital Ramon y Cajal | Servicio de Microbiologia Hospital Ramon y Cajal | Galan JC; Martinez L. Abreu M; Ponce M; y Gonzalez-Alba JM |
| EPI_ISL_2516600, EPI_ISL_2516705, EPI_ISL_2516706, EPI_ISL_2516707, EPI_ISL_2516708, EPI_ISL_2516725, EPI_ISL_2516752, EPI_ISL_2516755, EPI_ISL_2516859 | | | |
| see above | Servicio de Microbiología Clínica (Complejo Hospitalario de Navarra, Pamplona) | Centro de Secuenciación NASERTIC | Ana Miqueleiz; Ana Navascués; Carmen Ezpeleta Baquedano |
| EPI_ISL_2658213, EPI_ISL_2658235, EPI_ISL_2658236, EPI_ISL_2658240, EPI_ISL_2658243, EPI_ISL_2658245, EPI_ISL_2658252, EPI_ISL_2658253, EPI_ISL_2834008, EPI_ISL_2834010, EPI_ISL_2834014, EPI_ISL_2834017 | | | |
| see above | Servicio de Microbiología. Consorcio Hospital General Universitario de Valencia | SeqCOVID-SPAIN consortium/IBV(CSIC) | Begoña Fuster Escrivá; Carme Salvador García; Concepción Gimeno Cardona and SeqCOVID-SPAIN consortium; María Dolores Ocete; Rafael Medina González |
| EPI_ISL_2600911, EPI_ISL_2600917 | Servicio de Microbiología. Hospital Clínico Universitario de Valencia | SeqCOVID-SPAIN consortium/IBV(CSIC) | David Navarro Ortega; Eliseo Albert Vicent; Ignacio Torres and SeqCOVID-SPAIN consortium |
| EPI_ISL_2820638, EPI_ISL_2820644 | Servicio de Microbiología. Hospital Universitario Doctor Peset | SeqCOVID-SPAIN consortium/IBV(CSIC) | José Miguel Nogueira Coito and SeqCOVID-SPAIN consortium; Juan Alberola Enguídanos; Juan José Camarena Miñana; Rosa González Pellicer |
| EPI_ISL_5315473, EPI_ISL_5315475, EPI_ISL_5315476, EPI_ISL_5315477, EPI_ISL_5315478, EPI_ISL_5315479, EPI_ISL_5315482, EPI_ISL_5315488, EPI_ISL_5315489, EPI_ISL_5315495, EPI_ISL_5315496, EPI_ISL_5315501, EPI_ISL_5315503, EPI_ISL_5315504, EPI_ISL_5315507, EPI_ISL_5315510, EPI_ISL_5315512, EPI_ISL_5315517, EPI_ISL_5315518, EPI_ISL_5315519, EPI_ISL_5315521, EPI_ISL_5315524, EPI_ISL_5315525, EPI_ISL_5315527, EPI_ISL_5315528, EPI_ISL_5315529, EPI_ISL_5315530, EPI_ISL_5315533, EPI_ISL_5315534, EPI_ISL_5315535, EPI_ISL_5315537, EPI_ISL_5315542, EPI_ISL_6686901, EPI_ISL_6686902, EPI_ISL_6686908, EPI_ISL_6686909, EPI_ISL_6686922, EPI_ISL_6686936, EPI_ISL_6687689, EPI_ISL_6688104 | | | |
| see above | Shared Hospital Laboratory | Shared Hospital Laboratory | Christie Vermeiren; Finlay Maguire; Kevin Katz; Patryk Aftanas; Robert Kozak; Samira Mubareka |
| EPI_ISL_2712536, EPI_ISL_2712537, EPI_ISL_2712542, EPI_ISL_2712547 | Sharp HealthCare Laboratory | Andersen lab at Scripps Research | Art Mendoza; Cathy Woerle; Jacquelyn Berumen; Liam McGinnis; Omid Bakhtar; SEARCH Alliance San Diego with Aaron Harding |
| EPI_ISL_5517413 | Simple Laboratories | Gagnon Lab, Southern Illinois University | Keith Gagnon |
| EPI_ISL_2716912 | Southern Nevada Public Health Laboratory | Southern Nevada Public Health Laboratory | Michael Picker |
| EPI_ISL_2724799, EPI_ISL_2724815, EPI_ISL_2724834, EPI_ISL_2724844, EPI_ISL_2724855, EPI_ISL_2724856, EPI_ISL_2724873, EPI_ISL_2724876, EPI_ISL_2724885, EPI_ISL_2724909, EPI_ISL_3033102, EPI_ISL_3033111, EPI_ISL_3033190 | | | |
| see above | State Laboratories Division, Hawaii State Department of Health | State Laboratories Division, Hawaii State Department of Health | Ayana Garnet; Drew Kuwazaki; Edward Desmond; Pamela O'Brien; Razvan Sultana |
| EPI_ISL_2800567, EPI_ISL_2800568, EPI_ISL_2800569, EPI_ISL_2800570, EPI_ISL_2800571, EPI_ISL_2800572, EPI_ISL_2800573, EPI_ISL_2800577, EPI_ISL_2800578, EPI_ISL_2800579, EPI_ISL_2800581, EPI_ISL_2800582, EPI_ISL_2835227 | | | |
| see above | State Testing Facility | Altius Institute for Biomedical Research | Alex Isner; Alex Nguyen; Amanda Gale; Audra Johnson; Clem Green; Daniel Bates; Eric Thorland; Jacob Rodriguez; Jean Robinston; Jemma Nelson; Jessica Kunder; John Stamatoyannopoulos; Joshua Richards; Julia Wald; Kneshay Harper; Lauren Mitchell; Mark Frerker; Matt Hartman; Michael Buckley; Muhammad Halimun; Rebecca Bruders; Sadie Patraw; Sofia Olsson; Tobias Ragoczy |
| EPI_ISL_2618952, EPI_ISL_2618969, EPI_ISL_2619010, EPI_ISL_2619021, EPI_ISL_2619038, EPI_ISL_3998558, EPI_ISL_3998559, EPI_ISL_3998560, EPI_ISL_3998561, EPI_ISL_3998562, EPI_ISL_3998563, EPI_ISL_3998564, EPI_ISL_3998569, EPI_ISL_3998570, EPI_ISL_3998572, EPI_ISL_3998578, EPI_ISL_3998579, EPI_ISL_3998580, EPI_ISL_3998581, EPI_ISL_3998582, EPI_ISL_3998584, EPI_ISL_3998693 | | | |
| see above | Swedish national genomic surveillance program of SARS-CoV-2 | The Public Health Agency of Sweden | Alma Brolund; Maria Lind Karlberg; Maximilian Riess; Swedish national genomic surveillance program of SARS-CoV-2 |
| EPI_ISL_2868602, EPI_ISL_2868604, EPI_ISL_3370851 | TEMPUS LABS INC | Wadsworth Center, New York State Department of Health | Alexis Russell; Catharine Prussing; Daryl M. Lamson; Erasmus Schneider; Erica Lasek-Nesselquist; John Kelly; Jonathan Plitnick; Kirsten St. George; Matthew Shudt; Melissa A Leisner; Navjot Singh |

| | | | |
|---|---|---|---|
| EPI_ISL_2600471, EPI_ISL_2769191, EPI_ISL_2769210, EPI_ISL_2769249, EPI_ISL_2769316, EPI_ISL_2769327, EPI_ISL_2919353, EPI_ISL_2919452 | | | |
| see above | TGen North | TGen North | "Jolene Bowers; Ashlyn Pfeiffer; Brett Van Tassel; Chris French; Darrin Lemmer; Dave Engelthaler; Dave Engelthaler"; Hayley Yaglom; Heather Centner; The Arizona COVID Genomics Union (ACGU)" |
| EPI_ISL_2658964 | TXDSHS | TXDSHS | Anita Pokharel; Bonnie Oh; Chun Wang; Grace Kubin; Jenny Zhang; Karen Bobier; Lorraine Rodriguez; Maliha Rahman; Mayela Pedrueza; Myong Koag; Rachel Lee; Rashmi Tuladhar |
| EPI_ISL_2790540, EPI_ISL_2930464 | Tampa General Hospital Esoteric Lab | Tampa General Hospital Esoteric Research & Development Lab | Amorce Lima; Deanna Becker; Dominic Uy; Elaine Vendrone; Jon Faughn; Jorge Mecias-Frias; Suzane Silbert; Vicki Healer |
| EPI_ISL_2776071 | Texas Children's Hospital | Texas Children's Microbiome Center | Ila Singh; James Dunn; James Versalovic; Jennifer K. Spinler; Ruth Ann Luna |
| EPI_ISL_2756576, EPI_ISL_2756578, EPI_ISL_2756579, EPI_ISL_2756580, EPI_ISL_2756581, EPI_ISL_2756584 | The Caribbean Public Health Agency | Carrington Lab, Department of Preclinical Sciences, Faculty of Medical Sciences, The University of the West Indies, St Augustine Campus | Anushka Ramjag; Arianne Brown-Jordan; Avery Hinds; Christine V. F. Carrington; Gabriel Escobar; Nikita S. D. Sahadeo; Nuno Faria; Oliver Pybus; Risha Singh; Roshan Parasram; Sarah Hill; SueMin Nathaniel; Vernie Ramkissoon |
| EPI_ISL_2758892, EPI_ISL_2758897 | The Ohio State University Applied Microbiology Services Laboratory | The Ohio State University Applied Microbiology Services Laboratory | Seth A. Faith PhD |
| EPI_ISL_2620779 | Torrance Memorial Medical Center | Los Angeles County Public Health Laboratories | P. Hemarajata et al. |
| EPI_ISL_3215169, EPI_ISL_3215215, EPI_ISL_3215216, EPI_ISL_3215217, EPI_ISL_3215225 | U.O. Microbiologia Laboratorio Unico Centro Servizi - AUSL della Romagna | U.O. Microbiologia, Laboratorio Unico Centro Servizi - AUSL della Romagna | Giorgio Dirani |
| EPI_ISL_2625890 | UAB Diagnostikos laboratorija | Institute of Biotechnology, Life Sciences Center, Vilnius University | Emilija Vasiliunaite__ Milda Norkiene__ Danguole Ziogiene__ Albertas Timinskas__ Alma Gedvilaite |
| EPI_ISL_3912269, EPI_ISL_3912270 | UBS CAIC II | Analytical Competence Molecular Epidemiology Lab/ACME, Oswaldo Cruz Foundation, Ceara (FIOCRUZ CE) | Cleber Furtado Aksenen; Fabio Miyajima; Fernando Braga Stehling; Francisco Eder de Moura Lopes; Jamille Maria Mendes Bezerra; Joaquim Cesar do Nascimento Sousa Junior; Pedro Miguel Carneiro Jeronimo; Suzana Porto Almeida & Lucas Delerino on behalf of COVID-19 FIOCRUZ Genomic Network; Thais Ferreira de Oliveira; Thais de Oliveira Costa; Ticiane Cavalcante de Souza; Veridiana Pessoa Miyajima |
| EPI_ISL_5649591, EPI_ISL_5650358 | UBS DE MIRA ESTRELA | Instituto Butantan | Antonio Jorge Martins; Claudia Renata dos Santos Barros; David Schlesinger; Debora Botequio Moretti; Dimas Tadeu Covas; Elaine Cristina Marqueze; Elaine Vieira Santos; Evandra Strazza Rodrigues; Heidge Fukumasu; Jayme Augusto de Souza-Neto; José Salvatore Leister Patané; Luiz Alcantara; Luiz Lehmann Coutinho; Maria Carolina Elias; Maurício Lacerda Nogueira; Rafael dos Santos Bezerra; Raul Machado Neto; Rejane Maria Tommasini Grotto; Ricardo Haddad; Sandra Coccuzzo Sampaio Vessoni; Simone Kashima; Svetoslav Nanev Slavov; Vincent Louis Viala |
| EPI_ISL_5646304, EPI_ISL_5646309, EPI_ISL_5646319, EPI_ISL_5646320, EPI_ISL_5646323, EPI_ISL_5646332, EPI_ISL_5646345, EPI_ISL_5646363, EPI_ISL_5646369, EPI_ISL_5646382, EPI_ISL_5646383 | | | |
| see above | UBS DE TABAPUA | Instituto Butantan | Antonio Jorge Martins; Claudia Renata dos Santos Barros; David Schlesinger; Debora Botequio Moretti; Dimas Tadeu Covas; Elaine Cristina Marqueze; Elaine Vieira Santos; Evandra Strazza Rodrigues; Heidge Fukumasu; Jayme Augusto de Souza-Neto; José Salvatore Leister Patané; Luiz Alcantara; Luiz Lehmann Coutinho; Maria Carolina Elias; Maurício Lacerda Nogueira; Rafael dos Santos Bezerra; Raul Machado Neto; Rejane Maria Tommasini Grotto; Ricardo Haddad; Sandra Coccuzzo Sampaio Vessoni; Simone Kashima; Svetoslav Nanev Slavov; Vincent Louis Viala |
| EPI_ISL_3912280 | UBS DO BAIRRO SAO JOSE | Analytical Competence Molecular Epidemiology Lab/ACME, Oswaldo Cruz Foundation, Ceara (FIOCRUZ CE) | Cleber Furtado Aksenen; Fabio Miyajima; Fernando Braga Stehling; Francisco Eder de Moura Lopes; Jamille Maria Mendes Bezerra; Joaquim Cesar do Nascimento Sousa Junior; Pedro Miguel Carneiro Jeronimo; Suzana Porto Almeida & Lucas Delerino on behalf of COVID-19 FIOCRUZ Genomic Network; Thais Ferreira de Oliveira; Thais de Oliveira Costa; Ticiane Cavalcante de Souza; Veridiana Pessoa Miyajima |
| EPI_ISL_5645582 | UBS DR ALFREDO DANTAS DE SOUZA UMUARAMA | Instituto Butantan | Antonio Jorge Martins; Claudia Renata dos Santos Barros; David Schlesinger; Debora Botequio Moretti; Dimas Tadeu Covas; Elaine Cristina Marqueze; Elaine Vieira Santos; Evandra Strazza Rodrigues; Heidge Fukumasu; Jayme Augusto de Souza-Neto; José Salvatore Leister Patané; Luiz Alcantara; Luiz Lehmann Coutinho; Maria Carolina Elias; Maurício Lacerda Nogueira; Rafael dos Santos Bezerra; Raul Machado Neto; Rejane Maria Tommasini Grotto; Ricardo Haddad; Sandra Coccuzzo Sampaio Vessoni; Simone Kashima; Svetoslav Nanev Slavov; Vincent Louis Viala |
| EPI_ISL_5645739, EPI_ISL_5645762, EPI_ISL_5645824, EPI_ISL_5645936, EPI_ISL_5645947 | UBS II DE PIRAPOZINHO C SERVICO DE EACS E ESF | Instituto Butantan | Antonio Jorge Martins; Claudia Renata dos Santos Barros; David Schlesinger; Debora Botequio Moretti; Dimas Tadeu Covas; Elaine Cristina Marqueze; Elaine Vieira Santos; Evandra Strazza Rodrigues; Heidge Fukumasu; Jayme Augusto de Souza-Neto; José Salvatore Leister Patané; Luiz Alcantara; Luiz Lehmann Coutinho; Maria Carolina Elias; Maurício Lacerda Nogueira; Rafael dos Santos Bezerra; Raul Machado Neto; Rejane Maria Tommasini Grotto; Ricardo Haddad; Sandra Coccuzzo Sampaio Vessoni; Simone Kashima; Svetoslav Nanev Slavov; Vincent Louis Viala |
| EPI_ISL_5645917 | UBS II DE TACIBA | Instituto Butantan | Antonio Jorge Martins; Claudia Renata dos Santos Barros; David Schlesinger; Debora Botequio Moretti; Dimas Tadeu Covas; Elaine Cristina Marqueze; Elaine Vieira Santos; Evandra Strazza Rodrigues; Heidge Fukumasu; Jayme Augusto de Souza-Neto; José Salvatore Leister Patané; Luiz Alcantara; Luiz Lehmann Coutinho; Maria Carolina Elias; Maurício Lacerda Nogueira; Rafael dos Santos Bezerra; Raul Machado Neto; Rejane Maria Tommasini Grotto; Ricardo Haddad; Sandra Coccuzzo Sampaio Vessoni; Simone Kashima; Svetoslav Nanev Slavov; Vincent Louis Viala |
| EPI_ISL_5647123, EPI_ISL_5647133, EPI_ISL_5647136, EPI_ISL_5647153, EPI_ISL_5647154, EPI_ISL_5647168, EPI_ISL_5647169, EPI_ISL_5647189, EPI_ISL_5647190, EPI_ISL_5647191, EPI_ISL_5647193 | | | |
| see above | UBS II DE TANABI MILTON MARTINS PERCHES | Instituto Butantan | Antonio Jorge Martins; Claudia Renata dos Santos Barros; David Schlesinger; Debora Botequio Moretti; Dimas Tadeu Covas; Elaine Cristina Marqueze; Elaine Vieira Santos; Evandra Strazza Rodrigues; Heidge Fukumasu; Jayme Augusto de Souza-Neto; José Salvatore Leister Patané; Luiz Alcantara; Luiz Lehmann Coutinho; Maria Carolina Elias; Maurício Lacerda Nogueira; Rafael dos Santos Bezerra; Raul Machado Neto; Rejane Maria Tommasini Grotto; Ricardo Haddad; Sandra Coccuzzo Sampaio Vessoni; Simone Kashima; Svetoslav Nanev Slavov; Vincent Louis Viala |
| EPI_ISL_5645893 | UBS II DR JOSE CARLOS MACUCO JANINI | Instituto Butantan | Antonio Jorge Martins; Claudia Renata dos Santos Barros; David Schlesinger; Debora Botequio Moretti; Dimas Tadeu Covas; Elaine Cristina Marqueze; Elaine Vieira Santos; Evandra Strazza Rodrigues; Heidge Fukumasu; Jayme Augusto de Souza-Neto; José Salvatore Leister Patané; Luiz Alcantara; Luiz Lehmann Coutinho; Maria Carolina Elias; Maurício Lacerda Nogueira; Rafael dos Santos Bezerra; Raul Machado Neto; Rejane Maria Tommasini Grotto; Ricardo Haddad; Sandra Coccuzzo Sampaio Vessoni; Simone Kashima; Svetoslav Nanev Slavov; Vincent Louis Viala |
| EPI_ISL_5645715, EPI_ISL_5645718, EPI_ISL_5645755, EPI_ISL_5645810, EPI_ISL_5645825, EPI_ISL_5645910, EPI_ISL_5645928 | | | |
| see above | UBS III DE RANCHARIA | Instituto Butantan | Antonio Jorge Martins; Claudia Renata dos Santos Barros; David Schlesinger; Debora Botequio Moretti; Dimas Tadeu Covas; Elaine Cristina Marqueze; Elaine Vieira Santos; Evandra Strazza Rodrigues; Heidge Fukumasu; Jayme Augusto de Souza-Neto; José Salvatore Leister Patané; Luiz Alcantara; Luiz Lehmann Coutinho; Maria Carolina Elias; Maurício Lacerda Nogueira; Rafael dos Santos Bezerra; Raul Machado Neto; Rejane Maria Tommasini Grotto; Ricardo Haddad; Sandra Coccuzzo Sampaio Vessoni; Simone Kashima; Svetoslav Nanev Slavov; Vincent Louis Viala |
| EPI_ISL_5649589, EPI_ISL_5650344 | UBS LUIS FACHIN IPIGUA | Instituto Butantan | Antonio Jorge Martins; Claudia Renata dos Santos Barros; David Schlesinger; Debora Botequio Moretti; Dimas Tadeu Covas; Elaine Cristina Marqueze; Elaine Vieira Santos; Evandra Strazza Rodrigues; Heidge Fukumasu; Jayme Augusto de Souza-Neto; José Salvatore Leister Patané; Luiz Alcantara; Luiz Lehmann Coutinho; Maria Carolina Elias; Maurício Lacerda Nogueira; Rafael dos Santos Bezerra; Raul Machado Neto; Rejane Maria Tommasini Grotto; Ricardo Haddad; Sandra Coccuzzo Sampaio Vessoni; Simone Kashima; Svetoslav Nanev Slavov; Vincent Louis Viala |
| EPI_ISL_5647173 | UBS TEREZA GALLO UCHOA | Instituto Butantan | Antonio Jorge Martins; Claudia Renata dos Santos Barros; David Schlesinger; Debora Botequio Moretti; Dimas Tadeu Covas; Elaine Cristina Marqueze; Elaine Vieira Santos; Evandra Strazza Rodrigues; Heidge Fukumasu; Jayme Augusto de Souza-Neto; José Salvatore Leister Patané; Luiz Alcantara; Luiz Lehmann Coutinho; Maria Carolina Elias; Maurício Lacerda Nogueira; Rafael dos Santos Bezerra; Raul Machado Neto; Rejane Maria Tommasini Grotto; Ricardo Haddad; Sandra Coccuzzo Sampaio Vessoni; Simone Kashima; Svetoslav Nanev Slavov; Vincent Louis Viala |
| EPI_ISL_2960100 | UCC (Universidad Central del Caribe) | Grubaugh Lab - Yale School of Public Health | Anderson Brito; Andrea Firpo; Annie Watkins; Chaney Kalinich; Chantal Vogels; David Rivera Aponte; Ernesto Vazquez; Fabiola Cruz-Lopez; Fabiola Fontanet; Isabel Ott; Jessica Rothman; Joseph Fauver; Karolane Gonzalez; Kendall Billig; Mallery Breban; Mary Petrone; Natalie Machargo; Nathan Grubaugh; Patricia Serrano; Tania Mitwalli; Tara Alpert; Tobias Koch; Vianca Aponte |
| EPI_ISL_2772516, EPI_ISL_2772521 | UCLA Clinical Micro Lab | Los Angeles County Public Health Laboratories | P. Hemarajata et al. |
| EPI_ISL_2661544, EPI_ISL_2661545, EPI_ISL_2661546, EPI_ISL_2661547 | UHTL, University Hospitals | UHTL, University Hospitals | Alouani, D.; Sadri, N.; Song, X. |
| EPI_ISL_2695325, EPI_ISL_2695326 | ULSM - Matosinhos | Instituto Nacional de Saude (INSA) | Borges et al |
| EPI_ISL_3409334 | UMass Memorial Medical Center | Center for Microbiome Research | Doyle V. Ward; Karl J. Simon; Richard T. Ellison III |
| EPI_ISL_5646371 | UNIDADE BASICA DE SAUDE CIDINHA LEITE | Instituto Butantan | Antonio Jorge Martins; Claudia Renata dos Santos Barros; David Schlesinger; Debora Botequio Moretti; Dimas Tadeu Covas; Elaine Cristina Marqueze; Elaine Vieira Santos; Evandra Strazza Rodrigues; Heidge Fukumasu; Jayme Augusto de Souza-Neto; José Salvatore Leister Patané; Luiz Alcantara; Luiz Lehmann Coutinho; Maria Carolina Elias; Maurício Lacerda Nogueira; Rafael dos Santos Bezerra; Raul Machado Neto; Rejane Maria Tommasini Grotto; Ricardo Haddad; Sandra Coccuzzo Sampaio Vessoni; Simone Kashima; Svetoslav Nanev Slavov; Vincent Louis Viala |
| EPI_ISL_5650388 | UNIDADE BASICA DE SAUDE DA FAMILIA BILAC | Instituto Butantan | Antonio Jorge Martins; Claudia Renata dos Santos Barros; David Schlesinger; Debora Botequio Moretti; Dimas Tadeu Covas; Elaine Cristina Marqueze; Elaine Vieira Santos; Evandra Strazza Rodrigues; Heidge Fukumasu; Jayme Augusto de Souza-Neto; José Salvatore Leister Patané; Luiz Alcantara; Luiz Lehmann Coutinho; Maria Carolina Elias; Maurício Lacerda Nogueira; Rafael dos Santos Bezerra; Raul Machado Neto; Rejane Maria Tommasini Grotto; Ricardo Haddad; Sandra Coccuzzo Sampaio Vessoni; Simone Kashima; Svetoslav Nanev Slavov; Vincent Louis Viala |
| EPI_ISL_3912293 | UNIDADE BASICA DE SAUDE DA GIRILANDIA | Analytical Competence Molecular Epidemiology Lab/ACME, Oswaldo Cruz Foundation, Ceara (FIOCRUZ CE) | Cleber Furtado Aksenen; Fabio Miyajima; Fernando Braga Stehling; Francisco Eder de Moura Lopes; Jamille Maria Mendes Bezerra; Joaquim Cesar do Nascimento Sousa Junior; Pedro Miguel Carneiro Jeronimo; Suzana Porto Almeida & Lucas Delerino on behalf of COVID-19 FIOCRUZ Genomic Network; Thais Ferreira de Oliveira; Thais de Oliveira Costa; Ticiane Cavalcante de Souza; Veridiana Pessoa Miyajima |
| EPI_ISL_3912294, EPI_ISL_3912295 | UNIDADE BASICA DE SAUDE DA SEDE | Analytical Competence Molecular Epidemiology Lab/ACME, Oswaldo Cruz Foundation, Ceara (FIOCRUZ CE) | Cleber Furtado Aksenen; Fabio Miyajima; Fernando Braga Stehling; Francisco Eder de Moura Lopes; Jamille Maria Mendes Bezerra; Joaquim Cesar do Nascimento Sousa Junior; Pedro Miguel Carneiro Jeronimo; Suzana Porto Almeida & Lucas Delerino on behalf of COVID-19 FIOCRUZ Genomic Network; Thais Ferreira de Oliveira; Thais de Oliveira Costa; Ticiane Cavalcante de Souza; Veridiana Pessoa Miyajima |
| EPI_ISL_3912296 | UNIDADE BASICA DE SAUDE DE BOA AGUA | Analytical Competence Molecular Epidemiology Lab/ACME, Oswaldo Cruz Foundation, Ceara (FIOCRUZ CE) | Cleber Furtado Aksenen; Fabio Miyajima; Fernando Braga Stehling; Francisco Eder de Moura Lopes; Jamille Maria Mendes Bezerra; Joaquim Cesar do Nascimento Sousa Junior; Pedro Miguel Carneiro Jeronimo; Suzana Porto Almeida & Lucas Delerino on behalf of COVID-19 FIOCRUZ Genomic Network; Thais Ferreira de Oliveira; Thais de Oliveira Costa; Ticiane Cavalcante de Souza; Veridiana Pessoa Miyajima |
| EPI_ISL_5645572, EPI_ISL_5649614, EPI_ISL_5649633, EPI_ISL_5650202, EPI_ISL_5650215, EPI_ISL_5650223, EPI_ISL_5650330, EPI_ISL_5650339, EPI_ISL_5650351, EPI_ISL_5650360, EPI_ISL_5650379 | | | |
| see above | UNIDADE BASICA DE SAUDE DE TARUMA | Instituto Butantan | Antonio Jorge Martins; Claudia Renata dos Santos Barros; David Schlesinger; Debora Botequio Moretti; Dimas Tadeu Covas; Elaine Cristina Marqueze; Elaine Vieira Santos; Evandra Strazza Rodrigues; Heidge Fukumasu; Jayme Augusto de Souza-Neto; José Salvatore Leister Patané; Luiz Alcantara; Luiz Lehmann Coutinho; Maria Carolina Elias; Maurício Lacerda Nogueira; Rafael dos Santos Bezerra; Raul Machado Neto; Rejane Maria Tommasini Grotto; Ricardo Haddad; Sandra Coccuzzo Sampaio Vessoni; Simone Kashima; Svetoslav Nanev Slavov; Vincent Louis Viala |
| EPI_ISL_3912299 | UNIDADE BASICA DE SAUDE DO CAMPO DE AVIACAO | Analytical Competence Molecular Epidemiology Lab/ACME, Oswaldo Cruz Foundation, Ceara (FIOCRUZ CE) | Cleber Furtado Aksenen; Fabio Miyajima; Fernando Braga Stehling; Francisco Eder de Moura Lopes; Jamille Maria Mendes Bezerra; Joaquim Cesar do Nascimento Sousa Junior; Pedro Miguel Carneiro Jeronimo; Suzana Porto Almeida & Lucas Delerino on behalf of COVID-19 FIOCRUZ Genomic Network; Thais Ferreira de Oliveira; Thais de Oliveira Costa; Ticiane Cavalcante de Souza; Veridiana Pessoa Miyajima |

| | | | |
|---|---|---|---|
| EPI_ISL_5647151, EPI_ISL_5647178 | UNIDADE BASICA DE SAUDE DR MATHEUS GABRIEL BONASSA | Instituto Butantan | Antonio Jorge Martins; Claudia Renata dos Santos Barros; David Schlesinger; Debora Botequio Moretti; Dimas Tadeu Covas; Elaine Cristina Marqueze; Elaine Vieira Santos; Evandra Strazza Rodrigues; Heidge Fukumasu; Jayme Augusto de Souza-Neto; José Salvatore Leister Patané; Luiz Alcantara; Luiz Lehmann Coutinho; Maria Carolina Elias; Maurício Lacerda Nogueira; Rafael dos Santos Bezerra; Raul Machado Neto; Rejane Maria Tommasini Grotto; Ricardo Haddad; Sandra Coccuzzo Sampaio Vessoni; Simone Kashima; Svetoslav Nanev Slavov; Vincent Louis Viala |
| EPI_ISL_5645754, EPI_ISL_5645877, EPI_ISL_5645886 | UNIDADE BASICA DE SAUDE II CAIABU | Instituto Butantan | Antonio Jorge Martins; Claudia Renata dos Santos Barros; David Schlesinger; Debora Botequio Moretti; Dimas Tadeu Covas; Elaine Cristina Marqueze; Elaine Vieira Santos; Evandra Strazza Rodrigues; Heidge Fukumasu; Jayme Augusto de Souza-Neto; José Salvatore Leister Patané; Luiz Alcantara; Luiz Lehmann Coutinho; Maria Carolina Elias; Maurício Lacerda Nogueira; Rafael dos Santos Bezerra; Raul Machado Neto; Rejane Maria Tommasini Grotto; Ricardo Haddad; Sandra Coccuzzo Sampaio Vessoni; Simone Kashima; Svetoslav Nanev Slavov; Vincent Louis Viala |
| EPI_ISL_5645571, EPI_ISL_5645574, EPI_ISL_5645576, EPI_ISL_5649592, EPI_ISL_5650333, EPI_ISL_5650335, EPI_ISL_5650337, EPI_ISL_5650341, EPI_ISL_5650352, EPI_ISL_5650353, EPI_ISL_5650356, EPI_ISL_5650359, EPI_ISL_5650374, EPI_ISL_5650377, EPI_ISL_5650381 | | | |
| see above | UNIDADE BASICA DE SSAUDE DE POLONI | Instituto Butantan | Antonio Jorge Martins; Claudia Renata dos Santos Barros; David Schlesinger; Debora Botequio Moretti; Dimas Tadeu Covas; Elaine Cristina Marqueze; Elaine Vieira Santos; Evandra Strazza Rodrigues; Heidge Fukumasu; Jayme Augusto de Souza-Neto; José Salvatore Leister Patané; Luiz Alcantara; Luiz Lehmann Coutinho; Maria Carolina Elias; Maurício Lacerda Nogueira; Rafael dos Santos Bezerra; Raul Machado Neto; Rejane Maria Tommasini Grotto; Ricardo Haddad; Sandra Coccuzzo Sampaio Vessoni; Simone Kashima; Svetoslav Nanev Slavov; Vincent Louis Viala |
| EPI_ISL_5646347, EPI_ISL_5646353, EPI_ISL_5646372 | UNIDADE DE PRONTO ATENDIMENTO UPA 24 HORAS | Instituto Butantan | Antonio Jorge Martins; Claudia Renata dos Santos Barros; David Schlesinger; Debora Botequio Moretti; Dimas Tadeu Covas; Elaine Cristina Marqueze; Elaine Vieira Santos; Evandra Strazza Rodrigues; Heidge Fukumasu; Jayme Augusto de Souza-Neto; José Salvatore Leister Patané; Luiz Alcantara; Luiz Lehmann Coutinho; Maria Carolina Elias; Maurício Lacerda Nogueira; Rafael dos Santos Bezerra; Raul Machado Neto; Rejane Maria Tommasini Grotto; Ricardo Haddad; Sandra Coccuzzo Sampaio Vessoni; Simone Kashima; Svetoslav Nanev Slavov; Vincent Louis Viala |
| EPI_ISL_5646781 | UNIDADE DE VIGILANCIA EPIDEMIOLOGICA | Instituto Butantan | Antonio Jorge Martins; Claudia Renata dos Santos Barros; David Schlesinger; Debora Botequio Moretti; Dimas Tadeu Covas; Elaine Cristina Marqueze; Elaine Vieira Santos; Evandra Strazza Rodrigues; Heidge Fukumasu; Jayme Augusto de Souza-Neto; José Salvatore Leister Patané; Luiz Alcantara; Luiz Lehmann Coutinho; Maria Carolina Elias; Maurício Lacerda Nogueira; Rafael dos Santos Bezerra; Raul Machado Neto; Rejane Maria Tommasini Grotto; Ricardo Haddad; Sandra Coccuzzo Sampaio Vessoni; Simone Kashima; Svetoslav Nanev Slavov; Vincent Louis Viala |
| EPI_ISL_5646167, EPI_ISL_5646174 | UNIDADE MISTA ARACOIABA DA SERRA | Instituto Butantan | Antonio Jorge Martins; Claudia Renata dos Santos Barros; David Schlesinger; Debora Botequio Moretti; Dimas Tadeu Covas; Elaine Cristina Marqueze; Elaine Vieira Santos; Evandra Strazza Rodrigues; Heidge Fukumasu; Jayme Augusto de Souza-Neto; José Salvatore Leister Patané; Luiz Alcantara; Luiz Lehmann Coutinho; Maria Carolina Elias; Maurício Lacerda Nogueira; Rafael dos Santos Bezerra; Raul Machado Neto; Rejane Maria Tommasini Grotto; Ricardo Haddad; Sandra Coccuzzo Sampaio Vessoni; Simone Kashima; Svetoslav Nanev Slavov; Vincent Louis Viala |
| EPI_ISL_5645747 | UNIDADE MISTA DE SAUDE CAPELA DO ALTO | Instituto Butantan | Antonio Jorge Martins; Claudia Renata dos Santos Barros; David Schlesinger; Debora Botequio Moretti; Dimas Tadeu Covas; Elaine Cristina Marqueze; Elaine Vieira Santos; Evandra Strazza Rodrigues; Heidge Fukumasu; Jayme Augusto de Souza-Neto; José Salvatore Leister Patané; Luiz Alcantara; Luiz Lehmann Coutinho; Maria Carolina Elias; Maurício Lacerda Nogueira; Rafael dos Santos Bezerra; Raul Machado Neto; Rejane Maria Tommasini Grotto; Ricardo Haddad; Sandra Coccuzzo Sampaio Vessoni; Simone Kashima; Svetoslav Nanev Slavov; Vincent Louis Viala |
| EPI_ISL_5645795 | UNIDADE MISTA DE SAUDE DE SARAPUI | Instituto Butantan | Antonio Jorge Martins; Claudia Renata dos Santos Barros; David Schlesinger; Debora Botequio Moretti; Dimas Tadeu Covas; Elaine Cristina Marqueze; Elaine Vieira Santos; Evandra Strazza Rodrigues; Heidge Fukumasu; Jayme Augusto de Souza-Neto; José Salvatore Leister Patané; Luiz Alcantara; Luiz Lehmann Coutinho; Maria Carolina Elias; Maurício Lacerda Nogueira; Rafael dos Santos Bezerra; Raul Machado Neto; Rejane Maria Tommasini Grotto; Ricardo Haddad; Sandra Coccuzzo Sampaio Vessoni; Simone Kashima; Svetoslav Nanev Slavov; Vincent Louis Viala |
| EPI_ISL_5645743, EPI_ISL_5645770, EPI_ISL_5645909, EPI_ISL_5646147 | UNIDADE PSF TAQUARIVAI | Instituto Butantan | Antonio Jorge Martins; Claudia Renata dos Santos Barros; David Schlesinger; Debora Botequio Moretti; Dimas Tadeu Covas; Elaine Cristina Marqueze; Elaine Vieira Santos; Evandra Strazza Rodrigues; Heidge Fukumasu; Jayme Augusto de Souza-Neto; José Salvatore Leister Patané; Luiz Alcantara; Luiz Lehmann Coutinho; Maria Carolina Elias; Maurício Lacerda Nogueira; Rafael dos Santos Bezerra; Raul Machado Neto; Rejane Maria Tommasini Grotto; Ricardo Haddad; Sandra Coccuzzo Sampaio Vessoni; Simone Kashima; Svetoslav Nanev Slavov; Vincent Louis Viala |
| EPI_ISL_3912018, EPI_ISL_3912074, EPI_ISL_3912088, EPI_ISL_3912094, EPI_ISL_3912370, EPI_ISL_3912371, EPI_ISL_3912372, EPI_ISL_3912373, EPI_ISL_3912374, EPI_ISL_3912375, EPI_ISL_3912376, EPI_ISL_3912377, EPI_ISL_3912378, EPI_ISL_3912379, EPI_ISL_3912380 | | | |
| see above | UNIDADE REFERENCIA COVID 19 | Analytical Competence Molecular Epidemiology Lab/ACME, Oswaldo Cruz Foundation, Ceara (FIOCRUZ CE) | Cleber Furtado Aksenen; Fabio Miyajima; Fernando Braga Stehling; Francisco Eder de Moura Lopes; Jamille Maria Mendes Bezerra; Joaquim Cesar do Nascimento Sousa Junior; Pedro Miguel Carneiro Jeronimo; Suzana Porto Almeida & Lucas Delerino on behalf of COVID-19 FIOCRUZ Genomic Network; Thais Ferreira de Oliveira; Thais de Oliveira Costa; Ticiane Cavalcante de Souza; Veridiana Pessoa Miyajima |
| EPI_ISL_5645693, EPI_ISL_5645779, EPI_ISL_5645908, EPI_ISL_5646122, EPI_ISL_5646156, EPI_ISL_5646184 | UNIDADE SENTINELA COVID19 | Instituto Butantan | Antonio Jorge Martins; Claudia Renata dos Santos Barros; David Schlesinger; Debora Botequio Moretti; Dimas Tadeu Covas; Elaine Cristina Marqueze; Elaine Vieira Santos; Evandra Strazza Rodrigues; Heidge Fukumasu; Jayme Augusto de Souza-Neto; José Salvatore Leister Patané; Luiz Alcantara; Luiz Lehmann Coutinho; Maria Carolina Elias; Maurício Lacerda Nogueira; Rafael dos Santos Bezerra; Raul Machado Neto; Rejane Maria Tommasini Grotto; Ricardo Haddad; Sandra Coccuzzo Sampaio Vessoni; Simone Kashima; Svetoslav Nanev Slavov; Vincent Louis Viala |
| EPI_ISL_5645899 | UNIDADE SENTINELA VILA APARECIDA | Instituto Butantan | Antonio Jorge Martins; Claudia Renata dos Santos Barros; David Schlesinger; Debora Botequio Moretti; Dimas Tadeu Covas; Elaine Cristina Marqueze; Elaine Vieira Santos; Evandra Strazza Rodrigues; Heidge Fukumasu; Jayme Augusto de Souza-Neto; José Salvatore Leister Patané; Luiz Alcantara; Luiz Lehmann Coutinho; Maria Carolina Elias; Maurício Lacerda Nogueira; Rafael dos Santos Bezerra; Raul Machado Neto; Rejane Maria Tommasini Grotto; Ricardo Haddad; Sandra Coccuzzo Sampaio Vessoni; Simone Kashima; Svetoslav Nanev Slavov; Vincent Louis Viala |
| EPI_ISL_2628629, EPI_ISL_2628640, EPI_ISL_2628659, EPI_ISL_2628680, EPI_ISL_2628717, EPI_ISL_2628825, EPI_ISL_2645941, EPI_ISL_2645991, EPI_ISL_2645995 | | | |
| see above | UNILABS | Instituto Nacional de Saude (INSA) | Borges et al |
| EPI_ISL_5645612, EPI_ISL_5645619, EPI_ISL_5645623, EPI_ISL_5645690, EPI_ISL_5645726, EPI_ISL_5645805, EPI_ISL_5645811, EPI_ISL_5645826, EPI_ISL_5645827, EPI_ISL_5645828, EPI_ISL_5645859, EPI_ISL_5645887 | | | |
| see above | UPA DR AKIRA TADA | Instituto Butantan | Antonio Jorge Martins; Claudia Renata dos Santos Barros; David Schlesinger; Debora Botequio Moretti; Dimas Tadeu Covas; Elaine Cristina Marqueze; Elaine Vieira Santos; Evandra Strazza Rodrigues; Heidge Fukumasu; Jayme Augusto de Souza-Neto; José Salvatore Leister Patané; Luiz Alcantara; Luiz Lehmann Coutinho; Maria Carolina Elias; Maurício Lacerda Nogueira; Rafael dos Santos Bezerra; Raul Machado Neto; Rejane Maria Tommasini Grotto; Ricardo Haddad; Sandra Coccuzzo Sampaio Vessoni; Simone Kashima; Svetoslav Nanev Slavov; Vincent Louis Viala |
| EPI_ISL_5647225, EPI_ISL_5647252 | UPA GEISEL | Instituto Butantan | Antonio Jorge Martins; Claudia Renata dos Santos Barros; David Schlesinger; Debora Botequio Moretti; Dimas Tadeu Covas; Elaine Cristina Marqueze; Elaine Vieira Santos; Evandra Strazza Rodrigues; Heidge Fukumasu; Jayme Augusto de Souza-Neto; José Salvatore Leister Patané; Luiz Alcantara; Luiz Lehmann Coutinho; Maria Carolina Elias; Maurício Lacerda Nogueira; Rafael dos Santos Bezerra; Raul Machado Neto; Rejane Maria Tommasini Grotto; Ricardo Haddad; Sandra Coccuzzo Sampaio Vessoni; Simone Kashima; Svetoslav Nanev Slavov; Vincent Louis Viala |
| EPI_ISL_5645847, EPI_ISL_5645848, EPI_ISL_5645879 | UPA VILA SANTA CATARINA | Instituto Butantan | Antonio Jorge Martins; Claudia Renata dos Santos Barros; David Schlesinger; Debora Botequio Moretti; Dimas Tadeu Covas; Elaine Cristina Marqueze; Elaine Vieira Santos; Evandra Strazza Rodrigues; Heidge Fukumasu; Jayme Augusto de Souza-Neto; José Salvatore Leister Patané; Luiz Alcantara; Luiz Lehmann Coutinho; Maria Carolina Elias; Maurício Lacerda Nogueira; Rafael dos Santos Bezerra; Raul Machado Neto; Rejane Maria Tommasini Grotto; Ricardo Haddad; Sandra Coccuzzo Sampaio Vessoni; Simone Kashima; Svetoslav Nanev Slavov; Vincent Louis Viala |
| EPI_ISL_4293475 | US Air Force School of Aerospace Medicine | US Air Force School of Aerospace Medicine | Anthony Fries; Carol Garrett; Clarise Starr; Deanna Muehleman; Elizabeth Macias; Jennifer Meyer; Kelsey Lanter; Sarah Purves; William Gruner |
| EPI_ISL_5649670, EPI_ISL_5650362, EPI_ISL_5650367, EPI_ISL_5650370 | USF DE INUBIA PAULISTA | Instituto Butantan | Antonio Jorge Martins; Claudia Renata dos Santos Barros; David Schlesinger; Debora Botequio Moretti; Dimas Tadeu Covas; Elaine Cristina Marqueze; Elaine Vieira Santos; Evandra Strazza Rodrigues; Heidge Fukumasu; Jayme Augusto de Souza-Neto; José Salvatore Leister Patané; Luiz Alcantara; Luiz Lehmann Coutinho; Maria Carolina Elias; Maurício Lacerda Nogueira; Rafael dos Santos Bezerra; Raul Machado Neto; Rejane Maria Tommasini Grotto; Ricardo Haddad; Sandra Coccuzzo Sampaio Vessoni; Simone Kashima; Svetoslav Nanev Slavov; Vincent Louis Viala |
| EPI_ISL_2501396, EPI_ISL_2501397, EPI_ISL_2501470, EPI_ISL_2501472, EPI_ISL_2501473, EPI_ISL_2501474, EPI_ISL_2501475, EPI_ISL_2501476, EPI_ISL_2501477, EPI_ISL_2501478, EPI_ISL_2501479, EPI_ISL_2501480, EPI_ISL_2501481, EPI_ISL_2501482, EPI_ISL_2501483, EPI_ISL_2501484, EPI_ISL_2501485, EPI_ISL_2501486, EPI_ISL_2501487, EPI_ISL_2501488, EPI_ISL_2501489, EPI_ISL_2501490, EPI_ISL_2501491, EPI_ISL_2501492, EPI_ISL_2501493, EPI_ISL_2501494, EPI_ISL_2501495, EPI_ISL_2501496, EPI_ISL_2544982, EPI_ISL_2549409, EPI_ISL_2549432, EPI_ISL_2549440, EPI_ISL_2549442, EPI_ISL_2549447, EPI_ISL_2549450, EPI_ISL_2549451, EPI_ISL_2549458, EPI_ISL_2549471, EPI_ISL_2549476, EPI_ISL_2549478, EPI_ISL_2549479, EPI_ISL_2549481, EPI_ISL_2549493, EPI_ISL_2549495, EPI_ISL_2549496, EPI_ISL_2549499, EPI_ISL_2549504, EPI_ISL_2549507, EPI_ISL_2549509, EPI_ISL_2549519, EPI_ISL_2549520, EPI_ISL_2549523, EPI_ISL_2549525, EPI_ISL_2549532, EPI_ISL_2549540, EPI_ISL_2549543, EPI_ISL_2549548, EPI_ISL_2549553, EPI_ISL_2549555, EPI_ISL_2549556, EPI_ISL_2549557, EPI_ISL_2549558, EPI_ISL_2549559, EPI_ISL_2549561, EPI_ISL_2617987, EPI_ISL_2693781, EPI_ISL_2693801, EPI_ISL_2693813, EPI_ISL_2716479, EPI_ISL_2716488 | | | |
| see above | UW Virology Lab | UW Virology Lab | Alexander Greninger; Hong Xie; Keith R Jerome; Lasata Shrestha; Meei-Li Huang; Michelle Lin; Nathan Breit; Noah R. Baker; Patrick Mathias; Pavitra Roychoudhury; Ricardo Perez; Robert J. Livingston; Sean Ellis; Shah Mohamed Bakhash; Tien V. Nguyen |
| EPI_ISL_2681123, EPI_ISL_2681209, EPI_ISL_2681293 | Unidad de Investigación Biomédica de Zacatecas (UIBZ) | Instituto de Biotecnología de la UNAM | ; Alejandra García-Gasca; Alejandra Hernández-Terán; Alejandro Sánchez-Flores; Alfredo Herrera-Estrella; Alicia Ocaña-Mondragón; Andreu Comas-García; Angel Gustavo Salas-Lais; Antonio Loza Román; Bernardo Martínez-Miguel; Blanca Taboada; Brenda Irasema Maldonado-Meza; Bruno Gómez-Gil; Carla Ivón Herrera-Najera; Carlos F. Arias; Celia Boukadida; Clara Esperanza Santacruz-Tinoco; Concepción Grajales-Muñiz; Consorcio Mexicano de Vigilancia Genómica (CoViGen-Mex). Authors (in alphabetical order): Julio Elias Alvarado-Yaah; Cristóbal Cháidez-Quiróz; Célida Duque Molina; Célida Martínez- Rodríguez; Daniel Fregoso-Rueda; Daniel Lira Morales; Eduardo Becerril-Vargas; Fernando Fontove-Herrera; Fidencio Mejía-Nepomuceno; Francisco Pulido; Gloria Elena Espinosa-Ayala; Gloria María Molina-Salinas; Gloria Vazquez; Hector Esteban Paz-Juárez; Hector Montoya-Fuentes; Helen Haydee Fernanda Ramírez-Plascencia; Irvin González-López; Jean Pierre González; Jesús Hernández; Joel Armando Vázquez-Pérez.; Jorge Salas-Hernández; José Antonio Enciso-Moreno; José Arturo Martínez-Orozco; José Esteban Muñoz-Medina; José de Jesús Nuñez-Contreras; Juan Bautista Chale-Dzul; Julissa Enciso-Ibarra; Luis Alberto Ochoa-Carrera; Margarita Matías-Florentino; Mario Mújica-Sánchez; Marissa Perez-Garcia; María Guadalupe Santiago-Mauricio; María Guadalupe de Jesús Mireles-Rivera; Nelly Sélem-Mojica; Pavel Isa; Ricardo Ciria Merce; Ricardo Grande; Rosa María Gutiérrez Rios; Santiago Ávila-Ríos; Selene Zárate; Susana Lopez; Verónica Mata-Haro; Victor Eduardo García-Arias; Víctor Hugo Borja-Aburto |
| EPI_ISL_2681067, EPI_ISL_2681074, EPI_ISL_2681075, EPI_ISL_2681080, EPI_ISL_2681082, EPI_ISL_2681085, EPI_ISL_2681089, EPI_ISL_2681091, EPI_ISL_2681100, EPI_ISL_2681101, EPI_ISL_2681107, EPI_ISL_2681109, EPI_ISL_2681114, EPI_ISL_2681116, EPI_ISL_2681117, EPI_ISL_2681141, EPI_ISL_2681211, EPI_ISL_2681232, EPI_ISL_2681241, EPI_ISL_2681261, EPI_ISL_2681295, EPI_ISL_2681298, EPI_ISL_2681319, EPI_ISL_2681320, EPI_ISL_2681322, EPI_ISL_2681325, EPI_ISL_2681327, EPI_ISL_2681333, EPI_ISL_2681342, EPI_ISL_2681345, EPI_ISL_2681356, EPI_ISL_2681360, EPI_ISL_2681361, EPI_ISL_2681364, EPI_ISL_2681365, EPI_ISL_2681366, EPI_ISL_2681377, EPI_ISL_2681379, EPI_ISL_2681383, EPI_ISL_2681384, EPI_ISL_2801565, EPI_ISL_2801569, EPI_ISL_2801570, EPI_ISL_2801571, EPI_ISL_2801572, EPI_ISL_2801573, EPI_ISL_2801574, EPI_ISL_2801575, EPI_ISL_2801576, EPI_ISL_2801580, EPI_ISL_2801581, EPI_ISL_2801586, EPI_ISL_2801587, EPI_ISL_2801588, EPI_ISL_2801589, EPI_ISL_2801590, EPI_ISL_2801591, EPI_ISL_2801596, EPI_ISL_2801597, EPI_ISL_2801599, EPI_ISL_2801608, EPI_ISL_2801609, EPI_ISL_2801610, EPI_ISL_2801611, EPI_ISL_2801613, EPI_ISL_2801614, EPI_ISL_2801617, EPI_ISL_2801618, EPI_ISL_2801621, EPI_ISL_2801636, EPI_ISL_2801637, EPI_ISL_2801638, EPI_ISL_2801642 | | | |
| see above | Unidad de Investigación Médica de Yucatán (UIMY) | Instituto de Biotecnología de la UNAM | ; Alejandra García-Gasca; Alejandra Hernández-Terán; Alejandro Sánchez-Flores; Alfredo Herrera-Estrella; Alicia Ocaña-Mondragón; Andreu Comas-García; Angel Gustavo Salas-Lais; Antonio Loza Román; Bernardo Martínez-Miguel; Blanca Taboada; Brenda Irasema Maldonado-Meza; Bruno Gómez-Gil; Carla Ivón Herrera-Najera; Carlos F. Arias; Celia Boukadida; Clara Esperanza Santacruz-Tinoco; Concepción Grajales-Muñiz; Consorcio Mexicano de Vigilancia Genómica (CoViGen-Mex). Authors (in alphabetical order): Julio Elias Alvarado-Yaah; Cristóbal Cháidez-Quiróz; Célida Duque Molina; Célida Martínez- Rodríguez; Daniel Fregoso-Rueda; Daniel Lira Morales; Eduardo Becerril-Vargas; Fernando Fontove-Herrera; Fidencio Mejía-Nepomuceno; Francisco Pulido; Gloria Elena Espinosa-Ayala; Gloria María Molina-Salinas; Gloria Vazquez; Hector Esteban Paz-Juárez; Hector Montoya-Fuentes; Helen Haydee Fernanda Ramírez-Plascencia; Irvin González-López; Jean Pierre González; Jesús Hernández; Joel Armando Vázquez-Pérez.; Jorge Salas-Hernández; José Antonio Enciso-Moreno; José Arturo Martínez-Orozco; José Esteban Muñoz-Medina; José de Jesús Nuñez-Contreras; Juan Bautista Chale-Dzul; Julissa Enciso-Ibarra; Luis Alberto Ochoa-Carrera; Margarita Matías-Florentino; Mario Mújica-Sánchez; Marissa Perez-Garcia; María Guadalupe Santiago-Mauricio; María Guadalupe de Jesús Mireles-Rivera; Nelly Sélem-Mojica; Pavel Isa; Ricardo Ciria Merce; Ricardo Grande; Rosa María Gutiérrez Rios; Santiago Ávila-Ríos; Selene Zárate; Susana Lopez; Verónica Mata-Haro; Victor Eduardo García-Arias; Víctor Hugo Borja-Aburto |
| EPI_ISL_2691570, EPI_ISL_2691571, EPI_ISL_2691572, EPI_ISL_2691573, EPI_ISL_2691642 | Unidade de apoio ao diagnostico da COVID - UNADIG | Bioinformatics Laboratory / LNCC | Alessandra P Lamarca; Alexandra L Gerber; Amilcar Tanuri; Ana Paula de C Guimaraes; Ana Tereza R Vasconcelos; Andrea Cony Cavalcanti; Caio Luiz Pereira Ribeiro; Cassia Alves; Cintia Policarpo; Claudia Maria Braga de Mello; Cristiane Gomes da Silva; Diana Mariani; Douglas Terra Machado; Flavio Dias da Silva; Gleidson da Silva de Oliveira; Leandro Magalhaes de Souza; Liliane Cavalcante; Luiz G P de Almeida; Marcio Henrique de Oliveira Garcia; Mario Sergio Ribeiro; Ronaldo da Silva F Jr; Silvia Carvalho |
| EPI_ISL_2625638 | University Hospital Hradec Kralove | University Hospital Hradec Kralove | Helena Kovarikova; Marketa Gancarcikova |
| EPI_ISL_2483946, EPI_ISL_2483948, EPI_ISL_2483952 | University of Liège COVID-19 testing center | GIGA Medical Genomics | Bouchra Boujemla; Cécile Meex; Fabrice Bureau; Keith Durkin; Laurent Gillet; Maria Artesi; Marie-Pierre Hayette; Nathalie Renotte; Sébastien Bontems; Vincent Bours; Wouter Coppieters |
| EPI_ISL_2578648, EPI_ISL_2833782 | University of Wisconsin-Madison AIDS Vaccine Research Laboratories | University of Wisconsin-Madison AIDS Vaccine Research Laboratories | Gage Moreno; Katarina Braun; et al. AIDS Vaccine Research Laboratories |
| EPI_ISL_3014244 | Università Federico II - Dipartimento di scienze mediche traslazionali - Napoli | TIGEM | Antonio Grimaldi Patrizia Annunziata Francesco Panariello Teresa Giuliano Michele Cennamo Valentina Bouche Chiara Colantuono Lucio Di Filippo Mariano Fiorenza Anna Manfredi Marcello Salvi Giuseppe Portella Andrea Ballabio Davide Cacchiarelli |
| EPI_ISL_2600737, EPI_ISL_2600741, EPI_ISL_2600742, EPI_ISL_2600778 | Università degli Studi di Perugia | Istituto Zooprofilattico Sperimentale dell'Abruzzo e Molise "G. Caporale" | Ancora M; Biagetti M; Calistri P; Camilloni B; Cammà C; Curini V; Delli Compagni E; Di Domenico M; Di Pasquale A; Giammarioli M; Lorusso A; Mangone I; Marcacci M; Mencacci A; Puglia I; Rinaldi A; Savini G; Scialabba S |
| EPI_ISL_2856304, EPI_ISL_2856315, EPI_ISL_2856629, EPI_ISL_2856649, EPI_ISL_2856677, EPI_ISL_2856702, EPI_ISL_2856711, EPI_ISL_2856721, EPI_ISL_2856739, EPI_ISL_2856886, EPI_ISL_2856888, EPI_ISL_2856909, EPI_ISL_2856910 | | | |

| | | | |
|---|---|---|---|
| see above | Utah Public Health Laboratory | Utah Public Health Laboratory | Erin L. Young; Kelly F. Oakeson; Olinto Linares-Perdomo; Tara Gallagher |
| EPI_ISL_3355143 | VA-Division of Consolidated Laboratory Services | Centers for Disease Control and Prevention Division of Viral Diseases, Pathogen Discovery | Alex Burgin; Ben Rambo-Martin; Clinton Paden; Dakota Howard; Dave Wentworth; Dhwani Batra; Jasmine Padilla; Justin Lee; Krista Queen; Kristen Knipe; Kristine Lacek; Mark Burroughs; Matthew Schmerer; Meghan Bentz; Mili Sheth; Peter Cook; Sam Shepard; Sarah Nobles; Suxiang Tong; Vivien Dugan; Yvette Unoarumhi |
| EPI_ISL_5645570, EPI_ISL_5649575, EPI_ISL_5649705, EPI_ISL_5650331, EPI_ISL_5650338, EPI_ISL_5650345, EPI_ISL_5650346, EPI_ISL_5650350, EPI_ISL_5650373, EPI_ISL_5650391 | | | |
| see above | VIGILANCIA EM SAUDE SAE VIGILANCIA EPIDEMIOLOGICA | Instituto Butantan | Antonio Jorge Martins; Claudia Renata dos Santos Barros; David Schlesinger; Debora Botequio Moretti; Dimas Tadeu Covas; Elaine Cristina Marqueze; Elaine Vieira Santos; Evandra Strazza Rodrigues; Heidge Fukumasu; Jayme Augusto de Souza-Neto; José Salvatore Leister Patané; Luiz Alcantara; Luiz Lehmann Coutinho; Maria Carolina Elias; Maurício Lacerda Nogueira; Rafael dos Santos Bezerra; Raul Machado Neto; Rejane Maria Tommasini Grotto; Ricardo Haddad; Sandra Coccuzzo Sampaio Vessoni; Simone Kashima; Svetoslav Nanev Slavov; Vincent Louis Viala |
| EPI_ISL_5647165 | VIGILANCIA EPIDEMIOLOGICA DE LEME | Instituto Butantan | Antonio Jorge Martins; Claudia Renata dos Santos Barros; David Schlesinger; Debora Botequio Moretti; Dimas Tadeu Covas; Elaine Cristina Marqueze; Elaine Vieira Santos; Evandra Strazza Rodrigues; Heidge Fukumasu; Jayme Augusto de Souza-Neto; José Salvatore Leister Patané; Luiz Alcantara; Luiz Lehmann Coutinho; Maria Carolina Elias; Maurício Lacerda Nogueira; Rafael dos Santos Bezerra; Raul Machado Neto; Rejane Maria Tommasini Grotto; Ricardo Haddad; Sandra Coccuzzo Sampaio Vessoni; Simone Kashima; Svetoslav Nanev Slavov; Vincent Louis Viala |
| EPI_ISL_5647148, EPI_ISL_5647164 | VIGILANCIA EPIDEMIOLOGICA E CONTROLE DE VETORES PIRASSUNUN | Instituto Butantan | Antonio Jorge Martins; Claudia Renata dos Santos Barros; David Schlesinger; Debora Botequio Moretti; Dimas Tadeu Covas; Elaine Cristina Marqueze; Elaine Vieira Santos; Evandra Strazza Rodrigues; Heidge Fukumasu; Jayme Augusto de Souza-Neto; José Salvatore Leister Patané; Luiz Alcantara; Luiz Lehmann Coutinho; Maria Carolina Elias; Maurício Lacerda Nogueira; Rafael dos Santos Bezerra; Raul Machado Neto; Rejane Maria Tommasini Grotto; Ricardo Haddad; Sandra Coccuzzo Sampaio Vessoni; Simone Kashima; Svetoslav Nanev Slavov; Vincent Louis Viala |
| EPI_ISL_5645647, EPI_ISL_5646151 | VIGILANCIA EPIDIMIOLOGICA DE ARACARIGUAMA | Instituto Butantan | Antonio Jorge Martins; Claudia Renata dos Santos Barros; David Schlesinger; Debora Botequio Moretti; Dimas Tadeu Covas; Elaine Cristina Marqueze; Elaine Vieira Santos; Evandra Strazza Rodrigues; Heidge Fukumasu; Jayme Augusto de Souza-Neto; José Salvatore Leister Patané; Luiz Alcantara; Luiz Lehmann Coutinho; Maria Carolina Elias; Maurício Lacerda Nogueira; Rafael dos Santos Bezerra; Raul Machado Neto; Rejane Maria Tommasini Grotto; Ricardo Haddad; Sandra Coccuzzo Sampaio Vessoni; Simone Kashima; Svetoslav Nanev Slavov; Vincent Louis Viala |
| EPI_ISL_2544405, EPI_ISL_2611074, EPI_ISL_2662403 | Viollier AG | Department of Biosystems Science and Engineering, ETH Zürich | Andrea Patrignani; Andrea Patrizia Salzmann; Andreia Cabral de Gouvea; Catharine Aquino; Chaoran Chen; Christian Beisel; Christiane Beckmann; Christoph Noppen; Daniel Ehrsam; Doris Popovic; Elodie Burcklen; Griffin White; Henriette Kurth; Ina Nissen; Isabel Stürmer; Ivan Topolsky; Jay Tracy; Kim Philipp Jablonski; Lara Fuhrmann; Laura Neff; Lennart Opitz; Maria Domenica Moccia; Maurice Redondo; Mirjam Feldkamp; Natascha Santacroce; Niko Beerenwinkel; Olivier Kobel; Ralph Schlapbach; Rebecca Denes; Sarah Nadeau; Simon Grüter; Tanja Stadler; Timothy Sykes |
| EPI_ISL_2663473, EPI_ISL_2776043 | Virginia Division of Consolidated Laboratory Services | Virginia Division of Consolidated Laboratory Services | Virginia Division of Consolidated Laboratory Services |
| EPI_ISL_2884586, EPI_ISL_2884605 | Virology Laboratory, Scientific Department, Army Medical Center | Virology Laboratory, Scientific Department, Army Medical Center | Anella Monte; Anna Anselmo; Antonella Fortunato; Filippo Molinari; Florigio Lista; Francesco Giordani; Giancarlo Petralito; Giandomenico Cerreto; Giulia Campoli; Lucia Nicosia; Marzia Cavalli; Riccardo De Sanctis; Rossella Brandi; Silvia Fillo; Vanessa Vera Fain |
| EPI_ISL_2690098, EPI_ISL_2690108 | WA State Department of Health | Centers for Disease Control and Prevention Division of Viral Diseases, Pathogen Discovery | Alison Laufer Halpin; Ben L. Rambo-Martin; Clinton R. Paden; Dakota Howard; Darlene Wagner; Dave Wentworth; Dhwani Batra; Jasmine Padilla; Justin Lee; Katie Dillon; Krista Queen; Kristen Knipe; Kristine Lacek; Mark Burroughs; Matthew Schmerer; Mili Sheth; Peter Cook; Sam Shepard; Sarah Nobles; Shoshona Le; Suxiang Tong; Vivien Dugan; Yvette Unoarumhi |
| EPI_ISL_3031274 | WESTCHESTER MEDICAL CENTER | Wadsworth Center, New York State Department of Health | Alexis Russell; Catharine Prussing; Daryl M. Lamson; Erasmus Schneider; Erica Lasek-Nesselquist; John Kelly; Jonathan Plitnick; Kirsten St. George; Matthew Shudt; Melissa A Leisner; Navjot Singh |
| EPI_ISL_2787585 | WI State Laboratory of Hygiene | Centers for Disease Control and Prevention Division of Viral Diseases, Pathogen Discovery | Alison Laufer Halpin; Ben L. Rambo-Martin; Clinton R. Paden; Dakota Howard; Darlene Wagner; Dave Wentworth; Dhwani Batra; Jasmine Padilla; Justin Lee; Katie Dillon; Krista Queen; Kristen Knipe; Kristine Lacek; Mark Burroughs; Matthew Schmerer; Mili Sheth; Peter Cook; Sam Shepard; Sarah Nobles; Shoshona Le; Suxiang Tong; Vivien Dugan; Yvette Unoarumhi |
| EPI_ISL_2644654 | WSSE w Krakowie | 1. Academic Center for Pathomorphological and Genetic-Molecular Diagnostics ltd, Bialystok, Poland 2. National Institute of Public Health - National Institute of Hygiene, Warsaw, Poland | Anetta Sulewska; Jacek Nikliński; Janusz Dzięcioł; Joanna Kiśluk; Katarzyna Zacharczuk; Konrad Raczkowski; Magdalena Nowakowska; Małgorzata Sadkowska-Todys; Piotr Karabowicz; Piotr Majewski; Przemysław Biecek. Joanna Reszeć; Radosław Charkiewicz; Tomasz Wołkowicz |
| EPI_ISL_2557058, EPI_ISL_2557063, EPI_ISL_2557071, EPI_ISL_2557077, EPI_ISL_2651036, EPI_ISL_2651054, EPI_ISL_2651062, EPI_ISL_2651064, EPI_ISL_2651066, EPI_ISL_2651086, EPI_ISL_2651090, EPI_ISL_2716463, EPI_ISL_2716466, EPI_ISL_2834708 | | | |
| see above | Washington State Department of Health Public Health Laboratories | Washington State Department of Health Public Health Laboratories | Avi Singh; Darren Lucas; Denny Russell; Drew MacKellar; Geoff Melly; Hannah Gray; Joenice Gonzalez; JohnAric Peterson; Philip Dykema; Rebecca Cao; Vanessa De Los Santos |
| EPI_ISL_2466754, EPI_ISL_2466757, EPI_ISL_2499954 | Wichita State University - Molecular Diagnostics Lab | Kansas Health and Environmental Lab | Ben Olsen; Jonathan Barnell; Katherine Wiggins; Mike Grose; and Phil Adam |
| EPI_ISL_2648143 | Willis-Knighton Medical Center Hospital Laboratory | LSUHS Emerging Viral Threat Laboratory | Alexander Mijalis; Andrew D. Yurochko; April N. Johnson; Christopher G. Kevil; Gregory L. Ware; Jennifer L. Carroll; Jeremy P. Kamil; John A. Vanchiere; Joseph A. Bocchini; Lorie M. Atkins; Maarten Van Diest; Rona S. Scott |
| EPI_ISL_2727687, EPI_ISL_2727792 | Wisconsin State Laboratory of Hygiene Communicable Disease Division | Wisconsin State Laboratory of Hygiene Communicable Disease Division | Abigail C. Shockey; Alicia J. Mooney; Erika M. Hanson; Kelsey R. Florek; Richard Griesser; Sara Wagner; Tonya Danz |
| EPI_ISL_2560426, EPI_ISL_2712919, EPI_ISL_2712921, EPI_ISL_2713010, EPI_ISL_2726826, EPI_ISL_2801550, EPI_ISL_2876545, EPI_ISL_2876548 | | | |
| see above | Wyoming Public Health Laboratory | Wyoming Public Health Laboratory | Ashley Norberg; Brian Dominguez; Cari Sloma; Channing Weber; Chayse Rowley; Elliot Thomasson; Jim Mildenberger; Marley Goetz; Robert Petit; Sam Britz; Taylor Fearing; and Rob Christensen |
| EPI_ISL_2716245 | Yale Clinical Virology Lab | Grubaugh Lab - Yale School of Public Health | Anderson Brito; Annie Watkins; Chaney Kalinich; Chantal Vogels; Isabel Ott; Jessica Rothman; Joseph Fauver; Mallery Breban; Marie L. Landry; Mary Petrone; Nathan Grubaugh; Tara Alpert |
| EPI_ISL_2462974, EPI_ISL_2462976 | ZOL | Jessa | Berden et al. on behalf of the Jessa_cmdLab |
| EPI_ISL_2632267 | amedes MVZ für Laboratoriumsmedizin Fürstenfeldbruck | Robert Koch Institute | |
| EPI_ISL_5863413, EPI_ISL_5863414 | unknown | Noblis | Katharine Jennings; Lauren Brinkac; Lee F. Kolakowski; Minh Tran; Shane Mitchell; Shiela Diepold; William M. Nelson |
