## Supplement 3 for "Rapid and Accurate Identification of SARS-CoV-2 Omicron Variants Using Droplet Digital PCR (RT-ddPCR)": All_0607-0613.pdf

We gratefully acknowledge the following Authors from the Originating laboratories responsible for obtaining the specimens, as well as the Submitting laboratories where the genome data were generated and shared via GISAID, on which this research is based.

All Submitters of data may be contacted directly via www.gisaid.org

Authors are sorted alphabetically.

| Accession ID | Originating Laboratory | Submitting Laboratory | Authors |
|---|---|---|---|
| EPI_ISL_2987114 | "CDPH, Viral and Rickettsial Disease Laboratory" | Centers for Disease Control and Prevention Division of Viral Diseases, Pathogen Discovery | Alison Laufer Halpin; Ben L. Rambo-Martin; Clinton R. Paden; Dakota Howard; Darlene Wagner; Dave Wentworth; Dhwani Batra; Jasmine Padilla; Justin Lee; Katie Dillon; Krista Queen; Kristen Knipe; Kristine Lacek; Mark Burroughs; Matthew Schmerer; Mili Sheth; Peter Cook; Sam Shepard; Sarah Nobles; Shoshona Le; Suxiang Tong; Vivien Dugan; Yvette Unoarumhi |
| EPI_ISL_3276540, EPI_ISL_3276545 | ADILAB | Universidad Nacional de Colombia - Laboratorio Genómico One Health | Andres F. Cardona-Rios; Carlos Franco-Muñoz; Carolina Muñoz-Arango; Celeny Ortiz; Daniel O. Maldonado-Perez; Diego A. Álvarez-Díaz; Hector Alejandro Ruiz-Moreno; Idabely Betancur Ortiz; Jorge E. Osorio; Juan P. Hernandez-Ortiz; Karl A Ciuoderis; Katherine Laiton-Donato; Laura Silvana Perez; Lina M. Hurtado; Marcela Mercado-Reyes; Maria Angélica Maya; Maria Stella López; Rita Almanza Payares; Sandra Ines Cano; Simón Villegas Velásquez |
| EPI_ISL_5915159, EPI_ISL_5915161, EPI_ISL_5915171, EPI_ISL_5915215, EPI_ISL_5915227, EPI_ISL_5915228, EPI_ISL_5915234, EPI_ISL_5915268, EPI_ISL_5915286, EPI_ISL_5915297, EPI_ISL_5915301, EPI_ISL_5915308, EPI_ISL_6573819 | | | |
| see above | AMALAB/FACISA/UFRN | WallauLab on behalf of Fiocruz COVID-19 Genomic Surveillance Network | Alexandre Freitas da Silva; Allan Roberto Dias Nunes; Antonio Marinho da Silva Neto; Cassia Docena; Constância Flávia Junqueira Ayres; Filipe Zimmer Dezordi; Gabriel Luz Wallau; Gustavo Barbosa de Lima; Joana Cristina Medeiros Tavares Marques; Katya Anaya Jacinto; Lais Ceschini Machado; Lilian Caroliny Amorim Silva; Marcelo Henrique dos Santos Paiva; Mariane dos Santos Duarte; Matheus Filgueira Bezerra; Sinval Pinto Brandão Filho |
| EPI_ISL_5647224, EPI_ISL_5647258, EPI_ISL_5647289 | AMBULATORIO MEDICO MUNICIPAL DE AGUDOS | Instituto Butantan | Antonio Jorge Martins; Claudia Renata dos Santos Barros; David Schlesinger; Debora Botequio Moretti; Dimas Tadeu Covas; Elaine Cristina Marqueze; Elaine Vieira Santos; Evandra Strazza Rodrigues; Heidge Fukumasu; Jayme Augusto de Souza-Neto; José Salvatore Leister Patané; Luiz Alcantara; Luiz Lehmann Coutinho; Maria Carolina Elias; Maurício Lacerda Nogueira; Rafael dos Santos Bezerra; Raul Machado Neto; Rejane Maria Tommasini Grotto; Ricardo Haddad; Sandra Coccuzzo Sampaio Vessoni; Simone Kashima; Svetoslav Nanev Slavov; Vincent Louis Viala |
| EPI_ISL_2828002 | AREA DE SALUD GOICOECHEA 2 - CLINICA DR. JIMENEZ NUÑEZ | Inciensa, Instituto Costarricense de Investigación y Enseñanza en Nutrición y Salud | Adriana Godínez; Claudio Soto-Garita; Estela Cordero; Francisco Duarte; Hebleen Porras; Joselyn Prado & Juan Carlos Cartes; José Luis Vargas; Mariela Gutiérrez; Melany Calderón |
| EPI_ISL_2815338 | ASL BA | University of Bari Biomedical Sciences and Human Oncology | Maria Chironna |
| EPI_ISL_2815341 | ASL BARI | University of Bari Biomedical Sciences and Human Oncology | Maria Chironna |
| EPI_ISL_2963446, EPI_ISL_2963450 | AULSS 2 Marca Trevigiana | Istituto Zooprofilattico Sperimentale delle Venezie | Adelaide Milani; Alessia Schivo; Alice Fusaro; Ambra Pastori; Annalisa Salviato; Antonia Ricci; Calogero Terregino; Edoardo Giussani; Elisa Palumbo; Erika Giorgia Quaranta; Isabella Monne; Luca Tassoni |
| EPI_ISL_2925860 | AULSS 5 Polesana | Istituto Zooprofilattico Sperimentale delle Venezie | Adelaide Milani; Alessia Schivo; Alice Fusaro; Ambra Pastori; Annalisa Salviato; Antonia Ricci; Calogero Terregino; Edoardo Giussani; Elisa Palumbo; Erika Giorgia Quaranta; Isabella Monne; Luca Tassoni |
| EPI_ISL_4850554 | AYUDAS DIAGNOSTICAS SURA | Instituto Nacional de Salud- Dirección de Investigación en Salud Pública | Carlos Franco-Muñoz; Carmen Osorio; Diana Malo; Diego A. Álvarez-Díaz; Diego Andrés Prada; Gerardo Santamaría; Hector Alejandro Ruiz-Moreno; Jhonnatan Reales-González; Jorge Rivera; Juan Camilo Martinez; Julian Naizaque; Katherine Laiton-Donato; Lisseth Pardo; Magdalena Wiesner; Marcela Mercado-Reyes; Maria T. Herrera-Sepúlveda; Marta Lopez Blanco; Martha Lucia Ospina Martinez; Paola Rojas; Sergio Gomez; Sheryll Corchuelo; Ángela Alarcon Cruz |
| EPI_ISL_2652157 | AZ Klina | AZ Klina | Carl Vael - Lynsey Berckmans |
| EPI_ISL_2625749, EPI_ISL_2625753 | AZDelta | AZ Delta Medical Laboratories in Roeselare, Belgium | Dieter De Smet; Geert Martens; Merijn Vanhee; on behalf of AZ Delta COVID-19 Genomics core (member of Genomic surveillance of SARS-CoV-2 in Belgium network) |
| EPI_ISL_2686977, EPI_ISL_2686981, EPI_ISL_2686982, EPI_ISL_2686984, EPI_ISL_2686985, EPI_ISL_2686989, EPI_ISL_2686997, EPI_ISL_2687069, EPI_ISL_2687098, EPI_ISL_2687120, EPI_ISL_2687121, EPI_ISL_2687136, EPI_ISL_2687137, EPI_ISL_2687145, EPI_ISL_2687162, EPI_ISL_2687165, EPI_ISL_2687179, EPI_ISL_2687186, EPI_ISL_2687205, EPI_ISL_2687212, EPI_ISL_2687223, EPI_ISL_2687226, EPI_ISL_2687229, EPI_ISL_2687242, EPI_ISL_2687243, EPI_ISL_2687248, EPI_ISL_2687253, EPI_ISL_2687265, EPI_ISL_2687276, EPI_ISL_2687287, EPI_ISL_2687292, EPI_ISL_2687301, EPI_ISL_2687303, EPI_ISL_2687308, EPI_ISL_2687315, EPI_ISL_2687326, EPI_ISL_2687338, EPI_ISL_2687349, EPI_ISL_2687356, EPI_ISL_2687378, EPI_ISL_2687379, EPI_ISL_2687380, EPI_ISL_2687381, EPI_ISL_2687388, EPI_ISL_2687393, EPI_ISL_2687403, EPI_ISL_2687413, EPI_ISL_2687414, EPI_ISL_2687416, EPI_ISL_2687419, EPI_ISL_2687428, EPI_ISL_2687444, EPI_ISL_2687455, EPI_ISL_2687461, EPI_ISL_2687483, EPI_ISL_2687488, EPI_ISL_2687489, EPI_ISL_2687499, EPI_ISL_2687503, EPI_ISL_2687508, EPI_ISL_2687533, EPI_ISL_2687534, EPI_ISL_2687551, EPI_ISL_2687554, EPI_ISL_2687558, EPI_ISL_2687559, EPI_ISL_2687578, EPI_ISL_2709541, EPI_ISL_2709543, EPI_ISL_2709546, EPI_ISL_2709547, EPI_ISL_2785475, EPI_ISL_2785476, EPI_ISL_2785478, EPI_ISL_2785484, EPI_ISL_2785485, EPI_ISL_2785487, EPI_ISL_2785495, EPI_ISL_2785516, EPI_ISL_2832579, EPI_ISL_3220387, EPI_ISL_4364602 | | | |
| see above | Aegis Sciences Corporation | Centers for Disease Control and Prevention Division of Viral Diseases, Pathogen Discovery | Adrian Paskey; Alec Vest; Benjamin Rambo-Martin; Christopher Gulvick; Clinton Paden; Clinton R. Paden; Cyndi Clark; Dakota Howard; Darlene Wagner; Dhwani Batra; Dillon Nall; Duncan MacCannell; Erisa Sula; Ethan Sanders; Holly Houdeshell; Jason Caravas; Kara Moser; Kristine Lacek; Matthew Hardison; Matthew Schmerer; Ola Kvalvaag; Patrick Campbell; Peter Cook; Peter W. Cook; Rob Case; Scott Sammons; Shatavia Morrison; Shaun Westlund; Tymeckia Kendall; Victoria Caban Figueroa; Vikramsinha Ghorpade; Yvette Unoarumhi |
| EPI_ISL_2953286 | Ages | Elling and Cochella laboratories, IMBA/IMP | Alexander Stark; Ezgi Oezkan; Luisa Cochella and Ulrich Elling; Marcus Strobl; Olga Frank; Ramesh Yelagandula; Tanino Albanese |
| EPI_ISL_4219172, EPI_ISL_4219174, EPI_ISL_4219175 | Alameda County Public Health Department | Alameda County Public Health Department | Maria Elma; Vici Varghese |
| EPI_ISL_2626722, EPI_ISL_2626723, EPI_ISL_2626727, EPI_ISL_2626735, EPI_ISL_2626737, EPI_ISL_2626738, EPI_ISL_2626743, EPI_ISL_2626745, EPI_ISL_2893332 | | | |
| see above | Alaska State Virology Laboratory | Alaska State Virology Laboratory | Elva House; Jack Chen; Jacob Zidek; Lisa Smith; Ph.D.; Stephanie DeRonde |
| EPI_ISL_3030251 | Ancona | Microbiology University Politecnica delle Marche | Anna Valenza; Carla Acciarri; Katia Marinelli; Monica Lucia Ferreri; Patrizia Bagnarelli; Roberta Longo; Sara Caucci; Stefano Menzo |
| EPI_ISL_2716405, EPI_ISL_2716406, EPI_ISL_2716407, EPI_ISL_2716411 | Arizona State Public Health Laboratory | Arizona State Public Health Laboratory | Jessica Escobar; Katherine Fullerton; Linda Getsinger; Matthew Contursi; Nobuko Fukushima; Stacy White; Trung Huynh; Victor Waddell |
| EPI_ISL_3151264 | Arkansas Children's Hospital | Center for Global Health, University of New Mexico Health Sciences Center | Ashton Ingold; Bobby Boyanton; Darrell Dinwiddie; Daryl Dommman; Joshua L. Kennedy; Kurt Schwalm; Valerie Morley |
| EPI_ISL_2736280, EPI_ISL_2736298, EPI_ISL_3015721, EPI_ISL_3015722, EPI_ISL_3015723, EPI_ISL_3015725 | Atlas Genomics | Seattle Flu Study | Amanda Adler; Barry R. Lutz; Benjamin Pelle; Brian Hiatt; Caitlin R. Wolf; Chris D. Frazar; Deborah A. Nickerson; Elisabeth Brandstetter; Erica Ryke; Geoff Melly; Helen Y. Chu; Janet A. Englund; Jay Shendure; Jover Lee; Kairsten Fay; Kirsten Lacombe; Lea M. Starita; Mark J. Rieder; Matthew Richardson; Matthew Thompson; Melissa Truong; Michael Boeckh; Michael Famulare; Misja Ilcisin; Peter D. Han; Philip Dykema; Romesh Gautom; Scott Lindquist; Thomas R. Sibley; Trevor Bedford |
| EPI_ISL_2758010, EPI_ISL_2758011, EPI_ISL_2758012, EPI_ISL_2758013 | Austrian Agency for Health and Food Safety (AGES) | Bergthaler laboratory, CeMM Research Center for Molecular Medicine of the Austrian Academy of Sciences | Andreas Bergthaler; Anna Schedl; Bekir Erguner; Benedikt Agerer; Christoph Bock; Fabian Amman; Jan Laine; Lukas Endler; Maelle Le Moing; Martin Senekowitsch; Matthew Thornton; Michael Schuster; Petr Triska; Thomas Penz |
| EPI_ISL_3231359 | Azarian Lab at Burnett School of Biomedical Sciences | Azarian Lab at Burnett School of Biomedical Sciences | Catherine Johnston; Eleonora Cella; Taj Azarian |
| EPI_ISL_2622162 | Azienda Ospedaliera Pugliese Ciaccio di Catanzaro SOC Microbiologia e Virologia | Azienda Ospedaliera Pugliese Ciaccio di Catanzaro SOC Microbiologia e Virologia | Rossana Tallerico Federica Pasceri Marco De Fazio Ilenia Talotta Giuseppina Panduri Pasquale Minchella |
| EPI_ISL_2680975 | Azienda Sanitaria dell'Alto Adige - Laboratorio Aziendale di Microbiologia e Virologia | Azienda Sanitaria dell'Alto Adige | Irene Bianconi |
| EPI_ISL_2637954 | BIO ARD'AISNE | Department of Virology, Henri Mondor University Hospital, Assistance Publique Hôpitaux de Paris, Université Paris-Est Créteil, INSERM U955 | Alexandre Soulier; Christophe Rodriguez; Elisabeth Trawinski; Guillaume Gricourt; Jean-Michel Pawlotsky; Melissa N'Debi; Slim Fourati; Vanessa Demontant |
| EPI_ISL_5647010, EPI_ISL_5647011, EPI_ISL_5647012, EPI_ISL_5647013, EPI_ISL_5647015, EPI_ISL_5647019, EPI_ISL_5647020, EPI_ISL_5647021, EPI_ISL_5647025, EPI_ISL_5647026, EPI_ISL_5647031, EPI_ISL_5647032, EPI_ISL_5647033, EPI_ISL_5647035, EPI_ISL_5647036, EPI_ISL_5647037, EPI_ISL_5647038, EPI_ISL_5647039, EPI_ISL_5647040, EPI_ISL_5647041, EPI_ISL_5647042, EPI_ISL_5647043, EPI_ISL_5647047, EPI_ISL_5647048, EPI_ISL_5647050, EPI_ISL_5647051, EPI_ISL_5647054, EPI_ISL_5647055, EPI_ISL_5647056, EPI_ISL_5647059, EPI_ISL_5647068, EPI_ISL_5647069, EPI_ISL_5647070, EPI_ISL_5647071, EPI_ISL_5647072, EPI_ISL_5647073, EPI_ISL_5647076, EPI_ISL_5647077, EPI_ISL_5647078, EPI_ISL_5647080, EPI_ISL_5647081, EPI_ISL_5647082, EPI_ISL_5647083, EPI_ISL_5647084, EPI_ISL_5647086, EPI_ISL_5647087, EPI_ISL_5647088, EPI_ISL_5647090, EPI_ISL_5647093, EPI_ISL_5647094, EPI_ISL_5647095, EPI_ISL_5647096, EPI_ISL_5647097, EPI_ISL_5647099, EPI_ISL_5647100, EPI_ISL_5647101, EPI_ISL_5647102, EPI_ISL_5647103, EPI_ISL_5647104, EPI_ISL_5647959, EPI_ISL_5647960, EPI_ISL_5647964 | | | |
| see above | BIOFAST | Instituto Butantan | Antonio Jorge Martins; Claudia Renata dos Santos Barros; David Schlesinger; Debora Botequio Moretti; Dimas Tadeu Covas; Elaine Cristina Marqueze; Elaine Vieira Santos; Evandra Strazza Rodrigues; Heidge Fukumasu; Jayme Augusto de Souza-Neto; José Salvatore Leister Patané; Luiz Alcantara; Luiz Lehmann Coutinho; Maria Carolina Elias; Maurício Lacerda Nogueira; Rafael dos Santos Bezerra; Raul Machado Neto; Rejane Maria Tommasini Grotto; Ricardo Haddad; Sandra Coccuzzo Sampaio Vessoni; Simone Kashima; Svetoslav Nanev Slavov; Vincent Louis Viala |
| EPI_ISL_7672487 | Bambino Gesù Pediatric Hospital | Microbiology and Immunology Diagnosis Bambino Gesù Pediatric Hospital | Carlo Federico Perno; Claudia Alteri; Luna Colagrossi; Rossana Scutari; Valentino Costabile |
| EPI_ISL_3504653 | Baptist Health Medical Center | Center for Global Health, University of New Mexico Health Sciences Center | Amanda Novack; Darrell Dinwiddie; Daryl Domman; Dirk Haselow; Joshua L. Kennedy; Kurt Schwalm; Valerie Morley |

| | | | |
|---|---|---|---|
| EPI_ISL_2658950 | Basurto University Hospital: Clinical Microbiology Laboratory | Biocruces Bizkaia | Ana de la Hoz; Estibaliz Ugalde Zarraga; José Luis Díaz de Tuesta del Arco; Mikel Gallego Rodrigo; Mikel Urrutikoetxea-Gutiérrez; Mª Carmen Nieto Toboso |
| EPI_ISL_2626831 | Baylor Scott & White-Temple | Baylor Scott & White-Temple | Ari Rao; Kimberly Walker; Linden Morales; Marcus Volz; Shelby Johnson |
| EPI_ISL_2638007 | Biogroup Bio Lam-LCD Saint-Denis | Department of Virology, Henri Mondor University Hospital, Assistance Publique Hôpitaux de Paris, Université Paris-Est Créteil, INSERM U955 | Alexandre Soulier; Christophe Rodriguez; Elisabeth Trawinski; Guillaume Gricourt; Jean-Michel Pawlotsky; Melissa N'Debi; Slim Fourati; Vanessa Demontant |
| EPI_ISL_3145907, EPI_ISL_3145908, EPI_ISL_3145909 | BioneXt Lab | Laboratoire national de sante, Microbiology, Microbial Genomics Platform | Anke Wienecke-Baldacchino; Catherine Ragimbeau; Elodie Solarino; Fatu Djabi; Jessica Tapp; Lise Pignon; Raoul Salmon; Tamir Abdelrahman; Thibault Ferrandon; Virginie Jover |
| EPI_ISL_3479165, EPI_ISL_3479167, EPI_ISL_3479173, EPI_ISL_3479177, EPI_ISL_3490454, EPI_ISL_3490475, EPI_ISL_3490477, EPI_ISL_3490478, EPI_ISL_3490479, EPI_ISL_3490480, EPI_ISL_3490482, EPI_ISL_3490484, EPI_ISL_3490485, EPI_ISL_3490492, EPI_ISL_3490493, EPI_ISL_3490495, EPI_ISL_3490496, EPI_ISL_3490500, EPI_ISL_3490502, EPI_ISL_3490504, EPI_ISL_3490505, EPI_ISL_3490506, EPI_ISL_3490507, EPI_ISL_3490519, EPI_ISL_3490521, EPI_ISL_3490523, EPI_ISL_3490567, EPI_ISL_3490597, EPI_ISL_3490601, EPI_ISL_3490610, EPI_ISL_3490615, EPI_ISL_3490617, EPI_ISL_3490623, EPI_ISL_3490625, EPI_ISL_3490632, EPI_ISL_3490634, EPI_ISL_3490639, EPI_ISL_3490642, EPI_ISL_3490644, EPI_ISL_3490649, EPI_ISL_3490651, EPI_ISL_3490652, EPI_ISL_3490653, EPI_ISL_3490654, EPI_ISL_3490655, EPI_ISL_3490656, EPI_ISL_3490657, EPI_ISL_3490667, EPI_ISL_3490668, EPI_ISL_3490671, EPI_ISL_3490673, EPI_ISL_3490680, EPI_ISL_3490687, EPI_ISL_3490689, EPI_ISL_3490690, EPI_ISL_3490691, EPI_ISL_3490693, EPI_ISL_3490696, EPI_ISL_3490697, EPI_ISL_3490698, EPI_ISL_3490699, EPI_ISL_3490700, EPI_ISL_3490702, EPI_ISL_3490703, EPI_ISL_3490706, EPI_ISL_3490708, EPI_ISL_3490721, EPI_ISL_3490725, EPI_ISL_3490730, EPI_ISL_3490731, EPI_ISL_3490740, EPI_ISL_3490741, EPI_ISL_3490742, EPI_ISL_3490743, EPI_ISL_3490744, EPI_ISL_3490747, EPI_ISL_3490751, EPI_ISL_3490753, EPI_ISL_3490754, EPI_ISL_3490756, EPI_ISL_3490762, EPI_ISL_3490773, EPI_ISL_3490774, EPI_ISL_3490775, EPI_ISL_3490777, EPI_ISL_3490778, EPI_ISL_3490783, EPI_ISL_3490784, EPI_ISL_3490785, EPI_ISL_3490788, EPI_ISL_3490792, EPI_ISL_3490796, EPI_ISL_3490797, EPI_ISL_3490800, EPI_ISL_3490801, EPI_ISL_3490802, EPI_ISL_3490805, EPI_ISL_3490807, EPI_ISL_3490808, EPI_ISL_3490811, EPI_ISL_3490813, EPI_ISL_3490814, EPI_ISL_3490816, EPI_ISL_3490817, EPI_ISL_3490823, EPI_ISL_3490825, EPI_ISL_3490828, EPI_ISL_3490831, EPI_ISL_3490833, EPI_ISL_3490836, EPI_ISL_3490838, EPI_ISL_3490839, EPI_ISL_3490844, EPI_ISL_3490845, EPI_ISL_3490847, EPI_ISL_3490848, EPI_ISL_3490849, EPI_ISL_3490850, EPI_ISL_3490851, EPI_ISL_3490852, EPI_ISL_3490854, EPI_ISL_3490855, EPI_ISL_3490856, EPI_ISL_3490857, EPI_ISL_3490858, EPI_ISL_3490859, EPI_ISL_3490862, EPI_ISL_3490863, EPI_ISL_3490865, EPI_ISL_3490867, EPI_ISL_3490869, EPI_ISL_3490872, EPI_ISL_3490874, EPI_ISL_3490875, EPI_ISL_3490876, EPI_ISL_3490877, EPI_ISL_3490879, EPI_ISL_3490880, EPI_ISL_3490886, EPI_ISL_3490888, EPI_ISL_3490889, EPI_ISL_3490891, EPI_ISL_3490892, EPI_ISL_3490893, EPI_ISL_3490894, EPI_ISL_3490895, EPI_ISL_3490897, EPI_ISL_3490899, EPI_ISL_3490900, EPI_ISL_3490902, EPI_ISL_3490905, EPI_ISL_3490906, EPI_ISL_3490909, EPI_ISL_3490912, EPI_ISL_3490914, EPI_ISL_3490915, EPI_ISL_3490918, EPI_ISL_3490920, EPI_ISL_3490922, EPI_ISL_3490923, EPI_ISL_3490924, EPI_ISL_3490926, EPI_ISL_3490927, EPI_ISL_3490928, EPI_ISL_3490929, EPI_ISL_3490931, EPI_ISL_3490933, EPI_ISL_3490934, EPI_ISL_3490935, EPI_ISL_3490936, EPI_ISL_3490937, EPI_ISL_3490938, EPI_ISL_3490950, EPI_ISL_3490953, EPI_ISL_3490954, EPI_ISL_3490957, EPI_ISL_3490958, EPI_ISL_3490959, EPI_ISL_3490960, EPI_ISL_3490965, EPI_ISL_3490967, EPI_ISL_3490982, EPI_ISL_3490984, EPI_ISL_3490987, EPI_ISL_3490989, EPI_ISL_3490998, EPI_ISL_3491002, EPI_ISL_3491003, EPI_ISL_3491004, EPI_ISL_3491005, EPI_ISL_3491007, EPI_ISL_3491008, EPI_ISL_3491009, EPI_ISL_3491010, EPI_ISL_3491011, EPI_ISL_3491013, EPI_ISL_3491015, EPI_ISL_3491017, EPI_ISL_3491024, EPI_ISL_3491027, EPI_ISL_3491030, EPI_ISL_3491033, EPI_ISL_3491035, EPI_ISL_3491041, EPI_ISL_3491048, EPI_ISL_3491055, EPI_ISL_3491060, EPI_ISL_3491064, EPI_ISL_3491067, EPI_ISL_3491072, EPI_ISL_3491077, EPI_ISL_3491079, EPI_ISL_3491081, EPI_ISL_3491083, EPI_ISL_3491085, EPI_ISL_3491094, EPI_ISL_3491097, EPI_ISL_3491105, EPI_ISL_3491106, EPI_ISL_3491107, EPI_ISL_3491109, EPI_ISL_3491114, EPI_ISL_3491115, EPI_ISL_3491116, EPI_ISL_3491118, EPI_ISL_3491119, EPI_ISL_3491122, EPI_ISL_3491125, EPI_ISL_3491126, EPI_ISL_3491127, EPI_ISL_3491128, EPI_ISL_3491130, EPI_ISL_3491132, EPI_ISL_3491140, EPI_ISL_3491141, EPI_ISL_3491148, EPI_ISL_3491149, EPI_ISL_3491153, EPI_ISL_3491155, EPI_ISL_3491158, EPI_ISL_3491159, EPI_ISL_3491210, EPI_ISL_3491211, EPI_ISL_3491212, EPI_ISL_3491213, EPI_ISL_3491214, EPI_ISL_3491218, EPI_ISL_3491219 | | | |
| see above | British Columbia Centre For Disease Control | BCCDC Public Health Laboratory | Ana Pacagnella; Corrinne Ng; Dan Fornika; John Tyson; Kim Macdonald; Kimia Kamelian; Linda Hoang; Loretta Janz; Mel Krajden; Prystajecky Natalie; Robert Azana; Shannon Russell |
| EPI_ISL_2991515, EPI_ISL_2991536, EPI_ISL_2991615, EPI_ISL_2991624, EPI_ISL_2991644, EPI_ISL_2991648, EPI_ISL_2991673, EPI_ISL_2991711, EPI_ISL_2991713, EPI_ISL_2991742, EPI_ISL_2991845, EPI_ISL_2991871, EPI_ISL_2991908 | | | |
| see above | Broad Institute Clinical Research Sequencing Platform | Infectious Disease Program, Broad Institute of Harvard and MIT | A.E.; Adams, G.; Anahtar, M.; B.L.; B.W.; Bauer, M.; Birren; Branda, J.; Carter, A.; Cerrato, F.; Chaluvadi, S.; Chapman; Cusick, C.; D.J.; DeRuff, K.; Flowers, K.; Gallagher, G.; Gladden-Young, A.; Gnirke, A.; Harris, J.; J.E.; K.J.; LaRocque, R.; Lagerborg, K.; Lemieux; Lin; Loreth, C.; MacInnis; Neumann, A.; Normandin, E.; P.C.; Park; Pierce, V.; Reilly, S.; Rosenberg, E.; Rudy, M.; Ryan, E.; S.B.; Sabeti; Shaw, B.; Siddle; Slater, D.; Smole, S.; Tomkins-Tinch, C.; Turbett, S. |
| EPI_ISL_2921885, EPI_ISL_2922118, EPI_ISL_2923304, EPI_ISL_2923562, EPI_ISL_2923671, EPI_ISL_3669444, EPI_ISL_3669509, EPI_ISL_3669565, EPI_ISL_3669566, EPI_ISL_3669567, EPI_ISL_3669568, EPI_ISL_3669569, EPI_ISL_3669570, EPI_ISL_3669571, EPI_ISL_3669572, EPI_ISL_3669573, EPI_ISL_3669574, EPI_ISL_3669575, EPI_ISL_3669576, EPI_ISL_3669577, EPI_ISL_3669578, EPI_ISL_3669579, EPI_ISL_3669580, EPI_ISL_3669581, EPI_ISL_3669582, EPI_ISL_3669583, EPI_ISL_3669584, EPI_ISL_3669585, EPI_ISL_3669586, EPI_ISL_3669587, EPI_ISL_3669588, EPI_ISL_3669589, EPI_ISL_3669590, EPI_ISL_3669591, EPI_ISL_3669592, EPI_ISL_3669593, EPI_ISL_3669594, EPI_ISL_3669595, EPI_ISL_3669597, EPI_ISL_3669638, EPI_ISL_3669660 | | | |
| see above | CDPH VBL | California Department of Public Health | CDPH-COVIDNet; Emily Smith on behalf of CDPH-COVIDNet and UCSD EXCITE lab |
| EPI_ISL_2761751, EPI_ISL_2761840 | CENTOGENE Frankfurt Laboratory: Niederlassung Industriepark Höchst | Robert Koch Institute | |
| EPI_ISL_3102507 | CENTRO DE ATENDIMENTO PARA ENFRENTAMENTO AO COVID 19 | Analytical Competence Molecular Epidemiology Lab/ACME, Oswaldo Cruz Foundation, Ceara (FIOCRUZ CE) | Cleber Furtado Aksenen; Fabio Miyajima; Fernando Braga Stehling; Francisco Eder de Moura Lopes; Jamille Maria Mendes Bezerra; Joaquim César do Nascimento Sousa Junior; Pedro Miguel Carneiro Jeronimo; Suzana Porto Almeida e Lucas Delerino; Thais Ferreira de Oliveira; Thais de Oliveira Costa; Ticiane Cavalcante de Souza; Veridiana Pessoa Miyajima |
| EPI_ISL_5647016, EPI_ISL_5647049, EPI_ISL_5647067, EPI_ISL_5647079 | CENTRO DE GENOMAS | Instituto Butantan | Antonio Jorge Martins; Claudia Renata dos Santos Barros; David Schlesinger; Debora Botequio Moretti; Dimas Tadeu Covas; Elaine Cristina Marqueze; Elaine Vieira Santos; Evandra Strazza Rodrigues; Heidge Fukumasu; Jayme Augusto de Souza-Neto; José Salvatore Leister Patané; Luiz Alcantara; Luiz Lehmann Coutinho; Maria Carolina Elias; Maurício Lacerda Nogueira; Rafael dos Santos Bezerra; Raul Machado Neto; Rejane Maria Tommasini Grotto; Ricardo Haddad; Sandra Coccuzzo Sampaio Vessoni; Simone Kashima; Svetoslav Nanev Slavov; Vincent Louis Viala |
| EPI_ISL_5647941 | CENTRO DE SAUDE DE ANALANDIA | Instituto Butantan | Antonio Jorge Martins; Claudia Renata dos Santos Barros; David Schlesinger; Debora Botequio Moretti; Dimas Tadeu Covas; Elaine Cristina Marqueze; Elaine Vieira Santos; Evandra Strazza Rodrigues; Heidge Fukumasu; Jayme Augusto de Souza-Neto; José Salvatore Leister Patané; Luiz Alcantara; Luiz Lehmann Coutinho; Maria Carolina Elias; Maurício Lacerda Nogueira; Rafael dos Santos Bezerra; Raul Machado Neto; Rejane Maria Tommasini Grotto; Ricardo Haddad; Sandra Coccuzzo Sampaio Vessoni; Simone Kashima; Svetoslav Nanev Slavov; Vincent Louis Viala |
| EPI_ISL_5646464, EPI_ISL_5647242, EPI_ISL_5647277 | CENTRO DE SAUDE DE MARACAI | Instituto Butantan | Antonio Jorge Martins; Claudia Renata dos Santos Barros; David Schlesinger; Debora Botequio Moretti; Dimas Tadeu Covas; Elaine Cristina Marqueze; Elaine Vieira Santos; Evandra Strazza Rodrigues; Heidge Fukumasu; Jayme Augusto de Souza-Neto; José Salvatore Leister Patané; Luiz Alcantara; Luiz Lehmann Coutinho; Maria Carolina Elias; Maurício Lacerda Nogueira; Rafael dos Santos Bezerra; Raul Machado Neto; Rejane Maria Tommasini Grotto; Ricardo Haddad; Sandra Coccuzzo Sampaio Vessoni; Simone Kashima; Svetoslav Nanev Slavov; Vincent Louis Viala |
| EPI_ISL_5646674 | CENTRO DE SAUDE DE RIBEIRAO DO SUL | Instituto Butantan | Antonio Jorge Martins; Claudia Renata dos Santos Barros; David Schlesinger; Debora Botequio Moretti; Dimas Tadeu Covas; Elaine Cristina Marqueze; Elaine Vieira Santos; Evandra Strazza Rodrigues; Heidge Fukumasu; Jayme Augusto de Souza-Neto; José Salvatore Leister Patané; Luiz Alcantara; Luiz Lehmann Coutinho; Maria Carolina Elias; Maurício Lacerda Nogueira; Rafael dos Santos Bezerra; Raul Machado Neto; Rejane Maria Tommasini Grotto; Ricardo Haddad; Sandra Coccuzzo Sampaio Vessoni; Simone Kashima; Svetoslav Nanev Slavov; Vincent Louis Viala |
| EPI_ISL_5646523, EPI_ISL_5646615, EPI_ISL_5646656, EPI_ISL_5646698, EPI_ISL_5647231, EPI_ISL_5647278 | CENTRO DE SAUDE II DE BARIRI | Instituto Butantan | Antonio Jorge Martins; Claudia Renata dos Santos Barros; David Schlesinger; Debora Botequio Moretti; Dimas Tadeu Covas; Elaine Cristina Marqueze; Elaine Vieira Santos; Evandra Strazza Rodrigues; Heidge Fukumasu; Jayme Augusto de Souza-Neto; José Salvatore Leister Patané; Luiz Alcantara; Luiz Lehmann Coutinho; Maria Carolina Elias; Maurício Lacerda Nogueira; Rafael dos Santos Bezerra; Raul Machado Neto; Rejane Maria Tommasini Grotto; Ricardo Haddad; Sandra Coccuzzo Sampaio Vessoni; Simone Kashima; Svetoslav Nanev Slavov; Vincent Louis Viala |
| EPI_ISL_5646881, EPI_ISL_5646897, EPI_ISL_5646901, EPI_ISL_5646902, EPI_ISL_5646916 | CENTRO DE SAUDE II DE NHANDEARA | Instituto Butantan | Antonio Jorge Martins; Claudia Renata dos Santos Barros; David Schlesinger; Debora Botequio Moretti; Dimas Tadeu Covas; Elaine Cristina Marqueze; Elaine Vieira Santos; Evandra Strazza Rodrigues; Heidge Fukumasu; Jayme Augusto de Souza-Neto; José Salvatore Leister Patané; Luiz Alcantara; Luiz Lehmann Coutinho; Maria Carolina Elias; Maurício Lacerda Nogueira; Rafael dos Santos Bezerra; Raul Machado Neto; Rejane Maria Tommasini Grotto; Ricardo Haddad; Sandra Coccuzzo Sampaio Vessoni; Simone Kashima; Svetoslav Nanev Slavov; Vincent Louis Viala |
| EPI_ISL_5646936, EPI_ISL_5646937 | CENTRO DE SAUDE II DR ALCIDES FACUNDO ARROYO | Instituto Butantan | Antonio Jorge Martins; Claudia Renata dos Santos Barros; David Schlesinger; Debora Botequio Moretti; Dimas Tadeu Covas; Elaine Cristina Marqueze; Elaine Vieira Santos; Evandra Strazza Rodrigues; Heidge Fukumasu; Jayme Augusto de Souza-Neto; José Salvatore Leister Patané; Luiz Alcantara; Luiz Lehmann Coutinho; Maria Carolina Elias; Maurício Lacerda Nogueira; Rafael dos Santos Bezerra; Raul Machado Neto; Rejane Maria Tommasini Grotto; Ricardo Haddad; Sandra Coccuzzo Sampaio Vessoni; Simone Kashima; Svetoslav Nanev Slavov; Vincent Louis Viala |
| EPI_ISL_5646504, EPI_ISL_5646513, EPI_ISL_5646672, EPI_ISL_5646681, EPI_ISL_5646719, EPI_ISL_5646782, EPI_ISL_5646784, EPI_ISL_5646821, EPI_ISL_5646829 | | | |
| see above | CENTRO DE SAUDE II DR JOSE PAIONE MOCOCA | Instituto Butantan | Antonio Jorge Martins; Claudia Renata dos Santos Barros; David Schlesinger; Debora Botequio Moretti; Dimas Tadeu Covas; Elaine Cristina Marqueze; Elaine Vieira Santos; Evandra Strazza Rodrigues; Heidge Fukumasu; Jayme Augusto de Souza-Neto; José Salvatore Leister Patané; Luiz Alcantara; Luiz Lehmann Coutinho; Maria Carolina Elias; Maurício Lacerda Nogueira; Rafael dos Santos Bezerra; Raul Machado Neto; Rejane Maria Tommasini Grotto; Ricardo Haddad; Sandra Coccuzzo Sampaio Vessoni; Simone Kashima; Svetoslav Nanev Slavov; Vincent Louis Viala |
| EPI_ISL_5647920 | CENTRO DE SAUDE III DE DIVINOLANDIA | Instituto Butantan | Antonio Jorge Martins; Claudia Renata dos Santos Barros; David Schlesinger; Debora Botequio Moretti; Dimas Tadeu Covas; Elaine Cristina Marqueze; Elaine Vieira Santos; Evandra Strazza Rodrigues; Heidge Fukumasu; Jayme Augusto de Souza-Neto; José Salvatore Leister Patané; Luiz Alcantara; Luiz Lehmann Coutinho; Maria Carolina Elias; Maurício Lacerda Nogueira; Rafael dos Santos Bezerra; Raul Machado Neto; Rejane Maria Tommasini Grotto; Ricardo Haddad; Sandra Coccuzzo Sampaio Vessoni; Simone Kashima; Svetoslav Nanev Slavov; Vincent Louis Viala |
| EPI_ISL_5646401, EPI_ISL_5647274 | CENTRO DE SAUDE III GLICERIO GIMAIEL ELIAS GIMAIEL | Instituto Butantan | Antonio Jorge Martins; Claudia Renata dos Santos Barros; David Schlesinger; Debora Botequio Moretti; Dimas Tadeu Covas; Elaine Cristina Marqueze; Elaine Vieira Santos; Evandra Strazza Rodrigues; Heidge Fukumasu; Jayme Augusto de Souza-Neto; José Salvatore Leister Patané; Luiz Alcantara; Luiz Lehmann Coutinho; Maria Carolina Elias; Maurício Lacerda Nogueira; Rafael dos Santos Bezerra; Raul Machado Neto; Rejane Maria Tommasini Grotto; Ricardo Haddad; Sandra Coccuzzo Sampaio Vessoni; Simone Kashima; Svetoslav Nanev Slavov; Vincent Louis Viala |
| EPI_ISL_5646886, EPI_ISL_5646892 | CENTRO DE SAUDE III RINCAO | Instituto Butantan | Antonio Jorge Martins; Claudia Renata dos Santos Barros; David Schlesinger; Debora Botequio Moretti; Dimas Tadeu Covas; Elaine Cristina Marqueze; Elaine Vieira Santos; Evandra Strazza Rodrigues; Heidge Fukumasu; Jayme Augusto de Souza-Neto; José Salvatore Leister Patané; Luiz Alcantara; Luiz Lehmann Coutinho; Maria Carolina Elias; Maurício Lacerda Nogueira; Rafael dos Santos Bezerra; Raul Machado Neto; Rejane Maria Tommasini Grotto; Ricardo Haddad; Sandra Coccuzzo Sampaio Vessoni; Simone Kashima; Svetoslav Nanev Slavov; Vincent Louis Viala |
| EPI_ISL_5646423, EPI_ISL_5646468 | CENTRO DE SAUDE III TABATINGA | Instituto Butantan | Antonio Jorge Martins; Claudia Renata dos Santos Barros; David Schlesinger; Debora Botequio Moretti; Dimas Tadeu Covas; Elaine Cristina Marqueze; Elaine Vieira Santos; Evandra Strazza Rodrigues; Heidge Fukumasu; Jayme Augusto de Souza-Neto; José Salvatore Leister Patané; Luiz Alcantara; Luiz Lehmann Coutinho; Maria Carolina Elias; Maurício Lacerda Nogueira; Rafael dos Santos Bezerra; Raul Machado Neto; Rejane Maria Tommasini Grotto; Ricardo Haddad; Sandra Coccuzzo Sampaio Vessoni; Simone Kashima; Svetoslav Nanev Slavov; Vincent Louis Viala |
| EPI_ISL_5646550, EPI_ISL_5646572, EPI_ISL_5646607, EPI_ISL_5646627, EPI_ISL_5646631, EPI_ISL_5646650, EPI_ISL_5646668, EPI_ISL_5646693, EPI_ISL_5646694, EPI_ISL_5646702, EPI_ISL_5646712, EPI_ISL_5646732, EPI_ISL_5646746, EPI_ISL_5646747, EPI_ISL_5646770, EPI_ISL_5646771, EPI_ISL_5646772, EPI_ISL_5646802, EPI_ISL_5646816, EPI_ISL_5646817, EPI_ISL_5646819, EPI_ISL_5646825, EPI_ISL_5646834, EPI_ISL_5646841 | | | |
| see above | CENTRO DE TRIAGEM COVID 19 | Instituto Butantan | Antonio Jorge Martins; Claudia Renata dos Santos Barros; David Schlesinger; Debora Botequio Moretti; Dimas Tadeu Covas; Elaine Cristina Marqueze; Elaine Vieira Santos; Evandra Strazza Rodrigues; Heidge Fukumasu; Jayme Augusto de Souza-Neto; José Salvatore Leister Patané; Luiz Alcantara; Luiz Lehmann Coutinho; Maria Carolina Elias; Maurício Lacerda Nogueira; Rafael dos Santos Bezerra; Raul Machado Neto; Rejane Maria Tommasini Grotto; Ricardo Haddad; Sandra Coccuzzo Sampaio Vessoni; Simone Kashima; Svetoslav Nanev Slavov; Vincent Louis Viala |
| EPI_ISL_5646505, EPI_ISL_5646508, EPI_ISL_5646512, EPI_ISL_5646520, EPI_ISL_5646530, EPI_ISL_5646545, EPI_ISL_5646552, EPI_ISL_5646555, EPI_ISL_5646563, EPI_ISL_5646573, EPI_ISL_5646610, EPI_ISL_5646616, EPI_ISL_5646619, EPI_ISL_5646625, EPI_ISL_5646645, EPI_ISL_5646665, EPI_ISL_5646736, EPI_ISL_5646760, EPI_ISL_5646767, EPI_ISL_5646844 | | | |
| see above | CENTRO INTEGRADO DE SAUDE | Instituto Butantan | Antonio Jorge Martins; Claudia Renata dos Santos Barros; David Schlesinger; Debora Botequio Moretti; Dimas Tadeu Covas; Elaine Cristina Marqueze; Elaine Vieira Santos; Evandra Strazza Rodrigues; Heidge Fukumasu; Jayme Augusto de Souza-Neto; José Salvatore Leister Patané; Luiz Alcantara; Luiz Lehmann Coutinho; Maria Carolina Elias; Maurício Lacerda Nogueira; Rafael dos Santos Bezerra; Raul Machado Neto; Rejane Maria Tommasini Grotto; Ricardo Haddad; Sandra Coccuzzo Sampaio Vessoni; Simone Kashima; Svetoslav Nanev Slavov; Vincent Louis Viala |
| EPI_ISL_5647930 | CENTRO MEDICO DR NELSON SALOME DE CONCHAL | Instituto Butantan | Antonio Jorge Martins; Claudia Renata dos Santos Barros; David Schlesinger; Debora Botequio Moretti; Dimas Tadeu Covas; Elaine Cristina Marqueze; Elaine Vieira Santos; Evandra Strazza Rodrigues; Heidge Fukumasu; Jayme Augusto de Souza-Neto; José Salvatore Leister Patané; Luiz Alcantara; Luiz Lehmann Coutinho; Maria Carolina Elias; Maurício Lacerda Nogueira; Rafael dos Santos Bezerra; Raul Machado Neto; Rejane Maria Tommasini Grotto; Ricardo Haddad; Sandra Coccuzzo Sampaio Vessoni; Simone Kashima; Svetoslav Nanev Slavov; Vincent Louis Viala |
| EPI_ISL_2943231 | CENTRO MEDICO NAVAL SEMAR LAB BIOLOGIA MOLECULAR | CENTRO MEDICO NAVAL SEMAR LAB PATOLOGIA MOLECULAR | MEIXUEIRO CALDERON CLAUDIA. MORALES RAMIREZ OMAR. BALTIERRA URIBE SHANTAL. ZUNO CORONADO TEUTLI. GARCIA ALEGRIA IRMA.ALVAREZ JIMENEZ VIOLETA DEJANIRA. LOPEZ BUSTOS ABRAHAM. CABRERA GONZALEZ MOISES. ROMERO ROMAN EDGAR. LIEVANO ESQUINCA JAZMIN. FIGUEROA ANTONIO VICTOR. DIAZ PONCE MEDRANO JUAN ALBERTO. |
| EPI_ISL_2637834 | CERBALLIANCE Valmy | Department of Virology, Henri Mondor University Hospital, Assistance Publique Hôpitaux de Paris, Université Paris-Est Créteil, INSERM U955 | Alexandre Soulier; Christophe Rodriguez; Elisabeth Trawinski; Guillaume Gricourt; Jean-Michel Pawlotsky; Melissa N'Debi; Slim Fourati; Vanessa Demontant |
| EPI_ISL_2695390, EPI_ISL_2695391 | CH Tondela Viseu | Instituto Nacional de Saude (INSA) | Borges et al |
| EPI_ISL_2646209, EPI_ISL_2646210, EPI_ISL_2646212, EPI_ISL_2887728, | CH Valenciennes | CHU Lille - Laboratoire de Virologie | AIT YAHYA Emilie; ALIDJINOU Enagnon Kazali; BOCKET Laurence; CREPIN Michel; DEMAY Christophe; ENGELMANN Ilka; GEFFROY Sandrine; GUIGON Aurélie; LAMBERT Valérie; LAZREK Mouna; NOBILLIAUX Florian; PREVOST Brigitte; TCHANTCHOU NJOSSE YANICK; THUILLIER Caroline; TINEZ Claire |

| | | | |
|---|---|---|---|
| EPI_ISL_2887729 | | | |
| EPI_ISL_2629232, EPI_ISL_2629233, EPI_ISL_2629234, EPI_ISL_2835977 | CHC Andrée Rosemon | Institut Pasteur de la Guyane | Anne Lavergne; Antoine Enfissi; Arielle Salmier; Dominique Rousset |
| EPI_ISL_2894567 | CIAD Hermosillo | CIAD Hermosillo | ; Alejandra García-Gasca; Alejandra Hernández-Terán; Alejandro Sánchez-Flores; Alfredo Herrera-Estrella; Alicia Ocaña-Mondragón; Andreu Comas-García; Angel Gustavo Salas-Lais; Antonio Loza Román; Bernardo Martínez-Miguel; Blanca Taboada; Brenda Irasema Maldonado-Meza; Bruno Gómez-Gil; Carla Ivón Herrera-Najera; Carlos F. Arias; Celia Boukadida; Clara Esperanza Santacruz-Tinoco; Concepción Grajales-Muñiz; Consorcio Mexicano de Vigilancia Genómica (CoViGen-Mex). Authors (in alphabetical order): Julio Elias Alvarado-Yaah; Cristóbal Cháidez-Quiróz; Célida Duque Molina; Célida Martínez- Rodríguez; Daniel Fregoso-Rueda; Daniel Lira Morales; Eduardo Becerril-Vargas; Fernando Fontove-Herrera; Fidencio Mejía-Nepomuceno; Francisco Pulido; Gloria Elena Espinosa-Ayala; Gloria María Molina-Salinas; Gloria Vazquez; Hector Esteban Paz-Juárez; Hector Montoya-Fuentes; Helen Haydee Fernanda Ramírez-Plascencia; Irvin González-López; Jean Pierre González; Jesús Hernández; Joel Armando Vázquez-Pérez.; Jorge Salas-Hernández; José Antonio Enciso-Moreno; José Arturo Martínez-Orozco; José Esteban Muñoz-Medina; José de Jesús Nuñez-Contreras; Juan Bautista Chale-Dzul; Julissa Enciso-Ibarra; Luis Alberto Ochoa-Carrera; Margarita Matías-Florentino; Mario Mújica-Sánchez; Marissa Perez-Garcia; María Guadalupe Santiago-Mauricio; María Guadalupe de Jesús Mireles-Rivera; Nelly Sélem-Mojica; Pavel Isa; Ricardo Ciria Merce; Ricardo Grande; Rosa María Gutiérrez Rios; Santiago Ávila-Ríos; Selene Zárate; Susana Lopez; Verónica Mata-Haro; Victor Eduardo García-Arias; Víctor Hugo Borja-Aburto |
| EPI_ISL_2629235, EPI_ISL_2629236, EPI_ISL_2629237, EPI_ISL_2629238, EPI_ISL_2629239 | CNR Institut Pasteur de la Guyane | Institut Pasteur de la Guyane | Anne Lavergne; Antoine Enfissi; Arielle Salmier; Dominique Rousset |
| EPI_ISL_5646532, EPI_ISL_5646553, EPI_ISL_5646576, EPI_ISL_5646626, EPI_ISL_5646678, EPI_ISL_5646701, EPI_ISL_5646709, EPI_ISL_5646716, EPI_ISL_5646761 | | | |
| see above | COORDENADORIA MUNICIPAL DE SAUDE DE IRACEMAPOLIS | Instituto Butantan | Antonio Jorge Martins; Claudia Renata dos Santos Barros; David Schlesinger; Debora Botequio Moretti; Dimas Tadeu Covas; Elaine Cristina Marqueze; Elaine Vieira Santos; Evandra Strazza Rodrigues; Heidge Fukumasu; Jayme Augusto de Souza-Neto; José Salvatore Leister Patané; Luiz Alcantara; Luiz Lehmann Coutinho; Maria Carolina Elias; Maurício Lacerda Nogueira; Rafael dos Santos Bezerra; Raul Machado Neto; Rejane Maria Tommasini Grotto; Ricardo Haddad; Sandra Coccuzzo Sampaio Vessoni; Simone Kashima; Svetoslav Nanev Slavov; Vincent Louis Viala |
| EPI_ISL_5647271 | CS DE BALSAMO | Instituto Butantan | Antonio Jorge Martins; Claudia Renata dos Santos Barros; David Schlesinger; Debora Botequio Moretti; Dimas Tadeu Covas; Elaine Cristina Marqueze; Elaine Vieira Santos; Evandra Strazza Rodrigues; Heidge Fukumasu; Jayme Augusto de Souza-Neto; José Salvatore Leister Patané; Luiz Alcantara; Luiz Lehmann Coutinho; Maria Carolina Elias; Maurício Lacerda Nogueira; Rafael dos Santos Bezerra; Raul Machado Neto; Rejane Maria Tommasini Grotto; Ricardo Haddad; Sandra Coccuzzo Sampaio Vessoni; Simone Kashima; Svetoslav Nanev Slavov; Vincent Louis Viala |
| EPI_ISL_5647213, EPI_ISL_5647219, EPI_ISL_5647220, EPI_ISL_5647260, EPI_ISL_5647261, EPI_ISL_5647272 | CS DE CATIGUA | Instituto Butantan | Antonio Jorge Martins; Claudia Renata dos Santos Barros; David Schlesinger; Debora Botequio Moretti; Dimas Tadeu Covas; Elaine Cristina Marqueze; Elaine Vieira Santos; Evandra Strazza Rodrigues; Heidge Fukumasu; Jayme Augusto de Souza-Neto; José Salvatore Leister Patané; Luiz Alcantara; Luiz Lehmann Coutinho; Maria Carolina Elias; Maurício Lacerda Nogueira; Rafael dos Santos Bezerra; Raul Machado Neto; Rejane Maria Tommasini Grotto; Ricardo Haddad; Sandra Coccuzzo Sampaio Vessoni; Simone Kashima; Svetoslav Nanev Slavov; Vincent Louis Viala |
| EPI_ISL_5646913, EPI_ISL_5646914, EPI_ISL_5646930 | CS DE ICEM | Instituto Butantan | Antonio Jorge Martins; Claudia Renata dos Santos Barros; David Schlesinger; Debora Botequio Moretti; Dimas Tadeu Covas; Elaine Cristina Marqueze; Elaine Vieira Santos; Evandra Strazza Rodrigues; Heidge Fukumasu; Jayme Augusto de Souza-Neto; José Salvatore Leister Patané; Luiz Alcantara; Luiz Lehmann Coutinho; Maria Carolina Elias; Maurício Lacerda Nogueira; Rafael dos Santos Bezerra; Raul Machado Neto; Rejane Maria Tommasini Grotto; Ricardo Haddad; Sandra Coccuzzo Sampaio Vessoni; Simone Kashima; Svetoslav Nanev Slavov; Vincent Louis Viala |
| EPI_ISL_5647268 | CS DE JACI | Instituto Butantan | Antonio Jorge Martins; Claudia Renata dos Santos Barros; David Schlesinger; Debora Botequio Moretti; Dimas Tadeu Covas; Elaine Cristina Marqueze; Elaine Vieira Santos; Evandra Strazza Rodrigues; Heidge Fukumasu; Jayme Augusto de Souza-Neto; José Salvatore Leister Patané; Luiz Alcantara; Luiz Lehmann Coutinho; Maria Carolina Elias; Maurício Lacerda Nogueira; Rafael dos Santos Bezerra; Raul Machado Neto; Rejane Maria Tommasini Grotto; Ricardo Haddad; Sandra Coccuzzo Sampaio Vessoni; Simone Kashima; Svetoslav Nanev Slavov; Vincent Louis Viala |
| EPI_ISL_5647209, EPI_ISL_5647244, EPI_ISL_5647247 | CS DE PALMEIRA DOESTE | Instituto Butantan | Antonio Jorge Martins; Claudia Renata dos Santos Barros; David Schlesinger; Debora Botequio Moretti; Dimas Tadeu Covas; Elaine Cristina Marqueze; Elaine Vieira Santos; Evandra Strazza Rodrigues; Heidge Fukumasu; Jayme Augusto de Souza-Neto; José Salvatore Leister Patané; Luiz Alcantara; Luiz Lehmann Coutinho; Maria Carolina Elias; Maurício Lacerda Nogueira; Rafael dos Santos Bezerra; Raul Machado Neto; Rejane Maria Tommasini Grotto; Ricardo Haddad; Sandra Coccuzzo Sampaio Vessoni; Simone Kashima; Svetoslav Nanev Slavov; Vincent Louis Viala |
| EPI_ISL_5647212 | CS DE SANTA ALBERTINA | Instituto Butantan | Antonio Jorge Martins; Claudia Renata dos Santos Barros; David Schlesinger; Debora Botequio Moretti; Dimas Tadeu Covas; Elaine Cristina Marqueze; Elaine Vieira Santos; Evandra Strazza Rodrigues; Heidge Fukumasu; Jayme Augusto de Souza-Neto; José Salvatore Leister Patané; Luiz Alcantara; Luiz Lehmann Coutinho; Maria Carolina Elias; Maurício Lacerda Nogueira; Rafael dos Santos Bezerra; Raul Machado Neto; Rejane Maria Tommasini Grotto; Ricardo Haddad; Sandra Coccuzzo Sampaio Vessoni; Simone Kashima; Svetoslav Nanev Slavov; Vincent Louis Viala |
| EPI_ISL_5646822 | CS II DR JAHYR DE PAULA RIBEIRO GUARA | Instituto Butantan | Antonio Jorge Martins; Claudia Renata dos Santos Barros; David Schlesinger; Debora Botequio Moretti; Dimas Tadeu Covas; Elaine Cristina Marqueze; Elaine Vieira Santos; Evandra Strazza Rodrigues; Heidge Fukumasu; Jayme Augusto de Souza-Neto; José Salvatore Leister Patané; Luiz Alcantara; Luiz Lehmann Coutinho; Maria Carolina Elias; Maurício Lacerda Nogueira; Rafael dos Santos Bezerra; Raul Machado Neto; Rejane Maria Tommasini Grotto; Ricardo Haddad; Sandra Coccuzzo Sampaio Vessoni; Simone Kashima; Svetoslav Nanev Slavov; Vincent Louis Viala |
| EPI_ISL_5646679 | CS II DR JOSE CARQUEIJO SCRPARDO | Instituto Butantan | Antonio Jorge Martins; Claudia Renata dos Santos Barros; David Schlesinger; Debora Botequio Moretti; Dimas Tadeu Covas; Elaine Cristina Marqueze; Elaine Vieira Santos; Evandra Strazza Rodrigues; Heidge Fukumasu; Jayme Augusto de Souza-Neto; José Salvatore Leister Patané; Luiz Alcantara; Luiz Lehmann Coutinho; Maria Carolina Elias; Maurício Lacerda Nogueira; Rafael dos Santos Bezerra; Raul Machado Neto; Rejane Maria Tommasini Grotto; Ricardo Haddad; Sandra Coccuzzo Sampaio Vessoni; Simone Kashima; Svetoslav Nanev Slavov; Vincent Louis Viala |
| EPI_ISL_5646872, EPI_ISL_5646898, EPI_ISL_5646905, EPI_ISL_5646924 | CS II EGIDIO BRUNHARA MORRO AGUDO | Instituto Butantan | Antonio Jorge Martins; Claudia Renata dos Santos Barros; David Schlesinger; Debora Botequio Moretti; Dimas Tadeu Covas; Elaine Cristina Marqueze; Elaine Vieira Santos; Evandra Strazza Rodrigues; Heidge Fukumasu; Jayme Augusto de Souza-Neto; José Salvatore Leister Patané; Luiz Alcantara; Luiz Lehmann Coutinho; Maria Carolina Elias; Maurício Lacerda Nogueira; Rafael dos Santos Bezerra; Raul Machado Neto; Rejane Maria Tommasini Grotto; Ricardo Haddad; Sandra Coccuzzo Sampaio Vessoni; Simone Kashima; Svetoslav Nanev Slavov; Vincent Louis Viala |
| EPI_ISL_5646559, EPI_ISL_5646791, EPI_ISL_5646809 | CS III DR ANTONIO ANADAO SSGRAMA | Instituto Butantan | Antonio Jorge Martins; Claudia Renata dos Santos Barros; David Schlesinger; Debora Botequio Moretti; Dimas Tadeu Covas; Elaine Cristina Marqueze; Elaine Vieira Santos; Evandra Strazza Rodrigues; Heidge Fukumasu; Jayme Augusto de Souza-Neto; José Salvatore Leister Patané; Luiz Alcantara; Luiz Lehmann Coutinho; Maria Carolina Elias; Maurício Lacerda Nogueira; Rafael dos Santos Bezerra; Raul Machado Neto; Rejane Maria Tommasini Grotto; Ricardo Haddad; Sandra Coccuzzo Sampaio Vessoni; Simone Kashima; Svetoslav Nanev Slavov; Vincent Louis Viala |
| EPI_ISL_5646440, EPI_ISL_5647217 | CS III SANTOPOLIS DO AGUAPEI | Instituto Butantan | Antonio Jorge Martins; Claudia Renata dos Santos Barros; David Schlesinger; Debora Botequio Moretti; Dimas Tadeu Covas; Elaine Cristina Marqueze; Elaine Vieira Santos; Evandra Strazza Rodrigues; Heidge Fukumasu; Jayme Augusto de Souza-Neto; José Salvatore Leister Patané; Luiz Alcantara; Luiz Lehmann Coutinho; Maria Carolina Elias; Maurício Lacerda Nogueira; Rafael dos Santos Bezerra; Raul Machado Neto; Rejane Maria Tommasini Grotto; Ricardo Haddad; Sandra Coccuzzo Sampaio Vessoni; Simone Kashima; Svetoslav Nanev Slavov; Vincent Louis Viala |
| EPI_ISL_3912044, EPI_ISL_3912066, EPI_ISL_3912151, EPI_ISL_3912152 | CSF ARACATIACU | Analytical Competence Molecular Epidemiology Lab/ACME, Oswaldo Cruz Foundation, Ceara (FIOCRUZ CE) | Cleber Furtado Aksenen; Fabio Miyajima; Fernando Braga Stehling; Francisco Eder de Moura Lopes; Jamille Maria Mendes Bezerra; Joaquim Cesar do Nascimento Sousa Junior; Pedro Miguel Carneiro Jeronimo; Suzana Porto Almeida & Lucas Delerino on behalf of COVID-19 FIOCRUZ Genomic Network; Thais Ferreira de Oliveira; Thais de Oliveira Costa; Ticiane Cavalcante de Souza; Veridiana Pessoa Miyajima |
| EPI_ISL_3912153 | CSF COELCE | Analytical Competence Molecular Epidemiology Lab/ACME, Oswaldo Cruz Foundation, Ceara (FIOCRUZ CE) | Cleber Furtado Aksenen; Fabio Miyajima; Fernando Braga Stehling; Francisco Eder de Moura Lopes; Jamille Maria Mendes Bezerra; Joaquim Cesar do Nascimento Sousa Junior; Pedro Miguel Carneiro Jeronimo; Suzana Porto Almeida & Lucas Delerino on behalf of COVID-19 FIOCRUZ Genomic Network; Thais Ferreira de Oliveira; Thais de Oliveira Costa; Ticiane Cavalcante de Souza; Veridiana Pessoa Miyajima |
| EPI_ISL_3912155 | CSF JUNCO | Analytical Competence Molecular Epidemiology Lab/ACME, Oswaldo Cruz Foundation, Ceara (FIOCRUZ CE) | Cleber Furtado Aksenen; Fabio Miyajima; Fernando Braga Stehling; Francisco Eder de Moura Lopes; Jamille Maria Mendes Bezerra; Joaquim Cesar do Nascimento Sousa Junior; Pedro Miguel Carneiro Jeronimo; Suzana Porto Almeida & Lucas Delerino on behalf of COVID-19 FIOCRUZ Genomic Network; Thais Ferreira de Oliveira; Thais de Oliveira Costa; Ticiane Cavalcante de Souza; Veridiana Pessoa Miyajima |
| EPI_ISL_3912158 | CSF TERRENOS NOVOS 1 | Analytical Competence Molecular Epidemiology Lab/ACME, Oswaldo Cruz Foundation, Ceara (FIOCRUZ CE) | Cleber Furtado Aksenen; Fabio Miyajima; Fernando Braga Stehling; Francisco Eder de Moura Lopes; Jamille Maria Mendes Bezerra; Joaquim Cesar do Nascimento Sousa Junior; Pedro Miguel Carneiro Jeronimo; Suzana Porto Almeida & Lucas Delerino on behalf of COVID-19 FIOCRUZ Genomic Network; Thais Ferreira de Oliveira; Thais de Oliveira Costa; Ticiane Cavalcante de Souza; Veridiana Pessoa Miyajima |
| EPI_ISL_3023769 | CT Department of Public Health | CT Department of Public Health | Claire_Pearson; Tu_N_Nguyen |
| EPI_ISL_2892554, EPI_ISL_3536735, EPI_ISL_3536935, EPI_ISL_4005571, EPI_ISL_4330734 | California Department of Public Health | California Department of Public Health | CDPH IDLB COVIDNet et al |
| EPI_ISL_2772502 | California Hospital Medical Center | Los Angeles County Public Health Laboratories | P. Hemarajata et al. |
| EPI_ISL_2574068, EPI_ISL_2574072 | Center for Laboratory Medicine | Center for Laboratory Medicine | Yannick Gerth |
| EPI_ISL_3462528, EPI_ISL_3462529, EPI_ISL_3462543, EPI_ISL_3462559, EPI_ISL_3462560, EPI_ISL_3462563, EPI_ISL_3462580, EPI_ISL_3462656, EPI_ISL_3462657, EPI_ISL_3462658, EPI_ISL_3464728 | | | |
| see above | Central Laboratory, Bureau of Public Health (BOG) and Academic Hospital Paramaribo | Erasmus Medical Center | Bas B Oude Munnink; Cherise Beek; Consuella Partowidjojo; Dion Gajadin; Ed PF IJzerman; Emmanuelle Munger; Gary Gummels; Ingrid SK Krishnadath; Lycke Woittiez; Marion PG Koopmans; Mireille Van de Veer; Phyllis Pinas; Princes Wongsowidjojo; Radjesh Ori; Ranisha Doerbalie; Rohma Banwari; Soeradj Harkisoen; Stephen Vreden; Tilotmadebie Ramlal; Verne Nanhoe |
| EPI_ISL_3255279 | Central Public Health Laboratory - LACEN - Bahia, Salvador, Brazil | Central Public Health Laboratory - LACEN -Bahia, Salvador, Brazil | Arabela Leal; Breno Dominguez; Felicidade Pereira; Jaqueline Gomes; Luciana Oliveira; Luiz Alcantara; Marcela Gómez; Marta Giovanetti; Patrícia Cajado; Stephane Tosta; Vagner Fonseca; Vanessa Nardy |
| EPI_ISL_2842782, EPI_ISL_2842804 | Centro Medico ABC | Instituto Nacional de Medicina Genomica | Cedro-Tanda A; Cesar Lara; Daniel Aguirre; Escobar-Arrazola; Gonzalez-Barrera D; Herrera-Montalvo LA.; Hidalgo-Miranda A; Jacomine Reyes-Carrasco; M; Mendoza-Vargas A; Munguia-Garza P; Ramirez-Vega O; Rangel-DeLeon D; Reyes-Grajeda JP; Roxana Trejo-González |
| EPI_ISL_5926827, EPI_ISL_5926829, EPI_ISL_5926830 | Centro de Infectologia Charles Mérieux/ Laboratório Rodolphe Mérieux, FUNDHACRE | Bioinformatics Laboratory / LNCC | Alessandra P Lamarca; Alexandra L Gerber; Aline de Freitas Souza; Ana Paula de C Guimarães; Ana Tereza R Vasconcelos; Andreas Stocker; Cirley Maria de Oliveira Lobato; Douglas Terra Machado; Janaína Mazaro; Janete Taynã Nascimento Rodrigues; Luiz Fellype Alves de Souza; Luiz G P de Almeida; Ronaldo da Silva F Jr; Rutilene B. Souza |
| EPI_ISL_2942411 | Centro de Investigacion Biomedica de Occidente (CIBO) | Unidad de Genomica Avanzada | ; Alejandra Garcia-Gasca; Alejandra Hernandez-Teran; Alejandro Sanchez-Flores; Alfredo Herrera-Estrella; Alicia Ocaña-Mondragon; Andreu Comas-Garcia; Angel Gustavo Salas-Lais; Antonio Loza Roman; Bernardo Martinez-Miguel; Blanca Taboada; Brenda Irasema Maldonado-Meza; Bruno Gomez-Gil; Carla Ivon Herrera-Najera; Carlos F. Arias; Celia Boukadida; Celida Duque Molina; Celida Martinez- Rodriguez; Clara Esperanza Santacruz-Tinoco; Concepcion Grajales-Muñiz; Consorcio Mexicano de Vigilancia Genomica (CoViGen-Mex). Authors (in alphabetical order): Julio Elias Alvarado-Yaah; Cristobal Chaidez-Quiroz; Daniel Fregoso-Rueda; Daniel Lira Morales; Eduardo Becerril-Vargas; Fernando Fontove-Herrera; Fidencio Mejia-Nepomuceno; Francisco Pulido; Gloria Elena Espinosa-Ayala; Gloria Maria Molina-Salinas; Gloria Vazquez; Hector Esteban Paz-Juarez; Hector Montoya-Fuentes; Helen Haydee Fernanda Ramirez-Plascencia; Irvin Gonzalez-Lopez; Jean Pierre Gonzalez; Jesus Hernandez; Joel Armando Vazquez-Perez.; Jorge Salas-Hernandez; Jose Antonio Enciso-Moreno; Jose Arturo Martinez-Orozco; Jose Esteban Muñoz-Medina; Jose de Jesus Nuñez-Contreras; Juan Bautista Chale-Dzul; Julissa Enciso-Ibarra; Luis Alberto Ochoa-Carrera; Margarita Matias-Florentino; Maria Guadalupe Santiago-Mauricio; Maria Guadalupe de Jesus Mireles-Rivera; Mario Mujica-Sanchez; Marissa Perez-Garcia; Nelly Selem-Mojica; Pavel Isa; Ricardo Ciria Merce; Ricardo Grande; Rosa Maria Gutierrez Rios; Santiago avila-Rios; Selene Zarate; Susana Lopez; Veronica Mata-Haro; Victor Eduardo Garcia-Arias; Victor Hugo Borja-Aburto |
| EPI_ISL_2942484 | Centro de Investigacion Biomedica del Noreste (CIBIN) | Unidad de Genomica Avanzada | ; Alejandra Garcia-Gasca; Alejandra Hernandez-Teran; Alejandro Sanchez-Flores; Alfredo Herrera-Estrella; Alicia Ocaña-Mondragon; Andreu Comas-Garcia; Angel Gustavo Salas-Lais; Antonio Loza Roman; Bernardo Martinez-Miguel; Blanca Taboada; Brenda Irasema Maldonado-Meza; Bruno Gomez-Gil; Carla Ivon Herrera-Najera; Carlos F. Arias; Celia Boukadida; Celida Duque Molina; Celida Martinez- Rodriguez; Clara Esperanza Santacruz-Tinoco; Concepcion Grajales-Muñiz; Consorcio Mexicano de Vigilancia Genomica (CoViGen-Mex). Authors (in alphabetical order): Julio Elias Alvarado-Yaah; Cristobal |

| | | | |
|---|---|---|---|
| | | | Chaidez-Quiroz; Daniel Fregoso-Rueda; Daniel Lira Morales; Eduardo Becerril-Vargas; Fernando Fontove-Herrera; Fidencio Mejia-Nepomuceno; Francisco Pulido; Gloria Elena Espinosa-Ayala; Gloria Maria Molina-Salinas; Gloria Vazquez; Hector Esteban Paz-Juarez; Hector Montoya-Fuentes; Helen Haydee Fernanda Ramirez-Plascencia; Irvin Gonzalez-Lopez; Jean Pierre Gonzalez; Jesus Hernandez; Joel Armando Vazquez-Perez.; Jorge Salas-Hernandez; Jose Antonio Enciso-Moreno; Jose Arturo Martinez-Orozco; Jose Esteban Muñoz-Medina; Jose de Jesus Nuñez-Contreras; Juan Bautista Chale-Dzul; Julissa Enciso-Ibarra; Luis Alberto Ochoa-Carrera; Margarita Matias-Florentino; Maria Guadalupe Santiago-Mauricio; Maria Guadalupe de Jesus Mireles-Rivera; Mario Mujica-Sanchez; Marissa Perez-Garcia; Nelly Selem-Mojica; Pavel Isa; Ricardo Ciria Merce; Ricardo Grande; Rosa Maria Gutierrez Rios; Santiago avila-Rios; Selene Zarate; Susana Lopez; Veronica Mata-Haro; Victor Eduardo Garcia-Arias; Victor Hugo Borja-Aburto |
| EPI_ISL_4405422 | Centro de Investigaciones Básicas y Aplicadas, UNNOBA | Área de Secuenciación del Laboratorio de Virología del Hospital de Niños Dr. Ricardo Gutierrez on behalf of 'Proyecto Argentino Interinstitucional de genomica de SARS-CoV-2' (PAIS Consortium) | A; Acuña; Alaniz; Bagnis; Barbero; Bonadeo; Brandone; C; Cassarini; Castro; Chimento; Cristina; D; Demarchi; Español; F; Fernández; G; Goya; Gracia Balbi; Hernández del Pin; I; Icardi; L; LE; Lusso; M; MI; MS; Menite; Moroni; Morro; N; Nabaes Jodar; Natale; P; Palumbo; Pasquinelli; Perrone; R; Romano; Rosales; S; Sevic; Spinelli; V; Valinotto; Valla; Viegas, M.; Villafañe; Vitale |
| EPI_ISL_2801736, EPI_ISL_2801741, EPI_ISL_2801746, EPI_ISL_2801749, EPI_ISL_2801750, EPI_ISL_2801751, EPI_ISL_2801752 | | | |
| see above | Centro de Investigación Biomédica de Occidente (CIBO) | Instituto de Biotecnología de la UNAM | ; Alejandra García-Gasca; Alejandra Hernández-Terán; Alejandro Sánchez-Flores; Alfredo Herrera-Estrella; Alicia Ocaña-Mondragón; Andreu Comas-García; Angel Gustavo Salas-Lais; Antonio Loza Román; Bernardo Martínez-Miguel; Blanca Taboada; Brenda Irasema Maldonado-Meza; Bruno Gómez-Gil; Carla Ivón Herrera-Najera; Carlos F. Arias; Celia Boukadida; Clara Esperanza Santacruz-Tinoco; Concepción Grajales-Muñiz; Consorcio Mexicano de Vigilancia Genómica (CoViGen-Mex). Authors (in alphabetical order): Julio Elias Alvarado-Yaah; Cristóbal Cháidez-Quiróz; Célida Duque Molina; Célida Martínez- Rodríguez; Daniel Fregoso-Rueda; Daniel Lira Morales; Eduardo Becerril-Vargas; Fernando Fontove-Herrera; Fidencio Mejía-Nepomuceno; Francisco Pulido; Gloria Elena Espinosa-Ayala; Gloria María Molina-Salinas; Gloria Vazquez; Hector Esteban Paz-Juárez; Hector Montoya-Fuentes; Helen Haydee Fernanda Ramírez-Plascencia; Irvin González-López; Jean Pierre González; Jesús Hernández; Joel Armando Vázquez-Pérez.; Jorge Salas-Hernández; José Antonio Enciso-Moreno; José Arturo Martínez-Orozco; José Esteban Muñoz-Medina; José de Jesús Nuñez-Contreras; Juan Bautista Chale-Dzul; Julissa Enciso-Ibarra; Luis Alberto Ochoa-Carrera; Margarita Matías-Florentino; Mario Mújica-Sánchez; Marissa Perez-Garcia; María Guadalupe Santiago-Mauricio; María Guadalupe de Jesús Mireles-Rivera; Nelly Sélem-Mojica; Pavel Isa; Ricardo Ciria Merce; Ricardo Grande; Rosa María Gutiérrez Ríos; Santiago Ávila-Ríos; Selene Zárate; Susana Lopez; Verónica Mata-Haro; Víctor Eduardo García-Arias; Víctor Hugo Borja-Aburto |
| EPI_ISL_2801669, EPI_ISL_2801671, EPI_ISL_2801674, EPI_ISL_2801680, EPI_ISL_2801681, EPI_ISL_2801682, EPI_ISL_2801683, EPI_ISL_2801684, EPI_ISL_2801689, EPI_ISL_2801690, EPI_ISL_2801691 | | | |
| see above | Centro de Investigación Biomédica del Noreste (CIBIN) | Instituto de Biotecnología de la UNAM | ; Alejandra García-Gasca; Alejandra Hernández-Terán; Alejandro Sánchez-Flores; Alfredo Herrera-Estrella; Alicia Ocaña-Mondragón; Andreu Comas-García; Angel Gustavo Salas-Lais; Antonio Loza Román; Bernardo Martínez-Miguel; Blanca Taboada; Brenda Irasema Maldonado-Meza; Bruno Gómez-Gil; Carla Ivón Herrera-Najera; Carlos F. Arias; Celia Boukadida; Clara Esperanza Santacruz-Tinoco; Concepción Grajales-Muñiz; Consorcio Mexicano de Vigilancia Genómica (CoViGen-Mex). Authors (in alphabetical order): Julio Elias Alvarado-Yaah; Cristóbal Cháidez-Quiróz; Célida Duque Molina; Célida Martínez- Rodríguez; Daniel Fregoso-Rueda; Daniel Lira Morales; Eduardo Becerril-Vargas; Fernando Fontove-Herrera; Fidencio Mejía-Nepomuceno; Francisco Pulido; Gloria Elena Espinosa-Ayala; Gloria María Molina-Salinas; Gloria Vazquez; Hector Esteban Paz-Juárez; Hector Montoya-Fuentes; Helen Haydee Fernanda Ramírez-Plascencia; Irvin González-López; Jean Pierre González; Jesús Hernández; Joel Armando Vázquez-Pérez.; Jorge Salas-Hernández; José Antonio Enciso-Moreno; José Arturo Martínez-Orozco; José Esteban Muñoz-Medina; José de Jesús Nuñez-Contreras; Juan Bautista Chale-Dzul; Julissa Enciso-Ibarra; Luis Alberto Ochoa-Carrera; Margarita Matías-Florentino; Mario Mújica-Sánchez; Marissa Perez-Garcia; María Guadalupe Santiago-Mauricio; María Guadalupe de Jesús Mireles-Rivera; Nelly Sélem-Mojica; Pavel Isa; Ricardo Ciria Merce; Ricardo Grande; Rosa María Gutiérrez Ríos; Santiago Ávila-Ríos; Selene Zárate; Susana Lopez; Verónica Mata-Haro; Víctor Eduardo García-Arias; Víctor Hugo Borja-Aburto |
| EPI_ISL_4261889, EPI_ISL_4261892, EPI_ISL_4261916, EPI_ISL_4261917, EPI_ISL_4261918, EPI_ISL_4261919, EPI_ISL_4261920, EPI_ISL_4261924, EPI_ISL_4261953, EPI_ISL_4262028, EPI_ISL_4262038, EPI_ISL_4262059 | | | |
| see above | Chiu Laboratory, University of California, San Francisco | Chiu Laboratory, University of California, San Francisco | Alicia Sotomayor-Gonzalez; Baolin Wang; Candace Wang; Charles Chiu; Daniel Wong; Dustin Glasner; Venice Servellita; Xianding (Wayne) Deng; Yueyuan Zhang |
| EPI_ISL_2876281 | City of Milwaukee Health Department Laboratory | City of Milwaukee Health Department Laboratory | Amy Bauer; Jennifer Lentz; Manjeet Khubbar; Samantha Scott; Sanjib Bhattacharyya |
| EPI_ISL_3242490, EPI_ISL_3242503, EPI_ISL_3242504, EPI_ISL_3242509, EPI_ISL_3242520, EPI_ISL_3242521, EPI_ISL_3242524, EPI_ISL_3242539, EPI_ISL_3242552, EPI_ISL_3242579, EPI_ISL_3242593, EPI_ISL_3242636, EPI_ISL_3242682, EPI_ISL_3242683, EPI_ISL_3242684, EPI_ISL_3242685, EPI_ISL_3242686, EPI_ISL_3242687, EPI_ISL_3242688, EPI_ISL_3242689, EPI_ISL_3242690, EPI_ISL_3242691, EPI_ISL_3242692, EPI_ISL_3242693, EPI_ISL_3242694, EPI_ISL_3242695, EPI_ISL_3242696, EPI_ISL_3242697, EPI_ISL_3242698, EPI_ISL_3242699, EPI_ISL_3242700, EPI_ISL_3242701, EPI_ISL_3242743 | | | |
| see above | Clinica INDISA | "Facultad de Ciencias de la Vida, UNAB" | "Claudio Meneses; Ariel Orellana"; Claudio Olmos; Daniel Leon; Dayan Sanhueza; Eduardo Castro; Gonzalo Campaña; Macarena Bastias; Paola Pidal; Ricardo Yusta; Sebastian Wolter; Susana Saez; Víctor Monreal; Waldo Diaz |
| EPI_ISL_2932516 | Clinical Hospital of Infectious Diseases „Sfanta Parascheva" Iasi | National Institute of Infectious Diseases-Prof. Dr. Matei Bals Molecular Diagnostics Laboratory | Corina Casangiu; Dan Otelea; Leontina Banica; Marius Surleac; Petre Milu; Robert Hohan; Simona Paraschiv |
| EPI_ISL_2626011, EPI_ISL_2626014, EPI_ISL_2626019, EPI_ISL_2626029, EPI_ISL_2626043, EPI_ISL_2626082 | Cliniques universitaires Saint-Luc | UCLouvain/IREC/MBLG | Benoit Kabamba Mukadi; Bertrand Bearzatto; Jean Ruelle |
| EPI_ISL_4300436, EPI_ISL_4300441, EPI_ISL_4300442, EPI_ISL_4300445 | Color Genomics | Chiu Laboratory, University of California, San Francisco | Alicia Sotomayor-Gonzalez; Alicia Zhou; Baolin Wang; Charles Chiu; Daniel Wong; Erika Torres; Madyson Wanner; Noah Brazer; Scott Topper; Venice Servellita; Yueyuan Zhang |
| EPI_ISL_3149485, EPI_ISL_3403805, EPI_ISL_3403806, EPI_ISL_3403810, EPI_ISL_3403880, EPI_ISL_3403947 | Colorado Department of Public Health and Environment | Colorado Department of Public Health and Environment | Alexandria Rossheim; Diana Ir; Emily A. Travanty; Laura Bankers; Mandy Waters; Michael A. Martin; Michael Martin; Molly C. Hetherington-Rauth; Sarah Elizabeth Totten; Shannon R. Matzinger |
| EPI_ISL_2783655, EPI_ISL_2783658, EPI_ISL_2783660, EPI_ISL_2783663, EPI_ISL_2783975, EPI_ISL_2783978, EPI_ISL_2784218, EPI_ISL_2784222, EPI_ISL_2784224, EPI_ISL_2784227, EPI_ISL_2784312, EPI_ISL_2784314, EPI_ISL_2784315, EPI_ISL_2839689, EPI_ISL_2839726, EPI_ISL_2839730, EPI_ISL_2839733, EPI_ISL_2839753, EPI_ISL_2839781, EPI_ISL_2839838, EPI_ISL_2839842, EPI_ISL_2839845, EPI_ISL_2839868, EPI_ISL_2839872, EPI_ISL_2839887, EPI_ISL_2839902, EPI_ISL_2839913, EPI_ISL_2839941, EPI_ISL_2839945, EPI_ISL_3064617, EPI_ISL_3270301, EPI_ISL_3273356 | | | |
| see above | Curative Labs | Curative Labs | Elias L. Salfati; Eugenia Khorosheva; George Way; J.Cesar Ignacio-Espinoza; Janet Chen; Mikhail Hanewich-Hollatz; Nabjot Sandhu; Sophia Quasem; Vladimir Slepnev; Zhiyi Xie |
| EPI_ISL_2832469 | Curative Labs | New Mexico Department of Health Scientific Laboratory | Anastacia Griego-Fisher; D'eldra Malone; Ellie Johnson; Jennifer Benoit; Keila Gutierrez; Linda Salazar; Ratheesh Rajan |
| EPI_ISL_4415767, EPI_ISL_4415768, EPI_ISL_4415770, EPI_ISL_4415772, EPI_ISL_4415775, EPI_ISL_4415790, EPI_ISL_4415791, EPI_ISL_4415794, EPI_ISL_4415799, EPI_ISL_4415800, EPI_ISL_4415801, EPI_ISL_4415802, EPI_ISL_4415803, EPI_ISL_4415804, EPI_ISL_4415805, EPI_ISL_4415807, EPI_ISL_4415809, EPI_ISL_4415812, EPI_ISL_4415813, EPI_ISL_4415817, EPI_ISL_4415819, EPI_ISL_4415820, EPI_ISL_4415824, EPI_ISL_4415830, EPI_ISL_4415835, EPI_ISL_4415836, EPI_ISL_4415847, EPI_ISL_4415851, EPI_ISL_4415852, EPI_ISL_4415854, EPI_ISL_4415855, EPI_ISL_4415860, EPI_ISL_4415862, EPI_ISL_4415876, EPI_ISL_4415879, EPI_ISL_4415880, EPI_ISL_4415886, EPI_ISL_4415887, EPI_ISL_4415888, EPI_ISL_4415889, EPI_ISL_4415890, EPI_ISL_4415891, EPI_ISL_4415892, EPI_ISL_4415893, EPI_ISL_4415894, EPI_ISL_4415897, EPI_ISL_4415898, EPI_ISL_4415905, EPI_ISL_4415906, EPI_ISL_4415914, EPI_ISL_4415915, EPI_ISL_4415916, EPI_ISL_4415917, EPI_ISL_4415918, EPI_ISL_4415938, EPI_ISL_4415939, EPI_ISL_4415940, EPI_ISL_4415941, EPI_ISL_4415945, EPI_ISL_4415946, EPI_ISL_4415953, EPI_ISL_4415956, EPI_ISL_4415957, EPI_ISL_4415958, EPI_ISL_4415959, EPI_ISL_4415960, EPI_ISL_4415961, EPI_ISL_4415962, EPI_ISL_4415963, EPI_ISL_4415964, EPI_ISL_4415966, EPI_ISL_4415968, EPI_ISL_4415969, EPI_ISL_4415970, EPI_ISL_4415971, EPI_ISL_4415972, EPI_ISL_4415973, EPI_ISL_4415974, EPI_ISL_4415975, EPI_ISL_4415977, EPI_ISL_4415979, EPI_ISL_4415980, EPI_ISL_4415982, EPI_ISL_4415983, EPI_ISL_4415984, EPI_ISL_4415985, EPI_ISL_4415986, EPI_ISL_4415987, EPI_ISL_4415988, EPI_ISL_4415989, EPI_ISL_4415990, EPI_ISL_4415992, EPI_ISL_4415993, EPI_ISL_4415994, EPI_ISL_4415995, EPI_ISL_4415996, EPI_ISL_4415997, EPI_ISL_4415998, EPI_ISL_4415999, EPI_ISL_4416000, EPI_ISL_4416005, EPI_ISL_4416006, EPI_ISL_4416007, EPI_ISL_4416008, EPI_ISL_4416009, EPI_ISL_4416010, EPI_ISL_4416014, EPI_ISL_4416018, EPI_ISL_4416019, EPI_ISL_4416022, EPI_ISL_4416023, EPI_ISL_4416025, EPI_ISL_4416026, EPI_ISL_4416027, EPI_ISL_4416031, EPI_ISL_4416032, EPI_ISL_4416034, EPI_ISL_4416035, EPI_ISL_4416036, EPI_ISL_4416037, EPI_ISL_4416042, EPI_ISL_4416045, EPI_ISL_4416047, EPI_ISL_4416049, EPI_ISL_4416050, EPI_ISL_4416053, EPI_ISL_4416054, EPI_ISL_4416057, EPI_ISL_4416062, EPI_ISL_4416064, EPI_ISL_4416066, EPI_ISL_4416067, EPI_ISL_4416069, EPI_ISL_4416071, EPI_ISL_4416072, EPI_ISL_4416075, EPI_ISL_4416076, EPI_ISL_4416077, EPI_ISL_4416078, EPI_ISL_4416079, EPI_ISL_4416082, EPI_ISL_4416084, EPI_ISL_4416088, EPI_ISL_4416091, EPI_ISL_4416092, EPI_ISL_4416093, EPI_ISL_4416096, EPI_ISL_4416098, EPI_ISL_4416099, EPI_ISL_4422495, EPI_ISL_4422501, EPI_ISL_4422504, EPI_ISL_4422507, EPI_ISL_4422509, EPI_ISL_4422512, EPI_ISL_4422535, EPI_ISL_4422540, EPI_ISL_4422542, EPI_ISL_4422580, EPI_ISL_4422584, EPI_ISL_4422586, EPI_ISL_4422591, EPI_ISL_4422607, EPI_ISL_4422615, EPI_ISL_4422618, EPI_ISL_4422631, EPI_ISL_4422636, EPI_ISL_4422657, EPI_ISL_4422666, EPI_ISL_4422668, EPI_ISL_4422674, EPI_ISL_4422676, EPI_ISL_4422678, EPI_ISL_4422680, EPI_ISL_4422682, EPI_ISL_4422687, EPI_ISL_4422689, EPI_ISL_4422691, EPI_ISL_4422693, EPI_ISL_4422695, EPI_ISL_4422696, EPI_ISL_4422712, EPI_ISL_4422714, EPI_ISL_4422717, EPI_ISL_4422719, EPI_ISL_4422722, EPI_ISL_4422724, EPI_ISL_4422731, EPI_ISL_4422733, EPI_ISL_4422735, EPI_ISL_4422738, EPI_ISL_4422743, EPI_ISL_4422753, EPI_ISL_4422756, EPI_ISL_4422758, EPI_ISL_4422760, EPI_ISL_4422763, EPI_ISL_4422765, EPI_ISL_4422767, EPI_ISL_4422779, EPI_ISL_4422806, EPI_ISL_4422808, EPI_ISL_4422810, EPI_ISL_4422814, EPI_ISL_4422816, EPI_ISL_4422820, EPI_ISL_4422825, EPI_ISL_4422831, EPI_ISL_4422834, EPI_ISL_4422839, EPI_ISL_4422844, EPI_ISL_4422846, EPI_ISL_4422848, EPI_ISL_4422850, EPI_ISL_4422853, EPI_ISL_4422867, EPI_ISL_4422876, EPI_ISL_4422886, EPI_ISL_4422897, EPI_ISL_4422900, EPI_ISL_4422905, EPI_ISL_4422910, EPI_ISL_4422914, EPI_ISL_4422918, EPI_ISL_4422920, EPI_ISL_4422922, EPI_ISL_4422928, EPI_ISL_4422930, EPI_ISL_4422934, EPI_ISL_4422939, EPI_ISL_4422944, EPI_ISL_4422950, EPI_ISL_4422952, EPI_ISL_4422954, EPI_ISL_4422956, EPI_ISL_4422959, EPI_ISL_4422961, EPI_ISL_4422963, EPI_ISL_4422965, EPI_ISL_4422966, EPI_ISL_4422968, EPI_ISL_4422971, EPI_ISL_4422990, EPI_ISL_4422993, EPI_ISL_4422999, EPI_ISL_4423000, EPI_ISL_4423016, EPI_ISL_4423029, EPI_ISL_4423031, EPI_ISL_4423032, EPI_ISL_4423036, EPI_ISL_4423037, EPI_ISL_4423061, EPI_ISL_4423063, EPI_ISL_4423066, EPI_ISL_4423068, EPI_ISL_4423070, EPI_ISL_4423073, EPI_ISL_4423075, EPI_ISL_4423077, EPI_ISL_4423078, EPI_ISL_4423086, EPI_ISL_4423088, EPI_ISL_4423089, EPI_ISL_4423093, EPI_ISL_4423096, EPI_ISL_4423104, EPI_ISL_4423111, EPI_ISL_4423115, EPI_ISL_4423118, EPI_ISL_4423122, EPI_ISL_4423124, EPI_ISL_4423127, EPI_ISL_4423131, EPI_ISL_4423132, EPI_ISL_4423134, EPI_ISL_4423142, EPI_ISL_4423143, EPI_ISL_4423145, EPI_ISL_4423262, EPI_ISL_4423276, EPI_ISL_4423281, EPI_ISL_4423294, EPI_ISL_4423303, EPI_ISL_4423313, EPI_ISL_4423315, EPI_ISL_4423338, EPI_ISL_4423361, EPI_ISL_4423362, EPI_ISL_4423364, EPI_ISL_4423366, EPI_ISL_4423368, EPI_ISL_4423369, EPI_ISL_4423371, EPI_ISL_4423373, EPI_ISL_4423375, EPI_ISL_4423376, EPI_ISL_4423378, EPI_ISL_4423380, EPI_ISL_4423382, EPI_ISL_4423384, EPI_ISL_4423518, EPI_ISL_4423519, EPI_ISL_4423521, EPI_ISL_4423523, EPI_ISL_4423525, EPI_ISL_4426672, EPI_ISL_4426678, EPI_ISL_4426681, EPI_ISL_4426684, EPI_ISL_4426708, EPI_ISL_4426715, EPI_ISL_4426723, EPI_ISL_4426729, EPI_ISL_4426732, EPI_ISL_4426743, EPI_ISL_4426771, EPI_ISL_4426779, EPI_ISL_4426782, EPI_ISL_4426791, EPI_ISL_4426800, EPI_ISL_4426834, EPI_ISL_4426852, EPI_ISL_4427118, EPI_ISL_4427121, EPI_ISL_4427125, EPI_ISL_4427128, EPI_ISL_4427131, EPI_ISL_4427134, EPI_ISL_4427137, EPI_ISL_4427140, EPI_ISL_4427143, EPI_ISL_4427146, EPI_ISL_4427149, EPI_ISL_4427152, EPI_ISL_4427155, EPI_ISL_4427158, EPI_ISL_4427161, EPI_ISL_4427164, EPI_ISL_4427167, EPI_ISL_4427170, EPI_ISL_4427173, EPI_ISL_4427175, EPI_ISL_4427179, EPI_ISL_4427180, EPI_ISL_4427182, EPI_ISL_4427184, EPI_ISL_4427186, EPI_ISL_4427188, EPI_ISL_4427190, EPI_ISL_4427191, EPI_ISL_4427193, EPI_ISL_4427195, EPI_ISL_4427196, EPI_ISL_4427200, EPI_ISL_4427203, EPI_ISL_4427205, EPI_ISL_4427206, EPI_ISL_4427210, EPI_ISL_4427213, EPI_ISL_4427214, EPI_ISL_4427216, EPI_ISL_4427220, EPI_ISL_4427222, EPI_ISL_4427224, EPI_ISL_4427226, EPI_ISL_4427229, EPI_ISL_4427230, EPI_ISL_4427232, EPI_ISL_4427234, EPI_ISL_4427236, EPI_ISL_4427238, EPI_ISL_4427240, EPI_ISL_4427243, EPI_ISL_4427245, EPI_ISL_4427246, EPI_ISL_4427250, EPI_ISL_4427252, EPI_ISL_4427254, EPI_ISL_4427255, EPI_ISL_4427257, EPI_ISL_4427259, EPI_ISL_4427262, EPI_ISL_4427265, EPI_ISL_4427267, EPI_ISL_4427268, EPI_ISL_4427270, EPI_ISL_4427272, EPI_ISL_4427274, EPI_ISL_4427276, EPI_ISL_4427277, EPI_ISL_4427279, EPI_ISL_4427281, EPI_ISL_4427283, EPI_ISL_4427284, EPI_ISL_4427286, EPI_ISL_4427290, EPI_ISL_4427291, EPI_ISL_4427292, EPI_ISL_4427293, EPI_ISL_4427294, EPI_ISL_4427296, EPI_ISL_4427297, EPI_ISL_4427299, EPI_ISL_4427301, EPI_ISL_4427303, EPI_ISL_4427304, EPI_ISL_4427306, EPI_ISL_4427308, EPI_ISL_4427309, EPI_ISL_4427311, EPI_ISL_4427313, EPI_ISL_4427315, EPI_ISL_4427317, EPI_ISL_4427318, EPI_ISL_4427320, EPI_ISL_4427322, EPI_ISL_4427324, EPI_ISL_4427326, EPI_ISL_4427327, EPI_ISL_4427329, EPI_ISL_4427332, EPI_ISL_4427333, EPI_ISL_4427335, EPI_ISL_4427339, EPI_ISL_4427343, EPI_ISL_4427345, EPI_ISL_4427346, EPI_ISL_4427351, EPI_ISL_4427352, EPI_ISL_4427354, EPI_ISL_4427357, EPI_ISL_4427360, EPI_ISL_4427363, EPI_ISL_4427364, EPI_ISL_4427366, EPI_ISL_4427368, EPI_ISL_4427370, EPI_ISL_4427372, EPI_ISL_4427375, EPI_ISL_4427377, EPI_ISL_4427381, EPI_ISL_4427383, EPI_ISL_4427385, EPI_ISL_4427388, EPI_ISL_4427390, EPI_ISL_4427392, EPI_ISL_4427394, EPI_ISL_4427396, EPI_ISL_4427398, EPI_ISL_4427399, EPI_ISL_4427401, EPI_ISL_4427403, EPI_ISL_4427404, EPI_ISL_4427406, EPI_ISL_4427408, EPI_ISL_4427410, EPI_ISL_4427411, EPI_ISL_4427413, EPI_ISL_4427416, EPI_ISL_4427418, EPI_ISL_4427420, EPI_ISL_4427421, EPI_ISL_4427423, EPI_ISL_4427425, EPI_ISL_4427427, EPI_ISL_4427430, EPI_ISL_4427432, EPI_ISL_4427434, EPI_ISL_4427435, EPI_ISL_4427437, EPI_ISL_4427439, EPI_ISL_4427442, EPI_ISL_4427445, EPI_ISL_4427447, EPI_ISL_4427448, EPI_ISL_4427450, EPI_ISL_4427452, EPI_ISL_4427455, EPI_ISL_4427457, EPI_ISL_4427459, EPI_ISL_4427461, EPI_ISL_4427465, EPI_ISL_4427467, EPI_ISL_4427470, EPI_ISL_4427472, EPI_ISL_4427474, EPI_ISL_4427476, EPI_ISL_4427478, EPI_ISL_4427481, EPI_ISL_4427483, EPI_ISL_4427484, EPI_ISL_4427486, EPI_ISL_4427488, EPI_ISL_4427490, EPI_ISL_4427492, EPI_ISL_4427495, EPI_ISL_4427496, EPI_ISL_4427498, EPI_ISL_4427500, EPI_ISL_4427502, EPI_ISL_4427503, EPI_ISL_4427505, EPI_ISL_4427507, EPI_ISL_4427509, EPI_ISL_4427511, EPI_ISL_4427514, EPI_ISL_4427517, EPI_ISL_4427519, EPI_ISL_4427521, EPI_ISL_4427524, EPI_ISL_4427526, EPI_ISL_4427528, EPI_ISL_4427532, EPI_ISL_4427533, EPI_ISL_4427535, EPI_ISL_4427537, EPI_ISL_4427539, EPI_ISL_4427540, EPI_ISL_4427542, EPI_ISL_4427544, EPI_ISL_4427545, EPI_ISL_4427547, EPI_ISL_4427549, EPI_ISL_4428159, EPI_ISL_4428179, EPI_ISL_4428182, EPI_ISL_4428185, EPI_ISL_4428196, EPI_ISL_4428210, EPI_ISL_4428219, EPI_ISL_4428222, EPI_ISL_4428248, EPI_ISL_4428272, EPI_ISL_4428278, EPI_ISL_4428281, EPI_ISL_4428284, EPI_ISL_4428287, EPI_ISL_4428290, EPI_ISL_4428294, EPI_ISL_4428296, EPI_ISL_4428299, EPI_ISL_7743907, EPI_ISL_7743913, EPI_ISL_7743914, EPI_ISL_7743915, EPI_ISL_7743917, EPI_ISL_7743920 | | | |
| see above | DASA | DASA | Adriano Bonaldi; Angelica Hristov; Annelise Lopes; Bianca Cota; Cristina Oliveira; Jose Levi; Lidia Yamamoto; Paulo Pierry; Rodrigo Guarischi; Rodrigo Salazar |
| EPI_ISL_5646537, EPI_ISL_5646609 | DIRETORIA DE SAUDE | Instituto Butantan | Antonio Jorge Martins; Claudia Renata dos Santos Barros; David Schlesinger; Debora Botequio Moretti; Dimas Tadeu Covas; Elaine Cristina Marqueze; Elaine Vieira Santos; Evandra Strazza Rodrigues; Heidge Fukumasu; Jayme Augusto de Souza-Neto; José Salvatore Leister Patané; Luiz Alcantara; Luiz Lehmann Coutinho; Maria Carolina Elias; Maurício Lacerda Nogueira; Rafael dos Santos Bezerra; Raul Machado Neto; Rejane Maria Tommasini Grotto; Ricardo Haddad; Sandra Coccuzzo Sampaio Vessoni; Simone Kashima; Svetoslav Nanev Slavov; Vincent Louis Viala |
| EPI_ISL_5646718, EPI_ISL_5646794, EPI_ISL_5646824 | DIRETORIA MUNICIPAL DE SAUDE DE ENGENHEIRO COELHO | Instituto Butantan | Antonio Jorge Martins; Claudia Renata dos Santos Barros; David Schlesinger; Debora Botequio Moretti; Dimas Tadeu Covas; Elaine Cristina Marqueze; Elaine Vieira Santos; Evandra Strazza Rodrigues; Heidge Fukumasu; Jayme Augusto de Souza-Neto; José Salvatore Leister Patané; Luiz Alcantara; Luiz Lehmann Coutinho; Maria Carolina Elias; Maurício Lacerda Nogueira; Rafael dos Santos Bezerra; Raul Machado Neto; Rejane Maria Tommasini Grotto; Ricardo Haddad; Sandra Coccuzzo Sampaio Vessoni; Simone Kashima; Svetoslav Nanev Slavov; Vincent Louis Viala |
| EPI_ISL_2612553, EPI_ISL_2612554, EPI_ISL_2612555, EPI_ISL_2612556 | DOHMH Corona | New York City Public Health Laboratory | Jade Wang; et al. |
| EPI_ISL_3692595, EPI_ISL_3692598, EPI_ISL_3692632 | DPHL | Delaware Public Health Lab | Rebecca Savage |
| EPI_ISL_2637980 | DYNALAB | Department of Virology, Henri Mondor University Hospital, Assistance Publique Hôpitaux de Paris, Université Paris-Est Créteil, INSERM U955 | Alexandre Soulier; Christophe Rodriguez; Elisabeth Trawinski; Guillaume Gricourt; Jean-Michel Pawlotsky; Melissa N'Debi; Slim Fourati; Vanessa Demontant |

| | | | |
|---|---|---|---|
| EPI_ISL_2676629 | DYOMEDEA-LABORATOIRE DE LA SAUVEGARDE | CNR Virus des Infections Respiratoires - France SUD | Antonin Bal; Bruno Lina; Gregory Destras; Gwendolyne Burfin; Hadrien Regue; Laurence Josset; Martine Valette; Quentin Semanas |
| EPI_ISL_2669458 | Department of Bacteria, Parasites and Fungi, Statens Serum Institut, Copenhagen, Denmark | Statens Serum Institut Bioinformatics and Microbial Genomics | Danish Covid-19 Genome Consortium |
| EPI_ISL_2612782 | Department of Clinical Microbiology | GIGA Medical Genomics | Bouchra Boujemla; Cécile Meex; Keith Durkin; Maria Artesi; Marie-Pierre Hayette; Nathalie Renotte; Pierrette Melin; Raphaël Boreux; Sébastien Bontems; Vincent Bours |
| EPI_ISL_2673516, EPI_ISL_2673518 | Department of Virology, Istituto Zooprofilattico Sperimentale del Lazio e della Toscana (IZSLT) | Department of General Diagnostics; Department of Virology; Istituto Zooprofilattico Sperimentale del Lazio e della Toscana (IZSLT) | Alessia Franco; Antonella Cersini; Antonio Battisti.; Elena L. Diaconu; Fabiola Feltrin; Giuseppe Manna; Patricia Alba; Raffaela Conti; Teresa Scicluna; Virginia Carfora |
| EPI_ISL_2762312, EPI_ISL_2762314, EPI_ISL_2762356, EPI_ISL_2762365, EPI_ISL_2762395, EPI_ISL_2762431 | Diagnosticum - Labor Neukirchen | Robert Koch Institute | |
| EPI_ISL_3050695, EPI_ISL_3050697, EPI_ISL_3050700, EPI_ISL_3050701, EPI_ISL_3050704, EPI_ISL_3050718, EPI_ISL_3050720, EPI_ISL_3050723 | | | |
| see above | Dirección regional de salud del Callao (DIRESA-CALLAO) | Centro de Investigaciones Tecnológicas, Biomédicas y Medioambientales (CITBM) | B; Huaman; J. Alarcon; M. Cuellar; M. Ramirez; M. Sovero |
| EPI_ISL_3933507 | Division of Emerging Infectious Diseases, Bureau of Infectious Diseases Diagnosis Control, Korea Disease Control and Prevention Agency | Division of Emerging Infectious Diseases, Bureau of Infectious Diseases Diagnosis Control, Korea Disease Control and Prevention Agency | Ae Kyung Park; Chae Young Lee; Eun-Jin Kim; Heui Man Kim; Il-Hwan Kim; Jeong-Ah Kim |
| EPI_ISL_2644758, EPI_ISL_3474306 | Dutch COVID-19 response team | Medical Microbiology, Maastricht University Medical Centre | Brian van der Veer*; Carmen Reumkens; Christian Hoebe; Erik Beuken; Jozef Dingemans*; Lieke van Alphen; Paul Savelkoul |
| EPI_ISL_2610763, EPI_ISL_2672934, EPI_ISL_2672978, EPI_ISL_2673074, EPI_ISL_2673075, EPI_ISL_2673170, EPI_ISL_2673214, EPI_ISL_2673225, EPI_ISL_2673243, EPI_ISL_2673275, EPI_ISL_2787886, EPI_ISL_2787888, EPI_ISL_2787889, EPI_ISL_2787899, EPI_ISL_2787918 | | | |
| see above | Dutch COVID-19 response team | National Institute for Public Health and the Environment (RIVM) | Adam Meijer; AnneMarie van den Brandt; Annelies Kroneman; Bas van der Veer; Chantal Reusken; Dennis Schmitz; Dirk Eggink; Euníce Then; Florian Zwagemaker; Harry Vennema; Jeroen Cremer; Karim Hajji; Kim Freriks; Linda van de Nes; Lisa Wijsman; Lynn Aarts; Melissa van Tuil; Ryanne Jaarsma; Sanne Bos; Sharon van den Brink; on behalf of the national COVID-19 response team |
| EPI_ISL_4059814, EPI_ISL_4059816, EPI_ISL_4059822, EPI_ISL_4060091, EPI_ISL_4060109, EPI_ISL_4060110 | Dynacare Manitoba | Manitoba Cadham Provincial Laboratory | David Alexander; Hilary Racher; Hongbin Li; Melissa Desaulnier; Tintu Abraham |
| EPI_ISL_5647292 | EMERGENCIA RESPIRATORIA DE NOVA GRANADA | Instituto Butantan | Antonio Jorge Martins; Claudia Renata dos Santos Barros; David Schlesinger; Debora Botequio Moretti; Dimas Tadeu Covas; Elaine Cristina Marqueze; Elaine Vieira Santos; Evandra Strazza Rodrigues; Heidge Fukumasu; Jayme Augusto de Souza-Neto; José Salvatore Leister Patané; Luiz Alcantara; Luiz Lehmann Coutinho; Maria Carolina Elias; Maurício Lacerda Nogueira; Rafael dos Santos Bezerra; Raul Machado Neto; Rejane Maria Tommasini Grotto; Ricardo Haddad; Sandra Coccuzzo Sampaio Vessoni; Simone Kashima; Svetoslav Nanev Slavov; Vincent Louis Viala |
| EPI_ISL_5646877 | ESF NOVA TANABI II | Instituto Butantan | Antonio Jorge Martins; Claudia Renata dos Santos Barros; David Schlesinger; Debora Botequio Moretti; Dimas Tadeu Covas; Elaine Cristina Marqueze; Elaine Vieira Santos; Evandra Strazza Rodrigues; Heidge Fukumasu; Jayme Augusto de Souza-Neto; José Salvatore Leister Patané; Luiz Alcantara; Luiz Lehmann Coutinho; Maria Carolina Elias; Maurício Lacerda Nogueira; Rafael dos Santos Bezerra; Raul Machado Neto; Rejane Maria Tommasini Grotto; Ricardo Haddad; Sandra Coccuzzo Sampaio Vessoni; Simone Kashima; Svetoslav Nanev Slavov; Vincent Louis Viala |
| EPI_ISL_3215302, EPI_ISL_3215303 | EXCITE Lab | Andersen lab at Scripps Research | Abigail Schnapper; Angela Scioscia; Chip Schooley; David Pride; Helena Tubb; Natasha Martin Cheryl Anderson; Sawyer Farmer; Sharon Reed; Tommy Valles + SEARCH |
| EPI_ISL_2938523, EPI_ISL_2938537, EPI_ISL_2938538, EPI_ISL_2938551, EPI_ISL_2938557, EPI_ISL_2938560, EPI_ISL_2938587, EPI_ISL_2938589, EPI_ISL_2938613, EPI_ISL_2938614, EPI_ISL_2938615, EPI_ISL_2938616, EPI_ISL_2938631, EPI_ISL_2938633, EPI_ISL_2938652, EPI_ISL_2938653, EPI_ISL_2938654, EPI_ISL_2938656, EPI_ISL_2938845, EPI_ISL_2938868, EPI_ISL_2938893, EPI_ISL_2938909, EPI_ISL_3116500, EPI_ISL_3116505, EPI_ISL_3116506, EPI_ISL_3116883, EPI_ISL_3116884, EPI_ISL_3116887, EPI_ISL_3116941, EPI_ISL_3117263, EPI_ISL_3117264, EPI_ISL_3117265, EPI_ISL_3117272, EPI_ISL_3117396, EPI_ISL_3117397, EPI_ISL_3117398, EPI_ISL_3117404, EPI_ISL_3117407, EPI_ISL_3117420, EPI_ISL_3117421, EPI_ISL_3117422, EPI_ISL_3910229 | | | |
| see above | Edmonton Provincial Lab | Public Health Agency of Canada (PHAC) National Microbiology Laboratory | Buss; Croxen M; Deo A; Dieu P; E; Ferrato C; Gill K; Khan F; Koleva P; Li V; Lloyd C; Lynch T; Ma R; Murphy S; Pabbaraju K; Shokoples S; Thayer J; Tipples G; Whitehouse M; Wong A; Yu C; Zelyas N |
| EPI_ISL_3246274 | Eurofins Diatherix | Hudsonalpha Genome Sequencing Center | Ada Stewart; Jane Grimwood; Jenell Webber; Jennifer Cart; John Lovell; Joshua Stough; Leslie Malone; Lori H. Handley; Melissa Williams; Stefan Brzezinski; Teresa Jones; and Jeremy Schmutz |
| EPI_ISL_5646522, EPI_ISL_5646579, EPI_ISL_5646605, EPI_ISL_5646705, EPI_ISL_5646715, EPI_ISL_5646780 | FACULDADE DE MEDICINA ABC | Instituto Butantan | Antonio Jorge Martins; Claudia Renata dos Santos Barros; David Schlesinger; Debora Botequio Moretti; Dimas Tadeu Covas; Elaine Cristina Marqueze; Elaine Vieira Santos; Evandra Strazza Rodrigues; Heidge Fukumasu; Jayme Augusto de Souza-Neto; José Salvatore Leister Patané; Luiz Alcantara; Luiz Lehmann Coutinho; Maria Carolina Elias; Maurício Lacerda Nogueira; Rafael dos Santos Bezerra; Raul Machado Neto; Rejane Maria Tommasini Grotto; Ricardo Haddad; Sandra Coccuzzo Sampaio Vessoni; Simone Kashima; Svetoslav Nanev Slavov; Vincent Louis Viala |
| EPI_ISL_3066766 | FUNDACION VALLE DE LILI | Instituto Nacional de Salud | Carlos Franco-Muñoz; Carmen Osorio; Diana Malo; Diego A. Álvarez-Díaz; Diego Andrés Prada; Gerardo Santamaría; Hector Alejandro Ruiz-Moreno; Jhonnatan Reales-González; Jorge Rivera; Juan Camilo Martinez; Julian Naizaque; Katherine Laiton-Donato; Lisseth Pardo; Magdalena Wiesner; Marcela Mercado-Reyes; Maria T.Herrera-Sepúlveda; Marta Lopez Blanco; Martha Lucia Ospina Martinez; Paola Rojas; Sergio Gomez; Sheryll Corchuelo; Ángela Alarcon Cruz |
| EPI_ISL_2919141 | Fakultní nemocnice Hradec Králové | Fakultní nemocnice Hradec Králové | Helena Kovarikova; Marketa Gancarcikova |
| EPI_ISL_2836643, EPI_ISL_2836725, EPI_ISL_2836728, EPI_ISL_2836729, EPI_ISL_2836732, EPI_ISL_2836733, EPI_ISL_2836736, EPI_ISL_2836738, EPI_ISL_2836743, EPI_ISL_2836794, EPI_ISL_2836834, EPI_ISL_2836841, EPI_ISL_2836844, EPI_ISL_2920491, EPI_ISL_2920502, EPI_ISL_2920513, EPI_ISL_2920527, EPI_ISL_2920528, EPI_ISL_2920529, EPI_ISL_2920531, EPI_ISL_2920533, EPI_ISL_2920536, EPI_ISL_2920537, EPI_ISL_2920540, EPI_ISL_2920541, EPI_ISL_2920542, EPI_ISL_2920544, EPI_ISL_2920550, EPI_ISL_2920560, EPI_ISL_2920564, EPI_ISL_3100753, EPI_ISL_3100754, EPI_ISL_3100755, EPI_ISL_3100756, EPI_ISL_3100757, EPI_ISL_3100776, EPI_ISL_3100779, EPI_ISL_5542090, EPI_ISL_5542841, EPI_ISL_5866265, EPI_ISL_5866393, EPI_ISL_6050076, EPI_ISL_6050214 | | | |
| see above | Florida Bureau of Public Health Laboratories | Florida Bureau of Public Health Laboratories | Jason Blanton; Namratha Tarigopula; Sarah Schmedes; Tiffany Splatt |
| EPI_ISL_2663253, EPI_ISL_2663254 | Fondazione IRCCS Ca' Granda Ospedale Maggiore Policlinico | Fondazione IRCCS Ca' Granda Ospedale Maggiore Policlinico | Ferruccio Ceriotti; Sara Uceda Renteria |
| EPI_ISL_2598700, EPI_ISL_2598701, EPI_ISL_2598702, EPI_ISL_2598703, EPI_ISL_2598704, EPI_ISL_2598705, EPI_ISL_2598724, EPI_ISL_2598725, EPI_ISL_2598726, EPI_ISL_2598729, EPI_ISL_2598733, EPI_ISL_2598734, EPI_ISL_2598738, EPI_ISL_2598742, EPI_ISL_2598744, EPI_ISL_2598757, EPI_ISL_2598764, EPI_ISL_2598778, EPI_ISL_2598786, EPI_ISL_2598789, EPI_ISL_2598791, EPI_ISL_2686408, EPI_ISL_2686410, EPI_ISL_2686420, EPI_ISL_2686427, EPI_ISL_2686430, EPI_ISL_2686431, EPI_ISL_2686436, EPI_ISL_2686442, EPI_ISL_2686448, EPI_ISL_2686555, EPI_ISL_2686561, EPI_ISL_2686573, EPI_ISL_2686587, EPI_ISL_2686609, EPI_ISL_2686623, EPI_ISL_2686651, EPI_ISL_2686717, EPI_ISL_2686720, EPI_ISL_2686732, EPI_ISL_2686762, EPI_ISL_2686764, EPI_ISL_2686768, EPI_ISL_2686795, EPI_ISL_2785548, EPI_ISL_4369909 | | | |
| see above | Fulgent Genetics | Centers for Disease Control and Prevention Division of Viral Diseases, Pathogen Discovery | Adrian Paskey; Becky Tsai; Benafsh Sapra; Benjamin Rambo-Martin; Christopher Gulvick; Clinton Paden; Clinton R. Paden; Dakota Howard; Darlene Wagner; Dhwani Batra; Doreen Ng; Duncan MacCannell; Erisa Sula; Harry Gao; James Xie; Jason Caravas; John Gao; Joseph Fierro; Kara Moser; Kristine Lacek; Matthew Schmerer; Mickey Li; Peter Cook; Peter W. Cook; Scott Sammons; Shatavia Morrison; Tymeckia Kendall; Victoria Caban Figueroa; Yan Meng; Yvette Unoarumhi |
| EPI_ISL_5646399, EPI_ISL_5646457 | GRIPARIO FALCAO | Instituto Butantan | Antonio Jorge Martins; Claudia Renata dos Santos Barros; David Schlesinger; Debora Botequio Moretti; Dimas Tadeu Covas; Elaine Cristina Marqueze; Elaine Vieira Santos; Evandra Strazza Rodrigues; Heidge Fukumasu; Jayme Augusto de Souza-Neto; José Salvatore Leister Patané; Luiz Alcantara; Luiz Lehmann Coutinho; Maria Carolina Elias; Maurício Lacerda Nogueira; Rafael dos Santos Bezerra; Raul Machado Neto; Rejane Maria Tommasini Grotto; Ricardo Haddad; Sandra Coccuzzo Sampaio Vessoni; Simone Kashima; Svetoslav Nanev Slavov; Vincent Louis Viala |
| EPI_ISL_5646406, EPI_ISL_5646926 | GRUPO TECNICO DE VIGILANCIA SANITARIA E EPIDEMIOLOGICA | Instituto Butantan | Antonio Jorge Martins; Claudia Renata dos Santos Barros; David Schlesinger; Debora Botequio Moretti; Dimas Tadeu Covas; Elaine Cristina Marqueze; Elaine Vieira Santos; Evandra Strazza Rodrigues; Heidge Fukumasu; Jayme Augusto de Souza-Neto; José Salvatore Leister Patané; Luiz Alcantara; Luiz Lehmann Coutinho; Maria Carolina Elias; Maurício Lacerda Nogueira; Rafael dos Santos Bezerra; Raul Machado Neto; Rejane Maria Tommasini Grotto; Ricardo Haddad; Sandra Coccuzzo Sampaio Vessoni; Simone Kashima; Svetoslav Nanev Slavov; Vincent Louis Viala |
| EPI_ISL_2756071, EPI_ISL_2756072, EPI_ISL_2756073, EPI_ISL_2756077, EPI_ISL_2756078, EPI_ISL_2756079, EPI_ISL_2756080, EPI_ISL_2756081, EPI_ISL_2756082, EPI_ISL_2756085, EPI_ISL_2756088, EPI_ISL_2756092, EPI_ISL_2756094, EPI_ISL_2756095, EPI_ISL_2756097, EPI_ISL_2756098, EPI_ISL_2756100, EPI_ISL_2756101, EPI_ISL_2756104, EPI_ISL_2756105, EPI_ISL_2756106, EPI_ISL_2756108, EPI_ISL_2756109, EPI_ISL_2756110, EPI_ISL_2756112, EPI_ISL_2756115, EPI_ISL_2756116, EPI_ISL_2756119, EPI_ISL_2756121, EPI_ISL_2756122, EPI_ISL_2756124, EPI_ISL_2756125, EPI_ISL_2756126, EPI_ISL_2756134, EPI_ISL_2756137, EPI_ISL_2756138, EPI_ISL_2756139, EPI_ISL_2756140, EPI_ISL_2756142, EPI_ISL_2756143, EPI_ISL_2756144, EPI_ISL_2756150, EPI_ISL_2756151, EPI_ISL_2756152, EPI_ISL_2756153, EPI_ISL_2756158, EPI_ISL_2756171, EPI_ISL_2756173, EPI_ISL_2756178, EPI_ISL_2756179, EPI_ISL_2756180, EPI_ISL_2756181, EPI_ISL_2756182, EPI_ISL_2756183, EPI_ISL_2756184, EPI_ISL_2756185, EPI_ISL_2756186, EPI_ISL_2756187, EPI_ISL_2756188, EPI_ISL_2756189, EPI_ISL_2756190, EPI_ISL_2756191, EPI_ISL_2756193, EPI_ISL_2756194, EPI_ISL_2756221, EPI_ISL_2756222, EPI_ISL_2756223, EPI_ISL_2756224, EPI_ISL_2756226, EPI_ISL_2756228, EPI_ISL_2756229, EPI_ISL_2756230, EPI_ISL_2756233, EPI_ISL_2756234, EPI_ISL_2756236, EPI_ISL_2756237, EPI_ISL_2756240, EPI_ISL_2756241, EPI_ISL_2756244, EPI_ISL_2756245, EPI_ISL_2756252, EPI_ISL_2756253, EPI_ISL_2756255, EPI_ISL_2756256, EPI_ISL_2756257, EPI_ISL_2756260, EPI_ISL_2756264, EPI_ISL_2756269, EPI_ISL_2756272, EPI_ISL_2756274, EPI_ISL_2756311, EPI_ISL_2756317, EPI_ISL_2756319, EPI_ISL_2756335, EPI_ISL_2756338, EPI_ISL_2892616, EPI_ISL_2892619, EPI_ISL_2892629, EPI_ISL_2892664, EPI_ISL_2892665, EPI_ISL_2892667, EPI_ISL_2892668, EPI_ISL_2892669, EPI_ISL_2892671, EPI_ISL_2892672, EPI_ISL_2892673, EPI_ISL_2892674, EPI_ISL_2892675, EPI_ISL_2892676, EPI_ISL_2892677, EPI_ISL_2892678, EPI_ISL_2892679, EPI_ISL_2892718, EPI_ISL_2892720, EPI_ISL_2892721, EPI_ISL_2892722, EPI_ISL_2892808, EPI_ISL_2892809, EPI_ISL_2892812, EPI_ISL_2892813, EPI_ISL_2892815, EPI_ISL_2892816, EPI_ISL_2892817, EPI_ISL_2892818, EPI_ISL_2892820, EPI_ISL_2892828, EPI_ISL_2892867, EPI_ISL_2892870, EPI_ISL_2892871, EPI_ISL_2892872, EPI_ISL_2892873, EPI_ISL_2892874, EPI_ISL_2892875, EPI_ISL_2892877, EPI_ISL_2892878, EPI_ISL_2892879, EPI_ISL_2892895, EPI_ISL_2892900, EPI_ISL_2892919, EPI_ISL_2892920, EPI_ISL_2892929, EPI_ISL_2892930, EPI_ISL_2892931, EPI_ISL_2892932, EPI_ISL_2892934, EPI_ISL_2892936, EPI_ISL_2892961, EPI_ISL_2892963, EPI_ISL_2892967, EPI_ISL_2892968, EPI_ISL_2892969, EPI_ISL_2892970, EPI_ISL_2892971, EPI_ISL_2892972, EPI_ISL_2892973, EPI_ISL_2892980, EPI_ISL_2893023, EPI_ISL_2893024, EPI_ISL_2893025, EPI_ISL_2893026, EPI_ISL_2893027, EPI_ISL_2893028, EPI_ISL_2893029, EPI_ISL_2893030, EPI_ISL_3185615 | | | |
| see above | Genetica Molecular and Subdepartamento de Virologia ISP Chile | Instituto de Salud Publica de Chile | Andres Castillo; Barbara Parra; Constanza Campano; Gisselle Barra; Javier Tognarelli; Jorge Fernandez; Karen Orostica; Loredana Arata; Patricia Bustos; Rodrigo Fasce; Soledad Ulloa |
| EPI_ISL_2894597 | Genolife | Genolife | Yenelli Cedano-Thomas |
| EPI_ISL_2621556 | Genomics, Poplar Healthcare | Genomics, Poplar Healthcare | Truong, M. |
| EPI_ISL_2854970, EPI_ISL_2985620, EPI_ISL_2985641, EPI_ISL_2985642, EPI_ISL_2985650, EPI_ISL_2985689, EPI_ISL_2985697, EPI_ISL_2985736 | | | |
| see above | Gravity Diagnostics, LLC | Gravity Diagnostics, LLC | Gravity Diagnostics |
| EPI_ISL_3536210 | HEMOCE CENTRO DE HEMATOLOGIA E HEMOTERAPIA DO CEARA | Analytical Competence Molecular Epidemiology Lab/ACME, Oswaldo Cruz Foundation, Ceara (FIOCRUZ CE) | Cleber Furtado Aksenen; Fabio Miyajima; Fernando Braga Stehling; Francisco Eder de Moura Lopes; Jamille Maria Mendes Bezerra; Joaquim César do Nascimento Sousa Junior; Pedro Miguel Carneiro Jeronimo; Suzana Porto Almeida e Lucas Delerino; Thais Ferreira de Oliveira; Thais de Oliveira Costa; Ticiane Cavalcante de Souza; Veridiana Pessoa Miyajima |
| EPI_ISL_3102337, EPI_ISL_3536220 | HIAS HOSPITAL INFANTIL ALBERT SABIN | Analytical Competence Molecular Epidemiology Lab/ACME, Oswaldo | Cleber Furtado Aksenen; Fabio Miyajima; Fernando Braga Stehling; Francisco Eder de Moura Lopes; Jamille Maria Mendes Bezerra; Joaquim César do Nascimento Sousa Junior; Pedro Miguel Carneiro Jeronimo; Suzana Porto Almeida e Lucas Delerino; Thais Ferreira de Oliveira; Thais de Oliveira Costa; Ticiane Cavalcante de Souza; Veridiana Pessoa Miyajima |

| | | | |
|---|---|---|---|
| | | Cruz Foundation, Ceara (FIOCRUZ CE) | |
| EPI_ISL_2617605, EPI_ISL_2617606, EPI_ISL_2617622, EPI_ISL_2617623, EPI_ISL_2617624, EPI_ISL_2617625, EPI_ISL_2680902, EPI_ISL_2680903, EPI_ISL_2680904, EPI_ISL_2680905, EPI_ISL_2680906, EPI_ISL_2680907, EPI_ISL_2680908, EPI_ISL_2680909, EPI_ISL_2680910, EPI_ISL_2680911, EPI_ISL_2680912, EPI_ISL_2921538, EPI_ISL_2921539, EPI_ISL_2921540, EPI_ISL_2921541, EPI_ISL_2921542, EPI_ISL_2921543, EPI_ISL_2921544, EPI_ISL_2921545, EPI_ISL_2921546, EPI_ISL_2921547, EPI_ISL_2921548, EPI_ISL_2921551, EPI_ISL_2921559, EPI_ISL_2921560, EPI_ISL_2921571, EPI_ISL_2921574, EPI_ISL_2921575, EPI_ISL_2921604, EPI_ISL_3254318, EPI_ISL_3254319, EPI_ISL_3254330, EPI_ISL_3254331, EPI_ISL_3254332, EPI_ISL_3254333, EPI_ISL_3254418, EPI_ISL_3254419, EPI_ISL_3254420, EPI_ISL_3254421, EPI_ISL_3254422, EPI_ISL_3254423, EPI_ISL_3254424, EPI_ISL_3254425, EPI_ISL_3254426, EPI_ISL_3254427, EPI_ISL_3254428, EPI_ISL_3254429, EPI_ISL_3254430, EPI_ISL_3254431, EPI_ISL_3254432, EPI_ISL_3254433, EPI_ISL_3254434, EPI_ISL_3254435, EPI_ISL_3254436, EPI_ISL_3254437, EPI_ISL_3254438, EPI_ISL_3254439, EPI_ISL_3254440, EPI_ISL_3254441, EPI_ISL_3254442, EPI_ISL_3254443, EPI_ISL_3254444, EPI_ISL_3254445, EPI_ISL_3254446, EPI_ISL_3254447, EPI_ISL_3254448, EPI_ISL_3254449, EPI_ISL_3254450, EPI_ISL_3254451, EPI_ISL_3254452, EPI_ISL_3254453, EPI_ISL_3254454, EPI_ISL_3254455, EPI_ISL_3254456, EPI_ISL_3254457, EPI_ISL_3254458, EPI_ISL_3254459, EPI_ISL_3254460, EPI_ISL_3254461, EPI_ISL_3254462, EPI_ISL_3254463, EPI_ISL_3254464, EPI_ISL_3254466, EPI_ISL_3254467, EPI_ISL_3254468, EPI_ISL_3254469, EPI_ISL_3254470, EPI_ISL_3254471, EPI_ISL_3254472, EPI_ISL_3254473, EPI_ISL_3254474, EPI_ISL_3254475, EPI_ISL_3254476, EPI_ISL_3254477, EPI_ISL_3254478, EPI_ISL_3254479, EPI_ISL_3254480, EPI_ISL_3254481, EPI_ISL_3254482, EPI_ISL_3254483, EPI_ISL_3254537, EPI_ISL_3299423, EPI_ISL_3299428, EPI_ISL_4061628, EPI_ISL_4061648, EPI_ISL_4061649, EPI_ISL_4890091, EPI_ISL_5438925, EPI_ISL_5438926, EPI_ISL_5438937, EPI_ISL_5438944 | | | |
| see above | HLAGYN - Laboratorio de Imunologia de Transplantes de Goias | HLAGYN - Laboratorio de Imunologia de Transplantes de Goias | Alessandro Leonardo Alvares Magalhaes; Daniel Ferreira de Sousa; Danielle de Paiva Rezende; Erika Lopes Rocha Batista; Fernando Antonio Vinhal dos Santos; Frederico Rodrigues Vinhal; Kamila Oliveira Reis De Freitas.; Lucas Carlos Gomes Pereira; Sabrina Sara Moreira Duarte |
| EPI_ISL_3102453, EPI_ISL_3536205, EPI_ISL_3536232, EPI_ISL_3536233, EPI_ISL_3536307 | HM HOSPITAL DE MESSEJANA DR CARLOS ALBERTO STUDART GOMES | Analytical Competence Molecular Epidemiology Lab/ACME, Oswaldo Cruz Foundation, Ceara (FIOCRUZ CE) | Cleber Furtado Aksenen; Fabio Miyajima; Fernando Braga Stehling; Francisco Eder de Moura Lopes; Jamille Maria Mendes Bezerra; Joaquim César do Nascimento Sousa Junior; Pedro Miguel Carneiro Jeronimo; Suzana Porto Almeida e Lucas Delerino; Thais Ferreira de Oliveira; Thais de Oliveira Costa; Ticiane Cavalcante de Souza; Veridiana Pessoa Miyajima |
| EPI_ISL_3912191 | HOSP MUN ABELARDO GADELHA DA ROCHA | Analytical Competence Molecular Epidemiology Lab/ACME, Oswaldo Cruz Foundation, Ceara (FIOCRUZ CE) | Cleber Furtado Aksenen; Fabio Miyajima; Fernando Braga Stehling; Francisco Eder de Moura Lopes; Jamille Maria Mendes Bezerra; Joaquim Cesar do Nascimento Sousa Junior; Pedro Miguel Carneiro Jeronimo; Suzana Porto Almeida & Lucas Delerino on behalf of COVID-19 FIOCRUZ Genomic Network; Thais Ferreira de Oliveira; Thais de Oliveira Costa; Ticiane Cavalcante de Souza; Veridiana Pessoa Miyajima |
| EPI_ISL_5646868, EPI_ISL_5646870, EPI_ISL_5646935 | HOSP MUN JOSANIAS CASTANHA BRAGA | Instituto Butantan | Antonio Jorge Martins; Claudia Renata dos Santos Barros; David Schlesinger; Debora Botequio Moretti; Dimas Tadeu Covas; Elaine Cristina Marqueze; Elaine Vieira Santos; Evandra Strazza Rodrigues; Heidge Fukumasu; Jayme Augusto de Souza-Neto; José Salvatore Leister Patané; Luiz Alcantara; Luiz Lehmann Coutinho; Maria Carolina Elias; Maurício Lacerda Nogueira; Rafael dos Santos Bezerra; Raul Machado Neto; Rejane Maria Tommasini Grotto; Ricardo Haddad; Sandra Coccuzzo Sampaio Vessoni; Simone Kashima; Svetoslav Nanev Slavov; Vincent Louis Viala |
| EPI_ISL_3102273, EPI_ISL_3912020 | HOSPITAL CURA DARS | Analytical Competence Molecular Epidemiology Lab/ACME, Oswaldo Cruz Foundation, Ceara (FIOCRUZ CE) | Cleber Furtado Aksenen; Fabio Miyajima; Fernando Braga Stehling; Francisco Eder de Moura Lopes; Jamille Maria Mendes Bezerra; Joaquim Cesar do Nascimento Sousa Junior; Joaquim César do Nascimento Sousa Junior; Pedro Miguel Carneiro Jeronimo; Suzana Porto Almeida & Lucas Delerino on behalf of COVID-19 FIOCRUZ Genomic Network; Suzana Porto Almeida e Lucas Delerino; Thais Ferreira de Oliveira; Thais de Oliveira Costa; Ticiane Cavalcante de Souza; Veridiana Pessoa Miyajima |
| EPI_ISL_5646395, EPI_ISL_5646405, EPI_ISL_5646437, EPI_ISL_5646444, EPI_ISL_5646452, EPI_ISL_5646454, EPI_ISL_5646463 | | | |
| see above | HOSPITAL DE CAMPANHA COVID 19 DE MAIRIPORA | Instituto Butantan | Antonio Jorge Martins; Claudia Renata dos Santos Barros; David Schlesinger; Debora Botequio Moretti; Dimas Tadeu Covas; Elaine Cristina Marqueze; Elaine Vieira Santos; Evandra Strazza Rodrigues; Heidge Fukumasu; Jayme Augusto de Souza-Neto; José Salvatore Leister Patané; Luiz Alcantara; Luiz Lehmann Coutinho; Maria Carolina Elias; Maurício Lacerda Nogueira; Rafael dos Santos Bezerra; Raul Machado Neto; Rejane Maria Tommasini Grotto; Ricardo Haddad; Sandra Coccuzzo Sampaio Vessoni; Simone Kashima; Svetoslav Nanev Slavov; Vincent Louis Viala |
| EPI_ISL_5646762 | HOSPITAL DE CAMPANHA COVID 19 MUNICIPIO DE TAUBATE | Instituto Butantan | Antonio Jorge Martins; Claudia Renata dos Santos Barros; David Schlesinger; Debora Botequio Moretti; Dimas Tadeu Covas; Elaine Cristina Marqueze; Elaine Vieira Santos; Evandra Strazza Rodrigues; Heidge Fukumasu; Jayme Augusto de Souza-Neto; José Salvatore Leister Patané; Luiz Alcantara; Luiz Lehmann Coutinho; Maria Carolina Elias; Maurício Lacerda Nogueira; Rafael dos Santos Bezerra; Raul Machado Neto; Rejane Maria Tommasini Grotto; Ricardo Haddad; Sandra Coccuzzo Sampaio Vessoni; Simone Kashima; Svetoslav Nanev Slavov; Vincent Louis Viala |
| EPI_ISL_4601283 | HOSPITAL DE SANTA BARBARA | Instituto de Salud Carlos III | A. Monzón; ALEJANDRA; F. Casas; I. ASENJO VILLAMAYOR; I. Jiménez; M. Camarero; P. Zaballos; S. Cuesta; S. Iglesias-Caballero; S. Pozo; S. Varona; Sandonís; V. Vázquez-Morón |
| EPI_ISL_2828010 | HOSPITAL DR. RAFAEL A. CALDERON GUARDIA | Inciensa, Instituto Costarricense de Investigación y Enseñanza en Nutrición y Salud | Adriana Godínez; Claudio Soto-Garita; Estela Cordero; Francisco Duarte; Hebleen Porras; Joselyn Prado & Fabian Salas; José Luis Vargas; Mariela Gutiérrez; Melany Calderón |
| EPI_ISL_3536265, EPI_ISL_3536293 | HOSPITAL E MATERNIDADE DRA ZILDA ARNS NEUMANN | Analytical Competence Molecular Epidemiology Lab/ACME, Oswaldo Cruz Foundation, Ceara (FIOCRUZ CE) | Cleber Furtado Aksenen; Fabio Miyajima; Fernando Braga Stehling; Francisco Eder de Moura Lopes; Jamille Maria Mendes Bezerra; Joaquim César do Nascimento Sousa Junior; Pedro Miguel Carneiro Jeronimo; Suzana Porto Almeida e Lucas Delerino; Thais Ferreira de Oliveira; Thais de Oliveira Costa; Ticiane Cavalcante de Souza; Veridiana Pessoa Miyajima |
| EPI_ISL_5646708 | HOSPITAL ESTADUAL DE VILA ALPINA ORG SOCIAL SECONCI SAO PAUL | Instituto Butantan | Antonio Jorge Martins; Claudia Renata dos Santos Barros; David Schlesinger; Debora Botequio Moretti; Dimas Tadeu Covas; Elaine Cristina Marqueze; Elaine Vieira Santos; Evandra Strazza Rodrigues; Heidge Fukumasu; Jayme Augusto de Souza-Neto; José Salvatore Leister Patané; Luiz Alcantara; Luiz Lehmann Coutinho; Maria Carolina Elias; Maurício Lacerda Nogueira; Rafael dos Santos Bezerra; Raul Machado Neto; Rejane Maria Tommasini Grotto; Ricardo Haddad; Sandra Coccuzzo Sampaio Vessoni; Simone Kashima; Svetoslav Nanev Slavov; Vincent Louis Viala |
| EPI_ISL_3102275, EPI_ISL_3102291, EPI_ISL_3102360, EPI_ISL_3102386, EPI_ISL_3102393, EPI_ISL_3536149, EPI_ISL_3536169, EPI_ISL_3536184, EPI_ISL_3536271, EPI_ISL_3536274, EPI_ISL_3536283, EPI_ISL_3536299 | | | |
| see above | HOSPITAL ESTADUAL LEONARDO DA VINCI | Analytical Competence Molecular Epidemiology Lab/ACME, Oswaldo Cruz Foundation, Ceara (FIOCRUZ CE) | Cleber Furtado Aksenen; Fabio Miyajima; Fernando Braga Stehling; Francisco Eder de Moura Lopes; Jamille Maria Mendes Bezerra; Joaquim César do Nascimento Sousa Junior; Pedro Miguel Carneiro Jeronimo; Suzana Porto Almeida e Lucas Delerino; Thais Ferreira de Oliveira; Thais de Oliveira Costa; Ticiane Cavalcante de Souza; Veridiana Pessoa Miyajima |
| EPI_ISL_4601276 | HOSPITAL GENERAL UNIVERSITARIO DE ALBACETE | Instituto de Salud Carlos III | A. Monzón; CARIDAD; F. Casas; I. Jiménez; I.SAINZ DE BARANDA CAMINO; M. Sandonís; P. Zaballos; S. Cuesta; S. Iglesias-Caballero; S. Pozo; S. Varona; V. Camarero; Vázquez-Morón |
| EPI_ISL_4425234 | HOSPITAL GENERAL UNIVERSITARIO DE GUADALAJARA | Instituto de Salud Carlos III | A. Monzón; ALEJANDRO; F. Casas; I. Jiménez; I.GONZALEZ PRAETORIUS; Iglesias-Caballero; M. Sandonís; P. Zaballos; S. Camarero; S. Cuesta; S. Pozo; S. Varona; V. Vázquez-Morón |
| EPI_ISL_4601278, EPI_ISL_4601288 | HOSPITAL GENERAL UNIVERSITARIO REINA SOFIA | Instituto de Salud Carlos III | A. Monzón; F. Casas; I. Jiménez; I.NUÑEZ TRIGUEROS; M. Sandonís; Mª LUZ; P. Zaballos; S. Cuesta; S. Iglesias-Caballero; S. Pozo; S. Varona; V. Camarero; Vázquez-Morón |
| EPI_ISL_4425247, EPI_ISL_4425249 | HOSPITAL GENERAL UNIVERSITARIO SANTA LUCIA | Instituto de Salud Carlos III | A. Monzón; ANA; F. Casas; I. Jiménez; I.BLAZQUEZ ABELLAN; Iglesias-Caballero; M. Sandonís; P. Zaballos; S. Camarero; S. Cuesta; S. Pozo; S. Varona; V. Vázquez-Morón |
| EPI_ISL_4601282 | HOSPITAL GENERAL VIRGEN DE LA LUZ | Instituto de Salud Carlos III | A. Monzón; F. Casas; I. Jiménez; I. RODRIGUEZ ESCUDERO; M. Camarero; MARIA JOSE; P. Zaballos; S. Cuesta; S. Iglesias-Caballero; S. Pozo; S. Varona; Sandonís; V. Vázquez-Morón |
| EPI_ISL_5646927 | HOSPITAL GERAL JESUS TEIXEIRA DA COSTA GUAIANASES SAO PAULO | Instituto Butantan | Antonio Jorge Martins; Claudia Renata dos Santos Barros; David Schlesinger; Debora Botequio Moretti; Dimas Tadeu Covas; Elaine Cristina Marqueze; Elaine Vieira Santos; Evandra Strazza Rodrigues; Heidge Fukumasu; Jayme Augusto de Souza-Neto; José Salvatore Leister Patané; Luiz Alcantara; Luiz Lehmann Coutinho; Maria Carolina Elias; Maurício Lacerda Nogueira; Rafael dos Santos Bezerra; Raul Machado Neto; Rejane Maria Tommasini Grotto; Ricardo Haddad; Sandra Coccuzzo Sampaio Vessoni; Simone Kashima; Svetoslav Nanev Slavov; Vincent Louis Viala |
| EPI_ISL_5646419, EPI_ISL_5646633 | HOSPITAL HELIOPOLIS UNIDADE DE GESTAO ASSISTENCIAL I SP | Instituto Butantan | Antonio Jorge Martins; Claudia Renata dos Santos Barros; David Schlesinger; Debora Botequio Moretti; Dimas Tadeu Covas; Elaine Cristina Marqueze; Elaine Vieira Santos; Evandra Strazza Rodrigues; Heidge Fukumasu; Jayme Augusto de Souza-Neto; José Salvatore Leister Patané; Luiz Alcantara; Luiz Lehmann Coutinho; Maria Carolina Elias; Maurício Lacerda Nogueira; Rafael dos Santos Bezerra; Raul Machado Neto; Rejane Maria Tommasini Grotto; Ricardo Haddad; Sandra Coccuzzo Sampaio Vessoni; Simone Kashima; Svetoslav Nanev Slavov; Vincent Louis Viala |
| EPI_ISL_4601279 | HOSPITAL LOS ARCOS | Instituto de Salud Carlos III | A. Monzón; F. Casas; I. Jiménez; I.CAMARA SIMON; M. Sandonís; MARGARITA; P. Zaballos; S. Cuesta; S. Iglesias-Caballero; S. Pozo; S. Varona; V. Camarero; Vázquez-Morón |
| EPI_ISL_2827991, EPI_ISL_2827992 | HOSPITAL MEXICO | Inciensa, Instituto Costarricense de Investigación y Enseñanza en Nutrición y Salud | Adriana Godínez; Claudio Soto-Garita; Estela Cordero; Francisco Duarte; Hebleen Porras; Joselyn Prado & Juan Carlos Villalobos; José Luis Vargas; Mariela Gutiérrez; Melany Calderón |
| EPI_ISL_2828014 | HOSPITAL MONSEÑOR SANABRIA | Inciensa, Instituto Costarricense de Investigación y Enseñanza en Nutrición y Salud | Adriana Godínez; Claudio Soto-Garita; Estela Cordero; Francisco Duarte; Hebleen Porras; Joselyn Prado & María José Gómez; José Luis Vargas; Mariela Gutiérrez; Melany Calderón |
| EPI_ISL_5646517 | HOSPITAL MUNICIPAL DR TABAJARA RAMOS | Instituto Butantan | Antonio Jorge Martins; Claudia Renata dos Santos Barros; David Schlesinger; Debora Botequio Moretti; Dimas Tadeu Covas; Elaine Cristina Marqueze; Elaine Vieira Santos; Evandra Strazza Rodrigues; Heidge Fukumasu; Jayme Augusto de Souza-Neto; José Salvatore Leister Patané; Luiz Alcantara; Luiz Lehmann Coutinho; Maria Carolina Elias; Maurício Lacerda Nogueira; Rafael dos Santos Bezerra; Raul Machado Neto; Rejane Maria Tommasini Grotto; Ricardo Haddad; Sandra Coccuzzo Sampaio Vessoni; Simone Kashima; Svetoslav Nanev Slavov; Vincent Louis Viala |
| EPI_ISL_3102508 | HOSPITAL MUNICIPAL NOSSA SENHORA DOS MILAGRES | Analytical Competence Molecular Epidemiology Lab/ACME, Oswaldo Cruz Foundation, Ceara (FIOCRUZ CE) | Cleber Furtado Aksenen; Fabio Miyajima; Fernando Braga Stehling; Francisco Eder de Moura Lopes; Jamille Maria Mendes Bezerra; Joaquim César do Nascimento Sousa Junior; Pedro Miguel Carneiro Jeronimo; Suzana Porto Almeida e Lucas Delerino; Thais Ferreira de Oliveira; Thais de Oliveira Costa; Ticiane Cavalcante de Souza; Veridiana Pessoa Miyajima |
| EPI_ISL_3102313 | HOSPITAL OTOCLINICA | Analytical Competence Molecular Epidemiology Lab/ACME, Oswaldo Cruz Foundation, Ceara (FIOCRUZ CE) | Cleber Furtado Aksenen; Fabio Miyajima; Fernando Braga Stehling; Francisco Eder de Moura Lopes; Jamille Maria Mendes Bezerra; Joaquim César do Nascimento Sousa Junior; Pedro Miguel Carneiro Jeronimo; Suzana Porto Almeida e Lucas Delerino; Thais Ferreira de Oliveira; Thais de Oliveira Costa; Ticiane Cavalcante de Souza; Veridiana Pessoa Miyajima |
| EPI_ISL_2834732 | HOSPITAL REGIONAL DE ZIPAQUIRA | Instituto Nacional de Salud- Dirección de Investigación en Salud Pública | Carlos Franco-Muñoz; Carmen Osorio; Diana Malo; Diego A. Álvarez-Díaz; Diego Andrés Prada; Gerardo Santamaría; Hector Alejandro Ruiz-Moreno; Jhonnatan Reales-González; Jorge Rivera; Juan Camilo Martinez; Julian Naizaque; Katherine Laiton-Donato; Lisseth Pardo; Magdalena Wiesner; Marcela Mercado-Reyes; Maria T. Herrera-Sepúlveda; Marta Lopez Blanco; Martha Lucia Ospina Martinez; Paola Rojas; Sergio Gomez; Sheryll Corchuelo; Ángela Alarcon Cruz |
| EPI_ISL_2626145, EPI_ISL_2626160 | HOSPITAL UNIVERSITARIO SON ESPASES | HOSPITAL UNIVERSITARIO SON ESPASES | Hospital Universitario Son Espases; Servicio de Microbiología |
| EPI_ISL_3102435 | HOSPITAL UNIVERSITARIO WALTER CANTIDIO | Analytical Competence Molecular Epidemiology Lab/ACME, Oswaldo Cruz Foundation, Ceara (FIOCRUZ CE) | Cleber Furtado Aksenen; Fabio Miyajima; Fernando Braga Stehling; Francisco Eder de Moura Lopes; Jamille Maria Mendes Bezerra; Joaquim César do Nascimento Sousa Junior; Pedro Miguel Carneiro Jeronimo; Suzana Porto Almeida e Lucas Delerino; Thais Ferreira de Oliveira; Thais de Oliveira Costa; Ticiane Cavalcante de Souza; Veridiana Pessoa Miyajima |
| EPI_ISL_4601263, EPI_ISL_4601284, EPI_ISL_4601285 | HOSPITAL VIRGEN DE LA SALUD DE TOLEDO | Instituto de Salud Carlos III | A. Monzón; EVA; F. Casas; I. HEREDERO GALVEZ; I. Jiménez; M. Camarero; P. Zaballos; S. Cuesta; S. Iglesias-Caballero; S. Pozo; S. Varona; Sandonís; V. Vázquez-Morón |
| EPI_ISL_4743414 | HPTU | Laboratorio Departamental de Salud Publica de Antioquia | Andres F. Cardona-Rios; Cristian Arbey Velarde Hoyos; Gloria Isabel Escobar; Idabely Betancur Ortiz; Juan P. Hernandez-Ortiz; Maria Stella López |
| EPI_ISL_3388834 | Hosp. Pediatrico Dr. Avelino Castelán | Laboratorio de Biología Molecular, Instituto de Medicina Regional on behalf of 'Proyecto Argentino Interinstitucional de genomica de SARS-CoV-2' (PAIS Consortium) | Bettina Brusés; Gioia Lucia Marino; Griselda Oria; Horacio Lucero.; Javier Mussin; Julieta Ronchi; Laura Formichelli; María Andrea Gili; María Cecilia López; Melina Lorenzini Campos; Raúl Maximiliano Acevedo |

| | | | |
|---|---|---|---|
| EPI_ISL_3076926 | Hospital Carmen y Severo Ochoa | Laboratorio de Virología HUCA | Abreu F; Alvarez-Arguelles ME; Boga JA; Castelló C; Costales I; Coto E; Gómez de Oña J; Martín-Rodríguez G; Melón S; Perez-Martínez Z; Rojo S; Sandoval M |
| EPI_ISL_3147545 | Hospital Center Luxembourg | Laboratoire national de sante, Microbiology, Microbial Genomics Platform | Anke Wienecke-Baldacchino; Catherine Ragimbeau; Elodie Solarino; Fatu Djabi; Jean-Hugues Francois; Jessica Tapp; Lise Pignon; Michel Kohnen; Raoul Salmon; Tamir Abdelrahman; Virginie Jover |
| EPI_ISL_4946231, EPI_ISL_4946232 | Hospital Civil Ipiales | Instituto Nacional de Salud- Dirección de Investigación en Salud Pública | Carlos Franco-Muñoz; Carmen Osorio; Diana Malo; Diego A. Álvarez-Díaz; Diego Andrés Prada; Gerardo Santamaría; Hector Alejandro Ruiz-Moreno; Jhonnatan Reales-González; Jorge Rivera; Juan Camilo Martinez; Julian Naizaque; Katherine Laiton-Donato; Lisseth Pardo; Magdalena Wiesner; Marcela Mercado-Reyes; Maria T. Herrera-Sepúlveda; Marta Lopez Blanco; Martha Lucia Ospina Martinez; Paola Rojas; Sergio Gomez; Sheryll Corchuelo; Ángela Alarcon Cruz |
| EPI_ISL_2828327 | Hospital Clínico Universitario Virgen de la Arrixaca | Hospital Clínico Universitario Virgen de la Arrixaca | Marina Simón Páez y Laura Moreno Parrado |
| EPI_ISL_2601693, EPI_ISL_2601701, EPI_ISL_2601706, EPI_ISL_2601714, EPI_ISL_2601724 | Hospital General Universitario Gregorio Marañón | Hospital General Universitario Gregorio Marañón | Cristina Rodriguez-Grande; Darío García de Viedma; Julia Suárez; Laura Pérez-Lago; Marta Herranz Martin; Patricia Muñoz; Pedro Sola Campoy; Pilar Catalán; Sergio Buenestado Serrano; Victor Manuel de la Cueva |
| EPI_ISL_3216412 | Hospital General Universitario de Alicante - Instituto de Investigación Sanitaria y Biomédica de Alicante | SeqCOVID-SPAIN consortium/IBV(CSIC) | Carmen Molina Pardines and SeqCOVID-SPAIN consortium; Maripaz Ventero Martín |
| EPI_ISL_7740275 | Hospital Herrera Llerandi | Asociación de Salud Integral / Clínica Familiar "Luis Ángel García" | Ana S. Gonzalez-Reiche; Claudia Rangel; Danicela Mercado; Eduardo Arathon; Hilda Ruiz; Luis Aguirre; Luis Rivas; Narda Medina; Oscar Bonilla; Osmar Gamboa. |
| EPI_ISL_3033083, EPI_ISL_3033085, EPI_ISL_3260094 | Hospital Jaime Ferre - SAMCO Rafaela | Grupo de Genómica y Bioinformática del Instituto de Investigación de la Cadena Láctea CONICET-INTA on behalf of 'Proyecto Argentino Interinstitucional de genomica de SARS-CoV-2' (PAIS Consortium) | AF; Aliprandi D; Amadio; C; Eberhardt; Irazoqui; Isaia; JF; JM; MF; Pandolfi; Quaranta; Soratti R; V |
| EPI_ISL_2894556, EPI_ISL_2894560 | Hospital Margarita Maza de Juárez | Microbial Genomics Laboratory | ; Alejandra García-Gasca; Alejandra Hernández-Terán; Alejandro Sánchez-Flores; Alfredo Herrera-Estrella; Alicia Ocaña-Mondragón; Andreu Comas-García; Angel Gustavo Salas-Lais; Antonio Loza Román; Bernardo Martínez-Miguel; Blanca Taboada; Brenda Irasema Maldonado-Meza; Bruno Gómez-Gil; Carla Ivón Herrera-Najera; Carlos F. Arias; Celia Boukadida; Clara Esperanza Santacruz-Tinoco; Concepción Grajales-Muñiz; Consorcio Mexicano de Vigilancia Genómica (CoViGen-Mex). Authors (in alphabetical order): Julio Elias Alvarado-Yaah; Cristóbal Cháidez-Quiróz; Célida Duque Molina; Célida Martínez- Rodríguez; Daniel Fregoso-Rueda; Daniel Lira Morales; Eduardo Becerril-Vargas; Fernando Fontove-Herrera; Fidencio Mejía-Nepomuceno; Francisco Pulido; Gloria Elena Espinosa-Ayala; Gloria María Molina-Salinas; Gloria Vazquez; Hector Esteban Paz-Juárez; Hector Montoya-Fuentes; Helen Haydee Fernanda Ramírez-Plascencia; Irvin González-López; Jean Pierre González; Jesús Hernández; Joel Armando Vázquez-Pérez.; Jorge Salas-Hernández; José Antonio Enciso-Moreno; José Arturo Martínez-Orozco; José Esteban Muñoz-Medina; José de Jesús Nuñez-Contreras; Juan Bautista Chale-Dzul; Julissa Enciso-Ibarra; Luis Alberto Ochoa-Carrera; Margarita Matías-Florentino; Mario Mújica-Sánchez; Marissa Perez-Garcia; María Guadalupe Santiago-Mauricio; María Guadalupe de Jesús Mireles-Rivera; Nelly Sélem-Mojica; Pavel Isa; Ricardo Ciria Merce; Ricardo Grande; Rosa María Gutiérrez Rios; Santiago Ávila-Ríos; Selene Zárate; Susana Lopez; Verónica Mata-Haro; Victor Eduardo García-Arias; Víctor Hugo Borja-Aburto |
| EPI_ISL_2894565, EPI_ISL_2987715 | Hospital Sharp | Microbial Genomics Laboratory | ; Alejandra García-Gasca; Alejandra Hernández-Terán; Alejandro Sánchez-Flores; Alfredo Herrera-Estrella; Alicia Ocaña-Mondragón; Andreu Comas-García; Angel Gustavo Salas-Lais; Antonio Loza Román; Bernardo Martínez-Miguel; Blanca Taboada; Brenda Irasema Maldonado-Meza; Bruno Gómez-Gil; Carla Ivón Herrera-Najera; Carlos F. Arias; Celia Boukadida; Clara Esperanza Santacruz-Tinoco; Concepción Grajales-Muñiz; Consorcio Mexicano de Vigilancia Genómica (CoViGen-Mex). Authors (in alphabetical order): Julio Elias Alvarado-Yaah; Cristóbal Cháidez-Quiróz; Célida Duque Molina; Célida Martínez- Rodríguez; Daniel Fregoso-Rueda; Daniel Lira Morales; Eduardo Becerril-Vargas; Fernando Fontove-Herrera; Fidencio Mejía-Nepomuceno; Francisco Pulido; Gloria Elena Espinosa-Ayala; Gloria María Molina-Salinas; Gloria Vazquez; Hector Esteban Paz-Juárez; Hector Montoya-Fuentes; Helen Haydee Fernanda Ramírez-Plascencia; Irvin González-López; Jean Pierre González; Jesús Hernández; Joel Armando Vázquez-Pérez.; Jorge Salas-Hernández; José Antonio Enciso-Moreno; José Arturo Martínez-Orozco; José Esteban Muñoz-Medina; José de Jesús Nuñez-Contreras; Juan Bautista Chale-Dzul; Julissa Enciso-Ibarra; Luis Alberto Ochoa-Carrera; Margarita Matías-Florentino; Mario Mújica-Sánchez; Marissa Perez-Garcia; María Guadalupe Santiago-Mauricio; María Guadalupe de Jesús Mireles-Rivera; Nelly Sélem-Mojica; Pavel Isa; Ricardo Ciria Merce; Ricardo Grande; Rosa María Gutiérrez Rios; Santiago Ávila-Ríos; Selene Zárate; Susana Lopez; Verónica Mata-Haro; Victor Eduardo García-Arias; Víctor Hugo Borja-Aburto |
| EPI_ISL_2524776, EPI_ISL_2628167 | Hospital Universitari Arnau de Vilanova | Hospital Universitari Vall d'Hebron - Vall d'Hebron Institut de Recerca | Alejandra González-Sánchez; Andrés Antón; Ariadna Rando; Carla Castillo; Cristina Andrés; Damir Garcia-Cehic; Josep Quer; Juliana Esperalba; Maria Carmen Martin; Maria Gema Codina; Maria Piñana; Tomàs Pumarola |
| EPI_ISL_2637324 | Hospital Universitari Bellvitge | Microbiology Department | Aida Gonzalez-Diaz; Carmen Ardanuy; Jordi Camara; Jordi Niubó; Laura Calatayud; M Angeles Domínguez; Miguel Fernandez-Huerta; Sara Marti |
| EPI_ISL_2775952, EPI_ISL_2775959, EPI_ISL_2775966, EPI_ISL_2775970, EPI_ISL_2775971 | Hospital Universitari i Politècnic La Fe de València | SeqCOVID-SPAIN consortium/IBV(CSIC) | Ana Gil Brusola; Eva González Barberá; José Luis López Hontangas and SeqCOVID-SPAIN consortium; María Dolores Gómez Ruiz; Salvador Giner Almaraz |
| EPI_ISL_2620966 | Hospital Universitario Virgen de las Nieves de Granada-SAS | SeqCOVID-SPAIN consortium/IBV(CSIC) | Irene Pedrosa Corral; José M. Navarro-Marí and SeqCOVID-SPAIN consortium; Sara Sanbonmatsu Gámez |
| EPI_ISL_2886021, EPI_ISL_2886028 | Hospital Universitario de Guadalajara | SeqCOVID-SPAIN consortium/IBV(CSIC) | Alejandro González Praetorius y Mariela Martínez Ramírez and SeqCOVID-SPAIN consortium |
| EPI_ISL_5195508, EPI_ISL_5195510, EPI_ISL_5195531 | Hospital da Baleia | Instituto René Rachou / Fiocruz Minas | Alana Oliveira; Anna Salim; Camila Corsini; Daniel Miranda; Gabriel Fernandes; Mozar de Castro; Nathalie Almeida; Pedro Alves; Priscilla Filgueiras; Rafaella Fortini; Raphael Silva; Raquel Vilela; Rubens do Monte Neto; Sarah Gomes; Thaís Silva; Wander Jeremias |
| EPI_ISL_6513725 | Hospital da Clinicas - UNICAMP | Laboratorio de Estudos de Virus Emergentes | Bruno B. Benites; Daniel A. Toledo-Teixeira; Dewton Moraes Vasconcelos; Ester C. Sabino; Fabiana Granja; Fernando R. Spilki; Janine Schincariol Sabino; José Luiz Proenca-Modena; Julia Forato; Lia Furlaneto Marega; Luciana Souto Mofatto; Maria Marluce dos Santos Vilela; Mariene Ribeiro Amorim; Nuno R. Faria; Raquel Silveira Bello Stucchi; William M. de Souza |
| EPI_ISL_3023329, EPI_ISL_3023330 | Hospital of the University of Pennsylvania Molecular Pathology Lab | Bushman Lab - University of Pennsylvania | Aoife M. Roche; Arupa Ganguly; Ayannah S. Fitzgerald; Brendan Kelly; Jevon Graham-Wooten; John K. Everett; Kyle Rodino; Layla A. Khatib; Mike Feldman; Ronald G. Collman and Frederic Bushan; Samantha A. Whiteside; Scott Sherrill-Mix; Shantan Reddy; Young Hwang |
| EPI_ISL_5072860, EPI_ISL_5072878, EPI_ISL_5072879, EPI_ISL_5072881, EPI_ISL_5072889, EPI_ISL_5072910, EPI_ISL_5072950 | | | |
| see above | Houston Methodist Hospital | Houston Methodist Hospital | Ilya J. Finkelstein; James J. Davis; Jessica Cambric; Jimmy Gollihar; Kristina Reppond; Layne Pruitt; Madison N. Shyer; Marcus Nguyen; Matthew Ojeda Saavedra; Paul A. Christensen; Prasanti Yerramilli; Randall J. Olsen; Robert Olson; Ryan Gadd; S. Wesley Long; Sishir Subedi; and James M. Musser |
| EPI_ISL_2787364 | IA State Hygienic Laboratory | Centers for Disease Control and Prevention Division of Viral Diseases, Pathogen Discovery | Alison Laufer Halpin; Ben L. Rambo-Martin; Clinton R. Paden; Dakota Howard; Darlene Wagner; Dave Wentworth; Dhwani Batra; Jasmine Padilla; Justin Lee; Katie Dillon; Krista Queen; Kristen Knipe; Kristine Lacek; Mark Burroughs; Matthew Schmerer; Mili Sheth; Peter Cook; Sam Shepard; Sarah Nobles; Shoshona Le; Suxiang Tong; Vivien Dugan; Yvette Unoarumhi |
| EPI_ISL_3276639 | ICMT-Apartado | Universidad Nacional de Colombia - Laboratorio Genómico One Health | Andres F. Cardona-Rios; Carlos Franco-Muñoz; Carolina Muñoz-Arango; Celeny Ortiz; Daniel O. Maldonado-Perez; Diego A. Álvarez-Díaz; Hector Alejandro Ruiz-Moreno; Idabely Betancur Ortiz; Jorge E. Osorio; Juan P. Hernandez-Ortiz; Karl A Ciuoderis; Katherine Laiton-Donato; Laura Silvana Perez; Lina M. Hurtado; Marcela Mercado-Reyes; Maria Angélica Maya; Maria Stella López; Rita Almanza Payares; Sandra Ines Cano; Simón Villegas Velásquez |
| EPI_ISL_2840317, EPI_ISL_2840320, EPI_ISL_2987054 | ID Bureau of Laboratories | Centers for Disease Control and Prevention Division of Viral Diseases, Pathogen Discovery | Alison Laufer Halpin; Ben L. Rambo-Martin; Clinton R. Paden; Dakota Howard; Darlene Wagner; Dave Wentworth; Dhwani Batra; Jasmine Padilla; Justin Lee; Katie Dillon; Krista Queen; Kristen Knipe; Kristine Lacek; Mark Burroughs; Matthew Schmerer; Mili Sheth; Peter Cook; Sam Shepard; Sarah Nobles; Shoshona Le; Suxiang Tong; Vivien Dugan; Yvette Unoarumhi |
| EPI_ISL_2672096, EPI_ISL_2672099, EPI_ISL_2672108, EPI_ISL_2688187, EPI_ISL_2688230, EPI_ISL_2725059, EPI_ISL_2725060 | | | |
| see above | IN State Department of Health Laboratory Services | IN State Department of Health Laboratory Services | Brian Pope; Cassandra Campion; Jamie Yeadon; Kyle Brownlee; Lixia Liu; Mark Glazier; Melissa Hindenlang |
| EPI_ISL_2665397 | INMI Lazzaro Spallanzani IRCCS | National Institute for Infectious Diseases (INMI) L. Spallanzani I.R.C.C.S | A Di Caro; B Bartolini; CEM Gruber; E Giombini; F Messina; F Santini; G Bonfiglio; M Rueca; MR Capobianchi; O Butera |
| EPI_ISL_5646498, EPI_ISL_5646519, EPI_ISL_5646524, EPI_ISL_5646542, EPI_ISL_5646549, EPI_ISL_5646557, EPI_ISL_5646569, EPI_ISL_5646571, EPI_ISL_5646580, EPI_ISL_5646583, EPI_ISL_5646590, EPI_ISL_5646598, EPI_ISL_5646603, EPI_ISL_5646604, EPI_ISL_5646608, EPI_ISL_5646618, EPI_ISL_5646621, EPI_ISL_5646628, EPI_ISL_5646629, EPI_ISL_5646632, EPI_ISL_5646634, EPI_ISL_5646635, EPI_ISL_5646639, EPI_ISL_5646641, EPI_ISL_5646642, EPI_ISL_5646648, EPI_ISL_5646651, EPI_ISL_5646652, EPI_ISL_5646653, EPI_ISL_5646663, EPI_ISL_5646664, EPI_ISL_5646671, EPI_ISL_5646682, EPI_ISL_5646683, EPI_ISL_5646684, EPI_ISL_5646685, EPI_ISL_5646695, EPI_ISL_5646703, EPI_ISL_5646704, EPI_ISL_5646713, EPI_ISL_5646714, EPI_ISL_5646723, EPI_ISL_5646724, EPI_ISL_5646733, EPI_ISL_5646734, EPI_ISL_5646740, EPI_ISL_5646741, EPI_ISL_5646742, EPI_ISL_5646749, EPI_ISL_5646750, EPI_ISL_5646757, EPI_ISL_5646764, EPI_ISL_5646765, EPI_ISL_5646766, EPI_ISL_5646773, EPI_ISL_5646778, EPI_ISL_5646779, EPI_ISL_5646796, EPI_ISL_5646797, EPI_ISL_5646804, EPI_ISL_5646806, EPI_ISL_5646811, EPI_ISL_5646813, EPI_ISL_5646827, EPI_ISL_5646836, EPI_ISL_5646839, EPI_ISL_5646855, EPI_ISL_5647014, EPI_ISL_5647018, EPI_ISL_5647022, EPI_ISL_5647023, EPI_ISL_5647024, EPI_ISL_5647027, EPI_ISL_5647028, EPI_ISL_5647029, EPI_ISL_5647030, EPI_ISL_5647052, EPI_ISL_5647057, EPI_ISL_5647058, EPI_ISL_5647074, EPI_ISL_5647075, EPI_ISL_5647085, EPI_ISL_5647092, EPI_ISL_5647321, EPI_ISL_5647430, EPI_ISL_5647437, EPI_ISL_5647468, EPI_ISL_5647509, EPI_ISL_5647530, EPI_ISL_5647593, EPI_ISL_5647629 | | | |
| see above | INSIDE DIAGNOSTICOS | Instituto Butantan | Antonio Jorge Martins; Claudia Renata dos Santos Barros; David Schlesinger; Debora Botequio Moretti; Dimas Tadeu Covas; Elaine Cristina Marqueze; Elaine Vieira Santos; Evandra Strazza Rodrigues; Heidge Fukumasu; Jayme Augusto de Souza-Neto; José Salvatore Leister Patané; Luiz Alcantara; Luiz Lehmann Coutinho; Maria Carolina Elias; Maurício Lacerda Nogueira; Rafael dos Santos Bezerra; Raul Machado Neto; Rejane Maria Tommasini Grotto; Ricardo Haddad; Sandra Coccuzzo Sampaio Vessoni; Simone Kashima; Svetoslav Nanev Slavov; Vincent Louis Viala |
| EPI_ISL_2893556, EPI_ISL_2893596, EPI_ISL_2893599, EPI_ISL_2893600, EPI_ISL_2893601, EPI_ISL_2893604, EPI_ISL_2897544 | | | |
| see above | INSPI-CRN DE INFLUENZA Y OTROS VIRUS RESPIRATORIOS | NIC-INSPI | Alfredo Bruno; Domenica de Mora.; Jimmy Garcés; Johanna Laines; Lizbeth Patiño; Manuel Gonzalez; Maritza Olmedo; Michelle Páez |
| EPI_ISL_2604875, EPI_ISL_2604876, EPI_ISL_2604877 | INTERLAB | Omics Sciences Laboratory | Darlyn Amaya; Derly Andrade Molina; Gabriel Morey León; Juan Carlos Fernández Cadena; Rubén Armas González |
| EPI_ISL_2953137 | IPF (Institut fr Pathologie/Landeskrankenhaus Feldkirch) | Elling and Cochella laboratories, IMBA/IMP | Alexander Stark; Ezgi Oezkan; Luisa Cochella and Ulrich Elling; Marcus Strobl; Olga Frank; Ramesh Yelagandula; Tanino Albanese |
| EPI_ISL_2774175, EPI_ISL_2774176, EPI_ISL_2774179, EPI_ISL_2774182, EPI_ISL_2774188, EPI_ISL_2774190, EPI_ISL_2774194, EPI_ISL_2774199, EPI_ISL_2774200, EPI_ISL_2774201, EPI_ISL_2774202, EPI_ISL_2774207, EPI_ISL_2774208, EPI_ISL_2774209, EPI_ISL_2774210, EPI_ISL_2923024, EPI_ISL_2923025 | | | |
| see above | IRCCS San Gallicano Dermatological Institute | IRCCS Regina Elena National Cancer Institute | Aldo Morrone; Alice Massacci; Andrea Cazzani; Eleonora Sperandio; Elisabetta Trento; Fabrizio Ensoli; Francesca De Nicola; Francesca Maione; Francesca Sivori; Francisco Obregon; Frauke Goeman; Fulvia Fraticelli; Fulvia Pimpinelli; Gennaro Ciliberto; Giovanni Blandino; Giulia Orlandi; Grazia Prignano; Ilaria Cavallo; Ludovica Ciuffreda; Martina Pontone; Matteo Pallocca; Maurizio Fanciulli; Sabrina Strano; Sara Donzelli; Sara Petrolo; Serena Salvo; Valentina Ricca; di Domenico Enea Gino |
| EPI_ISL_2726940, EPI_ISL_2726941 | ISDH | Quantigen Biosciences | Paul J. Childress |
| EPI_ISL_3066924, EPI_ISL_3066972 | Idaho Bureau of Laboratories | Center for Global Health, University of New Mexico Health Sciences Center | Christopher Ball; Darrell Dinwiddie; Daryl Domman; Kurt Schwalm; Matthew Burns; Robert Voermans; Valerie Morley |

| | | | |
|---|---|---|---|
| EPI_ISL_2800749, EPI_ISL_2800761, EPI_ISL_2800764, EPI_ISL_2800828, EPI_ISL_2800846, EPI_ISL_2800851 | Illinois Department of Public Health | Illinois Department of Public Health - Chicago Lab | Ira Heimler; Joel Price; Vineet K. Dhiman |
| EPI_ISL_2598241, EPI_ISL_2598244, EPI_ISL_2598245, EPI_ISL_2774312, EPI_ISL_2774316, EPI_ISL_2774317, EPI_ISL_2774320, EPI_ISL_2774333, EPI_ISL_2774334, EPI_ISL_2774346, EPI_ISL_2774353, EPI_ISL_2774367, EPI_ISL_2774369, EPI_ISL_2774370 | | | |
| see above | Illinois Department of Public Health - Springfield Lab | Illinois Department of Public Health - Springfield Lab | Bryan Sim; Gordon McCall |
| EPI_ISL_3015619, EPI_ISL_3015626, EPI_ISL_3015628, EPI_ISL_3015629 | Incyte Diagnostics Spokane | Seattle Flu Study | Amanda Adler; Barry R. Lutz; Benjamin Pelle; Brian Hiatt; Caitlin R. Wolf; Chris D. Frazar; Deborah A. Nickerson; Elisabeth Brandstetter; Erica Ryke; Geoff Melly; Helen Y. Chu; Janet A. Englund; Jay Shendure; Jover Lee; Kairsten Fay; Kirsten Lacombe; Lea M. Starita; Mark J. Rieder; Matthew Richardson; Matthew Thompson; Melissa Truong; Michael Boeckh; Michael Famulare; Misja Ilcisin; Peter D. Han; Philip Dykema; Romesh Gautom; Scott Lindquist; Thomas R. Sibley; Trevor Bedford |
| EPI_ISL_2686856, EPI_ISL_2686857, EPI_ISL_2686871, EPI_ISL_2686883, EPI_ISL_2786635, EPI_ISL_2786653, EPI_ISL_2786659, EPI_ISL_2786668, EPI_ISL_2786679, EPI_ISL_2786690, EPI_ISL_2786699, EPI_ISL_4383691 | | | |
| see above | Infinity Biologix | Centers for Disease Control and Prevention Division of Viral Diseases, Pathogen Discovery | Adrian Paskey; Benjamin Rambo-Martin; Chirayu Goswami; Christian Bixby; Christopher Gulvick; Clinton Paden; Clinton R. Paden; Dakota Howard; Darlene Wagner; Dhwani Batra; Duncan MacCannell; Erisa Sula; Jason Caravas; Jonathan Schultz; Kara Moser; Kristine Lacek; Matthew Schmerer; Peter Cook; Peter W. Cook; Robin Grimwood; Russ Hager; Scott Sammons; Shatavia Morrison; Tymeckia Kendall; Victoria Caban Figueroa; Yihe Wang; Yvette Unoarumhi |
| EPI_ISL_2708978, EPI_ISL_2708979 | Institut de Virologie du CHU de Strasbourg | Institut de Virologie du CHU de Strasbourg | Fafi-Kremer Samira; Gallais Floriane; Gantner Pierre; Laugel Elodie; Solis Morgane; Velay Aurélie; Wendling Marie-Josée |
| EPI_ISL_3316202, EPI_ISL_3316242 | Instituto Adolfo Lutz - Regional de Bauru | Instituto Adolfo Lutz, Interdiciplinary Procedures Center, Strategic Laboratory | Caio Vinicius Dias Lopes; Claudia Regina Gonçalves; Claudio Tavares Sacchi; Karoline Rodrigues Campos; Leonardo Tadeu de Araujo; Marlon Benedito Nascomento Santos |
| EPI_ISL_2919263, EPI_ISL_3259691, EPI_ISL_3259692, EPI_ISL_3259693, EPI_ISL_3259694, EPI_ISL_3259695, EPI_ISL_3259697, EPI_ISL_3259698 | | | |
| see above | Instituto Adolfo Lutz - Regional de Campinas | Instituto Adolfo Lutz, Interdiciplinary Procedures Center, Strategic Laboratory | Caio Vinicius Dias Lopes; Claudia Regina Gonçalves; Claudio Tavares Sacchi; Erica Valessa Ramos Gomes; Karoline Rodrigues Campos; Leonardo Tadeu de Araujo; Marlon Benedito Nascomento Santos |
| EPI_ISL_3717853, EPI_ISL_3717854, EPI_ISL_3717855, EPI_ISL_3717856, EPI_ISL_3717857, EPI_ISL_3717858, EPI_ISL_3717859, EPI_ISL_3717860, EPI_ISL_3717861 | | | |
| see above | Instituto Adolfo Lutz - Regional de Marilia | Instituto Adolfo Lutz, Interdiciplinary Procedures Center, Strategic Laboratory | Caio Vinicius Dias Lopes; Claudia Regina Gonçalves; Claudio Tavares Sacchi; Karoline Rodrigues Campos; Leonardo Tadeu de Araujo; Marlon Benedito Nascimento Santos |
| EPI_ISL_3671866, EPI_ISL_3671867, EPI_ISL_3671868 | Instituto Adolfo Lutz - Regional de Presidente Prudente | Instituto Adolfo Lutz, Interdisciplinary Procedures Center, Strategic Laboratory | Caio Vinicius Dias Lopes; Claudia Regina Gonçalves; Claudio Tavares Sacchi; Karoline Rodrigues Campos; Leonardo Tadeu de Araujo; Marlon Benedito Nascomento Santos |
| EPI_ISL_5640218 | Instituto Adolfo Lutz - Regional de Rio Claro | Instituto Adolfo Lutz, Interdiciplinary Procedures Center, Strategic Laboratory | Caio Vinicius Dias Lopes; Claudia Regina Gonçalves; Claudio Tavares Sacchi; Karoline Rodrigues Campos; Leonardo Tadeu de Araujo; Marlon Benedito Nascimento Santos |
| EPI_ISL_3671879, EPI_ISL_3671880, EPI_ISL_3671881, EPI_ISL_3671882, EPI_ISL_3671883, EPI_ISL_3671884, EPI_ISL_3671885, EPI_ISL_3671886, EPI_ISL_3671887, EPI_ISL_3671888, EPI_ISL_3671889 | | | |
| see above | Instituto Adolfo Lutz - Regional de Santos | Instituto Adolfo Lutz, Interdiciplinary Procedures Center, Strategic Laboratory | Caio Vinicius Dias Lopes; Claudia Regina Gonçalves; Claudio Tavares Sacchi; Karoline Rodrigues Campos; Leonardo Tadeu de Araujo; Marlon Benedito Nascomento Santos |
| EPI_ISL_3259719, EPI_ISL_3259721, EPI_ISL_3259722 | Instituto Adolfo Lutz - Regional de Sao Jose do Rio Preto | Instituto Adolfo Lutz, Interdiciplinary Procedures Center, Strategic Laboratory | Caio Vinicius Dias Lopes; Claudia Regina Gonçalves; Claudio Tavares Sacchi; Erica Valessa Ramos Gomes; Karoline Rodrigues Campos; Leonardo Tadeu de Araujo; Marlon Benedito Nascomento Santos |
| EPI_ISL_3259677, EPI_ISL_3259678, EPI_ISL_3259679, EPI_ISL_3259680, EPI_ISL_3259681, EPI_ISL_3316161, EPI_ISL_3316165, EPI_ISL_3316199, EPI_ISL_3316237, EPI_ISL_3946605, EPI_ISL_3946607, EPI_ISL_3946608 | | | |
| see above | Instituto Adolfo Lutz - Regional de Sorocaba | Instituto Adolfo Lutz, Interdiciplinary Procedures Center, Strategic Laboratory | Caio Vinicius Dias Lopes; Claudia Regina Gonçalves; Claudio Tavares Sacchi; Erica Valessa Ramos Gomes; Karoline Rodrigues Campos; Leonardo Tadeu de Araujo; Marlon Benedito Nascomento Santos |
| EPI_ISL_2919233, EPI_ISL_2919234, EPI_ISL_2919268, EPI_ISL_2919269, EPI_ISL_2919270, EPI_ISL_2919271, EPI_ISL_2919272, EPI_ISL_2919273, EPI_ISL_2919274, EPI_ISL_2919275, EPI_ISL_2919276, EPI_ISL_2919277, EPI_ISL_2919278, EPI_ISL_2919279, EPI_ISL_2919280, EPI_ISL_2919281, EPI_ISL_2919282, EPI_ISL_2919283, EPI_ISL_2919284, EPI_ISL_2919285, EPI_ISL_2919286, EPI_ISL_2919290, EPI_ISL_2919291, EPI_ISL_2919292, EPI_ISL_2919293, EPI_ISL_3545803, EPI_ISL_4299349, EPI_ISL_4299350 | | | |
| see above | Instituto Adolfo Lutz Central | Instituto Adolfo Lutz, Interdiciplinary Procedures Center, Strategic Laboratory | Caio Vinicius Dias Lopes; Claudia Regina Gonçalves; Claudio Tavares Sacchi; Erica Valessa Ramos Gomes; Karoline Rodrigues Campos; Leonardo Tadeu de Araujo; Marlon Benedito Nascimento Santos; Marlon Benedito Nascomento Santos |
| EPI_ISL_2777223, EPI_ISL_2777227 | Instituto Nacional de Cancerología | Centro de Investigación en Enfermedades Infecciosas (CIENI), Instituto Nacional de Enfermedades Respiratorias (INER) | Alejandra Garcia-Gasca; Alejandra Hernández-Terán; Alejandro Sanchez-Flores; Alfredo Herrera-Estrella; Alicia Ocaña-Mondragón; Andreu Comas-García; Angel Gustavo Salas-Lais; Antonio Loza Román; Bernardo Martínez-Miguel; Blanca Taboada; Brenda Irasema Maldonado-Meza; Bruno Gomez-Gil; Carla Ivón Herrera-Najera; Carlos F. Arias; Celia Boukadida; Clara Esperanza Santacruz-Tinoco; Concepción Grajales-Muñiz; Consorcio Mexicano de Vigilancia Genómica (CoViGen-Mex). Authors (in alphabetical order): Julio Elias Alvarado-Yaah; Cristóbal Cháidez-Quiróz; Célida Duque Molina; Célida Martínez- Rodríguez; Daniel Fregoso-Rueda; Daniel Lira Morales; Eduardo Becerril-Vargas; Fernando Fontove-Herrera; Fidencio Mejía-Nepomuceno; Francisco Pulido; Gloria Elena Espinosa-Ayala; Gloria María Molina-Salinas; Gloria Vazquez; Hector Esteban Paz-Juárez; Hector Montoya-Fuentes; Helen Haydee Fernanda Ramírez-Plascencia; Irvin González-López; Jean Pierre González; Joel Armando Vázquez-Pérez.; Jorge Salas-Hernández; José Antonio Enciso-Moreno; José Arturo Martínez-Orozco; José Esteban Muñoz-Medina; José de Jesús Nuñez-Contreras; Juan Bautista Chale-Dzul; Julissa Enciso-Ibarra; Kathia Elizabeth Tapia-Diaz; Luis Alberto Ochoa-Carrera; Margarita Matías-Florentino; Mario Mújica-Sánchez; Marissa Pérez-García; María Guadalupe Santiago-Mauricio; María Guadalupe de Jesús Mireles-Rivera; Nelly Sélem-Mojica; Pavel Isa; Ricardo Ciria Merce; Ricardo Grande; Rosa María Gutierrez Rios; Santiago Ávila-Ríos; Selene Zárate; Susana Lopez; Victor Eduardo Garcia-Arias; Víctor Hugo Borja-Aburto |
| EPI_ISL_3983492 | Instituto Nacional de Cardiología (INC) | Centro de Investigación en Enfermedades Infecciosas (CIENI), Instituto Nacional de Enfermedades Respiratorias (INER) | Alejandra García-Gasca; Alejandra Hernández-Terán; Alejandro Sánchez-Flores; Alfredo Herrera-Estrella; Alicia Ocaña-Mondragón; Andreu Comas-García; Angel Gustavo Salas-Lais; Antonio Loza Román; Bernardo Martínez-Miguel; Blanca Taboada; Brenda Irasema Maldonado-Meza; Bruno Gómez-Gil; Carla Ivón Herrera-Najera; Carlos F. Arias; Celia Boukadida; Clara Esperanza Santacruz-Tinoco; Concepción Grajales-Muñiz; Consorcio Mexicano de Vigilancia Genómica (CoViGen-Mex). Authors (in alphabetical order): Julio Elias Alvarado-Yaah; Cristóbal Cháidez-Quiróz; Célida Duque Molina; Célida Martínez-Rodríguez; Daniel Fregoso-Rueda; Daniel Lira Morales; Eduardo Becerril-Vargas; Eduardo Rivera-Martínez; Fernando Fontove-Herrera; Fidencio Mejía-Nepomuceno; Francisco Pulido; Gabriel Chavira-Trujillo; Gloria Elena Espinosa-Ayala; Gloria María Molina-Salinas; Gloria Vazquez; Hector Montoya-Fuentes; Helen Haydee Fernanda Ramírez-Plascencia; Irvin González-López; Jean Pierre González; Jesús Hernández; Joel Armando Vázquez-Pérez).; Jorge Salas-Hernández; José Antonio Enciso-Moreno; José Arturo Martínez-Orozco; José Esteban Muñoz-Medina; José de Jesús Nuñez-Contreras; Juan Bautista Chale-Dzul; Julissa Enciso-Ibarra; Kathia Elizabeth Tapia-Diaz; Luis Alberto Ochoa-Carrera; Margarita Matías-Florentino; Mario Mújica-Sánchez; Marissa Perez-Garcia; María Eugenia Jiménez-Corona; María Guadalupe Santiago-Mauricio; María Guadalupe de Jesús Mireles-Rivera; Nelly Sélem-Mojica; Pavel Isa; Ricardo Ciria Merce; Ricardo Grande; Rosa María Gutiérrez Rios; Rosario Vazquez-Larios; Santiago Ávila-Ríos; Selene Zárate; Susana Lopez; Verónica Mata-Haro; Victor Eduardo García-Arias; Víctor Hugo Borja-Aburto |
| EPI_ISL_2777220 | Instituto Nacional de Enfermedades Respiratorias (INER) | Centro de Investigación en Enfermedades Infecciosas (CIENI), Instituto Nacional de Enfermedades Respiratorias (INER) | Alejandra Garcia-Gasca; Alejandra Hernández-Terán; Alejandro Sanchez-Flores; Alfredo Herrera-Estrella; Alicia Ocaña-Mondragón; Andreu Comas-García; Angel Gustavo Salas-Lais; Antonio Loza Román; Bernardo Martínez-Miguel; Blanca Taboada; Brenda Irasema Maldonado-Meza; Bruno Gomez-Gil; Carla Ivón Herrera-Najera; Carlos F. Arias; Celia Boukadida; Clara Esperanza Santacruz-Tinoco; Concepción Grajales-Muñiz; Consorcio Mexicano de Vigilancia Genómica (CoViGen-Mex). Authors (in alphabetical order): Julio Elias Alvarado-Yaah; Cristóbal Cháidez-Quiróz; Célida Duque Molina; Célida Martínez- Rodríguez; Daniel Fregoso-Rueda; Daniel Lira Morales; Eduardo Becerril-Vargas; Fernando Fontove-Herrera; Fidencio Mejía-Nepomuceno; Francisco Pulido; Gloria Elena Espinosa-Ayala; Gloria María Molina-Salinas; Gloria Vazquez; Hector Esteban Paz-Juárez; Hector Montoya-Fuentes; Helen Haydee Fernanda Ramírez-Plascencia; Irvin González-López; Jean Pierre González; Joel Armando Vázquez-Pérez.; Jorge Salas-Hernández; José Antonio Enciso-Moreno; José Arturo Martínez-Orozco; José Esteban Muñoz-Medina; José de Jesús Nuñez-Contreras; Juan Bautista Chale-Dzul; Julissa Enciso-Ibarra; Kathia Elizabeth Tapia-Diaz; Luis Alberto Ochoa-Carrera; Margarita Matías-Florentino; Mario Mújica-Sánchez; Marissa Pérez-García; María Guadalupe Santiago-Mauricio; María Guadalupe de Jesús Mireles-Rivera; Nelly Sélem-Mojica; Pavel Isa; Ricardo Ciria Merce; Ricardo Grande; Rosa María Gutierrez Rios; Santiago Ávila-Ríos; Selene Zárate; Susana Lopez; Victor Eduardo Garcia-Arias; Víctor Hugo Borja-Aburto |
| EPI_ISL_2844057, EPI_ISL_2844058, EPI_ISL_2844059, EPI_ISL_2844060, EPI_ISL_2844061, EPI_ISL_2844062, EPI_ISL_2844063, EPI_ISL_2844064, EPI_ISL_2844065, EPI_ISL_2844066, EPI_ISL_2844067, EPI_ISL_2844068, EPI_ISL_2844069, EPI_ISL_2844070, EPI_ISL_2844071, EPI_ISL_2844072, EPI_ISL_2844073, EPI_ISL_2844074, EPI_ISL_2844075, EPI_ISL_2844076, EPI_ISL_2844077, EPI_ISL_2844078, EPI_ISL_2844079, EPI_ISL_2844080, EPI_ISL_2844081, EPI_ISL_2844082, EPI_ISL_2844083, EPI_ISL_2844084, EPI_ISL_2844085, EPI_ISL_2844086, EPI_ISL_2844087, EPI_ISL_2844088, EPI_ISL_2844089, EPI_ISL_2844090, EPI_ISL_2844091, EPI_ISL_2844092, EPI_ISL_2844093, EPI_ISL_2844095, EPI_ISL_2844096, EPI_ISL_2844097, EPI_ISL_2844098, EPI_ISL_2844099, EPI_ISL_2844100, EPI_ISL_2844101, EPI_ISL_2844102, EPI_ISL_2844103, EPI_ISL_2844104, EPI_ISL_2844105, EPI_ISL_2844106, EPI_ISL_2844107, EPI_ISL_2844108, EPI_ISL_2844109, EPI_ISL_2844110, EPI_ISL_2844111, EPI_ISL_2844112, EPI_ISL_2844113, EPI_ISL_2844114, EPI_ISL_2844115, EPI_ISL_2844116, EPI_ISL_2844117, EPI_ISL_2844118, EPI_ISL_2844119, EPI_ISL_2844120, EPI_ISL_2844121, EPI_ISL_2844122, EPI_ISL_2844123, EPI_ISL_2844124, EPI_ISL_2844125, EPI_ISL_2844126, EPI_ISL_2844127, EPI_ISL_2844128, EPI_ISL_2844129, EPI_ISL_2844130, EPI_ISL_2844131, EPI_ISL_2844132, EPI_ISL_2844133, EPI_ISL_2844134, EPI_ISL_2844135, EPI_ISL_2844136, EPI_ISL_2844137, EPI_ISL_2844138, EPI_ISL_2844139, EPI_ISL_2844140, EPI_ISL_2844141, EPI_ISL_2844142, EPI_ISL_2844143, EPI_ISL_2844144, EPI_ISL_2844145, EPI_ISL_2844146, EPI_ISL_2844147, EPI_ISL_2844148, EPI_ISL_2844149, EPI_ISL_2844150, EPI_ISL_2844151, EPI_ISL_2844152, EPI_ISL_2844153, EPI_ISL_2844154, EPI_ISL_2844155, EPI_ISL_2844156, EPI_ISL_2844157, EPI_ISL_2844158, EPI_ISL_2844159, EPI_ISL_2844160, EPI_ISL_2844161, EPI_ISL_2844162, EPI_ISL_2844163, EPI_ISL_2844164, EPI_ISL_2844165, EPI_ISL_2844166, EPI_ISL_2844167, EPI_ISL_2844168, EPI_ISL_2844169, EPI_ISL_2844170, EPI_ISL_2844172, EPI_ISL_2844173, EPI_ISL_2844174, EPI_ISL_2844175, EPI_ISL_2844176, EPI_ISL_2844177, EPI_ISL_2844178, EPI_ISL_2844179, EPI_ISL_2844180, EPI_ISL_2844181, EPI_ISL_2844182, EPI_ISL_2844183, EPI_ISL_2844184, EPI_ISL_2844185, EPI_ISL_2844186, EPI_ISL_2844187, EPI_ISL_2844188, EPI_ISL_2844189, EPI_ISL_2844190, EPI_ISL_2844191, EPI_ISL_2844192, EPI_ISL_2844193, EPI_ISL_2844194, EPI_ISL_2844195, EPI_ISL_3030615, EPI_ISL_3118891, EPI_ISL_3118894, EPI_ISL_3118899, EPI_ISL_3118901, EPI_ISL_3118902, EPI_ISL_3118905, EPI_ISL_3118924, EPI_ISL_3118936, EPI_ISL_3118937, EPI_ISL_3244565, EPI_ISL_3244569, EPI_ISL_3244570, EPI_ISL_3244571, EPI_ISL_3244572, EPI_ISL_3244573, EPI_ISL_3244574, EPI_ISL_3244575, EPI_ISL_3244576, EPI_ISL_3244578, EPI_ISL_3244579, EPI_ISL_3244580, EPI_ISL_3244581, EPI_ISL_3244582, EPI_ISL_3244583, EPI_ISL_3244584, EPI_ISL_3244585, EPI_ISL_3244586, EPI_ISL_3244587, EPI_ISL_3244588, EPI_ISL_3244589, EPI_ISL_3244590, EPI_ISL_3244591, EPI_ISL_3244592, EPI_ISL_3244593, EPI_ISL_3244594 | | | |
| see above | Instituto de Biotecnologia - UNESP-Botucatu-SP | Instituto de Biotecnologia - UNESP-Botucatu-SP | Cecília Artico Banho; Cíntia Bittar; Fábio Sossai Possebon; Guilherme Campos; Helena Lage Ferreira; Jorge A. Petroli Marchesi; João Pessoa Araújo Jr.; Leila Sabrina Ullmann; Lívia Sacchetto; Maisa C. Pereira Parra; Marília Moraes; Maurício L. Nogueira; Paula Rahal; Paulo Inacio da Costa |
| EPI_ISL_3020245 | Instituto de Medicina Tropical Alexander Von Humboldt, Sede Cusco | Laboratorio de Genómica Microbiana, Universidad Peruana Cayetano Heredia | Alejandra Dávila-Barclay; Diego Cuicapuza; Guillermo Salvatierra; Janet Huancachoque; Luis González; Pablo Tsukayama; Pedro E. Romero; Pool Marcos |
| EPI_ISL_3347102 | Interpath Laboratory | Seattle Flu Study | Amanda Adler; Barry R. Lutz; Benjamin Pelle; Brian Hiatt; Caitlin R. Wolf; Chris D. Frazar; Deborah A. Nickerson; Elisabeth Brandstetter; Erica Ryke; Geoff Melly; Helen Y. Chu; Janet A. Englund; Jay Shendure; Jover Lee; Kairsten Fay; Kirsten Lacombe; Lea M. Starita; Mark J. Rieder; Matthew Richardson; Matthew Thompson; Melissa Truong; Michael Boeckh; Michael Famulare; Misja Ilcisin; Peter D. Han; Philip Dykema; Romesh Gautom; Scott Lindquist; Thomas R. Sibley; Trevor Bedford |
| EPI_ISL_3279318, EPI_ISL_3279332, EPI_ISL_3279359 | Israel Central Virology laboratory | Israel National Consortium for SARS-CoV-2 sequencing | Dana Bar-Ilan; Danit Sofer; Efrat Dahan Bucris; Efrat Glick-Saar; Ella Mendelson; Gideon Rechavi; Michal Mandelboim; Miranda Geva; Neta Zuckerman; Omer Asraf; Omri Nayshool; Oran Erster; Orna Mor |
| EPI_ISL_2641782, EPI_ISL_2641789, EPI_ISL_2641793, EPI_ISL_2641794, EPI_ISL_2641800, EPI_ISL_2641824, EPI_ISL_2641829, EPI_ISL_2641862, EPI_ISL_2641865, EPI_ISL_2641868, EPI_ISL_2641884, EPI_ISL_2641889, EPI_ISL_2641906, EPI_ISL_2641907, EPI_ISL_2641926, EPI_ISL_2641938, EPI_ISL_2642007, EPI_ISL_2642011, EPI_ISL_2642040 | | | |
| see above | Istituto Zooprofilattico Sperimentale del Mezzogiorno | Telethon Institute of Genetics and Medicine (TIGEM) | Antonio Grimaldi Patrizia Annunziata Francesco Panariello Biancamaria Pierri Claudia Tiberio Teresa Giuliano Valentina Bouche Chiara Colantuono Maria Concetta Cuomo Denise Di Concilio Lucio Di Filippo Anna Manfredi Marcello Salvi Antonio Limone Luigi Atripaldi Pellegrino Cerino Andrea Ballabio Davide Cacchiarelli |
| EPI_ISL_2601973, EPI_ISL_2601975, EPI_ISL_2601979 | Jessa | Jessa | Berden et al. on behalf of the Jessa_cmdLab |
| EPI_ISL_2931123 | John H Stroger Jr Hospital of Cook County | RIPHL at Rush University Medical Center | Ah-Young Sung; Felix Araujo Perez; Kevin Kunstman; Laura Furtado; Marieta Hyde; Max Kolton; Sharon Welbel; Stefan Green |

| | | | |
|---|---|---|---|
| EPI_ISL_2886367 | KU Leuven, Rega Institute, Clinical and Epidemiological Virology | KU Leuven, Rega Institute, Clinical and Epidemiological Virology | Bert Vanmechelen; Joan Marti-Carerras; Piet Maes; Tony Wawina-Bokalanga |
| EPI_ISL_3561203, EPI_ISL_3561592, EPI_ISL_3561701, EPI_ISL_3561878, EPI_ISL_3562071, EPI_ISL_3562092, EPI_ISL_3562117, EPI_ISL_3939799, EPI_ISL_3939805, EPI_ISL_3939807, EPI_ISL_3939816, EPI_ISL_3939818, EPI_ISL_3939825, EPI_ISL_3939826, EPI_ISL_3939831, EPI_ISL_3939836, EPI_ISL_3940788, EPI_ISL_3940804, EPI_ISL_3940806, EPI_ISL_3940810, EPI_ISL_3940823, EPI_ISL_3940837, EPI_ISL_3940884, EPI_ISL_3940885, EPI_ISL_3940886, EPI_ISL_3940889, EPI_ISL_3940890, EPI_ISL_3940904, EPI_ISL_3940920, EPI_ISL_3940922, EPI_ISL_3940924, EPI_ISL_3940932, EPI_ISL_3941010, EPI_ISL_3941019, EPI_ISL_3941020, EPI_ISL_3941026, EPI_ISL_3941029, EPI_ISL_3941037, EPI_ISL_3941042, EPI_ISL_3941060, EPI_ISL_3941063, EPI_ISL_3941068, EPI_ISL_4005463, EPI_ISL_4005472, EPI_ISL_4102671, EPI_ISL_4821413, EPI_ISL_4821427 | | | |
| see above | Kaiser Permanente NW Reginal Lab | OHSU MM Lab | Amber Halse; Jeannine Lama; Xuan Qin; Yun Wu |
| EPI_ISL_2933991 | Kaiser Regional | Santa Clara County Public Health Laboratory | Santa Clara County Public Health Department |
| EPI_ISL_2558702, EPI_ISL_2833925, EPI_ISL_2842072 | Kansas Health and Environmental Lab | Kansas Health and Environmental Lab | Ben Olsen; Jonathan Barnell; Katherine Wiggins; Mike Grose; and Phil Adam |
| EPI_ISL_4459181, EPI_ISL_4459183, EPI_ISL_4459186, EPI_ISL_4459188 | Kentucky State Public Health Lab | Kentucky State Public Health Lab | Anjayya Tummala; Chitralipi Kattunga; Joshua Tobias; Karim George; Matthew Johnson; Rachel Zinner; Rhonda Lucas; Vaneet Arora; William Grooms |
| EPI_ISL_5646588, EPI_ISL_5646646, EPI_ISL_5646661, EPI_ISL_5646783 | LABORATORIO DE FRANCA | Instituto Butantan | Antonio Jorge Martins; Claudia Renata dos Santos Barros; David Schlesinger; Debora Botequio Moretti; Dimas Tadeu Covas; Elaine Cristina Marqueze; Elaine Vieira Santos; Evandra Strazza Rodrigues; Heidge Fukumasu; Jayme Augusto de Souza-Neto; José Salvatore Leister Patané; Luiz Alcantara; Luiz Lehmann Coutinho; Maria Carolina Elias; Maurício Lacerda Nogueira; Rafael dos Santos Bezerra; Raul Machado Neto; Rejane Maria Tommasini Grotto; Ricardo Haddad; Sandra Coccuzzo Sampaio Vessoni; Simone Kashima; Svetoslav Nanev Slavov; Vincent Louis Viala |
| EPI_ISL_4743415 | LABORATORIO ECHAVARRIA | Laboratorio Departamental de Salud Publica de Antioquia | Andres F. Cardona-Rios; Cristian Arbey Velarde Hoyos; Gloria Isabel Escobar; Idabely Betancur Ortiz; Juan P. Hernandez-Ortiz; Maria Stella López |
| EPI_ISL_5646643 | LABORATORIO MUNICIPAL DE ANALISES CLINICAS DE ITANHAEM | Instituto Butantan | Antonio Jorge Martins; Claudia Renata dos Santos Barros; David Schlesinger; Debora Botequio Moretti; Dimas Tadeu Covas; Elaine Cristina Marqueze; Elaine Vieira Santos; Evandra Strazza Rodrigues; Heidge Fukumasu; Jayme Augusto de Souza-Neto; José Salvatore Leister Patané; Luiz Alcantara; Luiz Lehmann Coutinho; Maria Carolina Elias; Maurício Lacerda Nogueira; Rafael dos Santos Bezerra; Raul Machado Neto; Rejane Maria Tommasini Grotto; Ricardo Haddad; Sandra Coccuzzo Sampaio Vessoni; Simone Kashima; Svetoslav Nanev Slavov; Vincent Louis Viala |
| EPI_ISL_5646502, EPI_ISL_5646543, EPI_ISL_5646680, EPI_ISL_5646756, EPI_ISL_5646799, EPI_ISL_5646833, EPI_ISL_5646837, EPI_ISL_5647878 | | | |
| see above | LABORATORIO MUNICIPAL DE ANALISES CLINICAS DE RIO CLARO | Instituto Butantan | Antonio Jorge Martins; Claudia Renata dos Santos Barros; David Schlesinger; Debora Botequio Moretti; Dimas Tadeu Covas; Elaine Cristina Marqueze; Elaine Vieira Santos; Evandra Strazza Rodrigues; Heidge Fukumasu; Jayme Augusto de Souza-Neto; José Salvatore Leister Patané; Luiz Alcantara; Luiz Lehmann Coutinho; Maria Carolina Elias; Maurício Lacerda Nogueira; Rafael dos Santos Bezerra; Raul Machado Neto; Rejane Maria Tommasini Grotto; Ricardo Haddad; Sandra Coccuzzo Sampaio Vessoni; Simone Kashima; Svetoslav Nanev Slavov; Vincent Louis Viala |
| EPI_ISL_5646536, EPI_ISL_5646655, EPI_ISL_5646670, EPI_ISL_5646689, EPI_ISL_5646692, EPI_ISL_5646729, EPI_ISL_5647595, EPI_ISL_5647609, EPI_ISL_5647625 | | | |
| see above | LABORATORIO MUNICIPAL DE PIRACICABA | Instituto Butantan | Antonio Jorge Martins; Claudia Renata dos Santos Barros; David Schlesinger; Debora Botequio Moretti; Dimas Tadeu Covas; Elaine Cristina Marqueze; Elaine Vieira Santos; Evandra Strazza Rodrigues; Heidge Fukumasu; Jayme Augusto de Souza-Neto; José Salvatore Leister Patané; Luiz Alcantara; Luiz Lehmann Coutinho; Maria Carolina Elias; Maurício Lacerda Nogueira; Rafael dos Santos Bezerra; Raul Machado Neto; Rejane Maria Tommasini Grotto; Ricardo Haddad; Sandra Coccuzzo Sampaio Vessoni; Simone Kashima; Svetoslav Nanev Slavov; Vincent Louis Viala |
| EPI_ISL_4395874 | LABSAUDE | LGBio (Laboratorio de Genetica & Biodiversidade) | Alex Honda Bernardes; Amanda Alves de Melo; Aparecido Divino da Cruz; Cintia Pelegrineti Targueta de Azevedo Brito; Daniela de Melo e Silva; Elisangela de Paula Silveira Lacerda; Juliana Santana de Curcio; Luiz Augusto Pereira; Marc Alexandre Duarte Gigonzac; Mariana Pires de Campos Telles; Ramilla dos Santos Braga; Renata de Oliveira Dias; Rhewter Nunes; Thais Cidália Vieira Gigonzac; Thais Guimarães Castro; Thays Millena Alves Pedroso |
| EPI_ISL_3404856, EPI_ISL_3404857, EPI_ISL_3404858, EPI_ISL_3404859, EPI_ISL_3404861, EPI_ISL_3404862, EPI_ISL_3404863, EPI_ISL_3404864, EPI_ISL_3404865, EPI_ISL_3404866, EPI_ISL_3404867, EPI_ISL_3404868, EPI_ISL_3404869, EPI_ISL_3404870, EPI_ISL_3404871, EPI_ISL_3404872, EPI_ISL_3404873, EPI_ISL_3404874, EPI_ISL_3404875, EPI_ISL_3404876, EPI_ISL_3404877, EPI_ISL_3404878 | | | |
| see above | LACEN | LGBio (Laboratorio de Genetica & Biodiversidade) | Alex Honda Bernardes; Amanda Alves de Melo; Aparecido Divino da Cruz; Cintia Pelegrineti Targueta de Azevedo Brito; Daniela de Melo e Silva; Elisangela de Paula Silveira Lacerda; Juliana Santana de Curcio; Luiz Augusto Pereira; Marc Alexandre Duarte Gigonzac; Mariana Pires de Campos Telles; Ramilla dos Santos Braga; Renata de Oliveira Dias; Rhewter Nunes; Thais Cidália Vieira Gigonzac; Thais Guimarães Castro; Thays Millena Alves Pedroso |
| EPI_ISL_6721443, EPI_ISL_6721799 | LACEN - Laboratório Central de Saúde Pública de Pernambuco | Evandro Chagas Institute | Delana Andreza Melo Bezerra; Dielle Monteiro Teixeira; Jedson Ferreira Cardoso; Jessylene de Almeida Ferreira; Kenny da Costa Pinheiro; Luana Soares Barbagelata; Luana da Silva Soares; Mirleide Cordeiro dos Santos; Patrícia dos Santos Lobo; Rayssa Layna da Silva Bedran; Sandro Patroca da Silva |
| EPI_ISL_6721679, EPI_ISL_6721710 | LACEN - Laboratório Central de Saúde Pública do Amazonas | Evandro Chagas Institute | Delana Andreza Melo Bezerra; Dielle Monteiro Teixeira; Jedson Ferreira Cardoso; Jessylene de Almeida Ferreira; Kenny da Costa Pinheiro; Luana Soares Barbagelata; Luana da Silva Soares; Mirleide Cordeiro dos Santos; Patrícia dos Santos Lobo; Rayssa Layna da Silva Bedran; Sandro Patroca da Silva |
| EPI_ISL_6721376 | LACEN - Laboratório Central de Saúde Pública do Rio Grande do Norte | Evandro Chagas Institute | Delana Andreza Melo Bezerra; Dielle Monteiro Teixeira; Jedson Ferreira Cardoso; Jessylene de Almeida Ferreira; Kenny da Costa Pinheiro; Luana Soares Barbagelata; Luana da Silva Soares; Mirleide Cordeiro dos Santos; Patrícia dos Santos Lobo; Rayssa Layna da Silva Bedran; Sandro Patroca da Silva |
| EPI_ISL_6840913, EPI_ISL_6840917, EPI_ISL_6840926, EPI_ISL_6840934, EPI_ISL_6840944 | LACEN do Estado de Rondonia | Instituto Adolfo Lutz, Interdiciplinary Procedures Center, Strategic Laboratory | Ariadne Ferreira Amarante; Caio Vinicius Dias Lopes; Claudia Regina Gonçalves; Claudio Tavares Sacchi; Karoline Rodrigues Campos; Marlon Benedito Nascimento Santos |
| EPI_ISL_4206971 | LACEN do Estado do Piaui | Instituto Adolfo Lutz, Interdiciplinary Procedures Center, Strategic Laboratory | Caio Vinicius Dias Lopes; Claudia Regina Gonçalves; Claudio Tavares Sacchi; Karoline Rodrigues Campos; Marlon Benedito Nascimento Santos |
| EPI_ISL_4487753 | LACEN/PI | WallauLab on behalf of Fiocruz COVID-19 Genomic Surveillance Network | Adelino Soares Lima Neto; Alexandre Freitas da Silva; Antônio Marinho; Cassia Docena; Constância Flávia Junqueira Ayres; Filipe Zimmer Dezordi; Gabriel Luz Wallau; Gustavo Barbosa de Lima; Hellen de Oliveira Amaral; Jacenir Reis dos Santos Mallet; Joana Carolina Viana Lima; Lais Ceschini Machado; Leandro de Mattos; Lilian Caroliny Amorim Silva; Marcela de Lacerda Valença Queiroz; Marcelo Adriano da Cunha e Silva Vieira; Marcelo Henrique dos Santos Paiva; Matheus Filgueira Bezerra; Sinval Pinto Brandão Filho; Túlio de Lima Campos; Vladimir Costa Silva; Walterlene de Carvalho Goçalves |
| EPI_ISL_5193458, EPI_ISL_5193459, EPI_ISL_5193535, EPI_ISL_5193536, EPI_ISL_5193537, EPI_ISL_5193538, EPI_ISL_5193539, EPI_ISL_5193540, EPI_ISL_5193541, EPI_ISL_5193542, EPI_ISL_5193593, EPI_ISL_5193632, EPI_ISL_5193633 | | | |
| see above | LACEN/RR | Laboratorio de Ecologia de Doencas Transmissiveis na Amazonia, Instituto Leonidas e Maria Deane - Fiocruz Amazonia | André Corado; Cátia Alexandra Ribeiro Meneses; Felipe Naveca; Fernanda Nascimento; George Silva; Ilma de Aguiar Antony; Janis Cassimiro Lopes; Luciana Gonçalves; Maria Júlia Brandão; Matilde Mejía; Pricila Carlos Veloso; Rodrigo Melo Maito; Valdinete Nascimento; Victor Souza |
| EPI_ISL_2629291, EPI_ISL_2629301 | LAM GEN-BIO GRAVANCHES | CNR Virus des Infections Respiratoires - France SUD | Antonin Bal; Bruno Lina; Bruno Simon; Gregory Destras; Gwendolyne Burfin; Hadrien Regue; Laurence Josset; Martine Valette; Quentin Semanas |
| EPI_ISL_3010760, EPI_ISL_3010763, EPI_ISL_3010781, EPI_ISL_3010798, EPI_ISL_3010799, EPI_ISL_3010816, EPI_ISL_3010835, EPI_ISL_3010840, EPI_ISL_3010845, EPI_ISL_3010853, EPI_ISL_3010854, EPI_ISL_3010868, EPI_ISL_3010874, EPI_ISL_3010877, EPI_ISL_3010894, EPI_ISL_3010895, EPI_ISL_3010898, EPI_ISL_3010901, EPI_ISL_3010913, EPI_ISL_3010921, EPI_ISL_3010932, EPI_ISL_3020988, EPI_ISL_3020990, EPI_ISL_3047744, EPI_ISL_3261063, EPI_ISL_3261071, EPI_ISL_3261073, EPI_ISL_3261076, EPI_ISL_3261077, EPI_ISL_3261080, EPI_ISL_3261081, EPI_ISL_3261086, EPI_ISL_3261092, EPI_ISL_3261093, EPI_ISL_3261097, EPI_ISL_3261099, EPI_ISL_3261102, EPI_ISL_3261106, EPI_ISL_3261114, EPI_ISL_3267958, EPI_ISL_3267959, EPI_ISL_3268018, EPI_ISL_3268019, EPI_ISL_3298303 | | | |
| see above | LATE - Laboratório de Técnicas Especiais - Hospital Israelita Albert Einstein | LATE - Laboratório de Técnicas Especiais - Hospital Israelita Albert Einstein | Alexandre Hideaki Takara; Ana Paula Moreira Salles; Anelisie da Silva Santos; Deyvid Amgarten; Erick Gustavo Dorlass; Fernanda de Mello Malta; João Renato Rebello Pinho; Marcio Anunciacao Menezes; Pedro Henrique Sebe Rodrigues; Raquel Riyuzo |
| EPI_ISL_2827783 | LDSP - CAQUETA HOSPITAL MALVINAS | Instituto Nacional de Salud- Dirección de Investigación en Salud Pública | Carlos Franco-Muñoz; Carmen Osorio; Diana Malo; Diego A. Álvarez-Díaz; Diego Andrés Prada; Gerardo Santamaría; Hector Alejandro Ruiz-Moreno; Jhonnatan Reales-González; Jorge Rivera; Juan Camilo Martinez; Julian Naizaque; Katherine Laiton-Donato; Lisseth Pardo; Magdalena Wiesner; Marcela Mercado-Reyes; Maria T. Herrera-Sepúlveda; Marta Lopez Blanco; Martha Lucia Ospina Martinez; Paola Rojas; Sergio Gomez; Sheryll Corchuelo; Ángela Alarcon Cruz |
| EPI_ISL_2827779 | LDSP - CAQUETA HOSPITAL MARIA INMACULADA | Instituto Nacional de Salud- Dirección de Investigación en Salud Pública | Carlos Franco-Muñoz; Carmen Osorio; Diana Malo; Diego A. Álvarez-Díaz; Diego Andrés Prada; Gerardo Santamaría; Hector Alejandro Ruiz-Moreno; Jhonnatan Reales-González; Jorge Rivera; Juan Camilo Martinez; Julian Naizaque; Katherine Laiton-Donato; Lisseth Pardo; Magdalena Wiesner; Marcela Mercado-Reyes; Maria T. Herrera-Sepúlveda; Marta Lopez Blanco; Martha Lucia Ospina Martinez; Paola Rojas; Sergio Gomez; Sheryll Corchuelo; Ángela Alarcon Cruz |
| EPI_ISL_3007486 | LDSP VILLAVICENCIO - CLINICA PRIMAVERA DE VILLAVICENCIO | Instituto Nacional de Salud- Dirección de Investigación en Salud Pública | Carlos Franco-Muñoz; Carmen Osorio; Diana Malo; Diego A. Álvarez-Díaz; Diego Andrés Prada; Gerardo Santamaría; Hector Alejandro Ruiz-Moreno; Jhonnatan Reales-González; Jorge Rivera; Juan Camilo Martinez; Julian Naizaque; Katherine Laiton-Donato; Lisseth Pardo; Magdalena Wiesner; Marcela Mercado-Reyes; Maria T. Herrera-Sepúlveda; Marta Lopez Blanco; Martha Lucia Ospina Martinez; Paola Rojas; Sergio Gomez; Sheryll Corchuelo; Ángela Alarcon Cruz |
| EPI_ISL_4743383 | LDSP de Antioquia | Laboratorio Departamental de Salud Publica de Antioquia | Andres F. Cardona-Rios; Cristian Arbey Velarde Hoyos; Gloria Isabel Escobar; Idabely Betancur Ortiz; Juan P. Hernandez-Ortiz; Maria Stella López |
| EPI_ISL_2779059 | LESP Campeche | Instituto de Diagnostico y Referencia Epidemiologicos (INDRE) | Abril Rodriguez-Maldonado; Ariadna Medina-Benitez; Claudia Wong-Arambula; Ernesto Ramirez-Gonzalez.; Gisela Barrera-Badillo; Irma Lopez-Martinez; Joaquin Quiroz-Mercado; Lucia Hernandez-Rivas; Maribel Gonzalez-Villa; Natividad Cruz-Ortiz; Sergio Rangel-Guerrero; Tatiana Nunez-Garcia; Vanessa Rivero-Arredondo |
| EPI_ISL_2859008, EPI_ISL_2859009, EPI_ISL_2859010, EPI_ISL_2859011 | LESP Chiapas | Instituto de Diagnostico y Referencia Epidemiologicos (INDRE) | Abril Rodriguez-Maldonado; Ariadna Medina-Benitez; Claudia Wong-Arambula; Ernesto Ramirez-Gonzalez.; Gisela Barrera-Badillo; Irma Lopez-Martinez; Joaquin Quiroz-Mercado; Lucia Hernandez-Rivas; Maribel Gonzalez-Villa; Natividad Cruz-Ortiz; Sergio Rangel-Guerrero; Tatiana Nunez-Garcia; Vanessa Rivero-Arredondo |
| EPI_ISL_2937769 | LESP Ciudad de Mexico | Instituto de Diagnostico y Referencia Epidemiologicos (INDRE) | Abril Rodriguez-Maldonado; Ariadna Medina-Benitez; Claudia Wong-Arambula; Ernesto Ramirez-Gonzalez.; Gisela Barrera-Badillo; Irma Lopez-Martinez; Joaquin Quiroz-Mercado; Lucia Hernandez-Rivas; Maribel Gonzalez-Villa; Natividad Cruz-Ortiz; Sergio Rangel-Guerrero; Tatiana Nunez-Garcia; Vanessa Rivero-Arredondo |
| EPI_ISL_2937771 | LESP Estado de Mexico | Instituto de Diagnostico y Referencia Epidemiologicos (INDRE) | Abril Rodriguez-Maldonado; Ariadna Medina-Benitez; Claudia Wong-Arambula; Ernesto Ramirez-Gonzalez.; Gisela Barrera-Badillo; Irma Lopez-Martinez; Joaquin Quiroz-Mercado; Lucia Hernandez-Rivas; Maribel Gonzalez-Villa; Natividad Cruz-Ortiz; Sergio Rangel-Guerrero; Tatiana Nunez-Garcia; Vanessa Rivero-Arredondo |
| EPI_ISL_2779186, EPI_ISL_2779187, EPI_ISL_2779188, EPI_ISL_2779189, EPI_ISL_2779190, EPI_ISL_2779191, EPI_ISL_2779192, EPI_ISL_2779193, EPI_ISL_2800913, EPI_ISL_2800914, EPI_ISL_2800915, EPI_ISL_2800924, EPI_ISL_2858991, EPI_ISL_2858992, EPI_ISL_2858993 | | | |
| see above | LESP Guanajuato | Instituto de Diagnostico y Referencia Epidemiologicos (INDRE) | Abril Rodriguez-Maldonado; Ariadna Medina-Benitez; Claudia Wong-Arambula; Ernesto Ramirez-Gonzalez.; Gisela Barrera-Badillo; Irma Lopez-Martinez; Joaquin Quiroz-Mercado; Lucia Hernandez-Rivas; Maribel Gonzalez-Villa; Natividad Cruz-Ortiz; Sergio Rangel-Guerrero; Tatiana Nunez-Garcia; Vanessa Rivero-Arredondo |
| EPI_ISL_2674682, EPI_ISL_2859012 | LESP Hidalgo | Instituto de Diagnostico y Referencia Epidemiologicos (INDRE) | Abril Rodriguez-Maldonado; Ariadna Medina-Benitez; Claudia Wong-Arambula; Ernesto Ramirez-Gonzalez.; Gisela Barrera-Badillo; Irma Lopez-Martinez; Joaquin Quiroz-Mercado; Lucia Hernandez-Rivas; Maribel Gonzalez-Villa; Natividad Cruz-Ortiz; Sergio Rangel-Guerrero; Tatiana Nunez-Garcia; Vanessa Rivero-Arredondo |
| EPI_ISL_2779175, EPI_ISL_2858994, EPI_ISL_2858995, EPI_ISL_2937757 | LESP Jalisco | Instituto de Diagnostico y Referencia Epidemiologicos (INDRE) | Abril Rodriguez-Maldonado; Ariadna Medina-Benitez; Claudia Wong-Arambula; Ernesto Ramirez-Gonzalez.; Gisela Barrera-Badillo; Irma Lopez-Martinez; Joaquin Quiroz-Mercado; Lucia Hernandez-Rivas; Maribel Gonzalez-Villa; Natividad Cruz-Ortiz; Sergio Rangel-Guerrero; Tatiana Nunez-Garcia; Vanessa Rivero-Arredondo |
| EPI_ISL_2937756, EPI_ISL_3033596, EPI_ISL_3033598 | LESP Puebla | Instituto de Diagnostico y Referencia Epidemiologicos (INDRE) | Abril Rodriguez-Maldonado; Ariadna Medina-Benitez; Claudia Wong-Arambula; Ernesto Ramirez-Gonzalez.; Gisela Barrera-Badillo; Irma Lopez-Martinez; Joaquin Quiroz-Mercado; Lucia Hernandez-Rivas; Maribel Gonzalez-Villa; Natividad Cruz-Ortiz; Sergio Rangel-Guerrero; Tatiana Nunez-Garcia; Vanessa Rivero-Arredondo |

| | | | |
|---|---|---|---|
| EPI_ISL_2779179, EPI_ISL_2937758, EPI_ISL_2937759, EPI_ISL_2937760, EPI_ISL_2937761, EPI_ISL_2937762, EPI_ISL_2937763, EPI_ISL_2937764, EPI_ISL_2937766, EPI_ISL_2937798, EPI_ISL_2937799 | | | |
| see above | LESP Quintana Roo | Instituto de Diagnostico y Referencia Epidemiologicos (INDRE) | Abril Rodriguez-Maldonado; Ariadna Medina-Benitez; Claudia Wong-Arambula; Ernesto Ramirez-Gonzalez.; Gisela Barrera-Badillo; Irma Lopez-Martinez; Joaquin Quiroz-Mercado; Lucia Hernandez-Rivas; Maribel Gonzalez-Villa; Natividad Cruz-Ortiz; Sergio Rangel-Guerrero; Tatiana Nunez-Garcia; Vanessa Rivero-Arredondo |
| EPI_ISL_2779159, EPI_ISL_2937773 | LESP San Luis Potosi | Instituto de Diagnostico y Referencia Epidemiologicos (INDRE) | Abril Rodriguez-Maldonado; Ariadna Medina-Benitez; Claudia Wong-Arambula; Ernesto Ramirez-Gonzalez.; Gisela Barrera-Badillo; Irma Lopez-Martinez; Joaquin Quiroz-Mercado; Lucia Hernandez-Rivas; Maribel Gonzalez-Villa; Natividad Cruz-Ortiz; Sergio Rangel-Guerrero; Tatiana Nunez-Garcia; Vanessa Rivero-Arredondo |
| EPI_ISL_2779185, EPI_ISL_2937767 | LESP Sonora | Instituto de Diagnostico y Referencia Epidemiologicos (INDRE) | Abril Rodriguez-Maldonado; Ariadna Medina-Benitez; Claudia Wong-Arambula; Ernesto Ramirez-Gonzalez.; Gisela Barrera-Badillo; Irma Lopez-Martinez; Joaquin Quiroz-Mercado; Lucia Hernandez-Rivas; Maribel Gonzalez-Villa; Natividad Cruz-Ortiz; Sergio Rangel-Guerrero; Tatiana Nunez-Garcia; Vanessa Rivero-Arredondo |
| EPI_ISL_2674638, EPI_ISL_2779053, EPI_ISL_2779054, EPI_ISL_2779160, EPI_ISL_2779161, EPI_ISL_2779162, EPI_ISL_2779163, EPI_ISL_2779164, EPI_ISL_2779165, EPI_ISL_2779166, EPI_ISL_2779167, EPI_ISL_2779168, EPI_ISL_2779169, EPI_ISL_2779170, EPI_ISL_2779194, EPI_ISL_2859022, EPI_ISL_2937753, EPI_ISL_2937754, EPI_ISL_2937755 | | | |
| see above | LESP Tabasco | Instituto de Diagnostico y Referencia Epidemiologicos (INDRE) | Abril Rodriguez-Maldonado; Ariadna Medina-Benitez; Claudia Wong-Arambula; Ernesto Ramirez-Gonzalez.; Gisela Barrera-Badillo; Irma Lopez-Martinez; Joaquin Quiroz-Mercado; Lucia Hernandez-Rivas; Maribel Gonzalez-Villa; Natividad Cruz-Ortiz; Sergio Rangel-Guerrero; Tatiana Nunez-Garcia; Vanessa Rivero-Arredondo |
| EPI_ISL_2779061, EPI_ISL_2858996, EPI_ISL_2858998, EPI_ISL_2937776, EPI_ISL_2937777 | LESP Tamaulipas | Instituto de Diagnostico y Referencia Epidemiologicos (INDRE) | Abril Rodriguez-Maldonado; Ariadna Medina-Benitez; Claudia Wong-Arambula; Ernesto Ramirez-Gonzalez.; Gisela Barrera-Badillo; Irma Lopez-Martinez; Joaquin Quiroz-Mercado; Lucia Hernandez-Rivas; Maribel Gonzalez-Villa; Natividad Cruz-Ortiz; Sergio Rangel-Guerrero; Tatiana Nunez-Garcia; Vanessa Rivero-Arredondo |
| EPI_ISL_2859000, EPI_ISL_2937793 | LESP Veracruz | Instituto de Diagnostico y Referencia Epidemiologicos (INDRE) | Abril Rodriguez-Maldonado; Ariadna Medina-Benitez; Claudia Wong-Arambula; Ernesto Ramirez-Gonzalez.; Gisela Barrera-Badillo; Irma Lopez-Martinez; Joaquin Quiroz-Mercado; Lucia Hernandez-Rivas; Maribel Gonzalez-Villa; Natividad Cruz-Ortiz; Sergio Rangel-Guerrero; Tatiana Nunez-Garcia; Vanessa Rivero-Arredondo |
| EPI_ISL_2779114, EPI_ISL_2779115, EPI_ISL_2779116, EPI_ISL_2779117, EPI_ISL_2779118, EPI_ISL_2779119, EPI_ISL_2779122, EPI_ISL_2779123, EPI_ISL_2779124, EPI_ISL_2779125, EPI_ISL_2779126, EPI_ISL_2779127, EPI_ISL_2779128, EPI_ISL_2779129, EPI_ISL_2779130, EPI_ISL_2779131, EPI_ISL_2779132, EPI_ISL_2779133, EPI_ISL_2779134, EPI_ISL_2779135, EPI_ISL_2779136, EPI_ISL_2779137, EPI_ISL_2779138, EPI_ISL_2779139, EPI_ISL_2779140, EPI_ISL_2779141, EPI_ISL_2779142, EPI_ISL_2779143, EPI_ISL_2779144, EPI_ISL_2779145, EPI_ISL_2779146, EPI_ISL_2779147, EPI_ISL_2779148, EPI_ISL_2779149, EPI_ISL_2779150, EPI_ISL_2779151, EPI_ISL_2779152, EPI_ISL_2779153, EPI_ISL_2800916, EPI_ISL_2800918, EPI_ISL_2800919, EPI_ISL_2800920, EPI_ISL_2800921, EPI_ISL_2800922, EPI_ISL_2800923, EPI_ISL_3033479, EPI_ISL_3033480 | | | |
| see above | LESP Yucatan | Instituto de Diagnostico y Referencia Epidemiologicos (INDRE) | Abril Rodriguez-Maldonado; Ariadna Medina-Benitez; Claudia Wong-Arambula; Ernesto Ramirez-Gonzalez.; Gisela Barrera-Badillo; Irma Lopez-Martinez; Joaquin Quiroz-Mercado; Lucia Hernandez-Rivas; Maribel Gonzalez-Villa; Natividad Cruz-Ortiz; Sergio Rangel-Guerrero; Tatiana Nunez-Garcia; Vanessa Rivero-Arredondo |
| EPI_ISL_2605559, EPI_ISL_2605562, EPI_ISL_2605616, EPI_ISL_2605641, EPI_ISL_2861383, EPI_ISL_2861397, EPI_ISL_2861417, EPI_ISL_2861453, EPI_ISL_2861454 | | | |
| see above | Lab voor klinische biologie | Lab voor klinische biologie | Bruno Verhasselt; Hannelore Hamerlinck; Marija Janevska |
| EPI_ISL_2725206, EPI_ISL_2725216, EPI_ISL_2725223, EPI_ISL_2725229, EPI_ISL_2725238, EPI_ISL_2725239, EPI_ISL_2725245, EPI_ISL_2725248, EPI_ISL_2725249 | | | |
| see above | Lab. Microbiologia e Virologia Cotugno A.O. dei Colli - Istituto Zooprofilattico Sperimentale del Mezzogiorno | Telethon Institute of Genetics and Medicine (TIGEM) | Antonio Grimaldi Patrizia Annunziata Francesco Panariello Biancamaria Pierri Claudia Tiberio Teresa Giuliano Valentina Bouche Chiara Colantuono Maria Concetta Cuomo Denise Di Concilio Lucio Di Filippo Anna Manfredi Marcello Salvi Antonio Limone Luigi Atripaldi Pellegrino Cerino Andrea Ballabio Davide Cacchiarelli |
| EPI_ISL_2835851, EPI_ISL_2835852, EPI_ISL_2835854, EPI_ISL_2835856 | Labo Analyses Med | National Reference Center for Viruses of Respiratory Infections, Institut Pasteur, Paris | Angela Brisebarre; Camille Capel; Christophe Malabat; Corinne Maufrais; Etienne Simon-Lorière; Frédéric Lemoine; Louise Lefrançois; Marion Barbet; Maud Vanpeene; Méline Bizard; Sabine Trombert; Sylvie Behillil; Sylvie Van der Werf; Vincent Enouf |
| EPI_ISL_2709300 | Labo Analyses med | National Reference Center for Viruses of Respiratory Infections, Institut Pasteur, Paris | Angela Brisebarre; Camille Capel; Christophe Malabat; Corinne Maufrais; Damien Mornico; Etienne Simon-Lorière; Frédéric Lemoine; Louise Lefrançois; Marion Barbet; Maud Vanpeene; Méline Bizard; OphéLie Said-Delattre; Sylvie Behillil; Sylvie Van der Werf; Vincent Enouf |
| EPI_ISL_2694805, EPI_ISL_2694811, EPI_ISL_2694857 | Labor Berlin Charité Vivantes GmbH / Institut für Virologie | Charité Universitätsmedizin Berlin, Institut für Virologie/Labor Berlin | Barbara Mühlemann; Christian Drosten; Christine Stephan; Peter Menzel; Rolf Schwarzer; Terry Jones; Victor M Corman |
| EPI_ISL_2635148 | Labor Prof. Dr. G. Enders MVZ GbR | Robert Koch Institute | |
| EPI_ISL_2678754, EPI_ISL_2678763, EPI_ISL_2678774, EPI_ISL_2678786, EPI_ISL_2678801 | Labor ZOTZ\|KLIMAS; MVZ Düsseldorf-Centrum | Robert Koch Institute | |
| EPI_ISL_2835989, EPI_ISL_2835990 | Laboratoire Carage | Institut Pasteur de la Guyane | Anne Lavergne; Antoine Enfissi; Arielle Salmier; Dominique Rousset |
| EPI_ISL_2637629, EPI_ISL_2637761 | Laboratoire LENYS | Department of Virology, Henri Mondor University Hospital, Assistance Publique Hôpitaux de Paris, Université Paris-Est Créteil, INSERM U955 | Alexandre Soulier; Christophe Rodriguez; Elisabeth Trawinski; Guillaume Gricourt; Jean-Michel Pawlotsky; Melissa N'Debi; Slim Fourati; Vanessa Demontant |
| EPI_ISL_3458875, EPI_ISL_3458889, EPI_ISL_3458906, EPI_ISL_3458921, EPI_ISL_3458971, EPI_ISL_3458986 | Laboratoire de santé publique du Québec | Laboratoire de santé publique du Québec | Guillaume Bourque; Ioannis Ragoussis; Jesse Shapiro; Mark Lathrop and Michel Roger on behalf of the CoVSeQ research group; Sandrine Moreira |
| EPI_ISL_3148153 | Laboratoires d'analyses medicales - Ketterthill | Laboratoire national de sante, Microbiology, Microbial Genomics Platform | Anke Wienecke-Baldacchino; Caroline Scheiber; Catherine Ragimbeau; Elodie Solarino; Fatu Djabi; Jessica Tapp; Lise Pignon; Raoul Salmon; Serge Vedy; Tamir Abdelrahman; Virginie Jover |
| EPI_ISL_3906438 | Laboratori de Referència de Catalunya | SeqCOVID-SPAIN consortium/IBV(CSIC) | Aida Esperanza Ramírez Marinero; Alba Vilas Basil; Eduardo Padilla León; Mireia Canal Aranda and SeqCOVID-SPAIN consortium |
| EPI_ISL_2983184, EPI_ISL_2983186 | Laboratorio Central de Saude Publica do Estado de Goias (LACEN/GO) | Laboratory of Respiratory Viruses and Measles, Oswaldo Cruz Institute, FIOCRUZ | Agatha Cristinne Prudencio; Alice Sampaio Rocha; Ana Carolina Mendonca; Ana Flavia Mendonça; Anna Carolina Paixao; Carmen Helena Ramos; Cassiane Casanova; Elisa Cavalcante Pereira; Fernando Motta; Flavia Pereira Amorim da Silva; Ighor Leonardo Arantes Gomes; Luciana Appolinario; Luiz Augusto Pereira; Marilda Siqueira on behalf of the Fiocruz COVID-19 Genomic Surveillance Network; Paola Resende; Rafael Souza Guedes; Renata Serrano Lopes; Taina Venas; Vinicius Lemes da Silva |
| EPI_ISL_3539750, EPI_ISL_3539751 | Laboratorio Central de Saude Publica do Estado de Minas Gerais (LACEN/MG) | Laboratory of Respiratory Viruses and Measles, Oswaldo Cruz Institute, FIOCRUZ | Alice Sampaio Rocha; Ana Carolina Mendonca; Anna Carolina Paixao; Elisa Cavalcante Pereira; Fernando Motta; Luciana Appolinario; Marilda Siqueira on behalf of the Fiocruz COVID-19 Genomic Surveillance Network; Paola Resende; Renata Serrano Lopes; Rubens Pasa; Taina Venas |
| EPI_ISL_2731453, EPI_ISL_2731454, EPI_ISL_2731455, EPI_ISL_2731456, EPI_ISL_2863578, EPI_ISL_2863580, EPI_ISL_2863582, EPI_ISL_2863706, EPI_ISL_2863707, EPI_ISL_2863709, EPI_ISL_2863711, EPI_ISL_2863801, EPI_ISL_2863802, EPI_ISL_2863803, EPI_ISL_2863804, EPI_ISL_2863805, EPI_ISL_2863806, EPI_ISL_2863807, EPI_ISL_2863808, EPI_ISL_2863809, EPI_ISL_2863810, EPI_ISL_2863811, EPI_ISL_2863812, EPI_ISL_2863813, EPI_ISL_2863814, EPI_ISL_2863815, EPI_ISL_2863816, EPI_ISL_2863817, EPI_ISL_2863818, EPI_ISL_2863819, EPI_ISL_2863820, EPI_ISL_2863821, EPI_ISL_2863822, EPI_ISL_2863823, EPI_ISL_2863824, EPI_ISL_2863825, EPI_ISL_2863826, EPI_ISL_2863827, EPI_ISL_2863828, EPI_ISL_2863829, EPI_ISL_2863830, EPI_ISL_2863831, EPI_ISL_2863832, EPI_ISL_2863833, EPI_ISL_2863834, EPI_ISL_2863835, EPI_ISL_2863836, EPI_ISL_2863837, EPI_ISL_2863838, EPI_ISL_2863839, EPI_ISL_2863840, EPI_ISL_2863841, EPI_ISL_2863842, EPI_ISL_2863843, EPI_ISL_2863844, EPI_ISL_2863845, EPI_ISL_2863846, EPI_ISL_2863847, EPI_ISL_2863848, EPI_ISL_2863849, EPI_ISL_2863850, EPI_ISL_2863851, EPI_ISL_2863852, EPI_ISL_2863853, EPI_ISL_2863854, EPI_ISL_2863892, EPI_ISL_2863894 | | | |
| see above | Laboratorio Central de Saude Publica do Estado do Para (LACEN/PA) | Laboratory of Respiratory Viruses and Measles, Oswaldo Cruz Institute, FIOCRUZ | Agatha Cristinne Prudencio; Agatha Cristinne Prudencio Soares; Alice Sampaio Rocha; Ana Carolina Mendonca; Anna Carolina Paixao; Elisa Cavalcante Pereira; Fernando Motta; Ighor Leonardo Arantes Gomes; Luciana Appolinario; Marilda Siqueira on behalf of the Fiocruz COVID-19 Genomic Surveillance Network; Paola Resende; Renata Serrano Lopes; Taina Venas; Valnete Andrade |
| EPI_ISL_2686056, EPI_ISL_2686057, EPI_ISL_2686058, EPI_ISL_2686059 | Laboratorio Analisi Osp. Città di Castello - Azienda USL Umbria1 | Istituto Zooprofilattico Sperimentale dell'Abruzzo e Molise "G. Caporale" | Ancora M; Calistri P; Cammà C; Curini V; Delli Compagni E; Di Domenico M; Di Pasquale A; Lorusso A; Malagigi V; Mangone I; Marcacci M; Puglia I; Rinaldi A; Savini G; Scialabba S; Taccani P |
| EPI_ISL_2836972, EPI_ISL_2836975, EPI_ISL_2836976, EPI_ISL_2836977, EPI_ISL_2836978, EPI_ISL_2836979, EPI_ISL_2836980, EPI_ISL_2836981, EPI_ISL_2836982, EPI_ISL_2836983, EPI_ISL_2836984, EPI_ISL_2836985, EPI_ISL_2836986, EPI_ISL_2836987, EPI_ISL_2836988, EPI_ISL_2836989, EPI_ISL_2836990, EPI_ISL_2836991, EPI_ISL_2836992, EPI_ISL_2836993, EPI_ISL_2836996, EPI_ISL_2836998, EPI_ISL_2836999, EPI_ISL_2837000, EPI_ISL_2837001, EPI_ISL_2837002, EPI_ISL_2837003, EPI_ISL_2837004, EPI_ISL_2837005, EPI_ISL_2837006, EPI_ISL_2837007, EPI_ISL_2837008, EPI_ISL_2837009, EPI_ISL_2837010, EPI_ISL_2837011, EPI_ISL_2837012, EPI_ISL_2837013, EPI_ISL_2837014, EPI_ISL_2837015, EPI_ISL_2837016, EPI_ISL_2837017, EPI_ISL_2837018, EPI_ISL_2837019, EPI_ISL_2837020, EPI_ISL_2837021, EPI_ISL_2837022, EPI_ISL_2837024, EPI_ISL_2837026, EPI_ISL_2837027, EPI_ISL_2837028, EPI_ISL_2837029, EPI_ISL_2837030, EPI_ISL_2837031, EPI_ISL_2837032, EPI_ISL_2837033, EPI_ISL_2837034, EPI_ISL_2837035, EPI_ISL_2837036, EPI_ISL_2837037, EPI_ISL_2837038, EPI_ISL_2837039, EPI_ISL_2837040, EPI_ISL_2837042, EPI_ISL_2837044, EPI_ISL_2837045, EPI_ISL_2837046, EPI_ISL_2837047, EPI_ISL_2837048, EPI_ISL_2837049, EPI_ISL_2837050, EPI_ISL_2837051, EPI_ISL_2837052, EPI_ISL_2837054, EPI_ISL_2837055, EPI_ISL_2837056, EPI_ISL_2837057, EPI_ISL_2837058, EPI_ISL_2837059, EPI_ISL_2837060, EPI_ISL_2837061, EPI_ISL_2837062, EPI_ISL_2837063, EPI_ISL_2837064, EPI_ISL_2837065, EPI_ISL_2837066, EPI_ISL_2837067, EPI_ISL_2837068, EPI_ISL_2837069, EPI_ISL_2837070, EPI_ISL_2837071, EPI_ISL_2837072, EPI_ISL_2837073, EPI_ISL_2837074, EPI_ISL_2837075, EPI_ISL_2837076, EPI_ISL_2837077, EPI_ISL_2837078, EPI_ISL_2837079, EPI_ISL_2837080, EPI_ISL_2837081, EPI_ISL_2837082, EPI_ISL_2837083, EPI_ISL_2837084, EPI_ISL_2837085, EPI_ISL_2837086, EPI_ISL_2837087, EPI_ISL_2837088, EPI_ISL_2837089, EPI_ISL_2837090, EPI_ISL_2837091, EPI_ISL_2837092, EPI_ISL_2837093, EPI_ISL_2837094, EPI_ISL_2837095, EPI_ISL_2837096, EPI_ISL_2837097, EPI_ISL_2837098, EPI_ISL_2837099, EPI_ISL_2837100, EPI_ISL_2837101, EPI_ISL_2837102, EPI_ISL_2837103, EPI_ISL_2837104, EPI_ISL_2837105, EPI_ISL_2837106, EPI_ISL_2837107, EPI_ISL_2837108, EPI_ISL_2837109, EPI_ISL_2837110, EPI_ISL_2837111, EPI_ISL_2837112, EPI_ISL_2837113, EPI_ISL_2837114, EPI_ISL_2837123, EPI_ISL_2837124, EPI_ISL_2837125, EPI_ISL_2837126, EPI_ISL_2837127, EPI_ISL_2837128, EPI_ISL_2837129, EPI_ISL_2837130, EPI_ISL_2837131, EPI_ISL_2837132, EPI_ISL_2837139, EPI_ISL_2837140, EPI_ISL_2837141, EPI_ISL_2837142, EPI_ISL_2837143, EPI_ISL_2837144, EPI_ISL_2837145, EPI_ISL_2837146, EPI_ISL_2837147, EPI_ISL_2837150, EPI_ISL_2837153, EPI_ISL_2837154, EPI_ISL_2837155, EPI_ISL_2837156, EPI_ISL_2837157, EPI_ISL_2837158, EPI_ISL_2837159, EPI_ISL_2837160, EPI_ISL_2837161, EPI_ISL_2837162, EPI_ISL_2837163, EPI_ISL_2837173, EPI_ISL_2837174, EPI_ISL_2837175, EPI_ISL_2837177, EPI_ISL_2837189, EPI_ISL_2837190, EPI_ISL_2837191, EPI_ISL_2837192 | | | |
| see above | Laboratorio Central Noel Nutels | Bioinformatics Laboratory / LNCC | Alessandra P Lamarca; Alexandra L Gerber; Amilcar Tanuri; Ana Paula de C Guimaraes; Ana Tereza R Vasconcelos; Andrea Cony Cavalcanti; Caio Luiz Pereira Ribeiro; Cassia Alves; Cintia Policarpo; Claudia Maria Braga de Mello; Cristiane Gomes da Silva; Diana Mariani; Douglas Terra Machado; Flavio Dias da Silva; Gleidson da Silva de Oliveira; Leandro Magalhaes de Souza; Liliane Cavalcante; Luiz G P de Almeida; Marcio Henrique de Oliveira Garcia; Mario Sergio Ribeiro; Ronaldo da Silva F Jr; Silvia Carvalho |
| EPI_ISL_2942513, EPI_ISL_2942581, EPI_ISL_2942589, EPI_ISL_2942602, EPI_ISL_2942605, EPI_ISL_2942606, EPI_ISL_2942617 | | | |
| see above | Laboratorio Central de Epidemiologia (LCE) | Unidad de Genomica Avanzada | ; Alejandra Garcia-Gasca; Alejandra Hernandez-Teran; Alejandro Sanchez-Flores; Alfredo Herrera-Estrella; Alicia Ocaña-Mondragon; Andreu Comas-Garcia; Angel Gustavo Salas-Lais; Antonio Loza Roman; Bernardo Martinez-Miguel; Blanca Taboada; Brenda Irasema Maldonado-Meza; Bruno Gomez-Gil; Carla Ivon Herrera-Najera; Carlos F. Arias; Celia Boukadida; Celida Duque Molina; Celida Martinez- Rodriguez; Clara Esperanza Santacruz-Tinoco; Concepcion Grajales-Muñiz; Consorcio Mexicano de Vigilancia Genomica (CoViGen-Mex). Authors (in alphabetical order): Julio Elias Alvarado-Yaah; Cristobal Chaidez-Quiroz; Daniel Fregoso-Rueda; Daniel Lira Morales; Eduardo Becerril-Vargas; Fernando Fontove-Herrera; Fidencio Mejia-Nepomuceno; Francisco Pulido; Gloria Elena Espinosa-Ayala; Gloria Maria Molina-Salinas; Gloria Vazquez; Hector Esteban Paz-Juarez; Hector Montoya-Fuentes; Helen Haydee Fernanda Ramirez-Plascencia; Irvin Gonzalez-Lopez; Jean Pierre Gonzalez; Jesus Hernandez; Joel Armando Vazquez-Perez.; Jorge Salas-Hernandez; Jose Antonio Enciso-Moreno; Jose Arturo Martinez-Orozco; Jose Esteban Muñoz-Medina; Jose de Jesus Nuñez-Contreras; Juan Bautista Chale-Dzul; Julissa Enciso-Ibarra; Luis Alberto Ochoa-Carrera; Margarita Matias-Florentino; Maria Guadalupe Santiago-Mauricio; Maria Guadalupe de Jesus Mireles-Rivera; Mario Mujica-Sanchez; Marissa Perez-Garcia; Nelly Selem-Mojica; Pavel Isa; Ricardo Ciria Merce; Ricardo Grande; Rosa Maria Gutierrez Rios; Santiago avila-Rios; Selene Zarate; Susana Lopez; Veronica Mata-Haro; Victor Eduardo Garcia-Arias; Victor Hugo Borja-Aburto |

| | | | |
|---|---|---|---|
| EPI_ISL_2801857, EPI_ISL_2801858, EPI_ISL_2801860, EPI_ISL_2801861, EPI_ISL_2801862, EPI_ISL_2801867, EPI_ISL_2801876, EPI_ISL_2801877, EPI_ISL_2801880, EPI_ISL_2801883, EPI_ISL_2801884, EPI_ISL_2801889 | | | |
| see above | Laboratorio Central de Epidemiología (LCE) | Instituto de Biotecnología de la UNAM | ; Alejandra García-Gasca; Alejandra Hernández-Terán; Alejandro Sánchez-Flores; Alfredo Herrera-Estrella; Alicia Ocaña-Mondragón; Andreu Comas-García; Angel Gustavo Salas-Lais; Antonio Loza Román; Bernardo Martínez-Miguel; Blanca Taboada; Brenda Irasema Maldonado-Meza; Bruno Gómez-Gil; Carla Ivón Herrera-Najera; Carlos F. Arias; Celia Boukadida; Clara Esperanza Santacruz-Tinoco; Concepción Grajales-Muñiz; Consorcio Mexicano de Vigilancia Genómica (CoViGen-Mex). Authors (in alphabetical order): Julio Elias Alvarado-Yaah; Cristóbal Cháidez-Quiróz; Célida Duque Molina; Célida Martínez- Rodríguez; Daniel Fregoso-Rueda; Daniel Lira Morales; Eduardo Becerril-Vargas; Fernando Fontove-Herrera; Fidencio Mejía-Nepomuceno; Francisco Pulido; Gloria Elena Espinosa-Ayala; Gloria María Molina-Salinas; Gloria Vazquez; Hector Esteban Paz-Juárez; Hector Montoya-Fuentes; Helen Haydee Fernanda Ramírez-Plascencia; Irvin González-López; Jean Pierre González; Jesús Hernández; Joel Armando Vázquez-Pérez.; Jorge Salas-Hernández; José Antonio Enciso-Moreno; José Arturo Martínez-Orozco; José Esteban Muñoz-Medina; José de Jesús Nuñez-Contreras; Juan Bautista Chale-Dzul; Julissa Enciso-Ibarra; Luis Alberto Ochoa-Carrera; Margarita Matías-Florentino; Mario Mújica-Sánchez; Marissa Perez-Garcia; María Guadalupe Santiago-Mauricio; María Guadalupe de Jesús Mireles-Rivera; Nelly Sélem-Mojica; Pavel Isa; Ricardo Ciria Merce; Ricardo Grande; Rosa María Gutiérrez Rios; Santiago Ávila-Ríos; Selene Zárate; Susana Lopez; Verónica Mata-Haro; Victor Eduardo García-Arias; Víctor Hugo Borja-Aburto |
| EPI_ISL_6944557, EPI_ISL_6944562, EPI_ISL_6944563, EPI_ISL_6944564, EPI_ISL_6944571, EPI_ISL_6944574, EPI_ISL_6944580, EPI_ISL_6944582, EPI_ISL_6944592, EPI_ISL_6944604, EPI_ISL_6944605, EPI_ISL_6944611, EPI_ISL_6944612 | | | |
| see above | Laboratorio Central de Salud Publica | Viral Special Pathogens Branch, Centers for Disease Control and Prevention | Andrea Gómez de la Fuente; Cynthia Vázquez; Emir Talundzic; Joel Montgomery; John Klena; Juan Torales; Justin Lee; María José Ortega; María Liz Gamarra; Shannon Whitmer; Shirley Villalba |
| EPI_ISL_4518744, EPI_ISL_4518746 | Laboratorio Central de Salud Publica de Paraguay | Fundação Ezequiel Dias | Andre Leal; Andrea Gómez de la Fuente; Cynthia Vazquez; Elaine Cristina; Felipe Iani; Flavia Aburjaile; Gislene Garcia de Castro Lichs; Glauco Carvalho; Hegger Fritsch; Joilson Xavier; Juan Torales; Luiz Alcantara.; Luiz Henrique Ferraz Demarchi; Luiz Takao Watanabe; Marina Castilhos Souza Umaki Zardin; Marta Giovanetti; María José Ortega; María Liz Gamarra; Natalia Guimaraes; Raquel da Silva Ferreira; Shirley Villalba; Talita Adelino; Vagner Fonseca; de Oliveira |
| EPI_ISL_2863589, EPI_ISL_2863591, EPI_ISL_2863593 | Laboratorio Central de Saude Publica do Estado da Bahia (LACEN/BA) | Laboratory of Respiratory Viruses and Measles, Oswaldo Cruz Institute, FIOCRUZ | Alice Sampaio Rocha; Ana Carolina Mendonca; Anna Carolina Paixao; Elisa Cavalcante Pereira; Felicidade Pereira; Fernando Motta; Luciana Appolinario; Marilda Siqueira on behalf of the Fiocruz COVID-19 Genomic Surveillance Network; Paola Resende; Renata Serrano Lopes; Taina Venas |
| EPI_ISL_3190174, EPI_ISL_3190175, EPI_ISL_3190177, EPI_ISL_3190179, EPI_ISL_3190180, EPI_ISL_3190181, EPI_ISL_3190182, EPI_ISL_3190183, EPI_ISL_3190184, EPI_ISL_3190185, EPI_ISL_3190186, EPI_ISL_3190187, EPI_ISL_3190188, EPI_ISL_3434905 | | | |
| see above | Laboratorio Central de Saude Publica do Estado da Paraiba (LACEN/PB) | Laboratory of Respiratory Viruses and Measles, Oswaldo Cruz Institute, FIOCRUZ | Alice Sampaio Rocha; Ana Carolina Mendonca; Anna Carolina Paixao; Dalane Loudal Florentino Teixeira; Elisa Cavalcante Pereira; Fernando Motta; Joao Felipe Bezerra; Luciana Appolinario; Marilda Siqueira on behalf of the Fiocruz COVID-19 Genomic Surveillance Network; Paola Resende; Renata Serrano Lopes; Taina Venas |
| EPI_ISL_2983076, EPI_ISL_2983077, EPI_ISL_2983078, EPI_ISL_2983079, EPI_ISL_2983080, EPI_ISL_2983081, EPI_ISL_2983082, EPI_ISL_2983083, EPI_ISL_2983084, EPI_ISL_2983085, EPI_ISL_3190309, EPI_ISL_3190310, EPI_ISL_3190311, EPI_ISL_3190312, EPI_ISL_3190334, EPI_ISL_3190335, EPI_ISL_3190337, EPI_ISL_3190345 | | | |
| see above | Laboratorio Central de Saude Publica do Estado de Alagoas (LACEN/AL) | Laboratory of Respiratory Viruses and Measles, Oswaldo Cruz Institute, FIOCRUZ | Agatha Cristinne Prudencio; Alice Sampaio Rocha; Ana Carolina Mendonca; Anderson Brandao Leite; Anna Carolina Paixao; Elisa Cavalcante Pereira; Fernando Motta; Ighor Leonardo Arantes Gomes; Luciana Appolinario; Marilda Siqueira on behalf of the Fiocruz COVID-19 Genomic Surveillance Network; Paola Resende; Renata Serrano Lopes; Taina Venas |
| EPI_ISL_2983349, EPI_ISL_2983350, EPI_ISL_2983359, EPI_ISL_2983363 | Laboratorio Central de Saude Publica do Estado de Santa Catarina (LACEN-SC) | Laboratory of Respiratory Viruses and Measles, Oswaldo Cruz Institute, FIOCRUZ | Alice Sampaio Rocha; Ana Carolina Mendonca; Anna Carolina Paixao; Darcita Buerger Rovaris; Elisa Cavalcante Pereira; Fernando Motta; Luciana Appolinario; Marilda Siqueira on behalf of the Fiocruz COVID-19 Genomic Surveillance Network; Paola Resende; Renata Serrano Lopes; Sandra Bianchini Fernandes; Taina Venas |
| EPI_ISL_2983383, EPI_ISL_2983384, EPI_ISL_2983386, EPI_ISL_2983387, EPI_ISL_2983388, EPI_ISL_2983389, EPI_ISL_2983390, EPI_ISL_2983391, EPI_ISL_2983392, EPI_ISL_2983393, EPI_ISL_2983394, EPI_ISL_2983395, EPI_ISL_2983396, EPI_ISL_2983397, EPI_ISL_2983398, EPI_ISL_2983405, EPI_ISL_2983410, EPI_ISL_2983412, EPI_ISL_3235301 | | | |
| see above | Laboratorio Central de Saude Publica do Estado de Santa Catarina (LACEN/SC) | Laboratory of Respiratory Viruses and Measles, Oswaldo Cruz Institute, FIOCRUZ | Alice Sampaio Rocha; Ana Carolina Mendonca; Anna Carolina Paixao; Darcita Buerger Rovaris; Elisa Cavalcante Pereira; Fernando Motta; Luciana Appolinario; Marilda Siqueira on behalf of the Fiocruz COVID-19 Genomic Surveillance Network; Paola Resende; Renata Serrano Lopes; Sandra Bianchini Fernandes; Taina Venas |
| EPI_ISL_2863639, EPI_ISL_2863641, EPI_ISL_2863645, EPI_ISL_2863719 | Laboratorio Central de Saude Publica do Estado de Sergipe (LACEN/SE) | Laboratory of Respiratory Viruses and Measles, Oswaldo Cruz Institute, FIOCRUZ | Alice Sampaio Rocha; Ana Carolina Mendonca; Anna Carolina Paixao; Cliomar Alves dos Santos; Elisa Cavalcante Pereira; Fernando Motta; Luciana Appolinario; Marilda Siqueira on behalf of the Fiocruz COVID-19 Genomic Surveillance Network; Paola Resende; Renata Serrano Lopes; Tainá Moreira Martins Venas |
| EPI_ISL_2983426, EPI_ISL_2983427, EPI_ISL_2983445, EPI_ISL_2983446, EPI_ISL_3190190, EPI_ISL_3190191, EPI_ISL_3190192, EPI_ISL_3190266, EPI_ISL_3190268, EPI_ISL_3435048 | | | |
| see above | Laboratorio Central de Saude Publica do Estado do Parana (LACEN/PR) | Laboratory of Respiratory Viruses and Measles, Oswaldo Cruz Institute, FIOCRUZ | Agatha Soares; Alice Sampaio Rocha; Ana Carolina Mendonca; Anna Carolina Paixao; Elisa Cavalcante Pereira; Fernando Motta; Ighor Leonardo Arantes; Irina Riediger; Luciana Appolinario; Marilda Siqueira on behalf of the Fiocruz COVID-19 Genomic Surveillance Network; Paola Resende; Renata Serrano Lopes; Taina Venas |
| EPI_ISL_2982768, EPI_ISL_2982781, EPI_ISL_2982786, EPI_ISL_2982788, EPI_ISL_2982792, EPI_ISL_2982796, EPI_ISL_2982797, EPI_ISL_2982800, EPI_ISL_2982804, EPI_ISL_2982822 | | | |
| see above | Laboratorio Central de Saude Publica do Estado do Rio Grande do Sul (LACEN-RS) | Laboratory of Respiratory Viruses and Measles, Oswaldo Cruz Institute, FIOCRUZ | Alice Sampaio Rocha; Ana Carolina Mendonca; Anderson Brandao Leite; Anna Carolina Paixao; Elisa Cavalcante Pereira; Fernando Motta; Luciana Appolinario; Marilda Siqueira on behalf of the Fiocruz COVID-19 Genomic Surveillance Network; Paola Resende; Renata Serrano Lopes; Taina Venas |
| EPI_ISL_2863602, EPI_ISL_2863604, EPI_ISL_2863677, EPI_ISL_2863679, EPI_ISL_2863680, EPI_ISL_2863721, EPI_ISL_2863723 | | | |
| see above | Laboratorio Central de Saude Publica do Estado do Rio de Janeiro (LACEN-RJ) | Laboratory of Respiratory Viruses and Measles, Oswaldo Cruz Institute, FIOCRUZ | Agatha Cristinne Prudencio; Alice Sampaio Rocha; Ana Carolina Mendonca; Andrea Cony Cavalcanti; Anna Carolina Paixao; Elisa Cavalcante Pereira; Fernando Motta; Ighor Leonardo Arantes Gomes; Luciana Appolinario; Marilda Siqueira on behalf of the Fiocruz COVID-19 Genomic Surveillance Network; Paola Resende; Renata Serrano Lopes; Taina Venas |
| EPI_ISL_4196880 | Laboratorio Clínico Analizar | Gencore - Universidad de los Andes | Cristian Barrera; Gabriela Ariza; Luisa Sacristan; Marcela Guevara; Marcela Mercado; Silvia Restrepo |
| EPI_ISL_6121558, EPI_ISL_6121566 | Laboratorio de Alta Complejidad (LACMI), Instituto de Genética Humana (IgeHM) Parque de la Salud de la Provincia de Misiones, Universidad Nacional de Misiones | Grupo de Genómica y Bioinformática del Instituto de Investigación de la Cadena Láctea CONICET-INTA on behalf of 'Proyecto Argentino Interinstitucional de genomica de SARS-CoV-2' (PAIS Consortium) | AF; Aguirre; Amadio; Benitez; C; D; Eberhardt; Fontana; G; Gamarra; Gonzales; Irazoqui; JM; K; Ludojoski; M; MF; R; Salvatierra; Silva; TA; Urquiza; Vogel |
| EPI_ISL_4405311, EPI_ISL_4405631 | Laboratorio de Biología Molecular Bolívar | Área de Secuenciación del Laboratorio de Virología del Hospital de Niños Dr. Ricardo Gutierrez on behalf of 'Proyecto Argentino Interinstitucional de genomica de SARS-CoV-2' (PAIS Consortium) | Acuña; Cabrerizo; Caviglia; D; Di Claudio; D'Andrea; F; Goya; Jofré; LE; Larreche; Lescano; Lusso; M; ME; MI; ML; MR; MS; Martínez; Nabaes Jodar; Natale; R; S; Ticeira; Valinotto; Viegas, M. |
| EPI_ISL_4405363 | Laboratorio de Biología Molecular. Hospital Dr. Héctor Cura | Área de Secuenciación del Laboratorio de Virología del Hospital de Niños Dr. Ricardo Gutierrez on behalf of 'Proyecto Argentino Interinstitucional de genomica de SARS-CoV-2' (PAIS Consortium) | Acuña; D; Ghiano; Goya; J; LE; Lusso; MB; MI; MS; N; Nabaes Jodar; Natale; R; S; Spina; Turrina; Valinotto; Viegas, M.; Zaffanella |
| EPI_ISL_7406423 | Laboratorio de Biología molecular del Hospital General de Agudos Dr. Carlos G. Durand | Área de Secuenciación del Laboratorio de Virología del Hospital de Niños Dr. Ricardo Gutierrez on behalf of 'Proyecto Argentino Interinstitucional de genomica de SARS-CoV-2' (PAIS Consortium) | Acuña; Adriana Rozo; Ailin Domínguez; Albano Costa; Anisa Marchissio; Belen Warszatska; Camila Elsegood; Cecilia Irrazábal; Clara Theaux; Constanza Marina; D; Estela Dima; Eugenia Langan; Flavia Rivarola; Goya; Guillermo Notaristéfano; LE; Laura Jurado; Laura Álvarez; Lucas Dahinten; Lusso; MI; MS; Maricel Leivas; Martín Rodríguez Saá; Maryury Colina; María Belén Rodríguez Cardozo; María Fernanda Yaunguzian; Nabaes Jodar; Natale; Natalia Chiussi; Natasha Frisone; Noelia Anzorena; Noelia Narduzzi; Nuria Cañellas Walter Loayza; Rocío Carrón; Romina Gatica; S; Silvia Matillas; Valinotto; Viegas, M.; Yesica Castro. Promoción y Protección de la Salud: Miriam Bruno |
| EPI_ISL_4030317, EPI_ISL_4030326, EPI_ISL_4030345, EPI_ISL_4030348 | Laboratorio de Ecologia de Doencas Transmissiveis na Amazonia, Instituto Leonidas e Maria Deane - Fiocruz Amazonia | Laboratorio de Ecologia de Doencas Transmissiveis na Amazonia, Instituto Leonidas e Maria Deane - Fiocruz Amazonia | Alex Martins; André Corado; Debora Duarte; Fabiola Mendonça da Silva Chui; Felipe Naveca; Fernanda Nascimento; Fernando Fonseca de Almeida Val; George Silva; Gisely Cardoso de Melo; Karina Pessoa; Luciana Gonçalves; Marcus Vinicius Guimarães de Lacerda; Maria Júlia Brandão; Maria Paula Gomes Mourão; Mariana Xavier Simão; Matilde Mejía; Michele Jesus; Valdinete Nascimento; Vanderson de Souza Sampaio; Victor Souza; Ágatha Costa |
| EPI_ISL_3642592, EPI_ISL_3642594, EPI_ISL_3642595 | Laboratorio de Infectologia y Virologia Molecular | Laboratory of Molecular Virology, School of Medicine, Pontificia Universidad Catolica de Chile | Ana Maria Contreras; Andres E. Munoz-Marcos; Carlos Palma; Catalina Pardo-Roa; Constanza Maldonado; Constanza Martinez-Valdevenito; Eileen Serrano; Erick Salinas; Estefany Poblete; Francisco Melo; Jennifer Angulo; Jorge Levican; Leonardo I. Almonacid; M. Belen Leyton; Marcela Ferres; Maria Jose Avendano; Rafael A. Medina; Tamara Garcia-Salum |
| EPI_ISL_3048998, EPI_ISL_3048999, EPI_ISL_3049000, EPI_ISL_3761761 | Laboratorio de Pesquisa em Virologia, FAMERP, SJRP | Laboratorio de Pesquisa em Virologia, FAMERP, SJRP | Beatriz de Carvalho Marques; Cecília Artico Banho; Cíntia Bittar; Fábio Sossai Possebon; Guilherme Campos; Helena Lage Ferreira; Jorge A. Petroli Marchesi; João Pessoa Araújo Jr.; Leila Sabrina Ullmann; Lívia Sacchetto; Maisa C. Pereira Parra; Marília Moraes; Maurício L. Nogueira.; Paula Rahal; Paulo Inacio da Costa |
| EPI_ISL_3547035, EPI_ISL_3672407, EPI_ISL_3672408, EPI_ISL_3672409, EPI_ISL_3672411, EPI_ISL_3672424, EPI_ISL_3672427, EPI_ISL_3672431, EPI_ISL_3672432, EPI_ISL_3672433, EPI_ISL_3672434, EPI_ISL_3672439, EPI_ISL_3826389, EPI_ISL_3826390, EPI_ISL_3826391, EPI_ISL_3826412, EPI_ISL_3826425, EPI_ISL_3826426, EPI_ISL_3826432, EPI_ISL_3826433, EPI_ISL_3826443, EPI_ISL_3826444, EPI_ISL_3991472, EPI_ISL_3991475, EPI_ISL_3991478, EPI_ISL_3991481, EPI_ISL_3991485, EPI_ISL_3991488, EPI_ISL_3991491, EPI_ISL_3991494, EPI_ISL_3991497, EPI_ISL_3991501, EPI_ISL_3991633, EPI_ISL_4052705, EPI_ISL_4052743, EPI_ISL_4052750, EPI_ISL_4052758, EPI_ISL_4052762, EPI_ISL_4080441, EPI_ISL_4080442, EPI_ISL_4080448, EPI_ISL_4080450, EPI_ISL_4080456, EPI_ISL_4080461, EPI_ISL_4080462, EPI_ISL_4080463, EPI_ISL_4080464, EPI_ISL_4080465, EPI_ISL_4080466, EPI_ISL_4080467, EPI_ISL_4080468, EPI_ISL_4080473, EPI_ISL_4080474, EPI_ISL_4080475, EPI_ISL_4080517, EPI_ISL_4080521, EPI_ISL_4080522, EPI_ISL_4080527, EPI_ISL_4080541, EPI_ISL_4172674, EPI_ISL_4172677, EPI_ISL_4172691, EPI_ISL_4172695, EPI_ISL_4172697, EPI_ISL_4172699, EPI_ISL_4172700, EPI_ISL_4172704, EPI_ISL_4172708, EPI_ISL_4172714, EPI_ISL_4172719, EPI_ISL_4172721, EPI_ISL_4172723, EPI_ISL_4172906, EPI_ISL_4172913 | | | |
| see above | Laboratorio de Referencia Nacional de Virus Respiratorios. Centro Nacional de Salud Publica. Instituto Nacional de Salud Peru. | Laboratorio de Referencia Nacional de Virus Respiratorios. Centro Nacional de Salud Publica. Instituto Nacional de Salud Peru. | Carlos Padilla Rojas; Henri Bailon Calderon; Iris Silva Molina; Joseph Huayra Niquen; Lely Solari Zerpa; Luis Barcena Flores; Marco Galarza Perez; Nancy Rojas Serrano; Nieves Sevilla Castañeda; Omar Caceres Rey; Orson Mestanza Millones; Princesa Medrano Alhuay; Priscila Lope Pari; Sandra Morales Ruiz; Sara Gordillo Vilchez; Steve Acedo Lazo; Veronica Hurtado Vela; Victor Jimenez Vasquez; Wendy Lizarraga Olivares |
| EPI_ISL_2925741 | Laboratorio de Virología HUCA | Laboratorio de Virología HUCA | Abreu F; Alvarez-Arguelles ME; Boga JA; Castelló C; Costales I; Coto E; Gómez de Oña J; Martín-Rodríguez G; Melón S; Perez-Martínez Z; Rojo S; Sandoval M |
| EPI_ISL_4405138 | Laboratorio de Virología del Hospital de Niños Dr. Ricardo Gutierrez | Área de Secuenciación del Laboratorio de Virología del Hospital de Niños Dr. Ricardo Gutierrez on | AS; Acevedo; Acuña; Alexay; Alvarez Lopez; C; D; E; Goya; Gravis; Jacques; LE; Lusso; ME; MI; MS; Mistchenko; Nabaes Jodar; Natale; O; S; Valinotto; Viegas, M. |

| | | | |
|---|---|---|---|
| | | behalf of 'Proyecto Argentino Interinstitucional de genomica de SARS-CoV-2' (PAIS Consortium) | |
| EPI_ISL_4405637 | Laboratorio de genética y biología molecular del Hospital de trauma y emergencia Dr Federico Abete | Área de Secuenciación del Laboratorio de Virología del Hospital de Niños Dr. Ricardo Gutierrez on behalf of 'Proyecto Argentino Interinstitucional de genomica de SARS-CoV-2' (PAIS Consortium) | Acuña; D; Gallino; Goya; I; LE; Lusso; MI; MS; Nabaes Jodar; Natale; Primost; S; Valinotto; Viegas, M. |
| EPI_ISL_7620385 | Laboratorio del Hospital Regional Ushuaia Gdor. Ernesto Campos | Nodo de Secuenciación Tierra del Fuego - Hospital Regional Ushuaia - Centro Austral De Investigaciones Científicas - Universidad Nacional De Tierra Del Fuego on behalf PAIS project and ANLIS-Malbran | AE; Boutureira, MF.; CA; CB; CF; Castro; Ceballos; Cáceres; De Roccis; F; G; Gallego; Gramundi; ID; Nardi; Rojas; SB; SG; Yulan |
| EPI_ISL_2987691, EPI_ISL_2987692 | Laboratorios Delia Barraza | Laboratorio nacional-LANIIA-CIAD | ; Alejandra García-Gasca; Alejandra Hernández-Terán; Alejandro Sánchez-Flores; Alfredo Herrera-Estrella; Alicia Ocaña-Mondragón; Andreu Comas-García; Angel Gustavo Salas-Lais; Antonio Loza Román; Bernardo Martínez-Miguel; Blanca Taboada; Brenda Irasema Maldonado-Meza; Bruno Gómez-Gil; Carla Ivón Herrera-Najera; Carlos F. Arias; Celia Boukadida; Clara Esperanza Santacruz-Tinoco; Claudia Soto Félix; Concepción Grajales-Muñiz; Consorcio Mexicano de Vigilancia Genómica (CoViGen-Mex). Authors (in alphabetical order): Julio Elias Alvarado-Yaah; Cristóbal Cháidez-Quiróz; Célida Duque Molina; Célida Martínez- Rodríguez; Daniel Fregoso-Rueda; Daniel Lira Morales; Eduardo Becerril-Vargas; Fernando Fontove-Herrera; Fidencio Mejía-Nepomuceno; Francisco Pulido; Gloria Elena Espinosa-Ayala; Gloria María Molina-Salinas; Gloria Vazquez; Hector Esteban Paz-Juárez; Hector Montoya-Fuentes; Helen Haydee Fernanda Ramírez-Plascencia; Irvin González-López; Jean Pierre González; Jesús Hernández; Joel Armando Vázquez-Pérez. Laboratorios Delia Barraza: María Delia Barraza Sámano; Jorge Salas-Hernández; José Antonio Enciso-Moreno; José Arturo Martínez-Orozco; José Esteban Muñoz-Medina; José de Jesús Nuñez-Contreras; Juan Bautista Chale-Dzul; Julissa Enciso-Ibarra; Luis Alberto Ochoa-Carrera; Margarita Matías-Florentino; Mario Mújica-Sánchez; Marissa Perez-Garcia; María Guadalupe Santiago-Mauricio; María Guadalupe de Jesús Mireles-Rivera; Melissa García Angulo.; Nelly Sélem-Mojica; Pavel Isa; Ricardo Ciria Merce; Ricardo Grande; Rosa María Gutiérrez Rios; Santiago Ávila-Ríos; Selene Zárate; Susana Lopez; Verónica Mata-Haro; Victor Eduardo García-Arias; Víctor Hugo Borja-Aburto |
| EPI_ISL_2611384, EPI_ISL_2611392, EPI_ISL_2611397, EPI_ISL_2611398, EPI_ISL_2611413, EPI_ISL_2611418, EPI_ISL_2611428, EPI_ISL_2611812, EPI_ISL_2611823, EPI_ISL_2872165, EPI_ISL_2872473, EPI_ISL_2872474, EPI_ISL_2873122, EPI_ISL_2873124, EPI_ISL_2873127, EPI_ISL_4372568, EPI_ISL_4372572, EPI_ISL_4372611, EPI_ISL_4372658, EPI_ISL_4372751, EPI_ISL_4372818, EPI_ISL_4372883, EPI_ISL_4372890, EPI_ISL_4372929, EPI_ISL_4373051, EPI_ISL_4373137, EPI_ISL_4373266, EPI_ISL_4373307, EPI_ISL_4373338, EPI_ISL_4373466, EPI_ISL_4373493, EPI_ISL_4373684, EPI_ISL_4373919, EPI_ISL_4373925, EPI_ISL_4373963, EPI_ISL_4374052, EPI_ISL_4374134, EPI_ISL_4374149, EPI_ISL_4374153, EPI_ISL_4374177, EPI_ISL_4374226, EPI_ISL_4374238, EPI_ISL_4374329, EPI_ISL_4374432, EPI_ISL_4374494, EPI_ISL_4374525, EPI_ISL_4374612, EPI_ISL_4374629, EPI_ISL_4374640, EPI_ISL_4374739, EPI_ISL_4374764, EPI_ISL_4374810, EPI_ISL_4374836, EPI_ISL_4374920, EPI_ISL_4374964, EPI_ISL_4374998, EPI_ISL_4375011, EPI_ISL_4375016, EPI_ISL_4375026, EPI_ISL_4375061, EPI_ISL_4375113, EPI_ISL_4375146, EPI_ISL_4375284, EPI_ISL_4375556, EPI_ISL_4375571, EPI_ISL_4375667, EPI_ISL_4375728, EPI_ISL_4375884, EPI_ISL_4376077, EPI_ISL_4376095, EPI_ISL_4376359, EPI_ISL_4376361, EPI_ISL_4376372, EPI_ISL_4376376, EPI_ISL_4376474, EPI_ISL_4376485, EPI_ISL_4376564, EPI_ISL_4376592, EPI_ISL_4376607, EPI_ISL_4376814, EPI_ISL_4376853, EPI_ISL_4376867, EPI_ISL_4376911, EPI_ISL_4377144, EPI_ISL_4377390, EPI_ISL_4377590, EPI_ISL_4377689, EPI_ISL_4377727, EPI_ISL_4377742, EPI_ISL_4377776, EPI_ISL_4377778, EPI_ISL_4377866, EPI_ISL_4377891, EPI_ISL_4378089, EPI_ISL_4378140, EPI_ISL_4378253, EPI_ISL_4378336, EPI_ISL_4378355, EPI_ISL_4378448, EPI_ISL_4378575, EPI_ISL_4378601, EPI_ISL_4378671, EPI_ISL_4378673, EPI_ISL_4378750, EPI_ISL_4378764, EPI_ISL_4378844, EPI_ISL_4378947, EPI_ISL_4379000, EPI_ISL_4379053, EPI_ISL_4379156, EPI_ISL_4379275, EPI_ISL_4379296, EPI_ISL_4379369, EPI_ISL_4379381, EPI_ISL_4379385, EPI_ISL_4379395, EPI_ISL_4379448, EPI_ISL_4379452, EPI_ISL_4379569, EPI_ISL_4379703, EPI_ISL_4379752, EPI_ISL_4379927, EPI_ISL_4380135, EPI_ISL_4380152, EPI_ISL_4380214, EPI_ISL_4380269, EPI_ISL_4380517, EPI_ISL_4380676, EPI_ISL_4380691, EPI_ISL_4380850, EPI_ISL_4380979, EPI_ISL_4381007, EPI_ISL_4381047, EPI_ISL_4381071, EPI_ISL_4381192, EPI_ISL_4381258, EPI_ISL_4381314, EPI_ISL_4381388, EPI_ISL_4381530, EPI_ISL_4381793, EPI_ISL_4381866, EPI_ISL_4381979, EPI_ISL_4382149, EPI_ISL_4382213, EPI_ISL_4382344, EPI_ISL_4382382, EPI_ISL_4382422, EPI_ISL_4382531 | | | |
| see above | Laboratory Corporation of America | Centers for Disease Control and Prevention Division of Viral Diseases, Pathogen Discovery | Adrian Paskey; Amanda Douglas; Amanda Suchanek; Andrea Throop; Ayla Burns; Benjamin Rambo-Martin; Bobbi Croy; Brian Krueger; Brian Norvell; Christopher Gulvick; Christos Petropoulos; Clinton Paden; Clinton R. Paden; Craig Lukasik; Dakota Howard; Darlene Wagner; Debbie Boles; Dhwani Batra; Duncan MacCannell; Eyad Almasri; Goran Stevovic; Howard Engler; Hrushikesh Deshmukh; Jake Humphrey; Jana Schroth; Jason Caravas; Joe Voshell; John Pruitt; Jonathan Meltzer; Jonathan Williams; Kara Moser; Kimberly Wagner; Kristine Lacek; Lax Iyer; Lisa Pfefferle; Lyndon Tilson; Manoj Jain; Marcia Eisenberg; Mary Ann Cristobal; Mary Cristobal; Mary Williamson; Matthew Robinson; Matthew Schmerer; Michael Levandoski; Mike Sapeta; Mindy Nye; Minoo Agarwal; Mohan Kolli; Nuthawin Charoensri; Oren Cohen; Peter Cook; Peter W. Cook; Prashant Gupta; Qian Zeng; Rama Ghatti; Scott Parker; Scott Ryan; Scott Sammons; Shatavia Morrison; Stanley Letovsky; Steven Ragan; Suresh Babu Selvaraju; Suresh Selvaraju; Susan Countryman; Susan Hicks; Suzanne Dale; Thomas Urban; Tim Kuphal; Tricia Zwiefelhofer; Tymeckia Kendall; Victoria Caban Figueroa; Vincent Drouillon; Yvette Unoarumhi |
| EPI_ISL_2557409, EPI_ISL_2557410, EPI_ISL_2645506, EPI_ISL_2982720, EPI_ISL_2982721, EPI_ISL_2982722, EPI_ISL_2982723, EPI_ISL_2982724, EPI_ISL_2982725, EPI_ISL_2982726, EPI_ISL_2982727, EPI_ISL_2982728, EPI_ISL_2982729, EPI_ISL_3045382, EPI_ISL_3045468, EPI_ISL_3045469, EPI_ISL_3045470, EPI_ISL_3045472, EPI_ISL_3045473, EPI_ISL_3045474, EPI_ISL_3045475, EPI_ISL_3045476, EPI_ISL_3045477, EPI_ISL_3045478, EPI_ISL_3045495, EPI_ISL_3045553, EPI_ISL_5158276, EPI_ISL_5158277, EPI_ISL_6943987, EPI_ISL_6943988 | | | |
| see above | Laboratory of Respiratory Viruses and Measles, Oswaldo Cruz Institute, FIOCRUZ | Laboratory of Respiratory Viruses and Measles, Oswaldo Cruz Institute, FIOCRUZ | Agatha Cristinne Prudencio; Alice Sampaio Rocha; Ana Carolina Mendonca; Anna Carolina Paixao; Bruna Mendonça da Silva; Elisa Cavalcante Pereira; Fernando Motta; Ighor Arantes; Ighor Leonardo Arantes Gomes; Jéssica Graça Macedo de Carvalho; Larissa Macedo Pinto; Luciana Appolinario; Marilda Siqueira on behalf of the Fiocruz COVID-19 Genomic Surveillance Network; Paola Resende; Patricia Brasil; Renata Serrano Lopes; Taina Venas; Thais Aguiar; Victor Guimaraes |
| EPI_ISL_3025635, EPI_ISL_3461195, EPI_ISL_3461216, EPI_ISL_3461217, EPI_ISL_3461225, EPI_ISL_3461263, EPI_ISL_3461269, EPI_ISL_3461306 | | | |
| see above | Laboratory, Gravity Diagnostics | Laboratory, Gravity Diagnostics | Diagnostics, G. |
| EPI_ISL_4600475 | Laboratório Central de Saúde Pública Profº Gonçalo Moniz | Coordenação Geral de Laboratórios de Saúde Pública (CGLAB/DAEVS/SVS/MS) | Vagner Fonseca; et al. |
| EPI_ISL_4600521, EPI_ISL_4600522 | Laboratório Central de Saúde Pública de Pernambuco | Coordenação Geral de Laboratórios de Saúde Pública (CGLAB/DAEVS/SVS/MS) | Vagner Fonseca; et al. |
| EPI_ISL_3050337, EPI_ISL_3050338, EPI_ISL_3050339, EPI_ISL_3050340, EPI_ISL_3050341, EPI_ISL_3050342, EPI_ISL_3050343, EPI_ISL_3050344, EPI_ISL_3050345, EPI_ISL_3050346, EPI_ISL_3050347, EPI_ISL_3050348, EPI_ISL_3050349, EPI_ISL_3050350, EPI_ISL_3050351, EPI_ISL_3050352, EPI_ISL_3050353, EPI_ISL_3050354, EPI_ISL_3050355, EPI_ISL_3050356, EPI_ISL_3050358, EPI_ISL_3050359, EPI_ISL_3050360, EPI_ISL_3050361, EPI_ISL_3050362, EPI_ISL_3050363, EPI_ISL_3050364, EPI_ISL_3050365, EPI_ISL_3050366, EPI_ISL_3050367, EPI_ISL_3050368, EPI_ISL_3050369, EPI_ISL_3050370, EPI_ISL_3050371, EPI_ISL_3050372, EPI_ISL_3050373, EPI_ISL_3050374, EPI_ISL_3050375, EPI_ISL_3050376, EPI_ISL_3050377, EPI_ISL_3050378, EPI_ISL_3050379, EPI_ISL_3050380, EPI_ISL_3050381, EPI_ISL_3050382, EPI_ISL_3050383, EPI_ISL_3050384, EPI_ISL_3050385, EPI_ISL_3050386, EPI_ISL_3050387, EPI_ISL_3050388, EPI_ISL_3050389, EPI_ISL_3050390, EPI_ISL_3050391, EPI_ISL_3050392, EPI_ISL_3050393, EPI_ISL_3050394 | | | |
| see above | Laboratório Central de Saúde Pública do Amazonas - LACEN-AM | Laboratorio de Ecologia de Doencas Transmissiveis na Amazonia, Instituto Leonidas e Maria Deane - Fiocruz Amazonia | André Corado; Felipe Naveca; Fernanda Nascimento; George Silva; Karina Pessoa; Luciana Gonçalves; Maria Júlia Brandão; Matilde Mejía; Valdinete Nascimento; Victor Souza; Ágatha Costa |
| EPI_ISL_4170378, EPI_ISL_4170379, EPI_ISL_4170388, EPI_ISL_4170390, EPI_ISL_4212830, EPI_ISL_4212831, EPI_ISL_4212832, EPI_ISL_4212833, EPI_ISL_4220302, EPI_ISL_4220303, EPI_ISL_4220304, EPI_ISL_4220305, EPI_ISL_4220306, EPI_ISL_4220307, EPI_ISL_4220308, EPI_ISL_4220309, EPI_ISL_4220310, EPI_ISL_4220311, EPI_ISL_4220312, EPI_ISL_4220313, EPI_ISL_4220314, EPI_ISL_4220315, EPI_ISL_4220316, EPI_ISL_4220317, EPI_ISL_4220318, EPI_ISL_4271141, EPI_ISL_4271142, EPI_ISL_4271143, EPI_ISL_4271144, EPI_ISL_4271145, EPI_ISL_4271147, EPI_ISL_4271148, EPI_ISL_4271149, EPI_ISL_4271150, EPI_ISL_4271151, EPI_ISL_4271152, EPI_ISL_4271153, EPI_ISL_4271154, EPI_ISL_4271155, EPI_ISL_4271156, EPI_ISL_4271157, EPI_ISL_4271158, EPI_ISL_4271159, EPI_ISL_4271160, EPI_ISL_4271161, EPI_ISL_4271162, EPI_ISL_4271163, EPI_ISL_4271164, EPI_ISL_4271165, EPI_ISL_4271166, EPI_ISL_4271167, EPI_ISL_4271168, EPI_ISL_4271169, EPI_ISL_4271170, EPI_ISL_4271171, EPI_ISL_4271172, EPI_ISL_4271173, EPI_ISL_4271174, EPI_ISL_4271175, EPI_ISL_4271176, EPI_ISL_4271177, EPI_ISL_4271178, EPI_ISL_4271179, EPI_ISL_4271180, EPI_ISL_4271181, EPI_ISL_4271182, EPI_ISL_4271183, EPI_ISL_4271184, EPI_ISL_4271185, EPI_ISL_4271187, EPI_ISL_4271193, EPI_ISL_4271195, EPI_ISL_4271196, EPI_ISL_4271199, EPI_ISL_4271200, EPI_ISL_4271204, EPI_ISL_4271212, EPI_ISL_4271218, EPI_ISL_4271219, EPI_ISL_4271222, EPI_ISL_4271224, EPI_ISL_4271226, EPI_ISL_4271227, EPI_ISL_4271230, EPI_ISL_4271231, EPI_ISL_4271232, EPI_ISL_4271233, EPI_ISL_4271234, EPI_ISL_4271236, EPI_ISL_4271238, EPI_ISL_4271239, EPI_ISL_4271241, EPI_ISL_4271244, EPI_ISL_4271246, EPI_ISL_4271249, EPI_ISL_4271250, EPI_ISL_4271255, EPI_ISL_4414931, EPI_ISL_4414932, EPI_ISL_4414933, EPI_ISL_4414934, EPI_ISL_4414935, EPI_ISL_4414936, EPI_ISL_4414937, EPI_ISL_4414938, EPI_ISL_4414939, EPI_ISL_4414940, EPI_ISL_4414941, EPI_ISL_4414942, EPI_ISL_4414943, EPI_ISL_4414944, EPI_ISL_4414945, EPI_ISL_4414946, EPI_ISL_4414947, EPI_ISL_4414948, EPI_ISL_4414949, EPI_ISL_4414950, EPI_ISL_4414951, EPI_ISL_4414952, EPI_ISL_4414953, EPI_ISL_4414954, EPI_ISL_4414955, EPI_ISL_4414956, EPI_ISL_4414957, EPI_ISL_4414958, EPI_ISL_4418879, EPI_ISL_4543260, EPI_ISL_4543266, EPI_ISL_4543267, EPI_ISL_4543268, EPI_ISL_4543269, EPI_ISL_4543270, EPI_ISL_4543271, EPI_ISL_4543272, EPI_ISL_4543273, EPI_ISL_4543274, EPI_ISL_4543275, EPI_ISL_4543276, EPI_ISL_4543277, EPI_ISL_4543278, EPI_ISL_4543279, EPI_ISL_4543280, EPI_ISL_4543281, EPI_ISL_4543282, EPI_ISL_4543283, EPI_ISL_4543284, EPI_ISL_4543285, EPI_ISL_4543306, EPI_ISL_4543307, EPI_ISL_4543310, EPI_ISL_4543312, EPI_ISL_4543314, EPI_ISL_4543315, EPI_ISL_4543317, EPI_ISL_4543319, EPI_ISL_4543320, EPI_ISL_4543322, EPI_ISL_4543323, EPI_ISL_4543324, EPI_ISL_4543325, EPI_ISL_4543326, EPI_ISL_4543327, EPI_ISL_4543330, EPI_ISL_4543331, EPI_ISL_4543332, EPI_ISL_4543336, EPI_ISL_4543337, EPI_ISL_4543339, EPI_ISL_4543340, EPI_ISL_4543341, EPI_ISL_4543343, EPI_ISL_4543349, EPI_ISL_4543350, EPI_ISL_4543351, EPI_ISL_4543352, EPI_ISL_4543354, EPI_ISL_4543355, EPI_ISL_4543356, EPI_ISL_4543359, EPI_ISL_4543361, EPI_ISL_4543362, EPI_ISL_4543368, EPI_ISL_4543371 | | | |
| see above | Laboratório de Biotecnologia Aplicada (LBA) - Laboratório de Biologia Molecular - Hospital das Clínicas, Faculdade de Medicina de Botucatu, Departamento de Bioprocessos e Biotecnologia - Faculdade de Ciências Agronômicas, UNESP – Botucatu/SP | Laboratory of Respiratory Viruses and Measles, Oswaldo Cruz Institute, FIOCRUZ | Alice Sampaio Rocha; Ana Carolina Mendonca; Anna Carolina Paixao; Elisa Cavalcante Pereira; Felipe Allan da Silva Costa; Fernando Motta; Jayme Augusto de Souza Neto; Leonardo Nazario de Moraes; Luciana Appolinario; Marilda Siqueira on behalf of the Fiocruz COVID-19 Genomic Surveillance Network; Paola Resende; Patricia Akemi Assato; Rejane Maria Tommasini; Renata Serrano Lopes; Taina Venas |
| EPI_ISL_4520558, EPI_ISL_4520559, EPI_ISL_4520560, EPI_ISL_4520561, EPI_ISL_4520562, EPI_ISL_4520563, EPI_ISL_4520576, EPI_ISL_4520577, EPI_ISL_4520578, EPI_ISL_4520579, EPI_ISL_4520580, EPI_ISL_4520581, EPI_ISL_4520598, EPI_ISL_4520599 | | | |
| see above | Laboratório de Virologia e Cultivo Celular - LABVICC | Laboratório Baculovírus | Aline Belmok; Bergmann Morais Ribeiro; Clayton Luiz Borges; Déborah Anjos; Elen Regozino Muniz; Fabiola Souza Fiaccadori; Fernando L Melo; Gabriela Silvério Bazilio; Juliana Alves Parente Rocha; Keila Correia de Alcântara; Menira Souza; Nathânia Dábilla; Valéria Christina de Rezende Feres |
| EPI_ISL_2863595, EPI_ISL_2863600, EPI_ISL_2983419, EPI_ISL_2983420 | Labortorio Central de Saude Publica do Estado do Tocantins (LACEN/TO) | Laboratory of Respiratory Viruses and Measles, Oswaldo Cruz Institute, FIOCRUZ | Alice Sampaio Rocha; Ana Carolina Mendonca; Anna Carolina Paixao; Elisa Cavalcante Pereira; Fernando Motta; Jucimaria Dantas Galvao; Luciana Appolinario; Marilda Siqueira on behalf of the Fiocruz COVID-19 Genomic Surveillance Network; Paola Resende; Renata Serrano Lopes; Taina Venas |
| EPI_ISL_2554494, EPI_ISL_2568788, EPI_ISL_2640893 | Lighthouse Lab in Milton Keynes | Wellcome Sanger Institute for the COVID-19 Genomics UK (COG-UK) Consortium | Cordelia Langford; David K. Jackson; Dominic Kwiatkowski; Ewan Harrison; Ian Johnston; Jeffrey Barrett; John Sillitoe on behalf of the Wellcome Sanger Institute COVID-19 Surveillance Team; Roberto Amato; Sonia Goncalves; The Lighthouse Lab in Milton Keynes and Alex Alderton |
| EPI_ISL_2842582 | Lurie Children's Hospital of Chicago | Northwestern University - Ozer Lab | Egon A. Ozer; Judd F. Hultquist; Lacy M. Simons; Larry K. Kociolek; Michael G. Ison; Ramon Lorenzo-Redondo; Taylor J. Dean; William J. Muller; Xiaotian; Zheng |
| EPI_ISL_4063287 | M Health Fairview | Minnesota Department of Health, Public Health Laboratory | Alexandra Lorentz; Jacob Garfin; Matt Plumb; and Xiong Wang |
| EPI_ISL_5646832 | MARIA TEREZINHA BOGNAR CHECHETO | Instituto Butantan | Antonio Jorge Martins; Claudia Renata dos Santos Barros; David Schlesinger; Debora Botequio Moretti; Dimas Tadeu Covas; Elaine Cristina Marqueze; Elaine Vieira Santos; Evandra Strazza Rodrigues; Heidge Fukumasu; Jayme Augusto de Souza-Neto; José Salvatore Leister Patané; Luiz Alcantara; Luiz Lehmann Coutinho; Maria Carolina Elias; Maurício Lacerda Nogueira; Rafael dos Santos Bezerra; Raul Machado Neto; Rejane Maria Tommasini Grotto; Ricardo Haddad; Sandra Coccuzzo Sampaio Vessoni; Simone Kashima; Svetoslav Nanev Slavov; Vincent Louis Viala |
| EPI_ISL_4494427 | MB-Cadham Provincial laboratory | Manitoba Cadham Provincial Laboratory | Anna Majer; Anneliese Landgraff; David Alexander; Elsie Grudeski; Gary Van Domselaar; Grace Seo; Jared Bullard; Jennifer Tanner; Kerry Dust; Morag Graham; Nat; Natalie Knox; Paul Van Caeseele; Philip Mabon; Rhiannon Huzarewich; Russell Mandes; Shari Tyson |
| EPI_ISL_2659675 | MEPHI, Aix Marseille University | MEPHI, Aix Marseille University | Anthony LEVASSEUR |
| EPI_ISL_2646337, EPI_ISL_2646340, EPI_ISL_2690270 | MS Public Health Laboratory | University of Mississippi Medical Center, Molecular and Genomics Core Facility | Ashley C. Johnson; D. Ashley Robinson; Ithiel J. Frame; Krishna K. Ayyalasomayajula; Michael R. Garrett; Wenjie Wu |
| EPI_ISL_6492832 | MSHS Clinical Microbiology Laboratories | MSHS Pathogen Surveillance Program | Adolfo García-Sastre; Adriana van de Guchte; Ajay Obla; Alberto Paniz-Mondolfi; Ana S. Gonzalez-Reiche; Angela Amoako; Ashley Salimbangon; Betsaida Salom Melo; Bremy Alburquerque; Brianne Ciferri; Charles Gleason; Daniel Floda; Deena R. Altman; Denise Jurczyszak; Emilia Mia Sordillo; Gintaras Deikus; Giulio Kleiner; Gopi Patel; Hala Alshammary; Harm van Bakel; Irina Oussenko; Jayeeta Dutta; Juan Soto; Julia Matthews; Katherine Beach; Kathryn Twyman; Kayla Russo; Komal Srivastava; Levy Sominsky; Mahmoud Awawda; Marta Luksza; Matthew M. Hernandez; Melissa Gitman; Michael D. Nowak; Mitchell J. Sullivan; Nancy Francoeur; Robert Sebra; Sarah Schaefer; Shelcie Fabre; Shwetha Hara Sridhar; Viviana Simon; Ying-Chih Wang; Zain Khalil; Zenab Khan |
| EPI_ISL_2635865, EPI_ISL_2635867, EPI_ISL_2635874, EPI_ISL_2635888, EPI_ISL_2678714 | MVZ Labor Dr. Limbach & Kollegen GbR | Robert Koch Institute | |
| EPI_ISL_2635823 | MVZ Labor Dr. Reising-Ackermann und | Robert Koch Institute | |

| | | | |
|---|---|---|---|
| | Kollegen | | |
| EPI_ISL_2635480 | MVZ für Laboratoriumsmedizin und Mikrobiologie Würzburg | Robert Koch Institute | |
| EPI_ISL_3085974, EPI_ISL_4083936, EPI_ISL_4083937, EPI_ISL_4083938, EPI_ISL_4083940, EPI_ISL_4083942, EPI_ISL_4083945 | | | |
| see above | Maine Health and Environmental Testing Laboratory | Tewhey Lab, The Jackson Laboratory | Barter, M.; Dewey, H.; H. and Tewhey, R.; Iosue, F.; Lynch, R.; Matluk, N.; Munger |
| EPI_ISL_2786155, EPI_ISL_2786162, EPI_ISL_2786179, EPI_ISL_2786233, EPI_ISL_2786251, EPI_ISL_2786262, EPI_ISL_2786283, EPI_ISL_2786312, EPI_ISL_2786316, EPI_ISL_2786322, EPI_ISL_2786329, EPI_ISL_2786370, EPI_ISL_2786442, EPI_ISL_2875281, EPI_ISL_2875282, EPI_ISL_2875290, EPI_ISL_2875306, EPI_ISL_2875368 | | | |
| see above | Mako Medical | Centers for Disease Control and Prevention Division of Viral Diseases, Pathogen Discovery | Adrian Paskey; Benjamin Rambo-Martin; Christopher Gulvick; Clinton R. Paden; Dakota Howard; Darlene Wagner; Dhwani Batra; Duncan MacCannell; Jason Caravas; Kara Moser; Lauren Moon; Matthew Schmerer; Matthew Tugwell; Peter W. Cook; Scott Sammons; Shatavia Morrison; Yvette Unoarumhi |
| EPI_ISL_6666000 | Mass General Brigham | Mass General Brigham | A.E.; Adams, G.; Anahtar, M.; B.L.; B.W.; Bauer, M.; Birren; Branda, J.; Carter, A.; Cerrato, F.; Chaluvadi, S.; Chapman; Cusick, C.; D.J.; DeRuff, K.; E. and Sabeti; Flowers, K.; Gallagher, G.; Gladden-Young, A.; Gnirke, A.; Harris, J.; J.E.; K.J.; LaRocque, R.; Lagerborg, K.; Lemieux; Lin; Loreth, C.; MacInnis; Neumann, A.; Normandin, E.; P.C.; Park; Pierce, V.; Reilly, S.; Rosenberg; Rudy, M.; Ryan, E.; S.B.; Shaw, B.; Siddle; Slater, D.; Smole, S.; Tomkins-Tinch, C.; Turbett, S. |
| EPI_ISL_2920258, EPI_ISL_2920271 | Massachusetts State Public Health Laboratory | Massachusetts State Public Health Laboratory | Andrew Lang; Glen Gallagher; Sandra Smole; Timelia Fink |
| EPI_ISL_2772701 | Max von Pettenkofer Institute, Virology, National Reference Center for Retroviruses, LMU Munich | Laboratory for Functional Genome Analysis; Dept. Genomics; Gene Center of the LMU Munich | Alexander Graf; Helmut Blum; Marcel Stern; Oliver Keppler; Stefan Krebs |
| EPI_ISL_2663232, EPI_ISL_2663238 | Mayo Clinic & Mayo Clinic Laboratories | Minnesota Department of Health, Public Health Laboratory | Alexandra Lorentz; Jacob Garfin; Matt Plumb; and Xiong Wang |
| EPI_ISL_2544257 | Medica | Institute of Medical Virology | Alexandra Trkola; Annette Audigé; Cyril Shah; Gabriela Ziltener; Guido Bloemberg; Jon Huder; Jürg Böni; Kevin Steiner; Maria Grünberg; Maryam Zaheri; Michael Huber; Riccarda Capaul; Stefan Schmutz; Verena Kufner |
| EPI_ISL_2636815 | Medizinisch-Diagnostisches Labor Kempten allgäulab | Robert Koch Institute | |
| EPI_ISL_3047540 | Michael E. DeBakey VA Medical Center | Genomics and Discovery, Respiratory Viruses Branch, Division of Viral Diseases, Centers for Disease Control and Prevention | Adam Retchless; Anna Kelleher; Anna Uehara; Brian Lynch; Clinton R. Paden; Dhwani Batra; Haibin Wang; Han Jia Justin Ng; Jasmine Padilla; Jing Zhang; Justin Lee; Mark Burroughs; Mili Sheth; Morgan Davis; Peter Cook; Rachel Marine; Sarah Nobles; Suxiang Tong; Tara Coalter; Yan Li; Ying Tao |
| EPI_ISL_3030616 | Microbiology Department & Molecular Biology CORE \| CDB \| Hospital Clinic \| Barcelona | Microbiology Department & Molecular Biology CORE \| CDB \| Hospital Clinic \| Barcelona | Andrea Vergara Gomez; Jose Luis Villanueva Cañas; Miguel Julian Martínez Yoldi; Mª Angeles Marcos Maeso; Mª del Mar Mosquera Gutierrez |
| EPI_ISL_2967893 | Microbiology Department. Complexo Hospitalario Universitario de Vigo | Microbiology Department. Complexo Hospitalario Universitario de Vigo | Alfaya N; Alonso I; Alvarez M; Cabrera JJ; Carballo R; Cores O; Cortizo S; Davina C; Martinez L; Mediero G; Perez S; Potel C; Regueiro B; Rey S; Vasallo FJ; del-Campo V |
| EPI_ISL_3063340, EPI_ISL_3063345, EPI_ISL_3063349, EPI_ISL_3063351, EPI_ISL_3063361, EPI_ISL_3063365, EPI_ISL_3063369, EPI_ISL_3066744, EPI_ISL_3066745, EPI_ISL_3066751 | | | |
| see above | Ministry of Health Turkey | Ministry of Health Turkey | Fatma Bayrakdar; Gulay Korukluoglu; Suleyman Yalcin; Yasemin Cosgun |
| EPI_ISL_2598160, EPI_ISL_2790207 | Minnesota Department of Health, Public Health Laboratory | Minnesota Department of Health, Public Health Laboratory | Alexandra Lorentz; Jacob Garfin; Matt Plumb; and Xiong Wang |
| EPI_ISL_2861116 | Mobile County Health Department | Synergy Laboratories | Megan Cornwell |
| EPI_ISL_3046585 | Molecular Biology Laboratory, Westchester Medical Center | Genomics Core Laboratory, New York Medical College | Changhong Yin; Jian Zhuge; Utsav Pandey; Weihua Huang |
| EPI_ISL_2537374 | Montana Public Health Laboratory | Montana Public Health Laboratory | Carrie Biskupiak; Deborah Gibson; Joy Ritter; Michael Dills; Michelle Mozer |
| EPI_ISL_5681864 | NAMRU-6 | NAMRU-6 | Cristhopher Cruz; Eugenio Abente; Gilda Troncos; Marita Silva; Paul Graf.; Sonia Ampuero; Sonia Florez-Lucana; Victoria Espejo; Yeny Tinoco |
| EPI_ISL_2657668 | NCSLPH | NCSLPH | Chase K; Glover W; Greene S; Miller MC |
| EPI_ISL_2787857 | NJ Public Health and Environmental Laboratories | Centers for Disease Control and Prevention Division of Viral Diseases, Pathogen Discovery | Alison Laufer Halpin; Ben L. Rambo-Martin; Clinton R. Paden; Dakota Howard; Darlene Wagner; Dave Wentworth; Dhwani Batra; Jasmine Padilla; Justin Lee; Katie Dillon; Krista Queen; Kristen Knipe; Kristine Lacek; Mark Burroughs; Matthew Schmerer; Mili Sheth; Peter Cook; Sam Shepard; Sarah Nobles; Shoshona Le; Suxiang Tong; Vivien Dugan; Yvette Unoarumhi |
| EPI_ISL_2557148, EPI_ISL_2557157, EPI_ISL_2649881, EPI_ISL_2842678, EPI_ISL_2940311 | NJDOH, Public Health and Environmental Laboratories | NJ_PHEL | Allison Roder; Byeong Jeong; Chelsea San Filippo; Dana Woell; Jacquelyn Deverell; Lindsey Bodnar; Ryan Pachucki; Shiv K. Verma |
| EPI_ISL_3150980 | NL-Dr. Leonard A. Miller Centre for Health Services | National Microbiology Laboratory (NML) | Anna Majer; Anneliese Landgraff; CanCOGeN's metadata curation team; Darian Hole; Elsie Grudeski; Gary Van Domselaar; George Zahariadis; Grace Seo; Jennifer Tanner; Kerri Smith; Kirsten Biggar; Laura Gilbert; Madison Chapel; Morag Graham; Natalie Knox; Nathalie Bastien; Philip Mabon; Public Health Agency of Canada CanCOGeN team; Rhiannon Huzarewich; Robert Needle; Russell Mandes; Shari Tyson; Timothy Booth; Yan Li; Yang Yu |
| EPI_ISL_5646511, EPI_ISL_5646526, EPI_ISL_5646528, EPI_ISL_5646560, EPI_ISL_5646582, EPI_ISL_5646591, EPI_ISL_5646606, EPI_ISL_5646622, EPI_ISL_5646636, EPI_ISL_5646673, EPI_ISL_5646696, EPI_ISL_5646699, EPI_ISL_5646711, EPI_ISL_5646743, EPI_ISL_5646848, EPI_ISL_5647283, EPI_ISL_5647286 | | | |
| see above | NUCLEO DE SAUDE MARY DOTA DE BAURU | Instituto Butantan | Antonio Jorge Martins; Claudia Renata dos Santos Barros; David Schlesinger; Debora Botequio Moretti; Dimas Tadeu Covas; Elaine Cristina Marqueze; Elaine Vieira Santos; Evandra Strazza Rodrigues; Heidge Fukumasu; Jayme Augusto de Souza-Neto; José Salvatore Leister Patané; Luiz Alcantara; Luiz Lehmann Coutinho; Maria Carolina Elias; Maurício Lacerda Nogueira; Rafael dos Santos Bezerra; Raul Machado Neto; Rejane Maria Tommasini Grotto; Ricardo Haddad; Sandra Coccuzzo Sampaio Vessoni; Simone Kashima; Svetoslav Nanev Slavov; Vincent Louis Viala |
| EPI_ISL_5646534, EPI_ISL_5646554, EPI_ISL_5646676, EPI_ISL_5646706, EPI_ISL_5646845, EPI_ISL_5646852 | NUCLEO DE SAUDE VILA FALCAO DE BAURU | Instituto Butantan | Antonio Jorge Martins; Claudia Renata dos Santos Barros; David Schlesinger; Debora Botequio Moretti; Dimas Tadeu Covas; Elaine Cristina Marqueze; Elaine Vieira Santos; Evandra Strazza Rodrigues; Heidge Fukumasu; Jayme Augusto de Souza-Neto; José Salvatore Leister Patané; Luiz Alcantara; Luiz Lehmann Coutinho; Maria Carolina Elias; Maurício Lacerda Nogueira; Rafael dos Santos Bezerra; Raul Machado Neto; Rejane Maria Tommasini Grotto; Ricardo Haddad; Sandra Coccuzzo Sampaio Vessoni; Simone Kashima; Svetoslav Nanev Slavov; Vincent Louis Viala |
| EPI_ISL_2753941 | NYU Langone Health | Departments of Pathology and Medicine, New York University School of Medicine | Adriana Heguy; Christian Marier; Dacia Dimartino; Emily Guzman; Gael Westby; Guiqing Wang; Paolo Cotzia; Paul Zappile; Peter Meyn; Sitharam Ramaswami; Yutong Zhang |
| EPI_ISL_3058135, EPI_ISL_3058136 | Nastavni zavod za javno zdravstvo Splitsko- Dalmatinske Županije | Hrvatski zavod za javno zdravstvo | Irena Tabain; Ivana Ferenčak |
| EPI_ISL_2661571 | National Institute of Public Health | National Institute of Public Health | Helena Jirincova et al |
| EPI_ISL_2545436, EPI_ISL_2545443, EPI_ISL_2545444, EPI_ISL_2545445, EPI_ISL_2545446, EPI_ISL_2545451, EPI_ISL_2687961, EPI_ISL_2687962, EPI_ISL_2687972, EPI_ISL_2687976, EPI_ISL_2756196, EPI_ISL_2756197, EPI_ISL_2756199, EPI_ISL_2756200, EPI_ISL_2756203, EPI_ISL_2756212, EPI_ISL_2895656, EPI_ISL_2895657 | | | |
| see above | National Platform bis UMONS/Jolimont | National Platform bis UMONS/Jolimont | Eric Tarantino; Florian Juszczak; François Dufrasne; Gautier Detry; Guillaume Bayon-Vicente; Ruddy Wattiez |
| EPI_ISL_2621962, EPI_ISL_2621963 | National Public Health Laboratory, National Centre for Infectious Diseases | National Public Health Laboratory, National Centre for Infectious Diseases | Lin Cui; Raymond Tzer Pin Lin; Royce Ang; Tze Minn Mak; Zhenyang Zhou |
| EPI_ISL_2466748, EPI_ISL_2534347, EPI_ISL_2689682 | Nebraska Public Health Laboratory | NPHL COVID-19 Response Team | NPHL COVID-19 Response Team |
| EPI_ISL_2612507, EPI_ISL_4877707, EPI_ISL_4877714 | Nevada State Public Health Laboratory | Nevada State Public Health Laboratory | Andrew Gorzalski; Mark Pandori |
| EPI_ISL_2715250 | North Dakota Department of Health, Public Health Laboratory | North Dakota Department of Health, Public Health Laboratory | Lisa Wingerter |
| EPI_ISL_3015666, EPI_ISL_3015668, EPI_ISL_3015671, EPI_ISL_3015675, EPI_ISL_3015681, EPI_ISL_3015686, EPI_ISL_3015690, EPI_ISL_3015691, EPI_ISL_3015701, EPI_ISL_3015753 | | | |
| see above | Northwest Laboratory | Seattle Flu Study | Amanda Adler; Barry R. Lutz; Benjamin Pelle; Brian Hiatt; Caitlin R. Wolf; Chris D. Frazar; Deborah A. Nickerson; Elisabeth Brandstetter; Erica Ryke; Geoff Melly; Helen Y. Chu; Janet A. Englund; Jay Shendure; Jover Lee; Kairsten Fay; Kirsten Lacombe; Lea M. Starita; Mark J. Rieder; Matthew Richardson; Matthew Thompson; Melissa Truong; Michael Boeckh; Michael Famulare; Misja Ilcisin; Peter D. Han; Philip Dykema; Romesh Gautom; Scott Lindquist; Thomas R. Sibley; Trevor Bedford |
| EPI_ISL_2842612 | Northwestern Memorial Hospital | Northwestern University - Ozer Lab | Chad J. Achenbach; Chao Qi; Egon A. Ozer; Judd F. Hultquist; Lacy M. Simons; Lawrence J. Jennings; Michael G. Ison; Ramon Lorenzo-Redondo; Taylor J. Dean |
| EPI_ISL_2931120 | Northwestern Memorial Hospital | RIPHL at Rush University Medical Center | Amber Kimble; Chao Qi; Felix Araujo Perez; Kevin Kunstman; Laura Furtado; Marieta Hyde; Max Kolton; Stefan Green |
| EPI_ISL_2953665, EPI_ISL_2953697, EPI_ISL_2953775 | Novogenia GmbH | Elling and Cochella laboratories, IMBA/IMP | Alexander Stark; Ezgi Oezkan; Luisa Cochella and Ulrich Elling; Marcus Strobl; Olga Frank; Ramesh Yelagandula; Tanino Albanese |
| EPI_ISL_2838732, EPI_ISL_2838883 | OHSU Lab Services Molecular Microbiology Lab | Oregon SARS-CoV-2 Genome Sequencing Center | '[UsersoroakBox SyncPHI_SARS-COV-2_Sample_Sheet_infoGISAID_Submission_Infometa_master_2021.xlsx]release20_final'!D:AC; 26; FALSE); VLOOKUP(C547; VLOOKUP(C549 |
| EPI_ISL_3395825 | OLVZ Aalst | OLVZ Aalst | Astrid Holderbeke |

| | | | |
|---|---|---|---|
| EPI_ISL_2787436, EPI_ISL_2840415 | OR State PHL-Virology/Immunology Section | Centers for Disease Control and Prevention Division of Viral Diseases, Pathogen Discovery | Alison Laufer Halpin; Ben L. Rambo-Martin; Clinton R. Paden; Dakota Howard; Darlene Wagner; Dave Wentworth; Dhwani Batra; Jasmine Padilla; Justin Lee; Katie Dillon; Krista Queen; Kristen Knipe; Kristine Lacek; Mark Burroughs; Matthew Schmerer; Mili Sheth; Peter Cook; Sam Shepard; Sarah Nobles; Shoshona Le; Suxiang Tong; Vivien Dugan; Yvette Unoarumhi |
| EPI_ISL_2604871 | Omics Sciences Laboratory | Omics Sciences Laboratory | Darlyn Amaya; Derly Andrade Molina; Gabriel Morey León; Juan Carlos Fernández Cadena; Rubén Armas González |
| EPI_ISL_2928167, EPI_ISL_2928169, EPI_ISL_2928172, EPI_ISL_2928173, EPI_ISL_2928174, EPI_ISL_2928176 | Orange County Public Health Laboratory | Orange County Public Health Laboratory | Angelica Torres; Julia Wolfe |
| EPI_ISL_2604209, EPI_ISL_2612540, EPI_ISL_2612545, EPI_ISL_2631155 | Oregon State Public Health Laboratory | Oregon State Public Health Laboratory | Eugene Yeboah; John Fontana and Shane Sevey; Laura Tsaknaridis; Rafia Razzaque; Vanda Makris |
| EPI_ISL_2795670 | Ospedale S.M. Goretti di Latina | National Institute for Infectious Diseases (INMI) L. Spallanzani I.R.C.C.S | A Di Caro; B Bartolini; CEM Gruber; E Giombini; F Messina; F Santini; G Bonfiglio; M Rueca; MR Capobianchi; O Butera |
| EPI_ISL_5646769 | PAS AGROVILA 3 DE CAIUA | Instituto Butantan | Antonio Jorge Martins; Claudia Renata dos Santos Barros; David Schlesinger; Debora Botequio Moretti; Dimas Tadeu Covas; Elaine Cristina Marqueze; Elaine Vieira Santos; Evandra Strazza Rodrigues; Heidge Fukumasu; Jayme Augusto de Souza-Neto; José Salvatore Leister Patané; Luiz Alcantara; Luiz Lehmann Coutinho; Maria Carolina Elias; Maurício Lacerda Nogueira; Rafael dos Santos Bezerra; Raul Machado Neto; Rejane Maria Tommasini Grotto; Ricardo Haddad; Sandra Coccuzzo Sampaio Vessoni; Simone Kashima; Svetoslav Nanev Slavov; Vincent Louis Viala |
| EPI_ISL_5646596 | PAS JOAO ANTONIO DO NASCIMENTO | Instituto Butantan | Antonio Jorge Martins; Claudia Renata dos Santos Barros; David Schlesinger; Debora Botequio Moretti; Dimas Tadeu Covas; Elaine Cristina Marqueze; Elaine Vieira Santos; Evandra Strazza Rodrigues; Heidge Fukumasu; Jayme Augusto de Souza-Neto; José Salvatore Leister Patané; Luiz Alcantara; Luiz Lehmann Coutinho; Maria Carolina Elias; Maurício Lacerda Nogueira; Rafael dos Santos Bezerra; Raul Machado Neto; Rejane Maria Tommasini Grotto; Ricardo Haddad; Sandra Coccuzzo Sampaio Vessoni; Simone Kashima; Svetoslav Nanev Slavov; Vincent Louis Viala |
| EPI_ISL_5646567 | POSTO DE SAUDE BENVIRA E S F 4 | Instituto Butantan | Antonio Jorge Martins; Claudia Renata dos Santos Barros; David Schlesinger; Debora Botequio Moretti; Dimas Tadeu Covas; Elaine Cristina Marqueze; Elaine Vieira Santos; Evandra Strazza Rodrigues; Heidge Fukumasu; Jayme Augusto de Souza-Neto; José Salvatore Leister Patané; Luiz Alcantara; Luiz Lehmann Coutinho; Maria Carolina Elias; Maurício Lacerda Nogueira; Rafael dos Santos Bezerra; Raul Machado Neto; Rejane Maria Tommasini Grotto; Ricardo Haddad; Sandra Coccuzzo Sampaio Vessoni; Simone Kashima; Svetoslav Nanev Slavov; Vincent Louis Viala |
| EPI_ISL_5646801 | POSTO DE SAUDE DRCRISTOVAN RUTHILLER T MACIEL | Instituto Butantan | Antonio Jorge Martins; Claudia Renata dos Santos Barros; David Schlesinger; Debora Botequio Moretti; Dimas Tadeu Covas; Elaine Cristina Marqueze; Elaine Vieira Santos; Evandra Strazza Rodrigues; Heidge Fukumasu; Jayme Augusto de Souza-Neto; José Salvatore Leister Patané; Luiz Alcantara; Luiz Lehmann Coutinho; Maria Carolina Elias; Maurício Lacerda Nogueira; Rafael dos Santos Bezerra; Raul Machado Neto; Rejane Maria Tommasini Grotto; Ricardo Haddad; Sandra Coccuzzo Sampaio Vessoni; Simone Kashima; Svetoslav Nanev Slavov; Vincent Louis Viala |
| EPI_ISL_5646730 | PROGRAMA SAUDE DA FAMILIA I IEPE | Instituto Butantan | Antonio Jorge Martins; Claudia Renata dos Santos Barros; David Schlesinger; Debora Botequio Moretti; Dimas Tadeu Covas; Elaine Cristina Marqueze; Elaine Vieira Santos; Evandra Strazza Rodrigues; Heidge Fukumasu; Jayme Augusto de Souza-Neto; José Salvatore Leister Patané; Luiz Alcantara; Luiz Lehmann Coutinho; Maria Carolina Elias; Maurício Lacerda Nogueira; Rafael dos Santos Bezerra; Raul Machado Neto; Rejane Maria Tommasini Grotto; Ricardo Haddad; Sandra Coccuzzo Sampaio Vessoni; Simone Kashima; Svetoslav Nanev Slavov; Vincent Louis Viala |
| EPI_ISL_5646391, EPI_ISL_5646394, EPI_ISL_5646414, EPI_ISL_5646420, EPI_ISL_5646424, EPI_ISL_5646428, EPI_ISL_5646436, EPI_ISL_5646438, EPI_ISL_5646442, EPI_ISL_5646443, EPI_ISL_5646449, EPI_ISL_5646453, EPI_ISL_5646455, EPI_ISL_5646462, EPI_ISL_5646465, EPI_ISL_5646474, EPI_ISL_5646475, EPI_ISL_5646481 | | | |
| see above | PRONTO ATENDIMENTO DE VARGEM GRANDE PAULISTA | Instituto Butantan | Antonio Jorge Martins; Claudia Renata dos Santos Barros; David Schlesinger; Debora Botequio Moretti; Dimas Tadeu Covas; Elaine Cristina Marqueze; Elaine Vieira Santos; Evandra Strazza Rodrigues; Heidge Fukumasu; Jayme Augusto de Souza-Neto; José Salvatore Leister Patané; Luiz Alcantara; Luiz Lehmann Coutinho; Maria Carolina Elias; Maurício Lacerda Nogueira; Rafael dos Santos Bezerra; Raul Machado Neto; Rejane Maria Tommasini Grotto; Ricardo Haddad; Sandra Coccuzzo Sampaio Vessoni; Simone Kashima; Svetoslav Nanev Slavov; Vincent Louis Viala |
| EPI_ISL_5646482, EPI_ISL_5646483 | PRONTO ATENDIMENTO MUNICIPAL | Instituto Butantan | Antonio Jorge Martins; Claudia Renata dos Santos Barros; David Schlesinger; Debora Botequio Moretti; Dimas Tadeu Covas; Elaine Cristina Marqueze; Elaine Vieira Santos; Evandra Strazza Rodrigues; Heidge Fukumasu; Jayme Augusto de Souza-Neto; José Salvatore Leister Patané; Luiz Alcantara; Luiz Lehmann Coutinho; Maria Carolina Elias; Maurício Lacerda Nogueira; Rafael dos Santos Bezerra; Raul Machado Neto; Rejane Maria Tommasini Grotto; Ricardo Haddad; Sandra Coccuzzo Sampaio Vessoni; Simone Kashima; Svetoslav Nanev Slavov; Vincent Louis Viala |
| EPI_ISL_5646874 | PRONTO ATENDIMENTO MUNICIPAL IRIO TAINO | Instituto Butantan | Antonio Jorge Martins; Claudia Renata dos Santos Barros; David Schlesinger; Debora Botequio Moretti; Dimas Tadeu Covas; Elaine Cristina Marqueze; Elaine Vieira Santos; Evandra Strazza Rodrigues; Heidge Fukumasu; Jayme Augusto de Souza-Neto; José Salvatore Leister Patané; Luiz Alcantara; Luiz Lehmann Coutinho; Maria Carolina Elias; Maurício Lacerda Nogueira; Rafael dos Santos Bezerra; Raul Machado Neto; Rejane Maria Tommasini Grotto; Ricardo Haddad; Sandra Coccuzzo Sampaio Vessoni; Simone Kashima; Svetoslav Nanev Slavov; Vincent Louis Viala |
| EPI_ISL_5646871 | PSF DE IDA IOLANDA NHANDEARA | Instituto Butantan | Antonio Jorge Martins; Claudia Renata dos Santos Barros; David Schlesinger; Debora Botequio Moretti; Dimas Tadeu Covas; Elaine Cristina Marqueze; Elaine Vieira Santos; Evandra Strazza Rodrigues; Heidge Fukumasu; Jayme Augusto de Souza-Neto; José Salvatore Leister Patané; Luiz Alcantara; Luiz Lehmann Coutinho; Maria Carolina Elias; Maurício Lacerda Nogueira; Rafael dos Santos Bezerra; Raul Machado Neto; Rejane Maria Tommasini Grotto; Ricardo Haddad; Sandra Coccuzzo Sampaio Vessoni; Simone Kashima; Svetoslav Nanev Slavov; Vincent Louis Viala |
| EPI_ISL_5646647 | PSF DO PARQUE DOS PINHEIROS | Instituto Butantan | Antonio Jorge Martins; Claudia Renata dos Santos Barros; David Schlesinger; Debora Botequio Moretti; Dimas Tadeu Covas; Elaine Cristina Marqueze; Elaine Vieira Santos; Evandra Strazza Rodrigues; Heidge Fukumasu; Jayme Augusto de Souza-Neto; José Salvatore Leister Patané; Luiz Alcantara; Luiz Lehmann Coutinho; Maria Carolina Elias; Maurício Lacerda Nogueira; Rafael dos Santos Bezerra; Raul Machado Neto; Rejane Maria Tommasini Grotto; Ricardo Haddad; Sandra Coccuzzo Sampaio Vessoni; Simone Kashima; Svetoslav Nanev Slavov; Vincent Louis Viala |
| EPI_ISL_2631397, EPI_ISL_2631407, EPI_ISL_2634916, EPI_ISL_2634943, EPI_ISL_2634974, EPI_ISL_4096684 | Pandemic Response Lab - NYC | Pandemic Response Lab, R&D | Cybill del Castillo; Dylan Law; Haiping Hao; Henry Lee; Jon Laurent; Katharine Nelson; Melissa Hopkins; Michael Hammerling; Pradeep Bugga; Shinyoung Clair Kang; Sol Rey; William Ward |
| EPI_ISL_3050146 | Pathology, UAB | Pathology, UAB | E.J.; Hendrickson; Hernandez, J.; Leal; Lefkowitz; Moates, D.; R.C.; S.M. |
| EPI_ISL_5565845 | Philippine Red Cross - Clark Molecular Laboratory | Philippine Genome Center | Alethea R. de Guzman; Alyssa Joyce E. Telles; Anna Ong-Lim; Arianne A. Zamora; Benedict A. Maralit; Carlo M. Lapid; Celia Carlos; Cynthia P. Saloma; Devon Ray Pacial; Diomedes A. Cariño; Edsel Maurice Salvana; El King D. Morado; Elcid Aaron R. Pangilinan; Eva Maria Cutiongco-de la Paz; Francis A. Tablizo; Henrietta Marie Rodriguez; Jaime C. Montoya; Jan Michael C. Yap; Jarvin E. Nipales; Jo-Hannah S. Llames; John Michael Egana; John Q. Wong; Joshua Gregor A. Dizon; Joshua Jose Endozo; Juan Antonio R. Magalang; Karol Sophia Agape R. Padilla; Kris P. Punayan; Kristina Patriz Dela Cruz; Lindsay Clare D.L. Carandang; Ma. Exanil Plantig; Marc Edsel C. Ayes; Maria Rosario Singh-Vergeire; Maria Sofia L. Yangzon; Marielle M. Gamboa; Marissa Alejandria; Niña Francesca Bustamante; Razel Nikka M. Hao; Renato Jacinto Q. Mantaring; Rianna Patricia S. Cruz; Shiela Mae M. Araiza; Yvonne Valerie Austria; Zipporah Mariebelle R. Enriquez; Zyrel V. Mollejon |
| EPI_ISL_4405591 | Plataforma de Servicios Biotecnólogicos: UTTIPP/PSB , Universidad Nacional de Quilmes. | Área de Secuenciación del Laboratorio de Virología del Hospital de Niños Dr. Ricardo Gutierrez on behalf of 'Proyecto Argentino Interinstitucional de genomica de SARS-CoV-2' (PAIS Consortium) | A; Acuña; Cardama; Castello; D; Farina; G; Goya; Goñi; H; LE; Lusso; MI; MS; Nabaes Jodar; Natale; S; Valinotto; Viegas, M. |
| EPI_ISL_6907440, EPI_ISL_6907447, EPI_ISL_6907997, EPI_ISL_6908006, EPI_ISL_6908014 | Plataforma de Vigilancia Molecular (PVM) - FIOCRUZ/BA | Plataforma de Vigilancia Molecular (PVM) - FIOCRUZ/BA | Bruno Bezerril Andrade; Camila I. de Oliveira; Clarissa Araújo Gurgel; Eduardo Oyama; Francys A Rangel; Laila A da Silva; Laise de Moraes; Leonardo Paiva Farias; Marina Cucco; Pedro Silveira; Ricardo Khouri on behalf of the Fiocruz COVID-19 Genomic Surveillance Network.; Tiago Graf |
| EPI_ISL_2600926, EPI_ISL_2600943, EPI_ISL_2600944, EPI_ISL_2600948, EPI_ISL_3061935 | Plateforme de testing Namuroise | Plateforme de testing Namuroise | Degosserie Jonathan; Demars Aurore; Denis Olivier; Lesly NYINKEU KEMAMEN; Lesly Nyinkeu Kemamen; Maschietto Céline; Mullier François; Nicolas GILLIARD; Nobis Chloé; Otto Gaetan |
| EPI_ISL_2545610, EPI_ISL_2545636, EPI_ISL_2626852, EPI_ISL_2626853, EPI_ISL_2626882, EPI_ISL_2626883, EPI_ISL_2626884, EPI_ISL_2628061, EPI_ISL_2628062, EPI_ISL_2628063, EPI_ISL_2628064, EPI_ISL_2628065, EPI_ISL_2714770, EPI_ISL_2861572, EPI_ISL_2861631 | | | |
| see above | Platform BIS UZA/UAntwerpen | Labo Klinische Biologie, UZA | Basil Britto Xavier; Christine Lammens; Herman Goossens; Ines Verbesselt; Jasmine Coppens; Kathleen Holemans; Marie Le Mercier; Veerle Matheeussen |
| EPI_ISL_3120103 | Policlinico San Donato | Laboratory of Clinical Microbiology, Virology and Bioemergencies, ASST Fatebenefratelli Sacco - Sacco Hospital | Valeria Micheli |
| EPI_ISL_7406392, EPI_ISL_7406394, EPI_ISL_7406397, EPI_ISL_7406414 | Programa Laboratorio de Salud Pública "Dr Dalmiro Pérez Laborda" | Área de Secuenciación del Laboratorio de Virología del Hospital de Niños Dr. Ricardo Gutierrez on behalf of 'Proyecto Argentino Interinstitucional de genomica de SARS-CoV-2' (PAIS Consortium) | A; AM; Acuña; Aguilera; BH; Bohn; Cabral Bombardieri; Campos; Carrizo; D; E; Esteves; F; G; Goya; J; JM; JP; Jofré; L; LE; Lacaze; Lusso; M; MA; MI; MS; Mastrodonato; Molina; Nabaes Jodar; Natale; Olivera; Perez Diaz; Peñalva; Quijano; Rivero; Rosales; S; Talia; Valinotto; Viegas, M. |
| EPI_ISL_2709231 | Public Health Laboratory, Minnesota Department of Health | University of Minnesota Genomics Center | Corbin Dirkx; Daryl M. Gohl; Jaquelyn Kuriger-Laber; John Garbe; and Sean Wang |
| EPI_ISL_2686359, EPI_ISL_2686360, EPI_ISL_2686364, EPI_ISL_2686365, EPI_ISL_2686369, EPI_ISL_2686370, EPI_ISL_2686374, EPI_ISL_2686375, EPI_ISL_2686376, EPI_ISL_2686377, EPI_ISL_2686380, EPI_ISL_2686381, EPI_ISL_2686382, EPI_ISL_2686383, EPI_ISL_2686384, EPI_ISL_2686385, EPI_ISL_2686386, EPI_ISL_2686387, EPI_ISL_2686388, EPI_ISL_2872074, EPI_ISL_2876105, EPI_ISL_2876106, EPI_ISL_2876107, EPI_ISL_2876108, EPI_ISL_2876109, EPI_ISL_2876110, EPI_ISL_2876111, EPI_ISL_2876112, EPI_ISL_2876113, EPI_ISL_2876114, EPI_ISL_2876115, EPI_ISL_2876116, EPI_ISL_2876117, EPI_ISL_2876118, EPI_ISL_2876119, EPI_ISL_2876120, EPI_ISL_2876121, EPI_ISL_2876122, EPI_ISL_2876123, EPI_ISL_2876124, EPI_ISL_2876125, EPI_ISL_2876126, EPI_ISL_2876127, EPI_ISL_2876128, EPI_ISL_2876129, EPI_ISL_2876131, EPI_ISL_2876132, EPI_ISL_2876133, EPI_ISL_2876134, EPI_ISL_2876135, EPI_ISL_2876136, EPI_ISL_2876137, EPI_ISL_2876138, EPI_ISL_2876139, EPI_ISL_2876140, EPI_ISL_2876141, EPI_ISL_2876144, EPI_ISL_2876145, EPI_ISL_2876146, EPI_ISL_2876147, EPI_ISL_2876148, EPI_ISL_2876149, EPI_ISL_2876150, EPI_ISL_2876151, EPI_ISL_2876152, EPI_ISL_2876153, EPI_ISL_2876154, EPI_ISL_2876155, EPI_ISL_2876156, EPI_ISL_2876157, EPI_ISL_2876158, EPI_ISL_2876159, EPI_ISL_2876160, EPI_ISL_2876161, EPI_ISL_2876162, EPI_ISL_2876163, EPI_ISL_2876164, EPI_ISL_2876165, EPI_ISL_2876166, EPI_ISL_2876167, EPI_ISL_2876168, EPI_ISL_2876169, EPI_ISL_2876170, EPI_ISL_2876616, EPI_ISL_2876617, EPI_ISL_2876618, EPI_ISL_2876619, EPI_ISL_2876620, EPI_ISL_2876621, EPI_ISL_2876623, EPI_ISL_2876624, EPI_ISL_2876625, EPI_ISL_2876626, EPI_ISL_2876628, EPI_ISL_2876630, EPI_ISL_2876631, EPI_ISL_2876632, EPI_ISL_2876633, EPI_ISL_2876634, EPI_ISL_2876635, EPI_ISL_2876636, EPI_ISL_2876637, EPI_ISL_2876638, EPI_ISL_2876640, EPI_ISL_2876643, EPI_ISL_2876644, EPI_ISL_2876649, EPI_ISL_2876650, EPI_ISL_2876651, EPI_ISL_2876653, EPI_ISL_2876654, EPI_ISL_2876655, EPI_ISL_2876656, EPI_ISL_2876657, EPI_ISL_2876661, EPI_ISL_2876662, EPI_ISL_2876663, EPI_ISL_2876665, EPI_ISL_2876668, EPI_ISL_2876672, EPI_ISL_2876673, EPI_ISL_2876676, EPI_ISL_2876677, EPI_ISL_2876690, EPI_ISL_4368466, EPI_ISL_4368658, EPI_ISL_4369219, EPI_ISL_4370640, EPI_ISL_4371475, EPI_ISL_4372912 | | | |
| see above | Quest Diagnostics Incorporated | Centers for Disease Control and Prevention Division of Viral Diseases, Pathogen Discovery | A. Gerasimova; A. Perez; Adrian Paskey; B. Anderson; Benjamin Rambo-Martin; Christopher Gulvick; Clinton Paden; Clinton R. Paden; Dakota Howard; Darlene Wagner; Dhwani Batra; Duncan MacCannell; Erisa Sula; F. Lacbawan; I. A. Shlyakhter; I. Shlyakhter; Jason Caravas; K. Livingston; K.E. Livingston; Kara Moser; Kristine Lacek; L. Bernstein; L.E. Bernstein; M. Hua; Matthew Schmerer; P. Tanpaiboon; Peter Cook; Peter W. Cook; R. Kagan; R. M. Kagan; R. Owen; R. Rolando; R. V. Rolando; S. H. Rosenthal; S. Rosenthal; Scott Sammons; Shatavia Morrison; Tymeckia Kendall; Victoria Caban Figueroa; Y. Liu; Yvette Unoarumhi |
| EPI_ISL_2840374 | RI State Health Laboratories | Centers for Disease Control and Prevention Division of Viral Diseases, Pathogen Discovery | Alison Laufer Halpin; Ben L. Rambo-Martin; Clinton R. Paden; Dakota Howard; Darlene Wagner; Dave Wentworth; Dhwani Batra; Jasmine Padilla; Justin Lee; Katie Dillon; Krista Queen; Kristen Knipe; Kristine Lacek; Mark Burroughs; Matthew Schmerer; Mili Sheth; Peter Cook; Sam Shepard; Sarah Nobles; Shoshona Le; Suxiang Tong; Vivien Dugan; Yvette Unoarumhi |
| EPI_ISL_2795664, EPI_ISL_2795667 | RM4 | National Institute for Infectious Diseases (INMI) L. Spallanzani I.R.C.C.S | A Di Caro; B Bartolini; CEM Gruber; E Giombini; F Messina; F Santini; G Bonfiglio; M Rueca; MR Capobianchi; O Butera |
| EPI_ISL_2904313 | Respiratory Virus Unit, Microbiology Services Colindale, Public Health England | COVID-19 Genomics UK (COG-UK) Consortium | PHE Covid Sequencing Team |

| | | | |
|---|---|---|---|
| EPI_ISL_3091694 | Respiratory Viruses Branch, Centers for Disease Control and Prevention | Respiratory Viruses Branch, Centers for Disease Control and Prevention | Anderson, B.; Batra, D.; Bernstein, L.; C.R.; Caravas, J.; Cook; Gerasimova, A.; Gulvick, C.; Howard, D.; Hua, M.; I.A.; Kagan; Lacbawan, F.; Liu, Y.; Livingston, K.; MacCannell, D.; Morrison, S.; Moser, K.; Owen, R.; P.W.; Paden; Paskey, A.; Perez, A.; R.M.; R.V.; Rambo-Martin, B.; Rolando; Rosenthal; S.H.; Sammons, S.; Schmerer, M.; Shlyakhter; Tanpaiboon, P.; Unoarumhi, Y.; Wagner, D. |
| EPI_ISL_2725593 | Rhode Island State Health Laboratory | Rhode Island State Health Laboratory | Ewa King; Kristin Carpenter-Azevedo; Richard C. Huard |
| EPI_ISL_2840624 | S.C. Laboratorio Analisi, ASL 3 Liguria | U.O. Igiene, Ospedale Policlinico San Martino | Bruzzone Bianca; Caligiuri Patrizia; De Pace Vanessa; Domnich Alexander; Icardi Giancarlo on behalf of SARS-CoV-2 ITALIAN RESEARCH ENTERPRISE-(SCIRE) Collaborative Group; Orsi Andrea; Ricucci Valentina; Spitaleri Antonino |
| EPI_ISL_5647230, EPI_ISL_5647265, EPI_ISL_5647295 | SANTA CASA DE MISERICORDIA DE BRAGANCA PAULISTA | Instituto Butantan | Antonio Jorge Martins; Claudia Renata dos Santos Barros; David Schlesinger; Debora Botequio Moretti; Dimas Tadeu Covas; Elaine Cristina Marqueze; Elaine Vieira Santos; Evandra Strazza Rodrigues; Heidge Fukumasu; Jayme Augusto de Souza-Neto; José Salvatore Leister Patané; Luiz Alcantara; Luiz Lehmann Coutinho; Maria Carolina Elias; Maurício Lacerda Nogueira; Rafael dos Santos Bezerra; Raul Machado Neto; Rejane Maria Tommasini Grotto; Ricardo Haddad; Sandra Coccuzzo Sampaio Vessoni; Simone Kashima; Svetoslav Nanev Slavov; Vincent Louis Viala |
| EPI_ISL_5646548, EPI_ISL_5646578, EPI_ISL_5646775 | SANTA CASA DE MISERICORDIA DE UBATUBA | Instituto Butantan | Antonio Jorge Martins; Claudia Renata dos Santos Barros; David Schlesinger; Debora Botequio Moretti; Dimas Tadeu Covas; Elaine Cristina Marqueze; Elaine Vieira Santos; Evandra Strazza Rodrigues; Heidge Fukumasu; Jayme Augusto de Souza-Neto; José Salvatore Leister Patané; Luiz Alcantara; Luiz Lehmann Coutinho; Maria Carolina Elias; Maurício Lacerda Nogueira; Rafael dos Santos Bezerra; Raul Machado Neto; Rejane Maria Tommasini Grotto; Ricardo Haddad; Sandra Coccuzzo Sampaio Vessoni; Simone Kashima; Svetoslav Nanev Slavov; Vincent Louis Viala |
| EPI_ISL_5646597 | SANTA CASA DE PIRATININGA | Instituto Butantan | Antonio Jorge Martins; Claudia Renata dos Santos Barros; David Schlesinger; Debora Botequio Moretti; Dimas Tadeu Covas; Elaine Cristina Marqueze; Elaine Vieira Santos; Evandra Strazza Rodrigues; Heidge Fukumasu; Jayme Augusto de Souza-Neto; José Salvatore Leister Patané; Luiz Alcantara; Luiz Lehmann Coutinho; Maria Carolina Elias; Maurício Lacerda Nogueira; Rafael dos Santos Bezerra; Raul Machado Neto; Rejane Maria Tommasini Grotto; Ricardo Haddad; Sandra Coccuzzo Sampaio Vessoni; Simone Kashima; Svetoslav Nanev Slavov; Vincent Louis Viala |
| EPI_ISL_2986110 | SARS-CoV-2 Sequencing Castilla y Leon-Spain Consortium | SARS-CoV-2 Sequencing Castilla y Leon-Spain Consortium | Antonio Orduña-Domingo; Carlos Fuster Foz; Carmen Aldea-Mansilla; Carmen Gimeno Crespo; David Abad; Gabriel March Rossello; Gregoria Megías Lobón; Jose María Eiros Bouza; M. Isabel Fernandez-Natal; Marta Dominguez-Gil; Marta Hernandez; María Antonia García Castro; Mª Fe Brezmes-Valdivieso; Noelia Arenal Andrés; Silvia Rojo; Sonsoles Garcinuño Pérez |
| EPI_ISL_2621772 | SARS-CoV-2 testing team, National Institute of Infectious Diseases | Pathogen Genomics Center, National Institute of Infectious Diseases | Hazuka Y Furihata; Hiromizu Takahashi; Kentaro Itokawa; Makoto Kuroda; Masanori Hashino; Masumichi Saito; Naomi Nojiri; Nozomu Hanaoka; Rina Tanaka; Sana Uchikoba; Tsuguto Fujimoto; Tsuyoshi Sekizuka |
| EPI_ISL_2828404 | SC (UCO) Igiene e Sanità Pubblica, ASUGI, Trieste and SC Microbiologia e Virologia, ASFO, Pordenone | ARGO Laboratorio Genomica ed Epigenomica | Barbone F; Breda C; D'Agaro P; Dal Monego S; De Rosa R; Degasperi M; Licastro D; Marcello A; Segat L |
| EPI_ISL_5646396, EPI_ISL_5646397, EPI_ISL_5646400, EPI_ISL_5646409, EPI_ISL_5646411, EPI_ISL_5646412, EPI_ISL_5646413, EPI_ISL_5646421, EPI_ISL_5646426, EPI_ISL_5646435, EPI_ISL_5646448, EPI_ISL_5646458, EPI_ISL_5646472 | | | |
| see above | SECRETARIA DA SAUDE DE MAIRIPORA | Instituto Butantan | Antonio Jorge Martins; Claudia Renata dos Santos Barros; David Schlesinger; Debora Botequio Moretti; Dimas Tadeu Covas; Elaine Cristina Marqueze; Elaine Vieira Santos; Evandra Strazza Rodrigues; Heidge Fukumasu; Jayme Augusto de Souza-Neto; José Salvatore Leister Patané; Luiz Alcantara; Luiz Lehmann Coutinho; Maria Carolina Elias; Maurício Lacerda Nogueira; Rafael dos Santos Bezerra; Raul Machado Neto; Rejane Maria Tommasini Grotto; Ricardo Haddad; Sandra Coccuzzo Sampaio Vessoni; Simone Kashima; Svetoslav Nanev Slavov; Vincent Louis Viala |
| EPI_ISL_5646613, EPI_ISL_5646755, EPI_ISL_5646842 | SECRETARIA DE SAUDE | Instituto Butantan | Antonio Jorge Martins; Claudia Renata dos Santos Barros; David Schlesinger; Debora Botequio Moretti; Dimas Tadeu Covas; Elaine Cristina Marqueze; Elaine Vieira Santos; Evandra Strazza Rodrigues; Heidge Fukumasu; Jayme Augusto de Souza-Neto; José Salvatore Leister Patané; Luiz Alcantara; Luiz Lehmann Coutinho; Maria Carolina Elias; Maurício Lacerda Nogueira; Rafael dos Santos Bezerra; Raul Machado Neto; Rejane Maria Tommasini Grotto; Ricardo Haddad; Sandra Coccuzzo Sampaio Vessoni; Simone Kashima; Svetoslav Nanev Slavov; Vincent Louis Viala |
| EPI_ISL_5646850 | SECRETARIA DE SAUDE CABREUVA | Instituto Butantan | Antonio Jorge Martins; Claudia Renata dos Santos Barros; David Schlesinger; Debora Botequio Moretti; Dimas Tadeu Covas; Elaine Cristina Marqueze; Elaine Vieira Santos; Evandra Strazza Rodrigues; Heidge Fukumasu; Jayme Augusto de Souza-Neto; José Salvatore Leister Patané; Luiz Alcantara; Luiz Lehmann Coutinho; Maria Carolina Elias; Maurício Lacerda Nogueira; Rafael dos Santos Bezerra; Raul Machado Neto; Rejane Maria Tommasini Grotto; Ricardo Haddad; Sandra Coccuzzo Sampaio Vessoni; Simone Kashima; Svetoslav Nanev Slavov; Vincent Louis Viala |
| EPI_ISL_5647913 | SECRETARIA DE SAUDE DE SANTA GERTRUDES | Instituto Butantan | Antonio Jorge Martins; Claudia Renata dos Santos Barros; David Schlesinger; Debora Botequio Moretti; Dimas Tadeu Covas; Elaine Cristina Marqueze; Elaine Vieira Santos; Evandra Strazza Rodrigues; Heidge Fukumasu; Jayme Augusto de Souza-Neto; José Salvatore Leister Patané; Luiz Alcantara; Luiz Lehmann Coutinho; Maria Carolina Elias; Maurício Lacerda Nogueira; Rafael dos Santos Bezerra; Raul Machado Neto; Rejane Maria Tommasini Grotto; Ricardo Haddad; Sandra Coccuzzo Sampaio Vessoni; Simone Kashima; Svetoslav Nanev Slavov; Vincent Louis Viala |
| EPI_ISL_5647922 | SECRETARIA DE SAUDE DE SAO PEDRO | Instituto Butantan | Antonio Jorge Martins; Claudia Renata dos Santos Barros; David Schlesinger; Debora Botequio Moretti; Dimas Tadeu Covas; Elaine Cristina Marqueze; Elaine Vieira Santos; Evandra Strazza Rodrigues; Heidge Fukumasu; Jayme Augusto de Souza-Neto; José Salvatore Leister Patané; Luiz Alcantara; Luiz Lehmann Coutinho; Maria Carolina Elias; Maurício Lacerda Nogueira; Rafael dos Santos Bezerra; Raul Machado Neto; Rejane Maria Tommasini Grotto; Ricardo Haddad; Sandra Coccuzzo Sampaio Vessoni; Simone Kashima; Svetoslav Nanev Slavov; Vincent Louis Viala |
| EPI_ISL_5646566, EPI_ISL_5646620 | SECRETARIA MUNICIPAL DA SAUDE DE GUARATINGUETA | Instituto Butantan | Antonio Jorge Martins; Claudia Renata dos Santos Barros; David Schlesinger; Debora Botequio Moretti; Dimas Tadeu Covas; Elaine Cristina Marqueze; Elaine Vieira Santos; Evandra Strazza Rodrigues; Heidge Fukumasu; Jayme Augusto de Souza-Neto; José Salvatore Leister Patané; Luiz Alcantara; Luiz Lehmann Coutinho; Maria Carolina Elias; Maurício Lacerda Nogueira; Rafael dos Santos Bezerra; Raul Machado Neto; Rejane Maria Tommasini Grotto; Ricardo Haddad; Sandra Coccuzzo Sampaio Vessoni; Simone Kashima; Svetoslav Nanev Slavov; Vincent Louis Viala |
| EPI_ISL_5646744, EPI_ISL_5646854 | SECRETARIA MUNICIPAL DA SAUDE DE GUARIBA | Instituto Butantan | Antonio Jorge Martins; Claudia Renata dos Santos Barros; David Schlesinger; Debora Botequio Moretti; Dimas Tadeu Covas; Elaine Cristina Marqueze; Elaine Vieira Santos; Evandra Strazza Rodrigues; Heidge Fukumasu; Jayme Augusto de Souza-Neto; José Salvatore Leister Patané; Luiz Alcantara; Luiz Lehmann Coutinho; Maria Carolina Elias; Maurício Lacerda Nogueira; Rafael dos Santos Bezerra; Raul Machado Neto; Rejane Maria Tommasini Grotto; Ricardo Haddad; Sandra Coccuzzo Sampaio Vessoni; Simone Kashima; Svetoslav Nanev Slavov; Vincent Louis Viala |
| EPI_ISL_5646592 | SECRETARIA MUNICIPAL DE SAUDE | Instituto Butantan | Antonio Jorge Martins; Claudia Renata dos Santos Barros; David Schlesinger; Debora Botequio Moretti; Dimas Tadeu Covas; Elaine Cristina Marqueze; Elaine Vieira Santos; Evandra Strazza Rodrigues; Heidge Fukumasu; Jayme Augusto de Souza-Neto; José Salvatore Leister Patané; Luiz Alcantara; Luiz Lehmann Coutinho; Maria Carolina Elias; Maurício Lacerda Nogueira; Rafael dos Santos Bezerra; Raul Machado Neto; Rejane Maria Tommasini Grotto; Ricardo Haddad; Sandra Coccuzzo Sampaio Vessoni; Simone Kashima; Svetoslav Nanev Slavov; Vincent Louis Viala |
| EPI_ISL_5646617 | SECRETARIA MUNICIPAL DE SAUDE DE ANDRADINA | Instituto Butantan | Antonio Jorge Martins; Claudia Renata dos Santos Barros; David Schlesinger; Debora Botequio Moretti; Dimas Tadeu Covas; Elaine Cristina Marqueze; Elaine Vieira Santos; Evandra Strazza Rodrigues; Heidge Fukumasu; Jayme Augusto de Souza-Neto; José Salvatore Leister Patané; Luiz Alcantara; Luiz Lehmann Coutinho; Maria Carolina Elias; Maurício Lacerda Nogueira; Rafael dos Santos Bezerra; Raul Machado Neto; Rejane Maria Tommasini Grotto; Ricardo Haddad; Sandra Coccuzzo Sampaio Vessoni; Simone Kashima; Svetoslav Nanev Slavov; Vincent Louis Viala |
| EPI_ISL_5646503 | SECRETARIA MUNICIPAL DE SAUDE DE PRES BERNARDES SP | Instituto Butantan | Antonio Jorge Martins; Claudia Renata dos Santos Barros; David Schlesinger; Debora Botequio Moretti; Dimas Tadeu Covas; Elaine Cristina Marqueze; Elaine Vieira Santos; Evandra Strazza Rodrigues; Heidge Fukumasu; Jayme Augusto de Souza-Neto; José Salvatore Leister Patané; Luiz Alcantara; Luiz Lehmann Coutinho; Maria Carolina Elias; Maurício Lacerda Nogueira; Rafael dos Santos Bezerra; Raul Machado Neto; Rejane Maria Tommasini Grotto; Ricardo Haddad; Sandra Coccuzzo Sampaio Vessoni; Simone Kashima; Svetoslav Nanev Slavov; Vincent Louis Viala |
| EPI_ISL_3912245 | SECRETARIA MUNICIPAL DE SAUDE DE TIANGUA | Analytical Competence Molecular Epidemiology Lab/ACME, Oswaldo Cruz Foundation, Ceara (FIOCRUZ CE) | Cleber Furtado Aksenen; Fabio Miyajima; Fernando Braga Stehling; Francisco Eder de Moura Lopes; Jamille Maria Mendes Bezerra; Joaquim Cesar do Nascimento Sousa Junior; Pedro Miguel Carneiro Jeronimo; Suzana Porto Almeida & Lucas Delerino on behalf of COVID-19 FIOCRUZ Genomic Network; Thais Ferreira de Oliveira; Thais de Oliveira Costa; Ticiane Cavalcante de Souza; Veridiana Pessoa Miyajima |
| EPI_ISL_5648062, EPI_ISL_5648063, EPI_ISL_5648078, EPI_ISL_5648084, EPI_ISL_5648085, EPI_ISL_5648087, EPI_ISL_5648121 | | | |
| see above | SECRETARIA MUNICIPAL DE SAUDE DE TORRINHA | Instituto Butantan | Antonio Jorge Martins; Claudia Renata dos Santos Barros; David Schlesinger; Debora Botequio Moretti; Dimas Tadeu Covas; Elaine Cristina Marqueze; Elaine Vieira Santos; Evandra Strazza Rodrigues; Heidge Fukumasu; Jayme Augusto de Souza-Neto; José Salvatore Leister Patané; Luiz Alcantara; Luiz Lehmann Coutinho; Maria Carolina Elias; Maurício Lacerda Nogueira; Rafael dos Santos Bezerra; Raul Machado Neto; Rejane Maria Tommasini Grotto; Ricardo Haddad; Sandra Coccuzzo Sampaio Vessoni; Simone Kashima; Svetoslav Nanev Slavov; Vincent Louis Viala |
| EPI_ISL_5646510, EPI_ISL_5646518, EPI_ISL_5646521, EPI_ISL_5646538, EPI_ISL_5646540, EPI_ISL_5646556, EPI_ISL_5646574, EPI_ISL_5646577, EPI_ISL_5646587, EPI_ISL_5646593, EPI_ISL_5646595, EPI_ISL_5646601, EPI_ISL_5646630, EPI_ISL_5646637, EPI_ISL_5646644, EPI_ISL_5646662, EPI_ISL_5646666, EPI_ISL_5646667, EPI_ISL_5646675, EPI_ISL_5646690, EPI_ISL_5646691, EPI_ISL_5646700, EPI_ISL_5646710, EPI_ISL_5646720, EPI_ISL_5646728, EPI_ISL_5646737, EPI_ISL_5646738, EPI_ISL_5646745, EPI_ISL_5646754, EPI_ISL_5646774, EPI_ISL_5646776, EPI_ISL_5646792, EPI_ISL_5646793, EPI_ISL_5646795, EPI_ISL_5646800, EPI_ISL_5646810, EPI_ISL_5646835, EPI_ISL_5646847 | | | |
| see above | SECRETARIA MUNICIPAL DE SAUDE SOROCABA | Instituto Butantan | Antonio Jorge Martins; Claudia Renata dos Santos Barros; David Schlesinger; Debora Botequio Moretti; Dimas Tadeu Covas; Elaine Cristina Marqueze; Elaine Vieira Santos; Evandra Strazza Rodrigues; Heidge Fukumasu; Jayme Augusto de Souza-Neto; José Salvatore Leister Patané; Luiz Alcantara; Luiz Lehmann Coutinho; Maria Carolina Elias; Maurício Lacerda Nogueira; Rafael dos Santos Bezerra; Raul Machado Neto; Rejane Maria Tommasini Grotto; Ricardo Haddad; Sandra Coccuzzo Sampaio Vessoni; Simone Kashima; Svetoslav Nanev Slavov; Vincent Louis Viala |
| EPI_ISL_2673672 | SIESP CHIETI - DRIVE IN ORTONA | Istituto Zooprofilattico Sperimentale dell'Abruzzo e Molise "G. Caporale" | Ancora M; Calistri P; Cammà C; Caporale M; Curini V; Delli Compagni E; Di Domenico M; Di Lollo Valeria; Di Pasquale A; Lorusso A; Mangone I; Marcacci M; Puglia I; Rinaldi A; Savini G; Scialabba S |
| EPI_ISL_2889816, EPI_ISL_2889822, EPI_ISL_3255546, EPI_ISL_3255549, EPI_ISL_3255554, EPI_ISL_3255557, EPI_ISL_3255568, EPI_ISL_3255979 | | | |
| see above | SK-Roy Romanow Provincial Laboratory | National Microbiology Laboratory (NML) | Alanna Senecal; Amanda Lang; Anna Majer; Anneliese Landgraff; CanCOGeN's metadata curation team; Darian Hole; Elsie Grudeski; Gary Van Domselaar; Grace Seo; Jennifer Tanner; Jessica Minion; Kara Loos; Keith MacKenzie; Kirsten Biggar; Madison Chapel; Meredith Faires; Morag Graham; Natalie Knox; Nathalie Bastien; Philip Mabon; Public Health Agency of Canada CanCOGeN team; Rachel DePaulo; Rhiannon Huzarewich; Russell Mandes; Ryan McDonald; Shari Tyson; Timothy Booth; Yan Li |
| EPI_ISL_5844812, EPI_ISL_5844818, EPI_ISL_5844821, EPI_ISL_5844822, EPI_ISL_5844825, EPI_ISL_5844830, EPI_ISL_5844832, EPI_ISL_5844833, EPI_ISL_5844837, EPI_ISL_5844843, EPI_ISL_5844850, EPI_ISL_5844852, EPI_ISL_5844853, EPI_ISL_5844857, EPI_ISL_5844861, EPI_ISL_5844864, EPI_ISL_5844865, EPI_ISL_5844869, EPI_ISL_5844883, EPI_ISL_5844889, EPI_ISL_5844896, EPI_ISL_5844904, EPI_ISL_5844913, EPI_ISL_5844914, EPI_ISL_5844916, EPI_ISL_5844921, EPI_ISL_5844924, EPI_ISL_5844925, EPI_ISL_5844931, EPI_ISL_5844955, EPI_ISL_5844966, EPI_ISL_5844967, EPI_ISL_5844968, EPI_ISL_5844969, EPI_ISL_5844975, EPI_ISL_5844976, EPI_ISL_5844978, EPI_ISL_5844979, EPI_ISL_5844982, EPI_ISL_5844986, EPI_ISL_5844988, EPI_ISL_5844993, EPI_ISL_5845005, EPI_ISL_5845015, EPI_ISL_5845032, EPI_ISL_5845034, EPI_ISL_5845037, EPI_ISL_5845040, EPI_ISL_5845052 | | | |
| see above | SK-Roy Romanow Provincial Laboratory | Saskatchewan - Roy Romanow Provincial Laboratory (RRPL) | Alanna Senecal; Amanda Lang; Jessica Minion; Kara Loos; Keith MacKenzie; Meredith Faires; Rachel DePaulo; Roy Romanow Provincial Laboratory - Molecular Diagnostics; Ryan McDonald |
| EPI_ISL_2868260, EPI_ISL_2868261 | SUNY UPSTATE MEDICAL UNIVERSITY | Wadsworth Center, New York State Department of Health | Alexis Russell; Catharine Prussing; Daryl M. Lamson; Erasmus Schneider; Erica Lasek-Nesselquist; John Kelly; Jonathan Plitnick; Kirsten St. George; Matthew Shudt; Melissa A Leisner; Navjot Singh |
| EPI_ISL_4878290 | SURA Bogota | Instituto Nacional de Salud- Dirección de Investigación en Salud Pública | Carlos Franco-Muñoz; Carmen Osorio; Diana Malo; Diego A. Álvarez-Díaz; Diego Andrés Prada; Gerardo Santamaría; Hector Alejandro Ruiz-Moreno; Jhonnatan Reales-González; Jorge Rivera; Juan Camilo Martinez; Julian Naizaque; Katherine Laiton-Donato; Lisseth Pardo; Magdalena Wiesner; Marcela Mercado-Reyes; Maria T. Herrera-Sepúlveda; Marta Lopez Blanco; Martha Lucia Ospina Martinez; Paola Rojas; Sergio Gomez; Sheryll Corchuelo; Ángela Alarcon Cruz |
| EPI_ISL_2612781, EPI_ISL_2612783, EPI_ISL_2612784, EPI_ISL_2612802, EPI_ISL_2612803, EPI_ISL_2612833, EPI_ISL_2685855 | | | |
| see above | SYNLAB | GIGA Medical Genomics | Bouchra Boujemla; Cécile Meex; Keith Durkin; Maria Artesi; Marie-Pierre Hayette; Nathalie Renotte; Pierrette Melin; Raphaël Boreux; Sébastien Bontems; Vincent Bours |
| EPI_ISL_2633059, EPI_ISL_2635424, EPI_ISL_2635454 | SYNLAB MVZ Heidelberg | Robert Koch Institute | |
| EPI_ISL_2633321 | SYNLAB MVZ Leinfelden-Echterdingen | Robert Koch Institute | |
| EPI_ISL_2633310, EPI_ISL_2635200, EPI_ISL_2635291, EPI_ISL_2635292, EPI_ISL_2635317 | SYNLAB MVZ Leverkusen | Robert Koch Institute | |
| EPI_ISL_2633330 | SYNLAB MVZ Trier | Robert Koch Institute | |
| EPI_ISL_2633150, EPI_ISL_2635446 | SYNLAB MVZ Weiden | Robert Koch Institute | |
| EPI_ISL_2692551, EPI_ISL_2692552, EPI_ISL_2692558, EPI_ISL_2692564, EPI_ISL_2692565, EPI_ISL_2692566, EPI_ISL_2692573, EPI_ISL_2692580, EPI_ISL_2692582, EPI_ISL_2692588, EPI_ISL_2692594, EPI_ISL_2692595, EPI_ISL_2692608, EPI_ISL_2692609, EPI_ISL_2692615, EPI_ISL_2692616, EPI_ISL_2692624, EPI_ISL_2692625, EPI_ISL_2692626, EPI_ISL_2692628, EPI_ISL_2692629, EPI_ISL_2692632, EPI_ISL_2692634, EPI_ISL_2692637, EPI_ISL_2692638, EPI_ISL_2692639, EPI_ISL_2692640, EPI_ISL_2692643, EPI_ISL_2692644, EPI_ISL_2692646, EPI_ISL_2692647, EPI_ISL_2692648, EPI_ISL_2692650, EPI_ISL_2692652, EPI_ISL_2692653, EPI_ISL_2692654, EPI_ISL_2692655, EPI_ISL_2692656, EPI_ISL_2692658, EPI_ISL_2692659, EPI_ISL_2692661, EPI_ISL_2692664, EPI_ISL_2692666, EPI_ISL_2692667, EPI_ISL_2692668, EPI_ISL_2692669, EPI_ISL_2692670, EPI_ISL_2692671, EPI_ISL_2692672, EPI_ISL_2692673, EPI_ISL_2692675, EPI_ISL_2692677, EPI_ISL_2692678, EPI_ISL_2692679, EPI_ISL_2692680, EPI_ISL_2692683, EPI_ISL_2692685, EPI_ISL_2692686, EPI_ISL_2692687, EPI_ISL_2692688, EPI_ISL_2692690, EPI_ISL_2692691, EPI_ISL_2692692, EPI_ISL_2692693, EPI_ISL_2692694, EPI_ISL_2692695, EPI_ISL_2692698, EPI_ISL_2692699, EPI_ISL_2692700, EPI_ISL_2692701, EPI_ISL_2692781, EPI_ISL_2692782, EPI_ISL_2810102, EPI_ISL_2810114, EPI_ISL_2810264, EPI_ISL_2810270, EPI_ISL_2810271, EPI_ISL_2810284, EPI_ISL_2810285, EPI_ISL_2810292, EPI_ISL_2810310, EPI_ISL_2810311, EPI_ISL_2810318, EPI_ISL_2810320, EPI_ISL_2810325, EPI_ISL_2894155, EPI_ISL_2894161, EPI_ISL_2894162, EPI_ISL_2894165, EPI_ISL_2894168, EPI_ISL_2894169, EPI_ISL_2894170, EPI_ISL_2894174, EPI_ISL_2894175, EPI_ISL_2894177, EPI_ISL_2894178, EPI_ISL_2894180, EPI_ISL_2894181, EPI_ISL_2894182, EPI_ISL_2894188, EPI_ISL_2894189, EPI_ISL_2894190, EPI_ISL_2894193, EPI_ISL_2894195, EPI_ISL_2894196, EPI_ISL_2894197, EPI_ISL_2894200, EPI_ISL_2894204, EPI_ISL_2894205, EPI_ISL_2894207, EPI_ISL_2894212, EPI_ISL_2894213, EPI_ISL_2894214, EPI_ISL_2894220, EPI_ISL_2894221, EPI_ISL_2894222, EPI_ISL_2894223, EPI_ISL_2894225, EPI_ISL_2894226, EPI_ISL_2894228, EPI_ISL_2894229, EPI_ISL_2894231, EPI_ISL_2894232, EPI_ISL_2894233, EPI_ISL_2894234, EPI_ISL_2894235, EPI_ISL_2894331, EPI_ISL_2894332, EPI_ISL_2894333, EPI_ISL_2894335, EPI_ISL_2894337, EPI_ISL_2894338, EPI_ISL_2894339, EPI_ISL_2894341, EPI_ISL_2894342, EPI_ISL_2894344, EPI_ISL_2894345, EPI_ISL_2894347, EPI_ISL_2894348, EPI_ISL_2894349, EPI_ISL_2894350, EPI_ISL_2894351, EPI_ISL_2894352, EPI_ISL_2894353, EPI_ISL_2894354, EPI_ISL_2894356, EPI_ISL_2894358, EPI_ISL_2894359, EPI_ISL_2894361, EPI_ISL_2894362, EPI_ISL_2894365, EPI_ISL_2894378, EPI_ISL_2894379, EPI_ISL_2894381, EPI_ISL_2894390, EPI_ISL_2970378, EPI_ISL_6253799 | | | |
| see above | Salud Digna | Instituto Nacional de Medicina Genomica | Abraham Campos-Romero; Blancas S; Cedro-Tanda A; Cisneros-Villanueva M; Cruz-Islas Jazmin; Escobar-Arrazola; Gonzalez-Barrera D; Herrera-Montalvo LA.; Hidalgo-Miranda A; Luna-Ruiz Marco; M; Mendoza-Vargas A; Moreno-Camacho José Luis; Munguia-Garza P; Orjuela-Rodríguez M; Ramirez-Vega O; Rangel-DeLeon D; Reyes-Grajeda JP; Rodriguez-Gallegos Jorge |
| EPI_ISL_2841534, | San Diego County Public Health | San Diego County Public Health | Ashleigh Murphy Schafer; Brett Austin; San Diego County Public Health Laboratory |

| | | | |
|---|---|---|---|
| EPI_ISL_2841536, EPI_ISL_2841614, EPI_ISL_2873837 | Laboratory | Laboratory | |
| EPI_ISL_2774614, EPI_ISL_2934006, EPI_ISL_2934043 | Santa Clara Valley Medical Center | Santa Clara County Public Health Laboratory | Santa Clara County Public Health Department |
| EPI_ISL_3015602 | Seattle Flu Study | Seattle Flu Study | Amanda Adler; Barry R. Lutz; Benjamin Pelle; Caitlin R. Wolf; Chris D. Frazar; Deborah A. Nickerson; Elisabeth Brandstetter; Erica Ryke; Helen Y. Chu; Janet A. Englund; Jay Shendure; Jeff Duchin; Jover Lee; Kairsten Fay; Karen Cowgill; Kirsten Lacombe; Lea M. Starita; Mark J. Rieder; Matthew Richardson; Matthew Thompson; Melissa Truong; Michael Boeckh; Michael Famulare; Misja Ilcisin; Peter D. Han; Stephanie Schrag; Thomas R. Sibley; Trevor Bedford |
| EPI_ISL_4082396, EPI_ISL_4082509, EPI_ISL_4082511, EPI_ISL_4082512, EPI_ISL_4082532, EPI_ISL_4082574, EPI_ISL_4082575, EPI_ISL_4082579, EPI_ISL_4082586, EPI_ISL_4082609, EPI_ISL_4082629, EPI_ISL_4082632, EPI_ISL_4082647, EPI_ISL_4082654, EPI_ISL_4082662, EPI_ISL_4082664, EPI_ISL_4082665, EPI_ISL_4082681, EPI_ISL_4082682, EPI_ISL_4082684, EPI_ISL_4082686, EPI_ISL_4082687, EPI_ISL_4082688, EPI_ISL_4082691, EPI_ISL_4082710, EPI_ISL_4082711, EPI_ISL_4082712, EPI_ISL_4082713, EPI_ISL_4082715, EPI_ISL_4082717, EPI_ISL_4082724, EPI_ISL_4082728, EPI_ISL_4082729, EPI_ISL_4082731, EPI_ISL_4082736, EPI_ISL_4082737, EPI_ISL_4082739, EPI_ISL_4082740, EPI_ISL_4082741, EPI_ISL_4082742, EPI_ISL_4082743, EPI_ISL_4082745, EPI_ISL_4082746, EPI_ISL_4082747, EPI_ISL_4082748, EPI_ISL_4082749, EPI_ISL_4082750, EPI_ISL_4082754, EPI_ISL_4082756, EPI_ISL_4082757, EPI_ISL_4082758, EPI_ISL_4082761, EPI_ISL_4082773, EPI_ISL_4082775, EPI_ISL_4082789, EPI_ISL_4082792, EPI_ISL_4082793, EPI_ISL_4082803, EPI_ISL_4082804, EPI_ISL_4082805, EPI_ISL_4082806, EPI_ISL_4082807, EPI_ISL_4082808, EPI_ISL_4082810, EPI_ISL_4082811, EPI_ISL_4082812, EPI_ISL_4082813, EPI_ISL_4082830, EPI_ISL_4082832, EPI_ISL_4082833, EPI_ISL_4082834, EPI_ISL_4082843, EPI_ISL_4082844, EPI_ISL_4082852, EPI_ISL_4082877, EPI_ISL_4082885, EPI_ISL_4082897, EPI_ISL_4082930, EPI_ISL_4082991, EPI_ISL_4082997, EPI_ISL_4083021, EPI_ISL_4083022, EPI_ISL_4083030, EPI_ISL_4083031, EPI_ISL_4083099, EPI_ISL_4083114, EPI_ISL_4083118, EPI_ISL_4083165, EPI_ISL_4083204, EPI_ISL_4083323, EPI_ISL_4083324, EPI_ISL_5158411, EPI_ISL_5158419, EPI_ISL_5158421, EPI_ISL_5158423, EPI_ISL_5158427, EPI_ISL_5158428, EPI_ISL_5158429, EPI_ISL_6694378, EPI_ISL_6694438 | | | |
| see above | Servicio Virosis Respiratorias-Departamento Virología-INEI | Instituto Nacional Enfermedades Infecciosas C.G.Malbran | Avaro M.; Baumeister E.; Benedetti E.; Campos J.; Cisterna D.; Dattero ME; De Belder D.; Haim MS.; Lorenzo F.; Molina V.; Perandones C.; Poklepovich T.; Pontoriero A.; Russo M.; Sanchez Loria J.; Tuduri E. |
| EPI_ISL_3149734 | Servicio de Inmunología del Hospital Julio Perrando | Laboratorio de Biología Molecular, Instituto de Medicina Regional on behalf of 'Proyecto Argentino Interinstitucional de genomica de SARS-CoV-2' (PAIS Consortium) | Bettina Brusés; Florencia Vallejos Schulze; Griselda Oria; Horacio Lucero.; Javier Mussin; Laura Formichelli; María Delia Foussal; Melina Lorenzini Campos; Raúl Maximiliano Acevedo |
| EPI_ISL_2689709, EPI_ISL_2689710, EPI_ISL_2758565, EPI_ISL_2758594 | Servicio de Microbiologia Hospital Ramon y Cajal | Servicio de Microbiologia Hospital Ramon y Cajal | Galan JC; Martinez L. Abreu M; Ponce M; y Gonzalez-Alba JM |
| EPI_ISL_2834019, EPI_ISL_2834022, EPI_ISL_2834024, EPI_ISL_2834030, EPI_ISL_2834037, EPI_ISL_2834040 | Servicio de Microbiología. Consorcio Hospital General Universitario de Valencia | SeqCOVID-SPAIN consortium/IBV(CSIC) | Begoña Fuster Escrivá; Carme Salvador García; Concepción Gimeno Cardona and SeqCOVID-SPAIN consortium; María Dolores Ocete; Rafael Medina González |
| EPI_ISL_6686928, EPI_ISL_6686929, EPI_ISL_6687797, EPI_ISL_6688001 | Shared Hospital Laboratory | Shared Hospital Laboratory | Christie Vermeiren; Finlay Maguire; Kevin Katz; Patryk Aftanas; Robert Kozak; Samira Mubareka |
| EPI_ISL_2716924 | Southern Nevada Public Health Laboratory | Southern Nevada Public Health Laboratory | Michael Picker |
| EPI_ISL_2724792, EPI_ISL_2724857, EPI_ISL_2724858, EPI_ISL_2724862, EPI_ISL_2724869, EPI_ISL_2724880, EPI_ISL_2724889, EPI_ISL_2724891, EPI_ISL_2724899, EPI_ISL_2724900, EPI_ISL_2724901, EPI_ISL_2724902, EPI_ISL_3033089, EPI_ISL_3033092, EPI_ISL_3033093, EPI_ISL_3033095, EPI_ISL_3033103, EPI_ISL_3033104, EPI_ISL_3033105, EPI_ISL_3033106, EPI_ISL_3033107, EPI_ISL_3033108, EPI_ISL_3033115, EPI_ISL_3033120, EPI_ISL_3033122, EPI_ISL_3033123, EPI_ISL_3033133, EPI_ISL_3033158 | | | |
| see above | State Laboratories Division, Hawaii State Department of Health | State Laboratories Division, Hawaii State Department of Health | Ayana Garnet; Drew Kuwazaki; Edward Desmond; Pamela O'Brien; Razvan Sultana |
| EPI_ISL_3049994, EPI_ISL_3050004 | State Testing Facility | Altius Institute for Biomedical Research | Alex Isner; Alex Nguyen; Amanda Gale; Audra Johnson; Clem Green; Daniel Bates; Eric Thorland; Jacob Rodriguez; Jean Robinston; Jemma Nelson; Jessica Kunder; John Stamatoyannopoulos; Joshua Richards; Julia Wald; Kneshay Harper; Lauren Mitchell; Mark Frerker; Matt Hartman; Michael Buckley; Muhammad Halimun; Rebecca Bruders; Tobias Ragoczy |
| EPI_ISL_6794411 | State of New Hampshire Public Health Laboratories | State of New Hampshire Public Health Laboratories | Caitlin Mercier; Chris Benton; Jinfeng Li; Juan Bolanos; Xinglu Zhang |
| EPI_ISL_2792714, EPI_ISL_2792769, EPI_ISL_2792804, EPI_ISL_3998699, EPI_ISL_3998702, EPI_ISL_3998705, EPI_ISL_3998869 | | | |
| see above | Swedish national genomic surveillance program of SARS-CoV-2 | The Public Health Agency of Sweden | Alma Brolund; Maria Lind Karlberg; Maximilian Riess; Swedish national genomic surveillance program of SARS-CoV-2 |
| EPI_ISL_2633033, EPI_ISL_2633345 | Synlab MVZ Augsburg | Robert Koch Institute | |
| EPI_ISL_2868607 | TEMPUS LABS INC | Wadsworth Center, New York State Department of Health | Alexis Russell; Catharine Prussing; Daryl M. Lamson; Erasmus Schneider; Erica Lasek-Nesselquist; John Kelly; Jonathan Plitnick; Kirsten St. George; Matthew Shudt; Melissa A Leisner; Navjot Singh |
| EPI_ISL_2919322, EPI_ISL_2919491, EPI_ISL_2919521, EPI_ISL_2919662, EPI_ISL_3009255, EPI_ISL_3326813, EPI_ISL_4059572, EPI_ISL_6253861 | | | |
| see above | TGen North | TGen North | "Jolene Bowers; Brett Van Tassel; Chris French; Darrin Lemmer; Dave Engelthaler; Dave Engelthaler"; Hayley Yaglom; Heather Centner; Jolene Bowers |
| EPI_ISL_2658975, EPI_ISL_2712996, EPI_ISL_2983489 | TXDSHS | TXDSHS | Anita Pokharel; Bonnie Oh; Chun Wang; Grace Kubin; Jenny Zhang; Karen Bobier; Lorraine Rodriguez; Maliha Rahman; Mayela Pedrueza; Myong Koag; Rachel Lee; Rashmi Tuladhar |
| EPI_ISL_2649825, EPI_ISL_2649826, EPI_ISL_2800903, EPI_ISL_2833809, EPI_ISL_2930470 | Tampa General Hospital Esoteric Lab | Tampa General Hospital Esoteric Research & Development Lab | Amorce Lima; Deanna Becker; Dominic Uy; Elaine Vendrone; Jon Faughn; Jorge Mecias-Frias; Suzane Silbert; Vicki Healer |
| EPI_ISL_2716573, EPI_ISL_2716574, EPI_ISL_2716579, EPI_ISL_2716580, EPI_ISL_2716583, EPI_ISL_2716614 | The Caribbean Public Health Agency | Carrington Lab, Department of Preclinical Sciences, Faculty of Medical Sciences, The University of the West Indies, St Augustine Campus | Anushka Ramjag; Arianne Brown-Jordan; Avery Hinds; Christine V. F. Carrington; Gabriel Escobar; Nikita S. D. Sahadeo; Nuno Faria; Oliver Pybus; Risha Singh; Roshan Parasram; Sarah Hill; SueMin Nathaniel; Vernie Ramkissoon |
| EPI_ISL_2758914, EPI_ISL_2758925, EPI_ISL_2758927 | The Ohio State University Applied Microbiology Services Laboratory | The Ohio State University Applied Microbiology Services Laboratory | Seth A. Faith PhD |
| EPI_ISL_3152401 | TriCore Reference Laboratories | Center for Global Health, University of New Mexico Health Sciences Center | Cecilia Thompson; Darrell Dinwiddie; Daryl Domman; Karissa Culbreath; Kendra Pesko; Kurt Schwalm; Valerie Morley |
| EPI_ISL_2840625 | U.O. Igiene, Ospedale Policlinico San Martino IRCCS Genova | U.O. Igiene, Ospedale Policlinico San Martino | Bruzzone Bianca; Caligiuri Patrizia; De Pace Vanessa; Domnich Alexander; Icardi Giancarlo on behalf of SARS-CoV-2 ITALIAN RESEARCH ENTERPRISE-(SCIRE) Collaborative Group; Orsi Andrea; Ricucci Valentina |
| EPI_ISL_3215235, EPI_ISL_3215236 | U.O. Microbiologia Laboratorio Unico Centro Servizi - AUSL della Romagna | U.O. Microbiologia, Laboratorio Unico Centro Servizi - AUSL della Romagna | Giorgio Dirani |
| EPI_ISL_5646768 | UBS DR ABELARDO PINHEIRO GUIMARAES SCRPARDO | Instituto Butantan | Antonio Jorge Martins; Claudia Renata dos Santos Barros; David Schlesinger; Debora Botequio Moretti; Dimas Tadeu Covas; Elaine Cristina Marqueze; Elaine Vieira Santos; Evandra Strazza Rodrigues; Heidge Fukumasu; Jayme Augusto de Souza-Neto; José Salvatore Leister Patané; Luiz Alcantara; Luiz Lehmann Coutinho; Maria Carolina Elias; Maurício Lacerda Nogueira; Rafael dos Santos Bezerra; Raul Machado Neto; Rejane Maria Tommasini Grotto; Ricardo Haddad; Sandra Coccuzzo Sampaio Vessoni; Simone Kashima; Svetoslav Nanev Slavov; Vincent Louis Viala |
| EPI_ISL_5647237 | UBS DR ALFREDO DANTAS DE SOUZA UMUARAMA | Instituto Butantan | Antonio Jorge Martins; Claudia Renata dos Santos Barros; David Schlesinger; Debora Botequio Moretti; Dimas Tadeu Covas; Elaine Cristina Marqueze; Elaine Vieira Santos; Evandra Strazza Rodrigues; Heidge Fukumasu; Jayme Augusto de Souza-Neto; José Salvatore Leister Patané; Luiz Alcantara; Luiz Lehmann Coutinho; Maria Carolina Elias; Maurício Lacerda Nogueira; Rafael dos Santos Bezerra; Raul Machado Neto; Rejane Maria Tommasini Grotto; Ricardo Haddad; Sandra Coccuzzo Sampaio Vessoni; Simone Kashima; Svetoslav Nanev Slavov; Vincent Louis Viala |
| EPI_ISL_5646887, EPI_ISL_5646891, EPI_ISL_5646912, EPI_ISL_5646920, EPI_ISL_5646929 | UBS GETULINO JOSE DIAS | Instituto Butantan | Antonio Jorge Martins; Claudia Renata dos Santos Barros; David Schlesinger; Debora Botequio Moretti; Dimas Tadeu Covas; Elaine Cristina Marqueze; Elaine Vieira Santos; Evandra Strazza Rodrigues; Heidge Fukumasu; Jayme Augusto de Souza-Neto; José Salvatore Leister Patané; Luiz Alcantara; Luiz Lehmann Coutinho; Maria Carolina Elias; Maurício Lacerda Nogueira; Rafael dos Santos Bezerra; Raul Machado Neto; Rejane Maria Tommasini Grotto; Ricardo Haddad; Sandra Coccuzzo Sampaio Vessoni; Simone Kashima; Svetoslav Nanev Slavov; Vincent Louis Viala |
| EPI_ISL_5646388, EPI_ISL_5646445, EPI_ISL_5646470 | UBS II DA FAMILIA SEGISFREDO MAMEDE PINTO LOURDES | Instituto Butantan | Antonio Jorge Martins; Claudia Renata dos Santos Barros; David Schlesinger; Debora Botequio Moretti; Dimas Tadeu Covas; Elaine Cristina Marqueze; Elaine Vieira Santos; Evandra Strazza Rodrigues; Heidge Fukumasu; Jayme Augusto de Souza-Neto; José Salvatore Leister Patané; Luiz Alcantara; Luiz Lehmann Coutinho; Maria Carolina Elias; Maurício Lacerda Nogueira; Rafael dos Santos Bezerra; Raul Machado Neto; Rejane Maria Tommasini Grotto; Ricardo Haddad; Sandra Coccuzzo Sampaio Vessoni; Simone Kashima; Svetoslav Nanev Slavov; Vincent Louis Viala |
| EPI_ISL_5646547, EPI_ISL_5646669, EPI_ISL_5646748, EPI_ISL_5646818 | UBS II DE ALVARES MACHADO | Instituto Butantan | Antonio Jorge Martins; Claudia Renata dos Santos Barros; David Schlesinger; Debora Botequio Moretti; Dimas Tadeu Covas; Elaine Cristina Marqueze; Elaine Vieira Santos; Evandra Strazza Rodrigues; Heidge Fukumasu; Jayme Augusto de Souza-Neto; José Salvatore Leister Patané; Luiz Alcantara; Luiz Lehmann Coutinho; Maria Carolina Elias; Maurício Lacerda Nogueira; Rafael dos Santos Bezerra; Raul Machado Neto; Rejane Maria Tommasini Grotto; Ricardo Haddad; Sandra Coccuzzo Sampaio Vessoni; Simone Kashima; Svetoslav Nanev Slavov; Vincent Louis Viala |
| EPI_ISL_5646614, EPI_ISL_5646838 | UBS II DE PIRAPOZINHO C SERVICO DE EACS E ESF | Instituto Butantan | Antonio Jorge Martins; Claudia Renata dos Santos Barros; David Schlesinger; Debora Botequio Moretti; Dimas Tadeu Covas; Elaine Cristina Marqueze; Elaine Vieira Santos; Evandra Strazza Rodrigues; Heidge Fukumasu; Jayme Augusto de Souza-Neto; José Salvatore Leister Patané; Luiz Alcantara; Luiz Lehmann Coutinho; Maria Carolina Elias; Maurício Lacerda Nogueira; Rafael dos Santos Bezerra; Raul Machado Neto; Rejane Maria Tommasini Grotto; Ricardo Haddad; Sandra Coccuzzo Sampaio Vessoni; Simone Kashima; Svetoslav Nanev Slavov; Vincent Louis Viala |
| EPI_ISL_5646875, EPI_ISL_5646882, EPI_ISL_5646884, EPI_ISL_5646885, EPI_ISL_5646888, EPI_ISL_5646889, EPI_ISL_5646910, EPI_ISL_5646917, EPI_ISL_5646921, EPI_ISL_5646931, EPI_ISL_5646932, EPI_ISL_5646933 | | | |

| | | | |
|---|---|---|---|
| see above | UBS II DE TANABI MILTON MARTINS PERCHES | Instituto Butantan | Antonio Jorge Martins; Claudia Renata dos Santos Barros; David Schlesinger; Debora Botequio Moretti; Dimas Tadeu Covas; Elaine Cristina Marqueze; Elaine Vieira Santos; Evandra Strazza Rodrigues; Heidge Fukumasu; Jayme Augusto de Souza-Neto; José Salvatore Leister Patané; Luiz Alcantara; Luiz Lehmann Coutinho; Maria Carolina Elias; Maurício Lacerda Nogueira; Rafael dos Santos Bezerra; Raul Machado Neto; Rejane Maria Tommasini Grotto; Ricardo Haddad; Sandra Coccuzzo Sampaio Vessoni; Simone Kashima; Svetoslav Nanev Slavov; Vincent Louis Viala |
| EPI_ISL_5646638, EPI_ISL_5646697, EPI_ISL_5646739, EPI_ISL_5646846 | UBS II DR EXPEDITO SHIZUO KUROCE | Instituto Butantan | Antonio Jorge Martins; Claudia Renata dos Santos Barros; David Schlesinger; Debora Botequio Moretti; Dimas Tadeu Covas; Elaine Cristina Marqueze; Elaine Vieira Santos; Evandra Strazza Rodrigues; Heidge Fukumasu; Jayme Augusto de Souza-Neto; José Salvatore Leister Patané; Luiz Alcantara; Luiz Lehmann Coutinho; Maria Carolina Elias; Maurício Lacerda Nogueira; Rafael dos Santos Bezerra; Raul Machado Neto; Rejane Maria Tommasini Grotto; Ricardo Haddad; Sandra Coccuzzo Sampaio Vessoni; Simone Kashima; Svetoslav Nanev Slavov; Vincent Louis Viala |
| EPI_ISL_5646515, EPI_ISL_5646527, EPI_ISL_5646731 | UBS II DR JOSE CARLOS MACUCO JANINI | Instituto Butantan | Antonio Jorge Martins; Claudia Renata dos Santos Barros; David Schlesinger; Debora Botequio Moretti; Dimas Tadeu Covas; Elaine Cristina Marqueze; Elaine Vieira Santos; Evandra Strazza Rodrigues; Heidge Fukumasu; Jayme Augusto de Souza-Neto; José Salvatore Leister Patané; Luiz Alcantara; Luiz Lehmann Coutinho; Maria Carolina Elias; Maurício Lacerda Nogueira; Rafael dos Santos Bezerra; Raul Machado Neto; Rejane Maria Tommasini Grotto; Ricardo Haddad; Sandra Coccuzzo Sampaio Vessoni; Simone Kashima; Svetoslav Nanev Slavov; Vincent Louis Viala |
| EPI_ISL_5646500 | UBS III DE EUCLIDES DA CUNHA PAULISTA | Instituto Butantan | Antonio Jorge Martins; Claudia Renata dos Santos Barros; David Schlesinger; Debora Botequio Moretti; Dimas Tadeu Covas; Elaine Cristina Marqueze; Elaine Vieira Santos; Evandra Strazza Rodrigues; Heidge Fukumasu; Jayme Augusto de Souza-Neto; José Salvatore Leister Patané; Luiz Alcantara; Luiz Lehmann Coutinho; Maria Carolina Elias; Maurício Lacerda Nogueira; Rafael dos Santos Bezerra; Raul Machado Neto; Rejane Maria Tommasini Grotto; Ricardo Haddad; Sandra Coccuzzo Sampaio Vessoni; Simone Kashima; Svetoslav Nanev Slavov; Vincent Louis Viala |
| EPI_ISL_5646876, EPI_ISL_5646880, EPI_ISL_5646894, EPI_ISL_5646896, EPI_ISL_5646907, EPI_ISL_5646915 | UBS LUIS FACHIN IPIGUA | Instituto Butantan | Antonio Jorge Martins; Claudia Renata dos Santos Barros; David Schlesinger; Debora Botequio Moretti; Dimas Tadeu Covas; Elaine Cristina Marqueze; Elaine Vieira Santos; Evandra Strazza Rodrigues; Heidge Fukumasu; Jayme Augusto de Souza-Neto; José Salvatore Leister Patané; Luiz Alcantara; Luiz Lehmann Coutinho; Maria Carolina Elias; Maurício Lacerda Nogueira; Rafael dos Santos Bezerra; Raul Machado Neto; Rejane Maria Tommasini Grotto; Ricardo Haddad; Sandra Coccuzzo Sampaio Vessoni; Simone Kashima; Svetoslav Nanev Slavov; Vincent Louis Viala |
| EPI_ISL_5646722 | UBS MARCIA CRISTIANE DA SILVA DE OURO VERDE | Instituto Butantan | Antonio Jorge Martins; Claudia Renata dos Santos Barros; David Schlesinger; Debora Botequio Moretti; Dimas Tadeu Covas; Elaine Cristina Marqueze; Elaine Vieira Santos; Evandra Strazza Rodrigues; Heidge Fukumasu; Jayme Augusto de Souza-Neto; José Salvatore Leister Patané; Luiz Alcantara; Luiz Lehmann Coutinho; Maria Carolina Elias; Maurício Lacerda Nogueira; Rafael dos Santos Bezerra; Raul Machado Neto; Rejane Maria Tommasini Grotto; Ricardo Haddad; Sandra Coccuzzo Sampaio Vessoni; Simone Kashima; Svetoslav Nanev Slavov; Vincent Louis Viala |
| EPI_ISL_5646551 | UBS PARQUE CECAP | Instituto Butantan | Antonio Jorge Martins; Claudia Renata dos Santos Barros; David Schlesinger; Debora Botequio Moretti; Dimas Tadeu Covas; Elaine Cristina Marqueze; Elaine Vieira Santos; Evandra Strazza Rodrigues; Heidge Fukumasu; Jayme Augusto de Souza-Neto; José Salvatore Leister Patané; Luiz Alcantara; Luiz Lehmann Coutinho; Maria Carolina Elias; Maurício Lacerda Nogueira; Rafael dos Santos Bezerra; Raul Machado Neto; Rejane Maria Tommasini Grotto; Ricardo Haddad; Sandra Coccuzzo Sampaio Vessoni; Simone Kashima; Svetoslav Nanev Slavov; Vincent Louis Viala |
| EPI_ISL_5646893, EPI_ISL_5646903, EPI_ISL_5646904, EPI_ISL_5646911 | UBS RIBEIRAO CORRENTE | Instituto Butantan | Antonio Jorge Martins; Claudia Renata dos Santos Barros; David Schlesinger; Debora Botequio Moretti; Dimas Tadeu Covas; Elaine Cristina Marqueze; Elaine Vieira Santos; Evandra Strazza Rodrigues; Heidge Fukumasu; Jayme Augusto de Souza-Neto; José Salvatore Leister Patané; Luiz Alcantara; Luiz Lehmann Coutinho; Maria Carolina Elias; Maurício Lacerda Nogueira; Rafael dos Santos Bezerra; Raul Machado Neto; Rejane Maria Tommasini Grotto; Ricardo Haddad; Sandra Coccuzzo Sampaio Vessoni; Simone Kashima; Svetoslav Nanev Slavov; Vincent Louis Viala |
| EPI_ISL_5646925 | UBS SANTA MARIA GORETTI | Instituto Butantan | Antonio Jorge Martins; Claudia Renata dos Santos Barros; David Schlesinger; Debora Botequio Moretti; Dimas Tadeu Covas; Elaine Cristina Marqueze; Elaine Vieira Santos; Evandra Strazza Rodrigues; Heidge Fukumasu; Jayme Augusto de Souza-Neto; José Salvatore Leister Patané; Luiz Alcantara; Luiz Lehmann Coutinho; Maria Carolina Elias; Maurício Lacerda Nogueira; Rafael dos Santos Bezerra; Raul Machado Neto; Rejane Maria Tommasini Grotto; Ricardo Haddad; Sandra Coccuzzo Sampaio Vessoni; Simone Kashima; Svetoslav Nanev Slavov; Vincent Louis Viala |
| EPI_ISL_5646390, EPI_ISL_5646427, EPI_ISL_5646429, EPI_ISL_5646447, EPI_ISL_5646461 | UBS VILA RIBEIRO | Instituto Butantan | Antonio Jorge Martins; Claudia Renata dos Santos Barros; David Schlesinger; Debora Botequio Moretti; Dimas Tadeu Covas; Elaine Cristina Marqueze; Elaine Vieira Santos; Evandra Strazza Rodrigues; Heidge Fukumasu; Jayme Augusto de Souza-Neto; José Salvatore Leister Patané; Luiz Alcantara; Luiz Lehmann Coutinho; Maria Carolina Elias; Maurício Lacerda Nogueira; Rafael dos Santos Bezerra; Raul Machado Neto; Rejane Maria Tommasini Grotto; Ricardo Haddad; Sandra Coccuzzo Sampaio Vessoni; Simone Kashima; Svetoslav Nanev Slavov; Vincent Louis Viala |
| EPI_ISL_3067016, EPI_ISL_3067048 | UC-Christus Clinical Hospital Laboratory | Laboratory of Molecular Virology, School of Medicine, Pontificia Universidad Catolica de Chile | Ana Maira Guzman; Andres E. Munoz-Marcos; Catalina Pardo-Roa; Eileen Serrano; Erick Salinas; Estefany Poblete; Francisco Melo; Jorge Levican; Leonardo I. Almonacid; Maria Jose Avendano; Maria Patricia Vega; Rafael A. Medina; Ricardo Enrique de la Barra; Tamara Garcia-Salum |
| EPI_ISL_2960102, EPI_ISL_3025381, EPI_ISL_3025382, EPI_ISL_3025383, EPI_ISL_3025388, EPI_ISL_3025390, EPI_ISL_3025394 | | | |
| see above | UCC (Universidad Central del Caribe) | Grubaugh Lab - Yale School of Public Health | Anderson Brito; Andrea Firpo; Annie Watkins; Chaney Kalinich; Chantal Vogels; David Rivera Aponte; Ernesto Vazquez; Fabiola Cruz-Lopez; Fabiola Fontanet; Isabel Ott; Jessica Rothman; Joseph Fauver; Karolane Gonzalez; Kendall Billig; Mallery Breban; Mary Petrone; Natalie Machargo; Nathan Grubaugh; Patricia Serrano; Tania Mitwalli; Tara Alpert; Tobias Koch; Vianca Aponte |
| EPI_ISL_2661538 | UHTL, University Hospitals | UHTL, University Hospitals | Alouani, D.; Sadri, N.; Song, X. |
| EPI_ISL_2709012 | UMC Groningen, Clinical Virology, Department of Medical Microbiology and Infection Prevention | UMC Groningen, Clinical Virology, Department of Medical Microbiology and Infection Prevention | Alexander Friedrich; Coretta Van Leer-Buter; Erley Lizarazo-Forero; Hubert Niesters; Lilli Gard; Marjolein Knoester; Monika Fliss; Sigrid Rosema; Xuewei Zhou |
| EPI_ISL_2658343 | UMC Utrecht | UMC Utrecht | Annemarie Wensing; Anniek Tanja; Pascal Mantiri; Rob Schuurman |
| EPI_ISL_3066764 | UNIDAD DE DIAGNOSTICO HEMATO ONCOLOGICA | Instituto Nacional de Salud | Carlos Franco-Muñoz; Carmen Osorio; Diana Malo; Diego A. Álvarez-Díaz; Diego Andrés Prada; Gerardo Santamaría; Hector Alejandro Ruiz-Moreno; Jhonnatan Reales-González; Jorge Rivera; Juan Camilo Martinez; Julian Naizaque; Katherine Laiton-Donato; Lisseth Pardo; Magdalena Wiesner; Marcela Mercado-Reyes; Maria T.Herrera-Sepúlveda; Marta Lopez Blanco; Martha Lucia Ospina Martinez; Paola Rojas; Sergio Gomez; Sheryll Corchuelo; Ángela Alarcon Cruz |
| EPI_ISL_5646533, EPI_ISL_5646558, EPI_ISL_5646763, EPI_ISL_5646820 | UNIDADE BASICA DE SAUDE DE IPEUNA | Instituto Butantan | Antonio Jorge Martins; Claudia Renata dos Santos Barros; David Schlesinger; Debora Botequio Moretti; Dimas Tadeu Covas; Elaine Cristina Marqueze; Elaine Vieira Santos; Evandra Strazza Rodrigues; Heidge Fukumasu; Jayme Augusto de Souza-Neto; José Salvatore Leister Patané; Luiz Alcantara; Luiz Lehmann Coutinho; Maria Carolina Elias; Maurício Lacerda Nogueira; Rafael dos Santos Bezerra; Raul Machado Neto; Rejane Maria Tommasini Grotto; Ricardo Haddad; Sandra Coccuzzo Sampaio Vessoni; Simone Kashima; Svetoslav Nanev Slavov; Vincent Louis Viala |
| EPI_ISL_5647210, EPI_ISL_5647214 | UNIDADE BASICA DE SSAUDE DE POLONI | Instituto Butantan | Antonio Jorge Martins; Claudia Renata dos Santos Barros; David Schlesinger; Debora Botequio Moretti; Dimas Tadeu Covas; Elaine Cristina Marqueze; Elaine Vieira Santos; Evandra Strazza Rodrigues; Heidge Fukumasu; Jayme Augusto de Souza-Neto; José Salvatore Leister Patané; Luiz Alcantara; Luiz Lehmann Coutinho; Maria Carolina Elias; Maurício Lacerda Nogueira; Rafael dos Santos Bezerra; Raul Machado Neto; Rejane Maria Tommasini Grotto; Ricardo Haddad; Sandra Coccuzzo Sampaio Vessoni; Simone Kashima; Svetoslav Nanev Slavov; Vincent Louis Viala |
| EPI_ISL_5646514, EPI_ISL_5646535, EPI_ISL_5646721, EPI_ISL_5646823, EPI_ISL_5646831 | UNIDADE DE PRONTO ATENDIMENTO CENTRAL DE CARAGUATATUBA | Instituto Butantan | Antonio Jorge Martins; Claudia Renata dos Santos Barros; David Schlesinger; Debora Botequio Moretti; Dimas Tadeu Covas; Elaine Cristina Marqueze; Elaine Vieira Santos; Evandra Strazza Rodrigues; Heidge Fukumasu; Jayme Augusto de Souza-Neto; José Salvatore Leister Patané; Luiz Alcantara; Luiz Lehmann Coutinho; Maria Carolina Elias; Maurício Lacerda Nogueira; Rafael dos Santos Bezerra; Raul Machado Neto; Rejane Maria Tommasini Grotto; Ricardo Haddad; Sandra Coccuzzo Sampaio Vessoni; Simone Kashima; Svetoslav Nanev Slavov; Vincent Louis Viala |
| EPI_ISL_5646398, EPI_ISL_5646451, EPI_ISL_5646469 | UNIDADE DE PRONTO ATENDIMENTO TERRA PRETA | Instituto Butantan | Antonio Jorge Martins; Claudia Renata dos Santos Barros; David Schlesinger; Debora Botequio Moretti; Dimas Tadeu Covas; Elaine Cristina Marqueze; Elaine Vieira Santos; Evandra Strazza Rodrigues; Heidge Fukumasu; Jayme Augusto de Souza-Neto; José Salvatore Leister Patané; Luiz Alcantara; Luiz Lehmann Coutinho; Maria Carolina Elias; Maurício Lacerda Nogueira; Rafael dos Santos Bezerra; Raul Machado Neto; Rejane Maria Tommasini Grotto; Ricardo Haddad; Sandra Coccuzzo Sampaio Vessoni; Simone Kashima; Svetoslav Nanev Slavov; Vincent Louis Viala |
| EPI_ISL_5646544, EPI_ISL_5646600 | UNIDADE DE PRONTO ATENDIMENTO UPA SUL CARAGUATATUBA | Instituto Butantan | Antonio Jorge Martins; Claudia Renata dos Santos Barros; David Schlesinger; Debora Botequio Moretti; Dimas Tadeu Covas; Elaine Cristina Marqueze; Elaine Vieira Santos; Evandra Strazza Rodrigues; Heidge Fukumasu; Jayme Augusto de Souza-Neto; José Salvatore Leister Patané; Luiz Alcantara; Luiz Lehmann Coutinho; Maria Carolina Elias; Maurício Lacerda Nogueira; Rafael dos Santos Bezerra; Raul Machado Neto; Rejane Maria Tommasini Grotto; Ricardo Haddad; Sandra Coccuzzo Sampaio Vessoni; Simone Kashima; Svetoslav Nanev Slavov; Vincent Louis Viala |
| EPI_ISL_5646541, EPI_ISL_5646575, EPI_ISL_5646584, EPI_ISL_5646599, EPI_ISL_5646677, EPI_ISL_5646798 | UNIDADE DE SAUDE DA FAMILIA DR AMELIO TANGANELLI | Instituto Butantan | Antonio Jorge Martins; Claudia Renata dos Santos Barros; David Schlesinger; Debora Botequio Moretti; Dimas Tadeu Covas; Elaine Cristina Marqueze; Elaine Vieira Santos; Evandra Strazza Rodrigues; Heidge Fukumasu; Jayme Augusto de Souza-Neto; José Salvatore Leister Patané; Luiz Alcantara; Luiz Lehmann Coutinho; Maria Carolina Elias; Maurício Lacerda Nogueira; Rafael dos Santos Bezerra; Raul Machado Neto; Rejane Maria Tommasini Grotto; Ricardo Haddad; Sandra Coccuzzo Sampaio Vessoni; Simone Kashima; Svetoslav Nanev Slavov; Vincent Louis Viala |
| EPI_ISL_5647901 | UNIDADE DE VIGILANCIA EPIDEMIOLOGICA | Instituto Butantan | Antonio Jorge Martins; Claudia Renata dos Santos Barros; David Schlesinger; Debora Botequio Moretti; Dimas Tadeu Covas; Elaine Cristina Marqueze; Elaine Vieira Santos; Evandra Strazza Rodrigues; Heidge Fukumasu; Jayme Augusto de Souza-Neto; José Salvatore Leister Patané; Luiz Alcantara; Luiz Lehmann Coutinho; Maria Carolina Elias; Maurício Lacerda Nogueira; Rafael dos Santos Bezerra; Raul Machado Neto; Rejane Maria Tommasini Grotto; Ricardo Haddad; Sandra Coccuzzo Sampaio Vessoni; Simone Kashima; Svetoslav Nanev Slavov; Vincent Louis Viala |
| EPI_ISL_5646909 | UNIDADE MISTA DE SAUDE DE DUMONT | Instituto Butantan | Antonio Jorge Martins; Claudia Renata dos Santos Barros; David Schlesinger; Debora Botequio Moretti; Dimas Tadeu Covas; Elaine Cristina Marqueze; Elaine Vieira Santos; Evandra Strazza Rodrigues; Heidge Fukumasu; Jayme Augusto de Souza-Neto; José Salvatore Leister Patané; Luiz Alcantara; Luiz Lehmann Coutinho; Maria Carolina Elias; Maurício Lacerda Nogueira; Rafael dos Santos Bezerra; Raul Machado Neto; Rejane Maria Tommasini Grotto; Ricardo Haddad; Sandra Coccuzzo Sampaio Vessoni; Simone Kashima; Svetoslav Nanev Slavov; Vincent Louis Viala |
| EPI_ISL_3912320 | UNIDADE PRONTO ATENDIMENTO AUTRAN NUNES | Analytical Competence Molecular Epidemiology Lab/ACME, Oswaldo Cruz Foundation, Ceara (FIOCRUZ CE) | Cleber Furtado Aksenen; Fabio Miyajima; Fernando Braga Stehling; Francisco Eder de Moura Lopes; Jamille Maria Mendes Bezerra; Joaquim Cesar do Nascimento Sousa Junior; Pedro Miguel Carneiro Jeronimo; Suzana Porto Almeida & Lucas Delerino on behalf of COVID-19 FIOCRUZ Genomic Network; Thais Ferreira de Oliveira; Thais de Oliveira Costa; Ticiane Cavalcante de Souza; Veridiana Pessoa Miyajima |
| EPI_ISL_3912087, EPI_ISL_3912356, EPI_ISL_3912357, EPI_ISL_3912358, EPI_ISL_3912359, EPI_ISL_3912360, EPI_ISL_3912361, EPI_ISL_3912362, EPI_ISL_3912363 | | | |
| see above | UNIDADE REFERENCIA COVID 19 | Analytical Competence Molecular Epidemiology Lab/ACME, Oswaldo Cruz Foundation, Ceara (FIOCRUZ CE) | Cleber Furtado Aksenen; Fabio Miyajima; Fernando Braga Stehling; Francisco Eder de Moura Lopes; Jamille Maria Mendes Bezerra; Joaquim Cesar do Nascimento Sousa Junior; Pedro Miguel Carneiro Jeronimo; Suzana Porto Almeida & Lucas Delerino on behalf of COVID-19 FIOCRUZ Genomic Network; Thais Ferreira de Oliveira; Thais de Oliveira Costa; Ticiane Cavalcante de Souza; Veridiana Pessoa Miyajima |
| EPI_ISL_5646402, EPI_ISL_5646430, EPI_ISL_5646467 | UNIDADE REFERENCIAL SUDOESTE | Instituto Butantan | Antonio Jorge Martins; Claudia Renata dos Santos Barros; David Schlesinger; Debora Botequio Moretti; Dimas Tadeu Covas; Elaine Cristina Marqueze; Elaine Vieira Santos; Evandra Strazza Rodrigues; Heidge Fukumasu; Jayme Augusto de Souza-Neto; José Salvatore Leister Patané; Luiz Alcantara; Luiz Lehmann Coutinho; Maria Carolina Elias; Maurício Lacerda Nogueira; Rafael dos Santos Bezerra; Raul Machado Neto; Rejane Maria Tommasini Grotto; Ricardo Haddad; Sandra Coccuzzo Sampaio Vessoni; Simone Kashima; Svetoslav Nanev Slavov; Vincent Louis Viala |
| EPI_ISL_5646570, EPI_ISL_5646585, EPI_ISL_5646707, EPI_ISL_5646815 | UNIDADE SAUDE FAMILIA DR PIERRE ANTONIO COPPIETERS AREALVA | Instituto Butantan | Antonio Jorge Martins; Claudia Renata dos Santos Barros; David Schlesinger; Debora Botequio Moretti; Dimas Tadeu Covas; Elaine Cristina Marqueze; Elaine Vieira Santos; Evandra Strazza Rodrigues; Heidge Fukumasu; Jayme Augusto de Souza-Neto; José Salvatore Leister Patané; Luiz Alcantara; Luiz Lehmann Coutinho; Maria Carolina Elias; Maurício Lacerda Nogueira; Rafael dos Santos Bezerra; Raul Machado Neto; Rejane Maria Tommasini Grotto; Ricardo Haddad; Sandra Coccuzzo Sampaio Vessoni; Simone Kashima; Svetoslav Nanev Slavov; Vincent Louis Viala |
| EPI_ISL_2628990, EPI_ISL_2646099 | UNILABS | Instituto Nacional de Saude (INSA) | Borges et al |
| EPI_ISL_2796048 | UNILABS | Instituto Nacional de Saude (INSA) and Instituto Gulbenkian de Ciencia (IGC) | Borges et al |
| EPI_ISL_5651180, EPI_ISL_5651199, EPI_ISL_5653851 | UPA CAMPO DOS ALEMAES | Instituto Butantan | Antonio Jorge Martins; Claudia Renata dos Santos Barros; David Schlesinger; Debora Botequio Moretti; Dimas Tadeu Covas; Elaine Cristina Marqueze; Elaine Vieira Santos; Evandra Strazza Rodrigues; Heidge Fukumasu; Jayme Augusto de Souza-Neto; José Salvatore Leister Patané; Luiz Alcantara; Luiz Lehmann Coutinho; Maria Carolina Elias; Maurício Lacerda Nogueira; Rafael dos Santos Bezerra; Raul Machado Neto; Rejane Maria Tommasini Grotto; Ricardo Haddad; Sandra Coccuzzo Sampaio Vessoni; Simone Kashima; Svetoslav Nanev Slavov; Vincent Louis Viala |

| | | | |
|---|---|---|---|
| EPI_ISL_5646404, EPI_ISL_5646466, EPI_ISL_5646471, EPI_ISL_5646849 | UPA DE BEBEDOURO | Instituto Butantan | Antonio Jorge Martins; Claudia Renata dos Santos Barros; David Schlesinger; Debora Botequio Moretti; Dimas Tadeu Covas; Elaine Cristina Marqueze; Elaine Vieira Santos; Evandra Strazza Rodrigues; Heidge Fukumasu; Jayme Augusto de Souza-Neto; José Salvatore Leister Patané; Luiz Alcantara; Luiz Lehmann Coutinho; Maria Carolina Elias; Maurício Lacerda Nogueira; Rafael dos Santos Bezerra; Raul Machado Neto; Rejane Maria Tommasini Grotto; Ricardo Haddad; Sandra Coccuzzo Sampaio Vessoni; Simone Kashima; Svetoslav Nanev Slavov; Vincent Louis Viala |
| EPI_ISL_5646546, EPI_ISL_5646568, EPI_ISL_5646586, EPI_ISL_5646612, EPI_ISL_5646640, EPI_ISL_5646753, EPI_ISL_5646867, EPI_ISL_5646869, EPI_ISL_5646919, EPI_ISL_5646923, EPI_ISL_5646934 | | | |
| see above | UPA DR FRANCO DA ROCHA | Instituto Butantan | Antonio Jorge Martins; Claudia Renata dos Santos Barros; David Schlesinger; Debora Botequio Moretti; Dimas Tadeu Covas; Elaine Cristina Marqueze; Elaine Vieira Santos; Evandra Strazza Rodrigues; Heidge Fukumasu; Jayme Augusto de Souza-Neto; José Salvatore Leister Patané; Luiz Alcantara; Luiz Lehmann Coutinho; Maria Carolina Elias; Maurício Lacerda Nogueira; Rafael dos Santos Bezerra; Raul Machado Neto; Rejane Maria Tommasini Grotto; Ricardo Haddad; Sandra Coccuzzo Sampaio Vessoni; Simone Kashima; Svetoslav Nanev Slavov; Vincent Louis Viala |
| EPI_ISL_5647215 | UPA GEISEL | Instituto Butantan | Antonio Jorge Martins; Claudia Renata dos Santos Barros; David Schlesinger; Debora Botequio Moretti; Dimas Tadeu Covas; Elaine Cristina Marqueze; Elaine Vieira Santos; Evandra Strazza Rodrigues; Heidge Fukumasu; Jayme Augusto de Souza-Neto; José Salvatore Leister Patané; Luiz Alcantara; Luiz Lehmann Coutinho; Maria Carolina Elias; Maurício Lacerda Nogueira; Rafael dos Santos Bezerra; Raul Machado Neto; Rejane Maria Tommasini Grotto; Ricardo Haddad; Sandra Coccuzzo Sampaio Vessoni; Simone Kashima; Svetoslav Nanev Slavov; Vincent Louis Viala |
| EPI_ISL_5647248, EPI_ISL_5647291 | USF DE SAGRES | Instituto Butantan | Antonio Jorge Martins; Claudia Renata dos Santos Barros; David Schlesinger; Debora Botequio Moretti; Dimas Tadeu Covas; Elaine Cristina Marqueze; Elaine Vieira Santos; Evandra Strazza Rodrigues; Heidge Fukumasu; Jayme Augusto de Souza-Neto; José Salvatore Leister Patané; Luiz Alcantara; Luiz Lehmann Coutinho; Maria Carolina Elias; Maurício Lacerda Nogueira; Rafael dos Santos Bezerra; Raul Machado Neto; Rejane Maria Tommasini Grotto; Ricardo Haddad; Sandra Coccuzzo Sampaio Vessoni; Simone Kashima; Svetoslav Nanev Slavov; Vincent Louis Viala |
| EPI_ISL_5646602, EPI_ISL_5646803, EPI_ISL_5646840 | USF II ALADIA ARAN RODRIGUES | Instituto Butantan | Antonio Jorge Martins; Claudia Renata dos Santos Barros; David Schlesinger; Debora Botequio Moretti; Dimas Tadeu Covas; Elaine Cristina Marqueze; Elaine Vieira Santos; Evandra Strazza Rodrigues; Heidge Fukumasu; Jayme Augusto de Souza-Neto; José Salvatore Leister Patané; Luiz Alcantara; Luiz Lehmann Coutinho; Maria Carolina Elias; Maurício Lacerda Nogueira; Rafael dos Santos Bezerra; Raul Machado Neto; Rejane Maria Tommasini Grotto; Ricardo Haddad; Sandra Coccuzzo Sampaio Vessoni; Simone Kashima; Svetoslav Nanev Slavov; Vincent Louis Viala |
| EPI_ISL_2549337, EPI_ISL_2549343, EPI_ISL_2549346, EPI_ISL_2549354, EPI_ISL_2549359, EPI_ISL_2549366, EPI_ISL_2549371, EPI_ISL_2549378, EPI_ISL_2549382, EPI_ISL_2549385, EPI_ISL_2617628, EPI_ISL_2617970, EPI_ISL_2617975, EPI_ISL_2618038, EPI_ISL_2618039, EPI_ISL_2618047, EPI_ISL_2618049, EPI_ISL_2618053, EPI_ISL_2618056, EPI_ISL_2618070, EPI_ISL_2618085, EPI_ISL_2618088, EPI_ISL_2618098, EPI_ISL_2693129, EPI_ISL_2693139, EPI_ISL_2693299, EPI_ISL_2693301, EPI_ISL_2693306, EPI_ISL_2693307, EPI_ISL_2693310, EPI_ISL_2693312, EPI_ISL_2693684, EPI_ISL_2693702, EPI_ISL_2693738, EPI_ISL_2693739, EPI_ISL_2693746, EPI_ISL_2693752, EPI_ISL_2693753, EPI_ISL_2693763 | | | |
| see above | UW Virology Lab | UW Virology Lab | Alexander Greninger; Hong Xie; Keith R Jerome; Lasata Shrestha; Meei-Li Huang; Michelle Lin; Nathan Breit; Noah R. Baker; Patrick Mathias; Pavitra Roychoudhury; Ricardo Perez; Robert J. Livingston; Sean Ellis; Shah Mohamed Bakhash; Tien V. Nguyen |
| EPI_ISL_2801698 | Unidad de Investigación Biomédica de Zacatecas (UIBZ) | Instituto de Biotecnología de la UNAM | ; Alejandra García-Gasca; Alejandra Hernández-Terán; Alejandro Sánchez-Flores; Alfredo Herrera-Estrella; Alicia Ocaña-Mondragón; Andreu Comas-García; Angel Gustavo Salas-Lais; Antonio Loza Román; Bernardo Martínez-Miguel; Blanca Taboada; Brenda Irasema Maldonado-Meza; Bruno Gómez-Gil; Carla Ivón Herrera-Najera; Carlos F. Arias; Celia Boukadida; Clara Esperanza Santacruz-Tinoco; Concepción Grajales-Muñiz; Consorcio Mexicano de Vigilancia Genómica (CoViGen-Mex). Authors (in alphabetical order): Julio Elias Alvarado-Yaah; Cristóbal Cháidez-Quiróz; Célida Duque Molina; Célida Martínez- Rodríguez; Daniel Fregoso-Rueda; Daniel Lira Morales; Eduardo Becerril-Vargas; Fernando Fontove-Herrera; Fidencio Mejía-Nepomuceno; Francisco Pulido; Gloria Elena Espinosa-Ayala; Gloria María Molina-Salinas; Gloria Vazquez; Hector Esteban Paz-Juárez; Hector Montoya-Fuentes; Helen Haydee Fernanda Ramírez-Plascencia; Irvin González-López; Jean Pierre González; Jesús Hernández; Joel Armando Vázquez-Pérez.; Jorge Salas-Hernández; José Antonio Enciso-Moreno; José Arturo Martínez-Orozco; José Esteban Muñoz-Medina; José de Jesús Nuñez-Contreras; Juan Bautista Chale-Dzul; Julissa Enciso-Ibarra; Luis Alberto Ochoa-Carrera; Margarita Matías-Florentino; Mario Mújica-Sánchez; Marissa Perez-Garcia; María Guadalupe Santiago-Mauricio; María Guadalupe de Jesús Mireles-Rivera; Nelly Sélem-Mojica; Pavel Isa; Ricardo Ciria Merce; Ricardo Grande; Rosa María Gutiérrez Rios; Santiago Ávila-Ríos; Selene Zárate; Susana Lopez; Verónica Mata-Haro; Victor Eduardo García-Arias; Víctor Hugo Borja-Aburto |
| EPI_ISL_2801603, EPI_ISL_2801604, EPI_ISL_2801605, EPI_ISL_2801606, EPI_ISL_2801607, EPI_ISL_2801620, EPI_ISL_2801622, EPI_ISL_2801624, EPI_ISL_2801625, EPI_ISL_2801627, EPI_ISL_2801628, EPI_ISL_2801629, EPI_ISL_2801639, EPI_ISL_2801640, EPI_ISL_2801644, EPI_ISL_2801650, EPI_ISL_2801654, EPI_ISL_2801656, EPI_ISL_2801657 | | | |
| see above | Unidad de Investigación Médica de Yucatán (UIMY) | Instituto de Biotecnología de la UNAM | ; Alejandra García-Gasca; Alejandra Hernández-Terán; Alejandro Sánchez-Flores; Alfredo Herrera-Estrella; Alicia Ocaña-Mondragón; Andreu Comas-García; Angel Gustavo Salas-Lais; Antonio Loza Román; Bernardo Martínez-Miguel; Blanca Taboada; Brenda Irasema Maldonado-Meza; Bruno Gómez-Gil; Carla Ivón Herrera-Najera; Carlos F. Arias; Celia Boukadida; Clara Esperanza Santacruz-Tinoco; Concepción Grajales-Muñiz; Consorcio Mexicano de Vigilancia Genómica (CoViGen-Mex). Authors (in alphabetical order): Julio Elias Alvarado-Yaah; Cristóbal Cháidez-Quiróz; Célida Duque Molina; Célida Martínez- Rodríguez; Daniel Fregoso-Rueda; Daniel Lira Morales; Eduardo Becerril-Vargas; Fernando Fontove-Herrera; Fidencio Mejía-Nepomuceno; Francisco Pulido; Gloria Elena Espinosa-Ayala; Gloria María Molina-Salinas; Gloria Vazquez; Hector Esteban Paz-Juárez; Hector Montoya-Fuentes; Helen Haydee Fernanda Ramírez-Plascencia; Irvin González-López; Jean Pierre González; Jesús Hernández; Joel Armando Vázquez-Pérez.; Jorge Salas-Hernández; José Antonio Enciso-Moreno; José Arturo Martínez-Orozco; José Esteban Muñoz-Medina; José de Jesús Nuñez-Contreras; Juan Bautista Chale-Dzul; Julissa Enciso-Ibarra; Luis Alberto Ochoa-Carrera; Margarita Matías-Florentino; Mario Mújica-Sánchez; Marissa Perez-Garcia; María Guadalupe Santiago-Mauricio; María Guadalupe de Jesús Mireles-Rivera; Nelly Sélem-Mojica; Pavel Isa; Ricardo Ciria Merce; Ricardo Grande; Rosa María Gutiérrez Rios; Santiago Ávila-Ríos; Selene Zárate; Susana Lopez; Verónica Mata-Haro; Victor Eduardo García-Arias; Víctor Hugo Borja-Aburto |
| EPI_ISL_2837233, EPI_ISL_2837285, EPI_ISL_2837286, EPI_ISL_2837307 | Unidade de apoio ao diagnostico da COVID - UNADIG | Bioinformatics Laboratory / LNCC | Alessandra P Lamarca; Alexandra L Gerber; Amilcar Tanuri; Ana Paula de C Guimaraes; Ana Tereza R Vasconcelos; Andrea Cony Cavalcanti; Caio Luiz Pereira Ribeiro; Cassia Alves; Cintia Policarpo; Claudia Maria Braga de Mello; Cristiane Gomes da Silva; Diana Mariani; Douglas Terra Machado; Flavio Dias da Silva; Gleidson da Silva de Oliveira; Leandro Magalhaes de Souza; Liliane Cavalcante; Luiz G P de Almeida; Marcio Henrique de Oliveira Garcia; Mario Sergio Ribeiro; Ronaldo da Silva F Jr; Silvia Carvalho |
| EPI_ISL_2899164 | Unilabs | Karolinska University Hospital | Annelie Bjerkner; Isak Sylvin; Jan Albert; Karolina Ininbergs; Lina Guerra Blomqvist; Lynda Eneh; Martin Ekman; Martina Wahlund; Robert Dyrdak; Sandra Broddesson; Tanja Normark; Tobias Allander; Valtteri Wirta; Zhibing Yun |
| EPI_ISL_2776218 | Universidad Central del Caribe (UCC) | Grubaugh Lab - Yale School of Public Health | Anderson Brito; Andrea Firpo; Annie Watkins; Chaney Kalinich; Chantal Vogels; David Rivera Aponte; Ernesto Vazquez; Fabiola Cruz-Lopez; Fabiola Fontanet; Isabel Ott; Jessica Rothman; Joseph Fauver; Kendall Billig; Mallery Breban; Mary Petrone; Natalie Machargo; Nathan Grubaugh; Patricia Serrano; Tania Mitwalli; Tara Alpert; Tobias Koch; Vianca Aponte |
| EPI_ISL_5239242 | Universidad de Caldas | Instituto Nacional de Salud- Dirección de Investigación en Salud Pública | Beatriz de Arco; Carlos Franco-Muñoz; Carmen Osorio; Diana Malo; Diego A. Álvarez-Díaz; Diego Andrés Prada; Gerardo Santamaría; Hector Alejandro Ruiz-Moreno; Jhonnatan Reales-González; Jorge Rivera; Juan Camilo Martinez; Julian Naizaque; Katherine Laiton-Donato; Lisseth Pardo; Magdalena Wiesner; Marcela Mercado-Reyes; Maria T. Herrera-Sepúlveda; Marta Lopez Blanco; Martha Lucia Ospina Martinez; Paola Rojas; Sergio Gomez; Sheryll Corchuelo; Tatiana Cobos; Ángela Alarcon Cruz |
| EPI_ISL_2517694 | University College London, Great Ormond Street Hospital for Children NHS Foundation Trust, Imperial College Healthcare NHS Trust | COVID-19 Genomics UK (COG-UK) Consortium | Alison Holmes; Charlotte Williams; Helena Tutill; Jacqueline Findlay; James Price; Judith Breuer; Julianne Brown; Kathryn Harris; Leysa Forrest; Mark Kristiansen; Paola Niola; Paola Resende Silva; Patricia Dyal; Paul Randell; Rachel Williams; Samuel Weeks; Sergi Castellano; Sunando Roy; Tony Brooks; Yasmin Panchbhaya |
| EPI_ISL_2681457 | University Hospitals of Geneva, Laboratory of Virology | HUG, Laboratory of Virology and the Health2030 Genome Center | Ana Rita Goncalves; Deborah Penet; Emmanouil Dermitzakis; Henri Pegeot; Ioannis Xenarios; Keith Harshman; Laurent Kaiser; Lorenzo Cerutti; Melyssa Elies; Samuel Cordey |
| EPI_ISL_2500003, EPI_ISL_2500004, EPI_ISL_2500005 | University of Liège COVID-19 testing center | GIGA Medical Genomics | Bouchra Boujemla; Cécile Meex; Fabrice Bureau; Keith Durkin; Laurent Gillet; Maria Artesi; Marie-Pierre Hayette; Nathalie Renotte; Sébastien Bontems; Vincent Bours; Wouter Coppieters |
| EPI_ISL_2646329 | University of Mississippi Medical Center, Department of Pathology | University of Mississippi Medical Center, Molecular and Genomics Core Facility | Ashley C. Johnson; D. Ashley Robinson; Ithiel J. Frame; Krishna K. Ayyalasomayajula; Michael R. Garrett; Wenjie Wu |
| EPI_ISL_7635977, EPI_ISL_7635978, EPI_ISL_7635979 | University of Rome Tor Vergata: Departm Experim Medicine Chair of Virology | University of Rome Tor Vergata: Departm Experim Medicine Chair of Virology | Francesca Ceccherini-Silberstein; Loredana Sarmati; Lorenzo Piermatteo; Luca Carioti; Marco Iannetta; Maria Concetta Bellocchi; Massimo Andreoni |
| EPI_ISL_2686028, EPI_ISL_2686030, EPI_ISL_2686033, EPI_ISL_2686039, EPI_ISL_2686043, EPI_ISL_2686045 | Università degli Studi di Perugia | Istituto Zooprofilattico Sperimentale dell'Abruzzo e Molise “G. Caporale” | Ancora M; Biagetti M; Calistri P; Camilloni B; Cammà C; Curini V; Delli Compagni E; Di Domenico M; Di Pasquale A; Giammarioli M; Lorusso A; Mangone I; Marcacci M; Mencacci A; Puglia I; Rinaldi A; Savini G; Scialabba S |
| EPI_ISL_3030246 | Urbino | Microbiology University Politecnica delle Marche | Anna Valenza; Carla Acciarri; Katia Marinelli; Monica Lucia Ferreri; Patrizia Bagnarelli; Roberta Longo; Sara Caucci; Stefano Menzo |
| EPI_ISL_2856512, EPI_ISL_2856520, EPI_ISL_2856556, EPI_ISL_2856563, EPI_ISL_2856580, EPI_ISL_2856805, EPI_ISL_2856811, EPI_ISL_2856814, EPI_ISL_2856820, EPI_ISL_2856931, EPI_ISL_2857250, EPI_ISL_2857251, EPI_ISL_2857257, EPI_ISL_3132731, EPI_ISL_3132736, EPI_ISL_3132778, EPI_ISL_3132958, EPI_ISL_3132964, EPI_ISL_3132965, EPI_ISL_3132977, EPI_ISL_3133017 | | | |
| see above | Utah Public Health Laboratory | Utah Public Health Laboratory | Erin L. Young; Kelly F. Oakeson; Olinto Linares-Perdomo; Tara Gallagher |
| EPI_ISL_3526021 | Utah Public Health Laboratory, Utah Public Health Laboratory Infectious Disease submission group | Utah Public Health Laboratory, Utah Public Health Laboratory Infectious Disease submission group | Linares-Perdomo, O.; Oakeson, K.; Young, E. |
| EPI_ISL_3064772 | VA-Division of Consolidated Laboratory Services | Centers for Disease Control and Prevention Division of Viral Diseases, Pathogen Discovery | Alex Burgin; Ben L. Rambo-Martin; Clinton R. Paden; Dakota Howard; Dave Wentworth; Dhwani Batra; Jasmine Padilla; Justin Lee; Krista Queen; Kristen Knipe; Kristine Lacek; Mark Burroughs; Matthew Schmerer; Meghan Bentz; Mili Sheth; Peter Cook; Sam Shepard; Sarah Nobles; Suxiang Tong; Vivien Dugan; Yvette Unoarumhi |
| EPI_ISL_5646389, EPI_ISL_5646408, EPI_ISL_5646425, EPI_ISL_5646441, EPI_ISL_5646446, EPI_ISL_5646456 | VIGILANCIA EM SAUDE SAE VIGILANCIA EPIDEMIOLOGICA | Instituto Butantan | Antonio Jorge Martins; Claudia Renata dos Santos Barros; David Schlesinger; Debora Botequio Moretti; Dimas Tadeu Covas; Elaine Cristina Marqueze; Elaine Vieira Santos; Evandra Strazza Rodrigues; Heidge Fukumasu; Jayme Augusto de Souza-Neto; José Salvatore Leister Patané; Luiz Alcantara; Luiz Lehmann Coutinho; Maria Carolina Elias; Maurício Lacerda Nogueira; Rafael dos Santos Bezerra; Raul Machado Neto; Rejane Maria Tommasini Grotto; Ricardo Haddad; Sandra Coccuzzo Sampaio Vessoni; Simone Kashima; Svetoslav Nanev Slavov; Vincent Louis Viala |
| EPI_ISL_5647898, EPI_ISL_5647936 | VIGILANCIA EPIDEMIOLOGICA DE LEME | Instituto Butantan | Antonio Jorge Martins; Claudia Renata dos Santos Barros; David Schlesinger; Debora Botequio Moretti; Dimas Tadeu Covas; Elaine Cristina Marqueze; Elaine Vieira Santos; Evandra Strazza Rodrigues; Heidge Fukumasu; Jayme Augusto de Souza-Neto; José Salvatore Leister Patané; Luiz Alcantara; Luiz Lehmann Coutinho; Maria Carolina Elias; Maurício Lacerda Nogueira; Rafael dos Santos Bezerra; Raul Machado Neto; Rejane Maria Tommasini Grotto; Ricardo Haddad; Sandra Coccuzzo Sampaio Vessoni; Simone Kashima; Svetoslav Nanev Slavov; Vincent Louis Viala |
| EPI_ISL_5646393, EPI_ISL_5646407, EPI_ISL_5646422, EPI_ISL_5646432, EPI_ISL_5646433, EPI_ISL_5646434, EPI_ISL_5646450, EPI_ISL_5646459, EPI_ISL_5646460, EPI_ISL_5646478, EPI_ISL_5646507, EPI_ISL_5646938 | | | |
| see above | VIGILANCIA SANITARIA E VIG EPIDEMIOLOGICA DE ITAPEVI | Instituto Butantan | Antonio Jorge Martins; Claudia Renata dos Santos Barros; David Schlesinger; Debora Botequio Moretti; Dimas Tadeu Covas; Elaine Cristina Marqueze; Elaine Vieira Santos; Evandra Strazza Rodrigues; Heidge Fukumasu; Jayme Augusto de Souza-Neto; José Salvatore Leister Patané; Luiz Alcantara; Luiz Lehmann Coutinho; Maria Carolina Elias; Maurício Lacerda Nogueira; Rafael dos Santos Bezerra; Raul Machado Neto; Rejane Maria Tommasini Grotto; Ricardo Haddad; Sandra Coccuzzo Sampaio Vessoni; Simone Kashima; Svetoslav Nanev Slavov; Vincent Louis Viala |
| EPI_ISL_2622070 | Valais Hospital, Central Institute | Valais Hospital, Central Institute | Alexis Dumoulin; Deborah Penet; Emmanouil Dermitzakis; Henri Pegeot; Ioannis Xenarios; Keith Harshman; Lorenzo Cerutti; Melyssa Elies |
| EPI_ISL_2611109, EPI_ISL_2724503 | Viollier AG | Department of Biosystems Science and Engineering, ETH Zürich | Chaoran Chen; Christian Beisel; Christiane Beckmann; Christoph Noppen; Elodie Burcklen; Ina Nissen; Ivan Topolsky; Kim Philipp Jablonski; Lara Fuhrmann; Maurice Redondo; Mirjam Feldkamp; Natascha Santacroce; Niko Beerenwinkel; Olivier Kobel; Rebecca Denes; Sarah Nadeau; Tanja Stadler |
| EPI_ISL_2776051, EPI_ISL_2776054, EPI_ISL_2982646 | Virginia Division of Consolidated Laboratory Services | Virginia Division of Consolidated Laboratory Services | Virginia Division of Consolidated Laboratory Services |
| EPI_ISL_2884548 | Virology Laboratory, Scientific Department, Army Medical Center | Virology Laboratory, Scientific Department, Army Medical Center | Anella Monte; Anna Anselmo; Antonella Fortunato; Filippo Molinari; Florigio Lista; Francesco Giordani; Giancarlo Petralito; Giandomenico Cerreto; Giulia Campoli; Lucia Nicosia; Marzia Cavalli; Riccardo De Sanctis; Rossella Brandi; Silvia Fillo; Vanessa Vera Fain |
| EPI_ISL_3184305 | Virology Unit, AOUP | Virology Unit, AOUP | Annalisa Capria; Francesca Pasqualetti; Mauro Pistello; Pietro Giorgio Spezia; Susanna Cappellini |
| EPI_ISL_2788021, EPI_ISL_2788026 | WA State Department of Health | Centers for Disease Control and Prevention Division of Viral Diseases, Pathogen Discovery | Alison Laufer Halpin; Ben L. Rambo-Martin; Clinton R. Paden; Dakota Howard; Darlene Wagner; Dave Wentworth; Dhwani Batra; Jasmine Padilla; Justin Lee; Katie Dillon; Krista Queen; Kristen Knipe; Kristine Lacek; Mark Burroughs; Matthew Schmerer; Mili Sheth; Peter Cook; Sam Shepard; Sarah Nobles; Shoshona Le; Suxiang Tong; Vivien Dugan; Yvette Unoarumhi |
| EPI_ISL_3031272 | WESTCHESTER MEDICAL CENTER | Wadsworth Center, New York State | Alexis Russell; Catharine Prussing; Daryl M. Lamson; Erasmus Schneider; Erica Lasek-Nesselquist; John Kelly; Jonathan Plitnick; Kirsten St. George; Matthew Shudt; Melissa A Leisner; Navjot Singh |

| | | | |
|---|---|---|---|
| | | Department of Health | |
| EPI_ISL_2597899 | WSSE w Gdańsku | 1. Tricity SARS-CoV-2 sequencing consortium: University of Gdansk, Medical University of Gdansk, Vaxican Ltd., Invicta Ltd. 2. National Institute of Public Health - National Institute of Hygiene, Warsaw, Poland | Celina Cybulska; Karolina Gackowska; Katarzyna Groth; Katarzyna Zacharczuk; Krystyna Bienkowska Szewczyk; Lukasz Rabalski; Maciej Grzybek; Maciej Kosinski; Magdalena Nowakowska; Marcin Lubocki; Małgorzata Sadkowska-Todys; Tomasz Wołkowicz |
| EPI_ISL_2651083, EPI_ISL_2651084, EPI_ISL_2651097, EPI_ISL_2651105, EPI_ISL_2716464 | Washington State Department of Health Public Health Laboratories | Washington State Department of Health Public Health Laboratories | Avi Singh; Darren Lucas; Denny Russell; Drew MacKellar; Geoff Melly; Hannah Gray; Joenice Gonzalez; JohnAric Peterson; Philip Dykema; Rebecca Cao; Vanessa De Los Santos |
| EPI_ISL_3235365 | Weill Cornell Medicine | New York Genome Center | Andre Corvelo; Arryn Craney; Chris Mason; Dayna M. Oschwald; Hanna Rennert; Lars F Westblade; Margaret Elizabeth Ross; Melissa Cushing; Michael Zody; Olivier Elemento; Priya Velu; Samantha Fennessey; Tom Maniatis |
| EPI_ISL_2497908, EPI_ISL_2497922, EPI_ISL_2499942, EPI_ISL_2499943, EPI_ISL_2534256, EPI_ISL_2534262, EPI_ISL_2612191, EPI_ISL_2612195, EPI_ISL_2675193 | | | |
| see above | Wichita State University - Molecular Diagnostics Lab | Kansas Health and Environmental Lab | Ben Olsen; Jonathan Barnell; Katherine Wiggins; Mike Grose; and Phil Adam |
| EPI_ISL_2727766, EPI_ISL_2727767, EPI_ISL_2727775, EPI_ISL_2727782, EPI_ISL_3131920 | Wisconsin State Laboratory of Hygiene Communicable Disease Division | Wisconsin State Laboratory of Hygiene Communicable Disease Division | Abigail C. Shockey; Alicia J. Mooney; Erika M. Hanson; Kelsey R. Florek; Richard Griesser; Sara Wagner; Tonya Danz |
| EPI_ISL_2726819, EPI_ISL_2754064, EPI_ISL_2833845, EPI_ISL_2834976, EPI_ISL_2834977, EPI_ISL_2894510, EPI_ISL_2894513 | | | |
| see above | Wyoming Public Health Laboratory | Wyoming Public Health Laboratory | Ashley Norberg; Brian Dominguez; Cari Sloma; Channing Weber; Chayse Rowley; Elliot Thomasson; Jim Mildenberger; Marley Goetz; Robert Petit; Sam Britz; Taylor Fearing; and Rob Christensen |
| EPI_ISL_3115746, EPI_ISL_3115747 | ZOTZ KLIMAS MVZ Düsseldorf-Centrum GbR ÜBAG für Labormedizin, Genetik, Zytologie, Pathologie | Center of Medical Microbiology, Virology, and Hospital Hygiene, University of Duesseldorf | Alexander Dilthey; Andreas Walker; Daniel Strelow; Jessica Nicolai; Jörg Timm; Katrin Hoffmann; Klaus Pfeffer; Lisanna Hülse; Malte Kohns Vasconcelos; Maximilian Damagnez; Nadine Lübke; Patrick Finzer; Rainer Zotz; Tobias Wienemann; Torsten Houwaart |
| EPI_ISL_2726773 | Zakład Bakteriologii Szpital Matki Bożej Nieustającej Pomocy | 1. National Institute of Public Health - National Institute of Hygiene, Warsaw, Poland 2. Biobank Lab, University of Lodz 3. Laboratory of Respiratory Viruses, Teaching and Clinical Center of the Medical University of Lodz | Dominik Strapagiel; Izabela Dróżdż; Jakub Lach; Katarzyna Zacharczuk; Klaudyna Królikowska; Maciej Borowiec; Magdalena Nowakowska; Magdalena Traczyk-Borszyńska; Marcin Słomka; Marta Sobalska-Kwapis; Małgorzata Sadkowska-Todys; Tomasz Płoszaj; Tomasz Wołkowicz |
| EPI_ISL_2635609 | amedes MVZ Zentrum für Pathologie und Zytodiagnostik GmbH (Köln) | Robert Koch Institute | |
| EPI_ISL_2637965 | laboratoire Belle Epine | Department of Virology, Henri Mondor University Hospital, Assistance Publique Hôpitaux de Paris, Université Paris-Est Créteil, INSERM U955 | Alexandre Soulier; Christophe Rodriguez; Elisabeth Trawinski; Guillaume Gricourt; Jean-Michel Pawlotsky; Melissa N'Debi; Slim Fourati; Vanessa Demontant |
