## Supplement 3 for "Rapid and Accurate Identification of SARS-CoV-2 Omicron Variants Using Droplet Digital PCR (RT-ddPCR)": All_0621-0627.pdf

We gratefully acknowledge the following Authors from the Originating laboratories responsible for obtaining the specimens, as well as the Submitting laboratories where the genome data were generated and shared via GISAID, on which this research is based.

All Submitters of data may be contacted directly via www.gisaid.org

Authors are sorted alphabetically.

| Accession ID | Originating Laboratory | Submitting Laboratory | Authors |
| --- | --- | --- | --- |
| EPI_ISL_3128131, EPI_ISL_3128132, EPI_ISL_3128136 | "AZ SPHL, Arizona Department of Health Services" | Centers for Disease Control and Prevention Division of Viral Diseases, Pathogen Discovery | Alex Burgin; Ben L. Rambo-Martin; Clinton R. Paden; Dakota Howard; Dave Wentworth; Dhwani Batra; Jasmine Padilla; Justin Lee; Krista Queen; Kristen Knipe; Kristine Lacek; Mark Burroughs; Matthew Schmerer; Meghan Bentz; Mili Sheth; Peter Cook; Sam Shepard; Sarah Nobles; Suxiang Tong; Vivien Dugan; Yvette Unoarumhi |
| EPI_ISL_3355284 | "NM Dept. Health, Scientific Laboratory Division " | Centers for Disease Control and Prevention Division of Viral Diseases, Pathogen Discovery | Alex Burgin; Ben Rambo-Martin; Clinton Paden; Dakota Howard; Dave Wentworth; Dhwani Batra; Jasmine Padilla; Justin Lee; Krista Queen; Kristen Knipe; Kristine Lacek; Mark Burroughs; Matthew Schmerer; Meghan Bentz; Mili Sheth; Peter Cook; Sam Shepard; Sarah Nobles; Suxiang Tong; Vivien Dugan; Yvette Unoarumhi |
| EPI_ISL_3355430 | "NYSDOH Wadsworth Center, Virology Lab" | Centers for Disease Control and Prevention Division of Viral Diseases, Pathogen Discovery | Alex Burgin; Ben Rambo-Martin; Clinton Paden; Dakota Howard; Dave Wentworth; Dhwani Batra; Jasmine Padilla; Justin Lee; Krista Queen; Kristen Knipe; Kristine Lacek; Mark Burroughs; Matthew Schmerer; Meghan Bentz; Mili Sheth; Peter Cook; Sam Shepard; Sarah Nobles; Suxiang Tong; Vivien Dugan; Yvette Unoarumhi |
| EPI_ISL_5648755, EPI_ISL_5648782, EPI_ISL_5648783, EPI_ISL_5648784, EPI_ISL_5648791, EPI_ISL_5648792, EPI_ISL_5648793, EPI_ISL_5648801, EPI_ISL_5648816, EPI_ISL_5648821, EPI_ISL_5648828, EPI_ISL_5648841, EPI_ISL_5648842, EPI_ISL_5648844, EPI_ISL_5648854, EPI_ISL_5648862, EPI_ISL_5648863, EPI_ISL_5648864, EPI_ISL_5648866, EPI_ISL_5648867, EPI_ISL_5648869, EPI_ISL_5648874, EPI_ISL_5648888, EPI_ISL_5648894, EPI_ISL_5648911, EPI_ISL_5648917, EPI_ISL_5648933, EPI_ISL_5648949, EPI_ISL_5648950, EPI_ISL_5648953, EPI_ISL_5648955, EPI_ISL_5648958, EPI_ISL_5648962, EPI_ISL_5648963, EPI_ISL_5648968, EPI_ISL_5648976, EPI_ISL_5648977, EPI_ISL_5648984, EPI_ISL_5648985, EPI_ISL_5648988, EPI_ISL_5648989, EPI_ISL_5648990, EPI_ISL_5648996, EPI_ISL_5648997, EPI_ISL_5649001, EPI_ISL_5649002, EPI_ISL_5649003, EPI_ISL_5649004, EPI_ISL_5649012, EPI_ISL_5649013, EPI_ISL_5649025, EPI_ISL_5649043, EPI_ISL_5649049, EPI_ISL_5649050, EPI_ISL_5649059, EPI_ISL_5649060, EPI_ISL_5649064, EPI_ISL_5649065, EPI_ISL_5649066, EPI_ISL_5649071, EPI_ISL_5649072, EPI_ISL_5649078, EPI_ISL_5649079, EPI_ISL_5649088, EPI_ISL_5649089, EPI_ISL_5649098, EPI_ISL_5649099, EPI_ISL_5649116, EPI_ISL_5649117, EPI_ISL_5649123, EPI_ISL_5649128, EPI_ISL_5649130, EPI_ISL_5649132, EPI_ISL_5649138, EPI_ISL_5649141, EPI_ISL_5649143 | | | |
| see above | AFIP | Instituto Butantan | Antonio Jorge Martins; Claudia Renata dos Santos Barros; David Schlesinger; Debora Botequio Moretti; Dimas Tadeu Covas; Elaine Cristina Marqueze; Elaine Vieira Santos; Evandra Strazza Rodrigues; Heidge Fukumasu; Jayme Augusto de Souza-Neto; José Salvatore Leister Patané; Luiz Alcantara; Luiz Lehmann Coutinho; Maria Carolina Elias; Maurício Lacerda Nogueira; Rafael dos Santos Bezerra; Raul Machado Neto; Rejane Maria Tommasini Grotto; Ricardo Haddad; Sandra Coccuzzo Sampaio Vessoni; Simone Kashima; Svetoslav Nanev Slavov; Vincent Louis Viala |
| EPI_ISL_3031195 | ALBANY MEDICAL CENTER | Wadsworth Center, New York State Department of Health | Alexis Russell; Catharine Prussing; Daryl M. Lamson; Erasmus Schneider; Erica Lasek-Nesselquist; John Kelly; Jonathan Plitnick; Kirsten St. George; Matthew Shudt; Melissa A Leisner; Navjot Singh |
| EPI_ISL_5915131, EPI_ISL_5915173, EPI_ISL_5915189, EPI_ISL_5915217, EPI_ISL_5915231, EPI_ISL_5915236, EPI_ISL_5915239, EPI_ISL_5915274, EPI_ISL_5915275, EPI_ISL_5915276, EPI_ISL_5915277, EPI_ISL_5915302, EPI_ISL_5915316, EPI_ISL_5915317, EPI_ISL_6573786 | | | |
| see above | AMALAB/FACISA/UFRN | WallauLab on behalf of Fiocruz COVID-19 Genomic Surveillance Network | Alexandre Freitas da Silva; Allan Roberto Dias Nunes; Antonio Marinho da Silva Neto; Cassia Docena; Constância Flávia Junqueira Ayres; Filipe Zimmer Dezordi; Gabriel Luz Wallau; Gustavo Barbosa de Lima; Joana Cristina Medeiros Tavares Marques; Katya Anaya Jacinto; Lais Ceschini Machado; Lilian Caroliny Amorim Silva; Marcelo Henrique dos Santos Paiva; Mariane dos Santos Duarte; Matheus Filgueira Bezerra; Sinval Pinto Brandão Filho |
| EPI_ISL_5648546, EPI_ISL_5659598 | AMB MEDICO DR FRANCISCO FELIX DE MENDONCA SAUDADE | Instituto Butantan | Antonio Jorge Martins; Claudia Renata dos Santos Barros; David Schlesinger; Debora Botequio Moretti; Dimas Tadeu Covas; Elaine Cristina Marqueze; Elaine Vieira Santos; Evandra Strazza Rodrigues; Heidge Fukumasu; Jayme Augusto de Souza-Neto; José Salvatore Leister Patané; Luiz Alcantara; Luiz Lehmann Coutinho; Maria Carolina Elias; Maurício Lacerda Nogueira; Rafael dos Santos Bezerra; Raul Machado Neto; Rejane Maria Tommasini Grotto; Ricardo Haddad; Sandra Coccuzzo Sampaio Vessoni; Simone Kashima; Svetoslav Nanev Slavov; Vincent Louis Viala |
| EPI_ISL_5649369, EPI_ISL_5649383, EPI_ISL_5649418, EPI_ISL_5649430, EPI_ISL_5649478, EPI_ISL_5649479, EPI_ISL_5659526, EPI_ISL_5659527 | | | |
| see above | AMBULATORIO DE ESPECIALIDADE V E MOGI MIRIM | Instituto Butantan | Antonio Jorge Martins; Claudia Renata dos Santos Barros; David Schlesinger; Debora Botequio Moretti; Dimas Tadeu Covas; Elaine Cristina Marqueze; Elaine Vieira Santos; Evandra Strazza Rodrigues; Heidge Fukumasu; Jayme Augusto de Souza-Neto; José Salvatore Leister Patané; Luiz Alcantara; Luiz Lehmann Coutinho; Maria Carolina Elias; Maurício Lacerda Nogueira; Rafael dos Santos Bezerra; Raul Machado Neto; Rejane Maria Tommasini Grotto; Ricardo Haddad; Sandra Coccuzzo Sampaio Vessoni; Simone Kashima; Svetoslav Nanev Slavov; Vincent Louis Viala |
| EPI_ISL_5648438, EPI_ISL_5648484, EPI_ISL_5648503, EPI_ISL_5648570, EPI_ISL_5648623, EPI_ISL_5648722 | AMBULATORIO DR JOAO LANIA CENTRO JOSE BONIFACIO | Instituto Butantan | Antonio Jorge Martins; Claudia Renata dos Santos Barros; David Schlesinger; Debora Botequio Moretti; Dimas Tadeu Covas; Elaine Cristina Marqueze; Elaine Vieira Santos; Evandra Strazza Rodrigues; Heidge Fukumasu; Jayme Augusto de Souza-Neto; José Salvatore Leister Patané; Luiz Alcantara; Luiz Lehmann Coutinho; Maria Carolina Elias; Maurício Lacerda Nogueira; Rafael dos Santos Bezerra; Raul Machado Neto; Rejane Maria Tommasini Grotto; Ricardo Haddad; Sandra Coccuzzo Sampaio Vessoni; Simone Kashima; Svetoslav Nanev Slavov; Vincent Louis Viala |
| EPI_ISL_5649422, EPI_ISL_5649425, EPI_ISL_5649431, EPI_ISL_5649434, EPI_ISL_5649444, EPI_ISL_5649470, EPI_ISL_5649486, EPI_ISL_5659535 | | | |
| see above | AMBULATORIO IB | Instituto Butantan | Antonio Jorge Martins; Claudia Renata dos Santos Barros; David Schlesinger; Debora Botequio Moretti; Dimas Tadeu Covas; Elaine Cristina Marqueze; Elaine Vieira Santos; Evandra Strazza Rodrigues; Heidge Fukumasu; Jayme Augusto de Souza-Neto; José Salvatore Leister Patané; Luiz Alcantara; Luiz Lehmann Coutinho; Maria Carolina Elias; Maurício Lacerda Nogueira; Rafael dos Santos Bezerra; Raul Machado Neto; Rejane Maria Tommasini Grotto; Ricardo Haddad; Sandra Coccuzzo Sampaio Vessoni; Simone Kashima; Svetoslav Nanev Slavov; Vincent Louis Viala |
| EPI_ISL_5648583, EPI_ISL_5648707, EPI_ISL_5648739, EPI_ISL_5649343, EPI_ISL_5649379, EPI_ISL_5649387 | AMBULATORIO MEDICO DE ESPECIALIDADES DE PERUIBE | Instituto Butantan | Antonio Jorge Martins; Claudia Renata dos Santos Barros; David Schlesinger; Debora Botequio Moretti; Dimas Tadeu Covas; Elaine Cristina Marqueze; Elaine Vieira Santos; Evandra Strazza Rodrigues; Heidge Fukumasu; Jayme Augusto de Souza-Neto; José Salvatore Leister Patané; Luiz Alcantara; Luiz Lehmann Coutinho; Maria Carolina Elias; Maurício Lacerda Nogueira; Rafael dos Santos Bezerra; Raul Machado Neto; Rejane Maria Tommasini Grotto; Ricardo Haddad; Sandra Coccuzzo Sampaio Vessoni; Simone Kashima; Svetoslav Nanev Slavov; Vincent Louis Viala |
| EPI_ISL_5649270 | AMBULATORIO MEDICO E ODONTOLOGICO MUNICIPAL DE PIEDADE | Instituto Butantan | Antonio Jorge Martins; Claudia Renata dos Santos Barros; David Schlesinger; Debora Botequio Moretti; Dimas Tadeu Covas; Elaine Cristina Marqueze; Elaine Vieira Santos; Evandra Strazza Rodrigues; Heidge Fukumasu; Jayme Augusto de Souza-Neto; José Salvatore Leister Patané; Luiz Alcantara; Luiz Lehmann Coutinho; Maria Carolina Elias; Maurício Lacerda Nogueira; Rafael dos Santos Bezerra; Raul Machado Neto; Rejane Maria Tommasini Grotto; Ricardo Haddad; Sandra Coccuzzo Sampaio Vessoni; Simone Kashima; Svetoslav Nanev Slavov; Vincent Louis Viala |
| EPI_ISL_2963425 | AOPD | Istituto Zooprofilattico Sperimentale delle Venezie | Adelaide Milani; Alessia Schivo; Alice Fusaro; Ambra Pastori; Annalisa Salviato; Antonia Ricci; Calogero Terregino; Edoardo Giussani; Elisa Palumbo; Erika Giorgia Quaranta; Isabella Monne; Luca Tassoni |
| EPI_ISL_3037817 | AREA DE SALUD BELEN-FLORES - CLINICA DR. JORGE VOLIO | Inciensa, Instituto Costarricense de Investigación y Enseñanza en Nutrición y Salud | Adriana Godínez; Caterina Guzmán; Claudio Soto-Garita; Estela Cordero; Francisco Duarte; Hebleen Porras; José Luis Vargas; Mariela Gutiérrez Joselyn Prado; Melany Calderón; Nazareth Ruiz & Sandra Mora Leitón |
| EPI_ISL_3037821 | AREA DE SALUD COTO BRUS | Inciensa, Instituto Costarricense de Investigación y Enseñanza en Nutrición y Salud | Adriana Godínez; Caterina Guzmán; Claudio Soto-Garita; Estela Cordero; Francisco Duarte; Hebleen Porras; José Luis Vargas; Mariela Gutiérrez Joselyn Prado; Melany Calderón; Nazareth Ruiz & María Fernanda Matamoros |
| EPI_ISL_3298313, EPI_ISL_3298314 | AREA DE SALUD MATA REDONDA-HOSPITAL - CLINICA DR. MORENO CAÑAS | Inciensa, Instituto Costarricense de Investigación y Enseñanza en Nutrición y Salud | Adriana Godínez; Claudio Soto-Garita; Estela Cordero; Francisco Duarte; Hebleen Porras; Joselyn Prado & Ricardo González; José Luis Vargas; Mariela Gutiérrez; Melany Calderón |
| EPI_ISL_2716544, EPI_ISL_2716545, EPI_ISL_2716548, EPI_ISL_2716550, EPI_ISL_2716558, EPI_ISL_2716560, EPI_ISL_2716561, EPI_ISL_2716563, EPI_ISL_2812655, EPI_ISL_2812658, EPI_ISL_2812667, EPI_ISL_2812672, EPI_ISL_2812681, EPI_ISL_2812683, EPI_ISL_2812686 | | | |
| see above | ASL Napoli 1 Centro | AMES Centro Polidiagnostico Strumentale S.r.l. | "Giovanni Savarese; Antonella Di Carlo; Antonio Fico"; Eloisa Evangelista; Luigi D'Amore; Luisa Circelli; Maurizio D'Amora; Maurizio D'Amora Antonio Fico"; Monica Ianniello; Nadia Petrillo; Raffaella Ruggiero; Roberto Sirica |
| EPI_ISL_3122709 | ASST GOM NIGUARDA | ASST Grande ospedale Metropolitano Niguarda | Alice Nava |
| EPI_ISL_3028238 | AULSS 2 Marca Trevigiana | Istituto Zooprofilattico Sperimentale delle Venezie | Adelaide Milani; Alessia Schivo; Alice Fusaro; Ambra Pastori; Annalisa Salviato; Antonia Ricci; Calogero Terregino; Edoardo Giussani; Elisa Palumbo; Erika Giorgia Quaranta; Isabella Monne; Luca Tassoni |
| EPI_ISL_2963435, EPI_ISL_2963436, EPI_ISL_2963439 | AULSS 3 Venezia | Istituto Zooprofilattico Sperimentale delle Venezie | Adelaide Milani; Alessia Schivo; Alice Fusaro; Ambra Pastori; Annalisa Salviato; Antonia Ricci; Calogero Terregino; Edoardo Giussani; Elisa Palumbo; Erika Giorgia Quaranta; Isabella Monne; Luca Tassoni |
| EPI_ISL_3007277, EPI_ISL_3007279 | AULSS 3 Venezia | UOSD Genetica e Citogenetica - Azienda ULSS 3 Serenissima; Istituto Zooprofilattico Sperimentale delle Venezie | Adelaide Milani; Alessia Schivo; Alice Fusaro; Ambra Pastori; Annalisa Salviato; Antonia Ricci; Calogero Terregino; Claudia Perini; Edoardo Giussani; Elisa Palumbo; Elisa Squarcina; Erika Giorgia Quaranta; Isabella Monne; Laura Bevilacqua; Laura Squarzon; Luca Tassoni; Mosé Favarato |
| EPI_ISL_2975135, EPI_ISL_2975136, EPI_ISL_2975138, EPI_ISL_2975222, EPI_ISL_2975226, EPI_ISL_2975234, EPI_ISL_2975236 | | | |
| see above | AULSS 6 Euganea | Istituto Zooprofilattico Sperimentale delle Venezie | Adelaide Milani; Alessia Schivo; Alice Fusaro; Ambra Pastori; Annalisa Salviato; Antonia Ricci; Calogero Terregino; Edoardo Giussani; Elisa Palumbo; Erika Giorgia Quaranta; Isabella Monne; Luca Tassoni |
| EPI_ISL_2963476, EPI_ISL_2975154 | AULSS 8 Berica | Istituto Zooprofilattico Sperimentale delle Venezie | Adelaide Milani; Alessia Schivo; Alice Fusaro; Ambra Pastori; Annalisa Salviato; Antonia Ricci; Calogero Terregino; Edoardo Giussani; Elisa Palumbo; Erika Giorgia Quaranta; Isabella Monne; Luca Tassoni |
| EPI_ISL_2927999 | AULSS 9 Scaligera | Istituto Zooprofilattico Sperimentale delle Venezie | Adelaide Milani; Alessia Schivo; Alice Fusaro; Ambra Pastori; Annalisa Salviato; Antonia Ricci; Calogero Terregino; Edoardo Giussani; Elisa Palumbo; Erika Giorgia Quaranta; Isabella Monne; Luca Tassoni |
| EPI_ISL_2725502 | AZDelta | AZ Delta Medical Laboratories in Roeselare, Belgium | Dieter De Smet; Geert Martens; Merijn Vanhee; on behalf of AZ Delta COVID-19 Genomics core (member of Genomic surveillance of SARS-CoV-2 in Belgium network) |
| EPI_ISL_2784730, EPI_ISL_2784748, EPI_ISL_2784763, EPI_ISL_2784764, EPI_ISL_2784773, EPI_ISL_2784776, EPI_ISL_2784784, EPI_ISL_2784785, EPI_ISL_2784805, EPI_ISL_2784806, EPI_ISL_2784812, EPI_ISL_2784827, EPI_ISL_2784828, EPI_ISL_2784835, EPI_ISL_2784838, EPI_ISL_2785032, EPI_ISL_2785040, EPI_ISL_2785048, EPI_ISL_2785049, EPI_ISL_2832522, EPI_ISL_2832534, EPI_ISL_2832535, EPI_ISL_2875541, EPI_ISL_2875542, EPI_ISL_2875550, EPI_ISL_2875576, EPI_ISL_2875590, EPI_ISL_2875595, EPI_ISL_2875612, EPI_ISL_2875615, EPI_ISL_2875624, EPI_ISL_2875626, EPI_ISL_2875658, EPI_ISL_2875659, EPI_ISL_2875667, EPI_ISL_2875677, EPI_ISL_2875678, EPI_ISL_2875683, EPI_ISL_2875685, EPI_ISL_2875687, EPI_ISL_2875691, EPI_ISL_2875697, EPI_ISL_2875698, EPI_ISL_2875699, EPI_ISL_2875748, EPI_ISL_2875750, EPI_ISL_2875767, EPI_ISL_2875785, EPI_ISL_2875789, EPI_ISL_2875832, EPI_ISL_2875835, EPI_ISL_2875837, EPI_ISL_2875839, EPI_ISL_2875840, EPI_ISL_2875842, EPI_ISL_2875872, EPI_ISL_2875893, EPI_ISL_2875894, EPI_ISL_2875913, EPI_ISL_2875956, EPI_ISL_2877655, EPI_ISL_2877668, EPI_ISL_2877669, EPI_ISL_2877671, EPI_ISL_2877692, EPI_ISL_2877694, EPI_ISL_3218130, EPI_ISL_3305316, EPI_ISL_4361035, EPI_ISL_4361558, EPI_ISL_4364782, EPI_ISL_4365371, EPI_ISL_4365460 | | | |
| see above | Aegis Sciences Corporation | Centers for Disease Control and Prevention Division of Viral Diseases, Pathogen Discovery | Adrian Paskey; Alec Vest; Benjamin Rambo-Martin; Christopher Gulvick; Clinton Paden; Clinton R. Paden; Cyndi Clark; Dakota Howard; Darlene Wagner; Dhwani Batra; Dillon Nall; Duncan MacCannell; Erisa Sula; Ethan Sanders; Holly Houdeshell; Jason Caravas; Kara Moser; Kristine Lacek; Matthew Hardison; Matthew Schmerer; Ola Kvalvaag; Patrick Campbell; Peter Cook; Peter W. Cook; Rob Case; Scott Sammons; Shatavia Morrison; Shaun Westlund; Tymeckia Kendall; Victoria Caban Figueroa; Vikramsinha Ghorpade; Yvette Unoarumhi |

| | | | |
|---|---|---|---|
| EPI_ISL_4219157, EPI_ISL_4219158 | Alameda County Public Health Department | Alameda County Public Health Department | Maria Elma; Vici Varghese |
| EPI_ISL_2893331, EPI_ISL_2893375, EPI_ISL_2893376, EPI_ISL_2966433 | Alaska State Virology Laboratory | Alaska State Virology Laboratory | Elva House; Jack Chen; Jacob Zidek; Lisa Smith; Ph.D.; Stephanie DeRonde |
| EPI_ISL_3388711 | Alverno Labs | RIPHL at Rush University Medical Center | Edith Perez; Felix Araujo Perez; Kevin Kunstman; Laura Furtado; Marieta Hyde; Max Kolton; Melissa Mace; Stefan Green |
| EPI_ISL_2754301, EPI_ISL_2854198, EPI_ISL_2854199, EPI_ISL_2854200, EPI_ISL_2854201, EPI_ISL_2919071, EPI_ISL_2919072, EPI_ISL_2919073, EPI_ISL_2919074, EPI_ISL_2919075, EPI_ISL_2919076, EPI_ISL_2919093, EPI_ISL_2919101, EPI_ISL_2943052 | | | |
| see above | Arizona State Public Health Laboratory | Arizona State Public Health Laboratory | Jessica Escobar; Katherine Fullerton; Linda Getsinger; Matthew Contursi; Nobuko Fukushima; Stacy White; Trung Huynh; Victor Waddell |
| EPI_ISL_2839103, EPI_ISL_2839106, EPI_ISL_2863915, EPI_ISL_2863917, EPI_ISL_2863919, EPI_ISL_2863920, EPI_ISL_2863922 | | | |
| see above | Arizona State University | Arizona State University | Efrem S. Lim; Joshua LaBaer; Joy M. Blain; LaRinda A. Holland; Matthew F. Smith; Nicholas J. Mellor; Peter T. Skidmore; Rabia Maqsood; Valerie Harris; Vel Murugan |
| EPI_ISL_3504828, EPI_ISL_3504839, EPI_ISL_3504855 | Arkansas Children's Hospital | Center for Global Health, University of New Mexico Health Sciences Center | Ashton Ingold; Bobby Boyanton; Darrell Dinwiddie; Daryl Domman; Joshua L. Kennedy; Kurt Schwalm; Valerie Morley |
| EPI_ISL_3015774, EPI_ISL_3015776, EPI_ISL_3104892 | Atlas Genomics | Seattle Flu Study | Amanda Adler; Barry R. Lutz; Benjamin Pelle; Brian Hiatt; Caitlin R. Wolf; Chris D. Frazar; Deborah A. Nickerson; Elisabeth Brandstetter; Erica Ryke; Geoff Melly; Helen Y. Chu; Janet A. Englund; Jay Shendure; Jover Lee; Kairsten Fay; Kirsten Lacombe; Lea M. Starita; Mark J. Rieder; Matthew Richardson; Matthew Thompson; Melissa Truong; Michael Boeckh; Michael Famulare; Misja Ilcisin; Peter D. Han; Philip Dykema; Romesh Gautom; Scott Lindquist; Thomas R. Sibley; Trevor Bedford |
| EPI_ISL_2787073 | Azienda Ospedaliera San Camillo Forlanini | INMI Lazzaro Spallanzani IRCCS | B Bartolini; CEM Gruber; D Cerini; D Di Fusco; E Sperandio; F Santini; G Bonfiglio; L Ciuffreda; O Butera |
| EPI_ISL_2790288, EPI_ISL_2840656, EPI_ISL_2861280 | Azienda Ospedaliera Terni | Istituto Zooprofilattico Sperimentale dell’Abruzzo e Molise “G. Caporale” | Ancora M; Calistri P; Cammà C; Curini V; Delli Compagni E; Di Domenico M; Di Pasquale A; Lorusso A; Mangone I; Marcacci M; Palumbo M; Puglia I; Rinaldi A; Savini G; Scaccetti A; Scialabba S |
| EPI_ISL_2820998, EPI_ISL_2820999, EPI_ISL_2821000, EPI_ISL_2821002, EPI_ISL_2821004, EPI_ISL_2821005, EPI_ISL_2821006 | | | |
| see above | Azienda Ospedaliero - Universitaria di Modena Policlinico - Virologia e Microbiologia Molecolare | Istituto Zooprofilattico Sperimentale della Lombardia e dell'Emilia Romagna (IZSLER), Risk Analysis and Genomic Epidemiology Unit | Erika Scaltriti; Giulia Fregni Serpini; Ilaria Menozzi; Marina Morganti; Monica Pecorari; Stefano Pongolini; William Gennari |
| EPI_ISL_2787070, EPI_ISL_2787071, EPI_ISL_2787072 | Azienda Sanitaria Locale Roma 5 | INMI Lazzaro Spallanzani IRCCS | A Massacci; B Bartolini; CEM Gruber; D Cerini; D Di Fusco; E Giombini; E Sperandio; F De Nicola; F Messina; F Santini; G Bonfiglio; G Orlandi; GF Inger; M Rueca; O Butera |
| EPI_ISL_2790277, EPI_ISL_2790278, EPI_ISL_2840657, EPI_ISL_2840658, EPI_ISL_2840659, EPI_ISL_2840660, EPI_ISL_2840661, EPI_ISL_2840662, EPI_ISL_2840663, EPI_ISL_2840664, EPI_ISL_2840665, EPI_ISL_2840666, EPI_ISL_2840667, EPI_ISL_2840668, EPI_ISL_2840669 | | | |
| see above | Azienda USL Umbria 2 | Istituto Zooprofilattico Sperimentale dell’Abruzzo e Molise “G. Caporale” | Ancora M; Calistri P; Cammà C; Curini V; Delli Compagni E; Di Domenico M; Di Pasquale A; Lorusso A; Mangone I; Marcacci M; Pistoni E; Proietti A; Puglia I; Rinaldi A; Savini G; Scialabba S |
| EPI_ISL_2795913 | BIO67-BIOSPHERE | Department of Virology, Henri Mondor University Hospital, Assistance Publique Hôpitaux de Paris, Université Paris-Est Créteil, INSERM U955 | Alexandre Soulier; Christophe Rodriguez; Elisabeth Trawinski; Guillaume Gricourt; Jean-Michel Pawlotsky; Melissa N'Debi; Slim Fourati; Vanessa Demontant |
| EPI_ISL_5648312, EPI_ISL_5648324, EPI_ISL_5648345, EPI_ISL_5648367, EPI_ISL_5648401, EPI_ISL_5648432, EPI_ISL_5648435, EPI_ISL_5648444, EPI_ISL_5648451, EPI_ISL_5648453, EPI_ISL_5648455, EPI_ISL_5648457, EPI_ISL_5648459, EPI_ISL_5648464, EPI_ISL_5648465, EPI_ISL_5648466, EPI_ISL_5648470, EPI_ISL_5648471, EPI_ISL_5648475, EPI_ISL_5648476, EPI_ISL_5648480, EPI_ISL_5648481, EPI_ISL_5648487, EPI_ISL_5648490, EPI_ISL_5648491, EPI_ISL_5648493, EPI_ISL_5648495, EPI_ISL_5648496, EPI_ISL_5648497, EPI_ISL_5648506, EPI_ISL_5648511, EPI_ISL_5648518, EPI_ISL_5648522, EPI_ISL_5648523, EPI_ISL_5648536, EPI_ISL_5648543, EPI_ISL_5648547, EPI_ISL_5648556, EPI_ISL_5648557, EPI_ISL_5648558, EPI_ISL_5648559, EPI_ISL_5648561, EPI_ISL_5648562, EPI_ISL_5648563, EPI_ISL_5648566, EPI_ISL_5648568, EPI_ISL_5648569, EPI_ISL_5648578, EPI_ISL_5648584, EPI_ISL_5648587, EPI_ISL_5648593, EPI_ISL_5648597, EPI_ISL_5648599, EPI_ISL_5648609, EPI_ISL_5648611, EPI_ISL_5648613, EPI_ISL_5648614, EPI_ISL_5648618, EPI_ISL_5648619, EPI_ISL_5648620, EPI_ISL_5648629, EPI_ISL_5648630, EPI_ISL_5648641, EPI_ISL_5648642, EPI_ISL_5648643, EPI_ISL_5648649, EPI_ISL_5648650, EPI_ISL_5648651, EPI_ISL_5648660, EPI_ISL_5648666, EPI_ISL_5648671, EPI_ISL_5648672, EPI_ISL_5648673, EPI_ISL_5648674, EPI_ISL_5648679, EPI_ISL_5648680, EPI_ISL_5648685, EPI_ISL_5648697, EPI_ISL_5648698, EPI_ISL_5648705, EPI_ISL_5648706, EPI_ISL_5648710, EPI_ISL_5648716, EPI_ISL_5648717, EPI_ISL_5648718, EPI_ISL_5648719, EPI_ISL_5648720, EPI_ISL_5648721, EPI_ISL_5648724, EPI_ISL_5648727, EPI_ISL_5648742 | | | |
| see above | BIOFAST | Instituto Butantan | Antonio Jorge Martins; Claudia Renata dos Santos Barros; David Schlesinger; Debora Botequio Moretti; Dimas Tadeu Covas; Elaine Cristina Marqueze; Elaine Vieira Santos; Evandra Strazza Rodrigues; Heidge Fukumasu; Jayme Augusto de Souza-Neto; José Salvatore Leister Patané; Luiz Alcantara; Luiz Lehmann Coutinho; Maria Carolina Elias; Maurício Lacerda Nogueira; Rafael dos Santos Bezerra; Raul Machado Neto; Rejane Maria Tommasini Grotto; Ricardo Haddad; Sandra Coccuzzo Sampaio Vessoni; Simone Kashima; Svetoslav Nanev Slavov; Vincent Louis Viala |
| EPI_ISL_2933132, EPI_ISL_2933133, EPI_ISL_2933135, EPI_ISL_2933139, EPI_ISL_2933144, EPI_ISL_2933146, EPI_ISL_2933152, EPI_ISL_2933153, EPI_ISL_2933154, EPI_ISL_2933159, EPI_ISL_2933168, EPI_ISL_2933171, EPI_ISL_2933172, EPI_ISL_2933175, EPI_ISL_2933176, EPI_ISL_2933177, EPI_ISL_2933182, EPI_ISL_2933187, EPI_ISL_2933190, EPI_ISL_2933191, EPI_ISL_2933194, EPI_ISL_2933199, EPI_ISL_2933201, EPI_ISL_2933203, EPI_ISL_2933350, EPI_ISL_2933351, EPI_ISL_2933352, EPI_ISL_2933354, EPI_ISL_2933355, EPI_ISL_2933356, EPI_ISL_2933357, EPI_ISL_2933359, EPI_ISL_2933360, EPI_ISL_2933361, EPI_ISL_2933362, EPI_ISL_2933363, EPI_ISL_2933365, EPI_ISL_2933377, EPI_ISL_2933378, EPI_ISL_2933379, EPI_ISL_2933381, EPI_ISL_2933383, EPI_ISL_2933384, EPI_ISL_2933385, EPI_ISL_2933386, EPI_ISL_2933388, EPI_ISL_2933389, EPI_ISL_2933390, EPI_ISL_2933391, EPI_ISL_2933392, EPI_ISL_2933395, EPI_ISL_2933396, EPI_ISL_2933397, EPI_ISL_2933400, EPI_ISL_2933401, EPI_ISL_2933402, EPI_ISL_2933403, EPI_ISL_2933405, EPI_ISL_2933406, EPI_ISL_2933408, EPI_ISL_2933409, EPI_ISL_2933410, EPI_ISL_2933411, EPI_ISL_2933412, EPI_ISL_2933413, EPI_ISL_2933414, EPI_ISL_2933415, EPI_ISL_2933416, EPI_ISL_2933417, EPI_ISL_2933418, EPI_ISL_2933420, EPI_ISL_2933421, EPI_ISL_2933422, EPI_ISL_2933423, EPI_ISL_2933424, EPI_ISL_2933425, EPI_ISL_2933426, EPI_ISL_2933427, EPI_ISL_2933428, EPI_ISL_2933430, EPI_ISL_2933431, EPI_ISL_2933432, EPI_ISL_2933433, EPI_ISL_2933434, EPI_ISL_2933435, EPI_ISL_2933436, EPI_ISL_2933437, EPI_ISL_2933438, EPI_ISL_2933439, EPI_ISL_2933440, EPI_ISL_2933441, EPI_ISL_2933442, EPI_ISL_3130099, EPI_ISL_3130100 | | | |
| see above | BIOMNIS LYON | CNR Virus des Infections Respiratoires - France SUD | Antonin Bal; Bruno Lina; Gregory Destras; Gwendolyne Burfin; Hadrien Regue; Laurence Josset; Martine Valette; Quentin Semanas |
| EPI_ISL_2834162 | Baylor Scott & White-Temple | Baylor Scott & White-Temple | Ari Rao; Kimberly Walker; Linden Morales; Marcus Volz; Shelby Johnson |
| EPI_ISL_2928138, EPI_ISL_2928297, EPI_ISL_2928298, EPI_ISL_2928299, EPI_ISL_2928303, EPI_ISL_2928305 | Biomédicos de Mérida | Biomédicos de Mérida | CANCHE-PECH JOSE REYES; CAUICH-VERA KRISTIAN; CEBALLOS-LOPEZ ADRIAN; CHIN-HAU LETICIA; GARCIA-GONZALEZ IGRID; LOPEZ-NOVELO MARIA E; MENDOZA-OLIVEROS TAHALI; SAURI-MEDINA EMILY |
| EPI_ISL_3145910, EPI_ISL_3145911, EPI_ISL_3145912, EPI_ISL_3145913, EPI_ISL_3145914, EPI_ISL_3145915, EPI_ISL_3145916, EPI_ISL_3145917, EPI_ISL_3145918, EPI_ISL_3145919, EPI_ISL_3145920, EPI_ISL_3145921, EPI_ISL_3145922, EPI_ISL_3146548 | | | |
| see above | BioneXt Lab | Laboratoire national de sante, Microbiology, Microbial Genomics Platform | Anke Wienecke-Baldacchino; Catherine Ragimbeau; Elodie Solarino; Fatu Djabi; Jessica Tapp; Lise Pignon; Raoul Salmon; Tamir Abdelrahman; Thibault Ferrandon; Virginie Jover |
| EPI_ISL_3047315 | Brentwood Biomedical Research | Genomics and Discovery, Respiratory Viruses Branch, Division of Viral Diseases, Centers for Disease Control and Prevention | Adam Retchless; Anna Kelleher; Anna Uehara; Brian Lynch; Clinton R. Paden; Dhwani Batra; Haibin Wang; Han Jia Justin Ng; Jasmine Padilla; Jing Zhang; Justin Lee; Mark Burroughs; Mili Sheth; Morgan Davis; Peter Cook; Rachel Marine; Sarah Nobles; Suxiang Tong; Tara Coalter; Yan Li; Ying Tao |
| EPI_ISL_3596588, EPI_ISL_3596604, EPI_ISL_3602125, EPI_ISL_3602127, EPI_ISL_3602128, EPI_ISL_3602131, EPI_ISL_3602132, EPI_ISL_3602133, EPI_ISL_3602137, EPI_ISL_3602139, EPI_ISL_3602140, EPI_ISL_3602142, EPI_ISL_3602143, EPI_ISL_3602144, EPI_ISL_3602145, EPI_ISL_3602214, EPI_ISL_3602216, EPI_ISL_3602217, EPI_ISL_3602219, EPI_ISL_3602221, EPI_ISL_3602222, EPI_ISL_3602223, EPI_ISL_3602224, EPI_ISL_3602225, EPI_ISL_3602226, EPI_ISL_3602228, EPI_ISL_3602229, EPI_ISL_3602230, EPI_ISL_3602232, EPI_ISL_3602237, EPI_ISL_3602239, EPI_ISL_3602240, EPI_ISL_3602243, EPI_ISL_3602246, EPI_ISL_3602247, EPI_ISL_3602248, EPI_ISL_3602261, EPI_ISL_3602263, EPI_ISL_3602264, EPI_ISL_3602267, EPI_ISL_3602270, EPI_ISL_3602271, EPI_ISL_3602272, EPI_ISL_3602273, EPI_ISL_3602274, EPI_ISL_3602275, EPI_ISL_3602276, EPI_ISL_3602277, EPI_ISL_3602279, EPI_ISL_3602282, EPI_ISL_3602283, EPI_ISL_3602284, EPI_ISL_3602285, EPI_ISL_3602287, EPI_ISL_3602288, EPI_ISL_3602290, EPI_ISL_3602292, EPI_ISL_3602294, EPI_ISL_3602297, EPI_ISL_3602298, EPI_ISL_3602299, EPI_ISL_3602301, EPI_ISL_3602302, EPI_ISL_3602305, EPI_ISL_3602306, EPI_ISL_3602308, EPI_ISL_3602310, EPI_ISL_3602311, EPI_ISL_3602312, EPI_ISL_3602314, EPI_ISL_3602317, EPI_ISL_3602318, EPI_ISL_3602322, EPI_ISL_3602324, EPI_ISL_3602327, EPI_ISL_3602329, EPI_ISL_3602330, EPI_ISL_3602331, EPI_ISL_3602333, EPI_ISL_3602334, EPI_ISL_3602335, EPI_ISL_3602338, EPI_ISL_3602339, EPI_ISL_3602342, EPI_ISL_3602343, EPI_ISL_3602346, EPI_ISL_3602355, EPI_ISL_3602356, EPI_ISL_3602369, EPI_ISL_3602375, EPI_ISL_3602381, EPI_ISL_3602382, EPI_ISL_3602386, EPI_ISL_3602387, EPI_ISL_3602388, EPI_ISL_3602391, EPI_ISL_3602392, EPI_ISL_3602404, EPI_ISL_3602405, EPI_ISL_3602409, EPI_ISL_3602410, EPI_ISL_3602411, EPI_ISL_3602413, EPI_ISL_3602414, EPI_ISL_4423886, EPI_ISL_4423896, EPI_ISL_4423899, EPI_ISL_4423902, EPI_ISL_4423904 | | | |
| see above | British Columbia Centre For Disease Control | BCCDC Public Health Laboratory | Ana Pacagnella; Corrinne Ng; Dan Fornika; John Tyson; Kim Macdonald; Kimia Kamelian; Linda Hoang; Loretta Janz; Mel Krajden; Prystajecky Natalie; Robert Azana; Shannon Russell |
| EPI_ISL_3007349, EPI_ISL_3007351, EPI_ISL_3007406, EPI_ISL_3007483 | Broad Institute Clinical Research Sequencing Platform | Infectious Disease Program, Broad Institute of Harvard and MIT | Adams, G.; B.L.; B.W.; Bauer, M.; Birren; Blumenstiel, B.; Brown, C.; Carter, A.; Chaluvadi, S.; D.J.; DeFelice, M.; DeRuff, K.; Dodge, S.; Gabriel, S.; Gallagher, G.; Gladden-Young, A.; Granger, B.; J.E.; K.J.; Lagerborg, K.; Larkin, K.; Lee, M.; Lemieux; Lennon, N.; Loreth, C.; Madoff, L.; McGovern, S.; Meldrim, J.; Normandin, E.; P.C.; Park; Pearlman, L.; Reilly, S.; Rudy, M.; Sabeti; Siddle; Smole, S.; Tomkins-Tinch, C.; Vicente, G.; and MacInnis |
| EPI_ISL_5648305, EPI_ISL_5648498, EPI_ISL_5648505, EPI_ISL_5648728, EPI_ISL_5648733, EPI_ISL_5648750, EPI_ISL_5649433, EPI_ISL_5649457, EPI_ISL_5649465, EPI_ISL_5659539, EPI_ISL_5659541, EPI_ISL_5659554, EPI_ISL_5659622, EPI_ISL_5659631 | | | |
| see above | CADIP VILA REGINA FERNANDOPOLIS | Instituto Butantan | Antonio Jorge Martins; Claudia Renata dos Santos Barros; David Schlesinger; Debora Botequio Moretti; Dimas Tadeu Covas; Elaine Cristina Marqueze; Elaine Vieira Santos; Evandra Strazza Rodrigues; Heidge Fukumasu; Jayme Augusto de Souza-Neto; José Salvatore Leister Patané; Luiz Alcantara; Luiz Lehmann Coutinho; Maria Carolina Elias; Maurício Lacerda Nogueira; Rafael dos Santos Bezerra; Raul Machado Neto; Rejane Maria Tommasini Grotto; Ricardo Haddad; Sandra Coccuzzo Sampaio Vessoni; Simone Kashima; Svetoslav Nanev Slavov; Vincent Louis Viala |
| EPI_ISL_5649111 | CAIC CENTRO DE ATENDIMENTO INTEGRAL DA CRIANCAITAQUA | Instituto Butantan | Antonio Jorge Martins; Claudia Renata dos Santos Barros; David Schlesinger; Debora Botequio Moretti; Dimas Tadeu Covas; Elaine Cristina Marqueze; Elaine Vieira Santos; Evandra Strazza Rodrigues; Heidge Fukumasu; Jayme Augusto de Souza-Neto; José Salvatore Leister Patané; Luiz Alcantara; Luiz Lehmann Coutinho; Maria Carolina Elias; Maurício Lacerda Nogueira; Rafael dos Santos Bezerra; Raul Machado Neto; Rejane Maria Tommasini Grotto; Ricardo Haddad; Sandra Coccuzzo Sampaio Vessoni; Simone Kashima; Svetoslav Nanev Slavov; Vincent Louis Viala |
| EPI_ISL_3099595 | CAP Manlleu | Banc de Sang i Teixits | Carlos Hobeich; Francisco Vidal; Irene Corrales; Lorena Ramírez; Maria Glòria Soria; Natàlia Comes; Nina Borràs; Noemí Gonzalez; Silvia Sauleda |
| EPI_ISL_5659557 | CASA DE SAUDE DE INDIAPORA | Instituto Butantan | Antonio Jorge Martins; Claudia Renata dos Santos Barros; David Schlesinger; Debora Botequio Moretti; Dimas Tadeu Covas; Elaine Cristina Marqueze; Elaine Vieira Santos; Evandra Strazza Rodrigues; Heidge Fukumasu; Jayme Augusto de Souza-Neto; José Salvatore Leister Patané; Luiz Alcantara; Luiz Lehmann Coutinho; Maria Carolina Elias; Maurício Lacerda Nogueira; Rafael dos Santos Bezerra; Raul Machado Neto; Rejane Maria Tommasini Grotto; Ricardo Haddad; Sandra Coccuzzo Sampaio Vessoni; Simone Kashima; Svetoslav Nanev Slavov; Vincent Louis Viala |
| EPI_ISL_4491388, EPI_ISL_4492437, EPI_ISL_4492438, EPI_ISL_4492444, EPI_ISL_4508811, EPI_ISL_4509581, EPI_ISL_4510024, EPI_ISL_4510425, EPI_ISL_4511819, EPI_ISL_4511827 | | | |
| see above | CDPH VBL | California Department of Public Health | Emily Smith on behalf of CDPH-COVIDNet |
| EPI_ISL_2933353 | CEDIBIO-UNILABS-TOUNIS | CNR Virus des Infections Respiratoires - France SUD | Antonin Bal; Bruno Lina; Gregory Destras; Gwendolyne Burfin; Hadrien Regue; Laurence Josset; Martine Valette; Quentin Semanas |
| EPI_ISL_2845480 | CENTOGENE Frankfurt Laboratory: Niederlassung Industriepark Höchst | Robert Koch Institute | |

| | | | |
|---|---|---|---|
| EPI_ISL_5659581, EPI_ISL_5659639 | CENTRO DE ATENDIMENTO COVID 19 | Instituto Butantan | Antonio Jorge Martins; Claudia Renata dos Santos Barros; David Schlesinger; Debora Botequio Moretti; Dimas Tadeu Covas; Elaine Cristina Marqueze; Elaine Vieira Santos; Evandra Strazza Rodrigues; Heidge Fukumasu; Jayme Augusto de Souza-Neto; José Salvatore Leister Patané; Luiz Alcantara; Luiz Lehmann Coutinho; Maria Carolina Elias; Maurício Lacerda Nogueira; Rafael dos Santos Bezerra; Raul Machado Neto; Rejane Maria Tommasini Grotto; Ricardo Haddad; Sandra Coccuzzo Sampaio Vessoni; Simone Kashima; Svetoslav Nanev Slavov; Vincent Louis Viala |
| EPI_ISL_3912140, EPI_ISL_3912141 | CENTRO DE ATENDIMENTO PARA ENFRENTAMENTO AO COVID 19 | Analytical Competence Molecular Epidemiology Lab/ACME, Oswaldo Cruz Foundation, Ceara (FIOCRUZ CE) | Cleber Furtado Aksenen; Fabio Miyajima; Fernando Braga Stehling; Francisco Eder de Moura Lopes; Jamille Maria Mendes Bezerra; Joaquim Cesar do Nascimento Sousa Junior; Pedro Miguel Carneiro Jeronimo; Suzana Porto Almeida & Lucas Delerino on behalf of COVID-19 FIOCRUZ Genomic Network; Thais Ferreira de Oliveira; Thais de Oliveira Costa; Ticiane Cavalcante de Souza; Veridiana Pessoa Miyajima |
| EPI_ISL_5649236 | CENTRO DE DETENCAO PROVISORIA TACITO AP SANTANA DE CAIUA | Instituto Butantan | Antonio Jorge Martins; Claudia Renata dos Santos Barros; David Schlesinger; Debora Botequio Moretti; Dimas Tadeu Covas; Elaine Cristina Marqueze; Elaine Vieira Santos; Evandra Strazza Rodrigues; Heidge Fukumasu; Jayme Augusto de Souza-Neto; José Salvatore Leister Patané; Luiz Alcantara; Luiz Lehmann Coutinho; Maria Carolina Elias; Maurício Lacerda Nogueira; Rafael dos Santos Bezerra; Raul Machado Neto; Rejane Maria Tommasini Grotto; Ricardo Haddad; Sandra Coccuzzo Sampaio Vessoni; Simone Kashima; Svetoslav Nanev Slavov; Vincent Louis Viala |
| EPI_ISL_5649365 | CENTRO DE ESPECIALIDADES DE AGUAI AGUAI | Instituto Butantan | Antonio Jorge Martins; Claudia Renata dos Santos Barros; David Schlesinger; Debora Botequio Moretti; Dimas Tadeu Covas; Elaine Cristina Marqueze; Elaine Vieira Santos; Evandra Strazza Rodrigues; Heidge Fukumasu; Jayme Augusto de Souza-Neto; José Salvatore Leister Patané; Luiz Alcantara; Luiz Lehmann Coutinho; Maria Carolina Elias; Maurício Lacerda Nogueira; Rafael dos Santos Bezerra; Raul Machado Neto; Rejane Maria Tommasini Grotto; Ricardo Haddad; Sandra Coccuzzo Sampaio Vessoni; Simone Kashima; Svetoslav Nanev Slavov; Vincent Louis Viala |
| EPI_ISL_5648779, EPI_ISL_5648861 | CENTRO DE ESPECIALIDADES MARIAS DA GLORIA | Instituto Butantan | Antonio Jorge Martins; Claudia Renata dos Santos Barros; David Schlesinger; Debora Botequio Moretti; Dimas Tadeu Covas; Elaine Cristina Marqueze; Elaine Vieira Santos; Evandra Strazza Rodrigues; Heidge Fukumasu; Jayme Augusto de Souza-Neto; José Salvatore Leister Patané; Luiz Alcantara; Luiz Lehmann Coutinho; Maria Carolina Elias; Maurício Lacerda Nogueira; Rafael dos Santos Bezerra; Raul Machado Neto; Rejane Maria Tommasini Grotto; Ricardo Haddad; Sandra Coccuzzo Sampaio Vessoni; Simone Kashima; Svetoslav Nanev Slavov; Vincent Louis Viala |
| EPI_ISL_5648384, EPI_ISL_5648442, EPI_ISL_5648550, EPI_ISL_5648625, EPI_ISL_5648639, EPI_ISL_5648731 | CENTRO DE GENOMAS | Instituto Butantan | Antonio Jorge Martins; Claudia Renata dos Santos Barros; David Schlesinger; Debora Botequio Moretti; Dimas Tadeu Covas; Elaine Cristina Marqueze; Elaine Vieira Santos; Evandra Strazza Rodrigues; Heidge Fukumasu; Jayme Augusto de Souza-Neto; José Salvatore Leister Patané; Luiz Alcantara; Luiz Lehmann Coutinho; Maria Carolina Elias; Maurício Lacerda Nogueira; Rafael dos Santos Bezerra; Raul Machado Neto; Rejane Maria Tommasini Grotto; Ricardo Haddad; Sandra Coccuzzo Sampaio Vessoni; Simone Kashima; Svetoslav Nanev Slavov; Vincent Louis Viala |
| EPI_ISL_5648407, EPI_ISL_5648483, EPI_ISL_5648626, EPI_ISL_5649456, EPI_ISL_5659528 | CENTRO DE SAUDE DE ADAMANTINA | Instituto Butantan | Antonio Jorge Martins; Claudia Renata dos Santos Barros; David Schlesinger; Debora Botequio Moretti; Dimas Tadeu Covas; Elaine Cristina Marqueze; Elaine Vieira Santos; Evandra Strazza Rodrigues; Heidge Fukumasu; Jayme Augusto de Souza-Neto; José Salvatore Leister Patané; Luiz Alcantara; Luiz Lehmann Coutinho; Maria Carolina Elias; Maurício Lacerda Nogueira; Rafael dos Santos Bezerra; Raul Machado Neto; Rejane Maria Tommasini Grotto; Ricardo Haddad; Sandra Coccuzzo Sampaio Vessoni; Simone Kashima; Svetoslav Nanev Slavov; Vincent Louis Viala |
| EPI_ISL_5649217, EPI_ISL_5665746 | CENTRO DE SAUDE DE ANALANDIA | Instituto Butantan | Antonio Jorge Martins; Claudia Renata dos Santos Barros; David Schlesinger; Debora Botequio Moretti; Dimas Tadeu Covas; Elaine Cristina Marqueze; Elaine Vieira Santos; Evandra Strazza Rodrigues; Heidge Fukumasu; Jayme Augusto de Souza-Neto; José Salvatore Leister Patané; Luiz Alcantara; Luiz Lehmann Coutinho; Maria Carolina Elias; Maurício Lacerda Nogueira; Rafael dos Santos Bezerra; Raul Machado Neto; Rejane Maria Tommasini Grotto; Ricardo Haddad; Sandra Coccuzzo Sampaio Vessoni; Simone Kashima; Svetoslav Nanev Slavov; Vincent Louis Viala |
| EPI_ISL_5648889, EPI_ISL_5648923 | CENTRO DE SAUDE DE CHAVANTES | Instituto Butantan | Antonio Jorge Martins; Claudia Renata dos Santos Barros; David Schlesinger; Debora Botequio Moretti; Dimas Tadeu Covas; Elaine Cristina Marqueze; Elaine Vieira Santos; Evandra Strazza Rodrigues; Heidge Fukumasu; Jayme Augusto de Souza-Neto; José Salvatore Leister Patané; Luiz Alcantara; Luiz Lehmann Coutinho; Maria Carolina Elias; Maurício Lacerda Nogueira; Rafael dos Santos Bezerra; Raul Machado Neto; Rejane Maria Tommasini Grotto; Ricardo Haddad; Sandra Coccuzzo Sampaio Vessoni; Simone Kashima; Svetoslav Nanev Slavov; Vincent Louis Viala |
| EPI_ISL_5648825 | CENTRO DE SAUDE DE FLORIDA PAULISTA | Instituto Butantan | Antonio Jorge Martins; Claudia Renata dos Santos Barros; David Schlesinger; Debora Botequio Moretti; Dimas Tadeu Covas; Elaine Cristina Marqueze; Elaine Vieira Santos; Evandra Strazza Rodrigues; Heidge Fukumasu; Jayme Augusto de Souza-Neto; José Salvatore Leister Patané; Luiz Alcantara; Luiz Lehmann Coutinho; Maria Carolina Elias; Maurício Lacerda Nogueira; Rafael dos Santos Bezerra; Raul Machado Neto; Rejane Maria Tommasini Grotto; Ricardo Haddad; Sandra Coccuzzo Sampaio Vessoni; Simone Kashima; Svetoslav Nanev Slavov; Vincent Louis Viala |
| EPI_ISL_5648714 | CENTRO DE SAUDE DE IPAUCU | Instituto Butantan | Antonio Jorge Martins; Claudia Renata dos Santos Barros; David Schlesinger; Debora Botequio Moretti; Dimas Tadeu Covas; Elaine Cristina Marqueze; Elaine Vieira Santos; Evandra Strazza Rodrigues; Heidge Fukumasu; Jayme Augusto de Souza-Neto; José Salvatore Leister Patané; Luiz Alcantara; Luiz Lehmann Coutinho; Maria Carolina Elias; Maurício Lacerda Nogueira; Rafael dos Santos Bezerra; Raul Machado Neto; Rejane Maria Tommasini Grotto; Ricardo Haddad; Sandra Coccuzzo Sampaio Vessoni; Simone Kashima; Svetoslav Nanev Slavov; Vincent Louis Viala |
| EPI_ISL_5648657 | CENTRO DE SAUDE DE LUCELIA | Instituto Butantan | Antonio Jorge Martins; Claudia Renata dos Santos Barros; David Schlesinger; Debora Botequio Moretti; Dimas Tadeu Covas; Elaine Cristina Marqueze; Elaine Vieira Santos; Evandra Strazza Rodrigues; Heidge Fukumasu; Jayme Augusto de Souza-Neto; José Salvatore Leister Patané; Luiz Alcantara; Luiz Lehmann Coutinho; Maria Carolina Elias; Maurício Lacerda Nogueira; Rafael dos Santos Bezerra; Raul Machado Neto; Rejane Maria Tommasini Grotto; Ricardo Haddad; Sandra Coccuzzo Sampaio Vessoni; Simone Kashima; Svetoslav Nanev Slavov; Vincent Louis Viala |
| EPI_ISL_5648555 | CENTRO DE SAUDE DE MARACAI | Instituto Butantan | Antonio Jorge Martins; Claudia Renata dos Santos Barros; David Schlesinger; Debora Botequio Moretti; Dimas Tadeu Covas; Elaine Cristina Marqueze; Elaine Vieira Santos; Evandra Strazza Rodrigues; Heidge Fukumasu; Jayme Augusto de Souza-Neto; José Salvatore Leister Patané; Luiz Alcantara; Luiz Lehmann Coutinho; Maria Carolina Elias; Maurício Lacerda Nogueira; Rafael dos Santos Bezerra; Raul Machado Neto; Rejane Maria Tommasini Grotto; Ricardo Haddad; Sandra Coccuzzo Sampaio Vessoni; Simone Kashima; Svetoslav Nanev Slavov; Vincent Louis Viala |
| EPI_ISL_5648802, EPI_ISL_5648942, EPI_ISL_5649045, EPI_ISL_5649084 | CENTRO DE SAUDE DE OSVALDO CRUZ | Instituto Butantan | Antonio Jorge Martins; Claudia Renata dos Santos Barros; David Schlesinger; Debora Botequio Moretti; Dimas Tadeu Covas; Elaine Cristina Marqueze; Elaine Vieira Santos; Evandra Strazza Rodrigues; Heidge Fukumasu; Jayme Augusto de Souza-Neto; José Salvatore Leister Patané; Luiz Alcantara; Luiz Lehmann Coutinho; Maria Carolina Elias; Maurício Lacerda Nogueira; Rafael dos Santos Bezerra; Raul Machado Neto; Rejane Maria Tommasini Grotto; Ricardo Haddad; Sandra Coccuzzo Sampaio Vessoni; Simone Kashima; Svetoslav Nanev Slavov; Vincent Louis Viala |
| EPI_ISL_5649267, EPI_ISL_5649290 | CENTRO DE SAUDE DE PEREIRAS | Instituto Butantan | Antonio Jorge Martins; Claudia Renata dos Santos Barros; David Schlesinger; Debora Botequio Moretti; Dimas Tadeu Covas; Elaine Cristina Marqueze; Elaine Vieira Santos; Evandra Strazza Rodrigues; Heidge Fukumasu; Jayme Augusto de Souza-Neto; José Salvatore Leister Patané; Luiz Alcantara; Luiz Lehmann Coutinho; Maria Carolina Elias; Maurício Lacerda Nogueira; Rafael dos Santos Bezerra; Raul Machado Neto; Rejane Maria Tommasini Grotto; Ricardo Haddad; Sandra Coccuzzo Sampaio Vessoni; Simone Kashima; Svetoslav Nanev Slavov; Vincent Louis Viala |
| EPI_ISL_5649186, EPI_ISL_5649201 | CENTRO DE SAUDE DE PIRAJUI | Instituto Butantan | Antonio Jorge Martins; Claudia Renata dos Santos Barros; David Schlesinger; Debora Botequio Moretti; Dimas Tadeu Covas; Elaine Cristina Marqueze; Elaine Vieira Santos; Evandra Strazza Rodrigues; Heidge Fukumasu; Jayme Augusto de Souza-Neto; José Salvatore Leister Patané; Luiz Alcantara; Luiz Lehmann Coutinho; Maria Carolina Elias; Maurício Lacerda Nogueira; Rafael dos Santos Bezerra; Raul Machado Neto; Rejane Maria Tommasini Grotto; Ricardo Haddad; Sandra Coccuzzo Sampaio Vessoni; Simone Kashima; Svetoslav Nanev Slavov; Vincent Louis Viala |
| EPI_ISL_5648676 | CENTRO DE SAUDE DE PLATINA | Instituto Butantan | Antonio Jorge Martins; Claudia Renata dos Santos Barros; David Schlesinger; Debora Botequio Moretti; Dimas Tadeu Covas; Elaine Cristina Marqueze; Elaine Vieira Santos; Evandra Strazza Rodrigues; Heidge Fukumasu; Jayme Augusto de Souza-Neto; José Salvatore Leister Patané; Luiz Alcantara; Luiz Lehmann Coutinho; Maria Carolina Elias; Maurício Lacerda Nogueira; Rafael dos Santos Bezerra; Raul Machado Neto; Rejane Maria Tommasini Grotto; Ricardo Haddad; Sandra Coccuzzo Sampaio Vessoni; Simone Kashima; Svetoslav Nanev Slavov; Vincent Louis Viala |
| EPI_ISL_5648467, EPI_ISL_5648539 | CENTRO DE SAUDE DE RIBEIRAO DO SUL | Instituto Butantan | Antonio Jorge Martins; Claudia Renata dos Santos Barros; David Schlesinger; Debora Botequio Moretti; Dimas Tadeu Covas; Elaine Cristina Marqueze; Elaine Vieira Santos; Evandra Strazza Rodrigues; Heidge Fukumasu; Jayme Augusto de Souza-Neto; José Salvatore Leister Patané; Luiz Alcantara; Luiz Lehmann Coutinho; Maria Carolina Elias; Maurício Lacerda Nogueira; Rafael dos Santos Bezerra; Raul Machado Neto; Rejane Maria Tommasini Grotto; Ricardo Haddad; Sandra Coccuzzo Sampaio Vessoni; Simone Kashima; Svetoslav Nanev Slavov; Vincent Louis Viala |
| EPI_ISL_5648289, EPI_ISL_5648460, EPI_ISL_5648502, EPI_ISL_5648524, EPI_ISL_5648540, EPI_ISL_5648549, EPI_ISL_5648677 | | | |
| see above | CENTRO DE SAUDE DE SPEDRO DO TURVO | Instituto Butantan | Antonio Jorge Martins; Claudia Renata dos Santos Barros; David Schlesinger; Debora Botequio Moretti; Dimas Tadeu Covas; Elaine Cristina Marqueze; Elaine Vieira Santos; Evandra Strazza Rodrigues; Heidge Fukumasu; Jayme Augusto de Souza-Neto; José Salvatore Leister Patané; Luiz Alcantara; Luiz Lehmann Coutinho; Maria Carolina Elias; Maurício Lacerda Nogueira; Rafael dos Santos Bezerra; Raul Machado Neto; Rejane Maria Tommasini Grotto; Ricardo Haddad; Sandra Coccuzzo Sampaio Vessoni; Simone Kashima; Svetoslav Nanev Slavov; Vincent Louis Viala |
| EPI_ISL_3536339, EPI_ISL_3536340 | CENTRO DE SAUDE DONA TEREZA HOLANDA DE OLIVEIRA | Analytical Competence Molecular Epidemiology Lab/ACME, Oswaldo Cruz Foundation, Ceara (FIOCRUZ CE) | Cleber Furtado Aksenen; Fabio Miyajima; Fernando Braga Stehling; Francisco Eder de Moura Lopes; Jamille Maria Mendes Bezerra; Joaquim César do Nascimento Sousa Junior; Pedro Miguel Carneiro Jeronimo; Suzana Porto Almeida e Lucas Delerino; Thais Ferreira de Oliveira; Thais de Oliveira Costa; Ticiane Cavalcante de Souza; Veridiana Pessoa Miyajima |
| EPI_ISL_5649206, EPI_ISL_5649209 | CENTRO DE SAUDE DR JOSE AGOSTINHO NOGUEIRA PAULICEIA | Instituto Butantan | Antonio Jorge Martins; Claudia Renata dos Santos Barros; David Schlesinger; Debora Botequio Moretti; Dimas Tadeu Covas; Elaine Cristina Marqueze; Elaine Vieira Santos; Evandra Strazza Rodrigues; Heidge Fukumasu; Jayme Augusto de Souza-Neto; José Salvatore Leister Patané; Luiz Alcantara; Luiz Lehmann Coutinho; Maria Carolina Elias; Maurício Lacerda Nogueira; Rafael dos Santos Bezerra; Raul Machado Neto; Rejane Maria Tommasini Grotto; Ricardo Haddad; Sandra Coccuzzo Sampaio Vessoni; Simone Kashima; Svetoslav Nanev Slavov; Vincent Louis Viala |
| EPI_ISL_5649229, EPI_ISL_5665732 | CENTRO DE SAUDE DR MARIO DIAS DE AGUIAR CAPIVARI | Instituto Butantan | Antonio Jorge Martins; Claudia Renata dos Santos Barros; David Schlesinger; Debora Botequio Moretti; Dimas Tadeu Covas; Elaine Cristina Marqueze; Elaine Vieira Santos; Evandra Strazza Rodrigues; Heidge Fukumasu; Jayme Augusto de Souza-Neto; José Salvatore Leister Patané; Luiz Alcantara; Luiz Lehmann Coutinho; Maria Carolina Elias; Maurício Lacerda Nogueira; Rafael dos Santos Bezerra; Raul Machado Neto; Rejane Maria Tommasini Grotto; Ricardo Haddad; Sandra Coccuzzo Sampaio Vessoni; Simone Kashima; Svetoslav Nanev Slavov; Vincent Louis Viala |
| EPI_ISL_5648922 | CENTRO DE SAUDE DR NELSON DA CUNHA BASTOS | Instituto Butantan | Antonio Jorge Martins; Claudia Renata dos Santos Barros; David Schlesinger; Debora Botequio Moretti; Dimas Tadeu Covas; Elaine Cristina Marqueze; Elaine Vieira Santos; Evandra Strazza Rodrigues; Heidge Fukumasu; Jayme Augusto de Souza-Neto; José Salvatore Leister Patané; Luiz Alcantara; Luiz Lehmann Coutinho; Maria Carolina Elias; Maurício Lacerda Nogueira; Rafael dos Santos Bezerra; Raul Machado Neto; Rejane Maria Tommasini Grotto; Ricardo Haddad; Sandra Coccuzzo Sampaio Vessoni; Simone Kashima; Svetoslav Nanev Slavov; Vincent Louis Viala |
| EPI_ISL_5649442 | CENTRO DE SAUDE ESF IV ZONA RURAL DOMINGOS DE S SJRIOPARDO | Instituto Butantan | Antonio Jorge Martins; Claudia Renata dos Santos Barros; David Schlesinger; Debora Botequio Moretti; Dimas Tadeu Covas; Elaine Cristina Marqueze; Elaine Vieira Santos; Evandra Strazza Rodrigues; Heidge Fukumasu; Jayme Augusto de Souza-Neto; José Salvatore Leister Patané; Luiz Alcantara; Luiz Lehmann Coutinho; Maria Carolina Elias; Maurício Lacerda Nogueira; Rafael dos Santos Bezerra; Raul Machado Neto; Rejane Maria Tommasini Grotto; Ricardo Haddad; Sandra Coccuzzo Sampaio Vessoni; Simone Kashima; Svetoslav Nanev Slavov; Vincent Louis Viala |
| EPI_ISL_5649166, EPI_ISL_5649168, EPI_ISL_5649183 | CENTRO DE SAUDE I TACITO LEITE DE CARVALHO E SILVA | Instituto Butantan | Antonio Jorge Martins; Claudia Renata dos Santos Barros; David Schlesinger; Debora Botequio Moretti; Dimas Tadeu Covas; Elaine Cristina Marqueze; Elaine Vieira Santos; Evandra Strazza Rodrigues; Heidge Fukumasu; Jayme Augusto de Souza-Neto; José Salvatore Leister Patané; Luiz Alcantara; Luiz Lehmann Coutinho; Maria Carolina Elias; Maurício Lacerda Nogueira; Rafael dos Santos Bezerra; Raul Machado Neto; Rejane Maria Tommasini Grotto; Ricardo Haddad; Sandra Coccuzzo Sampaio Vessoni; Simone Kashima; Svetoslav Nanev Slavov; Vincent Louis Viala |
| EPI_ISL_5649164, EPI_ISL_5649179, EPI_ISL_5649213, EPI_ISL_5649218, EPI_ISL_5649220 | CENTRO DE SAUDE II | Instituto Butantan | Antonio Jorge Martins; Claudia Renata dos Santos Barros; David Schlesinger; Debora Botequio Moretti; Dimas Tadeu Covas; Elaine Cristina Marqueze; Elaine Vieira Santos; Evandra Strazza Rodrigues; Heidge Fukumasu; Jayme Augusto de Souza-Neto; José Salvatore Leister Patané; Luiz Alcantara; Luiz Lehmann Coutinho; Maria Carolina Elias; Maurício Lacerda Nogueira; Rafael dos Santos Bezerra; Raul Machado Neto; Rejane Maria Tommasini Grotto; Ricardo Haddad; Sandra Coccuzzo Sampaio Vessoni; Simone Kashima; Svetoslav Nanev Slavov; Vincent Louis Viala |
| EPI_ISL_5649172, EPI_ISL_5649174, EPI_ISL_5649239, EPI_ISL_5649339, EPI_ISL_5649342, EPI_ISL_5649344, EPI_ISL_5649347, EPI_ISL_5649366, EPI_ISL_5649371, EPI_ISL_5649373, EPI_ISL_5649374 | | | |
| see above | CENTRO DE SAUDE II BARRA BONITA | Instituto Butantan | Antonio Jorge Martins; Claudia Renata dos Santos Barros; David Schlesinger; Debora Botequio Moretti; Dimas Tadeu Covas; Elaine Cristina Marqueze; Elaine Vieira Santos; Evandra Strazza Rodrigues; Heidge Fukumasu; Jayme Augusto de Souza-Neto; José Salvatore Leister Patané; Luiz Alcantara; Luiz Lehmann Coutinho; Maria Carolina Elias; Maurício Lacerda Nogueira; Rafael dos Santos Bezerra; Raul Machado Neto; Rejane Maria Tommasini Grotto; Ricardo Haddad; Sandra Coccuzzo Sampaio Vessoni; Simone Kashima; Svetoslav Nanev Slavov; Vincent Louis Viala |
| EPI_ISL_5649198, EPI_ISL_5649240, EPI_ISL_5665761 | CENTRO DE SAUDE II DR JOSE PAIONE MOCOCA | Instituto Butantan | Antonio Jorge Martins; Claudia Renata dos Santos Barros; David Schlesinger; Debora Botequio Moretti; Dimas Tadeu Covas; Elaine Cristina Marqueze; Elaine Vieira Santos; Evandra Strazza Rodrigues; Heidge Fukumasu; Jayme Augusto de Souza-Neto; José Salvatore Leister Patané; Luiz Alcantara; Luiz Lehmann Coutinho; Maria Carolina Elias; Maurício Lacerda Nogueira; Rafael dos Santos Bezerra; Raul Machado Neto; Rejane Maria Tommasini Grotto; Ricardo Haddad; Sandra Coccuzzo Sampaio Vessoni; Simone Kashima; Svetoslav Nanev Slavov; Vincent Louis Viala |
| EPI_ISL_5649279 | CENTRO DE SAUDE II MAIRINQUE MAIRINQUE | Instituto Butantan | Antonio Jorge Martins; Claudia Renata dos Santos Barros; David Schlesinger; Debora Botequio Moretti; Dimas Tadeu Covas; Elaine Cristina Marqueze; Elaine Vieira Santos; Evandra Strazza Rodrigues; Heidge Fukumasu; Jayme Augusto de Souza-Neto; José Salvatore Leister Patané; Luiz Alcantara; Luiz Lehmann Coutinho; Maria Carolina Elias; Maurício Lacerda Nogueira; Rafael dos Santos Bezerra; Raul Machado Neto; Rejane Maria Tommasini Grotto; Ricardo Haddad; Sandra Coccuzzo Sampaio Vessoni; Simone Kashima; Svetoslav Nanev Slavov; Vincent Louis Viala |
| EPI_ISL_5649221, EPI_ISL_5665738 | CENTRO DE SAUDE III DE AGUAS DE SAO PEDRO | Instituto Butantan | Antonio Jorge Martins; Claudia Renata dos Santos Barros; David Schlesinger; Debora Botequio Moretti; Dimas Tadeu Covas; Elaine Cristina Marqueze; Elaine Vieira Santos; Evandra Strazza Rodrigues; Heidge Fukumasu; Jayme Augusto de Souza-Neto; José Salvatore Leister Patané; Luiz Alcantara; Luiz Lehmann Coutinho; Maria Carolina Elias; Maurício Lacerda Nogueira; Rafael dos Santos Bezerra; Raul Machado Neto; Rejane Maria Tommasini Grotto; Ricardo Haddad; Sandra Coccuzzo Sampaio Vessoni; Simone Kashima; Svetoslav Nanev Slavov; Vincent Louis Viala |
| EPI_ISL_5649453 | CENTRO DE SAUDE III SALES OLIVEIRA | Instituto Butantan | Antonio Jorge Martins; Claudia Renata dos Santos Barros; David Schlesinger; Debora Botequio Moretti; Dimas Tadeu Covas; Elaine Cristina Marqueze; Elaine Vieira Santos; Evandra Strazza Rodrigues; Heidge Fukumasu; Jayme Augusto de Souza-Neto; José Salvatore Leister Patané; Luiz Alcantara; Luiz Lehmann Coutinho; Maria Carolina Elias; Maurício Lacerda Nogueira; Rafael dos Santos Bezerra; Raul Machado Neto; Rejane Maria Tommasini Grotto; Ricardo Haddad; Sandra Coccuzzo Sampaio Vessoni; Simone Kashima; Svetoslav Nanev Slavov; Vincent Louis Viala |
| EPI_ISL_5649149, EPI_ISL_5649150, EPI_ISL_5649153, EPI_ISL_5649162, EPI_ISL_5649176, EPI_ISL_5649182, EPI_ISL_5649195, EPI_ISL_5649332 | | | |
| see above | CENTRO INTEGRADO DE SAUDE | Instituto Butantan | Antonio Jorge Martins; Claudia Renata dos Santos Barros; David Schlesinger; Debora Botequio Moretti; Dimas Tadeu Covas; Elaine Cristina Marqueze; Elaine Vieira Santos; Evandra Strazza Rodrigues; Heidge Fukumasu; Jayme Augusto de Souza-Neto; José Salvatore Leister Patané; Luiz Alcantara; Luiz Lehmann Coutinho; Maria Carolina Elias; Maurício Lacerda Nogueira; Rafael dos Santos Bezerra; Raul Machado Neto; Rejane Maria Tommasini Grotto; Ricardo Haddad; Sandra Coccuzzo Sampaio Vessoni; Simone Kashima; Svetoslav Nanev Slavov; Vincent Louis Viala |
| EPI_ISL_5649197 | CENTRO MEDICO DR NELSON SALOME DE CONCHAL | Instituto Butantan | Antonio Jorge Martins; Claudia Renata dos Santos Barros; David Schlesinger; Debora Botequio Moretti; Dimas Tadeu Covas; Elaine Cristina Marqueze; Elaine Vieira Santos; Evandra Strazza Rodrigues; Heidge Fukumasu; Jayme Augusto de Souza-Neto; José Salvatore Leister Patané; Luiz Alcantara; Luiz Lehmann Coutinho; Maria Carolina Elias; Maurício Lacerda Nogueira; Rafael dos Santos Bezerra; Raul Machado Neto; Rejane Maria Tommasini Grotto; Ricardo Haddad; Sandra Coccuzzo Sampaio Vessoni; Simone Kashima; Svetoslav Nanev Slavov; Vincent Louis Viala |
| EPI_ISL_2943233 | CENTRO MEDICO NAVAL SEMAR LAB BIOLOGIA MOLECULAR | CENTRO MEDICO NAVAL SEMAR LABORATORIO DE PATOLOGIA MOLECULAR | MEIXUEIRO CALDERON CLAUDIA. MORALES RAMIREZ OMAR. BALTIERRA URIBE SHANTAL. ZUNO CORONADO TEUTLI. GARCIA ALEGRIA IRMA.ALVAREZ JIMENEZ VIOLETA DEJANIRA. LOPEZ BUSTOS ABRAHAM. CABRERA GONZALEZ MOISES. ROMERO ROMAN EDGAR. LIEVANO ESQUINCA JAZMIN. FIGUEROA ANTONIO VICTOR. DIAZ PONCE MEDRANO JUAN ALBERTO. |
| EPI_ISL_5649194 | CENTRO SAUDE III DR JOSE TADEU MOURAO | Instituto Butantan | Antonio Jorge Martins; Claudia Renata dos Santos Barros; David Schlesinger; Debora Botequio Moretti; Dimas Tadeu Covas; Elaine Cristina Marqueze; Elaine Vieira Santos; Evandra Strazza Rodrigues; Heidge Fukumasu; Jayme Augusto de Souza-Neto; José Salvatore Leister Patané; Luiz Alcantara; Luiz Lehmann Coutinho; Maria Carolina Elias; Maurício Lacerda Nogueira; Rafael dos Santos Bezerra; Raul Machado Neto; Rejane Maria Tommasini Grotto; Ricardo Haddad; Sandra Coccuzzo Sampaio Vessoni; Simone Kashima; Svetoslav Nanev Slavov; Vincent Louis Viala |
| EPI_ISL_3046523 | CH ROUBAIX | CHU Lille - Laboratoire de Virologie | AIT YAHYA Emilie; ALIDJINOU Enagnon Kazali; BOCKET Laurence; CREPIN Michel; DEMAY Christophe; ENGELMANN Ilka; GEFFROY Sandrine; GUIGON Aurélie; LAMBERT Valérie; LAZREK Mouna; NOBILLIAUX Florian; PREVOST Brigitte; TCHANTCHOU NJOSSE YANICK; THUILLIER Caroline; TINEZ Claire |

| | | | |
|---|---|---|---|
| EPI_ISL_2983479 | CHC Andrée Rosemon | Institut Pasteur de la Guyane | Anne Lavergne; Antoine Enfissi; Arielle Salmier; Dominique Rousset |
| EPI_ISL_5648851, EPI_ISL_5648899, EPI_ISL_5648934, EPI_ISL_5648995, EPI_ISL_5649113 | COMPLEXO HOSPITALAR PADRE BENTO DE GUARULHOS | Instituto Butantan | Antonio Jorge Martins; Claudia Renata dos Santos Barros; David Schlesinger; Debora Botequio Moretti; Dimas Tadeu Covas; Elaine Cristina Marqueze; Elaine Vieira Santos; Evandra Strazza Rodrigues; Heidge Fukumasu; Jayme Augusto de Souza-Neto; José Salvatore Leister Patané; Luiz Alcantara; Luiz Lehmann Coutinho; Maria Carolina Elias; Maurício Lacerda Nogueira; Rafael dos Santos Bezerra; Raul Machado Neto; Rejane Maria Tommasini Grotto; Ricardo Haddad; Sandra Coccuzzo Sampaio Vessoni; Simone Kashima; Svetoslav Nanev Slavov; Vincent Louis Viala |
| EPI_ISL_5649184, EPI_ISL_5649203 | COORDENADORIA MUNICIPAL DE SAUDE DE IRACEMAPOLIS | Instituto Butantan | Antonio Jorge Martins; Claudia Renata dos Santos Barros; David Schlesinger; Debora Botequio Moretti; Dimas Tadeu Covas; Elaine Cristina Marqueze; Elaine Vieira Santos; Evandra Strazza Rodrigues; Heidge Fukumasu; Jayme Augusto de Souza-Neto; José Salvatore Leister Patané; Luiz Alcantara; Luiz Lehmann Coutinho; Maria Carolina Elias; Maurício Lacerda Nogueira; Rafael dos Santos Bezerra; Raul Machado Neto; Rejane Maria Tommasini Grotto; Ricardo Haddad; Sandra Coccuzzo Sampaio Vessoni; Simone Kashima; Svetoslav Nanev Slavov; Vincent Louis Viala |
| EPI_ISL_5648492, EPI_ISL_5649429 | CS ALVINLANDIA | Instituto Butantan | Antonio Jorge Martins; Claudia Renata dos Santos Barros; David Schlesinger; Debora Botequio Moretti; Dimas Tadeu Covas; Elaine Cristina Marqueze; Elaine Vieira Santos; Evandra Strazza Rodrigues; Heidge Fukumasu; Jayme Augusto de Souza-Neto; José Salvatore Leister Patané; Luiz Alcantara; Luiz Lehmann Coutinho; Maria Carolina Elias; Maurício Lacerda Nogueira; Rafael dos Santos Bezerra; Raul Machado Neto; Rejane Maria Tommasini Grotto; Ricardo Haddad; Sandra Coccuzzo Sampaio Vessoni; Simone Kashima; Svetoslav Nanev Slavov; Vincent Louis Viala |
| EPI_ISL_5659566 | CS DE APARECIDA DOESTE | Instituto Butantan | Antonio Jorge Martins; Claudia Renata dos Santos Barros; David Schlesinger; Debora Botequio Moretti; Dimas Tadeu Covas; Elaine Cristina Marqueze; Elaine Vieira Santos; Evandra Strazza Rodrigues; Heidge Fukumasu; Jayme Augusto de Souza-Neto; José Salvatore Leister Patané; Luiz Alcantara; Luiz Lehmann Coutinho; Maria Carolina Elias; Maurício Lacerda Nogueira; Rafael dos Santos Bezerra; Raul Machado Neto; Rejane Maria Tommasini Grotto; Ricardo Haddad; Sandra Coccuzzo Sampaio Vessoni; Simone Kashima; Svetoslav Nanev Slavov; Vincent Louis Viala |
| EPI_ISL_5659552, EPI_ISL_5659553, EPI_ISL_5659594, EPI_ISL_5659599, EPI_ISL_5659606 | CS DE COSMORAMA | Instituto Butantan | Antonio Jorge Martins; Claudia Renata dos Santos Barros; David Schlesinger; Debora Botequio Moretti; Dimas Tadeu Covas; Elaine Cristina Marqueze; Elaine Vieira Santos; Evandra Strazza Rodrigues; Heidge Fukumasu; Jayme Augusto de Souza-Neto; José Salvatore Leister Patané; Luiz Alcantara; Luiz Lehmann Coutinho; Maria Carolina Elias; Maurício Lacerda Nogueira; Rafael dos Santos Bezerra; Raul Machado Neto; Rejane Maria Tommasini Grotto; Ricardo Haddad; Sandra Coccuzzo Sampaio Vessoni; Simone Kashima; Svetoslav Nanev Slavov; Vincent Louis Viala |
| EPI_ISL_5648624, EPI_ISL_5659556, EPI_ISL_5659576, EPI_ISL_5659578, EPI_ISL_5659608 | CS DE ICEM | Instituto Butantan | Antonio Jorge Martins; Claudia Renata dos Santos Barros; David Schlesinger; Debora Botequio Moretti; Dimas Tadeu Covas; Elaine Cristina Marqueze; Elaine Vieira Santos; Evandra Strazza Rodrigues; Heidge Fukumasu; Jayme Augusto de Souza-Neto; José Salvatore Leister Patané; Luiz Alcantara; Luiz Lehmann Coutinho; Maria Carolina Elias; Maurício Lacerda Nogueira; Rafael dos Santos Bezerra; Raul Machado Neto; Rejane Maria Tommasini Grotto; Ricardo Haddad; Sandra Coccuzzo Sampaio Vessoni; Simone Kashima; Svetoslav Nanev Slavov; Vincent Louis Viala |
| EPI_ISL_5649464, EPI_ISL_5659564, EPI_ISL_5659580, EPI_ISL_5659596, EPI_ISL_5659618 | CS DE JACI | Instituto Butantan | Antonio Jorge Martins; Claudia Renata dos Santos Barros; David Schlesinger; Debora Botequio Moretti; Dimas Tadeu Covas; Elaine Cristina Marqueze; Elaine Vieira Santos; Evandra Strazza Rodrigues; Heidge Fukumasu; Jayme Augusto de Souza-Neto; José Salvatore Leister Patané; Luiz Alcantara; Luiz Lehmann Coutinho; Maria Carolina Elias; Maurício Lacerda Nogueira; Rafael dos Santos Bezerra; Raul Machado Neto; Rejane Maria Tommasini Grotto; Ricardo Haddad; Sandra Coccuzzo Sampaio Vessoni; Simone Kashima; Svetoslav Nanev Slavov; Vincent Louis Viala |
| EPI_ISL_5648964 | CS DE MIRASSOLANDIA | Instituto Butantan | Antonio Jorge Martins; Claudia Renata dos Santos Barros; David Schlesinger; Debora Botequio Moretti; Dimas Tadeu Covas; Elaine Cristina Marqueze; Elaine Vieira Santos; Evandra Strazza Rodrigues; Heidge Fukumasu; Jayme Augusto de Souza-Neto; José Salvatore Leister Patané; Luiz Alcantara; Luiz Lehmann Coutinho; Maria Carolina Elias; Maurício Lacerda Nogueira; Rafael dos Santos Bezerra; Raul Machado Neto; Rejane Maria Tommasini Grotto; Ricardo Haddad; Sandra Coccuzzo Sampaio Vessoni; Simone Kashima; Svetoslav Nanev Slavov; Vincent Louis Viala |
| EPI_ISL_5648655, EPI_ISL_5648746, EPI_ISL_5659543, EPI_ISL_5659545, EPI_ISL_5659555, EPI_ISL_5659562, EPI_ISL_5659573, EPI_ISL_5659583, EPI_ISL_5659590, EPI_ISL_5659591, EPI_ISL_5659597, EPI_ISL_5659601, EPI_ISL_5659609, EPI_ISL_5659612, EPI_ISL_5659616, EPI_ISL_5659627, EPI_ISL_5659635 | | | |
| see above | CS DE MONTE APRAZIVEL | Instituto Butantan | Antonio Jorge Martins; Claudia Renata dos Santos Barros; David Schlesinger; Debora Botequio Moretti; Dimas Tadeu Covas; Elaine Cristina Marqueze; Elaine Vieira Santos; Evandra Strazza Rodrigues; Heidge Fukumasu; Jayme Augusto de Souza-Neto; José Salvatore Leister Patané; Luiz Alcantara; Luiz Lehmann Coutinho; Maria Carolina Elias; Maurício Lacerda Nogueira; Rafael dos Santos Bezerra; Raul Machado Neto; Rejane Maria Tommasini Grotto; Ricardo Haddad; Sandra Coccuzzo Sampaio Vessoni; Simone Kashima; Svetoslav Nanev Slavov; Vincent Louis Viala |
| EPI_ISL_5648699, EPI_ISL_5659551, EPI_ISL_5659569, EPI_ISL_5659587 | CS DE NEVES PAULISTA | Instituto Butantan | Antonio Jorge Martins; Claudia Renata dos Santos Barros; David Schlesinger; Debora Botequio Moretti; Dimas Tadeu Covas; Elaine Cristina Marqueze; Elaine Vieira Santos; Evandra Strazza Rodrigues; Heidge Fukumasu; Jayme Augusto de Souza-Neto; José Salvatore Leister Patané; Luiz Alcantara; Luiz Lehmann Coutinho; Maria Carolina Elias; Maurício Lacerda Nogueira; Rafael dos Santos Bezerra; Raul Machado Neto; Rejane Maria Tommasini Grotto; Ricardo Haddad; Sandra Coccuzzo Sampaio Vessoni; Simone Kashima; Svetoslav Nanev Slavov; Vincent Louis Viala |
| EPI_ISL_5648907, EPI_ISL_5648943, EPI_ISL_5649039, EPI_ISL_5649052, EPI_ISL_5649083 | CS DE NOVO HORIZONTE | Instituto Butantan | Antonio Jorge Martins; Claudia Renata dos Santos Barros; David Schlesinger; Debora Botequio Moretti; Dimas Tadeu Covas; Elaine Cristina Marqueze; Elaine Vieira Santos; Evandra Strazza Rodrigues; Heidge Fukumasu; Jayme Augusto de Souza-Neto; José Salvatore Leister Patané; Luiz Alcantara; Luiz Lehmann Coutinho; Maria Carolina Elias; Maurício Lacerda Nogueira; Rafael dos Santos Bezerra; Raul Machado Neto; Rejane Maria Tommasini Grotto; Ricardo Haddad; Sandra Coccuzzo Sampaio Vessoni; Simone Kashima; Svetoslav Nanev Slavov; Vincent Louis Viala |
| EPI_ISL_5659572, EPI_ISL_5659585, EPI_ISL_5659626 | CS DE ONDA VERDE | Instituto Butantan | Antonio Jorge Martins; Claudia Renata dos Santos Barros; David Schlesinger; Debora Botequio Moretti; Dimas Tadeu Covas; Elaine Cristina Marqueze; Elaine Vieira Santos; Evandra Strazza Rodrigues; Heidge Fukumasu; Jayme Augusto de Souza-Neto; José Salvatore Leister Patané; Luiz Alcantara; Luiz Lehmann Coutinho; Maria Carolina Elias; Maurício Lacerda Nogueira; Rafael dos Santos Bezerra; Raul Machado Neto; Rejane Maria Tommasini Grotto; Ricardo Haddad; Sandra Coccuzzo Sampaio Vessoni; Simone Kashima; Svetoslav Nanev Slavov; Vincent Louis Viala |
| EPI_ISL_5648845 | CS DE OSCAR BRESSANE PSF | Instituto Butantan | Antonio Jorge Martins; Claudia Renata dos Santos Barros; David Schlesinger; Debora Botequio Moretti; Dimas Tadeu Covas; Elaine Cristina Marqueze; Elaine Vieira Santos; Evandra Strazza Rodrigues; Heidge Fukumasu; Jayme Augusto de Souza-Neto; José Salvatore Leister Patané; Luiz Alcantara; Luiz Lehmann Coutinho; Maria Carolina Elias; Maurício Lacerda Nogueira; Rafael dos Santos Bezerra; Raul Machado Neto; Rejane Maria Tommasini Grotto; Ricardo Haddad; Sandra Coccuzzo Sampaio Vessoni; Simone Kashima; Svetoslav Nanev Slavov; Vincent Louis Viala |
| EPI_ISL_5648809, EPI_ISL_5648839, EPI_ISL_5648872, EPI_ISL_5648972, EPI_ISL_5649015 | CS DE PALESTINA | Instituto Butantan | Antonio Jorge Martins; Claudia Renata dos Santos Barros; David Schlesinger; Debora Botequio Moretti; Dimas Tadeu Covas; Elaine Cristina Marqueze; Elaine Vieira Santos; Evandra Strazza Rodrigues; Heidge Fukumasu; Jayme Augusto de Souza-Neto; José Salvatore Leister Patané; Luiz Alcantara; Luiz Lehmann Coutinho; Maria Carolina Elias; Maurício Lacerda Nogueira; Rafael dos Santos Bezerra; Raul Machado Neto; Rejane Maria Tommasini Grotto; Ricardo Haddad; Sandra Coccuzzo Sampaio Vessoni; Simone Kashima; Svetoslav Nanev Slavov; Vincent Louis Viala |
| EPI_ISL_5648691, EPI_ISL_5659560, EPI_ISL_5659568 | CS DE PALMARES PAULISTA | Instituto Butantan | Antonio Jorge Martins; Claudia Renata dos Santos Barros; David Schlesinger; Debora Botequio Moretti; Dimas Tadeu Covas; Elaine Cristina Marqueze; Elaine Vieira Santos; Evandra Strazza Rodrigues; Heidge Fukumasu; Jayme Augusto de Souza-Neto; José Salvatore Leister Patané; Luiz Alcantara; Luiz Lehmann Coutinho; Maria Carolina Elias; Maurício Lacerda Nogueira; Rafael dos Santos Bezerra; Raul Machado Neto; Rejane Maria Tommasini Grotto; Ricardo Haddad; Sandra Coccuzzo Sampaio Vessoni; Simone Kashima; Svetoslav Nanev Slavov; Vincent Louis Viala |
| EPI_ISL_5648800 | CS DE POPULINA | Instituto Butantan | Antonio Jorge Martins; Claudia Renata dos Santos Barros; David Schlesinger; Debora Botequio Moretti; Dimas Tadeu Covas; Elaine Cristina Marqueze; Elaine Vieira Santos; Evandra Strazza Rodrigues; Heidge Fukumasu; Jayme Augusto de Souza-Neto; José Salvatore Leister Patané; Luiz Alcantara; Luiz Lehmann Coutinho; Maria Carolina Elias; Maurício Lacerda Nogueira; Rafael dos Santos Bezerra; Raul Machado Neto; Rejane Maria Tommasini Grotto; Ricardo Haddad; Sandra Coccuzzo Sampaio Vessoni; Simone Kashima; Svetoslav Nanev Slavov; Vincent Louis Viala |
| EPI_ISL_5649151 | CS DE ROSANA | Instituto Butantan | Antonio Jorge Martins; Claudia Renata dos Santos Barros; David Schlesinger; Debora Botequio Moretti; Dimas Tadeu Covas; Elaine Cristina Marqueze; Elaine Vieira Santos; Evandra Strazza Rodrigues; Heidge Fukumasu; Jayme Augusto de Souza-Neto; José Salvatore Leister Patané; Luiz Alcantara; Luiz Lehmann Coutinho; Maria Carolina Elias; Maurício Lacerda Nogueira; Rafael dos Santos Bezerra; Raul Machado Neto; Rejane Maria Tommasini Grotto; Ricardo Haddad; Sandra Coccuzzo Sampaio Vessoni; Simone Kashima; Svetoslav Nanev Slavov; Vincent Louis Viala |
| EPI_ISL_5659610, EPI_ISL_5659634 | CS DE SANTA ALBERTINA | Instituto Butantan | Antonio Jorge Martins; Claudia Renata dos Santos Barros; David Schlesinger; Debora Botequio Moretti; Dimas Tadeu Covas; Elaine Cristina Marqueze; Elaine Vieira Santos; Evandra Strazza Rodrigues; Heidge Fukumasu; Jayme Augusto de Souza-Neto; José Salvatore Leister Patané; Luiz Alcantara; Luiz Lehmann Coutinho; Maria Carolina Elias; Maurício Lacerda Nogueira; Rafael dos Santos Bezerra; Raul Machado Neto; Rejane Maria Tommasini Grotto; Ricardo Haddad; Sandra Coccuzzo Sampaio Vessoni; Simone Kashima; Svetoslav Nanev Slavov; Vincent Louis Viala |
| EPI_ISL_5648980, EPI_ISL_5649031, EPI_ISL_5649135 | CS DE SEBASTIANOPOLIS DO SUL | Instituto Butantan | Antonio Jorge Martins; Claudia Renata dos Santos Barros; David Schlesinger; Debora Botequio Moretti; Dimas Tadeu Covas; Elaine Cristina Marqueze; Elaine Vieira Santos; Evandra Strazza Rodrigues; Heidge Fukumasu; Jayme Augusto de Souza-Neto; José Salvatore Leister Patané; Luiz Alcantara; Luiz Lehmann Coutinho; Maria Carolina Elias; Maurício Lacerda Nogueira; Rafael dos Santos Bezerra; Raul Machado Neto; Rejane Maria Tommasini Grotto; Ricardo Haddad; Sandra Coccuzzo Sampaio Vessoni; Simone Kashima; Svetoslav Nanev Slavov; Vincent Louis Viala |
| EPI_ISL_5649445, EPI_ISL_5659621 | CS DE TURMALINA | Instituto Butantan | Antonio Jorge Martins; Claudia Renata dos Santos Barros; David Schlesinger; Debora Botequio Moretti; Dimas Tadeu Covas; Elaine Cristina Marqueze; Elaine Vieira Santos; Evandra Strazza Rodrigues; Heidge Fukumasu; Jayme Augusto de Souza-Neto; José Salvatore Leister Patané; Luiz Alcantara; Luiz Lehmann Coutinho; Maria Carolina Elias; Maurício Lacerda Nogueira; Rafael dos Santos Bezerra; Raul Machado Neto; Rejane Maria Tommasini Grotto; Ricardo Haddad; Sandra Coccuzzo Sampaio Vessoni; Simone Kashima; Svetoslav Nanev Slavov; Vincent Louis Viala |
| EPI_ISL_5648266, EPI_ISL_5648488, EPI_ISL_5659548, EPI_ISL_5659550, EPI_ISL_5659575, EPI_ISL_5659592, EPI_ISL_5659605, EPI_ISL_5659611 | | | |
| see above | CS DE VALENTIM GENTIL | Instituto Butantan | Antonio Jorge Martins; Claudia Renata dos Santos Barros; David Schlesinger; Debora Botequio Moretti; Dimas Tadeu Covas; Elaine Cristina Marqueze; Elaine Vieira Santos; Evandra Strazza Rodrigues; Heidge Fukumasu; Jayme Augusto de Souza-Neto; José Salvatore Leister Patané; Luiz Alcantara; Luiz Lehmann Coutinho; Maria Carolina Elias; Maurício Lacerda Nogueira; Rafael dos Santos Bezerra; Raul Machado Neto; Rejane Maria Tommasini Grotto; Ricardo Haddad; Sandra Coccuzzo Sampaio Vessoni; Simone Kashima; Svetoslav Nanev Slavov; Vincent Louis Viala |
| EPI_ISL_5649118 | CS II DR JOSE CARQUEIJO SCRPARDO | Instituto Butantan | Antonio Jorge Martins; Claudia Renata dos Santos Barros; David Schlesinger; Debora Botequio Moretti; Dimas Tadeu Covas; Elaine Cristina Marqueze; Elaine Vieira Santos; Evandra Strazza Rodrigues; Heidge Fukumasu; Jayme Augusto de Souza-Neto; José Salvatore Leister Patané; Luiz Alcantara; Luiz Lehmann Coutinho; Maria Carolina Elias; Maurício Lacerda Nogueira; Rafael dos Santos Bezerra; Raul Machado Neto; Rejane Maria Tommasini Grotto; Ricardo Haddad; Sandra Coccuzzo Sampaio Vessoni; Simone Kashima; Svetoslav Nanev Slavov; Vincent Louis Viala |
| EPI_ISL_5649173, EPI_ISL_5649185 | CS III DR ANTONIO ANADAO SSGRAMA | Instituto Butantan | Antonio Jorge Martins; Claudia Renata dos Santos Barros; David Schlesinger; Debora Botequio Moretti; Dimas Tadeu Covas; Elaine Cristina Marqueze; Elaine Vieira Santos; Evandra Strazza Rodrigues; Heidge Fukumasu; Jayme Augusto de Souza-Neto; José Salvatore Leister Patané; Luiz Alcantara; Luiz Lehmann Coutinho; Maria Carolina Elias; Maurício Lacerda Nogueira; Rafael dos Santos Bezerra; Raul Machado Neto; Rejane Maria Tommasini Grotto; Ricardo Haddad; Sandra Coccuzzo Sampaio Vessoni; Simone Kashima; Svetoslav Nanev Slavov; Vincent Louis Viala |
| EPI_ISL_4005587, EPI_ISL_4005588, EPI_ISL_4005589, EPI_ISL_4005595 | California Department of Public Health | California Department of Public Health | CDPH IDLB COVIDNet et al |
| EPI_ISL_7507368 | Cedars-Sinai Medical Center, Molecular Pathology Laboratory of Department of Pathology & Laboratory Medicine and Genomic Core | Cedars-Sinai Medical Center, Molecular Pathology Laboratory of Department of Pathology & Laboratory Medicine and Genomic Core | Brian Davis; Eric Vail; Jasmine T Plummer; Jorge Mario Sincuir Martinez; Stephanie Chen; Wenjuan Zhang |
| EPI_ISL_3462533, EPI_ISL_3462536, EPI_ISL_3462550, EPI_ISL_3462569, EPI_ISL_3462572, EPI_ISL_3462573, EPI_ISL_3462576, EPI_ISL_3462579, EPI_ISL_3462582, EPI_ISL_3462583, EPI_ISL_3462591, EPI_ISL_3462604, EPI_ISL_3462605, EPI_ISL_3462614, EPI_ISL_3462616, EPI_ISL_3462617, EPI_ISL_3462618, EPI_ISL_3462619, EPI_ISL_3462664, EPI_ISL_3462665, EPI_ISL_3462666, EPI_ISL_3462667, EPI_ISL_3462668, EPI_ISL_3462669, EPI_ISL_3462670, EPI_ISL_3462671, EPI_ISL_3462672, EPI_ISL_3462673, EPI_ISL_3462674, EPI_ISL_3462675, EPI_ISL_3462676, EPI_ISL_3462677, EPI_ISL_3464723, EPI_ISL_3464727 | | | |
| see above | Central Laboratory, Bureau of Public Health (BOG) and Academic Hospital Paramaribo | Erasmus Medical Center | Bas B Oude Munnink; Cherise Beek; Consuella Partowidjojo; Dion Gajadin; Ed PF IJzerman; Emmanuelle Munger; Gary Gummels; Ingrid SK Krishnadath; Lycke Woittiez; Marion PG Koopmans; Mireille Van de Veer; Phyllis Pinas; Princes Wongsowidjojo; Radjesh Ori; Ranisha Doerbalie; Rohma Banwari; Soeradj Harkisoen; Stephen Vreden; Tilotmadebie Ramlal; Verne Nanhoe |
| EPI_ISL_4296379, EPI_ISL_4296427 | Central Medical Laboratory | Baylor College of Medicine | Adrianna Maliga; Alexander Kneubehl; Allison Lino; Gerhaldine Morazan; Kristy Murray; Russell Manzanero; Sarah Gunter; Sarah Strobel; Shannon Ronca |
| EPI_ISL_3255211, EPI_ISL_3255215, EPI_ISL_3255222, EPI_ISL_3255224, EPI_ISL_3255228, EPI_ISL_3255229, EPI_ISL_3255234, EPI_ISL_3255235, EPI_ISL_3255236, EPI_ISL_3255238, EPI_ISL_3255239, EPI_ISL_3255240, EPI_ISL_3255242, EPI_ISL_3255243, EPI_ISL_3255247, EPI_ISL_3255249, EPI_ISL_3255251, EPI_ISL_3255254, EPI_ISL_3255257, EPI_ISL_3255259, EPI_ISL_3255261, EPI_ISL_3255263, EPI_ISL_3255264, EPI_ISL_3255270, EPI_ISL_3255274, EPI_ISL_3255276, EPI_ISL_3255278, EPI_ISL_3266072, EPI_ISL_3266073, EPI_ISL_4417758 | | | |
| see above | Central Public Health Laboratory - LACEN - Bahia, Salvador, Brazil | Central Public Health Laboratory - LACEN -Bahia, Salvador, Brazil | Arabela Leal; Breno Dominguez; Felicidade Pereira; Gabriela Menezes; Jaqueline Gomes; Lenisa Dandara; Luciana Oliveira; Luiz Alcantara; Marcela Gómez; Marta Giovanetti; Patrícia Cajado; Stephane Tosta; Vagner Fonseca; Vanessa Nardy |
| EPI_ISL_3147561 | Centre Hospitalier du Nord | Laboratoire national de sante, Microbiology, Microbial Genomics Platform | Anke Wienecke-Baldacchino; Catherine Ragimbeau; Elodie Solarino; Fatiha Boulmerka; Fatu Djabi; Jacqueline Parmentier; Jessica Tapp; Lise Pignon; Raoul Salmon; Tamir Abdelrahman; Virginie Jover |

| | | | |
|---|---|---|---|
| EPI_ISL_5926834, EPI_ISL_5926835 | Centro de Infectologia Charles Mérieux/ Laboratório Rodolphe Mérieux, FUNDHACRE | Bioinformatics Laboratory / LNCC | Alessandra P Lamarca; Alexandra L Gerber; Aline de Freitas Souza; Ana Paula de C Guimarães; Ana Tereza R Vasconcelos; Andreas Stocker; Cirley Maria de Oliveira Lobato; Douglas Terra Machado; Janaína Mazaro; Janete Taynã Nascimento Rodrigues; Luiz Fellype Alves de Souza; Luiz G P de Almeida; Ronaldo da Silva F Jr; Rutilene B. Souza |
| EPI_ISL_2942438, EPI_ISL_2942440, EPI_ISL_2942450, EPI_ISL_2942460, EPI_ISL_2942463 | Centro de Investigacion Biomedica de Occidente (CIBO) | Unidad de Genomica Avanzada | ; Alejandra Garcia-Gasca; Alejandra Hernandez-Teran; Alejandro Sanchez-Flores; Alfredo Herrera-Estrella; Alicia Ocaña-Mondragon; Andreu Comas-Garcia; Angel Gustavo Salas-Lais; Antonio Loza Roman; Bernardo Martinez-Miguel; Blanca Taboada; Brenda Irasema Maldonado-Meza; Bruno Gomez-Gil; Carla Ivon Herrera-Najera; Carlos F. Arias; Celia Boukadida; Celida Duque Molina; Celida Martinez- Rodriguez; Clara Esperanza Santacruz-Tinoco; Concepcion Grajales-Muñiz; Consorcio Mexicano de Vigilancia Genomica (CoViGen-Mex). Authors (in alphabetical order): Julio Elias Alvarado-Yaah; Cristobal Chaidez-Quiroz; Daniel Fregoso-Rueda; Daniel Lira Morales; Eduardo Becerril-Vargas; Fernando Fontove-Herrera; Fidencio Mejia-Nepomuceno; Francisco Pulido; Gloria Elena Espinosa-Ayala; Gloria Maria Molina-Salinas; Gloria Vazquez; Hector Esteban Paz-Juarez; Hector Montoya-Fuentes; Helen Haydee Fernanda Ramirez-Plascencia; Irvin Gonzalez-Lopez; Jean Pierre Gonzalez; Jesus Hernandez; Joel Armando Vazquez-Perez.; Jorge Salas-Hernandez; Jose Antonio Enciso-Moreno; Jose Arturo Martinez-Orozco; Jose Esteban Muñoz-Medina; Jose de Jesus Nuñez-Contreras; Juan Bautista Chale-Dzul; Julissa Enciso-Ibarra; Luis Alberto Ochoa-Carrera; Margarita Matias-Florentino; Maria Guadalupe Santiago-Mauricio; Maria Guadalupe de Jesus Mireles-Rivera; Mario Mujica-Sanchez; Marissa Perez-Garcia; Nelly Selem-Mojica; Pavel Isa; Ricardo Ciria Merce; Ricardo Grande; Rosa Maria Gutierrez Rios; Santiago avila-Rios; Selene Zarate; Susana Lopez; Veronica Mata-Haro; Victor Eduardo Garcia-Arias; Victor Hugo Borja-Aburto |
| EPI_ISL_2942510, EPI_ISL_2942756 | Centro de Investigacion Biomedica del Noreste (CIBIN) | Unidad de Genomica Avanzada | ; Alejandra Garcia-Gasca; Alejandra Hernandez-Teran; Alejandro Sanchez-Flores; Alfredo Herrera-Estrella; Alicia Ocaña-Mondragon; Andreu Comas-Garcia; Angel Gustavo Salas-Lais; Antonio Loza Roman; Bernardo Martinez-Miguel; Blanca Taboada; Brenda Irasema Maldonado-Meza; Bruno Gomez-Gil; Carla Ivon Herrera-Najera; Carlos F. Arias; Celia Boukadida; Celida Duque Molina; Celida Martinez- Rodriguez; Clara Esperanza Santacruz-Tinoco; Concepcion Grajales-Muñiz; Consorcio Mexicano de Vigilancia Genomica (CoViGen-Mex). Authors (in alphabetical order): Julio Elias Alvarado-Yaah; Cristobal Chaidez-Quiroz; Daniel Fregoso-Rueda; Daniel Lira Morales; Eduardo Becerril-Vargas; Fernando Fontove-Herrera; Fidencio Mejia-Nepomuceno; Francisco Pulido; Gloria Elena Espinosa-Ayala; Gloria Maria Molina-Salinas; Gloria Vazquez; Hector Esteban Paz-Juarez; Hector Montoya-Fuentes; Helen Haydee Fernanda Ramirez-Plascencia; Irvin Gonzalez-Lopez; Jean Pierre Gonzalez; Jesus Hernandez; Joel Armando Vazquez-Perez.; Jorge Salas-Hernandez; Jose Antonio Enciso-Moreno; Jose Arturo Martinez-Orozco; Jose Esteban Muñoz-Medina; Jose de Jesus Nuñez-Contreras; Juan Bautista Chale-Dzul; Julissa Enciso-Ibarra; Luis Alberto Ochoa-Carrera; Margarita Matias-Florentino; Maria Guadalupe Santiago-Mauricio; Maria Guadalupe de Jesus Mireles-Rivera; Mario Mujica-Sanchez; Marissa Perez-Garcia; Nelly Selem-Mojica; Pavel Isa; Ricardo Ciria Merce; Ricardo Grande; Rosa Maria Gutierrez Rios; Santiago avila-Rios; Selene Zarate; Susana Lopez; Veronica Mata-Haro; Victor Eduardo Garcia-Arias; Victor Hugo Borja-Aburto |
| EPI_ISL_4405204 | Centro de Investigaciones Básicas y Aplicadas, UNNOBA | Área de Secuenciación del Laboratorio de Virología del Hospital de Niños Dr. Ricardo Gutierrez on behalf of 'Proyecto Argentino Interinstitucional de genomica de SARS-CoV-2' (PAIS Consortium) | A; Acuña; Alaniz; Bagnis; Barbero; Bonadeo; Brandone; C; Cassarini; Castro; Chimento; Cristina; D; Demarchi; Español; F; Fernández; G; Goya; Gracia Balbi; Hernández del Pin; I; Icardi; L; LE; Lusso; M; MI; MS; Menite; Moroni; Morro; N; Nabaes Jodar; Natale; P; Palumbo; Pasquinelli; Perrone; R; Romano; Rosales; S; Sevic; Spinelli; V; Valinotto; Valla; Viegas, M.; Villafañe; Vitale |
| EPI_ISL_4498596 | Children's Hospital of Philadelphia | Planet | Ahmed M. Moustafa; Alex Arvanitis; Azad Ahmed; Brandy Neide; Colleen Bianco; Josh Chang Mell; Lidiya Denu; Paul J. Planet; Rebecca M. Harris; Susan Coffin; Swetha Rajagopal |
| EPI_ISL_3242501, EPI_ISL_3242527, EPI_ISL_3242528, EPI_ISL_3242551, EPI_ISL_3242709 | Clinica INDISA | "Facultad de Ciencias de la Vida, UNAB" | "Claudio Meneses; Ariel Orellana"; Claudio Olmos; Daniel Leon; Dayan Sanhueza; Eduardo Castro; Gonzalo Campaña; Macarena Bastias; Paola Pidal; Ricardo Yusta; Sebastian Wolter; Susana Saez; Víctor Monreal; Waldo Diaz |
| EPI_ISL_2833668 | Clinical Virology | Clinical Bacteriology | Adrian Egli; Alfredo Mari; Fanny Wegner; Hans Hirsch; Helena MB Seth-Smith; Julia Bielicki; Karoline Leuzinger; Manuel Battegay; Tim Roloff |
| EPI_ISL_4300470, EPI_ISL_4300484 | Color Genomics | Chiu Laboratory, University of California, San Francisco | Alicia Sotomayor-Gonzalez; Alicia Zhou; Baolin Wang; Charles Chiu; Daniel Wong; Erika Torres; Madyson Wanner; Noah Brazer; Scott Topper; Venice Servellita; Yueyuan Zhang |
| EPI_ISL_3404035, EPI_ISL_3537230, EPI_ISL_3583184, EPI_ISL_3583794 | Colorado Department of Public Health and Environment | Colorado Department of Public Health and Environment | Alexandria Rossheim; Diana Ir; Emily A. Travanty; Laura Bankers; Mandy Waters; Michael A. Martin; Michael Martin; Molly C. Hetherington-Rauth; Sarah Elizabeth Totten; Shannon R. Matzinger |
| EPI_ISL_2930322, EPI_ISL_2930325, EPI_ISL_2930327, EPI_ISL_2930330, EPI_ISL_2930338 | Cotugno | TIGEM | Antonio Grimaldi Patrizia Annunziata Francesco Panariello Biancamaria Pierri Claudia Tiberio Teresa Giuliano Valentina Bouche Chiara Colantuono Maria Concetta Cuomo Denise Di Concilio Lucio Di Filippo Anna Manfredi Marcello Salvi Antonio Limone Luigi Atripaldi Pellegrino Cerino Andrea Ballabio Davide Cacchiarelli |
| EPI_ISL_2839697, EPI_ISL_2839737, EPI_ISL_2839756, EPI_ISL_2839867, EPI_ISL_2839904, EPI_ISL_2839964, EPI_ISL_2839969, EPI_ISL_2839971, EPI_ISL_2839973, EPI_ISL_2839977, EPI_ISL_2941919, EPI_ISL_2941926, EPI_ISL_2942040, EPI_ISL_3064415, EPI_ISL_3064421, EPI_ISL_3064475, EPI_ISL_3064478, EPI_ISL_3064483, EPI_ISL_3064662, EPI_ISL_3064663, EPI_ISL_3270123, EPI_ISL_3270296, EPI_ISL_3270323 | | | |
| see above | Curative Labs | Curative Labs | Elias L. Salfati; Eugenia Khorosheva; George Way; J.Cesar Ignacio-Espinoza; Janet Chen; Mikhail Hanewich-Hollatz; Nabjot Sandhu; Sophia Quasem; Vladimir Slepnev; Zhiyi Xie |
| EPI_ISL_2941662, EPI_ISL_2941681, EPI_ISL_2941683, EPI_ISL_2941687, EPI_ISL_3086146 | Curative Labs | New Mexico Department of Health Scientific Laboratory | ; Anastacia Griego-Fisher; D'eldra Malone; Ellie Johnson; Jennifer Benoit; Keila Gutierrez; Linda Salazar; Ratheesh Rajan |
| EPI_ISL_3664274, EPI_ISL_4422976, EPI_ISL_4422977 | DASA | DASA | Adriano Bonaldi; Angelica Hristov; Annelise Lopes; Bianca Cota; Cristina Oliveira; Jose Levi; Lidia Yamamoto; Paulo Pierry; Rodrigo Guarischi; Rodrigo Salazar |
| EPI_ISL_2876488, EPI_ISL_2876490 | DC Public Health Lab/ Dept. of Forensic Sciences | DC Public Health Lab/ Dept. of Forensic Sciences | Brittany Hamilton; Connie Maza; David Payne; Elizabeth Zelaya; Janis Doss; Jocelyn Hauser; Monica Mann; Sarah Scott; Scott Nguyen |
| EPI_ISL_5648573, EPI_ISL_5648667, EPI_ISL_5648668, EPI_ISL_5648913 | DEPARTAMENTO MUNICIPAL DE SAUDE DE PARAGUACU PAULISTA | Instituto Butantan | Antonio Jorge Martins; Claudia Renata dos Santos Barros; David Schlesinger; Debora Botequio Moretti; Dimas Tadeu Covas; Elaine Cristina Marqueze; Elaine Vieira Santos; Evandra Strazza Rodrigues; Heidge Fukumasu; Jayme Augusto de Souza-Neto; José Salvatore Leister Patané; Luiz Alcantara; Luiz Lehmann Coutinho; Maria Carolina Elias; Maurício Lacerda Nogueira; Rafael dos Santos Bezerra; Raul Machado Neto; Rejane Maria Tommasini Grotto; Ricardo Haddad; Sandra Coccuzzo Sampaio Vessoni; Simone Kashima; Svetoslav Nanev Slavov; Vincent Louis Viala |
| EPI_ISL_5648374, EPI_ISL_5648474 | DIRETORIA MUNICIPAL DE SAUDE | Instituto Butantan | Antonio Jorge Martins; Claudia Renata dos Santos Barros; David Schlesinger; Debora Botequio Moretti; Dimas Tadeu Covas; Elaine Cristina Marqueze; Elaine Vieira Santos; Evandra Strazza Rodrigues; Heidge Fukumasu; Jayme Augusto de Souza-Neto; José Salvatore Leister Patané; Luiz Alcantara; Luiz Lehmann Coutinho; Maria Carolina Elias; Maurício Lacerda Nogueira; Rafael dos Santos Bezerra; Raul Machado Neto; Rejane Maria Tommasini Grotto; Ricardo Haddad; Sandra Coccuzzo Sampaio Vessoni; Simone Kashima; Svetoslav Nanev Slavov; Vincent Louis Viala |
| EPI_ISL_3404551, EPI_ISL_3404552 | DNAGYN | LGBio (Laboratorio de Genetica & Biodiversidade) | Alex Honda Bernardes; Amanda Alves de Melo; Aparecido Divino da Cruz; Cintia Pelegrineti Targueta de Azevedo Brito; Daniela de Melo e Silva; Elisangela de Paula Silveira Lacerda; Juliana Santana de Curcio; Luiz Augusto Pereira; Marc Alexandre Duarte Gigonzac; Mariana Pires de Campos Telles; Ramilla dos Santos Braga; Renata de Oliveira Dias; Rhewter Nunes; Thais Cidalia Vieira Gigonzac; Thais Guimarães Castro; Thays Millena Alves Pedroso |
| EPI_ISL_2747517, EPI_ISL_2748057, EPI_ISL_2748372, EPI_ISL_2784627 | Department of Bacteria, Parasites and Fungi, Statens Serum Institut, Copenhagen, Denmark | Statens Serum Institut Bioinformatics and Microbial Genomics | Danish Covid-19 Genome Consortium |
| EPI_ISL_2709754, EPI_ISL_2832360 | Department of Virology, Istituto Zooprofilattico Sperimentale del Lazio e della Toscana (IZSLT) | Department of General Diagnostics; Department of Virology; Istituto Zooprofilattico Sperimentale del Lazio e della Toscana (IZSLT) | Alessia Franco; Antonella Cersini; Antonio Battisti.; Elena L. Diaconu; Fabiola Feltrin; Giuseppe Manna; Patricia Alba; Raffaela Conti; Teresa Scicluna; Virginia Carfora |
| EPI_ISL_3237628 | Diagnosticum - Labor Neukirchen | Robert Koch Institute | |
| EPI_ISL_2862759, EPI_ISL_2862767, EPI_ISL_2981439, EPI_ISL_2981449, EPI_ISL_3056731, EPI_ISL_3056773, EPI_ISL_3136569 | | | |
| see above | Dutch COVID-19 response team | National Institute for Public Health and the Environment (RIVM) | Adam Meijer; AnneMarie van den Brandt; Annelies Kroneman; Bas van der Veer; Chantal Reusken; Dennis Schmitz; Dirk Eggink; Euníce Then; Florian Zwagemaker; Harry Vennema; Jeroen Cremer; Karim Hajji; Kim Freriks; Lisa Wijsman; Lynn Aarts; Melissa van Tuil; Ryanne Jaarsma; Sanne Bos; Sharon van den Brink; on behalf of the national COVID-19 response team |
| EPI_ISL_4059815, EPI_ISL_4059818, EPI_ISL_4060093 | Dynacare Manitoba | Manitoba Cadham Provincial Laboratory | David Alexander; Hilary Racher; Hongbin Li; Melissa Desaulnier; Tintu Abraham |
| EPI_ISL_5659586 | E S F DR EDSON GIANETTO JD CARLOS CASSETARI | Instituto Butantan | Antonio Jorge Martins; Claudia Renata dos Santos Barros; David Schlesinger; Debora Botequio Moretti; Dimas Tadeu Covas; Elaine Cristina Marqueze; Elaine Vieira Santos; Evandra Strazza Rodrigues; Heidge Fukumasu; Jayme Augusto de Souza-Neto; José Salvatore Leister Patané; Luiz Alcantara; Luiz Lehmann Coutinho; Maria Carolina Elias; Maurício Lacerda Nogueira; Rafael dos Santos Bezerra; Raul Machado Neto; Rejane Maria Tommasini Grotto; Ricardo Haddad; Sandra Coccuzzo Sampaio Vessoni; Simone Kashima; Svetoslav Nanev Slavov; Vincent Louis Viala |
| EPI_ISL_5648278, EPI_ISL_5648461, EPI_ISL_5648595 | EMERGENCIA RESPIRATORIA DE NOVA GRANADA | Instituto Butantan | Antonio Jorge Martins; Claudia Renata dos Santos Barros; David Schlesinger; Debora Botequio Moretti; Dimas Tadeu Covas; Elaine Cristina Marqueze; Elaine Vieira Santos; Evandra Strazza Rodrigues; Heidge Fukumasu; Jayme Augusto de Souza-Neto; José Salvatore Leister Patané; Luiz Alcantara; Luiz Lehmann Coutinho; Maria Carolina Elias; Maurício Lacerda Nogueira; Rafael dos Santos Bezerra; Raul Machado Neto; Rejane Maria Tommasini Grotto; Ricardo Haddad; Sandra Coccuzzo Sampaio Vessoni; Simone Kashima; Svetoslav Nanev Slavov; Vincent Louis Viala |
| EPI_ISL_5648489, EPI_ISL_5648690, EPI_ISL_5648725 | ESF COHAB IV | Instituto Butantan | Antonio Jorge Martins; Claudia Renata dos Santos Barros; David Schlesinger; Debora Botequio Moretti; Dimas Tadeu Covas; Elaine Cristina Marqueze; Elaine Vieira Santos; Evandra Strazza Rodrigues; Heidge Fukumasu; Jayme Augusto de Souza-Neto; José Salvatore Leister Patané; Luiz Alcantara; Luiz Lehmann Coutinho; Maria Carolina Elias; Maurício Lacerda Nogueira; Rafael dos Santos Bezerra; Raul Machado Neto; Rejane Maria Tommasini Grotto; Ricardo Haddad; Sandra Coccuzzo Sampaio Vessoni; Simone Kashima; Svetoslav Nanev Slavov; Vincent Louis Viala |
| EPI_ISL_5648772, EPI_ISL_5649026 | ESF NOVA TANABI II | Instituto Butantan | Antonio Jorge Martins; Claudia Renata dos Santos Barros; David Schlesinger; Debora Botequio Moretti; Dimas Tadeu Covas; Elaine Cristina Marqueze; Elaine Vieira Santos; Evandra Strazza Rodrigues; Heidge Fukumasu; Jayme Augusto de Souza-Neto; José Salvatore Leister Patané; Luiz Alcantara; Luiz Lehmann Coutinho; Maria Carolina Elias; Maurício Lacerda Nogueira; Rafael dos Santos Bezerra; Raul Machado Neto; Rejane Maria Tommasini Grotto; Ricardo Haddad; Sandra Coccuzzo Sampaio Vessoni; Simone Kashima; Svetoslav Nanev Slavov; Vincent Louis Viala |
| EPI_ISL_5648347 | ESF PQ UNIVERSITARIO DE ASSIS | Instituto Butantan | Antonio Jorge Martins; Claudia Renata dos Santos Barros; David Schlesinger; Debora Botequio Moretti; Dimas Tadeu Covas; Elaine Cristina Marqueze; Elaine Vieira Santos; Evandra Strazza Rodrigues; Heidge Fukumasu; Jayme Augusto de Souza-Neto; José Salvatore Leister Patané; Luiz Alcantara; Luiz Lehmann Coutinho; Maria Carolina Elias; Maurício Lacerda Nogueira; Rafael dos Santos Bezerra; Raul Machado Neto; Rejane Maria Tommasini Grotto; Ricardo Haddad; Sandra Coccuzzo Sampaio Vessoni; Simone Kashima; Svetoslav Nanev Slavov; Vincent Louis Viala |
| EPI_ISL_3215815, EPI_ISL_3475847, EPI_ISL_3475851 | EXCITE Lab | Andersen lab at Scripps Research | Abigail Schnapper; Angela Scioscia; Araz Majnoonian; Ashkan Hassani; Chip Schooley; David Pride; Fitri Carrissa Wijaya; Greg Humphrey; Helena Tubb; Laura Kohn; Marlene Flores; Megan Nguyen; Natasha Martin Cheryl Anderson; Patricia Gonzalez-Zuniga; Rebecca Fielding-Miller; Rodolfo Salido; Sawyer Farmer; Sharon Reed; Smruthi Karthikeyan; Tommy Valles + SEARCH; Victor Cantu; Vinton Omaleki |
| EPI_ISL_2939547, EPI_ISL_2939557, EPI_ISL_2939613, EPI_ISL_2939623, EPI_ISL_2939625, EPI_ISL_2939627, EPI_ISL_2939636, EPI_ISL_2939647, EPI_ISL_2939684, EPI_ISL_2939690, EPI_ISL_2939705, EPI_ISL_2939785, EPI_ISL_2939802, EPI_ISL_2939803, EPI_ISL_2939809, EPI_ISL_2939817, EPI_ISL_2939829, EPI_ISL_2939832, EPI_ISL_2939834, EPI_ISL_2939843, EPI_ISL_2939848, EPI_ISL_2939857, EPI_ISL_2939858, EPI_ISL_2939891, EPI_ISL_2939904, EPI_ISL_2939913, EPI_ISL_2939924, EPI_ISL_2939930, EPI_ISL_2939950, EPI_ISL_2939966, EPI_ISL_2939992, EPI_ISL_2940007, EPI_ISL_2940017, EPI_ISL_2940018, EPI_ISL_2940019, EPI_ISL_2940026, EPI_ISL_2940030, EPI_ISL_2940038, EPI_ISL_2940039, EPI_ISL_2940040, EPI_ISL_3116502, EPI_ISL_3116511, EPI_ISL_3116966, EPI_ISL_3116967, EPI_ISL_3116968, EPI_ISL_3116969, EPI_ISL_3116970, EPI_ISL_3116971, EPI_ISL_3116972, EPI_ISL_3116973, EPI_ISL_3116974, EPI_ISL_3116975, EPI_ISL_3116976, EPI_ISL_3116977, EPI_ISL_3116978, EPI_ISL_3117380 | | | |
| see above | Edmonton Provincial Lab | Public Health Agency of Canada (PHAC) National Microbiology Laboratory | Buss; Croxen M; Deo A; Dieu P; E; Ferrato C; Gill K; Khan F; Koleva P; Li V; Lloyd C; Lynch T; Ma R; Murphy S; Pabbaraju K; Shokoples S; Thayer J; Tipples G; Whitehouse M; Wong A; Yu C; Zelyas N |
| EPI_ISL_3246302 | Eurofins Diatherix | Hudsonalpha Genome Sequencing | Ada Stewart; Jane Grimwood; Jenell Webber; Jennifer Cart; John Lovell; Joshua Stough; Leslie Malone; Lori H. Handley; Melissa Williams; Stefan Brzezinski; Teresa Jones; and Jeremy Schmutz |

| | | | |
|---|---|---|---|
| | | Center | |
| EPI_ISL_5648811, EPI_ISL_5648820, EPI_ISL_5648850, EPI_ISL_5648852, EPI_ISL_5648875, EPI_ISL_5648905, EPI_ISL_5648969, EPI_ISL_5648970, EPI_ISL_5648991, EPI_ISL_5649036, EPI_ISL_5649080, EPI_ISL_5649124, EPI_ISL_5649131 | | | |
| see above | FACULDADE DE MEDICINA ABC | Instituto Butantan | Antonio Jorge Martins; Claudia Renata dos Santos Barros; David Schlesinger; Debora Botequio Moretti; Dimas Tadeu Covas; Elaine Cristina Marqueze; Elaine Vieira Santos; Evandra Strazza Rodrigues; Heidge Fukumasu; Jayme Augusto de Souza-Neto; José Salvatore Leister Patané; Luiz Alcantara; Luiz Lehmann Coutinho; Maria Carolina Elias; Maurício Lacerda Nogueira; Rafael dos Santos Bezerra; Raul Machado Neto; Rejane Maria Tommasini Grotto; Ricardo Haddad; Sandra Coccuzzo Sampaio Vessoni; Simone Kashima; Svetoslav Nanev Slavov; Vincent Louis Viala |
| EPI_ISL_3355340, EPI_ISL_3355380 | FL Bureau of Public Health Laboratories-Tampa | Centers for Disease Control and Prevention Division of Viral Diseases, Pathogen Discovery | Alex Burgin; Ben Rambo-Martin; Clinton Paden; Dakota Howard; Dave Wentworth; Dhwani Batra; Jasmine Padilla; Justin Lee; Krista Queen; Kristen Knipe; Kristine Lacek; Mark Burroughs; Matthew Schmerer; Meghan Bentz; Mili Sheth; Peter Cook; Sam Shepard; Sarah Nobles; Suxiang Tong; Vivien Dugan; Yvette Unoarumhi |
| EPI_ISL_2920620, EPI_ISL_2920621, EPI_ISL_2920622, EPI_ISL_2920623, EPI_ISL_2920624, EPI_ISL_2920625, EPI_ISL_2920627, EPI_ISL_2920628, EPI_ISL_2920629, EPI_ISL_2920630, EPI_ISL_2920631, EPI_ISL_2920632, EPI_ISL_2920633, EPI_ISL_2920634, EPI_ISL_2920635, EPI_ISL_2920636, EPI_ISL_2920637, EPI_ISL_2920638, EPI_ISL_2920640, EPI_ISL_2920641, EPI_ISL_2920642, EPI_ISL_2920643, EPI_ISL_2920644, EPI_ISL_2920645, EPI_ISL_2920646, EPI_ISL_2920650, EPI_ISL_2920668, EPI_ISL_2920676, EPI_ISL_3100864, EPI_ISL_3100867, EPI_ISL_3100873, EPI_ISL_3100881, EPI_ISL_3100882, EPI_ISL_3100883, EPI_ISL_3100887, EPI_ISL_3100889, EPI_ISL_5321920, EPI_ISL_5321940, EPI_ISL_5866271, EPI_ISL_5866272, EPI_ISL_5866273, EPI_ISL_5866274, EPI_ISL_5866275, EPI_ISL_5866276, EPI_ISL_5866277, EPI_ISL_5866322, EPI_ISL_5866655, EPI_ISL_5866713, EPI_ISL_5924954, EPI_ISL_5925178, EPI_ISL_5925179, EPI_ISL_5925180, EPI_ISL_6050080, EPI_ISL_6050081, EPI_ISL_6050082, EPI_ISL_6050083, EPI_ISL_6050084, EPI_ISL_6050200 | | | |
| see above | Florida Bureau of Public Health Laboratories | Florida Bureau of Public Health Laboratories | Jason Blanton; Namratha Tarigopula; Sarah Schmedes; Tiffany Splatt |
| EPI_ISL_2787052 | Fondazione Policlinico Universitario "A. Gemelli" IRCCS | INMI Lazzaro Spallanzani IRCCS | CEM Gruber; D Cerini; D Di Fusco; E Giombini; E Sperandio; F Santini; G Bonfiglio; O Butera; S Donzelli |
| EPI_ISL_2785737, EPI_ISL_2785761, EPI_ISL_2785773, EPI_ISL_2785777, EPI_ISL_2785779, EPI_ISL_2785803, EPI_ISL_2785844, EPI_ISL_2785856, EPI_ISL_2785857, EPI_ISL_2785858, EPI_ISL_2785859, EPI_ISL_2785889, EPI_ISL_2785891, EPI_ISL_2785912, EPI_ISL_2785916, EPI_ISL_2785919, EPI_ISL_2785932, EPI_ISL_2785940, EPI_ISL_2785953, EPI_ISL_2785954, EPI_ISL_2785960, EPI_ISL_2873401, EPI_ISL_2873415, EPI_ISL_2873426, EPI_ISL_2873427, EPI_ISL_2873442, EPI_ISL_2873443, EPI_ISL_2873458, EPI_ISL_2873461, EPI_ISL_2873463, EPI_ISL_2873470, EPI_ISL_2873497, EPI_ISL_2873794, EPI_ISL_2873796, EPI_ISL_2873801, EPI_ISL_2873802, EPI_ISL_2873808, EPI_ISL_2873815, EPI_ISL_2929136, EPI_ISL_2929138, EPI_ISL_2929150, EPI_ISL_2929152, EPI_ISL_2929153, EPI_ISL_2929158, EPI_ISL_2929162 | | | |
| see above | Fulgent Genetics | Centers for Disease Control and Prevention Division of Viral Diseases, Pathogen Discovery | Adrian Paskey; Becky Tsai; Benafsh Sapra; Benjamin Rambo-Martin; Christopher Gulvick; Clinton R. Paden; Dakota Howard; Darlene Wagner; Dhwani Batra; Doreen Ng; Duncan MacCannell; Harry Gao; James Xie; Jason Caravas; John Gao; Joseph Fierro; Kara Moser; Matthew Schmerer; Mickey Li; Peter W. Cook; Scott Sammons; Shatavia Morrison; Yan Meng; Yvette Unoarumhi |
| EPI_ISL_5648361 | GRIPARIO FALCAO | Instituto Butantan | Antonio Jorge Martins; Claudia Renata dos Santos Barros; David Schlesinger; Debora Botequio Moretti; Dimas Tadeu Covas; Elaine Cristina Marqueze; Elaine Vieira Santos; Evandra Strazza Rodrigues; Heidge Fukumasu; Jayme Augusto de Souza-Neto; José Salvatore Leister Patané; Luiz Alcantara; Luiz Lehmann Coutinho; Maria Carolina Elias; Maurício Lacerda Nogueira; Rafael dos Santos Bezerra; Raul Machado Neto; Rejane Maria Tommasini Grotto; Ricardo Haddad; Sandra Coccuzzo Sampaio Vessoni; Simone Kashima; Svetoslav Nanev Slavov; Vincent Louis Viala |
| EPI_ISL_2892598, EPI_ISL_2892612, EPI_ISL_2892660, EPI_ISL_2892661, EPI_ISL_2892685, EPI_ISL_2892686, EPI_ISL_2892687, EPI_ISL_2892692, EPI_ISL_2892693, EPI_ISL_2892694, EPI_ISL_2892698, EPI_ISL_2892709, EPI_ISL_2892710, EPI_ISL_2892711, EPI_ISL_2892715, EPI_ISL_2892724, EPI_ISL_2892725, EPI_ISL_2892743, EPI_ISL_2892744, EPI_ISL_2892745, EPI_ISL_2892746, EPI_ISL_2892747, EPI_ISL_2892755, EPI_ISL_2892757, EPI_ISL_2892761, EPI_ISL_2892762, EPI_ISL_2892767, EPI_ISL_2892780, EPI_ISL_2892786, EPI_ISL_2892789, EPI_ISL_2892790, EPI_ISL_2892791, EPI_ISL_2892823, EPI_ISL_2892825, EPI_ISL_2892826, EPI_ISL_2892827, EPI_ISL_2892829, EPI_ISL_2892831, EPI_ISL_2892832, EPI_ISL_2892836, EPI_ISL_2892837, EPI_ISL_2892838, EPI_ISL_2892839, EPI_ISL_2892840, EPI_ISL_2892841, EPI_ISL_2892842, EPI_ISL_2892843, EPI_ISL_2892844, EPI_ISL_2892845, EPI_ISL_2892846, EPI_ISL_2892853, EPI_ISL_2892854, EPI_ISL_2892855, EPI_ISL_2892856, EPI_ISL_2892857, EPI_ISL_2892858, EPI_ISL_2892859, EPI_ISL_2892863, EPI_ISL_2892864, EPI_ISL_2892865, EPI_ISL_2892868, EPI_ISL_2892893, EPI_ISL_2892894, EPI_ISL_2892898, EPI_ISL_2892905, EPI_ISL_2892909, EPI_ISL_2892912, EPI_ISL_2892951, EPI_ISL_2892959, EPI_ISL_2892978, EPI_ISL_2892981, EPI_ISL_2892982, EPI_ISL_2892983, EPI_ISL_2892985, EPI_ISL_2892986, EPI_ISL_2892987, EPI_ISL_2892988, EPI_ISL_2893006, EPI_ISL_2893014, EPI_ISL_2893019, EPI_ISL_2893020, EPI_ISL_2894891, EPI_ISL_2894892, EPI_ISL_2894911, EPI_ISL_2894914, EPI_ISL_2894915, EPI_ISL_2894916, EPI_ISL_2894917, EPI_ISL_2894918, EPI_ISL_2894919, EPI_ISL_2894921, EPI_ISL_2894923, EPI_ISL_2894924, EPI_ISL_2894929, EPI_ISL_2894930, EPI_ISL_2894959, EPI_ISL_2894960, EPI_ISL_2894963, EPI_ISL_2894964, EPI_ISL_2894966, EPI_ISL_2894967, EPI_ISL_2894970, EPI_ISL_2894971, EPI_ISL_2896155, EPI_ISL_2896156, EPI_ISL_2896159, EPI_ISL_2896160, EPI_ISL_2896161, EPI_ISL_2896162, EPI_ISL_2896163, EPI_ISL_2896164, EPI_ISL_2896165, EPI_ISL_2896166, EPI_ISL_2896167, EPI_ISL_2896168, EPI_ISL_2896169, EPI_ISL_2896170, EPI_ISL_2896171, EPI_ISL_2896172, EPI_ISL_2930511, EPI_ISL_2930512, EPI_ISL_2930513, EPI_ISL_2930514, EPI_ISL_2930515, EPI_ISL_2930516, EPI_ISL_2930517, EPI_ISL_2930518, EPI_ISL_2930536, EPI_ISL_2930539, EPI_ISL_2930542, EPI_ISL_2930545, EPI_ISL_2930546, EPI_ISL_2930547, EPI_ISL_2930548, EPI_ISL_3132197, EPI_ISL_3132198, EPI_ISL_3132199, EPI_ISL_3132200, EPI_ISL_3132201, EPI_ISL_3132202, EPI_ISL_3132203, EPI_ISL_3132204, EPI_ISL_3132205, EPI_ISL_3132206, EPI_ISL_3132207, EPI_ISL_3132208, EPI_ISL_3132209, EPI_ISL_3132210, EPI_ISL_3132211, EPI_ISL_3132212, EPI_ISL_3132213, EPI_ISL_3144908, EPI_ISL_3144911, EPI_ISL_3144912, EPI_ISL_3185385, EPI_ISL_3185386, EPI_ISL_3185387, EPI_ISL_3185388, EPI_ISL_3185389, EPI_ISL_3185390, EPI_ISL_3185408, EPI_ISL_3220038, EPI_ISL_3220040, EPI_ISL_3220042, EPI_ISL_3220043, EPI_ISL_3220045, EPI_ISL_3220047, EPI_ISL_3220051, EPI_ISL_3220053, EPI_ISL_3220054, EPI_ISL_3220056, EPI_ISL_3220058, EPI_ISL_3220060, EPI_ISL_3220061, EPI_ISL_3220063, EPI_ISL_3220065, EPI_ISL_3220067, EPI_ISL_3220068, EPI_ISL_3220070, EPI_ISL_3220072, EPI_ISL_3220074, EPI_ISL_3220075, EPI_ISL_3220077, EPI_ISL_3220079, EPI_ISL_3220085, EPI_ISL_3220087, EPI_ISL_3220088, EPI_ISL_3220090, EPI_ISL_3220092, EPI_ISL_3220096, EPI_ISL_3220098, EPI_ISL_3220101, EPI_ISL_3220120, EPI_ISL_3220129, EPI_ISL_3220133, EPI_ISL_3220136, EPI_ISL_3220138, EPI_ISL_3220145 | | | |
| see above | Genetica Molecular and Subdepartamento de Virologia ISP Chile | Instituto de Salud Publica de Chile | Andres Castillo; Barbara Parra; Constanza Campano; Gisselle Barra; Javier Tognarelli; Jorge Fernandez; Karen Orostica; Loredana Arata; Patricia Bustos; Rodrigo Fasce; Soledad Ulloa |
| EPI_ISL_2985664, EPI_ISL_2985665, EPI_ISL_2985668, EPI_ISL_2985674, EPI_ISL_2985675, EPI_ISL_2985714, EPI_ISL_2985751 | | | |
| see above | Gravity Diagnostics, LLC | Gravity Diagnostics, LLC | Gravity Diagnostics |
| EPI_ISL_7476174, EPI_ISL_7476661 | Grupo de Investigación en Enfermedades Tropicales del Ejército (GINETEJ), Laboratorio de Referencia e Investigación, Dirección de Sanidad Ejército, Bogotá, Colombia | Centro de Investigaciones en Microbiología y Biotecnología-UR (CIMBIUR), Facultad de Ciencias Naturales, Universidad del Rosario, Bogotá, Colombia | Alberto E. Paniz-Mondolfi; Angie L. Ramírez; Camilo Correa-Cardenas; Claudia Mendez; Enzo Guerrero-Araya; Juan David Ramírez; Julie Perez; Luz H. Patiño; Maria Clara Duque; Marina Muñoz; Nathalia Ballesteros; Nicolás Luna; Sergio Castañeda |
| EPI_ISL_3102352 | HGCC HOSPITAL GERAL DR CESAR CALS | Analytical Competence Molecular Epidemiology Lab/ACME, Oswaldo Cruz Foundation, Ceara (FIOCRUZ CE) | Cleber Furtado Aksenen; Fabio Miyajima; Fernando Braga Stehling; Francisco Eder de Moura Lopes; Jamille Maria Mendes Bezerra; Joaquim César do Nascimento Sousa Junior; Pedro Miguel Carneiro Jeronimo; Suzana Porto Almeida e Lucas Delerino; Thais Ferreira de Oliveira; Thais de Oliveira Costa; Ticiane Cavalcante de Souza; Veridiana Pessoa Miyajima |
| EPI_ISL_3102357, EPI_ISL_3102361, EPI_ISL_3102363 | HIAS HOSPITAL INFANTIL ALBERT SABIN | Analytical Competence Molecular Epidemiology Lab/ACME, Oswaldo Cruz Foundation, Ceara (FIOCRUZ CE) | Cleber Furtado Aksenen; Fabio Miyajima; Fernando Braga Stehling; Francisco Eder de Moura Lopes; Jamille Maria Mendes Bezerra; Joaquim César do Nascimento Sousa Junior; Pedro Miguel Carneiro Jeronimo; Suzana Porto Almeida e Lucas Delerino; Thais Ferreira de Oliveira; Thais de Oliveira Costa; Ticiane Cavalcante de Souza; Veridiana Pessoa Miyajima |
| EPI_ISL_2921569, EPI_ISL_2921570, EPI_ISL_2921580, EPI_ISL_2921581, EPI_ISL_2921582, EPI_ISL_2921583, EPI_ISL_2921584, EPI_ISL_2921585, EPI_ISL_2921586, EPI_ISL_2921587, EPI_ISL_2921588, EPI_ISL_2921589, EPI_ISL_2921590, EPI_ISL_2921591, EPI_ISL_2921592, EPI_ISL_2921593, EPI_ISL_2921594, EPI_ISL_2921595, EPI_ISL_2921596, EPI_ISL_2921597, EPI_ISL_2921598, EPI_ISL_2921599, EPI_ISL_2921600, EPI_ISL_2921601, EPI_ISL_3254565, EPI_ISL_3254566, EPI_ISL_3254567, EPI_ISL_3254568, EPI_ISL_3254569, EPI_ISL_3254570, EPI_ISL_3254571, EPI_ISL_3254572, EPI_ISL_3254573, EPI_ISL_3254574, EPI_ISL_3254575, EPI_ISL_3254576, EPI_ISL_3254577, EPI_ISL_3254578, EPI_ISL_3254579, EPI_ISL_3254580, EPI_ISL_3254581, EPI_ISL_3254582, EPI_ISL_3254583, EPI_ISL_3254584, EPI_ISL_3254585, EPI_ISL_3254586, EPI_ISL_3254587, EPI_ISL_3254588, EPI_ISL_3254589, EPI_ISL_3254590, EPI_ISL_3254591, EPI_ISL_3254592, EPI_ISL_3254593, EPI_ISL_3254594, EPI_ISL_3254595, EPI_ISL_3254596, EPI_ISL_3254597, EPI_ISL_3254598, EPI_ISL_3254599, EPI_ISL_3254600, EPI_ISL_3254601, EPI_ISL_3254602, EPI_ISL_3254603, EPI_ISL_3254604, EPI_ISL_3254605, EPI_ISL_3254606, EPI_ISL_3254607, EPI_ISL_3254608, EPI_ISL_3254609, EPI_ISL_3254610, EPI_ISL_3254611, EPI_ISL_3254612, EPI_ISL_3254613, EPI_ISL_3254614, EPI_ISL_3254615, EPI_ISL_3254616, EPI_ISL_3254617, EPI_ISL_3254618, EPI_ISL_3254619, EPI_ISL_3254620, EPI_ISL_3254621, EPI_ISL_3254622, EPI_ISL_3254623, EPI_ISL_3254624, EPI_ISL_3254625, EPI_ISL_3254626, EPI_ISL_3254627, EPI_ISL_3254628, EPI_ISL_3254629, EPI_ISL_3254630, EPI_ISL_3254631, EPI_ISL_3254632, EPI_ISL_3254633, EPI_ISL_3254634, EPI_ISL_3254635, EPI_ISL_3254674, EPI_ISL_3254675, EPI_ISL_3254676, EPI_ISL_3254677, EPI_ISL_3299426, EPI_ISL_3299427, EPI_ISL_3299429, EPI_ISL_4061652, EPI_ISL_4061655 | | | |
| see above | HLAGYN - Laboratorio de Imunologia de Transplantes de Goias | HLAGYN - Laboratorio de Imunologia de Transplantes de Goias | Alessandro Leonardo Alvares Magalhaes; Daniel Ferreira de Sousa; Erika Lopes Rocha Batista; Fernando Antonio Vinhal dos Santos; Frederico Rodrigues Vinhal; Kamila Oliveira Reis De Freitas.; Lucas Carlos Gomes Pereira; Sabrina Sara Moreira Duarte |
| EPI_ISL_3102431, EPI_ISL_3102440, EPI_ISL_3102462, EPI_ISL_3102474, EPI_ISL_3536224, EPI_ISL_3536225, EPI_ISL_3536234, EPI_ISL_3536241, EPI_ISL_3536253, EPI_ISL_3536255, EPI_ISL_3536256, EPI_ISL_3536257, EPI_ISL_3912173 | | | |
| see above | HM HOSPITAL DE MESSEJANA DR CARLOS ALBERTO STUDART GOMES | Analytical Competence Molecular Epidemiology Lab/ACME, Oswaldo Cruz Foundation, Ceara (FIOCRUZ CE) | Cleber Furtado Aksenen; Fabio Miyajima; Fernando Braga Stehling; Francisco Eder de Moura Lopes; Jamille Maria Mendes Bezerra; Joaquim Cesar do Nascimento Sousa Junior; Joaquim César do Nascimento Sousa Junior; Pedro Miguel Carneiro Jeronimo; Suzana Porto Almeida & Lucas Delerino on behalf of COVID-19 FIOCRUZ Genomic Network; Suzana Porto Almeida e Lucas Delerino; Thais Ferreira de Oliveira; Thais de Oliveira Costa; Ticiane Cavalcante de Souza; Veridiana Pessoa Miyajima |
| EPI_ISL_5649251, EPI_ISL_5649273 | HOSP E MATERNIDADE MUNICIPAL N SRA MONTE SERRAT | Instituto Butantan | Antonio Jorge Martins; Claudia Renata dos Santos Barros; David Schlesinger; Debora Botequio Moretti; Dimas Tadeu Covas; Elaine Cristina Marqueze; Elaine Vieira Santos; Evandra Strazza Rodrigues; Heidge Fukumasu; Jayme Augusto de Souza-Neto; José Salvatore Leister Patané; Luiz Alcantara; Luiz Lehmann Coutinho; Maria Carolina Elias; Maurício Lacerda Nogueira; Rafael dos Santos Bezerra; Raul Machado Neto; Rejane Maria Tommasini Grotto; Ricardo Haddad; Sandra Coccuzzo Sampaio Vessoni; Simone Kashima; Svetoslav Nanev Slavov; Vincent Louis Viala |
| EPI_ISL_5648948 | HOSP MAT SAO MIGUEL | Instituto Butantan | Antonio Jorge Martins; Claudia Renata dos Santos Barros; David Schlesinger; Debora Botequio Moretti; Dimas Tadeu Covas; Elaine Cristina Marqueze; Elaine Vieira Santos; Evandra Strazza Rodrigues; Heidge Fukumasu; Jayme Augusto de Souza-Neto; José Salvatore Leister Patané; Luiz Alcantara; Luiz Lehmann Coutinho; Maria Carolina Elias; Maurício Lacerda Nogueira; Rafael dos Santos Bezerra; Raul Machado Neto; Rejane Maria Tommasini Grotto; Ricardo Haddad; Sandra Coccuzzo Sampaio Vessoni; Simone Kashima; Svetoslav Nanev Slavov; Vincent Louis Viala |
| EPI_ISL_2812743 | HOSPITAL CAN MISSES | HOSPITAL UNIVERSITARIO SON ESPASES | Hospital Universitario Son Espases; Servicio de Microbiología |
| EPI_ISL_5649246 | HOSPITAL DE CAMPANHA COVID 19 CAIEIRAS | Instituto Butantan | Antonio Jorge Martins; Claudia Renata dos Santos Barros; David Schlesinger; Debora Botequio Moretti; Dimas Tadeu Covas; Elaine Cristina Marqueze; Elaine Vieira Santos; Evandra Strazza Rodrigues; Heidge Fukumasu; Jayme Augusto de Souza-Neto; José Salvatore Leister Patané; Luiz Alcantara; Luiz Lehmann Coutinho; Maria Carolina Elias; Maurício Lacerda Nogueira; Rafael dos Santos Bezerra; Raul Machado Neto; Rejane Maria Tommasini Grotto; Ricardo Haddad; Sandra Coccuzzo Sampaio Vessoni; Simone Kashima; Svetoslav Nanev Slavov; Vincent Louis Viala |
| EPI_ISL_5649447 | HOSPITAL DE CAMPANHA COVID 19 MUNICIPIO DE TAUBATE | Instituto Butantan | Antonio Jorge Martins; Claudia Renata dos Santos Barros; David Schlesinger; Debora Botequio Moretti; Dimas Tadeu Covas; Elaine Cristina Marqueze; Elaine Vieira Santos; Evandra Strazza Rodrigues; Heidge Fukumasu; Jayme Augusto de Souza-Neto; José Salvatore Leister Patané; Luiz Alcantara; Luiz Lehmann Coutinho; Maria Carolina Elias; Maurício Lacerda Nogueira; Rafael dos Santos Bezerra; Raul Machado Neto; Rejane Maria Tommasini Grotto; Ricardo Haddad; Sandra Coccuzzo Sampaio Vessoni; Simone Kashima; Svetoslav Nanev Slavov; Vincent Louis Viala |
| EPI_ISL_3298306 | HOSPITAL DE LAS MUJERES DR. ADOLFO CARIT | Inciensa, Instituto Costarricense de Investigación y Enseñanza en Nutrición y Salud | Adriana Godínez; Claudio Soto-Garita; Estela Cordero; Francisco Duarte; Hebleen Porras; Joselyn Prado & Karla Moreno; José Luis Vargas; Mariela Gutiérrez; Melany Calderón |
| EPI_ISL_3459378, EPI_ISL_3459388 | HOSPITAL DEPARTAMENTAL DE VILLAVICENCIO | Instituto Nacional de Salud | Carlos Franco-Muñoz; Carmen Osorio; Diana Malo; Diego A. Álvarez-Díaz; Diego Andrés Prada; Gerardo Santamaría; Hector Alejandro Ruiz-Moreno; Jhonnatan Reales-González; Jorge Rivera; Juan Camilo Martinez; Julian Naizaque; Katherine Laiton-Donato; Lisseth Pardo; Magdalena Wiesner; Marcela Mercado-Reyes; Maria T. Herrera-Sepúlveda; Marta Lopez Blanco; Martha Lucia Ospina Martinez; Paola Rojas; Sergio Gomez; Sheryll Corchuelo; Ángela Alarcon Cruz |
| EPI_ISL_3037815 | HOSPITAL DR. RAFAEL A. CALDERON GUARDIA | Inciensa, Instituto Costarricense de Investigación y Enseñanza en Nutrición y Salud | Adriana Godínez; Caterina Guzmán; Claudio Soto-Garita; Estela Cordero; Francisco Duarte; Hebleen Porras; José Luis Vargas; Mariela Gutiérrez Joselyn Prado; Melany Calderón; Nazareth Ruiz & Fabián Salas |
| EPI_ISL_3102334, EPI_ISL_3102353, EPI_ISL_3102399, EPI_ISL_3536151, EPI_ISL_3536154, EPI_ISL_3536155, EPI_ISL_3536170, EPI_ISL_3536188, EPI_ISL_3536266, EPI_ISL_3536279, EPI_ISL_3536280, EPI_ISL_3536286, EPI_ISL_3536350, EPI_ISL_3536359, EPI_ISL_3536364, EPI_ISL_3536366, EPI_ISL_3536398, EPI_ISL_3912120, EPI_ISL_3912206 | | | |
| see above | HOSPITAL ESTADUAL LEONARDO DA VINCI | Analytical Competence Molecular Epidemiology Lab/ACME, Oswaldo Cruz Foundation, Ceara (FIOCRUZ CE) | Cleber Furtado Aksenen; Fabio Miyajima; Fernando Braga Stehling; Francisco Eder de Moura Lopes; Jamille Maria Mendes Bezerra; Joaquim Cesar do Nascimento Sousa Junior; Joaquim César do Nascimento Sousa Junior; Pedro Miguel Carneiro Jeronimo; Suzana Porto Almeida & Lucas Delerino on behalf of COVID-19 FIOCRUZ Genomic Network; Suzana Porto Almeida e Lucas Delerino; Thais Ferreira de Oliveira; Thais de Oliveira Costa; Ticiane Cavalcante de Souza; Veridiana Pessoa Miyajima |
| EPI_ISL_3298312 | HOSPITAL MEXICO | Inciensa, Instituto Costarricense de Investigación y Enseñanza en Nutrición y Salud | Adriana Godínez; Claudio Soto-Garita; Estela Cordero; Francisco Duarte; Hebleen Porras; Joselyn Prado & Juan Carlos Villalobos; José Luis Vargas; Mariela Gutiérrez; Melany Calderón |
| EPI_ISL_5648930, EPI_ISL_5649033 | HOSPITAL MUN DR GUIDO GUIDA | Instituto Butantan | Antonio Jorge Martins; Claudia Renata dos Santos Barros; David Schlesinger; Debora Botequio Moretti; Dimas Tadeu Covas; Elaine Cristina Marqueze; Elaine Vieira Santos; Evandra Strazza Rodrigues; Heidge Fukumasu; Jayme Augusto de Souza-Neto; José Salvatore Leister Patané; Luiz Alcantara; Luiz Lehmann Coutinho; Maria Carolina Elias; Maurício Lacerda Nogueira; Rafael dos Santos Bezerra; Raul Machado Neto; Rejane Maria Tommasini Grotto; Ricardo Haddad; Sandra Coccuzzo Sampaio Vessoni; Simone Kashima; Svetoslav Nanev Slavov; Vincent Louis Viala |
| EPI_ISL_5649357, EPI_ISL_5649420, EPI_ISL_5649426, EPI_ISL_5649446 | HOSPITAL MUNICIPAL DE ITAPIRA | Instituto Butantan | Antonio Jorge Martins; Claudia Renata dos Santos Barros; David Schlesinger; Debora Botequio Moretti; Dimas Tadeu Covas; Elaine Cristina Marqueze; Elaine Vieira Santos; Evandra Strazza Rodrigues; Heidge Fukumasu; Jayme Augusto de Souza-Neto; José Salvatore Leister Patané; Luiz Alcantara; Luiz Lehmann Coutinho; Maria Carolina Elias; Maurício Lacerda Nogueira; Rafael dos Santos Bezerra; Raul Machado Neto; Rejane Maria Tommasini Grotto; Ricardo Haddad; Sandra Coccuzzo Sampaio Vessoni; Simone Kashima; Svetoslav Nanev Slavov; Vincent Louis Viala |
| EPI_ISL_5648515, EPI_ISL_5649324, EPI_ISL_5649333, EPI_ISL_5649370, EPI_ISL_5649380, EPI_ISL_5649389, EPI_ISL_5649396, EPI_ISL_5649397, EPI_ISL_5649401, EPI_ISL_5649405, EPI_ISL_5649407, EPI_ISL_5649409 | | | |
| see above | HOSPITAL MUNICIPAL DE SAO VICENTE | Instituto Butantan | Antonio Jorge Martins; Claudia Renata dos Santos Barros; David Schlesinger; Debora Botequio Moretti; Dimas Tadeu Covas; Elaine Cristina Marqueze; Elaine Vieira Santos; Evandra Strazza Rodrigues; Heidge Fukumasu; Jayme Augusto de Souza-Neto; José Salvatore Leister Patané; Luiz Alcantara; Luiz Lehmann Coutinho; Maria Carolina Elias; Maurício Lacerda Nogueira; Rafael dos Santos Bezerra; Raul Machado Neto; Rejane Maria Tommasini Grotto; Ricardo Haddad; Sandra Coccuzzo Sampaio Vessoni; Simone Kashima; Svetoslav Nanev Slavov; Vincent Louis Viala |

| | | | |
|---|---|---|---|
| EPI_ISL_5648769, EPI_ISL_5648775, EPI_ISL_5648823, EPI_ISL_5648843, EPI_ISL_5648860, EPI_ISL_5648870, EPI_ISL_5648877, EPI_ISL_5648890, EPI_ISL_5648965, EPI_ISL_5648966, EPI_ISL_5648986, EPI_ISL_5648999, EPI_ISL_5649067, EPI_ISL_5649112 | | | |
| see above | HOSPITAL MUNICIPAL DR MARIO GATTI CAMPINAS | Instituto Butantan | Antonio Jorge Martins; Claudia Renata dos Santos Barros; David Schlesinger; Debora Botequio Moretti; Dimas Tadeu Covas; Elaine Cristina Marqueze; Elaine Vieira Santos; Evandra Strazza Rodrigues; Heidge Fukumasu; Jayme Augusto de Souza-Neto; José Salvatore Leister Patané; Luiz Alcantara; Luiz Lehmann Coutinho; Maria Carolina Elias; Maurício Lacerda Nogueira; Rafael dos Santos Bezerra; Raul Machado Neto; Rejane Maria Tommasini Grotto; Ricardo Haddad; Sandra Coccuzzo Sampaio Vessoni; Simone Kashima; Svetoslav Nanev Slavov; Vincent Louis Viala |
| EPI_ISL_5648368, EPI_ISL_5648445, EPI_ISL_5648499, EPI_ISL_5648525, EPI_ISL_5648544, EPI_ISL_5648553, EPI_ISL_5648560, EPI_ISL_5648582, EPI_ISL_5648596, EPI_ISL_5648607, EPI_ISL_5648608, EPI_ISL_5648683, EPI_ISL_5648700, EPI_ISL_5648701, EPI_ISL_5648729, EPI_ISL_5649428 | | | |
| see above | HOSPITAL MUNICIPAL DR TABAJARA RAMOS | Instituto Butantan | Antonio Jorge Martins; Claudia Renata dos Santos Barros; David Schlesinger; Debora Botequio Moretti; Dimas Tadeu Covas; Elaine Cristina Marqueze; Elaine Vieira Santos; Evandra Strazza Rodrigues; Heidge Fukumasu; Jayme Augusto de Souza-Neto; José Salvatore Leister Patané; Luiz Alcantara; Luiz Lehmann Coutinho; Maria Carolina Elias; Maurício Lacerda Nogueira; Rafael dos Santos Bezerra; Raul Machado Neto; Rejane Maria Tommasini Grotto; Ricardo Haddad; Sandra Coccuzzo Sampaio Vessoni; Simone Kashima; Svetoslav Nanev Slavov; Vincent Louis Viala |
| EPI_ISL_3912047, EPI_ISL_3912063, EPI_ISL_3912110, EPI_ISL_3912113, EPI_ISL_3912115, EPI_ISL_3912122, EPI_ISL_3912218, EPI_ISL_3912219 | | | |
| see above | HOSPITAL MUNICIPAL NOSSA SENHORA DOS MILAGRES | Analytical Competence Molecular Epidemiology Lab/ACME, Oswaldo Cruz Foundation, Ceara (FIOCRUZ CE) | Cleber Furtado Aksenen; Fabio Miyajima; Fernando Braga Stehling; Francisco Eder de Moura Lopes; Jamille Maria Mendes Bezerra; Joaquim Cesar do Nascimento Sousa Junior; Pedro Miguel Carneiro Jeronimo; Suzana Porto Almeida & Lucas Delerino on behalf of COVID-19 FIOCRUZ Genomic Network; Thais Ferreira de Oliveira; Thais de Oliveira Costa; Ticiane Cavalcante de Souza; Veridiana Pessoa Miyajima |
| EPI_ISL_5649459 | HOSPITAL MUNICIPAL REYNALDO GUERRA CAJATI | Instituto Butantan | Antonio Jorge Martins; Claudia Renata dos Santos Barros; David Schlesinger; Debora Botequio Moretti; Dimas Tadeu Covas; Elaine Cristina Marqueze; Elaine Vieira Santos; Evandra Strazza Rodrigues; Heidge Fukumasu; Jayme Augusto de Souza-Neto; José Salvatore Leister Patané; Luiz Alcantara; Luiz Lehmann Coutinho; Maria Carolina Elias; Maurício Lacerda Nogueira; Rafael dos Santos Bezerra; Raul Machado Neto; Rejane Maria Tommasini Grotto; Ricardo Haddad; Sandra Coccuzzo Sampaio Vessoni; Simone Kashima; Svetoslav Nanev Slavov; Vincent Louis Viala |
| EPI_ISL_5648929 | HOSPITAL REGIONAL DE COTIA | Instituto Butantan | Antonio Jorge Martins; Claudia Renata dos Santos Barros; David Schlesinger; Debora Botequio Moretti; Dimas Tadeu Covas; Elaine Cristina Marqueze; Elaine Vieira Santos; Evandra Strazza Rodrigues; Heidge Fukumasu; Jayme Augusto de Souza-Neto; José Salvatore Leister Patané; Luiz Alcantara; Luiz Lehmann Coutinho; Maria Carolina Elias; Maurício Lacerda Nogueira; Rafael dos Santos Bezerra; Raul Machado Neto; Rejane Maria Tommasini Grotto; Ricardo Haddad; Sandra Coccuzzo Sampaio Vessoni; Simone Kashima; Svetoslav Nanev Slavov; Vincent Louis Viala |
| EPI_ISL_3037811 | HOSPITAL SAN JUAN DE DIOS | Inciensa, Instituto Costarricense de Investigación y Enseñanza en Nutrición y Salud | Adriana Godínez; Caterina Guzmán; Claudio Soto-Garita; Estela Cordero; Francisco Duarte; Hebleen Porras; José Luis Vargas; Mariela Gutiérrez Joselyn Prado; Melany Calderón; Nazareth Ruiz & Juliana Mora |
| EPI_ISL_3102450, EPI_ISL_3102465 | HOSPITAL SAO JOSE DE DOENCAS INFECCIOSAS | Analytical Competence Molecular Epidemiology Lab/ACME, Oswaldo Cruz Foundation, Ceara (FIOCRUZ CE) | Cleber Furtado Aksenen; Fabio Miyajima; Fernando Braga Stehling; Francisco Eder de Moura Lopes; Jamille Maria Mendes Bezerra; Joaquim César do Nascimento Sousa Junior; Pedro Miguel Carneiro Jeronimo; Suzana Porto Almeida e Lucas Delerino; Thais Ferreira de Oliveira; Thais de Oliveira Costa; Ticiane Cavalcante de Souza; Veridiana Pessoa Miyajima |
| EPI_ISL_5649181, EPI_ISL_5665663 | HOSPITAL SAO JOSE ITIRAPINA | Instituto Butantan | Antonio Jorge Martins; Claudia Renata dos Santos Barros; David Schlesinger; Debora Botequio Moretti; Dimas Tadeu Covas; Elaine Cristina Marqueze; Elaine Vieira Santos; Evandra Strazza Rodrigues; Heidge Fukumasu; Jayme Augusto de Souza-Neto; José Salvatore Leister Patané; Luiz Alcantara; Luiz Lehmann Coutinho; Maria Carolina Elias; Maurício Lacerda Nogueira; Rafael dos Santos Bezerra; Raul Machado Neto; Rejane Maria Tommasini Grotto; Ricardo Haddad; Sandra Coccuzzo Sampaio Vessoni; Simone Kashima; Svetoslav Nanev Slavov; Vincent Louis Viala |
| EPI_ISL_4820601, EPI_ISL_4820605 | HPTU | Laboratorio Departamental de Salud Publica de Antioquia | Cristian Arbey Velarde Hoyos; Gloria Isabel Escobar; Idabely Betancur Ortiz; Juan P. Hernandez-Ortiz; Maria Stella López |
| EPI_ISL_6484016, EPI_ISL_6484073, EPI_ISL_6484075, EPI_ISL_6484076, EPI_ISL_6484082, EPI_ISL_6484090, EPI_ISL_6484104, EPI_ISL_6484106, EPI_ISL_6484384, EPI_ISL_6484385, EPI_ISL_6484401, EPI_ISL_6484403, EPI_ISL_6484413, EPI_ISL_6484422, EPI_ISL_6484448, EPI_ISL_6484459, EPI_ISL_6484473, EPI_ISL_6484480, EPI_ISL_6484503, EPI_ISL_6484573, EPI_ISL_6484583, EPI_ISL_6484588, EPI_ISL_6484594, EPI_ISL_6484598, EPI_ISL_6484605, EPI_ISL_6484609, EPI_ISL_6484611, EPI_ISL_6484642, EPI_ISL_6484647, EPI_ISL_6484702, EPI_ISL_6484709, EPI_ISL_6484711, EPI_ISL_6484734, EPI_ISL_6484764, EPI_ISL_6484782, EPI_ISL_6484792, EPI_ISL_6484793, EPI_ISL_6484797, EPI_ISL_6484798, EPI_ISL_6484806, EPI_ISL_6484822, EPI_ISL_6484823, EPI_ISL_6484824, EPI_ISL_6484831, EPI_ISL_6484844, EPI_ISL_6484854, EPI_ISL_6484862, EPI_ISL_6484866, EPI_ISL_6484870, EPI_ISL_6484876, EPI_ISL_6484877, EPI_ISL_6485011 | | | |
| see above | Helix | Centers for Disease Control and Prevention Division of Viral Diseases, Pathogen Discovery | Benjamin Rambo-Martin; Christopher Gulvick; Clinton Paden; Dakota Howard; Dhwani Batra; Duncan MacCannell; Erisa Sula; Helix CA; Jason Caravas; Kristine Lacek; Matthew Schmerer; Peter Cook; Scott Sammons; Shatavia Morrison; Tymeckia Kendall; Victoria Caban Figueroa; Yvette Unoarumhi |
| EPI_ISL_3825478 | Hosp. Pediatrico Dr. Avelino Castelán | Laboratorio de Biología Molecular, Instituto de Medicina Regional on behalf of 'Proyecto Argentino Interinstitucional de genomica de SARS-CoV-2' (PAIS Consortium) | Bettina Brusés; Gioia Lucia Marino; Griselda Oria; Horacio Lucero.; Javier Mussin; Julieta Ronchi; Laura Formichelli; María Andrea Gili; María Cecilia López; Melina Lorenzini Campos; Raúl Maximiliano Acevedo |
| EPI_ISL_3092579 | Hospital Alvarez Buylla | Laboratorio de Virología HUCA | Abreu F; Alvarez-Arguelles ME; Boga JA; Castelló C; Costales I; Coto E; Gómez de Oña J; Martín-Rodríguez G; Melón S; Perez-Martínez Z; Rojo S; Sandoval M |
| EPI_ISL_2782321, EPI_ISL_2782332, EPI_ISL_2782348, EPI_ISL_2782352, EPI_ISL_2782356 | Hospital General Universitario Gregorio Marañón | Hospital General Universitario Gregorio Marañón | Cristina Rodriguez-Grande; Darío García de Viedma; Julia Suárez; Laura Pérez-Lago; Marta Herranz Martin; Patricia Muñoz; Pedro Sola Campoy; Pilar Catalán; Sergio Buenestado Serrano; Victor Manuel de la Cueva |
| EPI_ISL_3925418, EPI_ISL_3925428 | Hospital General Universitario de Alicante - Instituto de Investigación Sanitaria y Biomédica de Alicante | SeqCOVID-SPAIN consortium/IBV(CSIC) | Carmen Molina Pardines and SeqCOVID-SPAIN consortium; Maripaz Ventero Martín |
| EPI_ISL_5926922 | Hospital Herrera Llerandi | Asociación de Salud Integral / Clínica Familiar "Luis Ángel García" | Ana S. Gonzalez-Reiche; Claudia Rangel; Danicela Mercado; Eduardo Arathon; Hilda Ruiz; Luis Aguirre; Luis Rivas; Narda Medina; Oscar Bonilla; Osmar Gamboa. |
| EPI_ISL_3260108 | Hospital Jaime Ferre - SAMCO Rafaela | Grupo de Genómica y Bioinformática del Instituto de Investigación de la Cadena Láctea CONICET-INTA on behalf of 'Proyecto Argentino Interinstitucional de genomica de SARS-CoV-2' (PAIS Consortium) | AF; Aliprandi D; Amadio; C; Eberhardt; Irazoqui; Isaia; JF; JM; MF; Pandolfi; Quaranta; Soratti R; V |
| EPI_ISL_2927978 | Hospital Margarita Maza de Juárez | Microbial Genomics Laboratory | ; Alejandra García-Gasca; Alejandra Hernández-Terán; Alejandro Sánchez-Flores; Alfredo Herrera-Estrella; Alicia Ocaña-Mondragón; Andreu Comas-García; Angel Gustavo Salas-Lais; Antonio Loza Román; Bernardo Martínez-Miguel; Blanca Taboada; Brenda Irasema Maldonado-Meza; Bruno Gómez-Gil; Carla Ivón Herrera-Najera; Carlos F. Arias; Celia Boukadida; Clara Esperanza Santacruz-Tinoco; Concepción Grajales-Muñiz; Consorcio Mexicano de Vigilancia Genómica (CoViGen-Mex). Authors (in alphabetical order): Julio Elias Alvarado-Yaah; Cristóbal Cháidez-Quiróz; Célida Duque Molina; Célida Martínez- Rodríguez; Daniel Fregoso-Rueda; Daniel Lira Morales; Eduardo Becerril-Vargas; Fernando Fontove-Herrera; Fidencio Mejía-Nepomuceno; Francisco Pulido; Gloria Elena Espinosa-Ayala; Gloria María Molina-Salinas; Gloria Vazquez; Hector Esteban Paz-Juárez; Hector Montoya-Fuentes; Helen Haydee Fernanda Ramírez-Plascencia; Irvin González-López; Jean Pierre González; Jesús Hernández; Joel Armando Vázquez-Pérez.; Jorge Salas-Hernández; José Antonio Enciso-Moreno; José Arturo Martínez-Orozco; José Esteban Muñoz-Medina; José de Jesús Nuñez-Contreras; Juan Bautista Chale-Dzul; Julissa Enciso-Ibarra; Luis Alberto Ochoa-Carrera; Margarita Matías-Florentino; Mario Mújica-Sánchez; Marissa Perez-Garcia; María Guadalupe Santiago-Mauricio; María Guadalupe de Jesús Mireles-Rivera; Nelly Sélem-Mojica; Pavel Isa; Ricardo Ciria Merce; Ricardo Grande; Rosa María Gutiérrez Rios; Santiago Ávila-Ríos; Selene Zárate; Susana Lopez; Verónica Mata-Haro; Victor Eduardo García-Arias; Víctor Hugo Borja-Aburto |
| EPI_ISL_2828226, EPI_ISL_2828227 | Hospital Universitari Arnau de Vilanova | Hospital Universitari Vall d'Hebron - Vall d'Hebron Institut de Recerca | Alejandra González-Sánchez; Andrés Antón; Ariadna Rando; Carla Castillo; Cristina Andrés; Damir Garcia-Cehic; Josep Quer; Juliana Esperalba; Maria Carmen Martin; Maria Gema Codina; Maria Piñana; Tomàs Pumarola |
| EPI_ISL_2861766 | Hospital Universitari Bellvitge | Microbiology Department | Aida Gonzalez-Diaz; Carmen Ardanuy; Jordi Camara; Jordi Niubó; Laura Calatayud; M Angeles Domínguez; Miguel Fernandez-Huerta; Sara Marti |
| EPI_ISL_2657359, EPI_ISL_2657366, EPI_ISL_2657367 | Hospital Universitari Joan XXIII | Hospital Universitari Joan XXIII | Carla Martín; Clara Benavent; Cristina Gutiérrez; Ester Picó; Gemma Recio; Margarida Terrón; Natalia Bastón |
| EPI_ISL_2828109, EPI_ISL_2828111, EPI_ISL_2828116, EPI_ISL_2828131, EPI_ISL_2828138, EPI_ISL_2828151, EPI_ISL_2828215, EPI_ISL_2828217 | | | |
| see above | Hospital Universitari Vall d'Hebron - Vall d'Hebron Institut de Recerca | Hospital Universitari Vall d'Hebron - Vall d'Hebron Institut de Recerca | Alejandra González-Sánchez; Andrés Antón; Ariadna Rando; Carla Castillo; Cristina Andrés; Damir Garcia-Cehic; Josep Quer; Juliana Esperalba; Maria Carmen Martin; Maria Gema Codina; Maria Piñana; Tomàs Pumarola |
| EPI_ISL_3008463, EPI_ISL_3008470, EPI_ISL_3008483, EPI_ISL_3008519 | Hospital Universitari i Politècnic La Fe de València | SeqCOVID-SPAIN consortium/IBV(CSIC) | Ana Gil Brusola; Eva González Barberá; José Luis López Hontangas and SeqCOVID-SPAIN consortium; María Dolores Gómez Ruiz; Salvador Giner Almaraz |
| EPI_ISL_7307244 | Hospital Universitario San Ignacio | Centro de Investigaciones en Microbiología y Biotecnología-UR (CIMBIUR), Facultad de Ciencias Naturales, Universidad del Rosario, Bogotá, Colombia | Alberto Paniz-Mondolfi; Angie Ramírez; Beatriz Ariza; Camilo A. Correa-Cárdenas; Carlos Gómez-Restrepo; Claudia Cardozo-Romero; Claudia Méndez; David-Santiago Quevedo; Guido España; Hernando Díaz; Juan David Ramírez; Juliana Cuervo-Rojas; Julie Pérez; Luz H. Patiño; Manuel-Antonio Franco; Maria-Clara Duque; Marina Muñoz; Nathalia Ballesteros; Nicolas Luna; Sergio Castañeda; Zulma M. Cucunubá |
| EPI_ISL_2968598 | Houston Health Dept. | Houston Health Dept. | Adolpho Lara; Pamela Brown; Ryker Penn; Yanlai Lai |
| EPI_ISL_5073252, EPI_ISL_5073281, EPI_ISL_5073295, EPI_ISL_5073301, EPI_ISL_5073302, EPI_ISL_5073316, EPI_ISL_5073338 | | | |
| see above | Houston Methodist Hospital | Houston Methodist Hospital | Ilya J. Finkelstein; James J. Davis; Jessica Cambric; Jimmy Gollihar; Kristina Reppond; Layne Pruitt; Madison N. Shyer; Marcus Nguyen; Matthew Ojeda Saavedra; Paul A. Christensen; Prasanti Yerramilli; Randall J. Olsen; Robert Olson; Ryan Gadd; S. Wesley Long; Sishir Subedi; and James M. Musser |
| EPI_ISL_3010281, EPI_ISL_3010291, EPI_ISL_3010292 | Humboldt County Public Health Laboratory | Humboldt County Public Health Laboratory | Jeremy Corrigan |
| EPI_ISL_3393360 | Hôpital Cochin | Department of Virology, Henri Mondor University Hospital, Assistance Publique Hôpitaux de Paris, Université Paris-Est Créteil, INSERM U955 | Alexandre Soulier; Christophe Rodriguez; Elisabeth Trawinski; Guillaume Gricourt; Jean-Michel Pawlotsky; Melissa N'Debi; Slim Fourati; Vanessa Demontant |
| EPI_ISL_3128024, EPI_ISL_3128028, EPI_ISL_3355267, EPI_ISL_3355326 | ID Bureau of Laboratories | Centers for Disease Control and Prevention Division of Viral Diseases, Pathogen Discovery | Alex Burgin; Ben L. Rambo-Martin; Ben Rambo-Martin; Clinton Paden; Clinton R. Paden; Dakota Howard; Dave Wentworth; Dhwani Batra; Jasmine Padilla; Justin Lee; Krista Queen; Kristen Knipe; Kristine Lacek; Mark Burroughs; Matthew Schmerer; Meghan Bentz; Mili Sheth; Peter Cook; Sam Shepard; Sarah Nobles; Suxiang Tong; Vivien Dugan; Yvette Unoarumhi |

| | | | |
|---|---|---|---|
| EPI_ISL_2787053, EPI_ISL_2787054, EPI_ISL_2787076, EPI_ISL_2787077, EPI_ISL_2787078 | IDI-IRCCS | INMI Lazzaro Spallanzani IRCCS | A Tamburro; B Bartolini; CEM Gruber; E Conti; E Giombini; E Sperandio; F De Nicola; F Messina; F Santini; G Bonfiglio; G Cappiello; G Tramini; GF Inger; L Ciuffreda; M Meledandri; M Pallocca; M Rueca; ML Schiavone; O Butera; R Longo; S Donzelli |
| EPI_ISL_2709474, EPI_ISL_2725041 | IN State Department of Health Laboratory Services | IN State Department of Health Laboratory Services | Brian Pope; Cassandra Campion; Jamie Yeadon; Kyle Brownlee; Lixia Liu; Mark Glazier; Melissa Hindenlang |
| EPI_ISL_2787019, EPI_ISL_2787026, EPI_ISL_2787066 | INMI Lazzaro Spallanzani IRCCS | INMI Lazzaro Spallanzani IRCCS | A Massacci; B Bartolini; CEM Gruber; D Cerini; D Di Fusco; E Giombini; E Sperandio; F Messina; F Santini; G Bonfiglio; GF Inger; L Ciuffreda; M Pallocca; M Rueca; O Butera; S Donzelli |
| EPI_ISL_5648267, EPI_ISL_5648304, EPI_ISL_5648344, EPI_ISL_5648443, EPI_ISL_5648450, EPI_ISL_5648454, EPI_ISL_5648458, EPI_ISL_5648468, EPI_ISL_5648469, EPI_ISL_5648479, EPI_ISL_5648509, EPI_ISL_5648521, EPI_ISL_5648534, EPI_ISL_5648535, EPI_ISL_5648541, EPI_ISL_5648571, EPI_ISL_5648576, EPI_ISL_5648577, EPI_ISL_5648586, EPI_ISL_5648602, EPI_ISL_5648605, EPI_ISL_5648627, EPI_ISL_5648628, EPI_ISL_5648632, EPI_ISL_5648633, EPI_ISL_5648634, EPI_ISL_5648640, EPI_ISL_5648648, EPI_ISL_5648658, EPI_ISL_5648659, EPI_ISL_5648665, EPI_ISL_5648678, EPI_ISL_5648684, EPI_ISL_5648688, EPI_ISL_5648694, EPI_ISL_5648695, EPI_ISL_5648703, EPI_ISL_5648704, EPI_ISL_5648708, EPI_ISL_5648711, EPI_ISL_5648712, EPI_ISL_5648715, EPI_ISL_5648726, EPI_ISL_5648730, EPI_ISL_5648749, EPI_ISL_5648753, EPI_ISL_5648773, EPI_ISL_5648777, EPI_ISL_5648781, EPI_ISL_5648786, EPI_ISL_5648788, EPI_ISL_5648798, EPI_ISL_5648822, EPI_ISL_5648831, EPI_ISL_5648832, EPI_ISL_5648835, EPI_ISL_5648840, EPI_ISL_5648849, EPI_ISL_5648859, EPI_ISL_5648887, EPI_ISL_5648903, EPI_ISL_5648904, EPI_ISL_5648909, EPI_ISL_5648910, EPI_ISL_5648925, EPI_ISL_5648926, EPI_ISL_5648931, EPI_ISL_5648940, EPI_ISL_5648961, EPI_ISL_5648982, EPI_ISL_5649000, EPI_ISL_5649011, EPI_ISL_5649017, EPI_ISL_5649024, EPI_ISL_5649035, EPI_ISL_5649048, EPI_ISL_5649058, EPI_ISL_5649063, EPI_ISL_5649086, EPI_ISL_5649087, EPI_ISL_5649095, EPI_ISL_5649096, EPI_ISL_5649097, EPI_ISL_5649105, EPI_ISL_5649106, EPI_ISL_5649109, EPI_ISL_5649110, EPI_ISL_5649115, EPI_ISL_5649121, EPI_ISL_5649122, EPI_ISL_5649127, EPI_ISL_5661382, EPI_ISL_5661485, EPI_ISL_5661647, EPI_ISL_5661762, EPI_ISL_5661892, EPI_ISL_5661936, EPI_ISL_5661945, EPI_ISL_5661952, EPI_ISL_5662201, EPI_ISL_5662235, EPI_ISL_5662310, EPI_ISL_5662318, EPI_ISL_5662337, EPI_ISL_5662348, EPI_ISL_5662376, EPI_ISL_5662382, EPI_ISL_5662440, EPI_ISL_5662445, EPI_ISL_5662453, EPI_ISL_5662472, EPI_ISL_5662578, EPI_ISL_5662601, EPI_ISL_5662764, EPI_ISL_5662809, EPI_ISL_5662877, EPI_ISL_5662889, EPI_ISL_5662913, EPI_ISL_5663081, EPI_ISL_5663229, EPI_ISL_5663292 | | | |
| see above | INSIDE DIAGNOSTICOS | Instituto Butantan | Antonio Jorge Martins; Claudia Renata dos Santos Barros; David Schlesinger; Debora Botequio Moretti; Dimas Tadeu Covas; Elaine Cristina Marqueze; Elaine Vieira Santos; Evandra Strazza Rodrigues; Heidge Fukumasu; Jayme Augusto de Souza-Neto; José Salvatore Leister Patané; Luiz Alcantara; Luiz Lehmann Coutinho; Maria Carolina Elias; Maurício Lacerda Nogueira; Rafael dos Santos Bezerra; Raul Machado Neto; Rejane Maria Tommasini Grotto; Ricardo Haddad; Sandra Coccuzzo Sampaio Vessoni; Simone Kashima; Svetoslav Nanev Slavov; Vincent Louis Viala |
| EPI_ISL_2893597, EPI_ISL_2988466, EPI_ISL_2988529, EPI_ISL_2988531, EPI_ISL_2988534, EPI_ISL_2988568 | INSPI-CRN DE INFLUENZA Y OTROS VIRUS RESPIRATORIOS | NIC-INSPI | Alfredo Bruno; Domenica de Mora.; Jimmy Garcés; Johanna Laines; Lizbeth Patiño; Manuel Gonzalez; Maritza Olmedo; Michelle Páez |
| EPI_ISL_3770695, EPI_ISL_3770702 | IZSM-Virologia | Istituto Zooprofilattico Sperimentale del Mezzogiorno - Unità Operativa Complessa di Virologia | Antonio Limone; Claudio de Martinis; Ester De Carlo; Giovanna Fusco; Loredana Cozzolino; Lorena Cardillo; Maurizio Viscardi |
| EPI_ISL_3090836, EPI_ISL_3090838, EPI_ISL_3090841 | Idaho Bureau of Laboratories | Boise VA Medical Center, PALMS | Aimee Ceniseros; Cheri Lamb McFarlane; Christian Loera; Dr. Christopher Ball; Emily Bartlette; James Razor; Matthew Burns; Ying Pei |
| EPI_ISL_3231919, EPI_ISL_3231920, EPI_ISL_3232018 | Illinois Department of Public Health | Illinois Department of Public Health - Chicago Lab | Ira Heimler; Joel Price; Vineet K. Dhiman |
| EPI_ISL_2885410, EPI_ISL_2885423, EPI_ISL_2885425, EPI_ISL_2983631 | Illinois Department of Public Health - Springfield Lab | Illinois Department of Public Health - Springfield Lab | Bryan Sim; Gordon McCall |
| EPI_ISL_3104970, EPI_ISL_3105030, EPI_ISL_3105047, EPI_ISL_3105050, EPI_ISL_3105051 | Incyte Diagnostics Spokane | Seattle Flu Study | Amanda Adler; Barry R. Lutz; Benjamin Pelle; Brian Hiatt; Caitlin R. Wolf; Chris D. Frazar; Deborah A. Nickerson; Elisabeth Brandstetter; Erica Ryke; Geoff Melly; Helen Y. Chu; Janet A. Englund; Jay Shendure; Jover Lee; Kairsten Fay; Kirsten Lacombe; Lea M. Starita; Mark J. Rieder; Matthew Richardson; Matthew Thompson; Melissa Truong; Michael Boeckh; Michael Famulare; Misja Ilcisin; Peter D. Han; Philip Dykema; Romesh Gautom; Scott Lindquist; Thomas R. Sibley; Trevor Bedford |
| EPI_ISL_2876051, EPI_ISL_3020041, EPI_ISL_3085867 | Infinity Biologix | Centers for Disease Control and Prevention Division of Viral Diseases, Pathogen Discovery | Adrian Paskey; Benjamin Rambo-Martin; Chirayu Goswami; Christian Bixby; Christopher Gulvick; Clinton R. Paden; Dakota Howard; Darlene Wagner; Dhwani Batra; Duncan MacCannell; Jason Caravas; Jonathan Schultz; Kara Moser; Matthew Schmerer; Peter W. Cook; Robin Grimwood; Russ Hager; Scott Sammons; Shatavia Morrison; Yihe Wang; Yvette Unoarumhi |
| EPI_ISL_2812179 | Institute for Infectious Diseases | Institute for Infectious Diseases | Alban Ramette; Christian Baumann; Cora SÃ¤gesser; Franziska Suter-Riniker; Miguel A Terrazos Miani; Pascal Bittel; Peter Keller; Stefan Neuenschwander; Stephen L Leib |
| EPI_ISL_2861149, EPI_ISL_3062504, EPI_ISL_3062505 | Institute of Microbiology, Universidad San Francisco de Quito | Institute of Microbiology, Universidad San Francisco de Quito | Alexandra Gonzalez; Belén Prado-Vivar; Bernardo Gutiérrez; Betty Angulo; Christian Zambrano; Danilo Granda-Moncayo; Erika Muñoz; Fernanda Zurita; Francisco Guerra; Gabriel Trueba; Hugo Vergara; Jeaninna Peña; Jiyoung Lim; Juan Carlos Zuñiga; Juan José Guadalupe; Luis Fuenmayor; Luz-Angelica Castillo; Marco Viteri Yanez; Michelle Grunauer; Michelle Jacome; Monica Becerra-Wong; Natalia Parra; Patricio Rojas-Silva; Paúl Cárdenas; Rommel Guevara; Ronny Javier Pibaque; Stephanie Espín-Arroba; Sully Márquez; Valeria Armijos; Verónica Barragán |
| EPI_ISL_3316162, EPI_ISL_5538368 | Instituto Adolfo Lutz - Regional de Bauru | Instituto Adolfo Lutz, Interdiciplinary Procedures Center, Strategic Laboratory | Caio Vinicius Dias Lopes; Claudia Regina Gonçalves; Claudio Tavares Sacchi; Karoline Rodrigues Campos; Leonardo Tadeu de Araujo; Marlon Benedito Nascimento Santos; Marlon Benedito Nascomento Santos |
| EPI_ISL_3259712, EPI_ISL_3259713, EPI_ISL_3259714, EPI_ISL_3259715, EPI_ISL_3259716, EPI_ISL_3259717 | Instituto Adolfo Lutz - Regional de Campinas | Instituto Adolfo Lutz, Interdiciplinary Procedures Center, Strategic Laboratory | Caio Vinicius Dias Lopes; Claudia Regina Gonçalves; Claudio Tavares Sacchi; Erica Valessa Ramos Gomes; Karoline Rodrigues Campos; Leonardo Tadeu de Araujo; Marlon Benedito Nascomento Santos |
| EPI_ISL_3717841, EPI_ISL_3717842, EPI_ISL_3717843, EPI_ISL_3717844, EPI_ISL_3717868, EPI_ISL_3717869, EPI_ISL_3717870, EPI_ISL_3717871, EPI_ISL_3717889 | | | |
| see above | Instituto Adolfo Lutz - Regional de Marilia | Instituto Adolfo Lutz, Interdiciplinary Procedures Center, Strategic Laboratory | Caio Vinicius Dias Lopes; Claudia Regina Gonçalves; Claudio Tavares Sacchi; Karoline Rodrigues Campos; Leonardo Tadeu de Araujo; Marlon Benedito Nascimento Santos |
| EPI_ISL_3671894 | Instituto Adolfo Lutz - Regional de Presidente Prudente | Instituto Adolfo Lutz, Interdiciplinary Procedures Center, Strategic Laboratory | Caio Vinicius Dias Lopes; Claudia Regina Gonçalves; Claudio Tavares Sacchi; Karoline Rodrigues Campos; Leonardo Tadeu de Araujo; Marlon Benedito Nascomento Santos |
| EPI_ISL_3671872, EPI_ISL_3671873, EPI_ISL_3671874, EPI_ISL_3671875, EPI_ISL_3671876 | Instituto Adolfo Lutz - Regional de Presidente Prudente | Instituto Adolfo Lutz, Interdisciplinary Procedures Center, Strategic Laboratory | Caio Vinicius Dias Lopes; Claudia Regina Gonçalves; Claudio Tavares Sacchi; Karoline Rodrigues Campos; Leonardo Tadeu de Araujo; Marlon Benedito Nascomento Santos |
| EPI_ISL_3316171, EPI_ISL_3316178, EPI_ISL_3316192, EPI_ISL_3316240, EPI_ISL_3316301, EPI_ISL_3316302, EPI_ISL_3316304, EPI_ISL_3316305 | | | |
| see above | Instituto Adolfo Lutz - Regional de Ribeirao Preto | Instituto Adolfo Lutz, Interdiciplinary Procedures Center, Strategic Laboratory | Caio Vinicius Dias Lopes; Claudia Regina Gonçalves; Claudio Tavares Sacchi; Karoline Rodrigues Campos; Leonardo Tadeu de Araujo; Marlon Benedito Nascomento Santos |
| EPI_ISL_3316293, EPI_ISL_3316294, EPI_ISL_5640214, EPI_ISL_5640219 | Instituto Adolfo Lutz - Regional de Rio Claro | Instituto Adolfo Lutz, Interdiciplinary Procedures Center, Strategic Laboratory | Caio Vinicius Dias Lopes; Claudia Regina Gonçalves; Claudio Tavares Sacchi; Karoline Rodrigues Campos; Leonardo Tadeu de Araujo; Marlon Benedito Nascimento Santos; Marlon Benedito Nascomento Santos |
| EPI_ISL_4056276, EPI_ISL_4056277, EPI_ISL_4056278, EPI_ISL_4056279, EPI_ISL_4056280, EPI_ISL_4056281, EPI_ISL_4056282, EPI_ISL_4056283, EPI_ISL_4056284, EPI_ISL_4056285 | | | |
| see above | Instituto Adolfo Lutz - Regional de Santo Andre | Instituto Adolfo Lutz, Interdiciplinary Procedures Center, Strategic Laboratory | Caio Vinicius Dias Lopes; Claudia Regina Gonçalves; Claudio Tavares Sacchi; Karoline Rodrigues Campos; Leonardo Tadeu de Araujo; Marlon Benedito Nascimento Santos |
| EPI_ISL_3259726, EPI_ISL_3259727, EPI_ISL_3259728, EPI_ISL_3316244 | Instituto Adolfo Lutz - Regional de Sao Jose do Rio Preto | Instituto Adolfo Lutz, Interdiciplinary Procedures Center, Strategic Laboratory | Caio Vinicius Dias Lopes; Claudia Regina Gonçalves; Claudio Tavares Sacchi; Erica Valessa Ramos Gomes; Karoline Rodrigues Campos; Leonardo Tadeu de Araujo; Marlon Benedito Nascomento Santos |
| EPI_ISL_3259684, EPI_ISL_3259685, EPI_ISL_3259686, EPI_ISL_3259687, EPI_ISL_3259688 | Instituto Adolfo Lutz - Regional de Sorocaba | Instituto Adolfo Lutz, Interdiciplinary Procedures Center, Strategic Laboratory | Caio Vinicius Dias Lopes; Claudia Regina Gonçalves; Claudio Tavares Sacchi; Erica Valessa Ramos Gomes; Karoline Rodrigues Campos; Leonardo Tadeu de Araujo; Marlon Benedito Nascomento Santos |
| EPI_ISL_3259743, EPI_ISL_3259744, EPI_ISL_3259745, EPI_ISL_3259746, EPI_ISL_3259747, EPI_ISL_3259748, EPI_ISL_3259750, EPI_ISL_3259757, EPI_ISL_3316249, EPI_ISL_3316250, EPI_ISL_3316251, EPI_ISL_3316252, EPI_ISL_3316253, EPI_ISL_3316254, EPI_ISL_3316255, EPI_ISL_3316256, EPI_ISL_3316257, EPI_ISL_3316258, EPI_ISL_3316259, EPI_ISL_3316260, EPI_ISL_3316261, EPI_ISL_3316262, EPI_ISL_3316266, EPI_ISL_3316267, EPI_ISL_3316268, EPI_ISL_3316269, EPI_ISL_3316270, EPI_ISL_3316271, EPI_ISL_3316272, EPI_ISL_3316273, EPI_ISL_3316274, EPI_ISL_3316276, EPI_ISL_3316277, EPI_ISL_3316278, EPI_ISL_3316279, EPI_ISL_3316283, EPI_ISL_3316284, EPI_ISL_3316286, EPI_ISL_3316289, EPI_ISL_3316290, EPI_ISL_3671895, EPI_ISL_3671896, EPI_ISL_3671897, EPI_ISL_3671898, EPI_ISL_3671899, EPI_ISL_3671900, EPI_ISL_3671901, EPI_ISL_3710516, EPI_ISL_3717835, EPI_ISL_3717836, EPI_ISL_3717837, EPI_ISL_3717838, EPI_ISL_4206883, EPI_ISL_4206922, EPI_ISL_4206949, EPI_ISL_4299351, EPI_ISL_4299352, EPI_ISL_4299353, EPI_ISL_4299354, EPI_ISL_4299357, EPI_ISL_4299358, EPI_ISL_4299360, EPI_ISL_4888052, EPI_ISL_4888053, EPI_ISL_4888058, EPI_ISL_5030013, EPI_ISL_5030015, EPI_ISL_5030021 | | | |
| see above | Instituto Adolfo Lutz Central | Instituto Adolfo Lutz, Interdiciplinary Procedures Center, Strategic Laboratory | Caio Vinicius Dias Lopes; Claudia Regina Goncalves; Claudia Regina Gonçalves; Claudio Tavares Sacchi; Erica Valessa Ramos Gomes; Karoline Rodrigues Campos; Leonardo Tadeu de Araujo; Marlon Benedito Nascimento Santos; Marlon Benedito Nascomento Santos |

| | | | |
|---|---|---|---|
| EPI_ISL_2894236, EPI_ISL_2894255, EPI_ISL_3067685, EPI_ISL_3067782 | Instituto Nacional de Medicina Genomica | Instituto Nacional de Medicina Genomica | Abraham Campos-Romero; Cedro-Tanda A; Cisneros- Villanueva M; Escobar-Arrazola; Gonzalez-Barrera D; Herrera-Montalvo LA.; Hidalgo-Miranda A; Luna-Ruiz Marco; M; Mendoza-Vargas A; Moreno-Camacho José Luis; Munguia-Garza P; Ramirez-Vega O; Rangel-DeLeon D; Reyes-Grajeda JP; Rodriguez-Gallegos Jorge |
| EPI_ISL_3118918, EPI_ISL_3118919, EPI_ISL_3118921 | Instituto de Biotecnologia - UNESP-Botucatu-SP | Instituto de Biotecnologia - UNESP-Botucatu-SP | Cecília Artico Banho; Cíntia Bittar; Fábio Sossai Possebon; Guilherme Campos; Helena Lage Ferreira; Jorge A. Petroli Marchesi; João Pessoa Araújo Jr.; Leila Sabrina Ullmann; Lívia Sacchetto; Maisa C. Pereira Parra; Marília Moraes; Maurício L. Nogueira; Paula Rahal; Paulo Inacio da Costa |
| EPI_ISL_3347092, EPI_ISL_3347099 | Interpath Laboratory | Seattle Flu Study | Amanda Adler; Barry R. Lutz; Benjamin Pelle; Brian Hiatt; Caitlin R. Wolf; Chris D. Frazar; Deborah A. Nickerson; Elisabeth Brandstetter; Erica Ryke; Geoff Melly; Helen Y. Chu; Janet A. Englund; Jay Shendure; Jover Lee; Kairsten Fay; Kirsten Lacombe; Lea M. Starita; Mark J. Rieder; Matthew Richardson; Matthew Thompson; Melissa Truong; Michael Boeckh; Michael Famulare; Misja Ilcisin; Peter D. Han; Philip Dykema; Romesh Gautom; Scott Lindquist; Thomas R. Sibley; Trevor Bedford |
| EPI_ISL_2844338, EPI_ISL_2844339, EPI_ISL_2844341, EPI_ISL_2844389, EPI_ISL_2844416, EPI_ISL_2844422, EPI_ISL_2844429, EPI_ISL_2844445, EPI_ISL_2844446, EPI_ISL_2844448, EPI_ISL_2844460, EPI_ISL_2844464, EPI_ISL_2844468, EPI_ISL_2844474, EPI_ISL_2844475, EPI_ISL_2844485 | | | |
| see above | Istituto Zooprofilattico Sperimentale del Mezzogiorno | TIGEM | Antonio Grimaldi Patrizia Annunziata Francesco Panariello Biancamaria Pierri Claudia Tiberio Teresa Giuliano Valentina Bouche Chiara Colantuono Maria Concetta Cuomo Denise Di Concilio Lucio Di Filippo Anna Manfredi Marcello Salvi Antonio Limone Luigi Atripaldi Pellegrino Cerino Andrea Ballabio Davide Cacchiarelli |
| EPI_ISL_2725136, EPI_ISL_2725137, EPI_ISL_2725362, EPI_ISL_2725363, EPI_ISL_2725385, EPI_ISL_2725393, EPI_ISL_2725394, EPI_ISL_2725407 | | | |
| see above | Istituto Zooprofilattico Sperimentale del Mezzogiorno | Telethon Institute of Genetics and Medicine (TIGEM) | Antonio Grimaldi Patrizia Annunziata Francesco Panariello Biancamaria Pierri Claudia Tiberio Teresa Giuliano Valentina Bouche Chiara Colantuono Maria Concetta Cuomo Denise Di Concilio Lucio Di Filippo Anna Manfredi Marcello Salvi Antonio Limone Luigi Atripaldi Pellegrino Cerino Andrea Ballabio Davide Cacchiarelli |
| EPI_ISL_2725315, EPI_ISL_2725316, EPI_ISL_2725319 | Istituto Zooprofilattico Sperimentale del Mezzogiorno - Azienda Ospedaliera "Pugliese Ciaccio" di Catanzaro | Telethon Institute of Genetics and Medicine (TIGEM) | Antonio Grimaldi Patrizia Annunziata Francesco Panariello Biancamaria Pierri Claudia Tiberio Teresa Giuliano Valentina Bouche Chiara Colantuono Maria Concetta Cuomo Denise Di Concilio Lucio Di Filippo Anna Manfredi Pasquale Minchella Marcello Salvi Antonio Limone Luigi Atripaldi Pellegrino Cerino Andrea Ballabio Davide Cacchiarelli |
| EPI_ISL_2885153 | Johns Hopkins Hospital Department of Pathology | Johns Hopkins Hospital Department of Pathology | Adannaya Amadi; C. Paul Morris; Chun Huai Luo; Heba H. Mostafa; Matthew Schwartz; Nicholas Gallagher |
| EPI_ISL_5404874 | KU Leuven, Rega Institute, Clinical and Epidemiological Virology | KU Leuven, Rega Institute, Clinical and Epidemiological Virology | Bert Vanmechelen; Joan Marti-Carerras; Piet Maes; Tony Wawina-Bokalanga |
| EPI_ISL_3561761, EPI_ISL_3561766, EPI_ISL_4102590, EPI_ISL_4102591, EPI_ISL_4102593, EPI_ISL_4102605, EPI_ISL_4102607, EPI_ISL_4102620, EPI_ISL_4102629, EPI_ISL_4102630, EPI_ISL_4102639, EPI_ISL_4102641, EPI_ISL_4102645, EPI_ISL_4102647, EPI_ISL_4102651, EPI_ISL_4102652, EPI_ISL_4102655, EPI_ISL_4102776, EPI_ISL_4102782, EPI_ISL_4636778, EPI_ISL_4636779 | | | |
| see above | Kaiser Permanente NW Reginal Lab | OHSU MM Lab | Amber Halse; Jeannine Lama; Xuan Qin; Yun Wu |
| EPI_ISL_2688221, EPI_ISL_2842083 | Kansas Health and Environmental Lab | Kansas Health and Environmental Lab | Ben Olsen; Jonathan Barnell; Katherine Wiggins; Mike Grose; and Phil Adam |
| EPI_ISL_4417734 | Kentucky State Public Health Lab | Kentucky State Public Health Lab | Anjayya Tummala; Chitralipi Kattunga; Joshua Tobias; Karim George; Matthew Johnson; Rachel Zinner; Rhonda Lucas; Vaneet Arora; William Grooms |
| EPI_ISL_3064741 | LA Office of Public Health Laboratories | Centers for Disease Control and Prevention Division of Viral Diseases, Pathogen Discovery | Alex Burgin; Ben L. Rambo-Martin; Clinton R. Paden; Dakota Howard; Dave Wentworth; Dhwani Batra; Jasmine Padilla; Justin Lee; Krista Queen; Kristen Knipe; Kristine Lacek; Mark Burroughs; Matthew Schmerer; Meghan Bentz; Mili Sheth; Peter Cook; Sam Shepard; Sarah Nobles; Suxiang Tong; Vivien Dugan; Yvette Unoarumhi |
| EPI_ISL_2933178 | LABORATOIRE ACCOLAB SUD OUEST | CNR Virus des Infections Respiratoires - France SUD | Antonin Bal; Bruno Lina; Gregory Destras; Gwendolyne Burfin; Hadrien Regue; Laurence Josset; Martine Valette; Quentin Semanas |
| EPI_ISL_5649363, EPI_ISL_5649375, EPI_ISL_5649381, EPI_ISL_5649398, EPI_ISL_5649435, EPI_ISL_5649452, EPI_ISL_5649471, EPI_ISL_5649481, EPI_ISL_5659518, EPI_ISL_5659540 | | | |
| see above | LABORATORIO DE FRANCA | Instituto Butantan | Antonio Jorge Martins; Claudia Renata dos Santos Barros; David Schlesinger; Debora Botequio Moretti; Dimas Tadeu Covas; Elaine Cristina Marqueze; Elaine Vieira Santos; Evandra Strazza Rodrigues; Heidge Fukumasu; Jayme Augusto de Souza-Neto; José Salvatore Leister Patané; Luiz Alcantara; Luiz Lehmann Coutinho; Maria Carolina Elias; Maurício Lacerda Nogueira; Rafael dos Santos Bezerra; Raul Machado Neto; Rejane Maria Tommasini Grotto; Ricardo Haddad; Sandra Coccuzzo Sampaio Vessoni; Simone Kashima; Svetoslav Nanev Slavov; Vincent Louis Viala |
| EPI_ISL_5648329 | LABORATORIO MUNICIPAL DE ANALISES CLINICAS DE ITANHAEM | Instituto Butantan | Antonio Jorge Martins; Claudia Renata dos Santos Barros; David Schlesinger; Debora Botequio Moretti; Dimas Tadeu Covas; Elaine Cristina Marqueze; Elaine Vieira Santos; Evandra Strazza Rodrigues; Heidge Fukumasu; Jayme Augusto de Souza-Neto; José Salvatore Leister Patané; Luiz Alcantara; Luiz Lehmann Coutinho; Maria Carolina Elias; Maurício Lacerda Nogueira; Rafael dos Santos Bezerra; Raul Machado Neto; Rejane Maria Tommasini Grotto; Ricardo Haddad; Sandra Coccuzzo Sampaio Vessoni; Simone Kashima; Svetoslav Nanev Slavov; Vincent Louis Viala |
| EPI_ISL_5649210, EPI_ISL_5665670 | LABORATORIO MUNICIPAL DE ANALISES CLINICAS DE RIO CLARO | Instituto Butantan | Antonio Jorge Martins; Claudia Renata dos Santos Barros; David Schlesinger; Debora Botequio Moretti; Dimas Tadeu Covas; Elaine Cristina Marqueze; Elaine Vieira Santos; Evandra Strazza Rodrigues; Heidge Fukumasu; Jayme Augusto de Souza-Neto; José Salvatore Leister Patané; Luiz Alcantara; Luiz Lehmann Coutinho; Maria Carolina Elias; Maurício Lacerda Nogueira; Rafael dos Santos Bezerra; Raul Machado Neto; Rejane Maria Tommasini Grotto; Ricardo Haddad; Sandra Coccuzzo Sampaio Vessoni; Simone Kashima; Svetoslav Nanev Slavov; Vincent Louis Viala |
| EPI_ISL_5648797, EPI_ISL_5648833, EPI_ISL_5648947, EPI_ISL_5648979, EPI_ISL_5649006, EPI_ISL_5649047, EPI_ISL_5649056, EPI_ISL_5649061, EPI_ISL_5649068, EPI_ISL_5649108, EPI_ISL_5663388, EPI_ISL_5663482, EPI_ISL_5665157 | | | |
| see above | LABORATORIO MUNICIPAL DE PIRACICABA | Instituto Butantan | Antonio Jorge Martins; Claudia Renata dos Santos Barros; David Schlesinger; Debora Botequio Moretti; Dimas Tadeu Covas; Elaine Cristina Marqueze; Elaine Vieira Santos; Evandra Strazza Rodrigues; Heidge Fukumasu; Jayme Augusto de Souza-Neto; José Salvatore Leister Patané; Luiz Alcantara; Luiz Lehmann Coutinho; Maria Carolina Elias; Maurício Lacerda Nogueira; Rafael dos Santos Bezerra; Raul Machado Neto; Rejane Maria Tommasini Grotto; Ricardo Haddad; Sandra Coccuzzo Sampaio Vessoni; Simone Kashima; Svetoslav Nanev Slavov; Vincent Louis Viala |
| EPI_ISL_3404588, EPI_ISL_3404589, EPI_ISL_3404592, EPI_ISL_3404593, EPI_ISL_3404594, EPI_ISL_3404595, EPI_ISL_3404596, EPI_ISL_3404597 | | | |
| see above | LACEN | LGBio (Laboratorio de Genetica & Biodiversidade) | Alex Honda Bernardes; Amanda Alves de Melo; Aparecido Divino da Cruz; Cintia Pelegrineti Targueta de Azevedo Brito; Daniela de Melo e Silva; Elisangela de Paula Silveira Lacerda; Juliana Santana de Curcio; Luiz Augusto Pereira; Marc Alexandre Duarte Gigonzac; Mariana Pires de Campos Telles; Ramilla dos Santos Braga; Renata de Oliveira Dias; Rhewter Nunes; Thais Cidalia Vieira Gigonzac; Thais Guimarães Castro; Thays Millena Alves Pedroso |
| EPI_ISL_6721535 | LACEN - Laboratório Central de Saúde Pública de Pernambuco | Evandro Chagas Institute | Delana Andreza Melo Bezerra; Dielle Monteiro Teixeira; Jedson Ferreira Cardoso; Jessylene de Almeida Ferreira; Kenny da Costa Pinheiro; Luana Soares Barbagelata; Luana da Silva Soares; Mirleide Cordeiro dos Santos; Patrícia dos Santos Lobo; Rayssa Layna da Silva Bedran; Sandro Patroca da Silva |
| EPI_ISL_6721705, EPI_ISL_6721707 | LACEN - Laboratório Central de Saúde Pública do Amazonas | Evandro Chagas Institute | Delana Andreza Melo Bezerra; Dielle Monteiro Teixeira; Jedson Ferreira Cardoso; Jessylene de Almeida Ferreira; Kenny da Costa Pinheiro; Luana Soares Barbagelata; Luana da Silva Soares; Mirleide Cordeiro dos Santos; Patrícia dos Santos Lobo; Rayssa Layna da Silva Bedran; Sandro Patroca da Silva |
| EPI_ISL_6721631 | LACEN - Laboratório Central de Saúde Pública do Ceara | Evandro Chagas Institute | Delana Andreza Melo Bezerra; Dielle Monteiro Teixeira; Jedson Ferreira Cardoso; Jessylene de Almeida Ferreira; Kenny da Costa Pinheiro; Luana Soares Barbagelata; Luana da Silva Soares; Mirleide Cordeiro dos Santos; Patrícia dos Santos Lobo; Rayssa Layna da Silva Bedran; Sandro Patroca da Silva |
| EPI_ISL_5538536, EPI_ISL_5538541, EPI_ISL_5538553, EPI_ISL_5538562, EPI_ISL_5538568, EPI_ISL_5538582, EPI_ISL_5538593, EPI_ISL_5538599, EPI_ISL_5538607, EPI_ISL_5538622, EPI_ISL_5538638 | | | |
| see above | LACEN do Distrito Federal | Instituto Adolfo Lutz, Interdiciplinary Procedures Center, Strategic Laboratory | Caio Vinicius Dias Lopes; Claudia Regina Gonçalves; Claudio Tavares Sacchi; Karoline Rodrigues Campos; Leonardo Tadeu de Araujo; Marlon Benedito Nascimento Santos |
| EPI_ISL_2894547, EPI_ISL_2894548, EPI_ISL_2896234, EPI_ISL_2896239, EPI_ISL_2896240, EPI_ISL_2896241 | LACEN do Estado de Mato Grosso | Instituto Adolfo Lutz, Interdiciplinary Procedures Center, Strategic Laboratory | Caio Vinicius Dias Lopes; Claudia Regina Gonçalves; Claudio Tavares Sacchi; Karoline Rodrigues Campos |
| EPI_ISL_4487679, EPI_ISL_4487688, EPI_ISL_4487702, EPI_ISL_4487711 | LACEN/PI | WallauLab on behalf of Fiocruz COVID-19 Genomic Surveillance Network | Adelino Soares Lima Neto; Alexandre Freitas da Silva; Antônio Marinho; Cassia Docena; Constância Flávia Junqueira Ayres; Filipe Zimmer Dezordi; Gabriel Luz Wallau; Gustavo Barbosa de Lima; Hellen de Oliveira Amaral; Jacenir Reis dos Santos Mallet; Joana Carolina Viana Lima; Lais Ceschini Machado; Leandro de Mattos; Lilian Caroliny Amorim Silva; Marcela de Lacerda Valença Queiroz; Marcelo Adriano da Cunha e Silva Vieira; Marcelo Henrique dos Santos Paiva; Matheus Filgueira Bezerra; Sinval Pinto Brandão Filho; Túlio de Lima Campos; Vladimir Costa Silva; Walterlene de Carvalho Goçalves |
| EPI_ISL_5193461, EPI_ISL_5193518, EPI_ISL_5193522, EPI_ISL_5193523, EPI_ISL_5193524, EPI_ISL_5193525, EPI_ISL_5193526, EPI_ISL_5193527, EPI_ISL_5193583, EPI_ISL_5193631 | | | |
| see above | LACEN/RR | Laboratorio de Ecologia de Doencas Transmissiveis na Amazonia, Instituto Leonidas e Maria Deane - Fiocruz Amazonia | André Corado; Cátia Alexandra Ribeiro Meneses; Felipe Naveca; Fernanda Nascimento; George Silva; Ilma de Aguiar Antony; Janis Cassimiro Lopes; Luciana Gonçalves; Maria Júlia Brandão; Matilde Mejía; Pricila Carlos Veloso; Rodrigo Melo Maito; Valdinete Nascimento; Victor Souza |
| EPI_ISL_3261109, EPI_ISL_3267946, EPI_ISL_3267947, EPI_ISL_3267948, EPI_ISL_3267949, EPI_ISL_3267950, EPI_ISL_3267951, EPI_ISL_3267952, EPI_ISL_3267953, EPI_ISL_3267954, EPI_ISL_3267955, EPI_ISL_3267956, EPI_ISL_3267968, EPI_ISL_3267969, EPI_ISL_3267970, EPI_ISL_3267973, EPI_ISL_3267974, EPI_ISL_3267975, EPI_ISL_3267976, EPI_ISL_3267978, EPI_ISL_3267979, EPI_ISL_3267980, EPI_ISL_3267981, EPI_ISL_3267982, EPI_ISL_3267983, EPI_ISL_3267984, EPI_ISL_3267985, EPI_ISL_3267986, EPI_ISL_3267987, EPI_ISL_3267988, EPI_ISL_3267989, EPI_ISL_3267990, EPI_ISL_3267991, EPI_ISL_3267992, EPI_ISL_3267993, EPI_ISL_3267994, EPI_ISL_3267995, EPI_ISL_3267996, EPI_ISL_3267997, EPI_ISL_3267998, EPI_ISL_3267999, EPI_ISL_3268000, EPI_ISL_3268001, EPI_ISL_3268002, EPI_ISL_3268003, EPI_ISL_3268004, EPI_ISL_3268005, EPI_ISL_3268006, EPI_ISL_3268007, EPI_ISL_3268020, EPI_ISL_3268022, EPI_ISL_3268023, EPI_ISL_3268025 | | | |
| see above | LATE - Laboratório de Técnicas Especiais - Hospital Israelita Albert Einstein | LATE - Laboratório de Técnicas Especiais - Hospital Israelita Albert Einstein | Alexandre Hideaki Takara; Ana Paula Moreira Salles; Anelisie da Silva Santos; Deyvid Amgarten; Erick Gustavo Dorlass; Fernanda de Mello Malta; João Renato Rebello Pinho; Marcio Anunciacao Menezes; Pedro Henrique Sebe Rodrigues; Raquel Riyuzo |
| EPI_ISL_4666435 | LDSP | Laboratorio Departamental de Salud Publica de Antioquia | Andres F. Cardona-Rios; Cristian Arbey Velarde Hoyos; Gloria Isabel Escobar; Idabely Betancur Ortiz; Juan P. Hernandez-Ortiz; Maria Stella López |
| EPI_ISL_3033577, EPI_ISL_3033578 | LESP Baja California Sur | Instituto de Diagnostico y Referencia Epidemiologicos (INDRE) | Abril Rodriguez-Maldonado; Ariadna Medina-Benitez; Claudia Wong-Arambula; Ernesto Ramirez-Gonzalez.; Gisela Barrera-Badillo; Irma Lopez-Martinez; Joaquin Quiroz-Mercado; Lucia Hernandez-Rivas; Maribel Gonzalez-Villa; Natividad Cruz-Ortiz; Sergio Rangel-Guerrero; Tatiana Nunez-Garcia; Vanessa Rivero-Arredondo |
| EPI_ISL_3033474, EPI_ISL_3033475, EPI_ISL_3033476 | LESP Campeche | Instituto de Diagnostico y Referencia Epidemiologicos (INDRE) | Abril Rodriguez-Maldonado; Ariadna Medina-Benitez; Claudia Wong-Arambula; Ernesto Ramirez-Gonzalez.; Gisela Barrera-Badillo; Irma Lopez-Martinez; Joaquin Quiroz-Mercado; Lucia Hernandez-Rivas; Maribel Gonzalez-Villa; Natividad Cruz-Ortiz; Sergio Rangel-Guerrero; Tatiana Nunez-Garcia; Vanessa Rivero-Arredondo |
| EPI_ISL_2859061, EPI_ISL_2859062, EPI_ISL_2937804, EPI_ISL_2937805 | LESP Ciudad de Mexico | Instituto de Diagnostico y Referencia Epidemiologicos (INDRE) | Abril Rodriguez-Maldonado; Ariadna Medina-Benitez; Claudia Wong-Arambula; Ernesto Ramirez-Gonzalez.; Gisela Barrera-Badillo; Irma Lopez-Martinez; Joaquin Quiroz-Mercado; Lucia Hernandez-Rivas; Maribel Gonzalez-Villa; Natividad Cruz-Ortiz; Sergio Rangel-Guerrero; Tatiana Nunez-Garcia; Vanessa Rivero-Arredondo |
| EPI_ISL_3033589 | LESP Guanajuato | Instituto de Diagnostico y Referencia Epidemiologicos (INDRE) | Abril Rodriguez-Maldonado; Ariadna Medina-Benitez; Claudia Wong-Arambula; Ernesto Ramirez-Gonzalez.; Gisela Barrera-Badillo; Irma Lopez-Martinez; Joaquin Quiroz-Mercado; Lucia Hernandez-Rivas; Maribel Gonzalez-Villa; Natividad Cruz-Ortiz; Sergio Rangel-Guerrero; Tatiana Nunez-Garcia; Vanessa Rivero-Arredondo |
| EPI_ISL_3033562, EPI_ISL_3033566 | LESP Hidalgo | Instituto de Diagnostico y Referencia Epidemiologicos (INDRE) | Abril Rodriguez-Maldonado; Ariadna Medina-Benitez; Claudia Wong-Arambula; Ernesto Ramirez-Gonzalez.; Gisela Barrera-Badillo; Irma Lopez-Martinez; Joaquin Quiroz-Mercado; Lucia Hernandez-Rivas; Maribel Gonzalez-Villa; Natividad Cruz-Ortiz; Sergio Rangel-Guerrero; Tatiana Nunez-Garcia; Vanessa Rivero-Arredondo |

| | | | |
|---|---|---|---|
| EPI_ISL_2937815 | LESP Jalisco | Instituto de Diagnostico y Referencia Epidemiologicos (INDRE) | Abril Rodriguez-Maldonado; Ariadna Medina-Benitez; Claudia Wong-Arambula; Ernesto Ramirez-Gonzalez.; Gisela Barrera-Badillo; Irma Lopez-Martinez; Joaquin Quiroz-Mercado; Lucia Hernandez-Rivas; Maribel Gonzalez-Villa; Natividad Cruz-Ortiz; Sergio Rangel-Guerrero; Tatiana Nunez-Garcia; Vanessa Rivero-Arredondo |
| EPI_ISL_2859059 | LESP San Luis Potosi | Instituto de Diagnostico y Referencia Epidemiologicos (INDRE) | Abril Rodriguez-Maldonado; Ariadna Medina-Benitez; Claudia Wong-Arambula; Ernesto Ramirez-Gonzalez.; Gisela Barrera-Badillo; Irma Lopez-Martinez; Joaquin Quiroz-Mercado; Lucia Hernandez-Rivas; Maribel Gonzalez-Villa; Natividad Cruz-Ortiz; Sergio Rangel-Guerrero; Tatiana Nunez-Garcia; Vanessa Rivero-Arredondo |
| EPI_ISL_3033574 | LESP Sinaloa | Instituto de Diagnostico y Referencia Epidemiologicos (INDRE) | Abril Rodriguez-Maldonado; Ariadna Medina-Benitez; Claudia Wong-Arambula; Ernesto Ramirez-Gonzalez.; Gisela Barrera-Badillo; Irma Lopez-Martinez; Joaquin Quiroz-Mercado; Lucia Hernandez-Rivas; Maribel Gonzalez-Villa; Natividad Cruz-Ortiz; Sergio Rangel-Guerrero; Tatiana Nunez-Garcia; Vanessa Rivero-Arredondo |
| EPI_ISL_2859051, EPI_ISL_3033579, EPI_ISL_3033580 | LESP Sonora | Instituto de Diagnostico y Referencia Epidemiologicos (INDRE) | Abril Rodriguez-Maldonado; Ariadna Medina-Benitez; Claudia Wong-Arambula; Ernesto Ramirez-Gonzalez.; Gisela Barrera-Badillo; Irma Lopez-Martinez; Joaquin Quiroz-Mercado; Lucia Hernandez-Rivas; Maribel Gonzalez-Villa; Natividad Cruz-Ortiz; Sergio Rangel-Guerrero; Tatiana Nunez-Garcia; Vanessa Rivero-Arredondo |
| EPI_ISL_2859040, EPI_ISL_2859046, EPI_ISL_2859047, EPI_ISL_2859048, EPI_ISL_2859055, EPI_ISL_2859056, EPI_ISL_2859057, EPI_ISL_2937747, EPI_ISL_2937809, EPI_ISL_2937810, EPI_ISL_2937811, EPI_ISL_2937812, EPI_ISL_2937813, EPI_ISL_2937819, EPI_ISL_2937820, EPI_ISL_2937821, EPI_ISL_2937822, EPI_ISL_2937823, EPI_ISL_2937824, EPI_ISL_2937825, EPI_ISL_2937826, EPI_ISL_3033553 | | | |
| see above | LESP Tabasco | Instituto de Diagnostico y Referencia Epidemiologicos (INDRE) | Abril Rodriguez-Maldonado; Ariadna Medina-Benitez; Claudia Wong-Arambula; Ernesto Ramirez-Gonzalez.; Gisela Barrera-Badillo; Irma Lopez-Martinez; Joaquin Quiroz-Mercado; Lucia Hernandez-Rivas; Maribel Gonzalez-Villa; Natividad Cruz-Ortiz; Sergio Rangel-Guerrero; Tatiana Nunez-Garcia; Vanessa Rivero-Arredondo |
| EPI_ISL_3033469 | LESP Tamaulipas | Instituto de Diagnostico y Referencia Epidemiologicos (INDRE) | Abril Rodriguez-Maldonado; Ariadna Medina-Benitez; Claudia Wong-Arambula; Ernesto Ramirez-Gonzalez.; Gisela Barrera-Badillo; Irma Lopez-Martinez; Joaquin Quiroz-Mercado; Lucia Hernandez-Rivas; Maribel Gonzalez-Villa; Natividad Cruz-Ortiz; Sergio Rangel-Guerrero; Tatiana Nunez-Garcia; Vanessa Rivero-Arredondo |
| EPI_ISL_2937779, EPI_ISL_2937780, EPI_ISL_2937781, EPI_ISL_2937782, EPI_ISL_2937783, EPI_ISL_3033498, EPI_ISL_3033518, EPI_ISL_3033519, EPI_ISL_3033520, EPI_ISL_3033522, EPI_ISL_3033523, EPI_ISL_3033524, EPI_ISL_3033525, EPI_ISL_3033526, EPI_ISL_3033528, EPI_ISL_3033529, EPI_ISL_3033530, EPI_ISL_3033531, EPI_ISL_3033533, EPI_ISL_3033534, EPI_ISL_3033535, EPI_ISL_3033536, EPI_ISL_3033537, EPI_ISL_3033538, EPI_ISL_3033539, EPI_ISL_3033540, EPI_ISL_3033541, EPI_ISL_3033543, EPI_ISL_3033590, EPI_ISL_3033591, EPI_ISL_3046102, EPI_ISL_3046103 | | | |
| see above | LESP Yucatan | Instituto de Diagnostico y Referencia Epidemiologicos (INDRE) | Abril Rodriguez-Maldonado; Ariadna Medina-Benitez; Claudia Wong-Arambula; Ernesto Ramirez-Gonzalez.; Gisela Barrera-Badillo; Irma Lopez-Martinez; Joaquin Quiroz-Mercado; Lucia Hernandez-Rivas; Maribel Gonzalez-Villa; Natividad Cruz-Ortiz; Sergio Rangel-Guerrero; Tatiana Nunez-Garcia; Vanessa Rivero-Arredondo |
| EPI_ISL_4743406 | LIME | Laboratorio Departamental de Salud Publica de Antioquia | Andres F. Cardona-Rios; Cristian Arbey Velarde Hoyos; Gloria Isabel Escobar; Idabely Betancur Ortiz; Juan P. Hernandez-Ortiz; Maria Stella López |
| EPI_ISL_2861479, EPI_ISL_2861480 | Lab voor klinische biologie | Lab voor klinische biologie | Bruno Verhasselt; Hannelore Hamerlinck; Marija Janevska |
| EPI_ISL_2844492, EPI_ISL_2844494 | Lab. Microbiologia e Virologia Cotugno A.O. dei Colli - Istituto Zooprofilattico Sperimentale del Mezzogiorno | TIGEM | Antonio Grimaldi Patrizia Annunziata Francesco Panariello Biancamaria Pierri Claudia Tiberio Teresa Giuliano Valentina Bouche Chiara Colantuono Maria Concetta Cuomo Denise Di Concilio Lucio Di Filippo Anna Manfredi Marcello Salvi Antonio Limone Luigi Atripaldi Pellegrino Cerino Andrea Ballabio Davide Cacchiarelli |
| EPI_ISL_2725152, EPI_ISL_2725178 | Lab. Microbiologia e Virologia Cotugno A.O. dei Colli - Istituto Zooprofilattico Sperimentale del Mezzogiorno | Telethon Institute of Genetics and Medicine (TIGEM) | Antonio Grimaldi Patrizia Annunziata Francesco Panariello Biancamaria Pierri Claudia Tiberio Teresa Giuliano Valentina Bouche Chiara Colantuono Maria Concetta Cuomo Denise Di Concilio Lucio Di Filippo Anna Manfredi Marcello Salvi Antonio Limone Luigi Atripaldi Pellegrino Cerino Andrea Ballabio Davide Cacchiarelli |
| EPI_ISL_2835744, EPI_ISL_2923134, EPI_ISL_2923135, EPI_ISL_2923176 | Labo Analyses Med | National Reference Center for Viruses of Respiratory Infections, Institut Pasteur, Paris | Angela Brisebarre; Camille Capel; Christophe Malabat; Corinne Maufrais; Damien Mornico; Etienne Simon-Lorière; Frédéric Lemoine; Gael Millot; Louise Lefrançois; Marion Barbet; Maud Vanpeene; Méline Bizard; Sabine Trombert; Sylvie Behillil; Sylvie Van der Werf; TROMBERT Sabine; Vincent Enouf |
| EPI_ISL_2845132 | Labor Blackholm MVZ | Robert Koch Institute | |
| EPI_ISL_2845261 | Labor Dr. Wisplinghoff - Köln | Robert Koch Institute | |
| EPI_ISL_2878457 | Laboratoire Ana-L | Department of Virology, Henri Mondor University Hospital, Assistance Publique Hôpitaux de Paris, Université Paris-Est Créteil, INSERM U955 | Alexandre Soulier; Christophe Rodriguez; Elisabeth Trawinski; Guillaume Gricourt; Jean-Michel Pawlotsky; Melissa N'Debi; Slim Fourati; Vanessa Demontant |
| EPI_ISL_5136039 | Laboratoire de Virologie – CHU François Mitterrand - Dijon – France | INSERM U1231 - Equipe GAD | Ali SI-MOHAMMED; Hélène GIRAUDON; Lionel PIROTH; Yannis DUFFOURD |
| EPI_ISL_3459046 | Laboratoire de santé publique du Québec | Laboratoire de santé publique du Québec | Guillaume Bourque; Ioannis Ragoussis; Jesse Shapiro; Mark Lathrop and Michel Roger on behalf of the CoVSeQ research group; Sandrine Moreira |
| EPI_ISL_3147805 | Laboratoires d'analyses medicales - Ketterthill | Laboratoire national de sante, Microbiology, Microbial Genomics Platform | Anke Wienecke-Baldacchino; Caroline Scheiber; Catherine Ragimbeau; Elodie Solarino; Fatu Djabi; Jessica Tapp; Lise Pignon; Raoul Salmon; Serge Vedy; Tamir Abdelrahman; Virginie Jover |
| EPI_ISL_2983234 | Laboratorio Central de Saude Publica do Estado de Goias (LACEN/GO) | Laboratory of Respiratory Viruses and Measles, Oswaldo Cruz Institute, FIOCRUZ | Agatha Cristinne Prudencio; Alice Sampaio Rocha; Ana Carolina Mendonca; Ana Flavia Mendonça; Anna Carolina Paixao; Carmen Helena Ramos; Cassiane Casanova; Elisa Cavalcante Pereira; Fernando Motta; Flavia Pereira Amorim da Silva; Ighor Leonardo Arantes Gomes; Luciana Appolinario; Luiz Augusto Pereira; Marilda Siqueira on behalf of the Fiocruz COVID-19 Genomic Surveillance Network; Paola Resende; Rafael Souza Guedes; Renata Serrano Lopes; Taina Venas; Vinicius Lemes da Silva |
| EPI_ISL_3539761, EPI_ISL_3539762, EPI_ISL_3539763, EPI_ISL_3539764 | Laboratorio Central de Saude Publica do Estado de Minas Gerais (LACEN/MG) | Laboratory of Respiratory Viruses and Measles, Oswaldo Cruz Institute, FIOCRUZ | Alice Sampaio Rocha; Ana Carolina Mendonca; Anna Carolina Paixao; Elisa Cavalcante Pereira; Fernando Motta; Luciana Appolinario; Marilda Siqueira on behalf of the Fiocruz COVID-19 Genomic Surveillance Network; Paola Resende; Renata Serrano Lopes; Rubens Pasa; Taina Venas |
| EPI_ISL_4080770 | Laboratorio Central de Saude Publica do Estado do Para (LACEN/PA) | Laboratory of Respiratory Viruses and Measles, Oswaldo Cruz Institute, FIOCRUZ | Agatha Soares; Alice Sampaio Rocha; Ana Carolina Mendonca; Anna Carolina Paixao; Elisa Cavalcante Pereira; Fernando Motta; Ighor Arantes; Luciana Appolinario; Marilda Siqueira on behalf of the Fiocruz COVID-19 Genomic Surveillance Network; Paola Resende; Renata Serrano Lopes; Taina Venas; Valnete Andrade |
| EPI_ISL_2840655 | Laboratorio Analisi Osp. Città di Castello - Azienda USL Umbria1 | Istituto Zooprofilattico Sperimentale dell'Abruzzo e Molise "G. Caporale" | Ancora M; Calistri P; Cammà C; Curini V; Delli Compagni E; Di Domenico M; Di Pasquale A; Lorusso A; Malagigi V; Mangone I; Marcacci M; Puglia I; Rinaldi A; Savini G; Scialabba S; Taccani P |
| EPI_ISL_2926010, EPI_ISL_2926053, EPI_ISL_2926054 | Laboratorio CQRC | CQRC_QUALITY CONTROL CHEMICAL BIOLOGICAL RISK_AOOR Villa Sofia Cervello Palermo | Brunacci G.; Contino F.; Di Gaudio F. |
| EPI_ISL_4196694, EPI_ISL_4196695, EPI_ISL_4196703, EPI_ISL_4196704 | Laboratorio Central Mg. Luis Alfredo Pianciola | Laboratorio Central Mg. Luis Alfredo Pianciola on behalf of 'Proyecto Argentino Interinstitucional de genomica de SARS-CoV-2' (PAIS Consortium) | C Pintos; C Rastellini.; C Ziehm; J Ousset; L Pianciola; M Fernandez; M Mazzeo |
| EPI_ISL_3245101, EPI_ISL_3245102, EPI_ISL_3245104, EPI_ISL_3245106, EPI_ISL_3245107, EPI_ISL_3245108, EPI_ISL_3245109, EPI_ISL_3245110, EPI_ISL_3245111, EPI_ISL_3245112, EPI_ISL_3245113, EPI_ISL_3245114, EPI_ISL_3245115, EPI_ISL_3245116, EPI_ISL_3245118, EPI_ISL_3245130, EPI_ISL_3245131, EPI_ISL_3245132, EPI_ISL_3245133, EPI_ISL_3245134, EPI_ISL_3245138, EPI_ISL_3245145, EPI_ISL_3245149, EPI_ISL_3245150, EPI_ISL_3245151, EPI_ISL_3245152, EPI_ISL_3245166, EPI_ISL_3245167, EPI_ISL_3245168, EPI_ISL_3245169, EPI_ISL_3245176, EPI_ISL_3245177, EPI_ISL_3245187, EPI_ISL_3245340 | | | |
| see above | Laboratorio Central Noel Nutels | Bioinformatics Laboratory / LNCC | Alessandra P Lamarca; Alexandra L Gerber; Amilcar Tanuri; Ana Paula de C Guimaraes; Ana Tereza R Vasconcelos; Andrea Cony Cavalcanti; Caio Luiz Pereira Ribeiro; Cintia Policarpo; Claudia Maria Braga de Mello; Cristiane Gomes da Silva; Douglas Terra Machado; Erica Ramos dos Santos Nascimento; Fernanda Leitao dos Santos; Flavio Dias da Silva; Gleidson da Silva de Oliveira; Leandro Magalhaes de Souza; Liliane Cavalcante; Luiz G P de Almeida; Marcio Henrique de Oliveira Garcia; Mario Sergio Ribeiro; Ricardo Jose Barbosa Salviano; Ronaldo da Silva F Jr; Silvia Carvalho |
| EPI_ISL_2942569, EPI_ISL_2942573, EPI_ISL_2942829, EPI_ISL_2969936, EPI_ISL_2970058 | Laboratorio Central de Epidemiologia (LCE) | Unidad de Genomica Avanzada | ; Alejandra Garcia-Gasca; Alejandra Hernandez-Teran; Alejandro Sanchez-Flores; Alfredo Herrera-Estrella; Alicia Ocaña-Mondragon; Andreu Comas-Garcia; Angel Gustavo Salas-Lais; Antonio Loza Roman; Bernardo Martinez-Miguel; Blanca Taboada; Brenda Irasema Maldonado-Meza; Bruno Gomez-Gil; Carla Ivon Herrera-Najera; Carlos F. Arias; Celia Boukadida; Celida Duque Molina; Celida Martinez- Rodriguez; Clara Esperanza Santacruz-Tinoco; Concepcion Grajales-Muñiz; Consorcio Mexicano de Vigilancia Genomica (CoViGen-Mex). Authors (in alphabetical order): Julio Elias Alvarado-Yaah; Cristobal Chaidez-Quiroz; Daniel Fregoso-Rueda; Daniel Lira Morales; Eduardo Becerril-Vargas; Fernando Fontove-Herrera; Fidencio Mejia-Nepomuceno; Francisco Pulido; Gloria Elena Espinosa-Ayala; Gloria Maria Molina-Salinas; Gloria Vazquez; Hector Esteban Paz-Juarez; Hector Montoya-Fuentes; Helen Haydee Fernanda Ramirez-Plascencia; Irvin Gonzalez-Lopez; Jean Pierre Gonzalez; Jesus Hernandez; Joel Armando Vazquez-Perez.; Jorge Salas-Hernandez; Jose Antonio Enciso-Moreno; Jose Arturo Martinez-Orozco; Jose Esteban Muñoz-Medina; Jose de Jesus Nuñez-Contreras; Juan Bautista Chale-Dzul; Julissa Enciso-Ibarra; Luis Alberto Ochoa-Carrera; Margarita Matias-Florentino; Maria Guadalupe Santiago-Mauricio; Maria Guadalupe de Jesus Mireles-Rivera; Mario Mujica-Sanchez; Marissa Perez-Garcia; Nelly Selem-Mojica; Pavel Isa; Ricardo Ciria Merce; Ricardo Grande; Rosa Maria Gutierrez Rios; Santiago avila-Rios; Selene Zarate; Susana Lopez; Veronica Mata-Haro; Victor Eduardo Garcia-Arias; Victor Hugo Borja-Aburto |
| EPI_ISL_6944558, EPI_ISL_6944607, EPI_ISL_6944608, EPI_ISL_6944610 | Laboratorio Central de Salud Publica | Viral Special Pathogens Branch, Centers for Disease Control and Prevention | Andrea Gómez de la Fuente; Cynthia Vázquez; Emir Talundzic; Joel Montgomery; John Klena; Juan Torales; Justin Lee; María José Ortega; María Liz Gamarra; Shannon Whitmer; Shirley Villalba |
| EPI_ISL_4030323, EPI_ISL_5488364 | Laboratorio Central de Saude Publica do Amazonas - LACEN-AM | Laboratorio de Ecologia de Doencas Transmissiveis na Amazonia, Instituto Leonidas e Maria Deane - Fiocruz Amazonia | André Corado; Arlesson Viana Da Silva; Debora Duarte; Felipe Naveca; Fernanda Nascimento; George Silva; Karina Pessoa; Luciana Gonçalves; Maria Júlia Brandão; Matilde Mejía; Michele Jesus; Valdinete Nascimento; Victor Souza; Ágatha Costa |
| EPI_ISL_2983374, EPI_ISL_2983375, EPI_ISL_2983376 | Laboratorio Central de Saude Publica do Estado da Bahia (LACEN/BA) | Laboratory of Respiratory Viruses and Measles, Oswaldo Cruz Institute, FIOCRUZ | Alice Sampaio Rocha; Ana Carolina Mendonca; Anna Carolina Paixao; Elisa Cavalcante Pereira; Felicidade Pereira; Fernando Motta; Luciana Appolinario; Marilda Siqueira on behalf of the Fiocruz COVID-19 Genomic Surveillance Network; Paola Resende; Renata Serrano Lopes; Taina Venas |
| EPI_ISL_3190228, EPI_ISL_3190229, EPI_ISL_3190230, EPI_ISL_3190231, EPI_ISL_3190232, EPI_ISL_3190233, EPI_ISL_3190234, EPI_ISL_3190235, EPI_ISL_3190236, EPI_ISL_3190325, EPI_ISL_3190327, EPI_ISL_3190328, EPI_ISL_3190329, EPI_ISL_3190330, EPI_ISL_3190331, EPI_ISL_3190332, EPI_ISL_3434829, EPI_ISL_3434851 | | | |
| see above | Laboratorio Central de Saude Publica do Estado de Alagoas (LACEN/AL) | Laboratory of Respiratory Viruses and Measles, Oswaldo Cruz Institute, FIOCRUZ | Agatha Cristinne Prudencio; Agatha Soares; Alice Sampaio Rocha; Ana Carolina Mendonca; Anderson Brandao Leite; Anna Carolina Paixao; Elisa Cavalcante Pereira; Fernando Motta; Ighor Arantes; Ighor Leonardo Arantes Gomes; Luciana Appolinario; Marilda Siqueira on behalf of the Fiocruz COVID-19 Genomic Surveillance Network; Paola Resende; Renata Serrano Lopes; Taina Venas |
| EPI_ISL_2983340, EPI_ISL_2983341, EPI_ISL_2983342, EPI_ISL_2983343, EPI_ISL_2983344, EPI_ISL_2983348, EPI_ISL_2983351, EPI_ISL_2983357, EPI_ISL_2983358, EPI_ISL_2983366, EPI_ISL_2983367, EPI_ISL_2983370, EPI_ISL_2983371 | | | |
| see above | Laboratorio Central de Saude Publica do Estado de Santa Catarina (LACEN-SC) | Laboratory of Respiratory Viruses and Measles, Oswaldo Cruz Institute, FIOCRUZ | Alice Sampaio Rocha; Ana Carolina Mendonca; Anna Carolina Paixao; Darcita Buerger Rovaris; Elisa Cavalcante Pereira; Fernando Motta; Luciana Appolinario; Marilda Siqueira on behalf of the Fiocruz COVID-19 Genomic Surveillance Network; Paola Resende; Renata Serrano Lopes; Sandra Bianchini Fernandes; Taina Venas |

| | | | |
|---|---|---|---|
| EPI_ISL_3235299, EPI_ISL_3235300, EPI_ISL_3434925, EPI_ISL_3434927 | Laboratorio Central de Saude Publica do Estado de Santa Catarina (LACEN/SC) | Laboratory of Respiratory Viruses and Measles, Oswaldo Cruz Institute, FIOCRUZ | Alice Sampaio Rocha; Ana Carolina Mendonca; Anna Carolina Paixao; Darcita Buerger Rovaris; Elisa Cavalcante Pereira; Fernando Motta; Luciana Appolinario; Marilda Siqueira on behalf of the Fiocruz COVID-19 Genomic Surveillance Network; Paola Resende; Renata Serrano Lopes; Sandra Bianchini Fernandes; Taina Venas |
| EPI_ISL_3803019 | Laboratorio Central de Saude Publica do Estado de Sergipe (LACEN/SE) | Laboratory of Respiratory Viruses and Measles, Oswaldo Cruz Institute, FIOCRUZ | Agatha Soares; Alice Sampaio Rocha; Ana Carolina Mendonca; Anna Carolina Paixao; Cliomar Alves dos Santos; Elisa Cavalcante Pereira; Fernando Motta; Ighor Arantes; Luciana Appolinario; Marilda Siqueira on behalf of the Fiocruz COVID-19 Genomic Surveillance Network; Paola Resende; Renata Serrano Lopes; Tainá Moreira Martins Venas |
| EPI_ISL_3434960, EPI_ISL_3434961, EPI_ISL_3434962, EPI_ISL_3434964, EPI_ISL_3434965, EPI_ISL_3434966, EPI_ISL_3434975, EPI_ISL_3434979, EPI_ISL_3434980, EPI_ISL_3539741, EPI_ISL_3539742, EPI_ISL_3539743, EPI_ISL_3539744, EPI_ISL_3539745, EPI_ISL_3539746, EPI_ISL_3539932 | | | |
| see above | Laboratorio Central de Saude Publica do Estado do Amapa (LACEN/AP) | Laboratory of Respiratory Viruses and Measles, Oswaldo Cruz Institute, FIOCRUZ | Agatha Cristinne Prudencio; Alice Sampaio Rocha; Ana Carolina Mendonca; Andreia Santos Costa; Anna Carolina Paixao; Anne Caroline da Silva Soledade; Elisa Cavalcante Pereira; Fernando Motta; Ighor Leonardo Arantes Gomes; Lindomar dos Anjos Silva; Luciana Appolinario; Marcia Socorro Pereira Cavalcante; Marilda Siqueira on behalf of the Fiocruz COVID-19 Genomic Surveillance Network; Paola Resende; Renata Serrano Lopes; Taina Venas |
| EPI_ISL_3045465, EPI_ISL_3190244, EPI_ISL_3190245, EPI_ISL_3190246, EPI_ISL_3190248, EPI_ISL_3827894, EPI_ISL_3827895, EPI_ISL_3827903, EPI_ISL_3827908, EPI_ISL_3827928, EPI_ISL_3827938, EPI_ISL_3827961, EPI_ISL_3827964, EPI_ISL_3827979, EPI_ISL_3827995, EPI_ISL_3828004, EPI_ISL_3828008, EPI_ISL_3828011 | | | |
| see above | Laboratorio Central de Saude Publica do Estado do Espirito Santo (LACEN/ES) | Laboratory of Respiratory Viruses and Measles, Oswaldo Cruz Institute, FIOCRUZ | Alice Sampaio Rocha; Ana Carolina Mendonca; Anna Carolina Paixao; Elisa Cavalcante Pereira; Fernando Motta; Luciana Appolinario; Marilda Siqueira on behalf of the Fiocruz COVID-19 Genomic Surveillance Network; Paola Resende; Renata Serrano Lopes; Rodrigo Ribeiro Rodrigues; Taina Venas |
| EPI_ISL_3914147 | Laboratorio Central de Saude Publica do Estado do Parana (Instituto de Biologia Molecular do Paraná (LACEN-PR) | Instituto Carlos Chagas - Fiocruz | Alessandra De Melo Aguiar; Andreia Akemi Suzukawa; Andréa Rodrigues Ávila; Bruno Dallagiovanna; Dalila Zanette; Eduardo Balsanelli; Emanuel Maltempi de Souza; Fabio Passetti; Fabricio Klerynton Marchini; Fábio de Oliveira Pedrosa; Guilherme Becker; Helisson Faoro; Hellen Geremias dos Santos; Irina Nastassja Riediger; Letusa Albrecht; Lucas Blanes; Luis Gustavo Morello; Lysangela Ronalte Alves; Maria do Carmo Debur; Mauro de Medeiros Oliveira; Michelle Orane Schemberger; Paola Cristina Resende; Sheila Cristina Nardeli; Tiago Gräf; Valter Antônio de Baura |
| EPI_ISL_3190265, EPI_ISL_3190367, EPI_ISL_3190368, EPI_ISL_3190370, EPI_ISL_3190371, EPI_ISL_3190372, EPI_ISL_3190373, EPI_ISL_3190374, EPI_ISL_3190375, EPI_ISL_3190376, EPI_ISL_3190377, EPI_ISL_3190378, EPI_ISL_3190379, EPI_ISL_3190380 | | | |
| see above | Laboratorio Central de Saude Publica do Estado do Parana (LACEN/PR) | Laboratory of Respiratory Viruses and Measles, Oswaldo Cruz Institute, FIOCRUZ | Agatha Soares; Alice Sampaio Rocha; Ana Carolina Mendonca; Anna Carolina Paixao; Elisa Cavalcante Pereira; Fernando Motta; Ighor Arantes; Irina Riediger; Luciana Appolinario; Marilda Siqueira on behalf of the Fiocruz COVID-19 Genomic Surveillance Network; Paola Resende; Renata Serrano Lopes; Taina Venas |
| EPI_ISL_4418875 | Laboratorio Central de Saude Publica do Estado do Rio Grande do Norte (LACEN/RN) | Laboratory of Respiratory Viruses and Measles, Oswaldo Cruz Institute, FIOCRUZ | Agatha Soares; Alice Sampaio Rocha; Ana Carolina Mendonca; Ana Paula Ferreira Costa; Anna Carolina Paixao; Antonnyo Palmielly Diogenes Lima; Aurélio de Oliveira Bento; Elisa Cavalcante Pereira; Fernando Motta; Gessika Brenna Costa Alves; Heglayne Pereira Vital da Silva; Iago de Souza Gomes; Ighor Arantes; Isabelle Cristina Clemente dos Santos; Janaina Sonale Cavalcante Nogueira de Oliveira; Jayra Juliana Paiva Alves Abrantes; Jonas José da Silva; Luciana Appolinario; Marilda Siqueira on behalf of the Fiocruz COVID-19 Genomic Surveillance Network; Paola Resende; Renata Serrano Lopes; Taina Venas; Themis Rocha de Souza; Vítor Gabriel Saldanha Fernandes |
| EPI_ISL_3539846, EPI_ISL_3539938, EPI_ISL_3539939, EPI_ISL_3539940, EPI_ISL_3539941, EPI_ISL_3539942, EPI_ISL_3539943, EPI_ISL_3539944, EPI_ISL_3539945, EPI_ISL_3539946, EPI_ISL_3539947, EPI_ISL_3539948, EPI_ISL_3539949, EPI_ISL_3539950, EPI_ISL_3539951, EPI_ISL_3539952, EPI_ISL_3539953, EPI_ISL_3539954, EPI_ISL_3539955, EPI_ISL_3539956, EPI_ISL_3539957, EPI_ISL_3539958, EPI_ISL_3539959, EPI_ISL_3539960, EPI_ISL_3539961, EPI_ISL_3539962, EPI_ISL_3539963, EPI_ISL_3539964, EPI_ISL_3539965, EPI_ISL_3539966, EPI_ISL_3539967, EPI_ISL_3539968, EPI_ISL_3539969, EPI_ISL_3539970, EPI_ISL_3539971, EPI_ISL_3539972, EPI_ISL_3539973, EPI_ISL_3539974, EPI_ISL_3539977, EPI_ISL_3539979, EPI_ISL_3539991, EPI_ISL_4061391 | | | |
| see above | Laboratorio Central de Saude Publica do Estado do Rio Grande do Sul (LACEN-RS) | Laboratory of Respiratory Viruses and Measles, Oswaldo Cruz Institute, FIOCRUZ | Agatha Cristinne Soares; Alice Sampaio Rocha; Ana Carolina Mendonca; Anderson Brandao Leite; Anna Carolina Paixao; Elisa Cavalcante Pereira; Fernando Motta; Ighor Leonardo Arantes; Luciana Appolinario; Marilda Siqueira on behalf of the Fiocruz COVID-19 Genomic Surveillance Network; Paola Resende; Renata Serrano Lopes; Richard Salvato; Taina Venas; Tatiana Schaffer Gregianini |
| EPI_ISL_2982742, EPI_ISL_2982744, EPI_ISL_2982745, EPI_ISL_2982746, EPI_ISL_2982747, EPI_ISL_2982748, EPI_ISL_2982749, EPI_ISL_2982750, EPI_ISL_2982751, EPI_ISL_2982752, EPI_ISL_2982753, EPI_ISL_2982754, EPI_ISL_2982755, EPI_ISL_2982756, EPI_ISL_2983214 | | | |
| see above | Laboratorio Central de Saude Publica do Estado do Rio de Janeiro (LACEN-RJ) | Laboratory of Respiratory Viruses and Measles, Oswaldo Cruz Institute, FIOCRUZ | Agatha Cristinne Prudencio; Alice Sampaio Rocha; Ana Carolina Mendonca; Andrea Cony Cavalcanti; Anna Carolina Paixao; Elisa Cavalcante Pereira; Fernando Motta; Ighor Leonardo Arantes Gomes; Luciana Appolinario; Marilda Siqueira on behalf of the Fiocruz COVID-19 Genomic Surveillance Network; Paola Resende; Renata Serrano Lopes; Taina Venas |
| EPI_ISL_3033079, EPI_ISL_3260098, EPI_ISL_3260099, EPI_ISL_3260100, EPI_ISL_3260101, EPI_ISL_3260104, EPI_ISL_3260105 | | | |
| see above | Laboratorio Central de la Ciudad de Santa Fe | Grupo de Genómica y Bioinformática del Instituto de Investigación de la Cadena Láctea CONICET-INTA on behalf of 'Proyecto Argentino Interinstitucional de genomica de SARS-CoV-2' (PAIS Consortium) | AF; Amadio; C; Eberhardt; G; Irazoqui; JM; MF; Mugna; Ojeda; Pastor; Rompato; V |
| EPI_ISL_2940272, EPI_ISL_2940274, EPI_ISL_2940275, EPI_ISL_2940277, EPI_ISL_2940278, EPI_ISL_2940285, EPI_ISL_2940288, EPI_ISL_2940290 | | | |
| see above | Laboratorio Central, Ministerio de Salud Córdoba | Instituto de Patologia Vegetal (CIAP-INTA) on behalf of 'Proyecto Argentino Interinstitucional de genomica de SARS-CoV-2' (PAIS Consortium) | Barbas, G.; Castro, G.; Debat, HJ.; FD; Fernández; M; M.B.; Marquez, N.; Pisano; Re, V. |
| EPI_ISL_3150217 | Laboratorio Clínico Gamma | "Unidad de Secuenciación y Genómica, USACH" | "Carlos A. Villarroel; Francisco A. Cubillos"; Kamila Urbina; Mara Cuevas; Marcelo Cortez-San Martin; Yesseny Vásquez-Martínez |
| EPI_ISL_3333564 | Laboratorio Clínico Gamma | MELISA Institute Genomics and Proteomics SpA | Cristian Vargas; Elard Koch; Francisco Alvarez; Guillermo Nourdin; Mauricio Hernandez |
| EPI_ISL_2787068, EPI_ISL_2787069 | Laboratorio Genzano - ASL RM 6 | INMI Lazzaro Spallanzani IRCCS | B Bartolini; CEM Gruber; E Conti; E Giombini; E Sperandio; F De Nicola; F Messina; G Bonfiglio; G Cappiello; G Tramini; L Ciuffreda; M Pallocca; M Rueca; O Butera; R Longo |
| EPI_ISL_3275241, EPI_ISL_5617486, EPI_ISL_5617949 | Laboratorio Nacional de Salud, Ministerio de Salud Publica y Asistencia Social | Genomics and Proteomics Departament, Gorgas Memorial Institute For Health Studies | Alexander A Martinez; Ambar Moreno; Claudia Estrada; Claudia Gonzalez V; César Roberto Conde Pereira; Jessica Gondola; Leyda Abrego; Marlene Castillo; Melissa Gaitan; Oris Chavarria |
| EPI_ISL_4578885 | Laboratorio de Biología Molecular Bolívar | Área de Secuenciación del Laboratorio de Virología del Hospital de Niños Dr. Ricardo Gutierrez on behalf of 'Proyecto Argentino Interinstitucional de genomica de SARS-CoV-2' (PAIS Consortium) | Acuña; Cabrerizo; Caviglia; D; Di Claudio; D'Andrea; F; Goya; Jofré; LE; Larreche; Lescano; Lusso; M; ME; MI; ML; MR; MS; Martínez; Nabaes Jodar; Natale; R; S; Ticeira; Valinotto; Viegas, M. |
| EPI_ISL_4405155, EPI_ISL_4405157 | Laboratorio de Biología molecular del Hospital General de Agudos Dr. Carlos G. Durand | Área de Secuenciación del Laboratorio de Virología del Hospital de Niños Dr. Ricardo Gutierrez on behalf of 'Proyecto Argentino Interinstitucional de genomica de SARS-CoV-2' (PAIS Consortium) | A; Acuña; Anzorena; B; C; Carrón; Castro; Cañellas; Chiussi; Colina; Costa; D; Dahinten; Dima; Domínguez; E; Elsegood; F; Frisone; G; Gatica; Goya; Irrazábal; Jurado; L; LE; Leivas; Loayza; Lusso; M; MB; MF; MI; MS; Marchissio; Marina; Matillas; N; Nabaes Jodar; Narduzzi; Natale; Notaristéfano; R; Rivarola; Rodríguez Cardozo; Rodríguez Saá; Rozo; S; Theaux; Valinotto; Viegas, M.; W; Warszatska; Y; Yaunguzian |
| EPI_ISL_3050515, EPI_ISL_3050516, EPI_ISL_3050517, EPI_ISL_4030315, EPI_ISL_4030342, EPI_ISL_4030343, EPI_ISL_4030347 | | | |
| see above | Laboratorio de Ecologia de Doencas Transmissiveis na Amazonia, Instituto Leonidas e Maria Deane - Fiocruz Amazonia | Laboratorio de Ecologia de Doencas Transmissiveis na Amazonia, Instituto Leonidas e Maria Deane - Fiocruz Amazonia | Alex Martins; André Corado; Debora Duarte; Fabiola Mendonça da Silva Chui; Felipe Naveca; Fernanda Nascimento; Fernando Fonseca de Almeida Val; George Silva; Gisely Cardoso de Melo; Karina Pessoa; Luciana Gonçalves; Marcus Vinicius Guimarães de Lacerda; Maria Júlia Brandão; Maria Paula Gomes Mourão; Mariana Xavier Simão; Matilde Mejía; Michele Jesus; Valdinete Nascimento; Vanderson de Souza Sampaio; Victor Souza; Ágatha Costa |
| EPI_ISL_3133683, EPI_ISL_5994528, EPI_ISL_5994539 | Laboratorio de Infectologia y Virologia Molecular | Laboratory of Molecular Virology, School of Medicine, Pontificia Universidad Catolica de Chile | Alejandro Bhrun; Ana Maria Contreras; Andres E. Munoz-Marcos; Carlos Palma; Catalina Pardo-Roa; Constanza Maldonado; Constanza Martinez-Valdevenito; Eileen Serrano; Erick Salinas; Estefany Poblete; Eugenia L. Fuentes Luppichini; Francisco Melo; Jennifer Angulo; Jorge Levican; Leonardo I. Almonacid; M. Belen Leyton; Magdalena Vera; Marcela Ferres; Maria Jose Avendano; Rafael A. Medina; Rafael A. Medina.; Tamara Garcia-Salum |
| EPI_ISL_2854181, EPI_ISL_2854182, EPI_ISL_2854183, EPI_ISL_2854184, EPI_ISL_2854185, EPI_ISL_2907497, EPI_ISL_2907498, EPI_ISL_2907501, EPI_ISL_2907504, EPI_ISL_2907505, EPI_ISL_2907506, EPI_ISL_2907508 | | | |
| see above | Laboratorio de Medicina Molecular, Universidad de Magallanes | Centro Asistencial Docente y de Investigacion, Universidad de Magallanes | Constanza Ceroni; Diego Alvarez; Inés Cid; Jacqueline Aldridge; Jorge González; Marcelo Navarrete; Roberto Uribe-Paredes |
| EPI_ISL_3049013, EPI_ISL_3049014, EPI_ISL_3049015, EPI_ISL_3049016, EPI_ISL_3049017, EPI_ISL_3049018, EPI_ISL_3049019, EPI_ISL_3049020, EPI_ISL_3055529, EPI_ISL_3055530, EPI_ISL_3761535, EPI_ISL_3761587, EPI_ISL_3761599, EPI_ISL_3761647, EPI_ISL_3761648, EPI_ISL_3761649, EPI_ISL_3761650, EPI_ISL_3761651, EPI_ISL_3761652, EPI_ISL_3761653, EPI_ISL_3761654, EPI_ISL_3761655, EPI_ISL_3761656, EPI_ISL_3761657, EPI_ISL_3761658, EPI_ISL_3761659, EPI_ISL_3761660, EPI_ISL_3761661, EPI_ISL_3761662, EPI_ISL_3761663, EPI_ISL_3761664, EPI_ISL_3761665, EPI_ISL_3761666, EPI_ISL_3761667, EPI_ISL_3761668, EPI_ISL_3761669, EPI_ISL_3761670, EPI_ISL_3761671, EPI_ISL_3761672, EPI_ISL_3761673, EPI_ISL_3761767 | | | |
| see above | Laboratorio de Pesquisa em Virologia, FAMERP, SJRP | Laboratorio de Pesquisa em Virologia, FAMERP, SJRP | Beatriz de Carvalho Marques; Cecília Artico Banho; Cíntia Bittar; Fábio Sossai Possebon; Guilherme Campos; Helena Lage Ferreira; Jorge A. Petroli Marchesi; João Pessoa Araújo Jr.; Leila Sabrina Ullmann; Lívia Sacchetto; Maisa C. Pereira Parra; Marília Moraes; Maurício L. Nogueira.; Paula Rahal; Paulo Inacio da Costa |
| EPI_ISL_3991552, EPI_ISL_3991678, EPI_ISL_4080476, EPI_ISL_4080482, EPI_ISL_4080483, EPI_ISL_4080484, EPI_ISL_4080485, EPI_ISL_4080486, EPI_ISL_4080487, EPI_ISL_4080488, EPI_ISL_4080490, EPI_ISL_4080491, EPI_ISL_4080492, EPI_ISL_4080495, EPI_ISL_4080502, EPI_ISL_4080510, EPI_ISL_4080511, EPI_ISL_4080512, EPI_ISL_4080524, EPI_ISL_4080525, EPI_ISL_4080526, EPI_ISL_4080528, EPI_ISL_4080534, EPI_ISL_4172705, EPI_ISL_4172710, EPI_ISL_4172726, EPI_ISL_4172728, EPI_ISL_4172744, EPI_ISL_4172746, EPI_ISL_4172748, EPI_ISL_4172896, EPI_ISL_4172898, EPI_ISL_4172910, EPI_ISL_4172912, EPI_ISL_4172960 | | | |
| see above | Laboratorio de Referencia Nacional de Virus Respiratorios. Centro Nacional de Salud Publica. Instituto Nacional de Salud Peru. | Laboratorio de Referencia Nacional de Virus Respiratorios. Centro Nacional de Salud Publica. Instituto Nacional de Salud Peru. | Carlos Padilla Rojas; Henri Bailon Calderon; Iris Silva Molina; Joseph Huayra Niquen; Lely Solari Zerpa; Luis Barcena Flores; Marco Galarza Perez; Nancy Rojas Serrano; Nieves Sevilla Castañeda; Omar Caceres Rey; Orson Mestanza Millones; Princesa Medrano Alhuay; Priscila Lope Pari; Sandra Morales Ruiz; Sara Gordillo Vilchez; Steve Acedo Lazo; Veronica Hurtado Vela; Victor Jimenez Vasquez; Wendy Lizarraga Olivares |
| EPI_ISL_4405152 | Laboratorio de Virología del Hospital de Niños Dr. Ricardo Gutierrez | Área de Secuenciación del Laboratorio de Virología del Hospital de Niños Dr. Ricardo Gutierrez on | AS; Acevedo; Acuña; Alexay; Alvarez Lopez; C; D; E; Goya; Gravis; Jacques; LE; Lusso; ME; MI; MS; Mistchenko; Nabaes Jodar; Natale; O; S; Valinotto; Viegas, M. |

| | | | |
|---|---|---|---|
| | | behalf of 'Proyecto Argentino Interinstitucional de genomica de SARS-CoV-2' (PAIS Consortium) | |
| EPI_ISL_7620354 | Laboratorio del Hospital Regional Ushuaia Gdor. Ernesto Campos | Nodo de Secuenciación Tierra del Fuego - Hospital Regional Ushuaia - Centro Austral De Investigaciones Científicas - Universidad Nacional De Tierra Del Fuego on behalf PAIS project and ANLIS-Malbran | AE; Boutureira, MF.; CA; CB; CF; Castro; Ceballos; Cáceres; De Roccis; F; G; Gallego; Gramundi; ID; Nardi; Rojas; SB; SG; Yulan |
| EPI_ISL_2708972 | Laboratorio di Microbiologia | Laboratorio di Microbiologia | Martinetti Lucchini Gladys; Valeria Spina |
| EPI_ISL_2765554, EPI_ISL_2765565 | Laboratorio di Riferimento Regionale della Sicilia Occidentale per l'Emergenza COVID-19 | Laboratorio di Riferimento Regionale della Sicilia Occidentale per l'Emergenza COVID-19 | Carmelo Massimo Maida; Claudio Costantino; Daniela Di Naro; Fabio Tramuto; Francesco Vitale; Giorgio Graziano; Giulia Randazzo; Vincenzo Restivo; Walter Mazzucco |
| EPI_ISL_2872588, EPI_ISL_2872651, EPI_ISL_2872677, EPI_ISL_2872710, EPI_ISL_2926226, EPI_ISL_2926255, EPI_ISL_2926257, EPI_ISL_2926272, EPI_ISL_2926296, EPI_ISL_2926339, EPI_ISL_2926341, EPI_ISL_2926360, EPI_ISL_2926368, EPI_ISL_2926369, EPI_ISL_2926376, EPI_ISL_2926396, EPI_ISL_2926410, EPI_ISL_2926415, EPI_ISL_2926429, EPI_ISL_2926495, EPI_ISL_2926628, EPI_ISL_2926676, EPI_ISL_2926732, EPI_ISL_2926733, EPI_ISL_2926745, EPI_ISL_2926769, EPI_ISL_2926772, EPI_ISL_2926778, EPI_ISL_2926790, EPI_ISL_2926796, EPI_ISL_2926813, EPI_ISL_2926831, EPI_ISL_2926847, EPI_ISL_2926857, EPI_ISL_2926874, EPI_ISL_2926903, EPI_ISL_2926917, EPI_ISL_2926955, EPI_ISL_2926963, EPI_ISL_2926966, EPI_ISL_2926978, EPI_ISL_2926979, EPI_ISL_2927001, EPI_ISL_2927007, EPI_ISL_2927123, EPI_ISL_2927307, EPI_ISL_2927312, EPI_ISL_2927314, EPI_ISL_2927327, EPI_ISL_2927332, EPI_ISL_2927333, EPI_ISL_2927334, EPI_ISL_2927360, EPI_ISL_2927367, EPI_ISL_2927402, EPI_ISL_2927441, EPI_ISL_2927486, EPI_ISL_2927489, EPI_ISL_2927548, EPI_ISL_2927550, EPI_ISL_2927588, EPI_ISL_2927599, EPI_ISL_2927617, EPI_ISL_2927619, EPI_ISL_2927626, EPI_ISL_2927633, EPI_ISL_2927646, EPI_ISL_2927676, EPI_ISL_2927694, EPI_ISL_3017014, EPI_ISL_3017017, EPI_ISL_3017019, EPI_ISL_3017026, EPI_ISL_3017221, EPI_ISL_3017242, EPI_ISL_3017279, EPI_ISL_3017665, EPI_ISL_3328481, EPI_ISL_4380479 | | | |
| see above | Laboratory Corporation of America | Centers for Disease Control and Prevention Division of Viral Diseases, Pathogen Discovery | Adrian Paskey; Amanda Douglas; Amanda Suchanek; Andrea Throop; Ayla Burns; Benjamin Rambo-Martin; Bobbi Croy; Brian Krueger; Brian Norvell; Christopher Gulvick; Christos Petropoulos; Clinton Paden; Clinton R. Paden; Craig Lukasik; Dakota Howard; Darlene Wagner; Debbie Boles; Dhwani Batra; Duncan MacCannell; Eyad Almasri; Goran Stevovic; Howard Engler; Hrushikesh Deshmukh; Jake Humphrey; Jana Schroth; Jason Caravas; Joe Voshell; John Pruitt; Jonathan Meltzer; Jonathan Williams; Kara Moser; Kimberly Wagner; Kristine Lacek; Lax Iyer; Lisa Pfefferle; Lyndon Tilson; Manoj Jain; Marcia Eisenberg; Mary Ann Cristobal; Mary Cristobal; Mary Williamson; Matthew Robinson; Matthew Schmerer; Michael Levandoski; Mike Sapeta; Mindy Nye; Minoo Agarwal; Mohan Kolli; Nuthawin Charoensri; Oren Cohen; Peter Cook; Peter W. Cook; Prashant Gupta; Qian Zeng; Rama Ghatti; Scott Parker; Scott Ryan; Scott Sammons; Shatavia Morrison; Stanley Letovsky; Steven Ragan; Suresh Babu Selvaraju; Suresh Selvaraju; Susan Countryman; Susan Hicks; Suzanne Dale; Thomas Urban; Tim Kuphal; Tricia Zwiefelhofer; Tymeckia Kendall; Victoria Caban Figueroa; Vincent Drouillon; Yvette Unoarumhi |
| EPI_ISL_2863689, EPI_ISL_2863691, EPI_ISL_2863693, EPI_ISL_2982730, EPI_ISL_2982731, EPI_ISL_2982732, EPI_ISL_2982733, EPI_ISL_2982734, EPI_ISL_2982736, EPI_ISL_2982737, EPI_ISL_2982738, EPI_ISL_2982739, EPI_ISL_2982740, EPI_ISL_2982741, EPI_ISL_2983379, EPI_ISL_2983380, EPI_ISL_2983381, EPI_ISL_2983382, EPI_ISL_3045514, EPI_ISL_3045515, EPI_ISL_3045516, EPI_ISL_3045517, EPI_ISL_3045518, EPI_ISL_3045519, EPI_ISL_3045520, EPI_ISL_3045521, EPI_ISL_3045522, EPI_ISL_3045523, EPI_ISL_3045524, EPI_ISL_3045525, EPI_ISL_3045526, EPI_ISL_3045527, EPI_ISL_3045528, EPI_ISL_3045529, EPI_ISL_3045530, EPI_ISL_3045531, EPI_ISL_3045532, EPI_ISL_3045533, EPI_ISL_3045534, EPI_ISL_3045535, EPI_ISL_3045536, EPI_ISL_3045537, EPI_ISL_3045538, EPI_ISL_3045539, EPI_ISL_3045540, EPI_ISL_3045541, EPI_ISL_3045542, EPI_ISL_3045543, EPI_ISL_3045544, EPI_ISL_3045545, EPI_ISL_3045546, EPI_ISL_3045547, EPI_ISL_3045548, EPI_ISL_3434952, EPI_ISL_3435049, EPI_ISL_3435050, EPI_ISL_3435051, EPI_ISL_3435052 | | | |
| see above | Laboratory of Respiratory Viruses and Measles, Oswaldo Cruz Institute, FIOCRUZ | Laboratory of Respiratory Viruses and Measles, Oswaldo Cruz Institute, FIOCRUZ | Agatha Cristinne Prudencio; Alice Sampaio Rocha; Ana Carolina Mendonca; Anna Carolina Paixao; Elisa Cavalcante Pereira; Fernando Motta; Ighor Leonardo Arantes Gomes; Luciana Appolinario; Marilda Siqueira on behalf of the Fiocruz COVID-19 Genomic Surveillance Network; Paola Resende; Renata Serrano Lopes; Taina Venas |
| EPI_ISL_3461239, EPI_ISL_3461240, EPI_ISL_3461243, EPI_ISL_3461249, EPI_ISL_3461250, EPI_ISL_3461285, EPI_ISL_3461318 | | | |
| see above | Laboratory, Gravity Diagnostics | Laboratory, Gravity Diagnostics | Diagnostics, G. |
| EPI_ISL_4600456, EPI_ISL_4600457, EPI_ISL_4600458, EPI_ISL_4600466, EPI_ISL_4600470, EPI_ISL_4600478, EPI_ISL_4600480 | | | |
| see above | Laboratório Central de Saúde Pública Profº Gonçalo Moniz | Coordenação Geral de Laboratórios de Saúde Pública (CGLAB/DAEVS/SVS/MS) | Vagner Fonseca; et al. |
| EPI_ISL_4600509, EPI_ISL_4600532 | Laboratório Central de Saúde Pública de Pernambuco | Coordenação Geral de Laboratórios de Saúde Pública (CGLAB/DAEVS/SVS/MS) | Vagner Fonseca; et al. |
| EPI_ISL_3050502, EPI_ISL_3050503, EPI_ISL_3050504, EPI_ISL_3050505, EPI_ISL_3050506, EPI_ISL_3050507, EPI_ISL_3050508, EPI_ISL_3050509, EPI_ISL_3050510, EPI_ISL_3050511, EPI_ISL_3050512, EPI_ISL_3050513, EPI_ISL_3050514, EPI_ISL_3050518, EPI_ISL_3050519, EPI_ISL_3050520, EPI_ISL_3050521, EPI_ISL_3050522, EPI_ISL_3050523, EPI_ISL_3050524, EPI_ISL_3050525, EPI_ISL_3050526, EPI_ISL_3050527, EPI_ISL_3050528, EPI_ISL_3050529, EPI_ISL_3050530, EPI_ISL_3050531, EPI_ISL_3050532, EPI_ISL_3050533, EPI_ISL_3050534, EPI_ISL_3050535, EPI_ISL_3050536, EPI_ISL_3050537, EPI_ISL_3050538, EPI_ISL_3050539, EPI_ISL_3050540, EPI_ISL_3050541, EPI_ISL_3050542, EPI_ISL_3050543, EPI_ISL_3050544, EPI_ISL_3050545, EPI_ISL_3050546, EPI_ISL_3050547, EPI_ISL_3050548, EPI_ISL_3050549, EPI_ISL_3050550, EPI_ISL_3050551, EPI_ISL_3050552, EPI_ISL_3050553, EPI_ISL_3050554, EPI_ISL_3050555, EPI_ISL_3050556, EPI_ISL_3050557, EPI_ISL_3050558, EPI_ISL_3050559, EPI_ISL_3050560, EPI_ISL_3050561, EPI_ISL_3050562, EPI_ISL_3050563, EPI_ISL_3050564, EPI_ISL_3050565, EPI_ISL_3050566, EPI_ISL_3050567, EPI_ISL_3050568, EPI_ISL_3050569, EPI_ISL_3050570, EPI_ISL_3050571 | | | |
| see above | Laboratório Central de Saúde Pública do Amazonas - LACEN-AM | Laboratorio de Ecologia de Doencas Transmissiveis na Amazonia, Instituto Leonidas e Maria Deane - Fiocruz Amazonia | André Corado; Felipe Naveca; Fernanda Nascimento; George Silva; Karina Pessoa; Luciana Gonçalves; Maria Júlia Brandão; Matilde Mejía; Valdinete Nascimento; Victor Souza; Ágatha Costa |
| EPI_ISL_6677481, EPI_ISL_6677505 | Laboratório Central de Saúde Pública do Estado do Tocantins (LACEN/TO) | Bioinformatics and Biotechnology Laboratory (Labinftec/Federal University of Tocantins, Campus of Gurupi) | Fabrício Souza Campos; Jucimária Dantas Galvão; Raíssa Nunes dos Santos; Ueric José Borges de Souza |
| EPI_ISL_5195470, EPI_ISL_5195471, EPI_ISL_5195473, EPI_ISL_5195474, EPI_ISL_5195475, EPI_ISL_5195476, EPI_ISL_5195482 | | | |
| see above | Laboratório Municipal de Biologia Molecular | Instituto René Rachou / Fiocruz Minas | André Menezes; Anna Salim; Eneida Oliveira; Gabriel Fernandes; Pedro Alves; Rubens do Monte Neto; Thaís Silva |
| EPI_ISL_4434601 | Laboratório de Baculovirus, Universidade de Brasília (UnB), Instituto de Ciências Biológicas (IB) | Laboratório de Virologia, Faculdade de Medicina, Universidade Federal de Mato Grosso (UFMT) | Bergman Morais Ribeiro; Fernando Lucas Melo; Francisco Scoffoni Kennedy de Azevedo; Gessica Fernanda Colnago de Lima; Renata Dezengrini Slhessarenko; Thais Campos Cruz |
| EPI_ISL_3539775, EPI_ISL_3539776, EPI_ISL_3539777, EPI_ISL_3539778, EPI_ISL_3539779, EPI_ISL_3539780, EPI_ISL_3539781, EPI_ISL_3539782, EPI_ISL_3539783, EPI_ISL_3539784, EPI_ISL_3539785, EPI_ISL_3539786, EPI_ISL_3539787, EPI_ISL_3539791, EPI_ISL_3539792, EPI_ISL_4170375, EPI_ISL_4170393, EPI_ISL_4170395 | | | |
| see above | Laboratório de Biotecnologia Aplicada (LBA) - Laboratório de Biologia Molecular - Hospital das Clínicas, Faculdade de Medicina de Botucatu, Departamento de Bioprocessos e Biotecnologia - Faculdade de Ciências Agronômicas, UNESP – Botucatu/SP | Laboratory of Respiratory Viruses and Measles, Oswaldo Cruz Institute, FIOCRUZ | Alice Sampaio Rocha; Ana Carolina Mendonca; Anna Carolina Paixao; Elisa Cavalcante Pereira; Felipe Allan da Silva Costa; Fernando Motta; Jayme Augusto de Souza Neto; Leonardo Nazario de Moraes; Luciana Appolinario; Marilda Siqueira on behalf of the Fiocruz COVID-19 Genomic Surveillance Network; Paola Resende; Patricia Akemi Assato; Rejane Maria Tommasini; Renata Serrano Lopes; Taina Venas |
| EPI_ISL_3456063, EPI_ISL_3873663, EPI_ISL_3873665, EPI_ISL_4051963 | Laboratório de Microbiologia Molecular - Universidade FEEVALE | Molecular Microbiology Laboratory | Alana Witt Hansen; Fernando Rosado Spilki; Fágner Henrique Heldt; Juliana Schons Gulart; Juliana Schons Gularte; Juliane Deise Fleck; Mariana Soares da Silva; Matheus Nunes Weber; Meriane Demoliner; Michele Filippi.; Micheli Filippi; Paula Rodrigues de Almeida; Vyctoria Malayhka de Abreu Góes Pereira. |
| EPI_ISL_3045379 | Labortorio Central de Saude Publica do Estado do Rio de Janeiro (LACEN/RJ) | Laboratory of Respiratory Viruses and Measles, Oswaldo Cruz Institute, FIOCRUZ | Agatha Cristinne Prudencio Soares; Alice Sampaio Rocha; Ana Carolina Mendonca; Andrea Cony Cavalcanti; Anna Carolina Paixao; Elisa Cavalcante Pereira; Fernando Motta; Ighor Leonardo Arantes Gomes; Luciana Appolinario; Marilda Siqueira on behalf of the Fiocruz COVID-19 Genomic Surveillance Network; Paola Resende; Renata Serrano Lopes; Taina Venas |
| EPI_ISL_2983227, EPI_ISL_2983229, EPI_ISL_3190409 | Labortorio Central de Saude Publica do Estado do Tocantins (LACEN/TO) | Laboratory of Respiratory Viruses and Measles, Oswaldo Cruz Institute, FIOCRUZ | Agatha Soares; Alice Sampaio Rocha; Ana Carolina Mendonca; Anna Carolina Paixao; Elisa Cavalcante Pereira; Fernando Motta; Ighor Arantes; Jucimaria Dantas Galvao; Luciana Appolinario; Marilda Siqueira on behalf of the Fiocruz COVID-19 Genomic Surveillance Network; Paola Resende; Renata Serrano Lopes; Taina Venas |
| EPI_ISL_3044943 | Landesgesundheitsamt Baden-Württemberg | Robert Koch Institute | |
| EPI_ISL_2809112 | Lighthouse Lab in Milton Keynes | Wellcome Sanger Institute for the COVID-19 Genomics UK (COG-UK) Consortium | Cordelia Langford; David K. Jackson; Dominic Kwiatkowski; Ewan Harrison; Ian Johnston; Jeffrey Barrett; John Sillitoe on behalf of the Wellcome Sanger Institute COVID-19 Surveillance Team; Roberto Amato; Sonia Goncalves; The Lighthouse Lab in Milton Keynes and Alex Alderton |
| EPI_ISL_2894574, EPI_ISL_2894575, EPI_ISL_2894576, EPI_ISL_2894577, EPI_ISL_2894578, EPI_ISL_2894584 | Los Angeles County Public Health Laboratories | Los Angeles County Public Health Laboratories | P. Hemarajata et al. |
| EPI_ISL_3102455 | MATERNIDADE ESCOLA ASSIS CHATEAUBRIAND | Analytical Competence Molecular Epidemiology Lab/ACME, Oswaldo Cruz Foundation, Ceara (FIOCRUZ CE) | Cleber Furtado Aksenen; Fabio Miyajima; Fernando Braga Stehling; Francisco Eder de Moura Lopes; Jamille Maria Mendes Bezerra; Joaquim César do Nascimento Sousa Junior; Pedro Miguel Carneiro Jeronimo; Suzana Porto Almeida e Lucas Delerino; Thais Ferreira de Oliveira; Thais de Oliveira Costa; Ticiane Cavalcante de Souza; Veridiana Pessoa Miyajima |
| EPI_ISL_5434252 | MB-Cadham Provincial laboratory | Manitoba Cadham Provincial Laboratory | Anna Majer; Anneliese Landgraff; David Alexander; Elsie Grudeski; Gary Van Domselaar; Grace Seo; Jared Bullard; Jennifer Tanner; Kerry Dust; Morag Graham; Nat; Natalie Knox; Paul Van Caeseele; Philip Mabon; Rhiannon Huzarewich; Russell Mandes; Shari Tyson |
| EPI_ISL_4494342, EPI_ISL_4494343 | MB-Cadham Provincial laboratory | National Microbiology Laboratory (NML) | Anna Majer; Anneliese Landgraff; David Alexander; Elsie Grudeski; Gary Van Domselaar; Grace Seo; Jared Bullard; Jennifer Tanner; Kerry Dust; Morag Graham; Nat; Natalie Knox; Paul Van Caeseele; Philip Mabon; Rhiannon Huzarewich; Russell Mandes; Shari Tyson |
| EPI_ISL_2790050, EPI_ISL_2861207, EPI_ISL_2862024, EPI_ISL_2955566, EPI_ISL_2955569 | MD PHL | MD PHL | Maryland Department of Health Laboratories Administration |
| EPI_ISL_2858795, EPI_ISL_2858801 | MS Public Health Laboratory | University of Mississippi Medical Center, Molecular and Genomics | Ashley C. Johnson; D. Ashley Robinson; Ithiel J. Frame; Krishna K. Ayyalasomayajula; Michael R. Garrett; Wenjie Wu |

| | | | |
|---|---|---|---|
| | | Core Facility | |
| EPI_ISL_6492855 | MSHS Clinical Microbiology Laboratories | MSHS Pathogen Surveillance Program | Adolfo García-Sastre; Adriana van de Guchte; Ajay Obla; Alberto Paniz-Mondolfi; Ana S. Gonzalez-Reiche; Angela Amoako; Ashley Salimbangon; Betsaida Salom Melo; Bremy Alburquerque; Brianne Ciferri; Charles Gleason; Daniel Floda; Deena R. Altman; Denise Jurczyszak; Emilia Mia Sordillo; Gintaras Deikus; Giulio Kleiner; Gopi Patel; Hala Alshammary; Harm van Bakel; Irina Oussenko; Jayeeta Dutta; Juan Soto; Julia Matthews; Katherine Beach; Kathryn Twyman; Kayla Russo; Komal Srivastava; Levy Sominsky; Mahmoud Awawda; Marta Luksza; Matthew M. Hernandez; Melissa Gitman; Michael D. Nowak; Mitchell J. Sullivan; Nancy Francoeur; Robert Sebra; Sarah Schaefer; Shelcie Fabre; Shwetha Hara Sridhar; Viviana Simon; Ying-Chih Wang; Zain Khalil; Zenab Khan |
| EPI_ISL_2762510, EPI_ISL_2845213 | MVZ Labor Dr. Limbach & Kollegen GbR | Robert Koch Institute | |
| EPI_ISL_3239233 | MVZ Labor Dr. Quade & Kollegen GmbH | Robert Koch Institute | |
| EPI_ISL_3030216 | Macerata/Civitanova Marche | Microbiology University Politecnica delle Marche | Anna Valenza; Carla Acciarri; Katia Marinelli; Monica Lucia Ferreri; Patrizia Bagnarelli; Roberta Longo; Sara Caucci; Stefano Menzo |
| EPI_ISL_2875397, EPI_ISL_2875402, EPI_ISL_2875438, EPI_ISL_2875503, EPI_ISL_3065124, EPI_ISL_3065150, EPI_ISL_3065203, EPI_ISL_3065210 | | | |
| see above | Mako Medical | Centers for Disease Control and Prevention Division of Viral Diseases, Pathogen Discovery | Adrian Paskey; Benjamin Rambo-Martin; Christopher Gulvick; Clinton R. Paden; Dakota Howard; Darlene Wagner; Dhwani Batra; Duncan MacCannell; Jason Caravas; Kara Moser; Lauren Moon; Matthew Schmerer; Matthew Tugwell; Peter W. Cook; Scott Sammons; Shatavia Morrison; Yvette Unoarumhi |
| EPI_ISL_3021463 | Marian Medical Center Lab | County of San Luis Obispo Public Health Laboratory | Frances Sidhu; Katrina Erwin |
| EPI_ISL_4200530 | Marshfield Clinic Res Found | Centers for Disease Control and Prevention Division of Viral Diseases, Pathogen Discovery | Alex Burgin; Ben Rambo-Martin; Clinton Paden; Dakota Howard; Dave Wentworth; Dhwani Batra; Jasmine Padilla; Justin Lee; Krista Queen; Kristen Knipe; Kristine Lacek; Mark Burroughs; Matthew Schmerer; Meghan Bentz; Mili Sheth; Peter Cook; Sam Shepard; Sarah Nobles; Suxiang Tong; Vivien Dugan; Yvette Unoarumhi |
| EPI_ISL_2894590 | Martin Luther King, Jr. Outpatient Center | Los Angeles County Public Health Laboratories | P. Hemarajata et al. |
| EPI_ISL_3030661, EPI_ISL_3030663, EPI_ISL_3030666 | Microbiology Department & Molecular Biology CORE \| CDB \| Hospital Clinic \| Barcelona | Microbiology Department & Molecular Biology CORE \| CDB \| Hospital Clinic \| Barcelona | Andrea Vergara Gomez; Jose Luis Villanueva Cañas; Miguel Julian Martínez Yoldi; Mª Angeles Marcos Maeso; Mª del Mar Mosquera Gutierrez |
| EPI_ISL_2812448, EPI_ISL_2812455, EPI_ISL_2812464, EPI_ISL_2812471, EPI_ISL_2812485, EPI_ISL_2812499, EPI_ISL_2812522, EPI_ISL_2812531, EPI_ISL_2812534, EPI_ISL_2832140, EPI_ISL_2832177, EPI_ISL_2832188, EPI_ISL_2832189, EPI_ISL_2832201, EPI_ISL_3320286 | | | |
| see above | Microbiology Department, Laboratori Clínic Metropolitana Nord. Hospital Universitari Germans Trias i Pujol | Can Ruti SARS-CoV-2 Sequencing Hub (HUGTiP/IrsiCaixa/IGTP) | Alba Sánchez; Alexia París; Anna Not; Antoni E Bordoy; Bonaventura Clotet; Cristina Casañ; David Panisello; Francesc Catala-Moll; Gemma Clara; Ignacio Blanco; Laia Soler; Lauro Sumoy; Marc Noguera-Julian; Maria Casadellà; Mariona Parera; Mercedes Guerrero; Montserrat Giménez; Pere-Joan Cardona; Pilar Armengol; Roger Paredes; Verónica Saludes; and Elisa Martró on behalf of the Can Ruti SARS-CoV-2 Sequencing Hub |
| EPI_ISL_3026873, EPI_ISL_3026893 | Microbiology Division, SC DHEC | Microbiology Division, SC DHEC | Freeman; J.S. |
| EPI_ISL_2958557, EPI_ISL_2987761, EPI_ISL_2987789, EPI_ISL_2987790, EPI_ISL_2987793, EPI_ISL_2987795, EPI_ISL_2987796, EPI_ISL_2987810, EPI_ISL_2987811, EPI_ISL_2987812, EPI_ISL_2987813, EPI_ISL_2987815, EPI_ISL_2987816, EPI_ISL_2987817, EPI_ISL_2987818, EPI_ISL_2987819, EPI_ISL_2987820, EPI_ISL_2987973, EPI_ISL_3032633, EPI_ISL_3032635, EPI_ISL_3032636, EPI_ISL_3032638, EPI_ISL_3032640, EPI_ISL_3032644, EPI_ISL_3032654, EPI_ISL_3404541 | | | |
| see above | Ministry of Health Turkey | Ministry of Health Turkey | Fatma Bayrakdar; Gulay Korukluoglu; Suleyman Yalcin; Yasemin Cosgun |
| EPI_ISL_2727839, EPI_ISL_2727855, EPI_ISL_2727856, EPI_ISL_4591919 | Minnesota Department of Health, Public Health Laboratory | Minnesota Department of Health, Public Health Laboratory | Alexandra Lorentz; Jacob Garfin; Matt Plumb; and Xiong Wang |
| EPI_ISL_2861081, EPI_ISL_2861082 | Mobile County Health Department | Synergy Laboratories | Megan Cornwell |
| EPI_ISL_3941248 | Molecular & Genomic Pathology Laboratory, Thomas Jefferson University Hospital | Molecular & Genomic Pathology Laboratory, Thomas Jefferson University Hospital | Nitika Badjatia; Shannon Mahoney; Zi-xuan Wang |
| EPI_ISL_2718543, EPI_ISL_2718544 | Molecular Diagnostics Pathology Department Mater Dei Hospital Malta | Molecular Diagnostics Pathology Department Mater Dei Hospital Malta | C Cilia; G Zahra; L Grech; M Briffa; R Borg |
| EPI_ISL_2710355, EPI_ISL_2731437 | Montana Public Health Laboratory | Montana Public Health Laboratory | Carrie Biskupiak; Deborah Gibson; Joy Ritter; Michael Dills; Michelle Mozer |
| EPI_ISL_4557559 | Monterey County Public Health Laboratory | Monterey County Public Health Laboratory | Monterey County Public Health Laboratory |
| EPI_ISL_2854241, EPI_ISL_2854242, EPI_ISL_2854245 | NCSLPH | NCSLPH | Chase K; Glover W; Greene S; Miller MC |
| EPI_ISL_2987004, EPI_ISL_2987005, EPI_ISL_2987007 | NE Public Health Laboratory | Centers for Disease Control and Prevention Division of Viral Diseases, Pathogen Discovery | Alison Laufer Halpin; Ben L. Rambo-Martin; Clinton R. Paden; Dakota Howard; Darlene Wagner; Dave Wentworth; Dhwani Batra; Jasmine Padilla; Justin Lee; Katie Dillon; Krista Queen; Kristen Knipe; Kristine Lacek; Mark Burroughs; Matthew Schmerer; Mili Sheth; Peter Cook; Sam Shepard; Sarah Nobles; Shoshona Le; Suxiang Tong; Vivien Dugan; Yvette Unoarumhi |
| EPI_ISL_2842649, EPI_ISL_2842684, EPI_ISL_2940366, EPI_ISL_2940368 | NJDOH, Public Health and Environmental Laboratories | NJ_PHEL | Allison Roder; Byeong Jeong; Chelsea San Filippo; Dana Woell; Jacquelyn Deverell; Lindsey Bodnar; Ryan Pachucki; Shiv K. Verma |
| EPI_ISL_3031060 | NORTHWELL HEALTH LABORATORIES | Wadsworth Center, New York State Department of Health | Alexis Russell; Catharine Prussing; Daryl M. Lamson; Erasmus Schneider; Erica Lasek-Nesselquist; John Kelly; Jonathan Plitnick; Kirsten St. George; Matthew Shudt; Melissa A Leisner; Navjot Singh |
| EPI_ISL_5649169, EPI_ISL_5649237, EPI_ISL_5649338, EPI_ISL_5649345, EPI_ISL_5649361, EPI_ISL_5649377, EPI_ISL_5649384, EPI_ISL_5649469 | | | |
| see above | NUCLEO DE SAUDE MARY DOTA DE BAURU | Instituto Butantan | Antonio Jorge Martins; Claudia Renata dos Santos Barros; David Schlesinger; Debora Botequio Moretti; Dimas Tadeu Covas; Elaine Cristina Marqueze; Elaine Vieira Santos; Evandra Strazza Rodrigues; Heidge Fukumasu; Jayme Augusto de Souza-Neto; José Salvatore Leister Patané; Luiz Alcantara; Luiz Lehmann Coutinho; Maria Carolina Elias; Maurício Lacerda Nogueira; Rafael dos Santos Bezerra; Raul Machado Neto; Rejane Maria Tommasini Grotto; Ricardo Haddad; Sandra Coccuzzo Sampaio Vessoni; Simone Kashima; Svetoslav Nanev Slavov; Vincent Louis Viala |
| EPI_ISL_5649415, EPI_ISL_5649419, EPI_ISL_5649424, EPI_ISL_5649436, EPI_ISL_5649437, EPI_ISL_5649440, EPI_ISL_5649441, EPI_ISL_5649449, EPI_ISL_5649450, EPI_ISL_5649451, EPI_ISL_5649455, EPI_ISL_5649460, EPI_ISL_5649461, EPI_ISL_5649463, EPI_ISL_5649466, EPI_ISL_5649468, EPI_ISL_5649472, EPI_ISL_5649477, EPI_ISL_5659519, EPI_ISL_5659523 | | | |
| see above | NUCLEO SAUDE III N HABITACIONAL PRESIDENTE GEISEL BAURU | Instituto Butantan | Antonio Jorge Martins; Claudia Renata dos Santos Barros; David Schlesinger; Debora Botequio Moretti; Dimas Tadeu Covas; Elaine Cristina Marqueze; Elaine Vieira Santos; Evandra Strazza Rodrigues; Heidge Fukumasu; Jayme Augusto de Souza-Neto; José Salvatore Leister Patané; Luiz Alcantara; Luiz Lehmann Coutinho; Maria Carolina Elias; Maurício Lacerda Nogueira; Rafael dos Santos Bezerra; Raul Machado Neto; Rejane Maria Tommasini Grotto; Ricardo Haddad; Sandra Coccuzzo Sampaio Vessoni; Simone Kashima; Svetoslav Nanev Slavov; Vincent Louis Viala |
| EPI_ISL_3063136 | National Institute of Public Health | National Institute of Public Health | Alexander Nagy; Dusan Trnka; Helena Jirincova; Jaromira Vecerova; Timotej Suri |
| EPI_ISL_2895635 | National Platform bis UMONS/Jolimont | National Platform bis UMONS/Jolimont | Eric Tarantino; Florian Juszczak; François Dufrasne; Gautier Detry; Guillaume Bayon-Vicente; Ruddy Wattiez |
| EPI_ISL_3011287 | National Virus Reference Laboratory | National Virus Reference Laboratory | Charlene Bennett; Cillian F De Gascun; Gabriel Gonzalez; Jonathan Dean; Michael Carr; Zoe Yandle |
| EPI_ISL_2689123, EPI_ISL_2754087, EPI_ISL_2754090, EPI_ISL_2754879, EPI_ISL_2754901, EPI_ISL_2774536, EPI_ISL_2774538, EPI_ISL_2774539, EPI_ISL_2774540, EPI_ISL_2774542, EPI_ISL_2774550, EPI_ISL_2774552 | | | |
| see above | Nebraska Public Health Laboratory | NPHL COVID-19 Response Team | NPHL COVID-19 Response Team |
| EPI_ISL_2942242 | Nevada State Public Health Laboratory | Nevada State Public Health Laboratory | Andrew Gorzalski; Mark Pandori |
| EPI_ISL_3127152, EPI_ISL_3127156 | New Mexico Department of Health Scientific Laboratory | New Mexico Department of Health Scientific Laboratory | Anastacia Griego-Fisher; D'eldra Malone; Ellie Johnson; Jennifer Benoit; Linda Salazar; Ratheesh Rajan; T. Mark Willmon |
| EPI_ISL_3015789, EPI_ISL_3015809, EPI_ISL_3015814, EPI_ISL_3015817, EPI_ISL_3015819, EPI_ISL_3015834, EPI_ISL_3015838, EPI_ISL_3015840, EPI_ISL_3015841, EPI_ISL_3015843 | | | |
| see above | Northwest Laboratory | Seattle Flu Study | Amanda Adler; Barry R. Lutz; Benjamin Pelle; Brian Hiatt; Caitlin R. Wolf; Chris D. Frazar; Deborah A. Nickerson; Elisabeth Brandstetter; Erica Ryke; Geoff Melly; Helen Y. Chu; Janet A. Englund; Jay Shendure; Jover Lee; Kairsten Fay; Kirsten Lacombe; Lea M. Starita; Mark J. Rieder; Matthew Richardson; Matthew Thompson; Melissa Truong; Michael Boeckh; Michael Famulare; Misja Ilcisin; Peter D. Han; Philip Dykema; Romesh Gautom; Scott Lindquist; Thomas R. Sibley; Trevor Bedford |
| EPI_ISL_3355222 | OR State PHL-Virology/Immunology Section | Centers for Disease Control and Prevention Division of Viral Diseases, Pathogen Discovery | Alex Burgin; Ben Rambo-Martin; Clinton Paden; Dakota Howard; Dave Wentworth; Dhwani Batra; Jasmine Padilla; Justin Lee; Krista Queen; Kristen Knipe; Kristine Lacek; Mark Burroughs; Matthew Schmerer; Meghan Bentz; Mili Sheth; Peter Cook; Sam Shepard; Sarah Nobles; Suxiang Tong; Vivien Dugan; Yvette Unoarumhi |
| EPI_ISL_2840686 | OSPEDALE CIVILE TERAMO - CENTRO TRASFUSIONALE | Istituto Zooprofilattico Sperimentale dell'Abruzzo e Molise "G. Caporale" | Ancora M; Calistri P; Cammà C; Caporale M; Curini V; Delli Compagni E; Di Domenico M; Di Lollo Valeria; Di Pasquale A; Lorusso A; Mangone I; Marcacci M; Puglia I; Rinaldi A; Savini G; Scialabba S |
| EPI_ISL_2928203 | Orange County Public Health Laboratory | Orange County Public Health Laboratory | Angelica Torres; Julia Wolfe |
| EPI_ISL_2727476, EPI_ISL_2790735, EPI_ISL_2790740, EPI_ISL_2790741, EPI_ISL_2790742, EPI_ISL_2790743, EPI_ISL_2790744, EPI_ISL_2790745, EPI_ISL_2790746, EPI_ISL_2893428, EPI_ISL_3090726 | | | |
| see above | Oregon State Public Health Laboratory | Oregon State Public Health Laboratory | Eugene Yeboah; John Fontana and Shane Sevey; Laura Tsaknaridis; Rafia Razzaque; Vanda Makris |
| EPI_ISL_2796514, | Ospedale Giovanni Paolo II Ragusa RG | AOU Polclinico "G Rodolico-S.Marco" | STEFANI AND SCALIA |

| | | | |
|---|---|---|---|
| EPI_ISL_2796516 | | PO Rodolico and BIOMETEC UNICT CATANIA | |
| EPI_ISL_2787025, EPI_ISL_2787063 | Ospedale S.M. Goretti | INMI Lazzaro Spallanzani IRCCS | A Tamburro; B Bartolini; CEM Gruber; E Sperandio; F Natoni; F Santini; G Bonfiglio; G Orlandi; G Pessina; M Meledandri; O Butera; S Donzelli |
| EPI_ISL_2787018, EPI_ISL_2787055, EPI_ISL_2787056, EPI_ISL_2787057, EPI_ISL_2787058, EPI_ISL_2787059, EPI_ISL_2787060, EPI_ISL_2787061, EPI_ISL_2787062 | | | |
| see above | Ospedale San Filippo Neri | INMI Lazzaro Spallanzani IRCCS | A Massacci; B Bartolini; CEM Gruber; D Cerini; D Di Fusco; E Giombini; E Sperandio; F De Nicola; F Messina; F Santini; G Bonfiglio; G Orlandi; GF Inger; L Ciuffreda; M Meledandri; M Pallocca; M Rueca; ML Schiavone; O Butera; S Donzelli |
| EPI_ISL_2787020, EPI_ISL_2787021, EPI_ISL_2787022, EPI_ISL_2787023, EPI_ISL_2787074 | Ospedale Sandro Pertini ASL Roma2 | INMI Lazzaro Spallanzani IRCCS | A Massacci; B Bartolini; CEM Gruber; D Cerini; D Di Fusco; E Giombini; E Sperandio; F Messina; F Natoni; F Santini; G Bonfiglio; G Cappiello; G Orlandi; G Pessina; L Ciuffreda; M Pallocca; M Rueca; O Butera; R Longo; S Donzelli |
| EPI_ISL_3105048, EPI_ISL_3105049 | Overlake Hospital | Seattle Flu Study | Amanda Adler; Barry R. Lutz; Benjamin Pelle; Brian Hiatt; Caitlin R. Wolf; Chris D. Frazar; Deborah A. Nickerson; Elisabeth Brandstetter; Erica Ryke; Geoff Melly; Helen Y. Chu; Janet A. Englund; Jay Shendure; Jover Lee; Kairsten Fay; Kirsten Lacombe; Lea M. Starita; Mark J. Rieder; Matthew Richardson; Matthew Thompson; Melissa Truong; Michael Boeckh; Michael Famulare; Misja Ilcisin; Peter D. Han; Philip Dykema; Romesh Gautom; Scott Lindquist; Thomas R. Sibley; Trevor Bedford |
| EPI_ISL_2903092 | Oxford Viromics, NDM, University of Oxford; Oxford University Hospitals; Basingstoke and North Hampshire Hospital | COVID-19 Genomics UK (COG-UK) Consortium | Alex Mobbs; Amy Trebes; Anita Justice; Catrin Moore; Christophe Fraser; David Bonsall; David Buck; Emma Wise; George Macintyre; Jessica Lynch; John Todd; Mariateresa de Cesare; Matilde Mori; Monique Andersson; Nathan Moore; Nick Cortes; Robert Shaw; Stephen Kidd; Tanya Golubchik; Timothy Peto |
| EPI_ISL_5648255, EPI_ISL_5648283, EPI_ISL_5648439, EPI_ISL_5648500 | PA MATAO | Instituto Butantan | Antonio Jorge Martins; Claudia Renata dos Santos Barros; David Schlesinger; Debora Botequio Moretti; Dimas Tadeu Covas; Elaine Cristina Marqueze; Elaine Vieira Santos; Evandra Strazza Rodrigues; Heidge Fukumasu; Jayme Augusto de Souza-Neto; José Salvatore Leister Patané; Luiz Alcantara; Luiz Lehmann Coutinho; Maria Carolina Elias; Maurício Lacerda Nogueira; Rafael dos Santos Bezerra; Raul Machado Neto; Rejane Maria Tommasini Grotto; Ricardo Haddad; Sandra Coccuzzo Sampaio Vessoni; Simone Kashima; Svetoslav Nanev Slavov; Vincent Louis Viala |
| EPI_ISL_5659602 | PACS JOAO FRANCISCO MARQUES SANTA LUZIA JOSE BONIFACIO | Instituto Butantan | Antonio Jorge Martins; Claudia Renata dos Santos Barros; David Schlesinger; Debora Botequio Moretti; Dimas Tadeu Covas; Elaine Cristina Marqueze; Elaine Vieira Santos; Evandra Strazza Rodrigues; Heidge Fukumasu; Jayme Augusto de Souza-Neto; José Salvatore Leister Patané; Luiz Alcantara; Luiz Lehmann Coutinho; Maria Carolina Elias; Maurício Lacerda Nogueira; Rafael dos Santos Bezerra; Raul Machado Neto; Rejane Maria Tommasini Grotto; Ricardo Haddad; Sandra Coccuzzo Sampaio Vessoni; Simone Kashima; Svetoslav Nanev Slavov; Vincent Louis Viala |
| EPI_ISL_5648456, EPI_ISL_5648486, EPI_ISL_5648615 | PENITENCIARIA DR JOSE AUGUSTO CESAR SALGADO IRT | Instituto Butantan | Antonio Jorge Martins; Claudia Renata dos Santos Barros; David Schlesinger; Debora Botequio Moretti; Dimas Tadeu Covas; Elaine Cristina Marqueze; Elaine Vieira Santos; Evandra Strazza Rodrigues; Heidge Fukumasu; Jayme Augusto de Souza-Neto; José Salvatore Leister Patané; Luiz Alcantara; Luiz Lehmann Coutinho; Maria Carolina Elias; Maurício Lacerda Nogueira; Rafael dos Santos Bezerra; Raul Machado Neto; Rejane Maria Tommasini Grotto; Ricardo Haddad; Sandra Coccuzzo Sampaio Vessoni; Simone Kashima; Svetoslav Nanev Slavov; Vincent Louis Viala |
| EPI_ISL_5649242 | POLICLINICA COVID 19 ITAPETININGA | Instituto Butantan | Antonio Jorge Martins; Claudia Renata dos Santos Barros; David Schlesinger; Debora Botequio Moretti; Dimas Tadeu Covas; Elaine Cristina Marqueze; Elaine Vieira Santos; Evandra Strazza Rodrigues; Heidge Fukumasu; Jayme Augusto de Souza-Neto; José Salvatore Leister Patané; Luiz Alcantara; Luiz Lehmann Coutinho; Maria Carolina Elias; Maurício Lacerda Nogueira; Rafael dos Santos Bezerra; Raul Machado Neto; Rejane Maria Tommasini Grotto; Ricardo Haddad; Sandra Coccuzzo Sampaio Vessoni; Simone Kashima; Svetoslav Nanev Slavov; Vincent Louis Viala |
| EPI_ISL_5648895, EPI_ISL_5649019 | POLICLINICA DONA LUIZA | Instituto Butantan | Antonio Jorge Martins; Claudia Renata dos Santos Barros; David Schlesinger; Debora Botequio Moretti; Dimas Tadeu Covas; Elaine Cristina Marqueze; Elaine Vieira Santos; Evandra Strazza Rodrigues; Heidge Fukumasu; Jayme Augusto de Souza-Neto; José Salvatore Leister Patané; Luiz Alcantara; Luiz Lehmann Coutinho; Maria Carolina Elias; Maurício Lacerda Nogueira; Rafael dos Santos Bezerra; Raul Machado Neto; Rejane Maria Tommasini Grotto; Ricardo Haddad; Sandra Coccuzzo Sampaio Vessoni; Simone Kashima; Svetoslav Nanev Slavov; Vincent Louis Viala |
| EPI_ISL_5648785, EPI_ISL_5648946 | POLICLINICA JARDIM PARAISO | Instituto Butantan | Antonio Jorge Martins; Claudia Renata dos Santos Barros; David Schlesinger; Debora Botequio Moretti; Dimas Tadeu Covas; Elaine Cristina Marqueze; Elaine Vieira Santos; Evandra Strazza Rodrigues; Heidge Fukumasu; Jayme Augusto de Souza-Neto; José Salvatore Leister Patané; Luiz Alcantara; Luiz Lehmann Coutinho; Maria Carolina Elias; Maurício Lacerda Nogueira; Rafael dos Santos Bezerra; Raul Machado Neto; Rejane Maria Tommasini Grotto; Ricardo Haddad; Sandra Coccuzzo Sampaio Vessoni; Simone Kashima; Svetoslav Nanev Slavov; Vincent Louis Viala |
| EPI_ISL_5648812, EPI_ISL_5648827, EPI_ISL_5648868 | POLICLINICA MARIA DIRCE | Instituto Butantan | Antonio Jorge Martins; Claudia Renata dos Santos Barros; David Schlesinger; Debora Botequio Moretti; Dimas Tadeu Covas; Elaine Cristina Marqueze; Elaine Vieira Santos; Evandra Strazza Rodrigues; Heidge Fukumasu; Jayme Augusto de Souza-Neto; José Salvatore Leister Patané; Luiz Alcantara; Luiz Lehmann Coutinho; Maria Carolina Elias; Maurício Lacerda Nogueira; Rafael dos Santos Bezerra; Raul Machado Neto; Rejane Maria Tommasini Grotto; Ricardo Haddad; Sandra Coccuzzo Sampaio Vessoni; Simone Kashima; Svetoslav Nanev Slavov; Vincent Louis Viala |
| EPI_ISL_5648272, EPI_ISL_5648737 | POLICLINICA MUNIC DA EST TURISTICA DE HOLAMBRA | Instituto Butantan | Antonio Jorge Martins; Claudia Renata dos Santos Barros; David Schlesinger; Debora Botequio Moretti; Dimas Tadeu Covas; Elaine Cristina Marqueze; Elaine Vieira Santos; Evandra Strazza Rodrigues; Heidge Fukumasu; Jayme Augusto de Souza-Neto; José Salvatore Leister Patané; Luiz Alcantara; Luiz Lehmann Coutinho; Maria Carolina Elias; Maurício Lacerda Nogueira; Rafael dos Santos Bezerra; Raul Machado Neto; Rejane Maria Tommasini Grotto; Ricardo Haddad; Sandra Coccuzzo Sampaio Vessoni; Simone Kashima; Svetoslav Nanev Slavov; Vincent Louis Viala |
| EPI_ISL_5648876 | POLICLINICA PARAVENTI | Instituto Butantan | Antonio Jorge Martins; Claudia Renata dos Santos Barros; David Schlesinger; Debora Botequio Moretti; Dimas Tadeu Covas; Elaine Cristina Marqueze; Elaine Vieira Santos; Evandra Strazza Rodrigues; Heidge Fukumasu; Jayme Augusto de Souza-Neto; José Salvatore Leister Patané; Luiz Alcantara; Luiz Lehmann Coutinho; Maria Carolina Elias; Maurício Lacerda Nogueira; Rafael dos Santos Bezerra; Raul Machado Neto; Rejane Maria Tommasini Grotto; Ricardo Haddad; Sandra Coccuzzo Sampaio Vessoni; Simone Kashima; Svetoslav Nanev Slavov; Vincent Louis Viala |
| EPI_ISL_3536291 | POLO BASE DO TRILHO | Analytical Competence Molecular Epidemiology Lab/ACME, Oswaldo Cruz Foundation, Ceara (FIOCRUZ CE) | Cleber Furtado Aksenen; Fabio Miyajima; Fernando Braga Stehling; Francisco Eder de Moura Lopes; Jamille Maria Mendes Bezerra; Joaquim César do Nascimento Sousa Junior; Pedro Miguel Carneiro Jeronimo; Suzana Porto Almeida e Lucas Delerino; Thais Ferreira de Oliveira; Thais de Oliveira Costa; Ticiane Cavalcante de Souza; Veridiana Pessoa Miyajima |
| EPI_ISL_3536146 | POSTO DE SAUDE DE SANTA FELICIA | Analytical Competence Molecular Epidemiology Lab/ACME, Oswaldo Cruz Foundation, Ceara (FIOCRUZ CE) | Cleber Furtado Aksenen; Fabio Miyajima; Fernando Braga Stehling; Francisco Eder de Moura Lopes; Jamille Maria Mendes Bezerra; Joaquim César do Nascimento Sousa Junior; Pedro Miguel Carneiro Jeronimo; Suzana Porto Almeida e Lucas Delerino; Thais Ferreira de Oliveira; Thais de Oliveira Costa; Ticiane Cavalcante de Souza; Veridiana Pessoa Miyajima |
| EPI_ISL_5648810, EPI_ISL_5648813, EPI_ISL_5648834, EPI_ISL_5648974 | PRONTO ATENDIMENTO DE VARGEM GRANDE PAULISTA | Instituto Butantan | Antonio Jorge Martins; Claudia Renata dos Santos Barros; David Schlesinger; Debora Botequio Moretti; Dimas Tadeu Covas; Elaine Cristina Marqueze; Elaine Vieira Santos; Evandra Strazza Rodrigues; Heidge Fukumasu; Jayme Augusto de Souza-Neto; José Salvatore Leister Patané; Luiz Alcantara; Luiz Lehmann Coutinho; Maria Carolina Elias; Maurício Lacerda Nogueira; Rafael dos Santos Bezerra; Raul Machado Neto; Rejane Maria Tommasini Grotto; Ricardo Haddad; Sandra Coccuzzo Sampaio Vessoni; Simone Kashima; Svetoslav Nanev Slavov; Vincent Louis Viala |
| EPI_ISL_5649073 | PRONTO SOCORRO CENTRAL | Instituto Butantan | Antonio Jorge Martins; Claudia Renata dos Santos Barros; David Schlesinger; Debora Botequio Moretti; Dimas Tadeu Covas; Elaine Cristina Marqueze; Elaine Vieira Santos; Evandra Strazza Rodrigues; Heidge Fukumasu; Jayme Augusto de Souza-Neto; José Salvatore Leister Patané; Luiz Alcantara; Luiz Lehmann Coutinho; Maria Carolina Elias; Maurício Lacerda Nogueira; Rafael dos Santos Bezerra; Raul Machado Neto; Rejane Maria Tommasini Grotto; Ricardo Haddad; Sandra Coccuzzo Sampaio Vessoni; Simone Kashima; Svetoslav Nanev Slavov; Vincent Louis Viala |
| EPI_ISL_5648795, EPI_ISL_5648901, EPI_ISL_5648924, EPI_ISL_5648936, EPI_ISL_5649077, EPI_ISL_5649092, EPI_ISL_5649104, EPI_ISL_5649142 | | | |
| see above | PRONTO SOCORRO VEREADOR LUIZ CARLOS MARQUES | Instituto Butantan | Antonio Jorge Martins; Claudia Renata dos Santos Barros; David Schlesinger; Debora Botequio Moretti; Dimas Tadeu Covas; Elaine Cristina Marqueze; Elaine Vieira Santos; Evandra Strazza Rodrigues; Heidge Fukumasu; Jayme Augusto de Souza-Neto; José Salvatore Leister Patané; Luiz Alcantara; Luiz Lehmann Coutinho; Maria Carolina Elias; Maurício Lacerda Nogueira; Rafael dos Santos Bezerra; Raul Machado Neto; Rejane Maria Tommasini Grotto; Ricardo Haddad; Sandra Coccuzzo Sampaio Vessoni; Simone Kashima; Svetoslav Nanev Slavov; Vincent Louis Viala |
| EPI_ISL_5649250, EPI_ISL_5649259, EPI_ISL_5649289, EPI_ISL_5649303, EPI_ISL_5649319 | PSF DR ANTONIO PIRES DE ALMEIDA PORTO FELIZ | Instituto Butantan | Antonio Jorge Martins; Claudia Renata dos Santos Barros; David Schlesinger; Debora Botequio Moretti; Dimas Tadeu Covas; Elaine Cristina Marqueze; Elaine Vieira Santos; Evandra Strazza Rodrigues; Heidge Fukumasu; Jayme Augusto de Souza-Neto; José Salvatore Leister Patané; Luiz Alcantara; Luiz Lehmann Coutinho; Maria Carolina Elias; Maurício Lacerda Nogueira; Rafael dos Santos Bezerra; Raul Machado Neto; Rejane Maria Tommasini Grotto; Ricardo Haddad; Sandra Coccuzzo Sampaio Vessoni; Simone Kashima; Svetoslav Nanev Slavov; Vincent Louis Viala |
| EPI_ISL_5649241, EPI_ISL_5649268 | PSF JARDIM PANORAMA ALVARES MACHADO | Instituto Butantan | Antonio Jorge Martins; Claudia Renata dos Santos Barros; David Schlesinger; Debora Botequio Moretti; Dimas Tadeu Covas; Elaine Cristina Marqueze; Elaine Vieira Santos; Evandra Strazza Rodrigues; Heidge Fukumasu; Jayme Augusto de Souza-Neto; José Salvatore Leister Patané; Luiz Alcantara; Luiz Lehmann Coutinho; Maria Carolina Elias; Maurício Lacerda Nogueira; Rafael dos Santos Bezerra; Raul Machado Neto; Rejane Maria Tommasini Grotto; Ricardo Haddad; Sandra Coccuzzo Sampaio Vessoni; Simone Kashima; Svetoslav Nanev Slavov; Vincent Louis Viala |
| EPI_ISL_3536147 | PSF RODOVIARIA | Analytical Competence Molecular Epidemiology Lab/ACME, Oswaldo Cruz Foundation, Ceara (FIOCRUZ CE) | Cleber Furtado Aksenen; Fabio Miyajima; Fernando Braga Stehling; Francisco Eder de Moura Lopes; Jamille Maria Mendes Bezerra; Joaquim César do Nascimento Sousa Junior; Pedro Miguel Carneiro Jeronimo; Suzana Porto Almeida e Lucas Delerino; Thais Ferreira de Oliveira; Thais de Oliveira Costa; Ticiane Cavalcante de Souza; Veridiana Pessoa Miyajima |
| EPI_ISL_5649392 | PSF VIRGINIO BASSO | Instituto Butantan | Antonio Jorge Martins; Claudia Renata dos Santos Barros; David Schlesinger; Debora Botequio Moretti; Dimas Tadeu Covas; Elaine Cristina Marqueze; Elaine Vieira Santos; Evandra Strazza Rodrigues; Heidge Fukumasu; Jayme Augusto de Souza-Neto; José Salvatore Leister Patané; Luiz Alcantara; Luiz Lehmann Coutinho; Maria Carolina Elias; Maurício Lacerda Nogueira; Rafael dos Santos Bezerra; Raul Machado Neto; Rejane Maria Tommasini Grotto; Ricardo Haddad; Sandra Coccuzzo Sampaio Vessoni; Simone Kashima; Svetoslav Nanev Slavov; Vincent Louis Viala |
| EPI_ISL_3462479 | Palmdale Regional Medical Center | Los Angeles County Public Health Laboratories | P. Hemarajata et al. |
| EPI_ISL_2827489, EPI_ISL_2827500 | Pandemic Response Lab - NYC | Pandemic Response Lab, R&D | Cybill del Castillo; Dylan Law; Haiping Hao; Henry Lee; Isabel Fernandez Escapa; Jon Laurent; Melissa Hopkins; Michael Hammerling; Pradeep Bugga; Shinyoung Clair Kang; Sol Rey; William Ward |
| EPI_ISL_3127929 | Philadelphia Department of Public Health | Centers for Disease Control and Prevention Division of Viral Diseases, Pathogen Discovery | Alex Burgin; Ben L. Rambo-Martin; Clinton R. Paden; Dakota Howard; Dave Wentworth; Dhwani Batra; Jasmine Padilla; Justin Lee; Krista Queen; Kristen Knipe; Kristine Lacek; Mark Burroughs; Matthew Schmerer; Meghan Bentz; Mili Sheth; Peter Cook; Sam Shepard; Sarah Nobles; Suxiang Tong; Vivien Dugan; Yvette Unoarumhi |
| EPI_ISL_4405600 | Plataforma de Servicios Biotecnólogicos: UTTIPP/PSB , Universidad Nacional de Quilmes. | Área de Secuenciación del Laboratorio de Virología del Hospital de Niños Dr. Ricardo Gutierrez on behalf of 'Proyecto Argentino Interinstitucional de genomica de SARS-CoV-2' (PAIS Consortium) | A; Acuña; Cardama; Castello; D; Farina; G; Goya; Goñi; H; LE; Lusso; MI; MS; Nabaes Jodar; Natale; S; Valinotto; Viegas, M. |
| EPI_ISL_6907491, EPI_ISL_6907492, EPI_ISL_6907493, EPI_ISL_6907494, EPI_ISL_6907495 | Plataforma de Vigilancia Molecular (PVM) - FIOCRUZ/BA | Plataforma de Vigilancia Molecular (PVM) - FIOCRUZ/BA | Bruno Bezerril Andrade; Camila I. de Oliveira; Clarissa Araújo Gurgel; Eduardo Oyama; Francys A Rangel; Laila A da Silva; Laise de Moraes; Leonardo Paiva Farias; Marina Cucco; Pedro Silveira; Ricardo Khouri on behalf of the Fiocruz COVID-19 Genomic Surveillance Network.; Tiago Graf |
| EPI_ISL_2832239 | Plateforme de testing Namuroise | Plateforme de testing Namuroise | Degosserie Jonathan; Demars Aurore; Denis Olivier; Lesly NYINKEU K.; Maschietto Céline; Mullier François; Nicolas GILLIARD; Nobis Chloé; Otto Gaetan |
| EPI_ISL_2787064, EPI_ISL_2787065 | Policlinico Universitario Campus Bio-Medico | INMI Lazzaro Spallanzani IRCCS | A Massacci; B Bartolini; CEM Gruber; D Cerini; D Di Fusco; E Sperandio; F De Nicola; F Messina; F Santini; G Bonfiglio; GF Inger; M Rueca; O Butera |
| EPI_ISL_2876239 | Public Health Laboratory, Minnesota Department of Health | University of Minnesota Genomics Center | Corbin Dirkx; Daryl M. Gohl; Jaquelyn Kuriger-Laber; John Garbe; and Sean Wang |
| EPI_ISL_2876740, EPI_ISL_2876743, EPI_ISL_2876750, EPI_ISL_2876751, EPI_ISL_2876753, EPI_ISL_2876755, EPI_ISL_2876762, EPI_ISL_2876764, EPI_ISL_2876776, EPI_ISL_2876781, EPI_ISL_2876788, EPI_ISL_2876789, EPI_ISL_2876790, EPI_ISL_2876797, EPI_ISL_2876799, EPI_ISL_2876801, EPI_ISL_2876804, EPI_ISL_2876806, EPI_ISL_2876807, EPI_ISL_2876810, EPI_ISL_2876811, EPI_ISL_2876816, EPI_ISL_2876822, EPI_ISL_2876824, EPI_ISL_2876826, EPI_ISL_2876827, EPI_ISL_2876828, EPI_ISL_2876830, EPI_ISL_2876831, EPI_ISL_2876832, EPI_ISL_2876833, EPI_ISL_2876835, EPI_ISL_2876836, EPI_ISL_2876839, EPI_ISL_2876840, EPI_ISL_2876842, EPI_ISL_2876843, EPI_ISL_2876844, EPI_ISL_2876846, EPI_ISL_2876847, EPI_ISL_2876848, EPI_ISL_2876849, EPI_ISL_2876850, EPI_ISL_2876851, EPI_ISL_2876852, EPI_ISL_2876853, EPI_ISL_2876854, EPI_ISL_2876855, EPI_ISL_2876856, EPI_ISL_2876858, EPI_ISL_2876859, EPI_ISL_2876861, EPI_ISL_2876862, EPI_ISL_2876863, EPI_ISL_2876864, EPI_ISL_2876865, EPI_ISL_2876866, EPI_ISL_2876867, EPI_ISL_2876868, EPI_ISL_2876869, EPI_ISL_2876870, EPI_ISL_2876871, EPI_ISL_2876872, EPI_ISL_2876873, EPI_ISL_2876875, EPI_ISL_2876876, | | | |

| | | | |
|---|---|---|---|
| EPI_ISL_2876877, EPI_ISL_2876878, EPI_ISL_2876879, EPI_ISL_2876880, EPI_ISL_2876881, EPI_ISL_2876882, EPI_ISL_2876883, EPI_ISL_2876884, EPI_ISL_2876886, EPI_ISL_2876887, EPI_ISL_2876888, EPI_ISL_2876889, EPI_ISL_2876890, EPI_ISL_2876891, EPI_ISL_2876892, EPI_ISL_2876893, EPI_ISL_2876894, EPI_ISL_2876896, EPI_ISL_2876897, EPI_ISL_2876898, EPI_ISL_2876899, EPI_ISL_2876901, EPI_ISL_2876903, EPI_ISL_2876904, EPI_ISL_2876905, EPI_ISL_2876906, EPI_ISL_2876907, EPI_ISL_2876908, EPI_ISL_2876909, EPI_ISL_2876910, EPI_ISL_2876911, EPI_ISL_2876912, EPI_ISL_2876913, EPI_ISL_2876914, EPI_ISL_2876915, EPI_ISL_2876916, EPI_ISL_2876917, EPI_ISL_2876918, EPI_ISL_2876919, EPI_ISL_2876921, EPI_ISL_2876922, EPI_ISL_2876923, EPI_ISL_2876924, EPI_ISL_2876925, EPI_ISL_2876926, EPI_ISL_2876927, EPI_ISL_2876928, EPI_ISL_2876929, EPI_ISL_2876930, EPI_ISL_2876931, EPI_ISL_2876932, EPI_ISL_2876933, EPI_ISL_2876934, EPI_ISL_3305010, EPI_ISL_3305014, EPI_ISL_3305015, EPI_ISL_3305018, EPI_ISL_3305022, EPI_ISL_3305032, EPI_ISL_3305033, EPI_ISL_3305037, EPI_ISL_3305039, EPI_ISL_3305051, EPI_ISL_3305055, EPI_ISL_3305056, EPI_ISL_3305058, EPI_ISL_3305067, EPI_ISL_3305072, EPI_ISL_3305073, EPI_ISL_3305087, EPI_ISL_3305093, EPI_ISL_3305096 | | | |
| see above | Quest Diagnostics Incorporated | Centers for Disease Control and Prevention Division of Viral Diseases, Pathogen Discovery | A. Gerasimova; A. Perez; Adrian Paskey; B. Anderson; Benjamin Rambo-Martin; Christopher Gulvick; Clinton R. Paden; Dakota Howard; Darlene Wagner; Dhwani Batra; Duncan MacCannell; F. Lacbawan; I. A. Shlyakhter; Jason Caravas; K.E. Livingston; Kara Moser; L.E. Bernstein; M. Hua; Matthew Schmerer; P. Tanpaiboon; Peter W. Cook; R. M. Kagan; R. Owen; R. V. Rolando; S. H. Rosenthal; Scott Sammons; Shatavia Morrison; Y. Liu; Yvette Unoarumhi |
| EPI_ISL_3355373 | RI State Health Laboratories | Centers for Disease Control and Prevention Division of Viral Diseases, Pathogen Discovery | Alex Burgin; Ben Rambo-Martin; Clinton Paden; Dakota Howard; Dave Wentworth; Dhwani Batra; Jasmine Padilla; Justin Lee; Krista Queen; Kristen Knipe; Kristine Lacek; Mark Burroughs; Matthew Schmerer; Meghan Bentz; Mili Sheth; Peter Cook; Sam Shepard; Sarah Nobles; Suxiang Tong; Vivien Dugan; Yvette Unoarumhi |
| EPI_ISL_2933972 | Royal GD | Royal GD | Remco Dijkman |
| EPI_ISL_3388714 | Rush University Medical Center | RIPHL at Rush University Medical Center | Diane Springer; Edith Perez; Felix Araujo Perez; Joyce Houlihan; Kevin Kunstman; Laura Furtado; Marieta Hyde; Mary Hayden; Max Kolton; Stefan Green |
| EPI_ISL_2840628 | S.C. Laboratorio Analisi, ASL 3 Liguria | U.O. Igiene, Ospedale Policlinico San Martino | Bruzzone Bianca; Caligiuri Patrizia; De Pace Vanessa; Domnich Alexander; Icardi Giancarlo on behalf of SARS-CoV-2 ITALIAN RESEARCH ENTERPRISE-(SCIRE) Collaborative Group; Orsi Andrea; Ricucci Valentina; Spitaleri Antonino |
| EPI_ISL_5649337 | SANTA CASA DE DOIS CORREGOS | Instituto Butantan | Antonio Jorge Martins; Claudia Renata dos Santos Barros; David Schlesinger; Debora Botequio Moretti; Dimas Tadeu Covas; Elaine Cristina Marqueze; Elaine Vieira Santos; Evandra Strazza Rodrigues; Heidge Fukumasu; Jayme Augusto de Souza-Neto; José Salvatore Leister Patané; Luiz Alcantara; Luiz Lehmann Coutinho; Maria Carolina Elias; Maurício Lacerda Nogueira; Rafael dos Santos Bezerra; Raul Machado Neto; Rejane Maria Tommasini Grotto; Ricardo Haddad; Sandra Coccuzzo Sampaio Vessoni; Simone Kashima; Svetoslav Nanev Slavov; Vincent Louis Viala |
| EPI_ISL_5648878, EPI_ISL_5648893, EPI_ISL_5648921, EPI_ISL_5648954, EPI_ISL_5649023 | SANTA CASA DE GUARAREMA | Instituto Butantan | Antonio Jorge Martins; Claudia Renata dos Santos Barros; David Schlesinger; Debora Botequio Moretti; Dimas Tadeu Covas; Elaine Cristina Marqueze; Elaine Vieira Santos; Evandra Strazza Rodrigues; Heidge Fukumasu; Jayme Augusto de Souza-Neto; José Salvatore Leister Patané; Luiz Alcantara; Luiz Lehmann Coutinho; Maria Carolina Elias; Maurício Lacerda Nogueira; Rafael dos Santos Bezerra; Raul Machado Neto; Rejane Maria Tommasini Grotto; Ricardo Haddad; Sandra Coccuzzo Sampaio Vessoni; Simone Kashima; Svetoslav Nanev Slavov; Vincent Louis Viala |
| EPI_ISL_5648381, EPI_ISL_5648449, EPI_ISL_5648670 | SANTA CASA DE IBIRA | Instituto Butantan | Antonio Jorge Martins; Claudia Renata dos Santos Barros; David Schlesinger; Debora Botequio Moretti; Dimas Tadeu Covas; Elaine Cristina Marqueze; Elaine Vieira Santos; Evandra Strazza Rodrigues; Heidge Fukumasu; Jayme Augusto de Souza-Neto; José Salvatore Leister Patané; Luiz Alcantara; Luiz Lehmann Coutinho; Maria Carolina Elias; Maurício Lacerda Nogueira; Rafael dos Santos Bezerra; Raul Machado Neto; Rejane Maria Tommasini Grotto; Ricardo Haddad; Sandra Coccuzzo Sampaio Vessoni; Simone Kashima; Svetoslav Nanev Slavov; Vincent Louis Viala |
| EPI_ISL_5648612 | SANTA CASA DE JOSE BONIFACIO | Instituto Butantan | Antonio Jorge Martins; Claudia Renata dos Santos Barros; David Schlesinger; Debora Botequio Moretti; Dimas Tadeu Covas; Elaine Cristina Marqueze; Elaine Vieira Santos; Evandra Strazza Rodrigues; Heidge Fukumasu; Jayme Augusto de Souza-Neto; José Salvatore Leister Patané; Luiz Alcantara; Luiz Lehmann Coutinho; Maria Carolina Elias; Maurício Lacerda Nogueira; Rafael dos Santos Bezerra; Raul Machado Neto; Rejane Maria Tommasini Grotto; Ricardo Haddad; Sandra Coccuzzo Sampaio Vessoni; Simone Kashima; Svetoslav Nanev Slavov; Vincent Louis Viala |
| EPI_ISL_5649276, EPI_ISL_5649294 | SANTA CASA DE MISERICORDIA DE TIETE | Instituto Butantan | Antonio Jorge Martins; Claudia Renata dos Santos Barros; David Schlesinger; Debora Botequio Moretti; Dimas Tadeu Covas; Elaine Cristina Marqueze; Elaine Vieira Santos; Evandra Strazza Rodrigues; Heidge Fukumasu; Jayme Augusto de Souza-Neto; José Salvatore Leister Patané; Luiz Alcantara; Luiz Lehmann Coutinho; Maria Carolina Elias; Maurício Lacerda Nogueira; Rafael dos Santos Bezerra; Raul Machado Neto; Rejane Maria Tommasini Grotto; Ricardo Haddad; Sandra Coccuzzo Sampaio Vessoni; Simone Kashima; Svetoslav Nanev Slavov; Vincent Louis Viala |
| EPI_ISL_5649421 | SANTA CASA DE MISERICORDIA DE UBATUBA | Instituto Butantan | Antonio Jorge Martins; Claudia Renata dos Santos Barros; David Schlesinger; Debora Botequio Moretti; Dimas Tadeu Covas; Elaine Cristina Marqueze; Elaine Vieira Santos; Evandra Strazza Rodrigues; Heidge Fukumasu; Jayme Augusto de Souza-Neto; José Salvatore Leister Patané; Luiz Alcantara; Luiz Lehmann Coutinho; Maria Carolina Elias; Maurício Lacerda Nogueira; Rafael dos Santos Bezerra; Raul Machado Neto; Rejane Maria Tommasini Grotto; Ricardo Haddad; Sandra Coccuzzo Sampaio Vessoni; Simone Kashima; Svetoslav Nanev Slavov; Vincent Louis Viala |
| EPI_ISL_5648898, EPI_ISL_5649101 | SANTA CASA DE SANTA ADELIA | Instituto Butantan | Antonio Jorge Martins; Claudia Renata dos Santos Barros; David Schlesinger; Debora Botequio Moretti; Dimas Tadeu Covas; Elaine Cristina Marqueze; Elaine Vieira Santos; Evandra Strazza Rodrigues; Heidge Fukumasu; Jayme Augusto de Souza-Neto; José Salvatore Leister Patané; Luiz Alcantara; Luiz Lehmann Coutinho; Maria Carolina Elias; Maurício Lacerda Nogueira; Rafael dos Santos Bezerra; Raul Machado Neto; Rejane Maria Tommasini Grotto; Ricardo Haddad; Sandra Coccuzzo Sampaio Vessoni; Simone Kashima; Svetoslav Nanev Slavov; Vincent Louis Viala |
| EPI_ISL_5649152 | SANTA CASA SANTA RITA DO PASSA QUATRO | Instituto Butantan | Antonio Jorge Martins; Claudia Renata dos Santos Barros; David Schlesinger; Debora Botequio Moretti; Dimas Tadeu Covas; Elaine Cristina Marqueze; Elaine Vieira Santos; Evandra Strazza Rodrigues; Heidge Fukumasu; Jayme Augusto de Souza-Neto; José Salvatore Leister Patané; Luiz Alcantara; Luiz Lehmann Coutinho; Maria Carolina Elias; Maurício Lacerda Nogueira; Rafael dos Santos Bezerra; Raul Machado Neto; Rejane Maria Tommasini Grotto; Ricardo Haddad; Sandra Coccuzzo Sampaio Vessoni; Simone Kashima; Svetoslav Nanev Slavov; Vincent Louis Viala |
| EPI_ISL_2986642, EPI_ISL_2986643, EPI_ISL_2986644 | SARS-CoV-2 Sequencing Castilla y Leon-Spain Consortium | SARS-CoV-2 Sequencing Castilla y Leon-Spain Consortium | Antonio Orduña-Domingo; Carlos Fuster Foz; Carmen Aldea-Mansilla; Carmen Gimeno Crespo; David Abad; Gabriel March Rossello; Gregoria Megías Lobón; Jose María Eiros Bouza; M. Isabel Fernandez-Natal; Marta Dominguez-Gil; Marta Hernandez; María Antonia García Castro; Mª Fe Brezmes-Valdivieso; Noelia Arenal Andrés; Silvia Rojo; Sonsoles Garcinuño Pérez |
| EPI_ISL_2828304 | SARS-CoV-2 testing team, National Institute of Infectious Diseases | Pathogen Genomics Center, National Institute of Infectious Diseases | Hazuka Y Furihata; Hiromizu Takahashi; Kentaro Itokawa; Makoto Kuroda; Masanori Hashino; Masumichi Saito; Naomi Nojiri; Nozomu Hanaoka; Rina Tanaka; Tsuguto Fujimoto; Tsuyoshi Sekizuka |
| EPI_ISL_2894084 | SCM B12 | CNR Virus des Infections Respiratoires - France SUD | Antonin Bal; Bruno Lina; Gregory Destras; Gwendolyne Burfin; Hadrien Regue; Laurence Josset; Martine Valette; Quentin Semanas |
| EPI_ISL_3066770 | SECRETARIA DE SALUD DE SAN ANDRES ISLA | Instituto Nacional de Salud | Carlos Franco-Muñoz; Carmen Osorio; Diana Malo; Diego A. Álvarez-Díaz; Diego Andrés Prada; Gerardo Santamaría; Hector Alejandro Ruiz-Moreno; Jhonnatan Reales-González; Jorge Rivera; Juan Camilo Martinez; Julian Naizaque; Katherine Laiton-Donato; Lisseth Pardo; Magdalena Wiesner; Marcela Mercado-Reyes; Maria T.Herrera-Sepúlveda; Marta Lopez Blanco; Martha Lucia Ospina Martinez; Paola Rojas; Sergio Gomez; Sheryll Corchuelo; Ángela Alarcon Cruz |
| EPI_ISL_5649225, EPI_ISL_5665682, EPI_ISL_5665692 | SECRETARIA DE SAUDE DE SANTA GERTRUDES | Instituto Butantan | Antonio Jorge Martins; Claudia Renata dos Santos Barros; David Schlesinger; Debora Botequio Moretti; Dimas Tadeu Covas; Elaine Cristina Marqueze; Elaine Vieira Santos; Evandra Strazza Rodrigues; Heidge Fukumasu; Jayme Augusto de Souza-Neto; José Salvatore Leister Patané; Luiz Alcantara; Luiz Lehmann Coutinho; Maria Carolina Elias; Maurício Lacerda Nogueira; Rafael dos Santos Bezerra; Raul Machado Neto; Rejane Maria Tommasini Grotto; Ricardo Haddad; Sandra Coccuzzo Sampaio Vessoni; Simone Kashima; Svetoslav Nanev Slavov; Vincent Louis Viala |
| EPI_ISL_5649171, EPI_ISL_5649224, EPI_ISL_5665701, EPI_ISL_5665711 | SECRETARIA DE SAUDE DE SAO PEDRO | Instituto Butantan | Antonio Jorge Martins; Claudia Renata dos Santos Barros; David Schlesinger; Debora Botequio Moretti; Dimas Tadeu Covas; Elaine Cristina Marqueze; Elaine Vieira Santos; Evandra Strazza Rodrigues; Heidge Fukumasu; Jayme Augusto de Souza-Neto; José Salvatore Leister Patané; Luiz Alcantara; Luiz Lehmann Coutinho; Maria Carolina Elias; Maurício Lacerda Nogueira; Rafael dos Santos Bezerra; Raul Machado Neto; Rejane Maria Tommasini Grotto; Ricardo Haddad; Sandra Coccuzzo Sampaio Vessoni; Simone Kashima; Svetoslav Nanev Slavov; Vincent Louis Viala |
| EPI_ISL_5648408, EPI_ISL_5648436, EPI_ISL_5648463, EPI_ISL_5648636 | SECRETARIA DE SAUDE PUBLICA DE PRAIA GRANDE | Instituto Butantan | Antonio Jorge Martins; Claudia Renata dos Santos Barros; David Schlesinger; Debora Botequio Moretti; Dimas Tadeu Covas; Elaine Cristina Marqueze; Elaine Vieira Santos; Evandra Strazza Rodrigues; Heidge Fukumasu; Jayme Augusto de Souza-Neto; José Salvatore Leister Patané; Luiz Alcantara; Luiz Lehmann Coutinho; Maria Carolina Elias; Maurício Lacerda Nogueira; Rafael dos Santos Bezerra; Raul Machado Neto; Rejane Maria Tommasini Grotto; Ricardo Haddad; Sandra Coccuzzo Sampaio Vessoni; Simone Kashima; Svetoslav Nanev Slavov; Vincent Louis Viala |
| EPI_ISL_5649055 | SECRETARIA MUNICIPAL DE SAUDE | Instituto Butantan | Antonio Jorge Martins; Claudia Renata dos Santos Barros; David Schlesinger; Debora Botequio Moretti; Dimas Tadeu Covas; Elaine Cristina Marqueze; Elaine Vieira Santos; Evandra Strazza Rodrigues; Heidge Fukumasu; Jayme Augusto de Souza-Neto; José Salvatore Leister Patané; Luiz Alcantara; Luiz Lehmann Coutinho; Maria Carolina Elias; Maurício Lacerda Nogueira; Rafael dos Santos Bezerra; Raul Machado Neto; Rejane Maria Tommasini Grotto; Ricardo Haddad; Sandra Coccuzzo Sampaio Vessoni; Simone Kashima; Svetoslav Nanev Slavov; Vincent Louis Viala |
| EPI_ISL_5649175, EPI_ISL_5649223, EPI_ISL_5649230, EPI_ISL_5665749, EPI_ISL_5665754 | SECRETARIA MUNICIPAL DE SAUDE DE CORDEIROPOLIS | Instituto Butantan | Antonio Jorge Martins; Claudia Renata dos Santos Barros; David Schlesinger; Debora Botequio Moretti; Dimas Tadeu Covas; Elaine Cristina Marqueze; Elaine Vieira Santos; Evandra Strazza Rodrigues; Heidge Fukumasu; Jayme Augusto de Souza-Neto; José Salvatore Leister Patané; Luiz Alcantara; Luiz Lehmann Coutinho; Maria Carolina Elias; Maurício Lacerda Nogueira; Rafael dos Santos Bezerra; Raul Machado Neto; Rejane Maria Tommasini Grotto; Ricardo Haddad; Sandra Coccuzzo Sampaio Vessoni; Simone Kashima; Svetoslav Nanev Slavov; Vincent Louis Viala |
| EPI_ISL_5649406 | SECRETARIA MUNICIPAL DE SAUDE DE JAU | Instituto Butantan | Antonio Jorge Martins; Claudia Renata dos Santos Barros; David Schlesinger; Debora Botequio Moretti; Dimas Tadeu Covas; Elaine Cristina Marqueze; Elaine Vieira Santos; Evandra Strazza Rodrigues; Heidge Fukumasu; Jayme Augusto de Souza-Neto; José Salvatore Leister Patané; Luiz Alcantara; Luiz Lehmann Coutinho; Maria Carolina Elias; Maurício Lacerda Nogueira; Rafael dos Santos Bezerra; Raul Machado Neto; Rejane Maria Tommasini Grotto; Ricardo Haddad; Sandra Coccuzzo Sampaio Vessoni; Simone Kashima; Svetoslav Nanev Slavov; Vincent Louis Viala |
| EPI_ISL_5649170, EPI_ISL_5649208, EPI_ISL_5649219, EPI_ISL_5649226, EPI_ISL_5649231, EPI_ISL_5649235 | SECRETARIA MUNICIPAL DE SAUDE DE PRES BERNARDES SP | Instituto Butantan | Antonio Jorge Martins; Claudia Renata dos Santos Barros; David Schlesinger; Debora Botequio Moretti; Dimas Tadeu Covas; Elaine Cristina Marqueze; Elaine Vieira Santos; Evandra Strazza Rodrigues; Heidge Fukumasu; Jayme Augusto de Souza-Neto; José Salvatore Leister Patané; Luiz Alcantara; Luiz Lehmann Coutinho; Maria Carolina Elias; Maurício Lacerda Nogueira; Rafael dos Santos Bezerra; Raul Machado Neto; Rejane Maria Tommasini Grotto; Ricardo Haddad; Sandra Coccuzzo Sampaio Vessoni; Simone Kashima; Svetoslav Nanev Slavov; Vincent Louis Viala |
| EPI_ISL_3912243, EPI_ISL_3912244 | SECRETARIA MUNICIPAL DE SAUDE DE TIANGUA | Analytical Competence Molecular Epidemiology Lab/ACME, Oswaldo Cruz Foundation, Ceara (FIOCRUZ CE) | Cleber Furtado Aksenen; Fabio Miyajima; Fernando Braga Stehling; Francisco Eder de Moura Lopes; Jamille Maria Mendes Bezerra; Joaquim Cesar do Nascimento Sousa Junior; Pedro Miguel Carneiro Jeronimo; Suzana Porto Almeida & Lucas Delerino on behalf of COVID-19 FIOCRUZ Genomic Network; Thais Ferreira de Oliveira; Thais de Oliveira Costa; Ticiane Cavalcante de Souza; Veridiana Pessoa Miyajima |
| EPI_ISL_5648462, EPI_ISL_5648687 | SECRETARIA MUNICIPAL DE SAUDE DE VALPARAISO SP | Instituto Butantan | Antonio Jorge Martins; Claudia Renata dos Santos Barros; David Schlesinger; Debora Botequio Moretti; Dimas Tadeu Covas; Elaine Cristina Marqueze; Elaine Vieira Santos; Evandra Strazza Rodrigues; Heidge Fukumasu; Jayme Augusto de Souza-Neto; José Salvatore Leister Patané; Luiz Alcantara; Luiz Lehmann Coutinho; Maria Carolina Elias; Maurício Lacerda Nogueira; Rafael dos Santos Bezerra; Raul Machado Neto; Rejane Maria Tommasini Grotto; Ricardo Haddad; Sandra Coccuzzo Sampaio Vessoni; Simone Kashima; Svetoslav Nanev Slavov; Vincent Louis Viala |
| EPI_ISL_5649253, EPI_ISL_5649255, EPI_ISL_5649261, EPI_ISL_5649275, EPI_ISL_5649287, EPI_ISL_5649293, EPI_ISL_5649306, EPI_ISL_5649317, EPI_ISL_5649321 | | | |
| see above | SECRETARIA MUNICIPAL DE SAUDE SOROCABA | Instituto Butantan | Antonio Jorge Martins; Claudia Renata dos Santos Barros; David Schlesinger; Debora Botequio Moretti; Dimas Tadeu Covas; Elaine Cristina Marqueze; Elaine Vieira Santos; Evandra Strazza Rodrigues; Heidge Fukumasu; Jayme Augusto de Souza-Neto; José Salvatore Leister Patané; Luiz Alcantara; Luiz Lehmann Coutinho; Maria Carolina Elias; Maurício Lacerda Nogueira; Rafael dos Santos Bezerra; Raul Machado Neto; Rejane Maria Tommasini Grotto; Ricardo Haddad; Sandra Coccuzzo Sampaio Vessoni; Simone Kashima; Svetoslav Nanev Slavov; Vincent Louis Viala |
| EPI_ISL_2795722, EPI_ISL_2795758 | SELAS MEDILYS | Department of Virology, Henri Mondor University Hospital, Assistance Publique Hôpitaux de Paris, Université Paris-Est Créteil, INSERM U955 | Alexandre Soulier; Christophe Rodriguez; Elisabeth Trawinski; Guillaume Gricourt; Jean-Michel Pawlotsky; Melissa N'Debi; Slim Fourati; Vanessa Demontant |
| EPI_ISL_2895384, EPI_ISL_2895389 | SESARAM | Instituto Nacional de Saude (INSA) | Borges et al |
| EPI_ISL_3255867, EPI_ISL_3255869, EPI_ISL_3255878, EPI_ISL_3255889, EPI_ISL_3255892, EPI_ISL_3255896, EPI_ISL_3255916, EPI_ISL_3255917, EPI_ISL_3255919, EPI_ISL_3255922, EPI_ISL_3255925, EPI_ISL_3255929, EPI_ISL_3255933, EPI_ISL_3255934, EPI_ISL_3255939, EPI_ISL_3255954 | | | |
| see above | SK-Roy Romanow Provincial Laboratory | National Microbiology Laboratory (NML) | Alanna Senecal; Amanda Lang; Anna Majer; Anneliese Landgraff; CanCOGeN's metadata curation team; Darian Hole; Elsie Grudeski; Gary Van Domselaar; Grace Seo; Jennifer Tanner; Jessica Minion; Kara Loos; Keith MacKenzie; Kirsten Biggar; Madison Chapel; Meredith Faires; Morag Graham; Natalie Knox; Nathalie Bastien; Philip Mabon; Public Health Agency of Canada CanCOGeN team; Rachel DePaulo; Rhiannon Huzarewich; Russell Mandes; Ryan McDonald; Shari Tyson; Timothy Booth; Yan Li |
| EPI_ISL_5780495, EPI_ISL_5780543, EPI_ISL_5780544, EPI_ISL_5845145, EPI_ISL_5845149, EPI_ISL_5845154, EPI_ISL_5845161, EPI_ISL_5845164, EPI_ISL_5845165, EPI_ISL_5845168, EPI_ISL_5845177, EPI_ISL_5845178 | | | |
| see above | SK-Roy Romanow Provincial Laboratory | Saskatchewan - Roy Romanow Provincial Laboratory (RRPL) | Alanna Senecal; Amanda Lang; Jessica Minion; Kara Loos; Keith MacKenzie; Meredith Faires; Rachel DePaulo; Roy Romanow Provincial Laboratory - Molecular Diagnostics; Ryan McDonald |
| EPI_ISL_5649316, EPI_ISL_5649322 | SMS SECRETARIA MUNICIPAL DE SAUDE DE BOITUVA | Instituto Butantan | Antonio Jorge Martins; Claudia Renata dos Santos Barros; David Schlesinger; Debora Botequio Moretti; Dimas Tadeu Covas; Elaine Cristina Marqueze; Elaine Vieira Santos; Evandra Strazza Rodrigues; Heidge Fukumasu; Jayme Augusto de Souza-Neto; José Salvatore Leister Patané; Luiz Alcantara; Luiz Lehmann Coutinho; Maria Carolina Elias; Maurício Lacerda Nogueira; Rafael dos Santos Bezerra; Raul Machado Neto; Rejane Maria Tommasini Grotto; Ricardo Haddad; Sandra Coccuzzo Sampaio Vessoni; Simone Kashima; Svetoslav Nanev Slavov; Vincent Louis Viala |

| | | | |
|---|---|---|---|
| EPI_ISL_3536156, EPI_ISL_3536157, EPI_ISL_3536158, EPI_ISL_3536159, EPI_ISL_3536160, EPI_ISL_3536161, EPI_ISL_3536162 | | | |
| see above | SOCIEDADE HOSPITALAR PE DIONISIO | Analytical Competence Molecular Epidemiology Lab/ACME, Oswaldo Cruz Foundation, Ceara (FIOCRUZ CE) | Cleber Furtado Aksenen; Fabio Miyajima; Fernando Braga Stehling; Francisco Eder de Moura Lopes; Jamille Maria Mendes Bezerra; Joaquim César do Nascimento Sousa Junior; Pedro Miguel Carneiro Jeronimo; Suzana Porto Almeida e Lucas Delerino; Thais Ferreira de Oliveira; Thais de Oliveira Costa; Ticiane Cavalcante de Souza; Veridiana Pessoa Miyajima |
| EPI_ISL_4666457 | SURA | Laboratorio Departamental de Salud Publica de Antioquia | Andres F. Cardona-Rios; Cristian Arbey Velarde Hoyos; Gloria Isabel Escobar; Idabely Betancur Ortiz; Juan P. Hernandez-Ortiz; Maria Stella López |
| EPI_ISL_2930646 | SYNLAB | GIGA Medical Genomics | Bouchra Boujemla; Claire Gourzonès; Cécile Meex; Keith Durkin; Laurent Gillet; Maria Artesi; Marie-Pierre Hayette; Nadine Cambisano; Nathalie Renotte; Olivier Ek; Sébastien Bontems; Vincent Bours |
| EPI_ISL_2933358 | SYNLAB GASCOGNE | CNR Virus des Infections Respiratoires - France SUD | Antonin Bal; Bruno Lina; Gregory Destras; Gwendolyne Burfin; Hadrien Regue; Laurence Josset; Martine Valette; Quentin Semanas |
| EPI_ISL_2845000 | SYNLAB MVZ Dachau | Robert Koch Institute | |
| EPI_ISL_2844917, EPI_ISL_2844919, EPI_ISL_2844947, EPI_ISL_2844950 | SYNLAB MVZ Heidelberg | Robert Koch Institute | |
| EPI_ISL_2760678 | SYNLAB MVZ Leinfelden-Echterdingen | Robert Koch Institute | |
| EPI_ISL_2760707, EPI_ISL_2760783, EPI_ISL_2844910 | SYNLAB MVZ Leverkusen | Robert Koch Institute | |
| EPI_ISL_2844926, EPI_ISL_2844964, EPI_ISL_2844971, EPI_ISL_2844982, EPI_ISL_2844991 | SYNLAB MVZ Weiden | Robert Koch Institute | |
| EPI_ISL_2810032, EPI_ISL_2810033, EPI_ISL_2810034, EPI_ISL_2810035, EPI_ISL_2810044, EPI_ISL_2810049, EPI_ISL_2810052, EPI_ISL_2810060, EPI_ISL_2810065, EPI_ISL_2810070, EPI_ISL_2810073, EPI_ISL_2810079, EPI_ISL_2810080, EPI_ISL_2810081, EPI_ISL_2810082, EPI_ISL_2810086, EPI_ISL_2810088, EPI_ISL_2810094, EPI_ISL_2810096, EPI_ISL_2810176, EPI_ISL_2810179, EPI_ISL_2810208, EPI_ISL_2810218, EPI_ISL_2810247, EPI_ISL_2810256, EPI_ISL_2810262, EPI_ISL_2810294, EPI_ISL_2810313, EPI_ISL_2928485, EPI_ISL_2978441, EPI_ISL_2978456, EPI_ISL_2978600, EPI_ISL_2978608, EPI_ISL_2978614, EPI_ISL_2978615, EPI_ISL_2978620, EPI_ISL_2978621, EPI_ISL_2978622, EPI_ISL_2978627, EPI_ISL_2978628, EPI_ISL_2978633, EPI_ISL_2978634, EPI_ISL_2978635, EPI_ISL_2978640, EPI_ISL_2978641, EPI_ISL_2978642, EPI_ISL_2978657, EPI_ISL_2978662, EPI_ISL_2978663, EPI_ISL_2978664, EPI_ISL_2978671, EPI_ISL_2978678, EPI_ISL_2978679, EPI_ISL_3067659, EPI_ISL_3067663, EPI_ISL_3067896, EPI_ISL_3067899, EPI_ISL_3067931, EPI_ISL_3067932, EPI_ISL_3067933, EPI_ISL_3067934, EPI_ISL_3067935, EPI_ISL_3067936, EPI_ISL_3067937 | | | |
| see above | Salud Digna | Instituto Nacional de Medicina Genomica | Abraham Campos-Romero; Blancas S; Cedro-Tanda A; Cisneros-Villanueva M; Escobar-Arrazola; Gonzalez-Barrera D; Herrera-Montalvo LA.; Hidalgo-Miranda A; Luna-Ruiz Marco; M; Mendoza-Vargas A; Moreno-Camacho José Luis; Munguia-Garza P; Orjuela-Rodríguez M; Ramirez-Vega O; Rangel-DeLeon D; Reyes-Grajeda JP; Rodriguez-Gallegos Jorge |
| EPI_ISL_3374060, EPI_ISL_3374063, EPI_ISL_3374113, EPI_ISL_3374114, EPI_ISL_3374121 | San Diego County Public Health Laboratory | Andersen lab at Scripps Research | Brett Austin; Jovan Shephard; SEARCH Alliance San Diego with Ashleigh Murphy |
| EPI_ISL_3275048 | San Diego County Public Health Laboratory | San Diego County Public Health Laboratory | Ashleigh Murphy Schafer; Brett Austin |
| EPI_ISL_2833117, EPI_ISL_2833119, EPI_ISL_2833129 | San Gallicano Dermatologic InstituteIRCCS | National Institute for Infectious Diseases (INMI) L. Spallanzani I.R.C.C.S | A Di Caro; B Bartolini; CEM Gruber; E Giombini; F Messina; F Santini; G Bonfiglio; M Rueca; MR Capobianchi; O Butera |
| EPI_ISL_2967940 | Santa Clara Valley Medical Center | Santa Clara County Public Health Laboratory | Santa Clara County Public Health Department |
| EPI_ISL_2787075 | Sant’Eugenio/CTO ASL Roma 2. | INMI Lazzaro Spallanzani IRCCS | A Massacci; A Tamburro; B Bartolini; CEM Gruber; F Messina; G Bonfiglio; L Ciuffreda; M Rueca; ML Schiavone |
| EPI_ISL_4082818, EPI_ISL_4082820, EPI_ISL_4082889, EPI_ISL_4082918, EPI_ISL_4082922, EPI_ISL_4082941, EPI_ISL_4082943, EPI_ISL_4082945, EPI_ISL_4082946, EPI_ISL_4082947, EPI_ISL_4082948, EPI_ISL_4082949, EPI_ISL_4082950, EPI_ISL_4082966, EPI_ISL_4082973, EPI_ISL_4082981, EPI_ISL_4082983, EPI_ISL_4082985, EPI_ISL_4082986, EPI_ISL_4083002, EPI_ISL_4083003, EPI_ISL_4083007, EPI_ISL_4083008, EPI_ISL_4083009, EPI_ISL_4083010, EPI_ISL_4083011, EPI_ISL_4083012, EPI_ISL_4083013, EPI_ISL_4083017, EPI_ISL_4083018, EPI_ISL_4083039, EPI_ISL_4083040, EPI_ISL_4083041, EPI_ISL_4083042, EPI_ISL_4083043, EPI_ISL_4083045, EPI_ISL_4083046, EPI_ISL_4083050, EPI_ISL_4083051, EPI_ISL_4083052, EPI_ISL_4083053, EPI_ISL_4083054, EPI_ISL_4083060, EPI_ISL_4083063, EPI_ISL_4083064, EPI_ISL_4083066, EPI_ISL_4083095, EPI_ISL_4083119, EPI_ISL_4083122, EPI_ISL_4083123, EPI_ISL_4083139, EPI_ISL_4083140, EPI_ISL_4083141, EPI_ISL_4083142, EPI_ISL_4083143, EPI_ISL_4083144, EPI_ISL_4083145, EPI_ISL_4083166, EPI_ISL_4083220, EPI_ISL_4083224, EPI_ISL_4083226, EPI_ISL_4083231, EPI_ISL_4083236, EPI_ISL_4083241, EPI_ISL_4083246, EPI_ISL_4083249, EPI_ISL_4083253, EPI_ISL_4083254, EPI_ISL_4083273, EPI_ISL_4083274, EPI_ISL_4083285, EPI_ISL_4083290, EPI_ISL_4083319, EPI_ISL_4083322, EPI_ISL_5158432, EPI_ISL_5158434, EPI_ISL_5158537, EPI_ISL_5158861, EPI_ISL_6694375, EPI_ISL_6694377, EPI_ISL_6694379, EPI_ISL_6694398 | | | |
| see above | Servicio Virosis Respiratorias-Departamento Virología-INEI | Instituto Nacional Enfermedades Infecciosas C.G.Malbran | Avaro M.; Baumeister E.; Benedetti E.; Campos J.; Cisterna D.; Dattero ME; De Belder D.; Haim MS.; Lorenzo F.; Molina V.; Perandones C.; Poklepovich T.; Pontoriero A.; Russo M.; Sanchez Loria J.; Tuduri E. |
| EPI_ISL_3060984, EPI_ISL_3060990 | Servicio de Microbiología. Consorcio Hospital General Universitario de Valencia | SeqCOVID-SPAIN consortium/IBV(CSIC) | Begoña Fuster Escrivá; Carme Salvador García; Concepción Gimeno Cardona and SeqCOVID-SPAIN consortium; María Dolores Ocete; Rafael Medina González |
| EPI_ISL_3008598 | Servicio de Microbiología. Hospital Arnau de Vilanova | SeqCOVID-SPAIN consortium/IBV(CSIC) | Amparo Farga and SeqCOVID-SPAIN consortium; Rocío Falcón; Victoria Domínguez |
| EPI_ISL_3008587 | Servicio de Microbiología. Hospital General Universitario de Castellón | SeqCOVID-SPAIN consortium/IBV(CSIC) | María Dolores Tirado Balaguer and SeqCOVID-SPAIN consortium; Rosario Moreno Muñoz |
| EPI_ISL_3907299, EPI_ISL_3907401 | Servicio de Microbiología. Hospital Universitario Doctor Peset | SeqCOVID-SPAIN consortium/IBV(CSIC) | José Miguel Nogueira Coito and SeqCOVID-SPAIN consortium; Juan Alberola Enguídanos; Juan José Camarena Miñana; Rosa González Pellicer |
| EPI_ISL_2988836 | Sharp HealthCare Laboratory | Andersen lab at Scripps Research | Art Mendoza; Cathy Woerle; Jacquelyn Berumen; Liam McGinnis; Omid Bakhtar; SEARCH Alliance San Diego with Aaron Harding |
| EPI_ISL_3254819 | Simple Laboratories | Gagnon Lab, Southern Illinois University | Keith Gagnon |
| EPI_ISL_3050186, EPI_ISL_4072051, EPI_ISL_4072052 | Stanford Health Care | Stanford University School of Medicine, Clinical Virology Laboratory | Becky Jiang; Bernadette Troung; Daniel Solis; Hannah Wang; James Zehnder; Malaya K. Sahoo; Mamdouh Sibai; Nathan Hammond; Selamawit Bihon; and Benjamin A. Pinsky |
| EPI_ISL_3033163, EPI_ISL_3033180, EPI_ISL_3033191, EPI_ISL_3033207, EPI_ISL_3033212, EPI_ISL_3033223, EPI_ISL_3033236, EPI_ISL_3033237, EPI_ISL_3033238, EPI_ISL_3033254 | | | |
| see above | State Laboratories Division, Hawaii State Department of Health | State Laboratories Division, Hawaii State Department of Health | Ayana Garnet; Drew Kuwazaki; Edward Desmond; Pamela O'Brien; Razvan Sultana |
| EPI_ISL_3050022 | State Testing Facility | Altius Institute for Biomedical Research | Alex Isner; Alex Nguyen; Amanda Gale; Audra Johnson; Clem Green; Daniel Bates; Eric Thorland; Jacob Rodriguez; Jean Robinston; Jemma Nelson; Jessica Kunder; John Stamatoyannopoulos; Joshua Richards; Julia Wald; Kneshay Harper; Lauren Mitchell; Mark Frerker; Matt Hartman; Michael Buckley; Muhammad Halimun; Rebecca Bruders; Tobias Ragoczy |
| EPI_ISL_2888088, EPI_ISL_2888091, EPI_ISL_2888115, EPI_ISL_2980941, EPI_ISL_3795575, EPI_ISL_3999023, EPI_ISL_6135045 | | | |
| see above | Swedish national genomic surveillance program of SARS-CoV-2 | The Public Health Agency of Sweden | Alma Brolund; Maria Lind Karlberg; Maximilian Riess; Swedish national genomic surveillance program of SARS-CoV-2 |
| EPI_ISL_3088564, EPI_ISL_3088596, EPI_ISL_3088611, EPI_ISL_3088804, EPI_ISL_3325788, EPI_ISL_3326304, EPI_ISL_3326506, EPI_ISL_3326646, EPI_ISL_3326659, EPI_ISL_3326689 | | | |
| see above | TGen North | TGen North | "Jolene Bowers; Brett Van Tassel; Chris French; Darrin Lemmer; Dave Engelthaler; Dave Engelthaler"; Hayley Yaglom; Heather Centner; Jolene Bowers |
| EPI_ISL_3695212, EPI_ISL_3695229, EPI_ISL_3695237, EPI_ISL_3695265, EPI_ISL_3695284, EPI_ISL_3695292, EPI_ISL_3695305, EPI_ISL_3695339, EPI_ISL_3695345, EPI_ISL_3695348, EPI_ISL_3717096 | | | |
| see above | TPMG Regional Laboratory | California Department of Public Health | Emily Smith on behalf of CDPH-COVIDNet and UCI Genome Sciences Center/GHTF |
| EPI_ISL_3102156, EPI_ISL_3151158 | TXDSHS | TXDSHS | Anita Pokharel; Bonnie Oh; Chun Wang; Grace Kubin; Jenny Zhang; Karen Bobier; Lorraine Rodriguez; Maliha Rahman; Mayela Pedrueza; Myong Koag; Rachel Lee; Rashmi Tuladhar |
| EPI_ISL_2800905 | Tampa General Hospital Esoteric Lab | Tampa General Hospital Esoteric Research & Development Lab | Amorce Lima; Deanna Becker; Dominic Uy; Elaine Vendrone; Jon Faughn; Jorge Mecias-Frias; Suzane Silbert; Vicki Healer |
| EPI_ISL_3926036 | The National University Hospital of Iceland | deCODE genetics | Agnar Helgason; Alma Moller; Arna B Agustsdottir; Arnaldur Gylfason; Asgeir Sigurdsson; Aslaug Jonasdottir; Berglind Eiriksdottir; Bjarni Thorbjornsson; Brynjar O Jensson; Daniel F Gudbjartsson; Droplaug N Magnusdottir; Elisabet E Gardarsdottir; Emil A Thorarensen; Gardar Sveinbjornsson; Gisli Masson; Gudmundur Georgsson; Gudmundur L Norddahl; Gudrun Sigmundsdottir; Hakon Jonsson; Hannes Eggertsson; Hilma Holm; Ingileif Jonsdottir; Jona Saemundsdottir; Kamilla S Josefsdottir; Kari Stefansson; Karl G Kristinsson; Kjartan R Gudmundsson; Kristin E Sveinsdottir; Kristjan E Hjorleifsson; Louise le Roux; Maney Sveinsdottir; Olafia S Gretarsdottir; Olafur T Magnusson; Pall Melsted; Patrick Sulem; Run Fridriksdottir; Solvi Rognvaldsson; Thora R Gunnarsdottir; Thordur Kristjansson; Thorolfur Gudnason; Unnur Thorsteinsdottir |
| EPI_ISL_3634444, EPI_ISL_3634445, EPI_ISL_3634447 | Trinidad Public Health Laboratory | Carrington Lab, Department of Preclinical Sciences, Faculty of Medical Sciences, The University of the West Indies, St Augustine Campus | Anushka Ramjag; Arianne Brown-Jordan; Avery Hinds; Christine V. F. Carrington; Christopher Oura; Gabriel Escobar; Nikita S. D. Sahadeo; Nuno Faria; Oliver Pybus; Risha Singh; Roshan Parasram; Sarah Hill; SueMin Nathaniel; Vernie Ramkissoon |
| EPI_ISL_3266267, EPI_ISL_3266291, EPI_ISL_3266294 | U.O. Microbiologia Laboratorio Unico Centro Servizi - AUSL della Romagna | U.O. Microbiologia, Laboratorio Unico Centro Servizi - AUSL della Romagna | Giorgio Dirani |
| EPI_ISL_7235741, EPI_ISL_7313855 | U.O. Microbiologia Laboratorio Unico Centro Servizi AUSL della Romagna | U.O. Microbiologia Laboratorio Unico Centro Servizi AUSL della Romagna | Francesca Taddei; Giorgio Dirani; Giulia Gatti; Ilaria Poggianti; Silvia Zannoli; Vittorio Sambri |

| | | | |
|---|---|---|---|
| EPI_ISL_5648452 | UBS 01 VILA BANDEIRANTES BIRIGUI | Instituto Butantan | Antonio Jorge Martins; Claudia Renata dos Santos Barros; David Schlesinger; Debora Botequio Moretti; Dimas Tadeu Covas; Elaine Cristina Marqueze; Elaine Vieira Santos; Evandra Strazza Rodrigues; Heidge Fukumasu; Jayme Augusto de Souza-Neto; José Salvatore Leister Patané; Luiz Alcantara; Luiz Lehmann Coutinho; Maria Carolina Elias; Maurício Lacerda Nogueira; Rafael dos Santos Bezerra; Raul Machado Neto; Rejane Maria Tommasini Grotto; Ricardo Haddad; Sandra Coccuzzo Sampaio Vessoni; Simone Kashima; Svetoslav Nanev Slavov; Vincent Louis Viala |
| EPI_ISL_5648565 | UBS 02 JARDIM TOSELAR BIRIGUI | Instituto Butantan | Antonio Jorge Martins; Claudia Renata dos Santos Barros; David Schlesinger; Debora Botequio Moretti; Dimas Tadeu Covas; Elaine Cristina Marqueze; Elaine Vieira Santos; Evandra Strazza Rodrigues; Heidge Fukumasu; Jayme Augusto de Souza-Neto; José Salvatore Leister Patané; Luiz Alcantara; Luiz Lehmann Coutinho; Maria Carolina Elias; Maurício Lacerda Nogueira; Rafael dos Santos Bezerra; Raul Machado Neto; Rejane Maria Tommasini Grotto; Ricardo Haddad; Sandra Coccuzzo Sampaio Vessoni; Simone Kashima; Svetoslav Nanev Slavov; Vincent Louis Viala |
| EPI_ISL_5648693 | UBS 06 PARQUE DAS NACOES BIRIGUI | Instituto Butantan | Antonio Jorge Martins; Claudia Renata dos Santos Barros; David Schlesinger; Debora Botequio Moretti; Dimas Tadeu Covas; Elaine Cristina Marqueze; Elaine Vieira Santos; Evandra Strazza Rodrigues; Heidge Fukumasu; Jayme Augusto de Souza-Neto; José Salvatore Leister Patané; Luiz Alcantara; Luiz Lehmann Coutinho; Maria Carolina Elias; Maurício Lacerda Nogueira; Rafael dos Santos Bezerra; Raul Machado Neto; Rejane Maria Tommasini Grotto; Ricardo Haddad; Sandra Coccuzzo Sampaio Vessoni; Simone Kashima; Svetoslav Nanev Slavov; Vincent Louis Viala |
| EPI_ISL_5648794 | UBS 10 MANOEL SANCHES GRANEIRO COLINAS BIRIGUI | Instituto Butantan | Antonio Jorge Martins; Claudia Renata dos Santos Barros; David Schlesinger; Debora Botequio Moretti; Dimas Tadeu Covas; Elaine Cristina Marqueze; Elaine Vieira Santos; Evandra Strazza Rodrigues; Heidge Fukumasu; Jayme Augusto de Souza-Neto; José Salvatore Leister Patané; Luiz Alcantara; Luiz Lehmann Coutinho; Maria Carolina Elias; Maurício Lacerda Nogueira; Rafael dos Santos Bezerra; Raul Machado Neto; Rejane Maria Tommasini Grotto; Ricardo Haddad; Sandra Coccuzzo Sampaio Vessoni; Simone Kashima; Svetoslav Nanev Slavov; Vincent Louis Viala |
| EPI_ISL_5648574 | UBS 11 DR SERGIO AUGUSTO CLARK XAVIER SOARES PORTAL II | Instituto Butantan | Antonio Jorge Martins; Claudia Renata dos Santos Barros; David Schlesinger; Debora Botequio Moretti; Dimas Tadeu Covas; Elaine Cristina Marqueze; Elaine Vieira Santos; Evandra Strazza Rodrigues; Heidge Fukumasu; Jayme Augusto de Souza-Neto; José Salvatore Leister Patané; Luiz Alcantara; Luiz Lehmann Coutinho; Maria Carolina Elias; Maurício Lacerda Nogueira; Rafael dos Santos Bezerra; Raul Machado Neto; Rejane Maria Tommasini Grotto; Ricardo Haddad; Sandra Coccuzzo Sampaio Vessoni; Simone Kashima; Svetoslav Nanev Slavov; Vincent Louis Viala |
| EPI_ISL_5649126 | UBS ARACILIA | Instituto Butantan | Antonio Jorge Martins; Claudia Renata dos Santos Barros; David Schlesinger; Debora Botequio Moretti; Dimas Tadeu Covas; Elaine Cristina Marqueze; Elaine Vieira Santos; Evandra Strazza Rodrigues; Heidge Fukumasu; Jayme Augusto de Souza-Neto; José Salvatore Leister Patané; Luiz Alcantara; Luiz Lehmann Coutinho; Maria Carolina Elias; Maurício Lacerda Nogueira; Rafael dos Santos Bezerra; Raul Machado Neto; Rejane Maria Tommasini Grotto; Ricardo Haddad; Sandra Coccuzzo Sampaio Vessoni; Simone Kashima; Svetoslav Nanev Slavov; Vincent Louis Viala |
| EPI_ISL_5648853, EPI_ISL_5648892 | UBS ATALAIA DR MARIO AGUIAR FILHO | Instituto Butantan | Antonio Jorge Martins; Claudia Renata dos Santos Barros; David Schlesinger; Debora Botequio Moretti; Dimas Tadeu Covas; Elaine Cristina Marqueze; Elaine Vieira Santos; Evandra Strazza Rodrigues; Heidge Fukumasu; Jayme Augusto de Souza-Neto; José Salvatore Leister Patané; Luiz Alcantara; Luiz Lehmann Coutinho; Maria Carolina Elias; Maurício Lacerda Nogueira; Rafael dos Santos Bezerra; Raul Machado Neto; Rejane Maria Tommasini Grotto; Ricardo Haddad; Sandra Coccuzzo Sampaio Vessoni; Simone Kashima; Svetoslav Nanev Slavov; Vincent Louis Viala |
| EPI_ISL_5648514, EPI_ISL_5648682 | UBS CAIC OURINHOS | Instituto Butantan | Antonio Jorge Martins; Claudia Renata dos Santos Barros; David Schlesinger; Debora Botequio Moretti; Dimas Tadeu Covas; Elaine Cristina Marqueze; Elaine Vieira Santos; Evandra Strazza Rodrigues; Heidge Fukumasu; Jayme Augusto de Souza-Neto; José Salvatore Leister Patané; Luiz Alcantara; Luiz Lehmann Coutinho; Maria Carolina Elias; Maurício Lacerda Nogueira; Rafael dos Santos Bezerra; Raul Machado Neto; Rejane Maria Tommasini Grotto; Ricardo Haddad; Sandra Coccuzzo Sampaio Vessoni; Simone Kashima; Svetoslav Nanev Slavov; Vincent Louis Viala |
| EPI_ISL_5659544, EPI_ISL_5659559, EPI_ISL_5659565, EPI_ISL_5659584, EPI_ISL_5659588, EPI_ISL_5659595, EPI_ISL_5659633 | | | |
| see above | UBS CELESTINO CARNIELO | Instituto Butantan | Antonio Jorge Martins; Claudia Renata dos Santos Barros; David Schlesinger; Debora Botequio Moretti; Dimas Tadeu Covas; Elaine Cristina Marqueze; Elaine Vieira Santos; Evandra Strazza Rodrigues; Heidge Fukumasu; Jayme Augusto de Souza-Neto; José Salvatore Leister Patané; Luiz Alcantara; Luiz Lehmann Coutinho; Maria Carolina Elias; Maurício Lacerda Nogueira; Rafael dos Santos Bezerra; Raul Machado Neto; Rejane Maria Tommasini Grotto; Ricardo Haddad; Sandra Coccuzzo Sampaio Vessoni; Simone Kashima; Svetoslav Nanev Slavov; Vincent Louis Viala |
| EPI_ISL_5648939 | UBS CIDADE MARTINS | Instituto Butantan | Antonio Jorge Martins; Claudia Renata dos Santos Barros; David Schlesinger; Debora Botequio Moretti; Dimas Tadeu Covas; Elaine Cristina Marqueze; Elaine Vieira Santos; Evandra Strazza Rodrigues; Heidge Fukumasu; Jayme Augusto de Souza-Neto; José Salvatore Leister Patané; Luiz Alcantara; Luiz Lehmann Coutinho; Maria Carolina Elias; Maurício Lacerda Nogueira; Rafael dos Santos Bezerra; Raul Machado Neto; Rejane Maria Tommasini Grotto; Ricardo Haddad; Sandra Coccuzzo Sampaio Vessoni; Simone Kashima; Svetoslav Nanev Slavov; Vincent Louis Viala |
| EPI_ISL_5648770 | UBS CUMBICA | Instituto Butantan | Antonio Jorge Martins; Claudia Renata dos Santos Barros; David Schlesinger; Debora Botequio Moretti; Dimas Tadeu Covas; Elaine Cristina Marqueze; Elaine Vieira Santos; Evandra Strazza Rodrigues; Heidge Fukumasu; Jayme Augusto de Souza-Neto; José Salvatore Leister Patané; Luiz Alcantara; Luiz Lehmann Coutinho; Maria Carolina Elias; Maurício Lacerda Nogueira; Rafael dos Santos Bezerra; Raul Machado Neto; Rejane Maria Tommasini Grotto; Ricardo Haddad; Sandra Coccuzzo Sampaio Vessoni; Simone Kashima; Svetoslav Nanev Slavov; Vincent Louis Viala |
| EPI_ISL_5648927 | UBS CUMMINS | Instituto Butantan | Antonio Jorge Martins; Claudia Renata dos Santos Barros; David Schlesinger; Debora Botequio Moretti; Dimas Tadeu Covas; Elaine Cristina Marqueze; Elaine Vieira Santos; Evandra Strazza Rodrigues; Heidge Fukumasu; Jayme Augusto de Souza-Neto; José Salvatore Leister Patané; Luiz Alcantara; Luiz Lehmann Coutinho; Maria Carolina Elias; Maurício Lacerda Nogueira; Rafael dos Santos Bezerra; Raul Machado Neto; Rejane Maria Tommasini Grotto; Ricardo Haddad; Sandra Coccuzzo Sampaio Vessoni; Simone Kashima; Svetoslav Nanev Slavov; Vincent Louis Viala |
| EPI_ISL_5648588, EPI_ISL_5648606, EPI_ISL_5659549, EPI_ISL_5659561, EPI_ISL_5659579, EPI_ISL_5659589 | UBS DE MACEDONIA | Instituto Butantan | Antonio Jorge Martins; Claudia Renata dos Santos Barros; David Schlesinger; Debora Botequio Moretti; Dimas Tadeu Covas; Elaine Cristina Marqueze; Elaine Vieira Santos; Evandra Strazza Rodrigues; Heidge Fukumasu; Jayme Augusto de Souza-Neto; José Salvatore Leister Patané; Luiz Alcantara; Luiz Lehmann Coutinho; Maria Carolina Elias; Maurício Lacerda Nogueira; Rafael dos Santos Bezerra; Raul Machado Neto; Rejane Maria Tommasini Grotto; Ricardo Haddad; Sandra Coccuzzo Sampaio Vessoni; Simone Kashima; Svetoslav Nanev Slavov; Vincent Louis Viala |
| EPI_ISL_3536373, EPI_ISL_3536374 | UBS DE MUFUMBAL | Analytical Competence Molecular Epidemiology Lab/ACME, Oswaldo Cruz Foundation, Ceara (FIOCRUZ CE) | Cleber Furtado Aksenen; Fabio Miyajima; Fernando Braga Stehling; Francisco Eder de Moura Lopes; Jamille Maria Mendes Bezerra; Joaquim César do Nascimento Sousa Junior; Pedro Miguel Carneiro Jeronimo; Suzana Porto Almeida e Lucas Delerino; Thais Ferreira de Oliveira; Thais de Oliveira Costa; Ticiane Cavalcante de Souza; Veridiana Pessoa Miyajima |
| EPI_ISL_5648938, EPI_ISL_5649136 | UBS DE PEDRANOPOLIS | Instituto Butantan | Antonio Jorge Martins; Claudia Renata dos Santos Barros; David Schlesinger; Debora Botequio Moretti; Dimas Tadeu Covas; Elaine Cristina Marqueze; Elaine Vieira Santos; Evandra Strazza Rodrigues; Heidge Fukumasu; Jayme Augusto de Souza-Neto; José Salvatore Leister Patané; Luiz Alcantara; Luiz Lehmann Coutinho; Maria Carolina Elias; Maurício Lacerda Nogueira; Rafael dos Santos Bezerra; Raul Machado Neto; Rejane Maria Tommasini Grotto; Ricardo Haddad; Sandra Coccuzzo Sampaio Vessoni; Simone Kashima; Svetoslav Nanev Slavov; Vincent Louis Viala |
| EPI_ISL_5648572, EPI_ISL_5659546, EPI_ISL_5659558, EPI_ISL_5659571, EPI_ISL_5659582, EPI_ISL_5659617 | UBS DE TABAPUA | Instituto Butantan | Antonio Jorge Martins; Claudia Renata dos Santos Barros; David Schlesinger; Debora Botequio Moretti; Dimas Tadeu Covas; Elaine Cristina Marqueze; Elaine Vieira Santos; Evandra Strazza Rodrigues; Heidge Fukumasu; Jayme Augusto de Souza-Neto; José Salvatore Leister Patané; Luiz Alcantara; Luiz Lehmann Coutinho; Maria Carolina Elias; Maurício Lacerda Nogueira; Rafael dos Santos Bezerra; Raul Machado Neto; Rejane Maria Tommasini Grotto; Ricardo Haddad; Sandra Coccuzzo Sampaio Vessoni; Simone Kashima; Svetoslav Nanev Slavov; Vincent Louis Viala |
| EPI_ISL_3536375 | UBS DO CAMPO LINDO | Analytical Competence Molecular Epidemiology Lab/ACME, Oswaldo Cruz Foundation, Ceara (FIOCRUZ CE) | Cleber Furtado Aksenen; Fabio Miyajima; Fernando Braga Stehling; Francisco Eder de Moura Lopes; Jamille Maria Mendes Bezerra; Joaquim César do Nascimento Sousa Junior; Pedro Miguel Carneiro Jeronimo; Suzana Porto Almeida e Lucas Delerino; Thais Ferreira de Oliveira; Thais de Oliveira Costa; Ticiane Cavalcante de Souza; Veridiana Pessoa Miyajima |
| EPI_ISL_5648390, EPI_ISL_5648494, EPI_ISL_5648554, EPI_ISL_5648617, EPI_ISL_5648621, EPI_ISL_5648631, EPI_ISL_5648664, EPI_ISL_5648736, EPI_ISL_5648778, EPI_ISL_5648838, EPI_ISL_5648858, EPI_ISL_5649091, EPI_ISL_5649262, EPI_ISL_5649308 | | | |
| see above | UBS DR ALFREDO DANTAS DE SOUZA UMUARAMA | Instituto Butantan | Antonio Jorge Martins; Claudia Renata dos Santos Barros; David Schlesinger; Debora Botequio Moretti; Dimas Tadeu Covas; Elaine Cristina Marqueze; Elaine Vieira Santos; Evandra Strazza Rodrigues; Heidge Fukumasu; Jayme Augusto de Souza-Neto; José Salvatore Leister Patané; Luiz Alcantara; Luiz Lehmann Coutinho; Maria Carolina Elias; Maurício Lacerda Nogueira; Rafael dos Santos Bezerra; Raul Machado Neto; Rejane Maria Tommasini Grotto; Ricardo Haddad; Sandra Coccuzzo Sampaio Vessoni; Simone Kashima; Svetoslav Nanev Slavov; Vincent Louis Viala |
| EPI_ISL_5649102 | UBS DR WALDOMIRO FERREIRA NEVES SCRPARDO | Instituto Butantan | Antonio Jorge Martins; Claudia Renata dos Santos Barros; David Schlesinger; Debora Botequio Moretti; Dimas Tadeu Covas; Elaine Cristina Marqueze; Elaine Vieira Santos; Evandra Strazza Rodrigues; Heidge Fukumasu; Jayme Augusto de Souza-Neto; José Salvatore Leister Patané; Luiz Alcantara; Luiz Lehmann Coutinho; Maria Carolina Elias; Maurício Lacerda Nogueira; Rafael dos Santos Bezerra; Raul Machado Neto; Rejane Maria Tommasini Grotto; Ricardo Haddad; Sandra Coccuzzo Sampaio Vessoni; Simone Kashima; Svetoslav Nanev Slavov; Vincent Louis Viala |
| EPI_ISL_5648282, EPI_ISL_5648517, EPI_ISL_5648527, EPI_ISL_5648545, EPI_ISL_5648600, EPI_ISL_5648738, EPI_ISL_5648747 | | | |
| see above | UBS GLENDA RODELLA DUMAS | Instituto Butantan | Antonio Jorge Martins; Claudia Renata dos Santos Barros; David Schlesinger; Debora Botequio Moretti; Dimas Tadeu Covas; Elaine Cristina Marqueze; Elaine Vieira Santos; Evandra Strazza Rodrigues; Heidge Fukumasu; Jayme Augusto de Souza-Neto; José Salvatore Leister Patané; Luiz Alcantara; Luiz Lehmann Coutinho; Maria Carolina Elias; Maurício Lacerda Nogueira; Rafael dos Santos Bezerra; Raul Machado Neto; Rejane Maria Tommasini Grotto; Ricardo Haddad; Sandra Coccuzzo Sampaio Vessoni; Simone Kashima; Svetoslav Nanev Slavov; Vincent Louis Viala |
| EPI_ISL_5649148, EPI_ISL_5649154, EPI_ISL_5649155, EPI_ISL_5649157, EPI_ISL_5649178, EPI_ISL_5649207, EPI_ISL_5649211, EPI_ISL_5649227 | | | |
| see above | UBS II DE ALVARES MACHADO | Instituto Butantan | Antonio Jorge Martins; Claudia Renata dos Santos Barros; David Schlesinger; Debora Botequio Moretti; Dimas Tadeu Covas; Elaine Cristina Marqueze; Elaine Vieira Santos; Evandra Strazza Rodrigues; Heidge Fukumasu; Jayme Augusto de Souza-Neto; José Salvatore Leister Patané; Luiz Alcantara; Luiz Lehmann Coutinho; Maria Carolina Elias; Maurício Lacerda Nogueira; Rafael dos Santos Bezerra; Raul Machado Neto; Rejane Maria Tommasini Grotto; Ricardo Haddad; Sandra Coccuzzo Sampaio Vessoni; Simone Kashima; Svetoslav Nanev Slavov; Vincent Louis Viala |
| EPI_ISL_5649156, EPI_ISL_5649161, EPI_ISL_5649252 | UBS II DE TACIBA | Instituto Butantan | Antonio Jorge Martins; Claudia Renata dos Santos Barros; David Schlesinger; Debora Botequio Moretti; Dimas Tadeu Covas; Elaine Cristina Marqueze; Elaine Vieira Santos; Evandra Strazza Rodrigues; Heidge Fukumasu; Jayme Augusto de Souza-Neto; José Salvatore Leister Patané; Luiz Alcantara; Luiz Lehmann Coutinho; Maria Carolina Elias; Maurício Lacerda Nogueira; Rafael dos Santos Bezerra; Raul Machado Neto; Rejane Maria Tommasini Grotto; Ricardo Haddad; Sandra Coccuzzo Sampaio Vessoni; Simone Kashima; Svetoslav Nanev Slavov; Vincent Louis Viala |
| EPI_ISL_5648971, EPI_ISL_5648978, EPI_ISL_5649005, EPI_ISL_5649038, EPI_ISL_5649051, EPI_ISL_5649139 | UBS II DE TANABI MILTON MARTINS PERCHES | Instituto Butantan | Antonio Jorge Martins; Claudia Renata dos Santos Barros; David Schlesinger; Debora Botequio Moretti; Dimas Tadeu Covas; Elaine Cristina Marqueze; Elaine Vieira Santos; Evandra Strazza Rodrigues; Heidge Fukumasu; Jayme Augusto de Souza-Neto; José Salvatore Leister Patané; Luiz Alcantara; Luiz Lehmann Coutinho; Maria Carolina Elias; Maurício Lacerda Nogueira; Rafael dos Santos Bezerra; Raul Machado Neto; Rejane Maria Tommasini Grotto; Ricardo Haddad; Sandra Coccuzzo Sampaio Vessoni; Simone Kashima; Svetoslav Nanev Slavov; Vincent Louis Viala |
| EPI_ISL_5648847 | UBS III DE BENTO DE ABREU | Instituto Butantan | Antonio Jorge Martins; Claudia Renata dos Santos Barros; David Schlesinger; Debora Botequio Moretti; Dimas Tadeu Covas; Elaine Cristina Marqueze; Elaine Vieira Santos; Evandra Strazza Rodrigues; Heidge Fukumasu; Jayme Augusto de Souza-Neto; José Salvatore Leister Patané; Luiz Alcantara; Luiz Lehmann Coutinho; Maria Carolina Elias; Maurício Lacerda Nogueira; Rafael dos Santos Bezerra; Raul Machado Neto; Rejane Maria Tommasini Grotto; Ricardo Haddad; Sandra Coccuzzo Sampaio Vessoni; Simone Kashima; Svetoslav Nanev Slavov; Vincent Louis Viala |
| EPI_ISL_5648448, EPI_ISL_5648744 | UBS III DE RUBIACEA | Instituto Butantan | Antonio Jorge Martins; Claudia Renata dos Santos Barros; David Schlesinger; Debora Botequio Moretti; Dimas Tadeu Covas; Elaine Cristina Marqueze; Elaine Vieira Santos; Evandra Strazza Rodrigues; Heidge Fukumasu; Jayme Augusto de Souza-Neto; José Salvatore Leister Patané; Luiz Alcantara; Luiz Lehmann Coutinho; Maria Carolina Elias; Maurício Lacerda Nogueira; Rafael dos Santos Bezerra; Raul Machado Neto; Rejane Maria Tommasini Grotto; Ricardo Haddad; Sandra Coccuzzo Sampaio Vessoni; Simone Kashima; Svetoslav Nanev Slavov; Vincent Louis Viala |
| EPI_ISL_5648817, EPI_ISL_5649040 | UBS JARDIM CABUCU | Instituto Butantan | Antonio Jorge Martins; Claudia Renata dos Santos Barros; David Schlesinger; Debora Botequio Moretti; Dimas Tadeu Covas; Elaine Cristina Marqueze; Elaine Vieira Santos; Evandra Strazza Rodrigues; Heidge Fukumasu; Jayme Augusto de Souza-Neto; José Salvatore Leister Patané; Luiz Alcantara; Luiz Lehmann Coutinho; Maria Carolina Elias; Maurício Lacerda Nogueira; Rafael dos Santos Bezerra; Raul Machado Neto; Rejane Maria Tommasini Grotto; Ricardo Haddad; Sandra Coccuzzo Sampaio Vessoni; Simone Kashima; Svetoslav Nanev Slavov; Vincent Louis Viala |
| EPI_ISL_5648846, EPI_ISL_5648865, EPI_ISL_5648891, EPI_ISL_5648918, EPI_ISL_5648981 | UBS JARDIM CAIUBY | Instituto Butantan | Antonio Jorge Martins; Claudia Renata dos Santos Barros; David Schlesinger; Debora Botequio Moretti; Dimas Tadeu Covas; Elaine Cristina Marqueze; Elaine Vieira Santos; Evandra Strazza Rodrigues; Heidge Fukumasu; Jayme Augusto de Souza-Neto; José Salvatore Leister Patané; Luiz Alcantara; Luiz Lehmann Coutinho; Maria Carolina Elias; Maurício Lacerda Nogueira; Rafael dos Santos Bezerra; Raul Machado Neto; Rejane Maria Tommasini Grotto; Ricardo Haddad; Sandra Coccuzzo Sampaio Vessoni; Simone Kashima; Svetoslav Nanev Slavov; Vincent Louis Viala |
| EPI_ISL_5649103 | UBS JARDIM FORTALEZA | Instituto Butantan | Antonio Jorge Martins; Claudia Renata dos Santos Barros; David Schlesinger; Debora Botequio Moretti; Dimas Tadeu Covas; Elaine Cristina Marqueze; Elaine Vieira Santos; Evandra Strazza Rodrigues; Heidge Fukumasu; Jayme Augusto de Souza-Neto; José Salvatore Leister Patané; Luiz Alcantara; Luiz Lehmann Coutinho; Maria Carolina Elias; Maurício Lacerda Nogueira; Rafael dos Santos Bezerra; Raul Machado Neto; Rejane Maria Tommasini Grotto; Ricardo Haddad; Sandra Coccuzzo Sampaio Vessoni; Simone Kashima; Svetoslav Nanev Slavov; Vincent Louis Viala |
| EPI_ISL_5649137 | UBS JARDIM PALMIRA | Instituto Butantan | Antonio Jorge Martins; Claudia Renata dos Santos Barros; David Schlesinger; Debora Botequio Moretti; Dimas Tadeu Covas; Elaine Cristina Marqueze; Elaine Vieira Santos; Evandra Strazza Rodrigues; Heidge Fukumasu; Jayme Augusto de Souza-Neto; José Salvatore Leister Patané; Luiz Alcantara; Luiz Lehmann Coutinho; Maria Carolina Elias; Maurício Lacerda Nogueira; Rafael dos Santos Bezerra; Raul Machado Neto; Rejane Maria Tommasini Grotto; Ricardo Haddad; Sandra Coccuzzo Sampaio Vessoni; Simone Kashima; Svetoslav Nanev Slavov; Vincent Louis Viala |
| EPI_ISL_5648531 | UBS JARDIM PARANA | Instituto Butantan | Antonio Jorge Martins; Claudia Renata dos Santos Barros; David Schlesinger; Debora Botequio Moretti; Dimas Tadeu Covas; Elaine Cristina Marqueze; Elaine Vieira Santos; Evandra Strazza Rodrigues; Heidge Fukumasu; Jayme Augusto de Souza-Neto; José Salvatore Leister Patané; Luiz Alcantara; Luiz Lehmann Coutinho; Maria Carolina Elias; Maurício Lacerda Nogueira; Rafael dos Santos Bezerra; Raul Machado Neto; Rejane Maria Tommasini Grotto; Ricardo Haddad; Sandra Coccuzzo Sampaio Vessoni; Simone Kashima; Svetoslav Nanev Slavov; Vincent Louis Viala |
| EPI_ISL_5649134 | UBS JARDIM PARAVENTI | Instituto Butantan | Antonio Jorge Martins; Claudia Renata dos Santos Barros; David Schlesinger; Debora Botequio Moretti; Dimas Tadeu Covas; Elaine Cristina Marqueze; Elaine Vieira Santos; Evandra Strazza Rodrigues; Heidge Fukumasu; Jayme Augusto de Souza-Neto; José Salvatore Leister Patané; Luiz Alcantara; Luiz Lehmann Coutinho; Maria Carolina Elias; Maurício Lacerda Nogueira; Rafael dos Santos Bezerra; Raul Machado Neto; Rejane Maria Tommasini Grotto; Ricardo Haddad; Sandra Coccuzzo Sampaio Vessoni; Simone Kashima; Svetoslav Nanev Slavov; Vincent Louis Viala |
| EPI_ISL_5648819 | UBS JARDIM PRIMAVERA | Instituto Butantan | Antonio Jorge Martins; Claudia Renata dos Santos Barros; David Schlesinger; Debora Botequio Moretti; Dimas Tadeu Covas; Elaine Cristina Marqueze; Elaine Vieira Santos; Evandra Strazza Rodrigues; Heidge Fukumasu; Jayme Augusto de Souza-Neto; José Salvatore Leister Patané; Luiz Alcantara; Luiz Lehmann Coutinho; Maria Carolina Elias; Maurício Lacerda Nogueira; Rafael dos Santos Bezerra; Raul Machado Neto; Rejane Maria Tommasini Grotto; Ricardo Haddad; Sandra Coccuzzo Sampaio Vessoni; Simone Kashima; Svetoslav Nanev Slavov; Vincent Louis Viala |
| EPI_ISL_5648879 | UBS JARDIM ROSA DE FRANCA | Instituto Butantan | Antonio Jorge Martins; Claudia Renata dos Santos Barros; David Schlesinger; Debora Botequio Moretti; Dimas Tadeu Covas; Elaine Cristina Marqueze; Elaine Vieira Santos; Evandra Strazza Rodrigues; Heidge Fukumasu; Jayme Augusto de Souza-Neto; José Salvatore Leister Patané; Luiz Alcantara; Luiz Lehmann Coutinho; Maria Carolina Elias; Maurício Lacerda Nogueira; Rafael dos Santos Bezerra; Raul Machado Neto; Rejane Maria Tommasini Grotto; Ricardo Haddad; Sandra Coccuzzo Sampaio Vessoni; Simone Kashima; Svetoslav Nanev Slavov; Vincent Louis Viala |
| EPI_ISL_5649309, EPI_ISL_5649320 | UBS JARDIM VITORINO IPERO | Instituto Butantan | Antonio Jorge Martins; Claudia Renata dos Santos Barros; David Schlesinger; Debora Botequio Moretti; Dimas Tadeu Covas; Elaine Cristina Marqueze; Elaine Vieira Santos; Evandra Strazza Rodrigues; Heidge Fukumasu; Jayme Augusto de Souza-Neto; José Salvatore Leister Patané; Luiz Alcantara; Luiz Lehmann Coutinho; Maria Carolina Elias; Maurício Lacerda Nogueira; Rafael dos Santos Bezerra; Raul Machado Neto; Rejane Maria Tommasini Grotto; Ricardo Haddad; Sandra Coccuzzo Sampaio Vessoni; Simone Kashima; Svetoslav Nanev Slavov; Vincent Louis Viala |
| EPI_ISL_5648815, | UBS LAVRAS | Instituto Butantan | Antonio Jorge Martins; Claudia Renata dos Santos Barros; David Schlesinger; Debora Botequio Moretti; Dimas Tadeu Covas; Elaine Cristina Marqueze; Elaine Vieira Santos; Evandra Strazza Rodrigues; Heidge Fukumasu; Jayme Augusto de Souza-Neto; José Salvatore Leister Patané; Luiz Alcantara; |

| | | | |
|---|---|---|---|
| EPI_ISL_5648920, EPI_ISL_5649046 | | | Luiz Lehmann Coutinho; Maria Carolina Elias; Maurício Lacerda Nogueira; Rafael dos Santos Bezerra; Raul Machado Neto; Rejane Maria Tommasini Grotto; Ricardo Haddad; Sandra Coccuzzo Sampaio Vessoni; Simone Kashima; Svetoslav Nanev Slavov; Vincent Louis Viala |
| EPI_ISL_5648776, EPI_ISL_5648837, EPI_ISL_5649032 | UBS LUIS FACHIN IPIGUA | Instituto Butantan | Antonio Jorge Martins; Claudia Renata dos Santos Barros; David Schlesinger; Debora Botequio Moretti; Dimas Tadeu Covas; Elaine Cristina Marqueze; Elaine Vieira Santos; Evandra Strazza Rodrigues; Heidge Fukumasu; Jayme Augusto de Souza-Neto; José Salvatore Leister Patané; Luiz Alcantara; Luiz Lehmann Coutinho; Maria Carolina Elias; Maurício Lacerda Nogueira; Rafael dos Santos Bezerra; Raul Machado Neto; Rejane Maria Tommasini Grotto; Ricardo Haddad; Sandra Coccuzzo Sampaio Vessoni; Simone Kashima; Svetoslav Nanev Slavov; Vincent Louis Viala |
| EPI_ISL_5649158, EPI_ISL_5649163, EPI_ISL_5649188 | UBS MARCIA CRISTIANE DA SILVA DE OURO VERDE | Instituto Butantan | Antonio Jorge Martins; Claudia Renata dos Santos Barros; David Schlesinger; Debora Botequio Moretti; Dimas Tadeu Covas; Elaine Cristina Marqueze; Elaine Vieira Santos; Evandra Strazza Rodrigues; Heidge Fukumasu; Jayme Augusto de Souza-Neto; José Salvatore Leister Patané; Luiz Alcantara; Luiz Lehmann Coutinho; Maria Carolina Elias; Maurício Lacerda Nogueira; Rafael dos Santos Bezerra; Raul Machado Neto; Rejane Maria Tommasini Grotto; Ricardo Haddad; Sandra Coccuzzo Sampaio Vessoni; Simone Kashima; Svetoslav Nanev Slavov; Vincent Louis Viala |
| EPI_ISL_5648766 | UBS MONTE BELO | Instituto Butantan | Antonio Jorge Martins; Claudia Renata dos Santos Barros; David Schlesinger; Debora Botequio Moretti; Dimas Tadeu Covas; Elaine Cristina Marqueze; Elaine Vieira Santos; Evandra Strazza Rodrigues; Heidge Fukumasu; Jayme Augusto de Souza-Neto; José Salvatore Leister Patané; Luiz Alcantara; Luiz Lehmann Coutinho; Maria Carolina Elias; Maurício Lacerda Nogueira; Rafael dos Santos Bezerra; Raul Machado Neto; Rejane Maria Tommasini Grotto; Ricardo Haddad; Sandra Coccuzzo Sampaio Vessoni; Simone Kashima; Svetoslav Nanev Slavov; Vincent Louis Viala |
| EPI_ISL_5649107 | UBS NOVA BONSUCESSO | Instituto Butantan | Antonio Jorge Martins; Claudia Renata dos Santos Barros; David Schlesinger; Debora Botequio Moretti; Dimas Tadeu Covas; Elaine Cristina Marqueze; Elaine Vieira Santos; Evandra Strazza Rodrigues; Heidge Fukumasu; Jayme Augusto de Souza-Neto; José Salvatore Leister Patané; Luiz Alcantara; Luiz Lehmann Coutinho; Maria Carolina Elias; Maurício Lacerda Nogueira; Rafael dos Santos Bezerra; Raul Machado Neto; Rejane Maria Tommasini Grotto; Ricardo Haddad; Sandra Coccuzzo Sampaio Vessoni; Simone Kashima; Svetoslav Nanev Slavov; Vincent Louis Viala |
| EPI_ISL_5649133 | UBS PARQUE JANDAIA | Instituto Butantan | Antonio Jorge Martins; Claudia Renata dos Santos Barros; David Schlesinger; Debora Botequio Moretti; Dimas Tadeu Covas; Elaine Cristina Marqueze; Elaine Vieira Santos; Evandra Strazza Rodrigues; Heidge Fukumasu; Jayme Augusto de Souza-Neto; José Salvatore Leister Patané; Luiz Alcantara; Luiz Lehmann Coutinho; Maria Carolina Elias; Maurício Lacerda Nogueira; Rafael dos Santos Bezerra; Raul Machado Neto; Rejane Maria Tommasini Grotto; Ricardo Haddad; Sandra Coccuzzo Sampaio Vessoni; Simone Kashima; Svetoslav Nanev Slavov; Vincent Louis Viala |
| EPI_ISL_5649022 | UBS PARQUE MARENGO | Instituto Butantan | Antonio Jorge Martins; Claudia Renata dos Santos Barros; David Schlesinger; Debora Botequio Moretti; Dimas Tadeu Covas; Elaine Cristina Marqueze; Elaine Vieira Santos; Evandra Strazza Rodrigues; Heidge Fukumasu; Jayme Augusto de Souza-Neto; José Salvatore Leister Patané; Luiz Alcantara; Luiz Lehmann Coutinho; Maria Carolina Elias; Maurício Lacerda Nogueira; Rafael dos Santos Bezerra; Raul Machado Neto; Rejane Maria Tommasini Grotto; Ricardo Haddad; Sandra Coccuzzo Sampaio Vessoni; Simone Kashima; Svetoslav Nanev Slavov; Vincent Louis Viala |
| EPI_ISL_5648434, EPI_ISL_5648437, EPI_ISL_5648580, EPI_ISL_5648598, EPI_ISL_5648732, EPI_ISL_5648741, EPI_ISL_5648745, EPI_ISL_5648752, EPI_ISL_5659537, EPI_ISL_5659538, EPI_ISL_5659574, EPI_ISL_5659577, EPI_ISL_5659604, EPI_ISL_5659619, EPI_ISL_5659623, EPI_ISL_5659628, EPI_ISL_5659629, EPI_ISL_5659636 | | | |
| see above | UBS PASTOR MARCELINO DEUNGARO RIO GRANDE | Instituto Butantan | Antonio Jorge Martins; Claudia Renata dos Santos Barros; David Schlesinger; Debora Botequio Moretti; Dimas Tadeu Covas; Elaine Cristina Marqueze; Elaine Vieira Santos; Evandra Strazza Rodrigues; Heidge Fukumasu; Jayme Augusto de Souza-Neto; José Salvatore Leister Patané; Luiz Alcantara; Luiz Lehmann Coutinho; Maria Carolina Elias; Maurício Lacerda Nogueira; Rafael dos Santos Bezerra; Raul Machado Neto; Rejane Maria Tommasini Grotto; Ricardo Haddad; Sandra Coccuzzo Sampaio Vessoni; Simone Kashima; Svetoslav Nanev Slavov; Vincent Louis Viala |
| EPI_ISL_5648944 | UBS PONTE ALTA | Instituto Butantan | Antonio Jorge Martins; Claudia Renata dos Santos Barros; David Schlesinger; Debora Botequio Moretti; Dimas Tadeu Covas; Elaine Cristina Marqueze; Elaine Vieira Santos; Evandra Strazza Rodrigues; Heidge Fukumasu; Jayme Augusto de Souza-Neto; José Salvatore Leister Patané; Luiz Alcantara; Luiz Lehmann Coutinho; Maria Carolina Elias; Maurício Lacerda Nogueira; Rafael dos Santos Bezerra; Raul Machado Neto; Rejane Maria Tommasini Grotto; Ricardo Haddad; Sandra Coccuzzo Sampaio Vessoni; Simone Kashima; Svetoslav Nanev Slavov; Vincent Louis Viala |
| EPI_ISL_5649054 | UBS PONTE GRANDE | Instituto Butantan | Antonio Jorge Martins; Claudia Renata dos Santos Barros; David Schlesinger; Debora Botequio Moretti; Dimas Tadeu Covas; Elaine Cristina Marqueze; Elaine Vieira Santos; Evandra Strazza Rodrigues; Heidge Fukumasu; Jayme Augusto de Souza-Neto; José Salvatore Leister Patané; Luiz Alcantara; Luiz Lehmann Coutinho; Maria Carolina Elias; Maurício Lacerda Nogueira; Rafael dos Santos Bezerra; Raul Machado Neto; Rejane Maria Tommasini Grotto; Ricardo Haddad; Sandra Coccuzzo Sampaio Vessoni; Simone Kashima; Svetoslav Nanev Slavov; Vincent Louis Viala |
| EPI_ISL_5648805 | UBS SOIMCO | Instituto Butantan | Antonio Jorge Martins; Claudia Renata dos Santos Barros; David Schlesinger; Debora Botequio Moretti; Dimas Tadeu Covas; Elaine Cristina Marqueze; Elaine Vieira Santos; Evandra Strazza Rodrigues; Heidge Fukumasu; Jayme Augusto de Souza-Neto; José Salvatore Leister Patané; Luiz Alcantara; Luiz Lehmann Coutinho; Maria Carolina Elias; Maurício Lacerda Nogueira; Rafael dos Santos Bezerra; Raul Machado Neto; Rejane Maria Tommasini Grotto; Ricardo Haddad; Sandra Coccuzzo Sampaio Vessoni; Simone Kashima; Svetoslav Nanev Slavov; Vincent Louis Viala |
| EPI_ISL_5648824 | UBS TABOAO | Instituto Butantan | Antonio Jorge Martins; Claudia Renata dos Santos Barros; David Schlesinger; Debora Botequio Moretti; Dimas Tadeu Covas; Elaine Cristina Marqueze; Elaine Vieira Santos; Evandra Strazza Rodrigues; Heidge Fukumasu; Jayme Augusto de Souza-Neto; José Salvatore Leister Patané; Luiz Alcantara; Luiz Lehmann Coutinho; Maria Carolina Elias; Maurício Lacerda Nogueira; Rafael dos Santos Bezerra; Raul Machado Neto; Rejane Maria Tommasini Grotto; Ricardo Haddad; Sandra Coccuzzo Sampaio Vessoni; Simone Kashima; Svetoslav Nanev Slavov; Vincent Louis Viala |
| EPI_ISL_5648388, EPI_ISL_5659547, EPI_ISL_5659570, EPI_ISL_5659600 | UBS TEREZA GALLO UCHOA | Instituto Butantan | Antonio Jorge Martins; Claudia Renata dos Santos Barros; David Schlesinger; Debora Botequio Moretti; Dimas Tadeu Covas; Elaine Cristina Marqueze; Elaine Vieira Santos; Evandra Strazza Rodrigues; Heidge Fukumasu; Jayme Augusto de Souza-Neto; José Salvatore Leister Patané; Luiz Alcantara; Luiz Lehmann Coutinho; Maria Carolina Elias; Maurício Lacerda Nogueira; Rafael dos Santos Bezerra; Raul Machado Neto; Rejane Maria Tommasini Grotto; Ricardo Haddad; Sandra Coccuzzo Sampaio Vessoni; Simone Kashima; Svetoslav Nanev Slavov; Vincent Louis Viala |
| EPI_ISL_5648957 | UBS VILA BARROS | Instituto Butantan | Antonio Jorge Martins; Claudia Renata dos Santos Barros; David Schlesinger; Debora Botequio Moretti; Dimas Tadeu Covas; Elaine Cristina Marqueze; Elaine Vieira Santos; Evandra Strazza Rodrigues; Heidge Fukumasu; Jayme Augusto de Souza-Neto; José Salvatore Leister Patané; Luiz Alcantara; Luiz Lehmann Coutinho; Maria Carolina Elias; Maurício Lacerda Nogueira; Rafael dos Santos Bezerra; Raul Machado Neto; Rejane Maria Tommasini Grotto; Ricardo Haddad; Sandra Coccuzzo Sampaio Vessoni; Simone Kashima; Svetoslav Nanev Slavov; Vincent Louis Viala |
| EPI_ISL_5648564 | UBS VILA MARGARIDA | Instituto Butantan | Antonio Jorge Martins; Claudia Renata dos Santos Barros; David Schlesinger; Debora Botequio Moretti; Dimas Tadeu Covas; Elaine Cristina Marqueze; Elaine Vieira Santos; Evandra Strazza Rodrigues; Heidge Fukumasu; Jayme Augusto de Souza-Neto; José Salvatore Leister Patané; Luiz Alcantara; Luiz Lehmann Coutinho; Maria Carolina Elias; Maurício Lacerda Nogueira; Rafael dos Santos Bezerra; Raul Machado Neto; Rejane Maria Tommasini Grotto; Ricardo Haddad; Sandra Coccuzzo Sampaio Vessoni; Simone Kashima; Svetoslav Nanev Slavov; Vincent Louis Viala |
| EPI_ISL_5649423 | UBS VILA RIBEIRO | Instituto Butantan | Antonio Jorge Martins; Claudia Renata dos Santos Barros; David Schlesinger; Debora Botequio Moretti; Dimas Tadeu Covas; Elaine Cristina Marqueze; Elaine Vieira Santos; Evandra Strazza Rodrigues; Heidge Fukumasu; Jayme Augusto de Souza-Neto; José Salvatore Leister Patané; Luiz Alcantara; Luiz Lehmann Coutinho; Maria Carolina Elias; Maurício Lacerda Nogueira; Rafael dos Santos Bezerra; Raul Machado Neto; Rejane Maria Tommasini Grotto; Ricardo Haddad; Sandra Coccuzzo Sampaio Vessoni; Simone Kashima; Svetoslav Nanev Slavov; Vincent Louis Viala |
| EPI_ISL_5648808 | UBS VILA RIO DE JANEIRO | Instituto Butantan | Antonio Jorge Martins; Claudia Renata dos Santos Barros; David Schlesinger; Debora Botequio Moretti; Dimas Tadeu Covas; Elaine Cristina Marqueze; Elaine Vieira Santos; Evandra Strazza Rodrigues; Heidge Fukumasu; Jayme Augusto de Souza-Neto; José Salvatore Leister Patané; Luiz Alcantara; Luiz Lehmann Coutinho; Maria Carolina Elias; Maurício Lacerda Nogueira; Rafael dos Santos Bezerra; Raul Machado Neto; Rejane Maria Tommasini Grotto; Ricardo Haddad; Sandra Coccuzzo Sampaio Vessoni; Simone Kashima; Svetoslav Nanev Slavov; Vincent Louis Viala |
| EPI_ISL_2860321 | UCC | Grubaugh Lab - Yale School of Public Health | Anderson Brito; Andrea Firpo; Annie Watkins; Chaney Kalinich; Chantal Vogels; David Rivera Aponte; Ernesto Vazquez; Fabiola Cruz-Lopez; Fabiola Fontanet; Isabel Ott; Jessica Rothman; Joseph Fauver; Karolane Gonzalez; Kendall Billig; Mallery Breban; Mary Petrone; Natalie Machargo; Nathan Grubaugh; Patricia Serrano; Tania Mitwalli; Tara Alpert; Tobias Koch; Vianca Aponte |
| EPI_ISL_3710096 | UCLA Clinical Micro Lab | Los Angeles County Public Health Laboratories | P. Hemarajata et al. |
| EPI_ISL_3462087, EPI_ISL_3462089 | UHTL, University Hospitals | UHTL, University Hospitals | Alouani, D.; Sadri, N.; Song, X. |
| EPI_ISL_5648796, EPI_ISL_5648873 | UMS PIRAPORA DO BOM JESUS | Instituto Butantan | Antonio Jorge Martins; Claudia Renata dos Santos Barros; David Schlesinger; Debora Botequio Moretti; Dimas Tadeu Covas; Elaine Cristina Marqueze; Elaine Vieira Santos; Evandra Strazza Rodrigues; Heidge Fukumasu; Jayme Augusto de Souza-Neto; José Salvatore Leister Patané; Luiz Alcantara; Luiz Lehmann Coutinho; Maria Carolina Elias; Maurício Lacerda Nogueira; Rafael dos Santos Bezerra; Raul Machado Neto; Rejane Maria Tommasini Grotto; Ricardo Haddad; Sandra Coccuzzo Sampaio Vessoni; Simone Kashima; Svetoslav Nanev Slavov; Vincent Louis Viala |
| EPI_ISL_3536379 | UNIDADE BASICA DE SAUDE ALTO BRILHANTE | Analytical Competence Molecular Epidemiology Lab/ACME, Oswaldo Cruz Foundation, Ceara (FIOCRUZ CE) | Cleber Furtado Aksenen; Fabio Miyajima; Fernando Braga Stehling; Francisco Eder de Moura Lopes; Jamille Maria Mendes Bezerra; Joaquim César do Nascimento Sousa Junior; Pedro Miguel Carneiro Jeronimo; Suzana Porto Almeida e Lucas Delerino; Thais Ferreira de Oliveira; Thais de Oliveira Costa; Ticiane Cavalcante de Souza; Veridiana Pessoa Miyajima |
| EPI_ISL_5648519 | UNIDADE BASICA DE SAUDE CIDINHA LEITE | Instituto Butantan | Antonio Jorge Martins; Claudia Renata dos Santos Barros; David Schlesinger; Debora Botequio Moretti; Dimas Tadeu Covas; Elaine Cristina Marqueze; Elaine Vieira Santos; Evandra Strazza Rodrigues; Heidge Fukumasu; Jayme Augusto de Souza-Neto; José Salvatore Leister Patané; Luiz Alcantara; Luiz Lehmann Coutinho; Maria Carolina Elias; Maurício Lacerda Nogueira; Rafael dos Santos Bezerra; Raul Machado Neto; Rejane Maria Tommasini Grotto; Ricardo Haddad; Sandra Coccuzzo Sampaio Vessoni; Simone Kashima; Svetoslav Nanev Slavov; Vincent Louis Viala |
| EPI_ISL_3536380 | UNIDADE BASICA DE SAUDE COLIBRIS | Analytical Competence Molecular Epidemiology Lab/ACME, Oswaldo Cruz Foundation, Ceara (FIOCRUZ CE) | Cleber Furtado Aksenen; Fabio Miyajima; Fernando Braga Stehling; Francisco Eder de Moura Lopes; Jamille Maria Mendes Bezerra; Joaquim César do Nascimento Sousa Junior; Pedro Miguel Carneiro Jeronimo; Suzana Porto Almeida e Lucas Delerino; Thais Ferreira de Oliveira; Thais de Oliveira Costa; Ticiane Cavalcante de Souza; Veridiana Pessoa Miyajima |
| EPI_ISL_5648806 | UNIDADE BASICA DE SAUDE DE EMBAUBA | Instituto Butantan | Antonio Jorge Martins; Claudia Renata dos Santos Barros; David Schlesinger; Debora Botequio Moretti; Dimas Tadeu Covas; Elaine Cristina Marqueze; Elaine Vieira Santos; Evandra Strazza Rodrigues; Heidge Fukumasu; Jayme Augusto de Souza-Neto; José Salvatore Leister Patané; Luiz Alcantara; Luiz Lehmann Coutinho; Maria Carolina Elias; Maurício Lacerda Nogueira; Rafael dos Santos Bezerra; Raul Machado Neto; Rejane Maria Tommasini Grotto; Ricardo Haddad; Sandra Coccuzzo Sampaio Vessoni; Simone Kashima; Svetoslav Nanev Slavov; Vincent Louis Viala |
| EPI_ISL_5649147 | UNIDADE BASICA DE SAUDE DE IPEUNA | Instituto Butantan | Antonio Jorge Martins; Claudia Renata dos Santos Barros; David Schlesinger; Debora Botequio Moretti; Dimas Tadeu Covas; Elaine Cristina Marqueze; Elaine Vieira Santos; Evandra Strazza Rodrigues; Heidge Fukumasu; Jayme Augusto de Souza-Neto; José Salvatore Leister Patané; Luiz Alcantara; Luiz Lehmann Coutinho; Maria Carolina Elias; Maurício Lacerda Nogueira; Rafael dos Santos Bezerra; Raul Machado Neto; Rejane Maria Tommasini Grotto; Ricardo Haddad; Sandra Coccuzzo Sampaio Vessoni; Simone Kashima; Svetoslav Nanev Slavov; Vincent Louis Viala |
| EPI_ISL_5649075 | UNIDADE BASICA DE SAUDE DE TARUMA | Instituto Butantan | Antonio Jorge Martins; Claudia Renata dos Santos Barros; David Schlesinger; Debora Botequio Moretti; Dimas Tadeu Covas; Elaine Cristina Marqueze; Elaine Vieira Santos; Evandra Strazza Rodrigues; Heidge Fukumasu; Jayme Augusto de Souza-Neto; José Salvatore Leister Patané; Luiz Alcantara; Luiz Lehmann Coutinho; Maria Carolina Elias; Maurício Lacerda Nogueira; Rafael dos Santos Bezerra; Raul Machado Neto; Rejane Maria Tommasini Grotto; Ricardo Haddad; Sandra Coccuzzo Sampaio Vessoni; Simone Kashima; Svetoslav Nanev Slavov; Vincent Louis Viala |
| EPI_ISL_5649187, EPI_ISL_5649215 | UNIDADE BASICA DE SAUDE DR MATHEUS GABRIEL BONASSA | Instituto Butantan | Antonio Jorge Martins; Claudia Renata dos Santos Barros; David Schlesinger; Debora Botequio Moretti; Dimas Tadeu Covas; Elaine Cristina Marqueze; Elaine Vieira Santos; Evandra Strazza Rodrigues; Heidge Fukumasu; Jayme Augusto de Souza-Neto; José Salvatore Leister Patané; Luiz Alcantara; Luiz Lehmann Coutinho; Maria Carolina Elias; Maurício Lacerda Nogueira; Rafael dos Santos Bezerra; Raul Machado Neto; Rejane Maria Tommasini Grotto; Ricardo Haddad; Sandra Coccuzzo Sampaio Vessoni; Simone Kashima; Svetoslav Nanev Slavov; Vincent Louis Viala |
| EPI_ISL_5649160, EPI_ISL_5649199, EPI_ISL_5649205, EPI_ISL_5649214 | UNIDADE BASICA DE SAUDE II SANDOVALINA | Instituto Butantan | Antonio Jorge Martins; Claudia Renata dos Santos Barros; David Schlesinger; Debora Botequio Moretti; Dimas Tadeu Covas; Elaine Cristina Marqueze; Elaine Vieira Santos; Evandra Strazza Rodrigues; Heidge Fukumasu; Jayme Augusto de Souza-Neto; José Salvatore Leister Patané; Luiz Alcantara; Luiz Lehmann Coutinho; Maria Carolina Elias; Maurício Lacerda Nogueira; Rafael dos Santos Bezerra; Raul Machado Neto; Rejane Maria Tommasini Grotto; Ricardo Haddad; Sandra Coccuzzo Sampaio Vessoni; Simone Kashima; Svetoslav Nanev Slavov; Vincent Louis Viala |
| EPI_ISL_5648581, EPI_ISL_5648592, EPI_ISL_5648663, EPI_ISL_5659567, EPI_ISL_5659620 | UNIDADE BASICA DE SAUDE PREFEITO JOSE OLIVEIRA DE SOUZA | Instituto Butantan | Antonio Jorge Martins; Claudia Renata dos Santos Barros; David Schlesinger; Debora Botequio Moretti; Dimas Tadeu Covas; Elaine Cristina Marqueze; Elaine Vieira Santos; Evandra Strazza Rodrigues; Heidge Fukumasu; Jayme Augusto de Souza-Neto; José Salvatore Leister Patané; Luiz Alcantara; Luiz Lehmann Coutinho; Maria Carolina Elias; Maurício Lacerda Nogueira; Rafael dos Santos Bezerra; Raul Machado Neto; Rejane Maria Tommasini Grotto; Ricardo Haddad; Sandra Coccuzzo Sampaio Vessoni; Simone Kashima; Svetoslav Nanev Slavov; Vincent Louis Viala |
| EPI_ISL_3536381 | UNIDADE BASICA DE SAUDE SANTA TEREZA | Analytical Competence Molecular Epidemiology Lab/ACME, Oswaldo Cruz Foundation, Ceara (FIOCRUZ CE) | Cleber Furtado Aksenen; Fabio Miyajima; Fernando Braga Stehling; Francisco Eder de Moura Lopes; Jamille Maria Mendes Bezerra; Joaquim César do Nascimento Sousa Junior; Pedro Miguel Carneiro Jeronimo; Suzana Porto Almeida e Lucas Delerino; Thais Ferreira de Oliveira; Thais de Oliveira Costa; Ticiane Cavalcante de Souza; Veridiana Pessoa Miyajima |
| EPI_ISL_3536382 | UNIDADE BASICA DE SAUDE SANTANA | Analytical Competence Molecular Epidemiology Lab/ACME, Oswaldo Cruz Foundation, Ceara (FIOCRUZ CE) | Cleber Furtado Aksenen; Fabio Miyajima; Fernando Braga Stehling; Francisco Eder de Moura Lopes; Jamille Maria Mendes Bezerra; Joaquim César do Nascimento Sousa Junior; Pedro Miguel Carneiro Jeronimo; Suzana Porto Almeida e Lucas Delerino; Thais Ferreira de Oliveira; Thais de Oliveira Costa; Ticiane Cavalcante de Souza; Veridiana Pessoa Miyajima |
| EPI_ISL_3536148 | UNIDADE BASICA DE SAUDE VILA COBAL | Analytical Competence Molecular Epidemiology Lab/ACME, Oswaldo Cruz Foundation, Ceara (FIOCRUZ CE) | Cleber Furtado Aksenen; Fabio Miyajima; Fernando Braga Stehling; Francisco Eder de Moura Lopes; Jamille Maria Mendes Bezerra; Joaquim César do Nascimento Sousa Junior; Pedro Miguel Carneiro Jeronimo; Suzana Porto Almeida e Lucas Delerino; Thais Ferreira de Oliveira; Thais de Oliveira Costa; Ticiane Cavalcante de Souza; Veridiana Pessoa Miyajima |
| EPI_ISL_5659603, EPI_ISL_5659607 | UNIDADE BASICA DE SSAUDE DE POLONI | Instituto Butantan | Antonio Jorge Martins; Claudia Renata dos Santos Barros; David Schlesinger; Debora Botequio Moretti; Dimas Tadeu Covas; Elaine Cristina Marqueze; Elaine Vieira Santos; Evandra Strazza Rodrigues; Heidge Fukumasu; Jayme Augusto de Souza-Neto; José Salvatore Leister Patané; Luiz Alcantara; Luiz Lehmann Coutinho; Maria Carolina Elias; Maurício Lacerda Nogueira; Rafael dos Santos Bezerra; Raul Machado Neto; Rejane Maria Tommasini Grotto; Ricardo Haddad; Sandra Coccuzzo Sampaio Vessoni; Simone Kashima; Svetoslav Nanev Slavov; Vincent Louis Viala |
| EPI_ISL_5648297, EPI_ISL_5648357, EPI_ISL_5648391, EPI_ISL_5648415, EPI_ISL_5648538, EPI_ISL_5648548, EPI_ISL_5648594, EPI_ISL_5648635, EPI_ISL_5648692, EPI_ISL_5648713, EPI_ISL_5649204 | | | |
| see above | UNIDADE DA SAUDE DO ADULTO CASA BRANCA PREFEITURA | Instituto Butantan | Antonio Jorge Martins; Claudia Renata dos Santos Barros; David Schlesinger; Debora Botequio Moretti; Dimas Tadeu Covas; Elaine Cristina Marqueze; Elaine Vieira Santos; Evandra Strazza Rodrigues; Heidge Fukumasu; Jayme Augusto de Souza-Neto; José Salvatore Leister Patané; Luiz Alcantara; Luiz Lehmann Coutinho; Maria Carolina Elias; Maurício Lacerda Nogueira; Rafael dos Santos Bezerra; Raul Machado Neto; Rejane Maria Tommasini Grotto; Ricardo Haddad; Sandra Coccuzzo Sampaio Vessoni; Simone Kashima; Svetoslav Nanev Slavov; Vincent Louis Viala |
| EPI_ISL_5648771, EPI_ISL_5648790, EPI_ISL_5648807, EPI_ISL_5648818, EPI_ISL_5648856, EPI_ISL_5648896, EPI_ISL_5648897, EPI_ISL_5648900, EPI_ISL_5648908, EPI_ISL_5648960, EPI_ISL_5648973, EPI_ISL_5649007, EPI_ISL_5649021, EPI_ISL_5649076, EPI_ISL_5649140 | | | |
| see above | UNIDADE DE PRONTO ATENDIMENTO DE | Instituto Butantan | Antonio Jorge Martins; Claudia Renata dos Santos Barros; David Schlesinger; Debora Botequio Moretti; Dimas Tadeu Covas; Elaine Cristina Marqueze; Elaine Vieira Santos; Evandra Strazza Rodrigues; Heidge Fukumasu; Jayme Augusto de Souza-Neto; José Salvatore Leister Patané; Luiz Alcantara; |

| | | | |
|---|---|---|---|
| | VARZEA PAULISTA UPA II | | Luiz Lehmann Coutinho; Maria Carolina Elias; Maurício Lacerda Nogueira; Rafael dos Santos Bezerra; Raul Machado Neto; Rejane Maria Tommasini Grotto; Ricardo Haddad; Sandra Coccuzzo Sampaio Vessoni; Simone Kashima; Svetoslav Nanev Slavov; Vincent Louis Viala |
| EPI_ISL_5649480 | UNIDADE DE PRONTO ATENDIMENTO UPA | Instituto Butantan | Antonio Jorge Martins; Claudia Renata dos Santos Barros; David Schlesinger; Debora Botequio Moretti; Dimas Tadeu Covas; Elaine Cristina Marqueze; Elaine Vieira Santos; Evandra Strazza Rodrigues; Heidge Fukumasu; Jayme Augusto de Souza-Neto; José Salvatore Leister Patané; Luiz Alcantara; Luiz Lehmann Coutinho; Maria Carolina Elias; Maurício Lacerda Nogueira; Rafael dos Santos Bezerra; Raul Machado Neto; Rejane Maria Tommasini Grotto; Ricardo Haddad; Sandra Coccuzzo Sampaio Vessoni; Simone Kashima; Svetoslav Nanev Slavov; Vincent Louis Viala |
| EPI_ISL_5648928 | UNIDADE DE SAUDE DA FAMILIA JARDIM AMERICA ITAQUAQUECETUBA | Instituto Butantan | Antonio Jorge Martins; Claudia Renata dos Santos Barros; David Schlesinger; Debora Botequio Moretti; Dimas Tadeu Covas; Elaine Cristina Marqueze; Elaine Vieira Santos; Evandra Strazza Rodrigues; Heidge Fukumasu; Jayme Augusto de Souza-Neto; José Salvatore Leister Patané; Luiz Alcantara; Luiz Lehmann Coutinho; Maria Carolina Elias; Maurício Lacerda Nogueira; Rafael dos Santos Bezerra; Raul Machado Neto; Rejane Maria Tommasini Grotto; Ricardo Haddad; Sandra Coccuzzo Sampaio Vessoni; Simone Kashima; Svetoslav Nanev Slavov; Vincent Louis Viala |
| EPI_ISL_5648951, EPI_ISL_5648959, EPI_ISL_5649016 | UNIDADE DE SAUDE DA FAMILIA NOVO HORIZONTE | Instituto Butantan | Antonio Jorge Martins; Claudia Renata dos Santos Barros; David Schlesinger; Debora Botequio Moretti; Dimas Tadeu Covas; Elaine Cristina Marqueze; Elaine Vieira Santos; Evandra Strazza Rodrigues; Heidge Fukumasu; Jayme Augusto de Souza-Neto; José Salvatore Leister Patané; Luiz Alcantara; Luiz Lehmann Coutinho; Maria Carolina Elias; Maurício Lacerda Nogueira; Rafael dos Santos Bezerra; Raul Machado Neto; Rejane Maria Tommasini Grotto; Ricardo Haddad; Sandra Coccuzzo Sampaio Vessoni; Simone Kashima; Svetoslav Nanev Slavov; Vincent Louis Viala |
| EPI_ISL_5649212, EPI_ISL_5665718, EPI_ISL_5665723, EPI_ISL_5665728 | UNIDADE DE VIGILANCIA EPIDEMIOLOGICA | Instituto Butantan | Antonio Jorge Martins; Claudia Renata dos Santos Barros; David Schlesinger; Debora Botequio Moretti; Dimas Tadeu Covas; Elaine Cristina Marqueze; Elaine Vieira Santos; Evandra Strazza Rodrigues; Heidge Fukumasu; Jayme Augusto de Souza-Neto; José Salvatore Leister Patané; Luiz Alcantara; Luiz Lehmann Coutinho; Maria Carolina Elias; Maurício Lacerda Nogueira; Rafael dos Santos Bezerra; Raul Machado Neto; Rejane Maria Tommasini Grotto; Ricardo Haddad; Sandra Coccuzzo Sampaio Vessoni; Simone Kashima; Svetoslav Nanev Slavov; Vincent Louis Viala |
| EPI_ISL_5648723 | UNIDADE MISTA DE SAUDE DE ORIENTE | Instituto Butantan | Antonio Jorge Martins; Claudia Renata dos Santos Barros; David Schlesinger; Debora Botequio Moretti; Dimas Tadeu Covas; Elaine Cristina Marqueze; Elaine Vieira Santos; Evandra Strazza Rodrigues; Heidge Fukumasu; Jayme Augusto de Souza-Neto; José Salvatore Leister Patané; Luiz Alcantara; Luiz Lehmann Coutinho; Maria Carolina Elias; Maurício Lacerda Nogueira; Rafael dos Santos Bezerra; Raul Machado Neto; Rejane Maria Tommasini Grotto; Ricardo Haddad; Sandra Coccuzzo Sampaio Vessoni; Simone Kashima; Svetoslav Nanev Slavov; Vincent Louis Viala |
| EPI_ISL_5649167 | UNIDADE MISTA DE SAUDE MOMBUCA | Instituto Butantan | Antonio Jorge Martins; Claudia Renata dos Santos Barros; David Schlesinger; Debora Botequio Moretti; Dimas Tadeu Covas; Elaine Cristina Marqueze; Elaine Vieira Santos; Evandra Strazza Rodrigues; Heidge Fukumasu; Jayme Augusto de Souza-Neto; José Salvatore Leister Patané; Luiz Alcantara; Luiz Lehmann Coutinho; Maria Carolina Elias; Maurício Lacerda Nogueira; Rafael dos Santos Bezerra; Raul Machado Neto; Rejane Maria Tommasini Grotto; Ricardo Haddad; Sandra Coccuzzo Sampaio Vessoni; Simone Kashima; Svetoslav Nanev Slavov; Vincent Louis Viala |
| EPI_ISL_5649222, EPI_ISL_5649234 | UNIDADE MISTA OSVALDO P CAMARGO ELIAS FAUSTO | Instituto Butantan | Antonio Jorge Martins; Claudia Renata dos Santos Barros; David Schlesinger; Debora Botequio Moretti; Dimas Tadeu Covas; Elaine Cristina Marqueze; Elaine Vieira Santos; Evandra Strazza Rodrigues; Heidge Fukumasu; Jayme Augusto de Souza-Neto; José Salvatore Leister Patané; Luiz Alcantara; Luiz Lehmann Coutinho; Maria Carolina Elias; Maurício Lacerda Nogueira; Rafael dos Santos Bezerra; Raul Machado Neto; Rejane Maria Tommasini Grotto; Ricardo Haddad; Sandra Coccuzzo Sampaio Vessoni; Simone Kashima; Svetoslav Nanev Slavov; Vincent Louis Viala |
| EPI_ISL_3536194, EPI_ISL_3536195, EPI_ISL_3536196, EPI_ISL_3536197, EPI_ISL_3536198, EPI_ISL_3536199, EPI_ISL_3536200, EPI_ISL_3536201, EPI_ISL_3536202, EPI_ISL_3536203, EPI_ISL_3912108, EPI_ISL_3912355 | | | |
| see above | UNIDADE REFERENCIA COVID 19 | Analytical Competence Molecular Epidemiology Lab/ACME, Oswaldo Cruz Foundation, Ceara (FIOCRUZ CE) | Cleber Furtado Aksenen; Fabio Miyajima; Fernando Braga Stehling; Francisco Eder de Moura Lopes; Jamille Maria Mendes Bezerra; Joaquim Cesar do Nascimento Sousa Junior; Joaquim César do Nascimento Sousa Junior; Pedro Miguel Carneiro Jeronimo; Suzana Porto Almeida & Lucas Delerino on behalf of COVID-19 FIOCRUZ Genomic Network; Suzana Porto Almeida e Lucas Delerino; Thais Ferreira de Oliveira; Thais de Oliveira Costa; Ticiane Cavalcante de Souza; Veridiana Pessoa Miyajima |
| EPI_ISL_5648748 | UNIDADE REFERENCIAL SUDOESTE | Instituto Butantan | Antonio Jorge Martins; Claudia Renata dos Santos Barros; David Schlesinger; Debora Botequio Moretti; Dimas Tadeu Covas; Elaine Cristina Marqueze; Elaine Vieira Santos; Evandra Strazza Rodrigues; Heidge Fukumasu; Jayme Augusto de Souza-Neto; José Salvatore Leister Patané; Luiz Alcantara; Luiz Lehmann Coutinho; Maria Carolina Elias; Maurício Lacerda Nogueira; Rafael dos Santos Bezerra; Raul Machado Neto; Rejane Maria Tommasini Grotto; Ricardo Haddad; Sandra Coccuzzo Sampaio Vessoni; Simone Kashima; Svetoslav Nanev Slavov; Vincent Louis Viala |
| EPI_ISL_5648645 | UNIDADE RESPIRATORIA NOVA HORTOLANDIA | Instituto Butantan | Antonio Jorge Martins; Claudia Renata dos Santos Barros; David Schlesinger; Debora Botequio Moretti; Dimas Tadeu Covas; Elaine Cristina Marqueze; Elaine Vieira Santos; Evandra Strazza Rodrigues; Heidge Fukumasu; Jayme Augusto de Souza-Neto; José Salvatore Leister Patané; Luiz Alcantara; Luiz Lehmann Coutinho; Maria Carolina Elias; Maurício Lacerda Nogueira; Rafael dos Santos Bezerra; Raul Machado Neto; Rejane Maria Tommasini Grotto; Ricardo Haddad; Sandra Coccuzzo Sampaio Vessoni; Simone Kashima; Svetoslav Nanev Slavov; Vincent Louis Viala |
| EPI_ISL_2810705 | UNILABS | Instituto Nacional de Saude (INSA) | Borges et al |
| EPI_ISL_5648799, EPI_ISL_5648855 | UPA CUMBICA | Instituto Butantan | Antonio Jorge Martins; Claudia Renata dos Santos Barros; David Schlesinger; Debora Botequio Moretti; Dimas Tadeu Covas; Elaine Cristina Marqueze; Elaine Vieira Santos; Evandra Strazza Rodrigues; Heidge Fukumasu; Jayme Augusto de Souza-Neto; José Salvatore Leister Patané; Luiz Alcantara; Luiz Lehmann Coutinho; Maria Carolina Elias; Maurício Lacerda Nogueira; Rafael dos Santos Bezerra; Raul Machado Neto; Rejane Maria Tommasini Grotto; Ricardo Haddad; Sandra Coccuzzo Sampaio Vessoni; Simone Kashima; Svetoslav Nanev Slavov; Vincent Louis Viala |
| EPI_ISL_5649247, EPI_ISL_5649288, EPI_ISL_5649300, EPI_ISL_5649307, EPI_ISL_5649314 | UPA DR FRANCO DA ROCHA | Instituto Butantan | Antonio Jorge Martins; Claudia Renata dos Santos Barros; David Schlesinger; Debora Botequio Moretti; Dimas Tadeu Covas; Elaine Cristina Marqueze; Elaine Vieira Santos; Evandra Strazza Rodrigues; Heidge Fukumasu; Jayme Augusto de Souza-Neto; José Salvatore Leister Patané; Luiz Alcantara; Luiz Lehmann Coutinho; Maria Carolina Elias; Maurício Lacerda Nogueira; Rafael dos Santos Bezerra; Raul Machado Neto; Rejane Maria Tommasini Grotto; Ricardo Haddad; Sandra Coccuzzo Sampaio Vessoni; Simone Kashima; Svetoslav Nanev Slavov; Vincent Louis Viala |
| EPI_ISL_5648836 | UPA PAULISTA | Instituto Butantan | Antonio Jorge Martins; Claudia Renata dos Santos Barros; David Schlesinger; Debora Botequio Moretti; Dimas Tadeu Covas; Elaine Cristina Marqueze; Elaine Vieira Santos; Evandra Strazza Rodrigues; Heidge Fukumasu; Jayme Augusto de Souza-Neto; José Salvatore Leister Patané; Luiz Alcantara; Luiz Lehmann Coutinho; Maria Carolina Elias; Maurício Lacerda Nogueira; Rafael dos Santos Bezerra; Raul Machado Neto; Rejane Maria Tommasini Grotto; Ricardo Haddad; Sandra Coccuzzo Sampaio Vessoni; Simone Kashima; Svetoslav Nanev Slavov; Vincent Louis Viala |
| EPI_ISL_5649020 | UPA SADAKO SEDOGUTI | Instituto Butantan | Antonio Jorge Martins; Claudia Renata dos Santos Barros; David Schlesinger; Debora Botequio Moretti; Dimas Tadeu Covas; Elaine Cristina Marqueze; Elaine Vieira Santos; Evandra Strazza Rodrigues; Heidge Fukumasu; Jayme Augusto de Souza-Neto; José Salvatore Leister Patané; Luiz Alcantara; Luiz Lehmann Coutinho; Maria Carolina Elias; Maurício Lacerda Nogueira; Rafael dos Santos Bezerra; Raul Machado Neto; Rejane Maria Tommasini Grotto; Ricardo Haddad; Sandra Coccuzzo Sampaio Vessoni; Simone Kashima; Svetoslav Nanev Slavov; Vincent Louis Viala |
| EPI_ISL_5649119 | UPA SAO JOAO LAVRAS | Instituto Butantan | Antonio Jorge Martins; Claudia Renata dos Santos Barros; David Schlesinger; Debora Botequio Moretti; Dimas Tadeu Covas; Elaine Cristina Marqueze; Elaine Vieira Santos; Evandra Strazza Rodrigues; Heidge Fukumasu; Jayme Augusto de Souza-Neto; José Salvatore Leister Patané; Luiz Alcantara; Luiz Lehmann Coutinho; Maria Carolina Elias; Maurício Lacerda Nogueira; Rafael dos Santos Bezerra; Raul Machado Neto; Rejane Maria Tommasini Grotto; Ricardo Haddad; Sandra Coccuzzo Sampaio Vessoni; Simone Kashima; Svetoslav Nanev Slavov; Vincent Louis Viala |
| EPI_ISL_5648260, EPI_ISL_5648478, EPI_ISL_5648526, EPI_ISL_5648530, EPI_ISL_5648616, EPI_ISL_5648656 | UPA UNIDADE DE PRONTO ATENDIMENTO 24 HORAS BOM JESUS | Instituto Butantan | Antonio Jorge Martins; Claudia Renata dos Santos Barros; David Schlesinger; Debora Botequio Moretti; Dimas Tadeu Covas; Elaine Cristina Marqueze; Elaine Vieira Santos; Evandra Strazza Rodrigues; Heidge Fukumasu; Jayme Augusto de Souza-Neto; José Salvatore Leister Patané; Luiz Alcantara; Luiz Lehmann Coutinho; Maria Carolina Elias; Maurício Lacerda Nogueira; Rafael dos Santos Bezerra; Raul Machado Neto; Rejane Maria Tommasini Grotto; Ricardo Haddad; Sandra Coccuzzo Sampaio Vessoni; Simone Kashima; Svetoslav Nanev Slavov; Vincent Louis Viala |
| EPI_ISL_5648774, EPI_ISL_5648848, EPI_ISL_5648871, EPI_ISL_5649057, EPI_ISL_5649069, EPI_ISL_5649070 | UPA VILA SANTA CATARINA | Instituto Butantan | Antonio Jorge Martins; Claudia Renata dos Santos Barros; David Schlesinger; Debora Botequio Moretti; Dimas Tadeu Covas; Elaine Cristina Marqueze; Elaine Vieira Santos; Evandra Strazza Rodrigues; Heidge Fukumasu; Jayme Augusto de Souza-Neto; José Salvatore Leister Patané; Luiz Alcantara; Luiz Lehmann Coutinho; Maria Carolina Elias; Maurício Lacerda Nogueira; Rafael dos Santos Bezerra; Raul Machado Neto; Rejane Maria Tommasini Grotto; Ricardo Haddad; Sandra Coccuzzo Sampaio Vessoni; Simone Kashima; Svetoslav Nanev Slavov; Vincent Louis Viala |
| EPI_ISL_5648335, EPI_ISL_5648740, EPI_ISL_5648751, EPI_ISL_5649372 | USAFA FORTE | Instituto Butantan | Antonio Jorge Martins; Claudia Renata dos Santos Barros; David Schlesinger; Debora Botequio Moretti; Dimas Tadeu Covas; Elaine Cristina Marqueze; Elaine Vieira Santos; Evandra Strazza Rodrigues; Heidge Fukumasu; Jayme Augusto de Souza-Neto; José Salvatore Leister Patané; Luiz Alcantara; Luiz Lehmann Coutinho; Maria Carolina Elias; Maurício Lacerda Nogueira; Rafael dos Santos Bezerra; Raul Machado Neto; Rejane Maria Tommasini Grotto; Ricardo Haddad; Sandra Coccuzzo Sampaio Vessoni; Simone Kashima; Svetoslav Nanev Slavov; Vincent Louis Viala |
| EPI_ISL_5649352 | USAFA GUARAMAR | Instituto Butantan | Antonio Jorge Martins; Claudia Renata dos Santos Barros; David Schlesinger; Debora Botequio Moretti; Dimas Tadeu Covas; Elaine Cristina Marqueze; Elaine Vieira Santos; Evandra Strazza Rodrigues; Heidge Fukumasu; Jayme Augusto de Souza-Neto; José Salvatore Leister Patané; Luiz Alcantara; Luiz Lehmann Coutinho; Maria Carolina Elias; Maurício Lacerda Nogueira; Rafael dos Santos Bezerra; Raul Machado Neto; Rejane Maria Tommasini Grotto; Ricardo Haddad; Sandra Coccuzzo Sampaio Vessoni; Simone Kashima; Svetoslav Nanev Slavov; Vincent Louis Viala |
| EPI_ISL_5649393 | USAFA MIRIM | Instituto Butantan | Antonio Jorge Martins; Claudia Renata dos Santos Barros; David Schlesinger; Debora Botequio Moretti; Dimas Tadeu Covas; Elaine Cristina Marqueze; Elaine Vieira Santos; Evandra Strazza Rodrigues; Heidge Fukumasu; Jayme Augusto de Souza-Neto; José Salvatore Leister Patané; Luiz Alcantara; Luiz Lehmann Coutinho; Maria Carolina Elias; Maurício Lacerda Nogueira; Rafael dos Santos Bezerra; Raul Machado Neto; Rejane Maria Tommasini Grotto; Ricardo Haddad; Sandra Coccuzzo Sampaio Vessoni; Simone Kashima; Svetoslav Nanev Slavov; Vincent Louis Viala |
| EPI_ISL_5648542, EPI_ISL_5649408 | USAFA SAMAMBAIA | Instituto Butantan | Antonio Jorge Martins; Claudia Renata dos Santos Barros; David Schlesinger; Debora Botequio Moretti; Dimas Tadeu Covas; Elaine Cristina Marqueze; Elaine Vieira Santos; Evandra Strazza Rodrigues; Heidge Fukumasu; Jayme Augusto de Souza-Neto; José Salvatore Leister Patané; Luiz Alcantara; Luiz Lehmann Coutinho; Maria Carolina Elias; Maurício Lacerda Nogueira; Rafael dos Santos Bezerra; Raul Machado Neto; Rejane Maria Tommasini Grotto; Ricardo Haddad; Sandra Coccuzzo Sampaio Vessoni; Simone Kashima; Svetoslav Nanev Slavov; Vincent Louis Viala |
| EPI_ISL_5648646, EPI_ISL_5649364 | USAFA TUDE BASTOS | Instituto Butantan | Antonio Jorge Martins; Claudia Renata dos Santos Barros; David Schlesinger; Debora Botequio Moretti; Dimas Tadeu Covas; Elaine Cristina Marqueze; Elaine Vieira Santos; Evandra Strazza Rodrigues; Heidge Fukumasu; Jayme Augusto de Souza-Neto; José Salvatore Leister Patané; Luiz Alcantara; Luiz Lehmann Coutinho; Maria Carolina Elias; Maurício Lacerda Nogueira; Rafael dos Santos Bezerra; Raul Machado Neto; Rejane Maria Tommasini Grotto; Ricardo Haddad; Sandra Coccuzzo Sampaio Vessoni; Simone Kashima; Svetoslav Nanev Slavov; Vincent Louis Viala |
| EPI_ISL_2894588 | USC Clinical Lab | Los Angeles County Public Health Laboratories | P. Hemarajata et al. |
| EPI_ISL_5648520 | USF DE INUBIA PAULISTA | Instituto Butantan | Antonio Jorge Martins; Claudia Renata dos Santos Barros; David Schlesinger; Debora Botequio Moretti; Dimas Tadeu Covas; Elaine Cristina Marqueze; Elaine Vieira Santos; Evandra Strazza Rodrigues; Heidge Fukumasu; Jayme Augusto de Souza-Neto; José Salvatore Leister Patané; Luiz Alcantara; Luiz Lehmann Coutinho; Maria Carolina Elias; Maurício Lacerda Nogueira; Rafael dos Santos Bezerra; Raul Machado Neto; Rejane Maria Tommasini Grotto; Ricardo Haddad; Sandra Coccuzzo Sampaio Vessoni; Simone Kashima; Svetoslav Nanev Slavov; Vincent Louis Viala |
| EPI_ISL_5648686, EPI_ISL_5648998 | USF DE SAGRES | Instituto Butantan | Antonio Jorge Martins; Claudia Renata dos Santos Barros; David Schlesinger; Debora Botequio Moretti; Dimas Tadeu Covas; Elaine Cristina Marqueze; Elaine Vieira Santos; Evandra Strazza Rodrigues; Heidge Fukumasu; Jayme Augusto de Souza-Neto; José Salvatore Leister Patané; Luiz Alcantara; Luiz Lehmann Coutinho; Maria Carolina Elias; Maurício Lacerda Nogueira; Rafael dos Santos Bezerra; Raul Machado Neto; Rejane Maria Tommasini Grotto; Ricardo Haddad; Sandra Coccuzzo Sampaio Vessoni; Simone Kashima; Svetoslav Nanev Slavov; Vincent Louis Viala |
| EPI_ISL_5648532 | USF DR PAULO MINAMI | Instituto Butantan | Antonio Jorge Martins; Claudia Renata dos Santos Barros; David Schlesinger; Debora Botequio Moretti; Dimas Tadeu Covas; Elaine Cristina Marqueze; Elaine Vieira Santos; Evandra Strazza Rodrigues; Heidge Fukumasu; Jayme Augusto de Souza-Neto; José Salvatore Leister Patané; Luiz Alcantara; Luiz Lehmann Coutinho; Maria Carolina Elias; Maurício Lacerda Nogueira; Rafael dos Santos Bezerra; Raul Machado Neto; Rejane Maria Tommasini Grotto; Ricardo Haddad; Sandra Coccuzzo Sampaio Vessoni; Simone Kashima; Svetoslav Nanev Slavov; Vincent Louis Viala |
| EPI_ISL_5648780 | USF DR SAMUEL MARTINS FIGUEIRA SCRPARDO | Instituto Butantan | Antonio Jorge Martins; Claudia Renata dos Santos Barros; David Schlesinger; Debora Botequio Moretti; Dimas Tadeu Covas; Elaine Cristina Marqueze; Elaine Vieira Santos; Evandra Strazza Rodrigues; Heidge Fukumasu; Jayme Augusto de Souza-Neto; José Salvatore Leister Patané; Luiz Alcantara; Luiz Lehmann Coutinho; Maria Carolina Elias; Maurício Lacerda Nogueira; Rafael dos Santos Bezerra; Raul Machado Neto; Rejane Maria Tommasini Grotto; Ricardo Haddad; Sandra Coccuzzo Sampaio Vessoni; Simone Kashima; Svetoslav Nanev Slavov; Vincent Louis Viala |
| EPI_ISL_5648826, EPI_ISL_5649018 | USF I DE GUAIMBE | Instituto Butantan | Antonio Jorge Martins; Claudia Renata dos Santos Barros; David Schlesinger; Debora Botequio Moretti; Dimas Tadeu Covas; Elaine Cristina Marqueze; Elaine Vieira Santos; Evandra Strazza Rodrigues; Heidge Fukumasu; Jayme Augusto de Souza-Neto; José Salvatore Leister Patané; Luiz Alcantara; Luiz Lehmann Coutinho; Maria Carolina Elias; Maurício Lacerda Nogueira; Rafael dos Santos Bezerra; Raul Machado Neto; Rejane Maria Tommasini Grotto; Ricardo Haddad; Sandra Coccuzzo Sampaio Vessoni; Simone Kashima; Svetoslav Nanev Slavov; Vincent Louis Viala |
| EPI_ISL_5648472 | USF JARDIM CAMANDUCAIA | Instituto Butantan | Antonio Jorge Martins; Claudia Renata dos Santos Barros; David Schlesinger; Debora Botequio Moretti; Dimas Tadeu Covas; Elaine Cristina Marqueze; Elaine Vieira Santos; Evandra Strazza Rodrigues; Heidge Fukumasu; Jayme Augusto de Souza-Neto; José Salvatore Leister Patané; Luiz Alcantara; Luiz Lehmann Coutinho; Maria Carolina Elias; Maurício Lacerda Nogueira; Rafael dos Santos Bezerra; Raul Machado Neto; Rejane Maria Tommasini Grotto; Ricardo Haddad; Sandra Coccuzzo Sampaio Vessoni; Simone Kashima; Svetoslav Nanev Slavov; Vincent Louis Viala |
| EPI_ISL_5648937, EPI_ISL_5649090 | USF JARDIM JOSEFINA | Instituto Butantan | Antonio Jorge Martins; Claudia Renata dos Santos Barros; David Schlesinger; Debora Botequio Moretti; Dimas Tadeu Covas; Elaine Cristina Marqueze; Elaine Vieira Santos; Evandra Strazza Rodrigues; Heidge Fukumasu; Jayme Augusto de Souza-Neto; José Salvatore Leister Patané; Luiz Alcantara; Luiz Lehmann Coutinho; Maria Carolina Elias; Maurício Lacerda Nogueira; Rafael dos Santos Bezerra; Raul Machado Neto; Rejane Maria Tommasini Grotto; Ricardo Haddad; Sandra Coccuzzo Sampaio Vessoni; Simone Kashima; Svetoslav Nanev Slavov; Vincent Louis Viala |
| EPI_ISL_5648967 | USF JARDIM JOSELY | Instituto Butantan | Antonio Jorge Martins; Claudia Renata dos Santos Barros; David Schlesinger; Debora Botequio Moretti; Dimas Tadeu Covas; Elaine Cristina Marqueze; Elaine Vieira Santos; Evandra Strazza Rodrigues; Heidge Fukumasu; Jayme Augusto de Souza-Neto; José Salvatore Leister Patané; Luiz Alcantara; Luiz Lehmann Coutinho; Maria Carolina Elias; Maurício Lacerda Nogueira; Rafael dos Santos Bezerra; Raul Machado Neto; Rejane Maria Tommasini Grotto; Ricardo Haddad; Sandra Coccuzzo Sampaio Vessoni; Simone Kashima; Svetoslav Nanev Slavov; Vincent Louis Viala |
| EPI_ISL_5648789, EPI_ISL_5648919 | USF JARDIM PIRATININGA ITAQUAQUECETUBA | Instituto Butantan | Antonio Jorge Martins; Claudia Renata dos Santos Barros; David Schlesinger; Debora Botequio Moretti; Dimas Tadeu Covas; Elaine Cristina Marqueze; Elaine Vieira Santos; Evandra Strazza Rodrigues; Heidge Fukumasu; Jayme Augusto de Souza-Neto; José Salvatore Leister Patané; Luiz Alcantara; Luiz Lehmann Coutinho; Maria Carolina Elias; Maurício Lacerda Nogueira; Rafael dos Santos Bezerra; Raul Machado Neto; Rejane Maria Tommasini Grotto; Ricardo Haddad; Sandra Coccuzzo Sampaio Vessoni; Simone Kashima; Svetoslav Nanev Slavov; Vincent Louis Viala |
| EPI_ISL_5649427 | USF JARDIM SAO CARLOS | Instituto Butantan | Antonio Jorge Martins; Claudia Renata dos Santos Barros; David Schlesinger; Debora Botequio Moretti; Dimas Tadeu Covas; Elaine Cristina Marqueze; Elaine Vieira Santos; Evandra Strazza Rodrigues; Heidge Fukumasu; Jayme Augusto de Souza-Neto; José Salvatore Leister Patané; Luiz Alcantara; Luiz Lehmann Coutinho; Maria Carolina Elias; Maurício Lacerda Nogueira; Rafael dos Santos Bezerra; Raul Machado Neto; Rejane Maria Tommasini Grotto; Ricardo Haddad; Sandra Coccuzzo Sampaio Vessoni; Simone Kashima; Svetoslav Nanev Slavov; Vincent Louis Viala |
| EPI_ISL_5648906 | USF NICEA LOUZADA | Instituto Butantan | Antonio Jorge Martins; Claudia Renata dos Santos Barros; David Schlesinger; Debora Botequio Moretti; Dimas Tadeu Covas; Elaine Cristina Marqueze; Elaine Vieira Santos; Evandra Strazza Rodrigues; Heidge Fukumasu; Jayme Augusto de Souza-Neto; José Salvatore Leister Patané; Luiz Alcantara; Luiz Lehmann Coutinho; Maria Carolina Elias; Maurício Lacerda Nogueira; Rafael dos Santos Bezerra; Raul Machado Neto; Rejane Maria Tommasini Grotto; Ricardo Haddad; Sandra Coccuzzo Sampaio Vessoni; Simone Kashima; Svetoslav Nanev Slavov; Vincent Louis Viala |
| EPI_ISL_5648508, EPI_ISL_5649484, EPI_ISL_5659536, EPI_ISL_5659624, EPI_ISL_5659625, EPI_ISL_5659630, EPI_ISL_5659632, EPI_ISL_5659637, EPI_ISL_5659638, EPI_ISL_5659640 | | | |
| see above | USF PAULISTA FERNANDOPOLIS ANTONIO PIVATO | Instituto Butantan | Antonio Jorge Martins; Claudia Renata dos Santos Barros; David Schlesinger; Debora Botequio Moretti; Dimas Tadeu Covas; Elaine Cristina Marqueze; Elaine Vieira Santos; Evandra Strazza Rodrigues; Heidge Fukumasu; Jayme Augusto de Souza-Neto; José Salvatore Leister Patané; Luiz Alcantara; Luiz Lehmann Coutinho; Maria Carolina Elias; Maurício Lacerda Nogueira; Rafael dos Santos Bezerra; Raul Machado Neto; Rejane Maria Tommasini Grotto; Ricardo Haddad; Sandra Coccuzzo Sampaio Vessoni; Simone Kashima; Svetoslav Nanev Slavov; Vincent Louis Viala |

| | | | |
|---|---|---|---|
| EPI_ISL_5648935, EPI_ISL_5648945, EPI_ISL_5649008, EPI_ISL_5649085 | USF PEQUENO CORACAO | Instituto Butantan | Antonio Jorge Martins; Claudia Renata dos Santos Barros; David Schlesinger; Debora Botequio Moretti; Dimas Tadeu Covas; Elaine Cristina Marqueze; Elaine Vieira Santos; Evandra Strazza Rodrigues; Heidge Fukumasu; Jayme Augusto de Souza-Neto; José Salvatore Leister Patané; Luiz Alcantara; Luiz Lehmann Coutinho; Maria Carolina Elias; Maurício Lacerda Nogueira; Rafael dos Santos Bezerra; Raul Machado Neto; Rejane Maria Tommasini Grotto; Ricardo Haddad; Sandra Coccuzzo Sampaio Vessoni; Simone Kashima; Svetoslav Nanev Slavov; Vincent Louis Viala |
| EPI_ISL_5648433, EPI_ISL_5648575, EPI_ISL_5648604, EPI_ISL_5648637, EPI_ISL_5648638, EPI_ISL_5648702, EPI_ISL_5648734, EPI_ISL_5648735, EPI_ISL_5649334 | | | |
| see above | USF SALERNO | Instituto Butantan | Antonio Jorge Martins; Claudia Renata dos Santos Barros; David Schlesinger; Debora Botequio Moretti; Dimas Tadeu Covas; Elaine Cristina Marqueze; Elaine Vieira Santos; Evandra Strazza Rodrigues; Heidge Fukumasu; Jayme Augusto de Souza-Neto; José Salvatore Leister Patané; Luiz Alcantara; Luiz Lehmann Coutinho; Maria Carolina Elias; Maurício Lacerda Nogueira; Rafael dos Santos Bezerra; Raul Machado Neto; Rejane Maria Tommasini Grotto; Ricardo Haddad; Sandra Coccuzzo Sampaio Vessoni; Simone Kashima; Svetoslav Nanev Slavov; Vincent Louis Viala |
| EPI_ISL_5648669, EPI_ISL_5649438 | USF SANTA CECILIA | Instituto Butantan | Antonio Jorge Martins; Claudia Renata dos Santos Barros; David Schlesinger; Debora Botequio Moretti; Dimas Tadeu Covas; Elaine Cristina Marqueze; Elaine Vieira Santos; Evandra Strazza Rodrigues; Heidge Fukumasu; Jayme Augusto de Souza-Neto; José Salvatore Leister Patané; Luiz Alcantara; Luiz Lehmann Coutinho; Maria Carolina Elias; Maurício Lacerda Nogueira; Rafael dos Santos Bezerra; Raul Machado Neto; Rejane Maria Tommasini Grotto; Ricardo Haddad; Sandra Coccuzzo Sampaio Vessoni; Simone Kashima; Svetoslav Nanev Slavov; Vincent Louis Viala |
| EPI_ISL_5648585, EPI_ISL_5648622 | USF SANTA TEREZINHA | Instituto Butantan | Antonio Jorge Martins; Claudia Renata dos Santos Barros; David Schlesinger; Debora Botequio Moretti; Dimas Tadeu Covas; Elaine Cristina Marqueze; Elaine Vieira Santos; Evandra Strazza Rodrigues; Heidge Fukumasu; Jayme Augusto de Souza-Neto; José Salvatore Leister Patané; Luiz Alcantara; Luiz Lehmann Coutinho; Maria Carolina Elias; Maurício Lacerda Nogueira; Rafael dos Santos Bezerra; Raul Machado Neto; Rejane Maria Tommasini Grotto; Ricardo Haddad; Sandra Coccuzzo Sampaio Vessoni; Simone Kashima; Svetoslav Nanev Slavov; Vincent Louis Viala |
| EPI_ISL_5659522 | USF SERROTE REGISTRO | Instituto Butantan | Antonio Jorge Martins; Claudia Renata dos Santos Barros; David Schlesinger; Debora Botequio Moretti; Dimas Tadeu Covas; Elaine Cristina Marqueze; Elaine Vieira Santos; Evandra Strazza Rodrigues; Heidge Fukumasu; Jayme Augusto de Souza-Neto; José Salvatore Leister Patané; Luiz Alcantara; Luiz Lehmann Coutinho; Maria Carolina Elias; Maurício Lacerda Nogueira; Rafael dos Santos Bezerra; Raul Machado Neto; Rejane Maria Tommasini Grotto; Ricardo Haddad; Sandra Coccuzzo Sampaio Vessoni; Simone Kashima; Svetoslav Nanev Slavov; Vincent Louis Viala |
| EPI_ISL_2802913, EPI_ISL_2802914, EPI_ISL_2802923, EPI_ISL_2802934, EPI_ISL_2802937, EPI_ISL_2802944, EPI_ISL_2802947, EPI_ISL_2802949, EPI_ISL_2802955, EPI_ISL_2802959, EPI_ISL_2802976, EPI_ISL_2802990, EPI_ISL_2802995, EPI_ISL_2802998, EPI_ISL_2803005, EPI_ISL_2803018, EPI_ISL_2803048, EPI_ISL_2803065, EPI_ISL_2803066, EPI_ISL_2803098, EPI_ISL_2803123, EPI_ISL_2803139, EPI_ISL_2803145, EPI_ISL_2803308, EPI_ISL_2803309, EPI_ISL_3032160, EPI_ISL_3032167, EPI_ISL_3946117 | | | |
| see above | UW Virology Lab | UW Virology Lab | Alexander Greninger; Hong Xie; Keith R Jerome; Lasata Shrestha; Maria Lukes; Meei-Li Huang; Michelle Lin; Nathan Breit; Noah R. Baker; Patrick Mathias; Pavitra Roychoudhury; Ricardo Perez; Robert J. Livingston; Savanna S. Carmack; Sean Ellis; Shah Mohamed Bakhash; Tien V. Nguyen |
| EPI_ISL_2970038, EPI_ISL_2970105 | Unidad de Investigacion Biomedica de Zacatecas (UIBZ) | Unidad de Genomica Avanzada | ; Alejandra Garcia-Gasca; Alejandra Hernandez-Teran; Alejandro Sanchez-Flores; Alfredo Herrera-Estrella; Alicia Ocaña-Mondragon; Andreu Comas-Garcia; Angel Gustavo Salas-Lais; Antonio Loza Roman; Bernardo Martinez-Miguel; Blanca Taboada; Brenda Irasema Maldonado-Meza; Bruno Gomez-Gil; Carla Ivon Herrera-Najera; Carlos F. Arias; Celia Boukadida; Celida Duque Molina; Celida Martinez- Rodriguez; Clara Esperanza Santacruz-Tinoco; Concepcion Grajales-Muñiz; Consorcio Mexicano de Vigilancia Genomica (CoViGen-Mex). Authors (in alphabetical order): Julio Elias Alvarado-Yaah; Cristobal Chaidez-Quiroz; Daniel Fregoso-Rueda; Daniel Lira Morales; Eduardo Becerril-Vargas; Fernando Fontove-Herrera; Fidencio Mejia-Nepomuceno; Francisco Pulido; Gloria Elena Espinosa-Ayala; Gloria Maria Molina-Salinas; Gloria Vazquez; Hector Esteban Paz-Juarez; Hector Montoya-Fuentes; Helen Haydee Fernanda Ramirez-Plascencia; Irvin Gonzalez-Lopez; Jean Pierre Gonzalez; Jesus Hernandez; Joel Armando Vazquez-Perez.; Jorge Salas-Hernandez; Jose Antonio Enciso-Moreno; Jose Arturo Martinez-Orozco; Jose Esteban Muñoz-Medina; Jose de Jesus Nuñez-Contreras; Juan Bautista Chale-Dzul; Julissa Enciso-Ibarra; Luis Alberto Ochoa-Carrera; Margarita Matias-Florentino; Maria Guadalupe Santiago-Mauricio; Maria Guadalupe de Jesus Mireles-Rivera; Mario Mujica-Sanchez; Marissa Perez-Garcia; Nelly Selem-Mojica; Pavel Isa; Ricardo Ciria Merce; Ricardo Grande; Rosa Maria Gutierrez Rios; Santiago avila-Rios; Selene Zarate; Susana Lopez; Veronica Mata-Haro; Victor Eduardo Garcia-Arias; Victor Hugo Borja-Aburto |
| EPI_ISL_2942356, EPI_ISL_2942357, EPI_ISL_2942358, EPI_ISL_2942359, EPI_ISL_2942362, EPI_ISL_2942363, EPI_ISL_2942371, EPI_ISL_2942379, EPI_ISL_2942380, EPI_ISL_2942381, EPI_ISL_2942382, EPI_ISL_2942385, EPI_ISL_2942387, EPI_ISL_2942390, EPI_ISL_2942394, EPI_ISL_2942395, EPI_ISL_2942399, EPI_ISL_2942400, EPI_ISL_2942402, EPI_ISL_2942405, EPI_ISL_2942407, EPI_ISL_2942626, EPI_ISL_2942627, EPI_ISL_2942628, EPI_ISL_2942629, EPI_ISL_2942630, EPI_ISL_2942631, EPI_ISL_2942633, EPI_ISL_2942637, EPI_ISL_2942640, EPI_ISL_2942641, EPI_ISL_2942643, EPI_ISL_2942645, EPI_ISL_2942646, EPI_ISL_2942647, EPI_ISL_2942648, EPI_ISL_2942653, EPI_ISL_2942654, EPI_ISL_2942655, EPI_ISL_2942657, EPI_ISL_2942662, EPI_ISL_2942669, EPI_ISL_2942670, EPI_ISL_2942681, EPI_ISL_2942691, EPI_ISL_2942699, EPI_ISL_2942700, EPI_ISL_2942707, EPI_ISL_2970032, EPI_ISL_2970080, EPI_ISL_2970081, EPI_ISL_2970084, EPI_ISL_2970085 | | | |
| see above | Unidad de Investigacion Medica de Yucatan (UIMY) | Unidad de Genomica Avanzada | ; Alejandra Garcia-Gasca; Alejandra Hernandez-Teran; Alejandro Sanchez-Flores; Alfredo Herrera-Estrella; Alicia Ocaña-Mondragon; Andreu Comas-Garcia; Angel Gustavo Salas-Lais; Antonio Loza Roman; Bernardo Martinez-Miguel; Blanca Taboada; Brenda Irasema Maldonado-Meza; Bruno Gomez-Gil; Carla Ivon Herrera-Najera; Carlos F. Arias; Celia Boukadida; Celida Duque Molina; Celida Martinez- Rodriguez; Clara Esperanza Santacruz-Tinoco; Concepcion Grajales-Muñiz; Consorcio Mexicano de Vigilancia Genomica (CoViGen-Mex). Authors (in alphabetical order): Julio Elias Alvarado-Yaah; Cristobal Chaidez-Quiroz; Daniel Fregoso-Rueda; Daniel Lira Morales; Eduardo Becerril-Vargas; Fernando Fontove-Herrera; Fidencio Mejia-Nepomuceno; Francisco Pulido; Gloria Elena Espinosa-Ayala; Gloria Maria Molina-Salinas; Gloria Vazquez; Hector Esteban Paz-Juarez; Hector Montoya-Fuentes; Helen Haydee Fernanda Ramirez-Plascencia; Irvin Gonzalez-Lopez; Jean Pierre Gonzalez; Jesus Hernandez; Joel Armando Vazquez-Perez.; Jorge Salas-Hernandez; Jose Antonio Enciso-Moreno; Jose Arturo Martinez-Orozco; Jose Esteban Muñoz-Medina; Jose de Jesus Nuñez-Contreras; Juan Bautista Chale-Dzul; Julissa Enciso-Ibarra; Luis Alberto Ochoa-Carrera; Margarita Matias-Florentino; Maria Guadalupe Santiago-Mauricio; Maria Guadalupe de Jesus Mireles-Rivera; Mario Mujica-Sanchez; Marissa Perez-Garcia; Nelly Selem-Mojica; Pavel Isa; Ricardo Ciria Merce; Ricardo Grande; Rosa Maria Gutierrez Rios; Santiago avila-Rios; Selene Zarate; Susana Lopez; Veronica Mata-Haro; Victor Eduardo Garcia-Arias; Victor Hugo Borja-Aburto |
| EPI_ISL_3072167, EPI_ISL_3072168, EPI_ISL_3072170, EPI_ISL_3072171, EPI_ISL_3072257, EPI_ISL_3072259, EPI_ISL_3072261, EPI_ISL_3072353, EPI_ISL_3072419, EPI_ISL_3072421, EPI_ISL_3072423, EPI_ISL_3072541 | | | |
| see above | Unidade de apoio ao diagnostico da COVID - UNADIG | Bioinformatics Laboratory / LNCC | Alessandra P Lamarca; Alexandra L Gerber; Amilcar Tanuri; Ana Paula de C Guimaraes; Ana Tereza R Vasconcelos; Andrea Cony Cavalcanti; Caio Luiz Pereira Ribeiro; Cassia Alves; Cintia Policarpo; Claudia Maria Braga de Mello; Cristiane Gomes da Silva; Diana Mariani; Douglas Terra Machado; Flavio Dias da Silva; Gleidson da Silva de Oliveira; Leandro Magalhaes de Souza; Liliane Cavalcante; Luiz G P de Almeida; Marcio Henrique de Oliveira Garcia; Mario Sergio Ribeiro; Ronaldo da Silva F Jr; Silvia Carvalho |
| EPI_ISL_2840091, EPI_ISL_2840156, EPI_ISL_2897535 | Unilabs | Karolinska University Hospital | Annelie Bjerkner; Isak Sylvin; Jan Albert; Karolina Ininbergs; Lina Guerra Blomqvist; Lynda Eneh; Martin Ekman; Martina Wahlund; Robert Dyrdak; Sandra Broddesson; Tanja Normark; Tobias Allander; Valtteri Wirta; Zhibing Lin |
| EPI_ISL_3320720, EPI_ISL_3320743, EPI_ISL_3320748, EPI_ISL_3320752 | Universidad del Quindio | Instituto Nacional de Salud | Carlos Franco-Muñoz; Carmen Osorio; Diana Malo; Diego A. Álvarez-Díaz; Diego Andrés Prada; Gerardo Santamaría; Hector Alejandro Ruiz-Moreno; Jhonnatan Reales-González; Jorge Rivera; Juan Camilo Martinez; Julian Naizaque; Katherine Laiton-Donato; Lisseth Pardo; Magdalena Wiesner; Marcela Mercado-Reyes; Maria T. Herrera-Sepúlveda; Marta Lopez Blanco; Martha Lucia Ospina Martinez; Paola Rojas; Sergio Gomez; Sheryll Corchuelo; Ángela Alarcon Cruz |
| EPI_ISL_2787067 | University Hospital Sant'Andrea-Sapienza | INMI Lazzaro Spallanzani IRCCS | D Cerini; D Di Fusco; E Giombini; F Messina; F Santini; G Orlandi; M Pallocca; M Rueca; O Butera |
| EPI_ISL_3388664 | University of Illinois at Chicago | RIPHL at Rush University Medical Center | Edith Perez; Felix Araujo Perez; Iuliana Bentea; Kevin Kunstman; Laura Furtado; Marieta Hyde; Max Kolton; Omar Perez; Stefan Green |
| EPI_ISL_2833993 | University of Wisconsin-Madison AIDS Vaccine Research Laboratories | University of Wisconsin-Madison AIDS Vaccine Research Laboratories | Gage Moreno; Katarina Braun; et al. AIDS Vaccine Research Laboratories |
| EPI_ISL_2790255, EPI_ISL_2840648 | Università degli Studi di Perugia | Istituto Zooprofilattico Sperimentale dell'Abruzzo e Molise "G. Caporale" | Ancora M; Biagetti M; Calistri P; Camilloni B; Cammà C; Curini V; Delli Compagni E; Di Domenico M; Di Pasquale A; Giammarioli M; Lorusso A; Mangone I; Marcacci M; Mencacci A; Puglia I; Rinaldi A; Savini G; Scialabba S |
| EPI_ISL_6510838, EPI_ISL_6510839, EPI_ISL_6510847, EPI_ISL_6510855 | Università degli Studi di Perugia, Perugia, Italy | Istituto Superiore di Sanità (ISS) | Alessandra Lo Presti; Angela Di Martino; Antonella Mencacci; Barbara Camilloni; Luigina Ambrosio; Manuela Marra; Marco Crescenzi; Maria Carollo; Paola Stefanelli |
| EPI_ISL_2845773 | Universitätsklinikum Heidelberg | Robert Koch Institute | |
| EPI_ISL_3030227 | Urbino | Microbiology University Politecnica delle Marche | Anna Valenza; Carla Acciarri; Katia Marinelli; Monica Lucia Ferreri; Patrizia Bagnarelli; Roberta Longo; Sara Caucci; Stefano Menzo |
| EPI_ISL_3506002, EPI_ISL_3506141 | Usansolo-Galdakao University Hospital | Cruces University Hospital | Ana Belén de la Hoz; Ana Belén de la Hz; Ana Gual-de-Torrella; Izaskun Alejo-Cancho; Mikel Gallego |
| EPI_ISL_2857704, EPI_ISL_2857706, EPI_ISL_2857746, EPI_ISL_2857747, EPI_ISL_2857767, EPI_ISL_3756201, EPI_ISL_3756245, EPI_ISL_3756247, EPI_ISL_3756248, EPI_ISL_3756249, EPI_ISL_3756250, EPI_ISL_3756252, EPI_ISL_3756253, EPI_ISL_3756256, EPI_ISL_3756257 | | | |
| see above | Utah Public Health Laboratory | Utah Public Health Laboratory | Erin L. Young; Kelly F. Oakeson; Olinto Linares-Perdomo; Tara Gallagher |
| EPI_ISL_5649034 | VIGILANCIA EPIDEMIOLOGICA DE FERRAZ DE VASCONCELOS | Instituto Butantan | Antonio Jorge Martins; Claudia Renata dos Santos Barros; David Schlesinger; Debora Botequio Moretti; Dimas Tadeu Covas; Elaine Cristina Marqueze; Elaine Vieira Santos; Evandra Strazza Rodrigues; Heidge Fukumasu; Jayme Augusto de Souza-Neto; José Salvatore Leister Patané; Luiz Alcantara; Luiz Lehmann Coutinho; Maria Carolina Elias; Maurício Lacerda Nogueira; Rafael dos Santos Bezerra; Raul Machado Neto; Rejane Maria Tommasini Grotto; Ricardo Haddad; Sandra Coccuzzo Sampaio Vessoni; Simone Kashima; Svetoslav Nanev Slavov; Vincent Louis Viala |
| EPI_ISL_5665775, EPI_ISL_5665778 | VIGILANCIA EPIDEMIOLOGICA DE LEME | Instituto Butantan | Antonio Jorge Martins; Claudia Renata dos Santos Barros; David Schlesinger; Debora Botequio Moretti; Dimas Tadeu Covas; Elaine Cristina Marqueze; Elaine Vieira Santos; Evandra Strazza Rodrigues; Heidge Fukumasu; Jayme Augusto de Souza-Neto; José Salvatore Leister Patané; Luiz Alcantara; Luiz Lehmann Coutinho; Maria Carolina Elias; Maurício Lacerda Nogueira; Rafael dos Santos Bezerra; Raul Machado Neto; Rejane Maria Tommasini Grotto; Ricardo Haddad; Sandra Coccuzzo Sampaio Vessoni; Simone Kashima; Svetoslav Nanev Slavov; Vincent Louis Viala |
| EPI_ISL_5648765, EPI_ISL_5648902, EPI_ISL_5649114 | VIGILANCIA EPIDEMIOLOGICA DE SALESOPOLIS | Instituto Butantan | Antonio Jorge Martins; Claudia Renata dos Santos Barros; David Schlesinger; Debora Botequio Moretti; Dimas Tadeu Covas; Elaine Cristina Marqueze; Elaine Vieira Santos; Evandra Strazza Rodrigues; Heidge Fukumasu; Jayme Augusto de Souza-Neto; José Salvatore Leister Patané; Luiz Alcantara; Luiz Lehmann Coutinho; Maria Carolina Elias; Maurício Lacerda Nogueira; Rafael dos Santos Bezerra; Raul Machado Neto; Rejane Maria Tommasini Grotto; Ricardo Haddad; Sandra Coccuzzo Sampaio Vessoni; Simone Kashima; Svetoslav Nanev Slavov; Vincent Louis Viala |
| EPI_ISL_5649180, EPI_ISL_5649202, EPI_ISL_5649233 | VIGILANCIA EPIDEMIOLOGICA E CONTROLE DE VETORES PIRASSUNUN | Instituto Butantan | Antonio Jorge Martins; Claudia Renata dos Santos Barros; David Schlesinger; Debora Botequio Moretti; Dimas Tadeu Covas; Elaine Cristina Marqueze; Elaine Vieira Santos; Evandra Strazza Rodrigues; Heidge Fukumasu; Jayme Augusto de Souza-Neto; José Salvatore Leister Patané; Luiz Alcantara; Luiz Lehmann Coutinho; Maria Carolina Elias; Maurício Lacerda Nogueira; Rafael dos Santos Bezerra; Raul Machado Neto; Rejane Maria Tommasini Grotto; Ricardo Haddad; Sandra Coccuzzo Sampaio Vessoni; Simone Kashima; Svetoslav Nanev Slavov; Vincent Louis Viala |
| EPI_ISL_5649269, EPI_ISL_5649280, EPI_ISL_5649285 | VIGILANCIA EPIDIMIOLOGICA DE ARACARIGUAMA | Instituto Butantan | Antonio Jorge Martins; Claudia Renata dos Santos Barros; David Schlesinger; Debora Botequio Moretti; Dimas Tadeu Covas; Elaine Cristina Marqueze; Elaine Vieira Santos; Evandra Strazza Rodrigues; Heidge Fukumasu; Jayme Augusto de Souza-Neto; José Salvatore Leister Patané; Luiz Alcantara; Luiz Lehmann Coutinho; Maria Carolina Elias; Maurício Lacerda Nogueira; Rafael dos Santos Bezerra; Raul Machado Neto; Rejane Maria Tommasini Grotto; Ricardo Haddad; Sandra Coccuzzo Sampaio Vessoni; Simone Kashima; Svetoslav Nanev Slavov; Vincent Louis Viala |
| EPI_ISL_5648916, EPI_ISL_5648952, EPI_ISL_5649009, EPI_ISL_5649010, EPI_ISL_5649041, EPI_ISL_5649062, EPI_ISL_5649093, EPI_ISL_5649094, EPI_ISL_5649129, EPI_ISL_5649295 | | | |
| see above | VIGILANCIA SANITARIA E VIG EPIDEMIOLOGICA DE ITAPEVI | Instituto Butantan | Antonio Jorge Martins; Claudia Renata dos Santos Barros; David Schlesinger; Debora Botequio Moretti; Dimas Tadeu Covas; Elaine Cristina Marqueze; Elaine Vieira Santos; Evandra Strazza Rodrigues; Heidge Fukumasu; Jayme Augusto de Souza-Neto; José Salvatore Leister Patané; Luiz Alcantara; Luiz Lehmann Coutinho; Maria Carolina Elias; Maurício Lacerda Nogueira; Rafael dos Santos Bezerra; Raul Machado Neto; Rejane Maria Tommasini Grotto; Ricardo Haddad; Sandra Coccuzzo Sampaio Vessoni; Simone Kashima; Svetoslav Nanev Slavov; Vincent Louis Viala |
| EPI_ISL_5779915 | VUMC Molecular Infectious Diseases Laboratory (MIDL) | Dr. Suman Das Lab - Vanderbilt University Medical Center (VUMC) | Bookyung Park; Grant Vestal; Helen Boone; Hunter Brown; Jonathan E. Schmitz; Meghan Shilts; Seesandra Rajagopala; Suman B. Pakala; Suman Das |
| EPI_ISL_3030121 | Viollier AG | Clinical Bacteriology | Adrian Egli; Alfredo Mari; Christiane Beckmann; Fanny Wegner; Hans Hirsch; Helena MB Seth-Smith; Julia Bielicki; Karoline Leuzinger; Manuel Battegay; Tim Roloff |
| EPI_ISL_2938392, EPI_ISL_2938447, EPI_ISL_3007610 | Viollier AG | Department of Biosystems Science and Engineering, ETH Zürich | Andrea Patrignani; Andrea Patrizia Salzmann; Andreia Cabral de Gouvea; Catharine Aquino; Chaoran Chen; Christiane Beckmann; Christoph Noppen; Daniel Ehrsam; Doris Popovic; Griffin White; Henriette Kurth; Isabel Stürmer; Ivan Topolsky; Jay Tracy; Kim Philipp Jablonski; Lara Fuhrmann; Laura Neff; Lennart Opitz; Maria Domenica Moccia; Maurice Redondo; Niko Beerenwinkel; Olivier Kobel; Ralph Schlapbach; Sarah Nadeau; Simon Grüter; Tanja Stadler; Timothy Sykes |
| EPI_ISL_2920813 | Virginia Division of Consolidated Laboratory Services | Virginia Division of Consolidated Laboratory Services | Virginia Division of Consolidated Laboratory Services |
| EPI_ISL_2759091 | Virology Unit, AOUP | Virology Unit | Marialinda Vatteroni; Mauro Pistello; Susi Frateschi |
| EPI_ISL_2759090 | Virology Unit, AOUP | Virology Unit, AOUP | Marialinda Vatteroni; Mauro Pistello; Susi Frateschi |
| EPI_ISL_2716469, EPI_ISL_2716470, EPI_ISL_2716471, EPI_ISL_2716472, EPI_ISL_3016350, EPI_ISL_3016356, EPI_ISL_3016357, EPI_ISL_3016389, EPI_ISL_4187760 | | | |
| see above | Washington State Department of Health Public Health Laboratories | Washington State Department of Health Public Health Laboratories | Avi Singh; Darren Lucas; Denny Russell; Drew MacKellar; Geoff Melly; Hannah Gray; Joenice Gonzalez; JohnAric Peterson; Philip Dykema; Rebecca Cao; Vanessa De Los Santos |
| EPI_ISL_2876174 | Wexner Medical Center | The Ohio State University College of Medicine | Koenig, S.; Seminetta, J. |
| EPI_ISL_2689761, EPI_ISL_2689762, EPI_ISL_2689763, EPI_ISL_2689768, EPI_ISL_2689769, EPI_ISL_2689770, EPI_ISL_2756526, EPI_ISL_2756532, EPI_ISL_2756534 | | | |
| see above | Wichita State University - Molecular | Kansas Health and Environmental | Ben Olsen; Jonathan Barnell; Katherine Wiggins; Mike Grose; and Phil Adam |

| | | | |
|---|---|---|---|
| | Diagnostics Lab | Lab | |
| EPI_ISL_3131949 | Wisconsin State Laboratory of Hygiene Communicable Disease Division | Wisconsin State Laboratory of Hygiene Communicable Disease Division | Abigail C. Shockey; Alicia J. Mooney; Erika M. Hanson; Kelsey R. Florek; Richard Griesser; Sara Wagner; Tonya Danz |
| EPI_ISL_2943171, EPI_ISL_2960293, EPI_ISL_3038891, EPI_ISL_3038893, EPI_ISL_3160613, EPI_ISL_3207429, EPI_ISL_4470576 | | | |
| see above | Wyoming Public Health Laboratory | Wyoming Public Health Laboratory | Ashley Norberg; Brian Dominguez; Cari Sloma; Channing Weber; Chayse Rowley; Elliot Thomasson; Jim Mildenberger; Marley Goetz; Robert Petit; Sam Britz; Taylor Fearing; and Rob Christensen |
