## Supplement 3 for "Rapid and Accurate Identification of SARS-CoV-2 Omicron Variants Using Droplet Digital PCR (RT-ddPCR)": All_0628-0704.pdf

We gratefully acknowledge the following Authors from the Originating laboratories responsible for obtaining the specimens, as well as the Submitting laboratories where the genome data were generated and shared via GISAID, on which this research is based.

All Submitters of data may be contacted directly via www.gisaid.org

Authors are sorted alphabetically.

| Accession ID | Originating Laboratory | Submitting Laboratory | Authors |
|---|---|---|---|
| EPI_ISL_5660054, EPI_ISL_5660190, EPI_ISL_5660322, EPI_ISL_5660358, EPI_ISL_5660400, EPI_ISL_5660407, EPI_ISL_5660462, EPI_ISL_5660467, EPI_ISL_5660576, EPI_ISL_5660603, EPI_ISL_5660648, EPI_ISL_5660656, EPI_ISL_5660739, EPI_ISL_5660766, EPI_ISL_5660769, EPI_ISL_5660772, EPI_ISL_5660889, EPI_ISL_5660897, EPI_ISL_5660899, EPI_ISL_5660933, EPI_ISL_5660940, EPI_ISL_5660966, EPI_ISL_5660987, EPI_ISL_5663724, EPI_ISL_5663736, EPI_ISL_5663800, EPI_ISL_5663808, EPI_ISL_5663834, EPI_ISL_5663852, EPI_ISL_5663911, EPI_ISL_5663935, EPI_ISL_5663971, EPI_ISL_5663990, EPI_ISL_5664007, EPI_ISL_5664009, EPI_ISL_5664048, EPI_ISL_5664056, EPI_ISL_5664096, EPI_ISL_5664200, EPI_ISL_5664372, EPI_ISL_5664396, EPI_ISL_5664412, EPI_ISL_5664417, EPI_ISL_5664490, EPI_ISL_5664516, EPI_ISL_5664536, EPI_ISL_5664562, EPI_ISL_5664656, EPI_ISL_5664687, EPI_ISL_5664696, EPI_ISL_5664704, EPI_ISL_5664728, EPI_ISL_5664734, EPI_ISL_5664741, EPI_ISL_5664784, EPI_ISL_5664794, EPI_ISL_5664803, EPI_ISL_5664874, EPI_ISL_5664879, EPI_ISL_5664888, EPI_ISL_5665005, EPI_ISL_5665041, EPI_ISL_5665045, EPI_ISL_5665052, EPI_ISL_5665057, EPI_ISL_5665078, EPI_ISL_5665087, EPI_ISL_5665098, EPI_ISL_5665142, EPI_ISL_5665149, EPI_ISL_5665199, EPI_ISL_5665209, EPI_ISL_5665217, EPI_ISL_5665239, EPI_ISL_5665264, EPI_ISL_5665306, EPI_ISL_5665314, EPI_ISL_5665318, EPI_ISL_5665365, EPI_ISL_5665380, EPI_ISL_5665389, EPI_ISL_5665526 | | | |
| see above | AFIP | Instituto Butantan | Antonio Jorge Martins; Claudia Renata dos Santos Barros; David Schlesinger; Debora Botequio Moretti; Dimas Tadeu Covas; Elaine Cristina Marqueze; Elaine Vieira Santos; Evandra Strazza Rodrigues; Heidge Fukumasu; Jayme Augusto de Souza-Neto; José Salvatore Leister Patané; Luiz Alcantara; Luiz Lehmann Coutinho; Maria Carolina Elias; Maurício Lacerda Nogueira; Rafael dos Santos Bezerra; Raul Machado Neto; Rejane Maria Tommasini Grotto; Ricardo Haddad; Sandra Coccuzzo Sampaio Vessoni; Simone Kashima; Svetoslav Nanev Slavov; Vincent Louis Viala |
| EPI_ISL_5915125, EPI_ISL_5915150, EPI_ISL_5915192, EPI_ISL_5915232, EPI_ISL_5915240, EPI_ISL_5915279, EPI_ISL_5915282, EPI_ISL_5915298, EPI_ISL_5915323 | | | |
| see above | AMALAB/FACISA/UFRN | WallauLab on behalf of Fiocruz COVID-19 Genomic Surveillance Network | Alexandre Freitas da Silva; Allan Roberto Dias Nunes; Antonio Marinho da Silva Neto; Cassia Docena; Constância Flávia Junqueira Ayres; Filipe Zimmer Dezordi; Gabriel Luz Wallau; Gustavo Barbosa de Lima; Joana Cristina Medeiros Tavares Marques; Katya Anaya Jacinto; Lais Ceschini Machado; Lilian Caroliny Amorim Silva; Marcelo Henrique dos Santos Paiva; Mariane dos Santos Duarte; Matheus Filgueira Bezerra; Sinval Pinto Brandão Filho |
| EPI_ISL_5667941, EPI_ISL_5668151 | AMB MEDICO DR FRANCISCO FELIX DE MENDONCA SAUDADE | Instituto Butantan | Antonio Jorge Martins; Claudia Renata dos Santos Barros; David Schlesinger; Debora Botequio Moretti; Dimas Tadeu Covas; Elaine Cristina Marqueze; Elaine Vieira Santos; Evandra Strazza Rodrigues; Heidge Fukumasu; Jayme Augusto de Souza-Neto; José Salvatore Leister Patané; Luiz Alcantara; Luiz Lehmann Coutinho; Maria Carolina Elias; Maurício Lacerda Nogueira; Rafael dos Santos Bezerra; Raul Machado Neto; Rejane Maria Tommasini Grotto; Ricardo Haddad; Sandra Coccuzzo Sampaio Vessoni; Simone Kashima; Svetoslav Nanev Slavov; Vincent Louis Viala |
| EPI_ISL_5669373, EPI_ISL_5669385, EPI_ISL_5669459, EPI_ISL_5669553, EPI_ISL_5669908, EPI_ISL_5670050, EPI_ISL_5670064, EPI_ISL_5670384 | | | |
| see above | AMBULATORIO CENTRAL DE ESPECIALIDADES ITATIBA | Instituto Butantan | Antonio Jorge Martins; Claudia Renata dos Santos Barros; David Schlesinger; Debora Botequio Moretti; Dimas Tadeu Covas; Elaine Cristina Marqueze; Elaine Vieira Santos; Evandra Strazza Rodrigues; Heidge Fukumasu; Jayme Augusto de Souza-Neto; José Salvatore Leister Patané; Luiz Alcantara; Luiz Lehmann Coutinho; Maria Carolina Elias; Maurício Lacerda Nogueira; Rafael dos Santos Bezerra; Raul Machado Neto; Rejane Maria Tommasini Grotto; Ricardo Haddad; Sandra Coccuzzo Sampaio Vessoni; Simone Kashima; Svetoslav Nanev Slavov; Vincent Louis Viala |
| EPI_ISL_5660372, EPI_ISL_5660508, EPI_ISL_5660922 | AMBULATORIO DE ESPECIALIDADE V E MOGI MIRIM | Instituto Butantan | Antonio Jorge Martins; Claudia Renata dos Santos Barros; David Schlesinger; Debora Botequio Moretti; Dimas Tadeu Covas; Elaine Cristina Marqueze; Elaine Vieira Santos; Evandra Strazza Rodrigues; Heidge Fukumasu; Jayme Augusto de Souza-Neto; José Salvatore Leister Patané; Luiz Alcantara; Luiz Lehmann Coutinho; Maria Carolina Elias; Maurício Lacerda Nogueira; Rafael dos Santos Bezerra; Raul Machado Neto; Rejane Maria Tommasini Grotto; Ricardo Haddad; Sandra Coccuzzo Sampaio Vessoni; Simone Kashima; Svetoslav Nanev Slavov; Vincent Louis Viala |
| EPI_ISL_5667803, EPI_ISL_5667887, EPI_ISL_5667902, EPI_ISL_5667983, EPI_ISL_5668145, EPI_ISL_5668253, EPI_ISL_5668359, EPI_ISL_5668375 | | | |
| see above | AMBULATORIO DR JOAO LANIA CENTRO JOSE BONIFACIO | Instituto Butantan | Antonio Jorge Martins; Claudia Renata dos Santos Barros; David Schlesinger; Debora Botequio Moretti; Dimas Tadeu Covas; Elaine Cristina Marqueze; Elaine Vieira Santos; Evandra Strazza Rodrigues; Heidge Fukumasu; Jayme Augusto de Souza-Neto; José Salvatore Leister Patané; Luiz Alcantara; Luiz Lehmann Coutinho; Maria Carolina Elias; Maurício Lacerda Nogueira; Rafael dos Santos Bezerra; Raul Machado Neto; Rejane Maria Tommasini Grotto; Ricardo Haddad; Sandra Coccuzzo Sampaio Vessoni; Simone Kashima; Svetoslav Nanev Slavov; Vincent Louis Viala |
| EPI_ISL_3298316 | AREA DE SALUD EL GUARCO | Inciensa, Instituto Costarricense de Investigación y Enseñanza en Nutrición y Salud | Adriana Godínez; Claudio Soto-Garita; Estela Cordero; Francisco Duarte; Hebleen Porras; Joselyn Prado & Mónica Charpentier; José Luis Vargas; Mariela Gutiérrez; Melany Calderón |
| EPI_ISL_3037822 | AREA DE SALUD PARAISO-CERVANTES | Inciensa, Instituto Costarricense de Investigación y Enseñanza en Nutrición y Salud | Adriana Godínez; Caterina Guzmán; Claudio Soto-Garita; Estela Cordero; Francisco Duarte; Hebleen Porras; José Luis Vargas; Mariela Gutiérrez Joselyn Prado; Melany Calderón; Nazareth Ruiz & Mónica Charpentier |
| EPI_ISL_2861866, EPI_ISL_2861882, EPI_ISL_2861888, EPI_ISL_2861892, EPI_ISL_2861896, EPI_ISL_2861902, EPI_ISL_3062912, EPI_ISL_3062917 | | | |
| see above | ASL Napoli 1 Centro | AMES Centro Polidiagnostico Strumentale S.r.l. | "Giovanni Savarese; Antonella Di Carlo; Antonio Fico; Eloisa Evangelista; Giovanni Savarese; Luigi D'Amore; Luisa Circelli; Maurizio D'Amora; Maurizio D'Amora Antonio Fico"; Monica Ianniello; Nadia Petrillo; Raffaella Ruggiero; Roberto Sirica |
| EPI_ISL_3006783 | AULSS 8 Berica | Istituto Zooprofilattico Sperimentale delle Venezie | Adelaide Milani; Alessia Schivo; Alice Fusaro; Ambra Pastori; Annalisa Salviato; Antonia Ricci; Calogero Terregino; Edoardo Giussani; Elisa Palumbo; Erika Giorgia Quaranta; Isabella Monne; Luca Tassoni |
| EPI_ISL_3039626 | AULSS 9 Scaligera - Legnago | Istituto Zooprofilattico Sperimentale delle Venezie | Adelaide Milani; Alessia Schivo; Alice Fusaro; Ambra Pastori; Annalisa Salviato; Antonia Ricci; Calogero Terregino; Edoardo Giussani; Elisa Palumbo; Erika Giorgia Quaranta; Isabella Monne; Luca Tassoni |
| EPI_ISL_2790111, EPI_ISL_2790112, EPI_ISL_2790130 | AZDelta | AZ Delta Medical Laboratories in Roeselare, Belgium | Dieter De Smet; Geert Martens; Merijn Vanhee; on behalf of AZ Delta COVID-19 Genomics core (member of Genomic surveillance of SARS-CoV-2 in Belgium network) |
| EPI_ISL_2876014, EPI_ISL_3217615, EPI_ISL_3217619, EPI_ISL_3220328, EPI_ISL_3220333, EPI_ISL_3220336, EPI_ISL_3220364, EPI_ISL_3220373, EPI_ISL_3220375, EPI_ISL_3220408, EPI_ISL_3220435, EPI_ISL_3220484, EPI_ISL_3220485, EPI_ISL_3220501, EPI_ISL_3220513, EPI_ISL_3220536, EPI_ISL_3220542, EPI_ISL_3220577, EPI_ISL_3220578, EPI_ISL_3220582, EPI_ISL_3220589, EPI_ISL_3220590, EPI_ISL_3220594, EPI_ISL_3220640, EPI_ISL_3220667, EPI_ISL_3220670, EPI_ISL_3220672, EPI_ISL_3220675, EPI_ISL_3220704, EPI_ISL_3220708, EPI_ISL_3220720, EPI_ISL_3220744, EPI_ISL_3220768, EPI_ISL_3220774, EPI_ISL_3220783, EPI_ISL_3220880, EPI_ISL_3220894, EPI_ISL_3220896, EPI_ISL_3220903, EPI_ISL_3220908, EPI_ISL_3220917, EPI_ISL_3220969, EPI_ISL_3220989, EPI_ISL_3221003, EPI_ISL_3221005, EPI_ISL_3221007, EPI_ISL_3221008, EPI_ISL_3221016, EPI_ISL_3221022, EPI_ISL_3221038, EPI_ISL_3221048, EPI_ISL_3305299, EPI_ISL_3305308, EPI_ISL_3305322, EPI_ISL_3305350, EPI_ISL_3321345, EPI_ISL_3321349, EPI_ISL_3321358, EPI_ISL_4362431, EPI_ISL_4366567 | | | |
| see above | Aegis Sciences Corporation | Centers for Disease Control and Prevention Division of Viral Diseases, Pathogen Discovery | Adrian Paskey; Alec Vest; Benjamin Rambo-Martin; Christopher Gulvick; Clinton Paden; Clinton R. Paden; Cyndi Clark; Dakota Howard; Darlene Wagner; Dhwani Batra; Dillon Nall; Duncan MacCannell; Erisa Sula; Ethan Sanders; Holly Houdeshell; Jason Caravas; Kara Moser; Kristine Lacek; Matthew Hardison; Matthew Schmerer; Ola Kvalvaag; Patrick Campbell; Peter Cook; Peter W. Cook; Rob Case; Scott Sammons; Shatavia Morrison; Shaun Westlund; Tymeckia Kendall; Victoria Caban Figueroa; Vikramsinha Ghorpade; Yvette Unoarumhi |
| EPI_ISL_4219144, EPI_ISL_4219151, EPI_ISL_4219154 | Alameda County Public Health Department | Alameda County Public Health Department | Maria Elma; Vici Varghese |
| EPI_ISL_2966419, EPI_ISL_2966439, EPI_ISL_2966445, EPI_ISL_3025339, EPI_ISL_3118590 | Alaska State Virology Laboratory | Alaska State Virology Laboratory | Elva House; Jack Chen; Jacob Zidek; Lisa Smith; Ph.D.; Stephanie DeRonde |
| EPI_ISL_7711714 | Area De Salud Colorado [Abangares/Guanacaste] | Universidad de Costa Rica e Instituto Costarricense de Investigacion y Ensenanza en Nutricion y Salud | Brenes Hebleen; Camacho Erwin; Campos-Sanchez Rebeca; Ceballos Ana; Cordero Estela; Cristancho Marco; Duarte Francisco; Fernandez-Do Porto Dario; Herrera-Estrella Alfredo; Jimenez-Moraila Beatriz; Kreuze Jan; Mireles-Rivera Guadalupe; Molina-Mora Jose; Munoz-Medina Jose Esteban; Negri Tatiana; Nunes Gisele; Oliveira Guilherme; Oliveira Renato; Reales-Gonzalez Jhonnatan; Remes-Lenicov Federico; Reyes Alejandro; Sosa Ezequiel; Soto Claudio; Tsukayama Pablo; Turjanski Adrian; and Zayat Jonathan |
| EPI_ISL_7711721 | Area De Salud Desamparados 1 - Clinica Dr. Marcial Fallas [Grifo Alto/Desamparados/San Jose] | Universidad de Costa Rica e Instituto Costarricense de Investigacion y Ensenanza en Nutricion y Salud | Brenes Hebleen; Camacho Erwin; Campos-Sanchez Rebeca; Ceballos Ana; Cordero Estela; Cristancho Marco; Duarte Francisco; Fernandez-Do Porto Dario; Herrera-Estrella Alfredo; Jimenez-Moraila Beatriz; Kreuze Jan; Mireles-Rivera Guadalupe; Molina-Mora Jose; Munoz-Medina Jose Esteban; Negri Tatiana; Nunes Gisele; Oliveira Guilherme; Oliveira Renato; Reales-Gonzalez Jhonnatan; Remes-Lenicov Federico; Reyes Alejandro; Sosa Ezequiel; Soto Claudio; Tsukayama Pablo; Turjanski Adrian; and Zayat Jonathan |
| EPI_ISL_7711737 | Area De Salud Desamparados 3 [Grifo Alto/Desamparados/San Jose] | Universidad de Costa Rica e Instituto Costarricense de Investigacion y Ensenanza en Nutricion y Salud | Brenes Hebleen; Camacho Erwin; Campos-Sanchez Rebeca; Ceballos Ana; Cordero Estela; Cristancho Marco; Duarte Francisco; Fernandez-Do Porto Dario; Herrera-Estrella Alfredo; Jimenez-Moraila Beatriz; Kreuze Jan; Mireles-Rivera Guadalupe; Molina-Mora Jose; Munoz-Medina Jose Esteban; Negri Tatiana; Nunes Gisele; Oliveira Guilherme; Oliveira Renato; Reales-Gonzalez Jhonnatan; Remes-Lenicov Federico; Reyes Alejandro; Sosa Ezequiel; Soto Claudio; Tsukayama Pablo; Turjanski Adrian; and Zayat Jonathan |
| EPI_ISL_7711722 | Area De Salud Esparza [Aguabuena/Esparza/Puntarenas] | Universidad de Costa Rica e Instituto Costarricense de Investigacion y Ensenanza en Nutricion y Salud | Brenes Hebleen; Camacho Erwin; Campos-Sanchez Rebeca; Ceballos Ana; Cordero Estela; Cristancho Marco; Duarte Francisco; Fernandez-Do Porto Dario; Herrera-Estrella Alfredo; Jimenez-Moraila Beatriz; Kreuze Jan; Mireles-Rivera Guadalupe; Molina-Mora Jose; Munoz-Medina Jose Esteban; Negri Tatiana; Nunes Gisele; Oliveira Guilherme; Oliveira Renato; Reales-Gonzalez Jhonnatan; Remes-Lenicov Federico; Reyes Alejandro; Sosa Ezequiel; Soto Claudio; Tsukayama Pablo; Turjanski Adrian; and Zayat Jonathan |
| EPI_ISL_7711724 | Area De Salud Fortuna [San Carlos/Alajuela] | Universidad de Costa Rica e Instituto Costarricense de Investigacion y Ensenanza en Nutricion y Salud | Brenes Hebleen; Camacho Erwin; Campos-Sanchez Rebeca; Ceballos Ana; Cordero Estela; Cristancho Marco; Duarte Francisco; Fernandez-Do Porto Dario; Herrera-Estrella Alfredo; Jimenez-Moraila Beatriz; Kreuze Jan; Mireles-Rivera Guadalupe; Molina-Mora Jose; Munoz-Medina Jose Esteban; Negri Tatiana; Nunes Gisele; Oliveira Guilherme; Oliveira Renato; Reales-Gonzalez Jhonnatan; Remes-Lenicov Federico; Reyes Alejandro; Sosa Ezequiel; Soto Claudio; Tsukayama Pablo; Turjanski Adrian; and Zayat Jonathan |
| EPI_ISL_7711656 | Area De Salud San Rafael [Puerto Jimenez/Puntarenas/Puntarenas] | Universidad de Costa Rica e Instituto Costarricense de Investigacion y Ensenanza en Nutricion y Salud | Brenes Hebleen; Camacho Erwin; Campos-Sanchez Rebeca; Ceballos Ana; Cordero Estela; Cristancho Marco; Duarte Francisco; Fernandez-Do Porto Dario; Herrera-Estrella Alfredo; Jimenez-Moraila Beatriz; Kreuze Jan; Mireles-Rivera Guadalupe; Molina-Mora Jose; Munoz-Medina Jose Esteban; Negri Tatiana; Nunes Gisele; Oliveira Guilherme; Oliveira Renato; Reales-Gonzalez Jhonnatan; Remes-Lenicov Federico; Reyes Alejandro; Sosa Ezequiel; Soto Claudio; Tsukayama Pablo; Turjanski Adrian; and Zayat Jonathan |
| EPI_ISL_7711720 | Area De Salud Santa Cruz | Universidad de Costa Rica e Instituto Costarricense de Investigacion y Ensenanza en Nutricion y Salud | Brenes Hebleen; Camacho Erwin; Campos-Sanchez Rebeca; Ceballos Ana; Cordero Estela; Cristancho Marco; Duarte Francisco; Fernandez-Do Porto Dario; Herrera-Estrella Alfredo; Jimenez-Moraila Beatriz; Kreuze Jan; Mireles-Rivera Guadalupe; Molina-Mora Jose; Munoz-Medina Jose Esteban; Negri Tatiana; Nunes Gisele; Oliveira Guilherme; Oliveira Renato; Reales-Gonzalez Jhonnatan; Remes-Lenicov Federico; Reyes Alejandro; Sosa Ezequiel; Soto Claudio; Tsukayama Pablo; Turjanski Adrian; and Zayat Jonathan |
| EPI_ISL_2919078, EPI_ISL_2919079, EPI_ISL_2919080, EPI_ISL_2919081, EPI_ISL_2919082, EPI_ISL_2919083, EPI_ISL_2919084, EPI_ISL_2919097, EPI_ISL_2943060 | | | |
| see above | Arizona State Public Health Laboratory | Arizona State Public Health Laboratory | Jessica Escobar; Katherine Fullerton; Linda Getsinger; Matthew Contursi; Nobuko Fukushima; Stacy White; Trung Huynh; Victor Waddell |
| EPI_ISL_2863900, EPI_ISL_2863925, EPI_ISL_2863926, EPI_ISL_2968651, | Arizona State University | Arizona State University | Efrem S. Lim; Joshua LaBaer; Joy M. Blain; LaRinda A. Holland; Matthew F. Smith; Nicholas J. Mellor; Peter T. Skidmore; Rabia Maqsood; Valerie Harris; Vel Murugan |

| | | | |
|---|---|---|---|
| EPI_ISL_2968652 | | | |
| EPI_ISL_3105039 | Atlas Genomics | Seattle Flu Study | Amanda Adler; Barry R. Lutz; Benjamin Pelle; Brian Hiatt; Caitlin R. Wolf; Chris D. Frazar; Deborah A. Nickerson; Elisabeth Brandstetter; Erica Ryke; Geoff Melly; Helen Y. Chu; Janet A. Englund; Jay Shendure; Jover Lee; Kairsten Fay; Kirsten Lacombe; Lea M. Starita; Mark J. Rieder; Matthew Richardson; Matthew Thompson; Melissa Truong; Michael Boeckh; Michael Famulare; Misja Ilcisin; Peter D. Han; Philip Dykema; Romesh Gautom; Scott Lindquist; Thomas R. Sibley; Trevor Bedford |
| EPI_ISL_2919122, EPI_ISL_2919123, EPI_ISL_2919125, EPI_ISL_2919126, EPI_ISL_2919127 | Azienda USL Umbria 2 | Istituto Zooprofilattico Sperimentale dell'Abruzzo e Molise "G. Caporale" | Ancora M; Calistri P; Cammà C; Curini V; Delli Compagni E; Di Domenico M; Di Pasquale A; Lorusso A; Mangone I; Marcacci M; Pistoni E; Proietti A; Puglia I; Rinaldi A; Savini G; Scialabba S |
| EPI_ISL_5659642, EPI_ISL_5659655, EPI_ISL_5659734, EPI_ISL_5659753, EPI_ISL_5659771, EPI_ISL_5659777, EPI_ISL_5659783, EPI_ISL_5659796, EPI_ISL_5659827, EPI_ISL_5659841, EPI_ISL_5659853, EPI_ISL_5659900, EPI_ISL_5659907, EPI_ISL_5659912, EPI_ISL_5659920, EPI_ISL_5659936, EPI_ISL_5659951, EPI_ISL_5659973, EPI_ISL_5659992, EPI_ISL_5660029, EPI_ISL_5660045, EPI_ISL_5660050, EPI_ISL_5660059, EPI_ISL_5660069, EPI_ISL_5660083, EPI_ISL_5660097, EPI_ISL_5660121, EPI_ISL_5660139, EPI_ISL_5660161, EPI_ISL_5660166, EPI_ISL_5660169, EPI_ISL_5660175, EPI_ISL_5660180, EPI_ISL_5660187, EPI_ISL_5660211, EPI_ISL_5660227, EPI_ISL_5660243, EPI_ISL_5660259, EPI_ISL_5660272, EPI_ISL_5660277, EPI_ISL_5660295, EPI_ISL_5660333, EPI_ISL_5660413, EPI_ISL_5660440, EPI_ISL_5660487, EPI_ISL_5660589, EPI_ISL_5660608, EPI_ISL_5660615, EPI_ISL_5660664, EPI_ISL_5660695, EPI_ISL_5660703, EPI_ISL_5660726, EPI_ISL_5660746, EPI_ISL_5660780, EPI_ISL_5660786, EPI_ISL_5660795, EPI_ISL_5660799, EPI_ISL_5660820, EPI_ISL_5660828, EPI_ISL_5660858, EPI_ISL_5660864, EPI_ISL_5660870, EPI_ISL_5660908, EPI_ISL_5660946, EPI_ISL_5660956, EPI_ISL_5660973, EPI_ISL_5660978, EPI_ISL_5660993, EPI_ISL_5660998, EPI_ISL_5661040, EPI_ISL_5661055, EPI_ISL_5661065, EPI_ISL_5661081, EPI_ISL_5661087, EPI_ISL_5661117, EPI_ISL_5661708, EPI_ISL_5661832, EPI_ISL_5661971, EPI_ISL_5662010, EPI_ISL_5662083, EPI_ISL_5662090, EPI_ISL_5662127, EPI_ISL_5662193, EPI_ISL_5662196, EPI_ISL_5662272, EPI_ISL_5662286, EPI_ISL_5662383, EPI_ISL_5662395, EPI_ISL_5662456, EPI_ISL_5662485, EPI_ISL_5662541, EPI_ISL_5662588, EPI_ISL_5662595, EPI_ISL_5662617, EPI_ISL_5662628, EPI_ISL_5662656, EPI_ISL_5662661, EPI_ISL_5662771, EPI_ISL_5662788, EPI_ISL_5662817, EPI_ISL_5662826, EPI_ISL_5662855, EPI_ISL_5662907, EPI_ISL_5662980, EPI_ISL_5663025, EPI_ISL_5663028, EPI_ISL_5663033, EPI_ISL_5663051, EPI_ISL_5663058, EPI_ISL_5663117, EPI_ISL_5663140, EPI_ISL_5663144, EPI_ISL_5663233, EPI_ISL_5663256, EPI_ISL_5663285 | | | |
| see above | BIOFAST | Instituto Butantan | Antonio Jorge Martins; Claudia Renata dos Santos Barros; David Schlesinger; Debora Botequio Moretti; Dimas Tadeu Covas; Elaine Cristina Marqueze; Elaine Vieira Santos; Evandra Strazza Rodrigues; Heidge Fukumasu; Jayme Augusto de Souza-Neto; José Salvatore Leister Patané; Luiz Alcantara; Luiz Lehmann Coutinho; Maria Carolina Elias; Maurício Lacerda Nogueira; Rafael dos Santos Bezerra; Raul Machado Neto; Rejane Maria Tommasini Grotto; Ricardo Haddad; Sandra Coccuzzo Sampaio Vessoni; Simone Kashima; Svetoslav Nanev Slavov; Vincent Louis Viala |
| EPI_ISL_2933141, EPI_ISL_2933143, EPI_ISL_2933184, EPI_ISL_2933186, EPI_ISL_2933364, EPI_ISL_2933366, EPI_ISL_2933367, EPI_ISL_2933368, EPI_ISL_2933369, EPI_ISL_2933370, EPI_ISL_2933371, EPI_ISL_2933372, EPI_ISL_2933373, EPI_ISL_2933374, EPI_ISL_2933375, EPI_ISL_2933376, EPI_ISL_2933387, EPI_ISL_2933398, EPI_ISL_2933399, EPI_ISL_2933404, EPI_ISL_2933429 | | | |
| see above | BIOMNIS LYON | CNR Virus des Infections Respiratoires - France SUD | Antonin Bal; Bruno Lina; Gregory Destras; Gwendolyne Burfin; Hadrien Regue; Laurence Josset; Martine Valette; Quentin Semanas |
| EPI_ISL_2957164 | Bayerisches Landesamt für Gesundheit und Lebensmittelsicherheit (LGL) | Robert Koch Institute | |
| EPI_ISL_2928245, EPI_ISL_2928247, EPI_ISL_2928257, EPI_ISL_3031587 | Baylor Scott & White-Temple | Baylor Scott & White-Temple | Ari Rao; Kimberly Walker; Linden Morales; Marcus Volz; Shelby Johnson |
| EPI_ISL_3145901, EPI_ISL_3145923, EPI_ISL_3145924, EPI_ISL_3145925, EPI_ISL_3145926, EPI_ISL_3145927, EPI_ISL_3145928, EPI_ISL_3145929, EPI_ISL_3145930, EPI_ISL_3145931, EPI_ISL_3145932, EPI_ISL_3145933, EPI_ISL_3145934, EPI_ISL_3145935, EPI_ISL_3145936, EPI_ISL_3145937, EPI_ISL_3145938, EPI_ISL_3145939, EPI_ISL_3145940, EPI_ISL_3145941, EPI_ISL_3145942, EPI_ISL_3145943, EPI_ISL_3145944, EPI_ISL_3145945, EPI_ISL_3145946, EPI_ISL_3145947, EPI_ISL_3145948, EPI_ISL_3145949, EPI_ISL_3145950, EPI_ISL_3145951, EPI_ISL_3145952, EPI_ISL_3145953, EPI_ISL_3145954, EPI_ISL_3145955, EPI_ISL_3145956, EPI_ISL_3145957, EPI_ISL_3145958, EPI_ISL_3145959, EPI_ISL_3145960, EPI_ISL_3145961, EPI_ISL_3145962, EPI_ISL_3145963, EPI_ISL_3145964, EPI_ISL_3145965, EPI_ISL_3145966, EPI_ISL_3145967, EPI_ISL_3145968, EPI_ISL_3145969, EPI_ISL_3145970, EPI_ISL_3145971, EPI_ISL_3145972, EPI_ISL_3145973, EPI_ISL_3145974, EPI_ISL_3145975, EPI_ISL_3145976, EPI_ISL_3145977, EPI_ISL_3145978, EPI_ISL_3145979, EPI_ISL_3145980, EPI_ISL_3145981, EPI_ISL_3145982, EPI_ISL_3145983, EPI_ISL_3145984, EPI_ISL_3145985, EPI_ISL_3145986, EPI_ISL_3145987, EPI_ISL_3145988, EPI_ISL_3145989, EPI_ISL_3145990, EPI_ISL_3145991, EPI_ISL_3145992, EPI_ISL_3145993, EPI_ISL_3145994, EPI_ISL_3145995, EPI_ISL_3145996, EPI_ISL_3145997, EPI_ISL_3145998, EPI_ISL_3145999, EPI_ISL_3146000, EPI_ISL_3146001, EPI_ISL_3146002, EPI_ISL_3146003, EPI_ISL_3146004, EPI_ISL_3146005, EPI_ISL_3146006, EPI_ISL_3146007, EPI_ISL_3146008, EPI_ISL_3146009, EPI_ISL_3146010, EPI_ISL_3146011, EPI_ISL_3146012, EPI_ISL_3146013, EPI_ISL_3146014, EPI_ISL_3146015, EPI_ISL_3146016, EPI_ISL_3146017, EPI_ISL_3146018, EPI_ISL_3146019, EPI_ISL_3146020, EPI_ISL_3146021, EPI_ISL_3146022, EPI_ISL_3146023, EPI_ISL_3146024, EPI_ISL_3146025, EPI_ISL_3146026, EPI_ISL_3146027, EPI_ISL_3146028, EPI_ISL_3146029, EPI_ISL_3146030, EPI_ISL_3146031, EPI_ISL_3146032, EPI_ISL_3146033, EPI_ISL_3146034, EPI_ISL_3146035, EPI_ISL_3146036, EPI_ISL_3146037, EPI_ISL_3146038, EPI_ISL_3146039, EPI_ISL_3146040, EPI_ISL_3146041, EPI_ISL_3146042, EPI_ISL_3146043, EPI_ISL_3146044, EPI_ISL_3146045, EPI_ISL_3146046, EPI_ISL_3146047, EPI_ISL_3146048, EPI_ISL_3146049, EPI_ISL_3146050, EPI_ISL_3146051, EPI_ISL_3146052, EPI_ISL_3146053, EPI_ISL_3146054, EPI_ISL_3146055, EPI_ISL_3146056, EPI_ISL_3146057, EPI_ISL_3146058, EPI_ISL_3146059, EPI_ISL_3146264, EPI_ISL_3146273, EPI_ISL_3146276, EPI_ISL_3146277, EPI_ISL_3146289, EPI_ISL_3146460, EPI_ISL_3146463, EPI_ISL_3146467, EPI_ISL_3146473, EPI_ISL_3146475, EPI_ISL_3146478, EPI_ISL_3146481, EPI_ISL_3146491, EPI_ISL_3146492, EPI_ISL_3146494, EPI_ISL_3146498, EPI_ISL_3146501, EPI_ISL_3146508, EPI_ISL_3146511, EPI_ISL_3146524, EPI_ISL_3146534, EPI_ISL_3146542, EPI_ISL_3146547 | | | |
| see above | BioneXt Lab | Laboratoire national de sante, Microbiology, Microbial Genomics Platform | Anke Wienecke-Baldacchino; Catherine Ragimbeau; Elodie Solarino; Fatu Djabi; Jessica Tapp; Lise Pignon; Raoul Salmon; Tamir Abdelrahman; Thibault Ferrandon; Virginie Jover |
| EPI_ISL_3596607, EPI_ISL_3596609, EPI_ISL_3596612, EPI_ISL_3596615, EPI_ISL_3596618, EPI_ISL_3596621, EPI_ISL_3596624, EPI_ISL_3596627, EPI_ISL_3602358, EPI_ISL_3602361, EPI_ISL_3602363, EPI_ISL_3602418, EPI_ISL_3602419, EPI_ISL_3602422, EPI_ISL_3602423, EPI_ISL_3602424, EPI_ISL_3602425, EPI_ISL_3602427, EPI_ISL_3602429, EPI_ISL_3602431, EPI_ISL_3602432, EPI_ISL_3602435, EPI_ISL_3602446, EPI_ISL_3602447, EPI_ISL_3602448, EPI_ISL_3602450, EPI_ISL_3602452, EPI_ISL_3602453, EPI_ISL_3602456, EPI_ISL_3602457, EPI_ISL_3602458, EPI_ISL_3602459, EPI_ISL_3602460, EPI_ISL_3602461, EPI_ISL_3602462, EPI_ISL_3602463, EPI_ISL_3602464, EPI_ISL_3602467, EPI_ISL_3602470, EPI_ISL_3602474, EPI_ISL_3602475, EPI_ISL_3602476, EPI_ISL_3602477, EPI_ISL_3602479, EPI_ISL_3602482, EPI_ISL_3602487, EPI_ISL_3602488, EPI_ISL_3602492, EPI_ISL_3602493, EPI_ISL_3602494, EPI_ISL_3602495, EPI_ISL_3602498, EPI_ISL_3602500, EPI_ISL_3602501, EPI_ISL_3602503, EPI_ISL_3602510, EPI_ISL_3602513, EPI_ISL_3602517, EPI_ISL_3602518, EPI_ISL_3602520, EPI_ISL_3602522, EPI_ISL_3602529, EPI_ISL_3602530, EPI_ISL_3602531, EPI_ISL_3602536, EPI_ISL_3602537, EPI_ISL_3602538, EPI_ISL_3602539, EPI_ISL_3602540, EPI_ISL_3602541, EPI_ISL_3602542, EPI_ISL_3602543, EPI_ISL_3602544, EPI_ISL_3602546, EPI_ISL_3602547, EPI_ISL_3602548, EPI_ISL_3602549, EPI_ISL_3602550, EPI_ISL_3602565, EPI_ISL_3602569, EPI_ISL_3602571, EPI_ISL_3602572, EPI_ISL_3602573, EPI_ISL_3602582, EPI_ISL_3602586, EPI_ISL_3602587, EPI_ISL_3602590, EPI_ISL_3602593, EPI_ISL_3602594, EPI_ISL_3602598, EPI_ISL_3602603, EPI_ISL_3602607, EPI_ISL_3602609, EPI_ISL_3602610, EPI_ISL_3602613, EPI_ISL_3602614, EPI_ISL_3602618, EPI_ISL_3602620, EPI_ISL_3602621, EPI_ISL_3602623, EPI_ISL_3602624, EPI_ISL_3602626, EPI_ISL_3602627, EPI_ISL_3602631, EPI_ISL_3602632, EPI_ISL_3602740, EPI_ISL_3602743 | | | |
| see above | British Columbia Centre For Disease Control | BCCDC Public Health Laboratory | Ana Pacagnella; Corrinne Ng; Dan Fornika; John Tyson; Kim Macdonald; Kimia Kamelian; Linda Hoang; Loretta Janz; Mel Krajden; Prystajecky Natalie; Robert Azana; Shannon Russell |
| EPI_ISL_3007332, EPI_ISL_3007337, EPI_ISL_3007356, EPI_ISL_3007358, EPI_ISL_3007381, EPI_ISL_3007411, EPI_ISL_3007427, EPI_ISL_3007453 | | | |
| see above | Broad Institute Clinical Research Sequencing Platform | Infectious Disease Program, Broad Institute of Harvard and MIT | Adams, G.; B.L.; B.W.; Bauer, M.; Birren; Blumenstiel, B.; Brown, C.; Carter, A.; Chaluvadi, S.; D.J.; DeFelice, M.; DeRuff, K.; Dodge, S.; Gabriel, S.; Gallagher, G.; Gladden-Young, A.; Granger, B.; J.E.; K.J.; Lagerborg, K.; Larkin, K.; Lee, M.; Lemieux; Lennon, N.; Loreth, C.; Madoff, L.; McGovern, S.; Meldrim, J.; Normandin, E.; P.C.; Park; Pearlman, L.; Reilly, S.; Rudy, M.; Sabeti; Siddle; Smole, S.; Tomkins-Tinch, C.; Vicente, G.; and MacInnis |
| EPI_ISL_5667268, EPI_ISL_5667282, EPI_ISL_5667490, EPI_ISL_5667495, EPI_ISL_5667698, EPI_ISL_5667723, EPI_ISL_5667776 | | | |
| see above | CADIP VILA REGINA FERNANDOPOLIS | Instituto Butantan | Antonio Jorge Martins; Claudia Renata dos Santos Barros; David Schlesinger; Debora Botequio Moretti; Dimas Tadeu Covas; Elaine Cristina Marqueze; Elaine Vieira Santos; Evandra Strazza Rodrigues; Heidge Fukumasu; Jayme Augusto de Souza-Neto; José Salvatore Leister Patané; Luiz Alcantara; Luiz Lehmann Coutinho; Maria Carolina Elias; Maurício Lacerda Nogueira; Rafael dos Santos Bezerra; Raul Machado Neto; Rejane Maria Tommasini Grotto; Ricardo Haddad; Sandra Coccuzzo Sampaio Vessoni; Simone Kashima; Svetoslav Nanev Slavov; Vincent Louis Viala |
| EPI_ISL_7673820, EPI_ISL_7673826, EPI_ISL_7673829, EPI_ISL_7673832, EPI_ISL_7673833, EPI_ISL_7673843, EPI_ISL_7673856 | | | |
| see above | CDPH VBL | California Department of Public Health | Emily Smith on behalf of CDPH-COVIDNet |
| EPI_ISL_3043910, EPI_ISL_3043944 | CENTOGENE Frankfurt Laboratory: Niederlassung Industriepark Höchst | Robert Koch Institute | |
| EPI_ISL_5659674, EPI_ISL_5660325, EPI_ISL_5660836, EPI_ISL_5661389, EPI_ISL_5661414 | CENTRO DE APOIO EPIDEMIOLOGICO | Instituto Butantan | Antonio Jorge Martins; Claudia Renata dos Santos Barros; David Schlesinger; Debora Botequio Moretti; Dimas Tadeu Covas; Elaine Cristina Marqueze; Elaine Vieira Santos; Evandra Strazza Rodrigues; Heidge Fukumasu; Jayme Augusto de Souza-Neto; José Salvatore Leister Patané; Luiz Alcantara; Luiz Lehmann Coutinho; Maria Carolina Elias; Maurício Lacerda Nogueira; Rafael dos Santos Bezerra; Raul Machado Neto; Rejane Maria Tommasini Grotto; Ricardo Haddad; Sandra Coccuzzo Sampaio Vessoni; Simone Kashima; Svetoslav Nanev Slavov; Vincent Louis Viala |
| EPI_ISL_5662089, EPI_ISL_5663064 | CENTRO DE ATENCAO AO IDOSO UNICO GALLAFRIO | Instituto Butantan | Antonio Jorge Martins; Claudia Renata dos Santos Barros; David Schlesinger; Debora Botequio Moretti; Dimas Tadeu Covas; Elaine Cristina Marqueze; Elaine Vieira Santos; Evandra Strazza Rodrigues; Heidge Fukumasu; Jayme Augusto de Souza-Neto; José Salvatore Leister Patané; Luiz Alcantara; Luiz Lehmann Coutinho; Maria Carolina Elias; Maurício Lacerda Nogueira; Rafael dos Santos Bezerra; Raul Machado Neto; Rejane Maria Tommasini Grotto; Ricardo Haddad; Sandra Coccuzzo Sampaio Vessoni; Simone Kashima; Svetoslav Nanev Slavov; Vincent Louis Viala |
| EPI_ISL_5666858, EPI_ISL_5666973, EPI_ISL_5667238 | CENTRO DE ATENDIMENTO PARA ENFRENTAMENTO A COVID 19 DE COSMO | Instituto Butantan | Antonio Jorge Martins; Claudia Renata dos Santos Barros; David Schlesinger; Debora Botequio Moretti; Dimas Tadeu Covas; Elaine Cristina Marqueze; Elaine Vieira Santos; Evandra Strazza Rodrigues; Heidge Fukumasu; Jayme Augusto de Souza-Neto; José Salvatore Leister Patané; Luiz Alcantara; Luiz Lehmann Coutinho; Maria Carolina Elias; Maurício Lacerda Nogueira; Rafael dos Santos Bezerra; Raul Machado Neto; Rejane Maria Tommasini Grotto; Ricardo Haddad; Sandra Coccuzzo Sampaio Vessoni; Simone Kashima; Svetoslav Nanev Slavov; Vincent Louis Viala |
| EPI_ISL_3912138, EPI_ISL_3912139 | CENTRO DE ATENDIMENTO PARA ENFRENTAMENTO AO COVID 19 | Analytical Competence Molecular Epidemiology Lab/ACME, Oswaldo Cruz Foundation, Ceara (FIOCRUZ CE) | Cleber Furtado Aksenen; Fabio Miyajima; Fernando Braga Stehling; Francisco Eder de Moura Lopes; Jamille Maria Mendes Bezerra; Joaquim Cesar do Nascimento Sousa Junior; Pedro Miguel Carneiro Jeronimo; Suzana Porto Almeida & Lucas Delerino on behalf of COVID-19 FIOCRUZ Genomic Network; Thais Ferreira de Oliveira; Thais de Oliveira Costa; Ticiane Cavalcante de Souza; Veridiana Pessoa Miyajima |
| EPI_ISL_5664195, EPI_ISL_5664773 | CENTRO DE ESPECIALIDADES DE PRIMAVERA | Instituto Butantan | Antonio Jorge Martins; Claudia Renata dos Santos Barros; David Schlesinger; Debora Botequio Moretti; Dimas Tadeu Covas; Elaine Cristina Marqueze; Elaine Vieira Santos; Evandra Strazza Rodrigues; Heidge Fukumasu; Jayme Augusto de Souza-Neto; José Salvatore Leister Patané; Luiz Alcantara; Luiz Lehmann Coutinho; Maria Carolina Elias; Maurício Lacerda Nogueira; Rafael dos Santos Bezerra; Raul Machado Neto; Rejane Maria Tommasini Grotto; Ricardo Haddad; Sandra Coccuzzo Sampaio Vessoni; Simone Kashima; Svetoslav Nanev Slavov; Vincent Louis Viala |
| EPI_ISL_5659686, EPI_ISL_5660203 | CENTRO DE ESPECIALIDADES MARIAS DA GLORIA | Instituto Butantan | Antonio Jorge Martins; Claudia Renata dos Santos Barros; David Schlesinger; Debora Botequio Moretti; Dimas Tadeu Covas; Elaine Cristina Marqueze; Elaine Vieira Santos; Evandra Strazza Rodrigues; Heidge Fukumasu; Jayme Augusto de Souza-Neto; José Salvatore Leister Patané; Luiz Alcantara; Luiz Lehmann Coutinho; Maria Carolina Elias; Maurício Lacerda Nogueira; Rafael dos Santos Bezerra; Raul Machado Neto; Rejane Maria Tommasini Grotto; Ricardo Haddad; Sandra Coccuzzo Sampaio Vessoni; Simone Kashima; Svetoslav Nanev Slavov; Vincent Louis Viala |
| EPI_ISL_5667307, EPI_ISL_5667331, EPI_ISL_5667452, EPI_ISL_5667467, EPI_ISL_5667661, EPI_ISL_5667684, EPI_ISL_5667686, EPI_ISL_5667692 | | | |
| see above | CENTRO DE ESPECIALIDADES MUNICIPAL CESAR TORREZAN R BONITO | Instituto Butantan | Antonio Jorge Martins; Claudia Renata dos Santos Barros; David Schlesinger; Debora Botequio Moretti; Dimas Tadeu Covas; Elaine Cristina Marqueze; Elaine Vieira Santos; Evandra Strazza Rodrigues; Heidge Fukumasu; Jayme Augusto de Souza-Neto; José Salvatore Leister Patané; Luiz Alcantara; Luiz Lehmann Coutinho; Maria Carolina Elias; Maurício Lacerda Nogueira; Rafael dos Santos Bezerra; Raul Machado Neto; Rejane Maria Tommasini Grotto; Ricardo Haddad; Sandra Coccuzzo Sampaio Vessoni; Simone Kashima; Svetoslav Nanev Slavov; Vincent Louis Viala |
| EPI_ISL_5661288, EPI_ISL_5661698, EPI_ISL_5661794, EPI_ISL_5662071, EPI_ISL_5662148, EPI_ISL_5662258, EPI_ISL_5662279, EPI_ISL_5662796, EPI_ISL_5662957, EPI_ISL_5663105, EPI_ISL_5663182 | | | |
| see above | CENTRO DE GENOMAS | Instituto Butantan | Antonio Jorge Martins; Claudia Renata dos Santos Barros; David Schlesinger; Debora Botequio Moretti; Dimas Tadeu Covas; Elaine Cristina Marqueze; Elaine Vieira Santos; Evandra Strazza Rodrigues; Heidge Fukumasu; Jayme Augusto de Souza-Neto; José Salvatore Leister Patané; Luiz Alcantara; Luiz Lehmann Coutinho; Maria Carolina Elias; Maurício Lacerda Nogueira; Rafael dos Santos Bezerra; Raul Machado Neto; Rejane Maria Tommasini Grotto; Ricardo Haddad; Sandra Coccuzzo Sampaio Vessoni; Simone Kashima; Svetoslav Nanev Slavov; Vincent Louis Viala |
| EPI_ISL_5666302, EPI_ISL_5666421 | CENTRO DE SAUDE DE ADAMANTINA | Instituto Butantan | Antonio Jorge Martins; Claudia Renata dos Santos Barros; David Schlesinger; Debora Botequio Moretti; Dimas Tadeu Covas; Elaine Cristina Marqueze; Elaine Vieira Santos; Evandra Strazza Rodrigues; Heidge Fukumasu; Jayme Augusto de Souza-Neto; José Salvatore Leister Patané; Luiz Alcantara; Luiz Lehmann Coutinho; Maria Carolina Elias; Maurício Lacerda Nogueira; Rafael dos Santos Bezerra; Raul Machado Neto; Rejane Maria Tommasini Grotto; Ricardo Haddad; Sandra Coccuzzo Sampaio Vessoni; Simone Kashima; Svetoslav Nanev Slavov; Vincent Louis Viala |
| EPI_ISL_5665870, EPI_ISL_5671382 | CENTRO DE SAUDE DE ANALANDIA | Instituto Butantan | Antonio Jorge Martins; Claudia Renata dos Santos Barros; David Schlesinger; Debora Botequio Moretti; Dimas Tadeu Covas; Elaine Cristina Marqueze; Elaine Vieira Santos; Evandra Strazza Rodrigues; Heidge Fukumasu; Jayme Augusto de Souza-Neto; José Salvatore Leister Patané; Luiz Alcantara; Luiz Lehmann Coutinho; Maria Carolina Elias; Maurício Lacerda Nogueira; Rafael dos Santos Bezerra; Raul Machado Neto; Rejane Maria Tommasini Grotto; Ricardo Haddad; Sandra Coccuzzo Sampaio Vessoni; Simone Kashima; Svetoslav Nanev Slavov; Vincent Louis Viala |
| EPI_ISL_5661373, EPI_ISL_5661655, EPI_ISL_5662598 | CENTRO DE SAUDE DE CHAVANTES | Instituto Butantan | Antonio Jorge Martins; Claudia Renata dos Santos Barros; David Schlesinger; Debora Botequio Moretti; Dimas Tadeu Covas; Elaine Cristina Marqueze; Elaine Vieira Santos; Evandra Strazza Rodrigues; Heidge Fukumasu; Jayme Augusto de Souza-Neto; José Salvatore Leister Patané; Luiz Alcantara; Luiz Lehmann Coutinho; Maria Carolina Elias; Maurício Lacerda Nogueira; Rafael dos Santos Bezerra; Raul Machado Neto; Rejane Maria Tommasini Grotto; Ricardo Haddad; Sandra Coccuzzo Sampaio Vessoni; Simone Kashima; Svetoslav Nanev Slavov; Vincent Louis Viala |
| EPI_ISL_5666398, EPI_ISL_5666522, EPI_ISL_5666527, EPI_ISL_5666538, EPI_ISL_5666545, EPI_ISL_5666592, EPI_ISL_5666613 | | | |
| see above | CENTRO DE SAUDE DE IPAUCU | Instituto Butantan | Antonio Jorge Martins; Claudia Renata dos Santos Barros; David Schlesinger; Debora Botequio Moretti; Dimas Tadeu Covas; Elaine Cristina Marqueze; Elaine Vieira Santos; Evandra Strazza Rodrigues; Heidge Fukumasu; Jayme Augusto de Souza-Neto; José Salvatore Leister Patané; Luiz Alcantara; Luiz Lehmann Coutinho; Maria Carolina Elias; Maurício Lacerda Nogueira; Rafael dos Santos Bezerra; Raul Machado Neto; Rejane Maria Tommasini Grotto; Ricardo Haddad; Sandra Coccuzzo Sampaio Vessoni; Simone Kashima; Svetoslav Nanev Slavov; Vincent Louis Viala |
| EPI_ISL_5661325, EPI_ISL_5661571, EPI_ISL_5661858, EPI_ISL_5662263 | CENTRO DE SAUDE DE MARACAI | Instituto Butantan | Antonio Jorge Martins; Claudia Renata dos Santos Barros; David Schlesinger; Debora Botequio Moretti; Dimas Tadeu Covas; Elaine Cristina Marqueze; Elaine Vieira Santos; Evandra Strazza Rodrigues; Heidge Fukumasu; Jayme Augusto de Souza-Neto; José Salvatore Leister Patané; Luiz Alcantara; Luiz Lehmann Coutinho; Maria Carolina Elias; Maurício Lacerda Nogueira; Rafael dos Santos Bezerra; Raul Machado Neto; Rejane Maria Tommasini Grotto; Ricardo Haddad; Sandra Coccuzzo Sampaio Vessoni; Simone Kashima; Svetoslav Nanev Slavov; Vincent Louis Viala |

| | | | |
|---|---|---|---|
| EPI_ISL_5663357 | CENTRO DE SAUDE DE MONTE CASTELO | Instituto Butantan | Antonio Jorge Martins; Claudia Renata dos Santos Barros; David Schlesinger; Debora Botequio Moretti; Dimas Tadeu Covas; Elaine Cristina Marqueze; Elaine Vieira Santos; Evandra Strazza Rodrigues; Heidge Fukumasu; Jayme Augusto de Souza-Neto; José Salvatore Leister Patané; Luiz Alcantara; Luiz Lehmann Coutinho; Maria Carolina Elias; Maurício Lacerda Nogueira; Rafael dos Santos Bezerra; Raul Machado Neto; Rejane Maria Tommasini Grotto; Ricardo Haddad; Sandra Coccuzzo Sampaio Vessoni; Simone Kashima; Svetoslav Nanev Slavov; Vincent Louis Viala |
| EPI_ISL_5661615, EPI_ISL_5661705 | CENTRO DE SAUDE DE OSVALDO CRUZ | Instituto Butantan | Antonio Jorge Martins; Claudia Renata dos Santos Barros; David Schlesinger; Debora Botequio Moretti; Dimas Tadeu Covas; Elaine Cristina Marqueze; Elaine Vieira Santos; Evandra Strazza Rodrigues; Heidge Fukumasu; Jayme Augusto de Souza-Neto; José Salvatore Leister Patané; Luiz Alcantara; Luiz Lehmann Coutinho; Maria Carolina Elias; Maurício Lacerda Nogueira; Rafael dos Santos Bezerra; Raul Machado Neto; Rejane Maria Tommasini Grotto; Ricardo Haddad; Sandra Coccuzzo Sampaio Vessoni; Simone Kashima; Svetoslav Nanev Slavov; Vincent Louis Viala |
| EPI_ISL_5666957, EPI_ISL_5667082 | CENTRO DE SAUDE DE PIRAJUI | Instituto Butantan | Antonio Jorge Martins; Claudia Renata dos Santos Barros; David Schlesinger; Debora Botequio Moretti; Dimas Tadeu Covas; Elaine Cristina Marqueze; Elaine Vieira Santos; Evandra Strazza Rodrigues; Heidge Fukumasu; Jayme Augusto de Souza-Neto; José Salvatore Leister Patané; Luiz Alcantara; Luiz Lehmann Coutinho; Maria Carolina Elias; Maurício Lacerda Nogueira; Rafael dos Santos Bezerra; Raul Machado Neto; Rejane Maria Tommasini Grotto; Ricardo Haddad; Sandra Coccuzzo Sampaio Vessoni; Simone Kashima; Svetoslav Nanev Slavov; Vincent Louis Viala |
| EPI_ISL_5661458 | CENTRO DE SAUDE DE PLATINA | Instituto Butantan | Antonio Jorge Martins; Claudia Renata dos Santos Barros; David Schlesinger; Debora Botequio Moretti; Dimas Tadeu Covas; Elaine Cristina Marqueze; Elaine Vieira Santos; Evandra Strazza Rodrigues; Heidge Fukumasu; Jayme Augusto de Souza-Neto; José Salvatore Leister Patané; Luiz Alcantara; Luiz Lehmann Coutinho; Maria Carolina Elias; Maurício Lacerda Nogueira; Rafael dos Santos Bezerra; Raul Machado Neto; Rejane Maria Tommasini Grotto; Ricardo Haddad; Sandra Coccuzzo Sampaio Vessoni; Simone Kashima; Svetoslav Nanev Slavov; Vincent Louis Viala |
| EPI_ISL_5666173 | CENTRO DE SAUDE DR MARIO DIAS DE AGUIAR CAPIVARI | Instituto Butantan | Antonio Jorge Martins; Claudia Renata dos Santos Barros; David Schlesinger; Debora Botequio Moretti; Dimas Tadeu Covas; Elaine Cristina Marqueze; Elaine Vieira Santos; Evandra Strazza Rodrigues; Heidge Fukumasu; Jayme Augusto de Souza-Neto; José Salvatore Leister Patané; Luiz Alcantara; Luiz Lehmann Coutinho; Maria Carolina Elias; Maurício Lacerda Nogueira; Rafael dos Santos Bezerra; Raul Machado Neto; Rejane Maria Tommasini Grotto; Ricardo Haddad; Sandra Coccuzzo Sampaio Vessoni; Simone Kashima; Svetoslav Nanev Slavov; Vincent Louis Viala |
| EPI_ISL_5663365, EPI_ISL_5663453, EPI_ISL_5663755, EPI_ISL_5664149 | CENTRO DE SAUDE II | Instituto Butantan | Antonio Jorge Martins; Claudia Renata dos Santos Barros; David Schlesinger; Debora Botequio Moretti; Dimas Tadeu Covas; Elaine Cristina Marqueze; Elaine Vieira Santos; Evandra Strazza Rodrigues; Heidge Fukumasu; Jayme Augusto de Souza-Neto; José Salvatore Leister Patané; Luiz Alcantara; Luiz Lehmann Coutinho; Maria Carolina Elias; Maurício Lacerda Nogueira; Rafael dos Santos Bezerra; Raul Machado Neto; Rejane Maria Tommasini Grotto; Ricardo Haddad; Sandra Coccuzzo Sampaio Vessoni; Simone Kashima; Svetoslav Nanev Slavov; Vincent Louis Viala |
| EPI_ISL_5666787, EPI_ISL_5666945, EPI_ISL_5667070, EPI_ISL_5667121, EPI_ISL_5667140 | CENTRO DE SAUDE II DE CAFELANDIA | Instituto Butantan | Antonio Jorge Martins; Claudia Renata dos Santos Barros; David Schlesinger; Debora Botequio Moretti; Dimas Tadeu Covas; Elaine Cristina Marqueze; Elaine Vieira Santos; Evandra Strazza Rodrigues; Heidge Fukumasu; Jayme Augusto de Souza-Neto; José Salvatore Leister Patané; Luiz Alcantara; Luiz Lehmann Coutinho; Maria Carolina Elias; Maurício Lacerda Nogueira; Rafael dos Santos Bezerra; Raul Machado Neto; Rejane Maria Tommasini Grotto; Ricardo Haddad; Sandra Coccuzzo Sampaio Vessoni; Simone Kashima; Svetoslav Nanev Slavov; Vincent Louis Viala |
| EPI_ISL_5667393, EPI_ISL_5667431, EPI_ISL_5667580, EPI_ISL_5667605, EPI_ISL_5667685, EPI_ISL_5668212 | CENTRO DE SAUDE II DE POTIRENDABA CESARINO BENFATTI | Instituto Butantan | Antonio Jorge Martins; Claudia Renata dos Santos Barros; David Schlesinger; Debora Botequio Moretti; Dimas Tadeu Covas; Elaine Cristina Marqueze; Elaine Vieira Santos; Evandra Strazza Rodrigues; Heidge Fukumasu; Jayme Augusto de Souza-Neto; José Salvatore Leister Patané; Luiz Alcantara; Luiz Lehmann Coutinho; Maria Carolina Elias; Maurício Lacerda Nogueira; Rafael dos Santos Bezerra; Raul Machado Neto; Rejane Maria Tommasini Grotto; Ricardo Haddad; Sandra Coccuzzo Sampaio Vessoni; Simone Kashima; Svetoslav Nanev Slavov; Vincent Louis Viala |
| EPI_ISL_5665816, EPI_ISL_5665844, EPI_ISL_5665927 | CENTRO DE SAUDE II DR JOSE PAIONE MOCOCA | Instituto Butantan | Antonio Jorge Martins; Claudia Renata dos Santos Barros; David Schlesinger; Debora Botequio Moretti; Dimas Tadeu Covas; Elaine Cristina Marqueze; Elaine Vieira Santos; Evandra Strazza Rodrigues; Heidge Fukumasu; Jayme Augusto de Souza-Neto; José Salvatore Leister Patané; Luiz Alcantara; Luiz Lehmann Coutinho; Maria Carolina Elias; Maurício Lacerda Nogueira; Rafael dos Santos Bezerra; Raul Machado Neto; Rejane Maria Tommasini Grotto; Ricardo Haddad; Sandra Coccuzzo Sampaio Vessoni; Simone Kashima; Svetoslav Nanev Slavov; Vincent Louis Viala |
| EPI_ISL_5671064 | CENTRO DE SAUDE III DE AGUAS DE SAO PEDRO | Instituto Butantan | Antonio Jorge Martins; Claudia Renata dos Santos Barros; David Schlesinger; Debora Botequio Moretti; Dimas Tadeu Covas; Elaine Cristina Marqueze; Elaine Vieira Santos; Evandra Strazza Rodrigues; Heidge Fukumasu; Jayme Augusto de Souza-Neto; José Salvatore Leister Patané; Luiz Alcantara; Luiz Lehmann Coutinho; Maria Carolina Elias; Maurício Lacerda Nogueira; Rafael dos Santos Bezerra; Raul Machado Neto; Rejane Maria Tommasini Grotto; Ricardo Haddad; Sandra Coccuzzo Sampaio Vessoni; Simone Kashima; Svetoslav Nanev Slavov; Vincent Louis Viala |
| EPI_ISL_5665858, EPI_ISL_5665863, EPI_ISL_5671192 | CENTRO MEDICO DR NELSON SALOME DE CONCHAL | Instituto Butantan | Antonio Jorge Martins; Claudia Renata dos Santos Barros; David Schlesinger; Debora Botequio Moretti; Dimas Tadeu Covas; Elaine Cristina Marqueze; Elaine Vieira Santos; Evandra Strazza Rodrigues; Heidge Fukumasu; Jayme Augusto de Souza-Neto; José Salvatore Leister Patané; Luiz Alcantara; Luiz Lehmann Coutinho; Maria Carolina Elias; Maurício Lacerda Nogueira; Rafael dos Santos Bezerra; Raul Machado Neto; Rejane Maria Tommasini Grotto; Ricardo Haddad; Sandra Coccuzzo Sampaio Vessoni; Simone Kashima; Svetoslav Nanev Slavov; Vincent Louis Viala |
| EPI_ISL_5665764 | CENTRO SAUDE III DR JOSE TADEU MOURAO | Instituto Butantan | Antonio Jorge Martins; Claudia Renata dos Santos Barros; David Schlesinger; Debora Botequio Moretti; Dimas Tadeu Covas; Elaine Cristina Marqueze; Elaine Vieira Santos; Evandra Strazza Rodrigues; Heidge Fukumasu; Jayme Augusto de Souza-Neto; José Salvatore Leister Patané; Luiz Alcantara; Luiz Lehmann Coutinho; Maria Carolina Elias; Maurício Lacerda Nogueira; Rafael dos Santos Bezerra; Raul Machado Neto; Rejane Maria Tommasini Grotto; Ricardo Haddad; Sandra Coccuzzo Sampaio Vessoni; Simone Kashima; Svetoslav Nanev Slavov; Vincent Louis Viala |
| EPI_ISL_2933185, EPI_ISL_2933380 | CERBALLIANCE MIDI-PYRENEES | CNR Virus des Infections Respiratoires - France SUD | Antonin Bal; Bruno Lina; Gregory Destras; Gwendolyne Burfin; Hadrien Regue; Laurence Josset; Martine Valette; Quentin Semanas |
| EPI_ISL_3710614 | CHU Purpan - Laboratoire de Virologie - Institut Fédératif de Biologie | CHU Purpan - Laboratoire de Virologie - Institut Fédératif de Biologie | Boyer P.; Carcenac R.; Ferrer V.; Harter A.; Izopet J.; Jeanne N.; Latour J.; Ranger N.; Tremeaux R. |
| EPI_ISL_2978237 | CHUV | Laboratory of genomics and metagenomics | Claire Bertelli; Damien Jacot; Gilbert Greub; SÃ©bastien Aeby; Trestan Pillonel |
| EPI_ISL_2957044 | CLILAB | Microbiology Department | Aida Gonzalez-Diaz; Carmen Ardanuy; Jordi Camara; Jordi Niubó; Laura Calatayud; M Angeles Domínguez; Miguel Fernandez-Huerta; Sara Marti |
| EPI_ISL_5150517 | CLINICA MEDIHELP SERVICES | UNIMOL | Doris Gómez-Camargo; Jaison Torres-Pacheco; Mashiel Fernández-Ruiz; Steev Loyola |
| EPI_ISL_2983480 | CNR Institut Pasteur de la Guyane | Institut Pasteur de la Guyane | Anne Lavergne; Antoine Enfissi; Arielle Salmier; Dominique Rousset |
| EPI_ISL_5665782, EPI_ISL_5666166 | COORDENADORIA MUNICIPAL DE SAUDE DE IRACEMAPOLIS | Instituto Butantan | Antonio Jorge Martins; Claudia Renata dos Santos Barros; David Schlesinger; Debora Botequio Moretti; Dimas Tadeu Covas; Elaine Cristina Marqueze; Elaine Vieira Santos; Evandra Strazza Rodrigues; Heidge Fukumasu; Jayme Augusto de Souza-Neto; José Salvatore Leister Patané; Luiz Alcantara; Luiz Lehmann Coutinho; Maria Carolina Elias; Maurício Lacerda Nogueira; Rafael dos Santos Bezerra; Raul Machado Neto; Rejane Maria Tommasini Grotto; Ricardo Haddad; Sandra Coccuzzo Sampaio Vessoni; Simone Kashima; Svetoslav Nanev Slavov; Vincent Louis Viala |
| EPI_ISL_5666317, EPI_ISL_5666367, EPI_ISL_5666384, EPI_ISL_5666446, EPI_ISL_5666462, EPI_ISL_5666552 | CS ALVINLANDIA | Instituto Butantan | Antonio Jorge Martins; Claudia Renata dos Santos Barros; David Schlesinger; Debora Botequio Moretti; Dimas Tadeu Covas; Elaine Cristina Marqueze; Elaine Vieira Santos; Evandra Strazza Rodrigues; Heidge Fukumasu; Jayme Augusto de Souza-Neto; José Salvatore Leister Patané; Luiz Alcantara; Luiz Lehmann Coutinho; Maria Carolina Elias; Maurício Lacerda Nogueira; Rafael dos Santos Bezerra; Raul Machado Neto; Rejane Maria Tommasini Grotto; Ricardo Haddad; Sandra Coccuzzo Sampaio Vessoni; Simone Kashima; Svetoslav Nanev Slavov; Vincent Louis Viala |
| EPI_ISL_5667445, EPI_ISL_5667654 | CS DE COSMORAMA | Instituto Butantan | Antonio Jorge Martins; Claudia Renata dos Santos Barros; David Schlesinger; Debora Botequio Moretti; Dimas Tadeu Covas; Elaine Cristina Marqueze; Elaine Vieira Santos; Evandra Strazza Rodrigues; Heidge Fukumasu; Jayme Augusto de Souza-Neto; José Salvatore Leister Patané; Luiz Alcantara; Luiz Lehmann Coutinho; Maria Carolina Elias; Maurício Lacerda Nogueira; Rafael dos Santos Bezerra; Raul Machado Neto; Rejane Maria Tommasini Grotto; Ricardo Haddad; Sandra Coccuzzo Sampaio Vessoni; Simone Kashima; Svetoslav Nanev Slavov; Vincent Louis Viala |
| EPI_ISL_5666312, EPI_ISL_5666403, EPI_ISL_5667325, EPI_ISL_5667457, EPI_ISL_5667507, EPI_ISL_5667543, EPI_ISL_5667652 | | | |
| see above | CS DE ESTRELA DOESTE | Instituto Butantan | Antonio Jorge Martins; Claudia Renata dos Santos Barros; David Schlesinger; Debora Botequio Moretti; Dimas Tadeu Covas; Elaine Cristina Marqueze; Elaine Vieira Santos; Evandra Strazza Rodrigues; Heidge Fukumasu; Jayme Augusto de Souza-Neto; José Salvatore Leister Patané; Luiz Alcantara; Luiz Lehmann Coutinho; Maria Carolina Elias; Maurício Lacerda Nogueira; Rafael dos Santos Bezerra; Raul Machado Neto; Rejane Maria Tommasini Grotto; Ricardo Haddad; Sandra Coccuzzo Sampaio Vessoni; Simone Kashima; Svetoslav Nanev Slavov; Vincent Louis Viala |
| EPI_ISL_5666322, EPI_ISL_5666517, EPI_ISL_5666619, EPI_ISL_5666629 | CS DE GENERAL SALGADO | Instituto Butantan | Antonio Jorge Martins; Claudia Renata dos Santos Barros; David Schlesinger; Debora Botequio Moretti; Dimas Tadeu Covas; Elaine Cristina Marqueze; Elaine Vieira Santos; Evandra Strazza Rodrigues; Heidge Fukumasu; Jayme Augusto de Souza-Neto; José Salvatore Leister Patané; Luiz Alcantara; Luiz Lehmann Coutinho; Maria Carolina Elias; Maurício Lacerda Nogueira; Rafael dos Santos Bezerra; Raul Machado Neto; Rejane Maria Tommasini Grotto; Ricardo Haddad; Sandra Coccuzzo Sampaio Vessoni; Simone Kashima; Svetoslav Nanev Slavov; Vincent Louis Viala |
| EPI_ISL_5660317, EPI_ISL_5660670, EPI_ISL_5661123, EPI_ISL_5667526, EPI_ISL_5667593 | CS DE ICEM | Instituto Butantan | Antonio Jorge Martins; Claudia Renata dos Santos Barros; David Schlesinger; Debora Botequio Moretti; Dimas Tadeu Covas; Elaine Cristina Marqueze; Elaine Vieira Santos; Evandra Strazza Rodrigues; Heidge Fukumasu; Jayme Augusto de Souza-Neto; José Salvatore Leister Patané; Luiz Alcantara; Luiz Lehmann Coutinho; Maria Carolina Elias; Maurício Lacerda Nogueira; Rafael dos Santos Bezerra; Raul Machado Neto; Rejane Maria Tommasini Grotto; Ricardo Haddad; Sandra Coccuzzo Sampaio Vessoni; Simone Kashima; Svetoslav Nanev Slavov; Vincent Louis Viala |
| EPI_ISL_5668402 | CS DE MARAPOAMA | Instituto Butantan | Antonio Jorge Martins; Claudia Renata dos Santos Barros; David Schlesinger; Debora Botequio Moretti; Dimas Tadeu Covas; Elaine Cristina Marqueze; Elaine Vieira Santos; Evandra Strazza Rodrigues; Heidge Fukumasu; Jayme Augusto de Souza-Neto; José Salvatore Leister Patané; Luiz Alcantara; Luiz Lehmann Coutinho; Maria Carolina Elias; Maurício Lacerda Nogueira; Rafael dos Santos Bezerra; Raul Machado Neto; Rejane Maria Tommasini Grotto; Ricardo Haddad; Sandra Coccuzzo Sampaio Vessoni; Simone Kashima; Svetoslav Nanev Slavov; Vincent Louis Viala |
| EPI_ISL_5668394 | CS DE MIRASSOLANDIA | Instituto Butantan | Antonio Jorge Martins; Claudia Renata dos Santos Barros; David Schlesinger; Debora Botequio Moretti; Dimas Tadeu Covas; Elaine Cristina Marqueze; Elaine Vieira Santos; Evandra Strazza Rodrigues; Heidge Fukumasu; Jayme Augusto de Souza-Neto; José Salvatore Leister Patané; Luiz Alcantara; Luiz Lehmann Coutinho; Maria Carolina Elias; Maurício Lacerda Nogueira; Rafael dos Santos Bezerra; Raul Machado Neto; Rejane Maria Tommasini Grotto; Ricardo Haddad; Sandra Coccuzzo Sampaio Vessoni; Simone Kashima; Svetoslav Nanev Slavov; Vincent Louis Viala |
| EPI_ISL_5660447, EPI_ISL_5666208, EPI_ISL_5666237, EPI_ISL_5666253, EPI_ISL_5666390, EPI_ISL_5666414, EPI_ISL_5666540, EPI_ISL_5666569, EPI_ISL_5666608, EPI_ISL_5667315 | | | |
| see above | CS DE MONTE APRAZIVEL | Instituto Butantan | Antonio Jorge Martins; Claudia Renata dos Santos Barros; David Schlesinger; Debora Botequio Moretti; Dimas Tadeu Covas; Elaine Cristina Marqueze; Elaine Vieira Santos; Evandra Strazza Rodrigues; Heidge Fukumasu; Jayme Augusto de Souza-Neto; José Salvatore Leister Patané; Luiz Alcantara; Luiz Lehmann Coutinho; Maria Carolina Elias; Maurício Lacerda Nogueira; Rafael dos Santos Bezerra; Raul Machado Neto; Rejane Maria Tommasini Grotto; Ricardo Haddad; Sandra Coccuzzo Sampaio Vessoni; Simone Kashima; Svetoslav Nanev Slavov; Vincent Louis Viala |
| EPI_ISL_5666184, EPI_ISL_5666189, EPI_ISL_5666274, EPI_ISL_5666281, EPI_ISL_5666469, EPI_ISL_5666479, EPI_ISL_5666494 | | | |
| see above | CS DE PALESTINA | Instituto Butantan | Antonio Jorge Martins; Claudia Renata dos Santos Barros; David Schlesinger; Debora Botequio Moretti; Dimas Tadeu Covas; Elaine Cristina Marqueze; Elaine Vieira Santos; Evandra Strazza Rodrigues; Heidge Fukumasu; Jayme Augusto de Souza-Neto; José Salvatore Leister Patané; Luiz Alcantara; Luiz Lehmann Coutinho; Maria Carolina Elias; Maurício Lacerda Nogueira; Rafael dos Santos Bezerra; Raul Machado Neto; Rejane Maria Tommasini Grotto; Ricardo Haddad; Sandra Coccuzzo Sampaio Vessoni; Simone Kashima; Svetoslav Nanev Slavov; Vincent Louis Viala |
| EPI_ISL_5664113, EPI_ISL_5664288, EPI_ISL_5664528, EPI_ISL_5664607, EPI_ISL_5665027 | CS DE ROSANA | Instituto Butantan | Antonio Jorge Martins; Claudia Renata dos Santos Barros; David Schlesinger; Debora Botequio Moretti; Dimas Tadeu Covas; Elaine Cristina Marqueze; Elaine Vieira Santos; Evandra Strazza Rodrigues; Heidge Fukumasu; Jayme Augusto de Souza-Neto; José Salvatore Leister Patané; Luiz Alcantara; Luiz Lehmann Coutinho; Maria Carolina Elias; Maurício Lacerda Nogueira; Rafael dos Santos Bezerra; Raul Machado Neto; Rejane Maria Tommasini Grotto; Ricardo Haddad; Sandra Coccuzzo Sampaio Vessoni; Simone Kashima; Svetoslav Nanev Slavov; Vincent Louis Viala |
| EPI_ISL_5666564, EPI_ISL_5666585 | CS DE SANTA ALBERTINA | Instituto Butantan | Antonio Jorge Martins; Claudia Renata dos Santos Barros; David Schlesinger; Debora Botequio Moretti; Dimas Tadeu Covas; Elaine Cristina Marqueze; Elaine Vieira Santos; Evandra Strazza Rodrigues; Heidge Fukumasu; Jayme Augusto de Souza-Neto; José Salvatore Leister Patané; Luiz Alcantara; Luiz Lehmann Coutinho; Maria Carolina Elias; Maurício Lacerda Nogueira; Rafael dos Santos Bezerra; Raul Machado Neto; Rejane Maria Tommasini Grotto; Ricardo Haddad; Sandra Coccuzzo Sampaio Vessoni; Simone Kashima; Svetoslav Nanev Slavov; Vincent Louis Viala |
| EPI_ISL_5666342 | CS DE TURMALINA | Instituto Butantan | Antonio Jorge Martins; Claudia Renata dos Santos Barros; David Schlesinger; Debora Botequio Moretti; Dimas Tadeu Covas; Elaine Cristina Marqueze; Elaine Vieira Santos; Evandra Strazza Rodrigues; Heidge Fukumasu; Jayme Augusto de Souza-Neto; José Salvatore Leister Patané; Luiz Alcantara; Luiz Lehmann Coutinho; Maria Carolina Elias; Maurício Lacerda Nogueira; Rafael dos Santos Bezerra; Raul Machado Neto; Rejane Maria Tommasini Grotto; Ricardo Haddad; Sandra Coccuzzo Sampaio Vessoni; Simone Kashima; Svetoslav Nanev Slavov; Vincent Louis Viala |
| EPI_ISL_5667687 | CS DE UNIAO PAULISTA | Instituto Butantan | Antonio Jorge Martins; Claudia Renata dos Santos Barros; David Schlesinger; Debora Botequio Moretti; Dimas Tadeu Covas; Elaine Cristina Marqueze; Elaine Vieira Santos; Evandra Strazza Rodrigues; Heidge Fukumasu; Jayme Augusto de Souza-Neto; José Salvatore Leister Patané; Luiz Alcantara; Luiz Lehmann Coutinho; Maria Carolina Elias; Maurício Lacerda Nogueira; Rafael dos Santos Bezerra; Raul Machado Neto; Rejane Maria Tommasini Grotto; Ricardo Haddad; Sandra Coccuzzo Sampaio Vessoni; Simone Kashima; Svetoslav Nanev Slavov; Vincent Louis Viala |
| EPI_ISL_5659660, EPI_ISL_5659819, EPI_ISL_5659939, EPI_ISL_5660106, EPI_ISL_5660147, EPI_ISL_5660631, EPI_ISL_5660878, EPI_ISL_5660981 | | | |
| see above | CS DE URANIA | Instituto Butantan | Antonio Jorge Martins; Claudia Renata dos Santos Barros; David Schlesinger; Debora Botequio Moretti; Dimas Tadeu Covas; Elaine Cristina Marqueze; Elaine Vieira Santos; Evandra Strazza Rodrigues; Heidge Fukumasu; Jayme Augusto de Souza-Neto; José Salvatore Leister Patané; Luiz Alcantara; Luiz Lehmann Coutinho; Maria Carolina Elias; Maurício Lacerda Nogueira; Rafael dos Santos Bezerra; Raul Machado Neto; Rejane Maria Tommasini Grotto; Ricardo Haddad; Sandra Coccuzzo Sampaio Vessoni; Simone Kashima; Svetoslav Nanev Slavov; Vincent Louis Viala |
| EPI_ISL_5666400, EPI_ISL_5666427, | CS DE VALENTIM GENTIL | Instituto Butantan | Antonio Jorge Martins; Claudia Renata dos Santos Barros; David Schlesinger; Debora Botequio Moretti; Dimas Tadeu Covas; Elaine Cristina Marqueze; Elaine Vieira Santos; Evandra Strazza Rodrigues; Heidge Fukumasu; Jayme Augusto de Souza-Neto; José Salvatore Leister Patané; Luiz Alcantara; Luiz Lehmann Coutinho; Maria Carolina Elias; Maurício Lacerda Nogueira; Rafael dos Santos Bezerra; Raul Machado Neto; Rejane Maria Tommasini Grotto; Ricardo Haddad; Sandra Coccuzzo Sampaio Vessoni; Simone Kashima; Svetoslav Nanev Slavov; Vincent Louis Viala |

| | | | |
|---|---|---|---|
| EPI_ISL_5666509, EPI_ISL_5667368 | | | |
| EPI_ISL_5666728, EPI_ISL_5666737, EPI_ISL_5666948 | CS II DR JAHYR DE PAULA RIBEIRO GUARA | Instituto Butantan | Antonio Jorge Martins; Claudia Renata dos Santos Barros; David Schlesinger; Debora Botequio Moretti; Dimas Tadeu Covas; Elaine Cristina Marqueze; Elaine Vieira Santos; Evandra Strazza Rodrigues; Heidge Fukumasu; Jayme Augusto de Souza-Neto; José Salvatore Leister Patané; Luiz Alcantara; Luiz Lehmann Coutinho; Maria Carolina Elias; Maurício Lacerda Nogueira; Rafael dos Santos Bezerra; Raul Machado Neto; Rejane Maria Tommasini Grotto; Ricardo Haddad; Sandra Coccuzzo Sampaio Vessoni; Simone Kashima; Svetoslav Nanev Slavov; Vincent Louis Viala |
| EPI_ISL_5662948 | CS II DR JOSE CARQUEIJO SCRPARDO | Instituto Butantan | Antonio Jorge Martins; Claudia Renata dos Santos Barros; David Schlesinger; Debora Botequio Moretti; Dimas Tadeu Covas; Elaine Cristina Marqueze; Elaine Vieira Santos; Evandra Strazza Rodrigues; Heidge Fukumasu; Jayme Augusto de Souza-Neto; José Salvatore Leister Patané; Luiz Alcantara; Luiz Lehmann Coutinho; Maria Carolina Elias; Maurício Lacerda Nogueira; Rafael dos Santos Bezerra; Raul Machado Neto; Rejane Maria Tommasini Grotto; Ricardo Haddad; Sandra Coccuzzo Sampaio Vessoni; Simone Kashima; Svetoslav Nanev Slavov; Vincent Louis Viala |
| EPI_ISL_5666678 | CS II DR MIGUEL VITALIANO ORLANDIA | Instituto Butantan | Antonio Jorge Martins; Claudia Renata dos Santos Barros; David Schlesinger; Debora Botequio Moretti; Dimas Tadeu Covas; Elaine Cristina Marqueze; Elaine Vieira Santos; Evandra Strazza Rodrigues; Heidge Fukumasu; Jayme Augusto de Souza-Neto; José Salvatore Leister Patané; Luiz Alcantara; Luiz Lehmann Coutinho; Maria Carolina Elias; Maurício Lacerda Nogueira; Rafael dos Santos Bezerra; Raul Machado Neto; Rejane Maria Tommasini Grotto; Ricardo Haddad; Sandra Coccuzzo Sampaio Vessoni; Simone Kashima; Svetoslav Nanev Slavov; Vincent Louis Viala |
| EPI_ISL_5665888 | CS III DR ANTONIO ANADAO SSGRAMA | Instituto Butantan | Antonio Jorge Martins; Claudia Renata dos Santos Barros; David Schlesinger; Debora Botequio Moretti; Dimas Tadeu Covas; Elaine Cristina Marqueze; Elaine Vieira Santos; Evandra Strazza Rodrigues; Heidge Fukumasu; Jayme Augusto de Souza-Neto; José Salvatore Leister Patané; Luiz Alcantara; Luiz Lehmann Coutinho; Maria Carolina Elias; Maurício Lacerda Nogueira; Rafael dos Santos Bezerra; Raul Machado Neto; Rejane Maria Tommasini Grotto; Ricardo Haddad; Sandra Coccuzzo Sampaio Vessoni; Simone Kashima; Svetoslav Nanev Slavov; Vincent Louis Viala |
| EPI_ISL_5669442 | CSA COMPLEXO DE SAUDE ANHANGUERA LTDA | Instituto Butantan | Antonio Jorge Martins; Claudia Renata dos Santos Barros; David Schlesinger; Debora Botequio Moretti; Dimas Tadeu Covas; Elaine Cristina Marqueze; Elaine Vieira Santos; Evandra Strazza Rodrigues; Heidge Fukumasu; Jayme Augusto de Souza-Neto; José Salvatore Leister Patané; Luiz Alcantara; Luiz Lehmann Coutinho; Maria Carolina Elias; Maurício Lacerda Nogueira; Rafael dos Santos Bezerra; Raul Machado Neto; Rejane Maria Tommasini Grotto; Ricardo Haddad; Sandra Coccuzzo Sampaio Vessoni; Simone Kashima; Svetoslav Nanev Slavov; Vincent Louis Viala |
| EPI_ISL_5663512, EPI_ISL_5669219, EPI_ISL_5669252, EPI_ISL_5669398, EPI_ISL_5669767, EPI_ISL_5670382, EPI_ISL_5670407, EPI_ISL_5670464, EPI_ISL_5670517 | | | |
| see above | CSA COMPLEXO DE SAUDE ANHANGUERA VILA JAGUARA | Instituto Butantan | Antonio Jorge Martins; Claudia Renata dos Santos Barros; David Schlesinger; Debora Botequio Moretti; Dimas Tadeu Covas; Elaine Cristina Marqueze; Elaine Vieira Santos; Evandra Strazza Rodrigues; Heidge Fukumasu; Jayme Augusto de Souza-Neto; José Salvatore Leister Patané; Luiz Alcantara; Luiz Lehmann Coutinho; Maria Carolina Elias; Maurício Lacerda Nogueira; Rafael dos Santos Bezerra; Raul Machado Neto; Rejane Maria Tommasini Grotto; Ricardo Haddad; Sandra Coccuzzo Sampaio Vessoni; Simone Kashima; Svetoslav Nanev Slavov; Vincent Louis Viala |
| EPI_ISL_5663443 | CSII DR JOSE FERREIRA TELLES | Instituto Butantan | Antonio Jorge Martins; Claudia Renata dos Santos Barros; David Schlesinger; Debora Botequio Moretti; Dimas Tadeu Covas; Elaine Cristina Marqueze; Elaine Vieira Santos; Evandra Strazza Rodrigues; Heidge Fukumasu; Jayme Augusto de Souza-Neto; José Salvatore Leister Patané; Luiz Alcantara; Luiz Lehmann Coutinho; Maria Carolina Elias; Maurício Lacerda Nogueira; Rafael dos Santos Bezerra; Raul Machado Neto; Rejane Maria Tommasini Grotto; Ricardo Haddad; Sandra Coccuzzo Sampaio Vessoni; Simone Kashima; Svetoslav Nanev Slavov; Vincent Louis Viala |
| EPI_ISL_3023785, EPI_ISL_3061869, EPI_ISL_3151561 | CT Department of Public Health | CT Department of Public Health | Claire_Pearson; Tu_N_Nguyen |
| EPI_ISL_4005606, EPI_ISL_4005607 | California Department of Public Health | California Department of Public Health | CDPH IDLB COVIDNet et al |
| EPI_ISL_3276680 | California Hospital Medical Center | Los Angeles County Public Health Laboratories | P. Hemarajata et al. |
| EPI_ISL_7507375 | Cedars-Sinai Medical Center, Molecular Pathology Laboratory of Department of Pathology & Laboratory Medicine and Genomic Core | Cedars-Sinai Medical Center, Molecular Pathology Laboratory of Department of Pathology & Laboratory Medicine and Genomic Core | Brian Davis; Eric Vail; Jasmine T Plummer; Jorge Mario Sincuir Martinez; Stephanie Chen; Wenjuan Zhang |
| EPI_ISL_2907558, EPI_ISL_3276763 | Centinela Hospital Medical Center | Los Angeles County Public Health Laboratories | P. Hemarajata et al. |
| EPI_ISL_2958976 | Centracare Laboratory Services | Minnesota Department of Health, Public Health Laboratory | Alexandra Lorentz; Jacob Garfin; Matt Plumb; and Xiong Wang |
| EPI_ISL_3462551, EPI_ISL_3462612, EPI_ISL_3462679, EPI_ISL_3462680, EPI_ISL_3462681, EPI_ISL_3462682, EPI_ISL_3462683, EPI_ISL_3462684, EPI_ISL_3462685, EPI_ISL_3462686, EPI_ISL_3464735 | | | |
| see above | Central Laboratory, Bureau of Public Health (BOG) and Academic Hospital Paramaribo | Erasmus Medical Center | Bas B Oude Munnink; Cherise Beek; Consuella Partowidjojo; Dion Gajadin; Ed PF IJzerman; Emmanuelle Munger; Gary Gummels; Ingrid SK Krishnadath; Lycke Woittiez; Marion PG Koopmans; Mireille Van de Veer; Phyllis Pinas; Princes Wongsowidjojo; Radjesh Ori; Ranisha Doerbalie; Rohma Banwari; Soeradj Harkisoen; Stephen Vreden; Tilotmadebie Ramlal; Verne Nanhoe |
| EPI_ISL_4296404, EPI_ISL_4296410, EPI_ISL_4296411 | Central Medical Laboratory | Baylor College of Medicine | Adrianna Maliga; Alexander Kneubehl; Allison Lino; Gerhaldine Morazan; Kristy Murray; Russell Manzanero; Sarah Gunter; Sarah Strobel; Shannon Ronca |
| EPI_ISL_3255231, EPI_ISL_3255241, EPI_ISL_3255245, EPI_ISL_3255246, EPI_ISL_3255252, EPI_ISL_3255258, EPI_ISL_3255260, EPI_ISL_3255262, EPI_ISL_3255266, EPI_ISL_3255268, EPI_ISL_3255269, EPI_ISL_3255271, EPI_ISL_3255272, EPI_ISL_3255273, EPI_ISL_3255275, EPI_ISL_3255280, EPI_ISL_4417773 | | | |
| see above | Central Public Health Laboratory - LACEN -Bahia, Salvador, Brazil | Central Public Health Laboratory - LACEN -Bahia, Salvador, Brazil | Arabela Leal; Breno Dominguez; Felicidade Pereira; Gabriela Menezes; Jaqueline Gomes; Lenisa Dandara; Luciana Oliveira; Luiz Alcantara; Marcela Gómez; Marta Giovanetti; Patrícia Cajado; Stephane Tosta; Vagner Fonseca; Vanessa Nardy |
| EPI_ISL_3832900 | Central Washington Hospital | Seattle Flu Study | Amanda Adler; Barry R. Lutz; Benjamin Pelle; Brian Hiatt; Caitlin R. Wolf; Chris D. Frazar; Deborah A. Nickerson; Elisabeth Brandstetter; Erica Ryke; Geoff Melly; Helen Y. Chu; Janet A. Englund; Jay Shendure; Jover Lee; Kairsten Fay; Kirsten Lacombe; Lea M. Starita; Mark J. Rieder; Matthew Richardson; Matthew Thompson; Melissa Truong; Michael Boeckh; Michael Famulare; Misja Ilcisin; Peter D. Han; Philip Dykema; Romesh Gautom; Scott Lindquist; Thomas R. Sibley; Trevor Bedford |
| EPI_ISL_3147587 | Centre Hospitalier du Nord | Laboratoire national de sante, Microbiology, Microbial Genomics Platform | Anke Wienecke-Baldacchino; Catherine Ragimbeau; Elodie Solarino; Fatiha Boulmerka; Fatu Djabi; Jacqueline Parmentier; Jessica Tapp; Lise Pignon; Raoul Salmon; Tamir Abdelrahman; Virginie Jover |
| EPI_ISL_3825488 | Centro de Estudios Moleculares | Laboratorio de Biología Molecular, Instituto de Medicina Regional on behalf of 'Proyecto Argentino Interinstitucional de genomica de SARS-CoV-2' (PAIS Consortium) | Alexia Del Puerto; Bettina Brusés; Cecilia Urquijo; Gerardo Deluca; Griselda Oria; Horacio Lucero.; Javier Mussin; Laura Formichelli; Melina Lorenzini Campos; Raúl Maximiliano Acevedo |
| EPI_ISL_2942732, EPI_ISL_2942737 | Centro de Investigacion Biomedica de Occidente (CIBO) | Unidad de Genomica Avanzada | ; Alejandra Garcia-Gasca; Alejandra Hernandez-Teran; Alejandro Sanchez-Flores; Alfredo Herrera-Estrella; Alicia Ocaña-Mondragon; Andreu Comas-Garcia; Angel Gustavo Salas-Lais; Antonio Loza Roman; Bernardo Martinez-Miguel; Blanca Taboada; Brenda Irasema Maldonado-Meza; Bruno Gomez-Gil; Carla Ivon Herrera-Najera; Carlos F. Arias; Celia Boukadida; Celida Duque Molina; Celia Martinez- Rodriguez; Clara Esperanza Santacruz-Tinoco; Concepcion Grajales-Muñiz; Consorcio Mexicano de Vigilancia Genomica (CoViGen-Mex). Authors (in alphabetical order): Julio Elias Alvarado-Yaah; Cristobal Chaidez-Quiroz; Daniel Fregoso-Rueda; Daniel Lira Morales; Eduardo Becerril-Vargas; Fernando Fontove-Herrera; Fidencio Mejia-Nepomuceno; Francisco Pulido; Gloria Elena Espinosa-Ayala; Gloria Maria Molina-Salinas; Gloria Vazquez; Hector Esteban Paz-Juarez; Hector Montoya-Fuentes; Helen Haydee Fernanda Ramirez-Plascencia; Irvin Gonzalez-Lopez; Jean Pierre Gonzalez; Jesus Hernandez; Joel Armando Vazquez-Perez.; Jorge Salas-Hernandez; Jose Antonio Enciso-Moreno; Jose Arturo Martinez-Orozco; Jose Esteban Muñoz-Medina; Jose de Jesus Nuñez-Contreras; Juan Bautista Chale-Dzul; Julissa Enciso-Ibarra; Luis Alberto Ochoa-Carrera; Margarita Matias-Florentino; Maria Guadalupe Santiago-Mauricio; Maria Guadalupe de Jesus Mireles-Rivera; Mario Mujica-Sanchez; Marissa Perez-Garcia; Nelly Selem-Mojica; Pavel Isa; Ricardo Ciria Merce; Ricardo Grande; Rosa Maria Gutierrez Rios; Santiago avila-Rios; Selene Zarate; Susana Lopez; Veronica Mata-Haro; Victor Eduardo Garcia-Arias; Victor Hugo Borja-Aburto |
| EPI_ISL_2942770 | Centro de Investigacion Biomedica del Noreste (CIBIN) | Unidad de Genomica Avanzada | ; Alejandra Garcia-Gasca; Alejandra Hernandez-Teran; Alejandro Sanchez-Flores; Alfredo Herrera-Estrella; Alicia Ocaña-Mondragon; Andreu Comas-Garcia; Angel Gustavo Salas-Lais; Antonio Loza Roman; Bernardo Martinez-Miguel; Blanca Taboada; Brenda Irasema Maldonado-Meza; Bruno Gomez-Gil; Carla Ivon Herrera-Najera; Carlos F. Arias; Celia Boukadida; Celida Duque Molina; Celia Martinez- Rodriguez; Clara Esperanza Santacruz-Tinoco; Concepcion Grajales-Muñiz; Consorcio Mexicano de Vigilancia Genomica (CoViGen-Mex). Authors (in alphabetical order): Julio Elias Alvarado-Yaah; Cristobal Chaidez-Quiroz; Daniel Fregoso-Rueda; Daniel Lira Morales; Eduardo Becerril-Vargas; Fernando Fontove-Herrera; Fidencio Mejia-Nepomuceno; Francisco Pulido; Gloria Elena Espinosa-Ayala; Gloria Maria Molina-Salinas; Gloria Vazquez; Hector Esteban Paz-Juarez; Hector Montoya-Fuentes; Helen Haydee Fernanda Ramirez-Plascencia; Irvin Gonzalez-Lopez; Jean Pierre Gonzalez; Jesus Hernandez; Joel Armando Vazquez-Perez.; Jorge Salas-Hernandez; Jose Antonio Enciso-Moreno; Jose Arturo Martinez-Orozco; Jose Esteban Muñoz-Medina; Jose de Jesus Nuñez-Contreras; Juan Bautista Chale-Dzul; Julissa Enciso-Ibarra; Luis Alberto Ochoa-Carrera; Margarita Matias-Florentino; Maria Guadalupe Santiago-Mauricio; Maria Guadalupe de Jesus Mireles-Rivera; Mario Mujica-Sanchez; Marissa Perez-Garcia; Nelly Selem-Mojica; Pavel Isa; Ricardo Ciria Merce; Ricardo Grande; Rosa Maria Gutierrez Rios; Santiago avila-Rios; Selene Zarate; Susana Lopez; Veronica Mata-Haro; Victor Eduardo Garcia-Arias; Victor Hugo Borja-Aburto |
| EPI_ISL_3019538 | Centro de Investigación Biomédica de La Rioja - Hospital San Pedro Logroño | SeqCOVID-SPAIN consortium/IBV(CSIC) | José Manuel Azcona Gutiérrez; María Pilar Bea Escudero; María de Toro; Miriam Blasco Alberdi and SeqCOVID-SPAIN consortium |
| EPI_ISL_3242529, EPI_ISL_3242550, EPI_ISL_3242711, EPI_ISL_3242712, EPI_ISL_3242713, EPI_ISL_3242726 | Clinica INDISA | "Facultad de Ciencias de la Vida, UNAB" | "Claudio Meneses; Ariel Orellana"; Claudio Olmos; Daniel Leon; Dayan Sanhueza; Eduardo Castro; Gonzalo Campaña; Macarena Bastias; Paola Pidal; Ricardo Yusta; Sebastian Wolter; Susana Saez; Víctor Monreal; Waldo Diaz |
| EPI_ISL_2885659 | Cliniques universitaires Saint-Luc | UCLouvain/IREC/MBLG | Benoit Kabamba Mukadi; Bertrand Bearzatto; Jean Ruelle |
| EPI_ISL_5323539 | Color Genomics | Chiu Laboratory, University of California, San Francisco | Alicia Sotomayor-Gonzalez; Alicia Zhou; Baolin Wang; Charles Chiu; Daniel Wong; Erika Torres; Noah Brazer; Scott Topper; Venice Servellita |
| EPI_ISL_3583410, EPI_ISL_3583883, EPI_ISL_3696525, EPI_ISL_3696528, EPI_ISL_3988632 | Colorado Department of Public Health and Environment | Colorado Department of Public Health and Environment | Alexandria Rossheim; Diana Ir; Emily A. Travanty; Laura Bankers; Mandy Waters; Michael A. Martin; Michael Martin; Molly C. Hetherington-Rauth; Sarah Elizabeth Totten; Shannon R. Matzinger |
| EPI_ISL_2930362, EPI_ISL_2930407, EPI_ISL_2930408, EPI_ISL_2930410, EPI_ISL_2930417, EPI_ISL_2930429, EPI_ISL_3014094, EPI_ISL_3014173, EPI_ISL_3014174, EPI_ISL_3014175 | | | |
| see above | Cotugno | TIGEM | Antonio Grimaldi Patrizia Annunziata Francesco Panariello Biancamaria Pierri Claudia Tiberio Teresa Giuliano Valentina Bouche Chiara Colantuono Maria Concetta Cuomo Denise Di Concilio Lucio Di Filippo Anna Manfredi Marcello Salvi Antonio Limone Luigi Atripaldi Pellegrino Cerino Andrea Ballabio Davide Cacchiarelli |
| EPI_ISL_2942034, EPI_ISL_2942086, EPI_ISL_3064523, EPI_ISL_3064537, EPI_ISL_3064539, EPI_ISL_3064540, EPI_ISL_3064667, EPI_ISL_3270308, EPI_ISL_3270395, EPI_ISL_3270418, EPI_ISL_3273397, EPI_ISL_3273444 | | | |
| see above | Curative Labs | Curative Labs | Elias L. Salfati; Eugenia Khorosheva; George Way; J.Cesar Ignacio-Espinoza; Janet Chen; Mikhail Hanewich-Hollatz; Nabjot Sandhu; Sophia Quasem; Vladimir Slepnev; Zhiyi Xie |
| EPI_ISL_4414243, EPI_ISL_4414245, EPI_ISL_4414251, EPI_ISL_4414252, EPI_ISL_4414263, EPI_ISL_4414267, EPI_ISL_4414288, EPI_ISL_4414289, EPI_ISL_4414292, EPI_ISL_4414293, EPI_ISL_4414298, EPI_ISL_4414300, EPI_ISL_4414317, EPI_ISL_4414319, EPI_ISL_4414323, EPI_ISL_4414333, EPI_ISL_4414336, EPI_ISL_4414337, EPI_ISL_4414356, EPI_ISL_4414364, EPI_ISL_4414374, EPI_ISL_4414382, EPI_ISL_4414386, EPI_ISL_4414388, EPI_ISL_4414390, EPI_ISL_4414392, EPI_ISL_4414406, EPI_ISL_4414407, EPI_ISL_4414414, EPI_ISL_4414417, EPI_ISL_4414421, EPI_ISL_4414443, EPI_ISL_4414446, EPI_ISL_4414451, EPI_ISL_4414460, EPI_ISL_4414465, EPI_ISL_4414498, EPI_ISL_4414499, EPI_ISL_4414500, EPI_ISL_4414501, EPI_ISL_4414502, EPI_ISL_4414503, EPI_ISL_4414504, EPI_ISL_4414505, EPI_ISL_4414506, EPI_ISL_4414507, EPI_ISL_4414508, EPI_ISL_4414520, EPI_ISL_4414521, EPI_ISL_4414525, EPI_ISL_4414527, EPI_ISL_4414528, EPI_ISL_4414532, EPI_ISL_4414533, EPI_ISL_4414543, EPI_ISL_4414544, EPI_ISL_4414564, EPI_ISL_4414566, EPI_ISL_4414574, EPI_ISL_4422554, EPI_ISL_4422981, EPI_ISL_4422982, EPI_ISL_4422984, EPI_ISL_4423809, EPI_ISL_4423814, EPI_ISL_4423817, EPI_ISL_4423819, EPI_ISL_4423822, EPI_ISL_4423846, EPI_ISL_4423849, EPI_ISL_4423852, EPI_ISL_4423875, EPI_ISL_4423885, EPI_ISL_4423930, EPI_ISL_4423933, EPI_ISL_4423936, EPI_ISL_4423939, EPI_ISL_4423941, EPI_ISL_4423944, EPI_ISL_4423983, EPI_ISL_4423985, EPI_ISL_4423987, EPI_ISL_4423998, EPI_ISL_4424005, EPI_ISL_4424026, EPI_ISL_4424052, EPI_ISL_4424077, EPI_ISL_4424079, EPI_ISL_4424081, EPI_ISL_4424084, EPI_ISL_4424086, EPI_ISL_4424097, EPI_ISL_4424098, EPI_ISL_4424117, EPI_ISL_4424119, EPI_ISL_4424121, EPI_ISL_4424123, EPI_ISL_4424126, EPI_ISL_4424128, EPI_ISL_4424130, EPI_ISL_4424131, EPI_ISL_4424298, EPI_ISL_4424307, EPI_ISL_4424313, EPI_ISL_4424329, EPI_ISL_4424331, EPI_ISL_4424332, EPI_ISL_4424334, EPI_ISL_4424379, EPI_ISL_4424393, EPI_ISL_4424401, EPI_ISL_4424402, EPI_ISL_4424404, EPI_ISL_4424416, EPI_ISL_4424422, EPI_ISL_4424424, EPI_ISL_4424425, EPI_ISL_4424427, EPI_ISL_4424429, EPI_ISL_4424430, EPI_ISL_4424432, EPI_ISL_4424434, EPI_ISL_4424436, EPI_ISL_4424437, EPI_ISL_4424439, EPI_ISL_4424441, EPI_ISL_4424443, EPI_ISL_4424445, EPI_ISL_4424446, EPI_ISL_4424448, EPI_ISL_4424450, EPI_ISL_4424451, EPI_ISL_4424453, EPI_ISL_4424455, EPI_ISL_4424456, EPI_ISL_4424458, EPI_ISL_4424460, EPI_ISL_4424462, EPI_ISL_4424463, EPI_ISL_4424465, EPI_ISL_4424467, EPI_ISL_4424468, EPI_ISL_4424470, EPI_ISL_4424472, EPI_ISL_4424474, EPI_ISL_4424476, EPI_ISL_4424477, EPI_ISL_4424479, EPI_ISL_4424481, EPI_ISL_4424483, EPI_ISL_4424485, EPI_ISL_4424487, EPI_ISL_4424489, EPI_ISL_4424491, | | | |

| | | | |
|---|---|---|---|
| EPI_ISL_4424493, EPI_ISL_4424495, EPI_ISL_4424496, EPI_ISL_4424498, EPI_ISL_4424500, EPI_ISL_4424501, EPI_ISL_4424503, EPI_ISL_4424505, EPI_ISL_4424507, EPI_ISL_4424508, EPI_ISL_4424509, EPI_ISL_4424775, EPI_ISL_4424777, EPI_ISL_4424779, EPI_ISL_4424783, EPI_ISL_4424790, EPI_ISL_7743827, EPI_ISL_7743828, EPI_ISL_7743831 | | | |
| see above | DASA | DASA | Adriano Bonaldi; Angelica Hristov; Annelise Lopes; Bianca Cota; Cristina Oliveira; Jose Levi; Lidia Yamamoto; Paulo Pierry; Rodrigo Guarischi; Rodrigo Salazar |
| EPI_ISL_5666759 | DEPARTAMENTO DE SAUDE MUNICIPAL SOCORRO SP | Instituto Butantan | Antonio Jorge Martins; Claudia Renata dos Santos Barros; David Schlesinger; Debora Botequio Moretti; Dimas Tadeu Covas; Elaine Cristina Marqueze; Elaine Vieira Santos; Evandra Strazza Rodrigues; Heidge Fukumasu; Jayme Augusto de Souza-Neto; José Salvatore Leister Patané; Luiz Alcantara; Luiz Lehmann Coutinho; Maria Carolina Elias; Maurício Lacerda Nogueira; Rafael dos Santos Bezerra; Raul Machado Neto; Rejane Maria Tommasini Grotto; Ricardo Haddad; Sandra Coccuzzo Sampaio Vessoni; Simone Kashima; Svetoslav Nanev Slavov; Vincent Louis Viala |
| EPI_ISL_5661495, EPI_ISL_5661869, EPI_ISL_5661982, EPI_ISL_5666262, EPI_ISL_5666297, EPI_ISL_5666359, EPI_ISL_5666577 | | | |
| see above | DEPARTAMENTO MUNICIPAL DE SAUDE DE PARAGUACU PAULISTA | Instituto Butantan | Antonio Jorge Martins; Claudia Renata dos Santos Barros; David Schlesinger; Debora Botequio Moretti; Dimas Tadeu Covas; Elaine Cristina Marqueze; Elaine Vieira Santos; Evandra Strazza Rodrigues; Heidge Fukumasu; Jayme Augusto de Souza-Neto; José Salvatore Leister Patané; Luiz Alcantara; Luiz Lehmann Coutinho; Maria Carolina Elias; Maurício Lacerda Nogueira; Rafael dos Santos Bezerra; Raul Machado Neto; Rejane Maria Tommasini Grotto; Ricardo Haddad; Sandra Coccuzzo Sampaio Vessoni; Simone Kashima; Svetoslav Nanev Slavov; Vincent Louis Viala |
| EPI_ISL_5670991 | DIRETORIA MUNICIPAL DE SAUDE DE ENGENHEIRO COELHO | Instituto Butantan | Antonio Jorge Martins; Claudia Renata dos Santos Barros; David Schlesinger; Debora Botequio Moretti; Dimas Tadeu Covas; Elaine Cristina Marqueze; Elaine Vieira Santos; Evandra Strazza Rodrigues; Heidge Fukumasu; Jayme Augusto de Souza-Neto; José Salvatore Leister Patané; Luiz Alcantara; Luiz Lehmann Coutinho; Maria Carolina Elias; Maurício Lacerda Nogueira; Rafael dos Santos Bezerra; Raul Machado Neto; Rejane Maria Tommasini Grotto; Ricardo Haddad; Sandra Coccuzzo Sampaio Vessoni; Simone Kashima; Svetoslav Nanev Slavov; Vincent Louis Viala |
| EPI_ISL_3062444 | DOHMH Morrisania | New York City Public Health Laboratory | Jade Wang; et al. |
| EPI_ISL_2819380, EPI_ISL_2862720, EPI_ISL_2896927 | Department of Bacteria, Parasites and Fungi, Statens Serum Institut, Copenhagen, Denmark | Statens Serum Institut Bioinformatics and Microbial Genomics | Danish Covid-19 Genome Consortium |
| EPI_ISL_2930770, EPI_ISL_2930772 | Department of Clinical Microbiology | GIGA Medical Genomics | Bouchra Boujemla; Claire Gourzonès; Cécile Meex; Keith Durkin; Laurent Gillet; Maria Artesi; Marie-Pierre Hayette; Nadine Cambisano; Nathalie Renotte; Olivier Ek; Sébastien Bontems; Vincent Bours |
| EPI_ISL_4093608, EPI_ISL_4093609, EPI_ISL_4093610, EPI_ISL_4093611, EPI_ISL_4093612 | Department of Virology and Immunology, University of Helsinki and Helsinki University Hospital, Huslab Finland | Department of Virology, Faculty of Medicine, University of Helsinki, Helsinki, Finland | Essi Korhonen; Hanna Jarva; Hanna Liimatainen; Hannimari Kallio-Kokko; Harri Kangas; Hussein Alburkat; Jenni Virtanen; Maija Lappalainen; Maija Suvanto; Olli Vapalahti; Pekka Ellonen; Phuoc Truong; Ravi Kant; Sari Hannula; Satu Kurkela; Teemu Smura |
| EPI_ISL_3372577, EPI_ISL_3372579, EPI_ISL_3372580 | Dirección regional de salud del Callao (DIRESA-CALLAO) | Centro de Investigaciones Tecnológicas, Biomédicas y Medioambientales (CITBM) | B; Huaman; J. Alarcon; M. Cuellar; M. Ramirez; M. Sovero |
| EPI_ISL_2967544, EPI_ISL_2967545 | Division of Emerging Infectious Diseases, Bureau of Infectious Diseases Diagnosis Control, Korea Disease Control and Prevention Agency | Division of Emerging Infectious Diseases, Bureau of Infectious Diseases Diagnosis Control, Korea Disease Control and Prevention Agency | Ae Kyung Park; Chae Young Lee; Eun-Jin Kim; Heui Man Kim; Il-Hwan Kim; Jeong-Ah Kim; Jeong-Min Kim |
| EPI_ISL_2981315, EPI_ISL_2981755, EPI_ISL_2981889, EPI_ISL_2981890, EPI_ISL_2981895, EPI_ISL_3057480, EPI_ISL_3136605, EPI_ISL_3136615 | | | |
| see above | Dutch COVID-19 response team | National Institute for Public Health and the Environment (RIVM) | Adam Meijer; AnneMarie van den Brandt; Annelies Kroneman; Bas van der Veer; Chantal Reusken; Dennis Schmitz; Dirk Eggink; Euníce Then; Florian Zwagemaker; Harry Vennema; Jeroen Cremer; Karim Hajji; Kim Freriks; Lisa Wijsman; Lynn Aarts; Melissa van Tuil; Ryanne Jaarsma; Sanne Bos; Sharon van den Brink; on behalf of the national COVID-19 response team |
| EPI_ISL_4059819, EPI_ISL_4059820, EPI_ISL_4059823, EPI_ISL_4060090, EPI_ISL_4060106, EPI_ISL_4060107 | Dynacare Manitoba | Manitoba Cadham Provincial Laboratory | David Alexander; Hilary Racher; Hongbin Li; Melissa Desaulnier; Tintu Abraham |
| EPI_ISL_5668123, EPI_ISL_5668347 | E S F DR EDSON GIANETTO JD CARLOS CASSETARI | Instituto Butantan | Antonio Jorge Martins; Claudia Renata dos Santos Barros; David Schlesinger; Debora Botequio Moretti; Dimas Tadeu Covas; Elaine Cristina Marqueze; Elaine Vieira Santos; Evandra Strazza Rodrigues; Heidge Fukumasu; Jayme Augusto de Souza-Neto; José Salvatore Leister Patané; Luiz Alcantara; Luiz Lehmann Coutinho; Maria Carolina Elias; Maurício Lacerda Nogueira; Rafael dos Santos Bezerra; Raul Machado Neto; Rejane Maria Tommasini Grotto; Ricardo Haddad; Sandra Coccuzzo Sampaio Vessoni; Simone Kashima; Svetoslav Nanev Slavov; Vincent Louis Viala |
| EPI_ISL_5662006 | ESF BONFIM | Instituto Butantan | Antonio Jorge Martins; Claudia Renata dos Santos Barros; David Schlesinger; Debora Botequio Moretti; Dimas Tadeu Covas; Elaine Cristina Marqueze; Elaine Vieira Santos; Evandra Strazza Rodrigues; Heidge Fukumasu; Jayme Augusto de Souza-Neto; José Salvatore Leister Patané; Luiz Alcantara; Luiz Lehmann Coutinho; Maria Carolina Elias; Maurício Lacerda Nogueira; Rafael dos Santos Bezerra; Raul Machado Neto; Rejane Maria Tommasini Grotto; Ricardo Haddad; Sandra Coccuzzo Sampaio Vessoni; Simone Kashima; Svetoslav Nanev Slavov; Vincent Louis Viala |
| EPI_ISL_5667423, EPI_ISL_5667562 | ESF NOVA TANABI II | Instituto Butantan | Antonio Jorge Martins; Claudia Renata dos Santos Barros; David Schlesinger; Debora Botequio Moretti; Dimas Tadeu Covas; Elaine Cristina Marqueze; Elaine Vieira Santos; Evandra Strazza Rodrigues; Heidge Fukumasu; Jayme Augusto de Souza-Neto; José Salvatore Leister Patané; Luiz Alcantara; Luiz Lehmann Coutinho; Maria Carolina Elias; Maurício Lacerda Nogueira; Rafael dos Santos Bezerra; Raul Machado Neto; Rejane Maria Tommasini Grotto; Ricardo Haddad; Sandra Coccuzzo Sampaio Vessoni; Simone Kashima; Svetoslav Nanev Slavov; Vincent Louis Viala |
| EPI_ISL_5661365, EPI_ISL_5662092 | ESF PARQUE COLINAS | Instituto Butantan | Antonio Jorge Martins; Claudia Renata dos Santos Barros; David Schlesinger; Debora Botequio Moretti; Dimas Tadeu Covas; Elaine Cristina Marqueze; Elaine Vieira Santos; Evandra Strazza Rodrigues; Heidge Fukumasu; Jayme Augusto de Souza-Neto; José Salvatore Leister Patané; Luiz Alcantara; Luiz Lehmann Coutinho; Maria Carolina Elias; Maurício Lacerda Nogueira; Rafael dos Santos Bezerra; Raul Machado Neto; Rejane Maria Tommasini Grotto; Ricardo Haddad; Sandra Coccuzzo Sampaio Vessoni; Simone Kashima; Svetoslav Nanev Slavov; Vincent Louis Viala |
| EPI_ISL_5661356 | ESF PQ UNIVERSITARIO DE ASSIS | Instituto Butantan | Antonio Jorge Martins; Claudia Renata dos Santos Barros; David Schlesinger; Debora Botequio Moretti; Dimas Tadeu Covas; Elaine Cristina Marqueze; Elaine Vieira Santos; Evandra Strazza Rodrigues; Heidge Fukumasu; Jayme Augusto de Souza-Neto; José Salvatore Leister Patané; Luiz Alcantara; Luiz Lehmann Coutinho; Maria Carolina Elias; Maurício Lacerda Nogueira; Rafael dos Santos Bezerra; Raul Machado Neto; Rejane Maria Tommasini Grotto; Ricardo Haddad; Sandra Coccuzzo Sampaio Vessoni; Simone Kashima; Svetoslav Nanev Slavov; Vincent Louis Viala |
| EPI_ISL_5661221, EPI_ISL_5661427 | ESF VILA GLORIA I | Instituto Butantan | Antonio Jorge Martins; Claudia Renata dos Santos Barros; David Schlesinger; Debora Botequio Moretti; Dimas Tadeu Covas; Elaine Cristina Marqueze; Elaine Vieira Santos; Evandra Strazza Rodrigues; Heidge Fukumasu; Jayme Augusto de Souza-Neto; José Salvatore Leister Patané; Luiz Alcantara; Luiz Lehmann Coutinho; Maria Carolina Elias; Maurício Lacerda Nogueira; Rafael dos Santos Bezerra; Raul Machado Neto; Rejane Maria Tommasini Grotto; Ricardo Haddad; Sandra Coccuzzo Sampaio Vessoni; Simone Kashima; Svetoslav Nanev Slavov; Vincent Louis Viala |
| EPI_ISL_3116503, EPI_ISL_3909771, EPI_ISL_3909772, EPI_ISL_3909773, EPI_ISL_3909775, EPI_ISL_3909776, EPI_ISL_3909801, EPI_ISL_3909857, EPI_ISL_3909858 | | | |
| see above | Edmonton Provincial Lab | Public Health Agency of Canada (PHAC) National Microbiology Laboratory | Buss; Croxen M; Deo A; Dieu P; E; Ferrato C; Gill K; Khan F; Koleva P; Li V; Lloyd C; Lynch T; Ma R; Murphy S; Pabbaraju K; Shokoples S; Thayer J; Tipples G; Whitehouse M; Wong A; Yu C; Zelyas N |
| EPI_ISL_3718929 | Emory Molecular Diagnostics Laboratory, Emory Healthcare | Piantadosi Lab, Emory Department of Pathology | Ahmed Babiker; Anne Piantadosi; Ludy Registre Carmola |
| EPI_ISL_5659659, EPI_ISL_5659866, EPI_ISL_5660101, EPI_ISL_5660337, EPI_ISL_5660340, EPI_ISL_5660365, EPI_ISL_5660566, EPI_ISL_5660708, EPI_ISL_5660875, EPI_ISL_5673287, EPI_ISL_5673831, EPI_ISL_5674833, EPI_ISL_5675177 | | | |
| see above | FACULDADE DE MEDICINA ABC | Instituto Butantan | Antonio Jorge Martins; Claudia Renata dos Santos Barros; David Schlesinger; Debora Botequio Moretti; Dimas Tadeu Covas; Elaine Cristina Marqueze; Elaine Vieira Santos; Evandra Strazza Rodrigues; Heidge Fukumasu; Jayme Augusto de Souza-Neto; José Salvatore Leister Patané; Luiz Alcantara; Luiz Lehmann Coutinho; Maria Carolina Elias; Maurício Lacerda Nogueira; Rafael dos Santos Bezerra; Raul Machado Neto; Rejane Maria Tommasini Grotto; Ricardo Haddad; Sandra Coccuzzo Sampaio Vessoni; Simone Kashima; Svetoslav Nanev Slavov; Vincent Louis Viala |
| EPI_ISL_3474183, EPI_ISL_3474184 | FYR Diagnostics | FYR Diagnostics | Blohm; Ghai; Tre; Vikas |
| EPI_ISL_3100829, EPI_ISL_3100840, EPI_ISL_3100855, EPI_ISL_3100865, EPI_ISL_3100900, EPI_ISL_3100901, EPI_ISL_3100902, EPI_ISL_3100904, EPI_ISL_3100905, EPI_ISL_3100909, EPI_ISL_3100910, EPI_ISL_3100945, EPI_ISL_3100952, EPI_ISL_3100955, EPI_ISL_3100959, EPI_ISL_3100969, EPI_ISL_3100971, EPI_ISL_5322010, EPI_ISL_5322086, EPI_ISL_5322092, EPI_ISL_5322288, EPI_ISL_5322292, EPI_ISL_5322301, EPI_ISL_5322423, EPI_ISL_5542091, EPI_ISL_5866262, EPI_ISL_5866278, EPI_ISL_5866279, EPI_ISL_5866308, EPI_ISL_5866402, EPI_ISL_5925181, EPI_ISL_5925183, EPI_ISL_5925184, EPI_ISL_5925185, EPI_ISL_5925186, EPI_ISL_5925187, EPI_ISL_5925198 | | | |
| see above | Florida Bureau of Public Health Laboratories | Florida Bureau of Public Health Laboratories | Jason Blanton; Namratha Tarigopula; Sarah Schmedes; Tiffany Splatt |
| EPI_ISL_2873568, EPI_ISL_2873575, EPI_ISL_2873586, EPI_ISL_2873587, EPI_ISL_2873601, EPI_ISL_2873631, EPI_ISL_2873648, EPI_ISL_2873681, EPI_ISL_2873692, EPI_ISL_2873726, EPI_ISL_2873739, EPI_ISL_2873762, EPI_ISL_2873765, EPI_ISL_2873771, EPI_ISL_2928551, EPI_ISL_2928558, EPI_ISL_2928567, EPI_ISL_2928639, EPI_ISL_2928640, EPI_ISL_2928678, EPI_ISL_2928681, EPI_ISL_2928702, EPI_ISL_2928717, EPI_ISL_2928729, EPI_ISL_2928777, EPI_ISL_2928778, EPI_ISL_2928803, EPI_ISL_2928820, EPI_ISL_2928855, EPI_ISL_2928863, EPI_ISL_2928886, EPI_ISL_2928893, EPI_ISL_2928987, EPI_ISL_2929052, EPI_ISL_2929063, EPI_ISL_3016704, EPI_ISL_3299787, EPI_ISL_3299974, EPI_ISL_3299991 | | | |
| see above | Fulgent Genetics | Centers for Disease Control and Prevention Division of Viral Diseases, Pathogen Discovery | Adrian Paskey; Becky Tsai; Benafsh Sapra; Benjamin Rambo-Martin; Christopher Gulvick; Clinton R. Paden; Dakota Howard; Darlene Wagner; Dhwani Batra; Doreen Ng; Duncan MacCannell; Harry Gao; James Xie; Jason Caravas; John Gao; Joseph Fierro; Kara Moser; Matthew Schmerer; Mickey Li; Peter W. Cook; Scott Sammons; Shatavia Morrison; Yan Meng; Yvette Unoarumhi |
| EPI_ISL_3099596 | Fundacio Althaia-Manresa | Banc de Sang i Teixits | Carlos Hobeich; Francisco Vidal; Irene Corrales; Lorena Ramírez; Maria Glòria Soria; Natália Comes; Nina Borràs; Noemí Gonzalez; Silvia Sauleda |
| EPI_ISL_5672475 | GRUPO TECNICO DE VIGILANCIA SANITARIA E EPIDEMIOLOGICA | Instituto Butantan | Antonio Jorge Martins; Claudia Renata dos Santos Barros; David Schlesinger; Debora Botequio Moretti; Dimas Tadeu Covas; Elaine Cristina Marqueze; Elaine Vieira Santos; Evandra Strazza Rodrigues; Heidge Fukumasu; Jayme Augusto de Souza-Neto; José Salvatore Leister Patané; Luiz Alcantara; Luiz Lehmann Coutinho; Maria Carolina Elias; Maurício Lacerda Nogueira; Rafael dos Santos Bezerra; Raul Machado Neto; Rejane Maria Tommasini Grotto; Ricardo Haddad; Sandra Coccuzzo Sampaio Vessoni; Simone Kashima; Svetoslav Nanev Slavov; Vincent Louis Viala |
| EPI_ISL_4198489, EPI_ISL_4198496, EPI_ISL_4740274 | Gencore - Universidad de los Andes | Gencore - Universidad de los Andes | Cristian Barrera; David González; Gabriela Ariza; Luisa Sacristan; Marcela Guevara; Silvia Restrepo |
| EPI_ISL_2894925, EPI_ISL_2894926, EPI_ISL_2894927, EPI_ISL_2894928, EPI_ISL_2894945, EPI_ISL_2894947, EPI_ISL_2894948, EPI_ISL_2894949, EPI_ISL_2894951, EPI_ISL_2894952, EPI_ISL_2894953, EPI_ISL_2894954, EPI_ISL_2894955, EPI_ISL_2894956, EPI_ISL_2896150, EPI_ISL_2896151, EPI_ISL_2896152, EPI_ISL_2896158, EPI_ISL_2896173, EPI_ISL_2896174, EPI_ISL_2896175, EPI_ISL_2896176, EPI_ISL_2896177, EPI_ISL_2896178, EPI_ISL_2896179, EPI_ISL_2896180, EPI_ISL_2896181, EPI_ISL_2896183, EPI_ISL_2930485, EPI_ISL_2930492, EPI_ISL_2930493, EPI_ISL_2930494, EPI_ISL_2930495, EPI_ISL_2930497, EPI_ISL_2930499, EPI_ISL_2930500, EPI_ISL_2930501, EPI_ISL_2930502, EPI_ISL_2930504, EPI_ISL_2930505, EPI_ISL_2930506, EPI_ISL_2930507, EPI_ISL_2930508, EPI_ISL_2930509, EPI_ISL_2930534, EPI_ISL_2930550, EPI_ISL_2930551, EPI_ISL_2930552, EPI_ISL_2930553, EPI_ISL_2930554, EPI_ISL_3132217, EPI_ISL_3132218, EPI_ISL_3132220, EPI_ISL_3132221, EPI_ISL_3132222, EPI_ISL_3132223, EPI_ISL_3132224, EPI_ISL_3132226, EPI_ISL_3132227, EPI_ISL_3132228, EPI_ISL_3132231, EPI_ISL_3132232, EPI_ISL_3132233, EPI_ISL_3132234, EPI_ISL_3132235, EPI_ISL_3132236, EPI_ISL_3132237, EPI_ISL_3132238, EPI_ISL_3132239, EPI_ISL_3132240, EPI_ISL_3132241, EPI_ISL_3132242, EPI_ISL_3132243, EPI_ISL_3132244, EPI_ISL_3132245, EPI_ISL_3132246, EPI_ISL_3132247, EPI_ISL_3132248, EPI_ISL_3132249, EPI_ISL_3132250, EPI_ISL_3132251, EPI_ISL_3132266, EPI_ISL_3132267, EPI_ISL_3132268, EPI_ISL_3132271, EPI_ISL_3132635, EPI_ISL_3132638, EPI_ISL_3132639, EPI_ISL_3132640, EPI_ISL_3132641, EPI_ISL_3132642, EPI_ISL_3132643, EPI_ISL_3132645, EPI_ISL_3132646, EPI_ISL_3132647, EPI_ISL_3132650, EPI_ISL_3132651, EPI_ISL_3132652, EPI_ISL_3132655, EPI_ISL_3132656, EPI_ISL_3132657, EPI_ISL_3132658, EPI_ISL_3132659, EPI_ISL_3132660, EPI_ISL_3132661, EPI_ISL_3132662, EPI_ISL_3132667, EPI_ISL_3132668, EPI_ISL_3132669, EPI_ISL_3132670, EPI_ISL_3132671, EPI_ISL_3132673, EPI_ISL_3144892, EPI_ISL_3144893, EPI_ISL_3144894, EPI_ISL_3144895, EPI_ISL_3144896, EPI_ISL_3144897, EPI_ISL_3144899, EPI_ISL_3144900, EPI_ISL_3144901, EPI_ISL_3144905, EPI_ISL_3144909, EPI_ISL_3144913, EPI_ISL_3144914, EPI_ISL_3144915, EPI_ISL_3144916, EPI_ISL_3144919, EPI_ISL_3144920, EPI_ISL_3144921, EPI_ISL_3144923, EPI_ISL_3144924, EPI_ISL_3144925, EPI_ISL_3144926, EPI_ISL_3144927, EPI_ISL_3144928, EPI_ISL_3144929, EPI_ISL_3144930, EPI_ISL_3144931, EPI_ISL_3144932, EPI_ISL_3149071, EPI_ISL_3149073, EPI_ISL_3149075, EPI_ISL_3149076, EPI_ISL_3149077, EPI_ISL_3149078, EPI_ISL_3149080, EPI_ISL_3149081, EPI_ISL_3149082, EPI_ISL_3149083, EPI_ISL_3149084, EPI_ISL_3149085, EPI_ISL_3149087, EPI_ISL_3149088, EPI_ISL_3149089, EPI_ISL_3149090, EPI_ISL_3149091, EPI_ISL_3149092, EPI_ISL_3149093, EPI_ISL_3149094, EPI_ISL_3149095, EPI_ISL_3149096, EPI_ISL_3149097, EPI_ISL_3149098, EPI_ISL_3149099, EPI_ISL_3149100, EPI_ISL_3149101, EPI_ISL_3149103, EPI_ISL_3149107, EPI_ISL_3149108, EPI_ISL_3149109, EPI_ISL_3149110, EPI_ISL_3149111, EPI_ISL_3149112, EPI_ISL_3149113, EPI_ISL_3149114, EPI_ISL_3149115, EPI_ISL_3149116, EPI_ISL_3149117, EPI_ISL_3149118, EPI_ISL_3149119, EPI_ISL_3149120, EPI_ISL_3149121, EPI_ISL_3149122, EPI_ISL_3149123, EPI_ISL_3149124, EPI_ISL_3149125, EPI_ISL_3149126, EPI_ISL_3149127, EPI_ISL_3149128, EPI_ISL_3149129, EPI_ISL_3185391, EPI_ISL_3185394, EPI_ISL_3185395, EPI_ISL_3185399, EPI_ISL_3185400, EPI_ISL_3185401, EPI_ISL_3185402, EPI_ISL_3185405, EPI_ISL_3185406, EPI_ISL_3185407, EPI_ISL_3185409, EPI_ISL_3185410, EPI_ISL_3185411, EPI_ISL_3185413, EPI_ISL_3185414, EPI_ISL_3185415, EPI_ISL_3185416, EPI_ISL_3185417, EPI_ISL_3185419, EPI_ISL_3185420, EPI_ISL_3185421, EPI_ISL_3185422, EPI_ISL_3185423, EPI_ISL_3185424, EPI_ISL_3185425, EPI_ISL_3185426, EPI_ISL_3185427, EPI_ISL_3185428, EPI_ISL_3185429, EPI_ISL_3185430, EPI_ISL_3185431, EPI_ISL_3185432, EPI_ISL_3185433, EPI_ISL_3185434, EPI_ISL_3185435, EPI_ISL_3185436, EPI_ISL_3185437, EPI_ISL_3185439, EPI_ISL_3185441, EPI_ISL_3185442, EPI_ISL_3185443, EPI_ISL_3185444, EPI_ISL_3185445, EPI_ISL_3185446, EPI_ISL_3185447, EPI_ISL_3185448, EPI_ISL_3185449, EPI_ISL_3185450, EPI_ISL_3185451, EPI_ISL_3185453, EPI_ISL_3185456, EPI_ISL_3185457, EPI_ISL_3185459, EPI_ISL_3185464, EPI_ISL_3185467, EPI_ISL_3185470, EPI_ISL_3185473, EPI_ISL_3185477, EPI_ISL_3185478, EPI_ISL_3185481, EPI_ISL_3185482, EPI_ISL_3185483, EPI_ISL_3185484, EPI_ISL_3185486, EPI_ISL_3185487, EPI_ISL_3185489, EPI_ISL_3185492, EPI_ISL_3185493, EPI_ISL_3185494, EPI_ISL_3185495, EPI_ISL_3185496, EPI_ISL_3185497, EPI_ISL_3185498, EPI_ISL_3185500, EPI_ISL_3185501, EPI_ISL_3185502, EPI_ISL_3185503, EPI_ISL_3185504, EPI_ISL_3185505, EPI_ISL_3185506, EPI_ISL_3185507, EPI_ISL_3185508, EPI_ISL_3185509, EPI_ISL_3185510, EPI_ISL_3185512, EPI_ISL_3185513, EPI_ISL_3185514, EPI_ISL_3185515, EPI_ISL_3185516, EPI_ISL_3185518, EPI_ISL_3185519, EPI_ISL_3185520, EPI_ISL_3185522, EPI_ISL_3185523, EPI_ISL_3185524, EPI_ISL_3185525, EPI_ISL_3185535, EPI_ISL_3185538, EPI_ISL_3185539, EPI_ISL_3185540, EPI_ISL_3185541, EPI_ISL_3185542, EPI_ISL_3185550, EPI_ISL_3185551, EPI_ISL_3185552, EPI_ISL_3185555, EPI_ISL_3185556, EPI_ISL_3185557, EPI_ISL_3185559, EPI_ISL_3185561, EPI_ISL_3185576, EPI_ISL_3185577, EPI_ISL_3185578, EPI_ISL_3185579, EPI_ISL_3185583, EPI_ISL_3185584, EPI_ISL_3220142 | | | |
| see above | Genetica Molecular and Subdepartamento de Virologia ISP Chile | Instituto de Salud Publica de Chile | Andres Castillo; Barbara Parra; Constanza Campano; Gisselle Barra; Javier Tognarelli; Jorge Fernandez; Karen Orostica; Loredana Arata; Patricia Bustos; Rodrigo Fasce; Soledad Ulloa |

| | | | |
|---|---|---|---|
| EPI_ISL_2894605, EPI_ISL_2894606 | Genolife | Genolife | Yenelli Cedano-Thomas |
| EPI_ISL_2967965 | Good Samaritan Hospital | Santa Clara County Public Health Laboratory | Santa Clara County Public Health Department |
| EPI_ISL_2985782, EPI_ISL_2985786 | Gravity Diagnostics, LLC | Gravity Diagnostics, LLC | Gravity Diagnostics |
| EPI_ISL_3536150 | H J M A HOSPITAL JOSE MARTINIANO DE ALENCAR | Analytical Competence Molecular Epidemiology Lab/ACME, Oswaldo Cruz Foundation, Ceara (FIOCRUZ CE) | Cleber Furtado Aksenen; Fabio Miyajima; Fernando Braga Stehling; Francisco Eder de Moura Lopes; Jamille Maria Mendes Bezerra; Joaquim César do Nascimento Sousa Junior; Pedro Miguel Carneiro Jeronimo; Suzana Porto Almeida e Lucas Delerino; Thais Ferreira de Oliveira; Thais de Oliveira Costa; Ticiane Cavalcante de Souza; Veridiana Pessoa Miyajima |
| EPI_ISL_3102382 | HGCC HOSPITAL GERAL DR CESAR CALS | Analytical Competence Molecular Epidemiology Lab/ACME, Oswaldo Cruz Foundation, Ceara (FIOCRUZ CE) | Cleber Furtado Aksenen; Fabio Miyajima; Fernando Braga Stehling; Francisco Eder de Moura Lopes; Jamille Maria Mendes Bezerra; Joaquim César do Nascimento Sousa Junior; Pedro Miguel Carneiro Jeronimo; Suzana Porto Almeida e Lucas Delerino; Thais Ferreira de Oliveira; Thais de Oliveira Costa; Ticiane Cavalcante de Souza; Veridiana Pessoa Miyajima |
| EPI_ISL_3536218 | HIAS HOSPITAL INFANTIL ALBERT SABIN | Analytical Competence Molecular Epidemiology Lab/ACME, Oswaldo Cruz Foundation, Ceara (FIOCRUZ CE) | Cleber Furtado Aksenen; Fabio Miyajima; Fernando Braga Stehling; Francisco Eder de Moura Lopes; Jamille Maria Mendes Bezerra; Joaquim César do Nascimento Sousa Junior; Pedro Miguel Carneiro Jeronimo; Suzana Porto Almeida e Lucas Delerino; Thais Ferreira de Oliveira; Thais de Oliveira Costa; Ticiane Cavalcante de Souza; Veridiana Pessoa Miyajima |
| EPI_ISL_3254636, EPI_ISL_3254637, EPI_ISL_3254638, EPI_ISL_3254639, EPI_ISL_3254640, EPI_ISL_3254641, EPI_ISL_3254642, EPI_ISL_3254643, EPI_ISL_3254644, EPI_ISL_3254645, EPI_ISL_3254646, EPI_ISL_3254647, EPI_ISL_3254648, EPI_ISL_3254649, EPI_ISL_3254650, EPI_ISL_3254651, EPI_ISL_3254652, EPI_ISL_3254653, EPI_ISL_3254654, EPI_ISL_3254684, EPI_ISL_3259352, EPI_ISL_3386153, EPI_ISL_4061651, EPI_ISL_4061657, EPI_ISL_4061659, EPI_ISL_4061660, EPI_ISL_4061661, EPI_ISL_4061673 | | | |
| see above | HLAGYN - Laboratorio de Imunologia de Transplantes de Goias | HLAGYN - Laboratorio de Imunologia de Transplantes de Goias | Alessandro Leonardo Alvares Magalhaes; Erika Lopes Rocha Batista; Fernando Antonio Vinhal dos Santos; Frederico Rodrigues Vinhal; Kamila Oliveira Reis De Freitas.; Lucas Carlos Gomes Pereira; Sabrina Sara Moreira Duarte |
| EPI_ISL_3102420, EPI_ISL_3102430, EPI_ISL_3102432, EPI_ISL_3102461, EPI_ISL_3536237, EPI_ISL_3536238, EPI_ISL_3536243, EPI_ISL_3536259 | | | |
| see above | HM HOSPITAL DE MESSEJANA DR CARLOS ALBERTO STUDART GOMES | Analytical Competence Molecular Epidemiology Lab/ACME, Oswaldo Cruz Foundation, Ceara (FIOCRUZ CE) | Cleber Furtado Aksenen; Fabio Miyajima; Fernando Braga Stehling; Francisco Eder de Moura Lopes; Jamille Maria Mendes Bezerra; Joaquim César do Nascimento Sousa Junior; Pedro Miguel Carneiro Jeronimo; Suzana Porto Almeida e Lucas Delerino; Thais Ferreira de Oliveira; Thais de Oliveira Costa; Ticiane Cavalcante de Souza; Veridiana Pessoa Miyajima |
| EPI_ISL_5669764 | HOSP MAT SAO MIGUEL | Instituto Butantan | Antonio Jorge Martins; Claudia Renata dos Santos Barros; David Schlesinger; Debora Botequio Moretti; Dimas Tadeu Covas; Elaine Cristina Marqueze; Elaine Vieira Santos; Evandra Strazza Rodrigues; Heidge Fukumasu; Jayme Augusto de Souza-Neto; José Salvatore Leister Patané; Luiz Alcantara; Luiz Lehmann Coutinho; Maria Carolina Elias; Maurício Lacerda Nogueira; Rafael dos Santos Bezerra; Raul Machado Neto; Rejane Maria Tommasini Grotto; Ricardo Haddad; Sandra Coccuzzo Sampaio Vessoni; Simone Kashima; Svetoslav Nanev Slavov; Vincent Louis Viala |
| EPI_ISL_5672103, EPI_ISL_5672194, EPI_ISL_5672260, EPI_ISL_5672294, EPI_ISL_5672342 | HOSP MUN CIDADE TIRADENTES CARMEN PRUDENTE | Instituto Butantan | Antonio Jorge Martins; Claudia Renata dos Santos Barros; David Schlesinger; Debora Botequio Moretti; Dimas Tadeu Covas; Elaine Cristina Marqueze; Elaine Vieira Santos; Evandra Strazza Rodrigues; Heidge Fukumasu; Jayme Augusto de Souza-Neto; José Salvatore Leister Patané; Luiz Alcantara; Luiz Lehmann Coutinho; Maria Carolina Elias; Maurício Lacerda Nogueira; Rafael dos Santos Bezerra; Raul Machado Neto; Rejane Maria Tommasini Grotto; Ricardo Haddad; Sandra Coccuzzo Sampaio Vessoni; Simone Kashima; Svetoslav Nanev Slavov; Vincent Louis Viala |
| EPI_ISL_5663769, EPI_ISL_5664868 | HOSP MUN DE MOGI DAS CRUZES PREF WALDEMAR COSTA FILHO | Instituto Butantan | Antonio Jorge Martins; Claudia Renata dos Santos Barros; David Schlesinger; Debora Botequio Moretti; Dimas Tadeu Covas; Elaine Cristina Marqueze; Elaine Vieira Santos; Evandra Strazza Rodrigues; Heidge Fukumasu; Jayme Augusto de Souza-Neto; José Salvatore Leister Patané; Luiz Alcantara; Luiz Lehmann Coutinho; Maria Carolina Elias; Maurício Lacerda Nogueira; Rafael dos Santos Bezerra; Raul Machado Neto; Rejane Maria Tommasini Grotto; Ricardo Haddad; Sandra Coccuzzo Sampaio Vessoni; Simone Kashima; Svetoslav Nanev Slavov; Vincent Louis Viala |
| EPI_ISL_5673337, EPI_ISL_5673413, EPI_ISL_5674920 | HOSP MUN JOSANIAS CASTANHA BRAGA | Instituto Butantan | Antonio Jorge Martins; Claudia Renata dos Santos Barros; David Schlesinger; Debora Botequio Moretti; Dimas Tadeu Covas; Elaine Cristina Marqueze; Elaine Vieira Santos; Evandra Strazza Rodrigues; Heidge Fukumasu; Jayme Augusto de Souza-Neto; José Salvatore Leister Patané; Luiz Alcantara; Luiz Lehmann Coutinho; Maria Carolina Elias; Maurício Lacerda Nogueira; Rafael dos Santos Bezerra; Raul Machado Neto; Rejane Maria Tommasini Grotto; Ricardo Haddad; Sandra Coccuzzo Sampaio Vessoni; Simone Kashima; Svetoslav Nanev Slavov; Vincent Louis Viala |
| EPI_ISL_5659891, EPI_ISL_5660065, EPI_ISL_5660130, EPI_ISL_5660962 | HOSPITAL DA COSTA SUL DE SAO SEBASTIAO | Instituto Butantan | Antonio Jorge Martins; Claudia Renata dos Santos Barros; David Schlesinger; Debora Botequio Moretti; Dimas Tadeu Covas; Elaine Cristina Marqueze; Elaine Vieira Santos; Evandra Strazza Rodrigues; Heidge Fukumasu; Jayme Augusto de Souza-Neto; José Salvatore Leister Patané; Luiz Alcantara; Luiz Lehmann Coutinho; Maria Carolina Elias; Maurício Lacerda Nogueira; Rafael dos Santos Bezerra; Raul Machado Neto; Rejane Maria Tommasini Grotto; Ricardo Haddad; Sandra Coccuzzo Sampaio Vessoni; Simone Kashima; Svetoslav Nanev Slavov; Vincent Louis Viala |
| EPI_ISL_5659835, EPI_ISL_5662548, EPI_ISL_5672037, EPI_ISL_5672073, EPI_ISL_5672308, EPI_ISL_5672560, EPI_ISL_5675338 | | | |
| see above | HOSPITAL DE CAMPANHA COVID 19 CAIEIRAS | Instituto Butantan | Antonio Jorge Martins; Claudia Renata dos Santos Barros; David Schlesinger; Debora Botequio Moretti; Dimas Tadeu Covas; Elaine Cristina Marqueze; Elaine Vieira Santos; Evandra Strazza Rodrigues; Heidge Fukumasu; Jayme Augusto de Souza-Neto; José Salvatore Leister Patané; Luiz Alcantara; Luiz Lehmann Coutinho; Maria Carolina Elias; Maurício Lacerda Nogueira; Rafael dos Santos Bezerra; Raul Machado Neto; Rejane Maria Tommasini Grotto; Ricardo Haddad; Sandra Coccuzzo Sampaio Vessoni; Simone Kashima; Svetoslav Nanev Slavov; Vincent Louis Viala |
| EPI_ISL_5661920, EPI_ISL_5662229, EPI_ISL_5662360, EPI_ISL_5662676, EPI_ISL_5663408, EPI_ISL_5663675, EPI_ISL_5663822, EPI_ISL_5664322, EPI_ISL_5664408, EPI_ISL_5664669, EPI_ISL_5664765, EPI_ISL_5665014, EPI_ISL_5665022, EPI_ISL_5665106, EPI_ISL_5665356, EPI_ISL_5665371, EPI_ISL_5665459 | | | |
| see above | HOSPITAL DE CAMPANHA COVID 19 MUNICIPIO DE TAUBATE | Instituto Butantan | Antonio Jorge Martins; Claudia Renata dos Santos Barros; David Schlesinger; Debora Botequio Moretti; Dimas Tadeu Covas; Elaine Cristina Marqueze; Elaine Vieira Santos; Evandra Strazza Rodrigues; Heidge Fukumasu; Jayme Augusto de Souza-Neto; José Salvatore Leister Patané; Luiz Alcantara; Luiz Lehmann Coutinho; Maria Carolina Elias; Maurício Lacerda Nogueira; Rafael dos Santos Bezerra; Raul Machado Neto; Rejane Maria Tommasini Grotto; Ricardo Haddad; Sandra Coccuzzo Sampaio Vessoni; Simone Kashima; Svetoslav Nanev Slavov; Vincent Louis Viala |
| EPI_ISL_3037826 | HOSPITAL DE NIÑOS DR. CARLOS SAENZ HERRERA | Inciensa, Instituto Costarricense de Investigación y Enseñanza en Nutrición y Salud | Adriana Godínez; Caterina Guzmán; Claudio Soto-Garita; Estela Cordero; Francisco Duarte; Hebleen Porras; José Luis Vargas; Mariela Gutiérrez Joselyn Prado; Melany Calderón; Nazareth Ruiz & Cristian Pérez |
| EPI_ISL_3912027 | HOSPITAL DONA HELENA | Analytical Competence Molecular Epidemiology Lab/ACME, Oswaldo Cruz Foundation, Ceara (FIOCRUZ CE) | Cleber Furtado Aksenen; Fabio Miyajima; Fernando Braga Stehling; Francisco Eder de Moura Lopes; Jamille Maria Mendes Bezerra; Joaquim Cesar do Nascimento Sousa Junior; Pedro Miguel Carneiro Jeronimo; Suzana Porto Almeida & Lucas Delerino on behalf of COVID-19 FIOCRUZ Genomic Network; Thais Ferreira de Oliveira; Thais de Oliveira Costa; Ticiane Cavalcante de Souza; Veridiana Pessoa Miyajima |
| EPI_ISL_5660216, EPI_ISL_5660806, EPI_ISL_5660841, EPI_ISL_5660886, EPI_ISL_5661334, EPI_ISL_5662099 | HOSPITAL E MATERNIDADE NOSSA SENHORA DA AJUDA | Instituto Butantan | Antonio Jorge Martins; Claudia Renata dos Santos Barros; David Schlesinger; Debora Botequio Moretti; Dimas Tadeu Covas; Elaine Cristina Marqueze; Elaine Vieira Santos; Evandra Strazza Rodrigues; Heidge Fukumasu; Jayme Augusto de Souza-Neto; José Salvatore Leister Patané; Luiz Alcantara; Luiz Lehmann Coutinho; Maria Carolina Elias; Maurício Lacerda Nogueira; Rafael dos Santos Bezerra; Raul Machado Neto; Rejane Maria Tommasini Grotto; Ricardo Haddad; Sandra Coccuzzo Sampaio Vessoni; Simone Kashima; Svetoslav Nanev Slavov; Vincent Louis Viala |
| EPI_ISL_5662605, EPI_ISL_5663088, EPI_ISL_5663456, EPI_ISL_5663721, EPI_ISL_5663814, EPI_ISL_5663953, EPI_ISL_5664173, EPI_ISL_5664584, EPI_ISL_5665224, EPI_ISL_5665268, EPI_ISL_5665338 | | | |
| see above | HOSPITAL E PRONTO SOCORRO PORTINARI | Instituto Butantan | Antonio Jorge Martins; Claudia Renata dos Santos Barros; David Schlesinger; Debora Botequio Moretti; Dimas Tadeu Covas; Elaine Cristina Marqueze; Elaine Vieira Santos; Evandra Strazza Rodrigues; Heidge Fukumasu; Jayme Augusto de Souza-Neto; José Salvatore Leister Patané; Luiz Alcantara; Luiz Lehmann Coutinho; Maria Carolina Elias; Maurício Lacerda Nogueira; Rafael dos Santos Bezerra; Raul Machado Neto; Rejane Maria Tommasini Grotto; Ricardo Haddad; Sandra Coccuzzo Sampaio Vessoni; Simone Kashima; Svetoslav Nanev Slavov; Vincent Louis Viala |
| EPI_ISL_5661012, EPI_ISL_5661200, EPI_ISL_5661229, EPI_ISL_5661270, EPI_ISL_5661317, EPI_ISL_5661379, EPI_ISL_5661439, EPI_ISL_5661446, EPI_ISL_5661548, EPI_ISL_5661585, EPI_ISL_5661591, EPI_ISL_5661596, EPI_ISL_5661599, EPI_ISL_5661624, EPI_ISL_5661629, EPI_ISL_5661717, EPI_ISL_5662105 | | | |
| see above | HOSPITAL ENF ANTONIO POLICARPO DE OLIVEIRA | Instituto Butantan | Antonio Jorge Martins; Claudia Renata dos Santos Barros; David Schlesinger; Debora Botequio Moretti; Dimas Tadeu Covas; Elaine Cristina Marqueze; Elaine Vieira Santos; Evandra Strazza Rodrigues; Heidge Fukumasu; Jayme Augusto de Souza-Neto; José Salvatore Leister Patané; Luiz Alcantara; Luiz Lehmann Coutinho; Maria Carolina Elias; Maurício Lacerda Nogueira; Rafael dos Santos Bezerra; Raul Machado Neto; Rejane Maria Tommasini Grotto; Ricardo Haddad; Sandra Coccuzzo Sampaio Vessoni; Simone Kashima; Svetoslav Nanev Slavov; Vincent Louis Viala |
| EPI_ISL_5661768 | HOSPITAL ESCOLA DA UNIVERSIDADE DE TAUBATE | Instituto Butantan | Antonio Jorge Martins; Claudia Renata dos Santos Barros; David Schlesinger; Debora Botequio Moretti; Dimas Tadeu Covas; Elaine Cristina Marqueze; Elaine Vieira Santos; Evandra Strazza Rodrigues; Heidge Fukumasu; Jayme Augusto de Souza-Neto; José Salvatore Leister Patané; Luiz Alcantara; Luiz Lehmann Coutinho; Maria Carolina Elias; Maurício Lacerda Nogueira; Rafael dos Santos Bezerra; Raul Machado Neto; Rejane Maria Tommasini Grotto; Ricardo Haddad; Sandra Coccuzzo Sampaio Vessoni; Simone Kashima; Svetoslav Nanev Slavov; Vincent Louis Viala |
| EPI_ISL_5669618 | HOSPITAL ESTADUAL DE SAPOPEMBA SAO PAULO | Instituto Butantan | Antonio Jorge Martins; Claudia Renata dos Santos Barros; David Schlesinger; Debora Botequio Moretti; Dimas Tadeu Covas; Elaine Cristina Marqueze; Elaine Vieira Santos; Evandra Strazza Rodrigues; Heidge Fukumasu; Jayme Augusto de Souza-Neto; José Salvatore Leister Patané; Luiz Alcantara; Luiz Lehmann Coutinho; Maria Carolina Elias; Maurício Lacerda Nogueira; Rafael dos Santos Bezerra; Raul Machado Neto; Rejane Maria Tommasini Grotto; Ricardo Haddad; Sandra Coccuzzo Sampaio Vessoni; Simone Kashima; Svetoslav Nanev Slavov; Vincent Louis Viala |
| EPI_ISL_3102336, EPI_ISL_3102372, EPI_ISL_3102374, EPI_ISL_3102403, EPI_ISL_3536152, EPI_ISL_3536175, EPI_ISL_3536176, EPI_ISL_3536284, EPI_ISL_3536313, EPI_ISL_3536358, EPI_ISL_3536372, EPI_ISL_3536376, EPI_ISL_3536377, EPI_ISL_3536395 | | | |
| see above | HOSPITAL ESTADUAL LEONARDO DA VINCI | Analytical Competence Molecular Epidemiology Lab/ACME, Oswaldo Cruz Foundation, Ceara (FIOCRUZ CE) | Cleber Furtado Aksenen; Fabio Miyajima; Fernando Braga Stehling; Francisco Eder de Moura Lopes; Jamille Maria Mendes Bezerra; Joaquim César do Nascimento Sousa Junior; Pedro Miguel Carneiro Jeronimo; Suzana Porto Almeida e Lucas Delerino; Thais Ferreira de Oliveira; Thais de Oliveira Costa; Ticiane Cavalcante de Souza; Veridiana Pessoa Miyajima |
| EPI_ISL_4799527 | HOSPITAL GENERAL LA MANCHA CENTRO | Instituto de Salud Carlos III | A. Monzón; F. Casas; I. GAITAN PITERA; I. Jiménez; JORGE; M. Camarero; P. Zaballos; S. Cuesta; S. Iglesias-Caballero; S. Pozo; S. Varona; Sandonís; V. Vázquez-Morón |
| EPI_ISL_4799130, EPI_ISL_4799136 | HOSPITAL GENERAL NUESTRA SEÑORA DEL PRADO | Instituto de Salud Carlos III | A. Monzón; ALICIA; F. Casas; I. Jiménez; I.BETETA LOPEZ; M. Sandonís; P. Zaballos; S. Cuesta; S. Iglesias-Caballero; S. Pozo; S. Varona; V. Camarero; Vázquez-Morón |
| EPI_ISL_4799446 | HOSPITAL GENERAL UNIVERSITARIO SANTA LUCIA | Instituto de Salud Carlos III | A. Monzón; ANA; F. Casas; I. BLAZQUEZ ABELLAN; I. Jiménez; M. Camarero; P. Zaballos; S. Cuesta; S. Iglesias-Caballero; S. Pozo; S. Varona; Sandonís; V. Vázquez-Morón |
| EPI_ISL_5667815 | HOSPITAL GERAL DE ITAPEVI | Instituto Butantan | Antonio Jorge Martins; Claudia Renata dos Santos Barros; David Schlesinger; Debora Botequio Moretti; Dimas Tadeu Covas; Elaine Cristina Marqueze; Elaine Vieira Santos; Evandra Strazza Rodrigues; Heidge Fukumasu; Jayme Augusto de Souza-Neto; José Salvatore Leister Patané; Luiz Alcantara; Luiz Lehmann Coutinho; Maria Carolina Elias; Maurício Lacerda Nogueira; Rafael dos Santos Bezerra; Raul Machado Neto; Rejane Maria Tommasini Grotto; Ricardo Haddad; Sandra Coccuzzo Sampaio Vessoni; Simone Kashima; Svetoslav Nanev Slavov; Vincent Louis Viala |
| EPI_ISL_5662754, EPI_ISL_5662760, EPI_ISL_5663170, EPI_ISL_5667920, EPI_ISL_5667957, EPI_ISL_5668026, EPI_ISL_5668035, EPI_ISL_5668040, EPI_ISL_5672358, EPI_ISL_5672469, EPI_ISL_5674072 | | | |
| see above | HOSPITAL GERAL JESUS TEIXEIRA DA COSTA GUAIANASES SAO PAULO | Instituto Butantan | Antonio Jorge Martins; Claudia Renata dos Santos Barros; David Schlesinger; Debora Botequio Moretti; Dimas Tadeu Covas; Elaine Cristina Marqueze; Elaine Vieira Santos; Evandra Strazza Rodrigues; Heidge Fukumasu; Jayme Augusto de Souza-Neto; José Salvatore Leister Patané; Luiz Alcantara; Luiz Lehmann Coutinho; Maria Carolina Elias; Maurício Lacerda Nogueira; Rafael dos Santos Bezerra; Raul Machado Neto; Rejane Maria Tommasini Grotto; Ricardo Haddad; Sandra Coccuzzo Sampaio Vessoni; Simone Kashima; Svetoslav Nanev Slavov; Vincent Louis Viala |
| EPI_ISL_5664264 | HOSPITAL GERAL SANTA MARCELINA DE ITAIM PAULISTA SAO PAULO | Instituto Butantan | Antonio Jorge Martins; Claudia Renata dos Santos Barros; David Schlesinger; Debora Botequio Moretti; Dimas Tadeu Covas; Elaine Cristina Marqueze; Elaine Vieira Santos; Evandra Strazza Rodrigues; Heidge Fukumasu; Jayme Augusto de Souza-Neto; José Salvatore Leister Patané; Luiz Alcantara; Luiz Lehmann Coutinho; Maria Carolina Elias; Maurício Lacerda Nogueira; Rafael dos Santos Bezerra; Raul Machado Neto; Rejane Maria Tommasini Grotto; Ricardo Haddad; Sandra Coccuzzo Sampaio Vessoni; Simone Kashima; Svetoslav Nanev Slavov; Vincent Louis Viala |

| | | | |
|---|---|---|---|
| EPI_ISL_5668735 | HOSPITAL MUN DR GUIDO GUIDA | Instituto Butantan | Antonio Jorge Martins; Claudia Renata dos Santos Barros; David Schlesinger; Debora Botequio Moretti; Dimas Tadeu Covas; Elaine Cristina Marqueze; Elaine Vieira Santos; Evandra Strazza Rodrigues; Heidge Fukumasu; Jayme Augusto de Souza-Neto; José Salvatore Leister Patané; Luiz Alcantara; Luiz Lehmann Coutinho; Maria Carolina Elias; Maurício Lacerda Nogueira; Rafael dos Santos Bezerra; Raul Machado Neto; Rejane Maria Tommasini Grotto; Ricardo Haddad; Sandra Coccuzzo Sampaio Vessoni; Simone Kashima; Svetoslav Nanev Slavov; Vincent Louis Viala |
| EPI_ISL_5669869 | HOSPITAL MUNICIPAL DR ACILIO CARREON GARCIA | Instituto Butantan | Antonio Jorge Martins; Claudia Renata dos Santos Barros; David Schlesinger; Debora Botequio Moretti; Dimas Tadeu Covas; Elaine Cristina Marqueze; Elaine Vieira Santos; Evandra Strazza Rodrigues; Heidge Fukumasu; Jayme Augusto de Souza-Neto; José Salvatore Leister Patané; Luiz Alcantara; Luiz Lehmann Coutinho; Maria Carolina Elias; Maurício Lacerda Nogueira; Rafael dos Santos Bezerra; Raul Machado Neto; Rejane Maria Tommasini Grotto; Ricardo Haddad; Sandra Coccuzzo Sampaio Vessoni; Simone Kashima; Svetoslav Nanev Slavov; Vincent Louis Viala |
| EPI_ISL_5659723, EPI_ISL_5659843, EPI_ISL_5659859, EPI_ISL_5660154, EPI_ISL_5660720, EPI_ISL_5668562, EPI_ISL_5668589, EPI_ISL_5668641, EPI_ISL_5668703, EPI_ISL_5668993, EPI_ISL_5669001, EPI_ISL_5669446, EPI_ISL_5669656, EPI_ISL_5669826, EPI_ISL_5669848, EPI_ISL_5669891, EPI_ISL_5670070, EPI_ISL_5670103, EPI_ISL_5670111, EPI_ISL_5670164, EPI_ISL_5670190, EPI_ISL_5670212, EPI_ISL_5670270, EPI_ISL_5670425 | | | |
| see above | HOSPITAL MUNICIPAL DR TABAJARA RAMOS | Instituto Butantan | Antonio Jorge Martins; Claudia Renata dos Santos Barros; David Schlesinger; Debora Botequio Moretti; Dimas Tadeu Covas; Elaine Cristina Marqueze; Elaine Vieira Santos; Evandra Strazza Rodrigues; Heidge Fukumasu; Jayme Augusto de Souza-Neto; José Salvatore Leister Patané; Luiz Alcantara; Luiz Lehmann Coutinho; Maria Carolina Elias; Maurício Lacerda Nogueira; Rafael dos Santos Bezerra; Raul Machado Neto; Rejane Maria Tommasini Grotto; Ricardo Haddad; Sandra Coccuzzo Sampaio Vessoni; Simone Kashima; Svetoslav Nanev Slavov; Vincent Louis Viala |
| EPI_ISL_3912125, EPI_ISL_3912216, EPI_ISL_3912217 | HOSPITAL MUNICIPAL NOSSA SENHORA DOS MILAGRES | Analytical Competence Molecular Epidemiology Lab/ACME, Oswaldo Cruz Foundation, Ceara (FIOCRUZ CE) | Cleber Furtado Aksenen; Fabio Miyajima; Fernando Braga Stehling; Francisco Eder de Moura Lopes; Jamille Maria Mendes Bezerra; Joaquim Cesar do Nascimento Sousa Junior; Pedro Miguel Carneiro Jeronimo; Suzana Porto Almeida & Lucas Delerino on behalf of COVID-19 FIOCRUZ Genomic Network; Thais Ferreira de Oliveira; Thais de Oliveira Costa; Ticiane Cavalcante de Souza; Veridiana Pessoa Miyajima |
| EPI_ISL_3761773, EPI_ISL_3761782 | HOSPITAL NACIONAL DE NIÑOS | Inciensa, Instituto Costarricense de Investigación y Enseñanza en Nutrición y Salud | Cristian Pérez-Corrales & Valeria Peralta-Barquero |
| EPI_ISL_5669035, EPI_ISL_5670229, EPI_ISL_5670557, EPI_ISL_5670562 | HOSPITAL REGIONAL DE REGISTRO REGISTRO | Instituto Butantan | Antonio Jorge Martins; Claudia Renata dos Santos Barros; David Schlesinger; Debora Botequio Moretti; Dimas Tadeu Covas; Elaine Cristina Marqueze; Elaine Vieira Santos; Evandra Strazza Rodrigues; Heidge Fukumasu; Jayme Augusto de Souza-Neto; José Salvatore Leister Patané; Luiz Alcantara; Luiz Lehmann Coutinho; Maria Carolina Elias; Maurício Lacerda Nogueira; Rafael dos Santos Bezerra; Raul Machado Neto; Rejane Maria Tommasini Grotto; Ricardo Haddad; Sandra Coccuzzo Sampaio Vessoni; Simone Kashima; Svetoslav Nanev Slavov; Vincent Louis Viala |
| EPI_ISL_3912226 | HOSPITAL REGIONAL UNIMED | Analytical Competence Molecular Epidemiology Lab/ACME, Oswaldo Cruz Foundation, Ceara (FIOCRUZ CE) | Cleber Furtado Aksenen; Fabio Miyajima; Fernando Braga Stehling; Francisco Eder de Moura Lopes; Jamille Maria Mendes Bezerra; Joaquim Cesar do Nascimento Sousa Junior; Pedro Miguel Carneiro Jeronimo; Suzana Porto Almeida & Lucas Delerino on behalf of COVID-19 FIOCRUZ Genomic Network; Thais Ferreira de Oliveira; Thais de Oliveira Costa; Ticiane Cavalcante de Souza; Veridiana Pessoa Miyajima |
| EPI_ISL_4799970 | HOSPITAL SAN PEDRO | Instituto de Salud Carlos III | A. Monzón; F. Casas; I. Jiménez; I.BLASCO ALBERDI; Iglesias-Caballero; M. Sandonís; MIRIAN; P. Zaballos; S. Camarero; S. Cuesta; S. Pozo; S. Varona; V. Vázquez-Morón |
| EPI_ISL_3102409, EPI_ISL_3536207 | HOSPITAL SAO JOSE DE DOENCAS INFECCIOSAS | Analytical Competence Molecular Epidemiology Lab/ACME, Oswaldo Cruz Foundation, Ceara (FIOCRUZ CE) | Cleber Furtado Aksenen; Fabio Miyajima; Fernando Braga Stehling; Francisco Eder de Moura Lopes; Jamille Maria Mendes Bezerra; Joaquim César do Nascimento Sousa Junior; Pedro Miguel Carneiro Jeronimo; Suzana Porto Almeida e Lucas Delerino; Thais Ferreira de Oliveira; Thais de Oliveira Costa; Ticiane Cavalcante de Souza; Veridiana Pessoa Miyajima |
| EPI_ISL_5665898 | HOSPITAL SAO JOSE ITIRAPINA | Instituto Butantan | Antonio Jorge Martins; Claudia Renata dos Santos Barros; David Schlesinger; Debora Botequio Moretti; Dimas Tadeu Covas; Elaine Cristina Marqueze; Elaine Vieira Santos; Evandra Strazza Rodrigues; Heidge Fukumasu; Jayme Augusto de Souza-Neto; José Salvatore Leister Patané; Luiz Alcantara; Luiz Lehmann Coutinho; Maria Carolina Elias; Maurício Lacerda Nogueira; Rafael dos Santos Bezerra; Raul Machado Neto; Rejane Maria Tommasini Grotto; Ricardo Haddad; Sandra Coccuzzo Sampaio Vessoni; Simone Kashima; Svetoslav Nanev Slavov; Vincent Louis Viala |
| EPI_ISL_3007689 | HOSPITAL UNIVERSITARIO SON LLATZER | HOSPITAL UNIVERSITARIO SON ESPASES | Hospital Universitario Son Espases; Servicio de Microbiología |
| EPI_ISL_4799365 | HOSPITAL VIRGEN DE LA SALUD DE TOLEDO | Instituto de Salud Carlos III | A. Monzón; EVA; F. Casas; I. Jiménez; I.HEREDERO GALVEZ; M. Sandonís; P. Zaballos; S. Cuesta; S. Iglesias-Caballero; S. Pozo; S. Varona; V. Camarero; Vázquez-Morón |
| EPI_ISL_3912085, EPI_ISL_3912229 | HPP LUIZ ROBERTO PESSOA AIRES | Analytical Competence Molecular Epidemiology Lab/ACME, Oswaldo Cruz Foundation, Ceara (FIOCRUZ CE) | Cleber Furtado Aksenen; Fabio Miyajima; Fernando Braga Stehling; Francisco Eder de Moura Lopes; Jamille Maria Mendes Bezerra; Joaquim Cesar do Nascimento Sousa Junior; Pedro Miguel Carneiro Jeronimo; Suzana Porto Almeida & Lucas Delerino on behalf of COVID-19 FIOCRUZ Genomic Network; Thais Ferreira de Oliveira; Thais de Oliveira Costa; Ticiane Cavalcante de Souza; Veridiana Pessoa Miyajima |
| EPI_ISL_3062488 | Health sciences Department, Universidad Técnica Particular de Loja | Institute of Microbiology, Universidad San Francisco de Quito | Belén Prado-Vivar; Bernardo Gutiérrez; Erika Muñoz; Fernanda Zurita; Fernando Serrano; Gabriel Trueba; Juan José Guadalupe; Katty Ojeda; Michelle Grunauer; Monica Becerra-Wong; Paola Dalgo; Patricio Rojas-Silva; Paúl Cárdenas; Sully Márquez; Verónica Barragán |
| EPI_ISL_6480794, EPI_ISL_6480800, EPI_ISL_6480813, EPI_ISL_6480819, EPI_ISL_6480820, EPI_ISL_6480835, EPI_ISL_6480836, EPI_ISL_6480856, EPI_ISL_6480880, EPI_ISL_6480944, EPI_ISL_6480946, EPI_ISL_6480953, EPI_ISL_6480989, EPI_ISL_6481064, EPI_ISL_6481101, EPI_ISL_6481105, EPI_ISL_6481109, EPI_ISL_6481126, EPI_ISL_6481155, EPI_ISL_6481172, EPI_ISL_6481249, EPI_ISL_6484125, EPI_ISL_6484176, EPI_ISL_6484198, EPI_ISL_6484902, EPI_ISL_6484965, EPI_ISL_6484999, EPI_ISL_6485014, EPI_ISL_6485048, EPI_ISL_6485073, EPI_ISL_6485085, EPI_ISL_6485091, EPI_ISL_6485100, EPI_ISL_6485119, EPI_ISL_6485123, EPI_ISL_6485129, EPI_ISL_6485131, EPI_ISL_6485140, EPI_ISL_6485149, EPI_ISL_6485158, EPI_ISL_6485171, EPI_ISL_6485185, EPI_ISL_6485189 | | | |
| see above | Helix | Centers for Disease Control and Prevention Division of Viral Diseases, Pathogen Discovery | Benjamin Rambo-Martin; Christopher Gulvick; Clinton Paden; Dakota Howard; Dhwani Batra; Duncan MacCannell; Erisa Sula; Helix CA; Jason Caravas; Kristine Lacek; Matthew Schmerer; Peter Cook; Scott Sammons; Shatavia Morrison; Tymeckia Kendall; Victoria Caban Figueroa; Yvette Unoarumhi |
| EPI_ISL_3825489 | Hosp. 4 de Junio Dr. Ramón Carrillo | Laboratorio de Biología Molecular, Instituto de Medicina Regional on behalf of 'Proyecto Argentino Interinstitucional de genomica de SARS-CoV-2' (PAIS Consortium) | Andrea Carolina Piedrabuena; Bettina Brusés; Diana Pérez Quintana; Gabriela Jurasek; Griselda Oria; Horacio Lucero.; Javier Mussin; Laura Formichelli; Mayra Mendoza; Melina Lorenzini Campos; Raúl Maximiliano Acevedo |
| EPI_ISL_2923267 | Hospital | National Reference Center for Viruses of Respiratory Infections, Institut Pasteur, Paris | Angela Brisebarre; Camille Capel; Christophe Malabat; Corinne Maufrais; Etienne Simon-Lorière; FARFOUR Eric; Frédéric Lemoine; Gael Millot; Louise Lefrançois; Marion Barbet; Maud Vanpeene; Méline Bizard; Sylvie Behillil; Sylvie Van der Werf; Vincent Enouf |
| EPI_ISL_3147295, EPI_ISL_3147332, EPI_ISL_3147478, EPI_ISL_3147480, EPI_ISL_3147496, EPI_ISL_3147521, EPI_ISL_3147554, EPI_ISL_3147557, EPI_ISL_3147567, EPI_ISL_3147568 | | | |
| see above | Hospital Center Emile Mayrisch | Laboratoire national de sante, Microbiology, Microbial Genomics Platform | Anke Wienecke-Baldacchino; Catherine Ragimbeau; Cynthia Oxacelay; Elodie Solarino; Fatu Djabi; Jessica Tapp; Lise Pignon; Raoul Salmon; Tamir Abdelrahman; Virginie Jover |
| EPI_ISL_3147302, EPI_ISL_3147536, EPI_ISL_3147571, EPI_ISL_3147580 | Hospital Center Luxembourg | Laboratoire national de sante, Microbiology, Microbial Genomics Platform | Anke Wienecke-Baldacchino; Catherine Ragimbeau; Elodie Solarino; Fatu Djabi; Jean-Hugues Francois; Jessica Tapp; Lise Pignon; Michel Kohnen; Raoul Salmon; Tamir Abdelrahman; Virginie Jover |
| EPI_ISL_3167156 | Hospital General Universitario Gregorio Marañón | Hospital General Universitario Gregorio Marañón | Cristina Rodriguez-Grande; Darío García de Viedma; Julia Suárez; Laura Pérez-Lago; Marta Herranz Martin; Patricia Muñoz; Pedro Sola Campoy; Pilar Catalán; Sergio Buenestado Serrano; Victor Manuel de la Cueva |
| EPI_ISL_6971766, EPI_ISL_6971767 | Hospital General de Luque | Laboratorio Central de Salud Pública | Andrea Gómez de la Fuente; Cesar Cantero; Cynthia Vazquez; Juan Torales; Julio Barrios; Maria Liz Gamarra; Sandra Gonzalez; Shirley Villalba; Tania Alfonzo |
| EPI_ISL_5926920, EPI_ISL_7740276, EPI_ISL_7740285 | Hospital Herrera Llerandi | Asociación de Salud Integral / Clínica Familiar "Luis Ángel García" | Ana S. Gonzalez-Reiche; Claudia Rangel; Danicela Mercado; Eduardo Arathon; Hilda Ruiz; Luis Aguirre; Luis Rivas; Narda Medina; Oscar Bonilla; Osmar Gamboa. |
| EPI_ISL_3260106, EPI_ISL_3260110 | Hospital Jaime Ferre - SAMCO Rafaela | Grupo de Genómica y Bioinformática del Instituto de Investigación de la Cadena Láctea CONICET-INTA on behalf of 'Proyecto Argentino Interinstitucional de genomica de SARS-CoV-2' (PAIS Consortium) | AF; Aliprandi D; Amadio; C; Eberhardt; Irazoqui; Isaia; JF; JM; MF; Pandolfi; Quaranta; Soratti R; V |
| EPI_ISL_7711723 | Hospital Monseñor Sanabria | Universidad de Costa Rica e Instituto Costarricense de Investigacion y Ensenanza en Nutricion y Salud | Brenes Hebleen; Camacho Erwin; Campos-Sanchez Rebeca; Ceballos Ana; Cordero Estela; Cristancho Marco; Duarte Francisco; Fernandez-Do Porto Dario; Herrera-Estrella Alfredo; Jimenez-Moraila Beatriz; Kreuze Jan; Mireles-Rivera Guadalupe; Molina-Mora Jose; Munoz-Medina Jose Esteban; Negri Tatiana; Nunes Gisele; Oliveira Guilherme; Oliveira Renato; Reales-Gonzalez Jhonnatan; Remes-Lenicov Federico; Reyes Alejandro; Sosa Ezequiel; Soto Claudio; Tsukayama Pablo; Turjanski Adrian; and Zayat Jonathan |
| EPI_ISL_2828233 | Hospital Universitari Arnau de Vilanova | Hospital Universitari Vall d'Hebron - Vall d'Hebron Institut de Recerca | Alejandra González-Sánchez; Andrés Antón; Ariadna Rando; Carla Castillo; Cristina Andrés; Damir Garcia-Cehic; Josep Quer; Juliana Esperalba; Maria Carmen Martin; Maria Gema Codina; Maria Piñana; Tomàs Pumarola |
| EPI_ISL_6718313, EPI_ISL_6718325, EPI_ISL_6718329, EPI_ISL_6718347, EPI_ISL_6718389, EPI_ISL_6718420, EPI_ISL_6718424, EPI_ISL_6718445, EPI_ISL_6718466 | | | |
| see above | Hospital Universitari Dr. Josep Trueta | Institut d'Investigació Biomèdica de Girona \| Hospital Universitari Dr. Josep Trueta | Bernat del Olmo; Mel·lina Pinsach; Meritxell Deulofeu; Nuria Esther Neto; Paula Costa |
| EPI_ISL_2828114, EPI_ISL_2885277, EPI_ISL_2885302, EPI_ISL_2885342, EPI_ISL_2938032, EPI_ISL_2938048, EPI_ISL_2938086, EPI_ISL_2982331, EPI_ISL_2982387 | | | |
| see above | Hospital Universitari Vall d'Hebron - Vall d'Hebron Institut de Recerca | Hospital Universitari Vall d'Hebron - Vall d'Hebron Institut de Recerca | Alejandra González-Sánchez; Andrés Antón; Ariadna Rando; Carla Castillo; Cristina Andrés; Damir Garcia-Cehic; Josep Quer; Juliana Esperalba; Karen García; Maria Carmen Martin; Maria Gema Codina; Maria Piñana; Rodrigo Vásquez; Tomàs Pumarola |
| EPI_ISL_3087929, EPI_ISL_3087940 | Hospital Universitari i Politècnic La Fe de València | SeqCOVID-SPAIN consortium/IBV(CSIC) | Ana Gil Brusola; Eva González Barberá; José Luis López Hontangas and SeqCOVID-SPAIN consortium; María Dolores Gómez Ruiz; Salvador Giner Almaraz |
| EPI_ISL_7307241 | Hospital Universitario San Ignacio | Centro de Investigaciones en Microbiología y Biotecnología-UR (CIMBIUR), Facultad de Ciencias Naturales, Universidad del | Alberto Paniz-Mondolfi; Angie Ramírez; Beatriz Ariza; Camilo A. Correa-Cárdenas; Carlos Gómez-Restrepo; Claudia Cardozo-Romero; Claudia Méndez; David-Santiago Quevedo; Guido España; Hernando Díaz; Juan David Ramírez; Juliana Cuervo-Rojas; Julie Pérez; Luz H. Patiño; Manuel-Antonio Franco; Maria-Clara Duque; Marina Muñoz; Nathalia Ballesteros; Nicolas Luna; Sergio Castañeda; Zulma M. Cucunubá |

| | | | |
|---|---|---|---|
| | | Rosario, Bogotá, Colombia | |
| EPI_ISL_5195518, EPI_ISL_5195519 | Hospital da Baleia | Instituto René Rachou / Fiocruz Minas | Alana Oliveira; Anna Salim; Camila Corsini; Daniel Miranda; Gabriel Fernandes; Mozar de Castro; Nathalie Almeida; Pedro Alves; Priscilla Filgueiras; Rafaella Fortini; Raphael Silva; Raquel Vilela; Rubens do Monte Neto; Sarah Gomes; Thaís Silva; Wander Jeremias |
| EPI_ISL_2968613, EPI_ISL_2968620 | Houston Health Dept. | Houston Health Dept. | Adolpho Lara; Pamela Brown; Ryker Penn; Yanlai Lai |
| EPI_ISL_5073348, EPI_ISL_5073368, EPI_ISL_5073396, EPI_ISL_5073399, EPI_ISL_5073420, EPI_ISL_5073434, EPI_ISL_5073460, EPI_ISL_5073473, EPI_ISL_5073474, EPI_ISL_5073507, EPI_ISL_5073518, EPI_ISL_5073525 | | | |
| see above | Houston Methodist Hospital | Houston Methodist Hospital | Ilya J. Finkelstein; James J. Davis; Jessica Cambric; Jimmy Gollihar; Kristina Reppond; Layne Pruitt; Madison N. Shyer; Marcus Nguyen; Matthew Ojeda Saavedra; Paul A. Christensen; Prasanti Yerramilli; Randall J. Olsen; Robert Olson; Ryan Gadd; S. Wesley Long; Sishir Subedi; and James M. Musser |
| EPI_ISL_3276689 | Huntington Memorial Hospital | Los Angeles County Public Health Laboratories | P. Hemarajata et al. |
| EPI_ISL_2878256 | Hôpital Cochin | Department of Virology, Henri Mondor University Hospital, Assistance Publique Hôpitaux de Paris, Université Paris-Est Créteil, INSERM U955 | Alexandre Soulier; Christophe Rodriguez; Elisabeth Trawinski; Guillaume Gricourt; Jean-Michel Pawlotsky; Melissa N'Debi; Slim Fourati; Vanessa Demontant |
| EPI_ISL_3147532, EPI_ISL_3147538, EPI_ISL_3147560 | H⌠pitaux Robert Schuman | Laboratoire national de sante, Microbiology, Microbial Genomics Platform | Alain Hakim; Anke Wienecke-Baldacchino; Catherine Ragimbeau; Elodie Solarino; Fatu Djabi; Jessica Tapp; Lise Pignon; Nawfal Faik; Raoul Salmon; Tamir Abdelrahman; Virginie Jover |
| EPI_ISL_3128023, EPI_ISL_3128032 | ID Bureau of Laboratories | Centers for Disease Control and Prevention Division of Viral Diseases, Pathogen Discovery | Alex Burgin; Ben L. Rambo-Martin; Clinton R. Paden; Dakota Howard; Dave Wentworth; Dhwani Batra; Jasmine Padilla; Justin Lee; Krista Queen; Kristen Knipe; Kristine Lacek; Mark Burroughs; Matthew Schmerer; Meghan Bentz; Mili Sheth; Peter Cook; Sam Shepard; Sarah Nobles; Suxiang Tong; Vivien Dugan; Yvette Unoarumhi |
| EPI_ISL_2812810 | IFO Regina Elena | National Institute for Infectious Diseases (INMI) L. Spallanzani I.R.C.C.S | A Di Caro; B Bartolini; CEM Gruber; E Giombini; F Messina; F Santini; G Bonfiglio; M Rueca; MR Capobianchi; O Butera |
| EPI_ISL_2789899, EPI_ISL_2860400, EPI_ISL_2967005 | IN State Department of Health Laboratory Services | IN State Department of Health Laboratory Services | Brian Pope; Cassandra Campion; Jamie Yeadon; Kyle Brownlee; Lixia Liu; Mark Glazier; Melissa Hindenlang |
| EPI_ISL_5661436, EPI_ISL_5661559, EPI_ISL_5661608, EPI_ISL_5661657, EPI_ISL_5661732, EPI_ISL_5661748, EPI_ISL_5661818, EPI_ISL_5661887, EPI_ISL_5661899, EPI_ISL_5661923, EPI_ISL_5661960, EPI_ISL_5661965, EPI_ISL_5661967, EPI_ISL_5661991, EPI_ISL_5661998, EPI_ISL_5662021, EPI_ISL_5662033, EPI_ISL_5662039, EPI_ISL_5662051, EPI_ISL_5662060, EPI_ISL_5662065, EPI_ISL_5662085, EPI_ISL_5662086, EPI_ISL_5662103, EPI_ISL_5662107, EPI_ISL_5662114, EPI_ISL_5662117, EPI_ISL_5662134, EPI_ISL_5662162, EPI_ISL_5662180, EPI_ISL_5662291, EPI_ISL_5662294, EPI_ISL_5662303, EPI_ISL_5662320, EPI_ISL_5662325, EPI_ISL_5662331, EPI_ISL_5662368, EPI_ISL_5662385, EPI_ISL_5662404, EPI_ISL_5662418, EPI_ISL_5662432, EPI_ISL_5662465, EPI_ISL_5662477, EPI_ISL_5662503, EPI_ISL_5662511, EPI_ISL_5662516, EPI_ISL_5662533, EPI_ISL_5662564, EPI_ISL_5662584, EPI_ISL_5662636, EPI_ISL_5662647, EPI_ISL_5662685, EPI_ISL_5662691, EPI_ISL_5662701, EPI_ISL_5662778, EPI_ISL_5662803, EPI_ISL_5662842, EPI_ISL_5662847, EPI_ISL_5662886, EPI_ISL_5662894, EPI_ISL_5662898, EPI_ISL_5662902, EPI_ISL_5662922, EPI_ISL_5662931, EPI_ISL_5662933, EPI_ISL_5662962, EPI_ISL_5662969, EPI_ISL_5662973, EPI_ISL_5662997, EPI_ISL_5663018, EPI_ISL_5663038, EPI_ISL_5663046, EPI_ISL_5663085, EPI_ISL_5663110, EPI_ISL_5663204, EPI_ISL_5663216, EPI_ISL_5663240, EPI_ISL_5663250, EPI_ISL_5663264, EPI_ISL_5663271, EPI_ISL_5669106, EPI_ISL_5669192, EPI_ISL_5669509, EPI_ISL_5670677, EPI_ISL_5673609, EPI_ISL_5675214 | | | |
| see above | INSIDE DIAGNOSTICOS | Instituto Butantan | Antonio Jorge Martins; Claudia Renata dos Santos Barros; David Schlesinger; Debora Botequio Moretti; Dimas Tadeu Covas; Elaine Cristina Marqueze; Elaine Vieira Santos; Evandra Strazza Rodrigues; Heidge Fukumasu; Jayme Augusto de Souza-Neto; José Salvatore Leister Patané; Luiz Alcantara; Luiz Lehmann Coutinho; Maria Carolina Elias; Maurício Lacerda Nogueira; Rafael dos Santos Bezerra; Raul Machado Neto; Rejane Maria Tommasini Grotto; Ricardo Haddad; Sandra Coccuzzo Sampaio Vessoni; Simone Kashima; Svetoslav Nanev Slavov; Vincent Louis Viala |
| EPI_ISL_2988448, EPI_ISL_2988462, EPI_ISL_2988463, EPI_ISL_2988495, EPI_ISL_2988498, EPI_ISL_2988501, EPI_ISL_2988502, EPI_ISL_2988526, EPI_ISL_2988552, EPI_ISL_2988560, EPI_ISL_2988561, EPI_ISL_2988577, EPI_ISL_2988581, EPI_ISL_3010437, EPI_ISL_3010458, EPI_ISL_3068147, EPI_ISL_3068193, EPI_ISL_3068207, EPI_ISL_3068233 | | | |
| see above | INSPI-CRN DE INFLUENZA Y OTROS VIRUS RESPIRATORIOS | NIC-INSPI | Alfredo Bruno; Daniel Ramos; Domenica de Mora.; Jimmy Garcés; Johanna Laines; Lizbeth Patiño; Manuel Gonzalez; Marcela Mejía; Maria Angelica Becerra; Maritza Olmedo; Mayra Wilca; Michelle Páez; Milton Medina |
| EPI_ISL_3088316, EPI_ISL_3088317, EPI_ISL_3088319 | INTERLAB | Omics Sciences Laboratory | Darlyn Amaya; Derly Andrade Molina; Gabriel Morey León; Juan Carlos Fernández Cadena; Rubén Armas González |
| EPI_ISL_2922996, EPI_ISL_2923003, EPI_ISL_2923016, EPI_ISL_2923022, EPI_ISL_2923028, EPI_ISL_2923039, EPI_ISL_2923047, EPI_ISL_2923063, EPI_ISL_2923075, EPI_ISL_2923085, EPI_ISL_2923093, EPI_ISL_2985074, EPI_ISL_2985343 | | | |
| see above | IRCCS San Gallicano Dermatological Institute | IRCCS Regina Elena National Cancer Institute | Aldo Morrone; Alessia Lauretti; Alice Massacci; Andrea Cazzani; Antonio Federico; Eleonora Sperandio; Elisabetta Trento; Fabrizio Ensoli; Francesca De Nicola; Frauke Goeman; Fulvia Pimpinelli; Gennaro Ciliberto; Giovanna D'agosto; Giovanni Blandino; Giulia Orlandi; Grazia Prignano; Ilaria Cavallo; Lorenzo Furzi; Ludovica Ciuffreda; Martina Diano; Martina Pontone; Matteo Pallocca; Maurizio Fanciulli; Sabrina Strano; Sara Donzelli; Serena Salvo |
| EPI_ISL_5661742, EPI_ISL_5662911 | IRMANDADE DA SANTA CASA DE MISERICORDIA LORENA | Instituto Butantan | Antonio Jorge Martins; Claudia Renata dos Santos Barros; David Schlesinger; Debora Botequio Moretti; Dimas Tadeu Covas; Elaine Cristina Marqueze; Elaine Vieira Santos; Evandra Strazza Rodrigues; Heidge Fukumasu; Jayme Augusto de Souza-Neto; José Salvatore Leister Patané; Luiz Alcantara; Luiz Lehmann Coutinho; Maria Carolina Elias; Maurício Lacerda Nogueira; Rafael dos Santos Bezerra; Raul Machado Neto; Rejane Maria Tommasini Grotto; Ricardo Haddad; Sandra Coccuzzo Sampaio Vessoni; Simone Kashima; Svetoslav Nanev Slavov; Vincent Louis Viala |
| EPI_ISL_3218885, EPI_ISL_3281530 | IVY3 Central Lab, Vanderbilt University Medical Center | Lauring Lab, University of Michigan, Department of Microbiology and Immunology | Gilbert; Valesano |
| EPI_ISL_3275132 | Idaho Bureau of Laboratories | Center for Global Health, University of New Mexico Health Sciences Center | Christopher Ball; Darrell Dinwiddie; Daryl Domman; Kurt Schwalm; Matthew Burns; Robert Voermans; Valerie Morley |
| EPI_ISL_3254717, EPI_ISL_3254817 | Illinois Department of Public Health | Gagnon Lab, Southern Illinois University | Keith Gagnon |
| EPI_ISL_3231908, EPI_ISL_3231999, EPI_ISL_3232014, EPI_ISL_3232020 | Illinois Department of Public Health | Illinois Department of Public Health - Chicago Lab | Ira Heimler; Joel Price; Vineet K. Dhiman |
| EPI_ISL_3088454, EPI_ISL_3133523 | Illinois Department of Public Health - Springfield Lab | Illinois Department of Public Health - Springfield Lab | Bryan Sim; Gordon McCall; Marshall Smock |
| EPI_ISL_3105052, EPI_ISL_3347000 | Incyte Diagnostics Spokane | Seattle Flu Study | Amanda Adler; Barry R. Lutz; Benjamin Pelle; Brian Hiatt; Caitlin R. Wolf; Chris D. Frazar; Deborah A. Nickerson; Elisabeth Brandstetter; Erica Ryke; Geoff Melly; Helen Y. Chu; Janet A. Englund; Jay Shendure; Jover Lee; Kairsten Fay; Kirsten Lacombe; Lea M. Starita; Mark J. Rieder; Matthew Richardson; Matthew Thompson; Melissa Truong; Michael Boeckh; Michael Famulare; Misja Ilcisin; Peter D. Han; Philip Dykema; Romesh Gautom; Scott Lindquist; Thomas R. Sibley; Trevor Bedford |
| EPI_ISL_3020018, EPI_ISL_3020027, EPI_ISL_3020043, EPI_ISL_4383484 | Infinity Biologix | Centers for Disease Control and Prevention Division of Viral Diseases, Pathogen Discovery | Adrian Paskey; Benjamin Rambo-Martin; Chirayu Goswami; Christian Bixby; Christopher Gulvick; Clinton Paden; Clinton R. Paden; Dakota Howard; Darlene Wagner; Dhwani Batra; Duncan MacCannell; Erisa Sula; Jason Caravas; Jonathan Schultz; Kara Moser; Kristine Lacek; Matthew Schmerer; Peter Cook; Peter W. Cook; Robin Grimwood; Russ Hager; Scott Sammons; Shatavia Morrison; Tymeckia Kendall; Victoria Caban Figueroa; Yihe Wang; Yvette Unoarumhi |
| EPI_ISL_3031143 | Innlandet Hospital Trust, Division Lillehammer, Department for Medical Microbiology | Norwegian Institute of Public Health, Department of Virology | Atiya R Ali; Debech Nadia; Engebretsen Serina Beate; Garcia Llorente Ignacio; Hilde Elshaug; Hilde Vollan; Jon Bråte; Kamilla Heddeland Instefjord; Karoline Bragstad; Kathrine Stene-Johansen; Line Victoria Moen; Marie Paulsen Madsen; Olav Hungnes; Pedersen Benedikte Nevjen; Rasmus Riis Kopperud |
| EPI_ISL_2956071, EPI_ISL_2956076 | Institut für Labormedizin Mikrobiologie und Hygiene | Robert Koch Institute | |
| EPI_ISL_2840487 | Institute for Infectious Diseases | Institute for Infectious Diseases | Alban Ramette; Christian Baumann; Cora SÃ¤gesser; Franziska Suter-Riniker; Miguel A Terrazos Miani; Pascal Bittel; Peter Keller; Stefan Neuenschwander; Stephen L Leib |
| EPI_ISL_3426469 | Institute of Microbiology and Immunology, Faculty of Medicine, University of Ljubljana | Institute of Microbiology and Immunology, Faculty of Medicine, University of Ljubljana | Alen Suljič; Andraž Celar; Dominika Šturm; Doroteja Vlaj; Mario Poljak; Matic Brvar; Miša Korva; Patricija Pozvek; Samo Zakotnik; Tatjana Avšič – Županc; Tomaž Mark Zorec; Špela Pleh |
| EPI_ISL_2861156, EPI_ISL_3062498, EPI_ISL_3062500 | Institute of Microbiology, Universidad San Francisco de Quito | Institute of Microbiology, Universidad San Francisco de Quito | Belén Prado-Vivar; Bernardo Darquea; Bernardo Gutiérrez; Christian Zambrano; Erika Muñoz; Fernanda Zurita; Gabriel Trueba; Juan Carlos Zuñiga; Juan José Guadalupe; Michelle Grunauer; Monica Becerra-Wong; Patricio Rojas-Silva; Paúl Cárdenas; Sully Márquez; Verónica Barragán |
| EPI_ISL_5538373, EPI_ISL_5538378, EPI_ISL_5538388, EPI_ISL_5538649 | Instituto Adolfo Lutz - Regional de Bauru | Instituto Adolfo Lutz, Interdiciplinary Procedures Center, Strategic Laboratory | Caio Vinicius Dias Lopes; Claudia Regina Gonçalves; Claudio Tavares Sacchi; Karoline Rodrigues Campos; Leonardo Tadeu de Araujo; Marlon Benedito Nascimento Santos |
| EPI_ISL_3545799 | Instituto Adolfo Lutz - Regional de Campinas | Instituto Adolfo Lutz, Interdiciplinary Procedures Center, Strategic Laboratory | Caio Vinicius Dias Lopes; Claudia Regina Gonçalves; Claudio Tavares Sacchi; Karoline Rodrigues Campos; Leonardo Tadeu de Araujo; Marlon Benedito Nascomento Santos |
| EPI_ISL_3717846, EPI_ISL_3717847, EPI_ISL_3717848, EPI_ISL_3717849, EPI_ISL_3717874, EPI_ISL_3717875 | Instituto Adolfo Lutz - Regional de Marilia | Instituto Adolfo Lutz, Interdiciplinary Procedures Center, Strategic Laboratory | Caio Vinicius Dias Lopes; Claudia Regina Gonçalves; Claudio Tavares Sacchi; Karoline Rodrigues Campos; Leonardo Tadeu de Araujo; Marlon Benedito Nascimento Santos |
| EPI_ISL_3316164, EPI_ISL_3316206, EPI_ISL_3316243, EPI_ISL_3316307, EPI_ISL_4299342, EPI_ISL_4299343, EPI_ISL_4299344, EPI_ISL_4299345 | | | |
| see above | Instituto Adolfo Lutz - Regional de Ribeirao Preto | Instituto Adolfo Lutz, Interdiciplinary Procedures Center, Strategic Laboratory | Caio Vinicius Dias Lopes; Claudia Regina Gonçalves; Claudio Tavares Sacchi; Karoline Rodrigues Campos; Leonardo Tadeu de Araujo; Marlon Benedito Nascimento Santos; Marlon Benedito Nascomento Santos |
| EPI_ISL_3671877 | Instituto Adolfo Lutz - Regional de Ribeirao Preto | Instituto Adolfo Lutz, Interdisciplinary Procedures | Caio Vinicius Dias Lopes; Claudia Regina Gonçalves; Claudio Tavares Sacchi; Karoline Rodrigues Campos; Leonardo Tadeu de Araujo; Marlon Benedito Nascomento Santos |

| | | | |
|---|---|---|---|
| | | Center, Strategic Laboratory | |
| EPI_ISL_3316296, EPI_ISL_3316297, EPI_ISL_3316299, EPI_ISL_5640215, EPI_ISL_5640216, EPI_ISL_5640220 | Instituto Adolfo Lutz - Regional de Rio Claro | Instituto Adolfo Lutz, Interdiciplinary Procedures Center, Strategic Laboratory | Caio Vinicius Dias Lopes; Claudia Regina Gonçalves; Claudio Tavares Sacchi; Karoline Rodrigues Campos; Leonardo Tadeu de Araujo; Marlon Benedito Nascimento Santos; Marlon Benedito Nascomento Santos |
| EPI_ISL_3717876, EPI_ISL_3717877, EPI_ISL_3717878, EPI_ISL_3717882, EPI_ISL_3717884, EPI_ISL_3717885, EPI_ISL_3717886, EPI_ISL_4056259, EPI_ISL_4056260, EPI_ISL_4056262, EPI_ISL_4056263, EPI_ISL_4056264, EPI_ISL_4056289, EPI_ISL_4056290, EPI_ISL_4056291, EPI_ISL_4056292, EPI_ISL_4056293, EPI_ISL_4056294, EPI_ISL_4056295 | | | |
| see above | Instituto Adolfo Lutz - Regional de Santo Andre | Instituto Adolfo Lutz, Interdiciplinary Procedures Center, Strategic Laboratory | Caio Vinicius Dias Lopes; Claudia Regina Gonçalves; Claudio Tavares Sacchi; Karoline Rodrigues Campos; Leonardo Tadeu de Araujo; Marlon Benedito Nascimento Santos |
| EPI_ISL_3671878 | Instituto Adolfo Lutz - Regional de Santo Andre | Instituto Adolfo Lutz, Interdisciplinary Procedures Center, Strategic Laboratory | Caio Vinicius Dias Lopes; Claudia Regina Gonçalves; Claudio Tavares Sacchi; Karoline Rodrigues Campos; Leonardo Tadeu de Araujo; Marlon Benedito Nascimento Santos |
| EPI_ISL_3259689, EPI_ISL_3259690 | Instituto Adolfo Lutz - Regional de Sorocaba | Instituto Adolfo Lutz, Interdiciplinary Procedures Center, Strategic Laboratory | Caio Vinicius Dias Lopes; Claudia Regina Gonçalves; Claudio Tavares Sacchi; Erica Valessa Ramos Gomes; Karoline Rodrigues Campos; Leonardo Tadeu de Araujo; Marlon Benedito Nascimento Santos |
| EPI_ISL_3316280, EPI_ISL_3316281, EPI_ISL_3316287, EPI_ISL_3316288, EPI_ISL_3316291, EPI_ISL_3316292, EPI_ISL_3545798, EPI_ISL_3671905, EPI_ISL_3691691, EPI_ISL_3691692, EPI_ISL_3691693, EPI_ISL_3691700, EPI_ISL_4206898, EPI_ISL_4206917, EPI_ISL_4206921, EPI_ISL_4206934, EPI_ISL_4206938, EPI_ISL_4206942, EPI_ISL_4206947, EPI_ISL_4206957, EPI_ISL_4206968, EPI_ISL_4299346, EPI_ISL_4299347, EPI_ISL_4299348, EPI_ISL_4888067, EPI_ISL_4888072, EPI_ISL_4888081, EPI_ISL_4888087, EPI_ISL_4888102, EPI_ISL_5030014, EPI_ISL_5030018 | | | |
| see above | Instituto Adolfo Lutz Central | Instituto Adolfo Lutz, Interdiciplinary Procedures Center, Strategic Laboratory | Caio Vinicius Dias Lopes; Claudia Regina Goncalves; Claudia Regina Gonçalves; Claudio Tavares Sacchi; Karoline Rodrigues Campos; Leonardo Tadeu de Araujo; Marlon Benedito Nascimento Santos; Marlon Benedito Nascomento Santos |
| EPI_ISL_3268375, EPI_ISL_3268376, EPI_ISL_3268377 | Instituto Nacional de Enfermedades Respiratorias (INER) | Centro de Investigación en Enfermedades Infecciosas (CIENI), Instituto Nacional de Enfermedades Respiratorias (INER) | Alejandra Garcia-Gasca; Alejandra Hernández-Terán; Alejandro Sanchez-Flores; Alfredo Herrera-Estrella; Alicia Ocaña-Mondragón; Andreu Comas-García; Angel Gustavo Salas-Lais; Antonio Loza Román; Bernardo Martínez-Miguel; Blanca Taboada; Brenda Irasema Maldonado-Meza; Bruno Gomez-Gil; Carla Ivón Herrera-Najera; Carlos F. Arias; Celia Boukadida; Clara Esperanza Santacruz-Tinoco; Concepción Grajales-Muñiz; Consorcio Mexicano de Vigilancia Genómica (CoViGen-Mex). Authors (in alphabetical order): Julio Elias Alvarado-Yaah; Cristóbal Cháidez-Quiróz; Célida Duque Molina; Célida Martínez- Rodríguez; Daniel Fregoso-Rueda; Daniel Lira Morales; Eduardo Becerril-Vargas; Fernando Fontove-Herrera; Fidencio Mejía-Nepomuceno; Francisco Pulido; Gloria Elena Espinosa-Ayala; Gloria María Molina-Salinas; Gloria Vazquez; Hector Esteban Paz-Juárez; Hector Montoya-Fuentes; Helen Haydee Fernanda Ramírez-Plascencia; Irvin González-López; Jean Pierre González; Joel Armando Vázquez-Pérez.; Jorge Salas-Hernández; José Antonio Enciso-Moreno; José Arturo Martínez-Orozco; José Esteban Muñoz-Medina; José de Jesús Nuñez-Contreras; Juan Bautista Chale-Dzul; Julissa Enciso-Ibarra; Kathia Elizabeth Tapia-Diaz; Luis Alberto Ochoa-Carrera; Margarita Matías-Florentino; Mario Mújica-Sánchez; Marissa Pérez-García; María Guadalupe Santiago-Mauricio; María Guadalupe de Jesús Mireles-Rivera; Nelly Sélem-Mojica; Pavel Isa; Ricardo Ciria Merce; Ricardo Grande; Rosa María Gutierrez Rios; Santiago Ávila-Ríos; Selene Zárate; Susana Lopez; Victor Eduardo Garcia-Arias; Víctor Hugo Borja-Aburto |
| EPI_ISL_2894284, EPI_ISL_2894306, EPI_ISL_2894311, EPI_ISL_2894326, EPI_ISL_2894329 | Instituto Nacional de Medicina Genomica | Instituto Nacional de Medicina Genomica | Cedro-Tanda A; Cisneros- Villanueva M; Escobar-Arrazola; Gonzalez-Barrera D; Herrera-Montalvo LA.; Hidalgo-Miranda A; M; Mendoza-Vargas A; Munguia-Garza P; Ramirez-Vega O; Rangel-DeLeon D; Reyes-Grajeda JP |
| EPI_ISL_2844449, EPI_ISL_2844450, EPI_ISL_2844455, EPI_ISL_2844471, EPI_ISL_2844472, EPI_ISL_2844476, EPI_ISL_2844477, EPI_ISL_2844479, EPI_ISL_2844480, EPI_ISL_2844484, EPI_ISL_2844486, EPI_ISL_2844487 | | | |
| see above | Istituto Zooprofilattico Sperimentale del Mezzogiorno | TIGEM | Antonio Grimaldi Patrizia Annunziata Francesco Panariello Biancamaria Pierri Claudia Tiberio Teresa Giuliano Valentina Bouche Chiara Colantuono Maria Concetta Cuomo Denise Di Concilio Lucio Di Filippo Anna Manfredi Marcello Salvi Antonio Limone Luigi Atripaldi Pellegrino Cerino Andrea Ballabio Davide Cacchiarelli |
| EPI_ISL_3825500, EPI_ISL_3825521, EPI_ISL_3825650, EPI_ISL_3825690, EPI_ISL_3825724, EPI_ISL_4102673, EPI_ISL_4102687, EPI_ISL_4102785, EPI_ISL_4102788, EPI_ISL_4260949, EPI_ISL_4261019, EPI_ISL_4261059, EPI_ISL_4261108, EPI_ISL_4261130, EPI_ISL_4261133 | | | |
| see above | Kaiser Permanente NW Reginal Lab | OHSU MM Lab | Amber Halse; Jeannine Lama; Xuan Qin; Yun Wu |
| EPI_ISL_4417750, EPI_ISL_4419028, EPI_ISL_4419035, EPI_ISL_4461962, EPI_ISL_4461963, EPI_ISL_4461969 | Kentucky State Public Health Lab | Kentucky State Public Health Lab | Anjayya Tummala; Chitralipi Kattunga; Joshua Tobias; Karim George; Matthew Johnson; Rachel Zinner; Rhonda Lucas; Vaneet Arora; William Grooms |
| EPI_ISL_3007740 | LABORATORIO DE BIOLOGIA MOLECULAR CENTRO MEDICO NAVAL SEMAR | LABORATORIO DE PATOLOGÍA MOLECULAR CENTRO MEDICO NAVAL SEMAR | ; Alvarez Jimenez Violeta Dejanira; Baltierra Uribe Shantal; Cabrera Gonzalez Moises; Figueroa Antonio Victor; Garcia Alegria Irma; Lievano Esquinca Jazmin; Lopez Bustos Abraham; Meixueiro Calderon Claudia; Morales Ramirez Omar; Zuno Coronado Teutli |
| EPI_ISL_5666879 | LABORATORIO DE FRANCA | Instituto Butantan | Antonio Jorge Martins; Claudia Renata dos Santos Barros; David Schlesinger; Debora Botequio Moretti; Dimas Tadeu Covas; Elaine Cristina Marqueze; Elaine Vieira Santos; Evandra Strazza Rodrigues; Heidge Fukumasu; Jayme Augusto de Souza-Neto; José Salvatore Leister Patané; Luiz Alcantara; Luiz Lehmann Coutinho; Maria Carolina Elias; Maurício Lacerda Nogueira; Rafael dos Santos Bezerra; Raul Machado Neto; Rejane Maria Tommasini Grotto; Ricardo Haddad; Sandra Coccuzzo Sampaio Vessoni; Simone Kashima; Svetoslav Nanev Slavov; Vincent Louis Viala |
| EPI_ISL_5662625, EPI_ISL_5663363, EPI_ISL_5663450, EPI_ISL_5663552, EPI_ISL_5664178, EPI_ISL_5664637 | LABORATORIO DR PAULO EMILIO DALESSANDRO PINDAMONHANGABA | Instituto Butantan | Antonio Jorge Martins; Claudia Renata dos Santos Barros; David Schlesinger; Debora Botequio Moretti; Dimas Tadeu Covas; Elaine Cristina Marqueze; Elaine Vieira Santos; Evandra Strazza Rodrigues; Heidge Fukumasu; Jayme Augusto de Souza-Neto; José Salvatore Leister Patané; Luiz Alcantara; Luiz Lehmann Coutinho; Maria Carolina Elias; Maurício Lacerda Nogueira; Rafael dos Santos Bezerra; Raul Machado Neto; Rejane Maria Tommasini Grotto; Ricardo Haddad; Sandra Coccuzzo Sampaio Vessoni; Simone Kashima; Svetoslav Nanev Slavov; Vincent Louis Viala |
| EPI_ISL_4743385 | LABORATORIO ECHAVARRIA | Laboratorio Departamental de Salud Publica de Antioquia | Andres F. Cardona-Rios; Cristian Arbey Velarde Hoyos; Gloria Isabel Escobar; Idabely Betancur Ortiz; Juan P. Hernandez-Ortiz; Maria Stella López |
| EPI_ISL_5665792, EPI_ISL_5665820, EPI_ISL_5671262, EPI_ISL_5671313 | LABORATORIO MUNICIPAL DE ANALISES CLINICAS DE RIO CLARO | Instituto Butantan | Antonio Jorge Martins; Claudia Renata dos Santos Barros; David Schlesinger; Debora Botequio Moretti; Dimas Tadeu Covas; Elaine Cristina Marqueze; Elaine Vieira Santos; Evandra Strazza Rodrigues; Heidge Fukumasu; Jayme Augusto de Souza-Neto; José Salvatore Leister Patané; Luiz Alcantara; Luiz Lehmann Coutinho; Maria Carolina Elias; Maurício Lacerda Nogueira; Rafael dos Santos Bezerra; Raul Machado Neto; Rejane Maria Tommasini Grotto; Ricardo Haddad; Sandra Coccuzzo Sampaio Vessoni; Simone Kashima; Svetoslav Nanev Slavov; Vincent Louis Viala |
| EPI_ISL_5663345, EPI_ISL_5663578, EPI_ISL_5663667, EPI_ISL_5664067, EPI_ISL_5664091, EPI_ISL_5664136, EPI_ISL_5664358, EPI_ISL_5664437, EPI_ISL_5664554, EPI_ISL_5664715, EPI_ISL_5664814, EPI_ISL_5664993, EPI_ISL_5665128, EPI_ISL_5665484, EPI_ISL_5665511, EPI_ISL_5665575, EPI_ISL_5673262, EPI_ISL_5673459, EPI_ISL_5673677, EPI_ISL_5673726, EPI_ISL_5674089, EPI_ISL_5674394, EPI_ISL_5674503, EPI_ISL_5674615, EPI_ISL_5674996 | | | |
| see above | LABORATORIO MUNICIPAL DE PIRACICABA | Instituto Butantan | Antonio Jorge Martins; Claudia Renata dos Santos Barros; David Schlesinger; Debora Botequio Moretti; Dimas Tadeu Covas; Elaine Cristina Marqueze; Elaine Vieira Santos; Evandra Strazza Rodrigues; Heidge Fukumasu; Jayme Augusto de Souza-Neto; José Salvatore Leister Patané; Luiz Alcantara; Luiz Lehmann Coutinho; Maria Carolina Elias; Maurício Lacerda Nogueira; Rafael dos Santos Bezerra; Raul Machado Neto; Rejane Maria Tommasini Grotto; Ricardo Haddad; Sandra Coccuzzo Sampaio Vessoni; Simone Kashima; Svetoslav Nanev Slavov; Vincent Louis Viala |
| EPI_ISL_5661862, EPI_ISL_5662528, EPI_ISL_5663163, EPI_ISL_5663336, EPI_ISL_5663464, EPI_ISL_5663476, EPI_ISL_5663509, EPI_ISL_5663517, EPI_ISL_5663532, EPI_ISL_5663661, EPI_ISL_5663717, EPI_ISL_5663722, EPI_ISL_5663742, EPI_ISL_5663887, EPI_ISL_5663921, EPI_ISL_5664107, EPI_ISL_5664278, EPI_ISL_5664331, EPI_ISL_5664338, EPI_ISL_5664506, EPI_ISL_5664599, EPI_ISL_5664711, EPI_ISL_5664753, EPI_ISL_5664860, EPI_ISL_5664864, EPI_ISL_5664983, EPI_ISL_5665072, EPI_ISL_5665103, EPI_ISL_5665249, EPI_ISL_5665253, EPI_ISL_5665393 | | | |
| see above | LABORATORIO MUNICIPAL DE SUZANO | Instituto Butantan | Antonio Jorge Martins; Claudia Renata dos Santos Barros; David Schlesinger; Debora Botequio Moretti; Dimas Tadeu Covas; Elaine Cristina Marqueze; Elaine Vieira Santos; Evandra Strazza Rodrigues; Heidge Fukumasu; Jayme Augusto de Souza-Neto; José Salvatore Leister Patané; Luiz Alcantara; Luiz Lehmann Coutinho; Maria Carolina Elias; Maurício Lacerda Nogueira; Rafael dos Santos Bezerra; Raul Machado Neto; Rejane Maria Tommasini Grotto; Ricardo Haddad; Sandra Coccuzzo Sampaio Vessoni; Simone Kashima; Svetoslav Nanev Slavov; Vincent Louis Viala |
| EPI_ISL_2895184, EPI_ISL_2895187 | LAC UCoimbra | Instituto Nacional de Saude (INSA) | Borges et al |
| EPI_ISL_3404563, EPI_ISL_3404570, EPI_ISL_3404590, EPI_ISL_3404598, EPI_ISL_3404599, EPI_ISL_3404600, EPI_ISL_3404601, EPI_ISL_3404602, EPI_ISL_3404606, EPI_ISL_4395149 | | | |
| see above | LACEN | LGBio (Laboratorio de Genetica & Biodiversidade) | Alex Honda Bernardes; Amanda Alves de Melo; Aparecido Divino da Cruz; Cintia Pelegrineti Targueta de Azevedo Brito; Daniela de Melo e Silva; Elisangela de Paula Silveira Lacerda; Juliana Santana de Curcio; Luiz Augusto Pereira; Marc Alexandre Duarte Gigonzac; Mariana Pires de Campos Telles; Ramilla dos Santos Braga; Renata de Oliveira Dias; Rhewter Nunes; Thais Cidalia Vieira Gigonzac; Thais Guimarães Castro; Thays Millena Alves Pedroso |
| EPI_ISL_6721520, EPI_ISL_6722289 | LACEN - Laboratório Central de Saúde Pública de Pernambuco | Evandro Chagas Institute | Delana Andreza Melo Bezerra; Dielle Monteiro Teixeira; Jedson Ferreira Cardoso; Jessylene de Almeida Ferreira; Kenny da Costa Pinheiro; Luana Soares Barbagelata; Luana da Silva Soares; Mirleide Cordeiro dos Santos; Patrícia dos Santos Lobo; Rayssa Layna da Silva Bedran; Sandro Patroca da Silva |
| EPI_ISL_6721266 | LACEN - Laboratório Central de Saúde Pública de Roraima | Evandro Chagas Institute | Delana Andreza Melo Bezerra; Dielle Monteiro Teixeira; Jedson Ferreira Cardoso; Jessylene de Almeida Ferreira; Kenny da Costa Pinheiro; Luana Soares Barbagelata; Luana da Silva Soares; Mirleide Cordeiro dos Santos; Patrícia dos Santos Lobo; Rayssa Layna da Silva Bedran; Sandro Patroca da Silva |
| EPI_ISL_6721784 | LACEN - Laboratório Central de Saúde Pública do Rio Grande do Norte | Evandro Chagas Institute | Delana Andreza Melo Bezerra; Dielle Monteiro Teixeira; Jedson Ferreira Cardoso; Jessylene de Almeida Ferreira; Kenny da Costa Pinheiro; Luana Soares Barbagelata; Luana da Silva Soares; Mirleide Cordeiro dos Santos; Patrícia dos Santos Lobo; Rayssa Layna da Silva Bedran; Sandro Patroca da Silva |
| EPI_ISL_5538625 | LACEN do Distrito Federal | Instituto Adolfo Lutz, Interdiciplinary Procedures Center, Strategic Laboratory | Caio Vinicius Dias Lopes; Claudia Regina Gonçalves; Claudio Tavares Sacchi; Karoline Rodrigues Campos; Leonardo Tadeu de Araujo; Marlon Benedito Nascimento Santos |
| EPI_ISL_4206887 | LACEN do Estado de Mato Grosso | Instituto Adolfo Lutz, Interdiciplinary Procedures Center, Strategic Laboratory | Caio Vinicius Dias Lopes; Claudia Regina Gonçalves; Claudio Tavares Sacchi; Karoline Rodrigues Campos; Marlon Benedito Nascimento Santos |
| EPI_ISL_3046265, EPI_ISL_3046266, EPI_ISL_3046267, EPI_ISL_3046268, EPI_ISL_3046269, EPI_ISL_3046270, EPI_ISL_3046271, EPI_ISL_3046272, EPI_ISL_3046273, EPI_ISL_3046274, EPI_ISL_3046275, EPI_ISL_3046276, EPI_ISL_3046277, EPI_ISL_3046278, EPI_ISL_3046279, EPI_ISL_3046280, EPI_ISL_3046281, EPI_ISL_3046282, EPI_ISL_3046283, EPI_ISL_3046284, EPI_ISL_3046285, EPI_ISL_3046286, EPI_ISL_3046287, EPI_ISL_3046288, EPI_ISL_3046289, EPI_ISL_3046290, EPI_ISL_3046291, EPI_ISL_3046292, EPI_ISL_3046293, EPI_ISL_3046294, EPI_ISL_3046295, EPI_ISL_3046296, EPI_ISL_3046297, EPI_ISL_3046298, EPI_ISL_3046299, EPI_ISL_3046300, EPI_ISL_3046301, EPI_ISL_3046302, EPI_ISL_3046303, EPI_ISL_3046304, EPI_ISL_3046305, EPI_ISL_3046306, EPI_ISL_3046307, EPI_ISL_3046308, EPI_ISL_3046309, EPI_ISL_3046310, EPI_ISL_3046311, EPI_ISL_3046312, EPI_ISL_3046313, EPI_ISL_3046314, EPI_ISL_3046315, EPI_ISL_3046316, EPI_ISL_3046317, EPI_ISL_3046318, EPI_ISL_3046319, EPI_ISL_3046320, EPI_ISL_3046321, EPI_ISL_3046322, EPI_ISL_3046323, EPI_ISL_3046324, EPI_ISL_3046325, EPI_ISL_3046326, EPI_ISL_3046331, EPI_ISL_3046332, EPI_ISL_3046333, EPI_ISL_3046334, EPI_ISL_3046335, EPI_ISL_3046336, EPI_ISL_3046337, EPI_ISL_3046338, EPI_ISL_3046339, EPI_ISL_3046340, EPI_ISL_3046341, EPI_ISL_3046342, EPI_ISL_3046343, EPI_ISL_3046344, EPI_ISL_3046345, EPI_ISL_3046346, EPI_ISL_3046347, EPI_ISL_3046348, EPI_ISL_3046349, EPI_ISL_3134664, EPI_ISL_3134665, EPI_ISL_3134691 | | | |
| see above | LACEN/PE | WallauLab on behalf of Fiocruz COVID-19 Genomic Surveillance Network | Alexandre Freitas da Silva; Cassia Docena; Constância Flávia Junqueira Ayres; Filipe Zimmer Dezordi; Gabriel Luz Wallau; Gustavo Barbosa de Lima; Lais Ceschini Machado; Lilian Caroliny Amorim Silva; Marcelo Henrique dos Santos Paiva; Matheus Filgueira Bezerra; Sinval Pinto Brandão Filho |

| | | | |
|---|---|---|---|
| EPI_ISL_4487617, EPI_ISL_4487635, EPI_ISL_4487646, EPI_ISL_4487657, EPI_ISL_4487668 | LACEN/PI | WallauLab on behalf of Fiocruz COVID-19 Genomic Surveillance Network | Adelino Soares Lima Neto; Alexandre Freitas da Silva; Antônio Marinho; Cassia Docena; Constância Flávia Junqueira Ayres; Filipe Zimmer Dezordi; Gabriel Luz Wallau; Gustavo Barbosa de Lima; Hellen de Oliveira Amaral; Jacenir Reis dos Santos Mallet; Joana Carolina Viana Lima; Lais Ceschini Machado; Leandro de Mattos; Lilian Caroliny Amorim Silva; Marcela de Lacerda Valença Queiroz; Marcelo Adriano da Cunha e Silva Vieira; Marcelo Henrique dos Santos Paiva; Matheus Filgueira Bezerra; Sinval Pinto Brandão Filho; Túlio de Lima Campos; Vladimir Costa Silva; Walterlene de Carvalho Goçalves |
| EPI_ISL_2956938 | LADR Zentrallabor DR. Kramer & Kollegen Geesthacht | Robert Koch Institute | |
| EPI_ISL_3047701, EPI_ISL_3047710, EPI_ISL_3047719, EPI_ISL_3047727, EPI_ISL_3047729, EPI_ISL_3047730, EPI_ISL_3047742, EPI_ISL_3134977, EPI_ISL_3246189, EPI_ISL_3267906, EPI_ISL_3267907, EPI_ISL_3267908, EPI_ISL_3267909, EPI_ISL_3267910, EPI_ISL_3267911, EPI_ISL_3267912, EPI_ISL_3267913, EPI_ISL_3267914, EPI_ISL_3267915, EPI_ISL_3267916, EPI_ISL_3267917, EPI_ISL_3267918, EPI_ISL_3267919, EPI_ISL_3267920, EPI_ISL_3267921, EPI_ISL_3267922, EPI_ISL_3267923, EPI_ISL_3267924, EPI_ISL_3267925, EPI_ISL_3267926, EPI_ISL_3267927, EPI_ISL_3267928, EPI_ISL_3267929, EPI_ISL_3267930, EPI_ISL_3267931, EPI_ISL_3267932, EPI_ISL_3267933, EPI_ISL_3267934, EPI_ISL_3267935, EPI_ISL_3267936, EPI_ISL_3267937, EPI_ISL_3267938, EPI_ISL_3267939, EPI_ISL_3267940, EPI_ISL_3267941, EPI_ISL_3267942, EPI_ISL_3267943, EPI_ISL_3267961, EPI_ISL_3267962, EPI_ISL_3267963, EPI_ISL_3267971, EPI_ISL_3267972, EPI_ISL_3304422 | | | |
| see above | LATE - Laboratório de Técnicas Especiais - Hospital Israelita Albert Einstein | LATE - Laboratório de Técnicas Especiais - Hospital Israelita Albert Einstein | Alexandre Hideaki Takara; Ana Paula Moreira Salles; Anelisie da Silva Santos; Deyvid Amgarten; Erick Gustavo Dorlass; Fernanda de Mello Malta; João Renato Rebello Pinho; Marcio Anunciacao Menezes; Pedro Henrique Sebe Rodrigues; Raquel Riyuzo |
| EPI_ISL_2841778, EPI_ISL_2841782, EPI_ISL_2841785, EPI_ISL_2841787 | LBM ALPHABIO, Marseille | LBM ALPHABIO, Marseille | Vincent GARCIA |
| EPI_ISL_3320731, EPI_ISL_3320739 | LDSP Cundinamarca | Instituto Nacional de Salud | Carlos Franco-Muñoz; Carmen Osorio; Diana Malo; Diego A. Álvarez-Díaz; Diego Andrés Prada; Gerardo Santamaría; Hector Alejandro Ruiz-Moreno; Jhonnatan Reales-González; Jorge Rivera; Juan Camilo Martinez; Julian Naizaque; Katherine Laiton-Donato; Lisseth Pardo; Magdalena Wiesner; Marcela Mercado-Reyes; Maria T. Herrera-Sepúlveda; Marta Lopez Blanco; Martha Lucia Ospina Martinez; Paola Rojas; Sergio Gomez; Sheryll Corchuelo; Ángela Alarcon Cruz |
| EPI_ISL_3265588 | LESP Aguascalientes | Instituto de Diagnostico y Referencia Epidemiologicos (INDRE) | Abril Rodriguez-Maldonado; Ariadna Medina-Benitez; Claudia Wong-Arambula; Ernesto Ramirez-Gonzalez.; Gisela Barrera-Badillo; Irma Lopez-Martinez; Joaquin Quiroz-Mercado; Lucia Hernandez-Rivas; Maribel Gonzalez-Villa; Natividad Cruz-Ortiz; Sergio Rangel-Guerrero; Tatiana Nunez-Garcia; Vanessa Rivero-Arredondo |
| EPI_ISL_3265589, EPI_ISL_3265590, EPI_ISL_3265591 | LESP Campeche | Instituto de Diagnostico y Referencia Epidemiologicos (INDRE) | Abril Rodriguez-Maldonado; Ariadna Medina-Benitez; Claudia Wong-Arambula; Ernesto Ramirez-Gonzalez.; Gisela Barrera-Badillo; Irma Lopez-Martinez; Joaquin Quiroz-Mercado; Lucia Hernandez-Rivas; Maribel Gonzalez-Villa; Natividad Cruz-Ortiz; Sergio Rangel-Guerrero; Tatiana Nunez-Garcia; Vanessa Rivero-Arredondo |
| EPI_ISL_3265585, EPI_ISL_3265586 | LESP Estado de Mexico | Instituto de Diagnostico y Referencia Epidemiologicos (INDRE) | Abril Rodriguez-Maldonado; Ariadna Medina-Benitez; Claudia Wong-Arambula; Ernesto Ramirez-Gonzalez.; Gisela Barrera-Badillo; Irma Lopez-Martinez; Joaquin Quiroz-Mercado; Lucia Hernandez-Rivas; Maribel Gonzalez-Villa; Natividad Cruz-Ortiz; Sergio Rangel-Guerrero; Tatiana Nunez-Garcia; Vanessa Rivero-Arredondo |
| EPI_ISL_3265578, EPI_ISL_3265597 | LESP Hidalgo | Instituto de Diagnostico y Referencia Epidemiologicos (INDRE) | Abril Rodriguez-Maldonado; Ariadna Medina-Benitez; Claudia Wong-Arambula; Ernesto Ramirez-Gonzalez.; Gisela Barrera-Badillo; Irma Lopez-Martinez; Joaquin Quiroz-Mercado; Lucia Hernandez-Rivas; Maribel Gonzalez-Villa; Natividad Cruz-Ortiz; Sergio Rangel-Guerrero; Tatiana Nunez-Garcia; Vanessa Rivero-Arredondo |
| EPI_ISL_3033592 | LESP Michoacan | Instituto de Diagnostico y Referencia Epidemiologicos (INDRE) | Abril Rodriguez-Maldonado; Ariadna Medina-Benitez; Claudia Wong-Arambula; Ernesto Ramirez-Gonzalez.; Gisela Barrera-Badillo; Irma Lopez-Martinez; Joaquin Quiroz-Mercado; Lucia Hernandez-Rivas; Maribel Gonzalez-Villa; Natividad Cruz-Ortiz; Sergio Rangel-Guerrero; Tatiana Nunez-Garcia; Vanessa Rivero-Arredondo |
| EPI_ISL_3278289 | LESP Nayarit | Instituto de Diagnostico y Referencia Epidemiologicos (INDRE) | Abril Rodriguez-Maldonado; Ariadna Medina-Benitez; Claudia Wong-Arambula; Ernesto Ramirez-Gonzalez.; Gisela Barrera-Badillo; Irma Lopez-Martinez; Joaquin Quiroz-Mercado; Lucia Hernandez-Rivas; Maribel Gonzalez-Villa; Natividad Cruz-Ortiz; Sergio Rangel-Guerrero; Tatiana Nunez-Garcia; Vanessa Rivero-Arredondo |
| EPI_ISL_3033595 | LESP Queretaro | Instituto de Diagnostico y Referencia Epidemiologicos (INDRE) | Abril Rodriguez-Maldonado; Ariadna Medina-Benitez; Claudia Wong-Arambula; Ernesto Ramirez-Gonzalez.; Gisela Barrera-Badillo; Irma Lopez-Martinez; Joaquin Quiroz-Mercado; Lucia Hernandez-Rivas; Maribel Gonzalez-Villa; Natividad Cruz-Ortiz; Sergio Rangel-Guerrero; Tatiana Nunez-Garcia; Vanessa Rivero-Arredondo |
| EPI_ISL_3265595, EPI_ISL_3265596, EPI_ISL_3278285 | LESP Quintana Roo | Instituto de Diagnostico y Referencia Epidemiologicos (INDRE) | Abril Rodriguez-Maldonado; Ariadna Medina-Benitez; Claudia Wong-Arambula; Ernesto Ramirez-Gonzalez.; Gisela Barrera-Badillo; Irma Lopez-Martinez; Joaquin Quiroz-Mercado; Lucia Hernandez-Rivas; Maribel Gonzalez-Villa; Natividad Cruz-Ortiz; Sergio Rangel-Guerrero; Tatiana Nunez-Garcia; Vanessa Rivero-Arredondo |
| EPI_ISL_2859060 | LESP San Luis Potosi | Instituto de Diagnostico y Referencia Epidemiologicos (INDRE) | Abril Rodriguez-Maldonado; Ariadna Medina-Benitez; Claudia Wong-Arambula; Ernesto Ramirez-Gonzalez.; Gisela Barrera-Badillo; Irma Lopez-Martinez; Joaquin Quiroz-Mercado; Lucia Hernandez-Rivas; Maribel Gonzalez-Villa; Natividad Cruz-Ortiz; Sergio Rangel-Guerrero; Tatiana Nunez-Garcia; Vanessa Rivero-Arredondo |
| EPI_ISL_3265592, EPI_ISL_3265593, EPI_ISL_3265594, EPI_ISL_3265614 | LESP Sonora | Instituto de Diagnostico y Referencia Epidemiologicos (INDRE) | Abril Rodriguez-Maldonado; Ariadna Medina-Benitez; Claudia Wong-Arambula; Ernesto Ramirez-Gonzalez.; Gisela Barrera-Badillo; Irma Lopez-Martinez; Joaquin Quiroz-Mercado; Lucia Hernandez-Rivas; Maribel Gonzalez-Villa; Natividad Cruz-Ortiz; Sergio Rangel-Guerrero; Tatiana Nunez-Garcia; Vanessa Rivero-Arredondo |
| EPI_ISL_3033548, EPI_ISL_3033549, EPI_ISL_3033550, EPI_ISL_3033581, EPI_ISL_3033582, EPI_ISL_3033583, EPI_ISL_3033584, EPI_ISL_3033585, EPI_ISL_3033586, EPI_ISL_3033587, EPI_ISL_3033588, EPI_ISL_3033599, EPI_ISL_3033600, EPI_ISL_3046106, EPI_ISL_3459950, EPI_ISL_3459951 | | | |
| see above | LESP Tabasco | Instituto de Diagnostico y Referencia Epidemiologicos (INDRE) | Abril Rodriguez-Maldonado; Ariadna Medina-Benitez; Claudia Wong-Arambula; Ernesto Ramirez-Gonzalez.; Gisela Barrera-Badillo; Irma Lopez-Martinez; Joaquin Quiroz-Mercado; Lucia Hernandez-Rivas; Maribel Gonzalez-Villa; Natividad Cruz-Ortiz; Sergio Rangel-Guerrero; Tatiana Nunez-Garcia; Vanessa Rivero-Arredondo |
| EPI_ISL_3265606, EPI_ISL_3265607 | LESP Tamaulipas | Instituto de Diagnostico y Referencia Epidemiologicos (INDRE) | Abril Rodriguez-Maldonado; Ariadna Medina-Benitez; Claudia Wong-Arambula; Ernesto Ramirez-Gonzalez.; Gisela Barrera-Badillo; Irma Lopez-Martinez; Joaquin Quiroz-Mercado; Lucia Hernandez-Rivas; Maribel Gonzalez-Villa; Natividad Cruz-Ortiz; Sergio Rangel-Guerrero; Tatiana Nunez-Garcia; Vanessa Rivero-Arredondo |
| EPI_ISL_3265602 | LESP Veracruz | Instituto de Diagnostico y Referencia Epidemiologicos (INDRE) | Abril Rodriguez-Maldonado; Ariadna Medina-Benitez; Claudia Wong-Arambula; Ernesto Ramirez-Gonzalez.; Gisela Barrera-Badillo; Irma Lopez-Martinez; Joaquin Quiroz-Mercado; Lucia Hernandez-Rivas; Maribel Gonzalez-Villa; Natividad Cruz-Ortiz; Sergio Rangel-Guerrero; Tatiana Nunez-Garcia; Vanessa Rivero-Arredondo |
| EPI_ISL_3459958 | LESP Yucatan | Instituto de Diagnostico y Referencia Epidemiologicos (INDRE) | Abril Rodriguez-Maldonado; Ariadna Medina-Benitez; Claudia Wong-Arambula; Ernesto Ramirez-Gonzalez.; Gisela Barrera-Badillo; Irma Lopez-Martinez; Joaquin Quiroz-Mercado; Lucia Hernandez-Rivas; Maribel Gonzalez-Villa; Natividad Cruz-Ortiz; Sergio Rangel-Guerrero; Tatiana Nunez-Garcia; Vanessa Rivero-Arredondo |
| EPI_ISL_4743407 | LIME | Laboratorio Departamental de Salud Publica de Antioquia | Andres F. Cardona-Rios; Cristian Arbey Velarde Hoyos; Gloria Isabel Escobar; Idabely Betancur Ortiz; Juan P. Hernandez-Ortiz; Maria Stella López |
| EPI_ISL_2873838 | LUM Laboratorio Unico Metropolitano - AUSL BOLOGNA | U.O. Microbiologia, Laboratorio Unico Centro Servizi - AUSL della Romagna | Giorgio Dirani |
| EPI_ISL_2844510, EPI_ISL_2844517 | Lab. Microbiologia e Virologia Cotugno A.O. dei Colli - Istituto Zooprofilattico Sperimentale del Mezzogiorno | TIGEM | Antonio Grimaldi Patrizia Annunziata Francesco Panariello Biancamaria Pierri Claudia Tiberio Teresa Giuliano Valentina Bouche Chiara Colantuono Maria Concetta Cuomo Denise Di Concilio Lucio Di Filippo Anna Manfredi Marcello Salvi Antonio Limone Luigi Atripaldi Pellegrino Cerino Andrea Ballabio Davide Cacchiarelli |
| EPI_ISL_2923244, EPI_ISL_2958763 | Labo Analyses Med | National Reference Center for Viruses of Respiratory Infections, Institut Pasteur, Paris | Angela Brisebarre; BESSON Jérôme; Camille Capel; Christophe Malabat; Corinne Maufrais; Etienne Simon-Lorière; Frédéric Lemoine; Hub Bioinformatique Biostatistiques; Louise Lefrançois; Marion Barbet; Maud Vanpeene; Méline Bizard; POTIRON Grégoire; Pierre Lechat; Sylvie Behillil; Sylvie Van der Werf; Vincent Enouf |
| EPI_ISL_3113180, EPI_ISL_3113202, EPI_ISL_3113213 | Labor Berlin Charité Vivantes GmbH / Institut für Virologie | Charité Universitätsmedizin Berlin, Institut für Virologie/Labor Berlin | Barbara Mühlemann; Christian Drosten; Christine Stephan; Peter Menzel; Rolf Schwarzer; Terry Jones; Victor M Corman |
| EPI_ISL_2878505, EPI_ISL_2878531 | Laboratoire ESPACEBIO | Department of Virology, Henri Mondor University Hospital, Assistance Publique Hôpitaux de Paris, Université Paris-Est Créteil, INSERM U955 | Alexandre Soulier; Christophe Rodriguez; Elisabeth Trawinski; Guillaume Gricourt; Jean-Michel Pawlotsky; Melissa N'Debi; Slim Fourati; Vanessa Demontant |
| EPI_ISL_4028959, EPI_ISL_4029006 | Laboratoire National de Santé Publique – LNSP | Genomics and Proteomics Departament, Gorgas Memorial Institute For Health Studies | Alexander A Martinez; Ambar Moreno; Claudia Estrada; Claudia Gonzalez V; César Roberto Conde Pereira; Jessica Gondola; Leyda Abrego; Marlene Castillo; Melissa Gaitan; Oris Chavarria |
| EPI_ISL_3459082, EPI_ISL_3459085, EPI_ISL_3459097, EPI_ISL_3459100 | Laboratoire de santé publique du Québec | Laboratoire de santé publique du Québec | Guillaume Bourque; Ioannis Ragoussis; Jesse Shapiro; Mark Lathrop and Michel Roger on behalf of the CoVSeQ research group; Sandrine Moreira |
| EPI_ISL_3147323, EPI_ISL_3147325, EPI_ISL_3147326, EPI_ISL_3147479, EPI_ISL_3147503, EPI_ISL_3147512, EPI_ISL_3147550, EPI_ISL_3147551, EPI_ISL_3147562, EPI_ISL_3147578, EPI_ISL_3147593 | | | |
| see above | Laboratoire national de sante, Microbiology, Virology | Laboratoire national de sante, Microbiology, Microbial Genomics Platform | Anke Wienecke-Baldacchino; Catherine Ragimbeau; Elodie Solarino; Fatu Djabi; Jessica Tapp; Lise Pignon; Raoul Salmon; Tamir Abdelrahman; Trung Nguyen Nguyen; Virginie Jover |
| EPI_ISL_3147179, EPI_ISL_3147180, EPI_ISL_3147185, EPI_ISL_3147192, EPI_ISL_3147193, EPI_ISL_3147194, EPI_ISL_3147202, EPI_ISL_3147204, EPI_ISL_3147207, EPI_ISL_3147211, EPI_ISL_3147216, EPI_ISL_3147231, EPI_ISL_3147241, EPI_ISL_3147246, EPI_ISL_3147263, EPI_ISL_3147269, EPI_ISL_3147272, EPI_ISL_3147284, EPI_ISL_3147290, EPI_ISL_3147301, EPI_ISL_3147310, EPI_ISL_3147313, EPI_ISL_3147409, EPI_ISL_3147413, EPI_ISL_3147424, EPI_ISL_3147442, EPI_ISL_3147444, EPI_ISL_3147451, EPI_ISL_3147458, EPI_ISL_3147459, EPI_ISL_3147461, EPI_ISL_3147471, EPI_ISL_3147476, EPI_ISL_3147493, EPI_ISL_3147494, EPI_ISL_3147497, EPI_ISL_3147507, EPI_ISL_3147510, EPI_ISL_3147517, EPI_ISL_3147520, EPI_ISL_3147522, EPI_ISL_3147523, EPI_ISL_3147535, EPI_ISL_3147556, | | | |

| | | | |
|---|---|---|---|
| EPI_ISL_3147559, EPI_ISL_3147565, EPI_ISL_3147572, EPI_ISL_3147588, EPI_ISL_3147590, EPI_ISL_3147594, EPI_ISL_3147596 | | | |
| see above | Laboratoires Reunis | Laboratoire national de sante, Microbiology, Microbial Genomics Platform | Anke Wienecke-Baldacchino; Bernard Weber; Catherine Ragimbeau; Elodie Solarino; Fatu Djabi; Jessica Tapp; Lise Pignon; Raoul Salmon; Tamir Abdelrahman; Virginie Jover |
| EPI_ISL_3147718, EPI_ISL_3147746, EPI_ISL_3147764, EPI_ISL_3147769, EPI_ISL_3147808, EPI_ISL_3147817, EPI_ISL_3147826, EPI_ISL_3147829, EPI_ISL_3147839, EPI_ISL_3147858, EPI_ISL_3148091, EPI_ISL_3148118, EPI_ISL_3148123, EPI_ISL_3148125, EPI_ISL_3148127, EPI_ISL_3148128, EPI_ISL_3148133, EPI_ISL_3148136, EPI_ISL_3148154, EPI_ISL_3148155, EPI_ISL_3148156, EPI_ISL_3148157, EPI_ISL_3148158, EPI_ISL_3148159, EPI_ISL_3148160, EPI_ISL_3148161, EPI_ISL_3148162, EPI_ISL_3148163, EPI_ISL_3148164, EPI_ISL_3148165, EPI_ISL_3148166, EPI_ISL_3148167, EPI_ISL_3148168, EPI_ISL_3148169, EPI_ISL_3148170, EPI_ISL_3148171, EPI_ISL_3148172, EPI_ISL_3148173, EPI_ISL_3148174, EPI_ISL_3148175, EPI_ISL_3148176, EPI_ISL_3148177, EPI_ISL_3148178, EPI_ISL_3148179, EPI_ISL_3148180, EPI_ISL_3148181, EPI_ISL_3148182, EPI_ISL_3148183, EPI_ISL_3148184, EPI_ISL_3148185, EPI_ISL_3148186, EPI_ISL_3148187, EPI_ISL_3148188, EPI_ISL_3148189, EPI_ISL_3148190, EPI_ISL_3148191, EPI_ISL_3148192, EPI_ISL_3148193, EPI_ISL_3148194, EPI_ISL_3148195, EPI_ISL_3148196, EPI_ISL_3148197, EPI_ISL_3148198, EPI_ISL_3148199, EPI_ISL_3148200, EPI_ISL_3148201, EPI_ISL_3148202, EPI_ISL_3148203, EPI_ISL_3148204, EPI_ISL_3148205, EPI_ISL_3148206, EPI_ISL_3148207, EPI_ISL_3148208, EPI_ISL_3148209, EPI_ISL_3148210, EPI_ISL_3148211, EPI_ISL_3148212, EPI_ISL_3148213, EPI_ISL_3148214, EPI_ISL_3148215, EPI_ISL_3148216, EPI_ISL_3148217, EPI_ISL_3148218 | | | |
| see above | Laboratoires d'analyses medicales - Ketterthill | Laboratoire national de sante, Microbiology, Microbial Genomics Platform | Anke Wienecke-Baldacchino; Caroline Scheiber; Catherine Ragimbeau; Elodie Solarino; Fatu Djabi; Jessica Tapp; Lise Pignon; Raoul Salmon; Serge Vedy; Tamir Abdelrahman; Virginie Jover |
| EPI_ISL_3906440, EPI_ISL_3906483 | Laboratori de Referència de Catalunya | SeqCOVID-SPAIN consortium/IBV(CSIC) | Aida Esperanza Ramírez Marinero; Alba Vilas Basil; Eduardo Padilla León; Mireia Canal Aranda and SeqCOVID-SPAIN consortium |
| EPI_ISL_3190352, EPI_ISL_3190353, EPI_ISL_3190355, EPI_ISL_3190356, EPI_ISL_3190358 | Laboratorio Central de Saude Publica do Estado de Goias (LACEN/GO) | Laboratory of Respiratory Viruses and Measles, Oswaldo Cruz Institute, FIOCRUZ | Agatha Cristinne Prudencio; Alice Sampaio Rocha; Ana Carolina Mendonca; Ana Flavia Mendonça; Anna Carolina Paixao; Carmen Helena Ramos; Cassiane Casanova; Elisa Cavalcante Pereira; Fernando Motta; Flavia Pereira Amorim da Silva; Ighor Leonardo Arantes Gomes; Luciana Appolinario; Luiz Augusto Pereira; Marilda Siqueira on behalf of the Fiocruz COVID-19 Genomic Surveillance Network; Paola Resende; Rafael Souza Guedes; Renata Serrano Lopes; Taina Venas; Vinicius Lemes da Silva |
| EPI_ISL_3539765 | Laboratorio Central de Saude Publica do Estado de Minas Gerais (LACEN/MG) | Laboratory of Respiratory Viruses and Measles, Oswaldo Cruz Institute, FIOCRUZ | Alice Sampaio Rocha; Ana Carolina Mendonca; Anna Carolina Paixao; Elisa Cavalcante Pereira; Fernando Motta; Luciana Appolinario; Marilda Siqueira on behalf of the Fiocruz COVID-19 Genomic Surveillance Network; Paola Resende; Renata Serrano Lopes; Rubens Pasa; Taina Venas |
| EPI_ISL_4080766 | Laboratorio Central de Saude Publica do Estado do Para (LACEN/PA) | Laboratory of Respiratory Viruses and Measles, Oswaldo Cruz Institute, FIOCRUZ | Agatha Soares; Alice Sampaio Rocha; Ana Carolina Mendonca; Anna Carolina Paixao; Elisa Cavalcante Pereira; Fernando Motta; Ighor Arantes; Luciana Appolinario; Marilda Siqueira on behalf of the Fiocruz COVID-19 Genomic Surveillance Network; Paola Resende; Renata Serrano Lopes; Taina Venas; Valnete Andrade |
| EPI_ISL_2925957 | Laboratorio CQRC | CQRC_QUALITY CONTROL CHEMICAL BIOLOGICAL RISK_AOOR Villa Sofia Cervello Palermo | Brunacci G.; Contino F.; Di Gaudio F. |
| EPI_ISL_3245105, EPI_ISL_3245121, EPI_ISL_3245122, EPI_ISL_3245125, EPI_ISL_3245126, EPI_ISL_3245127, EPI_ISL_3245128, EPI_ISL_3245129, EPI_ISL_3245135, EPI_ISL_3245136, EPI_ISL_3245137, EPI_ISL_3245141, EPI_ISL_3245142, EPI_ISL_3245143, EPI_ISL_3245144, EPI_ISL_3245146, EPI_ISL_3245147, EPI_ISL_3245148, EPI_ISL_3245153, EPI_ISL_3245154, EPI_ISL_3245155, EPI_ISL_3245156, EPI_ISL_3245157, EPI_ISL_3245158, EPI_ISL_3245159, EPI_ISL_3245160, EPI_ISL_3245161, EPI_ISL_3245162, EPI_ISL_3245163, EPI_ISL_3245164, EPI_ISL_3245165, EPI_ISL_3245170, EPI_ISL_3245171, EPI_ISL_3245172, EPI_ISL_3245174, EPI_ISL_3245175, EPI_ISL_3245179, EPI_ISL_3245180, EPI_ISL_3245181, EPI_ISL_3245182, EPI_ISL_3245183, EPI_ISL_3245184, EPI_ISL_3245185, EPI_ISL_3245186, EPI_ISL_3245188, EPI_ISL_3245189, EPI_ISL_3245190, EPI_ISL_3245191, EPI_ISL_3245192, EPI_ISL_3245193, EPI_ISL_3245194, EPI_ISL_3245195, EPI_ISL_3245196, EPI_ISL_3245197, EPI_ISL_3245198, EPI_ISL_3245199, EPI_ISL_3245200, EPI_ISL_3245201, EPI_ISL_3245203, EPI_ISL_3245204, EPI_ISL_3245206, EPI_ISL_3245207, EPI_ISL_3245208, EPI_ISL_3245209, EPI_ISL_3245210, EPI_ISL_3245211, EPI_ISL_3245212, EPI_ISL_3245213, EPI_ISL_3245214, EPI_ISL_3245215, EPI_ISL_3245217, EPI_ISL_3245218, EPI_ISL_3245220, EPI_ISL_3245221, EPI_ISL_3245222, EPI_ISL_3245234, EPI_ISL_3245235, EPI_ISL_3245237, EPI_ISL_3245238, EPI_ISL_3245341, EPI_ISL_3245343, EPI_ISL_3245344, EPI_ISL_3245345 | | | |
| see above | Laboratorio Central Noel Nutels | Bioinformatics Laboratory / LNCC | Alessandra P Lamarca; Alexandra L Gerber; Amilcar Tanuri; Ana Paula de C Guimaraes; Ana Tereza R Vasconcelos; Andrea Cony Cavalcanti; Caio Luiz Pereira Ribeiro; Cintia Policarpo; Claudia Maria Braga de Mello; Cristiane Gomes da Silva; Douglas Terra Machado; Erica Ramos dos Santos Nascimento; Fernanda Leitao dos Santos; Flavio Dias da Silva; Gleidson da Silva de Oliveira; Leandro Magalhaes de Souza; Liliane Cavalcante; Luiz G P de Almeida; Marcio Henrique de Oliveira Garcia; Mario Sergio Ribeiro; Ricardo Jose Barbosa Salviano; Ronaldo da Silva F Jr; Silvia Carvalho |
| EPI_ISL_2942866, EPI_ISL_2942893, EPI_ISL_2942903, EPI_ISL_2942904, EPI_ISL_2942908, EPI_ISL_2942909, EPI_ISL_2942921, EPI_ISL_2942923, EPI_ISL_2942924, EPI_ISL_2942926, EPI_ISL_2942930, EPI_ISL_2942933 | | | |
| see above | Laboratorio Central de Epidemiologia (LCE) | Unidad de Genomica Avanzada | ; Alejandra Garcia-Gasca; Alejandra Hernandez-Teran; Alejandro Sanchez-Flores; Alfredo Herrera-Estrella; Alicia Ocaña-Mondragon; Andreu Comas-Garcia; Angel Gustavo Salas-Lais; Antonio Loza Roman; Bernardo Martinez-Miguel; Blanca Taboada; Brenda Irasema Maldonado-Meza; Bruno Gomez-Gil; Carla Ivon Herrera-Najera; Carlos F. Arias; Celia Boukadida; Celida Duque Molina; Celida Martinez- Rodriguez; Clara Esperanza Santacruz-Tinoco; Concepcion Grajales-Muñiz; Consorcio Mexicano de Vigilancia Genomica (CoViGen-Mex). Authors (in alphabetical order): Julio Elias Alvarado-Yaah; Cristobal Chaidez-Quiroz; Daniel Fregoso-Rueda; Daniel Lira Morales; Eduardo Becerril-Vargas; Fernando Fontove-Herrera; Fidencio Mejia-Nepomuceno; Francisco Pulido; Gloria Elena Espinosa-Ayala; Gloria Maria Molina-Salinas; Gloria Vazquez; Hector Esteban Paz-Juarez; Hector Montoya-Fuentes; Helen Haydee Fernanda Ramirez-Plascencia; Irvin Gonzalez-Lopez; Jean Pierre Gonzalez; Jesus Hernandez; Joel Armando Vazquez-Perez.; Jorge Salas-Hernandez; Jose Antonio Enciso-Moreno; Jose Arturo Martinez-Orozco; Jose Esteban Muñoz-Medina; Jose de Jesus Nuñez-Contreras; Juan Bautista Chale-Dzul; Julissa Enciso-Ibarra; Luis Alberto Ochoa-Carrera; Margarita Matias-Florentino; Maria Guadalupe Santiago-Mauricio; Maria Guadalupe de Jesus Mireles-Rivera; Mario Mujica-Sanchez; Marissa Perez-Garcia; Nelly Selem-Mojica; Pavel Isa; Ricardo Ciria Merce; Ricardo Grande; Rosa Maria Gutierrez Rios; Santiago avila-Rios; Selene Zarate; Susana Lopez; Veronica Mata-Haro; Victor Eduardo Garcia-Arias; Victor Hugo Borja-Aburto |
| EPI_ISL_3155547, EPI_ISL_3155550, EPI_ISL_3155564, EPI_ISL_3155575, EPI_ISL_3155625 | Laboratorio Central de Epidemiología (LCE) | Centro de Investigación en Enfermedades Infecciosas (CIENI), Instituto Nacional de Enfermedades Respiratorias (INER) | ; Alejandra García-Gasca; Alejandra Hernández-Terán; Alejandro Sánchez-Flores; Alfredo Herrera-Estrella; Alicia Ocaña-Mondragón; Andreu Comas-García; Angel Gustavo Salas-Lais; Antonio Loza Román; Bernardo Martínez-Miguel; Blanca Taboada; Brenda Irasema Maldonado-Meza; Bruno Gómez-Gil; Carla Ivón Herrera-Najera; Carlos F. Arias; Celia Boukadida; Célida Duque Molina; Célida Martínez- Rodríguez; Clara Esperanza Santacruz-Tinoco; Concepción Grajales-Muñiz; Consorcio Mexicano de Vigilancia Genómica (CoViGen-Mex). Authors (in alphabetical order): Julio Elias Alvarado-Yaah; Cristóbal Cháidez-Quiróz; Célida Duque Molina; Célida Martínez- Rodríguez; Daniel Fregoso-Rueda; Daniel Lira Morales; Eduardo Becerril-Vargas; Fernando Fontove-Herrera; Fidencio Mejía-Nepomuceno; Francisco Pulido; Gloria Elena Espinosa-Ayala; Gloria María Molina-Salinas; Gloria Vazquez; Hector Esteban Paz-Juárez; Hector Montoya-Fuentes; Helen Haydee Fernanda Ramírez-Plascencia; Irvin González-López; Jean Pierre González; Jesús Hernández; Joel Armando Vázquez-Pérez.; Jorge Salas-Hernández; José Antonio Enciso-Moreno; José Arturo Martínez-Orozco; José Esteban Muñoz-Medina; José de Jesús Nuñez-Contreras; Juan Bautista Chale-Dzul; Julissa Enciso-Ibarra; Kathia Elizabeth Tapia-Diaz; Luis Alberto Ochoa-Carrera; Margarita Matías-Florentino; Mario Mújica-Sánchez; Marissa Perez-Garcia; María Guadalupe Santiago-Mauricio; María Guadalupe de Jesús Mireles-Rivera; Nelly Sélem-Mojica; Pavel Isa; Ricardo Ciria Merce; Ricardo Grande; Rosa María Gutiérrez Rios; Santiago Ávila-Ríos; Selene Zárate; Susana Lopez; Verónica Mata-Haro; Victor Eduardo García-Arias; Víctor Hugo Borja-Aburto |
| EPI_ISL_6944560, EPI_ISL_6944565, EPI_ISL_6944572, EPI_ISL_6944579, EPI_ISL_6944591, EPI_ISL_6944595, EPI_ISL_6944596, EPI_ISL_6944597, EPI_ISL_6944600, EPI_ISL_6944613, EPI_ISL_6944614, EPI_ISL_6944615, EPI_ISL_6944640 | | | |
| see above | Laboratorio Central de Salud Publica | Viral Special Pathogens Branch, Centers for Disease Control and Prevention | Andrea Gómez de la Fuente; Cynthia Vázquez; Emir Talundzic; Joel Montgomery; John Klena; Juan Torales; Justin Lee; María José Ortega; María Liz Gamarra; Shannon Whitmer; Shirley Villalba |
| EPI_ISL_3045424, EPI_ISL_3045425, EPI_ISL_3045426 | Laboratorio Central de Saude Publica do Estado da Bahia (LACEN/BA) | Laboratory of Respiratory Viruses and Measles, Oswaldo Cruz Institute, FIOCRUZ | Alice Sampaio Rocha; Ana Carolina Mendonca; Anna Carolina Paixao; Elisa Cavalcante Pereira; Felicidade Pereira; Fernando Motta; Luciana Appolinario; Marilda Siqueira on behalf of the Fiocruz COVID-19 Genomic Surveillance Network; Paola Resende; Renata Serrano Lopes; Taina Venas |
| EPI_ISL_3434828, EPI_ISL_3434830, EPI_ISL_3434831, EPI_ISL_3434832, EPI_ISL_3434833, EPI_ISL_3434834, EPI_ISL_3434835, EPI_ISL_3434836, EPI_ISL_3434849, EPI_ISL_3434850, EPI_ISL_3434852, EPI_ISL_3434853, EPI_ISL_3434854, EPI_ISL_3434855, EPI_ISL_3434856, EPI_ISL_3434857, EPI_ISL_3434858, EPI_ISL_3434859, EPI_ISL_3434860, EPI_ISL_3434861, EPI_ISL_3434862, EPI_ISL_3434863, EPI_ISL_3434864, EPI_ISL_3434865, EPI_ISL_3434866, EPI_ISL_3434867, EPI_ISL_3434868, EPI_ISL_3434869, EPI_ISL_3434870, EPI_ISL_3434871, EPI_ISL_3434872, EPI_ISL_3434897, EPI_ISL_3434939, EPI_ISL_3434942, EPI_ISL_3539926, EPI_ISL_3539927, EPI_ISL_3539928, EPI_ISL_3539930 | | | |
| see above | Laboratorio Central de Saude Publica do Estado de Alagoas (LACEN/AL) | Laboratory of Respiratory Viruses and Measles, Oswaldo Cruz Institute, FIOCRUZ | Agatha Soares; Alice Sampaio Rocha; Ana Carolina Mendonca; Anderson Brandao Leite; Anna Carolina Paixao; Elisa Cavalcante Pereira; Fernando Motta; Ighor Arantes; Luciana Appolinario; Marilda Siqueira on behalf of the Fiocruz COVID-19 Genomic Surveillance Network; Paola Resende; Renata Serrano Lopes; Taina Venas |
| EPI_ISL_3190219, EPI_ISL_3190222, EPI_ISL_3190223, EPI_ISL_3190224, EPI_ISL_3190225, EPI_ISL_3190226, EPI_ISL_3434908, EPI_ISL_3434909, EPI_ISL_3434928, EPI_ISL_3434929, EPI_ISL_3434930, EPI_ISL_3434931, EPI_ISL_3434932, EPI_ISL_3434933, EPI_ISL_3434934, EPI_ISL_3434935, EPI_ISL_3434936, EPI_ISL_3434937, EPI_ISL_3434940 | | | |
| see above | Laboratorio Central de Saude Publica do Estado de Santa Catarina (LACEN/SC) | Laboratory of Respiratory Viruses and Measles, Oswaldo Cruz Institute, FIOCRUZ | Alice Sampaio Rocha; Ana Carolina Mendonca; Anna Carolina Paixao; Darcita Buerger Rovaris; Elisa Cavalcante Pereira; Fernando Motta; Luciana Appolinario; Marilda Siqueira on behalf of the Fiocruz COVID-19 Genomic Surveillance Network; Paola Resende; Renata Serrano Lopes; Sandra Bianchini Fernandes; Taina Venas |
| EPI_ISL_3803004, EPI_ISL_3803007, EPI_ISL_3803010, EPI_ISL_3803011, EPI_ISL_3803014, EPI_ISL_3803021 | Laboratorio Central de Saude Publica do Estado de Sergipe (LACEN/SE) | Laboratory of Respiratory Viruses and Measles, Oswaldo Cruz Institute, FIOCRUZ | Agatha Soares; Alice Sampaio Rocha; Ana Carolina Mendonca; Anna Carolina Paixao; Cliomar Alves dos Santos; Elisa Cavalcante Pereira; Fernando Motta; Ighor Arantes; Luciana Appolinario; Marilda Siqueira on behalf of the Fiocruz COVID-19 Genomic Surveillance Network; Paola Resende; Renata Serrano Lopes; Tainá Moreira Martins Venas |
| EPI_ISL_3434968, EPI_ISL_3434969, EPI_ISL_3434970, EPI_ISL_3434976, EPI_ISL_3434978, EPI_ISL_3539748 | Laboratorio Central de Saude Publica do Estado do Amapa (LACEN/AP) | Laboratory of Respiratory Viruses and Measles, Oswaldo Cruz Institute, FIOCRUZ | Agatha Cristinne Prudencio; Alice Sampaio Rocha; Ana Carolina Mendonca; Andreia Santos Costa; Anna Carolina Paixao; Anne Caroline da Silva Soledade; Elisa Cavalcante Pereira; Fernando Motta; Ighor Leonardo Arantes Gomes; Lindomar dos Anjos Silva; Luciana Appolinario; Marcia Socorro Pereira Cavalcante; Marilda Siqueira on behalf of the Fiocruz COVID-19 Genomic Surveillance Network; Paola Resende; Renata Serrano Lopes; Taina Venas |
| EPI_ISL_3045463, EPI_ISL_3045464, EPI_ISL_3045467, EPI_ISL_3045554, EPI_ISL_3045555, EPI_ISL_3045556, EPI_ISL_3190249, EPI_ISL_3190250, EPI_ISL_3190251, EPI_ISL_3190252, EPI_ISL_3190253, EPI_ISL_3435024, EPI_ISL_3435045, EPI_ISL_3827880, EPI_ISL_3827911, EPI_ISL_3827918, EPI_ISL_3827934, EPI_ISL_3827999, EPI_ISL_3828002, EPI_ISL_3828003 | | | |
| see above | Laboratorio Central de Saude Publica do Estado do Espirito Santo (LACEN/ES) | Laboratory of Respiratory Viruses and Measles, Oswaldo Cruz Institute, FIOCRUZ | Alice Sampaio Rocha; Ana Carolina Mendonca; Anna Carolina Paixao; Eilisa Cavalcante Pereira; Elisa Cavalcante Pereira; Fernando Motta; Luciana Appolinario; Marilda Siqueira on behalf of the Fiocruz COVID-19 Genomic Surveillance Network; Paola Resende; Renata Serrano Lopes; Rodrigo Ribeiro Rodrigues; Taina Venas |
| EPI_ISL_4880295 | Laboratorio Central de Saude Publica do Estado do Maranhao (LACEN-MA) | Laboratory of Respiratory Viruses and Measles, Oswaldo Cruz Institute, FIOCRUZ | Agatha Soares; Alice Sampaio Rocha; Ana Carolina Mendonca; Anna Carolina Paixao; Fernando Motta; Ighor Arantes; Lidio Gonçalves Lima Neto; Luciana Appolinario; Marilda Siqueira on behalf of the Fiocruz COVID-19 Genomic Surveillance Network; Paola Resende; Renata Serrano Lopes |
| EPI_ISL_3914053, EPI_ISL_3914064, EPI_ISL_3914082, EPI_ISL_3914087, EPI_ISL_3914095, EPI_ISL_3914107, EPI_ISL_3914129, EPI_ISL_3914134, EPI_ISL_3914158, EPI_ISL_3914166, EPI_ISL_3914171, EPI_ISL_3914179, EPI_ISL_3914198, EPI_ISL_3914206, EPI_ISL_3914223, EPI_ISL_3914232, EPI_ISL_3914240, EPI_ISL_3914244, EPI_ISL_3914258, EPI_ISL_3914262, EPI_ISL_3914287, EPI_ISL_3914300, EPI_ISL_3914304, EPI_ISL_3914308, EPI_ISL_3914355, EPI_ISL_3914359, EPI_ISL_3914363, EPI_ISL_3914368, EPI_ISL_3914376, EPI_ISL_3914384, EPI_ISL_3914389, EPI_ISL_3914394, EPI_ISL_3914397, EPI_ISL_3914402, EPI_ISL_3914407, EPI_ISL_3914415, EPI_ISL_3914418, EPI_ISL_3914423, EPI_ISL_3914427, EPI_ISL_3914430, EPI_ISL_3914438 | | | |
| see above | Laboratorio Central de Saude Publica do Estado do Parana (Instituto de Biologia Molecular do Paraná (LACEN-PR) | Instituto Carlos Chagas - Fiocruz | Alessandra De Melo Aguiar; Andreia Akemi Suzukawa; Andréa Rodrigues Ávila; Bruno Dallagiovanna; Dalila Zanette; Eduardo Balsanelli; Emanuel Maltempi de Souza; Fabio Passetti; Fabricio Klerynton Marchini; Fábio de Oliveira Pedrosa; Guilherme Becker; Helisson Faoro; Hellen Geremias dos Santos; Irina Nastassja Riediger; Letusa Albrecht; Lucas Blanes; Luis Gustavo Morello; Lysangela Ronalte Alves; Maria do Carmo Debur; Mauro de Medeiros Oliveira; Michelle Orane Schemberger; Paola Cristina Resende; Sheila Cristina Nardeli; Tiago Gräf; Valter Antônio de Baura |
| EPI_ISL_3190381, EPI_ISL_3190382, EPI_ISL_3190384, EPI_ISL_3190385, EPI_ISL_3190386, EPI_ISL_3190387, EPI_ISL_3190388, EPI_ISL_3190389, EPI_ISL_3190390, EPI_ISL_3190391, EPI_ISL_4634020 | | | |
| see above | Laboratorio Central de Saude Publica do Estado do Parana (LACEN/PR) | Laboratory of Respiratory Viruses and Measles, Oswaldo Cruz Institute, FIOCRUZ | Agatha Soares; Alice Sampaio Rocha; Ana Carolina Mendonca; Anna Carolina Paixao; Elisa Cavalcante Pereira; Fernando Motta; Ighor Arantes; Irina Riediger; Luciana Appolinario; Marilda Siqueira on behalf of the Fiocruz COVID-19 Genomic Surveillance Network; Paola Resende; Renata Serrano Lopes; Taina Venas |
| EPI_ISL_3190392, EPI_ISL_3190393, EPI_ISL_3190395, EPI_ISL_3190396, EPI_ISL_3190397, EPI_ISL_3190398, EPI_ISL_3190399, EPI_ISL_3190400, EPI_ISL_3190401, EPI_ISL_3190402, EPI_ISL_3190404 | | | |
| see above | Laboratorio Central de Saude Publica do Estado do Rio Grande do Norte (LACEN/RN) | Laboratory of Respiratory Viruses and Measles, Oswaldo Cruz Institute, FIOCRUZ | Agatha Soares; Alice Sampaio Rocha; Ana Carolina Mendonca; Ana Paula Ferreira Costa; Anna Carolina Paixao; Antonnyo Palmielly Diogenes Lima; Aurélio de Oliveira Bento; Elisa Cavalcante Pereira; Fernando Motta; Gessika Brenna Costa Alves; Heglayne Pereira Vital da Silva; Iago de Souza Gomes; Ighor Arantes; Isabelle Cristina Clemente dos Santos; Janaina Sonale Cavalcante Nogueira de Oliveira; Jayra Juliana Paiva Alves Abrantes; Jonas José da Silva; Luciana Appolinario; Marilda Siqueira on behalf of the Fiocruz COVID-19 Genomic Surveillance Network; Paola Resende; Renata Serrano Lopes; Taina Venas; Themis Rocha de Souza; Vítor Gabriel Saldanha Fernandes |

| | | | |
|---|---|---|---|
| EPI_ISL_3539241, EPI_ISL_3539242, EPI_ISL_3539243, EPI_ISL_3539244, EPI_ISL_3539767, EPI_ISL_3539768, EPI_ISL_3539769, EPI_ISL_3539840, EPI_ISL_3539841, EPI_ISL_3539842, EPI_ISL_3539843, EPI_ISL_3539844, EPI_ISL_3539845, EPI_ISL_3539847, EPI_ISL_3539848, EPI_ISL_3539849, EPI_ISL_3539850, EPI_ISL_3539851, EPI_ISL_3539852, EPI_ISL_3539853, EPI_ISL_3539854, EPI_ISL_3539855, EPI_ISL_3539856, EPI_ISL_3539858, EPI_ISL_3539859, EPI_ISL_3539860, EPI_ISL_3539861, EPI_ISL_3539862, EPI_ISL_3539863, EPI_ISL_3539864, EPI_ISL_3539865, EPI_ISL_3539867, EPI_ISL_3539868, EPI_ISL_3539869, EPI_ISL_3539870, EPI_ISL_3539871, EPI_ISL_3539872, EPI_ISL_3539873, EPI_ISL_3539874, EPI_ISL_3539875, EPI_ISL_3539876, EPI_ISL_3539877, EPI_ISL_3539878, EPI_ISL_3539879, EPI_ISL_3539880, EPI_ISL_3539881, EPI_ISL_3539882, EPI_ISL_3539883, EPI_ISL_3539884, EPI_ISL_3539885, EPI_ISL_3539886, EPI_ISL_3539887, EPI_ISL_3539888, EPI_ISL_3539889, EPI_ISL_3539890, EPI_ISL_3539891, EPI_ISL_3539892, EPI_ISL_3539893, EPI_ISL_3539894, EPI_ISL_3539895, EPI_ISL_3539896, EPI_ISL_3539897, EPI_ISL_3539898, EPI_ISL_3539899, EPI_ISL_3539900, EPI_ISL_3539901, EPI_ISL_3539902, EPI_ISL_3539903, EPI_ISL_3539904, EPI_ISL_3539905, EPI_ISL_3539976, EPI_ISL_3539978, EPI_ISL_3539980, EPI_ISL_3539981, EPI_ISL_3539982, EPI_ISL_3539983, EPI_ISL_3539984, EPI_ISL_3539985, EPI_ISL_3539986, EPI_ISL_3539987, EPI_ISL_3539988, EPI_ISL_3539989, EPI_ISL_3539990, EPI_ISL_3539992, EPI_ISL_3539993, EPI_ISL_3539994, EPI_ISL_3539995, EPI_ISL_3539996, EPI_ISL_3539997, EPI_ISL_3832461 | | | |
| see above | Laboratorio Central de Saude Publica do Estado do Rio Grande do Sul (LACEN-RS) | Laboratory of Respiratory Viruses and Measles, Oswaldo Cruz Institute, FIOCRUZ | Agatha Cristinne Soares; Alice Sampaio Rocha; Ana Carolina Mendonca; Anderson Brandao Leite; Anna Carolina Paixao; Elisa Cavalcante Pereira; Fernando Motta; Ighor Leonardo Arantes; Luciana Appolinario; Marilda Siqueira on behalf of the Fiocruz COVID-19 Genomic Surveillance Network; Paola Resende; Renata Serrano Lopes; Richard Salvato; Taina Venas; Tatiana Schaffer Gregianini |
| EPI_ISL_4237303, EPI_ISL_4237304, EPI_ISL_4237313, EPI_ISL_4237318, EPI_ISL_4237323, EPI_ISL_4237324, EPI_ISL_4237325, EPI_ISL_4237326, EPI_ISL_4237327, EPI_ISL_4237328, EPI_ISL_4237339, EPI_ISL_4237353, EPI_ISL_4237362, EPI_ISL_4237409 | | | |
| see above | Laboratorio Central de Saude Publica do Estado do Rio de Janeiro (LACEN-RJ) | Laboratory of Respiratory Viruses and Measles, Oswaldo Cruz Institute, FIOCRUZ | Agatha Cristinne Prudencio; Alice Sampaio Rocha; Ana Carolina Mendonca; Andrea Cony Cavalcanti; Anna Carolina Paixao; Elisa Cavalcante Pereira; Fernando Motta; Ighor Leonardo Arantes Gomes; Luciana Appolinario; Marilda Siqueira on behalf of the Fiocruz COVID-19 Genomic Surveillance Network; Paola Resende; Renata Serrano Lopes; Taina Venas |
| EPI_ISL_3434906 | Laboratorio Central de Saude Publica do Estado do Rio de Janeiro (LACEN/RJ) | Laboratory of Respiratory Viruses and Measles, Oswaldo Cruz Institute, FIOCRUZ | Agatha Cristinne Prudencio Soares; Alice Sampaio Rocha; Ana Carolina Mendonca; Andrea Cony Cavalcanti; Anna Carolina Paixao; Elisa Cavalcante Pereira; Fernando Motta; Ighor Leonardo Arantes Gomes; Luciana Appolinario; Marilda Siqueira on behalf of the Fiocruz COVID-19 Genomic Surveillance Network; Paola Resende; Renata Serrano Lopes; Taina Venas |
| EPI_ISL_3235304, EPI_ISL_3235305, EPI_ISL_3235306, EPI_ISL_3235308, EPI_ISL_3235309, EPI_ISL_3235310, EPI_ISL_3235311, EPI_ISL_3235312, EPI_ISL_3235314 | | | |
| see above | Laboratorio Central de Saude Publica do Estado do Tocantins (LACEN/TO) | Laboratory of Respiratory Viruses and Measles, Oswaldo Cruz Institute, FIOCRUZ | Agatha Soares; Alice Sampaio Rocha; Ana Carolina Mendonca; Anna Carolina Paixao; Elisa Cavalcante Pereira; Fernando Motta; Ighor Arantes; Jucimaria Dantas Galvao; Luciana Appolinario; Marilda Siqueira on behalf of the Fiocruz COVID-19 Genomic Surveillance Network; Paola Resende; Renata Serrano Lopes; Taina Venas |
| EPI_ISL_4849750 | Laboratorio Central de la Ciudad de Santa Fe | Grupo de Genómica y Bioinformática del Instituto de Investigación de la Cadena Láctea CONICET-INTA on behalf of 'Proyecto Argentino Interinstitucional de genomica de SARS-CoV-2' (PAIS Consortium) | AF; Amadio; C; Eberhardt; G; Irazoqui; JM; MF; Mugna; Ojeda; Pastor; Rompato; V |
| EPI_ISL_3150214, EPI_ISL_3150215 | Laboratorio Clínico Gamma | "Unidad de Secuenciación y Genómica, USACH" | "Carlos A. Villarroel; Francisco A. Cubillos"; Kamila Urbina; Mara Cuevas; Marcelo Cortez-San Martin; Yesseny Vásquez-Martínez |
| EPI_ISL_3671038 | Laboratorio Las Americas | Universidad Nacional de Colombia - Laboratorio Genómico One Health | Andres F. Cardona-Rios; Carlos Franco-Muñoz; Carolina Muñoz-Arango; Celeny Ortiz; Daniel O. Maldonado-Perez; Diego A. Álvarez-Díaz; Hector Alejandro Ruiz-Moreno; Idabely Betancur Ortiz; Jorge E. Osorio; Juan P. Hernandez-Ortiz; Karl A Ciuoderis; Katherine Laiton-Donato; Laura Silvana Perez; Lina M. Hurtado; Marcela Mercado-Reyes; Maria Angélica Maya; Maria Stella López; Rita Almanza Payares; Sandra Ines Cano; Simón Villegas Velásquez |
| EPI_ISL_3527820, EPI_ISL_3527822, EPI_ISL_3835378 | Laboratorio Nacional de Salud | Asociación de Salud Integral / Clínica Familiar "Luis Ángel García" | Ana S. Gonzalez-Reiche; Claudia Rangel; Danicela Mercado; Eduardo Arathon; Hilda Ruiz; Luis Aguirre; Luis Rivas; Narda Medina; Oscar Bonilla; Osmar Gamboa. |
| EPI_ISL_5617616 | Laboratorio Nacional de Salud, Ministerio de Salud Publica y Asistencia Social | Genomics and Proteomics Departament, Gorgas Memorial Institute For Health Studies | Alexander A Martinez; Ambar Moreno; Claudia Estrada; Claudia Gonzalez V; César Roberto Conde Pereira; Jessica Gondola; Leyda Abrego; Marlene Castillo; Melissa Gaitan; Oris Chavarria |
| EPI_ISL_3278303 | Laboratorio Olimpus | Instituto Nacional de Salud | Carlos Franco-Muñoz; Carmen Osorio; Diana Malo; Diego A. Álvarez-Díaz; Diego Andrés Prada; Gerardo Santamaría; Hector Alejandro Ruiz-Moreno; Jhonnatan Reales-González; Jorge Rivera; Juan Camilo Martinez; Julian Naizaque; Katherine Laiton-Donato; Lisseth Pardo; Magdalena Wiesner; Marcela Mercado-Reyes; Maria T. Herrera-Sepúlveda; Marta Lopez Blanco; Martha Lucia Ospina Martinez; Paola Rojas; Sergio Gomez; Sheryll Corchuelo; Ángela Alarcon Cruz |
| EPI_ISL_3671019 | Laboratorio UNIGEM | Universidad Nacional de Colombia - Laboratorio Genómico One Health | Andres F. Cardona-Rios; Carlos Franco-Muñoz; Carolina Muñoz-Arango; Celeny Ortiz; Daniel O. Maldonado-Perez; Diego A. Álvarez-Díaz; Hector Alejandro Ruiz-Moreno; Idabely Betancur Ortiz; Jorge E. Osorio; Juan P. Hernandez-Ortiz; Karl A Ciuoderis; Katherine Laiton-Donato; Laura Silvana Perez; Lina M. Hurtado; Marcela Mercado-Reyes; Maria Angélica Maya; Maria Stella López; Rita Almanza Payares; Sandra Ines Cano; Simón Villegas Velásquez |
| EPI_ISL_5332830, EPI_ISL_5332835, EPI_ISL_6121555, EPI_ISL_6121574, EPI_ISL_6121575 | Laboratorio de Alta Complejidad (LACMI), Instituto de Genética Humana (IgeHM) Parque de la Salud de la Provincia de Misiones, Universidad Nacional de Misiones | Grupo de Genómica y Bioinformática del Instituto de Investigación de la Cadena Láctea CONICET-INTA on behalf of 'Proyecto Argentino Interinstitucional de genomica de SARS-CoV-2' (PAIS Consortium) | AF; Aguirre; Amadio; Benitez; C; D; Eberhardt; Fontana; G; Gamarra; Gonzales; Irazoqui; JM; K; Ludojoski; M; MF; R; Salvatierra; Silva; TA; Urquiza; Vogel |
| EPI_ISL_3050574, EPI_ISL_3050591, EPI_ISL_3050615, EPI_ISL_3050616, EPI_ISL_3050617, EPI_ISL_3050618, EPI_ISL_4030316, EPI_ISL_4030328, EPI_ISL_4030330, EPI_ISL_4030331, EPI_ISL_4030351, EPI_ISL_4030362, EPI_ISL_4030366 | | | |
| see above | Laboratorio de Ecologia de Doencas Transmissiveis na Amazonia, Instituto Leonidas e Maria Deane - Fiocruz Amazonia | Laboratorio de Ecologia de Doencas Transmissiveis na Amazonia, Instituto Leonidas e Maria Deane - Fiocruz Amazonia | Alex Martins; André Corado; Debora Duarte; Fabiola Mendonça da Silva Chui; Felipe Naveca; Fernanda Nascimento; Fernando Fonseca de Almeida Val; George Silva; Gisely Cardoso de Melo; Karina Pessoa; Luciana Gonçalves; Marcus Vinicius Guimarães de Lacerda; Maria Júlia Brandão; Maria Paula Gomes Mourão; Mariana Xavier Simão; Matilde Mejía; Michele Jesus; Valdinete Nascimento; Vanderson de Souza Sampaio; Victor Souza; Ágatha Costa |
| EPI_ISL_3370443, EPI_ISL_4890157, EPI_ISL_4890171, EPI_ISL_5994556, EPI_ISL_5994564, EPI_ISL_5994571 | Laboratorio de Infectologia y Virologia Molecular | Laboratory of Molecular Virology, School of Medicine, Pontificia Universidad Catolica de Chile | Alejandro Bhrun; Ana Maria Contreras; Andres E. Munoz-Marcos; Carlos Palma; Catalina Pardo-Roa; Constanza Maldonado; Constanza Martinez-Valdevenito; Eileen Serrano; Erick Salinas; Estefany Poblete; Eugenia L. Fuentes Luppichini; Francisco Melo; Jennifer Angulo; Jorge Levican; Leonardo I. Almonacid; M. Belen Leyton; Magdalena Vera; Marcela Ferres; Maria Jose Avendano; Rafael A. Medina; Rafael A. Medina.; Tamara Garcia-Salum |
| EPI_ISL_2907512, EPI_ISL_2907513, EPI_ISL_3134279 | Laboratorio de Medicina Molecular, Universidad de Magallanes | Centro Asistencial Docente y de Investigacion, Universidad de Magallanes | Carolina Pérez; Constanza Ceroni; Diego Alvarez; Inés Cid; Jacqueline Aldridge; Jorge González; Juan Ríos; Marcelo Navarrete; Roberto Uribe-Paredes |
| EPI_ISL_3049001, EPI_ISL_3049021, EPI_ISL_3049022, EPI_ISL_3049023, EPI_ISL_3049024, EPI_ISL_3049025, EPI_ISL_3049026, EPI_ISL_3049027, EPI_ISL_3049028, EPI_ISL_3049029, EPI_ISL_3049030, EPI_ISL_3055532, EPI_ISL_3055533, EPI_ISL_3761522, EPI_ISL_3761523, EPI_ISL_3761528, EPI_ISL_3761539, EPI_ISL_3761558, EPI_ISL_3761563, EPI_ISL_3761573, EPI_ISL_3761586, EPI_ISL_3761588, EPI_ISL_3761674, EPI_ISL_3761675, EPI_ISL_3761676, EPI_ISL_3761677, EPI_ISL_3761678, EPI_ISL_3761679, EPI_ISL_3761680, EPI_ISL_3761754, EPI_ISL_3761755, EPI_ISL_3761756, EPI_ISL_3761757, EPI_ISL_3761758, EPI_ISL_3761759 | | | |
| see above | Laboratorio de Pesquisa em Virologia, FAMERP, SJRP | Laboratorio de Pesquisa em Virologia, FAMERP, SJRP | Beatriz de Carvalho Marques; Cecília Artico Banho; Cíntia Bittar; Fábio Sossai Possebon; Guilherme Campos; Helena Lage Ferreira; Jorge A. Petroli Marchesi; João Pessoa Araújo Jr.; Leila Sabrina Ullmann; Lívia Sacchetto; Maisa C. Pereira Parra; Marília Moraes; Maurício L. Nogueira.; Paula Rahal; Paulo Inacio da Costa |
| EPI_ISL_3991711, EPI_ISL_4080481, EPI_ISL_4080489, EPI_ISL_4080493, EPI_ISL_4080494, EPI_ISL_4080529, EPI_ISL_4080530, EPI_ISL_4080535, EPI_ISL_4080536, EPI_ISL_4080538, EPI_ISL_4080539, EPI_ISL_4080540, EPI_ISL_4080542, EPI_ISL_4080543, EPI_ISL_4080544, EPI_ISL_4080555, EPI_ISL_4172730, EPI_ISL_4172749, EPI_ISL_4172751, EPI_ISL_4172877, EPI_ISL_4172884, EPI_ISL_4172917, EPI_ISL_4172969, EPI_ISL_4279977, EPI_ISL_4279978, EPI_ISL_4279999, EPI_ISL_4280021, EPI_ISL_4416866, EPI_ISL_4416869, EPI_ISL_4416887, EPI_ISL_4417092, EPI_ISL_4417094, EPI_ISL_4417105, EPI_ISL_4417106, EPI_ISL_4417120, EPI_ISL_4417134 | | | |
| see above | Laboratorio de Referencia Nacional de Virus Respiratorios. Centro Nacional de Salud Publica. Instituto Nacional de Salud Peru. | Laboratorio de Referencia Nacional de Virus Respiratorios. Centro Nacional de Salud Publica. Instituto Nacional de Salud Peru. | Carlos Padilla Rojas; Henri Bailon Calderon; Iris Silva Molina; Joseph Huayra Niquen; Lely Solari Zerpa; Luis Barcena Flores; Marco Galarza Perez; Nancy Rojas Serrano; Nieves Sevilla Castañeda; Omar Caceres Rey; Orson Mestanza Millones; Princesa Medrano Alhuay; Priscila Lope Pari; Sandra Morales Ruiz; Sara Gordillo Vilchez; Steve Acedo Lazo; Veronica Hurtado Vela; Victor Jimenez Vasquez; Wendy Lizarraga Olivares |
| EPI_ISL_7747304, EPI_ISL_7747358, EPI_ISL_7747369 | Laboratorio de Salud Pública de Bogotá | Gencore - Universidad de los Andes | Alejandro Gomez; Cristian Barrera; Gabriela Ariza; Gabriela Delgado; Johana Hernandez; Luisa Sacristan; Marcela Guevara; Silvia Restrepo |
| EPI_ISL_5771889, EPI_ISL_5771890 | Laboratorio del Hospital Regional Ushuaia Gdor. Ernesto Campos | Nodo de Secuenciación Tierra del Fuego - Hospital Regional Ushuaia - Centro Austral De Investigaciones Científicas - Universidad Nacional De Tierra Del Fuego | AE; Boutureira, MF.; CA; CB; CF; Castro; Ceballos; Cáceres; De Roccis; F; G; Gallego; Gramundi; ID; Nardi; Rojas; SB; SG; Yulan |
| EPI_ISL_7620356, EPI_ISL_7620388 | Laboratorio del Hospital Regional Ushuaia Gdor. Ernesto Campos | Nodo de Secuenciación Tierra del Fuego - Hospital Regional Ushuaia - Centro Austral De Investigaciones Científicas - Universidad Nacional De Tierra Del Fuego on behalf PAIS project and ANLIS-Malbran | AE; Boutureira, MF.; CA; CB; CF; Castro; Ceballos; Cáceres; De Roccis; F; G; Gallego; Gramundi; ID; Nardi; Rojas; SB; SG; Yulan |
| EPI_ISL_2918882 | Laboratorio di Microbiologia | Laboratorio di Microbiologia | Martinetti Lucchini Gladys; Valeria Spina |
| EPI_ISL_2975218, EPI_ISL_2975220, EPI_ISL_3026010, EPI_ISL_3128491 | Laboratorio di Riferimento Regionale della Sicilia Occidentale per l'Emergenza COVID-19 | Laboratorio di Riferimento Regionale della Sicilia Occidentale per l'Emergenza COVID-19 | Carmelo Massimo Maida; Claudio Costantino; Daniela Di Naro; Fabio Tramuto; Francesco Vitale; Giorgio Graziano; Giulia Randazzo; Vincenzo Restivo; Walter Mazzucco |
| EPI_ISL_2926523, EPI_ISL_2926529, EPI_ISL_2926530, EPI_ISL_2926531, EPI_ISL_2926532, EPI_ISL_2926537, EPI_ISL_2927715, EPI_ISL_2927719, EPI_ISL_2927729, EPI_ISL_3016825, EPI_ISL_3016829, EPI_ISL_3016879, EPI_ISL_3016882, EPI_ISL_3016915, EPI_ISL_3016930, EPI_ISL_3016931, EPI_ISL_3016948, EPI_ISL_3016967, EPI_ISL_3016975, EPI_ISL_3016982, EPI_ISL_3017094, EPI_ISL_3017147, EPI_ISL_3017174, EPI_ISL_3017176, EPI_ISL_3017297, EPI_ISL_3017327, EPI_ISL_3017348, EPI_ISL_3017349, EPI_ISL_3017430, EPI_ISL_3017512, EPI_ISL_3017519, EPI_ISL_3017520, EPI_ISL_3017521, EPI_ISL_3017533, EPI_ISL_3017697, EPI_ISL_3017717, EPI_ISL_3017719, EPI_ISL_3017731, EPI_ISL_3017743, EPI_ISL_3017744, EPI_ISL_3017764, EPI_ISL_3017776, EPI_ISL_3017801, EPI_ISL_3017802, | | | |

| | | | |
|---|---|---|---|
| EPI_ISL_3017811, EPI_ISL_3017812, EPI_ISL_3017833, EPI_ISL_3017857, EPI_ISL_3017887, EPI_ISL_3017905, EPI_ISL_3017910, EPI_ISL_3017911, EPI_ISL_3017938, EPI_ISL_3017950, EPI_ISL_3017969, EPI_ISL_3017992, EPI_ISL_3017994, EPI_ISL_3018005, EPI_ISL_3018006, EPI_ISL_3018052, EPI_ISL_3018059, EPI_ISL_3018061, EPI_ISL_3018062, EPI_ISL_3018064, EPI_ISL_3018095, EPI_ISL_3018106, EPI_ISL_3018151, EPI_ISL_3020069, EPI_ISL_3111278, EPI_ISL_3111330, EPI_ISL_3111339, EPI_ISL_3111358, EPI_ISL_3111848, EPI_ISL_3112180, EPI_ISL_3112199, EPI_ISL_3112200, EPI_ISL_3112232, EPI_ISL_3112282, EPI_ISL_3112292, EPI_ISL_3112302, EPI_ISL_3112304, EPI_ISL_3112314, EPI_ISL_3112315, EPI_ISL_3112413, EPI_ISL_3112416, EPI_ISL_3112989, EPI_ISL_3112991, EPI_ISL_3112995, EPI_ISL_3113000, EPI_ISL_3113033, EPI_ISL_3113034, EPI_ISL_3113038, EPI_ISL_3113101, EPI_ISL_3113102, EPI_ISL_3113103, EPI_ISL_3113118, EPI_ISL_3113134, EPI_ISL_3113387, EPI_ISL_3113388, EPI_ISL_3113497, EPI_ISL_3328496, EPI_ISL_3328507, EPI_ISL_3328514, EPI_ISL_3330633, EPI_ISL_3330636, EPI_ISL_3330637, EPI_ISL_4373728 | | | |
| see above | Laboratory Corporation of America | Centers for Disease Control and Prevention Division of Viral Diseases, Pathogen Discovery | Adrian Paskey; Amanda Douglas; Amanda Suchanek; Andrea Throop; Ayla Burns; Benjamin Rambo-Martin; Bobbi Croy; Brian Krueger; Brian Norvell; Christopher Gulvick; Christos Petropoulos; Clinton Paden; Clinton R. Paden; Craig Lukasik; Dakota Howard; Darlene Wagner; Debbie Boles; Dhwani Batra; Duncan MacCannell; Eyad Almasri; Goran Stevovic; Howard Engler; Hrushikesh Deshmukh; Jake Humphrey; Jana Schroth; Jason Caravas; Joe Voshell; John Pruitt; Jonathan Meltzer; Jonathan Williams; Kara Moser; Kimberly Wagner; Kristine Lacek; Lax Iyer; Lisa Pfefferle; Lyndon Tilson; Manoj Jain; Marcia Eisenberg; Mary Ann Cristobal; Mary Cristobal; Mary Williamson; Matthew Robinson; Matthew Schmerer; Michael Levandoski; Mike Sapeta; Mindy Nye; Minoo Agarwal; Mohan Kolli; Nuthawin Charoensri; Oren Cohen; Peter Cook; Peter W. Cook; Prashant Gupta; Qian Zeng; Rama Ghatti; Scott Parker; Scott Ryan; Scott Sammons; Shatavia Morrison; Stanley Letovsky; Steven Ragan; Suresh Babu Selvaraju; Suresh Selvaraju; Susan Countryman; Susan Hicks; Suzanne Dale; Thomas Urban; Tim Kuphal; Tricia Zwiefelhofer; Tymeckia Kendall; Victoria Caban Figueroa; Vincent Drouillon; Yvette Unoarumhi |
| EPI_ISL_2983187, EPI_ISL_2983236, EPI_ISL_2983237, EPI_ISL_3045373, EPI_ISL_3045374, EPI_ISL_3045375, EPI_ISL_3045376, EPI_ISL_3045377, EPI_ISL_3045549, EPI_ISL_3045550, EPI_ISL_3045551, EPI_ISL_3045552, EPI_ISL_3827873, EPI_ISL_4880263, EPI_ISL_4880264, EPI_ISL_5158271 | | | |
| see above | Laboratory of Respiratory Viruses and Measles, Oswaldo Cruz Institute, FIOCRUZ | Laboratory of Respiratory Viruses and Measles, Oswaldo Cruz Institute, FIOCRUZ | Agatha Cristinne Prudencio; Alice Sampaio Rocha; Ana Carolina Mendonca; Anna Carolina Paixao; Elisa Cavalcante Pereira; Fernando Motta; Ighor Leonardo Arantes Gomes; Luciana Appolinario; Marilda Siqueira on behalf of the Fiocruz COVID-19 Genomic Surveillance Network; Paola Resende; Patricia Brasil; Renata Serrano Lopes; Taina Venas; Thais Aguiar |
| EPI_ISL_3461348, EPI_ISL_3461352 | Laboratory, Gravity Diagnostics | Laboratory, Gravity Diagnostics | Diagnostics, G. |
| EPI_ISL_4600459, EPI_ISL_4600460, EPI_ISL_4600461, EPI_ISL_4600462, EPI_ISL_4600464, EPI_ISL_4600467, EPI_ISL_4600474, EPI_ISL_4600479 | | | |
| see above | Laboratório Central de Saúde Pública Profº Gonçalo Moniz | Coordenação Geral de Laboratórios de Saúde Pública (CGLAB/DAEVS/SVS/MS) | Vagner Fonseca; et al. |
| EPI_ISL_6568661, EPI_ISL_6568685 | Laboratório Central de Saúde Pública de Pernambuco | Coordenação Geral de Laboratórios de Saúde Pública (CGLAB/DAEVS/SVS/MS) | Vagner Fonseca; et al. |
| EPI_ISL_3050575, EPI_ISL_3050576, EPI_ISL_3050577, EPI_ISL_3050578, EPI_ISL_3050579, EPI_ISL_3050580, EPI_ISL_3050581, EPI_ISL_3050582, EPI_ISL_3050583, EPI_ISL_3050584, EPI_ISL_3050585, EPI_ISL_3050586, EPI_ISL_3050587, EPI_ISL_3050588, EPI_ISL_3050589, EPI_ISL_3050590, EPI_ISL_3050592, EPI_ISL_3050593, EPI_ISL_3050594, EPI_ISL_3050595, EPI_ISL_3050596, EPI_ISL_3050597, EPI_ISL_3050598, EPI_ISL_3050599, EPI_ISL_3050600, EPI_ISL_3050601, EPI_ISL_3050602, EPI_ISL_3050603, EPI_ISL_3050604, EPI_ISL_3050605, EPI_ISL_3050606, EPI_ISL_3050607, EPI_ISL_3050608, EPI_ISL_3050609, EPI_ISL_3050610, EPI_ISL_3050611, EPI_ISL_3050612, EPI_ISL_3050613, EPI_ISL_3050614, EPI_ISL_3050619, EPI_ISL_3050620, EPI_ISL_3050621, EPI_ISL_3050622, EPI_ISL_3050623, EPI_ISL_3050624, EPI_ISL_3050625, EPI_ISL_3050626, EPI_ISL_3050627, EPI_ISL_3050628, EPI_ISL_3050629, EPI_ISL_3050630, EPI_ISL_3050631, EPI_ISL_3050632, EPI_ISL_3050633, EPI_ISL_3050634, EPI_ISL_3050635, EPI_ISL_3050636, EPI_ISL_3050637, EPI_ISL_3050638, EPI_ISL_3050639, EPI_ISL_3050640, EPI_ISL_4520325, EPI_ISL_4520326, EPI_ISL_4520327, EPI_ISL_4520328, EPI_ISL_4520370, EPI_ISL_4520384 | | | |
| see above | Laboratório Central de Saúde Pública do Amazonas - LACEN-AM | Laboratorio de Ecologia de Doencas Transmissiveis na Amazonia, Instituto Leonidas e Maria Deane - Fiocruz Amazonia | André Corado; Felipe Naveca; Fernanda Nascimento; George Silva; Karina Pessoa; Luciana Gonçalves; Maria Júlia Brandão; Matilde Mejía; Valdinete Nascimento; Victor Souza; Ágatha Costa |
| EPI_ISL_6677476, EPI_ISL_6677480, EPI_ISL_6677590, EPI_ISL_6678411, EPI_ISL_6678511, EPI_ISL_6679928, EPI_ISL_6681050, EPI_ISL_6681796, EPI_ISL_6682191, EPI_ISL_6683276, EPI_ISL_6685073, EPI_ISL_6685077, EPI_ISL_6685140, EPI_ISL_6686428, EPI_ISL_7235716, EPI_ISL_7235914, EPI_ISL_7236461, EPI_ISL_7237031, EPI_ISL_7243779, EPI_ISL_7244508, EPI_ISL_7246824, EPI_ISL_7247859, EPI_ISL_7252530, EPI_ISL_7257409, EPI_ISL_7258225, EPI_ISL_7258784, EPI_ISL_7261516 | | | |
| see above | Laboratório Central de Saúde Pública do Estado do Tocantins (LACEN/TO) | Bioinformatics and Biotechnology Laboratory (Labinftec/Federal University of Tocantins, Campus of Gurupi) | Fabrício Souza Campos; Jucimária Dantas Galvão; Raíssa Nunes dos Santos; Ueric José Borges de Souza |
| EPI_ISL_5195461, EPI_ISL_5195477, EPI_ISL_5195479, EPI_ISL_5195480, EPI_ISL_5195483, EPI_ISL_5195484 | Laboratório Municipal de Biologia Molecular | Instituto René Rachou / Fiocruz Minas | André Menezes; Anna Salim; Eneida Oliveira; Gabriel Fernandes; Pedro Alves; Rubens do Monte Neto; Thaís Silva |
| EPI_ISL_3492497, EPI_ISL_4051958 | Laboratório de Microbiologia Molecular - Universidade FEEVALE | Molecular Microbiology Laboratory | Alana Witt Hansen; Fernando Rosado Spilki; Fágner Henrique Heldt; Juliana Schons Gulart; Juliana Schons Gularte; Juliane Deise Fleck; Mariana Soares da Silva; Matheus Nunes Weber; Meriane Demoliner; Michele Filippi.; Micheli Filippi; Paula Rodrigues de Almeida; Vyctoria Malayhka de Abreu Góes Pereira. |
| EPI_ISL_3045380, EPI_ISL_3190193, EPI_ISL_3190194, EPI_ISL_3190195, EPI_ISL_3190196, EPI_ISL_3190197, EPI_ISL_3190198, EPI_ISL_3190199, EPI_ISL_3190200, EPI_ISL_3190201, EPI_ISL_3190202, EPI_ISL_3190205, EPI_ISL_3190206, EPI_ISL_3190207, EPI_ISL_3190208, EPI_ISL_3190209, EPI_ISL_3190210, EPI_ISL_3190211, EPI_ISL_3190217, EPI_ISL_3190218 | | | |
| see above | Labortorio Central de Saude Publica do Estado do Rio de Janeiro (LACEN/RJ) | Laboratory of Respiratory Viruses and Measles, Oswaldo Cruz Institute, FIOCRUZ | Agatha Cristinne Prudencio Soares; Alice Sampaio Rocha; Ana Carolina Mendonca; Andrea Cony Cavalcanti; Anna Carolina Paixao; Elisa Cavalcante Pereira; Fernando Motta; Ighor Leonardo Arantes Gomes; Luciana Appolinario; Marilda Siqueira on behalf of the Fiocruz COVID-19 Genomic Surveillance Network; Paola Resende; Renata Serrano Lopes; Taina Venas |
| EPI_ISL_3190410, EPI_ISL_3190411 | Labortorio Central de Saude Publica do Estado do Tocantins (LACEN/TO) | Laboratory of Respiratory Viruses and Measles, Oswaldo Cruz Institute, FIOCRUZ | Agatha Soares; Alice Sampaio Rocha; Ana Carolina Mendonca; Anna Carolina Paixao; Elisa Cavalcante Pereira; Fernando Motta; Ighor Arantes; Jucimaria Dantas Galvao; Luciana Appolinario; Marilda Siqueira on behalf of the Fiocruz COVID-19 Genomic Surveillance Network; Paola Resende; Renata Serrano Lopes; Taina Venas |
| EPI_ISL_5434437 | MB-Cadham Provincial laboratory | Manitoba Cadham Provincial Laboratory | Anna Majer; Anneliese Landgraff; David Alexander; Elsie Grudeski; Gary Van Domselaar; Grace Seo; Jared Bullard; Jennifer Tanner; Kerry Dust; Morag Graham; Nat; Natalie Knox; Paul Van Caeseele; Philip Mabon; Rhiannon Huzarewich; Russell Mandes; Shari Tyson |
| EPI_ISL_2955561, EPI_ISL_2980037, EPI_ISL_3020114, EPI_ISL_3030729, EPI_ISL_3045923, EPI_ISL_3099906 | MD PHL | MD PHL | Maryland Department of Health Laboratories Administration |
| EPI_ISL_3235569, EPI_ISL_3235571 | MD PHL | Maryland Department of Health Laboratories Administration | Ami Patel; Eric N. Keller; Jillian Loomis; Kwang Low; Terence L. Moore; and Robert Myers |
| EPI_ISL_2957399 | MDI Limbach Berlin GmbH; MVZ Labor Berlin | Robert Koch Institute | |
| EPI_ISL_2843674 | MEPHI, Aix Marseille University | MEPHI, Aix Marseille University | Anthony LEVASSEUR |
| EPI_ISL_3355449 | MI - Michigan Department of Health and Human Services - Bureau of Laboratories | Centers for Disease Control and Prevention Division of Viral Diseases, Pathogen Discovery | Alex Burgin; Ben Rambo-Martin; Clinton Paden; Dakota Howard; Dave Wentworth; Dhwani Batra; Jasmine Padilla; Justin Lee; Krista Queen; Kristen Knipe; Kristine Lacek; Mark Burroughs; Matthew Schmerer; Meghan Bentz; Mili Sheth; Peter Cook; Sam Shepard; Sarah Nobles; Suxiang Tong; Vivien Dugan; Yvette Unoarumhi |
| EPI_ISL_6492870 | MSHS Clinical Microbiology Laboratories | MSHS Pathogen Surveillance Program | Adolfo García-Sastre; Adriana van de Guchte; Ajay Obla; Alberto Paniz-Mondolfi; Ana S. Gonzalez-Reiche; Angela Amoako; Ashley Salimbangon; Betsaida Salom Melo; Bremy Alburquerque; Brianne Ciferri; Charles Gleason; Daniel Floda; Deena R. Altman; Denise Jurczyszak; Emilia Mia Sordillo; Gintaras Deikus; Giulio Kleiner; Gopi Patel; Hala Alshammary; Harm van Bakel; Irina Oussenko; Jayeeta Dutta; Juan Soto; Julia Matthews; Katherine Beach; Kathryn Twyman; Kayla Russo; Komal Srivastava; Levy Sominsky; Mahmoud Awawda; Marta Luksza; Matthew M. Hernandez; Melissa Gitman; Michael D. Nowak; Mitchell J. Sullivan; Nancy Francoeur; Robert Sebra; Sarah Schaefer; Shelcie Fabre; Shwetha Hara Sridhar; Viviana Simon; Ying-Chih Wang; Zain Khalil; Zenab Khan |
| EPI_ISL_4281319 | MUSC Molecular Pathology Laboratory | MUSC Molecular Pathology Laboratory | Dariusz Pytel; Frederick S. Nolte; Jaclyn Dunne; Jim Madory; Julie W. Hirschhorn; Kristen Maurer; Scott Curry; W. Bailey Glen Jr |
| EPI_ISL_2845961, EPI_ISL_2956720 | MVZ Labor Dr. Limbach & Kollegen GbR | Robert Koch Institute | |
| EPI_ISL_2957367 | MVZ für Laboratoriumsmedizin und Mikrobiologie Koblenz-Mittelrhein (Labor Koblenz) | Robert Koch Institute | |
| EPI_ISL_3030184, EPI_ISL_3030197, EPI_ISL_3030202 | Macerata/Civitanova Marche | Microbiology University Politecnica delle Marche | Anna Valenza; Carla Acciarri; Katia Marinelli; Monica Lucia Ferreri; Patrizia Bagnarelli; Roberta Longo; Sara Caucci; Stefano Menzo |
| EPI_ISL_3086117, EPI_ISL_3186511, EPI_ISL_3186534, EPI_ISL_4083981 | Maine Health and Environmental Testing Laboratory | Tewhey Lab, The Jackson Laboratory | Barter, M.; Dewey, H.; H. and Tewhey, R.; Iosue, F.; Lynch, R.; Matluk, N.; Munger |
| EPI_ISL_3065230, EPI_ISL_3065282, EPI_ISL_3065306, EPI_ISL_3110617, EPI_ISL_3110729, EPI_ISL_3110748, EPI_ISL_3110749, EPI_ISL_3110831 | | | |
| see above | Mako Medical | Centers for Disease Control and Prevention Division of Viral Diseases, Pathogen Discovery | Adrian Paskey; Benjamin Rambo-Martin; Christopher Gulvick; Clinton R. Paden; Dakota Howard; Darlene Wagner; Dhwani Batra; Duncan MacCannell; Jason Caravas; Kara Moser; Lauren Moon; Matthew Schmerer; Matthew Tugwell; Peter W. Cook; Scott Sammons; Shatavia Morrison; Yvette Unoarumhi |
| EPI_ISL_3435357 | Maryland Genomics, Institute for Genome Sciences, University of Maryland School of Medicine | Maryland Genomics, Institute for Genome Sciences, University of Maryland School of Medicine | Claire M; Fraser; Hazen; Holly; Humphrys; Ivette; Jacques; Jonathan; Kranthi; Lim; Lisa D; Luke J; Mike; Ott; Ravel; Roussey; Sadzewicz; Sandra; Santana-Cruz; Tallon; Tracy; Vavikolanu |
| EPI_ISL_3024786 | Maryland Public Health Laboratory (MD PHL) | Maryland Public Health Laboratory (MD PHL) | Maryland Department of Health Laboratories Administration |
| EPI_ISL_6666023 | Mass General Brigham | Mass General Brigham | A.E.; Adams, G.; Anahtar, M.; B.L.; B.W.; Bauer, M.; Birren; Branda, J.; Carter, A.; Cerrato, F.; Chaluvadi, S.; Chapman; Cusick, C.; D.J.; DeRuff, K.; E. and Sabeti; Flowers, K.; Gallagher, G.; Gladden-Young, A.; Gnirke, A.; Harris, J.; J.E.; K.J.; LaRocque, R.; Lagerborg, K.; Lemieux; Lin; Loreth, C.; MacInnis; Neumann, A.; Normandin, E.; P.C.; Park; Pierce, V.; Reilly, S.; Rosenberg; Rudy, M.; Ryan, E.; S.B.; Shaw, B.; Siddle; Slater, D.; Smole, S.; Tomkins-Tinch, C.; Turbett, S. |
| EPI_ISL_2958947 | Mayo Clinic & Mayo Clinic | Minnesota Department of Health, | Alexandra Lorentz; Jacob Garfin; Matt Plumb; and Xiong Wang |

| | | | |
|---|---|---|---|
| | Laboratories | Public Health Laboratory | |
| EPI_ISL_2956095 | Medizinische Laboratorien Düsseldorf | Robert Koch Institute | |
| EPI_ISL_2955023, EPI_ISL_2955043, EPI_ISL_2955045, EPI_ISL_2955066, EPI_ISL_2955067, EPI_ISL_2955074, EPI_ISL_2955112, EPI_ISL_2955116 | | | |
| see above | Medizinische Universitaet Innsbruck Institut fuer Virologie | Elling and Cochella laboratories, IMBA/IMP | Alexander Stark; Ezgi Oezkan; Luisa Cochella and Ulrich Elling; Marcus Strobl; Olga Frank; Ramesh Yelagandula; Tanino Albanese |
| EPI_ISL_3908105 | Microbiology Department & Molecular Biology CORE \| CDB \| Hospital Clinic \| Barcelona | Microbiology Department & Molecular Biology CORE \| CDB \| Hospital Clinic \| Barcelona | Andrea Vergara Gomez \| Jose Luis Villanueva Canas \| Miguel Julian Martinez Yoldi \| M. Mar Mosquera Gutierrez \| M. Angeles Marcos Maeso |
| EPI_ISL_2897003, EPI_ISL_2897006, EPI_ISL_2897030, EPI_ISL_2897048, EPI_ISL_2926114, EPI_ISL_2926152, EPI_ISL_2926161, EPI_ISL_2926172 | | | |
| see above | Microbiology Department, Laboratori Clínic Metropolitana Nord. Hospital Universitari Germans Trias i Pujol | Can Ruti SARS-CoV-2 Sequencing Hub (HUGTiP/IrsiCaixa/IGTP) | Alba Sánchez; Alexia París; Anna Not; Antoni E Bordoy; Bonaventura Clotet; Cristina Casañ; David Panisello; Francesc Catala-Moll; Gemma Clara; Ignacio Blanco; Laia Soler; Lauro Sumoy; Marc Noguera-Julian; Maria Casadellà; Mariona Parera; Mercedes Guerrero; Montserrat Giménez; Pere-Joan Cardona; Pilar Armengol; Roger Paredes; Verónica Saludes; and Elisa Martró on behalf of the Can Ruti SARS-CoV-2 Sequencing Hub |
| EPI_ISL_2861303 | Microbiology Department. Complexo Hospitalario Universitario de Vigo | Microbiology Department. Complexo Hospitalario Universitario de Vigo | Alfaya N; Alonso I; Alvarez M; Cabrera JJ; Carballo R; Cores O; Cortizo S; Davina C; Martinez L; Mediero G; Perez S; Potel C; Regueiro B; Rey S; Vasallo FJ; del-Campo V |
| EPI_ISL_3026895, EPI_ISL_3026896 | Microbiology Division, SC DHEC | Microbiology Division, SC DHEC | Freeman; J.S. |
| EPI_ISL_2987923, EPI_ISL_2988020, EPI_ISL_2988026, EPI_ISL_3906776, EPI_ISL_3906902, EPI_ISL_3907001, EPI_ISL_3907002, EPI_ISL_3907003 | | | |
| see above | Ministry of Health Turkey | Ministry of Health Turkey | Fatma Bayrakdar; Gulay Korukluoglu; Gülay Korukluoğlu; Suleyman Yalcin; Süleyman Yalcin; Yasemin Cosgun; Yasemin Coşgun |
| EPI_ISL_2958932 | Minnesota Department of Health, Public Health Laboratory | Minnesota Department of Health, Public Health Laboratory | Alexandra Lorentz; Jacob Garfin; Matt Plumb; and Xiong Wang |
| EPI_ISL_6661731 | Molecular Biology Laboratory, Westchester Medical Center | Genomics Core Laboratory, New York Medical College | Brahmaraju Mopidevi; Humayun K. Islam; Jian Zhuge; Salomon Amar; Utsav Pandey; Weihua Huang |
| EPI_ISL_2941763, EPI_ISL_2943174, EPI_ISL_2943199, EPI_ISL_2943207, EPI_ISL_2943208, EPI_ISL_2943219, EPI_ISL_2943220 | | | |
| see above | Montana Public Health Laboratory | Montana Public Health Laboratory | Carrie Biskupiak; Deborah Gibson; Joy Ritter; Michael Dills; Michelle Mozer |
| EPI_ISL_3148964 | Montefiore Nyack Hospital Affiliated Laboratory | Wadsworth Center, New York State Department of Health | Alexis Russell; Catharine Prussing; Daryl M. Lamson; Erasmus Schneider; Erica Lasek-Nesselquist; John Kelly; Jonathan Plitnick; Kirsten St. George; Matthew Shudt; Melissa A Leisner; Navjot Singh |
| EPI_ISL_3242077 | ND Dept. of Health Laboratory Services-Microbiology | Centers for Disease Control and Prevention Division of Viral Diseases, Pathogen Discovery | Alex Burgin; Ben L. Rambo-Martin; Clinton R. Paden; Dakota Howard; Dave Wentworth; Dhwani Batra; Jasmine Padilla; Justin Lee; Krista Queen; Kristen Knipe; Kristine Lacek; Mark Burroughs; Matthew Schmerer; Meghan Bentz; Mili Sheth; Peter Cook; Sam Shepard; Sarah Nobles; Suxiang Tong; Vivien Dugan; Yvette Unoarumhi |
| EPI_ISL_3800472 | NJ Public Health and Environmental Laboratories | Centers for Disease Control and Prevention Division of Viral Diseases, Pathogen Discovery | Alex Burgin; Ben Rambo-Martin; Clinton Paden; Dakota Howard; Dave Wentworth; Dhwani Batra; Jasmine Padilla; Justin Lee; Krista Queen; Kristen Knipe; Kristine Lacek; Mark Burroughs; Matthew Schmerer; Meghan Bentz; Mili Sheth; Peter Cook; Sam Shepard; Sarah Nobles; Suxiang Tong; Vivien Dugan; Yvette Unoarumhi |
| EPI_ISL_2940348, EPI_ISL_2940365, EPI_ISL_2940369, EPI_ISL_3023719 | NJDOH, Public Health and Environmental Laboratories | NJ_PHEL | Allison Roder; Byeong Jeong; Chelsea San Filippo; Dana Woell; Jacquelyn Deverell; Lindsey Bodnar; Ryan Pachucki; Shiv K. Verma |
| EPI_ISL_3031066, EPI_ISL_3031067 | NORTHWELL HEALTH LABORATORIES | Wadsworth Center, New York State Department of Health | Alexis Russell; Catharine Prussing; Daryl M. Lamson; Erasmus Schneider; Erica Lasek-Nesselquist; John Kelly; Jonathan Plitnick; Kirsten St. George; Matthew Shudt; Melissa A Leisner; Navjot Singh |
| EPI_ISL_5666739, EPI_ISL_5666875, EPI_ISL_5666979, EPI_ISL_5667035, EPI_ISL_5667164 | NUCLEO DE ESPECIALIDADES EM SAUDE | Instituto Butantan | Antonio Jorge Martins; Claudia Renata dos Santos Barros; David Schlesinger; Debora Botequio Moretti; Dimas Tadeu Covas; Elaine Cristina Marqueze; Elaine Vieira Santos; Evandra Strazza Rodrigues; Heidge Fukumasu; Jayme Augusto de Souza-Neto; José Salvatore Leister Patané; Luiz Alcantara; Luiz Lehmann Coutinho; Maria Carolina Elias; Maurício Lacerda Nogueira; Rafael dos Santos Bezerra; Raul Machado Neto; Rejane Maria Tommasini Grotto; Ricardo Haddad; Sandra Coccuzzo Sampaio Vessoni; Simone Kashima; Svetoslav Nanev Slavov; Vincent Louis Viala |
| EPI_ISL_5663646, EPI_ISL_5663762, EPI_ISL_5663844, EPI_ISL_5664310, EPI_ISL_5664388, EPI_ISL_5664405, EPI_ISL_5664493, EPI_ISL_5664497, EPI_ISL_5664744, EPI_ISL_5664749, EPI_ISL_5664898, EPI_ISL_5665100, EPI_ISL_5665179, EPI_ISL_5667135 | | | |
| see above | NUCLEO DE SAUDE MARY DOTA DE BAURU | Instituto Butantan | Antonio Jorge Martins; Claudia Renata dos Santos Barros; David Schlesinger; Debora Botequio Moretti; Dimas Tadeu Covas; Elaine Cristina Marqueze; Elaine Vieira Santos; Evandra Strazza Rodrigues; Heidge Fukumasu; Jayme Augusto de Souza-Neto; José Salvatore Leister Patané; Luiz Alcantara; Luiz Lehmann Coutinho; Maria Carolina Elias; Maurício Lacerda Nogueira; Rafael dos Santos Bezerra; Raul Machado Neto; Rejane Maria Tommasini Grotto; Ricardo Haddad; Sandra Coccuzzo Sampaio Vessoni; Simone Kashima; Svetoslav Nanev Slavov; Vincent Louis Viala |
| EPI_ISL_5666720, EPI_ISL_5666757, EPI_ISL_5667054, EPI_ISL_5667195, EPI_ISL_5667228 | NUCLEO DE SAUDE VILA FALCAO DE BAURU | Instituto Butantan | Antonio Jorge Martins; Claudia Renata dos Santos Barros; David Schlesinger; Debora Botequio Moretti; Dimas Tadeu Covas; Elaine Cristina Marqueze; Elaine Vieira Santos; Evandra Strazza Rodrigues; Heidge Fukumasu; Jayme Augusto de Souza-Neto; José Salvatore Leister Patané; Luiz Alcantara; Luiz Lehmann Coutinho; Maria Carolina Elias; Maurício Lacerda Nogueira; Rafael dos Santos Bezerra; Raul Machado Neto; Rejane Maria Tommasini Grotto; Ricardo Haddad; Sandra Coccuzzo Sampaio Vessoni; Simone Kashima; Svetoslav Nanev Slavov; Vincent Louis Viala |
| EPI_ISL_5663626, EPI_ISL_5663979, EPI_ISL_5664822, EPI_ISL_5665174, EPI_ISL_5665324, EPI_ISL_5666637, EPI_ISL_5666712, EPI_ISL_5666768, EPI_ISL_5666815, EPI_ISL_5666843, EPI_ISL_5666852, EPI_ISL_5666866, EPI_ISL_5666909, EPI_ISL_5666919, EPI_ISL_5666962, EPI_ISL_5666967, EPI_ISL_5667074, EPI_ISL_5667156, EPI_ISL_5667208, EPI_ISL_5667216, EPI_ISL_5667223, EPI_ISL_5667247 | | | |
| see above | NUCLEO SAUDE III N HABITACIONAL PRESIDENTE GEISEL BAURU | Instituto Butantan | Antonio Jorge Martins; Claudia Renata dos Santos Barros; David Schlesinger; Debora Botequio Moretti; Dimas Tadeu Covas; Elaine Cristina Marqueze; Elaine Vieira Santos; Evandra Strazza Rodrigues; Heidge Fukumasu; Jayme Augusto de Souza-Neto; José Salvatore Leister Patané; Luiz Alcantara; Luiz Lehmann Coutinho; Maria Carolina Elias; Maurício Lacerda Nogueira; Rafael dos Santos Bezerra; Raul Machado Neto; Rejane Maria Tommasini Grotto; Ricardo Haddad; Sandra Coccuzzo Sampaio Vessoni; Simone Kashima; Svetoslav Nanev Slavov; Vincent Louis Viala |
| EPI_ISL_3134777, EPI_ISL_3134778, EPI_ISL_3134779, EPI_ISL_3134780, EPI_ISL_3134781, EPI_ISL_3134782, EPI_ISL_3134783, EPI_ISL_3134784, EPI_ISL_3134785, EPI_ISL_3134786, EPI_ISL_3134787, EPI_ISL_3134789, EPI_ISL_3447413, EPI_ISL_3447420, EPI_ISL_3447423, EPI_ISL_3447426, EPI_ISL_3447436, EPI_ISL_3447437, EPI_ISL_3447448, EPI_ISL_3447517, EPI_ISL_3447518, EPI_ISL_3447519, EPI_ISL_3447520, EPI_ISL_3447521, EPI_ISL_3447522, EPI_ISL_3447523, EPI_ISL_3447524, EPI_ISL_3447525, EPI_ISL_3447534, EPI_ISL_3447535, EPI_ISL_3447536, EPI_ISL_3447537, EPI_ISL_3447538, EPI_ISL_3447539, EPI_ISL_3447540, EPI_ISL_3447541, EPI_ISL_3447542, EPI_ISL_3447543, EPI_ISL_3447544, EPI_ISL_3447545, EPI_ISL_3447546, EPI_ISL_3447547, EPI_ISL_3447548, EPI_ISL_3447550, EPI_ISL_3447552, EPI_ISL_3447553, EPI_ISL_3447554, EPI_ISL_3447555, EPI_ISL_3447560, EPI_ISL_3447562, EPI_ISL_3447565, EPI_ISL_3447566, EPI_ISL_3447567, EPI_ISL_3447594, EPI_ISL_3447595, EPI_ISL_3447597, EPI_ISL_3703782, EPI_ISL_3703783, EPI_ISL_3703784, EPI_ISL_3703785, EPI_ISL_3703786, EPI_ISL_3703787, EPI_ISL_3703788, EPI_ISL_3703789, EPI_ISL_3703790, EPI_ISL_3703791, EPI_ISL_3703792, EPI_ISL_3703793, EPI_ISL_3703794, EPI_ISL_3703795, EPI_ISL_3703796, EPI_ISL_3703797, EPI_ISL_3703798, EPI_ISL_3703799, EPI_ISL_3703801, EPI_ISL_3703813, EPI_ISL_3703816, EPI_ISL_3703820, EPI_ISL_3703823, EPI_ISL_3703827, EPI_ISL_3703831, EPI_ISL_3703834, EPI_ISL_3703846, EPI_ISL_3703849, EPI_ISL_3703853, EPI_ISL_3703856, EPI_ISL_3703868, EPI_ISL_3703876, EPI_ISL_3703886, EPI_ISL_3703896, EPI_ISL_3703928 | | | |
| see above | NUPIT/UFPE | WallauLab on behalf of Fiocruz COVID-19 Genomic Surveillance Network | Alexandre Freitas da Silva; Cassia Docena; Constância Flávia Junqueira Ayres; Filipe Zimmer Dezordi; Gabriel Luz Wallau; Gustavo Barbosa de Lima; Lais Ceschini Machado; Lilian Caroliny Amorim Silva; Maira Galdino da Rocha Pitta; Marcelo Henrique dos Santos Paiva; Matheus Filgueira Bezerra; Michelly Cristiny Pereira; Rômulo Pessoa e Silva; Sinval Pinto Brandão Filho |
| EPI_ISL_3060095 | Nacionalinis maisto ir veterinarijos rizikos vertinimo institutas | Vilnius University Hospital Santaros Klinikos, Center of Laboratory Medicine | Daniel Naumovas; Dovile Juozapaite; Gytis Dudas; Ingrida Olendraite; Ligita Raugaite; Monika Katenaite; Rimvydas Norvilas |
| EPI_ISL_3151163, EPI_ISL_3151164 | Nastavni zavod za javno zdravstvo Splitsko- Dalmatinske Županije | Hrvatski zavod za javno zdravstvo | Irena Tabain; Ivana Ferenčak |
| EPI_ISL_2895644 | National Platform bis UMONS/Jolimont | National Platform bis UMONS/Jolimont | Eric Tarantino; Florian Juszczak; François Dufrasne; Gautier Detry; Guillaume Bayon-Vicente; Ruddy Wattiez |
| EPI_ISL_2966506, EPI_ISL_2966862 | Nebraska Public Health Laboratory | NPHL COVID-19 Response Team | NPHL COVID-19 Response Team |
| EPI_ISL_2942192, EPI_ISL_2942194, EPI_ISL_3021058 | Nevada State Public Health Laboratory | Nevada State Public Health Laboratory | Andrew Gorzalski; Mark Pandori |
| EPI_ISL_3045099, EPI_ISL_3045101 | New Mexico Department of Health Scientific Laboratory | New Mexico Department of Health Scientific Laboratory | Anastacia Griego-Fisher; D'eldra Malone; Ellie Johnson; Jennifer Benoit; Linda Salazar; Ratheesh Rajan |
| EPI_ISL_2956370 | Niedersächsisches Landesgesundheitsamt (NLGA) | Robert Koch Institute | |
| EPI_ISL_3104861, EPI_ISL_3104909, EPI_ISL_3104929, EPI_ISL_3104938, EPI_ISL_3105057 | Northwest Laboratory | Seattle Flu Study | Amanda Adler; Barry R. Lutz; Benjamin Pelle; Brian Hiatt; Caitlin R. Wolf; Chris D. Frazar; Deborah A. Nickerson; Elisabeth Brandstetter; Erica Ryke; Geoff Melly; Helen Y. Chu; Janet A. Englund; Jay Shendure; Jover Lee; Kairsten Fay; Kirsten Lacombe; Lea M. Starita; Mark J. Rieder; Matthew Richardson; Matthew Thompson; Melissa Truong; Michael Boeckh; Michael Famulare; Misja Ilcisin; Peter D. Han; Philip Dykema; Romesh Gautom; Scott Lindquist; Thomas R. Sibley; Trevor Bedford |
| EPI_ISL_3319698, EPI_ISL_3319703, EPI_ISL_3319706, EPI_ISL_3319707, EPI_ISL_3319708, EPI_ISL_3319712, EPI_ISL_3319713, EPI_ISL_3319716, EPI_ISL_3319717, EPI_ISL_3319722, EPI_ISL_3319728, EPI_ISL_3319729, EPI_ISL_3319731, EPI_ISL_3319733, EPI_ISL_3319734, EPI_ISL_3319735, EPI_ISL_3319743, EPI_ISL_3319747, EPI_ISL_3319752, EPI_ISL_3319754, EPI_ISL_3319757, EPI_ISL_3319760, EPI_ISL_3346519, EPI_ISL_3547394 | | | |
| see above | OHSU-MM Lab | OHSU-MM Lab | Jeannine Lama |
| EPI_ISL_2840716, | OSPEDALE CIVILE TERAMO - | Istituto Zooprofilattico | Ancora M; Calistri P; Cammà C; Caporale M; Curini V; Delli Compagni E; Di Domenico M; Di Lollo Valeria; Di Pasquale A; Lorusso A; Mangone I; Marcacci M; Puglia I; Rinaldi A; Savini G; Scialabba S |

| | | | |
|---|---|---|---|
| EPI_ISL_2921231 | CENTRO TRASFUSIONALE | Sperimentale dell'Abruzzo e Molise "G. Caporale" | |
| EPI_ISL_2966615 | Ochsner Health | Bioinfoexperts, LLC | Amy Feehan; David J. Nolan; Judy Crabtree; Julia-Garcia-Diaz; Lucio Miele; Rebecca Rose; Susanna L. Lamers; Tessa LaFleur |
| EPI_ISL_3010309, EPI_ISL_3010316 | Orange County Public Health Laboratory | Orange County Public Health Laboratory | Angelica Torres; Julia Wolfe |
| EPI_ISL_3189593 | Orebro University Hospital, Dept Laboratory Medicine, Clinical Microbiology | Orebro University Hospital | Sundqvist M et al |
| EPI_ISL_2893422, EPI_ISL_2893423, EPI_ISL_2893424, EPI_ISL_2893425, EPI_ISL_2893426, EPI_ISL_2893427, EPI_ISL_2893429, EPI_ISL_2893430, EPI_ISL_2893431, EPI_ISL_2893432, EPI_ISL_2893433, EPI_ISL_2893438, EPI_ISL_2933949, EPI_ISL_2955407, EPI_ISL_3010594, EPI_ISL_3067393 | | | |
| see above | Oregon State Public Health Laboratory | Oregon State Public Health Laboratory | Eugene Yeboah; John Fontana and Shane Sevey; Laura Tsaknaridis; Rafia Razzaque; Vanda Makris |
| EPI_ISL_2925727 | Osp. G.B.Grassi | National Institute for Infectious Diseases (INMI) L. Spallanzani I.R.C.C.S | A Di Caro; B Bartolini; CEM Gruber; E Giombini; F Messina; F Santini; G Bonfiglio; M Rueca; MR Capobianchi; O Butera |
| EPI_ISL_2925695 | Osp. S.Giovanni Evangelista | National Institute for Infectious Diseases (INMI) L. Spallanzani I.R.C.C.S | A Di Caro; B Bartolini; CEM Gruber; E Giombini; F Messina; F Santini; G Bonfiglio; M Rueca; MR Capobianchi; O Butera |
| EPI_ISL_2925724, EPI_ISL_2925725 | Osp. S.Giovanni Evengelista | National Institute for Infectious Diseases (INMI) L. Spallanzani I.R.C.C.S | A Di Caro; B Bartolini; CEM Gruber; E Giombini; F Messina; F Santini; G Bonfiglio; M Rueca; MR Capobianchi; O Butera |
| EPI_ISL_2888021, EPI_ISL_2888022, EPI_ISL_2888025, EPI_ISL_2888026, EPI_ISL_2888029, EPI_ISL_2888030, EPI_ISL_2888031, EPI_ISL_2888032, EPI_ISL_2888035, EPI_ISL_2888041 | | | |
| see above | Ospedale San Filippo Neri | Department of General Diagnostics; Department of Virology; Istituto Zooprofilattico Sperimentale del Lazio e della Toscana (IZSLT) | Alessia Franco; Antonella Cersini; Antonio Battisti.; Elena L. Diaconu; Fabiola Feltrin; Giuseppe Manna; Patricia Alba; Raffaela Conti; Teresa Scicluna; Virginia Carfora |
| EPI_ISL_2888012, EPI_ISL_2888013, EPI_ISL_2888014, EPI_ISL_2888015 | Ospedale Sandro Pertini ASL Roma2 | Department of General Diagnostics; Department of Virology; Istituto Zooprofilattico Sperimentale del Lazio e della Toscana (IZSLT) | Alessia Franco; Antonella Cersini; Antonio Battisti.; Elena L. Diaconu; Fabiola Feltrin; Giuseppe Manna; Patricia Alba; Raffaela Conti; Teresa Scicluna; Virginia Carfora |
| EPI_ISL_2925501 | Ospedale di Genzano | National Institute for Infectious Diseases (INMI) L. Spallanzani I.R.C.C.S | A Di Caro; B Bartolini; CEM Gruber; E Giombini; F Messina; F Santini; G Bonfiglio; M Rueca; MR Capobianchi; O Butera |
| EPI_ISL_3219518, EPI_ISL_3219519, EPI_ISL_3219520 | Outre mer | National Reference Center for Viruses of Respiratory Infections, Institut Pasteur, Paris | Angela Brisebarre; Camille Capel; Christophe Malabat; Corinne Maufrais; Dominique Rousset; Etienne Simon-Lorière; Frédéric Lemoine; Hub de Bioinformatique et Biostatistique; Louise Lefrançois; Marion Barbet; Maud Vanpeene; Méline Bizard; Sylvie Behillil; Sylvie Van der Werf; Vincent Enouf |
| EPI_ISL_3104900, EPI_ISL_3105053 | Overlake Hospital | Seattle Flu Study | Amanda Adler; Barry R. Lutz; Benjamin Pelle; Brian Hiatt; Caitlin R. Wolf; Chris D. Frazar; Deborah A. Nickerson; Elisabeth Brandstetter; Erica Ryke; Geoff Melly; Helen Y. Chu; Janet A. Englund; Jay Shendure; Jover Lee; Kairsten Fay; Kirsten Lacombe; Lea M. Starita; Mark J. Rieder; Matthew Richardson; Matthew Thompson; Melissa Truong; Michael Boechh; Michael Famulare; Misja Ilcisin; Peter D. Han; Philip Dykema; Romesh Gautom; Scott Lindquist; Thomas R. Sibley; Trevor Bedford |
| EPI_ISL_5667809 | PACS JOAO FRANCISCO MARQUES SANTA LUZIA JOSE BONIFACIO | Instituto Butantan | Antonio Jorge Martins; Claudia Renata dos Santos Barros; David Schlesinger; Debora Botequio Moretti; Dimas Tadeu Covas; Elaine Cristina Marqueze; Elaine Vieira Santos; Evandra Strazza Rodrigues; Heidge Fukumasu; Jayme Augusto de Souza-Neto; José Salvatore Leister Patané; Luiz Alcantara; Luiz Lehmann Coutinho; Maria Carolina Elias; Maurício Lacerda Nogueira; Rafael dos Santos Bezerra; Raul Machado Neto; Rejane Maria Tommasini Grotto; Ricardo Haddad; Sandra Coccuzzo Sampaio Vessoni; Simone Kashima; Svetoslav Nanev Slavov; Vincent Louis Viala |
| EPI_ISL_2967972 | PAMF Palo Alto | Santa Clara County Public Health Laboratory | Santa Clara County Public Health Department |
| EPI_ISL_4743409 | PECET UDEA | Laboratorio Departamental de Salud Publica de Antioquia | Andres F. Cardona-Rios; Cristian Arbey Velarde Hoyos; Gloria Isabel Escobar; Idabely Betancur Ortiz; Juan P. Hernandez-Ortiz; Maria Stella López |
| EPI_ISL_5663322 | POLICLINICA BONSUCESSO | Instituto Butantan | Antonio Jorge Martins; Claudia Renata dos Santos Barros; David Schlesinger; Debora Botequio Moretti; Dimas Tadeu Covas; Elaine Cristina Marqueze; Elaine Vieira Santos; Evandra Strazza Rodrigues; Heidge Fukumasu; Jayme Augusto de Souza-Neto; José Salvatore Leister Patané; Luiz Alcantara; Luiz Lehmann Coutinho; Maria Carolina Elias; Maurício Lacerda Nogueira; Rafael dos Santos Bezerra; Raul Machado Neto; Rejane Maria Tommasini Grotto; Ricardo Haddad; Sandra Coccuzzo Sampaio Vessoni; Simone Kashima; Svetoslav Nanev Slavov; Vincent Louis Viala |
| EPI_ISL_5671691 | POLICLINICA COVID 19 ITAPETININGA | Instituto Butantan | Antonio Jorge Martins; Claudia Renata dos Santos Barros; David Schlesinger; Debora Botequio Moretti; Dimas Tadeu Covas; Elaine Cristina Marqueze; Elaine Vieira Santos; Evandra Strazza Rodrigues; Heidge Fukumasu; Jayme Augusto de Souza-Neto; José Salvatore Leister Patané; Luiz Alcantara; Luiz Lehmann Coutinho; Maria Carolina Elias; Maurício Lacerda Nogueira; Rafael dos Santos Bezerra; Raul Machado Neto; Rejane Maria Tommasini Grotto; Ricardo Haddad; Sandra Coccuzzo Sampaio Vessoni; Simone Kashima; Svetoslav Nanev Slavov; Vincent Louis Viala |
| EPI_ISL_5664522 | POLICLINICA DONA LUIZA | Instituto Butantan | Antonio Jorge Martins; Claudia Renata dos Santos Barros; David Schlesinger; Debora Botequio Moretti; Dimas Tadeu Covas; Elaine Cristina Marqueze; Elaine Vieira Santos; Evandra Strazza Rodrigues; Heidge Fukumasu; Jayme Augusto de Souza-Neto; José Salvatore Leister Patané; Luiz Alcantara; Luiz Lehmann Coutinho; Maria Carolina Elias; Maurício Lacerda Nogueira; Rafael dos Santos Bezerra; Raul Machado Neto; Rejane Maria Tommasini Grotto; Ricardo Haddad; Sandra Coccuzzo Sampaio Vessoni; Simone Kashima; Svetoslav Nanev Slavov; Vincent Louis Viala |
| EPI_ISL_5659654, EPI_ISL_5659666, EPI_ISL_5659842, EPI_ISL_5659929, EPI_ISL_5659963, EPI_ISL_5659999, EPI_ISL_5660265, EPI_ISL_5660306, EPI_ISL_5660419, EPI_ISL_5660492, EPI_ISL_5660961, EPI_ISL_5668631, EPI_ISL_5668661, EPI_ISL_5668709, EPI_ISL_5668763, EPI_ISL_5668800, EPI_ISL_5668936, EPI_ISL_5669077, EPI_ISL_5669319, EPI_ISL_5669401, EPI_ISL_5669420, EPI_ISL_5669531, EPI_ISL_5669612, EPI_ISL_5669705, EPI_ISL_5669758, EPI_ISL_5669782, EPI_ISL_5669790, EPI_ISL_5669896, EPI_ISL_5669979, EPI_ISL_5670014, EPI_ISL_5670030, EPI_ISL_5670051, EPI_ISL_5670119, EPI_ISL_5670202, EPI_ISL_5670261, EPI_ISL_5670358, EPI_ISL_5670376, EPI_ISL_5670401, EPI_ISL_5670525, EPI_ISL_5670542, EPI_ISL_5670702, EPI_ISL_5670832 | | | |
| see above | POLICLINICA HORTOLANDIA | Instituto Butantan | Antonio Jorge Martins; Claudia Renata dos Santos Barros; David Schlesinger; Debora Botequio Moretti; Dimas Tadeu Covas; Elaine Cristina Marqueze; Elaine Vieira Santos; Evandra Strazza Rodrigues; Heidge Fukumasu; Jayme Augusto de Souza-Neto; José Salvatore Leister Patané; Luiz Alcantara; Luiz Lehmann Coutinho; Maria Carolina Elias; Maurício Lacerda Nogueira; Rafael dos Santos Bezerra; Raul Machado Neto; Rejane Maria Tommasini Grotto; Ricardo Haddad; Sandra Coccuzzo Sampaio Vessoni; Simone Kashima; Svetoslav Nanev Slavov; Vincent Louis Viala |
| EPI_ISL_5662495 | POLICLINICA MARIA DIRCE | Instituto Butantan | Antonio Jorge Martins; Claudia Renata dos Santos Barros; David Schlesinger; Debora Botequio Moretti; Dimas Tadeu Covas; Elaine Cristina Marqueze; Elaine Vieira Santos; Evandra Strazza Rodrigues; Heidge Fukumasu; Jayme Augusto de Souza-Neto; José Salvatore Leister Patané; Luiz Alcantara; Luiz Lehmann Coutinho; Maria Carolina Elias; Maurício Lacerda Nogueira; Rafael dos Santos Bezerra; Raul Machado Neto; Rejane Maria Tommasini Grotto; Ricardo Haddad; Sandra Coccuzzo Sampaio Vessoni; Simone Kashima; Svetoslav Nanev Slavov; Vincent Louis Viala |
| EPI_ISL_5663608, EPI_ISL_5664120 | POLICLINICA PARAVENTI | Instituto Butantan | Antonio Jorge Martins; Claudia Renata dos Santos Barros; David Schlesinger; Debora Botequio Moretti; Dimas Tadeu Covas; Elaine Cristina Marqueze; Elaine Vieira Santos; Evandra Strazza Rodrigues; Heidge Fukumasu; Jayme Augusto de Souza-Neto; José Salvatore Leister Patané; Luiz Alcantara; Luiz Lehmann Coutinho; Maria Carolina Elias; Maurício Lacerda Nogueira; Rafael dos Santos Bezerra; Raul Machado Neto; Rejane Maria Tommasini Grotto; Ricardo Haddad; Sandra Coccuzzo Sampaio Vessoni; Simone Kashima; Svetoslav Nanev Slavov; Vincent Louis Viala |
| EPI_ISL_5660501, EPI_ISL_5661519, EPI_ISL_5662165, EPI_ISL_5664341, EPI_ISL_5665412 | POSTO DE SAUDE BENVIRA E S F 4 | Instituto Butantan | Antonio Jorge Martins; Claudia Renata dos Santos Barros; David Schlesinger; Debora Botequio Moretti; Dimas Tadeu Covas; Elaine Cristina Marqueze; Elaine Vieira Santos; Evandra Strazza Rodrigues; Heidge Fukumasu; Jayme Augusto de Souza-Neto; José Salvatore Leister Patané; Luiz Alcantara; Luiz Lehmann Coutinho; Maria Carolina Elias; Maurício Lacerda Nogueira; Rafael dos Santos Bezerra; Raul Machado Neto; Rejane Maria Tommasini Grotto; Ricardo Haddad; Sandra Coccuzzo Sampaio Vessoni; Simone Kashima; Svetoslav Nanev Slavov; Vincent Louis Viala |
| EPI_ISL_5659810 | POSTO DE SAUDE DO PORTAO ATIBAIA | Instituto Butantan | Antonio Jorge Martins; Claudia Renata dos Santos Barros; David Schlesinger; Debora Botequio Moretti; Dimas Tadeu Covas; Elaine Cristina Marqueze; Elaine Vieira Santos; Evandra Strazza Rodrigues; Heidge Fukumasu; Jayme Augusto de Souza-Neto; José Salvatore Leister Patané; Luiz Alcantara; Luiz Lehmann Coutinho; Maria Carolina Elias; Maurício Lacerda Nogueira; Rafael dos Santos Bezerra; Raul Machado Neto; Rejane Maria Tommasini Grotto; Ricardo Haddad; Sandra Coccuzzo Sampaio Vessoni; Simone Kashima; Svetoslav Nanev Slavov; Vincent Louis Viala |
| EPI_ISL_5663826 | POSTO DE SAUDE DRCRISTOVAN RUTHILLER T MACIEL | Instituto Butantan | Antonio Jorge Martins; Claudia Renata dos Santos Barros; David Schlesinger; Debora Botequio Moretti; Dimas Tadeu Covas; Elaine Cristina Marqueze; Elaine Vieira Santos; Evandra Strazza Rodrigues; Heidge Fukumasu; Jayme Augusto de Souza-Neto; José Salvatore Leister Patané; Luiz Alcantara; Luiz Lehmann Coutinho; Maria Carolina Elias; Maurício Lacerda Nogueira; Rafael dos Santos Bezerra; Raul Machado Neto; Rejane Maria Tommasini Grotto; Ricardo Haddad; Sandra Coccuzzo Sampaio Vessoni; Simone Kashima; Svetoslav Nanev Slavov; Vincent Louis Viala |
| EPI_ISL_5661502, EPI_ISL_5670155 | POSTO MEDICO EUROPA | Instituto Butantan | Antonio Jorge Martins; Claudia Renata dos Santos Barros; David Schlesinger; Debora Botequio Moretti; Dimas Tadeu Covas; Elaine Cristina Marqueze; Elaine Vieira Santos; Evandra Strazza Rodrigues; Heidge Fukumasu; Jayme Augusto de Souza-Neto; José Salvatore Leister Patané; Luiz Alcantara; Luiz Lehmann Coutinho; Maria Carolina Elias; Maurício Lacerda Nogueira; Rafael dos Santos Bezerra; Raul Machado Neto; Rejane Maria Tommasini Grotto; Ricardo Haddad; Sandra Coccuzzo Sampaio Vessoni; Simone Kashima; Svetoslav Nanev Slavov; Vincent Louis Viala |
| EPI_ISL_5664856 | PROGRAMA SAUDE DA FAMILIA I IEPE | Instituto Butantan | Antonio Jorge Martins; Claudia Renata dos Santos Barros; David Schlesinger; Debora Botequio Moretti; Dimas Tadeu Covas; Elaine Cristina Marqueze; Elaine Vieira Santos; Evandra Strazza Rodrigues; Heidge Fukumasu; Jayme Augusto de Souza-Neto; José Salvatore Leister Patané; Luiz Alcantara; Luiz Lehmann Coutinho; Maria Carolina Elias; Maurício Lacerda Nogueira; Rafael dos Santos Bezerra; Raul Machado Neto; Rejane Maria Tommasini Grotto; Ricardo Haddad; Sandra Coccuzzo Sampaio Vessoni; Simone Kashima; Svetoslav Nanev Slavov; Vincent Louis Viala |
| EPI_ISL_5661277, EPI_ISL_5661545, EPI_ISL_5662865 | PRONTO ATENDIMENTO MUNICIPAL PAULO BARBOSA RANGEL TREMEMBE | Instituto Butantan | Antonio Jorge Martins; Claudia Renata dos Santos Barros; David Schlesinger; Debora Botequio Moretti; Dimas Tadeu Covas; Elaine Cristina Marqueze; Elaine Vieira Santos; Evandra Strazza Rodrigues; Heidge Fukumasu; Jayme Augusto de Souza-Neto; José Salvatore Leister Patané; Luiz Alcantara; Luiz Lehmann Coutinho; Maria Carolina Elias; Maurício Lacerda Nogueira; Rafael dos Santos Bezerra; Raul Machado Neto; Rejane Maria Tommasini Grotto; Ricardo Haddad; Sandra Coccuzzo Sampaio Vessoni; Simone Kashima; Svetoslav Nanev Slavov; Vincent Louis Viala |
| EPI_ISL_5664246, EPI_ISL_5664452 | PRONTO ATENDIMENTO PARQUE ALVORADA | Instituto Butantan | Antonio Jorge Martins; Claudia Renata dos Santos Barros; David Schlesinger; Debora Botequio Moretti; Dimas Tadeu Covas; Elaine Cristina Marqueze; Elaine Vieira Santos; Evandra Strazza Rodrigues; Heidge Fukumasu; Jayme Augusto de Souza-Neto; José Salvatore Leister Patané; Luiz Alcantara; Luiz Lehmann Coutinho; Maria Carolina Elias; Maurício Lacerda Nogueira; Rafael dos Santos Bezerra; Raul Machado Neto; Rejane Maria Tommasini Grotto; Ricardo Haddad; Sandra Coccuzzo Sampaio Vessoni; Simone Kashima; Svetoslav Nanev Slavov; Vincent Louis Viala |
| EPI_ISL_5667096 | PRONTO SOCORRO CENTRAL DE BAURU | Instituto Butantan | Antonio Jorge Martins; Claudia Renata dos Santos Barros; David Schlesinger; Debora Botequio Moretti; Dimas Tadeu Covas; Elaine Cristina Marqueze; Elaine Vieira Santos; Evandra Strazza Rodrigues; Heidge Fukumasu; Jayme Augusto de Souza-Neto; José Salvatore Leister Patané; Luiz Alcantara; Luiz Lehmann Coutinho; Maria Carolina Elias; Maurício Lacerda Nogueira; Rafael dos Santos Bezerra; Raul Machado Neto; Rejane Maria Tommasini Grotto; Ricardo Haddad; Sandra Coccuzzo Sampaio Vessoni; Simone Kashima; Svetoslav Nanev Slavov; Vincent Louis Viala |
| EPI_ISL_5661294 | PRONTO SOCORRO DR ANTONIO FLAVIO FRANCA | Instituto Butantan | Antonio Jorge Martins; Claudia Renata dos Santos Barros; David Schlesinger; Debora Botequio Moretti; Dimas Tadeu Covas; Elaine Cristina Marqueze; Elaine Vieira Santos; Evandra Strazza Rodrigues; Heidge Fukumasu; Jayme Augusto de Souza-Neto; José Salvatore Leister Patané; Luiz Alcantara; Luiz Lehmann Coutinho; Maria Carolina Elias; Maurício Lacerda Nogueira; Rafael dos Santos Bezerra; Raul Machado Neto; Rejane Maria Tommasini Grotto; Ricardo Haddad; Sandra Coccuzzo Sampaio Vessoni; Simone Kashima; Svetoslav Nanev Slavov; Vincent Louis Viala |
| EPI_ISL_5663514, EPI_ISL_5663709, EPI_ISL_5664424, EPI_ISL_5665422 | PRONTO SOCORRO PARQUE SAO GEORGE | Instituto Butantan | Antonio Jorge Martins; Claudia Renata dos Santos Barros; David Schlesinger; Debora Botequio Moretti; Dimas Tadeu Covas; Elaine Cristina Marqueze; Elaine Vieira Santos; Evandra Strazza Rodrigues; Heidge Fukumasu; Jayme Augusto de Souza-Neto; José Salvatore Leister Patané; Luiz Alcantara; Luiz Lehmann Coutinho; Maria Carolina Elias; Maurício Lacerda Nogueira; Rafael dos Santos Bezerra; Raul Machado Neto; Rejane Maria Tommasini Grotto; Ricardo Haddad; Sandra Coccuzzo Sampaio Vessoni; Simone Kashima; Svetoslav Nanev Slavov; Vincent Louis Viala |
| EPI_ISL_5665956, EPI_ISL_5665977, EPI_ISL_5665996, EPI_ISL_5666008, EPI_ISL_5666698, EPI_ISL_5666781, EPI_ISL_5667112 | | | |
| see above | PSF PARQUE EMILIA | Instituto Butantan | Antonio Jorge Martins; Claudia Renata dos Santos Barros; David Schlesinger; Debora Botequio Moretti; Dimas Tadeu Covas; Elaine Cristina Marqueze; Elaine Vieira Santos; Evandra Strazza Rodrigues; Heidge Fukumasu; Jayme Augusto de Souza-Neto; José Salvatore Leister Patané; Luiz Alcantara; Luiz Lehmann Coutinho; Maria Carolina Elias; Maurício Lacerda Nogueira; Rafael dos Santos Bezerra; Raul Machado Neto; Rejane Maria Tommasini Grotto; Ricardo Haddad; Sandra Coccuzzo Sampaio Vessoni; Simone Kashima; Svetoslav Nanev Slavov; Vincent Louis Viala |
| EPI_ISL_5665971, EPI_ISL_5665990, EPI_ISL_5666020, EPI_ISL_5666660 | PSF VIRGINIO BASSO | Instituto Butantan | Antonio Jorge Martins; Claudia Renata dos Santos Barros; David Schlesinger; Debora Botequio Moretti; Dimas Tadeu Covas; Elaine Cristina Marqueze; Elaine Vieira Santos; Evandra Strazza Rodrigues; Heidge Fukumasu; Jayme Augusto de Souza-Neto; José Salvatore Leister Patané; Luiz Alcantara; Luiz Lehmann Coutinho; Maria Carolina Elias; Maurício Lacerda Nogueira; Rafael dos Santos Bezerra; Raul Machado Neto; Rejane Maria Tommasini Grotto; Ricardo Haddad; Sandra Coccuzzo Sampaio Vessoni; Simone Kashima; Svetoslav Nanev Slavov; Vincent Louis Viala |

| | | | |
|---|---|---|---|
| EPI_ISL_2923912, EPI_ISL_2923915, EPI_ISL_2923943 | Pandemic Response Lab - NYC | Pandemic Response Lab, R&D | Cybill del Castillo; Dylan Law; Haiping Hao; Henry Lee; Isabel Fernandez Escapa; Jon Laurent; Melissa Hopkins; Michael Hammerling; Pradeep Bugga; Shinyoung Clair Kang; Sol Rey; William Ward |
| EPI_ISL_3398772 | Pathology, UAB | Pathology, UAB | E.J.; Hendrickson; Leal; Lefkowitz; Moates, D.; R.C.; S.M. |
| EPI_ISL_6907496, EPI_ISL_6907502, EPI_ISL_6907503, EPI_ISL_6907512, EPI_ISL_6907517, EPI_ISL_6907518, EPI_ISL_6907524, EPI_ISL_6907529, EPI_ISL_6907530, EPI_ISL_6907537, EPI_ISL_6908024, EPI_ISL_6908032, EPI_ISL_6908038 | | | |
| see above | Plataforma de Vigilancia Molecular (PVM) - FIOCRUZ/BA | Plataforma de Vigilancia Molecular (PVM) - FIOCRUZ/BA | Bruno Bezerril Andrade; Camila I. de Oliveira; Clarissa Araújo Gurgel; Eduardo Oyama; Francys A Rangel; Laila A da Silva; Laise de Moraes; Leonardo Paiva Farias; Marina Cucco; Pedro Silveira; Ricardo Khouri on behalf of the Fiocruz COVID-19 Genomic Surveillance Network.; Tiago Graf |
| EPI_ISL_2878401, EPI_ISL_2878468, EPI_ISL_2878493, EPI_ISL_2878533 | Plateau technique Biomer Metz Verriers | Department of Virology, Henri Mondor University Hospital, Assistance Publique Hôpitaux de Paris, Université Paris-Est Créteil, INSERM U955 | Alexandre Soulier; Christophe Rodriguez; Elisabeth Trawinski; Guillaume Gricourt; Jean-Michel Pawlotsky; Melissa N'Debi; Slim Fourati; Vanessa Demontant |
| EPI_ISL_3061989 | Plateforme de testing Namuroise | Plateforme de testing Namuroise | Degosserie Jonathan; Demars Aurore; Denis Olivier; Lesly Nyinkeu Kemamen; Maschietto Céline; Mullier François; Nobis Chloé; Otto Gaetan |
| EPI_ISL_2925488 | Policlinico Tor Vergata | National Institute for Infectious Diseases (INMI) L. Spallanzani I.R.C.C.S | A Di Caro; B Bartolini; CEM Gruber; E Giombini; F Messina; F Santini; G Bonfiglio; M Rueca; MR Capobianchi; O Butera |
| EPI_ISL_2810027, EPI_ISL_2820510 | Poliklinika Vitalis | Alea Genetic Centre | Dino Pecar; Ivana Ceko; Lana Salihefendic; Rijad Konjhodzic |
| EPI_ISL_2876895, EPI_ISL_2876902, EPI_ISL_3085828, EPI_ISL_3085829, EPI_ISL_3085830, EPI_ISL_3085831, EPI_ISL_3085832, EPI_ISL_3085833, EPI_ISL_3085834, EPI_ISL_3085835, EPI_ISL_3085836, EPI_ISL_3085837, EPI_ISL_3085838, EPI_ISL_3085839, EPI_ISL_3085840, EPI_ISL_3085841, EPI_ISL_3085842, EPI_ISL_3085843, EPI_ISL_3085844, EPI_ISL_3085845, EPI_ISL_3085847, EPI_ISL_3085848, EPI_ISL_3085849, EPI_ISL_3085850, EPI_ISL_3085851, EPI_ISL_3085852, EPI_ISL_3085853, EPI_ISL_3085854, EPI_ISL_3085855, EPI_ISL_3085856, EPI_ISL_3085857, EPI_ISL_3085858, EPI_ISL_3085859, EPI_ISL_3085860, EPI_ISL_3085861, EPI_ISL_3085862, EPI_ISL_3085863, EPI_ISL_3085864, EPI_ISL_3085865, EPI_ISL_3085866, EPI_ISL_3303842, EPI_ISL_3303849, EPI_ISL_3303851, EPI_ISL_3303853, EPI_ISL_3303854, EPI_ISL_3303855, EPI_ISL_3303857, EPI_ISL_3303858, EPI_ISL_3303859, EPI_ISL_3303861, EPI_ISL_3303863, EPI_ISL_3303864, EPI_ISL_3303866, EPI_ISL_3303867, EPI_ISL_3303868, EPI_ISL_3303870, EPI_ISL_3303873, EPI_ISL_3303875, EPI_ISL_3303879, EPI_ISL_3303880, EPI_ISL_3303883, EPI_ISL_3303885, EPI_ISL_3303887, EPI_ISL_3303901, EPI_ISL_3304999, EPI_ISL_3305012, EPI_ISL_3305023, EPI_ISL_3305026, EPI_ISL_3305030, EPI_ISL_3305034, EPI_ISL_3305040, EPI_ISL_3305062, EPI_ISL_3305070, EPI_ISL_3305074, EPI_ISL_3305081, EPI_ISL_3305100, EPI_ISL_3305106, EPI_ISL_3305108, EPI_ISL_3305111, EPI_ISL_3305115, EPI_ISL_3305119, EPI_ISL_3305121, EPI_ISL_3305124, EPI_ISL_3305131, EPI_ISL_3305133, EPI_ISL_3305140, EPI_ISL_3305141, EPI_ISL_3305148, EPI_ISL_3305151, EPI_ISL_3305154, EPI_ISL_3305155, EPI_ISL_3305156, EPI_ISL_3305157, EPI_ISL_3305159, EPI_ISL_3305165, EPI_ISL_3305166, EPI_ISL_3305170, EPI_ISL_3305171, EPI_ISL_3305183, EPI_ISL_3305195, EPI_ISL_4368326, EPI_ISL_4372039, EPI_ISL_4373090 | | | |
| see above | Quest Diagnostics Incorporated | Centers for Disease Control and Prevention Division of Viral Diseases, Pathogen Discovery | A. Gerasimova; A. Perez; Adrian Paskey; B. Anderson; Benjamin Rambo-Martin; Christopher Gulvick; Clinton Paden; Clinton R. Paden; Dakota Howard; Darlene Wagner; Dhwani Batra; Duncan MacCannell; Erisa Sula; F. Lacbawan; I. A. Shlyakhter; I. Shlyakhter; Jason Caravas; K. Livingston; K.E. Livingston; Kara Moser; Kristine Lacek; L. Bernstein; L.E. Bernstein; M. Hua; Matthew Schmerer; P. Tanpaiboon; Peter Cook; Peter W. Cook; R. Kagan; R. M. Kagan; R. Owen; R. Rolando; R. V. Rolando; S. H. Rosenthal; S. Rosenthal; Scott Sammons; Shatavia Morrison; Tymeckia Kendall; Victoria Caban Figueroa; Y. Liu; Yvette Unoarumhi |
| EPI_ISL_2812794 | RM1 | National Institute for Infectious Diseases (INMI) L. Spallanzani I.R.C.C.S | A Di Caro; B Bartolini; CEM Gruber; E Giombini; F Messina; F Santini; G Bonfiglio; M Rueca; MR Capobianchi; O Butera |
| EPI_ISL_2834961 | Regions Hospital | Minnesota Department of Health, Public Health Laboratory | Alexandra Lorentz; Jacob Garfin; Matt Plumb; and Xiong Wang |
| EPI_ISL_2904392, EPI_ISL_2904440 | Respiratory Virus Unit, Microbiology Services Colindale, Public Health England | COVID-19 Genomics UK (COG-UK) Consortium | PHE Covid Sequencing Team |
| EPI_ISL_3982730 | Ribeirao Preto | Ribeirao Preto | Antonio Jorge Martins; Claudia Renata dos Santos Barros; David Schlesinger; Debora Botequio Moretti; Dimas Tadeu Covas; Elaine Cristina Marqueze; Elaine Vieira Santos; Evandra Strazza Rodrigues; Heidge Fukumasu; Jayme Augusto de Souza-Neto; José Salvatore Leister Patané; Luiz Alcantara; Luiz Lehmann Coutinho; Maria Carolina Elias; Maurício Lacerda Nogueira; Rafael dos Santos Bezerra; Raul Machado Neto; Rejane Maria Tommasini Grotto; Ricardo Haddad; Sandra Coccuzzo Sampaio Vessoni; Simone Kashima; Svetoslav Nanev Slavov; Vincent Louis Viala |
| EPI_ISL_3982693, EPI_ISL_3982697, EPI_ISL_3982701, EPI_ISL_3982705, EPI_ISL_3982711 | Ribeirao Preto SP | Laboratorio de Biologia Molecular Hemocentro de Ribeirao Preto | Antonio Jorge Martins; Claudia Renata dos Santos Barros; David Schlesinger; Debora Botequio Moretti; Dimas Tadeu Covas; Elaine Cristina Marqueze; Elaine Vieira Santos; Evandra Strazza Rodrigues; Heidge Fukumasu; Jayme Augusto de Souza-Neto; José Salvatore Leister Patané; Luiz Alcantara; Luiz Lehmann Coutinho; Maria Carolina Elias; Maurício Lacerda Nogueira; Rafael dos Santos Bezerra; Raul Machado Neto; Rejane Maria Tommasini Grotto; Ricardo Haddad; Sandra Coccuzzo Sampaio Vessoni; Simone Kashima; Svetoslav Nanev Slavov; Vincent Louis Viala |
| EPI_ISL_3388715, EPI_ISL_3388808 | Rush University Medical Center | RIPHL at Rush University Medical Center | Diane Springer; Edith Perez; Felix Araujo Perez; Joyce Houlihan; Kevin Kunstman; Laura Furtado; Marieta Hyde; Mary Hayden; Max Kolton; Stefan Green |
| EPI_ISL_2784689, EPI_ISL_2784690 | S Filippo Neri | National Institute for Infectious Diseases (INMI) L. Spallanzani I.R.C.C.S | A Di Caro; B Bartolini; CEM Gruber; E Giombini; F Messina; F Santini; G Bonfiglio; M Rueca; MR Capobianchi; O Butera |
| EPI_ISL_5660005 | SANTA CASA | Instituto Butantan | Antonio Jorge Martins; Claudia Renata dos Santos Barros; David Schlesinger; Debora Botequio Moretti; Dimas Tadeu Covas; Elaine Cristina Marqueze; Elaine Vieira Santos; Evandra Strazza Rodrigues; Heidge Fukumasu; Jayme Augusto de Souza-Neto; José Salvatore Leister Patané; Luiz Alcantara; Luiz Lehmann Coutinho; Maria Carolina Elias; Maurício Lacerda Nogueira; Rafael dos Santos Bezerra; Raul Machado Neto; Rejane Maria Tommasini Grotto; Ricardo Haddad; Sandra Coccuzzo Sampaio Vessoni; Simone Kashima; Svetoslav Nanev Slavov; Vincent Louis Viala |
| EPI_ISL_5672115, EPI_ISL_5672250, EPI_ISL_5672301, EPI_ISL_5672391, EPI_ISL_5672416, EPI_ISL_5672452, EPI_ISL_5672456, EPI_ISL_5672463, EPI_ISL_5672520, EPI_ISL_5673408, EPI_ISL_5674024 | | | |
| see above | SANTA CASA DE GUARAREMA | Instituto Butantan | Antonio Jorge Martins; Claudia Renata dos Santos Barros; David Schlesinger; Debora Botequio Moretti; Dimas Tadeu Covas; Elaine Cristina Marqueze; Elaine Vieira Santos; Evandra Strazza Rodrigues; Heidge Fukumasu; Jayme Augusto de Souza-Neto; José Salvatore Leister Patané; Luiz Alcantara; Luiz Lehmann Coutinho; Maria Carolina Elias; Maurício Lacerda Nogueira; Rafael dos Santos Bezerra; Raul Machado Neto; Rejane Maria Tommasini Grotto; Ricardo Haddad; Sandra Coccuzzo Sampaio Vessoni; Simone Kashima; Svetoslav Nanev Slavov; Vincent Louis Viala |
| EPI_ISL_5667374, EPI_ISL_5667399, EPI_ISL_5667572, EPI_ISL_5667702 | SANTA CASA DE IBIRA | Instituto Butantan | Antonio Jorge Martins; Claudia Renata dos Santos Barros; David Schlesinger; Debora Botequio Moretti; Dimas Tadeu Covas; Elaine Cristina Marqueze; Elaine Vieira Santos; Evandra Strazza Rodrigues; Heidge Fukumasu; Jayme Augusto de Souza-Neto; José Salvatore Leister Patané; Luiz Alcantara; Luiz Lehmann Coutinho; Maria Carolina Elias; Maurício Lacerda Nogueira; Rafael dos Santos Bezerra; Raul Machado Neto; Rejane Maria Tommasini Grotto; Ricardo Haddad; Sandra Coccuzzo Sampaio Vessoni; Simone Kashima; Svetoslav Nanev Slavov; Vincent Louis Viala |
| EPI_ISL_5667949, EPI_ISL_5667962, EPI_ISL_5668300 | SANTA CASA DE JOSE BONIFACIO | Instituto Butantan | Antonio Jorge Martins; Claudia Renata dos Santos Barros; David Schlesinger; Debora Botequio Moretti; Dimas Tadeu Covas; Elaine Cristina Marqueze; Elaine Vieira Santos; Evandra Strazza Rodrigues; Heidge Fukumasu; Jayme Augusto de Souza-Neto; José Salvatore Leister Patané; Luiz Alcantara; Luiz Lehmann Coutinho; Maria Carolina Elias; Maurício Lacerda Nogueira; Rafael dos Santos Bezerra; Raul Machado Neto; Rejane Maria Tommasini Grotto; Ricardo Haddad; Sandra Coccuzzo Sampaio Vessoni; Simone Kashima; Svetoslav Nanev Slavov; Vincent Louis Viala |
| EPI_ISL_5667417, EPI_ISL_5667587, EPI_ISL_5667620 | SANTA CASA DE MISERICORDIA DE BRAGANCA PAULISTA | Instituto Butantan | Antonio Jorge Martins; Claudia Renata dos Santos Barros; David Schlesinger; Debora Botequio Moretti; Dimas Tadeu Covas; Elaine Cristina Marqueze; Elaine Vieira Santos; Evandra Strazza Rodrigues; Heidge Fukumasu; Jayme Augusto de Souza-Neto; José Salvatore Leister Patané; Luiz Alcantara; Luiz Lehmann Coutinho; Maria Carolina Elias; Maurício Lacerda Nogueira; Rafael dos Santos Bezerra; Raul Machado Neto; Rejane Maria Tommasini Grotto; Ricardo Haddad; Sandra Coccuzzo Sampaio Vessoni; Simone Kashima; Svetoslav Nanev Slavov; Vincent Louis Viala |
| EPI_ISL_5659696, EPI_ISL_5660280, EPI_ISL_5660634, EPI_ISL_5661420, EPI_ISL_5661579, EPI_ISL_5662140 | SANTA CASA DE MISERICORDIA DE UBATUBA | Instituto Butantan | Antonio Jorge Martins; Claudia Renata dos Santos Barros; David Schlesinger; Debora Botequio Moretti; Dimas Tadeu Covas; Elaine Cristina Marqueze; Elaine Vieira Santos; Evandra Strazza Rodrigues; Heidge Fukumasu; Jayme Augusto de Souza-Neto; José Salvatore Leister Patané; Luiz Alcantara; Luiz Lehmann Coutinho; Maria Carolina Elias; Maurício Lacerda Nogueira; Rafael dos Santos Bezerra; Raul Machado Neto; Rejane Maria Tommasini Grotto; Ricardo Haddad; Sandra Coccuzzo Sampaio Vessoni; Simone Kashima; Svetoslav Nanev Slavov; Vincent Louis Viala |
| EPI_ISL_5660451, EPI_ISL_5660560 | SANTA CASA DE MISERICORDIA SAO JOSE | Instituto Butantan | Antonio Jorge Martins; Claudia Renata dos Santos Barros; David Schlesinger; Debora Botequio Moretti; Dimas Tadeu Covas; Elaine Cristina Marqueze; Elaine Vieira Santos; Evandra Strazza Rodrigues; Heidge Fukumasu; Jayme Augusto de Souza-Neto; José Salvatore Leister Patané; Luiz Alcantara; Luiz Lehmann Coutinho; Maria Carolina Elias; Maurício Lacerda Nogueira; Rafael dos Santos Bezerra; Raul Machado Neto; Rejane Maria Tommasini Grotto; Ricardo Haddad; Sandra Coccuzzo Sampaio Vessoni; Simone Kashima; Svetoslav Nanev Slavov; Vincent Louis Viala |
| EPI_ISL_5665880 | SANTA CASA DE MOCOCA MOCOCA | Instituto Butantan | Antonio Jorge Martins; Claudia Renata dos Santos Barros; David Schlesinger; Debora Botequio Moretti; Dimas Tadeu Covas; Elaine Cristina Marqueze; Elaine Vieira Santos; Evandra Strazza Rodrigues; Heidge Fukumasu; Jayme Augusto de Souza-Neto; José Salvatore Leister Patané; Luiz Alcantara; Luiz Lehmann Coutinho; Maria Carolina Elias; Maurício Lacerda Nogueira; Rafael dos Santos Bezerra; Raul Machado Neto; Rejane Maria Tommasini Grotto; Ricardo Haddad; Sandra Coccuzzo Sampaio Vessoni; Simone Kashima; Svetoslav Nanev Slavov; Vincent Louis Viala |
| EPI_ISL_5660292, EPI_ISL_5660351 | SANTA CASA DE SANTA ADELIA | Instituto Butantan | Antonio Jorge Martins; Claudia Renata dos Santos Barros; David Schlesinger; Debora Botequio Moretti; Dimas Tadeu Covas; Elaine Cristina Marqueze; Elaine Vieira Santos; Evandra Strazza Rodrigues; Heidge Fukumasu; Jayme Augusto de Souza-Neto; José Salvatore Leister Patané; Luiz Alcantara; Luiz Lehmann Coutinho; Maria Carolina Elias; Maurício Lacerda Nogueira; Rafael dos Santos Bezerra; Raul Machado Neto; Rejane Maria Tommasini Grotto; Ricardo Haddad; Sandra Coccuzzo Sampaio Vessoni; Simone Kashima; Svetoslav Nanev Slavov; Vincent Louis Viala |
| EPI_ISL_3056256 | SARS-CoV-2 Sequencing Castilla y Leon-Spain Consortium | SARS-CoV-2 Sequencing Castilla y Leon-Spain Consortium | Antonio Orduña-Domingo; Carlos Fuster Foz; Carmen Aldea-Mansilla; Carmen Gimeno Crespo; David Abad; Gabriel March Rosello; Gregoria Megías Lobón; Jose María Eiros Bouza; M. Isabel Fernandez-Natal; Marta Dominguez-Gil; Marta Hernandez; María Antonia García Castro; Mª Fe Brezmes-Valdivieso; Noelia Arenal Andrés; Silvia Rojo; Sonsoles Garcinuño Pérez |
| EPI_ISL_2930565 | SARS-CoV-2 testing team, National Institute of Infectious Diseases | Pathogen Genomics Center, National Institute of Infectious Diseases | Hazuka Y Furihata; Hiromizu Takahashi; Kentaro Itokawa; Makoto Kuroda; Masanori Hashino; Masumichi Saito; Naomi Nojiri; Nozomu Hanaoka; Rina Tanaka; Tsuguto Fujimoto; Tsuyoshi Sekizuka |
| EPI_ISL_5663440, EPI_ISL_5663503, EPI_ISL_5664379, EPI_ISL_5664432, EPI_ISL_5664550, EPI_ISL_5665099, EPI_ISL_5665499, EPI_ISL_5665505 | | | |
| see above | SECAO CENTRO DE DIAGNOSTICO SECEDI | Instituto Butantan | Antonio Jorge Martins; Claudia Renata dos Santos Barros; David Schlesinger; Debora Botequio Moretti; Dimas Tadeu Covas; Elaine Cristina Marqueze; Elaine Vieira Santos; Evandra Strazza Rodrigues; Heidge Fukumasu; Jayme Augusto de Souza-Neto; José Salvatore Leister Patané; Luiz Alcantara; Luiz Lehmann Coutinho; Maria Carolina Elias; Maurício Lacerda Nogueira; Rafael dos Santos Bezerra; Raul Machado Neto; Rejane Maria Tommasini Grotto; Ricardo Haddad; Sandra Coccuzzo Sampaio Vessoni; Simone Kashima; Svetoslav Nanev Slavov; Vincent Louis Viala |
| EPI_ISL_5668138, EPI_ISL_5671995 | SECRETARIA DA SAUDE DE MAIRIPORA | Instituto Butantan | Antonio Jorge Martins; Claudia Renata dos Santos Barros; David Schlesinger; Debora Botequio Moretti; Dimas Tadeu Covas; Elaine Cristina Marqueze; Elaine Vieira Santos; Evandra Strazza Rodrigues; Heidge Fukumasu; Jayme Augusto de Souza-Neto; José Salvatore Leister Patané; Luiz Alcantara; Luiz Lehmann Coutinho; Maria Carolina Elias; Maurício Lacerda Nogueira; Rafael dos Santos Bezerra; Raul Machado Neto; Rejane Maria Tommasini Grotto; Ricardo Haddad; Sandra Coccuzzo Sampaio Vessoni; Simone Kashima; Svetoslav Nanev Slavov; Vincent Louis Viala |
| EPI_ISL_5659806, EPI_ISL_5661017, EPI_ISL_5661257, EPI_ISL_5661346, EPI_ISL_5661673 | SECRETARIA DE SAUDE | Instituto Butantan | Antonio Jorge Martins; Claudia Renata dos Santos Barros; David Schlesinger; Debora Botequio Moretti; Dimas Tadeu Covas; Elaine Cristina Marqueze; Elaine Vieira Santos; Evandra Strazza Rodrigues; Heidge Fukumasu; Jayme Augusto de Souza-Neto; José Salvatore Leister Patané; Luiz Alcantara; Luiz Lehmann Coutinho; Maria Carolina Elias; Maurício Lacerda Nogueira; Rafael dos Santos Bezerra; Raul Machado Neto; Rejane Maria Tommasini Grotto; Ricardo Haddad; Sandra Coccuzzo Sampaio Vessoni; Simone Kashima; Svetoslav Nanev Slavov; Vincent Louis Viala |
| EPI_ISL_5665902 | SECRETARIA DE SAUDE DE SANTA GERTRUDES | Instituto Butantan | Antonio Jorge Martins; Claudia Renata dos Santos Barros; David Schlesinger; Debora Botequio Moretti; Dimas Tadeu Covas; Elaine Cristina Marqueze; Elaine Vieira Santos; Evandra Strazza Rodrigues; Heidge Fukumasu; Jayme Augusto de Souza-Neto; José Salvatore Leister Patané; Luiz Alcantara; Luiz Lehmann Coutinho; Maria Carolina Elias; Maurício Lacerda Nogueira; Rafael dos Santos Bezerra; Raul Machado Neto; Rejane Maria Tommasini Grotto; Ricardo Haddad; Sandra Coccuzzo Sampaio Vessoni; Simone Kashima; Svetoslav Nanev Slavov; Vincent Louis Viala |
| EPI_ISL_5665855 | SECRETARIA DE SAUDE DE SAO PEDRO | Instituto Butantan | Antonio Jorge Martins; Claudia Renata dos Santos Barros; David Schlesinger; Debora Botequio Moretti; Dimas Tadeu Covas; Elaine Cristina Marqueze; Elaine Vieira Santos; Evandra Strazza Rodrigues; Heidge Fukumasu; Jayme Augusto de Souza-Neto; José Salvatore Leister Patané; Luiz Alcantara; Luiz Lehmann Coutinho; Maria Carolina Elias; Maurício Lacerda Nogueira; Rafael dos Santos Bezerra; Raul Machado Neto; Rejane Maria Tommasini Grotto; Ricardo Haddad; Sandra Coccuzzo Sampaio Vessoni; Simone Kashima; Svetoslav Nanev Slavov; Vincent Louis Viala |
| EPI_ISL_5661245, EPI_ISL_5661264, EPI_ISL_5661302, EPI_ISL_5661469, EPI_ISL_5661475, EPI_ISL_5661565, EPI_ISL_5661639, EPI_ISL_5661666, EPI_ISL_5661722, EPI_ISL_5661781, EPI_ISL_5661786, EPI_ISL_5661825, EPI_ISL_5661930, EPI_ISL_5663186, EPI_ISL_5663223, EPI_ISL_5668492, EPI_ISL_5668751, EPI_ISL_5668925, EPI_ISL_5669072, EPI_ISL_5669484, EPI_ISL_5669606, EPI_ISL_5669739, EPI_ISL_5669775, EPI_ISL_5669862, EPI_ISL_5669936, EPI_ISL_5669943, EPI_ISL_5670001, EPI_ISL_5670008, EPI_ISL_5670074, EPI_ISL_5670257, EPI_ISL_5670386, EPI_ISL_5670395, EPI_ISL_5670800 | | | |

| | | | |
|---|---|---|---|
| see above | SECRETARIA MUNCIPAL DE SAUDE DE SANTA BARBARA D OESTE | Instituto Butantan | Antonio Jorge Martins; Claudia Renata dos Santos Barros; David Schlesinger; Debora Botequio Moretti; Dimas Tadeu Covas; Elaine Cristina Marqueze; Elaine Vieira Santos; Evandra Strazza Rodrigues; Heidge Fukumasu; Jayme Augusto de Souza-Neto; José Salvatore Leister Patané; Luiz Alcantara; Luiz Lehmann Coutinho; Maria Carolina Elias; Maurício Lacerda Nogueira; Rafael dos Santos Bezerra; Raul Machado Neto; Rejane Maria Tommasini Grotto; Ricardo Haddad; Sandra Coccuzzo Sampaio Vessoni; Simone Kashima; Svetoslav Nanev Slavov; Vincent Louis Viala |
| EPI_ISL_5660115, EPI_ISL_5660428, EPI_ISL_5660733, EPI_ISL_5661095, EPI_ISL_5661282 | SECRETARIA MUNICIPAL DA SAUDE DE GUARATINGUETA | Instituto Butantan | Antonio Jorge Martins; Claudia Renata dos Santos Barros; David Schlesinger; Debora Botequio Moretti; Dimas Tadeu Covas; Elaine Cristina Marqueze; Elaine Vieira Santos; Evandra Strazza Rodrigues; Heidge Fukumasu; Jayme Augusto de Souza-Neto; José Salvatore Leister Patané; Luiz Alcantara; Luiz Lehmann Coutinho; Maria Carolina Elias; Maurício Lacerda Nogueira; Rafael dos Santos Bezerra; Raul Machado Neto; Rejane Maria Tommasini Grotto; Ricardo Haddad; Sandra Coccuzzo Sampaio Vessoni; Simone Kashima; Svetoslav Nanev Slavov; Vincent Louis Viala |
| EPI_ISL_5667483, EPI_ISL_5667820 | SECRETARIA MUNICIPAL DE SAUDE | Instituto Butantan | Antonio Jorge Martins; Claudia Renata dos Santos Barros; David Schlesinger; Debora Botequio Moretti; Dimas Tadeu Covas; Elaine Cristina Marqueze; Elaine Vieira Santos; Evandra Strazza Rodrigues; Heidge Fukumasu; Jayme Augusto de Souza-Neto; José Salvatore Leister Patané; Luiz Alcantara; Luiz Lehmann Coutinho; Maria Carolina Elias; Maurício Lacerda Nogueira; Rafael dos Santos Bezerra; Raul Machado Neto; Rejane Maria Tommasini Grotto; Ricardo Haddad; Sandra Coccuzzo Sampaio Vessoni; Simone Kashima; Svetoslav Nanev Slavov; Vincent Louis Viala |
| EPI_ISL_5665810, EPI_ISL_5665915, EPI_ISL_5671367 | SECRETARIA MUNICIPAL DE SAUDE DE CORDEIROPOLIS | Instituto Butantan | Antonio Jorge Martins; Claudia Renata dos Santos Barros; David Schlesinger; Debora Botequio Moretti; Dimas Tadeu Covas; Elaine Cristina Marqueze; Elaine Vieira Santos; Evandra Strazza Rodrigues; Heidge Fukumasu; Jayme Augusto de Souza-Neto; José Salvatore Leister Patané; Luiz Alcantara; Luiz Lehmann Coutinho; Maria Carolina Elias; Maurício Lacerda Nogueira; Rafael dos Santos Bezerra; Raul Machado Neto; Rejane Maria Tommasini Grotto; Ricardo Haddad; Sandra Coccuzzo Sampaio Vessoni; Simone Kashima; Svetoslav Nanev Slavov; Vincent Louis Viala |
| EPI_ISL_5662870 | SECRETARIA MUNICIPAL DE SAUDE DE PIRACAIA | Instituto Butantan | Antonio Jorge Martins; Claudia Renata dos Santos Barros; David Schlesinger; Debora Botequio Moretti; Dimas Tadeu Covas; Elaine Cristina Marqueze; Elaine Vieira Santos; Evandra Strazza Rodrigues; Heidge Fukumasu; Jayme Augusto de Souza-Neto; José Salvatore Leister Patané; Luiz Alcantara; Luiz Lehmann Coutinho; Maria Carolina Elias; Maurício Lacerda Nogueira; Rafael dos Santos Bezerra; Raul Machado Neto; Rejane Maria Tommasini Grotto; Ricardo Haddad; Sandra Coccuzzo Sampaio Vessoni; Simone Kashima; Svetoslav Nanev Slavov; Vincent Louis Viala |
| EPI_ISL_5663494 | SECRETARIA MUNICIPAL DE SAUDE DE PRES BERNARDES SP | Instituto Butantan | Antonio Jorge Martins; Claudia Renata dos Santos Barros; David Schlesinger; Debora Botequio Moretti; Dimas Tadeu Covas; Elaine Cristina Marqueze; Elaine Vieira Santos; Evandra Strazza Rodrigues; Heidge Fukumasu; Jayme Augusto de Souza-Neto; José Salvatore Leister Patané; Luiz Alcantara; Luiz Lehmann Coutinho; Maria Carolina Elias; Maurício Lacerda Nogueira; Rafael dos Santos Bezerra; Raul Machado Neto; Rejane Maria Tommasini Grotto; Ricardo Haddad; Sandra Coccuzzo Sampaio Vessoni; Simone Kashima; Svetoslav Nanev Slavov; Vincent Louis Viala |
| EPI_ISL_3912242 | SECRETARIA MUNICIPAL DE SAUDE DE TIANGUA | Analytical Competence Molecular Epidemiology Lab/ACME, Oswaldo Cruz Foundation, Ceara (FIOCRUZ CE) | Cleber Furtado Aksenen; Fabio Miyajima; Fernando Braga Stehling; Francisco Eder de Moura Lopes; Jamille Maria Mendes Bezerra; Joaquim Cesar do Nascimento Sousa Junior; Pedro Miguel Carneiro Jeronimo; Suzana Porto Almeida & Lucas Delerino on behalf of COVID-19 FIOCRUZ Genomic Network; Thais Ferreira de Oliveira; Thais de Oliveira Costa; Ticiane Cavalcante de Souza; Veridiana Pessoa Miyajima |
| EPI_ISL_5659925, EPI_ISL_5660241, EPI_ISL_5660311, EPI_ISL_5660377, EPI_ISL_5660480, EPI_ISL_5660528, EPI_ISL_5660547, EPI_ISL_5660964, EPI_ISL_5661010, EPI_ISL_5661209, EPI_ISL_5661452, EPI_ISL_5661661, EPI_ISL_5661810, EPI_ISL_5661837, EPI_ISL_5661974, EPI_ISL_5662015, EPI_ISL_5662084, EPI_ISL_5662088, EPI_ISL_5662156, EPI_ISL_5662188, EPI_ISL_5662242, EPI_ISL_5662252, EPI_ISL_5662343, EPI_ISL_5662384, EPI_ISL_5662428, EPI_ISL_5662459, EPI_ISL_5662567, EPI_ISL_5662631, EPI_ISL_5662670, EPI_ISL_5662681, EPI_ISL_5662747, EPI_ISL_5662836, EPI_ISL_5662862, EPI_ISL_5662988, EPI_ISL_5663015, EPI_ISL_5663101, EPI_ISL_5663130, EPI_ISL_5663152, EPI_ISL_5663298, EPI_ISL_5663304, EPI_ISL_5663316, EPI_ISL_5663326, EPI_ISL_5663430, EPI_ISL_5663521, EPI_ISL_5663540, EPI_ISL_5663571, EPI_ISL_5663595, EPI_ISL_5663700, EPI_ISL_5663704, EPI_ISL_5663725, EPI_ISL_5663792, EPI_ISL_5663796, EPI_ISL_5663868, EPI_ISL_5663945, EPI_ISL_5663964, EPI_ISL_5664013, EPI_ISL_5664062, EPI_ISL_5664104, EPI_ISL_5664129, EPI_ISL_5664156, EPI_ISL_5664182, EPI_ISL_5664235, EPI_ISL_5664259, EPI_ISL_5664316, EPI_ISL_5664348, EPI_ISL_5664470, EPI_ISL_5664578, EPI_ISL_5664594, EPI_ISL_5664759, EPI_ISL_5664829, EPI_ISL_5664844, EPI_ISL_5664951, EPI_ISL_5664962, EPI_ISL_5664967, EPI_ISL_5664980, EPI_ISL_5665097, EPI_ISL_5665118, EPI_ISL_5665230, EPI_ISL_5665244, EPI_ISL_5665254, EPI_ISL_5665255, EPI_ISL_5665275, EPI_ISL_5665285, EPI_ISL_5665291, EPI_ISL_5665299, EPI_ISL_5665332, EPI_ISL_5665351, EPI_ISL_5665398, EPI_ISL_5665403, EPI_ISL_5665497, EPI_ISL_5665535, EPI_ISL_5665539, EPI_ISL_5666671, EPI_ISL_5666685, EPI_ISL_5666706, EPI_ISL_5666794, EPI_ISL_5666799, EPI_ISL_5666893, EPI_ISL_5666903, EPI_ISL_5666995, EPI_ISL_5667018, EPI_ISL_5667061, EPI_ISL_5667125, EPI_ISL_5667171, EPI_ISL_5667177, EPI_ISL_5667192 | | | |
| see above | SECRETARIA MUNICIPAL DE SAUDE SOROCABA | Instituto Butantan | Antonio Jorge Martins; Claudia Renata dos Santos Barros; David Schlesinger; Debora Botequio Moretti; Dimas Tadeu Covas; Elaine Cristina Marqueze; Elaine Vieira Santos; Evandra Strazza Rodrigues; Heidge Fukumasu; Jayme Augusto de Souza-Neto; José Salvatore Leister Patané; Luiz Alcantara; Luiz Lehmann Coutinho; Maria Carolina Elias; Maurício Lacerda Nogueira; Rafael dos Santos Bezerra; Raul Machado Neto; Rejane Maria Tommasini Grotto; Ricardo Haddad; Sandra Coccuzzo Sampaio Vessoni; Simone Kashima; Svetoslav Nanev Slavov; Vincent Louis Viala |
| EPI_ISL_5664293 | SERV DE VIG SANITARIA EPIDEMIO E CTRL DE ZOONOZES GUARUJA | Instituto Butantan | Antonio Jorge Martins; Claudia Renata dos Santos Barros; David Schlesinger; Debora Botequio Moretti; Dimas Tadeu Covas; Elaine Cristina Marqueze; Elaine Vieira Santos; Evandra Strazza Rodrigues; Heidge Fukumasu; Jayme Augusto de Souza-Neto; José Salvatore Leister Patané; Luiz Alcantara; Luiz Lehmann Coutinho; Maria Carolina Elias; Maurício Lacerda Nogueira; Rafael dos Santos Bezerra; Raul Machado Neto; Rejane Maria Tommasini Grotto; Ricardo Haddad; Sandra Coccuzzo Sampaio Vessoni; Simone Kashima; Svetoslav Nanev Slavov; Vincent Louis Viala |
| EPI_ISL_5780514, EPI_ISL_5780516, EPI_ISL_5780530, EPI_ISL_5780531, EPI_ISL_5780533, EPI_ISL_5845183, EPI_ISL_5845204 | | | |
| see above | SK-Roy Romanow Provincial Laboratory | Saskatchewan - Roy Romanow Provincial Laboratory (RRPL) | Alanna Senecal; Amanda Lang; Jessica Minion; Kara Loos; Keith MacKenzie; Meredith Faires; Rachel DePaulo; Roy Romanow Provincial Laboratory - Molecular Diagnostics; Ryan McDonald |
| EPI_ISL_4743381 | SOMER | Laboratorio Departamental de Salud Publica de Antioquia | Andres F. Cardona-Rios; Cristian Arbey Velarde Hoyos; Gloria Isabel Escobar; Idabely Betancur Ortiz; Juan P. Hernandez-Ortiz; Maria Stella López |
| EPI_ISL_4984566, EPI_ISL_4984649 | SURA | Laboratorio Departamental de Salud Publica de Antioquia | Andres F. Cardona-Rios; Cristian Arbey Velarde Hoyos; Gloria Isabel Escobar; Idabely Betancur Ortiz; Juan P. Hernandez-Ortiz; Maria Stella López |
| EPI_ISL_2930809, EPI_ISL_2930810, EPI_ISL_2930811 | SYNLAB | GIGA Medical Genomics | Bouchra Boujemla; Claire Gourzonès; Cécile Meex; Keith Durkin; Laurent Gillet; Maria Artesi; Marie-Pierre Hayette; Nadine Cambisano; Nathalie Renotte; Olivier Ek; Sébastien Bontems; Vincent Bours |
| EPI_ISL_2844900 | SYNLAB MVZ Leverkusen | Robert Koch Institute | |
| EPI_ISL_2956172, EPI_ISL_2956175, EPI_ISL_2956231, EPI_ISL_2956242, EPI_ISL_2956246, EPI_ISL_2956251 | SYNLAB MVZ Trier | Robert Koch Institute | |
| EPI_ISL_2844907, EPI_ISL_2956139 | SYNLAB MVZ Weiden | Robert Koch Institute | |
| EPI_ISL_2894410, EPI_ISL_2978389, EPI_ISL_2978391, EPI_ISL_2978396, EPI_ISL_2978404, EPI_ISL_2978442, EPI_ISL_2978444, EPI_ISL_2978462, EPI_ISL_2978469, EPI_ISL_2978484, EPI_ISL_2978489, EPI_ISL_2978491, EPI_ISL_2978499, EPI_ISL_2978503, EPI_ISL_2978517, EPI_ISL_2978552, EPI_ISL_2978563, EPI_ISL_3067661, EPI_ISL_3067670, EPI_ISL_3067677, EPI_ISL_3067712, EPI_ISL_3067849, EPI_ISL_3067926, EPI_ISL_3160863, EPI_ISL_3277606 | | | |
| see above | Salud Digna | Instituto Nacional de Medicina Genomica | Abraham Campos-Romero; Cedro-Tanda A; Escobar-Arrazola; Escobar-Arrazola MA; Gonzalez-Barrera D; Herrera-Montalvo LA.; Hidalgo-Miranda A; Luna-Ruiz Marco; M; Mendoza-Vargas A; Moreno-Camacho José Luis; Munguia-Garza P; Ramirez-Vega O; Rangel-DeLeon D; Reyes-Grajeda JP; Rodriguez-Gallegos Jorge |
| EPI_ISL_3373975, EPI_ISL_3373977, EPI_ISL_3373979, EPI_ISL_3373982, EPI_ISL_3373984, EPI_ISL_3373986, EPI_ISL_3373994, EPI_ISL_3373996, EPI_ISL_3374000, EPI_ISL_3374006, EPI_ISL_3374014, EPI_ISL_3374015, EPI_ISL_3374019, EPI_ISL_3374020, EPI_ISL_3374024, EPI_ISL_3374025, EPI_ISL_3374026, EPI_ISL_3374031, EPI_ISL_3374036, EPI_ISL_3374037, EPI_ISL_3374038, EPI_ISL_3374039, EPI_ISL_4411237, EPI_ISL_4411285, EPI_ISL_4411361 | | | |
| see above | Salud Digna, A.C | Andersen lab at Scripps Research | Abraham Garcia Gil; Jose Luis Moreno Camacho; Marco Antonio Luna Ruiz-Esparza; Miguel A. Fernandez Rojas; SEARCH Alliance with Abraham Campos Romero |
| EPI_ISL_3346597, EPI_ISL_3346605, EPI_ISL_3388861, EPI_ISL_3388866 | San Diego County Public Health Laboratory | San Diego County Public Health Laboratory | Ashleigh Murphy Schafer; Brett Austin |
| EPI_ISL_3219168 | Santa Clara Valley Medical Center | Santa Clara County Public Health Laboratory | Santa Clara County Public Health Department |
| EPI_ISL_3373720, EPI_ISL_5570352 | Scripps Medical Laboratory | Andersen lab at Scripps Research | Ellen Stefanski; Ian Mchardy; SEARCH Alliance San Diego with Michael Quigley |
| EPI_ISL_3104831, EPI_ISL_4234768 | Seattle Flu Study | Seattle Flu Study | Amanda Adler; Barry R. Lutz; Benjamin Pelle; Caitlin R. Wolf; Chris D. Frazar; Deborah A. Nickerson; Elisabeth Brandstetter; Erica Ryke; Helen Y. Chu; Janet A. Englund; Jay Shendure; Jeff Duchin; Jover Lee; Kairsten Fay; Karen Cowgill; Kirsten Lacombe; Lea M. Starita; Mark J. Rieder; Matthew Richardson; Matthew Thompson; Melissa Truong; Michael Boeckh; Michael Famulare; Misja Ilcisin; Peter D. Han; Stephanie Schrag; Thomas R. Sibley; Trevor Bedford |
| EPI_ISL_3030174 | Senigallia | Microbiology University Politecnica delle Marche | Anna Valenza; Carla Acciarri; Katia Marinelli; Monica Lucia Ferreri; Patrizia Bagnarelli; Roberta Longo; Sara Caucci; Stefano Menzo |
| EPI_ISL_4082968, EPI_ISL_4082969, EPI_ISL_4082970, EPI_ISL_4082978, EPI_ISL_4082993, EPI_ISL_4083001, EPI_ISL_4083014, EPI_ISL_4083037, EPI_ISL_4083038, EPI_ISL_4083048, EPI_ISL_4083049, EPI_ISL_4083072, EPI_ISL_4083073, EPI_ISL_4083074, EPI_ISL_4083076, EPI_ISL_4083077, EPI_ISL_4083084, EPI_ISL_4083085, EPI_ISL_4083088, EPI_ISL_4083089, EPI_ISL_4083100, EPI_ISL_4083103, EPI_ISL_4083104, EPI_ISL_4083108, EPI_ISL_4083109, EPI_ISL_4083113, EPI_ISL_4083121, EPI_ISL_4083125, EPI_ISL_4083126, EPI_ISL_4083127, EPI_ISL_4083128, EPI_ISL_4083134, EPI_ISL_4083136, EPI_ISL_4083138, EPI_ISL_4083146, EPI_ISL_4083147, EPI_ISL_4083148, EPI_ISL_4083149, EPI_ISL_4083158, EPI_ISL_4083161, EPI_ISL_4083162, EPI_ISL_4083167, EPI_ISL_4083168, EPI_ISL_4083169, EPI_ISL_4083170, EPI_ISL_4083171, EPI_ISL_4083186, EPI_ISL_4083233, EPI_ISL_4083238, EPI_ISL_4083242, EPI_ISL_4083243, EPI_ISL_4083247, EPI_ISL_4083248, EPI_ISL_4083250, EPI_ISL_4083255, EPI_ISL_4083257, EPI_ISL_4083260, EPI_ISL_4083261, EPI_ISL_4083262, EPI_ISL_4083263, EPI_ISL_4083264, EPI_ISL_4083266, EPI_ISL_4083268, EPI_ISL_4083269, EPI_ISL_4083270, EPI_ISL_4083271, EPI_ISL_4083275, EPI_ISL_4083276, EPI_ISL_4083278, EPI_ISL_4083296, EPI_ISL_4083315, EPI_ISL_4083325, EPI_ISL_4083331, EPI_ISL_4083332, EPI_ISL_4083333, EPI_ISL_4083334, EPI_ISL_4083335, EPI_ISL_4083336, EPI_ISL_4083337, EPI_ISL_4083338, EPI_ISL_4083339, EPI_ISL_4083340, EPI_ISL_4083341, EPI_ISL_4083342, EPI_ISL_4083343, EPI_ISL_4083344, EPI_ISL_4083347, EPI_ISL_4083349, EPI_ISL_4083350, EPI_ISL_4083351, EPI_ISL_4083353, EPI_ISL_4083360, EPI_ISL_4083376, EPI_ISL_4083387, EPI_ISL_4083390, EPI_ISL_5158425, EPI_ISL_5158451, EPI_ISL_5158474, EPI_ISL_5158475, EPI_ISL_5158519, EPI_ISL_5158548, EPI_ISL_5158559, EPI_ISL_5158560, EPI_ISL_5158595, EPI_ISL_5158829, EPI_ISL_5656043, EPI_ISL_6694439 | | | |
| see above | Servicio Virosis Respiratorias-Departamento Virología-INEI | Instituto Nacional Enfermedades Infecciosas C.G.Malbran | Avaro M.; Baumeister E.; Benedetti E.; Campos J.; Cisterna D.; Dattero ME; De Belder D.; Haim MS.; Lorenzo F.; Molina V.; Perandones C.; Poklepovich T.; Pontoriero A.; Russo M.; Sanchez Loria J.; Tuduri E. |
| EPI_ISL_2960221 | Servicio de Microbiologia Hospital Ramon y Cajal | Servicio de Microbiologia Hospital Ramon y Cajal | Galan JC; Martinez L. Abreu M; Ponce M; y Gonzalez-Alba JM |
| EPI_ISL_3022017, EPI_ISL_3022215, EPI_ISL_3024223 | Shamir Medical Center (Asaf Harofe) | Shamir Medical Center (Asaf Harofe) | Abu Hamad Ramzia; Adina Bar Chaim; Anna Vishnevsky; Chen Weiner; Nir Rainy; Patricia Benveniste-Lekovitz; Reut Sorek Abramovich; Yevgeni Yegorov |
| EPI_ISL_3215949, EPI_ISL_3216035, EPI_ISL_3373810 | Sharp HealthCare Laboratory | Andersen lab at Scripps Research | Art Mendoza; Cathy Woerle; Jacquelyn Berumen; Liam McGinnis; Omid Bakhtar; SEARCH Alliance San Diego with Aaron Harding |
| EPI_ISL_3050210, EPI_ISL_3050257 | Stanford Health Care | Stanford University School of Medicine, Clinical Virology Laboratory | Becky Jiang; Bernadette Troung; Daniel Solis; James Zehnder; Malaya K. Sahoo; Mamdouh Sibai; Nathan Hammond; Selamawit Bihon; and Benjamin A. Pinsky |
| EPI_ISL_2724849, EPI_ISL_3033271, EPI_ISL_3189637, EPI_ISL_3189663 | State Laboratories Division, Hawaii State Department of Health | State Laboratories Division, Hawaii State Department of Health | Ayana Garnet; Daniel Strange; Drew Kuwazaki; Edward Desmond; Pamela O'Brien; Razvan Sultana |
| EPI_ISL_3049992, EPI_ISL_3050032, EPI_ISL_3050082, | State Testing Facility | Altius Institute for Biomedical Research | Alex Isner; Alex Nguyen; Amanda Gale; Audra Johnson; Clem Green; Daniel Bates; Eric Thorland; Jacob Rodriguez; Jean Robinston; Jemma Nelson; Jessica Kunder; John Stamatoyannopoulos; Joshua Richards; Julia Wald; Kneshay Harper; Lauren Mitchell; Mark Frerker; Matt Hartman; Michael Buckley; Muhammad Halimun; Rebecca Bruders; Tobias Ragoczy |

| | | | |
|---|---|---|---|
| EPI_ISL_3050093 | | | |
| EPI_ISL_2980167, EPI_ISL_3999054 | Swedish national genomic surveillance program of SARS-CoV-2 | The Public Health Agency of Sweden | Alma Brolund; Maria Lind Karlberg; Maximilian Riess; Swedish national genomic surveillance program of SARS-CoV-2 |
| EPI_ISL_3030108 | Synlab Suisse SA | Clinical Bacteriology | Adrian Egli; Alfredo Mari; Fanny Wegner; Hans Hirsch; Helena MB Seth-Smith; Julia Bielicki; Karoline Leuzinger; Manuel Battegay; Tim Roloff |
| EPI_ISL_3088801, EPI_ISL_3088803, EPI_ISL_3326711, EPI_ISL_3460368, EPI_ISL_3460398, EPI_ISL_3460406, EPI_ISL_3460420, EPI_ISL_3460423, EPI_ISL_3460427, EPI_ISL_3460463, EPI_ISL_3460472, EPI_ISL_3460511, EPI_ISL_3460515, EPI_ISL_3460516, EPI_ISL_3460517, EPI_ISL_3460522, EPI_ISL_3460579, EPI_ISL_3460611, EPI_ISL_3460614, EPI_ISL_3460660, EPI_ISL_3661760, EPI_ISL_3826236, EPI_ISL_3826274 | | | |
| see above | TGen North | TGen North | "Jolene Bowers; Brett Van Tassel; Chris French; Darrin Lemmer; Dave Engelthaler; Dave Engelthaler"; Hayley Yaglom; Heather Centner; Jolene Bowers |
| EPI_ISL_3695355, EPI_ISL_3695366, EPI_ISL_3695387, EPI_ISL_3695409, EPI_ISL_3695411, EPI_ISL_3695418, EPI_ISL_3695491, EPI_ISL_3695521, EPI_ISL_3695541, EPI_ISL_3695542, EPI_ISL_3695566, EPI_ISL_3695589 | | | |
| see above | TPMG Regional Laboratory | California Department of Public Health | Emily Smith on behalf of CDPH-COVIDNet and UCI Genome Sciences Center/GHTF |
| EPI_ISL_3048186 | Tampa General Hospital Esoteric Lab | Tampa General Hospital Esoteric Research & Development Lab | Amorce Lima; Deanna Becker; Dominic Uy; Elaine Vendrone; Jon Faughn; Jorge Mecias-Frias; Marci O'Driscoll; Suzane Silbert; Vicki Healer |
| EPI_ISL_3148993 | Texas Children's Hospital | Texas Children's Microbiome Center | Ila Singh; James Dunn; James Versalovic; Jennifer K. Spinler; Ruth Ann Luna |
| EPI_ISL_2967990, EPI_ISL_3610774, EPI_ISL_3610775, EPI_ISL_3642743 | The Caribbean Public Health Agency | Carrington Lab, Department of Preclinical Sciences, Faculty of Medical Sciences, The University of the West Indies, St Augustine Campus | Anushka Ramjag; Arianne Brown-Jordan; Avery Hinds; Christine V. F. Carrington; Christopher Oura; Gabriel Escobar; Kenneth George; Nikita S. D. Sahadeo; Nuno Faria; Oliver Pybus; Risha Singh; Roshan Parasram; Sarah Hill; SueMin Nathaniel; Vernie Ramkissoon |
| EPI_ISL_3943629 | Torrance Memorial Medical Center | Los Angeles County Public Health Laboratories | P. Hemarajata et al. |
| EPI_ISL_3610769, EPI_ISL_3610773 | Trinidad Public Health Laboratory | Carrington Lab, Department of Preclinical Sciences, Faculty of Medical Sciences, The University of the West Indies, St Augustine Campus | Anushka Ramjag; Arianne Brown-Jordan; Avery Hinds; Christine V. F. Carrington; Gabriel Escobar; Nikita S. D. Sahadeo; Nuno Faria; Oliver Pybus; Risha Singh; Roshan Parasram; Sarah Hill; SueMin Nathaniel; Vernie Ramkissoon |
| EPI_ISL_3266323 | U.O. Microbiologia Laboratorio Unico Centro Servizi - AUSL della Romagna | U.O. Microbiologia, Laboratorio Unico Centro Servizi - AUSL della Romagna | Giorgio Dirani |
| EPI_ISL_5666213, EPI_ISL_5666249, EPI_ISL_5666265, EPI_ISL_5666349, EPI_ISL_5666374, EPI_ISL_5666487, EPI_ISL_5666502, EPI_ISL_5666557 | | | |
| see above | UBS ALCIMINIO DE ASSIS LOURENCO BADY BASSITT | Instituto Butantan | Antonio Jorge Martins; Claudia Renata dos Santos Barros; David Schlesinger; Debora Botequio Moretti; Dimas Tadeu Covas; Elaine Cristina Marqueze; Elaine Vieira Santos; Evandra Strazza Rodrigues; Heidge Fukumasu; Jayme Augusto de Souza-Neto; José Salvatore Leister Patané; Luiz Alcantara; Luiz Lehmann Coutinho; Maria Carolina Elias; Maurício Lacerda Nogueira; Rafael dos Santos Bezerra; Raul Machado Neto; Rejane Maria Tommasini Grotto; Ricardo Haddad; Sandra Coccuzzo Sampaio Vessoni; Simone Kashima; Svetoslav Nanev Slavov; Vincent Louis Viala |
| EPI_ISL_5664272 | UBS ANTONIO NHAN | Instituto Butantan | Antonio Jorge Martins; Claudia Renata dos Santos Barros; David Schlesinger; Debora Botequio Moretti; Dimas Tadeu Covas; Elaine Cristina Marqueze; Elaine Vieira Santos; Evandra Strazza Rodrigues; Heidge Fukumasu; Jayme Augusto de Souza-Neto; José Salvatore Leister Patané; Luiz Alcantara; Luiz Lehmann Coutinho; Maria Carolina Elias; Maurício Lacerda Nogueira; Rafael dos Santos Bezerra; Raul Machado Neto; Rejane Maria Tommasini Grotto; Ricardo Haddad; Sandra Coccuzzo Sampaio Vessoni; Simone Kashima; Svetoslav Nanev Slavov; Vincent Louis Viala |
| EPI_ISL_5663312, EPI_ISL_5663567, EPI_ISL_5663657, EPI_ISL_5664189, EPI_ISL_5664297, EPI_ISL_5664955, EPI_ISL_5665115, EPI_ISL_5665520, EPI_ISL_5665566 | | | |
| see above | UBS ATALAIA DR MARIO AGUIAR FILHO | Instituto Butantan | Antonio Jorge Martins; Claudia Renata dos Santos Barros; David Schlesinger; Debora Botequio Moretti; Dimas Tadeu Covas; Elaine Cristina Marqueze; Elaine Vieira Santos; Evandra Strazza Rodrigues; Heidge Fukumasu; Jayme Augusto de Souza-Neto; José Salvatore Leister Patané; Luiz Alcantara; Luiz Lehmann Coutinho; Maria Carolina Elias; Maurício Lacerda Nogueira; Rafael dos Santos Bezerra; Raul Machado Neto; Rejane Maria Tommasini Grotto; Ricardo Haddad; Sandra Coccuzzo Sampaio Vessoni; Simone Kashima; Svetoslav Nanev Slavov; Vincent Louis Viala |
| EPI_ISL_5666199, EPI_ISL_5666329, EPI_ISL_5666434 | UBS ATILIO UNGARO | Instituto Butantan | Antonio Jorge Martins; Claudia Renata dos Santos Barros; David Schlesinger; Debora Botequio Moretti; Dimas Tadeu Covas; Elaine Cristina Marqueze; Elaine Vieira Santos; Evandra Strazza Rodrigues; Heidge Fukumasu; Jayme Augusto de Souza-Neto; José Salvatore Leister Patané; Luiz Alcantara; Luiz Lehmann Coutinho; Maria Carolina Elias; Maurício Lacerda Nogueira; Rafael dos Santos Bezerra; Raul Machado Neto; Rejane Maria Tommasini Grotto; Ricardo Haddad; Sandra Coccuzzo Sampaio Vessoni; Simone Kashima; Svetoslav Nanev Slavov; Vincent Louis Viala |
| EPI_ISL_5666140, EPI_ISL_5666150, EPI_ISL_5666158, EPI_ISL_5666195, EPI_ISL_5666288 | UBS CAIC OURINHOS | Instituto Butantan | Antonio Jorge Martins; Claudia Renata dos Santos Barros; David Schlesinger; Debora Botequio Moretti; Dimas Tadeu Covas; Elaine Cristina Marqueze; Elaine Vieira Santos; Evandra Strazza Rodrigues; Heidge Fukumasu; Jayme Augusto de Souza-Neto; José Salvatore Leister Patané; Luiz Alcantara; Luiz Lehmann Coutinho; Maria Carolina Elias; Maurício Lacerda Nogueira; Rafael dos Santos Bezerra; Raul Machado Neto; Rejane Maria Tommasini Grotto; Ricardo Haddad; Sandra Coccuzzo Sampaio Vessoni; Simone Kashima; Svetoslav Nanev Slavov; Vincent Louis Viala |
| EPI_ISL_5665091 | UBS CARMENO NAGGHY JARDIM DAVILA | Instituto Butantan | Antonio Jorge Martins; Claudia Renata dos Santos Barros; David Schlesinger; Debora Botequio Moretti; Dimas Tadeu Covas; Elaine Cristina Marqueze; Elaine Vieira Santos; Evandra Strazza Rodrigues; Heidge Fukumasu; Jayme Augusto de Souza-Neto; José Salvatore Leister Patané; Luiz Alcantara; Luiz Lehmann Coutinho; Maria Carolina Elias; Maurício Lacerda Nogueira; Rafael dos Santos Bezerra; Raul Machado Neto; Rejane Maria Tommasini Grotto; Ricardo Haddad; Sandra Coccuzzo Sampaio Vessoni; Simone Kashima; Svetoslav Nanev Slavov; Vincent Louis Viala |
| EPI_ISL_5664907 | UBS CAROLINA MARIA DE JESUS | Instituto Butantan | Antonio Jorge Martins; Claudia Renata dos Santos Barros; David Schlesinger; Debora Botequio Moretti; Dimas Tadeu Covas; Elaine Cristina Marqueze; Elaine Vieira Santos; Evandra Strazza Rodrigues; Heidge Fukumasu; Jayme Augusto de Souza-Neto; José Salvatore Leister Patané; Luiz Alcantara; Luiz Lehmann Coutinho; Maria Carolina Elias; Maurício Lacerda Nogueira; Rafael dos Santos Bezerra; Raul Machado Neto; Rejane Maria Tommasini Grotto; Ricardo Haddad; Sandra Coccuzzo Sampaio Vessoni; Simone Kashima; Svetoslav Nanev Slavov; Vincent Louis Viala |
| EPI_ISL_5667870, EPI_ISL_5667977, EPI_ISL_5668014, EPI_ISL_5668168, EPI_ISL_5668369, EPI_ISL_5668378 | UBS CELESTINO CARNIELO | Instituto Butantan | Antonio Jorge Martins; Claudia Renata dos Santos Barros; David Schlesinger; Debora Botequio Moretti; Dimas Tadeu Covas; Elaine Cristina Marqueze; Elaine Vieira Santos; Evandra Strazza Rodrigues; Heidge Fukumasu; Jayme Augusto de Souza-Neto; José Salvatore Leister Patané; Luiz Alcantara; Luiz Lehmann Coutinho; Maria Carolina Elias; Maurício Lacerda Nogueira; Rafael dos Santos Bezerra; Raul Machado Neto; Rejane Maria Tommasini Grotto; Ricardo Haddad; Sandra Coccuzzo Sampaio Vessoni; Simone Kashima; Svetoslav Nanev Slavov; Vincent Louis Viala |
| EPI_ISL_5663720, EPI_ISL_5664001, EPI_ISL_5664630 | UBS CUMBICA | Instituto Butantan | Antonio Jorge Martins; Claudia Renata dos Santos Barros; David Schlesinger; Debora Botequio Moretti; Dimas Tadeu Covas; Elaine Cristina Marqueze; Elaine Vieira Santos; Evandra Strazza Rodrigues; Heidge Fukumasu; Jayme Augusto de Souza-Neto; José Salvatore Leister Patané; Luiz Alcantara; Luiz Lehmann Coutinho; Maria Carolina Elias; Maurício Lacerda Nogueira; Rafael dos Santos Bezerra; Raul Machado Neto; Rejane Maria Tommasini Grotto; Ricardo Haddad; Sandra Coccuzzo Sampaio Vessoni; Simone Kashima; Svetoslav Nanev Slavov; Vincent Louis Viala |
| EPI_ISL_5660639, EPI_ISL_5661527 | UBS DARCY ALVES E ROBALINHO | Instituto Butantan | Antonio Jorge Martins; Claudia Renata dos Santos Barros; David Schlesinger; Debora Botequio Moretti; Dimas Tadeu Covas; Elaine Cristina Marqueze; Elaine Vieira Santos; Evandra Strazza Rodrigues; Heidge Fukumasu; Jayme Augusto de Souza-Neto; José Salvatore Leister Patané; Luiz Alcantara; Luiz Lehmann Coutinho; Maria Carolina Elias; Maurício Lacerda Nogueira; Rafael dos Santos Bezerra; Raul Machado Neto; Rejane Maria Tommasini Grotto; Ricardo Haddad; Sandra Coccuzzo Sampaio Vessoni; Simone Kashima; Svetoslav Nanev Slavov; Vincent Louis Viala |
| EPI_ISL_5660459 | UBS DE JAMBEIRO JOCELINA MEIRELES | Instituto Butantan | Antonio Jorge Martins; Claudia Renata dos Santos Barros; David Schlesinger; Debora Botequio Moretti; Dimas Tadeu Covas; Elaine Cristina Marqueze; Elaine Vieira Santos; Evandra Strazza Rodrigues; Heidge Fukumasu; Jayme Augusto de Souza-Neto; José Salvatore Leister Patané; Luiz Alcantara; Luiz Lehmann Coutinho; Maria Carolina Elias; Maurício Lacerda Nogueira; Rafael dos Santos Bezerra; Raul Machado Neto; Rejane Maria Tommasini Grotto; Ricardo Haddad; Sandra Coccuzzo Sampaio Vessoni; Simone Kashima; Svetoslav Nanev Slavov; Vincent Louis Viala |
| EPI_ISL_5666335, EPI_ISL_5666437, EPI_ISL_5666531, EPI_ISL_5666601 | UBS DE TABAPUA | Instituto Butantan | Antonio Jorge Martins; Claudia Renata dos Santos Barros; David Schlesinger; Debora Botequio Moretti; Dimas Tadeu Covas; Elaine Cristina Marqueze; Elaine Vieira Santos; Evandra Strazza Rodrigues; Heidge Fukumasu; Jayme Augusto de Souza-Neto; José Salvatore Leister Patané; Luiz Alcantara; Luiz Lehmann Coutinho; Maria Carolina Elias; Maurício Lacerda Nogueira; Rafael dos Santos Bezerra; Raul Machado Neto; Rejane Maria Tommasini Grotto; Ricardo Haddad; Sandra Coccuzzo Sampaio Vessoni; Simone Kashima; Svetoslav Nanev Slavov; Vincent Louis Viala |
| EPI_ISL_5661904, EPI_ISL_5663993 | UBS DINAMARCA | Instituto Butantan | Antonio Jorge Martins; Claudia Renata dos Santos Barros; David Schlesinger; Debora Botequio Moretti; Dimas Tadeu Covas; Elaine Cristina Marqueze; Elaine Vieira Santos; Evandra Strazza Rodrigues; Heidge Fukumasu; Jayme Augusto de Souza-Neto; José Salvatore Leister Patané; Luiz Alcantara; Luiz Lehmann Coutinho; Maria Carolina Elias; Maurício Lacerda Nogueira; Rafael dos Santos Bezerra; Raul Machado Neto; Rejane Maria Tommasini Grotto; Ricardo Haddad; Sandra Coccuzzo Sampaio Vessoni; Simone Kashima; Svetoslav Nanev Slavov; Vincent Louis Viala |
| EPI_ISL_5663005 | UBS DR ABELARDO PINHEIRO GUIMARAES SCRPARDO | Instituto Butantan | Antonio Jorge Martins; Claudia Renata dos Santos Barros; David Schlesinger; Debora Botequio Moretti; Dimas Tadeu Covas; Elaine Cristina Marqueze; Elaine Vieira Santos; Evandra Strazza Rodrigues; Heidge Fukumasu; Jayme Augusto de Souza-Neto; José Salvatore Leister Patané; Luiz Alcantara; Luiz Lehmann Coutinho; Maria Carolina Elias; Maurício Lacerda Nogueira; Rafael dos Santos Bezerra; Raul Machado Neto; Rejane Maria Tommasini Grotto; Ricardo Haddad; Sandra Coccuzzo Sampaio Vessoni; Simone Kashima; Svetoslav Nanev Slavov; Vincent Louis Viala |
| EPI_ISL_5664834 | UBS DR ADAUTO RIBEIRO | Instituto Butantan | Antonio Jorge Martins; Claudia Renata dos Santos Barros; David Schlesinger; Debora Botequio Moretti; Dimas Tadeu Covas; Elaine Cristina Marqueze; Elaine Vieira Santos; Evandra Strazza Rodrigues; Heidge Fukumasu; Jayme Augusto de Souza-Neto; José Salvatore Leister Patané; Luiz Alcantara; Luiz Lehmann Coutinho; Maria Carolina Elias; Maurício Lacerda Nogueira; Rafael dos Santos Bezerra; Raul Machado Neto; Rejane Maria Tommasini Grotto; Ricardo Haddad; Sandra Coccuzzo Sampaio Vessoni; Simone Kashima; Svetoslav Nanev Slavov; Vincent Louis Viala |
| EPI_ISL_5663917, EPI_ISL_5664543, EPI_ISL_5666689, EPI_ISL_5666750, EPI_ISL_5666821, EPI_ISL_5666839, EPI_ISL_5666886, EPI_ISL_5666934, EPI_ISL_5666939, EPI_ISL_5666986, EPI_ISL_5667001, EPI_ISL_5667010, EPI_ISL_5667027, EPI_ISL_5667044, EPI_ISL_5667103, EPI_ISL_5667147, EPI_ISL_5667184 | | | |
| see above | UBS DR ALFREDO DANTAS DE SOUZA UMUARAMA | Instituto Butantan | Antonio Jorge Martins; Claudia Renata dos Santos Barros; David Schlesinger; Debora Botequio Moretti; Dimas Tadeu Covas; Elaine Cristina Marqueze; Elaine Vieira Santos; Evandra Strazza Rodrigues; Heidge Fukumasu; Jayme Augusto de Souza-Neto; José Salvatore Leister Patané; Luiz Alcantara; Luiz Lehmann Coutinho; Maria Carolina Elias; Maurício Lacerda Nogueira; Rafael dos Santos Bezerra; Raul Machado Neto; Rejane Maria Tommasini Grotto; Ricardo Haddad; Sandra Coccuzzo Sampaio Vessoni; Simone Kashima; Svetoslav Nanev Slavov; Vincent Louis Viala |
| EPI_ISL_5666217 | UBS DR HELIO MIGLIARI | Instituto Butantan | Antonio Jorge Martins; Claudia Renata dos Santos Barros; David Schlesinger; Debora Botequio Moretti; Dimas Tadeu Covas; Elaine Cristina Marqueze; Elaine Vieira Santos; Evandra Strazza Rodrigues; Heidge Fukumasu; Jayme Augusto de Souza-Neto; José Salvatore Leister Patané; Luiz Alcantara; Luiz Lehmann Coutinho; Maria Carolina Elias; Maurício Lacerda Nogueira; Rafael dos Santos Bezerra; Raul Machado Neto; Rejane Maria Tommasini Grotto; Ricardo Haddad; Sandra Coccuzzo Sampaio Vessoni; Simone Kashima; Svetoslav Nanev Slavov; Vincent Louis Viala |
| EPI_ISL_5661909, EPI_ISL_5662831, EPI_ISL_5662938 | UBS DR WALDOMIRO FERREIRA NEVES SCRPARDO | Instituto Butantan | Antonio Jorge Martins; Claudia Renata dos Santos Barros; David Schlesinger; Debora Botequio Moretti; Dimas Tadeu Covas; Elaine Cristina Marqueze; Elaine Vieira Santos; Evandra Strazza Rodrigues; Heidge Fukumasu; Jayme Augusto de Souza-Neto; José Salvatore Leister Patané; Luiz Alcantara; Luiz Lehmann Coutinho; Maria Carolina Elias; Maurício Lacerda Nogueira; Rafael dos Santos Bezerra; Raul Machado Neto; Rejane Maria Tommasini Grotto; Ricardo Haddad; Sandra Coccuzzo Sampaio Vessoni; Simone Kashima; Svetoslav Nanev Slavov; Vincent Louis Viala |
| EPI_ISL_5661402, EPI_ISL_5661537 | UBS EMILIA COSME CERQUEIRA | Instituto Butantan | Antonio Jorge Martins; Claudia Renata dos Santos Barros; David Schlesinger; Debora Botequio Moretti; Dimas Tadeu Covas; Elaine Cristina Marqueze; Elaine Vieira Santos; Evandra Strazza Rodrigues; Heidge Fukumasu; Jayme Augusto de Souza-Neto; José Salvatore Leister Patané; Luiz Alcantara; Luiz Lehmann Coutinho; Maria Carolina Elias; Maurício Lacerda Nogueira; Rafael dos Santos Bezerra; Raul Machado Neto; Rejane Maria Tommasini Grotto; Ricardo Haddad; Sandra Coccuzzo Sampaio Vessoni; Simone Kashima; Svetoslav Nanev Slavov; Vincent Louis Viala |
| EPI_ISL_5661215, EPI_ISL_5662556 | UBS FRANCISCO DIAS DA SILVA | Instituto Butantan | Antonio Jorge Martins; Claudia Renata dos Santos Barros; David Schlesinger; Debora Botequio Moretti; Dimas Tadeu Covas; Elaine Cristina Marqueze; Elaine Vieira Santos; Evandra Strazza Rodrigues; Heidge Fukumasu; Jayme Augusto de Souza-Neto; José Salvatore Leister Patané; Luiz Alcantara; Luiz Lehmann Coutinho; Maria Carolina Elias; Maurício Lacerda Nogueira; Rafael dos Santos Bezerra; Raul Machado Neto; Rejane Maria Tommasini Grotto; Ricardo Haddad; Sandra Coccuzzo Sampaio Vessoni; Simone Kashima; Svetoslav Nanev Slavov; Vincent Louis Viala |
| EPI_ISL_5662219, EPI_ISL_5664223 | UBS GETULINO JOSE DIAS | Instituto Butantan | Antonio Jorge Martins; Claudia Renata dos Santos Barros; David Schlesinger; Debora Botequio Moretti; Dimas Tadeu Covas; Elaine Cristina Marqueze; Elaine Vieira Santos; Evandra Strazza Rodrigues; Heidge Fukumasu; Jayme Augusto de Souza-Neto; José Salvatore Leister Patané; Luiz Alcantara; Luiz Lehmann Coutinho; Maria Carolina Elias; Maurício Lacerda Nogueira; Rafael dos Santos Bezerra; Raul Machado Neto; Rejane Maria Tommasini Grotto; Ricardo Haddad; Sandra Coccuzzo Sampaio Vessoni; Simone Kashima; Svetoslav Nanev Slavov; Vincent Louis Viala |
| EPI_ISL_5662082 | UBS GUILHERMINA DE NOBREGA ABREU | Instituto Butantan | Antonio Jorge Martins; Claudia Renata dos Santos Barros; David Schlesinger; Debora Botequio Moretti; Dimas Tadeu Covas; Elaine Cristina Marqueze; Elaine Vieira Santos; Evandra Strazza Rodrigues; Heidge Fukumasu; Jayme Augusto de Souza-Neto; José Salvatore Leister Patané; Luiz Alcantara; Luiz Lehmann Coutinho; Maria Carolina Elias; Maurício Lacerda Nogueira; Rafael dos Santos Bezerra; Raul Machado Neto; Rejane Maria Tommasini Grotto; Ricardo Haddad; Sandra Coccuzzo Sampaio Vessoni; Simone Kashima; Svetoslav Nanev Slavov; Vincent Louis Viala |
| EPI_ISL_5661376 | UBS HELENA MARREY | Instituto Butantan | Antonio Jorge Martins; Claudia Renata dos Santos Barros; David Schlesinger; Debora Botequio Moretti; Dimas Tadeu Covas; Elaine Cristina Marqueze; Elaine Vieira Santos; Evandra Strazza Rodrigues; Heidge Fukumasu; Jayme Augusto de Souza-Neto; José Salvatore Leister Patané; Luiz Alcantara; Luiz Lehmann Coutinho; Maria Carolina Elias; Maurício Lacerda Nogueira; Rafael dos Santos Bezerra; Raul Machado Neto; Rejane Maria Tommasini Grotto; Ricardo Haddad; Sandra Coccuzzo Sampaio Vessoni; Simone Kashima; Svetoslav Nanev Slavov; Vincent Louis Viala |
| EPI_ISL_5665474, EPI_ISL_5665544 | UBS II DE NARANDIBA | Instituto Butantan | Antonio Jorge Martins; Claudia Renata dos Santos Barros; David Schlesinger; Debora Botequio Moretti; Dimas Tadeu Covas; Elaine Cristina Marqueze; Elaine Vieira Santos; Evandra Strazza Rodrigues; Heidge Fukumasu; Jayme Augusto de Souza-Neto; José Salvatore Leister Patané; Luiz Alcantara; Luiz Lehmann Coutinho; Maria Carolina Elias; Maurício Lacerda Nogueira; Rafael dos Santos Bezerra; Raul Machado Neto; Rejane Maria Tommasini Grotto; Ricardo Haddad; Sandra Coccuzzo Sampaio Vessoni; Simone Kashima; Svetoslav Nanev Slavov; Vincent Louis Viala |

| | | | |
|---|---|---|---|
| EPI_ISL_5663379, EPI_ISL_5664444, EPI_ISL_5665415 | UBS II DE PIRAPOZINHO C SERVICO DE EACS E ESF | Instituto Butantan | Antonio Jorge Martins; Claudia Renata dos Santos Barros; David Schlesinger; Debora Botequio Moretti; Dimas Tadeu Covas; Elaine Cristina Marqueze; Elaine Vieira Santos; Evandra Strazza Rodrigues; Heidge Fukumasu; Jayme Augusto de Souza-Neto; José Salvatore Leister Patané; Luiz Alcantara; Luiz Lehmann Coutinho; Maria Carolina Elias; Maurício Lacerda Nogueira; Rafael dos Santos Bezerra; Raul Machado Neto; Rejane Maria Tommasini Grotto; Ricardo Haddad; Sandra Coccuzzo Sampaio Vessoni; Simone Kashima; Svetoslav Nanev Slavov; Vincent Louis Viala |
| EPI_ISL_5663424, EPI_ISL_5663637, EPI_ISL_5664302 | UBS II DE TACIBA | Instituto Butantan | Antonio Jorge Martins; Claudia Renata dos Santos Barros; David Schlesinger; Debora Botequio Moretti; Dimas Tadeu Covas; Elaine Cristina Marqueze; Elaine Vieira Santos; Evandra Strazza Rodrigues; Heidge Fukumasu; Jayme Augusto de Souza-Neto; José Salvatore Leister Patané; Luiz Alcantara; Luiz Lehmann Coutinho; Maria Carolina Elias; Maurício Lacerda Nogueira; Rafael dos Santos Bezerra; Raul Machado Neto; Rejane Maria Tommasini Grotto; Ricardo Haddad; Sandra Coccuzzo Sampaio Vessoni; Simone Kashima; Svetoslav Nanev Slavov; Vincent Louis Viala |
| EPI_ISL_5663487, EPI_ISL_5663591, EPI_ISL_5663616, EPI_ISL_5663648, EPI_ISL_5663679, EPI_ISL_5663718, EPI_ISL_5664041, EPI_ISL_5665010, EPI_ISL_5665428 | | | |
| see above | UBS II DR EXPEDITO SHIZUO KUROCE | Instituto Butantan | Antonio Jorge Martins; Claudia Renata dos Santos Barros; David Schlesinger; Debora Botequio Moretti; Dimas Tadeu Covas; Elaine Cristina Marqueze; Elaine Vieira Santos; Evandra Strazza Rodrigues; Heidge Fukumasu; Jayme Augusto de Souza-Neto; José Salvatore Leister Patané; Luiz Alcantara; Luiz Lehmann Coutinho; Maria Carolina Elias; Maurício Lacerda Nogueira; Rafael dos Santos Bezerra; Raul Machado Neto; Rejane Maria Tommasini Grotto; Ricardo Haddad; Sandra Coccuzzo Sampaio Vessoni; Simone Kashima; Svetoslav Nanev Slavov; Vincent Louis Viala |
| EPI_ISL_5668688, EPI_ISL_5668727, EPI_ISL_5668948, EPI_ISL_5669522 | UBS III DE ELDORADO ELDORADO S P | Instituto Butantan | Antonio Jorge Martins; Claudia Renata dos Santos Barros; David Schlesinger; Debora Botequio Moretti; Dimas Tadeu Covas; Elaine Cristina Marqueze; Elaine Vieira Santos; Evandra Strazza Rodrigues; Heidge Fukumasu; Jayme Augusto de Souza-Neto; José Salvatore Leister Patané; Luiz Alcantara; Luiz Lehmann Coutinho; Maria Carolina Elias; Maurício Lacerda Nogueira; Rafael dos Santos Bezerra; Raul Machado Neto; Rejane Maria Tommasini Grotto; Ricardo Haddad; Sandra Coccuzzo Sampaio Vessoni; Simone Kashima; Svetoslav Nanev Slavov; Vincent Louis Viala |
| EPI_ISL_5663371 | UBS III DE EUCLIDES DA CUNHA PAULISTA | Instituto Butantan | Antonio Jorge Martins; Claudia Renata dos Santos Barros; David Schlesinger; Debora Botequio Moretti; Dimas Tadeu Covas; Elaine Cristina Marqueze; Elaine Vieira Santos; Evandra Strazza Rodrigues; Heidge Fukumasu; Jayme Augusto de Souza-Neto; José Salvatore Leister Patané; Luiz Alcantara; Luiz Lehmann Coutinho; Maria Carolina Elias; Maurício Lacerda Nogueira; Rafael dos Santos Bezerra; Raul Machado Neto; Rejane Maria Tommasini Grotto; Ricardo Haddad; Sandra Coccuzzo Sampaio Vessoni; Simone Kashima; Svetoslav Nanev Slavov; Vincent Louis Viala |
| EPI_ISL_5665434, EPI_ISL_5665450 | UBS III DE RANCHARIA | Instituto Butantan | Antonio Jorge Martins; Claudia Renata dos Santos Barros; David Schlesinger; Debora Botequio Moretti; Dimas Tadeu Covas; Elaine Cristina Marqueze; Elaine Vieira Santos; Evandra Strazza Rodrigues; Heidge Fukumasu; Jayme Augusto de Souza-Neto; José Salvatore Leister Patané; Luiz Alcantara; Luiz Lehmann Coutinho; Maria Carolina Elias; Maurício Lacerda Nogueira; Rafael dos Santos Bezerra; Raul Machado Neto; Rejane Maria Tommasini Grotto; Ricardo Haddad; Sandra Coccuzzo Sampaio Vessoni; Simone Kashima; Svetoslav Nanev Slavov; Vincent Louis Viala |
| EPI_ISL_5659730, EPI_ISL_5660093, EPI_ISL_5660301 | UBS JARDIM ALIANCA | Instituto Butantan | Antonio Jorge Martins; Claudia Renata dos Santos Barros; David Schlesinger; Debora Botequio Moretti; Dimas Tadeu Covas; Elaine Cristina Marqueze; Elaine Vieira Santos; Evandra Strazza Rodrigues; Heidge Fukumasu; Jayme Augusto de Souza-Neto; José Salvatore Leister Patané; Luiz Alcantara; Luiz Lehmann Coutinho; Maria Carolina Elias; Maurício Lacerda Nogueira; Rafael dos Santos Bezerra; Raul Machado Neto; Rejane Maria Tommasini Grotto; Ricardo Haddad; Sandra Coccuzzo Sampaio Vessoni; Simone Kashima; Svetoslav Nanev Slavov; Vincent Louis Viala |
| EPI_ISL_5663907 | UBS JARDIM FLOR DA MONTANHA | Instituto Butantan | Antonio Jorge Martins; Claudia Renata dos Santos Barros; David Schlesinger; Debora Botequio Moretti; Dimas Tadeu Covas; Elaine Cristina Marqueze; Elaine Vieira Santos; Evandra Strazza Rodrigues; Heidge Fukumasu; Jayme Augusto de Souza-Neto; José Salvatore Leister Patané; Luiz Alcantara; Luiz Lehmann Coutinho; Maria Carolina Elias; Maurício Lacerda Nogueira; Rafael dos Santos Bezerra; Raul Machado Neto; Rejane Maria Tommasini Grotto; Ricardo Haddad; Sandra Coccuzzo Sampaio Vessoni; Simone Kashima; Svetoslav Nanev Slavov; Vincent Louis Viala |
| EPI_ISL_5663558 | UBS JARDIM MUNHOZ | Instituto Butantan | Antonio Jorge Martins; Claudia Renata dos Santos Barros; David Schlesinger; Debora Botequio Moretti; Dimas Tadeu Covas; Elaine Cristina Marqueze; Elaine Vieira Santos; Evandra Strazza Rodrigues; Heidge Fukumasu; Jayme Augusto de Souza-Neto; José Salvatore Leister Patané; Luiz Alcantara; Luiz Lehmann Coutinho; Maria Carolina Elias; Maurício Lacerda Nogueira; Rafael dos Santos Bezerra; Raul Machado Neto; Rejane Maria Tommasini Grotto; Ricardo Haddad; Sandra Coccuzzo Sampaio Vessoni; Simone Kashima; Svetoslav Nanev Slavov; Vincent Louis Viala |
| EPI_ISL_5662706 | UBS JARDIM PALMIRA | Instituto Butantan | Antonio Jorge Martins; Claudia Renata dos Santos Barros; David Schlesinger; Debora Botequio Moretti; Dimas Tadeu Covas; Elaine Cristina Marqueze; Elaine Vieira Santos; Evandra Strazza Rodrigues; Heidge Fukumasu; Jayme Augusto de Souza-Neto; José Salvatore Leister Patané; Luiz Alcantara; Luiz Lehmann Coutinho; Maria Carolina Elias; Maurício Lacerda Nogueira; Rafael dos Santos Bezerra; Raul Machado Neto; Rejane Maria Tommasini Grotto; Ricardo Haddad; Sandra Coccuzzo Sampaio Vessoni; Simone Kashima; Svetoslav Nanev Slavov; Vincent Louis Viala |
| EPI_ISL_5663499 | UBS JARDIM ROSA DE FRANCA | Instituto Butantan | Antonio Jorge Martins; Claudia Renata dos Santos Barros; David Schlesinger; Debora Botequio Moretti; Dimas Tadeu Covas; Elaine Cristina Marqueze; Elaine Vieira Santos; Evandra Strazza Rodrigues; Heidge Fukumasu; Jayme Augusto de Souza-Neto; José Salvatore Leister Patané; Luiz Alcantara; Luiz Lehmann Coutinho; Maria Carolina Elias; Maurício Lacerda Nogueira; Rafael dos Santos Bezerra; Raul Machado Neto; Rejane Maria Tommasini Grotto; Ricardo Haddad; Sandra Coccuzzo Sampaio Vessoni; Simone Kashima; Svetoslav Nanev Slavov; Vincent Louis Viala |
| EPI_ISL_5664251 | UBS JARDIM TRANQUILIDADE | Instituto Butantan | Antonio Jorge Martins; Claudia Renata dos Santos Barros; David Schlesinger; Debora Botequio Moretti; Dimas Tadeu Covas; Elaine Cristina Marqueze; Elaine Vieira Santos; Evandra Strazza Rodrigues; Heidge Fukumasu; Jayme Augusto de Souza-Neto; José Salvatore Leister Patané; Luiz Alcantara; Luiz Lehmann Coutinho; Maria Carolina Elias; Maurício Lacerda Nogueira; Rafael dos Santos Bezerra; Raul Machado Neto; Rejane Maria Tommasini Grotto; Ricardo Haddad; Sandra Coccuzzo Sampaio Vessoni; Simone Kashima; Svetoslav Nanev Slavov; Vincent Louis Viala |
| EPI_ISL_5662210 | UBS JOSE GUIMARAES DE ABREU | Instituto Butantan | Antonio Jorge Martins; Claudia Renata dos Santos Barros; David Schlesinger; Debora Botequio Moretti; Dimas Tadeu Covas; Elaine Cristina Marqueze; Elaine Vieira Santos; Evandra Strazza Rodrigues; Heidge Fukumasu; Jayme Augusto de Souza-Neto; José Salvatore Leister Patané; Luiz Alcantara; Luiz Lehmann Coutinho; Maria Carolina Elias; Maurício Lacerda Nogueira; Rafael dos Santos Bezerra; Raul Machado Neto; Rejane Maria Tommasini Grotto; Ricardo Haddad; Sandra Coccuzzo Sampaio Vessoni; Simone Kashima; Svetoslav Nanev Slavov; Vincent Louis Viala |
| EPI_ISL_5661463 | UBS JOSE HILARIO DOS SANTOS | Instituto Butantan | Antonio Jorge Martins; Claudia Renata dos Santos Barros; David Schlesinger; Debora Botequio Moretti; Dimas Tadeu Covas; Elaine Cristina Marqueze; Elaine Vieira Santos; Evandra Strazza Rodrigues; Heidge Fukumasu; Jayme Augusto de Souza-Neto; José Salvatore Leister Patané; Luiz Alcantara; Luiz Lehmann Coutinho; Maria Carolina Elias; Maurício Lacerda Nogueira; Rafael dos Santos Bezerra; Raul Machado Neto; Rejane Maria Tommasini Grotto; Ricardo Haddad; Sandra Coccuzzo Sampaio Vessoni; Simone Kashima; Svetoslav Nanev Slavov; Vincent Louis Viala |
| EPI_ISL_5661877 | UBS JOSE SABINO FERREIRA | Instituto Butantan | Antonio Jorge Martins; Claudia Renata dos Santos Barros; David Schlesinger; Debora Botequio Moretti; Dimas Tadeu Covas; Elaine Cristina Marqueze; Elaine Vieira Santos; Evandra Strazza Rodrigues; Heidge Fukumasu; Jayme Augusto de Souza-Neto; José Salvatore Leister Patané; Luiz Alcantara; Luiz Lehmann Coutinho; Maria Carolina Elias; Maurício Lacerda Nogueira; Rafael dos Santos Bezerra; Raul Machado Neto; Rejane Maria Tommasini Grotto; Ricardo Haddad; Sandra Coccuzzo Sampaio Vessoni; Simone Kashima; Svetoslav Nanev Slavov; Vincent Louis Viala |
| EPI_ISL_5661799, EPI_ISL_5664485 | UBS JOVAIA | Instituto Butantan | Antonio Jorge Martins; Claudia Renata dos Santos Barros; David Schlesinger; Debora Botequio Moretti; Dimas Tadeu Covas; Elaine Cristina Marqueze; Elaine Vieira Santos; Evandra Strazza Rodrigues; Heidge Fukumasu; Jayme Augusto de Souza-Neto; José Salvatore Leister Patané; Luiz Alcantara; Luiz Lehmann Coutinho; Maria Carolina Elias; Maurício Lacerda Nogueira; Rafael dos Santos Bezerra; Raul Machado Neto; Rejane Maria Tommasini Grotto; Ricardo Haddad; Sandra Coccuzzo Sampaio Vessoni; Simone Kashima; Svetoslav Nanev Slavov; Vincent Louis Viala |
| EPI_ISL_5664207 | UBS JUREMA | Instituto Butantan | Antonio Jorge Martins; Claudia Renata dos Santos Barros; David Schlesinger; Debora Botequio Moretti; Dimas Tadeu Covas; Elaine Cristina Marqueze; Elaine Vieira Santos; Evandra Strazza Rodrigues; Heidge Fukumasu; Jayme Augusto de Souza-Neto; José Salvatore Leister Patané; Luiz Alcantara; Luiz Lehmann Coutinho; Maria Carolina Elias; Maurício Lacerda Nogueira; Rafael dos Santos Bezerra; Raul Machado Neto; Rejane Maria Tommasini Grotto; Ricardo Haddad; Sandra Coccuzzo Sampaio Vessoni; Simone Kashima; Svetoslav Nanev Slavov; Vincent Louis Viala |
| EPI_ISL_5662408, EPI_ISL_5662468, EPI_ISL_5662575, EPI_ISL_5662729, EPI_ISL_5663174, EPI_ISL_5664574 | UBS LIA BUARQUE DE MACEDO GASPARINI | Instituto Butantan | Antonio Jorge Martins; Claudia Renata dos Santos Barros; David Schlesinger; Debora Botequio Moretti; Dimas Tadeu Covas; Elaine Cristina Marqueze; Elaine Vieira Santos; Evandra Strazza Rodrigues; Heidge Fukumasu; Jayme Augusto de Souza-Neto; José Salvatore Leister Patané; Luiz Alcantara; Luiz Lehmann Coutinho; Maria Carolina Elias; Maurício Lacerda Nogueira; Rafael dos Santos Bezerra; Raul Machado Neto; Rejane Maria Tommasini Grotto; Ricardo Haddad; Sandra Coccuzzo Sampaio Vessoni; Simone Kashima; Svetoslav Nanev Slavov; Vincent Louis Viala |
| EPI_ISL_5661032, EPI_ISL_5661253 | UBS LUCIANO RODRIGUES COSTA | Instituto Butantan | Antonio Jorge Martins; Claudia Renata dos Santos Barros; David Schlesinger; Debora Botequio Moretti; Dimas Tadeu Covas; Elaine Cristina Marqueze; Elaine Vieira Santos; Evandra Strazza Rodrigues; Heidge Fukumasu; Jayme Augusto de Souza-Neto; José Salvatore Leister Patané; Luiz Alcantara; Luiz Lehmann Coutinho; Maria Carolina Elias; Maurício Lacerda Nogueira; Rafael dos Santos Bezerra; Raul Machado Neto; Rejane Maria Tommasini Grotto; Ricardo Haddad; Sandra Coccuzzo Sampaio Vessoni; Simone Kashima; Svetoslav Nanev Slavov; Vincent Louis Viala |
| EPI_ISL_5663075 | UBS MARIA GIRADE CURY | Instituto Butantan | Antonio Jorge Martins; Claudia Renata dos Santos Barros; David Schlesinger; Debora Botequio Moretti; Dimas Tadeu Covas; Elaine Cristina Marqueze; Elaine Vieira Santos; Evandra Strazza Rodrigues; Heidge Fukumasu; Jayme Augusto de Souza-Neto; José Salvatore Leister Patané; Luiz Alcantara; Luiz Lehmann Coutinho; Maria Carolina Elias; Maurício Lacerda Nogueira; Rafael dos Santos Bezerra; Raul Machado Neto; Rejane Maria Tommasini Grotto; Ricardo Haddad; Sandra Coccuzzo Sampaio Vessoni; Simone Kashima; Svetoslav Nanev Slavov; Vincent Louis Viala |
| EPI_ISL_5662610 | UBS MARIA PIA DE OLIVEIRA | Instituto Butantan | Antonio Jorge Martins; Claudia Renata dos Santos Barros; David Schlesinger; Debora Botequio Moretti; Dimas Tadeu Covas; Elaine Cristina Marqueze; Elaine Vieira Santos; Evandra Strazza Rodrigues; Heidge Fukumasu; Jayme Augusto de Souza-Neto; José Salvatore Leister Patané; Luiz Alcantara; Luiz Lehmann Coutinho; Maria Carolina Elias; Maurício Lacerda Nogueira; Rafael dos Santos Bezerra; Raul Machado Neto; Rejane Maria Tommasini Grotto; Ricardo Haddad; Sandra Coccuzzo Sampaio Vessoni; Simone Kashima; Svetoslav Nanev Slavov; Vincent Louis Viala |
| EPI_ISL_5659880 | UBS MARTINHO PRADO | Instituto Butantan | Antonio Jorge Martins; Claudia Renata dos Santos Barros; David Schlesinger; Debora Botequio Moretti; Dimas Tadeu Covas; Elaine Cristina Marqueze; Elaine Vieira Santos; Evandra Strazza Rodrigues; Heidge Fukumasu; Jayme Augusto de Souza-Neto; José Salvatore Leister Patané; Luiz Alcantara; Luiz Lehmann Coutinho; Maria Carolina Elias; Maurício Lacerda Nogueira; Rafael dos Santos Bezerra; Raul Machado Neto; Rejane Maria Tommasini Grotto; Ricardo Haddad; Sandra Coccuzzo Sampaio Vessoni; Simone Kashima; Svetoslav Nanev Slavov; Vincent Louis Viala |
| EPI_ISL_5663773 | UBS ODUVALDO MAGLIO | Instituto Butantan | Antonio Jorge Martins; Claudia Renata dos Santos Barros; David Schlesinger; Debora Botequio Moretti; Dimas Tadeu Covas; Elaine Cristina Marqueze; Elaine Vieira Santos; Evandra Strazza Rodrigues; Heidge Fukumasu; Jayme Augusto de Souza-Neto; José Salvatore Leister Patané; Luiz Alcantara; Luiz Lehmann Coutinho; Maria Carolina Elias; Maurício Lacerda Nogueira; Rafael dos Santos Bezerra; Raul Machado Neto; Rejane Maria Tommasini Grotto; Ricardo Haddad; Sandra Coccuzzo Sampaio Vessoni; Simone Kashima; Svetoslav Nanev Slavov; Vincent Louis Viala |
| EPI_ISL_5661552, EPI_ISL_5663402, EPI_ISL_5663651 | UBS OTACILIO FIRMINO LOPES | Instituto Butantan | Antonio Jorge Martins; Claudia Renata dos Santos Barros; David Schlesinger; Debora Botequio Moretti; Dimas Tadeu Covas; Elaine Cristina Marqueze; Elaine Vieira Santos; Evandra Strazza Rodrigues; Heidge Fukumasu; Jayme Augusto de Souza-Neto; José Salvatore Leister Patané; Luiz Alcantara; Luiz Lehmann Coutinho; Maria Carolina Elias; Maurício Lacerda Nogueira; Rafael dos Santos Bezerra; Raul Machado Neto; Rejane Maria Tommasini Grotto; Ricardo Haddad; Sandra Coccuzzo Sampaio Vessoni; Simone Kashima; Svetoslav Nanev Slavov; Vincent Louis Viala |
| EPI_ISL_5666244 | UBS PARQUE MINAS GERAIS | Instituto Butantan | Antonio Jorge Martins; Claudia Renata dos Santos Barros; David Schlesinger; Debora Botequio Moretti; Dimas Tadeu Covas; Elaine Cristina Marqueze; Elaine Vieira Santos; Evandra Strazza Rodrigues; Heidge Fukumasu; Jayme Augusto de Souza-Neto; José Salvatore Leister Patané; Luiz Alcantara; Luiz Lehmann Coutinho; Maria Carolina Elias; Maurício Lacerda Nogueira; Rafael dos Santos Bezerra; Raul Machado Neto; Rejane Maria Tommasini Grotto; Ricardo Haddad; Sandra Coccuzzo Sampaio Vessoni; Simone Kashima; Svetoslav Nanev Slavov; Vincent Louis Viala |
| EPI_ISL_5661618 | UBS PARQUE PALMARES | Instituto Butantan | Antonio Jorge Martins; Claudia Renata dos Santos Barros; David Schlesinger; Debora Botequio Moretti; Dimas Tadeu Covas; Elaine Cristina Marqueze; Elaine Vieira Santos; Evandra Strazza Rodrigues; Heidge Fukumasu; Jayme Augusto de Souza-Neto; José Salvatore Leister Patané; Luiz Alcantara; Luiz Lehmann Coutinho; Maria Carolina Elias; Maurício Lacerda Nogueira; Rafael dos Santos Bezerra; Raul Machado Neto; Rejane Maria Tommasini Grotto; Ricardo Haddad; Sandra Coccuzzo Sampaio Vessoni; Simone Kashima; Svetoslav Nanev Slavov; Vincent Louis Viala |
| EPI_ISL_5667318, EPI_ISL_5667387, EPI_ISL_5667708, EPI_ISL_5667783 | UBS PASTOR MARCELINO DEUNGARO RIO GRANDE | Instituto Butantan | Antonio Jorge Martins; Claudia Renata dos Santos Barros; David Schlesinger; Debora Botequio Moretti; Dimas Tadeu Covas; Elaine Cristina Marqueze; Elaine Vieira Santos; Evandra Strazza Rodrigues; Heidge Fukumasu; Jayme Augusto de Souza-Neto; José Salvatore Leister Patané; Luiz Alcantara; Luiz Lehmann Coutinho; Maria Carolina Elias; Maurício Lacerda Nogueira; Rafael dos Santos Bezerra; Raul Machado Neto; Rejane Maria Tommasini Grotto; Ricardo Haddad; Sandra Coccuzzo Sampaio Vessoni; Simone Kashima; Svetoslav Nanev Slavov; Vincent Louis Viala |
| EPI_ISL_5660622, EPI_ISL_5663726 | UBS PE GUERRINO RICIOTTI | Instituto Butantan | Antonio Jorge Martins; Claudia Renata dos Santos Barros; David Schlesinger; Debora Botequio Moretti; Dimas Tadeu Covas; Elaine Cristina Marqueze; Elaine Vieira Santos; Evandra Strazza Rodrigues; Heidge Fukumasu; Jayme Augusto de Souza-Neto; José Salvatore Leister Patané; Luiz Alcantara; Luiz Lehmann Coutinho; Maria Carolina Elias; Maurício Lacerda Nogueira; Rafael dos Santos Bezerra; Raul Machado Neto; Rejane Maria Tommasini Grotto; Ricardo Haddad; Sandra Coccuzzo Sampaio Vessoni; Simone Kashima; Svetoslav Nanev Slavov; Vincent Louis Viala |
| EPI_ISL_5666127, EPI_ISL_5666131 | UBS REGIAO OESTE DE OURINHOS | Instituto Butantan | Antonio Jorge Martins; Claudia Renata dos Santos Barros; David Schlesinger; Debora Botequio Moretti; Dimas Tadeu Covas; Elaine Cristina Marqueze; Elaine Vieira Santos; Evandra Strazza Rodrigues; Heidge Fukumasu; Jayme Augusto de Souza-Neto; José Salvatore Leister Patané; Luiz Alcantara; Luiz Lehmann Coutinho; Maria Carolina Elias; Maurício Lacerda Nogueira; Rafael dos Santos Bezerra; Raul Machado Neto; Rejane Maria Tommasini Grotto; Ricardo Haddad; Sandra Coccuzzo Sampaio Vessoni; Simone Kashima; Svetoslav Nanev Slavov; Vincent Louis Viala |
| EPI_ISL_5668262, EPI_ISL_5668354 | UBS RESSACA | Instituto Butantan | Antonio Jorge Martins; Claudia Renata dos Santos Barros; David Schlesinger; Debora Botequio Moretti; Dimas Tadeu Covas; Elaine Cristina Marqueze; Elaine Vieira Santos; Evandra Strazza Rodrigues; Heidge Fukumasu; Jayme Augusto de Souza-Neto; José Salvatore Leister Patané; Luiz Alcantara; Luiz Lehmann Coutinho; Maria Carolina Elias; Maurício Lacerda Nogueira; Rafael dos Santos Bezerra; Raul Machado Neto; Rejane Maria Tommasini Grotto; Ricardo Haddad; Sandra Coccuzzo Sampaio Vessoni; Simone Kashima; Svetoslav Nanev Slavov; Vincent Louis Viala |
| EPI_ISL_5666809 | UBS RIBEIRAO CORRENTE | Instituto Butantan | Antonio Jorge Martins; Claudia Renata dos Santos Barros; David Schlesinger; Debora Botequio Moretti; Dimas Tadeu Covas; Elaine Cristina Marqueze; Elaine Vieira Santos; Evandra Strazza Rodrigues; Heidge Fukumasu; Jayme Augusto de Souza-Neto; José Salvatore Leister Patané; Luiz Alcantara; Luiz Lehmann Coutinho; Maria Carolina Elias; Maurício Lacerda Nogueira; Rafael dos Santos Bezerra; Raul Machado Neto; Rejane Maria Tommasini Grotto; Ricardo Haddad; Sandra Coccuzzo Sampaio Vessoni; Simone Kashima; Svetoslav Nanev Slavov; Vincent Louis Viala |
| EPI_ISL_5663687 | UBS SANTA GEMA GALGANI | Instituto Butantan | Antonio Jorge Martins; Claudia Renata dos Santos Barros; David Schlesinger; Debora Botequio Moretti; Dimas Tadeu Covas; Elaine Cristina Marqueze; Elaine Vieira Santos; Evandra Strazza Rodrigues; Heidge Fukumasu; Jayme Augusto de Souza-Neto; José Salvatore Leister Patané; Luiz Alcantara; Luiz Lehmann Coutinho; Maria Carolina Elias; Maurício Lacerda Nogueira; Rafael dos Santos Bezerra; Raul Machado Neto; Rejane Maria Tommasini Grotto; Ricardo Haddad; Sandra Coccuzzo Sampaio Vessoni; Simone Kashima; Svetoslav Nanev Slavov; Vincent Louis Viala |
| EPI_ISL_5662524 | UBS SANTA MARIA | Instituto Butantan | Antonio Jorge Martins; Claudia Renata dos Santos Barros; David Schlesinger; Debora Botequio Moretti; Dimas Tadeu Covas; Elaine Cristina Marqueze; Elaine Vieira Santos; Evandra Strazza Rodrigues; Heidge Fukumasu; Jayme Augusto de Souza-Neto; José Salvatore Leister Patané; Luiz Alcantara; Luiz Lehmann Coutinho; Maria Carolina Elias; Maurício Lacerda Nogueira; Rafael dos Santos Bezerra; Raul Machado Neto; Rejane Maria Tommasini Grotto; Ricardo Haddad; Sandra Coccuzzo Sampaio Vessoni; Simone Kashima; Svetoslav Nanev Slavov; Vincent Louis Viala |
| EPI_ISL_5662350, EPI_ISL_5663137 | UBS SANTA MARIA GORETTI | Instituto Butantan | Antonio Jorge Martins; Claudia Renata dos Santos Barros; David Schlesinger; Debora Botequio Moretti; Dimas Tadeu Covas; Elaine Cristina Marqueze; Elaine Vieira Santos; Evandra Strazza Rodrigues; Heidge Fukumasu; Jayme Augusto de Souza-Neto; José Salvatore Leister Patané; Luiz Alcantara; Luiz Lehmann Coutinho; Maria Carolina Elias; Maurício Lacerda Nogueira; Rafael dos Santos Bezerra; Raul Machado Neto; Rejane Maria Tommasini Grotto; Ricardo Haddad; Sandra Coccuzzo Sampaio Vessoni; Simone Kashima; Svetoslav Nanev Slavov; Vincent Louis Viala |
| EPI_ISL_5663859, EPI_ISL_5663984 | UBS SOIMCO | Instituto Butantan | Antonio Jorge Martins; Claudia Renata dos Santos Barros; David Schlesinger; Debora Botequio Moretti; Dimas Tadeu Covas; Elaine Cristina Marqueze; Elaine Vieira Santos; Evandra Strazza Rodrigues; Heidge Fukumasu; Jayme Augusto de Souza-Neto; José Salvatore Leister Patané; Luiz Alcantara; Luiz Lehmann Coutinho; Maria Carolina Elias; Maurício Lacerda Nogueira; Rafael dos Santos Bezerra; Raul Machado Neto; Rejane Maria Tommasini Grotto; Ricardo Haddad; Sandra Coccuzzo Sampaio Vessoni; Simone Kashima; Svetoslav Nanev Slavov; Vincent Louis Viala |
| EPI_ISL_5661634 | UBS SYLVIO JOAO L DE LUCIA | Instituto Butantan | Antonio Jorge Martins; Claudia Renata dos Santos Barros; David Schlesinger; Debora Botequio Moretti; Dimas Tadeu Covas; Elaine Cristina Marqueze; Elaine Vieira Santos; Evandra Strazza Rodrigues; Heidge Fukumasu; Jayme Augusto de Souza-Neto; José Salvatore Leister Patané; Luiz Alcantara; Luiz Lehmann Coutinho; Maria Carolina Elias; Maurício Lacerda Nogueira; Rafael dos Santos Bezerra; Raul Machado Neto; Rejane Maria Tommasini Grotto; Ricardo Haddad; Sandra Coccuzzo Sampaio Vessoni; Simone Kashima; Svetoslav Nanev Slavov; Vincent Louis Viala |
| EPI_ISL_5667356, EPI_ISL_5667534, EPI_ISL_5667682 | UBS TEREZA GALLO UCHOA | Instituto Butantan | Antonio Jorge Martins; Claudia Renata dos Santos Barros; David Schlesinger; Debora Botequio Moretti; Dimas Tadeu Covas; Elaine Cristina Marqueze; Elaine Vieira Santos; Evandra Strazza Rodrigues; Heidge Fukumasu; Jayme Augusto de Souza-Neto; José Salvatore Leister Patané; Luiz Alcantara; Luiz Lehmann Coutinho; Maria Carolina Elias; Maurício Lacerda Nogueira; Rafael dos Santos Bezerra; Raul Machado Neto; Rejane Maria Tommasini Grotto; Ricardo Haddad; Sandra Coccuzzo Sampaio Vessoni; Simone Kashima; Svetoslav Nanev Slavov; Vincent Louis Viala |
| EPI_ISL_5662087 | UBS VALDIR ALVES PEREIRA | Instituto Butantan | Antonio Jorge Martins; Claudia Renata dos Santos Barros; David Schlesinger; Debora Botequio Moretti; Dimas Tadeu Covas; Elaine Cristina Marqueze; Elaine Vieira Santos; Evandra Strazza Rodrigues; Heidge Fukumasu; Jayme Augusto de Souza-Neto; José Salvatore Leister Patané; Luiz Alcantara; Luiz Lehmann Coutinho; Maria Carolina Elias; Maurício Lacerda Nogueira; Rafael dos Santos Bezerra; Raul Machado Neto; Rejane Maria Tommasini Grotto; Ricardo Haddad; Sandra Coccuzzo Sampaio Vessoni; Simone Kashima; Svetoslav Nanev Slavov; Vincent Louis Viala |
| EPI_ISL_5661754 | UBS VASCO DA ROCHA LEAO | Instituto Butantan | Antonio Jorge Martins; Claudia Renata dos Santos Barros; David Schlesinger; Debora Botequio Moretti; Dimas Tadeu Covas; Elaine Cristina Marqueze; Elaine Vieira Santos; Evandra Strazza Rodrigues; Heidge Fukumasu; Jayme Augusto de Souza-Neto; José Salvatore Leister Patané; Luiz Alcantara; Luiz Lehmann Coutinho; Maria Carolina Elias; Maurício Lacerda Nogueira; Rafael dos Santos Bezerra; Raul Machado Neto; Rejane Maria Tommasini Grotto; Ricardo Haddad; Sandra Coccuzzo Sampaio Vessoni; Simone Kashima; Svetoslav Nanev Slavov; Vincent Louis Viala |
| EPI_ISL_5662722, | UBS VILA BRASIL | Instituto Butantan | Antonio Jorge Martins; Claudia Renata dos Santos Barros; David Schlesinger; Debora Botequio Moretti; Dimas Tadeu Covas; Elaine Cristina Marqueze; Elaine Vieira Santos; Evandra Strazza Rodrigues; Heidge Fukumasu; Jayme Augusto de Souza-Neto; José Salvatore Leister Patané; Luiz Alcantara; Luiz Lehmann |

| | | | |
|---|---|---|---|
| EPI_ISL_5666143 | | | Coutinho; Maria Carolina Elias; Maurício Lacerda Nogueira; Rafael dos Santos Bezerra; Raul Machado Neto; Rejane Maria Tommasini Grotto; Ricardo Haddad; Sandra Coccuzzo Sampaio Vessoni; Simone Kashima; Svetoslav Nanev Slavov; Vincent Louis Viala |
| EPI_ISL_5664460 | UBS VILA CARMELA | Instituto Butantan | Antonio Jorge Martins; Claudia Renata dos Santos Barros; David Schlesinger; Debora Botequio Moretti; Dimas Tadeu Covas; Elaine Cristina Marqueze; Elaine Vieira Santos; Evandra Strazza Rodrigues; Heidge Fukumasu; Jayme Augusto de Souza-Neto; José Salvatore Leister Patané; Luiz Alcantara; Luiz Lehmann Coutinho; Maria Carolina Elias; Maurício Lacerda Nogueira; Rafael dos Santos Bezerra; Raul Machado Neto; Rejane Maria Tommasini Grotto; Ricardo Haddad; Sandra Coccuzzo Sampaio Vessoni; Simone Kashima; Svetoslav Nanev Slavov; Vincent Louis Viala |
| EPI_ISL_5661312, EPI_ISL_5661409 | UBS VILA RIBEIRO | Instituto Butantan | Antonio Jorge Martins; Claudia Renata dos Santos Barros; David Schlesinger; Debora Botequio Moretti; Dimas Tadeu Covas; Elaine Cristina Marqueze; Elaine Vieira Santos; Evandra Strazza Rodrigues; Heidge Fukumasu; Jayme Augusto de Souza-Neto; José Salvatore Leister Patané; Luiz Alcantara; Luiz Lehmann Coutinho; Maria Carolina Elias; Maurício Lacerda Nogueira; Rafael dos Santos Bezerra; Raul Machado Neto; Rejane Maria Tommasini Grotto; Ricardo Haddad; Sandra Coccuzzo Sampaio Vessoni; Simone Kashima; Svetoslav Nanev Slavov; Vincent Louis Viala |
| EPI_ISL_5664175 | UBS VILA RIO DE JANEIRO | Instituto Butantan | Antonio Jorge Martins; Claudia Renata dos Santos Barros; David Schlesinger; Debora Botequio Moretti; Dimas Tadeu Covas; Elaine Cristina Marqueze; Elaine Vieira Santos; Evandra Strazza Rodrigues; Heidge Fukumasu; Jayme Augusto de Souza-Neto; José Salvatore Leister Patané; Luiz Alcantara; Luiz Lehmann Coutinho; Maria Carolina Elias; Maurício Lacerda Nogueira; Rafael dos Santos Bezerra; Raul Machado Neto; Rejane Maria Tommasini Grotto; Ricardo Haddad; Sandra Coccuzzo Sampaio Vessoni; Simone Kashima; Svetoslav Nanev Slavov; Vincent Louis Viala |
| EPI_ISL_3419663 | UC Davis Genome Center and Healthy Davis Together | UC Davis Genome Center | UC Davis |
| EPI_ISL_3067002, EPI_ISL_3067008, EPI_ISL_3067012, EPI_ISL_3067038, EPI_ISL_3321103, EPI_ISL_3321292, EPI_ISL_3370428 | | | |
| see above | UC-Christus Clinical Hospital Laboratory | Laboratory of Molecular Virology, School of Medicine, Pontificia Universidad Catolica de Chile | Ana Maira Guzman; Andres E. Munoz-Marcos; Catalina Pardo-Roa; Eileen Serrano; Erick Salinas; Estefany Poblete; Francisco Melo; Jorge Levican; Leonardo I. Almonacid; Maria Jose Avendano; Maria Patricia Vega; Rafael A. Medina; Ricardo Enrique de la Barra; Tamara Garcia-Salum |
| EPI_ISL_3104705, EPI_ISL_3104709 | UCC (Universidad Central del Caribe) | Grubaugh Lab - Yale School of Public Health | Alejandaro Vallejo Degaudenzi; Anderson Brito; Annie Watkins; Chaney Kalinich; Chantal Vogels; Elisa Contreras; Esperanza Mendoza; Isabel Ott; Jessica Rothman; Joseph Fauver; Kendall Billig; Mallery Breban; Mary Petrone; Nathan Grubaugh; Robert Paulino-Ramirez; Tara Alpert; Tobias Koch; Victor Virgilio Calderon |
| EPI_ISL_3462088 | UHTL, University Hospitals | UHTL, University Hospitals | Alouani, D.; Sadri, N.; Song, X. |
| EPI_ISL_5666725, EPI_ISL_5667050, EPI_ISL_5667090 | UNID PROGRAMA SAUDE FAMILIA BACURITI VILA SIMOES CAFELANDIA | Instituto Butantan | Antonio Jorge Martins; Claudia Renata dos Santos Barros; David Schlesinger; Debora Botequio Moretti; Dimas Tadeu Covas; Elaine Cristina Marqueze; Elaine Vieira Santos; Evandra Strazza Rodrigues; Heidge Fukumasu; Jayme Augusto de Souza-Neto; José Salvatore Leister Patané; Luiz Alcantara; Luiz Lehmann Coutinho; Maria Carolina Elias; Maurício Lacerda Nogueira; Rafael dos Santos Bezerra; Raul Machado Neto; Rejane Maria Tommasini Grotto; Ricardo Haddad; Sandra Coccuzzo Sampaio Vessoni; Simone Kashima; Svetoslav Nanev Slavov; Vincent Louis Viala |
| EPI_ISL_5659661, EPI_ISL_5659823, EPI_ISL_5660197, EPI_ISL_5660515, EPI_ISL_5660581 | UNIDADE BASICA DE SAUDE DE POTIM | Instituto Butantan | Antonio Jorge Martins; Claudia Renata dos Santos Barros; David Schlesinger; Debora Botequio Moretti; Dimas Tadeu Covas; Elaine Cristina Marqueze; Elaine Vieira Santos; Evandra Strazza Rodrigues; Heidge Fukumasu; Jayme Augusto de Souza-Neto; José Salvatore Leister Patané; Luiz Alcantara; Luiz Lehmann Coutinho; Maria Carolina Elias; Maurício Lacerda Nogueira; Rafael dos Santos Bezerra; Raul Machado Neto; Rejane Maria Tommasini Grotto; Ricardo Haddad; Sandra Coccuzzo Sampaio Vessoni; Simone Kashima; Svetoslav Nanev Slavov; Vincent Louis Viala |
| EPI_ISL_5661692 | UNIDADE BASICA DE SAUDE DE TARUMA | Instituto Butantan | Antonio Jorge Martins; Claudia Renata dos Santos Barros; David Schlesinger; Debora Botequio Moretti; Dimas Tadeu Covas; Elaine Cristina Marqueze; Elaine Vieira Santos; Evandra Strazza Rodrigues; Heidge Fukumasu; Jayme Augusto de Souza-Neto; José Salvatore Leister Patané; Luiz Alcantara; Luiz Lehmann Coutinho; Maria Carolina Elias; Maurício Lacerda Nogueira; Rafael dos Santos Bezerra; Raul Machado Neto; Rejane Maria Tommasini Grotto; Ricardo Haddad; Sandra Coccuzzo Sampaio Vessoni; Simone Kashima; Svetoslav Nanev Slavov; Vincent Louis Viala |
| EPI_ISL_5671181 | UNIDADE BASICA DE SAUDE DR MATHEUS GABRIEL BONASSA | Instituto Butantan | Antonio Jorge Martins; Claudia Renata dos Santos Barros; David Schlesinger; Debora Botequio Moretti; Dimas Tadeu Covas; Elaine Cristina Marqueze; Elaine Vieira Santos; Evandra Strazza Rodrigues; Heidge Fukumasu; Jayme Augusto de Souza-Neto; José Salvatore Leister Patané; Luiz Alcantara; Luiz Lehmann Coutinho; Maria Carolina Elias; Maurício Lacerda Nogueira; Rafael dos Santos Bezerra; Raul Machado Neto; Rejane Maria Tommasini Grotto; Ricardo Haddad; Sandra Coccuzzo Sampaio Vessoni; Simone Kashima; Svetoslav Nanev Slavov; Vincent Louis Viala |
| EPI_ISL_5663899 | UNIDADE BASICA DE SAUDE II DE INDIANA | Instituto Butantan | Antonio Jorge Martins; Claudia Renata dos Santos Barros; David Schlesinger; Debora Botequio Moretti; Dimas Tadeu Covas; Elaine Cristina Marqueze; Elaine Vieira Santos; Evandra Strazza Rodrigues; Heidge Fukumasu; Jayme Augusto de Souza-Neto; José Salvatore Leister Patané; Luiz Alcantara; Luiz Lehmann Coutinho; Maria Carolina Elias; Maurício Lacerda Nogueira; Rafael dos Santos Bezerra; Raul Machado Neto; Rejane Maria Tommasini Grotto; Ricardo Haddad; Sandra Coccuzzo Sampaio Vessoni; Simone Kashima; Svetoslav Nanev Slavov; Vincent Louis Viala |
| EPI_ISL_5663419, EPI_ISL_5664642, EPI_ISL_5664943 | UNIDADE BASICA DE SAUDE II ESTRELA DO NORTE | Instituto Butantan | Antonio Jorge Martins; Claudia Renata dos Santos Barros; David Schlesinger; Debora Botequio Moretti; Dimas Tadeu Covas; Elaine Cristina Marqueze; Elaine Vieira Santos; Evandra Strazza Rodrigues; Heidge Fukumasu; Jayme Augusto de Souza-Neto; José Salvatore Leister Patané; Luiz Alcantara; Luiz Lehmann Coutinho; Maria Carolina Elias; Maurício Lacerda Nogueira; Rafael dos Santos Bezerra; Raul Machado Neto; Rejane Maria Tommasini Grotto; Ricardo Haddad; Sandra Coccuzzo Sampaio Vessoni; Simone Kashima; Svetoslav Nanev Slavov; Vincent Louis Viala |
| EPI_ISL_5665490, EPI_ISL_5665549 | UNIDADE BASICA DE SAUDE III DE ALFREDO MARCONDES | Instituto Butantan | Antonio Jorge Martins; Claudia Renata dos Santos Barros; David Schlesinger; Debora Botequio Moretti; Dimas Tadeu Covas; Elaine Cristina Marqueze; Elaine Vieira Santos; Evandra Strazza Rodrigues; Heidge Fukumasu; Jayme Augusto de Souza-Neto; José Salvatore Leister Patané; Luiz Alcantara; Luiz Lehmann Coutinho; Maria Carolina Elias; Maurício Lacerda Nogueira; Rafael dos Santos Bezerra; Raul Machado Neto; Rejane Maria Tommasini Grotto; Ricardo Haddad; Sandra Coccuzzo Sampaio Vessoni; Simone Kashima; Svetoslav Nanev Slavov; Vincent Louis Viala |
| EPI_ISL_5659656, EPI_ISL_5661059, EPI_ISL_5661108 | UNIDADE BASICA DE SAUDE PREFEITO JOSE OLIVEIRA DE SOUZA | Instituto Butantan | Antonio Jorge Martins; Claudia Renata dos Santos Barros; David Schlesinger; Debora Botequio Moretti; Dimas Tadeu Covas; Elaine Cristina Marqueze; Elaine Vieira Santos; Evandra Strazza Rodrigues; Heidge Fukumasu; Jayme Augusto de Souza-Neto; José Salvatore Leister Patané; Luiz Alcantara; Luiz Lehmann Coutinho; Maria Carolina Elias; Maurício Lacerda Nogueira; Rafael dos Santos Bezerra; Raul Machado Neto; Rejane Maria Tommasini Grotto; Ricardo Haddad; Sandra Coccuzzo Sampaio Vessoni; Simone Kashima; Svetoslav Nanev Slavov; Vincent Louis Viala |
| EPI_ISL_5660676 | UNIDADE DE PRONTO ATENDIMENTO CENTRAL DE CARAGUATATUBA | Instituto Butantan | Antonio Jorge Martins; Claudia Renata dos Santos Barros; David Schlesinger; Debora Botequio Moretti; Dimas Tadeu Covas; Elaine Cristina Marqueze; Elaine Vieira Santos; Evandra Strazza Rodrigues; Heidge Fukumasu; Jayme Augusto de Souza-Neto; José Salvatore Leister Patané; Luiz Alcantara; Luiz Lehmann Coutinho; Maria Carolina Elias; Maurício Lacerda Nogueira; Rafael dos Santos Bezerra; Raul Machado Neto; Rejane Maria Tommasini Grotto; Ricardo Haddad; Sandra Coccuzzo Sampaio Vessoni; Simone Kashima; Svetoslav Nanev Slavov; Vincent Louis Viala |
| EPI_ISL_3912311 | UNIDADE DE PRONTO ATENDIMENTO DA JUREMA | Analytical Competence Molecular Epidemiology Lab/ACME, Oswaldo Cruz Foundation, Ceara (FIOCRUZ CE) | Cleber Furtado Aksenen; Fabio Miyajima; Fernando Braga Stehling; Francisco Eder de Moura Lopes; Jamille Maria Mendes Bezerra; Joaquim Cesar do Nascimento Sousa Junior; Pedro Miguel Carneiro Jeronimo; Suzana Porto Almeida & Lucas Delerino on behalf of COVID-19 FIOCRUZ Genomic Network; Thais Ferreira de Oliveira; Thais de Oliveira Costa; Ticiane Cavalcante de Souza; Veridiana Pessoa Miyajima |
| EPI_ISL_5672017, EPI_ISL_5672021, EPI_ISL_5672069, EPI_ISL_5672096, EPI_ISL_5672150, EPI_ISL_5672198, EPI_ISL_5672240, EPI_ISL_5672267, EPI_ISL_5672378, EPI_ISL_5672553, EPI_ISL_5672566 | | | |
| see above | UNIDADE DE PRONTO ATENDIMENTO DE AGENOR DE CAMPOS | Instituto Butantan | Antonio Jorge Martins; Claudia Renata dos Santos Barros; David Schlesinger; Debora Botequio Moretti; Dimas Tadeu Covas; Elaine Cristina Marqueze; Elaine Vieira Santos; Evandra Strazza Rodrigues; Heidge Fukumasu; Jayme Augusto de Souza-Neto; José Salvatore Leister Patané; Luiz Alcantara; Luiz Lehmann Coutinho; Maria Carolina Elias; Maurício Lacerda Nogueira; Rafael dos Santos Bezerra; Raul Machado Neto; Rejane Maria Tommasini Grotto; Ricardo Haddad; Sandra Coccuzzo Sampaio Vessoni; Simone Kashima; Svetoslav Nanev Slavov; Vincent Louis Viala |
| EPI_ISL_5670286 | UNIDADE DE PRONTO ATENDIMENTO UPA | Instituto Butantan | Antonio Jorge Martins; Claudia Renata dos Santos Barros; David Schlesinger; Debora Botequio Moretti; Dimas Tadeu Covas; Elaine Cristina Marqueze; Elaine Vieira Santos; Evandra Strazza Rodrigues; Heidge Fukumasu; Jayme Augusto de Souza-Neto; José Salvatore Leister Patané; Luiz Alcantara; Luiz Lehmann Coutinho; Maria Carolina Elias; Maurício Lacerda Nogueira; Rafael dos Santos Bezerra; Raul Machado Neto; Rejane Maria Tommasini Grotto; Ricardo Haddad; Sandra Coccuzzo Sampaio Vessoni; Simone Kashima; Svetoslav Nanev Slavov; Vincent Louis Viala |
| EPI_ISL_5666031, EPI_ISL_5666043, EPI_ISL_5666061, EPI_ISL_5666072, EPI_ISL_5666076, EPI_ISL_5666082, EPI_ISL_5666094, EPI_ISL_5666104, EPI_ISL_5666118 | | | |
| see above | UNIDADE DE PRONTO ATENDIMENTO UPA 24 HORAS | Instituto Butantan | Antonio Jorge Martins; Claudia Renata dos Santos Barros; David Schlesinger; Debora Botequio Moretti; Dimas Tadeu Covas; Elaine Cristina Marqueze; Elaine Vieira Santos; Evandra Strazza Rodrigues; Heidge Fukumasu; Jayme Augusto de Souza-Neto; José Salvatore Leister Patané; Luiz Alcantara; Luiz Lehmann Coutinho; Maria Carolina Elias; Maurício Lacerda Nogueira; Rafael dos Santos Bezerra; Raul Machado Neto; Rejane Maria Tommasini Grotto; Ricardo Haddad; Sandra Coccuzzo Sampaio Vessoni; Simone Kashima; Svetoslav Nanev Slavov; Vincent Louis Viala |
| EPI_ISL_5659981, EPI_ISL_5660523, EPI_ISL_5660683, EPI_ISL_5660689 | UNIDADE DE PRONTO ATENDIMENTO UPA SUL CARAGUATATUBA | Instituto Butantan | Antonio Jorge Martins; Claudia Renata dos Santos Barros; David Schlesinger; Debora Botequio Moretti; Dimas Tadeu Covas; Elaine Cristina Marqueze; Elaine Vieira Santos; Evandra Strazza Rodrigues; Heidge Fukumasu; Jayme Augusto de Souza-Neto; José Salvatore Leister Patané; Luiz Alcantara; Luiz Lehmann Coutinho; Maria Carolina Elias; Maurício Lacerda Nogueira; Rafael dos Santos Bezerra; Raul Machado Neto; Rejane Maria Tommasini Grotto; Ricardo Haddad; Sandra Coccuzzo Sampaio Vessoni; Simone Kashima; Svetoslav Nanev Slavov; Vincent Louis Viala |
| EPI_ISL_5659789, EPI_ISL_5660075, EPI_ISL_5660088 | UNIDADE DE SAUDE DA FAMILIA AGNALDO SOARES BARBOSA | Instituto Butantan | Antonio Jorge Martins; Claudia Renata dos Santos Barros; David Schlesinger; Debora Botequio Moretti; Dimas Tadeu Covas; Elaine Cristina Marqueze; Elaine Vieira Santos; Evandra Strazza Rodrigues; Heidge Fukumasu; Jayme Augusto de Souza-Neto; José Salvatore Leister Patané; Luiz Alcantara; Luiz Lehmann Coutinho; Maria Carolina Elias; Maurício Lacerda Nogueira; Rafael dos Santos Bezerra; Raul Machado Neto; Rejane Maria Tommasini Grotto; Ricardo Haddad; Sandra Coccuzzo Sampaio Vessoni; Simone Kashima; Svetoslav Nanev Slavov; Vincent Louis Viala |
| EPI_ISL_5660020, EPI_ISL_5660125 | UNIDADE DE SAUDE DA FAMILIA DR JOSE VILELA JUNQUEIRA | Instituto Butantan | Antonio Jorge Martins; Claudia Renata dos Santos Barros; David Schlesinger; Debora Botequio Moretti; Dimas Tadeu Covas; Elaine Cristina Marqueze; Elaine Vieira Santos; Evandra Strazza Rodrigues; Heidge Fukumasu; Jayme Augusto de Souza-Neto; José Salvatore Leister Patané; Luiz Alcantara; Luiz Lehmann Coutinho; Maria Carolina Elias; Maurício Lacerda Nogueira; Rafael dos Santos Bezerra; Raul Machado Neto; Rejane Maria Tommasini Grotto; Ricardo Haddad; Sandra Coccuzzo Sampaio Vessoni; Simone Kashima; Svetoslav Nanev Slavov; Vincent Louis Viala |
| EPI_ISL_5659875 | UNIDADE DE SAUDE DA FAMILIA JOAO BOSCO LELLIS | Instituto Butantan | Antonio Jorge Martins; Claudia Renata dos Santos Barros; David Schlesinger; Debora Botequio Moretti; Dimas Tadeu Covas; Elaine Cristina Marqueze; Elaine Vieira Santos; Evandra Strazza Rodrigues; Heidge Fukumasu; Jayme Augusto de Souza-Neto; José Salvatore Leister Patané; Luiz Alcantara; Luiz Lehmann Coutinho; Maria Carolina Elias; Maurício Lacerda Nogueira; Rafael dos Santos Bezerra; Raul Machado Neto; Rejane Maria Tommasini Grotto; Ricardo Haddad; Sandra Coccuzzo Sampaio Vessoni; Simone Kashima; Svetoslav Nanev Slavov; Vincent Louis Viala |
| EPI_ISL_5659662, EPI_ISL_5659669, EPI_ISL_5659670, EPI_ISL_5659736, EPI_ISL_5659748, EPI_ISL_5659765, EPI_ISL_5659799, EPI_ISL_5659887, EPI_ISL_5659971, EPI_ISL_5660037, EPI_ISL_5660220, EPI_ISL_5660396, EPI_ISL_5660595, EPI_ISL_5660754 | | | |
| see above | UNIDADE DE SAUDE DA FAMILIA NOVO HORIZONTE | Instituto Butantan | Antonio Jorge Martins; Claudia Renata dos Santos Barros; David Schlesinger; Debora Botequio Moretti; Dimas Tadeu Covas; Elaine Cristina Marqueze; Elaine Vieira Santos; Evandra Strazza Rodrigues; Heidge Fukumasu; Jayme Augusto de Souza-Neto; José Salvatore Leister Patané; Luiz Alcantara; Luiz Lehmann Coutinho; Maria Carolina Elias; Maurício Lacerda Nogueira; Rafael dos Santos Bezerra; Raul Machado Neto; Rejane Maria Tommasini Grotto; Ricardo Haddad; Sandra Coccuzzo Sampaio Vessoni; Simone Kashima; Svetoslav Nanev Slavov; Vincent Louis Viala |
| EPI_ISL_5660383, EPI_ISL_5660834, EPI_ISL_5660913, EPI_ISL_5661050 | UNIDADE DE SAUDE DA FAMILIA TARCISIO BARINI | Instituto Butantan | Antonio Jorge Martins; Claudia Renata dos Santos Barros; David Schlesinger; Debora Botequio Moretti; Dimas Tadeu Covas; Elaine Cristina Marqueze; Elaine Vieira Santos; Evandra Strazza Rodrigues; Heidge Fukumasu; Jayme Augusto de Souza-Neto; José Salvatore Leister Patané; Luiz Alcantara; Luiz Lehmann Coutinho; Maria Carolina Elias; Maurício Lacerda Nogueira; Rafael dos Santos Bezerra; Raul Machado Neto; Rejane Maria Tommasini Grotto; Ricardo Haddad; Sandra Coccuzzo Sampaio Vessoni; Simone Kashima; Svetoslav Nanev Slavov; Vincent Louis Viala |
| EPI_ISL_5666915, EPI_ISL_5666927, EPI_ISL_5666989 | UNIDADE DE VIGILANCIA EM SAUDE | Instituto Butantan | Antonio Jorge Martins; Claudia Renata dos Santos Barros; David Schlesinger; Debora Botequio Moretti; Dimas Tadeu Covas; Elaine Cristina Marqueze; Elaine Vieira Santos; Evandra Strazza Rodrigues; Heidge Fukumasu; Jayme Augusto de Souza-Neto; José Salvatore Leister Patané; Luiz Alcantara; Luiz Lehmann Coutinho; Maria Carolina Elias; Maurício Lacerda Nogueira; Rafael dos Santos Bezerra; Raul Machado Neto; Rejane Maria Tommasini Grotto; Ricardo Haddad; Sandra Coccuzzo Sampaio Vessoni; Simone Kashima; Svetoslav Nanev Slavov; Vincent Louis Viala |
| EPI_ISL_5665827, EPI_ISL_5665832, EPI_ISL_5671144, EPI_ISL_5671202 | UNIDADE DE VIGILANCIA EPIDEMIOLOGICA | Instituto Butantan | Antonio Jorge Martins; Claudia Renata dos Santos Barros; David Schlesinger; Debora Botequio Moretti; Dimas Tadeu Covas; Elaine Cristina Marqueze; Elaine Vieira Santos; Evandra Strazza Rodrigues; Heidge Fukumasu; Jayme Augusto de Souza-Neto; José Salvatore Leister Patané; Luiz Alcantara; Luiz Lehmann Coutinho; Maria Carolina Elias; Maurício Lacerda Nogueira; Rafael dos Santos Bezerra; Raul Machado Neto; Rejane Maria Tommasini Grotto; Ricardo Haddad; Sandra Coccuzzo Sampaio Vessoni; Simone Kashima; Svetoslav Nanev Slavov; Vincent Louis Viala |
| EPI_ISL_5664921 | UNIDADE MISTA DE SAUDE MONS CID FRANCA SANTOS | Instituto Butantan | Antonio Jorge Martins; Claudia Renata dos Santos Barros; David Schlesinger; Debora Botequio Moretti; Dimas Tadeu Covas; Elaine Cristina Marqueze; Elaine Vieira Santos; Evandra Strazza Rodrigues; Heidge Fukumasu; Jayme Augusto de Souza-Neto; José Salvatore Leister Patané; Luiz Alcantara; Luiz Lehmann Coutinho; Maria Carolina Elias; Maurício Lacerda Nogueira; Rafael dos Santos Bezerra; Raul Machado Neto; Rejane Maria Tommasini Grotto; Ricardo Haddad; Sandra Coccuzzo Sampaio Vessoni; Simone Kashima; Svetoslav Nanev Slavov; Vincent Louis Viala |
| EPI_ISL_5659704, EPI_ISL_5660570, EPI_ISL_5660815, EPI_ISL_5663193 | UNIDADE MISTA DE SAUDE ROSEIRA DR PAULO GUIMARAES CASTRO | Instituto Butantan | Antonio Jorge Martins; Claudia Renata dos Santos Barros; David Schlesinger; Debora Botequio Moretti; Dimas Tadeu Covas; Elaine Cristina Marqueze; Elaine Vieira Santos; Evandra Strazza Rodrigues; Heidge Fukumasu; Jayme Augusto de Souza-Neto; José Salvatore Leister Patané; Luiz Alcantara; Luiz Lehmann Coutinho; Maria Carolina Elias; Maurício Lacerda Nogueira; Rafael dos Santos Bezerra; Raul Machado Neto; Rejane Maria Tommasini Grotto; Ricardo Haddad; Sandra Coccuzzo Sampaio Vessoni; Simone Kashima; Svetoslav Nanev Slavov; Vincent Louis Viala |
| EPI_ISL_5665868, EPI_ISL_5671113, EPI_ISL_5671238 | UNIDADE MISTA OSVALDO P CAMARGO ELIAS FAUSTO | Instituto Butantan | Antonio Jorge Martins; Claudia Renata dos Santos Barros; David Schlesinger; Debora Botequio Moretti; Dimas Tadeu Covas; Elaine Cristina Marqueze; Elaine Vieira Santos; Evandra Strazza Rodrigues; Heidge Fukumasu; Jayme Augusto de Souza-Neto; José Salvatore Leister Patané; Luiz Alcantara; Luiz Lehmann Coutinho; Maria Carolina Elias; Maurício Lacerda Nogueira; Rafael dos Santos Bezerra; Raul Machado Neto; Rejane Maria Tommasini Grotto; Ricardo Haddad; Sandra Coccuzzo Sampaio Vessoni; Simone Kashima; Svetoslav Nanev Slavov; Vincent Louis Viala |
| EPI_ISL_3912059, EPI_ISL_3912080, EPI_ISL_3912335, EPI_ISL_3912336, EPI_ISL_3912337, EPI_ISL_3912338, EPI_ISL_3912339, EPI_ISL_3912340, EPI_ISL_3912341, EPI_ISL_3912342, EPI_ISL_3912343, EPI_ISL_3912344, EPI_ISL_3912345, EPI_ISL_3912346, EPI_ISL_3912347, EPI_ISL_3912348, EPI_ISL_3912349, EPI_ISL_3912350, EPI_ISL_3912351, EPI_ISL_3912352, EPI_ISL_3912353 | | | |
| see above | UNIDADE REFERENCIA COVID 19 | Analytical Competence Molecular Epidemiology Lab/ACME, Oswaldo Cruz Foundation, Ceara (FIOCRUZ CE) | Cleber Furtado Aksenen; Fabio Miyajima; Fernando Braga Stehling; Francisco Eder de Moura Lopes; Jamille Maria Mendes Bezerra; Joaquim Cesar do Nascimento Sousa Junior; Pedro Miguel Carneiro Jeronimo; Suzana Porto Almeida & Lucas Delerino on behalf of COVID-19 FIOCRUZ Genomic Network; Thais Ferreira de Oliveira; Thais de Oliveira Costa; Ticiane Cavalcante de Souza; Veridiana Pessoa Miyajima |

| | | | |
|---|---|---|---|
| EPI_ISL_5661394, EPI_ISL_5661510, EPI_ISL_5661686, EPI_ISL_5662055 | UNIDADE REFERENCIAL SUDOESTE | Instituto Butantan | Antonio Jorge Martins; Claudia Renata dos Santos Barros; David Schlesinger; Debora Botequio Moretti; Dimas Tadeu Covas; Elaine Cristina Marqueze; Elaine Vieira Santos; Evandra Strazza Rodrigues; Heidge Fukumasu; Jayme Augusto de Souza-Neto; José Salvatore Leister Patané; Luiz Alcantara; Luiz Lehmann Coutinho; Maria Carolina Elias; Maurício Lacerda Nogueira; Rafael dos Santos Bezerra; Raul Machado Neto; Rejane Maria Tommasini Grotto; Ricardo Haddad; Sandra Coccuzzo Sampaio Vessoni; Simone Kashima; Svetoslav Nanev Slavov; Vincent Louis Viala |
| EPI_ISL_5668593, EPI_ISL_5668697, EPI_ISL_5669017, EPI_ISL_5669187, EPI_ISL_5669562, EPI_ISL_5669836, EPI_ISL_5670285, EPI_ISL_5670353, EPI_ISL_5670548, EPI_ISL_5670817 | | | |
| see above | UNIDADE RESPIRATORIA NOVA HORTOLANDIA | Instituto Butantan | Antonio Jorge Martins; Claudia Renata dos Santos Barros; David Schlesinger; Debora Botequio Moretti; Dimas Tadeu Covas; Elaine Cristina Marqueze; Elaine Vieira Santos; Evandra Strazza Rodrigues; Heidge Fukumasu; Jayme Augusto de Souza-Neto; José Salvatore Leister Patané; Luiz Alcantara; Luiz Lehmann Coutinho; Maria Carolina Elias; Maurício Lacerda Nogueira; Rafael dos Santos Bezerra; Raul Machado Neto; Rejane Maria Tommasini Grotto; Ricardo Haddad; Sandra Coccuzzo Sampaio Vessoni; Simone Kashima; Svetoslav Nanev Slavov; Vincent Louis Viala |
| EPI_ISL_5659667, EPI_ISL_5660475 | UPA 24 HORAS CENTRO | Instituto Butantan | Antonio Jorge Martins; Claudia Renata dos Santos Barros; David Schlesinger; Debora Botequio Moretti; Dimas Tadeu Covas; Elaine Cristina Marqueze; Elaine Vieira Santos; Evandra Strazza Rodrigues; Heidge Fukumasu; Jayme Augusto de Souza-Neto; José Salvatore Leister Patané; Luiz Alcantara; Luiz Lehmann Coutinho; Maria Carolina Elias; Maurício Lacerda Nogueira; Rafael dos Santos Bezerra; Raul Machado Neto; Rejane Maria Tommasini Grotto; Ricardo Haddad; Sandra Coccuzzo Sampaio Vessoni; Simone Kashima; Svetoslav Nanev Slavov; Vincent Louis Viala |
| EPI_ISL_5661850, EPI_ISL_5663155, EPI_ISL_5663414, EPI_ISL_5664849, EPI_ISL_5665344, EPI_ISL_5665442 | UPA ATALAIA DRA RITA DE CASSIA SORIO | Instituto Butantan | Antonio Jorge Martins; Claudia Renata dos Santos Barros; David Schlesinger; Debora Botequio Moretti; Dimas Tadeu Covas; Elaine Cristina Marqueze; Elaine Vieira Santos; Evandra Strazza Rodrigues; Heidge Fukumasu; Jayme Augusto de Souza-Neto; José Salvatore Leister Patané; Luiz Alcantara; Luiz Lehmann Coutinho; Maria Carolina Elias; Maurício Lacerda Nogueira; Rafael dos Santos Bezerra; Raul Machado Neto; Rejane Maria Tommasini Grotto; Ricardo Haddad; Sandra Coccuzzo Sampaio Vessoni; Simone Kashima; Svetoslav Nanev Slavov; Vincent Louis Viala |
| EPI_ISL_5660256, EPI_ISL_5661148, EPI_ISL_5661236, EPI_ISL_5661340, EPI_ISL_5661682, EPI_ISL_5662174, EPI_ISL_5663198, EPI_ISL_5663602, EPI_ISL_5663719, EPI_ISL_5663723, EPI_ISL_5663966, EPI_ISL_5664033, EPI_ISL_5664044, EPI_ISL_5664084, EPI_ISL_5664164, EPI_ISL_5664477, EPI_ISL_5664612, EPI_ISL_5664723, EPI_ISL_5664913, EPI_ISL_5664998, EPI_ISL_5665033, EPI_ISL_5665138, EPI_ISL_5665360, EPI_ISL_5672136, EPI_ISL_5672207, EPI_ISL_5672327, EPI_ISL_5672353, EPI_ISL_5672443, EPI_ISL_5672504, EPI_ISL_5672507, EPI_ISL_5673530 | | | |
| see above | UPA DR AKIRA TADA | Instituto Butantan | Antonio Jorge Martins; Claudia Renata dos Santos Barros; David Schlesinger; Debora Botequio Moretti; Dimas Tadeu Covas; Elaine Cristina Marqueze; Elaine Vieira Santos; Evandra Strazza Rodrigues; Heidge Fukumasu; Jayme Augusto de Souza-Neto; José Salvatore Leister Patané; Luiz Alcantara; Luiz Lehmann Coutinho; Maria Carolina Elias; Maurício Lacerda Nogueira; Rafael dos Santos Bezerra; Raul Machado Neto; Rejane Maria Tommasini Grotto; Ricardo Haddad; Sandra Coccuzzo Sampaio Vessoni; Simone Kashima; Svetoslav Nanev Slavov; Vincent Louis Viala |
| EPI_ISL_5659663, EPI_ISL_5660344, EPI_ISL_5660554, EPI_ISL_5660850, EPI_ISL_5660927, EPI_ISL_5661023, EPI_ISL_5661774, EPI_ISL_5663876, EPI_ISL_5663882, EPI_ISL_5664076, EPI_ISL_5664140, EPI_ISL_5664229, EPI_ISL_5664283, EPI_ISL_5664568, EPI_ISL_5664719, EPI_ISL_5664930, EPI_ISL_5664938, EPI_ISL_5672010, EPI_ISL_5672143, EPI_ISL_5672159, EPI_ISL_5672223, EPI_ISL_5672274, EPI_ISL_5672382, EPI_ISL_5672402, EPI_ISL_5672410, EPI_ISL_5672491, EPI_ISL_5672539, EPI_ISL_5673446 | | | |
| see above | UPA DR FRANCO DA ROCHA | Instituto Butantan | Antonio Jorge Martins; Claudia Renata dos Santos Barros; David Schlesinger; Debora Botequio Moretti; Dimas Tadeu Covas; Elaine Cristina Marqueze; Elaine Vieira Santos; Evandra Strazza Rodrigues; Heidge Fukumasu; Jayme Augusto de Souza-Neto; José Salvatore Leister Patané; Luiz Alcantara; Luiz Lehmann Coutinho; Maria Carolina Elias; Maurício Lacerda Nogueira; Rafael dos Santos Bezerra; Raul Machado Neto; Rejane Maria Tommasini Grotto; Ricardo Haddad; Sandra Coccuzzo Sampaio Vessoni; Simone Kashima; Svetoslav Nanev Slavov; Vincent Louis Viala |
| EPI_ISL_5661309 | UPA JOSE CAMPOS BARRETO ZEQUINHA BARRETO | Instituto Butantan | Antonio Jorge Martins; Claudia Renata dos Santos Barros; David Schlesinger; Debora Botequio Moretti; Dimas Tadeu Covas; Elaine Cristina Marqueze; Elaine Vieira Santos; Evandra Strazza Rodrigues; Heidge Fukumasu; Jayme Augusto de Souza-Neto; José Salvatore Leister Patané; Luiz Alcantara; Luiz Lehmann Coutinho; Maria Carolina Elias; Maurício Lacerda Nogueira; Rafael dos Santos Bezerra; Raul Machado Neto; Rejane Maria Tommasini Grotto; Ricardo Haddad; Sandra Coccuzzo Sampaio Vessoni; Simone Kashima; Svetoslav Nanev Slavov; Vincent Louis Viala |
| EPI_ISL_5662520, EPI_ISL_5662737, EPI_ISL_5664025, EPI_ISL_5670215, EPI_ISL_5670707 | UPA PAULISTA | Instituto Butantan | Antonio Jorge Martins; Claudia Renata dos Santos Barros; David Schlesinger; Debora Botequio Moretti; Dimas Tadeu Covas; Elaine Cristina Marqueze; Elaine Vieira Santos; Evandra Strazza Rodrigues; Heidge Fukumasu; Jayme Augusto de Souza-Neto; José Salvatore Leister Patané; Luiz Alcantara; Luiz Lehmann Coutinho; Maria Carolina Elias; Maurício Lacerda Nogueira; Rafael dos Santos Bezerra; Raul Machado Neto; Rejane Maria Tommasini Grotto; Ricardo Haddad; Sandra Coccuzzo Sampaio Vessoni; Simone Kashima; Svetoslav Nanev Slavov; Vincent Louis Viala |
| EPI_ISL_5663530, EPI_ISL_5663896, EPI_ISL_5664213 | UPA SAO JOAO LAVRAS | Instituto Butantan | Antonio Jorge Martins; Claudia Renata dos Santos Barros; David Schlesinger; Debora Botequio Moretti; Dimas Tadeu Covas; Elaine Cristina Marqueze; Elaine Vieira Santos; Evandra Strazza Rodrigues; Heidge Fukumasu; Jayme Augusto de Souza-Neto; José Salvatore Leister Patané; Luiz Alcantara; Luiz Lehmann Coutinho; Maria Carolina Elias; Maurício Lacerda Nogueira; Rafael dos Santos Bezerra; Raul Machado Neto; Rejane Maria Tommasini Grotto; Ricardo Haddad; Sandra Coccuzzo Sampaio Vessoni; Simone Kashima; Svetoslav Nanev Slavov; Vincent Louis Viala |
| EPI_ISL_5659665, EPI_ISL_5659956 | UPA UNIDADE DE PRONTO ATENDIMENTO 24 HORAS BOM JESUS | Instituto Butantan | Antonio Jorge Martins; Claudia Renata dos Santos Barros; David Schlesinger; Debora Botequio Moretti; Dimas Tadeu Covas; Elaine Cristina Marqueze; Elaine Vieira Santos; Evandra Strazza Rodrigues; Heidge Fukumasu; Jayme Augusto de Souza-Neto; José Salvatore Leister Patané; Luiz Alcantara; Luiz Lehmann Coutinho; Maria Carolina Elias; Maurício Lacerda Nogueira; Rafael dos Santos Bezerra; Raul Machado Neto; Rejane Maria Tommasini Grotto; Ricardo Haddad; Sandra Coccuzzo Sampaio Vessoni; Simone Kashima; Svetoslav Nanev Slavov; Vincent Louis Viala |
| EPI_ISL_5660711, EPI_ISL_5663398, EPI_ISL_5663543, EPI_ISL_5663583, EPI_ISL_5663746, EPI_ISL_5663776, EPI_ISL_5663785, EPI_ISL_5664364, EPI_ISL_5664392, EPI_ISL_5664430, EPI_ISL_5664647, EPI_ISL_5664682, EPI_ISL_5664776, EPI_ISL_5665125, EPI_ISL_5665189, EPI_ISL_5665256, EPI_ISL_5668073 | | | |
| see above | UPA VILA SANTA CATARINA | Instituto Butantan | Antonio Jorge Martins; Claudia Renata dos Santos Barros; David Schlesinger; Debora Botequio Moretti; Dimas Tadeu Covas; Elaine Cristina Marqueze; Elaine Vieira Santos; Evandra Strazza Rodrigues; Heidge Fukumasu; Jayme Augusto de Souza-Neto; José Salvatore Leister Patané; Luiz Alcantara; Luiz Lehmann Coutinho; Maria Carolina Elias; Maurício Lacerda Nogueira; Rafael dos Santos Bezerra; Raul Machado Neto; Rejane Maria Tommasini Grotto; Ricardo Haddad; Sandra Coccuzzo Sampaio Vessoni; Simone Kashima; Svetoslav Nanev Slavov; Vincent Louis Viala |
| EPI_ISL_3048152, EPI_ISL_3048160, EPI_ISL_3375809 | US Air Force School of Aerospace Medicine | US Air Force School of Aerospace Medicine | Amanda Javorina; Anthony Fries; Carol Garrett; Clarise Starr; Elizabeth Macias; Jennifer Meyer; Sarah Purves; William Gruner |
| EPI_ISL_5662713 | USF DR APARECIDO RODRIGUES MOUCO SCRPARDO | Instituto Butantan | Antonio Jorge Martins; Claudia Renata dos Santos Barros; David Schlesinger; Debora Botequio Moretti; Dimas Tadeu Covas; Elaine Cristina Marqueze; Elaine Vieira Santos; Evandra Strazza Rodrigues; Heidge Fukumasu; Jayme Augusto de Souza-Neto; José Salvatore Leister Patané; Luiz Alcantara; Luiz Lehmann Coutinho; Maria Carolina Elias; Maurício Lacerda Nogueira; Rafael dos Santos Bezerra; Raul Machado Neto; Rejane Maria Tommasini Grotto; Ricardo Haddad; Sandra Coccuzzo Sampaio Vessoni; Simone Kashima; Svetoslav Nanev Slavov; Vincent Louis Viala |
| EPI_ISL_5662277 | USF DR SAMUEL MARTINS FIGUEIRA SCRPARDO | Instituto Butantan | Antonio Jorge Martins; Claudia Renata dos Santos Barros; David Schlesinger; Debora Botequio Moretti; Dimas Tadeu Covas; Elaine Cristina Marqueze; Elaine Vieira Santos; Evandra Strazza Rodrigues; Heidge Fukumasu; Jayme Augusto de Souza-Neto; José Salvatore Leister Patané; Luiz Alcantara; Luiz Lehmann Coutinho; Maria Carolina Elias; Maurício Lacerda Nogueira; Rafael dos Santos Bezerra; Raul Machado Neto; Rejane Maria Tommasini Grotto; Ricardo Haddad; Sandra Coccuzzo Sampaio Vessoni; Simone Kashima; Svetoslav Nanev Slavov; Vincent Louis Viala |
| EPI_ISL_5660288, EPI_ISL_5661003 | USF EUCALIPTOS | Instituto Butantan | Antonio Jorge Martins; Claudia Renata dos Santos Barros; David Schlesinger; Debora Botequio Moretti; Dimas Tadeu Covas; Elaine Cristina Marqueze; Elaine Vieira Santos; Evandra Strazza Rodrigues; Heidge Fukumasu; Jayme Augusto de Souza-Neto; José Salvatore Leister Patané; Luiz Alcantara; Luiz Lehmann Coutinho; Maria Carolina Elias; Maurício Lacerda Nogueira; Rafael dos Santos Bezerra; Raul Machado Neto; Rejane Maria Tommasini Grotto; Ricardo Haddad; Sandra Coccuzzo Sampaio Vessoni; Simone Kashima; Svetoslav Nanev Slavov; Vincent Louis Viala |
| EPI_ISL_5666412 | USF JARDIM SAO CARLOS | Instituto Butantan | Antonio Jorge Martins; Claudia Renata dos Santos Barros; David Schlesinger; Debora Botequio Moretti; Dimas Tadeu Covas; Elaine Cristina Marqueze; Elaine Vieira Santos; Evandra Strazza Rodrigues; Heidge Fukumasu; Jayme Augusto de Souza-Neto; José Salvatore Leister Patané; Luiz Alcantara; Luiz Lehmann Coutinho; Maria Carolina Elias; Maurício Lacerda Nogueira; Rafael dos Santos Bezerra; Raul Machado Neto; Rejane Maria Tommasini Grotto; Ricardo Haddad; Sandra Coccuzzo Sampaio Vessoni; Simone Kashima; Svetoslav Nanev Slavov; Vincent Louis Viala |
| EPI_ISL_5662081 | USF JARDIM SAO JOAO SCRPARDO | Instituto Butantan | Antonio Jorge Martins; Claudia Renata dos Santos Barros; David Schlesinger; Debora Botequio Moretti; Dimas Tadeu Covas; Elaine Cristina Marqueze; Elaine Vieira Santos; Evandra Strazza Rodrigues; Heidge Fukumasu; Jayme Augusto de Souza-Neto; José Salvatore Leister Patané; Luiz Alcantara; Luiz Lehmann Coutinho; Maria Carolina Elias; Maurício Lacerda Nogueira; Rafael dos Santos Bezerra; Raul Machado Neto; Rejane Maria Tommasini Grotto; Ricardo Haddad; Sandra Coccuzzo Sampaio Vessoni; Simone Kashima; Svetoslav Nanev Slavov; Vincent Louis Viala |
| EPI_ISL_5662041 | USF JOAO NASCIMENTO TELLES | Instituto Butantan | Antonio Jorge Martins; Claudia Renata dos Santos Barros; David Schlesinger; Debora Botequio Moretti; Dimas Tadeu Covas; Elaine Cristina Marqueze; Elaine Vieira Santos; Evandra Strazza Rodrigues; Heidge Fukumasu; Jayme Augusto de Souza-Neto; José Salvatore Leister Patané; Luiz Alcantara; Luiz Lehmann Coutinho; Maria Carolina Elias; Maurício Lacerda Nogueira; Rafael dos Santos Bezerra; Raul Machado Neto; Rejane Maria Tommasini Grotto; Ricardo Haddad; Sandra Coccuzzo Sampaio Vessoni; Simone Kashima; Svetoslav Nanev Slavov; Vincent Louis Viala |
| EPI_ISL_5666232 | USF PACHECO CHAVES | Instituto Butantan | Antonio Jorge Martins; Claudia Renata dos Santos Barros; David Schlesinger; Debora Botequio Moretti; Dimas Tadeu Covas; Elaine Cristina Marqueze; Elaine Vieira Santos; Evandra Strazza Rodrigues; Heidge Fukumasu; Jayme Augusto de Souza-Neto; José Salvatore Leister Patané; Luiz Alcantara; Luiz Lehmann Coutinho; Maria Carolina Elias; Maurício Lacerda Nogueira; Rafael dos Santos Bezerra; Raul Machado Neto; Rejane Maria Tommasini Grotto; Ricardo Haddad; Sandra Coccuzzo Sampaio Vessoni; Simone Kashima; Svetoslav Nanev Slavov; Vincent Louis Viala |
| EPI_ISL_5667291, EPI_ISL_5667297, EPI_ISL_5667337, EPI_ISL_5667381, EPI_ISL_5667406, EPI_ISL_5667727, EPI_ISL_5667737, EPI_ISL_5667747, EPI_ISL_5667752, EPI_ISL_5667757, EPI_ISL_5667763, EPI_ISL_5667773, EPI_ISL_5667794 | | | |
| see above | USF PAULISTA FERNANDOPOLIS ANTONIO PIVATO | Instituto Butantan | Antonio Jorge Martins; Claudia Renata dos Santos Barros; David Schlesinger; Debora Botequio Moretti; Dimas Tadeu Covas; Elaine Cristina Marqueze; Elaine Vieira Santos; Evandra Strazza Rodrigues; Heidge Fukumasu; Jayme Augusto de Souza-Neto; José Salvatore Leister Patané; Luiz Alcantara; Luiz Lehmann Coutinho; Maria Carolina Elias; Maurício Lacerda Nogueira; Rafael dos Santos Bezerra; Raul Machado Neto; Rejane Maria Tommasini Grotto; Ricardo Haddad; Sandra Coccuzzo Sampaio Vessoni; Simone Kashima; Svetoslav Nanev Slavov; Vincent Louis Viala |
| EPI_ISL_5665929, EPI_ISL_5665934, EPI_ISL_5665942, EPI_ISL_5665952, EPI_ISL_5665963, EPI_ISL_5665986, EPI_ISL_5666824, EPI_ISL_5666833, EPI_ISL_5667068 | | | |
| see above | USF SALERNO | Instituto Butantan | Antonio Jorge Martins; Claudia Renata dos Santos Barros; David Schlesinger; Debora Botequio Moretti; Dimas Tadeu Covas; Elaine Cristina Marqueze; Elaine Vieira Santos; Evandra Strazza Rodrigues; Heidge Fukumasu; Jayme Augusto de Souza-Neto; José Salvatore Leister Patané; Luiz Alcantara; Luiz Lehmann Coutinho; Maria Carolina Elias; Maurício Lacerda Nogueira; Rafael dos Santos Bezerra; Raul Machado Neto; Rejane Maria Tommasini Grotto; Ricardo Haddad; Sandra Coccuzzo Sampaio Vessoni; Simone Kashima; Svetoslav Nanev Slavov; Vincent Louis Viala |
| EPI_ISL_5659658, EPI_ISL_5659947 | USF SUECIA | Instituto Butantan | Antonio Jorge Martins; Claudia Renata dos Santos Barros; David Schlesinger; Debora Botequio Moretti; Dimas Tadeu Covas; Elaine Cristina Marqueze; Elaine Vieira Santos; Evandra Strazza Rodrigues; Heidge Fukumasu; Jayme Augusto de Souza-Neto; José Salvatore Leister Patané; Luiz Alcantara; Luiz Lehmann Coutinho; Maria Carolina Elias; Maurício Lacerda Nogueira; Rafael dos Santos Bezerra; Raul Machado Neto; Rejane Maria Tommasini Grotto; Ricardo Haddad; Sandra Coccuzzo Sampaio Vessoni; Simone Kashima; Svetoslav Nanev Slavov; Vincent Louis Viala |
| EPI_ISL_2895012, EPI_ISL_2895022, EPI_ISL_2967732, EPI_ISL_2967766, EPI_ISL_2967768, EPI_ISL_3025927, EPI_ISL_3025931, EPI_ISL_3025955, EPI_ISL_3025960, EPI_ISL_3025967, EPI_ISL_3025987, EPI_ISL_3411453, EPI_ISL_3693077 | | | |
| see above | UW Virology Lab | UW Virology Lab | Alexander Greninger; Hong Xie; Keith R Jerome; Lasata Shrestha; Maria Lukes; Meei-Li Huang; Michelle Lin; Nathan Breit; Noah R. Baker; Patrick Mathias; Pavitra Roychoudhury; Ricardo Perez; Robert J. Livingston; Savanna S. Carmack; Sean Ellis; Shah Mohamed Bakhash; Tien V. Nguyen |
| EPI_ISL_2942751, EPI_ISL_2970106 | Unidad de Investigacion Biomedica de Zacatecas (UIBZ) | Unidad de Genomica Avanzada | ; Alejandra Garcia-Gasca; Alejandra Hernandez-Teran; Alejandro Sanchez-Flores; Alfredo Herrera-Estrella; Alicia Ocaña-Mondragon; Andreu Comas-Garcia; Angel Gustavo Salas-Lais; Antonio Loza Roman; Bernardo Martinez-Miguel; Blanca Taboada; Brenda Irasema Maldonado-Meza; Bruno Gomez-Gil; Carla Ivon Herrera-Najera; Carlos F. Arias; Celia Boukadida; Celida Duque Molina; Celida Martinez- Rodriguez; Clara Esperanza Santacruz-Tinoco; Concepcion Grajales-Muñiz; Consorcio Mexicano de Vigilancia Genomica (CoViGen-Mex). Authors (in alphabetical order): Julio Elias Alvarado-Yaah; Cristobal Chaidez-Quiroz; Daniel Fregoso-Rueda; Daniel Lira Morales; Eduardo Becerril-Vargas; Fernando Fontove-Herrera; Fidencio Mejia-Nepomuceno; Francisco Pulido; Gloria Elena Espinosa-Ayala; Gloria Maria Molina-Salinas; Gloria Vazquez; Hector Esteban Paz-Juarez; Hector Montoya-Fuentes; Helen Haydee Fernanda Ramirez-Plascencia; Irvin Gonzalez-Lopez; Jean Pierre Gonzalez; Jesus Hernandez; Joel Armando Vazquez-Perez.; Jorge Salas-Hernandez; Jose Antonio Enciso-Moreno; Jose Arturo Martinez-Orozco; Jose Esteban Muñoz-Medina; Jose de Jesus Nuñez-Contreras; Juan Bautista Chale-Dzul; Julissa Enciso-Ibarra; Luis Alberto Ochoa-Carrera; Margarita Matias-Florentino; Maria Guadalupe Santiago-Mauricio; Maria Guadalupe de Jesus Mireles-Rivera; Mario Mujica-Sanchez; Marissa Perez-Garcia; Nelly Selem-Mojica; Pavel Isa; Ricardo Ciria Merce; Ricardo Grande; Rosa Maria Gutierrez Rios; Santiago avila-Rios; Selene Zarate; Susana Lopez; Veronica Mata-Haro; Victor Eduardo Garcia-Arias; Victor Hugo Borja-Aburto |
| EPI_ISL_2942671, EPI_ISL_2942672, EPI_ISL_2942675, EPI_ISL_2942677, EPI_ISL_2942678, EPI_ISL_2942679, EPI_ISL_2942680, EPI_ISL_2942686, EPI_ISL_2942692, EPI_ISL_2942696, EPI_ISL_2942698, EPI_ISL_2942702, EPI_ISL_2942705, EPI_ISL_2942713, EPI_ISL_2942714, EPI_ISL_2942715, EPI_ISL_2942716, EPI_ISL_2942717, EPI_ISL_2942718, EPI_ISL_2942719 | | | |
| see above | Unidad de Investigacion Medica de Yucatan (UIMY) | Unidad de Genomica Avanzada | ; Alejandra Garcia-Gasca; Alejandra Hernandez-Teran; Alejandro Sanchez-Flores; Alfredo Herrera-Estrella; Alicia Ocaña-Mondragon; Andreu Comas-Garcia; Angel Gustavo Salas-Lais; Antonio Loza Roman; Bernardo Martinez-Miguel; Blanca Taboada; Brenda Irasema Maldonado-Meza; Bruno Gomez-Gil; Carla Ivon Herrera-Najera; Carlos F. Arias; Celia Boukadida; Celida Duque Molina; Celida Martinez- Rodriguez; Clara Esperanza Santacruz-Tinoco; Concepcion Grajales-Muñiz; Consorcio Mexicano de Vigilancia Genomica (CoViGen-Mex). Authors (in alphabetical order): Julio Elias Alvarado-Yaah; Cristobal Chaidez-Quiroz; Daniel Fregoso-Rueda; Daniel Lira Morales; Eduardo Becerril-Vargas; Fernando Fontove-Herrera; Fidencio Mejia-Nepomuceno; Francisco Pulido; Gloria Elena Espinosa-Ayala; Gloria Maria Molina-Salinas; Gloria Vazquez; Hector Esteban Paz-Juarez; Hector Montoya-Fuentes; Helen Haydee Fernanda Ramirez-Plascencia; Irvin Gonzalez-Lopez; Jean Pierre Gonzalez; Jesus Hernandez; Joel Armando Vazquez-Perez.; Jorge Salas-Hernandez; Jose Antonio Enciso-Moreno; Jose Arturo Martinez-Orozco; Jose Esteban Muñoz-Medina; Jose de Jesus Nuñez-Contreras; Juan Bautista Chale-Dzul; Julissa Enciso-Ibarra; Luis Alberto Ochoa-Carrera; Margarita Matias-Florentino; Maria Guadalupe Santiago-Mauricio; Maria Guadalupe de Jesus Mireles-Rivera; Mario Mujica-Sanchez; Marissa Perez-Garcia; Nelly Selem-Mojica; Pavel Isa; Ricardo Ciria Merce; Ricardo Grande; Rosa Maria Gutierrez Rios; Santiago avila-Rios; Selene Zarate; Susana Lopez; Veronica Mata-Haro; Victor Eduardo Garcia-Arias; Victor Hugo Borja-Aburto |
| EPI_ISL_3155343, EPI_ISL_3155347, EPI_ISL_3155348, EPI_ISL_3155353, EPI_ISL_3155355, EPI_ISL_3155356, EPI_ISL_3155359, EPI_ISL_3155362, EPI_ISL_3155364, EPI_ISL_3155365, EPI_ISL_3155366, EPI_ISL_3155380, EPI_ISL_7780467 | | | |
| see above | Unidad de Investigación Médica de Yucatán (UIMY) | Centro de Investigación en Enfermedades Infecciosas (CIENI), Instituto Nacional de Enfermedades Respiratorias (INER) | ; Alejandra García-Gasca; Alejandra Hernández-Terán; Alejandro Sánchez-Flores; Alfredo Herrera-Estrella; Alicia Ocaña-Mondragón; Andreu Comas-García; Angel Gustavo Salas-Lais; Antonio Loza Román; Bernardo Martínez-Miguel; Blanca Taboada; Brenda Irasema Maldonado-Meza; Bruno Gómez-Gil; Carla Ivón Herrera-Najera; Carlos F. Arias; Celia Boukadida; Clara Esperanza Santacruz-Tinoco; Concepción Grajales-Muñiz; Consorcio Mexicano de Vigilancia Genómica (CoViGen-Mex). Authors (in alphabetical order): Julio Elias Alvarado-Yaah; Cristóbal Cháidez-Quiróz; Célida Duque Molina; Célida Martínez- Rodríguez; Célida Martínez-Rodríguez; Daniel Fregoso-Rueda; Daniel Lira Morales; Eduardo Becerril-Vargas; Eduardo Rivera-Martínez; Fernando Fontove-Herrera; Fidencio Mejía-Nepomuceno; Francisco Pulido; Gabriel Chavira-Trujillo; Gloria Elena Espinosa-Ayala; Gloria María Molina-Salinas; Gloria Vazquez; Hector Esteban Paz-Juárez; Hector Montoya-Fuentes; Helen Haydee Fernanda Ramírez-Plascencia; Irvin González-López; Jean Pierre González; Jesús Hernández; Joel Armando Vázquez-Pérez.; Jorge Salas-Hernández; José Antonio Enciso-Moreno; José Arturo Martínez-Orozco; José Esteban Muñoz-Medina; José de Jesús Nuñez-Contreras; Juan Bautista Chale-Dzul; Julissa Enciso-Ibarra; Kathia Elizabeth Tapia-Diaz; Luis Alberto Ochoa-Carrera; Margarita Matías-Florentino; Mario Mújica-Sánchez; Marissa Perez-Garcia; María Eugenia Jiménez-Corona; María Guadalupe Santiago-Mauricio; María Guadalupe de Jesús Mireles-Rivera; Nelly Sélem-Mojica; Pavel Isa; Ricardo Ciria Merce; Ricardo Grande; Rosa María Gutiérrez Rios; Rosario Vazquez-Larios; Santiago Ávila-Ríos; Selene Zárate; Susana Lopez; Verónica Mata-Haro; Victor Eduardo García-Arias; Víctor Hugo Borja-Aburto |
| EPI_ISL_3072149, EPI_ISL_3072151, EPI_ISL_3072152, EPI_ISL_3072153, EPI_ISL_3072154, EPI_ISL_3072155, EPI_ISL_3072156, EPI_ISL_3072157, EPI_ISL_3072158, EPI_ISL_3072160, EPI_ISL_3072161, EPI_ISL_3072162, EPI_ISL_3072163, EPI_ISL_3072164, EPI_ISL_3072165, EPI_ISL_3072166, EPI_ISL_3072169, EPI_ISL_3072172, EPI_ISL_3072173, EPI_ISL_3072174, EPI_ISL_3072175, EPI_ISL_3072176, EPI_ISL_3072177, EPI_ISL_3072178, EPI_ISL_3072179, EPI_ISL_3072180, EPI_ISL_3072181, EPI_ISL_3072182, EPI_ISL_3072183, EPI_ISL_3072186, EPI_ISL_3072187, EPI_ISL_3072188, EPI_ISL_3072189, EPI_ISL_3072190, EPI_ISL_3072191, EPI_ISL_3072192, EPI_ISL_3072193, EPI_ISL_3072194, EPI_ISL_3072195, EPI_ISL_3072196, EPI_ISL_3072197, EPI_ISL_3072198, EPI_ISL_3072199, EPI_ISL_3072200, EPI_ISL_3072201, EPI_ISL_3072202, EPI_ISL_3072203, EPI_ISL_3072204, EPI_ISL_3072205, EPI_ISL_3072206, EPI_ISL_3072207, EPI_ISL_3072208, EPI_ISL_3072209, EPI_ISL_3072210, EPI_ISL_3072211, EPI_ISL_3072212, EPI_ISL_3072213, EPI_ISL_3072214, EPI_ISL_3072215, EPI_ISL_3072216, EPI_ISL_3072217, EPI_ISL_3072218, EPI_ISL_3072219, EPI_ISL_3072220, EPI_ISL_3072221, EPI_ISL_3072222, EPI_ISL_3072223, EPI_ISL_3072224, EPI_ISL_3072225, EPI_ISL_3072226, EPI_ISL_3072227, EPI_ISL_3072228, EPI_ISL_3072229, EPI_ISL_3072230, EPI_ISL_3072232, EPI_ISL_3072233, EPI_ISL_3072234, EPI_ISL_3072235, EPI_ISL_3072236, EPI_ISL_3072237, EPI_ISL_3072238, EPI_ISL_3072239, EPI_ISL_3072240, EPI_ISL_3072241, EPI_ISL_3072242, EPI_ISL_3072243, EPI_ISL_3072244, EPI_ISL_3072245, EPI_ISL_3072246, EPI_ISL_3072247, EPI_ISL_3072248, EPI_ISL_3072249, EPI_ISL_3072250, EPI_ISL_3072251, EPI_ISL_3072252, EPI_ISL_3072253, EPI_ISL_3072254, EPI_ISL_3072255, EPI_ISL_3072265, EPI_ISL_3072266, EPI_ISL_3072267, EPI_ISL_3072268, EPI_ISL_3072269, EPI_ISL_3072270, EPI_ISL_3072271, EPI_ISL_3072272, EPI_ISL_3072275, EPI_ISL_3072276, EPI_ISL_3072277, EPI_ISL_3072278, EPI_ISL_3072279, EPI_ISL_3072280, EPI_ISL_3072281, EPI_ISL_3072282, EPI_ISL_3072283, EPI_ISL_3072284, EPI_ISL_3072285, EPI_ISL_3072286, EPI_ISL_3072287, EPI_ISL_3072288, EPI_ISL_3072289, EPI_ISL_3072290, EPI_ISL_3072291, EPI_ISL_3072292, EPI_ISL_3072293, EPI_ISL_3072294, EPI_ISL_3072295, EPI_ISL_3072296, EPI_ISL_3072297, EPI_ISL_3072298, EPI_ISL_3072299, EPI_ISL_3072300, EPI_ISL_3072301, EPI_ISL_3072302, EPI_ISL_3072303, EPI_ISL_3072304, EPI_ISL_3072305, EPI_ISL_3072306, EPI_ISL_3072307, EPI_ISL_3072308, EPI_ISL_3072309, EPI_ISL_3072310, EPI_ISL_3072311, EPI_ISL_3072312, EPI_ISL_3072313, EPI_ISL_3072314, EPI_ISL_3072316, EPI_ISL_3072317, EPI_ISL_3072318, EPI_ISL_3072327, EPI_ISL_3072329, EPI_ISL_3072331, EPI_ISL_3072333, EPI_ISL_3072335, EPI_ISL_3072337, EPI_ISL_3072339, EPI_ISL_3072341, EPI_ISL_3072343, EPI_ISL_3072345, EPI_ISL_3072347, EPI_ISL_3072349, EPI_ISL_3072351, EPI_ISL_3072359, EPI_ISL_3072370, EPI_ISL_3072374, EPI_ISL_3072378, EPI_ISL_3072394, EPI_ISL_3072396, EPI_ISL_3072399, EPI_ISL_3072403, EPI_ISL_3072405, EPI_ISL_3072407, EPI_ISL_3072409, EPI_ISL_3072411, EPI_ISL_3072413, EPI_ISL_3072415, EPI_ISL_3072417, EPI_ISL_3072425, EPI_ISL_3072427, EPI_ISL_3072430, EPI_ISL_3072431, EPI_ISL_3072434, EPI_ISL_3072436, EPI_ISL_3072438, EPI_ISL_3072440, EPI_ISL_3072442, EPI_ISL_3072444, EPI_ISL_3072448, EPI_ISL_3072450, EPI_ISL_3072452, EPI_ISL_3072454, EPI_ISL_3072456, EPI_ISL_3072458, EPI_ISL_3072460, EPI_ISL_3072462, EPI_ISL_3072463, EPI_ISL_3072467, EPI_ISL_3072469, | | | |

| | | | |
|---|---|---|---|
| EPI_ISL_3072471, EPI_ISL_3072473, EPI_ISL_3072475, EPI_ISL_3072477, EPI_ISL_3072479, EPI_ISL_3072481, EPI_ISL_3072485, EPI_ISL_3072487, EPI_ISL_3072489, EPI_ISL_3072491, EPI_ISL_3072494, EPI_ISL_3072497, EPI_ISL_3072499, EPI_ISL_3072501, EPI_ISL_3072503, EPI_ISL_3072513, EPI_ISL_3072515, EPI_ISL_3072517, EPI_ISL_3072519, EPI_ISL_3072521, EPI_ISL_3072523, EPI_ISL_3072525, EPI_ISL_3072527, EPI_ISL_3072529, EPI_ISL_3072531, EPI_ISL_3072533, EPI_ISL_3072536, EPI_ISL_3072539, EPI_ISL_3072551, EPI_ISL_3072553, EPI_ISL_3072567, EPI_ISL_3072569, EPI_ISL_3072571, EPI_ISL_3072573, EPI_ISL_3072575, EPI_ISL_3072577, EPI_ISL_3072579, EPI_ISL_3072581, EPI_ISL_3072584, EPI_ISL_3072586, EPI_ISL_3072588, EPI_ISL_3072590, EPI_ISL_3072592, EPI_ISL_3072595, EPI_ISL_3072597, EPI_ISL_3072598, EPI_ISL_3072600, EPI_ISL_3072603, EPI_ISL_3072604, EPI_ISL_3072606, EPI_ISL_3072608, EPI_ISL_3072610, EPI_ISL_3072612, EPI_ISL_3072614, EPI_ISL_3072616, EPI_ISL_3072618, EPI_ISL_3072620, EPI_ISL_3072622, EPI_ISL_3072624, EPI_ISL_3072626 | | | |
| see above | Unidade de apoio ao diagnostico da COVID - UNADIG | Bioinformatics Laboratory / LNCC | Alessandra P Lamarca; Alexandra L Gerber; Amilcar Tanuri; Ana Paula de C Guimaraes; Ana Tereza R Vasconcelos; Andrea Cony Cavalcanti; Caio Luiz Pereira Ribeiro; Cassia Alves; Cintia Policarpo; Claudia Maria Braga de Mello; Cristiane Gomes da Silva; Diana Mariani; Douglas Terra Machado; Flavio Dias da Silva; Gleidson da Silva de Oliveira; Leandro Magalhaes de Souza; Liliane Cavalcante; Luiz G P de Almeida; Marcio Henrique de Oliveira Garcia; Mario Sergio Ribeiro; Ronaldo da Silva F Jr; Silvia Carvalho |
| EPI_ISL_3276651 | Universidad Nacional de Colombia - Laboratorio Genómico One Health | Universidad Nacional de Colombia - Laboratorio Genómico One Health | Andres F. Cardona-Rios; Carlos Franco-Muñoz; Carolina Muñoz-Arango; Celeny Ortiz; Daniel O. Maldonado-Perez; Diego A. Álvarez-Díaz; Hector Alejandro Ruiz-Moreno; Idabely Betancur Ortiz; Jorge E. Osorio; Juan P. Hernandez-Ortiz; Karl A Ciuoderis; Katherine Laiton-Donato; Laura Silvana Perez; Lina M. Hurtado; Marcela Mercado-Reyes; Maria Angélica Maya; Maria Stella López; Rita Almanza Payares; Sandra Ines Cano; Simón Villegas Velásquez |
| EPI_ISL_3907215 | Universidad de León | SeqCOVID-SPAIN consortium/IBV(CSIC) | Ana Carvajal; Antonio J. Molina and SeqCOVID-SPAIN consortium; Héctor Argüello; Juan M. Fregeneda; Tania Fernández-Villa; Vicente Martín |
| EPI_ISL_3320725 | Universidad del Quindio | Instituto Nacional de Salud | Carlos Franco-Muñoz; Carmen Osorio; Diana Malo; Diego A. Álvarez-Díaz; Diego Andrés Prada; Gerardo Santamaría; Hector Alejandro Ruiz-Moreno; Jhonnatan Reales-González; Jorge Rivera; Juan Camilo Martinez; Julian Naizaque; Katherine Laiton-Donato; Lisseth Pardo; Magdalena Wiesner; Marcela Mercado-Reyes; Maria T. Herrera-Sepúlveda; Marta Lopez Blanco; Martha Lucia Ospina Martinez; Paola Rojas; Sergio Gomez; Sheryll Corchuelo; Ángela Alarcon Cruz |
| EPI_ISL_2981937, EPI_ISL_2981938 | University Hospitals of Geneva, Laboratory of Virology | HUG, Laboratory of Virology and the Health2030 Genome Center | Ana Rita Goncalves; Deborah Penet; Emmanouil Dermitzakis; Henri Pegeot; Ioannis Xenarios; Keith Harshman; Laurent Kaiser; Lorenzo Cerutti; Melyssa Elies; Samuel Cordey |
| EPI_ISL_3388712 | University of Illinois at Chicago | RIPHL at Rush University Medical Center | Edith Perez; Felix Araujo Perez; Iuliana Bentea; Kevin Kunstman; Laura Furtado; Marieta Hyde; Max Kolton; Omar Perez; Stefan Green |
| EPI_ISL_2840651, EPI_ISL_2919108, EPI_ISL_2919111, EPI_ISL_3026911 | Università degli Studi di Perugia | Istituto Zooprofilattico Sperimentale dell'Abruzzo e Molise "G. Caporale" | Ancora M; Biagetti M; Calistri P; Camilloni B; Cammà C; Curini V; Delli Compagni E; Di Domenico M; Di Pasquale A; Giammarioli M; Lorusso A; Mangone I; Marcacci M; Mencacci A; Puglia I; Rinaldi A; Savini G; Scialabba S |
| EPI_ISL_3044277 | Universitätsklinikum des Saarlandes - Institut für Virologie | Robert Koch Institute | |
| EPI_ISL_3268526, EPI_ISL_3268532, EPI_ISL_3268537, EPI_ISL_3268552, EPI_ISL_3268559, EPI_ISL_3268601, EPI_ISL_3268609, EPI_ISL_3268615, EPI_ISL_3268701, EPI_ISL_3268835, EPI_ISL_3268861, EPI_ISL_3268866, EPI_ISL_3268907, EPI_ISL_3268913, EPI_ISL_3268916, EPI_ISL_3268936, EPI_ISL_3268945, EPI_ISL_3268952, EPI_ISL_3269897, EPI_ISL_3756246, EPI_ISL_3756251, EPI_ISL_3756254, EPI_ISL_3756255, EPI_ISL_3756258, EPI_ISL_3756259, EPI_ISL_3756260 | | | |
| see above | Utah Public Health Laboratory | Utah Public Health Laboratory | Erin L. Young; Kelly F. Oakeson; Olinto Linares-Perdomo; Tara Gallagher |
| EPI_ISL_2978251 | VIDYMED LAUSANNE | Laboratory of genomics and metagenomics | Claire Bertelli; Damien Jacot; Gilbert Greub; Sébastien Aeby; Trestan Pillonel |
| EPI_ISL_5664166, EPI_ISL_5672284 | VIGILANCIA EM SAUDE DE FCO MORATO | Instituto Butantan | Antonio Jorge Martins; Claudia Renata dos Santos Barros; David Schlesinger; Debora Botequio Moretti; Dimas Tadeu Covas; Elaine Cristina Marqueze; Elaine Vieira Santos; Evandra Strazza Rodrigues; Heidge Fukumasu; Jayme Augusto de Souza-Neto; José Salvatore Leister Patané; Luiz Alcantara; Luiz Lehmann Coutinho; Maria Carolina Elias; Maurício Lacerda Nogueira; Rafael dos Santos Bezerra; Raul Machado Neto; Rejane Maria Tommasini Grotto; Ricardo Haddad; Sandra Coccuzzo Sampaio Vessoni; Simone Kashima; Svetoslav Nanev Slavov; Vincent Louis Viala |
| EPI_ISL_5673512, EPI_ISL_5673517, EPI_ISL_5673615, EPI_ISL_5674312 | VIGILANCIA EPIDEMIOLOGICA DE IBATE | Instituto Butantan | Antonio Jorge Martins; Claudia Renata dos Santos Barros; David Schlesinger; Debora Botequio Moretti; Dimas Tadeu Covas; Elaine Cristina Marqueze; Elaine Vieira Santos; Evandra Strazza Rodrigues; Heidge Fukumasu; Jayme Augusto de Souza-Neto; José Salvatore Leister Patané; Luiz Alcantara; Luiz Lehmann Coutinho; Maria Carolina Elias; Maurício Lacerda Nogueira; Rafael dos Santos Bezerra; Raul Machado Neto; Rejane Maria Tommasini Grotto; Ricardo Haddad; Sandra Coccuzzo Sampaio Vessoni; Simone Kashima; Svetoslav Nanev Slavov; Vincent Louis Viala |
| EPI_ISL_5665849, EPI_ISL_5665894 | VIGILANCIA EPIDEMIOLOGICA DE LEME | Instituto Butantan | Antonio Jorge Martins; Claudia Renata dos Santos Barros; David Schlesinger; Debora Botequio Moretti; Dimas Tadeu Covas; Elaine Cristina Marqueze; Elaine Vieira Santos; Evandra Strazza Rodrigues; Heidge Fukumasu; Jayme Augusto de Souza-Neto; José Salvatore Leister Patané; Luiz Alcantara; Luiz Lehmann Coutinho; Maria Carolina Elias; Maurício Lacerda Nogueira; Rafael dos Santos Bezerra; Raul Machado Neto; Rejane Maria Tommasini Grotto; Ricardo Haddad; Sandra Coccuzzo Sampaio Vessoni; Simone Kashima; Svetoslav Nanev Slavov; Vincent Louis Viala |
| EPI_ISL_5665813, EPI_ISL_5665837, EPI_ISL_5665874, EPI_ISL_5665908 | VIGILANCIA EPIDEMIOLOGICA E CONTROLE DE VETORES PIRASSUNUN | Instituto Butantan | Antonio Jorge Martins; Claudia Renata dos Santos Barros; David Schlesinger; Debora Botequio Moretti; Dimas Tadeu Covas; Elaine Cristina Marqueze; Elaine Vieira Santos; Evandra Strazza Rodrigues; Heidge Fukumasu; Jayme Augusto de Souza-Neto; José Salvatore Leister Patané; Luiz Alcantara; Luiz Lehmann Coutinho; Maria Carolina Elias; Maurício Lacerda Nogueira; Rafael dos Santos Bezerra; Raul Machado Neto; Rejane Maria Tommasini Grotto; Ricardo Haddad; Sandra Coccuzzo Sampaio Vessoni; Simone Kashima; Svetoslav Nanev Slavov; Vincent Louis Viala |
| EPI_ISL_5661844, EPI_ISL_5662025, EPI_ISL_5662171, EPI_ISL_5662436, EPI_ISL_5662640, EPI_ISL_5662993, EPI_ISL_5663280, EPI_ISL_5667989, EPI_ISL_5668415, EPI_ISL_5672031, EPI_ISL_5672050, EPI_ISL_5672077, EPI_ISL_5672169, EPI_ISL_5672232, EPI_ISL_5672364, EPI_ISL_5672396, EPI_ISL_5673734, EPI_ISL_5674122, EPI_ISL_5674266, EPI_ISL_5674505 | | | |
| see above | VIGILANCIA SANITARIA E VIG EPIDEMIOLOGICA DE ITAPEVI | Instituto Butantan | Antonio Jorge Martins; Claudia Renata dos Santos Barros; David Schlesinger; Debora Botequio Moretti; Dimas Tadeu Covas; Elaine Cristina Marqueze; Elaine Vieira Santos; Evandra Strazza Rodrigues; Heidge Fukumasu; Jayme Augusto de Souza-Neto; José Salvatore Leister Patané; Luiz Alcantara; Luiz Lehmann Coutinho; Maria Carolina Elias; Maurício Lacerda Nogueira; Rafael dos Santos Bezerra; Raul Machado Neto; Rejane Maria Tommasini Grotto; Ricardo Haddad; Sandra Coccuzzo Sampaio Vessoni; Simone Kashima; Svetoslav Nanev Slavov; Vincent Louis Viala |
| EPI_ISL_2938364, EPI_ISL_3007545, EPI_ISL_3007621 | Viollier AG | Department of Biosystems Science and Engineering, ETH Zürich | Andrea Patrizia Salzmann; Chaoran Chen; Christian Beisel; Christiane Beckmann; Christoph Noppen; Elodie Burcklen; Henriette Kurth; Ina Nissen; Ivan Topolsky; Kim Philipp Jablonski; Lara Fuhrmann; Maurice Redondo; Mirjam Feldkamp; Natascha Santacroce; Niko Beerenwinkel; Olivier Kobel; Rebecca Denes; Sarah Nadeau; Tanja Stadler |
| EPI_ISL_3021034, EPI_ISL_3025865, EPI_ISL_3025885, EPI_ISL_3214945 | Virginia Division of Consolidated Laboratory Services | Virginia Division of Consolidated Laboratory Services | Virginia Division of Consolidated Laboratory Services |
| EPI_ISL_3031273 | WESTCHESTER MEDICAL CENTER | Wadsworth Center, New York State Department of Health | Alexis Russell; Catharine Prussing; Daryl M. Lamson; Erasmus Schneider; Erica Lasek-Nesselquist; John Kelly; Jonathan Plitnick; Kirsten St. George; Matthew Shudt; Melissa A Leisner; Navjot Singh |
| EPI_ISL_2834707, EPI_ISL_2859440, EPI_ISL_3016353, EPI_ISL_3016360, EPI_ISL_3216979 | Washington State Department of Health Public Health Laboratories | Washington State Department of Health Public Health Laboratories | Avi Singh; Darren Lucas; Denny Russell; Drew MacKellar; Geoff Melly; Hannah Gray; Joenice Gonzalez; JohnAric Peterson; Philip Dykema; Rebecca Cao; Vanessa De Los Santos |
| EPI_ISL_3246657 | Weill Cornell Medicine | New York Genome Center | Amy Baldwin; Andre Corvelo; Arryn Craney; Chris Mason; Dayna M. Oschwald; Hanna Rennert; Lars F Westblade; Margaret Elizabeth Ross; Melissa Cushing; Michael Zody; Olivier Elemento; Priya Velu; Samantha Fennessey; Tom Maniatis |
| EPI_ISL_2833947, EPI_ISL_2842052, EPI_ISL_2842056 | Wichita State University- Molecular Diagnostic Lab | Kansas Health and Environmental Lab | Ben Olsen; Jonathan Barnell; Katherine Wiggins; Mike Grose; and Phil Adam |
| EPI_ISL_3131960 | Wisconsin State Laboratory of Hygiene Communicable Disease Division | Wisconsin State Laboratory of Hygiene Communicable Disease Division | Abigail C. Shockey; Alicia J. Mooney; Erika M. Hanson; Kelsey R. Florek; Richard Griesser; Sara Wagner; Tonya Danz |
| EPI_ISL_3062475, EPI_ISL_3066886, EPI_ISL_3160600, EPI_ISL_3298293 | Wyoming Public Health Laboratory | Wyoming Public Health Laboratory | Ashley Norberg; Brian Dominguez; Cari Sloma; Channing Weber; Chayse Rowley; Elliot Thomasson; Jim Mildenberger; Marley Goetz; Robert Petit; Sam Britz; Taylor Fearing; and Rob Christensen |
| EPI_ISL_3025423 | Yale Clinical Virology Lab | Grubaugh Lab - Yale School of Public Health | Anderson Brito; Annie Watkins; Chaney Kalinich; Chantal Vogels; Isabel Ott; Jessica Rothman; Joseph Fauver; Kendall Billig; Mallery Breban; Marie L. Landry; Mary Petrone; Nathan Grubaugh; Tara Alpert; Tobias Koch |
