## Supplement 3 for "Rapid and Accurate Identification of SARS-CoV-2 Omicron Variants Using Droplet Digital PCR (RT-ddPCR)": All_0705-0711.pdf

We gratefully acknowledge the following Authors from the Originating laboratories responsible for obtaining the specimens, as well as the Submitting laboratories where the genome data were generated and shared via GISAID, on which this research is based.

All Submitters of data may be contacted directly via www.gisaid.org

Authors are sorted alphabetically.

| Accession ID | Originating Laboratory | Submitting Laboratory | Authors |
|---|---|---|---|
| EPI_ISL_4945663, EPI_ISL_4945664 | "Laboratorio de inmunología y Biologia Molecular, Universidad del Cauca" | "Laboratorio de biotecnología, Universidad Icesi" | "Maria I. Gutierrez Lopez; Diana M. Florez Galindo; Marcela Mercado; Maria F. Villegas Torres; Paola A. Caicedo Burbano"; Programa Nacional de Caracterización Genómica de SARS-CoV-2; Sara Gonzalez |
| EPI_ISL_6179011 | ADPH | ADPH | Keah Higgins |
| EPI_ISL_5671018, EPI_ISL_5671029, EPI_ISL_5671069, EPI_ISL_5671129, EPI_ISL_5671136, EPI_ISL_5671208, EPI_ISL_5671272, EPI_ISL_5671288, EPI_ISL_5671310, EPI_ISL_5671324, EPI_ISL_5671390, EPI_ISL_5671420, EPI_ISL_5673696, EPI_ISL_5673777, EPI_ISL_5673790, EPI_ISL_5673826, EPI_ISL_5673859, EPI_ISL_5673895, EPI_ISL_5673964, EPI_ISL_5673982, EPI_ISL_5673989, EPI_ISL_5673996, EPI_ISL_5674003, EPI_ISL_5674012, EPI_ISL_5674157, EPI_ISL_5674175, EPI_ISL_5674185, EPI_ISL_5674191, EPI_ISL_5674195, EPI_ISL_5674225, EPI_ISL_5674255, EPI_ISL_5674270, EPI_ISL_5674288, EPI_ISL_5674296, EPI_ISL_5674305, EPI_ISL_5674347, EPI_ISL_5674379, EPI_ISL_5674406, EPI_ISL_5674410, EPI_ISL_5674415, EPI_ISL_5674441, EPI_ISL_5674542, EPI_ISL_5674571, EPI_ISL_5674638, EPI_ISL_5674677, EPI_ISL_5674683, EPI_ISL_5674715, EPI_ISL_5674727, EPI_ISL_5674733, EPI_ISL_5674831, EPI_ISL_5674884, EPI_ISL_5674890, EPI_ISL_5674945, EPI_ISL_5675117, EPI_ISL_5675122, EPI_ISL_5675128, EPI_ISL_5675151, EPI_ISL_5675160, EPI_ISL_5675170, EPI_ISL_5675267, EPI_ISL_5675315, EPI_ISL_5675458, EPI_ISL_5675479, EPI_ISL_5675512 | | | |
| see above | AFIP | Instituto Butantan | Antonio Jorge Martins; Claudia Renata dos Santos Barros; David Schlesinger; Debora Botequio Moretti; Dimas Tadeu Covas; Elaine Cristina Marqueze; Elaine Vieira Santos; Evandra Strazza Rodrigues; Heidge Fukumasu; Jayme Augusto de Souza-Neto; José Salvatore Leister Patané; Luiz Alcantara; Luiz Lehmann Coutinho; Maria Carolina Elias; Maurício Lacerda Nogueira; Rafael dos Santos Bezerra; Raul Machado Neto; Rejane Maria Tommasini Grotto; Ricardo Haddad; Sandra Coccuzzo Sampaio Vessoni; Simone Kashima; Svetoslav Nanev Slavov; Vincent Louis Viala |
| EPI_ISL_5671812 | AMBULATORIO MEDICO E ODONTOLOGICO MUNICIPAL DE PIEDADE | Instituto Butantan | Antonio Jorge Martins; Claudia Renata dos Santos Barros; David Schlesinger; Debora Botequio Moretti; Dimas Tadeu Covas; Elaine Cristina Marqueze; Elaine Vieira Santos; Evandra Strazza Rodrigues; Heidge Fukumasu; Jayme Augusto de Souza-Neto; José Salvatore Leister Patané; Luiz Alcantara; Luiz Lehmann Coutinho; Maria Carolina Elias; Maurício Lacerda Nogueira; Rafael dos Santos Bezerra; Raul Machado Neto; Rejane Maria Tommasini Grotto; Ricardo Haddad; Sandra Coccuzzo Sampaio Vessoni; Simone Kashima; Svetoslav Nanev Slavov; Vincent Louis Viala |
| EPI_ISL_5672654, EPI_ISL_5672741, EPI_ISL_5672965, EPI_ISL_5673063 | AMBULATORIO MEDICO MUNICIPAL DE AGUDOS | Instituto Butantan | Antonio Jorge Martins; Claudia Renata dos Santos Barros; David Schlesinger; Debora Botequio Moretti; Dimas Tadeu Covas; Elaine Cristina Marqueze; Elaine Vieira Santos; Evandra Strazza Rodrigues; Heidge Fukumasu; Jayme Augusto de Souza-Neto; José Salvatore Leister Patané; Luiz Alcantara; Luiz Lehmann Coutinho; Maria Carolina Elias; Maurício Lacerda Nogueira; Rafael dos Santos Bezerra; Raul Machado Neto; Rejane Maria Tommasini Grotto; Ricardo Haddad; Sandra Coccuzzo Sampaio Vessoni; Simone Kashima; Svetoslav Nanev Slavov; Vincent Louis Viala |
| EPI_ISL_3298336 | AREA DE SALUD CARTAGO | Inciensa, Instituto Costarricense de Investigación y Enseñanza en Nutrición y Salud | Adriana Godínez; Claudio Soto-Garita; Estela Cordero; Francisco Duarte; Hebleen Porras; Joselyn Prado & Monserrat Segura; José Luis Vargas; Mariela Gutiérrez; Melany Calderón |
| EPI_ISL_3298363 | AREA DE SALUD COBANO | Inciensa, Instituto Costarricense de Investigación y Enseñanza en Nutrición y Salud | Adriana Godínez; Claudio Soto-Garita; Estela Cordero; Francisco Duarte; Hebleen Porras; Joselyn Prado & María José Gómez; José Luis Vargas; Mariela Gutiérrez; Melany Calderón |
| EPI_ISL_3062039, EPI_ISL_3062957, EPI_ISL_3063004, EPI_ISL_3063032, EPI_ISL_3063033, EPI_ISL_3063035 | ASL Napoli 1 Centro | AMES Centro Polidiagnostico Strumentale S.r.l. | "Giovanni Savarese; Antonella Di Carlo; Eloisa Evangelista; Giovanni Savarese; Luigi D'Amore; Luisa Circelli; Maurizio D'Amora Antonio Fico; Maurizio D'Amora Antonio Fico"; Raffaella Ruggiero; Roberto Sirica |
| EPI_ISL_3130797, EPI_ISL_3130800 | ASTRALAB TASSIGNY | CNR Virus des Infections Respiratoires - France SUD | Antonin Bal; Bruno Lina; Gregory Destras; Gwendolyne Burfin; Hadrien Regue; Laurence Josset; Martine Valette; Quentin Semanas |
| EPI_ISL_3039697, EPI_ISL_3039702, EPI_ISL_3385302 | AULSS 8 Berica | Istituto Zooprofilattico Sperimentale delle Venezie | Adelaide Milani; Alessia Schivo; Alice Fusaro; Ambra Pastori; Annalisa Salviato; Antonia Ricci; Calogero Terregino; Edoardo Giussani; Elisa Palumbo; Erika Giorgia Quaranta; Isabella Monne; Luca Tassoni |
| EPI_ISL_2861345, EPI_ISL_2861346, EPI_ISL_2861347, EPI_ISL_2861356, EPI_ISL_2861364, EPI_ISL_2921139, EPI_ISL_2921140, EPI_ISL_2921142, EPI_ISL_2921159, EPI_ISL_2921164 | | | |
| see above | AZDelta | AZ Delta Medical Laboratories in Roeselare, Belgium | Dieter De Smet; Geert Martens; Merijn Vanhee; on behalf of AZ Delta COVID-19 Genomics core (member of Genomic surveillance of SARS-CoV-2 in Belgium network) |
| EPI_ISL_3217299, EPI_ISL_3217365, EPI_ISL_3217383, EPI_ISL_3217395, EPI_ISL_3217474, EPI_ISL_3217512, EPI_ISL_3217548, EPI_ISL_3217581, EPI_ISL_3217626, EPI_ISL_3217639, EPI_ISL_3217643, EPI_ISL_3217664, EPI_ISL_3217679, EPI_ISL_3217705, EPI_ISL_3217741, EPI_ISL_3217760, EPI_ISL_3217767, EPI_ISL_3217792, EPI_ISL_3217827, EPI_ISL_3217844, EPI_ISL_3217875, EPI_ISL_3217930, EPI_ISL_3217987, EPI_ISL_3218007, EPI_ISL_3218066, EPI_ISL_3218101, EPI_ISL_3218139, EPI_ISL_3218200, EPI_ISL_3218202, EPI_ISL_3218239, EPI_ISL_3218298, EPI_ISL_3218299, EPI_ISL_3218385, EPI_ISL_3218395, EPI_ISL_3218412, EPI_ISL_3218448, EPI_ISL_3218501, EPI_ISL_3218502, EPI_ISL_3218504, EPI_ISL_3218511, EPI_ISL_3218512, EPI_ISL_3218513, EPI_ISL_3218514, EPI_ISL_3218520, EPI_ISL_3218549, EPI_ISL_3218615, EPI_ISL_3218678, EPI_ISL_3218708, EPI_ISL_3218723, EPI_ISL_3218734, EPI_ISL_3218817, EPI_ISL_3219935, EPI_ISL_3219936, EPI_ISL_3219958, EPI_ISL_3219960, EPI_ISL_3220021, EPI_ISL_3220023, EPI_ISL_3220024, EPI_ISL_3220055, EPI_ISL_3220128, EPI_ISL_3220173, EPI_ISL_3220183, EPI_ISL_3220252, EPI_ISL_3220840, EPI_ISL_3220849, EPI_ISL_3305340, EPI_ISL_3305372, EPI_ISL_3305390, EPI_ISL_3305408, EPI_ISL_3305433, EPI_ISL_3305444, EPI_ISL_3305461, EPI_ISL_3305499, EPI_ISL_3305534, EPI_ISL_3321441, EPI_ISL_3321445, EPI_ISL_3321480, EPI_ISL_3321481, EPI_ISL_3321906, EPI_ISL_3321977, EPI_ISL_3322441, EPI_ISL_3323016, EPI_ISL_3323038, EPI_ISL_3429980, EPI_ISL_4186564, EPI_ISL_4364420, EPI_ISL_4364735, EPI_ISL_4365012 | | | |
| see above | Aegis Sciences Corporation | Centers for Disease Control and Prevention Division of Viral Diseases, Pathogen Discovery | Adrian Paskey; Alec Vest; Benjamin Rambo-Martin; Christopher Gulvick; Clinton Paden; Clinton R. Paden; Cyndi Clark; Dakota Howard; Darlene Wagner; Dhwani Batra; Dillon Nall; Duncan MacCannell; Erisa Sula; Ethan Sanders; Holly Houdeshell; Jason Caravas; Kara Moser; Kristine Lacek; Matthew Hardison; Matthew Schmerer; Ola Kvalvaag; Patrick Campbell; Peter Cook; Peter W. Cook; Rob Case; Scott Sammons; Shatavia Morrison; Shaun Westlund; Tymeckia Kendall; Victoria Caban Figueroa; Vikramsinha Ghorpade; Yvette Unoarumhi |
| EPI_ISL_3253977 | Ages,Lifebrain COVID 19 Labor GmbH Klinik Penzing Pavillon 17 | Elling and Cochella laboratories, IMBA/IMP | Alexander Stark; Ezgi Oezkan; Luisa Cochella and Ulrich Elling; Marcus Strobl; Olga Frank; Ramesh Yelagandula; Tanino Albanese |
| EPI_ISL_2987162 | Alaska State Virology Laboratory | Alaska State Virology Laboratory | Elva House; Jack Chen; Jacob Zidek; Lisa Smith; Ph.D.; Stephanie DeRonde |
| EPI_ISL_3388719, EPI_ISL_3388803 | Alverno Labs | RIPHL at Rush University Medical Center | Edith Perez; Felix Araujo Perez; Kevin Kunstman; Laura Furtado; Marieta Hyde; Max Kolton; Melissa Mace; Stefan Green |
| EPI_ISL_3127443 | Applied DNA Sciences Inc | Applied DNA Sciences Inc | Huan Dong; Lucas Liu; Spencer William |
| EPI_ISL_7711725 | Area De Salud Aserri [Aserri/San Jose] | Universidad de Costa Rica e Instituto Costarricense de Investigacion y Ensenanza en Nutricion y Salud | Brenes Hebleen; Camacho Erwin; Campos-Sanchez Rebeca; Ceballos Ana; Cordero Estela; Cristancho Marco; Duarte Francisco; Fernandez-Do Porto Dario; Herrera-Estrella Alfredo; Jimenez-Moraila Beatriz; Kreuze Jan; Mireles-Rivera Guadalupe; Molina-Mora Jose; Munoz-Medina Jose Esteban; Negri Tatiana; Nunes Gisele; Oliveira Guilherme; Oliveira Renato; Reales-Gonzalez Jhonnatan; Remes-Lenicov Federico; Reyes Alejandro; Sosa Ezequiel; Soto Claudio; Tsukayama Pablo; Turjanski Adrian; and Zayat Jonathan |
| EPI_ISL_2981919, EPI_ISL_3089183, EPI_ISL_3089196, EPI_ISL_3090069, EPI_ISL_3217104 | Arizona State Public Health Laboratory | Arizona State Public Health Laboratory | Jessica Escobar; Katherine Fullerton; Linda Getsinger; Matthew Contursi; Nobuko Fukushima; Stacy White; Trung Huynh; Victor Waddell |
| EPI_ISL_2968644, EPI_ISL_2968657, EPI_ISL_3064318, EPI_ISL_3064331, EPI_ISL_3064355, EPI_ISL_3064357 | Arizona State University | Arizona State University | Ajeet Bains; Efrem S. Lim; Joshua LaBaer; Joy M. Blain; LaRinda A. Holland; Matthew F. Smith; Nicholas J. Mellor; Peter T. Skidmore; Rabia Maqsood; Valerie Harris; Vel Murugan |
| EPI_ISL_3553424, EPI_ISL_3553485 | Arkansas Children's Hospital | Center for Global Health, University of New Mexico Health Sciences Center | Ashton Ingold; Bobby Boyanton; Darrell Dinwiddie; Daryl Domman; Joshua L. Kennedy; Kurt Schwalm; Valerie Morley |
| EPI_ISL_3048647 | Arkansas Public Health Laboratory, Arkansas Department of Health | University of Minnesota Genomics Center | Corbin Dirkx; Daryl M. Gohl; Jaquelyn Kuriger-Laber; John Garbe |
| EPI_ISL_2932705 | Arlon | Plateforme de testing Namuroise | Degosserie Jonathan; Demars Aurore; Denis Olivier; Lesly Nyinkeu Kemamen; Maschietto Céline; Mullier François; Nicolas Gilliard; Nobis Chloé; Otto Gaetan |
| EPI_ISL_3307684 | Austrian Agency for Health and Food Safety (AGES) | Bergthaler laboratory, CeMM Research Center for Molecular Medicine of the Austrian Academy of Sciences | Andreas Bergthaler; Anna Schedl; Bekir Erguner; Benedikt Agerer; Christoph Bock; Fabian Amman; Jan Laine; Lukas Endler; Maelle Le Moing; Martin Senekowitsch; Matthew Thornton; Michael Schuster; Petr Triska; Thomas Penz |
| EPI_ISL_3030468 | Azienda Ospedaliera Terni | Istituto Zooprofilattico Sperimentale dell'Abruzzo e Molise "G. Caporale" | Ancora M; Calistri P; Cammà C; Curini V; Delli Compagni E; Di Domenico M; Di Pasquale A; Lorusso A; Mangone I; Marcacci M; Palumbo M; Puglia I; Rinaldi A; Savini G; Scaccetti A; Scialabba S |
| EPI_ISL_3393459 | BIO ARD'AISNE | Department of Virology, Henri Mondor University Hospital, Assistance Publique Hôpitaux de Paris, Université Paris-Est Créteil, INSERM U955 | Alexandre Soulier; Christophe Rodriguez; Elisabeth Trawinski; Guillaume Gricourt; Jean-Michel Pawlotsky; Melissa N'Debi; Slim Fourati; Vanessa Demontant |
| EPI_ISL_5673276, EPI_ISL_5673311, EPI_ISL_5673322, EPI_ISL_5673360, EPI_ISL_5673389, EPI_ISL_5673425, EPI_ISL_5673478, EPI_ISL_5673490, EPI_ISL_5673569, EPI_ISL_5673587, EPI_ISL_5673590, EPI_ISL_5673599, EPI_ISL_5673631, EPI_ISL_5673648, EPI_ISL_5673703, EPI_ISL_5673749, EPI_ISL_5673753, EPI_ISL_5673769, EPI_ISL_5673891, EPI_ISL_5673902, EPI_ISL_5673912, EPI_ISL_5673928, | | | |

| | | | |
|---|---|---|---|
| EPI_ISL_5673941, EPI_ISL_5674017, EPI_ISL_5674068, EPI_ISL_5674082, EPI_ISL_5674098, EPI_ISL_5674103, EPI_ISL_5674139, EPI_ISL_5674165, EPI_ISL_5674176, EPI_ISL_5674241, EPI_ISL_5674246, EPI_ISL_5674262, EPI_ISL_5674298, EPI_ISL_5674331, EPI_ISL_5674341, EPI_ISL_5674353, EPI_ISL_5674367, EPI_ISL_5674373, EPI_ISL_5674422, EPI_ISL_5674455, EPI_ISL_5674459, EPI_ISL_5674484, EPI_ISL_5674491, EPI_ISL_5674554, EPI_ISL_5674560, EPI_ISL_5674580, EPI_ISL_5674584, EPI_ISL_5674592, EPI_ISL_5674601, EPI_ISL_5674609, EPI_ISL_5674612, EPI_ISL_5674648, EPI_ISL_5674693, EPI_ISL_5674764, EPI_ISL_5674768, EPI_ISL_5674774, EPI_ISL_5674783, EPI_ISL_5674788, EPI_ISL_5674794, EPI_ISL_5674802, EPI_ISL_5674846, EPI_ISL_5674854, EPI_ISL_5674860, EPI_ISL_5674864, EPI_ISL_5674903, EPI_ISL_5674913, EPI_ISL_5674959, EPI_ISL_5674961, EPI_ISL_5674969, EPI_ISL_5674974, EPI_ISL_5675025, EPI_ISL_5675032, EPI_ISL_5675066, EPI_ISL_5675083, EPI_ISL_5675091, EPI_ISL_5675094, EPI_ISL_5675105, EPI_ISL_5675107, EPI_ISL_5675113, EPI_ISL_5675133, EPI_ISL_5675139, EPI_ISL_5675141, EPI_ISL_5675184, EPI_ISL_5675192, EPI_ISL_5675236, EPI_ISL_5675242, EPI_ISL_5675248, EPI_ISL_5675255, EPI_ISL_5675289, EPI_ISL_5675321, EPI_ISL_5675330, EPI_ISL_5675344, EPI_ISL_5675424, EPI_ISL_5677792 | | | |
| see above | BIOFAST | Instituto Butantan | Antonio Jorge Martins; Claudia Renata dos Santos Barros; David Schlesinger; Debora Botequio Moretti; Dimas Tadeu Covas; Elaine Cristina Marqueze; Elaine Vieira Santos; Evandra Strazza Rodrigues; Heidge Fukumasu; Jayme Augusto de Souza-Neto; José Salvatore Leister Patané; Luiz Alcantara; Luiz Lehmann Coutinho; Maria Carolina Elias; Maurício Lacerda Nogueira; Rafael dos Santos Bezerra; Raul Machado Neto; Rejane Maria Tommasini Grotto; Ricardo Haddad; Sandra Coccuzzo Sampaio Vessoni; Simone Kashima; Svetoslav Nanev Slavov; Vincent Louis Viala |
| EPI_ISL_3392198 | BROUSSAIS | Department of Virology, Henri Mondor University Hospital, Assistance Publique Hôpitaux de Paris, Université Paris-Est Créteil, INSERM U955 | Alexandre Soulier; Christophe Rodriguez; Elisabeth Trawinski; Guillaume Gricourt; Jean-Michel Pawlotsky; Melissa N'Debi; Slim Fourati; Vanessa Demontant |
| EPI_ISL_2984216 | Biogroup Bio Lam-LCD Saint-Denis | Department of Virology, Henri Mondor University Hospital, Assistance Publique Hôpitaux de Paris, Université Paris-Est Créteil, INSERM U955 | Alexandre Soulier; Christophe Rodriguez; Elisabeth Trawinski; Guillaume Gricourt; Jean-Michel Pawlotsky; Melissa N'Debi; Slim Fourati; Vanessa Demontant |
| EPI_ISL_3145701, EPI_ISL_3146060, EPI_ISL_3146061, EPI_ISL_3146062, EPI_ISL_3146063, EPI_ISL_3146064, EPI_ISL_3146065, EPI_ISL_3146066, EPI_ISL_3146067, EPI_ISL_3146068, EPI_ISL_3146069, EPI_ISL_3146070, EPI_ISL_3146071, EPI_ISL_3146072, EPI_ISL_3146073, EPI_ISL_3146074, EPI_ISL_3146075, EPI_ISL_3146076, EPI_ISL_3146077, EPI_ISL_3146078, EPI_ISL_3146079, EPI_ISL_3146080, EPI_ISL_3146081, EPI_ISL_3146082, EPI_ISL_3146083, EPI_ISL_3146084, EPI_ISL_3146085, EPI_ISL_3146086, EPI_ISL_3146087, EPI_ISL_3146088, EPI_ISL_3146089, EPI_ISL_3146090, EPI_ISL_3146091, EPI_ISL_3146092, EPI_ISL_3146093, EPI_ISL_3146094, EPI_ISL_3146095, EPI_ISL_3146096, EPI_ISL_3146097, EPI_ISL_3146098, EPI_ISL_3146099, EPI_ISL_3146100, EPI_ISL_3146101, EPI_ISL_3146102, EPI_ISL_3146103, EPI_ISL_3146104, EPI_ISL_3146105, EPI_ISL_3146106, EPI_ISL_3146107, EPI_ISL_3146108, EPI_ISL_3146109, EPI_ISL_3146110, EPI_ISL_3146111, EPI_ISL_3146112, EPI_ISL_3146113, EPI_ISL_3146114, EPI_ISL_3146115, EPI_ISL_3146116, EPI_ISL_3146117, EPI_ISL_3146118, EPI_ISL_3146119, EPI_ISL_3146120, EPI_ISL_3146121, EPI_ISL_3146122, EPI_ISL_3146123, EPI_ISL_3146124, EPI_ISL_3146125, EPI_ISL_3146126, EPI_ISL_3146127, EPI_ISL_3146128, EPI_ISL_3146129, EPI_ISL_3146130, EPI_ISL_3146131, EPI_ISL_3146132, EPI_ISL_3146133, EPI_ISL_3146134, EPI_ISL_3146135, EPI_ISL_3146136, EPI_ISL_3146137, EPI_ISL_3146138, EPI_ISL_3146139, EPI_ISL_3146140, EPI_ISL_3146141, EPI_ISL_3146142, EPI_ISL_3146143, EPI_ISL_3146144, EPI_ISL_3146145, EPI_ISL_3146146, EPI_ISL_3146147, EPI_ISL_3146148, EPI_ISL_3146149, EPI_ISL_3146150, EPI_ISL_3146151, EPI_ISL_3146152, EPI_ISL_3146153, EPI_ISL_3146154, EPI_ISL_3146155, EPI_ISL_3146156, EPI_ISL_3146157, EPI_ISL_3146158, EPI_ISL_3146159, EPI_ISL_3146160, EPI_ISL_3146161, EPI_ISL_3146162, EPI_ISL_3146163, EPI_ISL_3146164, EPI_ISL_3146165, EPI_ISL_3146166, EPI_ISL_3146167, EPI_ISL_3146168, EPI_ISL_3146169, EPI_ISL_3146170, EPI_ISL_3146171, EPI_ISL_3146172, EPI_ISL_3146173, EPI_ISL_3146174, EPI_ISL_3146175, EPI_ISL_3146176, EPI_ISL_3146177, EPI_ISL_3146178, EPI_ISL_3146179, EPI_ISL_3146180, EPI_ISL_3146181, EPI_ISL_3146182, EPI_ISL_3146183, EPI_ISL_3146184, EPI_ISL_3146185, EPI_ISL_3146186, EPI_ISL_3146187, EPI_ISL_3146188, EPI_ISL_3146189, EPI_ISL_3146190, EPI_ISL_3146191, EPI_ISL_3146192, EPI_ISL_3146193, EPI_ISL_3146194, EPI_ISL_3146195, EPI_ISL_3146196, EPI_ISL_3146197, EPI_ISL_3146198, EPI_ISL_3146199, EPI_ISL_3146200, EPI_ISL_3146201, EPI_ISL_3146202, EPI_ISL_3146268, EPI_ISL_3146269, EPI_ISL_3146272, EPI_ISL_3146278, EPI_ISL_3146425, EPI_ISL_3146468, EPI_ISL_3146469, EPI_ISL_3146483, EPI_ISL_3146484, EPI_ISL_3146486, EPI_ISL_3146488, EPI_ISL_3146489, EPI_ISL_3146490, EPI_ISL_3146517, EPI_ISL_3146521, EPI_ISL_3146536, EPI_ISL_3146537, EPI_ISL_3146543, EPI_ISL_3146550 | | | |
| see above | BioneXt Lab | Laboratoire national de sante, Microbiology, Microbial Genomics Platform | Anke Wienecke-Baldacchino; Catherine Ragimbeau; Elodie Solarino; Fatu Djabi; Jessica Tapp; Lise Pignon; Raoul Salmon; Tamir Abdelrahman; Thibault Ferrandon; Virginie Jover |
| EPI_ISL_3596630, EPI_ISL_3602553, EPI_ISL_3602555, EPI_ISL_3602558, EPI_ISL_3602560, EPI_ISL_3602561, EPI_ISL_3602563, EPI_ISL_3602564, EPI_ISL_3602636, EPI_ISL_3602638, EPI_ISL_3602655, EPI_ISL_3602657, EPI_ISL_3602659, EPI_ISL_3602660, EPI_ISL_3602662, EPI_ISL_3602663, EPI_ISL_3602665, EPI_ISL_3602667, EPI_ISL_3602670, EPI_ISL_3602683, EPI_ISL_3602685, EPI_ISL_3602686, EPI_ISL_3602687, EPI_ISL_3602688, EPI_ISL_3602689, EPI_ISL_3602690, EPI_ISL_3602691, EPI_ISL_3602692, EPI_ISL_3602693, EPI_ISL_3602694, EPI_ISL_3602700, EPI_ISL_3602702, EPI_ISL_3602705, EPI_ISL_3602706, EPI_ISL_3602707, EPI_ISL_3602708, EPI_ISL_3602710, EPI_ISL_3602711, EPI_ISL_3602713, EPI_ISL_3602714, EPI_ISL_3602715, EPI_ISL_3602716, EPI_ISL_3602717, EPI_ISL_3602718, EPI_ISL_3602723, EPI_ISL_3602731, EPI_ISL_3602735, EPI_ISL_3602737, EPI_ISL_3602739, EPI_ISL_3602744, EPI_ISL_3602755, EPI_ISL_3602758, EPI_ISL_3602759, EPI_ISL_3602760, EPI_ISL_3602761, EPI_ISL_3602762, EPI_ISL_3602764, EPI_ISL_3602776, EPI_ISL_3602792, EPI_ISL_3602793, EPI_ISL_3602809, EPI_ISL_3602810, EPI_ISL_3602811, EPI_ISL_3602812, EPI_ISL_3602813, EPI_ISL_3602815, EPI_ISL_3602821, EPI_ISL_3602822, EPI_ISL_3602823, EPI_ISL_3602824, EPI_ISL_3602826, EPI_ISL_3602827, EPI_ISL_3602828, EPI_ISL_3602829, EPI_ISL_3602830, EPI_ISL_3602833, EPI_ISL_3602834, EPI_ISL_3602837, EPI_ISL_3602839, EPI_ISL_3602841, EPI_ISL_3602843, EPI_ISL_3602844, EPI_ISL_3602845, EPI_ISL_3602847, EPI_ISL_3602849, EPI_ISL_3602850, EPI_ISL_3602851, EPI_ISL_3602854, EPI_ISL_3602860, EPI_ISL_3602861, EPI_ISL_3602862, EPI_ISL_3602864, EPI_ISL_3602865, EPI_ISL_3602870, EPI_ISL_3602871, EPI_ISL_3602873, EPI_ISL_3602875, EPI_ISL_3602877, EPI_ISL_3602880, EPI_ISL_3602884, EPI_ISL_3602903, EPI_ISL_3602911, EPI_ISL_3602921, EPI_ISL_3602922, EPI_ISL_3602923, EPI_ISL_3602925, EPI_ISL_3602933, EPI_ISL_4423672, EPI_ISL_4423674 | | | |
| see above | British Columbia Centre For Disease Control | BCCDC Public Health Laboratory | Ana Pacagnella; Corrinne Ng; Dan Fornika; John Tyson; Kim Macdonald; Kimia Kamelian; Linda Hoang; Loretta Janz; Mel Krajden; Prystajecky Natalie; Robert Azana; Shannon Russell |
| EPI_ISL_3007359, EPI_ISL_3007370, EPI_ISL_3007441, EPI_ISL_3007446 | Broad Institute Clinical Research Sequencing Platform | Infectious Disease Program, Broad Institute of Harvard and MIT | Adams, G.; B.L.; B.W.; Bauer, M.; Birren; Blumenstiel, B.; Brown, C.; Carter, A.; Chaluvadi, S.; D.J.; DeFelice, M.; DeRuff, K.; Dodge, S.; Gabriel, S.; Gallagher, G.; Gladden-Young, A.; Granger, B.; J.E.; K.J.; Lagerborg, K.; Larkin, K.; Lee, M.; Lemieux; Lennon, N.; Loreth, C.; Madoff, L.; McGovern, S.; Meldrim, J.; Normandin, E.; P.C.; Park; Pearlman, L.; Reilly, S.; Rudy, M.; Sabeti; Siddle; Smole, S.; Tomkins-Tinch, C.; Vicente, G.; and MacInnis |
| EPI_ISL_5675352, EPI_ISL_5675370, EPI_ISL_5675381, EPI_ISL_5675411 | CADIP VILA REGINA FERNANDOPOLIS | Instituto Butantan | Antonio Jorge Martins; Claudia Renata dos Santos Barros; David Schlesinger; Debora Botequio Moretti; Dimas Tadeu Covas; Elaine Cristina Marqueze; Elaine Vieira Santos; Evandra Strazza Rodrigues; Heidge Fukumasu; Jayme Augusto de Souza-Neto; José Salvatore Leister Patané; Luiz Alcantara; Luiz Lehmann Coutinho; Maria Carolina Elias; Maurício Lacerda Nogueira; Rafael dos Santos Bezerra; Raul Machado Neto; Rejane Maria Tommasini Grotto; Ricardo Haddad; Sandra Coccuzzo Sampaio Vessoni; Simone Kashima; Svetoslav Nanev Slavov; Vincent Louis Viala |
| EPI_ISL_3099737 | CAP Les Hortes | Banc de Sang i Teixits | Carlos Hobeich; Francisco Vidal; Irene Corrales; Lorena Ramírez; Maria Glòria Soria; Natália Comes; Nina Borràs; Noemí Gonzalez; Silvia Sauleda |
| EPI_ISL_5460218, EPI_ISL_5460260, EPI_ISL_5460303, EPI_ISL_5460306, EPI_ISL_7703902, EPI_ISL_7703912, EPI_ISL_7703921, EPI_ISL_7703922, EPI_ISL_7703923, EPI_ISL_7703928, EPI_ISL_7703972, EPI_ISL_7703975, EPI_ISL_7703981, EPI_ISL_7703986, EPI_ISL_7704013 | | | |
| see above | CDPH VBL | California Department of Public Health | Emily Smith on behalf of CDPH-COVIDNet |
| EPI_ISL_3043892, EPI_ISL_3043939 | CENTOGENE Frankfurt Laboratory: Niederlassung Industriepark Höchst | Robert Koch Institute | |
| EPI_ISL_5673904, EPI_ISL_5674040, EPI_ISL_5674708, EPI_ISL_5674881, EPI_ISL_5675430 | CENTRO DE ATENDIMENTO E ENFRENTAMENTO A COVID 19 | Instituto Butantan | Antonio Jorge Martins; Claudia Renata dos Santos Barros; David Schlesinger; Debora Botequio Moretti; Dimas Tadeu Covas; Elaine Cristina Marqueze; Elaine Vieira Santos; Evandra Strazza Rodrigues; Heidge Fukumasu; Jayme Augusto de Souza-Neto; José Salvatore Leister Patané; Luiz Alcantara; Luiz Lehmann Coutinho; Maria Carolina Elias; Maurício Lacerda Nogueira; Rafael dos Santos Bezerra; Raul Machado Neto; Rejane Maria Tommasini Grotto; Ricardo Haddad; Sandra Coccuzzo Sampaio Vessoni; Simone Kashima; Svetoslav Nanev Slavov; Vincent Louis Viala |
| EPI_ISL_5675387 | CENTRO DE ESPECIALIDADES MARIAS DA GLORIA | Instituto Butantan | Antonio Jorge Martins; Claudia Renata dos Santos Barros; David Schlesinger; Debora Botequio Moretti; Dimas Tadeu Covas; Elaine Cristina Marqueze; Elaine Vieira Santos; Evandra Strazza Rodrigues; Heidge Fukumasu; Jayme Augusto de Souza-Neto; José Salvatore Leister Patané; Luiz Alcantara; Luiz Lehmann Coutinho; Maria Carolina Elias; Maurício Lacerda Nogueira; Rafael dos Santos Bezerra; Raul Machado Neto; Rejane Maria Tommasini Grotto; Ricardo Haddad; Sandra Coccuzzo Sampaio Vessoni; Simone Kashima; Svetoslav Nanev Slavov; Vincent Louis Viala |
| EPI_ISL_5673320, EPI_ISL_5673594, EPI_ISL_5673783, EPI_ISL_5674465, EPI_ISL_5674520, EPI_ISL_5674663, EPI_ISL_5674823 | | | |
| see above | CENTRO DE ESPECIALIDADES MUNICIPAL CESAR TORREZAN R BONITO | Instituto Butantan | Antonio Jorge Martins; Claudia Renata dos Santos Barros; David Schlesinger; Debora Botequio Moretti; Dimas Tadeu Covas; Elaine Cristina Marqueze; Elaine Vieira Santos; Evandra Strazza Rodrigues; Heidge Fukumasu; Jayme Augusto de Souza-Neto; José Salvatore Leister Patané; Luiz Alcantara; Luiz Lehmann Coutinho; Maria Carolina Elias; Maurício Lacerda Nogueira; Rafael dos Santos Bezerra; Raul Machado Neto; Rejane Maria Tommasini Grotto; Ricardo Haddad; Sandra Coccuzzo Sampaio Vessoni; Simone Kashima; Svetoslav Nanev Slavov; Vincent Louis Viala |
| EPI_ISL_5674935, EPI_ISL_5675002, EPI_ISL_5675292 | CENTRO DE REFERENCIA DO IDOSO DR HUMBERTO MENDES DE CARVALHO | Instituto Butantan | Antonio Jorge Martins; Claudia Renata dos Santos Barros; David Schlesinger; Debora Botequio Moretti; Dimas Tadeu Covas; Elaine Cristina Marqueze; Elaine Vieira Santos; Evandra Strazza Rodrigues; Heidge Fukumasu; Jayme Augusto de Souza-Neto; José Salvatore Leister Patané; Luiz Alcantara; Luiz Lehmann Coutinho; Maria Carolina Elias; Maurício Lacerda Nogueira; Rafael dos Santos Bezerra; Raul Machado Neto; Rejane Maria Tommasini Grotto; Ricardo Haddad; Sandra Coccuzzo Sampaio Vessoni; Simone Kashima; Svetoslav Nanev Slavov; Vincent Louis Viala |
| EPI_ISL_5673013 | CENTRO DE SAUDE DE AVAI | Instituto Butantan | Antonio Jorge Martins; Claudia Renata dos Santos Barros; David Schlesinger; Debora Botequio Moretti; Dimas Tadeu Covas; Elaine Cristina Marqueze; Elaine Vieira Santos; Evandra Strazza Rodrigues; Heidge Fukumasu; Jayme Augusto de Souza-Neto; José Salvatore Leister Patané; Luiz Alcantara; Luiz Lehmann Coutinho; Maria Carolina Elias; Maurício Lacerda Nogueira; Rafael dos Santos Bezerra; Raul Machado Neto; Rejane Maria Tommasini Grotto; Ricardo Haddad; Sandra Coccuzzo Sampaio Vessoni; Simone Kashima; Svetoslav Nanev Slavov; Vincent Louis Viala |
| EPI_ISL_5673970 | CENTRO DE SAUDE DE CAJOBI | Instituto Butantan | Antonio Jorge Martins; Claudia Renata dos Santos Barros; David Schlesinger; Debora Botequio Moretti; Dimas Tadeu Covas; Elaine Cristina Marqueze; Elaine Vieira Santos; Evandra Strazza Rodrigues; Heidge Fukumasu; Jayme Augusto de Souza-Neto; José Salvatore Leister Patané; Luiz Alcantara; Luiz Lehmann Coutinho; Maria Carolina Elias; Maurício Lacerda Nogueira; Rafael dos Santos Bezerra; Raul Machado Neto; Rejane Maria Tommasini Grotto; Ricardo Haddad; Sandra Coccuzzo Sampaio Vessoni; Simone Kashima; Svetoslav Nanev Slavov; Vincent Louis Viala |
| EPI_ISL_5673220, EPI_ISL_5673626, EPI_ISL_5674229, EPI_ISL_5674654 | CENTRO DE SAUDE DE CHAVANTES | Instituto Butantan | Antonio Jorge Martins; Claudia Renata dos Santos Barros; David Schlesinger; Debora Botequio Moretti; Dimas Tadeu Covas; Elaine Cristina Marqueze; Elaine Vieira Santos; Evandra Strazza Rodrigues; Heidge Fukumasu; Jayme Augusto de Souza-Neto; José Salvatore Leister Patané; Luiz Alcantara; Luiz Lehmann Coutinho; Maria Carolina Elias; Maurício Lacerda Nogueira; Rafael dos Santos Bezerra; Raul Machado Neto; Rejane Maria Tommasini Grotto; Ricardo Haddad; Sandra Coccuzzo Sampaio Vessoni; Simone Kashima; Svetoslav Nanev Slavov; Vincent Louis Viala |
| EPI_ISL_5672966, EPI_ISL_5672967 | CENTRO DE SAUDE DE LUCIANOPOLIS | Instituto Butantan | Antonio Jorge Martins; Claudia Renata dos Santos Barros; David Schlesinger; Debora Botequio Moretti; Dimas Tadeu Covas; Elaine Cristina Marqueze; Elaine Vieira Santos; Evandra Strazza Rodrigues; Heidge Fukumasu; Jayme Augusto de Souza-Neto; José Salvatore Leister Patané; Luiz Alcantara; Luiz Lehmann Coutinho; Maria Carolina Elias; Maurício Lacerda Nogueira; Rafael dos Santos Bezerra; Raul Machado Neto; Rejane Maria Tommasini Grotto; Ricardo Haddad; Sandra Coccuzzo Sampaio Vessoni; Simone Kashima; Svetoslav Nanev Slavov; Vincent Louis Viala |
| EPI_ISL_5672650, EPI_ISL_5672669, EPI_ISL_5672890, EPI_ISL_5672976, EPI_ISL_5672986 | CENTRO DE SAUDE DE PIRAJUI | Instituto Butantan | Antonio Jorge Martins; Claudia Renata dos Santos Barros; David Schlesinger; Debora Botequio Moretti; Dimas Tadeu Covas; Elaine Cristina Marqueze; Elaine Vieira Santos; Evandra Strazza Rodrigues; Heidge Fukumasu; Jayme Augusto de Souza-Neto; José Salvatore Leister Patané; Luiz Alcantara; Luiz Lehmann Coutinho; Maria Carolina Elias; Maurício Lacerda Nogueira; Rafael dos Santos Bezerra; Raul Machado Neto; Rejane Maria Tommasini Grotto; Ricardo Haddad; Sandra Coccuzzo Sampaio Vessoni; Simone Kashima; Svetoslav Nanev Slavov; Vincent Louis Viala |
| EPI_ISL_5673118, EPI_ISL_5674387 | CENTRO DE SAUDE DR JOSE AGOSTINHO NOGUEIRA PAULICEIA | Instituto Butantan | Antonio Jorge Martins; Claudia Renata dos Santos Barros; David Schlesinger; Debora Botequio Moretti; Dimas Tadeu Covas; Elaine Cristina Marqueze; Elaine Vieira Santos; Evandra Strazza Rodrigues; Heidge Fukumasu; Jayme Augusto de Souza-Neto; José Salvatore Leister Patané; Luiz Alcantara; Luiz Lehmann Coutinho; Maria Carolina Elias; Maurício Lacerda Nogueira; Rafael dos Santos Bezerra; Raul Machado Neto; Rejane Maria Tommasini Grotto; Ricardo Haddad; Sandra Coccuzzo Sampaio Vessoni; Simone Kashima; Svetoslav Nanev Slavov; Vincent Louis Viala |
| EPI_ISL_5680779 | CENTRO DE SAUDE DR MARIO DIAS DE AGUIAR CAPIVARI | Instituto Butantan | Antonio Jorge Martins; Claudia Renata dos Santos Barros; David Schlesinger; Debora Botequio Moretti; Dimas Tadeu Covas; Elaine Cristina Marqueze; Elaine Vieira Santos; Evandra Strazza Rodrigues; Heidge Fukumasu; Jayme Augusto de Souza-Neto; José Salvatore Leister Patané; Luiz Alcantara; Luiz Lehmann Coutinho; Maria Carolina Elias; Maurício Lacerda Nogueira; Rafael dos Santos Bezerra; Raul Machado Neto; Rejane Maria Tommasini Grotto; Ricardo Haddad; Sandra Coccuzzo Sampaio Vessoni; Simone Kashima; Svetoslav Nanev Slavov; Vincent Louis Viala |
| EPI_ISL_5671744 | CENTRO DE SAUDE DR RENATO DE CARVALHO RIBEIRO ANGATUBA | Instituto Butantan | Antonio Jorge Martins; Claudia Renata dos Santos Barros; David Schlesinger; Debora Botequio Moretti; Dimas Tadeu Covas; Elaine Cristina Marqueze; Elaine Vieira Santos; Evandra Strazza Rodrigues; Heidge Fukumasu; Jayme Augusto de Souza-Neto; José Salvatore Leister Patané; Luiz Alcantara; Luiz Lehmann Coutinho; Maria Carolina Elias; Maurício Lacerda Nogueira; Rafael dos Santos Bezerra; Raul Machado Neto; Rejane Maria Tommasini Grotto; Ricardo Haddad; Sandra Coccuzzo Sampaio Vessoni; Simone Kashima; Svetoslav Nanev Slavov; Vincent Louis Viala |
| EPI_ISL_5671003 | CENTRO DE SAUDE DR YOSHITO KANZAWA | Instituto Butantan | Antonio Jorge Martins; Claudia Renata dos Santos Barros; David Schlesinger; Debora Botequio Moretti; Dimas Tadeu Covas; Elaine Cristina Marqueze; Elaine Vieira Santos; Evandra Strazza Rodrigues; Heidge Fukumasu; Jayme Augusto de Souza-Neto; José Salvatore Leister Patané; Luiz Alcantara; Luiz Lehmann Coutinho; Maria Carolina Elias; Maurício Lacerda Nogueira; Rafael dos Santos Bezerra; Raul Machado Neto; Rejane Maria Tommasini Grotto; Ricardo Haddad; Sandra Coccuzzo Sampaio Vessoni; Simone Kashima; Svetoslav Nanev Slavov; Vincent Louis Viala |
| EPI_ISL_5672937, EPI_ISL_5672992, EPI_ISL_5673034, EPI_ISL_5673056, EPI_ISL_5673102, EPI_ISL_5673112 | CENTRO DE SAUDE I TACITO LEITE DE CARVALHO E SILVA | Instituto Butantan | Antonio Jorge Martins; Claudia Renata dos Santos Barros; David Schlesinger; Debora Botequio Moretti; Dimas Tadeu Covas; Elaine Cristina Marqueze; Elaine Vieira Santos; Evandra Strazza Rodrigues; Heidge Fukumasu; Jayme Augusto de Souza-Neto; José Salvatore Leister Patané; Luiz Alcantara; Luiz Lehmann Coutinho; Maria Carolina Elias; Maurício Lacerda Nogueira; Rafael dos Santos Bezerra; Raul Machado Neto; Rejane Maria Tommasini Grotto; Ricardo Haddad; Sandra Coccuzzo Sampaio Vessoni; Simone Kashima; Svetoslav Nanev Slavov; Vincent Louis Viala |
| EPI_ISL_5672829, | CENTRO DE SAUDE II BARRA | Instituto Butantan | Antonio Jorge Martins; Claudia Renata dos Santos Barros; David Schlesinger; Debora Botequio Moretti; Dimas Tadeu Covas; Elaine Cristina Marqueze; Elaine Vieira Santos; Evandra Strazza Rodrigues; Heidge Fukumasu; Jayme Augusto de Souza-Neto; José Salvatore Leister Patané; Luiz Alcantara; Luiz Lehmann |

| | | | |
|---|---|---|---|
| EPI_ISL_5672866, EPI_ISL_5672912, EPI_ISL_5672917, EPI_ISL_5672925, EPI_ISL_5673089 | BONITA | | Coutinho; Maria Carolina Elias; Maurício Lacerda Nogueira; Rafael dos Santos Bezerra; Raul Machado Neto; Rejane Maria Tommasini Grotto; Ricardo Haddad; Sandra Coccuzzo Sampaio Vessoni; Simone Kashima; Svetoslav Nanev Slavov; Vincent Louis Viala |
| EPI_ISL_5672943, EPI_ISL_5672990, EPI_ISL_5673137 | CENTRO DE SAUDE II DE CAFELANDIA | Instituto Butantan | Antonio Jorge Martins; Claudia Renata dos Santos Barros; David Schlesinger; Debora Botequio Moretti; Dimas Tadeu Covas; Elaine Cristina Marqueze; Elaine Vieira Santos; Evandra Strazza Rodrigues; Heidge Fukumasu; Jayme Augusto de Souza-Neto; José Salvatore Leister Patané; Luiz Alcantara; Luiz Lehmann Coutinho; Maria Carolina Elias; Maurício Lacerda Nogueira; Rafael dos Santos Bezerra; Raul Machado Neto; Rejane Maria Tommasini Grotto; Ricardo Haddad; Sandra Coccuzzo Sampaio Vessoni; Simone Kashima; Svetoslav Nanev Slavov; Vincent Louis Viala |
| EPI_ISL_5670900, EPI_ISL_5670915, EPI_ISL_5670939, EPI_ISL_5670961, EPI_ISL_5670975, EPI_ISL_5671105 | CENTRO DE SAUDE II DR GABRIEL MESQUITA VARGEM GDE DO SUL | Instituto Butantan | Antonio Jorge Martins; Claudia Renata dos Santos Barros; David Schlesinger; Debora Botequio Moretti; Dimas Tadeu Covas; Elaine Cristina Marqueze; Elaine Vieira Santos; Evandra Strazza Rodrigues; Heidge Fukumasu; Jayme Augusto de Souza-Neto; José Salvatore Leister Patané; Luiz Alcantara; Luiz Lehmann Coutinho; Maria Carolina Elias; Maurício Lacerda Nogueira; Rafael dos Santos Bezerra; Raul Machado Neto; Rejane Maria Tommasini Grotto; Ricardo Haddad; Sandra Coccuzzo Sampaio Vessoni; Simone Kashima; Svetoslav Nanev Slavov; Vincent Louis Viala |
| EPI_ISL_5671160, EPI_ISL_5671400, EPI_ISL_5671436 | CENTRO DE SAUDE II DR JOSE PAIONE MOCOCA | Instituto Butantan | Antonio Jorge Martins; Claudia Renata dos Santos Barros; David Schlesinger; Debora Botequio Moretti; Dimas Tadeu Covas; Elaine Cristina Marqueze; Elaine Vieira Santos; Evandra Strazza Rodrigues; Heidge Fukumasu; Jayme Augusto de Souza-Neto; José Salvatore Leister Patané; Luiz Alcantara; Luiz Lehmann Coutinho; Maria Carolina Elias; Maurício Lacerda Nogueira; Rafael dos Santos Bezerra; Raul Machado Neto; Rejane Maria Tommasini Grotto; Ricardo Haddad; Sandra Coccuzzo Sampaio Vessoni; Simone Kashima; Svetoslav Nanev Slavov; Vincent Louis Viala |
| EPI_ISL_5671549, EPI_ISL_5671862 | CENTRO DE SAUDE II TIETE DR RUI SILVEIRA MELO | Instituto Butantan | Antonio Jorge Martins; Claudia Renata dos Santos Barros; David Schlesinger; Debora Botequio Moretti; Dimas Tadeu Covas; Elaine Cristina Marqueze; Elaine Vieira Santos; Evandra Strazza Rodrigues; Heidge Fukumasu; Jayme Augusto de Souza-Neto; José Salvatore Leister Patané; Luiz Alcantara; Luiz Lehmann Coutinho; Maria Carolina Elias; Maurício Lacerda Nogueira; Rafael dos Santos Bezerra; Raul Machado Neto; Rejane Maria Tommasini Grotto; Ricardo Haddad; Sandra Coccuzzo Sampaio Vessoni; Simone Kashima; Svetoslav Nanev Slavov; Vincent Louis Viala |
| EPI_ISL_5680816 | CENTRO DE SAUDE III DE AGUAS DE SAO PEDRO | Instituto Butantan | Antonio Jorge Martins; Claudia Renata dos Santos Barros; David Schlesinger; Debora Botequio Moretti; Dimas Tadeu Covas; Elaine Cristina Marqueze; Elaine Vieira Santos; Evandra Strazza Rodrigues; Heidge Fukumasu; Jayme Augusto de Souza-Neto; José Salvatore Leister Patané; Luiz Alcantara; Luiz Lehmann Coutinho; Maria Carolina Elias; Maurício Lacerda Nogueira; Rafael dos Santos Bezerra; Raul Machado Neto; Rejane Maria Tommasini Grotto; Ricardo Haddad; Sandra Coccuzzo Sampaio Vessoni; Simone Kashima; Svetoslav Nanev Slavov; Vincent Louis Viala |
| EPI_ISL_5672884, EPI_ISL_5672960, EPI_ISL_5673179 | CENTRO DE SAUDE III DE IGARACU DO TIETE | Instituto Butantan | Antonio Jorge Martins; Claudia Renata dos Santos Barros; David Schlesinger; Debora Botequio Moretti; Dimas Tadeu Covas; Elaine Cristina Marqueze; Elaine Vieira Santos; Evandra Strazza Rodrigues; Heidge Fukumasu; Jayme Augusto de Souza-Neto; José Salvatore Leister Patané; Luiz Alcantara; Luiz Lehmann Coutinho; Maria Carolina Elias; Maurício Lacerda Nogueira; Rafael dos Santos Bezerra; Raul Machado Neto; Rejane Maria Tommasini Grotto; Ricardo Haddad; Sandra Coccuzzo Sampaio Vessoni; Simone Kashima; Svetoslav Nanev Slavov; Vincent Louis Viala |
| EPI_ISL_5672843, EPI_ISL_5673081 | CENTRO DE SAUDE III DE PANORAMA | Instituto Butantan | Antonio Jorge Martins; Claudia Renata dos Santos Barros; David Schlesinger; Debora Botequio Moretti; Dimas Tadeu Covas; Elaine Cristina Marqueze; Elaine Vieira Santos; Evandra Strazza Rodrigues; Heidge Fukumasu; Jayme Augusto de Souza-Neto; José Salvatore Leister Patané; Luiz Alcantara; Luiz Lehmann Coutinho; Maria Carolina Elias; Maurício Lacerda Nogueira; Rafael dos Santos Bezerra; Raul Machado Neto; Rejane Maria Tommasini Grotto; Ricardo Haddad; Sandra Coccuzzo Sampaio Vessoni; Simone Kashima; Svetoslav Nanev Slavov; Vincent Louis Viala |
| EPI_ISL_5670909 | CENTRO DE SAUDE III SALES OLIVEIRA | Instituto Butantan | Antonio Jorge Martins; Claudia Renata dos Santos Barros; David Schlesinger; Debora Botequio Moretti; Dimas Tadeu Covas; Elaine Cristina Marqueze; Elaine Vieira Santos; Evandra Strazza Rodrigues; Heidge Fukumasu; Jayme Augusto de Souza-Neto; José Salvatore Leister Patané; Luiz Alcantara; Luiz Lehmann Coutinho; Maria Carolina Elias; Maurício Lacerda Nogueira; Rafael dos Santos Bezerra; Raul Machado Neto; Rejane Maria Tommasini Grotto; Ricardo Haddad; Sandra Coccuzzo Sampaio Vessoni; Simone Kashima; Svetoslav Nanev Slavov; Vincent Louis Viala |
| EPI_ISL_5672810, EPI_ISL_5673174 | CENTRO DE SAUDE III TABATINGA | Instituto Butantan | Antonio Jorge Martins; Claudia Renata dos Santos Barros; David Schlesinger; Debora Botequio Moretti; Dimas Tadeu Covas; Elaine Cristina Marqueze; Elaine Vieira Santos; Evandra Strazza Rodrigues; Heidge Fukumasu; Jayme Augusto de Souza-Neto; José Salvatore Leister Patané; Luiz Alcantara; Luiz Lehmann Coutinho; Maria Carolina Elias; Maurício Lacerda Nogueira; Rafael dos Santos Bezerra; Raul Machado Neto; Rejane Maria Tommasini Grotto; Ricardo Haddad; Sandra Coccuzzo Sampaio Vessoni; Simone Kashima; Svetoslav Nanev Slavov; Vincent Louis Viala |
| EPI_ISL_5671592, EPI_ISL_5671611, EPI_ISL_5671808, EPI_ISL_5671896 | CENTRO DE SAUDE SAO ROQUE DR JOSE CARVALHO BRITO | Instituto Butantan | Antonio Jorge Martins; Claudia Renata dos Santos Barros; David Schlesinger; Debora Botequio Moretti; Dimas Tadeu Covas; Elaine Cristina Marqueze; Elaine Vieira Santos; Evandra Strazza Rodrigues; Heidge Fukumasu; Jayme Augusto de Souza-Neto; José Salvatore Leister Patané; Luiz Alcantara; Luiz Lehmann Coutinho; Maria Carolina Elias; Maurício Lacerda Nogueira; Rafael dos Santos Bezerra; Raul Machado Neto; Rejane Maria Tommasini Grotto; Ricardo Haddad; Sandra Coccuzzo Sampaio Vessoni; Simone Kashima; Svetoslav Nanev Slavov; Vincent Louis Viala |
| EPI_ISL_5672837 | CENTRO INTEGRADO DE SAUDE | Instituto Butantan | Antonio Jorge Martins; Claudia Renata dos Santos Barros; David Schlesinger; Debora Botequio Moretti; Dimas Tadeu Covas; Elaine Cristina Marqueze; Elaine Vieira Santos; Evandra Strazza Rodrigues; Heidge Fukumasu; Jayme Augusto de Souza-Neto; José Salvatore Leister Patané; Luiz Alcantara; Luiz Lehmann Coutinho; Maria Carolina Elias; Maurício Lacerda Nogueira; Rafael dos Santos Bezerra; Raul Machado Neto; Rejane Maria Tommasini Grotto; Ricardo Haddad; Sandra Coccuzzo Sampaio Vessoni; Simone Kashima; Svetoslav Nanev Slavov; Vincent Louis Viala |
| EPI_ISL_5671169 | CENTRO SAUDE III DR JOSE TADEU MOURAO | Instituto Butantan | Antonio Jorge Martins; Claudia Renata dos Santos Barros; David Schlesinger; Debora Botequio Moretti; Dimas Tadeu Covas; Elaine Cristina Marqueze; Elaine Vieira Santos; Evandra Strazza Rodrigues; Heidge Fukumasu; Jayme Augusto de Souza-Neto; José Salvatore Leister Patané; Luiz Alcantara; Luiz Lehmann Coutinho; Maria Carolina Elias; Maurício Lacerda Nogueira; Rafael dos Santos Bezerra; Raul Machado Neto; Rejane Maria Tommasini Grotto; Ricardo Haddad; Sandra Coccuzzo Sampaio Vessoni; Simone Kashima; Svetoslav Nanev Slavov; Vincent Louis Viala |
| EPI_ISL_2989092 | CH Porto - H Sto Antonio | Instituto Nacional de Saude (INSA) | Borges et al |
| EPI_ISL_3234552, EPI_ISL_3234556, EPI_ISL_3234560, EPI_ISL_3234561, EPI_ISL_3234582 | CHU Purpan - Laboratoire de Virologie - Institut Fédératif de Biologie | CHU Purpan - Laboratoire de Virologie - Institut Fédératif de Biologie | Bulach T.; Donnadieu C.; Izopet J.; Latour J.; Milhes M.; Nicot F.; Ranger N.; Salin G.; Tremeaux P. |
| EPI_ISL_3061060 | CHUV | Laboratory of genomics and metagenomics | Claire Bertelli; Damien Jacot; Gilbert Greub; Sébastien Aeby; Trestan Pillonel |
| EPI_ISL_5675440 | CIAS DR JORGE BELOTTI PEDRINHAS PTA | Instituto Butantan | Antonio Jorge Martins; Claudia Renata dos Santos Barros; David Schlesinger; Debora Botequio Moretti; Dimas Tadeu Covas; Elaine Cristina Marqueze; Elaine Vieira Santos; Evandra Strazza Rodrigues; Heidge Fukumasu; Jayme Augusto de Souza-Neto; José Salvatore Leister Patané; Luiz Alcantara; Luiz Lehmann Coutinho; Maria Carolina Elias; Maurício Lacerda Nogueira; Rafael dos Santos Bezerra; Raul Machado Neto; Rejane Maria Tommasini Grotto; Ricardo Haddad; Sandra Coccuzzo Sampaio Vessoni; Simone Kashima; Svetoslav Nanev Slavov; Vincent Louis Viala |
| EPI_ISL_3030542, EPI_ISL_3030557 | CLILAB | Microbiology Department | Aida Gonzalez-Diaz; Carmen Ardanuy; Jordi Camara; Jordi Niubó; Laura Calatayud; M Angeles Domínguez; Sara Marti |
| EPI_ISL_3298334 | CLINICA BIBLICA | Inciensa, Instituto Costarricense de Investigación y Enseñanza en Nutrición y Salud | Adriana Godínez; Claudio Soto-Garita; Estela Cordero; Francisco Duarte; Hebleen Porras; Joselyn Prado & Karla Gutiérrez; José Luis Vargas; Mariela Gutiérrez; Melany Calderón |
| EPI_ISL_4960515 | CLINICA EL ROSARIO SEDE TESORO | Laboratorio Departamental de Salud Publica de Antioquia | Cristian Arbey Velarde Hoyos; Gloria Isabel Escobar; Idabely Betancur Ortiz; Juan P. Hernandez-Ortiz; Maria Stella López |
| EPI_ISL_5669053, EPI_ISL_5669151, EPI_ISL_5670505, EPI_ISL_5670839, EPI_ISL_5678132, EPI_ISL_5678411, EPI_ISL_5678488 | | | |
| see above | COMPLEXO HOSPITALAR PADRE BENTO DE GUARULHOS | Instituto Butantan | Antonio Jorge Martins; Claudia Renata dos Santos Barros; David Schlesinger; Debora Botequio Moretti; Dimas Tadeu Covas; Elaine Cristina Marqueze; Elaine Vieira Santos; Evandra Strazza Rodrigues; Heidge Fukumasu; Jayme Augusto de Souza-Neto; José Salvatore Leister Patané; Luiz Alcantara; Luiz Lehmann Coutinho; Maria Carolina Elias; Maurício Lacerda Nogueira; Rafael dos Santos Bezerra; Raul Machado Neto; Rejane Maria Tommasini Grotto; Ricardo Haddad; Sandra Coccuzzo Sampaio Vessoni; Simone Kashima; Svetoslav Nanev Slavov; Vincent Louis Viala |
| EPI_ISL_5673562, EPI_ISL_5673773, EPI_ISL_5674624, EPI_ISL_5674987 | CS DE GENERAL SALGADO | Instituto Butantan | Antonio Jorge Martins; Claudia Renata dos Santos Barros; David Schlesinger; Debora Botequio Moretti; Dimas Tadeu Covas; Elaine Cristina Marqueze; Elaine Vieira Santos; Evandra Strazza Rodrigues; Heidge Fukumasu; Jayme Augusto de Souza-Neto; José Salvatore Leister Patané; Luiz Alcantara; Luiz Lehmann Coutinho; Maria Carolina Elias; Maurício Lacerda Nogueira; Rafael dos Santos Bezerra; Raul Machado Neto; Rejane Maria Tommasini Grotto; Ricardo Haddad; Sandra Coccuzzo Sampaio Vessoni; Simone Kashima; Svetoslav Nanev Slavov; Vincent Louis Viala |
| EPI_ISL_5673888, EPI_ISL_5674112, EPI_ISL_5674177, EPI_ISL_5674220, EPI_ISL_5675204, EPI_ISL_5675365, EPI_ISL_5675394, EPI_ISL_5675399, EPI_ISL_5675464, EPI_ISL_5675487, EPI_ISL_5675494 | | | |
| see above | CS DE MONTE APRAZIVEL | Instituto Butantan | Antonio Jorge Martins; Claudia Renata dos Santos Barros; David Schlesinger; Debora Botequio Moretti; Dimas Tadeu Covas; Elaine Cristina Marqueze; Elaine Vieira Santos; Evandra Strazza Rodrigues; Heidge Fukumasu; Jayme Augusto de Souza-Neto; José Salvatore Leister Patané; Luiz Alcantara; Luiz Lehmann Coutinho; Maria Carolina Elias; Maurício Lacerda Nogueira; Rafael dos Santos Bezerra; Raul Machado Neto; Rejane Maria Tommasini Grotto; Ricardo Haddad; Sandra Coccuzzo Sampaio Vessoni; Simone Kashima; Svetoslav Nanev Slavov; Vincent Louis Viala |
| EPI_ISL_5673920 | CS DE NEVES PAULISTA | Instituto Butantan | Antonio Jorge Martins; Claudia Renata dos Santos Barros; David Schlesinger; Debora Botequio Moretti; Dimas Tadeu Covas; Elaine Cristina Marqueze; Elaine Vieira Santos; Evandra Strazza Rodrigues; Heidge Fukumasu; Jayme Augusto de Souza-Neto; José Salvatore Leister Patané; Luiz Alcantara; Luiz Lehmann Coutinho; Maria Carolina Elias; Maurício Lacerda Nogueira; Rafael dos Santos Bezerra; Raul Machado Neto; Rejane Maria Tommasini Grotto; Ricardo Haddad; Sandra Coccuzzo Sampaio Vessoni; Simone Kashima; Svetoslav Nanev Slavov; Vincent Louis Viala |
| EPI_ISL_5673352 | CS DE ORINDIUVA | Instituto Butantan | Antonio Jorge Martins; Claudia Renata dos Santos Barros; David Schlesinger; Debora Botequio Moretti; Dimas Tadeu Covas; Elaine Cristina Marqueze; Elaine Vieira Santos; Evandra Strazza Rodrigues; Heidge Fukumasu; Jayme Augusto de Souza-Neto; José Salvatore Leister Patané; Luiz Alcantara; Luiz Lehmann Coutinho; Maria Carolina Elias; Maurício Lacerda Nogueira; Rafael dos Santos Bezerra; Raul Machado Neto; Rejane Maria Tommasini Grotto; Ricardo Haddad; Sandra Coccuzzo Sampaio Vessoni; Simone Kashima; Svetoslav Nanev Slavov; Vincent Louis Viala |
| EPI_ISL_5673922, EPI_ISL_5674278 | CS DE SEBASTIANOPOLIS DO SUL | Instituto Butantan | Antonio Jorge Martins; Claudia Renata dos Santos Barros; David Schlesinger; Debora Botequio Moretti; Dimas Tadeu Covas; Elaine Cristina Marqueze; Elaine Vieira Santos; Evandra Strazza Rodrigues; Heidge Fukumasu; Jayme Augusto de Souza-Neto; José Salvatore Leister Patané; Luiz Alcantara; Luiz Lehmann Coutinho; Maria Carolina Elias; Maurício Lacerda Nogueira; Rafael dos Santos Bezerra; Raul Machado Neto; Rejane Maria Tommasini Grotto; Ricardo Haddad; Sandra Coccuzzo Sampaio Vessoni; Simone Kashima; Svetoslav Nanev Slavov; Vincent Louis Viala |
| EPI_ISL_5673372, EPI_ISL_5673393, EPI_ISL_5673429, EPI_ISL_5673464, EPI_ISL_5673654, EPI_ISL_5673714, EPI_ISL_5673742, EPI_ISL_5673758, EPI_ISL_5673807, EPI_ISL_5674515, EPI_ISL_5674874, EPI_ISL_5674983, EPI_ISL_5675454 | | | |
| see above | CS DE VALENTIM GENTIL | Instituto Butantan | Antonio Jorge Martins; Claudia Renata dos Santos Barros; David Schlesinger; Debora Botequio Moretti; Dimas Tadeu Covas; Elaine Cristina Marqueze; Elaine Vieira Santos; Evandra Strazza Rodrigues; Heidge Fukumasu; Jayme Augusto de Souza-Neto; José Salvatore Leister Patané; Luiz Alcantara; Luiz Lehmann Coutinho; Maria Carolina Elias; Maurício Lacerda Nogueira; Rafael dos Santos Bezerra; Raul Machado Neto; Rejane Maria Tommasini Grotto; Ricardo Haddad; Sandra Coccuzzo Sampaio Vessoni; Simone Kashima; Svetoslav Nanev Slavov; Vincent Louis Viala |
| EPI_ISL_5673542, EPI_ISL_5674817 | CS I DE OURINHOS | Instituto Butantan | Antonio Jorge Martins; Claudia Renata dos Santos Barros; David Schlesinger; Debora Botequio Moretti; Dimas Tadeu Covas; Elaine Cristina Marqueze; Elaine Vieira Santos; Evandra Strazza Rodrigues; Heidge Fukumasu; Jayme Augusto de Souza-Neto; José Salvatore Leister Patané; Luiz Alcantara; Luiz Lehmann Coutinho; Maria Carolina Elias; Maurício Lacerda Nogueira; Rafael dos Santos Bezerra; Raul Machado Neto; Rejane Maria Tommasini Grotto; Ricardo Haddad; Sandra Coccuzzo Sampaio Vessoni; Simone Kashima; Svetoslav Nanev Slavov; Vincent Louis Viala |
| EPI_ISL_5671038 | CS II DE MIGUELOPOLIS | Instituto Butantan | Antonio Jorge Martins; Claudia Renata dos Santos Barros; David Schlesinger; Debora Botequio Moretti; Dimas Tadeu Covas; Elaine Cristina Marqueze; Elaine Vieira Santos; Evandra Strazza Rodrigues; Heidge Fukumasu; Jayme Augusto de Souza-Neto; José Salvatore Leister Patané; Luiz Alcantara; Luiz Lehmann Coutinho; Maria Carolina Elias; Maurício Lacerda Nogueira; Rafael dos Santos Bezerra; Raul Machado Neto; Rejane Maria Tommasini Grotto; Ricardo Haddad; Sandra Coccuzzo Sampaio Vessoni; Simone Kashima; Svetoslav Nanev Slavov; Vincent Louis Viala |
| EPI_ISL_5671266, EPI_ISL_5671407 | CS III DR ANTONIO ANADAO SSGRAMA | Instituto Butantan | Antonio Jorge Martins; Claudia Renata dos Santos Barros; David Schlesinger; Debora Botequio Moretti; Dimas Tadeu Covas; Elaine Cristina Marqueze; Elaine Vieira Santos; Evandra Strazza Rodrigues; Heidge Fukumasu; Jayme Augusto de Souza-Neto; José Salvatore Leister Patané; Luiz Alcantara; Luiz Lehmann Coutinho; Maria Carolina Elias; Maurício Lacerda Nogueira; Rafael dos Santos Bezerra; Raul Machado Neto; Rejane Maria Tommasini Grotto; Ricardo Haddad; Sandra Coccuzzo Sampaio Vessoni; Simone Kashima; Svetoslav Nanev Slavov; Vincent Louis Viala |
| EPI_ISL_5673690, EPI_ISL_5673744, EPI_ISL_5674234 | CS III SANTOPOLIS DO AGUAPEI | Instituto Butantan | Antonio Jorge Martins; Claudia Renata dos Santos Barros; David Schlesinger; Debora Botequio Moretti; Dimas Tadeu Covas; Elaine Cristina Marqueze; Elaine Vieira Santos; Evandra Strazza Rodrigues; Heidge Fukumasu; Jayme Augusto de Souza-Neto; José Salvatore Leister Patané; Luiz Alcantara; Luiz Lehmann Coutinho; Maria Carolina Elias; Maurício Lacerda Nogueira; Rafael dos Santos Bezerra; Raul Machado Neto; Rejane Maria Tommasini Grotto; Ricardo Haddad; Sandra Coccuzzo Sampaio Vessoni; Simone Kashima; Svetoslav Nanev Slavov; Vincent Louis Viala |
| EPI_ISL_3151562, EPI_ISL_3320691 | CT Department of Public Health | CT Department of Public Health | Claire_Pearson; Tu_N_Nguyen |
| EPI_ISL_4005612 | California Department of Public Health | California Department of Public Health | CDPH IDLB COVIDNet et al |
| EPI_ISL_3276686 | California Hospital Medical Center | Los Angeles County Public Health Laboratories | P. Hemarajata et al. |
| EPI_ISL_3300259 | Centinela Hospital Medical Center | Los Angeles County Public Health Laboratories | P. Hemarajata et al. |
| EPI_ISL_3462537, EPI_ISL_3462542, EPI_ISL_3462554, EPI_ISL_3462555, EPI_ISL_3462598, EPI_ISL_3462601, EPI_ISL_3462608, EPI_ISL_3462620, EPI_ISL_3462621, EPI_ISL_3462687, EPI_ISL_3462688, EPI_ISL_3462689, EPI_ISL_3462690, EPI_ISL_3462691, EPI_ISL_3462692, EPI_ISL_3462693, EPI_ISL_3462694, EPI_ISL_3462695, EPI_ISL_3464722, EPI_ISL_3464732, EPI_ISL_3464733, EPI_ISL_3464734 | | | |

| | | | |
|---|---|---|---|
| see above | Central Laboratory, Bureau of Public Health (BOG) and Academic Hospital Paramaribo | Erasmus Medical Center | Bas B Oude Munnink; Cherise Beek; Consuella Partowidjojo; Dion Gajadin; Ed PF IJzerman; Emmanuelle Munger; Gary Gummels; Ingrid SK Krishnadath; Lycke Woittiez; Marion PG Koopmans; Mireille Van de Veer; Phyllis Pinas; Princes Wongsowidjojo; Radjesh Ori; Ranisha Doerbalie; Rohma Banwari; Soeradj Harkisoen; Stephen Vreden; Tilotmadebie Ramlal; Verne Nanhoe |
| EPI_ISL_3147224 | Centre Hospitalier du Nord | Laboratoire national de sante, Microbiology, Microbial Genomics Platform | Anke Wienecke-Baldacchino; Catherine Ragimbeau; Elodie Solarino; Fatiha Boulmerka; Fatu Djabi; Jacqueline Parmentier; Jessica Tapp; Lise Pignon; Raoul Salmon; Tamir Abdelrahman; Virginie Jover |
| EPI_ISL_3129919 | Centro de Investigación Biomédica de La Rioja - Hospital San Pedro Logroño | SeqCOVID-SPAIN consortium/IBV(CSIC) | José Manuel Azcona Gutiérrez; María Pilar Bea Escudero; María de Toro; Miriam Blasco Alberdi and SeqCOVID-SPAIN consortium |
| EPI_ISL_3155465, EPI_ISL_3155471, EPI_ISL_3155472, EPI_ISL_3155482, EPI_ISL_3155489, EPI_ISL_3155492, EPI_ISL_3155494, EPI_ISL_3155500 | | | |
| see above | Centro de Investigación Biomédica de Occidente (CIBO) | Centro de Investigación en Enfermedades Infecciosas (CIENI), Instituto Nacional de Enfermedades Respiratorias (INER) | ; Alejandra García-Gasca; Alejandra Hernández-Terán; Alejandro Sánchez-Flores; Alfredo Herrera-Estrella; Alicia Ocaña-Mondragón; Andreu Comas-García; Angel Gustavo Salas-Lais; Antonio Loza Román; Bernardo Martínez-Miguel; Blanca Taboada; Brenda Irasema Maldonado-Meza; Bruno Gómez-Gil; Carla Ivón Herrera-Najera; Carlos F. Arias; Celia Boukadida; Clara Esperanza Santacruz-Tinoco; Concepción Grajales-Muñiz; Consorcio Mexicano de Vigilancia Genómica (CoViGen-Mex). Authors (in alphabetical order): Julio Elias Alvarado-Yaah; Cristóbal Cháidez-Quiróz; Célida Duque Molina; Célida Martínez- Rodríguez; Daniel Fregoso-Rueda; Daniel Lira Morales; Eduardo Becerril-Vargas; Fernando Fontove-Herrera; Fidencio Mejía-Nepomuceno; Francisco Pulido; Gloria Elena Espinosa-Ayala; Gloria María Molina-Salinas; Gloria Vazquez; Hector Esteban Paz-Juárez; Hector Montoya-Fuentes; Helen Haydee Fernanda Ramírez-Plascencia; Irvin González-López; Jean Pierre González; Jesús Hernández; Joel Armando Vázquez-Pérez.; Jorge Salas-Hernández; José Antonio Enciso-Moreno; José Arturo Martínez-Orozco; José Esteban Muñoz-Medina; José de Jesús Nuñez-Contreras; Juan Bautista Chale-Dzul; Julissa Enciso-Ibarra; Kathia Elizabeth Tapia-Diaz; Luis Alberto Ochoa-Carrera; Margarita Matías-Florentino; Mario Mújica-Sánchez; Marissa Perez-Garcia; María Guadalupe Santiago-Mauricio; María Guadalupe de Jesús Mireles-Rivera; Nelly Sélem-Mojica; Pavel Isa; Ricardo Ciria Merce; Ricardo Grande; Rosa María Gutiérrez Rios; Santiago Ávila-Ríos; Selene Zárate; Susana Lopez; Verónica Mata-Haro; Victor Eduardo García-Arias; Víctor Hugo Borja-Aburto |
| EPI_ISL_3155438 | Centro de Investigación Biomédica del Noreste (CIBIN) | Centro de Investigación en Enfermedades Infecciosas (CIENI), Instituto Nacional de Enfermedades Respiratorias (INER) | ; Alejandra García-Gasca; Alejandra Hernández-Terán; Alejandro Sánchez-Flores; Alfredo Herrera-Estrella; Alicia Ocaña-Mondragón; Andreu Comas-García; Angel Gustavo Salas-Lais; Antonio Loza Román; Bernardo Martínez-Miguel; Blanca Taboada; Brenda Irasema Maldonado-Meza; Bruno Gómez-Gil; Carla Ivón Herrera-Najera; Carlos F. Arias; Celia Boukadida; Clara Esperanza Santacruz-Tinoco; Concepción Grajales-Muñiz; Consorcio Mexicano de Vigilancia Genómica (CoViGen-Mex). Authors (in alphabetical order): Julio Elias Alvarado-Yaah; Cristóbal Cháidez-Quiróz; Célida Duque Molina; Célida Martínez- Rodríguez; Daniel Fregoso-Rueda; Daniel Lira Morales; Eduardo Becerril-Vargas; Fernando Fontove-Herrera; Fidencio Mejía-Nepomuceno; Francisco Pulido; Gloria Elena Espinosa-Ayala; Gloria María Molina-Salinas; Gloria Vazquez; Hector Esteban Paz-Juárez; Hector Montoya-Fuentes; Helen Haydee Fernanda Ramírez-Plascencia; Irvin González-López; Jean Pierre González; Jesús Hernández; Joel Armando Vázquez-Pérez.; Jorge Salas-Hernández; José Antonio Enciso-Moreno; José Arturo Martínez-Orozco; José Esteban Muñoz-Medina; José de Jesús Nuñez-Contreras; Juan Bautista Chale-Dzul; Julissa Enciso-Ibarra; Kathia Elizabeth Tapia-Diaz; Luis Alberto Ochoa-Carrera; Margarita Matías-Florentino; Mario Mújica-Sánchez; Marissa Perez-Garcia; María Guadalupe Santiago-Mauricio; María Guadalupe de Jesús Mireles-Rivera; Nelly Sélem-Mojica; Pavel Isa; Ricardo Ciria Merce; Ricardo Grande; Rosa María Gutiérrez Rios; Santiago Ávila-Ríos; Selene Zárate; Susana Lopez; Verónica Mata-Haro; Victor Eduardo García-Arias; Víctor Hugo Borja-Aburto |
| EPI_ISL_2983969, EPI_ISL_2983976, EPI_ISL_2983990, EPI_ISL_2983995, EPI_ISL_2984003, EPI_ISL_2984028, EPI_ISL_2984038, EPI_ISL_2984044, EPI_ISL_2984083, EPI_ISL_2984187 | | | |
| see above | Cerballiance Paris IDF est Saint-Denis | Department of Virology, Henri Mondor University Hospital, Assistance Publique Hôpitaux de Paris, Université Paris-Est Créteil, INSERM U955 | Alexandre Soulier; Christophe Rodriguez; Elisabeth Trawinski; Guillaume Gricourt; Jean-Michel Pawlotsky; Melissa N'Debi; Slim Fourati; Vanessa Demontant |
| EPI_ISL_3242714, EPI_ISL_3242715, EPI_ISL_3242716, EPI_ISL_3242717, EPI_ISL_3242718, EPI_ISL_3242719, EPI_ISL_3242722, EPI_ISL_3242744 | | | |
| see above | Clinica INDISA | "Facultad de Ciencias de la Vida, UNAB" | "Claudio Meneses; Ariel Orellana"; Claudio Olmos; Daniel Leon; Dayan Sanhueza; Eduardo Castro; Gonzalo Campaña; Macarena Bastias; Paola Pidal; Ricardo Yusta; Sebastian Wolter; Susana Saez; Víctor Monreal; Waldo Diaz |
| EPI_ISL_2988680 | Clinical Microbiology, Infection Prevention and Control | Section for Molecular Diagnostics | Björn Hallström; Jonas Björkman |
| EPI_ISL_2966260, EPI_ISL_2966296, EPI_ISL_2966305, EPI_ISL_2966319, EPI_ISL_3089083 | Cliniques universitaires Saint-Luc | UCLouvain/IREC/MBLG | Benoit Kabamba Mukadi; Bertrand Bearzatto; Jean Ruelle |
| EPI_ISL_3760586, EPI_ISL_3842262, EPI_ISL_3843010, EPI_ISL_3843056, EPI_ISL_3869513, EPI_ISL_3916490, EPI_ISL_3916502, EPI_ISL_4059186, EPI_ISL_4059229 | | | |
| see above | Colorado Department of Public Health and Environment | Colorado Department of Public Health and Environment | Alexandria Rossheim; Diana Ir; Emily A. Travanty; Laura Bankers; Mandy Waters; Michael A. Martin; Michael Martin; Molly C. Hetherington-Rauth; Sarah Elizabeth Totten; Shannon R. Matzinger |
| EPI_ISL_3014120, EPI_ISL_3014123, EPI_ISL_3014156, EPI_ISL_3014157, EPI_ISL_3014158, EPI_ISL_3014159, EPI_ISL_3014160, EPI_ISL_3014161, EPI_ISL_3014162, EPI_ISL_3014163, EPI_ISL_3014164, EPI_ISL_3014165, EPI_ISL_3014166, EPI_ISL_3014167, EPI_ISL_3014168, EPI_ISL_3014169, EPI_ISL_3014170, EPI_ISL_3014171, EPI_ISL_3092133, EPI_ISL_3092134, EPI_ISL_3092210, EPI_ISL_3092226, EPI_ISL_3092419, EPI_ISL_3092420, EPI_ISL_3092421, EPI_ISL_3092422, EPI_ISL_3092423, EPI_ISL_3092424, EPI_ISL_3092425, EPI_ISL_3092426, EPI_ISL_3092427, EPI_ISL_3092428, EPI_ISL_3092429, EPI_ISL_3092430 | | | |
| see above | Cotugno | TIGEM | Antonio Grimaldi Patrizia Annunziata Francesco Panariello Biancamaria Pierri Claudia Tiberio Teresa Giuliano Valentina Bouche Chiara Colantuono Maria Concetta Cuomo Denise Di Concilio Lucio Di Filippo Anna Manfredi Marcello Salvi Antonio Limone Luigi Atripaldi Pellegrino Cerino Andrea Ballabio Davide Cacchiarelli |
| EPI_ISL_3064684, EPI_ISL_3270245, EPI_ISL_3273234, EPI_ISL_3273235, EPI_ISL_3273286, EPI_ISL_3273307, EPI_ISL_3273459, EPI_ISL_3273469, EPI_ISL_3273483, EPI_ISL_3273540, EPI_ISL_5522672, EPI_ISL_5522684, EPI_ISL_5522767, EPI_ISL_5522809 | | | |
| see above | Curative Labs | Curative Labs | Elias L. Salfati; Eugenia Khorosheva; George Way; J.Cesar Ignacio-Espinoza; Janet Chen; Mikhail Hanewich-Hollatz; Nabjot Sandhu; Sophia Quasem; Vladimir Slepnev; Zhiyi Xie |
| EPI_ISL_4414145, EPI_ISL_4414146, EPI_ISL_4414242, EPI_ISL_4414244, EPI_ISL_4414247, EPI_ISL_4414248, EPI_ISL_4414249, EPI_ISL_4414258, EPI_ISL_4414259, EPI_ISL_4414264, EPI_ISL_4414265, EPI_ISL_4414266, EPI_ISL_4414268, EPI_ISL_4414269, EPI_ISL_4414274, EPI_ISL_4414277, EPI_ISL_4414278, EPI_ISL_4414279, EPI_ISL_4414281, EPI_ISL_4414283, EPI_ISL_4414284, EPI_ISL_4414286, EPI_ISL_4414287, EPI_ISL_4414290, EPI_ISL_4414291, EPI_ISL_4414294, EPI_ISL_4414295, EPI_ISL_4414296, EPI_ISL_4414297, EPI_ISL_4414301, EPI_ISL_4414302, EPI_ISL_4414303, EPI_ISL_4414304, EPI_ISL_4414305, EPI_ISL_4414307, EPI_ISL_4414308, EPI_ISL_4414309, EPI_ISL_4414310, EPI_ISL_4414312, EPI_ISL_4414314, EPI_ISL_4414315, EPI_ISL_4414316, EPI_ISL_4414324, EPI_ISL_4414325, EPI_ISL_4414326, EPI_ISL_4414327, EPI_ISL_4414328, EPI_ISL_4414331, EPI_ISL_4414338, EPI_ISL_4414340, EPI_ISL_4414343, EPI_ISL_4414344, EPI_ISL_4414345, EPI_ISL_4414347, EPI_ISL_4414348, EPI_ISL_4414353, EPI_ISL_4414357, EPI_ISL_4414358, EPI_ISL_4414359, EPI_ISL_4414360, EPI_ISL_4414361, EPI_ISL_4414363, EPI_ISL_4414367, EPI_ISL_4414368, EPI_ISL_4414369, EPI_ISL_4414373, EPI_ISL_4414376, EPI_ISL_4414379, EPI_ISL_4414380, EPI_ISL_4414381, EPI_ISL_4414383, EPI_ISL_4414387, EPI_ISL_4414389, EPI_ISL_4414391, EPI_ISL_4414395, EPI_ISL_4414398, EPI_ISL_4414399, EPI_ISL_4414400, EPI_ISL_4414401, EPI_ISL_4414403, EPI_ISL_4414404, EPI_ISL_4414405, EPI_ISL_4414410, EPI_ISL_4414412, EPI_ISL_4414422, EPI_ISL_4414423, EPI_ISL_4414424, EPI_ISL_4414425, EPI_ISL_4414426, EPI_ISL_4414427, EPI_ISL_4414428, EPI_ISL_4414429, EPI_ISL_4414431, EPI_ISL_4414432, EPI_ISL_4414433, EPI_ISL_4414434, EPI_ISL_4414435, EPI_ISL_4414436, EPI_ISL_4414437, EPI_ISL_4414439, EPI_ISL_4414440, EPI_ISL_4414442, EPI_ISL_4414447, EPI_ISL_4414449, EPI_ISL_4414450, EPI_ISL_4414453, EPI_ISL_4414455, EPI_ISL_4414457, EPI_ISL_4414458, EPI_ISL_4414459, EPI_ISL_4414461, EPI_ISL_4414462, EPI_ISL_4414463, EPI_ISL_4414464, EPI_ISL_4414470, EPI_ISL_4414471, EPI_ISL_4414473, EPI_ISL_4414474, EPI_ISL_4414475, EPI_ISL_4414494, EPI_ISL_4414496, EPI_ISL_4414497, EPI_ISL_4414514, EPI_ISL_4414517, EPI_ISL_4414518, EPI_ISL_4414519, EPI_ISL_4414530, EPI_ISL_4414531, EPI_ISL_4414540, EPI_ISL_4414541, EPI_ISL_4414546, EPI_ISL_4414558, EPI_ISL_4414562, EPI_ISL_4414563, EPI_ISL_4414569, EPI_ISL_4414571, EPI_ISL_4414573, EPI_ISL_4414576, EPI_ISL_4414577, EPI_ISL_4414580, EPI_ISL_4422987, EPI_ISL_4423812, EPI_ISL_4423823, EPI_ISL_4423840, EPI_ISL_4423854, EPI_ISL_4423857, EPI_ISL_4423872, EPI_ISL_4423879, EPI_ISL_4423881, EPI_ISL_4423883, EPI_ISL_4423887, EPI_ISL_4423890, EPI_ISL_4423892, EPI_ISL_4423895, EPI_ISL_4423898, EPI_ISL_4423901, EPI_ISL_4423903, EPI_ISL_4423906, EPI_ISL_4423908, EPI_ISL_4423910, EPI_ISL_4423912, EPI_ISL_4423928, EPI_ISL_4423946, EPI_ISL_4423949, EPI_ISL_4423951, EPI_ISL_4423954, EPI_ISL_4423956, EPI_ISL_4423958, EPI_ISL_4423960, EPI_ISL_4423963, EPI_ISL_4423965, EPI_ISL_4423968, EPI_ISL_4423970, EPI_ISL_4423973, EPI_ISL_4423976, EPI_ISL_4423989, EPI_ISL_4423991, EPI_ISL_4424001, EPI_ISL_4424007, EPI_ISL_4424008, EPI_ISL_4424010, EPI_ISL_4424028, EPI_ISL_4424032, EPI_ISL_4424033, EPI_ISL_4424048, EPI_ISL_4424054, EPI_ISL_4424056, EPI_ISL_4424065, EPI_ISL_4424073, EPI_ISL_4424075, EPI_ISL_4424082, EPI_ISL_4424088, EPI_ISL_4424090, EPI_ISL_4424095, EPI_ISL_4424100, EPI_ISL_4424102, EPI_ISL_4424104, EPI_ISL_4424113, EPI_ISL_4424133, EPI_ISL_4424136, EPI_ISL_4424139, EPI_ISL_4424140, EPI_ISL_4424142, EPI_ISL_4424144, EPI_ISL_4424146, EPI_ISL_4424149, EPI_ISL_4424152, EPI_ISL_4424155, EPI_ISL_4424158, EPI_ISL_4424161, EPI_ISL_4424162, EPI_ISL_4424164, EPI_ISL_4424166, EPI_ISL_4424168, EPI_ISL_4424170, EPI_ISL_4424171, EPI_ISL_4424174, EPI_ISL_4424176, EPI_ISL_4424178, EPI_ISL_4424180, EPI_ISL_4424182, EPI_ISL_4424184, EPI_ISL_4424187, EPI_ISL_4424191, EPI_ISL_4424193, EPI_ISL_4424196, EPI_ISL_4424199, EPI_ISL_4424202, EPI_ISL_4424204, EPI_ISL_4424206, EPI_ISL_4424207, EPI_ISL_4424209, EPI_ISL_4424211, EPI_ISL_4424213, EPI_ISL_4424271, EPI_ISL_4424272, EPI_ISL_4424279, EPI_ISL_4424300, EPI_ISL_4424304, EPI_ISL_4424309, EPI_ISL_4424311, EPI_ISL_4424314, EPI_ISL_4424316, EPI_ISL_4424320, EPI_ISL_4424322, EPI_ISL_4424324, EPI_ISL_4424325, EPI_ISL_4424327, EPI_ISL_4424336, EPI_ISL_4424338, EPI_ISL_4424339, EPI_ISL_4424341, EPI_ISL_4424352, EPI_ISL_4424354, EPI_ISL_4424359, EPI_ISL_4424360, EPI_ISL_4424362, EPI_ISL_4424364, EPI_ISL_4424366, EPI_ISL_4424368, EPI_ISL_4424370, EPI_ISL_4424371, EPI_ISL_4424373, EPI_ISL_4424375, EPI_ISL_4424383, EPI_ISL_4424388, EPI_ISL_4424395, EPI_ISL_4424397, EPI_ISL_4424406, EPI_ISL_4424407, EPI_ISL_4424411, EPI_ISL_4424510, EPI_ISL_4424511, EPI_ISL_4424512, EPI_ISL_4424514, EPI_ISL_4424515, EPI_ISL_4424517, EPI_ISL_4424519, EPI_ISL_4424521, EPI_ISL_4424523, EPI_ISL_4424524, EPI_ISL_4424526, EPI_ISL_4424528, EPI_ISL_4424530, EPI_ISL_4424531, EPI_ISL_4424533, EPI_ISL_4424535, EPI_ISL_4424537, EPI_ISL_4424539, EPI_ISL_4424541, EPI_ISL_4424542, EPI_ISL_4424544, EPI_ISL_4424546, EPI_ISL_4424548, EPI_ISL_4424549, EPI_ISL_4424551, EPI_ISL_4424553, EPI_ISL_4424554, EPI_ISL_4424556, EPI_ISL_4424558, EPI_ISL_4424559, EPI_ISL_4424561, EPI_ISL_4424563, EPI_ISL_4424565, EPI_ISL_4424566, EPI_ISL_4424568, EPI_ISL_4424570, EPI_ISL_4424572, EPI_ISL_4424573, EPI_ISL_4424575, EPI_ISL_4424577, EPI_ISL_4424578, EPI_ISL_4424580, EPI_ISL_4424582, EPI_ISL_4424584, EPI_ISL_4424585, EPI_ISL_4424587, EPI_ISL_4424589, EPI_ISL_4424590, EPI_ISL_4424592, EPI_ISL_4424594, EPI_ISL_4424596, EPI_ISL_4424597, EPI_ISL_4424599, EPI_ISL_4424601, EPI_ISL_4424603, EPI_ISL_4424604, EPI_ISL_4424606, EPI_ISL_4424608, EPI_ISL_4424609, EPI_ISL_4424611, EPI_ISL_4424613, EPI_ISL_4424614, EPI_ISL_4424616, EPI_ISL_4424618, EPI_ISL_4424619, EPI_ISL_4424621, EPI_ISL_4424623, EPI_ISL_4424624, EPI_ISL_4424627, EPI_ISL_4424629, EPI_ISL_4424631, EPI_ISL_4424632, EPI_ISL_4424634, EPI_ISL_4424636, EPI_ISL_4424638, EPI_ISL_4424640, EPI_ISL_4424641, EPI_ISL_4424643, EPI_ISL_4424647, EPI_ISL_4424649, EPI_ISL_4424652, EPI_ISL_4424654, EPI_ISL_4424656, EPI_ISL_4424658, EPI_ISL_4424659, EPI_ISL_4424661, EPI_ISL_4424663, EPI_ISL_4424768, EPI_ISL_4424770, EPI_ISL_4424781, EPI_ISL_4424792, EPI_ISL_4424794, EPI_ISL_7697191, EPI_ISL_7743829, EPI_ISL_7743830, EPI_ISL_7743832 | | | |
| see above | DASA | DASA | Adriano Bonaldi; Angelica Hristov; Annelise Lopes; Bianca Cota; Camila Romano; Cristina Oliveira; Jose Levi; Lidia Yamamoto; Luciane Sussuchi; Paulo Pierry; Rodrigo Guarischi; Rodrigo Salazar |
| EPI_ISL_3152370 | DC Public Health Lab/ Dept. of Forensic Sciences | DC Public Health Lab/ Dept. of Forensic Sciences | Brittany Hamilton; Connie Maza; David Payne; Elizabeth Zelaya; Janis Doss; Jocelyn Hauser; Monica Mann; Sarah Scott; Scott Nguyen |
| EPI_ISL_5650807, EPI_ISL_5653846 | DEPARTAMENTO DE SAUDE | Instituto Butantan | Antonio Jorge Martins; Claudia Renata dos Santos Barros; David Schlesinger; Debora Botequio Moretti; Dimas Tadeu Covas; Elaine Cristina Marqueze; Elaine Vieira Santos; Evandra Strazza Rodrigues; Heidge Fukumasu; Jayme Augusto de Souza-Neto; José Salvatore Leister Patané; Luiz Alcantara; Luiz Lehmann Coutinho; Maria Carolina Elias; Maurício Lacerda Nogueira; Rafael dos Santos Bezerra; Raul Machado Neto; Rejane Maria Tommasini Grotto; Ricardo Haddad; Sandra Coccuzzo Sampaio Vessoni; Simone Kashima; Svetoslav Nanev Slavov; Vincent Louis Viala |
| EPI_ISL_5669414, EPI_ISL_5669701, EPI_ISL_5669983, EPI_ISL_5670760, EPI_ISL_5671582, EPI_ISL_5671595, EPI_ISL_5671675, EPI_ISL_5671697, EPI_ISL_5671955 | | | |
| see above | DEPARTAMENTO DE SAUDE COLETIVA | Instituto Butantan | Antonio Jorge Martins; Claudia Renata dos Santos Barros; David Schlesinger; Debora Botequio Moretti; Dimas Tadeu Covas; Elaine Cristina Marqueze; Elaine Vieira Santos; Evandra Strazza Rodrigues; Heidge Fukumasu; Jayme Augusto de Souza-Neto; José Salvatore Leister Patané; Luiz Alcantara; Luiz Lehmann Coutinho; Maria Carolina Elias; Maurício Lacerda Nogueira; Rafael dos Santos Bezerra; Raul Machado Neto; Rejane Maria Tommasini Grotto; Ricardo Haddad; Sandra Coccuzzo Sampaio Vessoni; Simone Kashima; Svetoslav Nanev Slavov; Vincent Louis Viala |
| EPI_ISL_3692660, EPI_ISL_3692719 | DPHL | Delaware Public Health Lab | Rebecca Savage |
| EPI_ISL_2896690, EPI_ISL_2896719, EPI_ISL_2971800 | Department of Bacteria, Parasites and Fungi, Statens Serum Institut, Copenhagen, Denmark | Statens Serum Institut Bioinformatics and Microbial Genomics | Danish Covid-19 Genome Consortium |
| EPI_ISL_2930887, EPI_ISL_2930891, EPI_ISL_2930897, EPI_ISL_2930957, EPI_ISL_2930958, EPI_ISL_2930959, EPI_ISL_3008888, EPI_ISL_3008892 | | | |
| see above | Department of Clinical Microbiology | GIGA Medical Genomics | Bouchra Boujemla; Claire Gourzonès; Cécile Meex; Keith Durkin; Laurent Gillet; Maria Artesi; Marie-Pierre Hayette; Nadine Cambisano; Nathalie Renotte; Olivier Ek; Sébastien Bontems; Vincent Bours |
| EPI_ISL_3372584, EPI_ISL_3372586 | Dirección regional de salud del Callao (DIRESA-CALLAO) | Centro de Investigaciones Tecnológicas, Biomédicas y Medioambientales (CITBM) | B; Huaman; J. Alarcon; M. Cuellar; M. Ramirez; M. Sovero |
| EPI_ISL_3056821, EPI_ISL_3057865, EPI_ISL_3136701, EPI_ISL_3137106 | Dutch COVID-19 response team | National Institute for Public Health and the Environment (RIVM) | Adam Meijer; AnneMarie van den Brandt; Annelies Kroneman; Bas van der Veer; Chantal Reusken; Dennis Schmitz; Dirk Eggink; Euníce Then; Florian Zwagemaker; Harry Vennema; Jeroen Cremer; Karim Hajji; Kim Freriks; Lisa Wijsman; Lynn Aarts; Melissa van Tuil; Ryanne Jaarsma; Sanne Bos; Sharon van den Brink; on behalf of the national COVID-19 response team |
| EPI_ISL_4059813, EPI_ISL_4059821, EPI_ISL_4059824, EPI_ISL_4060095 | Dynacare Manitoba | Manitoba Cadham Provincial Laboratory | David Alexander; Hilary Racher; Hongbin Li; Melissa Desaulnier; Tintu Abraham |
| EPI_ISL_5673342, EPI_ISL_5673399, EPI_ISL_5673454, EPI_ISL_5673583, EPI_ISL_5673637, EPI_ISL_5673679, EPI_ISL_5673686, EPI_ISL_5673883, EPI_ISL_5674434 | | | |
| see above | EMERGENCIA RESPIRATORIA | Instituto Butantan | Antonio Jorge Martins; Claudia Renata dos Santos Barros; David Schlesinger; Debora Botequio Moretti; Dimas Tadeu Covas; Elaine Cristina Marqueze; Elaine Vieira Santos; Evandra Strazza Rodrigues; Heidge Fukumasu; Jayme Augusto de Souza-Neto; José Salvatore Leister Patané; Luiz Alcantara; Luiz Lehmann |

| | | | |
|---|---|---|---|
| | DE NOVA GRANADA | | Coutinho; Maria Carolina Elias; Maurício Lacerda Nogueira; Rafael dos Santos Bezerra; Raul Machado Neto; Rejane Maria Tommasini Grotto; Ricardo Haddad; Sandra Coccuzzo Sampaio Vessoni; Simone Kashima; Svetoslav Nanev Slavov; Vincent Louis Viala |
| EPI_ISL_3215798 | EXCITE Lab | Andersen lab at Scripps Research | Abigail Schnapper; Angela Scioscia; Cheryl Anderson; Chip Schooley; Greg Humphrey; Helena Tubb; Natasha Martin; Sawyer Farmer; Smruthi Karthikeyan; Tommy Valles + SEARCH |
| EPI_ISL_3116504, EPI_ISL_3116508, EPI_ISL_3116509, EPI_ISL_3116510, EPI_ISL_3909785, EPI_ISL_3909959, EPI_ISL_3910250, EPI_ISL_3910251, EPI_ISL_3910283, EPI_ISL_3910458, EPI_ISL_3910557 | | | |
| see above | Edmonton Provincial Lab | Public Health Agency of Canada (PHAC) National Microbiology Laboratory | Buss; Croxen M; Deo A; Dieu P; E; Ferrato C; Gill K; Khan F; Koleva P; Li V; Lloyd C; Lynch T; Ma R; Murphy S; Pabbaraju K; Shokoples S; Thayer J; Tipples G; Whitehouse M; Wong A; Yu C; Zelyas N |
| EPI_ISL_5673932, EPI_ISL_5674180, EPI_ISL_5674475, EPI_ISL_5674578, EPI_ISL_5674739, EPI_ISL_5674953, EPI_ISL_5675042 | | | |
| see above | FACULDADE DE MEDICINA ABC | Instituto Butantan | Antonio Jorge Martins; Claudia Renata dos Santos Barros; David Schlesinger; Debora Botequio Moretti; Dimas Tadeu Covas; Elaine Cristina Marqueze; Elaine Vieira Santos; Evandra Strazza Rodrigues; Heidge Fukumasu; Jayme Augusto de Souza-Neto; José Salvatore Leister Patané; Luiz Alcantara; Luiz Lehmann Coutinho; Maria Carolina Elias; Maurício Lacerda Nogueira; Rafael dos Santos Bezerra; Raul Machado Neto; Rejane Maria Tommasini Grotto; Ricardo Haddad; Sandra Coccuzzo Sampaio Vessoni; Simone Kashima; Svetoslav Nanev Slavov; Vincent Louis Viala |
| EPI_ISL_3242064, EPI_ISL_3242067 | FL Bureau of Public Health Laboratories-Tampa | Centers for Disease Control and Prevention Division of Viral Diseases, Pathogen Discovery | Alex Burgin; Ben L. Rambo-Martin; Clinton R. Paden; Dakota Howard; Dave Wentworth; Dhwani Batra; Jasmine Padilla; Justin Lee; Krista Queen; Kristen Knipe; Kristine Lacek; Mark Burroughs; Matthew Schmerer; Meghan Bentz; Mili Sheth; Peter Cook; Sam Shepard; Sarah Nobles; Suxiang Tong; Vivien Dugan; Yvette Unoarumhi |
| EPI_ISL_3486967, EPI_ISL_3487331 | FYR Diagnostics | FYR Diagnostics | Blohm; Ghai; Tre; Vikas |
| EPI_ISL_3100847, EPI_ISL_3903285, EPI_ISL_3903476, EPI_ISL_3903535, EPI_ISL_5322158, EPI_ISL_5322332, EPI_ISL_5322362, EPI_ISL_5322438, EPI_ISL_5322488, EPI_ISL_5322845, EPI_ISL_5925189, EPI_ISL_5925190, EPI_ISL_5925191, EPI_ISL_5925192, EPI_ISL_5925197 | | | |
| see above | Florida Bureau of Public Health Laboratories | Florida Bureau of Public Health Laboratories | Jason Blanton; Jiaqi Li; Namratha Tarigopula; Sarah Schmedes; Tiffany Splatt |
| EPI_ISL_2928856, EPI_ISL_2928996, EPI_ISL_2929015, EPI_ISL_2929021, EPI_ISL_2929022, EPI_ISL_2929073, EPI_ISL_2929077, EPI_ISL_2929078, EPI_ISL_2929080, EPI_ISL_2929083, EPI_ISL_2929085, EPI_ISL_2929086, EPI_ISL_2929093, EPI_ISL_2929125, EPI_ISL_3016511, EPI_ISL_3016520, EPI_ISL_3016527, EPI_ISL_3016544, EPI_ISL_3016551, EPI_ISL_3016552, EPI_ISL_3016580, EPI_ISL_3016581, EPI_ISL_3016654, EPI_ISL_3016655, EPI_ISL_3016671, EPI_ISL_3016702, EPI_ISL_3064813, EPI_ISL_3065030, EPI_ISL_3208196, EPI_ISL_3208204, EPI_ISL_3208249, EPI_ISL_3208256, EPI_ISL_3208262, EPI_ISL_3208266, EPI_ISL_3208287, EPI_ISL_3208290, EPI_ISL_3208297, EPI_ISL_3208299, EPI_ISL_3208310, EPI_ISL_3208320, EPI_ISL_3208332, EPI_ISL_3208341, EPI_ISL_3208345, EPI_ISL_3208643, EPI_ISL_3208689, EPI_ISL_3208706, EPI_ISL_3208781, EPI_ISL_3208806, EPI_ISL_3208819, EPI_ISL_3210139, EPI_ISL_3299799, EPI_ISL_3299857, EPI_ISL_3299859, EPI_ISL_3299864, EPI_ISL_3299867, EPI_ISL_3299871, EPI_ISL_3299893, EPI_ISL_3299894, EPI_ISL_3299924, EPI_ISL_3299939, EPI_ISL_3299967, EPI_ISL_3299968, EPI_ISL_3299983, EPI_ISL_3300002, EPI_ISL_3300021, EPI_ISL_4367432 | | | |
| see above | Fulgent Genetics | Centers for Disease Control and Prevention Division of Viral Diseases, Pathogen Discovery | Adrian Paskey; Becky Tsai; Benafsh Sapra; Benjamin Rambo-Martin; Christopher Gulvick; Clinton Paden; Clinton R. Paden; Dakota Howard; Darlene Wagner; Dhwani Batra; Doreen Ng; Duncan MacCannell; Erisa Sula; Harry Gao; James Xie; Jason Caravas; John Gao; Joseph Fierro; Kara Moser; Kristine Lacek; Matthew Schmerer; Mickey Li; Peter Cook; Peter W. Cook; Scott Sammons; Shatavia Morrison; Tymeckia Kendall; Victoria Caban Figueroa; Yan Meng; Yvette Unoarumhi |
| EPI_ISL_3800483 | GA Department of Public Health Laboratory | Centers for Disease Control and Prevention Division of Viral Diseases, Pathogen Discovery | Alex Burgin; Ben Rambo-Martin; Clinton Paden; Dakota Howard; Dave Wentworth; Dhwani Batra; Jasmine Padilla; Justin Lee; Krista Queen; Kristen Knipe; Kristine Lacek; Mark Burroughs; Matthew Schmerer; Meghan Bentz; Mili Sheth; Peter Cook; Sam Shepard; Sarah Nobles; Suxiang Tong; Vivien Dugan; Yvette Unoarumhi |
| EPI_ISL_3300573 | Gandhi Hospital | CDFD | Ashwin Dalal; Asmita Gupta; Divya Vashisht; Murali Bashyam; Nagamani Kammili; Reelina Basu; Vinay Donipadi |
| EPI_ISL_3132173, EPI_ISL_3132174, EPI_ISL_3132175, EPI_ISL_3132176, EPI_ISL_3132177, EPI_ISL_3132179, EPI_ISL_3132181, EPI_ISL_3132182, EPI_ISL_3132183, EPI_ISL_3132184, EPI_ISL_3132185, EPI_ISL_3132186, EPI_ISL_3132188, EPI_ISL_3132270, EPI_ISL_3132678, EPI_ISL_3132679, EPI_ISL_3132680, EPI_ISL_3132681, EPI_ISL_3132682, EPI_ISL_3132683, EPI_ISL_3132684, EPI_ISL_3132685, EPI_ISL_3132687, EPI_ISL_3132688, EPI_ISL_3132689, EPI_ISL_3132691, EPI_ISL_3132692, EPI_ISL_3132693, EPI_ISL_3132695, EPI_ISL_3132696, EPI_ISL_3132698, EPI_ISL_3149072, EPI_ISL_3149074, EPI_ISL_3149102, EPI_ISL_3149104, EPI_ISL_3149105, EPI_ISL_3149106, EPI_ISL_3185458, EPI_ISL_3185469, EPI_ISL_3185471, EPI_ISL_3185472, EPI_ISL_3185476, EPI_ISL_3185490, EPI_ISL_3185526, EPI_ISL_3185527, EPI_ISL_3185528, EPI_ISL_3185529, EPI_ISL_3185530, EPI_ISL_3185531, EPI_ISL_3185545, EPI_ISL_3185546, EPI_ISL_3185547, EPI_ISL_3185548, EPI_ISL_3185549, EPI_ISL_3185560, EPI_ISL_3185562, EPI_ISL_3185564, EPI_ISL_3185566, EPI_ISL_3185567, EPI_ISL_3185568, EPI_ISL_3185569, EPI_ISL_3185570, EPI_ISL_3185572, EPI_ISL_3185587, EPI_ISL_3185588, EPI_ISL_3185591, EPI_ISL_3185592, EPI_ISL_3185593, EPI_ISL_3185594, EPI_ISL_3185595, EPI_ISL_3185596, EPI_ISL_3185601, EPI_ISL_3185602, EPI_ISL_3185606, EPI_ISL_3185607, EPI_ISL_3185608, EPI_ISL_3185609, EPI_ISL_3185610, EPI_ISL_3185611, EPI_ISL_3185612, EPI_ISL_3185613, EPI_ISL_3185614, EPI_ISL_3185616, EPI_ISL_3185619, EPI_ISL_3185620, EPI_ISL_3185621, EPI_ISL_3185624, EPI_ISL_3185625, EPI_ISL_3185627, EPI_ISL_3185629, EPI_ISL_3185630, EPI_ISL_3185631, EPI_ISL_3185632, EPI_ISL_3185633, EPI_ISL_3185634, EPI_ISL_3185635, EPI_ISL_3185636, EPI_ISL_3185637, EPI_ISL_3185639, EPI_ISL_3185640, EPI_ISL_3185641, EPI_ISL_3185642, EPI_ISL_3185643, EPI_ISL_3185644, EPI_ISL_3185645, EPI_ISL_3185647, EPI_ISL_3185648, EPI_ISL_3185649, EPI_ISL_3185658, EPI_ISL_3185662, EPI_ISL_3185664, EPI_ISL_3185665, EPI_ISL_3185666, EPI_ISL_3185667, EPI_ISL_3185673, EPI_ISL_3185674, EPI_ISL_3185675, EPI_ISL_3185676, EPI_ISL_3185678, EPI_ISL_3185679, EPI_ISL_3185680, EPI_ISL_3185682, EPI_ISL_3185684, EPI_ISL_3185685, EPI_ISL_3185691, EPI_ISL_3185692, EPI_ISL_3185693, EPI_ISL_3185694, EPI_ISL_3185695, EPI_ISL_3185696, EPI_ISL_3185697, EPI_ISL_3185698, EPI_ISL_3185699, EPI_ISL_3185700, EPI_ISL_3185703, EPI_ISL_3185705, EPI_ISL_3185706, EPI_ISL_3185708, EPI_ISL_3185710, EPI_ISL_3185711, EPI_ISL_3185712, EPI_ISL_3185713, EPI_ISL_3185715, EPI_ISL_3185716, EPI_ISL_3185717, EPI_ISL_3185718, EPI_ISL_3185719, EPI_ISL_3185720, EPI_ISL_3185721, EPI_ISL_3185722, EPI_ISL_3185723, EPI_ISL_3185724, EPI_ISL_3185726, EPI_ISL_3185727, EPI_ISL_3185728, EPI_ISL_3185730, EPI_ISL_3185731, EPI_ISL_3185734, EPI_ISL_3185754, EPI_ISL_3185764, EPI_ISL_3185766, EPI_ISL_3185767, EPI_ISL_3185770, EPI_ISL_3185795, EPI_ISL_3185806, EPI_ISL_3358533, EPI_ISL_3358534, EPI_ISL_3358535, EPI_ISL_3358536, EPI_ISL_3358538, EPI_ISL_3358540, EPI_ISL_3358541, EPI_ISL_3358548, EPI_ISL_3358558, EPI_ISL_3358559, EPI_ISL_3358562, EPI_ISL_3358566, EPI_ISL_3358567, EPI_ISL_3358568, EPI_ISL_3358570, EPI_ISL_3358571, EPI_ISL_3358572, EPI_ISL_3358573, EPI_ISL_3368809, EPI_ISL_3368853, EPI_ISL_3368854, EPI_ISL_3368855, EPI_ISL_3368856, EPI_ISL_3368857, EPI_ISL_3368858, EPI_ISL_3368885, EPI_ISL_3368886, EPI_ISL_3368887, EPI_ISL_3368888, EPI_ISL_3368889, EPI_ISL_3368890, EPI_ISL_3368891, EPI_ISL_3368892, EPI_ISL_3368893, EPI_ISL_3368894, EPI_ISL_3368896, EPI_ISL_3368897, EPI_ISL_3369339, EPI_ISL_3369340, EPI_ISL_3369360, EPI_ISL_3369377, EPI_ISL_3369378, EPI_ISL_3369381, EPI_ISL_3369382, EPI_ISL_3369383, EPI_ISL_3369384, EPI_ISL_3369385, EPI_ISL_3369386, EPI_ISL_3369391, EPI_ISL_3369403, EPI_ISL_3369404, EPI_ISL_3369408, EPI_ISL_3369409, EPI_ISL_3981972, EPI_ISL_3981976 | | | |
| see above | Genetica Molecular and Subdepartamento de Virologia ISP Chile | Instituto de Salud Publica de Chile | Andres Castillo; Barbara Parra; Constanza Campano; Gisselle Barra; Javier Tognarelli; Jorge Fernandez; Karen Orostica; Loredana Arata; Patricia Bustos; Rodrigo Fasce; Soledad Ulloa |
| EPI_ISL_7476570 | Grupo de Investigación en Enfermedades Tropicales del Ejército (GINETEJ), Laboratorio de Referencia e Investigación, Dirección de Sanidad Ejército, Bogotá, Colombia | Centro de Investigaciones en Microbiología y Biotecnología-UR (CIMBIUR), Facultad de Ciencias Naturales, Universidad del Rosario, Bogotá, Colombia | Alberto E. Paniz-Mondolfi; Angie L. Ramírez; Camilo Correa-Cardenas; Claudia Mendez; Enzo Guerrero-Araya; Juan David Ramírez; Julie Perez; Luz H. Patiño; Maria Clara Duque; Marina Muñoz; Nathalia Ballesteros; Nicolás Luna; Sergio Castañeda |
| EPI_ISL_3536305 | H J M A HOSPITAL JOSE MARTINIANO DE ALENCAR | Analytical Competence Molecular Epidemiology Lab/ACME, Oswaldo Cruz Foundation, Ceara (FIOCRUZ CE) | Cleber Furtado Aksenen; Fabio Miyajima; Fernando Braga Stehling; Francisco Eder de Moura Lopes; Jamille Maria Mendes Bezerra; Joaquim César do Nascimento Sousa Junior; Pedro Miguel Carneiro Jeronimo; Suzana Porto Almeida e Lucas Delerino; Thais Ferreira de Oliveira; Thais de Oliveira Costa; Ticiane Cavalcante de Souza; Veridiana Pessoa Miyajima |
| EPI_ISL_3254655, EPI_ISL_3254656, EPI_ISL_3254657, EPI_ISL_3254658, EPI_ISL_3254659, EPI_ISL_3254660, EPI_ISL_3254661, EPI_ISL_3254662, EPI_ISL_3254663, EPI_ISL_3254664, EPI_ISL_3254665, EPI_ISL_3254666, EPI_ISL_3254667, EPI_ISL_3254668, EPI_ISL_3254669, EPI_ISL_3254678, EPI_ISL_3254679, EPI_ISL_3254680, EPI_ISL_3254681, EPI_ISL_3254683, EPI_ISL_3259353, EPI_ISL_3259354, EPI_ISL_3259373, EPI_ISL_3274744, EPI_ISL_3274745, EPI_ISL_3274746, EPI_ISL_3299424, EPI_ISL_3386142, EPI_ISL_3386151, EPI_ISL_3386154, EPI_ISL_4061630, EPI_ISL_4061631, EPI_ISL_4061667, EPI_ISL_4061668, EPI_ISL_4554943 | | | |
| see above | HLAGYN - Laboratorio de Imunologia de Transplantes de Goias | HLAGYN - Laboratorio de Imunologia de Transplantes de Goias | Alessandro Leonardo Alvares Magalhaes; Erika Lopes Rocha Batista; Fernando Antonio Vinhal dos Santos; Frederico Rodrigues Vinhal; Kamila Oliveira Reis De Freitas.; Lucas Carlos Gomes Pereira; Sabrina Sara Moreira Duarte |
| EPI_ISL_3536222, EPI_ISL_3536226, EPI_ISL_3536240, EPI_ISL_3536246, EPI_ISL_3536251, EPI_ISL_3536298, EPI_ISL_3536306 | | | |
| see above | HM HOSPITAL DE MESSEJANA DR CARLOS ALBERTO STUDART GOMES | Analytical Competence Molecular Epidemiology Lab/ACME, Oswaldo Cruz Foundation, Ceara (FIOCRUZ CE) | Cleber Furtado Aksenen; Fabio Miyajima; Fernando Braga Stehling; Francisco Eder de Moura Lopes; Jamille Maria Mendes Bezerra; Joaquim César do Nascimento Sousa Junior; Pedro Miguel Carneiro Jeronimo; Suzana Porto Almeida e Lucas Delerino; Thais Ferreira de Oliveira; Thais de Oliveira Costa; Ticiane Cavalcante de Souza; Veridiana Pessoa Miyajima |
| EPI_ISL_5671563, EPI_ISL_5671616, EPI_ISL_5676472, EPI_ISL_5677711 | HOSP E MATERNIDADE MUNICIPAL N SRA MONTE SERRAT | Instituto Butantan | Antonio Jorge Martins; Claudia Renata dos Santos Barros; David Schlesinger; Debora Botequio Moretti; Dimas Tadeu Covas; Elaine Cristina Marqueze; Elaine Vieira Santos; Evandra Strazza Rodrigues; Heidge Fukumasu; Jayme Augusto de Souza-Neto; José Salvatore Leister Patané; Luiz Alcantara; Luiz Lehmann Coutinho; Maria Carolina Elias; Maurício Lacerda Nogueira; Rafael dos Santos Bezerra; Raul Machado Neto; Rejane Maria Tommasini Grotto; Ricardo Haddad; Sandra Coccuzzo Sampaio Vessoni; Simone Kashima; Svetoslav Nanev Slavov; Vincent Louis Viala |
| EPI_ISL_5670819, EPI_ISL_5672313 | HOSP MUN CIDADE TIRADENTES CARMEN PRUDENTE | Instituto Butantan | Antonio Jorge Martins; Claudia Renata dos Santos Barros; David Schlesinger; Debora Botequio Moretti; Dimas Tadeu Covas; Elaine Cristina Marqueze; Elaine Vieira Santos; Evandra Strazza Rodrigues; Heidge Fukumasu; Jayme Augusto de Souza-Neto; José Salvatore Leister Patané; Luiz Alcantara; Luiz Lehmann Coutinho; Maria Carolina Elias; Maurício Lacerda Nogueira; Rafael dos Santos Bezerra; Raul Machado Neto; Rejane Maria Tommasini Grotto; Ricardo Haddad; Sandra Coccuzzo Sampaio Vessoni; Simone Kashima; Svetoslav Nanev Slavov; Vincent Louis Viala |
| EPI_ISL_5668529, EPI_ISL_5669266, EPI_ISL_5669645, EPI_ISL_5670292, EPI_ISL_5670305, EPI_ISL_5670711, EPI_ISL_5670729 | | | |
| see above | HOSPITAL DE CAMPANHA COVID 19 CAIEIRAS | Instituto Butantan | Antonio Jorge Martins; Claudia Renata dos Santos Barros; David Schlesinger; Debora Botequio Moretti; Dimas Tadeu Covas; Elaine Cristina Marqueze; Elaine Vieira Santos; Evandra Strazza Rodrigues; Heidge Fukumasu; Jayme Augusto de Souza-Neto; José Salvatore Leister Patané; Luiz Alcantara; Luiz Lehmann Coutinho; Maria Carolina Elias; Maurício Lacerda Nogueira; Rafael dos Santos Bezerra; Raul Machado Neto; Rejane Maria Tommasini Grotto; Ricardo Haddad; Sandra Coccuzzo Sampaio Vessoni; Simone Kashima; Svetoslav Nanev Slavov; Vincent Louis Viala |
| EPI_ISL_5669683, EPI_ISL_5669914 | HOSPITAL DE CAMPANHA COVID 19 DE MAIRIPORA | Instituto Butantan | Antonio Jorge Martins; Claudia Renata dos Santos Barros; David Schlesinger; Debora Botequio Moretti; Dimas Tadeu Covas; Elaine Cristina Marqueze; Elaine Vieira Santos; Evandra Strazza Rodrigues; Heidge Fukumasu; Jayme Augusto de Souza-Neto; José Salvatore Leister Patané; Luiz Alcantara; Luiz Lehmann Coutinho; Maria Carolina Elias; Maurício Lacerda Nogueira; Rafael dos Santos Bezerra; Raul Machado Neto; Rejane Maria Tommasini Grotto; Ricardo Haddad; Sandra Coccuzzo Sampaio Vessoni; Simone Kashima; Svetoslav Nanev Slavov; Vincent Louis Viala |
| EPI_ISL_3464519 | HOSPITAL DR. FERNANDO ESCALANTE PRADILLA | Inciensa, Instituto Costarricense de Investigación y Enseñanza en Nutrición y Salud | Adriana Godínez; Claudio Soto-Garita; Estela Cordero; Francisco Duarte; Hebleen Porras; Joselyn Prado & Yendri Ramírez Alpízar; José Luis Vargas; Mariela Gutiérrez; Melany Calderón |
| EPI_ISL_3536297, EPI_ISL_3536301 | HOSPITAL E MATERNIDADE DRA ZILDA ARNS NEUMANN | Analytical Competence Molecular Epidemiology Lab/ACME, Oswaldo Cruz Foundation, Ceara (FIOCRUZ CE) | Cleber Furtado Aksenen; Fabio Miyajima; Fernando Braga Stehling; Francisco Eder de Moura Lopes; Jamille Maria Mendes Bezerra; Joaquim César do Nascimento Sousa Junior; Pedro Miguel Carneiro Jeronimo; Suzana Porto Almeida e Lucas Delerino; Thais Ferreira de Oliveira; Thais de Oliveira Costa; Ticiane Cavalcante de Souza; Veridiana Pessoa Miyajima |
| EPI_ISL_3536164, EPI_ISL_3536167, EPI_ISL_3536168, EPI_ISL_3536171, EPI_ISL_3536174, EPI_ISL_3536270, EPI_ISL_3536295, EPI_ISL_3536312, EPI_ISL_3536322, EPI_ISL_3536348, EPI_ISL_3536392, EPI_ISL_3536394, EPI_ISL_3536396, EPI_ISL_3912036, EPI_ISL_3912053, EPI_ISL_3912205 | | | |
| see above | HOSPITAL ESTADUAL LEONARDO DA VINCI | Analytical Competence Molecular Epidemiology Lab/ACME, Oswaldo Cruz Foundation, Ceara (FIOCRUZ CE) | Cleber Furtado Aksenen; Fabio Miyajima; Fernando Braga Stehling; Francisco Eder de Moura Lopes; Jamille Maria Mendes Bezerra; Joaquim Cesar do Nascimento Sousa Junior; Joaquim César do Nascimento Sousa Junior; Pedro Miguel Carneiro Jeronimo; Suzana Porto Almeida & Lucas Delerino on behalf of COVID-19 FIOCRUZ Genomic Network; Suzana Porto Almeida e Lucas Delerino; Thais Ferreira de Oliveira; Thais de Oliveira Costa; Ticiane Cavalcante de Souza; Veridiana Pessoa Miyajima |
| EPI_ISL_4919277 | HOSPITAL GENERAL UNIVERSITARIO DE GUADALAJARA | Instituto de Salud Carlos III | A. Monzón; ALEJANDRO; F. Casas; I. GONZALEZ PRAETORIUS; I. Jiménez; M. Camarero; P. Zaballos; S. Cuesta; S. Iglesias-Caballero; S. Pozo; S. Varona; Sandonís; V. Vázquez-Morón |
| EPI_ISL_5672004, EPI_ISL_5672063, EPI_ISL_5672108, EPI_ISL_5672503, EPI_ISL_5673951 | HOSPITAL GERAL JESUS TEIXEIRA DA COSTA GUAIANASES SAO PAULO | Instituto Butantan | Antonio Jorge Martins; Claudia Renata dos Santos Barros; David Schlesinger; Debora Botequio Moretti; Dimas Tadeu Covas; Elaine Cristina Marqueze; Elaine Vieira Santos; Evandra Strazza Rodrigues; Heidge Fukumasu; Jayme Augusto de Souza-Neto; José Salvatore Leister Patané; Luiz Alcantara; Luiz Lehmann Coutinho; Maria Carolina Elias; Maurício Lacerda Nogueira; Rafael dos Santos Bezerra; Raul Machado Neto; Rejane Maria Tommasini Grotto; Ricardo Haddad; Sandra Coccuzzo Sampaio Vessoni; Simone Kashima; Svetoslav Nanev Slavov; Vincent Louis Viala |
| EPI_ISL_5673001, EPI_ISL_5673025, | HOSPITAL MATERNIDADE SAO JOSE ITAPUI | Instituto Butantan | Antonio Jorge Martins; Claudia Renata dos Santos Barros; David Schlesinger; Debora Botequio Moretti; Dimas Tadeu Covas; Elaine Cristina Marqueze; Elaine Vieira Santos; Evandra Strazza Rodrigues; Heidge Fukumasu; Jayme Augusto de Souza-Neto; José Salvatore Leister Patané; Luiz Alcantara; Luiz Lehmann Coutinho; Maria Carolina Elias; Maurício Lacerda Nogueira; Rafael dos Santos Bezerra; Raul Machado Neto; Rejane Maria Tommasini Grotto; Ricardo Haddad; Sandra Coccuzzo Sampaio Vessoni; Simone Kashima; Svetoslav Nanev Slavov; Vincent Louis Viala |

| | | | |
|---|---|---|---|
| EPI_ISL_5673076, EPI_ISL_5673150, EPI_ISL_5673208 | | | |
| EPI_ISL_3298328 | HOSPITAL MEXICO | Inciensa, Instituto Costarricense de Investigación y Enseñanza en Nutrición y Salud | Adriana Godínez; Claudio Soto-Garita; Estela Cordero; Francisco Duarte; Hebleen Porras; Joselyn Prado & Juan Carlos Villalobos; José Luis Vargas; Mariela Gutiérrez; Melany Calderón |
| EPI_ISL_5669129, EPI_ISL_5669136, EPI_ISL_5670330, EPI_ISL_5670432, EPI_ISL_5670470 | HOSPITAL MUN DR GUIDO GUIDA | Instituto Butantan | Antonio Jorge Martins; Claudia Renata dos Santos Barros; David Schlesinger; Debora Botequio Moretti; Dimas Tadeu Covas; Elaine Cristina Marqueze; Elaine Vieira Santos; Evandra Strazza Rodrigues; Heidge Fukumasu; Jayme Augusto de Souza-Neto; José Salvatore Leister Patané; Luiz Alcantara; Luiz Lehmann Coutinho; Maria Carolina Elias; Maurício Lacerda Nogueira; Rafael dos Santos Bezerra; Raul Machado Neto; Rejane Maria Tommasini Grotto; Ricardo Haddad; Sandra Coccuzzo Sampaio Vessoni; Simone Kashima; Svetoslav Nanev Slavov; Vincent Louis Viala |
| EPI_ISL_5671601 | HOSPITAL MUNICIPAL DE IBIUNA IBIUNA SP | Instituto Butantan | Antonio Jorge Martins; Claudia Renata dos Santos Barros; David Schlesinger; Debora Botequio Moretti; Dimas Tadeu Covas; Elaine Cristina Marqueze; Elaine Vieira Santos; Evandra Strazza Rodrigues; Heidge Fukumasu; Jayme Augusto de Souza-Neto; José Salvatore Leister Patané; Luiz Alcantara; Luiz Lehmann Coutinho; Maria Carolina Elias; Maurício Lacerda Nogueira; Rafael dos Santos Bezerra; Raul Machado Neto; Rejane Maria Tommasini Grotto; Ricardo Haddad; Sandra Coccuzzo Sampaio Vessoni; Simone Kashima; Svetoslav Nanev Slavov; Vincent Louis Viala |
| EPI_ISL_5670952, EPI_ISL_5670956, EPI_ISL_5670982, EPI_ISL_5670997, EPI_ISL_5671173, EPI_ISL_5671319, EPI_ISL_5671448 | | | |
| see above | HOSPITAL MUNICIPAL DE ITAPIRA | Instituto Butantan | Antonio Jorge Martins; Claudia Renata dos Santos Barros; David Schlesinger; Debora Botequio Moretti; Dimas Tadeu Covas; Elaine Cristina Marqueze; Elaine Vieira Santos; Evandra Strazza Rodrigues; Heidge Fukumasu; Jayme Augusto de Souza-Neto; José Salvatore Leister Patané; Luiz Alcantara; Luiz Lehmann Coutinho; Maria Carolina Elias; Maurício Lacerda Nogueira; Rafael dos Santos Bezerra; Raul Machado Neto; Rejane Maria Tommasini Grotto; Ricardo Haddad; Sandra Coccuzzo Sampaio Vessoni; Simone Kashima; Svetoslav Nanev Slavov; Vincent Louis Viala |
| EPI_ISL_5670965, EPI_ISL_5671007, EPI_ISL_5671012, EPI_ISL_5671021, EPI_ISL_5671091 | HOSPITAL MUNICIPAL DR TABAJARA RAMOS | Instituto Butantan | Antonio Jorge Martins; Claudia Renata dos Santos Barros; David Schlesinger; Debora Botequio Moretti; Dimas Tadeu Covas; Elaine Cristina Marqueze; Elaine Vieira Santos; Evandra Strazza Rodrigues; Heidge Fukumasu; Jayme Augusto de Souza-Neto; José Salvatore Leister Patané; Luiz Alcantara; Luiz Lehmann Coutinho; Maria Carolina Elias; Maurício Lacerda Nogueira; Rafael dos Santos Bezerra; Raul Machado Neto; Rejane Maria Tommasini Grotto; Ricardo Haddad; Sandra Coccuzzo Sampaio Vessoni; Simone Kashima; Svetoslav Nanev Slavov; Vincent Louis Viala |
| EPI_ISL_5654086 | HOSPITAL MUNICIPAL LEONARDUS VAN MELLIS | Instituto Butantan | Antonio Jorge Martins; Claudia Renata dos Santos Barros; David Schlesinger; Debora Botequio Moretti; Dimas Tadeu Covas; Elaine Cristina Marqueze; Elaine Vieira Santos; Evandra Strazza Rodrigues; Heidge Fukumasu; Jayme Augusto de Souza-Neto; José Salvatore Leister Patané; Luiz Alcantara; Luiz Lehmann Coutinho; Maria Carolina Elias; Maurício Lacerda Nogueira; Rafael dos Santos Bezerra; Raul Machado Neto; Rejane Maria Tommasini Grotto; Ricardo Haddad; Sandra Coccuzzo Sampaio Vessoni; Simone Kashima; Svetoslav Nanev Slavov; Vincent Louis Viala |
| EPI_ISL_5668429 | HOSPITAL MUNICIPAL REYNALDO GUERRA CAJATI | Instituto Butantan | Antonio Jorge Martins; Claudia Renata dos Santos Barros; David Schlesinger; Debora Botequio Moretti; Dimas Tadeu Covas; Elaine Cristina Marqueze; Elaine Vieira Santos; Evandra Strazza Rodrigues; Heidge Fukumasu; Jayme Augusto de Souza-Neto; José Salvatore Leister Patané; Luiz Alcantara; Luiz Lehmann Coutinho; Maria Carolina Elias; Maurício Lacerda Nogueira; Rafael dos Santos Bezerra; Raul Machado Neto; Rejane Maria Tommasini Grotto; Ricardo Haddad; Sandra Coccuzzo Sampaio Vessoni; Simone Kashima; Svetoslav Nanev Slavov; Vincent Louis Viala |
| EPI_ISL_5668789, EPI_ISL_5669096, EPI_ISL_5669271 | HOSPITAL REGIONAL DE ITAPETININGA | Instituto Butantan | Antonio Jorge Martins; Claudia Renata dos Santos Barros; David Schlesinger; Debora Botequio Moretti; Dimas Tadeu Covas; Elaine Cristina Marqueze; Elaine Vieira Santos; Evandra Strazza Rodrigues; Heidge Fukumasu; Jayme Augusto de Souza-Neto; José Salvatore Leister Patané; Luiz Alcantara; Luiz Lehmann Coutinho; Maria Carolina Elias; Maurício Lacerda Nogueira; Rafael dos Santos Bezerra; Raul Machado Neto; Rejane Maria Tommasini Grotto; Ricardo Haddad; Sandra Coccuzzo Sampaio Vessoni; Simone Kashima; Svetoslav Nanev Slavov; Vincent Louis Viala |
| EPI_ISL_3912225 | HOSPITAL REGIONAL UNIMED | Analytical Competence Molecular Epidemiology Lab/ACME, Oswaldo Cruz Foundation, Ceara (FIOCRUZ CE) | Cleber Furtado Aksenen; Fabio Miyajima; Fernando Braga Stehling; Francisco Eder de Moura Lopes; Jamille Maria Mendes Bezerra; Joaquim Cesar do Nascimento Sousa Junior; Pedro Miguel Carneiro Jeronimo; Suzana Porto Almeida & Lucas Delerino on behalf of COVID-19 FIOCRUZ Genomic Network; Thais Ferreira de Oliveira; Thais de Oliveira Costa; Ticiane Cavalcante de Souza; Veridiana Pessoa Miyajima |
| EPI_ISL_5679492 | HOSPITAL SANTO ANTONIO DE JUQUIA JUQUIA | Instituto Butantan | Antonio Jorge Martins; Claudia Renata dos Santos Barros; David Schlesinger; Debora Botequio Moretti; Dimas Tadeu Covas; Elaine Cristina Marqueze; Elaine Vieira Santos; Evandra Strazza Rodrigues; Heidge Fukumasu; Jayme Augusto de Souza-Neto; José Salvatore Leister Patané; Luiz Alcantara; Luiz Lehmann Coutinho; Maria Carolina Elias; Maurício Lacerda Nogueira; Rafael dos Santos Bezerra; Raul Machado Neto; Rejane Maria Tommasini Grotto; Ricardo Haddad; Sandra Coccuzzo Sampaio Vessoni; Simone Kashima; Svetoslav Nanev Slavov; Vincent Louis Viala |
| EPI_ISL_5680799, EPI_ISL_5680851 | HOSPITAL SAO JOSE ITIRAPINA | Instituto Butantan | Antonio Jorge Martins; Claudia Renata dos Santos Barros; David Schlesinger; Debora Botequio Moretti; Dimas Tadeu Covas; Elaine Cristina Marqueze; Elaine Vieira Santos; Evandra Strazza Rodrigues; Heidge Fukumasu; Jayme Augusto de Souza-Neto; José Salvatore Leister Patané; Luiz Alcantara; Luiz Lehmann Coutinho; Maria Carolina Elias; Maurício Lacerda Nogueira; Rafael dos Santos Bezerra; Raul Machado Neto; Rejane Maria Tommasini Grotto; Ricardo Haddad; Sandra Coccuzzo Sampaio Vessoni; Simone Kashima; Svetoslav Nanev Slavov; Vincent Louis Viala |
| EPI_ISL_5515607 | HOSPITAL UNIVERSITARIO 12 DE OCTUBRE | HOSPITAL UNIVERSITARIO 12 DE OCTUBRE | Carmen Martín-Higuera; Esther Viedma; Irene Múñoz-Gallego; M.ª Dolores Folgueira; Mar Aguilera; Noelia Moral; Rafael Delgado; Sagrario Zurita |
| EPI_ISL_4799343, EPI_ISL_4919054, EPI_ISL_4919093 | HOSPITAL VIRGEN DE LA SALUD DE TOLEDO | Instituto de Salud Carlos III | A. Monzón; EVA; F. Casas; I. Jiménez; I.HEREDERO GALVEZ; M. Sandonís; P. Zaballos; S. Cuesta; S. Iglesias-Caballero; S. Pozo; S. Varona; V. Camarero; Vázquez-Morón |
| EPI_ISL_3696378 | Hackensack Medical Center | New York Genome Center | Amy Baldwin; Andre Corvelo; Barry Kreiswirth; David Perlin; Dayna M. Oschwald; Jose Mediavilla; Kaelea Composto; Kar Chow; Liang Chen; Marcus Cunningham; Michael Zody; Samantha Fennessey; Tom Maniatis |
| EPI_ISL_5144908, EPI_ISL_5144913, EPI_ISL_5144971, EPI_ISL_6481211, EPI_ISL_6481244, EPI_ISL_6481263, EPI_ISL_6481279, EPI_ISL_6481301, EPI_ISL_6481312, EPI_ISL_6481315, EPI_ISL_6481337, EPI_ISL_6481353, EPI_ISL_6481362, EPI_ISL_6481438, EPI_ISL_6481459, EPI_ISL_6481499, EPI_ISL_6481510, EPI_ISL_6481525, EPI_ISL_6481549, EPI_ISL_6481606, EPI_ISL_6481616, EPI_ISL_6481662, EPI_ISL_6481693, EPI_ISL_6481788, EPI_ISL_6481826, EPI_ISL_6481860, EPI_ISL_6481870, EPI_ISL_6481871, EPI_ISL_6483955, EPI_ISL_6484539, EPI_ISL_6484569 | | | |
| see above | Helix | Centers for Disease Control and Prevention Division of Viral Diseases, Pathogen Discovery | Benjamin Rambo-Martin; Christopher Gulvick; Clinton Paden; Dakota Howard; Dhwani Batra; Duncan MacCannell; Erisa Sula; Helix CA; Jason Caravas; Kristine Lacek; Matthew Schmerer; Peter Cook; Scott Sammons; Shatavia Morrison; Tymeckia Kendall; Victoria Caban Figueroa; Yvette Unoarumhi |
| EPI_ISL_3147484, EPI_ISL_3147492, EPI_ISL_3147566, EPI_ISL_3147584, EPI_ISL_3147595, EPI_ISL_3147599 | Hospital Center Emile Mayrisch | Laboratoire national de sante, Microbiology, Microbial Genomics Platform | Anke Wienecke-Baldacchino; Catherine Ragimbeau; Cynthia Oxacelay; Elodie Solarino; Fatu Djabi; Jessica Tapp; Lise Pignon; Raoul Salmon; Tamir Abdelrahman; Virginie Jover |
| EPI_ISL_3147485, EPI_ISL_3147498, EPI_ISL_3147508, EPI_ISL_3147542, EPI_ISL_3147544, EPI_ISL_3147547, EPI_ISL_3147548, EPI_ISL_3147552, EPI_ISL_3147564, EPI_ISL_3147573, EPI_ISL_3147574, EPI_ISL_3147575, EPI_ISL_3147582, EPI_ISL_3147597 | | | |
| see above | Hospital Center Luxembourg | Laboratoire national de sante, Microbiology, Microbial Genomics Platform | Anke Wienecke-Baldacchino; Catherine Ragimbeau; Elodie Solarino; Fatu Djabi; Jean-Hugues Francois; Jessica Tapp; Lise Pignon; Michel Kohnen; Raoul Salmon; Tamir Abdelrahman; Virginie Jover |
| EPI_ISL_5926919, EPI_ISL_7740274 | Hospital Herrera Llerandi | Asociación de Salud Integral / Clínica Familiar "Luis Ángel García" | Ana S. Gonzalez-Reiche; Claudia Rangel; Danicela Mercado; Eduardo Arathon; Hilda Ruiz; Luis Aguirre; Luis Rivas; Narda Medina; Oscar Bonilla; Osmar Gamboa. |
| EPI_ISL_3260111 | Hospital Jaime Ferre - SAMCO Rafaela | Grupo de Genómica y Bioinformática del Instituto de Investigación de la Cadena Láctea CONICET-INTA on behalf of 'Proyecto Argentino Interinstitucional de genomica de SARS-CoV-2' (PAIS Consortium) | AF; Aliprandi D; Amadio; C; Eberhardt; Irazoqui; Isaia; JF; JM; MF; Pandolfi; Quaranta; Soratti R; V |
| EPI_ISL_2987707 | Hospital Margarita Maza de Juárez | Microbial Genomics Laboratory | ; Alejandra García-Gasca; Alejandra Hernández-Terán; Alejandro Sánchez-Flores; Alfredo Herrera-Estrella; Alicia Ocaña-Mondragón; Andreu Comas-García; Angel Gustavo Salas-Lais; Antonio Loza Román; Bernardo Martínez-Miguel; Blanca Taboada; Brenda Irasema Maldonado-Meza; Bruno Gómez-Gil; Carla Ivón Herrera-Najera; Carlos F. Arias; Celia Boukadida; Clara Esperanza Santacruz-Tinoco; Concepción Grajales-Muñiz; Consorcio Mexicano de Vigilancia Genómica (CoViGen-Mex). Authors (in alphabetical order): Julio Elias Alvarado-Yaah; Cristóbal Cháidez-Quiróz; Célida Duque Molina; Célida Martínez- Rodríguez; Daniel Fregoso-Rueda; Daniel Lira Morales; Eduardo Becerril-Vargas; Fernando Fontove-Herrera; Fidencio Mejía-Nepomuceno; Francisco Pulido; Gloria Elena Espinosa-Ayala; Gloria María Molina-Salinas; Gloria Vazquez; Hector Esteban Paz-Juárez; Hector Montoya-Fuentes; Helen Haydee Fernanda Ramírez-Plascencia; Irvin González-López; Jean Pierre González; Jesús Hernández; Joel Armando Vázquez-Pérez.; Jorge Salas-Hernández; José Antonio Enciso-Moreno; José Arturo Martínez-Orozco; José Esteban Muñoz-Medina; José de Jesús Nuñez-Contreras; Juan Bautista Chale-Dzul; Julissa Enciso-Ibarra; Luis Alberto Ochoa-Carrera; Margarita Matías-Florentino; Mario Mújica-Sánchez; Marissa Perez-Garcia; María Guadalupe Santiago-Mauricio; María Guadalupe de Jesús Mireles-Rivera; Nelly Sélem-Mojica; Pavel Isa; Ricardo Ciria Merce; Ricardo Grande; Rosa María Gutiérrez Rios; Santiago Ávila-Ríos; Selene Zárate; Susana Lopez; Verónica Mata-Haro; Victor Eduardo García-Arias; Víctor Hugo Borja-Aburto |
| EPI_ISL_3030597 | Hospital Universitari Bellvitge | Microbiology Department | Aida Gonzalez-Diaz; Carmen Ardanuy; Jordi Camara; Jordi Niubó; Laura Calatayud; M Angeles Domínguez; Sara Marti |
| EPI_ISL_2982418, EPI_ISL_3009857, EPI_ISL_3009909, EPI_ISL_3009910, EPI_ISL_3009942, EPI_ISL_3009983, EPI_ISL_3984105, EPI_ISL_4348612 | | | |
| see above | Hospital Universitari Vall d'Hebron - Vall d'Hebron Institut de Recerca | Hospital Universitari Vall d'Hebron - Vall d'Hebron Institut de Recerca | Alejandra González-Sánchez; Andrés Antón; Ariadna Rando; Carla Castillo; Cristina Andrés; Damir Garcia-Cehic; Josep Quer; Juliana Esperalba; Karen García; Maria Carmen Martin; Maria Gema Codina; Maria Piñana; Rodrigo Vásquez; Tomàs Pumarola |
| EPI_ISL_3545794 | Hospital de Base de Sao Jose do Rio Preto | Instituto Adolfo Lutz, Interdiciplinary Procedures Center, Strategic Laboratory | Caio Vinicius Dias Lopes; Claudia Regina Gonçalves; Claudio Tavares Sacchi; Karoline Rodrigues Campos; Leonardo Tadeu de Araujo; Marlon Benedito Nascomento Santos |
| EPI_ISL_2968664, EPI_ISL_2968665, EPI_ISL_3398922, EPI_ISL_3398956 | Houston Health Dept. | Houston Health Dept. | Adolpho Lara; Pamela Brown; Ryker Penn; Yanlai Lai |
| EPI_ISL_5339234, EPI_ISL_5339250, EPI_ISL_5339256, EPI_ISL_5339302, EPI_ISL_5339407, EPI_ISL_5339409 | Houston Methodist Hospital | Houston Methodist Hospital | Ilya J. Finkelstein; James J. Davis; Jessica Cambric; Jimmy Gollihar; Kristina Reppond; Layne Pruitt; Madison N. Shyer; Marcus Nguyen; Matthew Ojeda Saavedra; Paul A. Christensen; Prasanti Yerramilli; Randall J. Olsen; Robert Olson; Ryan Gadd; S. Wesley Long; Sishir Subedi; and James M. Musser |
| EPI_ISL_3276715 | Huntington Memorial Hospital | Los Angeles County Public Health Laboratories | P. Hemarajata et al. |
| EPI_ISL_3912024 | III CENTRO DE SAUDE | Analytical Competence Molecular Epidemiology Lab/ACME, Oswaldo Cruz Foundation, Ceara (FIOCRUZ | Cleber Furtado Aksenen; Fabio Miyajima; Fernando Braga Stehling; Francisco Eder de Moura Lopes; Jamille Maria Mendes Bezerra; Joaquim Cesar do Nascimento Sousa Junior; Pedro Miguel Carneiro Jeronimo; Suzana Porto Almeida & Lucas Delerino on behalf of COVID-19 FIOCRUZ Genomic Network; Thais Ferreira de Oliveira; Thais de Oliveira Costa; Ticiane Cavalcante de Souza; Veridiana Pessoa Miyajima |

| | | | |
|---|---|---|---|
| | | CE) | |
| EPI_ISL_3030065, EPI_ISL_3030076 | IN State Department of Health Laboratory Services | IN State Department of Health Laboratory Services | Brian Pope; Cassandra Campion; Jamie Yeadon; Kyle Brownlee; Lixia Liu; Mark Glazier; Melissa Hindenlang |
| EPI_ISL_2925717, EPI_ISL_2925718 | INMI | National Institute for Infectious Diseases (INMI) L. Spallanzani I.R.C.C.S | A Di Caro; B Bartolini; CEM Gruber; E Giombini; F Messina; F Santini; G Bonfiglio; M Rueca; MR Capobianchi; O Butera |
| EPI_ISL_2925570, EPI_ISL_2925571 | INMI "L.Spallanzani" | National Institute for Infectious Diseases (INMI) L. Spallanzani I.R.C.C.S | A Di Caro; B Bartolini; CEM Gruber; E Giombini; F Messina; F Santini; G Bonfiglio; M Rueca; MR Capobianchi; O Butera |
| EPI_ISL_5668425, EPI_ISL_5668437, EPI_ISL_5668443, EPI_ISL_5668469, EPI_ISL_5668478, EPI_ISL_5668501, EPI_ISL_5668519, EPI_ISL_5668536, EPI_ISL_5668567, EPI_ISL_5668576, EPI_ISL_5668579, EPI_ISL_5668618, EPI_ISL_5668635, EPI_ISL_5668715, EPI_ISL_5668758, EPI_ISL_5668775, EPI_ISL_5668779, EPI_ISL_5668831, EPI_ISL_5668860, EPI_ISL_5668933, EPI_ISL_5668971, EPI_ISL_5668986, EPI_ISL_5669007, EPI_ISL_5669027, EPI_ISL_5669059, EPI_ISL_5669063, EPI_ISL_5669083, EPI_ISL_5669087, EPI_ISL_5669113, EPI_ISL_5669120, EPI_ISL_5669140, EPI_ISL_5669147, EPI_ISL_5669183, EPI_ISL_5669194, EPI_ISL_5669198, EPI_ISL_5669205, EPI_ISL_5669244, EPI_ISL_5669288, EPI_ISL_5669307, EPI_ISL_5669313, EPI_ISL_5669342, EPI_ISL_5669350, EPI_ISL_5669362, EPI_ISL_5669367, EPI_ISL_5669408, EPI_ISL_5669424, EPI_ISL_5669454, EPI_ISL_5669524, EPI_ISL_5669526, EPI_ISL_5669541, EPI_ISL_5669551, EPI_ISL_5669566, EPI_ISL_5669570, EPI_ISL_5669590, EPI_ISL_5669599, EPI_ISL_5669647, EPI_ISL_5669661, EPI_ISL_5669689, EPI_ISL_5669697, EPI_ISL_5669793, EPI_ISL_5669878, EPI_ISL_5669884, EPI_ISL_5669902, EPI_ISL_5669920, EPI_ISL_5669928, EPI_ISL_5669959, EPI_ISL_5669970, EPI_ISL_5669990, EPI_ISL_5670020, EPI_ISL_5670025, EPI_ISL_5670039, EPI_ISL_5670061, EPI_ISL_5670097, EPI_ISL_5670129, EPI_ISL_5670138, EPI_ISL_5670176, EPI_ISL_5670180, EPI_ISL_5670196, EPI_ISL_5670220, EPI_ISL_5670283, EPI_ISL_5670370, EPI_ISL_5670416, EPI_ISL_5670450, EPI_ISL_5670460, EPI_ISL_5670489, EPI_ISL_5670496, EPI_ISL_5670511, EPI_ISL_5670607, EPI_ISL_5670739, EPI_ISL_5670753, EPI_ISL_5670783, EPI_ISL_5670795, EPI_ISL_5670825, EPI_ISL_5670847, EPI_ISL_5670852, EPI_ISL_5673236, EPI_ISL_5673302, EPI_ISL_5673366, EPI_ISL_5673376, EPI_ISL_5673498, EPI_ISL_5673502, EPI_ISL_5673534, EPI_ISL_5673573, EPI_ISL_5673603, EPI_ISL_5673643, EPI_ISL_5673664, EPI_ISL_5673669, EPI_ISL_5673719, EPI_ISL_5673816, EPI_ISL_5673837, EPI_ISL_5673870, EPI_ISL_5673936, EPI_ISL_5673957, EPI_ISL_5674045, EPI_ISL_5674055, EPI_ISL_5674063, EPI_ISL_5674077, EPI_ISL_5674095, EPI_ISL_5674118, EPI_ISL_5674128, EPI_ISL_5674134, EPI_ISL_5674204, EPI_ISL_5674214, EPI_ISL_5674360, EPI_ISL_5674447, EPI_ISL_5674481, EPI_ISL_5674548, EPI_ISL_5674642, EPI_ISL_5674689, EPI_ISL_5674745, EPI_ISL_5674750, EPI_ISL_5674755, EPI_ISL_5674839, EPI_ISL_5674895, EPI_ISL_5675013, EPI_ISL_5675020, EPI_ISL_5675051, EPI_ISL_5675055, EPI_ISL_5675060, EPI_ISL_5675073, EPI_ISL_5675221, EPI_ISL_5675227, EPI_ISL_5675275, EPI_ISL_5675280, EPI_ISL_5675283, EPI_ISL_5675305, EPI_ISL_5675317, EPI_ISL_5675496, EPI_ISL_5675506, EPI_ISL_5675518, EPI_ISL_5678731, EPI_ISL_5678806, EPI_ISL_5678961, EPI_ISL_5679185, EPI_ISL_5679208, EPI_ISL_5679273, EPI_ISL_5679361, EPI_ISL_5679689, EPI_ISL_5679880, EPI_ISL_5680153 | | | |
| see above | INSIDE DIAGNOSTICOS | Instituto Butantan | Antonio Jorge Martins; Claudia Renata dos Santos Barros; David Schlesinger; Debora Botequio Moretti; Dimas Tadeu Covas; Elaine Cristina Marqueze; Elaine Vieira Santos; Evandra Strazza Rodrigues; Heidge Fukumasu; Jayme Augusto de Souza-Neto; José Salvatore Leister Patané; Luiz Alcantara; Luiz Lehmann Coutinho; Maria Carolina Elias; Maurício Lacerda Nogueira; Rafael dos Santos Bezerra; Raul Machado Neto; Rejane Maria Tommasini Grotto; Ricardo Haddad; Sandra Coccuzzo Sampaio Vessoni; Simone Kashima; Svetoslav Nanev Slavov; Vincent Louis Viala |
| EPI_ISL_2988479, EPI_ISL_2988484, EPI_ISL_2988486, EPI_ISL_2988506, EPI_ISL_2988519, EPI_ISL_2988528, EPI_ISL_2988554, EPI_ISL_2988556, EPI_ISL_2988573, EPI_ISL_3068144, EPI_ISL_3068146, EPI_ISL_3068149, EPI_ISL_3068157, EPI_ISL_3068158, EPI_ISL_3068159, EPI_ISL_3068164, EPI_ISL_3068166, EPI_ISL_3068169, EPI_ISL_3068194, EPI_ISL_3068195, EPI_ISL_3068199, EPI_ISL_3068208, EPI_ISL_3068209, EPI_ISL_3068213, EPI_ISL_3068218, EPI_ISL_3068220, EPI_ISL_3068223, EPI_ISL_3068228, EPI_ISL_3068231, EPI_ISL_3068236, EPI_ISL_3133207, EPI_ISL_3133221, EPI_ISL_3274398, EPI_ISL_3274440, EPI_ISL_3274462, EPI_ISL_3274492, EPI_ISL_3274503, EPI_ISL_3274512, EPI_ISL_3274524, EPI_ISL_3274534, EPI_ISL_3274535, EPI_ISL_3274539, EPI_ISL_3274540, EPI_ISL_3274584, EPI_ISL_3274593, EPI_ISL_3274599, EPI_ISL_3274604, EPI_ISL_4496947, EPI_ISL_4497097, EPI_ISL_4497174, EPI_ISL_4497278 | | | |
| see above | INSPI-CRN DE INFLUENZA Y OTROS VIRUS RESPIRATORIOS | NIC-INSPI | Alfredo Bruno; Carlos Chiluisa; Daniel Ramos; Domenica de Mora.; Jimmy Garcés; Johanna Laines; Lizbeth Patiño; Manuel Gonzalez; Marcela Mejía; Maria Angelica Becerra; Maritza Olmedo; Mayra Wilca; Michelle Páez; Milton Medina |
| EPI_ISL_2985018, EPI_ISL_2985077, EPI_ISL_2985083, EPI_ISL_2985115, EPI_ISL_2985121, EPI_ISL_2985145, EPI_ISL_2985147, EPI_ISL_2985160, EPI_ISL_2985167, EPI_ISL_2985177, EPI_ISL_2985220, EPI_ISL_2985222, EPI_ISL_2985242, EPI_ISL_2985278, EPI_ISL_2985289, EPI_ISL_2985290, EPI_ISL_2985302, EPI_ISL_2985337, EPI_ISL_2985344, EPI_ISL_2985346, EPI_ISL_3068250 | | | |
| see above | IRCCS San Gallicano Dermatological Institute | IRCCS Regina Elena National Cancer Institute | Aldo Morrone; Alessia Lauretti; Alice Massacci; Andrea Cazzani; Carla Mottini; Eleonora Sperandio; Elisabetta Trento; Fabrizio Ensoli; Francesca De Nicola; Francisco Obregon; Frauke Goeman; Fulvia Fraticelli; Fulvia Pimpinelli; Gennaro Ciliberto; Giovanna D'agosto; Giovanni Blandino; Giulia Orlandi; Ilaria Cavallo; Ilaria Celesti; Lorenzo Furzi; Ludovica Ciuffreda; Martina Diano; Martina Pontone; Matteo Pallocca; Maurizio Fanciulli; Sara Donzelli; Sara Petrolo; Serena Salvo; di Domenico Enea Gino |
| EPI_ISL_3281552, EPI_ISL_3547122 | IVY3 Central Lab, Vanderbilt University Medical Center | Lauring Lab, University of Michigan, Department of Microbiology and Immunology | Gilbert |
| EPI_ISL_3100649, EPI_ISL_3100653 | Idaho Bureau of Laboratories | Boise VA Medical Center, PALMS | Aimee Ceniseros; Cheri Lamb McFarlane; Christian Loera; Dr. Christopher Ball; Emily Bartlette; James Razor; Matthew Burns; Ying Pei |
| EPI_ISL_3088460 | Illinois Department of Public Health - Springfield Lab | Illinois Department of Public Health - Springfield Lab | Bryan Sim; Marshall Smock |
| EPI_ISL_3110995, EPI_ISL_3111004, EPI_ISL_3111032, EPI_ISL_3111082, EPI_ISL_3111088, EPI_ISL_4383726 | Infinity Biologix | Centers for Disease Control and Prevention Division of Viral Diseases, Pathogen Discovery | Adrian Paskey; Benjamin Rambo-Martin; Chirayu Goswami; Christian Bixby; Christopher Gulvick; Clinton Paden; Clinton R. Paden; Dakota Howard; Darlene Wagner; Dhwani Batra; Duncan MacCannell; Erisa Sula; Jason Caravas; Jonathan Schultz; Kara Moser; Kristine Lacek; Matthew Schmerer; Peter Cook; Peter W. Cook; Robin Grimwood; Russ Hager; Scott Sammons; Shatavia Morrison; Tymeckia Kendall; Victoria Caban Figueroa; Yihe Wang; Yvette Unoarumhi |
| EPI_ISL_3123354 | Institute for Water Quality and Resource Management, Technical University Vienna | Bergthaler laboratory, CeMM Research Center for Molecular Medicine of the Austrian Academy of Sciences | Andreas Bergthaler; Anna Schedl; Bekir Erguner; Benedikt Agerer; Christoph Bock; Fabian Amman; Jan Laine; Lukas Endler; Maelle Le Moing; Martin Senekowitsch; Matthew Thornton; Michael Schuster; Petr Triska; Thomas Penz |
| EPI_ISL_3453310, EPI_ISL_3504024 | Institute of Microbiology and Immunology, Faculty of Medicine, University of Ljubljana | Institute of Microbiology and Immunology, Faculty of Medicine, University of Ljubljana | Alen Suljič; Andraž Celar; Dominika Šturm; Doroteja Vlaj; Mario Poljak; Matic Brvar; Miša Korva; Patricija Pozvek; Samo Zakotnik; Tatjana Avšič – Županc; Tomaž Mark Zorec; Špela Pleh |
| EPI_ISL_3062492, EPI_ISL_3062494 | Institute of Microbiology, Universidad San Francisco de Quito | Institute of Microbiology, Universidad San Francisco de Quito | Andrea Pinos; Belén Prado-Vivar; Bernardo Gutiérrez; Erika Muñoz; Fernanda Zurita; Gabriel Trueba; Juan José Guadalupe; Michelle Grunauer; Monica Becerra-Wong; Patricio Rojas-Silva; Paúl Cárdenas; Sully Márquez; Verónica Barragán |
| EPI_ISL_5538395, EPI_ISL_5538400, EPI_ISL_5538410, EPI_ISL_5538415, EPI_ISL_5538423, EPI_ISL_5538431, EPI_ISL_5538452, EPI_ISL_5538464 | | | |
| see above | Instituto Adolfo Lutz - Regional de Bauru | Instituto Adolfo Lutz, Interdiciplinary Procedures Center, Strategic Laboratory | Caio Vinicius Dias Lopes; Claudia Regina Gonçalves; Claudio Tavares Sacchi; Karoline Rodrigues Campos; Leonardo Tadeu de Araujo; Marlon Benedito Nascimento Santos |
| EPI_ISL_4299301, EPI_ISL_4299306, EPI_ISL_4299313, EPI_ISL_4299314, EPI_ISL_4299315, EPI_ISL_4299316, EPI_ISL_4299317, EPI_ISL_4299320, EPI_ISL_4299321, EPI_ISL_4299323 | | | |
| see above | Instituto Adolfo Lutz - Regional de Campinas | Instituto Adolfo Lutz, Interdiciplinary Procedures Center, Strategic Laboratory | Caio Vinicius Dias Lopes; Claudia Regina Gonçalves; Claudio Tavares Sacchi; Karoline Rodrigues Campos; Marlon Benedito Nascimento Santos |
| EPI_ISL_3545788, EPI_ISL_3545789, EPI_ISL_3545795, EPI_ISL_3545796 | Instituto Adolfo Lutz - Regional de Marilia | Instituto Adolfo Lutz, Interdiciplinary Procedures Center, Strategic Laboratory | Caio Vinicius Dias Lopes; Claudia Regina Gonçalves; Claudio Tavares Sacchi; Karoline Rodrigues Campos; Leonardo Tadeu de Araujo; Marlon Benedito Nascomento Santos |
| EPI_ISL_3316167, EPI_ISL_3545793 | Instituto Adolfo Lutz - Regional de Ribeirao Preto | Instituto Adolfo Lutz, Interdiciplinary Procedures Center, Strategic Laboratory | Caio Vinicius Dias Lopes; Claudia Regina Gonçalves; Claudio Tavares Sacchi; Karoline Rodrigues Campos; Leonardo Tadeu de Araujo; Marlon Benedito Nascomento Santos |
| EPI_ISL_3316298, EPI_ISL_4877303, EPI_ISL_4877304, EPI_ISL_4877305, EPI_ISL_4877306, EPI_ISL_4877307, EPI_ISL_4877308, EPI_ISL_4877309 | | | |
| see above | Instituto Adolfo Lutz - Regional de Rio Claro | Instituto Adolfo Lutz, Interdiciplinary Procedures Center, Strategic Laboratory | Caio Vinicius Dias Lopes; Claudia Regina Gonçalves; Claudio Tavares Sacchi; Karoline Rodrigues Campos; Leonardo Tadeu de Araujo; Marlon Benedito Nascimento Santos; Marlon Benedito Nascomento Santos |
| EPI_ISL_3946610 | Instituto Adolfo Lutz - Regional de Santo Andre | Instituto Adolfo Lutz, Interdiciplinary Procedures Center, Strategic Laboratory | Claudio Tavares Sacchi; Karoline Rodrigues Campos |
| EPI_ISL_6840852 | Instituto Adolfo Lutz - Regional de Sao Jose do Rio Preto | Instituto Adolfo Lutz, Interdiciplinary Procedures Center, Strategic Laboratory | Ariadne Ferreira Amarante; Caio Vinicius Dias Lopes; Claudia Regina Gonçalves; Claudio Tavares Sacchi; Karoline Rodrigues Campos; Marlon Benedito Nascimento Santos |
| EPI_ISL_3545790, EPI_ISL_3545791, EPI_ISL_3545797 | Instituto Adolfo Lutz - Regional de Sorocaba | Instituto Adolfo Lutz, Interdiciplinary Procedures Center, Strategic Laboratory | Caio Vinicius Dias Lopes; Claudia Regina Gonçalves; Claudio Tavares Sacchi; Karoline Rodrigues Campos; Leonardo Tadeu de Araujo; Marlon Benedito Nascomento Santos |
| EPI_ISL_3545792, EPI_ISL_4081132, EPI_ISL_4206919, EPI_ISL_4206929, EPI_ISL_4206931, EPI_ISL_4206936, EPI_ISL_4206937, EPI_ISL_4206948, EPI_ISL_4888088, EPI_ISL_4888094, EPI_ISL_5030099, EPI_ISL_5030100, EPI_ISL_5640228 | | | |
| see above | Instituto Adolfo Lutz Central | Instituto Adolfo Lutz, Interdiciplinary Procedures Center, Strategic Laboratory | Caio Vinicius Dias Lopes; Claudia Regina Goncalves; Claudia Regina Gonçalves; Claudio Tavares Sacchi; Karoline Rodrigues Campos; Leonardo Tadeu de Araujo; Marlon Benedito Nascimento Santos; Marlon Benedito Nascomento Santos |
| EPI_ISL_3268378, EPI_ISL_3268379, EPI_ISL_3717565 | Instituto Nacional de Enfermedades Respiratorias (INER) | Centro de Investigación en Enfermedades Infecciosas (CIENI), Instituto Nacional de Enfermedades Respiratorias (INER) | Alejandra Garcia-Gasca; Alejandra Hernández-Terán; Alejandro Sanchez-Flores; Alfredo Herrera-Estrella; Alicia Ocaña-Mondragón; Andreu Comas-García; Angel Gustavo Salas-Lais; Antonio Loza Román; Bernardo Martínez-Miguel; Blanca Taboada; Brenda Irasema Maldonado-Meza; Bruno Gomez-Gil; Carla Ivón Herrera-Najera; Carlos F. Arias; Celia Boukadida; Clara Esperanza Santacruz-Tinoco; Concepción Grajales-Muñiz; Consorcio Mexicano de Vigilancia Genómica (CoViGen-Mex). Authors (in alphabetical order): Julio Elias Alvarado-Yaah; Cristóbal Cháidez-Quiróz; Célida Duque Molina; Célida Martínez- Rodríguez; Daniel Fregoso-Rueda; Daniel Lira Morales; Eduardo Becerril-Vargas; Fernando Fontove-Herrera; Fidencio Mejía-Nepomuceno; Francisco Pulido; Gloria Elena Espinosa-Ayala; Gloria María Molina-Salinas; Gloria Vazquez; Hector Esteban Paz-Juárez; Hector Montoya-Fuentes; Helen Haydee Fernanda Ramírez-Plascencia; Irvin González-López; Jean Pierre González; Joel Armando Vázquez-Pérez.; Jorge Salas-Hernández; José Antonio Enciso-Moreno; José Arturo Martínez-Orozco; José Esteban Muñoz-Medina; José de Jesús Nuñez-Contreras; Juan Bautista Chale-Dzul; Julissa Enciso-Ibarra; Kathia Elizabeth Tapia-Diaz; Luis Alberto Ochoa-Carrera; Margarita Matías-Florentino; Mario Mújica-Sánchez; Marissa Pérez-García; María Guadalupe Santiago-Mauricio; María Guadalupe de Jesús Mireles-Rivera; Nelly Sélem-Mojica; Pavel Isa; Ricardo Ciria Merce; Ricardo Grande; Rosa María Gutierrez Rios; Santiago Ávila-Ríos; Selene Zárate; Susana Lopez; Victor Eduardo Garcia-Arias; Víctor Hugo Borja-Aburto |
| EPI_ISL_3118923, EPI_ISL_3118925, EPI_ISL_3118926, EPI_ISL_3118927, EPI_ISL_3118928, EPI_ISL_3118929, EPI_ISL_3118930, EPI_ISL_3118931, EPI_ISL_3118932, EPI_ISL_3118938, EPI_ISL_3118939, EPI_ISL_3118940, EPI_ISL_3118941, EPI_ISL_3118942, EPI_ISL_3118943, EPI_ISL_3118944, EPI_ISL_3118945, EPI_ISL_3118946, EPI_ISL_3118947, EPI_ISL_3118948 | | | |
| see above | Instituto de Biotecnologia - UNESP-Botucatu-SP | Instituto de Biotecnologia - UNESP-Botucatu-SP | Cecília Artico Banho; Cíntia Bittar; Fábio Sossai Possebon; Guilherme Campos; Helena Lage Ferreira; Jorge A. Petroli Marchesi; João Pessoa Araújo Jr.; Leila Sabrina Ullmann; Lívia Sacchetto; Maisa C. Pereira Parra; Marília Moraes; Maurício L. Nogueira; Paula Rahal; Paulo Inacio da Costa |
| EPI_ISL_3347068, EPI_ISL_3347084 | Interpath Laboratory | Seattle Flu Study | Amanda Adler; Barry R. Lutz; Benjamin Pelle; Brian Hiatt; Caitlin R. Wolf; Chris D. Frazar; Deborah A. Nickerson; Elisabeth Brandstetter; Erica Ryke; Geoff Melly; Helen Y. Chu; Janet A. Englund; Jay Shendure; Jover Lee; Kairsten Fay; Kirsten Lacombe; Lea M. Starita; Mark J. Rieder; Matthew Richardson; Matthew Thompson; Melissa Truong; Michael Boeckh; Michael Famulare; Misja Ilcisin; Peter D. Han; Philip Dykema; Romesh Gautom; Scott Lindquist; Thomas R. Sibley; Trevor Bedford |

| | | | |
|---|---|---|---|
| EPI_ISL_3388727 | John H Stroger Jr Hospital of Cook County | RIPHL at Rush University Medical Center | Ah Young Sung; Edith Perez; Felix Araujo Perez; Kevin Kunstman; Laura Furtado; Marieta Hyde; Max Kolton; Sharon Welbel; Stefan Green |
| EPI_ISL_5405417, EPI_ISL_5405606, EPI_ISL_5405612, EPI_ISL_5405622, EPI_ISL_5405632, EPI_ISL_5414619, EPI_ISL_5414991, EPI_ISL_5424282 | | | |
| see above | KU Leuven, Rega Institute, Clinical and Epidemiological Virology | KU Leuven, Rega Institute, Clinical and Epidemiological Virology | Bert Vanmechelen; Joan Marti-Carerras; Piet Maes; Tony Wawina-Bokalanga |
| EPI_ISL_3825531, EPI_ISL_3825693, EPI_ISL_3825696, EPI_ISL_3825711, EPI_ISL_3825730, EPI_ISL_3825748, EPI_ISL_3825754, EPI_ISL_3825760, EPI_ISL_3825772, EPI_ISL_3825787, EPI_ISL_3825791, EPI_ISL_3825799, EPI_ISL_4260988, EPI_ISL_4261139 | | | |
| see above | Kaiser Permanente NW Reginal Lab | OHSU MM Lab | Amber Halse; Jeannine Lama; Xuan Qin; Yun Wu |
| EPI_ISL_2893679, EPI_ISL_2893680 | Kansas Health and Environmental Lab | Kansas Health and Environmental Lab | Ben Olsen; Jonathan Barnell; Katherine Wiggins; Mike Grose; and Phil Adam |
| EPI_ISL_4493230, EPI_ISL_4493262, EPI_ISL_4493290, EPI_ISL_4493291 | Kentucky State Public Health Lab | Kentucky State Public Health Lab | Anjayya Tummala; Chitralipi Kattunga; Joshua Tobias; Karim George; Matthew Johnson; Rachel Zinner; Rhonda Lucas; Vaneet Arora; William Grooms |
| EPI_ISL_3205704 | LABORATOIRE ALPIGENE | CNR Virus des Infections Respiratoires - France SUD | Antonin Bal; Bruno Lina; Gregory Destras; Gwendolyne Burfin; Hadrien Regue; Laurence Josset; Martine Valette; Quentin Semanas |
| EPI_ISL_3130541 | LABORATOIRE CERBALLIANCE PLT VILLON | CNR Virus des Infections Respiratoires - France SUD | Antonin Bal; Bruno Lina; Gregory Destras; Gwendolyne Burfin; Hadrien Regue; Laurence Josset; Martine Valette; Quentin Semanas |
| EPI_ISL_5670970, EPI_ISL_5671216, EPI_ISL_5671303, EPI_ISL_5671361, EPI_ISL_5671429 | LABORATORIO DE FRANCA | Instituto Butantan | Antonio Jorge Martins; Claudia Renata dos Santos Barros; David Schlesinger; Debora Botequio Moretti; Dimas Tadeu Covas; Elaine Cristina Marqueze; Elaine Vieira Santos; Evandra Strazza Rodrigues; Heidge Fukumasu; Jayme Augusto de Souza-Neto; José Salvatore Leister Patané; Luiz Alcantara; Luiz Lehmann Coutinho; Maria Carolina Elias; Maurício Lacerda Nogueira; Rafael dos Santos Bezerra; Raul Machado Neto; Rejane Maria Tommasini Grotto; Ricardo Haddad; Sandra Coccuzzo Sampaio Vessoni; Simone Kashima; Svetoslav Nanev Slavov; Vincent Louis Viala |
| EPI_ISL_5669952, EPI_ISL_5670625 | LABORATORIO LOCAL DE ITAPECERICA DA SERRA | Instituto Butantan | Antonio Jorge Martins; Claudia Renata dos Santos Barros; David Schlesinger; Debora Botequio Moretti; Dimas Tadeu Covas; Elaine Cristina Marqueze; Elaine Vieira Santos; Evandra Strazza Rodrigues; Heidge Fukumasu; Jayme Augusto de Souza-Neto; José Salvatore Leister Patané; Luiz Alcantara; Luiz Lehmann Coutinho; Maria Carolina Elias; Maurício Lacerda Nogueira; Rafael dos Santos Bezerra; Raul Machado Neto; Rejane Maria Tommasini Grotto; Ricardo Haddad; Sandra Coccuzzo Sampaio Vessoni; Simone Kashima; Svetoslav Nanev Slavov; Vincent Louis Viala |
| EPI_ISL_5670888, EPI_ISL_5680809, EPI_ISL_5680868, EPI_ISL_5681073 | LABORATORIO MUNICIPAL DE ANALISES CLINICAS DE RIO CLARO | Instituto Butantan | Antonio Jorge Martins; Claudia Renata dos Santos Barros; David Schlesinger; Debora Botequio Moretti; Dimas Tadeu Covas; Elaine Cristina Marqueze; Elaine Vieira Santos; Evandra Strazza Rodrigues; Heidge Fukumasu; Jayme Augusto de Souza-Neto; José Salvatore Leister Patané; Luiz Alcantara; Luiz Lehmann Coutinho; Maria Carolina Elias; Maurício Lacerda Nogueira; Rafael dos Santos Bezerra; Raul Machado Neto; Rejane Maria Tommasini Grotto; Ricardo Haddad; Sandra Coccuzzo Sampaio Vessoni; Simone Kashima; Svetoslav Nanev Slavov; Vincent Louis Viala |
| EPI_ISL_5673348, EPI_ISL_5673386, EPI_ISL_5673547, EPI_ISL_5674170, EPI_ISL_5674537, EPI_ISL_5675263, EPI_ISL_5675781, EPI_ISL_5676028, EPI_ISL_5676908, EPI_ISL_5677131 | | | |
| see above | LABORATORIO MUNICIPAL DE PIRACICABA | Instituto Butantan | Antonio Jorge Martins; Claudia Renata dos Santos Barros; David Schlesinger; Debora Botequio Moretti; Dimas Tadeu Covas; Elaine Cristina Marqueze; Elaine Vieira Santos; Evandra Strazza Rodrigues; Heidge Fukumasu; Jayme Augusto de Souza-Neto; José Salvatore Leister Patané; Luiz Alcantara; Luiz Lehmann Coutinho; Maria Carolina Elias; Maurício Lacerda Nogueira; Rafael dos Santos Bezerra; Raul Machado Neto; Rejane Maria Tommasini Grotto; Ricardo Haddad; Sandra Coccuzzo Sampaio Vessoni; Simone Kashima; Svetoslav Nanev Slavov; Vincent Louis Viala |
| EPI_ISL_3404554, EPI_ISL_3404555, EPI_ISL_3404556, EPI_ISL_3404557, EPI_ISL_3404558, EPI_ISL_3404559, EPI_ISL_3404560, EPI_ISL_3404561, EPI_ISL_3404562, EPI_ISL_3404564, EPI_ISL_3404565, EPI_ISL_3404566, EPI_ISL_3404567, EPI_ISL_3404568, EPI_ISL_3404569, EPI_ISL_3404571, EPI_ISL_3404572, EPI_ISL_3404573, EPI_ISL_3404575, EPI_ISL_3404576, EPI_ISL_3404577, EPI_ISL_3404578, EPI_ISL_3404579, EPI_ISL_3404580, EPI_ISL_3404581, EPI_ISL_3404582, EPI_ISL_3404583, EPI_ISL_3404584, EPI_ISL_3404585, EPI_ISL_3404586, EPI_ISL_3404603, EPI_ISL_3404604, EPI_ISL_3404605, EPI_ISL_3404607, EPI_ISL_3404608, EPI_ISL_4395148 | | | |
| see above | LACEN | LGBio (Laboratorio de Genetica & Biodiversidade) | Alex Honda Bernardes; Amanda Alves de Melo; Aparecido Divino da Cruz; Cintia Pelegrineti Targueta de Azevedo Brito; Daniela de Melo e Silva; Elisangela de Paula Silveira Lacerda; Juliana Santana de Curcio; Luiz Augusto Pereira; Marc Alexandre Duarte Gigonzac; Mariana Pires de Campos Telles; Ramilla dos Santos Braga; Renata de Oliveira Dias; Rhewter Nunes; Thais Cidalia Vieira Gigonzac; Thais Guimarães Castro; Thays Millena Alves Pedroso |
| EPI_ISL_6721437, EPI_ISL_6721475 | LACEN - Laboratório Central de Saúde Pública de Pernambuco | Evandro Chagas Institute | Delana Andreza Melo Bezerra; Dielle Monteiro Teixeira; Jedson Ferreira Cardoso; Jessylene de Almeida Ferreira; Kenny da Costa Pinheiro; Luana Soares Barbagelata; Luana da Silva Soares; Mirleide Cordeiro dos Santos; Patrícia dos Santos Lobo; Rayssa Layna da Silva Bedran; Sandro Patroca da Silva |
| EPI_ISL_6721313 | LACEN - Laboratório Central de Saúde Pública de Roraima | Evandro Chagas Institute | Delana Andreza Melo Bezerra; Dielle Monteiro Teixeira; Jedson Ferreira Cardoso; Jessylene de Almeida Ferreira; Kenny da Costa Pinheiro; Luana Soares Barbagelata; Luana da Silva Soares; Mirleide Cordeiro dos Santos; Patrícia dos Santos Lobo; Rayssa Layna da Silva Bedran; Sandro Patroca da Silva |
| EPI_ISL_6721582, EPI_ISL_6721608, EPI_ISL_6721612, EPI_ISL_6721675 | LACEN - Laboratório Central de Saúde Pública do Ceara | Evandro Chagas Institute | Delana Andreza Melo Bezerra; Dielle Monteiro Teixeira; Jedson Ferreira Cardoso; Jessylene de Almeida Ferreira; Kenny da Costa Pinheiro; Luana Soares Barbagelata; Luana da Silva Soares; Mirleide Cordeiro dos Santos; Patrícia dos Santos Lobo; Rayssa Layna da Silva Bedran; Sandro Patroca da Silva |
| EPI_ISL_6721336, EPI_ISL_6721339, EPI_ISL_6721355, EPI_ISL_6721419 | LACEN - Laboratório Central de Saúde Pública do Rio Grande do Norte | Evandro Chagas Institute | Delana Andreza Melo Bezerra; Dielle Monteiro Teixeira; Jedson Ferreira Cardoso; Jessylene de Almeida Ferreira; Kenny da Costa Pinheiro; Luana Soares Barbagelata; Luana da Silva Soares; Mirleide Cordeiro dos Santos; Patrícia dos Santos Lobo; Rayssa Layna da Silva Bedran; Sandro Patroca da Silva |
| EPI_ISL_3046327, EPI_ISL_3046328, EPI_ISL_3046329, EPI_ISL_3046330, EPI_ISL_3134666, EPI_ISL_3134667, EPI_ISL_3134668, EPI_ISL_3134669, EPI_ISL_3134670, EPI_ISL_3134671, EPI_ISL_3134672, EPI_ISL_3134673, EPI_ISL_3134674, EPI_ISL_3134675, EPI_ISL_3134676, EPI_ISL_3134677, EPI_ISL_3134678, EPI_ISL_3134679, EPI_ISL_3134680, EPI_ISL_3134681, EPI_ISL_3134682, EPI_ISL_3134683, EPI_ISL_3134684, EPI_ISL_3134685, EPI_ISL_3134686, EPI_ISL_3134687, EPI_ISL_3134688, EPI_ISL_3134689, EPI_ISL_3134690, EPI_ISL_3134692, EPI_ISL_3134693, EPI_ISL_3134694, EPI_ISL_3134695, EPI_ISL_3134696, EPI_ISL_3134697, EPI_ISL_3134698, EPI_ISL_3134699, EPI_ISL_3134700, EPI_ISL_3134701, EPI_ISL_3134702, EPI_ISL_3134703, EPI_ISL_3134704, EPI_ISL_3134705, EPI_ISL_3134706, EPI_ISL_3134707, EPI_ISL_3134708, EPI_ISL_3134709, EPI_ISL_3134710, EPI_ISL_3134711, EPI_ISL_3134712, EPI_ISL_3134713, EPI_ISL_3134714, EPI_ISL_3134715, EPI_ISL_3134716, EPI_ISL_3134717, EPI_ISL_3134718, EPI_ISL_3134719, EPI_ISL_3134720, EPI_ISL_3134721, EPI_ISL_3134722, EPI_ISL_3134723, EPI_ISL_3447414, EPI_ISL_3447415, EPI_ISL_3447417, EPI_ISL_3447422, EPI_ISL_3447427, EPI_ISL_3447429, EPI_ISL_3447438, EPI_ISL_3447442, EPI_ISL_3447443, EPI_ISL_3447446, EPI_ISL_3447455, EPI_ISL_3447464, EPI_ISL_3447467, EPI_ISL_3447469, EPI_ISL_3447470, EPI_ISL_3447472, EPI_ISL_3447475, EPI_ISL_3447478, EPI_ISL_3447479, EPI_ISL_3447487, EPI_ISL_3447488, EPI_ISL_3447600 | | | |
| see above | LACEN/PE | WallauLab on behalf of Fiocruz COVID-19 Genomic Surveillance Network | Alexandre Freitas da Silva; Cassia Docena; Constância Flávia Junqueira Ayres; Filipe Zimmer Dezordi; Gabriel Luz Wallau; Gustavo Barbosa de Lima; Lais Ceschini Machado; Lilian Caroliny Amorim Silva; Marcelo Henrique dos Santos Paiva; Matheus Filgueira Bezerra; Sinval Pinto Brandão Filho |
| EPI_ISL_5193507, EPI_ISL_5193508, EPI_ISL_5193509, EPI_ISL_5193510, EPI_ISL_5193511, EPI_ISL_5193512, EPI_ISL_5193513, EPI_ISL_5193515, EPI_ISL_5193516, EPI_ISL_5193517, EPI_ISL_5193629, EPI_ISL_5193630 | | | |
| see above | LACEN/RR | Laboratorio de Ecologia de Doencas Transmissiveis na Amazonia, Instituto Leonidas e Maria Deane - Fiocruz Amazonia | André Corado; Cátia Alexandra Ribeiro Meneses; Felipe Naveca; Fernanda Nascimento; George Silva; Ilma de Aguiar Antony; Janis Cassimiro Lopes; Luciana Gonçalves; Maria Júlia Brandão; Matilde Mejía; Pricila Carlos Veloso; Rodrigo Melo Maito; Valdinete Nascimento; Victor Souza |
| EPI_ISL_3130641 | LAM CERBALLIANCE | CNR Virus des Infections Respiratoires - France SUD | Antonin Bal; Bruno Lina; Gregory Destras; Gwendolyne Burfin; Hadrien Regue; Laurence Josset; Martine Valette; Quentin Semanas |
| EPI_ISL_3149318, EPI_ISL_3149319, EPI_ISL_3149320, EPI_ISL_3149321 | LANIIA | LANIIA-Nayarit | ; Alejandra García-Gasca; Alejandra Hernández-Terán; Alejandro Sánchez-Flores; Alfredo Herrera-Estrella; Alicia Ocaña-Mondragón; Andreu Comas-García; Angel Gustavo Salas-Lais; Antonio Loza Román; Bernardo Martínez-Miguel; Blanca Taboada; Brenda Irasema Maldonado-Meza; Bruno Gómez-Gil; Carla Ivón Herrera-Najera; Carlos Eduardo Covantes-Rosales; Carlos F. Arias; Celia Boukadida; Clara Esperanza Santacruz-Tinoco; Concepción Grajales-Muñiz; Consorcio Mexicano de Vigilancia Genómica (CoViGen-Mex). Authors (in alphabetical order): Julio Elias Alvarado-Yaah; Cristóbal Cháidez-Quiróz; Célida Duque Molina; Célida Martínez- Rodríguez; Daniel Alberto Girón Pérez; Daniel Fregoso-Rueda; Daniel Lira Morales; Eduardo Becerril-Vargas; Fernando Fontove-Herrera; Fidencio Mejía-Nepomuceno; Francisco Pulido; Gloria Elena Espinosa-Ayala; Gloria María Molina-Salinas; Gloria Vazquez; Hector Esteban Paz-Juárez; Hector Montoya-Fuentes; Helen Haydee Fernanda Ramírez-Plascencia; Irvin González-López; Jean Pierre González; Jesús Hernández; Joel Armando Vázquez-Pérez. LANNIA Nayarit: Manuel Ivan Girón Pérez; Jorge Salas-Hernández; José Antonio Enciso-Moreno; José Arturo Martínez-Orozco; José Esteban Muñoz-Medina; José de Jesús Nuñez-Contreras; Juan Bautista Chale-Dzul; Julissa Enciso-Ibarra; Luis Alberto Ochoa-Carrera; Margarita Matías-Florentino; Mario Mújica-Sánchez; Marissa Perez-Garcia; María Guadalupe Santiago-Mauricio; María Guadalupe de Jesús Mireles-Rivera; Nelly Sélem-Mojica; Pavel Isa; Ricardo Ciria Merce; Ricardo Grande; Rosa María Gutiérrez Rios; Santiago Ávila-Ríos; Selene Zárate; Susana Lopez; Ulises Mercado.; Verónica Mata-Haro; Victor Eduardo García-Arias; Víctor Hugo Borja-Aburto |
| EPI_ISL_3047702, EPI_ISL_3047704, EPI_ISL_3047706, EPI_ISL_3047707, EPI_ISL_3047708, EPI_ISL_3047709, EPI_ISL_3047711, EPI_ISL_3047712, EPI_ISL_3047713, EPI_ISL_3047714, EPI_ISL_3047715, EPI_ISL_3047716, EPI_ISL_3047718, EPI_ISL_3047720, EPI_ISL_3047721, EPI_ISL_3047723, EPI_ISL_3047724, EPI_ISL_3047726, EPI_ISL_3047728, EPI_ISL_3047731, EPI_ISL_3047733, EPI_ISL_3047734, EPI_ISL_3047735, EPI_ISL_3047736, EPI_ISL_3047737, EPI_ISL_3047738, EPI_ISL_3047741, EPI_ISL_3047743, EPI_ISL_3047745, EPI_ISL_3134921, EPI_ISL_3134922, EPI_ISL_3134927, EPI_ISL_3134929, EPI_ISL_3134930, EPI_ISL_3134931, EPI_ISL_3134933, EPI_ISL_3134934, EPI_ISL_3134939, EPI_ISL_3134949, EPI_ISL_3134950, EPI_ISL_3134955, EPI_ISL_3134958, EPI_ISL_3134960, EPI_ISL_3134965, EPI_ISL_3134967, EPI_ISL_3134974 | | | |
| see above | LATE - Laboratório de Técnicas Especiais - Hospital Israelita Albert Einstein | LATE - Laboratório de Técnicas Especiais - Hospital Israelita Albert Einstein | Alexandre Hideaki Takara; Ana Paula Moreira Salles; Anelisie da Silva Santos; Deyvid Amgarten; Erick Gustavo Dorlass; Fernanda de Mello Malta; João Renato Rebello Pinho; Marcio Anunciacao Menezes; Pedro Henrique Sebe Rodrigues; Raquel Riyuzo |
| EPI_ISL_3459381 | LDSP CUNDINAMARCA | Instituto Nacional de Salud | Carlos Franco-Muñoz; Carmen Osorio; Diana Malo; Diego A. Álvarez-Díaz; Diego Andrés Prada; Gerardo Santamaría; Hector Alejandro Ruiz-Moreno; Jhonnatan Reales-González; Jorge Rivera; Juan Camilo Martinez; Julian Naizaque; Katherine Laiton-Donato; Lisseth Pardo; Magdalena Wiesner; Marcela Mercado-Reyes; Maria T. Herrera-Sepúlveda; Marta Lopez Blanco; Martha Lucia Ospina Martinez; Paola Rojas; Sergio Gomez; Sheryll Corchuelo; Ángela Alarcon Cruz |
| EPI_ISL_3265598, EPI_ISL_3265599, EPI_ISL_3265600, EPI_ISL_3265601 | LESP Aguascalientes | Instituto de Diagnostico y Referencia Epidemiologicos (INDRE) | Abril Rodriguez-Maldonado; Ariadna Medina-Benitez; Claudia Wong-Arambula; Ernesto Ramirez-Gonzalez.; Gisela Barrera-Badillo; Irma Lopez-Martinez; Joaquin Quiroz-Mercado; Lucia Hernandez-Rivas; Maribel Gonzalez-Villa; Natividad Cruz-Ortiz; Sergio Rangel-Guerrero; Tatiana Nunez-Garcia; Vanessa Rivero-Arredondo |
| EPI_ISL_3265617, EPI_ISL_3278290 | LESP Baja California | Instituto de Diagnostico y Referencia Epidemiologicos (INDRE) | Abril Rodriguez-Maldonado; Ariadna Medina-Benitez; Claudia Wong-Arambula; Ernesto Ramirez-Gonzalez.; Gisela Barrera-Badillo; Irma Lopez-Martinez; Joaquin Quiroz-Mercado; Lucia Hernandez-Rivas; Maribel Gonzalez-Villa; Natividad Cruz-Ortiz; Sergio Rangel-Guerrero; Tatiana Nunez-Garcia; Vanessa Rivero-Arredondo |
| EPI_ISL_3265609, EPI_ISL_3265612, EPI_ISL_3265613, EPI_ISL_3265616, EPI_ISL_3265618 | LESP Campeche | Instituto de Diagnostico y Referencia Epidemiologicos (INDRE) | Abril Rodriguez-Maldonado; Ariadna Medina-Benitez; Claudia Wong-Arambula; Ernesto Ramirez-Gonzalez.; Gisela Barrera-Badillo; Irma Lopez-Martinez; Joaquin Quiroz-Mercado; Lucia Hernandez-Rivas; Maribel Gonzalez-Villa; Natividad Cruz-Ortiz; Sergio Rangel-Guerrero; Tatiana Nunez-Garcia; Vanessa Rivero-Arredondo |
| EPI_ISL_3265579, EPI_ISL_3265580, EPI_ISL_3265581, EPI_ISL_3265582, EPI_ISL_3265583, EPI_ISL_3265584, EPI_ISL_3278284 | | | |
| see above | LESP Hidalgo | Instituto de Diagnostico y Referencia Epidemiologicos (INDRE) | Abril Rodriguez-Maldonado; Ariadna Medina-Benitez; Claudia Wong-Arambula; Ernesto Ramirez-Gonzalez.; Gisela Barrera-Badillo; Irma Lopez-Martinez; Joaquin Quiroz-Mercado; Lucia Hernandez-Rivas; Maribel Gonzalez-Villa; Natividad Cruz-Ortiz; Sergio Rangel-Guerrero; Tatiana Nunez-Garcia; Vanessa Rivero-Arredondo |

| | | | |
|---|---|---|---|
| EPI_ISL_3459946, EPI_ISL_3459947, EPI_ISL_3459957, EPI_ISL_3459964 | LESP Jalisco | Instituto de Diagnostico y Referencia Epidemiologicos (INDRE) | Abril Rodriguez-Maldonado; Ariadna Medina-Benitez; Claudia Wong-Arambula; Ernesto Ramirez-Gonzalez.; Gisela Barrera-Badillo; Irma Lopez-Martinez; Joaquin Quiroz-Mercado; Lucia Hernandez-Rivas; Maribel Gonzalez-Villa; Natividad Cruz-Ortiz; Sergio Rangel-Guerrero; Tatiana Nunez-Garcia; Vanessa Rivero-Arredondo |
| EPI_ISL_3459943 | LESP Morelos | Instituto de Diagnostico y Referencia Epidemiologicos (INDRE) | Abril Rodriguez-Maldonado; Ariadna Medina-Benitez; Claudia Wong-Arambula; Ernesto Ramirez-Gonzalez.; Gisela Barrera-Badillo; Irma Lopez-Martinez; Joaquin Quiroz-Mercado; Lucia Hernandez-Rivas; Maribel Gonzalez-Villa; Natividad Cruz-Ortiz; Sergio Rangel-Guerrero; Tatiana Nunez-Garcia; Vanessa Rivero-Arredondo |
| EPI_ISL_3265605 | LESP Nayarit | Instituto de Diagnostico y Referencia Epidemiologicos (INDRE) | Abril Rodriguez-Maldonado; Ariadna Medina-Benitez; Claudia Wong-Arambula; Ernesto Ramirez-Gonzalez.; Gisela Barrera-Badillo; Irma Lopez-Martinez; Joaquin Quiroz-Mercado; Lucia Hernandez-Rivas; Maribel Gonzalez-Villa; Natividad Cruz-Ortiz; Sergio Rangel-Guerrero; Tatiana Nunez-Garcia; Vanessa Rivero-Arredondo |
| EPI_ISL_3459902 | LESP Queretaro | Instituto de Diagnostico y Referencia Epidemiologicos (INDRE) | Abril Rodriguez-Maldonado; Ariadna Medina-Benitez; Claudia Wong-Arambula; Ernesto Ramirez-Gonzalez.; Gisela Barrera-Badillo; Irma Lopez-Martinez; Joaquin Quiroz-Mercado; Lucia Hernandez-Rivas; Maribel Gonzalez-Villa; Natividad Cruz-Ortiz; Sergio Rangel-Guerrero; Tatiana Nunez-Garcia; Vanessa Rivero-Arredondo |
| EPI_ISL_3265608 | LESP Quintana Roo | Instituto de Diagnostico y Referencia Epidemiologicos (INDRE) | Abril Rodriguez-Maldonado; Ariadna Medina-Benitez; Claudia Wong-Arambula; Ernesto Ramirez-Gonzalez.; Gisela Barrera-Badillo; Irma Lopez-Martinez; Joaquin Quiroz-Mercado; Lucia Hernandez-Rivas; Maribel Gonzalez-Villa; Natividad Cruz-Ortiz; Sergio Rangel-Guerrero; Tatiana Nunez-Garcia; Vanessa Rivero-Arredondo |
| EPI_ISL_3265615, EPI_ISL_3459940 | LESP Sonora | Instituto de Diagnostico y Referencia Epidemiologicos (INDRE) | Abril Rodriguez-Maldonado; Ariadna Medina-Benitez; Claudia Wong-Arambula; Ernesto Ramirez-Gonzalez.; Gisela Barrera-Badillo; Irma Lopez-Martinez; Joaquin Quiroz-Mercado; Lucia Hernandez-Rivas; Maribel Gonzalez-Villa; Natividad Cruz-Ortiz; Sergio Rangel-Guerrero; Tatiana Nunez-Garcia; Vanessa Rivero-Arredondo |
| EPI_ISL_3459948, EPI_ISL_3459949 | LESP Tabasco | Instituto de Diagnostico y Referencia Epidemiologicos (INDRE) | Abril Rodriguez-Maldonado; Ariadna Medina-Benitez; Claudia Wong-Arambula; Ernesto Ramirez-Gonzalez.; Gisela Barrera-Badillo; Irma Lopez-Martinez; Joaquin Quiroz-Mercado; Lucia Hernandez-Rivas; Maribel Gonzalez-Villa; Natividad Cruz-Ortiz; Sergio Rangel-Guerrero; Tatiana Nunez-Garcia; Vanessa Rivero-Arredondo |
| EPI_ISL_3265603, EPI_ISL_3265604, EPI_ISL_3278286, EPI_ISL_3278287, EPI_ISL_3278288 | LESP Veracruz | Instituto de Diagnostico y Referencia Epidemiologicos (INDRE) | Abril Rodriguez-Maldonado; Ariadna Medina-Benitez; Claudia Wong-Arambula; Ernesto Ramirez-Gonzalez.; Gisela Barrera-Badillo; Irma Lopez-Martinez; Joaquin Quiroz-Mercado; Lucia Hernandez-Rivas; Maribel Gonzalez-Villa; Natividad Cruz-Ortiz; Sergio Rangel-Guerrero; Tatiana Nunez-Garcia; Vanessa Rivero-Arredondo |
| EPI_ISL_3868312 | Lab voor klinische biologie | Lab voor klinische biologie | Bruno Verhasselt; Hannelore Hamerlinck; Marija Janevska |
| EPI_ISL_3214387, EPI_ISL_3214389, EPI_ISL_3214390, EPI_ISL_3214391, EPI_ISL_3214392 | Lab. Microbiologia e Virologia Cotugno A.O. dei Colli | TIGEM | Antonio Grimaldi Patrizia Annunziata Francesco Panariello Biancamaria Pierri Claudia Tiberio Teresa Giuliano Valentina Bouche Chiara Colantuono Maria Concetta Cuomo Denise Di Concilio Lucio Di Filippo Anna Manfredi Marcello Salvi Antonio Limone Luigi Atripaldi Pellegrino Cerino Andrea Ballabio Davide Cacchiarelli |
| EPI_ISL_3216457, EPI_ISL_3216465 | LabQ Clinical Diagnostics LLC | New York City Public Health Laboratory | Jade Wang; et al. |
| EPI_ISL_3058210, EPI_ISL_3245972 | Labo Analyses Med | National Reference Center for Viruses of Respiratory Infections, Institut Pasteur, Paris | Angela Brisebarre; BéNéDicte Roquebert; Camille Capel; Christophe Malabat; Corinne Maufrais; Etienne Simon-Lorière; Frédéric Lemoine; Hub de bioinformatique et biostatistique; Louise Lefrançois; Marion Barbet; Maud Vanpeene; Méline Bizard; Sylvie Behillil; Sylvie Van der Werf; Victoire Baillet; Vincent Enouf |
| EPI_ISL_3239261 | Labor Becker & Kollegen (Standort München) | Robert Koch Institute | |
| EPI_ISL_3044365 | Labor Dr. Wisplinghoff - Köln | Robert Koch Institute | |
| EPI_ISL_2957378 | Labor Prof. Dr. G. Enders MVZ GbR | Robert Koch Institute | |
| EPI_ISL_3496212, EPI_ISL_3496219, EPI_ISL_3496227, EPI_ISL_3496250 | Labor ZOTZ\|KLIMAS; MVZ Düsseldorf-Centrum | Robert Koch Institute | |
| EPI_ISL_3129959 | Laboratoire Carage | Institut Pasteur de la Guyane | Anne Lavergne; Antoine Enfissi; Arielle Salmier; Dominique Rousset |
| EPI_ISL_3601599 | Laboratoire Hôpital Sainte Musse | Laboratoire Hôpital Sainte Musse | Lionel Chollet |
| EPI_ISL_3147215, EPI_ISL_3147236, EPI_ISL_3147321, EPI_ISL_3147462, EPI_ISL_3147483, EPI_ISL_3147489, EPI_ISL_3147499, EPI_ISL_3147506, EPI_ISL_3147511, EPI_ISL_3147518, EPI_ISL_3147525, EPI_ISL_3147527, EPI_ISL_3147546, EPI_ISL_3147576, EPI_ISL_3147585, EPI_ISL_3147589 | | | |
| see above | Laboratoire national de sante, Microbiology, Virology | Laboratoire national de sante, Microbiology, Microbial Genomics Platform | Anke Wienecke-Baldacchino; Catherine Ragimbeau; Elodie Solarino; Fatu Djabi; Jessica Tapp; Lise Pignon; Raoul Salmon; Tamir Abdelrahman; Trung Nguyen Nguyen; Virginie Jover |
| EPI_ISL_3147183, EPI_ISL_3147189, EPI_ISL_3147205, EPI_ISL_3147235, EPI_ISL_3147243, EPI_ISL_3147252, EPI_ISL_3147298, EPI_ISL_3147333, EPI_ISL_3147345, EPI_ISL_3147405, EPI_ISL_3147408, EPI_ISL_3147434, EPI_ISL_3147435, EPI_ISL_3147438, EPI_ISL_3147443, EPI_ISL_3147449, EPI_ISL_3147450, EPI_ISL_3147456, EPI_ISL_3147467, EPI_ISL_3147470, EPI_ISL_3147501, EPI_ISL_3147502, EPI_ISL_3147509, EPI_ISL_3147513, EPI_ISL_3147529, EPI_ISL_3147537, EPI_ISL_3147539, EPI_ISL_3147541, EPI_ISL_3147549, EPI_ISL_3147553, EPI_ISL_3147569, EPI_ISL_3147586, EPI_ISL_3147592 | | | |
| see above | Laboratoires Reunis | Laboratoire national de sante, Microbiology, Microbial Genomics Platform | Anke Wienecke-Baldacchino; Bernard Weber; Catherine Ragimbeau; Elodie Solarino; Fatu Djabi; Jessica Tapp; Lise Pignon; Raoul Salmon; Tamir Abdelrahman; Virginie Jover |
| EPI_ISL_3147720, EPI_ISL_3147772, EPI_ISL_3147777, EPI_ISL_3147806, EPI_ISL_3147809, EPI_ISL_3147816, EPI_ISL_3147830, EPI_ISL_3147840, EPI_ISL_3147850, EPI_ISL_3148087, EPI_ISL_3148219, EPI_ISL_3148220, EPI_ISL_3148221, EPI_ISL_3148222, EPI_ISL_3148223, EPI_ISL_3148224, EPI_ISL_3148225, EPI_ISL_3148226, EPI_ISL_3148227, EPI_ISL_3148228, EPI_ISL_3148229, EPI_ISL_3148230, EPI_ISL_3148231, EPI_ISL_3148232, EPI_ISL_3148233, EPI_ISL_3148234, EPI_ISL_3148235, EPI_ISL_3148236, EPI_ISL_3148237, EPI_ISL_3148238, EPI_ISL_3148239, EPI_ISL_3148240, EPI_ISL_3148241, EPI_ISL_3148242, EPI_ISL_3148243, EPI_ISL_3148244, EPI_ISL_3148245, EPI_ISL_3148246, EPI_ISL_3148247, EPI_ISL_3148248, EPI_ISL_3148249, EPI_ISL_3148250, EPI_ISL_3148251, EPI_ISL_3148252, EPI_ISL_3148253, EPI_ISL_3148254, EPI_ISL_3148255, EPI_ISL_3148256, EPI_ISL_3148257, EPI_ISL_3148258, EPI_ISL_3148259, EPI_ISL_3148260, EPI_ISL_3148261, EPI_ISL_3148262, EPI_ISL_3148263, EPI_ISL_3148264, EPI_ISL_3148265, EPI_ISL_3148298 | | | |
| see above | Laboratoires d'analyses medicales - Ketterthill | Laboratoire national de sante, Microbiology, Microbial Genomics Platform | Anke Wienecke-Baldacchino; Caroline Scheiber; Catherine Ragimbeau; Elodie Solarino; Fatu Djabi; Jessica Tapp; Lise Pignon; Raoul Salmon; Serge Vedy; Tamir Abdelrahman; Virginie Jover |
| EPI_ISL_3190354, EPI_ISL_3539229 | Laboratorio Central de Saude Publica do Estado de Goias (LACEN/GO) | Laboratory of Respiratory Viruses and Measles, Oswaldo Cruz Institute, FIOCRUZ | Agatha Cristinne Prudencio; Alice Sampaio Rocha; Ana Carolina Mendonca; Ana Flavia Mendonça; Anna Carolina Paixao; Carmen Helena Ramos; Cassiane Casanova; Elisa Cavalcante Pereira; Fernando Motta; Flavia Pereira Amorim da Silva; Ighor Leonardo Arantes Gomes; Luciana Appolinario; Luiz Augusto Pereira; Marilda Siqueira on behalf of the Fiocruz COVID-19 Genomic Surveillance Network; Paola Resende; Rafael Souza Guedes; Renata Serrano Lopes; Taina Venas; Vinicius Lemes da Silva |
| EPI_ISL_2925913, EPI_ISL_2993691, EPI_ISL_2993696, EPI_ISL_2993697, EPI_ISL_2993703, EPI_ISL_3376863 | Laboratorio CQRC | CQRC_QUALITY CONTROL CHEMICAL BIOLOGICAL RISK_AOOR Villa Sofia Cervello Palermo | Brunacci G.; Contino F.; Di Gaudio F. |
| EPI_ISL_3245216, EPI_ISL_3245223, EPI_ISL_3245224, EPI_ISL_3245225, EPI_ISL_3245229, EPI_ISL_3245230, EPI_ISL_3245231, EPI_ISL_3245232, EPI_ISL_3245233, EPI_ISL_3245239, EPI_ISL_3245278, EPI_ISL_3245300, EPI_ISL_3245326, EPI_ISL_3245331 | | | |
| see above | Laboratorio Central Noel Nutels | Bioinformatics Laboratory / LNCC | Alessandra P Lamarca; Alexandra L Gerber; Amilcar Tanuri; Ana Paula de C Guimaraes; Ana Tereza R Vasconcelos; Andrea Cony Cavalcanti; Caio Luiz Pereira Ribeiro; Cintia Policarpo; Claudia Maria Braga de Mello; Cristiane Gomes da Silva; Douglas Terra Machado; Erica Ramos dos Santos Nascimento; Fernanda Leitao dos Santos; Flavio Dias da Silva; Gleidson da Silva de Oliveira; Leandro Magalhaes de Souza; Liliane Cavalcante; Luiz G P de Almeida; Marcio Henrique de Oliveira Garcia; Mario Sergio Ribeiro; Ricardo Jose Barbosa Salviano; Ronaldo da Silva F Jr; Silvia Carvalho |
| EPI_ISL_3155578, EPI_ISL_3155580, EPI_ISL_3155589, EPI_ISL_3155602, EPI_ISL_3155611, EPI_ISL_3155623, EPI_ISL_3155626, EPI_ISL_3155642 | | | |
| see above | Laboratorio Central de Epidemiología (LCE) | Centro de Investigación en Enfermedades Infecciosas (CIENI), Instituto Nacional de Enfermedades Respiratorias (INER) | ; Alejandra García-Gasca; Alejandra Hernández-Terán; Alejandro Sánchez-Flores; Alfredo Herrera-Estrella; Alicia Ocaña-Mondragón; Andreu Comas-García; Angel Gustavo Salas-Lais; Antonio Loza Román; Bernardo Martínez-Miguel; Blanca Taboada; Brenda Irasema Maldonado-Meza; Bruno Gómez-Gil; Carla Ivón Herrera-Najera; Carlos F. Arias; Celia Boukadida; Clara Esperanza Santacruz-Tinoco; Concepción Grajales-Muñiz; Consorcio Mexicano de Vigilancia Genómica (CoViGen-Mex). Authors (in alphabetical order): Julio Elias Alvarado-Yaah; Cristóbal Cháidez-Quiróz; Célida Duque Molina; Célida Martínez- Rodríguez; Daniel Fregoso-Rueda; Daniel Lira Morales; Eduardo Becerril-Vargas; Fernando Fontove-Herrera; Fidencio Mejía-Nepomuceno; Francisco Pulido; Gloria Elena Espinosa-Ayala; Gloria María Molina-Salinas; Gloria Vazquez; Hector Esteban Paz-Juárez; Hector Montoya-Fuentes; Helen Haydee Fernanda Ramírez-Plascencia; Irvin González-López; Jean Pierre González; Jesús Hernández; Joel Armando Vázquez-Pérez.; Jorge Salas-Hernández; José Antonio Enciso-Moreno; José Arturo Martínez-Orozco; José Esteban Muñoz-Medina; José de Jesús Nuñez-Contreras; Juan Bautista Chale-Dzul; Julissa Enciso-Ibarra; Kathia Elizabeth Tapia-Diaz; Luis Alberto Ochoa-Carrera; Margarita Matías-Florentino; Mario Újica-Sánchez; Marissa Perez-Garcia; María Guadalupe Santiago-Mauricio; María Guadalupe de Jesús Mireles-Rivera; Nelly Sélem-Mojica; Pavel Isa; Ricardo Ciria Merce; Ricardo Grande; Rosa María Gutiérrez Rios; Santiago Ávila-Ríos; Selene Zárate; Susana Lopez; Verónica Mata-Haro; Victor Eduardo García-Arias; Víctor Hugo Borja-Aburto |
| EPI_ISL_6944556, EPI_ISL_6944559, EPI_ISL_6944561, EPI_ISL_6944566, EPI_ISL_6944573, EPI_ISL_6944587, EPI_ISL_6944589, EPI_ISL_6944590, EPI_ISL_6944601, EPI_ISL_6944609, EPI_ISL_6944618, EPI_ISL_6944620, EPI_ISL_6944622, EPI_ISL_6944624, EPI_ISL_6944625, EPI_ISL_6944626, EPI_ISL_6944630, EPI_ISL_6944631, EPI_ISL_6944632, EPI_ISL_6944633, EPI_ISL_6944636, EPI_ISL_6944637 | | | |
| see above | Laboratorio Central de Salud Publica | Viral Special Pathogens Branch, Centers for Disease Control and Prevention | Andrea Gómez de la Fuente; Cynthia Vázquez; Emir Talundzic; Joel Montgomery; John Klena; Juan Torales; Justin Lee; María José Ortega; María Liz Gamarra; Shannon Whitmer; Shirley Villalba |
| EPI_ISL_4518742, EPI_ISL_4518745, EPI_ISL_4518747, EPI_ISL_4518749 | Laboratorio Central de Salud Publica de Paraguay | Fundação Ezequiel Dias | Andre Leal; Andrea Gómez de la Fuente; Cynthia Vazquez; Elaine Cristina; Felipe Iani; Flavia Aburjaile; Gislene Garcia de Castro Lichs; Glauco Carvalho; Hegger Fritsch; Joilson Xavier; Juan Torales; Luiz Alcantara.; Luiz Henrique Ferraz Demarchi; Luiz Takao Watanabe; Marina Castilhos Souza Umaki Zardin; Marta Giovanetti; María José Ortega; María Liz Gamarra; Natalia Guimaraes; Raquel da Silva Ferreira; Shirley Villalba; Talita Adelino; Vagner Fonseca; de Oliveira |
| EPI_ISL_5490447 | Laboratorio Central de Saude Publica do Amazonas - LACEN-AM | Laboratorio de Ecologia de Doencas Transmissiveis na Amazonia, Instituto Leonidas e | André Corado; Arlesson Viana Da Silva; Felipe Naveca; Fernanda Nascimento; George Silva; Luciana Gonçalves; Maria Júlia Brandão; Matilde Mejía; Valdinete Nascimento; Victor Souza |

| | | | |
|---|---|---|---|
| | | Maria Deane - Fiocruz Amazonia | |
| EPI_ISL_3190169, EPI_ISL_3190170, EPI_ISL_3190171 | Laboratorio Central de Saude Publica do Estado da Bahia (LACEN/BA) | Laboratory of Respiratory Viruses and Measles, Oswaldo Cruz Institute, FIOCRUZ | Alice Sampaio Rocha; Ana Carolina Mendonca; Anna Carolina Paixao; Elisa Cavalcante Pereira; Felicidade Pereira; Fernando Motta; Luciana Appolinario; Marilda Siqueira on behalf of the Fiocruz COVID-19 Genomic Surveillance Network; Paola Resende; Renata Serrano Lopes; Taina Venas |
| EPI_ISL_3434759 | Laboratorio Central de Saude Publica do Estado da Paraiba (LACEN/PB) | Laboratory of Respiratory Viruses and Measles, Oswaldo Cruz Institute, FIOCRUZ | Alice Sampaio Rocha; Ana Carolina Mendonca; Anna Carolina Paixao; Dalane Loudal Florentino Teixeira; Elisa Cavalcante Pereira; Fernando Motta; Joao Felipe Bezerra; Luciana Appolinario; Marilda Siqueira on behalf of the Fiocruz COVID-19 Genomic Surveillance Network; Paola Resende; Renata Serrano Lopes; Taina Venas |
| EPI_ISL_3434837, EPI_ISL_3434838, EPI_ISL_3434839, EPI_ISL_3434840, EPI_ISL_3434841, EPI_ISL_3434842, EPI_ISL_3434843, EPI_ISL_3434873, EPI_ISL_3434874, EPI_ISL_3434875, EPI_ISL_3434876, EPI_ISL_3434877, EPI_ISL_3434878, EPI_ISL_3434879, EPI_ISL_3434880, EPI_ISL_3434881, EPI_ISL_3434882, EPI_ISL_3434883, EPI_ISL_3434884, EPI_ISL_3434885, EPI_ISL_3434886, EPI_ISL_3434887, EPI_ISL_3434888, EPI_ISL_3434889, EPI_ISL_3434890, EPI_ISL_3434891, EPI_ISL_3434892, EPI_ISL_3434893, EPI_ISL_3434895, EPI_ISL_3434938, EPI_ISL_3539925, EPI_ISL_3802933 | | | |
| see above | Laboratorio Central de Saude Publica do Estado de Alagoas (LACEN/AL) | Laboratory of Respiratory Viruses and Measles, Oswaldo Cruz Institute, FIOCRUZ | Agatha Soares; Alice Sampaio Rocha; Ana Carolina Mendonca; Anderson Brandao Leite; Anna Carolina Paixao; Elisa Cavalcante Pereira; Fernando Motta; Ighor Arantes; Luciana Appolinario; Marilda Siqueira on behalf of the Fiocruz COVID-19 Genomic Surveillance Network; Paola Resende; Renata Serrano Lopes; Taina Venas |
| EPI_ISL_3235294, EPI_ISL_3235295, EPI_ISL_3235296, EPI_ISL_3235297, EPI_ISL_3235298, EPI_ISL_3235303, EPI_ISL_3434821, EPI_ISL_3434822, EPI_ISL_3434823, EPI_ISL_3434910, EPI_ISL_3434911, EPI_ISL_3434912, EPI_ISL_3434913, EPI_ISL_3434914, EPI_ISL_3434915, EPI_ISL_3434916, EPI_ISL_3539248, EPI_ISL_3539770, EPI_ISL_3539771 | | | |
| see above | Laboratorio Central de Saude Publica do Estado de Santa Catarina (LACEN/SC) | Laboratory of Respiratory Viruses and Measles, Oswaldo Cruz Institute, FIOCRUZ | Alice Sampaio Rocha; Ana Carolina Mendonca; Anna Carolina Paixao; Darcita Buerger Rovaris; Elisa Cavalcante Pereira; Fernando Motta; Luciana Appolinario; Marilda Siqueira on behalf of the Fiocruz COVID-19 Genomic Surveillance Network; Paola Resende; Renata Serrano Lopes; Sandra Bianchini Fernandes; Taina Venas |
| EPI_ISL_3803006, EPI_ISL_3803008, EPI_ISL_3803009, EPI_ISL_3803012, EPI_ISL_3803015, EPI_ISL_3803016, EPI_ISL_3803018, EPI_ISL_3803020, EPI_ISL_3803022, EPI_ISL_3803023, EPI_ISL_3803024, EPI_ISL_3803025, EPI_ISL_3803026, EPI_ISL_4170319, EPI_ISL_4212839 | | | |
| see above | Laboratorio Central de Saude Publica do Estado de Sergipe (LACEN/SE) | Laboratory of Respiratory Viruses and Measles, Oswaldo Cruz Institute, FIOCRUZ | Agatha Soares; Alice Sampaio Rocha; Ana Carolina Mendonca; Anna Carolina Paixao; Cliomar Alves dos Santos; Elisa Cavalcante Pereira; Fernando Motta; Ighor Arantes; Luciana Appolinario; Marilda Siqueira on behalf of the Fiocruz COVID-19 Genomic Surveillance Network; Paola Resende; Renata Serrano Lopes; Tainá Moreira Martins Venas |
| EPI_ISL_3434946, EPI_ISL_3434948, EPI_ISL_3434958, EPI_ISL_3434967, EPI_ISL_3434971, EPI_ISL_3434972, EPI_ISL_3434973, EPI_ISL_3434974, EPI_ISL_3434981, EPI_ISL_3539794, EPI_ISL_3539795 | | | |
| see above | Laboratorio Central de Saude Publica do Estado do Amapa (LACEN/AP) | Laboratory of Respiratory Viruses and Measles, Oswaldo Cruz Institute, FIOCRUZ | Agatha Cristinne Prudencio; Alice Sampaio Rocha; Ana Carolina Mendonca; Andreia Santos Costa; Anna Carolina Paixao; Anne Caroline da Silva Soledade; Elisa Cavalcante Pereira; Fernando Motta; Ighor Leonardo Arantes Gomes; Lindomar dos Anjos Silva; Luciana Appolinario; Marcia Socorro Pereira Cavalcante; Marilda Siqueira on behalf of the Fiocruz COVID-19 Genomic Surveillance Network; Paola Resende; Renata Serrano Lopes; Taina Venas |
| EPI_ISL_3434746, EPI_ISL_3434987, EPI_ISL_3434988, EPI_ISL_3434995, EPI_ISL_3434996, EPI_ISL_3434997, EPI_ISL_3434998, EPI_ISL_3434999, EPI_ISL_3435008, EPI_ISL_3435011, EPI_ISL_3435013, EPI_ISL_3435014, EPI_ISL_3435015, EPI_ISL_3435016, EPI_ISL_3435019, EPI_ISL_3435025, EPI_ISL_3435026, EPI_ISL_3435029, EPI_ISL_3435030, EPI_ISL_3435031, EPI_ISL_3435035, EPI_ISL_3435037, EPI_ISL_3435038, EPI_ISL_3435039, EPI_ISL_3435040, EPI_ISL_3435041, EPI_ISL_3435043, EPI_ISL_3435044, EPI_ISL_3435046, EPI_ISL_3435065, EPI_ISL_3539934, EPI_ISL_3827884, EPI_ISL_3827927, EPI_ISL_3827973, EPI_ISL_3827984, EPI_ISL_3827985, EPI_ISL_3828000, EPI_ISL_3828005, EPI_ISL_3828010, EPI_ISL_4220161, EPI_ISL_5329028 | | | |
| see above | Laboratorio Central de Saude Publica do Estado do Espirito Santo (LACEN/ES) | Laboratory of Respiratory Viruses and Measles, Oswaldo Cruz Institute, FIOCRUZ | Agatha Soares; Alice Sampaio Rocha; Ana Carolina Mendonca; Anna Carolina Paixao; Elisa Cavalcante Pereira; Fernando Motta; Ighor Leonardo Arantes; Luciana Appolinario; Marilda Siqueira on behalf of the Fiocruz COVID-19 Genomic Surveillance Network; Paola Resende; Renata Serrano Lopes; Rodrigo Ribeiro Rodrigues; Taina Venas |
| EPI_ISL_4880275, EPI_ISL_4880277, EPI_ISL_4880278, EPI_ISL_4880279, EPI_ISL_4880280, EPI_ISL_4880294, EPI_ISL_4880298, EPI_ISL_4880311, EPI_ISL_4880312, EPI_ISL_4880313, EPI_ISL_4880314, EPI_ISL_4880319 | | | |
| see above | Laboratorio Central de Saude Publica do Estado do Maranhao (LACEN-MA) | Laboratory of Respiratory Viruses and Measles, Oswaldo Cruz Institute, FIOCRUZ | Agatha Soares; Alice Sampaio Rocha; Ana Carolina Mendonca; Anna Carolina Paixao; Fernando Motta; Ighor Arantes; Lidio Gonçalves Lima Neto; Luciana Appolinario; Marilda Siqueira on behalf of the Fiocruz COVID-19 Genomic Surveillance Network; Paola Resende; Renata Serrano Lopes |
| EPI_ISL_6173114, EPI_ISL_6173115, EPI_ISL_6173116 | Laboratorio Central de Saude Publica do Estado do Maranhao (LACEN/MA) | Laboratory of Respiratory Viruses and Measles, Oswaldo Cruz Institute, FIOCRUZ | Alice Sampaio Rocha; Bruna Mendonça da Silva; Elisa Cavalcante Pereira; Fernando Motta; Ighor Arantes; Jéssica Graça Macedo de Carvalho; Larissa Macedo Pinto; Lidio Gonçalves Lima Neto; Luciana Appolinario; Marilda Siqueira on behalf of the Fiocruz COVID-19 Genomic Surveillance Network; Paola Resende |
| EPI_ISL_3914073, EPI_ISL_3914090, EPI_ISL_3914103, EPI_ISL_3914111, EPI_ISL_3914119, EPI_ISL_3914124, EPI_ISL_3914138, EPI_ISL_3914150, EPI_ISL_3914155, EPI_ISL_3914163, EPI_ISL_3914175, EPI_ISL_3914184, EPI_ISL_3914193, EPI_ISL_3914211, EPI_ISL_3914228, EPI_ISL_3914236, EPI_ISL_3914271, EPI_ISL_3914275, EPI_ISL_3914279, EPI_ISL_3914282, EPI_ISL_3914296, EPI_ISL_3914317, EPI_ISL_3914326, EPI_ISL_3914342, EPI_ISL_3914347, EPI_ISL_3914351, EPI_ISL_3914372, EPI_ISL_3914381, EPI_ISL_3914410, EPI_ISL_3914434 | | | |
| see above | Laboratorio Central de Saude Publica do Estado do Parana (Instituto de Biologia Molecular do Paraná (LACEN-PR) | Instituto Carlos Chagas - Fiocruz | Alessandra De Melo Aguiar; Andreia Akemi Suzukawa; Andréa Rodrigues Ávila; Bruno Dallagiovanna; Dalila Zanette; Eduardo Balsanelli; Emanuel Maltempi de Souza; Fabio Passetti; Fabricio Klerynton Marchini; Fábio de Oliveira Pedrosa; Guilherme Becker; Helisson Faoro; Hellen Geremias dos Santos; Irina Nastassja Riediger; Letusa Albrecht; Lucas Blanes; Luis Gustavo Morello; Lysangela Ronalte Alves; Maria do Carmo Debur; Mauro de Medeiros Oliveira; Michelle Orane Schemberger; Paola Cristina Resende; Sheila Cristina Nardeli; Tiago Gräf; Valter Antônio de Baura |
| EPI_ISL_3235268, EPI_ISL_3235269, EPI_ISL_3235270, EPI_ISL_3235271, EPI_ISL_3235272, EPI_ISL_3235273, EPI_ISL_3235274, EPI_ISL_3235275 | | | |
| see above | Laboratorio Central de Saude Publica do Estado do Rio Grande do Norte (LACEN/RN) | Laboratory of Respiratory Viruses and Measles, Oswaldo Cruz Institute, FIOCRUZ | Agatha Soares; Alice Sampaio Rocha; Ana Carolina Mendonca; Ana Paula Ferreira Costa; Anna Carolina Paixao; Antonnyo Palmielly Diogenes Lima; Aurélio de Oliveira Bento; Elisa Cavalcante Pereira; Fernando Motta; Gessika Brenna Costa Alves; Heglayne Pereira Vital da Silva; Iago de Souza Gomes; Ighor Arantes; Isabelle Cristina Clemente dos Santos; Janaina Sonale Cavalcante Nogueira de Oliveira; Jayra Juliana Paiva Alves Abrantes; Jonas José da Silva; Luciana Appolinario; Marilda Siqueira on behalf of the Fiocruz COVID-19 Genomic Surveillance Network; Paola Resende; Renata Serrano Lopes; Taina Venas; Themis Rocha de Souza; Vítor Gabriel Saldanha Fernandes |
| EPI_ISL_3190303, EPI_ISL_3190305, EPI_ISL_3190306, EPI_ISL_3190406, EPI_ISL_3190407, EPI_ISL_3190408, EPI_ISL_3235276, EPI_ISL_3235277, EPI_ISL_3235278, EPI_ISL_3235280, EPI_ISL_3235281, EPI_ISL_3235282, EPI_ISL_3235283, EPI_ISL_3235284, EPI_ISL_3235285, EPI_ISL_3235289, EPI_ISL_3235290, EPI_ISL_3235292, EPI_ISL_3235293, EPI_ISL_3434907, EPI_ISL_3832410, EPI_ISL_3832411, EPI_ISL_3832412, EPI_ISL_3832414, EPI_ISL_3832415, EPI_ISL_3832416, EPI_ISL_4061392 | | | |
| see above | Laboratorio Central de Saude Publica do Estado do Rio Grande do Sul (LACEN-RS) | Laboratory of Respiratory Viruses and Measles, Oswaldo Cruz Institute, FIOCRUZ | Agatha Cristinne Soares; Alice Sampaio Rocha; Ana Carolina Mendonca; Anderson Brandao Leite; Anna Carolina Paixao; Elisa Cavalcante Pereira; Fernando Motta; Ighor Leonardo Arantes; Luciana Appolinario; Marilda Siqueira on behalf of the Fiocruz COVID-19 Genomic Surveillance Network; Paola Resende; Renata Serrano Lopes; Richard Salvato; Taina Venas; Tatiana Schaffer Gregianini |
| EPI_ISL_4237273, EPI_ISL_4237275, EPI_ISL_4237302, EPI_ISL_4237307, EPI_ISL_4237309, EPI_ISL_4237311, EPI_ISL_4237312, EPI_ISL_4237314, EPI_ISL_4237315, EPI_ISL_4237316, EPI_ISL_4237320, EPI_ISL_4237321, EPI_ISL_4237322, EPI_ISL_4237330, EPI_ISL_4237331, EPI_ISL_4237332, EPI_ISL_4237333, EPI_ISL_4237336, EPI_ISL_4237337, EPI_ISL_4237338, EPI_ISL_4237352, EPI_ISL_4237359 | | | |
| see above | Laboratorio Central de Saude Publica do Estado do Rio de Janeiro (LACEN-RJ) | Laboratory of Respiratory Viruses and Measles, Oswaldo Cruz Institute, FIOCRUZ | Agatha Cristinne Prudencio; Alice Sampaio Rocha; Ana Carolina Mendonca; Andrea Cony Cavalcanti; Anna Carolina Paixao; Elisa Cavalcante Pereira; Fernando Motta; Ighor Leonardo Arantes Gomes; Luciana Appolinario; Marilda Siqueira on behalf of the Fiocruz COVID-19 Genomic Surveillance Network; Paola Resende; Renata Serrano Lopes; Taina Venas |
| EPI_ISL_3235307, EPI_ISL_3235313 | Laboratorio Central de Saude Publica do Estado do Tocantins (LACEN/TO) | Laboratory of Respiratory Viruses and Measles, Oswaldo Cruz Institute, FIOCRUZ | Agatha Soares; Alice Sampaio Rocha; Ana Carolina Mendonca; Anna Carolina Paixao; Elisa Cavalcante Pereira; Fernando Motta; Ighor Arantes; Jucimaria Dantas Galvao; Luciana Appolinario; Marilda Siqueira on behalf of the Fiocruz COVID-19 Genomic Surveillance Network; Paola Resende; Renata Serrano Lopes; Taina Venas |
| EPI_ISL_4849752, EPI_ISL_4849753, EPI_ISL_4849754, EPI_ISL_4849755, EPI_ISL_4849756, EPI_ISL_4849770 | Laboratorio Central de la Ciudad de Santa Fe | Grupo de Genómica y Bioinformática del Instituto de Investigación de la Cadena Láctea CONICET-INTA on behalf of 'Proyecto Argentino Interinstitucional de genomica de SARS-CoV-2' (PAIS Consortium) | AF; Amadio; C; Eberhardt; G; Irazoqui; JM; MF; Mugna; Ojeda; Pastor; Rompato; V |
| EPI_ISL_4003466 | Laboratorio Central, Ministerio de Salud Córdoba | Instituto de Patologia Vegetal (CIAP-INTA) on behalf of 'Proyecto Argentino Interinstitucional de genomica de SARS-CoV-2' (PAIS Consortium) | Barbas, G.; Castro, G.; Debat, HJ.; FD; Fernández; M; M.B.; Marquez, N.; Pisano; Re, V. |
| EPI_ISL_3150223 | Laboratorio Clínico Gamma | "Unidad de Secuenciación y Genómica, USACH" | "Carlos A. Villarroel; Francisco A. Cubillos"; Kamila Urbina; Mara Cuevas; Marcelo Cortez-San Martin; Yesseny Vásquez-Martínez |
| EPI_ISL_3527791, EPI_ISL_3527796, EPI_ISL_3835365, EPI_ISL_3835379, EPI_ISL_3835385 | Laboratorio Nacional de Salud | Asociación de Salud Integral / Clínica Familiar "Luis Ángel García" | Ana S. Gonzalez-Reiche; Claudia Rangel; Danicela Mercado; Eduardo Arathon; Hilda Ruiz; Luis Aguirre; Luis Rivas; Narda Medina; Oscar Bonilla; Osmar Gamboa. |
| EPI_ISL_5332831, EPI_ISL_5332832, EPI_ISL_5332838, EPI_ISL_6121556, EPI_ISL_6121557, EPI_ISL_6121563, EPI_ISL_6121564, EPI_ISL_6121577, EPI_ISL_6121578 | | | |
| see above | Laboratorio de Alta Complejidad (LACMI), Instituto de Genética Humana (IgeHM) Parque de la Salud de la Provincia de Misiones, Universidad Nacional de Misiones | Grupo de Genómica y Bioinformática del Instituto de Investigación de la Cadena Láctea CONICET-INTA on behalf of 'Proyecto Argentino Interinstitucional de genomica de SARS-CoV-2' (PAIS Consortium) | AF; Aguirre; Amadio; Benitez; C; D; Eberhardt; Fontana; G; Gamarra; Gonzales; Irazoqui; JM; K; Ludojoski; M; MF; R; Salvatierra; Silva; TA; Urquiza; Vogel |
| EPI_ISL_3050641, EPI_ISL_4030314, EPI_ISL_5490803 | Laboratorio de Ecologia de Doencas Transmissiveis na Amazonia, Instituto Leonidas e Maria Deane - Fiocruz Amazonia | Laboratorio de Ecologia de Doencas Transmissiveis na Amazonia, Instituto Leonidas e Maria Deane - Fiocruz Amazonia | Alex Martins; André Corado; Debora Duarte; Fabiola Mendonça da Silva Chui; Felipe Naveca; Fernanda Nascimento; Fernando Fonseca de Almeida Val; George Silva; Gisely Cardoso de Melo; Karina Pessoa; Luciana Gonçalves; Marcus Vinicius Guimarães de Lacerda; Maria Júlia Brandão; Maria Paula Gomes Mourão; Mariana Xavier Simão; Matilde Mejía; Michele Jesus; Valdinete Nascimento; Vanderson Sampaio; Vanderson de Souza Sampaio; Victor Souza; Ágatha Costa |
| EPI_ISL_3370435, EPI_ISL_3370436, EPI_ISL_3370437, EPI_ISL_3370438, EPI_ISL_3370439, EPI_ISL_3370440, EPI_ISL_3370441, EPI_ISL_3370442, EPI_ISL_3370456, EPI_ISL_3535735, EPI_ISL_5994580 | | | |
| see above | Laboratorio de Infectologia y | Laboratory of Molecular Virology, | Alejandro Bhrun; Ana Maria Contreras; Andres E. Munoz-Marcos; Carlos Palma; Catalina Pardo-Roa; Constanza Maldonado; Constanza Martinez-Valdevenito; Eileen Serrano; Erick Salinas; Estefany Poblete; Eugenia L. Fuentes Luppichini; Francisco Melo; Jennifer Angulo; Jorge Levican; Leonardo I. Almonacid; M. Belen |

| | | | |
|---|---|---|---|
| | Virologia Molecular | School of Medicine, Pontificia Universidad Catolica de Chile | Leyton; Magdalena Vera; Marcela Ferres; Maria Jose Avendano; Rafael A. Medina; Rafael A. Medina.; Tamara Garcia-Salum |
| EPI_ISL_4889739 | Laboratorio de Infectología, Servicio de Infectología, Hospital Universitario Dr. José Eleuterio González - Universidad Autónoma de Nuevo León | Centro de Investigación e Innovación en Virología Médica, Departamento de Bioquímica y Medicina Molecular,Facultad de Medicina - Universidad Autónoma de Nuevo León | Adrian Camacho-Ortiz; Ana M. Rivas-Estilla; Daniel Arellanos-Soto; Daniel Salas-Treviño; Eduardo Perez-Alba; Javier Ramos-Jiménez; Kame A. Galán-Huerta; Laura Nuzzolo-Shihadeh; Paola Bocanegra-Ibarias; Samantha M. Flores-Treviño; Sonia A. Lozano-Sepúlveda |
| EPI_ISL_3134248, EPI_ISL_3134251, EPI_ISL_3134252, EPI_ISL_3134255, EPI_ISL_3134256, EPI_ISL_3134257, EPI_ISL_3134258, EPI_ISL_3134259, EPI_ISL_3134262, EPI_ISL_3134263, EPI_ISL_3134265 | | | |
| see above | Laboratorio de Medicina Molecular, Universidad de Magallanes | Centro Asistencial Docente y de Investigacion, Universidad de Magallanes | Carolina Pérez; Constanza Ceroni; Diego Alvarez; Inés Cid; Jacqueline Aldridge; Jorge González; Juan Ríos; Marcelo Navarrete |
| EPI_ISL_3761530, EPI_ISL_3761537, EPI_ISL_3761544, EPI_ISL_3761545, EPI_ISL_3761546, EPI_ISL_3761547, EPI_ISL_3761554, EPI_ISL_3761556, EPI_ISL_3761561, EPI_ISL_3761567, EPI_ISL_3761569, EPI_ISL_3761589, EPI_ISL_3761597, EPI_ISL_3761600, EPI_ISL_3761605, EPI_ISL_3761607, EPI_ISL_3761608, EPI_ISL_3761614, EPI_ISL_3761681, EPI_ISL_3761682, EPI_ISL_3761683, EPI_ISL_3761684, EPI_ISL_3761685, EPI_ISL_3761686, EPI_ISL_3761687, EPI_ISL_3761688, EPI_ISL_3761689, EPI_ISL_3761690, EPI_ISL_3761691, EPI_ISL_3761692, EPI_ISL_3761693, EPI_ISL_3761694, EPI_ISL_3761695, EPI_ISL_3761696, EPI_ISL_3761697, EPI_ISL_3761698, EPI_ISL_3761699 | | | |
| see above | Laboratorio de Pesquisa em Virologia, FAMERP, SJRP | Laboratorio de Pesquisa em Virologia, FAMERP, SJRP | Beatriz de Carvalho Marques; Cecília Artico Banho; Cíntia Bittar; Fábio Sossai Possebon; Guilherme Campos; Helena Lage Ferreira; Jorge A. Petroli Marchesi; João Pessoa Araújo Jr.; Leila Sabrina Ullmann; Lívia Sacchetto; Maisa C. Pereira Parra; Marília Moraes; Maurício L. Nogueira.; Paula Rahal; Paulo Inacio da Costa |
| EPI_ISL_3991562, EPI_ISL_3991565, EPI_ISL_3991568, EPI_ISL_3991571, EPI_ISL_3991574, EPI_ISL_3991576, EPI_ISL_3991579, EPI_ISL_3991582, EPI_ISL_3991586, EPI_ISL_3991589, EPI_ISL_3991593, EPI_ISL_3991596, EPI_ISL_3991599, EPI_ISL_3991602, EPI_ISL_3991629, EPI_ISL_3991636, EPI_ISL_3991640, EPI_ISL_3991643, EPI_ISL_3991646, EPI_ISL_3991651, EPI_ISL_3991658, EPI_ISL_3991669, EPI_ISL_3991682, EPI_ISL_3991698, EPI_ISL_3991701, EPI_ISL_3991705, EPI_ISL_3991708, EPI_ISL_3991715, EPI_ISL_3991718, EPI_ISL_3991722, EPI_ISL_3991725, EPI_ISL_4080532, EPI_ISL_4080537, EPI_ISL_4080545, EPI_ISL_4080548, EPI_ISL_4080549, EPI_ISL_4172712, EPI_ISL_4172716, EPI_ISL_4172755, EPI_ISL_4172756, EPI_ISL_4172758, EPI_ISL_4172760, EPI_ISL_4172762, EPI_ISL_4172765, EPI_ISL_4172767, EPI_ISL_4172769, EPI_ISL_4172771, EPI_ISL_4172772, EPI_ISL_4172774, EPI_ISL_4172919, EPI_ISL_4172921, EPI_ISL_4172922, EPI_ISL_4172924, EPI_ISL_4172926, EPI_ISL_4172928, EPI_ISL_4279962, EPI_ISL_4279965, EPI_ISL_4279979, EPI_ISL_4279980, EPI_ISL_4279981, EPI_ISL_4279982, EPI_ISL_4279983, EPI_ISL_4279989, EPI_ISL_4279995, EPI_ISL_4280000, EPI_ISL_4280001, EPI_ISL_4280002, EPI_ISL_4280003, EPI_ISL_4280025, EPI_ISL_4416856, EPI_ISL_4416867, EPI_ISL_4416871, EPI_ISL_4416877, EPI_ISL_4416881, EPI_ISL_4416882, EPI_ISL_4416890, EPI_ISL_4416891, EPI_ISL_4416892, EPI_ISL_4416902, EPI_ISL_4416907, EPI_ISL_4416908, EPI_ISL_4416909, EPI_ISL_4416910, EPI_ISL_4416947, EPI_ISL_4417070, EPI_ISL_4417078, EPI_ISL_4417081, EPI_ISL_4417082, EPI_ISL_4417083, EPI_ISL_4417084, EPI_ISL_4417085, EPI_ISL_4417095, EPI_ISL_4417096, EPI_ISL_4417103, EPI_ISL_4417111, EPI_ISL_4417112, EPI_ISL_4417113 | | | |
| see above | Laboratorio de Referencia Nacional de Virus Respiratorios. Centro Nacional de Salud Publica. Instituto Nacional de Salud Peru. | Laboratorio de Referencia Nacional de Virus Respiratorios. Centro Nacional de Salud Publica. Instituto Nacional de Salud Peru. | Carlos Padilla Rojas; Henri Bailon Calderon; Iris Silva Molina; Joseph Huayra Niquen; Lely Solari Zerpa; Luis Barcena Flores; Marco Galarza Perez; Nancy Rojas Serrano; Nieves Sevilla Castañeda; Omar Caceres Rey; Orson Mestanza Millones; Princesa Medrano Alhuay; Priscila Lope Pari; Sandra Morales Ruiz; Sara Gordillo Vilchez; Steve Acedo Lazo; Veronica Hurtado Vela; Victor Jimenez Vasquez; Wendy Lizarraga Olivares |
| EPI_ISL_7747309, EPI_ISL_7747310, EPI_ISL_7747311, EPI_ISL_7747325, EPI_ISL_7747384 | Laboratorio de Salud Pública de Bogotá | Gencore - Universidad de los Andes | Alejandro Gomez; Cristian Barrera; Gabriela Ariza; Gabriela Delgado; Johana Hernandez; Luisa Sacristan; Marcela Guevara; Silvia Restrepo |
| EPI_ISL_3055559 | Laboratorio de salud publica Puebla | LABOPAT | Cynthia Penaloza; Luis Mendoza; Silvia Montilla |
| EPI_ISL_5771891 | Laboratorio del Hospital Regional Ushuaia Gdor. Ernesto Campos | Nodo de Secuenciación Tierra del Fuego - Hospital Regional Ushuaia - Centro Austral De Investigaciones Científicas - Universidad Nacional De Tierra Del Fuego | AE; Boutureira, MF.; CA; CB; CF; Castro; Ceballos; Cáceres; De Roccis; F; G; Gallego; Gramundi; ID; Nardi; Rojas; SB; SG; Yulan |
| EPI_ISL_7620357, EPI_ISL_7620358, EPI_ISL_7620390, EPI_ISL_7620391, EPI_ISL_7620392 | Laboratorio del Hospital Regional Ushuaia Gdor. Ernesto Campos | Nodo de Secuenciación Tierra del Fuego - Hospital Regional Ushuaia - Centro Austral De Investigaciones Científicas - Universidad Nacional De Tierra Del Fuego on behalf PAIS project and ANLIS-Malbran | AE; Boutureira, MF.; CA; CB; CF; Castro; Ceballos; Cáceres; De Roccis; F; G; Gallego; Gramundi; ID; Nardi; Rojas; SB; SG; Yulan |
| EPI_ISL_3128492 | Laboratorio di Riferimento Regionale della Sicilia Occidentale per l'Emergenza COVID-19 | Laboratorio di Riferimento Regionale della Sicilia Occidentale per l'Emergenza COVID-19 | Carmelo Massimo Maida; Claudio Costantino; Daniela Di Naro; Fabio Tramuto; Francesco Vitale; Giorgio Graziano; Giulia Randazzo; Vincenzo Restivo; Walter Mazzucco |
| EPI_ISL_3017548, EPI_ISL_3017557, EPI_ISL_3111442, EPI_ISL_3111457, EPI_ISL_3111471, EPI_ISL_3111489, EPI_ISL_3111491, EPI_ISL_3111492, EPI_ISL_3111576, EPI_ISL_3111586, EPI_ISL_3111598, EPI_ISL_3111601, EPI_ISL_3111605, EPI_ISL_3111677, EPI_ISL_3111699, EPI_ISL_3111704, EPI_ISL_3111761, EPI_ISL_3111779, EPI_ISL_3111811, EPI_ISL_3111814, EPI_ISL_3111941, EPI_ISL_3111969, EPI_ISL_3112402, EPI_ISL_3112437, EPI_ISL_3112459, EPI_ISL_3112478, EPI_ISL_3112540, EPI_ISL_3112569, EPI_ISL_3112652, EPI_ISL_3112656, EPI_ISL_3112705, EPI_ISL_3112706, EPI_ISL_3112727, EPI_ISL_3112749, EPI_ISL_3112764, EPI_ISL_3112866, EPI_ISL_3112884, EPI_ISL_3112892, EPI_ISL_3112926, EPI_ISL_3112956, EPI_ISL_3112957, EPI_ISL_3113219, EPI_ISL_3113280, EPI_ISL_3113287, EPI_ISL_3113321, EPI_ISL_3113359, EPI_ISL_3113366, EPI_ISL_3113408, EPI_ISL_3113495, EPI_ISL_3113502, EPI_ISL_3113522, EPI_ISL_3113550, EPI_ISL_3113568, EPI_ISL_3113578, EPI_ISL_3113579, EPI_ISL_3113608, EPI_ISL_3113617, EPI_ISL_3113622, EPI_ISL_3113681, EPI_ISL_3113726, EPI_ISL_3325548, EPI_ISL_3325577, EPI_ISL_3325611, EPI_ISL_3325649, EPI_ISL_3325732, EPI_ISL_3325734, EPI_ISL_3325864, EPI_ISL_3325895, EPI_ISL_3326058, EPI_ISL_3326343, EPI_ISL_3326376, EPI_ISL_3326389, EPI_ISL_3326419, EPI_ISL_3326531, EPI_ISL_3326546, EPI_ISL_3326561, EPI_ISL_3326562, EPI_ISL_3326570, EPI_ISL_3326572, EPI_ISL_3326649, EPI_ISL_3326658, EPI_ISL_3326664, EPI_ISL_3326671, EPI_ISL_3326736, EPI_ISL_3326907, EPI_ISL_3326911, EPI_ISL_3327047, EPI_ISL_3328025, EPI_ISL_3328026, EPI_ISL_3328088, EPI_ISL_3328532, EPI_ISL_3328657, EPI_ISL_3328756, EPI_ISL_3329154, EPI_ISL_3330638, EPI_ISL_3330657, EPI_ISL_3330704, EPI_ISL_3330706, EPI_ISL_4158934 | | | |
| see above | Laboratory Corporation of America | Centers for Disease Control and Prevention Division of Viral Diseases, Pathogen Discovery | Adrian Paskey; Amanda Douglas; Amanda Suchanek; Andrea Throop; Ayla Burns; Benjamin Rambo-Martin; Bobbi Croy; Brian Krueger; Brian Norvell; Christopher Gulvick; Christos Petropoulos; Clinton Paden; Clinton R. Paden; Craig Lukasik; Dakota Howard; Darlene Wagner; Debbie Boles; Dhwani Batra; Duncan MacCannell; Eyad Almasri; Goran Stevovic; Howard Engler; Hrushikesh Deshmukh; Jake Humphrey; Jana Schroth; Jason Caravas; Joe Voshell; John Pruitt; Jonathan Meltzer; Jonathan Williams; Kara Moser; Kimberly Wagner; Lax Iyer; Lisa Pfefferle; Lyndon Tilson; Manoj Jain; Marcia Eisenberg; Mary Ann Cristobal; Mary Cristobal; Mary Williamson; Matthew Robinson; Matthew Schmerer; Michael Levandoski; Mike Sapeta; Mindy Nye; Minoo Agarwal; Mohan Kolli; Nuthawin Charoensri; Oren Cohen; Peter Cook; Peter W. Cook; Prashant Gupta; Qian Zeng; Rama Ghatti; Scott Parker; Scott Ryan; Scott Sammons; Shatavia Morrison; Stanley Letovsky; Steven Ragan; Suresh Babu Selvaraju; Suresh Selvaraju; Susan Countryman; Susan Hicks; Suzanne Dale; Thomas Urban; Tim Kuphal; Tricia Zwiefelhofer; Vincent Drouillon; Yvette Unoarumhi |
| EPI_ISL_5160551 | Laboratory Medicine | Department of Laboratory Medicine, Lin-Kou Chang Gung Memorial Hospital, Taoyuan, Taiwan | Cheng-Hsun Chiu; Cheng-Ta Yang; Chung-Guei Huang; Guang-Wu Chen; Kuo-Chien Tsao; Kuo-Ming Lee; Mei-Jen Hsiao; Peng-Nien Huang; Po-Wei Huang; Shin-Ru Shih; Shu-Li Yang; Yi-Chun Liu; Yu-Nong Gong |
| EPI_ISL_3931694 | Laboratory of Microbiology and Virology, Ospedale Amedeo di Savoia, ASL "Città di Torino" | Laboratory of Microbiology and Virology, Ospedale Amedeo di Savoia, ASL "Città di Torino" | Debora Fiorito; Elisa Burdino; Francesco Cerutti; Gabriella Gregori; Maria Grazia Milia; Tiziano Allice; Valeria Ghisetti |
| EPI_ISL_3045423, EPI_ISL_3190168, EPI_ISL_3235267, EPI_ISL_3539998, EPI_ISL_3539999, EPI_ISL_3540000, EPI_ISL_4880265, EPI_ISL_4880266, EPI_ISL_6943989 | | | |
| see above | Laboratory of Respiratory Viruses and Measles, Oswaldo Cruz Institute, FIOCRUZ | Laboratory of Respiratory Viruses and Measles, Oswaldo Cruz Institute, FIOCRUZ | Agatha Cristinne Prudencio; Alice Sampaio Rocha; Ana Carolina Mendonca; Anna Carolina Paixao; Bruna Mendonça da Silva; Elisa Cavalcante Pereira; Fernando Motta; Ighor Arantes; Ighor Leonardo Arantes Gomes; Jéssica Graça Macedo de Carvalho; Larissa Macedo Pinto; Luciana Appolinario; Marilda Siqueira on behalf of the Fiocruz COVID-19 Genomic Surveillance Network; Paola Resende; Patricia Brasil; Renata Serrano Lopes; Taina Venas; Thais Aguiar; Victor Guimaraes |
| EPI_ISL_4600481, EPI_ISL_4600482 | Laboratório Central de Saúde Pública Profº Gonçalo Moniz | Coordenação Geral de Laboratórios de Saúde Pública (CGLAB/DAEVS/SVS/MS) | Vagner Fonseca; et al. |
| EPI_ISL_4600510, EPI_ISL_4600535 | Laboratório Central de Saúde Pública de Pernambuco | Coordenação Geral de Laboratórios de Saúde Pública (CGLAB/DAEVS/SVS/MS) | Vagner Fonseca; et al. |
| EPI_ISL_3050642, EPI_ISL_3050643, EPI_ISL_3050644, EPI_ISL_3050645, EPI_ISL_3050646, EPI_ISL_3050647, EPI_ISL_3050648, EPI_ISL_3050649, EPI_ISL_3050650, EPI_ISL_4520305, EPI_ISL_4520306, EPI_ISL_4520307, EPI_ISL_4520308, EPI_ISL_4520309, EPI_ISL_4520310, EPI_ISL_4520311, EPI_ISL_4520315, EPI_ISL_4520316, EPI_ISL_4520317, EPI_ISL_4520319, EPI_ISL_4520320, EPI_ISL_4520321, EPI_ISL_4520322, EPI_ISL_4520323, EPI_ISL_4520324, EPI_ISL_4520338, EPI_ISL_4520339, EPI_ISL_4520340, EPI_ISL_4520341, EPI_ISL_4520353, EPI_ISL_4520356, EPI_ISL_4520357, EPI_ISL_4520358, EPI_ISL_4520360, EPI_ISL_4520361, EPI_ISL_4520362, EPI_ISL_4520363, EPI_ISL_4520364, EPI_ISL_4520365, EPI_ISL_4520367, EPI_ISL_4520368, EPI_ISL_4520369, EPI_ISL_4520371, EPI_ISL_4520373, EPI_ISL_4520374, EPI_ISL_4520375, EPI_ISL_4520376, EPI_ISL_4520377, EPI_ISL_4520386, EPI_ISL_4520387, EPI_ISL_4520388, EPI_ISL_4520389, EPI_ISL_4520392, EPI_ISL_4520393, EPI_ISL_4520394, EPI_ISL_4520395, EPI_ISL_4520396, EPI_ISL_4520397, EPI_ISL_4520405, EPI_ISL_4520406, EPI_ISL_4520407, EPI_ISL_4520408, EPI_ISL_4520409, EPI_ISL_4520410, EPI_ISL_4520411, EPI_ISL_4520412, EPI_ISL_4520413, EPI_ISL_4520414, EPI_ISL_4520415, EPI_ISL_4520416, EPI_ISL_4520417, EPI_ISL_4520426, EPI_ISL_4520427, EPI_ISL_4520430, EPI_ISL_4520431, EPI_ISL_4520434, EPI_ISL_4520435, EPI_ISL_4520436 | | | |
| see above | Laboratório Central de Saúde Pública do Amazonas - LACEN-AM | Laboratorio de Ecologia de Doencas Transmissiveis na Amazonia, Instituto Leonidas e Maria Deane - Fiocruz Amazonia | André Corado; Felipe Naveca; Fernanda Nascimento; George Silva; Karina Pessoa; Luciana Gonçalves; Maria Júlia Brandão; Matilde Mejía; Valdinete Nascimento; Victor Souza; Ágatha Costa |
| EPI_ISL_7248142, EPI_ISL_7248428, EPI_ISL_7249088, EPI_ISL_7249880, EPI_ISL_7252071 | Laboratório Central de Saúde Pública do Estado do Tocantins (LACEN/TO) | Bioinformatics and Biotechnology Laboratory (Labinftec/Federal University of Tocantins, Campus of Gurupi) | Fabrício Souza Campos; Jucimária Dantas Galvão; Raíssa Nunes dos Santos; Ueric José Borges de Souza |
| EPI_ISL_5195464, EPI_ISL_5195478, EPI_ISL_5195481, EPI_ISL_5195485 | Laboratório Municipal de Biologia Molecular | Instituto René Rachou / Fiocruz Minas | André Menezes; Anna Salim; Eneida Oliveira; Gabriel Fernandes; Pedro Alves; Rubens do Monte Neto; Thaís Silva |
| EPI_ISL_5658382, EPI_ISL_5658389, EPI_ISL_5658390, EPI_ISL_5658396, EPI_ISL_5658401 | Laboratório Translacional, Centro de Pesquisa em Medicina Tropical, CEPEM/RO | Laboratório de Ecologia de Doenças Transmissíveis na Amazônia, Instituto Leônidas e Maria Deane - Fiocruz Amazônia | André Corado; Dhelio Pereira; Felipe Naveca on behalf of the Fiocruz COVID-19 Genomic Surveillance Network; Fernanda Nascimento; George Silva; Luciana Gonçalves; Maria Júlia Brandão; Matilde Mejía; Michelle Oliveira-Silva; Valdinete Nascimento; Victor Souza |

| | | | |
|---|---|---|---|
| EPI_ISL_3492499, EPI_ISL_3492505, EPI_ISL_4051961 | Laboratório de Microbiologia Molecular - Universidade FEEVALE | Molecular Microbiology Laboratory | Alana Witt Hansen; Fernando Rosado Spilki; Fágner Henrique Heldt; Juliana Schons Gulart; Juliana Schons Gularte; Juliane Deise Fleck; Mariana Soares da Silva; Matheus Nunes Weber; Meriane Demoliner; Michele Filippi.; Micheli Filippi; Paula Rodrigues de Almeida; Vyctoria Malayhka de Abreu Góes Pereira. |
| EPI_ISL_3190203, EPI_ISL_3190204, EPI_ISL_3190212, EPI_ISL_3190213, EPI_ISL_3190214, EPI_ISL_3190215, EPI_ISL_3190216, EPI_ISL_3190296 | | | |
| see above | Labortorio Central de Saude Publica do Estado do Rio de Janeiro (LACEN/RJ) | Laboratory of Respiratory Viruses and Measles, Oswaldo Cruz Institute, FIOCRUZ | Agatha Cristinne Prudencio Soares; Alice Sampaio Rocha; Ana Carolina Mendonca; Andrea Cony Cavalcanti; Anna Carolina Paixao; Elisa Cavalcante Pereira; Fernando Motta; Ighor Leonardo Arantes Gomes; Luciana Appolinario; Marilda Siqueira on behalf of the Fiocruz COVID-19 Genomic Surveillance Network; Paola Resende; Renata Serrano Lopes; Taina Venas |
| EPI_ISL_3044946 | Landesgesundheitsamt Baden-Württemberg | Robert Koch Institute | |
| EPI_ISL_3045051 | Limbach - MVZ Labor Ludwigsburg | Robert Koch Institute | |
| EPI_ISL_3099901 | MD PHL | MD PHL | Maryland Department of Health Laboratories Administration |
| EPI_ISL_3152288, EPI_ISL_3152291 | MD PHL | Maryland Department of Health Laboratories Administration | Ami Patel; Eric N. Keller; Jillian Loomis; Kwang Low; Terence L. Moore; and Robert Myers |
| EPI_ISL_2956711, EPI_ISL_3044167, EPI_ISL_3044168, EPI_ISL_3044171 | MVZ Labor Dr. Limbach & Kollegen GbR | Robert Koch Institute | |
| EPI_ISL_2957228 | MVZ für Laboratoriumsmedizin und Mikrobiologie Koblenz-Mittelrhein (Labor Koblenz) | Robert Koch Institute | |
| EPI_ISL_3065479, EPI_ISL_3065499, EPI_ISL_3110708, EPI_ISL_3110717, EPI_ISL_3110726, EPI_ISL_3110821, EPI_ISL_3110860, EPI_ISL_3110893, EPI_ISL_3110919, EPI_ISL_3110978 | | | |
| see above | Mako Medical | Centers for Disease Control and Prevention Division of Viral Diseases, Pathogen Discovery | Adrian Paskey; Benjamin Rambo-Martin; Christopher Gulvick; Clinton R. Paden; Dakota Howard; Darlene Wagner; Dhwani Batra; Duncan MacCannell; Jason Caravas; Kara Moser; Lauren Moon; Matthew Schmerer; Matthew Tugwell; Peter W. Cook; Scott Sammons; Shatavia Morrison; Yvette Unoarumhi |
| EPI_ISL_3276780, EPI_ISL_3276836 | Martin Luther King, Jr. Outpatient Center | Los Angeles County Public Health Laboratories | P. Hemarajata et al. |
| EPI_ISL_3435301, EPI_ISL_3435355, EPI_ISL_3435358, EPI_ISL_3435359 | Maryland Genomics, Institute for Genome Sciences, University of Maryland School of Medicine | Maryland Genomics, Institute for Genome Sciences, University of Maryland School of Medicine | Claire M; Fraser; Hazen; Holly; Humphrys; Ivette; Jacques; Jonathan; Kranthi; Lim; Lisa D; Luke J; Mike; Ott; Ravel; Roussey; Sadzewicz; Sandra; Santana-Cruz; Tallon; Tracy; Vavikolanu |
| EPI_ISL_2998619 | Microbiologia CATLAB | Can Ruti SARS-CoV-2 Sequencing Hub (HUGTiP/IrsiCaixa/IGTP) | Alba Sánchez; Alexia París; Anna Not; Antoni E Bordoy; Bonaventura Clotet; Cristina Casañ; David Panisello; Francesc Catala-Moll; Gemma Clara; Ignacio Blanco; Laia Soler; Lauro Sumoy; Marc Noguera-Julian; Maria Casadellà; Mariona Parera; Mercedes Guerrero; Montserrat Giménez; Pere-Joan Cardona; Pilar Armengol; Roger Paredes; Verónica Saludes; and Elisa Martró on behalf of the Can Ruti SARS-CoV-2 Sequencing Hub |
| EPI_ISL_3027975, EPI_ISL_3163990, EPI_ISL_3320201 | Microbiology Department, Laboratori Clínic Metropolitana Nord. Hospital Universitari Germans Trias i Pujol | Can Ruti SARS-CoV-2 Sequencing Hub (HUGTiP/IrsiCaixa/IGTP) | Alba Sánchez; Alexia París; Anna Not; Antoni E Bordoy; Bonaventura Clotet; Cristina Casañ; David Panisello; Francesc Catala-Moll; Gemma Clara; Ignacio Blanco; Laia Soler; Lauro Sumoy; Marc Noguera-Julian; Maria Casadellà; Mariona Parera; Mercedes Guerrero; Montserrat Giménez; Pere-Joan Cardona; Pilar Armengol; Roger Paredes; Verónica Saludes; and Elisa Martró on behalf of the Can Ruti SARS-CoV-2 Sequencing Hub |
| EPI_ISL_3316395 | Ministry of Health Turkey | Ministry of Health Turkey | Fatma Bayrakdar; Gülay Korukluoğlu; Süleyman Yalcin; Yasemin Coşgun |
| EPI_ISL_2958934, EPI_ISL_2958944 | Minnesota Department of Health, Public Health Laboratory | Minnesota Department of Health, Public Health Laboratory | Alexandra Lorentz; Jacob Garfin; Matt Plumb; and Xiong Wang |
| EPI_ISL_3161282 | Missouri State Public Health Laboratory | Missouri State Public Health Laboratory | Ashley New; Joshua Barry; Matthew Sinn |
| EPI_ISL_6174367 | Molecular Biology Laboratory, Vidant Medical Center | Brody Integrative Genomics Core, East Carolina University | Changhong Yin; Heather Duncan; John T. Fallon; Kimberly P. Briley; Weihua Huang |
| EPI_ISL_4688551 | Mon Health Medical Center | WVU and Marshall University Combined Genomics Core Facilities | James Denvir; Peter Perrotta; Peter Stoilov; Ryan Percifield; Wesley Kimble |
| EPI_ISL_2982696, EPI_ISL_2982701, EPI_ISL_2982709 | Montana Public Health Laboratory | Montana Public Health Laboratory | Carrie Biskupiak; Deborah Gibson; Joy Ritter; Michael Dills; Michelle Mozer |
| EPI_ISL_3242078 | ND Dept. of Health Laboratory Services-Microbiology | Centers for Disease Control and Prevention Division of Viral Diseases, Pathogen Discovery | Alex Burgin; Ben L. Rambo-Martin; Clinton R. Paden; Dakota Howard; Dave Wentworth; Dhwani Batra; Jasmine Padilla; Justin Lee; Krista Queen; Kristen Knipe; Kristine Lacek; Mark Burroughs; Matthew Schmerer; Meghan Bentz; Mili Sheth; Peter Cook; Sam Shepard; Sarah Nobles; Suxiang Tong; Vivien Dugan; Yvette Unoarumhi |
| EPI_ISL_4193683 | NJ Public Health and Environmental Laboratories | Centers for Disease Control and Prevention Division of Viral Diseases, Pathogen Discovery | Alex Burgin; Ben Rambo-Martin; Clinton Paden; Dakota Howard; Dave Wentworth; Dhwani Batra; Jasmine Padilla; Justin Lee; Krista Queen; Kristen Knipe; Kristine Lacek; Mark Burroughs; Matthew Schmerer; Meghan Bentz; Mili Sheth; Peter Cook; Sam Shepard; Sarah Nobles; Suxiang Tong; Vivien Dugan; Yvette Unoarumhi |
| EPI_ISL_3023739, EPI_ISL_3023754, EPI_ISL_3129978, EPI_ISL_3129992 | NJDOH, Public Health and Environmental Laboratories | NJ_PHEL | Allison Roder; Byeong Jeong; Chelsea San Filippo; Dana Woell; Jacquelyn Deverell; Lindsey Bodnar; Ryan Pachucki; Shiv K. Verma |
| EPI_ISL_5675198 | NS JARDIM MATILDE | Instituto Butantan | Antonio Jorge Martins; Claudia Renata dos Santos Barros; David Schlesinger; Debora Botequio Moretti; Dimas Tadeu Covas; Elaine Cristina Marqueze; Elaine Vieira Santos; Evandra Strazza Rodrigues; Heidge Fukumasu; Jayme Augusto de Souza-Neto; José Salvatore Leister Patané; Luiz Alcantara; Luiz Lehmann Coutinho; Maria Carolina Elias; Maurício Lacerda Nogueira; Rafael dos Santos Bezerra; Raul Machado Neto; Rejane Maria Tommasini Grotto; Ricardo Haddad; Sandra Coccuzzo Sampaio Vessoni; Simone Kashima; Svetoslav Nanev Slavov; Vincent Louis Viala |
| EPI_ISL_5672815, EPI_ISL_5672968 | NUCLEO DE SAUDE MARY DOTA DE BAURU | Instituto Butantan | Antonio Jorge Martins; Claudia Renata dos Santos Barros; David Schlesinger; Debora Botequio Moretti; Dimas Tadeu Covas; Elaine Cristina Marqueze; Elaine Vieira Santos; Evandra Strazza Rodrigues; Heidge Fukumasu; Jayme Augusto de Souza-Neto; José Salvatore Leister Patané; Luiz Alcantara; Luiz Lehmann Coutinho; Maria Carolina Elias; Maurício Lacerda Nogueira; Rafael dos Santos Bezerra; Raul Machado Neto; Rejane Maria Tommasini Grotto; Ricardo Haddad; Sandra Coccuzzo Sampaio Vessoni; Simone Kashima; Svetoslav Nanev Slavov; Vincent Louis Viala |
| EPI_ISL_5672616, EPI_ISL_5672693, EPI_ISL_5672705, EPI_ISL_5672714, EPI_ISL_5672746, EPI_ISL_5672755 | NUCLEO DE SAUDE VILA FALCAO DE BAURU | Instituto Butantan | Antonio Jorge Martins; Claudia Renata dos Santos Barros; David Schlesinger; Debora Botequio Moretti; Dimas Tadeu Covas; Elaine Cristina Marqueze; Elaine Vieira Santos; Evandra Strazza Rodrigues; Heidge Fukumasu; Jayme Augusto de Souza-Neto; José Salvatore Leister Patané; Luiz Alcantara; Luiz Lehmann Coutinho; Maria Carolina Elias; Maurício Lacerda Nogueira; Rafael dos Santos Bezerra; Raul Machado Neto; Rejane Maria Tommasini Grotto; Ricardo Haddad; Sandra Coccuzzo Sampaio Vessoni; Simone Kashima; Svetoslav Nanev Slavov; Vincent Louis Viala |
| EPI_ISL_5672580, EPI_ISL_5672585, EPI_ISL_5672590, EPI_ISL_5672596, EPI_ISL_5672624, EPI_ISL_5672628, EPI_ISL_5672635, EPI_ISL_5672644, EPI_ISL_5672661, EPI_ISL_5672664, EPI_ISL_5672678, EPI_ISL_5672710, EPI_ISL_5672723, EPI_ISL_5672895, EPI_ISL_5673007, EPI_ISL_5673156, EPI_ISL_5673185, EPI_ISL_5673215 | | | |
| see above | NUCLEO SAUDE III N HABITACIONAL PRESIDENTE GEISEL BAURU | Instituto Butantan | Antonio Jorge Martins; Claudia Renata dos Santos Barros; David Schlesinger; Debora Botequio Moretti; Dimas Tadeu Covas; Elaine Cristina Marqueze; Elaine Vieira Santos; Evandra Strazza Rodrigues; Heidge Fukumasu; Jayme Augusto de Souza-Neto; José Salvatore Leister Patané; Luiz Alcantara; Luiz Lehmann Coutinho; Maria Carolina Elias; Maurício Lacerda Nogueira; Rafael dos Santos Bezerra; Raul Machado Neto; Rejane Maria Tommasini Grotto; Ricardo Haddad; Sandra Coccuzzo Sampaio Vessoni; Simone Kashima; Svetoslav Nanev Slavov; Vincent Louis Viala |
| EPI_ISL_3134788, EPI_ISL_3134790, EPI_ISL_3134791, EPI_ISL_3134792, EPI_ISL_3134793, EPI_ISL_3134794, EPI_ISL_3134795, EPI_ISL_3134796, EPI_ISL_3134797, EPI_ISL_3134798, EPI_ISL_3134799, EPI_ISL_3134800, EPI_ISL_3134801, EPI_ISL_3134802, EPI_ISL_3134803, EPI_ISL_3134804, EPI_ISL_3134805, EPI_ISL_3134806, EPI_ISL_3134807, EPI_ISL_3134808, EPI_ISL_3134809, EPI_ISL_3134810, EPI_ISL_3134811, EPI_ISL_3134812, EPI_ISL_3134813, EPI_ISL_3134814, EPI_ISL_3447428, EPI_ISL_3447439, EPI_ISL_3447444, EPI_ISL_3447458, EPI_ISL_3447490, EPI_ISL_3447491, EPI_ISL_3447492, EPI_ISL_3447493, EPI_ISL_3447494, EPI_ISL_3447495, EPI_ISL_3447496, EPI_ISL_3447497, EPI_ISL_3447498, EPI_ISL_3447499, EPI_ISL_3447500, EPI_ISL_3447501, EPI_ISL_3447502, EPI_ISL_3447503, EPI_ISL_3447504, EPI_ISL_3447505, EPI_ISL_3447506, EPI_ISL_3447507, EPI_ISL_3447508, EPI_ISL_3447509, EPI_ISL_3447510, EPI_ISL_3447511, EPI_ISL_3447512, EPI_ISL_3447513, EPI_ISL_3447514, EPI_ISL_3447515, EPI_ISL_3447516, EPI_ISL_3447526, EPI_ISL_3447527, EPI_ISL_3447528, EPI_ISL_3447529, EPI_ISL_3447530, EPI_ISL_3447531, EPI_ISL_3447532, EPI_ISL_3447533, EPI_ISL_3447549, EPI_ISL_3447551, EPI_ISL_3447556, EPI_ISL_3447557, EPI_ISL_3447558, EPI_ISL_3447559, EPI_ISL_3447561, EPI_ISL_3447563, EPI_ISL_3447564, EPI_ISL_3447568, EPI_ISL_3447596, EPI_ISL_3703905, EPI_ISL_3703909, EPI_ISL_3703912, EPI_ISL_3703917, EPI_ISL_3703938, EPI_ISL_3703948, EPI_ISL_3703958, EPI_ISL_3703962, EPI_ISL_3703965, EPI_ISL_3703969, EPI_ISL_3703972, EPI_ISL_3703976, EPI_ISL_3703980, EPI_ISL_3703991, EPI_ISL_3704002, EPI_ISL_3704012, EPI_ISL_3704016, EPI_ISL_3704020, EPI_ISL_3704024, EPI_ISL_3704035, EPI_ISL_3704039, EPI_ISL_3704043, EPI_ISL_3704046, EPI_ISL_3704056, EPI_ISL_3704066, EPI_ISL_3704070, EPI_ISL_3704074, EPI_ISL_3704078, EPI_ISL_3704279, EPI_ISL_3704282, EPI_ISL_3704286 | | | |
| see above | NUPIT/UFPE | WallauLab on behalf of Fiocruz COVID-19 Genomic Surveillance Network | Alexandre Freitas da Silva; Cassia Docena; Constância Flávia Junqueira Ayres; Filipe Zimmer Dezordi; Gabriel Luz Wallau; Gustavo Barbosa de Lima; Lais Ceschini Machado; Lilian Caroliny Amorim Silva; Maira Galdino da Rocha Pitta; Marcelo Henrique dos Santos Paiva; Matheus Filgueira Bezerra; Michelly Cristiny Pereira; Rômulo Pessoa e Silva; Sinval Pinto Brandão Filho |
| EPI_ISL_3245519 | NYU Langone Health | Departments of Pathology and Medicine, New York University School of Medicine | Adriana Heguy; Christian Marier; Dacia Dimartino; Emily Guzman; Gael Westby; Guiqing Wang; Paolo Cotzia; Paul Zappile; Peter Meyn; Sitharam Ramaswami; Yutong Zhang |
| EPI_ISL_6814076, EPI_ISL_6814077 | National Platform bis COVID ULB-IBC | National Platform bis COVID ULB-IBC | Arnaud Marchant; Benoit Haerlingen; Coralie Henin; Marie-Luce Delforge; Ricardo De Mendonça |
| EPI_ISL_2966468, EPI_ISL_2966473, EPI_ISL_2966502 | Nebraska Public Health Laboratory | NPHL COVID-19 Response Team | NPHL COVID-19 Response Team |
| EPI_ISL_3534306 | Nebraska Public Health Laboratory | UNMC COVID-19 Response Team | UNMC COVID-19 Response Team |

| | | | |
|---|---|---|---|
| EPI_ISL_3021171, EPI_ISL_3021207, EPI_ISL_3372271, EPI_ISL_3372272 | Nevada State Public Health Laboratory | Nevada State Public Health Laboratory | Andrew Gorzalski; Mark Pandori |
| EPI_ISL_3086158, EPI_ISL_3127141, EPI_ISL_3127195 | New Mexico Department of Health Scientific Laboratory | New Mexico Department of Health Scientific Laboratory | Anastacia Griego-Fisher; D'eldra Malone; Ellie Johnson; Jennifer Benoit; Linda Salazar; Ratheesh Rajan; T. Mark Willmon |
| EPI_ISL_3105056, EPI_ISL_3105058 | Northwest Laboratory | Seattle Flu Study | Amanda Adler; Barry R. Lutz; Benjamin Pelle; Brian Hiatt; Caitlin R. Wolf; Chris D. Frazar; Deborah A. Nickerson; Elisabeth Brandstetter; Erica Ryke; Geoff Melly; Helen Y. Chu; Janet A. Englund; Jay Shendure; Jover Lee; Kairsten Fay; Kirsten Lacombe; Lea M. Starita; Mark J. Rieder; Matthew Richardson; Matthew Thompson; Melissa Truong; Michael Boeckh; Michael Famulare; Misja Ilcisin; Peter D. Han; Philip Dykema; Romesh Gautom; Scott Lindquist; Thomas R. Sibley; Trevor Bedford |
| EPI_ISL_3710095, EPI_ISL_3943699 | Olive View Medical Center | Los Angeles County Public Health Laboratories | P. Hemarajata et al. |
| EPI_ISL_3063046, EPI_ISL_3063059, EPI_ISL_3099977 | Orange County Public Health Laboratory | Orange County Public Health Laboratory | Angelica Torres; Julia Wolfe |
| EPI_ISL_2928905, EPI_ISL_3010589, EPI_ISL_3048118, EPI_ISL_3048120 | Oregon State Public Health Laboratory | Oregon State Public Health Laboratory | Eugene Yeboah; John Fontana and Shane Sevey; Laura Tsaknaridis; Rafia Razzaque; Vanda Makris |
| EPI_ISL_2985352, EPI_ISL_2985361 | Ospedale San Filippo Neri | Department of General Diagnostics; Department of Virology; Istituto Zooprofilattico Sperimentale del Lazio e della Toscana (IZSLT) | Alessia Franco; Antonella Cersini; Antonio Battisti.; Elena L. Diaconu; Fabiola Feltrin; Giuseppe Manna; Patricia Alba; Raffaela Conti; Teresa Scicluna; Virginia Carfora |
| EPI_ISL_3219521, EPI_ISL_3219522, EPI_ISL_3219523 | Outre mer | National Reference Center for Viruses of Respiratory Infections, Institut Pasteur, Paris | Angela Brisebarre; Camille Capel; Christophe Malabat; Corinne Maufrais; Dominique Rousset; Etienne Simon-Lorière; Frédéric Lemoine; Hub de Bioinformatique et Biostatistique; Louise Lefrançois; Marion Barbet; Maud Vanpeene; Méline Bizard; Sylvie Behillil; Sylvie Van der Werf; Vincent Enouf |
| EPI_ISL_5672826, EPI_ISL_5672858, EPI_ISL_5672931, EPI_ISL_5672954, EPI_ISL_5673051, EPI_ISL_5673143, EPI_ISL_5673162, EPI_ISL_5673193 | | | |
| see above | PAS JOAO ANTONIO DO NASCIMENTO | Instituto Butantan | Antonio Jorge Martins; Claudia Renata dos Santos Barros; David Schlesinger; Debora Botequio Moretti; Dimas Tadeu Covas; Elaine Cristina Marqueze; Elaine Vieira Santos; Evandra Strazza Rodrigues; Heidge Fukumasu; Jayme Augusto de Souza-Neto; José Salvatore Leister Patané; Luiz Alcantara; Luiz Lehmann Coutinho; Maria Carolina Elias; Maurício Lacerda Nogueira; Rafael dos Santos Bezerra; Raul Machado Neto; Rejane Maria Tommasini Grotto; Ricardo Haddad; Sandra Coccuzzo Sampaio Vessoni; Simone Kashima; Svetoslav Nanev Slavov; Vincent Louis Viala |
| EPI_ISL_5672770, EPI_ISL_5673131 | POLICLINICA ALCIA LINEIA VITTI REJANI | Instituto Butantan | Antonio Jorge Martins; Claudia Renata dos Santos Barros; David Schlesinger; Debora Botequio Moretti; Dimas Tadeu Covas; Elaine Cristina Marqueze; Elaine Vieira Santos; Evandra Strazza Rodrigues; Heidge Fukumasu; Jayme Augusto de Souza-Neto; José Salvatore Leister Patané; Luiz Alcantara; Luiz Lehmann Coutinho; Maria Carolina Elias; Maurício Lacerda Nogueira; Rafael dos Santos Bezerra; Raul Machado Neto; Rejane Maria Tommasini Grotto; Ricardo Haddad; Sandra Coccuzzo Sampaio Vessoni; Simone Kashima; Svetoslav Nanev Slavov; Vincent Louis Viala |
| EPI_ISL_5668487, EPI_ISL_5668503, EPI_ISL_5668843, EPI_ISL_5669233, EPI_ISL_5669258, EPI_ISL_5669466, EPI_ISL_5669583, EPI_ISL_5671557, EPI_ISL_5671636, EPI_ISL_5671802, EPI_ISL_5671881, EPI_ISL_5671901, EPI_ISL_5671972 | | | |
| see above | POLICLINICA COVID 19 ITAPETININGA | Instituto Butantan | Antonio Jorge Martins; Claudia Renata dos Santos Barros; David Schlesinger; Debora Botequio Moretti; Dimas Tadeu Covas; Elaine Cristina Marqueze; Elaine Vieira Santos; Evandra Strazza Rodrigues; Heidge Fukumasu; Jayme Augusto de Souza-Neto; José Salvatore Leister Patané; Luiz Alcantara; Luiz Lehmann Coutinho; Maria Carolina Elias; Maurício Lacerda Nogueira; Rafael dos Santos Bezerra; Raul Machado Neto; Rejane Maria Tommasini Grotto; Ricardo Haddad; Sandra Coccuzzo Sampaio Vessoni; Simone Kashima; Svetoslav Nanev Slavov; Vincent Louis Viala |
| EPI_ISL_5670225 | POLICLINICA JARDIM PARAISO | Instituto Butantan | Antonio Jorge Martins; Claudia Renata dos Santos Barros; David Schlesinger; Debora Botequio Moretti; Dimas Tadeu Covas; Elaine Cristina Marqueze; Elaine Vieira Santos; Evandra Strazza Rodrigues; Heidge Fukumasu; Jayme Augusto de Souza-Neto; José Salvatore Leister Patané; Luiz Alcantara; Luiz Lehmann Coutinho; Maria Carolina Elias; Maurício Lacerda Nogueira; Rafael dos Santos Bezerra; Raul Machado Neto; Rejane Maria Tommasini Grotto; Ricardo Haddad; Sandra Coccuzzo Sampaio Vessoni; Simone Kashima; Svetoslav Nanev Slavov; Vincent Louis Viala |
| EPI_ISL_5679466 | POLICLINICA MARIA DIRCE | Instituto Butantan | Antonio Jorge Martins; Claudia Renata dos Santos Barros; David Schlesinger; Debora Botequio Moretti; Dimas Tadeu Covas; Elaine Cristina Marqueze; Elaine Vieira Santos; Evandra Strazza Rodrigues; Heidge Fukumasu; Jayme Augusto de Souza-Neto; José Salvatore Leister Patané; Luiz Alcantara; Luiz Lehmann Coutinho; Maria Carolina Elias; Maurício Lacerda Nogueira; Rafael dos Santos Bezerra; Raul Machado Neto; Rejane Maria Tommasini Grotto; Ricardo Haddad; Sandra Coccuzzo Sampaio Vessoni; Simone Kashima; Svetoslav Nanev Slavov; Vincent Louis Viala |
| EPI_ISL_5676527 | PRONTO ATENDIMENTO MUNICIPAL ALUMINIO | Instituto Butantan | Antonio Jorge Martins; Claudia Renata dos Santos Barros; David Schlesinger; Debora Botequio Moretti; Dimas Tadeu Covas; Elaine Cristina Marqueze; Elaine Vieira Santos; Evandra Strazza Rodrigues; Heidge Fukumasu; Jayme Augusto de Souza-Neto; José Salvatore Leister Patané; Luiz Alcantara; Luiz Lehmann Coutinho; Maria Carolina Elias; Maurício Lacerda Nogueira; Rafael dos Santos Bezerra; Raul Machado Neto; Rejane Maria Tommasini Grotto; Ricardo Haddad; Sandra Coccuzzo Sampaio Vessoni; Simone Kashima; Svetoslav Nanev Slavov; Vincent Louis Viala |
| EPI_ISL_5670321 | PRONTO ATENDIMENTO MUNICIPAL DE JACUPIRANGA | Instituto Butantan | Antonio Jorge Martins; Claudia Renata dos Santos Barros; David Schlesinger; Debora Botequio Moretti; Dimas Tadeu Covas; Elaine Cristina Marqueze; Elaine Vieira Santos; Evandra Strazza Rodrigues; Heidge Fukumasu; Jayme Augusto de Souza-Neto; José Salvatore Leister Patané; Luiz Alcantara; Luiz Lehmann Coutinho; Maria Carolina Elias; Maurício Lacerda Nogueira; Rafael dos Santos Bezerra; Raul Machado Neto; Rejane Maria Tommasini Grotto; Ricardo Haddad; Sandra Coccuzzo Sampaio Vessoni; Simone Kashima; Svetoslav Nanev Slavov; Vincent Louis Viala |
| EPI_ISL_5668942, EPI_ISL_5669173, EPI_ISL_5669516, EPI_ISL_5669635, EPI_ISL_5670774 | PRONTO ATENDIMENTO MUNICIPAL ITALO SANTUCCI | Instituto Butantan | Antonio Jorge Martins; Claudia Renata dos Santos Barros; David Schlesinger; Debora Botequio Moretti; Dimas Tadeu Covas; Elaine Cristina Marqueze; Elaine Vieira Santos; Evandra Strazza Rodrigues; Heidge Fukumasu; Jayme Augusto de Souza-Neto; José Salvatore Leister Patané; Luiz Alcantara; Luiz Lehmann Coutinho; Maria Carolina Elias; Maurício Lacerda Nogueira; Rafael dos Santos Bezerra; Raul Machado Neto; Rejane Maria Tommasini Grotto; Ricardo Haddad; Sandra Coccuzzo Sampaio Vessoni; Simone Kashima; Svetoslav Nanev Slavov; Vincent Louis Viala |
| EPI_ISL_5670444 | PRONTO SOCORRO JARDIM JACIRA | Instituto Butantan | Antonio Jorge Martins; Claudia Renata dos Santos Barros; David Schlesinger; Debora Botequio Moretti; Dimas Tadeu Covas; Elaine Cristina Marqueze; Elaine Vieira Santos; Evandra Strazza Rodrigues; Heidge Fukumasu; Jayme Augusto de Souza-Neto; José Salvatore Leister Patané; Luiz Alcantara; Luiz Lehmann Coutinho; Maria Carolina Elias; Maurício Lacerda Nogueira; Rafael dos Santos Bezerra; Raul Machado Neto; Rejane Maria Tommasini Grotto; Ricardo Haddad; Sandra Coccuzzo Sampaio Vessoni; Simone Kashima; Svetoslav Nanev Slavov; Vincent Louis Viala |
| EPI_ISL_5669235, EPI_ISL_5670054 | PS E AMBULATORIO ENGENHEIRO CARDOSO | Instituto Butantan | Antonio Jorge Martins; Claudia Renata dos Santos Barros; David Schlesinger; Debora Botequio Moretti; Dimas Tadeu Covas; Elaine Cristina Marqueze; Elaine Vieira Santos; Evandra Strazza Rodrigues; Heidge Fukumasu; Jayme Augusto de Souza-Neto; José Salvatore Leister Patané; Luiz Alcantara; Luiz Lehmann Coutinho; Maria Carolina Elias; Maurício Lacerda Nogueira; Rafael dos Santos Bezerra; Raul Machado Neto; Rejane Maria Tommasini Grotto; Ricardo Haddad; Sandra Coccuzzo Sampaio Vessoni; Simone Kashima; Svetoslav Nanev Slavov; Vincent Louis Viala |
| EPI_ISL_5668551, EPI_ISL_5668721, EPI_ISL_5668743, EPI_ISL_5668807, EPI_ISL_5668853, EPI_ISL_5668873, EPI_ISL_5669159, EPI_ISL_5669277, EPI_ISL_5669536, EPI_ISL_5670769, EPI_ISL_5670790, EPI_ISL_5671508, EPI_ISL_5671643, EPI_ISL_5671737, EPI_ISL_5671819, EPI_ISL_5671841, EPI_ISL_5671962 | | | |
| see above | PSF DR ANTONIO PIRES DE ALMEIDA PORTO FELIZ | Instituto Butantan | Antonio Jorge Martins; Claudia Renata dos Santos Barros; David Schlesinger; Debora Botequio Moretti; Dimas Tadeu Covas; Elaine Cristina Marqueze; Elaine Vieira Santos; Evandra Strazza Rodrigues; Heidge Fukumasu; Jayme Augusto de Souza-Neto; José Salvatore Leister Patané; Luiz Alcantara; Luiz Lehmann Coutinho; Maria Carolina Elias; Maurício Lacerda Nogueira; Rafael dos Santos Bezerra; Raul Machado Neto; Rejane Maria Tommasini Grotto; Ricardo Haddad; Sandra Coccuzzo Sampaio Vessoni; Simone Kashima; Svetoslav Nanev Slavov; Vincent Louis Viala |
| EPI_ISL_5673522 | PSF III | Instituto Butantan | Antonio Jorge Martins; Claudia Renata dos Santos Barros; David Schlesinger; Debora Botequio Moretti; Dimas Tadeu Covas; Elaine Cristina Marqueze; Elaine Vieira Santos; Evandra Strazza Rodrigues; Heidge Fukumasu; Jayme Augusto de Souza-Neto; José Salvatore Leister Patané; Luiz Alcantara; Luiz Lehmann Coutinho; Maria Carolina Elias; Maurício Lacerda Nogueira; Rafael dos Santos Bezerra; Raul Machado Neto; Rejane Maria Tommasini Grotto; Ricardo Haddad; Sandra Coccuzzo Sampaio Vessoni; Simone Kashima; Svetoslav Nanev Slavov; Vincent Louis Viala |
| EPI_ISL_2923945, EPI_ISL_3010632, EPI_ISL_3010646, EPI_ISL_3010648, EPI_ISL_3010649, EPI_ISL_3010653, EPI_ISL_3010670, EPI_ISL_3010678, EPI_ISL_3010693, EPI_ISL_3010754, EPI_ISL_3010757 | | | |
| see above | Pandemic Response Lab - NYC | Pandemic Response Lab, R&D | Cybill del Castillo; Dylan Law; Haiping Hao; Henry Lee; Isabel Fernandez Escapa; Jon Laurent; Melissa Hopkins; Michael Hammerling; Pradeep Bugga; Shinyoung Clair Kang; Sol Rey; William Ward |
| EPI_ISL_3371092 | Pennsylvania Department of Health Bureau of Laboratories | Pennsylvania Department of Health Bureau of Laboratories | Dongxiang Xia |
| EPI_ISL_2925556 | Pertini | National Institute for Infectious Diseases (INMI) L. Spallanzani I.R.C.C.S | A Di Caro; B Bartolini; CEM Gruber; E Giombini; F Messina; F Santini; G Bonfiglio; M Rueca; MR Capobianchi; O Butera |
| EPI_ISL_6907545, EPI_ISL_6907550, EPI_ISL_6907551, EPI_ISL_6908039, EPI_ISL_6908045, EPI_ISL_6908049, EPI_ISL_6908057 | | | |
| see above | Plataforma de Vigilancia Molecular (PVM) - FIOCRUZ/BA | Plataforma de Vigilancia Molecular (PVM) - FIOCRUZ/BA | Bruno Bezerril Andrade; Camila I. de Oliveira; Clarissa Araújo Gurgel; Eduardo Oyama; Francys A Rangel; Laila A da Silva; Laise de Moraes; Leonardo Paiva Farias; Marina Cucco; Pedro Silveira; Ricardo Khouri on behalf of the Fiocruz COVID-19 Genomic Surveillance Network.; Tiago Graf |
| EPI_ISL_3391110 | Plateau technique Biomer Metz Verriers | Department of Virology, Henri Mondor University Hospital, Assistance Publique Hôpitaux de Paris, Université Paris-Est Créteil, INSERM U955 | Alexandre Soulier; Christophe Rodriguez; Elisabeth Trawinski; Guillaume Gricourt; Jean-Michel Pawlotsky; Melissa N'Debi; Slim Fourati; Vanessa Demontant |
| EPI_ISL_2932694 | Plateforme de testing Namuroise | Plateforme de testing Namuroise | Degosserie Jonathan; Demars Aurore; Denis Olivier; Lesly Nyinkeu Kemamen; Maschietto Céline; Mullier François; Nicolas Gilliard; Nobis Chloé; Otto Gaetan |
| EPI_ISL_2988282, EPI_ISL_3008658, EPI_ISL_3008660 | Platform BIS UZA/UAntwerpen | Labo Klinische Biologie, UZA | Basil Britto Xavier; Christine Lammens; Herman Goossens; Ines Verbesselt; Jasmine Coppens; Kathleen Holemans; Marie Le Mercier; Veerle Matheeussen |
| EPI_ISL_2925711 | Policlinico Tor Vergata | National Institute for Infectious Diseases (INMI) L. Spallanzani I.R.C.C.S | A Di Caro; B Bartolini; CEM Gruber; E Giombini; F Messina; F Santini; G Bonfiglio; M Rueca; MR Capobianchi; O Butera |
| EPI_ISL_3127425 | Presidio Ospedaliero "Madonna delle Grazie" di Matera | ASM - P.O. Madonna delle Grazie - Matera | Massimo Dell'Edera |
| EPI_ISL_3276769 | Providence Saint John's Health Center | Los Angeles County Public Health Laboratories | P. Hemarajata et al. |
| EPI_ISL_3214755 | Quest Diagnostics | Minnesota Department of Health, Public Health Laboratory | Alexandra Lorentz; Jacob Garfin; Matt Plumb; and Xiong Wang |
| EPI_ISL_3303838, EPI_ISL_3303839, EPI_ISL_3303840, EPI_ISL_3303841, EPI_ISL_3303843, EPI_ISL_3303844, EPI_ISL_3303845, EPI_ISL_3303846, EPI_ISL_3303847, EPI_ISL_3303848, EPI_ISL_3303852, EPI_ISL_3303856, EPI_ISL_3303860, EPI_ISL_3303862, EPI_ISL_3303869, EPI_ISL_3303871, EPI_ISL_3303872, EPI_ISL_3303874, EPI_ISL_3303877, EPI_ISL_3303878, EPI_ISL_3303881, EPI_ISL_3303882, EPI_ISL_3303884, EPI_ISL_3303886, EPI_ISL_3303888, EPI_ISL_3303889, EPI_ISL_3303890, EPI_ISL_3303891, EPI_ISL_3303892, EPI_ISL_3303893, EPI_ISL_3303894, EPI_ISL_3303895, EPI_ISL_3303896, EPI_ISL_3303897, EPI_ISL_3303898, EPI_ISL_3303899, EPI_ISL_3303900, EPI_ISL_3303902, EPI_ISL_3303903, EPI_ISL_3303904, EPI_ISL_3303905, EPI_ISL_3303906, EPI_ISL_3303907, EPI_ISL_3303908, EPI_ISL_3303909, EPI_ISL_3303910, EPI_ISL_3303912, EPI_ISL_3303913, EPI_ISL_3303914, EPI_ISL_3303915, EPI_ISL_3303916, EPI_ISL_3303917, EPI_ISL_3303918, EPI_ISL_3303922, EPI_ISL_3303924, EPI_ISL_3303925, EPI_ISL_4196476, EPI_ISL_4196477, EPI_ISL_4196480, EPI_ISL_4196483, EPI_ISL_4196484, EPI_ISL_4196492, EPI_ISL_4196497, EPI_ISL_4196505, EPI_ISL_4196511, EPI_ISL_4196516, EPI_ISL_4196517, EPI_ISL_4196518, EPI_ISL_4196527, EPI_ISL_4196529, EPI_ISL_4196532, EPI_ISL_4196537, EPI_ISL_4196539, EPI_ISL_4196543, EPI_ISL_4196548, EPI_ISL_4196550, EPI_ISL_4196556, EPI_ISL_4196559, EPI_ISL_4196566, EPI_ISL_4196567, EPI_ISL_4196569, EPI_ISL_4196572, EPI_ISL_4196575, EPI_ISL_4196579, EPI_ISL_4196593, EPI_ISL_4196601, EPI_ISL_4196602 | | | |
| see above | Quest Diagnostics | Centers for Disease Control and | A. Gerasimova; A. Perez; Adrian Paskey; B. Anderson; Benjamin Rambo-Martin; Christopher Gulvick; Clinton Paden; Clinton R. Paden; Dakota Howard; Darlene Wagner; Dhwani Batra; Duncan MacCannell; Erisa Sula; F. Lacbawan; I. A. Shlyakhter; I. Shlyakhter; Jason Caravas; K. Livingston; K.E. Livingston; Kara Moser; |

| | | | |
|---|---|---|---|
| | Incorporated | Prevention Division of Viral Diseases, Pathogen Discovery | Kristine Lacek; L. Bernstein; L.E. Bernstein; M. Hua; Matthew Schmerer; P. Tanpaiboon; Peter Cook; Peter W. Cook; R. Kagan; R. M. Kagan; R. Owen; R. Rolando; R. V. Rolando; S. H. Rosenthal; S. Rosenthal; Scott Sammons; Shatavia Morrison; Tymeckia Kendall; Victoria Caban Figueroa; Y. Liu; Yvette Unoarumhi |
| EPI_ISL_3004254 | Respiratory Virus Unit, Microbiology Services Colindale, Public Health England | COVID-19 Genomics UK (COG-UK) Consortium | PHE Covid Sequencing Team |
| EPI_ISL_5669165 | SAE SERVICO DE ASSISTENCIA ESPECIALIZADA M I ITAPEVA | Instituto Butantan | Antonio Jorge Martins; Claudia Renata dos Santos Barros; David Schlesinger; Debora Botequio Moretti; Dimas Tadeu Covas; Elaine Cristina Marqueze; Elaine Vieira Santos; Evandra Strazza Rodrigues; Heidge Fukumasu; Jayme Augusto de Souza-Neto; José Salvatore Leister Patané; Luiz Alcantara; Luiz Lehmann Coutinho; Maria Carolina Elias; Maurício Lacerda Nogueira; Rafael dos Santos Bezerra; Raul Machado Neto; Rejane Maria Tommasini Grotto; Ricardo Haddad; Sandra Coccuzzo Sampaio Vessoni; Simone Kashima; Svetoslav Nanev Slavov; Vincent Louis Viala |
| EPI_ISL_5673487 | SAE SERVICO DE ATENDIMENTO ESPECIALIZADO | Instituto Butantan | Antonio Jorge Martins; Claudia Renata dos Santos Barros; David Schlesinger; Debora Botequio Moretti; Dimas Tadeu Covas; Elaine Cristina Marqueze; Elaine Vieira Santos; Evandra Strazza Rodrigues; Heidge Fukumasu; Jayme Augusto de Souza-Neto; José Salvatore Leister Patané; Luiz Alcantara; Luiz Lehmann Coutinho; Maria Carolina Elias; Maurício Lacerda Nogueira; Rafael dos Santos Bezerra; Raul Machado Neto; Rejane Maria Tommasini Grotto; Ricardo Haddad; Sandra Coccuzzo Sampaio Vessoni; Simone Kashima; Svetoslav Nanev Slavov; Vincent Louis Viala |
| EPI_ISL_5651170 | SANTA CASA DE MISERICORDIA DE SAO JOSE DOS CAMPOS | Instituto Butantan | Antonio Jorge Martins; Claudia Renata dos Santos Barros; David Schlesinger; Debora Botequio Moretti; Dimas Tadeu Covas; Elaine Cristina Marqueze; Elaine Vieira Santos; Evandra Strazza Rodrigues; Heidge Fukumasu; Jayme Augusto de Souza-Neto; José Salvatore Leister Patané; Luiz Alcantara; Luiz Lehmann Coutinho; Maria Carolina Elias; Maurício Lacerda Nogueira; Rafael dos Santos Bezerra; Raul Machado Neto; Rejane Maria Tommasini Grotto; Ricardo Haddad; Sandra Coccuzzo Sampaio Vessoni; Simone Kashima; Svetoslav Nanev Slavov; Vincent Louis Viala |
| EPI_ISL_5671671 | SANTA CASA DE MISERICORDIA DE TIETE | Instituto Butantan | Antonio Jorge Martins; Claudia Renata dos Santos Barros; David Schlesinger; Debora Botequio Moretti; Dimas Tadeu Covas; Elaine Cristina Marqueze; Elaine Vieira Santos; Evandra Strazza Rodrigues; Heidge Fukumasu; Jayme Augusto de Souza-Neto; José Salvatore Leister Patané; Luiz Alcantara; Luiz Lehmann Coutinho; Maria Carolina Elias; Maurício Lacerda Nogueira; Rafael dos Santos Bezerra; Raul Machado Neto; Rejane Maria Tommasini Grotto; Ricardo Haddad; Sandra Coccuzzo Sampaio Vessoni; Simone Kashima; Svetoslav Nanev Slavov; Vincent Louis Viala |
| EPI_ISL_5668885, EPI_ISL_5671681, EPI_ISL_5675744, EPI_ISL_5677076 | SANTA CASA DE PORTO FELIZ | Instituto Butantan | Antonio Jorge Martins; Claudia Renata dos Santos Barros; David Schlesinger; Debora Botequio Moretti; Dimas Tadeu Covas; Elaine Cristina Marqueze; Elaine Vieira Santos; Evandra Strazza Rodrigues; Heidge Fukumasu; Jayme Augusto de Souza-Neto; José Salvatore Leister Patané; Luiz Alcantara; Luiz Lehmann Coutinho; Maria Carolina Elias; Maurício Lacerda Nogueira; Rafael dos Santos Bezerra; Raul Machado Neto; Rejane Maria Tommasini Grotto; Ricardo Haddad; Sandra Coccuzzo Sampaio Vessoni; Simone Kashima; Svetoslav Nanev Slavov; Vincent Louis Viala |
| EPI_ISL_5674152 | SANTA CASA MISER STACRUZ RIO PARDO | Instituto Butantan | Antonio Jorge Martins; Claudia Renata dos Santos Barros; David Schlesinger; Debora Botequio Moretti; Dimas Tadeu Covas; Elaine Cristina Marqueze; Elaine Vieira Santos; Evandra Strazza Rodrigues; Heidge Fukumasu; Jayme Augusto de Souza-Neto; José Salvatore Leister Patané; Luiz Alcantara; Luiz Lehmann Coutinho; Maria Carolina Elias; Maurício Lacerda Nogueira; Rafael dos Santos Bezerra; Raul Machado Neto; Rejane Maria Tommasini Grotto; Ricardo Haddad; Sandra Coccuzzo Sampaio Vessoni; Simone Kashima; Svetoslav Nanev Slavov; Vincent Louis Viala |
| EPI_ISL_3587525 | SARS-CoV-2 Sequencing Castilla y Leon-Spain Consortium | SARS-CoV-2 Sequencing Castilla y Leon-Spain Consortium | Antonio Orduña-Domingo; Carlos Fuster Foz; Carmen Aldea-Mansilla; Carmen Gimeno Crespo; David Abad; Gabriel March Rosello; Gregoria Megías Lobón; Jose María Eiros Bouza; M. Isabel Fernandez-Natal; Marta Dominguez-Gil; Marta Hernandez; María Antonia García Castro; Mª Fe Brezmes-Valdivieso; Noelia Arenal Andrés; Silvia Rojo; Sonsoles Garcinuño Pérez |
| EPI_ISL_3130809 | SCM B12 | CNR Virus des Infections Respiratoires - France SUD | Antonin Bal; Bruno Lina; Gregory Destras; Gwendolyne Burfin; Hadrien Regue; Laurence Josset; Martine Valette; Quentin Semanas |
| EPI_ISL_5675859, EPI_ISL_5675866, EPI_ISL_5675920, EPI_ISL_5677010, EPI_ISL_5677423, EPI_ISL_5677440, EPI_ISL_5677674 | | | |
| see above | SECAO CENTRO DE DIAGNOSTICO SECEDI | Instituto Butantan | Antonio Jorge Martins; Claudia Renata dos Santos Barros; David Schlesinger; Debora Botequio Moretti; Dimas Tadeu Covas; Elaine Cristina Marqueze; Elaine Vieira Santos; Evandra Strazza Rodrigues; Heidge Fukumasu; Jayme Augusto de Souza-Neto; José Salvatore Leister Patané; Luiz Alcantara; Luiz Lehmann Coutinho; Maria Carolina Elias; Maurício Lacerda Nogueira; Rafael dos Santos Bezerra; Raul Machado Neto; Rejane Maria Tommasini Grotto; Ricardo Haddad; Sandra Coccuzzo Sampaio Vessoni; Simone Kashima; Svetoslav Nanev Slavov; Vincent Louis Viala |
| EPI_ISL_5669667, EPI_ISL_5670085, EPI_ISL_5670480, EPI_ISL_5670861 | SECRETARIA DA SAUDE DE MAIRIPORA | Instituto Butantan | Antonio Jorge Martins; Claudia Renata dos Santos Barros; David Schlesinger; Debora Botequio Moretti; Dimas Tadeu Covas; Elaine Cristina Marqueze; Elaine Vieira Santos; Evandra Strazza Rodrigues; Heidge Fukumasu; Jayme Augusto de Souza-Neto; José Salvatore Leister Patané; Luiz Alcantara; Luiz Lehmann Coutinho; Maria Carolina Elias; Maurício Lacerda Nogueira; Rafael dos Santos Bezerra; Raul Machado Neto; Rejane Maria Tommasini Grotto; Ricardo Haddad; Sandra Coccuzzo Sampaio Vessoni; Simone Kashima; Svetoslav Nanev Slavov; Vincent Louis Viala |
| EPI_ISL_5680432 | SECRETARIA DE SAUDE DE SANTA GERTRUDES | Instituto Butantan | Antonio Jorge Martins; Claudia Renata dos Santos Barros; David Schlesinger; Debora Botequio Moretti; Dimas Tadeu Covas; Elaine Cristina Marqueze; Elaine Vieira Santos; Evandra Strazza Rodrigues; Heidge Fukumasu; Jayme Augusto de Souza-Neto; José Salvatore Leister Patané; Luiz Alcantara; Luiz Lehmann Coutinho; Maria Carolina Elias; Maurício Lacerda Nogueira; Rafael dos Santos Bezerra; Raul Machado Neto; Rejane Maria Tommasini Grotto; Ricardo Haddad; Sandra Coccuzzo Sampaio Vessoni; Simone Kashima; Svetoslav Nanev Slavov; Vincent Louis Viala |
| EPI_ISL_5671058, EPI_ISL_5671344 | SECRETARIA DE SAUDE DE SAO PEDRO | Instituto Butantan | Antonio Jorge Martins; Claudia Renata dos Santos Barros; David Schlesinger; Debora Botequio Moretti; Dimas Tadeu Covas; Elaine Cristina Marqueze; Elaine Vieira Santos; Evandra Strazza Rodrigues; Heidge Fukumasu; Jayme Augusto de Souza-Neto; José Salvatore Leister Patané; Luiz Alcantara; Luiz Lehmann Coutinho; Maria Carolina Elias; Maurício Lacerda Nogueira; Rafael dos Santos Bezerra; Raul Machado Neto; Rejane Maria Tommasini Grotto; Ricardo Haddad; Sandra Coccuzzo Sampaio Vessoni; Simone Kashima; Svetoslav Nanev Slavov; Vincent Louis Viala |
| EPI_ISL_5676487, EPI_ISL_5677867, EPI_ISL_5678140 | SECRETARIA DE SAUDE PUBLICA DE PRAIA GRANDE | Instituto Butantan | Antonio Jorge Martins; Claudia Renata dos Santos Barros; David Schlesinger; Debora Botequio Moretti; Dimas Tadeu Covas; Elaine Cristina Marqueze; Elaine Vieira Santos; Evandra Strazza Rodrigues; Heidge Fukumasu; Jayme Augusto de Souza-Neto; José Salvatore Leister Patané; Luiz Alcantara; Luiz Lehmann Coutinho; Maria Carolina Elias; Maurício Lacerda Nogueira; Rafael dos Santos Bezerra; Raul Machado Neto; Rejane Maria Tommasini Grotto; Ricardo Haddad; Sandra Coccuzzo Sampaio Vessoni; Simone Kashima; Svetoslav Nanev Slavov; Vincent Louis Viala |
| EPI_ISL_5668656, EPI_ISL_5668837 | SECRETARIA MUNICIPAL DA SAUDE DE ITAPEVA SP | Instituto Butantan | Antonio Jorge Martins; Claudia Renata dos Santos Barros; David Schlesinger; Debora Botequio Moretti; Dimas Tadeu Covas; Elaine Cristina Marqueze; Elaine Vieira Santos; Evandra Strazza Rodrigues; Heidge Fukumasu; Jayme Augusto de Souza-Neto; José Salvatore Leister Patané; Luiz Alcantara; Luiz Lehmann Coutinho; Maria Carolina Elias; Maurício Lacerda Nogueira; Rafael dos Santos Bezerra; Raul Machado Neto; Rejane Maria Tommasini Grotto; Ricardo Haddad; Sandra Coccuzzo Sampaio Vessoni; Simone Kashima; Svetoslav Nanev Slavov; Vincent Louis Viala |
| EPI_ISL_5671539 | SECRETARIA MUNICIPAL DA SAUDE DE ITARARE | Instituto Butantan | Antonio Jorge Martins; Claudia Renata dos Santos Barros; David Schlesinger; Debora Botequio Moretti; Dimas Tadeu Covas; Elaine Cristina Marqueze; Elaine Vieira Santos; Evandra Strazza Rodrigues; Heidge Fukumasu; Jayme Augusto de Souza-Neto; José Salvatore Leister Patané; Luiz Alcantara; Luiz Lehmann Coutinho; Maria Carolina Elias; Maurício Lacerda Nogueira; Rafael dos Santos Bezerra; Raul Machado Neto; Rejane Maria Tommasini Grotto; Ricardo Haddad; Sandra Coccuzzo Sampaio Vessoni; Simone Kashima; Svetoslav Nanev Slavov; Vincent Louis Viala |
| EPI_ISL_5674328, EPI_ISL_5674632 | SECRETARIA MUNICIPAL DE SAUDE DE ANDRADINA | Instituto Butantan | Antonio Jorge Martins; Claudia Renata dos Santos Barros; David Schlesinger; Debora Botequio Moretti; Dimas Tadeu Covas; Elaine Cristina Marqueze; Elaine Vieira Santos; Evandra Strazza Rodrigues; Heidge Fukumasu; Jayme Augusto de Souza-Neto; José Salvatore Leister Patané; Luiz Alcantara; Luiz Lehmann Coutinho; Maria Carolina Elias; Maurício Lacerda Nogueira; Rafael dos Santos Bezerra; Raul Machado Neto; Rejane Maria Tommasini Grotto; Ricardo Haddad; Sandra Coccuzzo Sampaio Vessoni; Simone Kashima; Svetoslav Nanev Slavov; Vincent Louis Viala |
| EPI_ISL_5671052, EPI_ISL_5680535, EPI_ISL_5681025 | SECRETARIA MUNICIPAL DE SAUDE DE CORDEIROPOLIS | Instituto Butantan | Antonio Jorge Martins; Claudia Renata dos Santos Barros; David Schlesinger; Debora Botequio Moretti; Dimas Tadeu Covas; Elaine Cristina Marqueze; Elaine Vieira Santos; Evandra Strazza Rodrigues; Heidge Fukumasu; Jayme Augusto de Souza-Neto; José Salvatore Leister Patané; Luiz Alcantara; Luiz Lehmann Coutinho; Maria Carolina Elias; Maurício Lacerda Nogueira; Rafael dos Santos Bezerra; Raul Machado Neto; Rejane Maria Tommasini Grotto; Ricardo Haddad; Sandra Coccuzzo Sampaio Vessoni; Simone Kashima; Svetoslav Nanev Slavov; Vincent Louis Viala |
| EPI_ISL_5672997, EPI_ISL_5673036, EPI_ISL_5673069, EPI_ISL_5673168, EPI_ISL_5673203 | SECRETARIA MUNICIPAL DE SAUDE DE JAU | Instituto Butantan | Antonio Jorge Martins; Claudia Renata dos Santos Barros; David Schlesinger; Debora Botequio Moretti; Dimas Tadeu Covas; Elaine Cristina Marqueze; Elaine Vieira Santos; Evandra Strazza Rodrigues; Heidge Fukumasu; Jayme Augusto de Souza-Neto; José Salvatore Leister Patané; Luiz Alcantara; Luiz Lehmann Coutinho; Maria Carolina Elias; Maurício Lacerda Nogueira; Rafael dos Santos Bezerra; Raul Machado Neto; Rejane Maria Tommasini Grotto; Ricardo Haddad; Sandra Coccuzzo Sampaio Vessoni; Simone Kashima; Svetoslav Nanev Slavov; Vincent Louis Viala |
| EPI_ISL_3912072, EPI_ISL_3912105 | SECRETARIA MUNICIPAL DE SAUDE DE TIANGUA | Analytical Competence Molecular Epidemiology Lab/ACME, Oswaldo Cruz Foundation, Ceara (FIOCRUZ CE) | Cleber Furtado Aksenen; Fabio Miyajima; Fernando Braga Stehling; Francisco Eder de Moura Lopes; Jamille Maria Mendes Bezerra; Joaquim Cesar do Nascimento Sousa Junior; Pedro Miguel Carneiro Jeronimo; Suzana Porto Almeida & Lucas Delerino on behalf of COVID-19 FIOCRUZ Genomic Network; Thais Ferreira de Oliveira; Thais de Oliveira Costa; Ticiane Cavalcante de Souza; Veridiana Pessoa Miyajima |
| EPI_ISL_5672849, EPI_ISL_5672874, EPI_ISL_5672902, EPI_ISL_5672949 | SECRETARIA MUNICIPAL DE SAUDE DE TUPI PAULISTA | Instituto Butantan | Antonio Jorge Martins; Claudia Renata dos Santos Barros; David Schlesinger; Debora Botequio Moretti; Dimas Tadeu Covas; Elaine Cristina Marqueze; Elaine Vieira Santos; Evandra Strazza Rodrigues; Heidge Fukumasu; Jayme Augusto de Souza-Neto; José Salvatore Leister Patané; Luiz Alcantara; Luiz Lehmann Coutinho; Maria Carolina Elias; Maurício Lacerda Nogueira; Rafael dos Santos Bezerra; Raul Machado Neto; Rejane Maria Tommasini Grotto; Ricardo Haddad; Sandra Coccuzzo Sampaio Vessoni; Simone Kashima; Svetoslav Nanev Slavov; Vincent Louis Viala |
| EPI_ISL_5671077 | SECRETARIA MUNICIPAL DE SAUDE DE VALPARAISO SP | Instituto Butantan | Antonio Jorge Martins; Claudia Renata dos Santos Barros; David Schlesinger; Debora Botequio Moretti; Dimas Tadeu Covas; Elaine Cristina Marqueze; Elaine Vieira Santos; Evandra Strazza Rodrigues; Heidge Fukumasu; Jayme Augusto de Souza-Neto; José Salvatore Leister Patané; Luiz Alcantara; Luiz Lehmann Coutinho; Maria Carolina Elias; Maurício Lacerda Nogueira; Rafael dos Santos Bezerra; Raul Machado Neto; Rejane Maria Tommasini Grotto; Ricardo Haddad; Sandra Coccuzzo Sampaio Vessoni; Simone Kashima; Svetoslav Nanev Slavov; Vincent Louis Viala |
| EPI_ISL_5668547, EPI_ISL_5668599, EPI_ISL_5668611, EPI_ISL_5668648, EPI_ISL_5668674, EPI_ISL_5668730, EPI_ISL_5668793, EPI_ISL_5668801, EPI_ISL_5668815, EPI_ISL_5668824, EPI_ISL_5668849, EPI_ISL_5668880, EPI_ISL_5668890, EPI_ISL_5668894, EPI_ISL_5668906, EPI_ISL_5668914, EPI_ISL_5668952, EPI_ISL_5668980, EPI_ISL_5669099, EPI_ISL_5669213, EPI_ISL_5669293, EPI_ISL_5669299, EPI_ISL_5669355, EPI_ISL_5669392, EPI_ISL_5669476, EPI_ISL_5669602, EPI_ISL_5669625, EPI_ISL_5669674, EPI_ISL_5669721, EPI_ISL_5669833, EPI_ISL_5671494, EPI_ISL_5671500, EPI_ISL_5671516, EPI_ISL_5671523, EPI_ISL_5671529, EPI_ISL_5671532, EPI_ISL_5671541, EPI_ISL_5671569, EPI_ISL_5671571, EPI_ISL_5671578, EPI_ISL_5671590, EPI_ISL_5671630, EPI_ISL_5671642, EPI_ISL_5671649, EPI_ISL_5671656, EPI_ISL_5671664, EPI_ISL_5671709, EPI_ISL_5671728, EPI_ISL_5671760, EPI_ISL_5671775, EPI_ISL_5671779, EPI_ISL_5671796, EPI_ISL_5671825, EPI_ISL_5671834, EPI_ISL_5671850, EPI_ISL_5671855, EPI_ISL_5671872, EPI_ISL_5671887, EPI_ISL_5671922, EPI_ISL_5671929, EPI_ISL_5671938, EPI_ISL_5671944, EPI_ISL_5671968, EPI_ISL_5671984, EPI_ISL_5671986, EPI_ISL_5672602, EPI_ISL_5672607, EPI_ISL_5672619, EPI_ISL_5672647, EPI_ISL_5672659, EPI_ISL_5672680, EPI_ISL_5672687, EPI_ISL_5672716, EPI_ISL_5672764, EPI_ISL_5675681, EPI_ISL_5675701, EPI_ISL_5675721, EPI_ISL_5675816, EPI_ISL_5675833, EPI_ISL_5675975, EPI_ISL_5676053, EPI_ISL_5676079, EPI_ISL_5676088, EPI_ISL_5676244, EPI_ISL_5676424, EPI_ISL_5676763, EPI_ISL_5676933, EPI_ISL_5676975, EPI_ISL_5676982, EPI_ISL_5677182, EPI_ISL_5677246, EPI_ISL_5677548, EPI_ISL_5677626, EPI_ISL_5677652, EPI_ISL_5677775, EPI_ISL_5677892, EPI_ISL_5677967, EPI_ISL_5678091 | | | |
| see above | SECRETARIA MUNICIPAL DE SAUDE SOROCABA | Instituto Butantan | Antonio Jorge Martins; Claudia Renata dos Santos Barros; David Schlesinger; Debora Botequio Moretti; Dimas Tadeu Covas; Elaine Cristina Marqueze; Elaine Vieira Santos; Evandra Strazza Rodrigues; Heidge Fukumasu; Jayme Augusto de Souza-Neto; José Salvatore Leister Patané; Luiz Alcantara; Luiz Lehmann Coutinho; Maria Carolina Elias; Maurício Lacerda Nogueira; Rafael dos Santos Bezerra; Raul Machado Neto; Rejane Maria Tommasini Grotto; Ricardo Haddad; Sandra Coccuzzo Sampaio Vessoni; Simone Kashima; Svetoslav Nanev Slavov; Vincent Louis Viala |
| EPI_ISL_2984109, EPI_ISL_3391234, EPI_ISL_3391251, EPI_ISL_3393387 | SELAS MEDILYS | Department of Virology, Henri Mondor University Hospital, Assistance Publique Hôpitaux de Paris, Université Paris-Est Créteil, INSERM U955 | Alexandre Soulier; Christophe Rodriguez; Elisabeth Trawinski; Guillaume Gricourt; Jean-Michel Pawlotsky; Melissa N'Debi; Slim Fourati; Vanessa Demontant |
| EPI_ISL_3534684, EPI_ISL_3534748 | SK-Roy Romanow Provincial Laboratory | National Microbiology Laboratory (NML) | Anna Majer; Anneliese Landgraff; Elsie Grudeski; Gary Van Domselaar; Grace Seo; Jennifer Tanner; Kara Loos; Keith MacKenzie; Meredith Faires; Morag Graham; Natalie Knox; Philip Mabon; Rachel; Rhiannon Huzarewich; Russell Mandes; Ryan McDonald; Shari Tyson |
| EPI_ISL_5668516, EPI_ISL_5668768, EPI_ISL_5668956, EPI_ISL_5668965 | SMS IPERO | Instituto Butantan | Antonio Jorge Martins; Claudia Renata dos Santos Barros; David Schlesinger; Debora Botequio Moretti; Dimas Tadeu Covas; Elaine Cristina Marqueze; Elaine Vieira Santos; Evandra Strazza Rodrigues; Heidge Fukumasu; Jayme Augusto de Souza-Neto; José Salvatore Leister Patané; Luiz Alcantara; Luiz Lehmann Coutinho; Maria Carolina Elias; Maurício Lacerda Nogueira; Rafael dos Santos Bezerra; Raul Machado Neto; Rejane Maria Tommasini Grotto; Ricardo Haddad; Sandra Coccuzzo Sampaio Vessoni; Simone Kashima; Svetoslav Nanev Slavov; Vincent Louis Viala |
| EPI_ISL_5671949, EPI_ISL_5671977 | SMS SECRETARIA MUNICIPAL DE SAUDE DE BOITUVA | Instituto Butantan | Antonio Jorge Martins; Claudia Renata dos Santos Barros; David Schlesinger; Debora Botequio Moretti; Dimas Tadeu Covas; Elaine Cristina Marqueze; Elaine Vieira Santos; Evandra Strazza Rodrigues; Heidge Fukumasu; Jayme Augusto de Souza-Neto; José Salvatore Leister Patané; Luiz Alcantara; Luiz Lehmann Coutinho; Maria Carolina Elias; Maurício Lacerda Nogueira; Rafael dos Santos Bezerra; Raul Machado Neto; Rejane Maria Tommasini Grotto; Ricardo Haddad; Sandra Coccuzzo Sampaio Vessoni; Simone Kashima; Svetoslav Nanev Slavov; Vincent Louis Viala |
| EPI_ISL_2930835, EPI_ISL_3008873, EPI_ISL_3087264 | SYNLAB | GIGA Medical Genomics | Bouchra Boujemla; Claire Gourzonès; Cécile Meex; Keith Durkin; Laurent Gillet; Maria Artesi; Marie-Pierre Hayette; Nadine Cambisano; Nathalie Renotte; Olivier Ek; Sébastien Bontems; Vincent Bours |
| EPI_ISL_3044006 | SYNLAB Labor München Zentrum LMZ | Robert Koch Institute | |
| EPI_ISL_3044040 | SYNLAB MVZ Leinfelden-Echterdingen | Robert Koch Institute | |
| EPI_ISL_2956160, EPI_ISL_3043979, EPI_ISL_3043991, | SYNLAB MVZ Leverkusen | Robert Koch Institute | |

| | | | |
|---|---|---|---|
| EPI_ISL_3044087 | | | |
| EPI_ISL_2956145, EPI_ISL_2956263, EPI_ISL_2956273, EPI_ISL_3043969, EPI_ISL_3044002, EPI_ISL_3044016, EPI_ISL_3044022, EPI_ISL_3044044, EPI_ISL_3044063, EPI_ISL_3044079, EPI_ISL_3044093, EPI_ISL_3044098, EPI_ISL_3044546, EPI_ISL_3044554, EPI_ISL_3044620, EPI_ISL_3044674, EPI_ISL_3044748 | | | |
| see above | SYNLAB MVZ Trier | Robert Koch Institute | |
| EPI_ISL_3276655, EPI_ISL_3276656 | Saint Francis Medical Center | Los Angeles County Public Health Laboratories | P. Hemarajata et al. |
| EPI_ISL_3160968, EPI_ISL_3160969, EPI_ISL_3277614, EPI_ISL_3503562, EPI_ISL_3721739, EPI_ISL_3721740, EPI_ISL_3721810, EPI_ISL_3721852, EPI_ISL_3721879, EPI_ISL_3721904, EPI_ISL_3721929, EPI_ISL_3721931, EPI_ISL_3721950, EPI_ISL_3721955, EPI_ISL_3721960, EPI_ISL_3905846, EPI_ISL_3905847, EPI_ISL_3905850, EPI_ISL_3905866, EPI_ISL_3905882, EPI_ISL_3905898, EPI_ISL_3905904, EPI_ISL_3905925, EPI_ISL_3905940, EPI_ISL_3905941, EPI_ISL_3943414, EPI_ISL_3943416, EPI_ISL_3943515, EPI_ISL_3943539, EPI_ISL_3943557, EPI_ISL_3943559 | | | |
| see above | Salud Digna | Instituto Nacional de Medicina Genomica | Abraham Campos-Romero; Cedro-Tanda A; Escobar-Arrazola MA; Herrera-Montalvo LA.; Hidalgo-Miranda A; Luna-Ruiz Marco; Mendoza-Vargas A; Moreno-Camacho José Luis; Munguia-Garza P; Ramirez-Vega O; Rangel-DeLeon D; Reyes-Grajeda JP; Rodriguez-Gallegos Jorge; Yair Alfaro-Mora |
| EPI_ISL_3374016, EPI_ISL_3374023, EPI_ISL_3374030, EPI_ISL_3912490, EPI_ISL_3912512, EPI_ISL_4411700 | Salud Digna, A.C | Andersen lab at Scripps Research | Abraham Garcia Gil; Jose Luis Moreno Camacho; Marco Antonio Luna Ruiz-Esparza; Miguel A. Fernandez Rojas; SEARCH Alliance with Abraham Campos Romero |
| EPI_ISL_3374068, EPI_ISL_3374069, EPI_ISL_3374071, EPI_ISL_3912959 | San Diego County Public Health Laboratory | Andersen lab at Scripps Research | Brett Austin; Jovan Shephard; SEARCH Alliance San Diego with Ashleigh Murphy; SEARCH Alliance San Diego with Ashleigh Murphy and Brett Austin |
| EPI_ISL_3388867 | San Diego County Public Health Laboratory | San Diego County Public Health Laboratory | Ashleigh Murphy Schafer; Brett Austin |
| EPI_ISL_3048920, EPI_ISL_3067573, EPI_ISL_3219167 | Santa Clara Valley Medical Center | Santa Clara County Public Health Laboratory | Santa Clara County Public Health Department |
| EPI_ISL_4601678 | Servicio Microbiología H.U. Dr. Negrín | Servicio Microbiología H.U. Dr. Negrín | Ana Bordes Benítez; Bartolomé Gómez Arroyo; Francisco Javier Chamizo López |
| EPI_ISL_4083071, EPI_ISL_4083177, EPI_ISL_4083178, EPI_ISL_4083181, EPI_ISL_4083182, EPI_ISL_4083185, EPI_ISL_4083187, EPI_ISL_4083188, EPI_ISL_4083256, EPI_ISL_4083279, EPI_ISL_4083292, EPI_ISL_4083294, EPI_ISL_4083297, EPI_ISL_4083298, EPI_ISL_4083300, EPI_ISL_4083301, EPI_ISL_4083303, EPI_ISL_4083304, EPI_ISL_4083305, EPI_ISL_4083306, EPI_ISL_4083308, EPI_ISL_4083309, EPI_ISL_4083310, EPI_ISL_4083312, EPI_ISL_4083317, EPI_ISL_4083318, EPI_ISL_4083326, EPI_ISL_4083345, EPI_ISL_4083354, EPI_ISL_4083355, EPI_ISL_4083357, EPI_ISL_4083358, EPI_ISL_4083359, EPI_ISL_4083367, EPI_ISL_4083368, EPI_ISL_4083375, EPI_ISL_4083379, EPI_ISL_4083380, EPI_ISL_4083386, EPI_ISL_4083389, EPI_ISL_4083396, EPI_ISL_4083397, EPI_ISL_4083398, EPI_ISL_4083399, EPI_ISL_4083400, EPI_ISL_4083411, EPI_ISL_4083412, EPI_ISL_4083426, EPI_ISL_4083427, EPI_ISL_4083429, EPI_ISL_4083430, EPI_ISL_4083431, EPI_ISL_4083432, EPI_ISL_4083434, EPI_ISL_4083435, EPI_ISL_4083436, EPI_ISL_4083438, EPI_ISL_4083439, EPI_ISL_4083440, EPI_ISL_4083441, EPI_ISL_4083453, EPI_ISL_4083454, EPI_ISL_4083455, EPI_ISL_4083456, EPI_ISL_4083457, EPI_ISL_4083460, EPI_ISL_4083461, EPI_ISL_4083462, EPI_ISL_4083463, EPI_ISL_4083464, EPI_ISL_4083465, EPI_ISL_4083467, EPI_ISL_4083468, EPI_ISL_4083469, EPI_ISL_4083470, EPI_ISL_4083471, EPI_ISL_4083472, EPI_ISL_4083473, EPI_ISL_4083474, EPI_ISL_4083498, EPI_ISL_4083499, EPI_ISL_4083510, EPI_ISL_4083512, EPI_ISL_5158435, EPI_ISL_5158473, EPI_ISL_5158478, EPI_ISL_5158488, EPI_ISL_5158512, EPI_ISL_5158520, EPI_ISL_5158550, EPI_ISL_5158553, EPI_ISL_5158554, EPI_ISL_5158558, EPI_ISL_5158561, EPI_ISL_5158597, EPI_ISL_5158735, EPI_ISL_5158736, EPI_ISL_5158740, EPI_ISL_5158741, EPI_ISL_5158776, EPI_ISL_5158814, EPI_ISL_5158816, EPI_ISL_5158817, EPI_ISL_5158819, EPI_ISL_5158828, EPI_ISL_5158837, EPI_ISL_5158860, EPI_ISL_5158863, EPI_ISL_6694516 | | | |
| see above | Servicio Virosis Respiratorias-Departamento Virología-INEI | Instituto Nacional Enfermedades Infecciosas C.G.Malbran | Avaro M.; Baumeister E.; Benedetti E.; Campos J.; Cisterna D.; Dattero ME; De Belder D.; Haim MS.; Lorenzo F.; Molina V.; Perandones C.; Poklepovich T.; Pontoriero A.; Russo M.; Sanchez Loria J.; Tuduri E. |
| EPI_ISL_3825494 | Servicio de Inmunología del Hospital Julio Perrando | Laboratorio de Biología Molecular, Instituto de Medicina Regional on behalf of 'Proyecto Argentino Interinstitucional de genomica de SARS-CoV-2' (PAIS Consortium) | Bettina Brusés; Florencia Vallejos Schulze; Griselda Oria; Horacio Lucero.; Javier Mussin; Laura Formichelli; María Delia Foussal; Melina Lorenzini Campos; Raúl Maximiliano Acevedo |
| EPI_ISL_3219108, EPI_ISL_3219110, EPI_ISL_3219120 | Servicio de Microbiologia Hospital Ramon y Cajal | Servicio de Microbiologia Hospital Ramon y Cajal | Galan JC; Martinez L. Abreu M; Ponce M; y Gonzalez-Alba JM |
| EPI_ISL_3024407, EPI_ISL_3024497 | Shamir Medical Center (Asaf Harofe) | Shamir Medical Center (Asaf Harofe) | Abu Hamad Ramzia; Adina Bar Chaim; Anna Vishnevsky; Chen Weiner; Nir Rainy; Patricia Benveniste-Lekovitz; Reut Sorek Abramovich; Yevgeni Yegorov |
| EPI_ISL_3373819, EPI_ISL_3373825 | Sharp HealthCare Laboratory | Andersen lab at Scripps Research | Art Mendoza; Cathy Woerle; Jacquelyn Berumen; Liam McGinnis; Omid Bakhtar; SEARCH Alliance San Diego with Aaron Harding |
| EPI_ISL_4194204 | Southern Nevada Public Health Laboratory | Southern Nevada Public Health Laboratory | Michael Picker |
| EPI_ISL_3050249, EPI_ISL_3236132 | Stanford Health Care | Stanford University School of Medicine, Clinical Virology Laboratory | Becky Jiang; Bernadette Troung; Daniel Solis; James Zehnder; Malaya K. Sahoo; Mamdouh Sibai; Nathan Hammond; Selamawit Bihon; and Benjamin A. Pinsky |
| EPI_ISL_3419018, EPI_ISL_3419023 | StarMed Healthcare | UNC Charlotte Environmental Monitoring Laboratory | Adam France; Cynthia Gibas; Erin Stiers; Jannatul Ferdous; Jessica Schlueter; Kevin Lambirth; Sam Kunkleman |
| EPI_ISL_3464209, EPI_ISL_3537437 | State Hygienic Laboratory at the University of Iowa | State Hygienic Laboratory at the University of Iowa | Alankar Kampoowale; Anna Yakos; Erik Twait; Kris Eveland; Kristen Zanon; Mariah Knutson; Valerie Reeb; Wes Hottel |
| EPI_ISL_3189705, EPI_ISL_3307817, EPI_ISL_3307957, EPI_ISL_3932795 | State Laboratories Division, Hawaii State Department of Health | State Laboratories Division, Hawaii State Department of Health | Ayana Garnet; Daniel Strange; Drew Kuwazaki; Edward Desmond; Pamela O'Brien; Razvan Sultana |
| EPI_ISL_3050050, EPI_ISL_3050096, EPI_ISL_3050121, EPI_ISL_3050131 | State Testing Facility | Altius Institute for Biomedical Research | Alex Isner; Alex Nguyen; Amanda Gale; Audra Johnson; Clem Green; Daniel Bates; Eric Thorland; Jacob Rodriguez; Jean Robinston; Jemma Nelson; Jessica Kunder; John Stamatoyannopoulos; Joshua Richards; Julia Wald; Kneshay Harper; Lauren Mitchell; Mark Frerker; Matt Hartman; Michael Buckley; Muhammad Halimun; Rebecca Bruders; Tobias Ragoczy |
| EPI_ISL_6794539 | State of New Hampshire Public Health Laboratories | State of New Hampshire Public Health Laboratories | Caitlin Mercier; Chris Benton; Jinfeng Li; Juan Bolanos; Xinglu Zhang |
| EPI_ISL_3157564 | Swedish national genomic surveillance program of SARS-CoV-2 | The Public Health Agency of Sweden | Alma Brolund; Maria Lind Karlberg; Maximilian Riess; Swedish national genomic surveillance program of SARS-CoV-2 |
| EPI_ISL_3325722, EPI_ISL_3325735, EPI_ISL_3325849, EPI_ISL_3325887, EPI_ISL_3325903, EPI_ISL_3325976, EPI_ISL_3326107, EPI_ISL_3326351, EPI_ISL_3326362, EPI_ISL_3326371, EPI_ISL_3326397, EPI_ISL_3326399, EPI_ISL_3326417, EPI_ISL_3326437, EPI_ISL_3326524, EPI_ISL_3326541, EPI_ISL_3326554, EPI_ISL_3326560, EPI_ISL_3326693, EPI_ISL_3326706, EPI_ISL_3326748, EPI_ISL_3460411, EPI_ISL_3460429, EPI_ISL_3460452, EPI_ISL_3460469, EPI_ISL_3460490, EPI_ISL_3460532, EPI_ISL_3460556, EPI_ISL_3460615, EPI_ISL_3460664, EPI_ISL_3661518, EPI_ISL_3661578, EPI_ISL_3661650, EPI_ISL_3661658, EPI_ISL_3661819, EPI_ISL_3661970 | | | |
| see above | TGen North | TGen North | "Jolene Bowers; Brett Van Tassel; Chris French; Darrin Lemmer; Dave Engelthaler; Dave Engelthaler"; Hayley Yaglom; Heather Centner; Jolene Bowers |
| EPI_ISL_3695610, EPI_ISL_3695731, EPI_ISL_3695767, EPI_ISL_3695799, EPI_ISL_3695801, EPI_ISL_3722892 | TPMG Regional Laboratory | California Department of Public Health | Emily Smith on behalf of CDPH-COVIDNet and UCI Genome Sciences Center/GHTF |
| EPI_ISL_3242037 | TX-Houston Health Department Laboratory | Centers for Disease Control and Prevention Division of Viral Diseases, Pathogen Discovery | Alex Burgin; Ben L. Rambo-Martin; Clinton R. Paden; Dakota Howard; Dave Wentworth; Dhwani Batra; Jasmine Padilla; Justin Lee; Krista Queen; Kristen Knipe; Kristine Lacek; Mark Burroughs; Matthew Schmerer; Meghan Bentz; Mili Sheth; Peter Cook; Sam Shepard; Sarah Nobles; Suxiang Tong; Vivien Dugan; Yvette Unoarumhi |
| EPI_ISL_3067396, EPI_ISL_3267887 | TXDSHS | TXDSHS | Anita Pokharel; Bonnie Oh; Chun Wang; Grace Kubin; Jenny Zhang; Karen Bobier; Lorraine Rodriguez; Maliha Rahman; Mayela Pedrueza; Myong Koag; Rachel Lee; Rashmi Tuladhar |
| EPI_ISL_2966844, EPI_ISL_2966850, EPI_ISL_2988616, EPI_ISL_3020314 | Tampa General Hospital Esoteric Lab | Tampa General Hospital Esoteric Research & Development Lab | Amorce Lima; Deanna Becker; Dominic Uy; Elaine Vendrone; Jon Faughn; Jorge Mecias-Frias; Suzane Silbert; Vicki Healer |
| EPI_ISL_3149042 | Texas Children's Hospital | Texas Children's Microbiome Center | Ila Singh; James Dunn; James Versalovic; Jennifer K. Spinler; Ruth Ann Luna |
| EPI_ISL_3642761 | The Caribbean Public Health Agency | Carrington Lab, Department of Preclinical Sciences, Faculty of Medical Sciences, The University of the West Indies, St Augustine Campus | Anushka Ramjag; Arianne Brown-Jordan; Avery Hinds; Christine V. F. Carrington; Christopher Oura; Gabriel Escobar; Nikita S. D. Sahadeo; Nuno Faria; Oliver Pybus; Risha Singh; Sarah Hill; Sharra Greenaway-Duberry; SueMin Nathaniel; Vernie Ramkissoon |
| EPI_ISL_3926065 | The National University Hospital of Iceland | deCODE genetics | Agnar Helgason; Alma Moller; Arna B Agustsdottir; Arnaldur Gylfason; Asgeir Sigurdsson; Aslaug Jonasdottir; Berglind Eiriksdottir; Bjarni Thorbjornsson; Brynjar O Jensson; Daniel F Gudbjartsson; Droplaug N Magnusdottir; Elisabet E Gardarsdottir; Emil A Thorarensen; Gardar Sveinbjornsson; Gisli Masson; Gudmundur Georgsson; Gudmundur L Norddahl; Gudrun Sigmundsdottir; Hakon Jonsson; Hannes Eggertsson; Hilma Holm; Ingileif Jonsdottir; Jona Saemundsdottir; Kamilla S Josefsdottir; Kari Stefansson; Karl G Kristinsson; Kjartan R Gudmundsson; Kristin E Sveinsdottir; Kristjan E Hjorleifsson; Louise le Roux; Maney Sveinsdottir; Olafia S Gretarsdottir; Olafur T Magnusson; Pall Melsted; Patrick Sulem; Run Fridriksdottir; Solvi Rognvaldsson; Thora R Gunnarsdottir; Thordur Kristjansson; Thorolfur Gudnason; Unnur Thorsteinsdottir |

| | | | |
|---|---|---|---|
| EPI_ISL_3276692, EPI_ISL_3276693, EPI_ISL_3276747, EPI_ISL_3276749 | Torrance Memorial Medical Center | Los Angeles County Public Health Laboratories | P. Hemarajata et al. |
| EPI_ISL_3610776, EPI_ISL_3610778, EPI_ISL_3642762, EPI_ISL_3642763 | Trinidad Public Health Laboratory | Carrington Lab, Department of Preclinical Sciences, Faculty of Medical Sciences, The University of the West Indies, St Augustine Campus | Anushka Ramjag; Arianne Brown-Jordan; Avery Hinds; Christine V. F. Carrington; Christopher Oura; Gabriel Escobar; Nikita S. D. Sahadeo; Nuno Faria; Oliver Pybus; Risha Singh; Roshan Parasram; Sarah Hill; SueMin Nathaniel; Vernie Ramkissoon |
| EPI_ISL_5671622, EPI_ISL_5671920 | UBS ALAMBARI | Instituto Butantan | Antonio Jorge Martins; Claudia Renata dos Santos Barros; David Schlesinger; Debora Botequio Moretti; Dimas Tadeu Covas; Elaine Cristina Marqueze; Elaine Vieira Santos; Evandra Strazza Rodrigues; Heidge Fukumasu; Jayme Augusto de Souza-Neto; José Salvatore Leister Patané; Luiz Alcantara; Luiz Lehmann Coutinho; Maria Carolina Elias; Maurício Lacerda Nogueira; Rafael dos Santos Bezerra; Raul Machado Neto; Rejane Maria Tommasini Grotto; Ricardo Haddad; Sandra Coccuzzo Sampaio Vessoni; Simone Kashima; Svetoslav Nanev Slavov; Vincent Louis Viala |
| EPI_ISL_5672422, EPI_ISL_5672485, EPI_ISL_5672514 | UBS CELESTINO CARNIELO | Instituto Butantan | Antonio Jorge Martins; Claudia Renata dos Santos Barros; David Schlesinger; Debora Botequio Moretti; Dimas Tadeu Covas; Elaine Cristina Marqueze; Elaine Vieira Santos; Evandra Strazza Rodrigues; Heidge Fukumasu; Jayme Augusto de Souza-Neto; José Salvatore Leister Patané; Luiz Alcantara; Luiz Lehmann Coutinho; Maria Carolina Elias; Maurício Lacerda Nogueira; Rafael dos Santos Bezerra; Raul Machado Neto; Rejane Maria Tommasini Grotto; Ricardo Haddad; Sandra Coccuzzo Sampaio Vessoni; Simone Kashima; Svetoslav Nanev Slavov; Vincent Louis Viala |
| EPI_ISL_5675359 | UBS DE MACEDONIA | Instituto Butantan | Antonio Jorge Martins; Claudia Renata dos Santos Barros; David Schlesinger; Debora Botequio Moretti; Dimas Tadeu Covas; Elaine Cristina Marqueze; Elaine Vieira Santos; Evandra Strazza Rodrigues; Heidge Fukumasu; Jayme Augusto de Souza-Neto; José Salvatore Leister Patané; Luiz Alcantara; Luiz Lehmann Coutinho; Maria Carolina Elias; Maurício Lacerda Nogueira; Rafael dos Santos Bezerra; Raul Machado Neto; Rejane Maria Tommasini Grotto; Ricardo Haddad; Sandra Coccuzzo Sampaio Vessoni; Simone Kashima; Svetoslav Nanev Slavov; Vincent Louis Viala |
| EPI_ISL_5673293, EPI_ISL_5673420, EPI_ISL_5673867, EPI_ISL_5674030, EPI_ISL_5674086, EPI_ISL_5674809 | UBS DE MIRA ESTRELA | Instituto Butantan | Antonio Jorge Martins; Claudia Renata dos Santos Barros; David Schlesinger; Debora Botequio Moretti; Dimas Tadeu Covas; Elaine Cristina Marqueze; Elaine Vieira Santos; Evandra Strazza Rodrigues; Heidge Fukumasu; Jayme Augusto de Souza-Neto; José Salvatore Leister Patané; Luiz Alcantara; Luiz Lehmann Coutinho; Maria Carolina Elias; Maurício Lacerda Nogueira; Rafael dos Santos Bezerra; Raul Machado Neto; Rejane Maria Tommasini Grotto; Ricardo Haddad; Sandra Coccuzzo Sampaio Vessoni; Simone Kashima; Svetoslav Nanev Slavov; Vincent Louis Viala |
| EPI_ISL_5674630 | UBS DE PEDRANOPOLIS | Instituto Butantan | Antonio Jorge Martins; Claudia Renata dos Santos Barros; David Schlesinger; Debora Botequio Moretti; Dimas Tadeu Covas; Elaine Cristina Marqueze; Elaine Vieira Santos; Evandra Strazza Rodrigues; Heidge Fukumasu; Jayme Augusto de Souza-Neto; José Salvatore Leister Patané; Luiz Alcantara; Luiz Lehmann Coutinho; Maria Carolina Elias; Maurício Lacerda Nogueira; Rafael dos Santos Bezerra; Raul Machado Neto; Rejane Maria Tommasini Grotto; Ricardo Haddad; Sandra Coccuzzo Sampaio Vessoni; Simone Kashima; Svetoslav Nanev Slavov; Vincent Louis Viala |
| EPI_ISL_5674869, EPI_ISL_5675408 | UBS DE TABAPUA | Instituto Butantan | Antonio Jorge Martins; Claudia Renata dos Santos Barros; David Schlesinger; Debora Botequio Moretti; Dimas Tadeu Covas; Elaine Cristina Marqueze; Elaine Vieira Santos; Evandra Strazza Rodrigues; Heidge Fukumasu; Jayme Augusto de Souza-Neto; José Salvatore Leister Patané; Luiz Alcantara; Luiz Lehmann Coutinho; Maria Carolina Elias; Maurício Lacerda Nogueira; Rafael dos Santos Bezerra; Raul Machado Neto; Rejane Maria Tommasini Grotto; Ricardo Haddad; Sandra Coccuzzo Sampaio Vessoni; Simone Kashima; Svetoslav Nanev Slavov; Vincent Louis Viala |
| EPI_ISL_5673250, EPI_ISL_5673660, EPI_ISL_5673798 | UBS DR ABELARDO PINHEIRO GUIMARAES SCRPARDO | Instituto Butantan | Antonio Jorge Martins; Claudia Renata dos Santos Barros; David Schlesinger; Debora Botequio Moretti; Dimas Tadeu Covas; Elaine Cristina Marqueze; Elaine Vieira Santos; Evandra Strazza Rodrigues; Heidge Fukumasu; Jayme Augusto de Souza-Neto; José Salvatore Leister Patané; Luiz Alcantara; Luiz Lehmann Coutinho; Maria Carolina Elias; Maurício Lacerda Nogueira; Rafael dos Santos Bezerra; Raul Machado Neto; Rejane Maria Tommasini Grotto; Ricardo Haddad; Sandra Coccuzzo Sampaio Vessoni; Simone Kashima; Svetoslav Nanev Slavov; Vincent Louis Viala |
| EPI_ISL_5670896, EPI_ISL_5670923, EPI_ISL_5671100, EPI_ISL_5671151, EPI_ISL_5671185, EPI_ISL_5671281, EPI_ISL_5671375, EPI_ISL_5671395, EPI_ISL_5671454, EPI_ISL_5673269, EPI_ISL_5673436, EPI_ISL_5674319, EPI_ISL_5674427, EPI_ISL_5674702, EPI_ISL_5675039, EPI_ISL_5675435 | | | |
| see above | UBS DR ALFREDO DANTAS DE SOUZA UMUARAMA | Instituto Butantan | Antonio Jorge Martins; Claudia Renata dos Santos Barros; David Schlesinger; Debora Botequio Moretti; Dimas Tadeu Covas; Elaine Cristina Marqueze; Elaine Vieira Santos; Evandra Strazza Rodrigues; Heidge Fukumasu; Jayme Augusto de Souza-Neto; José Salvatore Leister Patané; Luiz Alcantara; Luiz Lehmann Coutinho; Maria Carolina Elias; Maurício Lacerda Nogueira; Rafael dos Santos Bezerra; Raul Machado Neto; Rejane Maria Tommasini Grotto; Ricardo Haddad; Sandra Coccuzzo Sampaio Vessoni; Simone Kashima; Svetoslav Nanev Slavov; Vincent Louis Viala |
| EPI_ISL_5673553, EPI_ISL_5673842, EPI_ISL_5674530, EPI_ISL_5675006 | UBS DR WALDOMIRO FERREIRA NEVES SCRPARDO | Instituto Butantan | Antonio Jorge Martins; Claudia Renata dos Santos Barros; David Schlesinger; Debora Botequio Moretti; Dimas Tadeu Covas; Elaine Cristina Marqueze; Elaine Vieira Santos; Evandra Strazza Rodrigues; Heidge Fukumasu; Jayme Augusto de Souza-Neto; José Salvatore Leister Patané; Luiz Alcantara; Luiz Lehmann Coutinho; Maria Carolina Elias; Maurício Lacerda Nogueira; Rafael dos Santos Bezerra; Raul Machado Neto; Rejane Maria Tommasini Grotto; Ricardo Haddad; Sandra Coccuzzo Sampaio Vessoni; Simone Kashima; Svetoslav Nanev Slavov; Vincent Louis Viala |
| EPI_ISL_5673877, EPI_ISL_5673978, EPI_ISL_5674567 | UBS ESPIRITO SANTO DO TURVO | Instituto Butantan | Antonio Jorge Martins; Claudia Renata dos Santos Barros; David Schlesinger; Debora Botequio Moretti; Dimas Tadeu Covas; Elaine Cristina Marqueze; Elaine Vieira Santos; Evandra Strazza Rodrigues; Heidge Fukumasu; Jayme Augusto de Souza-Neto; José Salvatore Leister Patané; Luiz Alcantara; Luiz Lehmann Coutinho; Maria Carolina Elias; Maurício Lacerda Nogueira; Rafael dos Santos Bezerra; Raul Machado Neto; Rejane Maria Tommasini Grotto; Ricardo Haddad; Sandra Coccuzzo Sampaio Vessoni; Simone Kashima; Svetoslav Nanev Slavov; Vincent Louis Viala |
| EPI_ISL_5672428 | UBS EUFRASIO PEREIRA COSTA | Instituto Butantan | Antonio Jorge Martins; Claudia Renata dos Santos Barros; David Schlesinger; Debora Botequio Moretti; Dimas Tadeu Covas; Elaine Cristina Marqueze; Elaine Vieira Santos; Evandra Strazza Rodrigues; Heidge Fukumasu; Jayme Augusto de Souza-Neto; José Salvatore Leister Patané; Luiz Alcantara; Luiz Lehmann Coutinho; Maria Carolina Elias; Maurício Lacerda Nogueira; Rafael dos Santos Bezerra; Raul Machado Neto; Rejane Maria Tommasini Grotto; Ricardo Haddad; Sandra Coccuzzo Sampaio Vessoni; Simone Kashima; Svetoslav Nanev Slavov; Vincent Louis Viala |
| EPI_ISL_5673020, EPI_ISL_5673041 | UBS II DE TACIBA | Instituto Butantan | Antonio Jorge Martins; Claudia Renata dos Santos Barros; David Schlesinger; Debora Botequio Moretti; Dimas Tadeu Covas; Elaine Cristina Marqueze; Elaine Vieira Santos; Evandra Strazza Rodrigues; Heidge Fukumasu; Jayme Augusto de Souza-Neto; José Salvatore Leister Patané; Luiz Alcantara; Luiz Lehmann Coutinho; Maria Carolina Elias; Maurício Lacerda Nogueira; Rafael dos Santos Bezerra; Raul Machado Neto; Rejane Maria Tommasini Grotto; Ricardo Haddad; Sandra Coccuzzo Sampaio Vessoni; Simone Kashima; Svetoslav Nanev Slavov; Vincent Louis Viala |
| EPI_ISL_5671353 | UBS III DE BENTO DE ABREU | Instituto Butantan | Antonio Jorge Martins; Claudia Renata dos Santos Barros; David Schlesinger; Debora Botequio Moretti; Dimas Tadeu Covas; Elaine Cristina Marqueze; Elaine Vieira Santos; Evandra Strazza Rodrigues; Heidge Fukumasu; Jayme Augusto de Souza-Neto; José Salvatore Leister Patané; Luiz Alcantara; Luiz Lehmann Coutinho; Maria Carolina Elias; Maurício Lacerda Nogueira; Rafael dos Santos Bezerra; Raul Machado Neto; Rejane Maria Tommasini Grotto; Ricardo Haddad; Sandra Coccuzzo Sampaio Vessoni; Simone Kashima; Svetoslav Nanev Slavov; Vincent Louis Viala |
| EPI_ISL_5670659, EPI_ISL_5670671 | UBS III DE ELDORADO ELDORADO S P | Instituto Butantan | Antonio Jorge Martins; Claudia Renata dos Santos Barros; David Schlesinger; Debora Botequio Moretti; Dimas Tadeu Covas; Elaine Cristina Marqueze; Elaine Vieira Santos; Evandra Strazza Rodrigues; Heidge Fukumasu; Jayme Augusto de Souza-Neto; José Salvatore Leister Patané; Luiz Alcantara; Luiz Lehmann Coutinho; Maria Carolina Elias; Maurício Lacerda Nogueira; Rafael dos Santos Bezerra; Raul Machado Neto; Rejane Maria Tommasini Grotto; Ricardo Haddad; Sandra Coccuzzo Sampaio Vessoni; Simone Kashima; Svetoslav Nanev Slavov; Vincent Louis Viala |
| EPI_ISL_5673098 | UBS III DE EUCLIDES DA CUNHA PAULISTA | Instituto Butantan | Antonio Jorge Martins; Claudia Renata dos Santos Barros; David Schlesinger; Debora Botequio Moretti; Dimas Tadeu Covas; Elaine Cristina Marqueze; Elaine Vieira Santos; Evandra Strazza Rodrigues; Heidge Fukumasu; Jayme Augusto de Souza-Neto; José Salvatore Leister Patané; Luiz Alcantara; Luiz Lehmann Coutinho; Maria Carolina Elias; Maurício Lacerda Nogueira; Rafael dos Santos Bezerra; Raul Machado Neto; Rejane Maria Tommasini Grotto; Ricardo Haddad; Sandra Coccuzzo Sampaio Vessoni; Simone Kashima; Svetoslav Nanev Slavov; Vincent Louis Viala |
| EPI_ISL_5668586, EPI_ISL_5671769 | UBS JARDIM VITORINO IPERO | Instituto Butantan | Antonio Jorge Martins; Claudia Renata dos Santos Barros; David Schlesinger; Debora Botequio Moretti; Dimas Tadeu Covas; Elaine Cristina Marqueze; Elaine Vieira Santos; Evandra Strazza Rodrigues; Heidge Fukumasu; Jayme Augusto de Souza-Neto; José Salvatore Leister Patané; Luiz Alcantara; Luiz Lehmann Coutinho; Maria Carolina Elias; Maurício Lacerda Nogueira; Rafael dos Santos Bezerra; Raul Machado Neto; Rejane Maria Tommasini Grotto; Ricardo Haddad; Sandra Coccuzzo Sampaio Vessoni; Simone Kashima; Svetoslav Nanev Slavov; Vincent Louis Viala |
| EPI_ISL_5669345 | UBS JOSE AUGUSTO BATISTA FILHO MENEGUETI | Instituto Butantan | Antonio Jorge Martins; Claudia Renata dos Santos Barros; David Schlesinger; Debora Botequio Moretti; Dimas Tadeu Covas; Elaine Cristina Marqueze; Elaine Vieira Santos; Evandra Strazza Rodrigues; Heidge Fukumasu; Jayme Augusto de Souza-Neto; José Salvatore Leister Patané; Luiz Alcantara; Luiz Lehmann Coutinho; Maria Carolina Elias; Maurício Lacerda Nogueira; Rafael dos Santos Bezerra; Raul Machado Neto; Rejane Maria Tommasini Grotto; Ricardo Haddad; Sandra Coccuzzo Sampaio Vessoni; Simone Kashima; Svetoslav Nanev Slavov; Vincent Louis Viala |
| EPI_ISL_5672042, EPI_ISL_5672056, EPI_ISL_5672085, EPI_ISL_5672129, EPI_ISL_5672174, EPI_ISL_5672180, EPI_ISL_5672289, EPI_ISL_5672334, EPI_ISL_5672497, EPI_ISL_5672502, EPI_ISL_5672530, EPI_ISL_5672550, EPI_ISL_5675415, EPI_ISL_5675470 | | | |
| see above | UBS PASTOR MARCELINO DEUNGARO RIO GRANDE | Instituto Butantan | Antonio Jorge Martins; Claudia Renata dos Santos Barros; David Schlesinger; Debora Botequio Moretti; Dimas Tadeu Covas; Elaine Cristina Marqueze; Elaine Vieira Santos; Evandra Strazza Rodrigues; Heidge Fukumasu; Jayme Augusto de Souza-Neto; José Salvatore Leister Patané; Luiz Alcantara; Luiz Lehmann Coutinho; Maria Carolina Elias; Maurício Lacerda Nogueira; Rafael dos Santos Bezerra; Raul Machado Neto; Rejane Maria Tommasini Grotto; Ricardo Haddad; Sandra Coccuzzo Sampaio Vessoni; Simone Kashima; Svetoslav Nanev Slavov; Vincent Louis Viala |
| EPI_ISL_5672164 | UBS PINHEIRINHO | Instituto Butantan | Antonio Jorge Martins; Claudia Renata dos Santos Barros; David Schlesinger; Debora Botequio Moretti; Dimas Tadeu Covas; Elaine Cristina Marqueze; Elaine Vieira Santos; Evandra Strazza Rodrigues; Heidge Fukumasu; Jayme Augusto de Souza-Neto; José Salvatore Leister Patané; Luiz Alcantara; Luiz Lehmann Coutinho; Maria Carolina Elias; Maurício Lacerda Nogueira; Rafael dos Santos Bezerra; Raul Machado Neto; Rejane Maria Tommasini Grotto; Ricardo Haddad; Sandra Coccuzzo Sampaio Vessoni; Simone Kashima; Svetoslav Nanev Slavov; Vincent Louis Viala |
| EPI_ISL_5671035 | UBS RACLE BARRETO DA SILVA | Instituto Butantan | Antonio Jorge Martins; Claudia Renata dos Santos Barros; David Schlesinger; Debora Botequio Moretti; Dimas Tadeu Covas; Elaine Cristina Marqueze; Elaine Vieira Santos; Evandra Strazza Rodrigues; Heidge Fukumasu; Jayme Augusto de Souza-Neto; José Salvatore Leister Patané; Luiz Alcantara; Luiz Lehmann Coutinho; Maria Carolina Elias; Maurício Lacerda Nogueira; Rafael dos Santos Bezerra; Raul Machado Neto; Rejane Maria Tommasini Grotto; Ricardo Haddad; Sandra Coccuzzo Sampaio Vessoni; Simone Kashima; Svetoslav Nanev Slavov; Vincent Louis Viala |
| EPI_ISL_5671718 | UBS SALTO DE SAO JOSE | Instituto Butantan | Antonio Jorge Martins; Claudia Renata dos Santos Barros; David Schlesinger; Debora Botequio Moretti; Dimas Tadeu Covas; Elaine Cristina Marqueze; Elaine Vieira Santos; Evandra Strazza Rodrigues; Heidge Fukumasu; Jayme Augusto de Souza-Neto; José Salvatore Leister Patané; Luiz Alcantara; Luiz Lehmann Coutinho; Maria Carolina Elias; Maurício Lacerda Nogueira; Rafael dos Santos Bezerra; Raul Machado Neto; Rejane Maria Tommasini Grotto; Ricardo Haddad; Sandra Coccuzzo Sampaio Vessoni; Simone Kashima; Svetoslav Nanev Slavov; Vincent Louis Viala |
| EPI_ISL_5674400 | UBS TEREZA GALLO UCHOA | Instituto Butantan | Antonio Jorge Martins; Claudia Renata dos Santos Barros; David Schlesinger; Debora Botequio Moretti; Dimas Tadeu Covas; Elaine Cristina Marqueze; Elaine Vieira Santos; Evandra Strazza Rodrigues; Heidge Fukumasu; Jayme Augusto de Souza-Neto; José Salvatore Leister Patané; Luiz Alcantara; Luiz Lehmann Coutinho; Maria Carolina Elias; Maurício Lacerda Nogueira; Rafael dos Santos Bezerra; Raul Machado Neto; Rejane Maria Tommasini Grotto; Ricardo Haddad; Sandra Coccuzzo Sampaio Vessoni; Simone Kashima; Svetoslav Nanev Slavov; Vincent Louis Viala |
| EPI_ISL_5670945 | UBS ZONA NORTE | Instituto Butantan | Antonio Jorge Martins; Claudia Renata dos Santos Barros; David Schlesinger; Debora Botequio Moretti; Dimas Tadeu Covas; Elaine Cristina Marqueze; Elaine Vieira Santos; Evandra Strazza Rodrigues; Heidge Fukumasu; Jayme Augusto de Souza-Neto; José Salvatore Leister Patané; Luiz Alcantara; Luiz Lehmann Coutinho; Maria Carolina Elias; Maurício Lacerda Nogueira; Rafael dos Santos Bezerra; Raul Machado Neto; Rejane Maria Tommasini Grotto; Ricardo Haddad; Sandra Coccuzzo Sampaio Vessoni; Simone Kashima; Svetoslav Nanev Slavov; Vincent Louis Viala |
| EPI_ISL_3419683, EPI_ISL_3419684, EPI_ISL_3419690 | UC Davis Genome Center and Healthy Davis Together | UC Davis Genome Center | UC Davis |
| EPI_ISL_3067040, EPI_ISL_3067050 | UC-Christus Clinical Hospital Laboratory | Laboratory of Molecular Virology, School of Medicine, Pontificia Universidad Catolica de Chile | Ana Maira Guzman; Andres E. Munoz-Marcos; Catalina Pardo-Roa; Eileen Serrano; Erick Salinas; Estefany Poblete; Francisco Melo; Jorge Levican; Leonardo I. Almonacid; Maria Jose Avendano; Maria Patricia Vega; Rafael A. Medina; Ricardo Enrique de la Barra; Tamara Garcia-Salum |
| EPI_ISL_3104704, EPI_ISL_3236554 | UCC (Universidad Central del Caribe) | Grubaugh Lab - Yale School of Public Health | Alejandaro Vallejo Degaudenzi; Anderson Brito; Andrea Firpo; Annie Watkins; Chaney Kalinich; Chantal Vogels; David Rivera Aponte; Elisa Contreras; Ernesto Vazquez; Esperanza Mendoza; Fabiola Cruz-Lopez; Fabiola Fontanet; Isabel Ott; Jessica Rothman; Joseph Fauver; Karolane Gonzalez; Kendall Billig; Mallery Breban; Mary Petrone; Natalie Machargo; Nathan Grubaugh; Patricia Serrano; Robert Paulino-Ramirez; Tania Mitwalli; Tara Alpert; Tobias Koch; Vianca Aponte; Victor Virgilio Calderon |
| EPI_ISL_5673121 | UNIDADE BASICA DE SAUDE II DE TARABAI | Instituto Butantan | Antonio Jorge Martins; Claudia Renata dos Santos Barros; David Schlesinger; Debora Botequio Moretti; Dimas Tadeu Covas; Elaine Cristina Marqueze; Elaine Vieira Santos; Evandra Strazza Rodrigues; Heidge Fukumasu; Jayme Augusto de Souza-Neto; José Salvatore Leister Patané; Luiz Alcantara; Luiz Lehmann Coutinho; Maria Carolina Elias; Maurício Lacerda Nogueira; Rafael dos Santos Bezerra; Raul Machado Neto; Rejane Maria Tommasini Grotto; Ricardo Haddad; Sandra Coccuzzo Sampaio Vessoni; Simone Kashima; Svetoslav Nanev Slavov; Vincent Louis Viala |
| EPI_ISL_5673763, EPI_ISL_5674148 | UNIDADE BASICA DE SAUDE PREFEITO JOSE OLIVEIRA DE SOUZA | Instituto Butantan | Antonio Jorge Martins; Claudia Renata dos Santos Barros; David Schlesinger; Debora Botequio Moretti; Dimas Tadeu Covas; Elaine Cristina Marqueze; Elaine Vieira Santos; Evandra Strazza Rodrigues; Heidge Fukumasu; Jayme Augusto de Souza-Neto; José Salvatore Leister Patané; Luiz Alcantara; Luiz Lehmann Coutinho; Maria Carolina Elias; Maurício Lacerda Nogueira; Rafael dos Santos Bezerra; Raul Machado Neto; Rejane Maria Tommasini Grotto; Ricardo Haddad; Sandra Coccuzzo Sampaio Vessoni; Simone Kashima; Svetoslav Nanev Slavov; Vincent Louis Viala |
| EPI_ISL_5675524 | UNIDADE DE SAUDE DR PHEBO DE OLIVEIRA ROGE FERREIRA | Instituto Butantan | Antonio Jorge Martins; Claudia Renata dos Santos Barros; David Schlesinger; Debora Botequio Moretti; Dimas Tadeu Covas; Elaine Cristina Marqueze; Elaine Vieira Santos; Evandra Strazza Rodrigues; Heidge Fukumasu; Jayme Augusto de Souza-Neto; José Salvatore Leister Patané; Luiz Alcantara; Luiz Lehmann Coutinho; Maria Carolina Elias; Maurício Lacerda Nogueira; Rafael dos Santos Bezerra; Raul Machado Neto; Rejane Maria Tommasini Grotto; Ricardo Haddad; Sandra Coccuzzo Sampaio Vessoni; Simone Kashima; Svetoslav Nanev Slavov; Vincent Louis Viala |
| EPI_ISL_5670931, EPI_ISL_5671122, EPI_ISL_5671252 | UNIDADE DE VIGILANCIA EM SAUDE | Instituto Butantan | Antonio Jorge Martins; Claudia Renata dos Santos Barros; David Schlesinger; Debora Botequio Moretti; Dimas Tadeu Covas; Elaine Cristina Marqueze; Elaine Vieira Santos; Evandra Strazza Rodrigues; Heidge Fukumasu; Jayme Augusto de Souza-Neto; José Salvatore Leister Patané; Luiz Alcantara; Luiz Lehmann Coutinho; Maria Carolina Elias; Maurício Lacerda Nogueira; Rafael dos Santos Bezerra; Raul Machado Neto; Rejane Maria Tommasini Grotto; Ricardo Haddad; Sandra Coccuzzo Sampaio Vessoni; Simone Kashima; Svetoslav Nanev Slavov; Vincent Louis Viala |
| EPI_ISL_5671248, EPI_ISL_5671413 | UNIDADE DE VIGILANCIA EPIDEMIOLOGICA | Instituto Butantan | Antonio Jorge Martins; Claudia Renata dos Santos Barros; David Schlesinger; Debora Botequio Moretti; Dimas Tadeu Covas; Elaine Cristina Marqueze; Elaine Vieira Santos; Evandra Strazza Rodrigues; Heidge Fukumasu; Jayme Augusto de Souza-Neto; José Salvatore Leister Patané; Luiz Alcantara; Luiz Lehmann Coutinho; Maria Carolina Elias; Maurício Lacerda Nogueira; Rafael dos Santos Bezerra; Raul Machado Neto; Rejane Maria Tommasini Grotto; Ricardo Haddad; Sandra Coccuzzo Sampaio Vessoni; Simone Kashima; Svetoslav Nanev Slavov; Vincent Louis Viala |
| EPI_ISL_5671753, EPI_ISL_5671785, EPI_ISL_5671891 | UNIDADE MISTA ARACOIABA DA SERRA | Instituto Butantan | Antonio Jorge Martins; Claudia Renata dos Santos Barros; David Schlesinger; Debora Botequio Moretti; Dimas Tadeu Covas; Elaine Cristina Marqueze; Elaine Vieira Santos; Evandra Strazza Rodrigues; Heidge Fukumasu; Jayme Augusto de Souza-Neto; José Salvatore Leister Patané; Luiz Alcantara; Luiz Lehmann Coutinho; Maria Carolina Elias; Maurício Lacerda Nogueira; Rafael dos Santos Bezerra; Raul Machado Neto; Rejane Maria Tommasini Grotto; Ricardo Haddad; Sandra Coccuzzo Sampaio Vessoni; Simone Kashima; Svetoslav Nanev Slavov; Vincent Louis Viala |
| EPI_ISL_5669714 | UNIDADE MISTA DE SAUDE DE SARAPUI | Instituto Butantan | Antonio Jorge Martins; Claudia Renata dos Santos Barros; David Schlesinger; Debora Botequio Moretti; Dimas Tadeu Covas; Elaine Cristina Marqueze; Elaine Vieira Santos; Evandra Strazza Rodrigues; Heidge Fukumasu; Jayme Augusto de Souza-Neto; José Salvatore Leister Patané; Luiz Alcantara; Luiz Lehmann Coutinho; Maria Carolina Elias; Maurício Lacerda Nogueira; Rafael dos Santos Bezerra; Raul Machado Neto; Rejane Maria Tommasini Grotto; Ricardo Haddad; Sandra Coccuzzo Sampaio Vessoni; Simone Kashima; Svetoslav Nanev Slavov; Vincent Louis Viala |
| EPI_ISL_5671223 | UNIDADE MISTA OSVALDO P | Instituto Butantan | Antonio Jorge Martins; Claudia Renata dos Santos Barros; David Schlesinger; Debora Botequio Moretti; Dimas Tadeu Covas; Elaine Cristina Marqueze; Elaine Vieira Santos; Evandra Strazza Rodrigues; Heidge Fukumasu; Jayme Augusto de Souza-Neto; José Salvatore Leister Patané; Luiz Alcantara; Luiz Lehmann |

| | | | |
|---|---|---|---|
| | CAMARGO ELIAS FAUSTO | | Coutinho; Maria Carolina Elias; Maurício Lacerda Nogueira; Rafael dos Santos Bezerra; Raul Machado Neto; Rejane Maria Tommasini Grotto; Ricardo Haddad; Sandra Coccuzzo Sampaio Vessoni; Simone Kashima; Svetoslav Nanev Slavov; Vincent Louis Viala |
| EPI_ISL_5671627, EPI_ISL_5671703, EPI_ISL_5671791 | UNIDADE SENTINELA COVID19 | Instituto Butantan | Antonio Jorge Martins; Claudia Renata dos Santos Barros; David Schlesinger; Debora Botequio Moretti; Dimas Tadeu Covas; Elaine Cristina Marqueze; Elaine Vieira Santos; Evandra Strazza Rodrigues; Heidge Fukumasu; Jayme Augusto de Souza-Neto; José Salvatore Leister Patané; Luiz Alcantara; Luiz Lehmann Coutinho; Maria Carolina Elias; Maurício Lacerda Nogueira; Rafael dos Santos Bezerra; Raul Machado Neto; Rejane Maria Tommasini Grotto; Ricardo Haddad; Sandra Coccuzzo Sampaio Vessoni; Simone Kashima; Svetoslav Nanev Slavov; Vincent Louis Viala |
| EPI_ISL_5668866 | UNIDADE SENTINELA VILA APARECIDA | Instituto Butantan | Antonio Jorge Martins; Claudia Renata dos Santos Barros; David Schlesinger; Debora Botequio Moretti; Dimas Tadeu Covas; Elaine Cristina Marqueze; Elaine Vieira Santos; Evandra Strazza Rodrigues; Heidge Fukumasu; Jayme Augusto de Souza-Neto; José Salvatore Leister Patané; Luiz Alcantara; Luiz Lehmann Coutinho; Maria Carolina Elias; Maurício Lacerda Nogueira; Rafael dos Santos Bezerra; Raul Machado Neto; Rejane Maria Tommasini Grotto; Ricardo Haddad; Sandra Coccuzzo Sampaio Vessoni; Simone Kashima; Svetoslav Nanev Slavov; Vincent Louis Viala |
| EPI_ISL_5678127 | UPA CUMBICA | Instituto Butantan | Antonio Jorge Martins; Claudia Renata dos Santos Barros; David Schlesinger; Debora Botequio Moretti; Dimas Tadeu Covas; Elaine Cristina Marqueze; Elaine Vieira Santos; Evandra Strazza Rodrigues; Heidge Fukumasu; Jayme Augusto de Souza-Neto; José Salvatore Leister Patané; Luiz Alcantara; Luiz Lehmann Coutinho; Maria Carolina Elias; Maurício Lacerda Nogueira; Rafael dos Santos Bezerra; Raul Machado Neto; Rejane Maria Tommasini Grotto; Ricardo Haddad; Sandra Coccuzzo Sampaio Vessoni; Simone Kashima; Svetoslav Nanev Slavov; Vincent Louis Viala |
| EPI_ISL_5673330, EPI_ISL_5673382, EPI_ISL_5673474, EPI_ISL_5673852, EPI_ISL_5674928 | UPA DE BEBEDOURO | Instituto Butantan | Antonio Jorge Martins; Claudia Renata dos Santos Barros; David Schlesinger; Debora Botequio Moretti; Dimas Tadeu Covas; Elaine Cristina Marqueze; Elaine Vieira Santos; Evandra Strazza Rodrigues; Heidge Fukumasu; Jayme Augusto de Souza-Neto; José Salvatore Leister Patané; Luiz Alcantara; Luiz Lehmann Coutinho; Maria Carolina Elias; Maurício Lacerda Nogueira; Rafael dos Santos Bezerra; Raul Machado Neto; Rejane Maria Tommasini Grotto; Ricardo Haddad; Sandra Coccuzzo Sampaio Vessoni; Simone Kashima; Svetoslav Nanev Slavov; Vincent Louis Viala |
| EPI_ISL_5672217 | UPA DR AKIRA TADA | Instituto Butantan | Antonio Jorge Martins; Claudia Renata dos Santos Barros; David Schlesinger; Debora Botequio Moretti; Dimas Tadeu Covas; Elaine Cristina Marqueze; Elaine Vieira Santos; Evandra Strazza Rodrigues; Heidge Fukumasu; Jayme Augusto de Souza-Neto; José Salvatore Leister Patané; Luiz Alcantara; Luiz Lehmann Coutinho; Maria Carolina Elias; Maurício Lacerda Nogueira; Rafael dos Santos Bezerra; Raul Machado Neto; Rejane Maria Tommasini Grotto; Ricardo Haddad; Sandra Coccuzzo Sampaio Vessoni; Simone Kashima; Svetoslav Nanev Slavov; Vincent Louis Viala |
| EPI_ISL_5668671, EPI_ISL_5669012, EPI_ISL_5669047, EPI_ISL_5669325, EPI_ISL_5669500, EPI_ISL_5669594, EPI_ISL_5669799, EPI_ISL_5670033, EPI_ISL_5670079, EPI_ISL_5670173, EPI_ISL_5670298, EPI_ISL_5670311, EPI_ISL_5670361, EPI_ISL_5670581, EPI_ISL_5670592, EPI_ISL_5670601, EPI_ISL_5670620, EPI_ISL_5670692, EPI_ISL_5670732 | | | |
| see above | UPA DR FRANCO DA ROCHA | Instituto Butantan | Antonio Jorge Martins; Claudia Renata dos Santos Barros; David Schlesinger; Debora Botequio Moretti; Dimas Tadeu Covas; Elaine Cristina Marqueze; Elaine Vieira Santos; Evandra Strazza Rodrigues; Heidge Fukumasu; Jayme Augusto de Souza-Neto; José Salvatore Leister Patané; Luiz Alcantara; Luiz Lehmann Coutinho; Maria Carolina Elias; Maurício Lacerda Nogueira; Rafael dos Santos Bezerra; Raul Machado Neto; Rejane Maria Tommasini Grotto; Ricardo Haddad; Sandra Coccuzzo Sampaio Vessoni; Simone Kashima; Svetoslav Nanev Slavov; Vincent Louis Viala |
| EPI_ISL_5672277 | UPA GEISEL | Instituto Butantan | Antonio Jorge Martins; Claudia Renata dos Santos Barros; David Schlesinger; Debora Botequio Moretti; Dimas Tadeu Covas; Elaine Cristina Marqueze; Elaine Vieira Santos; Evandra Strazza Rodrigues; Heidge Fukumasu; Jayme Augusto de Souza-Neto; José Salvatore Leister Patané; Luiz Alcantara; Luiz Lehmann Coutinho; Maria Carolina Elias; Maurício Lacerda Nogueira; Rafael dos Santos Bezerra; Raul Machado Neto; Rejane Maria Tommasini Grotto; Ricardo Haddad; Sandra Coccuzzo Sampaio Vessoni; Simone Kashima; Svetoslav Nanev Slavov; Vincent Louis Viala |
| EPI_ISL_5668542, EPI_ISL_5669439, EPI_ISL_5669577, EPI_ISL_5670236, EPI_ISL_5670248, EPI_ISL_5670279, EPI_ISL_5670500, EPI_ISL_5670573, EPI_ISL_5670715, EPI_ISL_5670737 | | | |
| see above | UPA VILA SANTA CATARINA | Instituto Butantan | Antonio Jorge Martins; Claudia Renata dos Santos Barros; David Schlesinger; Debora Botequio Moretti; Dimas Tadeu Covas; Elaine Cristina Marqueze; Elaine Vieira Santos; Evandra Strazza Rodrigues; Heidge Fukumasu; Jayme Augusto de Souza-Neto; José Salvatore Leister Patané; Luiz Alcantara; Luiz Lehmann Coutinho; Maria Carolina Elias; Maurício Lacerda Nogueira; Rafael dos Santos Bezerra; Raul Machado Neto; Rejane Maria Tommasini Grotto; Ricardo Haddad; Sandra Coccuzzo Sampaio Vessoni; Simone Kashima; Svetoslav Nanev Slavov; Vincent Louis Viala |
| EPI_ISL_3375839, EPI_ISL_3375846, EPI_ISL_4078216 | US Air Force School of Aerospace Medicine | US Air Force School of Aerospace Medicine | Amanda Javorina; Anthony Fries; Carol Garrett; Clarise Starr; Cole Anderson; Elizabeth Macias; Fritz Castillo; Jennifer Black; Jennifer Meyer; Sarah Purves; William Gruner |
| EPI_ISL_3276671 | USC Clinical Lab | Los Angeles County Public Health Laboratories | P. Hemarajata et al. |
| EPI_ISL_5673623 | USF DR APARECIDO RODRIGUES MOUCO SCRPARDO | Instituto Butantan | Antonio Jorge Martins; Claudia Renata dos Santos Barros; David Schlesinger; Debora Botequio Moretti; Dimas Tadeu Covas; Elaine Cristina Marqueze; Elaine Vieira Santos; Evandra Strazza Rodrigues; Heidge Fukumasu; Jayme Augusto de Souza-Neto; José Salvatore Leister Patané; Luiz Alcantara; Luiz Lehmann Coutinho; Maria Carolina Elias; Maurício Lacerda Nogueira; Rafael dos Santos Bezerra; Raul Machado Neto; Rejane Maria Tommasini Grotto; Ricardo Haddad; Sandra Coccuzzo Sampaio Vessoni; Simone Kashima; Svetoslav Nanev Slavov; Vincent Louis Viala |
| EPI_ISL_5673709 | USF DR SAMUEL MARTINS FIGUEIRA SCRPARDO | Instituto Butantan | Antonio Jorge Martins; Claudia Renata dos Santos Barros; David Schlesinger; Debora Botequio Moretti; Dimas Tadeu Covas; Elaine Cristina Marqueze; Elaine Vieira Santos; Evandra Strazza Rodrigues; Heidge Fukumasu; Jayme Augusto de Souza-Neto; José Salvatore Leister Patané; Luiz Alcantara; Luiz Lehmann Coutinho; Maria Carolina Elias; Maurício Lacerda Nogueira; Rafael dos Santos Bezerra; Raul Machado Neto; Rejane Maria Tommasini Grotto; Ricardo Haddad; Sandra Coccuzzo Sampaio Vessoni; Simone Kashima; Svetoslav Nanev Slavov; Vincent Louis Viala |
| EPI_ISL_5672090, EPI_ISL_5672120, EPI_ISL_5672134, EPI_ISL_5672185, EPI_ISL_5672319, EPI_ISL_5672370 | USF PAULISTA FERNANDOPOLIS ANTONIO PIVATO | Instituto Butantan | Antonio Jorge Martins; Claudia Renata dos Santos Barros; David Schlesinger; Debora Botequio Moretti; Dimas Tadeu Covas; Elaine Cristina Marqueze; Elaine Vieira Santos; Evandra Strazza Rodrigues; Heidge Fukumasu; Jayme Augusto de Souza-Neto; José Salvatore Leister Patané; Luiz Alcantara; Luiz Lehmann Coutinho; Maria Carolina Elias; Maurício Lacerda Nogueira; Rafael dos Santos Bezerra; Raul Machado Neto; Rejane Maria Tommasini Grotto; Ricardo Haddad; Sandra Coccuzzo Sampaio Vessoni; Simone Kashima; Svetoslav Nanev Slavov; Vincent Louis Viala |
| EPI_ISL_2895003, EPI_ISL_2967608, EPI_ISL_2967614, EPI_ISL_2967625, EPI_ISL_2967658, EPI_ISL_2967672, EPI_ISL_2967691, EPI_ISL_2967692, EPI_ISL_2967707, EPI_ISL_2967725, EPI_ISL_3063503, EPI_ISL_3063518, EPI_ISL_3063560, EPI_ISL_3063563, EPI_ISL_3063607, EPI_ISL_3063609, EPI_ISL_3063657, EPI_ISL_3063680, EPI_ISL_3063684, EPI_ISL_3063726, EPI_ISL_3133329 | | | |
| see above | UW Virology Lab | UW Virology Lab | Alexander Greninger; Hong Xie; Keith R Jerome; Lasata Shrestha; Maria Lukes; Meei-Li Huang; Michelle Lin; Nathan Breit; Noah R. Baker; Patrick Mathias; Pavitra Roychoudhury; Ricardo Perez; Robert J. Livingston; Sean Ellis; Shah Mohamed Bakhash; Tien V. Nguyen |
| EPI_ISL_3155357, EPI_ISL_3155368, EPI_ISL_3155369, EPI_ISL_3155371, EPI_ISL_3155373, EPI_ISL_3155374, EPI_ISL_3155376, EPI_ISL_3155381, EPI_ISL_3155382, EPI_ISL_3155386, EPI_ISL_3155388, EPI_ISL_3155389, EPI_ISL_3155392, EPI_ISL_3155393, EPI_ISL_7780469 | | | |
| see above | Unidad de Investigación Médica de Yucatán (UIMY) | Centro de Investigación en Enfermedades Infecciosas (CIENI), Instituto Nacional de Enfermedades Respiratorias (INER) | ; Alejandra García-Gasca; Alejandra Hernández-Terán; Alejandro Sánchez-Flores; Alfredo Herrera-Estrella; Alicia Ocaña-Mondragón; Andreu Comas-García; Angel Gustavo Salas-Lais; Antonio Loza Román; Bernardo Martínez-Miguel; Blanca Taboada; Brenda Irasema Maldonado-Meza; Bruno Gómez-Gil; Carla Ivón Herrera-Najera; Carlos F. Arias; Celia Boukadida; Clara Esperanza Santacruz-Tinoco; Concepción Grajales-Muñiz; Consorcio Mexicano de Vigilancia Genómica (CoViGen-Mex). Authors (in alphabetical order): Julio Elias Alvarado-Yaah; Cristóbal Cháidez-Quiróz; Célida Duque Molina; Célida Martínez- Rodríguez; Célida Martínez-Rodríguez; Daniel Fregoso-Rueda; Daniel Lira Morales; Eduardo Becerril-Vargas; Eduardo Rivera-Martínez; Fernando Fontove-Herrera; Fidencio Mejía-Nepomuceno; Francisco Pulido; Gabriel Chavira-Trujillo; Gloria Elena Espinosa-Ayala; Gloria María Molina-Salinas; Gloria Vazquez; Hector Esteban Paz-Juárez; Hector Montoya-Fuentes; Helen Haydee Fernanda Ramírez-Plascencia; Irvin González-López; Jean Pierre González; Jesús Hernández; Joel Armando Vázquez-Pérez.; Jorge Salas-Hernández; José Antonio Enciso-Moreno; José Arturo Martínez-Orozco; José Esteban Muñoz-Medina; José de Jesús Nuñez-Contreras; Juan Bautista Chale-Dzul; Julissa Enciso-Ibarra; Kathia Elizabeth Tapia-Diaz; Luis Alberto Ochoa-Carrera; Margarita Matías-Florentino; Mario Mújica-Sánchez; Marissa Perez-Garcia; María Eugenia Jiménez-Corona; María Guadalupe Santiago-Mauricio; María Guadalupe de Jesús Mireles-Rivera; Nelly Sélem-Mojica; Pavel Isa; Ricardo Ciria Merce; Ricardo Grande; Rosa María Gutiérrez Rios; Rosario Vazquez-Larios; Santiago Ávila-Ríos; Selene Zárate; Susana Lopez; Verónica Mata-Haro; Victor Eduardo García-Arias; Víctor Hugo Borja-Aburto |
| EPI_ISL_3347533, EPI_ISL_3347534 | Unidad de Investigación Médica de Yucatán (UIMY) | Instituto de Biotecnología de la UNAM | ; Alejandra García-Gasca; Alejandra Hernández-Terán; Alejandro Sánchez-Flores; Alfredo Herrera-Estrella; Alicia Ocaña-Mondragón; Andreu Comas-García; Angel Gustavo Salas-Lais; Antonio Loza Román; Bernardo Martínez-Miguel; Blanca Taboada; Brenda Irasema Maldonado-Meza; Bruno Gómez-Gil; Carla Ivón Herrera-Najera; Carlos F. Arias; Celia Boukadida; Clara Esperanza Santacruz-Tinoco; Concepción Grajales-Muñiz; Consorcio Mexicano de Vigilancia Genómica (CoViGen-Mex). Authors (in alphabetical order): Julio Elias Alvarado-Yaah; Cristóbal Cháidez-Quiróz; Célida Duque Molina; Célida Martínez- Rodríguez; Daniel Fregoso-Rueda; Daniel Lira Morales; Eduardo Becerril-Vargas; Fernando Fontove-Herrera; Fidencio Mejía-Nepomuceno; Francisco Pulido; Gloria Elena Espinosa-Ayala; Gloria María Molina-Salinas; Gloria Vazquez; Hector Esteban Paz-Juárez; Hector Montoya-Fuentes; Helen Haydee Fernanda Ramírez-Plascencia; Irvin González-López; Jean Pierre González; Jesús Hernández; Joel Armando Vázquez-Pérez).; Jorge Salas-Hernández; José Antonio Enciso-Moreno; José Arturo Martínez-Orozco; José Esteban Muñoz-Medina; José de Jesús Nuñez-Contreras; Juan Bautista Chale-Dzul; Julissa Enciso-Ibarra; Kathia Elizabeth Tapia-Diaz; Luis Alberto Ochoa-Carrera; Margarita Matías-Florentino; Mario Mújica-Sánchez; Marissa Perez-Garcia; María Guadalupe Santiago-Mauricio; María Guadalupe de Jesús Mireles-Rivera; Nelly Sélem-Mojica; Pavel Isa; Ricardo Ciria Merce; Ricardo Grande; Rosa María Gutiérrez Rios; Santiago Ávila-Ríos; Selene Zárate; Susana Lopez; Verónica Mata-Haro; Victor Eduardo García-Arias; Víctor Hugo Borja-Aburto |
| EPI_ISL_3245362, EPI_ISL_3245370, EPI_ISL_3245374, EPI_ISL_3245380, EPI_ISL_3245381, EPI_ISL_3245382, EPI_ISL_3245383, EPI_ISL_3245395, EPI_ISL_3245396, EPI_ISL_3245409, EPI_ISL_3245411, EPI_ISL_3245412 | | | |
| see above | Unidade de apoio ao diagnostico da COVID - UNADIG | Bioinformatics Laboratory / LNCC | Alessandra P Lamarca; Alexandra L Gerber; Amilcar Tanuri; Ana Paula de C Guimaraes; Ana Tereza R Vasconcelos; Andrea Cony Cavalcanti; Caio Luiz Pereira Ribeiro; Cintia Policarpo; Claudia Maria Braga de Mello; Cristiane Gomes da Silva; Douglas Terra Machado; Erica Ramos dos Santos Nascimento; Fernanda Leitao dos Santos; Flavio Dias da Silva; Gleidson da Silva de Oliveira; Leandro Magalhaes de Souza; Liliane Cavalcante; Luiz G P de Almeida; Marcio Henrique de Oliveira Garcia; Mario Sergio Ribeiro; Ricardo Jose Barbosa Salviano; Ronaldo da Silva F Jr; Silvia Carvalho |
| EPI_ISL_3721601 | Universidad de Caldas | Universidad Nacional de Colombia - Laboratorio Genómico One Health | Andres F. Cardona-Rios; Carlos Franco-Muñoz; Carolina Muñoz-Arango; Celeny Ortiz; Daniel O. Maldonado-Perez; Diego A. Álvarez-Díaz; Hector Alejandro Ruiz-Moreno; Idabely Betancur Ortiz; Jorge E. Osorio; Juan P. Hernandez-Ortiz; Karl A Ciuoderis; Katherine Laiton-Donato; Laura Silvana Perez; Lina M. Hurtado; Marcela Mercado-Reyes; María Angélica Maya; Maria Stella López; Rita Almanza Payares; Sandra Ines Cano; Simón Villegas Velásquez |
| EPI_ISL_3076939 | University Clinical Hospital of Mostar, Department of Microbiology and Molecular Diagnostics | University of Sarajevo, Veterinary Faculty, Laboratory for Molecular Diagnostic and Research Laboratory | Goletic S; Goletic T; Hodzic A.; Jazic A.; Nicevic M.; Ostojic M; Sabic E.; Softic A; Terzic I. |
| EPI_ISL_3537377, EPI_ISL_3537381 | University of Iowa Hospitals and Clinics | State Hygienic Laboratory at the University of Iowa | Alankar Kampoowale; Anna Yakos; Erik Twait; Kris Eveland; Mariah Knutson; Valerie Reeb; Wes Hottel |
| EPI_ISL_3032251, EPI_ISL_3032262, EPI_ISL_3087324 | University of Liège COVID-19 testing center | GIGA Medical Genomics | Bouchra Boujemla; Claire Gourzonès; Cécile Meex; Keith Durkin; Laurent Gillet; Maria Artesi; Marie-Pierre Hayette; Nadine Cambisano; Nathalie Renotte; Olivier Ek; Sébastien Bontems; Vincent Bours |
| EPI_ISL_3087570 | University of Wisconsin-Madison AIDS Vaccine Research Laboratories | University of Wisconsin-Madison AIDS Vaccine Research Laboratories | Gage Moreno; Katarina Braun; et al. AIDS Vaccine Research Laboratories |
| EPI_ISL_3144682, EPI_ISL_3263861, EPI_ISL_3268740, EPI_ISL_3268786, EPI_ISL_3745387, EPI_ISL_3745388, EPI_ISL_3745390, EPI_ISL_3745392, EPI_ISL_3745393, EPI_ISL_3745397, EPI_ISL_3745398, EPI_ISL_3745400 | | | |
| see above | Utah Public Health Laboratory | Utah Public Health Laboratory | Erin L. Young; Kelly F. Oakeson; Olinto Linares-Perdomo; Pooja Gupta |
| EPI_ISL_2978252 | VIDYMED LAUSANNE | Laboratory of genomics and metagenomics | Claire Bertelli; Damien Jacot; Gilbert Greub; Sébastien Aeby; Trestan Pillonel |
| EPI_ISL_5673133 | VIGILANCIA EM SAUDE SAE VIGILANCIA EPIDEMIOLOGICA | Instituto Butantan | Antonio Jorge Martins; Claudia Renata dos Santos Barros; David Schlesinger; Debora Botequio Moretti; Dimas Tadeu Covas; Elaine Cristina Marqueze; Elaine Vieira Santos; Evandra Strazza Rodrigues; Heidge Fukumasu; Jayme Augusto de Souza-Neto; José Salvatore Leister Patané; Luiz Alcantara; Luiz Lehmann Coutinho; Maria Carolina Elias; Maurício Lacerda Nogueira; Rafael dos Santos Bezerra; Raul Machado Neto; Rejane Maria Tommasini Grotto; Ricardo Haddad; Sandra Coccuzzo Sampaio Vessoni; Simone Kashima; Svetoslav Nanev Slavov; Vincent Louis Viala |
| EPI_ISL_5671301, EPI_ISL_5671340, EPI_ISL_5671465, EPI_ISL_5680582, EPI_ISL_5680702 | VIGILANCIA EPIDEMIOLOGICA DE LEME | Instituto Butantan | Antonio Jorge Martins; Claudia Renata dos Santos Barros; David Schlesinger; Debora Botequio Moretti; Dimas Tadeu Covas; Elaine Cristina Marqueze; Elaine Vieira Santos; Evandra Strazza Rodrigues; Heidge Fukumasu; Jayme Augusto de Souza-Neto; José Salvatore Leister Patané; Luiz Alcantara; Luiz Lehmann Coutinho; Maria Carolina Elias; Maurício Lacerda Nogueira; Rafael dos Santos Bezerra; Raul Machado Neto; Rejane Maria Tommasini Grotto; Ricardo Haddad; Sandra Coccuzzo Sampaio Vessoni; Simone Kashima; Svetoslav Nanev Slavov; Vincent Louis Viala |
| EPI_ISL_5670869, EPI_ISL_5671297, EPI_ISL_5671444 | VIGILANCIA EPIDEMIOLOGICA E CONTROLE DE VETORES PIRASSUNUNGA | Instituto Butantan | Antonio Jorge Martins; Claudia Renata dos Santos Barros; David Schlesinger; Debora Botequio Moretti; Dimas Tadeu Covas; Elaine Cristina Marqueze; Elaine Vieira Santos; Evandra Strazza Rodrigues; Heidge Fukumasu; Jayme Augusto de Souza-Neto; José Salvatore Leister Patané; Luiz Alcantara; Luiz Lehmann Coutinho; Maria Carolina Elias; Maurício Lacerda Nogueira; Rafael dos Santos Bezerra; Raul Machado Neto; Rejane Maria Tommasini Grotto; Ricardo Haddad; Sandra Coccuzzo Sampaio Vessoni; Simone Kashima; Svetoslav Nanev Slavov; Vincent Louis Viala |
| EPI_ISL_5671606 | VIGILANCIA EPIDIMIOLOGICA DE ARACARIGUAMA | Instituto Butantan | Antonio Jorge Martins; Claudia Renata dos Santos Barros; David Schlesinger; Debora Botequio Moretti; Dimas Tadeu Covas; Elaine Cristina Marqueze; Elaine Vieira Santos; Evandra Strazza Rodrigues; Heidge Fukumasu; Jayme Augusto de Souza-Neto; José Salvatore Leister Patané; Luiz Alcantara; Luiz Lehmann Coutinho; Maria Carolina Elias; Maurício Lacerda Nogueira; Rafael dos Santos Bezerra; Raul Machado Neto; Rejane Maria Tommasini Grotto; Ricardo Haddad; Sandra Coccuzzo Sampaio Vessoni; Simone Kashima; Svetoslav Nanev Slavov; Vincent Louis Viala |
| EPI_ISL_5668464, EPI_ISL_5668683, EPI_ISL_5669226, EPI_ISL_5669433, EPI_ISL_5669545, EPI_ISL_5669813, EPI_ISL_5669854, EPI_ISL_5670436, EPI_ISL_5670587, EPI_ISL_5670722, EPI_ISL_5670763, EPI_ISL_5672439, EPI_ISL_5672542 | | | |
| see above | VIGILANCIA SANITARIA E VIG EPIDEMIOLOGICA DE ITAPEVI | Instituto Butantan | Antonio Jorge Martins; Claudia Renata dos Santos Barros; David Schlesinger; Debora Botequio Moretti; Dimas Tadeu Covas; Elaine Cristina Marqueze; Elaine Vieira Santos; Evandra Strazza Rodrigues; Heidge Fukumasu; Jayme Augusto de Souza-Neto; José Salvatore Leister Patané; Luiz Alcantara; Luiz Lehmann Coutinho; Maria Carolina Elias; Maurício Lacerda Nogueira; Rafael dos Santos Bezerra; Raul Machado Neto; Rejane Maria Tommasini Grotto; Ricardo Haddad; Sandra Coccuzzo Sampaio Vessoni; Simone Kashima; Svetoslav Nanev Slavov; Vincent Louis Viala |
| EPI_ISL_5779998 | VUMC Molecular Infectious Diseases Laboratory (MIDL) | Dr. Suman Das Lab - Vanderbilt University Medical Center (VUMC) | Bookyung Park; Grant Vestal; Helen Boone; Hunter Brown; Jonathan E. Schmitz; Meghan Shilts; Seesandra Rajagopala; Suman B. Pakala; Suman Das |

| | | | |
|---|---|---|---|
| EPI_ISL_2938458 | Viollier AG | Department of Biosystems Science and Engineering, ETH Zürich | Chaoran Chen; Christian Beisel; Christiane Beckmann; Christoph Noppen; Elodie Burcklen; Ina Nissen; Ivan Topolsky; Kim Philipp Jablonski; Lara Fuhrmann; Maurice Redondo; Mirjam Feldkamp; Natascha Santacroce; Niko Beerenwinkel; Olivier Kobel; Rebecca Denes; Sarah Nadeau; Tanja Stadler |
| EPI_ISL_3021028, EPI_ISL_3025879, EPI_ISL_3048085, EPI_ISL_3048097, EPI_ISL_3048111, EPI_ISL_3123430, EPI_ISL_3123435, EPI_ISL_3214960 | | | |
| see above | Virginia Division of Consolidated Laboratory Services | Virginia Division of Consolidated Laboratory Services | Virginia Division of Consolidated Laboratory Services |
| EPI_ISL_3303929 | WA State Department of Health | Centers for Disease Control and Prevention Division of Viral Diseases, Pathogen Discovery | Alex Burgin; Ben L. Rambo-Martin; Clinton R. Paden; Dakota Howard; Dave Wentworth; Dhwani Batra; Jasmine Padilla; Justin Lee; Krista Queen; Kristen Knipe; Kristine Lacek; Mark Burroughs; Matthew Schmerer; Meghan Bentz; Mili Sheth; Peter Cook; Sam Shepard; Sarah Nobles; Suxiang Tong; Vivien Dugan; Yvette Unoarumhi |
| EPI_ISL_3009183 | WSSE w Katowicach | 1. Academic Center for Pathomorphological and Genetic-Molecular Diagnostics ltd, Bialystok, Poland 2. National Institute of Public Health - National Institute of Hygiene, Warsaw, Poland | Anetta Sulewska; Jacek Nikliński; Janusz Dzięcioł; Joanna Kiśluk; Katarzyna Zacharczuk; Konrad Raczkowski; Magdalena Nowakowska; Małgorzata Sadkowska-Todys; Piotr Karabowicz; Piotr Majewski; Przemysław Biecek. Joanna Reszeć; Radosław Charkiewicz; Tomasz Wołkowicz |
| EPI_ISL_3016339, EPI_ISL_3216969, EPI_ISL_3216996, EPI_ISL_3217046 | Washington State Department of Health Public Health Laboratories | Washington State Department of Health Public Health Laboratories | Avi Singh; Darren Lucas; Denny Russell; Drew MacKellar; Geoff Melly; Hannah Gray; Joenice Gonzalez; JohnAric Peterson; Philip Dykema; Rebecca Cao; Vanessa De Los Santos |
| EPI_ISL_2885897 | Wichita State University-Molecular Diagnostic Lab | Kansas Health and Environmental Lab | Ben Olsen; Jonathan Barnell; Katherine Wiggins; Mike Grose; and Phil Adam |
| EPI_ISL_3234718, EPI_ISL_3234720 | Wisconsin State Laboratory of Hygiene Communicable Disease Division | Wisconsin State Laboratory of Hygiene Communicable Disease Division | Abigail C. Shockey; Alicia J. Mooney; Erika M. Hanson; Kelsey R. Florek; Richard Griesser; Sara Wagner; Tonya Danz |
| EPI_ISL_3032386 | Wojskowy Instytut Medycyny Lotniczej Warszawa | National Institute of Public Health NIH - National Research Institute | Gierczyński Rafał; Sadkowska-Todys Małgorzata; Wołkowicz Tomasz; Zacharczuk Katarzyna |
| EPI_ISL_3092082, EPI_ISL_3160630, EPI_ISL_6582679 | Wyoming Public Health Laboratory | Wyoming Public Health Laboratory | Ashley Norberg; Brian Dominguez; Cari Sloma; Channing Weber; Chayse Rowley; Elliot Thomasson; Jim Mildenberger; Marley Goetz; Robert Petit; Sam Britz; Taylor Fearing; and Rob Christensen |
| EPI_ISL_3025425, EPI_ISL_3104692, EPI_ISL_3104693, EPI_ISL_3104694, EPI_ISL_3188566 | Yale Clinical Virology Lab | Grubaugh Lab - Yale School of Public Health | Anderson Brito; Annie Watkins; Chaney Kalinich; Chantal Vogels; Isabel Ott; Jessica Rothman; Joseph Fauver; Kendall Billig; Mallery Breban; Marie L. Landry; Mary Petrone; Nathan Grubaugh; Tara Alpert; Tobias Koch |
| EPI_ISL_3117461, EPI_ISL_3117498 | ZOTZ KLIMAS MVZ Düsseldorf-Centrum GbR ÜBAG für Labormedizin, Genetik, Zytologie, Pathologie | Center of Medical Microbiology, Virology, and Hospital Hygiene, University of Duesseldorf | Alexander Dilthey; Andreas Walker; Daniel Strelow; Jessica Nicolai; Jörg Timm; Katrin Hoffmann; Klaus Pfeffer; Lisanna Hülse; Malte Kohns Vasconcelos; Maximilian Damagnez; Nadine Lübke; Patrick Finzer; Rainer Zotz; Tobias Wienemann; Torsten Houwaart |
